# Molecular signatures of chronic antibody-mediated rejection in human liver transplants

**DOI:** 10.1101/2024.02.08.24302515

**Authors:** Bastian Engel, Ahmed Alaswad, Alejandro Campos-Murguia, Martijn Zoodsma, Anne Klingbeil, Pablo Ruiz, Gonzalo Crespo, Kinan Chihab, Emily A. Bosselmann, Sophia Heinrich, Björn Hartleben, Danny D. Jonigk, Alba Diaz, Murielle Verboom, Michael Hallensleben, Robert Geffers, Christine S Falk, Sergej Ruff, Chris Lauber, Emilie Toft Skovgaard, Morten Karsdal, Diana Julie Leeming, Heiner Wedemeyer, Cheng-Jian Xu, Jordi Colmenero, Elmar Jaeckel, Yang Li, Richard Taubert

## Abstract

**Background&Aims:** The role of antibody-mediated rejection (ABMR) after liver transplantation (LT) remains controversial. Chronic ABMR (cABMR) is often subclinical and potentially missed without surveillance biopsies (svLbx). Transcriptome analysis previously characterized molecular changes in T cell-mediated rejection (TCMR) after solid organ transplantation. We aimed to identify molecular cABMR signatures after LT.

**Methods:** Indication and svLbx from two prospective institutional biorepositories were screened. We performed RNA-sequencing on liver biopsies (discovery cohort: n=71; Hannover validation cohort: n=58; Barcelona validation cohort: n=29). Downstream analyses explored the molecular features of cABMR, clinical (clinTCMR), subclinical TCMR (subTCMR) and chronic ductopenic rejection compared to no histological rejection (NHR).

**Results:** Nineteen percent of LT recipients with donor-specific antibodies had cABMR in the training cohort. Fifty seven percent of cABMR patients had normal/near normal liver enzymes, recognized only by svLbx. CABMR was associated with subsequent cirrhosis development in 40-50% and displayed differentially expressed genes (DEGs) uniquely enriched in fibrogenesis-, complement activation- and TNFα-signaling-related pathways. ClinTCMR showed no such recurrent cirrhosis and DEGs uniquely enriched in antigen presentation-, interferon-signaling-, and T cell receptor-signaling-related pathways. SubTCMR was molecularly almost indistinguishable from NHR. There was a high degree of correlation in the DEGs between the training and two independent validation cohorts.

**Conclusions:** We report transcriptome features of cABMR that is a unique molecular identity associated with inflammation and fibrogenesis as described in other solid organ transplantations and being linked to a high rate of liver graft failure.

**Impact and Implications:**

- Chronic antibody-mediated rejection (ABMR) is a recognized relevant cause of graft injury and unfavorable patient outcomes after kidney, heart and lung transplantation but the role of chronic ABMR after liver transplantation (LT) remains controversial, despite a recurrent cirrhosis rate of 40-50% in the present study. Therefore, we aimed to characterize rejection phenotypes after LT on a molecular basis, as done in other solids organ transplants, to identify unique features of chronic ABMR.
- Our findings highlight chronic ABMR to be a molecularly distinct clinical phenotype of rejection after LT that often presents with normal liver enzymes validated in three independent cohorts.
- Therefore, our work emphasizes the usage of surveillance graft biopsies for the detection of a fibrosis associated phenotype of chronic ABMR. The molecular signature may be used in the future by researchers and clinicians to discriminate chronic ABMR from graft injury of other causes.

## 1. Introduction

Liver transplantation (LT) is a life-saving treatment for end-stage liver disease, but long-term outcomes have not improved in recent decades[1]. Chronic morbidity and mortality are mainly due to immunosuppressive toxicity and graft dysfunction[1]. Surveillance liver biopsies (svLBx) reveal advanced fibrosis in 20–60% of recipientsL[2,3].

The role of antibody-mediated rejection (ABMR) in ABO-compatible LT has long been debated. Donor-specific antibodies (DSAs) are linked to graft injury, fibrosis, TCMR-related gene expression, and reduced graft and patient survivalL[4,5]. DSA prevalence increases over time, affecting up to 50% of recipients[2]. In 2016, the Banff Foundation published the first criteria for acute and chronic ABMR (cABMR) in LT[6], but its low incidence (<10–15%) and frequent absence of enzyme elevation hinder recognition without svLBx[7]. Thus, true prevalence and clinical impact remain unclear.

Gene expression profiling has clarified ABMR signatures in kidney, heart, and lung transplantsL[8,9], with comparable risk of graft loss irrespective of clinical differences[10]. This two-center study aimed to define the molecular signature of cABMR in LT and distinguish it from clinTCMR, subTCMR, and chronic ductopenic rejection (cDR).

## 2. Patients and methods

### 2.1 Patient cohort

Patient samples for discovery and internal validation cohorts were retrieved from Hannover’s ongoing prospective biorepository. The biorepository includes LTR that undergo liver biopsy (LBx) either for the work-up of elevated liver enzymes, liver fibrosis or graft surveillance to individualize immunosuppression[11]. Samples for external validation were retrieved from a local biorepository at Hospital Clinic Barcelona, Spain. All patients provided written consent.

The study was approved by respective local Ethics Committees (protocol number 933 for project Z2 of the comprehensive research center 738; MHH Ethikkommission, Hannover, Germany; HCB/2018/0911, Medicine Research Ethics Committee of Hospital Clinic, University of Barcelona, Barcelona, Spain) and conducted according to the ethical guidelines of the Declarations of Helsinki and Istanbul.

### 2.2 Liver biopsies, rejection phenotypes, histological grading and staging

Relevant obstructive cholestasis was excluded via ultrasound or MRCP. Biopsies from patients with infections or vascular complications were omitted. Viral sequences were screened in transcriptome data (Suppl. Method Section 1.4), but only sporadic, inconclusive hits were detected (Suppl. Figure 1). Histological evaluation, blinded and performed by expert liver pathologists, included RAI, inflammation grade, fibrosis staging (mHAI), central perivenulitis, portal microvasculitis, ductular reaction, steatosis, and total liver allograft fibrosis (LAF) scoring[6,12]. Significant fibrosis was defined as periportal fibrosis (Ishak F) ≥2 and/or any LAF component ≥2. Definitions for subTCMR and clinTCMR followed recent criteria[13]: subTCMR required RAI≥1+1+1 with liver enzymes <2×ULN; clinTCMR required the same RAI threshold with AST, ALT and/or AP >2×ULN. Possible cABMR was defined per the 2016 Banff guidelines[6] by presence of DSA (MFI>1000), moderate fibrosis (Ishak F≥2 and/or any LAF component ≥2), and compatible histological findings (e.g., RAI P≥1 and/orRAI D≥1 or Ishak A≥1 or central perivenulitis≥1 and/or RAI B≥1), excluding other injury causes (Figure 1A, Suppl. Table 1). Patients with no histological signs of rejection (NHR) had to have normal liver enzymes and RAI ≤1 without any relevant inflammation or fibrosis (Ishak F ≤1, each LAF score component ≤1) on histological examination. C4d staining was performed on FFPE sections (2 µm) according to local standard operating procedures using the SP91 antibody (Roche) on the Ventana BenchMark Ultra with CC1 protocol (64 min antigen retrieval, 32 min antibody incubation), detected via the ultraView Universal DAB kit (Roche). Sample selection and biomaterial availability are detailed in Suppl. Figure 2. CDR was defined per the 2006 Banff consensus[14]; selection criteria are shown in Suppl. Figure 11.

**Figure 1:**
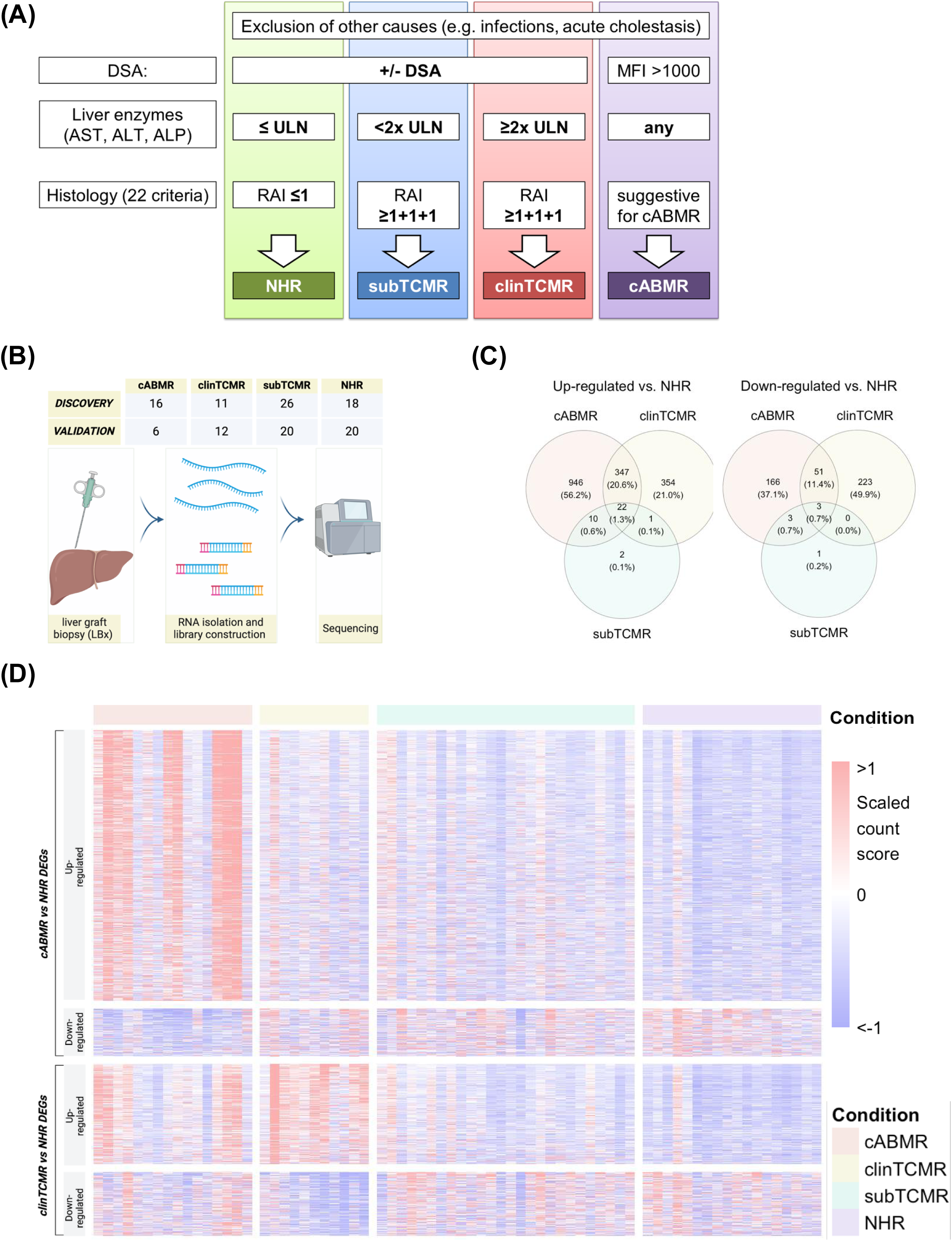
Study design and molecular rejection profiles. **(A)** Diagnostic flowchart for phenotypic rejection characterization. **(B)** Study design. **(C)** Venn diagram of up-/downregulated DEGs per rejection type vs. NHR. **(D)** Heatmap of unique DEGs in cABMR and clinTCMR. Columns: samples; rows: DEGs. *clinTCMR–clinical T cell-mediated rejection; cABMR–chronic antibody-mediated rejection; subTCMR–subclinical TCMR; NHR–no histological rejection; DEGs– differentially expressed genes; RAI-rejection activity index, DSA-donor specific antibodies*.

### 2.3 RNA isolation and sequencing

For the training cohort, total RNA was extracted from Allprotect-preserved biopsies using the AllPrep DNA/RNA/Protein Mini Kit (ID: 80004, Qiagen, The Netherlands) per manufacturer’s instructions. For the validation cohort, RNA was isolated from FFPE samples using the RNeasy FFPE Kit (ID: 73504, Qiagen, The Netherlands). RNA was stored at −80°C until use. RNA quality and integrity were assessed using the Agilent 2100 Bioanalyzer (Agilent Technologies, Germany). Library preparation and sequencing were performed at HZI Braunschweig (Germany) (training) and BGI Genomics (Hongkong, China) (validation), following local protocols.

In the training cohort, mRNA was purified from 500 ng total RNA using the Dynabeads® mRNA DIRECT™ Micro Kit (ID: 61021, Thermo Fisher, USA), followed by library prep with the ScriptSeq v2 RNA-Seq Kit (ID: SSV21106, Epicentre/Illumina, USA). Sequencing was performed on an Illumina HiSeq2500 (50-cycle, single-end; TruSeq SBS Kit v3-HS), yielding ∼30 million reads/sample. The validation cohort was sequenced on BGI’s DNBSEQ™ platform, producing ∼2×100 million paired-end reads per sample.

Eligibility for sequencing required a minimum RIN of 2 and DV200 >30–50% for FFPE-derived RNA. Median RIN values were 8.3 (6.6–9.5) for the training cohort, 2.4 (2.1–2.8) for the internal, and 2.2 (2.2–2.3) for the external validation cohort.

### 2.4 Gene expression data processing and analysis

Raw sequencing data were processed using the nf-core (v3.9) implemented in Nextflow[15,16]. Length-scaled gene counts were imported into DESeq2 (v1.40.1) via tximport (v1.28.0) for differential expression analysis[17,18]. Genes with |log2 fold change|>1 and FDR-adjusted pL<L0.05 were considered differentially expressed. Time from transplantation to biopsy, age, and sex were included as covariates. Functional enrichment of DEGs was performed using the over-representation analysis tool of the ConsensusPathDB-human platform[19].

Further details on materials and methods are available in the Supplementary CTAT Table and Supplementary Information file.

## 3. Results

### 3.1 Characterization of the training cohort

Liver biopsies classified as clinTCMR, subTCMR, cABMR, or NHR were selected from a cohort of over 700 biopsies collected over seven years (Figure 1A, Suppl. Figure 2A). The prevalence of cABMR was 19% in DSA-positive biopsies (18/97) and 19% (18/94) in representative biopsies with advanced fibrosis. Samples were grouped by clinical phenotype (Table 1, Suppl. Table 1, Suppl. Figure 4A). No significant age differences were observed between groups (pL=L0.30), though sex distribution differed (pL=L0.04). Time since liver transplantation did not differ significantly in post-hoc analysis (Suppl. Figure 4A). As defined, patients with NHR and subTCMR showed no liver enzyme elevation, while those with clinTCMR did. Applying the same threshold (≤2×ULN for AST, ALT, AP), 81% of cABMR cases were subclinical and likely detectable only by svLBx. Histological inflammation and fibrosis were absent in NHR but present in all other phenotypes, with inflammation highest in clinTCMR and fibrosis most pronounced in cABMR, which also showed the highest DSA frequency. Notably, 56% of cABMR biopsies displayed TCMR features (RAI≥1+1+1), resembling ABMR patterns in other solid organ transplants (SOT)[20]. Use of cyclosporine and tacrolimus differed significantly between groups (pL=L0.022 and pL=L0.006, respectively), whereas other immunosuppressive regimens did not differ.

**Table 1.** Continuous variables reported as median (range), categorical variables as number (frequency). Kruskal-Wallis test was used for the comparison of the four groups.

| **Table 1** | **Overall** | **NHR** | **subTCM**<br>**R** | **cABMR** | **clinTCM**<br>**R** | **p** |
| --- | --- | --- | --- | --- | --- | --- |
| sample number | 71 | 18 | 26 | 16 | 11 |  |
| age [years] | 52 (18–76) | 50 (20–69) | 52 (21–67) | 59 (22–67) | 47 (18–76) | 0.30 |
| female sex | 24 (33.8) | 8 (44.4) | 5 (19.2) | 4 (25.0) | 7 (63.6) | 0.04 |
| time after LT [months] | 13 (2–298) | 10 (5–60) | 19 (5–116) | 32 (6–298) | 7 (2–152) | 0.04 |
| AST [xULN] | 0.8 (0.3–11.9) (n = 70) | 0.6 (0.4–0.8) (n=17) | 0.7 (0.3–1.2) | 0.9 (0.5–1.6) | 3 (1.5–11.9) | < 0.001 |
| ALT [xULN] | 0.5 (0.1–17.2) | 0.4 (0.2–0.6) | 0.5 (0.1–1.5) | 0.5 (0.3–2.1) | 4.1 (2.4–17.2) | < 0.001 |
| AP [xULN] | 0.9 (0.2–3.9) (n = 62) | 0.7 (0.2–1.1) (n=14) | 0.8 (0.2–1.3) | 1.2 (0.7–3.1) | 1.8 (0.8–3.9) | < 0.001 |
| bilirubin [xULN] | 0.5 (0.2–2.5) | 0.5 (0.2–2.5) | 0.5 (0.2–1.5) | 0.6 (0.2–2.2) | 0.9 (0.3–2.5) | 0.02 |
| RAI | 3 (0–8) | 0 (0–1) | 3.5 (3–6) | 3.5 (1–8) | 5 (3–7) | < 0.001 |
| mHAI | 3 (0–8) | 1 (0–2) | 2 (0–8) | 5 (2–7) | 4 (2–6) | <<br>0.001 |
| Ishak fibrosis | 1 (0–6) | 0 (0–1) | 1 (0–4) | 2.5 (1–6) | 1 (0–2) | <<br>0.001 |
| LAF | 1 (0–8) | 0 (0–1) | 1 (0–5) | 4 (2–8) | 1 (0–3) | <<br>0.001 |
| DSA positive | 28<br>(39.4) | 1 (5.6) | 7 (26.9) | 16 (100) | 4 (36.4) | <<br>0.001 |
| TAC | 25<br>(35.2) | 10<br>(55.6) | 10 (38.5) | 0 (0.0) | 5 (45.5) | 0.006 |
| CsA | 44<br>(62.0) | 8 (44.4) | 15 (57.7) | 15<br>(93.8) | 6 (54.5) | 0.022 |
| Sirolimus | 1 (1.4) | 0 (0.0) | 0 (0.0) | 1 (6.3) | 0 (0.0) | 0.32 |
| Prednisolone | 57<br>(80.3) | 13<br>(72.2) | 21 (80.8) | 12<br>(75.0) | 1 (100) | 0.29 |
| Mycophenolate-mofetil | 63<br>(88.7) | 15<br>(83.3) | 24 (92.3) | 15<br>(93.8) | 9 (81.8) | 0.62 |
| Azathioprine | 1 (1.4) | 0 (0.0) | 0 (0.0) | 0 (0.0) | 1 (9.1) | 0.14 |

### 3.2 Signature genes of cABMR and clinTCMR are associated with distinct biological processes

The study workflow is summarized in Figure 1B. To assess transcriptomic changes in rejection phenotypes (cABMR, clinTCMR, subTCMR), differential expression analysis versus NHR was conducted in the training cohort (Suppl. Figure 5; Suppl. Table 2). The numbers of up- and down-regulated genes for each group (Suppl. Table 4) are shown in Figures 1C and 1D. In cABMR, up-regulated DEGs were enriched in fibrogenesis and TNF-signaling pathways (FDR-adjusted p<0.05, hypergeometric test), whereas in clinTCMR, DEGs were associated with allograft rejection and cytokine signaling, particularly interferon pathways (Suppl. Figure 5A, B; Suppl. Table 3). SubTCMR exhibited minimal transcriptomic changes compared to NHR, with DEGs linked to antigen presentation, IL-12 production, and dendritic cell differentiation (Suppl. Figure 5C; Suppl. Table 3).

We then identified DEGs unique to cABMR and clinTCMR. cABMR-specific DEGs were enriched in fibrogenesis, cell adhesion, and NF-κB signaling (Figure 2A, B; Suppl. Table 5), while clinTCMR-specific DEGs were associated with allograft rejection, Th1/Th2/Th17 differentiation, antigen processing, NK cell cytotoxicity, and interferon signaling (Figure 2C, D; Suppl. Table 5). The 369 DEGs up-regulated in both clinTCMR and cABMR pointed to shared activation of non-canonical TNF receptor/NF-κB signaling, chemokine signaling, and cytokine responses (Suppl. Figure 6; Suppl. Tables 6, 7).

**Figure 2:**
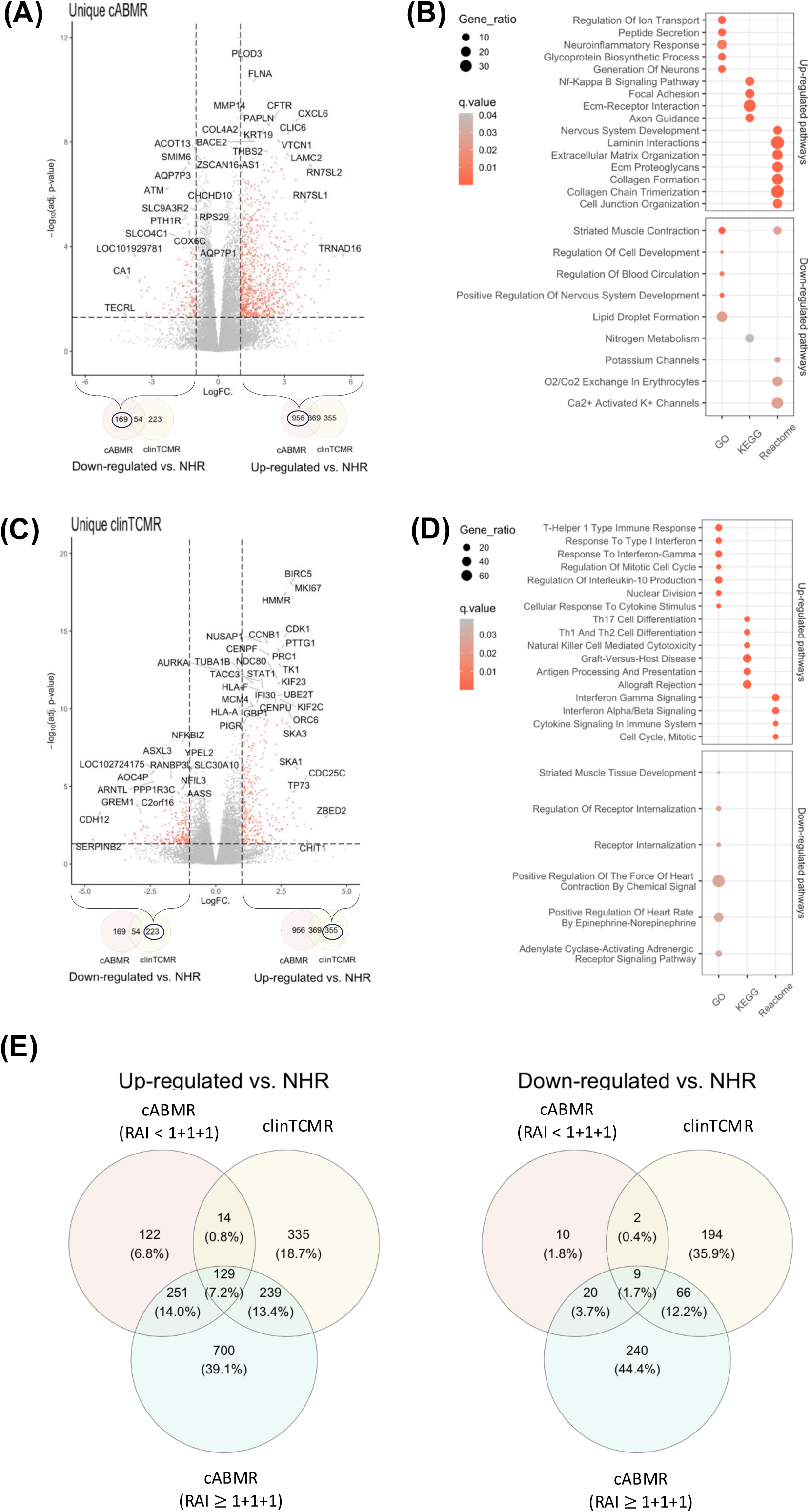
Molecular signatures and enriched pathways of cABMR and clinTCMR. **(A, C)** Volcano plots: unique DEGs of cABMR and clinTCMR vs. NHR (FDR<0.05, |log_₂_FC|>1). **(B, D)** Enriched pathways from DEGs in A and C. **(E)** Overlap of DEGs in cABMR with/without clinTCMR features and clinTCMR. *clinTCMR–clinical T cell-mediated rejection; cABMR–chronic antibody-mediated rejection; NHR–no histological rejection; DEGs–differentially expressed genes*.

Finally, cABMR biopsies with and without histologic features of TCMR (RAI≥1+1+1) were compared to clinTCMR (Figure 2E). Although overlap was greater between cABMR with TCMR features and clinTCMR, the majority of cABMR DEGs remained distinct.

### 3.3 Validation of cABMR and clinTCMR gene signature in an independent internal validation cohort

Clinical characteristics of the internal validation cohort were largely consistent with the training cohort (Table 2). Notably, the time from LT to biopsy was longer across all phenotypes, which was adjusted for in the statistical analysis (Methods 2.4; Suppl. Figure 4B). Unlike the training cohort, immunosuppressive regimens were comparable between groups due to the routine use of tacrolimus as the primary calcineurin inhibitor. The prevalence of cABMR was 13% (7/54) in DSA-positive biopsies and 5% (7/135) in biopsies with advanced fibrosis (Suppl. Figure 2B). As in the training cohort, 50% of cABMR cases exhibited liver enzyme levels below 2×ULN, making them detectable primarily via surveillance biopsies.

**Table 2.**
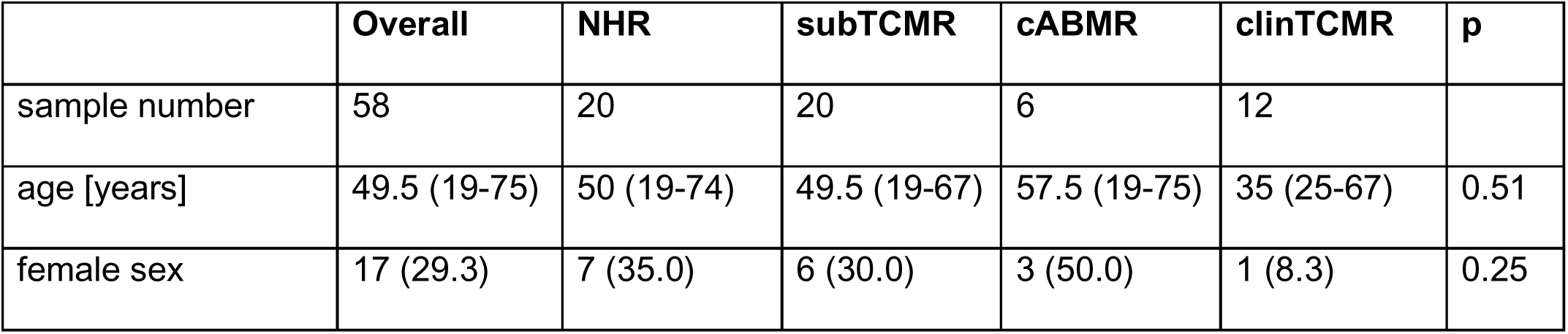
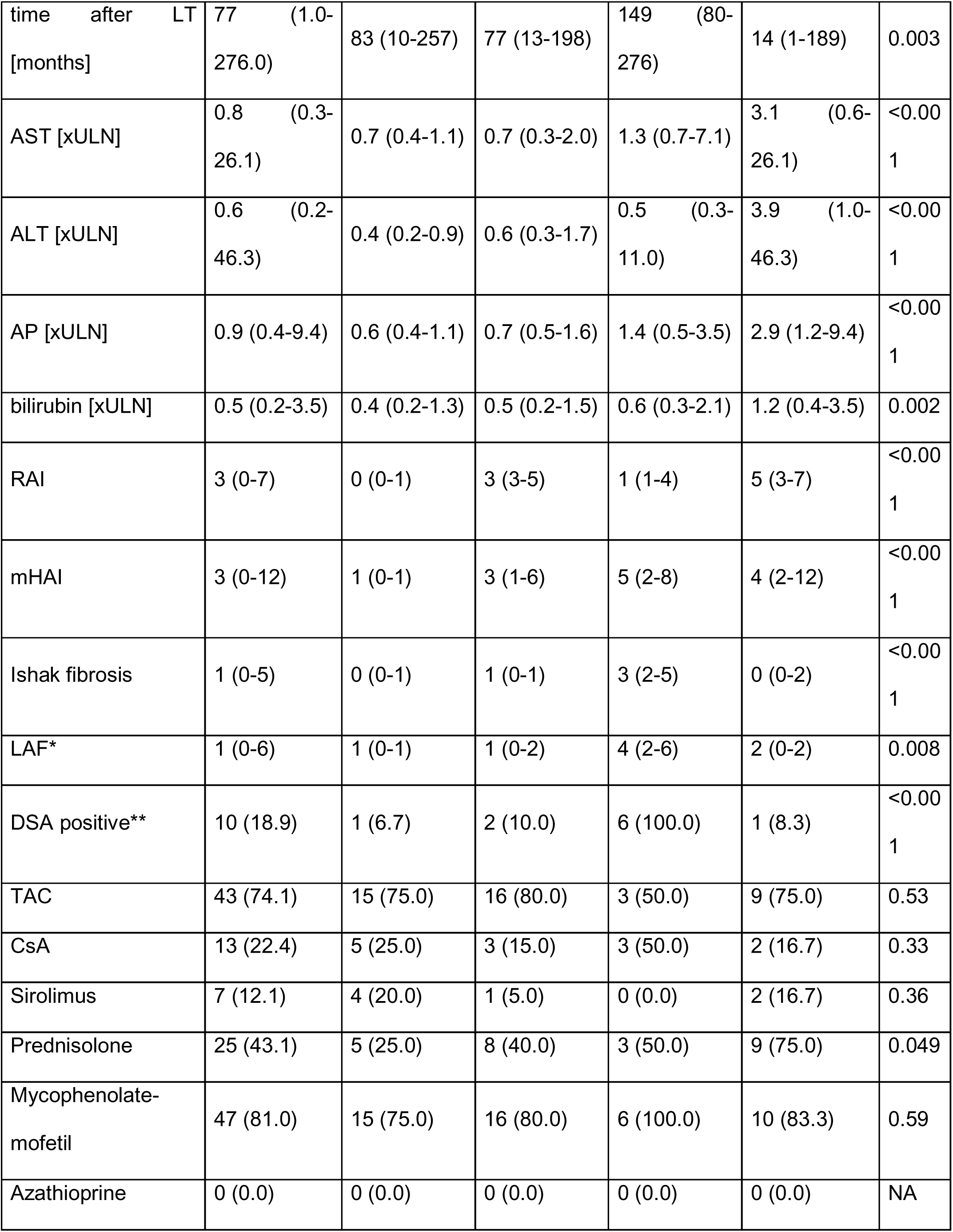
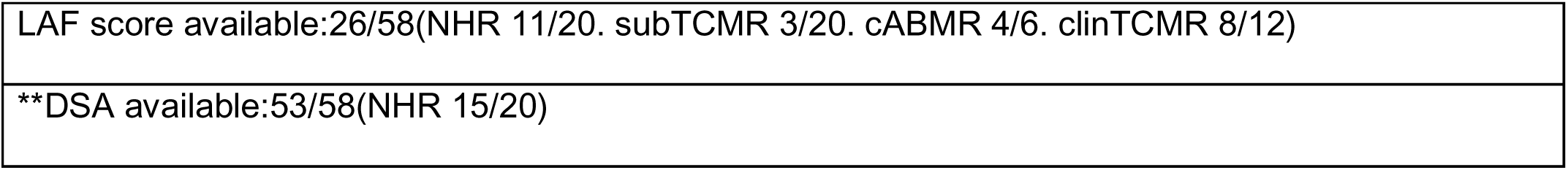
Continuous variables reported as median (range), categorical variables as number (frequency). Kruskal-Wallis test was used for the comparison of the four groups.

Gene expression profiles and pathway enrichment patterns for cABMR, clinTCMR, and subTCMR showed high concordance between cohorts (Figure 3A+B; Suppl. Figures 7A–C, 8A, 10A–B; Suppl. Tables 8, 9, 12, 13). Validation rates were 87% for cABMR DEGs and 93% for clinTCMR DEGs (Figure 3A). Immunofluorescence confirmed increased fibrogenesis and matrix remodeling in cABMR via COL14A1 and MMP2 staining (Suppl. Figure 9). cABMR-specific DEGs were enriched in fibrogenesis, cell adhesion, TNF signaling (via physiological ligands), TNFR2 non-canonical NF-κB signaling, and complement/coagulation cascades (Suppl. Figure 8B; Suppl. Tables 10, 11). ClinTCMR-specific DEGs were enriched in interferon signaling, antigen processing, Th1/2/17 differentiation, adaptive immunity, T cell receptor and IL-2 signaling, and other T cell-associated pathways (Suppl. Figure 8C; Suppl. Tables 10, 11), replicating findings from the training cohort.

**Figure 3:**
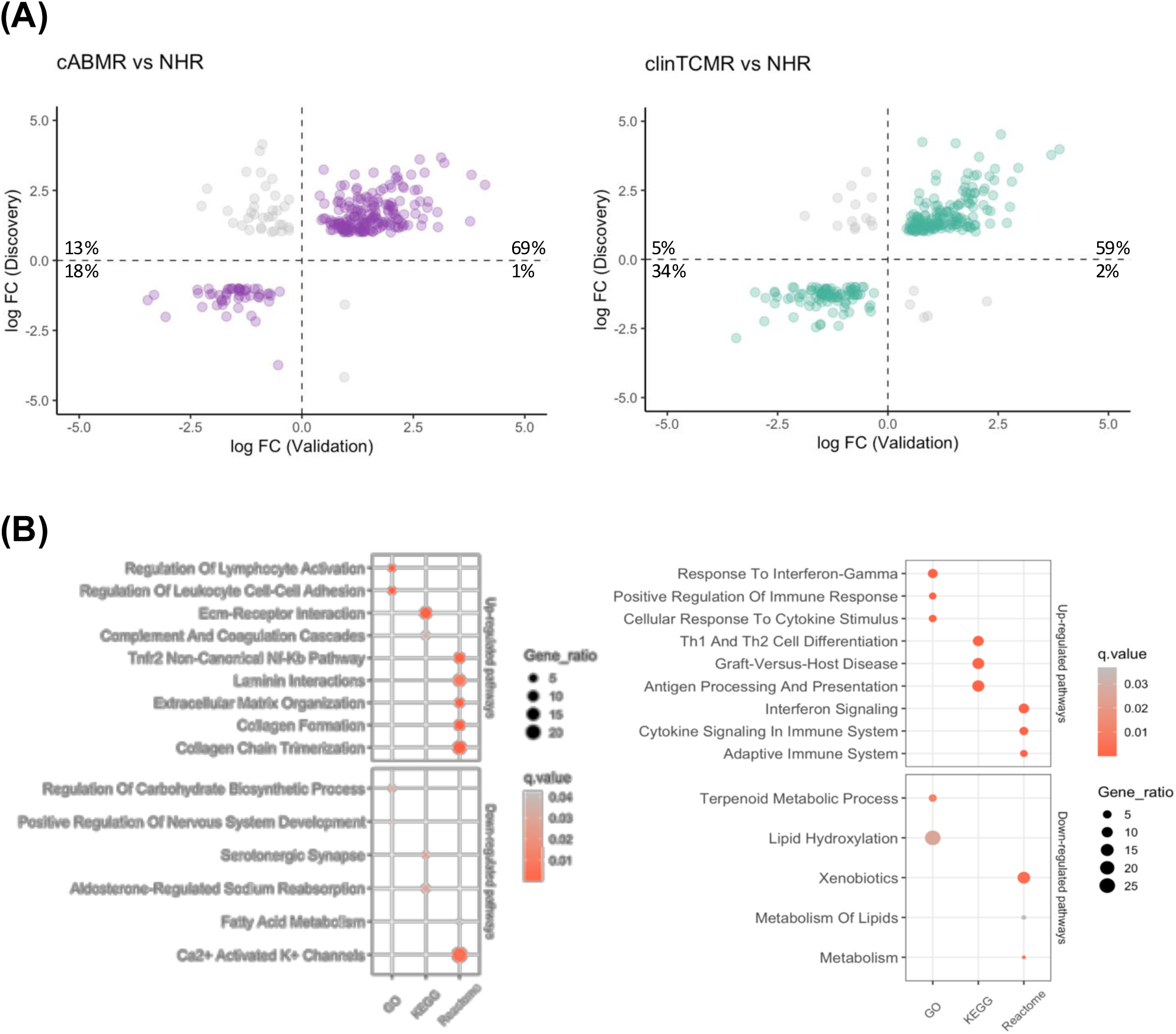
Internal validation of DEGs and pathways. **(A)** Validation of rejection vs. NHR DEGs (p < 0.05; Wald test). **(B)** Enriched pathways from validated DEGs. *clinTCMR–clinical T cell-mediated rejection; cABMR–chronic antibody-mediated rejection; NHR–no histological rejection; DEGs–differentially expressed genes*.

All available FFPE samples from patients with cDR (nL=L4) were included to assess overlap with cABMR-associated transcriptional changes (Suppl. Figure 11; Suppl. Tables 14, 15). Only a minority of DEGs were shared across cABMR, clinTCMR, and cDR (Suppl. Figure 12A). Notably, only cABMR DEGs—but not those of cDR—were enriched in complement and TNF signaling pathways (Suppl. Figure 12B; Figure 3B; Suppl. Table 16).

### 3.4 Validation of cABMR and clinTCMR gene signature in an external cohort

Thirty-two samples from Hospital Clinic Barcelona were available for external validation. Three cABMR cases were excluded for not meeting Banff diagnostic criteria. Among the remaining eight cABMR cases, 50% were classified as probable ABMR based on positive C4d staining[6]. Age (p=0.11) and sex (p=0.94) did not differ between groups; however, biopsies for cABMR were performed later post-transplant compared to clinTCMR and NHR. Use of tacrolimus and cyclosporine was comparable across groups. Additional clinical characteristics are summarized in Table 3.

**Table 3.** Continuous variables reported as median (range), categorical variables as number (frequency). Kruskal-Wallis test was used for the comparison of the four groups.

| **Table 3** | **NHR** | **clinTCMR** | **cABMR** | **p** |
| --- | --- | --- | --- | --- |
| Sample number | 7 | 14 | 8 |  |
| Age [years] | 58 [42-68] | 64 [38-78] | 51 [25-67] | 0.11 |
| female sex | 2 (28.6) | 3 (21.4) | 2 (25.0) | 0.94 |
| Time after LT [months] | 3.6 [2.7-4.3] | 33.8 [0.2-220.9] | 205.2 [16.2-330.2] | <0.001 |
| AST [xULN] | 0.4 [0.4-0.8] | 3.4 [0.8-14.2] | 1.4 [0.7-4.4] | <0.001 |
| ALT [xULN] | 0.5 [0.4-0.7] | 3.7 [1.3-18.6] | 1.4 [0.6-5.6] | <0.001 |
| AP [xULN] | 0.7 [0.4-1.5] | 12.0 [1.6-51.6] | 6.6 [1.1-29.7] | <0.001 |
| bilirubin [xULN] | 0.8 [0.4-1.0] | 1.7 [0.2-9.0] | 0.9 [0.5-1.3] | 0.041 |
| RAI | n/a | 5.0 [3.0-7.0] | 1.5 [1.0-4.0] | 0.001 |
| mHAI | 1.0 [0.0-2.0] | 3.5 [1.0-9.0] | 4.5 [2.0-9.0] | 0.001 |
| Ishak fibrosis | 0.0 [0.0-0.0] | 0.0 [0.0-1.0] | 3.0 [2.0-5.0] | <0.001 |
| LAF | 0.0 [0.0-0.0] | 0.0 [0.0-1.0] | 3.0 [2.0-4.0] | <0.001 |
| TAC | 4 (57.1) | 10 (71.4) | 7 (87.5) | 0.42 |
| CsA | 3 (42.9) | 2 (14.3) | 1 (12.5) | 0.25 |
| Everolimus | 0 (0.0) | 1 (7.1) | 0 (0.0) | 0.57 |
| Prednisolone | 7 (100.0) | 7 (50.0) | 1 (12.5) | 0.003 |
| Mycophenolate-mofetil | 1 (14.3) | 9 (64.3) | 5 (62.5) | 0.08 |
| DSA | 0 (0.0) | 0 (0.0) | 8 (100.0) | 0.001 |

Transcriptomic consistency with the discovery cohort was high: 79% of cABMR and 93% of clinTCMR DEGs were validated (Figure 4A; Suppl. Table 17). In cABMR, upregulated DEGs were associated with humoral immune mechanisms, complement activation, and fibrogenesis, whereas clinTCMR DEGs were associated with adaptive immune responses. Pathway analysis showed that cABMR DEGs were enriched in fibrogenesis and complement cascades, while clinTCMR DEGs were enriched in allograft rejection, interferon signaling, and adaptive immunity pathways (Figure 4B; Suppl. Table 18).

**Figure 4:**
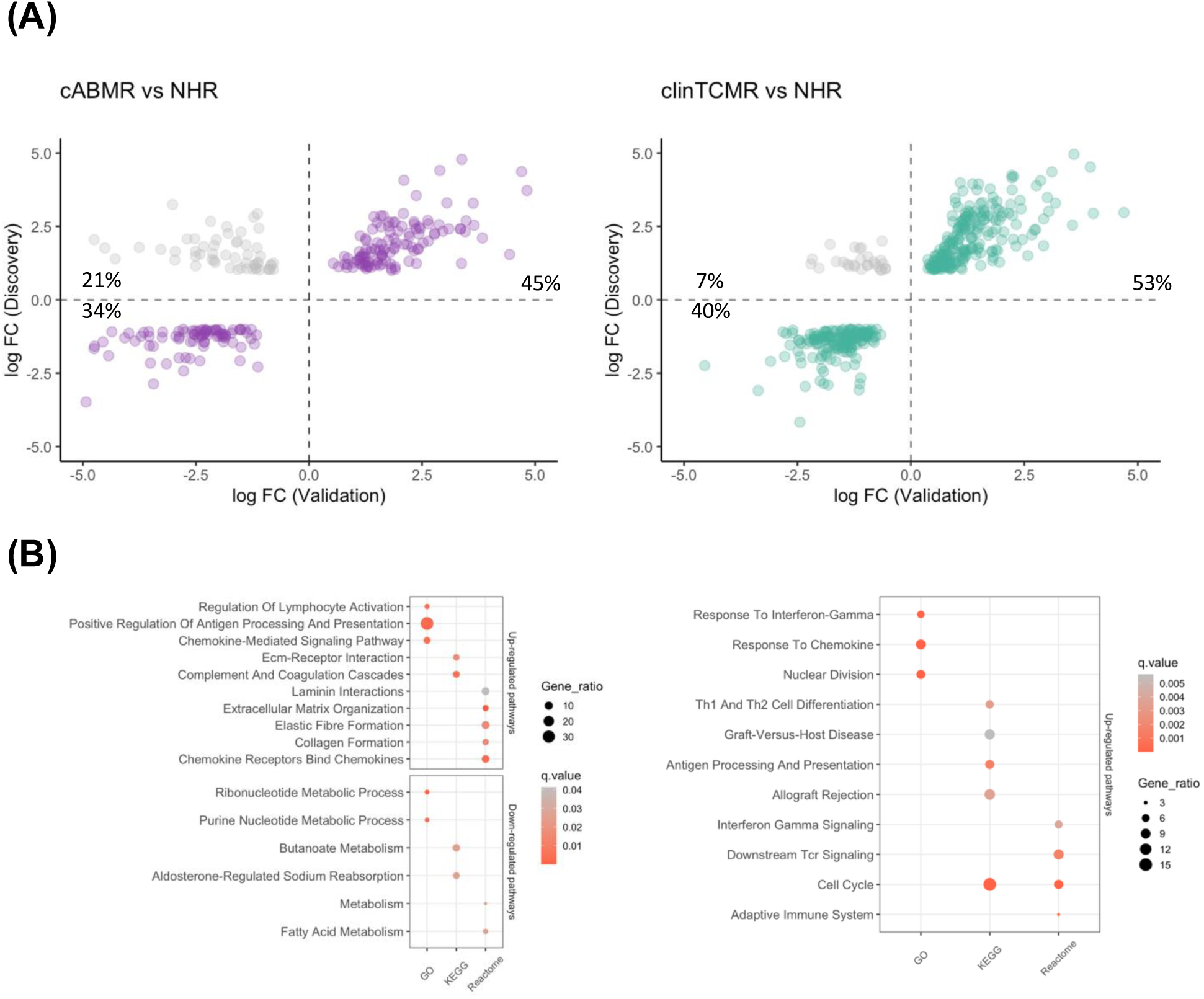
External validation of DEGs and pathways. **(A)** External cohort validation of rejection vs. NHR DEGs (p<0.05; Wald test). **(B)** Enriched pathways from DEGs in A. *clinTCMR–clinical T cell-mediated rejection; cABMR–chronic antibody-mediated rejection; NHR–no histological rejection; DEGs–differentially expressed genes*.

### 3.5 Plasma-soluble immune mediators and **s**erological neoepitope markers of extracellular matrix remodeling

We measured cytokines, complement factors, and extracellular matrix (ECM) remodeling markers in paired plasma samples (95/121) from the Hannover cohorts (Suppl. Figure 13). Plasma levels of 48 cytokines showed no significant differences between groups (Suppl. Figures 14A–E) and did not correlate with corresponding gene expression levels (Suppl. Figure 15). Among ECM markers, only C3M and C4M were elevated in cABMR compared to NHR (p=0.02 and p<0.01, respectively; Suppl. Figure 16). PRO-C3 levels were significantly higher in cABMR and clinTCMR versus subTCMR and NHR (Suppl. Figure 16F). Median plasma levels of complement factors C3c and C4 did not differ between groups (Suppl. Figure 17).

### 3.6 Outcomes

In the Hannover cohorts, follow-up periods did not differ significantly among groups: NHR (median 59 months, range 5–178), subTCMR (60 [6–164]), clinTCMR (44 [23–170]), cABMR (74 [24–170]), and cDR (34 [6–167]; p=0.67). At last follow-up, cirrhosis developed in none of the NHR patients, 12% of subTCMR, and 5% of clinTCMR cases. Cirrhosis prevalence was 50% in cDR and 40% in cABMR, with significant differences compared to NHR (p=0.03 and p=0.01, respectively). No significant differences were observed in survival or retransplantation rates (Log-Rank p=0.77). Clinical outcomes in the Barcelona cohort were similar; 50% of cABMR patients developed recurrent cirrhosis, while none of the controls or clinTCMR patients did.

## 4. Discussion

This study characterized the molecular signature of cABMR after LT, distinguishing it from clinTCMR, subTCMR, and cDR, using whole-transcriptome sequencing of mRNA from fresh-frozen and FFPE liver biopsies across three independent cohorts from two transplant centers. A substantial overlap (79–93%) in rejection-associated transcripts was observed across all cohorts.

Given the liver’s immunoprivileged status—reflected in low rates of acute cellular rejection[6], and rejection-related graft loss—many liver transplant centers initially questioned the relevance of ABMR despite its accepted relevance in other SOTs[21]. Subsequent findings, including reduced graft survival in DSA-positive LTR and associations between DSAs and fibrosis, challenged the notion of hepatic resistance to humoral injury[22,23]. The 2016 Banff criteria for acute and chronic ABMR enabled standardized diagnosis[6]. Although cABMR is less frequent after LT than in other SOTs, its prognosis—graft survival <50% at 5–10 years—parallels outcomes seen in other organs[7,24].

This two-center study confirms the poor prognosis of cABMR, with cirrhosis recurrence in 40–50% and re-listing in 18–25% of cases. Early detection remains difficult, as cABMR diagnosis requires at least moderate fibrosis histologically[6]. However, svLBx programs can identify cABMR before liver function tests deteriorate. A longitudinal pediatric study reported progression from DSA positivity to graft inflammation and eventually to graft fibrosis on paired svLBx[25]. The observed 13– 19% prevalence of cABMR among DSA-positive LT recipients is notable, given that up to 50% develop DSAs five years post-LT[2]. Moreover, 5–19% of those with advanced fibrosis had cABMR.

Clinical assessment of cABMR is challenging, as it requires DSA testing. Without svLBx, its prevalence is likely underestimated[21]. This may explain why ABMR-specific molecular signatures were undetected in the INTERLIVER study, which analyzed 235 prospectively collected biopsies, but included only 15% svLBx[26]. In the present study, 30 cABMR cases were identified from over 1,000 biopsies across 14 years at two high-volume centers. Notably, 57% of cABMR cases were subclinical and diagnosed exclusively via svLBx.

Although cABMR is associated with poor outcomes, the histologic criteria defined in 2016 remain debated. This study presents a validated, distinct molecular signature of cABMR consistent with those observed in other SOTs, reinforcing the clinical relevance of the 2016 criteria[6]. The findings support the concept that cABMR is pathophysiologically distinct from clinTCMR, driven by pro-fibrotic pathways, TNF–NF-κB signaling, and complement activation. The cABMR signature after LT showed enrichment in complement and humoral immune-related DEGs, including B-cell and immunoglobulin regulation (suppl. Table 10). In rat models, TNF–NF-κB-related DEGs were similarly elevated in ABMR after LT[29]. Despite structural and immunological differences across SOTs, cABMR signatures converge on similar pathways[8,9,27,28]. In contrast, clinTCMR was associated with DEGs reflecting T cell–mediated signaling, offering a molecular basis for further studies.

Several studies have sought to define a gene signature for clinTCMR in LTRs using microarray-based approaches[4,30,31]. In this study, 10 of 11 previously reported DEGs were significantly upregulated in clinTCMR versus NHR in the discovery cohort. Eight of the top 10 clinTCMR-associated genes from the INTERLIVER study were also overexpressed in both our discovery and internal validation cohorts. Notably, two genes—*GBP1* and *PSMB9*—were uniquely associated with clinTCMR in our cohort and are both interferon-inducible. Prior studies on clinTCMR signatures did not include any cABMR, thus classifying transcripts as TCMR-specific that were identified as transcripts being shared by cABMR and clinTCMR in the present study. In contrast, we observed overlap between clinTCMR and cABMR. Discrepancies in DEG profiles likely reflect differences in gene expression methodologies: microarrays in INTERLIVER and studies from the Sánchez-Fueyo group[26,30] vs. whole-transcriptome sequencing in our study. While platform differences influence gene counts, biological processes and diagnostic classifier performance remain largely consistent across technologies in KT cohorts[32]. Importantly, RNA-seq analyses show high concordance in mapped reads from fresh-frozen and FFPE tissue, despite RNA degradation during FFPE processing[32–35].

A limitation of this study is the predominantly negative C4d staining in both discovery and internal validation cohorts. According to the 2016 Banff Consensus, absence or

failure of C4d staining reduces diagnostic certainty, categorizing cABMR as possible rather than probable[6]. C4d assay performance varies by technique and center protocol[6,36]. While C4d staining on FFPE liver tissue performs acceptably in acute ABMR compared to immunofluorescence on frozen tissue, data on its use in cABMR are limited[36]. Sensitivity of C4d staining for cABMR is generally low[6]. This limitation was mitigated by the external Barcelona cohort, where 50% of cases were C4d-positive, potentially reflecting a more severe clinical phenotype (87% elevated liver enzymes in Barcelona vs. 27% in Hannover). Notably, molecular signature overlap between validation cohorts with and without C4d deposition ranged from 79-87%. Furthermore, cABMR diagnosis without C4d is well established and carries similar prognostic significance as C4d-positive ABMR in kidney and lung transplantation[37,38]. These findings align with current pathophysiological concepts of ABMR from other SOTs and reinforce evidence for cABMR in LT[28].

The three cohorts showed additional heterogeneity, including higher cyclosporine use in the older discovery cohort (2008–2016), variable liver enzyme elevations, later cABMR diagnosis post-LT in validation cohorts, and differing biopsy preservation methods (fresh frozen in discovery vs. FFPE in validation). Despite these differences, molecular rejection signatures overlapped 79–93%, indicating that cABMR and clinTCMR signatures are consistent regardless of center, immunosuppressive regimen, time post-LT, or sample type. The strong concordance with cABMR signatures from other SOTs further supports the study’s conclusions[27,28].

The molecular signature of cABMR complements prior studies linking TCMR-associated transcripts to liver graft injury and fibrosis progression in svLBx[4]. TCMR signatures have aided the development of clinical decision tools using non-invasive tests such as liver function tests, DSA, and liver stiffness measurements[30]. That study proposed that classical light microscopy injury patterns might overestimate

TCMR risk at the molecular level in svLBx. These patterns were used to screen immunosuppression minimization strategies in a prospective multicenter trial[30]. Thus, combining cABMR and TCMR signatures could guide and monitor personalized immunosuppression in both clinical and research settings, an approach already applied in kidney transplantation[39].

Except for extracellular matrix remodeling markers linked to liver graft fibrosis[40], neither complement nor cytokine levels in peripheral blood reflected liver graft injury in cABMR. However, blood samples did not adequately represent the liver’s T cell compartment[41,42].Currently, the only well-validated non-invasive screening tests for cABMR are DSA and liver stiffness measurement. Abnormal results from these tests may prompt a svLbx[2,43].

This study provides further evidence that cABMR progresses to end-stage liver graft failure in 40-50% of patients, despite diagnosis during a predominantly subclinical phase. The molecular signatures defining histological cABMR differ from clinTCMR, are externally validated, and resemble those seen in other SOTs. Identifying patients at high risk of graft failure due to cABMR is critical for personalizing future therapies. This is imperative to ensure the exclusion of a relevant rejection risk in the majority of LT recipients, with the objective of reducing immunosuppression and, by extension, the toxicity of long-term immunosuppressive treatments in LTR.

## Data Availability

The data that support the plots within this paper and other findings of this study are available from the corresponding author upon reasonable request. Representative histopathological whole slide images, the raw gene expression data, including a sample-data relationship file, as well as the count tables are stored conformant with the MINSEQE reporting standard (Version 1.0, June 2012, accessible at https://zenodo.org/doi/10.5281/zenodo.5706411) at MHH RepoMed repository https://doi.org/10.26068/mhhrpm/20240207-000.
The manuscript was published as a preprint at medRxiv on February 8th 2024 and updated with the revised versions on October 15th 2024 and May 15th 2025. The preprint can be accessed at medrxiv.org by entering the manuscript ID MEDRXIV/2024/302515.

## Acknowledgements

We thank Szilvia Ziegert and Konstantinos Iordanidis from the “immune tolerance working group” at Hannover Medical School for technical assistance. We would like to thank Raquel García (advanced practice nurse in liver transplantation), Ana Dura (secretary), Javier Muñoz (biotechnician) and Christian Rodríguez (research nurse) for their assistance with sample collection and processing. We thank Esma Turlak for assistance with immunofluorescent staining. We thank Stefan Huebscher, already retired from his work in the pathology department of the University of Birmingham/UK, for his advice in initiating this project and for his training of the pathologists at Hannover Medical School during a fruitful workshop in September 2016.

## Author contributions to manuscript

Study concept and design: BE, EJ, RT. Acquisition of data: BE (overall data management), ACM (validation cohort), AK and KC (training cohort), PR, GC, AD, JC (external validation cohort), BH & DDJ (histopathology), ETS (ECM markers), CSF (cytokine measurements). Analysis and interpretation of data: BE and ACM (overall data analysis and interpretation), AA (bioinformatics), SR, CL, CSF, ETS, DJL, EJ, YL, RT. Drafting of the manuscript: BE, AA, ACM, RT. Critical revision of the manuscript for important intellectual content: All authors. Statistical analysis: BE, AA, ACM, CJX, YL. Obtained funding: BE, ACM, KC, JC, GC, PR, CL, EJ, YL, RT. Administrative, technical or material support: All authors. Study supervision: CJX, EJ, YL, RT.

## Data availability statement

The data that support the plots within this paper and other findings of this study are available from the corresponding author upon reasonable request. Representative histopathological whole slide images, the raw gene expression data, including a sample-data relationship file, as well as the count tables are stored conformant with the MINSEQE reporting standard (Version 1.0, June 2012, accessible at https://zenodo.org/doi/10.5281/zenodo.5706411) at MHH RepoMed repository https://doi.org/10.26068/mhhrpm/20240207-000.

The manuscript was published as a preprint at medRxiv on February 8th 2024 and updated with the revised versions on October 15th 2024 and May 15th 2025. The preprint can be accessed at medrxiv.org by entering the manuscript ID MEDRXIV/2024/302515.

## Declaration of generative AI and AI-assisted technologies in the writing process

During the preparation of this work the author(s) used [DeepL and ChatGPT] in order to improve the usage of the English language. After using this tool/service, the author(s) reviewed and edited the content as needed and take(s) full responsibility for the content of the publication.

## Abbreviations

ABMR: antibody-mediated rejection
cABMR: chronic antibody-mediated rejection
cDR: chronic ductopenic rejection
CsA: cyclosporine A
clinTCMR: clinically overt T cell-mediated rejection
DEGs: differentially expressed genes
DSA: donor-specific anti-HLA antibodies
FDR: false-discovery rate
FFPE: formalin-fixed paraffin-embedded
KT: kidney transplantation
LAF: liver allograft fibrosis
LBx: liver graft biopsy
LT: liver transplantation
LTR: liver transplant recipient
NHR: no histological rejection
mHAI: modified histological activity index
RAI: rejection activity index
RIN: RNA integrity number
SOT: solid organ transplantation
subTCMR: subclinical T cell-mediated rejection
svLBx: surveillance liver graft biopsies
TAC: tacrolimus
TCMR: T cell-mediated rejection
ULN: upper limit of normal
xULN: times the upper limit of normal.

**Bold designates equally contributing authors in the references.**

