## Supplemental file for "Molecular signatures of chronic antibody-mediated rejection in human liver transplants"

Robert Geffers: 0000-0003-4409-016X

Martijn Zoodsma: 0000-0003-3636-2209

Murielle Verboom: 0000-0001-8260-2045

Cheng-Jian Xu: 0000-0003-1586-4672

Sergej Ruff: 0009-0000-8264-6347

Chris Lauber: 0000-0002-2265-2953

Christine S Falk: 0000-0003-1376-7318

Emilie Toft Skovgaard: 0009-0002-3875-488X

Morten Karsdal: 0000-0001-5026-8740

Diana Julie Leeming: 0000-0002-4256-140X

Jordi Colmenero: 0000-0001-6024-8479

Pablo Ruiz: 0000-0002-5810-8802

Gonzalo Crespo: 0000-0002-1178-4897

Alba Diaz: 0000-0002-0456-7085

[1]Department of Gastroenterology, Hepatology, Infectious Diseases and Endocrinology, Hannover Medical School, Hannover, Germany

[2]PRACTIS & CORE100Pilot Clinician Scientist Program Clinician Scientist Program, Dean's Office for Academic Career Development, Hannover Medical School, Germany

[3] MHH Transplant Center, Hannover Medical School, Germany

[4] Department of Computational Biology for Individualised Medicine, Centre for Individualised Infection Medicine (CiiM), a joint venture between the Helmholtz-Centre for Infection Research (HZI) and the Hannover Medical School (MHH), Hannover, Germany

[5]TWINCORE, a joint venture between the Helmholtz-Centre for Infection Research (HZI) and the Hannover Medical School (MHH), Hannover, Germany

[6] Liver Transplantation, Liver Unit. Hospital Clínic, Biomedical Research Institute August Pi i Sunyer (IDIBAPS), Biomedical Research Networking Center (CIBER), University of Barcelona, Spain

[7]Hospital Clínic, IDIBAPS - Biomedical Research Institute August Pi i Sunyer, CIBER - Biomedical Research Networking Center, University of Barcelona, Spain

[8]Institute for Pathology, Hannover Medical School, Hannover, Germany

[9] Institute of Pathology, RWTH University of Aachen, Aachen, Germany.

[10] Department of Pathology. Hospital Clínic, Biomedical Research Institute August Pi i Sunyer (IDIBAPS), Biomedical Research Networking Center (CIBER), University of Barcelona, Spain

[11] Institute for Transfusion Medicine and Transplant Engineering, Hannover Medical School, Hannover, Germany

[12] Research Group Genome Analytics, Helmholtz Centre for Infection Research, Braunschweig, Germany

[13] Institute of Transplant Immunology, Hannover Medical School (MHH), Hannover, Germany; German Centre for Infection Research, TTU-IICH Hannover, Hannover, Germany

[14] Institute for Experimental Virology, TWINCORE Centre for Experimental and Clinical Infection Research, a joint venture between the Hannover Medical School (MHH) and the Helmholtz Centre for Infection Research (HZI), Hannover, Germany

[15] Cluster of Excellence 2155 RESIST, Hannover, Germany

[16] Nordic Bioscience A/S, Herlev, Denmark

[17] Department of Internal Medicine and Radboud Institute for Molecular Life Sciences, Radboud University Medical Center, Nijmegen, the Netherlands

[18] Cluster of Excellence RESIST (EXC 2155), Hannover Medical School, Hannover, Germany

[19] Lower Saxony center for artificial intelligence and causal methods in medicine (CAIMed), Hannover, Germany

[&] Biomedical Research in End-stage and Obstructive Lung Disease Hannover (BREATH) of the German Center for Lung Research (DZL), Hannover, Germany.

#Current address: Ajmera Transplant Centre, Toronto General Hospital, United Health Network, University of Toronto, Toronto, Canada

**Table of contents**


| | |
|---|---|
| **Supplementary Methods & Patients** | **3** |
| **Supplementary Figures** | **9** |
| **Supplementary Tables** | **28** |
| **References** | **3594** |


#### Supplementary Methods & Patients

##### 1.1 Patient cohort

For the training cohort, only patients with available freshly conserved liver tissue from LBx were eligible to guarantee the highest quality of RNA for downstream analyses as previously published by our group[1]. Samples had to be obtained between December 2008 and October 2016 to be eligible for inclusion in the training cohort (Suppl. Figure 2A). Patients with possible cABMR were selected based on the Banff criteria of 2016[2]. Clinical management of LTR was changed to a risk-stratified approach guided by svLBx which was fully established in 2018 as published[3]. Hence, for the validation cohort, samples that were collected between 2018 and 2022 were eligible for study inclusion. Storage of freshly conserved liver tissue was not mandatory for inclusion into the validation cohort as formalin-fixed and paraffin-embedded (FFPE) liver tissue was more accessible. The selection is outlined in suppl. Figure 2B.

##### 1.2 Biochemical measurements

Laboratory parameters were obtained from patients' records and were measured in clinical routine using high throughput techniques. C3c and C4 complement were measured in EDTA-plasma samples that were collected at the time of biopsy and were stored at -80°C. Measurements were performed in the central laboratory facility of Hannover Medical School on a Cobas 8000 (Roche, Switzerland).

**1.3 Liver biopsies, rejection phenotypes, histological grading and staging**

LBx were performed percutaneously with local anesthesia. A 5 mm section of the biopsy cylinder was immediately preserved in Allprotect Tissue Reagent (ID: 76405, Qiagen N.V., Venlo, The Netherlands) and then stored at -80 °C. The remaining biopsy cylinder was fixed in 4% neutral buffered formalin, embedded in paraffin wax and subsequently used for routine histopathological evaluation. The majority of DSA positive biopsies (>98%) were negative for C4d deposition with our protocol and an external protocol used by Prof. Huebscher in Birmingham, UK and those that were positive did not show a typical staining pattern[1,4]. Therefore, C4d positive staining was not mandatory for the diagnosis of "possible" cABMR, in line with the current Banff consensus[2], in contrast to C4d-positive "probable" cABMR to be included as cABMR. In the following manuscript, cABMR reflects "possible" chronic ABMR. In line with the Banff consensus, features of TCMR may co-exist with cABMR[2]. No histological rejection (NHR) was defined as RAI ≤1 without any relevant inflammation or fibrosis (Ishak F ≤1, each LAF score component ≤1). Patients had to have liver enzymes within the normal range for their biopsy to be classified as NHR. Exemplary stains of each phenotype are provided in suppl. Figure 3 and distribution of histopathological scores within groups are summarized in suppl. table 1. In addition, representative whole slide images were uploaded to the repository along with the sequencing data. The repository can be assessed at: https://doi.org/10.26068/mhhrpm/20240207-000.

#### 1.4 Virus discovery in raw read sequencing data

Raw-read sequencing data was analyzed for the presence of sequences of viral origin using a Data-driven Virus Discovery approach[5]. The sequencing experiments were subjected to a profile Hidden Markov Model (pHMM)-based sequence homology search using multiple amino acid sequence alignments of viral marker genes for various eukaryotic DNA and RNA virus families. For the hits obtained we performed a seed-based, targeted viral genome assembly. The computational workflow is described in previous studies[6-8] and an implementation is provided by the Virushunter and Virusgatherer tools, which are available at https://github.com/lauberlab/VirusHunterGatherer. Visualization of the virus discovery results was done using Virusparies, which is available at https://github.com/SergejRuff/Virusparies.

#### 1.5 Cytokines and chemokines quantification

Forty-eight cytokines and chemokines were quantified for 95 of 129 patients from the training and validation cohorts using the Luminex-based multiplex technology (human 27-Plex, human 21- Plex, ICAM-1, VCAM-1, Bio-Rad Kits, #M50-OKCAF0Y, #MF0-005KMII, #171- B6009M, #171-B6022M) according to the manufacturer's instructions as published recently[9].

#### 1.6 Serological neoepitope markers of extracellular matrix remodeling

The biomarkers assessed in blood samples reflect extracellular matrix remodeling of type III, IV, and VI collagen (nordicPRO-C3™[10], nordicPRO-C4™[11], and nordicPRO-C6™[12]) and the degradation of type III and IV collagen generated by matrix metalloproteinase (MMP) cleavage (nordicC3M™[13] and nordicC4M™[14]).

Immunoregulatory-related biomarkers reflecting macrophage activity assessed by an MMP and citrullinated vimentin fragment (nordicVICM™[15]) and T-cell activity assessed by a granzyme B-generated type IV collagen neoepitope (nordicC4G™[16]) were also measured. The biomarkers were assessed using validated enzyme-linked immunosorbent assays (ELISAs) and high-sensitivity automated sandwich chemiluminescence assays (IDS-i10) developed by Nordic Bioscience A/S (Herlev, Denmark). Biomarker levels were based on a calibrator standard curve using SoftMax Pro v7.1 or the IDS-i10 software.

**1.7 Immunofluorescence Staining**

The deparaffinization process was conducted using xylene for a duration of two eight-minute intervals, followed by an ethanol dilution series (100%, 90%, 80%, 70%) with a five-minute duration each, concluding with distilled water and TBST rinses. To restore antigenicity the slides were subjected to a three-minute heating process in a pressure cooker. The blocking process was conducted with a 10-minute incubation period in a solution of 5% goat serum in TBST, followed by a washing step. The staining process entailed the application of the primary antibody (MMP2 (ABIN6143911) and Col14A1 (ABIN7183371) at a 1:50 dilution with overnight incubation at 4 °C, followed by a washing step. Subsequently, the secondary antibody (goat-anti-rabbit AF555; Invitrogen, A32732, California, USA) at a dilution of 1:500 was added for 60 minutes, after which a nuclear staining solution (Flouoromount-G, with DAPI (Invitrogen, REF 00-4959-52, California, USA) was applied. Samples were scanned using the PhenoImager® Fusion Whole Slide Scanner (Akoya Biosciences, Massachusetts, USA).

**1.8 Statistical analysis**

Statistical analyses were performed in IBM SPSS Statistics (version 27, Spss, Inc., Chicago, IL) and R Statistical Software (version 4.1.2, R Core Team). The Wilcoxon-Mann-Whitney U test or Student's t-test were used to compare quantitative data between two independent groups as applicable and the Kruskal-Wallis test with Dunn's multiple comparison post-hoc test was used for the comparison between more than two groups. The $\chi^2$ test was used to compare contingency tables with more than two groups.

#### Supplementary Figures

##### Supplementary Figure 1

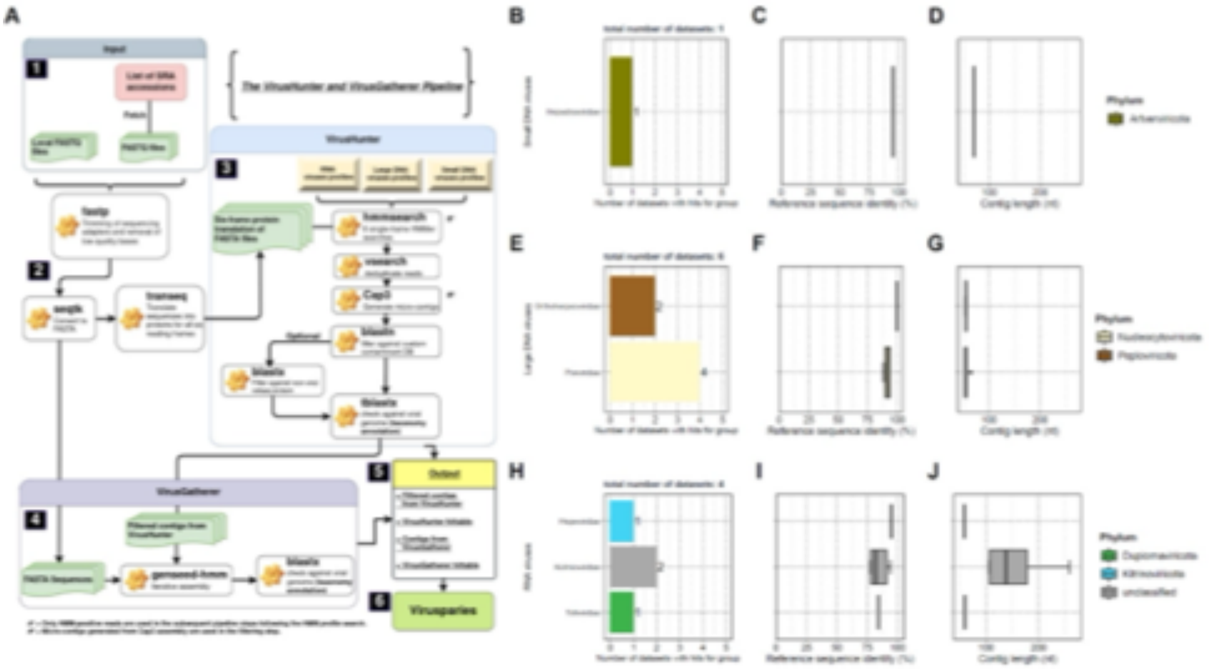


**Supplementary Figure 1. Virus discovery in raw sequencing data. (A)** Workflows of the Virushunter and Virusgatherer computational pipelines. The pipelines can process local FASTQ files or download sequencing data from the NCBI SRA database using SRA accessions (step 1). For both, the FASTQ files are converted into FASTA format. For Virushunter, the nucleotide sequences are *in silico* translated into amino acid sequences in all six reading frames (step 2). A profile Hidden Markov model-based homology search,with profiles for either DNA or RNA viruses, is employed to detect viral sequences. After deduplicating the detected viral reads, they are assembled into micro-contigs. Custom contaminants and, optionally, non-viral reference sequences are filtered out. Finally, taxonomic annotation is performed by pairwise aligning the remaining viral contigs against a viral reference database (step 3). The contigs from the Virushunter output are used for iterative, targeted assembly of longer viral contigs that may correspond to complete viral genomes or genome fragments. A final taxonomic annotation is then performed for these longer contigs (step 4). The results from Virushunter and Virusgatherer include contigs in Fasta format and hit tables in tab-separated format (step 5). Hit tables from Virushunter and Virusgatherer are used in the Virusparies R package for data processing, summary statistics, and visualizing results (step 6). **(B-J)** Shown are (from left to right) the number of datasets with viral hits, the amino acid sequence identity of these hits against the closest viral reference sequence, and the length of the viral contig sequences for hits against small DNA viruses **(B,C,D),** large DNA viruses **(E,F,G)**, and RNA viruses **(H,I,J).** Virus taxonomic phyla are discriminated by color.

#### Supplementary Figure 2

(A)

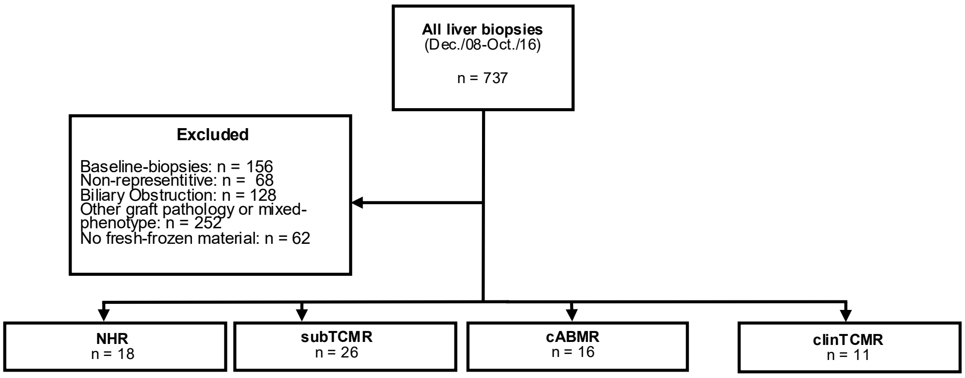


(B)

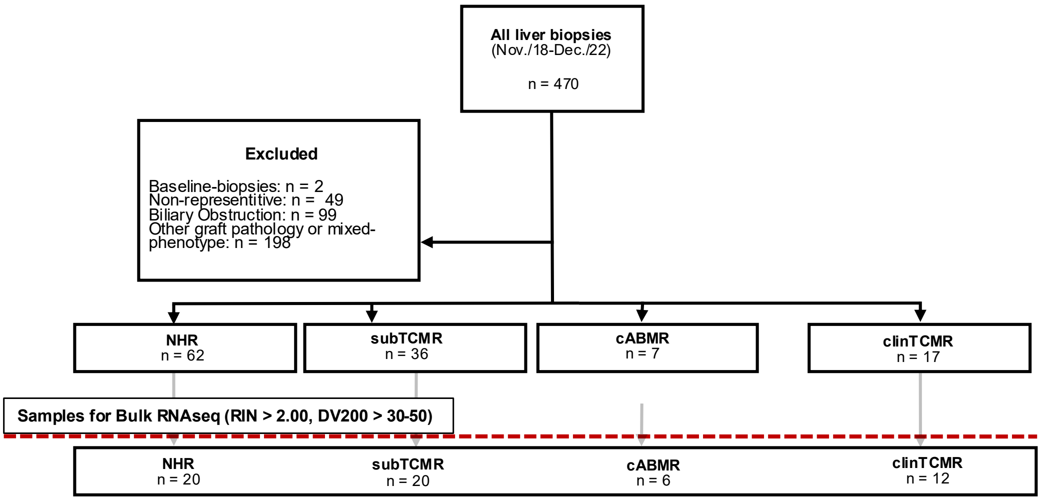


**Supp. Figure 2: Sample selection flow chart**
**(A)** Flow Chart highlighting sample selection for the discovery cohort (**B)** Flow Chart highlighting sample selection for the validation cohort. NHR and subTCMR were matched for age, sex and time after transplantation to clinTCMR and cABMR as far as possible in both cohorts.
RIN: RNA integrity number; DV200: RNA quality metric that assesses the frequency of RNA fragments > 200 nucleotides. clinTCMR: clinical T cell-mediated rejection; cABMR: chronic antibody-mediated rejection; subTCMR: subclinical T cell-mediated rejection; NHR: no histological rejection.

**Supplementary Figure 3**

**(A)**

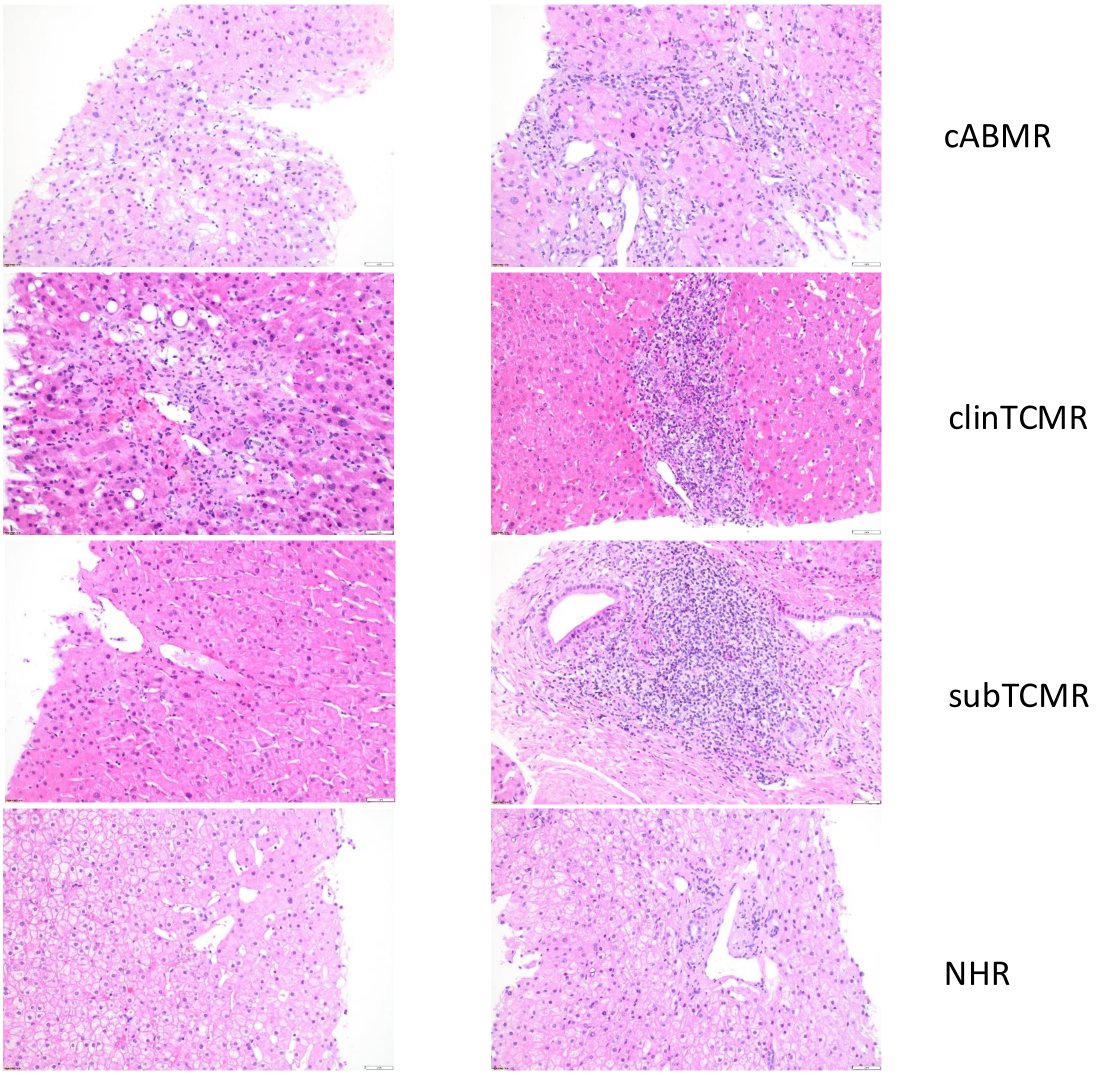


**Supp. Figure 3: Diagnostic scheme for phenotype classification and exemplary H&E stains**
**(A)** Exemplary hematoxylin & eosin stains for each phenotype with a focus on the lobular area on the left-sided sections and on the portal area on the right-sided sections. clinTCMR: clinical T cell-mediated rejection; cABMR: chronic antibody-mediated rejection; subTCMR: subclinical T cell-mediated rejection; NHR: no histological rejection, ULN: upper limit of normal, RAI: rejection activity index, MFI: mean fluorescence intensity, DSA: donor specific anti-HLA antibodies. Representative whole slide images were uploaded to the repository along with the sequencing data. The repository can be assessed at: https://doi.org/10.26068/mhhrpm/20240207-000.

#### Supplementary Figure 4

**(A)**

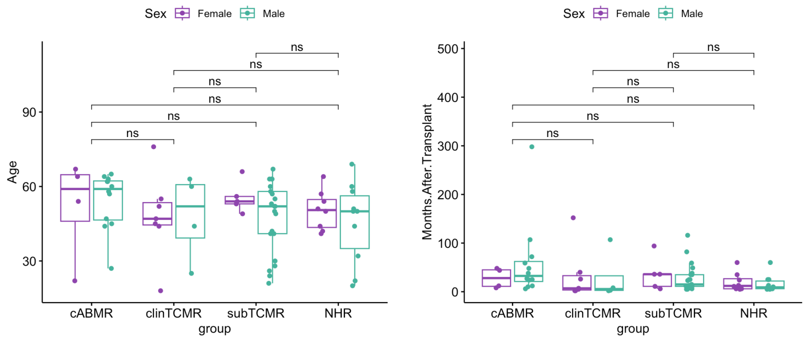


**(B)**

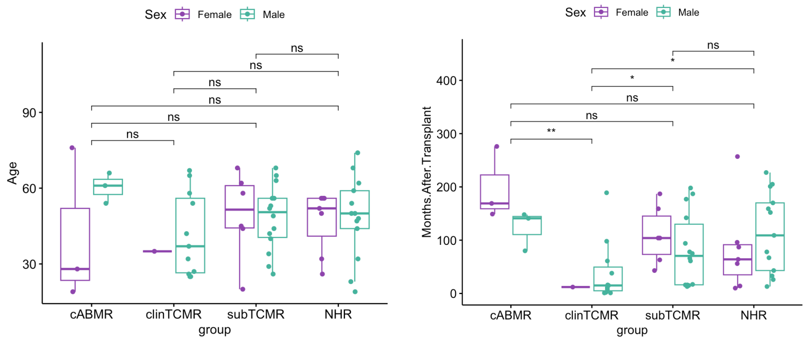


**Supp. Figure 4: Demographic data of the discovery and validation cohorts**

**(A)** Demographic data of the discovery cohort. (**B)** Demographic data of the validation cohort. clinTCMR: clinical T cell-mediated rejection; cABMR: chronic antibody-mediated rejection; subTCMR: subclinical T cell-mediated rejection; NHR: no histological rejection, ns: not significant. *: $p < 0.05$; **: $p < 0.01$. Kruskal-Wallis test with Dunn‘s post-hoc test.

#### Supplementary Figure 5

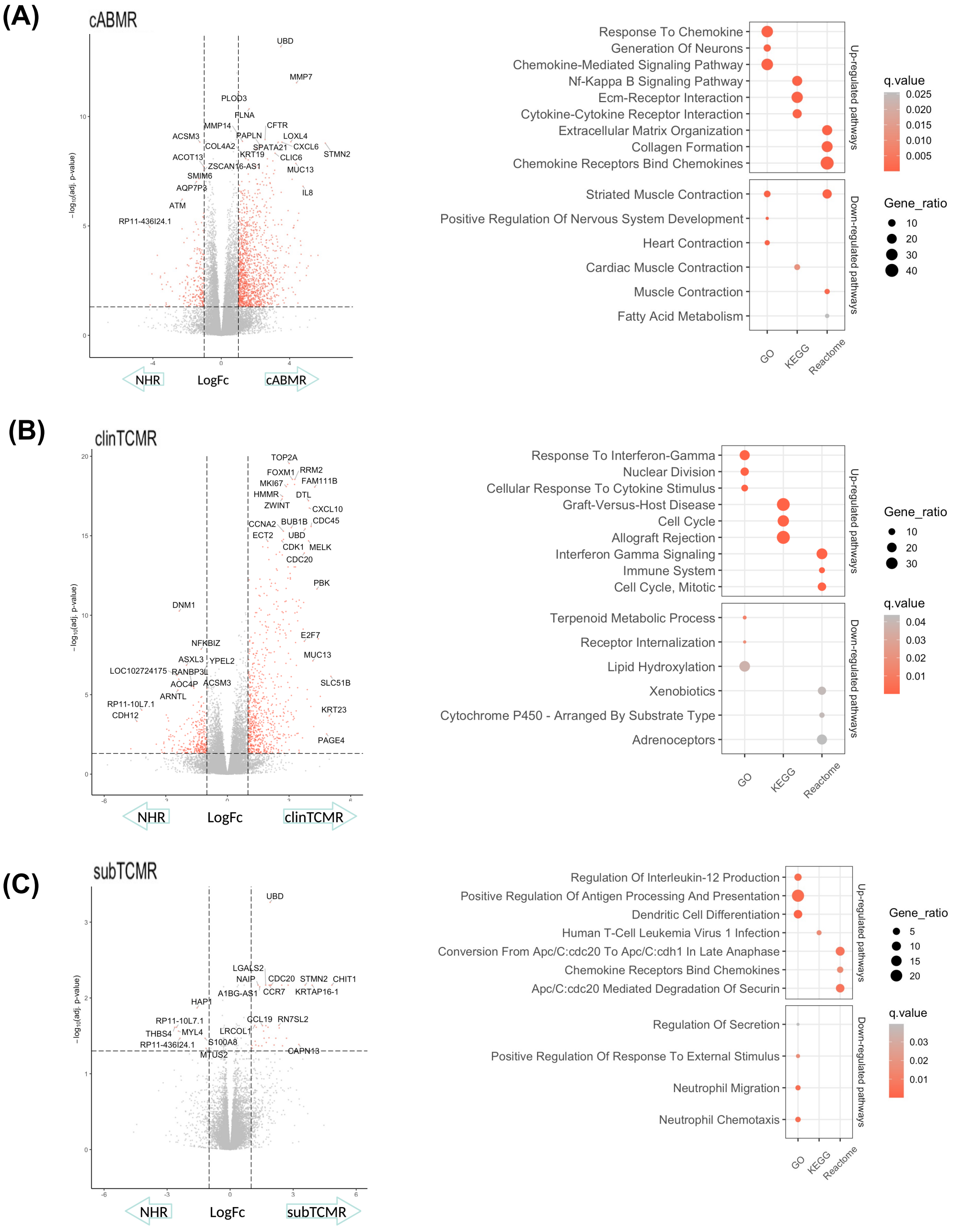


**Supp. Figure 5: Molecular signature and functional enrichment of DEGs of cABMR, clinTCMR and subTCMR in the discovery cohort**

Volcano plots showing cABMR vs NHR DEGs (**A**), clinTCMR vs NHR DEGs (**B**), subTCMR vs NHR DEGs (**C**) from the discovery cohort, using false discovery rate (FDR) < 0.05 and absolute log fold change (LogFc) > 1, with Over-represented pathways out of these DEGs. clinTCMR: clinical T cell-mediated rejection; cABMR: chronic antibody-mediated rejection; subTCMR: subclinical T cell-mediated rejection; NHR: no histological rejection, DEGs: differentially expressed genes.

#### Supplementary Figure 6

**(A)**

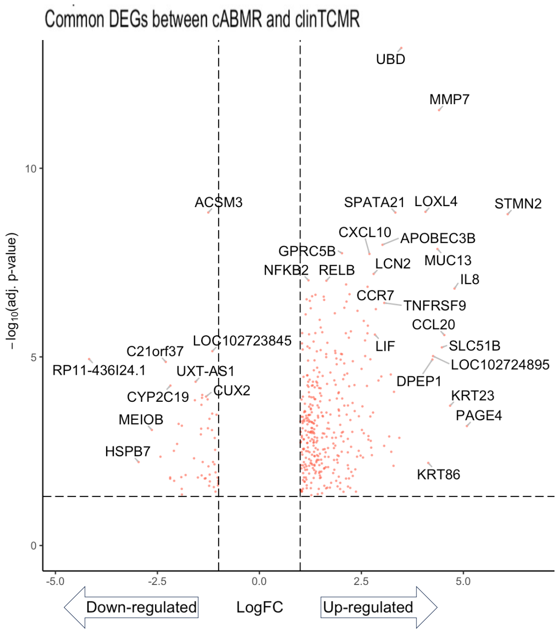


**(B)**

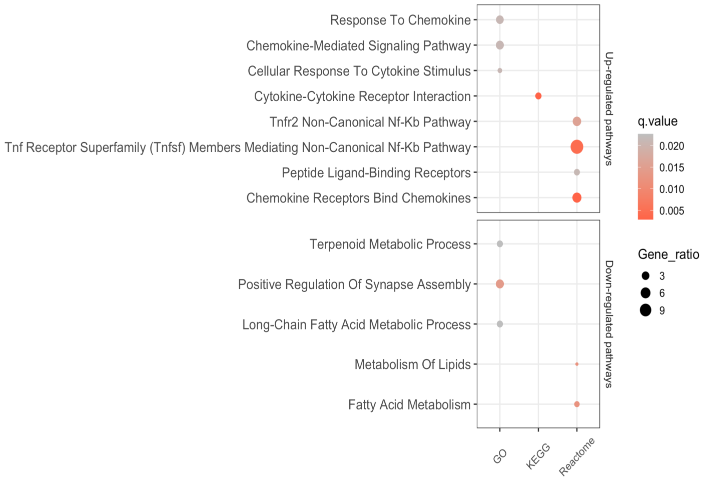


**Supp. Figure 6: Common features between cABMR and clinTCMR in the discovery cohort**

**(A)** Volcano plot showing the common features between cABMR and clinTCMR vs NHR DEGs, using false discovery rate (FDR) < 0.05 and absolute log fold change (LogFc) > 1. (**B**) Over-represented pathways out of DEGs in panel A. clinTCMR: clinical T cell-mediated rejection; cABMR: chronic antibody-mediated rejection; subTCMR: subclinical T cell-mediated rejection; NHR: no histological rejection, DEGs: differentially expressed genes.

#### Supplementary Figure 7

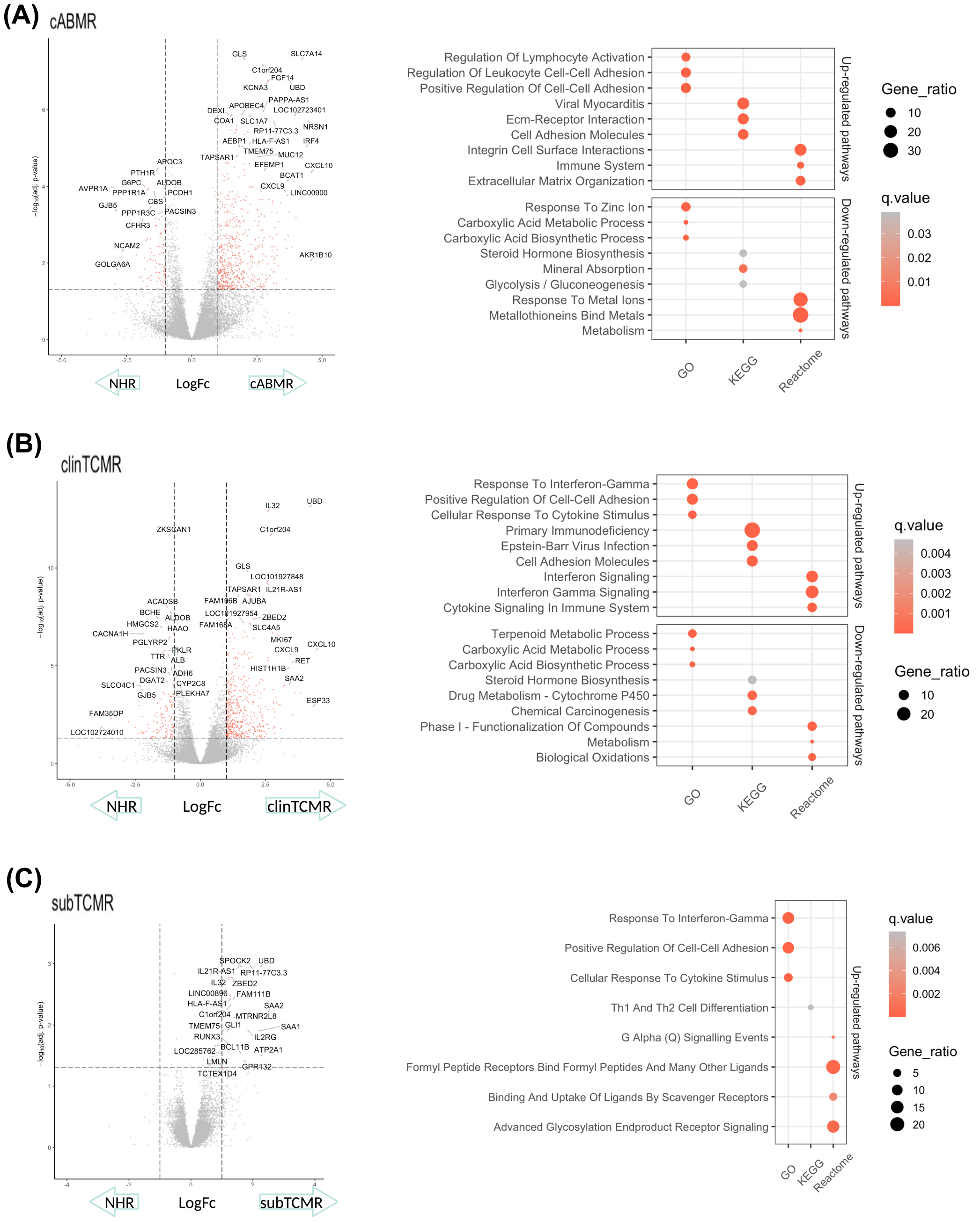


**Suppl. Figure 7: Molecular signature and functional enrichment of DEGs of cABMR, clinTCMR and subTCMR in the validation cohort**

Volcano plots showing cABMR vs NHR DEGs (**A**), clinTCMR vs NHR DEGs (**B**), subTCMR vs NHR DEGs (**C**) from the validation cohort, using false discovery rate (FDR) < 0.05 and absolute log fold change (LogFc) > 1, with Over-represented pathways out of these DEGs. clinTCMR: clinical T cell-mediated rejection; cABMR: chronic antibody-mediated rejection; subTCMR: subclinical T cell-mediated rejection; NHR: no histological rejection, DEGs: differentially expressed genes.

#### Supplementary Figure 8

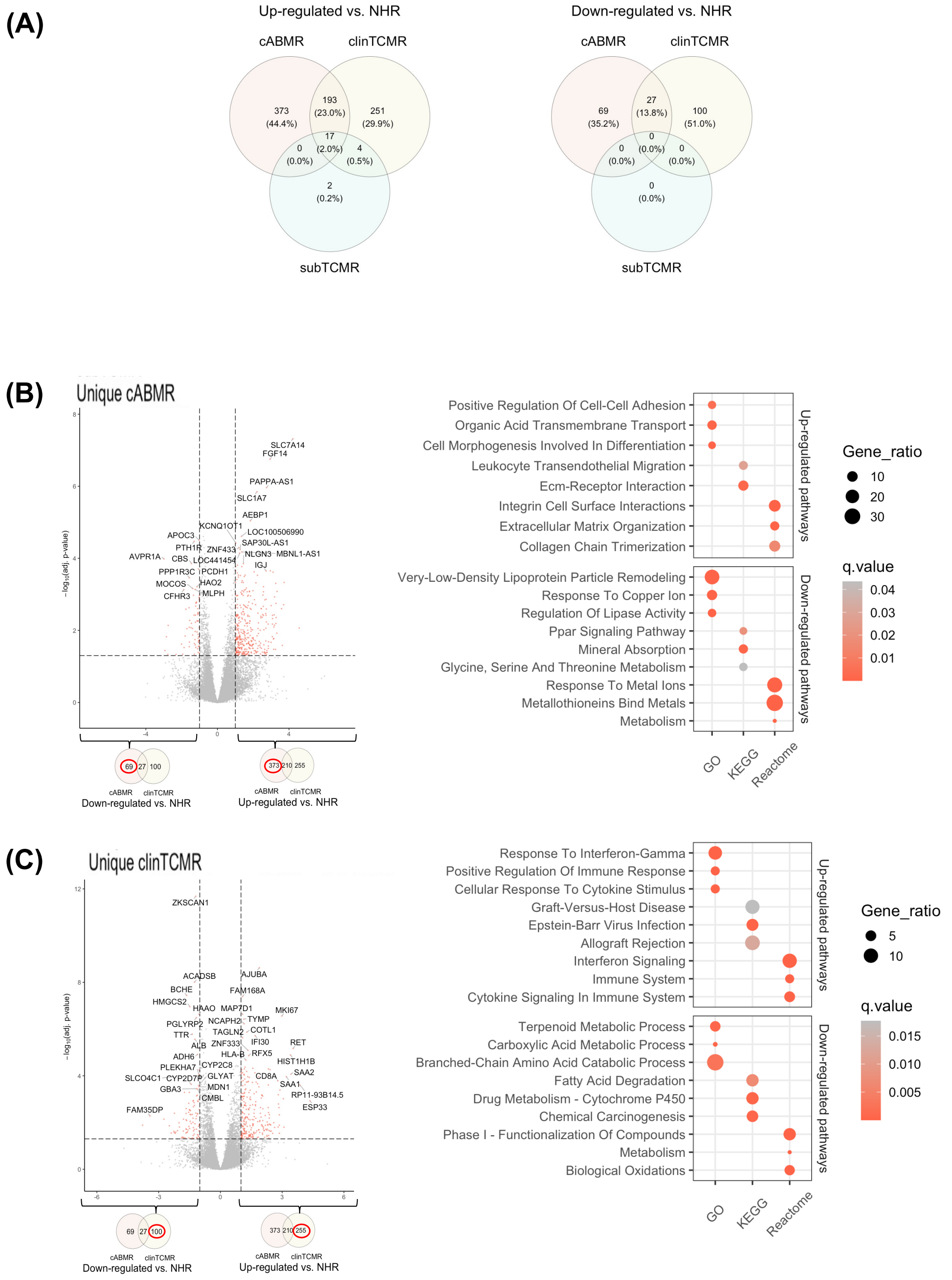


**Suppl. Figure 8: Molecular signature and functional enrichment of unique DEGs of cABMR and clinTCMR in the validation cohort.**

**(A)** Venn diagram showing the numbers of the up and down-regulated DEGs in each rejection group compared to NHR from validation cohort. Volcano plot showing the unshared (unique) cABMR vs NHR **(B)** and unshared (unique) clinTCMR vs NHR **(C)** DEGs from the validation cohort, using false discovery rate (FDR) < 0.05 and absolute log fold change (LogFc) > 1 with Over-represented pathways out of these. clinTCMR: clinical T cell-mediated rejection; cABMR: chronic antibody-mediated rejection; subTCMR: subclinical T cell-mediated rejection; NHR: no histological rejection, DEGs: differentially expressed genes.

**Supplementary Figure 9**

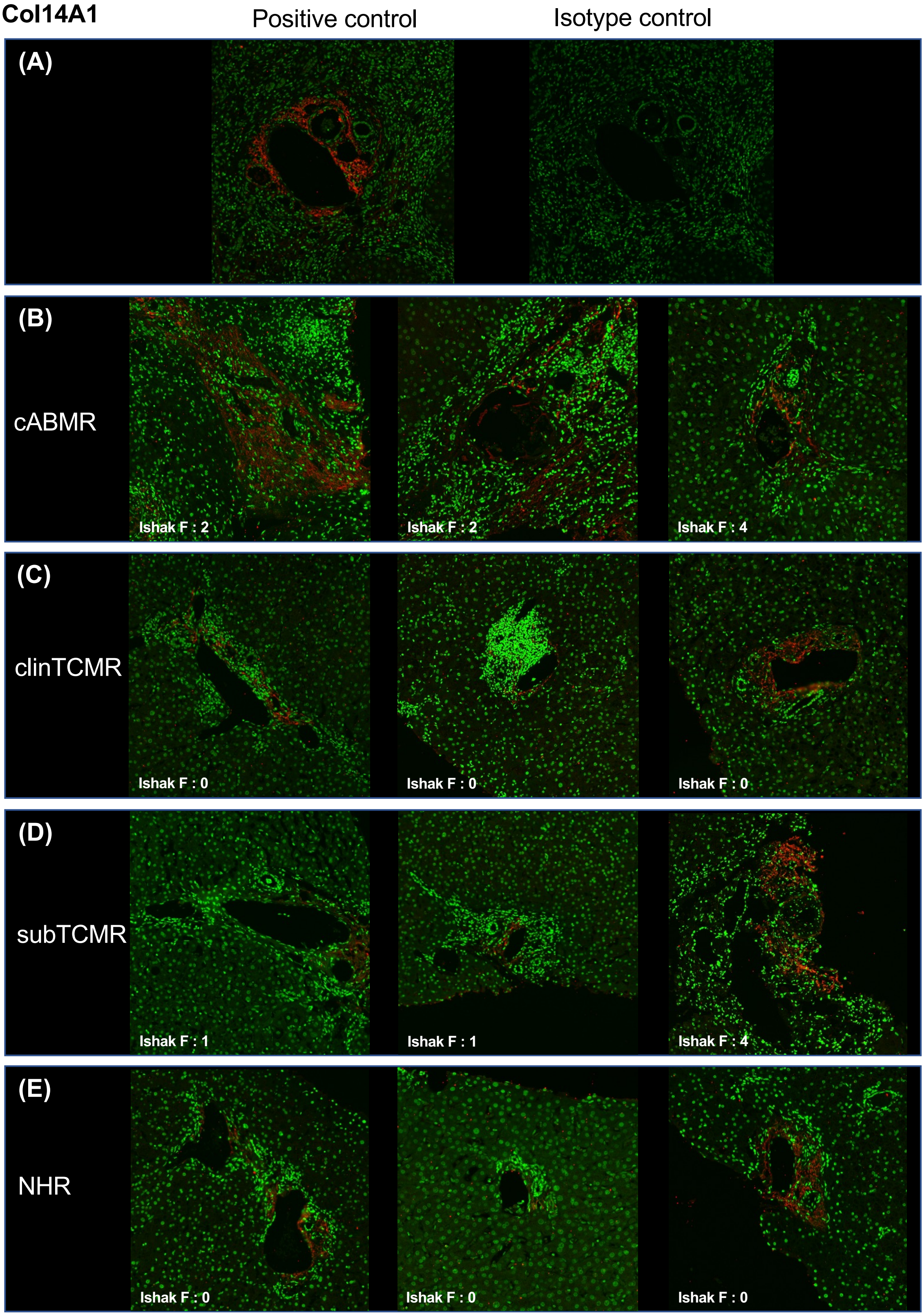

**Supplementary Figure 9**

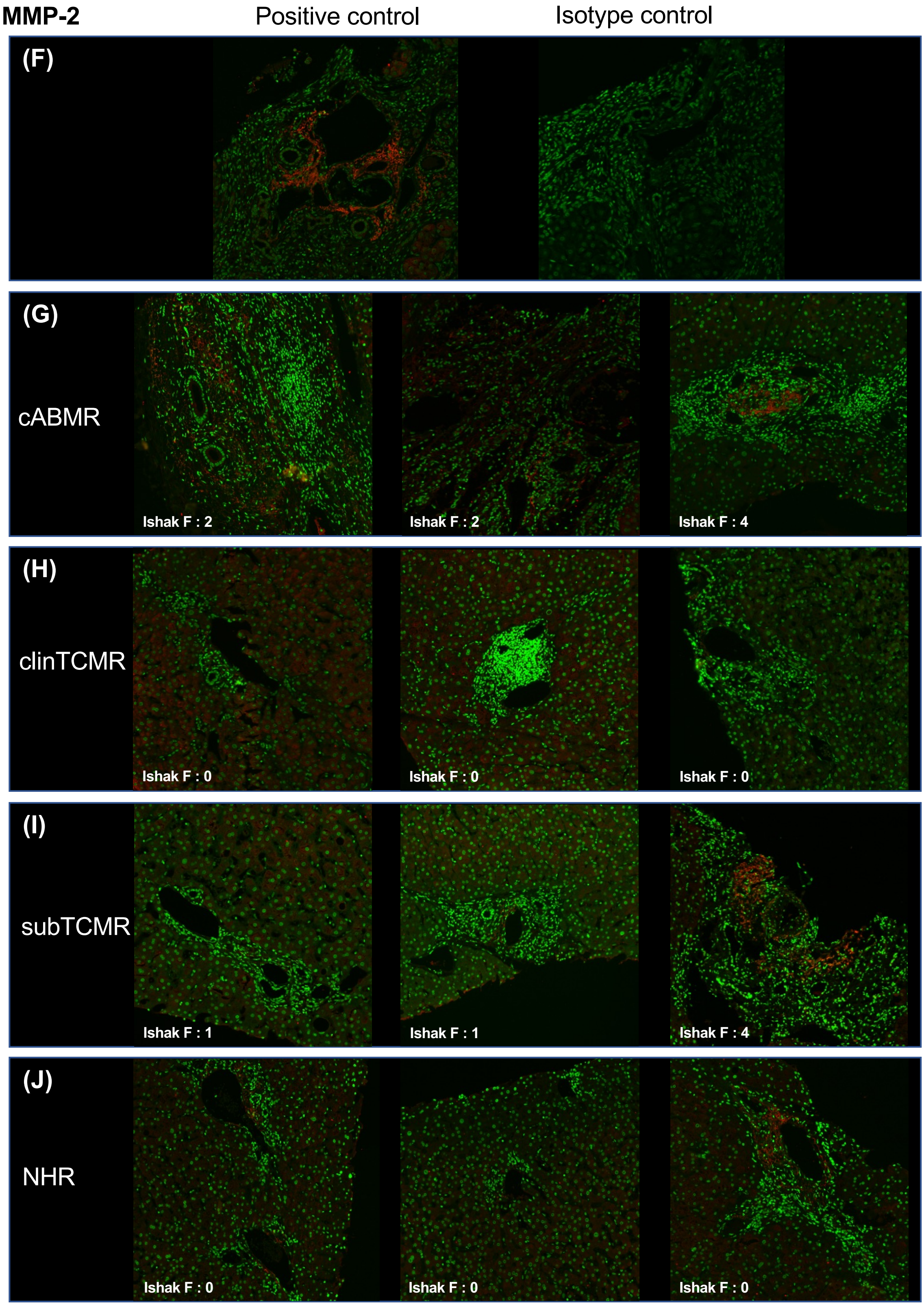

**Suppl. Figure 9: Immunofluorescence**
Exemplary immunofluorescence stains. **(A)** Positive and isotype control for Col14A1 **(B)** cABMR samples stained with Col14A1. **(C)** clinTCMR samples stained with Col14A1. **(D)** subTCMR samples stained with Col14A1. **(F)** NHR samples stained with Col14A1. **(F)** Positive and isotype control for MMP-2. **(G)** cABMR samples stained with MMP-2. **(H)** clinTCMR samples stained with MMP-2. **(I)** subTCMR samples stained with MMP-2. **(J)** NHR samples stained with MMP-2.
clinTCMR: clinical T cell-mediated rejection; cABMR: chronic antibody-mediated rejection; Col14A1: collagen Type XIV alpha 1 Chain; subTCMR: subclinical T cell-mediated rejection; MMP-2: matrix metalloproteinase-2; NHR: no histological rejection.

#### Supplementary Figure 10

(A)

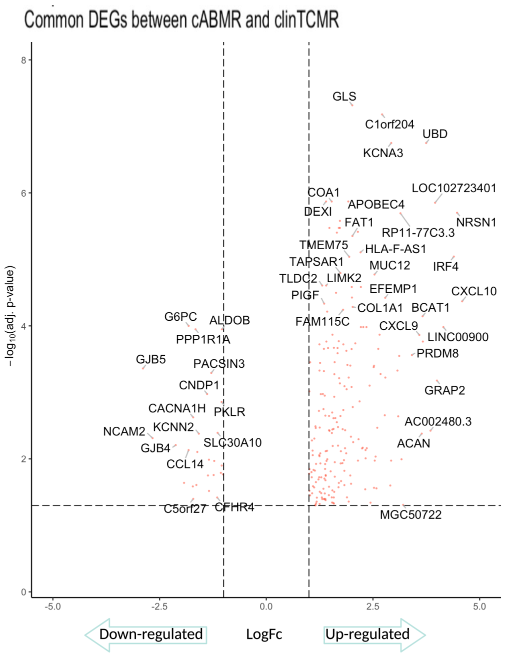


(B)

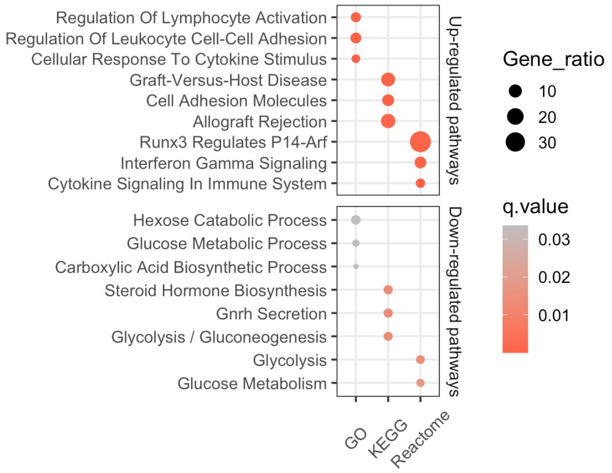


**Suppl. Figure 10: Common features between cABMR and clinTCMR in the validation cohort**

**(A)** Volcano plot showing the common features between cABMR and clinTCMR vs NHR DEGs from the validation cohort, using false discovery rate (FDR) < 0.05 and absolute log fold change (LogFc) > 1. (**B)** Over-represented pathways out of DEGs in panel A. clinTCMR: clinical T cell-mediated rejection; cABMR: chronic antibody-mediated rejection; subTCMR: subclinical T cell-mediated rejection; NHR: no histological rejection, DEGs: differentially expressed genes.

#### Supplementary Figure 11

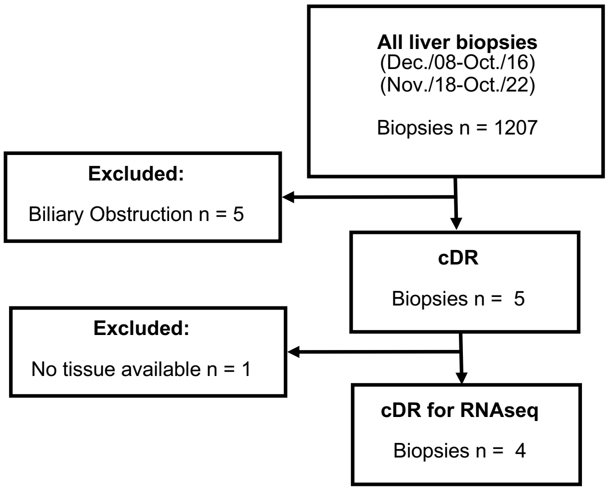


**Suppl. Figure 11: Flow chart showing selection of samples with chronic ductopenic rejection**
cDR: chronic ductopenic rejection

#### Supplementary Figure 12

**A**

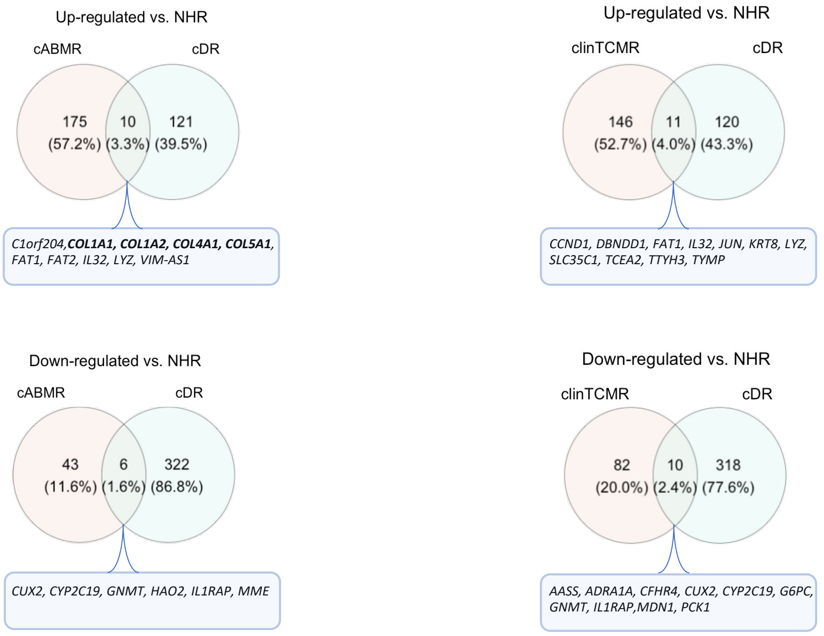


**B**

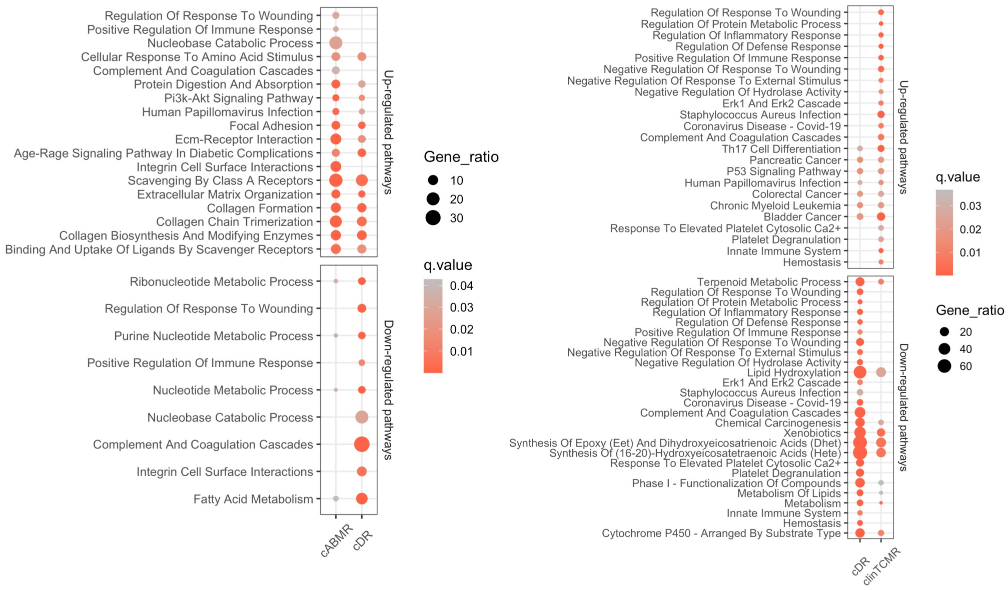


**Suppl. Figure 12: Transcriptome-unique features of chronic ductopenic rejection when compared to cABMR and clinTCMR**

**A.** Venn diagram showing the numbers of the up and down-regulated DEGs in each rejection group compared to NHR. Validated cABMR and cTCMR DEGs were used. **B.** Dotplot of commonly enriched pathways between cABMR and CDR (left), and cTCMR and CDR (right)

#### Supplementary Figure 13

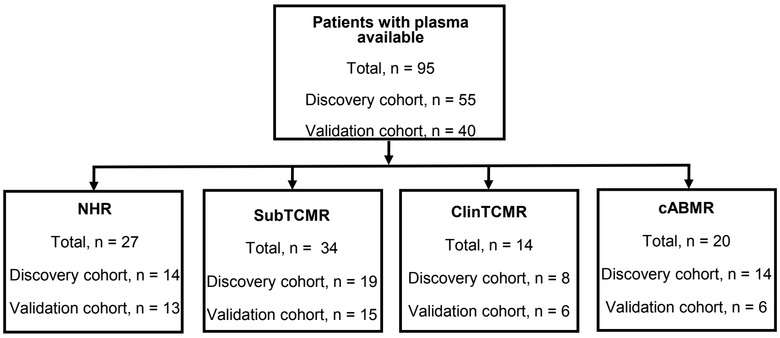


**Suppl. Figure 13: Flow chart showing selection of plasma samples**

cABMR: chronic antibody-mediated rejection; subTCMR: subclinical T cell-mediated rejection; NHR: no histological rejection.

#### Supplementary Figure 14

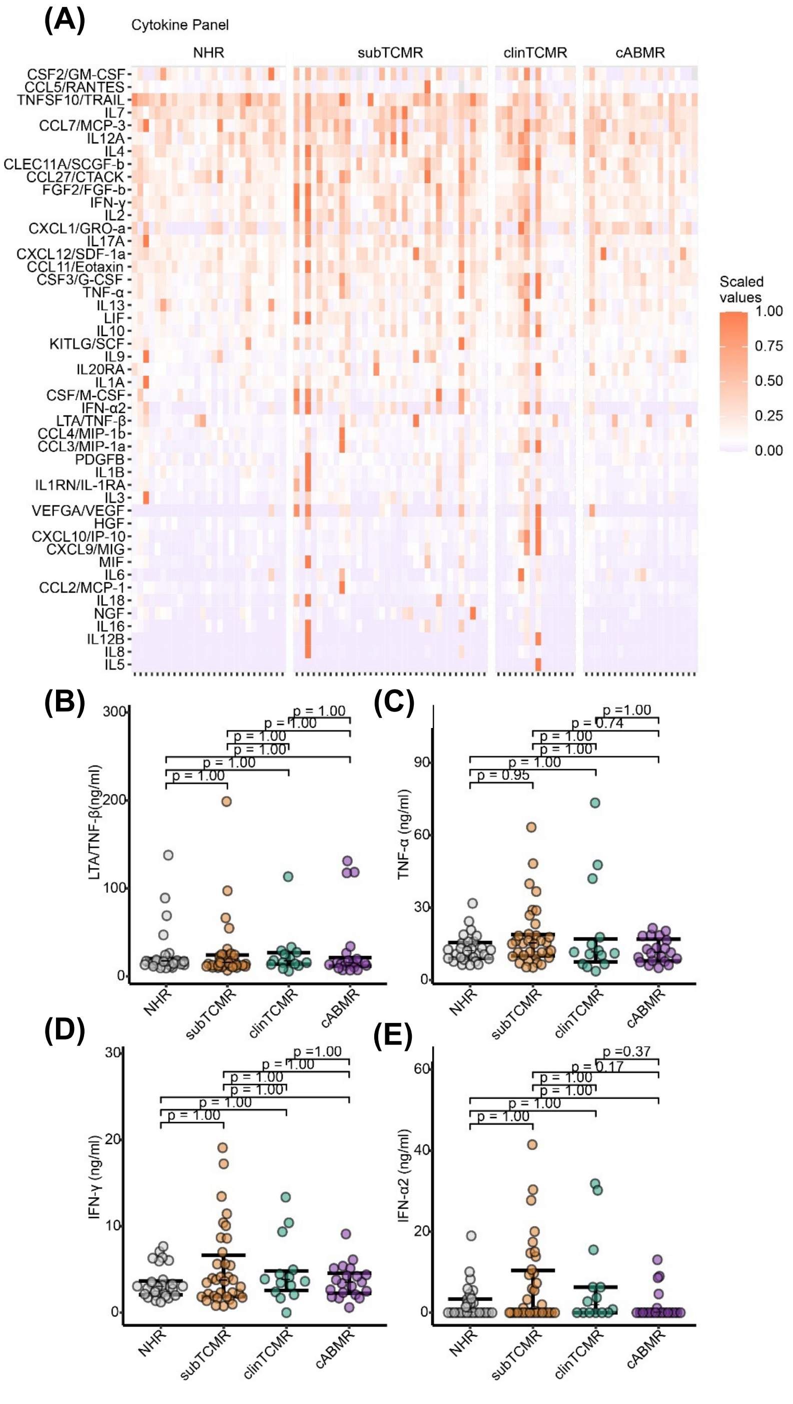


**Suppl. Figure 14: Distribution of plasma cytokine levels between the groups**
**(A)** The heatmap illustrates the scaled values of plasma cytokine levels, with each row representing a cytokine and each column representing a single sample. The samples are clustered according to their respective group, as indicated. **(B-E)** Dot plots indicate the distribution of IFN and TNF levels between the different groups. The median and interquartile range are shown with bars and bold lines inside the bars. The Kruskal-Wallis test and Dunn's post-hoc test were used to compute the p-value, a p-value < 0.05 is considered significant. cABMR: chronic antibody-mediated rejection; subTCMR: subclinical T cell-mediated rejection; NHR: no histological rejection.

#### Supplementary Figure 15

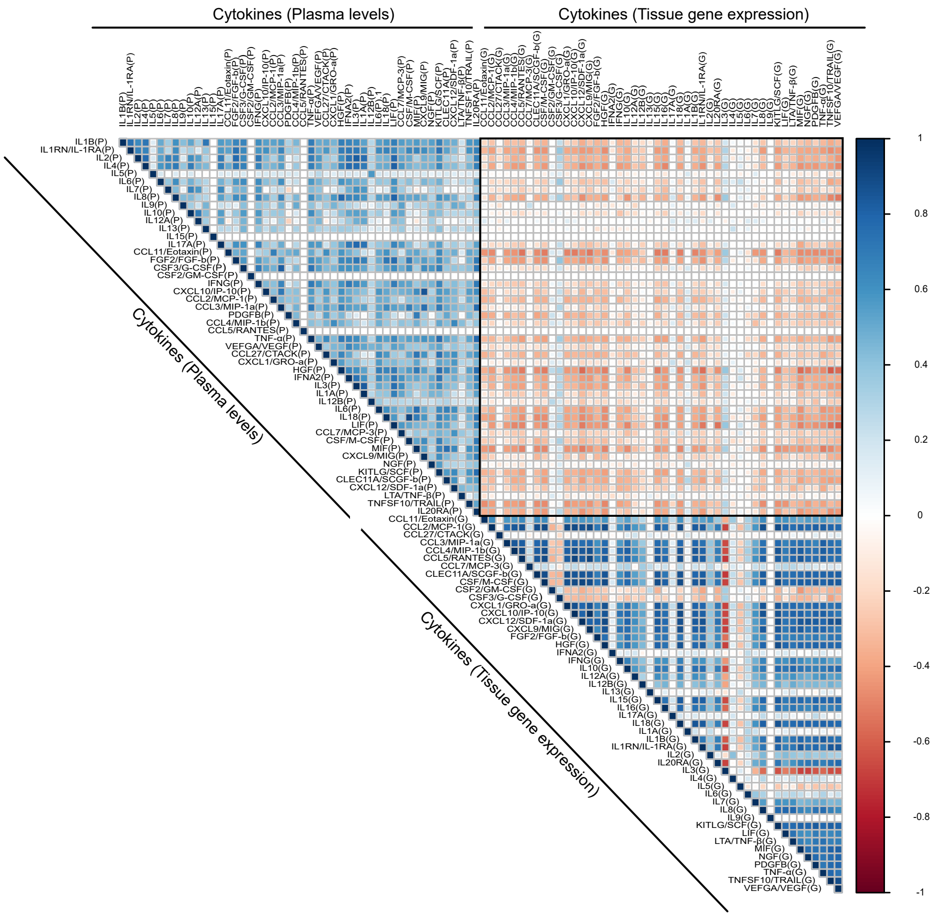


**Suppl. Figure 15: Correlation matrix of plasma cytokine levels with tissue gene expression of corresponding cytokines**

Spearman correlation plot illustrating the relationship between plasma cytokine levels (input as ng/ml) and graft gene expression of those same cytokines (input as gene counts). Correlation coefficients are indicated with a color gradient, where red values approach -1, indicating a strong negative correlation, and blue values approach 1, indicating a strong positive correlation. Values near 0 (neutral) are depicted in white. This plot visually represents the strength and direction of the monotonic relationships between the variables analyzed.

#### Supplementary Figure 16

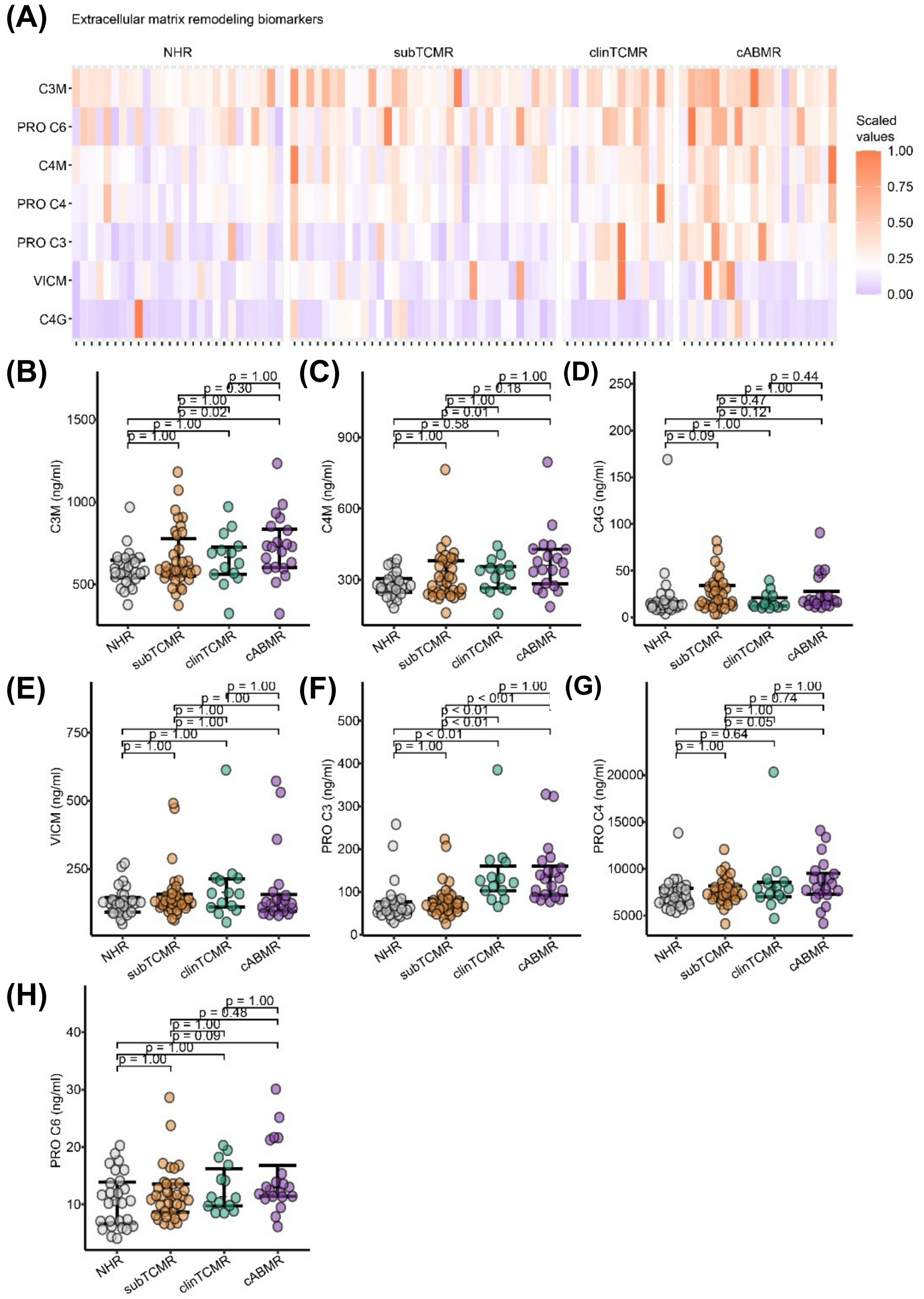


**Suppl. Figure 16: Distribution of extracellular matrix marker levels between the groups**
**(A)** Heatmap indicating scaled values of plasma extracellular matrix marker levels with each row indicating an extracellular matrix marker as indicated and each column representing a single sample. Samples are clustered according to their group as indicated. **(B-H)** Dot plots indicating distribution of respective extracellular matrix marker levels between the different groups. Median and interquartile range are shown with bars and bold lines inside the bars. Kruskal-Wallis Test and Dunns‘ post-hoc test were used for the p-value computation, a p-value y 0.05 is considered significant. cABMR: chronic antibody-mediated rejection; subTCMR: subclinical T cell-mediated rejection; NHR: no histological rejection.

#### Supplementary Figure 17

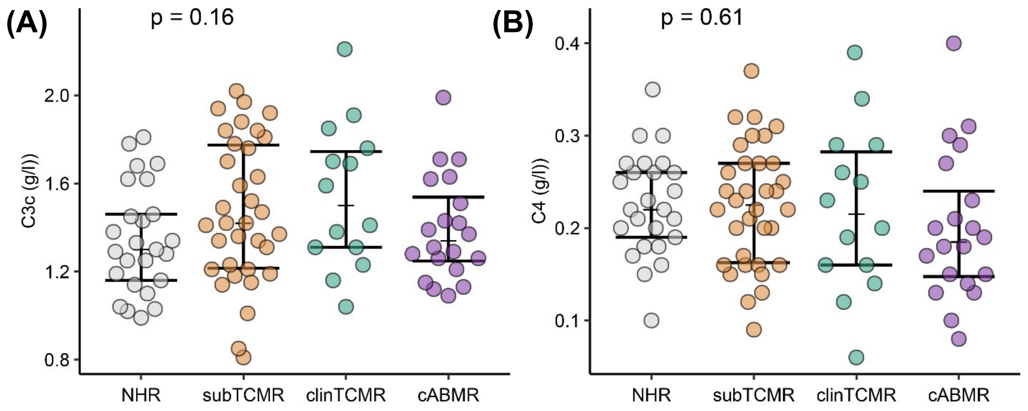


**Suppl. Figure 17: Median levels of complement factors measured from plasma of patients after liver transplantation**

Plasma levels of complement C3c **(A)** and C4 **(B).** Median and interquartile range are shown with bars and bold lines inside the bars. ANOVA was used for statistical comparison, a pvalue < 0.05 is considered significant. clinTCMR: clinical T cell-mediated rejection; cABMR: chronic antibody-mediated rejection; subTCMR: subclinical T cell-mediated rejection; NHR: no histological rejection.

### Supplementary tables

Suppl. Table 1

| Histological Assessment | No. | Median | Range | Score points 0 | 1 | 2 | 3 | 4 | 5 | 6 | 7 | 8 | 9 | 10 | 11 | 12 |
|---|---|---|---|---|---|---|---|---|---|---|---|---|---|---|---|---|
| **NHR** | | | | | | | | | | | | | | | | |
| Ishak_A | 38 | 0 | 0-0 | 100 | 0 | 0 | 0 | 0 | | | | | | | | |
| Ishak_B | 38 | 0 | 0-0 | 100 | 0 | 0 | 0 | 0 | 0 | 0 | | | | | | |
| Ishak_C | 38 | 0 | 0-1 | 79 | 21 | 0 | 0 | 0 | | | | | | | | |
| Ishak_D | 38 | 0 | 0-1 | 58 | 42 | 0 | 0 | 0 | | | | | | | | |
| Ishak_HAI | 38 | 1 | 0-2 | 42 | 53 | 5 | 0 | 0 | 0 | 0 | 0 | 0 | 0 | 0 | 0 | 0 |
| Ishak_F | 38 | 0 | 0-1 | 71 | 29 | 0 | 0 | 0 | 0 | 0 | | | | | | |
| LAF_portal | 29 | 0 | 0-1 | 69 | 31 | 0 | 0 | | | | | | | | | |
| LAF_sinusoidal | 29 | 0 | 0-0 | 100 | 0 | 0 | 0 | | | | | | | | | |
| LAF_perivenulary | 29 | 0 | 0-1 | 90 | 10 | 0 | 0 | | | | | | | | | |
| LAF_Total | 29 | 0 | 0-1 | 59 | 41 | 0 | 0 | 0 | 0 | 0 | 0 | 0 | 0 | | | |
| RAI_portal | 38 | 0 | 0-1 | 58 | 42 | 0 | 0 | | | | | | | | | |
| RAI_biliary | 38 | 0 | 0-1 | 97 | 3 | 0 | 0 | | | | | | | | | |
| RAI_venous_endothelial | 38 | 0 | 0-0 | 100 | 0 | 0 | 0 | | | | | | | | | |
| RAI_Total | 38 | 0 | 0-1 | 55 | 45 | 0 | 0 | 0 | 0 | 0 | 0 | 0 | 0 | | | |
| **subTCMR** | | | | | | | | | | | | | | | | |
| Ishak_A | 46 | 0 | 0-3 | 63 | 26 | 9 | 2 | 0 | | | | | | | | |
| Ishak_B | 46 | 0 | 0-2 | 93 | 4 | 2 | 0 | 0 | 0 | 0 | | | | | | |
| Ishak_C | 46 | 1 | 0-2 | 37 | 41 | 22 | 0 | 0 | | | | | | | | |
| Ishak_D | 46 | 2 | 0-4 | 2 | 46 | 46 | 4 | 2 | | | | | | | | |
| Ishak_HAI | 46 | 3 | 0-8 | 4 | 17 | 22 | 26 | 11 | 7 | 9 | 2 | 2 | 0 | 0 | 0 | 0 |
| Ishak_F | 46 | 1 | 0-4 | 39 | 52 | 2 | 4 | 2 | 0 | 0 | | | | | | |
| LAF_portal | 29 | 1 | 0-3 | 31 | 52 | 10 | 7 | | | | | | | | | |
| LAF_sinusoidal | 29 | 0 | 0-1 | 79 | 21 | 0 | 0 | | | | | | | | | |
| LAF_perivenulary | 29 | 0 | 0-1 | 79 | 21 | 0 | 0 | | | | | | | | | |
| LAF_Total | 29 | 1 | 0-5 | 31 | 31 | 21 | 10 | 3 | 3 | 0 | 0 | 0 | 0 | | | |
| RAI_portal | 46 | 1 | 1-3 | 0 | 63 | 35 | 2 | | | | | | | | | |
| RAI_biliary | 46 | 1 | 1-2 | 0 | 80 | 20 | 0 | | | | | | | | | |
| RAI_venous_endothelial | 46 | 1 | 1-2 | 0 | 91 | 9 | 0 | | | | | | | | | |
| RAI_Total | 46 | 3 | 3-6 | 0 | 0 | 0 | 59 | 20 | 17 | 4 | 0 | 0 | 0 | | | |
| **clinTCMR** | | | | | | | | | | | | | | | | |
| Ishak_A | 23 | 1 | 0-3 | 26 | 61 | 9 | 4 | 0 | | | | | | | | |
| Ishak_B | 23 | 0 | 0-3 | 96 | 0 | 0 | 4 | 0 | 0 | 0 | | | | | | |
| Ishak_C | 23 | 2 | 0-3 | 4 | 43 | 43 | 9 | 0 | | | | | | | | |
| Ishak_D | 23 | 2 | 1-3 | 0 | 43 | 43 | 13 | 0 | | | | | | | | |
| Ishak_HAI | 23 | 4 | 2-12 | 0 | 0 | 9 | 26 | 35 | 13 | 13 | 0 | 0 | 0 | 0 | 0 | 4 |
| Ishak_F | 23 | 1 | 0-2 | 48 | 43 | 9 | 0 | 0 | 0 | 0 | | | | | | |
| LAF_portal | 19 | 1 | 0-2 | 37 | 53 | 11 | 0 | | | | | | | | | |
| LAF_sinusoidal | 19 | 0 | 0-1 | 95 | 5 | 0 | 0 | | | | | | | | | |
| LAF_perivenulary | 19 | 0 | 0-1 | 74 | 26 | 0 | 0 | | | | | | | | | |
| LAF_Total | 19 | 1 | 0-3 | 32 | 37 | 26 | 5 | 0 | 0 | 0 | 0 | 0 | 0 | | | |
| RAI_portal | 23 | 2 | 1-3 | 0 | 35 | 57 | 9 | | | | | | | | | |
| RAI_biliary | 23 | 2 | 1-2 | 0 | 39 | 61 | 0 | | | | | | | | | |
| RAI_venous_endothelial | 23 | 1 | 1-3 | 0 | 74 | 22 | 4 | | | | | | | | | |
| RAI_Total | 23 | 5 | 3-7 | 0 | 0 | 0 | 26 | 22 | 26 | 13 | 13 | 0 | 0 | | | |
| **cABMR** | | | | | | | | | | | | | | | | |
| Ishak_A | 22 | 1,5 | 0-2 | 9 | 41 | 50 | 0 | 0 | | | | | | | | |
| Ishak_B | 22 | 0 | 0-2 | 95 | 0 | 5 | 0 | 0 | 0 | 0 | | | | | | |
| Ishak_C | 22 | 1 | 0-2 | 5 | 73 | 23 | 0 | 0 | | | | | | | | |
| Ishak_D | 22 | 2 | 1-4 | 0 | 23 | 50 | 23 | 5 | | | | | | | | |
| Ishak_HAI | 22 | 5 | 2-8 | 0 | 0 | 9 | 14 | 14 | 32 | 23 | 5 | 5 | 0 | 0 | 0 | 0 |
| Ishak_F | 22 | 2,5 | 1-6 | 0 | 5 | 45 | 14 | 27 | 5 | 5 | | | | | | |
| LAF_portal | 20 | 2 | 1-3 | 0 | 10 | 55 | 35 | | | | | | | | | |
| LAF_sinusoidal | 20 | 1 | 0-2 | 45 | 35 | 20 | 0 | | | | | | | | | |
| LAF_perivenulary | 20 | 1 | 0-3 | 40 | 45 | 5 | 10 | | | | | | | | | |
| LAF_Total | 20 | 4 | 2-8 | 0 | 0 | 30 | 15 | 25 | 15 | 5 | 5 | 5 | 0 | | | |
| RAI_portal | 22 | 2 | 1-3 | 0 | 41 | 50 | 9 | | | | | | | | | |
| RAI_biliary | 22 | 1 | 0-3 | 32 | 50 | 14 | 5 | | | | | | | | | |
| RAI_venous_endothelial | 22 | 0,5 | 0-2 | 50 | 36 | 14 | 0 | | | | | | | | | |
| RAI_Total | 22 | 3 | 1-8 | 0 | 27 | 14 | 14 | 23 | 9 | 9 | 0 | 5 | | | | |
| **CDR** | | | | | | | | | | | | | | | | |
| Ishak_A | 4 | 0 | 0-1 | 75 | 25 | 0 | 0 | 0 | | | | | | | | |
| Ishak_B | 4 | 0 | 0-0 | 100 | 0 | 0 | 0 | 0 | 0 | 0 | | | | | | |
| Ishak_C | 4 | 0,5 | 0-1 | 50 | 50 | 0 | 0 | 0 | | | | | | | | |
| Ishak_D | 4 | 1 | 0-2 | 25 | 50 | 25 | 0 | 0 | | | | | | | | |
| Ishak_HAI | 4 | 1,5 | 0-4 | 25 | 25 | 25 | 0 | 25 | 0 | 0 | 0 | 0 | 0 | 0 | 0 | 0 |
| Ishak_F | 4 | 2 | 0-4 | 25 | 25 | 0 | 25 | 25 | 0 | 0 | | | | | | |
| LAF_portal | 2 | 1 | 0-2 | 50 | 0 | 50 | 0 | | | | | | | | | |
| LAF_sinusoidal | 2 | 0,5 | 0-1 | 50 | 50 | 0 | 0 | | | | | | | | | |
| LAF_perivenulary | 2 | 1 | 1-1 | 0 | 100 | 0 | 0 | | | | | | | | | |
| LAF_Total | 2 | 2,5 | 1-4 | 0 | 50 | 0 | 0 | 50 | 0 | 0 | 0 | 0 | | | | |
| RAI_portal | 4 | 1,5 | 1-2 | 0 | 50 | 50 | 0 | | | | | | | | | |
| RAI_biliary | 4 | 1,5 | 1-3 | 0 | 50 | 25 | 25 | | | | | | | | | |
| RAI_venous_endothelial | 4 | 0,5 | 0-2 | 50 | 25 | 25 | 0 | | | | | | | | | |
| RAI_Total | 4 | 4 | 2-6 | 0 | 0 | 25 | 0 | 50 | 0 | 25 | 0 | 0 | | | | |

**Suppl. Table 1. Frequency table of histological scores.**
Frequency table summarizing histological scores, showing the percentage distribution of each histological score value within the dataset. The number of samples, the median and range are also shown. Color intensity ranges from red to green, where higher percentages are represented in shades of red, indicating higher prevalence, and lower percentages are represented in shades of green, indicating lower prevalence. cABMR: chronic antibody mediated rejection; cDR: chronic ductopenic rejection; HAI, histological activity index; subTCMR: subclinical T cell-mediated rejection; NHR: no histological rejection; RAI: rejection activity index; LAF: liver allograft fibrosis.

**Suppl. Table 2**

cABMR vs. NHR

| log2FoldChange | padj | gene_names | significance | direction |
|---|---|---|---|---|
| 3,478071266 | 6,29438E-14 | UBD | TRUE | Upregulated |
| 0,918935902 | 9,91942E-13 | MYH9 | FALSE | Upregulated |
| 4,403777448 | 2,80093E-12 | MMP7 | TRUE | Upregulated |
| 1,028804069 | 7,40951E-12 | PLOD3 | TRUE | Upregulated |
| 1,631709102 | 4,34351E-11 | FLNA | TRUE | Upregulated |
| 0,951716567 | 2,82535E-10 | SPTAN1 | FALSE | Upregulated |
| 0,568634231 | 4,53428E-10 | ARHGEF18 | FALSE | Upregulated |
| 0,869570076 | 1,10167E-09 | AGRN | FALSE | Upregulated |
| 2,562824093 | 1,10167E-09 | CFTR | TRUE | Upregulated |
| 1,242538694 | 1,32729E-09 | MMP14 | TRUE | Upregulated |
| -0,90029584 | 1,35116E-09 | COX7C | FALSE | Downregulated |
| 4,070652531 | 1,39048E-09 | LOXL4 | TRUE | Upregulated |
| -1,252803454 | 1,44719E-09 | ACSM3 | TRUE | Downregulated |
| 3,552175429 | 1,44719E-09 | CXCL6 | TRUE | Upregulated |
| 0,569801008 | 1,45183E-09 | FLII | FALSE | Upregulated |
| 3,332557182 | 1,45183E-09 | SPATA21 | TRUE | Upregulated |
| 6,086352207 | 1,60189E-09 | STMN2 | TRUE | Upregulated |
| 2,309907267 | 2,21808E-09 | PAPLN | TRUE | Upregulated |
| 2,034886752 | 2,96993E-09 | KRT19 | TRUE | Upregulated |
| 0,711609684 | 3,97326E-09 | MYO1C | FALSE | Upregulated |
| 3,146085976 | 4,61834E-09 | CLIC6 | TRUE | Upregulated |
| 1,43061686 | 8,16685E-09 | COL4A2 | TRUE | Upregulated |
| -0,468343756 | 8,32979E-09 | FAM204A | FALSE | Downregulated |
| 2,700316486 | 8,32979E-09 | VTCN1 | TRUE | Upregulated |
| 1,576336175 | 9,35002E-09 | BACE2 | TRUE | Upregulated |
| 3,017573296 | 1,03952E-08 | APOBEC3B | TRUE | Upregulated |
| 0,895074482 | 1,03952E-08 | USP11 | FALSE | Upregulated |
| 4,364393334 | 1,34524E-08 | MUC13 | TRUE | Upregulated |
| 0,584422829 | 1,50942E-08 | CHERP | FALSE | Upregulated |
| 2,024271343 | 1,73955E-08 | GPRC5B | TRUE | Upregulated |
| 2,69763015 | 1,81502E-08 | CXCL10 | TRUE | Upregulated |
| -1,033921301 | 1,8504E-08 | ACOT13 | TRUE | Downregulated |
| 2,179027599 | 2,16129E-08 | THBS2 | TRUE | Upregulated |
| 0,635791441 | 2,75402E-08 | SEPT9 | FALSE | Upregulated |
| -1,009926822 | 2,76644E-08 | ZSCAN16-AS1 | TRUE | Downregulated |
| 3,125265825 | 2,7787E-08 | LAMC2 | TRUE | Upregulated |
| 2,421754133 | 3,65955E-08 | FTH1P3 | TRUE | Upregulated |
| 3,066675927 | 3,99767E-08 | MTRNR2L8 | TRUE | Upregulated |
| 2,643692181 | 4,14407E-08 | EFEMP1 | TRUE | Upregulated |
| -0,637005175 | 4,14407E-08 | LOC100507217 | FALSE | Downregulated |
| -0,604124735 | 4,14407E-08 | NACA | FALSE | Downregulated |
| 0,769510152 | 4,14407E-08 | PYGB | FALSE | Upregulated |
| 1,579405329 | 4,14407E-08 | SLC12A2 | TRUE | Upregulated |
| -0,958631261 | 4,89456E-08 | TBCA | FALSE | Downregulated |
| 2,586219739 | 6,15533E-08 | ADRA2A | TRUE | Upregulated |
| 2,800194736 | 6,15533E-08 | LCN2 | TRUE | Upregulated |
| -1,290107322 | 6,73692E-08 | SMIM6 | TRUE | Downregulated |
| 4,146749971 | 7,21791E-08 | RN7SL2 | TRUE | Upregulated |

| | | | | |
|---|---|---|---|---|
| 2,042623439 | 7,2481E-08 | ITGA3 | TRUE | Upregulated |
| -0,900794413 | 8,95667E-08 | LINC00467 | FALSE | Downregulated |
| 1,202682533 | 9,09875E-08 | NFKB2 | TRUE | Upregulated |
| -0,717090582 | 9,15553E-08 | AC007246.3 | FALSE | Downregulated |
| -1,448747804 | 9,15553E-08 | AQP7P3 | TRUE | Downregulated |
| 2,404435315 | 9,15553E-08 | CLDN11 | TRUE | Upregulated |
| -0,895831852 | 9,31485E-08 | ATP5E | FALSE | Downregulated |
| 0,518134378 | 9,31485E-08 | CLCN7 | FALSE | Upregulated |
| -0,659867709 | 9,31485E-08 | LYRM5 | FALSE | Downregulated |
| 1,641532968 | 9,31485E-08 | RELB | TRUE | Upregulated |
| -0,678685021 | 1,03282E-07 | NDUFS4 | FALSE | Downregulated |
| 2,851023441 | 1,0331E-07 | CDH6 | TRUE | Upregulated |
| 2,861949251 | 1,12572E-07 | RGS4 | TRUE | Upregulated |
| 2,096268181 | 1,17097E-07 | PMEPA1 | TRUE | Upregulated |
| 2,125389619 | 1,17097E-07 | RGCC | TRUE | Upregulated |
| 2,277627629 | 1,2321E-07 | ITGB8 | TRUE | Upregulated |
| -0,654244236 | 1,2671E-07 | RPL22 | FALSE | Downregulated |
| 0,686012306 | 1,26796E-07 | TLN1 | FALSE | Upregulated |
| 2,485558807 | 1,30775E-07 | SPINT1 | TRUE | Upregulated |
| 1,065748276 | 1,33846E-07 | RHBDF2 | TRUE | Upregulated |
| 2,651033603 | 1,34957E-07 | CCR7 | TRUE | Upregulated |
| 4,782511474 | 1,5033E-07 | IL8 | TRUE | Upregulated |
| 0,811705282 | 1,71634E-07 | INF2 | FALSE | Upregulated |
| 2,937813294 | 1,71634E-07 | MOXD1 | TRUE | Upregulated |
| -0,521093068 | 1,71634E-07 | MTIF3 | FALSE | Downregulated |
| -0,82155919 | 1,97019E-07 | F7 | FALSE | Downregulated |
| 0,73917308 | 2,05879E-07 | TMEM184B | FALSE | Upregulated |
| -0,529160502 | 2,11197E-07 | ARV1 | FALSE | Downregulated |
| 2,209051365 | 2,12474E-07 | HLA-G | TRUE | Upregulated |
| 1,374586356 | 2,20082E-07 | SELM | TRUE | Upregulated |
| 2,134695373 | 2,20399E-07 | PTK7 | TRUE | Upregulated |
| 2,986686056 | 2,40494E-07 | FLJ44635 | TRUE | Upregulated |
| 1,29174774 | 2,40494E-07 | MCM5 | TRUE | Upregulated |
| 1,576832626 | 2,4664E-07 | BIRC3 | TRUE | Upregulated |
| -0,869128963 | 2,50495E-07 | UBE2D4 | FALSE | Downregulated |
| 1,073937536 | 2,50943E-07 | SLC25A24 | TRUE | Upregulated |
| 1,165400211 | 2,56718E-07 | KIRREL | TRUE | Upregulated |
| -0,537394183 | 2,83479E-07 | CEBPZ-AS1 | FALSE | Downregulated |
| 3,506019858 | 2,83479E-07 | FAM133CP | TRUE | Upregulated |
| 1,157001962 | 2,89735E-07 | TTYH3 | TRUE | Upregulated |
| 0,908646151 | 2,97244E-07 | VASH1 | FALSE | Upregulated |
| 1,586860013 | 3,00355E-07 | NFE2L3 | TRUE | Upregulated |
| 0,875512617 | 3,02077E-07 | PIEZO1 | FALSE | Upregulated |
| 2,263570453 | 3,07012E-07 | LAMC3 | TRUE | Upregulated |
| 0,803776265 | 3,07012E-07 | SLC12A4 | FALSE | Upregulated |
| -0,481291621 | 3,18148E-07 | FRA10AC1 | FALSE | Downregulated |
| 1,235585475 | 3,18148E-07 | MVP | TRUE | Upregulated |
| 0,710061804 | 3,18148E-07 | TNFRSF1B | FALSE | Upregulated |
| 1,367368625 | 3,21127E-07 | COL4A1 | TRUE | Upregulated |
| 1,979368227 | 3,21127E-07 | WISP1 | TRUE | Upregulated |
| 1,222719879 | 3,25463E-07 | CLEC2D | TRUE | Upregulated |

| | | | | |
|---|---|---|---|---|
| 2,134975442 | 3,33019E-07 | CCL2 | TRUE | Upregulated |
| -0,459694277 | 3,54759E-07 | MRPS14 | FALSE | Downregulated |
| 3,061093096 | 3,61164E-07 | TNFRSF9 | TRUE | Upregulated |
| 0,723783642 | 3,81643E-07 | GPI | FALSE | Upregulated |
| 1,275254944 | 3,82347E-07 | MRC2 | TRUE | Upregulated |
| -0,616278953 | 3,82347E-07 | OCIAD1 | FALSE | Downregulated |
| 1,370485077 | 3,95807E-07 | ATP2A3 | TRUE | Upregulated |
| 0,899643464 | 3,97607E-07 | ZNF319 | FALSE | Upregulated |
| 1,654657944 | 4,09126E-07 | ARNT2 | TRUE | Upregulated |
| -0,950579909 | 4,22923E-07 | CNBP | FALSE | Downregulated |
| 1,502866365 | 4,47423E-07 | LXN | TRUE | Upregulated |
| 0,663741349 | 4,7845E-07 | CD59 | FALSE | Upregulated |
| 1,824442014 | 4,84118E-07 | ANXA3 | TRUE | Upregulated |
| -0,776074292 | 4,84586E-07 | F9 | FALSE | Downregulated |
| 2,081769698 | 4,85788E-07 | FSTL3 | TRUE | Upregulated |
| 1,506822436 | 4,98298E-07 | PODXL | TRUE | Upregulated |
| -0,936715326 | 4,98298E-07 | SEC61A2 | FALSE | Downregulated |
| 1,258118187 | 5,00489E-07 | CORO1A | TRUE | Upregulated |
| 2,854327165 | 5,28243E-07 | KIAA1644 | TRUE | Upregulated |
| -2,283681997 | 5,85467E-07 | ATM | TRUE | Downregulated |
| 2,404634977 | 5,85467E-07 | DTL | TRUE | Upregulated |
| 1,307493 | 5,85467E-07 | GPR56 | TRUE | Upregulated |
| -0,64409707 | 6,02093E-07 | PEX7 | FALSE | Downregulated |
| 1,656239365 | 6,07194E-07 | ITGB4 | TRUE | Upregulated |
| 1,848734859 | 6,79363E-07 | PFKP | TRUE | Upregulated |
| 1,86706092 | 6,97634E-07 | WNK2 | TRUE | Upregulated |
| -0,701119121 | 6,98984E-07 | RUNDC3B | FALSE | Downregulated |
| -0,703822145 | 7,03998E-07 | PAIP2B | FALSE | Downregulated |
| 1,220094284 | 7,03998E-07 | VIM | TRUE | Upregulated |
| 1,133626738 | 7,26827E-07 | ITGA6 | TRUE | Upregulated |
| 0,701810993 | 7,52062E-07 | HIP1R | FALSE | Upregulated |
| 2,633050793 | 7,78544E-07 | MEX3B | TRUE | Upregulated |
| 1,314070991 | 9,43973E-07 | FMNL1 | TRUE | Upregulated |
| -0,472815463 | 9,64439E-07 | SNAP29 | FALSE | Downregulated |
| 0,615606771 | 9,80063E-07 | PLCG1 | FALSE | Upregulated |
| 0,943925564 | 9,86751E-07 | ITGAV | FALSE | Upregulated |
| 0,917652328 | 9,86751E-07 | PRKX | FALSE | Upregulated |
| -0,541117125 | 1,01328E-06 | EBAG9 | FALSE | Downregulated |
| 1,129527889 | 1,01328E-06 | LBH | TRUE | Upregulated |
| 1,477703316 | 1,01328E-06 | MICAL1 | TRUE | Upregulated |
| 1,934165444 | 1,03249E-06 | SOX4 | TRUE | Upregulated |
| 2,558102848 | 1,08297E-06 | CDC20 | TRUE | Upregulated |
| 0,936485707 | 1,09807E-06 | POLD1 | FALSE | Upregulated |
| 2,097733954 | 1,22558E-06 | IGSF3 | TRUE | Upregulated |
| 2,084741318 | 1,22558E-06 | THY1 | TRUE | Upregulated |
| 0,623256718 | 1,24417E-06 | CLIC1 | FALSE | Upregulated |
| -0,99998576 | 1,24417E-06 | POLR3GL | FALSE | Downregulated |
| -0,759501158 | 1,24417E-06 | RPL26 | FALSE | Downregulated |
| 2,507514177 | 1,25589E-06 | ITGA2 | TRUE | Upregulated |
| 1,486980856 | 1,33742E-06 | EHF | TRUE | Upregulated |
| 2,910921621 | 1,35053E-06 | LOC102723722 | TRUE | Upregulated |

| | | | | |
|---|---|---|---|---|
| -0,70823007 | 1,37144E-06 | NDUFB1 | FALSE | Downregulated |
| 1,317551704 | 1,3781E-06 | AFAP1 | TRUE | Upregulated |
| 0,966604594 | 1,3781E-06 | HSPG2 | FALSE | Upregulated |
| -0,516270383 | 1,43997E-06 | MRFAP1L1 | FALSE | Downregulated |
| -0,735435147 | 1,44903E-06 | ATP5J | FALSE | Downregulated |
| 2,658179516 | 1,44903E-06 | PCDP1 | TRUE | Upregulated |
| 1,497358501 | 1,44903E-06 | SVEP1 | TRUE | Upregulated |
| 2,044600275 | 1,48182E-06 | SLC34A2 | TRUE | Upregulated |
| 0,902945905 | 1,48182E-06 | TNFAIP3 | FALSE | Upregulated |
| 1,343372063 | 1,533E-06 | RHBDF1 | TRUE | Upregulated |
| 0,95546026 | 1,54378E-06 | FAM129B | FALSE | Upregulated |
| 1,780623492 | 1,59319E-06 | AEBP1 | TRUE | Upregulated |
| 1,502772216 | 1,62127E-06 | LSP1 | TRUE | Upregulated |
| 0,532249074 | 1,64275E-06 | LASP1 | FALSE | Upregulated |
| -0,454064658 | 1,68922E-06 | CDC26 | FALSE | Downregulated |
| 1,033258572 | 1,69221E-06 | ZYX | TRUE | Upregulated |
| -0,804353364 | 1,71238E-06 | RPL13A | FALSE | Downregulated |
| 0,39787462 | 1,78847E-06 | BAG6 | FALSE | Upregulated |
| -0,337074886 | 1,78847E-06 | FOPNL | FALSE | Downregulated |
| -0,434024429 | 1,78847E-06 | UBE2D3 | FALSE | Downregulated |
| -0,823704968 | 1,80155E-06 | MIR4458HG | FALSE | Downregulated |
| 1,21336714 | 1,8149E-06 | ITGAM | TRUE | Upregulated |
| 1,887633448 | 1,91889E-06 | BICC1 | TRUE | Upregulated |
| -0,74149525 | 1,91889E-06 | MRPS33 | FALSE | Downregulated |
| 0,391165592 | 1,94632E-06 | AP1B1 | FALSE | Upregulated |
| 0,502479487 | 1,94632E-06 | ARFIP1 | FALSE | Upregulated |
| 3,910546107 | 1,94632E-06 | RN7SL1 | TRUE | Upregulated |
| 1,692025503 | 2,02528E-06 | DUSP4 | TRUE | Upregulated |
| 2,241861021 | 2,04847E-06 | CXCL11 | TRUE | Upregulated |
| 0,919361291 | 2,07309E-06 | CASZ1 | FALSE | Upregulated |
| 2,274039904 | 2,07309E-06 | TACSTD2 | TRUE | Upregulated |
| 2,147510669 | 2,10309E-06 | TGFB2 | TRUE | Upregulated |
| -0,384921351 | 2,12932E-06 | NDUFA5 | FALSE | Downregulated |
| 1,499078765 | 2,14102E-06 | MOB3B | TRUE | Upregulated |
| 1,540881942 | 2,16057E-06 | SPON2 | TRUE | Upregulated |
| 0,547202125 | 2,1818E-06 | INTS1 | FALSE | Upregulated |
| -0,553422762 | 2,1818E-06 | TMEM41B | FALSE | Downregulated |
| -0,607804322 | 2,22537E-06 | ABHD10 | FALSE | Downregulated |
| 0,649934159 | 2,22537E-06 | LMNA | FALSE | Upregulated |
| 1,095374292 | 2,27527E-06 | NFATC1 | TRUE | Upregulated |
| 1,37058197 | 2,2775E-06 | JUN | TRUE | Upregulated |
| 1,048565738 | 2,2775E-06 | TAP1 | TRUE | Upregulated |
| 2,118633124 | 2,29946E-06 | IGJ | TRUE | Upregulated |
| 2,648406022 | 2,31026E-06 | SHISA3 | TRUE | Upregulated |
| 2,318222735 | 2,32348E-06 | SYT13 | TRUE | Upregulated |
| 1,363940394 | 2,44869E-06 | MCAM | TRUE | Upregulated |
| -0,346072117 | 2,44869E-06 | NRD1 | FALSE | Downregulated |
| 1,217031116 | 2,45498E-06 | PRDM1 | TRUE | Upregulated |
| 1,874006957 | 2,47107E-06 | SOX9 | TRUE | Upregulated |
| -0,494685169 | 2,50046E-06 | AMZ2 | FALSE | Downregulated |
| 2,831463267 | 2,50046E-06 | LIF | TRUE | Upregulated |

| | | | | |
|---|---|---|---|---|
| 0,453817062 | 2,51094E-06 | CAPN1 | FALSE | Upregulated |
| -0,633813385 | 2,51094E-06 | HAO1 | FALSE | Downregulated |
| 0,422101139 | 2,51094E-06 | UBE2Z | FALSE | Upregulated |
| -1,022940748 | 2,54232E-06 | CHCHD10 | TRUE | Downregulated |
| 4,532267875 | 2,56854E-06 | CCL20 | TRUE | Upregulated |
| -0,535212115 | 2,56854E-06 | RNF7 | FALSE | Downregulated |
| 1,489024364 | 2,70377E-06 | GRAMD1B | TRUE | Upregulated |
| -0,550907245 | 2,8164E-06 | NUCKS1 | FALSE | Downregulated |
| -0,469091926 | 2,84419E-06 | OMA1 | FALSE | Downregulated |
| -0,601384354 | 2,9255E-06 | MTERFD3 | FALSE | Downregulated |
| -0,591483321 | 2,9255E-06 | TXN2 | FALSE | Downregulated |
| 2,193183529 | 3,02335E-06 | CXCR3 | TRUE | Upregulated |
| 0,369721844 | 3,02335E-06 | PRRC2A | FALSE | Upregulated |
| 0,48889641 | 3,03965E-06 | RNF19A | FALSE | Upregulated |
| -0,415571849 | 3,06749E-06 | GPN1 | FALSE | Downregulated |
| 0,845622905 | 3,24252E-06 | RBM38 | FALSE | Upregulated |
| 2,048803947 | 3,27685E-06 | MALAT1 | TRUE | Upregulated |
| 1,004855701 | 3,31577E-06 | ARL4C | TRUE | Upregulated |
| 0,965237754 | 3,33079E-06 | MAFK | FALSE | Upregulated |
| -0,669424222 | 3,33079E-06 | PCCB | FALSE | Downregulated |
| 0,625460101 | 3,39048E-06 | MAPRE1 | FALSE | Upregulated |
| -0,53093412 | 3,43955E-06 | WIBG | FALSE | Downregulated |
| -0,796002321 | 3,56283E-06 | AR | FALSE | Downregulated |
| 2,58936578 | 3,56554E-06 | LOC102723407 | TRUE | Upregulated |
| 0,849585079 | 3,57977E-06 | C15orf39 | FALSE | Upregulated |
| 0,510901787 | 3,64259E-06 | VAT1 | FALSE | Upregulated |
| -1,070814224 | 3,72027E-06 | EPB41L4A-AS1 | TRUE | Downregulated |
| 2,241983531 | 3,72027E-06 | LOXL1 | TRUE | Upregulated |
| 1,840036223 | 3,72252E-06 | COL16A1 | TRUE | Upregulated |
| 1,32794301 | 3,82169E-06 | ANXA2 | TRUE | Upregulated |
| 0,365914584 | 3,85666E-06 | CSNK1D | FALSE | Upregulated |
| 1,841544763 | 3,85666E-06 | KIAA1211 | TRUE | Upregulated |
| -0,867284162 | 3,85666E-06 | MAP2K1 | FALSE | Downregulated |
| -0,734881778 | 3,99717E-06 | MOCS2 | FALSE | Downregulated |
| -1,01855785 | 4,00019E-06 | SLC9A3R2 | TRUE | Downregulated |
| 1,665623356 | 4,11962E-06 | NCAPH | TRUE | Upregulated |
| 1,761816929 | 4,11962E-06 | SGPP2 | TRUE | Upregulated |
| 1,200177854 | 4,22531E-06 | PTPRH | TRUE | Upregulated |
| -0,492125604 | 4,23952E-06 | DCAF6 | FALSE | Downregulated |
| 1,043625783 | 4,26858E-06 | DRAM1 | TRUE | Upregulated |
| -0,585997189 | 4,47087E-06 | CPB2 | FALSE | Downregulated |
| 0,491613632 | 4,75846E-06 | ATG7 | FALSE | Upregulated |
| -0,460940368 | 5,06678E-06 | TRAPPC6B | FALSE | Downregulated |
| -0,504596152 | 5,06678E-06 | ZBTB8OS | FALSE | Downregulated |
| 1,666835372 | 5,14581E-06 | CYR61 | TRUE | Upregulated |
| 1,17803861 | 5,15685E-06 | ECT2 | TRUE | Upregulated |
| 0,44096962 | 5,15685E-06 | TMEM43 | FALSE | Upregulated |
| 3,521765983 | 5,16937E-06 | TRPC4 | TRUE | Upregulated |
| 1,040652709 | 5,23821E-06 | IER5 | TRUE | Upregulated |
| 3,040702696 | 5,3044E-06 | ANXA2P1 | TRUE | Upregulated |
| 0,741443323 | 5,3044E-06 | CYTH2 | FALSE | Upregulated |

| | | | | |
|---|---|---|---|---|
| 2,610332881 | 5,39369E-06 | MTRNR2L6 | TRUE | Upregulated |
| 0,536609296 | 5,45318E-06 | ITGB5 | FALSE | Upregulated |
| 4,472658997 | 5,45318E-06 | SLC51B | TRUE | Upregulated |
| 1,331573497 | 5,55184E-06 | ARRDC2 | TRUE | Upregulated |
| 1,235992666 | 5,58262E-06 | COL3A1 | TRUE | Upregulated |
| -0,731041045 | 5,68123E-06 | HPX | FALSE | Downregulated |
| 1,229263804 | 5,68123E-06 | TYMP | TRUE | Upregulated |
| -0,355257033 | 5,88033E-06 | PPIL4 | FALSE | Downregulated |
| -0,503946801 | 5,89938E-06 | VTI1B | FALSE | Downregulated |
| 0,780698253 | 6,0547E-06 | NUP93 | FALSE | Upregulated |
| -0,620243701 | 6,13225E-06 | HRSP12 | FALSE | Downregulated |
| -0,763246481 | 6,15074E-06 | HSD17B6 | FALSE | Downregulated |
| 1,582582892 | 6,20094E-06 | HOPX | TRUE | Upregulated |
| 1,060631236 | 6,20094E-06 | STK17B | TRUE | Upregulated |
| -0,520255555 | 6,37869E-06 | PYURF | FALSE | Downregulated |
| 1,500920693 | 6,43294E-06 | CXCR4 | TRUE | Upregulated |
| -0,459200601 | 6,44073E-06 | KRR1 | FALSE | Downregulated |
| 1,932207805 | 6,4597E-06 | A1BG-AS1 | TRUE | Upregulated |
| 0,529058366 | 6,4597E-06 | ATP6V0A1 | FALSE | Upregulated |
| 0,944403909 | 6,4597E-06 | INPP5D | FALSE | Upregulated |
| -0,629935943 | 6,49611E-06 | PEBP1 | FALSE | Downregulated |
| 1,122073423 | 6,49611E-06 | TUBB6 | TRUE | Upregulated |
| -1,506100866 | 6,50402E-06 | DKFZP434I0714 | TRUE | Downregulated |
| 0,452630243 | 6,67476E-06 | MTOR | FALSE | Upregulated |
| 0,989971843 | 6,72623E-06 | MFSD10 | FALSE | Upregulated |
| 1,856414402 | 6,73125E-06 | GPR171 | TRUE | Upregulated |
| -0,416484773 | 6,74091E-06 | ZHX1 | FALSE | Downregulated |
| 2,534002559 | 6,83835E-06 | LOC101928318 | TRUE | Upregulated |
| -0,58129158 | 6,83835E-06 | PCBD2 | FALSE | Downregulated |
| -0,650408002 | 6,85214E-06 | HNMT | FALSE | Downregulated |
| 0,78130824 | 6,86696E-06 | ANKRD44 | FALSE | Upregulated |
| -0,427580381 | 6,86696E-06 | BABAM1 | FALSE | Downregulated |
| -1,153436192 | 6,86696E-06 | LOC102723845 | TRUE | Downregulated |
| 1,51852781 | 6,9058E-06 | AQP1 | TRUE | Upregulated |
| 2,093881384 | 6,9058E-06 | LTBP2 | TRUE | Upregulated |
| -0,9276795 | 6,94107E-06 | TMEM256 | FALSE | Downregulated |
| 1,407229245 | 6,94467E-06 | ITGAX | TRUE | Upregulated |
| 1,438969019 | 6,99486E-06 | EPHA3 | TRUE | Upregulated |
| 0,718669604 | 7,08934E-06 | ANXA4 | FALSE | Upregulated |
| 0,846705856 | 7,3527E-06 | ARHGAP31 | FALSE | Upregulated |
| 1,863153157 | 7,70835E-06 | IL32 | TRUE | Upregulated |
| 1,157166937 | 7,75896E-06 | DGKA | TRUE | Upregulated |
| 0,641405567 | 7,80634E-06 | ENG | FALSE | Upregulated |
| 0,914541689 | 7,93777E-06 | CHST2 | FALSE | Upregulated |
| 1,220330591 | 7,93777E-06 | ITPR3 | TRUE | Upregulated |
| 1,498638847 | 7,93777E-06 | MSC | TRUE | Upregulated |
| 1,156624529 | 7,93777E-06 | SCRN1 | TRUE | Upregulated |
| 2,000975308 | 8,11698E-06 | LEF1 | TRUE | Upregulated |
| -0,472479309 | 8,14605E-06 | OARD1 | FALSE | Downregulated |
| -1,454992741 | 8,14605E-06 | PTH1R | TRUE | Downregulated |
| 1,523584389 | 8,19339E-06 | CTNND2 | TRUE | Upregulated |

| | | | | |
|---|---|---|---|---|
| 1,636917277 | 8,20754E-06 | TYMS | TRUE | Upregulated |
| 2,024659887 | 8,27202E-06 | KLHL29 | TRUE | Upregulated |
| 3,341925304 | 8,30114E-06 | CXCL5 | TRUE | Upregulated |
| -0,864936048 | 8,45107E-06 | TMEM223 | FALSE | Downregulated |
| 0,436075064 | 8,50724E-06 | CTNNA1 | FALSE | Upregulated |
| 0,575375687 | 8,51098E-06 | PIAS3 | FALSE | Upregulated |
| 3,537697346 | 8,51098E-06 | RHOV | TRUE | Upregulated |
| -0,728911148 | 8,56904E-06 | PLA2G12A | FALSE | Downregulated |
| 1,259649761 | 8,64768E-06 | SERPINE2 | TRUE | Upregulated |
| 1,043338562 | 8,69448E-06 | CAPN2 | TRUE | Upregulated |
| -0,572466283 | 8,86163E-06 | COQ3 | FALSE | Downregulated |
| -0,487509759 | 8,87262E-06 | MRPS22 | FALSE | Downregulated |
| 3,13429163 | 8,87865E-06 | VSIG2 | TRUE | Upregulated |
| 0,893513933 | 8,98532E-06 | GMIP | FALSE | Upregulated |
| 1,490703255 | 9,2398E-06 | TAGLN | TRUE | Upregulated |
| 2,592264873 | 9,25483E-06 | DBNDD1 | TRUE | Upregulated |
| -0,517221811 | 9,27708E-06 | APOPT1 | FALSE | Downregulated |
| 1,172963937 | 9,27708E-06 | C1orf198 | TRUE | Upregulated |
| 4,256684595 | 9,27708E-06 | LOC102724895 | TRUE | Upregulated |
| 1,564481638 | 9,27708E-06 | PARM1 | TRUE | Upregulated |
| 0,786090412 | 9,27708E-06 | RFX5 | FALSE | Upregulated |
| 2,589454244 | 9,27708E-06 | SEZ6L2 | TRUE | Upregulated |
| 1,345608486 | 9,3917E-06 | SLCO2A1 | TRUE | Upregulated |
| 1,697627487 | 9,41905E-06 | FZD7 | TRUE | Upregulated |
| -0,680736365 | 9,56653E-06 | ACADM | FALSE | Downregulated |
| -0,305572995 | 9,56653E-06 | ATMIN | FALSE | Downregulated |
| 1,76908547 | 9,56653E-06 | NAIP | TRUE | Upregulated |
| 0,510858285 | 9,56653E-06 | PCBP1 | FALSE | Upregulated |
| 0,550821084 | 9,56653E-06 | RNPEPL1 | FALSE | Upregulated |
| -1,012351478 | 9,56653E-06 | RPS29 | TRUE | Downregulated |
| 0,478248707 | 9,67098E-06 | HGS | FALSE | Upregulated |
| -0,601345801 | 9,67098E-06 | SMIM11 | FALSE | Downregulated |
| 1,63710553 | 9,88357E-06 | FAT1 | TRUE | Upregulated |
| 1,50795545 | 9,96409E-06 | COL1A2 | TRUE | Upregulated |
| 2,046389135 | 1,03646E-05 | PAPPA | TRUE | Upregulated |
| 0,429114041 | 1,03646E-05 | URGCP | FALSE | Upregulated |
| 1,416523121 | 1,05317E-05 | SLFN13 | TRUE | Upregulated |
| -0,440229221 | 1,06446E-05 | C1orf115 | FALSE | Downregulated |
| 1,887027511 | 1,07396E-05 | CCL21 | TRUE | Upregulated |
| 0,932686336 | 1,08469E-05 | COL6A2 | FALSE | Upregulated |
| 2,247892286 | 1,08579E-05 | SPIB | TRUE | Upregulated |
| 0,7077904 | 1,09777E-05 | LRP10 | FALSE | Upregulated |
| -0,890809497 | 1,1031E-05 | ADK | FALSE | Downregulated |
| -4,174521499 | 1,11227E-05 | RP11-436I24.1 | TRUE | Downregulated |
| 2,719603704 | 1,12796E-05 | GDF15 | TRUE | Upregulated |
| 0,787920088 | 1,1298E-05 | RNF213 | FALSE | Upregulated |
| -0,388312518 | 1,1382E-05 | EMC4 | FALSE | Downregulated |
| 4,239925311 | 1,14569E-05 | DPEP1 | TRUE | Upregulated |
| 1,991011097 | 1,15612E-05 | SPP1 | TRUE | Upregulated |
| 0,691915818 | 1,15955E-05 | CTSD | FALSE | Upregulated |
| 0,751078551 | 1,16133E-05 | ALYREF | FALSE | Upregulated |

| | | | | |
|---|---|---|---|---|
| -0,980429529 | 1,18204E-05 | PALM3 | FALSE | Downregulated |
| 0,458067802 | 1,18204E-05 | UNC13B | FALSE | Upregulated |
| -0,650508579 | 1,18983E-05 | SMARCA1 | FALSE | Downregulated |
| -0,456556383 | 1,19297E-05 | C1orf27 | FALSE | Downregulated |
| 1,350376628 | 1,20282E-05 | CD2 | TRUE | Upregulated |
| 4,207054855 | 1,20282E-05 | MTRNR2L5 | TRUE | Upregulated |
| 0,43248125 | 1,20282E-05 | RNF10 | FALSE | Upregulated |
| 0,892972352 | 1,20282E-05 | SH2D3C | FALSE | Upregulated |
| -0,407909438 | 1,21463E-05 | NSMCE4A | FALSE | Downregulated |
| 0,883560757 | 1,21565E-05 | CCND2 | FALSE | Upregulated |
| 1,192826325 | 1,21904E-05 | CD96 | TRUE | Upregulated |
| -0,389517755 | 1,25804E-05 | UQCRB | FALSE | Downregulated |
| 2,28991136 | 1,26949E-05 | CDC45 | TRUE | Upregulated |
| -0,991256369 | 1,26949E-05 | LINC00478 | FALSE | Downregulated |
| 0,456743703 | 1,27334E-05 | CC2D1A | FALSE | Upregulated |
| 0,535576383 | 1,31416E-05 | VGLL4 | FALSE | Upregulated |
| -0,536526635 | 1,31508E-05 | FBXL12 | FALSE | Downregulated |
| -2,295855993 | 1,32173E-05 | C21orf37 | TRUE | Downregulated |
| 1,381107007 | 1,32173E-05 | GLIS2 | TRUE | Upregulated |
| 1,406782551 | 1,32173E-05 | PLVAP | TRUE | Upregulated |
| -1,354766789 | 1,36377E-05 | FITM1 | TRUE | Downregulated |
| 1,31104968 | 1,36516E-05 | COL5A1 | TRUE | Upregulated |
| 0,508430543 | 1,37601E-05 | H2AFY | FALSE | Upregulated |
| 0,872153705 | 1,38696E-05 | TEAD2 | FALSE | Upregulated |
| -0,533031714 | 1,40758E-05 | COA6 | FALSE | Downregulated |
| 1,077561071 | 1,42152E-05 | COL6A3 | TRUE | Upregulated |
| 0,737339559 | 1,43883E-05 | NCKAP5L | FALSE | Upregulated |
| 1,749795172 | 1,43883E-05 | PTGDS | TRUE | Upregulated |
| 2,002240324 | 1,44928E-05 | SEMA7A | TRUE | Upregulated |
| 2,079291061 | 1,453E-05 | FCAMR | TRUE | Upregulated |
| 0,801835895 | 1,47686E-05 | CD93 | FALSE | Upregulated |
| 0,616098946 | 1,47686E-05 | DGKZ | FALSE | Upregulated |
| 1,082461622 | 1,50154E-05 | PDIA3P1 | TRUE | Upregulated |
| 0,932111884 | 1,51222E-05 | PACS1 | FALSE | Upregulated |
| 0,990249165 | 1,51222E-05 | PNMA1 | FALSE | Upregulated |
| -0,45525117 | 1,51229E-05 | OSBPL9 | FALSE | Downregulated |
| 1,894568301 | 1,52616E-05 | PDGFA | TRUE | Upregulated |
| 0,454512627 | 1,5437E-05 | ATP1A1 | FALSE | Upregulated |
| 1,795585657 | 1,5437E-05 | CPZ | TRUE | Upregulated |
| 0,745464601 | 1,55322E-05 | RAB11FIP1 | FALSE | Upregulated |
| -0,603506353 | 1,55336E-05 | TMEM18 | FALSE | Downregulated |
| 0,639838877 | 1,55911E-05 | NEO1 | FALSE | Upregulated |
| 2,237407184 | 1,56908E-05 | LOC440311 | TRUE | Upregulated |
| 2,523276058 | 1,57996E-05 | KRT7 | TRUE | Upregulated |
| -0,94574142 | 1,58607E-05 | DNAJC25 | FALSE | Downregulated |
| -1,066097346 | 1,5962E-05 | DHRS4L2 | TRUE | Downregulated |
| -0,929788519 | 1,61271E-05 | CCL14 | FALSE | Downregulated |
| 2,079106077 | 1,61271E-05 | PABPC3 | TRUE | Upregulated |
| 1,897493561 | 1,64764E-05 | CD24 | TRUE | Upregulated |
| 1,27439523 | 1,64764E-05 | JAG1 | TRUE | Upregulated |
| -0,779812615 | 1,64764E-05 | NUPR1 | FALSE | Downregulated |

| | | | | |
|---|---|---|---|---|
| 1,333014516 | 1,67036E-05 | LAMA2 | TRUE | Upregulated |
| -0,35229824 | 1,67358E-05 | ADIPOR1 | FALSE | Downregulated |
| -0,434685165 | 1,71021E-05 | NDUFAF7 | FALSE | Downregulated |
| -0,721430717 | 1,71754E-05 | RWDD4 | FALSE | Downregulated |
| 0,894486141 | 1,73017E-05 | EMP3 | FALSE | Upregulated |
| -0,496727885 | 1,74576E-05 | CDADC1 | FALSE | Downregulated |
| 1,461646647 | 1,78837E-05 | LUM | TRUE | Upregulated |
| 0,879879763 | 1,78837E-05 | SLC27A4 | FALSE | Upregulated |
| -0,314403388 | 1,78837E-05 | WAC | FALSE | Downregulated |
| -0,636061181 | 1,80919E-05 | SERPINF1 | FALSE | Downregulated |
| -0,511184001 | 1,8254E-05 | CYB5D1 | FALSE | Downregulated |
| -1,619880141 | 1,82857E-05 | LOC102724655 | TRUE | Downregulated |
| -0,704755933 | 1,84627E-05 | RCAN1 | FALSE | Downregulated |
| 0,677245367 | 1,85872E-05 | TP53BP2 | FALSE | Upregulated |
| 1,415764295 | 1,88416E-05 | RHOF | TRUE | Upregulated |
| 1,541168976 | 1,9092E-05 | CACNB3 | TRUE | Upregulated |
| 0,720409936 | 1,93344E-05 | RANGAP1 | FALSE | Upregulated |
| -1,011430586 | 1,93553E-05 | TCP10L | TRUE | Downregulated |
| -0,482837397 | 1,94884E-05 | TMCO1 | FALSE | Downregulated |
| 1,907740023 | 2,01378E-05 | SPON1 | TRUE | Upregulated |
| -0,606578135 | 2,0274E-05 | APOC1 | FALSE | Downregulated |
| 2,504023698 | 2,0274E-05 | TNFRSF12A | TRUE | Upregulated |
| 0,983922737 | 2,02794E-05 | ARHGAP27 | FALSE | Upregulated |
| 1,344886068 | 2,02794E-05 | PLK3 | TRUE | Upregulated |
| -0,655702434 | 2,05053E-05 | POLR2J | FALSE | Downregulated |
| -0,313614236 | 2,05226E-05 | CREBL2 | FALSE | Downregulated |
| -0,681703849 | 2,05226E-05 | GRTP1 | FALSE | Downregulated |
| 1,088869688 | 2,05226E-05 | KCTD12 | TRUE | Upregulated |
| 0,739756501 | 2,05226E-05 | SERPINH1 | FALSE | Upregulated |
| 1,030304009 | 2,05226E-05 | SLFN5 | TRUE | Upregulated |
| -0,611452853 | 2,06988E-05 | AZGP1 | FALSE | Downregulated |
| 0,911440587 | 2,15254E-05 | ANXA1 | FALSE | Upregulated |
| -0,498483059 | 2,15254E-05 | HMGCL | FALSE | Downregulated |
| 2,102417037 | 2,1658E-05 | COL15A1 | TRUE | Upregulated |
| -0,797550937 | 2,1658E-05 | EXOSC6 | FALSE | Downregulated |
| 0,989885677 | 2,1658E-05 | HLA-F | FALSE | Upregulated |
| 0,610596465 | 2,16595E-05 | PKN1 | FALSE | Upregulated |
| 1,499482208 | 2,18694E-05 | PDGFD | TRUE | Upregulated |
| 0,926140976 | 2,19565E-05 | NFKBIE | FALSE | Upregulated |
| 1,334105103 | 2,20606E-05 | CD3E | TRUE | Upregulated |
| 1,722455358 | 2,20606E-05 | GEM | TRUE | Upregulated |
| 0,95294213 | 2,20606E-05 | IKZF1 | FALSE | Upregulated |
| 1,159257446 | 2,28486E-05 | SLC37A1 | TRUE | Upregulated |
| 0,650663413 | 2,3103E-05 | VPS37C | FALSE | Upregulated |
| -0,55640525 | 2,3211E-05 | ELP6 | FALSE | Downregulated |
| -0,44357772 | 2,33009E-05 | MRP63 | FALSE | Downregulated |
| -0,319039717 | 2,33032E-05 | IARS2 | FALSE | Downregulated |
| -0,537854573 | 2,33032E-05 | PIK3C2G | FALSE | Downregulated |
| 0,818415265 | 2,34103E-05 | PITPNM1 | FALSE | Upregulated |
| 0,797974387 | 2,361E-05 | HEG1 | FALSE | Upregulated |
| 3,870602628 | 2,44162E-05 | LAMP5 | TRUE | Upregulated |

| | | | | |
|---|---|---|---|---|
| 0,995750143 | 2,4802E-05 | NDE1 | FALSE | Upregulated |
| 1,469450261 | 2,48258E-05 | SEMA4A | TRUE | Upregulated |
| 1,440703535 | 2,48432E-05 | FBLIM1 | TRUE | Upregulated |
| -0,345456429 | 2,49163E-05 | TPMT | FALSE | Downregulated |
| 2,415733468 | 2,49464E-05 | TMEM200A | TRUE | Upregulated |
| -1,319048847 | 2,51604E-05 | COX6C | TRUE | Downregulated |
| -0,768494979 | 2,52287E-05 | LSM3 | FALSE | Downregulated |
| 0,584968158 | 2,52548E-05 | ABCD1 | FALSE | Upregulated |
| -0,355058747 | 2,52548E-05 | TMEM248 | FALSE | Downregulated |
| 0,512236147 | 2,52922E-05 | MED16 | FALSE | Upregulated |
| -0,695649469 | 2,52922E-05 | NME4 | FALSE | Downregulated |
| 1,74020792 | 2,53016E-05 | ATP10A | TRUE | Upregulated |
| 0,883767386 | 2,53016E-05 | G6PD | FALSE | Upregulated |
| 1,532541131 | 2,53016E-05 | VCAN | TRUE | Upregulated |
| 0,744537049 | 2,59529E-05 | KIAA0556 | FALSE | Upregulated |
| 1,805730729 | 2,59529E-05 | ST14 | TRUE | Upregulated |
| -0,582169808 | 2,59567E-05 | GPX4 | FALSE | Downregulated |
| 2,943148662 | 2,59567E-05 | OLR1 | TRUE | Upregulated |
| 1,453961984 | 2,59567E-05 | SUSD2 | TRUE | Upregulated |
| 1,25691718 | 2,60775E-05 | CRISPLD2 | TRUE | Upregulated |
| 0,866440288 | 2,68042E-05 | HIC1 | FALSE | Upregulated |
| 2,152887251 | 2,69109E-05 | CCL19 | TRUE | Upregulated |
| 1,179572769 | 2,70769E-05 | FLRT2 | TRUE | Upregulated |
| -0,486281218 | 2,71656E-05 | OXA1L | FALSE | Downregulated |
| 1,147780485 | 2,71656E-05 | SAMD3 | TRUE | Upregulated |
| 0,706703755 | 2,72526E-05 | ADAM15 | FALSE | Upregulated |
| -0,358814844 | 2,72526E-05 | DYNLT3 | FALSE | Downregulated |
| 0,491060715 | 2,72881E-05 | STK24 | FALSE | Upregulated |
| 1,292809173 | 2,77508E-05 | NFASC | TRUE | Upregulated |
| 1,734347008 | 2,78683E-05 | MMP24 | TRUE | Upregulated |
| -0,708047368 | 2,79871E-05 | EI24 | FALSE | Downregulated |
| -0,477523961 | 2,8131E-05 | DNAJC19 | FALSE | Downregulated |
| 3,220455981 | 2,85415E-05 | HKDC1 | TRUE | Upregulated |
| -0,796232554 | 2,87923E-05 | FBXL15 | FALSE | Downregulated |
| 0,718627623 | 2,91406E-05 | RALGDS | FALSE | Upregulated |
| 2,084961018 | 2,93074E-05 | DCDC2 | TRUE | Upregulated |
| -0,683438932 | 2,93074E-05 | ST13 | FALSE | Downregulated |
| 1,167813493 | 3,00319E-05 | CIITA | TRUE | Upregulated |
| 1,179187812 | 3,00319E-05 | MCM6 | TRUE | Upregulated |
| 0,5661113 | 3,00319E-05 | SEPN1 | FALSE | Upregulated |
| 1,609750253 | 3,02722E-05 | F3 | TRUE | Upregulated |
| 2,327199269 | 3,02722E-05 | GABRP | TRUE | Upregulated |
| -0,40335943 | 3,02722E-05 | SAP18 | FALSE | Downregulated |
| 0,689368003 | 3,08677E-05 | CNN2 | FALSE | Upregulated |
| 1,473978614 | 3,11586E-05 | MAP1B | TRUE | Upregulated |
| 1,550878988 | 3,11586E-05 | MTHFD2 | TRUE | Upregulated |
| -0,595814652 | 3,2175E-05 | CISD3 | FALSE | Downregulated |
| -1,014510196 | 3,22074E-05 | AQP7P1 | TRUE | Downregulated |
| 0,580157881 | 3,22074E-05 | BSG | FALSE | Upregulated |
| 2,988464598 | 3,22074E-05 | TNFRSF17 | TRUE | Upregulated |
| 1,432370331 | 3,28615E-05 | GZMK | TRUE | Upregulated |

| | | | | |
|---|---|---|---|---|
| 0,864866197 | 3,28615E-05 | TAGLN2 | FALSE | Upregulated |
| 1,081493757 | 3,29241E-05 | LIMD2 | TRUE | Upregulated |
| -0,404242032 | 3,35407E-05 | NDUFS2 | FALSE | Downregulated |
| -0,604793347 | 3,38375E-05 | ALB | FALSE | Downregulated |
| -0,514745411 | 3,38375E-05 | DCAF11 | FALSE | Downregulated |
| -0,49584524 | 3,38375E-05 | LIPT1 | FALSE | Downregulated |
| 1,227624525 | 3,45486E-05 | COL4A4 | TRUE | Upregulated |
| 0,417379851 | 3,50453E-05 | TBCD | FALSE | Upregulated |
| 0,854596884 | 3,52028E-05 | SLC1A5 | FALSE | Upregulated |
| 1,828794393 | 3,5387E-05 | FGF7 | TRUE | Upregulated |
| 0,645482682 | 3,54289E-05 | MAP4 | FALSE | Upregulated |
| 4,295774175 | 3,54289E-05 | SHISA9 | TRUE | Upregulated |
| -2,003915565 | 3,54289E-05 | SLCO4C1 | TRUE | Downregulated |
| -0,538436864 | 3,64822E-05 | SERF2-C15ORF63 | FALSE | Downregulated |
| 0,947420802 | 3,6488E-05 | TAP2 | FALSE | Upregulated |
| 1,103282199 | 3,73299E-05 | TNFRSF21 | TRUE | Upregulated |
| 2,05345095 | 3,79274E-05 | FAM111B | TRUE | Upregulated |
| 0,93978909 | 3,81355E-05 | SASH3 | FALSE | Upregulated |
| 0,817371106 | 3,82155E-05 | TCIRG1 | FALSE | Upregulated |
| 1,231351079 | 3,91424E-05 | PTPRCAP | TRUE | Upregulated |
| 0,438406487 | 3,94215E-05 | RPL36A-HNRNPH2 | FALSE | Upregulated |
| 0,494736702 | 3,94389E-05 | ESYT1 | FALSE | Upregulated |
| -0,43051805 | 3,94389E-05 | TCEB3 | FALSE | Downregulated |
| 1,18000022 | 3,94455E-05 | DBN1 | TRUE | Upregulated |
| 3,261181685 | 3,94935E-05 | GPRIN2 | TRUE | Upregulated |
| 2,333592375 | 4,01549E-05 | TMEM132A | TRUE | Upregulated |
| 0,875607134 | 4,02346E-05 | ZNF710 | FALSE | Upregulated |
| 1,348134412 | 4,02523E-05 | GPR176 | TRUE | Upregulated |
| 0,994109402 | 4,02523E-05 | RAB34 | FALSE | Upregulated |
| 1,836436636 | 4,08205E-05 | PHLDA3 | TRUE | Upregulated |
| 1,081531246 | 4,11827E-05 | BTG2 | TRUE | Upregulated |
| 2,918133004 | 4,11827E-05 | CTSE | TRUE | Upregulated |
| 0,678737535 | 4,11827E-05 | EMILIN1 | FALSE | Upregulated |
| 1,381745759 | 4,13843E-05 | ADAMTS10 | TRUE | Upregulated |
| 0,378836067 | 4,16117E-05 | HTT | FALSE | Upregulated |
| -0,362653102 | 4,16117E-05 | REEP5 | FALSE | Downregulated |
| 2,654596614 | 4,26138E-05 | LRRN4 | TRUE | Upregulated |
| -0,837344783 | 4,27605E-05 | DNASE1L3 | FALSE | Downregulated |
| 0,398331244 | 4,30176E-05 | TRIM28 | FALSE | Upregulated |
| 0,680427335 | 4,31653E-05 | GIPC1 | FALSE | Upregulated |
| -0,563216425 | 4,31653E-05 | MRPS18C | FALSE | Downregulated |
| 1,753845339 | 4,31653E-05 | TMEM136 | TRUE | Upregulated |
| -0,810623508 | 4,3869E-05 | PLGLB2 | FALSE | Downregulated |
| 1,394342038 | 4,40333E-05 | MYO1G | TRUE | Upregulated |
| 2,620103346 | 4,43989E-05 | HSP90AB4P | TRUE | Upregulated |
| 1,312105451 | 4,43989E-05 | MCM2 | TRUE | Upregulated |
| 1,197459506 | 4,44245E-05 | CD28 | TRUE | Upregulated |
| 2,815544135 | 4,44245E-05 | LOC100272216 | TRUE | Upregulated |
| 0,494438167 | 4,44245E-05 | PARVA | FALSE | Upregulated |
| -1,564271843 | 4,44245E-05 | UXT-AS1 | TRUE | Downregulated |
| -0,474046567 | 4,46409E-05 | CCNH | FALSE | Downregulated |

| | | | | |
|---|---|---|---|---|
| 1,049171949 | 4,65751E-05 | BTG3 | TRUE | Upregulated |
| 0,801525501 | 4,65751E-05 | SERINC2 | FALSE | Upregulated |
| 1,028838668 | 4,65751E-05 | SYT11 | TRUE | Upregulated |
| 1,491010008 | 4,71134E-05 | ZNF469 | TRUE | Upregulated |
| -0,842528218 | 4,74667E-05 | ANGPTL6 | FALSE | Downregulated |
| 0,706401547 | 4,74667E-05 | ATP9A | FALSE | Upregulated |
| 0,939410192 | 4,74667E-05 | FMNL3 | FALSE | Upregulated |
| 1,41674545 | 4,74667E-05 | GPRIN3 | TRUE | Upregulated |
| -0,989344807 | 4,74908E-05 | HMGCS2 | FALSE | Downregulated |
| 4,634040808 | 4,77017E-05 | KRTAP16-1 | TRUE | Upregulated |
| 0,722475341 | 4,87241E-05 | MAP7D1 | FALSE | Upregulated |
| 0,957570353 | 4,87241E-05 | NEDD4L | FALSE | Upregulated |
| -0,614846531 | 4,91491E-05 | DBI | FALSE | Downregulated |
| -0,707750028 | 4,93145E-05 | MCUR1 | FALSE | Downregulated |
| 1,658126936 | 5,06235E-05 | BUB1B | TRUE | Upregulated |
| 0,765486583 | 5,07551E-05 | ABR | FALSE | Upregulated |
| 0,845481884 | 5,14166E-05 | ADPRH | FALSE | Upregulated |
| 0,576980418 | 5,14166E-05 | HPS1 | FALSE | Upregulated |
| 1,207514838 | 5,14275E-05 | SLC45A4 | TRUE | Upregulated |
| -0,538580572 | 5,19711E-05 | BCKDHB | FALSE | Downregulated |
| 0,362512909 | 5,22306E-05 | BUB3 | FALSE | Upregulated |
| 0,54110521 | 5,27556E-05 | ZNF200 | FALSE | Upregulated |
| 3,2937323 | 5,32006E-05 | MTRNR2L7 | TRUE | Upregulated |
| -0,451985079 | 5,33796E-05 | COX17 | FALSE | Downregulated |
| 1,869457699 | 5,33796E-05 | SOX9-AS1 | TRUE | Upregulated |
| -0,436214697 | 5,37338E-05 | MRPS9 | FALSE | Downregulated |
| 0,661677353 | 5,37463E-05 | TPGS2 | FALSE | Upregulated |
| 1,523247658 | 5,42294E-05 | HNRNPA1P10 | TRUE | Upregulated |
| 2,32984681 | 5,43817E-05 | EYS | TRUE | Upregulated |
| 1,179551561 | 5,52617E-05 | CD37 | TRUE | Upregulated |
| 1,319611362 | 5,52617E-05 | LOXL2 | TRUE | Upregulated |
| 1,305776772 | 5,53004E-05 | MMP19 | TRUE | Upregulated |
| -0,593227887 | 5,58198E-05 | SPAG7 | FALSE | Downregulated |
| 1,419385072 | 5,60198E-05 | SPECC1 | TRUE | Upregulated |
| -0,699350226 | 5,60827E-05 | F11 | FALSE | Downregulated |
| -0,631693925 | 5,60827E-05 | PON3 | FALSE | Downregulated |
| -2,182207718 | 5,62142E-05 | CYP2C19 | TRUE | Downregulated |
| -0,733467439 | 5,62373E-05 | HFE2 | FALSE | Downregulated |
| -0,683723733 | 5,63833E-05 | ABHD14B | FALSE | Downregulated |
| -0,95995039 | 5,63833E-05 | GLOD5 | FALSE | Downregulated |
| 0,819226525 | 5,65039E-05 | WLS | FALSE | Upregulated |
| 1,28622218 | 5,65717E-05 | F2RL1 | TRUE | Upregulated |
| 1,07386391 | 5,71972E-05 | SLC6A6 | TRUE | Upregulated |
| -0,624004621 | 5,73593E-05 | ANGPTL3 | FALSE | Downregulated |
| 2,636342303 | 5,73593E-05 | NPTX2 | TRUE | Upregulated |
| 0,903139023 | 5,7643E-05 | GPR124 | FALSE | Upregulated |
| -0,46449228 | 5,7643E-05 | SEPHS2 | FALSE | Downregulated |
| 0,754708943 | 5,80177E-05 | GRAMD1A | FALSE | Upregulated |
| 2,200116207 | 5,85234E-05 | KCNJ5 | TRUE | Upregulated |
| 1,698378063 | 5,86134E-05 | GCNT4 | TRUE | Upregulated |
| -0,561240326 | 5,88497E-05 | ZCCHC9 | FALSE | Downregulated |

| | | | | |
|---|---|---|---|---|
| -0,822255133 | 5,91132E-05 | C16orf46 | FALSE | Downregulated |
| 0,62207046 | 5,92406E-05 | HMGN4 | FALSE | Upregulated |
| 0,646483487 | 5,9345E-05 | FGD6 | FALSE | Upregulated |
| 0,520213199 | 5,9389E-05 | CDIP1 | FALSE | Upregulated |
| -0,745557448 | 5,94311E-05 | CBWD5 | FALSE | Downregulated |
| 0,872276242 | 6,03715E-05 | C10orf54 | FALSE | Upregulated |
| 0,845956351 | 6,03715E-05 | ITGA4 | FALSE | Upregulated |
| 1,203407056 | 6,03832E-05 | SLC7A6 | TRUE | Upregulated |
| 0,753524269 | 6,0804E-05 | TRAF2 | FALSE | Upregulated |
| 2,991108246 | 6,11714E-05 | FAM150A | TRUE | Upregulated |
| -0,705773508 | 6,16884E-05 | HIRIP3 | FALSE | Downregulated |
| 1,57344842 | 6,23218E-05 | NOTCH3 | TRUE | Upregulated |
| 1,354669197 | 6,23383E-05 | SPOCK2 | TRUE | Upregulated |
| -0,581495111 | 6,24974E-05 | BLVRB | FALSE | Downregulated |
| 1,493262924 | 6,24974E-05 | COL1A1 | TRUE | Upregulated |
| -0,888577207 | 6,26079E-05 | CLEC4G | FALSE | Downregulated |
| -0,404170823 | 6,31342E-05 | HMGN3 | FALSE | Downregulated |
| -0,50409869 | 6,31342E-05 | RNF5 | FALSE | Downregulated |
| -0,532386273 | 6,44309E-05 | LIX1L | FALSE | Downregulated |
| -0,658483598 | 6,48684E-05 | SNRNP48 | FALSE | Downregulated |
| -0,41131454 | 6,50765E-05 | RPAIN | FALSE | Downregulated |
| 1,463177695 | 6,50817E-05 | CD5 | TRUE | Upregulated |
| 1,173500329 | 6,57758E-05 | ENTPD1 | TRUE | Upregulated |
| 0,433719372 | 6,57758E-05 | NUP188 | FALSE | Upregulated |
| 1,054104876 | 6,57758E-05 | RUNX3 | TRUE | Upregulated |
| 0,630978261 | 6,57758E-05 | SBNO2 | FALSE | Upregulated |
| -0,936040123 | 6,57758E-05 | SLC22A9 | FALSE | Downregulated |
| 1,500536344 | 6,6139E-05 | PTPN7 | TRUE | Upregulated |
| 0,96976879 | 6,6139E-05 | TRAF5 | FALSE | Upregulated |
| 1,167126987 | 6,64351E-05 | HUNK | TRUE | Upregulated |
| 0,993799067 | 6,66483E-05 | APOBEC3G | FALSE | Upregulated |
| 0,731191414 | 6,66833E-05 | TMSB10 | FALSE | Upregulated |
| -0,561957866 | 6,68013E-05 | C14orf2 | FALSE | Downregulated |
| 0,891357663 | 6,68013E-05 | HMHA1 | FALSE | Upregulated |
| 0,77578398 | 6,68013E-05 | PPP1R16B | FALSE | Upregulated |
| 3,026416582 | 6,68013E-05 | RP11-14D22.5 | TRUE | Upregulated |
| 0,45163804 | 6,68013E-05 | RPS6KA4 | FALSE | Upregulated |
| 0,776237114 | 6,68013E-05 | YWHAH | FALSE | Upregulated |
| 1,513878621 | 6,71237E-05 | LY9 | TRUE | Upregulated |
| -0,900197524 | 6,82623E-05 | RP11-317J10.2 | FALSE | Downregulated |
| -0,370947888 | 6,83749E-05 | USP47 | FALSE | Downregulated |
| 0,632478096 | 6,90145E-05 | DAPK3 | FALSE | Upregulated |
| 0,809816305 | 6,90145E-05 | LIMK1 | FALSE | Upregulated |
| -0,552125751 | 6,90145E-05 | NDUFB7 | FALSE | Downregulated |
| -0,40940516 | 6,90767E-05 | PARP16 | FALSE | Downregulated |
| -0,452535226 | 6,92525E-05 | UBB | FALSE | Downregulated |
| 2,407602973 | 7,06642E-05 | SCN7A | TRUE | Upregulated |
| 0,549953728 | 7,14867E-05 | MAP3K1 | FALSE | Upregulated |
| 0,826733024 | 7,24423E-05 | SMA5 | FALSE | Upregulated |
| 1,275059876 | 7,24968E-05 | SOGA2 | TRUE | Upregulated |
| 1,364923574 | 7,26948E-05 | ITIH5 | TRUE | Upregulated |

| | | | | |
|---|---|---|---|---|
| -0,980473436 | 7,26948E-05 | RAB11B-AS1 | FALSE | Downregulated |
| 0,740936306 | 7,29348E-05 | ZFP36 | FALSE | Upregulated |
| -0,751166354 | 7,29723E-05 | CDNF | FALSE | Downregulated |
| -0,983966742 | 7,32178E-05 | APOC4-APOC2 | FALSE | Downregulated |
| -0,658401123 | 7,36414E-05 | PTMS | FALSE | Downregulated |
| 0,48334851 | 7,36414E-05 | SMARCA4 | FALSE | Upregulated |
| -0,534885627 | 7,4565E-05 | NMRK1 | FALSE | Downregulated |
| 4,656332223 | 7,51871E-05 | MIR4426 | TRUE | Upregulated |
| 0,974316767 | 7,54178E-05 | MID1 | FALSE | Upregulated |
| -0,424228776 | 7,5815E-05 | NUDT6 | FALSE | Downregulated |
| -0,44722698 | 7,61712E-05 | ARL6IP4 | FALSE | Downregulated |
| -0,634935437 | 7,622E-05 | TRPT1 | FALSE | Downregulated |
| -0,440793615 | 7,62989E-05 | IRF2 | FALSE | Downregulated |
| -0,418046343 | 7,64733E-05 | COX20 | FALSE | Downregulated |
| 0,614840732 | 7,65019E-05 | ITPKC | FALSE | Upregulated |
| 0,533535915 | 7,69874E-05 | PNPLA6 | FALSE | Upregulated |
| 1,678954827 | 7,73644E-05 | NEURL3 | TRUE | Upregulated |
| 1,022608914 | 7,73644E-05 | UNC5B | TRUE | Upregulated |
| 0,601661517 | 7,73644E-05 | VOPP1 | FALSE | Upregulated |
| -0,526462719 | 7,73644E-05 | ZNF330 | FALSE | Downregulated |
| -0,540665972 | 7,73805E-05 | ACADL | FALSE | Downregulated |
| 0,379449596 | 7,79429E-05 | VARS | FALSE | Upregulated |
| -0,402256515 | 7,81115E-05 | ESD | FALSE | Downregulated |
| -0,421820417 | 7,81115E-05 | MPC2 | FALSE | Downregulated |
| 4,037343855 | 7,85144E-05 | OPRM1 | TRUE | Upregulated |
| -0,781825763 | 7,8863E-05 | DNAJC12 | FALSE | Downregulated |
| 1,247099084 | 7,89991E-05 | LFNG | TRUE | Upregulated |
| -0,580108391 | 7,89991E-05 | NSA2 | FALSE | Downregulated |
| 1,176607073 | 8,02835E-05 | C15orf52 | TRUE | Upregulated |
| -0,342055325 | 8,20662E-05 | BCCIP | FALSE | Downregulated |
| 1,220154135 | 8,21927E-05 | ACTA2 | TRUE | Upregulated |
| 3,298992774 | 8,21927E-05 | RAB3B | TRUE | Upregulated |
| -0,456544473 | 8,22639E-05 | TSR2 | FALSE | Downregulated |
| -0,730787198 | 8,28425E-05 | INO80B-WBP1 | FALSE | Downregulated |
| -0,769824193 | 8,34498E-05 | CLEC3B | FALSE | Downregulated |
| 2,554136956 | 8,4241E-05 | CLDN4 | TRUE | Upregulated |
| 0,330010872 | 8,42664E-05 | CCAR2 | FALSE | Upregulated |
| 1,213764253 | 8,42664E-05 | ISG20 | TRUE | Upregulated |
| 3,552700837 | 8,45779E-05 | FAXC | TRUE | Upregulated |
| 1,947113831 | 8,45896E-05 | EGR3 | TRUE | Upregulated |
| -0,452311796 | 8,51295E-05 | KRCC1 | FALSE | Downregulated |
| 1,269202283 | 8,54225E-05 | FBLN5 | TRUE | Upregulated |
| 2,599387844 | 8,64804E-05 | C1orf106 | TRUE | Upregulated |
| 0,770491474 | 8,65489E-05 | ARHGEF10 | FALSE | Upregulated |
| -0,316898863 | 8,66713E-05 | ATAD1 | FALSE | Downregulated |
| 0,436901204 | 8,67692E-05 | NSMAF | FALSE | Upregulated |
| -0,547704735 | 8,67692E-05 | SUGCT | FALSE | Downregulated |
| -0,532284227 | 8,82064E-05 | PIGV | FALSE | Downregulated |
| 0,792194723 | 8,84014E-05 | CRIP2 | FALSE | Upregulated |
| 0,864409311 | 8,93356E-05 | FAM60A | FALSE | Upregulated |
| 1,935568426 | 8,93356E-05 | LGALS2 | TRUE | Upregulated |

| | | | | |
|---|---|---|---|---|
| 0,898133471 | 8,93376E-05 | CD97 | FALSE | Upregulated |
| -0,335872566 | 8,93376E-05 | EIF2A | FALSE | Downregulated |
| 0,521005778 | 8,97037E-05 | TBC1D9 | FALSE | Upregulated |
| 1,487852767 | 8,9717E-05 | EPHB2 | TRUE | Upregulated |
| 1,96003539 | 8,9717E-05 | GLI2 | TRUE | Upregulated |
| -0,38573392 | 9,01233E-05 | SUOX | FALSE | Downregulated |
| -0,544099762 | 9,02403E-05 | GYS2 | FALSE | Downregulated |
| 0,308986685 | 9,045E-05 | DNM2 | FALSE | Upregulated |
| 1,280366927 | 9,045E-05 | IER3 | TRUE | Upregulated |
| -0,51080719 | 9,16898E-05 | FAM210B | FALSE | Downregulated |
| 0,546186927 | 9,22515E-05 | ILK | FALSE | Upregulated |
| 0,835969788 | 9,23879E-05 | TNFRSF11B | FALSE | Upregulated |
| -0,724394065 | 9,38864E-05 | ECH1 | FALSE | Downregulated |
| 0,720080247 | 9,41021E-05 | TGFB1 | FALSE | Upregulated |
| -0,624655619 | 9,41304E-05 | RAB30-AS1 | FALSE | Downregulated |
| 0,543814422 | 9,41304E-05 | SMPD1 | FALSE | Upregulated |
| 0,884491122 | 9,4419E-05 | ARHGEF3 | FALSE | Upregulated |
| 1,09296755 | 9,50113E-05 | CALHM2 | TRUE | Upregulated |
| -0,61925513 | 9,53397E-05 | C11orf1 | FALSE | Downregulated |
| -0,486741065 | 9,54783E-05 | RBM41 | FALSE | Downregulated |
| 0,810739206 | 9,574E-05 | STK17A | FALSE | Upregulated |
| -0,490697516 | 9,67056E-05 | C11orf74 | FALSE | Downregulated |
| 3,92617452 | 9,71974E-05 | PKP1 | TRUE | Upregulated |
| -0,717164008 | 9,71974E-05 | RWDD1 | FALSE | Downregulated |
| -0,452090592 | 9,75657E-05 | POLR2F | FALSE | Downregulated |
| 2,756050948 | 9,8516E-05 | PAPPA2 | TRUE | Upregulated |
| -0,458217028 | 9,86181E-05 | SAT1 | FALSE | Downregulated |
| -0,878317836 | 9,93308E-05 | CIDEB | FALSE | Downregulated |
| -0,448264906 | 9,94562E-05 | FBXO3 | FALSE | Downregulated |
| 0,294003196 | 9,94562E-05 | NKIRAS2 | FALSE | Upregulated |
| -1,401232076 | 9,94562E-05 | RP11-12A2.3 | TRUE | Downregulated |
| 0,587590976 | 9,94562E-05 | TNS3 | FALSE | Upregulated |
| 0,624789077 | 0,000100038 | GPX1 | FALSE | Upregulated |
| 2,659429244 | 0,0001004 | FAP | TRUE | Upregulated |
| 3,453203015 | 0,000100429 | PDZK1IP1 | TRUE | Upregulated |
| 1,207022019 | 0,000100456 | HEYL | TRUE | Upregulated |
| 0,420673631 | 0,000100568 | PRKCSH | FALSE | Upregulated |
| -0,785119364 | 0,000100568 | RPL17-C18orf32 | FALSE | Downregulated |
| -0,884645354 | 0,000100728 | ENDOG | FALSE | Downregulated |
| -0,667442734 | 0,000101277 | DDX60L | FALSE | Downregulated |
| 1,257817198 | 0,000101277 | KCNA3 | TRUE | Upregulated |
| 0,896677649 | 0,00010244 | PHLDB1 | FALSE | Upregulated |
| -0,628372473 | 0,000102856 | MBLAC2 | FALSE | Downregulated |
| 0,441252083 | 0,000102856 | NCOR2 | FALSE | Upregulated |
| 1,447356819 | 0,000103784 | PROM1 | TRUE | Upregulated |
| 1,568818923 | 0,000104602 | PAQR5 | TRUE | Upregulated |
| 1,514924682 | 0,000104602 | SLAMF8 | TRUE | Upregulated |
| -0,39261137 | 0,000105298 | GTF2H5 | FALSE | Downregulated |
| -0,825084229 | 0,000105408 | AQP7 | FALSE | Downregulated |
| 1,061947665 | 0,000105711 | ICAM1 | TRUE | Upregulated |
| 0,793238769 | 0,00010591 | COL6A1 | FALSE | Upregulated |

| | | | | |
|---|---|---|---|---|
| 0,266453676 | 0,000105964 | PDCD6IP | FALSE | Upregulated |
| -0,344663407 | 0,000106111 | ACP1 | FALSE | Downregulated |
| 1,732400982 | 0,000107716 | EPCAM | TRUE | Upregulated |
| -1,549151944 | 0,000107716 | MYB | TRUE | Downregulated |
| -1,307258666 | 0,000109582 | CUX2 | TRUE | Downregulated |
| 1,840715973 | 0,000109659 | ZNF578 | TRUE | Upregulated |
| -0,705471823 | 0,000109661 | ZNF771 | FALSE | Downregulated |
| 0,41723034 | 0,000110049 | STAT6 | FALSE | Upregulated |
| -1,241027209 | 0,000110145 | AC004862.6 | TRUE | Downregulated |
| 1,222157173 | 0,000110145 | IKBKE | TRUE | Upregulated |
| 1,066379769 | 0,000110208 | VAMP5 | TRUE | Upregulated |
| -0,535067166 | 0,00011044 | TYW3 | FALSE | Downregulated |
| 0,89685015 | 0,000111185 | SEL1L3 | FALSE | Upregulated |
| -0,488416373 | 0,000111871 | GJB1 | FALSE | Downregulated |
| -0,686239014 | 0,000111878 | C2orf72 | FALSE | Downregulated |
| 0,800023438 | 0,000111915 | ADCY3 | FALSE | Upregulated |
| -0,446281642 | 0,000112469 | ATP5H | FALSE | Downregulated |
| -0,438113098 | 0,000114482 | DPH5 | FALSE | Downregulated |
| -0,287408294 | 0,000114482 | FXR1 | FALSE | Downregulated |
| 1,325055657 | 0,000114482 | LAX1 | TRUE | Upregulated |
| 1,27311525 | 0,000114482 | RGS1 | TRUE | Upregulated |
| 0,995726411 | 0,000116712 | TRPV2 | FALSE | Upregulated |
| 2,005705108 | 0,00011675 | PPAP2C | TRUE | Upregulated |
| -0,502917213 | 0,000118318 | CDO1 | FALSE | Downregulated |
| 0,917308412 | 0,000118318 | EHD4 | FALSE | Upregulated |
| 0,93161439 | 0,000118318 | ITGB2 | FALSE | Upregulated |
| -1,406685485 | 0,000118318 | LINC01093 | TRUE | Downregulated |
| 0,542526699 | 0,000118796 | KCTD10 | FALSE | Upregulated |
| 1,528125887 | 0,000119891 | ELOVL7 | TRUE | Upregulated |
| -0,711919763 | 0,000119909 | EVI5 | FALSE | Downregulated |
| -0,378169809 | 0,000120863 | PINK1 | FALSE | Downregulated |
| 0,389976274 | 0,000121891 | ARF3 | FALSE | Upregulated |
| 0,766495101 | 0,000121891 | CLIP2 | FALSE | Upregulated |
| -0,51872699 | 0,000121891 | PET100 | FALSE | Downregulated |
| -0,410956873 | 0,000122131 | MTO1 | FALSE | Downregulated |
| -1,132548898 | 0,000122413 | C14orf37 | TRUE | Downregulated |
| -0,358252399 | 0,000122585 | CHMP2A | FALSE | Downregulated |
| 1,052709928 | 0,000123547 | B3GNT7 | TRUE | Upregulated |
| 2,029513645 | 0,000123675 | MELK | TRUE | Upregulated |
| 1,898123958 | 0,0001238 | DEPDC1B | TRUE | Upregulated |
| 0,99143722 | 0,000123813 | LIMK2 | FALSE | Upregulated |
| -0,811503587 | 0,000124215 | TMEM160 | FALSE | Downregulated |
| -0,668671337 | 0,000124215 | ZCWPW1 | FALSE | Downregulated |
| 1,348249855 | 0,000124397 | GSN | TRUE | Upregulated |
| 0,475021501 | 0,000124397 | NUMA1 | FALSE | Upregulated |
| -0,508163278 | 0,000125577 | ANAPC11 | FALSE | Downregulated |
| -0,538483122 | 0,000125899 | CMC4 | FALSE | Downregulated |
| 0,878639456 | 0,000126532 | PIM2 | FALSE | Upregulated |
| 1,446731932 | 0,000126746 | STMN3 | TRUE | Upregulated |
| 1,352597052 | 0,000127642 | CD69 | TRUE | Upregulated |
| 3,042582646 | 0,000127762 | EVPL | TRUE | Upregulated |

| | | | | |
|---|---|---|---|---|
| 1,664088424 | 0,000128341 | PLAU | TRUE | Upregulated |
| 0,86761082 | 0,000128341 | UNC13D | FALSE | Upregulated |
| -0,38120421 | 0,000128576 | MCCC2 | FALSE | Downregulated |
| 1,368915898 | 0,000128576 | SLC7A1 | TRUE | Upregulated |
| 0,904792503 | 0,000128576 | ZNF628 | FALSE | Upregulated |
| 3,480368044 | 0,000130068 | MUC16 | TRUE | Upregulated |
| -0,625345529 | 0,000130083 | C8A | FALSE | Downregulated |
| 0,731595295 | 0,000130083 | CCND3 | FALSE | Upregulated |
| -1,269266589 | 0,000130396 | RP11-458D21.1 | TRUE | Downregulated |
| -0,619832257 | 0,000130625 | UQCR11 | FALSE | Downregulated |
| -0,682981278 | 0,000130876 | C1orf21 | FALSE | Downregulated |
| 1,033036494 | 0,000131013 | HLA-DOA | TRUE | Upregulated |
| 0,786961849 | 0,000132101 | GLIS3 | FALSE | Upregulated |
| -0,29216629 | 0,000132591 | PPP2R5C | FALSE | Downregulated |
| 3,219650581 | 0,000132696 | C3orf52 | TRUE | Upregulated |
| -0,512438488 | 0,000133819 | AAMP | FALSE | Downregulated |
| -0,391532171 | 0,000134672 | NOL11 | FALSE | Downregulated |
| 2,703939762 | 0,000134682 | BDKRB2 | TRUE | Upregulated |
| 0,583343877 | 0,000135567 | ACOT9 | FALSE | Upregulated |
| -0,400103613 | 0,00013674 | ACAD10 | FALSE | Downregulated |
| -0,633580153 | 0,00013674 | HEBP2 | FALSE | Downregulated |
| 1,722679939 | 0,000137008 | CXCL9 | TRUE | Upregulated |
| -0,748108536 | 0,000137008 | EIF4E | FALSE | Downregulated |
| 3,143339271 | 0,000137008 | LRRIQ1 | TRUE | Upregulated |
| -1,581925439 | 0,00013739 | HSD17B14 | TRUE | Downregulated |
| 1,597962142 | 0,000138433 | C1orf116 | TRUE | Upregulated |
| 0,508248907 | 0,000141737 | BCL10 | FALSE | Upregulated |
| -0,801475948 | 0,000141737 | DCXR | FALSE | Downregulated |
| 1,383587644 | 0,000141895 | SCD5 | TRUE | Upregulated |
| 0,745369637 | 0,000143856 | MAP1S | FALSE | Upregulated |
| -0,343871006 | 0,000143856 | SRP14 | FALSE | Downregulated |
| 0,972632613 | 0,000146385 | RAP1GAP2 | FALSE | Upregulated |
| 1,490454095 | 0,000146805 | TLR7 | TRUE | Upregulated |
| -0,523110171 | 0,000148903 | RPL19 | FALSE | Downregulated |
| -0,75361421 | 0,000149837 | CALML4 | FALSE | Downregulated |
| 1,12272267 | 0,000149837 | PHACTR1 | TRUE | Upregulated |
| 1,72308786 | 0,000150257 | SLC1A7 | TRUE | Upregulated |
| 3,801309209 | 0,000150591 | RNU2-1 | TRUE | Upregulated |
| -0,868311142 | 0,000150629 | IGF2 | FALSE | Downregulated |
| -0,62031795 | 0,000151801 | ANKRA2 | FALSE | Downregulated |
| 0,572803757 | 0,000153037 | GPC1 | FALSE | Upregulated |
| 0,953835515 | 0,000153037 | GPX8 | FALSE | Upregulated |
| 0,404071645 | 0,000153037 | ITGB1 | FALSE | Upregulated |
| 0,522712685 | 0,000153037 | SYPL1 | FALSE | Upregulated |
| 0,451683413 | 0,000153037 | TNFRSF10B | FALSE | Upregulated |
| -1,195907544 | 0,000153127 | ASPDH | TRUE | Downregulated |
| 0,936921916 | 0,000153926 | TNFRSF10A | FALSE | Upregulated |
| 0,533517622 | 0,000154202 | GRN | FALSE | Upregulated |
| 1,098490331 | 0,000154202 | MYOF | TRUE | Upregulated |
| -0,634816494 | 0,000154202 | SMIM14 | FALSE | Downregulated |
| -0,244704483 | 0,000154417 | URI1 | FALSE | Downregulated |

| | | | | |
|---|---|---|---|---|
| 1,384560283 | 0,000157349 | PCNXL2 | TRUE | Upregulated |
| -0,53685662 | 0,000158644 | MRPS18B | FALSE | Downregulated |
| 0,861908679 | 0,000159167 | PIK3CD | FALSE | Upregulated |
| 1,403062541 | 0,00016048 | THBS1 | TRUE | Upregulated |
| 2,283768218 | 0,000161964 | MND1 | TRUE | Upregulated |
| -0,56370592 | 0,000162831 | TFB1M | FALSE | Downregulated |
| 0,344524295 | 0,000164656 | SF3B3 | FALSE | Upregulated |
| 0,353076017 | 0,000165451 | POLG | FALSE | Upregulated |
| -0,624314596 | 0,000166315 | ATP5G1 | FALSE | Downregulated |
| -0,480462527 | 0,000166888 | SLIRP | FALSE | Downregulated |
| 0,97377184 | 0,0001684 | SPINT2 | FALSE | Upregulated |
| 0,949537567 | 0,0001684 | STK39 | FALSE | Upregulated |
| 0,461333203 | 0,000168484 | STX6 | FALSE | Upregulated |
| 0,503775829 | 0,000169186 | SYNJ2 | FALSE | Upregulated |
| 0,386340596 | 0,000169289 | SPECC1L | FALSE | Upregulated |
| 1,114153119 | 0,000169646 | TCF19 | TRUE | Upregulated |
| 1,50417363 | 0,000170971 | LOX | TRUE | Upregulated |
| 0,501412688 | 0,000171784 | DCAF15 | FALSE | Upregulated |
| -0,472838509 | 0,000171853 | C1orf56 | FALSE | Downregulated |
| 1,435677261 | 0,000174344 | SULT1C4 | TRUE | Upregulated |
| 0,289328124 | 0,00017454 | RHOA | FALSE | Upregulated |
| 1,105164843 | 0,000176103 | MRVI1 | TRUE | Upregulated |
| 1,248203534 | 0,000176254 | THEMIS | TRUE | Upregulated |
| 1,059070151 | 0,000176878 | ADAMTS1 | TRUE | Upregulated |
| -1,207624731 | 0,000176878 | HM13-AS1 | TRUE | Downregulated |
| 0,417656498 | 0,000176902 | ATP13A1 | FALSE | Upregulated |
| 1,305852589 | 0,000176902 | BICD1 | TRUE | Upregulated |
| 3,955835136 | 0,000176902 | TUBB3 | TRUE | Upregulated |
| -0,458361953 | 0,000177238 | RPS27A | FALSE | Downregulated |
| -0,486038865 | 0,000178856 | DNAJB1 | FALSE | Downregulated |
| 0,762375214 | 0,000179448 | CYTH4 | FALSE | Upregulated |
| -0,30613032 | 0,00017986 | ZNF780A | FALSE | Downregulated |
| -0,469628144 | 0,000180348 | PIGY | FALSE | Downregulated |
| -1,421094224 | 0,000181756 | GPC5 | TRUE | Downregulated |
| 1,320463893 | 0,000182302 | APCDD1 | TRUE | Upregulated |
| -0,748228935 | 0,000182841 | FKBP2 | FALSE | Downregulated |
| 0,998788907 | 0,000183771 | DOK1 | FALSE | Upregulated |
| -0,641132785 | 0,00018457 | HCFC1R1 | FALSE | Downregulated |
| 1,481773471 | 0,000184699 | IL34 | TRUE | Upregulated |
| 1,670901979 | 0,000184699 | NALCN | TRUE | Upregulated |
| -0,389549039 | 0,000185366 | PET112 | FALSE | Downregulated |
| 0,860260863 | 0,000185873 | TNFAIP2 | FALSE | Upregulated |
| -0,338185273 | 0,000187226 | PROSC | FALSE | Downregulated |
| 0,985819837 | 0,000187464 | FGD3 | FALSE | Upregulated |
| -0,622752224 | 0,000187525 | PXMP2 | FALSE | Downregulated |
| -1,08901385 | 0,000187627 | C1orf173 | TRUE | Downregulated |
| 0,461219219 | 0,000187627 | RB1 | FALSE | Upregulated |
| 0,374693171 | 0,000187627 | WWP2 | FALSE | Upregulated |
| -0,412348101 | 0,000187627 | ZCRB1 | FALSE | Downregulated |
| -0,477652041 | 0,0001881 | TAF11 | FALSE | Downregulated |
| 0,728284226 | 0,000188236 | CORO7 | FALSE | Upregulated |

| | | | | |
|---|---|---|---|---|
| 0,632738808 | 0,000188236 | DSN1 | FALSE | Upregulated |
| 1,424599493 | 0,000188236 | IL21R | TRUE | Upregulated |
| 1,303215936 | 0,000188354 | HLA-H | TRUE | Upregulated |
| 2,170417598 | 0,00018886 | IGLL1 | TRUE | Upregulated |
| -0,840145154 | 0,000190145 | SPG20 | FALSE | Downregulated |
| 2,081300634 | 0,00019039 | DERL3 | TRUE | Upregulated |
| 0,532301989 | 0,000190624 | PTTG1IP | FALSE | Upregulated |
| -0,434572284 | 0,000190724 | F8A1 | FALSE | Downregulated |
| -0,773105225 | 0,000191376 | CROT | FALSE | Downregulated |
| 1,012325327 | 0,000191614 | KLC2 | TRUE | Upregulated |
| 4,674357245 | 0,000191614 | KRT23 | TRUE | Upregulated |
| 2,179430252 | 0,000192432 | CXCL1 | TRUE | Upregulated |
| -0,424494313 | 0,000192886 | NDUFA6 | FALSE | Downregulated |
| 1,345179509 | 0,000193544 | TNFSF11 | TRUE | Upregulated |
| 1,120523862 | 0,000196065 | FRMD6 | TRUE | Upregulated |
| 0,780866447 | 0,000196115 | PTRF | FALSE | Upregulated |
| 0,853027589 | 0,000196115 | SLC9A1 | FALSE | Upregulated |
| 0,775765696 | 0,000196373 | PIGR | FALSE | Upregulated |
| -0,493848791 | 0,000196556 | NUDT16 | FALSE | Downregulated |
| 1,945799345 | 0,000197156 | AP1M2 | TRUE | Upregulated |
| -0,589398877 | 0,00019757 | ARG1 | FALSE | Downregulated |
| 1,125598694 | 0,00019757 | CARD11 | TRUE | Upregulated |
| 0,573407833 | 0,000198671 | SLC39A13 | FALSE | Upregulated |
| 1,219759648 | 0,000199403 | ADAMTS12 | TRUE | Upregulated |
| -1,050272314 | 0,00019969 | CNGA1 | TRUE | Downregulated |
| 0,380927651 | 0,000199748 | ACTR2 | FALSE | Upregulated |
| -1,00209017 | 0,000199748 | EMBP1 | TRUE | Downregulated |
| 2,525030983 | 0,000199748 | FCRL5 | TRUE | Upregulated |
| 0,72605687 | 0,000199748 | WIPF1 | FALSE | Upregulated |
| -0,296889292 | 0,000202534 | WDR75 | FALSE | Downregulated |
| -0,488710003 | 0,00020256 | ALDH7A1 | FALSE | Downregulated |
| -0,608740999 | 0,000203471 | COBLL1 | FALSE | Downregulated |
| 0,631004379 | 0,000203496 | LAMP2 | FALSE | Upregulated |
| 1,218296987 | 0,000203496 | MTHFD1L | TRUE | Upregulated |
| 0,392133129 | 0,000203496 | ZFYVE26 | FALSE | Upregulated |
| -0,294738532 | 0,000203864 | ATPAF1 | FALSE | Downregulated |
| -0,344765733 | 0,000203999 | DAZAP2 | FALSE | Downregulated |
| -0,622206811 | 0,000204821 | ANAPC10 | FALSE | Downregulated |
| 0,644514803 | 0,000204821 | COTL1 | FALSE | Upregulated |
| -0,53449493 | 0,000204821 | MAF | FALSE | Downregulated |
| 0,45536867 | 0,000204821 | PRKD2 | FALSE | Upregulated |
| 0,383853397 | 0,000204821 | YWHAZ | FALSE | Upregulated |
| 1,056048587 | 0,000205706 | DTX3 | TRUE | Upregulated |
| 2,228068071 | 0,00020906 | ATP5EP2 | TRUE | Upregulated |
| 1,66856358 | 0,00020906 | NACA2 | TRUE | Upregulated |
| 1,579635906 | 0,000210146 | DPYSL3 | TRUE | Upregulated |
| 2,274687835 | 0,000211093 | MGAT3 | TRUE | Upregulated |
| 1,454664504 | 0,000214552 | CD8A | TRUE | Upregulated |
| -0,431918321 | 0,000214552 | NUDT5 | FALSE | Downregulated |
| 0,734644702 | 0,000214808 | ALDH3B1 | FALSE | Upregulated |
| 1,765797004 | 0,000214808 | NELL2 | TRUE | Upregulated |

| | | | | |
|---|---|---|---|---|
| 5,658307167 | 0,000214808 | TRNAD16 | TRUE | Upregulated |
| -3,739783506 | 0,000215225 | LOC101929781 | TRUE | Downregulated |
| -0,567849577 | 0,000218835 | DMGDH | FALSE | Downregulated |
| 0,959186927 | 0,000218835 | ZNF320 | FALSE | Upregulated |
| 0,622586326 | 0,000220224 | PLD2 | FALSE | Upregulated |
| 2,096511239 | 0,000221205 | OXTR | TRUE | Upregulated |
| -0,285861765 | 0,000223069 | CHMP5 | FALSE | Downregulated |
| 2,574744483 | 0,000223101 | SLC4A3 | TRUE | Upregulated |
| 0,427637969 | 0,000223101 | UNC45A | FALSE | Upregulated |
| 1,557160712 | 0,000223101 | ZNF208 | TRUE | Upregulated |
| 5,248271144 | 0,000224693 | RP11-551M15.1 | TRUE | Upregulated |
| 0,463960044 | 0,000225408 | ALG9 | FALSE | Upregulated |
| -0,536288896 | 0,000227301 | NIFK | FALSE | Downregulated |
| 1,259806195 | 0,000227301 | RASGRP1 | TRUE | Upregulated |
| 0,653596763 | 0,000227542 | MTMR2 | FALSE | Upregulated |
| 1,111112976 | 0,0002286 | WDFY4 | TRUE | Upregulated |
| 1,630502606 | 0,000228814 | LDLRAD2 | TRUE | Upregulated |
| -0,484417041 | 0,000230828 | IDH1 | FALSE | Downregulated |
| 0,66961269 | 0,000231256 | GALNT10 | FALSE | Upregulated |
| -0,470442257 | 0,000231256 | MRPS24 | FALSE | Downregulated |
| 0,735227778 | 0,000231256 | PDP1 | FALSE | Upregulated |
| -0,404382545 | 0,000231256 | PSMD8 | FALSE | Downregulated |
| 0,356989574 | 0,000231256 | RPTOR | FALSE | Upregulated |
| 0,827766919 | 0,000233088 | SH3RF3 | FALSE | Upregulated |
| 1,792299595 | 0,000235672 | MTRNR2L9 | TRUE | Upregulated |
| -0,566723289 | 0,000237916 | NFIA | FALSE | Downregulated |
| 0,831358988 | 0,000238534 | MFSD6 | FALSE | Upregulated |
| 1,262110558 | 0,000239622 | CD7 | TRUE | Upregulated |
| -0,356811897 | 0,000239648 | TEX2 | FALSE | Downregulated |
| 0,369202406 | 0,00024081 | MED15 | FALSE | Upregulated |
| 1,941551812 | 0,000241117 | NRIP2 | TRUE | Upregulated |
| 0,820901432 | 0,000241453 | SLC43A2 | FALSE | Upregulated |
| 0,511319386 | 0,000243329 | RAI1 | FALSE | Upregulated |
| 0,374344515 | 0,000245356 | LAMP1 | FALSE | Upregulated |
| -1,142822501 | 0,000247727 | LOC102724832 | TRUE | Downregulated |
| -0,448358691 | 0,000247727 | METTL7A | FALSE | Downregulated |
| 0,506863735 | 0,000248647 | CAPNS1 | FALSE | Upregulated |
| -0,626737178 | 0,00024958 | MARC2 | FALSE | Downregulated |
| 0,690452665 | 0,000249719 | PAPSS1 | FALSE | Upregulated |
| 2,124187132 | 0,000251658 | FAM227A | TRUE | Upregulated |
| 0,427127968 | 0,000253559 | ARPC5 | FALSE | Upregulated |
| 1,890530312 | 0,000255423 | CAPN6 | TRUE | Upregulated |
| -0,625949998 | 0,000255423 | FAM13A | FALSE | Downregulated |
| 0,428117121 | 0,000255423 | TP53 | FALSE | Upregulated |
| -0,721192746 | 0,000255554 | EMC6 | FALSE | Downregulated |
| 2,324235189 | 0,000256297 | UCA1 | TRUE | Upregulated |
| -0,49706745 | 0,000256567 | ENY2 | FALSE | Downregulated |
| -0,411529318 | 0,00025918 | IFT88 | FALSE | Downregulated |
| 1,950300057 | 0,000259696 | TTC9 | TRUE | Upregulated |
| -0,454515742 | 0,00026449 | GCSH | FALSE | Downregulated |
| 0,688109305 | 0,000264678 | DCHS1 | FALSE | Upregulated |

| | | | | |
|---|---|---|---|---|
| 1,512462667 | 0,000264678 | FCGBP | TRUE | Upregulated |
| 0,683548992 | 0,000265802 | GSTP1 | FALSE | Upregulated |
| 2,336043859 | 0,000265802 | MIR155HG | TRUE | Upregulated |
| 0,829772502 | 0,000265802 | NUSAP1 | FALSE | Upregulated |
| 1,000432519 | 0,000265802 | PALLD | TRUE | Upregulated |
| -0,535042303 | 0,000265802 | THUMPD2 | FALSE | Downregulated |
| 0,766987724 | 0,000266659 | FUT4 | FALSE | Upregulated |
| 1,110201122 | 0,000266659 | PLXNC1 | TRUE | Upregulated |
| 1,000018486 | 0,00026702 | BCL2 | TRUE | Upregulated |
| -0,430067617 | 0,000268712 | PLG | FALSE | Downregulated |
| -0,472145065 | 0,000270098 | PECR | FALSE | Downregulated |
| 1,121834786 | 0,000274317 | LYZ | TRUE | Upregulated |
| 0,253555873 | 0,000276005 | DHX9 | FALSE | Upregulated |
| 0,559065373 | 0,000276799 | ZNF219 | FALSE | Upregulated |
| 0,457981421 | 0,000277501 | PLEC | FALSE | Upregulated |
| 0,585526184 | 0,000277978 | TMEM185B | FALSE | Upregulated |
| 2,636778563 | 0,000277978 | WNT2B | TRUE | Upregulated |
| -0,412075774 | 0,000278492 | MRPL35 | FALSE | Downregulated |
| 3,302614986 | 0,000279886 | LOC102724241 | TRUE | Upregulated |
| 0,903632441 | 0,000279886 | PDE7A | FALSE | Upregulated |
| -0,314620543 | 0,000280584 | DDX47 | FALSE | Downregulated |
| 0,606788537 | 0,000280584 | TMBIM1 | FALSE | Upregulated |
| 0,673458048 | 0,000281277 | NOC4L | FALSE | Upregulated |
| 0,995580241 | 0,00028185 | GLUD2 | FALSE | Upregulated |
| 0,631027969 | 0,000283097 | APBB1 | FALSE | Upregulated |
| 0,80197369 | 0,000283306 | HLA-B | FALSE | Upregulated |
| 1,329059107 | 0,000283786 | CLHC1 | TRUE | Upregulated |
| -0,35352278 | 0,000283786 | DCUN1D1 | FALSE | Downregulated |
| 1,096153638 | 0,000285062 | KLHL6 | TRUE | Upregulated |
| -0,685468272 | 0,000285873 | CAT | FALSE | Downregulated |
| 0,678645515 | 0,000286287 | CASP4 | FALSE | Upregulated |
| 0,378892539 | 0,000286287 | CORO1B | FALSE | Upregulated |
| 1,29707245 | 0,000286287 | GAPT | TRUE | Upregulated |
| -1,005634818 | 0,000286287 | HAO2 | TRUE | Downregulated |
| 0,627066581 | 0,000286287 | IFNGR1 | FALSE | Upregulated |
| 1,283333526 | 0,000286287 | ITK | TRUE | Upregulated |
| -0,490649887 | 0,000286287 | PCTP | FALSE | Downregulated |
| 0,933079084 | 0,000286287 | SLC38A1 | FALSE | Upregulated |
| 0,993056344 | 0,000286648 | CD247 | FALSE | Upregulated |
| -0,740495492 | 0,000286648 | GSTZ1 | FALSE | Downregulated |
| -0,648304027 | 0,000287937 | FBXO4 | FALSE | Downregulated |
| -0,359009581 | 0,000288053 | SEPT10 | FALSE | Downregulated |
| -0,511871135 | 0,000288474 | RPL4 | FALSE | Downregulated |
| -0,606447939 | 0,000289213 | SEC62 | FALSE | Downregulated |
| 1,521714173 | 0,000289523 | CHEK1 | TRUE | Upregulated |
| 0,577721633 | 0,00029037 | TWSG1 | FALSE | Upregulated |
| 0,815124762 | 0,000290645 | RAC2 | FALSE | Upregulated |
| 0,886715049 | 0,000290896 | TBKBP1 | FALSE | Upregulated |
| 2,785653137 | 0,000291111 | ADAMDEC1 | TRUE | Upregulated |
| 0,461664483 | 0,000291111 | RECQL | FALSE | Upregulated |
| 0,520439645 | 0,000291111 | TRIM8 | FALSE | Upregulated |

| | | | | |
|---|---|---|---|---|
| 0,868311132 | 0,000291111 | ZNF462 | FALSE | Upregulated |
| -1,738724121 | 0,000291116 | RP11-423H2.3 | TRUE | Downregulated |
| 1,643068531 | 0,000291217 | IDO1 | TRUE | Upregulated |
| -0,442966283 | 0,000291248 | AHCY | FALSE | Downregulated |
| 0,312047961 | 0,000291823 | DNAJC5 | FALSE | Upregulated |
| 0,92470593 | 0,000294706 | PIK3CG | FALSE | Upregulated |
| 0,39451275 | 0,000296425 | RALB | FALSE | Upregulated |
| 0,894501637 | 0,000296571 | FERMT3 | FALSE | Upregulated |
| -0,509403302 | 0,000297343 | PCBD1 | FALSE | Downregulated |
| 0,535362552 | 0,000299404 | ANXA11 | FALSE | Upregulated |
| -0,612057999 | 0,000299428 | DNAJC28 | FALSE | Downregulated |
| 0,812545121 | 0,000300787 | ADCY7 | FALSE | Upregulated |
| 3,143972437 | 0,000300787 | RPL23AP32 | TRUE | Upregulated |
| -0,855128286 | 0,000300787 | SLC45A3 | FALSE | Downregulated |
| 1,629027499 | 0,000302173 | ITGB2-AS1 | TRUE | Upregulated |
| -1,815604035 | 0,000308801 | NEUROD2 | TRUE | Downregulated |
| 0,729831013 | 0,000308801 | STXBP1 | FALSE | Upregulated |
| 1,593878928 | 0,000310057 | ADORA1 | TRUE | Upregulated |
| -1,157359174 | 0,000310816 | RP11-242D8.1 | TRUE | Downregulated |
| -0,352433457 | 0,000310816 | SHFM1 | FALSE | Downregulated |
| 0,677300301 | 0,000312153 | CDC25B | FALSE | Upregulated |
| 0,82765067 | 0,000312817 | TNFAIP8 | FALSE | Upregulated |
| 0,740763891 | 0,000313146 | ACTG1 | FALSE | Upregulated |
| -1,182198009 | 0,000313146 | IGF1 | TRUE | Downregulated |
| -0,789522717 | 0,000313146 | NRN1 | FALSE | Downregulated |
| -0,396381822 | 0,000313146 | RPL5 | FALSE | Downregulated |
| 0,682931332 | 0,000313786 | PREX1 | FALSE | Upregulated |
| -0,55271828 | 0,000314286 | NAA38 | FALSE | Downregulated |
| -0,865331408 | 0,000314286 | TTC36 | FALSE | Downregulated |
| 1,075866617 | 0,000314445 | KLF5 | TRUE | Upregulated |
| -0,8949082 | 0,000316889 | LOC101929243 | FALSE | Downregulated |
| 0,479116235 | 0,000316889 | PIGS | FALSE | Upregulated |
| 0,58762847 | 0,000318697 | SGSH | FALSE | Upregulated |
| -0,531558387 | 0,000319696 | PPP2R3C | FALSE | Downregulated |
| -0,92306402 | 0,000321747 | MRPS11 | FALSE | Downregulated |
| 0,538897405 | 0,000321747 | TUBA1B | FALSE | Upregulated |
| -0,507043375 | 0,000322872 | EMC2 | FALSE | Downregulated |
| -0,38018487 | 0,000323877 | CEP70 | FALSE | Downregulated |
| 1,428532828 | 0,000324859 | EDARADD | TRUE | Upregulated |
| 1,740714248 | 0,000328086 | EGR2 | TRUE | Upregulated |
| 2,102626756 | 0,000329235 | ROR2 | TRUE | Upregulated |
| 0,44305744 | 0,000329358 | ATP6V0D1 | FALSE | Upregulated |
| 0,916750485 | 0,00032938 | ARHGAP4 | FALSE | Upregulated |
| -0,537896797 | 0,000329695 | NGRN | FALSE | Downregulated |
| 2,138915112 | 0,000330138 | IL4I1 | TRUE | Upregulated |
| 0,453191384 | 0,000330138 | MED12 | FALSE | Upregulated |
| 0,318065913 | 0,000330138 | WASF2 | FALSE | Upregulated |
| -0,622176044 | 0,00033093 | OST4 | FALSE | Downregulated |
| -0,495982693 | 0,000331236 | SLC16A2 | FALSE | Downregulated |
| -0,666227301 | 0,000332109 | GCHFR | FALSE | Downregulated |
| -1,051407577 | 0,000332259 | LOC101929633 | TRUE | Downregulated |

| | | | | |
|---|---|---|---|---|
| 1,242078617 | 0,000334771 | KDELR3 | TRUE | Upregulated |
| -0,540985882 | 0,000334771 | OXR1 | FALSE | Downregulated |
| -0,419430703 | 0,000337126 | ORAI3 | FALSE | Downregulated |
| 1,212123049 | 0,00033774 | HELLS | TRUE | Upregulated |
| -0,498071061 | 0,000338597 | MXI1 | FALSE | Downregulated |
| -0,380188272 | 0,000339936 | EIF5 | FALSE | Downregulated |
| 1,207401069 | 0,000339936 | TOP2A | TRUE | Upregulated |
| -0,53057493 | 0,000346281 | ETFDH | FALSE | Downregulated |
| 0,469009859 | 0,000351333 | YEATS2 | FALSE | Upregulated |
| 0,387881498 | 0,000352965 | DGCR2 | FALSE | Upregulated |
| 1,147301724 | 0,000353104 | LCK | TRUE | Upregulated |
| 0,487692814 | 0,000353722 | CEP170B | FALSE | Upregulated |
| 1,954399741 | 0,000353846 | IL1RL1 | TRUE | Upregulated |
| 1,434886296 | 0,000353846 | RLTPR | TRUE | Upregulated |
| 1,808655088 | 0,000355525 | MACC1 | TRUE | Upregulated |
| 1,547822605 | 0,000356835 | NETO2 | TRUE | Upregulated |
| 0,782968015 | 0,000358444 | FKBP10 | FALSE | Upregulated |
| 0,503015858 | 0,00035851 | ARHGEF1 | FALSE | Upregulated |
| 0,499797454 | 0,000360295 | POLR1A | FALSE | Upregulated |
| 1,055764131 | 0,000364338 | ABCC4 | TRUE | Upregulated |
| 0,58508498 | 0,000364845 | PRKCD | FALSE | Upregulated |
| 2,438371066 | 0,000364845 | RTKN2 | TRUE | Upregulated |
| -0,370314702 | 0,000366213 | TERF1 | FALSE | Downregulated |
| 0,739214862 | 0,000366562 | LPPR2 | FALSE | Upregulated |
| 1,557548262 | 0,000368668 | CD27 | TRUE | Upregulated |
| 0,460001632 | 0,000368668 | PPAP2A | FALSE | Upregulated |
| -0,566451372 | 0,000369289 | PLSCR4 | FALSE | Downregulated |
| 2,71913147 | 0,000369982 | KRT8P41 | TRUE | Upregulated |
| -0,663860996 | 0,000370607 | PPP1R11 | FALSE | Downregulated |
| 0,485876689 | 0,000372885 | VASP | FALSE | Upregulated |
| -0,780257706 | 0,000373678 | LINC00888 | FALSE | Downregulated |
| 1,024551293 | 0,000374781 | CCR5 | TRUE | Upregulated |
| 0,932252348 | 0,000375077 | NEDD9 | FALSE | Upregulated |
| 1,604794759 | 0,000375376 | KIAA1244 | TRUE | Upregulated |
| 0,787092516 | 0,000375376 | SLFN11 | FALSE | Upregulated |
| 1,063192705 | 0,00037939 | ARHGAP11A | TRUE | Upregulated |
| 1,438355947 | 0,000381097 | TNFRSF19 | TRUE | Upregulated |
| 0,59273335 | 0,000383234 | RHBDD2 | FALSE | Upregulated |
| -1,424504552 | 0,000384205 | LOC100509620 | TRUE | Downregulated |
| 0,467496557 | 0,000384312 | CALM2 | FALSE | Upregulated |
| 1,248995403 | 0,000386816 | PNMA2 | TRUE | Upregulated |
| -0,400258846 | 0,000387236 | MRPL46 | FALSE | Downregulated |
| 0,854085382 | 0,0003895 | PLEKHG2 | FALSE | Upregulated |
| 1,717166324 | 0,000389884 | CD200 | TRUE | Upregulated |
| -0,36354111 | 0,000390759 | CHRAC1 | FALSE | Downregulated |
| 1,440639258 | 0,000394608 | BTLA | TRUE | Upregulated |
| -0,344549891 | 0,000395511 | ATG4C | FALSE | Downregulated |
| -0,420410411 | 0,000395676 | RPL23 | FALSE | Downregulated |
| -0,568737844 | 0,000398272 | CITED2 | FALSE | Downregulated |
| 1,550760746 | 0,000399478 | ITGBL1 | TRUE | Upregulated |
| 1,177880353 | 0,00040347 | PSTPIP1 | TRUE | Upregulated |

| | | | | |
|---|---|---|---|---|
| 2,773927 | 0,000403978 | LOC100131257 | TRUE | Upregulated |
| 0,377413827 | 0,000405829 | MTA2 | FALSE | Upregulated |
| -0,541394503 | 0,000405829 | RBP5 | FALSE | Downregulated |
| 1,10615439 | 0,000407357 | CCL28 | TRUE | Upregulated |
| 0,594521271 | 0,000410292 | AKNA | FALSE | Upregulated |
| 0,871372801 | 0,000410292 | FAM78A | FALSE | Upregulated |
| -0,409336544 | 0,000410292 | KYNU | FALSE | Downregulated |
| 1,143165779 | 0,000413532 | ZWINT | TRUE | Upregulated |
| 0,654870836 | 0,000414783 | RHOC | FALSE | Upregulated |
| -0,501792226 | 0,000415999 | C1RL | FALSE | Downregulated |
| 0,243907577 | 0,000416626 | EFTUD2 | FALSE | Upregulated |
| 0,942025629 | 0,000417164 | EPDR1 | FALSE | Upregulated |
| 0,860371926 | 0,000418593 | CYTIP | FALSE | Upregulated |
| -0,338228527 | 0,000418746 | C1GALT1 | FALSE | Downregulated |
| 0,695854315 | 0,000418746 | QSOX1 | FALSE | Upregulated |
| -0,594420272 | 0,000423247 | TMEM176B | FALSE | Downregulated |
| -0,444792884 | 0,000424063 | HSPA9 | FALSE | Downregulated |
| 0,715131517 | 0,000424774 | ATP1B3 | FALSE | Upregulated |
| -0,864330243 | 0,000424788 | TMEM238 | FALSE | Downregulated |
| -1,224669344 | 0,000427014 | GNMT | TRUE | Downregulated |
| -0,650061616 | 0,000427014 | MBIP | FALSE | Downregulated |
| 0,939952369 | 0,000427014 | TGFB3 | FALSE | Upregulated |
| 0,387818189 | 0,000430907 | TNRC18 | FALSE | Upregulated |
| -0,601203676 | 0,000433571 | CBS | FALSE | Downregulated |
| -0,587780471 | 0,000436036 | TRMT10A | FALSE | Downregulated |
| 0,469986133 | 0,000436257 | UGCG | FALSE | Upregulated |
| 1,139363514 | 0,000437144 | FMN1 | TRUE | Upregulated |
| -0,595842375 | 0,000444558 | BEND7 | FALSE | Downregulated |
| 0,982954584 | 0,000447426 | LAIR1 | FALSE | Upregulated |
| 0,571547688 | 0,000448539 | B4GALT5 | FALSE | Upregulated |
| -0,370876849 | 0,000448539 | PSMD5 | FALSE | Downregulated |
| -0,280832916 | 0,000448539 | UBE2V1 | FALSE | Downregulated |
| -0,558330777 | 0,000449318 | SNHG16 | FALSE | Downregulated |
| 0,369800042 | 0,000449558 | PLEKHM2 | FALSE | Upregulated |
| 2,687205335 | 0,000450513 | CRMP1 | TRUE | Upregulated |
| 2,16483427 | 0,000450513 | PTGES | TRUE | Upregulated |
| 1,093656538 | 0,000450513 | SLC2A6 | TRUE | Upregulated |
| 0,632637553 | 0,000450591 | KIAA1522 | FALSE | Upregulated |
| 3,004870275 | 0,000450894 | MBOAT4 | TRUE | Upregulated |
| 1,20625989 | 0,000452105 | SLC35C1 | TRUE | Upregulated |
| 0,734642114 | 0,000452105 | VSIG10 | FALSE | Upregulated |
| -0,241612858 | 0,000456288 | UFSP2 | FALSE | Downregulated |
| 0,828195168 | 0,000457611 | CHST3 | FALSE | Upregulated |
| 2,229459455 | 0,000459528 | GDNF | TRUE | Upregulated |
| 0,733912682 | 0,000459695 | PTGFRN | FALSE | Upregulated |
| -0,569053119 | 0,000459923 | GPATCH1 | FALSE | Downregulated |
| -0,658749781 | 0,000460028 | ENPP1 | FALSE | Downregulated |
| 1,481574621 | 0,000462224 | GPSM1 | TRUE | Upregulated |
| -0,320740942 | 0,00046381 | SDHA | FALSE | Downregulated |
| 1,517433431 | 0,00046735 | TESC | TRUE | Upregulated |
| 0,746217803 | 0,000468076 | MSN | FALSE | Upregulated |

| | | | | |
|---|---|---|---|---|
| -0,456350423 | 0,00047097 | INVS | FALSE | Downregulated |
| 1,357432673 | 0,000471488 | RARRES1 | TRUE | Upregulated |
| 0,737457747 | 0,000472436 | LAPTM5 | FALSE | Upregulated |
| 4,590404645 | 0,000475421 | TFAMP1 | TRUE | Upregulated |
| 1,16691849 | 0,000478542 | FAM129A | TRUE | Upregulated |
| 0,362113108 | 0,000480174 | PCNXL3 | FALSE | Upregulated |
| 0,482883279 | 0,000482059 | FLNB | FALSE | Upregulated |
| -0,416983679 | 0,000483064 | PIPOX | FALSE | Downregulated |
| -0,998071336 | 0,000485116 | MTHFD2L | FALSE | Downregulated |
| -1,403172598 | 0,000485339 | FGF23 | TRUE | Downregulated |
| 1,22043654 | 0,000485339 | IER5L | TRUE | Upregulated |
| 1,905972111 | 0,000486443 | CDH11 | TRUE | Upregulated |
| 1,972863479 | 0,000486743 | TENM4 | TRUE | Upregulated |
| 2,886813079 | 0,00048926 | LINC00842 | TRUE | Upregulated |
| 2,3199636 | 0,000489566 | ST8SIA1 | TRUE | Upregulated |
| 0,423713843 | 0,000490014 | MAN2B1 | FALSE | Upregulated |
| 1,059549273 | 0,00049102 | AMICA1 | TRUE | Upregulated |
| 1,2272752 | 0,000493758 | C11orf63 | TRUE | Upregulated |
| -0,467251646 | 0,00049406 | CRB3 | FALSE | Downregulated |
| -0,92963472 | 0,000495044 | SH3BGR | FALSE | Downregulated |
| 2,933688636 | 0,000496754 | IGFBP7-AS1 | TRUE | Upregulated |
| -0,56196712 | 0,000496754 | L2HGDH | FALSE | Downregulated |
| 1,09716973 | 0,000496754 | MYC | TRUE | Upregulated |
| -0,345052853 | 0,000497798 | TM2D1 | FALSE | Downregulated |
| 1,242225461 | 0,000499293 | CRIP1 | TRUE | Upregulated |
| 0,931911353 | 0,000499293 | MICALL2 | FALSE | Upregulated |
| -0,440362935 | 0,000500834 | GDI2 | FALSE | Downregulated |
| 1,003216054 | 0,000500834 | KRT18 | TRUE | Upregulated |
| -1,466219265 | 0,000500834 | METRN | TRUE | Downregulated |
| 1,345737051 | 0,000500834 | MTRNR2L3 | TRUE | Upregulated |
| -1,331744031 | 0,000502814 | RND2 | TRUE | Downregulated |
| 0,682966206 | 0,000503565 | IL16 | FALSE | Upregulated |
| 0,514074118 | 0,000503653 | ERMP1 | FALSE | Upregulated |
| -0,392082611 | 0,000504873 | CCDC58 | FALSE | Downregulated |
| -0,517929916 | 0,000506164 | SLC41A2 | FALSE | Downregulated |
| 0,430429223 | 0,000508533 | RNF145 | FALSE | Upregulated |
| 1,21575287 | 0,000515156 | GPNMB | TRUE | Upregulated |
| 0,769822875 | 0,000516277 | ZNF101 | FALSE | Upregulated |
| -0,429517033 | 0,000517365 | EIF4B | FALSE | Downregulated |
| 1,284474867 | 0,000517413 | CDCP1 | TRUE | Upregulated |
| 1,942629977 | 0,000517413 | HS3ST1 | TRUE | Upregulated |
| 0,670423903 | 0,000517961 | LRRC16A | FALSE | Upregulated |
| 0,313796766 | 0,000520949 | RARS | FALSE | Upregulated |
| 0,647352812 | 0,000521615 | DSEL | FALSE | Upregulated |
| -0,295814765 | 0,000525258 | CNOT2 | FALSE | Downregulated |
| 1,046042126 | 0,000525258 | ENO2 | TRUE | Upregulated |
| 1,284361346 | 0,00052701 | CLDN7 | TRUE | Upregulated |
| 0,453038411 | 0,000527253 | GNA12 | FALSE | Upregulated |
| 0,559100065 | 0,000531755 | MR1 | FALSE | Upregulated |
| 1,097096138 | 0,000532442 | FILIP1L | TRUE | Upregulated |
| 0,418619093 | 0,000534854 | COLGALT1 | FALSE | Upregulated |

| | | | | |
|---|---|---|---|---|
| 0,548129347 | 0,000536499 | PEAK1 | FALSE | Upregulated |
| 0,734905715 | 0,000537161 | HLA-A | FALSE | Upregulated |
| 1,778739346 | 0,000537161 | STC1 | TRUE | Upregulated |
| -0,487429881 | 0,000538319 | A1CF | FALSE | Downregulated |
| 1,596062078 | 0,000542101 | EMP1 | TRUE | Upregulated |
| -0,850032864 | 0,000542101 | MTERFD2 | FALSE | Downregulated |
| -0,852189518 | 0,000543212 | ANKRD35 | FALSE | Downregulated |
| -0,977364886 | 0,000544218 | ACSM2A | FALSE | Downregulated |
| 1,687218343 | 0,000544936 | MTMR11 | TRUE | Upregulated |
| 2,68826403 | 0,000546132 | TMEM132C | TRUE | Upregulated |
| 1,059895908 | 0,000553231 | APOL3 | TRUE | Upregulated |
| 1,627009157 | 0,000553597 | C6orf132 | TRUE | Upregulated |
| 1,319302001 | 0,000555183 | RRM2 | TRUE | Upregulated |
| 0,684996124 | 0,000563422 | PRSS23 | FALSE | Upregulated |
| -0,441855536 | 0,000565008 | CDC40 | FALSE | Downregulated |
| 2,796739169 | 0,000565008 | SOGA3 | TRUE | Upregulated |
| -0,46146521 | 0,00056703 | GPATCH11 | FALSE | Downregulated |
| -0,406348337 | 0,00056703 | PLK1S1 | FALSE | Downregulated |
| 0,236750688 | 0,00056736 | EXT2 | FALSE | Upregulated |
| -1,212962589 | 0,00056736 | LOC101928384 | TRUE | Downregulated |
| 0,331673586 | 0,000567707 | ARPC2 | FALSE | Upregulated |
| 1,165938108 | 0,000567707 | MFI2 | TRUE | Upregulated |
| 1,33732172 | 0,000569602 | CD6 | TRUE | Upregulated |
| 1,056602752 | 0,000570643 | SLIT2 | TRUE | Upregulated |
| -0,322228454 | 0,000571681 | NEDD8 | FALSE | Downregulated |
| -0,796482259 | 0,000573052 | ORM2 | FALSE | Downregulated |
| 0,724899168 | 0,000576021 | CMTM3 | FALSE | Upregulated |
| -0,413457262 | 0,000578778 | ALDH8A1 | FALSE | Downregulated |
| 0,89244874 | 0,000581235 | EPHA2 | FALSE | Upregulated |
| -0,295797383 | 0,000581235 | SMEK1 | FALSE | Downregulated |
| -1,980038861 | 0,00058193 | TRHDE-AS1 | TRUE | Downregulated |
| -0,3604777 | 0,000582278 | NFU1 | FALSE | Downregulated |
| 1,07139242 | 0,000582278 | PTPN14 | TRUE | Upregulated |
| 1,095819518 | 0,000582344 | LRRC37A4P | TRUE | Upregulated |
| 2,476169317 | 0,000582344 | LYPD2 | TRUE | Upregulated |
| -0,346995385 | 0,000582657 | TIMM10B | FALSE | Downregulated |
| 0,383476414 | 0,000585678 | AMBRA1 | FALSE | Upregulated |
| 1,085809006 | 0,000585678 | DACT1 | TRUE | Upregulated |
| 0,695890019 | 0,000589272 | ANKRD13D | FALSE | Upregulated |
| -0,285034107 | 0,000590448 | ARFGAP2 | FALSE | Downregulated |
| 0,516703139 | 0,000590448 | MKL1 | FALSE | Upregulated |
| 0,471722096 | 0,000590448 | RPL28 | FALSE | Upregulated |
| 1,529715597 | 0,000590448 | SDK1 | TRUE | Upregulated |
| -0,28926586 | 0,000590448 | ZCCHC7 | FALSE | Downregulated |
| -0,393735134 | 0,00059121 | MLX | FALSE | Downregulated |
| -0,250130176 | 0,000593295 | SNW1 | FALSE | Downregulated |
| -0,35283901 | 0,000595147 | SEC63 | FALSE | Downregulated |
| 0,944435725 | 0,000595589 | PTPRU | FALSE | Upregulated |
| -0,390822124 | 0,000596197 | NDUFA1 | FALSE | Downregulated |
| 3,195731628 | 0,000596197 | SCN5A | TRUE | Upregulated |
| 1,177205814 | 0,000596801 | PLAUR | TRUE | Upregulated |

| | | | | |
|---|---|---|---|---|
| -0,312336283 | 0,000598454 | TAB3 | FALSE | Downregulated |
| -0,337060371 | 0,000603492 | C2orf47 | FALSE | Downregulated |
| 2,132073387 | 0,000603492 | KIAA1045 | TRUE | Upregulated |
| 0,516968965 | 0,000603492 | PIP5K1C | FALSE | Upregulated |
| 0,41638305 | 0,000605405 | JUP | FALSE | Upregulated |
| 1,046045477 | 0,000606882 | SRGAP1 | TRUE | Upregulated |
| -0,39981616 | 0,000607135 | EEF1D | FALSE | Downregulated |
| 3,722731658 | 0,00060954 | LINC00319 | TRUE | Upregulated |
| 0,476425682 | 0,000611563 | FOXK1 | FALSE | Upregulated |
| -0,486320209 | 0,000611563 | ISCU | FALSE | Downregulated |
| 0,656116604 | 0,000611563 | SLC15A3 | FALSE | Upregulated |
| -0,585933098 | 0,000614199 | SNAPC5 | FALSE | Downregulated |
| -0,488488741 | 0,000615876 | RAB7A | FALSE | Downregulated |
| 1,981224451 | 0,000616033 | E2F1 | TRUE | Upregulated |
| -0,465268387 | 0,000616033 | NFATC3 | FALSE | Downregulated |
| 0,943022957 | 0,000617139 | GLIPR2 | FALSE | Upregulated |
| -0,600400638 | 0,00061821 | CCDC104 | FALSE | Downregulated |
| 0,635074346 | 0,00061821 | CCDC146 | FALSE | Upregulated |
| 0,596455361 | 0,000621745 | STAP2 | FALSE | Upregulated |
| -0,432893686 | 0,000629111 | COX7A2L | FALSE | Downregulated |
| 0,784793117 | 0,000629111 | SEMA4D | FALSE | Upregulated |
| -0,585550459 | 0,000629243 | BIVM-ERCC5 | FALSE | Downregulated |
| -0,56110523 | 0,000629947 | ARMC5 | FALSE | Downregulated |
| -0,846868128 | 0,000632994 | LOC92249 | FALSE | Downregulated |
| -0,396526839 | 0,000636499 | STK11 | FALSE | Downregulated |
| 0,32859325 | 0,000636499 | USP5 | FALSE | Upregulated |
| 0,996043589 | 0,000639086 | TSPAN15 | FALSE | Upregulated |
| 0,782472483 | 0,000640235 | IL15RA | FALSE | Upregulated |
| -0,767758353 | 0,000640374 | DLEU1 | FALSE | Downregulated |
| 0,565460534 | 0,000641804 | TACC1 | FALSE | Upregulated |
| -0,620324515 | 0,000643105 | PXDC1 | FALSE | Downregulated |
| -0,449384657 | 0,000643785 | C19orf53 | FALSE | Downregulated |
| 3,785574656 | 0,000648295 | ARGFX | TRUE | Upregulated |
| 1,410853825 | 0,000648295 | UBASH3A | TRUE | Upregulated |
| -1,901151255 | 0,000650285 | HRASLS2 | TRUE | Downregulated |
| -0,413958937 | 0,000651584 | AKR1A1 | FALSE | Downregulated |
| 0,963094092 | 0,000651584 | CCDC88B | FALSE | Upregulated |
| 0,537788399 | 0,000651584 | FRY | FALSE | Upregulated |
| 0,449698437 | 0,000651584 | INTS7 | FALSE | Upregulated |
| -0,733836344 | 0,000651584 | IYD | FALSE | Downregulated |
| 2,954772139 | 0,000651584 | SCTR | TRUE | Upregulated |
| -0,395748558 | 0,00065245 | MRPL24 | FALSE | Downregulated |
| 0,390897833 | 0,000655389 | ZNF687 | FALSE | Upregulated |
| -0,272611386 | 0,000657886 | COPS7A | FALSE | Downregulated |
| -0,789892324 | 0,000657886 | FAM184A | FALSE | Downregulated |
| 1,634277186 | 0,000658104 | VANGL2 | TRUE | Upregulated |
| 5,086138253 | 0,000662871 | PAGE4 | TRUE | Upregulated |
| -0,461447584 | 0,000663157 | TAF15 | FALSE | Downregulated |
| 2,115573508 | 0,000664913 | DISC1FP1 | TRUE | Upregulated |
| 1,11802224 | 0,000667362 | PCSK5 | TRUE | Upregulated |
| 0,527889803 | 0,000667499 | TRIB2 | FALSE | Upregulated |

| | | | | |
|---|---|---|---|---|
| 1,282668339 | 0,000669999 | PLCE1 | TRUE | Upregulated |
| 0,485282733 | 0,000677469 | PSEN2 | FALSE | Upregulated |
| -0,742825156 | 0,000677876 | LOC100289361 | FALSE | Downregulated |
| 0,847031471 | 0,00067934 | PHF19 | FALSE | Upregulated |
| -0,575914638 | 0,000680039 | PHF3 | FALSE | Downregulated |
| 0,670050133 | 0,000680131 | PPA1 | FALSE | Upregulated |
| 0,94750026 | 0,000681311 | GALNT16 | FALSE | Upregulated |
| 0,818614939 | 0,000681469 | PDGFRB | FALSE | Upregulated |
| -0,424332444 | 0,000682148 | LAS1L | FALSE | Downregulated |
| 0,704984273 | 0,00068429 | SLCO3A1 | FALSE | Upregulated |
| 0,338542487 | 0,000684923 | DYNC1H1 | FALSE | Upregulated |
| -0,53097395 | 0,000685053 | NIPSNAP1 | FALSE | Downregulated |
| -0,832812482 | 0,00068639 | CYP4A11 | FALSE | Downregulated |
| 0,690054604 | 0,000687027 | CUEDC1 | FALSE | Upregulated |
| -0,62365225 | 0,000687027 | PLA2G16 | FALSE | Downregulated |
| -0,740799218 | 0,000687174 | APCS | FALSE | Downregulated |
| 2,050978667 | 0,000688722 | LSAMP | TRUE | Upregulated |
| 2,280975881 | 0,000693848 | CPNE7 | TRUE | Upregulated |
| 0,490691642 | 0,000694128 | SIPA1L3 | FALSE | Upregulated |
| -0,798354317 | 0,000705373 | GPR125 | FALSE | Downregulated |
| 0,899694879 | 0,000705914 | RCAN3 | FALSE | Upregulated |
| 0,682021646 | 0,000708299 | SERPINB1 | FALSE | Upregulated |
| -0,725038473 | 0,000710399 | APOC3 | FALSE | Downregulated |
| 1,029809863 | 0,000711253 | LOC115110 | TRUE | Upregulated |
| 0,897991603 | 0,000711422 | TNFSF13B | FALSE | Upregulated |
| 0,845186081 | 0,000713689 | KLF2 | FALSE | Upregulated |
| -0,454388864 | 0,000714475 | LIAS | FALSE | Downregulated |
| 0,456976102 | 0,000719693 | CREB3 | FALSE | Upregulated |
| -0,433791682 | 0,000722961 | ANAPC16 | FALSE | Downregulated |
| 0,772294834 | 0,000723104 | MAML2 | FALSE | Upregulated |
| 0,377271211 | 0,00072402 | CFL1 | FALSE | Upregulated |
| 0,767736118 | 0,0007242 | KIAA0513 | FALSE | Upregulated |
| 2,689730092 | 0,000725016 | POTEM | TRUE | Upregulated |
| 0,407207874 | 0,000725273 | PIK3CB | FALSE | Upregulated |
| 0,928371719 | 0,000725395 | KPNA2 | FALSE | Upregulated |
| 0,794888131 | 0,000725395 | PAQR8 | FALSE | Upregulated |
| -0,508168486 | 0,000725558 | RPL27 | FALSE | Downregulated |
| -0,48776853 | 0,000727991 | LRRFIP2 | FALSE | Downregulated |
| 0,694574712 | 0,000727991 | RARG | FALSE | Upregulated |
| 0,312862319 | 0,000727991 | SF3A1 | FALSE | Upregulated |
| 0,758845212 | 0,000729754 | RPS6KA1 | FALSE | Upregulated |
| 0,756060278 | 0,000729754 | TTC7A | FALSE | Upregulated |
| 3,926629321 | 0,000731557 | CAPN13 | TRUE | Upregulated |
| 1,397121073 | 0,000733101 | FAM99B | TRUE | Upregulated |
| 1,635786358 | 0,00073424 | NAV3 | TRUE | Upregulated |
| 0,297040846 | 0,000735975 | NFKB1 | FALSE | Upregulated |
| 0,547909267 | 0,000735975 | PARP8 | FALSE | Upregulated |
| -0,5731185 | 0,000738111 | RAB33B | FALSE | Downregulated |
| 0,506113154 | 0,000739332 | CLSTN1 | FALSE | Upregulated |
| 0,823490828 | 0,00074131 | LOC101928465 | FALSE | Upregulated |
| -0,60815813 | 0,00074131 | LSM5 | FALSE | Downregulated |

| | | | | |
|---|---|---|---|---|
| 4,4144693 | 0,000745838 | ADCY8 | TRUE | Upregulated |
| 1,083144051 | 0,000745838 | FAM57A | TRUE | Upregulated |
| -0,583460846 | 0,000745851 | ANAPC15 | FALSE | Downregulated |
| 1,27580658 | 0,000745851 | FOXM1 | TRUE | Upregulated |
| -0,815910913 | 0,000746015 | RDH16 | FALSE | Downregulated |
| -0,427596161 | 0,000746945 | ESRP2 | FALSE | Downregulated |
| 3,057113076 | 0,000748556 | FOXJ1 | TRUE | Upregulated |
| 1,294154661 | 0,000752413 | MARCH3 | TRUE | Upregulated |
| 0,8970707 | 0,000752713 | OSMR | FALSE | Upregulated |
| -0,909397561 | 0,000752713 | RP11-178C3.2 | FALSE | Downregulated |
| 3,670869289 | 0,000753051 | ACAN | TRUE | Upregulated |
| -0,420212522 | 0,000753051 | SNX2 | FALSE | Downregulated |
| -0,449719104 | 0,000753051 | ST3GAL1 | FALSE | Downregulated |
| -1,262190265 | 0,000758884 | IL1RAP | TRUE | Downregulated |
| -0,452403387 | 0,000758884 | NDUFS5 | FALSE | Downregulated |
| 1,058758426 | 0,000760174 | TP53I3 | TRUE | Upregulated |
| -0,250884996 | 0,000760885 | HNRNPK | FALSE | Downregulated |
| 1,193285839 | 0,000765257 | PCDH17 | TRUE | Upregulated |
| 0,828701708 | 0,000769056 | FMOD | FALSE | Upregulated |
| -0,377826483 | 0,000769811 | KIAA0391 | FALSE | Downregulated |
| 1,662786175 | 0,000769811 | UBXN10 | TRUE | Upregulated |
| 0,712578958 | 0,000770917 | E2F3 | FALSE | Upregulated |
| 0,372927503 | 0,000771528 | ANAPC2 | FALSE | Upregulated |
| 0,869519245 | 0,000777132 | FAM110A | FALSE | Upregulated |
| -1,502001762 | 0,000777796 | MPL | TRUE | Downregulated |
| -0,609892771 | 0,000778462 | AFM | FALSE | Downregulated |
| 0,8215948 | 0,000778462 | UAP1L1 | FALSE | Upregulated |
| 0,830861255 | 0,000779484 | ABCA3 | FALSE | Upregulated |
| -1,428187261 | 0,000779484 | ENHO | TRUE | Downregulated |
| 0,351462226 | 0,000781745 | EMC1 | FALSE | Upregulated |
| -0,715316214 | 0,00078557 | CCT6B | FALSE | Downregulated |
| 1,708975549 | 0,00078557 | HAPLN3 | TRUE | Upregulated |
| -0,463086527 | 0,00078557 | NDUFA7 | FALSE | Downregulated |
| 0,87639607 | 0,00078557 | RASAL3 | FALSE | Upregulated |
| 0,588647084 | 0,000786121 | LMF1 | FALSE | Upregulated |
| -0,421732008 | 0,000786121 | TARSL2 | FALSE | Downregulated |
| 0,779808422 | 0,000786121 | ZNF669 | FALSE | Upregulated |
| 0,508802382 | 0,000786299 | EML2 | FALSE | Upregulated |
| 2,487283143 | 0,000791809 | TRIM67 | TRUE | Upregulated |
| -0,32454853 | 0,000796255 | PEX19 | FALSE | Downregulated |
| -0,665441948 | 0,000798978 | RBM47 | FALSE | Downregulated |
| 0,411228352 | 0,000801346 | CC2D1B | FALSE | Upregulated |
| -0,638299856 | 0,000803641 | HAAO | FALSE | Downregulated |
| 0,892081439 | 0,000804448 | SRPX2 | FALSE | Upregulated |
| 0,628465535 | 0,000804448 | ZBTB45 | FALSE | Upregulated |
| 1,344150371 | 0,000805826 | CAPG | TRUE | Upregulated |
| -0,436242554 | 0,000805826 | ZNF696 | FALSE | Downregulated |
| 2,252872601 | 0,000806523 | ZPLD1 | TRUE | Upregulated |
| -0,547084498 | 0,000810845 | BTD | FALSE | Downregulated |
| -0,505468262 | 0,000810845 | EBNA1BP2 | FALSE | Downregulated |
| -0,475559312 | 0,000812521 | AIMP1 | FALSE | Downregulated |

| | | | | |
|---|---|---|---|---|
| 3,214724338 | 0,000812521 | MTRNR2L1 | TRUE | Upregulated |
| -0,577045369 | 0,000812521 | PBLD | FALSE | Downregulated |
| -0,348352356 | 0,000812521 | REEP3 | FALSE | Downregulated |
| 5,002368023 | 0,000816088 | LOC100508046 | TRUE | Upregulated |
| 2,701916304 | 0,000819505 | SLC35F3 | TRUE | Upregulated |
| 0,353833483 | 0,000821381 | RHOU | FALSE | Upregulated |
| 3,081217241 | 0,000825841 | SLC6A20 | TRUE | Upregulated |
| -0,32309735 | 0,000834784 | TCAIM | FALSE | Downregulated |
| 0,475729153 | 0,000837833 | SIPA1 | FALSE | Upregulated |
| 0,618379078 | 0,000838796 | PLEKHO1 | FALSE | Upregulated |
| 2,512552798 | 0,000841687 | C9orf172 | TRUE | Upregulated |
| 0,308539731 | 0,000842139 | DBNL | FALSE | Upregulated |
| 1,317074058 | 0,000842139 | KCNN3 | TRUE | Upregulated |
| -2,636448986 | 0,000842139 | MEIOB | TRUE | Downregulated |
| -0,30971383 | 0,000842763 | ZZZ3 | FALSE | Downregulated |
| -0,590532404 | 0,000843158 | TIMMDC1 | FALSE | Downregulated |
| -1,162358059 | 0,000849039 | DNM1 | TRUE | Downregulated |
| -0,275569832 | 0,000850774 | PEX13 | FALSE | Downregulated |
| 2,275193316 | 0,000852845 | CH25H | TRUE | Upregulated |
| -0,332498177 | 0,000854367 | MRPL37 | FALSE | Downregulated |
| 0,53574028 | 0,000854367 | NPDC1 | FALSE | Upregulated |
| -0,469381681 | 0,000859836 | PAH | FALSE | Downregulated |
| 0,888567943 | 0,000863107 | EVI2A | FALSE | Upregulated |
| 0,491654149 | 0,000863605 | DPAGT1 | FALSE | Upregulated |
| 0,622791508 | 0,000866583 | PPP1R18 | FALSE | Upregulated |
| 0,857532957 | 0,00086947 | AEN | FALSE | Upregulated |
| 0,479494183 | 0,00086947 | HDAC6 | FALSE | Upregulated |
| 0,583125648 | 0,000871872 | TPM4 | FALSE | Upregulated |
| -0,36357582 | 0,000874003 | ALKBH5 | FALSE | Downregulated |
| -1,261038469 | 0,000874313 | BRCA1 | TRUE | Downregulated |
| -0,513346694 | 0,000877109 | GOLIM4 | FALSE | Downregulated |
| 0,841449924 | 0,000878282 | ADAMTSL2 | FALSE | Upregulated |
| 1,131035347 | 0,000878282 | ARNTL2 | TRUE | Upregulated |
| 0,460757295 | 0,000878282 | SPTBN1 | FALSE | Upregulated |
| 0,282826686 | 0,000878282 | TINF2 | FALSE | Upregulated |
| -0,425442781 | 0,000878563 | TBL1X | FALSE | Downregulated |
| -0,356949163 | 0,00088263 | COA5 | FALSE | Downregulated |
| 1,504221965 | 0,000882645 | CACNA1C | TRUE | Upregulated |
| -0,478448329 | 0,000882645 | MRRF | FALSE | Downregulated |
| -0,272591605 | 0,000883764 | MBD2 | FALSE | Downregulated |
| 0,475433711 | 0,000885685 | ACTB | FALSE | Upregulated |
| -0,534477058 | 0,000887316 | TMEM53 | FALSE | Downregulated |
| 0,718121238 | 0,000893656 | GBP4 | FALSE | Upregulated |
| 0,355262019 | 0,000893656 | MARK2 | FALSE | Upregulated |
| 0,852858188 | 0,000893656 | SRC | FALSE | Upregulated |
| -0,442194725 | 0,000894789 | BLOC1S1 | FALSE | Downregulated |
| 0,30866086 | 0,000894789 | SF1 | FALSE | Upregulated |
| 0,965580487 | 0,000895945 | PTGER4 | FALSE | Upregulated |
| -0,515656611 | 0,00089662 | MPLKIP | FALSE | Downregulated |
| -0,712259865 | 0,000896846 | ACADSB | FALSE | Downregulated |
| 0,764394559 | 0,000896846 | HCP5 | FALSE | Upregulated |

| | | | | |
|---|---|---|---|---|
| -1,013602757 | 0,000896846 | HSPB9 | TRUE | Downregulated |
| -0,523113857 | 0,000896846 | PRDX6 | FALSE | Downregulated |
| 0,768131085 | 0,000896846 | TUBA1A | FALSE | Upregulated |
| 0,472252143 | 0,000897847 | DDX39A | FALSE | Upregulated |
| -0,30347289 | 0,000902173 | DLD | FALSE | Downregulated |
| -0,722886241 | 0,000902394 | SLC39A4 | FALSE | Downregulated |
| 1,61482571 | 0,000906054 | MTRNR2L10 | TRUE | Upregulated |
| -0,46131585 | 0,000906054 | PHF14 | FALSE | Downregulated |
| -0,563233075 | 0,000913371 | APRT | FALSE | Downregulated |
| -0,485521779 | 0,000914694 | MUT | FALSE | Downregulated |
| 1,016578891 | 0,000914694 | TC2N | TRUE | Upregulated |
| 0,88625212 | 0,000915566 | CSF2RA | FALSE | Upregulated |
| 1,2988547 | 0,000915566 | LAMA3 | TRUE | Upregulated |
| 0,514096252 | 0,000915566 | VCL | FALSE | Upregulated |
| 0,950094012 | 0,000915566 | ZNF607 | FALSE | Upregulated |
| 1,934962954 | 0,000922108 | DNAJC6 | TRUE | Upregulated |
| 1,188620631 | 0,000923642 | ADAM28 | TRUE | Upregulated |
| -1,057630568 | 0,000923642 | LOC101928230 | TRUE | Downregulated |
| 0,723246834 | 0,000923642 | PLEKHO2 | FALSE | Upregulated |
| 1,265096619 | 0,000923642 | PLK1 | TRUE | Upregulated |
| 0,532171313 | 0,000924017 | PPIC | FALSE | Upregulated |
| -0,474783379 | 0,000924894 | USP15 | FALSE | Downregulated |
| -0,437803239 | 0,000933168 | NKAP | FALSE | Downregulated |
| -0,81838872 | 0,000935577 | CYP2C8 | FALSE | Downregulated |
| 0,694675284 | 0,000938534 | ITPKB | FALSE | Upregulated |
| -0,285190863 | 0,000939165 | TALDO1 | FALSE | Downregulated |
| 0,478183401 | 0,000942163 | GNPDA1 | FALSE | Upregulated |
| 2,592708495 | 0,00094268 | CTAGE1 | TRUE | Upregulated |
| 0,485114644 | 0,00094268 | LAMB2 | FALSE | Upregulated |
| 1,121244132 | 0,000944477 | SH2D3A | TRUE | Upregulated |
| 2,048764824 | 0,000946001 | FAM19A5 | TRUE | Upregulated |
| 0,387293223 | 0,000946645 | CYB5R3 | FALSE | Upregulated |
| 2,277048981 | 0,000946645 | LOC642131 | TRUE | Upregulated |
| 0,519895323 | 0,000946645 | TARBP1 | FALSE | Upregulated |
| 1,195948953 | 0,000947108 | SLAMF7 | TRUE | Upregulated |
| 1,012665659 | 0,000947313 | LILRB3 | TRUE | Upregulated |
| -0,405808015 | 0,000947313 | MED11 | FALSE | Downregulated |
| -0,744908725 | 0,000947313 | PLD6 | FALSE | Downregulated |
| -0,433570944 | 0,000950524 | HINT1 | FALSE | Downregulated |
| 0,9728584 | 0,00095696 | MRAS | FALSE | Upregulated |
| -0,818975803 | 0,00095914 | IL6R | FALSE | Downregulated |
| 0,607876864 | 0,00095914 | IQGAP1 | FALSE | Upregulated |
| -0,36474543 | 0,000962178 | COMMD1 | FALSE | Downregulated |
| -0,47819434 | 0,000962178 | QDPR | FALSE | Downregulated |
| 0,884322308 | 0,000962414 | FANCA | FALSE | Upregulated |
| -0,39948905 | 0,000962414 | MRPL20 | FALSE | Downregulated |
| 1,252501709 | 0,00096315 | SNX20 | TRUE | Upregulated |
| 0,40294848 | 0,000963321 | MOV10 | FALSE | Upregulated |
| 0,603058997 | 0,000964008 | BYSL | FALSE | Upregulated |
| 0,45672895 | 0,000964008 | NFATC2 | FALSE | Upregulated |
| -0,39717109 | 0,000964008 | PIGB | FALSE | Downregulated |

| | | | | |
|---|---|---|---|---|
| -0,470495737 | 0,000973148 | COX14 | FALSE | Downregulated |
| 1,327200959 | 0,000973148 | FAM169A | TRUE | Upregulated |
| 0,367570932 | 0,000973148 | PPP2CB | FALSE | Upregulated |
| 1,140594835 | 0,0009804 | C7 | TRUE | Upregulated |
| 1,243498604 | 0,000981505 | TRPM2 | TRUE | Upregulated |
| -0,556162799 | 0,00098175 | SLC47A1 | FALSE | Downregulated |
| -0,392278272 | 0,000982385 | C5orf22 | FALSE | Downregulated |
| -0,484287619 | 0,000984704 | CTNNBL1 | FALSE | Downregulated |
| 1,828894912 | 0,000987783 | CPNE5 | TRUE | Upregulated |
| 0,389469391 | 0,000988607 | ARHGDIA | FALSE | Upregulated |
| 0,438489016 | 0,00099183 | THADA | FALSE | Upregulated |
| 2,020748503 | 0,000992003 | LINC00892 | TRUE | Upregulated |
| 0,461276581 | 0,000995416 | ABCA2 | FALSE | Upregulated |
| 0,407514407 | 0,000995416 | POMGNT1 | FALSE | Upregulated |
| -0,636903818 | 0,001005368 | ABCA5 | FALSE | Downregulated |
| 0,357594655 | 0,001006902 | DENND5A | FALSE | Upregulated |
| -0,343562933 | 0,001012011 | PRDX5 | FALSE | Downregulated |
| 1,177476046 | 0,001012854 | ITGA11 | TRUE | Upregulated |
| 1,289505323 | 0,001015834 | NCAPG | TRUE | Upregulated |
| -0,907225712 | 0,001019187 | IFITM10 | FALSE | Downregulated |
| -0,559363532 | 0,001019723 | MINPP1 | FALSE | Downregulated |
| 0,43794543 | 0,001019824 | PAK4 | FALSE | Upregulated |
| 2,55871891 | 0,001030208 | DUSP8 | TRUE | Upregulated |
| 0,45603682 | 0,001032565 | CDCA7L | FALSE | Upregulated |
| 0,797264155 | 0,001032565 | CEP128 | FALSE | Upregulated |
| -0,438476421 | 0,001033301 | STX17 | FALSE | Downregulated |
| -0,816311518 | 0,001033747 | SULT2A1 | FALSE | Downregulated |
| 1,553730809 | 0,001034846 | CD79A | TRUE | Upregulated |
| -0,465038727 | 0,001034954 | RTN4 | FALSE | Downregulated |
| 1,015261821 | 0,001035653 | ANKRD19P | TRUE | Upregulated |
| -0,586437905 | 0,001035924 | HSBP1L1 | FALSE | Downregulated |
| 0,854372011 | 0,001036377 | NIPAL2 | FALSE | Upregulated |
| 0,827317063 | 0,001038782 | RNASE6 | FALSE | Upregulated |
| -0,685849688 | 0,001041319 | ADAMTS13 | FALSE | Downregulated |
| 0,821205265 | 0,001041319 | PARVG | FALSE | Upregulated |
| 1,342231518 | 0,001041319 | SAMD11 | TRUE | Upregulated |
| -0,592776225 | 0,001043188 | CCDC82 | FALSE | Downregulated |
| 2,778690724 | 0,00104485 | MT1HL1 | TRUE | Upregulated |
| -0,363298393 | 0,001045437 | RPF2 | FALSE | Downregulated |
| 0,790651784 | 0,001045924 | DOCK8 | FALSE | Upregulated |
| 2,083738493 | 0,001051205 | MYOCD | TRUE | Upregulated |
| 1,139053229 | 0,001052224 | PLEKHB1 | TRUE | Upregulated |
| 0,864124147 | 0,001055148 | DDR1 | FALSE | Upregulated |
| 3,127568711 | 0,001055148 | DUOX1 | TRUE | Upregulated |
| -0,538944432 | 0,001056886 | RAB40B | FALSE | Downregulated |
| -0,381492321 | 0,001056886 | TM2D2 | FALSE | Downregulated |
| 0,471274155 | 0,001057003 | KIAA0930 | FALSE | Upregulated |
| 1,220072036 | 0,00105782 | HNRNPA3P1 | TRUE | Upregulated |
| 1,590021553 | 0,001057979 | HLA-DOB | TRUE | Upregulated |
| -0,518044103 | 0,001062498 | C8orf82 | FALSE | Downregulated |
| -0,464248518 | 0,001065739 | CFH | FALSE | Downregulated |

| | | | | |
|---|---|---|---|---|
| 1,425534967 | 0,001065739 | LPAR2 | TRUE | Upregulated |
| 1,268387036 | 0,001069565 | PLCXD3 | TRUE | Upregulated |
| -0,370141249 | 0,001072578 | NDUFV2 | FALSE | Downregulated |
| 0,64272419 | 0,001073366 | GLS | FALSE | Upregulated |
| -0,643893581 | 0,001073607 | GLYAT | FALSE | Downregulated |
| 2,10396965 | 0,001081933 | WISP2 | TRUE | Upregulated |
| 0,771284203 | 0,001084944 | FAIM | FALSE | Upregulated |
| 0,52904331 | 0,001088107 | NOMO2 | FALSE | Upregulated |
| 1,240988169 | 0,001088107 | ZIK1 | TRUE | Upregulated |
| -0,377653834 | 0,001089114 | CTSL | FALSE | Downregulated |
| -0,453024679 | 0,001089114 | ECHS1 | FALSE | Downregulated |
| 0,380507872 | 0,001093185 | NACC2 | FALSE | Upregulated |
| 0,319918589 | 0,001094412 | GNA13 | FALSE | Upregulated |
| -0,406743557 | 0,001094412 | ZBTB10 | FALSE | Downregulated |
| 0,869185742 | 0,001097016 | KRT8 | FALSE | Upregulated |
| -1,434993678 | 0,001104253 | AC097468.4 | TRUE | Downregulated |
| 0,771345306 | 0,001104253 | ODF3B | FALSE | Upregulated |
| 2,623709533 | 0,00111351 | APLP1 | TRUE | Upregulated |
| -0,471921521 | 0,00111351 | CMTM8 | FALSE | Downregulated |
| -0,473692521 | 0,001113873 | OXNAD1 | FALSE | Downregulated |
| -0,366511728 | 0,001113873 | TSHZ1 | FALSE | Downregulated |
| -0,350990664 | 0,001114606 | DCAF8 | FALSE | Downregulated |
| -0,606488598 | 0,001115117 | FABP1 | FALSE | Downregulated |
| 0,427783498 | 0,001116145 | CDC42SE1 | FALSE | Upregulated |
| 0,372951879 | 0,001116145 | RNASEK | FALSE | Upregulated |
| 0,349795409 | 0,001116145 | TAPBP | FALSE | Upregulated |
| -0,910118681 | 0,001116145 | TIMM13 | FALSE | Downregulated |
| 0,381000063 | 0,001116351 | STX4 | FALSE | Upregulated |
| 0,717824741 | 0,001122682 | CRIM1 | FALSE | Upregulated |
| 0,285318353 | 0,001124199 | GLYR1 | FALSE | Upregulated |
| 0,706602129 | 0,001125011 | ANKS6 | FALSE | Upregulated |
| -0,616034782 | 0,001128158 | LIMCH1 | FALSE | Downregulated |
| -0,497416406 | 0,001135454 | RPL36AL | FALSE | Downregulated |
| -0,536648287 | 0,00113686 | FMO3 | FALSE | Downregulated |
| 0,855686702 | 0,001137361 | STAT1 | FALSE | Upregulated |
| -0,50291799 | 0,001141678 | ZNF429 | FALSE | Downregulated |
| -0,428243389 | 0,001149077 | GCNT2 | FALSE | Downregulated |
| 0,549526949 | 0,001149077 | RCN2 | FALSE | Upregulated |
| -0,972039584 | 0,001153071 | C19orf18 | FALSE | Downregulated |
| 1,160907358 | 0,001153071 | TESPA1 | TRUE | Upregulated |
| -0,521154331 | 0,001155509 | NET1 | FALSE | Downregulated |
| -0,254995018 | 0,001160211 | ARPP19 | FALSE | Downregulated |
| -0,312364799 | 0,001166136 | CCDC90B | FALSE | Downregulated |
| -0,616382135 | 0,001166136 | MRPS25 | FALSE | Downregulated |
| -0,647541064 | 0,001169028 | COQ10A | FALSE | Downregulated |
| 1,184958025 | 0,001171632 | DAPK2 | TRUE | Upregulated |
| -0,325387042 | 0,001182749 | LOC100129361 | FALSE | Downregulated |
| -0,364745365 | 0,001185132 | HNRNPC | FALSE | Downregulated |
| -0,281669438 | 0,00118581 | C9orf78 | FALSE | Downregulated |
| 1,894717253 | 0,001187247 | ILDR1 | TRUE | Upregulated |
| -1,472279363 | 0,001187254 | LOC102723782 | TRUE | Downregulated |

| | | | | |
|---|---|---|---|---|
| -0,909240565 | 0,00119425 | HSD17B7 | FALSE | Downregulated |
| 2,334952058 | 0,0011959 | CTHRC1 | TRUE | Upregulated |
| 0,783491513 | 0,001197068 | HLA-DRA | FALSE | Upregulated |
| 0,787016004 | 0,001197348 | TRIM47 | FALSE | Upregulated |
| -0,370746193 | 0,001199591 | NAP1L4 | FALSE | Downregulated |
| 1,039676376 | 0,00120249 | RHOH | TRUE | Upregulated |
| -0,445276242 | 0,001204298 | ZNHIT1 | FALSE | Downregulated |
| 0,435521585 | 0,001220599 | CCT6A | FALSE | Upregulated |
| 2,672829145 | 0,001221666 | SEPT5-GP1BB | TRUE | Upregulated |
| 0,490222681 | 0,001224702 | FOCAD | FALSE | Upregulated |
| -0,725446922 | 0,0012255 | ALDOB | FALSE | Downregulated |
| 0,308795084 | 0,001230176 | ETV3 | FALSE | Upregulated |
| 2,000944891 | 0,001231679 | CCL18 | TRUE | Upregulated |
| -0,396729139 | 0,001238117 | ATP5G3 | FALSE | Downregulated |
| -0,377348405 | 0,001239716 | CCDC47 | FALSE | Downregulated |
| -0,430000897 | 0,001249915 | ALKBH8 | FALSE | Downregulated |
| 3,181691996 | 0,001252014 | MYEOV | TRUE | Upregulated |
| -0,250349666 | 0,001252689 | FAF1 | FALSE | Downregulated |
| -0,359027742 | 0,00125269 | ORC5 | FALSE | Downregulated |
| 0,46108054 | 0,001254245 | SLC12A9 | FALSE | Upregulated |
| -0,685860416 | 0,001255353 | C9 | FALSE | Downregulated |
| -0,615555311 | 0,00125832 | HOMER2 | FALSE | Downregulated |
| 0,678282146 | 0,001261336 | LYL1 | FALSE | Upregulated |
| 0,680159927 | 0,001261799 | RARB | FALSE | Upregulated |
| 0,652526915 | 0,001263692 | DAB2IP | FALSE | Upregulated |
| 0,919466294 | 0,001263692 | EMB | FALSE | Upregulated |
| -0,397986046 | 0,001265641 | FERMT2 | FALSE | Downregulated |
| -0,371867168 | 0,001267123 | UBE2L3 | FALSE | Downregulated |
| 1,647105075 | 0,001272953 | POU2AF1 | TRUE | Upregulated |
| 0,296741811 | 0,001272953 | WDR1 | FALSE | Upregulated |
| -0,605935562 | 0,001275448 | ACSM2B | FALSE | Downregulated |
| 1,490276577 | 0,001275448 | BLOC1S4 | TRUE | Upregulated |
| -0,304134858 | 0,001280702 | EPS15 | FALSE | Downregulated |
| 0,996731649 | 0,001283231 | BRCA2 | FALSE | Upregulated |
| 2,566215671 | 0,001283573 | CD207 | TRUE | Upregulated |
| -0,775486484 | 0,001286531 | LOC728290 | FALSE | Downregulated |
| 1,136567681 | 0,001291153 | MFSD7 | TRUE | Upregulated |
| 0,682513817 | 0,001291673 | TSSC4 | FALSE | Upregulated |
| 1,429032582 | 0,001298724 | SLC6A9 | TRUE | Upregulated |
| -0,297656398 | 0,001302062 | EXOSC10 | FALSE | Downregulated |
| 1,355003603 | 0,001303126 | HRH2 | TRUE | Upregulated |
| -0,696598172 | 0,001312889 | MARC1 | FALSE | Downregulated |
| 0,784646177 | 0,001315109 | APOBEC3C | FALSE | Upregulated |
| 0,370510697 | 0,001315503 | GNB2 | FALSE | Upregulated |
| 0,910193996 | 0,001319326 | OSBPL3 | FALSE | Upregulated |
| -0,459188945 | 0,001321959 | EIF4E2 | FALSE | Downregulated |
| 1,441784414 | 0,001323528 | BCL11A | TRUE | Upregulated |
| 2,777387704 | 0,001323528 | MTRNR2L4 | TRUE | Upregulated |
| 1,389002214 | 0,001325316 | MB21D2 | TRUE | Upregulated |
| 0,547885811 | 0,001328409 | ALDH16A1 | FALSE | Upregulated |
| -0,447091776 | 0,001328409 | DNAJC17 | FALSE | Downregulated |

| | | | | |
|---|---|---|---|---|
| 1,040295914 | 0,001328409 | PARPBP | TRUE | Upregulated |
| 0,568209299 | 0,001330836 | ACTN1 | FALSE | Upregulated |
| -0,264591109 | 0,001334354 | COPS5 | FALSE | Downregulated |
| 0,828889184 | 0,001339202 | TSHZ3 | FALSE | Upregulated |
| -0,347913683 | 0,001340389 | BCAS2 | FALSE | Downregulated |
| -0,857656208 | 0,001341571 | AASS | FALSE | Downregulated |
| -0,457414568 | 0,001341571 | ATP5D | FALSE | Downregulated |
| 0,437414432 | 0,001341571 | CHPF2 | FALSE | Upregulated |
| 0,866922514 | 0,001342929 | ACKR2 | FALSE | Upregulated |
| 4,322651328 | 0,001342929 | KRT17 | TRUE | Upregulated |
| -0,717413229 | 0,001342929 | PEX11A | FALSE | Downregulated |
| -0,344491182 | 0,001345041 | ERMARD | FALSE | Downregulated |
| 1,205704468 | 0,001345041 | KIFC1 | TRUE | Upregulated |
| -0,969553824 | 0,001345909 | LOC102724261 | FALSE | Downregulated |
| 1,536663625 | 0,001345909 | ZBP1 | TRUE | Upregulated |
| 0,365889749 | 0,001349152 | AGFG1 | FALSE | Upregulated |
| 0,192726995 | 0,001350244 | AP2M1 | FALSE | Upregulated |
| 3,701897731 | 0,001350244 | LOC101928262 | TRUE | Upregulated |
| 1,874431216 | 0,001350244 | MYO7B | TRUE | Upregulated |
| 1,112241705 | 0,001350936 | THSD1 | TRUE | Upregulated |
| -0,379800451 | 0,001354536 | DDX23 | FALSE | Downregulated |
| 0,782880287 | 0,001359939 | BCL6B | FALSE | Upregulated |
| -0,274855468 | 0,001359939 | CSNK1G3 | FALSE | Downregulated |
| 1,414305446 | 0,001359939 | GRIN2D | TRUE | Upregulated |
| 1,876741129 | 0,001359939 | VWA2 | TRUE | Upregulated |
| 1,78489187 | 0,001360947 | E2F8 | TRUE | Upregulated |
| 3,429708596 | 0,001363518 | LOC101929475 | TRUE | Upregulated |
| -0,764165695 | 0,001365904 | EMP2 | FALSE | Downregulated |
| -0,432646593 | 0,001365904 | TFPI | FALSE | Downregulated |
| 0,744206049 | 0,001365904 | TRAF1 | FALSE | Upregulated |
| 0,433747155 | 0,001367081 | XPO1 | FALSE | Upregulated |
| 1,096416169 | 0,001369331 | ENPP7 | TRUE | Upregulated |
| 0,531810574 | 0,001369626 | C18orf32 | FALSE | Upregulated |
| -1,095493239 | 0,001373166 | DCDC5 | TRUE | Downregulated |
| 0,947104661 | 0,001373166 | LGALS3 | FALSE | Upregulated |
| 0,253809378 | 0,001376796 | WDTC1 | FALSE | Upregulated |
| -0,342153703 | 0,001376796 | ZNF770 | FALSE | Downregulated |
| 0,576477244 | 0,00137913 | C12orf49 | FALSE | Upregulated |
| 3,111079304 | 0,00137913 | PTPRR | TRUE | Upregulated |
| 1,15589229 | 0,001381004 | ARHGEF16 | TRUE | Upregulated |
| 0,802392811 | 0,001384595 | RENBP | FALSE | Upregulated |
| -0,631912197 | 0,001388669 | PTGR1 | FALSE | Downregulated |
| 0,705869523 | 0,00139038 | DEF6 | FALSE | Upregulated |
| -3,312084197 | 0,001396431 | FBXO40 | TRUE | Downregulated |
| 1,018069289 | 0,001396431 | KCNAB2 | TRUE | Upregulated |
| 1,747168696 | 0,001396431 | TMPRSS13 | TRUE | Upregulated |
| 2,232705373 | 0,001397656 | ANKRD20A5P | TRUE | Upregulated |
| 0,459914556 | 0,001399673 | PPP1R15A | FALSE | Upregulated |
| 0,927097648 | 0,001410757 | ADAP1 | FALSE | Upregulated |
| 0,420551048 | 0,001411524 | ENTPD6 | FALSE | Upregulated |
| 0,405919702 | 0,001413888 | FAM98B | FALSE | Upregulated |

| | | | | |
|---|---|---|---|---|
| 1,260063936 | 0,001413888 | PROM2 | TRUE | Upregulated |
| -0,312382436 | 0,001413888 | TMEM254 | FALSE | Downregulated |
| 0,442908894 | 0,001413952 | RCBTB1 | FALSE | Upregulated |
| 0,43824828 | 0,001416964 | RCC2 | FALSE | Upregulated |
| 1,057046294 | 0,001420676 | SGK223 | TRUE | Upregulated |
| -0,459563608 | 0,001421567 | APITD1 | FALSE | Downregulated |
| 0,836320924 | 0,001422968 | GALNT3 | FALSE | Upregulated |
| -0,472079801 | 0,001422968 | HSD17B4 | FALSE | Downregulated |
| 2,463816713 | 0,001422968 | SPRED3 | TRUE | Upregulated |
| 0,862361958 | 0,001423488 | COL14A1 | FALSE | Upregulated |
| -0,264712179 | 0,001424113 | APEX1 | FALSE | Downregulated |
| 0,744598404 | 0,001425273 | KIFC3 | FALSE | Upregulated |
| -0,519154116 | 0,001430845 | MAGI2-AS3 | FALSE | Downregulated |
| -0,418906979 | 0,001432031 | PDAP1 | FALSE | Downregulated |
| 0,687233707 | 0,00143487 | TMC6 | FALSE | Upregulated |
| -0,724175711 | 0,001449005 | LRRC3DN | FALSE | Downregulated |
| 2,167807975 | 0,001449739 | ERCC6L | TRUE | Upregulated |
| -0,496164225 | 0,001449739 | PDIA5 | FALSE | Downregulated |
| -0,529783466 | 0,001453098 | TSR3 | FALSE | Downregulated |
| -0,349558706 | 0,00145782 | PNPLA8 | FALSE | Downregulated |
| -0,486192495 | 0,001464164 | TCEAL4 | FALSE | Downregulated |
| 0,253232097 | 0,001464364 | NUP205 | FALSE | Upregulated |
| 1,286508827 | 0,001470213 | BATF | TRUE | Upregulated |
| 0,745022668 | 0,001470591 | CD53 | FALSE | Upregulated |
| 0,350331784 | 0,001471635 | PFKL | FALSE | Upregulated |
| -0,343344479 | 0,00147476 | BBS4 | FALSE | Downregulated |
| -0,665951688 | 0,00147476 | TXLNG | FALSE | Downregulated |
| 0,564493269 | 0,001475407 | MAP3K14 | FALSE | Upregulated |
| -0,362455676 | 0,001477328 | RSAD1 | FALSE | Downregulated |
| 0,46891524 | 0,00148081 | SCFD2 | FALSE | Upregulated |
| 0,708462371 | 0,001481555 | ZNF668 | FALSE | Upregulated |
| 0,918123801 | 0,001486869 | TMEM164 | FALSE | Upregulated |
| -0,276191397 | 0,001487027 | PRPF31 | FALSE | Downregulated |
| 1,086804541 | 0,001490149 | CCRL2 | TRUE | Upregulated |
| -0,438466436 | 0,001490149 | NDUFA3 | FALSE | Downregulated |
| 0,580281337 | 0,001492074 | ZNF836 | FALSE | Upregulated |
| 0,957898834 | 0,001492225 | ARHGAP44 | FALSE | Upregulated |
| 1,08771587 | 0,001492225 | CCL5 | TRUE | Upregulated |
| -0,962668173 | 0,001497427 | RP5-1024G6.5 | FALSE | Downregulated |
| -0,527422907 | 0,001497427 | TCEA3 | FALSE | Downregulated |
| -0,413107961 | 0,001498093 | GNAI1 | FALSE | Downregulated |
| -0,852433672 | 0,001498093 | PBDC1 | FALSE | Downregulated |
| -0,778678559 | 0,001498093 | TTR | FALSE | Downregulated |
| 0,521000951 | 0,001499388 | SLC44A2 | FALSE | Upregulated |
| -0,666457031 | 0,001500773 | FAXDC2 | FALSE | Downregulated |
| 1,015584677 | 0,001502476 | PDZRN3 | TRUE | Upregulated |
| -0,370498095 | 0,001502476 | SLC25A11 | FALSE | Downregulated |
| 0,953058933 | 0,001507592 | PLA2G4C | FALSE | Upregulated |
| 0,626171577 | 0,001508736 | AZI1 | FALSE | Upregulated |
| 0,358606312 | 0,001518028 | GNS | FALSE | Upregulated |
| -0,762163782 | 0,001518843 | LOC149351 | FALSE | Downregulated |

| | | | | |
|---|---|---|---|---|
| -4,058187821 | 0,001520586 | CA1 | TRUE | Downregulated |
| 2,671674541 | 0,001520586 | RASGRF1 | TRUE | Upregulated |
| -0,747681989 | 0,001524198 | SIGIRR | FALSE | Downregulated |
| 0,520402184 | 0,00153215 | FAM160B2 | FALSE | Upregulated |
| -0,286409248 | 0,001534209 | UBA2 | FALSE | Downregulated |
| 0,904524778 | 0,0015423 | LRRK1 | FALSE | Upregulated |
| -0,359719796 | 0,001549108 | TRAP1 | FALSE | Downregulated |
| -1,014979303 | 0,001566841 | ARID3C | TRUE | Downregulated |
| 1,376366647 | 0,00156777 | CREB5 | TRUE | Upregulated |
| 1,329008444 | 0,001574009 | EMILIN2 | TRUE | Upregulated |
| -0,346300862 | 0,001574009 | MRPS5 | FALSE | Downregulated |
| -0,270056842 | 0,001574009 | PPIE | FALSE | Downregulated |
| -0,429072376 | 0,00157771 | CCS | FALSE | Downregulated |
| 0,675509616 | 0,00157868 | ZBTB46 | FALSE | Upregulated |
| 1,165894372 | 0,001589798 | JAG2 | TRUE | Upregulated |
| 2,313762681 | 0,001603818 | ANGPTL7 | TRUE | Upregulated |
| 1,471446019 | 0,001606469 | LAYN | TRUE | Upregulated |
| -0,305325807 | 0,001606469 | MRPS31 | FALSE | Downregulated |
| 0,788106051 | 0,001606469 | RAI2 | FALSE | Upregulated |
| 0,410703396 | 0,001614646 | LANCL2 | FALSE | Upregulated |
| -0,472012671 | 0,001619203 | C9orf41 | FALSE | Downregulated |
| 1,418012076 | 0,001624101 | RAB7B | TRUE | Upregulated |
| 1,05523646 | 0,001624134 | CDKN2B | TRUE | Upregulated |
| 0,354753071 | 0,001624134 | SHANK2 | FALSE | Upregulated |
| -0,293783636 | 0,0016266 | TMEM14C | FALSE | Downregulated |
| -0,876516961 | 0,00162923 | DAB1 | FALSE | Downregulated |
| -0,552752067 | 0,001631037 | SURF6 | FALSE | Downregulated |
| -0,518018195 | 0,001631798 | BPHL | FALSE | Downregulated |
| 2,729358638 | 0,001638989 | C15orf48 | TRUE | Upregulated |
| -0,614968586 | 0,001638989 | RSRC2 | FALSE | Downregulated |
| -0,758222777 | 0,001640103 | SLC25A23 | FALSE | Downregulated |
| 1,098670559 | 0,001640103 | TBC1D10C | TRUE | Upregulated |
| -0,436394441 | 0,001644195 | HECTD1 | FALSE | Downregulated |
| -0,321863849 | 0,001653814 | TSPAN6 | FALSE | Downregulated |
| 0,592180157 | 0,001654014 | ANXA6 | FALSE | Upregulated |
| 1,247861857 | 0,001655932 | TRAT1 | TRUE | Upregulated |
| 1,586100583 | 0,001661509 | ADAMTS15 | TRUE | Upregulated |
| -0,429470099 | 0,001678547 | TNFAIP8L1 | FALSE | Downregulated |
| 0,473422149 | 0,001678547 | UBE2B | FALSE | Upregulated |
| 1,255650194 | 0,001684127 | ENPP5 | TRUE | Upregulated |
| -0,490276528 | 0,001689373 | MFSD9 | FALSE | Downregulated |
| 0,310149462 | 0,001690106 | SCAF1 | FALSE | Upregulated |
| -0,290426984 | 0,001692289 | SUPV3L1 | FALSE | Downregulated |
| 0,539527207 | 0,001693093 | C15orf41 | FALSE | Upregulated |
| 2,901317552 | 0,001694652 | ANO4 | TRUE | Upregulated |
| 1,377157347 | 0,001699027 | TNXB | TRUE | Upregulated |
| -0,268360385 | 0,001699352 | BACE1 | FALSE | Downregulated |
| -0,545705746 | 0,001700876 | IFT46 | FALSE | Downregulated |
| 2,061429282 | 0,001702762 | HAR1A | TRUE | Upregulated |
| 0,971923533 | 0,001704416 | PDE1A | FALSE | Upregulated |
| -0,331167111 | 0,001707329 | DCTN3 | FALSE | Downregulated |

| | | | | |
|---|---|---|---|---|
| -0,274423115 | 0,001715843 | CDC123 | FALSE | Downregulated |
| 1,017216219 | 0,001715843 | PTPN22 | TRUE | Upregulated |
| 1,000807256 | 0,001721183 | CCDC102B | TRUE | Upregulated |
| -0,428023828 | 0,001723952 | GC | FALSE | Downregulated |
| -0,358574474 | 0,001723952 | IFT74 | FALSE | Downregulated |
| 0,643807721 | 0,001726704 | RASA3 | FALSE | Upregulated |
| 1,27134571 | 0,001728148 | ALDH1L2 | TRUE | Upregulated |
| 2,26183532 | 0,001730079 | LOC102724835 | TRUE | Upregulated |
| 0,768707881 | 0,001731074 | NCOA7 | FALSE | Upregulated |
| -0,342155606 | 0,00174362 | RBM8A | FALSE | Downregulated |
| 0,491899983 | 0,00174676 | TMEM63A | FALSE | Upregulated |
| 0,480122263 | 0,001747118 | DTNBP1 | FALSE | Upregulated |
| 0,56454428 | 0,001754485 | NEURL1B | FALSE | Upregulated |
| 1,08512443 | 0,001756508 | CHST9 | TRUE | Upregulated |
| 1,407368355 | 0,001756508 | GRHL2 | TRUE | Upregulated |
| 0,389263639 | 0,001756508 | SPTLC2 | FALSE | Upregulated |
| 0,573015907 | 0,001763338 | INO80C | FALSE | Upregulated |
| -0,445126117 | 0,001763375 | LACTB2 | FALSE | Downregulated |
| 0,593637352 | 0,001764616 | HK1 | FALSE | Upregulated |
| -0,462497934 | 0,001772168 | CCDC25 | FALSE | Downregulated |
| 1,333724748 | 0,001772888 | KIF20A | TRUE | Upregulated |
| 0,434762165 | 0,001776543 | LEPROTL1 | FALSE | Upregulated |
| -0,415282974 | 0,001783299 | TSFM | FALSE | Downregulated |
| -0,690121856 | 0,001783912 | RP9 | FALSE | Downregulated |
| 1,539709639 | 0,001785866 | LRRC1 | TRUE | Upregulated |
| -0,388044339 | 0,001786303 | CFDP1 | FALSE | Downregulated |
| 0,236896151 | 0,001788903 | POLDIP3 | FALSE | Upregulated |
| -0,342399075 | 0,00179313 | PDSS2 | FALSE | Downregulated |
| 2,56249534 | 0,00179313 | PTCHD4 | TRUE | Upregulated |
| 0,897935961 | 0,001793176 | ARHGEF25 | FALSE | Upregulated |
| 0,786618784 | 0,001793176 | PTGIS | FALSE | Upregulated |
| 4,974868077 | 0,001793463 | LINC00550 | TRUE | Upregulated |
| -0,329913626 | 0,001793753 | CASC4 | FALSE | Downregulated |
| 1,381610757 | 0,001793753 | FAM150B | TRUE | Upregulated |
| 1,395762684 | 0,001799597 | AURKB | TRUE | Upregulated |
| 0,317218103 | 0,001800606 | RAB35 | FALSE | Upregulated |
| -0,229207795 | 0,001800606 | SERBP1 | FALSE | Downregulated |
| 0,497806167 | 0,001800748 | YAP1 | FALSE | Upregulated |
| -0,631416946 | 0,00180132 | SNHG11 | FALSE | Downregulated |
| -0,272169931 | 0,001801607 | IMPACT | FALSE | Downregulated |
| -0,43961532 | 0,001801607 | R3HCC1 | FALSE | Downregulated |
| -0,540262525 | 0,001803751 | GLRX | FALSE | Downregulated |
| -0,264953199 | 0,001803751 | PITHD1 | FALSE | Downregulated |
| -0,307716753 | 0,001804577 | NFYA | FALSE | Downregulated |
| -0,394752807 | 0,001812583 | C11orf57 | FALSE | Downregulated |
| -0,231258077 | 0,001812996 | CUL4B | FALSE | Downregulated |
| 0,421619319 | 0,001815069 | HSDL1 | FALSE | Upregulated |
| 0,345566065 | 0,001815069 | ITFG3 | FALSE | Upregulated |
| -0,323392529 | 0,001816494 | KNG1 | FALSE | Downregulated |
| 0,663395095 | 0,001816912 | TES | FALSE | Upregulated |
| 0,509960366 | 0,00182448 | P4HTM | FALSE | Upregulated |

| | | | | |
|---|---|---|---|---|
| 1,006298497 | 0,001824627 | GAS7 | TRUE | Upregulated |
| 2,765221676 | 0,001830635 | CNGB1 | TRUE | Upregulated |
| 0,658273817 | 0,001830635 | NAV1 | FALSE | Upregulated |
| -0,448800805 | 0,001833187 | THAP5 | FALSE | Downregulated |
| -0,580494779 | 0,001836008 | DDX19B | FALSE | Downregulated |
| 2,878339713 | 0,001836008 | RPL29P2 | TRUE | Upregulated |
| 0,827925298 | 0,001837198 | MBOAT1 | FALSE | Upregulated |
| 1,326496672 | 0,001841833 | COL8A2 | TRUE | Upregulated |
| -0,31018864 | 0,001841833 | DNAJC8 | FALSE | Downregulated |
| 1,007019503 | 0,001841833 | XYLT1 | TRUE | Upregulated |
| 2,217639063 | 0,001844132 | CCL22 | TRUE | Upregulated |
| 1,426851231 | 0,001845865 | RPL23P8 | TRUE | Upregulated |
| 0,824476919 | 0,001850579 | ITGB3 | FALSE | Upregulated |
| 0,709665245 | 0,001851231 | GALNT6 | FALSE | Upregulated |
| 1,175492003 | 0,001851231 | TRIM59 | TRUE | Upregulated |
| 0,62745478 | 0,001852542 | ARID5B | FALSE | Upregulated |
| 0,913593769 | 0,001852542 | NRM | FALSE | Upregulated |
| -0,271820582 | 0,001858457 | PRR13 | FALSE | Downregulated |
| 1,243662375 | 0,001859751 | ANKRD36C | TRUE | Upregulated |
| -0,348101335 | 0,001861101 | CDC37L1 | FALSE | Downregulated |
| -0,314797758 | 0,001874796 | EFCAB14 | FALSE | Downregulated |
| 1,169750927 | 0,001876145 | GINS4 | TRUE | Upregulated |
| 1,645067696 | 0,001879141 | HLA-J | TRUE | Upregulated |
| 2,930907929 | 0,001879141 | RALGAPA1P | TRUE | Upregulated |
| 0,951421357 | 0,00189292 | GPR132 | FALSE | Upregulated |
| 0,655230649 | 0,001896654 | HLA-DMB | FALSE | Upregulated |
| -0,360864291 | 0,00190753 | AUH | FALSE | Downregulated |
| 0,351380615 | 0,001909478 | SCRIB | FALSE | Upregulated |
| -0,476222192 | 0,001911696 | SMDT1 | FALSE | Downregulated |
| -0,322112356 | 0,001913646 | DHX33 | FALSE | Downregulated |
| 0,301888066 | 0,001916329 | ARHGEF11 | FALSE | Upregulated |
| 1,505101645 | 0,00191984 | ID4 | TRUE | Upregulated |
| -1,276457213 | 0,00191984 | LOC102723583 | TRUE | Downregulated |
| 0,817074342 | 0,001926944 | PDGFRA | FALSE | Upregulated |
| 0,668367591 | 0,001930837 | ORAI2 | FALSE | Upregulated |
| 1,812644366 | 0,001931503 | MZB1 | TRUE | Upregulated |
| -0,543220593 | 0,001932184 | PEMT | FALSE | Downregulated |
| -0,23955519 | 0,001934835 | EIF2AK1 | FALSE | Downregulated |
| -0,571792287 | 0,00193532 | LOC643072 | FALSE | Downregulated |
| 1,197978126 | 0,00193532 | RCAN2 | TRUE | Upregulated |
| -3,552961188 | 0,00193532 | TCAP | TRUE | Downregulated |
| -0,381950259 | 0,001952689 | KTN1 | FALSE | Downregulated |
| -0,402395253 | 0,0019649 | RNMTL1 | FALSE | Downregulated |
| 0,659600206 | 0,001969405 | CTTNBP2NL | FALSE | Upregulated |
| 0,673621011 | 0,001969405 | FAR1 | FALSE | Upregulated |
| 0,674416071 | 0,001971938 | ROBO1 | FALSE | Upregulated |
| 0,494741992 | 0,001975224 | ZNFX1 | FALSE | Upregulated |
| -2,017602023 | 0,001976532 | NCAM2 | TRUE | Downregulated |
| 0,519702858 | 0,001977306 | PXDN | FALSE | Upregulated |
| 0,384532952 | 0,001982805 | AAGAB | FALSE | Upregulated |
| -0,552216252 | 0,001983097 | COX8A | FALSE | Downregulated |

| | | | | |
|---|---|---|---|---|
| -0,382332061 | 0,00198378 | SBDS | FALSE | Downregulated |
| 1,428696592 | 0,0019881 | GFPT2 | TRUE | Upregulated |
| -0,790849086 | 0,001999486 | C1orf53 | FALSE | Downregulated |
| -0,478560769 | 0,001999486 | MPDZ | FALSE | Downregulated |
| 0,646701145 | 0,002009228 | CLTCL1 | FALSE | Upregulated |
| -0,240083734 | 0,002009228 | RAB5A | FALSE | Downregulated |
| 0,345063907 | 0,002009566 | PTBP3 | FALSE | Upregulated |
| 2,279927695 | 0,002009566 | SLC52A3 | TRUE | Upregulated |
| 0,738179176 | 0,002013335 | NCF2 | FALSE | Upregulated |
| 0,464541589 | 0,002023174 | GPD2 | FALSE | Upregulated |
| -0,25079452 | 0,002028662 | PPCS | FALSE | Downregulated |
| 1,14573928 | 0,002030491 | GAREML | TRUE | Upregulated |
| 0,78973295 | 0,00203082 | TRIM22 | FALSE | Upregulated |
| 0,644854068 | 0,002034143 | CHD3 | FALSE | Upregulated |
| 1,033493376 | 0,0020413 | MFGE8 | TRUE | Upregulated |
| -0,447532409 | 0,002042068 | MBNL3 | FALSE | Downregulated |
| 0,801187076 | 0,00204807 | EVA1B | FALSE | Upregulated |
| -0,468590654 | 0,002054689 | RMDN2 | FALSE | Downregulated |
| -0,299581173 | 0,002055956 | SERINC3 | FALSE | Downregulated |
| 0,646447555 | 0,002056966 | TCF7 | FALSE | Upregulated |
| -0,353581872 | 0,00206067 | CCDC126 | FALSE | Downregulated |
| -0,736614667 | 0,002069441 | PNPLA3 | FALSE | Downregulated |
| 0,546564695 | 0,002070178 | PML | FALSE | Upregulated |
| 0,477985452 | 0,002074104 | FBXL14 | FALSE | Upregulated |
| 0,286058064 | 0,002074104 | STARD3 | FALSE | Upregulated |
| -0,358623133 | 0,002086627 | LRIF1 | FALSE | Downregulated |
| -0,262763508 | 0,002091854 | DCAF5 | FALSE | Downregulated |
| -0,498450932 | 0,002099206 | NAAA | FALSE | Downregulated |
| -0,503347375 | 0,002099206 | PSMG4 | FALSE | Downregulated |
| -0,284623377 | 0,002105847 | MTIF2 | FALSE | Downregulated |
| -0,287122579 | 0,002105847 | RMND1 | FALSE | Downregulated |
| 0,750709613 | 0,002122173 | NBEAL2 | FALSE | Upregulated |
| -1,352743434 | 0,002122479 | MTUS2 | TRUE | Downregulated |
| 1,227498819 | 0,002122479 | SLC44A3 | TRUE | Upregulated |
| 1,046469094 | 0,002128346 | PLXNA3 | TRUE | Upregulated |
| -0,433875758 | 0,002130599 | DNAAF2 | FALSE | Downregulated |
| -0,412882307 | 0,002132393 | DTD2 | FALSE | Downregulated |
| -0,527835479 | 0,0021404 | GCDH | FALSE | Downregulated |
| 0,439286079 | 0,002142177 | GRK6 | FALSE | Upregulated |
| 1,507246783 | 0,002146379 | SEMA3G | TRUE | Upregulated |
| -0,589106734 | 0,002148368 | MOSPD3 | FALSE | Downregulated |
| 0,538043801 | 0,002158232 | ABCC10 | FALSE | Upregulated |
| 0,473635023 | 0,002161727 | STARD3NL | FALSE | Upregulated |
| 0,489832612 | 0,002163963 | KIF2A | FALSE | Upregulated |
| -0,584283096 | 0,002164188 | GPHN | FALSE | Downregulated |
| 0,387469837 | 0,002164188 | ZMIZ1 | FALSE | Upregulated |
| -0,412017709 | 0,002166391 | TAB2 | FALSE | Downregulated |
| 1,507610123 | 0,002167798 | FFAR4 | TRUE | Upregulated |
| 0,921221627 | 0,002169195 | CD52 | FALSE | Upregulated |
| -0,393095291 | 0,002169195 | STYX | FALSE | Downregulated |
| 0,493505751 | 0,002173865 | AAK1 | FALSE | Upregulated |

| | | | | |
|---|---|---|---|---|
| 0,338058223 | 0,002178263 | SBF1 | FALSE | Upregulated |
| -0,312225297 | 0,00218531 | ELF2 | FALSE | Downregulated |
| -1,9755578 | 0,002186331 | SLITRK3 | TRUE | Downregulated |
| -1,034092804 | 0,002190546 | MME | TRUE | Downregulated |
| 0,340531981 | 0,002190553 | ZNF562 | FALSE | Upregulated |
| 1,860342568 | 0,002191105 | BCAS1 | TRUE | Upregulated |
| -0,349805352 | 0,002191123 | CHURC1 | FALSE | Downregulated |
| 1,327448492 | 0,002192011 | FCN1 | TRUE | Upregulated |
| -0,361337505 | 0,002192011 | PTPN18 | FALSE | Downregulated |
| -0,344063604 | 0,002192011 | RANBP1 | FALSE | Downregulated |
| 0,257040375 | 0,002194791 | ATP2C1 | FALSE | Upregulated |
| 3,492979762 | 0,002197059 | DNAAF1 | TRUE | Upregulated |
| -0,487736206 | 0,002197059 | METTL22 | FALSE | Downregulated |
| -0,431700643 | 0,002197672 | CXADR | FALSE | Downregulated |
| -1,01595978 | 0,002204831 | SULT1B1 | TRUE | Downregulated |
| 0,584456558 | 0,002211983 | FLI1 | FALSE | Upregulated |
| -0,375109535 | 0,002212635 | APOE | FALSE | Downregulated |
| -0,728972652 | 0,002216298 | ZRSR2 | FALSE | Downregulated |
| 0,657739899 | 0,002220849 | SLC26A2 | FALSE | Upregulated |
| 3,057945093 | 0,002221252 | AKR1B10 | TRUE | Upregulated |
| -0,937061638 | 0,00222626 | LOC102723766 | FALSE | Downregulated |
| -0,611201891 | 0,00222626 | TUBE1 | FALSE | Downregulated |
| -1,18883002 | 0,002243849 | KCNN2 | TRUE | Downregulated |
| -0,405226736 | 0,002244577 | ERP29 | FALSE | Downregulated |
| -0,249850378 | 0,002261504 | PTP4A2 | FALSE | Downregulated |
| -0,574574791 | 0,002266972 | CEP63 | FALSE | Downregulated |
| -0,568721489 | 0,002269535 | PARP2 | FALSE | Downregulated |
| 0,282291644 | 0,002269535 | SNRK | FALSE | Upregulated |
| 0,970104912 | 0,002272267 | GPC4 | FALSE | Upregulated |
| -0,354389138 | 0,002272267 | ZADH2 | FALSE | Downregulated |
| 1,24236134 | 0,002273608 | CHST4 | TRUE | Upregulated |
| -0,368930563 | 0,002273608 | ISY1 | FALSE | Downregulated |
| 0,597054494 | 0,002273608 | ITGAL | FALSE | Upregulated |
| 0,319184084 | 0,002273608 | TRAF7 | FALSE | Upregulated |
| -0,351345254 | 0,002273608 | TTC38 | FALSE | Downregulated |
| -3,562318573 | 0,002274299 | MYL3 | TRUE | Downregulated |
| 0,559551538 | 0,002281624 | ARHGEF2 | FALSE | Upregulated |
| -0,38206659 | 0,002286243 | UFM1 | FALSE | Downregulated |
| 0,318270939 | 0,002299818 | AP2A1 | FALSE | Upregulated |
| -0,334483223 | 0,002299818 | CLPX | FALSE | Downregulated |
| -0,531929456 | 0,002306202 | ACAA1 | FALSE | Downregulated |
| -0,436814594 | 0,002306202 | AMBP | FALSE | Downregulated |
| -0,645088818 | 0,002306202 | RAB13 | FALSE | Downregulated |
| -1,201561013 | 0,002311286 | LOC101927331 | TRUE | Downregulated |
| 0,493081938 | 0,002314337 | GAS2L1 | FALSE | Upregulated |
| 0,306433585 | 0,002314337 | SEC24C | FALSE | Upregulated |
| 3,0241031 | 0,0023181 | ANXA8L1 | TRUE | Upregulated |
| -0,514495113 | 0,0023181 | C11orf54 | FALSE | Downregulated |
| -1,614627594 | 0,002319913 | ATP5L2 | TRUE | Downregulated |
| 0,30306506 | 0,002325659 | CTDNEP1 | FALSE | Upregulated |
| -0,335639864 | 0,002329214 | PPID | FALSE | Downregulated |

| | | | | |
|---|---|---|---|---|
| 0,356731552 | 0,002329214 | PRR12 | FALSE | Upregulated |
| 0,599959433 | 0,002341564 | ABCC1 | FALSE | Upregulated |
| -0,679888915 | 0,002341564 | ADH4 | FALSE | Downregulated |
| 1,929763021 | 0,002341564 | RPL31P11 | TRUE | Upregulated |
| 0,421225699 | 0,002349792 | CABIN1 | FALSE | Upregulated |
| -0,267851018 | 0,002349792 | PDHX | FALSE | Downregulated |
| 2,247104622 | 0,002349792 | RP11-18H21.1 | TRUE | Upregulated |
| 0,337449403 | 0,002349792 | SLC15A4 | FALSE | Upregulated |
| -0,567306068 | 0,002349792 | SNRNP27 | FALSE | Downregulated |
| -0,376105643 | 0,002350816 | ZNF638 | FALSE | Downregulated |
| 1,21055827 | 0,002354148 | DPT | TRUE | Upregulated |
| 0,82467794 | 0,002354148 | LAMA5 | FALSE | Upregulated |
| -0,292136777 | 0,002354148 | PSMB7 | FALSE | Downregulated |
| 2,246931061 | 0,002355296 | ZNF541 | TRUE | Upregulated |
| -0,345727455 | 0,002358389 | ASB8 | FALSE | Downregulated |
| -0,462612949 | 0,002365645 | TMEM101 | FALSE | Downregulated |
| 0,665782901 | 0,002367635 | MAP3K12 | FALSE | Upregulated |
| 1,302642741 | 0,002367635 | POF1B | TRUE | Upregulated |
| -0,483987203 | 0,002382149 | C22orf46 | FALSE | Downregulated |
| -0,361895159 | 0,002382149 | FUNDC2 | FALSE | Downregulated |
| -0,589280789 | 0,002382281 | EBPL | FALSE | Downregulated |
| 0,582001902 | 0,002384521 | HLA-DMA | FALSE | Upregulated |
| 0,990663387 | 0,002386064 | RNF150 | FALSE | Upregulated |
| -0,376442372 | 0,002390288 | PIN4 | FALSE | Downregulated |
| 0,976397815 | 0,002399811 | FANCD2 | FALSE | Upregulated |
| 2,522565926 | 0,002407598 | TTLL6 | TRUE | Upregulated |
| 0,7591802 | 0,002410025 | PTPRC | FALSE | Upregulated |
| 0,267515688 | 0,002422587 | VPS4A | FALSE | Upregulated |
| -0,326525591 | 0,002425004 | AHCTF1 | FALSE | Downregulated |
| 0,891800048 | 0,002430152 | SAMD14 | FALSE | Upregulated |
| -0,885153876 | 0,002432344 | ZG16 | FALSE | Downregulated |
| -1,227113644 | 0,002433291 | AC074289.1 | TRUE | Downregulated |
| 1,187642232 | 0,002437762 | BCAT1 | TRUE | Upregulated |
| 2,311463452 | 0,002439309 | TPTE2P1 | TRUE | Upregulated |
| -0,370674229 | 0,002443401 | RBM26 | FALSE | Downregulated |
| -0,587597721 | 0,002447932 | AGMAT | FALSE | Downregulated |
| 0,780325531 | 0,002447932 | TJP3 | FALSE | Upregulated |
| 0,988669491 | 0,002461197 | PRC1 | FALSE | Upregulated |
| 0,665777685 | 0,002461822 | DOK2 | FALSE | Upregulated |
| 2,064887924 | 0,002462143 | LOC102724737 | TRUE | Upregulated |
| -0,397383705 | 0,002468735 | MRPL43 | FALSE | Downregulated |
| -0,230703284 | 0,002474072 | SENP6 | FALSE | Downregulated |
| 2,740016361 | 0,002475898 | LOC100126784 | TRUE | Upregulated |
| -0,604080614 | 0,002479652 | GCSHP3 | FALSE | Downregulated |
| -0,344262855 | 0,002481407 | PRKAA1 | FALSE | Downregulated |
| 0,995684759 | 0,002481837 | ZNF831 | FALSE | Upregulated |
| 1,150503157 | 0,002488685 | CCDC8 | TRUE | Upregulated |
| -0,336042914 | 0,002499357 | LOC101928527 | FALSE | Downregulated |
| -0,351773177 | 0,002504054 | RWDD2B | FALSE | Downregulated |
| -0,410974205 | 0,002511342 | RNF130 | FALSE | Downregulated |
| 1,298046863 | 0,002514751 | SEMA3E | TRUE | Upregulated |

| | | | | |
|---|---|---|---|---|
| -0,215114823 | 0,002518293 | RSL1D1 | FALSE | Downregulated |
| 0,60023856 | 0,002518293 | SFXN3 | FALSE | Upregulated |
| -0,379239012 | 0,002524332 | GLYATL1 | FALSE | Downregulated |
| 0,374398198 | 0,002535249 | KLHL5 | FALSE | Upregulated |
| 1,682220803 | 0,002535249 | PLA2G2D | TRUE | Upregulated |
| -0,895249412 | 0,002535249 | TEN1-CDK3 | FALSE | Downregulated |
| -1,154397735 | 0,002538086 | LOC102724493 | TRUE | Downregulated |
| -0,680870409 | 0,002538838 | LIPC | FALSE | Downregulated |
| -0,829047035 | 0,002539047 | CD82 | FALSE | Downregulated |
| -0,374859762 | 0,00254103 | RMDN1 | FALSE | Downregulated |
| 0,888371833 | 0,00254103 | RTN1 | FALSE | Upregulated |
| 0,991530809 | 0,002544144 | SLA2 | FALSE | Upregulated |
| 1,82755965 | 0,002545298 | TRABD2A | TRUE | Upregulated |
| 1,377639244 | 0,002550184 | BAIAP2L2 | TRUE | Upregulated |
| -0,396848112 | 0,002555767 | PDHB | FALSE | Downregulated |
| -0,338307026 | 0,002559377 | YES1 | FALSE | Downregulated |
| 0,401317425 | 0,002563433 | SF3A2 | FALSE | Upregulated |
| 1,759550304 | 0,002566702 | RERGL | TRUE | Upregulated |
| 1,008120038 | 0,0025716 | PNMAL2 | TRUE | Upregulated |
| -0,784358715 | 0,0025737 | LIPG | FALSE | Downregulated |
| -0,371545043 | 0,002576959 | PDHA1 | FALSE | Downregulated |
| 0,708121374 | 0,002577491 | ARHGEF17 | FALSE | Upregulated |
| 3,286886903 | 0,002580076 | MAGED4 | TRUE | Upregulated |
| -0,597179243 | 0,002585213 | IL17RC | FALSE | Downregulated |
| -0,368284591 | 0,002585213 | RPS20 | FALSE | Downregulated |
| 0,349332777 | 0,002596543 | ANKRD52 | FALSE | Upregulated |
| 0,484167696 | 0,002596543 | PRIM2 | FALSE | Upregulated |
| 3,411496401 | 0,002596543 | SCRT2 | TRUE | Upregulated |
| 0,82146666 | 0,002596543 | ZMAT3 | FALSE | Upregulated |
| 0,450632556 | 0,002596761 | PI4K2A | FALSE | Upregulated |
| 0,489522189 | 0,002599983 | CSNK1E | FALSE | Upregulated |
| -0,672910008 | 0,002599983 | NFIC | FALSE | Downregulated |
| -0,25432868 | 0,002603079 | HMGB1 | FALSE | Downregulated |
| -0,949390424 | 0,0026172 | ENPP3 | FALSE | Downregulated |
| -0,323071153 | 0,0026172 | NDUFA12 | FALSE | Downregulated |
| -0,762064602 | 0,0026172 | REPS1 | FALSE | Downregulated |
| -0,482090836 | 0,002618828 | APOA2 | FALSE | Downregulated |
| -0,292573006 | 0,002624061 | RPUSD4 | FALSE | Downregulated |
| -0,642347998 | 0,002630489 | ZCCHC6 | FALSE | Downregulated |
| -0,456006804 | 0,002633891 | TRAM2-AS1 | FALSE | Downregulated |
| 0,296106647 | 0,00263571 | DENND1A | FALSE | Upregulated |
| 1,091793905 | 0,002642897 | CD3G | TRUE | Upregulated |
| 3,605872748 | 0,002645777 | LOC102723603 | TRUE | Upregulated |
| -0,318815368 | 0,002645851 | CDK7 | FALSE | Downregulated |
| 1,178349592 | 0,002646147 | FHL2 | TRUE | Upregulated |
| 2,77013783 | 0,002655678 | COL22A1 | TRUE | Upregulated |
| 0,474945093 | 0,002657001 | CTC1 | FALSE | Upregulated |
| -1,74497687 | 0,00265873 | AC009501.4 | TRUE | Downregulated |
| -0,612424602 | 0,002671762 | SRRM1 | FALSE | Downregulated |
| 2,461670485 | 0,002672571 | BAI2 | TRUE | Upregulated |
| -0,488781929 | 0,002672571 | SLC27A2 | FALSE | Downregulated |

| | | | | |
|---|---|---|---|---|
| -0,594071811 | 0,002673542 | B3GNT1 | FALSE | Downregulated |
| 1,070137425 | 0,002679641 | FABP5 | TRUE | Upregulated |
| 3,032686429 | 0,002680573 | LOC101928466 | TRUE | Upregulated |
| -0,275451566 | 0,002680573 | ZNF33A | FALSE | Downregulated |
| -0,204166758 | 0,002682151 | SETD3 | FALSE | Downregulated |
| -0,506106617 | 0,002684769 | PABPN1 | FALSE | Downregulated |
| 0,303009239 | 0,002684769 | URB1 | FALSE | Upregulated |
| 0,320949995 | 0,002691978 | FBXW5 | FALSE | Upregulated |
| -0,422705603 | 0,002698673 | COQ9 | FALSE | Downregulated |
| -0,804280219 | 0,002699917 | LOC102724761 | FALSE | Downregulated |
| 0,989571355 | 0,002699917 | PAQR4 | FALSE | Upregulated |
| -0,400803904 | 0,002700949 | NIT2 | FALSE | Downregulated |
| -0,773992065 | 0,00270117 | CMBL | FALSE | Downregulated |
| -0,339593289 | 0,002702335 | ZNF271 | FALSE | Downregulated |
| -0,306953186 | 0,002705441 | ATP5J2 | FALSE | Downregulated |
| 1,889810657 | 0,002707836 | ACHE | TRUE | Upregulated |
| 0,726078626 | 0,002707836 | ZDHHC13 | FALSE | Upregulated |
| 0,830501016 | 0,002715834 | CCDC109B | FALSE | Upregulated |
| 1,599361631 | 0,002717123 | MYBL2 | TRUE | Upregulated |
| 1,154072391 | 0,002718504 | ANKRD33B | TRUE | Upregulated |
| -0,514019299 | 0,002718504 | LARP1B | FALSE | Downregulated |
| 0,462766926 | 0,002720894 | ATG2A | FALSE | Upregulated |
| 0,93478409 | 0,002725319 | LEPREL2 | FALSE | Upregulated |
| -0,408412299 | 0,002727699 | ADI1 | FALSE | Downregulated |
| -0,717198586 | 0,002735646 | KMT2E-AS1 | FALSE | Downregulated |
| 0,334777678 | 0,002739013 | ANAPC7 | FALSE | Upregulated |
| -0,49046459 | 0,002746182 | POLE3 | FALSE | Downregulated |
| 1,082892784 | 0,002763254 | CCNA2 | TRUE | Upregulated |
| -0,285012656 | 0,00276567 | BAG1 | FALSE | Downregulated |
| 0,401497964 | 0,00276567 | STAT5A | FALSE | Upregulated |
| 1,06703187 | 0,002768866 | FBLN2 | TRUE | Upregulated |
| 1,35644514 | 0,002771691 | CXorf36 | TRUE | Upregulated |
| 2,025673838 | 0,002773443 | NXPH3 | TRUE | Upregulated |
| -0,653237792 | 0,002778497 | LOC100506730 | FALSE | Downregulated |
| 2,496346597 | 0,002780048 | ANP32AP1 | TRUE | Upregulated |
| 1,562060769 | 0,002780048 | MCOLN2 | TRUE | Upregulated |
| -0,352738566 | 0,002780068 | EPB41L5 | FALSE | Downregulated |
| 0,586095105 | 0,002780068 | IRF8 | FALSE | Upregulated |
| -0,656282721 | 0,002780878 | LINC01184 | FALSE | Downregulated |
| 0,957896877 | 0,002782647 | RNF24 | FALSE | Upregulated |
| 0,476199387 | 0,002784645 | FRMD8 | FALSE | Upregulated |
| -0,446122998 | 0,002788754 | ALDH2 | FALSE | Downregulated |
| -0,487452261 | 0,002789162 | PALM2 | FALSE | Downregulated |
| -0,353020793 | 0,002789162 | ZNF569 | FALSE | Downregulated |
| 1,430754998 | 0,002801825 | ABCD2 | TRUE | Upregulated |
| 1,862952701 | 0,002804628 | H3F3C | TRUE | Upregulated |
| 0,30573521 | 0,002814273 | ADAM10 | FALSE | Upregulated |
| 2,162434414 | 0,002814273 | ADRA2C | TRUE | Upregulated |
| -0,630211421 | 0,002814739 | LOC257396 | FALSE | Downregulated |
| 0,959840488 | 0,002828491 | LAT | FALSE | Upregulated |
| -1,085633802 | 0,002836321 | LOC101927595 | TRUE | Downregulated |

| | | | | |
|---|---|---|---|---|
| -0,263243424 | 0,002836687 | HDHD2 | FALSE | Downregulated |
| -1,005851902 | 0,002836687 | KCNMA1 | TRUE | Downregulated |
| 1,427580882 | 0,002840812 | SLC22A15 | TRUE | Upregulated |
| -0,301065528 | 0,002847002 | ZNF830 | FALSE | Downregulated |
| -0,452288263 | 0,002850707 | ACAT1 | FALSE | Downregulated |
| -0,235861759 | 0,00285349 | LYRM7 | FALSE | Downregulated |
| 0,832593388 | 0,002861221 | TCEA2 | FALSE | Upregulated |
| 1,017954486 | 0,002861665 | FAM109B | TRUE | Upregulated |
| 0,39540182 | 0,002867324 | LEPRE1 | FALSE | Upregulated |
| -0,239262976 | 0,002869669 | GPBP1L1 | FALSE | Downregulated |
| -0,190990826 | 0,002869669 | PAPOLA | FALSE | Downregulated |
| 0,55526693 | 0,002870706 | MANEAL | FALSE | Upregulated |
| 0,902328719 | 0,00288659 | IKZF3 | FALSE | Upregulated |
| -0,381233502 | 0,002892304 | TFDP2 | FALSE | Downregulated |
| 1,879234023 | 0,002892488 | LOC101926970 | TRUE | Upregulated |
| -0,520698805 | 0,002895833 | C10orf11 | FALSE | Downregulated |
| 0,729727621 | 0,002899342 | PMP22 | FALSE | Upregulated |
| 0,692349221 | 0,002900675 | OSBPL5 | FALSE | Upregulated |
| -0,887889963 | 0,002905621 | SLC16A10 | FALSE | Downregulated |
| -0,562594739 | 0,002910959 | RNR2 | FALSE | Downregulated |
| 0,908792802 | 0,002910959 | ZNF486 | FALSE | Upregulated |
| -0,507615167 | 0,002916655 | APOH | FALSE | Downregulated |
| 2,502856704 | 0,002926722 | CASC15 | TRUE | Upregulated |
| 0,309846037 | 0,002933914 | BTBD2 | FALSE | Upregulated |
| -0,410875218 | 0,002936128 | NDUFAF2 | FALSE | Downregulated |
| 0,653672668 | 0,002936477 | LRRC32 | FALSE | Upregulated |
| 0,696034385 | 0,002974094 | PCED1B | FALSE | Upregulated |
| -0,332950199 | 0,002975316 | HIBADH | FALSE | Downregulated |
| 2,578558832 | 0,002975316 | MDFI | TRUE | Upregulated |
| -0,313662389 | 0,002978471 | PARD3 | FALSE | Downregulated |
| -0,506011244 | 0,002981147 | ZMAT2 | FALSE | Downregulated |
| 0,523647423 | 0,002984386 | ZFPM1 | FALSE | Upregulated |
| 0,413928825 | 0,002986951 | PSMB10 | FALSE | Upregulated |
| 0,974790774 | 0,002988159 | EMR2 | FALSE | Upregulated |
| -1,520949425 | 0,002994059 | FAM124B | TRUE | Downregulated |
| -0,608370848 | 0,002994059 | ROMO1 | FALSE | Downregulated |
| -0,792479457 | 0,002997125 | BCHE | FALSE | Downregulated |
| 0,92977548 | 0,002997125 | GFI1 | FALSE | Upregulated |
| -0,644194574 | 0,002997125 | LOC100507412 | FALSE | Downregulated |
| -0,394365201 | 0,002997125 | MRPL40 | FALSE | Downregulated |
| 2,434702248 | 0,003009775 | NRIP3 | TRUE | Upregulated |
| 0,908643335 | 0,003011335 | ALOX5AP | FALSE | Upregulated |
| -2,096148677 | 0,003011335 | LOC149684 | TRUE | Downregulated |
| 1,135489606 | 0,003011335 | MMP2 | TRUE | Upregulated |
| 0,743528345 | 0,003011335 | TAX1BP3 | FALSE | Upregulated |
| 0,76260351 | 0,003028921 | PLEKHF1 | FALSE | Upregulated |
| -0,492380539 | 0,003046008 | NDUFA2 | FALSE | Downregulated |
| 0,247203088 | 0,003049839 | PLXNB2 | FALSE | Upregulated |
| 1,14543027 | 0,003062455 | GINS2 | TRUE | Upregulated |
| -1,581430709 | 0,003062455 | KCNE1 | TRUE | Downregulated |
| 0,515528008 | 0,003074264 | FOSL2 | FALSE | Upregulated |

| | | | | |
|---|---|---|---|---|
| 0,785153326 | 0,003074622 | PKHD1 | FALSE | Upregulated |
| 1,784157496 | 0,003078407 | GOLGA7B | TRUE | Upregulated |
| 0,923346774 | 0,003078604 | CDC42BPG | FALSE | Upregulated |
| 0,445923967 | 0,00308881 | CLN6 | FALSE | Upregulated |
| 1,189751156 | 0,00309299 | HLA-L | TRUE | Upregulated |
| 4,600519014 | 0,00309299 | LOC101928510 | TRUE | Upregulated |
| 0,643733115 | 0,003103043 | WARS | FALSE | Upregulated |
| -0,272338346 | 0,003103709 | C4orf3 | FALSE | Downregulated |
| -0,461934705 | 0,003104767 | ING2 | FALSE | Downregulated |
| -0,454203553 | 0,003117573 | AKR1C4 | FALSE | Downregulated |
| -0,819383535 | 0,003117573 | AKR1D1 | FALSE | Downregulated |
| -0,56377129 | 0,003117573 | LOC730102 | FALSE | Downregulated |
| -0,661545798 | 0,003126658 | C7orf55 | FALSE | Downregulated |
| -0,257112857 | 0,00313082 | PDS5A | FALSE | Downregulated |
| -0,475214836 | 0,003133905 | CBX3 | FALSE | Downregulated |
| -0,396235644 | 0,003135265 | TSPAN12 | FALSE | Downregulated |
| 3,239046677 | 0,003143872 | FAT2 | TRUE | Upregulated |
| 0,245128383 | 0,003149818 | GRB2 | FALSE | Upregulated |
| -0,403574234 | 0,003155959 | CEP120 | FALSE | Downregulated |
| -0,546614049 | 0,00315717 | GYG2 | FALSE | Downregulated |
| 0,532131015 | 0,003158483 | MFSD12 | FALSE | Upregulated |
| -1,337511082 | 0,003160966 | DPPA4 | TRUE | Downregulated |
| -0,495618716 | 0,003167878 | LOC101928433 | FALSE | Downregulated |
| -1,574512705 | 0,003168475 | TRNV | TRUE | Downregulated |
| 0,788844537 | 0,003175837 | NFAM1 | FALSE | Upregulated |
| 0,997954799 | 0,003177273 | HSPA12B | FALSE | Upregulated |
| 2,017135323 | 0,003179216 | E2F7 | TRUE | Upregulated |
| 2,164456125 | 0,003179216 | LOC101927902 | TRUE | Upregulated |
| 2,204975561 | 0,003179216 | LYPD1 | TRUE | Upregulated |
| 0,399244351 | 0,003182613 | CHSY1 | FALSE | Upregulated |
| -0,889286052 | 0,003184239 | TMEM52 | FALSE | Downregulated |
| 0,889759098 | 0,003184988 | SPSB1 | FALSE | Upregulated |
| -0,538831984 | 0,003190437 | CYP27A1 | FALSE | Downregulated |
| 3,232069012 | 0,003193952 | LPAR3 | TRUE | Upregulated |
| 2,940115518 | 0,00319962 | DSC3 | TRUE | Upregulated |
| 1,33932407 | 0,00319962 | ETV7 | TRUE | Upregulated |
| -0,54070549 | 0,00319962 | SORD | FALSE | Downregulated |
| 1,135395005 | 0,00319962 | WTIP | TRUE | Upregulated |
| 1,587927393 | 0,003201576 | TRPV6 | TRUE | Upregulated |
| -0,865231371 | 0,003204868 | ATP6AP1L | FALSE | Downregulated |
| -0,508843191 | 0,003208278 | MRPL53 | FALSE | Downregulated |
| 0,984304959 | 0,003208278 | TAGAP | FALSE | Upregulated |
| 0,773126701 | 0,003208343 | NLRC5 | FALSE | Upregulated |
| 0,822885286 | 0,003208439 | CSRNP1 | FALSE | Upregulated |
| 2,416224395 | 0,00321065 | GPR141 | TRUE | Upregulated |
| 0,6414857 | 0,003216822 | KNTC1 | FALSE | Upregulated |
| 0,499076366 | 0,00321849 | LPXN | FALSE | Upregulated |
| 1,035689664 | 0,003239694 | IL18 | TRUE | Upregulated |
| 1,44658273 | 0,003241402 | FAM155A | TRUE | Upregulated |
| 0,529322216 | 0,003241402 | SFI1 | FALSE | Upregulated |
| -1,578949627 | 0,003242049 | LOC100996723 | TRUE | Downregulated |

| | | | | |
|---|---|---|---|---|
| -0,177656663 | 0,003242049 | PAIP2 | FALSE | Downregulated |
| -0,314209279 | 0,00324439 | CHCHD2 | FALSE | Downregulated |
| 0,416013639 | 0,003245531 | ABTB1 | FALSE | Upregulated |
| -0,257396629 | 0,003245875 | MAPK9 | FALSE | Downregulated |
| -0,292531457 | 0,003247822 | GHITM | FALSE | Downregulated |
| 0,684767797 | 0,003247822 | KIAA1462 | FALSE | Upregulated |
| 0,858123861 | 0,003247822 | P2RY1 | FALSE | Upregulated |
| 0,868738951 | 0,003247822 | SH2D1A | FALSE | Upregulated |
| -0,297369966 | 0,003247822 | WWP1 | FALSE | Downregulated |
| 0,36754128 | 0,003247822 | ZFYVE27 | FALSE | Upregulated |
| 0,910542575 | 0,003248119 | LPAR1 | FALSE | Upregulated |
| 0,34670147 | 0,003253428 | ATP6AP2 | FALSE | Upregulated |
| 3,162498857 | 0,003263776 | C9orf129 | TRUE | Upregulated |
| 0,307184351 | 0,003272216 | STX7 | FALSE | Upregulated |
| -0,369154514 | 0,003273344 | HLTF | FALSE | Downregulated |
| 0,649272282 | 0,00327592 | KCNQ1 | FALSE | Upregulated |
| 1,229765875 | 0,003276712 | C16orf54 | TRUE | Upregulated |
| 0,491749845 | 0,003276712 | FBXL18 | FALSE | Upregulated |
| 1,482239028 | 0,003278257 | KIAA1211L | TRUE | Upregulated |
| 1,92763321 | 0,003284719 | LEFTY1 | TRUE | Upregulated |
| 1,669685984 | 0,003285474 | FCER1A | TRUE | Upregulated |
| 1,115090036 | 0,003291898 | EVC2 | TRUE | Upregulated |
| 1,622613838 | 0,003300554 | ICOS | TRUE | Upregulated |
| -0,369749177 | 0,003305167 | SFT2D3 | FALSE | Downregulated |
| 2,167527347 | 0,003305495 | LOC101060038 | TRUE | Upregulated |
| 0,669817168 | 0,003307761 | TMPRSS2 | FALSE | Upregulated |
| -0,53566585 | 0,003320042 | APLF | FALSE | Downregulated |
| 0,532058962 | 0,003322203 | SH3BGRL3 | FALSE | Upregulated |
| 0,622577032 | 0,003372937 | METRNL | FALSE | Upregulated |
| 1,342702979 | 0,003376052 | SPHK1 | TRUE | Upregulated |
| 0,547090436 | 0,003388641 | PCNA | FALSE | Upregulated |
| 0,88205455 | 0,003389356 | TFPI2 | FALSE | Upregulated |
| -0,436891987 | 0,003391474 | ECSIT | FALSE | Downregulated |
| -0,624152607 | 0,003391474 | MYL5 | FALSE | Downregulated |
| 1,091927 | 0,003399219 | FJX1 | TRUE | Upregulated |
| -0,363205598 | 0,003405676 | MRPS15 | FALSE | Downregulated |
| -0,223150989 | 0,003409659 | UBE2R2 | FALSE | Downregulated |
| 0,436935684 | 0,003417956 | MCM7 | FALSE | Upregulated |
| -0,252797509 | 0,003427094 | TRAPPC11 | FALSE | Downregulated |
| 1,403126072 | 0,003433296 | CNNM1 | TRUE | Upregulated |
| -0,340270712 | 0,003433296 | NMD3 | FALSE | Downregulated |
| -0,303667483 | 0,003433513 | ZNF181 | FALSE | Downregulated |
| 0,236053501 | 0,003446003 | PRPF8 | FALSE | Upregulated |
| -0,407410042 | 0,003449221 | STX18 | FALSE | Downregulated |
| -0,649851453 | 0,003454067 | FXYD1 | FALSE | Downregulated |
| 0,726277194 | 0,003457862 | MOGAT3 | FALSE | Upregulated |
| 0,541645875 | 0,003471313 | NOD1 | FALSE | Upregulated |
| -0,463768666 | 0,003475626 | CFHR2 | FALSE | Downregulated |
| -0,402120858 | 0,003487427 | TRAPPC2L | FALSE | Downregulated |
| 0,26840695 | 0,00349249 | ENOPH1 | FALSE | Upregulated |
| -0,271836451 | 0,00349249 | ISG20L2 | FALSE | Downregulated |

| | | | | |
|---|---|---|---|---|
| -0,280273082 | 0,00349249 | LSM12 | FALSE | Downregulated |
| -0,419210873 | 0,003492887 | NUDT16P1 | FALSE | Downregulated |
| -0,564199769 | 0,003501887 | SEPT7P2 | FALSE | Downregulated |
| -0,572378549 | 0,003503823 | PDE3B | FALSE | Downregulated |
| -0,385011243 | 0,003507512 | OSTC | FALSE | Downregulated |
| 0,424052841 | 0,00352172 | PEA15 | FALSE | Upregulated |
| -0,324756957 | 0,00352641 | ATP5O | FALSE | Downregulated |
| -0,345495826 | 0,00352641 | PEX1 | FALSE | Downregulated |
| -3,130759144 | 0,003529064 | CPNE6 | TRUE | Downregulated |
| 3,411769219 | 0,003529064 | PLEKHS1 | TRUE | Upregulated |
| -0,451621043 | 0,003551741 | SNRPD1 | FALSE | Downregulated |
| -0,319576374 | 0,003573263 | C1S | FALSE | Downregulated |
| -0,392569079 | 0,003586358 | MRPS34 | FALSE | Downregulated |
| 1,213051228 | 0,003591878 | TMEM171 | TRUE | Upregulated |
| -0,716494344 | 0,003599797 | ABCC9 | FALSE | Downregulated |
| -0,219650704 | 0,003599797 | ARL5A | FALSE | Downregulated |
| 1,167147847 | 0,003599797 | IL3RA | TRUE | Upregulated |
| 1,483193474 | 0,003599797 | TUBB2B | TRUE | Upregulated |
| 0,898687842 | 0,003603875 | CSGALNACT1 | FALSE | Upregulated |
| 0,614127972 | 0,003610052 | C14orf169 | FALSE | Upregulated |
| 0,775601338 | 0,003620565 | CHTF18 | FALSE | Upregulated |
| 0,349468479 | 0,00363892 | CD63 | FALSE | Upregulated |
| -0,317781437 | 0,00363892 | UHRF1BP1L | FALSE | Downregulated |
| 0,919121283 | 0,003653418 | CNTNAP1 | FALSE | Upregulated |
| 0,828644901 | 0,003653418 | IL2RG | FALSE | Upregulated |
| 0,405950497 | 0,00365388 | GRK5 | FALSE | Upregulated |
| -0,416879369 | 0,003662854 | RPL37 | FALSE | Downregulated |
| -2,280600633 | 0,00366608 | GPR112 | TRUE | Downregulated |
| 0,530847334 | 0,003675184 | LMNB2 | FALSE | Upregulated |
| -0,571759073 | 0,003694654 | SULT1A2 | FALSE | Downregulated |
| -0,322246689 | 0,003701249 | KLHDC2 | FALSE | Downregulated |
| -0,354128259 | 0,003714154 | CCDC59 | FALSE | Downregulated |
| -0,308541151 | 0,003717451 | DERA | FALSE | Downregulated |
| -0,361508467 | 0,003717451 | FAM210A | FALSE | Downregulated |
| -0,347408176 | 0,003717451 | SCRN3 | FALSE | Downregulated |
| 1,03647231 | 0,003717737 | RP11-367J11.2 | TRUE | Upregulated |
| 0,808741994 | 0,003720671 | FCHO1 | FALSE | Upregulated |
| 0,884664525 | 0,003720671 | MYRF | FALSE | Upregulated |
| 0,282251879 | 0,003722349 | CCDC97 | FALSE | Upregulated |
| 0,521513413 | 0,003722349 | PECAM1 | FALSE | Upregulated |
| 0,743269467 | 0,003725959 | GBP1 | FALSE | Upregulated |
| -0,463443368 | 0,00373443 | GALT | FALSE | Downregulated |
| -0,425512943 | 0,00373443 | UFL1 | FALSE | Downregulated |
| 0,413880235 | 0,003746876 | APAF1 | FALSE | Upregulated |
| 0,293210033 | 0,003746876 | DCTN2 | FALSE | Upregulated |
| -0,506585874 | 0,003746876 | NUDT4 | FALSE | Downregulated |
| -1,95504721 | 0,003746876 | PF4 | TRUE | Downregulated |
| 0,756462085 | 0,003746876 | RGS10 | FALSE | Upregulated |
| 1,225937468 | 0,003762962 | NPNT | TRUE | Upregulated |
| -0,647876176 | 0,003766213 | ACVR1C | FALSE | Downregulated |
| 1,490907714 | 0,003766213 | PIWIL4 | TRUE | Upregulated |

| | | | | |
|---|---|---|---|---|
| 0,278419797 | 0,003771716 | YWHAQ | FALSE | Upregulated |
| 0,509557806 | 0,003773428 | TWF2 | FALSE | Upregulated |
| 1,038077166 | 0,003785513 | TLR10 | TRUE | Upregulated |
| 1,052531768 | 0,00379284 | SIGLEC10 | TRUE | Upregulated |
| -0,681666169 | 0,003793053 | ANXA10 | FALSE | Downregulated |
| -0,46442332 | 0,00379683 | SLC10A1 | FALSE | Downregulated |
| 0,822599668 | 0,003805909 | LINGO1 | FALSE | Upregulated |
| 1,394969032 | 0,003808428 | IRF4 | TRUE | Upregulated |
| 0,185826307 | 0,003812788 | PAFAH1B1 | FALSE | Upregulated |
| 0,704754611 | 0,003823263 | NUDT18 | FALSE | Upregulated |
| -0,336336528 | 0,003824463 | AIG1 | FALSE | Downregulated |
| 1,289927374 | 0,003824463 | CLSPN | TRUE | Upregulated |
| -0,653176258 | 0,003831094 | LOC101928061 | FALSE | Downregulated |
| 0,794913504 | 0,00383683 | APBB3 | FALSE | Upregulated |
| 0,624146318 | 0,003841675 | INTS9 | FALSE | Upregulated |
| -0,741677598 | 0,00384257 | BCL2L2-PABPN1 | FALSE | Downregulated |
| -0,384205924 | 0,00384257 | SYAP1 | FALSE | Downregulated |
| -0,524820498 | 0,003845863 | APOF | FALSE | Downregulated |
| -0,194372271 | 0,003845863 | SNRPD3 | FALSE | Downregulated |
| -0,606608939 | 0,003846484 | KCNE3 | FALSE | Downregulated |
| -0,53838229 | 0,003846484 | PIR | FALSE | Downregulated |
| -0,356426514 | 0,003846484 | PRDX1 | FALSE | Downregulated |
| -0,23469004 | 0,003851256 | SMEK2 | FALSE | Downregulated |
| -0,704718147 | 0,003866611 | LOC102724392 | FALSE | Downregulated |
| -0,50933995 | 0,003867483 | MPHOSPH10 | FALSE | Downregulated |
| 0,266808851 | 0,003867483 | SGPL1 | FALSE | Upregulated |
| -0,474512047 | 0,003871742 | C16orf13 | FALSE | Downregulated |
| 1,916358824 | 0,003871742 | SBSPON | TRUE | Upregulated |
| 0,739024248 | 0,003883606 | SPN | FALSE | Upregulated |
| 1,371716853 | 0,003892434 | PIPSL | TRUE | Upregulated |
| -0,65626859 | 0,003899695 | LOC100289097 | FALSE | Downregulated |
| 0,571275616 | 0,003899695 | MARCKS | FALSE | Upregulated |
| 2,198364189 | 0,003902855 | LMTK3 | TRUE | Upregulated |
| -0,341895061 | 0,003907398 | RBM23 | FALSE | Downregulated |
| -0,544234701 | 0,003907768 | C12orf43 | FALSE | Downregulated |
| 0,553849343 | 0,003908149 | PLXNA1 | FALSE | Upregulated |
| -0,697309895 | 0,003908149 | PPM1K | FALSE | Downregulated |
| -0,279861526 | 0,003908149 | RTCA | FALSE | Downregulated |
| -0,191736093 | 0,003910468 | COIL | FALSE | Downregulated |
| 1,411681544 | 0,003913039 | METTL24 | TRUE | Upregulated |
| -0,346017402 | 0,003914816 | RPL34 | FALSE | Downregulated |
| -0,473034817 | 0,003916427 | RPS24 | FALSE | Downregulated |
| -1,016671611 | 0,003916427 | THRB-AS1 | TRUE | Downregulated |
| 2,757175331 | 0,003922336 | LOC101059954 | TRUE | Upregulated |
| 0,402077031 | 0,003922451 | CLN3 | FALSE | Upregulated |
| -0,284549413 | 0,00395214 | SPAG16 | FALSE | Downregulated |
| -0,403878764 | 0,00395214 | TMEM57 | FALSE | Downregulated |
| 0,473990723 | 0,003954931 | ZBTB9 | FALSE | Upregulated |
| 0,421690585 | 0,003961087 | TBL3 | FALSE | Upregulated |
| 1,914959145 | 0,003965317 | PBK | TRUE | Upregulated |
| 0,474791656 | 0,003965317 | PRKAB1 | FALSE | Upregulated |

| | | | | |
|---|---|---|---|---|
| 1,396698367 | 0,003965317 | STK33 | TRUE | Upregulated |
| 0,62556415 | 0,003965317 | TRIM11 | FALSE | Upregulated |
| 0,389738103 | 0,004011583 | PLXND1 | FALSE | Upregulated |
| 0,460165822 | 0,00401679 | ROBO4 | FALSE | Upregulated |
| -0,357593731 | 0,004021038 | SNRPF | FALSE | Downregulated |
| 0,36347345 | 0,004027645 | TNFRSF1A | FALSE | Upregulated |
| 2,48045141 | 0,004028432 | CILP2 | TRUE | Upregulated |
| 0,370748537 | 0,00403522 | ETS1 | FALSE | Upregulated |
| -0,397909933 | 0,004046809 | SYF2 | FALSE | Downregulated |
| 1,133279951 | 0,004050986 | GATA2 | TRUE | Upregulated |
| -0,279236449 | 0,00406775 | THOC1 | FALSE | Downregulated |
| 0,737559943 | 0,004076546 | DLL1 | FALSE | Upregulated |
| 1,31026796 | 0,004081031 | NKD2 | TRUE | Upregulated |
| 0,39584513 | 0,004081031 | PDLIM1 | FALSE | Upregulated |
| -0,494412292 | 0,004083547 | ZFP36L1 | FALSE | Downregulated |
| 0,695059101 | 0,004083772 | ZDHHC1 | FALSE | Upregulated |
| 0,319914446 | 0,004084484 | CDC42 | FALSE | Upregulated |
| -0,442709956 | 0,004091321 | SLC37A4 | FALSE | Downregulated |
| -0,326536561 | 0,004093973 | ETFB | FALSE | Downregulated |
| 0,803876366 | 0,004094523 | TUSC3 | FALSE | Upregulated |
| -1,114481526 | 0,004106608 | LOC101927765 | TRUE | Downregulated |
| 0,525001863 | 0,004108614 | ERI1 | FALSE | Upregulated |
| 2,777601322 | 0,004108614 | FOXF2 | TRUE | Upregulated |
| 0,825526016 | 0,004108614 | RIMS3 | FALSE | Upregulated |
| 1,504513487 | 0,004108614 | TRIM29 | TRUE | Upregulated |
| 0,407349586 | 0,004113507 | ACOT7 | FALSE | Upregulated |
| 0,841540697 | 0,004113507 | FLJ32255 | FALSE | Upregulated |
| 1,348371347 | 0,004113507 | PANDAR | TRUE | Upregulated |
| -0,303447917 | 0,004121311 | CSNK2A2 | FALSE | Downregulated |
| 1,796626951 | 0,004121311 | FER1L4 | TRUE | Upregulated |
| 1,943466965 | 0,00414248 | PDCD1 | TRUE | Upregulated |
| 0,350046428 | 0,004143194 | SEPT8 | FALSE | Upregulated |
| 1,106583022 | 0,004148077 | GP1BA | TRUE | Upregulated |
| 1,441091171 | 0,00415419 | DUSP2 | TRUE | Upregulated |
| -0,440154906 | 0,00415682 | NUFIP1 | FALSE | Downregulated |
| 0,503710326 | 0,004162385 | POR | FALSE | Upregulated |
| -0,552743298 | 0,004170005 | IDNK | FALSE | Downregulated |
| 0,769664936 | 0,004170005 | RNASE1 | FALSE | Upregulated |
| 2,931612347 | 0,004182858 | C2orf91 | TRUE | Upregulated |
| 1,425833656 | 0,004191336 | CASKIN1 | TRUE | Upregulated |
| 0,316835496 | 0,004191336 | TOMM34 | FALSE | Upregulated |
| 1,353544111 | 0,004193687 | MNS1 | TRUE | Upregulated |
| -1,018223899 | 0,00419556 | LOC101929427 | TRUE | Downregulated |
| -0,655082365 | 0,004198559 | CYP2E1 | FALSE | Downregulated |
| 0,799386674 | 0,004198559 | GZMA | FALSE | Upregulated |
| 0,340979982 | 0,004224553 | ATP11A | FALSE | Upregulated |
| -0,305836073 | 0,004224553 | GFM1 | FALSE | Downregulated |
| 0,489909211 | 0,004225244 | LZTS2 | FALSE | Upregulated |
| 0,826611928 | 0,004236181 | CDR2L | FALSE | Upregulated |
| 1,074940686 | 0,00424455 | HSPA2 | TRUE | Upregulated |
| 0,444110524 | 0,004247246 | FUCA2 | FALSE | Upregulated |

| | | | | |
|---|---|---|---|---|
| -1,973084694 | 0,004248598 | COL28A1 | TRUE | Downregulated |
| -0,42762829 | 0,004264117 | TOMM5 | FALSE | Downregulated |
| -0,284344578 | 0,004272806 | CLDN12 | FALSE | Downregulated |
| 1,385688597 | 0,004277258 | WNT4 | TRUE | Upregulated |
| 0,935895251 | 0,004286068 | SERPINB8 | FALSE | Upregulated |
| 0,564727737 | 0,004299801 | TGIF1 | FALSE | Upregulated |
| 2,393332412 | 0,004304372 | TRIM31 | TRUE | Upregulated |
| 2,498816227 | 0,00430901 | SLC45A1 | TRUE | Upregulated |
| 1,525407323 | 0,004314503 | FLJ22184 | TRUE | Upregulated |
| -2,42560413 | 0,004314503 | LOC101927170 | TRUE | Downregulated |
| -0,617525999 | 0,004317186 | LNP1 | FALSE | Downregulated |
| -0,521390687 | 0,004317186 | PRKAB2 | FALSE | Downregulated |
| 1,606537507 | 0,004328729 | FZD2 | TRUE | Upregulated |
| -0,344406139 | 0,004329469 | ING1 | FALSE | Downregulated |
| 0,971649674 | 0,004329469 | LINC00857 | FALSE | Upregulated |
| 0,337948615 | 0,004329469 | PPM1F | FALSE | Upregulated |
| 0,436441886 | 0,004336441 | LAP3 | FALSE | Upregulated |
| 0,515315235 | 0,004343675 | PLEKHG1 | FALSE | Upregulated |
| -0,425391419 | 0,004348168 | TIMM8B | FALSE | Downregulated |
| 0,76199543 | 0,004352176 | CXorf21 | FALSE | Upregulated |
| 0,392818356 | 0,004354866 | ATP9B | FALSE | Upregulated |
| 0,295210946 | 0,004356201 | ZC3H4 | FALSE | Upregulated |
| 1,694151048 | 0,004356465 | DEPDC1 | TRUE | Upregulated |
| 1,303363123 | 0,004356465 | SBK1 | TRUE | Upregulated |
| -0,270498412 | 0,004356468 | FUNDC1 | FALSE | Downregulated |
| -0,36684429 | 0,004356468 | GIN1 | FALSE | Downregulated |
| 2,584259684 | 0,004361956 | CCNI2 | TRUE | Upregulated |
| 0,420644634 | 0,004365767 | CBX6 | FALSE | Upregulated |
| -0,63188378 | 0,004374406 | LOC100506603 | FALSE | Downregulated |
| -0,575113884 | 0,004376189 | RAD50 | FALSE | Downregulated |
| 0,727841284 | 0,004379816 | CDC42EP3 | FALSE | Upregulated |
| -0,381012093 | 0,004383426 | SLC25A13 | FALSE | Downregulated |
| -0,396991957 | 0,004383426 | USF2 | FALSE | Downregulated |
| 1,027804972 | 0,004397025 | FBXO41 | TRUE | Upregulated |
| 0,635694409 | 0,004397025 | PIK3R5 | FALSE | Upregulated |
| 0,467576225 | 0,004397025 | RHOG | FALSE | Upregulated |
| 1,08800887 | 0,004401898 | DAGLA | TRUE | Upregulated |
| 2,966301349 | 0,004403265 | PDE6A | TRUE | Upregulated |
| 1,451602612 | 0,00440444 | GPC3 | TRUE | Upregulated |
| 0,324989658 | 0,00440561 | DSC2 | FALSE | Upregulated |
| -0,214335213 | 0,004412712 | SLC30A9 | FALSE | Downregulated |
| 0,334436609 | 0,004420049 | FBXW11 | FALSE | Upregulated |
| -0,249256519 | 0,004420049 | GXYLT1 | FALSE | Downregulated |
| 0,473703703 | 0,004431672 | CTBP2 | FALSE | Upregulated |
| -0,401273686 | 0,004436195 | ZNF18 | FALSE | Downregulated |
| 0,28531433 | 0,004440373 | IGF2R | FALSE | Upregulated |
| -0,510172207 | 0,00444965 | PAXIP1-AS1 | FALSE | Downregulated |
| 0,760870276 | 0,004460947 | JAM3 | FALSE | Upregulated |
| 0,664552492 | 0,004461177 | IMPDH1 | FALSE | Upregulated |
| 0,728187679 | 0,004461177 | TRPV4 | FALSE | Upregulated |
| 0,423124718 | 0,004469178 | EHD1 | FALSE | Upregulated |

| | | | | |
|---|---|---|---|---|
| 1,093287816 | 0,004469178 | EVC | TRUE | Upregulated |
| 1,249158421 | 0,004470204 | GAS6-AS1 | TRUE | Upregulated |
| 0,356468683 | 0,004471061 | MPRIP | FALSE | Upregulated |
| -0,383508135 | 0,004471544 | RBKS | FALSE | Downregulated |
| 2,327586171 | 0,004494172 | KRT80 | TRUE | Upregulated |
| 3,255087302 | 0,004494172 | LOC650157 | TRUE | Upregulated |
| 0,22241296 | 0,004494172 | STAM2 | FALSE | Upregulated |
| -0,508681085 | 0,004494172 | ZFP1 | FALSE | Downregulated |
| 0,861968205 | 0,004496357 | CERCAM | FALSE | Upregulated |
| 0,858467591 | 0,004501697 | MAP9 | FALSE | Upregulated |
| -0,376685186 | 0,004501724 | RPL24 | FALSE | Downregulated |
| 0,559524808 | 0,004501724 | TMEM165 | FALSE | Upregulated |
| 0,561647562 | 0,004513801 | HIGD1A | FALSE | Upregulated |
| 2,808221595 | 0,004527631 | AACSP1 | TRUE | Upregulated |
| 0,969919549 | 0,004527631 | CD300C | FALSE | Upregulated |
| -0,416583122 | 0,004528304 | MTRF1L | FALSE | Downregulated |
| 2,539505634 | 0,004531003 | CD19 | TRUE | Upregulated |
| 0,519327184 | 0,004538655 | IFT52 | FALSE | Upregulated |
| 0,429439499 | 0,004549619 | SNRNP40 | FALSE | Upregulated |
| 0,687012596 | 0,004551183 | HID1 | FALSE | Upregulated |
| 0,909624688 | 0,004553917 | HLA-DQA1 | FALSE | Upregulated |
| -0,391143055 | 0,004553917 | HSD11B1 | FALSE | Downregulated |
| 0,770357544 | 0,004553917 | KCTD17 | FALSE | Upregulated |
| 1,929951971 | 0,004555012 | LINC01057 | TRUE | Upregulated |
| -3,48199845 | 0,004562123 | KLHL41 | TRUE | Downregulated |
| -0,453274699 | 0,004570765 | WDR19 | FALSE | Downregulated |
| 0,578158048 | 0,004574754 | ARHGEF5 | FALSE | Upregulated |
| 0,430539411 | 0,004575187 | MICALL1 | FALSE | Upregulated |
| 1,624402403 | 0,004575479 | BMP8B | TRUE | Upregulated |
| 2,08295918 | 0,004579686 | DLGAP1 | TRUE | Upregulated |
| 2,234786694 | 0,004579686 | PODNL1 | TRUE | Upregulated |
| -0,362446165 | 0,004593707 | TTC3 | FALSE | Downregulated |
| 1,139616928 | 0,004613792 | BCL2A1 | TRUE | Upregulated |
| -0,311969984 | 0,004613792 | CALM1 | FALSE | Downregulated |
| 0,332460986 | 0,00463459 | TSC22D2 | FALSE | Upregulated |
| 0,27813629 | 0,004642269 | PRKDC | FALSE | Upregulated |
| 0,796304496 | 0,004652716 | CYP11A1 | FALSE | Upregulated |
| 1,611457303 | 0,004667829 | FBXL16 | TRUE | Upregulated |
| -0,558998807 | 0,004671374 | FAM160A1 | FALSE | Downregulated |
| 0,923610376 | 0,004671374 | RELT | FALSE | Upregulated |
| 1,782148492 | 0,004671374 | SRSF12 | TRUE | Upregulated |
| 1,49268907 | 0,00467294 | LINC01094 | TRUE | Upregulated |
| 1,694220374 | 0,004688894 | B4GALNT3 | TRUE | Upregulated |
| -0,560491851 | 0,004688894 | PFDN1 | FALSE | Downregulated |
| 0,927880749 | 0,004690122 | ARMC9 | FALSE | Upregulated |
| 0,477815085 | 0,0046911 | CSF1 | FALSE | Upregulated |
| 2,994102947 | 0,004691939 | IGSF9B | TRUE | Upregulated |
| -3,210073347 | 0,004708255 | CA3 | TRUE | Downregulated |
| 0,569353335 | 0,004713375 | RAB11FIP4 | FALSE | Upregulated |
| -0,872585028 | 0,004713894 | LOC101930644 | FALSE | Downregulated |
| -1,20018549 | 0,004738018 | TRNS1 | TRUE | Downregulated |

| | | | | |
|---|---|---|---|---|
| 0,739531857 | 0,004754805 | ALOX5 | FALSE | Upregulated |
| 2,048236371 | 0,004754805 | LOC644936 | TRUE | Upregulated |
| -1,602180085 | 0,004754805 | TRHDE | TRUE | Downregulated |
| 0,346274385 | 0,004755013 | DSG2 | FALSE | Upregulated |
| 0,285237471 | 0,00476235 | KLC1 | FALSE | Upregulated |
| 1,579797072 | 0,004770329 | GABRB3 | TRUE | Upregulated |
| 0,469383707 | 0,004770992 | CARD10 | FALSE | Upregulated |
| -1,96544814 | 0,004777241 | LOC101929134 | TRUE | Downregulated |
| 0,361454597 | 0,00478914 | RFWD3 | FALSE | Upregulated |
| -0,846253318 | 0,00482671 | GRIA3 | FALSE | Downregulated |
| 0,445800254 | 0,00482671 | MGAT5 | FALSE | Upregulated |
| 0,871197888 | 0,004830214 | FAM83G | FALSE | Upregulated |
| -0,351048913 | 0,004833502 | SOD1 | FALSE | Downregulated |
| -0,886032066 | 0,004834669 | RP11-205M3.3 | FALSE | Downregulated |
| -0,493274086 | 0,004837936 | PROX1-AS1 | FALSE | Downregulated |
| 0,354237475 | 0,004847245 | CD2BP2 | FALSE | Upregulated |
| -0,66053428 | 0,004847245 | TRIM16L | FALSE | Downregulated |
| -0,349068577 | 0,004850255 | ICT1 | FALSE | Downregulated |
| -0,684209689 | 0,004850255 | SLC22A1 | FALSE | Downregulated |
| 1,029590608 | 0,004850255 | SLC35F2 | TRUE | Upregulated |
| -0,414183325 | 0,004850255 | TMEM141 | FALSE | Downregulated |
| 0,236817045 | 0,004850255 | ZC3H18 | FALSE | Upregulated |
| 0,965684641 | 0,004858581 | STXBP6 | FALSE | Upregulated |
| -0,683456438 | 0,004893244 | RP11-328K4.1 | FALSE | Downregulated |
| 0,633378214 | 0,004900685 | CMTM7 | FALSE | Upregulated |
| -0,437070103 | 0,004906871 | F13B | FALSE | Downregulated |
| 0,953711273 | 0,004920558 | CTSK | FALSE | Upregulated |
| -0,330013148 | 0,004924202 | HAT1 | FALSE | Downregulated |
| -0,355972602 | 0,004938925 | MRPL48 | FALSE | Downregulated |
| 3,009842014 | 0,004983163 | CACNA1I | TRUE | Upregulated |
| 2,442723559 | 0,00498502 | CRB2 | TRUE | Upregulated |
| 1,219307011 | 0,004996385 | SYTL1 | TRUE | Upregulated |
| -0,348752619 | 0,005006129 | SETD6 | FALSE | Downregulated |
| 1,333432572 | 0,005017512 | LOC102724312 | TRUE | Upregulated |
| 0,668653492 | 0,005022996 | ELF4 | FALSE | Upregulated |
| 0,576561143 | 0,005043322 | GAS6 | FALSE | Upregulated |
| 0,409581472 | 0,005046925 | KCTD11 | FALSE | Upregulated |
| -0,162366624 | 0,005047836 | ZNF146 | FALSE | Downregulated |
| 0,47041905 | 0,005051786 | BCL2L1 | FALSE | Upregulated |
| -0,244957852 | 0,005057787 | C14orf166 | FALSE | Downregulated |
| 0,496029251 | 0,005057787 | GPR137B | FALSE | Upregulated |
| 0,351818299 | 0,005064603 | PRPS2 | FALSE | Upregulated |
| -0,817382823 | 0,005100193 | SH3RF2 | FALSE | Downregulated |
| -0,323387723 | 0,005108878 | HACL1 | FALSE | Downregulated |
| -0,452104646 | 0,00511565 | MTHFS | FALSE | Downregulated |
| 0,395636548 | 0,005127031 | FUS | FALSE | Upregulated |
| 1,353484522 | 0,005127031 | GABRE | TRUE | Upregulated |
| -0,618564971 | 0,005127031 | GHR | FALSE | Downregulated |
| 0,267912056 | 0,005127031 | KIAA0247 | FALSE | Upregulated |
| 1,083176026 | 0,005127031 | MYEF2 | TRUE | Upregulated |
| 1,088615082 | 0,005127031 | ST6GALNAC4 | TRUE | Upregulated |

| | | | | |
|---|---|---|---|---|
| 1,125560929 | 0,005127031 | TNC | TRUE | Upregulated |
| 0,313535345 | 0,005163177 | CD47 | FALSE | Upregulated |
| 0,182391197 | 0,005169257 | KHDRBS1 | FALSE | Upregulated |
| 1,51298807 | 0,005170565 | C5orf20 | TRUE | Upregulated |
| 0,67396942 | 0,005170565 | DENND1C | FALSE | Upregulated |
| -0,346581403 | 0,005174431 | NDUFAF1 | FALSE | Downregulated |
| 0,511624225 | 0,005179884 | ELK3 | FALSE | Upregulated |
| 1,902051987 | 0,005181892 | LRRC43 | TRUE | Upregulated |
| 2,15423137 | 0,005183743 | CXADRP2 | TRUE | Upregulated |
| 0,637171447 | 0,005192968 | EFEMP2 | FALSE | Upregulated |
| 1,416618539 | 0,005192968 | ORC1 | TRUE | Upregulated |
| -0,32318818 | 0,00519378 | AKAP1 | FALSE | Downregulated |
| 0,254332824 | 0,005196439 | FIG4 | FALSE | Upregulated |
| 0,389449224 | 0,005206277 | FAM122B | FALSE | Upregulated |
| -0,341711235 | 0,00521446 | CEP57 | FALSE | Downregulated |
| -0,29995896 | 0,00522189 | SDHAF2 | FALSE | Downregulated |
| 1,594311423 | 0,00523061 | CCL4L1 | TRUE | Upregulated |
| -0,585089267 | 0,00523061 | ZNF658B | FALSE | Downregulated |
| 0,459808685 | 0,005231565 | PPAN | FALSE | Upregulated |
| -0,359213396 | 0,005233128 | IK | FALSE | Downregulated |
| -0,664687347 | 0,005244088 | HP | FALSE | Downregulated |
| 0,253500904 | 0,005247304 | VEZF1 | FALSE | Upregulated |
| 1,45220738 | 0,005248192 | KIF18A | TRUE | Upregulated |
| 0,505766501 | 0,005258423 | FAM117B | FALSE | Upregulated |
| 1,829732184 | 0,005265089 | IQGAP3 | TRUE | Upregulated |
| -0,202688541 | 0,005272606 | GTPBP10 | FALSE | Downregulated |
| 0,551560522 | 0,005282672 | FBXL19 | FALSE | Upregulated |
| 0,299458858 | 0,005282672 | ZNF394 | FALSE | Upregulated |
| -0,318462718 | 0,005287114 | PLA2G12B | FALSE | Downregulated |
| 0,37407319 | 0,005288107 | SLC12A7 | FALSE | Upregulated |
| -0,475962134 | 0,005288107 | XRCC5 | FALSE | Downregulated |
| 0,47042275 | 0,005295053 | IQCK | FALSE | Upregulated |
| 0,631789043 | 0,005295053 | IRF1 | FALSE | Upregulated |
| -0,290514938 | 0,005295053 | YBX1 | FALSE | Downregulated |
| 1,037643472 | 0,005303106 | SEMA3B | TRUE | Upregulated |
| 0,384611866 | 0,005303906 | AKIRIN1 | FALSE | Upregulated |
| 0,732310804 | 0,005303906 | GVINP1 | FALSE | Upregulated |
| 1,664672926 | 0,005303906 | SMOC2 | TRUE | Upregulated |
| -0,650530547 | 0,005303906 | ST3GAL6 | FALSE | Downregulated |
| -0,261211787 | 0,00531505 | CRKL | FALSE | Downregulated |
| -0,896602688 | 0,00531505 | SNHG19 | FALSE | Downregulated |
| 1,032760305 | 0,0053225 | CCR2 | TRUE | Upregulated |
| -0,937437286 | 0,00534341 | HMGN5 | FALSE | Downregulated |
| -0,246925336 | 0,00534341 | TADA2B | FALSE | Downregulated |
| -1,380918643 | 0,00534922 | LOC101927284 | TRUE | Downregulated |
| -0,30029647 | 0,005354153 | CREG1 | FALSE | Downregulated |
| -0,959103857 | 0,005365226 | SNORA8 | FALSE | Downregulated |
| -1,0875037 | 0,005382128 | KCNJ3 | TRUE | Downregulated |
| 0,630217727 | 0,005382128 | KLF6 | FALSE | Upregulated |
| 0,413355006 | 0,005386514 | SWAP70 | FALSE | Upregulated |
| -0,396440559 | 0,005391043 | AKAP8 | FALSE | Downregulated |

| | | | | |
|---|---|---|---|---|
| -0,333885698 | 0,005391043 | C8orf59 | FALSE | Downregulated |
| 2,578609692 | 0,005391043 | SLC44A4 | TRUE | Upregulated |
| 0,727715999 | 0,00540305 | HDX | FALSE | Upregulated |
| -0,696836075 | 0,005416555 | MIR22HG | FALSE | Downregulated |
| 0,329235193 | 0,00541725 | ACSF3 | FALSE | Upregulated |
| 0,474027166 | 0,005431817 | HLA-C | FALSE | Upregulated |
| 0,314427015 | 0,005446051 | PTPN21 | FALSE | Upregulated |
| 2,580547845 | 0,005450806 | KRTAP5-AS1 | TRUE | Upregulated |
| 0,312002342 | 0,005450806 | SHC1 | FALSE | Upregulated |
| 0,865846693 | 0,005455795 | PRICKLE1 | FALSE | Upregulated |
| 0,521664308 | 0,005469471 | AMOTL2 | FALSE | Upregulated |
| -0,716239269 | 0,005470276 | PEG3 | FALSE | Downregulated |
| -0,38294338 | 0,005473131 | TFR2 | FALSE | Downregulated |
| 0,577063801 | 0,005482864 | TACC3 | FALSE | Upregulated |
| -0,791152753 | 0,005495975 | GSTA1 | FALSE | Downregulated |
| -0,895659261 | 0,005495975 | TIPARP-AS1 | FALSE | Downregulated |
| 1,263557124 | 0,005495975 | TTK | TRUE | Upregulated |
| -0,267210748 | 0,005501513 | DNAJC7 | FALSE | Downregulated |
| -0,29082396 | 0,005507855 | MAPKAP1 | FALSE | Downregulated |
| 0,6947285 | 0,005508181 | CHST11 | FALSE | Upregulated |
| -0,538518419 | 0,005508181 | DHX35 | FALSE | Downregulated |
| -1,424615795 | 0,005508181 | HAP1 | TRUE | Downregulated |
| -0,354743843 | 0,005521577 | H3F3A | FALSE | Downregulated |
| 3,033513489 | 0,00553288 | CACNA1G | TRUE | Upregulated |
| 0,323331547 | 0,00553288 | FYCO1 | FALSE | Upregulated |
| -0,440023792 | 0,00553288 | METAP2 | FALSE | Downregulated |
| 0,489795191 | 0,00553288 | RASSF3 | FALSE | Upregulated |
| -0,413822061 | 0,005539376 | CDC34 | FALSE | Downregulated |
| -0,433962886 | 0,00554122 | CWC22 | FALSE | Downregulated |
| 0,266419854 | 0,00554122 | GBA | FALSE | Upregulated |
| 1,401316638 | 0,00554122 | RPS6KL1 | TRUE | Upregulated |
| 1,712166738 | 0,005542535 | TIAM2 | TRUE | Upregulated |
| 2,708018122 | 0,005542535 | TMEM52B | TRUE | Upregulated |
| 0,991569978 | 0,005548912 | CHAF1B | FALSE | Upregulated |
| 0,615900325 | 0,005556189 | ANXA5 | FALSE | Upregulated |
| 0,310123488 | 0,005556189 | UBE2A | FALSE | Upregulated |
| 0,840304039 | 0,005557557 | GPX7 | FALSE | Upregulated |
| 0,395086952 | 0,005557971 | WWC3 | FALSE | Upregulated |
| 1,420822877 | 0,005567366 | KLRC1 | TRUE | Upregulated |
| 0,636273008 | 0,005594626 | DOCK2 | FALSE | Upregulated |
| 0,386087079 | 0,005594626 | PELO | FALSE | Upregulated |
| 1,941286622 | 0,005599571 | KCNH8 | TRUE | Upregulated |
| 1,004818907 | 0,005603352 | CD22 | TRUE | Upregulated |
| -0,201071381 | 0,005604146 | OPA1 | FALSE | Downregulated |
| 1,890526687 | 0,00560662 | TMEM92 | TRUE | Upregulated |
| 0,710184707 | 0,005607228 | TIMP1 | FALSE | Upregulated |
| 2,357727997 | 0,005614543 | LINC00673 | TRUE | Upregulated |
| -0,397578407 | 0,005642351 | TTC39C | FALSE | Downregulated |
| 0,232227508 | 0,005670311 | POLR3C | FALSE | Upregulated |
| 0,618214204 | 0,005684605 | RASL11A | FALSE | Upregulated |
| 0,350586896 | 0,00569481 | NINJ1 | FALSE | Upregulated |

| | | | | |
|---|---|---|---|---|
| 2,652115876 | 0,005706399 | GPR84 | TRUE | Upregulated |
| -0,501169079 | 0,005706399 | KLKB1 | FALSE | Downregulated |
| 0,320909084 | 0,005711129 | CAPZA1 | FALSE | Upregulated |
| 0,733083096 | 0,005724796 | JUNB | FALSE | Upregulated |
| 0,304352788 | 0,005736997 | ZBED4 | FALSE | Upregulated |
| 0,677313977 | 0,005759119 | SLA | FALSE | Upregulated |
| 0,502932601 | 0,005759119 | SPATS2 | FALSE | Upregulated |
| 0,694763613 | 0,005783578 | ATP8B2 | FALSE | Upregulated |
| 1,990045156 | 0,005783578 | PDX1 | TRUE | Upregulated |
| -0,483058331 | 0,005783578 | PSENEN | FALSE | Downregulated |
| 0,867231084 | 0,005784076 | BAX | FALSE | Upregulated |
| 1,226279474 | 0,005787754 | LOC101927183 | TRUE | Upregulated |
| -0,25740127 | 0,005787754 | PDCD2 | FALSE | Downregulated |
| -0,39057305 | 0,005787754 | PET117 | FALSE | Downregulated |
| -0,694918942 | 0,005788195 | LINC00910 | FALSE | Downregulated |
| 0,311776281 | 0,005800137 | GAA | FALSE | Upregulated |
| 0,739533892 | 0,00580465 | NCEH1 | FALSE | Upregulated |
| 0,949863199 | 0,005818705 | KLHL25 | FALSE | Upregulated |
| 0,485358038 | 0,005834345 | FLYWCH1 | FALSE | Upregulated |
| -0,377531492 | 0,005834345 | MTMR4 | FALSE | Downregulated |
| -0,26888879 | 0,005834345 | PTER | FALSE | Downregulated |
| 1,533490273 | 0,005851334 | RPL21P44 | TRUE | Upregulated |
| 0,616310388 | 0,005855936 | ITPRIP | FALSE | Upregulated |
| 1,309117743 | 0,00593503 | FNDC1 | TRUE | Upregulated |
| 1,387910503 | 0,005953969 | IGSF21 | TRUE | Upregulated |
| -1,137079031 | 0,005955513 | EFCAB1 | TRUE | Downregulated |
| -1,390481467 | 0,00595807 | BCRP3 | TRUE | Downregulated |
| -2,961954778 | 0,005965794 | HSPB7 | TRUE | Downregulated |
| -0,409437341 | 0,005970721 | RPS4X | FALSE | Downregulated |
| 1,439824491 | 0,005981724 | FBXL2 | TRUE | Upregulated |
| -0,642805478 | 0,005985204 | LOC102724975 | FALSE | Downregulated |
| 0,569715898 | 0,005987955 | CTSC | FALSE | Upregulated |
| -0,341776169 | 0,005989289 | HSDL2 | FALSE | Downregulated |
| 1,083893968 | 0,005989289 | LOC729683 | TRUE | Upregulated |
| -0,177876747 | 0,005998043 | KLHL12 | FALSE | Downregulated |
| 1,089717595 | 0,005998043 | SLAMF1 | TRUE | Upregulated |
| 0,760077643 | 0,006003919 | PALD1 | FALSE | Upregulated |
| -0,316606768 | 0,006003919 | ZRANB1 | FALSE | Downregulated |
| 1,395474038 | 0,006028052 | CDT1 | TRUE | Upregulated |
| -0,407672823 | 0,006046492 | HADH | FALSE | Downregulated |
| 2,078035165 | 0,006047882 | SERPINA12 | TRUE | Upregulated |
| -0,496001977 | 0,00605948 | DNAJC21 | FALSE | Downregulated |
| -0,445943436 | 0,00605948 | TMEM120A | FALSE | Downregulated |
| 0,67920556 | 0,006060104 | ZNF532 | FALSE | Upregulated |
| -0,594151432 | 0,006073597 | ABCC6P1 | FALSE | Downregulated |
| -0,402147422 | 0,006077185 | PCK2 | FALSE | Downregulated |
| 2,438202753 | 0,006078718 | ASAP1-IT2 | TRUE | Upregulated |
| 0,367558003 | 0,006078718 | PIP4K2A | FALSE | Upregulated |
| -1,804652257 | 0,006113914 | LOC101927652 | TRUE | Downregulated |
| -0,542318173 | 0,006123558 | CDA | FALSE | Downregulated |
| 1,005402914 | 0,006123558 | CDC42EP5 | TRUE | Upregulated |

| | | | | |
|---|---|---|---|---|
| -2,082034066 | 0,006123558 | LRRC39 | TRUE | Downregulated |
| 0,297639936 | 0,006123558 | LYPLA2 | FALSE | Upregulated |
| 0,27492167 | 0,006123558 | MYH14 | FALSE | Upregulated |
| -0,275618937 | 0,006138574 | TPI1 | FALSE | Downregulated |
| 1,695025823 | 0,006141266 | UNC13A | TRUE | Upregulated |
| -0,504580335 | 0,006145554 | TMEM25 | FALSE | Downregulated |
| 0,454493642 | 0,006150095 | CRLF3 | FALSE | Upregulated |
| 1,63521627 | 0,006168008 | MST1R | TRUE | Upregulated |
| -0,441685424 | 0,006178435 | RNF144B | FALSE | Downregulated |
| 1,918303229 | 0,006196499 | FBN3 | TRUE | Upregulated |
| -0,523618756 | 0,006210071 | ADH6 | FALSE | Downregulated |
| 2,288943507 | 0,006211931 | KIAA1549L | TRUE | Upregulated |
| 0,615488138 | 0,006214525 | GJD3 | FALSE | Upregulated |
| -0,176706177 | 0,006218439 | TMEM30A | FALSE | Downregulated |
| 1,261694653 | 0,006245325 | PTGS2 | TRUE | Upregulated |
| 0,403849655 | 0,006276745 | ATP6V1H | FALSE | Upregulated |
| -0,750474138 | 0,006279714 | GPAM | FALSE | Downregulated |
| 2,564851289 | 0,006286636 | TMEM130 | TRUE | Upregulated |
| -0,743552079 | 0,006286636 | WNK3 | FALSE | Downregulated |
| 0,295708404 | 0,006303099 | F11R | FALSE | Upregulated |
| -0,300657553 | 0,006303099 | NDUFV1 | FALSE | Downregulated |
| -0,372550595 | 0,006304838 | BDH1 | FALSE | Downregulated |
| -1,280984479 | 0,006304838 | HMGCS1 | TRUE | Downregulated |
| 4,138966052 | 0,006304838 | KRT86 | TRUE | Upregulated |
| 0,212184306 | 0,006323254 | NSFL1C | FALSE | Upregulated |
| 0,415853045 | 0,006339937 | SLC41A3 | FALSE | Upregulated |
| -0,727007364 | 0,006357089 | RFESD | FALSE | Downregulated |
| 1,252242481 | 0,006372158 | ASF1B | TRUE | Upregulated |
| 0,463822054 | 0,006387413 | RAB8B | FALSE | Upregulated |
| -0,761795266 | 0,006390587 | EGFLAM | FALSE | Downregulated |
| 0,386640085 | 0,006397095 | ELOVL1 | FALSE | Upregulated |
| 0,693439879 | 0,006397095 | PLEKHB2 | FALSE | Upregulated |
| -0,328301421 | 0,006397095 | ZNF747 | FALSE | Downregulated |
| 1,025202185 | 0,006401706 | SEMA3D | TRUE | Upregulated |
| 1,198379363 | 0,006406386 | B3GALT2 | TRUE | Upregulated |
| 0,975979384 | 0,006423724 | RP11-696N14.1 | FALSE | Upregulated |
| 0,282816897 | 0,006476167 | POLR3A | FALSE | Upregulated |
| 0,452250473 | 0,006477568 | RAP2B | FALSE | Upregulated |
| -0,989407274 | 0,006477705 | CLEC4M | FALSE | Downregulated |
| 1,530546114 | 0,006502351 | UBE2C | TRUE | Upregulated |
| -0,740477019 | 0,006509713 | LOC100133985 | FALSE | Downregulated |
| 3,044783486 | 0,006514419 | LOC643387 | TRUE | Upregulated |
| 0,827948796 | 0,006527425 | KIAA1324L | FALSE | Upregulated |
| 1,275491034 | 0,006527425 | SIT1 | TRUE | Upregulated |
| -0,512423805 | 0,006553269 | IWS1 | FALSE | Downregulated |
| 0,380340972 | 0,006554539 | MPZL1 | FALSE | Upregulated |
| 0,573335766 | 0,006564465 | P4HA2 | FALSE | Upregulated |
| -0,2690879 | 0,006587236 | C9orf156 | FALSE | Downregulated |
| -0,987826914 | 0,006593639 | C17orf97 | FALSE | Downregulated |
| -0,328358475 | 0,006593639 | CENPBD1 | FALSE | Downregulated |
| 0,393341609 | 0,006593639 | NOL6 | FALSE | Upregulated |

| | | | | |
|---|---|---|---|---|
| 0,508817936 | 0,006603103 | CYBA | FALSE | Upregulated |
| 0,572163557 | 0,006612245 | FAM102B | FALSE | Upregulated |
| -0,452619506 | 0,006614056 | C4orf19 | FALSE | Downregulated |
| 0,433654685 | 0,006614056 | IQCE | FALSE | Upregulated |
| 0,418274184 | 0,006633039 | RELL1 | FALSE | Upregulated |
| 1,075017934 | 0,006633463 | LOC100128494 | TRUE | Upregulated |
| 0,850175819 | 0,006645995 | ANTXR1 | FALSE | Upregulated |
| 0,25431218 | 0,006683102 | SYMPK | FALSE | Upregulated |
| 0,474937813 | 0,006687549 | FAM189B | FALSE | Upregulated |
| 0,956777404 | 0,006691848 | ZNF681 | FALSE | Upregulated |
| -0,534746114 | 0,006707598 | LBX2-AS1 | FALSE | Downregulated |
| -0,831189394 | 0,006723952 | ADRA1A | FALSE | Downregulated |
| 3,578206876 | 0,006723952 | CEACAM6 | TRUE | Upregulated |
| 0,693534048 | 0,006730653 | FDXR | FALSE | Upregulated |
| -0,315704509 | 0,006734214 | CAAP1 | FALSE | Downregulated |
| 1,19592481 | 0,006740092 | CLDN10 | TRUE | Upregulated |
| 0,720793722 | 0,006765415 | TMC8 | FALSE | Upregulated |
| -0,644620161 | 0,006771697 | NPIPA1 | FALSE | Downregulated |
| 0,491686922 | 0,006776115 | SPARC | FALSE | Upregulated |
| 0,324598771 | 0,006777866 | CYLD | FALSE | Upregulated |
| -0,341848495 | 0,006789296 | FBXL3 | FALSE | Downregulated |
| 0,263031325 | 0,006789296 | KEAP1 | FALSE | Upregulated |
| 2,411658501 | 0,006789296 | PGAM4 | TRUE | Upregulated |
| 0,69008074 | 0,006789688 | ZNF367 | FALSE | Upregulated |
| 0,877685815 | 0,006820612 | CD83 | FALSE | Upregulated |
| -0,297285776 | 0,006820612 | SAP30BP | FALSE | Downregulated |
| -0,361519446 | 0,006820612 | SMARCE1 | FALSE | Downregulated |
| -0,347072589 | 0,006832349 | B3GNT2 | FALSE | Downregulated |
| -0,538494819 | 0,006832349 | DHRS4-AS1 | FALSE | Downregulated |
| 0,401810782 | 0,006832349 | GTPBP2 | FALSE | Upregulated |
| 1,292399991 | 0,00683613 | PRKAA2 | TRUE | Upregulated |
| -0,32073498 | 0,00683613 | SIRT3 | FALSE | Downregulated |
| -1,101753495 | 0,006859866 | SCG5 | TRUE | Downregulated |
| 2,290636164 | 0,006860753 | CDH3 | TRUE | Upregulated |
| 1,350782932 | 0,006860753 | SLC28A3 | TRUE | Upregulated |
| -0,418091821 | 0,006868219 | XPNPEP3 | FALSE | Downregulated |
| 0,691086919 | 0,006871188 | PTPRE | FALSE | Upregulated |
| -0,320998646 | 0,006878841 | MAX | FALSE | Downregulated |
| 0,433416139 | 0,006878841 | SARS2 | FALSE | Upregulated |
| 1,074259901 | 0,006882316 | DKK3 | TRUE | Upregulated |
| -0,295983657 | 0,006884536 | THAP1 | FALSE | Downregulated |
| 3,937420011 | 0,006901171 | HCN4 | TRUE | Upregulated |
| 1,125357337 | 0,006904907 | GPR18 | TRUE | Upregulated |
| 1,952096203 | 0,006904907 | KLC3 | TRUE | Upregulated |
| 1,528022763 | 0,006908904 | POM121L9P | TRUE | Upregulated |
| -0,22075726 | 0,006916932 | CHMP7 | FALSE | Downregulated |
| -0,345294118 | 0,006927644 | POFUT1 | FALSE | Downregulated |
| 0,523316712 | 0,006940495 | HM13 | FALSE | Upregulated |
| 0,212573856 | 0,006961069 | RNF40 | FALSE | Upregulated |
| -0,551911177 | 0,006980929 | C7orf63 | FALSE | Downregulated |
| -0,323795497 | 0,006982456 | SOWAHC | FALSE | Downregulated |

| | | | | |
|---|---|---|---|---|
| -0,261963392 | 0,006984243 | LINC00674 | FALSE | Downregulated |
| 1,321281155 | 0,006984243 | ROBO2 | TRUE | Upregulated |
| 0,40197533 | 0,007002208 | DDOST | FALSE | Upregulated |
| -0,35395609 | 0,007002208 | PSME1 | FALSE | Downregulated |
| 1,841086247 | 0,007002208 | TDRD1 | TRUE | Upregulated |
| -0,437762193 | 0,007006379 | ZBTB39 | FALSE | Downregulated |
| 1,297196102 | 0,007012811 | TRNP1 | TRUE | Upregulated |
| 1,098555466 | 0,007021961 | VIM-AS1 | TRUE | Upregulated |
| 0,22399508 | 0,007031616 | PTBP1 | FALSE | Upregulated |
| 0,248099237 | 0,007032974 | GTF3C1 | FALSE | Upregulated |
| 0,735511608 | 0,00705301 | SEPT1 | FALSE | Upregulated |
| 1,348481732 | 0,007056478 | GNB1L | TRUE | Upregulated |
| 0,380633421 | 0,007056478 | OGFR | FALSE | Upregulated |
| 0,485451664 | 0,007056478 | TUBB4B | FALSE | Upregulated |
| 1,01260012 | 0,007062815 | CD1C | TRUE | Upregulated |
| 0,338254674 | 0,007066069 | ADCY9 | FALSE | Upregulated |
| 0,278766036 | 0,007098207 | WDR26 | FALSE | Upregulated |
| -0,388919043 | 0,007111581 | APOOL | FALSE | Downregulated |
| -0,426671202 | 0,007115012 | LHPP | FALSE | Downregulated |
| 0,656129242 | 0,00712168 | CYBRD1 | FALSE | Upregulated |
| 0,30540029 | 0,007133971 | HS1BP3 | FALSE | Upregulated |
| -0,799108902 | 0,007136561 | FAM129C | FALSE | Downregulated |
| 0,415722912 | 0,007150871 | BCAR1 | FALSE | Upregulated |
| 0,390292078 | 0,00715217 | MAPK3 | FALSE | Upregulated |
| 2,450494478 | 0,007159377 | DGCR9 | TRUE | Upregulated |
| -0,79140697 | 0,007160705 | POLN | FALSE | Downregulated |
| 2,415929307 | 0,007162311 | KIAA1199 | TRUE | Upregulated |
| 0,657006286 | 0,007164915 | ZNF273 | FALSE | Upregulated |
| 0,507282725 | 0,007190025 | ERAP1 | FALSE | Upregulated |
| -0,430468144 | 0,007201775 | CBWD2 | FALSE | Downregulated |
| -2,086510684 | 0,007201775 | RPS14P3 | TRUE | Downregulated |
| -0,617698352 | 0,007205566 | LOC285074 | FALSE | Downregulated |
| 1,468701055 | 0,007209603 | RMI2 | TRUE | Upregulated |
| 0,921981758 | 0,007220477 | CCND1 | FALSE | Upregulated |
| 0,324568375 | 0,007234041 | ICMT | FALSE | Upregulated |
| 0,338362677 | 0,007234041 | MAP1LC3B | FALSE | Upregulated |
| -0,344668953 | 0,007234041 | UBE3A | FALSE | Downregulated |
| 0,694818763 | 0,007235956 | LPCAT4 | FALSE | Upregulated |
| 1,017893794 | 0,007246637 | CXCR6 | TRUE | Upregulated |
| -0,318281109 | 0,007256856 | HNRNPH3 | FALSE | Downregulated |
| 2,167255691 | 0,007259352 | YPEL4 | TRUE | Upregulated |
| 0,436389503 | 0,007275378 | SNX29 | FALSE | Upregulated |
| 0,571098826 | 0,00728597 | RASSF2 | FALSE | Upregulated |
| 1,065561232 | 0,007300456 | CCL4 | TRUE | Upregulated |
| -1,243843569 | 0,007312611 | SNHG9 | TRUE | Downregulated |
| 0,769892272 | 0,007366236 | IL2RB | FALSE | Upregulated |
| -0,226714181 | 0,007378133 | UBTF | FALSE | Downregulated |
| 0,34844097 | 0,007397374 | SOCS4 | FALSE | Upregulated |
| -0,665122263 | 0,007412359 | ADPRM | FALSE | Downregulated |
| 0,479675553 | 0,007416012 | MYO19 | FALSE | Upregulated |
| -0,598802645 | 0,007424177 | ACMSD | FALSE | Downregulated |

| | | | | |
|---|---|---|---|---|
| 0,519124017 | 0,007436676 | ATP8B1 | FALSE | Upregulated |
| 0,449681994 | 0,007437725 | PRKCH | FALSE | Upregulated |
| 1,351171912 | 0,007450496 | CENPK | TRUE | Upregulated |
| 0,271522627 | 0,007450496 | XPNPEP1 | FALSE | Upregulated |
| 0,381914942 | 0,00747783 | NOTCH1 | FALSE | Upregulated |
| 0,203555107 | 0,00747783 | TRPC4AP | FALSE | Upregulated |
| -0,413008782 | 0,007488281 | MINOS1 | FALSE | Downregulated |
| -0,391994361 | 0,007495742 | ALDH6A1 | FALSE | Downregulated |
| -0,453024344 | 0,007495742 | CRLS1 | FALSE | Downregulated |
| 0,331988885 | 0,007495742 | FBXO31 | FALSE | Upregulated |
| -0,386443891 | 0,007497194 | LOC102288414 | FALSE | Downregulated |
| 1,941684894 | 0,007503594 | DTNA | TRUE | Upregulated |
| 1,219276462 | 0,007503594 | UHRF1 | TRUE | Upregulated |
| 3,287736488 | 0,007517124 | LINC00313 | TRUE | Upregulated |
| 2,953422437 | 0,007517124 | NTM | TRUE | Upregulated |
| 0,739199105 | 0,007541195 | TGFBI | FALSE | Upregulated |
| -0,860250978 | 0,007552295 | LOC101927509 | FALSE | Downregulated |
| 0,854769668 | 0,007560358 | WWC1 | FALSE | Upregulated |
| -0,286345046 | 0,007564555 | FCGRT | FALSE | Downregulated |
| -0,362678835 | 0,007595678 | COX5B | FALSE | Downregulated |
| -0,307284073 | 0,007598453 | BCL2L2 | FALSE | Downregulated |
| 0,801836257 | 0,007620463 | STON1 | FALSE | Upregulated |
| 0,617739041 | 0,007622925 | LPCAT1 | FALSE | Upregulated |
| 0,755962694 | 0,007626114 | KIF3C | FALSE | Upregulated |
| 0,757320526 | 0,007627445 | AJUBA | FALSE | Upregulated |
| -0,859177042 | 0,007627445 | GSTM4 | FALSE | Downregulated |
| -3,442119055 | 0,007627674 | MB | TRUE | Downregulated |
| -0,510817327 | 0,007627674 | PANK3 | FALSE | Downregulated |
| 0,864429811 | 0,007666939 | ST13P4 | FALSE | Upregulated |
| 1,593909835 | 0,00766855 | MYH11 | TRUE | Upregulated |
| 0,239751378 | 0,007673202 | PSMD3 | FALSE | Upregulated |
| 1,397832164 | 0,007676262 | CRTAC1 | TRUE | Upregulated |
| -0,369467394 | 0,007685512 | GS1-251I9.4 | FALSE | Downregulated |
| -0,195275964 | 0,007685512 | RAD17 | FALSE | Downregulated |
| 0,297867145 | 0,007690174 | EPAS1 | FALSE | Upregulated |
| -0,232809634 | 0,007690174 | YWHAE | FALSE | Downregulated |
| -0,355411678 | 0,007693833 | AIFM1 | FALSE | Downregulated |
| -0,375479884 | 0,007693833 | MRPS2 | FALSE | Downregulated |
| -0,378416132 | 0,007693833 | SMN1 | FALSE | Downregulated |
| 1,496105828 | 0,007738604 | LOC153684 | TRUE | Upregulated |
| 0,329882003 | 0,007745716 | ZBTB4 | FALSE | Upregulated |
| 1,153432963 | 0,007745972 | LINC00152 | TRUE | Upregulated |
| -0,293701858 | 0,007745972 | RAB8A | FALSE | Downregulated |
| -1,24901151 | 0,007771971 | FAM9B | TRUE | Downregulated |
| 1,789945191 | 0,007771971 | NACAD | TRUE | Upregulated |
| 0,6397861 | 0,00779506 | CDC42EP1 | FALSE | Upregulated |
| 0,343944527 | 0,00779881 | IP6K1 | FALSE | Upregulated |
| 2,109969478 | 0,007807999 | DOK7 | TRUE | Upregulated |
| -0,312809072 | 0,007807999 | TMEM192 | FALSE | Downregulated |
| -0,552356533 | 0,007811628 | IMPA2 | FALSE | Downregulated |
| -0,192065221 | 0,007826488 | CSNK1A1 | FALSE | Downregulated |

| | | | | |
|---|---|---|---|---|
| -0,33496858 | 0,007840767 | PCGF5 | FALSE | Downregulated |
| -0,368487339 | 0,007846786 | BHMT2 | FALSE | Downregulated |
| 0,358405272 | 0,007870907 | MYO1D | FALSE | Upregulated |
| -0,276645035 | 0,007870907 | SRP9 | FALSE | Downregulated |
| 0,331216572 | 0,007871449 | RBM42 | FALSE | Upregulated |
| 2,870372324 | 0,007910324 | CXADRP3 | TRUE | Upregulated |
| -0,324973997 | 0,007924775 | KIF1C | FALSE | Downregulated |
| 0,269503408 | 0,007963563 | KIAA0947 | FALSE | Upregulated |
| 0,391682549 | 0,00797256 | APEX2 | FALSE | Upregulated |
| 1,22016655 | 0,007985105 | C12orf75 | TRUE | Upregulated |
| 0,77256177 | 0,007989902 | AC009948.5 | FALSE | Upregulated |
| 0,56757273 | 0,007989902 | TTF2 | FALSE | Upregulated |
| -0,323619555 | 0,007996235 | TFB2M | FALSE | Downregulated |
| 0,269799577 | 0,007998512 | DHX8 | FALSE | Upregulated |
| -0,283560904 | 0,007999098 | POLR2D | FALSE | Downregulated |
| 0,563762351 | 0,008031308 | KIAA1841 | FALSE | Upregulated |
| 0,300659947 | 0,008031308 | TMEM87B | FALSE | Upregulated |
| -0,32533203 | 0,00803636 | FBXO9 | FALSE | Downregulated |
| -0,474134547 | 0,008050797 | SH3BGRL2 | FALSE | Downregulated |
| -0,321683567 | 0,008078398 | RPS7 | FALSE | Downregulated |
| 0,61090041 | 0,00807946 | CD151 | FALSE | Upregulated |
| -0,464700932 | 0,008088094 | ZSWIM7 | FALSE | Downregulated |
| 1,035884365 | 0,00810091 | VWF | TRUE | Upregulated |
| -0,318410522 | 0,008109747 | SSBP1 | FALSE | Downregulated |
| 1,684802359 | 0,008130487 | NRXN2 | TRUE | Upregulated |
| -0,33095834 | 0,008133183 | CLYBL | FALSE | Downregulated |
| -0,377386553 | 0,008141499 | KLHL24 | FALSE | Downregulated |
| -0,514028549 | 0,008141499 | SRSF10 | FALSE | Downregulated |
| -0,345929341 | 0,008152863 | C14orf28 | FALSE | Downregulated |
| -0,30710043 | 0,008152863 | VAMP4 | FALSE | Downregulated |
| -0,391393906 | 0,008152863 | ZNF445 | FALSE | Downregulated |
| 1,627385644 | 0,008175807 | LINC00925 | TRUE | Upregulated |
| 0,626026812 | 0,008186476 | PALM | FALSE | Upregulated |
| -1,182968173 | 0,008206985 | LOC101927609 | TRUE | Downregulated |
| -0,2515629 | 0,008214452 | MRPL39 | FALSE | Downregulated |
| 2,3379019 | 0,008232755 | MIR635 | TRUE | Upregulated |
| -0,535832784 | 0,008232755 | TRIM52 | FALSE | Downregulated |
| 3,648667979 | 0,008236269 | RP11-394O9.1 | TRUE | Upregulated |
| 0,549628044 | 0,008241313 | GUCY1A3 | FALSE | Upregulated |
| -0,614368176 | 0,008243313 | ZNF829 | FALSE | Downregulated |
| 2,276491275 | 0,008253433 | OR2A9P | TRUE | Upregulated |
| -0,327745886 | 0,008263896 | CDK14 | FALSE | Downregulated |
| 0,492936363 | 0,008269284 | STK10 | FALSE | Upregulated |
| 0,335034747 | 0,008287318 | IDH3G | FALSE | Upregulated |
| 0,293369666 | 0,00828795 | SUFU | FALSE | Upregulated |
| -0,455487252 | 0,00829451 | ZNF688 | FALSE | Downregulated |
| 0,515409045 | 0,008302275 | GNG11 | FALSE | Upregulated |
| -1,485672256 | 0,008302275 | LOC101929561 | TRUE | Downregulated |
| -0,325370304 | 0,008302275 | XPA | FALSE | Downregulated |
| -0,467732808 | 0,008310659 | HECTD3 | FALSE | Downregulated |
| -0,324574531 | 0,008310659 | SNX13 | FALSE | Downregulated |

| | | | | |
|---|---|---|---|---|
| 1,381310538 | 0,008311632 | FAM167A | TRUE | Upregulated |
| 3,6042025 | 0,008314436 | MGC50722 | TRUE | Upregulated |
| 1,101571121 | 0,008314827 | P2RY10 | TRUE | Upregulated |
| 0,386861993 | 0,008316479 | TRANK1 | FALSE | Upregulated |
| -0,96579421 | 0,008359349 | ASXL3 | FALSE | Downregulated |
| -0,292089355 | 0,008359349 | DIMT1 | FALSE | Downregulated |
| -0,705259732 | 0,008359349 | HPGD | FALSE | Downregulated |
| -0,46986725 | 0,008360734 | C19orf25 | FALSE | Downregulated |
| 1,634201004 | 0,008362752 | PITRM1-AS1 | TRUE | Upregulated |
| 0,538676557 | 0,008364184 | PIK3IP1 | FALSE | Upregulated |
| 0,243306952 | 0,00838022 | CLTC | FALSE | Upregulated |
| 0,77103051 | 0,008384927 | GATS | FALSE | Upregulated |
| -1,210131191 | 0,008387415 | INTS6-AS1 | TRUE | Downregulated |
| -0,46098727 | 0,008387415 | MGMT | FALSE | Downregulated |
| 0,371641845 | 0,008413848 | ELMO2 | FALSE | Upregulated |
| -0,304977286 | 0,008420613 | SENP2 | FALSE | Downregulated |
| -0,350206967 | 0,008431368 | SMARCD2 | FALSE | Downregulated |
| 0,747840827 | 0,008443286 | RAMP1 | FALSE | Upregulated |
| -0,299995667 | 0,00845113 | KCTD3 | FALSE | Downregulated |
| 0,745362488 | 0,008453634 | LTBP4 | FALSE | Upregulated |
| -0,456564282 | 0,008453634 | SRD5A1 | FALSE | Downregulated |
| 0,410269116 | 0,008454576 | TMEM2 | FALSE | Upregulated |
| -0,372597595 | 0,008485939 | ZNF254 | FALSE | Downregulated |
| -0,326518696 | 0,008501179 | PPA2 | FALSE | Downregulated |
| -0,518488512 | 0,008505521 | LINC00667 | FALSE | Downregulated |
| -0,509006996 | 0,008505552 | C2orf88 | FALSE | Downregulated |
| -0,222707984 | 0,008511857 | AFG3L2 | FALSE | Downregulated |
| 1,026224225 | 0,008535678 | LOC101927903 | TRUE | Upregulated |
| 0,663312806 | 0,008552505 | ZNF28 | FALSE | Upregulated |
| 0,292341276 | 0,008585958 | APEH | FALSE | Upregulated |
| 0,287668583 | 0,008586769 | MAEA | FALSE | Upregulated |
| -0,681816679 | 0,008589171 | NSDHL | FALSE | Downregulated |
| -0,376645234 | 0,008598837 | ABCA6 | FALSE | Downregulated |
| -1,884899152 | 0,008605656 | EYA4 | TRUE | Downregulated |
| -1,159416821 | 0,008613869 | LINC01176 | TRUE | Downregulated |
| 2,745729218 | 0,008622087 | MKRN9P | TRUE | Upregulated |
| -1,333509579 | 0,008622087 | SQLE | TRUE | Downregulated |
| 1,311248597 | 0,008629092 | CDKN3 | TRUE | Upregulated |
| 0,322690962 | 0,008660401 | CORO1C | FALSE | Upregulated |
| 1,593601458 | 0,008660401 | RASD1 | TRUE | Upregulated |
| 1,009789187 | 0,008667679 | RASGRF2 | TRUE | Upregulated |
| -0,4793892 | 0,008669118 | CCDC69 | FALSE | Downregulated |
| 0,693031867 | 0,008669118 | GAS5 | FALSE | Upregulated |
| 0,94729156 | 0,008669118 | GBP3 | FALSE | Upregulated |
| 0,408478927 | 0,008669118 | PFKFB2 | FALSE | Upregulated |
| 0,242702338 | 0,008669118 | RALGAPB | FALSE | Upregulated |
| 2,079801069 | 0,008669118 | TLDC2 | TRUE | Upregulated |
| 0,923979691 | 0,008670338 | CENPU | FALSE | Upregulated |
| 0,925339566 | 0,00867842 | CCDC102A | FALSE | Upregulated |
| -0,410799179 | 0,00867842 | TMEM170B | FALSE | Downregulated |
| -0,306168609 | 0,00867842 | TOLLIP | FALSE | Downregulated |

| | | | | |
|---|---|---|---|---|
| 1,133972766 | 0,008699281 | LRRC70 | TRUE | Upregulated |
| 0,598594631 | 0,008705286 | RHOQ | FALSE | Upregulated |
| 0,717372427 | 0,008718401 | FANCI | FALSE | Upregulated |
| 1,556737885 | 0,008718401 | TTC16 | TRUE | Upregulated |
| 2,166009399 | 0,008730245 | FCRL2 | TRUE | Upregulated |
| 0,219861678 | 0,00876899 | ARF6 | FALSE | Upregulated |
| 1,028930835 | 0,008792854 | LRCOL1 | TRUE | Upregulated |
| 0,313422809 | 0,008793335 | STX12 | FALSE | Upregulated |
| -0,364544015 | 0,008799472 | JTB | FALSE | Downregulated |
| 0,460479847 | 0,008806001 | NAA40 | FALSE | Upregulated |
| -0,591368534 | 0,008806001 | PSMD7 | FALSE | Downregulated |
| -0,969746462 | 0,008826081 | LAMTOR5-AS1 | FALSE | Downregulated |
| 0,598889486 | 0,008831625 | AKR1B1 | FALSE | Upregulated |
| 0,78957209 | 0,008831625 | SLAMF6 | FALSE | Upregulated |
| 0,65876565 | 0,008848415 | ARHGAP9 | FALSE | Upregulated |
| 0,540733906 | 0,008848415 | ATP13A2 | FALSE | Upregulated |
| -0,810000663 | 0,00886162 | LINC00844 | FALSE | Downregulated |
| 0,748984262 | 0,008908364 | PDGFB | FALSE | Upregulated |
| 2,178902456 | 0,008911988 | EPHX3 | TRUE | Upregulated |
| -0,529351481 | 0,008911988 | RPL39 | FALSE | Downregulated |
| -0,292176159 | 0,008913765 | ADSL | FALSE | Downregulated |
| 1,584935585 | 0,008928381 | OIP5 | TRUE | Upregulated |
| -1,440930074 | 0,008928381 | TRNA | TRUE | Downregulated |
| -0,416398275 | 0,008939455 | THUMPD1 | FALSE | Downregulated |
| -0,941290391 | 0,008952199 | MT2A | FALSE | Downregulated |
| -0,329694085 | 0,008952199 | TADA1 | FALSE | Downregulated |
| -0,347221127 | 0,009025872 | CAMK2G | FALSE | Downregulated |
| 2,491810673 | 0,009041882 | SNORA24 | TRUE | Upregulated |
| 0,709378397 | 0,009068949 | KIF21B | FALSE | Upregulated |
| 0,677840955 | 0,009071171 | MATK | FALSE | Upregulated |
| 2,501598346 | 0,009098809 | CAPN11 | TRUE | Upregulated |
| -1,062707005 | 0,009124584 | LINC00238 | TRUE | Downregulated |
| -0,325017385 | 0,009139437 | MED28 | FALSE | Downregulated |
| 2,029802814 | 0,009154237 | VASH2 | TRUE | Upregulated |
| -0,511636108 | 0,009174669 | RP11-983P16.4 | FALSE | Downregulated |
| 0,345378426 | 0,009177747 | RRBP1 | FALSE | Upregulated |
| 0,296972298 | 0,009196272 | MOB4 | FALSE | Upregulated |
| -0,542924016 | 0,009244567 | FAM107B | FALSE | Downregulated |
| 2,114984324 | 0,0092539 | LOC158434 | TRUE | Upregulated |
| 0,433074872 | 0,009255013 | ASCC3 | FALSE | Upregulated |
| -1,132482203 | 0,009255013 | LOC102724559 | TRUE | Downregulated |
| 0,312556678 | 0,009269477 | PCSK7 | FALSE | Upregulated |
| -0,309103428 | 0,009277503 | ACOT8 | FALSE | Downregulated |
| 2,843007113 | 0,009281141 | CHGB | TRUE | Upregulated |
| 2,123848548 | 0,009281141 | WFDC2 | TRUE | Upregulated |
| -0,377661743 | 0,009336193 | LYRM1 | FALSE | Downregulated |
| 2,351124467 | 0,009336193 | RIBC2 | TRUE | Upregulated |
| 0,339899975 | 0,009342233 | SHKBP1 | FALSE | Upregulated |
| 0,878455869 | 0,00935292 | FBN1 | FALSE | Upregulated |
| 0,519970166 | 0,00935292 | IFI16 | FALSE | Upregulated |
| -0,296893755 | 0,00935292 | PSMA2 | FALSE | Downregulated |

| | | | | |
|---|---|---|---|---|
| 0,635022631 | 0,00935292 | UBASH3B | FALSE | Upregulated |
| -2,863486667 | 0,009355277 | MURC | TRUE | Downregulated |
| 0,947115693 | 0,009361541 | CDK1 | FALSE | Upregulated |
| -0,594339247 | 0,009361541 | TMEM161B-AS1 | FALSE | Downregulated |
| 1,303528007 | 0,009399799 | LOC644961 | TRUE | Upregulated |
| 0,564340678 | 0,009424911 | TMEM102 | FALSE | Upregulated |
| 0,642407608 | 0,009432456 | TTC21A | FALSE | Upregulated |
| -0,36963156 | 0,009436016 | KDELC1 | FALSE | Downregulated |
| 1,760593658 | 0,0094948 | FAM155B | TRUE | Upregulated |
| 1,167357999 | 0,009496755 | HJURP | TRUE | Upregulated |
| 0,797063674 | 0,00950059 | DBNDD2 | FALSE | Upregulated |
| -0,564386853 | 0,00950059 | VIPR1 | FALSE | Downregulated |
| 0,583025997 | 0,009525412 | PLTP | FALSE | Upregulated |
| 0,638845079 | 0,009525456 | ARHGAP23 | FALSE | Upregulated |
| 0,65216008 | 0,009527276 | NLRC3 | FALSE | Upregulated |
| -0,671802544 | 0,009552795 | ETNK2 | FALSE | Downregulated |
| -0,285418537 | 0,009552795 | HDGF | FALSE | Downregulated |
| 1,798099558 | 0,009552795 | PTX3 | TRUE | Upregulated |
| 1,24440767 | 0,009554574 | COLCA2 | TRUE | Upregulated |
| 0,522086747 | 0,009554574 | KIAA0754 | FALSE | Upregulated |
| 1,836418149 | 0,009554678 | GPRIN1 | TRUE | Upregulated |
| -0,411676038 | 0,009581954 | MANEA | FALSE | Downregulated |
| 1,944830215 | 0,009588159 | CPXM1 | TRUE | Upregulated |
| -0,249277247 | 0,009588159 | PTEN | FALSE | Downregulated |
| 0,215082896 | 0,009597621 | PPP6R1 | FALSE | Upregulated |
| -0,337378443 | 0,009597621 | SLC25A26 | FALSE | Downregulated |
| 0,262336149 | 0,009599546 | SLC4A2 | FALSE | Upregulated |
| 0,299437327 | 0,009599546 | TRRAP | FALSE | Upregulated |
| -0,576073885 | 0,009630795 | BMP4 | FALSE | Downregulated |
| 1,123437922 | 0,009630795 | MS4A1 | TRUE | Upregulated |
| -0,268858877 | 0,009630795 | NARS2 | FALSE | Downregulated |
| 1,027801076 | 0,009645321 | TVP23A | TRUE | Upregulated |
| -0,594818673 | 0,009683135 | PACSIN3 | FALSE | Downregulated |
| -0,334547066 | 0,009698111 | FAM120C | FALSE | Downregulated |
| 1,342189182 | 0,009698429 | ADD2 | TRUE | Upregulated |
| 0,51405177 | 0,009698429 | RAB3IP | FALSE | Upregulated |
| -0,469358615 | 0,009698429 | USP13 | FALSE | Downregulated |
| -0,737689267 | 0,009718661 | GADD45A | FALSE | Downregulated |
| 1,168919235 | 0,009730787 | NTRK2 | TRUE | Upregulated |
| 0,894568997 | 0,009730787 | PARP15 | FALSE | Upregulated |
| -0,308041716 | 0,009730787 | SLC35D1 | FALSE | Downregulated |
| 0,687190725 | 0,009738336 | SH3YL1 | FALSE | Upregulated |
| -0,605068451 | 0,009740237 | ZNF444 | FALSE | Downregulated |
| -0,525524402 | 0,009750359 | UBL7-AS1 | FALSE | Downregulated |
| 0,709967239 | 0,009750495 | ZNF43 | FALSE | Upregulated |
| 0,960334688 | 0,009750752 | GRAP2 | FALSE | Upregulated |
| 2,526723838 | 0,009810899 | CDC14C | TRUE | Upregulated |
| 0,897284073 | 0,009811167 | NHS | FALSE | Upregulated |
| -0,246595584 | 0,009829216 | REXO4 | FALSE | Downregulated |
| -0,279788374 | 0,009837388 | MRPL42 | FALSE | Downregulated |
| -0,45802817 | 0,009839446 | ELL2 | FALSE | Downregulated |

| | | | | |
|---|---|---|---|---|
| 0,804790519 | 0,009847571 | CLIP3 | FALSE | Upregulated |
| -0,267261128 | 0,009855365 | TMEM5 | FALSE | Downregulated |
| -0,342418724 | 0,009868684 | SNAPC1 | FALSE | Downregulated |
| -0,288527636 | 0,009960306 | RBP4 | FALSE | Downregulated |
| 0,785829768 | 0,009960306 | RNF122 | FALSE | Upregulated |
| 0,976425912 | 0,009964533 | ITM2A | FALSE | Upregulated |
| 0,264629136 | 0,009964533 | UBIAD1 | FALSE | Upregulated |
| 0,464126097 | 0,009979739 | ZNF765 | FALSE | Upregulated |
| 1,907681844 | 0,009980334 | CXCL3 | TRUE | Upregulated |
| 0,595170479 | 0,009980334 | UST | FALSE | Upregulated |
| 0,555019488 | 0,009993097 | FER | FALSE | Upregulated |
| 1,369305535 | 0,01002171 | SNAP25 | TRUE | Upregulated |
| 1,912339614 | 0,010022804 | FBXL13 | TRUE | Upregulated |
| -0,327758427 | 0,010022804 | WAC-AS1 | FALSE | Downregulated |
| 0,282530192 | 0,010024592 | GATAD2A | FALSE | Upregulated |
| -0,242053749 | 0,010034338 | MRS2 | FALSE | Downregulated |
| 0,224555909 | 0,010060161 | UBA1 | FALSE | Upregulated |
| 2,328532003 | 0,0101005 | POTEE | TRUE | Upregulated |
| 0,722761626 | 0,010107242 | MSRB3 | FALSE | Upregulated |
| -0,679165778 | 0,010114655 | SYT1 | FALSE | Downregulated |
| 1,394838295 | 0,010126399 | GAL3ST4 | TRUE | Upregulated |
| 0,7770414 | 0,01014037 | FUT6 | FALSE | Upregulated |
| 0,431926341 | 0,010141185 | BRMS1 | FALSE | Upregulated |
| 0,706745899 | 0,010153035 | PHLDA1 | FALSE | Upregulated |
| 0,309286073 | 0,010165587 | NOMO1 | FALSE | Upregulated |
| -0,311756863 | 0,010165603 | CYB5A | FALSE | Downregulated |
| -0,657201806 | 0,010184932 | ARHGAP20 | FALSE | Downregulated |
| -0,439191451 | 0,010186627 | C19orf66 | FALSE | Downregulated |
| -0,370966798 | 0,010187744 | RPL38 | FALSE | Downregulated |
| -0,44044899 | 0,01021201 | SMIM4 | FALSE | Downregulated |
| 0,444959652 | 0,01023414 | GNG2 | FALSE | Upregulated |
| -0,329683114 | 0,01023414 | PHLDB2 | FALSE | Downregulated |
| -0,384684361 | 0,01023414 | PTK2B | FALSE | Downregulated |
| -0,321829803 | 0,010259719 | RNF168 | FALSE | Downregulated |
| 0,64600208 | 0,010259719 | SEMA3F | FALSE | Upregulated |
| 4,21210722 | 0,010272218 | FGF19 | TRUE | Upregulated |
| 2,278687715 | 0,010285903 | GALNT5 | TRUE | Upregulated |
| -0,241696726 | 0,010286498 | SAR1B | FALSE | Downregulated |
| 0,525194337 | 0,010312552 | GLIPR1 | FALSE | Upregulated |
| -0,854731193 | 0,010314846 | DHRS4L1 | FALSE | Downregulated |
| -0,319102055 | 0,010314846 | GGNBP2 | FALSE | Downregulated |
| 0,299079693 | 0,010315864 | CAMSAP1 | FALSE | Upregulated |
| -1,099460009 | 0,010323613 | LOC100505985 | TRUE | Downregulated |
| 0,355343029 | 0,010324811 | TRAF3 | FALSE | Upregulated |
| 0,738830388 | 0,010333529 | ZNF439 | FALSE | Upregulated |
| -0,289675884 | 0,01035866 | CTAGE5 | FALSE | Downregulated |
| 0,319079602 | 0,010361381 | CCT2 | FALSE | Upregulated |
| 0,565336407 | 0,010361381 | MEGF6 | FALSE | Upregulated |
| 0,36731217 | 0,010371835 | WWTR1 | FALSE | Upregulated |
| -0,353761999 | 0,010376965 | THRAP3 | FALSE | Downregulated |
| -0,505299193 | 0,010382815 | SYDE2 | FALSE | Downregulated |

| | | | | |
|---|---|---|---|---|
| 0,970207971 | 0,010420495 | ACAP1 | FALSE | Upregulated |
| 0,46362009 | 0,010420495 | AKAP7 | FALSE | Upregulated |
| 0,492104516 | 0,010420495 | GNPDA2 | FALSE | Upregulated |
| -0,967189971 | 0,010420495 | SULT1E1 | FALSE | Downregulated |
| 0,415734807 | 0,010442625 | GORAB | FALSE | Upregulated |
| -0,614071537 | 0,010442625 | PPP1R1C | FALSE | Downregulated |
| 0,776505845 | 0,010450008 | BBC3 | FALSE | Upregulated |
| 0,352913471 | 0,010470726 | ZNF646 | FALSE | Upregulated |
| 0,261970935 | 0,010480961 | TAF6 | FALSE | Upregulated |
| 1,84743823 | 0,010483894 | SETSIP | TRUE | Upregulated |
| -0,26144595 | 0,010493705 | NDFIP1 | FALSE | Downregulated |
| -0,377241141 | 0,01050327 | C1R | FALSE | Downregulated |
| 0,242960715 | 0,01050327 | FKBP1A | FALSE | Upregulated |
| 1,745070519 | 0,01051201 | TFF3 | TRUE | Upregulated |
| 1,016063127 | 0,010514607 | ARL10 | TRUE | Upregulated |
| 0,316398799 | 0,010514607 | GTPBP1 | FALSE | Upregulated |
| -0,279977218 | 0,010514607 | RRP8 | FALSE | Downregulated |
| -0,321074813 | 0,010540916 | CCDC51 | FALSE | Downregulated |
| -0,419526326 | 0,010581782 | FAM214A | FALSE | Downregulated |
| 1,481419977 | 0,010616656 | NRCAM | TRUE | Upregulated |
| -0,474937131 | 0,010629971 | CD55 | FALSE | Downregulated |
| 1,069042317 | 0,010636057 | DOC2B | TRUE | Upregulated |
| 0,540974683 | 0,010636057 | MMD | FALSE | Upregulated |
| 0,905996884 | 0,010654837 | LZTS1 | FALSE | Upregulated |
| 1,385417658 | 0,010654837 | SPC25 | TRUE | Upregulated |
| 0,263815787 | 0,010663739 | WDFY1 | FALSE | Upregulated |
| 1,861562201 | 0,01066797 | CDKN2A | TRUE | Upregulated |
| 2,010984854 | 0,01066797 | MGAT4C | TRUE | Upregulated |
| -1,20202331 | 0,010669934 | LINC01171 | TRUE | Downregulated |
| -0,311116251 | 0,010693134 | PSMA7 | FALSE | Downregulated |
| 0,19249535 | 0,010701784 | PHRF1 | FALSE | Upregulated |
| 1,516260228 | 0,010707686 | C16orf59 | TRUE | Upregulated |
| 0,599277658 | 0,010719733 | TMEM107 | FALSE | Upregulated |
| 0,305887583 | 0,010732206 | IKBKB | FALSE | Upregulated |
| -0,657675542 | 0,010732206 | MGC72080 | FALSE | Downregulated |
| 0,777190741 | 0,010734948 | LOXL3 | FALSE | Upregulated |
| -0,301054549 | 0,010734948 | PCF11 | FALSE | Downregulated |
| 0,169115216 | 0,010747945 | GRSF1 | FALSE | Upregulated |
| 0,537212871 | 0,010747945 | HSPB1 | FALSE | Upregulated |
| 0,277510944 | 0,010747945 | NIPA2 | FALSE | Upregulated |
| -0,391575297 | 0,010768143 | DNPH1 | FALSE | Downregulated |
| -0,311885563 | 0,010778673 | FAM185A | FALSE | Downregulated |
| 1,474845684 | 0,010792522 | EXO1 | TRUE | Upregulated |
| 0,427803246 | 0,010792522 | HYAL2 | FALSE | Upregulated |
| 0,391423012 | 0,010801472 | SEC16A | FALSE | Upregulated |
| -0,534257747 | 0,010804149 | ANP32B | FALSE | Downregulated |
| 0,319698233 | 0,010842626 | MMGT1 | FALSE | Upregulated |
| -0,548099397 | 0,010850956 | GLDC | FALSE | Downregulated |
| -0,251915221 | 0,010853906 | AUP1 | FALSE | Downregulated |
| -0,269875686 | 0,010853906 | AZIN1 | FALSE | Downregulated |
| 2,745662061 | 0,010853906 | HOXD-AS1 | TRUE | Upregulated |

| | | | | |
|---|---|---|---|---|
| 0,773817356 | 0,010853906 | KIAA0226L | FALSE | Upregulated |
| 0,752851819 | 0,010853906 | TTLL1 | FALSE | Upregulated |
| -0,441267885 | 0,010853906 | UGT2B15 | FALSE | Downregulated |
| 2,013021576 | 0,010853906 | WBP11P1 | TRUE | Upregulated |
| 0,713989822 | 0,010853906 | WDHD1 | FALSE | Upregulated |
| -0,644716407 | 0,010853906 | XRCC6BP1 | FALSE | Downregulated |
| 0,294254885 | 0,010861043 | NOTCH2 | FALSE | Upregulated |
| 0,414845107 | 0,010861043 | PJA1 | FALSE | Upregulated |
| -0,215689068 | 0,010861043 | RPP14 | FALSE | Downregulated |
| 0,965708943 | 0,010884456 | RECQL4 | FALSE | Upregulated |
| -0,226022221 | 0,01089621 | SCO1 | FALSE | Downregulated |
| 0,692385718 | 0,010912453 | KITLG | FALSE | Upregulated |
| 1,04670814 | 0,010915867 | KIAA1377 | TRUE | Upregulated |
| 2,253176576 | 0,010922279 | PHLDA2 | TRUE | Upregulated |
| -0,831563792 | 0,010942978 | PPP1R3C | FALSE | Downregulated |
| 0,418899301 | 0,010947363 | RASSF4 | FALSE | Upregulated |
| 1,188304407 | 0,010948632 | ATL1 | TRUE | Upregulated |
| -1,121359342 | 0,010948632 | LRRTM2 | TRUE | Downregulated |
| 3,066665312 | 0,010963895 | HAPLN2 | TRUE | Upregulated |
| 0,31293812 | 0,010963895 | PREP | FALSE | Upregulated |
| 0,364093297 | 0,010966009 | COPG2 | FALSE | Upregulated |
| 0,992308638 | 0,010966009 | ELF3 | FALSE | Upregulated |
| 0,265675394 | 0,010966009 | PPP2R1A | FALSE | Upregulated |
| 2,000305262 | 0,010977101 | PABPC1P2 | TRUE | Upregulated |
| -1,015029734 | 0,010977101 | TIMD4 | TRUE | Downregulated |
| 0,635095809 | 0,010992134 | CELF2 | FALSE | Upregulated |
| -0,315254722 | 0,010992134 | SMS | FALSE | Downregulated |
| -0,38746603 | 0,010993074 | C1orf168 | FALSE | Downregulated |
| 1,098045942 | 0,011017723 | BHLHE22 | TRUE | Upregulated |
| -0,433252381 | 0,011062231 | RBFOX2 | FALSE | Downregulated |
| -2,078912321 | 0,011062344 | ANKRD23 | TRUE | Downregulated |
| -0,233896985 | 0,01108333 | YY1AP1 | FALSE | Downregulated |
| 0,235250474 | 0,011110986 | RCOR1 | FALSE | Upregulated |
| -0,390067845 | 0,011113285 | EHHADH | FALSE | Downregulated |
| -0,445166089 | 0,011113285 | ZFAND2A | FALSE | Downregulated |
| -0,89866103 | 0,011149588 | FABP3 | FALSE | Downregulated |
| 1,623916742 | 0,011152998 | SPDYE5 | TRUE | Upregulated |
| 2,001365872 | 0,011157586 | FOXL1 | TRUE | Upregulated |
| -0,365932401 | 0,011159319 | GKAP1 | FALSE | Downregulated |
| -0,282671574 | 0,011163029 | CDKN2AIP | FALSE | Downregulated |
| 0,576559041 | 0,011169386 | MCM4 | FALSE | Upregulated |
| 0,230922982 | 0,011179778 | DAP | FALSE | Upregulated |
| 0,922609743 | 0,011186518 | ZAP70 | FALSE | Upregulated |
| -0,25002338 | 0,011214366 | TMTC4 | FALSE | Downregulated |
| -1,074801146 | 0,01121742 | RP11-75N4.2 | TRUE | Downregulated |
| -0,392366751 | 0,011223989 | SMARCA2 | FALSE | Downregulated |
| 0,199067714 | 0,011256748 | GAK | FALSE | Upregulated |
| 0,901632733 | 0,011256748 | LOC100093631 | FALSE | Upregulated |
| 0,394959654 | 0,011256748 | ZNF286A | FALSE | Upregulated |
| 1,07623985 | 0,011289473 | TSPAN2 | TRUE | Upregulated |
| 0,852501838 | 0,011298182 | TIGIT | FALSE | Upregulated |

| | | | | |
|---|---|---|---|---|
| 1,339855018 | 0,011305073 | B3GNT3 | TRUE | Upregulated |
| 0,576103174 | 0,011305073 | FBXL7 | FALSE | Upregulated |
| 0,460990494 | 0,011305073 | ZNF235 | FALSE | Upregulated |
| -0,231246992 | 0,011308147 | C5orf15 | FALSE | Downregulated |
| -0,909021559 | 0,011309601 | RP11-279F6.1 | FALSE | Downregulated |
| 0,485889619 | 0,011313051 | NIN | FALSE | Upregulated |
| 2,111897098 | 0,011314602 | YWHAEP1 | TRUE | Upregulated |
| 0,694773387 | 0,011335758 | ATF3 | FALSE | Upregulated |
| 0,441244881 | 0,011349945 | PAPOLG | FALSE | Upregulated |
| 0,381245139 | 0,01135519 | ATR | FALSE | Upregulated |
| 1,005517736 | 0,01136436 | GBGT1 | TRUE | Upregulated |
| -0,361675693 | 0,01136436 | PQLC1 | FALSE | Downregulated |
| 1,317785783 | 0,011402576 | CDH13 | TRUE | Upregulated |
| -0,303652882 | 0,011417223 | SETD8 | FALSE | Downregulated |
| -0,367893231 | 0,011424176 | HDAC8 | FALSE | Downregulated |
| -0,369428696 | 0,011425166 | CCDC121 | FALSE | Downregulated |
| 0,457226235 | 0,011455394 | PARP12 | FALSE | Upregulated |
| -0,342221265 | 0,011461321 | PRPF40A | FALSE | Downregulated |
| 1,860158697 | 0,011466596 | PIFO | TRUE | Upregulated |
| -0,712312191 | 0,011469061 | PKLR | FALSE | Downregulated |
| 0,596182313 | 0,01148974 | RIPK4 | FALSE | Upregulated |
| 0,700384362 | 0,011501218 | KRBA1 | FALSE | Upregulated |
| 0,638926262 | 0,011501218 | TUBB2A | FALSE | Upregulated |
| 0,278987249 | 0,011512654 | GPS1 | FALSE | Upregulated |
| 0,225201232 | 0,011512654 | NRAS | FALSE | Upregulated |
| 2,272078967 | 0,011523233 | LOC102723559 | TRUE | Upregulated |
| -0,3808992 | 0,011523233 | LYAR | FALSE | Downregulated |
| -0,243556844 | 0,011533504 | MRPL3 | FALSE | Downregulated |
| 1,898166123 | 0,011566423 | PLEKHN1 | TRUE | Upregulated |
| -0,652912942 | 0,011587608 | LINC01018 | FALSE | Downregulated |
| 0,774486479 | 0,011617357 | MCTP1 | FALSE | Upregulated |
| 0,587022077 | 0,011617357 | MEAF6 | FALSE | Upregulated |
| -0,333000349 | 0,011621188 | THAP9 | FALSE | Downregulated |
| -0,299406794 | 0,011630301 | LSM6 | FALSE | Downregulated |
| 0,351156784 | 0,011651144 | AP5B1 | FALSE | Upregulated |
| 0,652632887 | 0,011651144 | FGGY | FALSE | Upregulated |
| 0,487743783 | 0,011651144 | RIC8B | FALSE | Upregulated |
| -0,261180327 | 0,011660445 | DPY30 | FALSE | Downregulated |
| 1,46582539 | 0,011681674 | MCM10 | TRUE | Upregulated |
| -0,385071234 | 0,011681674 | TLK1 | FALSE | Downregulated |
| 0,849396304 | 0,011687821 | MXRA8 | FALSE | Upregulated |
| 0,813030717 | 0,011700156 | MALL | FALSE | Upregulated |
| 1,808006911 | 0,01170482 | FAM154B | TRUE | Upregulated |
| 0,37599114 | 0,011721653 | SPATA2 | FALSE | Upregulated |
| 0,433720155 | 0,011741894 | DGKQ | FALSE | Upregulated |
| 0,335699775 | 0,011746632 | SETD1B | FALSE | Upregulated |
| 1,07201637 | 0,011746632 | SHCBP1 | TRUE | Upregulated |
| 0,784975099 | 0,011746632 | ZNF888 | FALSE | Upregulated |
| -0,829751479 | 0,011760766 | AC144449.1 | FALSE | Downregulated |
| -0,825834135 | 0,011760766 | CNDP1 | FALSE | Downregulated |
| 0,881226638 | 0,011781579 | MAP4K1 | FALSE | Upregulated |

| | | | | |
|---|---|---|---|---|
| 0,727206635 | 0,011789974 | LDOC1 | FALSE | Upregulated |
| 1,001941517 | 0,011795402 | ATP13A4 | TRUE | Upregulated |
| 1,561939611 | 0,011795402 | CNN1 | TRUE | Upregulated |
| 1,29138737 | 0,011804133 | RPL13AP6 | TRUE | Upregulated |
| -0,403837383 | 0,011806921 | UBL5 | FALSE | Downregulated |
| -0,709391252 | 0,011814367 | PITPNM3 | FALSE | Downregulated |
| -0,895808182 | 0,011819476 | LOC102546229 | FALSE | Downregulated |
| 0,732080469 | 0,011829857 | FIBIN | FALSE | Upregulated |
| -0,41575398 | 0,011838071 | KIAA1143 | FALSE | Downregulated |
| 0,261809359 | 0,011839038 | PXN | FALSE | Upregulated |
| 0,49433463 | 0,011844339 | ZNF563 | FALSE | Upregulated |
| 1,370923614 | 0,011864442 | POLQ | TRUE | Upregulated |
| -0,668355703 | 0,01186813 | GBA3 | FALSE | Downregulated |
| 0,300354718 | 0,01188466 | NUPL1 | FALSE | Upregulated |
| -0,457091879 | 0,011889979 | SLC2A8 | FALSE | Downregulated |
| 1,800932357 | 0,011905003 | N4BP3 | TRUE | Upregulated |
| 0,297191246 | 0,011913096 | VPS18 | FALSE | Upregulated |
| 2,126319299 | 0,011917552 | MMEL1 | TRUE | Upregulated |
| -0,187835645 | 0,011919054 | AP3S1 | FALSE | Downregulated |
| 0,626179038 | 0,011919054 | SFMBT2 | FALSE | Upregulated |
| 0,42132796 | 0,011935241 | PLXNA2 | FALSE | Upregulated |
| 0,316000767 | 0,011946628 | SRSF2 | FALSE | Upregulated |
| 0,342647405 | 0,012012335 | CNN3 | FALSE | Upregulated |
| -0,456583731 | 0,012015274 | GBE1 | FALSE | Downregulated |
| 0,498601863 | 0,012036106 | CTSS | FALSE | Upregulated |
| 1,21900781 | 0,01205585 | SNAP25-AS1 | TRUE | Upregulated |
| 2,738450964 | 0,012068736 | GSTM2P1 | TRUE | Upregulated |
| -0,285270124 | 0,012068736 | PGRMC2 | FALSE | Downregulated |
| 0,298358765 | 0,012068736 | ZFP36L2 | FALSE | Upregulated |
| 0,726813777 | 0,012072819 | ISYNA1 | FALSE | Upregulated |
| -0,636585258 | 0,012081347 | SC5D | FALSE | Downregulated |
| 1,362289978 | 0,012081347 | TMED10P1 | TRUE | Upregulated |
| -0,534358034 | 0,012081347 | TP53TG1 | FALSE | Downregulated |
| 0,783018331 | 0,012095133 | MICB | FALSE | Upregulated |
| 0,41672005 | 0,012100407 | ACLY | FALSE | Upregulated |
| -0,431434061 | 0,012150194 | PHACTR4 | FALSE | Downregulated |
| 1,764343978 | 0,012162536 | GRIN2A | TRUE | Upregulated |
| -1,086523658 | 0,012162536 | LOC100506675 | TRUE | Downregulated |
| -0,252040501 | 0,012164056 | EIF2B5 | FALSE | Downregulated |
| -0,38453499 | 0,012187005 | CNNM2 | FALSE | Downregulated |
| -0,521584392 | 0,012187005 | FCGR2B | FALSE | Downregulated |
| -0,330936714 | 0,012187005 | MSRB2 | FALSE | Downregulated |
| -0,906973382 | 0,012187005 | OAT | FALSE | Downregulated |
| -0,38412241 | 0,012187005 | OCEL1 | FALSE | Downregulated |
| 0,421826706 | 0,012196411 | SSBP3 | FALSE | Upregulated |
| -0,289999737 | 0,012212964 | AKT1S1 | FALSE | Downregulated |
| 0,589721639 | 0,012214694 | GBP2 | FALSE | Upregulated |
| 1,23295169 | 0,012214694 | IL1B | TRUE | Upregulated |
| -0,231816438 | 0,012232677 | LANCL1 | FALSE | Downregulated |
| -0,245967206 | 0,012232677 | USP48 | FALSE | Downregulated |
| -0,334497371 | 0,012248123 | RPL37A | FALSE | Downregulated |

| | | | | |
|---|---|---|---|---|
| -0,5033376 | 0,012268655 | SKAP1 | FALSE | Downregulated |
| 0,620328542 | 0,01228782 | TMOD2 | FALSE | Upregulated |
| 0,681947943 | 0,012303627 | TPBG | FALSE | Upregulated |
| 0,939734341 | 0,012321523 | CD3D | FALSE | Upregulated |
| -0,373122261 | 0,012334356 | RPS15 | FALSE | Downregulated |
| -0,288078492 | 0,012354332 | ATP5C1 | FALSE | Downregulated |
| -1,173172791 | 0,012358109 | TENM3 | TRUE | Downregulated |
| 0,287982176 | 0,012389472 | C22orf29 | FALSE | Upregulated |
| -0,308699901 | 0,012424632 | LDHD | FALSE | Downregulated |
| 2,336220231 | 0,012455905 | GXYLT2 | TRUE | Upregulated |
| -0,324394881 | 0,012463549 | SPOP | FALSE | Downregulated |
| -0,610702545 | 0,012471589 | FAM134B | FALSE | Downregulated |
| -0,473681874 | 0,012485265 | GPR146 | FALSE | Downregulated |
| 0,306131852 | 0,012503675 | ASNA1 | FALSE | Upregulated |
| -0,265180198 | 0,012516326 | METTL5 | FALSE | Downregulated |
| 1,319887451 | 0,012551174 | SEPT5 | TRUE | Upregulated |
| -0,627915709 | 0,012551676 | STAG3L4 | FALSE | Downregulated |
| 0,962004038 | 0,012551676 | TAF4B | FALSE | Upregulated |
| -0,376506836 | 0,01255757 | C6 | FALSE | Downregulated |
| 0,949744 | 0,012565211 | ZNF702P | FALSE | Upregulated |
| -0,41522512 | 0,012586989 | RBBP5 | FALSE | Downregulated |
| 0,89143274 | 0,01259715 | THBD | FALSE | Upregulated |
| -0,983680241 | 0,012601897 | ZDHHC11B | FALSE | Downregulated |
| -0,950695121 | 0,012602521 | PPM1E | FALSE | Downregulated |
| 0,970418168 | 0,012619596 | TMEM246 | FALSE | Upregulated |
| -0,280115972 | 0,012666658 | BRPF3 | FALSE | Downregulated |
| -0,296274435 | 0,012666658 | KLHL20 | FALSE | Downregulated |
| 0,371609935 | 0,012666658 | PHF5A | FALSE | Upregulated |
| 0,386360124 | 0,012676978 | MBOAT7 | FALSE | Upregulated |
| 2,641339062 | 0,01268217 | GABBR2 | TRUE | Upregulated |
| 1,122980867 | 0,012698212 | LOC389641 | TRUE | Upregulated |
| 2,238717811 | 0,01272989 | LOC102723542 | TRUE | Upregulated |
| -0,397256561 | 0,012772571 | EPT1 | FALSE | Downregulated |
| -0,311935799 | 0,012772571 | HSCB | FALSE | Downregulated |
| 0,82646255 | 0,012772571 | IL12RB1 | FALSE | Upregulated |
| 0,364293209 | 0,012772571 | PPRC1 | FALSE | Upregulated |
| -0,293123821 | 0,012772571 | RPS23 | FALSE | Downregulated |
| 0,675487584 | 0,012772571 | TLR2 | FALSE | Upregulated |
| -0,244488156 | 0,012788833 | NDEL1 | FALSE | Downregulated |
| -1,664285398 | 0,012792888 | C5orf27 | TRUE | Downregulated |
| -0,242320762 | 0,012865153 | NVL | FALSE | Downregulated |
| -1,219049654 | 0,012867888 | MYH3 | TRUE | Downregulated |
| -0,653071126 | 0,012889302 | G0S2 | FALSE | Downregulated |
| -0,46122386 | 0,012895648 | N4BP2L1 | FALSE | Downregulated |
| 0,60572217 | 0,012895648 | TMEM201 | FALSE | Upregulated |
| 0,739147166 | 0,012897204 | FPR3 | FALSE | Upregulated |
| 0,980379292 | 0,012900562 | CLDN2 | FALSE | Upregulated |
| -1,131890163 | 0,012922009 | TRNL1 | TRUE | Downregulated |
| 0,496223327 | 0,012924686 | BTN2A2 | FALSE | Upregulated |
| 0,472323844 | 0,012925532 | MMP15 | FALSE | Upregulated |
| 0,429505013 | 0,012942787 | APOL2 | FALSE | Upregulated |

| | | | | |
|---|---|---|---|---|
| -0,553476861 | 0,012942787 | BMPER | FALSE | Downregulated |
| -0,378956001 | 0,012942787 | SERTAD2 | FALSE | Downregulated |
| -1,613220397 | 0,012948134 | RET | TRUE | Downregulated |
| -0,357419285 | 0,01297928 | CRADD | FALSE | Downregulated |
| -0,513599491 | 0,013017917 | SERPING1 | FALSE | Downregulated |
| -0,37486291 | 0,013064734 | ATF7IP2 | FALSE | Downregulated |
| 0,477036251 | 0,01307725 | SLC38A2 | FALSE | Upregulated |
| -1,236335093 | 0,01310109 | EN1 | TRUE | Downregulated |
| -0,376159995 | 0,013140968 | DIABLO | FALSE | Downregulated |
| -1,808812813 | 0,013202697 | LOC102724208 | TRUE | Downregulated |
| -0,294918155 | 0,013219437 | ATPIF1 | FALSE | Downregulated |
| -0,368360485 | 0,013220471 | MRPS17 | FALSE | Downregulated |
| 0,639328201 | 0,013237135 | PKM | FALSE | Upregulated |
| 0,302840322 | 0,013238437 | MEX3C | FALSE | Upregulated |
| -0,638562681 | 0,013238437 | RNASEH1-AS1 | FALSE | Downregulated |
| 0,327371166 | 0,013241058 | HNRNPLL | FALSE | Upregulated |
| -0,963230439 | 0,013241058 | NBPF8 | FALSE | Downregulated |
| 0,730779748 | 0,013252363 | TLR5 | FALSE | Upregulated |
| 0,603559933 | 0,013269539 | MYO5A | FALSE | Upregulated |
| -0,61034492 | 0,013289567 | LOC101927600 | FALSE | Downregulated |
| 0,881755525 | 0,013324749 | TRO | FALSE | Upregulated |
| -0,220050055 | 0,013349059 | UBE2E3 | FALSE | Downregulated |
| 4,335278314 | 0,013362293 | PADI6 | TRUE | Upregulated |
| 0,461700759 | 0,013391537 | GPCPD1 | FALSE | Upregulated |
| -0,257393562 | 0,013433658 | UTP6 | FALSE | Downregulated |
| 0,812245604 | 0,013452294 | CENPE | FALSE | Upregulated |
| 0,618346027 | 0,013459345 | SRPX | FALSE | Upregulated |
| -1,415923821 | 0,013493644 | LGI1 | TRUE | Downregulated |
| 0,87388577 | 0,01350973 | CSF2RB | FALSE | Upregulated |
| 2,668102599 | 0,013510889 | LOC101929732 | TRUE | Upregulated |
| -0,214069103 | 0,013514969 | NDUFA9 | FALSE | Downregulated |
| 0,800601869 | 0,013514969 | SLC16A7 | FALSE | Upregulated |
| -0,4696787 | 0,013538723 | YBEY | FALSE | Downregulated |
| -0,541158931 | 0,013554933 | TCF25 | FALSE | Downregulated |
| 0,354720173 | 0,013554933 | TRAF4 | FALSE | Upregulated |
| -0,27727841 | 0,013568601 | COG6 | FALSE | Downregulated |
| 1,310824287 | 0,013568601 | CTBP1-AS | TRUE | Upregulated |
| 1,148516218 | 0,013568601 | MYOM1 | TRUE | Upregulated |
| 0,542405307 | 0,01357363 | CALCRL | FALSE | Upregulated |
| 1,939041893 | 0,013580912 | ANP32C | TRUE | Upregulated |
| 1,482694937 | 0,013590963 | GZMM | TRUE | Upregulated |
| -0,525168893 | 0,013618746 | RP5-886K2.3 | FALSE | Downregulated |
| 0,433935102 | 0,013643065 | ANKRD50 | FALSE | Upregulated |
| 2,159746935 | 0,013650701 | ELFN2 | TRUE | Upregulated |
| 0,258244352 | 0,013650701 | ZBTB17 | FALSE | Upregulated |
| 0,62681042 | 0,013699085 | SRGN | FALSE | Upregulated |
| 1,570273812 | 0,013699345 | CTAGE7P | TRUE | Upregulated |
| 0,804588226 | 0,013723518 | CDCA8 | FALSE | Upregulated |
| -1,013039259 | 0,013726755 | BBOX1 | TRUE | Downregulated |
| -0,356419802 | 0,013727375 | ABAT | FALSE | Downregulated |
| 0,750826947 | 0,013733377 | FAIM3 | FALSE | Upregulated |

| | | | | |
|---|---|---|---|---|
| 0,231017391 | 0,013740392 | EXOC8 | FALSE | Upregulated |
| 1,474361245 | 0,013760935 | LOC100996746 | TRUE | Upregulated |
| 0,477436656 | 0,013772318 | NXPE3 | FALSE | Upregulated |
| -0,224028238 | 0,0138442 | TEX261 | FALSE | Downregulated |
| -1,179244841 | 0,013851179 | MAN1B1-AS1 | TRUE | Downregulated |
| -0,511343186 | 0,013851179 | SLC24A1 | FALSE | Downregulated |
| 0,211504634 | 0,013856489 | KIF13B | FALSE | Upregulated |
| 0,744719837 | 0,013856489 | RAPGEF3 | FALSE | Upregulated |
| -0,386538135 | 0,013856489 | SNRPD2 | FALSE | Downregulated |
| -1,440976106 | 0,01391874 | LOC101929625 | TRUE | Downregulated |
| 0,323112591 | 0,013922613 | SMG9 | FALSE | Upregulated |
| 2,288976043 | 0,013925402 | AC020571.3 | TRUE | Upregulated |
| -0,600096812 | 0,013965423 | GLS2 | FALSE | Downregulated |
| 3,243304666 | 0,013974618 | LOC101928552 | TRUE | Upregulated |
| 2,047963117 | 0,013982293 | ZG16B | TRUE | Upregulated |
| 0,874013648 | 0,014007895 | LOC100507507 | FALSE | Upregulated |
| -0,331245349 | 0,014007895 | SREK1IP1 | FALSE | Downregulated |
| 0,584271227 | 0,014007895 | SYNGAP1 | FALSE | Upregulated |
| -0,752477144 | 0,01401155 | LOC113230 | FALSE | Downregulated |
| -0,792979572 | 0,014013155 | PFN2 | FALSE | Downregulated |
| -0,348759485 | 0,014041511 | RBMX2 | FALSE | Downregulated |
| 0,619178935 | 0,014048737 | PTAFR | FALSE | Upregulated |
| -0,313227517 | 0,014049108 | KLHL8 | FALSE | Downregulated |
| 0,832600261 | 0,014053841 | NOTUM | FALSE | Upregulated |
| 3,259155979 | 0,014060913 | LY6K | TRUE | Upregulated |
| 0,71793807 | 0,014061511 | FAR2 | FALSE | Upregulated |
| 1,057545142 | 0,014118926 | BANK1 | TRUE | Upregulated |
| -0,31392356 | 0,014121417 | GNAI2 | FALSE | Downregulated |
| 0,233233536 | 0,014121417 | PSMF1 | FALSE | Upregulated |
| 0,354683886 | 0,014126778 | IRAK1 | FALSE | Upregulated |
| 3,134101974 | 0,014126778 | PART1 | TRUE | Upregulated |
| 1,71256554 | 0,014135597 | ZNF683 | TRUE | Upregulated |
| -0,324195772 | 0,014136472 | MRPS6 | FALSE | Downregulated |
| 0,539761546 | 0,014136472 | RAB11FIP5 | FALSE | Upregulated |
| -0,390703107 | 0,014136472 | TMEM208 | FALSE | Downregulated |
| 0,699394965 | 0,014137792 | DUSP18 | FALSE | Upregulated |
| 0,380902705 | 0,014137792 | RHBDD3 | FALSE | Upregulated |
| -0,531383659 | 0,014144313 | ACSL1 | FALSE | Downregulated |
| 0,442014361 | 0,014148489 | CBL | FALSE | Upregulated |
| 0,680977623 | 0,01415492 | AOAH | FALSE | Upregulated |
| 2,506694195 | 0,014238307 | AREG | TRUE | Upregulated |
| 0,42405352 | 0,014238307 | CEP41 | FALSE | Upregulated |
| -0,696223879 | 0,014258446 | ASPA | FALSE | Downregulated |
| 0,299361362 | 0,014270227 | MTF1 | FALSE | Upregulated |
| -0,90942792 | 0,014273258 | LOC101927393 | FALSE | Downregulated |
| -0,509510909 | 0,014277508 | EGFR | FALSE | Downregulated |
| 2,024811655 | 0,014280356 | FA2H | TRUE | Upregulated |
| 2,117405277 | 0,014280812 | TMEM163 | TRUE | Upregulated |
| -0,307191364 | 0,014319803 | CPNE1 | FALSE | Downregulated |
| -0,308866376 | 0,014328528 | ACBD5 | FALSE | Downregulated |
| 0,239887492 | 0,014336376 | DCTN4 | FALSE | Upregulated |

| | | | | |
|---|---|---|---|---|
| 0,692561702 | 0,014396827 | LOC100132891 | FALSE | Upregulated |
| -0,426635674 | 0,014400126 | DDX10 | FALSE | Downregulated |
| 1,431972299 | 0,014420613 | DNAH7 | TRUE | Upregulated |
| -0,276919543 | 0,014420613 | UBXN1 | FALSE | Downregulated |
| 0,311108884 | 0,014422367 | FAM53B | FALSE | Upregulated |
| -0,565218496 | 0,014422367 | IGHMBP2 | FALSE | Downregulated |
| -0,408067472 | 0,014439867 | AOX1 | FALSE | Downregulated |
| 1,482143373 | 0,014439867 | C5orf66 | TRUE | Upregulated |
| 0,370751138 | 0,014444794 | DPYSL2 | FALSE | Upregulated |
| -0,348466653 | 0,014463988 | TMTC3 | FALSE | Downregulated |
| 0,261289519 | 0,01447242 | BCAP31 | FALSE | Upregulated |
| 0,461869385 | 0,01447242 | LAT2 | FALSE | Upregulated |
| -0,271963789 | 0,01447242 | TIMM21 | FALSE | Downregulated |
| 0,845718389 | 0,014477765 | GNA15 | FALSE | Upregulated |
| 0,860921714 | 0,014508292 | TRG-AS1 | FALSE | Upregulated |
| -0,265350804 | 0,014528245 | ACBD6 | FALSE | Downregulated |
| 1,679302304 | 0,014528245 | LOC102724248 | TRUE | Upregulated |
| 0,303145025 | 0,014537241 | SND1 | FALSE | Upregulated |
| -0,725812168 | 0,014537241 | SRD5A2 | FALSE | Downregulated |
| -0,375810777 | 0,014553921 | ATP5G2 | FALSE | Downregulated |
| 1,586127425 | 0,014553921 | FAM43B | TRUE | Upregulated |
| -0,344098302 | 0,014553921 | MPHOSPH9 | FALSE | Downregulated |
| 1,945627715 | 0,014588786 | MMP9 | TRUE | Upregulated |
| 0,936750504 | 0,014597292 | EGLN3 | FALSE | Upregulated |
| 0,818616761 | 0,014620046 | GJC1 | FALSE | Upregulated |
| -0,185958052 | 0,014634194 | PRPF38A | FALSE | Downregulated |
| 1,486520905 | 0,014658687 | LOC101927792 | TRUE | Upregulated |
| 0,405856767 | 0,01470302 | AMOT | FALSE | Upregulated |
| -0,299442959 | 0,014729043 | DHX36 | FALSE | Downregulated |
| -0,306208098 | 0,014748208 | ATF4 | FALSE | Downregulated |
| 0,44803927 | 0,014758836 | RBM3 | FALSE | Upregulated |
| -0,303438493 | 0,014767033 | HABP2 | FALSE | Downregulated |
| 0,384814994 | 0,014767033 | ZNF627 | FALSE | Upregulated |
| -0,421333969 | 0,014792381 | GOLGA4 | FALSE | Downregulated |
| 1,362639422 | 0,014824623 | CD1E | TRUE | Upregulated |
| 0,488855692 | 0,014826995 | LCP1 | FALSE | Upregulated |
| 0,475072062 | 0,014842868 | TLDC1 | FALSE | Upregulated |
| -0,336467786 | 0,014846442 | FAM172A | FALSE | Downregulated |
| -0,673322601 | 0,014863264 | HSD17B13 | FALSE | Downregulated |
| -0,316847148 | 0,014870441 | MRPL2 | FALSE | Downregulated |
| 0,762991124 | 0,014884385 | TMEM156 | FALSE | Upregulated |
| 0,534296136 | 0,014884559 | MYO1F | FALSE | Upregulated |
| 0,984537399 | 0,014914294 | EPHB3 | FALSE | Upregulated |
| -0,283626835 | 0,014935206 | CMTM6 | FALSE | Downregulated |
| 0,468120259 | 0,014942774 | SLC36A1 | FALSE | Upregulated |
| -0,21633267 | 0,014962714 | EIF1B | FALSE | Downregulated |
| 3,354358855 | 0,015013789 | DCC | TRUE | Upregulated |
| 1,94794147 | 0,015013789 | HIST2H2BC | TRUE | Upregulated |
| 0,692099635 | 0,015013789 | LRRC25 | FALSE | Upregulated |
| 2,67940834 | 0,015013789 | SFTA2 | TRUE | Upregulated |
| -1,914648 | 0,015018695 | RFPL2 | TRUE | Downregulated |

| | | | | |
|---|---|---|---|---|
| 0,727659611 | 0,015052691 | LILRA1 | FALSE | Upregulated |
| -0,538783578 | 0,015060919 | HYKK | FALSE | Downregulated |
| -0,746170616 | 0,015060919 | LOC100289495 | FALSE | Downregulated |
| 0,193682547 | 0,015083972 | AP2B1 | FALSE | Upregulated |
| 0,662591487 | 0,015083972 | DENND6B | FALSE | Upregulated |
| 0,734811918 | 0,015083972 | PRF1 | FALSE | Upregulated |
| 0,519273888 | 0,015096337 | TANC2 | FALSE | Upregulated |
| 2,018287719 | 0,015101961 | ATG9B | TRUE | Upregulated |
| -0,450042252 | 0,015116905 | LOC100506634 | FALSE | Downregulated |
| 0,578386674 | 0,015139796 | ADRBK2 | FALSE | Upregulated |
| -0,483372092 | 0,015139796 | NCL | FALSE | Downregulated |
| -0,551755517 | 0,015179899 | CLRN3 | FALSE | Downregulated |
| 1,761662285 | 0,015179899 | MMP11 | TRUE | Upregulated |
| -0,418301834 | 0,015179899 | NEU3 | FALSE | Downregulated |
| -0,536450793 | 0,015204863 | ASCC1 | FALSE | Downregulated |
| 0,475986156 | 0,015248798 | CYTH3 | FALSE | Upregulated |
| 1,093073287 | 0,015259733 | ANLN | TRUE | Upregulated |
| -2,187106206 | 0,015259733 | TNNT2 | TRUE | Downregulated |
| 0,300875059 | 0,015260772 | ZNF623 | FALSE | Upregulated |
| -0,467563756 | 0,015263893 | C1orf35 | FALSE | Downregulated |
| -0,45507493 | 0,01527681 | DPYS | FALSE | Downregulated |
| 0,625359157 | 0,015278658 | STPG1 | FALSE | Upregulated |
| 0,623642137 | 0,015278658 | TRERF1 | FALSE | Upregulated |
| -0,860059057 | 0,015304037 | JAKMIP2 | FALSE | Downregulated |
| 1,505295817 | 0,015308566 | CENPI | TRUE | Upregulated |
| 1,261478675 | 0,015314188 | EPS8L1 | TRUE | Upregulated |
| 1,025658009 | 0,015320437 | SFRP5 | TRUE | Upregulated |
| 1,13841905 | 0,015334399 | ASPHD2 | TRUE | Upregulated |
| -1,508257939 | 0,015339764 | DIO3OS | TRUE | Downregulated |
| -0,298126609 | 0,015374478 | CRTC2 | FALSE | Downregulated |
| 0,436035756 | 0,015380573 | ALG1 | FALSE | Upregulated |
| -0,51132446 | 0,015380573 | ARMCX5-GPRASP2 | FALSE | Downregulated |
| 0,918129663 | 0,015380573 | GLI3 | FALSE | Upregulated |
| -1,025575792 | 0,015399723 | LOC101928816 | TRUE | Downregulated |
| 0,322112066 | 0,015399723 | USP46 | FALSE | Upregulated |
| -0,344240447 | 0,015441444 | MRGBP | FALSE | Downregulated |
| -1,307452813 | 0,015497263 | RP11-539L10.2 | TRUE | Downregulated |
| 1,660137314 | 0,015507104 | GPR83 | TRUE | Upregulated |
| 0,693604411 | 0,015524105 | EML5 | FALSE | Upregulated |
| -2,424857011 | 0,015526105 | PGAM2 | TRUE | Downregulated |
| 1,167081001 | 0,015532129 | LOC440300 | TRUE | Upregulated |
| 0,285626237 | 0,015570205 | B4GALT3 | FALSE | Upregulated |
| -0,44211009 | 0,015623799 | ZMYM4 | FALSE | Downregulated |
| 2,544759476 | 0,015624977 | EPHA8 | TRUE | Upregulated |
| 0,329446694 | 0,015624977 | SIN3B | FALSE | Upregulated |
| 0,520567953 | 0,015624977 | SLC20A1 | FALSE | Upregulated |
| -0,457665891 | 0,015654495 | KIAA2018 | FALSE | Downregulated |
| -0,270827628 | 0,015670877 | NONO | FALSE | Downregulated |
| -0,33197574 | 0,015697815 | HTATSF1 | FALSE | Downregulated |
| -0,3184439 | 0,015713553 | SMIM20 | FALSE | Downregulated |
| -0,326330939 | 0,015729844 | CLU | FALSE | Downregulated |

| | | | | |
|---|---|---|---|---|
| 1,330620051 | 0,015770183 | CCNB2 | TRUE | Upregulated |
| 0,71519561 | 0,015775958 | PTGIR | FALSE | Upregulated |
| -0,315795132 | 0,01579186 | MRPL13 | FALSE | Downregulated |
| 1,394540679 | 0,015807656 | ZSCAN23 | TRUE | Upregulated |
| 0,262677059 | 0,015812759 | PPP4C | FALSE | Upregulated |
| 1,386493643 | 0,01582667 | FAM179A | TRUE | Upregulated |
| 1,23609523 | 0,01582667 | RAB38 | TRUE | Upregulated |
| 0,395787916 | 0,015855608 | SPRED1 | FALSE | Upregulated |
| 0,267530957 | 0,015855608 | TM9SF1 | FALSE | Upregulated |
| 3,101648752 | 0,015862467 | LOC101928722 | TRUE | Upregulated |
| 1,360515446 | 0,015882703 | NCF1B | TRUE | Upregulated |
| 1,00496464 | 0,015908265 | FRZB | TRUE | Upregulated |
| 0,652466357 | 0,015908265 | SOX18 | FALSE | Upregulated |
| 1,483467601 | 0,015910348 | LTA | TRUE | Upregulated |
| 0,344518916 | 0,015910658 | XYLT2 | FALSE | Upregulated |
| 0,165408897 | 0,015912859 | WDR82 | FALSE | Upregulated |
| 0,97478811 | 0,015930864 | MAFF | FALSE | Upregulated |
| -0,245308047 | 0,015983531 | NOB1 | FALSE | Downregulated |
| 1,314505101 | 0,016005972 | KCNK13 | TRUE | Upregulated |
| 0,260274809 | 0,016038143 | IGSF8 | FALSE | Upregulated |
| 2,521792495 | 0,016042204 | KCNJ4 | TRUE | Upregulated |
| -1,318601237 | 0,016062605 | MGAM | TRUE | Downregulated |
| -0,268844464 | 0,016095443 | HTRA2 | FALSE | Downregulated |
| 0,658031135 | 0,0161103 | CCNB1 | FALSE | Upregulated |
| -0,749799382 | 0,01614259 | AFP | FALSE | Downregulated |
| 0,84913154 | 0,01614259 | WDR76 | FALSE | Upregulated |
| -0,412634847 | 0,01614259 | ZHX3 | FALSE | Downregulated |
| 0,844617158 | 0,016149586 | C11orf80 | FALSE | Upregulated |
| 0,263181115 | 0,016197058 | CSNK1G1 | FALSE | Upregulated |
| -0,215009744 | 0,016197058 | PIK3C2A | FALSE | Downregulated |
| -0,634546257 | 0,016216373 | FUZ | FALSE | Downregulated |
| -0,580315506 | 0,016216373 | TM6SF2 | FALSE | Downregulated |
| 1,064817041 | 0,016251908 | GUSBP2 | TRUE | Upregulated |
| -0,627315896 | 0,01625686 | ACAT2 | FALSE | Downregulated |
| 2,195799522 | 0,01625686 | CCL3L3 | TRUE | Upregulated |
| 0,643675076 | 0,01625686 | CD44 | FALSE | Upregulated |
| -0,520066464 | 0,01625686 | XYLB | FALSE | Downregulated |
| 0,270456194 | 0,016286945 | FAM83H | FALSE | Upregulated |
| -0,649365715 | 0,016286945 | RP11-96H19.1 | FALSE | Downregulated |
| 0,460301445 | 0,016302452 | C11orf49 | FALSE | Upregulated |
| 0,391712136 | 0,016302452 | WDR35 | FALSE | Upregulated |
| 0,340703841 | 0,01631152 | DNAJC10 | FALSE | Upregulated |
| 0,47285669 | 0,016326934 | SAMHD1 | FALSE | Upregulated |
| 0,837174881 | 0,016326934 | SLC2A1 | FALSE | Upregulated |
| -0,429473767 | 0,016332281 | CCL16 | FALSE | Downregulated |
| -0,318746846 | 0,016335334 | NDUFB9 | FALSE | Downregulated |
| 0,380180256 | 0,016349045 | N6AMT1 | FALSE | Upregulated |
| 0,225228422 | 0,016349045 | PRRC2B | FALSE | Upregulated |
| 2,123451841 | 0,016363807 | NEIL3 | TRUE | Upregulated |
| 0,89981562 | 0,016369161 | MAB21L2 | FALSE | Upregulated |
| -1,340455452 | 0,016372342 | CYP1A1 | TRUE | Downregulated |

| | | | | |
|---|---|---|---|---|
| -0,353391323 | 0,01637312 | APOM | FALSE | Downregulated |
| 1,615095511 | 0,01637312 | LAMA1 | TRUE | Upregulated |
| 2,435566231 | 0,01637402 | LOC101927374 | TRUE | Upregulated |
| 0,315676444 | 0,016384325 | ZNF282 | FALSE | Upregulated |
| 0,902760305 | 0,016393907 | FRAS1 | FALSE | Upregulated |
| -0,843203295 | 0,016485288 | RNF152 | FALSE | Downregulated |
| 0,272418769 | 0,016505576 | CAPRIN1 | FALSE | Upregulated |
| 0,228872015 | 0,01650863 | UBTD2 | FALSE | Upregulated |
| 2,911970261 | 0,016516057 | CDC42P3 | TRUE | Upregulated |
| -0,343140079 | 0,016519552 | C12orf45 | FALSE | Downregulated |
| -0,22415453 | 0,016519552 | WDR41 | FALSE | Downregulated |
| -2,18718257 | 0,016526147 | DKFZp434J0226 | TRUE | Downregulated |
| 0,251839824 | 0,016526147 | LMF2 | FALSE | Upregulated |
| 0,761317577 | 0,016551847 | ABCB4 | FALSE | Upregulated |
| -0,344154532 | 0,01657079 | N4BP2 | FALSE | Downregulated |
| -0,813023497 | 0,016576655 | LOC100129781 | FALSE | Downregulated |
| 0,462740178 | 0,016587461 | ADAMTSL4 | FALSE | Upregulated |
| -0,432715121 | 0,016587461 | DPM3 | FALSE | Downregulated |
| 0,745947084 | 0,016605861 | PTGFR | FALSE | Upregulated |
| 0,884639606 | 0,0166132 | IL7R | FALSE | Upregulated |
| 2,145904828 | 0,01662761 | FUT2 | TRUE | Upregulated |
| 0,330136511 | 0,016656633 | C7orf26 | FALSE | Upregulated |
| 0,465726049 | 0,016664248 | ERBB2 | FALSE | Upregulated |
| -0,350115184 | 0,016664422 | SBDSP1 | FALSE | Downregulated |
| -0,266735311 | 0,016667145 | CCDC92 | FALSE | Downregulated |
| -0,319343401 | 0,016682252 | LINC00493 | FALSE | Downregulated |
| 0,381074354 | 0,016682252 | PTCHD3P1 | FALSE | Upregulated |
| 0,707936564 | 0,016683123 | TGM2 | FALSE | Upregulated |
| 0,293702548 | 0,016688993 | CDK5RAP2 | FALSE | Upregulated |
| 2,033406329 | 0,016690084 | LOC729739 | TRUE | Upregulated |
| -0,264741966 | 0,01670158 | R3HDM2 | FALSE | Downregulated |
| -0,582518345 | 0,01675279 | CD4 | FALSE | Downregulated |
| 0,85758934 | 0,01675279 | OSCAR | FALSE | Upregulated |
| 1,242676985 | 0,01675279 | SKP1P2 | TRUE | Upregulated |
| -0,332294866 | 0,01675279 | WDR59 | FALSE | Downregulated |
| 1,162872324 | 0,01675682 | CNIH3 | TRUE | Upregulated |
| 0,340850497 | 0,01675682 | RAVER1 | FALSE | Upregulated |
| 0,278222997 | 0,016776393 | ADO | FALSE | Upregulated |
| -0,6386248 | 0,016793522 | C19orf54 | FALSE | Downregulated |
| -0,401217048 | 0,016793522 | EYA3 | FALSE | Downregulated |
| -2,230982459 | 0,016825117 | ART5 | TRUE | Downregulated |
| 0,642283975 | 0,016825117 | FITM2 | FALSE | Upregulated |
| -1,040581169 | 0,016825117 | HS3ST3A1 | TRUE | Downregulated |
| 0,580271371 | 0,016835064 | ARMCX2 | FALSE | Upregulated |
| -0,468425469 | 0,016835064 | DUSP19 | FALSE | Downregulated |
| 0,520589469 | 0,016862755 | SYK | FALSE | Upregulated |
| 1,310166794 | 0,016878854 | WIPF3 | TRUE | Upregulated |
| -1,640621611 | 0,01694028 | RP11-43N5.1 | TRUE | Downregulated |
| 0,534661105 | 0,016955152 | AP001258.4 | FALSE | Upregulated |
| -0,372390639 | 0,016955152 | APMAP | FALSE | Downregulated |
| 0,433653736 | 0,016955152 | LACC1 | FALSE | Upregulated |

| | | | | |
|---|---|---|---|---|
| 0,534936822 | 0,016955152 | SIRPA | FALSE | Upregulated |
| 1,717588282 | 0,016960979 | NKAIN2 | TRUE | Upregulated |
| 0,672221668 | 0,016978793 | LPAR5 | FALSE | Upregulated |
| -0,427174271 | 0,016999875 | ANP32E | FALSE | Downregulated |
| 0,528113997 | 0,017002433 | ANKRD12 | FALSE | Upregulated |
| 0,698973672 | 0,017006398 | TNFRSF11A | FALSE | Upregulated |
| 0,779121794 | 0,017025551 | ROBO3 | FALSE | Upregulated |
| 0,296426843 | 0,017030039 | NUP155 | FALSE | Upregulated |
| -0,211556852 | 0,017030039 | SDCCAG8 | FALSE | Downregulated |
| 0,453644244 | 0,017047809 | IKBKG | FALSE | Upregulated |
| -0,430408838 | 0,017049966 | SLC25A37 | FALSE | Downregulated |
| 0,697853753 | 0,017054495 | FGR | FALSE | Upregulated |
| -0,334111843 | 0,017054495 | NDUFA11 | FALSE | Downregulated |
| 1,053090041 | 0,017085137 | SLIT3 | TRUE | Upregulated |
| -0,417582995 | 0,017085137 | TANGO6 | FALSE | Downregulated |
| -0,289132105 | 0,017090476 | TOM1L1 | FALSE | Downregulated |
| 0,431665481 | 0,017119655 | RAPGEF6 | FALSE | Upregulated |
| -0,57170353 | 0,017124137 | AMOTL1 | FALSE | Downregulated |
| -1,093649495 | 0,017124137 | LOC643327 | TRUE | Downregulated |
| -0,225697152 | 0,017124137 | NFE2L1 | FALSE | Downregulated |
| 3,162209137 | 0,017128198 | RBP2 | TRUE | Upregulated |
| 0,947381397 | 0,01712901 | SH2B2 | FALSE | Upregulated |
| -0,409500196 | 0,017146068 | HIBCH | FALSE | Downregulated |
| 0,843978894 | 0,017163794 | PIK3R6 | FALSE | Upregulated |
| 1,607045428 | 0,017182047 | PRKY | TRUE | Upregulated |
| -0,843658922 | 0,017183882 | PPP1R1A | FALSE | Downregulated |
| -0,323606312 | 0,017183882 | TOMM6 | FALSE | Downregulated |
| 0,959281959 | 0,017196787 | LOC101928429 | FALSE | Upregulated |
| 0,294564934 | 0,01720886 | ASPSCR1 | FALSE | Upregulated |
| -1,666069863 | 0,01720886 | LOC101927727 | TRUE | Downregulated |
| 1,155598937 | 0,017208907 | TNFRSF10C | TRUE | Upregulated |
| 0,336388884 | 0,017243279 | DMWD | FALSE | Upregulated |
| 0,864307752 | 0,017246583 | A4GALT | FALSE | Upregulated |
| -0,351264444 | 0,017246583 | ZFYVE19 | FALSE | Downregulated |
| -0,179039327 | 0,017248374 | MRPL16 | FALSE | Downregulated |
| -0,569391759 | 0,017310569 | RCL1 | FALSE | Downregulated |
| -0,682213319 | 0,017315082 | LOC100507577 | FALSE | Downregulated |
| 0,335785773 | 0,017315082 | POLA1 | FALSE | Upregulated |
| -1,552687194 | 0,017318011 | PROL1 | TRUE | Downregulated |
| 0,293787471 | 0,017318011 | SRSF3 | FALSE | Upregulated |
| -0,515058097 | 0,017318393 | HHLA3 | FALSE | Downregulated |
| 0,858551936 | 0,017351576 | FASLG | FALSE | Upregulated |
| 1,151436927 | 0,017354265 | DLGAP5 | TRUE | Upregulated |
| -0,470609779 | 0,017380885 | NPY1R | FALSE | Downregulated |
| -0,313167827 | 0,017380885 | PRNP | FALSE | Downregulated |
| -0,344276403 | 0,01739273 | TSPAN3 | FALSE | Downregulated |
| -0,3129883 | 0,017398396 | RARRES2 | FALSE | Downregulated |
| 2,00609475 | 0,017401124 | RBM5-AS1 | TRUE | Upregulated |
| -0,355164354 | 0,017405501 | NAT9 | FALSE | Downregulated |
| -0,20683082 | 0,017417111 | MAPKAPK5 | FALSE | Downregulated |
| -0,571208095 | 0,017417111 | UPF3A | FALSE | Downregulated |

| | | | | |
|---|---|---|---|---|
| 0,47386918 | 0,017427607 | IFT80 | FALSE | Upregulated |
| -0,301172239 | 0,017428852 | MRPL34 | FALSE | Downregulated |
| 0,903234462 | 0,017428852 | PPP2R2B | FALSE | Upregulated |
| 0,346676953 | 0,017428852 | SRGAP2 | FALSE | Upregulated |
| -0,902578918 | 0,017428852 | TMED6 | FALSE | Downregulated |
| 0,943307127 | 0,017428852 | ZNF665 | FALSE | Upregulated |
| -0,504724544 | 0,017481741 | AKAP6 | FALSE | Downregulated |
| -0,857560844 | 0,01748322 | RP5-1013A22.2 | FALSE | Downregulated |
| 0,36131928 | 0,017502352 | TDRD7 | FALSE | Upregulated |
| -0,230135282 | 0,017518228 | NBR1 | FALSE | Downregulated |
| -0,418440448 | 0,017520055 | ACAA2 | FALSE | Downregulated |
| 0,312445541 | 0,017520055 | KCTD2 | FALSE | Upregulated |
| 0,311156275 | 0,017520055 | TMUB2 | FALSE | Upregulated |
| -0,296653336 | 0,017538844 | IRGQ | FALSE | Downregulated |
| 0,440070384 | 0,017548505 | CDK18 | FALSE | Upregulated |
| 1,144198324 | 0,017556452 | CLSTN2 | TRUE | Upregulated |
| -1,514173106 | 0,017615733 | S100A1 | TRUE | Downregulated |
| 2,305518424 | 0,017615733 | SNORA62 | TRUE | Upregulated |
| -1,036682392 | 0,017615733 | TRNI | TRUE | Downregulated |
| -0,288773465 | 0,017615733 | UBR3 | FALSE | Downregulated |
| 0,594430472 | 0,017641409 | VEGFC | FALSE | Upregulated |
| -0,50415231 | 0,017670971 | SIVA1 | FALSE | Downregulated |
| 0,634478482 | 0,017698845 | ISLR | FALSE | Upregulated |
| -1,23338728 | 0,017716044 | LOC101927880 | TRUE | Downregulated |
| -0,344958644 | 0,017737635 | RABGEF1 | FALSE | Downregulated |
| -0,334732213 | 0,017745784 | FADD | FALSE | Downregulated |
| -0,293019765 | 0,017748855 | PLEKHA5 | FALSE | Downregulated |
| 0,350561542 | 0,017776845 | HLA-E | FALSE | Upregulated |
| 1,531726943 | 0,017782977 | PLD4 | TRUE | Upregulated |
| 0,67449322 | 0,01778998 | BAMBI | FALSE | Upregulated |
| 0,617763065 | 0,017824822 | STBD1 | FALSE | Upregulated |
| -0,382190636 | 0,017826828 | FAM117A | FALSE | Downregulated |
| -0,501332644 | 0,017826828 | TNFSF10 | FALSE | Downregulated |
| -1,636882287 | 0,01784431 | TRNW | TRUE | Downregulated |
| -0,288454501 | 0,017845484 | CPPED1 | FALSE | Downregulated |
| -0,333392551 | 0,017904843 | MAPRE3 | FALSE | Downregulated |
| -0,273208384 | 0,017923924 | CALCOCO2 | FALSE | Downregulated |
| 0,641760699 | 0,017959876 | ARSJ | FALSE | Upregulated |
| 0,65909869 | 0,017959876 | CDYL2 | FALSE | Upregulated |
| 0,202446318 | 0,017960544 | RANBP6 | FALSE | Upregulated |
| -0,213677851 | 0,017976276 | KLHDC3 | FALSE | Downregulated |
| 1,070320452 | 0,017976276 | SOCS1 | TRUE | Upregulated |
| 0,726444564 | 0,017982588 | PYHIN1 | FALSE | Upregulated |
| 0,295505072 | 0,01802541 | PTPN23 | FALSE | Upregulated |
| -0,340604555 | 0,018028293 | AP3S2 | FALSE | Downregulated |
| -0,206798296 | 0,018028293 | INTS10 | FALSE | Downregulated |
| -0,321740838 | 0,018028293 | LETMD1 | FALSE | Downregulated |
| -0,600146833 | 0,018046836 | CHN2 | FALSE | Downregulated |
| -0,549252564 | 0,01806219 | TSPAN7 | FALSE | Downregulated |
| 0,28958704 | 0,018084225 | INPP5K | FALSE | Upregulated |
| 0,771425486 | 0,01808497 | PDLIM7 | FALSE | Upregulated |

| | | | | |
|---|---|---|---|---|
| -0,293148125 | 0,018108748 | ARHGEF9 | FALSE | Downregulated |
| -0,332229252 | 0,018181752 | PCYOX1 | FALSE | Downregulated |
| -0,425490601 | 0,01818964 | MAT1A | FALSE | Downregulated |
| 0,307764099 | 0,018217651 | DOCK9 | FALSE | Upregulated |
| 0,341295498 | 0,018217651 | USP20 | FALSE | Upregulated |
| -0,501308085 | 0,018222468 | MFSD3 | FALSE | Downregulated |
| 0,50574005 | 0,018231561 | SYDE1 | FALSE | Upregulated |
| -1,282939103 | 0,018241573 | SYCE3 | TRUE | Downregulated |
| 2,223870778 | 0,018248919 | OTOGL | TRUE | Upregulated |
| -0,332040017 | 0,018284717 | UPF2 | FALSE | Downregulated |
| 0,729298861 | 0,018308263 | CST7 | FALSE | Upregulated |
| -0,225241547 | 0,018308263 | HBS1L | FALSE | Downregulated |
| -0,68793763 | 0,018313588 | LOC102723908 | FALSE | Downregulated |
| 0,213000165 | 0,018313588 | ZNF629 | FALSE | Upregulated |
| 0,440517259 | 0,018331072 | ESAM | FALSE | Upregulated |
| 0,86423389 | 0,018383823 | EVA1C | FALSE | Upregulated |
| 0,548240421 | 0,018394412 | CD33 | FALSE | Upregulated |
| -1,672915705 | 0,018394412 | LOC100128276 | TRUE | Downregulated |
| 1,592898602 | 0,018404106 | CASR | TRUE | Upregulated |
| 0,916473327 | 0,018429664 | BCL11B | FALSE | Upregulated |
| -0,347857122 | 0,018460409 | SIGMAR1 | FALSE | Downregulated |
| 0,91969924 | 0,018467142 | CD300LF | FALSE | Upregulated |
| -0,3903774 | 0,018467142 | SLC39A5 | FALSE | Downregulated |
| 0,962128525 | 0,018475652 | NBPF25P | FALSE | Upregulated |
| -0,262385595 | 0,01849283 | BNIP3 | FALSE | Downregulated |
| -0,345882002 | 0,018495913 | SMIM19 | FALSE | Downregulated |
| 0,206098925 | 0,018510047 | NDNL2 | FALSE | Upregulated |
| 0,339510579 | 0,018510047 | TDG | FALSE | Upregulated |
| 0,350882938 | 0,018519167 | USP6NL | FALSE | Upregulated |
| -0,705283382 | 0,018536586 | FNIP2 | FALSE | Downregulated |
| 0,253880888 | 0,018536586 | SMARCAL1 | FALSE | Upregulated |
| -1,229682168 | 0,018575083 | SIGLEC16 | TRUE | Downregulated |
| -0,241243318 | 0,018577256 | NIF3L1 | FALSE | Downregulated |
| 1,065284428 | 0,018587331 | FRMPD1 | TRUE | Upregulated |
| 0,491997994 | 0,018593175 | ITM2C | FALSE | Upregulated |
| 0,317470671 | 0,018606766 | TEP1 | FALSE | Upregulated |
| 0,631041756 | 0,018614869 | TUB | FALSE | Upregulated |
| 1,242342242 | 0,018646057 | RGL4 | TRUE | Upregulated |
| -0,218472476 | 0,018646057 | ZNF3 | FALSE | Downregulated |
| -0,390356835 | 0,018708022 | METAP1D | FALSE | Downregulated |
| 0,373206454 | 0,018751151 | DIS3L2 | FALSE | Upregulated |
| -1,084944172 | 0,018751151 | GLYATL3 | TRUE | Downregulated |
| -0,300281606 | 0,018754397 | ELOF1 | FALSE | Downregulated |
| -0,574207203 | 0,018754397 | LUC7L2 | FALSE | Downregulated |
| 1,882648509 | 0,018781187 | HS3ST2 | TRUE | Upregulated |
| 0,894694027 | 0,018781187 | SLC16A3 | FALSE | Upregulated |
| -0,318634574 | 0,018796309 | HSD17B10 | FALSE | Downregulated |
| 0,516046961 | 0,018796309 | UBE2L6 | FALSE | Upregulated |
| -2,089750337 | 0,018798889 | FAM9A | TRUE | Downregulated |
| 3,767947356 | 0,018843678 | LINC00966 | TRUE | Upregulated |
| -1,011113638 | 0,018861014 | RP11-15A1.7 | TRUE | Downregulated |

| | | | | |
|---|---|---|---|---|
| 0,223296077 | 0,018900666 | PPP2R5E | FALSE | Upregulated |
| -0,870459175 | 0,018900743 | MGC27382 | FALSE | Downregulated |
| -0,365438899 | 0,018951171 | RPL36A | FALSE | Downregulated |
| 0,306825907 | 0,018998599 | FIBP | FALSE | Upregulated |
| 0,220903651 | 0,018999387 | RALY | FALSE | Upregulated |
| 1,278843093 | 0,018999387 | SPTBN5 | TRUE | Upregulated |
| 0,916165237 | 0,019051549 | ANKDD1A | FALSE | Upregulated |
| -0,624683288 | 0,01915354 | LOC101927974 | FALSE | Downregulated |
| 0,83622938 | 0,019209969 | CDKN1A | FALSE | Upregulated |
| 0,284332908 | 0,019211019 | SSH3 | FALSE | Upregulated |
| 2,670358463 | 0,019241616 | GUCA2A | TRUE | Upregulated |
| -0,874380284 | 0,019247243 | HNF1A-AS1 | FALSE | Downregulated |
| 0,374915231 | 0,019258935 | RNASEL | FALSE | Upregulated |
| 1,28499627 | 0,019328964 | DDX12P | TRUE | Upregulated |
| 2,17220312 | 0,019334362 | COL10A1 | TRUE | Upregulated |
| 1,905182111 | 0,0193359 | FLJ44511 | TRUE | Upregulated |
| 0,653312435 | 0,019360178 | SLC51A | FALSE | Upregulated |
| 0,589212115 | 0,019400387 | HAVCR2 | FALSE | Upregulated |
| 2,958434778 | 0,019409284 | NPTX1 | TRUE | Upregulated |
| 1,385282361 | 0,019455115 | ENTPD2 | TRUE | Upregulated |
| -0,320469084 | 0,019482454 | PKIG | FALSE | Downregulated |
| 0,185477078 | 0,019482454 | RAB1B | FALSE | Upregulated |
| 0,468048669 | 0,019487376 | ATP2B4 | FALSE | Upregulated |
| -1,215345651 | 0,019487376 | LOC102724603 | TRUE | Downregulated |
| -0,672248238 | 0,019504522 | SAMD5 | FALSE | Downregulated |
| 0,359021199 | 0,019531795 | C1QTNF1 | FALSE | Upregulated |
| 0,317493622 | 0,019553041 | TNFRSF14 | FALSE | Upregulated |
| 0,378429596 | 0,019554348 | RFX1 | FALSE | Upregulated |
| 1,837839798 | 0,019595483 | PRSS16 | TRUE | Upregulated |
| -0,955816456 | 0,019602008 | OGFR-AS1 | FALSE | Downregulated |
| -0,36443601 | 0,019609993 | TTPA | FALSE | Downregulated |
| 0,288401152 | 0,01961155 | AXIN1 | FALSE | Upregulated |
| 0,634819846 | 0,019664585 | FLJ23867 | FALSE | Upregulated |
| -0,247983038 | 0,019665332 | PAXIP1 | FALSE | Downregulated |
| 0,940354881 | 0,019734875 | CTGF | FALSE | Upregulated |
| 2,710935523 | 0,019747006 | CDH16 | TRUE | Upregulated |
| 0,270732488 | 0,019747006 | SH3RF1 | FALSE | Upregulated |
| 1,104604373 | 0,019782465 | BUB1 | TRUE | Upregulated |
| -0,882037925 | 0,019782465 | GNN | FALSE | Downregulated |
| 1,108957119 | 0,019782465 | NEK2 | TRUE | Upregulated |
| -0,335251084 | 0,019782465 | SLC25A4 | FALSE | Downregulated |
| 1,187589253 | 0,019790769 | P2RX1 | TRUE | Upregulated |
| -0,25198153 | 0,019810072 | DHFRL1 | FALSE | Downregulated |
| -0,912570712 | 0,01982146 | PODXL2 | FALSE | Downregulated |
| -0,393241341 | 0,019829606 | BLOC1S3 | FALSE | Downregulated |
| 0,394793102 | 0,019839813 | TSPAN4 | FALSE | Upregulated |
| 0,742527315 | 0,019870777 | CLIP4 | FALSE | Upregulated |
| 0,552769737 | 0,0199118 | NOS3 | FALSE | Upregulated |
| 0,667991376 | 0,019918332 | C8orf47 | FALSE | Upregulated |
| 0,345240403 | 0,019918332 | GLTSCR1 | FALSE | Upregulated |
| 2,196781 | 0,019918332 | LOC101928152 | TRUE | Upregulated |

| | | | | |
|---|---|---|---|---|
| -0,451184588 | 0,019918332 | ZKSCAN1 | FALSE | Downregulated |
| 0,572947004 | 0,019938963 | TTC22 | FALSE | Upregulated |
| 1,241037016 | 0,019956068 | F13A1 | TRUE | Upregulated |
| 0,339339572 | 0,01995824 | EMC8 | FALSE | Upregulated |
| 2,902709962 | 0,019985082 | KCNQ3 | TRUE | Upregulated |
| 0,282932932 | 0,019989977 | TMEM62 | FALSE | Upregulated |
| -1,146046027 | 0,019992482 | OLFM2 | TRUE | Downregulated |
| 0,485991873 | 0,020109066 | MOSPD1 | FALSE | Upregulated |
| 0,241883053 | 0,020118857 | RAP1A | FALSE | Upregulated |
| 0,397980066 | 0,020118857 | SMYD2 | FALSE | Upregulated |
| -0,79860008 | 0,020163456 | MT1E | FALSE | Downregulated |
| -0,30002538 | 0,020167936 | ATP5L | FALSE | Downregulated |
| 1,034823093 | 0,020168918 | CUBN | TRUE | Upregulated |
| 0,30560071 | 0,020204995 | MAP3K11 | FALSE | Upregulated |
| -0,955044905 | 0,020208797 | FLJ36777 | FALSE | Downregulated |
| 0,836392987 | 0,020218573 | CGREF1 | FALSE | Upregulated |
| 0,408192255 | 0,020218573 | RHOT2 | FALSE | Upregulated |
| 0,973056414 | 0,020218573 | ZNF827 | FALSE | Upregulated |
| 2,722337255 | 0,020220764 | EGOT | TRUE | Upregulated |
| 0,268367063 | 0,020242055 | RBM12 | FALSE | Upregulated |
| 1,065211491 | 0,020242055 | SNCAIP | TRUE | Upregulated |
| -0,423826517 | 0,020247977 | PHYHD1 | FALSE | Downregulated |
| -0,217021525 | 0,020272008 | MAGEF1 | FALSE | Downregulated |
| 0,177393025 | 0,020343793 | EXOC7 | FALSE | Upregulated |
| 0,550546434 | 0,020368468 | SLC25A35 | FALSE | Upregulated |
| 0,241176391 | 0,020382039 | VPS45 | FALSE | Upregulated |
| -0,228124255 | 0,0203844 | ZNF480 | FALSE | Downregulated |
| 0,430789074 | 0,020389014 | BGN | FALSE | Upregulated |
| 1,789703398 | 0,020447116 | TKTL1 | TRUE | Upregulated |
| 0,305407262 | 0,020451182 | IARS | FALSE | Upregulated |
| 1,583865109 | 0,020451182 | PTPRQ | TRUE | Upregulated |
| -0,183086696 | 0,020454444 | RABEP1 | FALSE | Downregulated |
| -0,22785702 | 0,020483137 | ANKRD49 | FALSE | Downregulated |
| 0,234673904 | 0,020483137 | COPS8 | FALSE | Upregulated |
| 1,164119658 | 0,020483137 | MEIS3 | TRUE | Upregulated |
| -0,25670591 | 0,020483137 | WDR55 | FALSE | Downregulated |
| -0,301483928 | 0,020483137 | ZDHHC16 | FALSE | Downregulated |
| 0,470437982 | 0,020584585 | PPM1M | FALSE | Upregulated |
| 0,434835269 | 0,020584585 | PVRL2 | FALSE | Upregulated |
| 0,428464094 | 0,020584585 | WDR60 | FALSE | Upregulated |
| 0,333008448 | 0,020589253 | CXCL16 | FALSE | Upregulated |
| 1,18405577 | 0,020596835 | FGF12 | TRUE | Upregulated |
| 0,382219854 | 0,020603348 | MLH3 | FALSE | Upregulated |
| -0,379802166 | 0,020610985 | C21orf59 | FALSE | Downregulated |
| -0,700093343 | 0,020610985 | GPR98 | FALSE | Downregulated |
| -0,273438633 | 0,020618022 | DUSP3 | FALSE | Downregulated |
| 0,740224679 | 0,020619255 | LTB | FALSE | Upregulated |
| 0,774663507 | 0,020648186 | LOC102724592 | FALSE | Upregulated |
| -0,464782212 | 0,020648186 | PIK3R1 | FALSE | Downregulated |
| -0,346072368 | 0,020649451 | RBM34 | FALSE | Downregulated |
| 0,91223521 | 0,020667603 | DHRS13 | FALSE | Upregulated |

| | | | | |
|---|---|---|---|---|
| 0,486133987 | 0,020677614 | TRAPPC4 | FALSE | Upregulated |
| 0,632097478 | 0,020694758 | HPSE | FALSE | Upregulated |
| 0,677220808 | 0,020696094 | EIF5A2 | FALSE | Upregulated |
| 0,248836134 | 0,020696094 | TOR1A | FALSE | Upregulated |
| 2,822523766 | 0,020696094 | YBX3P1 | TRUE | Upregulated |
| -0,240656068 | 0,020740092 | ATP13A3 | FALSE | Downregulated |
| 0,594269017 | 0,020740092 | CD74 | FALSE | Upregulated |
| -0,322077317 | 0,020760126 | NDRG3 | FALSE | Downregulated |
| 1,55914596 | 0,020816427 | PPFIA2 | TRUE | Upregulated |
| -0,423579974 | 0,020829534 | CGN | FALSE | Downregulated |
| 1,161068879 | 0,020829534 | TIAF1 | TRUE | Upregulated |
| 0,162408504 | 0,020864935 | DHX30 | FALSE | Upregulated |
| 0,779728363 | 0,020892441 | ZNF14 | FALSE | Upregulated |
| -1,240497993 | 0,020916145 | SNTG1 | TRUE | Downregulated |
| 0,743480976 | 0,020951532 | FZD3 | FALSE | Upregulated |
| 0,606876565 | 0,020990583 | CD48 | FALSE | Upregulated |
| 0,484345813 | 0,020990613 | FZD6 | FALSE | Upregulated |
| -0,225219362 | 0,021073851 | ELP2 | FALSE | Downregulated |
| -0,244506789 | 0,021105333 | SAP30L | FALSE | Downregulated |
| -0,849963662 | 0,021137421 | LOC102724350 | FALSE | Downregulated |
| 1,033315988 | 0,021155641 | CHST10 | TRUE | Upregulated |
| 0,205897457 | 0,021155641 | SMARCD1 | FALSE | Upregulated |
| 0,328504579 | 0,021163318 | FAM49B | FALSE | Upregulated |
| 0,345258187 | 0,021176903 | UNC93B1 | FALSE | Upregulated |
| 1,270840532 | 0,021211065 | FCGR1A | TRUE | Upregulated |
| 1,758011282 | 0,021220733 | F2RL3 | TRUE | Upregulated |
| -0,658725197 | 0,021241697 | LYVE1 | FALSE | Downregulated |
| -0,610921204 | 0,021241697 | ZFYVE28 | FALSE | Downregulated |
| 1,981472737 | 0,021294071 | GSG2 | TRUE | Upregulated |
| -0,395421342 | 0,021337793 | SCML1 | FALSE | Downregulated |
| 0,602028416 | 0,021349095 | SAMSN1 | FALSE | Upregulated |
| -0,373387886 | 0,021378441 | ALAD | FALSE | Downregulated |
| 0,253041537 | 0,021387563 | NUPL2 | FALSE | Upregulated |
| 0,301027266 | 0,021402093 | HMGXB3 | FALSE | Upregulated |
| -0,321118742 | 0,021407776 | ZNF428 | FALSE | Downregulated |
| 0,592972399 | 0,021411915 | STAT4 | FALSE | Upregulated |
| 0,802072907 | 0,021424015 | PDZD2 | FALSE | Upregulated |
| -0,342720817 | 0,021424015 | TRPM7 | FALSE | Downregulated |
| 0,586840135 | 0,021439082 | FLT3LG | FALSE | Upregulated |
| -2,457128192 | 0,021442457 | BVES-AS1 | TRUE | Downregulated |
| 1,029823108 | 0,02145158 | LOC284454 | TRUE | Upregulated |
| 0,289808298 | 0,021454397 | ADNP2 | FALSE | Upregulated |
| -0,240291524 | 0,021456195 | LIN7C | FALSE | Downregulated |
| -0,488809571 | 0,021457581 | LIME1 | FALSE | Downregulated |
| 1,246431376 | 0,021463036 | SCART1 | TRUE | Upregulated |
| 1,250984056 | 0,021468605 | PLCB4 | TRUE | Upregulated |
| 2,231188982 | 0,021510426 | MESP2 | TRUE | Upregulated |
| -0,175498372 | 0,021510426 | PSMC1 | FALSE | Downregulated |
| 1,892410014 | 0,021576815 | LOC102724827 | TRUE | Upregulated |
| 2,414372884 | 0,021594564 | AIM1L | TRUE | Upregulated |
| 0,368803147 | 0,021594564 | DNAL4 | FALSE | Upregulated |

| | | | | |
|---|---|---|---|---|
| 0,28050995 | 0,021598928 | INPPL1 | FALSE | Upregulated |
| 1,314064125 | 0,02161852 | TMPRSS3 | TRUE | Upregulated |
| -0,223639728 | 0,021649717 | CSTF1 | FALSE | Downregulated |
| 0,353990766 | 0,021670295 | MAGED1 | FALSE | Upregulated |
| -1,012454752 | 0,021688607 | DOK6 | TRUE | Downregulated |
| -1,430320506 | 0,021688607 | EIF5AL1 | TRUE | Downregulated |
| 1,252802206 | 0,021717477 | KIF26B | TRUE | Upregulated |
| -0,267201865 | 0,021717477 | KLC4 | FALSE | Downregulated |
| -0,344625289 | 0,021717477 | QPRT | FALSE | Downregulated |
| -0,253903085 | 0,021717477 | ZC3H14 | FALSE | Downregulated |
| -0,439918009 | 0,021744068 | CCDC23 | FALSE | Downregulated |
| -0,457320021 | 0,021744068 | LOC100132352 | FALSE | Downregulated |
| -0,763614179 | 0,021744068 | LOC101928868 | FALSE | Downregulated |
| 0,400836114 | 0,021747487 | RHOBTB2 | FALSE | Upregulated |
| -0,578357516 | 0,021859479 | FCN2 | FALSE | Downregulated |
| 0,838387213 | 0,021887597 | PTTG1 | FALSE | Upregulated |
| 0,295187029 | 0,021896437 | PSME2 | FALSE | Upregulated |
| -0,199354417 | 0,021896564 | PHAX | FALSE | Downregulated |
| -0,269884477 | 0,021910111 | AP1AR | FALSE | Downregulated |
| 0,457134151 | 0,021913905 | DNMT1 | FALSE | Upregulated |
| 0,566472518 | 0,021920249 | CROCC | FALSE | Upregulated |
| -0,494414299 | 0,021945844 | CXCL14 | FALSE | Downregulated |
| -0,57411239 | 0,021945844 | SLC1A1 | FALSE | Downregulated |
| 0,288789842 | 0,021945844 | SMIM13 | FALSE | Upregulated |
| 0,316551386 | 0,021991433 | TTC13 | FALSE | Upregulated |
| 1,73868276 | 0,022011877 | LILRA4 | TRUE | Upregulated |
| 1,098850675 | 0,022011877 | LOC102724475 | TRUE | Upregulated |
| -0,201011248 | 0,022011877 | SLAIN2 | FALSE | Downregulated |
| -0,41912545 | 0,022011877 | SLC17A2 | FALSE | Downregulated |
| 0,845902684 | 0,022011877 | ZNF135 | FALSE | Upregulated |
| 0,349428852 | 0,022069284 | SQSTM1 | FALSE | Upregulated |
| 1,965763769 | 0,02208333 | LOC100288175 | TRUE | Upregulated |
| 0,665048327 | 0,0220997 | GPR161 | FALSE | Upregulated |
| 0,258390146 | 0,022103738 | ATP6AP1 | FALSE | Upregulated |
| 1,248973051 | 0,022105267 | SNAI3 | TRUE | Upregulated |
| -0,441799513 | 0,022138562 | SPRYD4 | FALSE | Downregulated |
| -0,263718757 | 0,022149276 | TTC19 | FALSE | Downregulated |
| 1,5743943 | 0,022159133 | MIR4697HG | TRUE | Upregulated |
| 0,367815734 | 0,022322155 | CDR2 | FALSE | Upregulated |
| 1,17404972 | 0,022322739 | TMEM51-AS1 | TRUE | Upregulated |
| 0,277898058 | 0,022394613 | NSD1 | FALSE | Upregulated |
| -0,216301224 | 0,022404555 | TATDN3 | FALSE | Downregulated |
| 0,953701813 | 0,022406471 | MDK | FALSE | Upregulated |
| -0,512911563 | 0,022445064 | ABCG2 | FALSE | Downregulated |
| -0,98630367 | 0,022462019 | IFI44L | FALSE | Downregulated |
| 0,819384288 | 0,022462019 | KCNJ15 | FALSE | Upregulated |
| 0,284975409 | 0,022487597 | GNG12 | FALSE | Upregulated |
| 0,852958075 | 0,022498029 | NPHP4 | FALSE | Upregulated |
| 0,803763413 | 0,022576307 | JAK3 | FALSE | Upregulated |
| 0,312364807 | 0,022607266 | FOXP4 | FALSE | Upregulated |
| 1,855056386 | 0,02263673 | SLC12A5 | TRUE | Upregulated |

| | | | | |
|---|---|---|---|---|
| -0,293577661 | 0,022655107 | ZNF326 | FALSE | Downregulated |
| 0,404765751 | 0,022675715 | SPRYD3 | FALSE | Upregulated |
| -0,380210906 | 0,022679894 | DECR1 | FALSE | Downregulated |
| -0,261011841 | 0,022679894 | NOP14 | FALSE | Downregulated |
| -0,177308041 | 0,022699766 | UBE2Q1 | FALSE | Downregulated |
| -0,863213845 | 0,022700881 | LOC101929107 | FALSE | Downregulated |
| 0,607247479 | 0,02276369 | NCS1 | FALSE | Upregulated |
| 0,429341135 | 0,02276732 | RAB3IL1 | FALSE | Upregulated |
| 1,76559692 | 0,022772397 | FRMD1 | TRUE | Upregulated |
| -0,317501342 | 0,022777505 | MKKS | FALSE | Downregulated |
| 0,269336934 | 0,022777505 | NR3C1 | FALSE | Upregulated |
| -0,662592087 | 0,022779548 | TMED8 | FALSE | Downregulated |
| -0,238570087 | 0,022785436 | TMBIM6 | FALSE | Downregulated |
| 0,210594885 | 0,022798428 | EIF4G1 | FALSE | Upregulated |
| 0,426926691 | 0,02282766 | IFFO2 | FALSE | Upregulated |
| -0,680476101 | 0,022855318 | ORM1 | FALSE | Downregulated |
| 0,261708251 | 0,022902668 | C2CD3 | FALSE | Upregulated |
| 0,236421211 | 0,022907644 | PI4KA | FALSE | Upregulated |
| -0,303185132 | 0,022920689 | PGM1 | FALSE | Downregulated |
| 1,02856623 | 0,022920689 | VEPH1 | TRUE | Upregulated |
| 0,193773899 | 0,022922854 | STXBP3 | FALSE | Upregulated |
| 1,646486251 | 0,022946297 | NOS2 | TRUE | Upregulated |
| 0,823352769 | 0,02295717 | TRPC1 | FALSE | Upregulated |
| 0,622227984 | 0,02296514 | LENG9 | FALSE | Upregulated |
| -1,832057571 | 0,022974708 | TNR | TRUE | Downregulated |
| -0,486470068 | 0,022989042 | SMC6 | FALSE | Downregulated |
| -0,282865588 | 0,023002829 | ZNF346 | FALSE | Downregulated |
| -0,455137192 | 0,023007036 | NAT2 | FALSE | Downregulated |
| -0,329099672 | 0,02301485 | CAPRIN2 | FALSE | Downregulated |
| -0,46928925 | 0,023062561 | CRHBP | FALSE | Downregulated |
| -0,268495096 | 0,023084295 | HSPB11 | FALSE | Downregulated |
| -0,272990377 | 0,023096672 | KBTBD6 | FALSE | Downregulated |
| 0,262748539 | 0,023101736 | SLC38A10 | FALSE | Upregulated |
| 2,371957678 | 0,023206245 | FRMPD3 | TRUE | Upregulated |
| -0,251872392 | 0,023254037 | C1QBP | FALSE | Downregulated |
| 0,406164674 | 0,023293443 | SARM1 | FALSE | Upregulated |
| 0,359897633 | 0,023302899 | RNF135 | FALSE | Upregulated |
| -0,476966424 | 0,023331091 | PISD | FALSE | Downregulated |
| 1,018326038 | 0,023331091 | RPSAP9 | TRUE | Upregulated |
| -0,199413408 | 0,023331091 | WDR20 | FALSE | Downregulated |
| 2,42443644 | 0,023347139 | WNT10A | TRUE | Upregulated |
| 0,289737963 | 0,023347485 | ATG13 | FALSE | Upregulated |
| 2,909846169 | 0,023349328 | LINC01186 | TRUE | Upregulated |
| 0,757585584 | 0,023391144 | NES | FALSE | Upregulated |
| -0,22222111 | 0,023391144 | ZC3H15 | FALSE | Downregulated |
| 0,378864559 | 0,023409143 | EFNB1 | FALSE | Upregulated |
| 0,450371687 | 0,023419012 | CSTB | FALSE | Upregulated |
| 0,735560064 | 0,023448441 | SNX22 | FALSE | Upregulated |
| 0,275718231 | 0,023470478 | FBXO42 | FALSE | Upregulated |
| 2,259637499 | 0,023472462 | ZNF365 | TRUE | Upregulated |
| 0,330764021 | 0,023485987 | NELFA | FALSE | Upregulated |

| | | | | |
|---|---|---|---|---|
| -0,264701452 | 0,023514109 | FAM120A | FALSE | Downregulated |
| -0,812262482 | 0,023514109 | NPIPB3 | FALSE | Downregulated |
| -0,476385636 | 0,023516059 | C20orf96 | FALSE | Downregulated |
| -0,996255343 | 0,023553549 | SCGB1B2P | FALSE | Downregulated |
| 1,094865392 | 0,023557748 | TMEM200C | TRUE | Upregulated |
| 0,533440615 | 0,023570436 | TMCC3 | FALSE | Upregulated |
| 1,145293522 | 0,02359268 | IGSF10 | TRUE | Upregulated |
| 0,496547019 | 0,023633474 | ZNF385A | FALSE | Upregulated |
| 0,668822509 | 0,023681841 | COL12A1 | FALSE | Upregulated |
| -0,724602923 | 0,023706289 | POMK | FALSE | Downregulated |
| -0,523833621 | 0,023742197 | MMAB | FALSE | Downregulated |
| 0,446139926 | 0,023743571 | MZT1 | FALSE | Upregulated |
| 0,285037937 | 0,023863264 | PDXK | FALSE | Upregulated |
| -0,699706943 | 0,023890771 | C19orf73 | FALSE | Downregulated |
| 0,826724421 | 0,023890771 | DGKH | FALSE | Upregulated |
| 0,561339593 | 0,023890771 | RRS1 | FALSE | Upregulated |
| 0,383671886 | 0,023917636 | SRSF7 | FALSE | Upregulated |
| -0,29711535 | 0,023917636 | ZNF226 | FALSE | Downregulated |
| -0,145636906 | 0,023948565 | CNOT7 | FALSE | Downregulated |
| -0,275278921 | 0,02396072 | PDCD5 | FALSE | Downregulated |
| 0,255012413 | 0,023974319 | USP19 | FALSE | Upregulated |
| -0,290692552 | 0,024005023 | NCBP2-AS2 | FALSE | Downregulated |
| -0,979949412 | 0,024009409 | FLJ30403 | FALSE | Downregulated |
| 0,914617937 | 0,024045907 | SLC4A8 | FALSE | Upregulated |
| 1,008795295 | 0,024048093 | CBR3 | TRUE | Upregulated |
| -0,331945325 | 0,024067285 | MTG2 | FALSE | Downregulated |
| -0,401320147 | 0,024086765 | GATM | FALSE | Downregulated |
| -0,603861191 | 0,024086765 | NPIPA8 | FALSE | Downregulated |
| 0,426905505 | 0,024104497 | C1orf216 | FALSE | Upregulated |
| 0,442169078 | 0,024104497 | SRM | FALSE | Upregulated |
| 0,331123855 | 0,0241078 | ZNF618 | FALSE | Upregulated |
| 0,445081786 | 0,024108798 | LIPA | FALSE | Upregulated |
| 2,499244658 | 0,024115726 | LOC285043 | TRUE | Upregulated |
| 0,801120545 | 0,024115726 | SIDT1 | FALSE | Upregulated |
| -0,312892195 | 0,024117349 | ISOC2 | FALSE | Downregulated |
| 0,328186177 | 0,024143024 | ACTN4 | FALSE | Upregulated |
| 0,490333141 | 0,024211413 | HLA-DPB1 | FALSE | Upregulated |
| -0,953790376 | 0,024211413 | LOC101928887 | FALSE | Downregulated |
| 0,309799429 | 0,024211413 | R3HDM4 | FALSE | Upregulated |
| -0,294669423 | 0,024211413 | SLC35A3 | FALSE | Downregulated |
| 0,897093385 | 0,024211413 | TNFSF15 | FALSE | Upregulated |
| -0,25769508 | 0,024214442 | ABHD17B | FALSE | Downregulated |
| 0,437185039 | 0,024214442 | FBXO44 | FALSE | Upregulated |
| 0,714723296 | 0,024214442 | LOC102723726 | FALSE | Upregulated |
| -0,246214201 | 0,024225997 | PIGN | FALSE | Downregulated |
| -1,517401343 | 0,024287013 | LOC102724402 | TRUE | Downregulated |
| -0,359604386 | 0,024308014 | MESDC2 | FALSE | Downregulated |
| 0,33885045 | 0,024317331 | RAP2A | FALSE | Upregulated |
| 1,68195362 | 0,024320553 | STC2 | TRUE | Upregulated |
| -0,234045917 | 0,024320995 | RHOT1 | FALSE | Downregulated |
| -0,559733883 | 0,024320995 | RP11-326F20.5 | FALSE | Downregulated |

| | | | | |
|---|---|---|---|---|
| -0,513208254 | 0,024324073 | FETUB | FALSE | Downregulated |
| -0,397312767 | 0,024324073 | IVNS1ABP | FALSE | Downregulated |
| 0,746880391 | 0,024331009 | BEST1 | FALSE | Upregulated |
| -0,564738819 | 0,024331009 | BTBD16 | FALSE | Downregulated |
| 1,168544372 | 0,024331009 | CCDC80 | TRUE | Upregulated |
| 0,564751453 | 0,024331009 | KLF4 | FALSE | Upregulated |
| 2,27250406 | 0,024335996 | SAA2-SAA4 | TRUE | Upregulated |
| -0,293658371 | 0,024423652 | COX19 | FALSE | Downregulated |
| -0,381139983 | 0,024423652 | FAM162A | FALSE | Downregulated |
| -0,23206116 | 0,02443136 | CCDC50 | FALSE | Downregulated |
| 0,836424216 | 0,024443187 | FXYD2 | FALSE | Upregulated |
| -0,294166271 | 0,024459685 | METTL18 | FALSE | Downregulated |
| -0,339079277 | 0,024459685 | ZCCHC17 | FALSE | Downregulated |
| 0,249351819 | 0,024509133 | C14orf159 | FALSE | Upregulated |
| 0,513247878 | 0,024519673 | TMEM106A | FALSE | Upregulated |
| -0,183019427 | 0,024519673 | VPS54 | FALSE | Downregulated |
| -0,294219673 | 0,024543224 | SLC9B2 | FALSE | Downregulated |
| 0,359034719 | 0,024569327 | DVL2 | FALSE | Upregulated |
| 0,762888949 | 0,024571254 | TSPAN11 | FALSE | Upregulated |
| -0,288765828 | 0,024584208 | RPS6KB2 | FALSE | Downregulated |
| -0,488555891 | 0,024610268 | FCN3 | FALSE | Downregulated |
| -0,36130835 | 0,024620643 | PON1 | FALSE | Downregulated |
| -0,317234548 | 0,024671558 | ARL3 | FALSE | Downregulated |
| -0,128842714 | 0,024682676 | SMU1 | FALSE | Downregulated |
| 1,080803582 | 0,024699977 | ESRP1 | TRUE | Upregulated |
| 1,160291779 | 0,024742508 | BHLHE41 | TRUE | Upregulated |
| 0,514619017 | 0,024781641 | IDUA | FALSE | Upregulated |
| -0,617176874 | 0,024790806 | TRAF3IP1 | FALSE | Downregulated |
| -0,367914481 | 0,024791765 | GIPC2 | FALSE | Downregulated |
| -0,555722714 | 0,024811198 | EIF5B | FALSE | Downregulated |
| 0,292196205 | 0,02481283 | PIGQ | FALSE | Upregulated |
| -0,329537787 | 0,024815797 | CCNYL1 | FALSE | Downregulated |
| -0,34448308 | 0,024825955 | ATP5I | FALSE | Downregulated |
| 0,367362289 | 0,024827602 | CLN8 | FALSE | Upregulated |
| -1,015622025 | 0,024863786 | LOC102724645 | TRUE | Downregulated |
| 1,197412063 | 0,024885388 | DND1 | TRUE | Upregulated |
| 0,238999438 | 0,02488872 | SUGP1 | FALSE | Upregulated |
| -0,523965482 | 0,024895244 | CBFA2T3 | FALSE | Downregulated |
| -0,306952786 | 0,02493037 | ITIH2 | FALSE | Downregulated |
| -0,235879039 | 0,024935545 | KRIT1 | FALSE | Downregulated |
| -0,316141225 | 0,024936067 | RPS28 | FALSE | Downregulated |
| -0,271137071 | 0,024965652 | ERLIN2 | FALSE | Downregulated |
| 1,691675559 | 0,024965652 | SIGLEC6 | TRUE | Upregulated |
| -0,423755249 | 0,02509124 | ITSN1 | FALSE | Downregulated |
| 1,670026188 | 0,025098188 | LIPH | TRUE | Upregulated |
| -0,423493518 | 0,025113465 | KBTBD11 | FALSE | Downregulated |
| 1,111194402 | 0,025115258 | BOLA3-AS1 | TRUE | Upregulated |
| -0,586025978 | 0,025115258 | RP11-60I3.5 | FALSE | Downregulated |
| 0,498526753 | 0,025147073 | DGCR14 | FALSE | Upregulated |
| 0,356941071 | 0,025153136 | TKT | FALSE | Upregulated |
| 0,807304013 | 0,025171563 | PLAT | FALSE | Upregulated |

| | | | | |
|---|---|---|---|---|
| -2,158182569 | 0,025180133 | NRAP | TRUE | Downregulated |
| 1,700428263 | 0,025201486 | FAM135B | TRUE | Upregulated |
| -2,106124932 | 0,025209421 | DCD | TRUE | Downregulated |
| -0,993165696 | 0,025213176 | LRRC16B | FALSE | Downregulated |
| 1,15164768 | 0,025213176 | RIN1 | TRUE | Upregulated |
| -0,142585061 | 0,025226065 | METAP1 | FALSE | Downregulated |
| 0,407898928 | 0,025226065 | PHF13 | FALSE | Upregulated |
| 1,757504356 | 0,025259917 | RNF219-AS1 | TRUE | Upregulated |
| 0,475081355 | 0,025271823 | HOMER3 | FALSE | Upregulated |
| -1,114391004 | 0,025278589 | USH2A | TRUE | Downregulated |
| 0,698914035 | 0,025278589 | ZC3H12A | FALSE | Upregulated |
| -1,177620019 | 0,025320236 | LINC00086 | TRUE | Downregulated |
| 0,248369697 | 0,025320236 | RBM4 | FALSE | Upregulated |
| -0,738178048 | 0,025326662 | RAB3A | FALSE | Downregulated |
| 0,285456137 | 0,025336796 | SF3B4 | FALSE | Upregulated |
| 0,336571889 | 0,025377165 | TCTN1 | FALSE | Upregulated |
| 1,590780044 | 0,025396676 | LOC100506098 | TRUE | Upregulated |
| 1,213241023 | 0,025413815 | SLC26A4 | TRUE | Upregulated |
| -0,225364442 | 0,025444965 | MAPK8 | FALSE | Downregulated |
| -1,232150571 | 0,025454844 | AVPR1A | TRUE | Downregulated |
| -0,273547339 | 0,025468375 | PEX16 | FALSE | Downregulated |
| 0,30670842 | 0,025523032 | CSRP1 | FALSE | Upregulated |
| 0,478882783 | 0,02552409 | HN1 | FALSE | Upregulated |
| 0,380962173 | 0,025530478 | EPS8L2 | FALSE | Upregulated |
| 0,265859808 | 0,025533233 | HMCES | FALSE | Upregulated |
| -0,315675989 | 0,025536203 | C4orf29 | FALSE | Downregulated |
| -0,212925528 | 0,025587527 | SECISBP2 | FALSE | Downregulated |
| 0,249123922 | 0,025587527 | TRAPPC12 | FALSE | Upregulated |
| -0,403997678 | 0,025620867 | TCEANC | FALSE | Downregulated |
| -0,435139746 | 0,025628999 | GRAMD3 | FALSE | Downregulated |
| -0,645509364 | 0,025628999 | PRG4 | FALSE | Downregulated |
| -0,782041307 | 0,025644143 | LOC730101 | FALSE | Downregulated |
| 2,145319099 | 0,025668012 | DNAH17 | TRUE | Upregulated |
| 0,519068815 | 0,025692545 | SLC25A36 | FALSE | Upregulated |
| -0,194124665 | 0,025692545 | SUMO1 | FALSE | Downregulated |
| -0,432594954 | 0,025747979 | SLC2A4RG | FALSE | Downregulated |
| 0,24814445 | 0,02576174 | CHMP1A | FALSE | Upregulated |
| 1,238899785 | 0,025767243 | GGT6 | TRUE | Upregulated |
| -0,26985463 | 0,02577709 | LINC00998 | FALSE | Downregulated |
| -0,727062745 | 0,025785664 | PDK4 | FALSE | Downregulated |
| 0,379742676 | 0,025801393 | MAP3K9 | FALSE | Upregulated |
| -0,439697672 | 0,025855072 | LTV1 | FALSE | Downregulated |
| 0,314634264 | 0,025919627 | ELMSAN1 | FALSE | Upregulated |
| -0,300903338 | 0,025919627 | MAP2K7 | FALSE | Downregulated |
| 1,48998852 | 0,025919627 | RASL12 | TRUE | Upregulated |
| 0,340845807 | 0,025931471 | PPP4R1 | FALSE | Upregulated |
| 2,451088809 | 0,025948783 | SMARCA5-AS1 | TRUE | Upregulated |
| 0,369805931 | 0,025963008 | RASAL2 | FALSE | Upregulated |
| 0,454285534 | 0,025983796 | PCGF2 | FALSE | Upregulated |
| 0,508403755 | 0,026012034 | ZC3H3 | FALSE | Upregulated |
| 0,457894099 | 0,02601749 | FBXL6 | FALSE | Upregulated |

| | | | | |
|---|---|---|---|---|
| -1,518001554 | 0,026077386 | CLGN | TRUE | Downregulated |
| 0,221523417 | 0,026096101 | ANKIB1 | FALSE | Upregulated |
| 0,292595255 | 0,026096101 | CCDC71 | FALSE | Upregulated |
| 0,593672645 | 0,026102979 | RPP25 | FALSE | Upregulated |
| 0,356971233 | 0,026106842 | RBM45 | FALSE | Upregulated |
| -0,538924496 | 0,026127138 | CKMT2-AS1 | FALSE | Downregulated |
| 0,714694079 | 0,026127138 | FAM212B | FALSE | Upregulated |
| 0,225614032 | 0,026143972 | SNX17 | FALSE | Upregulated |
| 0,768841334 | 0,026145293 | CDC7 | FALSE | Upregulated |
| 0,526629937 | 0,026147377 | HLA-DPA1 | FALSE | Upregulated |
| -0,335170399 | 0,026147377 | TMEM60 | FALSE | Downregulated |
| 0,262300472 | 0,026186186 | PIGG | FALSE | Upregulated |
| -1,80888265 | 0,026190879 | LOC101930100 | TRUE | Downregulated |
| 0,823819399 | 0,026223733 | DZIP1L | FALSE | Upregulated |
| 2,04073943 | 0,026229555 | RIMKLA | TRUE | Upregulated |
| -0,399627031 | 0,026233377 | C11orf71 | FALSE | Downregulated |
| -0,425198948 | 0,026233377 | FOLH1 | FALSE | Downregulated |
| 0,437637559 | 0,026233377 | HDAC7 | FALSE | Upregulated |
| 0,274873828 | 0,026233377 | STXBP2 | FALSE | Upregulated |
| 0,525885287 | 0,026233377 | WFS1 | FALSE | Upregulated |
| -0,166574518 | 0,026233377 | ZNF644 | FALSE | Downregulated |
| -0,336699254 | 0,026238575 | HERC3 | FALSE | Downregulated |
| -0,202308554 | 0,026238575 | NGDN | FALSE | Downregulated |
| 0,483480181 | 0,026268528 | NDOR1 | FALSE | Upregulated |
| 1,524204285 | 0,026268528 | NSG1 | TRUE | Upregulated |
| 0,270319393 | 0,026306516 | CLEC16A | FALSE | Upregulated |
| 0,393100679 | 0,026306516 | IRF9 | FALSE | Upregulated |
| 2,991471058 | 0,026306516 | SAA3P | TRUE | Upregulated |
| 0,192488088 | 0,026307851 | AMFR | FALSE | Upregulated |
| -0,330759875 | 0,026307851 | ASTE1 | FALSE | Downregulated |
| -0,341307295 | 0,026317955 | MAP10 | FALSE | Downregulated |
| 3,137450969 | 0,026317955 | SNORD3A | TRUE | Upregulated |
| 1,02840828 | 0,026340567 | GPR68 | TRUE | Upregulated |
| -0,322340579 | 0,02634581 | SETDB2 | FALSE | Downregulated |
| -0,281210072 | 0,026379125 | ELP4 | FALSE | Downregulated |
| 0,54670134 | 0,026379125 | ZFPM2 | FALSE | Upregulated |
| 1,273919541 | 0,026390963 | KIAA1614 | TRUE | Upregulated |
| 1,915461201 | 0,026390963 | STMND1 | TRUE | Upregulated |
| -0,281057403 | 0,026390963 | TOPORS | FALSE | Downregulated |
| 0,665899479 | 0,026390963 | TRAF3IP3 | FALSE | Upregulated |
| -0,259462193 | 0,026401612 | MAP3K2 | FALSE | Downregulated |
| -0,13675998 | 0,026401612 | SRSF1 | FALSE | Downregulated |
| 0,352588405 | 0,026415072 | RNFT1 | FALSE | Upregulated |
| -0,928575275 | 0,026461033 | HORMAD2 | FALSE | Downregulated |
| -0,358736321 | 0,026516335 | PTPLB | FALSE | Downregulated |
| 0,960090591 | 0,026533653 | FBXL19-AS1 | FALSE | Upregulated |
| 0,405246093 | 0,026538232 | ARSA | FALSE | Upregulated |
| 0,621403255 | 0,026551504 | ZNF763 | FALSE | Upregulated |
| -0,365789221 | 0,026554935 | IRS1 | FALSE | Downregulated |
| 0,636262975 | 0,02656572 | ME3 | FALSE | Upregulated |
| -0,716306889 | 0,02656572 | RP11-439L18.3 | FALSE | Downregulated |

| | | | | |
|---|---|---|---|---|
| 0,727387254 | 0,02656572 | RPSAP58 | FALSE | Upregulated |
| 0,252869126 | 0,02657671 | EPS8 | FALSE | Upregulated |
| 1,089023059 | 0,026588162 | LOC100506022 | TRUE | Upregulated |
| 0,594609605 | 0,026602988 | INMT | FALSE | Upregulated |
| 0,886250993 | 0,026602988 | LINC00342 | FALSE | Upregulated |
| 0,764560408 | 0,026602988 | MAD2L1 | FALSE | Upregulated |
| 0,19735211 | 0,026615893 | CLTA | FALSE | Upregulated |
| 0,244900841 | 0,026645403 | ACVR1 | FALSE | Upregulated |
| 2,171195806 | 0,026645403 | LOC102724385 | TRUE | Upregulated |
| 0,903360625 | 0,026678374 | SLC5A1 | FALSE | Upregulated |
| 0,468894878 | 0,026685858 | BAK1 | FALSE | Upregulated |
| -0,871822546 | 0,026696003 | NRG4 | FALSE | Downregulated |
| 0,983820975 | 0,026734854 | LRP8 | FALSE | Upregulated |
| 0,435374131 | 0,02676475 | GAS8 | FALSE | Upregulated |
| 0,174163143 | 0,026838628 | GCN1L1 | FALSE | Upregulated |
| 1,457845792 | 0,026888365 | EDA2R | TRUE | Upregulated |
| 0,591300212 | 0,026888365 | PRR26 | FALSE | Upregulated |
| -1,699050728 | 0,026888365 | PZP | TRUE | Downregulated |
| 0,729826343 | 0,026928594 | KLRD1 | FALSE | Upregulated |
| -0,563769335 | 0,026944791 | C3P1 | FALSE | Downregulated |
| 0,852717072 | 0,026996696 | MFAP4 | FALSE | Upregulated |
| -0,340113033 | 0,027019995 | DDIT3 | FALSE | Downregulated |
| -1,001731384 | 0,02702492 | XAGE3 | TRUE | Downregulated |
| -0,23482001 | 0,027047299 | FBXW2 | FALSE | Downregulated |
| 1,378475241 | 0,027047397 | LOC102723517 | TRUE | Upregulated |
| -0,579101434 | 0,027047397 | PID1 | FALSE | Downregulated |
| -0,304429893 | 0,027047397 | SFXN5 | FALSE | Downregulated |
| 0,357726214 | 0,027050462 | TTLL4 | FALSE | Upregulated |
| 1,05997802 | 0,027067731 | PLXDC1 | TRUE | Upregulated |
| 0,378417278 | 0,027106071 | CYYR1 | FALSE | Upregulated |
| -0,248929286 | 0,027110387 | ATG14 | FALSE | Downregulated |
| -0,184836677 | 0,027137888 | OS9 | FALSE | Downregulated |
| -0,550764794 | 0,027149058 | C1QTNF3 | FALSE | Downregulated |
| 0,79434656 | 0,027149058 | CCDC110 | FALSE | Upregulated |
| -0,2698175 | 0,027149058 | MSH3 | FALSE | Downregulated |
| -0,220607993 | 0,027149058 | ST6GAL1 | FALSE | Downregulated |
| 0,527314263 | 0,027166899 | RUNX1 | FALSE | Upregulated |
| -0,593082749 | 0,027192546 | RP11-141O11.2 | FALSE | Downregulated |
| -0,403392339 | 0,027192546 | SELENBP1 | FALSE | Downregulated |
| 0,793451722 | 0,027228477 | ABLIM2 | FALSE | Upregulated |
| 1,198848074 | 0,027359008 | TMEM158 | TRUE | Upregulated |
| 0,484174088 | 0,027392982 | MARVELD1 | FALSE | Upregulated |
| 0,542730777 | 0,027392982 | THEMIS2 | FALSE | Upregulated |
| -0,361449793 | 0,027401366 | CYP2C9 | FALSE | Downregulated |
| -0,415206204 | 0,027401366 | MITD1 | FALSE | Downregulated |
| 0,97143638 | 0,027428752 | FAM83B | FALSE | Upregulated |
| 0,450457903 | 0,027472836 | GPSM3 | FALSE | Upregulated |
| -0,289305021 | 0,027482148 | MTG1 | FALSE | Downregulated |
| 0,684339018 | 0,027508448 | HSH2D | FALSE | Upregulated |
| -0,257908626 | 0,027508448 | MYO1B | FALSE | Downregulated |
| -0,269989427 | 0,027529753 | SAYSD1 | FALSE | Downregulated |

| | | | | |
|---|---|---|---|---|
| 0,48577565 | 0,027535297 | GGT5 | FALSE | Upregulated |
| -0,704810281 | 0,027535297 | LOC553103 | FALSE | Downregulated |
| 2,262467626 | 0,027558517 | LOC102723996 | TRUE | Upregulated |
| -0,619447546 | 0,027565639 | LUC7L3 | FALSE | Downregulated |
| -1,117677519 | 0,027565639 | NPIPB6 | TRUE | Downregulated |
| 0,409082049 | 0,027568653 | TRIP6 | FALSE | Upregulated |
| 0,734909511 | 0,027593915 | CD34 | FALSE | Upregulated |
| 0,245557165 | 0,027627905 | ERH | FALSE | Upregulated |
| 0,305732692 | 0,027627905 | ZNF142 | FALSE | Upregulated |
| 2,332095826 | 0,027646874 | PCSK1N | TRUE | Upregulated |
| 0,90089641 | 0,027677574 | BATF3 | FALSE | Upregulated |
| 0,33451717 | 0,027677574 | UBL7 | FALSE | Upregulated |
| -1,104530189 | 0,027690962 | OR7E37P | TRUE | Downregulated |
| -0,304259564 | 0,027707693 | GLO1 | FALSE | Downregulated |
| 0,474348472 | 0,027742505 | CAMK1D | FALSE | Upregulated |
| -0,27593885 | 0,027828072 | APPL1 | FALSE | Downregulated |
| -0,205694894 | 0,02788628 | NOA1 | FALSE | Downregulated |
| 2,130028711 | 0,027904487 | HECW1 | TRUE | Upregulated |
| 0,308891074 | 0,027930118 | EGLN2 | FALSE | Upregulated |
| -0,263708229 | 0,027987933 | CACYBP | FALSE | Downregulated |
| -0,694117461 | 0,027987933 | POLR2A | FALSE | Downregulated |
| 0,279688282 | 0,027991901 | IFNAR1 | FALSE | Upregulated |
| 0,380134997 | 0,028007973 | COL5A2 | FALSE | Upregulated |
| 1,659363298 | 0,028121294 | KNDC1 | TRUE | Upregulated |
| -0,281762779 | 0,028140688 | ETFA | FALSE | Downregulated |
| 0,405366879 | 0,028147899 | C19orf44 | FALSE | Upregulated |
| -0,378725971 | 0,028149554 | ZYG11B | FALSE | Downregulated |
| 1,267963886 | 0,028204395 | UTS2B | TRUE | Upregulated |
| -1,049471958 | 0,028272387 | MT1G | TRUE | Downregulated |
| 2,207348529 | 0,028280998 | NRK | TRUE | Upregulated |
| 2,569448519 | 0,028304214 | CTXN1 | TRUE | Upregulated |
| 1,168541112 | 0,028304214 | MAP1A | TRUE | Upregulated |
| 0,529058969 | 0,028304214 | ZNF611 | FALSE | Upregulated |
| -0,471984904 | 0,028319787 | ADHFE1 | FALSE | Downregulated |
| 0,29085327 | 0,028401739 | NADSYN1 | FALSE | Upregulated |
| 0,670484567 | 0,0284354 | HSPA4L | FALSE | Upregulated |
| 1,729027308 | 0,02843855 | LINC00639 | TRUE | Upregulated |
| -0,231913813 | 0,028447162 | PROS1 | FALSE | Downregulated |
| -0,26657276 | 0,028508054 | VPS28 | FALSE | Downregulated |
| 2,108993844 | 0,028519591 | DCAF13P3 | TRUE | Upregulated |
| 1,662691507 | 0,028544422 | ANKRD36BP2 | TRUE | Upregulated |
| -0,323561294 | 0,028565745 | KCTD6 | FALSE | Downregulated |
| 1,335913987 | 0,028593687 | SCARNA9 | TRUE | Upregulated |
| 0,229370125 | 0,028616724 | ZFP64 | FALSE | Upregulated |
| 0,596313538 | 0,028656077 | EHD2 | FALSE | Upregulated |
| 0,459990712 | 0,028656077 | FBLN7 | FALSE | Upregulated |
| -0,435773729 | 0,028656077 | LOC101928319 | FALSE | Downregulated |
| 0,505644956 | 0,028656077 | LTBP3 | FALSE | Upregulated |
| -0,12918226 | 0,028656077 | MATR3 | FALSE | Downregulated |
| -0,166353805 | 0,028660422 | ARIH2 | FALSE | Downregulated |
| 0,981799499 | 0,028660422 | MIRLET7BHG | FALSE | Upregulated |

| | | | | |
|---|---|---|---|---|
| 0,682486611 | 0,028662209 | KCNC3 | FALSE | Upregulated |
| -0,328032809 | 0,028663983 | RPS15A | FALSE | Downregulated |
| -0,754410156 | 0,02866959 | COL11A2 | FALSE | Downregulated |
| -0,276309228 | 0,028677353 | RNF181 | FALSE | Downregulated |
| 0,347097404 | 0,028728392 | DHX57 | FALSE | Upregulated |
| 0,36036263 | 0,028728392 | LYN | FALSE | Upregulated |
| -0,61344586 | 0,028835825 | SYPL2 | FALSE | Downregulated |
| -0,397353837 | 0,028840354 | CD27-AS1 | FALSE | Downregulated |
| -0,334295481 | 0,02888229 | SFSWAP | FALSE | Downregulated |
| 1,082854453 | 0,028904338 | SIGLEC17P | TRUE | Upregulated |
| -0,209853595 | 0,028937279 | SYNJ2BP | FALSE | Downregulated |
| 0,317988872 | 0,028939209 | ABCB8 | FALSE | Upregulated |
| 2,032155424 | 0,028949106 | C18orf61 | TRUE | Upregulated |
| 2,226560143 | 0,028956116 | C2CD4A | TRUE | Upregulated |
| -0,495230953 | 0,028963585 | LIPT2 | FALSE | Downregulated |
| -0,199175518 | 0,028985503 | ASNSD1 | FALSE | Downregulated |
| 2,102117557 | 0,028985503 | UNC5A | TRUE | Upregulated |
| -0,84585786 | 0,028997212 | LRP2BP | FALSE | Downregulated |
| -0,471261745 | 0,029018233 | FLJ37035 | FALSE | Downregulated |
| 0,301040708 | 0,029021646 | PTPN6 | FALSE | Upregulated |
| -0,320587729 | 0,029024679 | RBM43 | FALSE | Downregulated |
| 0,63186811 | 0,029162238 | SGK1 | FALSE | Upregulated |
| -0,387933256 | 0,029168437 | NBR2 | FALSE | Downregulated |
| -0,655539895 | 0,029187856 | DDT | FALSE | Downregulated |
| -0,224374027 | 0,029188245 | MAF1 | FALSE | Downregulated |
| -0,250147608 | 0,029213604 | BAG4 | FALSE | Downregulated |
| 0,726593415 | 0,029213604 | HOXB5 | FALSE | Upregulated |
| 0,764052884 | 0,029213604 | MKI67 | FALSE | Upregulated |
| 0,462472265 | 0,029264975 | APH1B | FALSE | Upregulated |
| 0,247930563 | 0,029274738 | EXOC1 | FALSE | Upregulated |
| 0,474092084 | 0,029282483 | NCKAP1L | FALSE | Upregulated |
| 0,297138582 | 0,029282483 | VPS37B | FALSE | Upregulated |
| -0,682734385 | 0,029352304 | SMCO3 | FALSE | Downregulated |
| 1,193972992 | 0,029354402 | C1orf204 | TRUE | Upregulated |
| -0,212401574 | 0,029380136 | MRPL49 | FALSE | Downregulated |
| -0,871892544 | 0,029384069 | RP11-31F15.2 | FALSE | Downregulated |
| -0,288026634 | 0,029410945 | TUBD1 | FALSE | Downregulated |
| -0,785552574 | 0,029410945 | UBAP1L | FALSE | Downregulated |
| 0,233027544 | 0,0294356 | ARHGAP1 | FALSE | Upregulated |
| 0,658825048 | 0,0294356 | FAM43A | FALSE | Upregulated |
| -0,793227778 | 0,0294356 | RANBP3L | FALSE | Downregulated |
| 0,562118129 | 0,0294356 | SECTM1 | FALSE | Upregulated |
| -0,216212131 | 0,0294356 | ZNF143 | FALSE | Downregulated |
| -0,531956565 | 0,0294356 | ZNF652 | FALSE | Downregulated |
| 1,160171978 | 0,029447608 | SIGLEC8 | TRUE | Upregulated |
| 1,259967577 | 0,029455984 | IL2RA | TRUE | Upregulated |
| 0,96678351 | 0,029498188 | OMD | FALSE | Upregulated |
| -0,430632484 | 0,029498646 | LRP6 | FALSE | Downregulated |
| 0,286590263 | 0,029521475 | RFK | FALSE | Upregulated |
| 2,111077103 | 0,029558289 | GRID2IP | TRUE | Upregulated |
| -0,229728444 | 0,029567152 | EIF3E | FALSE | Downregulated |

| | | | | |
|---|---|---|---|---|
| 0,359042216 | 0,029578188 | C7orf50 | FALSE | Upregulated |
| 0,795691989 | 0,029630249 | PDZD4 | FALSE | Upregulated |
| -0,325023332 | 0,02966293 | OGG1 | FALSE | Downregulated |
| 2,520942201 | 0,029711917 | CD177 | TRUE | Upregulated |
| 1,003913423 | 0,029760734 | MAP1LC3B2 | TRUE | Upregulated |
| 0,964536958 | 0,029760734 | NOD2 | FALSE | Upregulated |
| 1,634771397 | 0,029788612 | CCAT1 | TRUE | Upregulated |
| 0,60833702 | 0,029792066 | ADA | FALSE | Upregulated |
| 0,223657501 | 0,029792066 | STRIP1 | FALSE | Upregulated |
| -0,420020741 | 0,029792066 | TMEM42 | FALSE | Downregulated |
| -0,238935002 | 0,029802757 | STRN3 | FALSE | Downregulated |
| 0,338828113 | 0,029808806 | DTX2 | FALSE | Upregulated |
| 0,270388598 | 0,029870863 | ARL2BP | FALSE | Upregulated |
| -0,851579771 | 0,029917202 | THRSP | FALSE | Downregulated |
| -0,330538248 | 0,030050441 | PDZD11 | FALSE | Downregulated |
| 1,50441791 | 0,030050677 | FAM189A2 | TRUE | Upregulated |
| 0,448723361 | 0,030050677 | SHB | FALSE | Upregulated |
| -0,40813413 | 0,030050677 | USP31 | FALSE | Downregulated |
| -0,54155924 | 0,030050677 | ZCCHC16 | FALSE | Downregulated |
| -0,230447073 | 0,030076218 | PATL1 | FALSE | Downregulated |
| 0,880834212 | 0,030076932 | GOLM1 | FALSE | Upregulated |
| -0,122077474 | 0,030098204 | USP7 | FALSE | Downregulated |
| -0,449289297 | 0,030103774 | MAP7D3 | FALSE | Downregulated |
| 0,807980101 | 0,030103774 | MARK1 | FALSE | Upregulated |
| -0,325699207 | 0,030138135 | NDUFA4 | FALSE | Downregulated |
| 0,274615399 | 0,030157563 | BBS1 | FALSE | Upregulated |
| 0,40758414 | 0,030191044 | CCDC120 | FALSE | Upregulated |
| 2,339034891 | 0,030208336 | BRD7P3 | TRUE | Upregulated |
| 0,736302833 | 0,030232261 | ROR1 | FALSE | Upregulated |
| 0,569060462 | 0,030302749 | LY96 | FALSE | Upregulated |
| 0,639600466 | 0,03030606 | TMEM229B | FALSE | Upregulated |
| -1,99379421 | 0,030330831 | LOC100996808 | TRUE | Downregulated |
| 0,472184463 | 0,03034276 | KSR1 | FALSE | Upregulated |
| 0,486649645 | 0,03035043 | ZNF506 | FALSE | Upregulated |
| 2,470452983 | 0,030362351 | LOC102724705 | TRUE | Upregulated |
| 0,196948034 | 0,030390257 | MCL1 | FALSE | Upregulated |
| 0,496553439 | 0,030418608 | AK1 | FALSE | Upregulated |
| -1,117215563 | 0,030418608 | LOC101928947 | TRUE | Downregulated |
| 2,709450435 | 0,030478907 | HES2 | TRUE | Upregulated |
| -0,231720657 | 0,030502437 | MPP5 | FALSE | Downregulated |
| -0,336511742 | 0,030502437 | NOP14-AS1 | FALSE | Downregulated |
| 0,417719734 | 0,030514855 | FAS | FALSE | Upregulated |
| 0,231077913 | 0,030514855 | ZNF609 | FALSE | Upregulated |
| 1,068073734 | 0,030568236 | OPRL1 | TRUE | Upregulated |
| -0,294297935 | 0,030570884 | RBM5 | FALSE | Downregulated |
| -0,220671311 | 0,030572195 | G3BP2 | FALSE | Downregulated |
| 1,733488414 | 0,030596517 | LOC101927481 | TRUE | Upregulated |
| -0,30583811 | 0,030615777 | UXT | FALSE | Downregulated |
| 0,668506409 | 0,030636362 | C3AR1 | FALSE | Upregulated |
| 0,428811516 | 0,030655073 | LOC642852 | FALSE | Upregulated |
| -0,228845605 | 0,030657128 | WASL | FALSE | Downregulated |

| | | | | |
|---|---|---|---|---|
| -0,249632269 | 0,03065729 | EDRF1 | FALSE | Downregulated |
| 0,260294547 | 0,030669023 | PIP5K1A | FALSE | Upregulated |
| -0,256608823 | 0,030669023 | SNX27 | FALSE | Downregulated |
| 0,563887418 | 0,030689834 | CLEC11A | FALSE | Upregulated |
| -0,410813607 | 0,030689834 | LOC220729 | FALSE | Downregulated |
| -1,903817428 | 0,030712949 | RP11-10L7.1 | TRUE | Downregulated |
| -0,26226502 | 0,030722383 | MTHFD1 | FALSE | Downregulated |
| 1,455360153 | 0,030754093 | FCRLA | TRUE | Upregulated |
| -0,382460377 | 0,030773231 | METTL20 | FALSE | Downregulated |
| 2,7124002 | 0,03077893 | HNRNPKP3 | TRUE | Upregulated |
| -0,17480261 | 0,03077893 | PANK2 | FALSE | Downregulated |
| -0,258051656 | 0,030836648 | EPM2AIP1 | FALSE | Downregulated |
| -0,289531934 | 0,030836648 | ZSCAN32 | FALSE | Downregulated |
| 0,353008403 | 0,030905193 | DLG5 | FALSE | Upregulated |
| -0,226375588 | 0,030905271 | SAFB | FALSE | Downregulated |
| 1,978567191 | 0,030922781 | CENPM | TRUE | Upregulated |
| 0,513857114 | 0,030942291 | GJA1 | FALSE | Upregulated |
| 1,000353971 | 0,030949009 | LGR6 | TRUE | Upregulated |
| 0,356120809 | 0,030989836 | LPGAT1 | FALSE | Upregulated |
| 0,625391329 | 0,031028068 | STARD9 | FALSE | Upregulated |
| 0,162288517 | 0,031058461 | CYFIP1 | FALSE | Upregulated |
| -0,503817489 | 0,031076213 | CCDC15 | FALSE | Downregulated |
| 0,543700028 | 0,031080921 | LILRB2 | FALSE | Upregulated |
| 0,355817384 | 0,031139569 | GNL3L | FALSE | Upregulated |
| 0,23571764 | 0,031139569 | TMEM209 | FALSE | Upregulated |
| 1,399366455 | 0,031143495 | LOC728026 | TRUE | Upregulated |
| -0,229408453 | 0,031168243 | IPO5 | FALSE | Downregulated |
| 1,074261285 | 0,031168436 | ETV4 | TRUE | Upregulated |
| -0,384570194 | 0,031168436 | HGSNAT | FALSE | Downregulated |
| 0,580233368 | 0,03118472 | FCHSD1 | FALSE | Upregulated |
| 0,383529868 | 0,03118472 | RRP7A | FALSE | Upregulated |
| 0,837593187 | 0,03118472 | XCL2 | FALSE | Upregulated |
| 0,458259743 | 0,031209598 | CYB561 | FALSE | Upregulated |
| -0,410638762 | 0,031209598 | UGT2B7 | FALSE | Downregulated |
| 0,686578191 | 0,031232712 | IL18BP | FALSE | Upregulated |
| -0,14638965 | 0,031248739 | C11orf58 | FALSE | Downregulated |
| 0,734712696 | 0,031248739 | PAFAH1B3 | FALSE | Upregulated |
| 0,237133177 | 0,031248739 | UBE2I | FALSE | Upregulated |
| -0,407292628 | 0,031283201 | PPFIBP1 | FALSE | Downregulated |
| 0,960806597 | 0,031283201 | TMEM159 | FALSE | Upregulated |
| -1,084519615 | 0,031342527 | LPA | TRUE | Downregulated |
| 0,223036877 | 0,031352792 | EIF4A3 | FALSE | Upregulated |
| 1,403667618 | 0,031370135 | CLCF1 | TRUE | Upregulated |
| -0,273044419 | 0,031384892 | APOB | FALSE | Downregulated |
| -0,372815219 | 0,031406764 | SEC16B | FALSE | Downregulated |
| -0,224037649 | 0,031407378 | NAE1 | FALSE | Downregulated |
| -0,195629999 | 0,031441557 | SNX5 | FALSE | Downregulated |
| -0,214050792 | 0,031460998 | HADHA | FALSE | Downregulated |
| 0,536654091 | 0,031492685 | DDB2 | FALSE | Upregulated |
| 0,227255827 | 0,031518617 | G3BP1 | FALSE | Upregulated |
| 1,743115946 | 0,031535212 | C12orf55 | TRUE | Upregulated |

| | | | | |
|---|---|---|---|---|
| 0,49696465 | 0,031580617 | TCF4 | FALSE | Upregulated |
| -0,391477169 | 0,031611517 | CPED1 | FALSE | Downregulated |
| 0,618544294 | 0,031611517 | NUAK2 | FALSE | Upregulated |
| -0,487199196 | 0,031611517 | PNPLA4 | FALSE | Downregulated |
| 0,752312041 | 0,031687328 | KCNQ1OT1 | FALSE | Upregulated |
| -0,206115692 | 0,031753082 | CDC5L | FALSE | Downregulated |
| 1,487159941 | 0,031753082 | LOC100996273 | TRUE | Upregulated |
| 0,317959854 | 0,031790217 | CBFB | FALSE | Upregulated |
| -0,447925593 | 0,031933181 | CHST7 | FALSE | Downregulated |
| 0,313369679 | 0,031933775 | PPP2R5B | FALSE | Upregulated |
| 0,639836945 | 0,032036969 | GAS2L3 | FALSE | Upregulated |
| 0,397766072 | 0,032038982 | FBXO10 | FALSE | Upregulated |
| 0,544983138 | 0,03206041 | PPP1R3D | FALSE | Upregulated |
| -0,691573884 | 0,03206041 | TMEM44-AS1 | FALSE | Downregulated |
| -0,950447203 | 0,032060502 | SEC14L3 | FALSE | Downregulated |
| -0,42010603 | 0,032126417 | SYTL5 | FALSE | Downregulated |
| 0,411340266 | 0,032126417 | TIMP2 | FALSE | Upregulated |
| -0,862846293 | 0,032155266 | SCARA5 | FALSE | Downregulated |
| -0,217289269 | 0,032155266 | ZSCAN21 | FALSE | Downregulated |
| 0,414422436 | 0,032168583 | WAS | FALSE | Upregulated |
| 1,53021706 | 0,032242603 | AC005594.3 | TRUE | Upregulated |
| 3,679846379 | 0,032265853 | CACNA1E | TRUE | Upregulated |
| 2,081397618 | 0,032273255 | LOC100335030 | TRUE | Upregulated |
| 0,336702493 | 0,032273255 | TNS1 | FALSE | Upregulated |
| -0,833430644 | 0,032308932 | SOAT2 | FALSE | Downregulated |
| -0,650852258 | 0,032311393 | CXXC4 | FALSE | Downregulated |
| -3,229933332 | 0,032311393 | TMOD4 | TRUE | Downregulated |
| -0,207977984 | 0,032312839 | AK6 | FALSE | Downregulated |
| 2,742892905 | 0,032416657 | FIGF | TRUE | Upregulated |
| -0,235917141 | 0,032419362 | PEX5 | FALSE | Downregulated |
| 1,225130636 | 0,032485438 | CLMP | TRUE | Upregulated |
| -0,269377049 | 0,032485438 | FIP1L1 | FALSE | Downregulated |
| 0,211154848 | 0,032517054 | NPLOC4 | FALSE | Upregulated |
| 0,691182381 | 0,03256711 | RERG | FALSE | Upregulated |
| 0,217180175 | 0,032571737 | ATXN2 | FALSE | Upregulated |
| -0,518770452 | 0,032571737 | COMTD1 | FALSE | Downregulated |
| 1,284412515 | 0,032571737 | MYO3A | TRUE | Upregulated |
| -0,290752377 | 0,032571737 | NR1H4 | FALSE | Downregulated |
| -0,380744903 | 0,032571737 | STK35 | FALSE | Downregulated |
| -0,2914845 | 0,032650515 | SDE2 | FALSE | Downregulated |
| 0,424450237 | 0,032694612 | CCDC88A | FALSE | Upregulated |
| -0,358767966 | 0,032717393 | AADAT | FALSE | Downregulated |
| 2,872223731 | 0,032717393 | GABRB2 | TRUE | Upregulated |
| 4,486336129 | 0,032717393 | PRODH | TRUE | Upregulated |
| -0,242879644 | 0,03272851 | CCDC12 | FALSE | Downregulated |
| -0,182386798 | 0,032747596 | GLOD4 | FALSE | Downregulated |
| 0,51237421 | 0,032800029 | RACGAP1 | FALSE | Upregulated |
| 1,00478598 | 0,032818265 | NUF2 | TRUE | Upregulated |
| -0,326018623 | 0,033115697 | NAGS | FALSE | Downregulated |
| -0,223488771 | 0,033119411 | PAPD5 | FALSE | Downregulated |
| 1,323399528 | 0,033142264 | CELF4 | TRUE | Upregulated |

| | | | | |
|---|---|---|---|---|
| -0,476040659 | 0,033142264 | TMEM220-AS1 | FALSE | Downregulated |
| -0,634679632 | 0,033169865 | IL27 | FALSE | Downregulated |
| 0,294977728 | 0,033190815 | C17orf70 | FALSE | Upregulated |
| 0,475712925 | 0,033194427 | HCK | FALSE | Upregulated |
| 1,303638872 | 0,03320399 | FRMD6-AS1 | TRUE | Upregulated |
| -0,396436245 | 0,033283387 | CLCN5 | FALSE | Downregulated |
| 1,149455936 | 0,033283387 | LMOD1 | TRUE | Upregulated |
| -0,708257832 | 0,033283387 | STAG3 | FALSE | Downregulated |
| 1,638588793 | 0,03331221 | SCN3A | TRUE | Upregulated |
| 2,98042721 | 0,033323585 | LINC00485 | TRUE | Upregulated |
| -0,162553329 | 0,033341688 | BRAP | FALSE | Downregulated |
| 0,436999983 | 0,033343269 | POLA2 | FALSE | Upregulated |
| 1,986523437 | 0,033351137 | MUC4 | TRUE | Upregulated |
| 1,0340911 | 0,033375663 | HS1BP3-IT1 | TRUE | Upregulated |
| 0,298938899 | 0,033395952 | SLCO2B1 | FALSE | Upregulated |
| -1,295699426 | 0,033439365 | CTC-558O2.1 | TRUE | Downregulated |
| 0,231295669 | 0,033533679 | ARL8A | FALSE | Upregulated |
| 1,450693006 | 0,033533679 | KCNK10 | TRUE | Upregulated |
| -0,326910063 | 0,033549242 | OBFC1 | FALSE | Downregulated |
| -0,237307827 | 0,033583374 | RBAK | FALSE | Downregulated |
| 2,56539963 | 0,03362417 | EPN3 | TRUE | Upregulated |
| 0,328082294 | 0,03362417 | TMSB4X | FALSE | Upregulated |
| -0,733806493 | 0,033713808 | HMGCR | FALSE | Downregulated |
| -0,496997586 | 0,033773999 | FAM76B | FALSE | Downregulated |
| 0,956527648 | 0,033773999 | GINS1 | FALSE | Upregulated |
| -0,224907373 | 0,033851126 | ATG4B | FALSE | Downregulated |
| 0,22185094 | 0,033863347 | CNDP2 | FALSE | Upregulated |
| 0,329561917 | 0,033877994 | CYP7B1 | FALSE | Upregulated |
| 1,245169939 | 0,033887832 | ARHGAP22 | TRUE | Upregulated |
| 1,596341121 | 0,033887832 | TRPC6 | TRUE | Upregulated |
| -0,369224435 | 0,033929738 | PUS3 | FALSE | Downregulated |
| 0,319385391 | 0,033953108 | PHPT1 | FALSE | Upregulated |
| -0,38651939 | 0,033955636 | AQP11 | FALSE | Downregulated |
| -0,796684616 | 0,03396024 | CYP1A2 | FALSE | Downregulated |
| -0,379443495 | 0,033998813 | MRPL41 | FALSE | Downregulated |
| -0,420625948 | 0,034047685 | FAM211B | FALSE | Downregulated |
| 0,351460753 | 0,034054549 | JADE2 | FALSE | Upregulated |
| 0,264364488 | 0,034057638 | CTSZ | FALSE | Upregulated |
| -0,857352997 | 0,034057638 | LGI4 | FALSE | Downregulated |
| 1,651238797 | 0,034057638 | LOC102724197 | TRUE | Upregulated |
| 0,222571821 | 0,034057638 | PIP4K2C | FALSE | Upregulated |
| 2,312635666 | 0,034057638 | SND1-IT1 | TRUE | Upregulated |
| -0,277584888 | 0,034057638 | SNRPA1 | FALSE | Downregulated |
| 1,797164802 | 0,034057638 | TSKS | TRUE | Upregulated |
| -0,494135112 | 0,034075908 | NFKBIZ | FALSE | Downregulated |
| -1,090673575 | 0,034105657 | LOC102724019 | TRUE | Downregulated |
| 0,361252573 | 0,034146828 | COMMD10 | FALSE | Upregulated |
| 2,333554115 | 0,034159532 | SERTAD4-AS1 | TRUE | Upregulated |
| 1,016116241 | 0,034193871 | ANKRD36 | TRUE | Upregulated |
| -0,494637242 | 0,034195008 | MDP1 | FALSE | Downregulated |
| 0,181058768 | 0,034196304 | MED24 | FALSE | Upregulated |

| | | | | |
|---|---|---|---|---|
| 0,271685678 | 0,034247608 | UNK | FALSE | Upregulated |
| -0,54242967 | 0,034289034 | NR1I2 | FALSE | Downregulated |
| 0,127032013 | 0,034304556 | XPO7 | FALSE | Upregulated |
| -0,947818297 | 0,034343059 | LOC102724889 | FALSE | Downregulated |
| -2,193689317 | 0,03450596 | UBE2DNL | TRUE | Downregulated |
| 0,212387262 | 0,034536357 | FAM53C | FALSE | Upregulated |
| 0,372718607 | 0,034536357 | MFSD5 | FALSE | Upregulated |
| 1,988838303 | 0,034536357 | SNORA81 | TRUE | Upregulated |
| 0,32408968 | 0,03455507 | CXorf38 | FALSE | Upregulated |
| -0,425506426 | 0,034598616 | C15orf61 | FALSE | Downregulated |
| 0,285825956 | 0,034630967 | METTL9 | FALSE | Upregulated |
| -0,206154794 | 0,034652799 | C6orf211 | FALSE | Downregulated |
| -0,525242275 | 0,034674175 | EBP | FALSE | Downregulated |
| -0,349580845 | 0,03471082 | DBT | FALSE | Downregulated |
| 0,286152559 | 0,034749474 | ARFGAP1 | FALSE | Upregulated |
| -0,395906695 | 0,034749474 | DPYD | FALSE | Downregulated |
| 1,358104412 | 0,034786403 | LOC101928105 | TRUE | Upregulated |
| 0,393631544 | 0,034787416 | TRABD | FALSE | Upregulated |
| 0,335502096 | 0,034890726 | SRA1 | FALSE | Upregulated |
| -1,790634542 | 0,034936879 | TRNT | TRUE | Downregulated |
| -0,304080501 | 0,035018968 | SRP54 | FALSE | Downregulated |
| 0,366234907 | 0,0350947 | IPPK | FALSE | Upregulated |
| 0,467245172 | 0,0350947 | TAPSAR1 | FALSE | Upregulated |
| 0,727037058 | 0,035147254 | HCG26 | FALSE | Upregulated |
| 0,332457613 | 0,035176302 | C15orf57 | FALSE | Upregulated |
| -0,29218764 | 0,035176302 | GPN3 | FALSE | Downregulated |
| 1,046255899 | 0,035176302 | SPATC1L | TRUE | Upregulated |
| 1,027675427 | 0,035267086 | RPL39L | TRUE | Upregulated |
| -0,252122899 | 0,035347732 | ARHGAP21 | FALSE | Downregulated |
| -0,19393516 | 0,035370281 | TLK2 | FALSE | Downregulated |
| -0,79453094 | 0,035424082 | IL17RE | FALSE | Downregulated |
| 0,313706435 | 0,035522223 | DPH1 | FALSE | Upregulated |
| -0,356795039 | 0,035531981 | SNTB1 | FALSE | Downregulated |
| -0,651655782 | 0,035533994 | TMTC1 | FALSE | Downregulated |
| 0,330017783 | 0,035558638 | GPN2 | FALSE | Upregulated |
| 0,973480612 | 0,035619602 | ARHGEF39 | FALSE | Upregulated |
| 0,285917981 | 0,035654557 | LRCH1 | FALSE | Upregulated |
| 1,675579761 | 0,035657024 | MMP23B | TRUE | Upregulated |
| -0,504404628 | 0,035681525 | YPEL2 | FALSE | Downregulated |
| 0,333498822 | 0,03572853 | MAST3 | FALSE | Upregulated |
| -0,332392711 | 0,035766035 | LINC00938 | FALSE | Downregulated |
| 1,04520871 | 0,035786415 | HSPA6 | TRUE | Upregulated |
| 0,350874087 | 0,035821322 | MSH2 | FALSE | Upregulated |
| 0,422858186 | 0,035821322 | SKAP2 | FALSE | Upregulated |
| 0,684181634 | 0,035839448 | C18orf54 | FALSE | Upregulated |
| -0,434354438 | 0,035917767 | SERPINA5 | FALSE | Downregulated |
| -1,225375931 | 0,035924394 | NEURL1 | TRUE | Downregulated |
| 0,161057897 | 0,035924394 | ZDHHC5 | FALSE | Upregulated |
| 0,220057992 | 0,035970005 | MTPN | FALSE | Upregulated |
| -0,262212214 | 0,035979593 | NT5DC1 | FALSE | Downregulated |
| -0,331558821 | 0,036007552 | SERPINA1 | FALSE | Downregulated |

| | | | | |
|---|---|---|---|---|
| 0,364839334 | 0,036010905 | ZNF217 | FALSE | Upregulated |
| -0,703794433 | 0,036041595 | ARRDC3 | FALSE | Downregulated |
| 0,382685924 | 0,036099279 | ITGA9 | FALSE | Upregulated |
| 0,351450697 | 0,03611487 | LITAF | FALSE | Upregulated |
| 0,503609481 | 0,036141578 | ZNF35 | FALSE | Upregulated |
| 0,518406859 | 0,036146382 | RP11-35G9.3 | FALSE | Upregulated |
| 1,458081429 | 0,03615759 | NMNAT2 | TRUE | Upregulated |
| -0,441085748 | 0,036178141 | LOC101928303 | FALSE | Downregulated |
| 0,571029483 | 0,036216953 | EZH2 | FALSE | Upregulated |
| 0,32567902 | 0,036233068 | BAIAP2L1 | FALSE | Upregulated |
| -0,382245015 | 0,036280611 | THAP9-AS1 | FALSE | Downregulated |
| -0,371733333 | 0,03631617 | GNL3 | FALSE | Downregulated |
| -1,07619641 | 0,036342068 | CALR3 | TRUE | Downregulated |
| 1,635916429 | 0,036342068 | MED12L | TRUE | Upregulated |
| 0,372824389 | 0,036372007 | FAM217B | FALSE | Upregulated |
| 1,035585131 | 0,036412332 | DIAPH3 | TRUE | Upregulated |
| -0,21245003 | 0,036423651 | FGFR1OP | FALSE | Downregulated |
| -0,697127788 | 0,036423651 | LOC100507316 | FALSE | Downregulated |
| 2,366546877 | 0,036429542 | C4orf6 | TRUE | Upregulated |
| 2,21603988 | 0,036429542 | DCX | TRUE | Upregulated |
| 0,84032557 | 0,03643312 | PRR15L | FALSE | Upregulated |
| -0,237545286 | 0,03643312 | RPL15 | FALSE | Downregulated |
| 1,331333261 | 0,036442253 | GAS6-AS2 | TRUE | Upregulated |
| 2,757460218 | 0,036446023 | UGT8 | TRUE | Upregulated |
| -0,439857675 | 0,036510006 | AMY2B | FALSE | Downregulated |
| -0,607002419 | 0,036510006 | FAM227B | FALSE | Downregulated |
| 0,453570351 | 0,036531681 | TMEM128 | FALSE | Upregulated |
| 1,346018299 | 0,036585955 | TMEM125 | TRUE | Upregulated |
| -0,330990974 | 0,036637817 | FAM96A | FALSE | Downregulated |
| 2,061259504 | 0,036656801 | POU5F1B | TRUE | Upregulated |
| 0,377608886 | 0,036677145 | CCDC130 | FALSE | Upregulated |
| 0,471398095 | 0,036677145 | WDR4 | FALSE | Upregulated |
| 2,811900452 | 0,036709669 | RP4-724E16.2 | TRUE | Upregulated |
| 0,288043488 | 0,03671471 | EIF5A | FALSE | Upregulated |
| -0,255192247 | 0,036729943 | PGPEP1 | FALSE | Downregulated |
| 2,190004182 | 0,036755455 | MCF2L2 | TRUE | Upregulated |
| 1,36369334 | 0,036825048 | CCDC141 | TRUE | Upregulated |
| 3,350446085 | 0,036832354 | LOC101927680 | TRUE | Upregulated |
| -0,35385744 | 0,03683739 | TP53INP1 | FALSE | Downregulated |
| 0,971570279 | 0,036861937 | GNAZ | FALSE | Upregulated |
| -0,414026901 | 0,036940991 | KAZN | FALSE | Downregulated |
| -0,356325817 | 0,036991028 | RALGPS2 | FALSE | Downregulated |
| -0,286123442 | 0,037003858 | HCFC2 | FALSE | Downregulated |
| -0,424348372 | 0,03704484 | ACP6 | FALSE | Downregulated |
| 0,791582359 | 0,037059584 | B3GALNT1 | FALSE | Upregulated |
| -1,043729543 | 0,037149642 | MPPED2 | TRUE | Downregulated |
| -0,498361832 | 0,037167352 | C15orf37 | FALSE | Downregulated |
| 1,724925486 | 0,037183707 | RASAL1 | TRUE | Upregulated |
| 0,225418757 | 0,037189624 | LEMD3 | FALSE | Upregulated |
| 0,228782948 | 0,037189624 | SUMF2 | FALSE | Upregulated |
| 0,385798682 | 0,037212648 | LRRC8B | FALSE | Upregulated |

| | | | | |
|---|---|---|---|---|
| 0,750648907 | 0,037300787 | CD300E | FALSE | Upregulated |
| 1,648203996 | 0,037300787 | MAL | TRUE | Upregulated |
| 0,175384924 | 0,037300787 | MAPK1IP1L | FALSE | Upregulated |
| 0,507952945 | 0,037314088 | ABCA7 | FALSE | Upregulated |
| -0,277098575 | 0,037314088 | MRPL33 | FALSE | Downregulated |
| 2,937339614 | 0,037338399 | SNORA44 | TRUE | Upregulated |
| -0,174390093 | 0,037338399 | UQCRC2 | FALSE | Downregulated |
| 0,290827585 | 0,03734124 | FBXO30 | FALSE | Upregulated |
| -0,738819941 | 0,037382895 | LOC101928424 | FALSE | Downregulated |
| -0,213116811 | 0,037382895 | N4BP2L2 | FALSE | Downregulated |
| -0,37311333 | 0,037382895 | OXLD1 | FALSE | Downregulated |
| 0,248786943 | 0,037382895 | SCAMP1 | FALSE | Upregulated |
| -0,231955871 | 0,037382895 | ZFP91 | FALSE | Downregulated |
| 1,008696595 | 0,037566292 | PLXNB3 | TRUE | Upregulated |
| -0,283359767 | 0,037596252 | USP8 | FALSE | Downregulated |
| 0,780836661 | 0,037629337 | AGAP2-AS1 | FALSE | Upregulated |
| -0,591544817 | 0,037629337 | ASCL2 | FALSE | Downregulated |
| -0,410187298 | 0,037629337 | HOGA1 | FALSE | Downregulated |
| -0,35129491 | 0,037629337 | SLC25A20 | FALSE | Downregulated |
| -0,40519 | 0,037688873 | RBM28 | FALSE | Downregulated |
| 1,29247558 | 0,037691861 | LOC101927337 | TRUE | Upregulated |
| -0,220164104 | 0,037708185 | ARF5 | FALSE | Downregulated |
| -0,228289382 | 0,037708185 | C19orf43 | FALSE | Downregulated |
| 1,13274798 | 0,037708185 | DNAH10 | TRUE | Upregulated |
| 0,748246442 | 0,037708185 | LDLRAD3 | FALSE | Upregulated |
| 0,954877402 | 0,037738462 | CD101 | FALSE | Upregulated |
| 0,26437817 | 0,037740712 | ADAT1 | FALSE | Upregulated |
| -0,364195254 | 0,037763796 | GLYCTK | FALSE | Downregulated |
| 1,117226016 | 0,037808486 | ADAMTS9-AS2 | TRUE | Upregulated |
| 0,909390981 | 0,037874119 | CREB3L1 | FALSE | Upregulated |
| 0,517778231 | 0,037908525 | HSPA12A | FALSE | Upregulated |
| -0,341377423 | 0,037965623 | ECM1 | FALSE | Downregulated |
| -1,571446488 | 0,038061893 | PADI2 | TRUE | Downregulated |
| 1,802037138 | 0,038076585 | LOC101928955 | TRUE | Upregulated |
| 1,521043231 | 0,038088535 | B3GALT5 | TRUE | Upregulated |
| -0,516556791 | 0,038088535 | RAPH1 | FALSE | Downregulated |
| -0,204385306 | 0,038113196 | METTL14 | FALSE | Downregulated |
| 0,653219824 | 0,038155178 | CD109 | FALSE | Upregulated |
| 0,565667391 | 0,038157876 | SLFN12 | FALSE | Upregulated |
| 0,804801998 | 0,038176713 | LOC102724535 | FALSE | Upregulated |
| -0,458579138 | 0,038184787 | AMDHD1 | FALSE | Downregulated |
| -0,215104344 | 0,038184787 | ARPC3 | FALSE | Downregulated |
| -3,216770871 | 0,038184787 | MYBPH | TRUE | Downregulated |
| 0,442502798 | 0,038251456 | ZNF48 | FALSE | Upregulated |
| -1,804584615 | 0,038266874 | MYL4 | TRUE | Downregulated |
| -0,487547069 | 0,038266874 | TBRG1 | FALSE | Downregulated |
| 1,72399711 | 0,038322445 | NCAN | TRUE | Upregulated |
| 0,522711966 | 0,038337017 | KIAA1456 | FALSE | Upregulated |
| 0,584132163 | 0,038388906 | MGC57346 | FALSE | Upregulated |
| -0,349538333 | 0,038449294 | LECT2 | FALSE | Downregulated |
| -0,825403435 | 0,038474329 | USP2-AS1 | FALSE | Downregulated |

| | | | | |
|---|---|---|---|---|
| 2,2875136 | 0,038479286 | SOX8 | TRUE | Upregulated |
| -0,491942655 | 0,038479286 | ZCWPW2 | FALSE | Downregulated |
| -0,229581539 | 0,038495142 | IPO8 | FALSE | Downregulated |
| -0,196504654 | 0,038499479 | MED21 | FALSE | Downregulated |
| -0,331767022 | 0,038508323 | DEK | FALSE | Downregulated |
| -0,588062794 | 0,03855232 | SOX7 | FALSE | Downregulated |
| 0,741535682 | 0,038653607 | SCARF2 | FALSE | Upregulated |
| 0,656099621 | 0,03868525 | ABCA4 | FALSE | Upregulated |
| 2,222675776 | 0,03868525 | CABYR | TRUE | Upregulated |
| 0,899581369 | 0,03868525 | SOD3 | FALSE | Upregulated |
| -0,259731373 | 0,038803509 | MPP6 | FALSE | Downregulated |
| -0,365829045 | 0,038813029 | CNST | FALSE | Downregulated |
| -0,248012563 | 0,038816396 | SUCLG2 | FALSE | Downregulated |
| 0,22913973 | 0,038906495 | CPSF2 | FALSE | Upregulated |
| 0,643367487 | 0,038953741 | NEK11 | FALSE | Upregulated |
| 0,326614889 | 0,03897731 | LDOC1L | FALSE | Upregulated |
| -0,200376 | 0,038999423 | MRPS30 | FALSE | Downregulated |
| 1,873504178 | 0,038999423 | TNS4 | TRUE | Upregulated |
| 3,266234892 | 0,039032281 | ABCA12 | TRUE | Upregulated |
| 1,135452946 | 0,039041532 | ANO9 | TRUE | Upregulated |
| 0,301472335 | 0,039041532 | KLHL21 | FALSE | Upregulated |
| -0,290274014 | 0,039099469 | RRNAD1 | FALSE | Downregulated |
| 0,645115244 | 0,039100375 | C1orf112 | FALSE | Upregulated |
| 0,261081052 | 0,039121311 | DNPEP | FALSE | Upregulated |
| 0,551279923 | 0,039121311 | IL17RB | FALSE | Upregulated |
| -0,265902471 | 0,039121311 | MLF2 | FALSE | Downregulated |
| 0,292426767 | 0,039121311 | NUDT15 | FALSE | Upregulated |
| -0,243946928 | 0,039127846 | TMEM11 | FALSE | Downregulated |
| 0,55763649 | 0,039150281 | LILRB4 | FALSE | Upregulated |
| 0,922449589 | 0,039175379 | LOC100506870 | FALSE | Upregulated |
| -0,626522272 | 0,039204195 | ADCY10 | FALSE | Downregulated |
| 0,573329443 | 0,039278295 | TBX2 | FALSE | Upregulated |
| 0,245924863 | 0,039339087 | MAD1L1 | FALSE | Upregulated |
| 0,168320384 | 0,039339087 | PPP1CA | FALSE | Upregulated |
| 0,131076217 | 0,039424719 | SZRD1 | FALSE | Upregulated |
| 0,247831026 | 0,039430652 | CS | FALSE | Upregulated |
| 0,394834366 | 0,039447157 | MICAL3 | FALSE | Upregulated |
| 0,544200673 | 0,039552817 | CECR1 | FALSE | Upregulated |
| 2,064538889 | 0,039612839 | LOC101929560 | TRUE | Upregulated |
| 0,250549126 | 0,039701985 | TESK1 | FALSE | Upregulated |
| 0,19416967 | 0,039729199 | SERPINB6 | FALSE | Upregulated |
| 0,645192055 | 0,039729465 | RALGPS1 | FALSE | Upregulated |
| 0,517076221 | 0,039735337 | ZNF185 | FALSE | Upregulated |
| 1,045247853 | 0,039760377 | LOC101927963 | TRUE | Upregulated |
| 0,302053257 | 0,039786594 | BAHD1 | FALSE | Upregulated |
| 1,54954919 | 0,039786594 | CPB2-AS1 | TRUE | Upregulated |
| -0,261518061 | 0,039786594 | NDUFS3 | FALSE | Downregulated |
| -4,164637545 | 0,039786594 | TECRL | TRUE | Downregulated |
| -0,189862384 | 0,039786594 | UBE2H | FALSE | Downregulated |
| -0,497565445 | 0,039893683 | LOC642441 | FALSE | Downregulated |
| 2,12377547 | 0,039911735 | RP11-103C16.2 | TRUE | Upregulated |

| | | | | |
|---|---|---|---|---|
| -0,487261459 | 0,039922225 | KLB | FALSE | Downregulated |
| 2,330553609 | 0,039972039 | ECT2L | TRUE | Upregulated |
| 0,552296477 | 0,039972039 | FHOD1 | FALSE | Upregulated |
| -0,224668943 | 0,039972039 | SLC25A32 | FALSE | Downregulated |
| 0,746420296 | 0,039997514 | LY75 | FALSE | Upregulated |
| 1,535750017 | 0,040017321 | TROAP | TRUE | Upregulated |
| 2,480208273 | 0,040071818 | AC003102.3 | TRUE | Upregulated |
| 1,595755625 | 0,040071818 | ACTG2 | TRUE | Upregulated |
| -0,606198458 | 0,040071818 | NPIPA2 | FALSE | Downregulated |
| 1,728642974 | 0,040083007 | ZCCHC18 | TRUE | Upregulated |
| 0,552726304 | 0,040113695 | RASIP1 | FALSE | Upregulated |
| 0,349294622 | 0,040118995 | ZBTB33 | FALSE | Upregulated |
| -0,504992936 | 0,040142544 | FDFT1 | FALSE | Downregulated |
| -0,268083316 | 0,040181412 | NAF1 | FALSE | Downregulated |
| -0,16378737 | 0,040211146 | CHMP2B | FALSE | Downregulated |
| 0,329084321 | 0,040211146 | ORAI1 | FALSE | Upregulated |
| 0,369920449 | 0,040211146 | SNAPC2 | FALSE | Upregulated |
| -0,265656009 | 0,040310352 | SMC3 | FALSE | Downregulated |
| -0,807329006 | 0,040332382 | LINC01144 | FALSE | Downregulated |
| -0,325395091 | 0,040384735 | THRB | FALSE | Downregulated |
| 0,565592598 | 0,040444645 | ZNF347 | FALSE | Upregulated |
| 1,548806184 | 0,040461793 | C14orf64 | TRUE | Upregulated |
| 0,337860853 | 0,040461793 | FAM219A | FALSE | Upregulated |
| 0,567674851 | 0,040461793 | RND1 | FALSE | Upregulated |
| -0,323033637 | 0,040461793 | UBE2W | FALSE | Downregulated |
| 0,194923354 | 0,040548239 | TCF20 | FALSE | Upregulated |
| 1,924482668 | 0,040565259 | LOC101927345 | TRUE | Upregulated |
| -0,37906775 | 0,040565259 | NABP2 | FALSE | Downregulated |
| -0,355363442 | 0,04058113 | PPM1H | FALSE | Downregulated |
| 1,37950397 | 0,040584579 | UGT2B28 | TRUE | Upregulated |
| 1,467137321 | 0,040586575 | SMG7-AS1 | TRUE | Upregulated |
| -0,435791561 | 0,040589537 | CFHR5 | FALSE | Downregulated |
| 0,472408805 | 0,040613049 | CKLF | FALSE | Upregulated |
| -0,296621415 | 0,040660328 | ERICH1 | FALSE | Downregulated |
| -1,006820145 | 0,040666229 | BRSK1 | TRUE | Downregulated |
| -1,451713321 | 0,040745938 | SPOCK3 | TRUE | Downregulated |
| 0,39725156 | 0,040745938 | TUBA1C | FALSE | Upregulated |
| 2,221207878 | 0,040807392 | KCNH4 | TRUE | Upregulated |
| -1,037039308 | 0,040846111 | FLVCR1-AS1 | TRUE | Downregulated |
| -0,192518968 | 0,040929425 | YTHDC1 | FALSE | Downregulated |
| -0,323569245 | 0,040930483 | ZRANB3 | FALSE | Downregulated |
| 0,725498582 | 0,041031664 | PLP2 | FALSE | Upregulated |
| 1,181559198 | 0,041039953 | RAB25 | TRUE | Upregulated |
| -0,568115597 | 0,041109373 | SLC22A25 | FALSE | Downregulated |
| -0,252812634 | 0,041200752 | POMP | FALSE | Downregulated |
| 0,192831237 | 0,041200752 | TAOK2 | FALSE | Upregulated |
| -0,53162497 | 0,041213731 | ASS1 | FALSE | Downregulated |
| -0,52793573 | 0,041213731 | TM7SF2 | FALSE | Downregulated |
| 1,575051475 | 0,041241672 | SCARNA7 | TRUE | Upregulated |
| -0,270516726 | 0,041268885 | RNF26 | FALSE | Downregulated |
| 1,194987819 | 0,041307454 | CLEC5A | TRUE | Upregulated |

| | | | | |
|---|---|---|---|---|
| -0,261402192 | 0,041331621 | KIN | FALSE | Downregulated |
| -0,619765776 | 0,041331943 | FLJ22763 | FALSE | Downregulated |
| 1,85274669 | 0,041347111 | LOC102724976 | TRUE | Upregulated |
| -0,21661227 | 0,041347111 | ZNF131 | FALSE | Downregulated |
| -0,37983738 | 0,041384533 | AK9 | FALSE | Downregulated |
| 0,25295137 | 0,04143253 | NCK1 | FALSE | Upregulated |
| 1,608635021 | 0,041438775 | GMCL1P1 | TRUE | Upregulated |
| 0,567211437 | 0,041462729 | FYB | FALSE | Upregulated |
| -0,393147414 | 0,041551204 | RBBP6 | FALSE | Downregulated |
| 0,438996052 | 0,041551204 | UBE2Q2 | FALSE | Upregulated |
| -0,207302504 | 0,041587337 | ABCE1 | FALSE | Downregulated |
| 0,308354494 | 0,041616637 | WSB2 | FALSE | Upregulated |
| 2,270861569 | 0,041738034 | ATCAY | TRUE | Upregulated |
| -0,843349179 | 0,041746887 | FAM151A | FALSE | Downregulated |
| 0,398943917 | 0,041746887 | TIMP3 | FALSE | Upregulated |
| 0,234748609 | 0,041798595 | OBSL1 | FALSE | Upregulated |
| 0,505411849 | 0,041869559 | SACS | FALSE | Upregulated |
| 0,887454886 | 0,041869559 | SNPH | FALSE | Upregulated |
| -0,461310143 | 0,041869559 | SRSF11 | FALSE | Downregulated |
| 0,315733978 | 0,041869559 | WDR13 | FALSE | Upregulated |
| -1,083599802 | 0,041931217 | KCNJ12 | TRUE | Downregulated |
| -0,409357646 | 0,041970331 | CYP8B1 | FALSE | Downregulated |
| 0,25637958 | 0,041973952 | MOGS | FALSE | Upregulated |
| -0,162090217 | 0,041973952 | SF3A3 | FALSE | Downregulated |
| -0,362225277 | 0,042040774 | LOC100130872 | FALSE | Downregulated |
| 0,776014352 | 0,042040774 | TMTC2 | FALSE | Upregulated |
| -0,162629033 | 0,042100906 | ATP6V1A | FALSE | Downregulated |
| 0,454488331 | 0,042168469 | C7orf43 | FALSE | Upregulated |
| -1,889630588 | 0,042231096 | MLANA | TRUE | Downregulated |
| 0,435309473 | 0,042274371 | ADAMTS2 | FALSE | Upregulated |
| 0,369632727 | 0,042288261 | C9orf142 | FALSE | Upregulated |
| 1,429903072 | 0,04229904 | SCIN | TRUE | Upregulated |
| -0,291606484 | 0,042330588 | RPLP1 | FALSE | Downregulated |
| -0,318419003 | 0,042385288 | NENF | FALSE | Downregulated |
| 1,275782875 | 0,042421825 | AC004019.13 | TRUE | Upregulated |
| -0,311350786 | 0,042459086 | CPSF4 | FALSE | Downregulated |
| -0,297970017 | 0,042459086 | RPL12 | FALSE | Downregulated |
| 0,155602154 | 0,042482732 | PUM1 | FALSE | Upregulated |
| 0,313925897 | 0,042544564 | BOP1 | FALSE | Upregulated |
| 0,171205408 | 0,042570678 | KIAA0100 | FALSE | Upregulated |
| 0,325937033 | 0,042570678 | STIP1 | FALSE | Upregulated |
| 0,466017704 | 0,042570678 | TTC39B | FALSE | Upregulated |
| 0,523960183 | 0,042577269 | TMEM243 | FALSE | Upregulated |
| 0,461417997 | 0,042590225 | PSMB9 | FALSE | Upregulated |
| 0,348925454 | 0,04264622 | ARL4A | FALSE | Upregulated |
| 0,513638814 | 0,042681714 | BATF2 | FALSE | Upregulated |
| 1,7007799 | 0,042758941 | KSR2 | TRUE | Upregulated |
| -0,468323655 | 0,042820812 | TPPP2 | FALSE | Downregulated |
| 0,258460891 | 0,042820812 | TSC22D4 | FALSE | Upregulated |
| -0,752359983 | 0,042822278 | LOC101929691 | FALSE | Downregulated |
| 0,730488935 | 0,042883198 | LOC100133445 | FALSE | Upregulated |

| | | | | |
|---|---|---|---|---|
| -0,267820007 | 0,042919808 | ZNF595 | FALSE | Downregulated |
| -0,185008253 | 0,042926979 | SUDS3 | FALSE | Downregulated |
| -0,326358897 | 0,042930024 | ALKBH7 | FALSE | Downregulated |
| -0,742377273 | 0,042932592 | RP11-372E1.4 | FALSE | Downregulated |
| 0,339911527 | 0,042956126 | APOL6 | FALSE | Upregulated |
| 0,370310251 | 0,042956126 | HAUS7 | FALSE | Upregulated |
| 0,282233647 | 0,043006437 | ABI2 | FALSE | Upregulated |
| 0,29725664 | 0,043014754 | METTL6 | FALSE | Upregulated |
| -0,304374405 | 0,043014754 | SDC2 | FALSE | Downregulated |
| 0,250784601 | 0,043037997 | ZFP90 | FALSE | Upregulated |
| 0,383306798 | 0,043094857 | DCLRE1B | FALSE | Upregulated |
| 1,708244081 | 0,043094857 | KCNH3 | TRUE | Upregulated |
| 0,962533133 | 0,043095893 | XCL1 | FALSE | Upregulated |
| 0,526172886 | 0,04313358 | CCDC112 | FALSE | Upregulated |
| -0,476488967 | 0,043154813 | P2RY13 | FALSE | Downregulated |
| 1,156059657 | 0,04318181 | LOC102724445 | TRUE | Upregulated |
| 1,051843993 | 0,043190164 | FBLN1 | TRUE | Upregulated |
| -0,308345114 | 0,043229823 | RPL30 | FALSE | Downregulated |
| -0,279209835 | 0,043275131 | PBX3 | FALSE | Downregulated |
| -0,246011968 | 0,043320846 | GTF2F2 | FALSE | Downregulated |
| 0,253788232 | 0,043320846 | ZNF134 | FALSE | Upregulated |
| -0,223844164 | 0,043372365 | RPRD1A | FALSE | Downregulated |
| -1,899286379 | 0,043443548 | THBS4 | TRUE | Downregulated |
| -0,280384734 | 0,043471546 | CFI | FALSE | Downregulated |
| -0,215908435 | 0,043471546 | EIF1AD | FALSE | Downregulated |
| -0,436326431 | 0,043471546 | TAMM41 | FALSE | Downregulated |
| -1,286169247 | 0,043471546 | TMEM257 | TRUE | Downregulated |
| 0,273994226 | 0,043479105 | ANAPC4 | FALSE | Upregulated |
| 1,447283676 | 0,043505344 | MMP16 | TRUE | Upregulated |
| 0,79302691 | 0,043520096 | CROCCP3 | FALSE | Upregulated |
| 0,555289077 | 0,043543964 | ZNF738 | FALSE | Upregulated |
| 0,382063153 | 0,043570577 | ZNF879 | FALSE | Upregulated |
| -0,506786149 | 0,043585682 | CFD | FALSE | Downregulated |
| -0,392915341 | 0,043624528 | AGL | FALSE | Downregulated |
| -0,562885077 | 0,043624528 | EXPH5 | FALSE | Downregulated |
| -0,482158537 | 0,043624528 | LOC102723701 | FALSE | Downregulated |
| -0,667933977 | 0,043625628 | PDE8B | FALSE | Downregulated |
| 0,215810746 | 0,043690402 | SNX12 | FALSE | Upregulated |
| 0,995301132 | 0,04371179 | LINC00920 | FALSE | Upregulated |
| -0,22472335 | 0,04371179 | RPL6 | FALSE | Downregulated |
| -0,318204627 | 0,04371179 | THNSL1 | FALSE | Downregulated |
| 0,152047305 | 0,043755963 | ARL8B | FALSE | Upregulated |
| 1,576455627 | 0,04377146 | C21orf88 | TRUE | Upregulated |
| 0,47393168 | 0,043787198 | GSKIP | FALSE | Upregulated |
| -1,406276427 | 0,043792369 | SLC4A1 | TRUE | Downregulated |
| -1,056214651 | 0,043817459 | LOC102724507 | TRUE | Downregulated |
| 0,643840826 | 0,043818177 | POC1A | FALSE | Upregulated |
| 0,716839945 | 0,043831166 | CCDC148 | FALSE | Upregulated |
| -0,585659609 | 0,043861552 | CTF1 | FALSE | Downregulated |
| 0,366432115 | 0,043861552 | GNB5 | FALSE | Upregulated |
| 1,297725269 | 0,04386716 | CXCR5 | TRUE | Upregulated |

| | | | | |
|---|---|---|---|---|
| 0,317759617 | 0,043868637 | KATNA1 | FALSE | Upregulated |
| 0,152344467 | 0,043880892 | USP4 | FALSE | Upregulated |
| -0,410706811 | 0,043881469 | TCERG1 | FALSE | Downregulated |
| -1,180715803 | 0,043903505 | HRC | TRUE | Downregulated |
| 0,327123066 | 0,043945266 | APBA3 | FALSE | Upregulated |
| 0,978603348 | 0,043953574 | MIAT | FALSE | Upregulated |
| 0,776422693 | 0,043956511 | SPARCL1 | FALSE | Upregulated |
| 0,308927047 | 0,043976139 | ARHGDIB | FALSE | Upregulated |
| -0,270568708 | 0,043976139 | CBR4 | FALSE | Downregulated |
| 0,262151984 | 0,043976139 | INIP | FALSE | Upregulated |
| 0,432673294 | 0,044006521 | SYTL2 | FALSE | Upregulated |
| -0,258374626 | 0,044051512 | TYSND1 | FALSE | Downregulated |
| 1,786797667 | 0,044115158 | LOC101929751 | TRUE | Upregulated |
| 0,35948943 | 0,044128851 | ZNF544 | FALSE | Upregulated |
| -0,375021684 | 0,044147527 | CCDC41 | FALSE | Downregulated |
| -0,724882787 | 0,044264738 | LOC101928645 | FALSE | Downregulated |
| 0,216266003 | 0,044285651 | COG1 | FALSE | Upregulated |
| -0,30691388 | 0,044285651 | NUDT22 | FALSE | Downregulated |
| -0,292763337 | 0,044285651 | PPP3CA | FALSE | Downregulated |
| 0,32542869 | 0,044344771 | ARL5B | FALSE | Upregulated |
| 1,722466564 | 0,044356621 | CA12 | TRUE | Upregulated |
| -3,025652024 | 0,044356621 | LOC102724480 | TRUE | Downregulated |
| -0,356590991 | 0,044395743 | SAT2 | FALSE | Downregulated |
| 0,260474227 | 0,04440524 | SCARB1 | FALSE | Upregulated |
| -0,499101711 | 0,044410601 | SAA4 | FALSE | Downregulated |
| -0,239447123 | 0,044430008 | RNF103 | FALSE | Downregulated |
| 0,365930045 | 0,04450708 | XRCC4 | FALSE | Upregulated |
| -0,333069239 | 0,044514085 | JMJD1C | FALSE | Downregulated |
| 1,090669156 | 0,044519065 | STK32A | TRUE | Upregulated |
| 1,057883844 | 0,04453075 | RYR2 | TRUE | Upregulated |
| -0,308906909 | 0,044543005 | NTAN1 | FALSE | Downregulated |
| 0,251390187 | 0,044589743 | HSPA4 | FALSE | Upregulated |
| 0,282542422 | 0,044592009 | CHDH | FALSE | Upregulated |
| 1,545882608 | 0,044646813 | DARC | TRUE | Upregulated |
| -0,357598559 | 0,044761396 | MRPL12 | FALSE | Downregulated |
| -0,286762065 | 0,044782506 | COMMD6 | FALSE | Downregulated |
| -0,344554364 | 0,044782506 | JADE1 | FALSE | Downregulated |
| 0,286981222 | 0,044927135 | FAM21A | FALSE | Upregulated |
| 1,274351588 | 0,044927135 | LOC101928149 | TRUE | Upregulated |
| -0,878304138 | 0,044927135 | SLC34A1 | FALSE | Downregulated |
| 2,203026785 | 0,044969243 | PDLIM4 | TRUE | Upregulated |
| -0,308434645 | 0,04497028 | MYD88 | FALSE | Downregulated |
| -0,361329164 | 0,045025724 | CIR1 | FALSE | Downregulated |
| 0,267082559 | 0,045025724 | DHX37 | FALSE | Upregulated |
| 0,921388483 | 0,045025724 | ORC6 | FALSE | Upregulated |
| -0,576788698 | 0,045073877 | IGLON5 | FALSE | Downregulated |
| 1,273824673 | 0,04507788 | IFNG | TRUE | Upregulated |
| 2,506126792 | 0,045125605 | LOC341056 | TRUE | Upregulated |
| -0,400350719 | 0,0451341 | TMEM99 | FALSE | Downregulated |
| 0,201918194 | 0,045166439 | KBTBD2 | FALSE | Upregulated |
| -3,262417896 | 0,045182915 | FAM157A | TRUE | Downregulated |

| | | | | |
|---|---|---|---|---|
| 0,432362464 | 0,045200594 | LHFPL2 | FALSE | Upregulated |
| -0,276955571 | 0,045227365 | ZMYM3 | FALSE | Downregulated |
| 0,397218067 | 0,045297567 | ZFP30 | FALSE | Upregulated |
| 0,497908343 | 0,045415731 | AXIN2 | FALSE | Upregulated |
| -0,27901659 | 0,045428277 | RPL21 | FALSE | Downregulated |
| -0,251130023 | 0,045445869 | SSRP1 | FALSE | Downregulated |
| -0,220657379 | 0,045458605 | CCDC28A | FALSE | Downregulated |
| 1,092879303 | 0,045464562 | TTYH1 | TRUE | Upregulated |
| 0,339734682 | 0,045472418 | PPP1R14B | FALSE | Upregulated |
| -0,412125878 | 0,045600155 | ATP11C | FALSE | Downregulated |
| 0,278402138 | 0,045637257 | PHYKPL | FALSE | Upregulated |
| -0,341334962 | 0,045645252 | ACOX2 | FALSE | Downregulated |
| 0,400501231 | 0,045752573 | MCM3 | FALSE | Upregulated |
| -0,355760004 | 0,045887373 | LCLAT1 | FALSE | Downregulated |
| -0,316369124 | 0,045910072 | TTC28-AS1 | FALSE | Downregulated |
| 1,455602628 | 0,045910072 | WBSCR17 | TRUE | Upregulated |
| -0,51600109 | 0,045917048 | STAB2 | FALSE | Downregulated |
| -0,150785758 | 0,04598372 | KHSRP | FALSE | Downregulated |
| -0,288688957 | 0,045986408 | RPL23A | FALSE | Downregulated |
| -0,218276406 | 0,046016216 | GABARAP | FALSE | Downregulated |
| 0,246141144 | 0,046038899 | TYK2 | FALSE | Upregulated |
| -0,227533216 | 0,046057326 | NAA35 | FALSE | Downregulated |
| 0,314380977 | 0,046072519 | CSRNP2 | FALSE | Upregulated |
| 0,336546788 | 0,046123802 | TBC1D22B | FALSE | Upregulated |
| 0,956412196 | 0,046152972 | LAMP3 | FALSE | Upregulated |
| 1,601110858 | 0,046152972 | TWIST2 | TRUE | Upregulated |
| 0,178880036 | 0,046182577 | VPS26A | FALSE | Upregulated |
| -0,853587888 | 0,046200557 | MAP2K6 | FALSE | Downregulated |
| -0,475129823 | 0,046202675 | ZGPAT | FALSE | Downregulated |
| 0,350239702 | 0,046278051 | C20orf194 | FALSE | Upregulated |
| 1,427484828 | 0,046306478 | CSNK2A3 | TRUE | Upregulated |
| -0,48746276 | 0,046321582 | LOC102724532 | FALSE | Downregulated |
| 2,221662045 | 0,046323601 | TTLL9 | TRUE | Upregulated |
| -0,618456578 | 0,046337424 | DIRAS3 | FALSE | Downregulated |
| -0,328816989 | 0,046378003 | ZMAT5 | FALSE | Downregulated |
| 0,450678551 | 0,046386161 | FLYWCH2 | FALSE | Upregulated |
| 0,687121928 | 0,046404384 | RPL19P12 | FALSE | Upregulated |
| -0,318458251 | 0,046404384 | SPTLC3 | FALSE | Downregulated |
| -0,345518319 | 0,046425471 | ABCA1 | FALSE | Downregulated |
| 0,299746359 | 0,046426212 | BTBD9 | FALSE | Upregulated |
| 2,071110298 | 0,046450357 | TAF1L | TRUE | Upregulated |
| 0,426734603 | 0,046453276 | BLVRA | FALSE | Upregulated |
| -0,623846686 | 0,046463218 | RNASEK-C17orf49 | FALSE | Downregulated |
| 0,410003986 | 0,046561725 | EHBP1L1 | FALSE | Upregulated |
| 0,454713325 | 0,046561725 | MAP2 | FALSE | Upregulated |
| 0,361756371 | 0,046567699 | ZNF408 | FALSE | Upregulated |
| -0,233765205 | 0,046633158 | ZNF302 | FALSE | Downregulated |
| 0,465000447 | 0,046638011 | ADM | FALSE | Upregulated |
| -0,712995488 | 0,046638011 | LOC100128531 | FALSE | Downregulated |
| -0,431022443 | 0,046658028 | ENTPD3-AS1 | FALSE | Downregulated |
| 1,477386953 | 0,046663051 | AC095067.1 | TRUE | Upregulated |

| | | | | |
|---|---|---|---|---|
| -0,152104176 | 0,046663051 | NRBP1 | FALSE | Downregulated |
| -0,6579613 | 0,046695633 | GOLGA8N | FALSE | Downregulated |
| -0,382371571 | 0,046715228 | TIAM1 | FALSE | Downregulated |
| -0,253134586 | 0,046743742 | CSE1L | FALSE | Downregulated |
| 1,998484287 | 0,046847874 | MADCAM1 | TRUE | Upregulated |
| -0,327739091 | 0,046865255 | UQCRH | FALSE | Downregulated |
| 1,852264712 | 0,046877074 | CHD5 | TRUE | Upregulated |
| -1,079616749 | 0,046877074 | IRX3 | TRUE | Downregulated |
| 0,39258069 | 0,046877074 | MROH1 | FALSE | Upregulated |
| -0,305246658 | 0,046887797 | TULP3 | FALSE | Downregulated |
| -0,212692042 | 0,046908024 | VBP1 | FALSE | Downregulated |
| -0,456758681 | 0,046908024 | ZNF415 | FALSE | Downregulated |
| 0,371806891 | 0,046970849 | ARPC1B | FALSE | Upregulated |
| -0,163342084 | 0,047002379 | DNTTIP2 | FALSE | Downregulated |
| 1,136317725 | 0,047023731 | BOC | TRUE | Upregulated |
| 0,341648748 | 0,047023731 | GOLT1B | FALSE | Upregulated |
| -3,10821698 | 0,047023731 | LOC101060569 | TRUE | Downregulated |
| 2,69637352 | 0,047057104 | CACNG4 | TRUE | Upregulated |
| 0,297906706 | 0,047120909 | UTRN | FALSE | Upregulated |
| 0,560226211 | 0,047124126 | C11orf84 | FALSE | Upregulated |
| -2,077189615 | 0,047124126 | DOCK9-AS1 | TRUE | Downregulated |
| 1,015406289 | 0,047124126 | LOC101929531 | TRUE | Upregulated |
| 0,247413381 | 0,047129606 | NIP7 | FALSE | Upregulated |
| -0,33317564 | 0,047150197 | EIF3J-AS1 | FALSE | Downregulated |
| -0,179987235 | 0,047150197 | PARK7 | FALSE | Downregulated |
| -0,27453432 | 0,047154123 | LAMTOR2 | FALSE | Downregulated |
| -0,294640416 | 0,047154123 | LZIC | FALSE | Downregulated |
| -0,457371182 | 0,047154123 | MOGAT2 | FALSE | Downregulated |
| 2,49307842 | 0,047154123 | TCN1 | TRUE | Upregulated |
| 0,94803805 | 0,047160873 | LGALS4 | FALSE | Upregulated |
| 2,397447589 | 0,047162688 | GJA3 | TRUE | Upregulated |
| -0,629370316 | 0,04716762 | ATF5 | FALSE | Downregulated |
| -0,461458069 | 0,04716762 | LCAT | FALSE | Downregulated |
| -0,207541742 | 0,04716762 | PDXDC1 | FALSE | Downregulated |
| 0,262681027 | 0,047176066 | ZNF574 | FALSE | Upregulated |
| 0,170399597 | 0,047261043 | KIAA0319L | FALSE | Upregulated |
| 1,963655446 | 0,047261043 | MIXL1 | TRUE | Upregulated |
| -0,509335864 | 0,047269885 | SLC17A1 | FALSE | Downregulated |
| 2,351311957 | 0,047338966 | C11orf70 | TRUE | Upregulated |
| -0,302287414 | 0,047338966 | RSRC1 | FALSE | Downregulated |
| -0,486817475 | 0,047338966 | SLC19A2 | FALSE | Downregulated |
| 0,678181884 | 0,047339319 | SLC16A6 | FALSE | Upregulated |
| 0,530595748 | 0,04734926 | ZDHHC24 | FALSE | Upregulated |
| -0,524696116 | 0,04746109 | CDKL3 | FALSE | Downregulated |
| 0,180473609 | 0,047483955 | DHX15 | FALSE | Upregulated |
| 0,34308387 | 0,047483955 | TJP1 | FALSE | Upregulated |
| -1,240814057 | 0,047506474 | PEG10 | TRUE | Downregulated |
| 1,197143474 | 0,047510968 | ANXA8 | TRUE | Upregulated |
| -0,772325374 | 0,047539078 | AGBL2 | FALSE | Downregulated |
| -0,964061873 | 0,047539078 | LOC101927226 | FALSE | Downregulated |
| 0,343601614 | 0,047539078 | ZNRF3 | FALSE | Upregulated |

| | | | | |
|---|---|---|---|---|
| 0,91164052 | 0,047573194 | KIF4A | FALSE | Upregulated |
| 0,891543752 | 0,04761194 | FOXC1 | FALSE | Upregulated |
| 1,836393269 | 0,047676982 | WSCD2 | TRUE | Upregulated |
| 0,782507727 | 0,047701291 | ZNF660 | FALSE | Upregulated |
| -0,162255812 | 0,047705088 | PSMD4 | FALSE | Downregulated |
| -0,456050654 | 0,047739628 | COX10-AS1 | FALSE | Downregulated |
| -0,374801319 | 0,047746481 | ARL16 | FALSE | Downregulated |
| 0,447112115 | 0,047746481 | KIF3A | FALSE | Upregulated |
| 0,326909959 | 0,047746481 | LRRC14 | FALSE | Upregulated |
| 0,379806644 | 0,047746481 | LRRC20 | FALSE | Upregulated |
| -0,226483319 | 0,047746481 | WDR12 | FALSE | Downregulated |
| -0,424180981 | 0,047778672 | ATXN7L1 | FALSE | Downregulated |
| 1,780473314 | 0,047792979 | ACTL10 | TRUE | Upregulated |
| 0,799883147 | 0,047805375 | PKNOX2 | FALSE | Upregulated |
| 0,232701014 | 0,047805375 | TSPAN17 | FALSE | Upregulated |
| -0,338407723 | 0,047805375 | YAF2 | FALSE | Downregulated |
| -0,242861987 | 0,047822648 | ATXN1 | FALSE | Downregulated |
| -0,216619773 | 0,047848571 | C16orf58 | FALSE | Downregulated |
| 0,348511749 | 0,047894299 | TANC1 | FALSE | Upregulated |
| -0,377628181 | 0,047915854 | NR3C2 | FALSE | Downregulated |
| 0,295221933 | 0,047915854 | RNASET2 | FALSE | Upregulated |
| 1,688079308 | 0,048030125 | LINC01152 | TRUE | Upregulated |
| 1,98968345 | 0,048044152 | ALS2CR11 | TRUE | Upregulated |
| 0,999026796 | 0,048044152 | AP000688.14 | FALSE | Upregulated |
| 0,776033477 | 0,048044152 | LINC01123 | FALSE | Upregulated |
| 1,078037593 | 0,048044152 | MDGA1 | TRUE | Upregulated |
| 1,005696901 | 0,048044152 | PPAPDC1A | TRUE | Upregulated |
| -0,216032517 | 0,048044152 | SDAD1 | FALSE | Downregulated |
| -0,296239681 | 0,048064728 | RPA3 | FALSE | Downregulated |
| 0,362619189 | 0,048123926 | LCMT1 | FALSE | Upregulated |
| -0,92486277 | 0,048161278 | COL9A3 | FALSE | Downregulated |
| -0,204859769 | 0,048161278 | PAICS | FALSE | Downregulated |
| -0,441563406 | 0,048184166 | AGXT2 | FALSE | Downregulated |
| 0,904774953 | 0,048184166 | FOS | FALSE | Upregulated |
| 1,24077289 | 0,048252539 | KCNN4 | TRUE | Upregulated |
| 0,284182292 | 0,048266371 | PLD3 | FALSE | Upregulated |
| -0,307109926 | 0,048305979 | DCAF4 | FALSE | Downregulated |
| 2,232263869 | 0,048305979 | SULT2B1 | TRUE | Upregulated |
| -0,189131558 | 0,048336791 | ORC4 | FALSE | Downregulated |
| 1,230784091 | 0,048371543 | HOXC4 | TRUE | Upregulated |
| 0,356565616 | 0,048470077 | SEMA4C | FALSE | Upregulated |
| 0,474052659 | 0,048511339 | FANCG | FALSE | Upregulated |
| -0,393144082 | 0,048519053 | GNL2 | FALSE | Downregulated |
| 0,713440094 | 0,048536744 | CD72 | FALSE | Upregulated |
| -0,616635572 | 0,048538419 | LINC01128 | FALSE | Downregulated |
| 0,540443513 | 0,048539065 | RNF207 | FALSE | Upregulated |
| -0,22094552 | 0,048562582 | CYP2J2 | FALSE | Downregulated |
| -0,338829691 | 0,048563043 | MAOA | FALSE | Downregulated |
| 0,326385701 | 0,048583848 | TMEM9 | FALSE | Upregulated |
| 0,289937668 | 0,048624527 | AGO2 | FALSE | Upregulated |
| -0,481156512 | 0,04873188 | GPER1 | FALSE | Downregulated |

| | | | | |
|---|---|---|---|---|
| -0,42334773 | 0,048838159 | CES3 | FALSE | Downregulated |
| 0,953352161 | 0,048859706 | CDH24 | FALSE | Upregulated |
| -0,446282687 | 0,048859706 | KHK | FALSE | Downregulated |
| 0,187547111 | 0,048864426 | ACTR3 | FALSE | Upregulated |
| -0,260311636 | 0,048864426 | MRPL51 | FALSE | Downregulated |
| 0,161238328 | 0,048864426 | MTMR14 | FALSE | Upregulated |
| 1,082097988 | 0,048864426 | NR4A1 | TRUE | Upregulated |
| -0,430192523 | 0,048931624 | HPR | FALSE | Downregulated |
| -1,260905421 | 0,049021796 | STAC3 | TRUE | Downregulated |
| -0,260115388 | 0,049041169 | ZFR | FALSE | Downregulated |
| -0,376669676 | 0,049047328 | TXNL4B | FALSE | Downregulated |
| -1,270416355 | 0,049053369 | LOC101929658 | TRUE | Downregulated |
| 0,200602188 | 0,049070686 | GSR | FALSE | Upregulated |
| -0,436719347 | 0,049070686 | LMLN | FALSE | Downregulated |
| -0,290023559 | 0,049110074 | ZC3H11A | FALSE | Downregulated |
| -0,364146619 | 0,049112093 | SAMD4A | FALSE | Downregulated |
| -0,245551096 | 0,049155013 | SBNO1 | FALSE | Downregulated |
| 2,683043538 | 0,049155013 | VGLL1 | TRUE | Upregulated |
| 0,420890414 | 0,04915813 | ZDHHC23 | FALSE | Upregulated |
| 0,430654825 | 0,049210205 | SRP14-AS1 | FALSE | Upregulated |
| 0,817680613 | 0,049218958 | SH2D2A | FALSE | Upregulated |
| 0,884222082 | 0,049278583 | CAP2 | FALSE | Upregulated |
| -0,27155975 | 0,049278583 | ZDHHC12 | FALSE | Downregulated |
| 0,375918107 | 0,049309514 | FUK | FALSE | Upregulated |
| -0,318238633 | 0,049309514 | PHF7 | FALSE | Downregulated |
| 0,493501794 | 0,049324973 | FAM105A | FALSE | Upregulated |
| 1,769443962 | 0,049324973 | RAB33A | TRUE | Upregulated |
| 0,445193821 | 0,049361102 | HTRA1 | FALSE | Upregulated |
| -0,39068577 | 0,049475365 | ANKRD39 | FALSE | Downregulated |
| 0,468138788 | 0,049550409 | SERPINB9 | FALSE | Upregulated |
| -0,466020583 | 0,049558988 | CCDC158 | FALSE | Downregulated |
| 0,545901275 | 0,04961029 | NRSN2 | FALSE | Upregulated |
| 0,53069034 | 0,049690707 | PRELP | FALSE | Upregulated |
| -0,512791521 | 0,04970942 | LOC102724250 | FALSE | Downregulated |
| 2,126762654 | 0,049730602 | APC2 | TRUE | Upregulated |
| 1,216382068 | 0,049757251 | ST8SIA3 | TRUE | Upregulated |
| 2,449486128 | 0,049770962 | HYDIN2 | TRUE | Upregulated |
| -0,296956427 | 0,049770962 | WDR5B | FALSE | Downregulated |
| 0,470978792 | 0,049774454 | DOCK11 | FALSE | Upregulated |
| -0,203005431 | 0,049774454 | TXNL1 | FALSE | Downregulated |
| 0,203469966 | 0,049795588 | CHFR | FALSE | Upregulated |
| 0,200800202 | 0,049912588 | ADRBK1 | FALSE | Upregulated |
| 1,825078615 | 0,049914506 | TPI1P3 | TRUE | Upregulated |
| 0,38720847 | 0,049928031 | CCNG2 | FALSE | Upregulated |
| 1,137155273 | 0,049946737 | FHAD1 | TRUE | Upregulated |
| 0,263078019 | 0,050030593 | TRAM1 | FALSE | Upregulated |
| 0,699543478 | 0,050044338 | WASH2P | FALSE | Upregulated |
| 0,887470622 | 0,050115517 | RAB36 | FALSE | Upregulated |
| 1,073295843 | 0,050119902 | CRNDE | FALSE | Upregulated |
| -0,307295578 | 0,050119902 | EXTL2 | FALSE | Downregulated |
| -0,353137481 | 0,050121391 | KBTBD3 | FALSE | Downregulated |

| | | | | |
|---|---|---|---|---|
| 0,737893755 | 0,050147284 | BIRC5 | FALSE | Upregulated |
| -0,266709793 | 0,050154783 | CDKN2AIPNL | FALSE | Downregulated |
| -0,634507541 | 0,050183918 | C19orf80 | FALSE | Downregulated |
| -0,484092846 | 0,050189752 | CASC10 | FALSE | Downregulated |
| 0,683558452 | 0,050191929 | ACE | FALSE | Upregulated |
| 2,225986585 | 0,050191929 | KCNF1 | FALSE | Upregulated |
| 0,252198393 | 0,050191929 | KDELR2 | FALSE | Upregulated |
| 0,878540581 | 0,050191929 | MYZAP | FALSE | Upregulated |
| 2,765854522 | 0,050191929 | SNORA6 | FALSE | Upregulated |
| -0,265126462 | 0,050327018 | SNRPA | FALSE | Downregulated |
| 1,125101417 | 0,050373087 | SUSD5 | FALSE | Upregulated |
| 0,486755081 | 0,050467456 | PROCR | FALSE | Upregulated |
| 0,179644027 | 0,050522412 | KLHL9 | FALSE | Upregulated |
| -0,384135941 | 0,050527074 | UBXN8 | FALSE | Downregulated |
| 1,511344165 | 0,050530869 | ANP32A-IT1 | FALSE | Upregulated |
| -0,984186534 | 0,050530869 | KCNH7 | FALSE | Downregulated |
| 0,35249889 | 0,050530869 | SDR42E1 | FALSE | Upregulated |
| -0,222709813 | 0,050590989 | MOCS1 | FALSE | Downregulated |
| -0,375536574 | 0,050613131 | COQ2 | FALSE | Downregulated |
| 0,252910352 | 0,050653749 | TRAPPC9 | FALSE | Upregulated |
| -0,810611528 | 0,050677626 | IL1RAPL2 | FALSE | Downregulated |
| 1,538927936 | 0,050732349 | PDPN | FALSE | Upregulated |
| -1,093515985 | 0,050753288 | ZBTB16 | FALSE | Downregulated |
| -0,354576667 | 0,050763572 | TMEM64 | FALSE | Downregulated |
| 0,583637768 | 0,050791716 | MTBP | FALSE | Upregulated |
| 1,598690632 | 0,050882023 | STAP1 | FALSE | Upregulated |
| 1,223618032 | 0,050882112 | CLEC4F | FALSE | Upregulated |
| 0,56757879 | 0,051000894 | TGFB1I1 | FALSE | Upregulated |
| -0,136786459 | 0,051001156 | RBM17 | FALSE | Downregulated |
| -0,637943225 | 0,051049487 | LOC80154 | FALSE | Downregulated |
| 0,387898353 | 0,051113642 | IFI27L1 | FALSE | Upregulated |
| 0,418720372 | 0,051113642 | LRRC59 | FALSE | Upregulated |
| -0,268573996 | 0,051113642 | REV1 | FALSE | Downregulated |
| 0,735272642 | 0,051142998 | AGAP2 | FALSE | Upregulated |
| -0,537085194 | 0,051142998 | LOC100505942 | FALSE | Downregulated |
| 0,643958127 | 0,051149395 | CENPW | FALSE | Upregulated |
| -0,391321407 | 0,05116849 | ANKRD37 | FALSE | Downregulated |
| -0,177728609 | 0,051175567 | BTBD7 | FALSE | Downregulated |
| 0,351794249 | 0,051175567 | PCBP4 | FALSE | Upregulated |
| -0,382268457 | 0,051175567 | WDR72 | FALSE | Downregulated |
| -0,227216454 | 0,05119518 | TCFL5 | FALSE | Downregulated |
| 0,569946277 | 0,051214381 | TNFSF8 | FALSE | Upregulated |
| 0,519652936 | 0,051236821 | ZNF430 | FALSE | Upregulated |
| 2,974620781 | 0,051274498 | NPHS1 | FALSE | Upregulated |
| 0,884426239 | 0,05127668 | CD8B | FALSE | Upregulated |
| 0,841178458 | 0,051374525 | LOC100506734 | FALSE | Upregulated |
| -0,162359413 | 0,051391112 | DCAF10 | FALSE | Downregulated |
| -0,882411238 | 0,051422827 | CHRNA4 | FALSE | Downregulated |
| 0,265595251 | 0,051425156 | LRP1 | FALSE | Upregulated |
| 0,326674247 | 0,051715907 | RND3 | FALSE | Upregulated |
| 1,798967119 | 0,051716685 | LOC101927254 | FALSE | Upregulated |

| | | | | |
|---|---|---|---|---|
| 2,479886883 | 0,051719071 | LOC102723913 | FALSE | Upregulated |
| 0,55829845 | 0,051723515 | PRKCB | FALSE | Upregulated |
| 1,21485504 | 0,05177556 | GLDN | FALSE | Upregulated |
| 0,444909036 | 0,051802793 | ACP5 | FALSE | Upregulated |
| -0,236984793 | 0,051822107 | PPP1R10 | FALSE | Downregulated |
| 2,016878348 | 0,051859023 | PSORS1C3 | FALSE | Upregulated |
| 0,39849551 | 0,051859023 | SLC29A3 | FALSE | Upregulated |
| -0,34811776 | 0,051871434 | ACADS | FALSE | Downregulated |
| -0,435664055 | 0,051889243 | DLGAP1-AS1 | FALSE | Downregulated |
| -0,341361227 | 0,051889243 | ZFAND4 | FALSE | Downregulated |
| -0,291981761 | 0,051976743 | POLR3H | FALSE | Downregulated |
| 0,788048601 | 0,051982901 | SOX17 | FALSE | Upregulated |
| -0,462089421 | 0,052035204 | CLEC1B | FALSE | Downregulated |
| 1,623640462 | 0,052253758 | CDKN2B-AS1 | FALSE | Upregulated |
| -0,205492496 | 0,05234053 | NDUFS1 | FALSE | Downregulated |
| 0,259381284 | 0,052482979 | SLC38A7 | FALSE | Upregulated |
| -0,386957952 | 0,052493706 | GNE | FALSE | Downregulated |
| 0,279139762 | 0,052493706 | PHIP | FALSE | Upregulated |
| -0,331473951 | 0,052516671 | CCSER2 | FALSE | Downregulated |
| 0,467012331 | 0,052529484 | ZNF341 | FALSE | Upregulated |
| -0,252142802 | 0,052541948 | SURF1 | FALSE | Downregulated |
| -0,377841709 | 0,052601853 | GPATCH2 | FALSE | Downregulated |
| 0,210171647 | 0,052601853 | HDGFRP2 | FALSE | Upregulated |
| -0,423014127 | 0,052620143 | PPIF | FALSE | Downregulated |
| -1,279765207 | 0,052627756 | ZBED3-AS1 | FALSE | Downregulated |
| 0,203045556 | 0,052701903 | KDM3B | FALSE | Upregulated |
| 0,674165286 | 0,052718916 | SNAI1 | FALSE | Upregulated |
| -0,305781406 | 0,05276524 | PMF1 | FALSE | Downregulated |
| -0,194705926 | 0,052850733 | LINC00657 | FALSE | Downregulated |
| -0,482301408 | 0,052850733 | LOC100507258 | FALSE | Downregulated |
| 0,192542544 | 0,052905628 | CHMP1B | FALSE | Upregulated |
| -0,272366197 | 0,052967748 | LMAN1 | FALSE | Downregulated |
| 1,56269437 | 0,052967748 | MMP28 | FALSE | Upregulated |
| -0,912650344 | 0,052985076 | LOC401127 | FALSE | Downregulated |
| 2,036499575 | 0,053018984 | ANKRD18B | FALSE | Upregulated |
| -0,341009438 | 0,053042082 | AGT | FALSE | Downregulated |
| -0,931234457 | 0,053042082 | LINC00685 | FALSE | Downregulated |
| -0,469011766 | 0,053091356 | ASB16-AS1 | FALSE | Downregulated |
| 0,225659175 | 0,053216276 | GMEB2 | FALSE | Upregulated |
| 0,252557904 | 0,053252116 | MTMR3 | FALSE | Upregulated |
| 1,80629647 | 0,053318036 | GPR110 | FALSE | Upregulated |
| -0,303402108 | 0,053322206 | MIB2 | FALSE | Downregulated |
| -0,398088941 | 0,053465131 | ABHD15 | FALSE | Downregulated |
| 2,18559524 | 0,053493518 | SRGAP2-AS1 | FALSE | Upregulated |
| 0,25384929 | 0,053505513 | C12orf10 | FALSE | Upregulated |
| 0,345769724 | 0,053561181 | NFKBIA | FALSE | Upregulated |
| -0,145497812 | 0,05362656 | SERINC1 | FALSE | Downregulated |
| -0,165623281 | 0,05363188 | FAF2 | FALSE | Downregulated |
| 0,664698116 | 0,05363188 | LOC100652768 | FALSE | Upregulated |
| -0,290347319 | 0,053654448 | IFT27 | FALSE | Downregulated |
| -0,38745928 | 0,053655967 | PNP | FALSE | Downregulated |

| | | | | |
|---|---|---|---|---|
| -0,228604001 | 0,053662048 | GNPNAT1 | FALSE | Downregulated |
| 0,230706977 | 0,053662048 | HEXA | FALSE | Upregulated |
| -0,672855653 | 0,053676187 | LOC102724833 | FALSE | Downregulated |
| -0,443544153 | 0,053676187 | RHOBTB3 | FALSE | Downregulated |
| 0,162115617 | 0,053676187 | UBQLN1 | FALSE | Upregulated |
| -0,215131203 | 0,053694019 | BCAP29 | FALSE | Downregulated |
| 0,650044471 | 0,053727108 | NBEA | FALSE | Upregulated |
| -0,356920883 | 0,053727108 | SMO | FALSE | Downregulated |
| 0,942635852 | 0,053744818 | LOC102723554 | FALSE | Upregulated |
| 2,843472377 | 0,053744818 | PPIAL4B | FALSE | Upregulated |
| 0,216035243 | 0,053831402 | GMCL1 | FALSE | Upregulated |
| 0,600267351 | 0,053840358 | AKAP5 | FALSE | Upregulated |
| -0,445221994 | 0,053840358 | BAAT | FALSE | Downregulated |
| 0,459778465 | 0,053840358 | LOC728743 | FALSE | Upregulated |
| 1,704887383 | 0,053844363 | CNFN | FALSE | Upregulated |
| 0,303500404 | 0,053967382 | ADCK2 | FALSE | Upregulated |
| -1,336465365 | 0,053986982 | NEUROD1 | FALSE | Downregulated |
| -0,531983503 | 0,054070693 | JPX | FALSE | Downregulated |
| 0,546139872 | 0,054081969 | SH3BP1 | FALSE | Upregulated |
| 0,269028332 | 0,05427164 | IL10RB | FALSE | Upregulated |
| -0,162762134 | 0,054343802 | UBE2K | FALSE | Downregulated |
| 0,159605013 | 0,054487484 | RBBP4 | FALSE | Upregulated |
| -0,190689482 | 0,054505446 | NOL10 | FALSE | Downregulated |
| 0,751206433 | 0,05452519 | C10orf35 | FALSE | Upregulated |
| -0,2960361 | 0,05452519 | MIOS | FALSE | Downregulated |
| 0,228013105 | 0,054526328 | RPN1 | FALSE | Upregulated |
| -0,352539474 | 0,05456843 | TF | FALSE | Downregulated |
| -0,213360404 | 0,054585587 | MED20 | FALSE | Downregulated |
| -0,226601327 | 0,054626762 | LARP7 | FALSE | Downregulated |
| 0,504476346 | 0,054646597 | TRIM62 | FALSE | Upregulated |
| -0,166891775 | 0,054735676 | COPS2 | FALSE | Downregulated |
| -0,402577213 | 0,054813532 | FAHD1 | FALSE | Downregulated |
| -0,229565357 | 0,054840584 | HDHD3 | FALSE | Downregulated |
| -0,312543591 | 0,054873559 | THAP7 | FALSE | Downregulated |
| -0,274008018 | 0,054881943 | SLC25A1 | FALSE | Downregulated |
| -0,302496387 | 0,05493057 | DGCR6L | FALSE | Downregulated |
| 1,247146664 | 0,05493057 | IGFBPL1 | FALSE | Upregulated |
| -0,182664134 | 0,05497995 | SNX3 | FALSE | Downregulated |
| 1,56520213 | 0,05497995 | TIFAB | FALSE | Upregulated |
| -1,611375437 | 0,055006557 | LOC102723643 | FALSE | Downregulated |
| -0,781253705 | 0,055024567 | LOC102723389 | FALSE | Downregulated |
| -1,047793411 | 0,055053771 | LOC101928810 | FALSE | Downregulated |
| 1,920603921 | 0,055100515 | LINC00854 | FALSE | Upregulated |
| -0,264563326 | 0,055216425 | ABHD17A | FALSE | Downregulated |
| 0,481441758 | 0,055389098 | PLK2 | FALSE | Upregulated |
| 1,716217727 | 0,055393037 | RPSAP52 | FALSE | Upregulated |
| -0,470178937 | 0,055503796 | RNPS1 | FALSE | Downregulated |
| 0,938966227 | 0,055514402 | CBX2 | FALSE | Upregulated |
| -0,87529172 | 0,055514402 | PKHD1L1 | FALSE | Downregulated |
| 0,483728533 | 0,055514402 | ZNF670 | FALSE | Upregulated |
| 0,22177232 | 0,055534192 | KAT5 | FALSE | Upregulated |

| | | | | |
|---|---|---|---|---|
| 0,883341853 | 0,055534192 | RHPN1 | FALSE | Upregulated |
| -0,352358449 | 0,055539272 | RPS27 | FALSE | Downregulated |
| -0,30549288 | 0,055539272 | THOC2 | FALSE | Downregulated |
| -0,290840346 | 0,055570451 | EDEM1 | FALSE | Downregulated |
| -0,218644727 | 0,055618596 | SRSF8 | FALSE | Downregulated |
| -0,2002037 | 0,055627341 | ABCD4 | FALSE | Downregulated |
| 0,71726632 | 0,055630249 | EEF1E1 | FALSE | Upregulated |
| 0,809269115 | 0,055630249 | LMCD1 | FALSE | Upregulated |
| 0,808978771 | 0,055630249 | P2RY14 | FALSE | Upregulated |
| 0,463203904 | 0,055678655 | H2AFX | FALSE | Upregulated |
| 0,542210354 | 0,055678655 | ZNF671 | FALSE | Upregulated |
| -0,254458876 | 0,055690024 | SDHB | FALSE | Downregulated |
| 0,376103706 | 0,055696666 | DISP1 | FALSE | Upregulated |
| 1,702732772 | 0,055696666 | LOC102724286 | FALSE | Upregulated |
| 0,149101447 | 0,055696666 | SMCR8 | FALSE | Upregulated |
| 0,262916606 | 0,05569958 | MCM9 | FALSE | Upregulated |
| -0,928794453 | 0,05572823 | LOC102723953 | FALSE | Downregulated |
| 0,274859201 | 0,055795901 | YIF1A | FALSE | Upregulated |
| -0,272129658 | 0,055813471 | PEX3 | FALSE | Downregulated |
| -0,321396359 | 0,055875278 | METTL21B | FALSE | Downregulated |
| 1,121055926 | 0,055876909 | LOXL1-AS1 | FALSE | Upregulated |
| 1,173243428 | 0,055889166 | LHX6 | FALSE | Upregulated |
| -0,361844522 | 0,055889166 | PRRG4 | FALSE | Downregulated |
| 0,254095557 | 0,05591372 | BRK1 | FALSE | Upregulated |
| 0,935090636 | 0,055934071 | ZNF625 | FALSE | Upregulated |
| 0,670151356 | 0,055979579 | HMMR | FALSE | Upregulated |
| 0,183192341 | 0,056106385 | HIF1AN | FALSE | Upregulated |
| 0,361108629 | 0,056111607 | PNKP | FALSE | Upregulated |
| -0,690585637 | 0,056142486 | LRP4 | FALSE | Downregulated |
| 0,710758467 | 0,056179918 | SPATA18 | FALSE | Upregulated |
| -0,263161547 | 0,05619217 | MRPL19 | FALSE | Downregulated |
| 0,681071945 | 0,056221189 | ANKRD29 | FALSE | Upregulated |
| -0,255719116 | 0,056238155 | FAU | FALSE | Downregulated |
| 0,142777149 | 0,056242618 | USP22 | FALSE | Upregulated |
| 0,828624267 | 0,056248495 | LINC00482 | FALSE | Upregulated |
| 0,396504488 | 0,056265171 | SPIRE2 | FALSE | Upregulated |
| -0,670808176 | 0,056320452 | ERRFI1 | FALSE | Downregulated |
| 0,279099769 | 0,056370499 | FCHO2 | FALSE | Upregulated |
| -1,358137358 | 0,056470054 | FAM85A | FALSE | Downregulated |
| -0,301080801 | 0,05648622 | NR1I3 | FALSE | Downregulated |
| -0,183142514 | 0,056486314 | VMA21 | FALSE | Downregulated |
| -0,176582083 | 0,056545627 | PDE8A | FALSE | Downregulated |
| 0,429324728 | 0,056582438 | BBS5 | FALSE | Upregulated |
| -0,667666245 | 0,056598129 | LOC101927559 | FALSE | Downregulated |
| -0,343223886 | 0,056619227 | MCC | FALSE | Downregulated |
| -0,238881661 | 0,056664801 | ALDH9A1 | FALSE | Downregulated |
| -1,555662644 | 0,056730731 | LRRC19 | FALSE | Downregulated |
| -0,188646967 | 0,056731194 | TM2D3 | FALSE | Downregulated |
| 0,49681155 | 0,056742624 | GALNT7 | FALSE | Upregulated |
| 1,977898182 | 0,056760408 | ZFY | FALSE | Upregulated |
| -0,41861451 | 0,056769031 | ZBED6 | FALSE | Downregulated |

| | | | | |
|---|---|---|---|---|
| -0,213451473 | 0,0568237 | SHARPIN | FALSE | Downregulated |
| 0,361858739 | 0,056848856 | PLIN3 | FALSE | Upregulated |
| 0,18858973 | 0,056877957 | MAPKAPK2 | FALSE | Upregulated |
| -0,236084068 | 0,056877957 | SLK | FALSE | Downregulated |
| 0,338347236 | 0,056915785 | CBR1 | FALSE | Upregulated |
| 0,303243811 | 0,056977922 | OSTM1 | FALSE | Upregulated |
| -0,96194842 | 0,057022765 | ATAD3C | FALSE | Downregulated |
| 0,282429353 | 0,05703687 | ACP2 | FALSE | Upregulated |
| 0,535209442 | 0,05704592 | CRELD2 | FALSE | Upregulated |
| -2,172454671 | 0,057065904 | FAM209A | FALSE | Downregulated |
| 0,771399497 | 0,057065904 | SNRK-AS1 | FALSE | Upregulated |
| 0,164643593 | 0,057089922 | NUP50 | FALSE | Upregulated |
| 0,451616546 | 0,057093716 | UBE2S | FALSE | Upregulated |
| -0,546712548 | 0,057110433 | FLJ38668 | FALSE | Downregulated |
| 0,7579052 | 0,057209954 | FAHD2CP | FALSE | Upregulated |
| 0,285540897 | 0,057324043 | GOLPH3 | FALSE | Upregulated |
| 1,023486475 | 0,05741325 | DNAJC5B | FALSE | Upregulated |
| 0,473561007 | 0,057508425 | BARD1 | FALSE | Upregulated |
| 0,218026964 | 0,057598588 | UTP14A | FALSE | Upregulated |
| -0,708515934 | 0,057733675 | ZBED3 | FALSE | Downregulated |
| 2,44929992 | 0,057740273 | FLJ41200 | FALSE | Upregulated |
| -0,263171918 | 0,057767713 | MED19 | FALSE | Downregulated |
| -0,281931669 | 0,057767713 | PROC | FALSE | Downregulated |
| 0,27604895 | 0,057819886 | PARD3B | FALSE | Upregulated |
| 2,23106352 | 0,057826193 | GSG1 | FALSE | Upregulated |
| -0,251981314 | 0,057932713 | CYP4F11 | FALSE | Downregulated |
| 0,180737596 | 0,058051277 | ZNF148 | FALSE | Upregulated |
| 0,94322182 | 0,058054363 | SEMA3A | FALSE | Upregulated |
| 1,542268014 | 0,058129349 | LOC102724740 | FALSE | Upregulated |
| 0,40102171 | 0,058134331 | KLF16 | FALSE | Upregulated |
| 0,491924593 | 0,058169723 | PDE4B | FALSE | Upregulated |
| -0,186160265 | 0,058238342 | BMPR2 | FALSE | Downregulated |
| 2,293132656 | 0,058266536 | LOC102723545 | FALSE | Upregulated |
| 0,767095335 | 0,05829355 | IGFBP2 | FALSE | Upregulated |
| 0,250548785 | 0,05829355 | PEX6 | FALSE | Upregulated |
| 0,72626742 | 0,058335655 | VWA7 | FALSE | Upregulated |
| -0,597473734 | 0,058377159 | RP11-342C23.4 | FALSE | Downregulated |
| 0,165963023 | 0,058432989 | EXOC4 | FALSE | Upregulated |
| 2,503163113 | 0,058493481 | SV2C | FALSE | Upregulated |
| 0,265324257 | 0,058509741 | ARHGAP26 | FALSE | Upregulated |
| 2,271771214 | 0,058509741 | TRPA1 | FALSE | Upregulated |
| -0,317899475 | 0,058524604 | SLC27A5 | FALSE | Downregulated |
| -0,263881595 | 0,05859394 | MNAT1 | FALSE | Downregulated |
| -0,416489905 | 0,058605752 | C2orf74 | FALSE | Downregulated |
| 0,195435846 | 0,058605752 | NECAP2 | FALSE | Upregulated |
| 0,279240592 | 0,058605752 | PSMA1 | FALSE | Upregulated |
| 1,468704864 | 0,058687311 | CCDC19 | FALSE | Upregulated |
| -0,313987232 | 0,058692183 | C10orf32 | FALSE | Downregulated |
| -0,195961402 | 0,058692183 | RNF185 | FALSE | Downregulated |
| 0,99405337 | 0,058706997 | SKA3 | FALSE | Upregulated |
| -0,323432755 | 0,05871756 | CYP4V2 | FALSE | Downregulated |

| | | | | |
|---|---|---|---|---|
| -0,72123001 | 0,05871756 | LOC101929046 | FALSE | Downregulated |
| -0,415036914 | 0,05871756 | NUDT3 | FALSE | Downregulated |
| 0,328705691 | 0,058763695 | GDA | FALSE | Upregulated |
| 0,314063945 | 0,058833468 | MAPK7 | FALSE | Upregulated |
| -0,520137862 | 0,059011313 | LOC101927809 | FALSE | Downregulated |
| -0,371985816 | 0,059042478 | RABEPK | FALSE | Downregulated |
| 1,158563539 | 0,059078999 | LOC102724428 | FALSE | Upregulated |
| 0,283332337 | 0,059081738 | PERP | FALSE | Upregulated |
| -0,424368029 | 0,059177884 | FGB | FALSE | Downregulated |
| -0,867964159 | 0,059187067 | PLIN1 | FALSE | Downregulated |
| -0,362106312 | 0,059207256 | FAM115A | FALSE | Downregulated |
| -0,309503735 | 0,05922586 | PAPD7 | FALSE | Downregulated |
| 0,968010728 | 0,059239882 | SH3TC2 | FALSE | Upregulated |
| -3,071103134 | 0,059263817 | LOC102723630 | FALSE | Downregulated |
| 0,553059794 | 0,059278728 | C5AR1 | FALSE | Upregulated |
| -0,317137602 | 0,059278728 | NDUFC2 | FALSE | Downregulated |
| -0,275722762 | 0,059278728 | RALA | FALSE | Downregulated |
| -0,33054135 | 0,059335025 | ACTR3B | FALSE | Downregulated |
| -1,228446685 | 0,059335025 | CMYA5 | FALSE | Downregulated |
| 0,240979459 | 0,059335025 | LDB1 | FALSE | Upregulated |
| 1,694773117 | 0,059335025 | OVOL2 | FALSE | Upregulated |
| 0,283786994 | 0,059403412 | AKIRIN2 | FALSE | Upregulated |
| -0,142654813 | 0,059403412 | ARPC1A | FALSE | Downregulated |
| -0,581022624 | 0,059404744 | HLA-F-AS1 | FALSE | Downregulated |
| 0,16305889 | 0,059404744 | TFIP11 | FALSE | Upregulated |
| 0,688992099 | 0,059481469 | ATP6V1C2 | FALSE | Upregulated |
| -2,061665427 | 0,059599466 | CDH12 | FALSE | Downregulated |
| 0,47865154 | 0,059624378 | ATAD3B | FALSE | Upregulated |
| 0,399911025 | 0,059768403 | LOC493754 | FALSE | Upregulated |
| 0,760576966 | 0,059768403 | ZNF714 | FALSE | Upregulated |
| -0,190122391 | 0,059807339 | STUB1 | FALSE | Downregulated |
| -0,766786328 | 0,059829879 | MANEA-AS1 | FALSE | Downregulated |
| 1,006178066 | 0,059859065 | GFM2 | FALSE | Upregulated |
| -0,40142753 | 0,059864331 | MGC27345 | FALSE | Downregulated |
| -1,016278816 | 0,059885257 | RFPL4A | FALSE | Downregulated |
| 0,378837451 | 0,059946202 | RRP12 | FALSE | Upregulated |
| -0,218558382 | 0,05999515 | PQBP1 | FALSE | Downregulated |
| -1,45353957 | 0,06000367 | LINC00890 | FALSE | Downregulated |
| 2,428317732 | 0,060046057 | EGFL6 | FALSE | Upregulated |
| -0,176363159 | 0,060075318 | THOC7 | FALSE | Downregulated |
| 0,900979687 | 0,06013567 | TMCC2 | FALSE | Upregulated |
| -0,256100067 | 0,060162787 | CEP57L1 | FALSE | Downregulated |
| 0,313313222 | 0,060235712 | FBXO25 | FALSE | Upregulated |
| 1,623891803 | 0,060331394 | CLEC4C | FALSE | Upregulated |
| 1,714466067 | 0,060331394 | SFN | FALSE | Upregulated |
| 0,861087127 | 0,06035453 | ERP27 | FALSE | Upregulated |
| -0,295274953 | 0,060369791 | MFAP3L | FALSE | Downregulated |
| 1,032295559 | 0,060369791 | SLMO2-ATP5E | FALSE | Upregulated |
| 1,140079068 | 0,060378463 | KLRC3 | FALSE | Upregulated |
| 0,180767025 | 0,060442161 | SPPL3 | FALSE | Upregulated |
| 0,996846071 | 0,060458914 | BCAS4 | FALSE | Upregulated |

| | | | | |
|---|---|---|---|---|
| 0,176063131 | 0,060458914 | VPS11 | FALSE | Upregulated |
| -0,19101368 | 0,060486557 | STK3 | FALSE | Downregulated |
| -0,280982287 | 0,060575967 | ANKS4B | FALSE | Downregulated |
| 0,16821933 | 0,060587077 | RIC8A | FALSE | Upregulated |
| 0,511400554 | 0,060601261 | STAMBPL1 | FALSE | Upregulated |
| 0,53638673 | 0,06063244 | BBS12 | FALSE | Upregulated |
| -0,284195676 | 0,060668183 | FAM104B | FALSE | Downregulated |
| 0,524210662 | 0,060690458 | PDE3A | FALSE | Upregulated |
| 0,472498099 | 0,060770204 | SLC2A3 | FALSE | Upregulated |
| 0,349183212 | 0,060840805 | GLMN | FALSE | Upregulated |
| -0,285318902 | 0,060872467 | GUCD1 | FALSE | Downregulated |
| 1,517335003 | 0,060872467 | HS6ST2 | FALSE | Upregulated |
| -0,278566729 | 0,060875185 | RTF1 | FALSE | Downregulated |
| -0,25620138 | 0,060888011 | INTS12 | FALSE | Downregulated |
| -0,270311589 | 0,060888898 | ADAL | FALSE | Downregulated |
| 0,303929986 | 0,061068298 | ORMDL2 | FALSE | Upregulated |
| -0,310380871 | 0,061107989 | TSTD1 | FALSE | Downregulated |
| 0,901979754 | 0,061131149 | LPHN3 | FALSE | Upregulated |
| 0,271255475 | 0,061131149 | MARS | FALSE | Upregulated |
| 0,645102551 | 0,061139327 | NLRP3 | FALSE | Upregulated |
| 1,656132845 | 0,061283902 | ESR2 | FALSE | Upregulated |
| 0,286449444 | 0,061283902 | IFIH1 | FALSE | Upregulated |
| 0,48640003 | 0,06133122 | FAM46C | FALSE | Upregulated |
| -0,31499419 | 0,061381026 | NFKBIL1 | FALSE | Downregulated |
| 0,334187833 | 0,061416187 | NCAPH2 | FALSE | Upregulated |
| 0,476910128 | 0,061425376 | GGT1 | FALSE | Upregulated |
| -0,196751033 | 0,061432912 | ARHGAP35 | FALSE | Downregulated |
| 1,452857585 | 0,061511845 | RUFY4 | FALSE | Upregulated |
| 0,239056532 | 0,061515031 | MYO9B | FALSE | Upregulated |
| 1,990261946 | 0,061566845 | CLK2P | FALSE | Upregulated |
| -0,307861189 | 0,061584335 | FXN | FALSE | Downregulated |
| 0,923379551 | 0,061584335 | OXCT2 | FALSE | Upregulated |
| 0,589240031 | 0,061584335 | THSD7A | FALSE | Upregulated |
| 0,961127429 | 0,061605808 | MAPK8IP2 | FALSE | Upregulated |
| 0,298788292 | 0,061674064 | C7orf49 | FALSE | Upregulated |
| -0,296351849 | 0,061674064 | CYB5D2 | FALSE | Downregulated |
| 0,32882521 | 0,061683818 | RIPK2 | FALSE | Upregulated |
| 0,358698555 | 0,061757795 | CCDC68 | FALSE | Upregulated |
| 0,663855562 | 0,061907783 | KLRB1 | FALSE | Upregulated |
| 0,610845063 | 0,061939958 | ST8SIA4 | FALSE | Upregulated |
| -0,693031149 | 0,061995246 | UBAC2-AS1 | FALSE | Downregulated |
| -0,247378591 | 0,06199887 | ERF | FALSE | Downregulated |
| 0,278211124 | 0,06201793 | RUFY2 | FALSE | Upregulated |
| 0,596395596 | 0,062065088 | PSIMCT-1 | FALSE | Upregulated |
| 2,16828881 | 0,062289947 | KLHL35 | FALSE | Upregulated |
| 1,466595142 | 0,062290578 | OPCML | FALSE | Upregulated |
| 0,549359979 | 0,062301808 | CENPF | FALSE | Upregulated |
| -0,56000997 | 0,062362889 | SHISA4 | FALSE | Downregulated |
| 0,216228233 | 0,062449912 | CCDC22 | FALSE | Upregulated |
| 0,252477568 | 0,062477764 | ITGA5 | FALSE | Upregulated |
| -0,212160444 | 0,062553841 | SPG7 | FALSE | Downregulated |

| | | | | |
|---|---|---|---|---|
| -0,243489974 | 0,062573733 | ALS2 | FALSE | Downregulated |
| 0,893359537 | 0,062577699 | GADD45G | FALSE | Upregulated |
| -0,3753225 | 0,062607522 | HDHD1 | FALSE | Downregulated |
| -0,592619044 | 0,062619814 | CYP51A1 | FALSE | Downregulated |
| -0,417459275 | 0,062694675 | ANKRD9 | FALSE | Downregulated |
| -0,350320423 | 0,062720407 | EIF4EBP1 | FALSE | Downregulated |
| -0,28241695 | 0,062769567 | BIVM | FALSE | Downregulated |
| -0,443439695 | 0,062805864 | LOC642361 | FALSE | Downregulated |
| 0,349588101 | 0,062905826 | ACAP3 | FALSE | Upregulated |
| 0,235557151 | 0,062928586 | PDCD6 | FALSE | Upregulated |
| -0,314836807 | 0,06295512 | IGFBP3 | FALSE | Downregulated |
| -0,660712269 | 0,06295512 | KCTD21-AS1 | FALSE | Downregulated |
| -0,282337097 | 0,063097002 | MRPS36 | FALSE | Downregulated |
| -2,067935284 | 0,063142138 | HIGD2B | FALSE | Downregulated |
| 0,934553758 | 0,063176058 | EBLN2 | FALSE | Upregulated |
| 0,468001154 | 0,063198191 | RBPMS2 | FALSE | Upregulated |
| -1,195702602 | 0,063238357 | ST18 | FALSE | Downregulated |
| -0,589625398 | 0,063250826 | FANCC | FALSE | Downregulated |
| 1,351971818 | 0,063280685 | DIRAS2 | FALSE | Upregulated |
| -0,228678952 | 0,063324884 | BCDIN3D | FALSE | Downregulated |
| 0,375473419 | 0,063324884 | TMCO3 | FALSE | Upregulated |
| -1,442624058 | 0,063400373 | HCG14 | FALSE | Downregulated |
| -0,281613623 | 0,063499078 | RPL17 | FALSE | Downregulated |
| 0,251475714 | 0,063530513 | E2F6 | FALSE | Upregulated |
| 1,203950251 | 0,063614095 | RP11-861A13.4 | FALSE | Upregulated |
| 0,494147235 | 0,063615558 | GPR65 | FALSE | Upregulated |
| -0,323956856 | 0,063666429 | TMEM116 | FALSE | Downregulated |
| 0,179124963 | 0,063691687 | LZTR1 | FALSE | Upregulated |
| 0,481171841 | 0,063728526 | SMPD2 | FALSE | Upregulated |
| 0,311992629 | 0,063732979 | H1FX | FALSE | Upregulated |
| 0,612646938 | 0,063740138 | STEAP4 | FALSE | Upregulated |
| -0,351248673 | 0,063740816 | F5 | FALSE | Downregulated |
| -0,200038568 | 0,063763471 | WBP2 | FALSE | Downregulated |
| -0,822235083 | 0,063811049 | LOC101929143 | FALSE | Downregulated |
| 0,535559742 | 0,063833821 | DSCC1 | FALSE | Upregulated |
| -0,746634577 | 0,06384707 | ZIC1 | FALSE | Downregulated |
| 0,308897385 | 0,063860953 | AGTRAP | FALSE | Upregulated |
| -0,354572305 | 0,063948846 | ARSK | FALSE | Downregulated |
| 0,287445853 | 0,063972996 | MSANTD4 | FALSE | Upregulated |
| 0,270551466 | 0,064042464 | YARS | FALSE | Upregulated |
| -0,152910722 | 0,064046088 | BUD13 | FALSE | Downregulated |
| -0,373213784 | 0,064093316 | TBC1D10B | FALSE | Downregulated |
| -0,163991398 | 0,064102681 | TRAPPC8 | FALSE | Downregulated |
| -0,197860456 | 0,064103032 | HBP1 | FALSE | Downregulated |
| 0,57451854 | 0,064194636 | AMPD3 | FALSE | Upregulated |
| 0,215156554 | 0,064309647 | PIGM | FALSE | Upregulated |
| 1,378348692 | 0,064332342 | TP73 | FALSE | Upregulated |
| -0,253212235 | 0,064537704 | HLCS | FALSE | Downregulated |
| 0,45205255 | 0,064553068 | NT5DC2 | FALSE | Upregulated |
| 0,247966102 | 0,064606742 | CUL7 | FALSE | Upregulated |
| -0,276254066 | 0,064660955 | CAMK1 | FALSE | Downregulated |

| | | | | |
|---|---|---|---|---|
| 0,81001894 | 0,064664482 | FSD1L | FALSE | Upregulated |
| 0,256965305 | 0,064664482 | RNF44 | FALSE | Upregulated |
| 1,243416646 | 0,064681632 | MFAP2 | FALSE | Upregulated |
| 0,244875586 | 0,064699708 | SPNS1 | FALSE | Upregulated |
| 1,033069377 | 0,064743254 | IQCD | FALSE | Upregulated |
| 1,192905143 | 0,064770616 | ROS1 | FALSE | Upregulated |
| -0,236172122 | 0,06481097 | MLLT10 | FALSE | Downregulated |
| 0,78959452 | 0,064824701 | ASNS | FALSE | Upregulated |
| -0,724982901 | 0,064824701 | HEXA-AS1 | FALSE | Downregulated |
| 0,375306997 | 0,064837472 | FKBPL | FALSE | Upregulated |
| 1,874997822 | 0,064840612 | LOC641746 | FALSE | Upregulated |
| 1,835361729 | 0,064850552 | LINC00900 | FALSE | Upregulated |
| -0,214103782 | 0,064866549 | RANBP2 | FALSE | Downregulated |
| -0,246578251 | 0,064905618 | TIPRL | FALSE | Downregulated |
| 0,301386935 | 0,064936162 | ZNF182 | FALSE | Upregulated |
| -0,285548883 | 0,064938573 | LMO7 | FALSE | Downregulated |
| -0,440932825 | 0,064983185 | ALDH1L1-AS2 | FALSE | Downregulated |
| 2,299105701 | 0,065042804 | FGF13-AS1 | FALSE | Upregulated |
| 0,209495516 | 0,065079563 | MAZ | FALSE | Upregulated |
| 0,191149339 | 0,065093151 | SORBS3 | FALSE | Upregulated |
| -0,454458156 | 0,065116627 | KNSTRN | FALSE | Downregulated |
| 0,304674651 | 0,065210123 | GMEB1 | FALSE | Upregulated |
| -0,385486431 | 0,065251596 | CXorf57 | FALSE | Downregulated |
| 1,759465603 | 0,065268197 | WTAPP1 | FALSE | Upregulated |
| 0,467466366 | 0,065272522 | GJA4 | FALSE | Upregulated |
| 2,550898814 | 0,065272522 | LOC643711 | FALSE | Upregulated |
| -0,247024346 | 0,065333858 | ZFAND1 | FALSE | Downregulated |
| 1,172414862 | 0,065363698 | LRFN4 | FALSE | Upregulated |
| -1,140795602 | 0,065372929 | GCK | FALSE | Downregulated |
| 0,234119636 | 0,065424789 | GTF2A2 | FALSE | Upregulated |
| 0,216850771 | 0,065501516 | GLRX3 | FALSE | Upregulated |
| 0,734512871 | 0,065501516 | HLA-DQB1 | FALSE | Upregulated |
| 1,655238058 | 0,065521602 | LOC101926943 | FALSE | Upregulated |
| 0,111347349 | 0,065521602 | TERF2IP | FALSE | Upregulated |
| 0,901620939 | 0,065533883 | VGLL3 | FALSE | Upregulated |
| 0,677982608 | 0,065559211 | AMIGO3 | FALSE | Upregulated |
| -0,217127356 | 0,065572877 | ERO1L | FALSE | Downregulated |
| -0,44624572 | 0,065607799 | MERTK | FALSE | Downregulated |
| -0,35987191 | 0,065858979 | MYCT1 | FALSE | Downregulated |
| 1,399413714 | 0,065858979 | RTEL1-TNFRSF6B | FALSE | Upregulated |
| -0,269371679 | 0,065861439 | FAM58A | FALSE | Downregulated |
| 0,489318971 | 0,065861439 | PLA2R1 | FALSE | Upregulated |
| -0,298111078 | 0,065864617 | ERBB2IP | FALSE | Downregulated |
| -0,263623341 | 0,066013903 | TMEM186 | FALSE | Downregulated |
| -0,221586859 | 0,066025647 | IKZF5 | FALSE | Downregulated |
| 1,500464786 | 0,066106687 | KCNA5 | FALSE | Upregulated |
| 2,196044685 | 0,066270481 | RP11-294O2.2 | FALSE | Upregulated |
| -0,301424349 | 0,066280003 | SMARCC1 | FALSE | Downregulated |
| -0,215032363 | 0,066328055 | IMMP2L | FALSE | Downregulated |
| 0,233011194 | 0,066363168 | ENTPD4 | FALSE | Upregulated |
| -2,069798051 | 0,066363168 | LOC101929051 | FALSE | Downregulated |

| | | | | |
|---|---|---|---|---|
| 1,063693506 | 0,066385147 | FGF11 | FALSE | Upregulated |
| 2,126616925 | 0,066390686 | PNOC | FALSE | Upregulated |
| 0,487039108 | 0,066437227 | VWA1 | FALSE | Upregulated |
| 0,667533549 | 0,066589352 | ZNF807 | FALSE | Upregulated |
| -0,196709187 | 0,066594453 | CASP6 | FALSE | Downregulated |
| -4,188494688 | 0,066649707 | GOLGA8J | FALSE | Downregulated |
| 0,237469513 | 0,066788163 | NFRKB | FALSE | Upregulated |
| -0,234066596 | 0,06680388 | CCDC174 | FALSE | Downregulated |
| 0,314134062 | 0,06680388 | DOCK6 | FALSE | Upregulated |
| 0,297438796 | 0,066886423 | TECPR1 | FALSE | Upregulated |
| 2,794578757 | 0,066958777 | CHIT1 | FALSE | Upregulated |
| 2,841693442 | 0,066969686 | SRD5A1P1 | FALSE | Upregulated |
| -0,593883955 | 0,06698779 | LOC100132111 | FALSE | Downregulated |
| -3,762330724 | 0,067005911 | SFRP2 | FALSE | Downregulated |
| -0,509891903 | 0,067053376 | PSMD5-AS1 | FALSE | Downregulated |
| -1,238901856 | 0,067191954 | LOC102723513 | FALSE | Downregulated |
| -0,548759428 | 0,067191954 | SSTR1 | FALSE | Downregulated |
| -0,31246034 | 0,067250751 | MTDH | FALSE | Downregulated |
| -0,754391715 | 0,067431733 | KCNK6 | FALSE | Downregulated |
| -0,395536086 | 0,067497351 | CWC25 | FALSE | Downregulated |
| -0,611924724 | 0,067506331 | SSTR2 | FALSE | Downregulated |
| -0,799301751 | 0,067519577 | LOC101927665 | FALSE | Downregulated |
| -0,278561504 | 0,067534832 | ZNF395 | FALSE | Downregulated |
| 0,440756047 | 0,067551107 | SELPLG | FALSE | Upregulated |
| 1,006183632 | 0,06756175 | ABCB9 | FALSE | Upregulated |
| 0,448097056 | 0,06756175 | APOLD1 | FALSE | Upregulated |
| 0,289980847 | 0,06756175 | C12orf4 | FALSE | Upregulated |
| 0,187400325 | 0,067581016 | NBAS | FALSE | Upregulated |
| 0,274673606 | 0,067609594 | UGGT1 | FALSE | Upregulated |
| -0,385862351 | 0,067620294 | NTHL1 | FALSE | Downregulated |
| -0,575164167 | 0,067689076 | ARL4D | FALSE | Downregulated |
| 2,257355001 | 0,067689076 | GPR3 | FALSE | Upregulated |
| -1,380296298 | 0,067689076 | LOC101929819 | FALSE | Downregulated |
| 0,18179263 | 0,067689076 | UBAP2 | FALSE | Upregulated |
| 0,616163695 | 0,067760653 | TMEM54 | FALSE | Upregulated |
| 0,198125739 | 0,067767075 | NELFCD | FALSE | Upregulated |
| 1,015522745 | 0,067767075 | RBMS3-AS3 | FALSE | Upregulated |
| -0,276536735 | 0,067768171 | TASP1 | FALSE | Downregulated |
| 0,921489712 | 0,067894133 | DMKN | FALSE | Upregulated |
| -0,574796492 | 0,067922311 | CETP | FALSE | Downregulated |
| -0,195689552 | 0,067992816 | GID4 | FALSE | Downregulated |
| -0,57236939 | 0,068013058 | UBXN4 | FALSE | Downregulated |
| -0,220249671 | 0,06805345 | TIMM50 | FALSE | Downregulated |
| -0,194400506 | 0,068065811 | PTDSS1 | FALSE | Downregulated |
| -0,447203566 | 0,068066111 | SMOC1 | FALSE | Downregulated |
| 2,368405356 | 0,068111168 | ST7-AS2 | FALSE | Upregulated |
| 2,423968908 | 0,068130073 | BARX2 | FALSE | Upregulated |
| -0,180223142 | 0,068160405 | TMED4 | FALSE | Downregulated |
| -0,364294446 | 0,068205865 | GABPB2 | FALSE | Downregulated |
| -0,27534582 | 0,068215208 | RBL2 | FALSE | Downregulated |
| -0,398072532 | 0,068252891 | SLCO1B1 | FALSE | Downregulated |

| | | | | |
|---|---|---|---|---|
| 0,248321296 | 0,068354624 | SLC35C2 | FALSE | Upregulated |
| 1,743970967 | 0,068412786 | LOC728739 | FALSE | Upregulated |
| -0,453837098 | 0,068415288 | SLC6A16 | FALSE | Downregulated |
| 1,737508492 | 0,068446145 | LOC102724448 | FALSE | Upregulated |
| -1,018514847 | 0,068476831 | PCP2 | FALSE | Downregulated |
| 0,618955209 | 0,068551182 | CORO2B | FALSE | Upregulated |
| -0,474705202 | 0,068551182 | HERC5 | FALSE | Downregulated |
| -0,281650111 | 0,068551182 | PRR3 | FALSE | Downregulated |
| 2,450323751 | 0,068551182 | RNF182 | FALSE | Upregulated |
| -0,222436924 | 0,068589631 | SDHD | FALSE | Downregulated |
| -0,245624071 | 0,068599292 | C6orf203 | FALSE | Downregulated |
| -0,220690463 | 0,068627279 | TMED1 | FALSE | Downregulated |
| -0,198797273 | 0,068656717 | DRG1 | FALSE | Downregulated |
| 0,675722534 | 0,068879648 | LINC00659 | FALSE | Upregulated |
| -0,259944413 | 0,068954385 | AK4 | FALSE | Downregulated |
| 2,54512917 | 0,068954385 | AKR1B15 | FALSE | Upregulated |
| -0,331558981 | 0,069006233 | NSRP1 | FALSE | Downregulated |
| -0,24129111 | 0,069035304 | PDCD4 | FALSE | Downregulated |
| 0,468457673 | 0,069043136 | MIIP | FALSE | Upregulated |
| 0,327916777 | 0,069069954 | ZNF414 | FALSE | Upregulated |
| 0,265972959 | 0,069143753 | VPS13B | FALSE | Upregulated |
| -0,32410394 | 0,069169765 | TTC1 | FALSE | Downregulated |
| 1,652430247 | 0,069313581 | RORB | FALSE | Upregulated |
| -0,200458014 | 0,069314971 | MTERFD1 | FALSE | Downregulated |
| -0,390468156 | 0,069314971 | NR2F1 | FALSE | Downregulated |
| 0,650708888 | 0,069317228 | CARD9 | FALSE | Upregulated |
| 0,44537879 | 0,069317228 | CYP2D6 | FALSE | Upregulated |
| 1,264532586 | 0,069321683 | LINC00996 | FALSE | Upregulated |
| -0,198225379 | 0,069323509 | CWC15 | FALSE | Downregulated |
| -0,241169254 | 0,069325759 | SIL1 | FALSE | Downregulated |
| -0,183506311 | 0,069376646 | ZNF252P | FALSE | Downregulated |
| -0,479030527 | 0,069421574 | VSNL1 | FALSE | Downregulated |
| 0,571903457 | 0,069458771 | CAMKK1 | FALSE | Upregulated |
| -0,268188455 | 0,069458771 | CEP95 | FALSE | Downregulated |
| -2,108579037 | 0,069458771 | LMOD3 | FALSE | Downregulated |
| 0,32433575 | 0,069507122 | RAB20 | FALSE | Upregulated |
| -0,231968804 | 0,069538009 | SCFD1 | FALSE | Downregulated |
| -0,157665828 | 0,069596098 | ANGEL2 | FALSE | Downregulated |
| 2,218395365 | 0,069630283 | LOC101928188 | FALSE | Upregulated |
| 0,265491868 | 0,069640759 | RFXANK | FALSE | Upregulated |
| 2,039513339 | 0,069784271 | CALB1 | FALSE | Upregulated |
| -0,163565575 | 0,069820056 | CASP3 | FALSE | Downregulated |
| -0,290300173 | 0,069970108 | NHLRC2 | FALSE | Downregulated |
| 0,645907876 | 0,070064066 | MAP6 | FALSE | Upregulated |
| 0,291992981 | 0,070189783 | NIPSNAP3A | FALSE | Upregulated |
| -0,509411377 | 0,070281351 | LINC00886 | FALSE | Downregulated |
| 1,879635592 | 0,070319436 | MSMP | FALSE | Upregulated |
| -2,193158027 | 0,070407399 | CRYBA4 | FALSE | Downregulated |
| -0,909765247 | 0,070473309 | NAPA-AS1 | FALSE | Downregulated |
| 0,147175066 | 0,070515855 | SART1 | FALSE | Upregulated |
| 0,453343488 | 0,070533106 | EPHB6 | FALSE | Upregulated |

| | | | | |
|---|---|---|---|---|
| -0,174597848 | 0,07058331 | RAD21 | FALSE | Downregulated |
| -0,45744805 | 0,070721227 | CATSPER2P1 | FALSE | Downregulated |
| -0,307068229 | 0,070721227 | DECR2 | FALSE | Downregulated |
| -0,470950838 | 0,070748375 | NEU4 | FALSE | Downregulated |
| 0,410495401 | 0,070979184 | ODC1 | FALSE | Upregulated |
| -2,617618816 | 0,071034726 | SMPX | FALSE | Downregulated |
| 2,863231458 | 0,071048403 | PPIAP30 | FALSE | Upregulated |
| 0,397254868 | 0,071114282 | HELZ2 | FALSE | Upregulated |
| -0,640800266 | 0,071135769 | RAB26 | FALSE | Downregulated |
| 0,24490817 | 0,07113622 | MYL6 | FALSE | Upregulated |
| -0,738357453 | 0,07113622 | POSTN | FALSE | Downregulated |
| -0,83819603 | 0,071236817 | PAX8-AS1 | FALSE | Downregulated |
| 0,615268059 | 0,071275483 | LARP6 | FALSE | Upregulated |
| -0,15120898 | 0,071419713 | MED29 | FALSE | Downregulated |
| -0,183513088 | 0,071440888 | FAM32A | FALSE | Downregulated |
| -1,416611471 | 0,071480439 | GREM1 | FALSE | Downregulated |
| 1,571859013 | 0,071627629 | PCBP2-OT1 | FALSE | Upregulated |
| 0,46961384 | 0,071671855 | ALDOA | FALSE | Upregulated |
| 1,516812288 | 0,071689027 | LOC101929523 | FALSE | Upregulated |
| 0,744501466 | 0,071707637 | LOC101928189 | FALSE | Upregulated |
| 0,209051994 | 0,071815627 | MAP3K13 | FALSE | Upregulated |
| -0,201667837 | 0,071864112 | NAA60 | FALSE | Downregulated |
| 0,38559901 | 0,071875693 | ZBTB25 | FALSE | Upregulated |
| -0,594701726 | 0,071879436 | REP15 | FALSE | Downregulated |
| 0,391023769 | 0,071906221 | FMNL2 | FALSE | Upregulated |
| 0,286535468 | 0,07192274 | MBD5 | FALSE | Upregulated |
| 0,274815012 | 0,07192274 | RPL23AP53 | FALSE | Upregulated |
| 0,311150837 | 0,071923345 | CEP78 | FALSE | Upregulated |
| -0,14651693 | 0,072032406 | MED4 | FALSE | Downregulated |
| -0,865320321 | 0,072077962 | LOC101927793 | FALSE | Downregulated |
| 0,229312864 | 0,072077962 | MAPK1 | FALSE | Upregulated |
| -0,832970189 | 0,072087335 | CXCR2 | FALSE | Downregulated |
| -1,597702087 | 0,072089515 | FAM41C | FALSE | Downregulated |
| 0,690990778 | 0,072089515 | KIF23 | FALSE | Upregulated |
| 0,314165075 | 0,072089515 | RAPGEF5 | FALSE | Upregulated |
| -1,701008432 | 0,072113791 | REG4 | FALSE | Downregulated |
| 0,269572707 | 0,072121741 | GIT1 | FALSE | Upregulated |
| 0,14643737 | 0,072149391 | PDCD7 | FALSE | Upregulated |
| 1,137782708 | 0,072151907 | ADAMTS3 | FALSE | Upregulated |
| -0,21812306 | 0,072151907 | ANKMY2 | FALSE | Downregulated |
| 1,093665138 | 0,072161495 | LYPD5 | FALSE | Upregulated |
| -0,631335251 | 0,072162209 | GNAO1 | FALSE | Downregulated |
| 0,178106307 | 0,072162209 | VAMP3 | FALSE | Upregulated |
| -0,282131092 | 0,072171741 | APOA1BP | FALSE | Downregulated |
| 0,205375632 | 0,072171741 | GLTPD1 | FALSE | Upregulated |
| 1,890317806 | 0,072218403 | NXPH4 | FALSE | Upregulated |
| -0,375526573 | 0,072228931 | THPO | FALSE | Downregulated |
| -0,407329379 | 0,072275123 | RP11-290F20.1 | FALSE | Downregulated |
| -0,15444465 | 0,07234761 | PIAS2 | FALSE | Downregulated |
| 0,27182307 | 0,072401433 | OGFOD3 | FALSE | Upregulated |
| 0,371450265 | 0,072416504 | LINC00963 | FALSE | Upregulated |

| | | | | |
|---|---|---|---|---|
| 1,022351537 | 0,072416817 | MS4A2 | FALSE | Upregulated |
| 0,961035295 | 0,072539749 | C22orf26 | FALSE | Upregulated |
| -0,213975655 | 0,072539749 | TRIM24 | FALSE | Downregulated |
| 0,286781391 | 0,072554014 | CLPB | FALSE | Upregulated |
| 2,374373039 | 0,07260634 | RPEL1 | FALSE | Upregulated |
| 0,189156666 | 0,072628588 | LRP11 | FALSE | Upregulated |
| 0,272858694 | 0,072642151 | MASTL | FALSE | Upregulated |
| -0,240207175 | 0,072775576 | DICER1 | FALSE | Downregulated |
| 0,27680842 | 0,072789275 | DIP2A | FALSE | Upregulated |
| -0,247621429 | 0,07281667 | SLC39A8 | FALSE | Downregulated |
| 0,338883399 | 0,072835794 | TRPM4 | FALSE | Upregulated |
| 0,400952507 | 0,072858677 | TCEAL3 | FALSE | Upregulated |
| 0,572615666 | 0,072873808 | PCED1B-AS1 | FALSE | Upregulated |
| -1,513422291 | 0,072974262 | KCNC2 | FALSE | Downregulated |
| 0,299277697 | 0,072984525 | MAP4K2 | FALSE | Upregulated |
| 0,957841836 | 0,072997454 | COL9A2 | FALSE | Upregulated |
| -0,468546466 | 0,072997454 | SLC25A47 | FALSE | Downregulated |
| 1,161443493 | 0,073100853 | C9orf153 | FALSE | Upregulated |
| 0,443470179 | 0,073100853 | CEP72 | FALSE | Upregulated |
| 0,976702313 | 0,073183892 | TMC7 | FALSE | Upregulated |
| -0,179007759 | 0,073207929 | LAPTM4A | FALSE | Downregulated |
| -0,270386259 | 0,073249827 | DCTPP1 | FALSE | Downregulated |
| 0,23337329 | 0,073277219 | WDR6 | FALSE | Upregulated |
| -0,224747185 | 0,073336972 | PTCD2 | FALSE | Downregulated |
| 0,650488988 | 0,073411449 | OSBP2 | FALSE | Upregulated |
| 0,488120316 | 0,073430382 | PDE4D | FALSE | Upregulated |
| -0,537253493 | 0,073459639 | CNKSR2 | FALSE | Downregulated |
| 0,559597716 | 0,073459639 | PVRIG | FALSE | Upregulated |
| 0,322432533 | 0,073516643 | ZNF79 | FALSE | Upregulated |
| 0,788734851 | 0,073518644 | SAMD15 | FALSE | Upregulated |
| 0,311688785 | 0,073572416 | CKAP4 | FALSE | Upregulated |
| -0,405273723 | 0,073572416 | FAM69B | FALSE | Downregulated |
| -0,2131281 | 0,073642419 | COL4A3BP | FALSE | Downregulated |
| -0,283271018 | 0,073642419 | SLTM | FALSE | Downregulated |
| 0,394941226 | 0,073738285 | ANGPTL2 | FALSE | Upregulated |
| -0,624266422 | 0,073797153 | NBPF20 | FALSE | Downregulated |
| 0,154357826 | 0,07384527 | UBE2V2 | FALSE | Upregulated |
| -0,39073381 | 0,073855078 | RTN4IP1 | FALSE | Downregulated |
| 1,800536277 | 0,073875069 | NRXN1 | FALSE | Upregulated |
| 0,454763466 | 0,073897894 | FCGR3A | FALSE | Upregulated |
| 0,255118107 | 0,073922544 | EIF2AK4 | FALSE | Upregulated |
| -0,72527728 | 0,073953393 | LOC102723501 | FALSE | Downregulated |
| 1,473631929 | 0,074046583 | C2orf81 | FALSE | Upregulated |
| -0,370159688 | 0,074060106 | ENDOV | FALSE | Downregulated |
| 1,445577251 | 0,074060106 | FOXS1 | FALSE | Upregulated |
| -0,761859752 | 0,074060106 | KIF6 | FALSE | Downregulated |
| -0,318016825 | 0,074098327 | BANF1 | FALSE | Downregulated |
| 0,174884701 | 0,074119511 | ACTR1A | FALSE | Upregulated |
| -1,95100289 | 0,074123679 | NEB | FALSE | Downregulated |
| -0,790200323 | 0,074171318 | NUDT9P1 | FALSE | Downregulated |
| 0,203263805 | 0,074202665 | DESI2 | FALSE | Upregulated |

| | | | | |
|---|---|---|---|---|
| 1,621177191 | 0,074208593 | RP11-353N14.5 | FALSE | Upregulated |
| 0,551710659 | 0,0742555 | BMP2 | FALSE | Upregulated |
| -0,225775965 | 0,074337995 | RXRA | FALSE | Downregulated |
| 0,469176164 | 0,074586413 | ZNF813 | FALSE | Upregulated |
| -0,517239788 | 0,07482317 | FAM99A | FALSE | Downregulated |
| -0,223591221 | 0,074844584 | SLC25A38 | FALSE | Downregulated |
| 2,047238853 | 0,074848407 | DLK1 | FALSE | Upregulated |
| -0,344981545 | 0,074848407 | ZNF554 | FALSE | Downregulated |
| -0,403650583 | 0,074957225 | LINC00261 | FALSE | Downregulated |
| -0,325829204 | 0,074972657 | C9orf91 | FALSE | Downregulated |
| 0,397026377 | 0,074972657 | ZMYND15 | FALSE | Upregulated |
| -1,140334191 | 0,074996864 | IP6K3 | FALSE | Downregulated |
| -0,223617781 | 0,074996864 | OPA3 | FALSE | Downregulated |
| -0,424940319 | 0,075124301 | RDX | FALSE | Downregulated |
| 0,41832774 | 0,075133129 | CD9 | FALSE | Upregulated |
| 1,488363324 | 0,075133129 | CLIC3 | FALSE | Upregulated |
| -0,386872074 | 0,075133129 | CYP39A1 | FALSE | Downregulated |
| 0,192402487 | 0,075133129 | TMEM115 | FALSE | Upregulated |
| 0,258730139 | 0,075133129 | TRIAP1 | FALSE | Upregulated |
| 0,546298523 | 0,075206738 | AP1S3 | FALSE | Upregulated |
| -0,656331625 | 0,075251303 | NBPF13P | FALSE | Downregulated |
| -0,371618969 | 0,075277174 | NIFK-AS1 | FALSE | Downregulated |
| -0,622827608 | 0,075309032 | LOC101928101 | FALSE | Downregulated |
| -0,2635311 | 0,075366466 | GPANK1 | FALSE | Downregulated |
| 0,672861017 | 0,075366466 | VNN2 | FALSE | Upregulated |
| 0,543872161 | 0,075370979 | CYSLTR1 | FALSE | Upregulated |
| 0,192604873 | 0,07537723 | WDR46 | FALSE | Upregulated |
| 0,350121321 | 0,075406575 | C10orf12 | FALSE | Upregulated |
| 0,228282622 | 0,075408255 | HMGN1 | FALSE | Upregulated |
| 0,245374888 | 0,075410616 | DALRD3 | FALSE | Upregulated |
| 0,934038852 | 0,075489256 | AUNIP | FALSE | Upregulated |
| 1,18156848 | 0,075798883 | RASL11B | FALSE | Upregulated |
| -0,234570411 | 0,075893639 | SLC46A1 | FALSE | Downregulated |
| 0,686829245 | 0,075997476 | GIMAP5 | FALSE | Upregulated |
| 0,869601691 | 0,075997476 | GULP1 | FALSE | Upregulated |
| 0,324553207 | 0,076189001 | RUSC2 | FALSE | Upregulated |
| -0,274165898 | 0,076228002 | RAB6A | FALSE | Downregulated |
| -0,26608488 | 0,076228002 | RPS17 | FALSE | Downregulated |
| -0,29678975 | 0,076228002 | WWOX | FALSE | Downregulated |
| 0,766178942 | 0,076326593 | ACKR3 | FALSE | Upregulated |
| 0,366416301 | 0,076326593 | CHPF | FALSE | Upregulated |
| -1,635525468 | 0,076326593 | CRYAB | FALSE | Downregulated |
| -0,543044612 | 0,076326593 | LOC646976 | FALSE | Downregulated |
| -0,214558451 | 0,076326593 | PIGH | FALSE | Downregulated |
| 0,512960811 | 0,076326593 | SLC52A2 | FALSE | Upregulated |
| -0,155207681 | 0,076360169 | TPP2 | FALSE | Downregulated |
| 0,311347396 | 0,076470013 | WDR18 | FALSE | Upregulated |
| 1,157310997 | 0,076473261 | LINC00243 | FALSE | Upregulated |
| 1,785969435 | 0,07650964 | KRT18P55 | FALSE | Upregulated |
| -1,556228986 | 0,07651817 | ATP6V1G2-DDX39B | FALSE | Downregulated |

| | | | | |
|---|---|---|---|---|
| 1,75210177 | 0,076522688 | COL6A5 | FALSE | Upregulated |
| -1,642727483 | 0,076591796 | INSC | FALSE | Downregulated |
| -0,233568094 | 0,076665579 | RNASEH2C | FALSE | Downregulated |
| -0,205753804 | 0,076683216 | CHIC2 | FALSE | Downregulated |
| 0,459151086 | 0,076725497 | ARL2 | FALSE | Upregulated |
| -0,400111152 | 0,076745795 | FLJ37453 | FALSE | Downregulated |
| -0,920080707 | 0,076801476 | LRRC3-AS1 | FALSE | Downregulated |
| 0,403717117 | 0,077019359 | SLC36A4 | FALSE | Upregulated |
| 0,228665194 | 0,077137865 | CASK | FALSE | Upregulated |
| 0,655219919 | 0,077185147 | BTC | FALSE | Upregulated |
| -0,249677113 | 0,077185147 | RWDD3 | FALSE | Downregulated |
| 0,688473642 | 0,07718535 | NUDT17 | FALSE | Upregulated |
| 0,480017644 | 0,077212245 | CRYM | FALSE | Upregulated |
| 0,200275381 | 0,077232944 | CALM3 | FALSE | Upregulated |
| -0,311414111 | 0,077232944 | HNRNPH2 | FALSE | Downregulated |
| -0,201694102 | 0,077371007 | GOLGA5 | FALSE | Downregulated |
| -0,327555734 | 0,077459966 | EIF3A | FALSE | Downregulated |
| -0,20818254 | 0,077536794 | DDX27 | FALSE | Downregulated |
| 0,187302123 | 0,077537373 | FARP1 | FALSE | Upregulated |
| -0,260273421 | 0,077537373 | TVP23B | FALSE | Downregulated |
| 1,186075792 | 0,077635481 | TNFRSF4 | FALSE | Upregulated |
| -0,422345153 | 0,077666161 | GPR89A | FALSE | Downregulated |
| -0,345283333 | 0,07767843 | SPATA5 | FALSE | Downregulated |
| -0,695455374 | 0,077681131 | FST | FALSE | Downregulated |
| -0,606636504 | 0,077701058 | MSMO1 | FALSE | Downregulated |
| 0,366315302 | 0,077793644 | SPNS2 | FALSE | Upregulated |
| -0,32510497 | 0,077821585 | GCFC2 | FALSE | Downregulated |
| -0,717229475 | 0,077861901 | CADM2 | FALSE | Downregulated |
| -0,411998391 | 0,077885143 | AKAP3 | FALSE | Downregulated |
| -0,229713563 | 0,077893091 | IL18R1 | FALSE | Downregulated |
| 0,58592165 | 0,077916876 | GDPGP1 | FALSE | Upregulated |
| 1,010275895 | 0,078000222 | RNF112 | FALSE | Upregulated |
| 0,686007314 | 0,078033146 | OLFML2B | FALSE | Upregulated |
| -1,888876853 | 0,078033146 | PCDH9-AS2 | FALSE | Downregulated |
| 0,196179765 | 0,078033146 | WIZ | FALSE | Upregulated |
| 0,931352362 | 0,078075612 | SPTSSB | FALSE | Upregulated |
| 0,176146806 | 0,078081648 | DENND6A | FALSE | Upregulated |
| 1,266451698 | 0,078145413 | LOC100505710 | FALSE | Upregulated |
| -0,748574257 | 0,078145413 | LRRC55 | FALSE | Downregulated |
| 1,784355869 | 0,078145413 | TCTEX1D4 | FALSE | Upregulated |
| -0,505235732 | 0,078167573 | GRHL1 | FALSE | Downregulated |
| -0,852010169 | 0,078167573 | STK31 | FALSE | Downregulated |
| 0,368636303 | 0,078179202 | RHNO1 | FALSE | Upregulated |
| -0,380268287 | 0,078179202 | RNF125 | FALSE | Downregulated |
| 0,298000195 | 0,078179202 | SGSM2 | FALSE | Upregulated |
| -0,251051386 | 0,078179202 | TOMM7 | FALSE | Downregulated |
| -0,270370147 | 0,078179202 | USP16 | FALSE | Downregulated |
| 1,245395523 | 0,078194581 | DOC2A | FALSE | Upregulated |
| -0,13397295 | 0,078194581 | TOMM70A | FALSE | Downregulated |
| -0,225026649 | 0,078258809 | GOT2 | FALSE | Downregulated |
| -0,191817951 | 0,078274936 | NSMCE1 | FALSE | Downregulated |

| | | | | |
|---|---|---|---|---|
| 0,26935005 | 0,078381969 | AHNAK | FALSE | Upregulated |
| -0,369904259 | 0,078381969 | FBXO5 | FALSE | Downregulated |
| -1,297602141 | 0,078381969 | LOC101927446 | FALSE | Downregulated |
| -0,197283082 | 0,078441543 | FKBP8 | FALSE | Downregulated |
| 0,178272111 | 0,078441543 | GALNT1 | FALSE | Upregulated |
| -0,429757749 | 0,078441543 | LINC00324 | FALSE | Downregulated |
| -0,353005181 | 0,078441543 | PMEL | FALSE | Downregulated |
| 0,964653864 | 0,078441543 | RP11-475O6.1 | FALSE | Upregulated |
| 0,514322545 | 0,078441543 | ZC2HC1A | FALSE | Upregulated |
| -0,277498306 | 0,078479324 | C19orf24 | FALSE | Downregulated |
| 0,172362381 | 0,078479324 | MINK1 | FALSE | Upregulated |
| -0,605051949 | 0,078522507 | RP5-914P20.5 | FALSE | Downregulated |
| 0,227374287 | 0,078522842 | PLEKHM1 | FALSE | Upregulated |
| -0,182284089 | 0,078620045 | PTPN4 | FALSE | Downregulated |
| 0,309551298 | 0,078620045 | RAD51D | FALSE | Upregulated |
| -0,159194036 | 0,078647908 | CLCN3 | FALSE | Downregulated |
| 0,740358983 | 0,078647908 | RP11-793H13.3 | FALSE | Upregulated |
| -0,203299328 | 0,078647908 | VPRBP | FALSE | Downregulated |
| -0,843596477 | 0,078653653 | TOLLIP-AS1 | FALSE | Downregulated |
| -0,420165105 | 0,078701673 | PLCXD1 | FALSE | Downregulated |
| 0,551111277 | 0,078716523 | ADAM8 | FALSE | Upregulated |
| -1,021404369 | 0,078778642 | LRIT3 | FALSE | Downregulated |
| 0,435942507 | 0,078784941 | TMEM86B | FALSE | Upregulated |
| 0,437859428 | 0,078837214 | CSF1R | FALSE | Upregulated |
| 0,154518863 | 0,078865287 | BSDC1 | FALSE | Upregulated |
| 1,961455692 | 0,078956124 | GPC2 | FALSE | Upregulated |
| -0,746483605 | 0,079199913 | GLIPR1L2 | FALSE | Downregulated |
| 1,448750859 | 0,079275981 | FAM64A | FALSE | Upregulated |
| 0,670929577 | 0,079275981 | PRMT6 | FALSE | Upregulated |
| -0,465488831 | 0,079280701 | ABLIM3 | FALSE | Downregulated |
| 0,665430029 | 0,079280701 | FAM171B | FALSE | Upregulated |
| -0,194654226 | 0,079302813 | COQ7 | FALSE | Downregulated |
| 0,444440332 | 0,079310176 | CCDC24 | FALSE | Upregulated |
| 0,127232641 | 0,079316831 | NCSTN | FALSE | Upregulated |
| -0,278983127 | 0,079377096 | KHDRBS3 | FALSE | Downregulated |
| -0,750522098 | 0,079377096 | LOC101927420 | FALSE | Downregulated |
| 0,471557787 | 0,079439007 | CD79B | FALSE | Upregulated |
| -0,371418634 | 0,079502021 | TAF13 | FALSE | Downregulated |
| 0,365032226 | 0,079568217 | TMEM234 | FALSE | Upregulated |
| 0,475052269 | 0,079569549 | PLEK | FALSE | Upregulated |
| 0,627385303 | 0,079632566 | SALL2 | FALSE | Upregulated |
| 2,259258962 | 0,079686445 | RP11-770E5.1 | FALSE | Upregulated |
| -0,251834645 | 0,079686579 | CD302 | FALSE | Downregulated |
| 0,231130778 | 0,079686579 | PDE12 | FALSE | Upregulated |
| -0,254603422 | 0,079697434 | ZNF654 | FALSE | Downregulated |
| 1,192944059 | 0,0797072 | CIB2 | FALSE | Upregulated |
| 1,377918554 | 0,079773013 | BIK | FALSE | Upregulated |
| -0,717457376 | 0,079773013 | CPE | FALSE | Downregulated |
| 1,918707156 | 0,079773013 | GRM6 | FALSE | Upregulated |
| 0,873593957 | 0,079773013 | GSTA5 | FALSE | Upregulated |
| -0,241949452 | 0,079773013 | LONP2 | FALSE | Downregulated |

| | | | | |
|---|---|---|---|---|
| 2,015260017 | 0,079910717 | LDLRAD1 | FALSE | Upregulated |
| 0,226153924 | 0,079976934 | IPO13 | FALSE | Upregulated |
| 1,244510654 | 0,079998161 | FDPSP2 | FALSE | Upregulated |
| -1,00193077 | 0,079998161 | ITLN2 | FALSE | Downregulated |
| -0,1995305 | 0,079998161 | LYPLA1 | FALSE | Downregulated |
| -0,224578353 | 0,079998161 | ZNF275 | FALSE | Downregulated |
| 0,389155175 | 0,080063656 | LCP2 | FALSE | Upregulated |
| 2,551230817 | 0,080116616 | ANKRD44-IT1 | FALSE | Upregulated |
| 1,555429462 | 0,080132574 | LOC401463 | FALSE | Upregulated |
| 1,153055452 | 0,080137297 | FAM66C | FALSE | Upregulated |
| -0,34886546 | 0,080137297 | RP11-392P7.6 | FALSE | Downregulated |
| 0,190046366 | 0,080137297 | SEC22A | FALSE | Upregulated |
| -0,939623075 | 0,080162443 | MT1F | FALSE | Downregulated |
| -0,186534338 | 0,080193867 | FAM179B | FALSE | Downregulated |
| -0,149837315 | 0,080219039 | B4GALT4 | FALSE | Downregulated |
| 0,23454005 | 0,080379321 | SLC25A40 | FALSE | Upregulated |
| 0,874415872 | 0,08040477 | GPR114 | FALSE | Upregulated |
| -0,196553785 | 0,080408695 | FOXO4 | FALSE | Downregulated |
| -0,265668176 | 0,080539142 | RPLP0 | FALSE | Downregulated |
| -0,464513602 | 0,080539142 | TMEM37 | FALSE | Downregulated |
| 0,199126809 | 0,08060175 | EDC4 | FALSE | Upregulated |
| -0,716260317 | 0,080601947 | PVALB | FALSE | Downregulated |
| 1,79389535 | 0,080682998 | TCL1A | FALSE | Upregulated |
| -0,987776788 | 0,080691836 | RP11-290F5.2 | FALSE | Downregulated |
| -0,322333628 | 0,080693502 | NOSTRIN | FALSE | Downregulated |
| 0,593946908 | 0,08070169 | ELN | FALSE | Upregulated |
| 0,13681575 | 0,080723629 | GOLGA7 | FALSE | Upregulated |
| -0,783057085 | 0,080785248 | FAM110D | FALSE | Downregulated |
| -0,184704127 | 0,080854508 | PPP1R15B | FALSE | Downregulated |
| 1,397679582 | 0,080942879 | LOC645752 | FALSE | Upregulated |
| -0,159505632 | 0,080942879 | PAFAH2 | FALSE | Downregulated |
| 1,737122709 | 0,080942879 | PROSER2-AS1 | FALSE | Upregulated |
| 0,433631787 | 0,080942879 | SPIN4 | FALSE | Upregulated |
| -0,208779845 | 0,080983161 | INTS6 | FALSE | Downregulated |
| 0,207678775 | 0,08098677 | DAGLB | FALSE | Upregulated |
| -0,324016598 | 0,081041065 | FRRS1 | FALSE | Downregulated |
| 1,734661517 | 0,081097211 | BEND3P3 | FALSE | Upregulated |
| 2,602018056 | 0,081174253 | TMEM212 | FALSE | Upregulated |
| 0,891259909 | 0,081184285 | KIAA1324 | FALSE | Upregulated |
| -0,174905193 | 0,081228807 | SNAP23 | FALSE | Downregulated |
| -0,993771451 | 0,081267628 | DGKB | FALSE | Downregulated |
| -0,287951955 | 0,081296784 | FOXJ3 | FALSE | Downregulated |
| 1,426326776 | 0,081308793 | LOC101928173 | FALSE | Upregulated |
| 1,30896012 | 0,081327898 | IGSF22 | FALSE | Upregulated |
| 0,536839366 | 0,081391705 | LOC102723521 | FALSE | Upregulated |
| -0,488901744 | 0,081433882 | PCDH9 | FALSE | Downregulated |
| -0,173112287 | 0,081444321 | SNUPN | FALSE | Downregulated |
| -0,743467736 | 0,081563102 | GUCA1B | FALSE | Downregulated |
| -0,955976125 | 0,081565909 | LOC101927635 | FALSE | Downregulated |
| -0,272506054 | 0,081565909 | SNRNP35 | FALSE | Downregulated |
| 0,699194833 | 0,081606764 | GJA5 | FALSE | Upregulated |

| | | | | |
|---|---|---|---|---|
| -0,267108165 | 0,081606764 | SETMAR | FALSE | Downregulated |
| 0,400974743 | 0,081642284 | SPI1 | FALSE | Upregulated |
| 0,185975337 | 0,08168568 | UBAP2L | FALSE | Upregulated |
| 1,548145499 | 0,081770165 | LINC00494 | FALSE | Upregulated |
| -0,601490955 | 0,081770165 | LINC00864 | FALSE | Downregulated |
| 0,230038966 | 0,081787557 | CCDC124 | FALSE | Upregulated |
| 0,199723074 | 0,081883715 | SMAD2 | FALSE | Upregulated |
| -0,503632446 | 0,081928159 | AC016629.8 | FALSE | Downregulated |
| -0,682675179 | 0,081951719 | CAPN3 | FALSE | Downregulated |
| 1,375078619 | 0,081965922 | TREM2 | FALSE | Upregulated |
| 0,478395369 | 0,081998181 | GAB3 | FALSE | Upregulated |
| 0,40205048 | 0,082003799 | METTL1 | FALSE | Upregulated |
| -0,257575681 | 0,082003799 | TRAM2 | FALSE | Downregulated |
| 1,317347087 | 0,082020499 | PRH2 | FALSE | Upregulated |
| -0,209471182 | 0,082121162 | STX8 | FALSE | Downregulated |
| -0,265704327 | 0,082158649 | STRADB | FALSE | Downregulated |
| -0,950532122 | 0,082211426 | CNTD2 | FALSE | Downregulated |
| -1,051730471 | 0,082221397 | LOC102467146 | FALSE | Downregulated |
| -0,463227269 | 0,082245724 | C17orf67 | FALSE | Downregulated |
| -0,241957845 | 0,082248377 | ING3 | FALSE | Downregulated |
| -0,163247447 | 0,082248377 | INTS8 | FALSE | Downregulated |
| 0,643151931 | 0,082308014 | FAM163B | FALSE | Upregulated |
| 0,656092958 | 0,082308014 | VLDLR | FALSE | Upregulated |
| 0,43524385 | 0,082351513 | MXRA5 | FALSE | Upregulated |
| 0,460223956 | 0,082386406 | ZNF785 | FALSE | Upregulated |
| -0,20734974 | 0,082388011 | CHCHD7 | FALSE | Downregulated |
| -0,282622728 | 0,082388011 | ITPA | FALSE | Downregulated |
| 0,309850498 | 0,082388011 | LIG1 | FALSE | Upregulated |
| 0,289531455 | 0,082457555 | ZNF624 | FALSE | Upregulated |
| 0,635961136 | 0,082524247 | TK1 | FALSE | Upregulated |
| -0,144972829 | 0,082625305 | CLDND1 | FALSE | Downregulated |
| -0,731946625 | 0,082711301 | CAMK2B | FALSE | Downregulated |
| -0,321821403 | 0,082771732 | H2AFJ | FALSE | Downregulated |
| 0,686923226 | 0,082796265 | KCNMB1 | FALSE | Upregulated |
| 1,625608661 | 0,082802032 | PTGER1 | FALSE | Upregulated |
| 1,798214988 | 0,08282352 | KDM5D | FALSE | Upregulated |
| 0,728849469 | 0,083007453 | NFATC4 | FALSE | Upregulated |
| 2,015313131 | 0,083007453 | NPAS1 | FALSE | Upregulated |
| -0,614860778 | 0,083010318 | RAPGEFL1 | FALSE | Downregulated |
| 0,189658012 | 0,083061019 | TMEM259 | FALSE | Upregulated |
| 0,322287065 | 0,083161528 | MAT2A | FALSE | Upregulated |
| 0,194268796 | 0,083161528 | SOCS7 | FALSE | Upregulated |
| -1,13297288 | 0,083259774 | TRPC3 | FALSE | Downregulated |
| -0,277817739 | 0,083310024 | ARVCF | FALSE | Downregulated |
| 0,643784472 | 0,083372087 | SHBG | FALSE | Upregulated |
| 0,695673291 | 0,083385837 | KAL1 | FALSE | Upregulated |
| -0,573128207 | 0,083385837 | LOC100505902 | FALSE | Downregulated |
| 1,371983595 | 0,083385837 | SPNS3 | FALSE | Upregulated |
| -0,448313523 | 0,083499278 | ACOT1 | FALSE | Downregulated |
| 0,387514992 | 0,083750664 | FES | FALSE | Upregulated |
| -0,195597044 | 0,083750664 | SSB | FALSE | Downregulated |

| | | | | |
|---|---|---|---|---|
| 0,159868646 | 0,083865621 | SPG11 | FALSE | Upregulated |
| 0,516518485 | 0,083918472 | HDAC9 | FALSE | Upregulated |
| 0,228715248 | 0,083953786 | RNF19B | FALSE | Upregulated |
| -0,255627727 | 0,083971173 | FAM174A | FALSE | Downregulated |
| 0,467742711 | 0,084079869 | DYNC2H1 | FALSE | Upregulated |
| -1,427385252 | 0,084089923 | TXLNB | FALSE | Downregulated |
| 0,204464035 | 0,08413226 | RBMXL1 | FALSE | Upregulated |
| -0,207978772 | 0,084157073 | PNKD | FALSE | Downregulated |
| 0,260820154 | 0,084157073 | TBC1D25 | FALSE | Upregulated |
| -0,220794229 | 0,084168125 | U2SURP | FALSE | Downregulated |
| 0,253732245 | 0,084183082 | RNF126 | FALSE | Upregulated |
| 0,348406352 | 0,084203903 | YPEL1 | FALSE | Upregulated |
| 0,308305568 | 0,084251756 | SYVN1 | FALSE | Upregulated |
| -0,421666916 | 0,084284463 | AMACR | FALSE | Downregulated |
| -0,266073121 | 0,084285907 | C5 | FALSE | Downregulated |
| 0,261670255 | 0,084308552 | C10orf88 | FALSE | Upregulated |
| -0,883598565 | 0,084308552 | RP11-261N11.8 | FALSE | Downregulated |
| 1,62001556 | 0,084308552 | TFAP2E | FALSE | Upregulated |
| -0,167668266 | 0,084309981 | PPWD1 | FALSE | Downregulated |
| 0,294846084 | 0,084336979 | RNF215 | FALSE | Upregulated |
| 1,165596804 | 0,084429402 | ADORA2B | FALSE | Upregulated |
| 0,834919848 | 0,084429402 | PROCA1 | FALSE | Upregulated |
| -0,146223351 | 0,084512611 | C1orf43 | FALSE | Downregulated |
| -0,413395658 | 0,084512611 | RWDD2A | FALSE | Downregulated |
| -0,145735557 | 0,084594257 | OSBPL2 | FALSE | Downregulated |
| 1,18907601 | 0,084795667 | LOC101927910 | FALSE | Upregulated |
| 0,541686627 | 0,084815541 | KLRK1 | FALSE | Upregulated |
| -0,559689354 | 0,084845857 | LOC100506286 | FALSE | Downregulated |
| -0,195603514 | 0,084895998 | ST7 | FALSE | Downregulated |
| 0,69875023 | 0,085007313 | P2RY6 | FALSE | Upregulated |
| 0,342079468 | 0,085007313 | SLC25A45 | FALSE | Upregulated |
| -0,29992015 | 0,085152977 | HELZ | FALSE | Downregulated |
| 0,174207178 | 0,085152977 | XRCC1 | FALSE | Upregulated |
| 1,038139763 | 0,085267768 | CMTM1 | FALSE | Upregulated |
| -0,733841606 | 0,085394178 | LOC613037 | FALSE | Downregulated |
| 0,269462677 | 0,08550157 | DCUN1D5 | FALSE | Upregulated |
| -0,184030256 | 0,085517103 | ZNF277 | FALSE | Downregulated |
| 1,040595213 | 0,08554186 | TRIP13 | FALSE | Upregulated |
| 0,209238236 | 0,085668455 | TUBB | FALSE | Upregulated |
| 0,336983348 | 0,085765518 | TRIM34 | FALSE | Upregulated |
| 0,384762107 | 0,085788391 | RARRES3 | FALSE | Upregulated |
| 0,247786307 | 0,085852847 | TRIO | FALSE | Upregulated |
| 0,753072138 | 0,085888659 | FLT3 | FALSE | Upregulated |
| -1,191589979 | 0,085925951 | LOC102723324 | FALSE | Downregulated |
| -1,657984096 | 0,085958073 | RYR1 | FALSE | Downregulated |
| -0,507259219 | 0,086085687 | ANKRD55 | FALSE | Downregulated |
| 1,468101215 | 0,086085687 | P2RX5 | FALSE | Upregulated |
| 0,934782958 | 0,086113795 | FAM188B | FALSE | Upregulated |
| 1,778308802 | 0,086113795 | LINC00506 | FALSE | Upregulated |
| 0,315286291 | 0,086162733 | B4GALT7 | FALSE | Upregulated |
| 0,231624793 | 0,086162733 | GLTP | FALSE | Upregulated |

| | | | | |
|---|---|---|---|---|
| 0,307258459 | 0,086254826 | BTN3A3 | FALSE | Upregulated |
| -0,209837505 | 0,086293303 | KIAA1586 | FALSE | Downregulated |
| -0,699739091 | 0,086358829 | PCK1 | FALSE | Downregulated |
| 0,196020794 | 0,086399778 | BRE | FALSE | Upregulated |
| 1,881455776 | 0,086399778 | CCL17 | FALSE | Upregulated |
| 2,376510278 | 0,086399778 | LOC102723371 | FALSE | Upregulated |
| 1,060656527 | 0,08658494 | CES1P2 | FALSE | Upregulated |
| -0,19641728 | 0,08658494 | H3F3AP4 | FALSE | Downregulated |
| 0,216859004 | 0,08658494 | PARP1 | FALSE | Upregulated |
| 0,226519446 | 0,086690514 | C15orf38 | FALSE | Upregulated |
| -0,353381506 | 0,086690514 | DEPDC7 | FALSE | Downregulated |
| 0,234217926 | 0,086690514 | DHDDS | FALSE | Upregulated |
| 1,804829987 | 0,086690514 | DUOX2 | FALSE | Upregulated |
| -0,144415105 | 0,086690514 | ITFG1 | FALSE | Downregulated |
| 0,207657259 | 0,086690514 | VPS8 | FALSE | Upregulated |
| -0,220705485 | 0,086756296 | PRKAR1A | FALSE | Downregulated |
| -1,475832955 | 0,086824788 | LOC102546298 | FALSE | Downregulated |
| 0,988991007 | 0,086897285 | MATN1-AS1 | FALSE | Upregulated |
| -1,327011307 | 0,086903192 | GALNT13 | FALSE | Downregulated |
| -0,260688476 | 0,086903192 | TMPRSS6 | FALSE | Downregulated |
| -0,517959403 | 0,086951364 | DNAJC27-AS1 | FALSE | Downregulated |
| -0,91801014 | 0,086951364 | NDNF | FALSE | Downregulated |
| 0,187971853 | 0,087001513 | M6PR | FALSE | Upregulated |
| 1,188135419 | 0,087042074 | CTAGE8 | FALSE | Upregulated |
| -0,290719188 | 0,087042074 | DCUN1D2 | FALSE | Downregulated |
| 0,408305053 | 0,087163937 | ID3 | FALSE | Upregulated |
| -0,214295686 | 0,087301335 | C2orf42 | FALSE | Downregulated |
| 0,11964512 | 0,087403055 | ADNP | FALSE | Upregulated |
| 0,183227388 | 0,087403055 | PHF2 | FALSE | Upregulated |
| 0,761037517 | 0,087558764 | AC011899.9 | FALSE | Upregulated |
| 0,185832785 | 0,087578756 | SENP3 | FALSE | Upregulated |
| 0,276502354 | 0,087681366 | POP1 | FALSE | Upregulated |
| 1,744278526 | 0,087697656 | DPP10-AS1 | FALSE | Upregulated |
| -0,241488432 | 0,087697656 | USO1 | FALSE | Downregulated |
| -0,256330395 | 0,087707382 | HAGH | FALSE | Downregulated |
| -0,518682426 | 0,087707382 | SMIM1 | FALSE | Downregulated |
| -0,310603338 | 0,087738223 | AMT | FALSE | Downregulated |
| 0,288950214 | 0,087743336 | EIF4E3 | FALSE | Upregulated |
| 0,90281693 | 0,087773431 | RASEF | FALSE | Upregulated |
| 0,232467491 | 0,087776301 | NIPAL3 | FALSE | Upregulated |
| -0,185952875 | 0,087842939 | NASP | FALSE | Downregulated |
| -0,165850977 | 0,087846626 | RABGGTB | FALSE | Downregulated |
| 0,508954211 | 0,087865301 | SOGA1 | FALSE | Upregulated |
| -0,32680506 | 0,087899915 | RNLS | FALSE | Downregulated |
| -0,179706971 | 0,087984316 | CCNY | FALSE | Downregulated |
| 0,161194801 | 0,087984316 | COG4 | FALSE | Upregulated |
| 0,885927807 | 0,087984316 | LOC100505727 | FALSE | Upregulated |
| 0,188590966 | 0,087984316 | PGK1 | FALSE | Upregulated |
| -0,268830835 | 0,087984316 | SKP2 | FALSE | Downregulated |
| -0,333033394 | 0,088039775 | SBF2-AS1 | FALSE | Downregulated |
| 0,132960032 | 0,088066205 | ILF3 | FALSE | Upregulated |

| | | | | |
|---|---|---|---|---|
| 0,94505151 | 0,088242859 | PRDM16 | FALSE | Upregulated |
| 0,26957715 | 0,088291478 | CABLES1 | FALSE | Upregulated |
| -1,025449241 | 0,088291478 | TYW1B | FALSE | Downregulated |
| 0,184244375 | 0,088311521 | OSER1 | FALSE | Upregulated |
| -1,164277378 | 0,088311521 | RBM20 | FALSE | Downregulated |
| -0,494876974 | 0,088311521 | RBM26-AS1 | FALSE | Downregulated |
| 1,218709794 | 0,088326648 | SKA1 | FALSE | Upregulated |
| -0,119654878 | 0,088353067 | CUL3 | FALSE | Downregulated |
| 0,366559642 | 0,08836063 | HECTD2 | FALSE | Upregulated |
| 0,613718029 | 0,088370089 | ASPM | FALSE | Upregulated |
| -0,264341992 | 0,08850429 | ILF3-AS1 | FALSE | Downregulated |
| -0,242624555 | 0,088510159 | COX16 | FALSE | Downregulated |
| -0,384296857 | 0,088520455 | FOXP2 | FALSE | Downregulated |
| -1,931805054 | 0,088669313 | LOC101926960 | FALSE | Downregulated |
| -0,366654718 | 0,088669313 | MTURN | FALSE | Downregulated |
| 1,011858777 | 0,088669313 | SEZ6L | FALSE | Upregulated |
| -0,532116773 | 0,088750579 | CD99P1 | FALSE | Downregulated |
| -0,518765476 | 0,088750579 | ONECUT1 | FALSE | Downregulated |
| -0,288611706 | 0,088750579 | RDH10 | FALSE | Downregulated |
| -0,190295215 | 0,088800403 | SLBP | FALSE | Downregulated |
| 1,557356852 | 0,088848546 | LOC102723735 | FALSE | Upregulated |
| 1,470562513 | 0,08894507 | LOC400548 | FALSE | Upregulated |
| -0,165601456 | 0,08894507 | MIER1 | FALSE | Downregulated |
| -0,212255991 | 0,088956539 | HOMEZ | FALSE | Downregulated |
| -1,09809355 | 0,088979574 | JAKMIP2-AS1 | FALSE | Downregulated |
| -0,132138078 | 0,08904759 | CUL5 | FALSE | Downregulated |
| 0,518180391 | 0,089099359 | S1PR2 | FALSE | Upregulated |
| 0,260031557 | 0,089116165 | DENND4B | FALSE | Upregulated |
| 1,635552887 | 0,0892456 | LOC101929469 | FALSE | Upregulated |
| 0,281097643 | 0,0892456 | ULK3 | FALSE | Upregulated |
| 0,281081934 | 0,089247827 | CAD | FALSE | Upregulated |
| 0,449669232 | 0,089436484 | GALNT4 | FALSE | Upregulated |
| -0,96932317 | 0,089436484 | LINC00398 | FALSE | Downregulated |
| 0,826166825 | 0,089553235 | COL5A3 | FALSE | Upregulated |
| 0,177694965 | 0,089642055 | DCAF12 | FALSE | Upregulated |
| 0,17244787 | 0,08971297 | EHMT2 | FALSE | Upregulated |
| -0,172814428 | 0,089753316 | ECD | FALSE | Downregulated |
| 0,147563726 | 0,089777907 | COPA | FALSE | Upregulated |
| 1,77755063 | 0,089777907 | SH3GL2 | FALSE | Upregulated |
| 0,215500847 | 0,089777907 | SZT2 | FALSE | Upregulated |
| -0,255937412 | 0,089790503 | TMEM14B | FALSE | Downregulated |
| -0,154492182 | 0,089893899 | GSK3B | FALSE | Downregulated |
| -0,212706265 | 0,089987122 | ZFYVE21 | FALSE | Downregulated |
| -0,152300126 | 0,090010741 | USP25 | FALSE | Downregulated |
| -0,171958538 | 0,090043935 | RFC1 | FALSE | Downregulated |
| 1,399852347 | 0,090088992 | CSPG5 | FALSE | Upregulated |
| -0,641870676 | 0,090135259 | PIWIL2 | FALSE | Downregulated |
| 0,234712604 | 0,090200364 | CTDP1 | FALSE | Upregulated |
| -0,510854773 | 0,09028451 | HIST1H2BC | FALSE | Downregulated |
| -0,215930343 | 0,090372202 | NOL8 | FALSE | Downregulated |
| -0,239972087 | 0,090432285 | SSSCA1 | FALSE | Downregulated |

| | | | | |
|---|---|---|---|---|
| 0,358525374 | 0,090455597 | MAP3K8 | FALSE | Upregulated |
| 0,322075285 | 0,090468652 | ST7L | FALSE | Upregulated |
| 2,364835653 | 0,090687907 | PTHLH | FALSE | Upregulated |
| -0,188650216 | 0,090707361 | NKIRAS1 | FALSE | Downregulated |
| 0,258014172 | 0,090738054 | CASP2 | FALSE | Upregulated |
| 1,038568077 | 0,09075208 | RGS9 | FALSE | Upregulated |
| -0,196530873 | 0,090779233 | DIS3L | FALSE | Downregulated |
| -0,411222895 | 0,090842545 | CRY1 | FALSE | Downregulated |
| -0,662534228 | 0,090842545 | RASSF6 | FALSE | Downregulated |
| 1,658029152 | 0,090880973 | AP000688.8 | FALSE | Upregulated |
| 0,399047741 | 0,090921573 | GTF2H2C | FALSE | Upregulated |
| -0,243986035 | 0,090954319 | TARS | FALSE | Downregulated |
| -0,41162483 | 0,091079653 | CBWD3 | FALSE | Downregulated |
| 0,606101406 | 0,091127089 | ADM2 | FALSE | Upregulated |
| -2,091571877 | 0,091127089 | GALP | FALSE | Downregulated |
| 1,188772654 | 0,091127089 | LRRC7 | FALSE | Upregulated |
| 0,258717847 | 0,091127089 | ZMYM5 | FALSE | Upregulated |
| 2,204351041 | 0,091129351 | GFI1B | FALSE | Upregulated |
| -0,342682075 | 0,091155205 | UGDH | FALSE | Downregulated |
| 0,723684046 | 0,091225304 | ANKRD13B | FALSE | Upregulated |
| 0,184363343 | 0,091225304 | FGD5-AS1 | FALSE | Upregulated |
| 0,403766417 | 0,091261458 | WRAP53 | FALSE | Upregulated |
| 2,435596429 | 0,091331501 | LOC102724971 | FALSE | Upregulated |
| -0,370009962 | 0,091331501 | MROH8 | FALSE | Downregulated |
| -2,185726208 | 0,091481112 | MYOM3 | FALSE | Downregulated |
| -0,15902788 | 0,091481112 | ZNF197 | FALSE | Downregulated |
| 1,000262275 | 0,091542983 | EME2 | FALSE | Upregulated |
| -0,155256166 | 0,091564236 | ALG13 | FALSE | Downregulated |
| -3,278174918 | 0,091576405 | KLHL40 | FALSE | Downregulated |
| -0,201529219 | 0,091624023 | HYAL1 | FALSE | Downregulated |
| 0,176266669 | 0,091624023 | PTPN1 | FALSE | Upregulated |
| 0,274918745 | 0,09165447 | ALPK1 | FALSE | Upregulated |
| 0,637596995 | 0,09165447 | TEAD4 | FALSE | Upregulated |
| 0,643957712 | 0,091683826 | S100A6 | FALSE | Upregulated |
| 0,30645524 | 0,091698108 | FUT11 | FALSE | Upregulated |
| 0,184565596 | 0,09171204 | EDC3 | FALSE | Upregulated |
| -0,312002817 | 0,091861261 | RNF180 | FALSE | Downregulated |
| 1,581692307 | 0,091979345 | MYO3B | FALSE | Upregulated |
| 0,434152748 | 0,091979724 | LINC00959 | FALSE | Upregulated |
| 0,907367334 | 0,091982673 | ADTRP | FALSE | Upregulated |
| 0,159074872 | 0,092017256 | CAP1 | FALSE | Upregulated |
| 0,490924054 | 0,092070363 | RTP4 | FALSE | Upregulated |
| -0,150715472 | 0,092085489 | IPO9 | FALSE | Downregulated |
| 2,320776958 | 0,092085489 | RP11-219G17.4 | FALSE | Upregulated |
| -1,645873491 | 0,092098482 | CASC9 | FALSE | Downregulated |
| -0,719798788 | 0,092098482 | YBX2 | FALSE | Downregulated |
| -0,428425419 | 0,092150383 | ZBTB7A | FALSE | Downregulated |
| -0,699046846 | 0,092182499 | LINC00263 | FALSE | Downregulated |
| -0,40163988 | 0,092203842 | ZNF550 | FALSE | Downregulated |
| -0,195083774 | 0,09226229 | DHTKD1 | FALSE | Downregulated |
| 1,908946114 | 0,09226229 | SEZ6 | FALSE | Upregulated |

| | | | | |
|---|---|---|---|---|
| 0,949169696 | 0,092328873 | CD80 | FALSE | Upregulated |
| 0,65164314 | 0,092397299 | CCDC64 | FALSE | Upregulated |
| 0,625619244 | 0,092397299 | NDC80 | FALSE | Upregulated |
| 0,372416187 | 0,092478497 | LLGL1 | FALSE | Upregulated |
| 1,936903068 | 0,09251261 | EPHA6 | FALSE | Upregulated |
| -0,641230308 | 0,09251261 | RP5-1159O4.2 | FALSE | Downregulated |
| -0,18593314 | 0,092594716 | UROD | FALSE | Downregulated |
| 1,056586182 | 0,09260098 | AFF2 | FALSE | Upregulated |
| -0,208394923 | 0,09260098 | ZMYM6NB | FALSE | Downregulated |
| -0,43614532 | 0,092631442 | SLC6A1 | FALSE | Downregulated |
| -0,17174705 | 0,092696108 | SETD2 | FALSE | Downregulated |
| 0,261594098 | 0,092696108 | ZNF92 | FALSE | Upregulated |
| 0,365266819 | 0,092720701 | AP4M1 | FALSE | Upregulated |
| 1,299976217 | 0,092770952 | KCNA6 | FALSE | Upregulated |
| 0,127916505 | 0,092783547 | RBMX | FALSE | Upregulated |
| -0,18815614 | 0,092783547 | TRIM41 | FALSE | Downregulated |
| 0,432466471 | 0,092876021 | HEPH | FALSE | Upregulated |
| -0,141053733 | 0,09287966 | DCBLD2 | FALSE | Downregulated |
| 0,721301591 | 0,092944772 | SSR4 | FALSE | Upregulated |
| 0,332453528 | 0,093002923 | PROSER2 | FALSE | Upregulated |
| -0,267242701 | 0,093066809 | C16orf87 | FALSE | Downregulated |
| 0,880336811 | 0,093066809 | CCR6 | FALSE | Upregulated |
| -0,177575779 | 0,093085486 | PSMA6 | FALSE | Downregulated |
| 1,019896384 | 0,093242823 | EPHA1-AS1 | FALSE | Upregulated |
| -1,894025032 | 0,093242823 | HIGD1C | FALSE | Downregulated |
| -0,789816399 | 0,093242823 | RP4-756H11.3 | FALSE | Downregulated |
| -0,243701796 | 0,093242823 | TMX4 | FALSE | Downregulated |
| 0,164340702 | 0,093320886 | ZNF317 | FALSE | Upregulated |
| 0,44075569 | 0,093436423 | PTRH1 | FALSE | Upregulated |
| 0,384956695 | 0,093463466 | HGF | FALSE | Upregulated |
| -1,441443873 | 0,09347659 | LOC646903 | FALSE | Downregulated |
| 2,312108096 | 0,093509091 | NEFL | FALSE | Upregulated |
| -0,378107466 | 0,09360265 | NOP58 | FALSE | Downregulated |
| -0,633375701 | 0,093610155 | PCOLCE2 | FALSE | Downregulated |
| -0,149263005 | 0,093610155 | SCARB2 | FALSE | Downregulated |
| 0,299092432 | 0,093610155 | TSEN34 | FALSE | Upregulated |
| -0,166984362 | 0,093610155 | VPS39 | FALSE | Downregulated |
| 1,634864064 | 0,093627629 | TNFRSF18 | FALSE | Upregulated |
| -0,59551484 | 0,093859742 | SNORA18 | FALSE | Downregulated |
| -0,287415511 | 0,093941811 | FRK | FALSE | Downregulated |
| -0,187417417 | 0,093941811 | HSF1 | FALSE | Downregulated |
| 0,803149465 | 0,094040811 | GTSE1 | FALSE | Upregulated |
| -0,296056919 | 0,094156448 | NBPF14 | FALSE | Downregulated |
| 2,398034574 | 0,094160034 | LOC101929227 | FALSE | Upregulated |
| 1,033188225 | 0,094203222 | CCDC85A | FALSE | Upregulated |
| 1,017040961 | 0,094207461 | LOC102723928 | FALSE | Upregulated |
| -0,257146974 | 0,094239961 | SMIM12 | FALSE | Downregulated |
| -1,520079272 | 0,094309219 | LINC00488 | FALSE | Downregulated |
| 0,263335037 | 0,094330682 | CEP170 | FALSE | Upregulated |
| 0,474066246 | 0,094330682 | PFKFB3 | FALSE | Upregulated |
| 0,231612596 | 0,094330682 | SLC35F6 | FALSE | Upregulated |

| | | | | |
|---|---|---|---|---|
| 0,376276238 | 0,094336298 | SIRT6 | FALSE | Upregulated |
| -0,731110301 | 0,094395124 | RGPD8 | FALSE | Downregulated |
| -0,742659769 | 0,094414368 | LOC100507395 | FALSE | Downregulated |
| -0,297457318 | 0,094498828 | RNASEH2B | FALSE | Downregulated |
| -0,271370047 | 0,094528494 | NR2C1 | FALSE | Downregulated |
| -1,359207395 | 0,094548156 | LINC00624 | FALSE | Downregulated |
| -0,174270216 | 0,094757996 | ALKBH3 | FALSE | Downregulated |
| 0,351129803 | 0,094757996 | ZNF579 | FALSE | Upregulated |
| 0,525906345 | 0,094763016 | PLAGL1 | FALSE | Upregulated |
| 0,295801699 | 0,094787769 | ASAP1 | FALSE | Upregulated |
| -0,341896647 | 0,094787769 | HEXIM2 | FALSE | Downregulated |
| 1,913602409 | 0,094814511 | ARL9 | FALSE | Upregulated |
| 0,643084386 | 0,094814511 | NRGN | FALSE | Upregulated |
| -0,291241101 | 0,094814511 | PHYH | FALSE | Downregulated |
| 0,265269125 | 0,094820554 | MBNL2 | FALSE | Upregulated |
| 0,177910324 | 0,094827358 | NECAP1 | FALSE | Upregulated |
| 0,489693281 | 0,094833588 | MARCH1 | FALSE | Upregulated |
| 0,391860031 | 0,094834345 | TLR1 | FALSE | Upregulated |
| -0,547801142 | 0,094841121 | FAM167B | FALSE | Downregulated |
| -0,158695998 | 0,09485351 | FECH | FALSE | Downregulated |
| 0,375420506 | 0,094916735 | CDKL5 | FALSE | Upregulated |
| -0,614528096 | 0,094940136 | PELI2 | FALSE | Downregulated |
| 0,162626446 | 0,094940136 | RPE | FALSE | Upregulated |
| -0,334553115 | 0,094972372 | CTSH | FALSE | Downregulated |
| -0,355769736 | 0,094995303 | KLF15 | FALSE | Downregulated |
| 0,261014673 | 0,095063818 | AGA | FALSE | Upregulated |
| 0,422245883 | 0,095419552 | LOC81691 | FALSE | Upregulated |
| 0,246475902 | 0,09551201 | ITGB1BP1 | FALSE | Upregulated |
| 1,002545422 | 0,09551905 | C16orf93 | FALSE | Upregulated |
| 1,754522209 | 0,095604958 | ZNF730 | FALSE | Upregulated |
| 0,532898725 | 0,095615273 | NKG7 | FALSE | Upregulated |
| 1,271443288 | 0,095647835 | NTS | FALSE | Upregulated |
| -0,248581255 | 0,095702234 | IQGAP2 | FALSE | Downregulated |
| -0,314741505 | 0,095705726 | SLC9A7 | FALSE | Downregulated |
| -0,217442133 | 0,095725498 | ICK | FALSE | Downregulated |
| 1,917425004 | 0,095756262 | TPSD1 | FALSE | Upregulated |
| 2,088852957 | 0,095773273 | LOC101929159 | FALSE | Upregulated |
| -0,247980503 | 0,095820712 | THTPA | FALSE | Downregulated |
| 0,217668629 | 0,095820712 | USP37 | FALSE | Upregulated |
| -0,608841462 | 0,095839034 | LOC102724481 | FALSE | Downregulated |
| 0,39242729 | 0,095918374 | TELO2 | FALSE | Upregulated |
| 0,319383559 | 0,095929766 | DTD1 | FALSE | Upregulated |
| -0,505987481 | 0,095933662 | HIST1H2BD | FALSE | Downregulated |
| -0,223189697 | 0,095938852 | COA3 | FALSE | Downregulated |
| 0,356621024 | 0,095938852 | TCP11L1 | FALSE | Upregulated |
| 1,414193929 | 0,095980363 | HAVCR1 | FALSE | Upregulated |
| -0,857395951 | 0,096002776 | FAM84A | FALSE | Downregulated |
| 0,125231764 | 0,096087901 | RTFDC1 | FALSE | Upregulated |
| -0,866184575 | 0,096245396 | GPR158 | FALSE | Downregulated |
| 0,357353773 | 0,096321261 | STX3 | FALSE | Upregulated |
| 0,407694485 | 0,096321261 | TTYH2 | FALSE | Upregulated |

| | | | | |
|---|---|---|---|---|
| -0,298771756 | 0,096332681 | CCNK | FALSE | Downregulated |
| 0,148772035 | 0,096343133 | C10orf76 | FALSE | Upregulated |
| -0,790954419 | 0,096343133 | CTNNA3 | FALSE | Downregulated |
| 0,941715191 | 0,096343133 | CYP2D7P | FALSE | Upregulated |
| -0,349505132 | 0,096343133 | DHRS4 | FALSE | Downregulated |
| 0,324262855 | 0,096343133 | PLEKHA8 | FALSE | Upregulated |
| 1,500940412 | 0,096343133 | PRC1-AS1 | FALSE | Upregulated |
| 0,352789883 | 0,096343133 | STK11IP | FALSE | Upregulated |
| -0,151685106 | 0,096366942 | DSTN | FALSE | Downregulated |
| -0,168474575 | 0,096398802 | NSUN2 | FALSE | Downregulated |
| 0,320430876 | 0,096477778 | HHAT | FALSE | Upregulated |
| -0,203304444 | 0,096477778 | MICU1 | FALSE | Downregulated |
| 1,304753666 | 0,096477778 | NOV | FALSE | Upregulated |
| 0,480522245 | 0,096501575 | SSBP2 | FALSE | Upregulated |
| -0,264661477 | 0,096524406 | XKR8 | FALSE | Downregulated |
| -0,500945118 | 0,096597464 | NDRG1 | FALSE | Downregulated |
| -0,599235824 | 0,096653722 | MDN1 | FALSE | Downregulated |
| -0,637221279 | 0,096653722 | PDXDC2P | FALSE | Downregulated |
| 0,369929279 | 0,096653722 | SUSD1 | FALSE | Upregulated |
| 0,443804149 | 0,096672509 | ABCC5 | FALSE | Upregulated |
| 1,431505845 | 0,096672509 | ARRDC5 | FALSE | Upregulated |
| 2,226778672 | 0,096672509 | ATP6V0D2 | FALSE | Upregulated |
| 1,57837485 | 0,096700239 | ARL14 | FALSE | Upregulated |
| -0,21273243 | 0,096822538 | ANP32A | FALSE | Downregulated |
| 1,507229777 | 0,096983438 | LOC729080 | FALSE | Upregulated |
| -0,189884185 | 0,096983438 | NUP88 | FALSE | Downregulated |
| -0,301826823 | 0,097014948 | PCED1A | FALSE | Downregulated |
| -0,361558807 | 0,097014948 | RBM6 | FALSE | Downregulated |
| 0,510753959 | 0,097149581 | ANGPT1 | FALSE | Upregulated |
| -1,798686841 | 0,097149581 | IL17D | FALSE | Downregulated |
| -0,370137791 | 0,097184239 | CCDC122 | FALSE | Downregulated |
| 0,229445736 | 0,097184239 | MFSD11 | FALSE | Upregulated |
| 0,183910918 | 0,097184239 | NELFB | FALSE | Upregulated |
| 0,333796537 | 0,097184239 | SLC18B1 | FALSE | Upregulated |
| 0,328359347 | 0,097212491 | MDM2 | FALSE | Upregulated |
| 1,211514819 | 0,097290036 | C19orf84 | FALSE | Upregulated |
| 0,278008124 | 0,097290036 | RRM2B | FALSE | Upregulated |
| -0,122938683 | 0,097342456 | AK2 | FALSE | Downregulated |
| -1,005705634 | 0,097397403 | CWH43 | FALSE | Downregulated |
| 1,206528794 | 0,097508531 | CACNG8 | FALSE | Upregulated |
| -0,763746626 | 0,097508531 | RPS18P9 | FALSE | Downregulated |
| -0,148018409 | 0,097632316 | COG2 | FALSE | Downregulated |
| -0,20279897 | 0,097798538 | TMED5 | FALSE | Downregulated |
| -0,473674531 | 0,09780505 | FTCDNL1 | FALSE | Downregulated |
| -0,666506813 | 0,097825084 | C2orf16 | FALSE | Downregulated |
| 1,262831535 | 0,097825084 | DLEC1 | FALSE | Upregulated |
| -0,197795008 | 0,097866464 | SEPT7 | FALSE | Downregulated |
| 1,93722134 | 0,098019438 | CSNK1A1L | FALSE | Upregulated |
| -0,222729108 | 0,098019438 | GNG5 | FALSE | Downregulated |
| 0,155354761 | 0,09807997 | TTC27 | FALSE | Upregulated |
| -0,33866665 | 0,098109255 | CCDC84 | FALSE | Downregulated |

| | | | | |
|---|---|---|---|---|
| -0,420659906 | 0,098109255 | CNTN3 | FALSE | Downregulated |
| -1,558329521 | 0,098109255 | SRL | FALSE | Downregulated |
| -0,177269182 | 0,098185803 | TSPAN31 | FALSE | Downregulated |
| 0,300883112 | 0,09819835 | SMCO4 | FALSE | Upregulated |
| 0,18519725 | 0,098230359 | MADD | FALSE | Upregulated |
| 0,980135305 | 0,098290095 | LOC100506621 | FALSE | Upregulated |
| 0,362103478 | 0,098317131 | B3GNT9 | FALSE | Upregulated |
| 0,297914768 | 0,098317131 | C17orf62 | FALSE | Upregulated |
| -0,643719063 | 0,098317131 | LRRIQ3 | FALSE | Downregulated |
| 0,398403686 | 0,098354807 | IFI27L2 | FALSE | Upregulated |
| 0,648455023 | 0,098354807 | NKD1 | FALSE | Upregulated |
| 0,423383156 | 0,098368106 | C5orf63 | FALSE | Upregulated |
| -0,4066337 | 0,09839245 | C6orf136 | FALSE | Downregulated |
| 0,188929528 | 0,098393905 | BTBD10 | FALSE | Upregulated |
| 1,017705356 | 0,09841607 | CDCA7 | FALSE | Upregulated |
| 0,605717596 | 0,098446572 | DUSP1 | FALSE | Upregulated |
| 1,304995752 | 0,098454784 | SLC26A10 | FALSE | Upregulated |
| 0,303985445 | 0,098600992 | GLA | FALSE | Upregulated |
| -0,262074087 | 0,099030237 | ZNF621 | FALSE | Downregulated |
| -0,262766245 | 0,099030938 | ITIH1 | FALSE | Downregulated |
| 1,058046109 | 0,099039827 | SNORA61 | FALSE | Upregulated |
| 0,728444328 | 0,099042106 | KCNJ16 | FALSE | Upregulated |
| 1,602805837 | 0,099042106 | RP11-16E12.2 | FALSE | Upregulated |
| 1,033176035 | 0,099129162 | NRG2 | FALSE | Upregulated |
| 0,5654096 | 0,099134003 | ZNF93 | FALSE | Upregulated |
| -0,191485535 | 0,099172564 | MED13L | FALSE | Downregulated |
| 0,256219574 | 0,099196825 | RPP30 | FALSE | Upregulated |
| -0,24060032 | 0,099197109 | ITCH | FALSE | Downregulated |
| 0,650140852 | 0,099213809 | LYPD6 | FALSE | Upregulated |
| -0,288321263 | 0,099229027 | GDF2 | FALSE | Downregulated |
| -0,903686164 | 0,099249768 | TMEM182 | FALSE | Downregulated |
| -0,396546476 | 0,09929684 | C14orf1 | FALSE | Downregulated |
| 0,304865831 | 0,099320891 | SNN | FALSE | Upregulated |
| 2,426451706 | 0,099356198 | SYT2 | FALSE | Upregulated |
| -0,161010492 | 0,099421617 | ABCB7 | FALSE | Downregulated |
| -1,360590491 | 0,099454694 | HAO2-IT1 | FALSE | Downregulated |
| 0,785890998 | 0,099454694 | SIGLEC5 | FALSE | Upregulated |
| 0,826060165 | 0,099454694 | ZACN | FALSE | Upregulated |
| 1,907647884 | 0,099464339 | CAPN14 | FALSE | Upregulated |
| -0,536794185 | 0,099475738 | LOC101927579 | FALSE | Downregulated |
| 0,186748768 | 0,099475738 | VPS33A | FALSE | Upregulated |
| -0,434991032 | 0,099494579 | ADAM1A | FALSE | Downregulated |
| 0,186485347 | 0,099517518 | RNF31 | FALSE | Upregulated |
| 0,336732161 | 0,09959492 | ZSCAN9 | FALSE | Upregulated |
| 0,534993968 | 0,099691219 | SP140 | FALSE | Upregulated |
| -0,625457804 | 0,099703123 | TUBB1 | FALSE | Downregulated |
| -1,363699783 | 0,09980523 | GTSCR1 | FALSE | Downregulated |
| -0,369908369 | 0,099812042 | LOC100288778 | FALSE | Downregulated |
| -0,274393689 | 0,099896309 | CPT2 | FALSE | Downregulated |
| -0,904909391 | 0,099896309 | HEATR4 | FALSE | Downregulated |
| 0,195295315 | 0,10002005 | UGGT2 | FALSE | Upregulated |

| | | | | |
|---|---|---|---|---|
| 0,752972416 | 0,100040138 | FGFBP2 | FALSE | Upregulated |
| -0,13591663 | 0,100164686 | ATXN7L3B | FALSE | Downregulated |
| -0,230578908 | 0,10017393 | C11orf31 | FALSE | Downregulated |
| 0,517960191 | 0,10017393 | TLR8 | FALSE | Upregulated |
| 0,373746505 | 0,100238529 | BCAM | FALSE | Upregulated |
| 0,344296415 | 0,100291703 | LOC102724002 | FALSE | Upregulated |
| 1,627980326 | 0,10029296 | PKP3 | FALSE | Upregulated |
| -0,238303691 | 0,100329117 | PRPSAP1 | FALSE | Downregulated |
| 2,660175907 | 0,100329117 | TCEB3B | FALSE | Upregulated |
| 0,222044251 | 0,100369443 | BMP1 | FALSE | Upregulated |
| 0,274550979 | 0,100531695 | BAD | FALSE | Upregulated |
| -0,447017384 | 0,100531695 | C14orf132 | FALSE | Downregulated |
| 2,163337497 | 0,10053177 | RP11-227H15.4 | FALSE | Upregulated |
| 1,624479043 | 0,100633279 | L1CAM | FALSE | Upregulated |
| 0,433800128 | 0,100633279 | LY86 | FALSE | Upregulated |
| 0,221640174 | 0,10087181 | HERC2 | FALSE | Upregulated |
| -0,128116474 | 0,100885611 | TMEM66 | FALSE | Downregulated |
| 2,353191744 | 0,10095894 | LOC101927757 | FALSE | Upregulated |
| 0,189318357 | 0,10108254 | LRRC42 | FALSE | Upregulated |
| 0,250638042 | 0,101093394 | DYRK2 | FALSE | Upregulated |
| -0,456371254 | 0,101123634 | GALK1 | FALSE | Downregulated |
| -0,339310982 | 0,101123634 | PAM16 | FALSE | Downregulated |
| 0,307377016 | 0,101131065 | ACSS3 | FALSE | Upregulated |
| -0,301439085 | 0,101131065 | LDHA | FALSE | Downregulated |
| 0,342586859 | 0,101131065 | LGALS9 | FALSE | Upregulated |
| -0,178968952 | 0,101421822 | FBXO38 | FALSE | Downregulated |
| 1,446022201 | 0,101445303 | TAAR3 | FALSE | Upregulated |
| -0,974588919 | 0,101477514 | LOC100996756 | FALSE | Downregulated |
| 0,223323071 | 0,101477514 | PTAR1 | FALSE | Upregulated |
| -0,298867127 | 0,101512858 | ADH1B | FALSE | Downregulated |
| 0,47724398 | 0,101512858 | INHBA | FALSE | Upregulated |
| 0,266045716 | 0,101512858 | TSHZ2 | FALSE | Upregulated |
| 0,930816659 | 0,101555429 | CELF5 | FALSE | Upregulated |
| -0,311622012 | 0,101654057 | NAT1 | FALSE | Downregulated |
| -0,548678008 | 0,10165774 | LOC101927340 | FALSE | Downregulated |
| 0,190926605 | 0,10165774 | MARCH5 | FALSE | Upregulated |
| 0,155076811 | 0,101715273 | CANT1 | FALSE | Upregulated |
| -0,275629267 | 0,101715273 | EEA1 | FALSE | Downregulated |
| 0,419781183 | 0,101715273 | HENMT1 | FALSE | Upregulated |
| -0,175180154 | 0,101715273 | MRPS10 | FALSE | Downregulated |
| -0,714685902 | 0,101727333 | ZNF667-AS1 | FALSE | Downregulated |
| 0,237808242 | 0,101805925 | FAM98A | FALSE | Upregulated |
| 0,439925307 | 0,101805925 | ZNF816 | FALSE | Upregulated |
| -0,951401956 | 0,101882811 | CBLN4 | FALSE | Downregulated |
| 0,708097276 | 0,102043999 | SOWAHD | FALSE | Upregulated |
| -0,181964356 | 0,102165385 | ANKRD28 | FALSE | Downregulated |
| 1,643712372 | 0,102165385 | GABRD | FALSE | Upregulated |
| 1,203273525 | 0,1022871 | LOC101927734 | FALSE | Upregulated |
| 1,085213751 | 0,102377357 | GDF10 | FALSE | Upregulated |
| -0,259689833 | 0,102489744 | DET1 | FALSE | Downregulated |
| -0,339055106 | 0,102693609 | RPP21 | FALSE | Downregulated |

| | | | | |
|---|---|---|---|---|
| 0,230939019 | 0,102703169 | AP5M1 | FALSE | Upregulated |
| -1,202537836 | 0,102703169 | LOC388780 | FALSE | Downregulated |
| -0,462426748 | 0,102703169 | TTC6 | FALSE | Downregulated |
| 0,19394842 | 0,102704296 | DAZAP1 | FALSE | Upregulated |
| 0,899201147 | 0,10279724 | FOXQ1 | FALSE | Upregulated |
| -0,677238231 | 0,10279724 | RP11-132A1.4 | FALSE | Downregulated |
| -0,282007913 | 0,102810741 | SIRT1 | FALSE | Downregulated |
| -0,49930324 | 0,102810741 | TPRG1 | FALSE | Downregulated |
| 0,281295506 | 0,102826758 | POGLUT1 | FALSE | Upregulated |
| 0,281479619 | 0,102826758 | TIFA | FALSE | Upregulated |
| -0,248079479 | 0,102869277 | REPS2 | FALSE | Downregulated |
| -0,292027736 | 0,102946573 | NUDT19 | FALSE | Downregulated |
| 0,203864619 | 0,103112037 | COPB1 | FALSE | Upregulated |
| 0,418363476 | 0,103112037 | PTPN13 | FALSE | Upregulated |
| -0,369127437 | 0,10332078 | LINC00339 | FALSE | Downregulated |
| -0,26244601 | 0,103408327 | FKBP7 | FALSE | Downregulated |
| -0,321724323 | 0,103408327 | NFKBIB | FALSE | Downregulated |
| -0,617856916 | 0,103415842 | LOC100507547 | FALSE | Downregulated |
| 1,625741863 | 0,103415842 | LOC102723769 | FALSE | Upregulated |
| 1,472782665 | 0,103449407 | LOC101929188 | FALSE | Upregulated |
| -0,469515463 | 0,103449903 | LOC100130093 | FALSE | Downregulated |
| 0,300039092 | 0,103661532 | CERS5 | FALSE | Upregulated |
| 1,726530712 | 0,103661532 | RPA4 | FALSE | Upregulated |
| 0,303890204 | 0,103661532 | ZNF416 | FALSE | Upregulated |
| 0,329328594 | 0,103673739 | FXYD5 | FALSE | Upregulated |
| -0,176100955 | 0,103684957 | SMAP2 | FALSE | Downregulated |
| -3,101036618 | 0,103726092 | MYO18B | FALSE | Downregulated |
| -0,67487407 | 0,103730961 | ENO3 | FALSE | Downregulated |
| -0,225933206 | 0,103760903 | FTSJ3 | FALSE | Downregulated |
| -0,14858377 | 0,103774927 | RMDN3 | FALSE | Downregulated |
| 1,374471522 | 0,103807203 | RP11-430B1.2 | FALSE | Upregulated |
| 1,062666819 | 0,103932711 | LOC646652 | FALSE | Upregulated |
| -0,209470834 | 0,103932711 | MTCH2 | FALSE | Downregulated |
| 0,22768849 | 0,1039359 | PPME1 | FALSE | Upregulated |
| 1,253335991 | 0,104110678 | APOBEC3H | FALSE | Upregulated |
| -1,007122674 | 0,104110678 | FAM27E3 | FALSE | Downregulated |
| -0,2077933 | 0,104110678 | FAM96B | FALSE | Downregulated |
| 0,175011192 | 0,104118373 | EPS15L1 | FALSE | Upregulated |
| 0,415500758 | 0,104203976 | SHROOM1 | FALSE | Upregulated |
| 0,130848323 | 0,104211887 | CSTF2T | FALSE | Upregulated |
| 2,025155462 | 0,104268412 | MSX2P1 | FALSE | Upregulated |
| -0,181349131 | 0,104290942 | ATG3 | FALSE | Downregulated |
| 0,579315676 | 0,104324184 | GLT8D2 | FALSE | Upregulated |
| 0,321797611 | 0,104334605 | PIK3R3 | FALSE | Upregulated |
| 0,339853633 | 0,104605291 | LOC101927371 | FALSE | Upregulated |
| 0,214596984 | 0,104746797 | C1GALT1C1 | FALSE | Upregulated |
| 0,60836884 | 0,104746797 | GBP5 | FALSE | Upregulated |
| 0,851230959 | 0,104746797 | GPAT2 | FALSE | Upregulated |
| 0,332042752 | 0,104770179 | CCNJ | FALSE | Upregulated |
| -0,332752965 | 0,104772883 | TMEM56 | FALSE | Downregulated |
| -0,498216679 | 0,104831569 | CLDN14 | FALSE | Downregulated |

| | | | | |
|---|---|---|---|---|
| 0,484214902 | 0,104846341 | INTU | FALSE | Upregulated |
| -0,222900388 | 0,104857344 | GPATCH8 | FALSE | Downregulated |
| 2,624705863 | 0,104857344 | NPSR1-AS1 | FALSE | Upregulated |
| 1,972512909 | 0,10485832 | ABCC13 | FALSE | Upregulated |
| 0,436158532 | 0,104920524 | ENC1 | FALSE | Upregulated |
| 1,24950023 | 0,104920524 | LOC100652736 | FALSE | Upregulated |
| -0,227704009 | 0,104920524 | R3HDM1 | FALSE | Downregulated |
| 0,678470659 | 0,104940577 | LOC100506473 | FALSE | Upregulated |
| -0,301250139 | 0,104972153 | PHLPP1 | FALSE | Downregulated |
| -0,882098464 | 0,104972688 | IFI6 | FALSE | Downregulated |
| -0,450037814 | 0,10500043 | NRL | FALSE | Downregulated |
| 0,758007978 | 0,105025704 | ANKRD20A9P | FALSE | Upregulated |
| 1,444363682 | 0,105026387 | MPP2 | FALSE | Upregulated |
| 0,757165837 | 0,105079302 | LOC100505564 | FALSE | Upregulated |
| -0,443967668 | 0,105115619 | SRRM5 | FALSE | Downregulated |
| 0,318408897 | 0,105152466 | LUZP1 | FALSE | Upregulated |
| 1,161751984 | 0,105242744 | VENTX | FALSE | Upregulated |
| 0,15208357 | 0,105242744 | YTHDF3 | FALSE | Upregulated |
| -0,148864667 | 0,105392475 | NUP160 | FALSE | Downregulated |
| -0,148096489 | 0,105418027 | DROSHA | FALSE | Downregulated |
| -0,75403648 | 0,105418027 | RP5-968D22.3 | FALSE | Downregulated |
| 0,965245804 | 0,105443417 | MIR4435-1HG | FALSE | Upregulated |
| 0,402105894 | 0,105443732 | SCNN1A | FALSE | Upregulated |
| 0,172723674 | 0,105460726 | PEX26 | FALSE | Upregulated |
| 0,281349209 | 0,10554221 | ZBTB20 | FALSE | Upregulated |
| 0,223581701 | 0,105635355 | NMI | FALSE | Upregulated |
| 0,286954941 | 0,105649514 | WDSUB1 | FALSE | Upregulated |
| 0,546766049 | 0,105650472 | GPR137C | FALSE | Upregulated |
| -0,261632807 | 0,105677528 | COLEC10 | FALSE | Downregulated |
| 1,490638932 | 0,105677528 | ZFP57 | FALSE | Upregulated |
| 1,004998532 | 0,10568523 | SEC14L6 | FALSE | Upregulated |
| -0,703897164 | 0,105963032 | HPGDS | FALSE | Downregulated |
| -0,220137035 | 0,105963032 | ZNF225 | FALSE | Downregulated |
| -0,903689204 | 0,106433801 | LOC102724329 | FALSE | Downregulated |
| 1,868066782 | 0,106524763 | SP6 | FALSE | Upregulated |
| 1,452861491 | 0,106563534 | LINC00877 | FALSE | Upregulated |
| 0,185075249 | 0,10660475 | PSMC3 | FALSE | Upregulated |
| 0,289869489 | 0,106704206 | EID2 | FALSE | Upregulated |
| -0,398934172 | 0,106704206 | RPS6KA6 | FALSE | Downregulated |
| -0,612625186 | 0,106857778 | SYNM | FALSE | Downregulated |
| 0,214078025 | 0,106857778 | VPS13C | FALSE | Upregulated |
| 1,507519697 | 0,106992052 | LOC101929950 | FALSE | Upregulated |
| 1,253150588 | 0,10701652 | WNT7B | FALSE | Upregulated |
| 0,37961023 | 0,107050485 | RAB31 | FALSE | Upregulated |
| 0,743399835 | 0,107152714 | CKAP2L | FALSE | Upregulated |
| 0,770897973 | 0,107152714 | LTF | FALSE | Upregulated |
| -0,746245588 | 0,107261914 | MT1X | FALSE | Downregulated |
| -0,17432952 | 0,107437917 | HNRNPM | FALSE | Downregulated |
| -0,257172415 | 0,107447223 | PRDX3 | FALSE | Downregulated |
| 0,213506886 | 0,107463518 | INSR | FALSE | Upregulated |
| -0,161262616 | 0,107463518 | MRPL44 | FALSE | Downregulated |

| | | | | |
|---|---|---|---|---|
| 0,148118053 | 0,107463518 | PANX1 | FALSE | Upregulated |
| 0,763401791 | 0,107463518 | RAB6B | FALSE | Upregulated |
| 0,217566667 | 0,107463518 | S100A16 | FALSE | Upregulated |
| -0,470176864 | 0,107463518 | TMEM255B | FALSE | Downregulated |
| -0,365491817 | 0,107493101 | MRI1 | FALSE | Downregulated |
| 0,348620412 | 0,107582067 | C14orf105 | FALSE | Upregulated |
| 0,295702838 | 0,107662258 | GALC | FALSE | Upregulated |
| 0,48106868 | 0,107840368 | CCDC40 | FALSE | Upregulated |
| 0,417221918 | 0,107840368 | CDCA4 | FALSE | Upregulated |
| -0,171542548 | 0,107840368 | NDUFB10 | FALSE | Downregulated |
| -0,231382189 | 0,107863819 | PRKACA | FALSE | Downregulated |
| 1,663502603 | 0,107945831 | B4GALNT4 | FALSE | Upregulated |
| 1,049439907 | 0,107970057 | HAS2 | FALSE | Upregulated |
| 0,231634707 | 0,108022783 | B3GALNT2 | FALSE | Upregulated |
| -0,265703739 | 0,108022783 | GSK3A | FALSE | Downregulated |
| 0,776758647 | 0,108022783 | IPO11-LRRC70 | FALSE | Upregulated |
| 0,295832835 | 0,108068902 | SLC38A6 | FALSE | Upregulated |
| -0,320555993 | 0,108076462 | GCAT | FALSE | Downregulated |
| -0,705387249 | 0,108110401 | SNAI3-AS1 | FALSE | Downregulated |
| 0,853211253 | 0,108310377 | PTP4A3 | FALSE | Upregulated |
| -0,494814579 | 0,108334027 | ANO3 | FALSE | Downregulated |
| 0,242583085 | 0,108450695 | TNFAIP1 | FALSE | Upregulated |
| -0,850407227 | 0,108590427 | CACNA1H | FALSE | Downregulated |
| 0,536245336 | 0,108706065 | ARHGAP39 | FALSE | Upregulated |
| 0,431084662 | 0,108737718 | CD244 | FALSE | Upregulated |
| -0,678510178 | 0,108792414 | SLC16A14 | FALSE | Downregulated |
| -0,564123789 | 0,108836845 | ATP1B2 | FALSE | Downregulated |
| -0,455983562 | 0,109000968 | IL13RA2 | FALSE | Downregulated |
| -0,286590222 | 0,109017648 | SH3BP5 | FALSE | Downregulated |
| 0,184919269 | 0,109017648 | SH3D19 | FALSE | Upregulated |
| -0,822420219 | 0,109017648 | TEX21P | FALSE | Downregulated |
| 0,713204959 | 0,109152811 | C8orf88 | FALSE | Upregulated |
| -0,189115633 | 0,109185114 | C17orf85 | FALSE | Downregulated |
| -0,522803463 | 0,109224932 | GPR182 | FALSE | Downregulated |
| 0,42223251 | 0,109245186 | FAM65B | FALSE | Upregulated |
| -0,192177385 | 0,109245186 | MTTP | FALSE | Downregulated |
| 0,253744191 | 0,109245186 | TSC22D1 | FALSE | Upregulated |
| -0,180120523 | 0,109280559 | KPNA1 | FALSE | Downregulated |
| 1,02092731 | 0,109280559 | MUC1 | FALSE | Upregulated |
| -0,334707963 | 0,109361576 | L3MBTL3 | FALSE | Downregulated |
| -0,334759265 | 0,109485927 | TSPAN33 | FALSE | Downregulated |
| -0,415293874 | 0,109559986 | RORC | FALSE | Downregulated |
| 0,231543493 | 0,109615271 | CSK | FALSE | Upregulated |
| 1,153310844 | 0,109889994 | ANKRD36BP1 | FALSE | Upregulated |
| 0,345599616 | 0,10995114 | ST3GAL4 | FALSE | Upregulated |
| -1,419407445 | 0,110007985 | LOC101928916 | FALSE | Downregulated |
| -1,256986089 | 0,110151496 | LOC102724265 | FALSE | Downregulated |
| -0,966431827 | 0,11023064 | FAM83A-AS1 | FALSE | Downregulated |
| 1,015886589 | 0,110249743 | TLR9 | FALSE | Upregulated |
| -0,229822812 | 0,110263771 | CTBP1-AS2 | FALSE | Downregulated |
| 0,268830455 | 0,110263771 | DYNLT1 | FALSE | Upregulated |

| | | | | |
|---|---|---|---|---|
| 0,428590607 | 0,110322606 | SLC25A29 | FALSE | Upregulated |
| 1,498703846 | 0,110523686 | CNR1 | FALSE | Upregulated |
| -0,271572531 | 0,110523686 | GATA4 | FALSE | Downregulated |
| -0,269529146 | 0,110542952 | NTN4 | FALSE | Downregulated |
| 0,186025901 | 0,110567472 | TRIM38 | FALSE | Upregulated |
| -0,363473065 | 0,110606833 | KCNT2 | FALSE | Downregulated |
| 1,637889681 | 0,110750185 | GALNT8 | FALSE | Upregulated |
| 0,545150871 | 0,110799621 | ARG2 | FALSE | Upregulated |
| -0,255978779 | 0,110799621 | METTL15 | FALSE | Downregulated |
| -0,190999087 | 0,110879149 | CDK4 | FALSE | Downregulated |
| -0,2915426 | 0,110879149 | SFPQ | FALSE | Downregulated |
| -0,747016176 | 0,110932867 | NOL4 | FALSE | Downregulated |
| 0,110417724 | 0,110932867 | QARS | FALSE | Upregulated |
| 0,156643951 | 0,110932867 | RNF4 | FALSE | Upregulated |
| 2,206330249 | 0,111069489 | C22orf31 | FALSE | Upregulated |
| -0,290661836 | 0,111161911 | RPS14 | FALSE | Downregulated |
| 0,475178579 | 0,111236958 | NUDT1 | FALSE | Upregulated |
| -0,261114787 | 0,111346663 | SEC14L2 | FALSE | Downregulated |
| -0,205954669 | 0,111365744 | NDUFB8 | FALSE | Downregulated |
| 0,694592608 | 0,111588134 | FGF13 | FALSE | Upregulated |
| 0,635550878 | 0,11163768 | ZNF382 | FALSE | Upregulated |
| -0,330624629 | 0,111665357 | SMAP1 | FALSE | Downregulated |
| 0,935295419 | 0,111710113 | LOC284837 | FALSE | Upregulated |
| -0,533488172 | 0,11175649 | HERC2P7 | FALSE | Downregulated |
| 1,10579403 | 0,11175649 | NECAB1 | FALSE | Upregulated |
| 1,418110651 | 0,111826667 | RN7SK | FALSE | Upregulated |
| 0,220315807 | 0,112025772 | ABHD4 | FALSE | Upregulated |
| 0,314522278 | 0,112031338 | NME1 | FALSE | Upregulated |
| -0,31715953 | 0,112070377 | NGFR | FALSE | Downregulated |
| -0,180060834 | 0,112070377 | TXNRD3 | FALSE | Downregulated |
| 0,216187975 | 0,112095962 | B4GALT1 | FALSE | Upregulated |
| 0,30695618 | 0,112095962 | CEP68 | FALSE | Upregulated |
| 0,270600182 | 0,112095962 | ERCC6L2 | FALSE | Upregulated |
| 1,470852427 | 0,112095962 | MYH15 | FALSE | Upregulated |
| -0,552624805 | 0,112097723 | CYP3A5 | FALSE | Downregulated |
| -0,183302623 | 0,112121237 | FRG1 | FALSE | Downregulated |
| 0,728368156 | 0,112129767 | NANOS1 | FALSE | Upregulated |
| 0,335309734 | 0,112156914 | PPP1R35 | FALSE | Upregulated |
| 0,598029775 | 0,112156914 | SIRPB2 | FALSE | Upregulated |
| 0,689019975 | 0,112249124 | DNAH2 | FALSE | Upregulated |
| 0,671172014 | 0,112249124 | FCRL6 | FALSE | Upregulated |
| -0,490769933 | 0,112249124 | PDE7B | FALSE | Downregulated |
| 0,93139318 | 0,11225931 | PTGDR2 | FALSE | Upregulated |
| 0,485049213 | 0,11225931 | RUNX1T1 | FALSE | Upregulated |
| 0,44322051 | 0,112320535 | NLRP1 | FALSE | Upregulated |
| 0,88958836 | 0,112322628 | LINC00926 | FALSE | Upregulated |
| -0,873285467 | 0,112375455 | ARNTL | FALSE | Downregulated |
| -0,213730794 | 0,112409023 | SCP2 | FALSE | Downregulated |
| 1,010783327 | 0,112477603 | ATP1A3 | FALSE | Upregulated |
| -0,267558571 | 0,112477603 | FUOM | FALSE | Downregulated |
| 0,519182315 | 0,112540116 | ZNF154 | FALSE | Upregulated |

| | | | | |
|---|---|---|---|---|
| -0,198486624 | 0,112634568 | CEP44 | FALSE | Downregulated |
| 0,25289827 | 0,112739078 | AMDHD2 | FALSE | Upregulated |
| 0,390774885 | 0,112813116 | AP1G2 | FALSE | Upregulated |
| -0,361743598 | 0,112820445 | FAM195A | FALSE | Downregulated |
| 0,68799761 | 0,112892273 | C9orf9 | FALSE | Upregulated |
| -0,669631522 | 0,112997454 | TAT | FALSE | Downregulated |
| -0,298542198 | 0,112999368 | MYCBP2 | FALSE | Downregulated |
| -0,34015085 | 0,113012663 | LOC101929291 | FALSE | Downregulated |
| 0,628334422 | 0,113023393 | LOC101929758 | FALSE | Upregulated |
| 0,437885133 | 0,113028091 | ZNF468 | FALSE | Upregulated |
| -0,28110942 | 0,113037159 | IL33 | FALSE | Downregulated |
| 0,143720446 | 0,113037159 | USP3 | FALSE | Upregulated |
| 0,219804197 | 0,113049811 | NCLN | FALSE | Upregulated |
| 1,828921905 | 0,113059473 | DNM1P41 | FALSE | Upregulated |
| -1,088089272 | 0,113059938 | LRRC37A11P | FALSE | Downregulated |
| 0,278160028 | 0,113131145 | ATAD3A | FALSE | Upregulated |
| -0,308298353 | 0,113193596 | LOC101929343 | FALSE | Downregulated |
| -0,313148538 | 0,113237883 | GLTPD2 | FALSE | Downregulated |
| -0,916627285 | 0,113313318 | LINC01132 | FALSE | Downregulated |
| -0,302759497 | 0,113500802 | PAXBP1 | FALSE | Downregulated |
| 1,605251933 | 0,113521859 | FAIM2 | FALSE | Upregulated |
| -0,475168795 | 0,113521859 | IQSEC2 | FALSE | Downregulated |
| 1,674437913 | 0,113521859 | SLC7A14 | FALSE | Upregulated |
| -0,269632968 | 0,113525659 | RDH11 | FALSE | Downregulated |
| -0,330882101 | 0,113583192 | GTF2IRD2 | FALSE | Downregulated |
| 0,508595303 | 0,113631821 | EFCAB4B | FALSE | Upregulated |
| 1,7544126 | 0,1136942 | ITGB6 | FALSE | Upregulated |
| 0,251205565 | 0,113697917 | CSTF2 | FALSE | Upregulated |
| -0,318712789 | 0,113733664 | DDC | FALSE | Downregulated |
| 0,413893703 | 0,113829424 | EXD3 | FALSE | Upregulated |
| 1,351525706 | 0,113829424 | LOC101929240 | FALSE | Upregulated |
| -0,166346524 | 0,114011012 | PPIP5K2 | FALSE | Downregulated |
| -0,290281441 | 0,114025088 | PPOX | FALSE | Downregulated |
| 0,308716795 | 0,114065164 | NR2C2AP | FALSE | Upregulated |
| 0,13871288 | 0,114076358 | ANKFY1 | FALSE | Upregulated |
| 0,207648347 | 0,114127174 | CASD1 | FALSE | Upregulated |
| 0,866670536 | 0,114238208 | BCL2L14 | FALSE | Upregulated |
| -0,421265799 | 0,11429968 | PPARGC1A | FALSE | Downregulated |
| -0,270017767 | 0,11429968 | RPSA | FALSE | Downregulated |
| 0,551465655 | 0,114313799 | DCLK2 | FALSE | Upregulated |
| -0,188154808 | 0,11448565 | CELF1 | FALSE | Downregulated |
| 0,368157415 | 0,114651459 | TXN | FALSE | Upregulated |
| 0,810535897 | 0,114737494 | FAM131B | FALSE | Upregulated |
| 0,171915964 | 0,114743039 | LUZP6 | FALSE | Upregulated |
| -0,509582998 | 0,114764222 | LOC101929959 | FALSE | Downregulated |
| 0,255977252 | 0,114809957 | NUP62 | FALSE | Upregulated |
| -0,254821367 | 0,114809957 | SDHAF1 | FALSE | Downregulated |
| -0,186491957 | 0,114809957 | SDHC | FALSE | Downregulated |
| 0,256694563 | 0,114819656 | IRAK4 | FALSE | Upregulated |
| 1,211112436 | 0,115007532 | PNMA3 | FALSE | Upregulated |
| -0,426927904 | 0,115007532 | PNN | FALSE | Downregulated |

| | | | | |
|---|---|---|---|---|
| -0,218636303 | 0,115021116 | RPL26L1 | FALSE | Downregulated |
| 0,298473457 | 0,115137629 | HMGA1 | FALSE | Upregulated |
| -0,233097701 | 0,115137629 | MRPL21 | FALSE | Downregulated |
| -0,447613762 | 0,115137629 | RP11-473M20.14 | FALSE | Downregulated |
| -0,166337911 | 0,115156947 | TTLL5 | FALSE | Downregulated |
| -0,912678362 | 0,115232578 | LOC101928891 | FALSE | Downregulated |
| 0,53409288 | 0,115378849 | FLRT3 | FALSE | Upregulated |
| 0,238342279 | 0,115442929 | ZNF234 | FALSE | Upregulated |
| -0,647341259 | 0,11548453 | EFHD1 | FALSE | Downregulated |
| 0,372163317 | 0,11548453 | LGALS3BP | FALSE | Upregulated |
| 0,512877753 | 0,11548453 | TSPYL5 | FALSE | Upregulated |
| -0,361500262 | 0,11548453 | VEGFA | FALSE | Downregulated |
| 0,232562926 | 0,115531111 | WDR77 | FALSE | Upregulated |
| -0,32041059 | 0,115596021 | PSMD1 | FALSE | Downregulated |
| 0,129575818 | 0,115720551 | GLG1 | FALSE | Upregulated |
| -0,638964614 | 0,115767752 | LOC102724782 | FALSE | Downregulated |
| 0,781486687 | 0,115767752 | USH1C | FALSE | Upregulated |
| 0,586240619 | 0,115819252 | LOC102724872 | FALSE | Upregulated |
| 0,563412897 | 0,115922872 | IGSF23 | FALSE | Upregulated |
| 0,959267596 | 0,115922872 | RASAL2-AS1 | FALSE | Upregulated |
| -0,61749073 | 0,115987936 | LOC101928047 | FALSE | Downregulated |
| -0,558099172 | 0,115987936 | ZNF774 | FALSE | Downregulated |
| -0,20663055 | 0,116000486 | ELP5 | FALSE | Downregulated |
| 1,378498648 | 0,116000486 | LOC101929494 | FALSE | Upregulated |
| 0,18098095 | 0,116024983 | ERGIC3 | FALSE | Upregulated |
| 2,081292971 | 0,116072791 | LINC00330 | FALSE | Upregulated |
| -0,172457978 | 0,116130095 | WDR3 | FALSE | Downregulated |
| 0,3214208 | 0,116139536 | ARHGAP15 | FALSE | Upregulated |
| 0,317444865 | 0,116166521 | CDPF1 | FALSE | Upregulated |
| 0,862922168 | 0,116259051 | RP11-65J3.1 | FALSE | Upregulated |
| 0,248153916 | 0,116261905 | CHD7 | FALSE | Upregulated |
| -0,345716415 | 0,116261905 | HNF4A-AS1 | FALSE | Downregulated |
| -0,192876255 | 0,116261905 | PCMT1 | FALSE | Downregulated |
| -2,113680473 | 0,116261905 | PPP1R27 | FALSE | Downregulated |
| -0,185508888 | 0,116261905 | SGMS1 | FALSE | Downregulated |
| -0,144340429 | 0,116261905 | USP9X | FALSE | Downregulated |
| 0,446890738 | 0,116276939 | PDE5A | FALSE | Upregulated |
| -1,768292846 | 0,11628122 | PKIA | FALSE | Downregulated |
| -0,297792672 | 0,116291666 | PKD2 | FALSE | Downregulated |
| -0,14552905 | 0,11630099 | SECISBP2L | FALSE | Downregulated |
| -0,323526386 | 0,116356002 | ECHDC2 | FALSE | Downregulated |
| 0,423069678 | 0,11641453 | ZNF83 | FALSE | Upregulated |
| -0,925146111 | 0,116429449 | DLK2 | FALSE | Downregulated |
| 0,852619673 | 0,116429449 | ZNF296 | FALSE | Upregulated |
| 0,280135983 | 0,116455325 | TP53BP1 | FALSE | Upregulated |
| 0,38495323 | 0,116479948 | KBTBD8 | FALSE | Upregulated |
| 0,860456905 | 0,116584356 | C11orf21 | FALSE | Upregulated |
| -0,933260797 | 0,116717848 | RASSF1-AS1 | FALSE | Downregulated |
| 0,23650413 | 0,116753534 | GSE1 | FALSE | Upregulated |
| 0,77806633 | 0,116753534 | VSTM2L | FALSE | Upregulated |
| 2,092545949 | 0,116766573 | CRLF1 | FALSE | Upregulated |

| | | | | |
|---|---|---|---|---|
| 0,351688147 | 0,116783744 | ZNF786 | FALSE | Upregulated |
| 1,601692602 | 0,116938314 | GUSBP3 | FALSE | Upregulated |
| -0,881527789 | 0,117061608 | AGBL4 | FALSE | Downregulated |
| 0,209030178 | 0,117061608 | INTS3 | FALSE | Upregulated |
| 0,18680809 | 0,117061608 | KIAA1279 | FALSE | Upregulated |
| -0,219829624 | 0,117061608 | NUDT12 | FALSE | Downregulated |
| 0,27551475 | 0,11714715 | PQLC3 | FALSE | Upregulated |
| -0,162794097 | 0,117275243 | FAM35A | FALSE | Downregulated |
| -0,178657136 | 0,11739458 | SORBS1 | FALSE | Downregulated |
| -0,746086269 | 0,11751988 | LOC100129518 | FALSE | Downregulated |
| 1,362546287 | 0,117544267 | RP11-27N21.3 | FALSE | Upregulated |
| -0,766285465 | 0,117544267 | TRAPPC3L | FALSE | Downregulated |
| 0,643766787 | 0,11757899 | C12orf5 | FALSE | Upregulated |
| -0,191634739 | 0,11757899 | ROGDI | FALSE | Downregulated |
| -0,319740759 | 0,117644882 | GPR126 | FALSE | Downregulated |
| -0,476104314 | 0,117644882 | LOC100652824 | FALSE | Downregulated |
| -0,391918379 | 0,117939761 | GPATCH4 | FALSE | Downregulated |
| 0,192867502 | 0,118057783 | ZNF672 | FALSE | Upregulated |
| 0,404999156 | 0,118202251 | BIN1 | FALSE | Upregulated |
| -0,261563456 | 0,118325465 | MBTD1 | FALSE | Downregulated |
| -0,342617695 | 0,118447819 | CEBPA | FALSE | Downregulated |
| 0,662179432 | 0,118447819 | CIT | FALSE | Upregulated |
| -0,24568763 | 0,118447819 | CTNNBIP1 | FALSE | Downregulated |
| -0,367666043 | 0,118447819 | LDLRAD4 | FALSE | Downregulated |
| 0,597833895 | 0,118447819 | PTGER2 | FALSE | Upregulated |
| -0,17064292 | 0,118473321 | CHCHD3 | FALSE | Downregulated |
| -0,190765858 | 0,118509865 | ETAA1 | FALSE | Downregulated |
| -0,396725556 | 0,118555395 | ADH1C | FALSE | Downregulated |
| -0,223156319 | 0,118737552 | C18orf21 | FALSE | Downregulated |
| -0,494938376 | 0,118737552 | LOC101928728 | FALSE | Downregulated |
| 0,37435148 | 0,118822324 | DOCK10 | FALSE | Upregulated |
| -0,402009647 | 0,118895169 | AP5S1 | FALSE | Downregulated |
| 1,13281466 | 0,118895169 | DCLK1 | FALSE | Upregulated |
| 0,237422604 | 0,118895169 | NIT1 | FALSE | Upregulated |
| -0,230438614 | 0,118895169 | OSBP | FALSE | Downregulated |
| 0,494025914 | 0,118895169 | ZNF525 | FALSE | Upregulated |
| -0,262180646 | 0,118936801 | RPL32 | FALSE | Downregulated |
| 0,228644055 | 0,118982847 | ABHD12 | FALSE | Upregulated |
| -0,211967282 | 0,118982847 | PDLIM2 | FALSE | Downregulated |
| -0,782910852 | 0,119017494 | LOC101928487 | FALSE | Downregulated |
| -0,662419871 | 0,119022264 | GOLGA6B | FALSE | Downregulated |
| 0,306736247 | 0,119158056 | DENND2D | FALSE | Upregulated |
| -0,171280984 | 0,119158056 | MGEA5 | FALSE | Downregulated |
| 0,206423831 | 0,119172877 | LATS2 | FALSE | Upregulated |
| -0,239999782 | 0,119213941 | NDRG2 | FALSE | Downregulated |
| 1,709276447 | 0,119213941 | RP11-397C18.2 | FALSE | Upregulated |
| 0,610600955 | 0,119221095 | IL18RAP | FALSE | Upregulated |
| -0,594335161 | 0,119223221 | PTPRT | FALSE | Downregulated |
| 0,514202036 | 0,1193265 | SLC5A6 | FALSE | Upregulated |
| -0,498528777 | 0,119393637 | SGCE | FALSE | Downregulated |
| 1,014601358 | 0,119414428 | PLCXD2 | FALSE | Upregulated |

| | | | | |
|---|---|---|---|---|
| 0,234319544 | 0,11961089 | ZBTB38 | FALSE | Upregulated |
| -0,415234178 | 0,119618747 | AGPAT5 | FALSE | Downregulated |
| 0,752586139 | 0,119707263 | CDHR1 | FALSE | Upregulated |
| -1,776818831 | 0,119762811 | LOC101927125 | FALSE | Downregulated |
| -0,2460115 | 0,119796818 | ARGLU1 | FALSE | Downregulated |
| -0,199141022 | 0,119840027 | HAX1 | FALSE | Downregulated |
| 2,682448788 | 0,119861554 | LOC730268 | FALSE | Upregulated |
| 0,409568357 | 0,119894743 | HK3 | FALSE | Upregulated |
| 1,715591864 | 0,119953742 | HIST1H4H | FALSE | Upregulated |
| -0,490598243 | 0,119953742 | IGFALS | FALSE | Downregulated |
| 0,538958455 | 0,119955671 | ZBTB7C | FALSE | Upregulated |
| 0,627796898 | 0,119967796 | AQP4 | FALSE | Upregulated |
| 1,389218543 | 0,119967796 | C22orf23 | FALSE | Upregulated |
| 1,115275717 | 0,119967796 | C9orf152 | FALSE | Upregulated |
| -0,182629926 | 0,119967796 | CHD1 | FALSE | Downregulated |
| -0,192243202 | 0,119967796 | SLC25A43 | FALSE | Downregulated |
| 0,795450087 | 0,119967796 | SSPN | FALSE | Upregulated |
| -0,313077881 | 0,120007954 | TTC14 | FALSE | Downregulated |
| 0,178342504 | 0,12015539 | CCDC137 | FALSE | Upregulated |
| 0,839875089 | 0,12015539 | LOC101927011 | FALSE | Upregulated |
| -0,458393088 | 0,12015539 | LOC143666 | FALSE | Downregulated |
| 0,25957933 | 0,120169293 | TBC1D16 | FALSE | Upregulated |
| 0,439059939 | 0,120202664 | CKS2 | FALSE | Upregulated |
| 0,34969509 | 0,120224563 | TMEM173 | FALSE | Upregulated |
| 1,423244793 | 0,120233069 | DKFZp686L13185 | FALSE | Upregulated |
| 0,991846921 | 0,12047233 | PVT1 | FALSE | Upregulated |
| 1,541656516 | 0,120547504 | ADAM32 | FALSE | Upregulated |
| -0,836079264 | 0,120563078 | LOC101927121 | FALSE | Downregulated |
| 0,162204452 | 0,120574062 | GGA3 | FALSE | Upregulated |
| -0,122867083 | 0,120637065 | ZFYVE9 | FALSE | Downregulated |
| -0,29951089 | 0,120740284 | MRPL23 | FALSE | Downregulated |
| 2,207617373 | 0,120779822 | GLRA1 | FALSE | Upregulated |
| 1,252412465 | 0,120859999 | LOC102724765 | FALSE | Upregulated |
| 0,300829418 | 0,120859999 | RASGRP3 | FALSE | Upregulated |
| 0,215627844 | 0,1208878 | TMEM9B | FALSE | Upregulated |
| 0,594022568 | 0,120909191 | KIF14 | FALSE | Upregulated |
| -0,178078923 | 0,120930835 | FOXN3 | FALSE | Downregulated |
| 0,253703516 | 0,120934439 | TBC1D10A | FALSE | Upregulated |
| -0,253865076 | 0,120978781 | GGCT | FALSE | Downregulated |
| 1,535238788 | 0,120978781 | LOC101927811 | FALSE | Upregulated |
| 0,705962891 | 0,120990644 | FMO2 | FALSE | Upregulated |
| -0,21786769 | 0,121016389 | CYTH1 | FALSE | Downregulated |
| 0,516908318 | 0,121016389 | HDGFRP3 | FALSE | Upregulated |
| 0,321945814 | 0,121016389 | IFITM3 | FALSE | Upregulated |
| 0,183865043 | 0,121016389 | SAE1 | FALSE | Upregulated |
| 0,263508268 | 0,121016389 | THG1L | FALSE | Upregulated |
| -0,293333956 | 0,12107699 | EPB41L4B | FALSE | Downregulated |
| 0,174879694 | 0,121088558 | ACO2 | FALSE | Upregulated |
| 0,615280088 | 0,12115265 | CPT1C | FALSE | Upregulated |
| 0,395096594 | 0,121220351 | KANK3 | FALSE | Upregulated |
| 0,289773775 | 0,121293781 | SPRY1 | FALSE | Upregulated |

| | | | | |
|---|---|---|---|---|
| 0,150153271 | 0,121387467 | RYBP | FALSE | Upregulated |
| -0,349105858 | 0,121393251 | UPF3B | FALSE | Downregulated |
| 0,217015137 | 0,121581453 | IFNAR2 | FALSE | Upregulated |
| 0,482898092 | 0,121596793 | LOC101928377 | FALSE | Upregulated |
| 0,2964594 | 0,121685947 | PPP2R1B | FALSE | Upregulated |
| -0,137735427 | 0,121745342 | ATP5F1 | FALSE | Downregulated |
| 0,37507041 | 0,121745342 | FAM47E-STBD1 | FALSE | Upregulated |
| -1,38945512 | 0,121745342 | LINC01118 | FALSE | Downregulated |
| -0,421215046 | 0,122088382 | ASB13 | FALSE | Downregulated |
| 0,232165044 | 0,122235479 | KIAA0226 | FALSE | Upregulated |
| -0,424635192 | 0,12223936 | HIST1H2AC | FALSE | Downregulated |
| 0,134296182 | 0,122436139 | DNAJC11 | FALSE | Upregulated |
| -0,210400719 | 0,122436139 | NT5C3B | FALSE | Downregulated |
| 0,139090552 | 0,122501052 | CHD8 | FALSE | Upregulated |
| -0,329982086 | 0,122517163 | DGAT1 | FALSE | Downregulated |
| 1,794609564 | 0,122549785 | TMEM252 | FALSE | Upregulated |
| -0,280230445 | 0,122611826 | SAFB2 | FALSE | Downregulated |
| 0,804661266 | 0,12267027 | CCL3 | FALSE | Upregulated |
| 0,432290085 | 0,12281488 | DFNB31 | FALSE | Upregulated |
| 1,495840895 | 0,123061992 | RPPH1 | FALSE | Upregulated |
| -0,332517148 | 0,123183649 | CEBPA-AS1 | FALSE | Downregulated |
| 0,418326853 | 0,123267946 | INPP5F | FALSE | Upregulated |
| -0,417123946 | 0,123405071 | LGALS1 | FALSE | Downregulated |
| 0,265322632 | 0,123405071 | TMEM104 | FALSE | Upregulated |
| -1,504581848 | 0,123405819 | HEMGN | FALSE | Downregulated |
| 0,398488917 | 0,123405819 | ZNF749 | FALSE | Upregulated |
| 0,703273786 | 0,123421494 | ZNF257 | FALSE | Upregulated |
| -0,362309531 | 0,12344153 | APOC4 | FALSE | Downregulated |
| 0,201649418 | 0,123504273 | POLRMT | FALSE | Upregulated |
| 0,381805642 | 0,123546019 | C1QTNF6 | FALSE | Upregulated |
| 1,714957724 | 0,123546019 | LMO3 | FALSE | Upregulated |
| -0,170760557 | 0,123576293 | RPS6KB1 | FALSE | Downregulated |
| 0,604626901 | 0,123720633 | WDR17 | FALSE | Upregulated |
| 0,629400938 | 0,12398821 | AC159540.1-2 | FALSE | Upregulated |
| 1,296640305 | 0,12399011 | TLL2 | FALSE | Upregulated |
| -0,889537858 | 0,12399645 | ACOT6 | FALSE | Downregulated |
| 2,054489729 | 0,124054723 | NLRP7 | FALSE | Upregulated |
| 1,353984551 | 0,124234827 | S100Z | FALSE | Upregulated |
| -0,173016351 | 0,124264261 | NFIB | FALSE | Downregulated |
| 0,295189756 | 0,124326163 | PLEKHG3 | FALSE | Upregulated |
| -0,379681756 | 0,124326163 | ZNF138 | FALSE | Downregulated |
| 0,505344751 | 0,124393859 | CENPH | FALSE | Upregulated |
| 0,233017232 | 0,124479633 | CXorf56 | FALSE | Upregulated |
| 0,364096222 | 0,124479633 | RGS3 | FALSE | Upregulated |
| 0,336805833 | 0,124509599 | RBL1 | FALSE | Upregulated |
| 1,710325892 | 0,124574449 | LIMS3-LOC440895 | FALSE | Upregulated |
| -1,28906063 | 0,124601444 | DNAI1 | FALSE | Downregulated |
| -1,170526486 | 0,124686253 | UGT2A2 | FALSE | Downregulated |
| -0,195995238 | 0,124785436 | RMND5B | FALSE | Downregulated |
| -0,273753149 | 0,124841988 | INPP5B | FALSE | Downregulated |
| 0,375278984 | 0,124853873 | RCSD1 | FALSE | Upregulated |

| | | | | |
|---|---|---|---|---|
| 0,191839152 | 0,124870988 | MAP3K3 | FALSE | Upregulated |
| -0,216443804 | 0,124870988 | RALGAPA2 | FALSE | Downregulated |
| 0,267152615 | 0,124982456 | FGD5 | FALSE | Upregulated |
| -0,299877221 | 0,125052975 | MFAP1 | FALSE | Downregulated |
| 0,27111156 | 0,125128023 | MIF4GD | FALSE | Upregulated |
| 0,518820501 | 0,125128023 | ZFP82 | FALSE | Upregulated |
| 0,194646099 | 0,125259458 | SLC25A3 | FALSE | Upregulated |
| -0,284793513 | 0,125409508 | ZNF680 | FALSE | Downregulated |
| 0,885370408 | 0,125422251 | GLI1 | FALSE | Upregulated |
| 1,462912434 | 0,125435407 | SNORA70 | FALSE | Upregulated |
| -0,232282107 | 0,125634192 | USP36 | FALSE | Downregulated |
| 0,158159022 | 0,12563948 | YIPF3 | FALSE | Upregulated |
| 0,921867633 | 0,125649199 | FLJ38576 | FALSE | Upregulated |
| 0,718637839 | 0,125656721 | ZBTB12 | FALSE | Upregulated |
| 0,858865293 | 0,125660072 | PKD1L1 | FALSE | Upregulated |
| 0,18388202 | 0,12571392 | ZBTB40 | FALSE | Upregulated |
| 0,598441717 | 0,125717309 | PYCR1 | FALSE | Upregulated |
| 0,364281974 | 0,125772469 | PSMC3IP | FALSE | Upregulated |
| 0,260344743 | 0,125832902 | RBM14 | FALSE | Upregulated |
| -0,644888548 | 0,125923859 | FAM198A | FALSE | Downregulated |
| 0,214315869 | 0,125923859 | NAGPA | FALSE | Upregulated |
| 0,449676834 | 0,126070154 | ZNF132 | FALSE | Upregulated |
| -0,462011578 | 0,12615902 | OSBPL6 | FALSE | Downregulated |
| 0,840797785 | 0,126215588 | ANKRD18A | FALSE | Upregulated |
| 0,362699084 | 0,126215588 | NID2 | FALSE | Upregulated |
| 0,207288653 | 0,126332645 | CCM2 | FALSE | Upregulated |
| 1,089105049 | 0,126367307 | FCRL1 | FALSE | Upregulated |
| 0,383907993 | 0,126394911 | ECSCR | FALSE | Upregulated |
| -2,138952817 | 0,126489085 | LOC285556 | FALSE | Downregulated |
| 0,253297373 | 0,126533643 | RUNDC1 | FALSE | Upregulated |
| -2,004662996 | 0,126630671 | LOC101060026 | FALSE | Downregulated |
| -1,583886337 | 0,126764716 | CDHR4 | FALSE | Downregulated |
| 1,59175655 | 0,126942744 | FXYD3 | FALSE | Upregulated |
| 0,307180674 | 0,127120277 | C10orf128 | FALSE | Upregulated |
| -0,157729166 | 0,127120277 | FTSJ2 | FALSE | Downregulated |
| 0,245467226 | 0,127120277 | ZNF555 | FALSE | Upregulated |
| -0,582655089 | 0,127205511 | LOC100506060 | FALSE | Downregulated |
| 0,186261006 | 0,127205511 | RBM19 | FALSE | Upregulated |
| 0,204419388 | 0,127267967 | COL18A1 | FALSE | Upregulated |
| 0,134166481 | 0,127293871 | STIM1 | FALSE | Upregulated |
| 0,441148413 | 0,127495925 | SOD2 | FALSE | Upregulated |
| 0,24094273 | 0,127527913 | RBM15 | FALSE | Upregulated |
| -0,358322372 | 0,127565609 | CPEB3 | FALSE | Downregulated |
| 0,419069463 | 0,127565609 | TMEM17 | FALSE | Upregulated |
| -0,406535408 | 0,127722295 | RORA | FALSE | Downregulated |
| 0,132064074 | 0,127942186 | FAM104A | FALSE | Upregulated |
| 0,496056023 | 0,127974027 | FRMD3 | FALSE | Upregulated |
| 0,293190478 | 0,127974027 | LINC00843 | FALSE | Upregulated |
| -0,16338305 | 0,127974027 | MED14 | FALSE | Downregulated |
| -0,235779564 | 0,127974027 | NPRL2 | FALSE | Downregulated |
| -0,287778387 | 0,127974027 | TGDS | FALSE | Downregulated |

| | | | | |
|---|---|---|---|---|
| -0,204072913 | 0,128208703 | LRRC40 | FALSE | Downregulated |
| 0,206450115 | 0,128317694 | DNAJB14 | FALSE | Upregulated |
| 0,334286091 | 0,128331052 | CYP1B1 | FALSE | Upregulated |
| 0,230783468 | 0,128478068 | ODF2 | FALSE | Upregulated |
| -0,309159872 | 0,128509499 | C9orf72 | FALSE | Downregulated |
| 0,380295467 | 0,12862041 | LYSMD2 | FALSE | Upregulated |
| -0,262767666 | 0,128648352 | AKAP9 | FALSE | Downregulated |
| 0,256157663 | 0,128701471 | ABLIM1 | FALSE | Upregulated |
| -1,361697494 | 0,128701471 | RP11-486A14.2 | FALSE | Downregulated |
| -0,875814815 | 0,128814503 | GPR88 | FALSE | Downregulated |
| -0,213812394 | 0,129162085 | IBA57 | FALSE | Downregulated |
| 0,162160888 | 0,129224118 | DYNLL2 | FALSE | Upregulated |
| -0,231239246 | 0,129258147 | SMYD3 | FALSE | Downregulated |
| 0,221269153 | 0,129300534 | TLE4 | FALSE | Upregulated |
| 1,23528453 | 0,129438698 | TEKT3 | FALSE | Upregulated |
| 0,153723479 | 0,129573366 | TFDP1 | FALSE | Upregulated |
| 0,380367329 | 0,129575948 | FAM198B | FALSE | Upregulated |
| 0,546552075 | 0,129689093 | LOC374443 | FALSE | Upregulated |
| -0,291851399 | 0,129703028 | FAM84B | FALSE | Downregulated |
| 4,362138994 | 0,129715002 | RNU1-4 | FALSE | Upregulated |
| -0,232845421 | 0,129899439 | MIA3 | FALSE | Downregulated |
| 0,596792534 | 0,129917464 | AIF1L | FALSE | Upregulated |
| 0,494204666 | 0,129917464 | EOMES | FALSE | Upregulated |
| 1,179071292 | 0,129927216 | XCR1 | FALSE | Upregulated |
| 2,230205133 | 0,130024432 | C6orf222 | FALSE | Upregulated |
| -2,027172063 | 0,130024432 | CASQ2 | FALSE | Downregulated |
| -1,516907759 | 0,130024432 | LOC102723842 | FALSE | Downregulated |
| -0,210680215 | 0,130024432 | PPM1B | FALSE | Downregulated |
| 0,184165054 | 0,130133367 | PTPRF | FALSE | Upregulated |
| 0,617423721 | 0,130231914 | MFSD2A | FALSE | Upregulated |
| 0,343152977 | 0,130231914 | PLSCR1 | FALSE | Upregulated |
| 1,287238705 | 0,1302978 | AK5 | FALSE | Upregulated |
| -0,315433167 | 0,130344909 | GFRA1 | FALSE | Downregulated |
| 0,097981279 | 0,130367472 | EIF4G2 | FALSE | Upregulated |
| 0,567444188 | 0,130714256 | HAS3 | FALSE | Upregulated |
| 0,146834614 | 0,130714256 | MED1 | FALSE | Upregulated |
| 0,531376785 | 0,131118528 | CLEC12A | FALSE | Upregulated |
| 1,257336848 | 0,131210959 | ATP8A2 | FALSE | Upregulated |
| 0,160607732 | 0,131210959 | ETF1 | FALSE | Upregulated |
| -0,173777364 | 0,131210959 | FAM200B | FALSE | Downregulated |
| 0,293742775 | 0,131210959 | OCLN | FALSE | Upregulated |
| 1,108748313 | 0,131329469 | CASP5 | FALSE | Upregulated |
| 0,563150249 | 0,131329469 | MCTP2 | FALSE | Upregulated |
| 0,480761241 | 0,131367137 | C10orf10 | FALSE | Upregulated |
| -0,257000572 | 0,131435566 | SERPINA4 | FALSE | Downregulated |
| -0,160293838 | 0,131520194 | SNX25 | FALSE | Downregulated |
| 0,360673848 | 0,131520194 | ZNF250 | FALSE | Upregulated |
| -0,210065992 | 0,13153652 | TMBIM4 | FALSE | Downregulated |
| -0,214534662 | 0,131551212 | GIGYF2 | FALSE | Downregulated |
| -1,346825668 | 0,13157807 | GSTT2B | FALSE | Downregulated |
| -0,195781916 | 0,13180108 | YOD1 | FALSE | Downregulated |

| | | | | |
|---|---|---|---|---|
| 1,129005557 | 0,131833053 | MORN3 | FALSE | Upregulated |
| -1,831592327 | 0,131854655 | LOC644086 | FALSE | Downregulated |
| 0,259510751 | 0,131929154 | BLZF1 | FALSE | Upregulated |
| -0,232243998 | 0,131940032 | BOLA1 | FALSE | Downregulated |
| -2,219831385 | 0,13197586 | LOC101928963 | FALSE | Downregulated |
| 0,185028878 | 0,13197586 | PPP2R4 | FALSE | Upregulated |
| -0,262509123 | 0,132054939 | DEXI | FALSE | Downregulated |
| 0,145015054 | 0,132158525 | TBC1D1 | FALSE | Upregulated |
| 0,361764371 | 0,132165192 | PILRA | FALSE | Upregulated |
| 1,689106244 | 0,132260626 | MRAP2 | FALSE | Upregulated |
| -0,12884484 | 0,132273722 | CLCC1 | FALSE | Downregulated |
| -0,721046694 | 0,132273722 | MSX1 | FALSE | Downregulated |
| -0,505916745 | 0,132346043 | DGAT2 | FALSE | Downregulated |
| -0,635297171 | 0,132468987 | TBC1D3 | FALSE | Downregulated |
| -0,810605988 | 0,13252126 | SCGB3A1 | FALSE | Downregulated |
| -0,183385979 | 0,13252126 | SRSF4 | FALSE | Downregulated |
| 0,72527496 | 0,132558247 | CHRDL2 | FALSE | Upregulated |
| 0,13642477 | 0,132558247 | TRA2B | FALSE | Upregulated |
| -0,161574628 | 0,13257842 | RNF170 | FALSE | Downregulated |
| 0,22075643 | 0,132582254 | HOOK3 | FALSE | Upregulated |
| 0,183667224 | 0,132582254 | RXRB | FALSE | Upregulated |
| -0,259304458 | 0,132582254 | TXNDC16 | FALSE | Downregulated |
| 0,220689253 | 0,132757215 | COQ10B | FALSE | Upregulated |
| 0,726648274 | 0,132757215 | TMEM45B | FALSE | Upregulated |
| -0,291571439 | 0,132803201 | SHMT1 | FALSE | Downregulated |
| 1,109027752 | 0,132860311 | SLC6A19 | FALSE | Upregulated |
| 0,330120557 | 0,132868409 | DENND3 | FALSE | Upregulated |
| -0,537396636 | 0,132881418 | CSRNP3 | FALSE | Downregulated |
| -0,442980924 | 0,132881418 | SOBP | FALSE | Downregulated |
| 0,283840023 | 0,132892413 | EPHA1 | FALSE | Upregulated |
| -0,307708781 | 0,132930252 | C11orf65 | FALSE | Downregulated |
| -0,313151138 | 0,132983266 | PIAS1 | FALSE | Downregulated |
| 1,414148737 | 0,133010961 | LOC102723775 | FALSE | Upregulated |
| -0,165130077 | 0,133100791 | BCL2L13 | FALSE | Downregulated |
| -0,215018293 | 0,133306488 | ZNF253 | FALSE | Downregulated |
| 0,449830979 | 0,133377131 | EXOC3L2 | FALSE | Upregulated |
| 1,114677171 | 0,133393727 | RASD2 | FALSE | Upregulated |
| -0,241053284 | 0,133449834 | MRE11A | FALSE | Downregulated |
| 0,969007346 | 0,133498467 | LOC101927681 | FALSE | Upregulated |
| -2,815324953 | 0,133533444 | C10orf71 | FALSE | Downregulated |
| 0,293082707 | 0,133577506 | C20orf112 | FALSE | Upregulated |
| -0,273707852 | 0,13365783 | ALKBH6 | FALSE | Downregulated |
| 0,359945378 | 0,133730243 | ZNF845 | FALSE | Upregulated |
| 1,799458945 | 0,133807503 | LY86-AS1 | FALSE | Upregulated |
| 0,908450965 | 0,133843938 | LOC102724436 | FALSE | Upregulated |
| -0,186672888 | 0,133843938 | NHLRC3 | FALSE | Downregulated |
| 0,229936969 | 0,133887868 | SLC25A28 | FALSE | Upregulated |
| -0,747059659 | 0,13420907 | G6PC | FALSE | Downregulated |
| 0,454242948 | 0,134235875 | LILRA2 | FALSE | Upregulated |
| 1,406090959 | 0,134243575 | CLNK | FALSE | Upregulated |
| 0,120495986 | 0,134291687 | TERF2 | FALSE | Upregulated |

| | | | | |
|---|---|---|---|---|
| -0,189874252 | 0,134409622 | C12orf65 | FALSE | Downregulated |
| 0,406362416 | 0,134461115 | AMIGO1 | FALSE | Upregulated |
| -0,157200044 | 0,134631468 | POLR2C | FALSE | Downregulated |
| 0,59820021 | 0,134883892 | ANXA2R | FALSE | Upregulated |
| -0,814212417 | 0,134883892 | GRM8 | FALSE | Downregulated |
| -0,703690335 | 0,134883892 | LOC728554 | FALSE | Downregulated |
| 1,592381929 | 0,134894955 | OLAH | FALSE | Upregulated |
| 0,251044166 | 0,134931864 | TMEM175 | FALSE | Upregulated |
| 1,636372224 | 0,13495803 | LOC101927085 | FALSE | Upregulated |
| -0,237395932 | 0,135056921 | EIF1AX | FALSE | Downregulated |
| 0,200527052 | 0,135056921 | PICK1 | FALSE | Upregulated |
| -0,21121844 | 0,135056921 | ZRANB2 | FALSE | Downregulated |
| 0,254792953 | 0,135130066 | DOLK | FALSE | Upregulated |
| 0,205061785 | 0,135139395 | RAN | FALSE | Upregulated |
| -0,987128075 | 0,135279279 | LOC101927369 | FALSE | Downregulated |
| 1,45383446 | 0,135306974 | LOC101927366 | FALSE | Upregulated |
| -0,157868352 | 0,135312857 | ATL2 | FALSE | Downregulated |
| 0,240651642 | 0,135384803 | P4HB | FALSE | Upregulated |
| -0,499875888 | 0,135426096 | NPIPA5 | FALSE | Downregulated |
| 2,02534005 | 0,135426096 | PRPH | FALSE | Upregulated |
| 2,191965455 | 0,135542455 | NCCRP1 | FALSE | Upregulated |
| 2,012278095 | 0,135766616 | UBE2MP1 | FALSE | Upregulated |
| -0,244803707 | 0,135808118 | GPS2 | FALSE | Downregulated |
| -0,319694939 | 0,135808118 | SKIDA1 | FALSE | Downregulated |
| 0,146043632 | 0,135819611 | UEVLD | FALSE | Upregulated |
| -0,249157409 | 0,135901261 | ALDH5A1 | FALSE | Downregulated |
| 0,282733859 | 0,135905429 | KIAA1598 | FALSE | Upregulated |
| -0,133342857 | 0,135905429 | TOM1L2 | FALSE | Downregulated |
| -0,848841043 | 0,135928429 | ATP6V1G2 | FALSE | Downregulated |
| -0,706548643 | 0,135950258 | EVPLL | FALSE | Downregulated |
| 0,722030762 | 0,136025549 | LRCH2 | FALSE | Upregulated |
| -0,185161355 | 0,136081418 | CETN2 | FALSE | Downregulated |
| 0,3729555 | 0,136101042 | CYBB | FALSE | Upregulated |
| -0,666079219 | 0,136163233 | HAPLN4 | FALSE | Downregulated |
| 1,037727443 | 0,136175299 | KEL | FALSE | Upregulated |
| -0,261247632 | 0,136194697 | GMFG | FALSE | Downregulated |
| 0,194270502 | 0,136244126 | ISOC1 | FALSE | Upregulated |
| 0,811966325 | 0,136277817 | NCR3LG1 | FALSE | Upregulated |
| 1,771338806 | 0,13629399 | DNAH9 | FALSE | Upregulated |
| 2,643095425 | 0,136317914 | RNU11 | FALSE | Upregulated |
| 0,173200102 | 0,136332077 | PRMT2 | FALSE | Upregulated |
| -0,163377314 | 0,136332136 | SPTY2D1 | FALSE | Downregulated |
| -0,187281959 | 0,136342789 | NRBF2 | FALSE | Downregulated |
| -0,494954888 | 0,136342789 | SYT7 | FALSE | Downregulated |
| 0,71475186 | 0,136357154 | HRCT1 | FALSE | Upregulated |
| 0,18188166 | 0,136548804 | ALCAM | FALSE | Upregulated |
| 0,483540056 | 0,136576143 | DAPP1 | FALSE | Upregulated |
| -1,02883664 | 0,136576143 | FKBP5 | FALSE | Downregulated |
| -0,143570953 | 0,136673718 | SART3 | FALSE | Downregulated |
| 0,253144998 | 0,136767375 | DRG2 | FALSE | Upregulated |
| -0,413031656 | 0,136798832 | RTTN | FALSE | Downregulated |

| | | | | |
|---|---|---|---|---|
| -0,584352612 | 0,136979071 | LOC102724870 | FALSE | Downregulated |
| 0,596905194 | 0,1371101 | DEFB1 | FALSE | Upregulated |
| 1,681605823 | 0,137179337 | LOC101060363 | FALSE | Upregulated |
| 0,226413955 | 0,137179337 | TP53RK | FALSE | Upregulated |
| -1,525716412 | 0,137254805 | LOC102724484 | FALSE | Downregulated |
| -0,302217921 | 0,13729481 | GRAMD1C | FALSE | Downregulated |
| -2,152858351 | 0,137328299 | NXPH1 | FALSE | Downregulated |
| -0,459555669 | 0,137375626 | RP5-968D22.1 | FALSE | Downregulated |
| 0,362083868 | 0,137376727 | TMED3 | FALSE | Upregulated |
| 0,163897832 | 0,137405909 | ARPC4 | FALSE | Upregulated |
| -0,17262017 | 0,137515861 | BIRC2 | FALSE | Downregulated |
| 0,183924687 | 0,137528231 | ABCF2 | FALSE | Upregulated |
| -0,16783264 | 0,137528231 | DDHD2 | FALSE | Downregulated |
| 0,188860985 | 0,137528231 | ZDHHC7 | FALSE | Upregulated |
| 0,146412534 | 0,137562889 | UTP18 | FALSE | Upregulated |
| 0,771644053 | 0,137590105 | LOC102724356 | FALSE | Upregulated |
| 0,130154053 | 0,137596468 | HEATR1 | FALSE | Upregulated |
| 0,107931239 | 0,137616915 | DDB1 | FALSE | Upregulated |
| -0,179829324 | 0,137775705 | NR5A2 | FALSE | Downregulated |
| -0,60673345 | 0,137797149 | LOC101927287 | FALSE | Downregulated |
| 1,178641508 | 0,137797149 | POM121L10P | FALSE | Upregulated |
| 0,631631456 | 0,137821997 | CCDC65 | FALSE | Upregulated |
| -0,172010546 | 0,137929977 | NPM1 | FALSE | Downregulated |
| 0,113418996 | 0,137974129 | GNB1 | FALSE | Upregulated |
| -1,311041651 | 0,137974129 | LOC102724336 | FALSE | Downregulated |
| 0,13789465 | 0,137974129 | SEPT2 | FALSE | Upregulated |
| -0,405736832 | 0,138115765 | C8orf49 | FALSE | Downregulated |
| 0,226890732 | 0,138115765 | MINA | FALSE | Upregulated |
| -0,678589324 | 0,13815786 | LINC00337 | FALSE | Downregulated |
| 0,319719847 | 0,138218428 | RBMS3 | FALSE | Upregulated |
| -2,058913529 | 0,138333073 | LOC101929008 | FALSE | Downregulated |
| -0,197972274 | 0,138334198 | ELMO1 | FALSE | Downregulated |
| 0,291769731 | 0,138396749 | C7orf60 | FALSE | Upregulated |
| 0,400426927 | 0,138631747 | NPR2 | FALSE | Upregulated |
| -0,130903729 | 0,138664041 | TAF5L | FALSE | Downregulated |
| -0,187451803 | 0,138664041 | WDR83OS | FALSE | Downregulated |
| 0,188001383 | 0,138737539 | RABGGTA | FALSE | Upregulated |
| -0,175256021 | 0,138737539 | ZNF441 | FALSE | Downregulated |
| 0,154957524 | 0,138815151 | CHTOP | FALSE | Upregulated |
| 0,199290535 | 0,13901758 | SMYD4 | FALSE | Upregulated |
| 1,35462177 | 0,139070757 | FAM182A | FALSE | Upregulated |
| -1,231787418 | 0,139188932 | TEKT4 | FALSE | Downregulated |
| 0,607770995 | 0,139193512 | LINC00861 | FALSE | Upregulated |
| 0,153218891 | 0,139193512 | TPD52L2 | FALSE | Upregulated |
| 1,830689549 | 0,139193521 | LOC100507387 | FALSE | Upregulated |
| -0,266019232 | 0,139261088 | NSUN6 | FALSE | Downregulated |
| 0,23124365 | 0,139292621 | ABL2 | FALSE | Upregulated |
| -0,274687661 | 0,139292621 | NAA16 | FALSE | Downregulated |
| 0,559859341 | 0,139314697 | LOC646021 | FALSE | Upregulated |
| 0,215143489 | 0,139352969 | ALG5 | FALSE | Upregulated |
| -0,145625319 | 0,139352969 | BRD7 | FALSE | Downregulated |

| | | | | |
|---|---|---|---|---|
| 0,205562869 | 0,139355804 | CHMP6 | FALSE | Upregulated |
| 2,281486381 | 0,139473994 | FSTL4 | FALSE | Upregulated |
| 1,762280965 | 0,139473994 | KCNJ13 | FALSE | Upregulated |
| -0,25478957 | 0,139473994 | SOWAHB | FALSE | Downregulated |
| -0,189234319 | 0,139473994 | VKORC1L1 | FALSE | Downregulated |
| -0,181730318 | 0,139473994 | ZNF721 | FALSE | Downregulated |
| -0,320572181 | 0,1394765 | DHX58 | FALSE | Downregulated |
| -0,421309435 | 0,1394765 | LPAL2 | FALSE | Downregulated |
| 0,639596905 | 0,1394765 | RAD51AP1 | FALSE | Upregulated |
| -1,029916191 | 0,139497377 | LINC00940 | FALSE | Downregulated |
| -0,187310607 | 0,139505588 | AK3 | FALSE | Downregulated |
| 0,160011027 | 0,139505588 | APPBP2 | FALSE | Upregulated |
| 0,240305461 | 0,139505588 | C9orf69 | FALSE | Upregulated |
| 0,209702299 | 0,139505588 | CRTC3 | FALSE | Upregulated |
| 0,110445909 | 0,139505588 | DCTN1 | FALSE | Upregulated |
| 0,215606819 | 0,139505588 | DPH3 | FALSE | Upregulated |
| -1,846323172 | 0,139505588 | LOC102724694 | FALSE | Downregulated |
| -0,187325676 | 0,139505588 | PIK3CA | FALSE | Downregulated |
| 0,245383691 | 0,139505588 | SEC22C | FALSE | Upregulated |
| -0,268261456 | 0,139505588 | WHAMM | FALSE | Downregulated |
| 0,949090203 | 0,139592627 | C16orf98 | FALSE | Upregulated |
| 0,609505145 | 0,139632143 | GPR143 | FALSE | Upregulated |
| -0,203056414 | 0,139681277 | AGTR1 | FALSE | Downregulated |
| -1,823604395 | 0,139904364 | LOC101928212 | FALSE | Downregulated |
| -0,20925753 | 0,139944203 | SEMA4G | FALSE | Downregulated |
| 1,177184153 | 0,139948703 | GK3P | FALSE | Upregulated |
| 0,24483778 | 0,139969108 | DAXX | FALSE | Upregulated |
| 0,633847629 | 0,139989994 | C15orf60 | FALSE | Upregulated |
| 0,185892493 | 0,140133087 | TGFBRAP1 | FALSE | Upregulated |
| -0,187375466 | 0,140171934 | WBSCR22 | FALSE | Downregulated |
| 0,632176662 | 0,140172348 | HMCN2 | FALSE | Upregulated |
| 0,406805044 | 0,140318211 | MXD3 | FALSE | Upregulated |
| 1,613170815 | 0,140319523 | TXLNG2P | FALSE | Upregulated |
| -0,141698302 | 0,140490502 | MAGOH | FALSE | Downregulated |
| 0,412781194 | 0,140508514 | TRIM69 | FALSE | Upregulated |
| -0,244634191 | 0,140547649 | ANGEL1 | FALSE | Downregulated |
| 0,331346187 | 0,140547649 | CCBE1 | FALSE | Upregulated |
| 1,16002578 | 0,140547649 | STYK1 | FALSE | Upregulated |
| -0,349589804 | 0,140560095 | U2AF1L4 | FALSE | Downregulated |
| -0,19108188 | 0,140820439 | PMM1 | FALSE | Downregulated |
| -0,162174642 | 0,140849334 | ENSA | FALSE | Downregulated |
| -0,211834693 | 0,140906357 | ZNF174 | FALSE | Downregulated |
| -0,155675434 | 0,141089986 | DCTN6 | FALSE | Downregulated |
| 0,372442335 | 0,141145337 | OPHN1 | FALSE | Upregulated |
| 0,148106268 | 0,14116625 | RNF6 | FALSE | Upregulated |
| 0,808556285 | 0,14124329 | LOC102724141 | FALSE | Upregulated |
| 0,332539098 | 0,141399119 | RPS27L | FALSE | Upregulated |
| -0,854667794 | 0,141447418 | CYP17A1 | FALSE | Downregulated |
| 1,181804195 | 0,141525436 | LOC101927070 | FALSE | Upregulated |
| 0,664343284 | 0,14164498 | EPPK1 | FALSE | Upregulated |
| -0,875988509 | 0,14164498 | UBQLNL | FALSE | Downregulated |

| | | | | |
|---|---|---|---|---|
| 0,323902196 | 0,141722599 | CTR9 | FALSE | Upregulated |
| 0,701002141 | 0,141810714 | CD40LG | FALSE | Upregulated |
| -1,569606334 | 0,142062954 | LOC101928430 | FALSE | Downregulated |
| 0,283424723 | 0,142081679 | ENPP4 | FALSE | Upregulated |
| 0,311720236 | 0,142116191 | UNC119 | FALSE | Upregulated |
| -0,144620111 | 0,142119722 | RAB2A | FALSE | Downregulated |
| -0,149045597 | 0,142189418 | SMARCB1 | FALSE | Downregulated |
| -0,190856523 | 0,142264535 | PIH1D1 | FALSE | Downregulated |
| 1,154843335 | 0,14227366 | CTLA4 | FALSE | Upregulated |
| 0,465772667 | 0,14227366 | ZSWIM4 | FALSE | Upregulated |
| -0,451558647 | 0,14236335 | SLC35E2 | FALSE | Downregulated |
| 2,873840821 | 0,142380159 | POTEF | FALSE | Upregulated |
| 0,203770823 | 0,142428126 | AGAP1 | FALSE | Upregulated |
| 0,804214265 | 0,142428126 | RAB39B | FALSE | Upregulated |
| 1,450666546 | 0,142462302 | AC131097.3 | FALSE | Upregulated |
| -0,163126596 | 0,142472648 | BCLAF1 | FALSE | Downregulated |
| 0,29175246 | 0,142489051 | OGFRL1 | FALSE | Upregulated |
| -0,438214033 | 0,142548467 | LOC102724795 | FALSE | Downregulated |
| 0,196964264 | 0,14260473 | PACS2 | FALSE | Upregulated |
| 0,238793328 | 0,142631537 | LAMC1 | FALSE | Upregulated |
| -1,625098691 | 0,142816292 | ABCA17P | FALSE | Downregulated |
| 0,270073106 | 0,142816292 | GUCY1B3 | FALSE | Upregulated |
| -0,251620411 | 0,142816292 | SLC23A1 | FALSE | Downregulated |
| 0,724600196 | 0,14282839 | FAM211A | FALSE | Upregulated |
| -0,227994635 | 0,142904945 | DYNC2LI1 | FALSE | Downregulated |
| 0,348061078 | 0,142904945 | SLC7A7 | FALSE | Upregulated |
| -1,100013241 | 0,142963295 | LOC101929070 | FALSE | Downregulated |
| 0,544479201 | 0,143382233 | C3orf14 | FALSE | Upregulated |
| -2,649939879 | 0,143509925 | CHRNA1 | FALSE | Downregulated |
| -0,291658283 | 0,143509925 | SLC22A7 | FALSE | Downregulated |
| 0,128973598 | 0,143570667 | RAB5C | FALSE | Upregulated |
| -0,3793299 | 0,143570667 | SLC29A1 | FALSE | Downregulated |
| 0,474672975 | 0,143570667 | TPM1 | FALSE | Upregulated |
| -0,248968361 | 0,143578055 | LOC148413 | FALSE | Downregulated |
| -0,304418053 | 0,143660921 | ARSD | FALSE | Downregulated |
| 2,516596846 | 0,143660921 | RP11-193H5.1 | FALSE | Upregulated |
| -0,75771502 | 0,143737391 | RP11-101E14.2 | FALSE | Downregulated |
| -0,637371553 | 0,143737391 | SERHL2 | FALSE | Downregulated |
| -0,70356605 | 0,14386647 | CAPN12 | FALSE | Downregulated |
| -0,457287881 | 0,14386647 | HYAL3 | FALSE | Downregulated |
| 0,433435817 | 0,144010168 | ZNF8 | FALSE | Upregulated |
| 1,288518643 | 0,144058131 | RIMBP3B | FALSE | Upregulated |
| 0,42265973 | 0,14415221 | FLVCR2 | FALSE | Upregulated |
| 0,164006679 | 0,144159897 | EED | FALSE | Upregulated |
| -1,062321252 | 0,144159897 | FAM69C | FALSE | Downregulated |
| 0,986151968 | 0,144159897 | FOSB | FALSE | Upregulated |
| 0,709804473 | 0,144363113 | BEAN1 | FALSE | Upregulated |
| 0,317967435 | 0,144383435 | FAM86C1 | FALSE | Upregulated |
| -0,330507028 | 0,144449558 | ACACB | FALSE | Downregulated |
| 0,977235588 | 0,144449558 | GPR35 | FALSE | Upregulated |
| -0,870860829 | 0,144557758 | UGT2B17 | FALSE | Downregulated |

| | | | | |
|---|---|---|---|---|
| -0,259498544 | 0,144643668 | OTC | FALSE | Downregulated |
| -0,329551251 | 0,14466433 | TYROBP | FALSE | Downregulated |
| -0,280035057 | 0,14468633 | CARS | FALSE | Downregulated |
| 2,076717497 | 0,144722389 | KCNA2 | FALSE | Upregulated |
| -0,157406112 | 0,144722389 | PEX14 | FALSE | Downregulated |
| -0,244297654 | 0,144822764 | ABHD8 | FALSE | Downregulated |
| 1,087591577 | 0,144960279 | SHC4 | FALSE | Upregulated |
| 0,277681314 | 0,145092122 | TTL | FALSE | Upregulated |
| -0,212820158 | 0,145154934 | DDX55 | FALSE | Downregulated |
| -3,270557923 | 0,145199513 | TRNAD14 | FALSE | Downregulated |
| 0,171121933 | 0,145270701 | TXNRD1 | FALSE | Upregulated |
| 0,801951295 | 0,145339995 | LOC102724792 | FALSE | Upregulated |
| -0,166761418 | 0,145341902 | TAPT1 | FALSE | Downregulated |
| -0,201397315 | 0,145472371 | CLP1 | FALSE | Downregulated |
| -0,580779546 | 0,145472371 | LINC00115 | FALSE | Downregulated |
| -1,163138543 | 0,145472371 | TRNL2 | FALSE | Downregulated |
| -1,844430889 | 0,145570865 | LOC101927999 | FALSE | Downregulated |
| -0,124488655 | 0,145585417 | CAPN7 | FALSE | Downregulated |
| -0,197813612 | 0,145585417 | ZNF805 | FALSE | Downregulated |
| 0,727945916 | 0,145591306 | JAKMIP1 | FALSE | Upregulated |
| -0,277147829 | 0,145591306 | SETD9 | FALSE | Downregulated |
| -0,480230225 | 0,145591306 | TRIM55 | FALSE | Downregulated |
| 0,36512459 | 0,145595028 | ALPL | FALSE | Upregulated |
| 0,26763189 | 0,145595028 | DSP | FALSE | Upregulated |
| -0,420790468 | 0,145595028 | SLC16A1 | FALSE | Downregulated |
| 0,39054038 | 0,145621661 | AURKA | FALSE | Upregulated |
| 0,176926439 | 0,145621661 | FAM13B | FALSE | Upregulated |
| -1,207528242 | 0,145621661 | LOC101927879 | FALSE | Downregulated |
| 0,956258882 | 0,145678432 | ISLR2 | FALSE | Upregulated |
| 0,621592129 | 0,145694709 | PLA2G5 | FALSE | Upregulated |
| 1,149845773 | 0,14571061 | LOC100287387 | FALSE | Upregulated |
| 0,621283778 | 0,145725182 | PMAIP1 | FALSE | Upregulated |
| -1,508033124 | 0,145830953 | C19orf77 | FALSE | Downregulated |
| 0,421699658 | 0,145830953 | CITED4 | FALSE | Upregulated |
| 0,308313496 | 0,145830953 | PPARD | FALSE | Upregulated |
| 0,364341258 | 0,145904773 | TMEM51 | FALSE | Upregulated |
| 2,246899247 | 0,145912468 | GRIN2C | FALSE | Upregulated |
| 0,117762432 | 0,146072252 | TRMT5 | FALSE | Upregulated |
| 0,200080195 | 0,146195585 | GRINA | FALSE | Upregulated |
| 0,524727787 | 0,146238521 | ZC3H12D | FALSE | Upregulated |
| -0,451487991 | 0,146265387 | APOA5 | FALSE | Downregulated |
| 0,445278954 | 0,146351735 | RPL13AP5 | FALSE | Upregulated |
| 0,257925162 | 0,146351735 | TIRAP | FALSE | Upregulated |
| -0,243286103 | 0,146437176 | KDM4B | FALSE | Downregulated |
| 0,209258335 | 0,146444256 | HEATR5A | FALSE | Upregulated |
| 0,763358548 | 0,146700918 | B3GNT5 | FALSE | Upregulated |
| 2,060548704 | 0,1468081 | CHRM3-AS2 | FALSE | Upregulated |
| 0,54899279 | 0,147000877 | EDN1 | FALSE | Upregulated |
| 0,276079716 | 0,147000877 | ZNF74 | FALSE | Upregulated |
| -0,267957131 | 0,147013951 | SEC24B | FALSE | Downregulated |
| -0,139432626 | 0,147071996 | PDS5B | FALSE | Downregulated |

| | | | | |
|---|---|---|---|---|
| 0,203565519 | 0,147206023 | MAGT1 | FALSE | Upregulated |
| -0,293867725 | 0,147206023 | UBALD1 | FALSE | Downregulated |
| 0,282719731 | 0,147218808 | B3GAT3 | FALSE | Upregulated |
| -0,415872369 | 0,147218808 | BLOC1S1-RDH5 | FALSE | Downregulated |
| -0,552757761 | 0,147428881 | HULC | FALSE | Downregulated |
| -0,134972871 | 0,147428881 | ZNF410 | FALSE | Downregulated |
| 0,871363248 | 0,147442381 | TMIGD2 | FALSE | Upregulated |
| -0,678423762 | 0,147447825 | LOC100288842 | FALSE | Downregulated |
| 0,150986914 | 0,147468522 | OTUB1 | FALSE | Upregulated |
| -0,317666881 | 0,147468562 | FBXO48 | FALSE | Downregulated |
| -0,181974454 | 0,147468562 | TAF5 | FALSE | Downregulated |
| 1,173712836 | 0,147478144 | IFNG-AS1 | FALSE | Upregulated |
| 0,651855719 | 0,147521239 | LOC101927263 | FALSE | Upregulated |
| 0,941524753 | 0,147672398 | LOC101929211 | FALSE | Upregulated |
| -0,336973911 | 0,147672398 | MREG | FALSE | Downregulated |
| -0,792069802 | 0,147687542 | RP11-406A20.4 | FALSE | Downregulated |
| 1,538037171 | 0,14779906 | VSIG1 | FALSE | Upregulated |
| -0,178052422 | 0,147945151 | RPL14 | FALSE | Downregulated |
| 0,312184458 | 0,148007482 | SRPRB | FALSE | Upregulated |
| 0,220481058 | 0,148149952 | QSOX2 | FALSE | Upregulated |
| -0,210974475 | 0,148149952 | TADA2A | FALSE | Downregulated |
| -1,023459779 | 0,148198629 | GPR123 | FALSE | Downregulated |
| 0,731675594 | 0,148321683 | LOC101928615 | FALSE | Upregulated |
| 0,252878159 | 0,148383258 | GIMAP2 | FALSE | Upregulated |
| -0,20094013 | 0,148383258 | KIAA1109 | FALSE | Downregulated |
| 0,255219108 | 0,148383258 | MECR | FALSE | Upregulated |
| -0,206230339 | 0,148492917 | TMEM161B | FALSE | Downregulated |
| -0,338564986 | 0,148501925 | LINC00649 | FALSE | Downregulated |
| -0,752217227 | 0,148501925 | LOC101928958 | FALSE | Downregulated |
| -0,166891756 | 0,148558453 | CYC1 | FALSE | Downregulated |
| -0,392803775 | 0,148576798 | HHIP | FALSE | Downregulated |
| -0,243855747 | 0,1487193 | ALG10B | FALSE | Downregulated |
| 0,258956764 | 0,148739475 | CA5B | FALSE | Upregulated |
| -0,55023746 | 0,148739475 | LOC102724105 | FALSE | Downregulated |
| -0,287478082 | 0,148816246 | TOB2 | FALSE | Downregulated |
| -0,431773396 | 0,148857281 | BHMT | FALSE | Downregulated |
| 1,661183077 | 0,148857281 | MYBPC3 | FALSE | Upregulated |
| -0,22809162 | 0,148861983 | PMVK | FALSE | Downregulated |
| -0,79464481 | 0,148893776 | LOC101928325 | FALSE | Downregulated |
| 0,161101889 | 0,148955511 | SGSM3 | FALSE | Upregulated |
| -1,82430646 | 0,149059433 | CCDC116 | FALSE | Downregulated |
| -0,275223588 | 0,149059433 | EPHX1 | FALSE | Downregulated |
| 0,180247038 | 0,149059433 | GALNT11 | FALSE | Upregulated |
| -0,121241646 | 0,149059433 | VAMP7 | FALSE | Downregulated |
| 0,54747419 | 0,149111706 | AP000472.2 | FALSE | Upregulated |
| 1,056852545 | 0,149114109 | CABP7 | FALSE | Upregulated |
| 1,704869025 | 0,149151879 | PTCHD1 | FALSE | Upregulated |
| 0,407142852 | 0,149302339 | C11orf96 | FALSE | Upregulated |
| -0,315666978 | 0,149408514 | GPR116 | FALSE | Downregulated |
| 0,47696333 | 0,149465956 | LOC389906 | FALSE | Upregulated |
| 0,77044712 | 0,149465956 | PIP5K1B | FALSE | Upregulated |

| | | | | |
|---|---|---|---|---|
| 0,127763454 | 0,149465956 | USP39 | FALSE | Upregulated |
| -0,20609376 | 0,149498253 | AATF | FALSE | Downregulated |
| -0,238043804 | 0,149498253 | FAM229B | FALSE | Downregulated |
| -0,267553952 | 0,149498253 | NFXL1 | FALSE | Downregulated |
| 0,375117917 | 0,149498253 | RGS19 | FALSE | Upregulated |
| -0,363119958 | 0,149498253 | SLC38A4 | FALSE | Downregulated |
| 0,218784135 | 0,149498253 | TMEM140 | FALSE | Upregulated |
| -0,389483934 | 0,149571972 | PPIG | FALSE | Downregulated |
| -0,270296689 | 0,149674445 | RSBN1L-AS1 | FALSE | Downregulated |
| -0,240115926 | 0,149718788 | DNAJC1 | FALSE | Downregulated |
| -0,253902451 | 0,149718788 | SAC3D1 | FALSE | Downregulated |
| -0,197732509 | 0,149751712 | ATP6V0E1 | FALSE | Downregulated |
| -2,056411253 | 0,149751712 | LOC101929085 | FALSE | Downregulated |
| 0,206604242 | 0,149769922 | UBXN2A | FALSE | Upregulated |
| -0,307382968 | 0,149830915 | GPD1 | FALSE | Downregulated |
| 0,167899617 | 0,149834955 | U2AF1 | FALSE | Upregulated |
| -0,180369169 | 0,149997979 | CARKD | FALSE | Downregulated |
| -0,708847736 | 0,150021184 | TGFBR3L | FALSE | Downregulated |
| 0,529927673 | 0,150145305 | MAGEE1 | FALSE | Upregulated |
| 1,643522405 | 0,150145305 | ULK4P3 | FALSE | Upregulated |
| 0,538852164 | 0,150163445 | VDR | FALSE | Upregulated |
| 0,679054758 | 0,150198807 | RAD51 | FALSE | Upregulated |
| 1,157402466 | 0,150225713 | TTTY15 | FALSE | Upregulated |
| -0,752257339 | 0,150341477 | SELE | FALSE | Downregulated |
| 0,260767546 | 0,150381327 | MYO7A | FALSE | Upregulated |
| 0,2331855 | 0,150381327 | NRF1 | FALSE | Upregulated |
| 1,674129224 | 0,150389433 | LOC494141 | FALSE | Upregulated |
| 0,910547064 | 0,15050404 | CSPG4 | FALSE | Upregulated |
| 0,621326647 | 0,150548906 | TBC1D30 | FALSE | Upregulated |
| -0,330556085 | 0,150636833 | DHFR | FALSE | Downregulated |
| -0,213954223 | 0,150873841 | ATL3 | FALSE | Downregulated |
| -0,217653063 | 0,150873841 | WHSC1L1 | FALSE | Downregulated |
| 3,217418283 | 0,15096958 | KRT81 | FALSE | Upregulated |
| 1,106717282 | 0,151017271 | ANKS1B | FALSE | Upregulated |
| -0,171418754 | 0,151028196 | DIAPH2 | FALSE | Downregulated |
| 0,714518694 | 0,15110116 | LOC102724451 | FALSE | Upregulated |
| 1,1132922 | 0,151139509 | LINC00853 | FALSE | Upregulated |
| 1,442453039 | 0,151310878 | MPP4 | FALSE | Upregulated |
| -0,905098853 | 0,151375272 | ITGB1BP2 | FALSE | Downregulated |
| -0,295494631 | 0,151423452 | PSMC4 | FALSE | Downregulated |
| 0,3281345150 | 0,151460062 | TBC1D7 | FALSE | Upregulated |
| 0,35043187 | 0,151460062 | XXYLT1 | FALSE | Upregulated |
| -0,108127512 | 0,151595822 | TSN | FALSE | Downregulated |
| 0,169149434 | 0,151595822 | WTAP | FALSE | Upregulated |
| 0,52703861 | 0,151604661 | OXCT1 | FALSE | Upregulated |
| -0,150590861 | 0,151700072 | SLC4A1AP | FALSE | Downregulated |
| 0,210963144 | 0,151807765 | TMCC1 | FALSE | Upregulated |
| -0,241322934 | 0,151978692 | LOC102723773 | FALSE | Downregulated |
| -0,214414169 | 0,152021318 | FAM21C | FALSE | Downregulated |
| 0,172493097 | 0,152049099 | EPG5 | FALSE | Upregulated |
| 0,764748417 | 0,152133394 | ULBP3 | FALSE | Upregulated |

| | | | | |
|---|---|---|---|---|
| -0,289336045 | 0,152165528 | HPRT1 | FALSE | Downregulated |
| -0,351873932 | 0,152172781 | BMP10 | FALSE | Downregulated |
| 0,640005361 | 0,152172781 | SERPINE1 | FALSE | Upregulated |
| -0,172997118 | 0,152269983 | KLHL2 | FALSE | Downregulated |
| -0,76138184 | 0,15232216 | POPDC2 | FALSE | Downregulated |
| -0,314785795 | 0,152381959 | GLCE | FALSE | Downregulated |
| -1,779099575 | 0,152381994 | LOC101928779 | FALSE | Downregulated |
| -0,303301245 | 0,152537248 | QPCTL | FALSE | Downregulated |
| 0,355105723 | 0,152583141 | DOHH | FALSE | Upregulated |
| -0,260557003 | 0,152647535 | GCKR | FALSE | Downregulated |
| 0,180222994 | 0,152647535 | NSF | FALSE | Upregulated |
| -0,290226334 | 0,152663985 | BAZ1B | FALSE | Downregulated |
| 0,147430978 | 0,152752298 | DCP1A | FALSE | Upregulated |
| 1,235869851 | 0,152752298 | LOC100129215 | FALSE | Upregulated |
| 0,40463061 | 0,152752298 | RAB43 | FALSE | Upregulated |
| -0,262910232 | 0,152752298 | TFAP4 | FALSE | Downregulated |
| -0,323424586 | 0,152858825 | ZNF585B | FALSE | Downregulated |
| 0,253720896 | 0,153170294 | VRK1 | FALSE | Upregulated |
| -0,544737045 | 0,153193452 | KCNMB4 | FALSE | Downregulated |
| -0,428431864 | 0,15322141 | CCDC150 | FALSE | Downregulated |
| 1,223757573 | 0,15322141 | CDC25C | FALSE | Upregulated |
| -0,103325613 | 0,15322141 | KCMF1 | FALSE | Downregulated |
| 0,879147803 | 0,15322141 | LOC339192 | FALSE | Upregulated |
| -0,156583077 | 0,15322141 | SSU72 | FALSE | Downregulated |
| -0,212309098 | 0,153280309 | GABPB1-AS1 | FALSE | Downregulated |
| 1,499489899 | 0,15330572 | NXF5 | FALSE | Upregulated |
| 0,844270091 | 0,153394299 | RPL13AP20 | FALSE | Upregulated |
| 0,307206701 | 0,153525805 | HABP4 | FALSE | Upregulated |
| 0,915995452 | 0,153525805 | LOC101928576 | FALSE | Upregulated |
| -0,173283689 | 0,153655007 | CPN2 | FALSE | Downregulated |
| -0,338524889 | 0,153655007 | TOR3A | FALSE | Downregulated |
| -0,885840336 | 0,15366446 | ABCA13 | FALSE | Downregulated |
| 2,007932 | 0,153773848 | PVRL4 | FALSE | Upregulated |
| -1,306026966 | 0,153915899 | EPHA7 | FALSE | Downregulated |
| 0,384787878 | 0,153915899 | NUMBL | FALSE | Upregulated |
| -1,314968969 | 0,153946008 | LOC101927702 | FALSE | Downregulated |
| 0,659055624 | 0,153946008 | ZNRF2P1 | FALSE | Upregulated |
| 2,054977511 | 0,153986276 | CCR8 | FALSE | Upregulated |
| -0,342995938 | 0,153986276 | ZNF467 | FALSE | Downregulated |
| -0,125635979 | 0,153989871 | CDC37 | FALSE | Downregulated |
| 0,375463704 | 0,153989871 | RRN3P3 | FALSE | Upregulated |
| -0,191127269 | 0,15403589 | KAT2B | FALSE | Downregulated |
| 0,287192408 | 0,15408915 | CXCL12 | FALSE | Upregulated |
| -0,20616344 | 0,154238659 | USP33 | FALSE | Downregulated |
| 0,498952484 | 0,154310656 | PLK4 | FALSE | Upregulated |
| 0,42541435 | 0,154409131 | PLA2G4A | FALSE | Upregulated |
| -0,180331425 | 0,154409131 | SLU7 | FALSE | Downregulated |
| -0,231013544 | 0,154409131 | SNRNP25 | FALSE | Downregulated |
| -0,200794961 | 0,154416784 | POP5 | FALSE | Downregulated |
| 2,557182479 | 0,154416784 | RP11-24J23.2 | FALSE | Upregulated |
| 1,865620455 | 0,154449371 | MYT1 | FALSE | Upregulated |

| | | | | |
|---|---|---|---|---|
| 0,147083938 | 0,154592144 | PSMC5 | FALSE | Upregulated |
| 0,318299962 | 0,154676986 | GCA | FALSE | Upregulated |
| 0,392807508 | 0,15476806 | RFX2 | FALSE | Upregulated |
| -0,168212228 | 0,154777883 | TRMT112 | FALSE | Downregulated |
| -0,144975587 | 0,154884721 | MEN1 | FALSE | Downregulated |
| 0,210755073 | 0,154884721 | TRAFD1 | FALSE | Upregulated |
| 0,873976197 | 0,154893809 | ATXN8OS | FALSE | Upregulated |
| 1,639517377 | 0,154963038 | CLEC17A | FALSE | Upregulated |
| -1,057370947 | 0,155048683 | RP11-575F12.1 | FALSE | Downregulated |
| 0,712121064 | 0,155193711 | BNC2 | FALSE | Upregulated |
| -1,063986935 | 0,155193711 | VWDE | FALSE | Downregulated |
| -0,154909762 | 0,155294107 | LOC100129034 | FALSE | Downregulated |
| -0,179467158 | 0,155536654 | BCL7B | FALSE | Downregulated |
| -0,337838625 | 0,15562733 | RP11-372E1.6 | FALSE | Downregulated |
| -0,235028427 | 0,155806139 | CLEC2B | FALSE | Downregulated |
| 0,878050299 | 0,155839377 | EGFR-AS1 | FALSE | Upregulated |
| -0,40798593 | 0,155989951 | FOLH1B | FALSE | Downregulated |
| -0,949187717 | 0,156065387 | SYCE1 | FALSE | Downregulated |
| 0,202422427 | 0,156122296 | BMPR1A | FALSE | Upregulated |
| 0,698869036 | 0,156273956 | SIRPG | FALSE | Upregulated |
| 0,783372544 | 0,156330997 | KIAA1656 | FALSE | Upregulated |
| -0,286859754 | 0,156376217 | HMBS | FALSE | Downregulated |
| -0,206889223 | 0,156376217 | TPD52L1 | FALSE | Downregulated |
| 0,190375002 | 0,156394435 | RPS6KA2 | FALSE | Upregulated |
| 0,592978035 | 0,156394435 | SGIP1 | FALSE | Upregulated |
| 0,437132963 | 0,156503783 | TEX9 | FALSE | Upregulated |
| -0,251231326 | 0,156584914 | GPT | FALSE | Downregulated |
| -0,18484491 | 0,156768151 | IGBP1 | FALSE | Downregulated |
| -0,416143685 | 0,156779709 | RP11-736K20.6 | FALSE | Downregulated |
| 0,329449178 | 0,156812989 | PARVB | FALSE | Upregulated |
| 1,608323342 | 0,156815122 | RBM11 | FALSE | Upregulated |
| 0,401206996 | 0,156924422 | KLF8 | FALSE | Upregulated |
| 0,853846714 | 0,156924422 | SAMD13 | FALSE | Upregulated |
| -0,174462554 | 0,156924422 | SUCLG1 | FALSE | Downregulated |
| 0,427907331 | 0,157034705 | EEPD1 | FALSE | Upregulated |
| 0,242870913 | 0,157067782 | GCLM | FALSE | Upregulated |
| 0,972653414 | 0,157104404 | CTCFL | FALSE | Upregulated |
| -0,258903222 | 0,157104404 | SORL1 | FALSE | Downregulated |
| 0,791764271 | 0,157182831 | LRRN1 | FALSE | Upregulated |
| 0,222664541 | 0,157186664 | GOLGA3 | FALSE | Upregulated |
| -0,369127291 | 0,15721867 | GLT1D1 | FALSE | Downregulated |
| -0,165873822 | 0,157296029 | SLC25A44 | FALSE | Downregulated |
| 1,967628919 | 0,157349637 | LINC00960 | FALSE | Upregulated |
| 0,2224858 | 0,157402287 | GTDC1 | FALSE | Upregulated |
| -0,595172839 | 0,157404975 | LOC101930307 | FALSE | Downregulated |
| -0,221871328 | 0,157410567 | ROCK1 | FALSE | Downregulated |
| 0,827151174 | 0,157472065 | SHC3 | FALSE | Upregulated |
| -0,20056145 | 0,157515421 | TDRP | FALSE | Downregulated |
| 0,486133661 | 0,157516379 | FBXW9 | FALSE | Upregulated |
| -0,314280343 | 0,157535488 | TOPORS-AS1 | FALSE | Downregulated |
| 0,392599606 | 0,157569438 | TSPO | FALSE | Upregulated |

| | | | | |
|---|---|---|---|---|
| 1,090977776 | 0,157594745 | LOC646127 | FALSE | Upregulated |
| 0,32416343 | 0,157675729 | ASAH2B | FALSE | Upregulated |
| 0,680852003 | 0,15772019 | RASGEF1A | FALSE | Upregulated |
| 0,675133395 | 0,157766445 | MINOS1-NBL1 | FALSE | Upregulated |
| -0,585415015 | 0,157766445 | MOGAT1 | FALSE | Downregulated |
| 0,227169856 | 0,157784017 | NME7 | FALSE | Upregulated |
| -0,18355601 | 0,1579389 | RNF214 | FALSE | Downregulated |
| -0,887366196 | 0,157960351 | MESP1 | FALSE | Downregulated |
| 0,538944673 | 0,158076344 | SLC35G2 | FALSE | Upregulated |
| -0,287926008 | 0,15821486 | TPCN2 | FALSE | Downregulated |
| 0,700718424 | 0,158256706 | ADARB2 | FALSE | Upregulated |
| -1,510630262 | 0,158256706 | LOC101927764 | FALSE | Downregulated |
| 0,625736272 | 0,158264352 | RP11-163N6.2 | FALSE | Upregulated |
| 0,3648048 | 0,158304121 | HDAC11 | FALSE | Upregulated |
| -0,353092691 | 0,158429079 | GPM6A | FALSE | Downregulated |
| 0,502193269 | 0,158463283 | EMR1 | FALSE | Upregulated |
| -0,300583428 | 0,158496876 | C9orf37 | FALSE | Downregulated |
| -0,234532864 | 0,158496876 | GRB14 | FALSE | Downregulated |
| -0,209795553 | 0,158496876 | NOC3L | FALSE | Downregulated |
| -0,120090656 | 0,158496876 | SS18 | FALSE | Downregulated |
| 0,360074921 | 0,158518043 | LINC01138 | FALSE | Upregulated |
| -0,435293342 | 0,158531097 | C1orf101 | FALSE | Downregulated |
| -0,151621721 | 0,158581496 | TTC17 | FALSE | Downregulated |
| -0,242234855 | 0,158687793 | FAM213A | FALSE | Downregulated |
| 0,202724902 | 0,158687793 | GMNN | FALSE | Upregulated |
| -0,193003868 | 0,158687793 | ORMDL3 | FALSE | Downregulated |
| 2,751938228 | 0,158866326 | C3orf56 | FALSE | Upregulated |
| -0,560834758 | 0,158892513 | PAX8 | FALSE | Downregulated |
| -0,712274722 | 0,15897449 | LOC101928347 | FALSE | Downregulated |
| -0,5809438 | 0,158976534 | LOC101927009 | FALSE | Downregulated |
| 0,907267706 | 0,158979699 | LOC100506411 | FALSE | Upregulated |
| -0,290470862 | 0,159141609 | ARID4A | FALSE | Downregulated |
| 0,337667963 | 0,159141609 | POLR2J4 | FALSE | Upregulated |
| 0,744636559 | 0,159147188 | LOC102060414 | FALSE | Upregulated |
| 0,198941402 | 0,159187909 | OIP5-AS1 | FALSE | Upregulated |
| -0,22646569 | 0,159201559 | DNAJC2 | FALSE | Downregulated |
| -0,146749366 | 0,159242335 | LARP1 | FALSE | Downregulated |
| -1,00367343 | 0,159242335 | LINC00907 | FALSE | Downregulated |
| 0,262322423 | 0,159270485 | INPP4A | FALSE | Upregulated |
| 0,681743363 | 0,159396132 | QPCT | FALSE | Upregulated |
| 1,973871852 | 0,159443158 | CXXC1P1 | FALSE | Upregulated |
| 0,438801479 | 0,159527264 | TRIM15 | FALSE | Upregulated |
| -0,316077861 | 0,159537367 | DUSP28 | FALSE | Downregulated |
| 0,208274797 | 0,159537367 | PPM1D | FALSE | Upregulated |
| 0,28548132 | 0,159693616 | ARHGEF6 | FALSE | Upregulated |
| 0,177652795 | 0,159693616 | LPP | FALSE | Upregulated |
| -0,279484954 | 0,159712141 | CCPG1 | FALSE | Downregulated |
| 1,199879503 | 0,159712141 | KIR3DL2 | FALSE | Upregulated |
| 0,570564396 | 0,15988211 | FANCE | FALSE | Upregulated |
| 2,279667609 | 0,15988211 | SNORA65 | FALSE | Upregulated |
| 0,203816163 | 0,159908622 | MYBBP1A | FALSE | Upregulated |

| | | | | |
|---|---|---|---|---|
| 0,487361968 | 0,160053654 | CCT6P1 | FALSE | Upregulated |
| 0,535385721 | 0,160062063 | FAM26F | FALSE | Upregulated |
| 1,729509235 | 0,160062063 | KCP | FALSE | Upregulated |
| 1,091756444 | 0,160096189 | E2F2 | FALSE | Upregulated |
| -0,458384097 | 0,160106039 | ARHGAP10 | FALSE | Downregulated |
| -0,278078442 | 0,160202933 | ZNF511 | FALSE | Downregulated |
| 0,245754581 | 0,160293943 | CRYBG3 | FALSE | Upregulated |
| 0,564574539 | 0,16055697 | ANKRD53 | FALSE | Upregulated |
| 0,450919451 | 0,16055697 | CHSY3 | FALSE | Upregulated |
| 0,331300008 | 0,160563098 | WDR25 | FALSE | Upregulated |
| -0,285526633 | 0,16067922 | CPS1 | FALSE | Downregulated |
| 0,85423204 | 0,16067922 | FAM83F | FALSE | Upregulated |
| 0,336229727 | 0,16067922 | HS6ST1 | FALSE | Upregulated |
| -0,361311909 | 0,16067922 | LHX2 | FALSE | Downregulated |
| -0,419924496 | 0,16067922 | RPS26P11 | FALSE | Downregulated |
| 0,956609104 | 0,160755031 | LOC645638 | FALSE | Upregulated |
| 0,747371281 | 0,16082518 | PTCH2 | FALSE | Upregulated |
| 0,526855753 | 0,16082518 | SDK2 | FALSE | Upregulated |
| 0,223060868 | 0,16082518 | SMURF2 | FALSE | Upregulated |
| 0,133791452 | 0,16095311 | KDM4A | FALSE | Upregulated |
| -0,219089991 | 0,160965236 | NDUFB3 | FALSE | Downregulated |
| -0,191454574 | 0,160983747 | EPB41 | FALSE | Downregulated |
| -1,750926785 | 0,16101404 | ANK1 | FALSE | Downregulated |
| 0,211189387 | 0,161069573 | VWA5A | FALSE | Upregulated |
| 0,183912258 | 0,161275794 | ALG12 | FALSE | Upregulated |
| -0,928569937 | 0,161382302 | LOC102724474 | FALSE | Downregulated |
| 0,226163175 | 0,161382302 | THEM6 | FALSE | Upregulated |
| 0,616138926 | 0,161407199 | LOC102724571 | FALSE | Upregulated |
| 0,208061109 | 0,161603301 | ASB1 | FALSE | Upregulated |
| 0,272115247 | 0,161644275 | METTL4 | FALSE | Upregulated |
| 0,415621436 | 0,161644275 | TMEM216 | FALSE | Upregulated |
| -0,131028357 | 0,161718025 | PRKAR2A | FALSE | Downregulated |
| -0,58709199 | 0,161775977 | C14orf180 | FALSE | Downregulated |
| -0,196183866 | 0,161775977 | DIEXF | FALSE | Downregulated |
| -0,430592847 | 0,161775977 | MROH6 | FALSE | Downregulated |
| -0,594033948 | 0,161775977 | NUDT10 | FALSE | Downregulated |
| 0,29357754 | 0,161775977 | PIK3C2B | FALSE | Upregulated |
| -0,216635351 | 0,161775977 | PVRL3 | FALSE | Downregulated |
| -0,172635072 | 0,161775977 | TMEM261 | FALSE | Downregulated |
| 2,264470076 | 0,161826963 | ADIPOQ-AS1 | FALSE | Upregulated |
| -0,279504196 | 0,161880653 | SPTBN2 | FALSE | Downregulated |
| -0,447117571 | 0,162200745 | FBXO15 | FALSE | Downregulated |
| -0,272753678 | 0,162228175 | CNTLN | FALSE | Downregulated |
| 0,13680862 | 0,162449629 | BROX | FALSE | Upregulated |
| -0,434433703 | 0,162490395 | LOC102723954 | FALSE | Downregulated |
| 0,11993322 | 0,162508474 | DVL3 | FALSE | Upregulated |
| 0,392918315 | 0,162508474 | RRP7B | FALSE | Upregulated |
| -0,449378118 | 0,162523209 | DBH-AS1 | FALSE | Downregulated |
| 0,190310955 | 0,162523209 | JOSD1 | FALSE | Upregulated |
| -0,656370393 | 0,162523209 | LOC102724935 | FALSE | Downregulated |
| 0,158835209 | 0,162523209 | PAWR | FALSE | Upregulated |

| | | | | |
|---|---|---|---|---|
| -0,130967854 | 0,162523209 | QRICH1 | FALSE | Downregulated |
| -0,185817697 | 0,162648013 | HPN | FALSE | Downregulated |
| 0,232777698 | 0,162946505 | NDUFB6 | FALSE | Upregulated |
| 0,24634416 | 0,162946505 | PPTC7 | FALSE | Upregulated |
| -4,063212924 | 0,163003646 | LOC100505841 | FALSE | Downregulated |
| 0,298483799 | 0,163067408 | RBP1 | FALSE | Upregulated |
| -0,780003592 | 0,163067592 | CYP3A43 | FALSE | Downregulated |
| -0,183245761 | 0,163163085 | PRDX2 | FALSE | Downregulated |
| -0,159138944 | 0,163180317 | FAM149B1 | FALSE | Downregulated |
| 0,28978389 | 0,163193943 | JRK | FALSE | Upregulated |
| -0,302686964 | 0,163288526 | ADCY1 | FALSE | Downregulated |
| -0,378280919 | 0,163299723 | PDE4DIP | FALSE | Downregulated |
| -0,217504567 | 0,163324008 | AFMID | FALSE | Downregulated |
| 0,563757301 | 0,163559412 | TRIM45 | FALSE | Upregulated |
| -0,134814279 | 0,163627042 | TRMT1L | FALSE | Downregulated |
| 0,154173383 | 0,163649179 | GPAA1 | FALSE | Upregulated |
| -0,151120798 | 0,163722252 | MIB1 | FALSE | Downregulated |
| 0,278892524 | 0,163790114 | CARD16 | FALSE | Upregulated |
| 1,591412233 | 0,163816233 | NYAP2 | FALSE | Upregulated |
| 0,169620216 | 0,163816233 | TMEM168 | FALSE | Upregulated |
| -0,951934506 | 0,163889498 | GUCY2C | FALSE | Downregulated |
| -0,152288159 | 0,163939509 | NDUFA10 | FALSE | Downregulated |
| 1,625134763 | 0,164052506 | LINC01181 | FALSE | Upregulated |
| 1,439856198 | 0,164057633 | LOC388282 | FALSE | Upregulated |
| 0,624504047 | 0,1641286 | CRTAM | FALSE | Upregulated |
| 0,24959559 | 0,1641286 | E4F1 | FALSE | Upregulated |
| 0,155011551 | 0,164190285 | AP1M1 | FALSE | Upregulated |
| 0,227465788 | 0,164190285 | CACUL1 | FALSE | Upregulated |
| -0,452301381 | 0,164190285 | DPF3 | FALSE | Downregulated |
| 0,246186496 | 0,164190285 | MDFIC | FALSE | Upregulated |
| 1,037321828 | 0,164190285 | PPIAL4G | FALSE | Upregulated |
| 0,656092925 | 0,164226359 | HES4 | FALSE | Upregulated |
| -1,491301486 | 0,164414454 | CR1L | FALSE | Downregulated |
| -0,134444902 | 0,164414454 | ZNF641 | FALSE | Downregulated |
| -1,540731441 | 0,164445372 | RSPH10B2 | FALSE | Downregulated |
| 2,034819869 | 0,164446451 | TFAP2C | FALSE | Upregulated |
| 0,219132174 | 0,164614888 | MARK4 | FALSE | Upregulated |
| -0,162763016 | 0,164614888 | MRPL45 | FALSE | Downregulated |
| 0,362530866 | 0,164624164 | SLC4A7 | FALSE | Upregulated |
| 0,539226853 | 0,164736957 | NOVA2 | FALSE | Upregulated |
| -0,539228264 | 0,164773689 | ASPRV1 | FALSE | Downregulated |
| -0,659101585 | 0,164773689 | GSTA3 | FALSE | Downregulated |
| 0,425842129 | 0,164773689 | PPP1R14A | FALSE | Upregulated |
| 0,259796584 | 0,164773689 | SERPINA7 | FALSE | Upregulated |
| -0,163364172 | 0,164908258 | COPS4 | FALSE | Downregulated |
| 0,691375324 | 0,164952336 | PIN4P1 | FALSE | Upregulated |
| -0,217536138 | 0,164952336 | TMA7 | FALSE | Downregulated |
| -0,268007465 | 0,165042157 | GLUD1 | FALSE | Downregulated |
| 0,474581764 | 0,165055119 | LURAP1L | FALSE | Upregulated |
| 0,146289912 | 0,165190078 | RASA1 | FALSE | Upregulated |
| -0,181055164 | 0,165195912 | MTRF1 | FALSE | Downregulated |

| | | | | |
|---|---|---|---|---|
| -0,19732639 | 0,165220014 | GEMIN6 | FALSE | Downregulated |
| 0,427021274 | 0,165220014 | P2RY8 | FALSE | Upregulated |
| 0,244607891 | 0,165220014 | SLC4A4 | FALSE | Upregulated |
| 0,885862801 | 0,165220014 | TG | FALSE | Upregulated |
| 0,214614181 | 0,16548227 | MORF4L2 | FALSE | Upregulated |
| -0,183454962 | 0,16548227 | RC3H1 | FALSE | Downregulated |
| -0,123297511 | 0,165484514 | PHKB | FALSE | Downregulated |
| 0,487263917 | 0,165484514 | ZNF718 | FALSE | Upregulated |
| 0,202464696 | 0,165596001 | SHROOM3 | FALSE | Upregulated |
| 0,177512479 | 0,165707135 | DDX5 | FALSE | Upregulated |
| 0,261147101 | 0,165808709 | JUND | FALSE | Upregulated |
| -2,223835535 | 0,16580908 | ZNF890P | FALSE | Downregulated |
| -0,241574231 | 0,165837727 | SLC40A1 | FALSE | Downregulated |
| -0,792314198 | 0,165876937 | LOC101926977 | FALSE | Downregulated |
| 0,299208678 | 0,165891858 | NAV2 | FALSE | Upregulated |
| 0,731161145 | 0,165940289 | LOC646278 | FALSE | Upregulated |
| 0,834527709 | 0,165961542 | IL23A | FALSE | Upregulated |
| -0,408469718 | 0,165971722 | C16orf95 | FALSE | Downregulated |
| 1,699475417 | 0,166122825 | LOC101927143 | FALSE | Upregulated |
| 0,264249247 | 0,166178243 | MFNG | FALSE | Upregulated |
| -0,776295837 | 0,166178243 | RP11-408H20.2 | FALSE | Downregulated |
| -0,213349642 | 0,166178243 | TPT1 | FALSE | Downregulated |
| 0,392073024 | 0,166178997 | ABHD17C | FALSE | Upregulated |
| 0,352226838 | 0,166395537 | CX3CL1 | FALSE | Upregulated |
| 0,19811469 | 0,166417799 | ALMS1 | FALSE | Upregulated |
| -0,152333075 | 0,166417799 | RSL24D1 | FALSE | Downregulated |
| -0,559555081 | 0,166588203 | LOC101929351 | FALSE | Downregulated |
| -1,578689882 | 0,166682068 | LOC100133182 | FALSE | Downregulated |
| 0,164378054 | 0,16668811 | PARP4 | FALSE | Upregulated |
| 0,487925434 | 0,166762348 | APBA2 | FALSE | Upregulated |
| 0,164495313 | 0,166762348 | CHUK | FALSE | Upregulated |
| 1,838364101 | 0,166762348 | LAMB4 | FALSE | Upregulated |
| -0,856647811 | 0,166762348 | LOC101929506 | FALSE | Downregulated |
| 0,375000594 | 0,166762348 | MAPK13 | FALSE | Upregulated |
| -1,156123922 | 0,166785882 | AC007879.2 | FALSE | Downregulated |
| 0,36881687 | 0,166801804 | FGD2 | FALSE | Upregulated |
| -1,197239143 | 0,166801804 | IL20RB | FALSE | Downregulated |
| 1,882447322 | 0,166801804 | NTNG1 | FALSE | Upregulated |
| 0,220420459 | 0,166805448 | CPNE2 | FALSE | Upregulated |
| -0,19313657 | 0,166843825 | LEO1 | FALSE | Downregulated |
| 0,3610169 | 0,166928232 | DLG3 | FALSE | Upregulated |
| -0,206987385 | 0,166987973 | C12orf76 | FALSE | Downregulated |
| -0,652787574 | 0,167007912 | WBP2NL | FALSE | Downregulated |
| -0,411835738 | 0,167119876 | MRO | FALSE | Downregulated |
| -0,338632528 | 0,16712301 | LOC652276 | FALSE | Downregulated |
| 0,401418189 | 0,167255167 | FCGR2A | FALSE | Upregulated |
| 1,024684546 | 0,167288903 | C3orf80 | FALSE | Upregulated |
| 0,266129554 | 0,167429419 | PPP1R13L | FALSE | Upregulated |
| -0,199491367 | 0,167435058 | HADHB | FALSE | Downregulated |
| 0,370552724 | 0,167476426 | EBF4 | FALSE | Upregulated |
| 1,521780959 | 0,167577012 | MAK | FALSE | Upregulated |

| | | | | |
|---|---|---|---|---|
| -0,241616084 | 0,167658436 | SEPT11 | FALSE | Downregulated |
| 0,592507841 | 0,167673031 | PNMA6A | FALSE | Upregulated |
| -0,646868915 | 0,167777823 | ASB9 | FALSE | Downregulated |
| -0,176839851 | 0,167923076 | SHMT2 | FALSE | Downregulated |
| 1,890271096 | 0,168064434 | IRG1 | FALSE | Upregulated |
| 1,152826586 | 0,168290846 | PRPH2 | FALSE | Upregulated |
| 0,144592376 | 0,168322856 | TBC1D5 | FALSE | Upregulated |
| 0,202457073 | 0,168383869 | CYSTM1 | FALSE | Upregulated |
| 0,233067608 | 0,168387106 | TAF6L | FALSE | Upregulated |
| 0,380310833 | 0,168402471 | SP5 | FALSE | Upregulated |
| -0,152034117 | 0,16851741 | GDAP2 | FALSE | Downregulated |
| 1,46597743 | 0,16851741 | MSTN | FALSE | Upregulated |
| 0,281547705 | 0,16851741 | PPP1R12B | FALSE | Upregulated |
| 0,12307971 | 0,16857912 | FAM168B | FALSE | Upregulated |
| 0,6268875 | 0,16869202 | LOC100131655 | FALSE | Upregulated |
| 0,986583151 | 0,168694206 | SLC29A4 | FALSE | Upregulated |
| 1,458305523 | 0,168694206 | SPINK1 | FALSE | Upregulated |
| 0,180377847 | 0,168847192 | PARS2 | FALSE | Upregulated |
| -0,116209983 | 0,168847192 | QKI | FALSE | Downregulated |
| -0,780816737 | 0,168847192 | SYCE1L | FALSE | Downregulated |
| 0,24738137 | 0,168848367 | FN3KRP | FALSE | Upregulated |
| -0,189515068 | 0,168868924 | PCGF6 | FALSE | Downregulated |
| -1,63024469 | 0,168884779 | ALAS2 | FALSE | Downregulated |
| -0,279332641 | 0,168972123 | TEK | FALSE | Downregulated |
| 0,170588491 | 0,168984997 | NCOA3 | FALSE | Upregulated |
| -0,52902991 | 0,169036764 | PPP4R4 | FALSE | Downregulated |
| 0,330856758 | 0,169053449 | BTK | FALSE | Upregulated |
| 0,362114601 | 0,169191078 | FGL2 | FALSE | Upregulated |
| 0,239958387 | 0,169244743 | NEDD4 | FALSE | Upregulated |
| 1,067203036 | 0,169344823 | CHI3L1 | FALSE | Upregulated |
| 1,551650523 | 0,16943007 | KCNK15 | FALSE | Upregulated |
| 0,516931337 | 0,16944265 | KLHL17 | FALSE | Upregulated |
| 0,192833684 | 0,169535541 | PRMT5 | FALSE | Upregulated |
| 0,120598098 | 0,169535541 | TRAK2 | FALSE | Upregulated |
| -0,336379842 | 0,169545014 | NUDT13 | FALSE | Downregulated |
| 0,672363797 | 0,169593525 | EME1 | FALSE | Upregulated |
| 0,466894548 | 0,169593525 | HYOU1 | FALSE | Upregulated |
| 1,254761325 | 0,169593525 | TMEM31 | FALSE | Upregulated |
| 0,944450374 | 0,169593525 | ULBP2 | FALSE | Upregulated |
| 0,169262268 | 0,169593525 | WDR37 | FALSE | Upregulated |
| 0,187529586 | 0,169593525 | ZBED5 | FALSE | Upregulated |
| 0,254024421 | 0,169631936 | BOK | FALSE | Upregulated |
| 0,711889576 | 0,169679265 | NPM2 | FALSE | Upregulated |
| -0,223897614 | 0,169679265 | TMEM126A | FALSE | Downregulated |
| -0,119712642 | 0,169679265 | YPEL5 | FALSE | Downregulated |
| 0,188554924 | 0,169684931 | TBPL1 | FALSE | Upregulated |
| 0,224469694 | 0,169859447 | KIAA0922 | FALSE | Upregulated |
| 0,228609451 | 0,169873307 | PHF23 | FALSE | Upregulated |
| -0,268436642 | 0,169873307 | SEC11C | FALSE | Downregulated |
| 0,260318382 | 0,169928713 | PPL | FALSE | Upregulated |
| -0,169731496 | 0,169954613 | DCAF17 | FALSE | Downregulated |

| | | | | |
|---|---|---|---|---|
| -0,388616976 | 0,169954613 | SMIM2-AS1 | FALSE | Downregulated |
| 0,40256932 | 0,169954613 | SYTL3 | FALSE | Upregulated |
| -0,137931892 | 0,169971448 | KIAA1429 | FALSE | Downregulated |
| 1,260194375 | 0,17012941 | LOC102724009 | FALSE | Upregulated |
| -0,662187366 | 0,17012941 | RP11-434D9.1 | FALSE | Downregulated |
| -0,54732488 | 0,170173602 | SLC18A2 | FALSE | Downregulated |
| -1,049422051 | 0,170185887 | NPPA-AS1 | FALSE | Downregulated |
| -0,190557752 | 0,170186889 | RSC1A1 | FALSE | Downregulated |
| 0,704961514 | 0,170310121 | C18orf56 | FALSE | Upregulated |
| -0,872644172 | 0,170310121 | LOC101927206 | FALSE | Downregulated |
| -0,341578185 | 0,17032068 | LRRC61 | FALSE | Downregulated |
| -0,559585074 | 0,170480161 | HESX1 | FALSE | Downregulated |
| -0,242171533 | 0,170624303 | ENPEP | FALSE | Downregulated |
| -0,255916505 | 0,170624303 | FOXA1 | FALSE | Downregulated |
| -0,253131723 | 0,170651014 | ZNF433 | FALSE | Downregulated |
| -0,257293785 | 0,170683591 | MYEOV2 | FALSE | Downregulated |
| 1,696550176 | 0,170721785 | HTR1F | FALSE | Upregulated |
| -0,13829352 | 0,170721785 | PSMD6 | FALSE | Downregulated |
| -0,203309854 | 0,170721785 | UQCR10 | FALSE | Downregulated |
| 1,647571462 | 0,170769623 | LOC650368 | FALSE | Upregulated |
| -0,13904773 | 0,170769623 | SLC25A46 | FALSE | Downregulated |
| -0,756326478 | 0,170793262 | MCM3AP-AS1 | FALSE | Downregulated |
| 1,09631061 | 0,170891273 | EDIL3 | FALSE | Upregulated |
| 0,520707612 | 0,171149903 | H2AFY2 | FALSE | Upregulated |
| 1,028953564 | 0,171318385 | AHNAK2 | FALSE | Upregulated |
| 0,283111198 | 0,171318385 | HNRNPA1L2 | FALSE | Upregulated |
| 0,170130367 | 0,171318385 | KLHL22 | FALSE | Upregulated |
| 0,92787637 | 0,17133582 | FCGR1B | FALSE | Upregulated |
| 0,144866309 | 0,171480062 | AP4E1 | FALSE | Upregulated |
| 0,448707997 | 0,171482384 | NEBL | FALSE | Upregulated |
| -0,583179113 | 0,171482384 | PPM1N | FALSE | Downregulated |
| 0,691375588 | 0,171517619 | BACE1-AS | FALSE | Upregulated |
| 0,22306572 | 0,171517619 | DPP7 | FALSE | Upregulated |
| 0,578002592 | 0,171517619 | EFNA3 | FALSE | Upregulated |
| 1,753085896 | 0,171517619 | KRT5 | FALSE | Upregulated |
| -0,430125387 | 0,171517619 | TSPYL2 | FALSE | Downregulated |
| -0,25503898 | 0,171544254 | LOC100506746 | FALSE | Downregulated |
| -0,204434752 | 0,171883333 | AASDH | FALSE | Downregulated |
| 0,137659212 | 0,171883333 | PSMD13 | FALSE | Upregulated |
| -0,892511741 | 0,171950612 | EXTL3-AS1 | FALSE | Downregulated |
| -0,556600909 | 0,171955257 | CFHR4 | FALSE | Downregulated |
| 0,240699308 | 0,171955257 | GIMAP1 | FALSE | Upregulated |
| 0,643998232 | 0,172162282 | LRRC49 | FALSE | Upregulated |
| 1,35449422 | 0,172201775 | IGSF5 | FALSE | Upregulated |
| -0,635048887 | 0,172236222 | LOC100288974 | FALSE | Downregulated |
| 0,13546019 | 0,172279036 | RNH1 | FALSE | Upregulated |
| 0,389051321 | 0,172381145 | MORC4 | FALSE | Upregulated |
| 0,235743639 | 0,172382285 | PIGW | FALSE | Upregulated |
| 0,128260998 | 0,172424009 | NCBP1 | FALSE | Upregulated |
| 1,70838911 | 0,172502921 | CALCA | FALSE | Upregulated |
| -0,572761823 | 0,172502921 | LINC01127 | FALSE | Downregulated |

| | | | | |
|---|---|---|---|---|
| 0,213730481 | 0,17252669 | BRAT1 | FALSE | Upregulated |
| -0,262034222 | 0,17252669 | RBM27 | FALSE | Downregulated |
| 1,352814005 | 0,17254038 | ICAM4 | FALSE | Upregulated |
| 0,574820525 | 0,172540504 | C8orf31 | FALSE | Upregulated |
| 0,51876591 | 0,172540504 | CCM2L | FALSE | Upregulated |
| 0,257533322 | 0,172540504 | ENDOD1 | FALSE | Upregulated |
| 2,509153803 | 0,172540504 | GLB1L3 | FALSE | Upregulated |
| -0,486801208 | 0,172540504 | INHBE | FALSE | Downregulated |
| 2,085975918 | 0,172540504 | LOC101928733 | FALSE | Upregulated |
| -0,311347345 | 0,172540504 | NAALADL2 | FALSE | Downregulated |
| 0,215582023 | 0,172540504 | OSGEP | FALSE | Upregulated |
| -0,191051114 | 0,172540504 | TBC1D4 | FALSE | Downregulated |
| 1,833690571 | 0,172838648 | ERC2 | FALSE | Upregulated |
| 0,535070086 | 0,17293621 | RPL13P5 | FALSE | Upregulated |
| 0,466822338 | 0,173000852 | CDK5R1 | FALSE | Upregulated |
| 2,459771162 | 0,173000852 | LOC643406 | FALSE | Upregulated |
| -0,272891095 | 0,173000852 | MS4A6A | FALSE | Downregulated |
| 1,214456502 | 0,173000852 | TEKT2 | FALSE | Upregulated |
| 0,743448147 | 0,173063489 | FAM83H-AS1 | FALSE | Upregulated |
| 0,187971152 | 0,173063489 | IPMK | FALSE | Upregulated |
| -0,700897382 | 0,173063489 | LOC100506014 | FALSE | Downregulated |
| -0,08795182 | 0,173294227 | RAB5B | FALSE | Downregulated |
| -0,314724697 | 0,17333772 | ECHDC3 | FALSE | Downregulated |
| -0,195152462 | 0,173491778 | NCOA6 | FALSE | Downregulated |
| 0,305989945 | 0,173491778 | PPAPDC1B | FALSE | Upregulated |
| -0,227424282 | 0,173491778 | PSMG1 | FALSE | Downregulated |
| 0,369155051 | 0,173508423 | SIGLEC9 | FALSE | Upregulated |
| 0,862962851 | 0,173508765 | FAM3B | FALSE | Upregulated |
| 0,136376774 | 0,173566195 | RNF114 | FALSE | Upregulated |
| 1,04069782 | 0,173582827 | SLCO4A1 | FALSE | Upregulated |
| 0,210603189 | 0,173615641 | B2M | FALSE | Upregulated |
| 0,17137657 | 0,173615641 | PSMD9 | FALSE | Upregulated |
| -0,207690303 | 0,173710525 | DNAJC22 | FALSE | Downregulated |
| 1,653655711 | 0,173724416 | LCN10 | FALSE | Upregulated |
| 1,303793561 | 0,173724416 | SLC2A1-AS1 | FALSE | Upregulated |
| 1,346938404 | 0,173764346 | FAM171A2 | FALSE | Upregulated |
| 1,569656335 | 0,173831676 | OSTCP2 | FALSE | Upregulated |
| 0,303008561 | 0,17403861 | ZNF500 | FALSE | Upregulated |
| 0,183297104 | 0,174099227 | CLUH | FALSE | Upregulated |
| 0,427618503 | 0,174189847 | ZNF713 | FALSE | Upregulated |
| -0,159480637 | 0,174204939 | AGO4 | FALSE | Downregulated |
| 1,185643731 | 0,174360352 | TMEM178B | FALSE | Upregulated |
| 0,163442996 | 0,174387816 | PTGES2 | FALSE | Upregulated |
| 0,310525626 | 0,174387816 | SESN1 | FALSE | Upregulated |
| 0,18264413 | 0,174392489 | VPS51 | FALSE | Upregulated |
| 0,517596532 | 0,174475631 | FOXD2 | FALSE | Upregulated |
| 0,154035675 | 0,174475631 | PGAM1 | FALSE | Upregulated |
| -0,218851769 | 0,174475964 | MTMR10 | FALSE | Downregulated |
| -0,997298518 | 0,174478604 | HYDIN | FALSE | Downregulated |
| -0,413594802 | 0,174478604 | LOC100272217 | FALSE | Downregulated |
| 0,253227457 | 0,174646165 | DCK | FALSE | Upregulated |

| | | | | |
|---|---|---|---|---|
| -0,202301377 | 0,174646165 | MASP2 | FALSE | Downregulated |
| 0,908492245 | 0,17465452 | LRRN2 | FALSE | Upregulated |
| 0,17141625 | 0,174671165 | CPNE8 | FALSE | Upregulated |
| -1,214901875 | 0,174671165 | LOC101927870 | FALSE | Downregulated |
| 1,328871876 | 0,174753398 | FLJ39095 | FALSE | Upregulated |
| -1,790582531 | 0,174791218 | IDI2 | FALSE | Downregulated |
| 1,781538075 | 0,174852721 | SERPINA9 | FALSE | Upregulated |
| 2,190201325 | 0,174893681 | GRIK3 | FALSE | Upregulated |
| 0,394989626 | 0,175037109 | SDF2L1 | FALSE | Upregulated |
| 1,429168488 | 0,175273021 | EFHC2 | FALSE | Upregulated |
| 0,593688116 | 0,175273021 | KIF2C | FALSE | Upregulated |
| 1,196866506 | 0,175273021 | SLC4A11 | FALSE | Upregulated |
| 0,296778894 | 0,175351868 | CNTN4 | FALSE | Upregulated |
| 0,330076773 | 0,175351868 | MITF | FALSE | Upregulated |
| -0,272016626 | 0,175357018 | C19orf68 | FALSE | Downregulated |
| -0,18010686 | 0,175357018 | MTFR1 | FALSE | Downregulated |
| 0,738993917 | 0,175361321 | LOC388210 | FALSE | Upregulated |
| 0,205331055 | 0,175394497 | LRRFIP1 | FALSE | Upregulated |
| 0,563970924 | 0,175408442 | NECAB2 | FALSE | Upregulated |
| -0,193903179 | 0,175410317 | STK16 | FALSE | Downregulated |
| 1,069641025 | 0,175415896 | ZNF233 | FALSE | Upregulated |
| 1,353384408 | 0,175459894 | SCG3 | FALSE | Upregulated |
| 0,471699373 | 0,175579295 | INPP4B | FALSE | Upregulated |
| 0,978809802 | 0,175599574 | GRIK4 | FALSE | Upregulated |
| 0,186041233 | 0,175647712 | CHD4 | FALSE | Upregulated |
| 1,402054043 | 0,175654566 | SLC10A6 | FALSE | Upregulated |
| 0,713237509 | 0,17569549 | RP11-84C10.4 | FALSE | Upregulated |
| 0,192056643 | 0,17569549 | ZNF76 | FALSE | Upregulated |
| 0,348963179 | 0,17570899 | IFIT3 | FALSE | Upregulated |
| 1,03303693 | 0,175737105 | NPBWR1 | FALSE | Upregulated |
| -0,172494625 | 0,175828549 | KLHDC10 | FALSE | Downregulated |
| 0,621238376 | 0,175830452 | FHL3 | FALSE | Upregulated |
| -1,055549317 | 0,175830452 | LOC101927135 | FALSE | Downregulated |
| 0,119708892 | 0,175830452 | RAP1B | FALSE | Upregulated |
| 0,731315566 | 0,175964036 | FLJ38717 | FALSE | Upregulated |
| 0,365416239 | 0,176012457 | LOC441155 | FALSE | Upregulated |
| -0,450418699 | 0,176040297 | IDI1 | FALSE | Downregulated |
| -0,538332058 | 0,176046863 | LOC102724800 | FALSE | Downregulated |
| 0,290178523 | 0,176078238 | ZNF548 | FALSE | Upregulated |
| -0,166774918 | 0,176193722 | ERI2 | FALSE | Downregulated |
| 0,315738552 | 0,176193722 | FAM122C | FALSE | Upregulated |
| 0,211660038 | 0,176268866 | DUS2 | FALSE | Upregulated |
| 1,954626312 | 0,176268866 | LOC102724865 | FALSE | Upregulated |
| 0,119674491 | 0,176383792 | ABCF3 | FALSE | Upregulated |
| 0,45328676 | 0,176383792 | NCF1C | FALSE | Upregulated |
| 0,240034844 | 0,176416552 | ZNF787 | FALSE | Upregulated |
| 1,253500856 | 0,176499512 | GGTLC1 | FALSE | Upregulated |
| -0,289539571 | 0,176499512 | LOC101929145 | FALSE | Downregulated |
| 0,265065468 | 0,176499512 | SLC26A11 | FALSE | Upregulated |
| 0,337829371 | 0,176712848 | TRAF3IP2 | FALSE | Upregulated |
| 0,280645629 | 0,176717965 | CCDC71L | FALSE | Upregulated |

| | | | | |
|---|---|---|---|---|
| -0,128556485 | 0,176853327 | GNAS | FALSE | Downregulated |
| 1,160437384 | 0,176853327 | MIR143HG | FALSE | Upregulated |
| 0,696686476 | 0,176853327 | SLC6A8 | FALSE | Upregulated |
| -0,350415012 | 0,17686703 | NIPAL1 | FALSE | Downregulated |
| -0,105847759 | 0,176967258 | ARL14EP | FALSE | Downregulated |
| -0,248000595 | 0,176967258 | CYP2C18 | FALSE | Downregulated |
| 0,144415383 | 0,177193881 | C2orf69 | FALSE | Upregulated |
| 0,168341163 | 0,17727523 | SDCBP | FALSE | Upregulated |
| -0,748880054 | 0,17727523 | TBX10 | FALSE | Downregulated |
| -0,142734641 | 0,177410993 | NUDT9 | FALSE | Downregulated |
| 2,058341221 | 0,177427006 | SLC13A1 | FALSE | Upregulated |
| 0,370338831 | 0,177436074 | RCN3 | FALSE | Upregulated |
| -0,243949126 | 0,177436074 | TMEM38B | FALSE | Downregulated |
| 1,734591843 | 0,17749123 | PSG4 | FALSE | Upregulated |
| -0,189746352 | 0,177546209 | IGFBP4 | FALSE | Downregulated |
| 0,242263245 | 0,177546209 | TLR3 | FALSE | Upregulated |
| -1,731177091 | 0,177663139 | LOC101927818 | FALSE | Downregulated |
| -0,800250471 | 0,177663139 | RP11-579D7.4 | FALSE | Downregulated |
| -0,134626313 | 0,17768006 | HELQ | FALSE | Downregulated |
| 0,673919103 | 0,17768006 | HOXA3 | FALSE | Upregulated |
| -0,775502841 | 0,17768006 | HSD17B7P2 | FALSE | Downregulated |
| -0,944618302 | 0,177884325 | C2orf78 | FALSE | Downregulated |
| -0,183912459 | 0,177884325 | SNHG6 | FALSE | Downregulated |
| -1,29578062 | 0,177898703 | LINC00563 | FALSE | Downregulated |
| -0,184157881 | 0,178007678 | KDM1B | FALSE | Downregulated |
| 0,166374973 | 0,178146736 | AGPAT6 | FALSE | Upregulated |
| 0,137979473 | 0,178357029 | CKAP5 | FALSE | Upregulated |
| -0,198050272 | 0,178370772 | TXNRD2 | FALSE | Downregulated |
| 0,154273868 | 0,178373931 | ZBTB6 | FALSE | Upregulated |
| -0,25075312 | 0,178373931 | ZNF490 | FALSE | Downregulated |
| -0,633858798 | 0,178463736 | LOC101928158 | FALSE | Downregulated |
| 1,055431056 | 0,178471707 | LOC389602 | FALSE | Upregulated |
| -0,289953208 | 0,17850545 | FAM76A | FALSE | Downregulated |
| 0,260389779 | 0,17850545 | LOC100862671 | FALSE | Upregulated |
| 0,248647311 | 0,17850545 | RPRD1B | FALSE | Upregulated |
| 0,273278796 | 0,17850545 | ZKSCAN3 | FALSE | Upregulated |
| 0,220446167 | 0,178508119 | EOGT | FALSE | Upregulated |
| -0,179175211 | 0,17877843 | METTL3 | FALSE | Downregulated |
| -0,218297759 | 0,178945526 | CTBS | FALSE | Downregulated |
| -0,240531373 | 0,178963767 | IL11RA | FALSE | Downregulated |
| -0,453668745 | 0,179008286 | LOC653160 | FALSE | Downregulated |
| 1,419081009 | 0,179008286 | PLA2G2A | FALSE | Upregulated |
| -0,527825608 | 0,179008286 | SLITRK6 | FALSE | Downregulated |
| -0,296203802 | 0,179038159 | LOC101928505 | FALSE | Downregulated |
| -0,200382823 | 0,179069931 | MED13 | FALSE | Downregulated |
| -0,121643228 | 0,179095402 | MIEF1 | FALSE | Downregulated |
| -0,534231061 | 0,179205425 | CACNA1A | FALSE | Downregulated |
| 0,235157468 | 0,179223483 | ZNF799 | FALSE | Upregulated |
| -0,559758484 | 0,179318782 | COPG2IT1 | FALSE | Downregulated |
| 0,270414186 | 0,179318782 | STK36 | FALSE | Upregulated |
| -0,184614467 | 0,179415497 | ABCC2 | FALSE | Downregulated |

| | | | | |
|---|---|---|---|---|
| 0,169567418 | 0,179464314 | MLLT6 | FALSE | Upregulated |
| 0,573692583 | 0,179464314 | PATL2 | FALSE | Upregulated |
| -0,148612336 | 0,179529987 | ERP44 | FALSE | Downregulated |
| 0,354941652 | 0,179529987 | IPCEF1 | FALSE | Upregulated |
| -0,256748006 | 0,179529987 | LIG4 | FALSE | Downregulated |
| -0,231857868 | 0,179529987 | MRPL55 | FALSE | Downregulated |
| 0,173895648 | 0,179529987 | PANK4 | FALSE | Upregulated |
| 0,117408602 | 0,179541094 | KPNA4 | FALSE | Upregulated |
| -0,188738927 | 0,179541094 | ZNF615 | FALSE | Downregulated |
| -0,181679374 | 0,179621753 | BMS1 | FALSE | Downregulated |
| -0,324815059 | 0,179707134 | SLC25A5-AS1 | FALSE | Downregulated |
| -0,174565104 | 0,179903588 | TRMT12 | FALSE | Downregulated |
| 0,252660765 | 0,179976552 | SLC25A22 | FALSE | Upregulated |
| 0,377614314 | 0,180160878 | EIF3C | FALSE | Upregulated |
| 0,15161708 | 0,180238417 | CCNI | FALSE | Upregulated |
| -0,246927085 | 0,180238417 | FAM114A1 | FALSE | Downregulated |
| 1,105290548 | 0,180238417 | GIPR | FALSE | Upregulated |
| 0,371528171 | 0,180238417 | LHFP | FALSE | Upregulated |
| 0,308094921 | 0,180238417 | TNFRSF10D | FALSE | Upregulated |
| -0,117875225 | 0,180238417 | UBE4A | FALSE | Downregulated |
| 1,102105194 | 0,180275562 | TSPAN10 | FALSE | Upregulated |
| 0,292562902 | 0,180418782 | MEIS3P1 | FALSE | Upregulated |
| -0,1711449 | 0,180418782 | ZNF281 | FALSE | Downregulated |
| 0,416182452 | 0,180448641 | ELOVL2 | FALSE | Upregulated |
| 0,587114064 | 0,180448641 | IL12RB2 | FALSE | Upregulated |
| 0,319031791 | 0,180485196 | AFAP1L2 | FALSE | Upregulated |
| 0,213360479 | 0,180510647 | RBX1 | FALSE | Upregulated |
| -0,75341941 | 0,180512904 | LOC101929289 | FALSE | Downregulated |
| -0,327490289 | 0,180559534 | LOC101927471 | FALSE | Downregulated |
| 0,190500976 | 0,180570185 | EP400 | FALSE | Upregulated |
| -0,173718743 | 0,180589816 | CTDSP2 | FALSE | Downregulated |
| 0,345386563 | 0,180643509 | ARHGAP30 | FALSE | Upregulated |
| -0,185102428 | 0,180657376 | MEMO1 | FALSE | Downregulated |
| 0,194031776 | 0,180737564 | ARHGAP18 | FALSE | Upregulated |
| 0,306093928 | 0,180779007 | FAM110C | FALSE | Upregulated |
| -0,181951422 | 0,180801083 | POLR3B | FALSE | Downregulated |
| 0,212741662 | 0,180827014 | HERC1 | FALSE | Upregulated |
| 0,377683166 | 0,180933973 | SLC25A19 | FALSE | Upregulated |
| 0,159557093 | 0,181022173 | NDUFS7 | FALSE | Upregulated |
| 1,119421368 | 0,181212298 | ZNF80 | FALSE | Upregulated |
| -0,257781368 | 0,181232549 | PDCL3 | FALSE | Downregulated |
| -0,379594303 | 0,18124883 | DLEU2 | FALSE | Downregulated |
| 1,238033911 | 0,181273105 | LOC151174 | FALSE | Upregulated |
| 0,419512393 | 0,181452336 | LOC102724168 | FALSE | Upregulated |
| 0,896731807 | 0,18167653 | LOC101927260 | FALSE | Upregulated |
| -1,170555346 | 0,18169608 | ELMOD1 | FALSE | Downregulated |
| 0,36628871 | 0,18179709 | TRMT61A | FALSE | Upregulated |
| 0,92668552 | 0,181847712 | LGI2 | FALSE | Upregulated |
| 0,312043338 | 0,181847712 | NLE1 | FALSE | Upregulated |
| -0,250320926 | 0,181871284 | SCAND1 | FALSE | Downregulated |
| -0,234365392 | 0,181924988 | HYLS1 | FALSE | Downregulated |

| | | | | |
|---|---|---|---|---|
| 0,658926887 | 0,181924988 | LOC101927069 | FALSE | Upregulated |
| 0,221762878 | 0,181929279 | SFR1 | FALSE | Upregulated |
| -0,259682859 | 0,181982481 | USP30 | FALSE | Downregulated |
| 0,395639731 | 0,182147407 | TYRO3 | FALSE | Upregulated |
| -0,634184631 | 0,182263346 | SCN2A | FALSE | Downregulated |
| -0,321765573 | 0,182275118 | HTR7P1 | FALSE | Downregulated |
| -0,30960326 | 0,182275118 | RBM25 | FALSE | Downregulated |
| 1,373996815 | 0,182286842 | FAM196B | FALSE | Upregulated |
| 0,691903361 | 0,182286842 | LOC101929574 | FALSE | Upregulated |
| -0,1939023 | 0,182286842 | MMP24-AS1 | FALSE | Downregulated |
| -0,182939415 | 0,1823529 | C17orf80 | FALSE | Downregulated |
| 0,365976713 | 0,1824071 | SYT15 | FALSE | Upregulated |
| -0,222361928 | 0,182447879 | MAGOHB | FALSE | Downregulated |
| -0,163610548 | 0,182617511 | SETD7 | FALSE | Downregulated |
| -0,229702056 | 0,182685341 | SMIM8 | FALSE | Downregulated |
| 0,217191311 | 0,182803757 | TOR2A | FALSE | Upregulated |
| 0,772319956 | 0,182823847 | CCDC89 | FALSE | Upregulated |
| 1,01935874 | 0,182864308 | EFCAB5 | FALSE | Upregulated |
| -0,119290392 | 0,182864308 | SMG7 | FALSE | Downregulated |
| 0,723657553 | 0,182944113 | LOC102724187 | FALSE | Upregulated |
| -0,166087331 | 0,183146869 | IBTK | FALSE | Downregulated |
| -0,333389715 | 0,183149793 | LOC728323 | FALSE | Downregulated |
| 0,170036191 | 0,183154022 | PRCP | FALSE | Upregulated |
| 0,258410944 | 0,183189192 | DDX28 | FALSE | Upregulated |
| 2,313243824 | 0,183189192 | ZCCHC12 | FALSE | Upregulated |
| -0,219009366 | 0,183206426 | ING5 | FALSE | Downregulated |
| 0,258583665 | 0,183412681 | ZFP69 | FALSE | Upregulated |
| -0,191389288 | 0,183428515 | BCS1L | FALSE | Downregulated |
| -0,815908856 | 0,183428515 | LOC101928462 | FALSE | Downregulated |
| -2,15089975 | 0,183514864 | GYPA | FALSE | Downregulated |
| 0,722070694 | 0,183514864 | LOC100996662 | FALSE | Upregulated |
| -0,362481819 | 0,183584296 | LOC102723992 | FALSE | Downregulated |
| 0,12764546 | 0,183826449 | TATDN2 | FALSE | Upregulated |
| 1,5228662 | 0,183896807 | LOC284009 | FALSE | Upregulated |
| -0,326458331 | 0,18400901 | MEX3A | FALSE | Downregulated |
| -0,381414699 | 0,184077942 | TBC1D2 | FALSE | Downregulated |
| -0,256834428 | 0,184110622 | MOCOS | FALSE | Downregulated |
| -0,262719424 | 0,184316041 | RAB28 | FALSE | Downregulated |
| -0,578296033 | 0,184339305 | LOC101928504 | FALSE | Downregulated |
| 0,163863446 | 0,184339305 | NOP56 | FALSE | Upregulated |
| -0,116719884 | 0,184339305 | TAF2 | FALSE | Downregulated |
| -0,491234437 | 0,184339305 | VNN3 | FALSE | Downregulated |
| 0,142010714 | 0,184344306 | METTL16 | FALSE | Upregulated |
| 1,494942141 | 0,184375667 | IGDCC4 | FALSE | Upregulated |
| -1,000885471 | 0,184456847 | RNA5-8S5 | FALSE | Downregulated |
| 0,592548846 | 0,184732062 | DNAJC18 | FALSE | Upregulated |
| 1,09764064 | 0,184866406 | FAM133DP | FALSE | Upregulated |
| 0,575774338 | 0,18489957 | ITPKA | FALSE | Upregulated |
| -0,212641941 | 0,185081285 | HNRNPUL2-BSCL2 | FALSE | Downregulated |
| 0,207744913 | 0,185215003 | LINC00997 | FALSE | Upregulated |
| 0,244048505 | 0,185220847 | SLC22A5 | FALSE | Upregulated |

| | | | | |
|---|---|---|---|---|
| -0,27325216 | 0,185220847 | TMEM39A | FALSE | Downregulated |
| -0,918611731 | 0,185304008 | KIAA1239 | FALSE | Downregulated |
| 0,263148631 | 0,185304008 | TMEM194B | FALSE | Upregulated |
| 2,255410423 | 0,185315033 | LOC101927131 | FALSE | Upregulated |
| 0,569751552 | 0,185315033 | OSBPL7 | FALSE | Upregulated |
| -0,622580833 | 0,185324624 | LINC00885 | FALSE | Downregulated |
| 2,221984085 | 0,18535044 | AGR2 | FALSE | Upregulated |
| -0,196163594 | 0,185387751 | GOLPH3L | FALSE | Downregulated |
| -0,162199491 | 0,185405677 | BET1L | FALSE | Downregulated |
| -0,213807413 | 0,185405677 | GNPAT | FALSE | Downregulated |
| -0,636798651 | 0,185405677 | LOC101926956 | FALSE | Downregulated |
| 0,376930605 | 0,18546946 | ARSB | FALSE | Upregulated |
| 0,128947445 | 0,18546946 | PLAA | FALSE | Upregulated |
| -0,495030908 | 0,185548599 | SUSD3 | FALSE | Downregulated |
| 0,553845391 | 0,185667081 | SPA17 | FALSE | Upregulated |
| 1,274743661 | 0,185681212 | CNTN2 | FALSE | Upregulated |
| -0,140144515 | 0,185681212 | VDAC2 | FALSE | Downregulated |
| -0,425112251 | 0,185752429 | SLC6A13 | FALSE | Downregulated |
| 0,570345596 | 0,185859277 | PINLYP | FALSE | Upregulated |
| -0,246065636 | 0,185934178 | SYT17 | FALSE | Downregulated |
| -0,663656408 | 0,186017143 | C2orf40 | FALSE | Downregulated |
| -0,286157369 | 0,186017143 | CMC2 | FALSE | Downregulated |
| -0,556244142 | 0,186118032 | CAND2 | FALSE | Downregulated |
| -0,392268547 | 0,186118032 | LOC100652758 | FALSE | Downregulated |
| -0,427345175 | 0,186118032 | RP5-1073O3.7 | FALSE | Downregulated |
| 0,187812617 | 0,186241564 | FKBP9 | FALSE | Upregulated |
| 0,165483902 | 0,186265591 | FAM188A | FALSE | Upregulated |
| 0,952313555 | 0,186291416 | LMCD1-AS1 | FALSE | Upregulated |
| -0,262919621 | 0,186504766 | LOC728769 | FALSE | Downregulated |
| 0,154969516 | 0,186704148 | PSMD12 | FALSE | Upregulated |
| -0,220129243 | 0,18672296 | UGP2 | FALSE | Downregulated |
| -0,551375141 | 0,186936698 | GOLGA6A | FALSE | Downregulated |
| 0,212163473 | 0,187036467 | RGP1 | FALSE | Upregulated |
| -0,149544988 | 0,187145443 | STAU1 | FALSE | Downregulated |
| 0,86266315 | 0,187147864 | NR4A3 | FALSE | Upregulated |
| -0,377997772 | 0,187147864 | PLAC8 | FALSE | Downregulated |
| -0,146560051 | 0,187147864 | PTPLAD1 | FALSE | Downregulated |
| -0,57050226 | 0,187219193 | ANGPT2 | FALSE | Downregulated |
| -0,814630378 | 0,187341167 | FHL1 | FALSE | Downregulated |
| 0,936751436 | 0,187341167 | IGF2-AS | FALSE | Upregulated |
| -0,161602118 | 0,187341167 | POT1 | FALSE | Downregulated |
| 1,195952763 | 0,187734686 | CRB1 | FALSE | Upregulated |
| 0,275803176 | 0,187961664 | SLC39A10 | FALSE | Upregulated |
| 0,504365133 | 0,187977922 | CNTD1 | FALSE | Upregulated |
| -0,211835223 | 0,187977922 | TMEM41A | FALSE | Downregulated |
| 0,401458903 | 0,188119078 | MECOM | FALSE | Upregulated |
| -0,58761141 | 0,188155858 | COL6A6 | FALSE | Downregulated |
| -0,983338023 | 0,188155858 | GOLGA6L10 | FALSE | Downregulated |
| -0,263253077 | 0,188155858 | PLD1 | FALSE | Downregulated |
| -0,172410837 | 0,188421991 | PARP3 | FALSE | Downregulated |
| -0,500755499 | 0,188652434 | ISM1 | FALSE | Downregulated |

| | | | | |
|---|---|---|---|---|
| 0,314963228 | 0,188652434 | ITPR1 | FALSE | Upregulated |
| 0,235355408 | 0,188662198 | RGS12 | FALSE | Upregulated |
| -0,515888106 | 0,188772766 | LOC102723323 | FALSE | Downregulated |
| -0,198828599 | 0,188868527 | PRR14 | FALSE | Downregulated |
| 0,51779982 | 0,188975616 | MB21D1 | FALSE | Upregulated |
| -0,187240758 | 0,188975616 | NAA15 | FALSE | Downregulated |
| -0,17166118 | 0,188975616 | SERF2 | FALSE | Downregulated |
| 0,147749471 | 0,189056428 | ANKH | FALSE | Upregulated |
| -0,755270979 | 0,189056428 | EFHB | FALSE | Downregulated |
| 0,370963222 | 0,189056428 | PDF | FALSE | Upregulated |
| -1,153966878 | 0,189084484 | LOC392364 | FALSE | Downregulated |
| -0,820537236 | 0,189084484 | VIL1 | FALSE | Downregulated |
| -0,265145003 | 0,189216365 | SMC1A | FALSE | Downregulated |
| 1,81979617 | 0,189253973 | ZBTB8B | FALSE | Upregulated |
| 0,54072765 | 0,1892817 | CCRN4L | FALSE | Upregulated |
| -0,128551233 | 0,189351799 | TMEM59 | FALSE | Downregulated |
| -0,216064054 | 0,189375207 | DDO | FALSE | Downregulated |
| 1,330952212 | 0,189375207 | PBX4 | FALSE | Upregulated |
| 1,829430534 | 0,189438972 | RP11-725G5.2 | FALSE | Upregulated |
| -0,297102135 | 0,189482371 | TMEM27 | FALSE | Downregulated |
| 1,037006598 | 0,189504397 | DNAAF3 | FALSE | Upregulated |
| -0,15315723 | 0,189763579 | GALM | FALSE | Downregulated |
| 0,77257917 | 0,189779918 | LOC101928153 | FALSE | Upregulated |
| 0,60200004 | 0,189928822 | MAP3K15 | FALSE | Upregulated |
| 0,290351951 | 0,190081527 | CEP112 | FALSE | Upregulated |
| -0,11332771 | 0,190205136 | PPHLN1 | FALSE | Downregulated |
| 0,19515168 | 0,190378567 | PFAS | FALSE | Upregulated |
| 0,789288464 | 0,190471553 | MKLN1-AS | FALSE | Upregulated |
| -0,322238751 | 0,190580114 | LINC01071 | FALSE | Downregulated |
| 1,69814535 | 0,190580114 | PTCHD2 | FALSE | Upregulated |
| 0,185784872 | 0,190605126 | IFT20 | FALSE | Upregulated |
| 0,290189972 | 0,190614885 | ZNF426 | FALSE | Upregulated |
| 1,499699971 | 0,190618386 | CAPN8 | FALSE | Upregulated |
| 0,322075447 | 0,190618386 | CDK2AP2 | FALSE | Upregulated |
| 0,272041318 | 0,190687428 | FSCN1 | FALSE | Upregulated |
| -0,253756804 | 0,19072288 | PAXIP1-AS2 | FALSE | Downregulated |
| 0,343725215 | 0,190787217 | ACSL6 | FALSE | Upregulated |
| -0,199617179 | 0,190787217 | FAM126A | FALSE | Downregulated |
| -0,188170704 | 0,190849715 | C1orf50 | FALSE | Downregulated |
| 0,990100526 | 0,190849715 | GFAP | FALSE | Upregulated |
| -1,370285211 | 0,190849715 | HCG17 | FALSE | Downregulated |
| -0,618252958 | 0,190849715 | KGFLP1 | FALSE | Downregulated |
| 1,543127161 | 0,190849715 | MAGEL2 | FALSE | Upregulated |
| 1,27752108 | 0,190849715 | SLC26A9 | FALSE | Upregulated |
| -1,041298954 | 0,190906985 | LOC101929555 | FALSE | Downregulated |
| -0,133853002 | 0,191094569 | PNPO | FALSE | Downregulated |
| 0,499181752 | 0,19113877 | LINC00883 | FALSE | Upregulated |
| 0,448720375 | 0,191176958 | MAPK11 | FALSE | Upregulated |
| 1,145963672 | 0,191191197 | GABRG1 | FALSE | Upregulated |
| -1,415895614 | 0,191191197 | LDHC | FALSE | Downregulated |
| 0,172723361 | 0,191203959 | EXOC2 | FALSE | Upregulated |

| | | | | |
|---|---|---|---|---|
| -0,206339291 | 0,191203959 | GNG7 | FALSE | Downregulated |
| -0,290779431 | 0,191203959 | LOC100132815 | FALSE | Downregulated |
| -0,1891019 | 0,191203959 | SS18L1 | FALSE | Downregulated |
| 0,126248968 | 0,191203959 | YWHAB | FALSE | Upregulated |
| 0,107295707 | 0,191274399 | PPP6C | FALSE | Upregulated |
| 0,217660441 | 0,191364575 | ARAP2 | FALSE | Upregulated |
| 0,186022029 | 0,191364575 | ZNF516 | FALSE | Upregulated |
| -0,19887069 | 0,191369574 | EEF1B2 | FALSE | Downregulated |
| -0,331536295 | 0,191438535 | DNAJC3-AS1 | FALSE | Downregulated |
| 0,198701556 | 0,191545111 | DLGAP4 | FALSE | Upregulated |
| 1,369327124 | 0,191545111 | UNC80 | FALSE | Upregulated |
| -1,215656451 | 0,191653274 | LOC101928263 | FALSE | Downregulated |
| 0,21319557 | 0,191684177 | HPS6 | FALSE | Upregulated |
| -0,196323678 | 0,191764469 | SERPIND1 | FALSE | Downregulated |
| -0,801064621 | 0,191825929 | LOC102723356 | FALSE | Downregulated |
| -0,095537857 | 0,191846908 | NUDT21 | FALSE | Downregulated |
| 0,475328764 | 0,192364218 | RP11-346I3.4 | FALSE | Upregulated |
| 1,220073598 | 0,192458816 | LOC101926889 | FALSE | Upregulated |
| 0,186936347 | 0,192509595 | ALG8 | FALSE | Upregulated |
| -0,224848229 | 0,192511751 | ZFP14 | FALSE | Downregulated |
| 1,096941589 | 0,192766966 | LOC102724006 | FALSE | Upregulated |
| 1,117666665 | 0,192781145 | WTH3DI | FALSE | Upregulated |
| 0,261826608 | 0,192781145 | ZBTB42 | FALSE | Upregulated |
| -0,099095608 | 0,192796073 | AIDA | FALSE | Downregulated |
| 0,250462711 | 0,192961246 | NUDT7 | FALSE | Upregulated |
| 0,11732753 | 0,192975383 | DERL1 | FALSE | Upregulated |
| 2,173789491 | 0,192975383 | SLC30A2 | FALSE | Upregulated |
| 0,512220878 | 0,193032846 | KIF7 | FALSE | Upregulated |
| 0,329013515 | 0,193032846 | ZC3H6 | FALSE | Upregulated |
| 0,894961255 | 0,193044357 | AJAP1 | FALSE | Upregulated |
| 0,497955488 | 0,193044357 | EID3 | FALSE | Upregulated |
| -0,254317619 | 0,193271244 | VEGFB | FALSE | Downregulated |
| -1,11453765 | 0,193271935 | LOC101927354 | FALSE | Downregulated |
| -0,194927375 | 0,193285887 | ZNF75D | FALSE | Downregulated |
| 0,208030829 | 0,193372641 | PIGU | FALSE | Upregulated |
| -0,104203415 | 0,193395971 | ATP5A1 | FALSE | Downregulated |
| 0,22837447 | 0,19346342 | HEATR2 | FALSE | Upregulated |
| -0,138470933 | 0,193499916 | TOMM22 | FALSE | Downregulated |
| 0,694683265 | 0,193588728 | MEI1 | FALSE | Upregulated |
| 0,3754222 | 0,193659843 | MSR1 | FALSE | Upregulated |
| 0,19022798 | 0,193836377 | BBX | FALSE | Upregulated |
| -0,195013173 | 0,193836377 | CCDC106 | FALSE | Downregulated |
| 0,218957244 | 0,193836377 | SLC35B2 | FALSE | Upregulated |
| -0,216238124 | 0,193904494 | ESPN | FALSE | Downregulated |
| 0,12316289 | 0,193904494 | RPA1 | FALSE | Upregulated |
| -0,841344861 | 0,193989762 | TFCP2L1 | FALSE | Downregulated |
| 0,143156776 | 0,193996735 | HCFC1 | FALSE | Upregulated |
| 0,328603343 | 0,194058387 | ENTHD2 | FALSE | Upregulated |
| -0,235533254 | 0,194145263 | HYI | FALSE | Downregulated |
| 0,3887658 | 0,194235516 | SMOX | FALSE | Upregulated |
| 0,301641224 | 0,194308234 | CHAF1A | FALSE | Upregulated |

| | | | | |
|---|---|---|---|---|
| -0,155601779 | 0,194315973 | BRMS1L | FALSE | Downregulated |
| 0,291122317 | 0,194315973 | CYB5RL | FALSE | Upregulated |
| -0,127194572 | 0,194315973 | GABPA | FALSE | Downregulated |
| 0,240749628 | 0,194315973 | GIMAP6 | FALSE | Upregulated |
| 1,230964363 | 0,194315973 | GSDMA | FALSE | Upregulated |
| 0,657874245 | 0,194315973 | MMS22L | FALSE | Upregulated |
| -0,262149608 | 0,194439552 | RASGEF1B | FALSE | Downregulated |
| 0,168525561 | 0,194460532 | NEDD1 | FALSE | Upregulated |
| 1,017196623 | 0,19446123 | LINC01021 | FALSE | Upregulated |
| 0,224834445 | 0,194473492 | CEP97 | FALSE | Upregulated |
| -0,22281431 | 0,194490832 | BDH2 | FALSE | Downregulated |
| -0,193873216 | 0,194495284 | TPST2 | FALSE | Downregulated |
| -0,279820748 | 0,194597297 | TMEM133 | FALSE | Downregulated |
| 0,301673004 | 0,19460069 | FAM213B | FALSE | Upregulated |
| -0,201280406 | 0,194694536 | OSGEPL1 | FALSE | Downregulated |
| -0,474802531 | 0,194744688 | NDST3 | FALSE | Downregulated |
| -0,365958261 | 0,194870261 | UGDH-AS1 | FALSE | Downregulated |
| -0,248567441 | 0,194903621 | TRAPPC6A | FALSE | Downregulated |
| 0,133457199 | 0,194973448 | SEP15 | FALSE | Upregulated |
| -0,118238264 | 0,194981045 | PRPF18 | FALSE | Downregulated |
| 0,397323608 | 0,195024278 | TFEC | FALSE | Upregulated |
| 1,371887336 | 0,195046319 | CHRNB2 | FALSE | Upregulated |
| 0,330478864 | 0,195059752 | MAGIX | FALSE | Upregulated |
| 1,65389026 | 0,195289005 | MATN3 | FALSE | Upregulated |
| 1,169271205 | 0,195375499 | PI15 | FALSE | Upregulated |
| 0,298095744 | 0,195375499 | PPDPF | FALSE | Upregulated |
| -0,183449246 | 0,195375499 | SEH1L | FALSE | Downregulated |
| 1,122171891 | 0,195744263 | DKK2 | FALSE | Upregulated |
| -0,318912452 | 0,196049017 | SREK1 | FALSE | Downregulated |
| 0,248178257 | 0,196049017 | ZNF333 | FALSE | Upregulated |
| 0,141580579 | 0,196309953 | FEM1C | FALSE | Upregulated |
| -0,231902624 | 0,196309953 | FLT4 | FALSE | Downregulated |
| 0,147053289 | 0,196309953 | HN1L | FALSE | Upregulated |
| 0,412601965 | 0,196309953 | PCYOX1L | FALSE | Upregulated |
| 0,538661036 | 0,196537672 | UBE2T | FALSE | Upregulated |
| -0,196309476 | 0,196784424 | LETM1 | FALSE | Downregulated |
| -0,362988493 | 0,196900044 | HCG18 | FALSE | Downregulated |
| -0,186031028 | 0,197030715 | MEGF8 | FALSE | Downregulated |
| 0,254463464 | 0,197098326 | C19orf10 | FALSE | Upregulated |
| -0,222527858 | 0,197098326 | CXXC5 | FALSE | Downregulated |
| 0,533745773 | 0,197124988 | NAPSB | FALSE | Upregulated |
| 0,410827692 | 0,197134706 | ADAMTS7 | FALSE | Upregulated |
| 0,640323272 | 0,197280861 | LOC102723926 | FALSE | Upregulated |
| 0,149812443 | 0,197293348 | TFE3 | FALSE | Upregulated |
| -0,366303949 | 0,197354143 | AC007551.3 | FALSE | Downregulated |
| 0,217063269 | 0,197354143 | PACSIN2 | FALSE | Upregulated |
| 0,113523065 | 0,197377617 | ELP3 | FALSE | Upregulated |
| 0,515233324 | 0,197377617 | SRGAP2C | FALSE | Upregulated |
| 0,220445013 | 0,197510326 | BRPF1 | FALSE | Upregulated |
| -0,185288678 | 0,197510326 | DDX3X | FALSE | Downregulated |
| 0,243744025 | 0,197667824 | RABEP2 | FALSE | Upregulated |

| | | | | |
|---|---|---|---|---|
| -0,333926682 | 0,19769804 | TLN2 | FALSE | Downregulated |
| -0,147383916 | 0,197716906 | C2CD5 | FALSE | Downregulated |
| -0,10693283 | 0,197717634 | LSM14A | FALSE | Downregulated |
| 0,234723873 | 0,197717634 | PNPLA2 | FALSE | Upregulated |
| -0,348020243 | 0,197717634 | SCN9A | FALSE | Downregulated |
| 0,151244254 | 0,197717634 | VCPIP1 | FALSE | Upregulated |
| 1,071693899 | 0,197779781 | CADM3 | FALSE | Upregulated |
| 1,298714347 | 0,197779781 | CC2D2B | FALSE | Upregulated |
| -0,172509572 | 0,197792043 | MAP7 | FALSE | Downregulated |
| -0,199903577 | 0,197814918 | ITPR2 | FALSE | Downregulated |
| 0,29196551 | 0,19784215 | BIN2 | FALSE | Upregulated |
| 0,237329807 | 0,197968164 | ACER3 | FALSE | Upregulated |
| 0,95044675 | 0,197968164 | HTR7 | FALSE | Upregulated |
| 0,530252057 | 0,198132151 | ZNF711 | FALSE | Upregulated |
| -0,258412278 | 0,19816411 | RHEBP2 | FALSE | Downregulated |
| 1,799440652 | 0,198184614 | LOC653786 | FALSE | Upregulated |
| 0,380422049 | 0,198196404 | TNRC6C-AS1 | FALSE | Upregulated |
| 0,124103862 | 0,198224798 | TSG101 | FALSE | Upregulated |
| 0,490178774 | 0,198268721 | CLDN9 | FALSE | Upregulated |
| 1,647848469 | 0,198268721 | ZNF716 | FALSE | Upregulated |
| -0,144723009 | 0,198324366 | TRIM44 | FALSE | Downregulated |
| -0,211266956 | 0,198394889 | C8G | FALSE | Downregulated |
| 0,098555666 | 0,19850509 | GPR107 | FALSE | Upregulated |
| -1,252328117 | 0,198520756 | LOC102724280 | FALSE | Downregulated |
| -0,237009131 | 0,198533939 | INHBC | FALSE | Downregulated |
| 0,157207207 | 0,198533939 | MTMR6 | FALSE | Upregulated |
| 0,25962363 | 0,198562962 | LRMP | FALSE | Upregulated |
| -0,136312233 | 0,198562962 | NCOA1 | FALSE | Downregulated |
| 0,815415868 | 0,1988436 | TREM1 | FALSE | Upregulated |
| 0,402581074 | 0,198932017 | SEMA4F | FALSE | Upregulated |
| -0,50738088 | 0,199246024 | HOTAIRM1 | FALSE | Downregulated |
| 0,987347043 | 0,19947505 | LOC101928884 | FALSE | Upregulated |
| -0,33400267 | 0,199515626 | SYNE4 | FALSE | Downregulated |
| -0,203855482 | 0,199665627 | OGDHL | FALSE | Downregulated |
| -0,240920223 | 0,199671098 | SH3BP2 | FALSE | Downregulated |
| 0,351834983 | 0,199693059 | WBP5 | FALSE | Upregulated |
| 0,801151477 | 0,199753398 | PAK3 | FALSE | Upregulated |
| 0,191007406 | 0,199798119 | LIMA1 | FALSE | Upregulated |
| -0,250182278 | 0,199875996 | SMPDL3A | FALSE | Downregulated |
| 0,972175774 | 0,199883439 | THSD7B | FALSE | Upregulated |
| -0,273527237 | 0,19994164 | LOC646719 | FALSE | Downregulated |
| 0,271363799 | 0,199950163 | DDHD1 | FALSE | Upregulated |
| -0,310092744 | 0,199976212 | SLC7A2 | FALSE | Downregulated |
| 0,334744712 | 0,199984864 | C1orf85 | FALSE | Upregulated |
| 0,34432541 | 0,200017728 | CNRIP1 | FALSE | Upregulated |
| -0,869204304 | 0,200017728 | EPO | FALSE | Downregulated |
| 0,515369932 | 0,200095968 | BAI3 | FALSE | Upregulated |
| -0,151112692 | 0,200169808 | SARS | FALSE | Downregulated |
| -0,256887426 | 0,200183679 | USMG5 | FALSE | Downregulated |
| -0,383044765 | 0,200204817 | KRTCAP3 | FALSE | Downregulated |
| -1,258107403 | 0,200335032 | RSPO4 | FALSE | Downregulated |

| | | | | |
|---|---|---|---|---|
| -0,354249901 | 0,200510172 | PACRG | FALSE | Downregulated |
| 0,883496551 | 0,200530773 | KLHDC7B | FALSE | Upregulated |
| 0,286758712 | 0,20062003 | FAM86A | FALSE | Upregulated |
| 1,055817602 | 0,200828716 | JPH3 | FALSE | Upregulated |
| -0,131648404 | 0,200828716 | ZNF398 | FALSE | Downregulated |
| 0,422340727 | 0,200839436 | CACHD1 | FALSE | Upregulated |
| 0,374683043 | 0,201020957 | CENPP | FALSE | Upregulated |
| 0,163860674 | 0,201020957 | MAPRE2 | FALSE | Upregulated |
| 0,190453459 | 0,201020957 | PAK1 | FALSE | Upregulated |
| 0,377029917 | 0,201062181 | TMEM206 | FALSE | Upregulated |
| 0,22429277 | 0,201397516 | BCL9L | FALSE | Upregulated |
| 0,170659702 | 0,201397516 | UBA6 | FALSE | Upregulated |
| -0,538469928 | 0,201425752 | TRNF | FALSE | Downregulated |
| -0,184710369 | 0,201478347 | KATNAL1 | FALSE | Downregulated |
| 0,134466618 | 0,201478347 | SUB1 | FALSE | Upregulated |
| 0,152566203 | 0,201630678 | ZNF227 | FALSE | Upregulated |
| 0,129171428 | 0,201640109 | SIDT2 | FALSE | Upregulated |
| 1,053675173 | 0,201662357 | AC104820.2 | FALSE | Upregulated |
| 0,980606171 | 0,201662357 | TMEM217 | FALSE | Upregulated |
| -0,31766773 | 0,201796725 | PUS7L | FALSE | Downregulated |
| 1,459914668 | 0,201817353 | RP11-154D17.1 | FALSE | Upregulated |
| -1,620479804 | 0,202037559 | LOC102723347 | FALSE | Downregulated |
| 0,984299394 | 0,202105468 | FAM19A1 | FALSE | Upregulated |
| 0,212120237 | 0,202132717 | HRAS | FALSE | Upregulated |
| 0,402580573 | 0,202217195 | XRCC3 | FALSE | Upregulated |
| -0,402593722 | 0,202277776 | BCL2L10 | FALSE | Downregulated |
| 0,285660993 | 0,202277776 | CEP135 | FALSE | Upregulated |
| 0,215584864 | 0,202277776 | HEXIM1 | FALSE | Upregulated |
| 1,493919738 | 0,202277776 | LOC441455 | FALSE | Upregulated |
| -0,178078506 | 0,202277776 | MOCS3 | FALSE | Downregulated |
| -0,196231174 | 0,202277776 | OAF | FALSE | Downregulated |
| -0,190723916 | 0,202277776 | TMEM134 | FALSE | Downregulated |
| -0,188866176 | 0,202277776 | UBN1 | FALSE | Downregulated |
| 0,248861258 | 0,202288086 | HIVEP2 | FALSE | Upregulated |
| -0,683336115 | 0,202288086 | LOC101929079 | FALSE | Downregulated |
| -0,152611629 | 0,202515775 | TAF12 | FALSE | Downregulated |
| 0,326325828 | 0,202559346 | AXL | FALSE | Upregulated |
| 0,250035712 | 0,202559346 | GSTCD | FALSE | Upregulated |
| -0,352003074 | 0,202559346 | HOXA4 | FALSE | Downregulated |
| -0,312585824 | 0,202559346 | IKZF4 | FALSE | Downregulated |
| -0,166354216 | 0,202559346 | MRPL18 | FALSE | Downregulated |
| 0,28535414 | 0,202559346 | PHC1 | FALSE | Upregulated |
| 1,401812901 | 0,202559346 | RUNDC3A | FALSE | Upregulated |
| 0,92505088 | 0,202660282 | CERKL | FALSE | Upregulated |
| 0,266274712 | 0,202660282 | URB2 | FALSE | Upregulated |
| -0,087897515 | 0,202849109 | API5 | FALSE | Downregulated |
| 0,313855806 | 0,202969977 | HILPDA | FALSE | Upregulated |
| -0,237248881 | 0,203056894 | C14orf93 | FALSE | Downregulated |
| 0,191988108 | 0,203170317 | PNRC1 | FALSE | Upregulated |
| 0,280696154 | 0,203237965 | RDH13 | FALSE | Upregulated |
| 0,77309401 | 0,203362706 | ANKRD22 | FALSE | Upregulated |

| | | | | |
|---|---|---|---|---|
| -0,647609871 | 0,20338067 | LOC101928794 | FALSE | Downregulated |
| 0,202127256 | 0,203385105 | ADARB1 | FALSE | Upregulated |
| 0,893100386 | 0,203385105 | LOC102724554 | FALSE | Upregulated |
| 2,568800391 | 0,203445244 | LOC285819 | FALSE | Upregulated |
| 1,067193145 | 0,203445244 | TPSB2 | FALSE | Upregulated |
| -0,254326739 | 0,203445244 | UBXN2B | FALSE | Downregulated |
| -0,691358628 | 0,203554933 | LOC102724184 | FALSE | Downregulated |
| 0,3179137 | 0,203612367 | PWWP2B | FALSE | Upregulated |
| 0,150174426 | 0,203698932 | CDK5RAP1 | FALSE | Upregulated |
| 0,383424701 | 0,203698932 | CX3CR1 | FALSE | Upregulated |
| -0,188067028 | 0,203698932 | SIRT7 | FALSE | Downregulated |
| 0,371790153 | 0,203698932 | SULF2 | FALSE | Upregulated |
| 0,120921365 | 0,203771128 | ARF1 | FALSE | Upregulated |
| 0,65014647 | 0,203925126 | LRRC37A6P | FALSE | Upregulated |
| 2,065681158 | 0,20407032 | CYP4F8 | FALSE | Upregulated |
| 1,686970714 | 0,204162005 | OR6W1P | FALSE | Upregulated |
| 1,391951102 | 0,204258895 | LOC101060153 | FALSE | Upregulated |
| -0,652693635 | 0,204258895 | PAQR6 | FALSE | Downregulated |
| -0,147364937 | 0,204272236 | DCAKD | FALSE | Downregulated |
| -0,199875327 | 0,204304388 | RPS10-NUDT3 | FALSE | Downregulated |
| -0,913158844 | 0,20445942 | MT1H | FALSE | Downregulated |
| -0,207371528 | 0,204703619 | MCFD2 | FALSE | Downregulated |
| -0,12831692 | 0,204819972 | FAM8A1 | FALSE | Downregulated |
| -0,297427759 | 0,204846671 | CECR2 | FALSE | Downregulated |
| -0,915369796 | 0,204846671 | LOC101928635 | FALSE | Downregulated |
| -0,266035429 | 0,204881804 | SLC48A1 | FALSE | Downregulated |
| 0,208577381 | 0,204935061 | GGA1 | FALSE | Upregulated |
| -0,315058465 | 0,205043506 | UPB1 | FALSE | Downregulated |
| -0,371578295 | 0,205061038 | PNISR | FALSE | Downregulated |
| -0,212108995 | 0,205061038 | UAP1 | FALSE | Downregulated |
| 0,309581467 | 0,205076594 | ARMCX1 | FALSE | Upregulated |
| 1,584343452 | 0,20516689 | RP11-588G21.2 | FALSE | Upregulated |
| -2,090818046 | 0,205167342 | LOC283683 | FALSE | Downregulated |
| 0,353207289 | 0,205257185 | PRR5L | FALSE | Upregulated |
| 0,362048074 | 0,205386083 | ZSCAN20 | FALSE | Upregulated |
| -1,703324044 | 0,205718083 | LOC101927721 | FALSE | Downregulated |
| -0,127509092 | 0,205792136 | BUD31 | FALSE | Downregulated |
| 0,272591697 | 0,205806454 | ZNF283 | FALSE | Upregulated |
| -0,195306888 | 0,205903825 | ACN9 | FALSE | Downregulated |
| -0,132319814 | 0,205903825 | LRPPRC | FALSE | Downregulated |
| -0,666790571 | 0,205903825 | RP11-475O6.1-2 | FALSE | Downregulated |
| -0,577745073 | 0,205942808 | LOC101928830 | FALSE | Downregulated |
| -0,237385349 | 0,205942808 | SEPP1 | FALSE | Downregulated |
| -0,12547685 | 0,205993876 | BRD2 | FALSE | Downregulated |
| 0,373200649 | 0,206418247 | ALS2CL | FALSE | Upregulated |
| -0,191086865 | 0,206418247 | BTRC | FALSE | Downregulated |
| -0,149102493 | 0,206418247 | CLINT1 | FALSE | Downregulated |
| 0,921351714 | 0,206431175 | LOC100134868 | FALSE | Upregulated |
| 0,181698925 | 0,206486121 | SMYD5 | FALSE | Upregulated |
| 0,207899888 | 0,20650159 | TRMT44 | FALSE | Upregulated |
| -0,367298701 | 0,206564015 | PPP1R3B | FALSE | Downregulated |

| | | | | |
|---|---|---|---|---|
| -0,214885199 | 0,206570645 | RGS14 | FALSE | Downregulated |
| -0,185711071 | 0,206843654 | MRPS28 | FALSE | Downregulated |
| 0,147838852 | 0,207173925 | KIF13A | FALSE | Upregulated |
| -0,474360486 | 0,207173925 | LOC101930114 | FALSE | Downregulated |
| -0,531830531 | 0,207173925 | TENM1 | FALSE | Downregulated |
| 2,068572284 | 0,207207918 | ECEL1 | FALSE | Upregulated |
| -0,228187067 | 0,207535104 | UXS1 | FALSE | Downregulated |
| -0,731387665 | 0,207575941 | ADPRHL1 | FALSE | Downregulated |
| 0,223729053 | 0,207586393 | RGL2 | FALSE | Upregulated |
| 0,080970562 | 0,207605998 | IST1 | FALSE | Upregulated |
| 0,692409569 | 0,207808615 | C1QTNF2 | FALSE | Upregulated |
| -0,186956891 | 0,207921922 | TMEM110 | FALSE | Downregulated |
| 0,674549823 | 0,207987148 | RP11-503C24.4 | FALSE | Upregulated |
| 0,975959644 | 0,208063248 | DNM1P46 | FALSE | Upregulated |
| 2,305554666 | 0,208063248 | HTR4 | FALSE | Upregulated |
| 1,618188997 | 0,208136644 | PTGER4P2-CDK2AP2P2 | FALSE | Upregulated |
| -0,248073638 | 0,208161059 | ZNF280C | FALSE | Downregulated |
| -0,248135107 | 0,208226549 | BMS1P20 | FALSE | Downregulated |
| -0,781018854 | 0,208226549 | DDX25 | FALSE | Downregulated |
| 0,597650983 | 0,208240686 | ADAMTS6 | FALSE | Upregulated |
| -0,171387307 | 0,208353092 | ZCCHC2 | FALSE | Downregulated |
| -0,193908808 | 0,208403916 | NR1H3 | FALSE | Downregulated |
| 0,581967692 | 0,208451902 | ARHGAP33 | FALSE | Upregulated |
| 0,442374902 | 0,20846372 | C3orf70 | FALSE | Upregulated |
| 0,694340384 | 0,208574808 | LOC102724851 | FALSE | Upregulated |
| 0,134995875 | 0,208574808 | PURB | FALSE | Upregulated |
| -0,723375248 | 0,208623738 | FADS1 | FALSE | Downregulated |
| -0,810089739 | 0,208634326 | COX6A2 | FALSE | Downregulated |
| -0,162997354 | 0,20871582 | EDF1 | FALSE | Downregulated |
| -0,258954905 | 0,208736615 | SLC39A6 | FALSE | Downregulated |
| -0,13141478 | 0,208736615 | WDR89 | FALSE | Downregulated |
| 1,756013637 | 0,208849517 | NETO1 | FALSE | Upregulated |
| 0,429307978 | 0,208864359 | RTN4RL1 | FALSE | Upregulated |
| 0,133991661 | 0,208903081 | AKT1 | FALSE | Upregulated |
| 0,564014856 | 0,208903081 | TMEM132E | FALSE | Upregulated |
| 1,238679538 | 0,208942554 | LOC101927931 | FALSE | Upregulated |
| -0,140037984 | 0,209261266 | EXOSC8 | FALSE | Downregulated |
| -0,250030893 | 0,209378648 | CHORDC1 | FALSE | Downregulated |
| 0,231059062 | 0,209387445 | CDH1 | FALSE | Upregulated |
| -0,782290867 | 0,209387445 | LOC339240 | FALSE | Downregulated |
| 0,692991504 | 0,209443183 | LOC101929545 | FALSE | Upregulated |
| -0,084459951 | 0,209461516 | EIF4H | FALSE | Downregulated |
| 0,381827974 | 0,209526974 | ASTN2 | FALSE | Upregulated |
| 0,201266414 | 0,209573671 | SLC6A12 | FALSE | Upregulated |
| -0,82006834 | 0,209629607 | LOC101929619 | FALSE | Downregulated |
| 0,201036559 | 0,209629607 | SCOC | FALSE | Upregulated |
| -0,134730028 | 0,209781438 | MRPS18A | FALSE | Downregulated |
| -0,861068947 | 0,209940019 | LOC100130987 | FALSE | Downregulated |
| 1,129536186 | 0,20995108 | RFPL1 | FALSE | Upregulated |
| 0,244082879 | 0,210181888 | ZNF432 | FALSE | Upregulated |
| 0,204224707 | 0,210238012 | SDC4 | FALSE | Upregulated |

| | | | | |
|---|---|---|---|---|
| -0,222866584 | 0,210356544 | CWC27 | FALSE | Downregulated |
| 0,315072328 | 0,210356544 | DNAH14 | FALSE | Upregulated |
| -1,338914297 | 0,210356544 | MFAP5 | FALSE | Downregulated |
| 0,141505759 | 0,210435296 | TNPO2 | FALSE | Upregulated |
| 0,728234181 | 0,210536524 | SEMA3C | FALSE | Upregulated |
| 0,635662561 | 0,210756038 | COL8A1 | FALSE | Upregulated |
| -0,156118575 | 0,210756038 | DHX29 | FALSE | Downregulated |
| 1,414047409 | 0,210756038 | FNDC8 | FALSE | Upregulated |
| 0,83449892 | 0,210802259 | LOC100996660 | FALSE | Upregulated |
| -0,428093315 | 0,210802259 | LOC102723864 | FALSE | Downregulated |
| -0,114421229 | 0,210843295 | UBE2G1 | FALSE | Downregulated |
| -0,119227355 | 0,21097658 | MRPS35 | FALSE | Downregulated |
| -0,452383983 | 0,211025574 | ANKHD1-EIF4EBP3 | FALSE | Downregulated |
| 0,897121254 | 0,211093416 | COL7A1 | FALSE | Upregulated |
| 1,046241516 | 0,211143847 | LOC100506901 | FALSE | Upregulated |
| -1,442206001 | 0,211238384 | RP1 | FALSE | Downregulated |
| -2,582247411 | 0,211239198 | RBFOX1 | FALSE | Downregulated |
| -0,672750573 | 0,211270622 | LOC102724766 | FALSE | Downregulated |
| -1,832429581 | 0,211403073 | LOC101927971 | FALSE | Downregulated |
| 0,711527865 | 0,2115643 | GATSL2 | FALSE | Upregulated |
| -0,354653512 | 0,211692989 | COL25A1 | FALSE | Downregulated |
| 0,572147128 | 0,211741011 | SH2D1B | FALSE | Upregulated |
| -0,195427298 | 0,211749768 | AP5Z1 | FALSE | Downregulated |
| 0,140674254 | 0,211749768 | BTBD3 | FALSE | Upregulated |
| 1,283071979 | 0,211749768 | LOC101928255 | FALSE | Upregulated |
| -0,250891426 | 0,211749768 | MAP4K4 | FALSE | Downregulated |
| -0,353429628 | 0,211750566 | ZKSCAN7 | FALSE | Downregulated |
| 1,025259126 | 0,211869553 | LOC102724281 | FALSE | Upregulated |
| -1,127411818 | 0,211924875 | KIAA1751 | FALSE | Downregulated |
| -0,60640057 | 0,212204514 | LOC102723566 | FALSE | Downregulated |
| 0,891648732 | 0,212263038 | ANKDD1B | FALSE | Upregulated |
| 0,233804003 | 0,212263038 | SLC10A3 | FALSE | Upregulated |
| -0,893823183 | 0,212382779 | LOC100996695 | FALSE | Downregulated |
| 0,232029254 | 0,212433841 | TBC1D31 | FALSE | Upregulated |
| -0,341389532 | 0,212549894 | PRLR | FALSE | Downregulated |
| 0,462176832 | 0,212658261 | LRRC4B | FALSE | Upregulated |
| 0,793384243 | 0,212658261 | PAK7 | FALSE | Upregulated |
| 1,252871728 | 0,212720091 | LHFPL3-AS2 | FALSE | Upregulated |
| -0,228605032 | 0,212856282 | NDUFV3 | FALSE | Downregulated |
| -0,240810379 | 0,212903133 | ZNF248 | FALSE | Downregulated |
| 1,46149617 | 0,212987306 | C4orf50 | FALSE | Upregulated |
| -0,174608374 | 0,213095181 | INTS2 | FALSE | Downregulated |
| -0,395989559 | 0,213248435 | C8orf48 | FALSE | Downregulated |
| -0,110972839 | 0,213248435 | MAP3K7 | FALSE | Downregulated |
| 1,530725499 | 0,213248435 | PLA2G4F | FALSE | Upregulated |
| 1,213072339 | 0,213248435 | TMEM151A | FALSE | Upregulated |
| 0,159303774 | 0,21335253 | KPNA3 | FALSE | Upregulated |
| 0,528533415 | 0,213462767 | CCDC157 | FALSE | Upregulated |
| -0,560567359 | 0,213561807 | PPP2R3A | FALSE | Downregulated |
| -0,184688985 | 0,213561807 | SLC11A2 | FALSE | Downregulated |

| | | | | |
|---|---|---|---|---|
| -0,170456098 | 0,213626015 | CISD1 | FALSE | Downregulated |
| 1,093333723 | 0,21365775 | SPATA9 | FALSE | Upregulated |
| 0,194441489 | 0,21367219 | SPRY2 | FALSE | Upregulated |
| 0,680714683 | 0,213812683 | PCBP3 | FALSE | Upregulated |
| 0,669129148 | 0,213927908 | C1QTNF9B-AS1 | FALSE | Upregulated |
| -0,348093719 | 0,213927908 | HGD | FALSE | Downregulated |
| 1,335792301 | 0,21405642 | HCAR1 | FALSE | Upregulated |
| -0,101925793 | 0,21405642 | MARCH7 | FALSE | Downregulated |
| -0,393724508 | 0,214114724 | COL21A1 | FALSE | Downregulated |
| 1,139749825 | 0,214221523 | AK8 | FALSE | Upregulated |
| 0,439171157 | 0,214362254 | DDX26B | FALSE | Upregulated |
| -0,799066824 | 0,214466209 | PPBP | FALSE | Downregulated |
| 1,015463945 | 0,214476028 | C10orf105 | FALSE | Upregulated |
| 0,370139261 | 0,214476028 | HSPA13 | FALSE | Upregulated |
| 0,524555553 | 0,214776898 | TOX3 | FALSE | Upregulated |
| -0,305544681 | 0,214967646 | PITPNA-AS1 | FALSE | Downregulated |
| 1,2664912 | 0,215005237 | CIDEC | FALSE | Upregulated |
| 0,203264218 | 0,215005237 | MVB12B | FALSE | Upregulated |
| 0,773119377 | 0,215056878 | ZNF483 | FALSE | Upregulated |
| 1,584670921 | 0,215134465 | LOC101929272 | FALSE | Upregulated |
| -0,322761206 | 0,215155728 | TMEM14A | FALSE | Downregulated |
| 0,597593907 | 0,215198473 | LRRC46 | FALSE | Upregulated |
| 0,239404949 | 0,215198473 | LSM10 | FALSE | Upregulated |
| 1,45042623 | 0,215284609 | LOC100996369 | FALSE | Upregulated |
| -0,234857632 | 0,215284609 | MAN1A1 | FALSE | Downregulated |
| -0,38983735 | 0,215284609 | OXER1 | FALSE | Downregulated |
| -0,106550203 | 0,215308379 | SCYL1 | FALSE | Downregulated |
| 0,344598161 | 0,215364011 | PRKRIP1 | FALSE | Upregulated |
| -0,335200908 | 0,215419839 | LONRF3 | FALSE | Downregulated |
| 0,708314035 | 0,215473113 | PABPC4L | FALSE | Upregulated |
| -0,364010624 | 0,215473113 | RGS5 | FALSE | Downregulated |
| -0,174960179 | 0,215626689 | EHMT1 | FALSE | Downregulated |
| -0,217173922 | 0,215655052 | MCEE | FALSE | Downregulated |
| -0,158349703 | 0,215683933 | METTL10 | FALSE | Downregulated |
| 0,747420643 | 0,215683933 | SCN4B | FALSE | Upregulated |
| -1,002541443 | 0,215723226 | LRRC9 | FALSE | Downregulated |
| -0,170687353 | 0,215886302 | C22orf39 | FALSE | Downregulated |
| -0,213415808 | 0,216000241 | ALG14 | FALSE | Downregulated |
| 0,205686267 | 0,216167662 | MFHAS1 | FALSE | Upregulated |
| 0,592577763 | 0,216225421 | DOPEY2 | FALSE | Upregulated |
| -0,229750061 | 0,216225421 | RNF217 | FALSE | Downregulated |
| 0,401728624 | 0,216243996 | HCST | FALSE | Upregulated |
| 0,835871687 | 0,216399027 | LINC00571 | FALSE | Upregulated |
| -0,66791082 | 0,216399027 | LINC00852 | FALSE | Downregulated |
| -0,56907614 | 0,216468382 | GSTA7P | FALSE | Downregulated |
| -0,138478844 | 0,216588009 | FBXL5 | FALSE | Downregulated |
| 1,078164477 | 0,216601208 | HLA-DPB2 | FALSE | Upregulated |
| 0,366895785 | 0,216601208 | KIF17 | FALSE | Upregulated |
| -2,170581573 | 0,216601208 | UNC45B | FALSE | Downregulated |
| 0,58564429 | 0,216727024 | ANKRD36B | FALSE | Upregulated |
| 1,163198794 | 0,216780617 | DPCR1 | FALSE | Upregulated |

| | | | | |
|---|---|---|---|---|
| -0,290362325 | 0,216780617 | SLCO1B3 | FALSE | Downregulated |
| -0,162185238 | 0,216780617 | WDR92 | FALSE | Downregulated |
| -0,1363989 | 0,216892141 | FUCA1 | FALSE | Downregulated |
| -0,57444825 | 0,216993513 | TRIM73 | FALSE | Downregulated |
| -0,436388724 | 0,217135687 | TRNM | FALSE | Downregulated |
| 0,160354945 | 0,217170779 | ABI1 | FALSE | Upregulated |
| 0,15766378 | 0,217258168 | ABHD16A | FALSE | Upregulated |
| 0,164613482 | 0,217419412 | CNP | FALSE | Upregulated |
| -0,30471429 | 0,217488187 | ZNF585A | FALSE | Downregulated |
| -0,726009705 | 0,217515742 | LOC101060553 | FALSE | Downregulated |
| -0,172450818 | 0,217544663 | FAM133B | FALSE | Downregulated |
| 0,295827545 | 0,217657082 | CAV1 | FALSE | Upregulated |
| -0,102181088 | 0,217657082 | GOPC | FALSE | Downregulated |
| 0,322046717 | 0,217657082 | TONSL | FALSE | Upregulated |
| -2,128090463 | 0,217657082 | UGT2A1 | FALSE | Downregulated |
| 0,176144018 | 0,217757421 | ERI3 | FALSE | Upregulated |
| 0,204999261 | 0,217838796 | TXNDC9 | FALSE | Upregulated |
| -1,450424686 | 0,218066836 | LOC102724593 | FALSE | Downregulated |
| -0,361915014 | 0,218097344 | CES4A | FALSE | Downregulated |
| 1,793033509 | 0,218097344 | KLK11 | FALSE | Upregulated |
| 0,238754592 | 0,218123234 | ASPN | FALSE | Upregulated |
| -0,127312079 | 0,218123234 | ING4 | FALSE | Downregulated |
| 1,20701216 | 0,218133128 | LOC101927596 | FALSE | Upregulated |
| -0,175884376 | 0,218156907 | RPP25L | FALSE | Downregulated |
| -0,17088451 | 0,218156907 | VARS2 | FALSE | Downregulated |
| -0,737873881 | 0,218238564 | PSRC1 | FALSE | Downregulated |
| -0,201961186 | 0,218320796 | COX6A1 | FALSE | Downregulated |
| 0,727821792 | 0,218393058 | COCH | FALSE | Upregulated |
| -0,428472916 | 0,218393293 | CYP3A4 | FALSE | Downregulated |
| 0,82256048 | 0,218409041 | EDN2 | FALSE | Upregulated |
| -0,144829409 | 0,218409041 | PPP2R5A | FALSE | Downregulated |
| -0,678978452 | 0,21845341 | PFN4 | FALSE | Downregulated |
| 1,972043936 | 0,21845341 | TMEM213 | FALSE | Upregulated |
| -0,492519246 | 0,21845341 | TSPAN8 | FALSE | Downregulated |
| 0,49118477 | 0,218702043 | ZNF300P1 | FALSE | Upregulated |
| -0,091589942 | 0,21873442 | DNAJC14 | FALSE | Downregulated |
| 0,208509515 | 0,218895034 | BTG1 | FALSE | Upregulated |
| -0,132496999 | 0,218899546 | TAF4 | FALSE | Downregulated |
| -0,192490472 | 0,218994489 | NR2F6 | FALSE | Downregulated |
| -1,053991149 | 0,219190546 | LOC102724539 | FALSE | Downregulated |
| 1,240168795 | 0,219216243 | LOC101928292 | FALSE | Upregulated |
| 0,474265894 | 0,219282462 | C15orf65 | FALSE | Upregulated |
| -0,247121499 | 0,219414852 | AADAC | FALSE | Downregulated |
| -0,267343064 | 0,219414852 | PDK3 | FALSE | Downregulated |
| -0,211347056 | 0,219448373 | PDZD8 | FALSE | Downregulated |
| 0,212235208 | 0,219526943 | CAMLG | FALSE | Upregulated |
| 0,690424529 | 0,219526943 | ESRRG | FALSE | Upregulated |
| -0,377666188 | 0,21957732 | SLC30A10 | FALSE | Downregulated |
| -0,596258845 | 0,219633448 | TTC40 | FALSE | Downregulated |
| 0,11808223 | 0,219720121 | PCYT1A | FALSE | Upregulated |
| 1,534416251 | 0,219740856 | CD1A | FALSE | Upregulated |

| | | | | |
|---|---|---|---|---|
| -0,208648593 | 0,219740856 | KLHDC8B | FALSE | Downregulated |
| 1,268842848 | 0,219876313 | FUT7 | FALSE | Upregulated |
| -0,239996237 | 0,219895732 | RAI14 | FALSE | Downregulated |
| -0,399877578 | 0,219919779 | NXN | FALSE | Downregulated |
| -0,121392185 | 0,219919779 | TARS2 | FALSE | Downregulated |
| -0,155890818 | 0,219935789 | PAAF1 | FALSE | Downregulated |
| -0,214303073 | 0,219972693 | PDDC1 | FALSE | Downregulated |
| 1,490291284 | 0,219972693 | POU3F2 | FALSE | Upregulated |
| 1,478656276 | 0,220001723 | FBLL1 | FALSE | Upregulated |
| 0,299949811 | 0,220039856 | CSRP2 | FALSE | Upregulated |
| -0,393293785 | 0,220039856 | FBXO32 | FALSE | Downregulated |
| -0,271739469 | 0,220039856 | NAT6 | FALSE | Downregulated |
| -0,186922841 | 0,220039944 | MGST1 | FALSE | Downregulated |
| -0,15565425 | 0,220192148 | F2 | FALSE | Downregulated |
| 0,205776282 | 0,220192148 | RNF149 | FALSE | Upregulated |
| 0,219963159 | 0,22032364 | C4orf32 | FALSE | Upregulated |
| 0,299864185 | 0,220466631 | CD320 | FALSE | Upregulated |
| 0,25626769 | 0,220466631 | CD58 | FALSE | Upregulated |
| 0,317887128 | 0,220466631 | CHST12 | FALSE | Upregulated |
| -0,490102431 | 0,220466631 | LOC100133331 | FALSE | Downregulated |
| 0,319190021 | 0,220495164 | ELMO3 | FALSE | Upregulated |
| 1,482306283 | 0,220543896 | AC226118.1 | FALSE | Upregulated |
| 0,247296025 | 0,220543896 | DPCD | FALSE | Upregulated |
| -0,33462617 | 0,220578138 | LOC729291 | FALSE | Downregulated |
| 1,105487266 | 0,220578138 | SAA1 | FALSE | Upregulated |
| 0,351310819 | 0,220578138 | TCTN2 | FALSE | Upregulated |
| 0,171964229 | 0,220578138 | ZSCAN25 | FALSE | Upregulated |
| 0,139406358 | 0,220594248 | TBC1D22A | FALSE | Upregulated |
| -0,141992819 | 0,220677862 | JADE3 | FALSE | Downregulated |
| 0,153000407 | 0,220677902 | PPP3CC | FALSE | Upregulated |
| -0,236629219 | 0,220813732 | CAMKMT | FALSE | Downregulated |
| -0,214422633 | 0,220850808 | CENPV | FALSE | Downregulated |
| -0,128502135 | 0,220910721 | CNOT4 | FALSE | Downregulated |
| -0,334525611 | 0,220910721 | NBPF10 | FALSE | Downregulated |
| -2,49407338 | 0,220910721 | SNORD3C | FALSE | Downregulated |
| -1,079153111 | 0,220931133 | LOC100506797 | FALSE | Downregulated |
| 0,160435063 | 0,220968546 | CDK2 | FALSE | Upregulated |
| -0,185158919 | 0,220968546 | TMEM219 | FALSE | Downregulated |
| 1,500632901 | 0,221015719 | LOC101060632 | FALSE | Upregulated |
| -0,205569326 | 0,221441237 | PFDN2 | FALSE | Downregulated |
| -0,160505166 | 0,221531095 | DESI1 | FALSE | Downregulated |
| -0,366655584 | 0,221531095 | THAP7-AS1 | FALSE | Downregulated |
| -0,758960725 | 0,221577467 | KLHL31 | FALSE | Downregulated |
| 1,126335135 | 0,221577467 | LOC101927119 | FALSE | Upregulated |
| -0,134841982 | 0,221577467 | PPP5C | FALSE | Downregulated |
| 0,814520773 | 0,221577467 | SLFN12L | FALSE | Upregulated |
| 3,180340882 | 0,221586673 | TMPRSS7 | FALSE | Upregulated |
| 0,230906308 | 0,221721602 | TRMT1 | FALSE | Upregulated |
| 2,143009755 | 0,221808745 | TRNAD8 | FALSE | Upregulated |
| 0,243605968 | 0,221824562 | SCPEP1 | FALSE | Upregulated |
| 0,752293537 | 0,221826008 | PMFBP1 | FALSE | Upregulated |

| | | | | |
|---|---|---|---|---|
| 0,230415554 | 0,221989116 | EPHB4 | FALSE | Upregulated |
| 0,359339573 | 0,221989116 | HHIPL1 | FALSE | Upregulated |
| -1,396547602 | 0,221989116 | LOC101928926 | FALSE | Downregulated |
| -0,167796344 | 0,221989116 | PAF1 | FALSE | Downregulated |
| 0,361845492 | 0,221989116 | PRKAG2-AS1 | FALSE | Upregulated |
| 0,173171586 | 0,221989116 | RIN2 | FALSE | Upregulated |
| 1,941556683 | 0,222160407 | SLC6A7 | FALSE | Upregulated |
| 0,820807339 | 0,222189697 | CBLN1 | FALSE | Upregulated |
| 0,174183174 | 0,222189697 | GTPBP6 | FALSE | Upregulated |
| 0,510014804 | 0,222231968 | OASL | FALSE | Upregulated |
| -0,338760228 | 0,222271549 | PANK1 | FALSE | Downregulated |
| -0,139571316 | 0,222293444 | HINT3 | FALSE | Downregulated |
| -0,79582579 | 0,222452574 | CCER2 | FALSE | Downregulated |
| 1,080157786 | 0,222455555 | ADAM11 | FALSE | Upregulated |
| -0,176428323 | 0,222607963 | ZKSCAN8 | FALSE | Downregulated |
| -0,170367646 | 0,222629244 | POLR1C | FALSE | Downregulated |
| 0,209758946 | 0,222665536 | PPP1R26 | FALSE | Upregulated |
| -0,167527436 | 0,222740041 | RAPGEF2 | FALSE | Downregulated |
| 0,561258787 | 0,22280103 | TNFAIP8L3 | FALSE | Upregulated |
| -0,409251852 | 0,22281645 | CES5A | FALSE | Downregulated |
| 1,077704232 | 0,222902104 | SUSD4 | FALSE | Upregulated |
| 1,302928274 | 0,222949526 | SMKR1 | FALSE | Upregulated |
| -0,167949338 | 0,222949526 | ZNF33B | FALSE | Downregulated |
| 0,279474969 | 0,222965184 | PAG1 | FALSE | Upregulated |
| -0,146755968 | 0,223011173 | DRAP1 | FALSE | Downregulated |
| -0,089321724 | 0,223077959 | STARD7 | FALSE | Downregulated |
| -0,158553542 | 0,223080803 | ABHD3 | FALSE | Downregulated |
| 1,710675529 | 0,223080803 | XRCC6P5 | FALSE | Upregulated |
| -0,312786675 | 0,223269355 | FAM47E | FALSE | Downregulated |
| -0,253943543 | 0,223298601 | ALG3 | FALSE | Downregulated |
| 1,420040556 | 0,223298601 | HHIPL2 | FALSE | Upregulated |
| -0,29940003 | 0,223298601 | LRTOMT | FALSE | Downregulated |
| 0,220674539 | 0,223298601 | SHROOM2 | FALSE | Upregulated |
| 0,919167052 | 0,223359813 | CDRT1 | FALSE | Upregulated |
| 2,083925135 | 0,223359813 | TBC1D27 | FALSE | Upregulated |
| 0,18442062 | 0,223589864 | PPP1R12C | FALSE | Upregulated |
| 0,765017838 | 0,223729985 | TNFRSF8 | FALSE | Upregulated |
| 0,327467398 | 0,223776576 | IFIT2 | FALSE | Upregulated |
| -0,184791754 | 0,223853127 | LNX2 | FALSE | Downregulated |
| 0,240544321 | 0,223921549 | RASSF5 | FALSE | Upregulated |
| 0,4917092 | 0,224119963 | CCNE1 | FALSE | Upregulated |
| -0,459061208 | 0,224201929 | RTN4R | FALSE | Downregulated |
| 0,882484323 | 0,224291566 | OR2A20P | FALSE | Upregulated |
| -0,316330137 | 0,224365182 | FAM26E | FALSE | Downregulated |
| 0,724278568 | 0,224415549 | NPEPL1 | FALSE | Upregulated |
| -0,205363485 | 0,224435286 | ACBD4 | FALSE | Downregulated |
| 0,545506552 | 0,224552004 | CDC25A | FALSE | Upregulated |
| -0,213107374 | 0,224558295 | ECM2 | FALSE | Downregulated |
| 0,782681671 | 0,22460347 | NRXN3 | FALSE | Upregulated |
| -0,214590344 | 0,224613492 | CDC14B | FALSE | Downregulated |
| -0,10861219 | 0,224827975 | PMPCB | FALSE | Downregulated |

| | | | | |
|---|---|---|---|---|
| -0,334674232 | 0,224922476 | PSAT1 | FALSE | Downregulated |
| -1,442373112 | 0,225005309 | LOC102724874 | FALSE | Downregulated |
| -0,779505008 | 0,225156373 | RUNX2 | FALSE | Downregulated |
| 1,073750923 | 0,225331432 | TPBGL | FALSE | Upregulated |
| -1,255220166 | 0,225525665 | RP1-69M21.2 | FALSE | Downregulated |
| -0,187470182 | 0,22583929 | GOLGA2 | FALSE | Downregulated |
| 0,136279432 | 0,225906148 | MXD4 | FALSE | Upregulated |
| -0,175916439 | 0,225929917 | COMMD2 | FALSE | Downregulated |
| 0,668260976 | 0,225976534 | LOC101929638 | FALSE | Upregulated |
| -0,184611361 | 0,226000402 | LINC00909 | FALSE | Downregulated |
| 0,178820583 | 0,226257502 | HSPD1 | FALSE | Upregulated |
| 1,111941305 | 0,226292033 | B4GALNT1 | FALSE | Upregulated |
| 0,110418641 | 0,226292033 | GNA11 | FALSE | Upregulated |
| 1,066844409 | 0,226385939 | CCDC36 | FALSE | Upregulated |
| 0,205162484 | 0,226430372 | AGTPBP1 | FALSE | Upregulated |
| 0,281640631 | 0,226632638 | MXRA7 | FALSE | Upregulated |
| 0,146748447 | 0,226632738 | RHBDD1 | FALSE | Upregulated |
| 0,558749997 | 0,226746181 | NYNRIN | FALSE | Upregulated |
| -0,24281395 | 0,227121993 | TTC30B | FALSE | Downregulated |
| 0,294048559 | 0,227186775 | MCF2L | FALSE | Upregulated |
| 0,458987295 | 0,227449101 | PLCD3 | FALSE | Upregulated |
| -0,32496467 | 0,227455814 | FGG | FALSE | Downregulated |
| 1,290664293 | 0,227513584 | FAM132B | FALSE | Upregulated |
| 0,238936139 | 0,227641001 | ZNF792 | FALSE | Upregulated |
| -0,14810758 | 0,227739655 | CREBBP | FALSE | Downregulated |
| -0,153138957 | 0,227739655 | PALB2 | FALSE | Downregulated |
| 0,343006769 | 0,227739655 | STIL | FALSE | Upregulated |
| -0,692655077 | 0,227841555 | GSTA2 | FALSE | Downregulated |
| -0,268068587 | 0,22792447 | MPND | FALSE | Downregulated |
| -0,209527984 | 0,227977911 | CCDC159 | FALSE | Downregulated |
| -0,138536564 | 0,227977911 | UIMC1 | FALSE | Downregulated |
| 0,922817552 | 0,228260945 | SYCP2L | FALSE | Upregulated |
| 1,325459787 | 0,228336542 | PCSK1 | FALSE | Upregulated |
| 0,802394355 | 0,228385564 | CYP27C1 | FALSE | Upregulated |
| -0,850949145 | 0,228412767 | LOC102724387 | FALSE | Downregulated |
| -0,242142664 | 0,228544491 | DHRS7 | FALSE | Downregulated |
| 0,287733629 | 0,228544491 | TTC3P1 | FALSE | Upregulated |
| -2,018049198 | 0,228574519 | LOC100996379 | FALSE | Downregulated |
| 1,208280528 | 0,228634101 | FAM178B | FALSE | Upregulated |
| 0,217130752 | 0,228646846 | MED22 | FALSE | Upregulated |
| 0,130148307 | 0,228657474 | COPS6 | FALSE | Upregulated |
| 0,294596174 | 0,228657474 | ETS2 | FALSE | Upregulated |
| -0,147011713 | 0,228657474 | ZNF318 | FALSE | Downregulated |
| -1,145881634 | 0,228766304 | LINC00277 | FALSE | Downregulated |
| 0,171289022 | 0,229144739 | ARHGAP17 | FALSE | Upregulated |
| 0,821865806 | 0,229144739 | LOC102723558 | FALSE | Upregulated |
| 0,955813364 | 0,229206183 | HPN-AS1 | FALSE | Upregulated |
| -0,507569709 | 0,229208274 | LOC101929664 | FALSE | Downregulated |
| -0,454593457 | 0,229400445 | MST1L | FALSE | Downregulated |
| 0,576584179 | 0,229466106 | NLGN3 | FALSE | Upregulated |
| 0,319973282 | 0,229637935 | IKZF2 | FALSE | Upregulated |

| | | | | |
|---|---|---|---|---|
| 0,27131745 | 0,229943916 | ECSCR-2 | FALSE | Upregulated |
| -0,26313467 | 0,229943916 | KLF12 | FALSE | Downregulated |
| 0,165314775 | 0,229943916 | RABIF | FALSE | Upregulated |
| 1,214858898 | 0,229943916 | SMC1B | FALSE | Upregulated |
| -0,160989551 | 0,229943916 | SNRPC | FALSE | Downregulated |
| 0,34855878 | 0,229943916 | ZFAND5 | FALSE | Upregulated |
| -0,36887373 | 0,229993303 | LINC00870 | FALSE | Downregulated |
| 0,223582337 | 0,230061865 | INCENP | FALSE | Upregulated |
| 0,188384036 | 0,230146582 | CBFA2T2 | FALSE | Upregulated |
| 0,262700054 | 0,230175351 | ETV5 | FALSE | Upregulated |
| 0,199596622 | 0,230437616 | CHML | FALSE | Upregulated |
| 0,636413448 | 0,230437616 | FAM225A | FALSE | Upregulated |
| -0,887221881 | 0,23045395 | AC002398.5 | FALSE | Downregulated |
| 1,761269642 | 0,230482987 | LIPN | FALSE | Upregulated |
| 0,127067543 | 0,230482987 | PSMB2 | FALSE | Upregulated |
| -0,192685805 | 0,23051007 | ACAD11 | FALSE | Downregulated |
| -0,255919602 | 0,23051007 | DHRS1 | FALSE | Downregulated |
| -0,39756585 | 0,23051007 | ZMAT1 | FALSE | Downregulated |
| 0,145557334 | 0,230515367 | CDK16 | FALSE | Upregulated |
| 0,110548688 | 0,230770634 | LEPROT | FALSE | Upregulated |
| -0,149145158 | 0,230770634 | ZW10 | FALSE | Downregulated |
| 0,755272071 | 0,230846881 | CEACAM22P | FALSE | Upregulated |
| -0,138428177 | 0,230846881 | CLDN1 | FALSE | Downregulated |
| 1,34908064 | 0,230905188 | ENTPD3 | FALSE | Upregulated |
| -0,403031051 | 0,231070253 | DFFB | FALSE | Downregulated |
| 1,491554358 | 0,231258052 | SMIM22 | FALSE | Upregulated |
| 1,071078083 | 0,231307153 | PHEX | FALSE | Upregulated |
| 0,294457418 | 0,231307153 | SMC2 | FALSE | Upregulated |
| 0,342936254 | 0,231328899 | NCF4 | FALSE | Upregulated |
| -0,213290736 | 0,231328899 | NGFRAP1 | FALSE | Downregulated |
| 0,238892556 | 0,231392863 | IFT43 | FALSE | Upregulated |
| -0,995460031 | 0,231392863 | PPFIA4 | FALSE | Downregulated |
| 1,419931245 | 0,231392863 | RAB6C | FALSE | Upregulated |
| -0,095560229 | 0,231392863 | TM9SF4 | FALSE | Downregulated |
| -0,680420898 | 0,231591098 | PCAT6 | FALSE | Downregulated |
| -0,126516921 | 0,23160387 | EXOSC2 | FALSE | Downregulated |
| -0,205608077 | 0,231627123 | SIK3 | FALSE | Downregulated |
| 0,637116696 | 0,231890878 | CDCA2 | FALSE | Upregulated |
| 0,172292197 | 0,231890878 | G6PC3 | FALSE | Upregulated |
| 0,905384372 | 0,231890878 | RP3-395M20.9 | FALSE | Upregulated |
| 0,746242122 | 0,231948252 | WT1 | FALSE | Upregulated |
| 1,293970648 | 0,232103484 | RGS17 | FALSE | Upregulated |
| 0,273514451 | 0,232183331 | DYNLL1-AS1 | FALSE | Upregulated |
| -0,648783986 | 0,232262071 | KCNK5 | FALSE | Downregulated |
| 0,147286984 | 0,232348528 | HSPA14 | FALSE | Upregulated |
| -0,306653093 | 0,232443478 | LINC01024 | FALSE | Downregulated |
| 0,515371464 | 0,232541588 | GRHL3 | FALSE | Upregulated |
| 0,317249838 | 0,232673784 | ZNF324B | FALSE | Upregulated |
| 0,440638098 | 0,233055616 | PRNCR1 | FALSE | Upregulated |
| 1,521520974 | 0,233089574 | NXPH2 | FALSE | Upregulated |
| 0,441846402 | 0,233211949 | TLCD1 | FALSE | Upregulated |

| | | | | |
|---|---|---|---|---|
| 0,57696895 | 0,23323982 | ZSCAN12P1 | FALSE | Upregulated |
| 0,216455987 | 0,233309977 | EXTL3 | FALSE | Upregulated |
| 1,035509933 | 0,233309977 | HIST2H2BF | FALSE | Upregulated |
| 1,169289318 | 0,233309977 | LINC00461 | FALSE | Upregulated |
| 0,405502967 | 0,233309977 | ZNF287 | FALSE | Upregulated |
| 1,472159531 | 0,233449002 | LOC100130298 | FALSE | Upregulated |
| 1,462731752 | 0,233465427 | RAB44 | FALSE | Upregulated |
| 1,675867116 | 0,233549496 | LOC102724424 | FALSE | Upregulated |
| -0,598383419 | 0,233614915 | F2RL2 | FALSE | Downregulated |
| 0,912116158 | 0,23368558 | FOSL1 | FALSE | Upregulated |
| -0,481176054 | 0,233733745 | CYP2B6 | FALSE | Downregulated |
| 0,247247395 | 0,23375967 | RAB27A | FALSE | Upregulated |
| 1,148312383 | 0,23377212 | C1QL1 | FALSE | Upregulated |
| 0,139495057 | 0,233833637 | CAPZA2 | FALSE | Upregulated |
| 0,345632013 | 0,233901087 | EXOC3L1 | FALSE | Upregulated |
| -0,398762835 | 0,233963974 | ZNF582-AS1 | FALSE | Downregulated |
| -0,297726239 | 0,234064411 | WDR78 | FALSE | Downregulated |
| 1,592052056 | 0,23407772 | CHRM5 | FALSE | Upregulated |
| -0,346324797 | 0,23407772 | TSNAXIP1 | FALSE | Downregulated |
| -0,219095202 | 0,234094965 | TIMM8A | FALSE | Downregulated |
| -0,313054541 | 0,234302718 | JOSD2 | FALSE | Downregulated |
| -0,139336414 | 0,234302718 | RPAP1 | FALSE | Downregulated |
| 0,223621075 | 0,234377089 | NUP210 | FALSE | Upregulated |
| -0,788543023 | 0,234390311 | PP13 | FALSE | Downregulated |
| 0,176312362 | 0,234440724 | NFE2L2 | FALSE | Upregulated |
| -0,147869516 | 0,234440724 | SLC31A1 | FALSE | Downregulated |
| 1,749025548 | 0,234440724 | SNORA14B | FALSE | Upregulated |
| -0,213227782 | 0,234512024 | BOLA3 | FALSE | Downregulated |
| 0,21940567 | 0,234512024 | IFNGR2 | FALSE | Upregulated |
| -1,013108621 | 0,234514074 | FAM83C | FALSE | Downregulated |
| -0,569775397 | 0,234573639 | LOC100505650 | FALSE | Downregulated |
| 1,091350236 | 0,234720334 | MAP1LC3C | FALSE | Upregulated |
| 0,247658411 | 0,234738921 | FTSJ1 | FALSE | Upregulated |
| -0,263943472 | 0,234811894 | DHRS11 | FALSE | Downregulated |
| 0,245006423 | 0,234811894 | IER2 | FALSE | Upregulated |
| -0,6208441 | 0,234914233 | IHH | FALSE | Downregulated |
| 0,998058894 | 0,23497392 | RP11-290F20.2 | FALSE | Upregulated |
| 1,106035716 | 0,235017729 | KCNIP1 | FALSE | Upregulated |
| -0,350787245 | 0,235071249 | DUSP10 | FALSE | Downregulated |
| 0,262543287 | 0,235071249 | FLVCR1 | FALSE | Upregulated |
| -1,352300236 | 0,235071249 | LOC100128374 | FALSE | Downregulated |
| 0,163285541 | 0,235071249 | TNIP2 | FALSE | Upregulated |
| 1,686034359 | 0,235073159 | GLIS1 | FALSE | Upregulated |
| 0,169203082 | 0,235326353 | EEFSEC | FALSE | Upregulated |
| 0,439712228 | 0,235336069 | FAM13C | FALSE | Upregulated |
| -0,532596687 | 0,235648194 | LOC727978 | FALSE | Downregulated |
| -0,23723574 | 0,235765985 | CHD1L | FALSE | Downregulated |
| 1,397677829 | 0,235765985 | RP11-673E1.1 | FALSE | Upregulated |
| -0,332310522 | 0,235765985 | ST20 | FALSE | Downregulated |
| 0,337901215 | 0,235765985 | ZNF837 | FALSE | Upregulated |
| -0,171918879 | 0,235857008 | C21orf33 | FALSE | Downregulated |

| | | | | |
|---|---|---|---|---|
| 1,391839507 | 0,235857008 | SPATC1 | FALSE | Upregulated |
| -0,389156201 | 0,23588882 | LPPR1 | FALSE | Downregulated |
| 0,178002386 | 0,235938113 | TRIM25 | FALSE | Upregulated |
| -0,172731269 | 0,236114281 | BPGM | FALSE | Downregulated |
| 0,100708277 | 0,236114281 | CAMTA2 | FALSE | Upregulated |
| 0,948205632 | 0,236114281 | RP11-344B5.2 | FALSE | Upregulated |
| 0,226508009 | 0,236135853 | PQLC2 | FALSE | Upregulated |
| -0,447356611 | 0,236164812 | LOC100287497 | FALSE | Downregulated |
| 0,202556903 | 0,236869779 | CYB5R4 | FALSE | Upregulated |
| -0,72430764 | 0,237118706 | LOC102723398 | FALSE | Downregulated |
| -0,35863559 | 0,237118706 | MPV17L | FALSE | Downregulated |
| 0,7772961 | 0,237118706 | POU5F1 | FALSE | Upregulated |
| 0,195237488 | 0,237118706 | RFT1 | FALSE | Upregulated |
| 0,275684142 | 0,237118706 | TOR4A | FALSE | Upregulated |
| 1,62183969 | 0,23712064 | ACOXL | FALSE | Upregulated |
| -1,059610507 | 0,23712064 | DDIT4L | FALSE | Downregulated |
| -0,142962179 | 0,23712064 | HEBP1 | FALSE | Downregulated |
| 1,692098744 | 0,23712064 | LOC100129473 | FALSE | Upregulated |
| 1,427867376 | 0,23712064 | LOC101928139 | FALSE | Upregulated |
| 0,87498563 | 0,23712064 | LOC102724031 | FALSE | Upregulated |
| 1,148375821 | 0,23712064 | OSTCP1 | FALSE | Upregulated |
| 0,193561715 | 0,23712064 | PPP1R37 | FALSE | Upregulated |
| 0,297659331 | 0,237133165 | FANCM | FALSE | Upregulated |
| -0,184641309 | 0,237147174 | GSDMD | FALSE | Downregulated |
| 0,170723732 | 0,237336929 | AARSD1 | FALSE | Upregulated |
| 0,12687825 | 0,237336929 | SETDB1 | FALSE | Upregulated |
| 0,126773228 | 0,237336929 | TMEM127 | FALSE | Upregulated |
| 0,651870468 | 0,237443398 | RP11-701P16.5 | FALSE | Upregulated |
| 0,222436026 | 0,237516439 | SMC4 | FALSE | Upregulated |
| -0,626582783 | 0,237546868 | LOC101929443 | FALSE | Downregulated |
| -0,706194082 | 0,237546868 | LOC102723622 | FALSE | Downregulated |
| -0,270933835 | 0,237565843 | TTC5 | FALSE | Downregulated |
| -0,177483337 | 0,237608297 | LAMTOR5 | FALSE | Downregulated |
| 0,291345955 | 0,237608297 | SPIN2B | FALSE | Upregulated |
| 1,488542127 | 0,237650563 | MEIS1-AS3 | FALSE | Upregulated |
| -0,164301419 | 0,237669243 | DAAM1 | FALSE | Downregulated |
| 0,1694851 | 0,237669243 | TEX10 | FALSE | Upregulated |
| -0,164248759 | 0,237715673 | HIGD2A | FALSE | Downregulated |
| 0,145408857 | 0,23774062 | IDH3A | FALSE | Upregulated |
| -0,161909062 | 0,23789348 | C9orf114 | FALSE | Downregulated |
| 0,397079712 | 0,237996474 | PHKG1 | FALSE | Upregulated |
| 0,162253472 | 0,23817828 | ATRN | FALSE | Upregulated |
| 0,138196617 | 0,238268247 | PIGO | FALSE | Upregulated |
| 0,137737872 | 0,238268247 | SKP1 | FALSE | Upregulated |
| 1,509583157 | 0,238268247 | VWA3A | FALSE | Upregulated |
| -0,31840134 | 0,238268247 | ZC3H13 | FALSE | Downregulated |
| 0,919636879 | 0,238499653 | OR2A7 | FALSE | Upregulated |
| -0,193744573 | 0,238525378 | NMRAL1 | FALSE | Downregulated |
| 0,119086712 | 0,238611018 | XPO5 | FALSE | Upregulated |
| 0,764263709 | 0,238703231 | LOC102724231 | FALSE | Upregulated |
| 1,548703664 | 0,238703231 | NKPD1 | FALSE | Upregulated |

| | | | | |
|---|---|---|---|---|
| -0,947065226 | 0,238703231 | PRAMENP | FALSE | Downregulated |
| -0,166420406 | 0,238761789 | GNB2L1 | FALSE | Downregulated |
| 0,420014601 | 0,238761789 | MBLAC1 | FALSE | Upregulated |
| -0,164228953 | 0,238761789 | SRRT | FALSE | Downregulated |
| -0,348617918 | 0,238803406 | NBPF19 | FALSE | Downregulated |
| -0,286660906 | 0,238803406 | SLC22A23 | FALSE | Downregulated |
| 0,320355201 | 0,238803406 | TRAPPC2P1 | FALSE | Upregulated |
| 3,131076613 | 0,239028496 | RNU1-28P | FALSE | Upregulated |
| 0,261728028 | 0,239102772 | ZNF213 | FALSE | Upregulated |
| 0,137813505 | 0,239140703 | MICU2 | FALSE | Upregulated |
| 0,447857834 | 0,239319812 | SREBF1 | FALSE | Upregulated |
| 0,337765662 | 0,239512952 | PDE1B | FALSE | Upregulated |
| 0,867957279 | 0,239713017 | RP11-7F17.5 | FALSE | Upregulated |
| 0,178762046 | 0,239833064 | FAM73A | FALSE | Upregulated |
| 0,28716154 | 0,239838901 | LAMB1 | FALSE | Upregulated |
| 1,392790961 | 0,239838901 | MLK7-AS1 | FALSE | Upregulated |
| 0,20355174 | 0,240056767 | PPT1 | FALSE | Upregulated |
| 0,883629929 | 0,240116346 | C19orf35 | FALSE | Upregulated |
| 0,206117976 | 0,240116346 | NOP2 | FALSE | Upregulated |
| 0,953056465 | 0,240243104 | BEND6 | FALSE | Upregulated |
| -0,350129297 | 0,240274535 | SLC31A2 | FALSE | Downregulated |
| 0,779316819 | 0,240276306 | ARSI | FALSE | Upregulated |
| 0,546457221 | 0,24028782 | ATHL1 | FALSE | Upregulated |
| -0,919219077 | 0,240320845 | AQP6 | FALSE | Downregulated |
| 0,912159863 | 0,240320845 | C1orf167 | FALSE | Upregulated |
| -0,872101515 | 0,24032991 | CYP7A1 | FALSE | Downregulated |
| 1,080108454 | 0,24047222 | LOC102724384 | FALSE | Upregulated |
| 2,10757063 | 0,240496704 | TRIM50 | FALSE | Upregulated |
| 1,338807071 | 0,240600548 | ZNF695 | FALSE | Upregulated |
| 0,172477679 | 0,240631066 | ATIC | FALSE | Upregulated |
| -0,428272359 | 0,240631066 | PRUNE2 | FALSE | Downregulated |
| 0,192760963 | 0,240764176 | ADPRHL2 | FALSE | Upregulated |
| -0,156609078 | 0,240809236 | AURKAIP1 | FALSE | Downregulated |
| -0,242822284 | 0,240809236 | CCDC66 | FALSE | Downregulated |
| -0,136378342 | 0,240950704 | KIAA1671 | FALSE | Downregulated |
| 0,778097169 | 0,240992836 | C1orf233 | FALSE | Upregulated |
| -0,223759648 | 0,24101264 | BAZ2B | FALSE | Downregulated |
| -0,164214521 | 0,24101264 | FAHD2A | FALSE | Downregulated |
| 0,525217844 | 0,241122417 | L3MBTL1 | FALSE | Upregulated |
| 0,220125468 | 0,241122417 | MPZL3 | FALSE | Upregulated |
| 0,15765617 | 0,241122417 | SLC35A2 | FALSE | Upregulated |
| 1,692204982 | 0,241122417 | ZBED2 | FALSE | Upregulated |
| 0,453625325 | 0,241218153 | ZNF610 | FALSE | Upregulated |
| 1,718305587 | 0,241250572 | FER1L6 | FALSE | Upregulated |
| -0,481740938 | 0,241314249 | LOC644794 | FALSE | Downregulated |
| 1,013153461 | 0,241335766 | INPP5J | FALSE | Upregulated |
| 0,191498616 | 0,24160903 | FADS3 | FALSE | Upregulated |
| -0,128847127 | 0,241668999 | CUTA | FALSE | Downregulated |
| -0,347483872 | 0,241805265 | ASPG | FALSE | Downregulated |
| -0,15028364 | 0,241805265 | GCLC | FALSE | Downregulated |
| 1,145726407 | 0,241805265 | LOC102724426 | FALSE | Upregulated |

| | | | | |
|---|---|---|---|---|
| -0,170263045 | 0,241976492 | ECI2 | FALSE | Downregulated |
| 0,224592086 | 0,241996378 | WRAP73 | FALSE | Upregulated |
| 0,200073266 | 0,242152181 | LONRF1 | FALSE | Upregulated |
| 1,736948054 | 0,242264503 | ZNF732 | FALSE | Upregulated |
| 0,16843404 | 0,242371415 | ADAM17 | FALSE | Upregulated |
| -0,36535624 | 0,242371415 | ADRA2B | FALSE | Downregulated |
| -1,441189752 | 0,242371415 | LRRC3B | FALSE | Downregulated |
| 0,31072088 | 0,242371415 | VSTM4 | FALSE | Upregulated |
| -0,239666732 | 0,242576851 | RITA1 | FALSE | Downregulated |
| 0,1726111 | 0,242652231 | MYO5B | FALSE | Upregulated |
| 0,093117654 | 0,242680352 | VWA9 | FALSE | Upregulated |
| 0,223441997 | 0,24268915 | NPM3 | FALSE | Upregulated |
| 1,152168801 | 0,242727134 | TTTY14 | FALSE | Upregulated |
| -0,748439146 | 0,242730947 | MYLK3 | FALSE | Downregulated |
| -0,347320459 | 0,242740353 | LCN12 | FALSE | Downregulated |
| -0,262462047 | 0,242740353 | ZNF121 | FALSE | Downregulated |
| 1,231309976 | 0,242792834 | MOG | FALSE | Upregulated |
| -0,140165065 | 0,243021286 | SNRPG | FALSE | Downregulated |
| -0,158434034 | 0,243026962 | RPL18 | FALSE | Downregulated |
| 0,286087208 | 0,243026962 | ZNF461 | FALSE | Upregulated |
| -0,320685995 | 0,243069287 | AP3D1 | FALSE | Downregulated |
| 0,309600021 | 0,243178168 | ZNF256 | FALSE | Upregulated |
| -0,881057366 | 0,243535193 | DEFB132 | FALSE | Downregulated |
| -0,125853933 | 0,243535193 | RNF141 | FALSE | Downregulated |
| -0,308887298 | 0,243574184 | KDR | FALSE | Downregulated |
| -0,288170589 | 0,243616867 | KRAS | FALSE | Downregulated |
| 0,151577754 | 0,243672683 | FAM219B | FALSE | Upregulated |
| 0,603461605 | 0,243672683 | MGP | FALSE | Upregulated |
| 0,757656282 | 0,243672683 | RRAD | FALSE | Upregulated |
| -0,190259359 | 0,243672683 | SASH1 | FALSE | Downregulated |
| 0,245974093 | 0,243740586 | C19orf47 | FALSE | Upregulated |
| 0,136596978 | 0,243740586 | TCP1 | FALSE | Upregulated |
| 0,456641164 | 0,243955405 | ENPP2 | FALSE | Upregulated |
| -0,324716923 | 0,244074476 | DGKE | FALSE | Downregulated |
| -0,516184335 | 0,244342302 | MYOM2 | FALSE | Downregulated |
| -0,997123036 | 0,244456768 | LOC102723997 | FALSE | Downregulated |
| 1,678385966 | 0,244485986 | CTGLF12P | FALSE | Upregulated |
| -0,236435371 | 0,244600712 | GPR180 | FALSE | Downregulated |
| 0,108786356 | 0,244630547 | PHC2 | FALSE | Upregulated |
| -1,695206571 | 0,244630547 | TUBA3E | FALSE | Downregulated |
| 0,189292595 | 0,244656983 | ANO10 | FALSE | Upregulated |
| -0,150472795 | 0,244798643 | CYB5B | FALSE | Downregulated |
| -0,12521732 | 0,244859945 | ADH5 | FALSE | Downregulated |
| 1,243636016 | 0,244905186 | CSTF3-AS1 | FALSE | Upregulated |
| -0,56965667 | 0,244961758 | APOC1P1 | FALSE | Downregulated |
| 0,232844253 | 0,244973179 | IL4R | FALSE | Upregulated |
| -0,789622331 | 0,244984066 | CNGA4 | FALSE | Downregulated |
| -0,149011709 | 0,244984066 | ZBTB14 | FALSE | Downregulated |
| 1,152763902 | 0,245014009 | LOC100507616 | FALSE | Upregulated |
| 0,424408744 | 0,245117122 | TXK | FALSE | Upregulated |
| -0,174918481 | 0,245141067 | SGMS2 | FALSE | Downregulated |

| | | | | |
|---|---|---|---|---|
| 0,46043547 | 0,245141067 | TBX21 | FALSE | Upregulated |
| -0,785674242 | 0,245173344 | HSPB6 | FALSE | Downregulated |
| -0,317586306 | 0,245176633 | PARK2 | FALSE | Downregulated |
| -0,104356083 | 0,24520985 | SF3B2 | FALSE | Downregulated |
| -0,744287918 | 0,245467272 | RP11-503N18.1 | FALSE | Downregulated |
| -0,137374779 | 0,245497559 | SPATS2L | FALSE | Downregulated |
| -0,186070922 | 0,245598205 | OCIAD2 | FALSE | Downregulated |
| -0,582195424 | 0,245603659 | LOC101926980 | FALSE | Downregulated |
| 0,15100659 | 0,245647546 | COMMD5 | FALSE | Upregulated |
| -0,172230396 | 0,245704944 | RPL3 | FALSE | Downregulated |
| -1,521353288 | 0,245859611 | SLC27A6 | FALSE | Downregulated |
| -0,774115687 | 0,246178254 | LOC101927382 | FALSE | Downregulated |
| 1,304431776 | 0,246178254 | MORC1 | FALSE | Upregulated |
| 1,457431734 | 0,246215054 | FAM212B-AS1 | FALSE | Upregulated |
| 0,177845676 | 0,246215054 | FARSA | FALSE | Upregulated |
| 0,269480033 | 0,246215054 | OGT | FALSE | Upregulated |
| 0,083198092 | 0,246215054 | SH3GLB1 | FALSE | Upregulated |
| 1,396410307 | 0,246260361 | PGM5-AS1 | FALSE | Upregulated |
| 0,204082514 | 0,246320551 | CADM1 | FALSE | Upregulated |
| 0,607630898 | 0,246320551 | XRCC2 | FALSE | Upregulated |
| -0,384377701 | 0,246502603 | GJC3 | FALSE | Downregulated |
| -0,119889012 | 0,246520149 | SUCLA2 | FALSE | Downregulated |
| 0,145186792 | 0,246634206 | ZBTB18 | FALSE | Upregulated |
| 0,244812011 | 0,246687178 | TM4SF1 | FALSE | Upregulated |
| 0,193503585 | 0,246745198 | ZNF337 | FALSE | Upregulated |
| 0,154043436 | 0,24675561 | KANSL2 | FALSE | Upregulated |
| -0,162191765 | 0,246758479 | KPNB1 | FALSE | Downregulated |
| -0,163871648 | 0,24678917 | UFD1L | FALSE | Downregulated |
| 0,100594847 | 0,246905139 | DFFA | FALSE | Upregulated |
| 0,217989269 | 0,247017476 | BTN3A1 | FALSE | Upregulated |
| -0,197885671 | 0,247017476 | NDUFS8 | FALSE | Downregulated |
| 0,112419484 | 0,247017476 | SDF4 | FALSE | Upregulated |
| 0,620663166 | 0,247017476 | SNORA4 | FALSE | Upregulated |
| 0,211480499 | 0,247146062 | ZBTB47 | FALSE | Upregulated |
| -0,431113086 | 0,247200371 | SH3PXD2A | FALSE | Downregulated |
| -0,649962688 | 0,247376774 | ADRB1 | FALSE | Downregulated |
| 0,675739206 | 0,247376774 | LOC101928667 | FALSE | Upregulated |
| -0,187271016 | 0,247376774 | RRAGD | FALSE | Downregulated |
| 0,480358132 | 0,24749713 | SH3D21 | FALSE | Upregulated |
| 1,175416035 | 0,247570598 | RP11-58E21.3 | FALSE | Upregulated |
| 0,303464064 | 0,247611223 | STMN1 | FALSE | Upregulated |
| 0,206767307 | 0,247624393 | ALG6 | FALSE | Upregulated |
| -0,311610501 | 0,247747407 | GCOM1 | FALSE | Downregulated |
| -0,87551218 | 0,247747407 | LOC100130539 | FALSE | Downregulated |
| 0,398689036 | 0,247747407 | LOC400043 | FALSE | Upregulated |
| -0,232483737 | 0,247818985 | C5orf51 | FALSE | Downregulated |
| 0,121104084 | 0,247928102 | ARAP1 | FALSE | Upregulated |
| 0,092607376 | 0,247928102 | GGA2 | FALSE | Upregulated |
| -0,584464917 | 0,24804765 | NIM1K | FALSE | Downregulated |
| 0,507052111 | 0,24818213 | KIF15 | FALSE | Upregulated |
| -0,289680654 | 0,248250558 | AS3MT | FALSE | Downregulated |

| | | | | |
|---|---|---|---|---|
| -0,126276224 | 0,248528981 | ZNF212 | FALSE | Downregulated |
| -0,315772027 | 0,248581024 | ENTPD8 | FALSE | Downregulated |
| 0,212427148 | 0,248581024 | SNX11 | FALSE | Upregulated |
| -0,108833133 | 0,248924212 | PHF10 | FALSE | Downregulated |
| 0,24142034 | 0,248924212 | SPDL1 | FALSE | Upregulated |
| -0,500779024 | 0,24898605 | TBX6 | FALSE | Downregulated |
| 0,596057544 | 0,249063588 | SCD | FALSE | Upregulated |
| -0,090769039 | 0,249162888 | NFX1 | FALSE | Downregulated |
| -0,162867771 | 0,249162888 | ZNF622 | FALSE | Downregulated |
| -0,908147277 | 0,249285819 | RP11-118B18.1 | FALSE | Downregulated |
| 1,899169583 | 0,249348484 | CSMD1 | FALSE | Upregulated |
| 0,276759159 | 0,249431505 | CASC5 | FALSE | Upregulated |
| -0,168262094 | 0,249617784 | RPAP2 | FALSE | Downregulated |
| -0,152097228 | 0,249672644 | MTFR1L | FALSE | Downregulated |
| -0,229447717 | 0,249821622 | HSD17B8 | FALSE | Downregulated |
| 0,181923335 | 0,249956071 | RPL8 | FALSE | Upregulated |
| -0,151626167 | 0,250097334 | PPP1R2 | FALSE | Downregulated |
| -0,474715362 | 0,250279489 | LRRC29 | FALSE | Downregulated |
| -1,018208281 | 0,250338501 | CTD-2337A12.1 | FALSE | Downregulated |
| -0,380808199 | 0,250468005 | BTNL9 | FALSE | Downregulated |
| 0,252987321 | 0,25051412 | CASP1 | FALSE | Upregulated |
| 0,54689644 | 0,25051412 | TNFRSF25 | FALSE | Upregulated |
| -2,261683618 | 0,250570209 | EGF | FALSE | Downregulated |
| 0,14578162 | 0,250570209 | MUL1 | FALSE | Upregulated |
| -0,284820395 | 0,25058383 | CDK19 | FALSE | Downregulated |
| -0,120269788 | 0,250770062 | RBPJ | FALSE | Downregulated |
| -0,102156587 | 0,251023522 | RLF | FALSE | Downregulated |
| 0,454481294 | 0,251088482 | EBI3 | FALSE | Upregulated |
| 0,103106685 | 0,251297709 | YWHAG | FALSE | Upregulated |
| -0,142929558 | 0,251368854 | UBXN6 | FALSE | Downregulated |
| 0,280065113 | 0,251397552 | C7orf31 | FALSE | Upregulated |
| -0,186299919 | 0,251535422 | ATG4A | FALSE | Downregulated |
| -0,129091115 | 0,251596244 | ZNF384 | FALSE | Downregulated |
| -0,144225383 | 0,251650914 | HDAC3 | FALSE | Downregulated |
| 0,237830338 | 0,251854439 | ELMOD3 | FALSE | Upregulated |
| -0,162190632 | 0,251857665 | STARD8 | FALSE | Downregulated |
| 0,861596784 | 0,2521214 | LOC101928660 | FALSE | Upregulated |
| 1,725952559 | 0,252171274 | NLGN1 | FALSE | Upregulated |
| 1,404993525 | 0,252213785 | AC007750.5 | FALSE | Upregulated |
| 0,463410422 | 0,252213785 | APOBEC3D | FALSE | Upregulated |
| 0,136535087 | 0,252236335 | NR2C2 | FALSE | Upregulated |
| 0,176016953 | 0,25224566 | CAMSAP3 | FALSE | Upregulated |
| 0,15483566 | 0,25224566 | FLOT1 | FALSE | Upregulated |
| -1,62708132 | 0,252376508 | TRIM63 | FALSE | Downregulated |
| -0,52519765 | 0,252573693 | TRIM58 | FALSE | Downregulated |
| 0,286697114 | 0,252813972 | ZNF124 | FALSE | Upregulated |
| 0,179048697 | 0,252832845 | CCNT2 | FALSE | Upregulated |
| 1,131285415 | 0,252912736 | SIRPD | FALSE | Upregulated |
| 0,115250634 | 0,252968463 | CDC42SE2 | FALSE | Upregulated |
| 0,115038653 | 0,253031193 | GART | FALSE | Upregulated |
| -0,1524488 | 0,253243373 | C7orf73 | FALSE | Downregulated |

| | | | | |
|---|---|---|---|---|
| -0,308194924 | 0,253243373 | RAD54B | FALSE | Downregulated |
| -0,252913097 | 0,253251772 | PPP1R3E | FALSE | Downregulated |
| -0,096645282 | 0,253432185 | ANKRD13C | FALSE | Downregulated |
| 0,497137406 | 0,253755284 | EMC3-AS1 | FALSE | Upregulated |
| -0,30902425 | 0,253774411 | AHDC1 | FALSE | Downregulated |
| 0,173809522 | 0,254003497 | A1BG | FALSE | Upregulated |
| 1,001991501 | 0,254003497 | GCNT3 | FALSE | Upregulated |
| -0,183742005 | 0,254003497 | THEM4 | FALSE | Downregulated |
| -0,140106045 | 0,254003497 | TMEM230 | FALSE | Downregulated |
| -0,145553955 | 0,254025517 | C2 | FALSE | Downregulated |
| -0,508636157 | 0,254086614 | LOC100133669 | FALSE | Downregulated |
| 1,358793447 | 0,254112302 | SLC12A3 | FALSE | Upregulated |
| -0,220436128 | 0,254166239 | POP7 | FALSE | Downregulated |
| 1,721664589 | 0,254287723 | CRYM-AS1 | FALSE | Upregulated |
| -0,186927419 | 0,254385824 | ACTL6A | FALSE | Downregulated |
| -0,415578084 | 0,25448396 | STK4-AS1 | FALSE | Downregulated |
| -0,402828877 | 0,254547376 | AD000090.2 | FALSE | Downregulated |
| -0,906622353 | 0,254595275 | PHOSPHO1 | FALSE | Downregulated |
| 0,452435607 | 0,254769411 | CLEC7A | FALSE | Upregulated |
| 0,249919853 | 0,254769411 | MIER2 | FALSE | Upregulated |
| -0,258800007 | 0,254769411 | ZNF527 | FALSE | Downregulated |
| 0,144958882 | 0,254784547 | DBR1 | FALSE | Upregulated |
| -0,273622909 | 0,254937221 | SNED1 | FALSE | Downregulated |
| 0,242644231 | 0,254952524 | MYLIP | FALSE | Upregulated |
| 0,331254254 | 0,255058872 | SH3PXD2B | FALSE | Upregulated |
| 1,976591592 | 0,255063105 | GAD1 | FALSE | Upregulated |
| 0,191725393 | 0,255063105 | IFT140 | FALSE | Upregulated |
| 0,156077247 | 0,255122448 | KATNB1 | FALSE | Upregulated |
| -0,111555751 | 0,255157516 | KIAA0368 | FALSE | Downregulated |
| -0,252287618 | 0,255160618 | RP11-81N13.1 | FALSE | Downregulated |
| 0,535218139 | 0,255235155 | DNAH11 | FALSE | Upregulated |
| 1,474098386 | 0,255335705 | LOC100996873 | FALSE | Upregulated |
| 0,162937646 | 0,255335705 | NPC1 | FALSE | Upregulated |
| -0,20968637 | 0,255420463 | BCAR3 | FALSE | Downregulated |
| 0,360781182 | 0,255420463 | MAPK10 | FALSE | Upregulated |
| -0,141700984 | 0,255420463 | RER1 | FALSE | Downregulated |
| 1,019531548 | 0,255420463 | STK32B | FALSE | Upregulated |
| 0,195174733 | 0,255450631 | TRIM21 | FALSE | Upregulated |
| 0,280506878 | 0,255454013 | EFCAB7 | FALSE | Upregulated |
| -0,099399805 | 0,255839707 | UBAP1 | FALSE | Downregulated |
| 0,540919988 | 0,255886256 | FAM156B | FALSE | Upregulated |
| 0,140409004 | 0,255886256 | PIAS4 | FALSE | Upregulated |
| 1,61030794 | 0,255925197 | XXYLT1-AS2 | FALSE | Upregulated |
| 0,730622571 | 0,255929989 | TIGD4 | FALSE | Upregulated |
| 0,524567717 | 0,255929989 | ZC3HAV1L | FALSE | Upregulated |
| 0,583012484 | 0,25598229 | AC097662.2 | FALSE | Upregulated |
| -0,802854074 | 0,255994501 | MEIG1 | FALSE | Downregulated |
| 1,639405846 | 0,256168317 | LOC101929173 | FALSE | Upregulated |
| -0,277731193 | 0,256202193 | DNALI1 | FALSE | Downregulated |
| 0,977659838 | 0,256206238 | HAGHL | FALSE | Upregulated |
| 0,162431582 | 0,256235151 | GTF3C4 | FALSE | Upregulated |

| | | | | |
|---|---|---|---|---|
| 0,396127445 | 0,256323641 | TRIM6 | FALSE | Upregulated |
| 0,375890023 | 0,256647244 | NPHP1 | FALSE | Upregulated |
| -1,499904337 | 0,256691531 | LOC101927486 | FALSE | Downregulated |
| -0,33565131 | 0,256691531 | TPM3 | FALSE | Downregulated |
| 0,212925541 | 0,256823681 | POC5 | FALSE | Upregulated |
| -0,337020325 | 0,256855392 | SESN3 | FALSE | Downregulated |
| 0,577645734 | 0,256922516 | PCLO | FALSE | Upregulated |
| -0,237099306 | 0,257034676 | NT5DC3 | FALSE | Downregulated |
| 0,267637952 | 0,257141462 | CLEC14A | FALSE | Upregulated |
| 0,076673182 | 0,257161857 | NEK9 | FALSE | Upregulated |
| -0,65244618 | 0,257161857 | POU6F2 | FALSE | Downregulated |
| 0,168904978 | 0,257207495 | KIAA0753 | FALSE | Upregulated |
| -0,177360505 | 0,257303808 | TNIP1 | FALSE | Downregulated |
| -0,166942121 | 0,257346416 | CENPBD1P1 | FALSE | Downregulated |
| 0,216205252 | 0,257346416 | DUS3L | FALSE | Upregulated |
| 0,782278666 | 0,257346416 | NOX4 | FALSE | Upregulated |
| 0,117528392 | 0,257383795 | SCAMP3 | FALSE | Upregulated |
| 0,320249296 | 0,257410009 | KCTD15 | FALSE | Upregulated |
| -0,192200304 | 0,257410009 | MRPL11 | FALSE | Downregulated |
| 0,107106005 | 0,257410009 | UBA3 | FALSE | Upregulated |
| 0,192958075 | 0,257455052 | POLD3 | FALSE | Upregulated |
| 0,279534691 | 0,257478229 | GAN | FALSE | Upregulated |
| 1,353711225 | 0,257518979 | FAM183B | FALSE | Upregulated |
| 0,122791947 | 0,257564522 | SNAPIN | FALSE | Upregulated |
| 0,170277675 | 0,257823658 | POMT2 | FALSE | Upregulated |
| -0,153658899 | 0,257825049 | RBM39 | FALSE | Downregulated |
| 0,552370212 | 0,257883823 | LINC00470 | FALSE | Upregulated |
| -0,129760343 | 0,257883823 | LRRC8D | FALSE | Downregulated |
| 1,673311899 | 0,257908114 | FGD5P1 | FALSE | Upregulated |
| -0,161602265 | 0,257992805 | EPM2A | FALSE | Downregulated |
| -0,134424906 | 0,257992805 | R3HCC1L | FALSE | Downregulated |
| 0,456899809 | 0,258095979 | A2M | FALSE | Upregulated |
| -0,19669575 | 0,258170135 | TMEM187 | FALSE | Downregulated |
| 0,189217671 | 0,258367343 | CENPN | FALSE | Upregulated |
| 0,111977075 | 0,258416154 | GLB1 | FALSE | Upregulated |
| -0,161418734 | 0,258416154 | PHF20L1 | FALSE | Downregulated |
| -0,295592621 | 0,258416154 | RP11-545I5.3 | FALSE | Downregulated |
| 0,284413696 | 0,258431213 | TLR6 | FALSE | Upregulated |
| -0,487486394 | 0,258448211 | LOC653513 | FALSE | Downregulated |
| 0,277159579 | 0,258477645 | ADAMTS9 | FALSE | Upregulated |
| -1,22559985 | 0,258477645 | CAMK2A | FALSE | Downregulated |
| 0,968179641 | 0,258527282 | BCL2L15 | FALSE | Upregulated |
| -0,220430366 | 0,258527282 | CAMK2N1 | FALSE | Downregulated |
| 0,163467197 | 0,258527282 | MNT | FALSE | Upregulated |
| 0,368174962 | 0,258527282 | NCAM1 | FALSE | Upregulated |
| 0,337034756 | 0,258527282 | TPRN | FALSE | Upregulated |
| -0,133281281 | 0,25876459 | FMR1 | FALSE | Downregulated |
| 0,743812389 | 0,258767138 | USP12-AS2 | FALSE | Upregulated |
| -0,232316848 | 0,258796626 | LOC101927752 | FALSE | Downregulated |
| 0,148464368 | 0,258796626 | XAB2 | FALSE | Upregulated |
| 0,186388 | 0,258796626 | ZNF761 | FALSE | Upregulated |

| | | | | |
|---|---|---|---|---|
| 0,137666352 | 0,258823611 | SIAH2 | FALSE | Upregulated |
| 0,114699857 | 0,25883801 | ELAC2 | FALSE | Upregulated |
| -0,257800037 | 0,25885799 | C19orf82 | FALSE | Downregulated |
| 0,526063459 | 0,258976007 | ZNF137P | FALSE | Upregulated |
| -0,085259367 | 0,259008205 | GPBP1 | FALSE | Downregulated |
| -0,168332135 | 0,25911715 | C1R-2 | FALSE | Downregulated |
| 0,346045384 | 0,259151562 | NBPF15 | FALSE | Upregulated |
| -0,271038098 | 0,259151562 | RSPO3 | FALSE | Downregulated |
| 0,166561303 | 0,259249127 | GARS | FALSE | Upregulated |
| -0,395194429 | 0,259249127 | LOC102723863 | FALSE | Downregulated |
| 1,260144024 | 0,259249127 | MEOX1 | FALSE | Upregulated |
| 0,407614214 | 0,259249127 | MRGPRF | FALSE | Upregulated |
| 0,412159814 | 0,259249127 | RIPK3 | FALSE | Upregulated |
| -0,354054314 | 0,259249127 | SAMD12 | FALSE | Downregulated |
| -0,387095535 | 0,259268026 | PHACTR3 | FALSE | Downregulated |
| 2,205439087 | 0,259560538 | SCARNA10 | FALSE | Upregulated |
| -0,222936775 | 0,259560538 | ZFHX4 | FALSE | Downregulated |
| 0,196363621 | 0,259578878 | FN1 | FALSE | Upregulated |
| -0,500999098 | 0,259578878 | RP11-314C16.1 | FALSE | Downregulated |
| 0,197335338 | 0,259734214 | DLAT | FALSE | Upregulated |
| 0,766810313 | 0,259872491 | CTSG | FALSE | Upregulated |
| -0,195168248 | 0,259901932 | CREB3L4 | FALSE | Downregulated |
| 0,97414958 | 0,259901932 | FANCB | FALSE | Upregulated |
| -0,316789467 | 0,259905923 | UVSSA | FALSE | Downregulated |
| -0,272322779 | 0,260052979 | CCDC134 | FALSE | Downregulated |
| 0,173422226 | 0,260214776 | MTMR12 | FALSE | Upregulated |
| -0,440569935 | 0,260214776 | SPIC | FALSE | Downregulated |
| 1,858553119 | 0,260327923 | PRND | FALSE | Upregulated |
| -0,305304231 | 0,260327923 | XRRA1 | FALSE | Downregulated |
| -0,294562397 | 0,260361253 | HSD11B1L | FALSE | Downregulated |
| -0,579069851 | 0,260413474 | RSPH4A | FALSE | Downregulated |
| 0,1519471 | 0,260747197 | MTX2 | FALSE | Upregulated |
| 0,371427076 | 0,260763001 | LRRC8E | FALSE | Upregulated |
| -0,700238168 | 0,260770431 | LOC102723461 | FALSE | Downregulated |
| 0,501104633 | 0,260770431 | PTGDR | FALSE | Upregulated |
| 1,292219565 | 0,260803 | SNORA55 | FALSE | Upregulated |
| 0,257563156 | 0,260838901 | LINC00526 | FALSE | Upregulated |
| 1,209858612 | 0,260864843 | LOC102723419 | FALSE | Upregulated |
| 1,640318361 | 0,260864843 | RNY4 | FALSE | Upregulated |
| 0,149293029 | 0,260881304 | NMT2 | FALSE | Upregulated |
| 0,701640409 | 0,260881304 | RPS15AP10 | FALSE | Upregulated |
| -0,129397951 | 0,260894107 | ATP6V1D | FALSE | Downregulated |
| 1,152129465 | 0,260894107 | NCRNA00185 | FALSE | Upregulated |
| 0,705678719 | 0,260946057 | IQUB | FALSE | Upregulated |
| 0,585133646 | 0,261005433 | NME5 | FALSE | Upregulated |
| -0,28760448 | 0,261014275 | KLF13 | FALSE | Downregulated |
| 0,510664546 | 0,261078104 | GALNT12 | FALSE | Upregulated |
| 0,466418503 | 0,261078104 | PGF | FALSE | Upregulated |
| 0,117197492 | 0,261078104 | ZDHHC9 | FALSE | Upregulated |
| -0,161689357 | 0,261174969 | CAMK2D | FALSE | Downregulated |
| -1,718845329 | 0,261174969 | CD81-AS1 | FALSE | Downregulated |

| | | | | |
|---|---|---|---|---|
| 1,651308383 | 0,261241192 | LINC00954 | FALSE | Upregulated |
| -0,593703516 | 0,261241192 | MT1DP | FALSE | Downregulated |
| -0,468285472 | 0,261308752 | PI4KAP1 | FALSE | Downregulated |
| 0,374374513 | 0,261322316 | KIF24 | FALSE | Upregulated |
| 1,20164078 | 0,261329138 | TSSK3 | FALSE | Upregulated |
| 1,123135242 | 0,261363706 | OR52N4 | FALSE | Upregulated |
| 1,618781189 | 0,261460631 | FRMD5 | FALSE | Upregulated |
| 1,639919097 | 0,261460631 | LOC101927954 | FALSE | Upregulated |
| -0,250770535 | 0,261493949 | ALKBH1 | FALSE | Downregulated |
| 0,343649068 | 0,261588167 | HSP90B2P | FALSE | Upregulated |
| 0,488402522 | 0,261610965 | ADC | FALSE | Upregulated |
| 1,008792986 | 0,261616206 | GPR20 | FALSE | Upregulated |
| 0,210841703 | 0,261616206 | SCLY | FALSE | Upregulated |
| 0,275897384 | 0,261639006 | FBF1 | FALSE | Upregulated |
| -0,359503419 | 0,261671632 | SLC25A34 | FALSE | Downregulated |
| 0,12940061 | 0,261698714 | NF1 | FALSE | Upregulated |
| -0,122333828 | 0,261707803 | MKLN1 | FALSE | Downregulated |
| 1,359446239 | 0,261795566 | CMA1 | FALSE | Upregulated |
| -0,106363338 | 0,261828418 | AGPAT3 | FALSE | Downregulated |
| 0,102085461 | 0,261999417 | KDM5A | FALSE | Upregulated |
| 0,164499149 | 0,262079032 | BID | FALSE | Upregulated |
| 0,535639136 | 0,262079032 | CDCA5 | FALSE | Upregulated |
| 0,318740126 | 0,262079032 | JAM2 | FALSE | Upregulated |
| -0,147158225 | 0,262079032 | TATDN1 | FALSE | Downregulated |
| -0,922886645 | 0,262221311 | LOC102724683 | FALSE | Downregulated |
| 0,142413204 | 0,262428902 | KIAA0195 | FALSE | Upregulated |
| -0,183590966 | 0,262450987 | SESN2 | FALSE | Downregulated |
| -0,448777364 | 0,262519708 | LEKR1 | FALSE | Downregulated |
| -0,143550006 | 0,262520406 | SSNA1 | FALSE | Downregulated |
| 0,383454254 | 0,26270208 | BSN | FALSE | Upregulated |
| 0,520655305 | 0,26270208 | LGR5 | FALSE | Upregulated |
| 0,322470367 | 0,26270208 | ZNF517 | FALSE | Upregulated |
| -0,17078555 | 0,262765041 | CMC1 | FALSE | Downregulated |
| 1,292276154 | 0,262833339 | RASA4B | FALSE | Upregulated |
| 0,115352521 | 0,262980026 | SMPD4 | FALSE | Upregulated |
| -0,119597647 | 0,263164379 | TOP2B | FALSE | Downregulated |
| 0,422953599 | 0,263272381 | DNMBP-AS1 | FALSE | Upregulated |
| 0,419316706 | 0,26341946 | LINC00240 | FALSE | Upregulated |
| -0,193188087 | 0,263460712 | SFXN4 | FALSE | Downregulated |
| 0,284087953 | 0,263529886 | ABCG1 | FALSE | Upregulated |
| 0,522026706 | 0,263812744 | ALS2CR12 | FALSE | Upregulated |
| 0,476182699 | 0,263812744 | C20orf196 | FALSE | Upregulated |
| -0,399314521 | 0,263892494 | BCO2 | FALSE | Downregulated |
| 1,004937986 | 0,263892494 | BDNF | FALSE | Upregulated |
| -0,824271143 | 0,263892494 | LOC101928264 | FALSE | Downregulated |
| 0,081251625 | 0,264131161 | GANAB | FALSE | Upregulated |
| 0,212298822 | 0,264152607 | ABCB11 | FALSE | Upregulated |
| -1,011541346 | 0,264152607 | PDE6C | FALSE | Downregulated |
| -0,321271727 | 0,264240662 | TRPV1 | FALSE | Downregulated |
| 0,238187113 | 0,264446955 | FGFR2 | FALSE | Upregulated |
| 0,22100939 | 0,264501005 | C20orf24 | FALSE | Upregulated |

| | | | | |
|---|---|---|---|---|
| 0,136784076 | 0,264564383 | CCT5 | FALSE | Upregulated |
| -0,421403145 | 0,264674132 | AIFM3 | FALSE | Downregulated |
| 0,522790144 | 0,264754086 | LOC101929356 | FALSE | Upregulated |
| 0,905268375 | 0,264764405 | HERC2P4 | FALSE | Upregulated |
| 1,35405005 | 0,264803041 | FAM27B | FALSE | Upregulated |
| -0,194349716 | 0,264946432 | RPS5 | FALSE | Downregulated |
| -0,223910178 | 0,264997284 | ZNF493 | FALSE | Downregulated |
| -0,220914228 | 0,265025727 | LOC101927045 | FALSE | Downregulated |
| 1,419591465 | 0,265103664 | LOC391722 | FALSE | Upregulated |
| -0,295563027 | 0,265113971 | FMO5 | FALSE | Downregulated |
| 1,816321478 | 0,265358789 | PGM5P2 | FALSE | Upregulated |
| 0,229277279 | 0,265560873 | SFT2D1 | FALSE | Upregulated |
| -0,156959451 | 0,265729415 | MRPL50 | FALSE | Downregulated |
| -1,187017913 | 0,265820163 | HCG23 | FALSE | Downregulated |
| 0,170923467 | 0,265863334 | IRF2BP2 | FALSE | Upregulated |
| 0,301265425 | 0,265958571 | RNASEH2A | FALSE | Upregulated |
| 1,578805838 | 0,266023306 | IL31RA | FALSE | Upregulated |
| 1,808283884 | 0,266132904 | BHLHE23 | FALSE | Upregulated |
| -0,117555572 | 0,266132904 | UTP3 | FALSE | Downregulated |
| 1,552565558 | 0,266470089 | IFNL1 | FALSE | Upregulated |
| 0,111912137 | 0,266605888 | SCAF11 | FALSE | Upregulated |
| 0,284390833 | 0,266608866 | PTGS1 | FALSE | Upregulated |
| -0,16165771 | 0,266608866 | RPS8 | FALSE | Downregulated |
| 0,212479462 | 0,266608866 | SGTB | FALSE | Upregulated |
| 0,45871899 | 0,266667168 | GBP1P1 | FALSE | Upregulated |
| -0,16054035 | 0,266726506 | LZTFL1 | FALSE | Downregulated |
| -0,484892615 | 0,266733144 | LOC285638 | FALSE | Downregulated |
| -0,233176429 | 0,266924519 | MRPS12 | FALSE | Downregulated |
| 0,74973027 | 0,267160432 | LOC101929887 | FALSE | Upregulated |
| 2,358330605 | 0,267179117 | CTAGE15 | FALSE | Upregulated |
| 0,877528596 | 0,267179117 | WDR93 | FALSE | Upregulated |
| -0,088027815 | 0,267179117 | ZFYVE20 | FALSE | Downregulated |
| -0,118501807 | 0,267195582 | WBP4 | FALSE | Downregulated |
| 0,705396854 | 0,267274612 | LINC00312 | FALSE | Upregulated |
| -0,379962088 | 0,267294043 | LOC101927506 | FALSE | Downregulated |
| 0,285608116 | 0,26744214 | ZEB1-AS1 | FALSE | Upregulated |
| 0,218164317 | 0,267584331 | DOCK5 | FALSE | Upregulated |
| -0,138103273 | 0,267641064 | TMEM69 | FALSE | Downregulated |
| -0,136996582 | 0,267650874 | C1D | FALSE | Downregulated |
| 0,179877667 | 0,267813591 | SH3GLB2 | FALSE | Upregulated |
| 0,791480738 | 0,267871485 | LOC102724611 | FALSE | Upregulated |
| -0,186763592 | 0,267871485 | NAPRT1 | FALSE | Downregulated |
| 0,502976619 | 0,267871485 | RP11-307E17.8 | FALSE | Upregulated |
| -0,102390268 | 0,26798398 | ATG12 | FALSE | Downregulated |
| 0,90221036 | 0,26798398 | CENPA | FALSE | Upregulated |
| -0,234060679 | 0,26798398 | CP | FALSE | Downregulated |
| -0,196619405 | 0,268079753 | RUFY3 | FALSE | Downregulated |
| -0,287761656 | 0,268153127 | NCKAP5 | FALSE | Downregulated |
| -0,135576783 | 0,268178151 | TJP2 | FALSE | Downregulated |
| 1,174701965 | 0,268199779 | ASPHD1 | FALSE | Upregulated |
| -0,105872019 | 0,268199779 | EIF2S1 | FALSE | Downregulated |

| | | | | |
|---|---|---|---|---|
| -0,165160887 | 0,268211928 | ABTB2 | FALSE | Downregulated |
| 0,586110904 | 0,268211928 | C17orf53 | FALSE | Upregulated |
| 0,097361452 | 0,268281133 | KCTD18 | FALSE | Upregulated |
| 0,421963938 | 0,268281133 | MRPL45P2 | FALSE | Upregulated |
| 0,202980071 | 0,268281133 | PHTF2 | FALSE | Upregulated |
| -1,558825962 | 0,268298151 | RP11-885B4.2 | FALSE | Downregulated |
| -0,313753638 | 0,268298151 | SLC10A5 | FALSE | Downregulated |
| 0,919125038 | 0,268330236 | TCEAL2 | FALSE | Upregulated |
| 0,14226536 | 0,268349174 | MOB1A | FALSE | Upregulated |
| 0,136946064 | 0,268349174 | YIPF2 | FALSE | Upregulated |
| -0,215481005 | 0,268448203 | ACSM5 | FALSE | Downregulated |
| -0,243679419 | 0,268562492 | ZNF684 | FALSE | Downregulated |
| -0,137830699 | 0,268912656 | GPALPP1 | FALSE | Downregulated |
| 0,493701151 | 0,268951005 | HAND2-AS1 | FALSE | Upregulated |
| 0,154768378 | 0,269066421 | BRWD1 | FALSE | Upregulated |
| -0,114910807 | 0,269066421 | FBXL17 | FALSE | Downregulated |
| 0,183652694 | 0,269116308 | PDIA6 | FALSE | Upregulated |
| 0,887554165 | 0,269116308 | RNA18S5 | FALSE | Upregulated |
| -0,108197986 | 0,269211021 | APH1A | FALSE | Downregulated |
| -0,380360714 | 0,269264296 | TNRC18P1 | FALSE | Downregulated |
| 0,209211557 | 0,269348676 | MEX3D | FALSE | Upregulated |
| 0,176082051 | 0,269468759 | WEE1 | FALSE | Upregulated |
| 0,237125373 | 0,269903497 | NR2F1-AS1 | FALSE | Upregulated |
| 0,355885167 | 0,269914984 | TECTA | FALSE | Upregulated |
| -0,307086248 | 0,270077683 | CBWD1 | FALSE | Downregulated |
| -0,341261943 | 0,270330774 | CCNB3 | FALSE | Downregulated |
| 0,144920951 | 0,270345349 | VMP1 | FALSE | Upregulated |
| -0,226551667 | 0,270400195 | LSM7 | FALSE | Downregulated |
| 0,226848809 | 0,270475582 | ARL6 | FALSE | Upregulated |
| 0,800020133 | 0,270654873 | FRY-AS1 | FALSE | Upregulated |
| -0,411541156 | 0,270749153 | SPTBN4 | FALSE | Downregulated |
| 1,298809077 | 0,270759181 | KLHL34 | FALSE | Upregulated |
| 0,753129979 | 0,270788175 | PKMYT1 | FALSE | Upregulated |
| 1,723959489 | 0,270824962 | SUMO1P1 | FALSE | Upregulated |
| 0,118298803 | 0,27097626 | TOM1 | FALSE | Upregulated |
| 0,135220539 | 0,271275921 | ZNF512 | FALSE | Upregulated |
| 0,67420014 | 0,271360566 | LOC102723465 | FALSE | Upregulated |
| -0,085485214 | 0,271526481 | PTCD3 | FALSE | Downregulated |
| 0,365925708 | 0,271591781 | C17orf96 | FALSE | Upregulated |
| -0,114848662 | 0,27165587 | POLDIP2 | FALSE | Downregulated |
| 0,219776316 | 0,27169501 | ZNF316 | FALSE | Upregulated |
| -0,228618912 | 0,271737847 | DUSP23 | FALSE | Downregulated |
| 0,621946957 | 0,272019137 | PLCH1 | FALSE | Upregulated |
| -0,202970658 | 0,272024477 | CCNB1IP1 | FALSE | Downregulated |
| -0,743644105 | 0,272094029 | FAM35BP | FALSE | Downregulated |
| -0,245643873 | 0,272094029 | SETBP1 | FALSE | Downregulated |
| -0,252452226 | 0,272221828 | PSIP1 | FALSE | Downregulated |
| 0,555946675 | 0,272364782 | EMR4P | FALSE | Upregulated |
| -0,147256575 | 0,272433408 | MTX1 | FALSE | Downregulated |
| 1,788369685 | 0,272545223 | ADGB | FALSE | Upregulated |
| -0,187344228 | 0,272545223 | ANKEF1 | FALSE | Downregulated |

| | | | | |
|---|---|---|---|---|
| 0,45598539 | 0,272777312 | EMID1 | FALSE | Upregulated |
| 1,130993078 | 0,272777312 | LOC102724685 | FALSE | Upregulated |
| -0,376579561 | 0,272777312 | LRRN3 | FALSE | Downregulated |
| -0,156782542 | 0,272777312 | MAN2B2 | FALSE | Downregulated |
| 0,693131878 | 0,272808437 | LYPD6B | FALSE | Upregulated |
| -0,105140311 | 0,272808437 | ZNF268 | FALSE | Downregulated |
| 0,489527409 | 0,272827539 | PLEKHM1P | FALSE | Upregulated |
| 1,448184294 | 0,273107704 | LOC100128164 | FALSE | Upregulated |
| -0,225047266 | 0,273107704 | THAP8 | FALSE | Downregulated |
| 0,198563997 | 0,273263613 | GAR1 | FALSE | Upregulated |
| 1,235033449 | 0,273576985 | ANTXRLP1 | FALSE | Upregulated |
| 0,26000479 | 0,273576985 | IL10RA | FALSE | Upregulated |
| 2,620300926 | 0,273597822 | CNTNAP5 | FALSE | Upregulated |
| 0,388565784 | 0,273764472 | TARP | FALSE | Upregulated |
| -0,161172305 | 0,274014593 | TMED2 | FALSE | Downregulated |
| 0,604329503 | 0,274538634 | LOC101928399 | FALSE | Upregulated |
| -0,2293223 | 0,274565449 | GAS2 | FALSE | Downregulated |
| 0,422197578 | 0,274648966 | PDZRN4 | FALSE | Upregulated |
| -0,067194164 | 0,274742591 | ARIH1 | FALSE | Downregulated |
| -0,279564575 | 0,274742591 | PTPRB | FALSE | Downregulated |
| 0,11048107 | 0,274742591 | SYNCRIP | FALSE | Upregulated |
| -0,517210301 | 0,274808952 | LOC101929140 | FALSE | Downregulated |
| 0,088563353 | 0,275332882 | TNPO1 | FALSE | Upregulated |
| 1,05158826 | 0,275375955 | LOC100289455 | FALSE | Upregulated |
| 0,853114017 | 0,27539892 | ROCK1P1 | FALSE | Upregulated |
| -0,124377663 | 0,275444054 | CENPB | FALSE | Downregulated |
| 0,242375979 | 0,275451201 | NOTCH4 | FALSE | Upregulated |
| 0,119752639 | 0,275451201 | RBM18 | FALSE | Upregulated |
| -0,326543451 | 0,275542937 | LOC645513 | FALSE | Downregulated |
| 0,096492713 | 0,275666071 | USP24 | FALSE | Upregulated |
| 0,173906736 | 0,275669828 | CDKAL1 | FALSE | Upregulated |
| 0,417878185 | 0,275680648 | CD163L1 | FALSE | Upregulated |
| -0,279789577 | 0,27568882 | SNHG7 | FALSE | Downregulated |
| 0,091809649 | 0,275858449 | KANSL3 | FALSE | Upregulated |
| -0,420271418 | 0,275866296 | LOC101927571 | FALSE | Downregulated |
| -0,141624271 | 0,275888408 | RPP38 | FALSE | Downregulated |
| -0,165475901 | 0,276009269 | STOM | FALSE | Downregulated |
| 0,31428612 | 0,276065979 | ZNF575 | FALSE | Upregulated |
| -0,184118022 | 0,276135687 | PRODH2 | FALSE | Downregulated |
| 0,114393719 | 0,276482603 | HIAT1 | FALSE | Upregulated |
| 0,960477119 | 0,276482603 | SLFNL1 | FALSE | Upregulated |
| 0,575374041 | 0,276596244 | RP11-611D20.2 | FALSE | Upregulated |
| -0,162841162 | 0,276835748 | CHD2 | FALSE | Downregulated |
| 0,956362193 | 0,276835748 | KIRREL3 | FALSE | Upregulated |
| 0,622028798 | 0,276964567 | FAM189A1 | FALSE | Upregulated |
| 0,298120929 | 0,277043873 | EPB41L3 | FALSE | Upregulated |
| 1,180086168 | 0,277061655 | GNG4 | FALSE | Upregulated |
| -0,232252033 | 0,277083229 | CD36 | FALSE | Downregulated |
| 1,452568846 | 0,277248091 | DLGAP1-AS3 | FALSE | Upregulated |
| 0,761878589 | 0,277276023 | CTRL | FALSE | Upregulated |
| -0,141067757 | 0,277276023 | KLF3 | FALSE | Downregulated |

| | | | | |
|---|---|---|---|---|
| 0,590349016 | 0,277361018 | LOC101928054 | FALSE | Upregulated |
| -0,106835347 | 0,277496389 | WDR5 | FALSE | Downregulated |
| -0,188007243 | 0,277525431 | GORASP2 | FALSE | Downregulated |
| -0,40259149 | 0,277538138 | LOC100996724 | FALSE | Downregulated |
| 1,459360802 | 0,27764997 | CTNNA2 | FALSE | Upregulated |
| 0,397580859 | 0,27764997 | SLC11A1 | FALSE | Upregulated |
| -0,170277065 | 0,27764997 | STARD10 | FALSE | Downregulated |
| -0,167554317 | 0,277793622 | NUP35 | FALSE | Downregulated |
| -0,597471312 | 0,27807617 | RP11-434C1.1 | FALSE | Downregulated |
| 0,898820078 | 0,278348764 | PRR16 | FALSE | Upregulated |
| 0,775705655 | 0,278498899 | DEGS2 | FALSE | Upregulated |
| 1,455347195 | 0,278527512 | PTPRZ1 | FALSE | Upregulated |
| -0,549417947 | 0,278906521 | FAR2P2 | FALSE | Downregulated |
| 0,748782587 | 0,27911151 | LOC101929381 | FALSE | Upregulated |
| 0,147075745 | 0,279113625 | FAM222B | FALSE | Upregulated |
| 0,160618766 | 0,279164565 | PRPSAP2 | FALSE | Upregulated |
| 0,08610157 | 0,279168386 | UPF1 | FALSE | Upregulated |
| -0,236711584 | 0,279194222 | RPL9 | FALSE | Downregulated |
| 1,21143985 | 0,279355962 | ABCC8 | FALSE | Upregulated |
| 1,282208905 | 0,279706886 | LOC728715 | FALSE | Upregulated |
| 0,126184944 | 0,279706886 | SOX13 | FALSE | Upregulated |
| 0,431511671 | 0,27977407 | CHRM2 | FALSE | Upregulated |
| -0,131870856 | 0,27977407 | IMPAD1 | FALSE | Downregulated |
| 0,290270218 | 0,27977407 | INPP5E | FALSE | Upregulated |
| -0,136139958 | 0,279774983 | HP1BP3 | FALSE | Downregulated |
| 1,347199432 | 0,279963937 | SNORD17 | FALSE | Upregulated |
| 0,164943782 | 0,279984224 | ASB6 | FALSE | Upregulated |
| 0,327684382 | 0,280183976 | FTX | FALSE | Upregulated |
| -0,268342373 | 0,280193353 | FBP1 | FALSE | Downregulated |
| -0,165313184 | 0,280193353 | TESK2 | FALSE | Downregulated |
| 0,21089185 | 0,280390228 | KLHL26 | FALSE | Upregulated |
| -0,190000316 | 0,280525299 | NDUFB2 | FALSE | Downregulated |
| -0,24225456 | 0,280587296 | ZNF846 | FALSE | Downregulated |
| -0,846982695 | 0,280628333 | LOC102723899 | FALSE | Downregulated |
| 1,599822745 | 0,280641252 | FLJ42393 | FALSE | Upregulated |
| 0,151975735 | 0,280641252 | INO80E | FALSE | Upregulated |
| -0,446076237 | 0,280641252 | SNHG18 | FALSE | Downregulated |
| 0,131591811 | 0,280642153 | PFN1 | FALSE | Upregulated |
| -0,246980412 | 0,280766426 | ZNHIT6 | FALSE | Downregulated |
| -0,977217374 | 0,28078145 | FOXH1 | FALSE | Downregulated |
| 0,25315915 | 0,28078145 | GEMIN7 | FALSE | Upregulated |
| -0,130201517 | 0,28078145 | PEX12 | FALSE | Downregulated |
| 1,444764001 | 0,28078499 | CD1B | FALSE | Upregulated |
| 0,257200958 | 0,281050574 | FUT8 | FALSE | Upregulated |
| 0,838399552 | 0,281135094 | C17orf72 | FALSE | Upregulated |
| -0,287561944 | 0,281135094 | SEPT4 | FALSE | Downregulated |
| -0,100083374 | 0,281262815 | SLC39A9 | FALSE | Downregulated |
| 0,858700536 | 0,281336013 | APOA4 | FALSE | Upregulated |
| 0,153206778 | 0,281336013 | MCOLN1 | FALSE | Upregulated |
| 0,165532512 | 0,281490705 | LIN7A | FALSE | Upregulated |
| 1,47707554 | 0,281490705 | LOC102467147 | FALSE | Upregulated |

| | | | | |
|---|---|---|---|---|
| 0,221722214 | 0,281906531 | CRELD1 | FALSE | Upregulated |
| -0,133033226 | 0,282041219 | COX18 | FALSE | Downregulated |
| 0,643511753 | 0,282041219 | LOC101930071 | FALSE | Upregulated |
| 0,50903092 | 0,28204172 | COL13A1 | FALSE | Upregulated |
| -0,073453344 | 0,28204172 | EIF2B1 | FALSE | Downregulated |
| 0,200322345 | 0,282094957 | SSBP4 | FALSE | Upregulated |
| 0,262688173 | 0,282094957 | TMEM231 | FALSE | Upregulated |
| 0,247639589 | 0,282139086 | STK19 | FALSE | Upregulated |
| 0,254974466 | 0,282149979 | ARAP3 | FALSE | Upregulated |
| 0,242655192 | 0,282149979 | BASP1 | FALSE | Upregulated |
| 0,593270293 | 0,282149979 | DTHD1 | FALSE | Upregulated |
| 0,208166467 | 0,282149979 | LIN52 | FALSE | Upregulated |
| -0,85070046 | 0,282149979 | RALYL | FALSE | Downregulated |
| -0,104336682 | 0,282336568 | CUEDC2 | FALSE | Downregulated |
| 0,149108409 | 0,282336568 | MCTS1 | FALSE | Upregulated |
| 0,184335864 | 0,282477709 | QTRT1 | FALSE | Upregulated |
| -0,290270105 | 0,282477709 | TPST1 | FALSE | Downregulated |
| 0,718887487 | 0,282545817 | GLB1L2 | FALSE | Upregulated |
| -0,103227313 | 0,282559745 | SMAD4 | FALSE | Downregulated |
| -0,228242065 | 0,282595716 | PSTK | FALSE | Downregulated |
| -0,13413234 | 0,282595716 | TIMM44 | FALSE | Downregulated |
| -0,27742625 | 0,282610914 | TMEM38A | FALSE | Downregulated |
| 0,157268856 | 0,28285536 | SHQ1 | FALSE | Upregulated |
| -0,154866602 | 0,282862589 | MKS1 | FALSE | Downregulated |
| 0,214802399 | 0,283050904 | SLC30A1 | FALSE | Upregulated |
| -0,297282872 | 0,283291968 | C9orf16 | FALSE | Downregulated |
| 0,156483815 | 0,283291968 | GPR108 | FALSE | Upregulated |
| -0,856150583 | 0,283514524 | AC002116.7 | FALSE | Downregulated |
| -0,249467041 | 0,283560852 | PAOX | FALSE | Downregulated |
| 0,886464394 | 0,284055301 | DENND5B-AS1 | FALSE | Upregulated |
| 0,170178538 | 0,284103041 | C19orf52 | FALSE | Upregulated |
| -0,428276245 | 0,284332817 | LOC100506688 | FALSE | Downregulated |
| 0,206211455 | 0,284445571 | EML1 | FALSE | Upregulated |
| -0,10227428 | 0,284591797 | KIAA1147 | FALSE | Downregulated |
| -0,223355447 | 0,284614568 | AKAP12 | FALSE | Downregulated |
| -0,157569389 | 0,284618405 | SUN2 | FALSE | Downregulated |
| -0,753361272 | 0,284641877 | LOC102723708 | FALSE | Downregulated |
| -0,238195868 | 0,284694268 | TBCEL | FALSE | Downregulated |
| 0,141628448 | 0,284709701 | TFG | FALSE | Upregulated |
| 0,255049997 | 0,284730776 | BMF | FALSE | Upregulated |
| 0,83387257 | 0,284737216 | LOC101928624 | FALSE | Upregulated |
| 0,17313847 | 0,284844989 | INPP1 | FALSE | Upregulated |
| -0,134395883 | 0,284846624 | CDK8 | FALSE | Downregulated |
| 0,462486805 | 0,284846624 | CYS1 | FALSE | Upregulated |
| 0,41385765 | 0,284846624 | FAM13A-AS1 | FALSE | Upregulated |
| 0,207722456 | 0,284846624 | NTMT1 | FALSE | Upregulated |
| -0,327342886 | 0,284876152 | GMDS | FALSE | Downregulated |
| -0,097533267 | 0,284878844 | GUF1 | FALSE | Downregulated |
| 0,531622804 | 0,284885582 | TMEM119 | FALSE | Upregulated |
| -1,175266979 | 0,284932946 | TPM2 | FALSE | Downregulated |
| -0,188541237 | 0,284989586 | ABCF1 | FALSE | Downregulated |

| | | | | |
|---|---|---|---|---|
| -1,578314922 | 0,284989586 | LOC101929826 | FALSE | Downregulated |
| -0,29829826 | 0,284989586 | MACROD2 | FALSE | Downregulated |
| -1,166550654 | 0,284989586 | RP11-381O7.3 | FALSE | Downregulated |
| 1,432117066 | 0,285007811 | DNAH3 | FALSE | Upregulated |
| 0,433984487 | 0,285224898 | EFNA5 | FALSE | Upregulated |
| 0,304609373 | 0,285224898 | LOC388692 | FALSE | Upregulated |
| -0,121086367 | 0,285238412 | PHB | FALSE | Downregulated |
| -0,238221311 | 0,285239875 | LOC101928499 | FALSE | Downregulated |
| -0,818762162 | 0,285248124 | GPR179 | FALSE | Downregulated |
| 1,410110947 | 0,285367987 | LOC100616530 | FALSE | Upregulated |
| 0,092702034 | 0,285636513 | ACTR1B | FALSE | Upregulated |
| 0,193961362 | 0,285741854 | ZSWIM1 | FALSE | Upregulated |
| 1,104463584 | 0,285750814 | UTY | FALSE | Upregulated |
| 0,605078137 | 0,285773664 | COL4A3 | FALSE | Upregulated |
| -0,515910851 | 0,285773664 | PRELID2 | FALSE | Downregulated |
| -0,163385524 | 0,285773664 | PTPN2 | FALSE | Downregulated |
| 0,172813896 | 0,285884622 | ADCK4 | FALSE | Upregulated |
| 0,732605505 | 0,285969422 | CPA3 | FALSE | Upregulated |
| 0,167234262 | 0,286209809 | SCMH1 | FALSE | Upregulated |
| 0,130008641 | 0,286358845 | EIF4A1 | FALSE | Upregulated |
| 0,087064341 | 0,286365079 | GTF2H1 | FALSE | Upregulated |
| 0,741945406 | 0,286365079 | LOC100506691 | FALSE | Upregulated |
| 0,170608423 | 0,286365079 | TBCCD1 | FALSE | Upregulated |
| -0,128732386 | 0,286400675 | CCDC43 | FALSE | Downregulated |
| 0,168968329 | 0,286573421 | ZNF764 | FALSE | Upregulated |
| -0,204199017 | 0,286724827 | MGST3 | FALSE | Downregulated |
| 0,392148468 | 0,286724827 | RASA4 | FALSE | Upregulated |
| -0,324443777 | 0,286767816 | IL1RL2 | FALSE | Downregulated |
| 1,024372438 | 0,286867151 | RGPD3 | FALSE | Upregulated |
| -0,138527038 | 0,286976532 | EIF4EBP2 | FALSE | Downregulated |
| 0,500194348 | 0,286987746 | KCNK1 | FALSE | Upregulated |
| 0,120237001 | 0,287177898 | FCHSD2 | FALSE | Upregulated |
| 0,194521872 | 0,287177898 | GOSR2 | FALSE | Upregulated |
| 1,88471179 | 0,287177898 | LOC102724393 | FALSE | Upregulated |
| 1,226908585 | 0,287181904 | CHRFAM7A | FALSE | Upregulated |
| -0,107043685 | 0,287181904 | DDX18 | FALSE | Downregulated |
| -0,317801526 | 0,287181904 | FADS6 | FALSE | Downregulated |
| 0,683769678 | 0,287181904 | LOC101928862 | FALSE | Upregulated |
| 0,134876941 | 0,287181904 | MPP1 | FALSE | Upregulated |
| -0,563808705 | 0,287181904 | PAGE5 | FALSE | Downregulated |
| -0,149089141 | 0,287181904 | PFDN5 | FALSE | Downregulated |
| -0,190678632 | 0,287181904 | ZNF546 | FALSE | Downregulated |
| 0,245748996 | 0,287381134 | RBPMS | FALSE | Upregulated |
| -0,101362625 | 0,287474202 | CRIPT | FALSE | Downregulated |
| -0,166899706 | 0,287566514 | CTSO | FALSE | Downregulated |
| 0,614364418 | 0,287566514 | TICAM2 | FALSE | Upregulated |
| -1,24333644 | 0,287600196 | RFPL3S | FALSE | Downregulated |
| -0,118327774 | 0,287681727 | STAM | FALSE | Downregulated |
| 0,475317336 | 0,287837583 | FAT3 | FALSE | Upregulated |
| -0,118143099 | 0,287923413 | FH | FALSE | Downregulated |
| -0,310957219 | 0,287923413 | SOX5 | FALSE | Downregulated |

| | | | | |
|---|---|---|---|---|
| 1,92493068 | 0,287944323 | ZNF295-AS1 | FALSE | Upregulated |
| 0,318417797 | 0,287996256 | NEK8 | FALSE | Upregulated |
| 0,863295725 | 0,287996256 | OSR1 | FALSE | Upregulated |
| 0,812805503 | 0,28807016 | AC005550.4 | FALSE | Upregulated |
| 1,452489717 | 0,288098778 | DNASE1L2 | FALSE | Upregulated |
| 0,147577447 | 0,288098778 | SOAT1 | FALSE | Upregulated |
| 0,313590244 | 0,288168275 | RPL32P3 | FALSE | Upregulated |
| -0,162902016 | 0,288261089 | MFSD8 | FALSE | Downregulated |
| 0,105095711 | 0,288383571 | LIMS1 | FALSE | Upregulated |
| 1,102732781 | 0,288433862 | ERVW-1 | FALSE | Upregulated |
| 0,432889979 | 0,288433862 | F11-AS1 | FALSE | Upregulated |
| -0,750128657 | 0,288433862 | SCXB | FALSE | Downregulated |
| 0,336149385 | 0,28852548 | TMOD1 | FALSE | Upregulated |
| 0,26659779 | 0,288550524 | PPP1R3F | FALSE | Upregulated |
| 1,290624893 | 0,288646698 | TMSB15B | FALSE | Upregulated |
| 0,60037335 | 0,288738455 | SSSCA1-AS1 | FALSE | Upregulated |
| 0,252066349 | 0,288738455 | ZXDA | FALSE | Upregulated |
| 1,523712816 | 0,288777514 | LOC100505685 | FALSE | Upregulated |
| 0,13581329 | 0,288808786 | TMA16 | FALSE | Upregulated |
| -0,130870422 | 0,288837125 | RSBN1 | FALSE | Downregulated |
| -0,119675222 | 0,288879042 | MANBA | FALSE | Downregulated |
| 0,609964196 | 0,288893136 | HOXA2 | FALSE | Upregulated |
| -0,156178101 | 0,289008076 | RNF139 | FALSE | Downregulated |
| 0,090987934 | 0,289090821 | KIAA0196 | FALSE | Upregulated |
| 0,144454332 | 0,289209602 | MAT2B | FALSE | Upregulated |
| 2,349780343 | 0,289210258 | LOC102723799 | FALSE | Upregulated |
| 0,118493094 | 0,289848692 | SIPA1L1 | FALSE | Upregulated |
| 0,161447458 | 0,289878173 | TMEM50B | FALSE | Upregulated |
| -0,189227983 | 0,289939825 | C12orf66 | FALSE | Downregulated |
| -0,658032921 | 0,289939825 | MIR6843 | FALSE | Downregulated |
| -0,25905563 | 0,289939825 | PTPRD | FALSE | Downregulated |
| -1,03404607 | 0,289939825 | SNX31 | FALSE | Downregulated |
| 0,162991151 | 0,290022354 | TMLHE | FALSE | Upregulated |
| -0,175950229 | 0,290030487 | PRDM10 | FALSE | Downregulated |
| 0,408453233 | 0,290423331 | EMCN | FALSE | Upregulated |
| 1,296489472 | 0,290528438 | LOC101929340 | FALSE | Upregulated |
| -0,281060594 | 0,29053289 | TAF3 | FALSE | Downregulated |
| 0,41832015 | 0,290824975 | SORCS2 | FALSE | Upregulated |
| -0,288352216 | 0,290837736 | PPP2R3B | FALSE | Downregulated |
| 1,462056918 | 0,290921623 | AC073283.4 | FALSE | Upregulated |
| 1,738957308 | 0,290952718 | ADAMTS8 | FALSE | Upregulated |
| -0,514034345 | 0,291032132 | TMEM249 | FALSE | Downregulated |
| 0,588599069 | 0,291074744 | AGPAT4 | FALSE | Upregulated |
| 1,06242138 | 0,291074744 | COL17A1 | FALSE | Upregulated |
| 0,134377399 | 0,291074744 | PPP1CB | FALSE | Upregulated |
| 0,134745131 | 0,291087583 | CTDSP1 | FALSE | Upregulated |
| -0,192781947 | 0,291112943 | DST | FALSE | Downregulated |
| 0,808406824 | 0,291112943 | RP11-682N22.1 | FALSE | Upregulated |
| -0,126227221 | 0,291112943 | SMIM7 | FALSE | Downregulated |
| 0,49157434 | 0,291115787 | CLVS1 | FALSE | Upregulated |
| 0,168663579 | 0,291145211 | AKAP13 | FALSE | Upregulated |

| | | | | |
|---|---|---|---|---|
| -0,194936714 | 0,291168864 | PHTF1 | FALSE | Downregulated |
| 0,558527979 | 0,29122854 | LOC101927610 | FALSE | Upregulated |
| -0,150381688 | 0,29122854 | LRRC58 | FALSE | Downregulated |
| 0,218183545 | 0,29129675 | ABCB1 | FALSE | Upregulated |
| 0,122902726 | 0,29129675 | CUL9 | FALSE | Upregulated |
| -0,165161119 | 0,29129675 | MRPL52 | FALSE | Downregulated |
| 1,169444415 | 0,29129675 | NXNL2 | FALSE | Upregulated |
| 0,290814733 | 0,291575703 | ASAP3 | FALSE | Upregulated |
| -0,165454162 | 0,291631271 | SPRED2 | FALSE | Downregulated |
| 0,893865753 | 0,291688587 | AC104655.2 | FALSE | Upregulated |
| -0,176675854 | 0,291718589 | C1orf131 | FALSE | Downregulated |
| -0,250773611 | 0,291718589 | CES1 | FALSE | Downregulated |
| -0,55336609 | 0,291718589 | LOC102724483 | FALSE | Downregulated |
| -0,214555792 | 0,291824344 | NME3 | FALSE | Downregulated |
| 0,161443627 | 0,291857421 | CRAMP1L | FALSE | Upregulated |
| 0,210977712 | 0,291857421 | PON2 | FALSE | Upregulated |
| -0,919824968 | 0,291876599 | NPY6R | FALSE | Downregulated |
| 0,734630847 | 0,291982961 | LOC102723683 | FALSE | Upregulated |
| 0,515906193 | 0,292056768 | PNMAL1 | FALSE | Upregulated |
| 0,094677605 | 0,292074406 | PICALM | FALSE | Upregulated |
| -0,211519964 | 0,292181114 | ARCN1 | FALSE | Downregulated |
| 0,136287775 | 0,292181114 | BCOR | FALSE | Upregulated |
| 0,158740653 | 0,292181114 | FAM220A | FALSE | Upregulated |
| 0,102235916 | 0,292181114 | IL17RA | FALSE | Upregulated |
| 1,059622243 | 0,292181114 | LINC00167 | FALSE | Upregulated |
| -0,443962659 | 0,292181114 | LOC100506023 | FALSE | Downregulated |
| 0,210074209 | 0,292181114 | PITPNM2 | FALSE | Upregulated |
| -0,870448517 | 0,292181114 | RP11-214C8.2 | FALSE | Downregulated |
| -1,091505874 | 0,292181114 | TRIM9 | FALSE | Downregulated |
| -0,508397284 | 0,292181114 | USP43 | FALSE | Downregulated |
| 0,874898625 | 0,292534988 | PF4V1 | FALSE | Upregulated |
| -0,182282396 | 0,292534988 | RPS10 | FALSE | Downregulated |
| 0,173406714 | 0,292651757 | TMEM245 | FALSE | Upregulated |
| -2,032010965 | 0,292743785 | LOC102723678 | FALSE | Downregulated |
| 1,640648556 | 0,292750288 | KCNT1 | FALSE | Upregulated |
| -0,18769546 | 0,292750288 | MAGI2 | FALSE | Downregulated |
| 1,618297243 | 0,292871951 | NCRUPAR | FALSE | Upregulated |
| -0,112946626 | 0,292871951 | QTRTD1 | FALSE | Downregulated |
| -1,444868131 | 0,293216266 | LOC101927814 | FALSE | Downregulated |
| 0,126563017 | 0,293241473 | FBXO34 | FALSE | Upregulated |
| -0,159607723 | 0,293241473 | ZNF16 | FALSE | Downregulated |
| 1,228519951 | 0,293429403 | LOC101927775 | FALSE | Upregulated |
| 0,229369878 | 0,293471457 | ATAT1 | FALSE | Upregulated |
| 1,751031432 | 0,293517848 | RBBP8NL | FALSE | Upregulated |
| -0,15122432 | 0,29354222 | HSF2 | FALSE | Downregulated |
| -0,362442695 | 0,29354222 | LOC441242 | FALSE | Downregulated |
| 0,422558213 | 0,293570259 | DGCR11 | FALSE | Upregulated |
| 1,449860258 | 0,293628784 | TMC1 | FALSE | Upregulated |
| 0,747632599 | 0,293663139 | CAMK1G | FALSE | Upregulated |
| 0,250140035 | 0,293721897 | RPUSD1 | FALSE | Upregulated |
| 0,590905813 | 0,293790468 | PRDM8 | FALSE | Upregulated |

| | | | | |
|---|---|---|---|---|
| 0,801461611 | 0,293790468 | RP11-760N9.1 | FALSE | Upregulated |
| 0,625673387 | 0,293809659 | SCGN | FALSE | Upregulated |
| -0,554620572 | 0,29388457 | LOC101927895 | FALSE | Downregulated |
| -1,602182064 | 0,29438526 | CIDEA | FALSE | Downregulated |
| 0,713722258 | 0,29438526 | ZNF114 | FALSE | Upregulated |
| 0,274258156 | 0,294468178 | MYL12A | FALSE | Upregulated |
| 0,163435228 | 0,294511307 | DPY19L3 | FALSE | Upregulated |
| -0,293736189 | 0,294511307 | FAM86C2P | FALSE | Downregulated |
| 1,101412086 | 0,294511307 | LOC100506731 | FALSE | Upregulated |
| 0,239731253 | 0,294600512 | RARA | FALSE | Upregulated |
| -0,178082098 | 0,294733732 | JHDM1D-AS1 | FALSE | Downregulated |
| -0,129862578 | 0,294733732 | WDR44 | FALSE | Downregulated |
| 0,900791243 | 0,295059144 | TBX1 | FALSE | Upregulated |
| 1,324693773 | 0,295226011 | C12orf68 | FALSE | Upregulated |
| 0,350369834 | 0,295226011 | RAVER2 | FALSE | Upregulated |
| 0,109280434 | 0,29539241 | SQRDL | FALSE | Upregulated |
| 0,258367835 | 0,295407773 | C8orf4 | FALSE | Upregulated |
| -0,118116101 | 0,295407773 | LLPH | FALSE | Downregulated |
| -0,132742924 | 0,295407773 | POLR1D | FALSE | Downregulated |
| 0,206143211 | 0,295407773 | WDR24 | FALSE | Upregulated |
| -0,136101953 | 0,295569006 | CDH2 | FALSE | Downregulated |
| -0,309073779 | 0,295586569 | C9orf43 | FALSE | Downregulated |
| 0,266324399 | 0,295586569 | CMKLR1 | FALSE | Upregulated |
| -0,33885896 | 0,295586569 | DLG2 | FALSE | Downregulated |
| -0,283268981 | 0,295586569 | LOC158960 | FALSE | Downregulated |
| 0,434547581 | 0,295586569 | PHYHIPL | FALSE | Upregulated |
| -0,577940103 | 0,295586569 | RP11-112L6.4 | FALSE | Downregulated |
| -0,216132444 | 0,295636123 | KIAA1217 | FALSE | Downregulated |
| -1,090668308 | 0,295668587 | LINC00348 | FALSE | Downregulated |
| -0,824813538 | 0,295677399 | LMX1A | FALSE | Downregulated |
| 0,817331928 | 0,296062497 | TAS2R14 | FALSE | Upregulated |
| 0,156828043 | 0,296195457 | RYK | FALSE | Upregulated |
| -0,182505335 | 0,296195457 | TMEM45A | FALSE | Downregulated |
| -0,168858841 | 0,296343416 | REXO1 | FALSE | Downregulated |
| 0,123446499 | 0,296538996 | NCOA5 | FALSE | Upregulated |
| -1,195788351 | 0,29665195 | RGMA | FALSE | Downregulated |
| -0,131377914 | 0,296660852 | TBCE | FALSE | Downregulated |
| 0,186611095 | 0,296660852 | ZSCAN30 | FALSE | Upregulated |
| 0,600102793 | 0,296690843 | LOC100507131 | FALSE | Upregulated |
| -0,15833588 | 0,296764585 | NDUFA13 | FALSE | Downregulated |
| 0,671951232 | 0,296824053 | ARMC4 | FALSE | Upregulated |
| -0,321365152 | 0,296871115 | FAM124A | FALSE | Downregulated |
| -0,169330968 | 0,296871115 | XPO4 | FALSE | Downregulated |
| 0,144975823 | 0,296918764 | KIF5B | FALSE | Upregulated |
| 0,635346694 | 0,296918764 | LOC102723989 | FALSE | Upregulated |
| 0,321296797 | 0,297060448 | CD84 | FALSE | Upregulated |
| 0,407897363 | 0,297211996 | PRTFDC1 | FALSE | Upregulated |
| -0,248550288 | 0,297278768 | SLC2A13 | FALSE | Downregulated |
| 0,535282737 | 0,297401859 | C21orf62 | FALSE | Upregulated |
| 0,188379253 | 0,297401859 | RCC1 | FALSE | Upregulated |
| 0,128493546 | 0,297401859 | SLC9A8 | FALSE | Upregulated |

| | | | | |
|---|---|---|---|---|
| -0,08472213 | 0,297401859 | ZDHHC3 | FALSE | Downregulated |
| 0,124771017 | 0,297441152 | SNRPB | FALSE | Upregulated |
| -0,105096241 | 0,297672079 | PPP2R2D | FALSE | Downregulated |
| -1,438532771 | 0,297743661 | ST20-MTHFS | FALSE | Downregulated |
| 1,677849145 | 0,297864326 | OVOL1 | FALSE | Upregulated |
| -0,173120608 | 0,298068612 | MCRS1 | FALSE | Downregulated |
| -1,139260117 | 0,298135745 | SPTY2D1-AS1 | FALSE | Downregulated |
| 0,339943923 | 0,298204406 | BCL3 | FALSE | Upregulated |
| 0,191156226 | 0,298443618 | EVA1A | FALSE | Upregulated |
| 0,130459803 | 0,298451563 | FASTKD5 | FALSE | Upregulated |
| -0,219338874 | 0,298456395 | ZNF106 | FALSE | Downregulated |
| 1,29249696 | 0,298525342 | FABP5P3 | FALSE | Upregulated |
| -0,297729802 | 0,298571421 | MLXIPL | FALSE | Downregulated |
| -0,150592841 | 0,298684471 | RNF219 | FALSE | Downregulated |
| -0,480783395 | 0,298787089 | LOC102546294 | FALSE | Downregulated |
| 0,179328608 | 0,298804454 | NLRX1 | FALSE | Upregulated |
| 0,278386101 | 0,299079155 | CDON | FALSE | Upregulated |
| 0,333746686 | 0,299079155 | ZNF285 | FALSE | Upregulated |
| -1,171063369 | 0,299311762 | LPPR4 | FALSE | Downregulated |
| 0,981850836 | 0,299346313 | LOC101929641 | FALSE | Upregulated |
| 0,217711331 | 0,299346313 | TMEM86A | FALSE | Upregulated |
| -0,180825661 | 0,299375697 | CNKSR3 | FALSE | Downregulated |
| -3,281447578 | 0,299375697 | DUSP27 | FALSE | Downregulated |
| 0,2468414 | 0,299384178 | ACYP1 | FALSE | Upregulated |
| -0,199209885 | 0,299384178 | BHLHE40 | FALSE | Downregulated |
| 0,713181252 | 0,299384178 | CHRDL1 | FALSE | Upregulated |
| -0,526282596 | 0,299384178 | RP1-232P20.1 | FALSE | Downregulated |
| -0,148601287 | 0,299384178 | SERPINC1 | FALSE | Downregulated |
| 0,428945577 | 0,299384178 | SLC24A4 | FALSE | Upregulated |
| -0,223889135 | 0,299469776 | VAMP2 | FALSE | Downregulated |
| -0,36818997 | 0,299662172 | IGF2BP3 | FALSE | Downregulated |
| -0,456264055 | 0,29969699 | NIPSNAP3B | FALSE | Downregulated |
| 0,21904731 | 0,29984249 | GATA6 | FALSE | Upregulated |
| -0,148581082 | 0,299876057 | BRD8 | FALSE | Downregulated |
| -0,123535081 | 0,300287945 | CCBL2 | FALSE | Downregulated |
| -0,258313824 | 0,300287945 | KCND3 | FALSE | Downregulated |
| -0,143496821 | 0,300299799 | COA4 | FALSE | Downregulated |
| -0,282864429 | 0,300335462 | FAM65A | FALSE | Downregulated |
| 1,034060791 | 0,300335462 | KIR2DL1 | FALSE | Upregulated |
| 0,558983458 | 0,300335462 | ZNF835 | FALSE | Upregulated |
| -0,210178548 | 0,300354328 | AIF1 | FALSE | Downregulated |
| 0,93629714 | 0,300363437 | HMGN2P46 | FALSE | Upregulated |
| -0,10384433 | 0,300366482 | WDR33 | FALSE | Downregulated |
| 0,195279574 | 0,300512983 | ACVRL1 | FALSE | Upregulated |
| 0,265267244 | 0,300512983 | GFPT1 | FALSE | Upregulated |
| 0,423338955 | 0,300564284 | PPP1R9A | FALSE | Upregulated |
| 0,377150817 | 0,300641566 | BZRAP1-AS1 | FALSE | Upregulated |
| 0,136292724 | 0,300641566 | CCP110 | FALSE | Upregulated |
| 0,409828634 | 0,300641566 | ELL3 | FALSE | Upregulated |
| -0,135930688 | 0,300641566 | IP6K2 | FALSE | Downregulated |
| 0,734282443 | 0,300641566 | LOC100507103 | FALSE | Upregulated |

| | | | | |
|---|---|---|---|---|
| 0,885925876 | 0,300641566 | LOC101928428 | FALSE | Upregulated |
| -0,842545773 | 0,300641566 | LOC101929814 | FALSE | Downregulated |
| -1,446733146 | 0,300641566 | LOC102723316 | FALSE | Downregulated |
| -0,446942283 | 0,300641566 | LY6G5C | FALSE | Downregulated |
| -0,101514072 | 0,300641566 | MAK16 | FALSE | Downregulated |
| 0,151874865 | 0,300641566 | SNX33 | FALSE | Upregulated |
| 0,243874813 | 0,300641566 | ZNF704 | FALSE | Upregulated |
| 0,776611861 | 0,300694174 | HSPA7 | FALSE | Upregulated |
| -0,185831144 | 0,300777118 | NUBPL | FALSE | Downregulated |
| -0,1317571 | 0,300863904 | SFXN1 | FALSE | Downregulated |
| 0,142840868 | 0,300863904 | SUN1 | FALSE | Upregulated |
| 1,026190463 | 0,30091438 | ALOX15B | FALSE | Upregulated |
| 1,672579104 | 0,30091438 | LOC390956 | FALSE | Upregulated |
| 0,175898633 | 0,30091438 | PP7080 | FALSE | Upregulated |
| 0,352872503 | 0,30091438 | TRDMT1 | FALSE | Upregulated |
| -0,220649332 | 0,300944167 | SPIN3 | FALSE | Downregulated |
| 0,242717611 | 0,30105927 | SYTL4 | FALSE | Upregulated |
| -0,160093304 | 0,301180546 | KIAA1009 | FALSE | Downregulated |
| -0,129552287 | 0,301209155 | PPFIA1 | FALSE | Downregulated |
| 0,832015296 | 0,301278042 | ALDH3A1 | FALSE | Upregulated |
| 0,360067492 | 0,301278042 | RAP1GAP | FALSE | Upregulated |
| 0,101042388 | 0,301297915 | KIAA0430 | FALSE | Upregulated |
| 0,155408977 | 0,301644029 | ETNK1 | FALSE | Upregulated |
| 0,573438007 | 0,301644029 | IQCA1 | FALSE | Upregulated |
| -1,138520194 | 0,301644029 | RBMXL2 | FALSE | Downregulated |
| -1,034181123 | 0,301842136 | LOC100128568 | FALSE | Downregulated |
| -0,505009801 | 0,301842136 | LOC100506985 | FALSE | Downregulated |
| 0,363025954 | 0,302193625 | WDR54 | FALSE | Upregulated |
| -1,356340824 | 0,30220024 | LOC102723950 | FALSE | Downregulated |
| 0,461617764 | 0,302274574 | SGOL1 | FALSE | Upregulated |
| 1,237973018 | 0,302358595 | ZNF724P | FALSE | Upregulated |
| 0,12924263 | 0,302437975 | GEMIN5 | FALSE | Upregulated |
| 0,182184834 | 0,302437975 | ZNF707 | FALSE | Upregulated |
| 0,103718945 | 0,302479407 | LTA4H | FALSE | Upregulated |
| 0,562866234 | 0,302568421 | TMEM154 | FALSE | Upregulated |
| 1,286151755 | 0,302638024 | NLGN4Y | FALSE | Upregulated |
| 1,651590848 | 0,302721382 | AGAP7 | FALSE | Upregulated |
| 0,128196485 | 0,302721382 | DNTTIP1 | FALSE | Upregulated |
| -0,789584051 | 0,302721382 | STPG2 | FALSE | Downregulated |
| 0,390300113 | 0,30280672 | ACVR2B-AS1 | FALSE | Upregulated |
| -0,530850497 | 0,302819119 | LOC200772 | FALSE | Downregulated |
| 0,222431562 | 0,302926845 | ABI3 | FALSE | Upregulated |
| -0,666347478 | 0,302926845 | NEK10 | FALSE | Downregulated |
| 1,015234537 | 0,302988813 | ANKRD45 | FALSE | Upregulated |
| -1,182999376 | 0,303089385 | LOC101929946 | FALSE | Downregulated |
| -0,500409105 | 0,303129194 | C2orf82 | FALSE | Downregulated |
| 2,324307996 | 0,303375471 | HCCAT5 | FALSE | Upregulated |
| 1,237225299 | 0,303375471 | ICAM5 | FALSE | Upregulated |
| -0,093113253 | 0,303375471 | UTP14C | FALSE | Downregulated |
| 1,391886749 | 0,303375471 | WNT10B | FALSE | Upregulated |
| -0,191853246 | 0,303404354 | DHX16 | FALSE | Downregulated |

| | | | | |
|---|---|---|---|---|
| -0,134393904 | 0,303404354 | UBAC1 | FALSE | Downregulated |
| -0,166581839 | 0,303501552 | RABGAP1L | FALSE | Downregulated |
| -0,204988547 | 0,303558325 | RABL6 | FALSE | Downregulated |
| -0,224566799 | 0,303578438 | KLHDC4 | FALSE | Downregulated |
| -1,085629369 | 0,303578438 | SLAMF9 | FALSE | Downregulated |
| -0,245015001 | 0,303578438 | SLC7A8 | FALSE | Downregulated |
| 0,268128523 | 0,303615512 | CBX8 | FALSE | Upregulated |
| -0,272481337 | 0,303615512 | ZNF850 | FALSE | Downregulated |
| 0,267708855 | 0,303804503 | CYGB | FALSE | Upregulated |
| 0,691441893 | 0,303870675 | ZNF804A | FALSE | Upregulated |
| 0,141856838 | 0,303878196 | SETX | FALSE | Upregulated |
| -0,59547644 | 0,303960783 | RNF186 | FALSE | Downregulated |
| 0,214124743 | 0,303964464 | NAP1L5 | FALSE | Upregulated |
| 0,211600274 | 0,303996001 | C19orf12 | FALSE | Upregulated |
| 0,265629618 | 0,303996001 | FAM109A | FALSE | Upregulated |
| -0,386618654 | 0,304140441 | FDPS | FALSE | Downregulated |
| 0,370822356 | 0,304313378 | APOL4 | FALSE | Upregulated |
| 1,094041201 | 0,304388415 | FLJ38379 | FALSE | Upregulated |
| 0,247380087 | 0,304388415 | PPP1R9B | FALSE | Upregulated |
| -0,180230372 | 0,304388415 | SLC10A7 | FALSE | Downregulated |
| 0,529331875 | 0,30454678 | LOC101928051 | FALSE | Upregulated |
| 0,57446932 | 0,304552773 | OLFML2A | FALSE | Upregulated |
| 0,568842584 | 0,304627604 | ABCA10 | FALSE | Upregulated |
| 0,569005572 | 0,304627604 | HCAR3 | FALSE | Upregulated |
| -0,183989314 | 0,304658861 | PRR5 | FALSE | Downregulated |
| -2,645797484 | 0,304869339 | ADCY2 | FALSE | Downregulated |
| -0,225784615 | 0,304903594 | SEMA6A | FALSE | Downregulated |
| -0,549432751 | 0,304964287 | LOC101929513 | FALSE | Downregulated |
| -0,304460696 | 0,304964287 | TTPAL | FALSE | Downregulated |
| 0,694154423 | 0,305275921 | RAD54L | FALSE | Upregulated |
| 0,090110833 | 0,305371678 | AFTPH | FALSE | Upregulated |
| -0,429122485 | 0,305371678 | PDE11A | FALSE | Downregulated |
| -1,0632135 | 0,305456515 | CBLN2 | FALSE | Downregulated |
| -0,391067537 | 0,305564256 | MYLK4 | FALSE | Downregulated |
| 0,152394475 | 0,305564256 | TMEM185A | FALSE | Upregulated |
| 1,004124214 | 0,305667648 | HIGD1B | FALSE | Upregulated |
| -1,433418563 | 0,305771107 | THEG | FALSE | Downregulated |
| 0,402449257 | 0,305809902 | KIAA0101 | FALSE | Upregulated |
| 0,10549349 | 0,305974081 | TXNDC15 | FALSE | Upregulated |
| 0,267220282 | 0,306033622 | RAPGEF4 | FALSE | Upregulated |
| -0,250617963 | 0,30613731 | C14orf79 | FALSE | Downregulated |
| 1,574364993 | 0,30613731 | C1orf200 | FALSE | Upregulated |
| -0,090660827 | 0,30613731 | DARS | FALSE | Downregulated |
| 0,928259255 | 0,30613731 | LOC101928801 | FALSE | Upregulated |
| 0,964805675 | 0,30613731 | LOXHD1 | FALSE | Upregulated |
| 0,732871792 | 0,30613731 | NTRK3 | FALSE | Upregulated |
| 0,384863423 | 0,30613731 | PBX1 | FALSE | Upregulated |
| 0,119636712 | 0,306160178 | BTBD1 | FALSE | Upregulated |
| 1,637243901 | 0,306333318 | BMP3 | FALSE | Upregulated |
| 1,322533431 | 0,306333318 | GABARAPL3 | FALSE | Upregulated |
| 1,052351305 | 0,306333318 | RP11-277P12.20 | FALSE | Upregulated |

| | | | | |
|---|---|---|---|---|
| -0,160488873 | 0,306509019 | MRPL14 | FALSE | Downregulated |
| -0,126860739 | 0,306510357 | TTI2 | FALSE | Downregulated |
| 0,210213604 | 0,306644991 | NCK2 | FALSE | Upregulated |
| -0,120619048 | 0,306644991 | STAG2 | FALSE | Downregulated |
| 0,261477718 | 0,306678807 | LPIN2 | FALSE | Upregulated |
| 0,15799687 | 0,306721853 | COPE | FALSE | Upregulated |
| -1,444301515 | 0,306729406 | LOC102723580 | FALSE | Downregulated |
| 0,693512504 | 0,306798987 | C6orf164 | FALSE | Upregulated |
| 0,323783792 | 0,306840624 | DGCR5 | FALSE | Upregulated |
| 0,646177272 | 0,306861233 | IGFBP1 | FALSE | Upregulated |
| -0,11711537 | 0,306861233 | OAZ1 | FALSE | Downregulated |
| 1,029817967 | 0,306903334 | HTRA4 | FALSE | Upregulated |
| 0,115445714 | 0,306906979 | AEBP2 | FALSE | Upregulated |
| 0,115771404 | 0,306906979 | KIAA1430 | FALSE | Upregulated |
| -0,14548105 | 0,306906979 | PSMA4 | FALSE | Downregulated |
| 0,516495199 | 0,306932158 | LOC728392 | FALSE | Upregulated |
| -0,120749566 | 0,306932158 | MRPS7 | FALSE | Downregulated |
| -0,137245799 | 0,306958146 | METTL17 | FALSE | Downregulated |
| 0,288999229 | 0,306964869 | C4orf21 | FALSE | Upregulated |
| -0,317992625 | 0,30701716 | LOC102724164 | FALSE | Downregulated |
| 0,277463078 | 0,30701716 | LOC102724200 | FALSE | Upregulated |
| 0,170066048 | 0,307046267 | ABCC3 | FALSE | Upregulated |
| -0,174104278 | 0,307046267 | TPR | FALSE | Downregulated |
| 0,148872214 | 0,307046267 | VPS41 | FALSE | Upregulated |
| 0,132362528 | 0,307143962 | ACAD8 | FALSE | Upregulated |
| 0,888731446 | 0,307200168 | SMIM17 | FALSE | Upregulated |
| 0,211070488 | 0,307227486 | DCN | FALSE | Upregulated |
| 0,193524706 | 0,307265546 | CABLES2 | FALSE | Upregulated |
| 0,417560446 | 0,307265546 | DACT3 | FALSE | Upregulated |
| 0,17426934 | 0,307350515 | DTNB | FALSE | Upregulated |
| -0,508052842 | 0,307384547 | LINC00535 | FALSE | Downregulated |
| 0,292523142 | 0,30747278 | LCA5 | FALSE | Upregulated |
| -0,506594641 | 0,307531239 | SNORA25 | FALSE | Downregulated |
| 1,222992214 | 0,30759231 | RNF157-AS1 | FALSE | Upregulated |
| 0,096690335 | 0,307626189 | ATP6V1C1 | FALSE | Upregulated |
| 0,431571291 | 0,307626189 | LINC00869 | FALSE | Upregulated |
| -0,398448359 | 0,307861849 | LOC101928042 | FALSE | Downregulated |
| 0,092059553 | 0,30788139 | CPSF3L | FALSE | Upregulated |
| -0,89291778 | 0,307920939 | LOC101927032 | FALSE | Downregulated |
| -0,301562515 | 0,307920939 | LOC101928707 | FALSE | Downregulated |
| -0,331406644 | 0,3084112 | CHAD | FALSE | Downregulated |
| 0,144185968 | 0,3084112 | HECA | FALSE | Upregulated |
| 1,434936028 | 0,308620891 | C3orf36 | FALSE | Upregulated |
| -0,254653627 | 0,308665229 | HECW2 | FALSE | Downregulated |
| 0,372736966 | 0,308824927 | EDNRA | FALSE | Upregulated |
| 0,102800054 | 0,308883691 | SH3GL1 | FALSE | Upregulated |
| 0,229486942 | 0,308944381 | OPLAH | FALSE | Upregulated |
| -0,321577798 | 0,308944381 | RHCE | FALSE | Downregulated |
| 1,303043836 | 0,309074434 | FAM106CP | FALSE | Upregulated |
| 0,26108139 | 0,309179458 | PKDCC | FALSE | Upregulated |
| 0,187540703 | 0,309650813 | CASP8 | FALSE | Upregulated |

| | | | | |
|---|---|---|---|---|
| 1,197564925 | 0,309650813 | CCL25 | FALSE | Upregulated |
| 0,247490311 | 0,309650813 | FAM63B | FALSE | Upregulated |
| 1,341317468 | 0,309650813 | IL24 | FALSE | Upregulated |
| -1,009539991 | 0,309650813 | LDLRAD4-AS1 | FALSE | Downregulated |
| 0,257651848 | 0,309650813 | SERPINI1 | FALSE | Upregulated |
| -0,194477792 | 0,309650813 | SLC2A11 | FALSE | Downregulated |
| 0,097887698 | 0,309650813 | USP1 | FALSE | Upregulated |
| 1,01720583 | 0,309679546 | TAS1R3 | FALSE | Upregulated |
| 0,118012457 | 0,309727082 | HMG20B | FALSE | Upregulated |
| -0,397406105 | 0,309727082 | LOC101928403 | FALSE | Downregulated |
| 0,150192445 | 0,309727082 | PCMTD2 | FALSE | Upregulated |
| -0,131907964 | 0,309842546 | GNG10 | FALSE | Downregulated |
| -0,203439442 | 0,309929193 | HOOK1 | FALSE | Downregulated |
| -0,156652046 | 0,309942387 | APOL1 | FALSE | Downregulated |
| 0,15714668 | 0,310028704 | C6orf47 | FALSE | Upregulated |
| 0,235810301 | 0,310067611 | TIA1 | FALSE | Upregulated |
| -0,257557867 | 0,310126513 | NEIL1 | FALSE | Downregulated |
| 0,750192439 | 0,310175472 | C4orf48 | FALSE | Upregulated |
| 0,635185562 | 0,3101867 | MED4-AS1 | FALSE | Upregulated |
| -0,124898954 | 0,310247242 | PSMB1 | FALSE | Downregulated |
| -1,032789789 | 0,310247242 | TRNK | FALSE | Downregulated |
| 0,126740284 | 0,310267645 | RAB1A | FALSE | Upregulated |
| -0,286294727 | 0,310306449 | BST2 | FALSE | Downregulated |
| 0,110284255 | 0,310466903 | CRK | FALSE | Upregulated |
| 0,820212273 | 0,310469102 | LINC00605 | FALSE | Upregulated |
| -0,187689395 | 0,310494822 | ADAMTSL3 | FALSE | Downregulated |
| 0,588916883 | 0,310568836 | LIPE-AS1 | FALSE | Upregulated |
| -0,384995265 | 0,31062753 | DNLZ | FALSE | Downregulated |
| 1,201529064 | 0,310636486 | RHOXF1 | FALSE | Upregulated |
| 0,115235523 | 0,310657597 | UBE2N | FALSE | Upregulated |
| -0,400895838 | 0,310789066 | RP11-333E1.1 | FALSE | Downregulated |
| -0,175545351 | 0,310957335 | FKBP4 | FALSE | Downregulated |
| 0,134365407 | 0,311092028 | PYGO2 | FALSE | Upregulated |
| -0,157189728 | 0,311092028 | YEATS4 | FALSE | Downregulated |
| -0,135297084 | 0,311098743 | COQ5 | FALSE | Downregulated |
| 0,278571718 | 0,311115651 | NABP1 | FALSE | Upregulated |
| 0,269985344 | 0,311128938 | AACS | FALSE | Upregulated |
| 0,837527208 | 0,311128938 | C19orf33 | FALSE | Upregulated |
| 0,949710553 | 0,311128938 | CCL8 | FALSE | Upregulated |
| -0,395135241 | 0,31168571 | GPIHBP1 | FALSE | Downregulated |
| -0,16151562 | 0,31168571 | PGAP2 | FALSE | Downregulated |
| 0,132656092 | 0,311822477 | NOC2L | FALSE | Upregulated |
| 1,13775515 | 0,311860227 | PCNA-AS1 | FALSE | Upregulated |
| 1,060761719 | 0,311860864 | ARHGEF34P | FALSE | Upregulated |
| 1,365861122 | 0,311860864 | SIX3 | FALSE | Upregulated |
| 0,225370072 | 0,311860864 | SPAG5 | FALSE | Upregulated |
| -0,585432333 | 0,311934609 | STX16-NPEPL1 | FALSE | Downregulated |
| -0,11257241 | 0,312033816 | KIAA1033 | FALSE | Downregulated |
| 0,132634164 | 0,312033816 | TAOK1 | FALSE | Upregulated |
| -0,301663642 | 0,312052943 | PMF1-BGLAP | FALSE | Downregulated |
| -0,10503673 | 0,312135872 | FGFR1OP2 | FALSE | Downregulated |

| | | | | |
|---|---|---|---|---|
| 0,25831509 | 0,312139969 | REL | FALSE | Upregulated |
| -0,24980201 | 0,312381872 | BRI3BP | FALSE | Downregulated |
| 1,31826144 | 0,312381872 | HIST4H4 | FALSE | Upregulated |
| 0,35332348 | 0,312381872 | LOC284889 | FALSE | Upregulated |
| 0,137481829 | 0,312381872 | MTA3 | FALSE | Upregulated |
| 0,245284688 | 0,312381872 | PRDM11 | FALSE | Upregulated |
| -0,392560746 | 0,312501291 | IZUMO4 | FALSE | Downregulated |
| -1,104153534 | 0,312890974 | CPA4 | FALSE | Downregulated |
| 1,18674595 | 0,312890974 | OR2A1 | FALSE | Upregulated |
| 0,241035024 | 0,313119133 | ATP2A2 | FALSE | Upregulated |
| 0,879184264 | 0,313119133 | PWP2 | FALSE | Upregulated |
| 0,358169238 | 0,31322548 | ZFP28 | FALSE | Upregulated |
| -0,087391393 | 0,313707913 | RAB14 | FALSE | Downregulated |
| -0,142464021 | 0,313861201 | ZNRF2 | FALSE | Downregulated |
| 0,391898752 | 0,314228372 | KIT | FALSE | Upregulated |
| 0,487291363 | 0,314228372 | SERINC4 | FALSE | Upregulated |
| -0,161091269 | 0,314243349 | GRHPR | FALSE | Downregulated |
| 0,152225163 | 0,314300291 | GIT2 | FALSE | Upregulated |
| 0,423233713 | 0,314300291 | RASL10B | FALSE | Upregulated |
| 1,76745256 | 0,314300291 | RP11-707A18.1 | FALSE | Upregulated |
| -0,180932246 | 0,314375952 | GFOD2 | FALSE | Downregulated |
| 0,12657047 | 0,314569664 | ELMOD2 | FALSE | Upregulated |
| 0,491520989 | 0,314569664 | HCG4 | FALSE | Upregulated |
| 0,24200662 | 0,314569664 | TIPIN | FALSE | Upregulated |
| 1,132306958 | 0,31469146 | FGF18 | FALSE | Upregulated |
| -0,21943174 | 0,314719578 | CYP4F2 | FALSE | Downregulated |
| 0,695965459 | 0,314882193 | STX19 | FALSE | Upregulated |
| -0,150637056 | 0,314935197 | NDUFB4 | FALSE | Downregulated |
| -0,299009983 | 0,314940559 | DAK | FALSE | Downregulated |
| 0,443322407 | 0,314940559 | FGF1 | FALSE | Upregulated |
| -1,43427245 | 0,314940559 | LOC101928976 | FALSE | Downregulated |
| -0,262528878 | 0,314940559 | PRPF38B | FALSE | Downregulated |
| 0,089392802 | 0,314940559 | RABL3 | FALSE | Upregulated |
| 0,153715664 | 0,314940559 | STK38L | FALSE | Upregulated |
| 0,243267883 | 0,314940559 | VAV3 | FALSE | Upregulated |
| -1,281822103 | 0,315016648 | ZIC4 | FALSE | Downregulated |
| -0,131936683 | 0,315128216 | TACO1 | FALSE | Downregulated |
| -0,319859516 | 0,315246602 | LINC01140 | FALSE | Downregulated |
| 0,86372926 | 0,315249713 | PRCD | FALSE | Upregulated |
| -0,484306328 | 0,315541648 | C17orf51 | FALSE | Downregulated |
| 0,612999482 | 0,315541648 | GPR55 | FALSE | Upregulated |
| 0,816211668 | 0,315541648 | KCNH2 | FALSE | Upregulated |
| -0,359093439 | 0,315541648 | RP11-480I12.7 | FALSE | Downregulated |
| 0,102135604 | 0,315541648 | WIPF2 | FALSE | Upregulated |
| -0,774823719 | 0,315710039 | DPP6 | FALSE | Downregulated |
| 0,118709669 | 0,315710039 | ELAVL1 | FALSE | Upregulated |
| 0,302628349 | 0,315710039 | EPB41L4A | FALSE | Upregulated |
| 0,239290941 | 0,315710039 | ZNF383 | FALSE | Upregulated |
| 1,189582909 | 0,315749602 | MSTO2P | FALSE | Upregulated |
| -0,145344692 | 0,315832837 | MPDU1 | FALSE | Downregulated |
| 1,497507559 | 0,316025783 | TRNAG4 | FALSE | Upregulated |

| | | | | |
|---|---|---|---|---|
| -0,123876929 | 0,316031419 | MIPEP | FALSE | Downregulated |
| 0,265762736 | 0,316135342 | PLCB2 | FALSE | Upregulated |
| 0,51628717 | 0,316214732 | KCNK3 | FALSE | Upregulated |
| 1,030958987 | 0,316261734 | AFAP1-AS1 | FALSE | Upregulated |
| -0,206379868 | 0,316274207 | PDCD2L | FALSE | Downregulated |
| 1,276906687 | 0,316388063 | AC098823.3 | FALSE | Upregulated |
| -0,249656416 | 0,316388063 | TSLP | FALSE | Downregulated |
| 0,183319603 | 0,316471634 | POLH | FALSE | Upregulated |
| 1,503712445 | 0,316475765 | DSC1 | FALSE | Upregulated |
| 0,462811624 | 0,316475765 | LINC00654 | FALSE | Upregulated |
| -1,201997799 | 0,31672096 | AC012309.5 | FALSE | Downregulated |
| 0,303028663 | 0,31672096 | FBXO2 | FALSE | Upregulated |
| 0,093792942 | 0,31672096 | OGDH | FALSE | Upregulated |
| -0,401938709 | 0,316739041 | LOC100506358 | FALSE | Downregulated |
| 1,106234636 | 0,316920003 | SLC7A10 | FALSE | Upregulated |
| -0,15669039 | 0,31706894 | CENPC | FALSE | Downregulated |
| 1,355632623 | 0,317156672 | DSCR9 | FALSE | Upregulated |
| -0,200092697 | 0,317156672 | WBP1L | FALSE | Downregulated |
| 1,304474552 | 0,317156672 | ZFR2 | FALSE | Upregulated |
| 0,521301773 | 0,317184815 | LOC101927929 | FALSE | Upregulated |
| 0,120477727 | 0,317208521 | SPOPL | FALSE | Upregulated |
| 0,641602855 | 0,317272704 | LOC101928070 | FALSE | Upregulated |
| 0,242735711 | 0,317359667 | ABHD11 | FALSE | Upregulated |
| 0,268701253 | 0,317359667 | POU6F1 | FALSE | Upregulated |
| 0,122092331 | 0,317359667 | SNIP1 | FALSE | Upregulated |
| 1,222959779 | 0,317550647 | CDK15 | FALSE | Upregulated |
| -0,113646523 | 0,317550647 | FAN1 | FALSE | Downregulated |
| -0,353970622 | 0,31759832 | GYLTL1B | FALSE | Downregulated |
| -1,511135948 | 0,317738288 | LOC102724914 | FALSE | Downregulated |
| 1,008354008 | 0,317738288 | PRSS27 | FALSE | Upregulated |
| 0,087516792 | 0,318045928 | CDV3 | FALSE | Upregulated |
| 0,503992155 | 0,318110359 | LOC441179 | FALSE | Upregulated |
| 0,173260941 | 0,318396471 | SEC14L1 | FALSE | Upregulated |
| 0,836857152 | 0,318891026 | TMEM132D | FALSE | Upregulated |
| 1,496287385 | 0,318891026 | ZNF664-FAM101A | FALSE | Upregulated |
| 0,166132619 | 0,31893001 | RFC2 | FALSE | Upregulated |
| 0,714444392 | 0,31896945 | CCR4 | FALSE | Upregulated |
| 0,511626835 | 0,31896945 | IGFBP6 | FALSE | Upregulated |
| 0,094669645 | 0,319011895 | RRAGA | FALSE | Upregulated |
| 0,489675147 | 0,319029477 | GNRHR2 | FALSE | Upregulated |
| 0,406144827 | 0,319178485 | STK32C | FALSE | Upregulated |
| -0,16579179 | 0,319335506 | CARM1 | FALSE | Downregulated |
| 0,125208518 | 0,319359124 | PLEKHM3 | FALSE | Upregulated |
| 0,109726977 | 0,319424325 | TMX1 | FALSE | Upregulated |
| 1,222819032 | 0,319455592 | ARHGAP11B | FALSE | Upregulated |
| 1,071014158 | 0,31953248 | LOC102723489 | FALSE | Upregulated |
| -1,327810904 | 0,319612785 | LOC100129617 | FALSE | Downregulated |
| -1,326256703 | 0,319805228 | LOC101927800 | FALSE | Downregulated |
| 1,032566247 | 0,319831945 | PDZD3 | FALSE | Upregulated |
| -0,315214839 | 0,31986958 | LOC100506157 | FALSE | Downregulated |
| 0,723340616 | 0,319933697 | HDC | FALSE | Upregulated |

| | | | | |
|---|---|---|---|---|
| 0,935195289 | 0,319933697 | LOC101930665 | FALSE | Upregulated |
| -0,218523358 | 0,319959032 | ENOSF1 | FALSE | Downregulated |
| -0,275401598 | 0,319989139 | KCNC4 | FALSE | Downregulated |
| 0,33009932 | 0,320022906 | USHBP1 | FALSE | Upregulated |
| -0,894379227 | 0,320051569 | DPEP3 | FALSE | Downregulated |
| 0,814357856 | 0,320051569 | GTSF1 | FALSE | Upregulated |
| 1,258599981 | 0,320051569 | PAPPA-AS1 | FALSE | Upregulated |
| -0,164162527 | 0,320051569 | RPS25 | FALSE | Downregulated |
| -0,250710248 | 0,320051569 | ZNF502 | FALSE | Downregulated |
| 0,381138847 | 0,32014813 | LIMD1-AS1 | FALSE | Upregulated |
| 1,275209858 | 0,320148285 | SLC35F1 | FALSE | Upregulated |
| -0,649084293 | 0,32033762 | AC092295.7 | FALSE | Downregulated |
| -0,132844944 | 0,32033762 | IMP3 | FALSE | Downregulated |
| -0,20213148 | 0,320381654 | POLR2M | FALSE | Downregulated |
| 0,135181953 | 0,320381654 | SOCS5 | FALSE | Upregulated |
| -0,127304366 | 0,320527811 | PRKCZ | FALSE | Downregulated |
| 0,168549671 | 0,320647267 | ARHGAP19 | FALSE | Upregulated |
| -0,232865782 | 0,320647267 | FAM221A | FALSE | Downregulated |
| -0,098635839 | 0,320647267 | ZDHHC6 | FALSE | Downregulated |
| -0,146620305 | 0,32073908 | LRRC28 | FALSE | Downregulated |
| -0,281753983 | 0,320752977 | TNFSF13 | FALSE | Downregulated |
| 0,553033051 | 0,320854315 | LOC102723365 | FALSE | Upregulated |
| 0,436523863 | 0,320858833 | SLC16A5 | FALSE | Upregulated |
| 0,445973221 | 0,320967848 | CHST1 | FALSE | Upregulated |
| 0,687346377 | 0,320989154 | MAMDC2 | FALSE | Upregulated |
| -0,695637304 | 0,321079186 | LRP2 | FALSE | Downregulated |
| -0,146936945 | 0,321275893 | C2orf49 | FALSE | Downregulated |
| -0,165923032 | 0,321324165 | PMS2P5 | FALSE | Downregulated |
| 0,19003206 | 0,321324165 | ZNF436 | FALSE | Upregulated |
| 0,136333663 | 0,321330912 | DYSF | FALSE | Upregulated |
| -0,148249265 | 0,32146749 | TMEM150A | FALSE | Downregulated |
| 0,232625554 | 0,321523647 | CCBL1 | FALSE | Upregulated |
| 0,146367918 | 0,321523647 | CRTC1 | FALSE | Upregulated |
| -0,18710652 | 0,321523647 | MLLT1 | FALSE | Downregulated |
| -0,226735092 | 0,321523647 | MPC1 | FALSE | Downregulated |
| -0,227137724 | 0,321523647 | ZNF580 | FALSE | Downregulated |
| 0,550382544 | 0,321604162 | MSH5-SAPCD1 | FALSE | Upregulated |
| 0,06910296 | 0,321751256 | ZC3H7A | FALSE | Upregulated |
| -0,219180945 | 0,321755638 | KIF20B | FALSE | Downregulated |
| 0,705894135 | 0,321755638 | SMPDL3B | FALSE | Upregulated |
| -0,234701642 | 0,321804542 | FAAH2 | FALSE | Downregulated |
| 0,161624675 | 0,321804542 | TIE1 | FALSE | Upregulated |
| 0,257649253 | 0,321804542 | YIF1B | FALSE | Upregulated |
| -0,234012234 | 0,321857917 | FBXO17 | FALSE | Downregulated |
| -0,686274424 | 0,321857917 | LOC101929482 | FALSE | Downregulated |
| 0,161497942 | 0,321857917 | TFPT | FALSE | Upregulated |
| -0,240916327 | 0,321928314 | C3orf62 | FALSE | Downregulated |
| 0,263276381 | 0,321934978 | WNT3 | FALSE | Upregulated |
| 0,135976759 | 0,321989176 | C2orf43 | FALSE | Upregulated |
| -0,110680229 | 0,321989176 | COASY | FALSE | Downregulated |
| 0,137478872 | 0,321989176 | SGPP1 | FALSE | Upregulated |

| | | | | |
|---|---|---|---|---|
| 0,859750756 | 0,321989176 | SOX15 | FALSE | Upregulated |
| 0,148442573 | 0,321997879 | IMP4 | FALSE | Upregulated |
| 0,157135595 | 0,321997879 | LDLRAP1 | FALSE | Upregulated |
| -1,714936129 | 0,321997879 | LOC284344 | FALSE | Downregulated |
| -0,371275568 | 0,321997879 | TMPPE | FALSE | Downregulated |
| -0,107849797 | 0,321997879 | UHMK1 | FALSE | Downregulated |
| 0,3433671 | 0,321997879 | USP6NL-IT1 | FALSE | Upregulated |
| -0,233396809 | 0,322069796 | ZNF529 | FALSE | Downregulated |
| 0,309507335 | 0,322076099 | SPSB2 | FALSE | Upregulated |
| 0,311807213 | 0,322127182 | RP11-521B24.3 | FALSE | Upregulated |
| -0,123824761 | 0,322133051 | NAA25 | FALSE | Downregulated |
| 0,535368094 | 0,322251974 | EHHADH-AS1 | FALSE | Upregulated |
| -0,116128522 | 0,322259696 | RPAP3 | FALSE | Downregulated |
| 0,18279288 | 0,322337812 | TMEM138 | FALSE | Upregulated |
| 0,869725553 | 0,322453638 | PKD2L2 | FALSE | Upregulated |
| 0,586035815 | 0,322463213 | KIF5C | FALSE | Upregulated |
| 0,762463035 | 0,322602464 | LOC101929074 | FALSE | Upregulated |
| -0,075931696 | 0,322930941 | PAIP1 | FALSE | Downregulated |
| 0,191867167 | 0,322930941 | SEC61A1 | FALSE | Upregulated |
| 0,074253058 | 0,322964992 | BAG5 | FALSE | Upregulated |
| -0,145187323 | 0,323171828 | NAT10 | FALSE | Downregulated |
| -0,216852949 | 0,323176562 | TMEM176A | FALSE | Downregulated |
| -1,320979472 | 0,323239602 | DKK4 | FALSE | Downregulated |
| 1,008950395 | 0,323239602 | USP9Y | FALSE | Upregulated |
| 0,125826411 | 0,323259698 | CWF19L2 | FALSE | Upregulated |
| -0,333311845 | 0,323696006 | C7orf13 | FALSE | Downregulated |
| -0,164562273 | 0,323759246 | GOLGB1 | FALSE | Downregulated |
| -1,3615555 | 0,323772451 | DOC2GP | FALSE | Downregulated |
| 0,42001478 | 0,32381992 | NRG1 | FALSE | Upregulated |
| 0,531802561 | 0,323825461 | ANKRD65 | FALSE | Upregulated |
| -0,153580935 | 0,323825461 | CFL2 | FALSE | Downregulated |
| -0,196430978 | 0,323825461 | FLJ31306 | FALSE | Downregulated |
| -0,180049917 | 0,323825461 | MACROD1 | FALSE | Downregulated |
| 0,204275035 | 0,323825461 | ZNF335 | FALSE | Upregulated |
| 0,246781633 | 0,323848775 | TAF8 | FALSE | Upregulated |
| 0,596753728 | 0,324134462 | LOC101928641 | FALSE | Upregulated |
| 0,642795209 | 0,324134462 | PIF1 | FALSE | Upregulated |
| 0,575780762 | 0,324281985 | IMPG2 | FALSE | Upregulated |
| 0,129687648 | 0,324294426 | EXOC6B | FALSE | Upregulated |
| 0,200174788 | 0,324341824 | AAMDC | FALSE | Upregulated |
| 0,19644115 | 0,324612841 | TEAD1 | FALSE | Upregulated |
| -0,144862869 | 0,324675339 | FARS2 | FALSE | Downregulated |
| 0,662780918 | 0,324752718 | SCUBE3 | FALSE | Upregulated |
| 1,312847812 | 0,324757557 | LOC101929491 | FALSE | Upregulated |
| 0,294103748 | 0,324837244 | STAG3L3 | FALSE | Upregulated |
| -0,425560301 | 0,325013574 | ADORA3 | FALSE | Downregulated |
| 0,146749588 | 0,325013574 | AKAP11 | FALSE | Upregulated |
| 0,333253105 | 0,325013574 | ATAD5 | FALSE | Upregulated |
| 1,077084396 | 0,325013574 | DLG1-AS1 | FALSE | Upregulated |
| -0,346584284 | 0,325013574 | EFCAB4A | FALSE | Downregulated |
| 0,313604833 | 0,325013574 | RHOB | FALSE | Upregulated |

| | | | | |
|---|---|---|---|---|
| -0,2102846 | 0,325130748 | HMGB3 | FALSE | Downregulated |
| 0,169866712 | 0,325228517 | ASAH1 | FALSE | Upregulated |
| 0,261052954 | 0,325320613 | NBPF11 | FALSE | Upregulated |
| 0,170647371 | 0,325419643 | CCDC61 | FALSE | Upregulated |
| -0,145927877 | 0,325450038 | ARMCX3 | FALSE | Downregulated |
| 0,151628386 | 0,325450038 | IPO4 | FALSE | Upregulated |
| 0,402070951 | 0,325450038 | TMEM55A | FALSE | Upregulated |
| 0,431859203 | 0,325741921 | LOC606724 | FALSE | Upregulated |
| 0,123575675 | 0,325741921 | MRPL28 | FALSE | Upregulated |
| -0,151038501 | 0,325916864 | WRN | FALSE | Downregulated |
| -0,259537419 | 0,325985415 | PLIN5 | FALSE | Downregulated |
| 1,029059417 | 0,326022299 | LOC102723443 | FALSE | Upregulated |
| -0,304016652 | 0,326022299 | RGS2 | FALSE | Downregulated |
| -1,429095763 | 0,326086424 | CXorf22 | FALSE | Downregulated |
| 0,104112504 | 0,326175813 | CAB39 | FALSE | Upregulated |
| 0,62724671 | 0,326175813 | SHANK1 | FALSE | Upregulated |
| -0,196851604 | 0,326312616 | C10orf118 | FALSE | Downregulated |
| 0,283739395 | 0,326419681 | SELP | FALSE | Upregulated |
| -0,414950078 | 0,32642759 | LINC01125 | FALSE | Downregulated |
| -1,372819694 | 0,32648362 | LOC101927012 | FALSE | Downregulated |
| -0,564358971 | 0,326621185 | KCNB1 | FALSE | Downregulated |
| -0,144922861 | 0,326984192 | ZC3H8 | FALSE | Downregulated |
| 0,133024447 | 0,327073013 | ST3GAL2 | FALSE | Upregulated |
| 1,351297976 | 0,32713255 | LOC101929500 | FALSE | Upregulated |
| -0,456010654 | 0,32713255 | MEGF10 | FALSE | Downregulated |
| -0,57591138 | 0,327228866 | KDM4A-AS1 | FALSE | Downregulated |
| 0,485738564 | 0,327489312 | KCNE4 | FALSE | Upregulated |
| -0,161798272 | 0,327528844 | TPD52 | FALSE | Downregulated |
| 0,096992403 | 0,327786846 | MORC2 | FALSE | Upregulated |
| -0,148632774 | 0,327828777 | NOP10 | FALSE | Downregulated |
| 0,841651434 | 0,327991127 | LOC101926916 | FALSE | Upregulated |
| 0,335609796 | 0,328120195 | FAM131A | FALSE | Upregulated |
| -0,165351702 | 0,328200191 | KMT2C | FALSE | Downregulated |
| -0,261342699 | 0,328300198 | C4orf33 | FALSE | Downregulated |
| 0,345141921 | 0,328324336 | LOC101927943 | FALSE | Upregulated |
| -0,245913239 | 0,328451721 | SOX12 | FALSE | Downregulated |
| 1,352574272 | 0,328471023 | WFIKKN2 | FALSE | Upregulated |
| 0,151238878 | 0,328578257 | MPG | FALSE | Upregulated |
| -1,080656743 | 0,328583422 | LOC101928261 | FALSE | Downregulated |
| -1,228138895 | 0,328690869 | CKMT2 | FALSE | Downregulated |
| 0,220439836 | 0,328722051 | DONSON | FALSE | Upregulated |
| 0,901998883 | 0,328722051 | LINC00539 | FALSE | Upregulated |
| -0,171792018 | 0,328722051 | PELI1 | FALSE | Downregulated |
| -0,103508402 | 0,328748729 | CHTF8 | FALSE | Downregulated |
| 0,211746016 | 0,328970079 | KCTD7 | FALSE | Upregulated |
| 0,545634181 | 0,329113054 | LOC102724607 | FALSE | Upregulated |
| 0,123926055 | 0,329124934 | KDELC2 | FALSE | Upregulated |
| -0,077026797 | 0,329124934 | ZNF140 | FALSE | Downregulated |
| -0,144545767 | 0,329152834 | ERCC5 | FALSE | Downregulated |
| -0,126146479 | 0,329406624 | TCTA | FALSE | Downregulated |
| 0,378177344 | 0,329411726 | HK2 | FALSE | Upregulated |

| | | | | |
|---|---|---|---|---|
| 0,246194042 | 0,329411726 | MLKL | FALSE | Upregulated |
| 0,630987602 | 0,329637449 | LOC101927886 | FALSE | Upregulated |
| 0,1137774 | 0,329662967 | ABL1 | FALSE | Upregulated |
| -0,096756456 | 0,329785506 | CAST | FALSE | Downregulated |
| 0,581919719 | 0,329911883 | SLMO1 | FALSE | Upregulated |
| 0,176298777 | 0,329946851 | IFIT5 | FALSE | Upregulated |
| 0,231795285 | 0,329994709 | CHST15 | FALSE | Upregulated |
| -0,144952612 | 0,329994709 | COPRS | FALSE | Downregulated |
| 0,338771693 | 0,329994709 | REC8 | FALSE | Upregulated |
| 0,349418611 | 0,330092374 | KIAA1524 | FALSE | Upregulated |
| -1,148074501 | 0,330092374 | SLC2A14 | FALSE | Downregulated |
| -0,479163619 | 0,330120549 | AGAP9 | FALSE | Downregulated |
| -0,265337452 | 0,330249247 | OVGP1 | FALSE | Downregulated |
| 0,12478235 | 0,330249247 | USF1 | FALSE | Upregulated |
| -0,136376117 | 0,330322496 | DLG1 | FALSE | Downregulated |
| -0,764033367 | 0,330337028 | LOC101928238 | FALSE | Downregulated |
| 0,156332608 | 0,330428468 | KLHL18 | FALSE | Upregulated |
| -0,215132342 | 0,330551922 | PHF8 | FALSE | Downregulated |
| 0,243701051 | 0,330551922 | ZNF587 | FALSE | Upregulated |
| -0,164538078 | 0,330632124 | MPST | FALSE | Downregulated |
| 0,335741329 | 0,330707335 | C3orf67 | FALSE | Upregulated |
| 0,1596344 | 0,330707335 | SPATA13 | FALSE | Upregulated |
| 1,548203897 | 0,330736053 | SNORD55 | FALSE | Upregulated |
| 1,008645261 | 0,330828942 | LOC100128653 | FALSE | Upregulated |
| 0,105226517 | 0,330908268 | KIFAP3 | FALSE | Upregulated |
| 1,243800866 | 0,331006683 | GYG2P1 | FALSE | Upregulated |
| -0,196047867 | 0,331072392 | SLX4IP | FALSE | Downregulated |
| 0,114555063 | 0,331105151 | RRAGC | FALSE | Upregulated |
| -0,200868103 | 0,331121096 | TNFSF12 | FALSE | Downregulated |
| 1,216755386 | 0,331262368 | NTSR1 | FALSE | Upregulated |
| 0,201930225 | 0,331383757 | PKD1 | FALSE | Upregulated |
| -0,296067262 | 0,33139253 | GATSL3 | FALSE | Downregulated |
| 0,406042086 | 0,331555395 | DHRS9 | FALSE | Upregulated |
| 0,351842637 | 0,331714127 | SLC22A17 | FALSE | Upregulated |
| 0,111461615 | 0,33176387 | KAT7 | FALSE | Upregulated |
| 0,401894499 | 0,331805867 | TOX | FALSE | Upregulated |
| -0,20249099 | 0,331977471 | TMEM150C | FALSE | Downregulated |
| -0,098821184 | 0,332107019 | PDIK1L | FALSE | Downregulated |
| 0,197698607 | 0,332179412 | DAB2 | FALSE | Upregulated |
| 0,115435994 | 0,332247301 | NFIX | FALSE | Upregulated |
| 1,164914581 | 0,332302607 | FLJ27354 | FALSE | Upregulated |
| 0,897554996 | 0,332302607 | TTC23L | FALSE | Upregulated |
| -0,443370441 | 0,332531655 | LOC101929264 | FALSE | Downregulated |
| -0,172149165 | 0,332545941 | LENG1 | FALSE | Downregulated |
| -0,61256767 | 0,332545941 | PTPN20B | FALSE | Downregulated |
| -0,093901766 | 0,332545941 | RAB11B | FALSE | Downregulated |
| 0,564690837 | 0,332545941 | STX1A | FALSE | Upregulated |
| 0,10564951 | 0,332567613 | MFSD1 | FALSE | Upregulated |
| -0,081866796 | 0,33264547 | FTO | FALSE | Downregulated |
| 0,75175339 | 0,33264547 | LOC101927512 | FALSE | Upregulated |
| 0,159150267 | 0,33264547 | NACC1 | FALSE | Upregulated |

| | | | | |
|---|---|---|---|---|
| 0,234872178 | 0,33264547 | TNFAIP8L2 | FALSE | Upregulated |
| -0,169622588 | 0,332813283 | TIMM17B | FALSE | Downregulated |
| -0,265617281 | 0,332873437 | PDP2 | FALSE | Downregulated |
| -0,102959235 | 0,332943776 | WDR43 | FALSE | Downregulated |
| -0,292742482 | 0,332991475 | RP4-639F20.1 | FALSE | Downregulated |
| 0,419475319 | 0,332991475 | ZSWIM8-AS1 | FALSE | Upregulated |
| 0,64717347 | 0,333005761 | RPS2P32 | FALSE | Upregulated |
| -0,511758317 | 0,333011245 | C6orf52 | FALSE | Downregulated |
| -0,512969832 | 0,333011245 | LINC01023 | FALSE | Downregulated |
| -0,398932254 | 0,333017562 | CYP4A22 | FALSE | Downregulated |
| 0,161049692 | 0,333017562 | SLC25A6 | FALSE | Upregulated |
| -0,181069393 | 0,333017562 | TGFBR3 | FALSE | Downregulated |
| -0,329758526 | 0,333028105 | ARHGEF26 | FALSE | Downregulated |
| -0,115376153 | 0,333028105 | PSMD11 | FALSE | Downregulated |
| 0,581775327 | 0,333058875 | MIR3064 | FALSE | Upregulated |
| -0,091073267 | 0,333186526 | KPNA6 | FALSE | Downregulated |
| 1,411196429 | 0,333369676 | C6orf223 | FALSE | Upregulated |
| -0,26286527 | 0,33339479 | SGK2 | FALSE | Downregulated |
| 1,682856799 | 0,333435993 | EREG | FALSE | Upregulated |
| 0,085253846 | 0,333435993 | FAM208A | FALSE | Upregulated |
| -0,587638664 | 0,333450067 | CTB-118N6.3 | FALSE | Downregulated |
| -0,117319565 | 0,333524318 | C1orf52 | FALSE | Downregulated |
| 0,212319106 | 0,333745805 | ZWILCH | FALSE | Upregulated |
| 0,097821492 | 0,333753525 | FKBP15 | FALSE | Upregulated |
| 1,742187714 | 0,333753525 | SH2D5 | FALSE | Upregulated |
| 1,28372234 | 0,333778457 | LOC102724149 | FALSE | Upregulated |
| 0,656437801 | 0,333784932 | FLJ31104 | FALSE | Upregulated |
| -0,423948466 | 0,333856186 | UNC93A | FALSE | Downregulated |
| -0,118306982 | 0,333944602 | SPAST | FALSE | Downregulated |
| -0,097567343 | 0,334089539 | NLK | FALSE | Downregulated |
| 0,824542804 | 0,33412936 | LOC90246 | FALSE | Upregulated |
| -0,131558659 | 0,334134088 | EXOSC9 | FALSE | Downregulated |
| 0,22137313 | 0,334168959 | BCL2L12 | FALSE | Upregulated |
| -0,239332761 | 0,334168959 | NLGN2 | FALSE | Downregulated |
| 0,557021897 | 0,334168959 | RGPD4 | FALSE | Upregulated |
| 0,495868185 | 0,334205365 | FBN2 | FALSE | Upregulated |
| -0,176509581 | 0,334243262 | ZNF619 | FALSE | Downregulated |
| -0,908877356 | 0,334289935 | MAPK4 | FALSE | Downregulated |
| 0,228438476 | 0,334310973 | IGF1R | FALSE | Upregulated |
| -0,149980935 | 0,334337751 | MRPL32 | FALSE | Downregulated |
| 0,300634211 | 0,334477689 | SLC13A3 | FALSE | Upregulated |
| -0,54470844 | 0,334538384 | C8orf44-SGK3 | FALSE | Downregulated |
| 0,349182192 | 0,334538384 | LOC101927151 | FALSE | Upregulated |
| -0,191269957 | 0,334538384 | PRADC1 | FALSE | Downregulated |
| 0,717448089 | 0,334690722 | LOC102723534 | FALSE | Upregulated |
| -0,308196186 | 0,334705079 | ARID3A | FALSE | Downregulated |
| -0,076729416 | 0,334708551 | NFYC | FALSE | Downregulated |
| -0,181822184 | 0,334748105 | STARD13 | FALSE | Downregulated |
| 0,139634115 | 0,335137182 | TAF9B | FALSE | Upregulated |
| 0,146670994 | 0,335244713 | KATNBL1 | FALSE | Upregulated |
| 0,447078168 | 0,335641587 | ALDH1A3 | FALSE | Upregulated |

| | | | | |
|---|---|---|---|---|
| 0,11356368 | 0,336398448 | UBC | FALSE | Upregulated |
| -0,202108664 | 0,336505113 | ATP6V0E2 | FALSE | Downregulated |
| -0,217079019 | 0,336505113 | CLK4 | FALSE | Downregulated |
| 1,592760411 | 0,336505113 | GPR17 | FALSE | Upregulated |
| -0,16588689 | 0,336505113 | RABAC1 | FALSE | Downregulated |
| -0,156148921 | 0,336505113 | ZNF605 | FALSE | Downregulated |
| 1,450869579 | 0,336523634 | LOC102724425 | FALSE | Upregulated |
| -0,097548796 | 0,336828823 | TMEM242 | FALSE | Downregulated |
| 0,342706585 | 0,33690694 | PTPLAD2 | FALSE | Upregulated |
| -1,614173446 | 0,33699271 | KLHL38 | FALSE | Downregulated |
| 1,1702586 | 0,337004155 | WEE2 | FALSE | Upregulated |
| -0,182910324 | 0,33707312 | MSRA | FALSE | Downregulated |
| -0,175986303 | 0,337155216 | C12orf23 | FALSE | Downregulated |
| 0,15274319 | 0,337165173 | HDAC5 | FALSE | Upregulated |
| -0,248577879 | 0,337467117 | LINC00663 | FALSE | Downregulated |
| 0,27463571 | 0,337562365 | ZFP3 | FALSE | Upregulated |
| -0,193920428 | 0,337659818 | MAML3 | FALSE | Downregulated |
| 0,373682041 | 0,337659818 | PGR | FALSE | Upregulated |
| 0,324666373 | 0,337716775 | TM6SF1 | FALSE | Upregulated |
| 0,646282247 | 0,337790085 | RCOR2 | FALSE | Upregulated |
| -0,119969842 | 0,337822297 | MLST8 | FALSE | Downregulated |
| 0,101603979 | 0,337910241 | TM9SF2 | FALSE | Upregulated |
| -0,272256685 | 0,338148715 | CCDC138 | FALSE | Downregulated |
| -0,145569033 | 0,338374649 | ABCC6 | FALSE | Downregulated |
| 0,657947819 | 0,338511197 | LOC441081 | FALSE | Upregulated |
| -0,597720501 | 0,338530741 | LOC101928895 | FALSE | Downregulated |
| -0,101120322 | 0,338591062 | AES | FALSE | Downregulated |
| 0,351305751 | 0,338591062 | CLEC4A | FALSE | Upregulated |
| 0,372430501 | 0,338591062 | FUT1 | FALSE | Upregulated |
| -0,629226052 | 0,338591062 | KLHL33 | FALSE | Downregulated |
| 0,133911253 | 0,338804506 | NUTF2 | FALSE | Upregulated |
| 0,397844173 | 0,338815514 | GPBAR1 | FALSE | Upregulated |
| 0,546573328 | 0,338815514 | ITPRIPL1 | FALSE | Upregulated |
| 1,835896282 | 0,338815514 | JPH4 | FALSE | Upregulated |
| 0,203460772 | 0,338815514 | SLX4 | FALSE | Upregulated |
| 0,649796925 | 0,338820328 | LOC102723311 | FALSE | Upregulated |
| 1,277545004 | 0,338902272 | LOC102467217 | FALSE | Upregulated |
| -0,909639121 | 0,338902272 | LOC102724682 | FALSE | Downregulated |
| 0,459950343 | 0,339113396 | ENTPD1-AS1 | FALSE | Upregulated |
| -0,611820167 | 0,339175562 | PRR4 | FALSE | Downregulated |
| -1,435138891 | 0,339345508 | LOC101928195 | FALSE | Downregulated |
| -0,529624508 | 0,339370764 | MT1M | FALSE | Downregulated |
| -0,175860232 | 0,339452474 | PIK3R4 | FALSE | Downregulated |
| 0,269809228 | 0,339462247 | CKAP2 | FALSE | Upregulated |
| -0,185062219 | 0,339462247 | SLC30A6 | FALSE | Downregulated |
| 0,112774346 | 0,339518847 | TNRC6B | FALSE | Upregulated |
| 0,788864913 | 0,339522711 | LOC101929333 | FALSE | Upregulated |
| -0,809392414 | 0,339715201 | C6orf7 | FALSE | Downregulated |
| 0,116024006 | 0,339715201 | CTIF | FALSE | Upregulated |
| -0,664031112 | 0,339757175 | HMGB3P1 | FALSE | Downregulated |
| 1,480330643 | 0,339810971 | LOC101927685 | FALSE | Upregulated |

| | | | | |
|---|---|---|---|---|
| -1,158323102 | 0,339876483 | GPR15 | FALSE | Downregulated |
| -0,118173352 | 0,339876483 | NFS1 | FALSE | Downregulated |
| 0,131370045 | 0,339993686 | TOP3A | FALSE | Upregulated |
| -0,135601184 | 0,340123644 | CARHSP1 | FALSE | Downregulated |
| -0,185673941 | 0,340440363 | DYNC1I2 | FALSE | Downregulated |
| 1,779112229 | 0,340440363 | LOC100996405 | FALSE | Upregulated |
| 0,590904508 | 0,34047159 | ARHGEF38 | FALSE | Upregulated |
| -0,1122311 | 0,34047159 | DDX56 | FALSE | Downregulated |
| -0,269224351 | 0,340484893 | WHAMMP1 | FALSE | Downregulated |
| 0,135903366 | 0,340688905 | VTI1A | FALSE | Upregulated |
| -0,262670266 | 0,340917554 | STARD5 | FALSE | Downregulated |
| 0,120040487 | 0,341138562 | GBF1 | FALSE | Upregulated |
| 0,281220093 | 0,341257381 | THUMPD3-AS1 | FALSE | Upregulated |
| 0,175381662 | 0,341257381 | WDFY2 | FALSE | Upregulated |
| 2,56111917 | 0,341292586 | SNORD3B-1 | FALSE | Upregulated |
| -1,579571856 | 0,341343163 | GUCY2EP | FALSE | Downregulated |
| -0,234917602 | 0,341590788 | POLR3G | FALSE | Downregulated |
| 0,341818369 | 0,341752206 | ARL11 | FALSE | Upregulated |
| 0,190862163 | 0,341756889 | ZNF81 | FALSE | Upregulated |
| 1,034232993 | 0,341810136 | SNORA3 | FALSE | Upregulated |
| 0,396160828 | 0,341813464 | TSPAN5 | FALSE | Upregulated |
| 1,518693605 | 0,341856024 | LINGO2 | FALSE | Upregulated |
| 0,198188644 | 0,341950914 | KIAA1919 | FALSE | Upregulated |
| -0,077869345 | 0,341950914 | RAB11A | FALSE | Downregulated |
| 0,613414855 | 0,342061454 | GSTM5 | FALSE | Upregulated |
| -0,460047674 | 0,342232341 | IDH1-AS1 | FALSE | Downregulated |
| -0,404057515 | 0,342232341 | RP5-1021I20.1 | FALSE | Downregulated |
| -0,185725987 | 0,342279278 | ZFX | FALSE | Downregulated |
| -0,086562953 | 0,342358536 | GSPT1 | FALSE | Downregulated |
| -0,102675522 | 0,342358536 | SLMAP | FALSE | Downregulated |
| 1,0244201 | 0,342480993 | NWD1 | FALSE | Upregulated |
| 0,208621282 | 0,342511695 | SCO2 | FALSE | Upregulated |
| -0,498353578 | 0,342540646 | DRD1 | FALSE | Downregulated |
| 0,656158328 | 0,342540646 | SNORD22 | FALSE | Upregulated |
| 0,114749868 | 0,342642887 | ZDHHC20 | FALSE | Upregulated |
| -0,097276766 | 0,342653486 | HDAC1 | FALSE | Downregulated |
| 0,138976118 | 0,342695327 | TIGD6 | FALSE | Upregulated |
| -0,144317918 | 0,342766866 | AMPD2 | FALSE | Downregulated |
| -0,056254447 | 0,34289226 | MKRN1 | FALSE | Downregulated |
| 0,858980824 | 0,342911642 | TAS2R5 | FALSE | Upregulated |
| 0,488705809 | 0,342953066 | CEP55 | FALSE | Upregulated |
| 0,2384261 | 0,342993752 | EPB41L2 | FALSE | Upregulated |
| 0,73238466 | 0,343055041 | CECR6 | FALSE | Upregulated |
| 0,126078478 | 0,343121608 | NKRF | FALSE | Upregulated |
| 1,234042746 | 0,343213189 | LOC100506526 | FALSE | Upregulated |
| 1,052475413 | 0,34323035 | LRRC71 | FALSE | Upregulated |
| -1,327840525 | 0,34323035 | PYGM | FALSE | Downregulated |
| 0,934238078 | 0,343330817 | C5orf64 | FALSE | Upregulated |
| 0,280113832 | 0,34356529 | E2F5 | FALSE | Upregulated |
| 0,173360208 | 0,34362686 | CSPP1 | FALSE | Upregulated |
| 0,146721061 | 0,343647674 | DMAP1 | FALSE | Upregulated |

| | | | | |
|---|---|---|---|---|
| 0,420692617 | 0,343647674 | PRKCQ | FALSE | Upregulated |
| -0,192118106 | 0,343660955 | C1orf86 | FALSE | Downregulated |
| -0,314130257 | 0,343660955 | HBCBP | FALSE | Downregulated |
| 0,347331593 | 0,343660955 | LINC00924 | FALSE | Upregulated |
| -0,10256361 | 0,343660955 | MARCH6 | FALSE | Downregulated |
| 1,452144231 | 0,343660955 | SOX11 | FALSE | Upregulated |
| -0,379360342 | 0,343946549 | CELF6 | FALSE | Downregulated |
| -0,320383001 | 0,343946549 | PROZ | FALSE | Downregulated |
| -1,118043894 | 0,343946549 | SYT4 | FALSE | Downregulated |
| 0,292605119 | 0,344019853 | GPR34 | FALSE | Upregulated |
| -0,760386406 | 0,344184865 | RP11-362F19.1 | FALSE | Downregulated |
| -0,11407787 | 0,344228018 | IL13RA1 | FALSE | Downregulated |
| 0,241438617 | 0,344261063 | NDN | FALSE | Upregulated |
| 1,279600834 | 0,344329097 | ANGPT4 | FALSE | Upregulated |
| -1,164661811 | 0,344329097 | BLOC1S5-TXNDC5 | FALSE | Downregulated |
| -0,232183753 | 0,344329097 | CTPS1 | FALSE | Downregulated |
| 1,243251338 | 0,344329097 | KATNBL1P6 | FALSE | Upregulated |
| -0,509486936 | 0,344329097 | LRRC24 | FALSE | Downregulated |
| 0,945925823 | 0,344414324 | SPEG | FALSE | Upregulated |
| 0,182428574 | 0,344511313 | VANGL1 | FALSE | Upregulated |
| -0,088149014 | 0,3448206 | TCEA1 | FALSE | Downregulated |
| 0,163380412 | 0,344822226 | HPCAL1 | FALSE | Upregulated |
| 0,128921601 | 0,344890521 | TGFBR2 | FALSE | Upregulated |
| -0,096828163 | 0,345017017 | ZBTB44 | FALSE | Downregulated |
| -0,140414125 | 0,345166024 | SPRTN | FALSE | Downregulated |
| 0,209890619 | 0,345178822 | HAUS4 | FALSE | Upregulated |
| 0,130483335 | 0,345249986 | ERGIC2 | FALSE | Upregulated |
| 0,095531852 | 0,345265376 | ADSS | FALSE | Upregulated |
| -0,082877638 | 0,345294137 | AP2A2 | FALSE | Downregulated |
| -0,771411325 | 0,345294137 | LOC102723972 | FALSE | Downregulated |
| 0,118066637 | 0,345294137 | MTERF | FALSE | Upregulated |
| 0,158064256 | 0,345348437 | INSIG2 | FALSE | Upregulated |
| 0,17236029 | 0,34551226 | SYNPO | FALSE | Upregulated |
| 0,538359002 | 0,34565285 | TWIST1 | FALSE | Upregulated |
| -0,137889785 | 0,345734556 | USPL1 | FALSE | Downregulated |
| 0,239433584 | 0,345812608 | BAG3 | FALSE | Upregulated |
| 1,558639386 | 0,34593615 | MMP23A | FALSE | Upregulated |
| -0,577119704 | 0,346063787 | RP11-637A17.2 | FALSE | Downregulated |
| 0,957524266 | 0,346278139 | GPR19 | FALSE | Upregulated |
| 0,538340577 | 0,346323964 | CDCA3 | FALSE | Upregulated |
| 0,237805565 | 0,346323964 | PDE10A | FALSE | Upregulated |
| -0,200441881 | 0,346423015 | SRP19 | FALSE | Downregulated |
| 0,393198337 | 0,346436307 | LRRC56 | FALSE | Upregulated |
| -0,379033698 | 0,346838762 | LOC100289098 | FALSE | Downregulated |
| -0,083028211 | 0,346841922 | C16orf72 | FALSE | Downregulated |
| -0,158513354 | 0,346949028 | CPOX | FALSE | Downregulated |
| 1,647197981 | 0,346952157 | HS6ST3 | FALSE | Upregulated |
| -0,06990322 | 0,347014539 | RBM22 | FALSE | Downregulated |
| 0,090937599 | 0,347087493 | CDK9 | FALSE | Upregulated |
| 0,892355082 | 0,347087493 | DNM1P35 | FALSE | Upregulated |
| 0,14174079 | 0,347149728 | ADAM9 | FALSE | Upregulated |

| | | | | |
|---|---|---|---|---|
| -0,299859033 | 0,347304285 | MYCL | FALSE | Downregulated |
| 0,236049829 | 0,347314306 | OSBPL10 | FALSE | Upregulated |
| 0,127914797 | 0,347417786 | DAG1 | FALSE | Upregulated |
| 0,207916405 | 0,347562421 | RANGRF | FALSE | Upregulated |
| 0,100629345 | 0,347666918 | HIRA | FALSE | Upregulated |
| 0,249555774 | 0,347808787 | LOC101927556 | FALSE | Upregulated |
| 0,126807001 | 0,347808787 | TXNDC12 | FALSE | Upregulated |
| 1,204773673 | 0,347879087 | LOC101929165 | FALSE | Upregulated |
| -0,223619887 | 0,347933757 | GIMAP1-GIMAP5 | FALSE | Downregulated |
| 1,636655337 | 0,347990766 | LOC101927333 | FALSE | Upregulated |
| -0,342243514 | 0,348073952 | TRAF3IP2-AS1 | FALSE | Downregulated |
| -0,155537909 | 0,348148786 | ZFAND2B | FALSE | Downregulated |
| 1,110010641 | 0,348164913 | BRE-AS1 | FALSE | Upregulated |
| 0,439600557 | 0,348182238 | BEND4 | FALSE | Upregulated |
| 0,281910477 | 0,348209332 | STAG3L2 | FALSE | Upregulated |
| -0,828633285 | 0,348345642 | LOC100996813 | FALSE | Downregulated |
| 0,161865093 | 0,348380977 | USP38 | FALSE | Upregulated |
| 0,329841677 | 0,348470456 | RP11-307C12.11 | FALSE | Upregulated |
| 0,244725515 | 0,348497668 | S100A11 | FALSE | Upregulated |
| 0,60081547 | 0,348606826 | FHDC1 | FALSE | Upregulated |
| -0,196941611 | 0,349060091 | ONECUT2 | FALSE | Downregulated |
| -0,215550618 | 0,349062306 | ABCA11P | FALSE | Downregulated |
| -1,881751066 | 0,349161428 | LOC102723828 | FALSE | Downregulated |
| 0,83063554 | 0,349268892 | LOC101927733 | FALSE | Upregulated |
| 0,349779484 | 0,349482376 | MGC16142 | FALSE | Upregulated |
| 1,329745536 | 0,349549295 | CITED1 | FALSE | Upregulated |
| -0,166036519 | 0,349549295 | THAP2 | FALSE | Downregulated |
| 0,104109489 | 0,34961327 | HUWE1 | FALSE | Upregulated |
| 0,100191109 | 0,349618632 | ATP6V1G1 | FALSE | Upregulated |
| -0,173798374 | 0,349669753 | DUS4L | FALSE | Downregulated |
| 0,767326405 | 0,349871298 | ERVMER34-1 | FALSE | Upregulated |
| -0,210221915 | 0,350151668 | TP53I13 | FALSE | Downregulated |
| 0,125373135 | 0,350212435 | ZNF473 | FALSE | Upregulated |
| -1,127316414 | 0,350645615 | RP4-794H19.4 | FALSE | Downregulated |
| 1,260618961 | 0,350712508 | OTX1 | FALSE | Upregulated |
| 0,362585115 | 0,351053209 | ANK2 | FALSE | Upregulated |
| -0,162477993 | 0,351053209 | BRD4 | FALSE | Downregulated |
| 0,066292854 | 0,351177856 | NUDCD3 | FALSE | Upregulated |
| 0,843369711 | 0,351306803 | LOC102724010 | FALSE | Upregulated |
| -0,514373731 | 0,351396183 | CHRNA10 | FALSE | Downregulated |
| 1,018300106 | 0,351417272 | PRSS22 | FALSE | Upregulated |
| -0,28327825 | 0,35146865 | MDM4 | FALSE | Downregulated |
| -0,154442366 | 0,35150169 | PPP6R2 | FALSE | Downregulated |
| -0,26902353 | 0,351582993 | SCN8A | FALSE | Downregulated |
| -0,136221813 | 0,351659203 | TBC1D12 | FALSE | Downregulated |
| 0,342302443 | 0,351659203 | ZNF71 | FALSE | Upregulated |
| -0,7690452 | 0,351666216 | FLJ20712 | FALSE | Downregulated |
| -0,106887103 | 0,351737299 | PBXIP1 | FALSE | Downregulated |
| -0,157139874 | 0,351763709 | OXSM | FALSE | Downregulated |
| -0,168122307 | 0,351767544 | C4BPA | FALSE | Downregulated |
| -0,104916044 | 0,351850172 | GID8 | FALSE | Downregulated |

| | | | | |
|---|---|---|---|---|
| 1,525854572 | 0,351850172 | RNF17 | FALSE | Upregulated |
| 0,184796534 | 0,352308069 | TM4SF4 | FALSE | Upregulated |
| -1,371176583 | 0,352432078 | NUTM2G | FALSE | Downregulated |
| -0,462169585 | 0,352477133 | RP11-379B18.5 | FALSE | Downregulated |
| 0,231378587 | 0,35250186 | SEPW1 | FALSE | Upregulated |
| 0,249895406 | 0,352553008 | RPGRIP1L | FALSE | Upregulated |
| 0,283808412 | 0,352610766 | SDHAP2 | FALSE | Upregulated |
| 0,196868584 | 0,352615437 | SPAG1 | FALSE | Upregulated |
| 1,122078555 | 0,352824721 | LOC102724292 | FALSE | Upregulated |
| 0,091455278 | 0,353012831 | STRAP | FALSE | Upregulated |
| 0,126451616 | 0,353022895 | TUBGCP5 | FALSE | Upregulated |
| 0,628162585 | 0,353049762 | ITGA8 | FALSE | Upregulated |
| 1,127867769 | 0,353049762 | LOC102723754 | FALSE | Upregulated |
| 0,727894143 | 0,353186214 | NAV2-AS4 | FALSE | Upregulated |
| 0,291095617 | 0,353186214 | PTPRO | FALSE | Upregulated |
| 0,090798032 | 0,353243542 | CDK13 | FALSE | Upregulated |
| -0,27215691 | 0,353265427 | SRRM2 | FALSE | Downregulated |
| 1,351649263 | 0,353321022 | C17orf66 | FALSE | Upregulated |
| 0,477293658 | 0,353519819 | LOC100289230 | FALSE | Upregulated |
| 1,377850929 | 0,3535204 | LOC102724642 | FALSE | Upregulated |
| -0,122856291 | 0,353798996 | SSFA2 | FALSE | Downregulated |
| 0,665123496 | 0,353829984 | B3GAT2 | FALSE | Upregulated |
| 0,13178 | 0,353829984 | BRF1 | FALSE | Upregulated |
| 0,218181227 | 0,353829984 | CCDC3 | FALSE | Upregulated |
| 0,118455365 | 0,353829984 | GORASP1 | FALSE | Upregulated |
| -0,259030283 | 0,353829984 | IFIT1 | FALSE | Downregulated |
| -0,297994484 | 0,353829984 | NRTN | FALSE | Downregulated |
| -0,54095463 | 0,353829984 | SLC22A10 | FALSE | Downregulated |
| -0,587277508 | 0,353829984 | TNKS2-AS1 | FALSE | Downregulated |
| -0,423775499 | 0,353992809 | MIR6723 | FALSE | Downregulated |
| -0,152795576 | 0,353992809 | UCK1 | FALSE | Downregulated |
| -0,166169375 | 0,354097087 | CTNNAL1 | FALSE | Downregulated |
| 0,52467474 | 0,354097087 | HSF2BP | FALSE | Upregulated |
| -0,384790375 | 0,354097087 | IRAK1BP1 | FALSE | Downregulated |
| 0,867998724 | 0,354097087 | SAA2 | FALSE | Upregulated |
| -0,201758834 | 0,354162583 | CCDC167 | FALSE | Downregulated |
| -0,443832884 | 0,354299468 | ASB4 | FALSE | Downregulated |
| -0,409355746 | 0,354299468 | TSPAN13 | FALSE | Downregulated |
| 0,211172452 | 0,354344051 | ARF4 | FALSE | Upregulated |
| 0,38072781 | 0,354344051 | LOC102723342 | FALSE | Upregulated |
| 1,482491028 | 0,354344051 | MAP2K4P1 | FALSE | Upregulated |
| 0,498017712 | 0,354515348 | BEX2 | FALSE | Upregulated |
| 0,116889263 | 0,354602966 | TMEM50A | FALSE | Upregulated |
| -0,311142025 | 0,354671512 | SUCNR1 | FALSE | Downregulated |
| -0,273307921 | 0,354819382 | FLJ46906 | FALSE | Downregulated |
| 0,181219597 | 0,354819382 | PPT2 | FALSE | Upregulated |
| 0,16499304 | 0,354819382 | TAF9 | FALSE | Upregulated |
| -1,248138549 | 0,354854591 | LOC102723838 | FALSE | Downregulated |
| -0,132057286 | 0,354869515 | SCAPER | FALSE | Downregulated |
| -0,142678687 | 0,354917776 | PPARA | FALSE | Downregulated |
| -0,329849158 | 0,354917776 | TBX15 | FALSE | Downregulated |

| | | | | |
|---|---|---|---|---|
| 0,259292275 | 0,355063301 | LOC102723553 | FALSE | Upregulated |
| -0,154083399 | 0,355171844 | USP53 | FALSE | Downregulated |
| -0,126063258 | 0,355280388 | EMC7 | FALSE | Downregulated |
| 0,145256748 | 0,35540511 | KMT2B | FALSE | Upregulated |
| -0,526740462 | 0,35543089 | LOC101928113 | FALSE | Downregulated |
| -0,422180515 | 0,35543089 | MBL1P | FALSE | Downregulated |
| -0,140323542 | 0,355546641 | RAE1 | FALSE | Downregulated |
| 0,47168908 | 0,355662435 | LOC101929223 | FALSE | Upregulated |
| 0,08574634 | 0,355700466 | SP1 | FALSE | Upregulated |
| -0,52835511 | 0,355715424 | GRAPL | FALSE | Downregulated |
| 0,090170115 | 0,355715424 | GTF2I | FALSE | Upregulated |
| -0,51369165 | 0,355715424 | LOC102724296 | FALSE | Downregulated |
| 0,143095809 | 0,355715424 | RNF166 | FALSE | Upregulated |
| -0,352067532 | 0,355715424 | SLC22A18AS | FALSE | Downregulated |
| -0,113680226 | 0,35575099 | TIMM22 | FALSE | Downregulated |
| -0,205359992 | 0,355805156 | TP53INP2 | FALSE | Downregulated |
| 0,090892822 | 0,355807306 | BICD2 | FALSE | Upregulated |
| 0,108138478 | 0,355807306 | E2F4 | FALSE | Upregulated |
| 1,268352457 | 0,355844251 | DHDH | FALSE | Upregulated |
| -0,159870058 | 0,355844251 | KIAA1467 | FALSE | Downregulated |
| -0,127692089 | 0,355844251 | SPRYD7 | FALSE | Downregulated |
| -0,090649725 | 0,356084619 | MIER3 | FALSE | Downregulated |
| 0,302217458 | 0,356167716 | CD38 | FALSE | Upregulated |
| -0,172556667 | 0,356268981 | SLC39A7 | FALSE | Downregulated |
| 0,460784026 | 0,356281604 | GLRB | FALSE | Upregulated |
| -0,099771199 | 0,356339547 | HNRNPD | FALSE | Downregulated |
| -0,508997982 | 0,356347513 | LOC102723602 | FALSE | Downregulated |
| 0,456594106 | 0,356401455 | GUSBP5 | FALSE | Upregulated |
| -0,10858036 | 0,356482198 | SUPT4H1 | FALSE | Downregulated |
| 0,128519304 | 0,356519852 | PITPNA | FALSE | Upregulated |
| 0,202761613 | 0,356567651 | ZSCAN18 | FALSE | Upregulated |
| -0,227794676 | 0,356627939 | PELP1 | FALSE | Downregulated |
| -0,205659799 | 0,357517793 | PHGDH | FALSE | Downregulated |
| -0,378287722 | 0,357584407 | ACR | FALSE | Downregulated |
| -0,787366713 | 0,357745322 | DNMT3L | FALSE | Downregulated |
| -0,198493894 | 0,357745322 | TTC7B | FALSE | Downregulated |
| -0,42952016 | 0,357867067 | KCNIP3 | FALSE | Downregulated |
| -0,257017534 | 0,357966125 | SLC17A4 | FALSE | Downregulated |
| -0,088422257 | 0,358036742 | ARMC10 | FALSE | Downregulated |
| -0,555018214 | 0,358200894 | LOC101927124 | FALSE | Downregulated |
| 0,316641561 | 0,358242998 | DZIP1 | FALSE | Upregulated |
| -0,148854179 | 0,3583943 | POLL | FALSE | Downregulated |
| 0,819250072 | 0,3583943 | ZFP91-CNTF | FALSE | Upregulated |
| -0,936300897 | 0,358700112 | RP11-286E11.1 | FALSE | Downregulated |
| 0,129242464 | 0,358700112 | ZBTB2 | FALSE | Upregulated |
| -0,247072882 | 0,358704907 | CLCN2 | FALSE | Downregulated |
| -0,490985442 | 0,358819551 | LOC101928143 | FALSE | Downregulated |
| -1,585204287 | 0,358819551 | LOC102724689 | FALSE | Downregulated |
| 0,39313003 | 0,358834353 | GABBR1 | FALSE | Upregulated |
| 0,096381465 | 0,358834353 | TRIOBP | FALSE | Upregulated |
| 1,671508892 | 0,358886598 | LOC100996517 | FALSE | Upregulated |

| | | | | |
|---|---|---|---|---|
| 0,110005659 | 0,358986999 | UBE2O | FALSE | Upregulated |
| 0,131206861 | 0,359003264 | AMD1 | FALSE | Upregulated |
| -0,126180302 | 0,359003264 | NDUFS6 | FALSE | Downregulated |
| 0,320399856 | 0,359028396 | SCML4 | FALSE | Upregulated |
| 0,214911256 | 0,359028396 | ZSWIM3 | FALSE | Upregulated |
| 0,098913534 | 0,359140555 | IKBKAP | FALSE | Upregulated |
| 0,140766075 | 0,359161907 | PMS2 | FALSE | Upregulated |
| 0,151927954 | 0,359258989 | MSI2 | FALSE | Upregulated |
| -1,181585497 | 0,359345463 | TMC3 | FALSE | Downregulated |
| 0,634242563 | 0,359532587 | BHLHA15 | FALSE | Upregulated |
| 0,200270319 | 0,359909135 | TOMM40L | FALSE | Upregulated |
| 0,206976276 | 0,359925585 | CASKIN2 | FALSE | Upregulated |
| -0,739206888 | 0,359925585 | LOC100506753 | FALSE | Downregulated |
| -0,122398536 | 0,359925585 | SRPK1 | FALSE | Downregulated |
| -0,419085143 | 0,360015057 | FBXL22 | FALSE | Downregulated |
| 0,534030664 | 0,360090322 | RFPL1S | FALSE | Upregulated |
| -1,289021584 | 0,360119127 | LCE2D | FALSE | Downregulated |
| -0,160545504 | 0,360213396 | IRF6 | FALSE | Downregulated |
| -0,194602405 | 0,36025631 | TMEM110-MUSTN1 | FALSE | Downregulated |
| 0,126470645 | 0,36044041 | DEPDC5 | FALSE | Upregulated |
| 0,102735228 | 0,360538036 | MAML1 | FALSE | Upregulated |
| -0,189836053 | 0,360950775 | C10orf2 | FALSE | Downregulated |
| 1,141841873 | 0,361023497 | SYT6 | FALSE | Upregulated |
| 0,636653566 | 0,361066936 | CAMK2N2 | FALSE | Upregulated |
| -0,123659933 | 0,361222687 | LCORL | FALSE | Downregulated |
| -0,117199904 | 0,361222687 | MAOB | FALSE | Downregulated |
| 0,133874629 | 0,361222687 | SLC9A6 | FALSE | Upregulated |
| 0,487497677 | 0,361222687 | TMEM150B | FALSE | Upregulated |
| -1,145304346 | 0,361470715 | ANKK1 | FALSE | Downregulated |
| 0,161888331 | 0,361470715 | TRIM68 | FALSE | Upregulated |
| 0,165238511 | 0,361470715 | TTLL12 | FALSE | Upregulated |
| -0,457323421 | 0,361501189 | MVD | FALSE | Downregulated |
| 0,142230309 | 0,361501189 | RRP15 | FALSE | Upregulated |
| -0,405764241 | 0,361519038 | LSMEM1 | FALSE | Downregulated |
| 0,1214927 | 0,361648594 | DSTYK | FALSE | Upregulated |
| 0,155070612 | 0,361706301 | IRF3 | FALSE | Upregulated |
| -0,271804429 | 0,361743508 | HSD11B2 | FALSE | Downregulated |
| 1,04252512 | 0,36191293 | RP11-475N22.4 | FALSE | Upregulated |
| 1,463313474 | 0,361918795 | FLJ35934 | FALSE | Upregulated |
| -0,105016758 | 0,361918795 | RSPRY1 | FALSE | Downregulated |
| 0,371969924 | 0,362056439 | LOC101929251 | FALSE | Upregulated |
| 0,100576859 | 0,362068167 | NIPA1 | FALSE | Upregulated |
| -0,133359758 | 0,362086469 | UTP15 | FALSE | Downregulated |
| -0,141357787 | 0,36221122 | KIAA0907 | FALSE | Downregulated |
| -0,287176682 | 0,362232258 | CCDC73 | FALSE | Downregulated |
| -0,182153429 | 0,362232258 | EPHX2 | FALSE | Downregulated |
| -0,518465804 | 0,362232258 | MYH7B | FALSE | Downregulated |
| 0,136656531 | 0,362232258 | POLE | FALSE | Upregulated |
| 0,205803664 | 0,362354601 | BCL7C | FALSE | Upregulated |
| 0,14614861 | 0,362406786 | DNAL1 | FALSE | Upregulated |

| | | | | |
|---|---|---|---|---|
| 0,096926154 | 0,362463203 | CD46 | FALSE | Upregulated |
| -0,15180005 | 0,362646519 | APOC2 | FALSE | Downregulated |
| -0,458805149 | 0,362646519 | CYP2B7P | FALSE | Downregulated |
| -0,282289929 | 0,362646519 | NTF3 | FALSE | Downregulated |
| 0,382471903 | 0,362720744 | CKB | FALSE | Upregulated |
| 0,341271393 | 0,362786924 | C11orf82 | FALSE | Upregulated |
| -0,15725447 | 0,362997666 | AGXT | FALSE | Downregulated |
| -0,175537316 | 0,363162393 | DPP9 | FALSE | Downregulated |
| -0,335530096 | 0,363366852 | C1orf228 | FALSE | Downregulated |
| -1,10658967 | 0,363366852 | CEP170P1 | FALSE | Downregulated |
| -1,031642942 | 0,363366852 | MYCBP2-AS1 | FALSE | Downregulated |
| 0,125704147 | 0,363376486 | BBS10 | FALSE | Upregulated |
| 0,820780658 | 0,363376486 | COX4I2 | FALSE | Upregulated |
| 0,0813527 | 0,363376486 | MOAP1 | FALSE | Upregulated |
| 0,105722699 | 0,363376486 | TMEM33 | FALSE | Upregulated |
| 0,145327355 | 0,363376486 | TRIM32 | FALSE | Upregulated |
| -0,117022798 | 0,363376486 | XPOT | FALSE | Downregulated |
| -0,129429632 | 0,363393536 | RUVBL1 | FALSE | Downregulated |
| -0,541195647 | 0,363394632 | C1orf213 | FALSE | Downregulated |
| -0,129563847 | 0,363394632 | RPS16 | FALSE | Downregulated |
| 1,292297388 | 0,36347246 | PPP1R2P3 | FALSE | Upregulated |
| -0,155053861 | 0,363514869 | PTRH2 | FALSE | Downregulated |
| -0,208735156 | 0,363815362 | UQCRBP1 | FALSE | Downregulated |
| -0,167563989 | 0,363996773 | PDE2A | FALSE | Downregulated |
| -0,194104956 | 0,364108485 | STX2 | FALSE | Downregulated |
| 0,555091677 | 0,36418313 | LOC90834 | FALSE | Upregulated |
| -0,916383232 | 0,36418313 | RP11-166O4.5 | FALSE | Downregulated |
| 0,557939429 | 0,364352615 | LOC646471 | FALSE | Upregulated |
| 0,283459583 | 0,36441274 | ALAS1 | FALSE | Upregulated |
| 0,214876731 | 0,36441274 | KIAA0040 | FALSE | Upregulated |
| 0,284282664 | 0,36441274 | NLRC4 | FALSE | Upregulated |
| 0,494754804 | 0,36441274 | PRDM6 | FALSE | Upregulated |
| 0,382273662 | 0,364434986 | LOC100499489 | FALSE | Upregulated |
| -0,231292515 | 0,364548812 | RP5-1033H22.2 | FALSE | Downregulated |
| 0,200693788 | 0,364549256 | PVR | FALSE | Upregulated |
| 0,891043683 | 0,364668276 | CCDC181 | FALSE | Upregulated |
| -0,171100774 | 0,364931494 | MTUS1 | FALSE | Downregulated |
| -0,135724355 | 0,365005374 | DDX46 | FALSE | Downregulated |
| -0,351847625 | 0,36516192 | LGSN | FALSE | Downregulated |
| 0,50487065 | 0,365299307 | SNORA67 | FALSE | Upregulated |
| 0,205968437 | 0,365328623 | HARBI1 | FALSE | Upregulated |
| 0,266328137 | 0,365483582 | PLIN4 | FALSE | Upregulated |
| 0,296968052 | 0,365554303 | RIMKLB | FALSE | Upregulated |
| 0,201553688 | 0,365752963 | ZBTB48 | FALSE | Upregulated |
| -0,505239936 | 0,365838539 | EPHB1 | FALSE | Downregulated |
| 0,157037292 | 0,365838539 | NDST2 | FALSE | Upregulated |
| 0,997685439 | 0,366041536 | PRR19 | FALSE | Upregulated |
| -0,184832368 | 0,366041536 | UACA | FALSE | Downregulated |
| 0,313532478 | 0,366279464 | UNC5C | FALSE | Upregulated |
| 1,045564904 | 0,366366588 | LOC101928027 | FALSE | Upregulated |
| 0,735461548 | 0,366722657 | INHBA-AS1 | FALSE | Upregulated |

| | | | | |
|---|---|---|---|---|
| -0,104520934 | 0,366775698 | CDS2 | FALSE | Downregulated |
| 0,388223177 | 0,36682716 | ZSWIM5 | FALSE | Upregulated |
| 1,117904615 | 0,366851027 | OR51E1 | FALSE | Upregulated |
| -0,283728388 | 0,366921609 | ALDH1L1 | FALSE | Downregulated |
| 0,211381465 | 0,367340427 | DHX34 | FALSE | Upregulated |
| -0,158894845 | 0,367394805 | ACSF2 | FALSE | Downregulated |
| 0,157140697 | 0,367394805 | FZR1 | FALSE | Upregulated |
| -0,156991012 | 0,367394805 | MZT2A | FALSE | Downregulated |
| -0,101529335 | 0,367404794 | WDR70 | FALSE | Downregulated |
| 0,12685592 | 0,367413163 | AKIP1 | FALSE | Upregulated |
| 0,878462218 | 0,367413163 | LOC101929609 | FALSE | Upregulated |
| 1,077195462 | 0,367505082 | CA4 | FALSE | Upregulated |
| -0,11124574 | 0,367505082 | GATAD1 | FALSE | Downregulated |
| -0,309515183 | 0,367707388 | SOWAHA | FALSE | Downregulated |
| -0,100801228 | 0,367991061 | FAM114A2 | FALSE | Downregulated |
| 0,569573762 | 0,368191664 | CCDC74A | FALSE | Upregulated |
| 0,244757984 | 0,36830415 | PGP | FALSE | Upregulated |
| -0,151130789 | 0,368318673 | RPL13 | FALSE | Downregulated |
| -0,185187059 | 0,368362337 | FLT1 | FALSE | Downregulated |
| -1,104120323 | 0,368362337 | GJB3 | FALSE | Downregulated |
| 0,104499185 | 0,368362337 | MFF | FALSE | Upregulated |
| 1,268861601 | 0,368362337 | SCGB2B3P | FALSE | Upregulated |
| 0,173996313 | 0,368362337 | TSEN2 | FALSE | Upregulated |
| -0,207814981 | 0,368362337 | ZNF141 | FALSE | Downregulated |
| 0,928376051 | 0,368373979 | CCDC155 | FALSE | Upregulated |
| 0,400138597 | 0,368393075 | GNAL | FALSE | Upregulated |
| 1,274077676 | 0,36840384 | TPTE2 | FALSE | Upregulated |
| -0,574935442 | 0,368905092 | CCDC38 | FALSE | Downregulated |
| -1,121118356 | 0,368905092 | PRH1 | FALSE | Downregulated |
| 0,569493953 | 0,368913718 | LOC100130899 | FALSE | Upregulated |
| 0,160918531 | 0,368972596 | RPS3 | FALSE | Upregulated |
| -0,143153697 | 0,368982583 | ZNF865 | FALSE | Downregulated |
| -0,113072374 | 0,369090306 | PTGR2 | FALSE | Downregulated |
| 0,173816643 | 0,369102793 | C11orf68 | FALSE | Upregulated |
| -1,355303933 | 0,369102793 | HMGA2 | FALSE | Downregulated |
| 0,092961891 | 0,369413116 | KIAA0141 | FALSE | Upregulated |
| 0,959350907 | 0,369544158 | LOC101059906 | FALSE | Upregulated |
| 1,101747133 | 0,369710645 | ARRDC3-AS1 | FALSE | Upregulated |
| -1,479640817 | 0,369710645 | RP11-231I16.1 | FALSE | Downregulated |
| 1,453916678 | 0,369767475 | C12orf42 | FALSE | Upregulated |
| 0,192467113 | 0,369816277 | HOOK2 | FALSE | Upregulated |
| 0,113942313 | 0,36984081 | STRN4 | FALSE | Upregulated |
| 0,172361742 | 0,369889246 | SCRN2 | FALSE | Upregulated |
| 2,217745567 | 0,369889246 | TRNAD15 | FALSE | Upregulated |
| -0,167630101 | 0,369958801 | AGPAT1 | FALSE | Downregulated |
| 0,146298073 | 0,370042483 | PARP10 | FALSE | Upregulated |
| -1,275821743 | 0,370050092 | RP11-1079K10.5 | FALSE | Downregulated |
| 0,110335901 | 0,370059568 | TIMM23 | FALSE | Upregulated |
| -0,843537959 | 0,370221889 | PM20D1 | FALSE | Downregulated |
| 1,215881072 | 0,370350142 | PCDH8 | FALSE | Upregulated |
| 0,086775208 | 0,370681235 | MBD1 | FALSE | Upregulated |

| | | | | |
|---|---|---|---|---|
| 0,700888039 | 0,370947265 | LOC100129516 | FALSE | Upregulated |
| -0,127230091 | 0,371166444 | CRTAP | FALSE | Downregulated |
| 1,046506571 | 0,371166444 | RP11-175K6.1 | FALSE | Upregulated |
| 1,047993571 | 0,371290343 | HCN2 | FALSE | Upregulated |
| -0,3704384 | 0,371457509 | C9orf96 | FALSE | Downregulated |
| -1,399209771 | 0,371468706 | FAM66D | FALSE | Downregulated |
| -0,328480073 | 0,371588769 | SHD | FALSE | Downregulated |
| 0,295817077 | 0,371633856 | ZNF773 | FALSE | Upregulated |
| 0,136729012 | 0,371649884 | FOXP1 | FALSE | Upregulated |
| 0,135888526 | 0,37167285 | LCMT2 | FALSE | Upregulated |
| 0,538304258 | 0,372092123 | DACT2 | FALSE | Upregulated |
| 1,493128826 | 0,37210229 | SRRM3 | FALSE | Upregulated |
| -0,274461101 | 0,372290937 | NOXA1 | FALSE | Downregulated |
| 1,015306738 | 0,372377745 | C20orf197 | FALSE | Upregulated |
| -0,325581742 | 0,372450628 | AK7 | FALSE | Downregulated |
| 0,303030638 | 0,372450628 | PPFIA3 | FALSE | Upregulated |
| -0,348608718 | 0,372450628 | RXFP1 | FALSE | Downregulated |
| -1,045141203 | 0,372630437 | ASB11 | FALSE | Downregulated |
| 0,184553674 | 0,372630437 | DCAF16 | FALSE | Upregulated |
| -0,099751548 | 0,372630437 | GTF2B | FALSE | Downregulated |
| -0,157670482 | 0,372630437 | KIAA1958 | FALSE | Downregulated |
| -0,604826271 | 0,372630437 | LOC100268168 | FALSE | Downregulated |
| -0,586674835 | 0,372630437 | LOC101929846 | FALSE | Downregulated |
| 0,086481983 | 0,372630437 | ZKSCAN5 | FALSE | Upregulated |
| -0,586292447 | 0,372835061 | LOC101929065 | FALSE | Downregulated |
| -0,385322632 | 0,3728381 | LRP5L | FALSE | Downregulated |
| -0,224140908 | 0,372917227 | MATN2 | FALSE | Downregulated |
| 0,213779411 | 0,372923549 | SEC23B | FALSE | Upregulated |
| -0,150487818 | 0,373070381 | ERLEC1 | FALSE | Downregulated |
| -0,750814897 | 0,37320259 | LOC100287934 | FALSE | Downregulated |
| -0,09775521 | 0,373274646 | SOS2 | FALSE | Downregulated |
| -0,253406132 | 0,373300407 | CD180 | FALSE | Downregulated |
| -0,119542153 | 0,373384299 | EMG1 | FALSE | Downregulated |
| 0,086896068 | 0,373391769 | ERCC3 | FALSE | Upregulated |
| 0,690270163 | 0,373391769 | FAM95C | FALSE | Upregulated |
| -1,254970723 | 0,373391769 | LOC101928814 | FALSE | Downregulated |
| -0,092533122 | 0,373522095 | VDAC3 | FALSE | Downregulated |
| 1,270225639 | 0,373630344 | LOC100506859 | FALSE | Upregulated |
| 0,541131103 | 0,373958088 | VIP | FALSE | Upregulated |
| 0,081462274 | 0,374277077 | EXOC3 | FALSE | Upregulated |
| 1,11805413 | 0,374277077 | LINC00982 | FALSE | Upregulated |
| -0,228086799 | 0,37433368 | SMUG1 | FALSE | Downregulated |
| -0,314482502 | 0,374362881 | ARL17A | FALSE | Downregulated |
| -0,536462619 | 0,374773819 | CXorf66 | FALSE | Downregulated |
| -0,51045462 | 0,374773819 | RP11-685M7.3 | FALSE | Downregulated |
| 0,790304788 | 0,374822207 | SNORD58C | FALSE | Upregulated |
| -0,68782816 | 0,374925565 | CALML6 | FALSE | Downregulated |
| -0,165454957 | 0,374925565 | PITPNC1 | FALSE | Downregulated |
| -0,702749791 | 0,374929408 | RGSL1 | FALSE | Downregulated |
| -0,116023637 | 0,374957492 | MAPK6 | FALSE | Downregulated |
| -0,699285136 | 0,374957492 | RP11-298I3.1 | FALSE | Downregulated |

| | | | | |
|---|---|---|---|---|
| -0,089589797 | 0,374978722 | FAM105B | FALSE | Downregulated |
| 0,58898373 | 0,375246468 | LOC102724040 | FALSE | Upregulated |
| -0,141279521 | 0,375408235 | FBXO46 | FALSE | Downregulated |
| 1,010893349 | 0,375408235 | SERTAD4 | FALSE | Upregulated |
| 0,246116776 | 0,375410693 | EFHC1 | FALSE | Upregulated |
| 0,589990766 | 0,375588554 | AC104134.2 | FALSE | Upregulated |
| 0,280153656 | 0,375640176 | KPTN | FALSE | Upregulated |
| 0,48808758 | 0,375660223 | TMPRSS9 | FALSE | Upregulated |
| 0,296505756 | 0,375700987 | DTX1 | FALSE | Upregulated |
| 0,260347083 | 0,37573761 | NRROS | FALSE | Upregulated |
| -0,125979811 | 0,376085533 | WASF1 | FALSE | Downregulated |
| -0,224793673 | 0,376091626 | GPR128 | FALSE | Downregulated |
| 0,102444909 | 0,376187163 | RAP1GDS1 | FALSE | Upregulated |
| 0,160617218 | 0,376243716 | PDXP | FALSE | Upregulated |
| 0,48031213 | 0,376285265 | ACSL4 | FALSE | Upregulated |
| 0,391795931 | 0,376285265 | BSPRY | FALSE | Upregulated |
| 1,107109772 | 0,376285265 | HOXA7 | FALSE | Upregulated |
| -0,643276446 | 0,376436779 | FLJ21408 | FALSE | Downregulated |
| -0,141099005 | 0,376589455 | MEST | FALSE | Downregulated |
| 1,152232926 | 0,376589455 | WDR16 | FALSE | Upregulated |
| -0,17038672 | 0,376600783 | C10orf25 | FALSE | Downregulated |
| 1,294242549 | 0,376869787 | RNU6-31P | FALSE | Upregulated |
| -0,192396254 | 0,376883853 | ANO8 | FALSE | Downregulated |
| 0,288343038 | 0,3769472 | S100A4 | FALSE | Upregulated |
| 0,712183098 | 0,376974295 | CLIC5 | FALSE | Upregulated |
| 0,31130418 | 0,377318947 | CNTNAP2 | FALSE | Upregulated |
| -0,68478332 | 0,377318947 | LOC102724256 | FALSE | Downregulated |
| -0,385169312 | 0,377318947 | PRSS12 | FALSE | Downregulated |
| -0,318925083 | 0,377318947 | RP11-695J4.2 | FALSE | Downregulated |
| -0,694281958 | 0,377318947 | SLC25A48 | FALSE | Downregulated |
| -0,207214589 | 0,377318947 | ZNF77 | FALSE | Downregulated |
| 0,308352998 | 0,37733907 | ADAMTSL5 | FALSE | Upregulated |
| -0,070066716 | 0,37733907 | GLE1 | FALSE | Downregulated |
| 0,512009127 | 0,37733907 | LOC440157 | FALSE | Upregulated |
| -0,12568934 | 0,377355222 | ERLIN1 | FALSE | Downregulated |
| 0,111999218 | 0,37739572 | SLC35D2 | FALSE | Upregulated |
| 0,261144331 | 0,377428055 | SAMD9L | FALSE | Upregulated |
| 0,207708652 | 0,377448814 | NUTM2A | FALSE | Upregulated |
| -0,518682074 | 0,377940317 | CYP2A7 | FALSE | Downregulated |
| -0,784207982 | 0,377940317 | IL5RA | FALSE | Downregulated |
| -0,127990381 | 0,377994262 | ISY1-RAB43 | FALSE | Downregulated |
| 0,134381418 | 0,378005231 | EIF3M | FALSE | Upregulated |
| 0,154380478 | 0,378192021 | ZNF551 | FALSE | Upregulated |
| -0,104380666 | 0,378249262 | ZFAND6 | FALSE | Downregulated |
| -0,317779947 | 0,378374203 | TRPC5 | FALSE | Downregulated |
| 0,143391078 | 0,378396631 | FKRP | FALSE | Upregulated |
| -0,142222016 | 0,378479213 | CIPC | FALSE | Downregulated |
| 0,122247678 | 0,378686342 | HEIH | FALSE | Upregulated |
| -0,426623482 | 0,378752059 | RP11-61I13.3 | FALSE | Downregulated |
| -1,70055796 | 0,37876737 | KBTBD12 | FALSE | Downregulated |
| 1,169017406 | 0,378855716 | ANKRD62 | FALSE | Upregulated |

| | | | | |
|---|---|---|---|---|
| 0,115514401 | 0,378855716 | FBRS | FALSE | Upregulated |
| -1,852177381 | 0,378855716 | SLC5A12 | FALSE | Downregulated |
| 0,078550965 | 0,378855716 | WDR48 | FALSE | Upregulated |
| 0,116251026 | 0,379034991 | CNNM3 | FALSE | Upregulated |
| 1,44970063 | 0,379034991 | LUCAT1 | FALSE | Upregulated |
| 0,413275088 | 0,379034991 | MTMR8 | FALSE | Upregulated |
| 0,203005225 | 0,379034991 | TMEM184A | FALSE | Upregulated |
| -1,349585892 | 0,379121273 | LOC101059936 | FALSE | Downregulated |
| 0,715865152 | 0,379121273 | MCOLN3 | FALSE | Upregulated |
| -0,175959166 | 0,379121273 | TSC1 | FALSE | Downregulated |
| -0,144799525 | 0,379222817 | ANKZF1 | FALSE | Downregulated |
| -0,122990242 | 0,379435455 | ERMAP | FALSE | Downregulated |
| 0,236846862 | 0,379435455 | MTFP1 | FALSE | Upregulated |
| -0,166897125 | 0,379617053 | ZNF542 | FALSE | Downregulated |
| -0,905763824 | 0,379653757 | RBAK-RBAKDN | FALSE | Downregulated |
| 0,966226727 | 0,380001888 | TMEM221 | FALSE | Upregulated |
| 1,036448193 | 0,380166696 | MUC5B | FALSE | Upregulated |
| 0,138395573 | 0,380166696 | SUMF1 | FALSE | Upregulated |
| 0,316925707 | 0,380248188 | FAM127C | FALSE | Upregulated |
| 0,178011963 | 0,380248188 | TMEM237 | FALSE | Upregulated |
| -0,850622971 | 0,380376279 | LOC102724285 | FALSE | Downregulated |
| -0,08876129 | 0,380410593 | RAD1 | FALSE | Downregulated |
| 0,468244348 | 0,380444547 | PLCH2 | FALSE | Upregulated |
| -0,319933481 | 0,380467906 | COL27A1 | FALSE | Downregulated |
| 1,587989703 | 0,380529525 | PHOSPHO2-KLHL23 | FALSE | Upregulated |
| -0,158131172 | 0,380763559 | L3HYPDH | FALSE | Downregulated |
| -0,115817476 | 0,380774022 | NDFIP2 | FALSE | Downregulated |
| 0,189341121 | 0,381095423 | ZC2HC1C | FALSE | Upregulated |
| -0,103975762 | 0,381102161 | ANKRD17 | FALSE | Downregulated |
| -0,372566183 | 0,381318965 | TEKT4P2 | FALSE | Downregulated |
| -0,126497198 | 0,381322372 | CCDC53 | FALSE | Downregulated |
| -0,160534677 | 0,381332784 | CYP4F3 | FALSE | Downregulated |
| 1,138093667 | 0,381412442 | DPYSL4 | FALSE | Upregulated |
| -0,612449851 | 0,381412442 | GUCA2B | FALSE | Downregulated |
| 0,537123869 | 0,381566242 | CCDC87 | FALSE | Upregulated |
| -0,528893992 | 0,38166879 | LOC728989 | FALSE | Downregulated |
| 1,052190554 | 0,381675642 | POU3F1 | FALSE | Upregulated |
| 0,305013971 | 0,381675642 | RAD51B | FALSE | Upregulated |
| -0,109424517 | 0,381982647 | KCTD5 | FALSE | Downregulated |
| -0,360222605 | 0,381982647 | LOC100506083 | FALSE | Downregulated |
| 1,719360014 | 0,382043455 | FABP6 | FALSE | Upregulated |
| 0,12388693 | 0,382043455 | ITPK1 | FALSE | Upregulated |
| -0,116466228 | 0,382043455 | MCPH1 | FALSE | Downregulated |
| 0,170245639 | 0,382072124 | DDX58 | FALSE | Upregulated |
| 0,739989571 | 0,382253044 | FLJ45139 | FALSE | Upregulated |
| -0,359499054 | 0,382253044 | TUBA8 | FALSE | Downregulated |
| -0,151682298 | 0,382299784 | TINAGL1 | FALSE | Downregulated |
| -0,122614469 | 0,382502335 | SRP72 | FALSE | Downregulated |
| 0,15413107 | 0,38251947 | CPEB4 | FALSE | Upregulated |
| 0,207391177 | 0,38251947 | GOT1 | FALSE | Upregulated |
| 0,398930638 | 0,38251947 | USP27X-AS1 | FALSE | Upregulated |

| | | | | |
|---|---|---|---|---|
| -0,124147229 | 0,38300605 | PLS3 | FALSE | Downregulated |
| 0,867964862 | 0,383127669 | KIR3DL1 | FALSE | Upregulated |
| 0,288932951 | 0,38317349 | CCDC170 | FALSE | Upregulated |
| 0,739001019 | 0,38317349 | SLC22A20 | FALSE | Upregulated |
| 0,145326357 | 0,383186296 | HACE1 | FALSE | Upregulated |
| 0,741595115 | 0,383393391 | KIF18B | FALSE | Upregulated |
| 0,526523267 | 0,383507549 | NR1D1 | FALSE | Upregulated |
| -0,72094909 | 0,383748222 | VWA3B | FALSE | Downregulated |
| -0,470727653 | 0,383934622 | LOC102724175 | FALSE | Downregulated |
| -1,237053285 | 0,384253217 | PIR-FIGF | FALSE | Downregulated |
| 0,83276917 | 0,384331213 | CADPS | FALSE | Upregulated |
| 0,172253747 | 0,384331213 | LRFN3 | FALSE | Upregulated |
| 0,338282715 | 0,384331213 | NHSL2 | FALSE | Upregulated |
| -0,135791492 | 0,384553248 | MKNK2 | FALSE | Downregulated |
| 1,329686354 | 0,38478543 | PRR15 | FALSE | Upregulated |
| -0,715915229 | 0,384789721 | CACNA2D3 | FALSE | Downregulated |
| 1,507265415 | 0,38481855 | CACNA1B | FALSE | Upregulated |
| -0,169742733 | 0,38481855 | OFD1 | FALSE | Downregulated |
| -0,178147483 | 0,384868974 | CST3 | FALSE | Downregulated |
| 1,340830273 | 0,384868974 | LGI3 | FALSE | Upregulated |
| -0,088785214 | 0,384868974 | UBE2J2 | FALSE | Downregulated |
| 0,501107501 | 0,384869673 | PYGO1 | FALSE | Upregulated |
| 0,32637957 | 0,384926594 | ZNF775 | FALSE | Upregulated |
| -0,122419766 | 0,385192525 | ESCO1 | FALSE | Downregulated |
| -1,719976703 | 0,385398083 | HOXC8 | FALSE | Downregulated |
| -0,648324943 | 0,385414948 | DPY19L2P3 | FALSE | Downregulated |
| 1,039983329 | 0,385414948 | PWRN1 | FALSE | Upregulated |
| 0,677535744 | 0,385430264 | CYP26A1 | FALSE | Upregulated |
| 1,346253545 | 0,385557391 | DGCR10 | FALSE | Upregulated |
| -0,585138373 | 0,385631875 | LOC101928946 | FALSE | Downregulated |
| 0,256695408 | 0,385631875 | NBPF3 | FALSE | Upregulated |
| 0,205834141 | 0,385637652 | ZNF211 | FALSE | Upregulated |
| -0,115922724 | 0,385701756 | CHP1 | FALSE | Downregulated |
| 0,169303663 | 0,38589766 | ZNF354A | FALSE | Upregulated |
| 0,603153747 | 0,386030597 | APLN | FALSE | Upregulated |
| 0,164055212 | 0,386030597 | TRNT1 | FALSE | Upregulated |
| 0,230336726 | 0,386610449 | WSCD1 | FALSE | Upregulated |
| -0,178786079 | 0,386618324 | RPL11 | FALSE | Downregulated |
| -0,560573563 | 0,386618324 | RPL34-AS1 | FALSE | Downregulated |
| 0,38546046 | 0,386665382 | TMEM71 | FALSE | Upregulated |
| 0,202973265 | 0,386675958 | FZD8 | FALSE | Upregulated |
| 0,537793888 | 0,386676398 | ZNF648 | FALSE | Upregulated |
| -0,964241671 | 0,386740487 | C9orf84 | FALSE | Downregulated |
| 0,117582787 | 0,386818924 | ELOVL5 | FALSE | Upregulated |
| 0,345645556 | 0,386876276 | FPR1 | FALSE | Upregulated |
| -0,336659943 | 0,386876276 | RP11-761I4.3 | FALSE | Downregulated |
| -0,12622781 | 0,3869696 | CCT3 | FALSE | Downregulated |
| -0,193443983 | 0,387045997 | TMEM258 | FALSE | Downregulated |
| -0,569018477 | 0,387129812 | LOC101928820 | FALSE | Downregulated |
| -0,648085175 | 0,387145118 | GGTLC2 | FALSE | Downregulated |
| 0,908424549 | 0,387145118 | LOC102723884 | FALSE | Upregulated |

| | | | | |
|---|---|---|---|---|
| 0,16969439 | 0,387145118 | MCM8 | FALSE | Upregulated |
| 0,10706035 | 0,387452025 | CWF19L1 | FALSE | Upregulated |
| 0,729397976 | 0,387681206 | AC010890.1 | FALSE | Upregulated |
| -0,173086363 | 0,387875421 | FBXO8 | FALSE | Downregulated |
| -0,112510823 | 0,387875421 | TMEM218 | FALSE | Downregulated |
| -0,148670083 | 0,388048437 | UGT2B4 | FALSE | Downregulated |
| -0,139119691 | 0,388090548 | TRIT1 | FALSE | Downregulated |
| 0,571175198 | 0,388178092 | ERMN | FALSE | Upregulated |
| 0,443932886 | 0,388178092 | HOXA1 | FALSE | Upregulated |
| 0,120973897 | 0,388178092 | TNKS | FALSE | Upregulated |
| 0,618971387 | 0,388220226 | RP11-1134I14.8 | FALSE | Upregulated |
| 0,26396115 | 0,388248742 | MUC20 | FALSE | Upregulated |
| 0,103238277 | 0,388411267 | C6orf62 | FALSE | Upregulated |
| -0,131707188 | 0,388411267 | KIAA1715 | FALSE | Downregulated |
| 0,225524505 | 0,388463998 | CCZ1 | FALSE | Upregulated |
| 0,145573209 | 0,388463998 | NPC2 | FALSE | Upregulated |
| -0,599996783 | 0,388601917 | DCLK3 | FALSE | Downregulated |
| -0,079082716 | 0,388794411 | GAPVD1 | FALSE | Downregulated |
| 0,153899682 | 0,388877405 | TCEB1 | FALSE | Upregulated |
| 0,34128176 | 0,38892634 | FAM86JP | FALSE | Upregulated |
| 1,274706819 | 0,38892634 | LOC644919 | FALSE | Upregulated |
| -0,149154574 | 0,38892634 | ZNF552 | FALSE | Downregulated |
| 0,086170524 | 0,388978735 | ATP11B | FALSE | Upregulated |
| -0,224126301 | 0,389079711 | AURKC | FALSE | Downregulated |
| -0,218794425 | 0,389079711 | CCZ1B | FALSE | Downregulated |
| -0,902714425 | 0,389079711 | LOC101060400 | FALSE | Downregulated |
| -0,811355159 | 0,389151153 | LOC345051 | FALSE | Downregulated |
| 0,182796082 | 0,389151153 | SMTN | FALSE | Upregulated |
| -0,491120011 | 0,389222057 | LOC101928548 | FALSE | Downregulated |
| 0,192524461 | 0,389370094 | SH3KBP1 | FALSE | Upregulated |
| 0,720460341 | 0,389456934 | LOC101929426 | FALSE | Upregulated |
| 0,093030087 | 0,389507961 | HNRNPL | FALSE | Upregulated |
| -0,10038149 | 0,389507961 | TRIM13 | FALSE | Downregulated |
| 1,065394674 | 0,389512423 | LOC100130075 | FALSE | Upregulated |
| -1,056357845 | 0,389512423 | RNU6-46P | FALSE | Downregulated |
| 0,107933741 | 0,389549141 | ORC3 | FALSE | Upregulated |
| -0,099014737 | 0,389549141 | STRBP | FALSE | Downregulated |
| 0,092541764 | 0,389672427 | PLOD1 | FALSE | Upregulated |
| 0,350041483 | 0,389718794 | FNDC5 | FALSE | Upregulated |
| -0,647463757 | 0,389718794 | XG | FALSE | Downregulated |
| -0,121599789 | 0,389817782 | KLHL15 | FALSE | Downregulated |
| 0,559856755 | 0,389881805 | LOC101929719 | FALSE | Upregulated |
| -0,942074951 | 0,389881805 | PGLYRP1 | FALSE | Downregulated |
| 0,066591104 | 0,389881805 | XRCC6 | FALSE | Upregulated |
| 0,103304678 | 0,389890698 | KHNYN | FALSE | Upregulated |
| -0,646343678 | 0,389897384 | CALN1 | FALSE | Downregulated |
| -0,188680194 | 0,38993012 | KIAA0895 | FALSE | Downregulated |
| 0,965617785 | 0,390021496 | CUZD1 | FALSE | Upregulated |
| -0,264073182 | 0,390149428 | LOC102724967 | FALSE | Downregulated |
| -0,992911992 | 0,390156007 | LOC102724681 | FALSE | Downregulated |
| 0,190148774 | 0,390180794 | SRXN1 | FALSE | Upregulated |

| | | | | |
|---|---|---|---|---|
| 0,132159414 | 0,390210899 | AGBL5 | FALSE | Upregulated |
| 0,8712831 | 0,390279521 | VAC14-AS1 | FALSE | Upregulated |
| -0,147175015 | 0,390289557 | CEPT1 | FALSE | Downregulated |
| -0,849309391 | 0,390289557 | FAM46B | FALSE | Downregulated |
| -0,148610384 | 0,390325862 | MCAT | FALSE | Downregulated |
| -0,161034901 | 0,390464469 | FBXW7 | FALSE | Downregulated |
| -0,529371193 | 0,390471327 | NOVA1 | FALSE | Downregulated |
| 0,103167664 | 0,390493161 | LOC101928170 | FALSE | Upregulated |
| 0,282034602 | 0,390510489 | GINS3 | FALSE | Upregulated |
| 0,919054486 | 0,390510489 | MYRFL | FALSE | Upregulated |
| -0,244887053 | 0,390659954 | RPGR | FALSE | Downregulated |
| 0,120350266 | 0,390664868 | TRMT10C | FALSE | Upregulated |
| -0,149635078 | 0,39078114 | SNTA1 | FALSE | Downregulated |
| 0,535072518 | 0,390841282 | RP5-1043L13.1-2 | FALSE | Upregulated |
| -0,176183744 | 0,390864213 | RPS26 | FALSE | Downregulated |
| 0,855222228 | 0,390909527 | FBXW4P1 | FALSE | Upregulated |
| -0,112812435 | 0,391152512 | TMEM183A | FALSE | Downregulated |
| -0,087494664 | 0,391502343 | MAP4K5 | FALSE | Downregulated |
| 0,275569868 | 0,391524773 | EAF2 | FALSE | Upregulated |
| -0,123890898 | 0,391678666 | C9orf85 | FALSE | Downregulated |
| -0,129144583 | 0,391678666 | NAPG | FALSE | Downregulated |
| 0,350720333 | 0,391699834 | LOC643733 | FALSE | Upregulated |
| -0,295882862 | 0,391822023 | NKAPL | FALSE | Downregulated |
| -0,164784732 | 0,391999898 | CAPN5 | FALSE | Downregulated |
| -0,607724213 | 0,392050127 | CXCR1 | FALSE | Downregulated |
| 0,538657685 | 0,392057068 | WDR86 | FALSE | Upregulated |
| 0,073235318 | 0,392109793 | MTCH1 | FALSE | Upregulated |
| 0,327646883 | 0,392172562 | GAS1 | FALSE | Upregulated |
| -0,103588831 | 0,392191445 | DKC1 | FALSE | Downregulated |
| 0,476150008 | 0,392191445 | PLAC9 | FALSE | Upregulated |
| 0,141393487 | 0,392363513 | ITPRIPL2 | FALSE | Upregulated |
| 0,354880199 | 0,392441841 | LINC01106 | FALSE | Upregulated |
| -0,218489815 | 0,39267201 | C16orf86 | FALSE | Downregulated |
| 0,293637111 | 0,392745033 | FOXO3B | FALSE | Upregulated |
| 0,26283699 | 0,392805934 | ANGPTL1 | FALSE | Upregulated |
| 0,100061961 | 0,392884211 | TRAPPC3 | FALSE | Upregulated |
| -0,178065445 | 0,392884211 | UROC1 | FALSE | Downregulated |
| 0,156934395 | 0,392902194 | ATP7A | FALSE | Upregulated |
| 1,665862427 | 0,392902194 | KRT72 | FALSE | Upregulated |
| 0,142566778 | 0,392902194 | TSPAN14 | FALSE | Upregulated |
| -0,13514238 | 0,393276422 | REST | FALSE | Downregulated |
| 0,219318213 | 0,393523197 | ATP6V1E2 | FALSE | Upregulated |
| -0,13818106 | 0,393573196 | CBX4 | FALSE | Downregulated |
| -0,337753385 | 0,393755277 | ACSS2 | FALSE | Downregulated |
| 0,209803113 | 0,393755277 | C17orf59 | FALSE | Upregulated |
| -0,904790835 | 0,393889537 | LOC101928841 | FALSE | Downregulated |
| -0,098173541 | 0,394108712 | TMED10 | FALSE | Downregulated |
| -0,091513715 | 0,394108712 | UBA5 | FALSE | Downregulated |
| 0,512540863 | 0,39427848 | MFI2-AS1 | FALSE | Upregulated |
| 0,123905098 | 0,394283238 | METTL21A | FALSE | Upregulated |
| -0,25006969 | 0,394286433 | TMEM97 | FALSE | Downregulated |

| | | | | |
|---|---|---|---|---|
| -0,512625699 | 0,394308129 | LOC100288102 | FALSE | Downregulated |
| -0,155773756 | 0,394344178 | ZSCAN22 | FALSE | Downregulated |
| -0,155638277 | 0,394619905 | SERPINA10 | FALSE | Downregulated |
| 0,59401017 | 0,394646792 | LOC101927837 | FALSE | Upregulated |
| -0,996006847 | 0,394674899 | SEMA3B-AS1 | FALSE | Downregulated |
| -0,086395613 | 0,395012112 | PEX11B | FALSE | Downregulated |
| -0,084381447 | 0,395314146 | FAM192A | FALSE | Downregulated |
| 0,865972672 | 0,395419158 | SMIM5 | FALSE | Upregulated |
| -0,27540163 | 0,395555361 | GREB1 | FALSE | Downregulated |
| -0,244862921 | 0,395594369 | LINC00999 | FALSE | Downregulated |
| -0,082761048 | 0,395594369 | MAPK14 | FALSE | Downregulated |
| 0,882864912 | 0,395594369 | OVCH1-AS1 | FALSE | Upregulated |
| -0,074521721 | 0,395655933 | KARS | FALSE | Downregulated |
| 0,165260254 | 0,395702661 | FBXO21 | FALSE | Upregulated |
| 0,542368079 | 0,395716534 | IL10 | FALSE | Upregulated |
| -0,69815332 | 0,395773214 | ALOX12 | FALSE | Downregulated |
| 0,141554938 | 0,395947109 | PACRGL | FALSE | Upregulated |
| -0,38958961 | 0,396352194 | LOC100270804 | FALSE | Downregulated |
| 0,186049834 | 0,396352194 | TOB1 | FALSE | Upregulated |
| -0,274953655 | 0,396386071 | ROM1 | FALSE | Downregulated |
| 0,14494343 | 0,396654114 | PFDN4 | FALSE | Upregulated |
| -0,077518909 | 0,39668126 | EIF2S3 | FALSE | Downregulated |
| -0,132766201 | 0,396768525 | NADK | FALSE | Downregulated |
| 0,28200596 | 0,396801233 | FAHD2B | FALSE | Upregulated |
| 0,844168357 | 0,396886285 | IL12A | FALSE | Upregulated |
| 0,108124603 | 0,396904497 | MSL3 | FALSE | Upregulated |
| -0,222030055 | 0,397083975 | LOC102723493 | FALSE | Downregulated |
| -0,665511361 | 0,397083975 | LRP1B | FALSE | Downregulated |
| -0,415255099 | 0,397369818 | FKBP1B | FALSE | Downregulated |
| -0,717692896 | 0,39747785 | MRAP | FALSE | Downregulated |
| -0,098230276 | 0,397492999 | SLC35A1 | FALSE | Downregulated |
| 0,146234854 | 0,397532531 | FAM3C | FALSE | Upregulated |
| 0,136346738 | 0,397532531 | GAB1 | FALSE | Upregulated |
| 1,493086513 | 0,397532531 | NLRP9 | FALSE | Upregulated |
| -0,090537961 | 0,39765048 | SKI | FALSE | Downregulated |
| 0,115828351 | 0,397797667 | LMTK2 | FALSE | Upregulated |
| -0,123365779 | 0,397842564 | AFF1 | FALSE | Downregulated |
| 0,180170204 | 0,397842564 | PAN3 | FALSE | Upregulated |
| -0,0938005 | 0,398001153 | TRIM33 | FALSE | Downregulated |
| -0,207760575 | 0,398182307 | ELFN1 | FALSE | Downregulated |
| 0,254076139 | 0,398347828 | PTN | FALSE | Upregulated |
| 0,087654381 | 0,398347858 | HMGN2 | FALSE | Upregulated |
| 0,270526491 | 0,398347858 | SHROOM4 | FALSE | Upregulated |
| 1,49758517 | 0,398408218 | FLJ45079 | FALSE | Upregulated |
| -0,22322613 | 0,398408218 | SEC14L4 | FALSE | Downregulated |
| 0,35346598 | 0,398453999 | SUMO1P3 | FALSE | Upregulated |
| 0,101570196 | 0,398508351 | ATG16L1 | FALSE | Upregulated |
| -0,174845028 | 0,398522842 | CD2AP | FALSE | Downregulated |
| -0,102737345 | 0,398522842 | LAMTOR1 | FALSE | Downregulated |
| 0,411498171 | 0,398584568 | GZMB | FALSE | Upregulated |
| -0,094292217 | 0,398584568 | TRUB1 | FALSE | Downregulated |

| | | | | |
|---|---|---|---|---|
| 0,422508852 | 0,398636795 | ATP8B3 | FALSE | Upregulated |
| 0,335416959 | 0,398636795 | LBP | FALSE | Upregulated |
| -0,958313603 | 0,398636795 | LOC100506082 | FALSE | Downregulated |
| 0,258076573 | 0,398732609 | ZNF431 | FALSE | Upregulated |
| 0,873656638 | 0,398734001 | CTAGE10P | FALSE | Upregulated |
| 0,127645032 | 0,398825847 | ADRM1 | FALSE | Upregulated |
| 1,101979264 | 0,398825847 | DKK1 | FALSE | Upregulated |
| -1,352654116 | 0,398825847 | LINC01139 | FALSE | Downregulated |
| 0,077379503 | 0,398825847 | NCBP2 | FALSE | Upregulated |
| 0,13659331 | 0,398825847 | SKIV2L | FALSE | Upregulated |
| 0,123891537 | 0,398840702 | WARS2 | FALSE | Upregulated |
| -0,079114447 | 0,398966861 | PPM1A | FALSE | Downregulated |
| -0,065370091 | 0,398982185 | KCTD20 | FALSE | Downregulated |
| -0,090900516 | 0,399055744 | HDLBP | FALSE | Downregulated |
| 0,109558714 | 0,399140426 | HFE | FALSE | Upregulated |
| 1,051205234 | 0,399140426 | SHISA2 | FALSE | Upregulated |
| 0,235350626 | 0,399140426 | WDYHV1 | FALSE | Upregulated |
| -0,10909477 | 0,399232965 | WWC2 | FALSE | Downregulated |
| -0,372697 | 0,399314926 | LOC100506235 | FALSE | Downregulated |
| -0,148729262 | 0,399314926 | PRPF3 | FALSE | Downregulated |
| -0,218878146 | 0,399314926 | TMEM98 | FALSE | Downregulated |
| 0,779854389 | 0,399392246 | DYDC2 | FALSE | Upregulated |
| 0,356268533 | 0,399392246 | TPX2 | FALSE | Upregulated |
| 0,211183299 | 0,399453072 | CENPQ | FALSE | Upregulated |
| 0,169913576 | 0,39950578 | MIS18BP1 | FALSE | Upregulated |
| 1,064842498 | 0,399512253 | SPDYE2 | FALSE | Upregulated |
| 0,118534934 | 0,399571936 | MTMR9 | FALSE | Upregulated |
| 1,128267479 | 0,399935413 | MSX2 | FALSE | Upregulated |
| -0,25924085 | 0,400045284 | WFDC1 | FALSE | Downregulated |
| 0,725185426 | 0,400110726 | CELP | FALSE | Upregulated |
| 0,73927015 | 0,400255508 | C1orf127 | FALSE | Upregulated |
| 0,136325286 | 0,400255508 | RC3H2 | FALSE | Upregulated |
| -0,090899186 | 0,400255508 | UBR7 | FALSE | Downregulated |
| 0,165097846 | 0,400361625 | C5orf28 | FALSE | Upregulated |
| -0,173728666 | 0,400361625 | CDK11A | FALSE | Downregulated |
| -0,231631339 | 0,400361625 | SYT9 | FALSE | Downregulated |
| -0,123664467 | 0,400374629 | ACVR2A | FALSE | Downregulated |
| 0,079651794 | 0,400374629 | VPS52 | FALSE | Upregulated |
| 0,12699904 | 0,400706811 | SAV1 | FALSE | Upregulated |
| 0,219012938 | 0,40084655 | SERTAD1 | FALSE | Upregulated |
| -0,0734726 | 0,40084945 | PPP1R7 | FALSE | Downregulated |
| 1,139121629 | 0,401015864 | RP11-138B4.1 | FALSE | Upregulated |
| 0,584504701 | 0,401193796 | LOC101928517 | FALSE | Upregulated |
| 0,118534867 | 0,401238031 | FUBP1 | FALSE | Upregulated |
| -0,162779115 | 0,401458445 | LRRCC1 | FALSE | Downregulated |
| 0,136659564 | 0,401458445 | ZNF720 | FALSE | Upregulated |
| 0,207314912 | 0,40153182 | FZD1 | FALSE | Upregulated |
| 0,102052123 | 0,401764602 | TWF1 | FALSE | Upregulated |
| 0,088567147 | 0,401931713 | SH3BP5L | FALSE | Upregulated |
| -0,776378809 | 0,40205915 | DYNLRB2 | FALSE | Downregulated |
| 0,459173872 | 0,402410889 | SPAG4 | FALSE | Upregulated |

| | | | | |
|---|---|---|---|---|
| 1,182382151 | 0,402507699 | HOXD8 | FALSE | Upregulated |
| 0,158923479 | 0,402507699 | TAF1B | FALSE | Upregulated |
| 1,446985974 | 0,402537328 | ATP10B | FALSE | Upregulated |
| -0,159018012 | 0,402537328 | TPP1 | FALSE | Downregulated |
| -0,267380124 | 0,402552603 | AKR7L | FALSE | Downregulated |
| 0,961844606 | 0,402552603 | LOC101928360 | FALSE | Upregulated |
| -0,71794757 | 0,402552603 | LOC102724092 | FALSE | Downregulated |
| 0,599769099 | 0,402552603 | LOC102724156 | FALSE | Upregulated |
| -0,196264852 | 0,402552603 | PHC3 | FALSE | Downregulated |
| -0,160519781 | 0,402601762 | STRA13 | FALSE | Downregulated |
| 0,748317034 | 0,402817264 | HIST2H4B | FALSE | Upregulated |
| -0,431676976 | 0,402998186 | NSUN7 | FALSE | Downregulated |
| -0,097734395 | 0,402998186 | SBF2 | FALSE | Downregulated |
| 0,139561434 | 0,402998186 | ZSWIM8 | FALSE | Upregulated |
| 0,661446939 | 0,403403546 | CLRN1-AS1 | FALSE | Upregulated |
| 0,762353047 | 0,403455101 | CCL24 | FALSE | Upregulated |
| 0,324307445 | 0,403533911 | COLQ | FALSE | Upregulated |
| -0,201602038 | 0,403533911 | RP11-305E17.6 | FALSE | Downregulated |
| -0,153350267 | 0,403533911 | SEC61B | FALSE | Downregulated |
| 0,146647092 | 0,403786366 | IMPDH2 | FALSE | Upregulated |
| 0,272503562 | 0,403786366 | PLEKHG6 | FALSE | Upregulated |
| 0,221121182 | 0,403802777 | GUCY1A2 | FALSE | Upregulated |
| 0,706847548 | 0,403821793 | SLC5A10 | FALSE | Upregulated |
| 0,828187069 | 0,403975186 | LOC102724024 | FALSE | Upregulated |
| 0,227514398 | 0,404066785 | CCNF | FALSE | Upregulated |
| 1,677380054 | 0,404116864 | OPRK1 | FALSE | Upregulated |
| -0,869430343 | 0,404116864 | TTLL13 | FALSE | Downregulated |
| 0,214838386 | 0,404243839 | GLUL | FALSE | Upregulated |
| 0,232926787 | 0,404247084 | CPVL | FALSE | Upregulated |
| 0,469943875 | 0,404247084 | EFR3B | FALSE | Upregulated |
| -0,576109966 | 0,404247084 | FAM133A | FALSE | Downregulated |
| -0,156532479 | 0,404247084 | TMEM177 | FALSE | Downregulated |
| -0,096945308 | 0,404247084 | UBR5 | FALSE | Downregulated |
| -0,392739616 | 0,40425193 | RAMP2-AS1 | FALSE | Downregulated |
| 1,086097095 | 0,404458371 | RLN2 | FALSE | Upregulated |
| 0,541285833 | 0,404481735 | DLEU7 | FALSE | Upregulated |
| 0,223403166 | 0,40457337 | LAMB3 | FALSE | Upregulated |
| -0,293340834 | 0,404686962 | LOC102724711 | FALSE | Downregulated |
| 0,141964349 | 0,404774224 | LCOR | FALSE | Upregulated |
| 1,238473246 | 0,404975406 | LOC400891 | FALSE | Upregulated |
| 0,205530386 | 0,404975406 | ZNF597 | FALSE | Upregulated |
| 1,651640923 | 0,405167327 | GPR156 | FALSE | Upregulated |
| -1,037233358 | 0,405235249 | LOC150051 | FALSE | Downregulated |
| -0,283546883 | 0,405291272 | LOC101929761 | FALSE | Downregulated |
| -0,108401256 | 0,40530445 | CRYL1 | FALSE | Downregulated |
| -0,077288376 | 0,40530445 | CTBP1 | FALSE | Downregulated |
| 0,204165499 | 0,40530445 | MAP3K10 | FALSE | Upregulated |
| 0,118707239 | 0,40530445 | ZFP41 | FALSE | Upregulated |
| -0,093792076 | 0,40533565 | MCCC1 | FALSE | Downregulated |
| 0,105099608 | 0,40539528 | SPPL2A | FALSE | Upregulated |
| 1,133126342 | 0,405638771 | LOC101929038 | FALSE | Upregulated |

| | | | | |
|---|---|---|---|---|
| -0,445327135 | 0,405651558 | DPY19L1P1 | FALSE | Downregulated |
| -0,661623031 | 0,405651558 | LINC00598 | FALSE | Downregulated |
| -0,211833035 | 0,405829134 | FNDC4 | FALSE | Downregulated |
| -0,637965673 | 0,405935809 | LOC284865 | FALSE | Downregulated |
| 0,131697749 | 0,406001606 | AIP | FALSE | Upregulated |
| -0,61620897 | 0,406133211 | COX7A1 | FALSE | Downregulated |
| -0,181547538 | 0,406564806 | C21orf2 | FALSE | Downregulated |
| -0,106150064 | 0,406564806 | CSNK1G2 | FALSE | Downregulated |
| -0,402678663 | 0,406564806 | GSTO2 | FALSE | Downregulated |
| -0,152793303 | 0,406564806 | SPSB3 | FALSE | Downregulated |
| 0,103549364 | 0,406569953 | USP34 | FALSE | Upregulated |
| -0,368854135 | 0,406738396 | ETV2 | FALSE | Downregulated |
| 0,163263709 | 0,406757237 | NAIF1 | FALSE | Upregulated |
| 0,179930629 | 0,406769194 | KIF16B | FALSE | Upregulated |
| -0,142605571 | 0,406786782 | DNAJC27 | FALSE | Downregulated |
| -0,561651808 | 0,406786782 | GRIP2 | FALSE | Downregulated |
| -0,126395046 | 0,406798276 | PES1 | FALSE | Downregulated |
| -0,669563691 | 0,406811169 | LOC101928083 | FALSE | Downregulated |
| 0,971000955 | 0,406811169 | TRPC2 | FALSE | Upregulated |
| 0,106951179 | 0,406988339 | VAMP8 | FALSE | Upregulated |
| 0,265921479 | 0,407097009 | LEPR | FALSE | Upregulated |
| -0,397252803 | 0,407098597 | LOC730183 | FALSE | Downregulated |
| -0,156940345 | 0,407127432 | SMLR1 | FALSE | Downregulated |
| 2,501394212 | 0,407175398 | TRNAK26 | FALSE | Upregulated |
| 0,279672421 | 0,40718593 | LRRC4C | FALSE | Upregulated |
| 0,182521687 | 0,407243311 | CCDC107 | FALSE | Upregulated |
| -0,203477857 | 0,407243311 | SPP2 | FALSE | Downregulated |
| 0,227388613 | 0,407561037 | ALG10 | FALSE | Upregulated |
| -1,036955179 | 0,407561037 | C15orf26 | FALSE | Downregulated |
| 0,133959418 | 0,407561037 | PLCD1 | FALSE | Upregulated |
| -0,405467183 | 0,407708661 | AMN | FALSE | Downregulated |
| -0,163681833 | 0,407954431 | DNAJC4 | FALSE | Downregulated |
| -0,378895966 | 0,408060359 | AC010226.4 | FALSE | Downregulated |
| 0,121715163 | 0,408060359 | CNNM4 | FALSE | Upregulated |
| 0,629739336 | 0,408193866 | LOC102724552 | FALSE | Upregulated |
| 0,088143253 | 0,408193866 | REPIN1 | FALSE | Upregulated |
| 0,119033933 | 0,408250316 | PSMG2 | FALSE | Upregulated |
| 0,205889245 | 0,408427726 | CBLC | FALSE | Upregulated |
| 0,200648217 | 0,408541599 | ANKAR | FALSE | Upregulated |
| 0,419997904 | 0,408541599 | LOC101928092 | FALSE | Upregulated |
| -0,108281441 | 0,408541599 | TMEM205 | FALSE | Downregulated |
| 0,193062837 | 0,408541599 | ZNF862 | FALSE | Upregulated |
| -0,160012733 | 0,408685696 | F8 | FALSE | Downregulated |
| 1,142560197 | 0,408756024 | PURG | FALSE | Upregulated |
| 0,133512754 | 0,408766081 | MYCBP | FALSE | Upregulated |
| 0,844967368 | 0,408806199 | RP11-37B2.1 | FALSE | Upregulated |
| -0,18260204 | 0,408850862 | LOC101060195 | FALSE | Downregulated |
| 1,016426711 | 0,408908717 | ZNRF3-AS1 | FALSE | Upregulated |
| -0,155839662 | 0,408962532 | ARID5A | FALSE | Downregulated |
| -0,082273174 | 0,408962532 | RNF121 | FALSE | Downregulated |
| 0,127493181 | 0,409034104 | ATP5SL | FALSE | Upregulated |

| | | | | |
|---|---|---|---|---|
| 1,289666572 | 0,409034104 | SFTPB | FALSE | Upregulated |
| 1,096343811 | 0,409099038 | KIAA0319 | FALSE | Upregulated |
| -0,141197552 | 0,409099038 | NDUFB11 | FALSE | Downregulated |
| -0,09769274 | 0,409099038 | PDCD10 | FALSE | Downregulated |
| -0,598820679 | 0,409307981 | KIF25 | FALSE | Downregulated |
| 0,116946053 | 0,409307981 | TTC4 | FALSE | Upregulated |
| 0,195965866 | 0,409322672 | FAM49A | FALSE | Upregulated |
| 0,98592597 | 0,409331982 | MIR181A2HG | FALSE | Upregulated |
| -0,165229629 | 0,409457964 | ARL13B | FALSE | Downregulated |
| -1,340863793 | 0,409457964 | GUCY1B2 | FALSE | Downregulated |
| 0,528223395 | 0,409501269 | LOC102723671 | FALSE | Upregulated |
| -0,747559247 | 0,409501269 | RP5-1004I9.1 | FALSE | Downregulated |
| 0,209163118 | 0,409529306 | FBXO36 | FALSE | Upregulated |
| 0,626402435 | 0,409529306 | LOC283693 | FALSE | Upregulated |
| 0,995038689 | 0,409641543 | FAM72A | FALSE | Upregulated |
| 0,098682084 | 0,409849431 | FRS2 | FALSE | Upregulated |
| -0,218188844 | 0,409849431 | RAC3 | FALSE | Downregulated |
| -0,555824135 | 0,409970951 | ANKLE1 | FALSE | Downregulated |
| 0,266396535 | 0,409970951 | ZNF701 | FALSE | Upregulated |
| 0,125906654 | 0,409992717 | CACFD1 | FALSE | Upregulated |
| -0,287579031 | 0,409992717 | CYP2A6 | FALSE | Downregulated |
| -0,443745195 | 0,409992717 | LOC100288637 | FALSE | Downregulated |
| 1,011870992 | 0,409992717 | SPRN | FALSE | Upregulated |
| -0,122768699 | 0,409992717 | TMEM189 | FALSE | Downregulated |
| -0,148448994 | 0,410266203 | ALDH1A1 | FALSE | Downregulated |
| -0,188088217 | 0,410266203 | ATN1 | FALSE | Downregulated |
| 0,439130694 | 0,410325245 | EBF1 | FALSE | Upregulated |
| -0,147563045 | 0,410436396 | FOXRED1 | FALSE | Downregulated |
| -0,259788822 | 0,410436396 | GRK4 | FALSE | Downregulated |
| 0,935452895 | 0,410436396 | SSUH2 | FALSE | Upregulated |
| -0,114211175 | 0,410447924 | ZMYND8 | FALSE | Downregulated |
| 1,353608946 | 0,410448259 | CDH22 | FALSE | Upregulated |
| 0,169479059 | 0,410448259 | MAST2 | FALSE | Upregulated |
| 0,119503578 | 0,410649965 | CACTIN | FALSE | Upregulated |
| 0,095037487 | 0,410679609 | CPSF6 | FALSE | Upregulated |
| 0,296340728 | 0,410692192 | NCF1 | FALSE | Upregulated |
| 0,342509999 | 0,410692192 | PRRT3 | FALSE | Upregulated |
| 0,14669709 | 0,410711022 | GADD45GIP1 | FALSE | Upregulated |
| 0,651164331 | 0,410867904 | ESYT3 | FALSE | Upregulated |
| -0,111733513 | 0,410867904 | PIGF | FALSE | Downregulated |
| 1,058139268 | 0,410895598 | FLRT1 | FALSE | Upregulated |
| 0,183843523 | 0,410952886 | PIK3AP1 | FALSE | Upregulated |
| 0,100299854 | 0,410952886 | SCAMP2 | FALSE | Upregulated |
| 1,143555213 | 0,411064299 | IGLL3P | FALSE | Upregulated |
| 0,251553967 | 0,411171419 | ZNF891 | FALSE | Upregulated |
| 0,210557656 | 0,411195252 | PEAR1 | FALSE | Upregulated |
| 0,522324973 | 0,411203262 | HIST3H2A | FALSE | Upregulated |
| 0,679673173 | 0,411263166 | FANK1 | FALSE | Upregulated |
| 0,257482847 | 0,411414968 | LRP12 | FALSE | Upregulated |
| 1,032959297 | 0,411544704 | THEM5 | FALSE | Upregulated |
| -0,189596598 | 0,411579154 | DYRK3 | FALSE | Downregulated |

| | | | | |
|---|---|---|---|---|
| -0,139440368 | 0,411855773 | METTL23 | FALSE | Downregulated |
| 0,116758191 | 0,411980219 | MBP | FALSE | Upregulated |
| 0,260637504 | 0,411980219 | ZNF366 | FALSE | Upregulated |
| -0,871926737 | 0,412036614 | C1orf180 | FALSE | Downregulated |
| 0,499781844 | 0,412119827 | GRIN3A | FALSE | Upregulated |
| -0,158047776 | 0,412153508 | THOP1 | FALSE | Downregulated |
| 0,778479206 | 0,412379821 | LAIR2 | FALSE | Upregulated |
| 0,16476905 | 0,412379821 | LDHB | FALSE | Upregulated |
| -0,972419429 | 0,412379821 | LUZP2 | FALSE | Downregulated |
| 0,284117434 | 0,412379821 | MILR1 | FALSE | Upregulated |
| -0,195256928 | 0,412379821 | PSD3 | FALSE | Downregulated |
| -0,16591721 | 0,412401552 | C12orf57 | FALSE | Downregulated |
| -0,36384598 | 0,412452535 | LOC101928236 | FALSE | Downregulated |
| 0,485152406 | 0,412473311 | LOC101928291 | FALSE | Upregulated |
| -0,17311231 | 0,412577206 | GBP7 | FALSE | Downregulated |
| -0,127740055 | 0,412904286 | PRPF4B | FALSE | Downregulated |
| -1,153246321 | 0,412955713 | LOC102724508 | FALSE | Downregulated |
| 0,120481849 | 0,412955713 | NPRL3 | FALSE | Upregulated |
| -0,127935975 | 0,412955713 | SCCPDH | FALSE | Downregulated |
| -0,097844919 | 0,41303779 | ARNT | FALSE | Downregulated |
| -0,637703876 | 0,413111694 | C9orf24 | FALSE | Downregulated |
| 0,494741072 | 0,413142985 | LOC100288911 | FALSE | Upregulated |
| 0,116527736 | 0,413142985 | SSH1 | FALSE | Upregulated |
| 0,258367469 | 0,413142985 | ZNF169 | FALSE | Upregulated |
| 0,21964838 | 0,413142985 | ZNF503-AS2 | FALSE | Upregulated |
| 0,319506038 | 0,413196828 | PFKFB4 | FALSE | Upregulated |
| 0,153070936 | 0,413442326 | NT5C3A | FALSE | Upregulated |
| 0,350890276 | 0,413452758 | PCP4L1 | FALSE | Upregulated |
| 0,243310321 | 0,413479408 | ENAH | FALSE | Upregulated |
| 0,466017556 | 0,413479408 | LINC00672 | FALSE | Upregulated |
| 0,511837513 | 0,413561835 | MIPEPP3 | FALSE | Upregulated |
| 0,218110575 | 0,413687395 | APOBEC3F | FALSE | Upregulated |
| -0,137594543 | 0,41378823 | BRWD3 | FALSE | Downregulated |
| -0,094954831 | 0,41378823 | C6orf120 | FALSE | Downregulated |
| -0,148750075 | 0,41378823 | CALD1 | FALSE | Downregulated |
| -0,193338472 | 0,41378823 | CCDC18 | FALSE | Downregulated |
| 0,744713207 | 0,41378823 | LOC102724601 | FALSE | Upregulated |
| 1,370919478 | 0,41378823 | NEXN-AS1 | FALSE | Upregulated |
| 0,082350439 | 0,413885183 | UBR1 | FALSE | Upregulated |
| -0,096684058 | 0,41399167 | PRKRA | FALSE | Downregulated |
| -1,275021757 | 0,414065951 | FLJ30375 | FALSE | Downregulated |
| 0,933666277 | 0,41413155 | LOC101928245 | FALSE | Upregulated |
| 0,086185365 | 0,414150822 | GPKOW | FALSE | Upregulated |
| 0,259043994 | 0,414150822 | LSM11 | FALSE | Upregulated |
| 0,508103236 | 0,414150822 | RGS16 | FALSE | Upregulated |
| -0,982201936 | 0,414165899 | LOC101927986 | FALSE | Downregulated |
| -0,189737165 | 0,414271916 | ZNF568 | FALSE | Downregulated |
| 0,086389164 | 0,414380087 | DCBLD1 | FALSE | Upregulated |
| 0,124476922 | 0,41440863 | SRRD | FALSE | Upregulated |
| 0,653776293 | 0,41476406 | LRRTM1 | FALSE | Upregulated |
| -0,152795261 | 0,414781366 | MTR | FALSE | Downregulated |

| | | | | |
|---|---|---|---|---|
| -0,247171714 | 0,415016755 | SNHG8 | FALSE | Downregulated |
| 0,675921463 | 0,415220362 | LOC102724908 | FALSE | Upregulated |
| 0,076538743 | 0,415341821 | AP3M1 | FALSE | Upregulated |
| -0,13865137 | 0,415466588 | SLC20A2 | FALSE | Downregulated |
| 0,288451371 | 0,415466588 | SYT12 | FALSE | Upregulated |
| 0,156034136 | 0,415466588 | TANGO2 | FALSE | Upregulated |
| -1,075956337 | 0,415587021 | CCDC162P | FALSE | Downregulated |
| 0,215727155 | 0,415690014 | HSPA1B | FALSE | Upregulated |
| 0,094876995 | 0,415690085 | SLC35A5 | FALSE | Upregulated |
| 0,95989448 | 0,415863876 | AC006145.4 | FALSE | Upregulated |
| 0,668811 | 0,415863876 | DISP2 | FALSE | Upregulated |
| -0,129791454 | 0,415863876 | NLN | FALSE | Downregulated |
| -0,10725343 | 0,415863876 | TIMM9 | FALSE | Downregulated |
| 0,133831463 | 0,415929105 | DOT1L | FALSE | Upregulated |
| 0,483814307 | 0,415929105 | S1PR4 | FALSE | Upregulated |
| 0,554610337 | 0,415934785 | FABP4 | FALSE | Upregulated |
| -0,225227637 | 0,416110475 | CTTNBP2 | FALSE | Downregulated |
| 0,191609384 | 0,416144828 | VPS53 | FALSE | Upregulated |
| -0,109536937 | 0,416179099 | FZD5 | FALSE | Downregulated |
| -0,827951621 | 0,416179099 | KIR2DS4 | FALSE | Downregulated |
| 0,718178103 | 0,416283137 | KCNE2 | FALSE | Upregulated |
| -1,306755004 | 0,416323341 | EPB42 | FALSE | Downregulated |
| -0,880193479 | 0,41660166 | LOC101929451 | FALSE | Downregulated |
| 0,128724261 | 0,416650702 | HIATL1 | FALSE | Upregulated |
| -0,865980228 | 0,416650702 | PROK2 | FALSE | Downregulated |
| 0,209121159 | 0,416858826 | HVCN1 | FALSE | Upregulated |
| -0,069842301 | 0,417096802 | ARFGEF1 | FALSE | Downregulated |
| 0,825279664 | 0,417102467 | BCORP1 | FALSE | Upregulated |
| 0,794461675 | 0,417102467 | RPH3A | FALSE | Upregulated |
| 0,399911431 | 0,417102467 | SNORA40 | FALSE | Upregulated |
| 1,008897027 | 0,4172007 | RIMS2 | FALSE | Upregulated |
| 0,275500255 | 0,417230754 | C19orf40 | FALSE | Upregulated |
| 0,167457205 | 0,417230754 | LYNX1 | FALSE | Upregulated |
| 0,153573676 | 0,417397738 | NUP43 | FALSE | Upregulated |
| 0,238338669 | 0,417535075 | LOC101928154 | FALSE | Upregulated |
| 0,244107982 | 0,417535075 | SPATA24 | FALSE | Upregulated |
| -0,222123811 | 0,417588389 | TMEM56-RWDD3 | FALSE | Downregulated |
| -0,166601701 | 0,417782812 | LIFR | FALSE | Downregulated |
| -0,859633756 | 0,418113099 | PI16 | FALSE | Downregulated |
| 0,234936083 | 0,41816551 | CEBPB | FALSE | Upregulated |
| 1,158888743 | 0,41816551 | SYN2 | FALSE | Upregulated |
| 0,146049439 | 0,418167656 | SKIL | FALSE | Upregulated |
| -0,61487659 | 0,418219733 | LOC400553 | FALSE | Downregulated |
| 0,268602596 | 0,418326916 | SMIM3 | FALSE | Upregulated |
| 0,312574327 | 0,418568519 | CYP4F22 | FALSE | Upregulated |
| 0,13769958 | 0,41861686 | AP1S1 | FALSE | Upregulated |
| -0,366542974 | 0,418640212 | LOC388906 | FALSE | Downregulated |
| 1,230314226 | 0,418640212 | SNORA23 | FALSE | Upregulated |
| 0,903978135 | 0,418695381 | AC007386.2 | FALSE | Upregulated |
| 0,261618448 | 0,418695381 | ARHGEF37 | FALSE | Upregulated |
| 1,788395529 | 0,418695381 | FAM226B | FALSE | Upregulated |

| | | | | |
|---|---|---|---|---|
| 0,21798638 | 0,418695381 | SAMD9 | FALSE | Upregulated |
| -0,079998484 | 0,418703394 | ACAD9 | FALSE | Downregulated |
| -0,110596073 | 0,418703394 | RNF216 | FALSE | Downregulated |
| -0,16115054 | 0,418783817 | RBM33 | FALSE | Downregulated |
| -0,684033174 | 0,418868513 | LOC102724824 | FALSE | Downregulated |
| -0,164426217 | 0,418933021 | GAB2 | FALSE | Downregulated |
| -0,364879291 | 0,418933021 | HCG8 | FALSE | Downregulated |
| 0,148070005 | 0,419012242 | PPIH | FALSE | Upregulated |
| -0,655666511 | 0,419077907 | LOC101927511 | FALSE | Downregulated |
| -0,097309195 | 0,419077907 | NADK2 | FALSE | Downregulated |
| 0,193879197 | 0,419077907 | TMEM44 | FALSE | Upregulated |
| 0,370800638 | 0,419107251 | MEFV | FALSE | Upregulated |
| 0,092197312 | 0,419141396 | CPSF3 | FALSE | Upregulated |
| 0,150293867 | 0,419141396 | FDX1L | FALSE | Upregulated |
| -0,315908759 | 0,419141396 | PPP1R32 | FALSE | Downregulated |
| 0,953770362 | 0,419174265 | C6orf99 | FALSE | Upregulated |
| 0,08684211 | 0,419281731 | DDX20 | FALSE | Upregulated |
| -0,089857473 | 0,419281731 | EAPP | FALSE | Downregulated |
| 0,378886482 | 0,419281731 | GALNT15 | FALSE | Upregulated |
| -0,45903888 | 0,419281731 | TUBA3FP | FALSE | Downregulated |
| 0,925923923 | 0,419503182 | TRNH | FALSE | Upregulated |
| -0,188039503 | 0,419540318 | ALDH18A1 | FALSE | Downregulated |
| 0,127738943 | 0,419627228 | PSKH1 | FALSE | Upregulated |
| 0,765371467 | 0,420272261 | RP1L1 | FALSE | Upregulated |
| 0,694028322 | 0,420404274 | AP000330.8 | FALSE | Upregulated |
| 0,644571733 | 0,420535792 | LOC100128108 | FALSE | Upregulated |
| -0,275116641 | 0,420553974 | TCHH | FALSE | Downregulated |
| 0,123863091 | 0,420691532 | GTF2E1 | FALSE | Upregulated |
| -0,476341418 | 0,420859595 | MKX | FALSE | Downregulated |
| -1,607881841 | 0,420859595 | MYOC | FALSE | Downregulated |
| 0,115639963 | 0,420859595 | RNF34 | FALSE | Upregulated |
| -0,158628903 | 0,420913982 | SLC25A30 | FALSE | Downregulated |
| -0,113288707 | 0,420962637 | CHMP4A | FALSE | Downregulated |
| 0,148986015 | 0,420962637 | MAFG | FALSE | Upregulated |
| -0,106627446 | 0,421400988 | PDCD11 | FALSE | Downregulated |
| -0,168719734 | 0,421912419 | MMACHC | FALSE | Downregulated |
| -0,189986477 | 0,421947409 | TRIM2 | FALSE | Downregulated |
| 0,850872591 | 0,422047557 | RP11-809M12.1 | FALSE | Upregulated |
| 1,372577646 | 0,42322127 | FCRL4 | FALSE | Upregulated |
| 0,564222649 | 0,423224787 | ACPP | FALSE | Upregulated |
| -0,144065894 | 0,423225951 | C11orf95 | FALSE | Downregulated |
| 0,100046329 | 0,423225951 | VPS13D | FALSE | Upregulated |
| 0,297652165 | 0,423335255 | LRRC31 | FALSE | Upregulated |
| -0,151607783 | 0,423404025 | RAB7L1 | FALSE | Downregulated |
| -0,132248545 | 0,423447368 | MAGI3 | FALSE | Downregulated |
| 0,995396501 | 0,423684906 | GGT2 | FALSE | Upregulated |
| -0,188050651 | 0,424061711 | POLR2J3 | FALSE | Downregulated |
| 0,835513394 | 0,424274042 | INTS4L1 | FALSE | Upregulated |
| 0,141468029 | 0,42433789 | ZNF32 | FALSE | Upregulated |
| 0,232123105 | 0,424382524 | IRF7 | FALSE | Upregulated |
| -0,454524471 | 0,4243945 | RP11-53O19.1 | FALSE | Downregulated |

| | | | | |
|---|---|---|---|---|
| 0,107488874 | 0,424404952 | DPY19L4 | FALSE | Upregulated |
| 0,095963534 | 0,424458945 | SAMD4B | FALSE | Upregulated |
| -0,247807729 | 0,424548811 | FAM174B | FALSE | Downregulated |
| -0,148920345 | 0,424573216 | RPL35 | FALSE | Downregulated |
| 0,458755672 | 0,424583997 | SLC1A3 | FALSE | Upregulated |
| 1,305227914 | 0,424999917 | CHRNA2 | FALSE | Upregulated |
| -0,112526002 | 0,424999917 | ZNF823 | FALSE | Downregulated |
| 0,572448744 | 0,425065491 | TMSB4Y | FALSE | Upregulated |
| -0,609726228 | 0,425065491 | YJEFN3 | FALSE | Downregulated |
| 0,780100456 | 0,425074725 | PDE1C | FALSE | Upregulated |
| -0,235854069 | 0,425169727 | ITGAE | FALSE | Downregulated |
| 0,369513687 | 0,425169727 | LINC01004 | FALSE | Upregulated |
| -0,139622821 | 0,425247301 | SLC12A6 | FALSE | Downregulated |
| 0,57684597 | 0,425305596 | LOC102723579 | FALSE | Upregulated |
| -0,388384415 | 0,425337765 | SLC25A27 | FALSE | Downregulated |
| -0,09502213 | 0,425417243 | RCN1 | FALSE | Downregulated |
| -0,17560839 | 0,425417243 | TAPBPL | FALSE | Downregulated |
| 0,141341974 | 0,425494721 | HNRNPAB | FALSE | Upregulated |
| -0,93346736 | 0,425494721 | LOC646513 | FALSE | Downregulated |
| 0,552654812 | 0,425660755 | LOC100287467 | FALSE | Upregulated |
| 0,299502849 | 0,425882162 | GPR160 | FALSE | Upregulated |
| -0,131930791 | 0,425961114 | RILPL2 | FALSE | Downregulated |
| 0,218544543 | 0,426029597 | RBM14-RBM4 | FALSE | Upregulated |
| 0,102320796 | 0,426047651 | TOP1MT | FALSE | Upregulated |
| 0,097185851 | 0,426053882 | TNKS2 | FALSE | Upregulated |
| -0,193736527 | 0,426134614 | SNX16 | FALSE | Downregulated |
| 0,094367927 | 0,426190447 | PARG | FALSE | Upregulated |
| -0,347979086 | 0,426313535 | GPR85 | FALSE | Downregulated |
| 0,696745166 | 0,426530042 | TSPY26P | FALSE | Upregulated |
| -0,150425864 | 0,426594429 | ADH1A | FALSE | Downregulated |
| -0,185506862 | 0,426594429 | SRD5A3 | FALSE | Downregulated |
| 0,792401419 | 0,426594429 | TPSAB1 | FALSE | Upregulated |
| -0,180220781 | 0,426707179 | C3orf18 | FALSE | Downregulated |
| 0,984230367 | 0,42693925 | AL109763.2 | FALSE | Upregulated |
| -0,632265954 | 0,42696347 | FBXW10 | FALSE | Downregulated |
| -0,797053329 | 0,427118801 | NPIPB15 | FALSE | Downregulated |
| 0,146080726 | 0,427263305 | MAN2C1 | FALSE | Upregulated |
| -1,392972529 | 0,427290384 | RYR3 | FALSE | Downregulated |
| 0,374568875 | 0,427334175 | CCDC28B | FALSE | Upregulated |
| 0,830549579 | 0,427334175 | LOC102723696 | FALSE | Upregulated |
| -0,134248734 | 0,427334175 | TMEM135 | FALSE | Downregulated |
| 0,132666036 | 0,427334175 | ZFYVE16 | FALSE | Upregulated |
| 0,356755427 | 0,427452364 | LINC00528 | FALSE | Upregulated |
| 0,278343524 | 0,427452364 | LOC100506713 | FALSE | Upregulated |
| 0,114852941 | 0,427541843 | KIAA1191 | FALSE | Upregulated |
| -0,405756593 | 0,427541843 | RP11-143J12.2 | FALSE | Downregulated |
| 0,747393764 | 0,427556709 | LOC100506497 | FALSE | Upregulated |
| 0,604674457 | 0,427715088 | LOC101928409 | FALSE | Upregulated |
| 0,098318887 | 0,427744089 | STK38 | FALSE | Upregulated |
| -0,18959211 | 0,427744089 | ZNF354B | FALSE | Downregulated |
| 0,881912844 | 0,427833147 | LOC101927116 | FALSE | Upregulated |

| | | | | |
|---|---|---|---|---|
| 0,540384061 | 0,427857305 | FLJ43681 | FALSE | Upregulated |
| -0,626534683 | 0,427857305 | TPSG1 | FALSE | Downregulated |
| 0,242141118 | 0,427914184 | DUSP6 | FALSE | Upregulated |
| 0,598309687 | 0,427914184 | LOC101930275 | FALSE | Upregulated |
| -1,257825861 | 0,427986062 | LOC102724720 | FALSE | Downregulated |
| 0,849313995 | 0,427986062 | TDRD15 | FALSE | Upregulated |
| -0,167346951 | 0,428024682 | SMAD7 | FALSE | Downregulated |
| 1,276466443 | 0,428177403 | LOC102724810 | FALSE | Upregulated |
| -0,176095819 | 0,428177403 | OIT3 | FALSE | Downregulated |
| 0,123637816 | 0,428177403 | RBM12B | FALSE | Upregulated |
| -0,20929547 | 0,428177403 | TMEM8B | FALSE | Downregulated |
| -0,10639251 | 0,428211181 | SDCCAG3 | FALSE | Downregulated |
| 0,398371941 | 0,428413653 | PRRT1 | FALSE | Upregulated |
| -0,151429535 | 0,428446887 | NDUFAF6 | FALSE | Downregulated |
| -0,263085498 | 0,428580282 | RP11-384K6.6 | FALSE | Downregulated |
| 0,401627412 | 0,428659126 | LOC100507054 | FALSE | Upregulated |
| 0,579729085 | 0,428681643 | KLRC2 | FALSE | Upregulated |
| 0,39091876 | 0,428681643 | LONRF2 | FALSE | Upregulated |
| 0,311890038 | 0,428754078 | RIPPLY1 | FALSE | Upregulated |
| -0,083926446 | 0,428839015 | BFAR | FALSE | Downregulated |
| 1,241151468 | 0,428839015 | LOC101928171 | FALSE | Upregulated |
| 0,175025118 | 0,428911369 | HCLS1 | FALSE | Upregulated |
| 0,196296711 | 0,428911369 | TEN1 | FALSE | Upregulated |
| -0,466255246 | 0,42892374 | TREH | FALSE | Downregulated |
| -0,34524431 | 0,42892374 | ZDHHC11 | FALSE | Downregulated |
| 1,123505162 | 0,429080869 | C19orf83 | FALSE | Upregulated |
| -0,219569707 | 0,429094009 | EPOR | FALSE | Downregulated |
| 0,847435679 | 0,429094009 | FAM90A1 | FALSE | Upregulated |
| 0,205951466 | 0,429094009 | NAB2 | FALSE | Upregulated |
| 0,457308831 | 0,429216494 | FUT8-AS1 | FALSE | Upregulated |
| -0,170581415 | 0,429336171 | DMRTA1 | FALSE | Downregulated |
| -0,197116183 | 0,429433434 | FGD4 | FALSE | Downregulated |
| -0,364433753 | 0,429542609 | TOB2P1 | FALSE | Downregulated |
| -0,141213187 | 0,429655172 | AGMO | FALSE | Downregulated |
| -1,172373379 | 0,429655172 | SDC4P | FALSE | Downregulated |
| 0,163696489 | 0,429655172 | STXBP5 | FALSE | Upregulated |
| 0,689489504 | 0,429757483 | SLC52A1 | FALSE | Upregulated |
| 0,54433418 | 0,429830396 | AC072062.1 | FALSE | Upregulated |
| 0,311446588 | 0,430088619 | MYO15B | FALSE | Upregulated |
| 0,175962826 | 0,430196488 | FBXO6 | FALSE | Upregulated |
| 0,113675345 | 0,430196488 | GTF3C3 | FALSE | Upregulated |
| 0,202659046 | 0,430196488 | ID2 | FALSE | Upregulated |
| 0,183913905 | 0,430196488 | RREB1 | FALSE | Upregulated |
| 0,214807022 | 0,43026894 | SLC37A2 | FALSE | Upregulated |
| 0,124211939 | 0,430781473 | STX10 | FALSE | Upregulated |
| 0,137837424 | 0,430899046 | H1F0 | FALSE | Upregulated |
| -0,222716372 | 0,430899046 | PLGLA | FALSE | Downregulated |
| 0,187735941 | 0,430929102 | GMFB | FALSE | Upregulated |
| 0,114143099 | 0,430932749 | PPAPDC2 | FALSE | Upregulated |
| 0,083295437 | 0,431003436 | RBM15B | FALSE | Upregulated |
| 0,408794835 | 0,431003436 | SV2A | FALSE | Upregulated |

| | | | | |
|---|---|---|---|---|
| 0,106858162 | 0,431133242 | RNF2 | FALSE | Upregulated |
| 0,159609206 | 0,43126789 | TPM3P9 | FALSE | Upregulated |
| 1,499954426 | 0,43145121 | RMRP | FALSE | Upregulated |
| 0,121598907 | 0,431458631 | BTN2A1 | FALSE | Upregulated |
| 1,016754783 | 0,431458631 | SOX2 | FALSE | Upregulated |
| -0,342410925 | 0,431640532 | LOC100129534 | FALSE | Downregulated |
| -0,173241634 | 0,431665003 | IRF5 | FALSE | Downregulated |
| 0,282351634 | 0,431706398 | C19orf55 | FALSE | Upregulated |
| -0,199781498 | 0,431706398 | SEMA6D | FALSE | Downregulated |
| 0,111652177 | 0,431722575 | TPCN1 | FALSE | Upregulated |
| 0,169686435 | 0,431843714 | KALRN | FALSE | Upregulated |
| -0,368313411 | 0,431843714 | LOC102724098 | FALSE | Downregulated |
| 0,202374576 | 0,432107151 | FDXACB1 | FALSE | Upregulated |
| 0,120718615 | 0,432290051 | ARL6IP5 | FALSE | Upregulated |
| -0,979156781 | 0,432390795 | PLN | FALSE | Downregulated |
| 0,326045763 | 0,432427201 | ADAM19 | FALSE | Upregulated |
| 0,567844043 | 0,432427201 | LOC102724026 | FALSE | Upregulated |
| 0,179806272 | 0,432427201 | MTHFSD | FALSE | Upregulated |
| -0,092769735 | 0,432427201 | SIN3A | FALSE | Downregulated |
| 2,385103713 | 0,432427201 | TRNAH5 | FALSE | Upregulated |
| 0,1145156 | 0,432427201 | ZKSCAN2 | FALSE | Upregulated |
| 0,561754767 | 0,43246636 | CEACAM3 | FALSE | Upregulated |
| -0,09729843 | 0,432482919 | CETN3 | FALSE | Downregulated |
| 0,125412014 | 0,432482919 | CPM | FALSE | Upregulated |
| 0,130634699 | 0,432482919 | ZNF17 | FALSE | Upregulated |
| 0,196862132 | 0,432615763 | PUSL1 | FALSE | Upregulated |
| -0,42391435 | 0,432825553 | LOC101928068 | FALSE | Downregulated |
| 0,465087511 | 0,432825553 | SLC9A7P1 | FALSE | Upregulated |
| 0,51857881 | 0,432984044 | HIF1A-AS2 | FALSE | Upregulated |
| 0,527860233 | 0,433065441 | TFAP2A | FALSE | Upregulated |
| 0,193298221 | 0,433353947 | UBE3D | FALSE | Upregulated |
| -0,166538215 | 0,433575918 | PAN2 | FALSE | Downregulated |
| 0,598457844 | 0,433657568 | SLC38A5 | FALSE | Upregulated |
| 0,105483118 | 0,433699743 | ARID2 | FALSE | Upregulated |
| 0,616657139 | 0,433699743 | ESPL1 | FALSE | Upregulated |
| 0,745609225 | 0,433699743 | LOC101928555 | FALSE | Upregulated |
| -0,089689999 | 0,433699743 | PRPS1 | FALSE | Downregulated |
| -0,486969166 | 0,434005523 | LOC101926907 | FALSE | Downregulated |
| -0,120061259 | 0,434213902 | SLC2A2 | FALSE | Downregulated |
| -0,170420101 | 0,43422171 | CNPY3 | FALSE | Downregulated |
| -0,274731725 | 0,434242674 | NUP62CL | FALSE | Downregulated |
| -0,10035513 | 0,434334338 | SRP68 | FALSE | Downregulated |
| 1,426235721 | 0,43440109 | MT1IP | FALSE | Upregulated |
| 0,601892937 | 0,434409741 | LOC101927789 | FALSE | Upregulated |
| -1,047731128 | 0,434409741 | LOC102724400 | FALSE | Downregulated |
| -0,30435006 | 0,434409741 | LOC728752 | FALSE | Downregulated |
| -1,200057104 | 0,434419101 | FLJ20444 | FALSE | Downregulated |
| 0,662876931 | 0,434423195 | AP4B1-AS1 | FALSE | Upregulated |
| 0,737639266 | 0,434442986 | LOC101928621 | FALSE | Upregulated |
| -0,145426112 | 0,434563656 | ERO1LB | FALSE | Downregulated |
| -0,104718342 | 0,43456691 | PCNXL4 | FALSE | Downregulated |

| | | | | |
|---|---|---|---|---|
| 0,106545231 | 0,434765919 | PHF20 | FALSE | Upregulated |
| 0,111002641 | 0,434831927 | AKTIP | FALSE | Upregulated |
| -0,296870944 | 0,434831927 | FGFBP3 | FALSE | Downregulated |
| 0,203075216 | 0,434968705 | RGL3 | FALSE | Upregulated |
| -0,599448685 | 0,435027366 | RP11-63P12.6 | FALSE | Downregulated |
| 0,502837842 | 0,435084063 | NACAP1 | FALSE | Upregulated |
| 0,447153906 | 0,435087302 | AC011747.4 | FALSE | Upregulated |
| -0,095697385 | 0,435173475 | GRPEL1 | FALSE | Downregulated |
| 0,199423144 | 0,435173475 | VCPKMT | FALSE | Upregulated |
| -0,283778042 | 0,435173855 | GOLGA2P5 | FALSE | Downregulated |
| -0,106356944 | 0,435234035 | CRAT | FALSE | Downregulated |
| -0,265817331 | 0,43542558 | ABCC11 | FALSE | Downregulated |
| -0,721167704 | 0,43542558 | CACNB1 | FALSE | Downregulated |
| 0,090336523 | 0,435506071 | RNF169 | FALSE | Upregulated |
| 0,316129564 | 0,43590407 | A2M-AS1 | FALSE | Upregulated |
| -0,133924717 | 0,436198017 | GLRX5 | FALSE | Downregulated |
| -0,274538043 | 0,436198017 | HAL | FALSE | Downregulated |
| 0,943735182 | 0,436254581 | RP11-538P18.2 | FALSE | Upregulated |
| 0,083655398 | 0,436361831 | GINM1 | FALSE | Upregulated |
| -0,404181241 | 0,436457441 | DLG5-AS1 | FALSE | Downregulated |
| 0,145556276 | 0,436542183 | PIGA | FALSE | Upregulated |
| -0,325720623 | 0,436542183 | SLC16A9 | FALSE | Downregulated |
| 0,160823128 | 0,436542183 | SLC26A1 | FALSE | Upregulated |
| 0,59003129 | 0,436609839 | FAM71F2 | FALSE | Upregulated |
| -0,119988943 | 0,436678802 | CEP104 | FALSE | Downregulated |
| 1,042578707 | 0,436688818 | C10orf95 | FALSE | Upregulated |
| 0,115262606 | 0,436688818 | FAM178A | FALSE | Upregulated |
| 0,409072942 | 0,436688818 | RGAG1 | FALSE | Upregulated |
| 0,220891315 | 0,436779458 | ZNF841 | FALSE | Upregulated |
| 1,327757947 | 0,43688596 | GRIA1 | FALSE | Upregulated |
| -0,100502858 | 0,437056649 | DYNLRB1 | FALSE | Downregulated |
| 1,059200041 | 0,437056649 | LINC00664 | FALSE | Upregulated |
| -0,100368947 | 0,437213885 | ECHDC1 | FALSE | Downregulated |
| 0,323415661 | 0,437266577 | ASRGL1 | FALSE | Upregulated |
| 0,53910612 | 0,437403876 | FAM223B | FALSE | Upregulated |
| 0,995844871 | 0,437438384 | ZNF488 | FALSE | Upregulated |
| 0,14986986 | 0,437440311 | SEPT6 | FALSE | Upregulated |
| -0,114333561 | 0,437565199 | EEF1G | FALSE | Downregulated |
| -0,612503108 | 0,437565199 | LOC102723331 | FALSE | Downregulated |
| -0,108289809 | 0,437770628 | SIRT2 | FALSE | Downregulated |
| 0,508417203 | 0,43781328 | FREM2 | FALSE | Upregulated |
| 0,113143347 | 0,437926539 | JARID2 | FALSE | Upregulated |
| 0,294745922 | 0,437926539 | ZNF85 | FALSE | Upregulated |
| -0,181125996 | 0,438050243 | PARP6 | FALSE | Downregulated |
| -0,124374381 | 0,438070566 | SWI5 | FALSE | Downregulated |
| 0,772114919 | 0,438076497 | RP11-219B17.1 | FALSE | Upregulated |
| -0,110624289 | 0,438181544 | WBSCR16 | FALSE | Downregulated |
| 0,236412325 | 0,438284402 | GPR157 | FALSE | Upregulated |
| -0,109725562 | 0,438308648 | ZNF606 | FALSE | Downregulated |
| -0,069711479 | 0,438417691 | IMMT | FALSE | Downregulated |
| 0,955405024 | 0,438417691 | SLCO5A1 | FALSE | Upregulated |

| | | | | |
|---|---|---|---|---|
| 0,436322748 | 0,438454975 | SRRM2-AS1 | FALSE | Upregulated |
| 0,148614762 | 0,438495811 | ABHD14A | FALSE | Upregulated |
| -0,119561143 | 0,438495811 | LTBR | FALSE | Downregulated |
| 0,100477305 | 0,438495811 | TRIM4 | FALSE | Upregulated |
| -0,08448364 | 0,438524795 | TRIM39 | FALSE | Downregulated |
| 0,113543575 | 0,438639729 | ATP2B1 | FALSE | Upregulated |
| -0,727082934 | 0,438639729 | MEG9 | FALSE | Downregulated |
| -0,089359733 | 0,438794926 | EXOSC3 | FALSE | Downregulated |
| 0,25010619 | 0,439130041 | ATP8A1 | FALSE | Upregulated |
| 0,34585909 | 0,439156177 | LILRA6 | FALSE | Upregulated |
| 0,083424603 | 0,439205935 | RNF41 | FALSE | Upregulated |
| -0,111563166 | 0,439292447 | ANAPC13 | FALSE | Downregulated |
| -0,183888461 | 0,439357357 | CA2 | FALSE | Downregulated |
| 0,827904931 | 0,439357357 | LOC101929125 | FALSE | Upregulated |
| 0,385323773 | 0,439474979 | CBWD6 | FALSE | Upregulated |
| -0,698039489 | 0,439621853 | LOC102723988 | FALSE | Downregulated |
| 1,291295185 | 0,439621853 | LOC102724348 | FALSE | Upregulated |
| 0,626747812 | 0,439621853 | RP11-276H19.1 | FALSE | Upregulated |
| -0,518656371 | 0,439804514 | JMJD1C-AS1 | FALSE | Downregulated |
| -1,959287946 | 0,440058536 | MYF6 | FALSE | Downregulated |
| 1,164066017 | 0,440062233 | TINCR | FALSE | Upregulated |
| -0,088850443 | 0,440137562 | BTF3 | FALSE | Downregulated |
| 0,196007688 | 0,440137562 | MSTO1 | FALSE | Upregulated |
| 0,809622406 | 0,440161165 | LOC100653133 | FALSE | Upregulated |
| 0,12574076 | 0,440292157 | ASGR1 | FALSE | Upregulated |
| -0,67691393 | 0,440325666 | PADI4 | FALSE | Downregulated |
| 0,359931602 | 0,440484014 | MTCP1 | FALSE | Upregulated |
| -0,088418077 | 0,440532257 | UBR2 | FALSE | Downregulated |
| 0,273412131 | 0,441084121 | BEND3 | FALSE | Upregulated |
| -0,15439078 | 0,441398488 | GPLD1 | FALSE | Downregulated |
| -0,718747031 | 0,441428264 | LINC01016 | FALSE | Downregulated |
| -0,386824073 | 0,441611479 | CREB3L3 | FALSE | Downregulated |
| 0,065228895 | 0,441740349 | INO80 | FALSE | Upregulated |
| 1,305257645 | 0,441740349 | KCNJ1 | FALSE | Upregulated |
| -0,559724126 | 0,441740349 | PRSS50 | FALSE | Downregulated |
| 0,083538813 | 0,441740349 | SEC23IP | FALSE | Upregulated |
| -1,09508155 | 0,441902688 | LOC102723694 | FALSE | Downregulated |
| -0,28872431 | 0,44208659 | STARD4 | FALSE | Downregulated |
| -0,195317062 | 0,442096995 | LRR1 | FALSE | Downregulated |
| -0,359545992 | 0,442172151 | SMAD5-AS1 | FALSE | Downregulated |
| -0,268258353 | 0,442365784 | AC009299.3 | FALSE | Downregulated |
| 0,251531123 | 0,442431412 | SRGAP3 | FALSE | Upregulated |
| 0,154818405 | 0,44248389 | HINFP | FALSE | Upregulated |
| -0,38476708 | 0,442642566 | AGAP5 | FALSE | Downregulated |
| -0,453282349 | 0,442642566 | BCYRN1 | FALSE | Downregulated |
| -0,124233819 | 0,442648191 | LMAN2L | FALSE | Downregulated |
| -0,820518957 | 0,442777562 | APOD | FALSE | Downregulated |
| 0,749751805 | 0,442821085 | LOC101928911 | FALSE | Upregulated |
| -0,210403993 | 0,442894687 | CYP4F12 | FALSE | Downregulated |
| -0,376265403 | 0,442894687 | RDH12 | FALSE | Downregulated |
| 0,113032639 | 0,442910686 | BIN3 | FALSE | Upregulated |

| | | | | |
|---|---|---|---|---|
| 1,23895598 | 0,442910686 | CPA1 | FALSE | Upregulated |
| 0,993451065 | 0,442910686 | RP11-379F4.4 | FALSE | Upregulated |
| -0,170306915 | 0,443033034 | FNBP4 | FALSE | Downregulated |
| -0,147867755 | 0,443095884 | MUTYH | FALSE | Downregulated |
| 0,554132612 | 0,443162611 | LOC100505501 | FALSE | Upregulated |
| -0,454699323 | 0,443253235 | C19orf71 | FALSE | Downregulated |
| -0,106771451 | 0,443253235 | PAGR1 | FALSE | Downregulated |
| 0,81304201 | 0,443516253 | TUBBP5 | FALSE | Upregulated |
| 0,671174564 | 0,443628653 | NPAP1 | FALSE | Upregulated |
| 0,099790058 | 0,443628653 | PSMD2 | FALSE | Upregulated |
| -0,096305773 | 0,443647483 | KANK1 | FALSE | Downregulated |
| 0,263558693 | 0,443832429 | ARHGAP6 | FALSE | Upregulated |
| -0,168515608 | 0,443975296 | C9orf3 | FALSE | Downregulated |
| 0,115638376 | 0,444101847 | MGAT4B | FALSE | Upregulated |
| 0,8354946 | 0,444188153 | LINC00271 | FALSE | Upregulated |
| 0,198285172 | 0,444403228 | FAM86DP | FALSE | Upregulated |
| 0,458862888 | 0,444403228 | P2RY11 | FALSE | Upregulated |
| -0,176186027 | 0,444492689 | RP11-85F14.5 | FALSE | Downregulated |
| -0,102209799 | 0,444771498 | DDX52 | FALSE | Downregulated |
| -0,264605605 | 0,444771498 | LOC101928076 | FALSE | Downregulated |
| -0,185848141 | 0,444771498 | RPS3A | FALSE | Downregulated |
| 0,118658721 | 0,444779162 | POLR2E | FALSE | Upregulated |
| -0,151864617 | 0,444863058 | BLNK | FALSE | Downregulated |
| -0,347402546 | 0,445031982 | AMHR2 | FALSE | Downregulated |
| 0,083662395 | 0,445031982 | KIDINS220 | FALSE | Upregulated |
| -0,156864438 | 0,445187759 | CASP9 | FALSE | Downregulated |
| 0,775217494 | 0,445299416 | LOC102723750 | FALSE | Upregulated |
| 0,18241278 | 0,445299416 | RFX3 | FALSE | Upregulated |
| 0,102765182 | 0,445348896 | EMD | FALSE | Upregulated |
| 0,102099303 | 0,445348896 | TMEM123 | FALSE | Upregulated |
| 0,177459762 | 0,445459886 | ADAP2 | FALSE | Upregulated |
| 0,951478562 | 0,445506409 | GGT8P | FALSE | Upregulated |
| 0,285795759 | 0,445506409 | SULF1 | FALSE | Upregulated |
| 0,872573625 | 0,44551958 | GRM7 | FALSE | Upregulated |
| 0,224263103 | 0,445607188 | TMEM79 | FALSE | Upregulated |
| -0,410608198 | 0,445702676 | LOC101929612 | FALSE | Downregulated |
| 0,203278419 | 0,445746821 | GRAMD4 | FALSE | Upregulated |
| 0,647934527 | 0,445918078 | TGM3 | FALSE | Upregulated |
| -0,137920124 | 0,445949922 | TOR1AIP2 | FALSE | Downregulated |
| 0,056876208 | 0,446153677 | SNX9 | FALSE | Upregulated |
| 0,80790784 | 0,446158967 | NDUFAF4P1 | FALSE | Upregulated |
| 0,126229959 | 0,446326719 | GON4L | FALSE | Upregulated |
| -0,98186788 | 0,446326719 | LINC00311 | FALSE | Downregulated |
| -0,256981584 | 0,446580983 | SDHAP1 | FALSE | Downregulated |
| 0,895381946 | 0,446764228 | DNAH8 | FALSE | Upregulated |
| -0,41516836 | 0,446885065 | FLJ37201 | FALSE | Downregulated |
| 0,404458288 | 0,446941749 | RAB39A | FALSE | Upregulated |
| -0,091243618 | 0,44697198 | EGLN1 | FALSE | Downregulated |
| 0,124263505 | 0,446983316 | TBL2 | FALSE | Upregulated |
| -0,878361912 | 0,447119616 | TRNAG20 | FALSE | Downregulated |
| 0,517163697 | 0,447217395 | EGR1 | FALSE | Upregulated |

| | | | | |
|---|---|---|---|---|
| 0,60666377 | 0,447358671 | ACTG1P4 | FALSE | Upregulated |
| -0,131271455 | 0,447584446 | ESF1 | FALSE | Downregulated |
| -0,176355826 | 0,447627596 | STX1B | FALSE | Downregulated |
| -0,330907697 | 0,447684672 | LOC100128288 | FALSE | Downregulated |
| 0,139462672 | 0,447741093 | LGALSL | FALSE | Upregulated |
| 0,091738198 | 0,447747243 | CLASP2 | FALSE | Upregulated |
| 0,113665236 | 0,447769098 | ATXN2L | FALSE | Upregulated |
| -0,307340776 | 0,447769098 | SH3BP5-AS1 | FALSE | Downregulated |
| 0,855960596 | 0,447769098 | SMCO2 | FALSE | Upregulated |
| -0,082648036 | 0,447769098 | SNX14 | FALSE | Downregulated |
| -0,187380331 | 0,447769098 | TREX1 | FALSE | Downregulated |
| -0,320800101 | 0,447822597 | TTLL7 | FALSE | Downregulated |
| 2,736789864 | 0,447921462 | C15orf38-AP3S2 | FALSE | Upregulated |
| -0,196651596 | 0,447921462 | NINL | FALSE | Downregulated |
| -0,298013137 | 0,447921462 | NPHP3-ACAD11 | FALSE | Downregulated |
| 0,181236749 | 0,447930571 | DMTF1 | FALSE | Upregulated |
| 0,77143815 | 0,447943604 | GJC2 | FALSE | Upregulated |
| -0,212140801 | 0,447966367 | ZNF814 | FALSE | Downregulated |
| 0,317211697 | 0,448259174 | CLUHP3 | FALSE | Upregulated |
| 0,617940381 | 0,448259174 | RP11-81H14.2 | FALSE | Upregulated |
| 1,011884171 | 0,448326029 | CDH17 | FALSE | Upregulated |
| 0,11525421 | 0,448326029 | PYGL | FALSE | Upregulated |
| -0,22126764 | 0,448357785 | HIST1H1C | FALSE | Downregulated |
| 0,408641083 | 0,448357785 | ST7-AS1 | FALSE | Upregulated |
| 0,141436589 | 0,448364256 | BBS7 | FALSE | Upregulated |
| 1,913623975 | 0,448676049 | CKMT1B | FALSE | Upregulated |
| -0,145454713 | 0,448904976 | LYSMD1 | FALSE | Downregulated |
| 1,166924389 | 0,448923782 | LOC494127 | FALSE | Upregulated |
| -1,141861279 | 0,449142871 | SCHIP1 | FALSE | Downregulated |
| -1,217016594 | 0,449400627 | C21orf128 | FALSE | Downregulated |
| -0,229432352 | 0,449417835 | LINC00087 | FALSE | Downregulated |
| 1,723833004 | 0,449417835 | LOC100289026 | FALSE | Upregulated |
| -0,099253417 | 0,449417835 | NDUFB5 | FALSE | Downregulated |
| 0,221527189 | 0,449417835 | WBP1 | FALSE | Upregulated |
| 0,408727645 | 0,449468224 | PPP1R3G | FALSE | Upregulated |
| -0,175716693 | 0,449665784 | LOC100506302 | FALSE | Downregulated |
| 0,176105743 | 0,449820639 | ZNF600 | FALSE | Upregulated |
| 0,086421878 | 0,449843913 | STAT5B | FALSE | Upregulated |
| -0,742619387 | 0,449945983 | SLC39A2 | FALSE | Downregulated |
| 0,847780213 | 0,450004608 | SCNN1B | FALSE | Upregulated |
| -0,120769238 | 0,45019241 | MIEN1 | FALSE | Downregulated |
| -0,440011429 | 0,450501044 | LOC102723467 | FALSE | Downregulated |
| 0,140958618 | 0,450526007 | TSTA3 | FALSE | Upregulated |
| 0,351193549 | 0,450788309 | SLC12A8 | FALSE | Upregulated |
| 1,061595343 | 0,450887593 | FAM92A1P2 | FALSE | Upregulated |
| -0,97264101 | 0,450943795 | LOC100506253 | FALSE | Downregulated |
| 0,997862281 | 0,450949707 | LOC100289333 | FALSE | Upregulated |
| -0,133612681 | 0,451143204 | SPAG9 | FALSE | Downregulated |
| 0,15481906 | 0,451200941 | JAK2 | FALSE | Upregulated |
| -0,366016572 | 0,451200941 | SLC2A4 | FALSE | Downregulated |
| -0,773303141 | 0,451638828 | C12orf39 | FALSE | Downregulated |

| | | | | |
|---|---|---|---|---|
| 0,178781388 | 0,451638828 | EFCAB11 | FALSE | Upregulated |
| 0,330531356 | 0,451638828 | GATA5 | FALSE | Upregulated |
| -0,356979774 | 0,451638828 | LOC100996763 | FALSE | Downregulated |
| 1,127083609 | 0,451660055 | MFSD6L | FALSE | Upregulated |
| -0,178289162 | 0,451690707 | ZNF417 | FALSE | Downregulated |
| 1,20477563 | 0,451729334 | GPR37L1 | FALSE | Upregulated |
| -0,13514296 | 0,451729334 | IVD | FALSE | Downregulated |
| 0,701330734 | 0,451729334 | KLHL14 | FALSE | Upregulated |
| -0,719213929 | 0,451729334 | LOC100499484-C9ORF174 | FALSE | Downregulated |
| 0,201682108 | 0,451729334 | ST6GAL2 | FALSE | Upregulated |
| 0,86632877 | 0,451729334 | TBL1Y | FALSE | Upregulated |
| 1,01174935 | 0,451755498 | ADAD2 | FALSE | Upregulated |
| 0,130213409 | 0,451795105 | PNO1 | FALSE | Upregulated |
| -0,364687842 | 0,451840395 | SYNPO2 | FALSE | Downregulated |
| 0,425916599 | 0,451850764 | COLCA1 | FALSE | Upregulated |
| -0,623912994 | 0,451850764 | POU6F2-AS1 | FALSE | Downregulated |
| -0,1357657 | 0,451850764 | TRMT2A | FALSE | Downregulated |
| 0,093708707 | 0,452045706 | AARS2 | FALSE | Upregulated |
| 0,317977506 | 0,452045706 | C16orf96 | FALSE | Upregulated |
| 0,191280945 | 0,452075965 | HPS3 | FALSE | Upregulated |
| -0,181685005 | 0,452075965 | ZNF385B | FALSE | Downregulated |
| -0,732429919 | 0,452092643 | LINC00525 | FALSE | Downregulated |
| 0,158145495 | 0,452092643 | TXNIP | FALSE | Upregulated |
| -0,1208455 | 0,452137244 | CEP350 | FALSE | Downregulated |
| -0,081977694 | 0,452137244 | SUV420H1 | FALSE | Downregulated |
| -0,156596392 | 0,452137244 | ZNF485 | FALSE | Downregulated |
| -0,612829651 | 0,452193301 | ESPNP | FALSE | Downregulated |
| 0,437630459 | 0,452202119 | ARPP21 | FALSE | Upregulated |
| -0,061501867 | 0,452202119 | FBXO11 | FALSE | Downregulated |
| -0,087417221 | 0,452202119 | SLC35B3 | FALSE | Downregulated |
| 0,118094959 | 0,452202119 | SMG6 | FALSE | Upregulated |
| -0,154341792 | 0,452202119 | UBXN7 | FALSE | Downregulated |
| 0,768115803 | 0,45226366 | LOC101928692 | FALSE | Upregulated |
| -0,538500213 | 0,452463312 | GPM6B | FALSE | Downregulated |
| 0,092670519 | 0,452526706 | SLC35E3 | FALSE | Upregulated |
| 0,164560852 | 0,452608453 | NCDN | FALSE | Upregulated |
| -0,127836956 | 0,452673601 | MED18 | FALSE | Downregulated |
| 0,120677018 | 0,452673957 | SEC22B | FALSE | Upregulated |
| -0,123944881 | 0,452715767 | ATG2B | FALSE | Downregulated |
| 0,106509836 | 0,452981562 | GTF2H3 | FALSE | Upregulated |
| 0,142538044 | 0,452981562 | MGAT4A | FALSE | Upregulated |
| -0,078268598 | 0,453017485 | ZHX2 | FALSE | Downregulated |
| 0,411139317 | 0,45305064 | RAB42 | FALSE | Upregulated |
| 0,244526767 | 0,453164434 | LOC102724064 | FALSE | Upregulated |
| -0,078848412 | 0,45324622 | BTF3L4 | FALSE | Downregulated |
| 0,196819978 | 0,453295845 | CDKN1C | FALSE | Upregulated |
| 0,277011246 | 0,453349699 | OAS3 | FALSE | Upregulated |
| -0,110756876 | 0,453349699 | TBC1D8 | FALSE | Downregulated |
| -0,181506192 | 0,453404299 | MID1IP1 | FALSE | Downregulated |
| 1,171977932 | 0,453559437 | CACNG3 | FALSE | Upregulated |

| | | | | |
|---|---|---|---|---|
| -0,570619766 | 0,453559437 | SIM1 | FALSE | Downregulated |
| -0,158216382 | 0,453559437 | TNK1 | FALSE | Downregulated |
| 0,103332554 | 0,453686056 | SNX7 | FALSE | Upregulated |
| 0,157515405 | 0,453835679 | FAM69A | FALSE | Upregulated |
| -1,043826612 | 0,45392113 | SNORD54 | FALSE | Downregulated |
| -0,226395222 | 0,453949235 | CSAD | FALSE | Downregulated |
| 0,856504428 | 0,454080169 | C3orf83 | FALSE | Upregulated |
| -0,476461642 | 0,454080169 | LOC101929977 | FALSE | Downregulated |
| 0,639009132 | 0,454080169 | TICRR | FALSE | Upregulated |
| 0,659823603 | 0,454080169 | ZNF99 | FALSE | Upregulated |
| 0,204499301 | 0,454143365 | GIPC3 | FALSE | Upregulated |
| -0,486858882 | 0,454143365 | GPR153 | FALSE | Downregulated |
| -0,183460725 | 0,454143365 | LY6E | FALSE | Downregulated |
| -0,091306315 | 0,454143365 | NUBP1 | FALSE | Downregulated |
| 0,157272905 | 0,454277589 | FAT4 | FALSE | Upregulated |
| -0,099578268 | 0,454378402 | TECPR2 | FALSE | Downregulated |
| 0,120476986 | 0,454443231 | NDC1 | FALSE | Upregulated |
| -0,081076452 | 0,454485082 | PIGK | FALSE | Downregulated |
| -0,690561205 | 0,454585856 | ANKRD18DP | FALSE | Downregulated |
| 0,246355168 | 0,454726331 | FAM228B | FALSE | Upregulated |
| -0,368371432 | 0,455028332 | LOC101928513 | FALSE | Downregulated |
| 0,157656614 | 0,455108933 | PAM | FALSE | Upregulated |
| 0,695814123 | 0,455208494 | AXDND1 | FALSE | Upregulated |
| 0,850743794 | 0,455350037 | CACNA1C-AS1 | FALSE | Upregulated |
| 0,541540104 | 0,455364372 | LOC101059949 | FALSE | Upregulated |
| 0,465512317 | 0,455491832 | MAP7D2 | FALSE | Upregulated |
| 0,947575518 | 0,455613494 | LOC102724614 | FALSE | Upregulated |
| 0,109505472 | 0,455613494 | MRPL47 | FALSE | Upregulated |
| -0,118780258 | 0,455613494 | RNF43 | FALSE | Downregulated |
| -0,226084621 | 0,455751341 | HCN3 | FALSE | Downregulated |
| -0,367957937 | 0,455751341 | ST3GAL6-AS1 | FALSE | Downregulated |
| -0,465829722 | 0,455761396 | PLXNA4 | FALSE | Downregulated |
| 0,413011723 | 0,455761396 | RSPH1 | FALSE | Upregulated |
| 0,11505536 | 0,455774434 | PLA2G15 | FALSE | Upregulated |
| 0,641029046 | 0,456059604 | LOC102724016 | FALSE | Upregulated |
| 0,147505728 | 0,45606489 | LINS | FALSE | Upregulated |
| 0,053726088 | 0,456073842 | MORF4L1 | FALSE | Upregulated |
| 0,099771777 | 0,456309281 | ZZEF1 | FALSE | Upregulated |
| -0,292377566 | 0,456391223 | MIF | FALSE | Downregulated |
| 0,53108549 | 0,456391223 | ULBP1 | FALSE | Upregulated |
| -0,31291839 | 0,456471187 | AC011747.7 | FALSE | Downregulated |
| 0,138792784 | 0,456471187 | CAV2 | FALSE | Upregulated |
| 0,112245348 | 0,456694003 | ABT1 | FALSE | Upregulated |
| -0,237462722 | 0,456996614 | CYB5R2 | FALSE | Downregulated |
| 0,179861618 | 0,457009668 | GDF11 | FALSE | Upregulated |
| -0,174013516 | 0,457009668 | RSBN1L | FALSE | Downregulated |
| -0,414429573 | 0,457207713 | PLA2G4B | FALSE | Downregulated |
| -0,225818191 | 0,457207713 | RHOD | FALSE | Downregulated |
| 0,216051404 | 0,457336816 | C1QTNF5 | FALSE | Upregulated |
| -0,11992558 | 0,45739874 | TBC1D8B | FALSE | Downregulated |
| 1,141144515 | 0,45759197 | AC104654.2 | FALSE | Upregulated |

| | | | | |
|---|---|---|---|---|
| -0,273256572 | 0,457627044 | NAMPT | FALSE | Downregulated |
| 0,99009721 | 0,457652854 | RP11-973H7.4 | FALSE | Upregulated |
| -0,096230696 | 0,457733901 | DMXL1 | FALSE | Downregulated |
| -0,449329684 | 0,457733901 | LOC101927894 | FALSE | Downregulated |
| -0,125498757 | 0,457752967 | HNF4A | FALSE | Downregulated |
| 1,375214174 | 0,457752967 | LANCL3 | FALSE | Upregulated |
| 0,245110514 | 0,457752967 | MORN4 | FALSE | Upregulated |
| 0,122504316 | 0,457768195 | CCDC115 | FALSE | Upregulated |
| 1,153721964 | 0,457768195 | MUC2 | FALSE | Upregulated |
| 0,122410916 | 0,457768195 | RRP1 | FALSE | Upregulated |
| 0,084449969 | 0,457822567 | MFAP3 | FALSE | Upregulated |
| 0,517699412 | 0,457881875 | CD300LB | FALSE | Upregulated |
| 0,105116538 | 0,457881875 | KIAA2026 | FALSE | Upregulated |
| -0,62981434 | 0,457890306 | TEKT5 | FALSE | Downregulated |
| -0,103214298 | 0,457914428 | PCM1 | FALSE | Downregulated |
| 0,194439246 | 0,458207899 | MYL9 | FALSE | Upregulated |
| -0,156456098 | 0,458212141 | RPL41 | FALSE | Downregulated |
| 0,267119946 | 0,458339886 | PLEKHH1 | FALSE | Upregulated |
| -0,349882867 | 0,45862239 | GADD45B | FALSE | Downregulated |
| -0,081428455 | 0,458726666 | MRPL38 | FALSE | Downregulated |
| -0,860298295 | 0,459126096 | ASCL1 | FALSE | Downregulated |
| -0,138775975 | 0,459207716 | SS18L2 | FALSE | Downregulated |
| 0,136351403 | 0,459417588 | ZMIZ2 | FALSE | Upregulated |
| 1,218109754 | 0,459541742 | IGBP1P1 | FALSE | Upregulated |
| 1,079068499 | 0,459541742 | LOC100505498 | FALSE | Upregulated |
| 0,127771364 | 0,459541742 | PCYT2 | FALSE | Upregulated |
| 0,982278949 | 0,459669478 | LOC102723867 | FALSE | Upregulated |
| -0,109424163 | 0,459929951 | RICTOR | FALSE | Downregulated |
| 0,471614957 | 0,459929951 | RP11-466F5.8 | FALSE | Upregulated |
| 2,209686951 | 0,459977304 | GABRA5 | FALSE | Upregulated |
| -0,687479696 | 0,459977304 | TRNS2 | FALSE | Downregulated |
| 0,124477482 | 0,460185651 | CPSF1 | FALSE | Upregulated |
| 0,178530943 | 0,460200248 | PRICKLE2 | FALSE | Upregulated |
| 1,09079273 | 0,460593162 | MAGED4B | FALSE | Upregulated |
| 0,426968821 | 0,46060266 | AC093391.2 | FALSE | Upregulated |
| -0,169061705 | 0,460676118 | AKAP17A | FALSE | Downregulated |
| 0,586917234 | 0,460676118 | LOC100505767 | FALSE | Upregulated |
| -0,483313251 | 0,460676118 | TUBB4A | FALSE | Downregulated |
| 0,270699411 | 0,460678204 | NR4A2 | FALSE | Upregulated |
| 0,198909259 | 0,460796617 | ARHGAP24 | FALSE | Upregulated |
| 0,076318523 | 0,460852338 | NPTN | FALSE | Upregulated |
| 0,110952672 | 0,461140368 | IPO11 | FALSE | Upregulated |
| 0,083600894 | 0,461175821 | CGGBP1 | FALSE | Upregulated |
| 0,839023534 | 0,461247372 | WDR63 | FALSE | Upregulated |
| -0,098387044 | 0,461346498 | CHCHD4 | FALSE | Downregulated |
| -0,182626525 | 0,461426406 | SFXN2 | FALSE | Downregulated |
| -0,35929584 | 0,461541649 | NS3BP | FALSE | Downregulated |
| -0,132440601 | 0,461584597 | ANKS1A | FALSE | Downregulated |
| -0,462709814 | 0,461584597 | C2orf27A | FALSE | Downregulated |
| -0,300166259 | 0,461584597 | SLC24A3 | FALSE | Downregulated |
| -0,143712043 | 0,461702077 | IQSEC1 | FALSE | Downregulated |

| | | | | |
|---|---|---|---|---|
| -0,300324397 | 0,461702077 | ZFP37 | FALSE | Downregulated |
| 0,142000332 | 0,461726976 | SLC35B1 | FALSE | Upregulated |
| -0,21153839 | 0,461740431 | SFRP1 | FALSE | Downregulated |
| -0,160663707 | 0,461865406 | LOC100505761 | FALSE | Downregulated |
| 0,243216898 | 0,461948233 | DOK3 | FALSE | Upregulated |
| -0,158292944 | 0,461948233 | KDM6A | FALSE | Downregulated |
| -0,084762753 | 0,461966517 | PIN1 | FALSE | Downregulated |
| -0,891166949 | 0,462000792 | LOC100996549 | FALSE | Downregulated |
| -0,73784079 | 0,462394992 | LOC102724209 | FALSE | Downregulated |
| -0,115613956 | 0,462405028 | C16orf80 | FALSE | Downregulated |
| 0,471019567 | 0,462405028 | LOC389831 | FALSE | Upregulated |
| 0,198568032 | 0,462567675 | HNF1B | FALSE | Upregulated |
| -0,136334385 | 0,462567675 | ZCCHC4 | FALSE | Downregulated |
| 0,12916149 | 0,462658574 | NDUFAF4 | FALSE | Upregulated |
| -0,161111339 | 0,462918427 | CD1D | FALSE | Downregulated |
| 0,523544622 | 0,462981234 | FMO1 | FALSE | Upregulated |
| -0,251397154 | 0,462981234 | TRNY | FALSE | Downregulated |
| 0,467820956 | 0,463250257 | LETM2 | FALSE | Upregulated |
| 0,212832317 | 0,463406761 | DCPS | FALSE | Upregulated |
| 0,234454431 | 0,46384405 | KIAA0825 | FALSE | Upregulated |
| 0,163249139 | 0,464027067 | CDHR5 | FALSE | Upregulated |
| -0,197480039 | 0,464027067 | CEP85L | FALSE | Downregulated |
| -0,141549624 | 0,464032788 | ZNF259 | FALSE | Downregulated |
| 0,964601257 | 0,464398222 | RP11-449D8.1 | FALSE | Upregulated |
| 0,153587987 | 0,464594339 | APIP | FALSE | Upregulated |
| 0,719625678 | 0,464599773 | LOC100130705 | FALSE | Upregulated |
| 0,06193391 | 0,464721393 | DCAF7 | FALSE | Upregulated |
| 0,287218983 | 0,464721393 | PPP4R1L | FALSE | Upregulated |
| 0,98553722 | 0,464721393 | PRSS21 | FALSE | Upregulated |
| -0,274257407 | 0,464721393 | SNHG17 | FALSE | Downregulated |
| -0,196049193 | 0,465024033 | IQCC | FALSE | Downregulated |
| -1,443536931 | 0,465062124 | C20orf26 | FALSE | Downregulated |
| -0,767538314 | 0,465062124 | LOC102724901 | FALSE | Downregulated |
| -0,196083455 | 0,465062124 | RGN | FALSE | Downregulated |
| -0,102707716 | 0,465073232 | DCLRE1A | FALSE | Downregulated |
| -0,230143323 | 0,465109388 | TP53I11 | FALSE | Downregulated |
| -0,103993494 | 0,465109388 | ZNF655 | FALSE | Downregulated |
| 1,220602415 | 0,465166214 | RP3-399L15.3 | FALSE | Upregulated |
| 0,368353135 | 0,465282256 | GNLY | FALSE | Upregulated |
| -0,455127336 | 0,465393896 | LOC101929824 | FALSE | Downregulated |
| 1,306435191 | 0,465426094 | FOXD4 | FALSE | Upregulated |
| 0,817472921 | 0,465567 | SLC12A1 | FALSE | Upregulated |
| -0,483869134 | 0,465712546 | CTSLP2 | FALSE | Downregulated |
| 0,114407214 | 0,465712546 | IPP | FALSE | Upregulated |
| 0,493889831 | 0,465712546 | ST6GALNAC1 | FALSE | Upregulated |
| -0,071224107 | 0,465725455 | ASH2L | FALSE | Downregulated |
| -0,104446499 | 0,465725455 | KAT6A | FALSE | Downregulated |
| 0,120843028 | 0,465725455 | PSMB8 | FALSE | Upregulated |
| -0,275490837 | 0,465725455 | TMEM232 | FALSE | Downregulated |
| -0,482450496 | 0,465753693 | RP3-322G13.5 | FALSE | Downregulated |
| 0,805236849 | 0,465833412 | C21orf140 | FALSE | Upregulated |

| | | | | |
|---|---|---|---|---|
| 0,115142947 | 0,465858412 | JKAMP | FALSE | Upregulated |
| 0,118398259 | 0,465858412 | RABL5 | FALSE | Upregulated |
| 0,076445734 | 0,465858412 | RBM10 | FALSE | Upregulated |
| 0,536158866 | 0,465937301 | FSIP1 | FALSE | Upregulated |
| 0,330448129 | 0,465937301 | LIPE | FALSE | Upregulated |
| 1,437015488 | 0,465937301 | MMP1 | FALSE | Upregulated |
| 0,77661935 | 0,466043021 | LOC101928323 | FALSE | Upregulated |
| 0,114982772 | 0,466369412 | POLR3D | FALSE | Upregulated |
| 2,270719403 | 0,466600987 | TRNAG30 | FALSE | Upregulated |
| 0,26140895 | 0,466619889 | SLC25A33 | FALSE | Upregulated |
| -0,134319866 | 0,466684741 | PRPF39 | FALSE | Downregulated |
| 0,084641991 | 0,466684741 | ZMYM2 | FALSE | Upregulated |
| 0,471845888 | 0,466955614 | GTF2IRD1P1 | FALSE | Upregulated |
| 0,34913545 | 0,466999827 | PA2G4P4 | FALSE | Upregulated |
| 0,244406174 | 0,467068706 | HCG11 | FALSE | Upregulated |
| -1,008771302 | 0,467125904 | ADAM21 | FALSE | Downregulated |
| 0,166927325 | 0,467299376 | DENND2A | FALSE | Upregulated |
| 1,952678486 | 0,467571903 | CKMT1A | FALSE | Upregulated |
| -0,160661962 | 0,467593243 | ZNF593 | FALSE | Downregulated |
| -0,099704935 | 0,467727998 | COX5A | FALSE | Downregulated |
| -0,143238278 | 0,467727998 | PRDX4 | FALSE | Downregulated |
| 0,139043641 | 0,467727998 | RBFA | FALSE | Upregulated |
| 0,069495048 | 0,467727998 | SLC35A4 | FALSE | Upregulated |
| -0,204696963 | 0,467977969 | MRC1 | FALSE | Downregulated |
| -0,588763204 | 0,467977969 | RP1-1J6.2 | FALSE | Downregulated |
| -0,132816082 | 0,46829661 | ST3GAL5 | FALSE | Downregulated |
| 0,187959172 | 0,46838203 | IL7 | FALSE | Upregulated |
| -0,548222632 | 0,46838203 | LOC102723428 | FALSE | Downregulated |
| 0,31131977 | 0,46838203 | LOC102724550 | FALSE | Upregulated |
| 0,492387103 | 0,46838203 | TNIP3 | FALSE | Upregulated |
| 0,100523893 | 0,468468848 | EIF3F | FALSE | Upregulated |
| 0,482206857 | 0,468468848 | HCG4B | FALSE | Upregulated |
| -0,667142438 | 0,468577928 | RNASE13 | FALSE | Downregulated |
| -0,067283706 | 0,468739414 | ANKRD40 | FALSE | Downregulated |
| -0,18960377 | 0,468739414 | C17orf107 | FALSE | Downregulated |
| -0,059778591 | 0,468739414 | CSNK2A1 | FALSE | Downregulated |
| -0,096624293 | 0,468739414 | DPM1 | FALSE | Downregulated |
| 0,644062651 | 0,468739414 | FHL5 | FALSE | Upregulated |
| 0,117795857 | 0,468739414 | NAGLU | FALSE | Upregulated |
| 0,482305256 | 0,468739414 | ZFHX4-AS1 | FALSE | Upregulated |
| 0,09513362 | 0,468739414 | ZNF592 | FALSE | Upregulated |
| 0,877322547 | 0,469031561 | PCYT1B | FALSE | Upregulated |
| -0,102926305 | 0,469031561 | SHPK | FALSE | Downregulated |
| 0,645002622 | 0,469262603 | HIATL2 | FALSE | Upregulated |
| -0,084699288 | 0,469262603 | UQCRC1 | FALSE | Downregulated |
| 0,157050827 | 0,46946658 | CPN1 | FALSE | Upregulated |
| 0,590786892 | 0,469566608 | DPY19L2P2 | FALSE | Upregulated |
| 0,153424681 | 0,469615459 | GNB4 | FALSE | Upregulated |
| 0,967926744 | 0,469615459 | TRPC5OS | FALSE | Upregulated |
| -0,217761327 | 0,469683347 | ACTR5 | FALSE | Downregulated |
| -0,901007762 | 0,469683347 | C1orf61 | FALSE | Downregulated |

| | | | | |
|---|---|---|---|---|
| -0,07673272 | 0,469683347 | DDX19A | FALSE | Downregulated |
| 0,161914488 | 0,469683347 | MYO10 | FALSE | Upregulated |
| 0,515350139 | 0,469700268 | C1orf95 | FALSE | Upregulated |
| 0,510094982 | 0,469700268 | SLIT1 | FALSE | Upregulated |
| 0,50146263 | 0,469736759 | LOC100506047 | FALSE | Upregulated |
| 0,132177021 | 0,469853811 | LINC00623 | FALSE | Upregulated |
| 0,393140722 | 0,470048822 | UQCRHL | FALSE | Upregulated |
| 0,088820789 | 0,470221844 | ADPGK | FALSE | Upregulated |
| 0,154872937 | 0,470221844 | NEURL4 | FALSE | Upregulated |
| -0,117918896 | 0,470241326 | CHRNB1 | FALSE | Downregulated |
| -0,11333959 | 0,470342548 | AIMP2 | FALSE | Downregulated |
| 0,86201481 | 0,470504077 | TTC24 | FALSE | Upregulated |
| -0,214076387 | 0,471044972 | ACOT2 | FALSE | Downregulated |
| 0,338630705 | 0,471150905 | SLC23A3 | FALSE | Upregulated |
| 0,097496191 | 0,471190168 | CTSB | FALSE | Upregulated |
| -0,28664468 | 0,471190168 | PREX2 | FALSE | Downregulated |
| 0,137595227 | 0,471353848 | ZNF112 | FALSE | Upregulated |
| -0,734298428 | 0,471469436 | LOC101060264 | FALSE | Downregulated |
| 0,112530956 | 0,471553485 | FAM103A1 | FALSE | Upregulated |
| 0,356395664 | 0,471553485 | LOC101928588 | FALSE | Upregulated |
| 0,580805211 | 0,471680883 | ADAM33 | FALSE | Upregulated |
| 1,113652893 | 0,471691154 | WNT9A | FALSE | Upregulated |
| 0,718875012 | 0,471790618 | PRICKLE2-AS1 | FALSE | Upregulated |
| -0,45709999 | 0,471790618 | RP11-452H21.4 | FALSE | Downregulated |
| 0,246675613 | 0,471799665 | LOC101929986 | FALSE | Upregulated |
| 1,121065671 | 0,471844274 | LOC101929536 | FALSE | Upregulated |
| 0,635757764 | 0,471914934 | FLJ33360 | FALSE | Upregulated |
| -0,628484381 | 0,471925208 | HSPB2-C11orf52 | FALSE | Downregulated |
| -0,078356148 | 0,471944379 | TIAL1 | FALSE | Downregulated |
| -0,246671262 | 0,471982314 | GOLGA6C | FALSE | Downregulated |
| 1,079503497 | 0,471982314 | LINC01150 | FALSE | Upregulated |
| -0,231291817 | 0,472190884 | TTC32 | FALSE | Downregulated |
| -0,103200344 | 0,472310532 | FBXO28 | FALSE | Downregulated |
| -0,318617695 | 0,472357214 | FOXN3-AS1 | FALSE | Downregulated |
| 0,330630317 | 0,472453615 | MSANTD3-TMEFF1 | FALSE | Upregulated |
| 0,2630873 | 0,472499292 | PER1 | FALSE | Upregulated |
| -0,139012782 | 0,472513814 | PROX1 | FALSE | Downregulated |
| -0,155273175 | 0,472900916 | AGO3 | FALSE | Downregulated |
| -0,253526022 | 0,472900916 | NR1D2 | FALSE | Downregulated |
| -0,107334166 | 0,472900916 | PRIMPOL | FALSE | Downregulated |
| 0,095803361 | 0,473553207 | ATF7IP | FALSE | Upregulated |
| 0,081743149 | 0,473553207 | C1orf174 | FALSE | Upregulated |
| -0,862775686 | 0,473553207 | LOC100506351 | FALSE | Downregulated |
| -0,488519403 | 0,473553207 | LOC102724791 | FALSE | Downregulated |
| -0,327531407 | 0,473553207 | LOC441124 | FALSE | Downregulated |
| 0,151786223 | 0,473553207 | PPIL3 | FALSE | Upregulated |
| 0,099139573 | 0,473572502 | NAA10 | FALSE | Upregulated |
| -0,072131572 | 0,473572502 | NGLY1 | FALSE | Downregulated |
| 0,266755381 | 0,473572502 | RAB27B | FALSE | Upregulated |
| -0,098826423 | 0,473572502 | RPUSD3 | FALSE | Downregulated |
| 0,138510116 | 0,473907962 | GLTSCR2 | FALSE | Upregulated |

| | | | | |
|---|---|---|---|---|
| -0,465946464 | 0,473981704 | C6orf141 | FALSE | Downregulated |
| 0,926914234 | 0,473981704 | FAM221B | FALSE | Upregulated |
| -0,24272574 | 0,473981704 | FAM71E1 | FALSE | Downregulated |
| 1,182167938 | 0,473981704 | PMP2 | FALSE | Upregulated |
| 0,106736971 | 0,473981704 | SPICE1 | FALSE | Upregulated |
| -0,329253062 | 0,474077244 | LOC101928913 | FALSE | Downregulated |
| -0,898061326 | 0,474081809 | LOC101928011 | FALSE | Downregulated |
| 0,113106111 | 0,474124179 | JMJD6 | FALSE | Upregulated |
| -0,134979531 | 0,4741543 | REEP6 | FALSE | Downregulated |
| 0,398214551 | 0,474548918 | ARPC4-TTLL3 | FALSE | Upregulated |
| 0,810248885 | 0,47462334 | RNFT2 | FALSE | Upregulated |
| -0,931564427 | 0,474700294 | C5orf60 | FALSE | Downregulated |
| -0,643933072 | 0,474700294 | LOC100132813 | FALSE | Downregulated |
| 0,082718499 | 0,474700294 | TMOD3 | FALSE | Upregulated |
| -0,080255379 | 0,474719076 | AQR | FALSE | Downregulated |
| -0,280197079 | 0,47481988 | CYP21A2 | FALSE | Downregulated |
| 1,192223921 | 0,47481988 | FLG2 | FALSE | Upregulated |
| -0,175216472 | 0,474996869 | LOC100129550 | FALSE | Downregulated |
| -0,914847615 | 0,475017902 | LINC01091 | FALSE | Downregulated |
| 0,438809323 | 0,475165148 | WDR86-AS1 | FALSE | Upregulated |
| 0,831663249 | 0,475439863 | LOC101929336 | FALSE | Upregulated |
| 0,175637199 | 0,475450142 | EXOC3L4 | FALSE | Upregulated |
| 0,810594174 | 0,475511492 | IL23R | FALSE | Upregulated |
| 0,283733226 | 0,475516098 | KCNJ2 | FALSE | Upregulated |
| 0,840071215 | 0,475516098 | LOC644172 | FALSE | Upregulated |
| -0,330537245 | 0,475648949 | RNF39 | FALSE | Downregulated |
| -0,142408351 | 0,475648949 | TOP1 | FALSE | Downregulated |
| -1,184802357 | 0,475648949 | ZBTB20-AS1 | FALSE | Downregulated |
| 0,660442071 | 0,475651534 | SNORA32 | FALSE | Upregulated |
| 0,118733188 | 0,47605709 | ADD3 | FALSE | Upregulated |
| -0,329827578 | 0,476181874 | LOC101928745 | FALSE | Downregulated |
| -0,981883462 | 0,476211965 | LOC102724196 | FALSE | Downregulated |
| -1,312770609 | 0,476217389 | CSAG3 | FALSE | Downregulated |
| -0,172274028 | 0,47634842 | FRG1B | FALSE | Downregulated |
| -0,150122198 | 0,476381023 | TNRC6C | FALSE | Downregulated |
| 0,122488526 | 0,476436593 | PTPRG | FALSE | Upregulated |
| -0,062870664 | 0,476693162 | CNOT8 | FALSE | Downregulated |
| 1,222443777 | 0,476763985 | GPR1 | FALSE | Upregulated |
| 0,355854015 | 0,477165805 | DISC1 | FALSE | Upregulated |
| 0,104155608 | 0,477165805 | ERCC2 | FALSE | Upregulated |
| -0,362436695 | 0,477165805 | LOC101060581 | FALSE | Downregulated |
| -0,20175289 | 0,477189901 | PMS2CL | FALSE | Downregulated |
| 0,113356203 | 0,477232587 | SPPL2B | FALSE | Upregulated |
| -0,189420677 | 0,477307361 | KLHDC1 | FALSE | Downregulated |
| 0,072263416 | 0,477373239 | CLPTM1 | FALSE | Upregulated |
| -0,171170945 | 0,477393713 | CLASRP | FALSE | Downregulated |
| 0,105525102 | 0,477543468 | B3GALT6 | FALSE | Upregulated |
| 0,256753045 | 0,477739737 | OLFM1 | FALSE | Upregulated |
| 0,676684723 | 0,477919902 | CCL13 | FALSE | Upregulated |
| 0,095861351 | 0,478077651 | RNF13 | FALSE | Upregulated |
| 1,00423535 | 0,478296174 | LOC102724377 | FALSE | Upregulated |

| | | | | |
|---|---|---|---|---|
| 0,219801233 | 0,478513638 | THAP10 | FALSE | Upregulated |
| 0,165501433 | 0,478690505 | CXorf40B | FALSE | Upregulated |
| -0,394845404 | 0,478690505 | LOC102724643 | FALSE | Downregulated |
| -0,219394403 | 0,478690505 | TPK1 | FALSE | Downregulated |
| -0,09732484 | 0,478847802 | SDF2 | FALSE | Downregulated |
| -0,933372212 | 0,478925035 | FSD2 | FALSE | Downregulated |
| 0,522406072 | 0,479180265 | LOC286190 | FALSE | Upregulated |
| 0,529605441 | 0,479180265 | LOC729737 | FALSE | Upregulated |
| -0,292211487 | 0,479397404 | DNAH6 | FALSE | Downregulated |
| 0,296173025 | 0,47952902 | SPECC1L-ADORA2A | FALSE | Upregulated |
| -0,476670889 | 0,479765521 | BNIPL | FALSE | Downregulated |
| 0,615857228 | 0,479765521 | PDZD7 | FALSE | Upregulated |
| 0,629160107 | 0,479807998 | LOC102724028 | FALSE | Upregulated |
| 0,731699053 | 0,479834079 | ENO1-AS1 | FALSE | Upregulated |
| -0,302539794 | 0,479834079 | LOC101927589 | FALSE | Downregulated |
| 0,15124789 | 0,479890048 | THAP3 | FALSE | Upregulated |
| -0,087582641 | 0,479993172 | MED10 | FALSE | Downregulated |
| 0,149416633 | 0,48020063 | MIEF2 | FALSE | Upregulated |
| 0,231263885 | 0,480204643 | ADCY4 | FALSE | Upregulated |
| 0,104167609 | 0,480204643 | CCDC101 | FALSE | Upregulated |
| 1,167927047 | 0,480204643 | KLK14 | FALSE | Upregulated |
| 0,782780216 | 0,480204643 | RAET1E-AS1 | FALSE | Upregulated |
| -0,088757274 | 0,480204643 | SENP1 | FALSE | Downregulated |
| 0,18596101 | 0,480283001 | NEDD8-MDP1 | FALSE | Upregulated |
| 0,43032233 | 0,480283001 | PKN3 | FALSE | Upregulated |
| 0,076046932 | 0,480283001 | SUMO2 | FALSE | Upregulated |
| -0,430052303 | 0,480617783 | MEI4 | FALSE | Downregulated |
| -1,138430565 | 0,480625813 | LOC101929157 | FALSE | Downregulated |
| -0,06228203 | 0,480625813 | ZNF664 | FALSE | Downregulated |
| 0,322302242 | 0,480648585 | FAM53A | FALSE | Upregulated |
| -0,742709601 | 0,480681966 | C10orf67 | FALSE | Downregulated |
| 0,152298121 | 0,480828356 | PLCG2 | FALSE | Upregulated |
| 0,474087104 | 0,481098879 | LOC100294362 | FALSE | Upregulated |
| -0,075284284 | 0,481098879 | RTN3 | FALSE | Downregulated |
| -0,109676011 | 0,481336929 | RNF128 | FALSE | Downregulated |
| -0,476407835 | 0,481350715 | MEP1B | FALSE | Downregulated |
| -0,248331979 | 0,481419681 | TRIM66 | FALSE | Downregulated |
| 0,453563519 | 0,48163005 | ZNF454 | FALSE | Upregulated |
| 0,161514687 | 0,481656846 | FAM211A-AS1 | FALSE | Upregulated |
| 0,892793906 | 0,481915768 | STAC2 | FALSE | Upregulated |
| 0,131829858 | 0,481930511 | RRP9 | FALSE | Upregulated |
| -0,106064925 | 0,482101933 | TMEM129 | FALSE | Downregulated |
| -0,159509385 | 0,482101933 | ZNF470 | FALSE | Downregulated |
| 0,130828013 | 0,482125532 | SMG5 | FALSE | Upregulated |
| -0,096326301 | 0,482156497 | ERBB3 | FALSE | Downregulated |
| 0,873877132 | 0,482217698 | FLJ41481 | FALSE | Upregulated |
| -0,421432143 | 0,482537535 | C11orf85 | FALSE | Downregulated |
| -0,283861594 | 0,482545372 | ZNF736 | FALSE | Downregulated |
| 2,74440709 | 0,48259914 | TRNAV19 | FALSE | Upregulated |
| -0,455537671 | 0,482607673 | NMB | FALSE | Downregulated |
| 0,474560243 | 0,48272928 | CCDC64B | FALSE | Upregulated |

| | | | | |
|---|---|---|---|---|
| -0,253288744 | 0,482801848 | HES6 | FALSE | Downregulated |
| -0,282098806 | 0,482801848 | ZNF165 | FALSE | Downregulated |
| 0,121022862 | 0,483079518 | RBCK1 | FALSE | Upregulated |
| -0,063255878 | 0,483110813 | GNL1 | FALSE | Downregulated |
| 0,80376059 | 0,483588865 | DYX1C1-CCPG1 | FALSE | Upregulated |
| -0,647951947 | 0,483616509 | LOC100630923 | FALSE | Downregulated |
| 0,220528712 | 0,483882989 | ITGB3BP | FALSE | Upregulated |
| -0,117089605 | 0,483984837 | SLC25A16 | FALSE | Downregulated |
| 0,885140889 | 0,484024682 | LOC102723814 | FALSE | Upregulated |
| -0,483029028 | 0,484024682 | LOC102724169 | FALSE | Downregulated |
| -0,93712806 | 0,484024682 | SLC39A12 | FALSE | Downregulated |
| 0,59161387 | 0,484097211 | C1orf177 | FALSE | Upregulated |
| 0,088400153 | 0,484351058 | SMARCA5 | FALSE | Upregulated |
| 0,819321521 | 0,48459944 | SMIM2 | FALSE | Upregulated |
| -0,146711417 | 0,48462212 | C5orf45 | FALSE | Downregulated |
| -0,492719921 | 0,484655266 | DDTL | FALSE | Downregulated |
| 0,142366961 | 0,484808994 | MED27 | FALSE | Upregulated |
| -0,43184138 | 0,484897746 | LOC101927377 | FALSE | Downregulated |
| 0,106528429 | 0,484902285 | MDH1 | FALSE | Upregulated |
| 0,804119607 | 0,48493951 | PCAT5 | FALSE | Upregulated |
| -0,147538652 | 0,48518522 | TEX30 | FALSE | Downregulated |
| 1,391421673 | 0,485393632 | EPS8L3 | FALSE | Upregulated |
| 0,445850001 | 0,485423874 | SIGLEC15 | FALSE | Upregulated |
| 0,839696875 | 0,485448513 | TCERG1L | FALSE | Upregulated |
| -0,560913057 | 0,485693135 | MUSTN1 | FALSE | Downregulated |
| -0,118257149 | 0,485875863 | PARD6B | FALSE | Downregulated |
| 0,430535521 | 0,486189283 | ADAM22 | FALSE | Upregulated |
| 0,140263339 | 0,486189283 | GAMT | FALSE | Upregulated |
| -1,017464207 | 0,486221654 | EGFL8 | FALSE | Downregulated |
| 0,345781463 | 0,486300533 | NPAS3 | FALSE | Upregulated |
| 0,554637386 | 0,486389185 | LOC728819 | FALSE | Upregulated |
| 0,377450025 | 0,486427571 | CCNT2-AS1 | FALSE | Upregulated |
| 0,368975154 | 0,486655466 | BMS1P6 | FALSE | Upregulated |
| -0,199465573 | 0,486655466 | EFNA4 | FALSE | Downregulated |
| -0,143380298 | 0,486655466 | XDH | FALSE | Downregulated |
| 0,101432709 | 0,486655466 | ZNF438 | FALSE | Upregulated |
| -0,158888809 | 0,486749254 | ZNF674 | FALSE | Downregulated |
| 1,114530517 | 0,486880009 | DRC1 | FALSE | Upregulated |
| 0,21225653 | 0,486880009 | ZNF788 | FALSE | Upregulated |
| 0,110762225 | 0,486895545 | GM2A | FALSE | Upregulated |
| 0,189986903 | 0,486895545 | TUBGCP6 | FALSE | Upregulated |
| -0,111381155 | 0,4870334 | HIPK2 | FALSE | Downregulated |
| -0,413678209 | 0,487111708 | HSD3B2 | FALSE | Downregulated |
| 0,35847269 | 0,487111708 | SCAMP5 | FALSE | Upregulated |
| 0,114200556 | 0,487111708 | ZNF136 | FALSE | Upregulated |
| 0,120536784 | 0,487334568 | MVB12A | FALSE | Upregulated |
| 0,13088702 | 0,48739727 | NXT2 | FALSE | Upregulated |
| 0,915745842 | 0,487462864 | LOC102723354 | FALSE | Upregulated |
| 0,597016048 | 0,487462864 | PLEKHG4B | FALSE | Upregulated |
| -0,236042837 | 0,487462864 | TRABD2B | FALSE | Downregulated |
| -0,133151913 | 0,487512917 | COX7A2 | FALSE | Downregulated |

| | | | | |
|---|---|---|---|---|
| 1,167075114 | 0,487512917 | EIF3IP1 | FALSE | Upregulated |
| -0,362820754 | 0,487512917 | RP3-400B16.1 | FALSE | Downregulated |
| -0,126565202 | 0,487927742 | HMGB2 | FALSE | Downregulated |
| 0,958706889 | 0,487927742 | LOC102723408 | FALSE | Upregulated |
| -0,0781188 | 0,487927742 | RNF187 | FALSE | Downregulated |
| 0,206732888 | 0,487927742 | SDSL | FALSE | Upregulated |
| -0,3086593 | 0,487927742 | WNT2 | FALSE | Downregulated |
| 1,648874152 | 0,488246041 | MUC5AC | FALSE | Upregulated |
| -0,364830654 | 0,488279724 | C10orf126 | FALSE | Downregulated |
| 1,115293321 | 0,488286488 | HSFX2 | FALSE | Upregulated |
| 1,050792657 | 0,488539271 | QRFPR | FALSE | Upregulated |
| 0,584424018 | 0,488581482 | HCG25 | FALSE | Upregulated |
| 0,737649007 | 0,488681803 | C16orf74 | FALSE | Upregulated |
| -0,138412307 | 0,488696854 | N6AMT2 | FALSE | Downregulated |
| 0,255600854 | 0,488951772 | HTR2B | FALSE | Upregulated |
| 0,808353365 | 0,489047005 | LOC102724081 | FALSE | Upregulated |
| 0,172981979 | 0,489057365 | CACNA1D | FALSE | Upregulated |
| -0,135961524 | 0,48923733 | AUTS2 | FALSE | Downregulated |
| -0,147464127 | 0,489284615 | ZC4H2 | FALSE | Downregulated |
| 0,159177127 | 0,489455731 | SP4 | FALSE | Upregulated |
| 0,142152545 | 0,489601669 | TENC1 | FALSE | Upregulated |
| 0,708326978 | 0,489611374 | ZNF280B | FALSE | Upregulated |
| 0,174639141 | 0,489707782 | CHRNE | FALSE | Upregulated |
| 0,286677733 | 0,489707782 | HBEGF | FALSE | Upregulated |
| 0,930417538 | 0,48971996 | RNF148 | FALSE | Upregulated |
| 0,121929255 | 0,489809885 | ATOX1 | FALSE | Upregulated |
| -0,80152483 | 0,489809885 | SLC5A4 | FALSE | Downregulated |
| 0,754126081 | 0,490112306 | LOC102723693 | FALSE | Upregulated |
| -0,137987659 | 0,490149722 | RFX7 | FALSE | Downregulated |
| 0,16187396 | 0,490229611 | ARHGAP25 | FALSE | Upregulated |
| -0,083249388 | 0,490229611 | EIF3I | FALSE | Downregulated |
| -0,830340995 | 0,490229611 | LOC100505625 | FALSE | Downregulated |
| -0,138661953 | 0,490390709 | CASP10 | FALSE | Downregulated |
| 0,452491722 | 0,490518437 | TNFSF14 | FALSE | Upregulated |
| 0,0788491 | 0,490565506 | CHAMP1 | FALSE | Upregulated |
| 0,720215694 | 0,49065081 | LOC102723746 | FALSE | Upregulated |
| -0,098119256 | 0,49065081 | THYN1 | FALSE | Downregulated |
| 0,640766742 | 0,490731542 | C16orf89 | FALSE | Upregulated |
| -0,146990891 | 0,490982444 | SATB1 | FALSE | Downregulated |
| 0,72340186 | 0,491101856 | LOC102723317 | FALSE | Upregulated |
| -0,092558156 | 0,491101856 | USP12 | FALSE | Downregulated |
| -0,342081539 | 0,491123955 | LINC00638 | FALSE | Downregulated |
| 0,103022734 | 0,491123955 | MRPS26 | FALSE | Upregulated |
| 0,705223971 | 0,49119835 | SLITRK2 | FALSE | Upregulated |
| 0,085977356 | 0,491337148 | XPO6 | FALSE | Upregulated |
| 0,106387456 | 0,491371815 | LRRC57 | FALSE | Upregulated |
| -0,561295183 | 0,491677678 | LINC01134 | FALSE | Downregulated |
| 0,241885008 | 0,49181228 | ZBTB26 | FALSE | Upregulated |
| -0,069680222 | 0,491912378 | RAB22A | FALSE | Downregulated |
| 0,54750044 | 0,492172825 | LOC102724037 | FALSE | Upregulated |
| 0,144965771 | 0,492172825 | ZNF513 | FALSE | Upregulated |

| | | | | |
|---|---|---|---|---|
| 1,616291072 | 0,492246167 | TRNAE19 | FALSE | Upregulated |
| -0,261241031 | 0,492246167 | UBE2Q2P1 | FALSE | Downregulated |
| 0,401899603 | 0,492587 | TGM1 | FALSE | Upregulated |
| 0,075242318 | 0,492688673 | ABHD13 | FALSE | Upregulated |
| -0,136018496 | 0,49269564 | ATRIP | FALSE | Downregulated |
| -0,883926675 | 0,492994054 | LOC101929149 | FALSE | Downregulated |
| -0,202353494 | 0,493473953 | GUSBP1 | FALSE | Downregulated |
| 0,223112945 | 0,493783444 | CCDC171 | FALSE | Upregulated |
| 0,114774075 | 0,493783444 | FAM89B | FALSE | Upregulated |
| -1,275512882 | 0,493783444 | RGS7 | FALSE | Downregulated |
| -0,101927246 | 0,494219573 | ZNF420 | FALSE | Downregulated |
| -0,103094109 | 0,494322118 | NDUFAB1 | FALSE | Downregulated |
| -0,961368817 | 0,494382251 | CECR5-AS1 | FALSE | Downregulated |
| 0,755053681 | 0,494423993 | GAP43 | FALSE | Upregulated |
| 0,328304274 | 0,494596701 | FCRL3 | FALSE | Upregulated |
| 0,082905281 | 0,494596701 | FOXK2 | FALSE | Upregulated |
| 0,169027465 | 0,494596701 | RFC4 | FALSE | Upregulated |
| 0,587580262 | 0,494596701 | ZMYND10 | FALSE | Upregulated |
| 0,464365482 | 0,49460775 | ADAM23 | FALSE | Upregulated |
| 0,382879877 | 0,49460775 | SIGLEC14 | FALSE | Upregulated |
| 2,44064235 | 0,494616702 | SIK1 | FALSE | Upregulated |
| 1,106934942 | 0,49486367 | KRT15 | FALSE | Upregulated |
| 0,180319021 | 0,494975163 | MAPK8IP1 | FALSE | Upregulated |
| -0,215051913 | 0,494975163 | SLC16A4 | FALSE | Downregulated |
| -0,498124854 | 0,494975163 | STRC | FALSE | Downregulated |
| -0,479333561 | 0,495026874 | RP11-432I5.1 | FALSE | Downregulated |
| 0,115444468 | 0,495098973 | COMMD8 | FALSE | Upregulated |
| -0,255586344 | 0,495098973 | LOC102724023 | FALSE | Downregulated |
| 0,215743853 | 0,495432645 | LRRC17 | FALSE | Upregulated |
| -0,342331557 | 0,495530721 | AGAP4 | FALSE | Downregulated |
| -0,585674099 | 0,495530721 | MROH7-TTC4 | FALSE | Downregulated |
| -0,08191805 | 0,495642056 | TANK | FALSE | Downregulated |
| 1,315461627 | 0,495755782 | TMEM72 | FALSE | Upregulated |
| 0,105650964 | 0,495914298 | PKNOX1 | FALSE | Upregulated |
| 0,192337089 | 0,495972675 | ZNF821 | FALSE | Upregulated |
| 0,859950538 | 0,496033016 | RP11-445H22.3 | FALSE | Upregulated |
| -0,128740396 | 0,496036343 | VPS29 | FALSE | Downregulated |
| -0,214435932 | 0,496234211 | LOC101928431 | FALSE | Downregulated |
| -0,207527277 | 0,496293318 | TAL1 | FALSE | Downregulated |
| -0,117698137 | 0,49629475 | PTPN11 | FALSE | Downregulated |
| 0,444911576 | 0,496324451 | C6orf201 | FALSE | Upregulated |
| -0,391657249 | 0,496840056 | LOC101928881 | FALSE | Downregulated |
| 0,129392887 | 0,496840056 | RGMB | FALSE | Upregulated |
| 0,052345784 | 0,496964496 | MBTPS1 | FALSE | Upregulated |
| -0,27918234 | 0,497153744 | RNF5P1 | FALSE | Downregulated |
| -0,126114128 | 0,49739896 | SYNE1 | FALSE | Downregulated |
| -0,058775534 | 0,497531812 | TFCP2 | FALSE | Downregulated |
| -0,111234601 | 0,497670005 | KIF21A | FALSE | Downregulated |
| -0,135752382 | 0,497670005 | RRAGB | FALSE | Downregulated |
| -0,362891203 | 0,497779711 | LOC101927407 | FALSE | Downregulated |
| 0,249295413 | 0,497887982 | BMP5 | FALSE | Upregulated |

| | | | | |
|---|---|---|---|---|
| 0,076978593 | 0,498034701 | ANAPC5 | FALSE | Upregulated |
| -0,098161633 | 0,498034701 | ILF2 | FALSE | Downregulated |
| -0,181256624 | 0,498034701 | LOC100506990 | FALSE | Downregulated |
| -0,081421927 | 0,498034701 | UTP11L | FALSE | Downregulated |
| 0,076745239 | 0,498056068 | ANKRD13A | FALSE | Upregulated |
| 0,353812592 | 0,498068471 | FAM212A | FALSE | Upregulated |
| -0,581684883 | 0,498581982 | C21orf90 | FALSE | Downregulated |
| -0,20497877 | 0,498581982 | USP51 | FALSE | Downregulated |
| -0,133444003 | 0,498922247 | NCALD | FALSE | Downregulated |
| 0,112725314 | 0,499002813 | CIC | FALSE | Upregulated |
| 0,956728776 | 0,499002813 | LOC100131825 | FALSE | Upregulated |
| 0,778346841 | 0,499002813 | P4HA2-AS1 | FALSE | Upregulated |
| -0,267234115 | 0,499326043 | AC022007.5 | FALSE | Downregulated |
| 1,004425751 | 0,499326043 | RNU6-26P | FALSE | Upregulated |
| -0,190082964 | 0,499326043 | SSH2 | FALSE | Downregulated |
| 0,189077689 | 0,49939363 | ADSSL1 | FALSE | Upregulated |
| -0,066591244 | 0,499421989 | RARS2 | FALSE | Downregulated |
| 0,574842027 | 0,499467489 | FAM228A | FALSE | Upregulated |
| -0,769070974 | 0,499485251 | LIMS3L | FALSE | Downregulated |
| 0,595606611 | 0,499500677 | CR2 | FALSE | Upregulated |
| 0,42786425 | 0,499500677 | HCG27 | FALSE | Upregulated |
| -0,134629716 | 0,499542076 | PEX11G | FALSE | Downregulated |
| -0,261424293 | 0,49955366 | SLC5A9 | FALSE | Downregulated |
| -0,069099011 | 0,499575529 | TMEM181 | FALSE | Downregulated |
| -0,265781668 | 0,499575529 | WDR52 | FALSE | Downregulated |
| -0,180776393 | 0,499691437 | DNMT3B | FALSE | Downregulated |
| 0,064518709 | 0,499691437 | RUFY1 | FALSE | Upregulated |
| 0,991083526 | 0,499871535 | RP11-158M2.5 | FALSE | Upregulated |
| -0,096915827 | 0,499918698 | ACSL3 | FALSE | Downregulated |
| -0,150911722 | 0,50009118 | AC074286.1 | FALSE | Downregulated |
| -0,268971085 | 0,500097635 | ZNRD1-AS1 | FALSE | Downregulated |
| 0,120325174 | 0,500613383 | RAD18 | FALSE | Upregulated |
| 0,06060177 | 0,500655188 | DNAJB12 | FALSE | Upregulated |
| 0,158360027 | 0,500655188 | RGAG4 | FALSE | Upregulated |
| 0,202374459 | 0,501133684 | IRAK3 | FALSE | Upregulated |
| 0,910133299 | 0,501133684 | TRIM46 | FALSE | Upregulated |
| -0,381165085 | 0,501230229 | LOC101928472 | FALSE | Downregulated |
| 0,763872884 | 0,501230229 | PLSCR3 | FALSE | Upregulated |
| 0,17108522 | 0,501253412 | FNDC3B | FALSE | Upregulated |
| 1,097784531 | 0,501342091 | RP11-887P2.5 | FALSE | Upregulated |
| -0,072043525 | 0,501410533 | SNX4 | FALSE | Downregulated |
| 0,139235447 | 0,501466114 | LIMS2 | FALSE | Upregulated |
| -0,148852159 | 0,501498068 | EFNA1 | FALSE | Downregulated |
| 1,123881991 | 0,501530156 | MRPL23-AS1 | FALSE | Upregulated |
| -0,092213504 | 0,501530156 | WDR7 | FALSE | Downregulated |
| -0,582725611 | 0,50158894 | LOC101929195 | FALSE | Downregulated |
| 0,466841439 | 0,501747329 | LOC101060351 | FALSE | Upregulated |
| -0,128623101 | 0,501861049 | FAM98C | FALSE | Downregulated |
| -0,080424434 | 0,501979315 | SLC39A11 | FALSE | Downregulated |
| 0,132001849 | 0,502045662 | ARHGEF40 | FALSE | Upregulated |
| 0,169101356 | 0,502109159 | RNF208 | FALSE | Upregulated |

| | | | | |
|---|---|---|---|---|
| -0,122321923 | 0,502172223 | MANSC1 | FALSE | Downregulated |
| -0,07226426 | 0,502187768 | TADA3 | FALSE | Downregulated |
| 0,417541623 | 0,502612258 | S100B | FALSE | Upregulated |
| -0,719054376 | 0,502740576 | MYT1L | FALSE | Downregulated |
| -0,127367457 | 0,502740576 | PGS1 | FALSE | Downregulated |
| -0,170908901 | 0,502740576 | UQCRQ | FALSE | Downregulated |
| 0,419796568 | 0,502919925 | ASTN1 | FALSE | Upregulated |
| -0,085267299 | 0,502919925 | FGFRL1 | FALSE | Downregulated |
| 0,231908869 | 0,502919925 | TET1 | FALSE | Upregulated |
| -0,141739292 | 0,502980613 | DPH6 | FALSE | Downregulated |
| 0,254235978 | 0,503086065 | ZNF521 | FALSE | Upregulated |
| 0,982212671 | 0,50310888 | LINC00889 | FALSE | Upregulated |
| 0,087671431 | 0,503450827 | TMEM87A | FALSE | Upregulated |
| 1,143647803 | 0,503471274 | TRNAK16 | FALSE | Upregulated |
| 0,772783567 | 0,503538031 | ANKRD61 | FALSE | Upregulated |
| 0,266464942 | 0,503538031 | MFSD4 | FALSE | Upregulated |
| 0,361509424 | 0,503538031 | TNF | FALSE | Upregulated |
| 0,133952908 | 0,503669011 | CEP89 | FALSE | Upregulated |
| -0,25245474 | 0,503698325 | P4HA3 | FALSE | Downregulated |
| 0,704981429 | 0,503741777 | RRH | FALSE | Upregulated |
| 0,097091739 | 0,503741777 | TNKS1BP1 | FALSE | Upregulated |
| -0,087862485 | 0,50389664 | MAL2 | FALSE | Downregulated |
| 0,127974194 | 0,50389664 | NSUN5 | FALSE | Upregulated |
| -0,093813475 | 0,50389664 | RPA3-AS1 | FALSE | Downregulated |
| 0,115488357 | 0,50389664 | ZBTB41 | FALSE | Upregulated |
| 0,160836976 | 0,503997211 | PGBD1 | FALSE | Upregulated |
| -0,172690895 | 0,50416888 | RP11-620J15.3 | FALSE | Downregulated |
| 0,791388341 | 0,504205047 | EDNRB-AS1 | FALSE | Upregulated |
| -0,129148112 | 0,504400641 | CARD8 | FALSE | Downregulated |
| 0,148505642 | 0,504400641 | IRAK2 | FALSE | Upregulated |
| 0,869120015 | 0,504400641 | SNORD76 | FALSE | Upregulated |
| 0,083247788 | 0,504622023 | EMC3 | FALSE | Upregulated |
| 1,844275268 | 0,504622023 | FAM226A | FALSE | Upregulated |
| 0,154617461 | 0,504623443 | PYCARD | FALSE | Upregulated |
| 0,152635018 | 0,504783228 | JMJD7 | FALSE | Upregulated |
| -1,134684181 | 0,504893949 | GOLGA8S | FALSE | Downregulated |
| 0,07378645 | 0,504893949 | PIP4K2B | FALSE | Upregulated |
| -0,132545506 | 0,504893949 | PTBP2 | FALSE | Downregulated |
| -0,394154352 | 0,50493681 | SOX2-OT | FALSE | Downregulated |
| -0,196092877 | 0,504988148 | KATNAL2 | FALSE | Downregulated |
| 0,104418896 | 0,505329622 | UCK2 | FALSE | Upregulated |
| 0,201434579 | 0,505392001 | AMIGO2 | FALSE | Upregulated |
| 0,179067397 | 0,505416247 | MYO5C | FALSE | Upregulated |
| -0,216039665 | 0,505452616 | CAPS | FALSE | Downregulated |
| 0,248623516 | 0,505452616 | MX2 | FALSE | Upregulated |
| -0,119145108 | 0,505452616 | NHSL1 | FALSE | Downregulated |
| -0,050076594 | 0,505687907 | DDX24 | FALSE | Downregulated |
| 0,068007839 | 0,505728077 | VIPAS39 | FALSE | Upregulated |
| 0,636767202 | 0,505743889 | C1orf229 | FALSE | Upregulated |
| 0,083908816 | 0,505743889 | MAN1B1 | FALSE | Upregulated |
| 1,109063974 | 0,505743889 | PRR5-ARHGAP8 | FALSE | Upregulated |

| | | | | |
|---|---|---|---|---|
| 0,508861124 | 0,505743889 | RNF212 | FALSE | Upregulated |
| 0,075883842 | 0,505744129 | ESYT2 | FALSE | Upregulated |
| 0,110516106 | 0,505744129 | FAM149A | FALSE | Upregulated |
| 0,092165284 | 0,505744129 | VAV2 | FALSE | Upregulated |
| 0,189090849 | 0,505836317 | HOXB2 | FALSE | Upregulated |
| -0,081223635 | 0,506160289 | GATAD2B | FALSE | Downregulated |
| 0,135563734 | 0,506166786 | ZSCAN12 | FALSE | Upregulated |
| 0,143932461 | 0,506254385 | RPS18 | FALSE | Upregulated |
| 0,265192168 | 0,506388187 | GFRA2 | FALSE | Upregulated |
| -0,794803239 | 0,506388187 | LOC100188947 | FALSE | Downregulated |
| -0,182854182 | 0,506422888 | MYO16 | FALSE | Downregulated |
| 0,128218997 | 0,506555097 | POMGNT2 | FALSE | Upregulated |
| 0,9532886 | 0,506756951 | SH3RF3-AS1 | FALSE | Upregulated |
| 0,087788141 | 0,506790064 | COPS3 | FALSE | Upregulated |
| 0,142956309 | 0,506833829 | ZNF587B | FALSE | Upregulated |
| 0,770281989 | 0,507032381 | CPS1-IT1 | FALSE | Upregulated |
| 0,504390211 | 0,507111127 | USP32P1 | FALSE | Upregulated |
| -0,223797666 | 0,507277062 | AZGP1P1 | FALSE | Downregulated |
| -0,164293364 | 0,507277062 | PRICKLE3 | FALSE | Downregulated |
| -0,154558489 | 0,507290896 | CLK1 | FALSE | Downregulated |
| 0,154645382 | 0,507290896 | LPAR6 | FALSE | Upregulated |
| 0,296571897 | 0,507290896 | LRG1 | FALSE | Upregulated |
| 0,132792624 | 0,507290896 | SUV39H2 | FALSE | Upregulated |
| -0,186336849 | 0,507421052 | FTCD | FALSE | Downregulated |
| 0,420548983 | 0,507421052 | NOMO3 | FALSE | Upregulated |
| 0,199012558 | 0,507473368 | CCT6P3 | FALSE | Upregulated |
| 1,009475761 | 0,507473368 | CHST5 | FALSE | Upregulated |
| -0,264122706 | 0,507473368 | IQCH | FALSE | Downregulated |
| 0,08747619 | 0,507473368 | MOB2 | FALSE | Upregulated |
| 0,178335719 | 0,507473368 | PTPRN2 | FALSE | Upregulated |
| 0,913869799 | 0,507473368 | RGS13 | FALSE | Upregulated |
| 0,079433032 | 0,507473368 | ZNF189 | FALSE | Upregulated |
| -0,129225481 | 0,507526672 | TBC1D24 | FALSE | Downregulated |
| 0,327507279 | 0,507819216 | TFRC | FALSE | Upregulated |
| 0,438186495 | 0,508075014 | CASC1 | FALSE | Upregulated |
| -0,354142823 | 0,508075014 | PLGLB1 | FALSE | Downregulated |
| 0,16055317 | 0,50839407 | GRB7 | FALSE | Upregulated |
| 0,064611372 | 0,50839407 | HMG20A | FALSE | Upregulated |
| -0,749454217 | 0,508560716 | LOC100132741 | FALSE | Downregulated |
| -0,775531754 | 0,508560716 | RAPSN | FALSE | Downregulated |
| -0,086645522 | 0,508594216 | GBA2 | FALSE | Downregulated |
| -0,087386394 | 0,508683701 | SWT1 | FALSE | Downregulated |
| -0,123402987 | 0,508874147 | ACBD3 | FALSE | Downregulated |
| -0,078621557 | 0,508874147 | GSPT2 | FALSE | Downregulated |
| -0,104156617 | 0,508874147 | PIGP | FALSE | Downregulated |
| -0,101459385 | 0,508874147 | ZNF343 | FALSE | Downregulated |
| -0,466157665 | 0,509023904 | PCDH20 | FALSE | Downregulated |
| 1,113057067 | 0,509190096 | ABCB5 | FALSE | Upregulated |
| 0,053970681 | 0,509190096 | SLC35E1 | FALSE | Upregulated |
| 0,104826491 | 0,509203742 | ANKRD27 | FALSE | Upregulated |
| -0,124499658 | 0,509203742 | C11orf30 | FALSE | Downregulated |

| | | | | |
|---|---|---|---|---|
| 0,655885567 | 0,509407928 | KRTAP5-6 | FALSE | Upregulated |
| 0,619507489 | 0,509407928 | LOC101930375 | FALSE | Upregulated |
| -0,898583705 | 0,50952795 | CMTM2 | FALSE | Downregulated |
| -0,217588489 | 0,509623312 | LOC100505771 | FALSE | Downregulated |
| 0,575829046 | 0,509639705 | CIRBP-AS1 | FALSE | Upregulated |
| 0,190481262 | 0,509806936 | AC004893.11 | FALSE | Upregulated |
| 0,579599448 | 0,509806936 | LOC102724476 | FALSE | Upregulated |
| -0,122393427 | 0,509827447 | CLOCK | FALSE | Downregulated |
| -0,185991166 | 0,509827447 | FEZ1 | FALSE | Downregulated |
| 1,197223221 | 0,509827447 | KLF14 | FALSE | Upregulated |
| 0,129320309 | 0,509827447 | LRIG3 | FALSE | Upregulated |
| 0,117605398 | 0,509840559 | SIX5 | FALSE | Upregulated |
| -0,599729782 | 0,509840559 | SNX29P1 | FALSE | Downregulated |
| 1,058082186 | 0,510038356 | EPHA10 | FALSE | Upregulated |
| 0,412687329 | 0,510346815 | NDUFB2-AS1 | FALSE | Upregulated |
| 0,966912577 | 0,510346815 | UPK1A | FALSE | Upregulated |
| -1,048984892 | 0,510395658 | LOC102723437 | FALSE | Downregulated |
| -0,393868761 | 0,510444717 | LOC101929265 | FALSE | Downregulated |
| -0,075171856 | 0,510631932 | EEF2 | FALSE | Downregulated |
| -0,284238031 | 0,510723887 | CFHR3 | FALSE | Downregulated |
| 0,244523915 | 0,510723887 | LOC100996740 | FALSE | Upregulated |
| 0,894586036 | 0,510723887 | LOC399815 | FALSE | Upregulated |
| 0,101341112 | 0,510723887 | MAPKAPK3 | FALSE | Upregulated |
| -0,863369087 | 0,510962796 | PPEF1 | FALSE | Downregulated |
| -0,073496722 | 0,511416794 | RAB3GAP1 | FALSE | Downregulated |
| 0,458234482 | 0,511493974 | AKR7A2P1 | FALSE | Upregulated |
| 0,967072333 | 0,511508797 | HRASLS5 | FALSE | Upregulated |
| -0,096289294 | 0,511508797 | RALGAPA1 | FALSE | Downregulated |
| -0,215020907 | 0,511641669 | TMEM198B | FALSE | Downregulated |
| -0,144641374 | 0,511699885 | LOH12CR1 | FALSE | Downregulated |
| -0,065517946 | 0,511832593 | CNOT1 | FALSE | Downregulated |
| -0,105671779 | 0,511861182 | RAB32 | FALSE | Downregulated |
| -0,177724163 | 0,511864458 | LEAP2 | FALSE | Downregulated |
| 0,89331082 | 0,511888216 | CCL11 | FALSE | Upregulated |
| -0,081700129 | 0,511888216 | MMAA | FALSE | Downregulated |
| 0,281259775 | 0,511888216 | TCTEX1D2 | FALSE | Upregulated |
| 0,894588343 | 0,511903775 | LOC102724376 | FALSE | Upregulated |
| 0,073832684 | 0,511949956 | GTF3C2 | FALSE | Upregulated |
| 0,103898341 | 0,512257689 | RHEB | FALSE | Upregulated |
| 0,966806219 | 0,512314762 | PDILT | FALSE | Upregulated |
| -1,235093205 | 0,512329691 | AMPD1 | FALSE | Downregulated |
| 0,115964649 | 0,512329691 | CDAN1 | FALSE | Upregulated |
| 0,61000563 | 0,512329691 | CXorf65 | FALSE | Upregulated |
| 0,221898109 | 0,512329691 | SLC35G1 | FALSE | Upregulated |
| -0,158331323 | 0,512329691 | TBXA2R | FALSE | Downregulated |
| 0,589725999 | 0,512835 | ARL17B | FALSE | Upregulated |
| 0,485991427 | 0,512835 | DLEU2L | FALSE | Upregulated |
| -0,231551871 | 0,512835 | IFITM1 | FALSE | Downregulated |
| -0,141325627 | 0,512835 | SLC13A5 | FALSE | Downregulated |
| 1,088775909 | 0,512835 | TBC1D3P1-DHX40P1 | FALSE | Upregulated |
| 0,905557482 | 0,51289848 | CAMP | FALSE | Upregulated |

| | | | | |
|---|---|---|---|---|
| 0,293876276 | 0,513171543 | BCDIN3D-AS1 | FALSE | Upregulated |
| 0,655522229 | 0,513290913 | ATP2C2 | FALSE | Upregulated |
| 0,117147704 | 0,513290913 | CAMSAP2 | FALSE | Upregulated |
| -0,55943067 | 0,513454491 | MUC6 | FALSE | Downregulated |
| 0,667727189 | 0,513715659 | GABRR2 | FALSE | Upregulated |
| 0,609448544 | 0,513723647 | ENO4 | FALSE | Upregulated |
| -0,064443624 | 0,514028665 | PCMTD1 | FALSE | Downregulated |
| -0,108009664 | 0,514028665 | ZSCAN26 | FALSE | Downregulated |
| -0,151397293 | 0,514042961 | HEXDC | FALSE | Downregulated |
| 0,137326314 | 0,514233117 | GRAP | FALSE | Upregulated |
| -0,642688824 | 0,514362485 | RBM24 | FALSE | Downregulated |
| 0,452796317 | 0,514362485 | SPINK5 | FALSE | Upregulated |
| -1,015555284 | 0,51472787 | SSTR5-AS1 | FALSE | Downregulated |
| 0,09600513 | 0,514781084 | AP3B1 | FALSE | Upregulated |
| -0,598939433 | 0,514781084 | C6orf25 | FALSE | Downregulated |
| 0,64384469 | 0,514781084 | CLEC12B | FALSE | Upregulated |
| -0,13840693 | 0,514781084 | HUS1 | FALSE | Downregulated |
| -0,099184956 | 0,514781084 | MUS81 | FALSE | Downregulated |
| 1,741457869 | 0,514781084 | PAPOLB | FALSE | Upregulated |
| 0,104906286 | 0,514781084 | RAB40C | FALSE | Upregulated |
| 0,164762387 | 0,514781084 | SPTSSA | FALSE | Upregulated |
| 0,148069548 | 0,514781084 | ZNF133 | FALSE | Upregulated |
| 0,692269699 | 0,514856665 | LOC388499 | FALSE | Upregulated |
| -0,069965288 | 0,515019126 | FAM134A | FALSE | Downregulated |
| 0,126083551 | 0,515269371 | BZW2 | FALSE | Upregulated |
| 1,045198565 | 0,515412949 | RP11-710F7.3 | FALSE | Upregulated |
| -0,572465321 | 0,515507218 | LOC101928102 | FALSE | Downregulated |
| 0,194294718 | 0,515608338 | C2orf76 | FALSE | Upregulated |
| 0,531853122 | 0,515608338 | PLD5 | FALSE | Upregulated |
| 1,086978288 | 0,515608338 | RPS4Y2 | FALSE | Upregulated |
| -0,102014898 | 0,515624579 | AKR7A2 | FALSE | Downregulated |
| -0,314661283 | 0,515724513 | P2RY12 | FALSE | Downregulated |
| 0,461932011 | 0,515816736 | GPR173 | FALSE | Upregulated |
| 0,608910776 | 0,515834216 | CCDC42B | FALSE | Upregulated |
| -0,783278183 | 0,5158742 | TMEM240 | FALSE | Downregulated |
| 0,552986704 | 0,515996092 | FSIP2 | FALSE | Upregulated |
| -0,117560242 | 0,516254205 | PEX10 | FALSE | Downregulated |
| 0,243507501 | 0,516378647 | DNAH1 | FALSE | Upregulated |
| 0,155443548 | 0,516378647 | HSPA1A | FALSE | Upregulated |
| -0,119425289 | 0,516378647 | TRAPPC5 | FALSE | Downregulated |
| 0,254578775 | 0,516521898 | LILRA5 | FALSE | Upregulated |
| 0,365122627 | 0,516824931 | SLC22A4 | FALSE | Upregulated |
| -0,081527619 | 0,516838491 | RNF113A | FALSE | Downregulated |
| -0,119257156 | 0,516970783 | PGD | FALSE | Downregulated |
| 0,171730472 | 0,517018765 | CA5BP1 | FALSE | Upregulated |
| 0,090859061 | 0,51704174 | CCNDBP1 | FALSE | Upregulated |
| 0,316369862 | 0,51704174 | FICD | FALSE | Upregulated |
| -0,101754381 | 0,51704174 | MORC3 | FALSE | Downregulated |
| 0,410198759 | 0,517133962 | LY6G5B | FALSE | Upregulated |
| 0,95428566 | 0,517172828 | LOC102723507 | FALSE | Upregulated |
| -0,310593541 | 0,517240127 | LOC101927105 | FALSE | Downregulated |

| | | | | |
|---|---|---|---|---|
| -0,114990128 | 0,517262259 | GEMIN4 | FALSE | Downregulated |
| 1,067030552 | 0,517262259 | LOC101929354 | FALSE | Upregulated |
| 0,884135534 | 0,517262259 | LOC646588 | FALSE | Upregulated |
| 0,176231414 | 0,517262259 | MAST4 | FALSE | Upregulated |
| 1,072215455 | 0,517262259 | MGAT5B | FALSE | Upregulated |
| 0,15268028 | 0,517262259 | TAF1A | FALSE | Upregulated |
| 0,166671244 | 0,517262259 | ZNF276 | FALSE | Upregulated |
| 0,111244729 | 0,51733299 | TBRG4 | FALSE | Upregulated |
| 0,072373826 | 0,517697606 | MAP4K3 | FALSE | Upregulated |
| 0,269880794 | 0,517802264 | ENKUR | FALSE | Upregulated |
| 0,641454114 | 0,517802753 | LOC102724661 | FALSE | Upregulated |
| -0,160903117 | 0,518088302 | HLF | FALSE | Downregulated |
| 0,441118009 | 0,518298028 | KLRC4 | FALSE | Upregulated |
| -0,107432766 | 0,518436738 | BET1 | FALSE | Downregulated |
| 0,196182783 | 0,518449679 | VAMP1 | FALSE | Upregulated |
| -0,566208822 | 0,518488931 | CPB1 | FALSE | Downregulated |
| 0,121103251 | 0,518592586 | YRDC | FALSE | Upregulated |
| 0,466625334 | 0,518652126 | LOC101929479 | FALSE | Upregulated |
| -0,402201434 | 0,518652126 | STAC | FALSE | Downregulated |
| 0,210445773 | 0,519079367 | LENG8 | FALSE | Upregulated |
| 0,098784544 | 0,519115149 | ZNHIT3 | FALSE | Upregulated |
| 0,302444296 | 0,519153942 | ATOH8 | FALSE | Upregulated |
| 0,146355594 | 0,519153942 | LRIG2 | FALSE | Upregulated |
| -0,313841916 | 0,519180244 | FAM19A2 | FALSE | Downregulated |
| -0,156447667 | 0,519206943 | GREB1L | FALSE | Downregulated |
| -0,107502203 | 0,519242434 | RNF216P1 | FALSE | Downregulated |
| 0,658674333 | 0,519289779 | LOC101928307 | FALSE | Upregulated |
| -0,810740611 | 0,519427429 | ATP1A2 | FALSE | Downregulated |
| 0,841786949 | 0,519427429 | FAM154A | FALSE | Upregulated |
| -0,267728896 | 0,519427429 | KANK4 | FALSE | Downregulated |
| -0,182376159 | 0,519427429 | LOC102723779 | FALSE | Downregulated |
| 0,16607832 | 0,519501561 | RFC3 | FALSE | Upregulated |
| -0,373142007 | 0,519586329 | LOC102724017 | FALSE | Downregulated |
| -0,067236207 | 0,519636665 | SUPT6H | FALSE | Downregulated |
| 0,161456769 | 0,519937264 | FLOT2 | FALSE | Upregulated |
| -0,121838073 | 0,520007418 | COMT | FALSE | Downregulated |
| 0,426179629 | 0,520129722 | COLEC12 | FALSE | Upregulated |
| -0,084057806 | 0,520321604 | FRYL | FALSE | Downregulated |
| -0,071233121 | 0,520344251 | BRIX1 | FALSE | Downregulated |
| -0,072815863 | 0,520433056 | PSMA5 | FALSE | Downregulated |
| 0,155044917 | 0,520708618 | SEMA5A | FALSE | Upregulated |
| -0,107764026 | 0,520750881 | DLC1 | FALSE | Downregulated |
| 0,73043851 | 0,521129591 | CLDN19 | FALSE | Upregulated |
| -0,751408325 | 0,521148166 | ENDOU | FALSE | Downregulated |
| -0,375475089 | 0,521269513 | ROPN1L | FALSE | Downregulated |
| 0,416883173 | 0,521285306 | FLG-AS1 | FALSE | Upregulated |
| -0,096106194 | 0,521376351 | COPB2 | FALSE | Downregulated |
| -0,484644604 | 0,521434372 | ALLC | FALSE | Downregulated |
| 0,080060746 | 0,521434372 | FYN | FALSE | Upregulated |
| -0,187391493 | 0,521452848 | FAM65C | FALSE | Downregulated |
| 0,126722679 | 0,521930374 | ART4 | FALSE | Upregulated |

| | | | | |
|---|---|---|---|---|
| 0,077225313 | 0,521938718 | FBXO22 | FALSE | Upregulated |
| -0,104631174 | 0,521952208 | LOC102724364 | FALSE | Downregulated |
| 0,081480784 | 0,522034131 | RNGTT | FALSE | Upregulated |
| 0,141917822 | 0,522128201 | C6orf48 | FALSE | Upregulated |
| -0,158870785 | 0,522128201 | YY2 | FALSE | Downregulated |
| -0,272393281 | 0,522161894 | RP11-622A1.2 | FALSE | Downregulated |
| 0,161896612 | 0,522165308 | LOC100506161 | FALSE | Upregulated |
| 1,090626557 | 0,522274442 | ANKRD30BP3 | FALSE | Upregulated |
| 0,220028073 | 0,52231922 | RPL21P28 | FALSE | Upregulated |
| -0,055777449 | 0,52233759 | MFN1 | FALSE | Downregulated |
| -0,110311949 | 0,522519596 | COX11 | FALSE | Downregulated |
| 0,140366456 | 0,523093402 | IFT81 | FALSE | Upregulated |
| -0,472982166 | 0,523093402 | LOC101928829 | FALSE | Downregulated |
| -0,16740499 | 0,523118204 | ERN1 | FALSE | Downregulated |
| -0,170564058 | 0,52327147 | ACVR2B | FALSE | Downregulated |
| 0,166637714 | 0,52327147 | STOML1 | FALSE | Upregulated |
| 0,532029988 | 0,523444789 | LOC100132356 | FALSE | Upregulated |
| 0,108574331 | 0,523444789 | LRCH3 | FALSE | Upregulated |
| 0,103856183 | 0,52345453 | MAD2L1BP | FALSE | Upregulated |
| 0,147220995 | 0,523456445 | SHC2 | FALSE | Upregulated |
| 0,067540382 | 0,523517275 | TMEM131 | FALSE | Upregulated |
| -0,116852045 | 0,523529808 | ZNF717 | FALSE | Downregulated |
| -0,049260912 | 0,523538275 | AHCYL1 | FALSE | Downregulated |
| -0,113069198 | 0,523556974 | CUTC | FALSE | Downregulated |
| 1,249977235 | 0,523556974 | SPATA31D1 | FALSE | Upregulated |
| 0,098026077 | 0,523660018 | PSMB4 | FALSE | Upregulated |
| 0,408860952 | 0,523666346 | RP5-855F14.1 | FALSE | Upregulated |
| 0,665523494 | 0,523746728 | ZNF20 | FALSE | Upregulated |
| -0,09426464 | 0,523782811 | TST | FALSE | Downregulated |
| 0,133716478 | 0,523809355 | TRIM3 | FALSE | Upregulated |
| -0,089332041 | 0,523826765 | LARGE | FALSE | Downregulated |
| 0,143072203 | 0,524125362 | ICAM2 | FALSE | Upregulated |
| 0,881223357 | 0,524184254 | SLC9C2 | FALSE | Upregulated |
| -0,094785002 | 0,524218529 | HMOX2 | FALSE | Downregulated |
| 0,942391111 | 0,524269316 | LOC101928349 | FALSE | Upregulated |
| 0,437607496 | 0,524279113 | ICA1L | FALSE | Upregulated |
| 0,627508846 | 0,524279113 | LOC101928665 | FALSE | Upregulated |
| -0,977143054 | 0,524279113 | LOC102724057 | FALSE | Downregulated |
| -0,078873691 | 0,524378919 | MBD3 | FALSE | Downregulated |
| 0,563566251 | 0,524450522 | CSMD2 | FALSE | Upregulated |
| 0,101107912 | 0,524450522 | IFRD2 | FALSE | Upregulated |
| 0,110058549 | 0,524450522 | MARCKSL1 | FALSE | Upregulated |
| -0,128803498 | 0,524570795 | NDUFAF3 | FALSE | Downregulated |
| 0,109609887 | 0,524652767 | BACH1 | FALSE | Upregulated |
| 0,122192763 | 0,524673652 | FAM208B | FALSE | Upregulated |
| 0,829655694 | 0,524673652 | LOC102724919 | FALSE | Upregulated |
| -0,076942316 | 0,524873297 | ACTR6 | FALSE | Downregulated |
| 0,17990486 | 0,524945433 | UNKL | FALSE | Upregulated |
| -0,375716159 | 0,525076704 | TRPM6 | FALSE | Downregulated |
| 0,382941125 | 0,525107832 | CERS6-AS1 | FALSE | Upregulated |
| 0,572548576 | 0,525402016 | LOC100506498 | FALSE | Upregulated |

| | | | | |
|---|---|---|---|---|
| 0,13728671 | 0,52558397 | ASAP2 | FALSE | Upregulated |
| -0,100073341 | 0,52558397 | FBRSL1 | FALSE | Downregulated |
| -0,24879374 | 0,52558397 | LSS | FALSE | Downregulated |
| 0,171746343 | 0,52558397 | SDC3 | FALSE | Upregulated |
| 1,253794751 | 0,52566762 | HNRNPCP5 | FALSE | Upregulated |
| 0,120954246 | 0,525779593 | CBLB | FALSE | Upregulated |
| -0,360168337 | 0,525779593 | LOC101929670 | FALSE | Downregulated |
| 0,872924336 | 0,525779593 | LOC642934 | FALSE | Upregulated |
| -0,232154176 | 0,525779593 | TMEM91 | FALSE | Downregulated |
| 0,265947834 | 0,525779593 | WASH5P | FALSE | Upregulated |
| -0,081163464 | 0,525817772 | DYNC1LI1 | FALSE | Downregulated |
| -0,097879646 | 0,525935751 | PIGT | FALSE | Downregulated |
| 0,264228444 | 0,525935751 | PRKCDBP | FALSE | Upregulated |
| 1,014668142 | 0,526243483 | C1orf110 | FALSE | Upregulated |
| 0,916375443 | 0,5262747 | FCGR1C | FALSE | Upregulated |
| 0,11518486 | 0,5262747 | YIPF1 | FALSE | Upregulated |
| 0,402353516 | 0,526446545 | RAB19 | FALSE | Upregulated |
| -0,175635888 | 0,526565757 | C1orf122 | FALSE | Downregulated |
| -0,056291871 | 0,526565757 | GRIPAP1 | FALSE | Downregulated |
| -0,292255428 | 0,526565757 | MIR210HG | FALSE | Downregulated |
| 0,115514892 | 0,526565757 | PUS1 | FALSE | Upregulated |
| -0,098452364 | 0,526574164 | ZEB1 | FALSE | Downregulated |
| 0,31089064 | 0,526600141 | MEF2B | FALSE | Upregulated |
| -0,091465511 | 0,526600141 | SNX30 | FALSE | Downregulated |
| 0,147284499 | 0,526693081 | LIMD1 | FALSE | Upregulated |
| -1,017159909 | 0,526693081 | LOC101928080 | FALSE | Downregulated |
| 0,7361565 | 0,526780723 | DDX3Y | FALSE | Upregulated |
| -1,484385521 | 0,527013615 | TRNAK3 | FALSE | Downregulated |
| 0,175676746 | 0,527168491 | ADCK5 | FALSE | Upregulated |
| 0,107869734 | 0,527332318 | C8orf76 | FALSE | Upregulated |
| -0,047280859 | 0,527332318 | TNPO3 | FALSE | Downregulated |
| 0,345701219 | 0,528038008 | ESCO2 | FALSE | Upregulated |
| 0,099677082 | 0,528038008 | MLLT4 | FALSE | Upregulated |
| -0,083023301 | 0,528038008 | SUPT20H | FALSE | Downregulated |
| 0,100249138 | 0,528038008 | TBCB | FALSE | Upregulated |
| 0,119289259 | 0,528038008 | UVRAG | FALSE | Upregulated |
| 0,212964534 | 0,528381568 | ITGB7 | FALSE | Upregulated |
| 0,148459681 | 0,528464032 | DDR2 | FALSE | Upregulated |
| -0,417642365 | 0,528509832 | HLA-DQA2 | FALSE | Downregulated |
| -0,106007499 | 0,528509832 | TARBP2 | FALSE | Downregulated |
| -0,168165809 | 0,528541532 | FAM180A | FALSE | Downregulated |
| -0,133429874 | 0,528541532 | KIAA1107 | FALSE | Downregulated |
| -0,077913685 | 0,528541532 | WDR61 | FALSE | Downregulated |
| 0,297897762 | 0,528701354 | SPDYE3 | FALSE | Upregulated |
| -0,098229445 | 0,528825146 | FARSB | FALSE | Downregulated |
| 0,115465884 | 0,528827322 | APOA1 | FALSE | Upregulated |
| 0,090245577 | 0,528844693 | NHP2 | FALSE | Upregulated |
| 0,061795324 | 0,529107691 | SNRPB2 | FALSE | Upregulated |
| 1,553650341 | 0,529114776 | TNFSF12-TNFSF13 | FALSE | Upregulated |
| -0,092164259 | 0,529618076 | NUDCD1 | FALSE | Downregulated |
| 0,214185099 | 0,529733102 | ZNF491 | FALSE | Upregulated |

| | | | | |
|---|---|---|---|---|
| 1,203047822 | 0,52975008 | COL2A1 | FALSE | Upregulated |
| 0,913542875 | 0,52976382 | LOC101929409 | FALSE | Upregulated |
| -0,330278586 | 0,52976382 | RP1-93I3.1 | FALSE | Downregulated |
| -0,764798095 | 0,529849538 | LOC102724135 | FALSE | Downregulated |
| -0,538546535 | 0,529856137 | ITGA2B | FALSE | Downregulated |
| 0,385555777 | 0,530177517 | MYO1A | FALSE | Upregulated |
| 0,299437979 | 0,530187252 | ZNF793 | FALSE | Upregulated |
| -0,084582426 | 0,530299281 | DNASE2 | FALSE | Downregulated |
| -0,126298072 | 0,53066778 | ASB3 | FALSE | Downregulated |
| 0,08724062 | 0,53066778 | CYP20A1 | FALSE | Upregulated |
| 0,080076362 | 0,53066778 | GABPB1 | FALSE | Upregulated |
| 0,082692887 | 0,53066778 | MEF2D | FALSE | Upregulated |
| 0,691191878 | 0,53066778 | PTCD1 | FALSE | Upregulated |
| 0,252876652 | 0,530766362 | BMS1P4 | FALSE | Upregulated |
| 0,585756255 | 0,530766362 | LOC100287808 | FALSE | Upregulated |
| -0,309084287 | 0,530766362 | SPACA6P | FALSE | Downregulated |
| 0,18629346 | 0,531244971 | ARMC2 | FALSE | Upregulated |
| -0,533993089 | 0,531352172 | ABO | FALSE | Downregulated |
| 0,076610333 | 0,531440486 | PGGT1B | FALSE | Upregulated |
| -0,096843525 | 0,531450395 | DYRK4 | FALSE | Downregulated |
| 0,294227464 | 0,531481976 | LOC100996573 | FALSE | Upregulated |
| 1,07914772 | 0,531913306 | OTUD6A | FALSE | Upregulated |
| -0,172227716 | 0,532081686 | FOXA2 | FALSE | Downregulated |
| 0,760255065 | 0,532152023 | KIAA2022 | FALSE | Upregulated |
| 0,485917737 | 0,532206738 | LOC102724235 | FALSE | Upregulated |
| -0,195471664 | 0,532206738 | RP6-109B7.3 | FALSE | Downregulated |
| -0,131088072 | 0,532206738 | ZNF224 | FALSE | Downregulated |
| 0,110396204 | 0,532328612 | KMT2D | FALSE | Upregulated |
| 0,085224434 | 0,532328612 | SLC30A7 | FALSE | Upregulated |
| 0,683191838 | 0,532349044 | POU2F3 | FALSE | Upregulated |
| -0,065712763 | 0,532356689 | CDC23 | FALSE | Downregulated |
| -0,058442837 | 0,532356689 | H2AFV | FALSE | Downregulated |
| 0,044723413 | 0,532687252 | HNRNPU | FALSE | Upregulated |
| 0,07209915 | 0,532921314 | LRP5 | FALSE | Upregulated |
| 0,887097321 | 0,532931756 | AGPAT4-IT1 | FALSE | Upregulated |
| 0,15458368 | 0,5329414 | ARMC7 | FALSE | Upregulated |
| -0,217734951 | 0,5329414 | EDA | FALSE | Downregulated |
| -0,186319553 | 0,5329414 | LRRC37A | FALSE | Downregulated |
| -0,093456596 | 0,5329414 | RRM1 | FALSE | Downregulated |
| -0,216416536 | 0,532954546 | LINC01089 | FALSE | Downregulated |
| -0,116194996 | 0,533069864 | ERCC8 | FALSE | Downregulated |
| -0,174415423 | 0,533083756 | CCDC14 | FALSE | Downregulated |
| -0,142752816 | 0,533320657 | C6orf57 | FALSE | Downregulated |
| 0,405279548 | 0,533320657 | ZNF90 | FALSE | Upregulated |
| 0,10068869 | 0,533517142 | KIF22 | FALSE | Upregulated |
| -0,288942489 | 0,533517142 | RP11-66A2.2 | FALSE | Downregulated |
| 0,125699093 | 0,533517142 | TMEM194A | FALSE | Upregulated |
| 0,106679255 | 0,533671301 | DDX51 | FALSE | Upregulated |
| -0,09996872 | 0,533671301 | LGMN | FALSE | Downregulated |
| 0,324323717 | 0,533671301 | RGS11 | FALSE | Upregulated |
| 0,401506741 | 0,533709178 | LOC102723544 | FALSE | Upregulated |

| | | | | |
|---|---|---|---|---|
| -0,701441987 | 0,534059236 | DLEU7-AS1 | FALSE | Downregulated |
| -0,337899236 | 0,534133817 | RP11-996F15.2 | FALSE | Downregulated |
| -0,872360792 | 0,534280561 | LOC151760 | FALSE | Downregulated |
| 0,441033284 | 0,534416366 | RP11-367N14.2 | FALSE | Upregulated |
| -0,119749091 | 0,534530445 | LGR4 | FALSE | Downregulated |
| -0,634147425 | 0,534530445 | RSPH10B | FALSE | Downregulated |
| -0,222272095 | 0,534530445 | TDO2 | FALSE | Downregulated |
| 0,328649052 | 0,534684787 | AMPH | FALSE | Upregulated |
| -0,674914811 | 0,534751156 | LOC101927673 | FALSE | Downregulated |
| 0,091229844 | 0,534972416 | N4BP1 | FALSE | Upregulated |
| 0,09891604 | 0,535102406 | RINT1 | FALSE | Upregulated |
| 0,109597329 | 0,535315246 | TOE1 | FALSE | Upregulated |
| -0,228154164 | 0,53545935 | WNT5B | FALSE | Downregulated |
| 0,671709721 | 0,535537464 | RP11-838N2.4 | FALSE | Upregulated |
| 0,312593258 | 0,535560394 | EFS | FALSE | Upregulated |
| 0,127848733 | 0,535880369 | MALT1 | FALSE | Upregulated |
| 0,297599189 | 0,535880369 | WDR65 | FALSE | Upregulated |
| 0,080804442 | 0,535988147 | PI4KB | FALSE | Upregulated |
| 0,16075443 | 0,536168546 | FAM89A | FALSE | Upregulated |
| 0,177575783 | 0,536168546 | KIF11 | FALSE | Upregulated |
| 0,219245214 | 0,536168546 | TMEM200B | FALSE | Upregulated |
| 0,089003114 | 0,536168546 | ZNF407 | FALSE | Upregulated |
| 0,124718729 | 0,536175286 | UROS | FALSE | Upregulated |
| -0,632319538 | 0,5363891 | PDLIM3 | FALSE | Downregulated |
| 0,082585246 | 0,5363891 | RRP36 | FALSE | Upregulated |
| 0,140612007 | 0,5363891 | ZNF780B | FALSE | Upregulated |
| 0,197079523 | 0,536784568 | GK | FALSE | Upregulated |
| -0,128187592 | 0,536784568 | MON1A | FALSE | Downregulated |
| 0,082848814 | 0,536904495 | CNOT6L | FALSE | Upregulated |
| 0,146060992 | 0,536949926 | PTCH1 | FALSE | Upregulated |
| -0,107421995 | 0,53710784 | AQP9 | FALSE | Downregulated |
| 0,13502486 | 0,537135387 | CEP290 | FALSE | Upregulated |
| 0,713932916 | 0,537135387 | LOC102723733 | FALSE | Upregulated |
| 0,335544181 | 0,537209385 | MEOX2 | FALSE | Upregulated |
| -0,514168432 | 0,537428723 | HP09025 | FALSE | Downregulated |
| -0,143252088 | 0,537656918 | HIST2H2BE | FALSE | Downregulated |
| 0,207463631 | 0,537656918 | IGSF6 | FALSE | Upregulated |
| 1,096491132 | 0,537656918 | RP11-542B15.1 | FALSE | Upregulated |
| -0,809893057 | 0,537760319 | C9orf170 | FALSE | Downregulated |
| 0,399045525 | 0,537760319 | FKBP1AP1 | FALSE | Upregulated |
| 0,200375751 | 0,537760319 | ZNF37BP | FALSE | Upregulated |
| -0,082814561 | 0,53787046 | MKL2 | FALSE | Downregulated |
| -0,306627969 | 0,53787046 | ZNF876P | FALSE | Downregulated |
| -0,10452851 | 0,538261961 | CALU | FALSE | Downregulated |
| -0,54145787 | 0,53833471 | LOC102467080 | FALSE | Downregulated |
| 0,489623381 | 0,538371904 | PEBP4 | FALSE | Upregulated |
| 0,447020197 | 0,53840344 | LINC00242 | FALSE | Upregulated |
| 0,169797046 | 0,538701043 | GSAP | FALSE | Upregulated |
| -0,08188444 | 0,538779551 | CD99L2 | FALSE | Downregulated |
| -0,087803109 | 0,538843688 | GUK1 | FALSE | Downregulated |
| -0,278136412 | 0,538914827 | C7orf55-LUC7L2 | FALSE | Downregulated |

| | | | | |
|---|---|---|---|---|
| -1,006410511 | 0,538914827 | INHA | FALSE | Downregulated |
| -0,479741223 | 0,538914827 | LOC102723927 | FALSE | Downregulated |
| 0,072268446 | 0,538914827 | PBX2 | FALSE | Upregulated |
| -0,105930362 | 0,538914827 | RPL29 | FALSE | Downregulated |
| 0,127134512 | 0,538914827 | TMEM106C | FALSE | Upregulated |
| -0,163961324 | 0,539098782 | FGA | FALSE | Downregulated |
| 0,138464819 | 0,539098782 | FGFR1 | FALSE | Upregulated |
| -0,117401432 | 0,539144358 | NNT-AS1 | FALSE | Downregulated |
| -0,081028133 | 0,539220992 | CDYL | FALSE | Downregulated |
| 0,112318676 | 0,539511895 | CGRRF1 | FALSE | Upregulated |
| 0,961736133 | 0,539533649 | ATP1A4 | FALSE | Upregulated |
| -0,512797651 | 0,539533649 | DCT | FALSE | Downregulated |
| 0,203517037 | 0,539578253 | AGBL3 | FALSE | Upregulated |
| 0,441069449 | 0,539801158 | LOC100128361 | FALSE | Upregulated |
| 0,079203099 | 0,539821995 | DDX39B | FALSE | Upregulated |
| 1,021443746 | 0,539957405 | LOC101929216 | FALSE | Upregulated |
| -1,169261662 | 0,540039541 | DUSP26 | FALSE | Downregulated |
| 0,428657495 | 0,540174655 | ARHGEF33 | FALSE | Upregulated |
| 0,059630965 | 0,540178431 | RBBP7 | FALSE | Upregulated |
| -0,144768227 | 0,54049837 | BAZ1A | FALSE | Downregulated |
| -0,0618704 | 0,540731236 | CCDC6 | FALSE | Downregulated |
| 0,07024846 | 0,540731236 | PCNP | FALSE | Upregulated |
| 0,044776709 | 0,540731236 | UBE3B | FALSE | Upregulated |
| -0,064825016 | 0,540772739 | CEBPZ | FALSE | Downregulated |
| 1,038941878 | 0,540772739 | IL9R | FALSE | Upregulated |
| 1,167894958 | 0,540772739 | TNFSF18 | FALSE | Upregulated |
| -0,148668319 | 0,540934305 | MYO9A | FALSE | Downregulated |
| -0,219591369 | 0,541006433 | CYP4X1 | FALSE | Downregulated |
| 0,145172748 | 0,541224618 | DAPK1 | FALSE | Upregulated |
| 0,130354419 | 0,541297542 | TUBG1 | FALSE | Upregulated |
| 0,922575705 | 0,541395615 | LOC101926896 | FALSE | Upregulated |
| -0,358011153 | 0,541395615 | NPIPB9 | FALSE | Downregulated |
| -0,080075397 | 0,541395615 | USP32 | FALSE | Downregulated |
| 0,100681722 | 0,541469007 | AHSA1 | FALSE | Upregulated |
| 0,193752191 | 0,541469007 | CCL15-CCL14 | FALSE | Upregulated |
| -0,06118629 | 0,541469007 | CUX1 | FALSE | Downregulated |
| 0,176409115 | 0,541469007 | DSTNP2 | FALSE | Upregulated |
| 0,079153921 | 0,541469007 | POMT1 | FALSE | Upregulated |
| 0,180820049 | 0,541508073 | IFI30 | FALSE | Upregulated |
| 0,065140296 | 0,541593615 | GNAI3 | FALSE | Upregulated |
| -0,314618513 | 0,541593615 | MTMR7 | FALSE | Downregulated |
| -0,073409669 | 0,541638884 | STT3B | FALSE | Downregulated |
| -1,226328203 | 0,541932044 | ARX | FALSE | Downregulated |
| 0,403519154 | 0,541932044 | FLJ39632 | FALSE | Upregulated |
| 0,308367202 | 0,541932044 | ITGA7 | FALSE | Upregulated |
| -0,406534151 | 0,541978404 | MIR564 | FALSE | Downregulated |
| -0,164321306 | 0,542246285 | BRD9 | FALSE | Downregulated |
| -0,164558331 | 0,542246285 | DLL4 | FALSE | Downregulated |
| -0,116084799 | 0,54233082 | TTLL11 | FALSE | Downregulated |
| 0,440855539 | 0,542644387 | LOC101926994 | FALSE | Upregulated |
| 0,926517859 | 0,542647931 | LOC101927799 | FALSE | Upregulated |

| | | | | |
|---|---|---|---|---|
| 0,17706407 | 0,542865096 | HMCN1 | FALSE | Upregulated |
| 0,740493492 | 0,542877738 | FAM71D | FALSE | Upregulated |
| 0,35951002 | 0,543076123 | ACBD7 | FALSE | Upregulated |
| 0,076411702 | 0,543279737 | PIGC | FALSE | Upregulated |
| 0,438662915 | 0,543398633 | COL4A6 | FALSE | Upregulated |
| 0,186146101 | 0,543398633 | GCNT1 | FALSE | Upregulated |
| -0,11607658 | 0,543403497 | HAUS8 | FALSE | Downregulated |
| 0,56924595 | 0,543403497 | LINC00092 | FALSE | Upregulated |
| -0,677826103 | 0,543551529 | LINC00707 | FALSE | Downregulated |
| 0,338790144 | 0,543583355 | GLUD1P3 | FALSE | Upregulated |
| -0,14184072 | 0,543649288 | HSP90B1 | FALSE | Downregulated |
| 0,108795591 | 0,543649288 | RPUSD2 | FALSE | Upregulated |
| 0,068837037 | 0,543871215 | YTHDF1 | FALSE | Upregulated |
| -0,517524729 | 0,544067662 | KIAA0408 | FALSE | Downregulated |
| -0,308480425 | 0,544487689 | SLC16A1-AS1 | FALSE | Downregulated |
| -0,159739506 | 0,544487689 | ZNF571 | FALSE | Downregulated |
| 0,109684503 | 0,544491154 | NAP1L1 | FALSE | Upregulated |
| 0,259600229 | 0,544602372 | METTL12 | FALSE | Upregulated |
| 0,102999261 | 0,544602372 | SLC38A3 | FALSE | Upregulated |
| -0,276506689 | 0,544675794 | SLC14A1 | FALSE | Downregulated |
| 0,365841128 | 0,544731128 | PRKXP1 | FALSE | Upregulated |
| 0,159110905 | 0,544824616 | NBPF1 | FALSE | Upregulated |
| 1,145681818 | 0,544837657 | DNER | FALSE | Upregulated |
| 0,335494674 | 0,544904948 | C1orf186 | FALSE | Upregulated |
| 1,037384793 | 0,544975838 | CYTL1 | FALSE | Upregulated |
| -0,138118918 | 0,544975838 | EIF3J | FALSE | Downregulated |
| -0,256012673 | 0,544975838 | TSKU | FALSE | Downregulated |
| -0,904370779 | 0,545161966 | CALB2 | FALSE | Downregulated |
| 0,112958541 | 0,545161966 | CDK10 | FALSE | Upregulated |
| -0,50183894 | 0,545165817 | FXYD7 | FALSE | Downregulated |
| 0,149525187 | 0,545165817 | GPRC5C | FALSE | Upregulated |
| 0,09767953 | 0,545165817 | PCBP2 | FALSE | Upregulated |
| -0,45685426 | 0,545165817 | TCAM1P | FALSE | Downregulated |
| -0,571696575 | 0,545277869 | ZNF385D | FALSE | Downregulated |
| -0,070811349 | 0,545290673 | PPIL2 | FALSE | Downregulated |
| -0,209217615 | 0,545333844 | RNF165 | FALSE | Downregulated |
| 0,085521881 | 0,545333844 | TRNAU1AP | FALSE | Upregulated |
| 0,174777542 | 0,5454815 | C6orf1 | FALSE | Upregulated |
| 0,170544645 | 0,5454815 | CHN1 | FALSE | Upregulated |
| -0,057810066 | 0,5454815 | CSDE1 | FALSE | Downregulated |
| -0,169244154 | 0,5454815 | ENTPD7 | FALSE | Downregulated |
| -0,195905178 | 0,545493374 | NEURL2 | FALSE | Downregulated |
| 0,258042152 | 0,54554632 | GDPD1 | FALSE | Upregulated |
| 0,146202685 | 0,545603606 | CLDN5 | FALSE | Upregulated |
| 0,086776593 | 0,54562976 | LRP3 | FALSE | Upregulated |
| -0,109525309 | 0,54562976 | PROSER1 | FALSE | Downregulated |
| 0,667457967 | 0,545987478 | LOC284648 | FALSE | Upregulated |
| -0,072837321 | 0,545987478 | PARL | FALSE | Downregulated |
| 0,828076756 | 0,546094064 | SH2D6 | FALSE | Upregulated |
| -0,165718693 | 0,546094064 | TRMT10B | FALSE | Downregulated |
| 0,387025388 | 0,546354273 | SLC26A5 | FALSE | Upregulated |

| | | | | |
|---|---|---|---|---|
| -0,343048308 | 0,546367162 | SV2B | FALSE | Downregulated |
| 0,147740215 | 0,546478632 | PKD1P1 | FALSE | Upregulated |
| -0,126439994 | 0,54661364 | C11orf52 | FALSE | Downregulated |
| -0,438194418 | 0,54661364 | DNAH12 | FALSE | Downregulated |
| 0,616691296 | 0,54661364 | RBFADN | FALSE | Upregulated |
| 0,072141052 | 0,54661364 | UBP1 | FALSE | Upregulated |
| 0,311127311 | 0,546645418 | LOC100506472 | FALSE | Upregulated |
| -0,174454148 | 0,546889719 | DLG4 | FALSE | Downregulated |
| 0,676418138 | 0,546889719 | FOXN4 | FALSE | Upregulated |
| 0,122737615 | 0,546889719 | OPTN | FALSE | Upregulated |
| -0,096926161 | 0,546889719 | PFDN6 | FALSE | Downregulated |
| 0,360158502 | 0,546929874 | SLC3A1 | FALSE | Upregulated |
| 0,06894064 | 0,547128297 | ATRAID | FALSE | Upregulated |
| -0,132691104 | 0,547355701 | FAM126B | FALSE | Downregulated |
| 0,226818347 | 0,547373747 | GUSBP11 | FALSE | Upregulated |
| 0,062704106 | 0,547373747 | RAB9A | FALSE | Upregulated |
| -0,062313746 | 0,547373747 | TARDBP | FALSE | Downregulated |
| 0,119592328 | 0,547442411 | CCDC85C | FALSE | Upregulated |
| 0,118048406 | 0,547442411 | LOC90784 | FALSE | Upregulated |
| 0,190279365 | 0,547570577 | HSPA5 | FALSE | Upregulated |
| -0,186075105 | 0,547642031 | LOC102724528 | FALSE | Downregulated |
| -0,15772269 | 0,547715075 | LOC100506538 | FALSE | Downregulated |
| -0,441001386 | 0,547715075 | NFE2 | FALSE | Downregulated |
| 0,331691817 | 0,547715075 | RP11-844P9.2 | FALSE | Upregulated |
| 0,090286149 | 0,547715075 | SLC3A2 | FALSE | Upregulated |
| 0,362839406 | 0,547715075 | TPPP3 | FALSE | Upregulated |
| -0,104852946 | 0,548256012 | MAN2A2 | FALSE | Downregulated |
| 0,077521821 | 0,548347783 | RPS6KC1 | FALSE | Upregulated |
| 0,135341373 | 0,548618202 | GTPBP3 | FALSE | Upregulated |
| 0,069207927 | 0,548828825 | ATG9A | FALSE | Upregulated |
| 0,30246437 | 0,548828825 | LOC102724783 | FALSE | Upregulated |
| -0,082300691 | 0,5489523 | RNF20 | FALSE | Downregulated |
| 0,069784002 | 0,549242759 | FAM91A1 | FALSE | Upregulated |
| 0,125194416 | 0,54963909 | EXO5 | FALSE | Upregulated |
| 0,146413581 | 0,54963909 | OLFML1 | FALSE | Upregulated |
| 2,570089585 | 0,54977597 | SYNPO2L | FALSE | Upregulated |
| -0,131627945 | 0,54977597 | TUSC1 | FALSE | Downregulated |
| 0,151048732 | 0,550048863 | NBPF9 | FALSE | Upregulated |
| -0,084399319 | 0,550072466 | AP4S1 | FALSE | Downregulated |
| -0,092078043 | 0,550077622 | RTKN | FALSE | Downregulated |
| 0,122306752 | 0,550148625 | DUSP5 | FALSE | Upregulated |
| -0,336675402 | 0,550148625 | GNRH1 | FALSE | Downregulated |
| 1,392478109 | 0,550151565 | HHLA2 | FALSE | Upregulated |
| -0,104614477 | 0,550151565 | SPEN | FALSE | Downregulated |
| 0,951955734 | 0,55023853 | IFITM4P | FALSE | Upregulated |
| 0,053671773 | 0,550784059 | RAD23B | FALSE | Upregulated |
| 0,649340574 | 0,550788059 | LOC100287072 | FALSE | Upregulated |
| 0,915841983 | 0,550806775 | FAM71F1 | FALSE | Upregulated |
| -0,198813016 | 0,550987789 | NR6A1 | FALSE | Downregulated |
| 0,398400713 | 0,550990361 | FBXO43 | FALSE | Upregulated |
| 0,116730337 | 0,550990361 | LINC01146 | FALSE | Upregulated |

| | | | | |
|---|---|---|---|---|
| -0,07432318 | 0,551191782 | DDX59 | FALSE | Downregulated |
| 0,777362056 | 0,551268967 | REEP2 | FALSE | Upregulated |
| 0,208537632 | 0,551478727 | CDC42EP2 | FALSE | Upregulated |
| 0,213635502 | 0,55181491 | MGC32805 | FALSE | Upregulated |
| 0,132242874 | 0,55181491 | TMEM170A | FALSE | Upregulated |
| -0,122726479 | 0,552037009 | CCSAP | FALSE | Downregulated |
| -0,132728236 | 0,55205974 | C8B | FALSE | Downregulated |
| -0,411743674 | 0,552081672 | HIST1H3E | FALSE | Downregulated |
| -1,52882782 | 0,552551874 | FBP2 | FALSE | Downregulated |
| 0,57199593 | 0,552571251 | HIST1H2BJ | FALSE | Upregulated |
| 0,213019172 | 0,552639035 | PLB1 | FALSE | Upregulated |
| 0,675649537 | 0,552800224 | LOC102724797 | FALSE | Upregulated |
| -0,120807207 | 0,552921177 | WDR31 | FALSE | Downregulated |
| 0,071533427 | 0,553276841 | CNPPD1 | FALSE | Upregulated |
| 0,079768608 | 0,553521726 | TTC23 | FALSE | Upregulated |
| -0,328765697 | 0,553592793 | LOC100506454 | FALSE | Downregulated |
| 0,078069287 | 0,553592793 | YARS2 | FALSE | Upregulated |
| -0,098922103 | 0,554075981 | STRN | FALSE | Downregulated |
| -0,108932086 | 0,554385925 | ZNF175 | FALSE | Downregulated |
| -0,659591545 | 0,554504744 | MEIS1-AS2 | FALSE | Downregulated |
| -0,308807367 | 0,554623976 | DBH | FALSE | Downregulated |
| 0,543668924 | 0,554674447 | LOC101928059 | FALSE | Upregulated |
| -0,075434162 | 0,554711265 | FNDC3A | FALSE | Downregulated |
| 0,2127851 | 0,554711265 | SNHG12 | FALSE | Upregulated |
| 0,285273731 | 0,55501443 | LOC101928595 | FALSE | Upregulated |
| -0,199394842 | 0,555059202 | DEPDC4 | FALSE | Downregulated |
| 0,877467856 | 0,555118729 | PROKR2 | FALSE | Upregulated |
| 0,313125603 | 0,555159192 | CCDC135 | FALSE | Upregulated |
| -0,510448309 | 0,555319231 | GS1-259H13.2 | FALSE | Downregulated |
| -0,079156607 | 0,555354108 | FNIP1 | FALSE | Downregulated |
| 0,786702528 | 0,555519871 | GRM2 | FALSE | Upregulated |
| 0,793662598 | 0,555519871 | HIST1H2AK | FALSE | Upregulated |
| 0,727355006 | 0,555519871 | PPT2-EGFL8 | FALSE | Upregulated |
| -0,747902309 | 0,555588776 | LOC100134368 | FALSE | Downregulated |
| 0,114019659 | 0,555693104 | AP4B1 | FALSE | Upregulated |
| -0,367210155 | 0,555693104 | C6orf163 | FALSE | Downregulated |
| -0,504155122 | 0,555693104 | CCDC39 | FALSE | Downregulated |
| -0,073842785 | 0,555693104 | CLN5 | FALSE | Downregulated |
| -0,719564445 | 0,555693104 | EMX2 | FALSE | Downregulated |
| 1,082531329 | 0,555693104 | ISL2 | FALSE | Upregulated |
| -0,119784429 | 0,555693104 | POLM | FALSE | Downregulated |
| 0,157832931 | 0,555693104 | RAD52 | FALSE | Upregulated |
| 0,103923753 | 0,555759671 | HSP90AB1 | FALSE | Upregulated |
| 0,103176541 | 0,555759671 | PTDSS2 | FALSE | Upregulated |
| 0,722189165 | 0,555759671 | SCN3B | FALSE | Upregulated |
| 1,131221498 | 0,555801815 | LOC102724621 | FALSE | Upregulated |
| -0,24184344 | 0,556002921 | GPR133 | FALSE | Downregulated |
| 0,379051836 | 0,556156919 | LOC728763 | FALSE | Upregulated |
| -0,166619552 | 0,556192935 | FGF14-AS2 | FALSE | Downregulated |
| -0,119617324 | 0,556193711 | DPM2 | FALSE | Downregulated |
| -0,178526536 | 0,556202611 | EZR | FALSE | Downregulated |

| | | | | |
|---|---|---|---|---|
| -0,43423749 | 0,556202611 | LINC00607 | FALSE | Downregulated |
| 0,080299622 | 0,556202611 | PEX2 | FALSE | Upregulated |
| 0,241077155 | 0,556202611 | PMS2P4 | FALSE | Upregulated |
| 0,579514778 | 0,556202611 | TMEM255A | FALSE | Upregulated |
| 0,097654251 | 0,556209406 | LOC100294145 | FALSE | Upregulated |
| -0,259909819 | 0,556246764 | TRNC | FALSE | Downregulated |
| 0,30298958 | 0,556346665 | GZMH | FALSE | Upregulated |
| 0,092102644 | 0,556378806 | MCU | FALSE | Upregulated |
| 0,114679543 | 0,55664188 | LRSAM1 | FALSE | Upregulated |
| -0,484333984 | 0,556790845 | COL6A4P2 | FALSE | Downregulated |
| -0,083839105 | 0,557013452 | TMF1 | FALSE | Downregulated |
| 0,134828971 | 0,557161141 | NSUN3 | FALSE | Upregulated |
| -0,089188059 | 0,557163689 | OCRL | FALSE | Downregulated |
| 0,133949698 | 0,557261893 | PMM2 | FALSE | Upregulated |
| -0,439722019 | 0,557268612 | LOC100131347 | FALSE | Downregulated |
| 0,074496252 | 0,557528722 | COMMD3-BMI1 | FALSE | Upregulated |
| 0,093612907 | 0,557528722 | GDI1 | FALSE | Upregulated |
| 1,084366542 | 0,557644713 | RNU4-1 | FALSE | Upregulated |
| 0,152610018 | 0,557783998 | USB1 | FALSE | Upregulated |
| -0,076727485 | 0,557985784 | ASGR2 | FALSE | Downregulated |
| 0,094415838 | 0,558035064 | DNAJC9 | FALSE | Upregulated |
| 0,934638442 | 0,558234231 | FZD10-AS1 | FALSE | Upregulated |
| -0,053306486 | 0,558254245 | IPO7 | FALSE | Downregulated |
| -0,23802889 | 0,558254245 | MCHR1 | FALSE | Downregulated |
| 0,081384618 | 0,558254245 | TGFBR1 | FALSE | Upregulated |
| -1,077942631 | 0,558404788 | LOC102723923 | FALSE | Downregulated |
| -0,117029763 | 0,558409383 | YLPM1 | FALSE | Downregulated |
| -0,19984499 | 0,558511897 | C15orf62 | FALSE | Downregulated |
| 0,117484486 | 0,558511897 | CREB3L2 | FALSE | Upregulated |
| 0,079110147 | 0,558511897 | FBL | FALSE | Upregulated |
| -0,402728894 | 0,558511897 | LOC100507670 | FALSE | Downregulated |
| 0,294178397 | 0,558511897 | MAB21L3 | FALSE | Upregulated |
| 0,467453001 | 0,558511897 | MYLK-AS1 | FALSE | Upregulated |
| 0,119783591 | 0,558511897 | PPARG | FALSE | Upregulated |
| 0,109331948 | 0,558511897 | TRIM65 | FALSE | Upregulated |
| -0,087380114 | 0,558538359 | RNF8 | FALSE | Downregulated |
| 0,271370186 | 0,558564013 | LOC101928192 | FALSE | Upregulated |
| -1,016477114 | 0,558636651 | LINC00515 | FALSE | Downregulated |
| -0,283668987 | 0,558801037 | ACE2 | FALSE | Downregulated |
| -0,880934026 | 0,55880706 | SPIN2A | FALSE | Downregulated |
| 0,216917408 | 0,55880706 | ZNF547 | FALSE | Upregulated |
| -0,193500339 | 0,558923575 | TTBK1 | FALSE | Downregulated |
| -0,904884175 | 0,559083415 | TFAP2A-AS1 | FALSE | Downregulated |
| 0,84414617 | 0,559103046 | DNM1P50 | FALSE | Upregulated |
| -0,245208905 | 0,559171509 | LOC101927989 | FALSE | Downregulated |
| -0,331513156 | 0,559331878 | LOC102723965 | FALSE | Downregulated |
| -0,190899276 | 0,559402927 | MVK | FALSE | Downregulated |
| -0,113349607 | 0,559457061 | CMSS1 | FALSE | Downregulated |
| 0,115621297 | 0,559457061 | GCGR | FALSE | Upregulated |
| 0,116996957 | 0,559457061 | MET | FALSE | Upregulated |
| -0,099018804 | 0,559457061 | PRDM2 | FALSE | Downregulated |

| | | | | |
|---|---|---|---|---|
| 0,134313005 | 0,559654428 | ZNF251 | FALSE | Upregulated |
| -0,124700449 | 0,559730425 | MED31 | FALSE | Downregulated |
| 0,372103828 | 0,559730425 | NCR3 | FALSE | Upregulated |
| 0,561155696 | 0,559760504 | ST7-OT4 | FALSE | Upregulated |
| 0,095342579 | 0,559802742 | RILP | FALSE | Upregulated |
| -0,097458873 | 0,559802742 | TTC31 | FALSE | Downregulated |
| 0,672633635 | 0,55989667 | C19orf59 | FALSE | Upregulated |
| -0,280926431 | 0,559980349 | LOC100126582 | FALSE | Downregulated |
| -0,282598338 | 0,560161657 | LOC101928249 | FALSE | Downregulated |
| -0,080245985 | 0,560194838 | KCTD21 | FALSE | Downregulated |
| 0,364129795 | 0,560219821 | SOCS2 | FALSE | Upregulated |
| -0,559759114 | 0,560947545 | COL19A1 | FALSE | Downregulated |
| -0,143863516 | 0,561100136 | KLHDC9 | FALSE | Downregulated |
| -0,11393845 | 0,561100136 | LINC00094 | FALSE | Downregulated |
| -0,115072123 | 0,561136641 | OVCA2 | FALSE | Downregulated |
| 0,143341599 | 0,561397859 | TUBG2 | FALSE | Upregulated |
| 0,216814191 | 0,561487773 | LOC102724129 | FALSE | Upregulated |
| 0,708917973 | 0,561819059 | SCRG1 | FALSE | Upregulated |
| 0,154078085 | 0,561885237 | ANK3 | FALSE | Upregulated |
| -0,171573022 | 0,561885237 | FAM35DP | FALSE | Downregulated |
| 0,253250651 | 0,561885237 | INHBB | FALSE | Upregulated |
| 0,416170745 | 0,561885237 | ZNF812 | FALSE | Upregulated |
| -0,198398005 | 0,562030679 | CA11 | FALSE | Downregulated |
| 0,112391579 | 0,56224858 | ABCB6 | FALSE | Upregulated |
| 0,073781945 | 0,56224858 | NNT | FALSE | Upregulated |
| -0,093353353 | 0,562358067 | PXMP4 | FALSE | Downregulated |
| 0,502133027 | 0,562409159 | SYCE2 | FALSE | Upregulated |
| -0,0821729 | 0,562648967 | PAPD4 | FALSE | Downregulated |
| 0,417023414 | 0,562652666 | RHBDL2 | FALSE | Upregulated |
| 0,479617024 | 0,562669732 | ZEB2-AS1 | FALSE | Upregulated |
| -0,066170764 | 0,563049708 | MYNN | FALSE | Downregulated |
| 0,069589817 | 0,563091325 | CBX5 | FALSE | Upregulated |
| -0,799714424 | 0,563123998 | LOC101928738 | FALSE | Downregulated |
| 0,475187064 | 0,563189892 | ZNF843 | FALSE | Upregulated |
| -0,15340529 | 0,563282971 | PLEKHH3 | FALSE | Downregulated |
| -0,1421595 | 0,563315834 | PRH1-PRR4 | FALSE | Downregulated |
| 0,136779323 | 0,563729975 | ATG4D | FALSE | Upregulated |
| 0,076603854 | 0,563729975 | PLCL2 | FALSE | Upregulated |
| 0,053783128 | 0,563729975 | STK25 | FALSE | Upregulated |
| -0,25666515 | 0,563956579 | KLHL4 | FALSE | Downregulated |
| 0,586945942 | 0,563956579 | LOC646670 | FALSE | Upregulated |
| 0,089305259 | 0,563956579 | PPIP5K1 | FALSE | Upregulated |
| -0,1622267 | 0,563956579 | RAD9A | FALSE | Downregulated |
| 0,0489687 | 0,563956579 | UBE4B | FALSE | Upregulated |
| -0,067194773 | 0,564080012 | TMEM184C | FALSE | Downregulated |
| -0,637113439 | 0,564187356 | NPIPB8 | FALSE | Downregulated |
| 0,095871829 | 0,564214981 | ABCD3 | FALSE | Upregulated |
| 0,929181463 | 0,564214981 | AC096559.1 | FALSE | Upregulated |
| -0,675410538 | 0,564214981 | CORO6 | FALSE | Downregulated |
| 0,099972114 | 0,564214981 | TRIQK | FALSE | Upregulated |
| -0,147705685 | 0,564214981 | ZNF214 | FALSE | Downregulated |

| | | | | |
|---|---|---|---|---|
| 0,540877117 | 0,564260409 | CATSPERB | FALSE | Upregulated |
| -0,113467167 | 0,564330986 | RMND5A | FALSE | Downregulated |
| 0,778353459 | 0,564383099 | GTSE1-AS1 | FALSE | Upregulated |
| 0,509901411 | 0,564438121 | MAGI1-IT1 | FALSE | Upregulated |
| 0,211899507 | 0,564438121 | PCSK9 | FALSE | Upregulated |
| 0,121028112 | 0,564438121 | RINL | FALSE | Upregulated |
| -0,104593032 | 0,564439563 | DDX17 | FALSE | Downregulated |
| -0,528958787 | 0,564439563 | INA | FALSE | Downregulated |
| -0,066807368 | 0,564439563 | RING1 | FALSE | Downregulated |
| -0,102436708 | 0,564659854 | FPGS | FALSE | Downregulated |
| -0,459541628 | 0,564659854 | USP32P2 | FALSE | Downregulated |
| -0,783438199 | 0,564724606 | MIR4767 | FALSE | Downregulated |
| 0,109294389 | 0,564754428 | MYH10 | FALSE | Upregulated |
| 0,626675931 | 0,564934809 | DUSP15 | FALSE | Upregulated |
| 0,605879747 | 0,564970906 | CHMP1B2P | FALSE | Upregulated |
| -0,06909426 | 0,565162951 | NOL7 | FALSE | Downregulated |
| 0,995914872 | 0,565256275 | LOC100507403 | FALSE | Upregulated |
| 0,138569257 | 0,565256275 | PSMG3 | FALSE | Upregulated |
| 0,144545818 | 0,565331706 | TMX3 | FALSE | Upregulated |
| 0,35580953 | 0,565431633 | C3orf35 | FALSE | Upregulated |
| 0,135203708 | 0,565431633 | GFOD1 | FALSE | Upregulated |
| 0,325685065 | 0,565431633 | POLR2J2 | FALSE | Upregulated |
| -0,102271392 | 0,565431633 | TPRA1 | FALSE | Downregulated |
| 0,177621658 | 0,565670442 | TRIM35 | FALSE | Upregulated |
| 0,106321802 | 0,56600297 | SNX21 | FALSE | Upregulated |
| -0,703915746 | 0,566056868 | LGALS8-AS1 | FALSE | Downregulated |
| 0,196796468 | 0,566121145 | CCDC152 | FALSE | Upregulated |
| 0,775405428 | 0,566347577 | LOC102723305 | FALSE | Upregulated |
| 0,071421278 | 0,566347577 | OTUD7B | FALSE | Upregulated |
| 0,10811345 | 0,566417831 | BCL7A | FALSE | Upregulated |
| 0,159588729 | 0,566417831 | GLIDR | FALSE | Upregulated |
| -0,260034084 | 0,566417831 | UBE2QL1 | FALSE | Downregulated |
| 0,085082744 | 0,566421615 | MID2 | FALSE | Upregulated |
| -0,064501105 | 0,566444451 | AKT2 | FALSE | Downregulated |
| 0,914945123 | 0,566444451 | MOBP | FALSE | Upregulated |
| -0,178372471 | 0,566444451 | RP11-95P2.1 | FALSE | Downregulated |
| -0,144997407 | 0,566454223 | BDNF-AS | FALSE | Downregulated |
| 0,126177088 | 0,566454223 | EMC9 | FALSE | Upregulated |
| 0,128529221 | 0,566454223 | FAM101B | FALSE | Upregulated |
| -0,533679438 | 0,566454223 | LOC101930310 | FALSE | Downregulated |
| 0,373187343 | 0,566454223 | LSMEM2 | FALSE | Upregulated |
| 0,521734234 | 0,566454223 | RAB3C | FALSE | Upregulated |
| 0,132633613 | 0,566509614 | NEK3 | FALSE | Upregulated |
| -0,105309325 | 0,566584696 | MDH2 | FALSE | Downregulated |
| -0,111813678 | 0,566813416 | ZSWIM6 | FALSE | Downregulated |
| 0,176397529 | 0,566927554 | GTF2IRD1 | FALSE | Upregulated |
| 0,81568953 | 0,567101828 | LOC102724875 | FALSE | Upregulated |
| 1,01907475 | 0,56734801 | FOXD1 | FALSE | Upregulated |
| -0,084559483 | 0,56734801 | PRMT3 | FALSE | Downregulated |
| 0,097994217 | 0,56813285 | ARFIP2 | FALSE | Upregulated |
| -0,199483834 | 0,56813285 | PAXBP1-AS1 | FALSE | Downregulated |

| | | | | |
|---|---|---|---|---|
| 0,191674937 | 0,56813285 | S100A13 | FALSE | Upregulated |
| 0,136948943 | 0,56823926 | TMEM117 | FALSE | Upregulated |
| -0,813944253 | 0,568325272 | LOC102724205 | FALSE | Downregulated |
| 0,070927503 | 0,568325272 | MCMBP | FALSE | Upregulated |
| -0,06680716 | 0,568358973 | FAM134C | FALSE | Downregulated |
| -0,595292387 | 0,568358973 | SPDYE1 | FALSE | Downregulated |
| 0,356272615 | 0,568538761 | SEC14L1P1 | FALSE | Upregulated |
| -0,163241308 | 0,568573796 | GAREM | FALSE | Downregulated |
| 0,126285665 | 0,568645098 | CYB561D2 | FALSE | Upregulated |
| 0,238992307 | 0,568645098 | LINC00865 | FALSE | Upregulated |
| -0,074461889 | 0,568672648 | KDM4C | FALSE | Downregulated |
| 0,396167136 | 0,569081889 | FAM81A | FALSE | Upregulated |
| 1,145501921 | 0,569081889 | FLJ36000 | FALSE | Upregulated |
| -0,144458142 | 0,569269344 | ABCA8 | FALSE | Downregulated |
| -0,080138999 | 0,569269344 | KDSR | FALSE | Downregulated |
| -0,122343585 | 0,569401343 | NCOR1 | FALSE | Downregulated |
| 0,298302264 | 0,569480066 | TSSK6 | FALSE | Upregulated |
| 0,16659119 | 0,569762615 | LAMA4 | FALSE | Upregulated |
| 0,182962537 | 0,569782879 | MBOAT2 | FALSE | Upregulated |
| 0,174254219 | 0,569782879 | PCDH18 | FALSE | Upregulated |
| 0,118795851 | 0,569782879 | TET2 | FALSE | Upregulated |
| 0,286494233 | 0,570006975 | IGSF9 | FALSE | Upregulated |
| 0,160567823 | 0,570687836 | LOC100505812 | FALSE | Upregulated |
| 0,074581852 | 0,570687836 | VPS25 | FALSE | Upregulated |
| -0,180926987 | 0,570967268 | ARHGAP5-AS1 | FALSE | Downregulated |
| -0,547708534 | 0,570967268 | KISS1 | FALSE | Downregulated |
| -0,141767497 | 0,571062741 | ELTD1 | FALSE | Downregulated |
| -0,148348934 | 0,571113581 | TUFT1 | FALSE | Downregulated |
| -0,07089975 | 0,571242805 | ILKAP | FALSE | Downregulated |
| 0,209601834 | 0,571453743 | SUV420H2 | FALSE | Upregulated |
| -0,454057594 | 0,571533592 | DLGAP3 | FALSE | Downregulated |
| -0,741381244 | 0,571621356 | LINC00944 | FALSE | Downregulated |
| 0,694140549 | 0,571705735 | TPH1 | FALSE | Upregulated |
| -0,242262542 | 0,572027782 | ACRC | FALSE | Downregulated |
| -0,135824337 | 0,572158423 | ARHGAP32 | FALSE | Downregulated |
| -1,154458701 | 0,572158423 | KCNA7 | FALSE | Downregulated |
| -0,097166806 | 0,572207596 | RCOR3 | FALSE | Downregulated |
| 0,364627317 | 0,572246967 | PDZD9 | FALSE | Upregulated |
| 0,127083733 | 0,572359216 | CCDC57 | FALSE | Upregulated |
| -0,333283542 | 0,572535048 | CACNB2 | FALSE | Downregulated |
| -0,096337699 | 0,572829271 | PCSK6 | FALSE | Downregulated |
| 0,235597111 | 0,572997237 | LOC100133091 | FALSE | Upregulated |
| 0,14300814 | 0,573385525 | ARHGEF15 | FALSE | Upregulated |
| 0,067460816 | 0,573499287 | STAU2 | FALSE | Upregulated |
| -0,055346742 | 0,573519025 | FEM1B | FALSE | Downregulated |
| 0,114602851 | 0,573521521 | NCAPG2 | FALSE | Upregulated |
| 0,273796219 | 0,573521521 | PODN | FALSE | Upregulated |
| 0,134093898 | 0,573521521 | SMG1 | FALSE | Upregulated |
| 0,232439515 | 0,573563706 | ADAMTSL1 | FALSE | Upregulated |
| -0,697927146 | 0,573577474 | SLC8A3 | FALSE | Downregulated |
| -0,068448792 | 0,573577474 | ZXDC | FALSE | Downregulated |

| | | | | |
|---|---|---|---|---|
| 0,109635264 | 0,57371654 | RECQL5 | FALSE | Upregulated |
| 0,184572576 | 0,57371654 | SLPI | FALSE | Upregulated |
| 0,06218697 | 0,573758369 | C8orf33 | FALSE | Upregulated |
| 0,324887261 | 0,573758369 | MMP25 | FALSE | Upregulated |
| -0,076662282 | 0,573818009 | ZNF274 | FALSE | Downregulated |
| -0,190944606 | 0,573966932 | CCDC34 | FALSE | Downregulated |
| 0,805977554 | 0,573966932 | GOLGA8K | FALSE | Upregulated |
| -0,14647314 | 0,573966932 | PLA1A | FALSE | Downregulated |
| 0,416722803 | 0,573966932 | STARD4-AS1 | FALSE | Upregulated |
| 0,111808837 | 0,573966932 | TOP3B | FALSE | Upregulated |
| 0,301091636 | 0,573987555 | ALPK3 | FALSE | Upregulated |
| 0,89834127 | 0,573987555 | HIPK4 | FALSE | Upregulated |
| 0,243617128 | 0,573987555 | NPY5R | FALSE | Upregulated |
| -0,074829362 | 0,573987555 | VRK3 | FALSE | Downregulated |
| 0,203678371 | 0,573987555 | ZNF514 | FALSE | Upregulated |
| 0,371130681 | 0,574004628 | LOC441666 | FALSE | Upregulated |
| -0,220764973 | 0,574004628 | PRR18 | FALSE | Downregulated |
| 0,420462267 | 0,574049946 | UGT3A2 | FALSE | Upregulated |
| -0,870480426 | 0,57419404 | LOC102724177 | FALSE | Downregulated |
| 0,107684013 | 0,57419404 | ZNF576 | FALSE | Upregulated |
| -0,154775722 | 0,574242252 | LOC100506518 | FALSE | Downregulated |
| 0,671957748 | 0,574248668 | LOC102723697 | FALSE | Upregulated |
| -0,913543117 | 0,574248668 | RCAN3AS | FALSE | Downregulated |
| -0,055318688 | 0,574248668 | SHOC2 | FALSE | Downregulated |
| -0,212989833 | 0,574248668 | ZDHHC19 | FALSE | Downregulated |
| 0,623219226 | 0,57429824 | HOXA10 | FALSE | Upregulated |
| 0,047006431 | 0,574306724 | RAB21 | FALSE | Upregulated |
| 0,109513046 | 0,574447942 | STK4 | FALSE | Upregulated |
| 0,094007512 | 0,574791206 | RBBP9 | FALSE | Upregulated |
| 0,169690321 | 0,574993736 | ATP2B2 | FALSE | Upregulated |
| -0,135126463 | 0,575073766 | SLC5A3 | FALSE | Downregulated |
| 0,458810145 | 0,57513202 | LOC729732 | FALSE | Upregulated |
| -0,412050994 | 0,57513264 | LOC101929953 | FALSE | Downregulated |
| 0,078676069 | 0,57513264 | MTMR1 | FALSE | Upregulated |
| 0,717926608 | 0,575191589 | NOG | FALSE | Upregulated |
| 0,116046669 | 0,575282167 | FANCL | FALSE | Upregulated |
| 0,728014739 | 0,57528753 | LOC91450 | FALSE | Upregulated |
| 0,144932878 | 0,575296634 | RTP3 | FALSE | Upregulated |
| 0,538469265 | 0,575311009 | CA9 | FALSE | Upregulated |
| -0,053627635 | 0,575570965 | VTA1 | FALSE | Downregulated |
| -0,059021853 | 0,575590469 | CCDC117 | FALSE | Downregulated |
| -0,316700976 | 0,575590469 | P2RX6 | FALSE | Downregulated |
| 0,151496442 | 0,575590469 | RHOJ | FALSE | Upregulated |
| -0,374219917 | 0,575600194 | AC009473.1 | FALSE | Downregulated |
| 0,511309323 | 0,575609337 | ST3GAL4-AS1 | FALSE | Upregulated |
| -0,077479737 | 0,575648562 | MEPCE | FALSE | Downregulated |
| 0,568194071 | 0,575774394 | PTPRG-AS1 | FALSE | Upregulated |
| 0,110857745 | 0,575822031 | HIF1A | FALSE | Upregulated |
| 0,351268653 | 0,575822031 | MTL5 | FALSE | Upregulated |
| -0,084097088 | 0,575850419 | ATXN3 | FALSE | Downregulated |
| -0,120984067 | 0,576120236 | TMEM143 | FALSE | Downregulated |

| | | | | |
|---|---|---|---|---|
| 0,085956171 | 0,576120236 | WDFY3 | FALSE | Upregulated |
| -0,314675181 | 0,576239556 | NXF3 | FALSE | Downregulated |
| -0,193000925 | 0,576352414 | NUCB2 | FALSE | Downregulated |
| -0,291506536 | 0,576380428 | FAM86HP | FALSE | Downregulated |
| 0,096791233 | 0,576531177 | PRMT7 | FALSE | Upregulated |
| -0,462262106 | 0,576602463 | LOC101928803 | FALSE | Downregulated |
| -0,218532632 | 0,576605462 | B9D2 | FALSE | Downregulated |
| -1,88266236 | 0,576620766 | LOC102724594 | FALSE | Downregulated |
| -0,049430458 | 0,576738705 | VPS37A | FALSE | Downregulated |
| 0,743083775 | 0,576760321 | GCSAML | FALSE | Upregulated |
| -0,247845595 | 0,576906482 | CCNJL | FALSE | Downregulated |
| 0,394926885 | 0,576906482 | FAM166B | FALSE | Upregulated |
| 0,198893297 | 0,576906482 | GALNT14 | FALSE | Upregulated |
| 0,121240908 | 0,576906482 | HAUS1 | FALSE | Upregulated |
| 0,357876234 | 0,576914584 | GPX2 | FALSE | Upregulated |
| 0,125806379 | 0,576988465 | NEIL2 | FALSE | Upregulated |
| 0,134883199 | 0,576988465 | RPL22L1 | FALSE | Upregulated |
| -0,802859262 | 0,576992311 | SCN2B | FALSE | Downregulated |
| -0,150070366 | 0,577063637 | LOC102606465 | FALSE | Downregulated |
| 0,812761135 | 0,577136643 | LOC102724342 | FALSE | Upregulated |
| -0,28977238 | 0,577306455 | RAD21-AS1 | FALSE | Downregulated |
| 0,242577005 | 0,577534798 | GCSAM | FALSE | Upregulated |
| 0,069216365 | 0,577534798 | GTF3C5 | FALSE | Upregulated |
| -0,135799288 | 0,577934133 | GET4 | FALSE | Downregulated |
| -0,061965071 | 0,577989183 | NAA20 | FALSE | Downregulated |
| 0,093370526 | 0,578017479 | MXD1 | FALSE | Upregulated |
| 0,2547516 | 0,578049652 | GPR37 | FALSE | Upregulated |
| -0,08962647 | 0,578049652 | KCNJ8 | FALSE | Downregulated |
| 0,138021362 | 0,578049652 | SNRNP70 | FALSE | Upregulated |
| 0,605246665 | 0,578098386 | TUBA4B | FALSE | Upregulated |
| -0,765314924 | 0,578241869 | LOC101928668 | FALSE | Downregulated |
| -0,30632616 | 0,578450905 | LOC101927057 | FALSE | Downregulated |
| 0,084807841 | 0,578732702 | RIF1 | FALSE | Upregulated |
| 0,275098443 | 0,578734345 | PRKCE | FALSE | Upregulated |
| -0,253796726 | 0,578735573 | SEMA6C | FALSE | Downregulated |
| -0,096799712 | 0,578898655 | INADL | FALSE | Downregulated |
| 0,741513769 | 0,579233084 | LOC730338 | FALSE | Upregulated |
| 0,687236783 | 0,579299023 | DNASE2B | FALSE | Upregulated |
| 0,11704176 | 0,579542191 | ZKSCAN4 | FALSE | Upregulated |
| 0,122901136 | 0,579680176 | STAT2 | FALSE | Upregulated |
| -0,071206166 | 0,57976407 | DNAJC24 | FALSE | Downregulated |
| -0,223964072 | 0,57976407 | NREP | FALSE | Downregulated |
| 0,386964348 | 0,580330329 | LOC102723328 | FALSE | Upregulated |
| 0,128723059 | 0,580410941 | SLC9A9 | FALSE | Upregulated |
| -0,105631936 | 0,58044962 | SLC25A10 | FALSE | Downregulated |
| -0,265637359 | 0,580608132 | NPIPB11 | FALSE | Downregulated |
| -0,425777142 | 0,580608132 | RP11-142C4.6 | FALSE | Downregulated |
| -0,363197937 | 0,580608132 | SPATA32 | FALSE | Downregulated |
| 0,082486446 | 0,580633794 | SORBS2 | FALSE | Upregulated |
| -0,085775358 | 0,580633794 | ZNF649 | FALSE | Downregulated |
| 0,11252775 | 0,581015416 | ANKRD6 | FALSE | Upregulated |

| | | | | |
|---|---|---|---|---|
| 0,347988078 | 0,581086057 | LOC101928806 | FALSE | Upregulated |
| 0,549899245 | 0,581108323 | C1orf185 | FALSE | Upregulated |
| -0,767146894 | 0,581108323 | GRM3 | FALSE | Downregulated |
| -0,104531984 | 0,581290964 | POLE4 | FALSE | Downregulated |
| 0,31328858 | 0,581290964 | SNHG5 | FALSE | Upregulated |
| 0,440851969 | 0,58131205 | ASIC1 | FALSE | Upregulated |
| -0,068979811 | 0,581406281 | RBM7 | FALSE | Downregulated |
| 0,554363269 | 0,58159541 | HIST1H2BG | FALSE | Upregulated |
| -0,346740397 | 0,581625823 | LOC100129722 | FALSE | Downregulated |
| -0,691237283 | 0,581625823 | LOC102723709 | FALSE | Downregulated |
| 2,159204081 | 0,581625823 | SLC6A11 | FALSE | Upregulated |
| -0,151252852 | 0,581740156 | ECE2 | FALSE | Downregulated |
| 0,160060422 | 0,581740156 | TSGA10 | FALSE | Upregulated |
| -0,07045953 | 0,581827854 | RALBP1 | FALSE | Downregulated |
| -0,894744431 | 0,581830732 | ZIM2 | FALSE | Downregulated |
| -1,006516886 | 0,582001642 | LEP | FALSE | Downregulated |
| -0,201507902 | 0,582023723 | CABP4 | FALSE | Downregulated |
| -0,139296598 | 0,582050765 | EIF2AK2 | FALSE | Downregulated |
| -0,069996203 | 0,582053118 | WRNIP1 | FALSE | Downregulated |
| 0,298393959 | 0,582158561 | BEND5 | FALSE | Upregulated |
| -0,138695478 | 0,582351127 | BAALC | FALSE | Downregulated |
| 0,105149909 | 0,582743023 | ZDHHC21 | FALSE | Upregulated |
| -0,201011829 | 0,582769839 | STL | FALSE | Downregulated |
| -0,412443837 | 0,582936199 | C19orf26 | FALSE | Downregulated |
| 0,649406897 | 0,582936199 | LOC100996696 | FALSE | Upregulated |
| -0,063031628 | 0,582936199 | PSME4 | FALSE | Downregulated |
| -0,266224261 | 0,582980085 | CLEC1A | FALSE | Downregulated |
| -0,815046153 | 0,582980085 | LOC401098 | FALSE | Downregulated |
| -0,141750615 | 0,583243566 | CRIPAK | FALSE | Downregulated |
| 0,989341208 | 0,583243566 | OR7D2 | FALSE | Upregulated |
| -0,279232879 | 0,583243566 | SNX29P2 | FALSE | Downregulated |
| 0,253569105 | 0,583267906 | ANO7 | FALSE | Upregulated |
| 0,066980694 | 0,583310624 | ZNF41 | FALSE | Upregulated |
| 0,476147407 | 0,583435018 | C9orf66 | FALSE | Upregulated |
| 0,472394101 | 0,583468733 | RNASE7 | FALSE | Upregulated |
| 0,486608481 | 0,583468733 | SLC26A3 | FALSE | Upregulated |
| 0,101669679 | 0,583497847 | CNOT6 | FALSE | Upregulated |
| -0,360936586 | 0,583512295 | RNA45S5 | FALSE | Downregulated |
| -1,570596932 | 0,583512295 | SMTNL1 | FALSE | Downregulated |
| 0,220770577 | 0,58366466 | CTSW | FALSE | Upregulated |
| -0,139703516 | 0,58366466 | DACH1 | FALSE | Downregulated |
| 0,066227962 | 0,58366466 | MKNK1 | FALSE | Upregulated |
| -0,082366137 | 0,58366466 | PDZK1 | FALSE | Downregulated |
| 0,545535381 | 0,583701495 | EEF1DP3 | FALSE | Upregulated |
| -0,091761838 | 0,583701495 | FURIN | FALSE | Downregulated |
| -0,728884085 | 0,583701495 | LOC102723561 | FALSE | Downregulated |
| -0,189229786 | 0,583701495 | MAN1C1 | FALSE | Downregulated |
| 0,525168623 | 0,583701495 | TMC5 | FALSE | Upregulated |
| 0,079637164 | 0,583701495 | TPRG1L | FALSE | Upregulated |
| -0,737128385 | 0,583825744 | NBEAP1 | FALSE | Downregulated |
| -0,106922514 | 0,583825744 | TACC2 | FALSE | Downregulated |

| | | | | |
|---|---|---|---|---|
| -0,096613722 | 0,583825744 | THAP4 | FALSE | Downregulated |
| -0,616270597 | 0,583839476 | WNK4 | FALSE | Downregulated |
| 0,1578409 | 0,583879248 | FOLR2 | FALSE | Upregulated |
| -0,138672476 | 0,583879248 | GSTA4 | FALSE | Downregulated |
| -0,28056246 | 0,583931361 | LOC102724345 | FALSE | Downregulated |
| 0,493376795 | 0,583931361 | ST7-OT3 | FALSE | Upregulated |
| 0,387420852 | 0,583975756 | LINC00965 | FALSE | Upregulated |
| 0,073235988 | 0,584000615 | C7orf25 | FALSE | Upregulated |
| 0,612066158 | 0,584132831 | RP11-840I19.3 | FALSE | Upregulated |
| -0,168436677 | 0,584271753 | IFI27 | FALSE | Downregulated |
| 0,564278451 | 0,584333937 | XKR6 | FALSE | Upregulated |
| 0,15278264 | 0,584346012 | ZSCAN5A | FALSE | Upregulated |
| 0,49370402 | 0,584441502 | ZNF215 | FALSE | Upregulated |
| 0,695495777 | 0,584489011 | GSDMC | FALSE | Upregulated |
| 0,106048373 | 0,584497636 | KDM2B | FALSE | Upregulated |
| 0,250818512 | 0,584866272 | FASN | FALSE | Upregulated |
| -0,251249195 | 0,584866272 | INSIG1 | FALSE | Downregulated |
| -0,14804023 | 0,585582163 | ANG | FALSE | Downregulated |
| 0,456254771 | 0,585597748 | CRABP2 | FALSE | Upregulated |
| 0,059772215 | 0,585765378 | RLIM | FALSE | Upregulated |
| -0,722606238 | 0,585812041 | RIMBP2 | FALSE | Downregulated |
| 0,335315872 | 0,585927485 | RP11-420A23.1 | FALSE | Upregulated |
| 0,142748279 | 0,5859563 | WSB1 | FALSE | Upregulated |
| 0,20286609 | 0,586884053 | CCDC96 | FALSE | Upregulated |
| -0,326007142 | 0,586884053 | NPW | FALSE | Downregulated |
| 0,067340878 | 0,587026747 | ADCY6 | FALSE | Upregulated |
| -0,177012808 | 0,587228077 | WEE2-AS1 | FALSE | Downregulated |
| -0,604742404 | 0,587396998 | TCTE1 | FALSE | Downregulated |
| -0,556060137 | 0,587435961 | LOC102723505 | FALSE | Downregulated |
| -0,107774247 | 0,58754614 | LDB2 | FALSE | Downregulated |
| 0,549484077 | 0,58754614 | TMEM72-AS1 | FALSE | Upregulated |
| 0,081160893 | 0,587581107 | TBCK | FALSE | Upregulated |
| -0,082121504 | 0,587630461 | FOXJ2 | FALSE | Downregulated |
| -0,176488338 | 0,587630461 | ZNF540 | FALSE | Downregulated |
| 0,719730881 | 0,587889588 | RP11-446H18.5 | FALSE | Upregulated |
| 0,06249156 | 0,587902151 | DEDD | FALSE | Upregulated |
| -0,263683691 | 0,587902151 | LOC101928274 | FALSE | Downregulated |
| 0,064334911 | 0,588022572 | APTX | FALSE | Upregulated |
| 0,111738319 | 0,588036078 | FAM20A | FALSE | Upregulated |
| 0,631573449 | 0,58808313 | PTPRVP | FALSE | Upregulated |
| -0,218429246 | 0,588479172 | GGACT | FALSE | Downregulated |
| 0,754249529 | 0,588479172 | KRT85 | FALSE | Upregulated |
| 0,423904956 | 0,588485851 | IDI2-AS1 | FALSE | Upregulated |
| -0,134490716 | 0,588770316 | GEN1 | FALSE | Downregulated |
| 0,581589818 | 0,588770316 | LOC100996557 | FALSE | Upregulated |
| -0,143454199 | 0,588770316 | MAP3K5 | FALSE | Downregulated |
| 0,153796789 | 0,588878764 | MOK | FALSE | Upregulated |
| -0,210895007 | 0,588878764 | MYL6B | FALSE | Downregulated |
| -0,073376096 | 0,588878764 | ZNF280D | FALSE | Downregulated |
| 0,089349658 | 0,589096699 | FLCN | FALSE | Upregulated |
| 0,521227115 | 0,589096699 | RAB9B | FALSE | Upregulated |

| | | | | |
|---|---|---|---|---|
| -0,060297865 | 0,589163134 | TMEM167A | FALSE | Downregulated |
| 0,120811695 | 0,589359053 | RPIA | FALSE | Upregulated |
| 0,822373256 | 0,589710512 | MMP8 | FALSE | Upregulated |
| -0,191384345 | 0,589710512 | SPIRE1 | FALSE | Downregulated |
| -0,076362896 | 0,589900493 | KIF1B | FALSE | Downregulated |
| -0,125727667 | 0,590027888 | IL1R2 | FALSE | Downregulated |
| 0,768589718 | 0,590058263 | LOC101927875 | FALSE | Upregulated |
| 0,777669137 | 0,590637242 | TM4SF19-AS1 | FALSE | Upregulated |
| -0,38675078 | 0,5906937 | CLEC4GP1 | FALSE | Downregulated |
| 0,094774451 | 0,5906937 | DUSP7 | FALSE | Upregulated |
| 0,066553277 | 0,5906937 | JMJD8 | FALSE | Upregulated |
| 0,093004273 | 0,5906937 | LEMD2 | FALSE | Upregulated |
| -0,093776709 | 0,590848841 | CLIC4 | FALSE | Downregulated |
| -0,108367874 | 0,590876084 | TEC | FALSE | Downregulated |
| -0,124218859 | 0,590911501 | ACOT4 | FALSE | Downregulated |
| 0,119665171 | 0,590911501 | ARFGAP3 | FALSE | Upregulated |
| -0,663738377 | 0,590911501 | BARX1 | FALSE | Downregulated |
| 0,554403426 | 0,590911501 | LOC101928562 | FALSE | Upregulated |
| 0,710530614 | 0,590974376 | RP11-508P1.2 | FALSE | Upregulated |
| -0,24074492 | 0,591446781 | ASAH2 | FALSE | Downregulated |
| 0,099247852 | 0,591446781 | PARP14 | FALSE | Upregulated |
| -0,104010578 | 0,591446781 | SGK3 | FALSE | Downregulated |
| 0,122326166 | 0,591662275 | C15orf40 | FALSE | Upregulated |
| -0,078274425 | 0,591662275 | PRRG1 | FALSE | Downregulated |
| -0,080587762 | 0,591674744 | ARHGAP29 | FALSE | Downregulated |
| 0,119300633 | 0,591674744 | GYS1 | FALSE | Upregulated |
| -0,21054735 | 0,591674744 | RP11-15I11.3 | FALSE | Downregulated |
| 0,154132368 | 0,591674744 | ZNF620 | FALSE | Upregulated |
| 0,948743886 | 0,592066055 | LOC100507156 | FALSE | Upregulated |
| 0,134910377 | 0,592288617 | ATP10D | FALSE | Upregulated |
| -0,529500294 | 0,592288617 | LOC101927044 | FALSE | Downregulated |
| -0,1062248 | 0,592304915 | LMBRD2 | FALSE | Downregulated |
| 0,585503364 | 0,592304915 | NHLRC4 | FALSE | Upregulated |
| 0,098857893 | 0,592657614 | C14orf142 | FALSE | Upregulated |
| 0,103473869 | 0,592752107 | GPATCH2L | FALSE | Upregulated |
| -0,113703179 | 0,592910448 | PHF21A | FALSE | Downregulated |
| 0,498158745 | 0,593196943 | LOC102723581 | FALSE | Upregulated |
| -0,837210739 | 0,593196943 | RGPD4-AS1 | FALSE | Downregulated |
| 0,75468711 | 0,593349019 | H2BFXP | FALSE | Upregulated |
| 0,51469614 | 0,593493301 | POM121L8P | FALSE | Upregulated |
| 0,132621798 | 0,593543139 | PYROXD1 | FALSE | Upregulated |
| -0,109277022 | 0,593550523 | FRAT2 | FALSE | Downregulated |
| -0,200445787 | 0,593550523 | LOC100129461 | FALSE | Downregulated |
| -0,418099933 | 0,593550523 | MEDAG | FALSE | Downregulated |
| -0,05595071 | 0,593550523 | POLR3E | FALSE | Downregulated |
| 0,110874968 | 0,593550523 | TLR4 | FALSE | Upregulated |
| -0,100458689 | 0,593550523 | TRIM26 | FALSE | Downregulated |
| 2,089099185 | 0,593732575 | CRYAA | FALSE | Upregulated |
| 1,136138613 | 0,594127665 | POU6F2-AS2 | FALSE | Upregulated |
| 0,29032588 | 0,594147881 | CCDC13 | FALSE | Upregulated |
| 0,108609799 | 0,594147881 | ZNF34 | FALSE | Upregulated |

| | | | | |
|---|---|---|---|---|
| 0,098118452 | 0,594399933 | EEF1A1 | FALSE | Upregulated |
| -0,7341979 | 0,594399933 | FSCN2 | FALSE | Downregulated |
| 0,061309885 | 0,5944768 | GPATCH3 | FALSE | Upregulated |
| -0,079785227 | 0,594581177 | ISCA1 | FALSE | Downregulated |
| 0,138143026 | 0,594619727 | ARMCX6 | FALSE | Upregulated |
| 0,744157945 | 0,594619727 | LINC00941 | FALSE | Upregulated |
| 0,813211578 | 0,594619727 | SLC5A5 | FALSE | Upregulated |
| 0,117552524 | 0,594619727 | SNAPC4 | FALSE | Upregulated |
| 0,136487825 | 0,594670529 | PELI3 | FALSE | Upregulated |
| -0,085121712 | 0,594677849 | JAGN1 | FALSE | Downregulated |
| 0,593598624 | 0,594677849 | RP11-661P17.1 | FALSE | Upregulated |
| -0,591953585 | 0,594803453 | DKFZP434K028 | FALSE | Downregulated |
| -0,373730476 | 0,594803453 | LOC152225 | FALSE | Downregulated |
| -0,081833876 | 0,594803453 | PHB2 | FALSE | Downregulated |
| 0,075411128 | 0,594803453 | SAMM50 | FALSE | Upregulated |
| -0,873181476 | 0,594824063 | TUBA3D | FALSE | Downregulated |
| -0,189572336 | 0,594841626 | OTUD3 | FALSE | Downregulated |
| -0,129244634 | 0,594915073 | NAPEPLD | FALSE | Downregulated |
| -0,103009213 | 0,595064637 | CCDC125 | FALSE | Downregulated |
| -0,072234829 | 0,595122478 | TMCO4 | FALSE | Downregulated |
| -0,086208109 | 0,595178694 | LYSMD3 | FALSE | Downregulated |
| -0,260460049 | 0,595178694 | UPK3B | FALSE | Downregulated |
| 0,155678414 | 0,595287718 | TMPO | FALSE | Upregulated |
| -0,09960528 | 0,595347355 | TTC30A | FALSE | Downregulated |
| 0,103728332 | 0,595377654 | EIF4G3 | FALSE | Upregulated |
| 0,760118664 | 0,59544376 | LOC102723714 | FALSE | Upregulated |
| -0,192534478 | 0,595536622 | TNN | FALSE | Downregulated |
| 0,087940314 | 0,595596291 | ENOX2 | FALSE | Upregulated |
| -0,150293237 | 0,595658106 | FCER1G | FALSE | Downregulated |
| -0,31979891 | 0,595658106 | LOC102723647 | FALSE | Downregulated |
| 0,227210066 | 0,595658106 | NRG3 | FALSE | Upregulated |
| 0,534555591 | 0,595658106 | TMPRSS4 | FALSE | Upregulated |
| -0,129866947 | 0,595663603 | USP27X | FALSE | Downregulated |
| -0,066481129 | 0,595870079 | ZCCHC10 | FALSE | Downregulated |
| 0,875949018 | 0,595977121 | SH3GL1P2 | FALSE | Upregulated |
| 0,573174196 | 0,595979738 | HSD3BP4 | FALSE | Upregulated |
| -0,056154771 | 0,596016587 | LRRC47 | FALSE | Downregulated |
| -0,858359432 | 0,596123169 | LOC101927582 | FALSE | Downregulated |
| -0,108792731 | 0,596123169 | NRIP1 | FALSE | Downregulated |
| -0,108086762 | 0,596565917 | ZNRD1 | FALSE | Downregulated |
| 0,26256554 | 0,596754891 | FLJ13197 | FALSE | Upregulated |
| -0,313994691 | 0,59709834 | GDF9 | FALSE | Downregulated |
| -0,324998464 | 0,597127 | MAP6D1 | FALSE | Downregulated |
| 0,106235542 | 0,597449489 | SDC1 | FALSE | Upregulated |
| 0,138482155 | 0,597556817 | PSTPIP2 | FALSE | Upregulated |
| 0,64653074 | 0,597577401 | MAG | FALSE | Upregulated |
| -0,86929779 | 0,597579685 | FAR2P3 | FALSE | Downregulated |
| -0,141118193 | 0,597582478 | KIAA1804 | FALSE | Downregulated |
| 0,394541227 | 0,597630243 | LOC102723640 | FALSE | Upregulated |
| 0,161329462 | 0,597630243 | MPP7 | FALSE | Upregulated |
| -0,095257212 | 0,597714772 | SMAD1 | FALSE | Downregulated |

| | | | | |
|---|---|---|---|---|
| 0,262534165 | 0,597887339 | COL26A1 | FALSE | Upregulated |
| -0,268492677 | 0,598094112 | LOC100506974 | FALSE | Downregulated |
| 0,156147414 | 0,598350104 | MRM1 | FALSE | Upregulated |
| -0,084559169 | 0,598362764 | TJAP1 | FALSE | Downregulated |
| 0,171080874 | 0,598591102 | CD226 | FALSE | Upregulated |
| -0,377202835 | 0,598742732 | LRRTM4 | FALSE | Downregulated |
| 0,736363697 | 0,5987743 | LOC101929151 | FALSE | Upregulated |
| -0,072223719 | 0,598882869 | HIPK3 | FALSE | Downregulated |
| 0,30910998 | 0,598882869 | SDS | FALSE | Upregulated |
| 0,049128635 | 0,59891659 | GTF2A1 | FALSE | Upregulated |
| 0,552160878 | 0,599223434 | LOC102723353 | FALSE | Upregulated |
| -0,442830698 | 0,599243351 | FAM66B | FALSE | Downregulated |
| -0,104262171 | 0,599243351 | ZNF358 | FALSE | Downregulated |
| -0,709231316 | 0,599332687 | LOC101928082 | FALSE | Downregulated |
| 0,69278514 | 0,599332687 | LOC102723811 | FALSE | Upregulated |
| -0,517411719 | 0,599354752 | C15orf59 | FALSE | Downregulated |
| -0,08115634 | 0,599354752 | XBP1 | FALSE | Downregulated |
| 0,316864315 | 0,599504424 | CNKSR1 | FALSE | Upregulated |
| -0,087856846 | 0,599504424 | SETD5 | FALSE | Downregulated |
| 0,18348205 | 0,599534509 | TSPAN1 | FALSE | Upregulated |
| 0,136109723 | 0,599596648 | GBAS | FALSE | Upregulated |
| -0,09933594 | 0,599756523 | RPS9 | FALSE | Downregulated |
| 0,394984914 | 0,599780892 | CCSER1 | FALSE | Upregulated |
| -0,085437588 | 0,599780892 | ZCCHC24 | FALSE | Downregulated |
| -0,119300356 | 0,600187857 | NANS | FALSE | Downregulated |
| 0,117092415 | 0,600187857 | ZNF589 | FALSE | Upregulated |
| 0,204346888 | 0,60031691 | IL27RA | FALSE | Upregulated |
| 0,39121338 | 0,60031691 | LOC101241902 | FALSE | Upregulated |
| -0,280845741 | 0,60031691 | LOC101927934 | FALSE | Downregulated |
| -0,074664691 | 0,60031691 | SRBD1 | FALSE | Downregulated |
| -0,986206419 | 0,600452239 | LOC102724703 | FALSE | Downregulated |
| -0,252587279 | 0,600480665 | GSTM2 | FALSE | Downregulated |
| 0,092291226 | 0,600495477 | PLGRKT | FALSE | Upregulated |
| -0,083713358 | 0,600613055 | ANKRD46 | FALSE | Downregulated |
| 0,51949765 | 0,600615858 | DPY19L2 | FALSE | Upregulated |
| 0,377812322 | 0,600615858 | EYA2 | FALSE | Upregulated |
| -0,178645166 | 0,600615858 | RP11-119F19.2 | FALSE | Downregulated |
| -0,089196955 | 0,600717946 | MRPL36 | FALSE | Downregulated |
| -0,092662904 | 0,600766936 | USP42 | FALSE | Downregulated |
| 0,215407827 | 0,600838616 | LOC102723983 | FALSE | Upregulated |
| -0,089355685 | 0,600888901 | LAMTOR4 | FALSE | Downregulated |
| -0,05812918 | 0,600888901 | PRKCI | FALSE | Downregulated |
| -0,197461761 | 0,601036028 | C17orf100 | FALSE | Downregulated |
| -0,186284674 | 0,601037796 | ATRNL1 | FALSE | Downregulated |
| 0,190115693 | 0,601037796 | CYFIP2 | FALSE | Upregulated |
| -0,25909174 | 0,601087812 | LOC101928069 | FALSE | Downregulated |
| -0,624617154 | 0,601087812 | MIR1247 | FALSE | Downregulated |
| -0,067144133 | 0,601087812 | TBL1XR1 | FALSE | Downregulated |
| -0,170909047 | 0,601135857 | RAB23 | FALSE | Downregulated |
| -0,226770153 | 0,601234998 | CPLX1 | FALSE | Downregulated |
| 0,115856889 | 0,601234998 | DBF4 | FALSE | Upregulated |

| | | | | |
|---|---|---|---|---|
| 0,189996461 | 0,601234998 | MNDA | FALSE | Upregulated |
| -0,427771138 | 0,601283378 | LOC100653233 | FALSE | Downregulated |
| -0,106593989 | 0,601607588 | CEP250 | FALSE | Downregulated |
| -0,701210657 | 0,601607588 | PCA3 | FALSE | Downregulated |
| 0,597421293 | 0,602024861 | LOC646938 | FALSE | Upregulated |
| -0,625865615 | 0,602024861 | LOC731157 | FALSE | Downregulated |
| -0,078743831 | 0,602126994 | ASPH | FALSE | Downregulated |
| 0,057781722 | 0,602285078 | PCID2 | FALSE | Upregulated |
| -0,078168574 | 0,602370534 | ZC3HC1 | FALSE | Downregulated |
| 0,177967325 | 0,602404202 | IL15 | FALSE | Upregulated |
| -0,087454035 | 0,602624164 | GTF3A | FALSE | Downregulated |
| -0,101489219 | 0,602624873 | POLD2 | FALSE | Downregulated |
| -0,500794638 | 0,602825697 | MIR4647 | FALSE | Downregulated |
| -0,793460875 | 0,603074404 | LOC285095 | FALSE | Downregulated |
| -1,229082402 | 0,603124035 | MIR5685 | FALSE | Downregulated |
| -0,066431112 | 0,603124035 | SLC25A5 | FALSE | Downregulated |
| 0,279229178 | 0,603147602 | BACH2 | FALSE | Upregulated |
| 0,07055818 | 0,603147602 | CDK17 | FALSE | Upregulated |
| 1,12495525 | 0,603147602 | TRIM40 | FALSE | Upregulated |
| -0,131885791 | 0,603346213 | ZDBF2 | FALSE | Downregulated |
| 0,699294701 | 0,60346672 | LINC00964 | FALSE | Upregulated |
| 0,110095335 | 0,603547283 | MICAL2 | FALSE | Upregulated |
| 0,288756357 | 0,603593032 | ZNF790-AS1 | FALSE | Upregulated |
| 0,955909422 | 0,603639793 | MYCNOS | FALSE | Upregulated |
| -0,265687089 | 0,603639793 | PILRB | FALSE | Downregulated |
| -0,839334397 | 0,603702579 | NUDT11 | FALSE | Downregulated |
| 0,45284636 | 0,603734154 | LOC102723610 | FALSE | Upregulated |
| 0,550093409 | 0,603763399 | LOC102724339 | FALSE | Upregulated |
| -0,280871817 | 0,603815866 | CRYGS | FALSE | Downregulated |
| -0,393827355 | 0,603825133 | LOC102724615 | FALSE | Downregulated |
| -0,06089308 | 0,603950429 | CERS2 | FALSE | Downregulated |
| -0,4858108 | 0,603950429 | LOC101929918 | FALSE | Downregulated |
| 0,158434985 | 0,604077732 | LPCAT2 | FALSE | Upregulated |
| -0,25765514 | 0,604077732 | PSMD6-AS2 | FALSE | Downregulated |
| -0,198766906 | 0,604123602 | DGKG | FALSE | Downregulated |
| 0,293217173 | 0,604138356 | CACNA2D1 | FALSE | Upregulated |
| -0,625880363 | 0,604138356 | LINC00652 | FALSE | Downregulated |
| -0,362692197 | 0,604138356 | MOV10L1 | FALSE | Downregulated |
| -0,242001886 | 0,604195981 | LOC102723804 | FALSE | Downregulated |
| -0,293622931 | 0,604220881 | DLGAP2 | FALSE | Downregulated |
| 0,341738982 | 0,604220881 | TMEM63C | FALSE | Upregulated |
| -0,596487775 | 0,604231431 | AC093375.1 | FALSE | Downregulated |
| 0,136035374 | 0,604231431 | CLDN16 | FALSE | Upregulated |
| -0,089801842 | 0,604403073 | COPZ2 | FALSE | Downregulated |
| -0,238293711 | 0,604403073 | KCNAB1 | FALSE | Downregulated |
| 0,085275023 | 0,604403073 | TMEM260 | FALSE | Upregulated |
| -0,264285591 | 0,604463904 | C6orf226 | FALSE | Downregulated |
| -0,191795235 | 0,604463904 | CD160 | FALSE | Downregulated |
| 0,087232887 | 0,604463904 | CLSTN3 | FALSE | Upregulated |
| 0,060574395 | 0,604463904 | FBXO33 | FALSE | Upregulated |
| 0,302071105 | 0,604463904 | LOC100507460 | FALSE | Upregulated |

| | | | | |
|---|---|---|---|---|
| 0,179764588 | 0,604463904 | SMAD9 | FALSE | Upregulated |
| -0,952192599 | 0,604463904 | TRNR | FALSE | Downregulated |
| 0,258861067 | 0,604649108 | FBXL8 | FALSE | Upregulated |
| -0,122897386 | 0,604926159 | HHEX | FALSE | Downregulated |
| 0,369399257 | 0,605096619 | MUSK | FALSE | Upregulated |
| -0,046484729 | 0,605121625 | AGGF1 | FALSE | Downregulated |
| 0,156995564 | 0,605121625 | LINC00987 | FALSE | Upregulated |
| 0,297972199 | 0,605175395 | GPR174 | FALSE | Upregulated |
| 0,665918579 | 0,605281678 | LOC101927594 | FALSE | Upregulated |
| -0,242301811 | 0,605554075 | CCDC168 | FALSE | Downregulated |
| 0,393006622 | 0,605928538 | LOC100507747 | FALSE | Upregulated |
| 0,104471888 | 0,605942963 | CHST14 | FALSE | Upregulated |
| -0,075447763 | 0,606361136 | AGO1 | FALSE | Downregulated |
| 0,061600547 | 0,606623057 | SCAP | FALSE | Upregulated |
| -0,160123673 | 0,606901292 | GANC | FALSE | Downregulated |
| -0,35855392 | 0,606920018 | HIST2H2AC | FALSE | Downregulated |
| 0,402266885 | 0,606920018 | SLC5A2 | FALSE | Upregulated |
| -0,164578706 | 0,60710268 | ZNF682 | FALSE | Downregulated |
| 0,129337845 | 0,607211891 | ADCY5 | FALSE | Upregulated |
| 0,063946431 | 0,607211891 | MLH1 | FALSE | Upregulated |
| -0,039841026 | 0,607234667 | DDX50 | FALSE | Downregulated |
| 0,157987884 | 0,607314376 | P2RX7 | FALSE | Upregulated |
| -0,082759014 | 0,607363258 | LSR | FALSE | Downregulated |
| 0,087862458 | 0,607646683 | LRRC45 | FALSE | Upregulated |
| 0,368410671 | 0,607646683 | SNORD89 | FALSE | Upregulated |
| 0,891026949 | 0,607684783 | CCDC177 | FALSE | Upregulated |
| 0,102114758 | 0,607684783 | MAPKBP1 | FALSE | Upregulated |
| 0,886854752 | 0,607707755 | BRSK2 | FALSE | Upregulated |
| 0,109538569 | 0,607707755 | FAM3A | FALSE | Upregulated |
| 0,300792433 | 0,607783862 | CYSLTR2 | FALSE | Upregulated |
| 0,801144897 | 0,607783862 | TTTY10 | FALSE | Upregulated |
| 0,505826353 | 0,607851986 | GRIK1-AS2 | FALSE | Upregulated |
| 0,049312463 | 0,607851986 | VAPA | FALSE | Upregulated |
| -0,35785368 | 0,607912653 | KLHL23 | FALSE | Downregulated |
| 0,816760784 | 0,607912653 | MIR2861 | FALSE | Upregulated |
| 0,467144209 | 0,607912653 | PTOV1-AS1 | FALSE | Upregulated |
| -0,260553234 | 0,608023887 | NUTM2B | FALSE | Downregulated |
| 0,050859929 | 0,608137458 | CTTN | FALSE | Upregulated |
| -0,068953665 | 0,608137458 | POM121 | FALSE | Downregulated |
| -0,117438545 | 0,608382514 | FLJ10038 | FALSE | Downregulated |
| -0,921117884 | 0,608382514 | LOC102724113 | FALSE | Downregulated |
| -0,07029082 | 0,608679924 | ALDH3A2 | FALSE | Downregulated |
| -0,103991929 | 0,608741425 | CD3EAP | FALSE | Downregulated |
| 0,236596801 | 0,608808822 | FHOD3 | FALSE | Upregulated |
| -0,078288169 | 0,608873698 | POLR2G | FALSE | Downregulated |
| 0,069090919 | 0,608925924 | MED9 | FALSE | Upregulated |
| 0,167941572 | 0,609013235 | C5orf56 | FALSE | Upregulated |
| -0,07417236 | 0,609013235 | POLR2K | FALSE | Downregulated |
| 0,71064461 | 0,609207125 | NR2E1 | FALSE | Upregulated |
| -0,061968709 | 0,609207125 | NSL1 | FALSE | Downregulated |
| 0,315369321 | 0,609250729 | LOC101927178 | FALSE | Upregulated |

| | | | | |
|---|---|---|---|---|
| -0,056103027 | 0,609250729 | SEC11A | FALSE | Downregulated |
| 0,605575489 | 0,609309791 | CPEB1 | FALSE | Upregulated |
| -0,13637404 | 0,609382852 | NPR1 | FALSE | Downregulated |
| -0,642628743 | 0,609408528 | FAM157C | FALSE | Downregulated |
| -1,005700599 | 0,609427542 | PAX7 | FALSE | Downregulated |
| 0,142958803 | 0,609433121 | MRPS31P5 | FALSE | Upregulated |
| 3,507054143 | 0,609444429 | H19 | FALSE | Upregulated |
| 0,684281967 | 0,609901565 | LOC101927375 | FALSE | Upregulated |
| -0,046049511 | 0,609901565 | MRFAP1 | FALSE | Downregulated |
| 0,069352941 | 0,609901565 | TMEM179B | FALSE | Upregulated |
| -0,265739935 | 0,610732715 | CYP46A1 | FALSE | Downregulated |
| 0,971541791 | 0,610732715 | PPP2R2C | FALSE | Upregulated |
| 0,134924149 | 0,610764813 | MPEG1 | FALSE | Upregulated |
| 0,389332854 | 0,610918946 | FOXP3 | FALSE | Upregulated |
| 0,636405385 | 0,611158856 | C1QL3 | FALSE | Upregulated |
| -0,22178709 | 0,611158856 | GLUD1P7 | FALSE | Downregulated |
| 0,662885375 | 0,611158856 | LINC00260 | FALSE | Upregulated |
| -0,484471193 | 0,611158856 | LOC101928875 | FALSE | Downregulated |
| 0,077429984 | 0,611158856 | SURF4 | FALSE | Upregulated |
| 0,426043877 | 0,611158856 | TSC22D1-AS1 | FALSE | Upregulated |
| -0,087950555 | 0,611158856 | UNG | FALSE | Downregulated |
| -0,677493414 | 0,611379392 | LINC00202-2 | FALSE | Downregulated |
| 0,115568737 | 0,61142765 | MYO6 | FALSE | Upregulated |
| 0,069188888 | 0,611579728 | NHP2L1 | FALSE | Upregulated |
| -0,406322748 | 0,611697868 | MMP21 | FALSE | Downregulated |
| -0,076652337 | 0,611797096 | SDR39U1 | FALSE | Downregulated |
| 0,260546838 | 0,611797096 | SMPD3 | FALSE | Upregulated |
| 0,231937176 | 0,611887921 | LOC101928657 | FALSE | Upregulated |
| -0,052643489 | 0,611999831 | ATP6V1B2 | FALSE | Downregulated |
| 0,078154904 | 0,612197726 | KLHL7 | FALSE | Upregulated |
| 0,323497392 | 0,612197726 | LOC101929115 | FALSE | Upregulated |
| 0,665101678 | 0,612197726 | SYNGR3 | FALSE | Upregulated |
| -0,107411728 | 0,612197726 | TBX3 | FALSE | Downregulated |
| 0,08245938 | 0,612314534 | C17orf103 | FALSE | Upregulated |
| 0,70464288 | 0,612325869 | SLC46A2 | FALSE | Upregulated |
| 0,068771713 | 0,612632068 | COG5 | FALSE | Upregulated |
| -0,127958898 | 0,612632068 | ZBTB49 | FALSE | Downregulated |
| -0,107516965 | 0,612651551 | MGST2 | FALSE | Downregulated |
| 0,471749117 | 0,612717119 | GP5 | FALSE | Upregulated |
| -0,106494728 | 0,612744217 | MIA2 | FALSE | Downregulated |
| -0,074295667 | 0,613042734 | ECE1 | FALSE | Downregulated |
| -0,198452982 | 0,613131777 | C1orf226 | FALSE | Downregulated |
| 0,143152455 | 0,613131777 | ENKD1 | FALSE | Upregulated |
| 0,090184391 | 0,613131777 | ULK2 | FALSE | Upregulated |
| 0,220858669 | 0,613196984 | BLM | FALSE | Upregulated |
| 0,069670108 | 0,61325918 | EMC10 | FALSE | Upregulated |
| -0,066286252 | 0,613269865 | DUSP11 | FALSE | Downregulated |
| -0,140681766 | 0,613269974 | CCDC176 | FALSE | Downregulated |
| -0,098065928 | 0,613269974 | KDM7A | FALSE | Downregulated |
| -0,281412192 | 0,61337326 | BLK | FALSE | Downregulated |
| -0,390361056 | 0,613437404 | GOLGA8I | FALSE | Downregulated |

| | | | | |
|---|---|---|---|---|
| -0,504004695 | 0,613468359 | LOC102724676 | FALSE | Downregulated |
| -0,407170828 | 0,613468359 | MAPT | FALSE | Downregulated |
| -0,114184012 | 0,613468359 | RPS19BP1 | FALSE | Downregulated |
| 0,140586367 | 0,613626805 | C8orf58 | FALSE | Upregulated |
| 0,227368995 | 0,613657884 | UGT2B11 | FALSE | Upregulated |
| 0,075464301 | 0,613847573 | NEU1 | FALSE | Upregulated |
| -0,052563761 | 0,613884097 | PWWP2A | FALSE | Downregulated |
| 0,114881466 | 0,614139312 | EVI5L | FALSE | Upregulated |
| -0,093381701 | 0,61424678 | FAM45A | FALSE | Downregulated |
| 0,053646152 | 0,614277305 | PPP2CA | FALSE | Upregulated |
| -0,064287697 | 0,614395682 | YIPF4 | FALSE | Downregulated |
| -0,070769674 | 0,614484209 | MTA1 | FALSE | Downregulated |
| -0,206947253 | 0,614484209 | TMIE | FALSE | Downregulated |
| 0,741427315 | 0,614491027 | AIPL1 | FALSE | Upregulated |
| 0,425726306 | 0,614491027 | CHRM3 | FALSE | Upregulated |
| -0,47922321 | 0,614509137 | RBM12B-AS1 | FALSE | Downregulated |
| 0,126892856 | 0,614782018 | NKX3-1 | FALSE | Upregulated |
| 0,079654373 | 0,614808658 | TM7SF3 | FALSE | Upregulated |
| 0,292408045 | 0,614859956 | LOC100996760 | FALSE | Upregulated |
| 0,83297167 | 0,614859956 | SLC18A1 | FALSE | Upregulated |
| -0,060027702 | 0,614859956 | STK40 | FALSE | Downregulated |
| -0,12489625 | 0,614859956 | THOC3 | FALSE | Downregulated |
| 0,50021349 | 0,61490892 | HSF5 | FALSE | Upregulated |
| -0,080152342 | 0,61490892 | SIAE | FALSE | Downregulated |
| -0,257295266 | 0,61490892 | TCEAL7 | FALSE | Downregulated |
| 0,39366449 | 0,61492103 | HSD3B1 | FALSE | Upregulated |
| 0,123054965 | 0,614925712 | ZBTB22 | FALSE | Upregulated |
| -0,066130762 | 0,614935665 | SMARCAD1 | FALSE | Downregulated |
| 0,792934036 | 0,615111074 | ANKRD30A | FALSE | Upregulated |
| 0,171153235 | 0,615140492 | CPT1A | FALSE | Upregulated |
| -0,387771901 | 0,615140492 | RP11-789C1.1 | FALSE | Downregulated |
| 0,877626089 | 0,615213827 | ZNF705E | FALSE | Upregulated |
| -0,435573069 | 0,615384617 | CISH | FALSE | Downregulated |
| 0,645039282 | 0,615432399 | AC079610.2 | FALSE | Upregulated |
| 0,060656792 | 0,615432399 | EIF3D | FALSE | Upregulated |
| -0,199738888 | 0,615453101 | KLHL3 | FALSE | Downregulated |
| 0,473339811 | 0,61548173 | PTCRA | FALSE | Upregulated |
| -0,123328951 | 0,615527112 | TTBK2 | FALSE | Downregulated |
| -0,074811661 | 0,615601653 | SERPINA6 | FALSE | Downregulated |
| -0,062982699 | 0,615601653 | TRUB2 | FALSE | Downregulated |
| 0,318336733 | 0,61568551 | FAM166A | FALSE | Upregulated |
| -0,298530222 | 0,61568551 | RUSC1-AS1 | FALSE | Downregulated |
| 0,272353802 | 0,615764567 | LOC440386 | FALSE | Upregulated |
| 0,093689533 | 0,615764567 | ZFHX3 | FALSE | Upregulated |
| 0,079904147 | 0,615783284 | PCNT | FALSE | Upregulated |
| 0,079810045 | 0,615834106 | DAD1 | FALSE | Upregulated |
| 0,076653494 | 0,615908538 | ST6GALNAC6 | FALSE | Upregulated |
| -0,10102788 | 0,615967415 | DOPEY1 | FALSE | Downregulated |
| 0,067200012 | 0,615967415 | PSMD10 | FALSE | Upregulated |
| 0,861286914 | 0,616017268 | MRPL42P5 | FALSE | Upregulated |
| 0,51140061 | 0,616082094 | CCDC144A | FALSE | Upregulated |

| | | | | |
|---|---|---|---|---|
| 0,075130547 | 0,616082094 | RFC5 | FALSE | Upregulated |
| 0,21036823 | 0,616082094 | VILL | FALSE | Upregulated |
| 0,116234979 | 0,616082094 | ZNF613 | FALSE | Upregulated |
| 0,152248943 | 0,616082094 | ZNF737 | FALSE | Upregulated |
| -0,077408533 | 0,616308587 | PSMB6 | FALSE | Downregulated |
| 0,580876706 | 0,61632248 | ADPGK-AS1 | FALSE | Upregulated |
| -0,073055258 | 0,616325815 | EIF3L | FALSE | Downregulated |
| 0,750855404 | 0,616325815 | SYNDIG1L | FALSE | Upregulated |
| -0,215271563 | 0,616574608 | PAN3-AS1 | FALSE | Downregulated |
| -0,113716314 | 0,616693141 | CCDC142 | FALSE | Downregulated |
| 0,130554407 | 0,616693141 | PLXDC2 | FALSE | Upregulated |
| -0,088466368 | 0,616693141 | TSC2 | FALSE | Downregulated |
| -0,085386522 | 0,616694954 | ANKHD1 | FALSE | Downregulated |
| -0,071404097 | 0,616694954 | EIF1 | FALSE | Downregulated |
| 0,08590607 | 0,616727431 | DCTD | FALSE | Upregulated |
| -0,445918137 | 0,616727431 | KCNG1 | FALSE | Downregulated |
| -0,674660934 | 0,616727431 | LOC100289092 | FALSE | Downregulated |
| 1,341619758 | 0,616727431 | RNU1-1 | FALSE | Upregulated |
| -0,074266757 | 0,616727431 | TRIM37 | FALSE | Downregulated |
| -0,333642413 | 0,616752689 | LOC149134 | FALSE | Downregulated |
| 0,138944148 | 0,616767967 | WDR90 | FALSE | Upregulated |
| 0,048023769 | 0,616946952 | PRKRIR | FALSE | Upregulated |
| -0,350879696 | 0,616946952 | RP11-21L23.3 | FALSE | Downregulated |
| 0,690067385 | 0,617014027 | LOC101929035 | FALSE | Upregulated |
| 0,481859267 | 0,617278111 | LOC101928117 | FALSE | Upregulated |
| 0,253719536 | 0,6173632 | TLE6 | FALSE | Upregulated |
| -0,094580725 | 0,61748915 | MLYCD | FALSE | Downregulated |
| -0,075147933 | 0,617583091 | PIGX | FALSE | Downregulated |
| 0,130837297 | 0,617911678 | FAM171A1 | FALSE | Upregulated |
| 0,374122587 | 0,617911678 | KCTD14 | FALSE | Upregulated |
| -0,432015256 | 0,617911678 | LOC102723582 | FALSE | Downregulated |
| -0,050142335 | 0,617911678 | SMNDC1 | FALSE | Downregulated |
| 0,864411484 | 0,618093895 | FSTL5 | FALSE | Upregulated |
| -0,216828651 | 0,618203497 | IQCH-AS1 | FALSE | Downregulated |
| 0,195237419 | 0,618203497 | RGS18 | FALSE | Upregulated |
| -0,151738537 | 0,618203497 | RNPC3 | FALSE | Downregulated |
| -0,045101573 | 0,618203497 | TUBGCP2 | FALSE | Downregulated |
| -0,141483622 | 0,618362298 | ERICH2 | FALSE | Downregulated |
| -0,723929368 | 0,618663914 | EBF2 | FALSE | Downregulated |
| 0,377852727 | 0,618747184 | LOC101928774 | FALSE | Upregulated |
| -0,206763054 | 0,618761345 | LOC102724005 | FALSE | Downregulated |
| 0,55333464 | 0,618761345 | SLC7A4 | FALSE | Upregulated |
| -0,071948107 | 0,618905 | MPI | FALSE | Downregulated |
| -0,038389666 | 0,618905 | POLR2B | FALSE | Downregulated |
| 0,087270865 | 0,618967657 | CERK | FALSE | Upregulated |
| -0,086365483 | 0,618983306 | BIRC6 | FALSE | Downregulated |
| 0,613205741 | 0,618983306 | GNB3 | FALSE | Upregulated |
| -0,061132391 | 0,618983306 | TBCC | FALSE | Downregulated |
| 0,08113574 | 0,619258121 | UBA52 | FALSE | Upregulated |
| -0,897145395 | 0,619389698 | ATP12A | FALSE | Downregulated |
| -0,239110092 | 0,619584756 | LOC101927181 | FALSE | Downregulated |

| | | | | |
|---|---|---|---|---|
| 0,115301275 | 0,619829518 | ABCG8 | FALSE | Upregulated |
| 0,088072512 | 0,619836629 | GLT8D1 | FALSE | Upregulated |
| 0,684826751 | 0,619842171 | GATA3-AS1 | FALSE | Upregulated |
| -0,059666623 | 0,619935959 | EFR3A | FALSE | Downregulated |
| -0,17199823 | 0,620098134 | ALDOC | FALSE | Downregulated |
| -0,344863386 | 0,620098134 | RP5-1050D4.5 | FALSE | Downregulated |
| -0,429643449 | 0,620208434 | ACSM1 | FALSE | Downregulated |
| 0,53049125 | 0,620208434 | GDNF-AS1 | FALSE | Upregulated |
| 0,151341681 | 0,620208434 | GPR4 | FALSE | Upregulated |
| -0,535587614 | 0,620208434 | KCNC4-AS1 | FALSE | Downregulated |
| 0,582232551 | 0,620208434 | LOC101928373 | FALSE | Upregulated |
| 0,063449776 | 0,620208434 | LPHN2 | FALSE | Upregulated |
| 0,169476073 | 0,620208434 | MICA | FALSE | Upregulated |
| -0,063479963 | 0,620208434 | NF2 | FALSE | Downregulated |
| 0,793491515 | 0,620519376 | CDCP2 | FALSE | Upregulated |
| -0,073720955 | 0,620695374 | MTM1 | FALSE | Downregulated |
| -0,413418239 | 0,620949099 | OBSCN | FALSE | Downregulated |
| 0,298019099 | 0,620949099 | PRTG | FALSE | Upregulated |
| 0,169089926 | 0,62173494 | RBMS2 | FALSE | Upregulated |
| 0,134930059 | 0,621770957 | SNX10 | FALSE | Upregulated |
| -0,158784161 | 0,621773101 | IFI44 | FALSE | Downregulated |
| 0,056557007 | 0,621773101 | TUFM | FALSE | Upregulated |
| -0,06036419 | 0,621773101 | ZNF766 | FALSE | Downregulated |
| 0,163190901 | 0,622237318 | CDKN2C | FALSE | Upregulated |
| -0,679867466 | 0,622481717 | TP53AIP1 | FALSE | Downregulated |
| 0,502490695 | 0,62265132 | CHST6 | FALSE | Upregulated |
| 0,392409675 | 0,622747827 | SLC44A5 | FALSE | Upregulated |
| 0,300619833 | 0,622747827 | ZNF192P1 | FALSE | Upregulated |
| -0,166316521 | 0,622857929 | C21orf119 | FALSE | Downregulated |
| 0,579609047 | 0,622857929 | MAMDC2-AS1 | FALSE | Upregulated |
| 0,091403566 | 0,622857929 | PGAM5 | FALSE | Upregulated |
| 0,597563213 | 0,622857929 | SNORA29 | FALSE | Upregulated |
| -0,08850085 | 0,622882452 | CDK2AP1 | FALSE | Downregulated |
| 0,090550822 | 0,622882452 | FAM161B | FALSE | Upregulated |
| -0,66923022 | 0,622882452 | LOC101926887 | FALSE | Downregulated |
| 0,079119446 | 0,622882452 | SFMBT1 | FALSE | Upregulated |
| 0,14466776 | 0,622882452 | UPP1 | FALSE | Upregulated |
| 0,328676549 | 0,622889597 | LOC642846 | FALSE | Upregulated |
| 0,596345537 | 0,623218152 | DPRXP4 | FALSE | Upregulated |
| -0,187215176 | 0,62347997 | KIF26A | FALSE | Downregulated |
| -0,250059198 | 0,62347997 | LOC643770 | FALSE | Downregulated |
| 0,080356254 | 0,62347997 | PTMA | FALSE | Upregulated |
| 0,632244738 | 0,62350727 | TNFRSF13C | FALSE | Upregulated |
| 0,521035082 | 0,623632296 | LOC101928659 | FALSE | Upregulated |
| 0,118423943 | 0,623884258 | DPH7 | FALSE | Upregulated |
| 0,103179739 | 0,624026643 | RPS19 | FALSE | Upregulated |
| 0,226011831 | 0,624050216 | LOC101927002 | FALSE | Upregulated |
| 0,521588582 | 0,624149361 | LGALS9C | FALSE | Upregulated |
| -0,077540116 | 0,624149361 | SELO | FALSE | Downregulated |
| -0,09173375 | 0,624487703 | ZNF155 | FALSE | Downregulated |
| -0,270253429 | 0,624546635 | IDO2 | FALSE | Downregulated |

| | | | | |
|---|---|---|---|---|
| 0,1016837 | 0,624546635 | RIN3 | FALSE | Upregulated |
| -0,153769178 | 0,624546635 | ZBTB21 | FALSE | Downregulated |
| 0,32991618 | 0,624634138 | CD300LG | FALSE | Upregulated |
| 0,501291633 | 0,62465801 | RFX8 | FALSE | Upregulated |
| -0,057881392 | 0,624886951 | ATF1 | FALSE | Downregulated |
| -0,159196717 | 0,624886951 | FRS3 | FALSE | Downregulated |
| -0,158945354 | 0,624886951 | NAT8 | FALSE | Downregulated |
| -0,054031023 | 0,624912915 | GOLGA1 | FALSE | Downregulated |
| 0,69820019 | 0,624939246 | PCDH15 | FALSE | Upregulated |
| -0,10934399 | 0,62504706 | HSD17B12 | FALSE | Downregulated |
| 0,114748452 | 0,625288424 | ALKBH4 | FALSE | Upregulated |
| -0,466375574 | 0,625407006 | SFRP4 | FALSE | Downregulated |
| 0,08756883 | 0,625628274 | PPAT | FALSE | Upregulated |
| -0,192236646 | 0,625791534 | TSTD3 | FALSE | Downregulated |
| 0,09008885 | 0,625791534 | UBE2D1 | FALSE | Upregulated |
| -0,240620294 | 0,6261165 | HSF4 | FALSE | Downregulated |
| -0,086214289 | 0,62625629 | BBS9 | FALSE | Downregulated |
| -0,417508793 | 0,62625629 | HIST1H3H | FALSE | Downregulated |
| -0,055848339 | 0,62625629 | ZNF800 | FALSE | Downregulated |
| 0,150572671 | 0,626416824 | TMEM144 | FALSE | Upregulated |
| 0,316381288 | 0,626467421 | LOC102723770 | FALSE | Upregulated |
| -0,407208745 | 0,626729176 | LOC101927646 | FALSE | Downregulated |
| -0,522802822 | 0,62674439 | LOC102723627 | FALSE | Downregulated |
| 0,139633631 | 0,626777611 | ODF2L | FALSE | Upregulated |
| 0,09125688 | 0,626777611 | TRMU | FALSE | Upregulated |
| -0,09299535 | 0,626898742 | DENND4A | FALSE | Downregulated |
| 0,096815897 | 0,626898742 | PC | FALSE | Upregulated |
| 0,160014893 | 0,627051412 | BCAT2 | FALSE | Upregulated |
| 0,177347764 | 0,627072035 | LINC00630 | FALSE | Upregulated |
| 0,218528916 | 0,627074865 | IL17RD | FALSE | Upregulated |
| 0,060810794 | 0,62712143 | CCDC127 | FALSE | Upregulated |
| -0,077901672 | 0,62712143 | ZNF75A | FALSE | Downregulated |
| 0,446714456 | 0,627267334 | OTOA | FALSE | Upregulated |
| -0,346706799 | 0,627284708 | RASL10A | FALSE | Downregulated |
| 2,288338257 | 0,627343146 | RPS4Y1 | FALSE | Upregulated |
| -0,886460515 | 0,627418577 | LRFN5 | FALSE | Downregulated |
| 0,500800459 | 0,627477261 | SRD5A3-AS1 | FALSE | Upregulated |
| -0,243869521 | 0,627546508 | ADAM12 | FALSE | Downregulated |
| -0,181077813 | 0,627546508 | KGFLP2 | FALSE | Downregulated |
| -0,109329079 | 0,627546508 | RAB37 | FALSE | Downregulated |
| -0,149421901 | 0,627546508 | ZNF577 | FALSE | Downregulated |
| 0,35103049 | 0,627554392 | PAK6 | FALSE | Upregulated |
| 0,27453814 | 0,627554392 | ZNF391 | FALSE | Upregulated |
| -0,07746423 | 0,627624768 | NARF | FALSE | Downregulated |
| -0,073490837 | 0,627717077 | DCP1B | FALSE | Downregulated |
| -0,714192222 | 0,627717077 | LOC100131315 | FALSE | Downregulated |
| 0,375920182 | 0,627717077 | MTRNR2L2 | FALSE | Upregulated |
| -0,201481617 | 0,627717077 | NFYC-AS1 | FALSE | Downregulated |
| 0,657124433 | 0,627770995 | CABP1 | FALSE | Upregulated |
| -0,054007665 | 0,627980684 | WAPAL | FALSE | Downregulated |
| -0,146143283 | 0,628107368 | CHKB | FALSE | Downregulated |

| | | | | |
|---|---|---|---|---|
| 0,17546508 | 0,628150046 | LRRC37A3 | FALSE | Upregulated |
| 0,860134553 | 0,628150046 | RP4-565E6.1 | FALSE | Upregulated |
| 0,074830926 | 0,628150046 | SLC38A9 | FALSE | Upregulated |
| -0,07260555 | 0,628449551 | ANTXR2 | FALSE | Downregulated |
| -0,095773403 | 0,628500886 | ZNF449 | FALSE | Downregulated |
| 0,54168982 | 0,628536026 | NME9 | FALSE | Upregulated |
| 0,514823927 | 0,628632282 | SNORD57 | FALSE | Upregulated |
| 0,096087221 | 0,62866479 | ETV6 | FALSE | Upregulated |
| -0,094827373 | 0,62866479 | PRELID1 | FALSE | Downregulated |
| -0,161187695 | 0,628818282 | ETV1 | FALSE | Downregulated |
| -0,723858067 | 0,628886815 | LOC101929540 | FALSE | Downregulated |
| 0,060320563 | 0,628886815 | PBRM1 | FALSE | Upregulated |
| 0,754706761 | 0,628886815 | PNMT | FALSE | Upregulated |
| 0,048615421 | 0,628886815 | PRPF6 | FALSE | Upregulated |
| -1,732981929 | 0,629028805 | ATP5J2-PTCD1 | FALSE | Downregulated |
| 0,062886904 | 0,629071078 | HCCS | FALSE | Upregulated |
| 0,387576032 | 0,629104448 | FCGR2C | FALSE | Upregulated |
| -0,088213793 | 0,629104448 | MAP2K2 | FALSE | Downregulated |
| 0,096795832 | 0,629104448 | TDP1 | FALSE | Upregulated |
| 0,141744806 | 0,629286839 | IKBIP | FALSE | Upregulated |
| -0,107173756 | 0,629624843 | SARDH | FALSE | Downregulated |
| -0,27019456 | 0,629709075 | FAM78B | FALSE | Downregulated |
| 0,439299484 | 0,62992121 | MMP17 | FALSE | Upregulated |
| -0,286045015 | 0,629965587 | BVES | FALSE | Downregulated |
| -0,288271654 | 0,629965587 | CTD-3080P12.3 | FALSE | Downregulated |
| 0,276752852 | 0,629965587 | LINC01011 | FALSE | Upregulated |
| 0,151361511 | 0,629965587 | ZNF418 | FALSE | Upregulated |
| 0,698424173 | 0,630055586 | LOC102724271 | FALSE | Upregulated |
| -0,455536649 | 0,630139243 | MGARP | FALSE | Downregulated |
| 0,082702716 | 0,630139243 | OGFOD1 | FALSE | Upregulated |
| 0,095834187 | 0,630139243 | SPATA7 | FALSE | Upregulated |
| 0,302647618 | 0,63025186 | ALPK2 | FALSE | Upregulated |
| 0,092207522 | 0,63025186 | BOD1L1 | FALSE | Upregulated |
| 0,173456238 | 0,63025186 | LINC01124 | FALSE | Upregulated |
| 0,103596369 | 0,630273146 | GLCCI1 | FALSE | Upregulated |
| -0,558201108 | 0,630390578 | EPGN | FALSE | Downregulated |
| 0,207644961 | 0,630541569 | LOC729603 | FALSE | Upregulated |
| 0,197814349 | 0,630702197 | HRH1 | FALSE | Upregulated |
| 1,228867725 | 0,630792029 | SNORD3B-2 | FALSE | Upregulated |
| -0,199405721 | 0,6308596 | LPIN3 | FALSE | Downregulated |
| -0,102980576 | 0,631128494 | SELK | FALSE | Downregulated |
| -0,324035029 | 0,631216478 | LOC101927372 | FALSE | Downregulated |
| 0,075137241 | 0,63136652 | ZNF791 | FALSE | Upregulated |
| -0,070731461 | 0,63143745 | EIF2D | FALSE | Downregulated |
| -0,356231519 | 0,63143745 | INE1 | FALSE | Downregulated |
| 0,607241452 | 0,63143745 | PROX2 | FALSE | Upregulated |
| -0,476793096 | 0,63156423 | TSHR | FALSE | Downregulated |
| -0,137726058 | 0,632007108 | DSG1 | FALSE | Downregulated |
| 0,720221824 | 0,632077228 | LOC102724440 | FALSE | Upregulated |
| 0,112846237 | 0,632165013 | DUSP12 | FALSE | Upregulated |
| -0,119995468 | 0,632319236 | AMMECR1 | FALSE | Downregulated |

| | | | | |
|---|---|---|---|---|
| 0,045910421 | 0,632319236 | DHX38 | FALSE | Upregulated |
| -1,116452224 | 0,632319236 | INTS4L2 | FALSE | Downregulated |
| 0,11196815 | 0,632319236 | PIM3 | FALSE | Upregulated |
| 0,35584866 | 0,632319236 | SCARNA12 | FALSE | Upregulated |
| -0,470142471 | 0,63277317 | LPL | FALSE | Downregulated |
| 0,248685676 | 0,632838688 | PABPC5 | FALSE | Upregulated |
| 0,059894318 | 0,632838688 | UNC50 | FALSE | Upregulated |
| 0,275034412 | 0,63293943 | SSR4P1 | FALSE | Upregulated |
| 0,051275027 | 0,633186108 | FXR2 | FALSE | Upregulated |
| 0,36726751 | 0,633186108 | LOC101928825 | FALSE | Upregulated |
| -0,075230937 | 0,633186108 | MAGI1 | FALSE | Downregulated |
| 0,066589957 | 0,633186108 | SF3B1 | FALSE | Upregulated |
| 0,312163631 | 0,633186108 | TMEM132B | FALSE | Upregulated |
| -0,689377327 | 0,633416716 | RAET1G | FALSE | Downregulated |
| -0,627464376 | 0,633469287 | ZNF474 | FALSE | Downregulated |
| 0,114898635 | 0,633529134 | SLC35E2B | FALSE | Upregulated |
| 0,616905739 | 0,633949245 | HERC2P10 | FALSE | Upregulated |
| 0,081142523 | 0,633949245 | IL1R1 | FALSE | Upregulated |
| 0,296245014 | 0,633949245 | LOC100507472 | FALSE | Upregulated |
| 0,764950473 | 0,633949245 | TAS2R60 | FALSE | Upregulated |
| -0,056281499 | 0,634036575 | HERC4 | FALSE | Downregulated |
| 0,590079144 | 0,6343129 | ADAMTS16 | FALSE | Upregulated |
| 0,439557165 | 0,634355107 | LOC101928474 | FALSE | Upregulated |
| 0,301911214 | 0,634591104 | WNT11 | FALSE | Upregulated |
| 0,414713596 | 0,634847235 | LOC102724379 | FALSE | Upregulated |
| -0,122780262 | 0,634878717 | PIGL | FALSE | Downregulated |
| -0,909394892 | 0,63505717 | AQP8 | FALSE | Downregulated |
| 0,101069799 | 0,635137075 | LEPREL4 | FALSE | Upregulated |
| -0,147390909 | 0,635220589 | RPL23AP7 | FALSE | Downregulated |
| -0,093211486 | 0,635426096 | DERL2 | FALSE | Downregulated |
| 0,339531062 | 0,635426096 | SPATA17 | FALSE | Upregulated |
| 0,094449003 | 0,63551327 | MTHFR | FALSE | Upregulated |
| 0,234430737 | 0,63551327 | ST8SIA6 | FALSE | Upregulated |
| -0,0521674 | 0,635704915 | LSM14B | FALSE | Downregulated |
| 0,068975746 | 0,635877662 | SHISA5 | FALSE | Upregulated |
| 0,18888336 | 0,63591192 | CCDC30 | FALSE | Upregulated |
| -0,154075544 | 0,635964111 | IMMP1L | FALSE | Downregulated |
| -0,081575323 | 0,636035309 | WRB | FALSE | Downregulated |
| 0,164141203 | 0,636065829 | BCL2L11 | FALSE | Upregulated |
| -0,138487273 | 0,636065829 | C12orf73 | FALSE | Downregulated |
| -0,983722625 | 0,636065829 | CLVS2 | FALSE | Downregulated |
| -0,25875867 | 0,636065829 | LINC00471 | FALSE | Downregulated |
| -0,242789109 | 0,636065829 | PRSS53 | FALSE | Downregulated |
| 0,057458624 | 0,636065829 | TBC1D9B | FALSE | Upregulated |
| 0,192055231 | 0,636065829 | ZNF853 | FALSE | Upregulated |
| -0,402074598 | 0,636123285 | LOC101927754 | FALSE | Downregulated |
| -0,287580042 | 0,636215195 | PRDM5 | FALSE | Downregulated |
| -0,276081988 | 0,636993282 | LOC102724306 | FALSE | Downregulated |
| 0,214582487 | 0,637048683 | LINC01197 | FALSE | Upregulated |
| 0,26736343 | 0,637048683 | LOC100288069 | FALSE | Upregulated |
| -0,10587098 | 0,637048683 | PGBD4 | FALSE | Downregulated |

| | | | | |
|---|---|---|---|---|
| 0,191969984 | 0,637092435 | GMPPB | FALSE | Upregulated |
| 0,386226501 | 0,637092435 | RBM46 | FALSE | Upregulated |
| -0,330301373 | 0,637384837 | ENPP6 | FALSE | Downregulated |
| -0,099267621 | 0,637384837 | POLK | FALSE | Downregulated |
| -0,095564583 | 0,6376222 | DNAJA1 | FALSE | Downregulated |
| 0,168305683 | 0,6376222 | DNASE1 | FALSE | Upregulated |
| 0,111919051 | 0,6376222 | LNX1 | FALSE | Upregulated |
| 0,116754375 | 0,6376222 | SCLT1 | FALSE | Upregulated |
| 0,785725456 | 0,6376222 | SHOX | FALSE | Upregulated |
| -0,184054277 | 0,63777437 | NINJ2 | FALSE | Downregulated |
| -0,08980276 | 0,637918788 | UBL3 | FALSE | Downregulated |
| -0,23348368 | 0,6380811 | C14orf182 | FALSE | Downregulated |
| -0,13795553 | 0,638607949 | CCDC7 | FALSE | Downregulated |
| -0,425110657 | 0,638607949 | ELF5 | FALSE | Downregulated |
| 0,073573148 | 0,638945085 | RERE | FALSE | Upregulated |
| -0,183909655 | 0,639053138 | ADCK1 | FALSE | Downregulated |
| -0,102715131 | 0,639053138 | C4BPB | FALSE | Downregulated |
| 0,065970295 | 0,639053138 | CMTR2 | FALSE | Upregulated |
| -0,06157206 | 0,639053138 | KIAA1468 | FALSE | Downregulated |
| 0,373963797 | 0,639053138 | LOC100996455 | FALSE | Upregulated |
| -0,05286073 | 0,639053138 | MGAT1 | FALSE | Downregulated |
| -0,244343592 | 0,639112274 | C5AR2 | FALSE | Downregulated |
| 0,083378398 | 0,639408175 | EML4 | FALSE | Upregulated |
| 0,088606188 | 0,639408175 | TEFM | FALSE | Upregulated |
| 0,093633893 | 0,639424925 | FAM135A | FALSE | Upregulated |
| -0,034773529 | 0,639624586 | DDX42 | FALSE | Downregulated |
| -0,753227037 | 0,63973653 | FAM86KP | FALSE | Downregulated |
| -0,141383135 | 0,639955056 | LOC101060179 | FALSE | Downregulated |
| -0,058669927 | 0,640172004 | HNRNPA3 | FALSE | Downregulated |
| -0,09496404 | 0,640306298 | ZNF524 | FALSE | Downregulated |
| -0,587177077 | 0,640359283 | COL18A1-AS2 | FALSE | Downregulated |
| 0,15551938 | 0,640359283 | HPS5 | FALSE | Upregulated |
| -0,104930574 | 0,640687425 | ZFAS1 | FALSE | Downregulated |
| -0,157802645 | 0,641237063 | OAS1 | FALSE | Downregulated |
| 0,089297112 | 0,641456988 | HSPBAP1 | FALSE | Upregulated |
| -0,210518911 | 0,64153286 | LOC101927759 | FALSE | Downregulated |
| -0,074735146 | 0,642027523 | ACY1 | FALSE | Downregulated |
| -0,437286387 | 0,642140418 | RP11-554F20.1 | FALSE | Downregulated |
| 0,665817668 | 0,642170373 | DPH3P1 | FALSE | Upregulated |
| 0,076950694 | 0,642378353 | YKT6 | FALSE | Upregulated |
| -0,591807201 | 0,64244066 | EBF3 | FALSE | Downregulated |
| 0,098497285 | 0,64244066 | INS-IGF2 | FALSE | Upregulated |
| 0,543003182 | 0,642627396 | CXorf58 | FALSE | Upregulated |
| 0,045744855 | 0,642883782 | ZNF24 | FALSE | Upregulated |
| 0,04746945 | 0,642887077 | RNF220 | FALSE | Upregulated |
| 0,978864739 | 0,642887077 | RP11-143M1.2 | FALSE | Upregulated |
| 0,188689303 | 0,642904535 | LPHN1 | FALSE | Upregulated |
| -0,048272829 | 0,643156605 | CCNC | FALSE | Downregulated |
| 0,333138418 | 0,643156605 | CNR2 | FALSE | Upregulated |
| -0,176537292 | 0,643161368 | CA14 | FALSE | Downregulated |
| -0,356289291 | 0,643184946 | METTL21EP | FALSE | Downregulated |

| | | | | |
|---|---|---|---|---|
| 0,190884635 | 0,643216081 | P2RX5-TAX1BP3 | FALSE | Upregulated |
| 0,505417586 | 0,643280025 | CDH15 | FALSE | Upregulated |
| -0,874339535 | 0,643280025 | SNORD45B | FALSE | Downregulated |
| 0,072347616 | 0,643324356 | C18orf8 | FALSE | Upregulated |
| -0,699315913 | 0,643324356 | RP3-476K8.3 | FALSE | Downregulated |
| 0,140933928 | 0,64345414 | CC2D2A | FALSE | Upregulated |
| -0,051322096 | 0,643558249 | MMS19 | FALSE | Downregulated |
| 0,434053468 | 0,643783559 | HEPN1 | FALSE | Upregulated |
| -0,402183029 | 0,643905734 | LOC400655 | FALSE | Downregulated |
| 0,308012397 | 0,643944938 | HR | FALSE | Upregulated |
| -0,099757121 | 0,644019238 | ZNF354C | FALSE | Downregulated |
| 0,751948446 | 0,644202376 | KRT36 | FALSE | Upregulated |
| 0,096203839 | 0,644202376 | PAQR3 | FALSE | Upregulated |
| -0,090602835 | 0,644400526 | DIO1 | FALSE | Downregulated |
| 0,067759457 | 0,644449262 | DPP8 | FALSE | Upregulated |
| -0,357986966 | 0,644449262 | GNG3 | FALSE | Downregulated |
| -0,099882908 | 0,644449262 | SRSF6 | FALSE | Downregulated |
| 0,298381156 | 0,644506203 | MPP3 | FALSE | Upregulated |
| 0,052820705 | 0,644537822 | LARP4B | FALSE | Upregulated |
| -0,637983744 | 0,644683192 | LOC101929726 | FALSE | Downregulated |
| -0,052744924 | 0,644691945 | USP40 | FALSE | Downregulated |
| 0,079375119 | 0,644691945 | ZBTB5 | FALSE | Upregulated |
| -0,056547461 | 0,644925285 | SLC30A5 | FALSE | Downregulated |
| 0,065402409 | 0,644933271 | MED23 | FALSE | Upregulated |
| 0,186082921 | 0,645012152 | KIFC2 | FALSE | Upregulated |
| -0,052238169 | 0,64506847 | WDR36 | FALSE | Downregulated |
| -0,058213276 | 0,645273575 | CECR5 | FALSE | Downregulated |
| -0,091599091 | 0,645307254 | LOC101927187 | FALSE | Downregulated |
| 0,079027734 | 0,645329146 | SESTD1 | FALSE | Upregulated |
| 0,231977284 | 0,645472179 | STAG3L5P-PVRIG2P-PILRB | FALSE | Upregulated |
| -0,301555415 | 0,645598565 | LOC101927686 | FALSE | Downregulated |
| 0,059481674 | 0,645598565 | NUFIP2 | FALSE | Upregulated |
| -0,232150365 | 0,645598565 | TRNN | FALSE | Downregulated |
| 0,115839703 | 0,645598565 | TSSC1 | FALSE | Upregulated |
| -0,077094476 | 0,645938184 | ASMTL | FALSE | Downregulated |
| -0,063296048 | 0,645938184 | DDA1 | FALSE | Downregulated |
| -0,053393789 | 0,645938184 | FAM160B1 | FALSE | Downregulated |
| 0,057034797 | 0,645938184 | MAGED2 | FALSE | Upregulated |
| -0,06459999 | 0,645938184 | SEPSECS | FALSE | Downregulated |
| 0,672502785 | 0,646122174 | LOC728637 | FALSE | Upregulated |
| 0,164604298 | 0,646156859 | CCDC88C | FALSE | Upregulated |
| 0,077763231 | 0,646156859 | DEGS1 | FALSE | Upregulated |
| -0,23721716 | 0,646156859 | TAS2R20 | FALSE | Downregulated |
| 0,173669401 | 0,646156859 | TRPS1 | FALSE | Upregulated |
| -0,736545361 | 0,646222132 | RP11-554A11.6 | FALSE | Downregulated |
| -0,048964761 | 0,646304835 | MED6 | FALSE | Downregulated |
| 0,050272882 | 0,646381726 | DDX6 | FALSE | Upregulated |
| 0,067132898 | 0,646381726 | LIG3 | FALSE | Upregulated |
| 0,106213221 | 0,646486432 | SRI | FALSE | Upregulated |
| -0,074830761 | 0,646702832 | C19orf60 | FALSE | Downregulated |

| | | | | |
|---|---|---|---|---|
| -0,101304572 | 0,646702832 | CLIC2 | FALSE | Downregulated |
| -0,076915307 | 0,646702832 | FRMD4B | FALSE | Downregulated |
| 0,826304909 | 0,646761357 | TMEM59L | FALSE | Upregulated |
| 0,307921146 | 0,646949571 | LOC101928584 | FALSE | Upregulated |
| -0,873710462 | 0,646961342 | B3GALT1 | FALSE | Downregulated |
| 0,390285813 | 0,647013574 | CLUL1 | FALSE | Upregulated |
| -0,133051754 | 0,647178634 | CLPTM1L | FALSE | Downregulated |
| 0,209316672 | 0,647178634 | RP6-206I17.2 | FALSE | Upregulated |
| 0,141333052 | 0,647178634 | TMEM67 | FALSE | Upregulated |
| 0,825559326 | 0,647335789 | GP2 | FALSE | Upregulated |
| 0,301190968 | 0,647848006 | ADAMTS4 | FALSE | Upregulated |
| 0,554471659 | 0,647892052 | KHDRBS2 | FALSE | Upregulated |
| 0,272327521 | 0,648128349 | PRSS35 | FALSE | Upregulated |
| 0,053734997 | 0,648147885 | DPF2 | FALSE | Upregulated |
| -0,251780018 | 0,648147885 | S100A8 | FALSE | Downregulated |
| 0,05231462 | 0,648147885 | USP14 | FALSE | Upregulated |
| 0,538969482 | 0,648385 | ZDHHC8P1 | FALSE | Upregulated |
| -0,096660476 | 0,648389539 | HERPUD1 | FALSE | Downregulated |
| -0,273368378 | 0,648403192 | LOC101927144 | FALSE | Downregulated |
| -0,29084709 | 0,648413953 | DCAF4L1 | FALSE | Downregulated |
| 0,467853168 | 0,648563243 | OCA2 | FALSE | Upregulated |
| 0,623755439 | 0,648628952 | FLJ38122 | FALSE | Upregulated |
| 0,04806438 | 0,648653134 | CCT7 | FALSE | Upregulated |
| 0,370299813 | 0,648824044 | LOC102724117 | FALSE | Upregulated |
| 0,09319858 | 0,648942995 | NISCH | FALSE | Upregulated |
| 0,094149392 | 0,648942995 | NOP9 | FALSE | Upregulated |
| -0,249759705 | 0,648942995 | PRSS36 | FALSE | Downregulated |
| 0,087768515 | 0,648972012 | SIK2 | FALSE | Upregulated |
| 0,608804158 | 0,649003311 | CATSPER1 | FALSE | Upregulated |
| 0,140478159 | 0,649049131 | CELSR1 | FALSE | Upregulated |
| 0,491995805 | 0,649049131 | CFL1P1 | FALSE | Upregulated |
| 0,059399902 | 0,649772684 | AIM1 | FALSE | Upregulated |
| 0,091868467 | 0,649772684 | TSPYL4 | FALSE | Upregulated |
| -0,121262742 | 0,649859951 | ALKBH2 | FALSE | Downregulated |
| 0,088889698 | 0,649978657 | TMEM39B | FALSE | Upregulated |
| -0,263427422 | 0,650142459 | CYP3A7 | FALSE | Downregulated |
| 0,220635051 | 0,650142459 | SMARCD3 | FALSE | Upregulated |
| -0,218680782 | 0,650270914 | ADCY10P1 | FALSE | Downregulated |
| -0,520066337 | 0,650270914 | LOC101928414 | FALSE | Downregulated |
| -0,409082778 | 0,650270914 | LOC102724921 | FALSE | Downregulated |
| 0,079905491 | 0,650270914 | RAB4A | FALSE | Upregulated |
| 0,30862675 | 0,650361511 | ZNF385C | FALSE | Upregulated |
| 0,08887697 | 0,650563274 | ARHGEF10L | FALSE | Upregulated |
| -0,137639751 | 0,650658661 | LRRK2 | FALSE | Downregulated |
| -0,153122846 | 0,650658661 | TVP23C | FALSE | Downregulated |
| -0,180243194 | 0,650678068 | AKR1C2 | FALSE | Downregulated |
| 0,27329927 | 0,650678068 | GHRLOS | FALSE | Upregulated |
| -0,330986085 | 0,650678068 | LINC00671 | FALSE | Downregulated |
| 0,126787723 | 0,650678068 | S100A10 | FALSE | Upregulated |
| -0,05224825 | 0,650678068 | ZBED1 | FALSE | Downregulated |
| -0,430262097 | 0,650702584 | SULT1C2 | FALSE | Downregulated |

| | | | | |
|---|---|---|---|---|
| 0,085420624 | 0,650949322 | CD40 | FALSE | Upregulated |
| 0,206788668 | 0,65101925 | DCST2 | FALSE | Upregulated |
| -0,072142341 | 0,651063057 | SMC5 | FALSE | Downregulated |
| 0,0869539 | 0,651091727 | LRCH4 | FALSE | Upregulated |
| -0,058625036 | 0,651091727 | MRPS23 | FALSE | Downregulated |
| -0,323930455 | 0,651135172 | B3GAT1 | FALSE | Downregulated |
| 0,176971496 | 0,651135172 | PDE6B | FALSE | Upregulated |
| -0,433748286 | 0,651351155 | GRID1 | FALSE | Downregulated |
| 0,403332992 | 0,651464106 | PLEKHG4 | FALSE | Upregulated |
| -0,045882161 | 0,651464106 | PLRG1 | FALSE | Downregulated |
| 0,17503525 | 0,651470928 | RTEL1 | FALSE | Upregulated |
| -0,315915274 | 0,651894293 | LOC102724751 | FALSE | Downregulated |
| 0,114121714 | 0,651894293 | SLC43A3 | FALSE | Upregulated |
| -0,677929203 | 0,651923305 | KRTAP5-2 | FALSE | Downregulated |
| 0,488412933 | 0,652650351 | SNHG4 | FALSE | Upregulated |
| -0,04416334 | 0,653071869 | OTUD5 | FALSE | Downregulated |
| -0,093868834 | 0,653126282 | DENND2C | FALSE | Downregulated |
| -0,073664279 | 0,653143794 | ARRDC1 | FALSE | Downregulated |
| 0,111769002 | 0,653143794 | CARD6 | FALSE | Upregulated |
| -0,13987006 | 0,653143794 | ENGASE | FALSE | Downregulated |
| 0,766511004 | 0,653143794 | KIF5A | FALSE | Upregulated |
| 0,059729621 | 0,653143794 | UBE2D2 | FALSE | Upregulated |
| -0,746893068 | 0,653371265 | FAM132A | FALSE | Downregulated |
| 0,503398802 | 0,653371265 | NPTXR | FALSE | Upregulated |
| 0,576158173 | 0,653371265 | SLC7A5P2 | FALSE | Upregulated |
| 0,368389517 | 0,653379641 | EPB41L4A-AS2 | FALSE | Upregulated |
| 0,057267737 | 0,653379641 | LAMTOR3 | FALSE | Upregulated |
| -0,599096527 | 0,653379641 | LOC101929462 | FALSE | Downregulated |
| 0,340211791 | 0,65352788 | IL20RA | FALSE | Upregulated |
| -0,387387071 | 0,65352788 | LOC101593348 | FALSE | Downregulated |
| 0,886762311 | 0,65352788 | LOC101929078 | FALSE | Upregulated |
| -0,385120607 | 0,65352788 | PLAC8L1 | FALSE | Downregulated |
| -0,069280977 | 0,653590545 | C2orf44 | FALSE | Downregulated |
| 0,311960243 | 0,653590545 | FGF21 | FALSE | Upregulated |
| 0,06743161 | 0,653590545 | TMEM161A | FALSE | Upregulated |
| 0,143874767 | 0,653607881 | DCLRE1C | FALSE | Upregulated |
| 0,204159283 | 0,653607881 | KDM4D | FALSE | Upregulated |
| -0,140527185 | 0,653683736 | AKR1C3 | FALSE | Downregulated |
| -0,391679232 | 0,653683736 | LOC101929675 | FALSE | Downregulated |
| -0,09314085 | 0,653929964 | BZW1 | FALSE | Downregulated |
| 0,101378139 | 0,654024538 | KLF7 | FALSE | Upregulated |
| 0,677143551 | 0,654024538 | LINC00278 | FALSE | Upregulated |
| -0,061938833 | 0,654024538 | RFWD2 | FALSE | Downregulated |
| -0,191356901 | 0,654024538 | TPGS1 | FALSE | Downregulated |
| -0,101892951 | 0,654024538 | UBR4 | FALSE | Downregulated |
| -0,667638038 | 0,654024538 | ZNF98 | FALSE | Downregulated |
| 0,060094284 | 0,654284806 | GMPS | FALSE | Upregulated |
| -0,520437604 | 0,654284806 | LHFPL5 | FALSE | Downregulated |
| -0,350601478 | 0,654284806 | LINC00862 | FALSE | Downregulated |
| -0,088283423 | 0,654432336 | RPL36 | FALSE | Downregulated |
| 0,072793114 | 0,654432336 | SEC23A | FALSE | Upregulated |

| | | | | |
|---|---|---|---|---|
| 0,084366594 | 0,654432336 | ZNF691 | FALSE | Upregulated |
| 0,055723932 | 0,654571656 | GTF2E2 | FALSE | Upregulated |
| -0,150018823 | 0,654726129 | SPATA6L | FALSE | Downregulated |
| 0,650796902 | 0,654728868 | KRTAP5-8 | FALSE | Upregulated |
| -0,064209378 | 0,654728868 | MRPS21 | FALSE | Downregulated |
| -0,80506374 | 0,654846676 | LOC102724143 | FALSE | Downregulated |
| 0,441154998 | 0,655148651 | LOC102723974 | FALSE | Upregulated |
| -0,087733139 | 0,655169113 | TCHP | FALSE | Downregulated |
| 0,245216404 | 0,655486777 | LINC00426 | FALSE | Upregulated |
| 0,712505995 | 0,655541914 | FAM153C | FALSE | Upregulated |
| 0,80122057 | 0,655541914 | TEPP | FALSE | Upregulated |
| 0,570814736 | 0,655726771 | LOC102723721 | FALSE | Upregulated |
| -0,344793553 | 0,655773991 | FAM229A | FALSE | Downregulated |
| 0,202725147 | 0,655978927 | TMEM88 | FALSE | Upregulated |
| 0,083143748 | 0,656071118 | HDAC4 | FALSE | Upregulated |
| 0,082302347 | 0,65611268 | CHD6 | FALSE | Upregulated |
| -0,071428779 | 0,65611268 | DTWD1 | FALSE | Downregulated |
| -0,11800714 | 0,65611268 | FBXO27 | FALSE | Downregulated |
| 0,290081533 | 0,65611268 | RP11-473M20.7 | FALSE | Upregulated |
| 0,081527526 | 0,65611268 | SLC39A3 | FALSE | Upregulated |
| 0,31293603 | 0,656134132 | NEGR1 | FALSE | Upregulated |
| -0,039589047 | 0,656321163 | SKIV2L2 | FALSE | Downregulated |
| 0,104181093 | 0,656322522 | ARMC6 | FALSE | Upregulated |
| -0,442589711 | 0,656322522 | LOC100996761 | FALSE | Downregulated |
| -0,172006481 | 0,656341729 | AC074138.3 | FALSE | Downregulated |
| 0,496276334 | 0,65647907 | LOC101929330 | FALSE | Upregulated |
| 0,176730436 | 0,656545592 | MYBL1 | FALSE | Upregulated |
| -0,775852445 | 0,656680458 | LOC102724004 | FALSE | Downregulated |
| 0,241330643 | 0,65676744 | PABPC1L | FALSE | Upregulated |
| 0,128669579 | 0,656840585 | LINC00574 | FALSE | Upregulated |
| 0,073256334 | 0,657021093 | NT5C | FALSE | Upregulated |
| -0,648373694 | 0,657124142 | FLJ31356 | FALSE | Downregulated |
| -0,067429768 | 0,657533131 | EIF6 | FALSE | Downregulated |
| 0,273931299 | 0,657616691 | RPL13AP3 | FALSE | Upregulated |
| 0,088963726 | 0,657641903 | MRPL54 | FALSE | Upregulated |
| -0,090542362 | 0,657674435 | KANSL1L | FALSE | Downregulated |
| 0,107776062 | 0,657706655 | ISPD | FALSE | Upregulated |
| -0,062509026 | 0,657729038 | MKRN2 | FALSE | Downregulated |
| 0,787435642 | 0,657729038 | UNQ9370 | FALSE | Upregulated |
| 0,146514895 | 0,65830735 | C5orf30 | FALSE | Upregulated |
| -0,130282701 | 0,65830735 | ICAM3 | FALSE | Downregulated |
| -0,083651439 | 0,658386238 | RSU1 | FALSE | Downregulated |
| -0,315418722 | 0,658425527 | NDUFA6-AS1 | FALSE | Downregulated |
| 0,102028097 | 0,658425527 | SUGP2 | FALSE | Upregulated |
| -0,458334776 | 0,658485741 | ABCG4 | FALSE | Downregulated |
| -0,119974922 | 0,658485741 | DANCR | FALSE | Downregulated |
| 0,061162434 | 0,658485741 | DHX40 | FALSE | Upregulated |
| -0,059240523 | 0,658485741 | IDS | FALSE | Downregulated |
| -0,238988846 | 0,658485741 | KIAA1875 | FALSE | Downregulated |
| -0,394654117 | 0,658485741 | LOC101928021 | FALSE | Downregulated |
| 0,201380209 | 0,658485741 | ZFP69B | FALSE | Upregulated |

| | | | | |
|---|---|---|---|---|
| 0,151714137 | 0,658855421 | HOXB4 | FALSE | Upregulated |
| -0,098287776 | 0,658855421 | SERPINF2 | FALSE | Downregulated |
| -0,272768131 | 0,658895986 | LRRC66 | FALSE | Downregulated |
| -0,234550629 | 0,658895986 | NLRP6 | FALSE | Downregulated |
| 0,716556455 | 0,658895986 | TRNAK6 | FALSE | Upregulated |
| -0,080207434 | 0,658908395 | TRAK1 | FALSE | Downregulated |
| 0,086284603 | 0,658933019 | CXorf23 | FALSE | Upregulated |
| 0,345405212 | 0,658958755 | RP11-274B18.2 | FALSE | Upregulated |
| 0,668100492 | 0,659049664 | MEP1A | FALSE | Upregulated |
| -0,066459664 | 0,659071299 | CSTF3 | FALSE | Downregulated |
| -0,120908872 | 0,659071299 | JAZF1 | FALSE | Downregulated |
| 0,070749461 | 0,659170838 | DDX49 | FALSE | Upregulated |
| -0,072719153 | 0,659170838 | PPP2R2A | FALSE | Downregulated |
| 0,097881752 | 0,659210656 | C3orf58 | FALSE | Upregulated |
| -0,148611695 | 0,659311342 | CHIC1 | FALSE | Downregulated |
| -0,079267512 | 0,659472771 | POLR1E | FALSE | Downregulated |
| -0,185377521 | 0,659481864 | C21orf58 | FALSE | Downregulated |
| 0,193800374 | 0,659481864 | SLC9B1 | FALSE | Upregulated |
| 0,638771387 | 0,659577014 | GPA33 | FALSE | Upregulated |
| -0,052136279 | 0,659633165 | SUMO3 | FALSE | Downregulated |
| 0,633952866 | 0,659637345 | CORIN | FALSE | Upregulated |
| -0,529547231 | 0,659637345 | MAB21L1 | FALSE | Downregulated |
| 0,271590003 | 0,659903403 | MPZ | FALSE | Upregulated |
| -0,083973739 | 0,659935325 | DMD | FALSE | Downregulated |
| 0,058173272 | 0,659935325 | FBXO45 | FALSE | Upregulated |
| -0,248905883 | 0,659935325 | WBSCR27 | FALSE | Downregulated |
| -0,119123021 | 0,659993097 | TUBA4A | FALSE | Downregulated |
| 0,221805589 | 0,660465467 | CYP26B1 | FALSE | Upregulated |
| -0,113108851 | 0,660465467 | UBN2 | FALSE | Downregulated |
| -0,276114798 | 0,660469471 | UNC79 | FALSE | Downregulated |
| 0,067750518 | 0,66074985 | ATXN7 | FALSE | Upregulated |
| 0,349134978 | 0,66074985 | GPR113 | FALSE | Upregulated |
| 0,232167033 | 0,66074985 | TDGF1 | FALSE | Upregulated |
| 0,083839254 | 0,66074985 | TGIF2 | FALSE | Upregulated |
| -0,436174025 | 0,660788535 | LOC102724262 | FALSE | Downregulated |
| 0,10300861 | 0,660987711 | ATP6V0B | FALSE | Upregulated |
| -0,542373587 | 0,660987711 | LOC101926936 | FALSE | Downregulated |
| -0,503868224 | 0,660987711 | LOC101930567 | FALSE | Downregulated |
| -0,069893147 | 0,661043461 | CYB5R1 | FALSE | Downregulated |
| 0,102435633 | 0,661143874 | TCN2 | FALSE | Upregulated |
| 0,525071879 | 0,661283769 | IGF2BP1 | FALSE | Upregulated |
| 0,875889368 | 0,661283769 | PIWIL1 | FALSE | Upregulated |
| 0,754569314 | 0,66149531 | CHRNA7 | FALSE | Upregulated |
| -0,104536572 | 0,661512047 | ZNF19 | FALSE | Downregulated |
| 0,239815358 | 0,661587987 | GPSM2 | FALSE | Upregulated |
| 0,043567521 | 0,661695391 | PDPK1 | FALSE | Upregulated |
| -0,048153905 | 0,661726578 | SNRNP200 | FALSE | Downregulated |
| -0,116631336 | 0,661786562 | PALMD | FALSE | Downregulated |
| 0,135019926 | 0,66181351 | SIGLEC1 | FALSE | Upregulated |
| 0,458383715 | 0,661824821 | VWA8-AS1 | FALSE | Upregulated |
| 0,054463961 | 0,661871668 | PHF12 | FALSE | Upregulated |

| | | | | |
|---|---|---|---|---|
| 0,229654288 | 0,661981381 | LOC101928415 | FALSE | Upregulated |
| 0,119263071 | 0,662037174 | B4GALT6 | FALSE | Upregulated |
| 0,409254849 | 0,662043234 | MID1-2 | FALSE | Upregulated |
| -0,275607012 | 0,662515818 | LOC101928768 | FALSE | Downregulated |
| 0,256784151 | 0,662515818 | MT1L | FALSE | Upregulated |
| -0,72384376 | 0,662878927 | FMR1NB | FALSE | Downregulated |
| 0,085015408 | 0,662895899 | RFNG | FALSE | Upregulated |
| -0,078521258 | 0,663003407 | EXOC6 | FALSE | Downregulated |
| -0,28494392 | 0,66308431 | CLDND2 | FALSE | Downregulated |
| 0,472931704 | 0,663128694 | MYCN | FALSE | Upregulated |
| -0,079908735 | 0,663153745 | PINX1 | FALSE | Downregulated |
| -0,670882034 | 0,663159615 | NCAM1-AS1 | FALSE | Downregulated |
| -0,174159833 | 0,66348985 | IQSEC3 | FALSE | Downregulated |
| 0,261710209 | 0,663660474 | ZNF781 | FALSE | Upregulated |
| 0,502526827 | 0,664169783 | HIST1H2AG | FALSE | Upregulated |
| -0,126043069 | 0,664233962 | RAD54L2 | FALSE | Downregulated |
| 0,525086662 | 0,664420323 | RP11-14N7.2 | FALSE | Upregulated |
| -0,107750563 | 0,664420323 | THSD4 | FALSE | Downregulated |
| -0,057576668 | 0,664829595 | DENND4C | FALSE | Downregulated |
| -0,735625126 | 0,664993003 | ANKRD20A3 | FALSE | Downregulated |
| -0,275693397 | 0,665005054 | CAHM | FALSE | Downregulated |
| 0,218747789 | 0,665005054 | LOC100288181 | FALSE | Upregulated |
| -0,045771629 | 0,665032503 | GDE1 | FALSE | Downregulated |
| 0,05034897 | 0,66510589 | PARN | FALSE | Upregulated |
| 0,101752282 | 0,665121708 | HAUS5 | FALSE | Upregulated |
| -0,126517517 | 0,665121708 | MZF1 | FALSE | Downregulated |
| -0,10170809 | 0,665272806 | MPZL2 | FALSE | Downregulated |
| -0,605349084 | 0,665342283 | AQPEP | FALSE | Downregulated |
| 0,734213251 | 0,665585773 | INGX | FALSE | Upregulated |
| 0,188007838 | 0,665585773 | SLC1A2 | FALSE | Upregulated |
| -0,246932275 | 0,665585773 | TMEM253 | FALSE | Downregulated |
| 0,662917199 | 0,665617337 | GABRA4 | FALSE | Upregulated |
| 0,060766758 | 0,665674249 | BCAS3 | FALSE | Upregulated |
| 0,609648276 | 0,665674249 | TBC1D29 | FALSE | Upregulated |
| -0,470689715 | 0,665843326 | SEC24B-AS1 | FALSE | Downregulated |
| -0,693727179 | 0,665852526 | CXorf64 | FALSE | Downregulated |
| -0,300045037 | 0,666205407 | LOC102723824 | FALSE | Downregulated |
| 0,057073149 | 0,666334076 | FNTA | FALSE | Upregulated |
| -0,18966496 | 0,666334076 | MEF2C | FALSE | Downregulated |
| 0,099367236 | 0,666425894 | F2R | FALSE | Upregulated |
| 0,11570016 | 0,666465618 | LUC7L | FALSE | Upregulated |
| 0,186311965 | 0,666686769 | ELOVL2-AS1 | FALSE | Upregulated |
| -0,377059457 | 0,666972777 | CRYBB3 | FALSE | Downregulated |
| -0,231565444 | 0,667041517 | LOC102723415 | FALSE | Downregulated |
| -0,049482791 | 0,667041517 | LSM8 | FALSE | Downregulated |
| 0,057948187 | 0,667041517 | SNRPE | FALSE | Upregulated |
| -0,500594815 | 0,667041517 | SPG20OS | FALSE | Downregulated |
| -0,276146594 | 0,667232522 | FGF14 | FALSE | Downregulated |
| 1,824908597 | 0,667243865 | EIF1AY | FALSE | Upregulated |
| -0,664416763 | 0,667550943 | SLC22A11 | FALSE | Downregulated |
| -0,110682425 | 0,667586341 | CFP | FALSE | Downregulated |

| | | | | |
|---|---|---|---|---|
| 0,066717923 | 0,667637947 | KDM3A | FALSE | Upregulated |
| 0,187311994 | 0,667647672 | SPSB4 | FALSE | Upregulated |
| -0,281329988 | 0,667650146 | LOC401286 | FALSE | Downregulated |
| 0,084140964 | 0,667892384 | PHKG2 | FALSE | Upregulated |
| 0,059874749 | 0,667916416 | APPL2 | FALSE | Upregulated |
| -0,291913828 | 0,667916416 | LOC646626 | FALSE | Downregulated |
| 0,044951071 | 0,667941686 | HNRNPUL2 | FALSE | Upregulated |
| -0,800428162 | 0,667998792 | CCDC79 | FALSE | Downregulated |
| 0,061689498 | 0,668017662 | PTOV1 | FALSE | Upregulated |
| -0,080263411 | 0,66806547 | CCNG1 | FALSE | Downregulated |
| 0,074548454 | 0,66806547 | RCBTB2 | FALSE | Upregulated |
| -0,117913776 | 0,668198414 | CRYBB2P1 | FALSE | Downregulated |
| 0,096149782 | 0,668212202 | GOLT1A | FALSE | Upregulated |
| -0,613062408 | 0,668212202 | LOC100505938 | FALSE | Downregulated |
| -0,510685009 | 0,668212202 | LOC102723870 | FALSE | Downregulated |
| 0,138289673 | 0,668212202 | MIR497HG | FALSE | Upregulated |
| -0,050889741 | 0,668212202 | NUP54 | FALSE | Downregulated |
| -0,275185142 | 0,668212202 | SFTPD | FALSE | Downregulated |
| 0,102441023 | 0,668212202 | TMEM80 | FALSE | Upregulated |
| 0,057156784 | 0,668212202 | TMX2 | FALSE | Upregulated |
| -0,057237956 | 0,668212202 | VWA8 | FALSE | Downregulated |
| -0,784773532 | 0,668212202 | XKRX | FALSE | Downregulated |
| 0,060317977 | 0,668274161 | INTS4 | FALSE | Upregulated |
| -0,095758216 | 0,668292742 | BSCL2 | FALSE | Downregulated |
| -0,189859187 | 0,668292742 | CCDC151 | FALSE | Downregulated |
| 0,170497397 | 0,668292742 | GRASP | FALSE | Upregulated |
| -0,201400638 | 0,668292742 | LOC101929072 | FALSE | Downregulated |
| 0,710258294 | 0,668416455 | BEST4 | FALSE | Upregulated |
| 0,619289257 | 0,668416455 | LOC101929447 | FALSE | Upregulated |
| -0,050486323 | 0,668428125 | KDELR1 | FALSE | Downregulated |
| 0,447019668 | 0,668428125 | RP11-173B14.5 | FALSE | Upregulated |
| -0,212705711 | 0,668539646 | COL4A5 | FALSE | Downregulated |
| 0,10658442 | 0,668548519 | AKR7A3 | FALSE | Upregulated |
| -0,342441594 | 0,66885659 | FAM163A | FALSE | Downregulated |
| 0,190443188 | 0,668877644 | BMS1P5 | FALSE | Upregulated |
| -0,503321526 | 0,669057906 | ADCYAP1R1 | FALSE | Downregulated |
| 0,485954957 | 0,66909482 | LOC102724930 | FALSE | Upregulated |
| 0,046254865 | 0,669167414 | VPS35 | FALSE | Upregulated |
| 0,141117128 | 0,669180445 | LINC00341 | FALSE | Upregulated |
| -0,786681954 | 0,669296359 | CHIA | FALSE | Downregulated |
| -0,276580116 | 0,669296359 | HEPACAM | FALSE | Downregulated |
| 0,41466345 | 0,669631964 | RNF138P1 | FALSE | Upregulated |
| 0,831157278 | 0,669647027 | ELOVL3 | FALSE | Upregulated |
| -0,067186006 | 0,669748862 | GGCX | FALSE | Downregulated |
| -0,062819513 | 0,669748862 | GRB10 | FALSE | Downregulated |
| -0,089400131 | 0,669748862 | KMT2E | FALSE | Downregulated |
| 0,255866068 | 0,669748862 | LINC00265 | FALSE | Upregulated |
| 0,074243372 | 0,669748862 | PHLDB3 | FALSE | Upregulated |
| -0,343851955 | 0,669903102 | RGMB-AS1 | FALSE | Downregulated |
| 0,052122461 | 0,670086712 | ASF1A | FALSE | Upregulated |
| 0,728919817 | 0,670086712 | FLJ45872 | FALSE | Upregulated |

| | | | | |
|---|---|---|---|---|
| -0,167195051 | 0,670108924 | C8orf44 | FALSE | Downregulated |
| 0,091977142 | 0,670108924 | STX16 | FALSE | Upregulated |
| 0,058519339 | 0,670108924 | VHL | FALSE | Upregulated |
| -0,709379633 | 0,670378389 | LOC101928231 | FALSE | Downregulated |
| -0,087576686 | 0,670378389 | NMNAT1 | FALSE | Downregulated |
| -0,497911179 | 0,670396148 | NCOR1P1 | FALSE | Downregulated |
| 0,112408608 | 0,670457657 | ZNF608 | FALSE | Upregulated |
| 0,051300076 | 0,670702569 | NUP98 | FALSE | Upregulated |
| 0,545571961 | 0,670755464 | LOC101928574 | FALSE | Upregulated |
| 0,294971454 | 0,670930654 | LOC399900 | FALSE | Upregulated |
| 0,123620732 | 0,67102078 | DYNLL1 | FALSE | Upregulated |
| 0,276640227 | 0,67102078 | LOC102723901 | FALSE | Upregulated |
| -0,057514187 | 0,67102078 | MRPL1 | FALSE | Downregulated |
| -0,66287809 | 0,67102078 | RP11-26J3.1 | FALSE | Downregulated |
| 0,79531022 | 0,671176729 | CLEC18C | FALSE | Upregulated |
| -0,058620951 | 0,671176729 | DPY19L1 | FALSE | Downregulated |
| -0,162303862 | 0,671176729 | IGFBP7 | FALSE | Downregulated |
| 0,05996769 | 0,671176729 | PIK3R2 | FALSE | Upregulated |
| 0,371335212 | 0,671176729 | TLL1 | FALSE | Upregulated |
| 0,229730048 | 0,671176729 | XK | FALSE | Upregulated |
| 0,324294762 | 0,671384096 | KIAA1755 | FALSE | Upregulated |
| -0,104846216 | 0,671384096 | MARCH9 | FALSE | Downregulated |
| -0,132500808 | 0,671384096 | NQO2 | FALSE | Downregulated |
| -0,339912758 | 0,671417787 | LOC102724126 | FALSE | Downregulated |
| 0,090340044 | 0,671417787 | LSM4 | FALSE | Upregulated |
| 0,147117527 | 0,671615353 | GPR183 | FALSE | Upregulated |
| -0,58732355 | 0,671615353 | PPAPDC3 | FALSE | Downregulated |
| 0,312649717 | 0,671615353 | SPATA25 | FALSE | Upregulated |
| 0,045802835 | 0,671639636 | AREL1 | FALSE | Upregulated |
| 0,109867579 | 0,671701748 | PTPRM | FALSE | Upregulated |
| 0,763042793 | 0,671890089 | DMRT2 | FALSE | Upregulated |
| -0,259459417 | 0,671890089 | SEC14L5 | FALSE | Downregulated |
| -0,124932727 | 0,671890089 | TIGD1 | FALSE | Downregulated |
| 0,109003265 | 0,67191922 | AAED1 | FALSE | Upregulated |
| 0,102760804 | 0,672045488 | IL22RA1 | FALSE | Upregulated |
| 0,450150694 | 0,672132388 | RP11-439L8.4 | FALSE | Upregulated |
| 0,137629207 | 0,672189925 | ADRA1B | FALSE | Upregulated |
| 0,572731121 | 0,672189925 | CACNA1F | FALSE | Upregulated |
| -0,075903166 | 0,672200664 | ATAD2B | FALSE | Downregulated |
| -0,48391464 | 0,672217181 | SEBOX | FALSE | Downregulated |
| -0,296381151 | 0,6722191 | CHAC1 | FALSE | Downregulated |
| 0,038338697 | 0,672358107 | UBE3C | FALSE | Upregulated |
| 0,558252722 | 0,672397885 | CSDC2 | FALSE | Upregulated |
| -0,107741893 | 0,672397885 | EGFL7 | FALSE | Downregulated |
| 0,352259707 | 0,672670041 | LINC00665 | FALSE | Upregulated |
| 0,107874659 | 0,672686333 | RASSF8 | FALSE | Upregulated |
| 0,209888408 | 0,672761062 | CCDC183-AS1 | FALSE | Upregulated |
| -0,541566832 | 0,672761062 | LOC102723656 | FALSE | Downregulated |
| 0,584613141 | 0,673032826 | JAZF1-AS1 | FALSE | Upregulated |
| 0,133252617 | 0,673206354 | ADAMTS17 | FALSE | Upregulated |
| 0,774540635 | 0,673206354 | ARHGAP40 | FALSE | Upregulated |

| | | | | |
|---|---|---|---|---|
| 0,07300094 | 0,673206354 | ATF6 | FALSE | Upregulated |
| -0,141771995 | 0,67341626 | GEMIN8P4 | FALSE | Downregulated |
| 0,952464229 | 0,67341626 | KCNQ2 | FALSE | Upregulated |
| 0,536340313 | 0,67341626 | MSH4 | FALSE | Upregulated |
| -0,130055328 | 0,67341626 | TMEM82 | FALSE | Downregulated |
| 0,17362443 | 0,673571067 | LOC100996255 | FALSE | Upregulated |
| -0,202599514 | 0,673571067 | LOC400927 | FALSE | Downregulated |
| -0,186105154 | 0,674159761 | PTGES2-AS1 | FALSE | Downregulated |
| 0,504990487 | 0,67417972 | GRM1 | FALSE | Upregulated |
| 0,378302881 | 0,67417972 | LOC729224 | FALSE | Upregulated |
| -0,129941278 | 0,67417972 | PAQR7 | FALSE | Downregulated |
| -0,32779904 | 0,67417972 | RHBDL1 | FALSE | Downregulated |
| 0,451275586 | 0,67417972 | ZNF571-AS1 | FALSE | Upregulated |
| -0,044598172 | 0,674209395 | IREB2 | FALSE | Downregulated |
| 0,544538348 | 0,674315315 | TNFRSF13B | FALSE | Upregulated |
| 0,480207744 | 0,674523454 | RP11-76N22.2 | FALSE | Upregulated |
| -1,335738884 | 0,674690102 | TRNAH6 | FALSE | Downregulated |
| -0,265070615 | 0,674691117 | LOC102724768 | FALSE | Downregulated |
| -0,585918532 | 0,674691117 | TEX22 | FALSE | Downregulated |
| -0,169520082 | 0,674950335 | HOXB3 | FALSE | Downregulated |
| -0,302030061 | 0,675051585 | GDPD3 | FALSE | Downregulated |
| -0,223443073 | 0,675051585 | GLP2R | FALSE | Downregulated |
| 0,462238621 | 0,675051585 | MEF2C-AS1 | FALSE | Upregulated |
| -0,318371269 | 0,675075476 | LOC100505915 | FALSE | Downregulated |
| -0,078912831 | 0,675319656 | SCNM1 | FALSE | Downregulated |
| -0,097840105 | 0,675319656 | SDPR | FALSE | Downregulated |
| -0,065403356 | 0,675552857 | HKR1 | FALSE | Downregulated |
| 0,098413989 | 0,675647301 | TRIM5 | FALSE | Upregulated |
| 0,041769154 | 0,675678887 | ARMC8 | FALSE | Upregulated |
| 0,121534469 | 0,675839157 | SLC8A1 | FALSE | Upregulated |
| 0,124816288 | 0,675839157 | ZNF284 | FALSE | Upregulated |
| 0,070259395 | 0,675876276 | DCAF13 | FALSE | Upregulated |
| -0,090184922 | 0,675914287 | MTSS1 | FALSE | Downregulated |
| 0,082319786 | 0,675914287 | ORMDL1 | FALSE | Upregulated |
| -0,106695269 | 0,676011967 | CDK20 | FALSE | Downregulated |
| -0,374809857 | 0,676011967 | NTN3 | FALSE | Downregulated |
| -0,053346547 | 0,676011967 | SCAF4 | FALSE | Downregulated |
| -0,109439503 | 0,676011967 | ZNF559 | FALSE | Downregulated |
| -0,128249929 | 0,67627076 | GIMAP4 | FALSE | Downregulated |
| 0,175339245 | 0,676307332 | TMEM198 | FALSE | Upregulated |
| -0,16978113 | 0,676445172 | ETNPPL | FALSE | Downregulated |
| 0,135097855 | 0,67701262 | FAM86EP | FALSE | Upregulated |
| 0,096731061 | 0,67708457 | LMNB1 | FALSE | Upregulated |
| -0,083030119 | 0,67708457 | RPL27A | FALSE | Downregulated |
| 0,137557622 | 0,677191607 | LINC01137 | FALSE | Upregulated |
| 0,472370346 | 0,677191607 | LOC100996506 | FALSE | Upregulated |
| -0,247263019 | 0,677191607 | LOC101926967 | FALSE | Downregulated |
| 0,124467978 | 0,677191607 | MS4A7 | FALSE | Upregulated |
| -0,10603871 | 0,677254685 | BLOC1S5 | FALSE | Downregulated |
| -0,102622416 | 0,677275147 | GARNL3 | FALSE | Downregulated |
| -0,069476182 | 0,677288827 | CBX7 | FALSE | Downregulated |

| | | | | |
|---|---|---|---|---|
| -0,177868442 | 0,677288827 | USP3-AS1 | FALSE | Downregulated |
| -0,582375529 | 0,677529308 | FAM156A | FALSE | Downregulated |
| 0,16800057 | 0,677529308 | SPATA6 | FALSE | Upregulated |
| 0,725115688 | 0,677634007 | ZBTB32 | FALSE | Upregulated |
| 0,04765891 | 0,677641226 | YIPF5 | FALSE | Upregulated |
| -0,823263908 | 0,678242872 | MYOZ3 | FALSE | Downregulated |
| 0,063875362 | 0,678242872 | PEF1 | FALSE | Upregulated |
| -0,068972408 | 0,678410637 | ITGA1 | FALSE | Downregulated |
| 0,825280868 | 0,678545945 | FAM106A | FALSE | Upregulated |
| 0,09652663 | 0,678545945 | GIGYF1 | FALSE | Upregulated |
| -0,382124517 | 0,678545945 | RP11-333A23.4 | FALSE | Downregulated |
| -0,118388621 | 0,678558179 | GK5 | FALSE | Downregulated |
| -0,069683469 | 0,678610895 | TRIP11 | FALSE | Downregulated |
| 0,223167968 | 0,678682277 | LOC101929178 | FALSE | Upregulated |
| 0,087321055 | 0,678845844 | JMJD4 | FALSE | Upregulated |
| -0,800103329 | 0,678909457 | ASB12 | FALSE | Downregulated |
| 0,069906421 | 0,678909457 | FNBP1L | FALSE | Upregulated |
| 0,054230161 | 0,678909457 | POGK | FALSE | Upregulated |
| 0,361535956 | 0,678909457 | USP44 | FALSE | Upregulated |
| 0,665540077 | 0,6789258 | LOC102724898 | FALSE | Upregulated |
| 0,395095928 | 0,678964785 | TSPAN32 | FALSE | Upregulated |
| 0,331732787 | 0,679034932 | CACNB4 | FALSE | Upregulated |
| -0,5912561 | 0,679034932 | FLNC | FALSE | Downregulated |
| 0,101563878 | 0,679397541 | CAB39L | FALSE | Upregulated |
| 0,064233168 | 0,679410107 | MAN2A1 | FALSE | Upregulated |
| 0,640255171 | 0,679471709 | WT1-AS | FALSE | Upregulated |
| -0,082687636 | 0,68009577 | CDK5 | FALSE | Downregulated |
| 0,0433195 | 0,68009577 | CUL2 | FALSE | Upregulated |
| -0,055734873 | 0,68009577 | GSTK1 | FALSE | Downregulated |
| 0,539099291 | 0,68009577 | LOC102724423 | FALSE | Upregulated |
| -0,636851587 | 0,680258106 | LOC102724119 | FALSE | Downregulated |
| 0,180443082 | 0,680258106 | SYBU | FALSE | Upregulated |
| -0,194889847 | 0,680333091 | PIH1D2 | FALSE | Downregulated |
| -0,09683353 | 0,680345804 | NHLRC1 | FALSE | Downregulated |
| -0,321437426 | 0,680397949 | FSBP | FALSE | Downregulated |
| 0,047406269 | 0,680427607 | CLK3 | FALSE | Upregulated |
| 0,500183061 | 0,680427607 | OTOF | FALSE | Upregulated |
| -0,625543271 | 0,680431368 | AC007966.1 | FALSE | Downregulated |
| -0,503919083 | 0,680698161 | LOC102724636 | FALSE | Downregulated |
| 0,565195933 | 0,680720712 | C1QTNF1-AS1 | FALSE | Upregulated |
| -0,219364155 | 0,680720712 | DNAJA4 | FALSE | Downregulated |
| -0,33534073 | 0,680720712 | HIST1H2AM | FALSE | Downregulated |
| 0,606478029 | 0,680720712 | LOC101929259 | FALSE | Upregulated |
| 0,075140881 | 0,680720712 | PSMD14 | FALSE | Upregulated |
| -0,056220157 | 0,680720712 | SIKE1 | FALSE | Downregulated |
| 0,612752764 | 0,680747821 | LINC01122 | FALSE | Upregulated |
| -0,079491463 | 0,681347027 | TMEM222 | FALSE | Downregulated |
| -0,048331057 | 0,681494657 | DYM | FALSE | Downregulated |
| -0,101456184 | 0,681533074 | PLEKHA7 | FALSE | Downregulated |
| 0,055061656 | 0,681665489 | SCAMP4 | FALSE | Upregulated |
| 0,070276064 | 0,681775522 | BCL9 | FALSE | Upregulated |

| | | | | |
|---|---|---|---|---|
| 0,04722272 | 0,681775522 | DR1 | FALSE | Upregulated |
| -0,666728504 | 0,681775522 | RGS9BP | FALSE | Downregulated |
| -0,128043105 | 0,681930016 | TMCC1-AS1 | FALSE | Downregulated |
| -0,049514725 | 0,682287312 | VPS72 | FALSE | Downregulated |
| 0,241163315 | 0,682287312 | ZNF833P | FALSE | Upregulated |
| -0,043128449 | 0,682326063 | RRN3 | FALSE | Downregulated |
| 0,208763448 | 0,682388892 | LOC101929177 | FALSE | Upregulated |
| 0,077470953 | 0,682590416 | ZNF782 | FALSE | Upregulated |
| 0,071611217 | 0,682682395 | IRF2BP1 | FALSE | Upregulated |
| -0,177218669 | 0,682719297 | SGOL2 | FALSE | Downregulated |
| -0,218814027 | 0,682916541 | HTRA3 | FALSE | Downregulated |
| 0,191025108 | 0,682916541 | SSC5D | FALSE | Upregulated |
| -0,04917446 | 0,682921156 | MAVS | FALSE | Downregulated |
| 0,317391507 | 0,682929249 | MUC3A | FALSE | Upregulated |
| 0,250785705 | 0,682978221 | TDRD9 | FALSE | Upregulated |
| -0,141848889 | 0,683438313 | DDX11 | FALSE | Downregulated |
| -0,076664859 | 0,683438313 | FAM207A | FALSE | Downregulated |
| -0,628423132 | 0,683438313 | LOC101928402 | FALSE | Downregulated |
| 0,491832284 | 0,683438313 | MYCBPAP | FALSE | Upregulated |
| 0,097219763 | 0,683547821 | PPIB | FALSE | Upregulated |
| 0,100748274 | 0,683562333 | IDH2 | FALSE | Upregulated |
| 0,088802323 | 0,683572713 | GLRX2 | FALSE | Upregulated |
| 0,159804909 | 0,683576507 | ARIH2OS | FALSE | Upregulated |
| 0,404535861 | 0,683576507 | FLJ38723 | FALSE | Upregulated |
| 0,170741801 | 0,683847658 | B3GALT4 | FALSE | Upregulated |
| 0,484053826 | 0,683847658 | CHKB-CPT1B | FALSE | Upregulated |
| 0,181033617 | 0,683847658 | CSTA | FALSE | Upregulated |
| 0,255779094 | 0,683847658 | H1FX-AS1 | FALSE | Upregulated |
| 0,38550093 | 0,683847658 | SNORA33 | FALSE | Upregulated |
| 0,071559511 | 0,683847658 | TRMT11 | FALSE | Upregulated |
| -0,07693354 | 0,683948069 | CERS6 | FALSE | Downregulated |
| 0,291346149 | 0,68411259 | EMILIN3 | FALSE | Upregulated |
| -0,043869766 | 0,684446171 | TK2 | FALSE | Downregulated |
| 0,046591317 | 0,684650934 | TBC1D20 | FALSE | Upregulated |
| 0,324497746 | 0,68484657 | C20orf202 | FALSE | Upregulated |
| -0,135568998 | 0,68484657 | GALNT18 | FALSE | Downregulated |
| -0,125109596 | 0,684927536 | MICALCL | FALSE | Downregulated |
| 0,09901465 | 0,684927536 | ZBED6CL | FALSE | Upregulated |
| -0,303107107 | 0,685051161 | CASP16 | FALSE | Downregulated |
| 0,099291148 | 0,685051161 | DMXL2 | FALSE | Upregulated |
| -0,077759726 | 0,685051161 | FAAH | FALSE | Downregulated |
| 0,082799082 | 0,685051161 | FAM127B | FALSE | Upregulated |
| -0,516658189 | 0,685051161 | IGSF1 | FALSE | Downregulated |
| 0,065303669 | 0,685051161 | LBR | FALSE | Upregulated |
| 0,165371441 | 0,685051161 | LDLR | FALSE | Upregulated |
| 0,069707918 | 0,685051161 | NCOA4 | FALSE | Upregulated |
| -0,581071004 | 0,685051161 | RP11-430H10.1 | FALSE | Downregulated |
| 0,054118384 | 0,685051161 | ZNF45 | FALSE | Upregulated |
| -0,094812364 | 0,685213193 | ADRB2 | FALSE | Downregulated |
| -0,098020715 | 0,685626383 | MICU3 | FALSE | Downregulated |
| -0,543843142 | 0,685732078 | LOC101929060 | FALSE | Downregulated |

| | | | | |
|---|---|---|---|---|
| -0,073458401 | 0,685846271 | TSEN15 | FALSE | Downregulated |
| -0,181199573 | 0,686190634 | RP1-27K12.2 | FALSE | Downregulated |
| 0,101700509 | 0,686216811 | MYRIP | FALSE | Upregulated |
| 0,376248888 | 0,686260999 | EMR3 | FALSE | Upregulated |
| -0,044106645 | 0,68628191 | THUMPD3 | FALSE | Downregulated |
| 0,124903973 | 0,68628191 | ZNF446 | FALSE | Upregulated |
| -0,06059728 | 0,686563303 | RNASEH1 | FALSE | Downregulated |
| -0,230254723 | 0,686653964 | CXCL2 | FALSE | Downregulated |
| 0,179046573 | 0,686653964 | GDF7 | FALSE | Upregulated |
| 0,551280755 | 0,686653964 | HMSD | FALSE | Upregulated |
| 0,07923712 | 0,686653964 | MOSPD2 | FALSE | Upregulated |
| -0,2618978 | 0,686653964 | SPESP1 | FALSE | Downregulated |
| 0,127873035 | 0,686653964 | STON2 | FALSE | Upregulated |
| 0,102241543 | 0,686653964 | TRIM16 | FALSE | Upregulated |
| -0,168070059 | 0,686656334 | ABRACL | FALSE | Downregulated |
| 0,179235205 | 0,686656334 | KIF9-AS1 | FALSE | Upregulated |
| -0,635348464 | 0,686656334 | LOC102723361 | FALSE | Downregulated |
| -0,05758215 | 0,686656334 | OSBPL8 | FALSE | Downregulated |
| -0,398404223 | 0,686771999 | BSN-AS2 | FALSE | Downregulated |
| 0,223429069 | 0,686854027 | ITGA10 | FALSE | Upregulated |
| -0,044314139 | 0,686854027 | KANSL1 | FALSE | Downregulated |
| -0,059894292 | 0,686854027 | SPIDR | FALSE | Downregulated |
| -0,116910096 | 0,686854027 | ZBTB11-AS1 | FALSE | Downregulated |
| -0,661398523 | 0,686862712 | LOC102724093 | FALSE | Downregulated |
| -0,081026504 | 0,686887287 | MARVELD3 | FALSE | Downregulated |
| 0,077085265 | 0,686887287 | NOM1 | FALSE | Upregulated |
| 0,654399513 | 0,686887287 | SLC22A14 | FALSE | Upregulated |
| 0,09590449 | 0,686887287 | ZNF784 | FALSE | Upregulated |
| -0,079095414 | 0,686911354 | ZNF30 | FALSE | Downregulated |
| 0,085606186 | 0,68693105 | TXNDC5 | FALSE | Upregulated |
| -0,38035723 | 0,686966757 | LOC101929533 | FALSE | Downregulated |
| 0,500964449 | 0,686971328 | MSANTD1 | FALSE | Upregulated |
| -0,190501959 | 0,686985483 | KDM8 | FALSE | Downregulated |
| 0,63450848 | 0,686985483 | LOC400499 | FALSE | Upregulated |
| 0,263646968 | 0,687039789 | SPAG8 | FALSE | Upregulated |
| -0,118577159 | 0,687402254 | EHD3 | FALSE | Downregulated |
| -1,052980517 | 0,687402254 | ZNF559-ZNF177 | FALSE | Downregulated |
| 0,363388025 | 0,687454361 | OSGEPL1-AS1 | FALSE | Upregulated |
| -0,190320342 | 0,68751337 | UCP2 | FALSE | Downregulated |
| -0,328798104 | 0,687523968 | RP11-421P23.1 | FALSE | Downregulated |
| 0,054593478 | 0,688280712 | EPN1 | FALSE | Upregulated |
| -0,407263609 | 0,688280712 | MAEL | FALSE | Downregulated |
| -0,062516103 | 0,688280712 | NAGA | FALSE | Downregulated |
| -0,120777441 | 0,688280712 | SCML2 | FALSE | Downregulated |
| 0,065469999 | 0,688280712 | ZBTB7B | FALSE | Upregulated |
| -0,062525402 | 0,688299686 | ANO6 | FALSE | Downregulated |
| 0,421333016 | 0,688299686 | FGF17 | FALSE | Upregulated |
| 0,106487772 | 0,688299686 | NRP1 | FALSE | Upregulated |
| -0,379799334 | 0,688299686 | TRAM1L1 | FALSE | Downregulated |
| 0,443131761 | 0,68840751 | COL11A1 | FALSE | Upregulated |
| 0,637426018 | 0,68840751 | MALRD1 | FALSE | Upregulated |

| | | | | |
|---|---|---|---|---|
| 0,069126788 | 0,688414346 | UHRF1BP1 | FALSE | Upregulated |
| -0,318595994 | 0,688735513 | C2CD4B | FALSE | Downregulated |
| 0,458597535 | 0,688735513 | LOC100127974 | FALSE | Upregulated |
| -0,056482842 | 0,688874961 | UBE2F | FALSE | Downregulated |
| 0,072464759 | 0,688996741 | RANBP10 | FALSE | Upregulated |
| 0,690624587 | 0,689129604 | SNORD58A | FALSE | Upregulated |
| 0,086269801 | 0,68914952 | RP11-890B15.2 | FALSE | Upregulated |
| -0,068062711 | 0,689201635 | DHCR24 | FALSE | Downregulated |
| -0,066490187 | 0,689354987 | HNF1A | FALSE | Downregulated |
| -0,879782457 | 0,689603892 | NT5C1B-RDH14 | FALSE | Downregulated |
| 0,399388613 | 0,689603892 | PGA4 | FALSE | Upregulated |
| -0,164444125 | 0,689603892 | THNSL2 | FALSE | Downregulated |
| -0,632613803 | 0,689987135 | JPH2 | FALSE | Downregulated |
| -0,182305354 | 0,689987135 | LINC00174 | FALSE | Downregulated |
| -0,07240324 | 0,690073428 | COMMD3 | FALSE | Downregulated |
| -0,710173146 | 0,690073428 | LOC729658 | FALSE | Downregulated |
| -0,134023936 | 0,690096197 | RPH3AL | FALSE | Downregulated |
| -0,553756827 | 0,690204036 | HSFX1 | FALSE | Downregulated |
| -0,153411503 | 0,690204036 | LOC100996492 | FALSE | Downregulated |
| 0,573165464 | 0,690204036 | LOC554223 | FALSE | Upregulated |
| 0,062538136 | 0,690526492 | BMP2K | FALSE | Upregulated |
| -0,170233947 | 0,690526492 | ICOSLG | FALSE | Downregulated |
| 0,468089213 | 0,690694042 | FAM72B | FALSE | Upregulated |
| -0,046709282 | 0,690694042 | TMEM68 | FALSE | Downregulated |
| -0,130620095 | 0,690703811 | CA5A | FALSE | Downregulated |
| 0,06641159 | 0,690703811 | CRYZ | FALSE | Upregulated |
| -0,137627458 | 0,690714552 | LOH12CR2 | FALSE | Downregulated |
| 0,104840427 | 0,690720403 | DTYMK | FALSE | Upregulated |
| 0,118553145 | 0,690720403 | TRIB3 | FALSE | Upregulated |
| 0,375223331 | 0,6908924 | CCDC114 | FALSE | Upregulated |
| 0,039696105 | 0,690893587 | TCF12 | FALSE | Upregulated |
| -0,151982008 | 0,690937594 | VMO1 | FALSE | Downregulated |
| 0,058970305 | 0,691370713 | ATP1B1 | FALSE | Upregulated |
| -0,075112525 | 0,691370713 | EDEM3 | FALSE | Downregulated |
| -0,165293286 | 0,691370713 | HNRNPU-AS1 | FALSE | Downregulated |
| 0,441388138 | 0,691370713 | KIR2DL4 | FALSE | Upregulated |
| -0,335950761 | 0,691370713 | LOC101929741 | FALSE | Downregulated |
| -0,739613645 | 0,691370713 | RHD | FALSE | Downregulated |
| -0,585765866 | 0,691428239 | ESRG | FALSE | Downregulated |
| 0,313927431 | 0,691518854 | CATSPER2 | FALSE | Upregulated |
| -0,076858784 | 0,69154071 | AOC3 | FALSE | Downregulated |
| -0,082593786 | 0,69154071 | CDC73 | FALSE | Downregulated |
| -0,077430472 | 0,691737948 | HSD17B11 | FALSE | Downregulated |
| -0,09374405 | 0,691767354 | HS3ST3B1 | FALSE | Downregulated |
| -0,11704391 | 0,691984704 | CA13 | FALSE | Downregulated |
| -0,352290466 | 0,691984704 | KCNJ11 | FALSE | Downregulated |
| 0,240385595 | 0,692185813 | SLC25A21 | FALSE | Upregulated |
| -0,053980901 | 0,692362319 | MANBAL | FALSE | Downregulated |
| -0,081401869 | 0,692364512 | LYRM9 | FALSE | Downregulated |
| 0,121859405 | 0,692751871 | ZNF425 | FALSE | Upregulated |
| 0,400259143 | 0,692835241 | FOLR1 | FALSE | Upregulated |

| | | | | |
|---|---|---|---|---|
| 0,054239658 | 0,693453923 | DHPS | FALSE | Upregulated |
| -0,064027011 | 0,693632624 | DEDD2 | FALSE | Downregulated |
| -0,042526808 | 0,693632624 | PRDM4 | FALSE | Downregulated |
| 0,561779794 | 0,69370476 | ANKRD30B | FALSE | Upregulated |
| -0,100232246 | 0,693824124 | SPEF2 | FALSE | Downregulated |
| 0,059835171 | 0,694247373 | PLS1 | FALSE | Upregulated |
| -0,161100887 | 0,694262948 | C1orf210 | FALSE | Downregulated |
| 0,106284002 | 0,694421238 | C3orf33 | FALSE | Upregulated |
| 0,698246681 | 0,694816537 | FBXL21 | FALSE | Upregulated |
| 0,053228259 | 0,69490269 | MAU2 | FALSE | Upregulated |
| -0,069242371 | 0,695174315 | EARS2 | FALSE | Downregulated |
| 0,07440495 | 0,695174315 | FLAD1 | FALSE | Upregulated |
| 0,044431354 | 0,695179507 | RAB18 | FALSE | Upregulated |
| 0,083840275 | 0,695356058 | L3MBTL4 | FALSE | Upregulated |
| -0,161088848 | 0,695381275 | LOC728024 | FALSE | Downregulated |
| -0,233037484 | 0,695395616 | BEX5 | FALSE | Downregulated |
| 0,222232567 | 0,695395616 | GDF6 | FALSE | Upregulated |
| 0,750806131 | 0,695395616 | LOC102724378 | FALSE | Upregulated |
| -0,344803026 | 0,695480204 | LOC102724965 | FALSE | Downregulated |
| -0,06048287 | 0,695664596 | DZIP3 | FALSE | Downregulated |
| -0,148579189 | 0,695692714 | GPR75 | FALSE | Downregulated |
| 0,086658631 | 0,6957129 | IFI35 | FALSE | Upregulated |
| -0,176977219 | 0,695841411 | DBP | FALSE | Downregulated |
| 0,608919255 | 0,696139004 | SLC8A2 | FALSE | Upregulated |
| 0,051548208 | 0,696142949 | UBLCP1 | FALSE | Upregulated |
| -0,136622809 | 0,696853147 | LOC100128398 | FALSE | Downregulated |
| 0,247255703 | 0,696853147 | UBE2Q2P3 | FALSE | Upregulated |
| 0,073242818 | 0,696913547 | PM20D2 | FALSE | Upregulated |
| -0,153825954 | 0,696925572 | RASGRP2 | FALSE | Downregulated |
| -0,493019828 | 0,697197374 | SERF1B | FALSE | Downregulated |
| 0,290934702 | 0,697239734 | KCNRG | FALSE | Upregulated |
| 0,068084438 | 0,697242583 | PARP11 | FALSE | Upregulated |
| 0,172245972 | 0,697242583 | ZNF599 | FALSE | Upregulated |
| 0,055951462 | 0,697380047 | PPP1CC | FALSE | Upregulated |
| 0,069521672 | 0,697456266 | PSME3 | FALSE | Upregulated |
| -0,104500213 | 0,697457983 | LINC00116 | FALSE | Downregulated |
| 0,181770548 | 0,697457983 | SNCG | FALSE | Upregulated |
| -0,18449502 | 0,697535492 | BCMO1 | FALSE | Downregulated |
| 0,177918107 | 0,697535492 | C11orf45 | FALSE | Upregulated |
| -0,566708558 | 0,697535492 | OMP | FALSE | Downregulated |
| 0,119742149 | 0,697535492 | OSCP1 | FALSE | Upregulated |
| 0,27326383 | 0,697535492 | ZNF727 | FALSE | Upregulated |
| 0,091012406 | 0,697586397 | ZNF267 | FALSE | Upregulated |
| -0,197854421 | 0,697874795 | LOC101926931 | FALSE | Downregulated |
| -0,058059801 | 0,697928756 | CNIH1 | FALSE | Downregulated |
| 0,254244987 | 0,698066027 | LOC441454 | FALSE | Upregulated |
| 0,056332313 | 0,698415653 | FKBP3 | FALSE | Upregulated |
| -0,06799915 | 0,698423497 | CHMP4C | FALSE | Downregulated |
| -0,201938693 | 0,698423497 | DNHD1 | FALSE | Downregulated |
| 0,042175879 | 0,698423497 | NUMB | FALSE | Upregulated |
| 0,141673104 | 0,698423497 | SCARA3 | FALSE | Upregulated |

| | | | | |
|---|---|---|---|---|
| -0,473375329 | 0,69847815 | LOC101927259 | FALSE | Downregulated |
| -0,153834438 | 0,698514039 | TTC18 | FALSE | Downregulated |
| 0,908887032 | 0,698643926 | ARTN | FALSE | Upregulated |
| 0,062217795 | 0,698713964 | NFATC2IP | FALSE | Upregulated |
| -0,708079916 | 0,69873355 | NUP210L | FALSE | Downregulated |
| 0,285258295 | 0,69873355 | RASA4CP | FALSE | Upregulated |
| -0,073222596 | 0,69873355 | TRMT13 | FALSE | Downregulated |
| 0,069728945 | 0,698876368 | BNIP1 | FALSE | Upregulated |
| -0,516237695 | 0,698876368 | CCR10 | FALSE | Downregulated |
| 0,375726163 | 0,698876368 | HOXB-AS1 | FALSE | Upregulated |
| 0,065345933 | 0,698876368 | LACTB | FALSE | Upregulated |
| -0,202524413 | 0,698876368 | OR7E14P | FALSE | Downregulated |
| -0,107170848 | 0,698876368 | OSBPL1A | FALSE | Downregulated |
| 0,187123014 | 0,698876368 | RASSF9 | FALSE | Upregulated |
| 0,234654973 | 0,698891908 | LOC554206 | FALSE | Upregulated |
| 0,696345176 | 0,699380088 | TAS1R1 | FALSE | Upregulated |
| 0,563536077 | 0,699423282 | LRRC36 | FALSE | Upregulated |
| -0,084501519 | 0,699535705 | MLEC | FALSE | Downregulated |
| -0,059487621 | 0,699540096 | SLC7A6OS | FALSE | Downregulated |
| -0,075614986 | 0,699563186 | ABCB10 | FALSE | Downregulated |
| -0,243345081 | 0,699975618 | LOC102724450 | FALSE | Downregulated |
| 0,252912764 | 0,699975618 | LOC102724811 | FALSE | Upregulated |
| -0,132677673 | 0,699975618 | RP11-418J17.1 | FALSE | Downregulated |
| -0,177582712 | 0,699975618 | RRN3P1 | FALSE | Downregulated |
| 0,06792922 | 0,699975618 | SRCAP | FALSE | Upregulated |
| 0,067849394 | 0,699975618 | TIGD2 | FALSE | Upregulated |
| 0,067202937 | 0,699975618 | TMEM47 | FALSE | Upregulated |
| 0,062084111 | 0,699975618 | ZNF263 | FALSE | Upregulated |
| 0,328038016 | 0,70015016 | LOC100287098 | FALSE | Upregulated |
| -0,15859566 | 0,70015016 | ULK4 | FALSE | Downregulated |
| -0,059982009 | 0,700152921 | HEXB | FALSE | Downregulated |
| 0,062721029 | 0,700152921 | ZNF84 | FALSE | Upregulated |
| 0,115929692 | 0,70024806 | BRIP1 | FALSE | Upregulated |
| 0,089986874 | 0,700646911 | LNPEP | FALSE | Upregulated |
| 0,121542397 | 0,700646911 | NPC1L1 | FALSE | Upregulated |
| -0,080814994 | 0,700646911 | PCGF3 | FALSE | Downregulated |
| -0,271452301 | 0,700646911 | PHYHIP | FALSE | Downregulated |
| -0,072089526 | 0,700646911 | ZNF25 | FALSE | Downregulated |
| -0,151712145 | 0,700743283 | VSIG4 | FALSE | Downregulated |
| 0,182418922 | 0,701060544 | LOC100499484 | FALSE | Upregulated |
| -0,20120783 | 0,701060544 | LOC101060376 | FALSE | Downregulated |
| 0,085894635 | 0,701060803 | NRDE2 | FALSE | Upregulated |
| -0,314685303 | 0,701261505 | LOC102724314 | FALSE | Downregulated |
| 0,087328331 | 0,701402395 | C8orf37 | FALSE | Upregulated |
| 0,555023714 | 0,70148262 | TTLL2 | FALSE | Upregulated |
| 0,038235507 | 0,701557575 | EWSR1 | FALSE | Upregulated |
| 0,557848961 | 0,70156223 | C10orf85 | FALSE | Upregulated |
| 0,04028401 | 0,70156223 | EXD2 | FALSE | Upregulated |
| 0,040131449 | 0,70156223 | FAM199X | FALSE | Upregulated |
| -0,304173749 | 0,70156223 | LINC00202-1 | FALSE | Downregulated |
| 0,15148996 | 0,70156223 | MAPK8IP3 | FALSE | Upregulated |

| | | | | |
|---|---|---|---|---|
| 0,089207501 | 0,70156223 | MMRN2 | FALSE | Upregulated |
| -0,066824374 | 0,70156223 | MTX3 | FALSE | Downregulated |
| 0,191747903 | 0,70156223 | RNF157 | FALSE | Upregulated |
| -0,184227812 | 0,701861385 | LOC101927054 | FALSE | Downregulated |
| -0,759056303 | 0,701864698 | MROH7 | FALSE | Downregulated |
| -0,391855176 | 0,701989203 | LOC100131496 | FALSE | Downregulated |
| -0,190711409 | 0,701989203 | SEC31B | FALSE | Downregulated |
| 0,123283502 | 0,702362642 | LIN9 | FALSE | Upregulated |
| 0,25475507 | 0,702424637 | DDIT4 | FALSE | Upregulated |
| 0,443955119 | 0,702973208 | LRRC2-AS1 | FALSE | Upregulated |
| 0,056890268 | 0,703013932 | MGA | FALSE | Upregulated |
| 0,167309614 | 0,703168834 | TRAIP | FALSE | Upregulated |
| -0,34278296 | 0,703342029 | LOC102723570 | FALSE | Downregulated |
| 0,229975534 | 0,703355455 | AHCTF1P1 | FALSE | Upregulated |
| 0,083202386 | 0,703355455 | GEMIN8 | FALSE | Upregulated |
| -0,07378621 | 0,703355455 | RPLP2 | FALSE | Downregulated |
| -0,077981865 | 0,703355455 | TOMM40 | FALSE | Downregulated |
| 0,051859389 | 0,703440954 | BRD1 | FALSE | Upregulated |
| -0,246629159 | 0,703700005 | LOC100505912 | FALSE | Downregulated |
| -0,24918872 | 0,703700005 | SCARNA17 | FALSE | Downregulated |
| 0,076938372 | 0,703833972 | CNIH4 | FALSE | Upregulated |
| -0,109112142 | 0,703833972 | GIMAP7 | FALSE | Downregulated |
| 0,038774034 | 0,703833972 | JAK1 | FALSE | Upregulated |
| 0,206921761 | 0,703833972 | NQO1 | FALSE | Upregulated |
| -0,564711847 | 0,703833972 | SIGLEC12 | FALSE | Downregulated |
| 0,163786244 | 0,703960999 | LOC100630918 | FALSE | Upregulated |
| 0,073611352 | 0,704057754 | TMED9 | FALSE | Upregulated |
| -0,080375915 | 0,704057754 | UQCC2 | FALSE | Downregulated |
| 0,299662012 | 0,704086383 | ELOVL4 | FALSE | Upregulated |
| 0,061624167 | 0,704086383 | TEAD3 | FALSE | Upregulated |
| -0,084610284 | 0,704298179 | KPNA5 | FALSE | Downregulated |
| -0,059280577 | 0,704380684 | DCUN1D4 | FALSE | Downregulated |
| -0,488651991 | 0,704380684 | LOC375196 | FALSE | Downregulated |
| -0,268779356 | 0,704412515 | LOC102724192 | FALSE | Downregulated |
| -0,046180122 | 0,70459526 | NCKAP1 | FALSE | Downregulated |
| 0,042589495 | 0,704626729 | ZNF207 | FALSE | Upregulated |
| -0,149504951 | 0,704758795 | XAF1 | FALSE | Downregulated |
| 0,074791517 | 0,704852152 | PLAGL2 | FALSE | Upregulated |
| -0,206815658 | 0,704896758 | ADIRF | FALSE | Downregulated |
| 0,241436698 | 0,704925095 | WDR62 | FALSE | Upregulated |
| 0,038447006 | 0,705083925 | DDX1 | FALSE | Upregulated |
| 0,325058224 | 0,705083925 | LOC645967 | FALSE | Upregulated |
| 0,632760685 | 0,705083925 | SORCS1 | FALSE | Upregulated |
| 0,107770937 | 0,705083925 | TPT1-AS1 | FALSE | Upregulated |
| -0,138881278 | 0,705237668 | ARHGEF35 | FALSE | Downregulated |
| 2,698991226 | 0,705237668 | LOC102724219 | FALSE | Upregulated |
| 0,054330875 | 0,705290975 | DIS3 | FALSE | Upregulated |
| 0,443102616 | 0,705293665 | LOC102724944 | FALSE | Upregulated |
| 0,067619117 | 0,705314405 | NANP | FALSE | Upregulated |
| 0,566475471 | 0,705319468 | CPLX4 | FALSE | Upregulated |
| 0,751227242 | 0,70592006 | SPEF1 | FALSE | Upregulated |

| | | | | |
|---|---|---|---|---|
| -0,317327028 | 0,705929498 | LOC101927104 | FALSE | Downregulated |
| 0,300293514 | 0,705929498 | LTC4S | FALSE | Upregulated |
| -0,66189863 | 0,705979751 | SNORD4A | FALSE | Downregulated |
| -0,110572018 | 0,705998975 | UGT2A3 | FALSE | Downregulated |
| -0,357132259 | 0,70603504 | LOC100506860 | FALSE | Downregulated |
| 0,241968618 | 0,706271782 | AC012314.8 | FALSE | Upregulated |
| -0,093901044 | 0,706271782 | CEP164 | FALSE | Downregulated |
| -0,058300673 | 0,706271782 | STOML2 | FALSE | Downregulated |
| -0,072596616 | 0,706271782 | YPEL3 | FALSE | Downregulated |
| 0,257862902 | 0,706297095 | SAP25 | FALSE | Upregulated |
| 0,043097868 | 0,706365728 | FUBP3 | FALSE | Upregulated |
| 0,430116543 | 0,706411036 | LOC100288748 | FALSE | Upregulated |
| 0,087607287 | 0,706411036 | NXF1 | FALSE | Upregulated |
| -0,68488258 | 0,706430652 | LOC102723310 | FALSE | Downregulated |
| 0,064615521 | 0,706489 | ACOX3 | FALSE | Upregulated |
| -0,065486319 | 0,706489 | BRCC3 | FALSE | Downregulated |
| 0,033638472 | 0,706489 | DDX41 | FALSE | Upregulated |
| 0,085644654 | 0,706489 | GCH1 | FALSE | Upregulated |
| -0,199346493 | 0,706489 | LOC101928235 | FALSE | Downregulated |
| -0,125575156 | 0,706489 | OTUB2 | FALSE | Downregulated |
| -0,098517219 | 0,706489 | TAF1C | FALSE | Downregulated |
| 0,55157742 | 0,706531697 | LOC100288162 | FALSE | Upregulated |
| 0,533101379 | 0,706531697 | LOC102724335 | FALSE | Upregulated |
| -0,110213639 | 0,706733239 | HLX | FALSE | Downregulated |
| 0,446044858 | 0,706815062 | EMX2OS | FALSE | Upregulated |
| -0,644012226 | 0,706815062 | LINC00629 | FALSE | Downregulated |
| 0,233006245 | 0,706815062 | LOC441204 | FALSE | Upregulated |
| -0,320960286 | 0,706815062 | OXT | FALSE | Downregulated |
| -0,040942556 | 0,70696182 | RNF111 | FALSE | Downregulated |
| -0,429511222 | 0,70696182 | ZAN | FALSE | Downregulated |
| -0,174910866 | 0,707087627 | SPATA1 | FALSE | Downregulated |
| 0,140534421 | 0,707286945 | CDH19 | FALSE | Upregulated |
| 0,862631447 | 0,707286945 | DAZ4 | FALSE | Upregulated |
| 0,149952593 | 0,707286945 | INCA1 | FALSE | Upregulated |
| -0,11901046 | 0,707286945 | INO80B | FALSE | Downregulated |
| 0,331677069 | 0,707286945 | LOC101926895 | FALSE | Upregulated |
| -0,314654004 | 0,707286945 | LOC101927252 | FALSE | Downregulated |
| 0,296024885 | 0,707286945 | LOC102724699 | FALSE | Upregulated |
| -0,487895573 | 0,707286945 | NR2E3 | FALSE | Downregulated |
| 0,151490947 | 0,707286945 | RFTN2 | FALSE | Upregulated |
| 0,103384598 | 0,707286945 | SATB2 | FALSE | Upregulated |
| -0,070753194 | 0,707286945 | TMEM63B | FALSE | Downregulated |
| -0,072750377 | 0,707420969 | GABARAPL1 | FALSE | Downregulated |
| -0,08060469 | 0,708108951 | KTI12 | FALSE | Downregulated |
| -0,208433103 | 0,708108951 | PAX5 | FALSE | Downregulated |
| 0,061674874 | 0,708159369 | TMEM30B | FALSE | Upregulated |
| -0,10393211 | 0,708227261 | G2E3 | FALSE | Downregulated |
| 0,245796074 | 0,708370269 | B3GNT8 | FALSE | Upregulated |
| 0,180300922 | 0,708370269 | LIFR-AS1 | FALSE | Upregulated |
| 0,751387735 | 0,708370269 | LOC101928910 | FALSE | Upregulated |
| -0,081800536 | 0,708608654 | DNAJB4 | FALSE | Downregulated |

| | | | | |
|---|---|---|---|---|
| 0,093559229 | 0,708608654 | SCN1B | FALSE | Upregulated |
| 0,126627237 | 0,708608654 | SLC2A12 | FALSE | Upregulated |
| 0,065113153 | 0,708799823 | SIAH1 | FALSE | Upregulated |
| 0,086533062 | 0,708799823 | ZNF614 | FALSE | Upregulated |
| -0,088616259 | 0,708951036 | SERINC5 | FALSE | Downregulated |
| 0,057964806 | 0,709006625 | DUSP22 | FALSE | Upregulated |
| 0,077662086 | 0,709195042 | UBOX5 | FALSE | Upregulated |
| 0,200299487 | 0,709519755 | PLCL1 | FALSE | Upregulated |
| -0,117197277 | 0,709626059 | FOXA3 | FALSE | Downregulated |
| -0,060301852 | 0,70970869 | SUV39H1 | FALSE | Downregulated |
| -0,166137572 | 0,709738248 | ALMS1-IT1 | FALSE | Downregulated |
| 0,473966964 | 0,709738248 | LOC101929431 | FALSE | Upregulated |
| 0,287520997 | 0,709738248 | RP1-257A7.4 | FALSE | Upregulated |
| 0,438189114 | 0,71086919 | KCNK2 | FALSE | Upregulated |
| -0,088728206 | 0,71086919 | MAP1LC3A | FALSE | Downregulated |
| -0,054387375 | 0,71086919 | SRPK2 | FALSE | Downregulated |
| 0,276464806 | 0,710888918 | LOC101929667 | FALSE | Upregulated |
| -0,178798024 | 0,710895898 | MEG3 | FALSE | Downregulated |
| 0,158084025 | 0,710895898 | RAD9B | FALSE | Upregulated |
| -0,064077857 | 0,710933305 | COX6B1 | FALSE | Downregulated |
| 0,40836035 | 0,710933305 | CXCL17 | FALSE | Upregulated |
| 0,074507518 | 0,710933305 | LATS1 | FALSE | Upregulated |
| 0,350614932 | 0,710933305 | LOC102723652 | FALSE | Upregulated |
| 0,071473222 | 0,710933305 | PDE6D | FALSE | Upregulated |
| 0,114012146 | 0,710933305 | ZDHHC2 | FALSE | Upregulated |
| 0,120374834 | 0,710960467 | C1QC | FALSE | Upregulated |
| 0,414283976 | 0,711043531 | LOC101927066 | FALSE | Upregulated |
| 0,549154487 | 0,711043531 | LOC101929490 | FALSE | Upregulated |
| -0,056367072 | 0,711043531 | TUBGCP4 | FALSE | Downregulated |
| 0,829527579 | 0,711044836 | KLRC4-KLRK1 | FALSE | Upregulated |
| -0,105133599 | 0,71109827 | LILRB5 | FALSE | Downregulated |
| 0,156706658 | 0,71109827 | RP11-15A1.3 | FALSE | Upregulated |
| -0,382992835 | 0,71109827 | UBE2Q2P2 | FALSE | Downregulated |
| -0,064355354 | 0,711128445 | XRN1 | FALSE | Downregulated |
| 0,143307842 | 0,711209214 | PHOSPHO2 | FALSE | Upregulated |
| 0,71320374 | 0,711426514 | CPEB2-AS1 | FALSE | Upregulated |
| 0,062287081 | 0,711426514 | ZNF23 | FALSE | Upregulated |
| 0,5821595 | 0,71155574 | TRIM17 | FALSE | Upregulated |
| -0,066690151 | 0,711606163 | GALK2 | FALSE | Downregulated |
| -0,056174502 | 0,711606163 | SLC43A1 | FALSE | Downregulated |
| -0,053197731 | 0,711791547 | AAAS | FALSE | Downregulated |
| 0,053988178 | 0,711791547 | PSMC2 | FALSE | Upregulated |
| -0,072150646 | 0,711905065 | KRBOX4 | FALSE | Downregulated |
| -0,201789851 | 0,711990146 | C22orf34 | FALSE | Downregulated |
| -0,139406727 | 0,711990146 | NFIL3 | FALSE | Downregulated |
| 0,036484643 | 0,711993269 | PSEN1 | FALSE | Upregulated |
| 0,038485013 | 0,711999183 | CAMKK2 | FALSE | Upregulated |
| -0,070162868 | 0,712424176 | KANK2 | FALSE | Downregulated |
| 0,045915901 | 0,712480131 | KBTBD4 | FALSE | Upregulated |
| -0,067985997 | 0,712480131 | TRIM27 | FALSE | Downregulated |
| 0,416202771 | 0,712564158 | PRR7 | FALSE | Upregulated |

| | | | | |
|---|---|---|---|---|
| 0,635987709 | 0,712647041 | FAM95A | FALSE | Upregulated |
| -0,309340908 | 0,712647041 | LOC101927842 | FALSE | Downregulated |
| -0,349397593 | 0,712647041 | SUGT1P3 | FALSE | Downregulated |
| -0,830554995 | 0,712664822 | LOC102723943 | FALSE | Downregulated |
| -0,141237065 | 0,712664822 | PER3 | FALSE | Downregulated |
| -0,059239369 | 0,712664822 | SIRT5 | FALSE | Downregulated |
| -0,053130058 | 0,712664822 | UQCC1 | FALSE | Downregulated |
| -0,092829704 | 0,713082123 | AP3M2 | FALSE | Downregulated |
| -0,06326967 | 0,713103004 | MEF2A | FALSE | Downregulated |
| -0,078656628 | 0,713791894 | WDR34 | FALSE | Downregulated |
| 0,091717016 | 0,714013325 | GTF2H4 | FALSE | Upregulated |
| 0,167852463 | 0,714013325 | KIF12 | FALSE | Upregulated |
| -0,15597263 | 0,714013325 | KRT222 | FALSE | Downregulated |
| -0,061079474 | 0,714122524 | ZMYM6 | FALSE | Downregulated |
| 0,134626869 | 0,714175624 | GPD1L | FALSE | Upregulated |
| 0,350969682 | 0,714207429 | LOC729652 | FALSE | Upregulated |
| 0,406313656 | 0,714315475 | LOC102724732 | FALSE | Upregulated |
| -0,244641703 | 0,714358758 | SCUBE1 | FALSE | Downregulated |
| -0,127832826 | 0,714485521 | RDH5 | FALSE | Downregulated |
| -0,469794517 | 0,714502337 | APOL5 | FALSE | Downregulated |
| -0,071838122 | 0,714502337 | ATP5S | FALSE | Downregulated |
| -0,077090426 | 0,714502337 | C1orf74 | FALSE | Downregulated |
| 0,349146134 | 0,714502337 | CRYBB1 | FALSE | Upregulated |
| -0,074852434 | 0,714502337 | GIMAP8 | FALSE | Downregulated |
| 0,490039366 | 0,714502337 | LOC101929375 | FALSE | Upregulated |
| -0,224184515 | 0,714597433 | AOC4P | FALSE | Downregulated |
| 0,160172376 | 0,714597433 | CIDECP | FALSE | Upregulated |
| 0,331079182 | 0,714597433 | FMR1-AS1 | FALSE | Upregulated |
| -0,502428455 | 0,714597433 | LINC00173 | FALSE | Downregulated |
| -0,13096177 | 0,714597433 | MAP3K7CL | FALSE | Downregulated |
| 0,19411962 | 0,714597433 | PRR22 | FALSE | Upregulated |
| 0,309568755 | 0,714597433 | PSD2 | FALSE | Upregulated |
| 0,08713726 | 0,714597433 | SLC25A14 | FALSE | Upregulated |
| 0,541949897 | 0,714597433 | SNORA14A | FALSE | Upregulated |
| -0,083009771 | 0,714597433 | ST3GAL3 | FALSE | Downregulated |
| 0,384467277 | 0,714597433 | TIMP4 | FALSE | Upregulated |
| 0,622183541 | 0,714718746 | CHST8 | FALSE | Upregulated |
| -0,194258675 | 0,714718746 | FCGR3B | FALSE | Downregulated |
| 0,144907239 | 0,714718746 | POU2F2 | FALSE | Upregulated |
| -0,068620132 | 0,714718746 | RPS12 | FALSE | Downregulated |
| -0,508922567 | 0,714718746 | SLC24A2 | FALSE | Downregulated |
| -0,209135008 | 0,714751718 | PCDH7 | FALSE | Downregulated |
| -0,132287836 | 0,714900874 | UGT3A1 | FALSE | Downregulated |
| -0,066103153 | 0,715176848 | POLR2I | FALSE | Downregulated |
| 0,058841838 | 0,71561101 | UBE2M | FALSE | Upregulated |
| 0,4201607 | 0,71561101 | VPREB3 | FALSE | Upregulated |
| -0,052465005 | 0,715753656 | ARRB2 | FALSE | Downregulated |
| -0,228991587 | 0,715826161 | AHRR | FALSE | Downregulated |
| -0,032521156 | 0,715826161 | BLOC1S6 | FALSE | Downregulated |
| 0,068865586 | 0,715826161 | GALNS | FALSE | Upregulated |
| -0,043513041 | 0,715826161 | HARS2 | FALSE | Downregulated |

| | | | | |
|---|---|---|---|---|
| -0,105047228 | 0,715826161 | IFNLR1 | FALSE | Downregulated |
| 0,14946829 | 0,715826161 | LOC102723919 | FALSE | Upregulated |
| 0,356277719 | 0,716003931 | CLDN22 | FALSE | Upregulated |
| -0,86526026 | 0,716003931 | LOC102723595 | FALSE | Downregulated |
| -0,092417846 | 0,716005612 | HSP90AA1 | FALSE | Downregulated |
| -0,072318753 | 0,716005612 | MED17 | FALSE | Downregulated |
| 0,113143186 | 0,716005612 | VNN1 | FALSE | Upregulated |
| 0,228618619 | 0,716024853 | GBP6 | FALSE | Upregulated |
| 0,063486816 | 0,716131628 | SYS1 | FALSE | Upregulated |
| 0,142651545 | 0,716180333 | RGPD1 | FALSE | Upregulated |
| 0,666212275 | 0,716477332 | ALK | FALSE | Upregulated |
| 0,110975644 | 0,716477332 | FAM173A | FALSE | Upregulated |
| 0,577221744 | 0,716477332 | LOC101929241 | FALSE | Upregulated |
| 0,04347579 | 0,716477332 | WNK1 | FALSE | Upregulated |
| -0,239831022 | 0,716604613 | GOLGA8A | FALSE | Downregulated |
| -0,062124264 | 0,716725832 | C12orf44 | FALSE | Downregulated |
| -0,093298002 | 0,716725832 | NT5E | FALSE | Downregulated |
| 0,044322541 | 0,716725832 | PYCR2 | FALSE | Upregulated |
| 0,041391471 | 0,716851264 | TMEM109 | FALSE | Upregulated |
| -0,310518003 | 0,717181773 | CYP2A13 | FALSE | Downregulated |
| 0,071309685 | 0,717201793 | ANKRD16 | FALSE | Upregulated |
| 0,621920725 | 0,717231585 | LOC101929937 | FALSE | Upregulated |
| 0,040628718 | 0,717231585 | TSTD2 | FALSE | Upregulated |
| -0,037910943 | 0,717820246 | ATXN10 | FALSE | Downregulated |
| -0,108247496 | 0,717960723 | ADORA2A-AS1 | FALSE | Downregulated |
| -0,070999099 | 0,7181414 | KNOP1 | FALSE | Downregulated |
| 0,07411568 | 0,718203615 | PDRG1 | FALSE | Upregulated |
| 0,088378708 | 0,718325703 | BTN3A2 | FALSE | Upregulated |
| 0,497123658 | 0,718325703 | ELAVL3 | FALSE | Upregulated |
| -0,044740706 | 0,718325703 | FAM168A | FALSE | Downregulated |
| 0,049489907 | 0,718325703 | VCP | FALSE | Upregulated |
| -0,08733137 | 0,718581729 | KMO | FALSE | Downregulated |
| 0,143452442 | 0,718581729 | PRICKLE4 | FALSE | Upregulated |
| -0,444436195 | 0,718581729 | XPNPEP2 | FALSE | Downregulated |
| 0,074272089 | 0,718581729 | ZDHHC14 | FALSE | Upregulated |
| 0,253693831 | 0,718626515 | ARHGAP28 | FALSE | Upregulated |
| 0,262706599 | 0,718648703 | LOC102723755 | FALSE | Upregulated |
| -0,052425315 | 0,718648703 | YAE1D1 | FALSE | Downregulated |
| -0,091089815 | 0,718671462 | CARF | FALSE | Downregulated |
| 0,054084807 | 0,718878472 | ZNF451 | FALSE | Upregulated |
| -0,440535773 | 0,719195256 | LOC101928730 | FALSE | Downregulated |
| -0,080273411 | 0,719255333 | HSPE1 | FALSE | Downregulated |
| 0,094798982 | 0,719309187 | FHIT | FALSE | Upregulated |
| 0,591813055 | 0,719357542 | DQX1 | FALSE | Upregulated |
| 0,078225013 | 0,719378464 | CPEB2 | FALSE | Upregulated |
| 0,247744116 | 0,719446047 | OR2A1-AS1 | FALSE | Upregulated |
| 0,285654748 | 0,719482675 | LOC642648 | FALSE | Upregulated |
| -0,808614491 | 0,719568816 | HHATL | FALSE | Downregulated |
| 0,562675491 | 0,719697461 | DGKI | FALSE | Upregulated |
| 0,230199808 | 0,719697461 | ITGAD | FALSE | Upregulated |
| -0,080257686 | 0,71981751 | PGAP1 | FALSE | Downregulated |

| | | | | |
|---|---|---|---|---|
| -0,240826455 | 0,719873919 | LINC00266-3 | FALSE | Downregulated |
| -0,059228638 | 0,719874199 | PPP3R1 | FALSE | Downregulated |
| -0,070350787 | 0,719940279 | ABHD6 | FALSE | Downregulated |
| -0,075079032 | 0,719996782 | ZNF396 | FALSE | Downregulated |
| 0,161825813 | 0,719997067 | PRKG1 | FALSE | Upregulated |
| 0,785387466 | 0,720076099 | OSR2 | FALSE | Upregulated |
| -0,08554704 | 0,720076099 | TSPAN18 | FALSE | Downregulated |
| -0,040926766 | 0,720303415 | TRIP12 | FALSE | Downregulated |
| 0,29102794 | 0,720375695 | AOX2P | FALSE | Upregulated |
| 0,249315895 | 0,720387626 | APOBEC3A | FALSE | Upregulated |
| -0,056819421 | 0,72043401 | COA7 | FALSE | Downregulated |
| 0,343131494 | 0,720558936 | FPGT-TNNI3K | FALSE | Upregulated |
| 0,283506714 | 0,720659053 | LOC101927469 | FALSE | Upregulated |
| -0,144717168 | 0,720659053 | LRAT | FALSE | Downregulated |
| 0,745714903 | 0,720803346 | CORT | FALSE | Upregulated |
| 0,256551423 | 0,720878443 | FLJ44313 | FALSE | Upregulated |
| -0,086850365 | 0,720878443 | ZUFSP | FALSE | Downregulated |
| -0,0690789 | 0,720883917 | PNPT1 | FALSE | Downregulated |
| 0,576129705 | 0,720931558 | LOC101927617 | FALSE | Upregulated |
| -0,384141104 | 0,720931558 | LOC101929463 | FALSE | Downregulated |
| -0,325241612 | 0,720931558 | PCAT18 | FALSE | Downregulated |
| -0,046421446 | 0,721205567 | CBLL1 | FALSE | Downregulated |
| 0,071678694 | 0,721372931 | FAM111A | FALSE | Upregulated |
| -0,172409369 | 0,721372931 | LOC100505555 | FALSE | Downregulated |
| 0,313807001 | 0,721372931 | LOC729083 | FALSE | Upregulated |
| -0,205696101 | 0,721372931 | SGCD | FALSE | Downregulated |
| -0,062002378 | 0,721459961 | LZTS3 | FALSE | Downregulated |
| -0,135225122 | 0,72153346 | RPS6KA5 | FALSE | Downregulated |
| -0,527107669 | 0,722195929 | LOC101927437 | FALSE | Downregulated |
| -0,307763091 | 0,722203901 | AC058791.1 | FALSE | Downregulated |
| -0,135852026 | 0,722204771 | FAM216A | FALSE | Downregulated |
| 0,090899977 | 0,722204771 | PLBD1 | FALSE | Upregulated |
| 0,04837578 | 0,722204771 | PTPN12 | FALSE | Upregulated |
| -1,068462166 | 0,722204771 | RNVU1-7 | FALSE | Downregulated |
| 0,217405686 | 0,722778037 | ID1 | FALSE | Upregulated |
| -0,561005477 | 0,722890887 | CTAGE4 | FALSE | Downregulated |
| -0,06146309 | 0,72308909 | FAH | FALSE | Downregulated |
| -0,08140565 | 0,723309896 | ANO1 | FALSE | Downregulated |
| 0,049843978 | 0,723801969 | AVL9 | FALSE | Upregulated |
| 0,656812054 | 0,72406104 | SCG2 | FALSE | Upregulated |
| -0,117599854 | 0,724110345 | LOC101929787 | FALSE | Downregulated |
| -0,285884616 | 0,724612319 | CBLN3 | FALSE | Downregulated |
| 0,203715278 | 0,724612319 | FAM86B3P | FALSE | Upregulated |
| 0,484081877 | 0,724612319 | LOC102724449 | FALSE | Upregulated |
| -0,074089861 | 0,724612319 | NECAB3 | FALSE | Downregulated |
| 0,117032277 | 0,724612319 | PVRL1 | FALSE | Upregulated |
| -0,051019782 | 0,724813755 | MRPL4 | FALSE | Downregulated |
| -0,083624632 | 0,724837339 | FAM50B | FALSE | Downregulated |
| 0,326878151 | 0,724880775 | KIAA1024 | FALSE | Upregulated |
| 0,490676818 | 0,724883544 | AMH | FALSE | Upregulated |
| 0,494315133 | 0,7250071 | LOC101928717 | FALSE | Upregulated |

| | | | | |
|---|---|---|---|---|
| -0,093287436 | 0,725095865 | LOC286437 | FALSE | Downregulated |
| 0,043355927 | 0,72510788 | PRPF4 | FALSE | Upregulated |
| 0,177027888 | 0,725112005 | CRHR1-IT1 | FALSE | Upregulated |
| -0,382928027 | 0,725119801 | FLJ34503 | FALSE | Downregulated |
| -0,065075063 | 0,725196572 | NDUFAF5 | FALSE | Downregulated |
| 0,036014529 | 0,725209012 | CAND1 | FALSE | Upregulated |
| -0,037066886 | 0,725209012 | MARCH8 | FALSE | Downregulated |
| -0,438391363 | 0,725209012 | TCP11 | FALSE | Downregulated |
| 0,711907314 | 0,725225111 | GSN-AS1 | FALSE | Upregulated |
| 0,199647039 | 0,725248526 | LOC100287015 | FALSE | Upregulated |
| 0,579441714 | 0,725248526 | RP11-106M7.1 | FALSE | Upregulated |
| 0,105292437 | 0,725248526 | SCIMP | FALSE | Upregulated |
| 0,038464243 | 0,725248526 | THOC5 | FALSE | Upregulated |
| -0,059341391 | 0,725485722 | TOR1AIP1 | FALSE | Downregulated |
| 0,140898326 | 0,725508326 | HMGCLL1 | FALSE | Upregulated |
| 0,089094393 | 0,725508326 | LST1 | FALSE | Upregulated |
| 0,424534951 | 0,725508326 | RAB40A | FALSE | Upregulated |
| -0,05017542 | 0,725508326 | SOCS6 | FALSE | Downregulated |
| 0,303207581 | 0,725588787 | ZNF860 | FALSE | Upregulated |
| -0,642687105 | 0,725628863 | IL36G | FALSE | Downregulated |
| 0,03401476 | 0,725732281 | ANXA7 | FALSE | Upregulated |
| -0,03799631 | 0,725732281 | CAPZB | FALSE | Downregulated |
| -0,251901962 | 0,725732281 | LOC101928706 | FALSE | Downregulated |
| -0,165755989 | 0,725732281 | LOC101929200 | FALSE | Downregulated |
| 0,061360773 | 0,72604322 | EXOSC1 | FALSE | Upregulated |
| -0,416567139 | 0,72604322 | L1TD1 | FALSE | Downregulated |
| -0,054693005 | 0,72604322 | RPL10 | FALSE | Downregulated |
| -0,047298557 | 0,72617747 | LIN54 | FALSE | Downregulated |
| 0,08895881 | 0,726226137 | PPIL1 | FALSE | Upregulated |
| 1,344690726 | 0,726489892 | TRNAG15 | FALSE | Upregulated |
| -0,597834615 | 0,726520105 | ZPBP | FALSE | Downregulated |
| 0,140746871 | 0,727443017 | ZNF573 | FALSE | Upregulated |
| 0,105895853 | 0,727562711 | OSGIN1 | FALSE | Upregulated |
| -0,058521034 | 0,727595283 | SLC27A1 | FALSE | Downregulated |
| -0,078582343 | 0,727595283 | ZNF484 | FALSE | Downregulated |
| -0,042494253 | 0,727759933 | SP100 | FALSE | Downregulated |
| -0,061259349 | 0,727828005 | FAM214B | FALSE | Downregulated |
| 0,04030096 | 0,727828005 | SUPT5H | FALSE | Upregulated |
| 0,130929546 | 0,728223832 | AC091729.9 | FALSE | Upregulated |
| -0,042809102 | 0,728541254 | SET | FALSE | Downregulated |
| -0,061924526 | 0,728706911 | JDP2 | FALSE | Downregulated |
| -0,074522086 | 0,728854065 | ACSL5 | FALSE | Downregulated |
| 0,738555522 | 0,729067358 | OPN1SW | FALSE | Upregulated |
| 0,340483895 | 0,729123071 | HSPB2 | FALSE | Upregulated |
| 0,07805332 | 0,72932035 | ZNF440 | FALSE | Upregulated |
| 0,078281085 | 0,729335268 | EHBP1 | FALSE | Upregulated |
| 0,237632043 | 0,729464454 | LOC100996246 | FALSE | Upregulated |
| 0,132781775 | 0,729464454 | TNIK | FALSE | Upregulated |
| 0,126486875 | 0,729516245 | PLA2G6 | FALSE | Upregulated |
| -0,045353906 | 0,729530615 | DNAJA3 | FALSE | Downregulated |
| -0,040366024 | 0,729536708 | PUF60 | FALSE | Downregulated |

| | | | | |
|---|---|---|---|---|
| -0,074301805 | 0,729928133 | DCUN1D3 | FALSE | Downregulated |
| -0,476559778 | 0,729928133 | LOC101929047 | FALSE | Downregulated |
| 0,073627024 | 0,729928133 | SALL1 | FALSE | Upregulated |
| 0,618127076 | 0,730138746 | OSM | FALSE | Upregulated |
| -0,060666216 | 0,730618191 | AFF4 | FALSE | Downregulated |
| -0,06573024 | 0,730618191 | ZNF596 | FALSE | Downregulated |
| -0,530830637 | 0,730736796 | TNMD | FALSE | Downregulated |
| -0,177381211 | 0,730784486 | RAD51AP2 | FALSE | Downregulated |
| 0,624178695 | 0,73087667 | LOC101927696 | FALSE | Upregulated |
| 0,170060532 | 0,73087667 | SIRPB1 | FALSE | Upregulated |
| 0,113912645 | 0,73087667 | SLC26A6 | FALSE | Upregulated |
| -0,053151958 | 0,73105941 | IER3IP1 | FALSE | Downregulated |
| 0,559694397 | 0,731076305 | LOC101928589 | FALSE | Upregulated |
| -0,074349478 | 0,731076305 | NOL9 | FALSE | Downregulated |
| -0,093422345 | 0,731406837 | FAM161A | FALSE | Downregulated |
| 0,067106314 | 0,731481754 | MEIS2 | FALSE | Upregulated |
| 1,133863749 | 0,731481754 | PRAMEF10 | FALSE | Upregulated |
| -0,081094913 | 0,731481754 | ZBTB24 | FALSE | Downregulated |
| 0,215271198 | 0,731497892 | LRRC34 | FALSE | Upregulated |
| -0,236555299 | 0,731497892 | SLC4A10 | FALSE | Downregulated |
| 0,551550042 | 0,731656027 | LOC101926997 | FALSE | Upregulated |
| 0,12039478 | 0,731674746 | CEP152 | FALSE | Upregulated |
| 0,045353892 | 0,732072746 | ADD1 | FALSE | Upregulated |
| 0,084599855 | 0,732072746 | CCDC85B | FALSE | Upregulated |
| 0,268529077 | 0,732085541 | CYP1B1-AS1 | FALSE | Upregulated |
| 0,643983569 | 0,732085541 | LOC101927262 | FALSE | Upregulated |
| 0,143550941 | 0,732085541 | LOC101927451 | FALSE | Upregulated |
| 0,231314381 | 0,732085541 | PACSIN1 | FALSE | Upregulated |
| -0,341398734 | 0,732085541 | RP3-510O8.4 | FALSE | Downregulated |
| 0,090907212 | 0,732085541 | WHAMMP3 | FALSE | Upregulated |
| 0,054875562 | 0,732085541 | ZNF561 | FALSE | Upregulated |
| 0,049811261 | 0,732422498 | UBAC2 | FALSE | Upregulated |
| -0,326913335 | 0,732463041 | TMPRSS5 | FALSE | Downregulated |
| 0,05038651 | 0,732534025 | ATP7B | FALSE | Upregulated |
| 0,041568528 | 0,732646424 | RNF11 | FALSE | Upregulated |
| -0,067625315 | 0,73278864 | LPCAT3 | FALSE | Downregulated |
| -0,046650937 | 0,732985223 | UBE2E1 | FALSE | Downregulated |
| 0,340907549 | 0,732985223 | ZNF286B | FALSE | Upregulated |
| -0,277219388 | 0,733107746 | LOC101927435 | FALSE | Downregulated |
| -0,042712525 | 0,733214084 | NOLC1 | FALSE | Downregulated |
| 0,591639914 | 0,733216891 | DUOXA1 | FALSE | Upregulated |
| 0,512983503 | 0,733216891 | TRNG | FALSE | Upregulated |
| 0,060873866 | 0,733219937 | PMPCA | FALSE | Upregulated |
| -0,231916332 | 0,733219937 | SALL4 | FALSE | Downregulated |
| -0,210356736 | 0,733224403 | LOC101927841 | FALSE | Downregulated |
| 0,099476539 | 0,733224403 | SH3BP4 | FALSE | Upregulated |
| 0,628702575 | 0,733224403 | WNT9B | FALSE | Upregulated |
| 0,084485151 | 0,733224403 | ZNF419 | FALSE | Upregulated |
| -0,232395007 | 0,733266245 | SERF1A | FALSE | Downregulated |
| -0,09289342 | 0,733555193 | NFAT5 | FALSE | Downregulated |
| -0,07192456 | 0,733582751 | AKAP2 | FALSE | Downregulated |

| | | | | |
|---|---|---|---|---|
| 0,050153215 | 0,733582751 | GPC6 | FALSE | Upregulated |
| -0,073220138 | 0,733582751 | ZMYM1 | FALSE | Downregulated |
| 0,096737613 | 0,733688726 | ACTRT3 | FALSE | Upregulated |
| -0,613399758 | 0,733688726 | FLJ30679 | FALSE | Downregulated |
| 0,163975941 | 0,733688726 | LOC102723897 | FALSE | Upregulated |
| 0,113384276 | 0,733756158 | LOC100506714 | FALSE | Upregulated |
| -0,28057906 | 0,733756158 | RP11-79E3.3 | FALSE | Downregulated |
| -0,198114654 | 0,733788185 | RP5-1125A11.1 | FALSE | Downregulated |
| 0,159503789 | 0,733803717 | FAM118A | FALSE | Upregulated |
| 0,079535681 | 0,733908914 | GGH | FALSE | Upregulated |
| 2,19962591 | 0,733931394 | EEF1A2 | FALSE | Upregulated |
| 0,05019035 | 0,733931394 | PRMT1 | FALSE | Upregulated |
| -0,035347593 | 0,733931394 | TTI1 | FALSE | Downregulated |
| 0,075486647 | 0,734294732 | ARRB1 | FALSE | Upregulated |
| -0,594603149 | 0,734509565 | AC005022.1 | FALSE | Downregulated |
| -0,138573597 | 0,734616274 | LINC01061 | FALSE | Downregulated |
| -0,057350322 | 0,734616274 | MDC1 | FALSE | Downregulated |
| 0,389155903 | 0,734628324 | PTH2R | FALSE | Upregulated |
| 0,351419399 | 0,734809071 | GPRC5A | FALSE | Upregulated |
| -0,055105247 | 0,734809071 | TMEM8A | FALSE | Downregulated |
| 0,161231256 | 0,734851261 | LOC150776 | FALSE | Upregulated |
| -0,63760423 | 0,734851261 | OR2C1 | FALSE | Downregulated |
| -0,428941213 | 0,734886895 | LOC101929396 | FALSE | Downregulated |
| -0,088236404 | 0,734987939 | ZMYND12 | FALSE | Downregulated |
| 0,048765911 | 0,735017942 | CPD | FALSE | Upregulated |
| 0,048995447 | 0,735017942 | DIP2B | FALSE | Upregulated |
| -0,089458757 | 0,735139002 | CLCN6 | FALSE | Downregulated |
| -0,381377495 | 0,735279163 | ASB14 | FALSE | Downregulated |
| 0,039095251 | 0,735375955 | CCT4 | FALSE | Upregulated |
| -0,053697688 | 0,735528344 | ACADVL | FALSE | Downregulated |
| -0,261458748 | 0,735604528 | LOC102724805 | FALSE | Downregulated |
| 0,43501605 | 0,735604528 | MINOS1P1 | FALSE | Upregulated |
| -0,061720967 | 0,735670561 | MGLL | FALSE | Downregulated |
| -0,379492626 | 0,735721709 | MLIP-IT1 | FALSE | Downregulated |
| 0,507272542 | 0,735876809 | LOC102724193 | FALSE | Upregulated |
| 0,067214465 | 0,735889928 | ANKRD54 | FALSE | Upregulated |
| 0,056267678 | 0,735889928 | EML3 | FALSE | Upregulated |
| -0,080041321 | 0,735889928 | MLPH | FALSE | Downregulated |
| 0,057708559 | 0,735889928 | TRMT6 | FALSE | Upregulated |
| -0,048832609 | 0,735889928 | ZFP62 | FALSE | Downregulated |
| 0,191634633 | 0,735909118 | HSD17B3 | FALSE | Upregulated |
| 0,056216123 | 0,735909118 | PRMT9 | FALSE | Upregulated |
| 0,038543398 | 0,735909118 | WDR11 | FALSE | Upregulated |
| -0,042475522 | 0,735941717 | CTNND1 | FALSE | Downregulated |
| 0,100915745 | 0,7360886 | SIGLEC11 | FALSE | Upregulated |
| 0,035713615 | 0,736144514 | DDX54 | FALSE | Upregulated |
| 0,482725718 | 0,736284629 | PAPL | FALSE | Upregulated |
| 0,19675001 | 0,736284629 | RP11-509J21.1 | FALSE | Upregulated |
| -0,085008779 | 0,736284629 | TMEM169 | FALSE | Downregulated |
| -0,051328527 | 0,736527653 | TUBGCP3 | FALSE | Downregulated |
| -0,064552323 | 0,736584995 | C16orf45 | FALSE | Downregulated |

| | | | | |
|---|---|---|---|---|
| -0,084576848 | 0,736584995 | PKP2 | FALSE | Downregulated |
| -0,044076795 | 0,736584995 | PPIA | FALSE | Downregulated |
| 0,063019752 | 0,736648267 | FAM195B | FALSE | Upregulated |
| -0,244862173 | 0,736772535 | C1orf192 | FALSE | Downregulated |
| 0,044603683 | 0,736847077 | EPRS | FALSE | Upregulated |
| -0,097656307 | 0,736847077 | TRIM52-AS1 | FALSE | Downregulated |
| 0,134428476 | 0,737098025 | LOC101929792 | FALSE | Upregulated |
| 0,092960961 | 0,7371471 | AIFM2 | FALSE | Upregulated |
| 0,061266915 | 0,7371471 | SNTB2 | FALSE | Upregulated |
| 0,525204282 | 0,73719116 | MAPK15 | FALSE | Upregulated |
| -0,04561923 | 0,73722832 | UTP20 | FALSE | Downregulated |
| -0,048942807 | 0,737332664 | FBXW4 | FALSE | Downregulated |
| 0,109676587 | 0,737510461 | ACSS1 | FALSE | Upregulated |
| 0,270350469 | 0,737510461 | CYP2G1P | FALSE | Upregulated |
| -0,086721677 | 0,737510461 | PHKA1 | FALSE | Downregulated |
| -0,120333542 | 0,737606507 | LOC731656 | FALSE | Downregulated |
| 0,604232018 | 0,737715467 | LOC441956 | FALSE | Upregulated |
| 0,110483416 | 0,737715467 | ZNF117 | FALSE | Upregulated |
| -0,509125567 | 0,737898099 | RP11-197N18.7 | FALSE | Downregulated |
| 0,062852046 | 0,738095672 | NSMCE2 | FALSE | Upregulated |
| -0,197101014 | 0,738132994 | SNCA | FALSE | Downregulated |
| 0,104907377 | 0,738362069 | ACOT12 | FALSE | Upregulated |
| 0,087190606 | 0,738362069 | AKT3 | FALSE | Upregulated |
| -0,052122475 | 0,738362069 | PUS10 | FALSE | Downregulated |
| -0,098748288 | 0,738763678 | C9orf89 | FALSE | Downregulated |
| -0,176973472 | 0,738763678 | FLJ44342 | FALSE | Downregulated |
| 0,054436845 | 0,738763678 | WDR45 | FALSE | Upregulated |
| -0,117080594 | 0,73898186 | CTU1 | FALSE | Downregulated |
| -0,465497234 | 0,739004638 | LOC286178 | FALSE | Downregulated |
| 0,047140123 | 0,739009808 | COX4I1 | FALSE | Upregulated |
| -0,310787817 | 0,739019641 | SRGAP2D | FALSE | Downregulated |
| -0,107190538 | 0,739019641 | STEAP1 | FALSE | Downregulated |
| -0,064778136 | 0,739078521 | CTDSPL2 | FALSE | Downregulated |
| -0,042641282 | 0,739078521 | RIOK1 | FALSE | Downregulated |
| 0,051200551 | 0,739078521 | SLC39A1 | FALSE | Upregulated |
| 0,04771718 | 0,739078521 | SRPR | FALSE | Upregulated |
| -0,03829849 | 0,739078521 | STIM2 | FALSE | Downregulated |
| 0,241014122 | 0,739253259 | CCDC153 | FALSE | Upregulated |
| 0,589451463 | 0,739253259 | EFCAB10 | FALSE | Upregulated |
| -0,038289817 | 0,739253259 | ZNF260 | FALSE | Downregulated |
| -0,059742394 | 0,739592497 | NUDT16L1 | FALSE | Downregulated |
| -0,371201669 | 0,739651345 | LOC440461 | FALSE | Downregulated |
| 0,124008859 | 0,739651345 | RP11-3P17.4 | FALSE | Upregulated |
| 0,329677098 | 0,739707824 | FCAR | FALSE | Upregulated |
| 0,265382134 | 0,739707824 | HSPB8 | FALSE | Upregulated |
| 0,136109018 | 0,739717764 | PRRG2 | FALSE | Upregulated |
| 0,105771398 | 0,739755816 | SGCB | FALSE | Upregulated |
| 0,063828254 | 0,739988937 | INTS5 | FALSE | Upregulated |
| -0,045112381 | 0,740094912 | ATF6B | FALSE | Downregulated |
| 0,066291605 | 0,740526809 | ESRRA | FALSE | Upregulated |
| 0,34139816 | 0,740895258 | LOC101928044 | FALSE | Upregulated |

| | | | | |
|---|---|---|---|---|
| 0,163058095 | 0,740895258 | SDHAP3 | FALSE | Upregulated |
| -0,116429023 | 0,740895258 | WHAMMP2 | FALSE | Downregulated |
| 0,984642114 | 0,741185323 | P2RX2 | FALSE | Upregulated |
| -0,086917759 | 0,741236923 | ZNF658 | FALSE | Downregulated |
| -0,448665094 | 0,741557229 | AC104777.2 | FALSE | Downregulated |
| 0,084162461 | 0,741719658 | ANKRD32 | FALSE | Upregulated |
| -0,044713006 | 0,741739792 | NIPBL | FALSE | Downregulated |
| -0,057747414 | 0,741783776 | USE1 | FALSE | Downregulated |
| -0,062260009 | 0,742538155 | HDDC3 | FALSE | Downregulated |
| 1,044129303 | 0,742666728 | RAD51L3-RFFL | FALSE | Upregulated |
| -0,564362365 | 0,742808733 | LOC101930026 | FALSE | Downregulated |
| -0,045556823 | 0,742808733 | MED8 | FALSE | Downregulated |
| -0,114776631 | 0,742989739 | GNG12-AS1 | FALSE | Downregulated |
| -0,103177835 | 0,742989739 | PXN-AS1 | FALSE | Downregulated |
| 0,078112193 | 0,743001677 | GDAP1 | FALSE | Upregulated |
| 0,069581059 | 0,743001677 | TMEM70 | FALSE | Upregulated |
| 0,534676079 | 0,743236125 | TEX11 | FALSE | Upregulated |
| -0,526431901 | 0,74326592 | TBC1D3H | FALSE | Downregulated |
| 0,37422857 | 0,743373325 | LOC101930452 | FALSE | Upregulated |
| -0,054431399 | 0,743377496 | FANCF | FALSE | Downregulated |
| 0,291354122 | 0,743377496 | LOC101929206 | FALSE | Upregulated |
| -0,058636404 | 0,743433512 | CLUAP1 | FALSE | Downregulated |
| 0,308268923 | 0,743433512 | DIO3 | FALSE | Upregulated |
| 0,061679797 | 0,743649439 | TMED7 | FALSE | Upregulated |
| 0,249192771 | 0,743994734 | FCER2 | FALSE | Upregulated |
| 0,236819905 | 0,743994734 | SEPT3 | FALSE | Upregulated |
| 0,242868164 | 0,744043127 | AC147651.4 | FALSE | Upregulated |
| 0,06613563 | 0,744265428 | METTL25 | FALSE | Upregulated |
| -0,047217484 | 0,744265428 | ZNF397 | FALSE | Downregulated |
| -0,073839789 | 0,744549011 | FN3K | FALSE | Downregulated |
| -0,1497365 | 0,744563717 | C14orf80 | FALSE | Downregulated |
| 0,189167578 | 0,744563717 | CELSR3 | FALSE | Upregulated |
| -2,390648973 | 0,744790888 | LOC101928874 | FALSE | Downregulated |
| -0,033830039 | 0,744995807 | PSMC6 | FALSE | Downregulated |
| 0,107732676 | 0,744995807 | TMEM204 | FALSE | Upregulated |
| 0,241229183 | 0,74513175 | DSCAML1 | FALSE | Upregulated |
| 0,146136894 | 0,745143973 | GNGT2 | FALSE | Upregulated |
| 0,394510099 | 0,745143973 | RP11-426L16.8 | FALSE | Upregulated |
| -0,062798314 | 0,745271411 | AGK | FALSE | Downregulated |
| -0,054837505 | 0,745285279 | TTC9C | FALSE | Downregulated |
| 0,188393922 | 0,745404365 | C1orf51 | FALSE | Upregulated |
| 0,232442239 | 0,745463264 | CPXM2 | FALSE | Upregulated |
| 0,521276534 | 0,745587202 | LOC101929270 | FALSE | Upregulated |
| -0,106349002 | 0,74566765 | CYP2S1 | FALSE | Downregulated |
| 0,260570326 | 0,745726064 | LOC100507336 | FALSE | Upregulated |
| 0,221569588 | 0,745726064 | LOC100507487 | FALSE | Upregulated |
| -0,181982019 | 0,745726064 | LOC101929552 | FALSE | Downregulated |
| -0,591810868 | 0,745726064 | MCCD1 | FALSE | Downregulated |
| 0,05698354 | 0,745726064 | URM1 | FALSE | Upregulated |
| 0,132077706 | 0,745726064 | ZNF229 | FALSE | Upregulated |
| -0,085315749 | 0,745873151 | BCORL1 | FALSE | Downregulated |

| | | | | |
|---|---|---|---|---|
| -0,064278736 | 0,745873151 | MST4 | FALSE | Downregulated |
| 0,049867549 | 0,746129565 | ERGIC1 | FALSE | Upregulated |
| 0,11464713 | 0,746252108 | KLLN | FALSE | Upregulated |
| 0,29252006 | 0,746321069 | CCDC62 | FALSE | Upregulated |
| -0,954892469 | 0,746458005 | TRNAH3 | FALSE | Downregulated |
| 0,050861702 | 0,746521081 | CIAPIN1 | FALSE | Upregulated |
| -0,137762827 | 0,746521081 | RSAD2 | FALSE | Downregulated |
| 0,458947245 | 0,746604238 | ANO2 | FALSE | Upregulated |
| -0,700766413 | 0,746604238 | SMTNL2 | FALSE | Downregulated |
| 0,151431055 | 0,746726767 | CCNE2 | FALSE | Upregulated |
| -0,433210879 | 0,746726767 | TSPEAR | FALSE | Downregulated |
| 0,385937101 | 0,746841723 | LOC101929422 | FALSE | Upregulated |
| 0,036172231 | 0,746841723 | MCM3AP | FALSE | Upregulated |
| -0,067352022 | 0,746841723 | METTL8 | FALSE | Downregulated |
| 0,209008206 | 0,746841723 | MLF1 | FALSE | Upregulated |
| -0,351047951 | 0,746841723 | PVRL3-AS1 | FALSE | Downregulated |
| -0,507692456 | 0,746993343 | LOC102723475 | FALSE | Downregulated |
| 0,428119499 | 0,747028143 | CEACAM20 | FALSE | Upregulated |
| 0,058979313 | 0,747028143 | HNF4G | FALSE | Upregulated |
| 0,080922681 | 0,747465748 | EXT1 | FALSE | Upregulated |
| -0,276149187 | 0,747465748 | LOC729162 | FALSE | Downregulated |
| 0,18728081 | 0,747578618 | PYROXD2 | FALSE | Upregulated |
| 0,084314393 | 0,747590472 | SCAND3 | FALSE | Upregulated |
| 0,077663135 | 0,747672345 | LTBP1 | FALSE | Upregulated |
| -0,058495709 | 0,747672345 | NCAPD3 | FALSE | Downregulated |
| 0,191633967 | 0,747672345 | SCN11A | FALSE | Upregulated |
| 0,023091521 | 0,747781677 | C6orf89 | FALSE | Upregulated |
| 0,245975639 | 0,747781677 | PKD1L3 | FALSE | Upregulated |
| 0,076303342 | 0,747948213 | PINK1-AS | FALSE | Upregulated |
| 0,370183329 | 0,748039272 | LOC102724176 | FALSE | Upregulated |
| -0,050842666 | 0,748043135 | C2CD2 | FALSE | Downregulated |
| 0,09924278 | 0,748043135 | DNAJB11 | FALSE | Upregulated |
| 0,069440538 | 0,748191342 | FXYD6 | FALSE | Upregulated |
| 0,562834103 | 0,748191342 | PCDH10 | FALSE | Upregulated |
| -0,087986382 | 0,748191342 | UGT2B10 | FALSE | Downregulated |
| 0,467449173 | 0,748335022 | LINC01013 | FALSE | Upregulated |
| -0,391680626 | 0,748640094 | AC007743.1 | FALSE | Downregulated |
| 0,077877403 | 0,748664826 | ULK1 | FALSE | Upregulated |
| -0,268506215 | 0,748827462 | CHRNA5 | FALSE | Downregulated |
| -0,271409022 | 0,748963251 | PANO | FALSE | Downregulated |
| -0,352617075 | 0,749105349 | TCTE3 | FALSE | Downregulated |
| -0,075282351 | 0,749369468 | THAP6 | FALSE | Downregulated |
| -0,103953146 | 0,749527983 | TICAM1 | FALSE | Downregulated |
| 0,299038655 | 0,749544786 | C4orf36 | FALSE | Upregulated |
| -0,217145436 | 0,749544786 | MAST1 | FALSE | Downregulated |
| 0,12718086 | 0,749544786 | ZFAT | FALSE | Upregulated |
| 0,488833913 | 0,749706467 | CSMD3 | FALSE | Upregulated |
| -0,112473567 | 0,749835295 | KLF9 | FALSE | Downregulated |
| 0,063246719 | 0,74990562 | ENTPD5 | FALSE | Upregulated |
| 0,105996393 | 0,74990562 | HMGN3-AS1 | FALSE | Upregulated |
| -0,385565406 | 0,74990562 | RNY5 | FALSE | Downregulated |

|  |  |  |  |  |
|---|---|---|---|---|
| 0,226037233 | 0,749906649 | C14orf164 | FALSE | Upregulated |
| 0,087021627 | 0,749906649 | MAP3K6 | FALSE | Upregulated |
| 0,09379784 | 0,749940438 | NLGN4X | FALSE | Upregulated |
| -0,204290438 | 0,750046611 | DKFZP434F142 | FALSE | Downregulated |
| 0,687827112 | 0,750046611 | LOC284600 | FALSE | Upregulated |
| -0,072876962 | 0,750290323 | ZNF69 | FALSE | Downregulated |
| 0,473817763 | 0,750397535 | LINC01108 | FALSE | Upregulated |
| -0,325136688 | 0,750397535 | LOC100130417 | FALSE | Downregulated |
| -0,152749201 | 0,750397535 | QRICH2 | FALSE | Downregulated |
| 0,46686838 | 0,750458803 | LOC101929521 | FALSE | Upregulated |
| -0,054309826 | 0,750458803 | MSH6 | FALSE | Downregulated |
| 0,391533826 | 0,750458803 | PLSCR2 | FALSE | Upregulated |
| 0,232649 | 0,750817713 | IL10RB-AS1 | FALSE | Upregulated |
| 0,043499093 | 0,750928075 | ITM2B | FALSE | Upregulated |
| -0,51442578 | 0,750928075 | LOC100422556 | FALSE | Downregulated |
| 0,481764944 | 0,750928075 | OR10R2 | FALSE | Upregulated |
| 0,098145159 | 0,750928075 | ZNF497 | FALSE | Upregulated |
| -0,446774394 | 0,75108298 | HRASLS | FALSE | Downregulated |
| 0,368066379 | 0,751110647 | SKOR1 | FALSE | Upregulated |
| 0,039631227 | 0,751128214 | CREB1 | FALSE | Upregulated |
| 0,033309046 | 0,751128214 | SPTLC1 | FALSE | Upregulated |
| 0,041559844 | 0,751152535 | ZBTB1 | FALSE | Upregulated |
| -0,094991398 | 0,751441395 | POMZP3 | FALSE | Downregulated |
| 0,527540901 | 0,75169551 | LOC100507346 | FALSE | Upregulated |
| -0,396349998 | 0,751947343 | FOXI2 | FALSE | Downregulated |
| -0,204643978 | 0,75221618 | GOLGA6L4 | FALSE | Downregulated |
| -0,062993582 | 0,75221618 | ISCA2 | FALSE | Downregulated |
| 0,123580732 | 0,75221618 | MPV17L2 | FALSE | Upregulated |
| -0,061694363 | 0,75221618 | NME6 | FALSE | Downregulated |
| 0,436100919 | 0,752270285 | LOC100132731 | FALSE | Upregulated |
| 0,314220214 | 0,75230129 | FLJ25363 | FALSE | Upregulated |
| -0,470451969 | 0,752386538 | RP11-260E18.1 | FALSE | Downregulated |
| -0,108602863 | 0,752677825 | SNAI2 | FALSE | Downregulated |
| 0,202817413 | 0,752860991 | HLA-DQB2 | FALSE | Upregulated |
| 0,065253926 | 0,752860991 | UBE2E2 | FALSE | Upregulated |
| -0,130954033 | 0,752965819 | ZNF204P | FALSE | Downregulated |
| 0,571757632 | 0,753065633 | LOC100130876 | FALSE | Upregulated |
| 0,17080107 | 0,753065633 | MIR198 | FALSE | Upregulated |
| 0,040400387 | 0,753248323 | PSAP | FALSE | Upregulated |
| 0,188182587 | 0,753343774 | ATE1-AS1 | FALSE | Upregulated |
| 0,059667058 | 0,753406242 | PCIF1 | FALSE | Upregulated |
| -0,05870802 | 0,753407161 | BANP | FALSE | Downregulated |
| -0,062593394 | 0,753408061 | HINT2 | FALSE | Downregulated |
| -0,093864673 | 0,753470045 | SPATA20 | FALSE | Downregulated |
| 0,180840993 | 0,753558271 | ZRANB2-AS1 | FALSE | Upregulated |
| -0,306844601 | 0,753879897 | SLC14A2 | FALSE | Downregulated |
| -0,098469498 | 0,753952158 | KIAA1328 | FALSE | Downregulated |
| 0,053197875 | 0,754069105 | ZNF350 | FALSE | Upregulated |
| 0,492683248 | 0,754149209 | SLC9A2 | FALSE | Upregulated |
| -0,043913901 | 0,75427645 | KAT6B | FALSE | Downregulated |
| -0,091336569 | 0,75428803 | BST1 | FALSE | Downregulated |

| | | | | |
|---|---|---|---|---|
| 0,150234523 | 0,75428803 | CD248 | FALSE | Upregulated |
| -0,182521718 | 0,75428803 | MIR4461 | FALSE | Downregulated |
| 0,088216994 | 0,75428803 | PGM3 | FALSE | Upregulated |
| -0,053758285 | 0,75428803 | POLR2H | FALSE | Downregulated |
| -0,124911137 | 0,75428803 | VCAM1 | FALSE | Downregulated |
| 0,089330488 | 0,754407647 | ACY3 | FALSE | Upregulated |
| -0,163365083 | 0,754407647 | DICER1-AS1 | FALSE | Downregulated |
| 0,860405047 | 0,754407647 | KCNJ9 | FALSE | Upregulated |
| 0,070690972 | 0,754660293 | COPG1 | FALSE | Upregulated |
| 0,069003635 | 0,754660293 | NME2 | FALSE | Upregulated |
| -0,122165109 | 0,75488059 | LIN7B | FALSE | Downregulated |
| 0,079123937 | 0,75488059 | MTSS1L | FALSE | Upregulated |
| 0,069574085 | 0,755068659 | AMER1 | FALSE | Upregulated |
| -0,037736603 | 0,755068659 | CMTR1 | FALSE | Downregulated |
| 0,066068376 | 0,755068659 | CNPY4 | FALSE | Upregulated |
| 0,208707002 | 0,755345698 | LOC100289561 | FALSE | Upregulated |
| 0,294795073 | 0,755345698 | PHKA2-AS1 | FALSE | Upregulated |
| -0,034826679 | 0,755476177 | SUPT7L | FALSE | Downregulated |
| -0,111428519 | 0,755522646 | BZRAP1 | FALSE | Downregulated |
| 0,103866086 | 0,755522646 | IGFBP5 | FALSE | Upregulated |
| 0,435210237 | 0,755522646 | LOC101927249 | FALSE | Upregulated |
| 0,260285521 | 0,755763806 | XKR9 | FALSE | Upregulated |
| 0,172114342 | 0,755944509 | C5orf34 | FALSE | Upregulated |
| 0,061295423 | 0,755944509 | KIAA1161 | FALSE | Upregulated |
| -0,069778465 | 0,756080291 | TAOK3 | FALSE | Downregulated |
| 0,133291692 | 0,756092258 | PIGZ | FALSE | Upregulated |
| 0,327222642 | 0,75618607 | C10orf107 | FALSE | Upregulated |
| -0,30629144 | 0,75618607 | LOC101929191 | FALSE | Downregulated |
| -0,032969723 | 0,75618607 | MARK3 | FALSE | Downregulated |
| -0,18534201 | 0,756378219 | C5orf49 | FALSE | Downregulated |
| 0,050819135 | 0,756378219 | HSPBP1 | FALSE | Upregulated |
| -0,155427739 | 0,756426546 | LOC101928370 | FALSE | Downregulated |
| 0,043095478 | 0,756426546 | MFN2 | FALSE | Upregulated |
| 0,069642452 | 0,756426546 | OGFOD2 | FALSE | Upregulated |
| 0,093088318 | 0,756426546 | TBC1D32 | FALSE | Upregulated |
| 0,321266792 | 0,756871965 | LINC01119 | FALSE | Upregulated |
| 0,033781744 | 0,756914388 | ATXN7L3 | FALSE | Upregulated |
| -0,058058888 | 0,756914388 | CREBRF | FALSE | Downregulated |
| 0,255490607 | 0,756914388 | LOC101927198 | FALSE | Upregulated |
| 0,52995548 | 0,756914388 | LOC101928671 | FALSE | Upregulated |
| -0,038233301 | 0,756914388 | TDP2 | FALSE | Downregulated |
| 0,057119668 | 0,756945861 | ARHGAP42 | FALSE | Upregulated |
| 0,055868388 | 0,756945861 | F10 | FALSE | Upregulated |
| 0,040037004 | 0,756945861 | TSR1 | FALSE | Upregulated |
| -0,214094578 | 0,756956847 | LOC102724316 | FALSE | Downregulated |
| 0,215922464 | 0,756996874 | BRINP2 | FALSE | Upregulated |
| 0,152398497 | 0,756996874 | CLEC10A | FALSE | Upregulated |
| -0,04809735 | 0,756996874 | CLNS1A | FALSE | Downregulated |
| 0,169692884 | 0,756996874 | LOC100506282 | FALSE | Upregulated |
| -0,542488286 | 0,756996874 | LOC101928445 | FALSE | Downregulated |
| 0,036293699 | 0,756996874 | LRPAP1 | FALSE | Upregulated |

| | | | | |
|---|---|---|---|---|
| -0,050333111 | 0,756996874 | LRRC37BP1 | FALSE | Downregulated |
| 0,050741724 | 0,756996874 | MRPL15 | FALSE | Upregulated |
| -0,104634403 | 0,756996874 | NXT1 | FALSE | Downregulated |
| 0,598966483 | 0,756996874 | OR51B5 | FALSE | Upregulated |
| -0,084581793 | 0,756996874 | PGLYRP2 | FALSE | Downregulated |
| 0,050847538 | 0,756996874 | RFFL | FALSE | Upregulated |
| -0,07227317 | 0,756996874 | RPL31 | FALSE | Downregulated |
| 0,037380895 | 0,756996874 | SCYL3 | FALSE | Upregulated |
| 0,456376581 | 0,756996874 | SPOCK1 | FALSE | Upregulated |
| 0,052836365 | 0,756996874 | SYNJ1 | FALSE | Upregulated |
| -0,209316151 | 0,756996874 | TMEM105 | FALSE | Downregulated |
| 0,038364899 | 0,756996874 | UBFD1 | FALSE | Upregulated |
| -0,077262528 | 0,756996874 | ZNF100 | FALSE | Downregulated |
| 0,099221686 | 0,75706739 | C4orf46 | FALSE | Upregulated |
| -0,083270273 | 0,75706739 | ZNF653 | FALSE | Downregulated |
| -0,057378571 | 0,757216019 | ZNF26 | FALSE | Downregulated |
| 0,308819122 | 0,757242736 | KIR3DX1 | FALSE | Upregulated |
| -0,036635706 | 0,757489859 | KIF3B | FALSE | Downregulated |
| -0,113659751 | 0,757504157 | TMEM120B | FALSE | Downregulated |
| 0,051776008 | 0,757819646 | ACTR8 | FALSE | Upregulated |
| 0,204987218 | 0,757850483 | LOC102723327 | FALSE | Upregulated |
| 0,31946013 | 0,75803266 | KCNJ2-AS1 | FALSE | Upregulated |
| -0,093305712 | 0,75803266 | LRIG1 | FALSE | Downregulated |
| 0,06108594 | 0,758070581 | DUSP14 | FALSE | Upregulated |
| 0,134559488 | 0,758079739 | VAT1L | FALSE | Upregulated |
| 0,040128876 | 0,758242747 | CNOT3 | FALSE | Upregulated |
| 0,052630764 | 0,758242747 | MED7 | FALSE | Upregulated |
| 0,413837011 | 0,758270185 | LOC256880 | FALSE | Upregulated |
| -0,335803516 | 0,758402458 | LOC101926940 | FALSE | Downregulated |
| -0,045835854 | 0,758402458 | POLR1B | FALSE | Downregulated |
| 0,280595286 | 0,758402458 | RP11-399K21.14 | FALSE | Upregulated |
| -0,059303736 | 0,758536444 | CYB561D1 | FALSE | Downregulated |
| 0,05338055 | 0,758536444 | LSM1 | FALSE | Upregulated |
| 0,105908446 | 0,758582211 | DHCR7 | FALSE | Upregulated |
| 0,226342352 | 0,75866264 | PLEKHG5 | FALSE | Upregulated |
| 0,45452002 | 0,758948072 | RP11-723O4.2 | FALSE | Upregulated |
| -0,437261227 | 0,758960287 | HIST1H1E | FALSE | Downregulated |
| -0,71893286 | 0,759030841 | KCNH6 | FALSE | Downregulated |
| 0,060596111 | 0,759047774 | DNM1L | FALSE | Upregulated |
| 0,606773512 | 0,759218409 | NOX5 | FALSE | Upregulated |
| 0,590868536 | 0,75938444 | SNORD74 | FALSE | Upregulated |
| 0,171991735 | 0,759388619 | LOC102724275 | FALSE | Upregulated |
| 0,278837029 | 0,759703674 | DKFZp667F0711 | FALSE | Upregulated |
| -0,271934264 | 0,759961567 | LOC102724372 | FALSE | Downregulated |
| 0,073545392 | 0,760051802 | PCDH12 | FALSE | Upregulated |
| -0,21967334 | 0,760075679 | C2orf54 | FALSE | Downregulated |
| 0,06089803 | 0,760130028 | HNRNPA1 | FALSE | Upregulated |
| 0,097214096 | 0,760172875 | THBS3 | FALSE | Upregulated |
| -0,136919323 | 0,760300117 | CCDC11 | FALSE | Downregulated |
| -0,233768501 | 0,760300117 | CCDC81 | FALSE | Downregulated |
| 0,321032128 | 0,760300117 | LOC100507530 | FALSE | Upregulated |

| | | | | |
|---|---|---|---|---|
| 0,437224181 | 0,760300117 | LOC257358 | FALSE | Upregulated |
| -0,736882067 | 0,760300117 | PTGES3L | FALSE | Downregulated |
| 0,076457553 | 0,760300117 | SCARF1 | FALSE | Upregulated |
| -0,137736334 | 0,760300117 | SLC25A12 | FALSE | Downregulated |
| 0,04747047 | 0,760300117 | TAF1 | FALSE | Upregulated |
| 0,121230024 | 0,760300117 | TTN-AS1 | FALSE | Upregulated |
| 0,047705834 | 0,760300117 | UNC119B | FALSE | Upregulated |
| 0,12246715 | 0,760358095 | PALM2-AKAP2 | FALSE | Upregulated |
| 0,148023436 | 0,76069579 | ZNF239 | FALSE | Upregulated |
| -0,042496261 | 0,76126434 | SEPHS1 | FALSE | Downregulated |
| -0,245595984 | 0,761357948 | TACR2 | FALSE | Downregulated |
| 0,155684072 | 0,761408593 | S100A9 | FALSE | Upregulated |
| -0,31680805 | 0,761408593 | SULT1A3 | FALSE | Downregulated |
| 0,147636243 | 0,76143516 | C1QTNF7 | FALSE | Upregulated |
| 0,032430836 | 0,76143516 | CIAO1 | FALSE | Upregulated |
| 0,386881008 | 0,76143516 | TACR1 | FALSE | Upregulated |
| -0,118824994 | 0,76143516 | ZFP2 | FALSE | Downregulated |
| -0,162378563 | 0,761554665 | GTF2H2 | FALSE | Downregulated |
| -0,672555692 | 0,761554665 | HIST2H2AA4 | FALSE | Downregulated |
| 0,109148859 | 0,761678056 | CEBPD | FALSE | Upregulated |
| 0,099978246 | 0,761765186 | APLNR | FALSE | Upregulated |
| -0,601480787 | 0,762713467 | LOC101928317 | FALSE | Downregulated |
| 0,043131803 | 0,76283312 | LARP4 | FALSE | Upregulated |
| -0,182952051 | 0,762898422 | LINC00346 | FALSE | Downregulated |
| -0,094469175 | 0,762898422 | SPRY4 | FALSE | Downregulated |
| -0,431885597 | 0,763031988 | HTR2A-AS1 | FALSE | Downregulated |
| 0,101335312 | 0,763247338 | DNAJB5 | FALSE | Upregulated |
| -0,331395022 | 0,763247338 | LOC100129316 | FALSE | Downregulated |
| -0,505815738 | 0,76325135 | ASB9P1 | FALSE | Downregulated |
| -0,493653768 | 0,76325135 | ASMT | FALSE | Downregulated |
| 0,100970422 | 0,76325135 | KCTD1 | FALSE | Upregulated |
| 0,34123129 | 0,76325135 | LOC100129138 | FALSE | Upregulated |
| 0,150601597 | 0,76325135 | NPAS2 | FALSE | Upregulated |
| 0,053928716 | 0,76325135 | PLXNB1 | FALSE | Upregulated |
| 0,044379541 | 0,763312936 | CD81 | FALSE | Upregulated |
| -0,156846675 | 0,763317219 | LOC100996419 | FALSE | Downregulated |
| -0,045807692 | 0,763317219 | REXO2 | FALSE | Downregulated |
| -0,04226693 | 0,763340234 | ATP6V0A2 | FALSE | Downregulated |
| -0,061320866 | 0,763340234 | CDC42BPA | FALSE | Downregulated |
| -0,065752406 | 0,763340234 | CYCS | FALSE | Downregulated |
| -0,389213712 | 0,763340234 | KCTD16 | FALSE | Downregulated |
| -0,094800492 | 0,763340234 | KLF11 | FALSE | Downregulated |
| 0,08219721 | 0,763340234 | LOC102723990 | FALSE | Upregulated |
| -0,11244129 | 0,763340234 | MGC70870 | FALSE | Downregulated |
| -0,063507296 | 0,763340234 | PEPD | FALSE | Downregulated |
| -0,029967838 | 0,763417305 | SAR1A | FALSE | Downregulated |
| -0,094333108 | 0,763531268 | PLCB1 | FALSE | Downregulated |
| -0,154854368 | 0,763604249 | RP1-234P15.4 | FALSE | Downregulated |
| 0,207598381 | 0,763739889 | CLEC4E | FALSE | Upregulated |
| -0,51129346 | 0,763755737 | DIO2 | FALSE | Downregulated |
| -0,176640491 | 0,763755737 | HSD17B1 | FALSE | Downregulated |

| | | | | |
|---|---|---|---|---|
| 0,078265077 | 0,763755737 | ITIH3 | FALSE | Upregulated |
| 0,052895273 | 0,763755737 | METTL2B | FALSE | Upregulated |
| -0,066129727 | 0,763755737 | PGBD2 | FALSE | Downregulated |
| -0,06762671 | 0,763755737 | RASA2 | FALSE | Downregulated |
| -0,44578053 | 0,763835856 | AC105053.3 | FALSE | Downregulated |
| 0,344822957 | 0,763897378 | FAM209B | FALSE | Upregulated |
| -0,067379999 | 0,763897378 | GMDS-AS1 | FALSE | Downregulated |
| 0,234211975 | 0,763897378 | MAP3K14-AS1 | FALSE | Upregulated |
| 0,661055619 | 0,763897378 | STEAP1B | FALSE | Upregulated |
| -0,099185091 | 0,763897378 | SULT1A1 | FALSE | Downregulated |
| -0,708953806 | 0,763897378 | TFF2 | FALSE | Downregulated |
| -0,600269827 | 0,763897378 | TULP2 | FALSE | Downregulated |
| -0,033526605 | 0,763897378 | U2AF2 | FALSE | Downregulated |
| -0,045036782 | 0,763929947 | CASC3 | FALSE | Downregulated |
| -0,070803129 | 0,763929947 | TCEB2 | FALSE | Downregulated |
| 0,047703125 | 0,76404652 | PKN2 | FALSE | Upregulated |
| -0,634169215 | 0,76404652 | TSSK4 | FALSE | Downregulated |
| 0,064076211 | 0,764153297 | FKBP14 | FALSE | Upregulated |
| -0,097027671 | 0,764423406 | FKBP11 | FALSE | Downregulated |
| -0,066567115 | 0,764423406 | LEPREL1 | FALSE | Downregulated |
| 0,117720473 | 0,764423406 | NAT14 | FALSE | Upregulated |
| -0,635464403 | 0,76444325 | C1QL2 | FALSE | Downregulated |
| 0,045965459 | 0,76444325 | CD99 | FALSE | Upregulated |
| 0,110136444 | 0,76444325 | LOC728613 | FALSE | Upregulated |
| 0,473971566 | 0,76444325 | RP11-864J10.4 | FALSE | Upregulated |
| -0,140794296 | 0,764456341 | LINC00641 | FALSE | Downregulated |
| -0,23997183 | 0,764579658 | KL | FALSE | Downregulated |
| -0,041858446 | 0,764579658 | ZNF689 | FALSE | Downregulated |
| -0,079603455 | 0,764605194 | ZNF772 | FALSE | Downregulated |
| 0,205605693 | 0,764931377 | LOC101929580 | FALSE | Upregulated |
| 0,445605945 | 0,765609413 | CTC-436P18.1 | FALSE | Upregulated |
| -0,044301392 | 0,765621429 | GTPBP4 | FALSE | Downregulated |
| -0,53058575 | 0,765621429 | LOC102467214 | FALSE | Downregulated |
| 0,296887877 | 0,765729467 | HOXB-AS3 | FALSE | Upregulated |
| 0,058595611 | 0,765842796 | BBIP1 | FALSE | Upregulated |
| 0,219795339 | 0,765842796 | LOC101927838 | FALSE | Upregulated |
| 0,052648027 | 0,765960776 | DSE | FALSE | Upregulated |
| 0,053033401 | 0,765960776 | KLHL28 | FALSE | Upregulated |
| 0,168339946 | 0,765960776 | ZNF789 | FALSE | Upregulated |
| 0,275998089 | 0,766005105 | LOC101929287 | FALSE | Upregulated |
| 0,04063343 | 0,766138823 | SNX8 | FALSE | Upregulated |
| 0,03667384 | 0,766284335 | NEK7 | FALSE | Upregulated |
| 0,417670891 | 0,766284335 | RASSF10 | FALSE | Upregulated |
| 0,065120165 | 0,766542484 | FAM175A | FALSE | Upregulated |
| -0,221870421 | 0,766542484 | TEX14 | FALSE | Downregulated |
| 0,235409774 | 0,766680611 | LOC100505570 | FALSE | Upregulated |
| 0,090067694 | 0,766680611 | PASK | FALSE | Upregulated |
| -0,034851955 | 0,766680611 | RAB3GAP2 | FALSE | Downregulated |
| 0,556773984 | 0,766680611 | RNY1 | FALSE | Upregulated |
| -0,174917139 | 0,766797983 | PDCL3P4 | FALSE | Downregulated |
| 0,037596684 | 0,766797983 | SNX6 | FALSE | Upregulated |

| | | | | |
|---|---|---|---|---|
| 0,56475271 | 0,766797983 | TRNAK9 | FALSE | Upregulated |
| -0,085828843 | 0,766825633 | KIF27 | FALSE | Downregulated |
| 0,31126267 | 0,766825633 | LOC101928334 | FALSE | Upregulated |
| 0,088919724 | 0,766825633 | PRKAR1B | FALSE | Upregulated |
| 0,152464957 | 0,766825633 | RP11-112H10.4 | FALSE | Upregulated |
| 0,091949435 | 0,766825633 | TGFA | FALSE | Upregulated |
| 0,382679585 | 0,766825633 | TPTE2P6 | FALSE | Upregulated |
| 0,356752827 | 0,766927767 | LOC101929661 | FALSE | Upregulated |
| -0,073813923 | 0,766942386 | RASSF7 | FALSE | Downregulated |
| -0,313315328 | 0,766942386 | RP11-426A6.5 | FALSE | Downregulated |
| -0,265665662 | 0,766942386 | RP5-977B1.7 | FALSE | Downregulated |
| 0,073879537 | 0,766942386 | ZNF443 | FALSE | Upregulated |
| 0,17796258 | 0,766942386 | ZNF783 | FALSE | Upregulated |
| -0,092366067 | 0,76713187 | CORO2A | FALSE | Downregulated |
| 0,272473368 | 0,767151923 | GPR97 | FALSE | Upregulated |
| 0,049813061 | 0,767151923 | RAP2C | FALSE | Upregulated |
| -0,087909116 | 0,767151923 | STARD7-AS1 | FALSE | Downregulated |
| -0,385632645 | 0,767202556 | LINC00479 | FALSE | Downregulated |
| -0,231492009 | 0,767238991 | PLA2G1B | FALSE | Downregulated |
| -0,125379529 | 0,767550223 | LINC01006 | FALSE | Downregulated |
| -0,25836551 | 0,76790436 | FLJ42258 | FALSE | Downregulated |
| 2,135894529 | 0,768021212 | ANKRD1 | FALSE | Upregulated |
| 0,068205517 | 0,768021212 | HSD17B2 | FALSE | Upregulated |
| -0,029880064 | 0,768021212 | PTK2 | FALSE | Downregulated |
| 0,040173729 | 0,768021212 | RAD23A | FALSE | Upregulated |
| 0,03448248 | 0,768021212 | RIOK3 | FALSE | Upregulated |
| 0,167911512 | 0,768118218 | PXDNL | FALSE | Upregulated |
| -0,227032881 | 0,768293278 | BTNL8 | FALSE | Downregulated |
| 0,095837215 | 0,768400789 | SLC25A51 | FALSE | Upregulated |
| -0,041988718 | 0,768441407 | DDRGK1 | FALSE | Downregulated |
| 0,168733458 | 0,768449821 | LYSMD4 | FALSE | Upregulated |
| 0,044678428 | 0,768449821 | SRF | FALSE | Upregulated |
| -0,075146959 | 0,768449821 | ZNF331 | FALSE | Downregulated |
| -0,071415258 | 0,768484935 | FTL | FALSE | Downregulated |
| -0,242037782 | 0,768484935 | SEC1P | FALSE | Downregulated |
| -0,161865514 | 0,768484935 | SGK494 | FALSE | Downregulated |
| -0,111431367 | 0,768553641 | APBA1 | FALSE | Downregulated |
| 0,177761723 | 0,768553641 | CXorf31 | FALSE | Upregulated |
| -0,244295805 | 0,768553641 | KLHL7-AS1 | FALSE | Downregulated |
| -0,221330855 | 0,76861034 | ATOH7 | FALSE | Downregulated |
| 0,223106428 | 0,76861034 | BNC1 | FALSE | Upregulated |
| 0,126607672 | 0,76861034 | CD5L | FALSE | Upregulated |
| 0,151184214 | 0,76861034 | LOC201651 | FALSE | Upregulated |
| 0,053839987 | 0,768622801 | FIS1 | FALSE | Upregulated |
| 0,111193047 | 0,768712086 | ABCA9 | FALSE | Upregulated |
| 0,087154961 | 0,768712086 | CRY2 | FALSE | Upregulated |
| 0,163159181 | 0,768712086 | GPRASP2 | FALSE | Upregulated |
| -0,494132813 | 0,768712086 | HIST1H2BO | FALSE | Downregulated |
| -0,026561435 | 0,768712086 | PTPRA | FALSE | Downregulated |
| 0,044923127 | 0,768712086 | RAB11FIP3 | FALSE | Upregulated |
| -0,036140533 | 0,768875877 | NUP107 | FALSE | Downregulated |

| | | | | |
|---|---|---|---|---|
| 0,705016351 | 0,769007315 | AL022476.2 | FALSE | Upregulated |
| -0,061862019 | 0,769007315 | TTC33 | FALSE | Downregulated |
| -0,114044588 | 0,769007315 | ZNF528 | FALSE | Downregulated |
| 0,047305303 | 0,769010637 | KLHL36 | FALSE | Upregulated |
| 0,379969245 | 0,769071773 | CYP4F35P | FALSE | Upregulated |
| 0,056823392 | 0,769071773 | FAM63A | FALSE | Upregulated |
| -0,736375929 | 0,769152326 | SNORD62A | FALSE | Downregulated |
| -0,09640735 | 0,769225577 | ARHGEF28 | FALSE | Downregulated |
| 0,053300466 | 0,769225577 | LACE1 | FALSE | Upregulated |
| 0,155376875 | 0,769304666 | LOC101928485 | FALSE | Upregulated |
| -0,131874101 | 0,769835011 | LOC102723666 | FALSE | Downregulated |
| 0,126181793 | 0,769880723 | RIC3 | FALSE | Upregulated |
| -0,053678123 | 0,769880723 | SRSF5 | FALSE | Downregulated |
| 0,166341617 | 0,76995339 | MTMR9LP | FALSE | Upregulated |
| -0,139205928 | 0,769958083 | EPB41L1 | FALSE | Downregulated |
| -0,048698439 | 0,769979717 | BLMH | FALSE | Downregulated |
| 0,029754508 | 0,769979717 | CTCF | FALSE | Upregulated |
| 0,22834681 | 0,769979717 | DYNC1I1 | FALSE | Upregulated |
| 0,301527677 | 0,769979717 | RGS6 | FALSE | Upregulated |
| 0,067177059 | 0,770060914 | SASS6 | FALSE | Upregulated |
| -0,040397916 | 0,770163775 | ARID1B | FALSE | Downregulated |
| -0,068709228 | 0,770297818 | ATAD2 | FALSE | Downregulated |
| 0,175961312 | 0,770297818 | GOLGA8R | FALSE | Upregulated |
| -0,186357241 | 0,770297818 | LOC284581 | FALSE | Downregulated |
| 0,600855011 | 0,770297818 | MEF2BNB-MEF2B | FALSE | Upregulated |
| 0,035663623 | 0,770444998 | COG7 | FALSE | Upregulated |
| 0,124830222 | 0,770524451 | CAMK4 | FALSE | Upregulated |
| 0,166371048 | 0,770757133 | EFCAB12 | FALSE | Upregulated |
| 0,049522152 | 0,770968876 | DIP2C | FALSE | Upregulated |
| 0,207128655 | 0,770970425 | ADAM20P1 | FALSE | Upregulated |
| -0,284278357 | 0,770970425 | DOK5 | FALSE | Downregulated |
| 0,463667175 | 0,770970425 | LINC00923 | FALSE | Upregulated |
| 0,081852952 | 0,77103354 | ANKMY1 | FALSE | Upregulated |
| 0,044557951 | 0,77103354 | DOCK7 | FALSE | Upregulated |
| 0,320812424 | 0,77103354 | LOC102724907 | FALSE | Upregulated |
| -0,083824567 | 0,77103354 | PMS2L2 | FALSE | Downregulated |
| 0,346415022 | 0,771251627 | PCAT1 | FALSE | Upregulated |
| -0,167051086 | 0,771320571 | LOC100506124 | FALSE | Downregulated |
| 0,226823497 | 0,771362358 | IAPP | FALSE | Upregulated |
| -0,070646588 | 0,771505624 | TIGD7 | FALSE | Downregulated |
| 0,130164148 | 0,771505624 | TRIL | FALSE | Upregulated |
| 0,039931934 | 0,771506325 | PKP4 | FALSE | Upregulated |
| 0,08922508 | 0,771656587 | PIDD | FALSE | Upregulated |
| 0,304303783 | 0,771686745 | CCDC42 | FALSE | Upregulated |
| -0,075699258 | 0,771769641 | PREB | FALSE | Downregulated |
| -0,038528226 | 0,772430816 | CRBN | FALSE | Downregulated |
| 0,247000094 | 0,772430816 | LOC101929647 | FALSE | Upregulated |
| -0,040296425 | 0,772689204 | ARFRP1 | FALSE | Downregulated |
| 0,383752584 | 0,772890197 | LOC102723510 | FALSE | Upregulated |
| 0,515890889 | 0,772906997 | AZU1 | FALSE | Upregulated |
| -0,074905684 | 0,772906997 | PRIM1 | FALSE | Downregulated |

| | | | | |
|---|---|---|---|---|
| -0,566608888 | 0,772931873 | TNFSF9 | FALSE | Downregulated |
| -0,103811303 | 0,773013116 | ZC3H12B | FALSE | Downregulated |
| 0,21449282 | 0,773127652 | LOC102723672 | FALSE | Upregulated |
| -0,407954994 | 0,773425155 | LOC102724322 | FALSE | Downregulated |
| 0,485090737 | 0,773591439 | LOC729468 | FALSE | Upregulated |
| 0,054257461 | 0,773689003 | CEACAM1 | FALSE | Upregulated |
| 0,0360792 | 0,773689003 | SP3 | FALSE | Upregulated |
| 0,097806212 | 0,773726915 | ARSG | FALSE | Upregulated |
| 0,200647341 | 0,773726915 | PDE4C | FALSE | Upregulated |
| 0,444895189 | 0,773797261 | LOC101060301 | FALSE | Upregulated |
| 0,175251515 | 0,773951274 | LOC155060 | FALSE | Upregulated |
| -0,107562773 | 0,77398507 | BMP6 | FALSE | Downregulated |
| 0,069315888 | 0,77398889 | BAIAP2 | FALSE | Upregulated |
| 0,352645074 | 0,77398889 | CCDC108 | FALSE | Upregulated |
| -0,089680126 | 0,77398889 | CREBZF | FALSE | Downregulated |
| 0,220793376 | 0,77398889 | LOC101929465 | FALSE | Upregulated |
| -0,300686729 | 0,77398889 | LOC644554 | FALSE | Downregulated |
| 0,483736784 | 0,77398889 | PRO2852 | FALSE | Upregulated |
| 0,466767766 | 0,77398889 | RAET1K | FALSE | Upregulated |
| 0,031552558 | 0,77398889 | RNMT | FALSE | Upregulated |
| 0,058374919 | 0,77398889 | SMURF1 | FALSE | Upregulated |
| 0,214417276 | 0,77398889 | ZDHHC15 | FALSE | Upregulated |
| -0,032575152 | 0,774069751 | EXOC5 | FALSE | Downregulated |
| -0,13726815 | 0,774182149 | SLC38A11 | FALSE | Downregulated |
| 0,10808927 | 0,774232799 | SIMC1 | FALSE | Upregulated |
| -0,05035971 | 0,774258485 | ELF1 | FALSE | Downregulated |
| -0,18704915 | 0,774281574 | LOC102723648 | FALSE | Downregulated |
| 0,111740019 | 0,774281574 | RAP2C-AS1 | FALSE | Upregulated |
| 0,069273896 | 0,77453071 | DNMT3A | FALSE | Upregulated |
| -0,07285611 | 0,77474854 | PTPN9 | FALSE | Downregulated |
| 0,181853363 | 0,774838783 | LOC101927330 | FALSE | Upregulated |
| -0,058433036 | 0,7751991 | CTSF | FALSE | Downregulated |
| 0,070379706 | 0,775266751 | HEY2 | FALSE | Upregulated |
| 0,467901723 | 0,775294586 | CERS1 | FALSE | Upregulated |
| -0,090542473 | 0,775385064 | RPS21 | FALSE | Downregulated |
| -0,078760144 | 0,775385064 | TOP1P1 | FALSE | Downregulated |
| 0,294640141 | 0,775516465 | LOC102724049 | FALSE | Upregulated |
| 0,358579231 | 0,775516465 | STON1-GTF2A1L | FALSE | Upregulated |
| -0,144543501 | 0,775731261 | LOC100996664 | FALSE | Downregulated |
| 0,075210138 | 0,775785478 | WDPCP | FALSE | Upregulated |
| 0,054238556 | 0,775869032 | BAG2 | FALSE | Upregulated |
| -0,085284377 | 0,776057947 | C1orf172 | FALSE | Downregulated |
| 0,54120266 | 0,776057947 | ODF3L1 | FALSE | Upregulated |
| -0,084673133 | 0,776125676 | LOC101929786 | FALSE | Downregulated |
| -0,034014093 | 0,776125676 | OSTF1 | FALSE | Downregulated |
| -0,119326439 | 0,776125676 | PLAG1 | FALSE | Downregulated |
| -0,22184058 | 0,776125676 | RP11-261C10.3 | FALSE | Downregulated |
| -0,038079901 | 0,776125676 | UQCRFS1 | FALSE | Downregulated |
| -0,051653595 | 0,776199328 | H6PD | FALSE | Downregulated |
| -0,476573848 | 0,776305085 | LOC101929692 | FALSE | Downregulated |
| -0,139327178 | 0,776746618 | CEACAM19 | FALSE | Downregulated |

| | | | | |
|---|---|---|---|---|
| -0,428532339 | 0,776746618 | RP5-1142A6.2 | FALSE | Downregulated |
| -0,079008197 | 0,776753133 | ANO5 | FALSE | Downregulated |
| -0,213045747 | 0,776753133 | GOLGA6L5P | FALSE | Downregulated |
| -0,111785645 | 0,776753133 | TCTEX1D1 | FALSE | Downregulated |
| -0,111108465 | 0,776770492 | ZNF692 | FALSE | Downregulated |
| -0,065035104 | 0,776826581 | NMNAT3 | FALSE | Downregulated |
| 0,992909994 | 0,776826581 | TRNAG27 | FALSE | Upregulated |
| -0,175957402 | 0,776947842 | TMEM74B | FALSE | Downregulated |
| 0,065254373 | 0,777073841 | BRI3 | FALSE | Upregulated |
| 0,22301098 | 0,777073841 | KCND2 | FALSE | Upregulated |
| -0,131461879 | 0,777667421 | LINC00961 | FALSE | Downregulated |
| 0,103484312 | 0,777680779 | RASGRP4 | FALSE | Upregulated |
| 0,462996232 | 0,777763023 | LOC101927031 | FALSE | Upregulated |
| 0,037031194 | 0,777835819 | L3MBTL2 | FALSE | Upregulated |
| 0,093484235 | 0,777835819 | SRCIN1 | FALSE | Upregulated |
| 0,294396839 | 0,778144055 | C10orf90 | FALSE | Upregulated |
| 0,503235511 | 0,778144055 | GRIK1-AS1 | FALSE | Upregulated |
| -0,357991404 | 0,778208024 | LOC441528 | FALSE | Downregulated |
| 0,141612134 | 0,778222503 | LOC102724112 | FALSE | Upregulated |
| -0,053263514 | 0,778239302 | APBB2 | FALSE | Downregulated |
| -0,192283711 | 0,778318758 | ATP6V0E2-AS1 | FALSE | Downregulated |
| -0,062910266 | 0,778318758 | DTX4 | FALSE | Downregulated |
| 0,141671846 | 0,778318758 | SLCO1B7 | FALSE | Upregulated |
| 0,076880477 | 0,778318758 | TDRKH | FALSE | Upregulated |
| -0,34920396 | 0,778318758 | WFIKKN1 | FALSE | Downregulated |
| 0,06941563 | 0,7785301 | ZNF222 | FALSE | Upregulated |
| 0,140941397 | 0,778701028 | REEP1 | FALSE | Upregulated |
| -0,286950598 | 0,778906747 | LINC00266-1 | FALSE | Downregulated |
| 0,467262284 | 0,779072082 | GRPR | FALSE | Upregulated |
| -0,207244987 | 0,779082343 | ZNF205-AS1 | FALSE | Downregulated |
| -0,254595019 | 0,779113818 | FAM218A | FALSE | Downregulated |
| 0,365640627 | 0,779113818 | LOC102723543 | FALSE | Upregulated |
| -0,049608001 | 0,77924544 | BAIAP2-AS1 | FALSE | Downregulated |
| -0,434759451 | 0,77924544 | C4orf26 | FALSE | Downregulated |
| -0,086591391 | 0,77924544 | CDH23 | FALSE | Downregulated |
| -0,041161339 | 0,77924544 | CXXC1 | FALSE | Downregulated |
| 0,120784871 | 0,77924544 | LOC101929747 | FALSE | Upregulated |
| -0,142959202 | 0,77924544 | NAALAD2 | FALSE | Downregulated |
| -0,055603135 | 0,779301425 | LOC145783 | FALSE | Downregulated |
| 0,034035609 | 0,779310494 | RAC1 | FALSE | Upregulated |
| -0,060323539 | 0,779370317 | SLC15A1 | FALSE | Downregulated |
| -0,121195385 | 0,779445911 | WDFY3-AS2 | FALSE | Downregulated |
| -0,065258203 | 0,779654224 | MYL12B | FALSE | Downregulated |
| -0,397855737 | 0,779708608 | RSPO1 | FALSE | Downregulated |
| 0,110764896 | 0,779708608 | ZNF677 | FALSE | Upregulated |
| 0,06382453 | 0,779758081 | ELAC1 | FALSE | Upregulated |
| 0,062211389 | 0,779758081 | FIZ1 | FALSE | Upregulated |
| 0,086325761 | 0,779758081 | GYG1 | FALSE | Upregulated |
| 0,111730939 | 0,779758081 | RTN4RL2 | FALSE | Upregulated |
| -0,037291181 | 0,779763056 | ACIN1 | FALSE | Downregulated |
| 0,309882334 | 0,779853919 | LOC102723917 | FALSE | Upregulated |

| | | | | |
|---|---|---|---|---|
| -0,113255856 | 0,779862762 | DNA2 | FALSE | Downregulated |
| 0,407577481 | 0,780072668 | FLJ16779 | FALSE | Upregulated |
| -0,290029724 | 0,780109134 | LINC00612 | FALSE | Downregulated |
| -0,127135785 | 0,780486046 | AVIL | FALSE | Downregulated |
| -0,052035401 | 0,780935309 | APP | FALSE | Downregulated |
| 0,288002535 | 0,780935309 | C10orf62 | FALSE | Upregulated |
| -0,035160014 | 0,780935309 | EPC1 | FALSE | Downregulated |
| -0,127065832 | 0,780935309 | EPSTI1 | FALSE | Downregulated |
| -0,034476937 | 0,780935309 | GNAQ | FALSE | Downregulated |
| 0,517527078 | 0,780935309 | HAS2-AS1 | FALSE | Upregulated |
| 0,122848474 | 0,780935309 | RP11-290F20.3 | FALSE | Upregulated |
| 0,040043553 | 0,780935309 | SENP5 | FALSE | Upregulated |
| -0,063978708 | 0,780935309 | SLC17A5 | FALSE | Downregulated |
| -0,054267434 | 0,780935309 | TTC28 | FALSE | Downregulated |
| -0,045968225 | 0,780992771 | CLTB | FALSE | Downregulated |
| 0,139906308 | 0,781080518 | LINC00921 | FALSE | Upregulated |
| 0,084171471 | 0,781127155 | ZNF487 | FALSE | Upregulated |
| 0,256762426 | 0,781313613 | VPS9D1-AS1 | FALSE | Upregulated |
| -0,051931313 | 0,781437253 | GSTO1 | FALSE | Downregulated |
| 0,130588065 | 0,781437253 | TMEM100 | FALSE | Upregulated |
| -0,038425747 | 0,781535141 | EID1 | FALSE | Downregulated |
| 0,037126609 | 0,781535141 | NHEJ1 | FALSE | Upregulated |
| -0,144184233 | 0,781535141 | TMEM74 | FALSE | Downregulated |
| 0,119863283 | 0,781551334 | SPAG5-AS1 | FALSE | Upregulated |
| 0,460165963 | 0,781562337 | LRRN4CL | FALSE | Upregulated |
| -0,080458697 | 0,781562337 | PI4KAP2 | FALSE | Downregulated |
| 0,055249701 | 0,781562337 | TRMT2B | FALSE | Upregulated |
| -0,05946402 | 0,781562337 | UBA6-AS1 | FALSE | Downregulated |
| -0,049848908 | 0,781736749 | CEP76 | FALSE | Downregulated |
| 0,232619332 | 0,781736749 | ELAVL4 | FALSE | Upregulated |
| 0,367659156 | 0,781736749 | RP1-90L6.2 | FALSE | Upregulated |
| -0,053646023 | 0,781736749 | SF3B5 | FALSE | Downregulated |
| -0,033799811 | 0,781743235 | CIZ1 | FALSE | Downregulated |
| 0,044002545 | 0,781743235 | DTWD2 | FALSE | Upregulated |
| -0,045893572 | 0,781743235 | GRWD1 | FALSE | Downregulated |
| -0,083551317 | 0,781743235 | SSX2IP | FALSE | Downregulated |
| 0,475345442 | 0,781743235 | SYCP1 | FALSE | Upregulated |
| 0,337818466 | 0,781743235 | TRPM1 | FALSE | Upregulated |
| 0,583703544 | 0,781755443 | AMY2A | FALSE | Upregulated |
| -0,027014313 | 0,781832089 | FAM20B | FALSE | Downregulated |
| 0,32885744 | 0,781832089 | WASH7P | FALSE | Upregulated |
| 0,044220471 | 0,781869023 | ZNF44 | FALSE | Upregulated |
| -0,051783441 | 0,782134047 | DGUOK | FALSE | Downregulated |
| -0,090230397 | 0,782134047 | IRS2 | FALSE | Downregulated |
| -0,241051886 | 0,782134047 | MSS51 | FALSE | Downregulated |
| 0,189134589 | 0,782134047 | PPM1J | FALSE | Upregulated |
| 0,142561706 | 0,7821367 | ST6GALNAC2 | FALSE | Upregulated |
| 0,14546675 | 0,782164176 | LOC202181 | FALSE | Upregulated |
| 0,042921749 | 0,782166388 | MAP2K5 | FALSE | Upregulated |
| 0,038783783 | 0,782172479 | C16orf62 | FALSE | Upregulated |
| 0,095767574 | 0,782172479 | LOC100506639 | FALSE | Upregulated |

| | | | | |
|---|---|---|---|---|
| -0,354122704 | 0,78217632 | GPR21 | FALSE | Downregulated |
| -0,20609533 | 0,782308561 | LOC101928182 | FALSE | Downregulated |
| -0,089891285 | 0,782308561 | RP1-249H1.4 | FALSE | Downregulated |
| 0,223695492 | 0,782459704 | CYP4F24P | FALSE | Upregulated |
| 0,063979311 | 0,782459704 | IMPA1 | FALSE | Upregulated |
| -0,634844862 | 0,782841254 | LOC101928961 | FALSE | Downregulated |
| 0,318187711 | 0,782841254 | PAX6 | FALSE | Upregulated |
| -0,204489832 | 0,782841254 | SLX1A-SULT1A3 | FALSE | Downregulated |
| -0,201669629 | 0,783222048 | RP11-356I2.4 | FALSE | Downregulated |
| -0,527675692 | 0,783246228 | RP11-341A22.2 | FALSE | Downregulated |
| -0,111360047 | 0,783246228 | ZFP92 | FALSE | Downregulated |
| 0,24374941 | 0,783319647 | LOC101928965 | FALSE | Upregulated |
| -0,041235232 | 0,783389321 | GRPEL2 | FALSE | Downregulated |
| -0,050843259 | 0,783658236 | TULP4 | FALSE | Downregulated |
| -0,131487929 | 0,783790862 | GSDMB | FALSE | Downregulated |
| -0,085600092 | 0,784019582 | UBTD1 | FALSE | Downregulated |
| -0,057823653 | 0,784063752 | DUSP16 | FALSE | Downregulated |
| -0,043093071 | 0,784063752 | MAP2K3 | FALSE | Downregulated |
| -0,155602071 | 0,78418377 | RP5-1103G7.4 | FALSE | Downregulated |
| 0,422441682 | 0,784311149 | C3orf20 | FALSE | Upregulated |
| 0,390268985 | 0,784363313 | BHLHE40-AS1 | FALSE | Upregulated |
| -0,514161489 | 0,784363313 | URGCP-MRPS24 | FALSE | Downregulated |
| -0,22970922 | 0,784487303 | POMC | FALSE | Downregulated |
| -0,223627597 | 0,784695071 | SMN2 | FALSE | Downregulated |
| 0,230526584 | 0,784836359 | OGN | FALSE | Upregulated |
| -0,047879416 | 0,785178067 | ARL6IP1 | FALSE | Downregulated |
| 0,14941764 | 0,785353706 | KLRF1 | FALSE | Upregulated |
| -0,057412442 | 0,785353706 | NAGK | FALSE | Downregulated |
| -0,268539528 | 0,78552302 | CBX3P2 | FALSE | Downregulated |
| 0,114409429 | 0,78564167 | SHH | FALSE | Upregulated |
| 0,429436327 | 0,785810409 | RLN1 | FALSE | Upregulated |
| -0,042222326 | 0,785827771 | RSPH3 | FALSE | Downregulated |
| -0,276306722 | 0,786209479 | LOC100996442 | FALSE | Downregulated |
| 0,141855571 | 0,786226543 | KIAA1549 | FALSE | Upregulated |
| 0,044774134 | 0,786272128 | PDGFC | FALSE | Upregulated |
| -0,042338549 | 0,786391992 | MSL1 | FALSE | Downregulated |
| 0,040421718 | 0,786478757 | C3orf38 | FALSE | Upregulated |
| 0,361302314 | 0,786478757 | LOC101928514 | FALSE | Upregulated |
| -0,150348093 | 0,786478757 | LOC102724229 | FALSE | Downregulated |
| 0,213508687 | 0,786478757 | MGC39372 | FALSE | Upregulated |
| 0,061900817 | 0,786478757 | STX5 | FALSE | Upregulated |
| 0,055837696 | 0,786478757 | ZSCAN2 | FALSE | Upregulated |
| -0,051416198 | 0,786535345 | DTX3L | FALSE | Downregulated |
| 0,053356483 | 0,786535345 | GAPDH | FALSE | Upregulated |
| -0,213376804 | 0,786535345 | LINGO4 | FALSE | Downregulated |
| 0,814262874 | 0,786535345 | RP4-669L17.10-2 | FALSE | Upregulated |
| 0,047015971 | 0,786535345 | RPRD2 | FALSE | Upregulated |
| -0,110099404 | 0,786955977 | ZNF221 | FALSE | Downregulated |
| -0,093580468 | 0,78721202 | SEMA6B | FALSE | Downregulated |
| 0,224277889 | 0,787292191 | LOC101928178 | FALSE | Upregulated |
| 0,083079028 | 0,787325182 | LRRC8C | FALSE | Upregulated |

| | | | | |
|---|---|---|---|---|
| -0,028443181 | 0,787482091 | RPL7L1 | FALSE | Downregulated |
| 0,126572391 | 0,787505642 | ZNF519 | FALSE | Upregulated |
| 0,337283807 | 0,787597928 | HCN1 | FALSE | Upregulated |
| -0,529057785 | 0,787597928 | RFPL4AL1 | FALSE | Downregulated |
| -0,064493426 | 0,787633341 | LRRC2 | FALSE | Downregulated |
| -0,451652458 | 0,787979857 | RP11-534N16.1 | FALSE | Downregulated |
| -0,055120674 | 0,787985154 | HRG | FALSE | Downregulated |
| 0,554728491 | 0,787985154 | PRSS42 | FALSE | Upregulated |
| 0,159648401 | 0,788158263 | EPHA4 | FALSE | Upregulated |
| -0,071547107 | 0,788158263 | PPARGC1B | FALSE | Downregulated |
| -0,14519572 | 0,78819594 | RTN2 | FALSE | Downregulated |
| -0,532338243 | 0,788324036 | LOC102723320 | FALSE | Downregulated |
| -0,037502939 | 0,788428368 | TOPBP1 | FALSE | Downregulated |
| 0,071600076 | 0,788526147 | C19orf70 | FALSE | Upregulated |
| -0,095974619 | 0,788694129 | ZNF334 | FALSE | Downregulated |
| -0,057429194 | 0,788694129 | ZNF518A | FALSE | Downregulated |
| 0,094894438 | 0,788748524 | ZNF844 | FALSE | Upregulated |
| 0,325057802 | 0,789006182 | LOC101930305 | FALSE | Upregulated |
| 0,022622711 | 0,789018202 | AP1G1 | FALSE | Upregulated |
| -0,087222385 | 0,789018202 | EIF4EBP3 | FALSE | Downregulated |
| 0,447801187 | 0,789058355 | GRID2 | FALSE | Upregulated |
| 0,055244749 | 0,789374315 | ME2 | FALSE | Upregulated |
| 0,49922385 | 0,789738567 | MFSD2B | FALSE | Upregulated |
| 0,180354778 | 0,789815278 | LOC101928055 | FALSE | Upregulated |
| -0,068355306 | 0,789845975 | DPH2 | FALSE | Downregulated |
| -0,466524214 | 0,789871969 | SCHLAP1 | FALSE | Downregulated |
| 0,04440542 | 0,790224806 | RAB30 | FALSE | Upregulated |
| 0,044863196 | 0,790272622 | DEAF1 | FALSE | Upregulated |
| -0,063415724 | 0,790357889 | MEIS1 | FALSE | Downregulated |
| -0,658603805 | 0,790357889 | SNORD62B | FALSE | Downregulated |
| 0,047763671 | 0,790357889 | VPS13A | FALSE | Upregulated |
| 0,102181279 | 0,790410687 | CCNL2 | FALSE | Upregulated |
| -0,070947073 | 0,79044382 | GPR89B | FALSE | Downregulated |
| -0,020429996 | 0,79044382 | HNRNPA2B1 | FALSE | Downregulated |
| 0,518532288 | 0,79044382 | LGALS9B | FALSE | Upregulated |
| -0,165245705 | 0,79044382 | LOC339874 | FALSE | Downregulated |
| 0,248919191 | 0,79044382 | LRRC15 | FALSE | Upregulated |
| -0,340892103 | 0,79044382 | MED15P9 | FALSE | Downregulated |
| 0,231340823 | 0,79044382 | NOXRED1 | FALSE | Upregulated |
| -0,034208524 | 0,79044382 | ZNF22 | FALSE | Downregulated |
| -0,307375582 | 0,790490924 | LOC101927896 | FALSE | Downregulated |
| -0,227329845 | 0,790490924 | ZNF676 | FALSE | Downregulated |
| -0,276588304 | 0,790694856 | MESTIT1 | FALSE | Downregulated |
| 0,055301092 | 0,790762199 | DPP3 | FALSE | Upregulated |
| -0,658218565 | 0,790762199 | PPP1R14C | FALSE | Downregulated |
| 0,045398881 | 0,79079056 | RIOK2 | FALSE | Upregulated |
| -0,192784309 | 0,791093078 | LOC100287175 | FALSE | Downregulated |
| 0,110388336 | 0,791118923 | ZNF815P | FALSE | Upregulated |
| 0,351246047 | 0,791176118 | AC097500.2 | FALSE | Upregulated |
| -0,275247359 | 0,791176118 | ITPR1-AS1 | FALSE | Downregulated |
| -0,069715639 | 0,791176118 | PPCDC | FALSE | Downregulated |

| | | | | |
|---|---|---|---|---|
| 0,048579299 | 0,791214178 | NOSIP | FALSE | Upregulated |
| 0,211570702 | 0,791508114 | CLECL1 | FALSE | Upregulated |
| -0,048016165 | 0,791508114 | FBXW8 | FALSE | Downregulated |
| 0,058008787 | 0,791508114 | MEF2BNB | FALSE | Upregulated |
| 0,06082291 | 0,791508114 | POFUT2 | FALSE | Upregulated |
| -0,188012464 | 0,791508114 | SMC5-AS1 | FALSE | Downregulated |
| 0,038051677 | 0,791508114 | TOR1B | FALSE | Upregulated |
| -0,035692296 | 0,791516305 | QRSL1 | FALSE | Downregulated |
| 0,464801347 | 0,791531717 | MIR6772 | FALSE | Upregulated |
| 0,312144979 | 0,791621868 | ADAMTS14 | FALSE | Upregulated |
| 0,261152152 | 0,791652081 | CNBD2 | FALSE | Upregulated |
| 0,042450215 | 0,791690204 | C4orf27 | FALSE | Upregulated |
| -1,628586381 | 0,791690204 | LOC102724828 | FALSE | Downregulated |
| 0,577751839 | 0,791690204 | LOC388242 | FALSE | Upregulated |
| -0,125841959 | 0,791788073 | BTBD8 | FALSE | Downregulated |
| 0,282152182 | 0,791788073 | TRNP | FALSE | Upregulated |
| -0,135726486 | 0,791791304 | ANKRD31 | FALSE | Downregulated |
| 0,120301469 | 0,791791304 | LINC00338 | FALSE | Upregulated |
| 0,166061456 | 0,791791304 | LOC102724852 | FALSE | Upregulated |
| 0,035500669 | 0,791967191 | ATE1 | FALSE | Upregulated |
| -0,145862922 | 0,791992763 | KCNK17 | FALSE | Downregulated |
| -0,228701327 | 0,792091456 | LOC101929796 | FALSE | Downregulated |
| 0,072301988 | 0,792219413 | ATP1A1OS | FALSE | Upregulated |
| -0,047487065 | 0,792403775 | CCAR1 | FALSE | Downregulated |
| -0,053468589 | 0,792403775 | CKS1B | FALSE | Downregulated |
| 0,428431081 | 0,792663701 | CECR7 | FALSE | Upregulated |
| -0,239908301 | 0,792668484 | CCR9 | FALSE | Downregulated |
| -0,052860789 | 0,79272137 | STT3A | FALSE | Downregulated |
| -0,050944021 | 0,79272137 | WDR83 | FALSE | Downregulated |
| -0,049508177 | 0,79277326 | HMGXB4 | FALSE | Downregulated |
| -0,138092336 | 0,79277326 | KCNJ14 | FALSE | Downregulated |
| -0,176374027 | 0,79277326 | LOC102724159 | FALSE | Downregulated |
| -0,146700842 | 0,792896463 | LOC101928434 | FALSE | Downregulated |
| 0,04414748 | 0,792896463 | NARFL | FALSE | Upregulated |
| -0,131319062 | 0,792929409 | KLHL32 | FALSE | Downregulated |
| 0,04779823 | 0,792931693 | GHDC | FALSE | Upregulated |
| -0,451302045 | 0,792940667 | HIST2H4A | FALSE | Downregulated |
| -0,043705299 | 0,792977432 | C11orf24 | FALSE | Downregulated |
| -0,044320875 | 0,792977432 | C17orf58 | FALSE | Downregulated |
| -0,136572928 | 0,792977432 | CPAMD8 | FALSE | Downregulated |
| 0,12619172 | 0,792977432 | RAD51-AS1 | FALSE | Upregulated |
| -0,16270168 | 0,793170068 | LOC102724455 | FALSE | Downregulated |
| -0,067059373 | 0,79321282 | SAP30 | FALSE | Downregulated |
| 0,41722474 | 0,793446332 | LOC101928439 | FALSE | Upregulated |
| 0,047180683 | 0,793494579 | MTRR | FALSE | Upregulated |
| -0,038102858 | 0,793780751 | PRKAG1 | FALSE | Downregulated |
| -0,376337058 | 0,793923274 | LOC101928484 | FALSE | Downregulated |
| -0,038275122 | 0,793923274 | VAPB | FALSE | Downregulated |
| -1,422400562 | 0,794178157 | TRNAK20 | FALSE | Downregulated |
| -0,181181691 | 0,794179769 | C15orf43 | FALSE | Downregulated |
| 0,101620388 | 0,794189257 | EFCC1 | FALSE | Upregulated |

| | | | | |
|---|---|---|---|---|
| -0,092503795 | 0,794189257 | LCA5L | FALSE | Downregulated |
| -0,149574566 | 0,794189257 | LOC102724517 | FALSE | Downregulated |
| -0,04729474 | 0,794189257 | SACM1L | FALSE | Downregulated |
| 0,197861978 | 0,794189257 | TRPM3 | FALSE | Upregulated |
| 0,041144968 | 0,794349052 | ZNF304 | FALSE | Upregulated |
| 0,055433569 | 0,794413583 | CMTM4 | FALSE | Upregulated |
| 0,061951092 | 0,794413583 | TFEB | FALSE | Upregulated |
| -0,158060017 | 0,794413583 | ZGLP1 | FALSE | Downregulated |
| -0,218985573 | 0,795173724 | AC018647.3 | FALSE | Downregulated |
| -0,042627494 | 0,795408794 | NTPCR | FALSE | Downregulated |
| -0,689391096 | 0,795408794 | PERM1 | FALSE | Downregulated |
| 0,047284471 | 0,795408794 | PXK | FALSE | Upregulated |
| 0,030008026 | 0,795408794 | RAF1 | FALSE | Upregulated |
| -0,221352984 | 0,795408794 | RBM44 | FALSE | Downregulated |
| -0,032827292 | 0,795408794 | TBC1D23 | FALSE | Downregulated |
| -0,027342861 | 0,795415264 | CDC27 | FALSE | Downregulated |
| -0,036500457 | 0,795415264 | RAB11FIP2 | FALSE | Downregulated |
| 0,323036656 | 0,795495512 | LOC100128882 | FALSE | Upregulated |
| 0,167655031 | 0,795954144 | CBR3-AS1 | FALSE | Upregulated |
| -0,145876148 | 0,796098722 | LOC102723616 | FALSE | Downregulated |
| 0,470124781 | 0,796098722 | RP11-170N11.1 | FALSE | Upregulated |
| 0,034623929 | 0,796098722 | ZNF362 | FALSE | Upregulated |
| 0,218643918 | 0,796190008 | FAM162B | FALSE | Upregulated |
| -0,138269413 | 0,796190008 | LINC00294 | FALSE | Downregulated |
| 0,092914231 | 0,796275241 | ZNF460 | FALSE | Upregulated |
| 0,18196304 | 0,79647772 | AC007038.7 | FALSE | Upregulated |
| 0,045572146 | 0,796616497 | CTSA | FALSE | Upregulated |
| 0,056594577 | 0,796658583 | C20orf27 | FALSE | Upregulated |
| -0,239435562 | 0,796722669 | LOC101928123 | FALSE | Downregulated |
| -0,145993408 | 0,796778526 | LOC101928826 | FALSE | Downregulated |
| -0,331011127 | 0,797314967 | RHBDL3 | FALSE | Downregulated |
| -0,122719399 | 0,79738836 | AGAP6 | FALSE | Downregulated |
| -0,248163517 | 0,797465013 | LOC101928764 | FALSE | Downregulated |
| 0,046817817 | 0,797759766 | RPL18A | FALSE | Upregulated |
| -0,044294004 | 0,797759766 | RQCD1 | FALSE | Downregulated |
| -0,056248349 | 0,797759766 | TIMM10 | FALSE | Downregulated |
| 0,185250608 | 0,797851857 | BMX | FALSE | Upregulated |
| 0,050403367 | 0,797851857 | EPN2 | FALSE | Upregulated |
| -0,064405108 | 0,797851857 | LOC100130744 | FALSE | Downregulated |
| -0,045252245 | 0,797851857 | SEC13 | FALSE | Downregulated |
| 0,079270675 | 0,797991098 | ZNF709 | FALSE | Upregulated |
| -0,039283254 | 0,798119846 | PMS2P1 | FALSE | Downregulated |
| -0,049625853 | 0,798236892 | CMAS | FALSE | Downregulated |
| 0,046140452 | 0,798236892 | FAM92A1 | FALSE | Upregulated |
| -0,34328155 | 0,798324277 | LOC101929837 | FALSE | Downregulated |
| 0,427520916 | 0,798341802 | KRT27 | FALSE | Upregulated |
| 0,275681365 | 0,798384058 | ASB2 | FALSE | Upregulated |
| -0,049491136 | 0,798413053 | ACOX1 | FALSE | Downregulated |
| 0,341581018 | 0,798413053 | C1orf145 | FALSE | Upregulated |
| -0,288810832 | 0,798413053 | LOC101926894 | FALSE | Downregulated |
| 0,057139819 | 0,798446931 | PLEKHJ1 | FALSE | Upregulated |

| | | | | |
|---|---|---|---|---|
| 0,03495315 | 0,798481281 | LTN1 | FALSE | Upregulated |
| 0,046666375 | 0,79855499 | MARVELD2 | FALSE | Upregulated |
| -0,20547849 | 0,798705239 | NEXN | FALSE | Downregulated |
| -0,164794113 | 0,798705239 | ROPN1B | FALSE | Downregulated |
| -0,34841689 | 0,79870823 | AC010127.3 | FALSE | Downregulated |
| 0,203163285 | 0,798794692 | BAI1 | FALSE | Upregulated |
| -0,033399916 | 0,798803449 | HERPUD2 | FALSE | Downregulated |
| 0,057575554 | 0,798829277 | WDR91 | FALSE | Upregulated |
| -0,190801511 | 0,798835201 | GOLGA2P7 | FALSE | Downregulated |
| 0,047476206 | 0,798930138 | AVEN | FALSE | Upregulated |
| 0,332690986 | 0,798931045 | MIR4442 | FALSE | Upregulated |
| 0,452772253 | 0,799003559 | LINC00968 | FALSE | Upregulated |
| 0,038792524 | 0,799048217 | BCKDK | FALSE | Upregulated |
| -0,080578039 | 0,799080663 | ORC2 | FALSE | Downregulated |
| -0,029689851 | 0,799080663 | SARNP | FALSE | Downregulated |
| -0,411372428 | 0,79917708 | LOC101928690 | FALSE | Downregulated |
| 0,056698225 | 0,79920684 | ZNF496 | FALSE | Upregulated |
| 0,379662222 | 0,799236627 | LOC101929017 | FALSE | Upregulated |
| -0,464011661 | 0,799431804 | C1QTNF4 | FALSE | Downregulated |
| 0,077079567 | 0,799431804 | PDE4A | FALSE | Upregulated |
| -0,078486526 | 0,799553192 | ZNF442 | FALSE | Downregulated |
| 0,29649821 | 0,799577957 | GYPE | FALSE | Upregulated |
| -0,03224 | 0,799771551 | BCR | FALSE | Downregulated |
| 0,057488023 | 0,799771551 | MLXIP | FALSE | Upregulated |
| 0,439556254 | 0,799771551 | MS4A3 | FALSE | Upregulated |
| 0,065330496 | 0,799771551 | PLEKHA8P1 | FALSE | Upregulated |
| -0,046818597 | 0,799771551 | PRRC1 | FALSE | Downregulated |
| -0,04705283 | 0,799771551 | ZNF510 | FALSE | Downregulated |
| -0,032439051 | 0,799795244 | FAM122A | FALSE | Downregulated |
| 0,09693837 | 0,799795244 | FOXO1 | FALSE | Upregulated |
| 0,07768854 | 0,799795244 | SLC2A10 | FALSE | Upregulated |
| 0,521870747 | 0,799795244 | SLCO1C1 | FALSE | Upregulated |
| -0,042599749 | 0,799819864 | AP2S1 | FALSE | Downregulated |
| -0,042117717 | 0,799902874 | ZDHHC18 | FALSE | Downregulated |
| 0,321754415 | 0,800110983 | FGF9 | FALSE | Upregulated |
| 0,39445493 | 0,800174396 | CNIH2 | FALSE | Upregulated |
| -0,224444243 | 0,800174396 | TECTB | FALSE | Downregulated |
| -0,121692195 | 0,800189477 | RP11-461O7.1 | FALSE | Downregulated |
| 0,080871271 | 0,800320296 | C1orf159 | FALSE | Upregulated |
| 0,040913081 | 0,800392195 | UFC1 | FALSE | Upregulated |
| 0,044578495 | 0,800712289 | EIF2B2 | FALSE | Upregulated |
| 0,328770891 | 0,800712289 | NHLH1 | FALSE | Upregulated |
| 0,066801077 | 0,800712289 | SP110 | FALSE | Upregulated |
| -0,044397432 | 0,800713786 | LGALS8 | FALSE | Downregulated |
| -0,148465325 | 0,800733341 | SLC13A2 | FALSE | Downregulated |
| 0,061224613 | 0,800816083 | MOB3A | FALSE | Upregulated |
| -0,059299193 | 0,801013176 | MZT2B | FALSE | Downregulated |
| 0,026907309 | 0,801168902 | ARHGAP12 | FALSE | Upregulated |
| 0,209704054 | 0,801168902 | BEGAIN | FALSE | Upregulated |
| 0,161819659 | 0,801168902 | TMEM183B | FALSE | Upregulated |
| -0,065143419 | 0,801418345 | TCF7L1 | FALSE | Downregulated |

| | | | | |
|---|---|---|---|---|
| 0,221775818 | 0,801899186 | PPP1R36 | FALSE | Upregulated |
| -0,040839678 | 0,802081313 | DENND5B | FALSE | Downregulated |
| 0,370161773 | 0,802090495 | SCGB2B2 | FALSE | Upregulated |
| 0,063440262 | 0,802371136 | NBEAL1 | FALSE | Upregulated |
| 0,051554482 | 0,802392609 | HEATR3 | FALSE | Upregulated |
| -0,133402906 | 0,802392609 | MIP | FALSE | Downregulated |
| -0,431691758 | 0,802392609 | TXNDC2 | FALSE | Downregulated |
| -0,129214431 | 0,802656604 | VN1R1 | FALSE | Downregulated |
| 0,142601115 | 0,802705188 | MN1 | FALSE | Upregulated |
| 0,236666391 | 0,802878556 | PPP5D1 | FALSE | Upregulated |
| 0,386854057 | 0,802953477 | LOC101928109 | FALSE | Upregulated |
| -0,080322537 | 0,803015827 | C1QA | FALSE | Downregulated |
| -0,110744992 | 0,803015827 | TNFSF4 | FALSE | Downregulated |
| -0,206221797 | 0,803181554 | FKBP9L | FALSE | Downregulated |
| -0,130776532 | 0,803187289 | MMRN1 | FALSE | Downregulated |
| 0,074246886 | 0,803207091 | DAAM2 | FALSE | Upregulated |
| -0,307808466 | 0,803207091 | DNAJC9-AS1 | FALSE | Downregulated |
| -0,037851893 | 0,803207091 | HNRNPDL | FALSE | Downregulated |
| 0,131390142 | 0,803207091 | RHBG | FALSE | Upregulated |
| -0,07529371 | 0,803207091 | SLC25A18 | FALSE | Downregulated |
| 0,401507193 | 0,803207091 | VSTM1 | FALSE | Upregulated |
| -0,408132119 | 0,803533446 | IGSF11 | FALSE | Downregulated |
| -0,17207527 | 0,804340862 | DDX43 | FALSE | Downregulated |
| -0,059078449 | 0,804575576 | NOTCH2NL | FALSE | Downregulated |
| -0,026772424 | 0,804588477 | RAB10 | FALSE | Downregulated |
| 0,243700278 | 0,804648174 | LOC101927268 | FALSE | Upregulated |
| -0,079387145 | 0,804680986 | C1orf162 | FALSE | Downregulated |
| 0,040041182 | 0,804680986 | CDKN1B | FALSE | Upregulated |
| 0,30270684 | 0,804680986 | CHODL | FALSE | Upregulated |
| -0,129557032 | 0,804680986 | DFNB59 | FALSE | Downregulated |
| -0,31204959 | 0,804680986 | GPR62 | FALSE | Downregulated |
| -0,379379751 | 0,804680986 | LOC100506114 | FALSE | Downregulated |
| 0,38993063 | 0,804680986 | LOC727710 | FALSE | Upregulated |
| -0,160802148 | 0,804680986 | NAGPA-AS1 | FALSE | Downregulated |
| -0,105328684 | 0,804680986 | NUDT4P1 | FALSE | Downregulated |
| 0,32971533 | 0,804680986 | TH | FALSE | Upregulated |
| -0,040829463 | 0,804680986 | TRMT61B | FALSE | Downregulated |
| -0,079465791 | 0,804680986 | UBXN11 | FALSE | Downregulated |
| 0,31783444 | 0,804680986 | WFDC3 | FALSE | Upregulated |
| 0,043775641 | 0,804727251 | LRRC27 | FALSE | Upregulated |
| 0,074113562 | 0,804742945 | GDPD5 | FALSE | Upregulated |
| 0,06536847 | 0,804773373 | CADM4 | FALSE | Upregulated |
| 0,035857682 | 0,80478851 | GCC1 | FALSE | Upregulated |
| 0,365402825 | 0,804955395 | C1orf189 | FALSE | Upregulated |
| 0,233051267 | 0,804955395 | LINC00441 | FALSE | Upregulated |
| 0,10828872 | 0,804955395 | TAPT1-AS1 | FALSE | Upregulated |
| 0,126581154 | 0,804961477 | BTN2A3P | FALSE | Upregulated |
| -0,02661613 | 0,804961477 | MAP2K4 | FALSE | Downregulated |
| -0,063790131 | 0,804961477 | SREBF2 | FALSE | Downregulated |
| -0,065368752 | 0,804961477 | SUPT3H | FALSE | Downregulated |
| -0,039351672 | 0,805455392 | UCHL3 | FALSE | Downregulated |

| | | | | |
|---|---|---|---|---|
| -0,057909323 | 0,805815578 | COX10 | FALSE | Downregulated |
| 0,1716485 | 0,805815578 | CYP3A7-CYP3AP1 | FALSE | Upregulated |
| 0,122472562 | 0,805815578 | FREM1 | FALSE | Upregulated |
| -0,251721959 | 0,805815578 | MIR29C | FALSE | Downregulated |
| -0,432788881 | 0,805815578 | RP11-829H16.3 | FALSE | Downregulated |
| 0,033669844 | 0,805815578 | YIPF6 | FALSE | Upregulated |
| 0,12835682 | 0,805989218 | CDHR2 | FALSE | Upregulated |
| -0,059997736 | 0,805989218 | GTF2IRD2B | FALSE | Downregulated |
| 0,152665269 | 0,805989218 | LOC101927060 | FALSE | Upregulated |
| -0,099457376 | 0,805989218 | PRKCQ-AS1 | FALSE | Downregulated |
| -0,032916247 | 0,805989218 | RNF38 | FALSE | Downregulated |
| 0,093136559 | 0,805989218 | TLE3 | FALSE | Upregulated |
| -0,168093498 | 0,806075233 | LINC00894 | FALSE | Downregulated |
| -0,221377265 | 0,806352984 | TENM2 | FALSE | Downregulated |
| 0,218863019 | 0,806479523 | SENCR | FALSE | Upregulated |
| 0,059344949 | 0,806533407 | MAGEH1 | FALSE | Upregulated |
| 0,040406966 | 0,80660711 | CIB1 | FALSE | Upregulated |
| 0,040450194 | 0,806649564 | NAA30 | FALSE | Upregulated |
| -0,387733511 | 0,807005803 | ST8SIA5 | FALSE | Downregulated |
| -0,089325741 | 0,807030743 | EVI2B | FALSE | Downregulated |
| -0,048911079 | 0,807244101 | ACD | FALSE | Downregulated |
| -0,235176131 | 0,807244101 | ARHGEF4 | FALSE | Downregulated |
| -0,282760117 | 0,807244101 | IL1RAPL1 | FALSE | Downregulated |
| -0,18302344 | 0,807244101 | PDGFRL | FALSE | Downregulated |
| 0,537965149 | 0,807244101 | SNORD56 | FALSE | Upregulated |
| -0,050838464 | 0,807244101 | TRAPPC1 | FALSE | Downregulated |
| 0,037372159 | 0,807259785 | KIAA1432 | FALSE | Upregulated |
| 0,019233192 | 0,807405225 | CUL4A | FALSE | Upregulated |
| -0,082538774 | 0,807405225 | LOC102724873 | FALSE | Downregulated |
| 0,157803621 | 0,807445592 | BTBD19 | FALSE | Upregulated |
| 0,281824449 | 0,80745878 | DOCK3 | FALSE | Upregulated |
| -0,102545621 | 0,807548882 | FAM83D | FALSE | Downregulated |
| 0,068334862 | 0,807826383 | ALDH1B1 | FALSE | Upregulated |
| -0,047785177 | 0,808057041 | COQ6 | FALSE | Downregulated |
| 0,367106052 | 0,808057041 | LOC101928279 | FALSE | Upregulated |
| -0,479419599 | 0,808137954 | POU4F1 | FALSE | Downregulated |
| -0,16208904 | 0,808230721 | MIR17HG | FALSE | Downregulated |
| -0,170757856 | 0,808672128 | LOC100507554 | FALSE | Downregulated |
| 0,047391026 | 0,808780794 | MIS12 | FALSE | Upregulated |
| -0,215257533 | 0,809178189 | AC007773.2 | FALSE | Downregulated |
| 0,077586103 | 0,809178189 | GLI4 | FALSE | Upregulated |
| -0,028614769 | 0,809178189 | PAK2 | FALSE | Downregulated |
| 0,054583249 | 0,809178189 | TMUB1 | FALSE | Upregulated |
| -0,074663731 | 0,80931068 | S1PR1 | FALSE | Downregulated |
| 0,182313315 | 0,809328745 | P2RX3 | FALSE | Upregulated |
| 0,072561095 | 0,809365776 | DDX60 | FALSE | Upregulated |
| 0,03350506 | 0,809365776 | EIF2S2 | FALSE | Upregulated |
| -0,363657433 | 0,809365776 | RDM1 | FALSE | Downregulated |
| -0,240436155 | 0,809365776 | TEX15 | FALSE | Downregulated |
| -0,356099413 | 0,809514094 | ACER1 | FALSE | Downregulated |
| -0,034020237 | 0,809514094 | OTUD4 | FALSE | Downregulated |

| | | | | |
|---|---|---|---|---|
| 0,197696007 | 0,809859582 | MROH2A | FALSE | Upregulated |
| 0,044733893 | 0,809887185 | ALG11 | FALSE | Upregulated |
| -0,033377208 | 0,809887185 | CMPK1 | FALSE | Downregulated |
| 0,22448744 | 0,809887185 | ERI3-IT1 | FALSE | Upregulated |
| -0,518162974 | 0,809887185 | FTH1P18 | FALSE | Downregulated |
| -0,117123041 | 0,809887185 | LOC101927216 | FALSE | Downregulated |
| -0,32336562 | 0,809887185 | LOC101927720 | FALSE | Downregulated |
| -0,433415593 | 0,809887185 | REXO1L1P | FALSE | Downregulated |
| -0,024488816 | 0,809887185 | VPS36 | FALSE | Downregulated |
| 0,049678328 | 0,809887185 | ZNF512B | FALSE | Upregulated |
| -0,152752742 | 0,809912019 | LOC400863 | FALSE | Downregulated |
| -0,049499219 | 0,809912019 | SNX24 | FALSE | Downregulated |
| -0,059105095 | 0,80998716 | PFKFB1 | FALSE | Downregulated |
| 0,049972065 | 0,809995124 | ACYP2 | FALSE | Upregulated |
| 0,03907934 | 0,810161868 | PPP1R13B | FALSE | Upregulated |
| 0,027932737 | 0,810493004 | HNRNPF | FALSE | Upregulated |
| -0,229118309 | 0,810576909 | ASIC4 | FALSE | Downregulated |
| -0,085596317 | 0,810576909 | IGF2BP2 | FALSE | Downregulated |
| 0,052857849 | 0,810583585 | STYXL1 | FALSE | Upregulated |
| 0,035129785 | 0,810683167 | PLEKHA1 | FALSE | Upregulated |
| 1,802271498 | 0,81069193 | IGFN1 | FALSE | Upregulated |
| -0,220978299 | 0,81069193 | SULT4A1 | FALSE | Downregulated |
| 0,038795805 | 0,81080458 | BBS2 | FALSE | Upregulated |
| -0,041855956 | 0,81080458 | LYST | FALSE | Downregulated |
| 0,036712888 | 0,811017633 | CNPY2 | FALSE | Upregulated |
| 0,030956357 | 0,811025282 | TRAF6 | FALSE | Upregulated |
| -0,265196229 | 0,811231736 | TIGD3 | FALSE | Downregulated |
| 0,048157986 | 0,811344969 | FBXL20 | FALSE | Upregulated |
| -0,098610853 | 0,811437197 | CNTFR | FALSE | Downregulated |
| 0,139000639 | 0,811437197 | FERMT1 | FALSE | Upregulated |
| 0,327562096 | 0,811437197 | LOC102724560 | FALSE | Upregulated |
| -0,04072149 | 0,811437197 | MEGF9 | FALSE | Downregulated |
| -0,044312953 | 0,811437197 | OLA1 | FALSE | Downregulated |
| -0,127167148 | 0,811437197 | YTHDF3-AS1 | FALSE | Downregulated |
| 0,040946121 | 0,811566431 | COMMD4 | FALSE | Upregulated |
| 0,053921079 | 0,811566431 | DGKD | FALSE | Upregulated |
| -0,323376208 | 0,811566431 | HFM1 | FALSE | Downregulated |
| -0,043736882 | 0,811566431 | INPP5A | FALSE | Downregulated |
| 0,196000745 | 0,811566431 | MTFR2 | FALSE | Upregulated |
| 0,34259924 | 0,811566431 | SGCZ | FALSE | Upregulated |
| 0,110297149 | 0,811566431 | ZNF300 | FALSE | Upregulated |
| -0,193990287 | 0,811638331 | LOC100507634 | FALSE | Downregulated |
| -0,033371412 | 0,811755272 | COMMD9 | FALSE | Downregulated |
| 0,302609083 | 0,811755272 | DYNAP | FALSE | Upregulated |
| 0,448172106 | 0,81193395 | LOC101927898 | FALSE | Upregulated |
| -0,309406102 | 0,812064017 | LOC101928967 | FALSE | Downregulated |
| -0,214168685 | 0,812189109 | LOC102724420 | FALSE | Downregulated |
| -0,06065197 | 0,812190094 | TPPP | FALSE | Downregulated |
| 0,378676164 | 0,812213989 | ARC | FALSE | Upregulated |
| 0,317547082 | 0,812213989 | MT1JP | FALSE | Upregulated |
| 0,532663754 | 0,812340203 | ADORA2A | FALSE | Upregulated |

| | | | | |
|---|---|---|---|---|
| 0,059053536 | 0,812426084 | PDPR | FALSE | Upregulated |
| -0,12625713 | 0,812462749 | CASC2 | FALSE | Downregulated |
| -0,225867152 | 0,812462749 | LOC102724850 | FALSE | Downregulated |
| -0,159162472 | 0,812508944 | ASIC3 | FALSE | Downregulated |
| 0,114200427 | 0,812508944 | CAPN10-AS1 | FALSE | Upregulated |
| -0,032656911 | 0,812524947 | CBX1 | FALSE | Downregulated |
| -0,035528428 | 0,812547718 | UCHL5 | FALSE | Downregulated |
| -1,622955377 | 0,812585321 | SLX1A | FALSE | Downregulated |
| -0,29672707 | 0,812592901 | LOC101928592 | FALSE | Downregulated |
| -1,785448277 | 0,812592901 | RP11-451G4.2 | FALSE | Downregulated |
| 0,110051603 | 0,812623279 | KLHL13 | FALSE | Upregulated |
| -0,095434924 | 0,812700724 | SLC17A9 | FALSE | Downregulated |
| -0,174736759 | 0,813012786 | HOXA-AS2 | FALSE | Downregulated |
| -0,062107217 | 0,813012786 | PCDH1 | FALSE | Downregulated |
| -0,156094243 | 0,813012786 | RD3L | FALSE | Downregulated |
| -0,057668245 | 0,813012786 | SH2D4A | FALSE | Downregulated |
| -0,082613659 | 0,813136305 | CAPS2 | FALSE | Downregulated |
| -0,056061632 | 0,813365294 | MBD6 | FALSE | Downregulated |
| 0,066663488 | 0,813543321 | C9orf40 | FALSE | Upregulated |
| -0,050649662 | 0,813601157 | TCEANC2 | FALSE | Downregulated |
| 0,030963808 | 0,813738548 | RPF1 | FALSE | Upregulated |
| -0,272894662 | 0,814166587 | AC068831.10 | FALSE | Downregulated |
| 0,124887362 | 0,814166587 | EML6 | FALSE | Upregulated |
| -0,262278734 | 0,814166587 | KCNIP2-AS1 | FALSE | Downregulated |
| 0,258582353 | 0,814339632 | KLHL10 | FALSE | Upregulated |
| 0,212032367 | 0,814339632 | LOC101928676 | FALSE | Upregulated |
| 0,147629061 | 0,814339632 | MLLT11 | FALSE | Upregulated |
| -0,04837629 | 0,814339632 | RP2 | FALSE | Downregulated |
| 0,390992779 | 0,81434995 | LOC102723446 | FALSE | Upregulated |
| 0,171336542 | 0,814618555 | PKD1L2 | FALSE | Upregulated |
| 0,04651064 | 0,814649704 | CALCOCO1 | FALSE | Upregulated |
| -0,212149462 | 0,814684936 | GNRH2 | FALSE | Downregulated |
| -0,191893262 | 0,814684936 | LINC00704 | FALSE | Downregulated |
| -0,303096047 | 0,814692771 | ANKRD20A12P | FALSE | Downregulated |
| 0,210937095 | 0,814692771 | RNASE2 | FALSE | Upregulated |
| 0,290786708 | 0,814757764 | NBPF7 | FALSE | Upregulated |
| 0,075080395 | 0,814791235 | LOC101060398 | FALSE | Upregulated |
| 0,331666646 | 0,81495939 | LOC102723959 | FALSE | Upregulated |
| -0,084288168 | 0,815004311 | TMEM254-AS1 | FALSE | Downregulated |
| 0,097115132 | 0,815006458 | NDUFA4L2 | FALSE | Upregulated |
| -0,383745903 | 0,815089248 | LOC102724462 | FALSE | Downregulated |
| 0,290980311 | 0,815183589 | SPDYE6 | FALSE | Upregulated |
| 0,15236767 | 0,815252242 | LINC00310 | FALSE | Upregulated |
| -0,424611442 | 0,815282638 | GABRA2 | FALSE | Downregulated |
| 0,091892997 | 0,815293904 | MGC12916 | FALSE | Upregulated |
| -0,041335059 | 0,815459971 | RPL35A | FALSE | Downregulated |
| 0,358039177 | 0,815496962 | LOC100507033 | FALSE | Upregulated |
| 0,066573391 | 0,815496962 | PIEZO2 | FALSE | Upregulated |
| -0,042826044 | 0,815496962 | PRKAG2 | FALSE | Downregulated |
| 0,0425249 | 0,815496962 | PTP4A1 | FALSE | Upregulated |
| -0,067997958 | 0,8155021 | METTL7B | FALSE | Downregulated |

| | | | | |
|---|---|---|---|---|
| 0,040804175 | 0,815543135 | C17orf75 | FALSE | Upregulated |
| -0,215645136 | 0,815572033 | KPNA7 | FALSE | Downregulated |
| 0,207990747 | 0,815572033 | LOC101928705 | FALSE | Upregulated |
| -0,050746909 | 0,815670961 | ATP8B4 | FALSE | Downregulated |
| -0,21501667 | 0,815670961 | LINC00880 | FALSE | Downregulated |
| 0,03138187 | 0,815670961 | PABPC1 | FALSE | Upregulated |
| 0,031317783 | 0,815670961 | PITRM1 | FALSE | Upregulated |
| -0,26102857 | 0,815670975 | LOC401010 | FALSE | Downregulated |
| 0,250843677 | 0,815842336 | ADM5 | FALSE | Upregulated |
| -1,748442959 | 0,81601674 | HYPK | FALSE | Downregulated |
| -0,062031467 | 0,816096775 | CLMN | FALSE | Downregulated |
| -0,031406232 | 0,816106639 | FCF1 | FALSE | Downregulated |
| 0,407848172 | 0,816106639 | LOC101930370 | FALSE | Upregulated |
| -0,251813888 | 0,816106639 | MSI1 | FALSE | Downregulated |
| -0,054470243 | 0,816106639 | SHF | FALSE | Downregulated |
| -0,036385647 | 0,816106639 | XRN2 | FALSE | Downregulated |
| -0,036823043 | 0,816106639 | ZNF768 | FALSE | Downregulated |
| 0,163158236 | 0,816205022 | DOCK9-AS2 | FALSE | Upregulated |
| 0,185085474 | 0,816205022 | SLC22A2 | FALSE | Upregulated |
| 0,214580579 | 0,816205022 | SPC24 | FALSE | Upregulated |
| -0,035988893 | 0,816205022 | TGOLN2 | FALSE | Downregulated |
| 0,106853511 | 0,816442205 | LOC283177 | FALSE | Upregulated |
| 0,401211166 | 0,816570649 | LCTL | FALSE | Upregulated |
| 0,030353344 | 0,816820977 | DUS1L | FALSE | Upregulated |
| 0,255792964 | 0,816820977 | LOC101928482 | FALSE | Upregulated |
| 0,410876879 | 0,817190013 | ATP2A1 | FALSE | Upregulated |
| -0,103183866 | 0,817324847 | WWC2-AS2 | FALSE | Downregulated |
| -0,33809773 | 0,817326019 | PCAT14 | FALSE | Downregulated |
| 0,043768256 | 0,817610476 | RRAS2 | FALSE | Upregulated |
| -0,086839305 | 0,817641627 | ME1 | FALSE | Downregulated |
| 0,473441124 | 0,817875176 | LOC645485 | FALSE | Upregulated |
| 0,178314515 | 0,817876762 | UCP3 | FALSE | Upregulated |
| -0,07723754 | 0,818070933 | C1QB | FALSE | Downregulated |
| 0,404188996 | 0,818197117 | LOC101927185 | FALSE | Upregulated |
| 0,027909761 | 0,818573739 | TCEAL8 | FALSE | Upregulated |
| 0,04805004 | 0,818645107 | CCDC86 | FALSE | Upregulated |
| 0,355493712 | 0,818674091 | RP11-280K24.4 | FALSE | Upregulated |
| 0,057634723 | 0,81884922 | KCTD13 | FALSE | Upregulated |
| 0,281032853 | 0,81884922 | LOC729041 | FALSE | Upregulated |
| 0,04968822 | 0,81884922 | PDIA3 | FALSE | Upregulated |
| -0,030627067 | 0,818907662 | NBN | FALSE | Downregulated |
| 0,100478724 | 0,81893391 | RPLP0P2 | FALSE | Upregulated |
| 0,079538497 | 0,818937863 | PKI55 | FALSE | Upregulated |
| 0,041062687 | 0,819124984 | ZDHHC4 | FALSE | Upregulated |
| -0,172534887 | 0,819162569 | LOC102724861 | FALSE | Downregulated |
| -0,037875377 | 0,819386486 | PCCA | FALSE | Downregulated |
| -0,069958895 | 0,819386486 | PDIA4 | FALSE | Downregulated |
| -0,417373651 | 0,819408942 | TDGF1P3 | FALSE | Downregulated |
| -0,057228458 | 0,819456681 | SLC9A3R1 | FALSE | Downregulated |
| -0,037138802 | 0,819993387 | CTPS2 | FALSE | Downregulated |
| -0,093558178 | 0,820057631 | ERVK13-1 | FALSE | Downregulated |

| | | | | |
|---|---|---|---|---|
| -0,302404636 | 0,820057631 | LINC00942 | FALSE | Downregulated |
| 0,188475121 | 0,820107185 | GOLGA6D | FALSE | Upregulated |
| 0,132524127 | 0,820107185 | NUTM2D | FALSE | Upregulated |
| -0,064949306 | 0,820107185 | SLC22A18 | FALSE | Downregulated |
| -0,077223741 | 0,820107185 | TAZ | FALSE | Downregulated |
| 0,410273915 | 0,820363508 | SNORA7B | FALSE | Upregulated |
| 0,241221003 | 0,820363508 | TMEM108 | FALSE | Upregulated |
| 0,067076542 | 0,820363508 | ZNF266 | FALSE | Upregulated |
| 0,386636726 | 0,820418058 | PNMA5 | FALSE | Upregulated |
| -0,390106845 | 0,820445073 | LOC101929634 | FALSE | Downregulated |
| 0,185944427 | 0,820456465 | C16orf71 | FALSE | Upregulated |
| -0,031754979 | 0,820456465 | POP4 | FALSE | Downregulated |
| -0,059234587 | 0,820485058 | SLC33A1 | FALSE | Downregulated |
| 0,140971872 | 0,820531231 | LOC102723780 | FALSE | Upregulated |
| 0,033137351 | 0,820531231 | SAMD8 | FALSE | Upregulated |
| -0,063541191 | 0,820549627 | ZNF675 | FALSE | Downregulated |
| 0,04978177 | 0,820604799 | FARP2 | FALSE | Upregulated |
| 0,041889334 | 0,820729539 | IFT57 | FALSE | Upregulated |
| 0,109962066 | 0,820803954 | LOC100129148 | FALSE | Upregulated |
| 0,138761801 | 0,820919719 | LOC643355 | FALSE | Upregulated |
| 0,104570031 | 0,821007947 | RP11-160O5.1 | FALSE | Upregulated |
| -0,024673095 | 0,821015176 | FAM175B | FALSE | Downregulated |
| 0,180566475 | 0,821015176 | GATM-AS1 | FALSE | Upregulated |
| -0,589684781 | 0,821015176 | LOC102723399 | FALSE | Downregulated |
| -0,037366762 | 0,82102214 | SNX18 | FALSE | Downregulated |
| -0,091454255 | 0,821406137 | TPTE2P5 | FALSE | Downregulated |
| 0,062648863 | 0,821406137 | ZNF662 | FALSE | Upregulated |
| 0,101649942 | 0,821485025 | NPIPB5 | FALSE | Upregulated |
| -0,20613549 | 0,821598123 | LINC00282 | FALSE | Downregulated |
| -0,080141047 | 0,821622481 | SIRT4 | FALSE | Downregulated |
| -0,037718523 | 0,821755473 | MDM1 | FALSE | Downregulated |
| 0,040341322 | 0,822136498 | TTC21B | FALSE | Upregulated |
| 0,041579954 | 0,822136498 | ZBTB34 | FALSE | Upregulated |
| -1,418629041 | 0,822344092 | EEF1E1-BLOC1S5 | FALSE | Downregulated |
| -0,035543331 | 0,822344092 | FDX1 | FALSE | Downregulated |
| 0,029924514 | 0,822510141 | DNMBP | FALSE | Upregulated |
| 0,164509395 | 0,822512946 | MAATS1 | FALSE | Upregulated |
| 0,252599251 | 0,822614965 | SLC5A11 | FALSE | Upregulated |
| -0,462574854 | 0,822630294 | LOC101929192 | FALSE | Downregulated |
| -0,029353333 | 0,822644687 | TRIP4 | FALSE | Downregulated |
| -0,030720605 | 0,822700322 | PJA2 | FALSE | Downregulated |
| 0,106225895 | 0,822851651 | ISG15 | FALSE | Upregulated |
| -0,077423 | 0,822947678 | LINC-PINT | FALSE | Downregulated |
| -0,136415251 | 0,822981326 | FAM222A-AS1 | FALSE | Downregulated |
| 0,101569651 | 0,822982001 | CENPL | FALSE | Upregulated |
| 0,213954902 | 0,822982001 | DNAH5 | FALSE | Upregulated |
| -0,092294311 | 0,822982001 | FGL1 | FALSE | Downregulated |
| 0,301780588 | 0,822982001 | MSH5 | FALSE | Upregulated |
| -0,116498404 | 0,823108677 | LOC100506388 | FALSE | Downregulated |
| 0,219891382 | 0,823108677 | TSACC | FALSE | Upregulated |
| 0,213079298 | 0,823151397 | DPP10 | FALSE | Upregulated |

| | | | | |
|---|---|---|---|---|
| -0,047905671 | 0,823233802 | MRPL27 | FALSE | Downregulated |
| -0,176789894 | 0,823279752 | LOC102723539 | FALSE | Downregulated |
| -0,055879645 | 0,823491515 | ANXA9 | FALSE | Downregulated |
| -0,058991661 | 0,823491515 | PLBD2 | FALSE | Downregulated |
| -0,041109194 | 0,82352963 | DKFZP586I1420 | FALSE | Downregulated |
| -0,133219764 | 0,82352963 | PRRT3-AS1 | FALSE | Downregulated |
| -0,255803626 | 0,823534096 | HMP19 | FALSE | Downregulated |
| 0,050478326 | 0,823557063 | MPHOSPH6 | FALSE | Upregulated |
| 0,446731774 | 0,823744111 | FOXD4L1 | FALSE | Upregulated |
| 0,055707443 | 0,82374916 | LSM2 | FALSE | Upregulated |
| -0,052286763 | 0,823919789 | LOC102723728 | FALSE | Downregulated |
| -0,093445537 | 0,823919789 | LRRC48 | FALSE | Downregulated |
| -0,021413279 | 0,823919789 | TSNAX | FALSE | Downregulated |
| -0,348218116 | 0,824131986 | LOC102724943 | FALSE | Downregulated |
| 0,047539407 | 0,824146734 | GNA14 | FALSE | Upregulated |
| 0,653518643 | 0,824146734 | LOC100288570 | FALSE | Upregulated |
| 0,093591697 | 0,824146734 | PTPRS | FALSE | Upregulated |
| -0,090021434 | 0,824146734 | WASH1 | FALSE | Downregulated |
| 0,045688378 | 0,824238378 | AFAP1L1 | FALSE | Upregulated |
| 0,035761695 | 0,824238378 | PLCB3 | FALSE | Upregulated |
| 0,06289307 | 0,824238378 | RAB3D | FALSE | Upregulated |
| 0,473886452 | 0,824249177 | LOC102724259 | FALSE | Upregulated |
| 0,040125623 | 0,824329612 | CYP2R1 | FALSE | Upregulated |
| 0,103429386 | 0,824329612 | USP2 | FALSE | Upregulated |
| 0,03040219 | 0,824370945 | LONP1 | FALSE | Upregulated |
| 0,398837005 | 0,824450686 | LOC102724246 | FALSE | Upregulated |
| -0,043963862 | 0,824537194 | SENP7 | FALSE | Downregulated |
| 0,168892079 | 0,824698632 | AC017060.1 | FALSE | Upregulated |
| -0,246699527 | 0,824773938 | LOC101928233 | FALSE | Downregulated |
| 0,051021492 | 0,824881948 | H2AFZ | FALSE | Upregulated |
| 0,031966037 | 0,824977166 | CDC42EP4 | FALSE | Upregulated |
| 0,0284085 | 0,825066662 | MOB1B | FALSE | Upregulated |
| 0,096600497 | 0,825066662 | NUDT8 | FALSE | Upregulated |
| 1,681987137 | 0,825066662 | RNU2-2P | FALSE | Upregulated |
| -0,399859241 | 0,825066662 | RP11-264C15.2 | FALSE | Downregulated |
| 0,157794309 | 0,825135444 | NPFF | FALSE | Upregulated |
| -0,122896933 | 0,825135444 | STAG3L5P | FALSE | Downregulated |
| 0,305473767 | 0,825213664 | IPO9-AS1 | FALSE | Upregulated |
| 0,076942752 | 0,825270872 | LOC102724905 | FALSE | Upregulated |
| -0,249496509 | 0,825283142 | AC079145.4 | FALSE | Downregulated |
| 0,082611653 | 0,825283142 | PRR11 | FALSE | Upregulated |
| -0,036434091 | 0,825312507 | PRUNE | FALSE | Downregulated |
| -0,119503467 | 0,825394103 | LOC100996457 | FALSE | Downregulated |
| 0,447753372 | 0,82573761 | FLJ45974 | FALSE | Upregulated |
| 0,052819049 | 0,825859617 | SLC28A1 | FALSE | Upregulated |
| 0,034387794 | 0,825867721 | TCTN3 | FALSE | Upregulated |
| 0,034495131 | 0,826079033 | ATP5B | FALSE | Upregulated |
| 0,03702274 | 0,826094989 | TBC1D17 | FALSE | Upregulated |
| -0,074212476 | 0,826414175 | SERPINA3 | FALSE | Downregulated |
| 0,030457197 | 0,826451368 | GATC | FALSE | Upregulated |
| -0,182779279 | 0,826451368 | RHEBL1 | FALSE | Downregulated |

| | | | | |
|---|---|---|---|---|
| 0,103076288 | 0,826451368 | SERP2 | FALSE | Upregulated |
| -0,062808974 | 0,826451368 | ZNF57 | FALSE | Downregulated |
| -0,225384366 | 0,826465011 | RP4-640H8.2 | FALSE | Downregulated |
| -0,107822404 | 0,82666195 | CATSPER3 | FALSE | Downregulated |
| -0,028330775 | 0,82666195 | CCDC93 | FALSE | Downregulated |
| -0,048432346 | 0,82666195 | CIRBP | FALSE | Downregulated |
| 0,031517908 | 0,82666195 | DNASE1L1 | FALSE | Upregulated |
| -0,126274794 | 0,82666195 | JAKMIP3 | FALSE | Downregulated |
| 0,299160959 | 0,82666195 | LOC101927212 | FALSE | Upregulated |
| 0,344277407 | 0,82666195 | LOC101929587 | FALSE | Upregulated |
| 0,173889106 | 0,82666195 | LOC101929882 | FALSE | Upregulated |
| 0,025140105 | 0,82666195 | RANBP3 | FALSE | Upregulated |
| -0,063232246 | 0,82666195 | RASSF8-AS1 | FALSE | Downregulated |
| -0,044016529 | 0,82666195 | RHPN2 | FALSE | Downregulated |
| 0,049801317 | 0,82666195 | SRR | FALSE | Upregulated |
| 0,038073218 | 0,82666195 | ZNRF1 | FALSE | Upregulated |
| -0,131061183 | 0,826763311 | MS4A14 | FALSE | Downregulated |
| -0,204499681 | 0,826777651 | CACTIN-AS1 | FALSE | Downregulated |
| -0,197566559 | 0,826777651 | LOC101559451 | FALSE | Downregulated |
| 0,045465115 | 0,826777651 | ZNF700 | FALSE | Upregulated |
| -0,244105986 | 0,826789404 | HORMAD1 | FALSE | Downregulated |
| -0,022263879 | 0,826977853 | ATG5 | FALSE | Downregulated |
| -0,163623188 | 0,827057359 | ZNF638-IT1 | FALSE | Downregulated |
| 0,296055089 | 0,827143667 | LOC101927854 | FALSE | Upregulated |
| 0,306972997 | 0,827502503 | ADAM20 | FALSE | Upregulated |
| -0,142178627 | 0,827644786 | SLC28A2 | FALSE | Downregulated |
| 0,06700191 | 0,827644786 | ZNF808 | FALSE | Upregulated |
| -0,188230831 | 0,827660494 | ZNF826P | FALSE | Downregulated |
| -0,038115215 | 0,82802714 | WDR53 | FALSE | Downregulated |
| -0,050160276 | 0,828077368 | APOO | FALSE | Downregulated |
| 0,070484157 | 0,828096313 | KIAA1407 | FALSE | Upregulated |
| 0,374389502 | 0,828096313 | LOC101927505 | FALSE | Upregulated |
| 0,118337899 | 0,828456681 | HOMER1 | FALSE | Upregulated |
| -0,071131946 | 0,828456681 | SCAI | FALSE | Downregulated |
| -0,379985032 | 0,828623511 | BREA2 | FALSE | Downregulated |
| -0,358361889 | 0,828623511 | LOC101928066 | FALSE | Downregulated |
| -0,029076232 | 0,828623511 | TYW5 | FALSE | Downregulated |
| 0,044959625 | 0,828861303 | CHCHD1 | FALSE | Upregulated |
| 0,130191165 | 0,828889853 | BOK-AS1 | FALSE | Upregulated |
| 0,381709704 | 0,828933972 | FAM186A | FALSE | Upregulated |
| -0,03023251 | 0,828954417 | HAUS6 | FALSE | Downregulated |
| -0,196308673 | 0,828982473 | RP11-135J2.4 | FALSE | Downregulated |
| 0,13446386 | 0,828989784 | DTX2P1-UPK3BP1-PMS2P11 | FALSE | Upregulated |
| -0,212663152 | 0,828989784 | LOC101929018 | FALSE | Downregulated |
| 0,040542097 | 0,829018563 | KIAA1731 | FALSE | Upregulated |
| -0,101386336 | 0,829181693 | RP11-115J16.1 | FALSE | Downregulated |
| -0,052510174 | 0,829265579 | KREMEN1 | FALSE | Downregulated |
| 0,344776873 | 0,829477352 | LOC729770 | FALSE | Upregulated |
| -0,373621542 | 0,829477352 | MIR5010 | FALSE | Downregulated |
| 0,026490594 | 0,829609865 | DNAJC16 | FALSE | Upregulated |

| | | | | |
|---|---|---|---|---|
| 0,066977363 | 0,829817117 | C4B | FALSE | Upregulated |
| 0,46816895 | 0,829817117 | NKX2-3 | FALSE | Upregulated |
| -0,038072365 | 0,829817117 | TMEM55B | FALSE | Downregulated |
| 0,047093724 | 0,829820106 | ZSCAN16 | FALSE | Upregulated |
| -0,044397332 | 0,829945135 | CCDC132 | FALSE | Downregulated |
| -0,17503039 | 0,829945135 | LINC01160 | FALSE | Downregulated |
| -0,053482979 | 0,829945135 | MIPOL1 | FALSE | Downregulated |
| -0,030671732 | 0,830073546 | C9orf64 | FALSE | Downregulated |
| -0,27627745 | 0,830073546 | CAPN8-2 | FALSE | Downregulated |
| -0,104273131 | 0,830198547 | LOC101928762 | FALSE | Downregulated |
| 0,443254675 | 0,830322044 | C1orf111 | FALSE | Upregulated |
| -0,205969063 | 0,830322044 | LOC101927885 | FALSE | Downregulated |
| -0,140703682 | 0,830322044 | NT5M | FALSE | Downregulated |
| -0,044782144 | 0,830409311 | POLI | FALSE | Downregulated |
| -0,14266259 | 0,830520024 | C4orf47 | FALSE | Downregulated |
| -0,345550672 | 0,830520024 | LOC101928477 | FALSE | Downregulated |
| 0,588543845 | 0,830547228 | TRNAK1 | FALSE | Upregulated |
| -0,158181216 | 0,830815113 | CARNS1 | FALSE | Downregulated |
| 0,125488082 | 0,830951796 | ARHGEF19 | FALSE | Upregulated |
| 0,137529848 | 0,831079075 | WDR64 | FALSE | Upregulated |
| 0,270400571 | 0,831117057 | ASIP | FALSE | Upregulated |
| 0,092942579 | 0,831117057 | DPEP2 | FALSE | Upregulated |
| -0,070095282 | 0,831117057 | LOC102724884 | FALSE | Downregulated |
| 0,290329403 | 0,831207152 | FLJ43315 | FALSE | Upregulated |
| -0,035990375 | 0,831377715 | CES2 | FALSE | Downregulated |
| 0,019502211 | 0,831678814 | VEZT | FALSE | Upregulated |
| -0,022804505 | 0,831791747 | DYNC1LI2 | FALSE | Downregulated |
| 0,193203459 | 0,832003262 | AGAP1-IT1 | FALSE | Upregulated |
| -0,027455241 | 0,832280468 | DNAJB2 | FALSE | Downregulated |
| 0,304805143 | 0,832280468 | RP11-134P9.3 | FALSE | Upregulated |
| -0,043262478 | 0,832348601 | BHLHB9 | FALSE | Downregulated |
| -0,0396573 | 0,832537995 | FBXL4 | FALSE | Downregulated |
| 0,348612092 | 0,832847845 | LOC102724848 | FALSE | Upregulated |
| 0,03178043 | 0,832893484 | CCDC94 | FALSE | Upregulated |
| -0,228638751 | 0,832966892 | LOC101928326 | FALSE | Downregulated |
| -0,301064285 | 0,833047288 | AOC1 | FALSE | Downregulated |
| 0,053784909 | 0,833047439 | D2HGDH | FALSE | Upregulated |
| -0,340728155 | 0,833202296 | C9orf173 | FALSE | Downregulated |
| -0,177340847 | 0,833202296 | SOCS2-AS1 | FALSE | Downregulated |
| 0,325252699 | 0,833526462 | CETN4P | FALSE | Upregulated |
| 0,085884395 | 0,833526462 | ZNF345 | FALSE | Upregulated |
| 0,283277037 | 0,833562658 | LOC101928355 | FALSE | Upregulated |
| -0,107891678 | 0,833562658 | LOC102724733 | FALSE | Downregulated |
| 0,411357729 | 0,833728783 | TRNAD17 | FALSE | Upregulated |
| -0,245210879 | 0,833863623 | FAM153A | FALSE | Downregulated |
| -0,036500094 | 0,833863623 | NUS1 | FALSE | Downregulated |
| 0,122374234 | 0,833863623 | OAZ3 | FALSE | Upregulated |
| 0,118872346 | 0,834026857 | SRCRB4D | FALSE | Upregulated |
| -0,074895606 | 0,834256998 | GATA3 | FALSE | Downregulated |
| 0,311838461 | 0,834260954 | AC090044.1 | FALSE | Upregulated |
| 0,043493383 | 0,834399015 | FAM118B | FALSE | Upregulated |

| | | | | |
|---|---|---|---|---|
| -0,155187328 | 0,834681785 | RSPO2 | FALSE | Downregulated |
| -0,039891244 | 0,834681785 | SRFBP1 | FALSE | Downregulated |
| -0,095121967 | 0,834768229 | EID2B | FALSE | Downregulated |
| 0,035393279 | 0,834768229 | WHSC1 | FALSE | Upregulated |
| -0,094783569 | 0,834832911 | LOC100996433 | FALSE | Downregulated |
| -0,167227981 | 0,834919555 | LOC102724569 | FALSE | Downregulated |
| -0,053275354 | 0,834919555 | STAT3 | FALSE | Downregulated |
| 0,07089952 | 0,834919555 | URAHP | FALSE | Upregulated |
| 0,065242913 | 0,835080993 | SMIM10 | FALSE | Upregulated |
| -0,189788384 | 0,835182434 | GRIK5 | FALSE | Downregulated |
| -0,038826988 | 0,83542512 | KBTBD7 | FALSE | Downregulated |
| 0,348229641 | 0,83542512 | LOC102723877 | FALSE | Upregulated |
| -0,022541593 | 0,835488208 | EIF3B | FALSE | Downregulated |
| -0,028992279 | 0,835488208 | KCTD9 | FALSE | Downregulated |
| -0,039367378 | 0,835488208 | P2RX4 | FALSE | Downregulated |
| -0,325968441 | 0,835497469 | LHFPL1 | FALSE | Downregulated |
| -0,2168438 | 0,835497469 | LOC101929615 | FALSE | Downregulated |
| -0,040781021 | 0,835497469 | UBE2J1 | FALSE | Downregulated |
| 0,036575445 | 0,835551283 | FTH1 | FALSE | Upregulated |
| -0,243219907 | 0,835551283 | TOMM20L | FALSE | Downregulated |
| 0,112366438 | 0,835645455 | FLJ35282 | FALSE | Upregulated |
| -0,320457665 | 0,835768173 | ZNF32-AS2 | FALSE | Downregulated |
| -0,376092385 | 0,836085491 | LOC101929866 | FALSE | Downregulated |
| 0,026178405 | 0,836127014 | PPP3CB | FALSE | Upregulated |
| -0,122897632 | 0,836204173 | COL23A1 | FALSE | Downregulated |
| 0,115882542 | 0,836376025 | ILDR2 | FALSE | Upregulated |
| -0,027120548 | 0,836376025 | ZC3H7B | FALSE | Downregulated |
| -0,266937055 | 0,8364567 | CCDC178 | FALSE | Downregulated |
| -0,021726104 | 0,836515505 | CCT8 | FALSE | Downregulated |
| -0,076375635 | 0,836691297 | RSG1 | FALSE | Downregulated |
| -0,238320605 | 0,836731665 | LOC102723840 | FALSE | Downregulated |
| 0,365381272 | 0,83673795 | LRRC3C | FALSE | Upregulated |
| -0,307401353 | 0,836757206 | LOC100271836 | FALSE | Downregulated |
| -0,02954832 | 0,836757206 | VRK2 | FALSE | Downregulated |
| -0,3025277 | 0,836783894 | A4GNT | FALSE | Downregulated |
| -0,032751405 | 0,836783894 | LOC100506710 | FALSE | Downregulated |
| -0,356399103 | 0,836783894 | RP11-593F23.1 | FALSE | Downregulated |
| -0,109335689 | 0,836785246 | ANKRD34A | FALSE | Downregulated |
| 0,057285458 | 0,836785246 | HIC2 | FALSE | Upregulated |
| 0,337116135 | 0,836785246 | HIST1H4C | FALSE | Upregulated |
| -0,03292463 | 0,836785246 | OAZ2 | FALSE | Downregulated |
| -0,2043063 | 0,836785246 | RP11-27K13.3 | FALSE | Downregulated |
| 0,04812478 | 0,836785246 | TSNARE1 | FALSE | Upregulated |
| 0,421521563 | 0,836999471 | CCDC74B | FALSE | Upregulated |
| 0,10804983 | 0,837283758 | TRIM61 | FALSE | Upregulated |
| 0,037030014 | 0,837339229 | COX15 | FALSE | Upregulated |
| -0,195996114 | 0,837873454 | LPAR4 | FALSE | Downregulated |
| 0,031136684 | 0,837976403 | FAM200A | FALSE | Upregulated |
| -0,0300056 | 0,838020095 | APC | FALSE | Downregulated |
| -0,447255834 | 0,838167992 | ART3 | FALSE | Downregulated |
| -0,049567706 | 0,838167992 | C16orf91 | FALSE | Downregulated |

| | | | | |
|---|---|---|---|---|
| -0,112745343 | 0,838309235 | CCL23 | FALSE | Downregulated |
| 0,045072802 | 0,838356728 | EXOSC5 | FALSE | Upregulated |
| 0,412196216 | 0,838369399 | RP3-522P13.3 | FALSE | Upregulated |
| -0,021356157 | 0,838377794 | TBK1 | FALSE | Downregulated |
| -0,080040203 | 0,838393198 | HERC2P9 | FALSE | Downregulated |
| -0,028120522 | 0,838448016 | KXD1 | FALSE | Downregulated |
| -0,052068337 | 0,838534253 | PPAP2B | FALSE | Downregulated |
| -0,092363801 | 0,838544588 | LOC101927663 | FALSE | Downregulated |
| 0,072925592 | 0,838982099 | SYNGR1 | FALSE | Upregulated |
| -0,059495473 | 0,839111551 | ANKRD24 | FALSE | Downregulated |
| -0,169794916 | 0,839313395 | S100A12 | FALSE | Downregulated |
| 0,276744941 | 0,839853966 | LOC344967 | FALSE | Upregulated |
| -0,026912425 | 0,840145596 | SMAD5 | FALSE | Downregulated |
| -0,045903176 | 0,840145596 | ZNF232 | FALSE | Downregulated |
| 0,050115897 | 0,840176548 | CHID1 | FALSE | Upregulated |
| -0,050570426 | 0,840250093 | AMN1 | FALSE | Downregulated |
| -0,147517389 | 0,840256618 | ASMTL-AS1 | FALSE | Downregulated |
| -0,066531891 | 0,840256618 | SDCBP2-AS1 | FALSE | Downregulated |
| 0,023930261 | 0,840257783 | TTC37 | FALSE | Upregulated |
| 0,20351629 | 0,840322988 | LOC102724957 | FALSE | Upregulated |
| 0,220577702 | 0,840366485 | C9orf163 | FALSE | Upregulated |
| 0,412901096 | 0,840366485 | LOC101927705 | FALSE | Upregulated |
| 0,212310624 | 0,840366485 | RP11-474D1.3 | FALSE | Upregulated |
| -0,025674989 | 0,840366485 | TUG1 | FALSE | Downregulated |
| -0,051114596 | 0,840420733 | LOC646762 | FALSE | Downregulated |
| -0,103934854 | 0,840420733 | MBNL1-AS1 | FALSE | Downregulated |
| -0,030468931 | 0,840448124 | NFYB | FALSE | Downregulated |
| 0,195228857 | 0,840448124 | SCN4A | FALSE | Upregulated |
| 0,094902394 | 0,840448124 | ZNF703 | FALSE | Upregulated |
| -0,052535296 | 0,840528688 | ZNF565 | FALSE | Downregulated |
| 0,094574938 | 0,840546732 | ZNF818P | FALSE | Upregulated |
| 0,09875591 | 0,840546895 | C1RL-AS1 | FALSE | Upregulated |
| -0,023470269 | 0,840747729 | BNIP2 | FALSE | Downregulated |
| -0,272885163 | 0,840879692 | FAM184B | FALSE | Downregulated |
| -0,084117774 | 0,841078489 | EIF1B-AS1 | FALSE | Downregulated |
| 0,097013487 | 0,841078489 | PLA2G7 | FALSE | Upregulated |
| 0,223295154 | 0,841091514 | CCDC180 | FALSE | Upregulated |
| -0,049355442 | 0,841143862 | HEMK1 | FALSE | Downregulated |
| 0,109235752 | 0,841264338 | CCDC103 | FALSE | Upregulated |
| -0,074817458 | 0,841303733 | MAMLD1 | FALSE | Downregulated |
| -0,036303905 | 0,841303733 | PREPL | FALSE | Downregulated |
| 0,153462492 | 0,841303733 | SUZ12P1 | FALSE | Upregulated |
| 0,275367353 | 0,841314512 | LOC102724545 | FALSE | Upregulated |
| -0,029762825 | 0,841353523 | SNF8 | FALSE | Downregulated |
| 0,043084445 | 0,84146273 | CTNS | FALSE | Upregulated |
| -0,06985241 | 0,84146273 | LOC100132832 | FALSE | Downregulated |
| -0,448410731 | 0,84146273 | LOC102724830 | FALSE | Downregulated |
| 0,043989204 | 0,84146273 | MALSU1 | FALSE | Upregulated |
| -0,044555395 | 0,84146273 | MORN2 | FALSE | Downregulated |
| 0,191775913 | 0,84146273 | SLC16A12 | FALSE | Upregulated |
| -0,028288801 | 0,84146273 | SLMO2 | FALSE | Downregulated |

| | | | | |
|---|---|---|---|---|
| 0,049114662 | 0,84146273 | SYNE2 | FALSE | Upregulated |
| -0,26936512 | 0,841598794 | LOC100130238 | FALSE | Downregulated |
| -0,048072393 | 0,841789829 | BDP1 | FALSE | Downregulated |
| -0,050665278 | 0,84186565 | C11orf83 | FALSE | Downregulated |
| -0,028859637 | 0,84186565 | NDUFA8 | FALSE | Downregulated |
| -0,113269447 | 0,841872896 | TMEM9B-AS1 | FALSE | Downregulated |
| -0,046352246 | 0,842463211 | CD14 | FALSE | Downregulated |
| 0,065012866 | 0,842664104 | ZBTB3 | FALSE | Upregulated |
| 0,045333084 | 0,842736789 | GLB1L | FALSE | Upregulated |
| 0,04092652 | 0,842736789 | MST1 | FALSE | Upregulated |
| -0,138726514 | 0,842872545 | PKD2L1 | FALSE | Downregulated |
| -0,04442355 | 0,842942142 | AQP3 | FALSE | Downregulated |
| -0,130198741 | 0,842942142 | RP11-500B12.1 | FALSE | Downregulated |
| 0,038747703 | 0,843280523 | FAM173B | FALSE | Upregulated |
| -0,093066179 | 0,843310364 | CATSPERG | FALSE | Downregulated |
| 0,026874049 | 0,843356314 | TGS1 | FALSE | Upregulated |
| -0,289467612 | 0,843397555 | LOC102724065 | FALSE | Downregulated |
| 0,080517669 | 0,843423419 | ACCS | FALSE | Upregulated |
| 0,089580791 | 0,843423419 | FNTB | FALSE | Upregulated |
| -0,022558449 | 0,843423419 | KDM1A | FALSE | Downregulated |
| -0,134891641 | 0,843423419 | LOC102724660 | FALSE | Downregulated |
| -0,129683813 | 0,843423419 | NNMT | FALSE | Downregulated |
| 0,095276416 | 0,843423419 | NR0B2 | FALSE | Upregulated |
| 0,047266631 | 0,843423419 | PRPF40B | FALSE | Upregulated |
| -0,033037588 | 0,843423419 | ZC3H10 | FALSE | Downregulated |
| 0,025730151 | 0,843492191 | CSRP2BP | FALSE | Upregulated |
| 0,041811983 | 0,843492191 | THOC6 | FALSE | Upregulated |
| 0,044149747 | 0,843791417 | TET3 | FALSE | Upregulated |
| -0,405284488 | 0,843791417 | VWA5B2 | FALSE | Downregulated |
| 0,03160531 | 0,843791417 | ZNF790 | FALSE | Upregulated |
| 0,044106988 | 0,84392687 | C21orf91 | FALSE | Upregulated |
| 0,034792173 | 0,843928011 | ZNF91 | FALSE | Upregulated |
| -0,119104643 | 0,844012551 | CDS1 | FALSE | Downregulated |
| -0,162236803 | 0,844300279 | MLC1 | FALSE | Downregulated |
| 0,128243747 | 0,844361273 | LOC102724687 | FALSE | Upregulated |
| 0,342990413 | 0,844361273 | LOC283731 | FALSE | Upregulated |
| -0,255043567 | 0,844361273 | PRSS45 | FALSE | Downregulated |
| 0,481100845 | 0,844361273 | RNU6-36P | FALSE | Upregulated |
| 0,106333087 | 0,844409399 | SYNC | FALSE | Upregulated |
| -0,262166971 | 0,844668984 | C1orf140 | FALSE | Downregulated |
| 0,402546379 | 0,844750103 | LOC101929790 | FALSE | Upregulated |
| 0,036013462 | 0,844937276 | VPS9D1 | FALSE | Upregulated |
| 0,038623056 | 0,845284369 | FAM203A | FALSE | Upregulated |
| 0,222154619 | 0,84537904 | RAB6C-AS1 | FALSE | Upregulated |
| -0,039618038 | 0,845402948 | ANKRD10 | FALSE | Downregulated |
| 0,102985061 | 0,845402948 | CASS4 | FALSE | Upregulated |
| 0,072322156 | 0,845402948 | CCR1 | FALSE | Upregulated |
| 0,312017837 | 0,845516044 | OLFM4 | FALSE | Upregulated |
| -0,175788707 | 0,845695784 | SPTA1 | FALSE | Downregulated |
| -0,036052659 | 0,845720053 | CASP7 | FALSE | Downregulated |
| -0,041508287 | 0,845720053 | PGM2 | FALSE | Downregulated |

| | | | | |
|---|---|---|---|---|
| 0,034617301 | 0,845744032 | ATP6V1F | FALSE | Upregulated |
| 0,290542321 | 0,845744032 | C16orf3 | FALSE | Upregulated |
| 0,473140127 | 0,845744032 | FRMPD4 | FALSE | Upregulated |
| -0,023919821 | 0,845744032 | GOSR1 | FALSE | Downregulated |
| 0,171432578 | 0,845744032 | LOC101927858 | FALSE | Upregulated |
| -0,02910885 | 0,845744032 | SOS1 | FALSE | Downregulated |
| 0,410457088 | 0,846022265 | NLRP8 | FALSE | Upregulated |
| 0,179847411 | 0,846104123 | DIRC3 | FALSE | Upregulated |
| 0,038025597 | 0,846154149 | SERGEF | FALSE | Upregulated |
| 0,112667747 | 0,84617679 | LOC101928673 | FALSE | Upregulated |
| 0,124065927 | 0,846250965 | CLEC9A | FALSE | Upregulated |
| -0,284856174 | 0,846447843 | DMBX1 | FALSE | Downregulated |
| -0,053754675 | 0,846447843 | SMAD6 | FALSE | Downregulated |
| 0,18982259 | 0,846553306 | LOC102724948 | FALSE | Upregulated |
| -0,040478399 | 0,846591947 | RBMS1 | FALSE | Downregulated |
| 0,247639727 | 0,846708727 | AC073133.1 | FALSE | Upregulated |
| 0,036112912 | 0,846708727 | ECI1 | FALSE | Upregulated |
| -0,122892485 | 0,846708727 | LOC100134822 | FALSE | Downregulated |
| 0,032847021 | 0,846708727 | MEA1 | FALSE | Upregulated |
| -0,190227739 | 0,846708727 | PSPN | FALSE | Downregulated |
| 0,030173321 | 0,846708727 | ZBTB11 | FALSE | Upregulated |
| 0,224637703 | 0,846714868 | BMP8A | FALSE | Upregulated |
| 0,043763041 | 0,846714868 | FRAT1 | FALSE | Upregulated |
| 0,194451187 | 0,846714868 | GOLGA8O | FALSE | Upregulated |
| -0,195228254 | 0,846714868 | LOC100996634 | FALSE | Downregulated |
| -0,144501588 | 0,846714868 | LOC101928058 | FALSE | Downregulated |
| -0,031767115 | 0,846714868 | MED26 | FALSE | Downregulated |
| -0,110612818 | 0,846714868 | RIBC1 | FALSE | Downregulated |
| 0,078177038 | 0,846735535 | ZNF471 | FALSE | Upregulated |
| 0,036643397 | 0,847044233 | KDM5B | FALSE | Upregulated |
| -0,289669406 | 0,847044233 | LINC01133 | FALSE | Downregulated |
| 0,123470952 | 0,847047044 | LINC00472 | FALSE | Upregulated |
| 0,196906806 | 0,847116251 | MAFG-AS1 | FALSE | Upregulated |
| -0,033090168 | 0,847183973 | DGCR8 | FALSE | Downregulated |
| -0,06125994 | 0,847183973 | PNPLA7 | FALSE | Downregulated |
| 0,181387891 | 0,847791823 | CCDC160 | FALSE | Upregulated |
| -0,084866864 | 0,847830797 | LOC101928451 | FALSE | Downregulated |
| -0,247923701 | 0,84798356 | LINC01010 | FALSE | Downregulated |
| -0,226324476 | 0,84798356 | SLC22A24 | FALSE | Downregulated |
| 0,033423008 | 0,848168162 | TIMELESS | FALSE | Upregulated |
| -0,06424937 | 0,848468618 | CHURC1-FNTB | FALSE | Downregulated |
| -0,037156247 | 0,848468618 | PAK1IP1 | FALSE | Downregulated |
| -0,376674523 | 0,848514499 | LOC102724106 | FALSE | Downregulated |
| -0,033896629 | 0,848514499 | SPCS2 | FALSE | Downregulated |
| 0,181394284 | 0,848526257 | ESM1 | FALSE | Upregulated |
| 0,068950985 | 0,848586251 | SLC25A53 | FALSE | Upregulated |
| -0,286495275 | 0,848661731 | FAM170A | FALSE | Downregulated |
| -0,034481941 | 0,848847673 | TMEM65 | FALSE | Downregulated |
| -0,257621334 | 0,848948074 | LOC101929566 | FALSE | Downregulated |
| -0,041376037 | 0,848996067 | SLC50A1 | FALSE | Downregulated |
| -0,049321559 | 0,848996067 | TMEM180 | FALSE | Downregulated |

| | | | | |
|---|---|---|---|---|
| 0,050212119 | 0,849402485 | ZNF570 | FALSE | Upregulated |
| -0,196040149 | 0,849421392 | TMC4 | FALSE | Downregulated |
| -0,235565943 | 0,849511479 | FAM95B1 | FALSE | Downregulated |
| -0,025019565 | 0,849511479 | RAPGEF1 | FALSE | Downregulated |
| -0,718216422 | 0,849511479 | TRNAH2 | FALSE | Downregulated |
| -0,036461015 | 0,849625628 | ADIPOR2 | FALSE | Downregulated |
| 0,416975352 | 0,849625628 | APOOP5 | FALSE | Upregulated |
| -0,328208561 | 0,849625628 | GALNT9 | FALSE | Downregulated |
| 0,033412868 | 0,849625628 | KIAA0586 | FALSE | Upregulated |
| -0,074161166 | 0,849625628 | LOC100996909 | FALSE | Downregulated |
| 0,119523913 | 0,849625628 | MT1A | FALSE | Upregulated |
| 0,1060875 | 0,849625628 | MX1 | FALSE | Upregulated |
| -0,249343456 | 0,849625628 | RNF144A-AS1 | FALSE | Downregulated |
| 0,186632416 | 0,849625628 | RP5-899E9.1 | FALSE | Upregulated |
| 0,038826207 | 0,849625628 | SCYL2 | FALSE | Upregulated |
| 0,457691425 | 0,849625628 | ST8SIA2 | FALSE | Upregulated |
| -0,525239143 | 0,849625628 | TNFRSF6B | FALSE | Downregulated |
| -0,039088002 | 0,849666483 | ERG | FALSE | Downregulated |
| 0,037442581 | 0,849695647 | TMEM126B | FALSE | Upregulated |
| 0,242037118 | 0,850113461 | TEX26 | FALSE | Upregulated |
| 0,448608206 | 0,850295356 | LOC100131626 | FALSE | Upregulated |
| 0,294638795 | 0,850352305 | LOC102723922 | FALSE | Upregulated |
| -0,229363727 | 0,850568046 | LOC102723366 | FALSE | Downregulated |
| 0,292996901 | 0,850596957 | FGF10 | FALSE | Upregulated |
| -0,184272575 | 0,85068455 | LOC102724806 | FALSE | Downregulated |
| -0,124338295 | 0,850800083 | FCRLB | FALSE | Downregulated |
| 0,078259003 | 0,850800083 | LINC00899 | FALSE | Upregulated |
| -0,056991421 | 0,850800083 | LRWD1 | FALSE | Downregulated |
| -0,039724856 | 0,850800083 | RPL23AP82 | FALSE | Downregulated |
| 0,066714441 | 0,850844902 | PRAM1 | FALSE | Upregulated |
| 0,353976848 | 0,850871886 | FGF20 | FALSE | Upregulated |
| -0,149676537 | 0,850871886 | LOC102723340 | FALSE | Downregulated |
| -0,065853897 | 0,850902175 | ABHD1 | FALSE | Downregulated |
| 0,095992069 | 0,850902175 | C2orf15 | FALSE | Upregulated |
| -0,365741093 | 0,850902175 | LOC100996716 | FALSE | Downregulated |
| 0,346357037 | 0,850931731 | FAM57B | FALSE | Upregulated |
| -0,136822126 | 0,850931731 | STRIP2 | FALSE | Downregulated |
| 0,040484308 | 0,850987881 | ZNF697 | FALSE | Upregulated |
| -0,09233911 | 0,8512744 | EP400NL | FALSE | Downregulated |
| 0,165452341 | 0,8512744 | LOC101928725 | FALSE | Upregulated |
| 0,162647812 | 0,8512744 | SATL1 | FALSE | Upregulated |
| -0,020345698 | 0,8512744 | SCAF8 | FALSE | Downregulated |
| 0,120455398 | 0,85143384 | AATK | FALSE | Upregulated |
| 0,073862969 | 0,85143384 | AC003665.1 | FALSE | Upregulated |
| 0,057155843 | 0,85143384 | CXorf40A | FALSE | Upregulated |
| -0,096878145 | 0,85143384 | HOXA5 | FALSE | Downregulated |
| -0,034066222 | 0,85143384 | KRTCAP2 | FALSE | Downregulated |
| -0,077274708 | 0,85143384 | NOS1AP | FALSE | Downregulated |
| 0,409388644 | 0,85143384 | SAG | FALSE | Upregulated |
| 0,158841735 | 0,85143384 | SGCA | FALSE | Upregulated |
| -0,024509451 | 0,851513834 | SPCS1 | FALSE | Downregulated |

| | | | | |
|---|---|---|---|---|
| 0,04342881 | 0,851552856 | MESDC1 | FALSE | Upregulated |
| -0,058037177 | 0,851552856 | STXBP4 | FALSE | Downregulated |
| -0,037228459 | 0,851556814 | HEATR5B | FALSE | Downregulated |
| 0,08213647 | 0,851556814 | LINC00957 | FALSE | Upregulated |
| 0,168020089 | 0,851570666 | CDH4 | FALSE | Upregulated |
| 0,383268651 | 0,851570666 | COMP | FALSE | Upregulated |
| -0,039941839 | 0,851570666 | THRA | FALSE | Downregulated |
| -0,590596143 | 0,851570666 | TRNAK34 | FALSE | Downregulated |
| -0,106549771 | 0,851650842 | CSNK1A1P1 | FALSE | Downregulated |
| -0,176384065 | 0,851650842 | REN | FALSE | Downregulated |
| -0,323537909 | 0,851811993 | C17orf104 | FALSE | Downregulated |
| -0,031699973 | 0,851811993 | MSRB1 | FALSE | Downregulated |
| -0,313136933 | 0,851832121 | LOC101059976 | FALSE | Downregulated |
| -0,195258284 | 0,852119047 | HIST1H4J | FALSE | Downregulated |
| -0,059515845 | 0,85216127 | AFG3L1P | FALSE | Downregulated |
| -0,030941824 | 0,852215521 | NICN1 | FALSE | Downregulated |
| 0,052811907 | 0,852523666 | SERP1 | FALSE | Upregulated |
| 0,047903297 | 0,852537334 | GPR39 | FALSE | Upregulated |
| -0,130118944 | 0,852537334 | LOC101926935 | FALSE | Downregulated |
| -0,051658143 | 0,852615654 | ABHD5 | FALSE | Downregulated |
| 0,07903871 | 0,852615654 | AGPAT9 | FALSE | Upregulated |
| 0,263148197 | 0,852615654 | LOC100130520 | FALSE | Upregulated |
| 0,103685127 | 0,852615654 | MIR503HG | FALSE | Upregulated |
| -0,033746929 | 0,852615654 | QSER1 | FALSE | Downregulated |
| -0,023382954 | 0,852626408 | EIF3H | FALSE | Downregulated |
| 0,028647287 | 0,85277666 | TUSC2 | FALSE | Upregulated |
| -0,331342755 | 0,852820637 | FAM177B | FALSE | Downregulated |
| 0,451040642 | 0,852820637 | FAM27C | FALSE | Upregulated |
| 0,350250609 | 0,852820637 | LOC102723363 | FALSE | Upregulated |
| -0,173405528 | 0,852820637 | LOC439994 | FALSE | Downregulated |
| 0,02219372 | 0,852820637 | NUP214 | FALSE | Upregulated |
| -0,031410658 | 0,852820637 | RMI1 | FALSE | Downregulated |
| -0,222776197 | 0,852820637 | RNU6ATAC | FALSE | Downregulated |
| 0,029310847 | 0,852820637 | ZNF616 | FALSE | Upregulated |
| 0,072592226 | 0,852911734 | SVIL | FALSE | Upregulated |
| 0,024191034 | 0,852974625 | CD164 | FALSE | Upregulated |
| -0,173131826 | 0,852982271 | C1QTNF9 | FALSE | Downregulated |
| -0,307210844 | 0,852982271 | LOC101928284 | FALSE | Downregulated |
| -0,099378674 | 0,853017007 | PPM1L | FALSE | Downregulated |
| 0,021963032 | 0,85309967 | RB1CC1 | FALSE | Upregulated |
| -0,168480708 | 0,853103395 | LOC101929713 | FALSE | Downregulated |
| -0,054454358 | 0,853138272 | C5orf55 | FALSE | Downregulated |
| 0,22150856 | 0,853138272 | C9orf47 | FALSE | Upregulated |
| 0,038187572 | 0,853138272 | DDX31 | FALSE | Upregulated |
| -0,017837587 | 0,853138272 | PRCC | FALSE | Downregulated |
| -0,060451611 | 0,853138272 | SLC17A3 | FALSE | Downregulated |
| 0,025160975 | 0,853138272 | SNX19 | FALSE | Upregulated |
| -0,051938771 | 0,853138272 | TRPM8 | FALSE | Downregulated |
| 0,333311065 | 0,853238008 | ANKRD20A8P | FALSE | Upregulated |
| 0,021940696 | 0,853753232 | AAR2 | FALSE | Upregulated |
| -0,068295845 | 0,853967147 | SLC25A25 | FALSE | Downregulated |

| | | | | |
|---|---|---|---|---|
| -0,08987085 | 0,853993913 | RELL2 | FALSE | Downregulated |
| 0,138373122 | 0,854180142 | COLGALT2 | FALSE | Upregulated |
| -0,043666352 | 0,854180142 | FAM20C | FALSE | Downregulated |
| -0,256855211 | 0,854180142 | KIAA1257 | FALSE | Downregulated |
| 0,358805086 | 0,854180142 | LINC00643 | FALSE | Upregulated |
| -0,147826312 | 0,854180142 | LOC101929571 | FALSE | Downregulated |
| -0,045925945 | 0,854180142 | MAFB | FALSE | Downregulated |
| -0,019072841 | 0,854180142 | PUM2 | FALSE | Downregulated |
| -0,094489662 | 0,854250314 | FAM151B | FALSE | Downregulated |
| 0,02142424 | 0,854250314 | NUP133 | FALSE | Upregulated |
| -0,043701251 | 0,854250314 | ZBED5-AS1 | FALSE | Downregulated |
| -0,097973353 | 0,85425098 | RNF32 | FALSE | Downregulated |
| -0,096558887 | 0,85433528 | AGER | FALSE | Downregulated |
| -0,110505982 | 0,854394307 | LOC101929055 | FALSE | Downregulated |
| 1,411451418 | 0,854526428 | MYBPC2 | FALSE | Upregulated |
| 0,062673327 | 0,854577374 | MSANTD2 | FALSE | Upregulated |
| -0,077587497 | 0,854633518 | NBPF26 | FALSE | Downregulated |
| -0,126163336 | 0,854633518 | ZMIZ1-AS1 | FALSE | Downregulated |
| -0,200042413 | 0,854641615 | LOC101929098 | FALSE | Downregulated |
| -0,03786136 | 0,854641615 | UBL4A | FALSE | Downregulated |
| 0,140892561 | 0,854679614 | LOC729887 | FALSE | Upregulated |
| 0,051069698 | 0,854699089 | GALNT2 | FALSE | Upregulated |
| 0,112439655 | 0,854699089 | HCAR2 | FALSE | Upregulated |
| -0,196498578 | 0,855058406 | SYN3 | FALSE | Downregulated |
| 0,03590533 | 0,855204748 | TXNL4A | FALSE | Upregulated |
| 0,033424112 | 0,855338626 | C12orf29 | FALSE | Upregulated |
| 0,399620468 | 0,855338626 | DKFZP434A062 | FALSE | Upregulated |
| 0,052890663 | 0,855603051 | ARRDC4 | FALSE | Upregulated |
| 0,065216827 | 0,855603051 | CSF3R | FALSE | Upregulated |
| -0,051321979 | 0,855603051 | KIF9 | FALSE | Downregulated |
| -0,250816809 | 0,855603051 | MROH2B | FALSE | Downregulated |
| 0,041326307 | 0,855692038 | HTATIP2 | FALSE | Upregulated |
| 0,31497806 | 0,855692038 | MAFA | FALSE | Upregulated |
| -0,017921141 | 0,855692038 | PNRC2 | FALSE | Downregulated |
| -0,300121863 | 0,855692038 | VIT | FALSE | Downregulated |
| -0,192330013 | 0,856284612 | C1orf105 | FALSE | Downregulated |
| -0,06055716 | 0,856342027 | DNAJB9 | FALSE | Downregulated |
| -0,032158852 | 0,85634825 | BOD1 | FALSE | Downregulated |
| 0,168272424 | 0,85634825 | LOC100293704 | FALSE | Upregulated |
| 0,03042994 | 0,85634825 | PI4K2B | FALSE | Upregulated |
| 0,059658627 | 0,856455832 | CHAC2 | FALSE | Upregulated |
| -0,032466388 | 0,856455832 | PSMA3 | FALSE | Downregulated |
| -0,032293123 | 0,856455832 | XPR1 | FALSE | Downregulated |
| 0,136367157 | 0,856681797 | LOC102724608 | FALSE | Upregulated |
| -0,038157906 | 0,856681797 | POLR2L | FALSE | Downregulated |
| -0,218649673 | 0,856681797 | RP5-1139I1.1 | FALSE | Downregulated |
| -0,022906424 | 0,856681797 | SON | FALSE | Downregulated |
| -0,104598001 | 0,857106405 | PTPLA | FALSE | Downregulated |
| -0,063279636 | 0,857106405 | RP11-752G15.6 | FALSE | Downregulated |
| -0,037709931 | 0,857399146 | FASTK | FALSE | Downregulated |
| 0,038871376 | 0,857435061 | TNRC6A | FALSE | Upregulated |

| | | | | |
|---|---|---|---|---|
| 0,29580421 | 0,85757053 | ATP6V1B1 | FALSE | Upregulated |
| 0,185474856 | 0,85757053 | LOC101927157 | FALSE | Upregulated |
| 0,298200367 | 0,85757053 | LOC101928632 | FALSE | Upregulated |
| -0,138910186 | 0,85757053 | SCUBE2 | FALSE | Downregulated |
| -0,025701977 | 0,85757053 | TAX1BP1 | FALSE | Downregulated |
| 1,287233029 | 0,857727784 | LDB3 | FALSE | Upregulated |
| 0,159224432 | 0,857753719 | LOC101926902 | FALSE | Upregulated |
| 0,411307578 | 0,857836963 | CYP26C1 | FALSE | Upregulated |
| 0,280782207 | 0,857841318 | CLCN1 | FALSE | Upregulated |
| -0,220446858 | 0,857841947 | ALG1L | FALSE | Downregulated |
| 0,050050182 | 0,857841947 | REEP4 | FALSE | Upregulated |
| 0,211517429 | 0,857855842 | INE2 | FALSE | Upregulated |
| 0,161457245 | 0,858071882 | LOC100131532 | FALSE | Upregulated |
| 0,097777959 | 0,858231969 | KIF19 | FALSE | Upregulated |
| 0,04164435 | 0,858231969 | ZCCHC14 | FALSE | Upregulated |
| -0,296820082 | 0,858238829 | LOC102723420 | FALSE | Downregulated |
| 0,030003299 | 0,858531072 | MRPL22 | FALSE | Upregulated |
| -0,030452486 | 0,858531072 | POC1B | FALSE | Downregulated |
| 0,168494427 | 0,858747731 | LOC101929687 | FALSE | Upregulated |
| 0,088075893 | 0,858747731 | RP1-60O19.1 | FALSE | Upregulated |
| -0,083870431 | 0,858805408 | DBF4B | FALSE | Downregulated |
| 0,03510964 | 0,858948795 | ZNF236 | FALSE | Upregulated |
| 0,31146448 | 0,859078612 | FMN2 | FALSE | Upregulated |
| 0,364325367 | 0,859078612 | LOC101927803 | FALSE | Upregulated |
| 0,03493227 | 0,859078612 | PHF6 | FALSE | Upregulated |
| 0,038228538 | 0,85928467 | SPCS3 | FALSE | Upregulated |
| -0,020927097 | 0,859434853 | ASB7 | FALSE | Downregulated |
| 0,155248798 | 0,859434853 | RP11-401P9.5 | FALSE | Upregulated |
| -0,11394101 | 0,859434853 | TP63 | FALSE | Downregulated |
| 0,082579778 | 0,859469444 | NKAPP1 | FALSE | Upregulated |
| 0,310993244 | 0,859469444 | UPK1B | FALSE | Upregulated |
| 0,028150176 | 0,859610684 | HEATR6 | FALSE | Upregulated |
| -0,182278517 | 0,859684029 | SNORD68 | FALSE | Downregulated |
| -0,032066331 | 0,859684029 | TCEAL1 | FALSE | Downregulated |
| 0,056634606 | 0,859733255 | AKR1C6P | FALSE | Upregulated |
| -0,260086194 | 0,859964451 | KCNH1 | FALSE | Downregulated |
| -0,234924959 | 0,859964451 | LOC101927725 | FALSE | Downregulated |
| -0,100869944 | 0,859980392 | KLRAP1 | FALSE | Downregulated |
| 0,293349559 | 0,859980392 | LOC102724501 | FALSE | Upregulated |
| -0,01906731 | 0,859980392 | RDH14 | FALSE | Downregulated |
| 0,043872614 | 0,859980392 | RPS11 | FALSE | Upregulated |
| 0,046267786 | 0,859980392 | ZNF584 | FALSE | Upregulated |
| -0,030611324 | 0,860253405 | RPS13 | FALSE | Downregulated |
| -0,179787454 | 0,860277031 | TMEM191C | FALSE | Downregulated |
| 0,026239392 | 0,860322347 | C18orf25 | FALSE | Upregulated |
| -0,157556996 | 0,860353192 | DBIL5P | FALSE | Downregulated |
| 0,028605812 | 0,860355677 | BTBD6 | FALSE | Upregulated |
| 0,033893188 | 0,860355677 | CLPP | FALSE | Upregulated |
| -0,040142745 | 0,860355677 | LIN37 | FALSE | Downregulated |
| -0,062359049 | 0,860355677 | ST6GALNAC3 | FALSE | Downregulated |
| -0,092712166 | 0,860424407 | GAS5-AS1 | FALSE | Downregulated |

| | | | | |
|---|---|---|---|---|
| 0,352135084 | 0,860424407 | LOC101928867 | FALSE | Upregulated |
| -0,060347571 | 0,860424407 | RP11-1072A3.2 | FALSE | Downregulated |
| -0,328113365 | 0,860424407 | RP11-133O22.6 | FALSE | Downregulated |
| -0,079090117 | 0,860584365 | GOLGA6L9 | FALSE | Downregulated |
| 0,032659452 | 0,86105745 | POLR3K | FALSE | Upregulated |
| -0,605346003 | 0,861089109 | LOC101060389 | FALSE | Downregulated |
| 0,085907169 | 0,861162257 | KIAA1683 | FALSE | Upregulated |
| -0,036428437 | 0,861162257 | SH3TC1 | FALSE | Downregulated |
| -0,070181344 | 0,861209988 | NBPF12 | FALSE | Downregulated |
| 0,176016123 | 0,861479987 | LOC101929004 | FALSE | Upregulated |
| 0,047050078 | 0,861479987 | NPL | FALSE | Upregulated |
| 0,284458908 | 0,861479987 | TAS2R31 | FALSE | Upregulated |
| 0,242655391 | 0,861479987 | TMEM191A | FALSE | Upregulated |
| 0,323860949 | 0,861872177 | CRYBB2 | FALSE | Upregulated |
| 0,047216401 | 0,861872177 | HSPH1 | FALSE | Upregulated |
| -0,106060533 | 0,861872177 | LOC100190986 | FALSE | Downregulated |
| 0,10706416 | 0,861872177 | TRIM36 | FALSE | Upregulated |
| -0,026345447 | 0,861939908 | CIRH1A | FALSE | Downregulated |
| 0,235662814 | 0,861939908 | PROK1 | FALSE | Upregulated |
| -0,036503901 | 0,862000145 | CLCN4 | FALSE | Downregulated |
| -0,042756914 | 0,862092792 | PYCRL | FALSE | Downregulated |
| 0,050782959 | 0,862241317 | EFNB2 | FALSE | Upregulated |
| 0,022213932 | 0,862349319 | BPNT1 | FALSE | Upregulated |
| 0,323454482 | 0,862349319 | LOC102724826 | FALSE | Upregulated |
| 0,028941154 | 0,862349319 | METTL2A | FALSE | Upregulated |
| -0,024837831 | 0,862349319 | NPEPPS | FALSE | Downregulated |
| -0,030710056 | 0,862349319 | NUDCD2 | FALSE | Downregulated |
| 0,087266389 | 0,862349319 | RGS7BP | FALSE | Upregulated |
| 0,074123009 | 0,862375348 | NMUR1 | FALSE | Upregulated |
| -0,068557662 | 0,86265864 | RP9P | FALSE | Downregulated |
| -0,043831199 | 0,862877287 | MYSM1 | FALSE | Downregulated |
| 0,159519295 | 0,862879234 | ARMC12 | FALSE | Upregulated |
| 0,040315487 | 0,86300104 | DOK4 | FALSE | Upregulated |
| 0,128273649 | 0,863037773 | CEACAM4 | FALSE | Upregulated |
| -0,021609278 | 0,863037773 | FYTTD1 | FALSE | Downregulated |
| 0,349414641 | 0,863049239 | RP11-1057B8.2 | FALSE | Upregulated |
| 0,034787387 | 0,86318055 | SP2 | FALSE | Upregulated |
| 0,134632672 | 0,863476433 | CDC6 | FALSE | Upregulated |
| -0,031935845 | 0,863617831 | DENND1B | FALSE | Downregulated |
| -0,048030693 | 0,863617831 | PCAT19 | FALSE | Downregulated |
| 0,044264187 | 0,863656067 | DAO | FALSE | Upregulated |
| -0,026768585 | 0,863656067 | KIAA2013 | FALSE | Downregulated |
| 0,028866263 | 0,863745101 | ALDH4A1 | FALSE | Upregulated |
| -0,316933638 | 0,863745101 | ASB16 | FALSE | Downregulated |
| 0,053645285 | 0,863745101 | CCNL1 | FALSE | Upregulated |
| 0,047520563 | 0,863745101 | CGNL1 | FALSE | Upregulated |
| -0,021925582 | 0,863745101 | USP28 | FALSE | Downregulated |
| 0,117168215 | 0,86395206 | LOC100240735 | FALSE | Upregulated |
| -0,04585402 | 0,864197089 | ZNF324 | FALSE | Downregulated |
| -0,020952859 | 0,864237644 | ARFGEF2 | FALSE | Downregulated |
| -0,031783164 | 0,864269821 | NID1 | FALSE | Downregulated |

| | | | | |
|---|---|---|---|---|
| -0,099951081 | 0,864317279 | LOC100130992 | FALSE | Downregulated |
| -0,082996659 | 0,864345024 | CCDC183 | FALSE | Downregulated |
| -0,107103319 | 0,864345024 | LOC100506036 | FALSE | Downregulated |
| -0,02860895 | 0,864443934 | BRF2 | FALSE | Downregulated |
| 0,041875058 | 0,864580075 | DHRS7B | FALSE | Upregulated |
| 0,284612542 | 0,86458171 | LOC102723382 | FALSE | Upregulated |
| -0,355968646 | 0,864614122 | LOC102724075 | FALSE | Downregulated |
| -0,050067788 | 0,864903457 | TCP11L2 | FALSE | Downregulated |
| 0,219899067 | 0,86511058 | GS1-122H1.2 | FALSE | Upregulated |
| 0,035359982 | 0,865146466 | SVIP | FALSE | Upregulated |
| 0,025265786 | 0,865162199 | ZCCHC8 | FALSE | Upregulated |
| 0,036450259 | 0,865346174 | SSR1 | FALSE | Upregulated |
| -0,062316328 | 0,86550064 | C6orf123 | FALSE | Downregulated |
| 0,221516396 | 0,865504126 | CHL1 | FALSE | Upregulated |
| 0,025653898 | 0,865504126 | GABARAPL2 | FALSE | Upregulated |
| 0,254573829 | 0,866115041 | LOC101928906 | FALSE | Upregulated |
| -0,024690222 | 0,866272646 | AHCYL2 | FALSE | Downregulated |
| -0,068796942 | 0,866372641 | ADAMTS5 | FALSE | Downregulated |
| 0,075030219 | 0,866372641 | C1orf63 | FALSE | Upregulated |
| -0,107076628 | 0,866372641 | CCDC17 | FALSE | Downregulated |
| -0,033628692 | 0,866372641 | EFHD2 | FALSE | Downregulated |
| -0,119300151 | 0,866372641 | FBXO39 | FALSE | Downregulated |
| 0,304453414 | 0,866372641 | FMO6P | FALSE | Upregulated |
| 0,26956845 | 0,866372641 | LINC00977 | FALSE | Upregulated |
| 0,308682398 | 0,866372641 | LOC102723729 | FALSE | Upregulated |
| -0,028252264 | 0,866372641 | NEK1 | FALSE | Downregulated |
| 0,175574109 | 0,866372641 | RP3-323P13.2 | FALSE | Upregulated |
| -0,033203835 | 0,866372641 | SEC24A | FALSE | Downregulated |
| 0,421406939 | 0,866372641 | SERPINB2 | FALSE | Upregulated |
| 0,320665069 | 0,866590978 | DEFA3 | FALSE | Upregulated |
| 0,25190461 | 0,866846533 | IL12B | FALSE | Upregulated |
| 0,063761828 | 0,866846533 | RBP7 | FALSE | Upregulated |
| 0,046904897 | 0,866846827 | ACPL2 | FALSE | Upregulated |
| 0,063149506 | 0,866846827 | C4A | FALSE | Upregulated |
| 0,153556471 | 0,866846827 | LOC101926922 | FALSE | Upregulated |
| -0,086381009 | 0,866846827 | NSUN5P2 | FALSE | Downregulated |
| 0,089293704 | 0,866846827 | SLC16A11 | FALSE | Upregulated |
| -0,025383818 | 0,866876148 | EPC2 | FALSE | Downregulated |
| -0,029800179 | 0,866876148 | IAH1 | FALSE | Downregulated |
| 0,122497155 | 0,86696186 | ACOT11 | FALSE | Upregulated |
| 0,167325381 | 0,866977133 | NOXO1 | FALSE | Upregulated |
| -0,116367282 | 0,867053731 | LOC102723576 | FALSE | Downregulated |
| 0,143763731 | 0,867053731 | TAS2R10 | FALSE | Upregulated |
| 0,288743658 | 0,867056661 | KIF28P | FALSE | Upregulated |
| 0,026892991 | 0,867056661 | LRBA | FALSE | Upregulated |
| 0,111740372 | 0,867056661 | TSC22D3 | FALSE | Upregulated |
| 0,139906226 | 0,867337015 | C9orf139 | FALSE | Upregulated |
| -0,022841098 | 0,867337015 | RANBP9 | FALSE | Downregulated |
| 0,076635236 | 0,867341742 | HHIP-AS1 | FALSE | Upregulated |
| 0,094421458 | 0,867341742 | LOC101928464 | FALSE | Upregulated |
| -0,307611536 | 0,867341742 | LOC102724731 | FALSE | Downregulated |

| | | | | |
|---|---|---|---|---|
| -0,07077466 | 0,867487245 | C7orf76 | FALSE | Downregulated |
| -0,02643027 | 0,867622948 | PSMB3 | FALSE | Downregulated |
| 0,062197207 | 0,867731369 | LOC102723665 | FALSE | Upregulated |
| 0,029334146 | 0,867830931 | DOLPP1 | FALSE | Upregulated |
| 0,109568015 | 0,86788441 | LOC440434 | FALSE | Upregulated |
| 0,396334085 | 0,867959885 | LINC00989 | FALSE | Upregulated |
| -0,325542546 | 0,867963366 | LOC100505874 | FALSE | Downregulated |
| -0,031775918 | 0,868070669 | OTUD6B | FALSE | Downregulated |
| -0,036244369 | 0,868070669 | SH2B3 | FALSE | Downregulated |
| -0,044689256 | 0,868269372 | PRAP1 | FALSE | Downregulated |
| -0,084173274 | 0,868393254 | CTC-338M12.4 | FALSE | Downregulated |
| -0,035393056 | 0,868393254 | SNAP47 | FALSE | Downregulated |
| -0,020990283 | 0,86856819 | ANKLE2 | FALSE | Downregulated |
| -0,203656522 | 0,86856819 | PLCD4 | FALSE | Downregulated |
| 0,253692618 | 0,868652157 | MIR6717 | FALSE | Upregulated |
| 0,04926137 | 0,868783947 | ANKS3 | FALSE | Upregulated |
| -0,264152346 | 0,868783947 | HIST1H3D | FALSE | Downregulated |
| 0,209927212 | 0,868783947 | KCNMB2 | FALSE | Upregulated |
| -0,023025965 | 0,868783947 | MBNL1 | FALSE | Downregulated |
| -0,488034566 | 0,869001584 | TRNAK22 | FALSE | Downregulated |
| -0,029596386 | 0,869001584 | TTC8 | FALSE | Downregulated |
| -0,034484799 | 0,869016526 | COG3 | FALSE | Downregulated |
| 0,051167687 | 0,869016526 | EFNB3 | FALSE | Upregulated |
| -0,419057763 | 0,869016526 | LOC440292 | FALSE | Downregulated |
| 0,079346858 | 0,8691793 | CACNA2D2 | FALSE | Upregulated |
| -0,26973267 | 0,869268021 | LOC102724003 | FALSE | Downregulated |
| 0,371924233 | 0,869268021 | SVILP1 | FALSE | Upregulated |
| -0,365612384 | 0,869268021 | TSNAX-DISC1 | FALSE | Downregulated |
| 0,040355766 | 0,86927361 | BAHCC1 | FALSE | Upregulated |
| 0,290843681 | 0,86927361 | GRIN2B | FALSE | Upregulated |
| 0,031656404 | 0,869403824 | ASXL1 | FALSE | Upregulated |
| -0,235021507 | 0,869629908 | C1QTNF9B | FALSE | Downregulated |
| 0,096364771 | 0,869629908 | RPL23AP64 | FALSE | Upregulated |
| 0,140776539 | 0,869652652 | GPR64 | FALSE | Upregulated |
| -0,098106735 | 0,869652652 | NLRP14 | FALSE | Downregulated |
| -0,18838775 | 0,869868427 | LOC100996643 | FALSE | Downregulated |
| 0,083440637 | 0,869868427 | NRARP | FALSE | Upregulated |
| -0,041288489 | 0,869868427 | SWSAP1 | FALSE | Downregulated |
| -0,047730721 | 0,869868427 | ZXDB | FALSE | Downregulated |
| 0,037948987 | 0,869872229 | CNEP1R1 | FALSE | Upregulated |
| -0,110483769 | 0,869872229 | VIPR2 | FALSE | Downregulated |
| -0,060271201 | 0,870069125 | FLJ20021 | FALSE | Downregulated |
| 0,037638123 | 0,870069125 | ZNF205 | FALSE | Upregulated |
| 0,185444106 | 0,870077385 | SNORD79 | FALSE | Upregulated |
| 0,05219525 | 0,870077385 | TOX2 | FALSE | Upregulated |
| 0,039884372 | 0,870098262 | CLDN23 | FALSE | Upregulated |
| -0,031881448 | 0,870455293 | ANKRD42 | FALSE | Downregulated |
| -0,045705845 | 0,870499986 | CD68 | FALSE | Downregulated |
| -0,092904302 | 0,870843223 | BMS1P2 | FALSE | Downregulated |
| 0,023827735 | 0,870891572 | FAM120B | FALSE | Upregulated |
| -0,112051317 | 0,87093879 | KIF1A | FALSE | Downregulated |

| | | | | |
|---|---|---|---|---|
| 0,069762138 | 0,871081619 | FGF2 | FALSE | Upregulated |
| 1,258404044 | 0,871667601 | LOC102724843 | FALSE | Upregulated |
| -0,029843316 | 0,871809406 | BLCAP | FALSE | Downregulated |
| 0,115226622 | 0,87184962 | MDH1B | FALSE | Upregulated |
| 0,163180804 | 0,87184962 | MPO | FALSE | Upregulated |
| 0,067858653 | 0,87184962 | PDE9A | FALSE | Upregulated |
| -0,05826038 | 0,87184962 | RAMP3 | FALSE | Downregulated |
| -0,026194212 | 0,87184962 | TBC1D14 | FALSE | Downregulated |
| -0,040583469 | 0,87184962 | TP73-AS1 | FALSE | Downregulated |
| -0,035226408 | 0,872022061 | ACTR3C | FALSE | Downregulated |
| -0,127913461 | 0,872022061 | GOLGA6L3 | FALSE | Downregulated |
| -0,224085742 | 0,872022061 | RP13-228J13.1 | FALSE | Downregulated |
| -0,017692924 | 0,872022061 | SNAPC3 | FALSE | Downregulated |
| -0,022596793 | 0,872022061 | TXNDC11 | FALSE | Downregulated |
| -0,353853917 | 0,872087774 | TRNAG22 | FALSE | Downregulated |
| 0,025749334 | 0,872242227 | DRAM2 | FALSE | Upregulated |
| -0,055502166 | 0,872242227 | MCMDC2 | FALSE | Downregulated |
| 0,030959598 | 0,872391994 | ZAK | FALSE | Upregulated |
| 0,324703735 | 0,872549762 | LINC00908 | FALSE | Upregulated |
| -0,021247954 | 0,872798975 | RNF146 | FALSE | Downregulated |
| 0,033165231 | 0,872871511 | KIAA1551 | FALSE | Upregulated |
| 0,08844228 | 0,872912256 | TMEM178A | FALSE | Upregulated |
| -0,036731163 | 0,873029006 | AKAP8L | FALSE | Downregulated |
| -0,035300881 | 0,873029006 | MGRN1 | FALSE | Downregulated |
| -0,032415625 | 0,873029006 | RAD51C | FALSE | Downregulated |
| -0,055472 | 0,873490436 | VSIG10L | FALSE | Downregulated |
| 0,084152739 | 0,87349294 | LOC100131564 | FALSE | Upregulated |
| -0,054070558 | 0,873761276 | ZNF594 | FALSE | Downregulated |
| -0,132215808 | 0,87382249 | CTD-2201I18.1 | FALSE | Downregulated |
| -0,052722576 | 0,87382249 | KRBA2 | FALSE | Downregulated |
| -0,060028352 | 0,873876493 | LAD1 | FALSE | Downregulated |
| -0,029886709 | 0,873890829 | GALE | FALSE | Downregulated |
| -0,038906439 | 0,873890829 | TRADD | FALSE | Downregulated |
| 0,061440942 | 0,874078167 | GATA6-AS1 | FALSE | Upregulated |
| -0,169048304 | 0,874078167 | REM2 | FALSE | Downregulated |
| 0,035032728 | 0,874122249 | ATRX | FALSE | Upregulated |
| -0,026260063 | 0,874474188 | PTPRK | FALSE | Downregulated |
| -0,033305314 | 0,874589173 | NOL12 | FALSE | Downregulated |
| -0,018181522 | 0,87483443 | NR1H2 | FALSE | Downregulated |
| 0,118039908 | 0,87483443 | SOCS3 | FALSE | Upregulated |
| -0,250603855 | 0,874846484 | LOC101930580 | FALSE | Downregulated |
| -0,048313114 | 0,875079944 | SNHG1 | FALSE | Downregulated |
| 0,041636618 | 0,875104995 | VASN | FALSE | Upregulated |
| 0,038130594 | 0,875143767 | DOCK4 | FALSE | Upregulated |
| -0,023141292 | 0,875230147 | NDST1 | FALSE | Downregulated |
| 0,027806612 | 0,875408333 | AARS | FALSE | Upregulated |
| 0,441598132 | 0,875408333 | DLX4 | FALSE | Upregulated |
| 0,064344389 | 0,875408333 | HOXB6 | FALSE | Upregulated |
| -0,022522502 | 0,875408333 | XPC | FALSE | Downregulated |
| 0,236844935 | 0,875498891 | LOC101929132 | FALSE | Upregulated |
| 0,068961886 | 0,875543641 | ALDH1A2 | FALSE | Upregulated |

| | | | | |
|---|---|---|---|---|
| -0,049093645 | 0,875817038 | STS | FALSE | Downregulated |
| -0,149002158 | 0,875831106 | FAM201A | FALSE | Downregulated |
| -0,030542214 | 0,875831106 | MON2 | FALSE | Downregulated |
| -0,449125711 | 0,875831106 | MYH4 | FALSE | Downregulated |
| -0,016942974 | 0,875831106 | RNF14 | FALSE | Downregulated |
| 0,061193859 | 0,875871453 | CLDN15 | FALSE | Upregulated |
| 0,023089743 | 0,875878196 | ZER1 | FALSE | Upregulated |
| 0,030375807 | 0,875964876 | CAPN15 | FALSE | Upregulated |
| 0,035727889 | 0,875964876 | CSGALNACT2 | FALSE | Upregulated |
| -0,063873853 | 0,876051275 | NKTR | FALSE | Downregulated |
| -0,038386321 | 0,876051275 | VIMP | FALSE | Downregulated |
| -0,017342237 | 0,876051275 | WIPI2 | FALSE | Downregulated |
| -0,161429144 | 0,876051275 | ZP3 | FALSE | Downregulated |
| -0,066288643 | 0,876388576 | CILP | FALSE | Downregulated |
| -0,032473934 | 0,876407876 | ANKRD11 | FALSE | Downregulated |
| 0,423989102 | 0,876407876 | GRM4 | FALSE | Upregulated |
| -0,071411679 | 0,876407876 | LOC102723405 | FALSE | Downregulated |
| -0,072998044 | 0,876435941 | PMS2P3 | FALSE | Downregulated |
| -0,104215564 | 0,876441392 | GOLGA8B | FALSE | Downregulated |
| -0,149798111 | 0,876701296 | HAR1B | FALSE | Downregulated |
| 0,016935268 | 0,876701296 | LYRM2 | FALSE | Upregulated |
| 0,030519557 | 0,876827081 | FRMD4A | FALSE | Upregulated |
| -0,06741042 | 0,876829729 | WDR27 | FALSE | Downregulated |
| -0,076046343 | 0,877109005 | LOC100129203 | FALSE | Downregulated |
| -0,020134402 | 0,877331579 | HNRNPUL1 | FALSE | Downregulated |
| -0,360670995 | 0,878006612 | LOC101929414 | FALSE | Downregulated |
| 0,02396608 | 0,878021131 | HIPK1 | FALSE | Upregulated |
| 0,027529581 | 0,878021131 | TECR | FALSE | Upregulated |
| 0,083589195 | 0,878021131 | TMPO-AS1 | FALSE | Upregulated |
| 0,239841872 | 0,878046715 | IL21R-AS1 | FALSE | Upregulated |
| -0,024055901 | 0,878201542 | ARHGEF12 | FALSE | Downregulated |
| 0,297248774 | 0,878208365 | TEX12 | FALSE | Upregulated |
| 0,271922937 | 0,8785152 | LRRC10B | FALSE | Upregulated |
| 0,171400215 | 0,878628195 | RNF175 | FALSE | Upregulated |
| -0,168773304 | 0,878744724 | AL132709.5 | FALSE | Downregulated |
| 0,034947457 | 0,878744724 | ARPC5L | FALSE | Upregulated |
| 0,030302507 | 0,878744724 | CASP8AP2 | FALSE | Upregulated |
| 0,036694737 | 0,878744724 | MAPKAPK5-AS1 | FALSE | Upregulated |
| -0,032148427 | 0,878744724 | WDR81 | FALSE | Downregulated |
| -0,113139833 | 0,878816757 | LOC101929112 | FALSE | Downregulated |
| -0,078753507 | 0,878816757 | LOC284023 | FALSE | Downregulated |
| 0,054257934 | 0,878816757 | TLE2 | FALSE | Upregulated |
| -0,016097628 | 0,87910806 | MRPL30 | FALSE | Downregulated |
| 0,083172897 | 0,879215109 | HOXB7 | FALSE | Upregulated |
| -0,029732385 | 0,879215109 | PSPC1 | FALSE | Downregulated |
| -0,021897199 | 0,879351304 | CRYZL1 | FALSE | Downregulated |
| 0,218007858 | 0,879484616 | LOC100506504 | FALSE | Upregulated |
| 0,085381528 | 0,879689158 | SLC17A8 | FALSE | Upregulated |
| -0,13887917 | 0,880107454 | LINC00222 | FALSE | Downregulated |
| -0,028098508 | 0,880107454 | NAPB | FALSE | Downregulated |
| 0,089772613 | 0,880130716 | C19orf57 | FALSE | Upregulated |

| | | | | |
|---|---|---|---|---|
| 0,019600458 | 0,880146696 | AASDHPPT | FALSE | Upregulated |
| 0,055553927 | 0,880699465 | LOC101928419 | FALSE | Upregulated |
| 0,126240051 | 0,880755008 | LOC102723981 | FALSE | Upregulated |
| -0,030070092 | 0,880755008 | POGZ | FALSE | Downregulated |
| -0,069215092 | 0,880755008 | SLC16A13 | FALSE | Downregulated |
| 0,064978996 | 0,881054454 | LOC100507642 | FALSE | Upregulated |
| 0,031344733 | 0,881073016 | ARID3B | FALSE | Upregulated |
| 0,021070298 | 0,881073016 | ATF2 | FALSE | Upregulated |
| 0,034353938 | 0,881073016 | DIXDC1 | FALSE | Upregulated |
| -0,025198746 | 0,881073016 | PLOD2 | FALSE | Downregulated |
| 0,032084706 | 0,881073016 | WDR73 | FALSE | Upregulated |
| -0,084181458 | 0,881127501 | GPR82 | FALSE | Downregulated |
| -0,027912765 | 0,881541652 | SLC25A17 | FALSE | Downregulated |
| 0,837588879 | 0,881541652 | TRNAK33 | FALSE | Upregulated |
| -0,027300315 | 0,881651555 | ZNF507 | FALSE | Downregulated |
| -0,025406231 | 0,881762965 | SEC31A | FALSE | Downregulated |
| -0,046223966 | 0,881826011 | RAG1 | FALSE | Downregulated |
| 0,283368904 | 0,882148614 | HPCAL4 | FALSE | Upregulated |
| -0,017802776 | 0,882148614 | OXSR1 | FALSE | Downregulated |
| 0,204048978 | 0,882179788 | C12orf74 | FALSE | Upregulated |
| 0,036462333 | 0,882370789 | SPATA2L | FALSE | Upregulated |
| 0,2178003 | 0,882545355 | ERBB4 | FALSE | Upregulated |
| 0,246558791 | 0,882579073 | TMEM40 | FALSE | Upregulated |
| -0,165891682 | 0,882591878 | KC6 | FALSE | Downregulated |
| 0,024821476 | 0,882591878 | UCKL1 | FALSE | Upregulated |
| -0,025721144 | 0,882870191 | FEZ2 | FALSE | Downregulated |
| -0,019723817 | 0,882870191 | NSUN4 | FALSE | Downregulated |
| 0,279972029 | 0,882910314 | KCNIP4 | FALSE | Upregulated |
| 0,124670697 | 0,882910314 | LOC102724788 | FALSE | Upregulated |
| -0,033563528 | 0,882979436 | RAB4B | FALSE | Downregulated |
| -0,056114547 | 0,883060769 | LINC00476 | FALSE | Downregulated |
| -0,141896739 | 0,883169097 | BPI | FALSE | Downregulated |
| -0,391721948 | 0,883249671 | RP11-513O17.2-2 | FALSE | Downregulated |
| 0,03041634 | 0,883369647 | NCKIPSD | FALSE | Upregulated |
| 0,035006891 | 0,883394518 | OLFML3 | FALSE | Upregulated |
| -0,023360826 | 0,883617828 | ZNF706 | FALSE | Downregulated |
| -0,098837827 | 0,883734732 | DSCAM | FALSE | Downregulated |
| -0,301547363 | 0,883734732 | LINC01141 | FALSE | Downregulated |
| 0,154692162 | 0,883734732 | LOC100288846 | FALSE | Upregulated |
| -0,096329477 | 0,883770014 | LOC101927300 | FALSE | Downregulated |
| -0,200404335 | 0,883819808 | PTCHD3 | FALSE | Downregulated |
| 0,053821213 | 0,883883476 | LRRC23 | FALSE | Upregulated |
| -0,02640232 | 0,883894023 | ILVBL | FALSE | Downregulated |
| 0,07630921 | 0,884089462 | LOC101927258 | FALSE | Upregulated |
| -0,042322932 | 0,884104746 | VKORC1 | FALSE | Downregulated |
| -0,018905437 | 0,884168482 | PPP6R3 | FALSE | Downregulated |
| 0,22262518 | 0,884355303 | CD70 | FALSE | Upregulated |
| -0,03415684 | 0,884355303 | CEP192 | FALSE | Downregulated |
| 0,052854713 | 0,884355303 | FOXF1 | FALSE | Upregulated |
| -0,020465794 | 0,884355303 | LARS2 | FALSE | Downregulated |
| 0,073646423 | 0,884355303 | SYCP2 | FALSE | Upregulated |

| | | | | |
|---|---|---|---|---|
| -0,040007114 | 0,88477106 | CREM | FALSE | Downregulated |
| 0,022206171 | 0,88487655 | ADAR | FALSE | Upregulated |
| -0,041456131 | 0,884929429 | LPP-AS2 | FALSE | Downregulated |
| 0,103570548 | 0,884929429 | YBX3 | FALSE | Upregulated |
| 0,328127515 | 0,884932859 | RP3-400N23.6 | FALSE | Upregulated |
| 0,084524342 | 0,885491723 | PTK6 | FALSE | Upregulated |
| -0,175917412 | 0,885749221 | KCNQ5 | FALSE | Downregulated |
| 0,036555813 | 0,886082467 | MPV17 | FALSE | Upregulated |
| 0,022872034 | 0,886211776 | FBXO7 | FALSE | Upregulated |
| 0,03771197 | 0,886211776 | IFT172 | FALSE | Upregulated |
| 0,265049806 | 0,886211776 | LOC101927848 | FALSE | Upregulated |
| -0,050753069 | 0,886211776 | MIR1181 | FALSE | Downregulated |
| 0,052355242 | 0,886516338 | LBX2 | FALSE | Upregulated |
| -0,221764811 | 0,88693714 | LOC100131395 | FALSE | Downregulated |
| -0,038864107 | 0,886969224 | FIGN | FALSE | Downregulated |
| 0,21130287 | 0,886969224 | SIM2 | FALSE | Upregulated |
| 0,123753251 | 0,887100297 | LOC101060341 | FALSE | Upregulated |
| -0,338372034 | 0,887226149 | CRISP3 | FALSE | Downregulated |
| -0,04303196 | 0,887559553 | CHKA | FALSE | Downregulated |
| -0,021903663 | 0,887559553 | EIF4A2 | FALSE | Downregulated |
| 0,033357966 | 0,887559553 | NOP16 | FALSE | Upregulated |
| -0,133984829 | 0,887866426 | LOC100996735 | FALSE | Downregulated |
| 0,225037014 | 0,888147579 | LOC102724351 | FALSE | Upregulated |
| 0,079444021 | 0,888237715 | GUSBP4 | FALSE | Upregulated |
| -0,244812617 | 0,888237715 | LOC102467073 | FALSE | Downregulated |
| -0,041849746 | 0,888237715 | WASH3P | FALSE | Downregulated |
| 0,093831595 | 0,888297614 | DNM3 | FALSE | Upregulated |
| -0,261430471 | 0,888306768 | KLHDC8A | FALSE | Downregulated |
| -0,20893488 | 0,888306768 | RD3 | FALSE | Downregulated |
| 0,018272092 | 0,888306768 | TWISTNB | FALSE | Upregulated |
| -0,143570115 | 0,888314197 | FLJ45513 | FALSE | Downregulated |
| 0,24901816 | 0,888319133 | SNORD16 | FALSE | Upregulated |
| -0,055319615 | 0,888479517 | STX11 | FALSE | Downregulated |
| -0,027769191 | 0,888618764 | COA1 | FALSE | Downregulated |
| 0,022362338 | 0,888622106 | DARS2 | FALSE | Upregulated |
| 0,021675479 | 0,888878758 | DIDO1 | FALSE | Upregulated |
| -0,061166725 | 0,888878758 | LOC101929704 | FALSE | Downregulated |
| -0,036883923 | 0,888878758 | RGL1 | FALSE | Downregulated |
| -0,030993783 | 0,888878758 | ZNF503 | FALSE | Downregulated |
| 0,043032684 | 0,888954947 | FIGNL1 | FALSE | Upregulated |
| -0,092643774 | 0,888994869 | C1orf220 | FALSE | Downregulated |
| 0,176432409 | 0,889093784 | LOC101928240 | FALSE | Upregulated |
| -0,102716507 | 0,889301697 | RP11-557H15.4 | FALSE | Downregulated |
| -0,033515124 | 0,889314875 | CENPT | FALSE | Downregulated |
| 0,240672037 | 0,889365497 | RP11-17A4.2 | FALSE | Upregulated |
| -0,034164776 | 0,889365497 | TTC12 | FALSE | Downregulated |
| -0,035562852 | 0,889365497 | USP21 | FALSE | Downregulated |
| 0,048565097 | 0,889396594 | C5orf54 | FALSE | Upregulated |
| 0,172302988 | 0,889420254 | LOC101928699 | FALSE | Upregulated |
| -0,021419444 | 0,889445867 | ZC3HAV1 | FALSE | Downregulated |
| 0,027323896 | 0,889460551 | TSPAN9 | FALSE | Upregulated |

| | | | | |
|---|---|---|---|---|
| -0,05614967 | 0,889562637 | CACNA2D4 | FALSE | Downregulated |
| -0,255435739 | 0,889569804 | SLC10A4 | FALSE | Downregulated |
| 0,093358083 | 0,889943676 | RANBP17 | FALSE | Upregulated |
| -0,040864594 | 0,89002233 | MBL2 | FALSE | Downregulated |
| 0,040671521 | 0,8901515 | NSMF | FALSE | Upregulated |
| 0,079357284 | 0,890277125 | LOC102724330 | FALSE | Upregulated |
| 0,031790187 | 0,89046615 | FKTN | FALSE | Upregulated |
| 0,055370553 | 0,89046615 | SOX6 | FALSE | Upregulated |
| -0,648797588 | 0,890620774 | CXCL13 | FALSE | Downregulated |
| 0,255122131 | 0,890663872 | HTR2A | FALSE | Upregulated |
| -0,017693092 | 0,890712176 | HSBP1 | FALSE | Downregulated |
| -0,131751482 | 0,890753674 | CEL | FALSE | Downregulated |
| 0,126142102 | 0,890753674 | ERVFRD-1 | FALSE | Upregulated |
| -0,211887768 | 0,89090952 | LURAP1 | FALSE | Downregulated |
| -0,030173141 | 0,89092157 | RBM48 | FALSE | Downregulated |
| -0,027245612 | 0,890978274 | ASL | FALSE | Downregulated |
| 0,020554985 | 0,891124133 | TBC1D13 | FALSE | Upregulated |
| -0,082669648 | 0,891172086 | LOC101928714 | FALSE | Downregulated |
| -0,023612541 | 0,891276433 | FAM206A | FALSE | Downregulated |
| 0,033488856 | 0,89130121 | S100PBP | FALSE | Upregulated |
| -0,1149502 | 0,891318409 | RP11-486O12.2 | FALSE | Downregulated |
| 0,229363661 | 0,891560004 | RP11-148L24.1 | FALSE | Upregulated |
| -0,029797102 | 0,891560004 | SLC46A3 | FALSE | Downregulated |
| -0,031628006 | 0,891625585 | ABHD2 | FALSE | Downregulated |
| 0,230541157 | 0,891625585 | LOC102723619 | FALSE | Upregulated |
| -0,1253933 | 0,891745426 | LAMB2P1 | FALSE | Downregulated |
| 0,03356234 | 0,89197233 | CDH5 | FALSE | Upregulated |
| -0,041730368 | 0,891999991 | IFRD1 | FALSE | Downregulated |
| 0,040419576 | 0,891999991 | SEMA4B | FALSE | Upregulated |
| -0,044836955 | 0,892192788 | RFXAP | FALSE | Downregulated |
| -0,155451805 | 0,892232127 | LOC101929346 | FALSE | Downregulated |
| -0,014964949 | 0,892258455 | CUL1 | FALSE | Downregulated |
| -0,12496907 | 0,892258455 | DDX11L2 | FALSE | Downregulated |
| -0,070146624 | 0,892258455 | LOC595101 | FALSE | Downregulated |
| -0,126706277 | 0,892296279 | LOC102724541 | FALSE | Downregulated |
| -0,090539862 | 0,892453508 | LTB4R2 | FALSE | Downregulated |
| 0,062577405 | 0,892587672 | ATG16L2 | FALSE | Upregulated |
| -0,020925776 | 0,892587672 | FPGT | FALSE | Downregulated |
| 0,029248025 | 0,892739405 | PLEKHA2 | FALSE | Upregulated |
| -0,019656559 | 0,892763074 | SPIN1 | FALSE | Downregulated |
| 0,064336155 | 0,893089659 | ACKR4 | FALSE | Upregulated |
| -0,085990067 | 0,893089659 | ALMS1P | FALSE | Downregulated |
| -0,030404207 | 0,893089659 | BAZ2A | FALSE | Downregulated |
| -0,025402497 | 0,893089659 | COPS7B | FALSE | Downregulated |
| -0,086069681 | 0,893089659 | ICA1 | FALSE | Downregulated |
| -0,02125694 | 0,893089659 | IDE | FALSE | Downregulated |
| -0,065685021 | 0,893089659 | LOC102724133 | FALSE | Downregulated |
| -0,115394544 | 0,893089659 | LOC102724970 | FALSE | Downregulated |
| 0,270855789 | 0,893089659 | PLP1 | FALSE | Upregulated |
| 0,021155056 | 0,893089659 | STRADA | FALSE | Upregulated |
| -0,170040858 | 0,893199939 | LOC102724230 | FALSE | Downregulated |

| | | | | |
|---|---|---|---|---|
| -1,072857916 | 0,893199939 | XIST | FALSE | Downregulated |
| -0,130178168 | 0,89326601 | ZNF177 | FALSE | Downregulated |
| -0,027364661 | 0,893572063 | ANPEP | FALSE | Downregulated |
| -0,060198069 | 0,893572063 | BFSP1 | FALSE | Downregulated |
| -0,263971778 | 0,893572063 | C19orf45 | FALSE | Downregulated |
| -0,033443919 | 0,893572063 | DCP2 | FALSE | Downregulated |
| -0,03783701 | 0,893572063 | GTF2H2B | FALSE | Downregulated |
| -0,061112523 | 0,893572063 | LOC100507373 | FALSE | Downregulated |
| -0,239605413 | 0,893572063 | LOC102723730 | FALSE | Downregulated |
| -0,020886952 | 0,893572063 | LSG1 | FALSE | Downregulated |
| -0,035057993 | 0,893572063 | MIDN | FALSE | Downregulated |
| -0,093871145 | 0,893572063 | MLIP | FALSE | Downregulated |
| -0,083323679 | 0,893572063 | MORC2-AS1 | FALSE | Downregulated |
| 0,049179086 | 0,893572063 | PLEKHA4 | FALSE | Upregulated |
| 0,034797544 | 0,893572063 | PRKCA | FALSE | Upregulated |
| -0,02949433 | 0,893572063 | PSMG3-AS1 | FALSE | Downregulated |
| -0,147237446 | 0,893572063 | SLC26A7 | FALSE | Downregulated |
| 0,275779923 | 0,893572063 | SPATA22 | FALSE | Upregulated |
| 0,094154581 | 0,893572063 | TMEM236 | FALSE | Upregulated |
| 0,049269166 | 0,893572063 | ZNF311 | FALSE | Upregulated |
| -0,018851137 | 0,893572063 | ZSCAN29 | FALSE | Downregulated |
| -0,238822153 | 0,893692164 | LINC00896 | FALSE | Downregulated |
| -0,069108925 | 0,893754403 | C21orf67 | FALSE | Downregulated |
| -0,065300334 | 0,893754403 | DMPK | FALSE | Downregulated |
| 0,083175797 | 0,893754403 | ESPNL | FALSE | Upregulated |
| -0,150814828 | 0,893754403 | LOC102724697 | FALSE | Downregulated |
| 0,114065906 | 0,893754403 | RBPMS-AS1 | FALSE | Upregulated |
| 0,092734934 | 0,893754403 | TTC39A | FALSE | Upregulated |
| 0,048063383 | 0,893754403 | ZNF107 | FALSE | Upregulated |
| -0,289796513 | 0,894037345 | LOC729506 | FALSE | Downregulated |
| 0,039020852 | 0,894354798 | DFNA5 | FALSE | Upregulated |
| 0,036052411 | 0,894395794 | DEPTOR | FALSE | Upregulated |
| 0,020124691 | 0,894395794 | EIF3K | FALSE | Upregulated |
| 0,053846923 | 0,894582699 | PCSK4 | FALSE | Upregulated |
| -0,018777738 | 0,894744434 | CHMP4B | FALSE | Downregulated |
| 0,131252591 | 0,894744434 | LINC00839 | FALSE | Upregulated |
| -0,145293326 | 0,89502513 | ADAMTSL4-AS1 | FALSE | Downregulated |
| -0,039837683 | 0,89502513 | LOC100507599 | FALSE | Downregulated |
| -0,035914091 | 0,895190168 | SLC30A4 | FALSE | Downregulated |
| -0,037937171 | 0,895340972 | OTUD1 | FALSE | Downregulated |
| 0,016189578 | 0,895458508 | EAF1 | FALSE | Upregulated |
| 0,13514161 | 0,895577554 | LOC102724583 | FALSE | Upregulated |
| -0,037431875 | 0,895596734 | NRP2 | FALSE | Downregulated |
| 0,047964402 | 0,895765156 | FSTL1 | FALSE | Upregulated |
| 0,016392268 | 0,895831471 | UBE2G2 | FALSE | Upregulated |
| -0,041090565 | 0,896087469 | AFF3 | FALSE | Downregulated |
| -0,061471385 | 0,896087469 | ANGPTL4 | FALSE | Downregulated |
| -0,021590428 | 0,896087469 | CARS2 | FALSE | Downregulated |
| 0,085214144 | 0,896145458 | ADAT3 | FALSE | Upregulated |
| 0,063066135 | 0,896286698 | AIM2 | FALSE | Upregulated |
| -0,040127617 | 0,896504505 | C17orf89 | FALSE | Downregulated |

| | | | | |
|---|---|---|---|---|
| 0,032097838 | 0,896504505 | CFB | FALSE | Upregulated |
| 0,018293671 | 0,896504505 | MASP1 | FALSE | Upregulated |
| -0,024207556 | 0,896504505 | PRR14L | FALSE | Downregulated |
| 0,024025654 | 0,89660169 | SLC37A3 | FALSE | Upregulated |
| -0,070527049 | 0,89689504 | RP11-677M14.7 | FALSE | Downregulated |
| -0,11700169 | 0,897047765 | SLC13A4 | FALSE | Downregulated |
| -0,160860491 | 0,897430193 | HAMP | FALSE | Downregulated |
| -0,10961602 | 0,897598634 | PTGER3 | FALSE | Downregulated |
| -0,203912414 | 0,897649952 | LOC285768 | FALSE | Downregulated |
| -0,014726945 | 0,897672865 | MTAP | FALSE | Downregulated |
| 0,032823766 | 0,898194622 | CADPS2 | FALSE | Upregulated |
| -0,025463802 | 0,898194622 | MUM1 | FALSE | Downregulated |
| 0,220974839 | 0,898194622 | RGS22 | FALSE | Upregulated |
| 0,023088471 | 0,898261623 | SH3BGRL | FALSE | Upregulated |
| 0,024431695 | 0,898261742 | C16orf52 | FALSE | Upregulated |
| -0,093092613 | 0,898261742 | LOC101060212 | FALSE | Downregulated |
| -0,018726119 | 0,898277355 | GUSB | FALSE | Downregulated |
| -0,026137146 | 0,898277355 | ZNF740 | FALSE | Downregulated |
| -0,02925828 | 0,898308378 | CCDC91 | FALSE | Downregulated |
| 0,108356818 | 0,898314132 | LOC101928612 | FALSE | Upregulated |
| -0,067132265 | 0,898357842 | LOC102723546 | FALSE | Downregulated |
| -0,058636715 | 0,898357842 | NFKBID | FALSE | Downregulated |
| -0,380287517 | 0,898567672 | TRNAG19 | FALSE | Downregulated |
| -0,082643954 | 0,898873255 | MORF4L2-AS1 | FALSE | Downregulated |
| -0,038633306 | 0,899101644 | IL1RN | FALSE | Downregulated |
| -0,027390061 | 0,899249669 | CPSF7 | FALSE | Downregulated |
| 0,035965406 | 0,899305966 | ARID4B | FALSE | Upregulated |
| -0,0170616 | 0,899305966 | CHMP3 | FALSE | Downregulated |
| 0,299566561 | 0,899305966 | LOC101929054 | FALSE | Upregulated |
| -0,149838564 | 0,899322186 | GNAT1 | FALSE | Downregulated |
| 0,017385317 | 0,89948654 | ARMC1 | FALSE | Upregulated |
| -0,064620322 | 0,89948654 | DKFZp779M0652 | FALSE | Downregulated |
| -0,03851197 | 0,89948654 | LOC389765 | FALSE | Downregulated |
| 0,230210336 | 0,899636528 | AC104024.1 | FALSE | Upregulated |
| -0,032151708 | 0,899675587 | CLK2 | FALSE | Downregulated |
| -0,238554923 | 0,899675587 | EMX1 | FALSE | Downregulated |
| -0,101684559 | 0,899685706 | PROB1 | FALSE | Downregulated |
| -0,03306532 | 0,899825614 | IQCG | FALSE | Downregulated |
| -0,064011643 | 0,899887069 | LOC100506100 | FALSE | Downregulated |
| 0,095222566 | 0,900050309 | AC010524.2 | FALSE | Upregulated |
| 0,02022497 | 0,900050309 | CD276 | FALSE | Upregulated |
| -0,107873326 | 0,900050309 | GPR135 | FALSE | Downregulated |
| -0,027983238 | 0,900050309 | MACF1 | FALSE | Downregulated |
| 0,064310718 | 0,900050309 | POLE2 | FALSE | Upregulated |
| 0,085248754 | 0,900050309 | SLC35E4 | FALSE | Upregulated |
| 0,014396105 | 0,900050309 | YME1L1 | FALSE | Upregulated |
| -0,047754731 | 0,900050309 | ZNF678 | FALSE | Downregulated |
| -0,150005939 | 0,900229741 | LOC101928344 | FALSE | Downregulated |
| -0,041320351 | 0,900606694 | PARD6G | FALSE | Downregulated |
| 0,250627096 | 0,900621024 | LINC00298 | FALSE | Upregulated |
| -0,0309384 | 0,900621024 | TRIM56 | FALSE | Downregulated |

| | | | | |
|---|---|---|---|---|
| -0,030896739 | 0,900688093 | ZNF626 | FALSE | Downregulated |
| -0,023494715 | 0,90074356 | CFLAR | FALSE | Downregulated |
| 0,017896618 | 0,90074356 | POM121C | FALSE | Upregulated |
| -0,180219451 | 0,900788353 | CES1P1 | FALSE | Downregulated |
| 0,043662659 | 0,900864403 | SLC1A4 | FALSE | Upregulated |
| 0,232814649 | 0,900867982 | LOC101928343 | FALSE | Upregulated |
| -0,04398622 | 0,900926904 | EXOG | FALSE | Downregulated |
| -0,126090527 | 0,900926904 | LOC101928558 | FALSE | Downregulated |
| -0,12159963 | 0,901027949 | FRMD7 | FALSE | Downregulated |
| 0,0663654 | 0,901346025 | LOC146880 | FALSE | Upregulated |
| -0,054228294 | 0,901433961 | CCL15 | FALSE | Downregulated |
| 0,13619272 | 0,901462451 | MS4A4E | FALSE | Upregulated |
| -0,028757933 | 0,901643785 | LINC00847 | FALSE | Downregulated |
| -0,032930761 | 0,901668348 | ZNF543 | FALSE | Downregulated |
| -0,023940018 | 0,901789982 | CTDSPL | FALSE | Downregulated |
| 0,036098697 | 0,901806795 | BORA | FALSE | Upregulated |
| 0,15849117 | 0,901806795 | RGPD5 | FALSE | Upregulated |
| 0,033148797 | 0,901950196 | EXOSC4 | FALSE | Upregulated |
| -0,044592743 | 0,901950196 | GREM2 | FALSE | Downregulated |
| 0,102612371 | 0,901950196 | LOC101927278 | FALSE | Upregulated |
| 0,410231984 | 0,902472436 | TRNAH10 | FALSE | Upregulated |
| 0,081891707 | 0,902614476 | DPH6-AS1 | FALSE | Upregulated |
| 0,225360148 | 0,902614476 | KCNK7 | FALSE | Upregulated |
| 0,25330662 | 0,902614476 | LGALS17A | FALSE | Upregulated |
| 0,225250506 | 0,902656319 | LOC101928716 | FALSE | Upregulated |
| -0,068061561 | 0,902904227 | LOC102724061 | FALSE | Downregulated |
| 0,204314524 | 0,903025401 | CEACAM8 | FALSE | Upregulated |
| -0,016395873 | 0,903028224 | CCDC9 | FALSE | Downregulated |
| -0,044455976 | 0,903041221 | OSBPL11 | FALSE | Downregulated |
| 0,057010846 | 0,903041221 | TLCD2 | FALSE | Upregulated |
| 0,047606198 | 0,903059736 | TMEM26 | FALSE | Upregulated |
| -0,048906903 | 0,903075783 | PRG2 | FALSE | Downregulated |
| -0,246706672 | 0,903125507 | DNAJB13 | FALSE | Downregulated |
| 0,024657043 | 0,903139506 | CLIP1 | FALSE | Upregulated |
| -0,017380307 | 0,903192629 | SAP130 | FALSE | Downregulated |
| 0,074033698 | 0,903192629 | ZNF667 | FALSE | Upregulated |
| -0,030607929 | 0,903217861 | SLC7A9 | FALSE | Downregulated |
| 0,02714197 | 0,903604689 | GGT7 | FALSE | Upregulated |
| -0,027252418 | 0,903738236 | PCGF1 | FALSE | Downregulated |
| -0,036058306 | 0,903746758 | USP18 | FALSE | Downregulated |
| -0,24446015 | 0,903784463 | MARCH4 | FALSE | Downregulated |
| -0,010157554 | 0,903934117 | BAP1 | FALSE | Downregulated |
| -0,05742023 | 0,903934117 | LOC401320 | FALSE | Downregulated |
| -0,024030196 | 0,903976105 | EFTUD1 | FALSE | Downregulated |
| -0,112855113 | 0,903976105 | LOC101927400 | FALSE | Downregulated |
| -0,053674454 | 0,904049284 | NPR3 | FALSE | Downregulated |
| 0,021285156 | 0,904049284 | NUDC | FALSE | Upregulated |
| -0,01814604 | 0,904049284 | SUZ12 | FALSE | Downregulated |
| 0,018667914 | 0,904201961 | ARAF | FALSE | Upregulated |
| -0,982786075 | 0,904358138 | MYL1 | FALSE | Downregulated |
| 0,052874075 | 0,904571489 | FAM193B | FALSE | Upregulated |

| | | | | |
|---|---|---|---|---|
| 0,20892721 | 0,904571489 | GAL3ST2 | FALSE | Upregulated |
| 0,162337571 | 0,904571489 | LOC102724537 | FALSE | Upregulated |
| 0,128496104 | 0,904571489 | TMEFF2 | FALSE | Upregulated |
| -0,06950862 | 0,904629723 | LOC100287042 | FALSE | Downregulated |
| -0,066362273 | 0,904629723 | LOC102723772 | FALSE | Downregulated |
| -0,132437597 | 0,904629723 | STAR | FALSE | Downregulated |
| -0,030772328 | 0,904704706 | HARS | FALSE | Downregulated |
| -0,024809361 | 0,904704706 | RPS2 | FALSE | Downregulated |
| -0,126312576 | 0,904946195 | FLJ22447 | FALSE | Downregulated |
| -0,059568629 | 0,90498944 | ZNF726 | FALSE | Downregulated |
| -0,036623554 | 0,905016275 | STAB1 | FALSE | Downregulated |
| -0,034295119 | 0,905027441 | LPIN1 | FALSE | Downregulated |
| 0,145567554 | 0,905296069 | CCDC41-AS1 | FALSE | Upregulated |
| 0,223895459 | 0,905296069 | MKRN3 | FALSE | Upregulated |
| 0,064470403 | 0,905296069 | NGF | FALSE | Upregulated |
| 0,087450559 | 0,905322429 | CCDC147 | FALSE | Upregulated |
| -0,022547675 | 0,905322429 | DEF8 | FALSE | Downregulated |
| -0,021716443 | 0,905322429 | DUT | FALSE | Downregulated |
| 0,043541961 | 0,905322429 | MS4A4A | FALSE | Upregulated |
| -0,02278074 | 0,905322429 | OPN3 | FALSE | Downregulated |
| -0,101212981 | 0,905322429 | SUMO4 | FALSE | Downregulated |
| 0,103373233 | 0,905500053 | STAG3L1 | FALSE | Upregulated |
| -0,022699165 | 0,905622051 | ATP6V1E1 | FALSE | Downregulated |
| -0,053556663 | 0,905642101 | FGD1 | FALSE | Downregulated |
| -0,205856402 | 0,905644973 | RP11-576I22.2 | FALSE | Downregulated |
| -0,031274396 | 0,905684191 | AMZ2P1 | FALSE | Downregulated |
| 0,017834143 | 0,905684191 | YY1 | FALSE | Upregulated |
| 0,021698062 | 0,905826365 | CRCP | FALSE | Upregulated |
| 0,124911278 | 0,906048027 | ALOX15 | FALSE | Upregulated |
| 0,041165187 | 0,906050604 | LINC00662 | FALSE | Upregulated |
| -0,04367542 | 0,906050604 | LOC100506127 | FALSE | Downregulated |
| 0,011345455 | 0,906050604 | PIK3C3 | FALSE | Upregulated |
| 0,153438673 | 0,906055784 | KLK10 | FALSE | Upregulated |
| 0,234003931 | 0,906171431 | BTNL3 | FALSE | Upregulated |
| -0,037653486 | 0,906171431 | CERS4 | FALSE | Downregulated |
| -0,025399857 | 0,906171431 | ERC1 | FALSE | Downregulated |
| -0,059955173 | 0,906171431 | LOC389834 | FALSE | Downregulated |
| -0,215348408 | 0,906171431 | NEFM | FALSE | Downregulated |
| -0,096027989 | 0,906171431 | PCOLCE-AS1 | FALSE | Downregulated |
| 0,096987616 | 0,906274631 | C12orf79 | FALSE | Upregulated |
| 0,057050787 | 0,906274631 | RP11-98I9.4 | FALSE | Upregulated |
| -0,197741503 | 0,906274631 | TRIM74 | FALSE | Downregulated |
| -0,035834115 | 0,906292597 | ZNF572 | FALSE | Downregulated |
| 0,016812922 | 0,906348822 | PA2G4 | FALSE | Upregulated |
| -0,192761145 | 0,906375721 | MT1B | FALSE | Downregulated |
| 0,153301821 | 0,90648646 | LOC102724328 | FALSE | Upregulated |
| 0,103745613 | 0,906562188 | LOC389607 | FALSE | Upregulated |
| 0,023049622 | 0,906685001 | DOCK1 | FALSE | Upregulated |
| 0,199525672 | 0,906685001 | NBL1 | FALSE | Upregulated |
| -0,027307974 | 0,906716877 | NTN1 | FALSE | Downregulated |
| -0,02330254 | 0,906716877 | NUBP2 | FALSE | Downregulated |

| | | | | |
|---|---|---|---|---|
| -0,243413448 | 0,906716877 | TUBB8 | FALSE | Downregulated |
| 0,013700143 | 0,906796953 | DENR | FALSE | Upregulated |
| -0,952876939 | 0,906796953 | MYOT | FALSE | Downregulated |
| -0,021983922 | 0,906796953 | PRKACB | FALSE | Downregulated |
| -0,024067748 | 0,906796953 | RNF25 | FALSE | Downregulated |
| 0,019092278 | 0,906796953 | ZNF37A | FALSE | Upregulated |
| -0,180531712 | 0,907251748 | LINC00032 | FALSE | Downregulated |
| -0,016590645 | 0,907275219 | WDR45B | FALSE | Downregulated |
| -0,029229731 | 0,907358389 | FASTKD1 | FALSE | Downregulated |
| 0,02966465 | 0,907477921 | VAC14 | FALSE | Upregulated |
| 0,011398746 | 0,907726712 | C3orf17 | FALSE | Upregulated |
| 0,043887896 | 0,907735561 | STOX2 | FALSE | Upregulated |
| 0,143328925 | 0,90780344 | CDKL2 | FALSE | Upregulated |
| 0,029754326 | 0,90780344 | F12 | FALSE | Upregulated |
| -0,070232759 | 0,90780344 | LOC102724238 | FALSE | Downregulated |
| 0,174321562 | 0,90780344 | SPAG17 | FALSE | Upregulated |
| 0,015034033 | 0,90780344 | STAMBP | FALSE | Upregulated |
| -0,074034205 | 0,907949197 | LOC646214 | FALSE | Downregulated |
| -0,042660096 | 0,908241747 | GPT2 | FALSE | Downregulated |
| 0,025209992 | 0,908328817 | NUP85 | FALSE | Upregulated |
| 0,08158205 | 0,908560138 | KAZALD1 | FALSE | Upregulated |
| -0,111178588 | 0,908560138 | LRRC69 | FALSE | Downregulated |
| -0,113517247 | 0,908593915 | C6orf165 | FALSE | Downregulated |
| 0,070961459 | 0,908593915 | CTAGE11P | FALSE | Upregulated |
| 0,028092381 | 0,908593915 | GCC2 | FALSE | Upregulated |
| -0,194895705 | 0,908593915 | LOC101926969 | FALSE | Downregulated |
| 0,170918717 | 0,908593915 | LOC101928062 | FALSE | Upregulated |
| 0,170853495 | 0,908593915 | PNLDC1 | FALSE | Upregulated |
| 0,01709482 | 0,908593915 | RRP1B | FALSE | Upregulated |
| -0,050297401 | 0,908593915 | SLC25A21-AS1 | FALSE | Downregulated |
| 0,016876857 | 0,908593915 | ZNF564 | FALSE | Upregulated |
| -0,042237559 | 0,908593915 | ZNF852 | FALSE | Downregulated |
| -0,021546145 | 0,908658333 | GFER | FALSE | Downregulated |
| -0,232427773 | 0,908658333 | LOC100134040 | FALSE | Downregulated |
| -0,068003457 | 0,908658333 | LOC102723564 | FALSE | Downregulated |
| 0,014822352 | 0,908658333 | LRRC41 | FALSE | Upregulated |
| -0,026992236 | 0,908658333 | MARS2 | FALSE | Downregulated |
| 0,019427056 | 0,908828587 | ARL1 | FALSE | Upregulated |
| 0,150883679 | 0,908828587 | RP11-483E7.1 | FALSE | Upregulated |
| 0,110489574 | 0,908828587 | RRN3P2 | FALSE | Upregulated |
| 0,023471997 | 0,908828587 | ZNF264 | FALSE | Upregulated |
| 0,016574638 | 0,908832891 | COMMD7 | FALSE | Upregulated |
| -0,078723984 | 0,908832891 | LOC100270746 | FALSE | Downregulated |
| -0,93900544 | 0,908832891 | RNA5S11 | FALSE | Downregulated |
| 0,030060253 | 0,908976117 | ZBTB8A | FALSE | Upregulated |
| 0,073888553 | 0,909543623 | SYCP3 | FALSE | Upregulated |
| 0,011440217 | 0,909588665 | VPS4B | FALSE | Upregulated |
| -0,308701924 | 0,909723484 | TNXA | FALSE | Downregulated |
| 0,132916098 | 0,909723484 | WWTR1-AS1 | FALSE | Upregulated |
| 0,152412985 | 0,909790103 | SLC7A11 | FALSE | Upregulated |
| 0,022744511 | 0,909835731 | CYB561A3 | FALSE | Upregulated |

| | | | | |
|---|---|---|---|---|
| 0,034223761 | 0,909911012 | SYNRG | FALSE | Upregulated |
| -0,221716677 | 0,909973815 | CCDC175 | FALSE | Downregulated |
| -0,086312442 | 0,909973815 | MAGOH2 | FALSE | Downregulated |
| -0,03275064 | 0,909973815 | NRBP2 | FALSE | Downregulated |
| -0,018264299 | 0,909973815 | PURA | FALSE | Downregulated |
| 0,02548434 | 0,910076759 | ZNF329 | FALSE | Upregulated |
| -0,148955071 | 0,91009063 | LINC00693 | FALSE | Downregulated |
| 0,02856934 | 0,910173646 | SLC25A42 | FALSE | Upregulated |
| -0,023789847 | 0,910209294 | FAM50A | FALSE | Downregulated |
| 0,014424557 | 0,91031744 | PCNX | FALSE | Upregulated |
| 0,046082945 | 0,910460204 | GPX3 | FALSE | Upregulated |
| 0,031735152 | 0,910525975 | BRD3 | FALSE | Upregulated |
| -0,213853928 | 0,910525975 | LOC642249 | FALSE | Downregulated |
| -0,9201944 | 0,910525975 | SLN | FALSE | Downregulated |
| -0,036105838 | 0,910636454 | AHI1 | FALSE | Downregulated |
| -0,169710406 | 0,910742995 | LOC102723390 | FALSE | Downregulated |
| 0,055744127 | 0,910864486 | LOC101929668 | FALSE | Upregulated |
| 0,019820783 | 0,910864486 | NAB1 | FALSE | Upregulated |
| 0,070773722 | 0,910901149 | LINC00884 | FALSE | Upregulated |
| 0,050644339 | 0,91109699 | CD274 | FALSE | Upregulated |
| 0,029761007 | 0,91109699 | DPP4 | FALSE | Upregulated |
| 0,031189692 | 0,911169393 | TBC1D2B | FALSE | Upregulated |
| 0,039280426 | 0,911343041 | C1orf54 | FALSE | Upregulated |
| 0,186250908 | 0,911343041 | HIST2H3D | FALSE | Upregulated |
| 0,01593668 | 0,91141015 | PPP4R2 | FALSE | Upregulated |
| 0,093390188 | 0,911541885 | NAALADL1 | FALSE | Upregulated |
| -0,051428636 | 0,911550628 | FAM186B | FALSE | Downregulated |
| 0,041768854 | 0,911550628 | SLC2A9 | FALSE | Upregulated |
| -0,088490258 | 0,911612852 | LOC101927520 | FALSE | Downregulated |
| 0,137942402 | 0,911612852 | SAP30L-AS1 | FALSE | Upregulated |
| -0,170153789 | 0,911674495 | C10orf111 | FALSE | Downregulated |
| 0,146841748 | 0,911674495 | CA8 | FALSE | Upregulated |
| 0,098525322 | 0,911674495 | STXBP5-AS1 | FALSE | Upregulated |
| -0,051267719 | 0,911674495 | UNC5CL | FALSE | Downregulated |
| -0,04521297 | 0,911674495 | WASF3 | FALSE | Downregulated |
| 0,031113398 | 0,911766641 | CAPN10 | FALSE | Upregulated |
| -0,021199603 | 0,911855141 | LLGL2 | FALSE | Downregulated |
| -0,061448893 | 0,912202028 | PRKD1 | FALSE | Downregulated |
| -0,039498039 | 0,912413207 | CD86 | FALSE | Downregulated |
| -0,163984019 | 0,912413207 | LRRC63 | FALSE | Downregulated |
| 0,035996059 | 0,912413207 | PGM2L1 | FALSE | Upregulated |
| 0,027772961 | 0,912413207 | PHF1 | FALSE | Upregulated |
| -0,136938505 | 0,912413207 | SLED1 | FALSE | Downregulated |
| 0,014142471 | 0,912619186 | UMPS | FALSE | Upregulated |
| 0,169333408 | 0,91276005 | LINC00675 | FALSE | Upregulated |
| 0,104518703 | 0,912764182 | UNQ6494 | FALSE | Upregulated |
| -0,034887569 | 0,912770072 | BCL6 | FALSE | Downregulated |
| 0,018463529 | 0,912770072 | IL6ST | FALSE | Upregulated |
| -0,023490572 | 0,912770072 | KRT10 | FALSE | Downregulated |
| -0,125601109 | 0,912770072 | LOC102724584 | FALSE | Downregulated |
| -0,017477212 | 0,912770072 | MTF2 | FALSE | Downregulated |

| | | | | |
|---|---|---|---|---|
| -0,15997722 | 0,912770072 | RCVRN | FALSE | Downregulated |
| 0,068407077 | 0,912770072 | SDCBP2 | FALSE | Upregulated |
| 0,024800678 | 0,912770072 | TXNDC17 | FALSE | Upregulated |
| -0,012662727 | 0,912770072 | WBP11 | FALSE | Downregulated |
| 0,064245082 | 0,912869 | LINC00969 | FALSE | Upregulated |
| 0,133111338 | 0,912869 | LINC01105 | FALSE | Upregulated |
| -0,035629296 | 0,913014295 | CTH | FALSE | Downregulated |
| -0,201733075 | 0,913014295 | DCST1 | FALSE | Downregulated |
| 0,236142884 | 0,913014295 | FUT9 | FALSE | Upregulated |
| 0,076498113 | 0,913014295 | IQCJ-SCHIP1 | FALSE | Upregulated |
| 0,09473602 | 0,913014295 | LOC654342 | FALSE | Upregulated |
| -0,019848883 | 0,913014295 | PTPMT1 | FALSE | Downregulated |
| 0,02184851 | 0,913014295 | TIPARP | FALSE | Upregulated |
| -0,022710774 | 0,913014295 | UBA7 | FALSE | Downregulated |
| 0,19359742 | 0,913014295 | ZNF492 | FALSE | Upregulated |
| -0,020420085 | 0,913020704 | EEF2K | FALSE | Downregulated |
| -0,024311114 | 0,913020704 | TEX264 | FALSE | Downregulated |
| 0,025165419 | 0,913084463 | DCTN5 | FALSE | Upregulated |
| -0,061009906 | 0,913113637 | LOC283788 | FALSE | Downregulated |
| -0,018114722 | 0,913118744 | TXLNA | FALSE | Downregulated |
| 0,031048024 | 0,913121228 | SLC22A3 | FALSE | Upregulated |
| -0,017120806 | 0,913124043 | C6orf106 | FALSE | Downregulated |
| -0,033297986 | 0,913124043 | FUT10 | FALSE | Downregulated |
| -0,086666742 | 0,913124043 | LOC102724940 | FALSE | Downregulated |
| 0,01743617 | 0,913213909 | MPPE1 | FALSE | Upregulated |
| -0,133613136 | 0,913261369 | LOC102723352 | FALSE | Downregulated |
| -0,029824686 | 0,913312349 | RUSC1 | FALSE | Downregulated |
| 0,204595852 | 0,913312349 | VENTXP7 | FALSE | Upregulated |
| -0,023461248 | 0,913316159 | JMY | FALSE | Downregulated |
| 0,177329068 | 0,913316159 | SLC6A2 | FALSE | Upregulated |
| 0,029833251 | 0,913316159 | ZNF518B | FALSE | Upregulated |
| 0,105191101 | 0,913325058 | TMEM233 | FALSE | Upregulated |
| 0,065849395 | 0,913420454 | LOC101928524 | FALSE | Upregulated |
| -0,022393556 | 0,913517138 | TAF10 | FALSE | Downregulated |
| -0,204411907 | 0,913774354 | LOC101927568 | FALSE | Downregulated |
| 0,256366538 | 0,913774354 | SYNGR4 | FALSE | Upregulated |
| 0,067296151 | 0,913875656 | WDR96 | FALSE | Upregulated |
| -0,064574931 | 0,914100245 | BMPR1B | FALSE | Downregulated |
| -0,018463773 | 0,914262794 | ZNF7 | FALSE | Downregulated |
| -0,022513614 | 0,914378185 | ERCC4 | FALSE | Downregulated |
| 0,060527649 | 0,914378185 | PDCD4-AS1 | FALSE | Upregulated |
| 0,030880844 | 0,914378185 | ZBTB37 | FALSE | Upregulated |
| 0,046240197 | 0,914497234 | IGFLR1 | FALSE | Upregulated |
| -0,02626609 | 0,914591711 | KDM6B | FALSE | Downregulated |
| 0,01875419 | 0,914591711 | NR2F2 | FALSE | Upregulated |
| 0,166288055 | 0,914830658 | CXorf30 | FALSE | Upregulated |
| -0,016487379 | 0,914873309 | EP300 | FALSE | Downregulated |
| 0,013449307 | 0,914873309 | HNRNPR | FALSE | Upregulated |
| 0,116558447 | 0,914873309 | LOC102723820 | FALSE | Upregulated |
| -0,056220426 | 0,914873309 | LRFN1 | FALSE | Downregulated |
| 0,049879151 | 0,914873309 | MGC16275 | FALSE | Upregulated |

| | | | | |
|---|---|---|---|---|
| -0,017874795 | 0,914873309 | OSGIN2 | FALSE | Downregulated |
| -0,043604124 | 0,914933964 | LOC101928126 | FALSE | Downregulated |
| -0,016798791 | 0,914933964 | NT5C2 | FALSE | Downregulated |
| 0,022441455 | 0,914933964 | PMS1 | FALSE | Upregulated |
| 0,872796623 | 0,914954633 | CACNA1S | FALSE | Upregulated |
| -0,022210203 | 0,914954633 | DVL1 | FALSE | Downregulated |
| -0,018356738 | 0,914954633 | ZNF180 | FALSE | Downregulated |
| -0,016557396 | 0,914980433 | GMPR2 | FALSE | Downregulated |
| -0,029076908 | 0,915034363 | FASTKD3 | FALSE | Downregulated |
| 0,073087922 | 0,915094224 | FAM115C | FALSE | Upregulated |
| 0,021580523 | 0,915094224 | GTPBP8 | FALSE | Upregulated |
| -0,225077524 | 0,915094224 | LOC102724536 | FALSE | Downregulated |
| 0,051607653 | 0,915094224 | PFKM | FALSE | Upregulated |
| -0,104989723 | 0,915112845 | NAT8B | FALSE | Downregulated |
| -0,053995662 | 0,915202588 | CMPK2 | FALSE | Downregulated |
| 0,018908589 | 0,915202588 | DDX21 | FALSE | Upregulated |
| 0,024897622 | 0,915202588 | FAM127A | FALSE | Upregulated |
| -0,081701269 | 0,915202588 | HTATSF1P2 | FALSE | Downregulated |
| 0,189975339 | 0,915202588 | LOC101928407 | FALSE | Upregulated |
| 0,179549319 | 0,915319793 | ODF3L2 | FALSE | Upregulated |
| -0,107568488 | 0,915419737 | SPTB | FALSE | Downregulated |
| -0,198934518 | 0,915438712 | RP11-82L2.1 | FALSE | Downregulated |
| 0,182466527 | 0,915438712 | SNORA41 | FALSE | Upregulated |
| -0,025453253 | 0,915464285 | LINC00984 | FALSE | Downregulated |
| -0,017064169 | 0,915788671 | RUVBL2 | FALSE | Downregulated |
| -0,041121213 | 0,915788671 | WNT5A | FALSE | Downregulated |
| 0,030912387 | 0,916107208 | CHCHD6 | FALSE | Upregulated |
| -0,076325685 | 0,916107208 | LOC102724190 | FALSE | Downregulated |
| 0,024309791 | 0,916176054 | NUGGC | FALSE | Upregulated |
| 0,084488064 | 0,916239072 | LOC101928882 | FALSE | Upregulated |
| -0,01769987 | 0,916589086 | AZI2 | FALSE | Downregulated |
| 0,115901277 | 0,916756779 | LOC285389 | FALSE | Upregulated |
| -0,774155648 | 0,916790268 | ATP1B4 | FALSE | Downregulated |
| -0,024190129 | 0,916790268 | TMEM251 | FALSE | Downregulated |
| 0,019615379 | 0,917291843 | ARID1A | FALSE | Upregulated |
| -0,164741331 | 0,917291843 | LOC102724883 | FALSE | Downregulated |
| 0,032463861 | 0,917291843 | TTC26 | FALSE | Upregulated |
| -0,035990757 | 0,917388293 | HMOX1 | FALSE | Downregulated |
| 0,025963732 | 0,917395075 | LOC550643 | FALSE | Upregulated |
| -0,097375199 | 0,917475952 | LOC101928281 | FALSE | Downregulated |
| -0,068386884 | 0,917528469 | LOC101927850 | FALSE | Downregulated |
| -0,131818731 | 0,917848888 | JPH1 | FALSE | Downregulated |
| -0,03480106 | 0,918016617 | SLC23A2 | FALSE | Downregulated |
| -0,101068765 | 0,918040585 | DCHS2 | FALSE | Downregulated |
| -0,07733748 | 0,918040585 | LOC101927086 | FALSE | Downregulated |
| 0,067199547 | 0,918040585 | PPIEL | FALSE | Upregulated |
| -0,021960771 | 0,918040585 | ZMYND19 | FALSE | Downregulated |
| -0,074896708 | 0,91821149 | LOC101929374 | FALSE | Downregulated |
| 0,037275468 | 0,918790223 | ARMCX4 | FALSE | Upregulated |
| 0,022764732 | 0,918790223 | CHCHD5 | FALSE | Upregulated |
| -0,018198687 | 0,918790223 | TTF1 | FALSE | Downregulated |

| | | | | |
|---|---|---|---|---|
| 0,03841119 | 0,918895759 | PRR24 | FALSE | Upregulated |
| -0,040468809 | 0,919076352 | NAP1L3 | FALSE | Downregulated |
| 0,075182858 | 0,91910016 | STOX1 | FALSE | Upregulated |
| -0,02011333 | 0,919218954 | DHRSX | FALSE | Downregulated |
| 0,154890268 | 0,919435213 | SCT | FALSE | Upregulated |
| -0,030740668 | 0,91958292 | GMPR | FALSE | Downregulated |
| 0,025564276 | 0,91958292 | GPR137 | FALSE | Upregulated |
| -0,042636523 | 0,919775475 | PDSS1 | FALSE | Downregulated |
| -0,017101989 | 0,919781837 | UHRF2 | FALSE | Downregulated |
| -0,03237811 | 0,919899566 | PGBD5 | FALSE | Downregulated |
| -0,278687463 | 0,91996706 | LOC102724956 | FALSE | Downregulated |
| -0,026746818 | 0,920073437 | FEM1A | FALSE | Downregulated |
| -0,209614972 | 0,92016626 | LOC101928921 | FALSE | Downregulated |
| 0,03480791 | 0,92016626 | LRRC4 | FALSE | Upregulated |
| 0,348898007 | 0,92016626 | NPTN-IT1 | FALSE | Upregulated |
| -0,074499776 | 0,920418402 | ATP8B5P | FALSE | Downregulated |
| -0,164185354 | 0,920418402 | LOC101928530 | FALSE | Downregulated |
| 0,019084891 | 0,920512633 | SPR | FALSE | Upregulated |
| 0,173228414 | 0,920707604 | LOC102723891 | FALSE | Upregulated |
| -0,143939872 | 0,920808966 | LOC101928765 | FALSE | Downregulated |
| -0,821061837 | 0,920845309 | MYH1 | FALSE | Downregulated |
| -0,06339405 | 0,920845309 | SYT10 | FALSE | Downregulated |
| -0,028648254 | 0,920882694 | CCDC149 | FALSE | Downregulated |
| 0,224038774 | 0,920882694 | RP11-536C5.7 | FALSE | Upregulated |
| -0,011206607 | 0,920882694 | ZFAND3 | FALSE | Downregulated |
| -0,823796103 | 0,921051246 | TRNAV4 | FALSE | Downregulated |
| -0,161186926 | 0,921142346 | LOC100505622 | FALSE | Downregulated |
| 0,143154261 | 0,921142346 | TNFAIP6 | FALSE | Upregulated |
| 0,176738282 | 0,92128414 | GALNTL6 | FALSE | Upregulated |
| 0,016458663 | 0,921384945 | MYO18A | FALSE | Upregulated |
| 0,133031788 | 0,921384945 | NOS1 | FALSE | Upregulated |
| -0,022700744 | 0,921384945 | SHPRH | FALSE | Downregulated |
| -0,053895398 | 0,921384945 | SPATA41 | FALSE | Downregulated |
| 0,341001344 | 0,921384945 | TRNAH4 | FALSE | Upregulated |
| -0,022148876 | 0,921384945 | TSEN54 | FALSE | Downregulated |
| -0,01678072 | 0,921384945 | ZFPL1 | FALSE | Downregulated |
| -0,021777837 | 0,921384945 | ZNF557 | FALSE | Downregulated |
| 0,119662971 | 0,921451715 | CNTN5 | FALSE | Upregulated |
| -0,017325282 | 0,92157816 | GTF3C6 | FALSE | Downregulated |
| -0,224923546 | 0,92157816 | LOC100996717 | FALSE | Downregulated |
| -0,053242246 | 0,92157816 | P4HA1 | FALSE | Downregulated |
| -0,028467197 | 0,921793251 | RHOBTB1 | FALSE | Downregulated |
| -0,025589319 | 0,921793251 | YDJC | FALSE | Downregulated |
| -0,034301393 | 0,921814316 | KLHL11 | FALSE | Downregulated |
| -0,033391136 | 0,922006386 | ZNHIT2 | FALSE | Downregulated |
| -0,036599686 | 0,922190461 | RAB15 | FALSE | Downregulated |
| -0,055447664 | 0,922195449 | CRIP3 | FALSE | Downregulated |
| 0,10506231 | 0,92231015 | NAPSA | FALSE | Upregulated |
| 0,024239863 | 0,92231015 | TLE1 | FALSE | Upregulated |
| -0,019456094 | 0,92231015 | ZNF184 | FALSE | Downregulated |
| -0,153118394 | 0,922353647 | ANKRD20A1 | FALSE | Downregulated |

| | | | | |
|---|---|---|---|---|
| 0,03359199 | 0,922353647 | C19orf48 | FALSE | Upregulated |
| -0,07418004 | 0,922353647 | CCDC173 | FALSE | Downregulated |
| 0,013742072 | 0,922353647 | CPNE3 | FALSE | Upregulated |
| -0,014332237 | 0,922353647 | ERAL1 | FALSE | Downregulated |
| 0,044426795 | 0,922353647 | LOC101927160 | FALSE | Upregulated |
| -0,642104655 | 0,922353647 | MIR1184-1 | FALSE | Downregulated |
| -0,021147468 | 0,922353647 | PGRMC1 | FALSE | Downregulated |
| -0,081468152 | 0,922353647 | PRKG2 | FALSE | Downregulated |
| 0,032186623 | 0,922353647 | SPATA33 | FALSE | Upregulated |
| 0,053856387 | 0,922353647 | TPI1P2 | FALSE | Upregulated |
| 0,024093917 | 0,922353647 | TRIM14 | FALSE | Upregulated |
| -0,016116745 | 0,922354551 | ACVR1B | FALSE | Downregulated |
| -0,0319717 | 0,922354551 | APBB1IP | FALSE | Downregulated |
| -0,022708409 | 0,922354551 | IRF2BPL | FALSE | Downregulated |
| 0,239337334 | 0,922354551 | KRT1 | FALSE | Upregulated |
| -0,071184089 | 0,922354551 | LOC100507656 | FALSE | Downregulated |
| 0,063373399 | 0,922354551 | LOC101928719 | FALSE | Upregulated |
| 0,02085981 | 0,922354551 | MRTO4 | FALSE | Upregulated |
| 0,102615718 | 0,922354551 | REM1 | FALSE | Upregulated |
| -0,023060187 | 0,922354551 | SP140L | FALSE | Downregulated |
| -0,111516308 | 0,922354551 | TRIM7 | FALSE | Downregulated |
| -0,014934125 | 0,922538045 | NCAPD2 | FALSE | Downregulated |
| 0,019420253 | 0,922564536 | C11orf73 | FALSE | Upregulated |
| -0,050479398 | 0,922564536 | KLRG1 | FALSE | Downregulated |
| -0,020495469 | 0,922564536 | PPM1G | FALSE | Downregulated |
| 0,045942722 | 0,922564536 | RP11-345L23.1 | FALSE | Upregulated |
| 0,168405352 | 0,922717654 | LINC00958 | FALSE | Upregulated |
| 0,048768677 | 0,923105604 | AHSA2 | FALSE | Upregulated |
| 0,015954201 | 0,923105604 | RAB2B | FALSE | Upregulated |
| 0,094973356 | 0,923122113 | LOC102723993 | FALSE | Upregulated |
| 0,072154367 | 0,923152464 | HCG16 | FALSE | Upregulated |
| -0,296149447 | 0,923203087 | GSTT2 | FALSE | Downregulated |
| 0,047098049 | 0,923203087 | LOC728407 | FALSE | Upregulated |
| 0,019884826 | 0,923203087 | ZNF292 | FALSE | Upregulated |
| 0,012650126 | 0,923280576 | DLST | FALSE | Upregulated |
| -0,016904675 | 0,923386203 | AGFG2 | FALSE | Downregulated |
| -0,068863463 | 0,923607096 | LOC101928111 | FALSE | Downregulated |
| -0,158573476 | 0,923922923 | SNORD6 | FALSE | Downregulated |
| 0,00994488 | 0,923976618 | BECN1 | FALSE | Upregulated |
| -0,080995299 | 0,923976618 | LOC102723597 | FALSE | Downregulated |
| 0,09471923 | 0,923976618 | LOC102723958 | FALSE | Upregulated |
| -0,032823696 | 0,923976618 | LTB4R | FALSE | Downregulated |
| 0,045967329 | 0,924035062 | RP11-425I13.3 | FALSE | Upregulated |
| -0,015465572 | 0,924170615 | CDC42BPB | FALSE | Downregulated |
| -0,04064619 | 0,924442055 | C11orf48 | FALSE | Downregulated |
| -0,025221096 | 0,92447071 | PUS7 | FALSE | Downregulated |
| 0,046457675 | 0,924709353 | FAM86FP | FALSE | Upregulated |
| 0,038459346 | 0,924760304 | RAMP2 | FALSE | Upregulated |
| -0,118720498 | 0,924829533 | TMEM179 | FALSE | Downregulated |
| 0,221773427 | 0,92485174 | ATP2B3 | FALSE | Upregulated |
| 0,107482369 | 0,925038709 | SLCO1A2 | FALSE | Upregulated |

| | | | | |
|---|---|---|---|---|
| -0,016189126 | 0,925085024 | CPQ | FALSE | Downregulated |
| 0,14678655 | 0,925264503 | FAM159A | FALSE | Upregulated |
| 0,019680349 | 0,925264503 | SMCHD1 | FALSE | Upregulated |
| 0,09424093 | 0,925264503 | SMG1P1 | FALSE | Upregulated |
| -0,059969479 | 0,925264503 | ZYG11A | FALSE | Downregulated |
| 0,015729174 | 0,925272805 | ELK4 | FALSE | Upregulated |
| 0,148722634 | 0,925272805 | LOC102724077 | FALSE | Upregulated |
| 0,209818431 | 0,925272805 | LOC102724488 | FALSE | Upregulated |
| -0,114909404 | 0,925272805 | OCLM | FALSE | Downregulated |
| 0,029613985 | 0,925289437 | GJB2 | FALSE | Upregulated |
| 0,202891508 | 0,925289437 | PI3 | FALSE | Upregulated |
| -0,015183874 | 0,925289437 | SMG8 | FALSE | Downregulated |
| -0,027353572 | 0,925456762 | TNK2 | FALSE | Downregulated |
| -0,019223448 | 0,925662411 | SYNGR2 | FALSE | Downregulated |
| -0,769570744 | 0,925674451 | GTF2IP1 | FALSE | Downregulated |
| 0,015604156 | 0,925688926 | DNAJC13 | FALSE | Upregulated |
| 0,037519715 | 0,925754461 | KLHDC7A | FALSE | Upregulated |
| -0,145230555 | 0,925780402 | LOC101928838 | FALSE | Downregulated |
| 0,017061014 | 0,925789486 | ZCCHC11 | FALSE | Upregulated |
| -0,012197052 | 0,925872435 | ASUN | FALSE | Downregulated |
| 0,013297498 | 0,926162282 | SNX1 | FALSE | Upregulated |
| 0,048447473 | 0,926327623 | FOXD2-AS1 | FALSE | Upregulated |
| 0,162446319 | 0,92647955 | LOC101928993 | FALSE | Upregulated |
| 0,049601668 | 0,926494172 | KCND1 | FALSE | Upregulated |
| 0,012435039 | 0,926624613 | RIPK1 | FALSE | Upregulated |
| 0,029413676 | 0,926624613 | TRIP10 | FALSE | Upregulated |
| -0,029160457 | 0,926712086 | AKR1C1 | FALSE | Downregulated |
| -0,106252343 | 0,926735859 | LOC100505534 | FALSE | Downregulated |
| -0,032925207 | 0,926863378 | SNHG3 | FALSE | Downregulated |
| -0,018111014 | 0,926958102 | AKAP10 | FALSE | Downregulated |
| 0,035288949 | 0,926958102 | KCNMB3 | FALSE | Upregulated |
| -0,046610363 | 0,926958102 | ZNF404 | FALSE | Downregulated |
| -0,042015225 | 0,926985388 | LOC101928000 | FALSE | Downregulated |
| -0,052862029 | 0,926988787 | LOC102724770 | FALSE | Downregulated |
| -0,761545399 | 0,927081435 | RNA5S17 | FALSE | Downregulated |
| 0,178512276 | 0,927081435 | RP11-319G6.1 | FALSE | Upregulated |
| -0,042787613 | 0,9271129 | CENPO | FALSE | Downregulated |
| 0,087647535 | 0,927306171 | C9orf169 | FALSE | Upregulated |
| 0,021519913 | 0,927306171 | IQCB1 | FALSE | Upregulated |
| -0,011876273 | 0,927306171 | TFAM | FALSE | Downregulated |
| -0,037702717 | 0,927306171 | ZNF501 | FALSE | Downregulated |
| 0,039525476 | 0,927307259 | DHODH | FALSE | Upregulated |
| -0,055837097 | 0,92745332 | ANXA13 | FALSE | Downregulated |
| 0,018306974 | 0,92745332 | RABL2B | FALSE | Upregulated |
| -0,037307702 | 0,927458902 | TMEM81 | FALSE | Downregulated |
| 0,020846874 | 0,927473097 | ASXL2 | FALSE | Upregulated |
| 0,155100799 | 0,927473097 | DMC1 | FALSE | Upregulated |
| 0,030994686 | 0,927473097 | HSPA1L | FALSE | Upregulated |
| 0,014756311 | 0,927473097 | KRI1 | FALSE | Upregulated |
| 0,033959822 | 0,927473097 | LAPTM4B | FALSE | Upregulated |
| 0,080388782 | 0,927473097 | LOC407835 | FALSE | Upregulated |

| | | | | |
|---|---|---|---|---|
| 0,022989328 | 0,927473097 | MPHOSPH8 | FALSE | Upregulated |
| 0,035288679 | 0,927473097 | PDCD1LG2 | FALSE | Upregulated |
| -0,01408294 | 0,927473097 | RNF167 | FALSE | Downregulated |
| -0,049531558 | 0,927473097 | TLX1 | FALSE | Downregulated |
| 0,013193619 | 0,927473097 | UBQLN4 | FALSE | Upregulated |
| -0,029364715 | 0,927473097 | ZNF10 | FALSE | Downregulated |
| -0,019877164 | 0,927473097 | ZNF230 | FALSE | Downregulated |
| 0,145896841 | 0,927509974 | SNORD35B | FALSE | Upregulated |
| 0,055101461 | 0,927524878 | FPR2 | FALSE | Upregulated |
| 0,069308553 | 0,927621587 | AGAP11 | FALSE | Upregulated |
| 0,016351477 | 0,92786855 | PGLS | FALSE | Upregulated |
| 0,656526227 | 0,927961314 | ANKRD2 | FALSE | Upregulated |
| 0,741385681 | 0,927962875 | HIST2H2AA3 | FALSE | Upregulated |
| -0,019771849 | 0,928105782 | NDUFC1 | FALSE | Downregulated |
| -0,019282973 | 0,92810705 | ATP6V0C | FALSE | Downregulated |
| -0,018382969 | 0,92810705 | IFFO1 | FALSE | Downregulated |
| -0,019610426 | 0,928135144 | CDK6 | FALSE | Downregulated |
| 0,043755268 | 0,928135144 | ZNF423 | FALSE | Upregulated |
| -0,116914949 | 0,928438196 | CALCR | FALSE | Downregulated |
| 0,082875559 | 0,928438196 | FAM45B | FALSE | Upregulated |
| -0,043722752 | 0,928438196 | MCF2L-AS1 | FALSE | Downregulated |
| -0,104616267 | 0,928438196 | RHCG | FALSE | Downregulated |
| 0,013820055 | 0,928440574 | ZNF12 | FALSE | Upregulated |
| 0,019564968 | 0,92857453 | C2orf68 | FALSE | Upregulated |
| 0,034613087 | 0,92857453 | FILIP1 | FALSE | Upregulated |
| 0,073006192 | 0,92857453 | FLJ42351 | FALSE | Upregulated |
| -0,063117193 | 0,928740461 | LINC00893 | FALSE | Downregulated |
| 0,150753477 | 0,928764365 | NALCN-AS1 | FALSE | Upregulated |
| 0,011930505 | 0,928860829 | VPS26B | FALSE | Upregulated |
| -0,215034347 | 0,929069651 | CLCNKA | FALSE | Downregulated |
| 0,05777703 | 0,929284347 | OTUD7A | FALSE | Upregulated |
| 0,015995056 | 0,929284347 | TCF3 | FALSE | Upregulated |
| -0,01449592 | 0,929335362 | EIF2B4 | FALSE | Downregulated |
| 0,08309121 | 0,929704876 | LOC102724748 | FALSE | Upregulated |
| 0,151953014 | 0,929834166 | AC010091.1 | FALSE | Upregulated |
| -0,019000149 | 0,929834166 | POLD4 | FALSE | Downregulated |
| 0,018744685 | 0,929895869 | NUP37 | FALSE | Upregulated |
| 0,013168847 | 0,929929779 | ATF7 | FALSE | Upregulated |
| 0,02770881 | 0,929929779 | PER2 | FALSE | Upregulated |
| 0,025639621 | 0,930128283 | HIST1H2BK | FALSE | Upregulated |
| -0,010503161 | 0,930450894 | IDH3B | FALSE | Downregulated |
| 0,012824716 | 0,930450894 | PPP1R21 | FALSE | Upregulated |
| -0,018233674 | 0,930450894 | TCF7L2 | FALSE | Downregulated |
| -0,012422405 | 0,930450894 | ZCCHC3 | FALSE | Downregulated |
| -0,024632528 | 0,930518834 | TM4SF18 | FALSE | Downregulated |
| 0,033233278 | 0,930533856 | WDR66 | FALSE | Upregulated |
| -0,037842241 | 0,930589368 | CRISPLD1 | FALSE | Downregulated |
| -0,085202837 | 0,930589368 | FBXO22-AS1 | FALSE | Downregulated |
| -0,120685448 | 0,930589368 | PRRT4 | FALSE | Downregulated |
| -0,146400146 | 0,930589368 | RP11-10A14.5 | FALSE | Downregulated |
| -0,061410692 | 0,930589368 | TPTEP1 | FALSE | Downregulated |

| | | | | |
|---|---|---|---|---|
| 0,087655664 | 0,93062685 | LOC100996425 | FALSE | Upregulated |
| 0,15963506 | 0,930639033 | ABHD12B | FALSE | Upregulated |
| -0,139446907 | 0,930967273 | DNAJB7 | FALSE | Downregulated |
| -0,025726585 | 0,930967273 | LOC100287896 | FALSE | Downregulated |
| -0,04171369 | 0,930967273 | LRRC37A2 | FALSE | Downregulated |
| 0,16986745 | 0,930967273 | RP11-544M22.1 | FALSE | Upregulated |
| 0,035290555 | 0,930967273 | USP54 | FALSE | Upregulated |
| 0,015210886 | 0,931027637 | CNOT10 | FALSE | Upregulated |
| 0,069045628 | 0,931028268 | KIAA1652 | FALSE | Upregulated |
| 0,058811398 | 0,931075278 | CHKB-AS1 | FALSE | Upregulated |
| 0,047521655 | 0,931126561 | FAM107A | FALSE | Upregulated |
| 0,11320972 | 0,931126561 | LOC101928583 | FALSE | Upregulated |
| 0,017305294 | 0,931126561 | RPL7 | FALSE | Upregulated |
| -0,71133232 | 0,931126561 | TRDN | FALSE | Downregulated |
| 0,071910937 | 0,931357064 | TMEM121 | FALSE | Upregulated |
| 0,121987344 | 0,931376893 | BCRP2 | FALSE | Upregulated |
| -0,167936297 | 0,931400187 | HAPLN1 | FALSE | Downregulated |
| -0,015998052 | 0,931400187 | LMAN2 | FALSE | Downregulated |
| 0,146529588 | 0,931400187 | LOC102724759 | FALSE | Upregulated |
| 0,042975676 | 0,931400187 | MANF | FALSE | Upregulated |
| 0,56625941 | 0,931400187 | MYLK2 | FALSE | Upregulated |
| 0,064216027 | 0,931400187 | NR2F2-AS1 | FALSE | Upregulated |
| -0,026006736 | 0,931400187 | PTPDC1 | FALSE | Downregulated |
| -0,027917145 | 0,931434674 | ZNF708 | FALSE | Downregulated |
| 0,031720039 | 0,931456435 | GMNC | FALSE | Upregulated |
| 0,097430721 | 0,931560184 | C2CD4C | FALSE | Upregulated |
| -0,029679014 | 0,931560184 | ERCC6-PGBD3 | FALSE | Downregulated |
| 0,019830398 | 0,931569082 | SUCO | FALSE | Upregulated |
| -0,020521079 | 0,931614365 | GEMIN2 | FALSE | Downregulated |
| -0,022432588 | 0,931614365 | HGFAC | FALSE | Downregulated |
| 0,171273091 | 0,931614365 | KCNE1L | FALSE | Upregulated |
| 0,1024848 | 0,931614365 | LOC102724012 | FALSE | Upregulated |
| -0,036793951 | 0,931638102 | CDKL1 | FALSE | Downregulated |
| 0,018277594 | 0,931779191 | CCHCR1 | FALSE | Upregulated |
| -0,635688084 | 0,931899197 | GSTM1 | FALSE | Downregulated |
| 0,182268423 | 0,932211509 | C20orf166-AS1 | FALSE | Upregulated |
| 0,021584941 | 0,932211509 | MYADM | FALSE | Upregulated |
| -0,016021697 | 0,932577772 | LOC339803 | FALSE | Downregulated |
| -0,03937972 | 0,933005297 | LILRB1 | FALSE | Downregulated |
| 0,015279142 | 0,933005297 | UPRT | FALSE | Upregulated |
| -0,037540453 | 0,933096758 | MARCO | FALSE | Downregulated |
| -0,03466783 | 0,933111977 | C19orf38 | FALSE | Downregulated |
| 0,126566972 | 0,933111977 | KIR2DL3 | FALSE | Upregulated |
| -0,020395027 | 0,933111977 | RILPL1 | FALSE | Downregulated |
| -0,030483625 | 0,933111977 | SAMD10 | FALSE | Downregulated |
| 0,061512561 | 0,933121414 | AKR1E2 | FALSE | Upregulated |
| -0,137535881 | 0,933123126 | AC006272.1 | FALSE | Downregulated |
| -0,03887795 | 0,933123126 | B3GNTL1 | FALSE | Downregulated |
| -0,150176247 | 0,933126575 | LOC102723458 | FALSE | Downregulated |
| -0,138981591 | 0,93338832 | EIF3CL | FALSE | Downregulated |
| -0,069526816 | 0,93338832 | RP5-1074L1.1 | FALSE | Downregulated |

| | | | | |
|---|---|---|---|---|
| -0,016190323 | 0,93338832 | SPATA5L1 | FALSE | Downregulated |
| -0,025751602 | 0,93338832 | TMCO6 | FALSE | Downregulated |
| 0,036527984 | 0,93338832 | TRNQ | FALSE | Upregulated |
| -0,153033326 | 0,933505556 | LOC102723837 | FALSE | Downregulated |
| 0,080188894 | 0,93356117 | PFN1P2 | FALSE | Upregulated |
| 0,244978675 | 0,933602511 | ANKRD20A2 | FALSE | Upregulated |
| 0,052615614 | 0,933602511 | LOC100507520 | FALSE | Upregulated |
| 0,117095651 | 0,933643973 | AURKAPS1 | FALSE | Upregulated |
| 0,017761324 | 0,933643973 | CMIP | FALSE | Upregulated |
| -0,018978645 | 0,933643973 | FAM177A1 | FALSE | Downregulated |
| -0,013997523 | 0,933643973 | GSS | FALSE | Downregulated |
| -0,015862461 | 0,933643973 | IFT122 | FALSE | Downregulated |
| 0,024987324 | 0,933643973 | KLF10 | FALSE | Upregulated |
| 0,140242971 | 0,933643973 | KRT42P | FALSE | Upregulated |
| -0,029469853 | 0,933643973 | LAGE3 | FALSE | Downregulated |
| -0,082605518 | 0,933643973 | LOC100653005 | FALSE | Downregulated |
| -0,07304979 | 0,933643973 | LOC101927932 | FALSE | Downregulated |
| -0,040111526 | 0,933643973 | LOC283070 | FALSE | Downregulated |
| 0,011882501 | 0,933643973 | MTPAP | FALSE | Upregulated |
| -0,027734577 | 0,933643973 | NUDT2 | FALSE | Downregulated |
| -0,030560581 | 0,933643973 | PCBP1-AS1 | FALSE | Downregulated |
| 0,021631571 | 0,933643973 | RASSF1 | FALSE | Upregulated |
| -0,688949703 | 0,933643973 | RNA5S1 | FALSE | Downregulated |
| -0,01780919 | 0,933643973 | RNASE4 | FALSE | Downregulated |
| -0,01623731 | 0,933643973 | RNF138 | FALSE | Downregulated |
| 0,052389719 | 0,933643973 | RP11-1223D19.1 | FALSE | Upregulated |
| -0,030578602 | 0,933643973 | SIGLEC7 | FALSE | Downregulated |
| -0,075386045 | 0,933643973 | SLC25A41 | FALSE | Downregulated |
| -0,015970175 | 0,933643973 | SPHK2 | FALSE | Downregulated |
| 0,236741554 | 0,933643973 | TMEM256-PLSCR3 | FALSE | Upregulated |
| -0,014600459 | 0,933643973 | ZNF526 | FALSE | Downregulated |
| -0,011169673 | 0,933643973 | ZNF777 | FALSE | Downregulated |
| 0,15130795 | 0,933740442 | LOC100653515 | FALSE | Upregulated |
| 0,094634042 | 0,933787252 | LOC101927479 | FALSE | Upregulated |
| -0,019334618 | 0,933787252 | SETD4 | FALSE | Downregulated |
| -0,096763511 | 0,933908501 | DSG4 | FALSE | Downregulated |
| -0,016343059 | 0,933923431 | DHRS3 | FALSE | Downregulated |
| -0,156481895 | 0,933982517 | LOC101928981 | FALSE | Downregulated |
| -0,070562288 | 0,933982517 | TEX29 | FALSE | Downregulated |
| -0,125191057 | 0,934176237 | CLEC4D | FALSE | Downregulated |
| -0,011762467 | 0,934176237 | EIF4ENIF1 | FALSE | Downregulated |
| 0,038955082 | 0,934176237 | KIAA0895L | FALSE | Upregulated |
| 0,136198405 | 0,934176237 | LEMD1-AS1 | FALSE | Upregulated |
| -0,11181389 | 0,934176237 | LOC101927308 | FALSE | Downregulated |
| 0,07358405 | 0,934176237 | LOC101927546 | FALSE | Upregulated |
| 0,03717734 | 0,934176237 | LOC728975 | FALSE | Upregulated |
| 0,029762443 | 0,934176237 | PORCN | FALSE | Upregulated |
| -0,016297774 | 0,934176237 | PSMB5 | FALSE | Downregulated |
| -0,09232258 | 0,934176237 | RP11-584P21.2 | FALSE | Downregulated |
| 0,675719846 | 0,934176237 | RPL3L | FALSE | Upregulated |
| 0,019184065 | 0,934176237 | RPS6 | FALSE | Upregulated |

| | | | | |
|---|---|---|---|---|
| 0,011037355 | 0,934176237 | UBQLN2 | FALSE | Upregulated |
| 0,067863659 | 0,934179155 | CHI3L2 | FALSE | Upregulated |
| 0,051709871 | 0,934179155 | KCNS3 | FALSE | Upregulated |
| 0,048392627 | 0,934179155 | LOC100233156 | FALSE | Upregulated |
| -0,072760353 | 0,934179155 | LOC102723589 | FALSE | Downregulated |
| -0,097070715 | 0,934179155 | NTN5 | FALSE | Downregulated |
| 0,015685265 | 0,934179155 | PTPRJ | FALSE | Upregulated |
| -0,040801084 | 0,934179155 | RPGRIP1 | FALSE | Downregulated |
| 0,024645063 | 0,93425564 | PRDM15 | FALSE | Upregulated |
| -0,068959878 | 0,93425564 | TSPEAR-AS1 | FALSE | Downregulated |
| 0,046553549 | 0,93429767 | PDE6G | FALSE | Upregulated |
| -0,020210708 | 0,934444389 | RPL10A | FALSE | Downregulated |
| -0,066660497 | 0,934445284 | LOC101929174 | FALSE | Downregulated |
| -0,058575363 | 0,93463728 | UFSP1 | FALSE | Downregulated |
| -0,100480903 | 0,934682506 | CALML3 | FALSE | Downregulated |
| -0,037070026 | 0,934682506 | CEACAM21 | FALSE | Downregulated |
| -0,035641505 | 0,934682506 | HEY1 | FALSE | Downregulated |
| -0,014187113 | 0,934682506 | LOC100506548 | FALSE | Downregulated |
| 0,054700907 | 0,934682506 | LOC101927322 | FALSE | Upregulated |
| 0,069146276 | 0,934682506 | LOC102724217 | FALSE | Upregulated |
| 0,051020028 | 0,934682506 | LOC102724814 | FALSE | Upregulated |
| -0,131580905 | 0,934682506 | RP11-374P20.4 | FALSE | Downregulated |
| 0,011501477 | 0,934682506 | TAB1 | FALSE | Upregulated |
| 0,014410462 | 0,934700633 | AGAP3 | FALSE | Upregulated |
| -0,019255651 | 0,934775429 | LMO2 | FALSE | Downregulated |
| -0,044102285 | 0,934782551 | KCNIP2 | FALSE | Downregulated |
| 0,010243123 | 0,934784373 | GTF2F1 | FALSE | Upregulated |
| -0,02939823 | 0,934784373 | NEFH | FALSE | Downregulated |
| 0,013884292 | 0,934784373 | SF3B14 | FALSE | Upregulated |
| -0,013387891 | 0,934784373 | SGTA | FALSE | Downregulated |
| 0,014679166 | 0,934879116 | CANX | FALSE | Upregulated |
| -0,012453435 | 0,934879116 | LMBRD1 | FALSE | Downregulated |
| -0,149592035 | 0,934879116 | RP11-492A10.1 | FALSE | Downregulated |
| -0,103360695 | 0,934889454 | ZNF878 | FALSE | Downregulated |
| 0,027305435 | 0,934949639 | TBC1D19 | FALSE | Upregulated |
| 0,039062128 | 0,934949639 | ZNF767 | FALSE | Upregulated |
| -0,017201495 | 0,935522591 | KLHL42 | FALSE | Downregulated |
| 0,035697508 | 0,935702879 | CMAHP | FALSE | Upregulated |
| 0,179898914 | 0,935702879 | FAM3D | FALSE | Upregulated |
| -0,023815914 | 0,935702879 | RNF144A | FALSE | Downregulated |
| -0,124407842 | 0,935702879 | SNORA7A | FALSE | Downregulated |
| -0,048092228 | 0,935714161 | LOC101929715 | FALSE | Downregulated |
| 0,655533915 | 0,935714161 | MYPN | FALSE | Upregulated |
| 0,032339007 | 0,935720394 | P2RY2 | FALSE | Upregulated |
| -0,024370309 | 0,935720394 | SFT2D2 | FALSE | Downregulated |
| 0,652474634 | 0,935842474 | CASQ1 | FALSE | Upregulated |
| 0,044638064 | 0,93589237 | KCNJ10 | FALSE | Upregulated |
| -0,063594859 | 0,93592441 | RNA28S5 | FALSE | Downregulated |
| -0,069613151 | 0,93592441 | TBX20 | FALSE | Downregulated |
| -0,045334272 | 0,935974418 | C21orf49 | FALSE | Downregulated |
| 0,057003023 | 0,935974418 | GSTM3 | FALSE | Upregulated |

| | | | | |
|---|---|---|---|---|
| 0,076598359 | 0,935974418 | LOC101926917 | FALSE | Upregulated |
| 0,145018872 | 0,935974418 | LOC102723724 | FALSE | Upregulated |
| -0,012230079 | 0,935974418 | MSL2 | FALSE | Downregulated |
| 0,04107949 | 0,935974418 | RP11-75C9.1 | FALSE | Upregulated |
| -0,011034377 | 0,936217622 | MAP3K4 | FALSE | Downregulated |
| -0,121796757 | 0,936303626 | LOC101928039 | FALSE | Downregulated |
| 0,030278214 | 0,9364827 | ELOVL6 | FALSE | Upregulated |
| -0,031239723 | 0,936489184 | CDKN2D | FALSE | Downregulated |
| 0,029542614 | 0,936489184 | SLC25A15 | FALSE | Upregulated |
| 0,019025818 | 0,936542838 | FAM110B | FALSE | Upregulated |
| -0,128788155 | 0,936619765 | RP11-383C5.3 | FALSE | Downregulated |
| 0,011312532 | 0,936668953 | LARS | FALSE | Upregulated |
| -0,011818414 | 0,936668953 | LMO4 | FALSE | Downregulated |
| -0,044931449 | 0,937144516 | LINC00939 | FALSE | Downregulated |
| 0,138394272 | 0,937158528 | LGALS12 | FALSE | Upregulated |
| -0,116369028 | 0,937213906 | LOC102724122 | FALSE | Downregulated |
| -0,451590048 | 0,937253964 | TRNAH7 | FALSE | Downregulated |
| 0,043973987 | 0,937419836 | LOC100506258 | FALSE | Upregulated |
| -0,10758762 | 0,937460009 | LOC100287166 | FALSE | Downregulated |
| -0,032747095 | 0,937647806 | CXCR2P1 | FALSE | Downregulated |
| 0,066833045 | 0,93772654 | TRPV3 | FALSE | Upregulated |
| 0,021272146 | 0,937933586 | SLC27A3 | FALSE | Upregulated |
| -0,059576807 | 0,937933586 | SUGT1P1 | FALSE | Downregulated |
| 0,072311823 | 0,937960094 | ZNF321P | FALSE | Upregulated |
| -0,019529348 | 0,938027067 | HDAC10 | FALSE | Downregulated |
| -0,104149164 | 0,938027067 | OVCH1 | FALSE | Downregulated |
| 0,116142749 | 0,938265607 | RADIL | FALSE | Upregulated |
| -0,018247076 | 0,938279654 | MYO1E | FALSE | Downregulated |
| 0,077846668 | 0,938279654 | RP11-57H12.3 | FALSE | Upregulated |
| -0,065835008 | 0,938291803 | HIF3A | FALSE | Downregulated |
| -0,06389749 | 0,938291803 | LOC102724163 | FALSE | Downregulated |
| 0,025035251 | 0,938291803 | LOC647859 | FALSE | Upregulated |
| 0,060045328 | 0,938669568 | ASH1L-AS1 | FALSE | Upregulated |
| 0,011603328 | 0,938734454 | TRIM23 | FALSE | Upregulated |
| 0,037121033 | 0,938845384 | VWCE | FALSE | Upregulated |
| 0,012303951 | 0,938954732 | CNTROB | FALSE | Upregulated |
| -0,013964096 | 0,938954732 | COQ4 | FALSE | Downregulated |
| 0,045273091 | 0,938980939 | CNTN1 | FALSE | Upregulated |
| 0,019591825 | 0,939091458 | AMMECR1L | FALSE | Upregulated |
| -0,437647718 | 0,939091458 | LOC102723360 | FALSE | Downregulated |
| 0,016214002 | 0,939091458 | MARCH2 | FALSE | Upregulated |
| -0,012784242 | 0,939091458 | NUB1 | FALSE | Downregulated |
| -0,170589707 | 0,939312989 | LOC101928628 | FALSE | Downregulated |
| 0,01550623 | 0,939312989 | THAP11 | FALSE | Upregulated |
| 0,027803818 | 0,939464236 | LRRC6 | FALSE | Upregulated |
| 0,198922671 | 0,939464236 | PCSK2 | FALSE | Upregulated |
| 0,032016196 | 0,93977826 | STEAP2 | FALSE | Upregulated |
| -0,016731221 | 0,939901035 | NELFE | FALSE | Downregulated |
| 0,050268836 | 0,939962205 | MST1P2 | FALSE | Upregulated |
| 0,054121542 | 0,939962205 | NEAT1 | FALSE | Upregulated |
| 0,01678737 | 0,939962205 | PPP1R16A | FALSE | Upregulated |

| | | | | |
|---|---|---|---|---|
| -0,013306007 | 0,939973533 | ADCK3 | FALSE | Downregulated |
| -0,069815408 | 0,939973533 | LOC101927192 | FALSE | Downregulated |
| 0,04710407 | 0,940069962 | STX18-AS1 | FALSE | Upregulated |
| 0,053776193 | 0,940104356 | C12orf61 | FALSE | Upregulated |
| -0,103317378 | 0,940104356 | DDX11-AS1 | FALSE | Downregulated |
| -0,048602026 | 0,940104356 | LINC00933 | FALSE | Downregulated |
| 0,018813168 | 0,940104356 | LRRC8A | FALSE | Upregulated |
| -0,013233006 | 0,940104356 | RNF123 | FALSE | Downregulated |
| -0,087948392 | 0,940104356 | SNORA60 | FALSE | Downregulated |
| -0,028369391 | 0,940104356 | SPRY3 | FALSE | Downregulated |
| 0,052973197 | 0,940216962 | SLC29A2 | FALSE | Upregulated |
| -0,062210731 | 0,940216962 | TAF7L | FALSE | Downregulated |
| 0,606804443 | 0,940412994 | LMOD2 | FALSE | Upregulated |
| -0,021452903 | 0,940412994 | PTRHD1 | FALSE | Downregulated |
| 0,009233845 | 0,940412994 | RAB12 | FALSE | Upregulated |
| -0,123036379 | 0,94050589 | SHOX2 | FALSE | Downregulated |
| 0,060484643 | 0,940619242 | NCR1 | FALSE | Upregulated |
| 0,023444338 | 0,940619242 | RECK | FALSE | Upregulated |
| 0,271761173 | 0,940619242 | SNORA59B | FALSE | Upregulated |
| 0,009598813 | 0,940641953 | DHX32 | FALSE | Upregulated |
| -0,026106382 | 0,940641953 | HERC6 | FALSE | Downregulated |
| 0,064832857 | 0,940641953 | NAT8L | FALSE | Upregulated |
| 0,014885899 | 0,940641953 | TCOF1 | FALSE | Upregulated |
| 0,009184683 | 0,940649315 | SETD1A | FALSE | Upregulated |
| -0,013144881 | 0,940852454 | EZH1 | FALSE | Downregulated |
| 0,05140133 | 0,940852454 | TTLL3 | FALSE | Upregulated |
| 0,013919978 | 0,941019957 | ASH1L | FALSE | Upregulated |
| 0,028413435 | 0,941019957 | SLC7A5 | FALSE | Upregulated |
| 0,053489655 | 0,941304727 | EDAR | FALSE | Upregulated |
| -0,012740131 | 0,941304727 | HAUS3 | FALSE | Downregulated |
| 0,036180702 | 0,941354716 | TBX19 | FALSE | Upregulated |
| -0,009177264 | 0,941721576 | DYRK1A | FALSE | Downregulated |
| 0,035577219 | 0,941721576 | LOC100379224 | FALSE | Upregulated |
| 0,122473678 | 0,941786527 | LOC102723902 | FALSE | Upregulated |
| 0,043020148 | 0,941793937 | PLEK2 | FALSE | Upregulated |
| -0,010821509 | 0,941961847 | SMARCC2 | FALSE | Downregulated |
| 0,011508202 | 0,942111884 | CBY1 | FALSE | Upregulated |
| -0,031900422 | 0,942327568 | CDHR3 | FALSE | Downregulated |
| 0,088903712 | 0,942327568 | CRP | FALSE | Upregulated |
| -0,089129429 | 0,942327568 | LOC101928669 | FALSE | Downregulated |
| 0,172322257 | 0,942327568 | LOC102724069 | FALSE | Upregulated |
| -0,017368128 | 0,942327568 | PLEKHA6 | FALSE | Downregulated |
| -0,046423177 | 0,9423929 | BTBD11 | FALSE | Downregulated |
| -0,063796698 | 0,942453025 | CDH26 | FALSE | Downregulated |
| -0,0183115 | 0,942453025 | UBALD2 | FALSE | Downregulated |
| -0,075000434 | 0,942681819 | ZRANB2-AS2 | FALSE | Downregulated |
| 0,061359927 | 0,942694151 | LOC644090 | FALSE | Upregulated |
| -0,078464427 | 0,943189616 | LOC101060494 | FALSE | Downregulated |
| -0,014981811 | 0,943189616 | RCCD1 | FALSE | Downregulated |
| -0,099254194 | 0,94327346 | KLHL30 | FALSE | Downregulated |
| -0,065802341 | 0,943361205 | LOC102723866 | FALSE | Downregulated |

| | | | | |
|---|---|---|---|---|
| -0,014166545 | 0,943361205 | MSANTD3 | FALSE | Downregulated |
| 0,025009518 | 0,943518093 | ABCC6P2 | FALSE | Upregulated |
| -0,164622672 | 0,943972369 | ABCC12 | FALSE | Downregulated |
| -0,012262985 | 0,943972369 | HPS4 | FALSE | Downregulated |
| -0,00950546 | 0,944027529 | MRPL9 | FALSE | Downregulated |
| 0,129900354 | 0,944069124 | DYX1C1 | FALSE | Upregulated |
| -0,020120718 | 0,944258185 | SLC39A14 | FALSE | Downregulated |
| 0,018370015 | 0,944470899 | MYPOP | FALSE | Upregulated |
| -0,036899932 | 0,944528738 | LAG3 | FALSE | Downregulated |
| -0,012713999 | 0,944841524 | TMEM167B | FALSE | Downregulated |
| -0,073709977 | 0,944912207 | DRD4 | FALSE | Downregulated |
| -0,014522942 | 0,945060663 | DNAJC30 | FALSE | Downregulated |
| 0,122667956 | 0,94506719 | LOC101930137 | FALSE | Upregulated |
| -0,153162504 | 0,94506719 | SLITRK5 | FALSE | Downregulated |
| -0,051896543 | 0,945210167 | LOC100288152 | FALSE | Downregulated |
| -0,038116263 | 0,945210167 | LOC154761 | FALSE | Downregulated |
| 0,011319481 | 0,945210167 | MON1B | FALSE | Upregulated |
| 0,01550383 | 0,945210167 | ZNF2 | FALSE | Upregulated |
| 0,118484037 | 0,945299975 | RASGEF1C | FALSE | Upregulated |
| 0,017422545 | 0,945416399 | TUT1 | FALSE | Upregulated |
| 0,075717272 | 0,945489736 | LOC101929724 | FALSE | Upregulated |
| 0,014234033 | 0,945509848 | DDAH2 | FALSE | Upregulated |
| -0,131375185 | 0,945541777 | BOLA2B | FALSE | Downregulated |
| 0,530587532 | 0,945541777 | MYOZ1 | FALSE | Upregulated |
| -0,0290668 | 0,945541777 | PLLP | FALSE | Downregulated |
| -0,031205865 | 0,945541777 | SLC19A1 | FALSE | Downregulated |
| 0,024022505 | 0,945541777 | SLC41A1 | FALSE | Upregulated |
| 0,024773464 | 0,945738981 | AHR | FALSE | Upregulated |
| -0,046868284 | 0,945744741 | FADS2 | FALSE | Downregulated |
| 0,010400415 | 0,945744741 | SELT | FALSE | Upregulated |
| 0,014423986 | 0,945748653 | KIAA0020 | FALSE | Upregulated |
| -0,013888469 | 0,945770381 | C3 | FALSE | Downregulated |
| -0,05345267 | 0,945770381 | CCDC136 | FALSE | Downregulated |
| -0,01657541 | 0,945770381 | SPG21 | FALSE | Downregulated |
| 0,520771389 | 0,945770381 | TNNI1 | FALSE | Upregulated |
| 0,073249958 | 0,946017693 | SIX4 | FALSE | Upregulated |
| 0,107835074 | 0,946243061 | LOC101927632 | FALSE | Upregulated |
| -0,010886652 | 0,946345028 | PHKA2 | FALSE | Downregulated |
| 0,557322815 | 0,946384176 | ACTN3 | FALSE | Upregulated |
| 0,553125089 | 0,946384176 | MYBPC1 | FALSE | Upregulated |
| 0,0096597 | 0,946384176 | ZFYVE1 | FALSE | Upregulated |
| -0,013622436 | 0,946387755 | TMEM199 | FALSE | Downregulated |
| 0,012084597 | 0,946387755 | TRA2A | FALSE | Upregulated |
| 0,109582679 | 0,946634945 | MIMT1 | FALSE | Upregulated |
| 0,057182671 | 0,946655479 | LOC101927645 | FALSE | Upregulated |
| 0,057466246 | 0,94670529 | LOC102723679 | FALSE | Upregulated |
| -0,057983205 | 0,94670529 | LOC102723980 | FALSE | Downregulated |
| 0,062641323 | 0,94670529 | PARD6G-AS1 | FALSE | Upregulated |
| -0,062837311 | 0,946739344 | RHPN1-AS1 | FALSE | Downregulated |
| 0,105363757 | 0,946739344 | ZNF32-AS1 | FALSE | Upregulated |
| 0,073679892 | 0,94685399 | FAM83E | FALSE | Upregulated |

| | | | | |
|---|---|---|---|---|
| -0,015359884 | 0,94685399 | RNPEP | FALSE | Downregulated |
| 0,015652502 | 0,947041119 | HECTD4 | FALSE | Upregulated |
| 0,08008697 | 0,94707462 | UCHL1 | FALSE | Upregulated |
| 0,095101023 | 0,94707462 | ZHX1-C8ORF76 | FALSE | Upregulated |
| 0,042833003 | 0,947333675 | GAL3ST1 | FALSE | Upregulated |
| 0,018268103 | 0,947333675 | RAB17 | FALSE | Upregulated |
| -0,293996155 | 0,947333675 | TRNAH1 | FALSE | Downregulated |
| 0,04515875 | 0,947485032 | CYP21A1P | FALSE | Upregulated |
| 0,073731272 | 0,947485032 | FOXO6 | FALSE | Upregulated |
| 0,013814445 | 0,947485032 | ZDHHC17 | FALSE | Upregulated |
| -0,010022957 | 0,947542 | FASTKD2 | FALSE | Downregulated |
| -0,067099505 | 0,947843267 | RP11-563J2.2 | FALSE | Downregulated |
| 0,012035508 | 0,948082078 | FAM73B | FALSE | Upregulated |
| -0,083623145 | 0,948082078 | PEG3-AS1 | FALSE | Downregulated |
| -0,012288515 | 0,948165016 | CDK11B | FALSE | Downregulated |
| 0,021371743 | 0,948177771 | HIVEP3 | FALSE | Upregulated |
| 0,095620715 | 0,948313587 | MIR3936 | FALSE | Upregulated |
| -0,008346404 | 0,94842847 | MECP2 | FALSE | Downregulated |
| -0,070270725 | 0,948507804 | LINC00299 | FALSE | Downregulated |
| 0,006788843 | 0,948522062 | FAM120AOS | FALSE | Upregulated |
| 0,008768292 | 0,948527333 | HIP1 | FALSE | Upregulated |
| -0,018107871 | 0,948527333 | PRKAR2B | FALSE | Downregulated |
| 0,009716452 | 0,948531928 | PABPC4 | FALSE | Upregulated |
| 0,071784256 | 0,948531928 | TRND | FALSE | Upregulated |
| -0,012932053 | 0,94874505 | KMT2A | FALSE | Downregulated |
| -0,119190153 | 0,948903621 | AMZ1 | FALSE | Downregulated |
| 0,026620167 | 0,948964592 | EFNA2 | FALSE | Upregulated |
| -0,015539171 | 0,948964592 | INO80D | FALSE | Downregulated |
| -0,01065632 | 0,949027524 | MBTPS2 | FALSE | Downregulated |
| 0,047828517 | 0,949043028 | BRICD5 | FALSE | Upregulated |
| 0,011091837 | 0,949057042 | SEL1L | FALSE | Upregulated |
| -0,053002929 | 0,949057042 | TREML2 | FALSE | Downregulated |
| -0,143136744 | 0,949057042 | ZNF625-ZNF20 | FALSE | Downregulated |
| 0,018293879 | 0,9491703 | HAND2 | FALSE | Upregulated |
| -0,011564839 | 0,949218815 | ATPAF2 | FALSE | Downregulated |
| -0,037918149 | 0,949218815 | CNTNAP3 | FALSE | Downregulated |
| 0,022812584 | 0,949218815 | DZANK1 | FALSE | Upregulated |
| 0,044744101 | 0,949218815 | ERAP2 | FALSE | Upregulated |
| 0,111853241 | 0,949218815 | FLJ31945 | FALSE | Upregulated |
| 0,110330274 | 0,949218815 | LINC00327 | FALSE | Upregulated |
| 0,107128888 | 0,949218815 | LINC00891 | FALSE | Upregulated |
| 0,107642853 | 0,949218815 | SNORA31 | FALSE | Upregulated |
| 0,118278303 | 0,949218815 | TREML1 | FALSE | Upregulated |
| 0,010571013 | 0,949218815 | VPS16 | FALSE | Upregulated |
| 0,017464038 | 0,949218815 | ZFC3H1 | FALSE | Upregulated |
| -0,010613569 | 0,949644698 | TROVE2 | FALSE | Downregulated |
| -0,109524193 | 0,949708653 | LOC101928045 | FALSE | Downregulated |
| 0,028950463 | 0,949708653 | LOC101929130 | FALSE | Upregulated |
| -0,010957656 | 0,949731467 | ACO1 | FALSE | Downregulated |
| 0,009863084 | 0,94974408 | DAP3 | FALSE | Upregulated |
| -0,012205861 | 0,949765658 | BPTF | FALSE | Downregulated |

| | | | | |
|---|---|---|---|---|
| -0,009456236 | 0,949765658 | COG8 | FALSE | Downregulated |
| -0,105658392 | 0,949765658 | LOC102724144 | FALSE | Downregulated |
| 0,060501981 | 0,949765658 | LOC102724153 | FALSE | Upregulated |
| 0,038606395 | 0,949765658 | MAMDC4 | FALSE | Upregulated |
| -0,00809187 | 0,949765658 | NEK6 | FALSE | Downregulated |
| 0,017648752 | 0,949765658 | PRX | FALSE | Upregulated |
| -0,01383911 | 0,949765658 | PSD4 | FALSE | Downregulated |
| 0,111919699 | 0,949765658 | WDR88 | FALSE | Upregulated |
| 0,018847297 | 0,949765658 | ZNF581 | FALSE | Upregulated |
| -0,020608977 | 0,949765658 | ZNF630 | FALSE | Downregulated |
| -0,014618189 | 0,950005408 | FOXN2 | FALSE | Downregulated |
| -0,010486313 | 0,950139062 | TBP | FALSE | Downregulated |
| 0,142283506 | 0,950194381 | MSLN | FALSE | Upregulated |
| -0,012539398 | 0,950257732 | SAAL1 | FALSE | Downregulated |
| -0,028192226 | 0,950261295 | SNHG10 | FALSE | Downregulated |
| -0,019178988 | 0,950278973 | TMEM214 | FALSE | Downregulated |
| 0,085170046 | 0,950407073 | LOC101928369 | FALSE | Upregulated |
| -0,041754368 | 0,950480191 | HIST1H2AE | FALSE | Downregulated |
| -0,013949979 | 0,950480191 | HPD | FALSE | Downregulated |
| 0,021527723 | 0,950480191 | MAFIP | FALSE | Upregulated |
| 0,015282994 | 0,950480191 | ZNF778 | FALSE | Upregulated |
| 0,009667603 | 0,950504644 | ELK1 | FALSE | Upregulated |
| -0,013027466 | 0,950593587 | PGAP3 | FALSE | Downregulated |
| -0,006582866 | 0,950602414 | DNAJA2 | FALSE | Downregulated |
| 0,050304109 | 0,950602414 | FSD1 | FALSE | Upregulated |
| 0,025311315 | 0,950602414 | SYNE3 | FALSE | Upregulated |
| -0,511690705 | 0,950602414 | TRNAV6 | FALSE | Downregulated |
| -0,046380275 | 0,950714947 | BRINP1 | FALSE | Downregulated |
| -0,017137176 | 0,950829066 | DDAH1 | FALSE | Downregulated |
| 0,042597607 | 0,950876752 | LOC100129917 | FALSE | Upregulated |
| -0,012052568 | 0,950876752 | RSF1 | FALSE | Downregulated |
| -0,053338262 | 0,950979425 | GBAP1 | FALSE | Downregulated |
| -0,0142106 | 0,951008353 | CISD2 | FALSE | Downregulated |
| -0,057638769 | 0,951121361 | GRIK2 | FALSE | Downregulated |
| -0,058433798 | 0,951121361 | LOC101927745 | FALSE | Downregulated |
| -0,02532996 | 0,951126492 | RPS10P7 | FALSE | Downregulated |
| -0,0144604 | 0,951126492 | ZNF567 | FALSE | Downregulated |
| -0,024615942 | 0,951126492 | ZNF674-AS1 | FALSE | Downregulated |
| 0,109191416 | 0,951181509 | NME8 | FALSE | Upregulated |
| -0,0086535 | 0,951376915 | SMIM15 | FALSE | Downregulated |
| -0,086854386 | 0,951382367 | OOEP | FALSE | Downregulated |
| 0,070532255 | 0,951382367 | TCP10L2 | FALSE | Upregulated |
| -0,365060671 | 0,951382367 | TNNI2 | FALSE | Downregulated |
| 0,011848832 | 0,951405001 | NARG2 | FALSE | Upregulated |
| -0,095974055 | 0,951405001 | RGPD2 | FALSE | Downregulated |
| 0,030970824 | 0,95147796 | FAM222A | FALSE | Upregulated |
| -0,017047408 | 0,95147796 | TMEM147 | FALSE | Downregulated |
| 0,017250552 | 0,951570196 | PTS | FALSE | Upregulated |
| 0,495952636 | 0,951783053 | ACTC1 | FALSE | Upregulated |
| -0,496009009 | 0,951812905 | MYL2 | FALSE | Downregulated |
| -0,060629952 | 0,952004791 | HRH4 | FALSE | Downregulated |

| | | | | |
|---|---|---|---|---|
| 0,035901567 | 0,952004791 | LOC101927402 | FALSE | Upregulated |
| -0,011264911 | 0,952004791 | ZNF598 | FALSE | Downregulated |
| -0,013626345 | 0,952075355 | ALG2 | FALSE | Downregulated |
| -0,046440284 | 0,952075355 | REREP3 | FALSE | Downregulated |
| 0,046264186 | 0,952177642 | ZNF883 | FALSE | Upregulated |
| -0,019917981 | 0,952313736 | ACACA | FALSE | Downregulated |
| -0,014716228 | 0,952313736 | GZF1 | FALSE | Downregulated |
| -0,099415996 | 0,952313736 | MIR142 | FALSE | Downregulated |
| 0,033724114 | 0,952738831 | CD209 | FALSE | Upregulated |
| -0,048812022 | 0,952738831 | SNORA63 | FALSE | Downregulated |
| 0,014698407 | 0,952763726 | CTU2 | FALSE | Upregulated |
| -0,034508504 | 0,952830726 | VPS37D | FALSE | Downregulated |
| -0,010392762 | 0,952889571 | COBL | FALSE | Downregulated |
| 0,068449377 | 0,952889571 | FAM169B | FALSE | Upregulated |
| 0,041869962 | 0,952889571 | NDRG4 | FALSE | Upregulated |
| 0,012509807 | 0,952889571 | SH2B1 | FALSE | Upregulated |
| 0,052274132 | 0,952889571 | TCF15 | FALSE | Upregulated |
| 0,064651768 | 0,953503645 | LOC101927190 | FALSE | Upregulated |
| -0,017873073 | 0,953515926 | WIPI1 | FALSE | Downregulated |
| -0,14660386 | 0,953544393 | GPR1-AS | FALSE | Downregulated |
| 0,064527524 | 0,953544393 | LOC100130285 | FALSE | Upregulated |
| -0,133142425 | 0,953544393 | MIA-RAB4B | FALSE | Downregulated |
| 0,014878978 | 0,953544393 | ORAOV1 | FALSE | Upregulated |
| 0,020414416 | 0,953544393 | USP30-AS1 | FALSE | Upregulated |
| -0,012726355 | 0,95359872 | FAM160A2 | FALSE | Downregulated |
| -0,015530875 | 0,953654613 | COX7B | FALSE | Downregulated |
| -0,008440672 | 0,953681971 | C5orf24 | FALSE | Downregulated |
| 0,020907874 | 0,953681971 | EFCAB2 | FALSE | Upregulated |
| -0,010596179 | 0,95394914 | MRPS16 | FALSE | Downregulated |
| -0,013269562 | 0,954165711 | ACAP2 | FALSE | Downregulated |
| -0,037196245 | 0,954165711 | C9orf116 | FALSE | Downregulated |
| -0,010940731 | 0,954165711 | ERCC1 | FALSE | Downregulated |
| -0,12711695 | 0,954291865 | LOC729222 | FALSE | Downregulated |
| -0,13938507 | 0,954316536 | NPB | FALSE | Downregulated |
| 0,010055225 | 0,95456929 | TRAPPC13 | FALSE | Upregulated |
| 0,467012458 | 0,954721657 | DES | FALSE | Upregulated |
| 0,07036772 | 0,954817054 | LOC101927051 | FALSE | Upregulated |
| -0,035739564 | 0,954822818 | LOC100996741 | FALSE | Downregulated |
| -0,017464847 | 0,954974828 | MIR100HG | FALSE | Downregulated |
| 0,097775524 | 0,955006438 | CCDC78 | FALSE | Upregulated |
| -0,023601309 | 0,955006438 | PPP1R26-AS1 | FALSE | Downregulated |
| 0,015000603 | 0,955171893 | USP46-AS1 | FALSE | Upregulated |
| 0,011223221 | 0,955193557 | ARMCX5 | FALSE | Upregulated |
| -0,051165268 | 0,955193557 | RIPPLY3 | FALSE | Downregulated |
| -0,011158055 | 0,955193557 | SMAD3 | FALSE | Downregulated |
| 0,101946781 | 0,955371858 | BEND2 | FALSE | Upregulated |
| -0,007078285 | 0,955371858 | GGPS1 | FALSE | Downregulated |
| 0,016092812 | 0,955371858 | TEF | FALSE | Upregulated |
| -0,01374985 | 0,955423113 | SHANK3 | FALSE | Downregulated |
| 0,016310284 | 0,955538555 | TRIM10 | FALSE | Upregulated |
| -0,019129062 | 0,955574902 | ZNF699 | FALSE | Downregulated |

| | | | | |
|---|---|---|---|---|
| -0,030867801 | 0,955599898 | TTC25 | FALSE | Downregulated |
| -0,070436293 | 0,956053991 | LOC101929295 | FALSE | Downregulated |
| 0,094099834 | 0,956053991 | NTRK1 | FALSE | Upregulated |
| 0,013254659 | 0,956053991 | PPP1R12A | FALSE | Upregulated |
| -0,012602723 | 0,956053991 | REV3L | FALSE | Downregulated |
| -0,080393549 | 0,956082631 | LOC440934 | FALSE | Downregulated |
| 0,00977807 | 0,956082631 | RETSAT | FALSE | Upregulated |
| -0,009029374 | 0,956150336 | PIKFYVE | FALSE | Downregulated |
| -0,011923836 | 0,956238493 | ZDHHC8 | FALSE | Downregulated |
| -0,00908001 | 0,956302421 | RPN2 | FALSE | Downregulated |
| -0,021568501 | 0,956420928 | AP1S2 | FALSE | Downregulated |
| -0,056363494 | 0,956441859 | CDK3 | FALSE | Downregulated |
| 0,006463015 | 0,956441859 | NAA50 | FALSE | Upregulated |
| 0,012751479 | 0,956462752 | KDM5C | FALSE | Upregulated |
| -0,027923053 | 0,956462752 | PLEKHH2 | FALSE | Downregulated |
| 0,023975512 | 0,956685726 | C11orf35 | FALSE | Upregulated |
| -0,02331297 | 0,956685726 | DNM3OS | FALSE | Downregulated |
| 0,016564759 | 0,956685726 | GMPPA | FALSE | Upregulated |
| -0,009811067 | 0,956685726 | HS2ST1 | FALSE | Downregulated |
| -0,031953318 | 0,956685726 | LOC100133315 | FALSE | Downregulated |
| -0,11264383 | 0,956685726 | LOC101929248 | FALSE | Downregulated |
| 0,062642162 | 0,956685726 | LOC101929666 | FALSE | Upregulated |
| 0,127604422 | 0,956685726 | MSMB | FALSE | Upregulated |
| -0,018067344 | 0,956685726 | NDUFC2-KCTD14 | FALSE | Downregulated |
| -0,018736357 | 0,956685726 | NOL3 | FALSE | Downregulated |
| -0,023879801 | 0,956685726 | PKIB | FALSE | Downregulated |
| -0,049871655 | 0,956685726 | SNORD104 | FALSE | Downregulated |
| -0,450474961 | 0,956685726 | TRNAK14 | FALSE | Downregulated |
| -0,020860447 | 0,956685726 | USP35 | FALSE | Downregulated |
| 0,010967495 | 0,956735024 | POU2F1 | FALSE | Upregulated |
| 0,398405754 | 0,956764117 | APOBEC2 | FALSE | Upregulated |
| 0,018213791 | 0,957049086 | NUDT14 | FALSE | Upregulated |
| -0,053771249 | 0,957049086 | SLC26A8 | FALSE | Downregulated |
| 0,017981445 | 0,957056836 | SAMD1 | FALSE | Upregulated |
| -0,146639224 | 0,957109368 | CNTNAP4 | FALSE | Downregulated |
| -0,030893994 | 0,957109368 | MLLT4-AS1 | FALSE | Downregulated |
| 0,018996801 | 0,957280203 | ERV3-1 | FALSE | Upregulated |
| 0,063631323 | 0,957280203 | LOC101929511 | FALSE | Upregulated |
| 0,063283728 | 0,957280203 | LOC400685 | FALSE | Upregulated |
| -0,006785381 | 0,957280203 | PPP2R5D | FALSE | Downregulated |
| 0,038459277 | 0,957280203 | PRSS3 | FALSE | Upregulated |
| -0,013593277 | 0,957313896 | ZNF558 | FALSE | Downregulated |
| 0,025925212 | 0,957323367 | ITLN1 | FALSE | Upregulated |
| 0,008036843 | 0,957461366 | ASCC2 | FALSE | Upregulated |
| 0,010824013 | 0,957461366 | CDK5RAP3 | FALSE | Upregulated |
| -0,020843239 | 0,957561151 | CLDN3 | FALSE | Downregulated |
| -0,008122802 | 0,957599669 | ELL | FALSE | Downregulated |
| -0,438845206 | 0,957728005 | TRNAV29 | FALSE | Downregulated |
| 0,03590595 | 0,957749141 | GPR162 | FALSE | Upregulated |
| -0,060580612 | 0,957749141 | GPRC5D | FALSE | Downregulated |
| 0,0160747 | 0,957753646 | ATG10 | FALSE | Upregulated |

| | | | | |
|---|---|---|---|---|
| -0,017426432 | 0,957753646 | CD300A | FALSE | Downregulated |
| 0,00691537 | 0,957753646 | GLTSCR1L | FALSE | Upregulated |
| 0,052900498 | 0,957753646 | MAMSTR | FALSE | Upregulated |
| 0,008686344 | 0,957753646 | NPAT | FALSE | Upregulated |
| -0,032305538 | 0,957753646 | RP5-1057J7.6 | FALSE | Downregulated |
| -0,435498759 | 0,957753646 | TRNAV-CAC | FALSE | Downregulated |
| -0,435498759 | 0,957753646 | TRNAV2 | FALSE | Downregulated |
| -0,435498759 | 0,957753646 | TRNAV22 | FALSE | Downregulated |
| -0,435498759 | 0,957753646 | TRNAV25 | FALSE | Downregulated |
| -0,007423512 | 0,957971786 | EIF3G | FALSE | Downregulated |
| -0,044603417 | 0,958102279 | DUSP9 | FALSE | Downregulated |
| 0,045494518 | 0,958102279 | LOC102724738 | FALSE | Upregulated |
| -0,020720365 | 0,95823478 | LIPJ | FALSE | Downregulated |
| 0,056699594 | 0,958243656 | ESRRB | FALSE | Upregulated |
| -0,056854269 | 0,958243656 | NTNG2 | FALSE | Downregulated |
| 0,013015261 | 0,958243656 | PAFAH1B2 | FALSE | Upregulated |
| -0,083453442 | 0,958243656 | PRSS30P | FALSE | Downregulated |
| 0,007852325 | 0,95833003 | ARHGEF7 | FALSE | Upregulated |
| -0,006997678 | 0,958398817 | BNIP3L | FALSE | Downregulated |
| 0,021866634 | 0,958399413 | CROCCP2 | FALSE | Upregulated |
| 0,019746055 | 0,958399413 | FEN1 | FALSE | Upregulated |
| -0,055081716 | 0,958399413 | SLC17A7 | FALSE | Downregulated |
| -0,009875525 | 0,958529829 | BLOC1S2 | FALSE | Downregulated |
| -0,072948822 | 0,958529829 | FAM83A | FALSE | Downregulated |
| -0,045769084 | 0,958555648 | LOC102724757 | FALSE | Downregulated |
| 0,023218955 | 0,958741245 | MPPED1 | FALSE | Upregulated |
| 0,012479788 | 0,958741245 | ZC3H12C | FALSE | Upregulated |
| 0,041703779 | 0,959128873 | C17orf49 | FALSE | Upregulated |
| -0,007351398 | 0,959174684 | ARHGAP5 | FALSE | Downregulated |
| 0,309821549 | 0,959285773 | MYH6 | FALSE | Upregulated |
| 0,016376787 | 0,959299301 | PGM5 | FALSE | Upregulated |
| 0,012084498 | 0,959640196 | PHF11 | FALSE | Upregulated |
| -0,060039083 | 0,959640196 | SNORD10 | FALSE | Downregulated |
| 0,01134691 | 0,959643612 | ITIH4 | FALSE | Upregulated |
| -0,02114873 | 0,959643612 | PRKAR2A-AS1 | FALSE | Downregulated |
| 0,028888251 | 0,959685101 | SPDYC | FALSE | Upregulated |
| 0,050506425 | 0,959685101 | UCKL1-AS1 | FALSE | Upregulated |
| -0,011693619 | 0,959979951 | AHSG | FALSE | Downregulated |
| -0,031210658 | 0,959979951 | SCNN1D | FALSE | Downregulated |
| -0,014390621 | 0,960273709 | ZEB2 | FALSE | Downregulated |
| -0,016482825 | 0,960273709 | ZNF70 | FALSE | Downregulated |
| 0,016649569 | 0,960304549 | LOC101060181 | FALSE | Upregulated |
| 0,093879071 | 0,960445396 | LOC100652930 | FALSE | Upregulated |
| -0,125170476 | 0,960452658 | RIMS1 | FALSE | Downregulated |
| -0,114702319 | 0,960452658 | RP11-320G24.1 | FALSE | Downregulated |
| 0,006618537 | 0,960486761 | CNOT11 | FALSE | Upregulated |
| 0,014390326 | 0,960625248 | ZNF586 | FALSE | Upregulated |
| -0,012025382 | 0,960630786 | KIAA0232 | FALSE | Downregulated |
| 0,053423441 | 0,960630786 | LINC00189 | FALSE | Upregulated |
| -0,065988803 | 0,960630786 | NME1-NME2 | FALSE | Downregulated |
| -0,009480276 | 0,960630786 | ZMPSTE24 | FALSE | Downregulated |

| | | | | |
|---|---|---|---|---|
| 0,011250651 | 0,961070834 | CCDC77 | FALSE | Upregulated |
| 0,082434497 | 0,961074444 | FIRRE | FALSE | Upregulated |
| -0,005432365 | 0,961074444 | H3F3B | FALSE | Downregulated |
| 0,012397973 | 0,961074444 | SERPINA11 | FALSE | Upregulated |
| 0,007701324 | 0,961074444 | SLC35F5 | FALSE | Upregulated |
| -0,018346789 | 0,96107638 | ADAT2 | FALSE | Downregulated |
| 0,027898646 | 0,96107638 | LINC00936 | FALSE | Upregulated |
| -0,036078894 | 0,961357574 | FAM86B1 | FALSE | Downregulated |
| -0,033084443 | 0,961624936 | FIGNL2 | FALSE | Downregulated |
| 0,008454871 | 0,961769683 | CEBPG | FALSE | Upregulated |
| 0,05085459 | 0,961951031 | HERC2P3 | FALSE | Upregulated |
| -0,024195796 | 0,961951031 | NLRP11 | FALSE | Downregulated |
| -0,038280041 | 0,961951031 | NYAP1 | FALSE | Downregulated |
| 0,012350358 | 0,962003328 | FMO4 | FALSE | Upregulated |
| -0,392149943 | 0,962011475 | MYH2 | FALSE | Downregulated |
| 0,015551414 | 0,962011475 | SENP8 | FALSE | Upregulated |
| -0,007183512 | 0,962354908 | C14orf119 | FALSE | Downregulated |
| 0,007750614 | 0,962354908 | LMBR1 | FALSE | Upregulated |
| -0,040159118 | 0,962354908 | LOC101928096 | FALSE | Downregulated |
| -0,012883914 | 0,962354908 | SSR2 | FALSE | Downregulated |
| -0,006092589 | 0,962354908 | TM9SF3 | FALSE | Downregulated |
| -0,008090478 | 0,962354908 | XIAP | FALSE | Downregulated |
| -0,062196978 | 0,962359487 | FAM74A1 | FALSE | Downregulated |
| 0,066650436 | 0,962393435 | TMED7-TICAM2 | FALSE | Upregulated |
| 0,06341989 | 0,962401283 | LINC01135 | FALSE | Upregulated |
| -0,00715939 | 0,962420462 | FAM136A | FALSE | Downregulated |
| 0,033587281 | 0,962420462 | LOC101927578 | FALSE | Upregulated |
| 0,042252766 | 0,962420462 | LOC101927699 | FALSE | Upregulated |
| -0,106271184 | 0,962420462 | LOC101929714 | FALSE | Downregulated |
| 0,010427231 | 0,962420462 | RAB24 | FALSE | Upregulated |
| -0,011705499 | 0,962420462 | ZNF839 | FALSE | Downregulated |
| -0,006610268 | 0,962477425 | MGME1 | FALSE | Downregulated |
| 0,008013973 | 0,962477425 | TAF1D | FALSE | Upregulated |
| 0,045560969 | 0,962591127 | HNRNPCL1 | FALSE | Upregulated |
| 0,04607847 | 0,962610314 | LOC101409256 | FALSE | Upregulated |
| -0,008276092 | 0,962626898 | EXOSC7 | FALSE | Downregulated |
| 0,383519234 | 0,962710309 | MYLPF | FALSE | Upregulated |
| 0,100260306 | 0,962943305 | TUSC5 | FALSE | Upregulated |
| 0,087069761 | 0,963228987 | LOC102724951 | FALSE | Upregulated |
| 0,014718305 | 0,963958011 | LMBR1L | FALSE | Upregulated |
| 0,029309755 | 0,963974159 | LOC102723794 | FALSE | Upregulated |
| -0,073009914 | 0,963974159 | NEK5 | FALSE | Downregulated |
| -0,011890721 | 0,963974159 | SLC35B4 | FALSE | Downregulated |
| 0,04692122 | 0,963987354 | APITD1-CORT | FALSE | Upregulated |
| 0,00772214 | 0,964195212 | YTHDC2 | FALSE | Upregulated |
| -0,012492287 | 0,964266293 | ESR1 | FALSE | Downregulated |
| 0,005855533 | 0,964267875 | CRNKL1 | FALSE | Upregulated |
| 0,008478076 | 0,96439655 | ZNF776 | FALSE | Upregulated |
| -0,165880917 | 0,964415056 | SNORD103B | FALSE | Downregulated |
| -0,053950396 | 0,96472703 | CTD-2258A20.5 | FALSE | Downregulated |
| -0,010713945 | 0,96472703 | CYP2U1 | FALSE | Downregulated |

| | | | | |
|---|---|---|---|---|
| -0,020483755 | 0,96472703 | PRORSD1P | FALSE | Downregulated |
| -0,01180358 | 0,96472703 | SERTAD3 | FALSE | Downregulated |
| -0,005326401 | 0,96472703 | ZNF322 | FALSE | Downregulated |
| -0,031159643 | 0,964773988 | KTN1-AS1 | FALSE | Downregulated |
| -0,020374322 | 0,964865427 | PARD6A | FALSE | Downregulated |
| 0,006337335 | 0,964865427 | PRKD3 | FALSE | Upregulated |
| 0,09925571 | 0,964870581 | BPIFB2 | FALSE | Upregulated |
| -0,006156152 | 0,964870581 | CSNK2B | FALSE | Downregulated |
| -0,025497492 | 0,964870581 | SEMA5B | FALSE | Downregulated |
| 0,021566184 | 0,964876833 | MUM1L1 | FALSE | Upregulated |
| 0,093066492 | 0,96503875 | ABHD16B | FALSE | Upregulated |
| 0,007970459 | 0,96503875 | HDDC2 | FALSE | Upregulated |
| 0,03503156 | 0,965224047 | SORT1 | FALSE | Upregulated |
| -0,047297129 | 0,965282761 | SLC6A4 | FALSE | Downregulated |
| 0,02682639 | 0,965537544 | LOC644656 | FALSE | Upregulated |
| -0,098218136 | 0,965541095 | STRA6 | FALSE | Downregulated |
| -0,016546575 | 0,965603349 | APOBR | FALSE | Downregulated |
| -0,108287018 | 0,96577403 | AC087859.1 | FALSE | Downregulated |
| -0,015973619 | 0,965809747 | GPRASP1 | FALSE | Downregulated |
| 0,091470589 | 0,965809747 | IFIT1B | FALSE | Upregulated |
| 0,046909781 | 0,965809747 | LOC102724634 | FALSE | Upregulated |
| 0,017380316 | 0,965840622 | SYP | FALSE | Upregulated |
| 0,034013098 | 0,96584817 | PRIMA1 | FALSE | Upregulated |
| -0,022053612 | 0,966117131 | NSUN5P1 | FALSE | Downregulated |
| -0,029630485 | 0,966153736 | LOC102724059 | FALSE | Downregulated |
| 0,038201179 | 0,966345734 | KRBOX1 | FALSE | Upregulated |
| 0,019612923 | 0,966498966 | SELL | FALSE | Upregulated |
| 0,015633358 | 0,966558068 | ARSE | FALSE | Upregulated |
| -0,007427555 | 0,966558068 | BCKDHA | FALSE | Downregulated |
| 0,006999062 | 0,966558068 | RNF115 | FALSE | Upregulated |
| -0,008641036 | 0,966558068 | TRAPPC10 | FALSE | Downregulated |
| -0,025548974 | 0,966563607 | FBXO24 | FALSE | Downregulated |
| 0,008400016 | 0,966664564 | MTFMT | FALSE | Upregulated |
| -0,072637221 | 0,96687814 | LOC100101148 | FALSE | Downregulated |
| -0,060371651 | 0,967478355 | LOC101929512 | FALSE | Downregulated |
| -0,07189891 | 0,967665731 | DIRAS1 | FALSE | Downregulated |
| -0,022113027 | 0,967688249 | CD200R1 | FALSE | Downregulated |
| -0,007015708 | 0,967688249 | RTCB | FALSE | Downregulated |
| -0,011038674 | 0,967792885 | ETHE1 | FALSE | Downregulated |
| 0,097225438 | 0,96787542 | LOC100128593 | FALSE | Upregulated |
| -0,007014867 | 0,967877735 | HDAC2 | FALSE | Downregulated |
| -0,011352708 | 0,96795294 | DNAJC3 | FALSE | Downregulated |
| -0,01246405 | 0,968157797 | TMEM220 | FALSE | Downregulated |
| 0,006929795 | 0,968161475 | RIT1 | FALSE | Upregulated |
| -0,012997364 | 0,968161475 | TMEM139 | FALSE | Downregulated |
| 0,057102827 | 0,968184386 | FLJ45825 | FALSE | Upregulated |
| -0,059101443 | 0,968184386 | USP50 | FALSE | Downregulated |
| 0,00631595 | 0,968184386 | UTP23 | FALSE | Upregulated |
| 0,00974469 | 0,968187726 | ZNF583 | FALSE | Upregulated |
| -0,006330435 | 0,968211706 | ZNF639 | FALSE | Downregulated |
| -0,005802491 | 0,96826844 | CYHR1 | FALSE | Downregulated |

| | | | | |
|---|---|---|---|---|
| -0,007297184 | 0,96829554 | ANAPC1 | FALSE | Downregulated |
| 0,011896977 | 0,96829554 | FOXRED2 | FALSE | Upregulated |
| 0,010592004 | 0,96829554 | HSPA8 | FALSE | Upregulated |
| -0,046239194 | 0,96829554 | LOC100507195 | FALSE | Downregulated |
| -0,016890372 | 0,96829554 | LOC100996497 | FALSE | Downregulated |
| 0,029234503 | 0,96829554 | LOC102723422 | FALSE | Upregulated |
| 0,011282943 | 0,96829554 | NGEF | FALSE | Upregulated |
| -0,127136806 | 0,96829554 | SNORD103A | FALSE | Downregulated |
| -0,022396335 | 0,968458267 | SLC4A5 | FALSE | Downregulated |
| -0,269867414 | 0,968462652 | TRNAG23 | FALSE | Downregulated |
| 0,016207607 | 0,968554627 | ACRBP | FALSE | Upregulated |
| -0,031808682 | 0,968577695 | LOC101926918 | FALSE | Downregulated |
| -0,019712916 | 0,968591765 | KANSL1-AS1 | FALSE | Downregulated |
| 0,036747271 | 0,968707377 | GRIK1 | FALSE | Upregulated |
| -0,025725852 | 0,968728932 | KCNQ4 | FALSE | Downregulated |
| -0,032062459 | 0,968728932 | LOC100652901 | FALSE | Downregulated |
| -0,055919851 | 0,968728932 | LOC101930531 | FALSE | Downregulated |
| 0,326892414 | 0,968857605 | BOLA2 | FALSE | Upregulated |
| -0,042535939 | 0,968857605 | CCDC154 | FALSE | Downregulated |
| -0,031813323 | 0,968857605 | CPT1B | FALSE | Downregulated |
| 0,04948992 | 0,968857605 | CYP27B1 | FALSE | Upregulated |
| 0,057236076 | 0,968857605 | LOC101927697 | FALSE | Upregulated |
| 0,021345788 | 0,968857605 | TOB1-AS1 | FALSE | Upregulated |
| -0,00823813 | 0,968857605 | ZNF202 | FALSE | Downregulated |
| 0,028456983 | 0,968936356 | LOC283887 | FALSE | Upregulated |
| 0,011129258 | 0,968992129 | DNAJC15 | FALSE | Upregulated |
| -0,058676164 | 0,968992129 | LOC101927326 | FALSE | Downregulated |
| 0,012305784 | 0,968992129 | MORN1 | FALSE | Upregulated |
| -0,009851765 | 0,969032473 | LRRC3 | FALSE | Downregulated |
| -0,008565309 | 0,969079998 | CEP85 | FALSE | Downregulated |
| 0,006519015 | 0,969079998 | PAPSS2 | FALSE | Upregulated |
| 0,01175911 | 0,969079998 | RRAS | FALSE | Upregulated |
| 0,044420512 | 0,969224147 | GPR63 | FALSE | Upregulated |
| -0,100055906 | 0,969272521 | GGN | FALSE | Downregulated |
| -0,010162897 | 0,969368409 | ITSN2 | FALSE | Downregulated |
| -0,007116941 | 0,969368409 | TMEM203 | FALSE | Downregulated |
| -0,005500251 | 0,969379269 | KDM2A | FALSE | Downregulated |
| -0,007606138 | 0,969629284 | SLC8B1 | FALSE | Downregulated |
| 0,008848841 | 0,969683766 | IGIP | FALSE | Upregulated |
| -0,058077924 | 0,969683766 | LOC102724349 | FALSE | Downregulated |
| -0,081904113 | 0,969683766 | NAP1L6 | FALSE | Downregulated |
| -0,027329663 | 0,969683766 | SLITRK4 | FALSE | Downregulated |
| 0,018634026 | 0,969683766 | SNHG15 | FALSE | Upregulated |
| 0,031615482 | 0,969742204 | SGSM1 | FALSE | Upregulated |
| 0,065578997 | 0,969834602 | C3orf27 | FALSE | Upregulated |
| 0,041556595 | 0,969834602 | RP11-182I10.3 | FALSE | Upregulated |
| -0,030982364 | 0,969979471 | AC159540.1 | FALSE | Downregulated |
| 0,006604735 | 0,969980502 | RPA2 | FALSE | Upregulated |
| 0,019177569 | 0,970055849 | SIPA1L2 | FALSE | Upregulated |
| -0,170196529 | 0,970941881 | RNU1-27P | FALSE | Downregulated |
| -0,048396082 | 0,971111322 | AC104653.1 | FALSE | Downregulated |

| | | | | |
|---|---|---|---|---|
| -0,036170625 | 0,971111322 | FLJ20464 | FALSE | Downregulated |
| -0,038727089 | 0,971111322 | FUT3 | FALSE | Downregulated |
| 0,006999857 | 0,971111322 | NEMF | FALSE | Upregulated |
| -0,008536965 | 0,971184056 | SURF2 | FALSE | Downregulated |
| 0,301888907 | 0,971248745 | ACTA1 | FALSE | Upregulated |
| 0,006620574 | 0,971380382 | NUP153 | FALSE | Upregulated |
| 0,021607871 | 0,971392466 | ARHGEF26-AS1 | FALSE | Upregulated |
| 0,041454455 | 0,971791101 | LOC101927026 | FALSE | Upregulated |
| 0,014498721 | 0,971791101 | PANX2 | FALSE | Upregulated |
| -0,047888761 | 0,971791101 | RP11-770J1.3 | FALSE | Downregulated |
| -0,052262882 | 0,97179197 | LOC100132354 | FALSE | Downregulated |
| -0,068004391 | 0,971825066 | LOC102724652 | FALSE | Downregulated |
| -0,012470577 | 0,97201511 | CEP19 | FALSE | Downregulated |
| -0,005424694 | 0,972055217 | SUGT1 | FALSE | Downregulated |
| -0,014425901 | 0,972065999 | RNR1 | FALSE | Downregulated |
| 0,010201619 | 0,972073337 | BMI1 | FALSE | Upregulated |
| 0,014626728 | 0,972079235 | PLIN2 | FALSE | Upregulated |
| -0,012544846 | 0,972080972 | C12orf60 | FALSE | Downregulated |
| -0,033955492 | 0,972339696 | TBX18 | FALSE | Downregulated |
| 0,011015768 | 0,972373204 | CENPJ | FALSE | Upregulated |
| 0,009858423 | 0,972373204 | FGFR3 | FALSE | Upregulated |
| -0,006748716 | 0,972373204 | MAN1A2 | FALSE | Downregulated |
| -0,013006638 | 0,972373204 | RARA-AS1 | FALSE | Downregulated |
| 0,004548414 | 0,972373204 | TSPYL1 | FALSE | Upregulated |
| 0,033524934 | 0,972410619 | LOC101928085 | FALSE | Upregulated |
| 0,009880725 | 0,972525918 | HIVEP1 | FALSE | Upregulated |
| -0,00720032 | 0,972545212 | PDLIM5 | FALSE | Downregulated |
| -0,019492426 | 0,972609865 | TDRD10 | FALSE | Downregulated |
| -0,021040947 | 0,972613431 | CNTNAP3B | FALSE | Downregulated |
| -0,007088091 | 0,972613431 | MMADHC | FALSE | Downregulated |
| 0,006361384 | 0,972637737 | ERCC6 | FALSE | Upregulated |
| -0,036245357 | 0,972658851 | CYP4Z1 | FALSE | Downregulated |
| 0,033209305 | 0,972658851 | LINC01126 | FALSE | Upregulated |
| -0,043239423 | 0,972658851 | LOC644249 | FALSE | Downregulated |
| 0,007423992 | 0,972658851 | MBD4 | FALSE | Upregulated |
| -0,024938632 | 0,972658851 | N4BP2L2-IT2 | FALSE | Downregulated |
| 0,008607762 | 0,972658851 | PIBF1 | FALSE | Upregulated |
| 0,012921061 | 0,972658851 | PIM1 | FALSE | Upregulated |
| 0,004105086 | 0,972658851 | PPP1R8 | FALSE | Upregulated |
| 0,032138833 | 0,972843679 | SLFNL1-AS1 | FALSE | Upregulated |
| 0,015241936 | 0,972857558 | LOC100129637 | FALSE | Upregulated |
| 0,041434798 | 0,972867021 | RNF139-AS1 | FALSE | Upregulated |
| 0,064578922 | 0,972885732 | LINC00593 | FALSE | Upregulated |
| 0,088569555 | 0,972885732 | PAX2 | FALSE | Upregulated |
| 0,008609644 | 0,972960243 | KIAA0355 | FALSE | Upregulated |
| 0,009250702 | 0,972960243 | PRAF2 | FALSE | Upregulated |
| -0,069813922 | 0,972960243 | TNFAIP8L2-SCNM1 | FALSE | Downregulated |
| -0,006343807 | 0,973114216 | EDEM2 | FALSE | Downregulated |
| 0,00834979 | 0,973149551 | EVL | FALSE | Upregulated |
| -0,008207008 | 0,973149551 | FNBP1 | FALSE | Downregulated |

| | | | | |
|---|---|---|---|---|
| -0,006650227 | 0,973192774 | CHPT1 | FALSE | Downregulated |
| 0,081997554 | 0,973192774 | LINC01087 | FALSE | Upregulated |
| -0,00874759 | 0,973192774 | TRAPPC2 | FALSE | Downregulated |
| -0,024089118 | 0,97323369 | LOC102724562 | FALSE | Downregulated |
| -0,280803431 | 0,97323369 | TRNAV9 | FALSE | Downregulated |
| 0,013923758 | 0,973252256 | KAT2A | FALSE | Upregulated |
| -0,010375057 | 0,973258882 | RBBP8 | FALSE | Downregulated |
| -0,006018463 | 0,973395852 | BRAF | FALSE | Downregulated |
| 0,018220015 | 0,973395852 | C8orf46 | FALSE | Upregulated |
| -0,012478619 | 0,973395852 | CFHR1 | FALSE | Downregulated |
| -0,026686722 | 0,973572294 | PCDH19 | FALSE | Downregulated |
| 0,012181781 | 0,973572294 | TMEM241 | FALSE | Upregulated |
| 0,006700277 | 0,973873283 | HNRNPH1 | FALSE | Upregulated |
| 0,034400543 | 0,973921049 | HLX-AS1 | FALSE | Upregulated |
| -0,011767566 | 0,973955703 | CCDC113 | FALSE | Downregulated |
| -0,033943138 | 0,973963838 | FLJ41309 | FALSE | Downregulated |
| -0,00614816 | 0,973987625 | MRPS27 | FALSE | Downregulated |
| 0,040588593 | 0,97418295 | GOLGA8H | FALSE | Upregulated |
| 0,046481125 | 0,974211907 | DGUOK-AS1 | FALSE | Upregulated |
| -0,00502793 | 0,974254116 | C1orf109 | FALSE | Downregulated |
| -0,03543335 | 0,974254116 | UCN | FALSE | Downregulated |
| 0,012678671 | 0,974268403 | CD163 | FALSE | Upregulated |
| -0,004026761 | 0,974313672 | FBXO18 | FALSE | Downregulated |
| 0,05206763 | 0,974313672 | LOC102724696 | FALSE | Upregulated |
| 0,061303325 | 0,974313672 | RP11-365O16.6 | FALSE | Upregulated |
| 0,063941331 | 0,974481621 | EGFEM1P | FALSE | Upregulated |
| 0,062750123 | 0,974481621 | LOC100505774 | FALSE | Upregulated |
| -0,030507818 | 0,974481621 | LOC101060626 | FALSE | Downregulated |
| 0,033960645 | 0,974481621 | LOC101927279 | FALSE | Upregulated |
| -0,016189366 | 0,974481621 | LOC101928498 | FALSE | Downregulated |
| -0,057824161 | 0,974481621 | LOC102724273 | FALSE | Downregulated |
| 0,008837237 | 0,974481621 | MED30 | FALSE | Upregulated |
| -0,007254108 | 0,974481621 | PTPN3 | FALSE | Downregulated |
| 0,005672456 | 0,974481621 | RCHY1 | FALSE | Upregulated |
| 0,033178263 | 0,974481621 | RP4-562J12.2 | FALSE | Upregulated |
| 0,038311687 | 0,974548421 | ARHGAP8 | FALSE | Upregulated |
| 0,035207832 | 0,974548421 | CARD14 | FALSE | Upregulated |
| -0,009060515 | 0,974548421 | HAUS2 | FALSE | Downregulated |
| -0,212628025 | 0,974548421 | SNORA59A | FALSE | Downregulated |
| 0,010533284 | 0,974603515 | MLLT3 | FALSE | Upregulated |
| 0,063756934 | 0,974699042 | PYY2 | FALSE | Upregulated |
| 0,006996143 | 0,974770097 | ITFG2 | FALSE | Upregulated |
| 0,025730169 | 0,974770097 | MC1R | FALSE | Upregulated |
| -0,037435356 | 0,974770097 | MUC12 | FALSE | Downregulated |
| 0,00464289 | 0,974770097 | PLEKHA3 | FALSE | Upregulated |
| -0,006455497 | 0,974804534 | EDNRB | FALSE | Downregulated |
| -0,259776807 | 0,974809308 | RNA5S2 | FALSE | Downregulated |
| 0,007224139 | 0,974809308 | ZNF566 | FALSE | Upregulated |
| -0,010707213 | 0,974943478 | FOXO3 | FALSE | Downregulated |
| -0,055636448 | 0,974943478 | LINC00487 | FALSE | Downregulated |
| 0,009669685 | 0,974943478 | MSL3P1 | FALSE | Upregulated |

| | | | | |
|---|---|---|---|---|
| -0,009491148 | 0,974943478 | ZNF582 | FALSE | Downregulated |
| 0,012996814 | 0,975115993 | LINC01000 | FALSE | Upregulated |
| -0,253737863 | 0,975115993 | TNNC2 | FALSE | Downregulated |
| -0,014040918 | 0,975185536 | LOC101927942 | FALSE | Downregulated |
| 0,045676187 | 0,97527014 | LINC00540 | FALSE | Upregulated |
| 0,026697732 | 0,975483953 | C18orf42 | FALSE | Upregulated |
| -0,017644188 | 0,975483953 | FBXO16 | FALSE | Downregulated |
| -0,003284371 | 0,975483953 | ZMYND11 | FALSE | Downregulated |
| 0,009063742 | 0,975647303 | RPP40 | FALSE | Upregulated |
| 0,06343316 | 0,975779082 | GRK5-IT1 | FALSE | Upregulated |
| 0,019418197 | 0,97585739 | SYN1 | FALSE | Upregulated |
| -0,038241987 | 0,976052169 | ADCYAP1 | FALSE | Downregulated |
| -0,004734669 | 0,976052169 | CDC16 | FALSE | Downregulated |
| -0,006793676 | 0,976052169 | DDI2 | FALSE | Downregulated |
| 0,005120689 | 0,976052169 | DNAJB6 | FALSE | Upregulated |
| 0,008922702 | 0,976052169 | SLC19A3 | FALSE | Upregulated |
| -0,1527631 | 0,976052169 | TRNAK2 | FALSE | Downregulated |
| -0,027536876 | 0,976270228 | CTSV | FALSE | Downregulated |
| -0,00813975 | 0,976307644 | SEC61G | FALSE | Downregulated |
| 0,008787885 | 0,976379293 | DMTN | FALSE | Upregulated |
| 0,024983755 | 0,976379293 | ZNF252P-AS1 | FALSE | Upregulated |
| -0,009869941 | 0,976631056 | DXO | FALSE | Downregulated |
| 0,036193737 | 0,976631056 | SPDYA | FALSE | Upregulated |
| -0,018200388 | 0,976765955 | LOC100507462 | FALSE | Downregulated |
| -0,065272845 | 0,976809541 | LOC102724108 | FALSE | Downregulated |
| 0,003681062 | 0,976953754 | RABGAP1 | FALSE | Upregulated |
| 0,00415297 | 0,977016647 | PTGES3 | FALSE | Upregulated |
| -0,030896485 | 0,977322995 | LRGUK | FALSE | Downregulated |
| -0,028374083 | 0,977366861 | SLC45A2 | FALSE | Downregulated |
| 0,014390478 | 0,97749655 | S1PR5 | FALSE | Upregulated |
| -0,212953583 | 0,977714623 | ACTN2 | FALSE | Downregulated |
| -0,005482639 | 0,977741172 | MYLK | FALSE | Downregulated |
| 0,030795164 | 0,978003292 | COL24A1 | FALSE | Upregulated |
| 0,004230649 | 0,978057202 | TYW1 | FALSE | Upregulated |
| 0,003989189 | 0,978115563 | ACTR10 | FALSE | Upregulated |
| -0,010785554 | 0,978224089 | S100A14 | FALSE | Downregulated |
| -0,013524995 | 0,978230758 | LOC101927126 | FALSE | Downregulated |
| -0,227824095 | 0,978369343 | RNVU1-18 | FALSE | Downregulated |
| -0,004256692 | 0,978407354 | POLR3F | FALSE | Downregulated |
| -0,028677646 | 0,978476846 | LOC101929280 | FALSE | Downregulated |
| -0,04245076 | 0,978712365 | CTD-2127H9.1 | FALSE | Downregulated |
| 0,003769014 | 0,978712365 | YTHDF2 | FALSE | Upregulated |
| -0,005577484 | 0,978712365 | ZNF195 | FALSE | Downregulated |
| 0,033754987 | 0,978792664 | TRNAE11 | FALSE | Upregulated |
| 0,056822109 | 0,978873586 | TRNAK13 | FALSE | Upregulated |
| -0,022836052 | 0,97904158 | RXRG | FALSE | Downregulated |
| 0,015129165 | 0,979056041 | CELSR2 | FALSE | Upregulated |
| 0,00733988 | 0,979056041 | DYRK1B | FALSE | Upregulated |
| -0,006120275 | 0,979056041 | STEAP3 | FALSE | Downregulated |
| 0,004229131 | 0,979072342 | NCOA2 | FALSE | Upregulated |
| 0,048919328 | 0,97912333 | LOC101929057 | FALSE | Upregulated |

| | | | | |
|---|---|---|---|---|
| 0,006013495 | 0,979205516 | USP45 | FALSE | Upregulated |
| -0,013932721 | 0,979458352 | LOC102724880 | FALSE | Downregulated |
| 0,032222692 | 0,979526094 | LOC101930332 | FALSE | Upregulated |
| 0,007490743 | 0,979592535 | SEC24D | FALSE | Upregulated |
| -0,035661725 | 0,97965407 | LOC730257 | FALSE | Downregulated |
| 0,029966126 | 0,97972015 | LOC102723894 | FALSE | Upregulated |
| -0,035884855 | 0,979816888 | LOC643802 | FALSE | Downregulated |
| -0,007741165 | 0,979816888 | PAQR9 | FALSE | Downregulated |
| -0,005777887 | 0,979816888 | PHLPP2 | FALSE | Downregulated |
| -0,013193119 | 0,979828644 | ENAM | FALSE | Downregulated |
| -0,019792921 | 0,979828644 | LOC101928852 | FALSE | Downregulated |
| 0,013556106 | 0,979828644 | LOC283922 | FALSE | Upregulated |
| -0,006305104 | 0,979840088 | AVPI1 | FALSE | Downregulated |
| -0,020332308 | 0,979932469 | FFAR2 | FALSE | Downregulated |
| -0,008473936 | 0,979951436 | VMAC | FALSE | Downregulated |
| 0,005412896 | 0,979987107 | CHD9 | FALSE | Upregulated |
| 0,068769821 | 0,98000561 | KCNS1 | FALSE | Upregulated |
| -0,023364723 | 0,980039344 | ADD3-AS1 | FALSE | Downregulated |
| 0,008105655 | 0,980039344 | COLEC11 | FALSE | Upregulated |
| -0,044427016 | 0,980039344 | ELANE | FALSE | Downregulated |
| 0,016306858 | 0,980039344 | LOC101927339 | FALSE | Upregulated |
| 0,030616451 | 0,980039344 | LRRC73 | FALSE | Upregulated |
| -0,039187704 | 0,980039344 | RNY3 | FALSE | Downregulated |
| -0,004380662 | 0,980042234 | SLC25A39 | FALSE | Downregulated |
| -0,0052392 | 0,980042234 | WDR74 | FALSE | Downregulated |
| -0,041032487 | 0,980112726 | LOC101927762 | FALSE | Downregulated |
| 0,017409076 | 0,980180934 | LYPD3 | FALSE | Upregulated |
| -0,004649425 | 0,980357559 | DIRC2 | FALSE | Downregulated |
| 0,00763579 | 0,980712726 | USP49 | FALSE | Upregulated |
| -0,005169867 | 0,980722255 | ARL15 | FALSE | Downregulated |
| 0,042431109 | 0,980783229 | GSG1L | FALSE | Upregulated |
| 0,004034165 | 0,980783229 | PATZ1 | FALSE | Upregulated |
| 0,023866093 | 0,980783229 | ZNF728 | FALSE | Upregulated |
| 0,018467211 | 0,981039283 | HNRNPA1P33 | FALSE | Upregulated |
| 0,005829651 | 0,981039283 | NUAK1 | FALSE | Upregulated |
| -0,003159101 | 0,981039283 | PRPF19 | FALSE | Downregulated |
| 0,00344108 | 0,981277496 | METTL13 | FALSE | Upregulated |
| 0,023172372 | 0,981593401 | CEMP1 | FALSE | Upregulated |
| 0,037319384 | 0,98161755 | LOC102723659 | FALSE | Upregulated |
| -0,004697926 | 0,981769355 | FZD4 | FALSE | Downregulated |
| -0,040211385 | 0,981769355 | LOC102724518 | FALSE | Downregulated |
| 0,006219159 | 0,981769355 | TRIB1 | FALSE | Upregulated |
| 0,006251944 | 0,981903178 | BTAF1 | FALSE | Upregulated |
| 0,008029682 | 0,981903178 | LRRC37B | FALSE | Upregulated |
| -0,015038958 | 0,981903178 | TDRD6 | FALSE | Downregulated |
| 0,004500934 | 0,981903178 | TMEM19 | FALSE | Upregulated |
| -0,027475296 | 0,981903178 | UPP2 | FALSE | Downregulated |
| 0,02073937 | 0,981985585 | C22orf43 | FALSE | Upregulated |
| -0,004550549 | 0,981985585 | MGAT2 | FALSE | Downregulated |
| 0,004280118 | 0,982096682 | FAM46A | FALSE | Upregulated |
| 0,010011748 | 0,982311596 | B9D1 | FALSE | Upregulated |

| | | | | |
|---|---|---|---|---|
| -0,041459198 | 0,982367908 | LOC100131289 | FALSE | Downregulated |
| -0,005788685 | 0,982367908 | PDK2 | FALSE | Downregulated |
| -0,00316809 | 0,982367908 | PITPNB | FALSE | Downregulated |
| -0,005994136 | 0,982367908 | POLB | FALSE | Downregulated |
| -0,013530113 | 0,982367908 | PSD | FALSE | Downregulated |
| 0,00585472 | 0,982367908 | SHE | FALSE | Upregulated |
| 0,013423955 | 0,982367908 | SLC15A2 | FALSE | Upregulated |
| -0,138440308 | 0,982367908 | SLX1B | FALSE | Downregulated |
| -0,00538161 | 0,982367908 | ZNF223 | FALSE | Downregulated |
| 0,016806363 | 0,982392957 | ADH7 | FALSE | Upregulated |
| -0,042375751 | 0,982392957 | PLCZ1 | FALSE | Downregulated |
| -0,004725165 | 0,982392957 | TPRKB | FALSE | Downregulated |
| -0,020412077 | 0,982479864 | ENOX1 | FALSE | Downregulated |
| 0,003602433 | 0,982506943 | ENO1 | FALSE | Upregulated |
| 0,022197368 | 0,982506943 | LOC101927458 | FALSE | Upregulated |
| 0,021130777 | 0,982506943 | LOC101928361 | FALSE | Upregulated |
| -0,033930926 | 0,982513202 | NANOG | FALSE | Downregulated |
| -0,181517625 | 0,982513202 | SMYD1 | FALSE | Downregulated |
| -0,099522995 | 0,982513202 | TRNAV11 | FALSE | Downregulated |
| -0,01226344 | 0,98270558 | CHRD | FALSE | Downregulated |
| 0,028961628 | 0,982811745 | LOC644662 | FALSE | Upregulated |
| -0,005763299 | 0,982811745 | MIS18A | FALSE | Downregulated |
| 0,008481167 | 0,982811745 | NPHP3 | FALSE | Upregulated |
| 0,005830082 | 0,982811745 | SSR3 | FALSE | Upregulated |
| 0,00424431 | 0,982811775 | SKA2 | FALSE | Upregulated |
| 0,015240124 | 0,982869381 | MYO15A | FALSE | Upregulated |
| 0,005614241 | 0,982960117 | ABCG5 | FALSE | Upregulated |
| -0,176952248 | 0,982960117 | TRNAV21 | FALSE | Downregulated |
| 0,004912502 | 0,982968875 | JRKL | FALSE | Upregulated |
| 0,03854557 | 0,983035719 | LOC100289511 | FALSE | Upregulated |
| 0,006243115 | 0,983096413 | TVP23C-CDRT4 | FALSE | Upregulated |
| 0,006446274 | 0,983096413 | ZNF549 | FALSE | Upregulated |
| -0,00908558 | 0,983129349 | NAP1L2 | FALSE | Downregulated |
| -0,17313036 | 0,983129349 | XIRP2 | FALSE | Downregulated |
| -0,003385286 | 0,983146166 | GNPTAB | FALSE | Downregulated |
| 0,008570074 | 0,983217843 | SMA4 | FALSE | Upregulated |
| 0,029846399 | 0,983245807 | PRRX1 | FALSE | Upregulated |
| -0,017368902 | 0,983279308 | GP6 | FALSE | Downregulated |
| -0,012092443 | 0,983279308 | RP4-782L23.1 | FALSE | Downregulated |
| 0,017419268 | 0,983365514 | GNAT2 | FALSE | Upregulated |
| 0,010370059 | 0,983365514 | LOC101929005 | FALSE | Upregulated |
| 0,003960354 | 0,983373705 | CAMTA1 | FALSE | Upregulated |
| 0,036375172 | 0,983439439 | LOC102723654 | FALSE | Upregulated |
| 0,004956229 | 0,983494036 | PDK1 | FALSE | Upregulated |
| -0,00299095 | 0,983514726 | C16orf70 | FALSE | Downregulated |
| 0,010364774 | 0,983514726 | CCDC163P | FALSE | Upregulated |
| -0,030850407 | 0,983514726 | LOC101929434 | FALSE | Downregulated |
| -0,013456421 | 0,983514726 | LOC648987 | FALSE | Downregulated |
| 0,010062135 | 0,983514726 | LTK | FALSE | Upregulated |
| -0,004901451 | 0,983514726 | TIMM17A | FALSE | Downregulated |
| -0,00219184 | 0,983514726 | VDAC1 | FALSE | Downregulated |

| | | | | |
|---|---|---|---|---|
| -0,003438139 | 0,983514726 | VTN | FALSE | Downregulated |
| -0,001902338 | 0,983521028 | PWP1 | FALSE | Downregulated |
| -0,005714607 | 0,983542212 | FAM102A | FALSE | Downregulated |
| -0,007529062 | 0,983542212 | HES1 | FALSE | Downregulated |
| 0,005069652 | 0,983542212 | RELN | FALSE | Upregulated |
| -0,004893007 | 0,983553952 | DHRS12 | FALSE | Downregulated |
| -0,013557517 | 0,983860229 | LOC101927763 | FALSE | Downregulated |
| 0,003241211 | 0,983860229 | RPS6KA3 | FALSE | Upregulated |
| 0,003215847 | 0,983991289 | ZBTB43 | FALSE | Upregulated |
| -0,002354232 | 0,984014821 | ATXN1L | FALSE | Downregulated |
| 0,010488373 | 0,984014821 | LOC102724252 | FALSE | Upregulated |
| 0,016630583 | 0,984014821 | LOC440895 | FALSE | Upregulated |
| -0,004942103 | 0,984014821 | LYRM4 | FALSE | Downregulated |
| 0,006957813 | 0,984014821 | RFTN1 | FALSE | Upregulated |
| -0,005964333 | 0,9840856 | PSPH | FALSE | Downregulated |
| 0,01011296 | 0,984173111 | HERC2P2 | FALSE | Upregulated |
| 0,022139363 | 0,984209503 | LOC440354 | FALSE | Upregulated |
| 0,031986039 | 0,984272528 | LOC102723801 | FALSE | Upregulated |
| 0,013233481 | 0,984272528 | PRRT2 | FALSE | Upregulated |
| -0,004324434 | 0,984576035 | C5orf42 | FALSE | Downregulated |
| -0,032133013 | 0,984576035 | FAM194A | FALSE | Downregulated |
| -0,015582578 | 0,984576035 | LINC00622 | FALSE | Downregulated |
| 0,0114245 | 0,984576035 | LOC642236 | FALSE | Upregulated |
| 0,013578144 | 0,984656906 | NLRP2 | FALSE | Upregulated |
| 0,016746814 | 0,984746151 | MIR1282 | FALSE | Upregulated |
| 0,02386234 | 0,984869509 | HIST1H2BN | FALSE | Upregulated |
| 0,012560309 | 0,984992764 | GRIP1 | FALSE | Upregulated |
| 0,023705288 | 0,985032323 | TMLHE-AS1 | FALSE | Upregulated |
| -0,007267829 | 0,985659208 | S1PR3 | FALSE | Downregulated |
| -0,009233582 | 0,985659208 | ZNF880 | FALSE | Downregulated |
| -0,002909558 | 0,985749901 | NMT1 | FALSE | Downregulated |
| 0,03186378 | 0,985758861 | LOC100128573 | FALSE | Upregulated |
| -0,002340155 | 0,985758861 | NAPA | FALSE | Downregulated |
| 0,037841143 | 0,985758861 | SPOCD1 | FALSE | Upregulated |
| 0,0023577 | 0,986142403 | SUPT16H | FALSE | Upregulated |
| -0,002530656 | 0,986181035 | APLP2 | FALSE | Downregulated |
| -0,004731209 | 0,986181035 | EIF2AK3 | FALSE | Downregulated |
| -0,005852515 | 0,986214593 | BAIAP3 | FALSE | Downregulated |
| 0,005130396 | 0,986314294 | GGTA1P | FALSE | Upregulated |
| 0,131243999 | 0,986314294 | TTN | FALSE | Upregulated |
| 0,00494085 | 0,986347492 | AGPAT2 | FALSE | Upregulated |
| -0,003212627 | 0,986347492 | PLEKHF2 | FALSE | Downregulated |
| 0,122208496 | 0,986444344 | TRIM54 | FALSE | Upregulated |
| -0,036860245 | 0,98657918 | CLEC19A | FALSE | Downregulated |
| -0,13652196 | 0,986642049 | MYOZ2 | FALSE | Downregulated |
| -0,001821244 | 0,986699512 | TBC1D15 | FALSE | Downregulated |
| -0,035423715 | 0,986776917 | GNAS-AS1 | FALSE | Downregulated |
| -0,008564901 | 0,986776917 | SEPSECS-AS1 | FALSE | Downregulated |
| 0,132642468 | 0,987027976 | LOC102724441 | FALSE | Upregulated |
| 0,031642904 | 0,987043649 | AC002480.3 | FALSE | Upregulated |
| -0,010183152 | 0,987043649 | AKR1CL1 | FALSE | Downregulated |

| | | | | |
|---|---|---|---|---|
| -0,00359448 | 0,987043649 | LYPLAL1 | FALSE | Downregulated |
| 0,019054066 | 0,987043649 | M1AP | FALSE | Upregulated |
| -0,130881403 | 0,987043649 | TNNC1 | FALSE | Downregulated |
| -0,062881106 | 0,987043649 | TSIX | FALSE | Downregulated |
| 0,018457093 | 0,987137493 | LOC102723756 | FALSE | Upregulated |
| -0,018642681 | 0,987137493 | MDGA2 | FALSE | Downregulated |
| 0,009191298 | 0,987137493 | NPIPB4 | FALSE | Upregulated |
| 0,018580965 | 0,987137493 | NRN1L | FALSE | Upregulated |
| 0,00767608 | 0,987137493 | PPIL6 | FALSE | Upregulated |
| -0,003647982 | 0,987137493 | WDR47 | FALSE | Downregulated |
| 0,024945159 | 0,987369775 | LOC102724543 | FALSE | Upregulated |
| -0,004731409 | 0,987369775 | SMAGP | FALSE | Downregulated |
| -0,010744959 | 0,987555809 | LOC101928767 | FALSE | Downregulated |
| -0,02776746 | 0,987555809 | RNF103-CHMP3 | FALSE | Downregulated |
| -0,042872755 | 0,987576417 | LOC613038 | FALSE | Downregulated |
| 0,001864923 | 0,987583972 | CTNNB1 | FALSE | Upregulated |
| -0,015197347 | 0,987583972 | LOC284412 | FALSE | Downregulated |
| -0,001986785 | 0,987583972 | STAG1 | FALSE | Downregulated |
| 0,008270528 | 0,987583972 | ZNF503-AS1 | FALSE | Upregulated |
| -0,012883396 | 0,987588343 | LOC102723641 | FALSE | Downregulated |
| 0,003182713 | 0,987621272 | CINP | FALSE | Upregulated |
| 0,00999663 | 0,987812711 | LOC101929385 | FALSE | Upregulated |
| 0,006035978 | 0,987812711 | PRSS8 | FALSE | Upregulated |
| -0,011692834 | 0,987949786 | LOC102724307 | FALSE | Downregulated |
| 0,010485102 | 0,988080085 | ARSF | FALSE | Upregulated |
| -0,029273758 | 0,988080085 | JSRP1 | FALSE | Downregulated |
| -0,002463697 | 0,988080085 | MRPL10 | FALSE | Downregulated |
| -0,017281625 | 0,988080085 | SLC9A5 | FALSE | Downregulated |
| -0,016775894 | 0,988243879 | LOC101928512 | FALSE | Downregulated |
| -0,020547232 | 0,988252492 | AC007405.4 | FALSE | Downregulated |
| -0,009529897 | 0,988252492 | CHEK2 | FALSE | Downregulated |
| 0,006937227 | 0,988252492 | ZFHX2 | FALSE | Upregulated |
| -0,006661364 | 0,988266322 | ACER2 | FALSE | Downregulated |
| 0,004530231 | 0,988266322 | IFITM2 | FALSE | Upregulated |
| 0,008468988 | 0,988266322 | LOC100507424 | FALSE | Upregulated |
| -0,0340971 | 0,988266322 | PXT1 | FALSE | Downregulated |
| -0,009614207 | 0,988289003 | USP6 | FALSE | Downregulated |
| 0,003121484 | 0,988305881 | CNTRL | FALSE | Upregulated |
| -0,017015886 | 0,988363892 | RP11-230B22.1 | FALSE | Downregulated |
| 0,004834071 | 0,988573158 | VAV1 | FALSE | Upregulated |
| -0,007715297 | 0,988615679 | CHADL | FALSE | Downregulated |
| -0,002459523 | 0,988615679 | TOMM20 | FALSE | Downregulated |
| -0,001764135 | 0,989126221 | TMEM106B | FALSE | Downregulated |
| 0,003045516 | 0,989149279 | LOC93622 | FALSE | Upregulated |
| -0,01261768 | 0,989149279 | SEPT14 | FALSE | Downregulated |
| -0,006277658 | 0,989330748 | CR1 | FALSE | Downregulated |
| 0,009602749 | 0,989330748 | LOC102724927 | FALSE | Upregulated |
| -0,021010764 | 0,989509054 | DPYSL5 | FALSE | Downregulated |
| -0,020690728 | 0,989529946 | MKRN7P | FALSE | Downregulated |
| 0,10393554 | 0,989602178 | PADI1 | FALSE | Upregulated |
| -0,016829208 | 0,989618333 | SOX30 | FALSE | Downregulated |

| | | | | |
|---|---|---|---|---|
| 0,002696746 | 0,989704823 | SERAC1 | FALSE | Upregulated |
| 0,005437668 | 0,990029428 | FENDRR | FALSE | Upregulated |
| 0,003753255 | 0,990029428 | ZSCAN31 | FALSE | Upregulated |
| -0,008578899 | 0,990123937 | LINC00570 | FALSE | Downregulated |
| -0,017371932 | 0,990123937 | SLC2A5 | FALSE | Downregulated |
| -0,031584925 | 0,990199443 | KY | FALSE | Downregulated |
| -0,00362795 | 0,990199443 | TBXAS1 | FALSE | Downregulated |
| 0,002445621 | 0,990199443 | VPS33B | FALSE | Upregulated |
| 0,015771314 | 0,990299141 | RP11-1080G15.1 | FALSE | Upregulated |
| -0,0147383 | 0,990299141 | S100P | FALSE | Downregulated |
| 0,003609887 | 0,99052963 | PAMR1 | FALSE | Upregulated |
| 0,003500096 | 0,990960751 | ARL6IP6 | FALSE | Upregulated |
| 0,002721863 | 0,990960751 | SLAIN1 | FALSE | Upregulated |
| 0,004388644 | 0,990960751 | TCF21 | FALSE | Upregulated |
| -0,01941782 | 0,991038753 | CLEC18A | FALSE | Downregulated |
| -0,003281439 | 0,991076805 | NUTM2A-AS1 | FALSE | Downregulated |
| 0,004064636 | 0,991076805 | TM4SF5 | FALSE | Upregulated |
| -0,004110244 | 0,991219207 | RP11-395P17.3 | FALSE | Downregulated |
| -0,008479972 | 0,991319805 | RP11-540A21.2 | FALSE | Downregulated |
| -0,011464132 | 0,991469451 | GHRL | FALSE | Downregulated |
| -0,002770358 | 0,991494756 | ANKRD26 | FALSE | Downregulated |
| -0,087008631 | 0,991494756 | CSRP3 | FALSE | Downregulated |
| 0,001922763 | 0,991494756 | ROCK2 | FALSE | Upregulated |
| 0,002575544 | 0,991494756 | ZNF160 | FALSE | Upregulated |
| 0,002035249 | 0,991519907 | MED25 | FALSE | Upregulated |
| -0,004318645 | 0,991612945 | LINC01003 | FALSE | Downregulated |
| -0,002655608 | 0,991975936 | CHST13 | FALSE | Downregulated |
| -0,003982876 | 0,992006284 | RCE1 | FALSE | Downregulated |
| -0,080556934 | 0,992006284 | RNU1-2 | FALSE | Downregulated |
| 0,020776765 | 0,992006284 | VAX2 | FALSE | Upregulated |
| 0,004367762 | 0,99236386 | ABI3BP | FALSE | Upregulated |
| 0,0772382 | 0,992662129 | TNNT1 | FALSE | Upregulated |
| 0,001575021 | 0,992732931 | C1orf123 | FALSE | Upregulated |
| 0,001098783 | 0,992732931 | DIAPH1 | FALSE | Upregulated |
| 0,001686387 | 0,992732931 | MOB3C | FALSE | Upregulated |
| -0,003026171 | 0,99278467 | FLJ42627 | FALSE | Downregulated |
| 0,001399738 | 0,99278467 | GNPTG | FALSE | Upregulated |
| 0,001334561 | 0,99278467 | HNRNPA0 | FALSE | Upregulated |
| 0,008808538 | 0,99278467 | LHX9 | FALSE | Upregulated |
| -0,01127341 | 0,99278467 | LINC00881 | FALSE | Downregulated |
| -0,000984488 | 0,99278467 | USP10 | FALSE | Downregulated |
| -0,003980833 | 0,992839444 | PCOLCE | FALSE | Downregulated |
| -0,001259034 | 0,993005691 | RELA | FALSE | Downregulated |
| 0,003788563 | 0,993120209 | SRGAP2B | FALSE | Upregulated |
| -0,001788623 | 0,993298537 | CDIPT | FALSE | Downregulated |
| -0,001577759 | 0,993298537 | KAT8 | FALSE | Downregulated |
| -0,003207129 | 0,993298537 | RABL2A | FALSE | Downregulated |
| -0,021723036 | 0,993298537 | RGPD6 | FALSE | Downregulated |
| -0,003452342 | 0,993325907 | TRNE | FALSE | Downregulated |
| -0,014544395 | 0,993385146 | LOC102724758 | FALSE | Downregulated |
| 0,002317949 | 0,993443311 | BEX4 | FALSE | Upregulated |

| | | | | |
|---|---|---|---|---|
| -0,014391579 | 0,993443311 | LINC00887 | FALSE | Downregulated |
| -0,012052901 | 0,993443311 | LOC102723761 | FALSE | Downregulated |
| 0,006819255 | 0,993443311 | PKDREJ | FALSE | Upregulated |
| -0,001075657 | 0,993518812 | SRSF9 | FALSE | Downregulated |
| 0,005267289 | 0,993665163 | KCNAB3 | FALSE | Upregulated |
| 0,001699997 | 0,993841034 | B3GALTL | FALSE | Upregulated |
| 0,014749088 | 0,993841034 | GLIPR1L1 | FALSE | Upregulated |
| -0,001858131 | 0,994271543 | PARP9 | FALSE | Downregulated |
| 0,007678566 | 0,994360951 | SH3GL1P1 | FALSE | Upregulated |
| -0,006760424 | 0,994511303 | FAM24B | FALSE | Downregulated |
| 0,001535886 | 0,994511303 | FGFR4 | FALSE | Upregulated |
| -0,003389723 | 0,994511303 | MIR600HG | FALSE | Downregulated |
| 0,001871245 | 0,994511303 | POLG2 | FALSE | Upregulated |
| -0,005409125 | 0,994511303 | SNX15 | FALSE | Downregulated |
| -0,002475252 | 0,994511303 | TIGD5 | FALSE | Downregulated |
| -0,056836978 | 0,994511303 | TNNT3 | FALSE | Downregulated |
| 0,047356893 | 0,994511303 | TRNAG21 | FALSE | Upregulated |
| 0,016066164 | 0,994511303 | WNT7A | FALSE | Upregulated |
| -0,001267297 | 0,9945795 | NUCB1 | FALSE | Downregulated |
| 0,008208291 | 0,994713535 | CAPZA3 | FALSE | Upregulated |
| -0,003815602 | 0,994713535 | LOC101928228 | FALSE | Downregulated |
| -0,010028413 | 0,994799701 | FUT5 | FALSE | Downregulated |
| -0,009041808 | 0,99482508 | A2MP1 | FALSE | Downregulated |
| 0,002326408 | 0,99482508 | NCMAP | FALSE | Upregulated |
| -0,001594788 | 0,99486856 | GYPC | FALSE | Downregulated |
| -0,006573565 | 0,99486856 | LINC00176 | FALSE | Downregulated |
| 0,010674269 | 0,994949759 | GRIN3B | FALSE | Upregulated |
| 0,005853916 | 0,994996653 | SAPCD2 | FALSE | Upregulated |
| -0,002607331 | 0,995125128 | HELB | FALSE | Downregulated |
| -0,003308792 | 0,99513702 | ALOX12P2 | FALSE | Downregulated |
| 0,002877998 | 0,99513702 | BEX1 | FALSE | Upregulated |
| -0,003076615 | 0,99513702 | EFCAB13 | FALSE | Downregulated |
| 0,001597 | 0,99513702 | HMBOX1 | FALSE | Upregulated |
| -0,002704672 | 0,99513702 | OAS2 | FALSE | Downregulated |
| -0,001850217 | 0,995258538 | SCAND2P | FALSE | Downregulated |
| -0,002127535 | 0,995348896 | EFCAB6 | FALSE | Downregulated |
| -0,003287504 | 0,995397767 | RP5-1180C10.2 | FALSE | Downregulated |
| 0,000806144 | 0,995546085 | COPZ1 | FALSE | Upregulated |
| 0,003141831 | 0,995546085 | GOLGA8M | FALSE | Upregulated |
| -0,005933386 | 0,995546085 | LOC286367 | FALSE | Downregulated |
| 0,001046972 | 0,995546085 | NEK4 | FALSE | Upregulated |
| -0,010036845 | 0,995546085 | NNAT | FALSE | Downregulated |
| 0,00098935 | 0,996232048 | RPL7A | FALSE | Upregulated |
| -0,008468519 | 0,996276836 | LOC102723682 | FALSE | Downregulated |
| -0,002602037 | 0,9963679 | AOC2 | FALSE | Downregulated |
| 0,03800308 | 0,996459152 | XIRP1 | FALSE | Upregulated |
| -0,037104911 | 0,996517913 | MYH7 | FALSE | Downregulated |
| -0,001023859 | 0,996647085 | PHACTR2 | FALSE | Downregulated |
| 0,004523078 | 0,996698005 | MCF2 | FALSE | Upregulated |
| -0,000850659 | 0,996698005 | PPFIBP2 | FALSE | Downregulated |
| -0,001568784 | 0,99684617 | RP11-513G11.3 | FALSE | Downregulated |

| | | | | |
|---|---|---|---|---|
| 0,002562522 | 0,997069247 | MAPK12 | FALSE | Upregulated |
| -0,00110823 | 0,997069247 | RBM4B | FALSE | Downregulated |
| 0,007556707 | 0,997069247 | TSPO2 | FALSE | Upregulated |
| -0,000661911 | 0,997090735 | EIF2B3 | FALSE | Downregulated |
| -0,002268835 | 0,997242078 | LYG1 | FALSE | Downregulated |
| 0,006133822 | 0,997329551 | LOC101928824 | FALSE | Upregulated |
| -0,000811921 | 0,997533561 | HSD3B7 | FALSE | Downregulated |
| 0,001678951 | 0,997533561 | LINC00950 | FALSE | Upregulated |
| -0,002145288 | 0,997645315 | CCDC144B | FALSE | Downregulated |
| 0,00538398 | 0,997645315 | LOC102723975 | FALSE | Upregulated |
| 0,000691925 | 0,997890332 | B4GALT2 | FALSE | Upregulated |
| -0,003984949 | 0,997955979 | CRX | FALSE | Downregulated |
| 0,001763688 | 0,99796857 | LOC727896 | FALSE | Upregulated |
| -0,004706092 | 0,998375571 | KRTAP5-9 | FALSE | Downregulated |
| -0,0007824 | 0,998461017 | MAD2L2 | FALSE | Downregulated |
| 0,00071475 | 0,9986995 | CALR | FALSE | Upregulated |
| 0,002161002 | 0,9986995 | CPO | FALSE | Upregulated |
| 0,000414599 | 0,9986995 | ZNF746 | FALSE | Upregulated |
| 0,001648187 | 0,99893396 | GDPD4 | FALSE | Upregulated |
| -0,001861727 | 0,998947728 | DHRS2 | FALSE | Downregulated |
| 0,000702185 | 0,998953833 | CDC14A | FALSE | Upregulated |
| -0,00129928 | 0,999100349 | SSPO | FALSE | Downregulated |
| -0,000291678 | 0,999129423 | ST5 | FALSE | Downregulated |
| 0,001750388 | 0,999167299 | ALG1L9P | FALSE | Upregulated |
| -0,000846004 | 0,999167299 | ATXN7L2 | FALSE | Downregulated |
| -0,000414468 | 0,999167299 | C2CD2L | FALSE | Downregulated |
| -0,00020264 | 0,999167299 | CCNT1 | FALSE | Downregulated |
| 0,001800147 | 0,999167299 | LOC101930129 | FALSE | Upregulated |
| 0,000236498 | 0,999167299 | NARS | FALSE | Upregulated |
| 0,0002379 | 0,999167299 | PDCL | FALSE | Upregulated |
| 0,000399984 | 0,999167299 | PRRC2C | FALSE | Upregulated |
| 0,00244783 | 0,999167299 | TAS2R4 | FALSE | Upregulated |
| -0,000269885 | 0,999186072 | AGPS | FALSE | Downregulated |
| 0,01138761 | 0,999186072 | CKM | FALSE | Upregulated |
| 0,000510871 | 0,999272873 | LOC101928120 | FALSE | Upregulated |
| -0,001748207 | 0,999275745 | WDR11-AS1 | FALSE | Downregulated |
| 0,001626055 | 0,999314894 | C21orf15 | FALSE | Upregulated |
| 0,00021577 | 0,999314894 | MRPL17 | FALSE | Upregulated |
| 0,000702144 | 0,999314894 | NLRP12 | FALSE | Upregulated |
| 0,005621964 | 0,999314894 | TRNAH9 | FALSE | Upregulated |
| 0,001628511 | 0,999427423 | AC104777.4 | FALSE | Upregulated |
| -0,000127702 | 0,999482121 | CDK12 | FALSE | Downregulated |
| 0,000441931 | 0,999482121 | LOC100506922 | FALSE | Upregulated |
| -0,000734122 | 0,999709126 | LOC101928560 | FALSE | Downregulated |
| -0,000178132 | 0,999709126 | OSER1-AS1 | FALSE | Downregulated |
| 0,000836585 | 0,999766275 | LOC102724812 | FALSE | Upregulated |
| -8,8435E-05 | 0,99980939 | CLASP1 | FALSE | Downregulated |
| 6,98677E-05 | 0,999895957 | CHM | FALSE | Upregulated |
| -1,85352E-05 | 1 | DSCR3 | FALSE | Downregulated |
| 3,79446E-05 | 1 | FAM193A | FALSE | Upregulated |
| -6,94391E-07 | 1 | GPR155 | FALSE | Downregulated |

| | | | | |
|---|---|---|---|---|
| -6,98676E-06 | 1 | PLEKHG7 | FALSE | Downregulated |
| 0 | 1 | RNA5S10 | FALSE | Upregulated |
| 0 | 1 | RNA5S3 | FALSE | Upregulated |
| 0 | 1 | RNA5S5 | FALSE | Upregulated |
| 1,94108E-05 | 1 | SLC44A1 | FALSE | Upregulated |
| -9,82808E-06 | 1 | TDRD3 | FALSE | Downregulated |
| 0,000107118 | 1 | ZNF530 | FALSE | Upregulated |
| | | RNA5S12 | | |
| | | RNA5S13 | | |
| | | RNA5S14 | | |
| | | RNA5S15 | | |
| | | RNA5S16 | | |
| | | RNA5S4 | | |
| | | RNA5S6 | | |
| | | RNA5S7 | | |
| | | RNA5S8 | | |

<u>ClinTCMR vs. NHR</u>

| log2FoldChange | padj | gene_names | significance | direction |
|---|---|---|---|---|
| 3,007207166 | 2,75925E-20 | TOP2A | TRUE | Upregulated |
| 3,166241378 | 2,78515E-19 | FOXM1 | TRUE | Upregulated |
| 3,279421268 | 2,78515E-19 | RRM2 | TRUE | Upregulated |
| 2,928404885 | 5,78565E-19 | BIRC5 | TRUE | Upregulated |
| 3,267074727 | 5,78565E-19 | NCAPG | TRUE | Upregulated |
| 2,864489382 | 7,91385E-19 | MKI67 | TRUE | Upregulated |
| 4,247248403 | 8,45992E-19 | FAM111B | TRUE | Upregulated |
| 2,683708758 | 3,29788E-18 | HMMR | TRUE | Upregulated |
| 2,66115725 | 4,72699E-18 | ZWINT | TRUE | Upregulated |
| 3,948238297 | 6,14311E-18 | DTL | TRUE | Upregulated |
| 3,984378996 | 1,7596E-17 | CXCL10 | TRUE | Upregulated |
| 4,071329182 | 2,57077E-16 | CDC45 | TRUE | Upregulated |
| 3,104666573 | 3,05676E-16 | BUB1B | TRUE | Upregulated |
| 3,781778143 | 3,60589E-16 | UBD | TRUE | Upregulated |
| 2,724742889 | 5,08252E-16 | CCNA2 | TRUE | Upregulated |
| 2,727014185 | 1,66285E-15 | FABP5 | TRUE | Upregulated |
| 2,647062583 | 2,0257E-15 | CDK1 | TRUE | Upregulated |
| 3,946220899 | 2,0257E-15 | MELK | TRUE | Upregulated |
| 1,965534688 | 2,17753E-15 | ECT2 | TRUE | Upregulated |
| 1,719797874 | 4,85211E-15 | NUSAP1 | TRUE | Upregulated |
| 1,9664764 | 5,40764E-15 | CCNB1 | TRUE | Upregulated |
| 2,541100153 | 1,04363E-14 | PTTG1 | TRUE | Upregulated |
| 3,744134277 | 1,18307E-14 | CDC20 | TRUE | Upregulated |
| 2,375606992 | 1,46334E-14 | PRC1 | TRUE | Upregulated |
| 2,670350465 | 1,58318E-14 | KIFC1 | TRUE | Upregulated |
| 2,017957016 | 3,1056E-14 | CENPF | TRUE | Upregulated |
| 3,311142266 | 8,9897E-14 | CXCL9 | TRUE | Upregulated |
| 3,248667632 | 8,9897E-14 | DLGAP5 | TRUE | Upregulated |
| 2,950604718 | 9,03806E-14 | KIF20A | TRUE | Upregulated |
| 2,384116838 | 1,19303E-13 | HELLS | TRUE | Upregulated |
| 2,423113276 | 1,44913E-13 | TK1 | TRUE | Upregulated |

| | | | | |
|---|---|---|---|---|
| 3,050269978 | 2,30306E-13 | NUF2 | TRUE | Upregulated |
| 3,062245659 | 4,30326E-13 | CLSPN | TRUE | Upregulated |
| 2,942842112 | 4,60657E-13 | HJURP | TRUE | Upregulated |
| 3,063826777 | 5,2132E-13 | UHRF1 | TRUE | Upregulated |
| 1,580904233 | 5,24129E-13 | TAP1 | TRUE | Upregulated |
| 2,066223433 | 5,56918E-13 | TCF19 | TRUE | Upregulated |
| 1,059011111 | 5,87302E-13 | TUBA1B | TRUE | Upregulated |
| 2,145375492 | 9,60161E-13 | APOL3 | TRUE | Upregulated |
| 2,329856827 | 9,60612E-13 | NDC80 | TRUE | Upregulated |
| 2,109878629 | 1,47077E-12 | PARPBP | TRUE | Upregulated |
| 1,631081244 | 1,57267E-12 | AURKA | TRUE | Upregulated |
| 2,856833543 | 2,0246E-12 | AURKB | TRUE | Upregulated |
| 1,388382255 | 2,05012E-12 | TACC3 | TRUE | Upregulated |
| 4,355382932 | 2,21926E-12 | PBK | TRUE | Upregulated |
| 2,378669061 | 2,41517E-12 | KIF23 | TRUE | Upregulated |
| 2,451206099 | 2,75527E-12 | NCAPH | TRUE | Upregulated |
| 1,786270044 | 4,18686E-12 | STAT1 | TRUE | Upregulated |
| 1,539182966 | 5,48416E-12 | TTYH3 | TRUE | Upregulated |
| 1,919433512 | 5,58778E-12 | MCM6 | TRUE | Upregulated |
| 1,58906853 | 6,12565E-12 | HLA-F | TRUE | Upregulated |
| 3,173533111 | 9,67143E-12 | CDKN3 | TRUE | Upregulated |
| 3,494683058 | 1,09853E-11 | UBE2C | TRUE | Upregulated |
| 2,856077879 | 1,25628E-11 | ANLN | TRUE | Upregulated |
| 3,352882106 | 1,33305E-11 | MYBL2 | TRUE | Upregulated |
| 2,801041096 | 1,36181E-11 | ASF1B | TRUE | Upregulated |
| 2,391028207 | 1,3795E-11 | UBE2T | TRUE | Upregulated |
| 3,709952716 | 1,54814E-11 | E2F1 | TRUE | Upregulated |
| 2,261902712 | 1,98841E-11 | CENPU | TRUE | Upregulated |
| 3,457196343 | 2,61673E-11 | APOBEC3B | TRUE | Upregulated |
| 2,463467482 | 2,64046E-11 | KIF2C | TRUE | Upregulated |
| 3,568281383 | 3,38838E-11 | EXO1 | TRUE | Upregulated |
| 1,559682512 | 3,62393E-11 | IFI30 | TRUE | Upregulated |
| 2,406058933 | 4,04841E-11 | TYMS | TRUE | Upregulated |
| 1,635851129 | 4,66755E-11 | MCM5 | TRUE | Upregulated |
| -2,345474466 | 5,38405E-11 | DNM1 | TRUE | Downregulated |
| 2,846715311 | 5,73789E-11 | BUB1 | TRUE | Upregulated |
| 2,661917634 | 5,9029E-11 | CHEK1 | TRUE | Upregulated |
| 3,38884905 | 6,36761E-11 | E2F8 | TRUE | Upregulated |
| 1,425312821 | 6,75031E-11 | MCM4 | TRUE | Upregulated |
| 1,367728088 | 6,78273E-11 | HLA-A | TRUE | Upregulated |
| 2,714105293 | 6,78273E-11 | ORC6 | TRUE | Upregulated |
| 3,285531296 | 7,0513E-11 | CCNB2 | TRUE | Upregulated |
| 0,88241728 | 7,08603E-11 | PSMB10 | FALSE | Upregulated |
| 2,051380644 | 8,83447E-11 | MCM2 | TRUE | Upregulated |
| 0,895925297 | 9,09514E-11 | B2M | FALSE | Upregulated |
| 1,964543731 | 9,63143E-11 | ISG20 | TRUE | Upregulated |
| 1,331090839 | 1,20323E-10 | PIGR | TRUE | Upregulated |
| 2,71656408 | 1,85355E-10 | NEK2 | TRUE | Upregulated |
| 0,914752399 | 2,10956E-10 | CENPN | FALSE | Upregulated |
| 3,421472716 | 2,20064E-10 | MCM10 | TRUE | Upregulated |
| 1,589934186 | 2,49122E-10 | GBP1 | TRUE | Upregulated |

| | | | | |
|---|---|---|---|---|
| 1,846695409 | 2,53733E-10 | ARHGAP11A | TRUE | Upregulated |
| 0,701179511 | 2,62532E-10 | LASP1 | FALSE | Upregulated |
| 2,273174059 | 2,63595E-10 | PLK1 | TRUE | Upregulated |
| 2,999463793 | 3,60391E-10 | CENPK | TRUE | Upregulated |
| 2,476904631 | 3,66622E-10 | SHCBP1 | TRUE | Upregulated |
| 1,694116428 | 3,96494E-10 | KPNA2 | TRUE | Upregulated |
| 2,563162007 | 3,97165E-10 | GTSE1 | TRUE | Upregulated |
| 1,860873023 | 4,63818E-10 | CDCA8 | TRUE | Upregulated |
| 1,905113576 | 5,3745E-10 | LYZ | TRUE | Upregulated |
| 2,509587681 | 5,52909E-10 | CKAP2L | TRUE | Upregulated |
| 1,995177426 | 5,52909E-10 | MAD2L1 | TRUE | Upregulated |
| 2,394996545 | 6,62754E-10 | SLAMF8 | TRUE | Upregulated |
| 3,685279833 | 7,34569E-10 | MND1 | TRUE | Upregulated |
| 2,558732725 | 7,93007E-10 | KIF4A | TRUE | Upregulated |
| 2,714008849 | 8,48899E-10 | CDCA2 | TRUE | Upregulated |
| 1,954650464 | 8,48904E-10 | PAQR4 | TRUE | Upregulated |
| 3,082214262 | 9,5561E-10 | SPC25 | TRUE | Upregulated |
| 0,833173977 | 9,97296E-10 | PSMB8 | FALSE | Upregulated |
| 2,340752623 | 1,13391E-09 | RAD51AP1 | TRUE | Upregulated |
| 1,892240329 | 1,21741E-09 | CENPE | TRUE | Upregulated |
| 1,33845002 | 1,28043E-09 | HLA-B | TRUE | Upregulated |
| 2,810352416 | 1,32053E-09 | SKA3 | TRUE | Upregulated |
| 2,553197489 | 1,44967E-09 | TTK | TRUE | Upregulated |
| 1,581093946 | 1,63261E-09 | FANCI | TRUE | Upregulated |
| 2,846042806 | 1,72991E-09 | CXCL11 | TRUE | Upregulated |
| 0,575154651 | 1,77016E-09 | ARPC2 | FALSE | Upregulated |
| 0,547449158 | 1,96414E-09 | WDR1 | FALSE | Upregulated |
| 0,669803347 | 2,23498E-09 | RPS6KA4 | FALSE | Upregulated |
| 1,149679229 | 2,34858E-09 | LGALS9 | TRUE | Upregulated |
| 2,848953206 | 2,63889E-09 | KIF18A | TRUE | Upregulated |
| 4,424231152 | 3,00318E-09 | ADAMDEC1 | TRUE | Upregulated |
| 1,912457766 | 3,00318E-09 | KIAA0101 | TRUE | Upregulated |
| 1,08695508 | 3,80111E-09 | TMSB10 | TRUE | Upregulated |
| 0,639060194 | 3,83507E-09 | VAMP8 | FALSE | Upregulated |
| 2,471562351 | 3,9888E-09 | IL32 | TRUE | Upregulated |
| 3,742960176 | 4,28895E-09 | E2F7 | TRUE | Upregulated |
| 1,081275471 | 4,49925E-09 | PCNA | TRUE | Upregulated |
| 3,316166043 | 5,14139E-09 | DEPDC1 | TRUE | Upregulated |
| 1,751314927 | 5,18504E-09 | CENPW | TRUE | Upregulated |
| 1,262194258 | 5,45497E-09 | PSMB9 | TRUE | Upregulated |
| 1,21966247 | 5,61408E-09 | HLA-DMB | TRUE | Upregulated |
| 0,723426037 | 6,39351E-09 | PSME2 | FALSE | Upregulated |
| 2,567143683 | 9,46099E-09 | DIAPH3 | TRUE | Upregulated |
| 1,859802119 | 1,12033E-08 | CDC7 | TRUE | Upregulated |
| 1,202984813 | 1,16115E-08 | TAGLN2 | TRUE | Upregulated |
| -1,280897771 | 1,34166E-08 | NFKBIZ | TRUE | Downregulated |
| 1,678360732 | 1,37152E-08 | LGALS3 | TRUE | Upregulated |
| 1,078975117 | 1,37394E-08 | SLC15A3 | TRUE | Upregulated |
| 1,934928522 | 1,42005E-08 | CD72 | TRUE | Upregulated |
| 1,927017058 | 1,49663E-08 | KIF14 | TRUE | Upregulated |
| 1,45297649 | 1,75283E-08 | EZH2 | TRUE | Upregulated |

| | | | | |
|---|---|---|---|---|
| 0,873584735 | 1,75845E-08 | PLOD3 | FALSE | Upregulated |
| 1,085211073 | 1,78913E-08 | CSTB | TRUE | Upregulated |
| -0,979965393 | 1,8327E-08 | NUDT4 | FALSE | Downregulated |
| 2,280056034 | 1,95875E-08 | CDCA5 | TRUE | Upregulated |
| 0,658899585 | 2,12933E-08 | ARPC5 | FALSE | Upregulated |
| 2,028554082 | 2,21146E-08 | SLAMF7 | TRUE | Upregulated |
| 2,11452043 | 2,27428E-08 | GINS2 | TRUE | Upregulated |
| 1,43054159 | 2,42139E-08 | CKS2 | TRUE | Upregulated |
| 2,970037435 | 2,4554E-08 | RMI2 | TRUE | Upregulated |
| 1,399222758 | 2,4554E-08 | ZNF367 | TRUE | Upregulated |
| 1,054412095 | 2,48036E-08 | CDC25B | TRUE | Upregulated |
| 1,278709499 | 2,71791E-08 | HCP5 | TRUE | Upregulated |
| 1,260869518 | 2,71791E-08 | KIF11 | TRUE | Upregulated |
| 0,998271985 | 2,77302E-08 | RFX5 | FALSE | Upregulated |
| 1,724613323 | 2,91402E-08 | BIRC3 | TRUE | Upregulated |
| 1,872332239 | 3,27989E-08 | ISG15 | TRUE | Upregulated |
| 1,369406998 | 3,42373E-08 | SECTM1 | TRUE | Upregulated |
| -0,59563116 | 4,49506E-08 | RBAK | FALSE | Downregulated |
| 1,338148619 | 4,67913E-08 | HAVCR2 | TRUE | Upregulated |
| 0,773218162 | 4,67913E-08 | TNFRSF1B | FALSE | Upregulated |
| 1,824789945 | 4,75866E-08 | CCRL2 | TRUE | Upregulated |
| 3,07129429 | 5,72378E-08 | OIP5 | TRUE | Upregulated |
| 0,610496643 | 5,83591E-08 | CFL1 | FALSE | Upregulated |
| 0,630995324 | 5,86626E-08 | ELOVL5 | FALSE | Upregulated |
| 1,041913454 | 6,05255E-08 | MCM3 | TRUE | Upregulated |
| 1,491897706 | 6,64418E-08 | TYMP | TRUE | Upregulated |
| 1,810035104 | 7,13513E-08 | ASPM | TRUE | Upregulated |
| 4,156401677 | 7,32968E-08 | MUC13 | TRUE | Upregulated |
| 0,967437261 | 7,37264E-08 | ARPC1B | FALSE | Upregulated |
| 0,764486158 | 7,55527E-08 | DDX39A | FALSE | Upregulated |
| 2,307025092 | 8,61815E-08 | CEP55 | TRUE | Upregulated |
| 0,880476543 | 9,6006E-08 | H2AFZ | FALSE | Upregulated |
| 0,748178589 | 9,99661E-08 | ABCD1 | FALSE | Upregulated |
| 1,301576275 | 1,02722E-07 | LILRB2 | TRUE | Upregulated |
| 2,848607542 | 1,04767E-07 | ICOS | TRUE | Upregulated |
| 1,246443875 | 1,06068E-07 | STMN1 | TRUE | Upregulated |
| 1,497648606 | 1,19137E-07 | LAIR1 | TRUE | Upregulated |
| 1,558219802 | 1,20976E-07 | ANXA2 | TRUE | Upregulated |
| 1,228040275 | 1,21232E-07 | SASH3 | TRUE | Upregulated |
| 0,442545527 | 1,38239E-07 | AP1B1 | FALSE | Upregulated |
| -1,996165453 | 1,38239E-07 | ASXL3 | TRUE | Downregulated |
| 0,889393279 | 1,38239E-07 | IFI35 | FALSE | Upregulated |
| 3,356636004 | 1,40624E-07 | ERCC6L | TRUE | Upregulated |
| 1,804376235 | 1,46265E-07 | WDR76 | TRUE | Upregulated |
| 0,920608436 | 1,68488E-07 | RASSF3 | FALSE | Upregulated |
| 1,714856191 | 1,79979E-07 | SERPINB8 | TRUE | Upregulated |
| 1,225272382 | 1,80633E-07 | TAP2 | TRUE | Upregulated |
| 1,352826546 | 2,32743E-07 | LILRB4 | TRUE | Upregulated |
| -0,413248735 | 2,41355E-07 | LYRM7 | FALSE | Downregulated |
| 1,181011153 | 2,62309E-07 | H2AFX | TRUE | Upregulated |
| 0,459888677 | 2,67723E-07 | CAP1 | FALSE | Upregulated |

| | | | | |
|---|---|---|---|---|
| 1,446028218 | 3,02706E-07 | VAMP5 | TRUE | Upregulated |
| 0,989154911 | 3,29321E-07 | HLA-DMA | FALSE | Upregulated |
| 1,611925756 | 3,39958E-07 | RELB | TRUE | Upregulated |
| 1,053450969 | 3,60828E-07 | LIMK1 | TRUE | Upregulated |
| 0,79282432 | 3,82591E-07 | TMEM2 | FALSE | Upregulated |
| -0,585391819 | 4,0372E-07 | PCNXL4 | FALSE | Downregulated |
| 1,539210853 | 4,15305E-07 | CD52 | TRUE | Upregulated |
| 1,003380944 | 4,17578E-07 | CTSS | TRUE | Upregulated |
| 0,567934627 | 4,17578E-07 | MPP1 | FALSE | Upregulated |
| 1,275227678 | 4,17578E-07 | PHF19 | TRUE | Upregulated |
| 0,668413225 | 4,62925E-07 | CLIC1 | FALSE | Upregulated |
| -1,1889795 | 4,70271E-07 | YPEL2 | TRUE | Downregulated |
| 1,063491261 | 4,94178E-07 | CHAF1A | TRUE | Upregulated |
| 2,455721395 | 4,94178E-07 | ORC1 | TRUE | Upregulated |
| 2,043234724 | 5,03166E-07 | CAPG | TRUE | Upregulated |
| 1,690282516 | 5,04492E-07 | RGS1 | TRUE | Upregulated |
| 0,616674568 | 5,44839E-07 | CORO1C | FALSE | Upregulated |
| 1,757235792 | 5,61283E-07 | CHAF1B | TRUE | Upregulated |
| 1,027015645 | 5,93096E-07 | SERPINB1 | TRUE | Upregulated |
| -2,458682195 | 5,98809E-07 | LOC102724175 | TRUE | Downregulated |
| 1,148519673 | 6,02297E-07 | RACGAP1 | TRUE | Upregulated |
| -0,595555685 | 6,0467E-07 | LOC100506548 | FALSE | Downregulated |
| 1,230436763 | 6,31191E-07 | SLA | TRUE | Upregulated |
| 2,619870288 | 6,46773E-07 | POLQ | TRUE | Upregulated |
| 1,318782607 | 6,53133E-07 | PRDM1 | TRUE | Upregulated |
| -0,811205641 | 6,81904E-07 | IRS1 | FALSE | Downregulated |
| 1,546739504 | 6,81904E-07 | PLXNC1 | TRUE | Upregulated |
| 5,034533437 | 6,81904E-07 | SLC51B | TRUE | Upregulated |
| 1,095913714 | 6,84429E-07 | GBP4 | TRUE | Upregulated |
| 0,775541412 | 7,20184E-07 | VOPP1 | FALSE | Upregulated |
| 3,104937404 | 7,23887E-07 | SKA1 | TRUE | Upregulated |
| 2,423722371 | 7,3311E-07 | CDT1 | TRUE | Upregulated |
| 0,597648105 | 7,39923E-07 | H2AFY | FALSE | Upregulated |
| 1,313839528 | 7,39923E-07 | MSR1 | TRUE | Upregulated |
| 1,078935497 | 7,88933E-07 | UBE2L6 | TRUE | Upregulated |
| 0,811003668 | 8,3816E-07 | LITAF | FALSE | Upregulated |
| 0,811361394 | 8,39923E-07 | MR1 | FALSE | Upregulated |
| 0,654464242 | 8,74489E-07 | RAB32 | FALSE | Upregulated |
| 2,136998952 | 8,85921E-07 | GINS1 | TRUE | Upregulated |
| 0,964993576 | 8,97419E-07 | CCND3 | FALSE | Upregulated |
| 0,848283063 | 9,21924E-07 | APOL2 | FALSE | Upregulated |
| 1,097512519 | 9,21924E-07 | UBE2S | TRUE | Upregulated |
| 1,843882274 | 9,37803E-07 | GINS4 | TRUE | Upregulated |
| 1,571136253 | 9,4108E-07 | TP53I3 | TRUE | Upregulated |
| -2,371318993 | 1,03141E-06 | AOC4P | TRUE | Downregulated |
| 0,682655839 | 1,04082E-06 | VASP | FALSE | Upregulated |
| -0,456382782 | 1,06084E-06 | ZNF33A | FALSE | Downregulated |
| 1,641929877 | 1,08516E-06 | SPON2 | TRUE | Upregulated |
| 1,269508136 | 1,08931E-06 | CD48 | TRUE | Upregulated |
| 1,219220922 | 1,1148E-06 | STIL | TRUE | Upregulated |
| 0,700416968 | 1,23834E-06 | BSG | FALSE | Upregulated |

| | | | | |
|---|---|---|---|---|
| -0,78727474 | 1,27218E-06 | JMJD1C | FALSE | Downregulated |
| 0,876187696 | 1,27235E-06 | PARP12 | FALSE | Upregulated |
| 0,78701675 | 1,42975E-06 | ZNFX1 | FALSE | Upregulated |
| 2,22385724 | 1,52165E-06 | IDO1 | TRUE | Upregulated |
| 0,916177103 | 1,52181E-06 | TUBA1C | FALSE | Upregulated |
| 1,087638099 | 1,53008E-06 | CMTM7 | TRUE | Upregulated |
| 0,503432705 | 1,58346E-06 | UBC | FALSE | Upregulated |
| 1,647860165 | 1,60926E-06 | CXCR4 | TRUE | Upregulated |
| 2,938293077 | 1,65667E-06 | TNFRSF9 | TRUE | Upregulated |
| 1,108457047 | 1,67969E-06 | RAC2 | TRUE | Upregulated |
| 1,723682057 | 1,71051E-06 | GPNMB | TRUE | Upregulated |
| 1,22681832 | 1,75446E-06 | APOBEC3G | TRUE | Upregulated |
| 1,115708991 | 1,75446E-06 | NFKB2 | TRUE | Upregulated |
| 1,486898765 | 1,94899E-06 | BRCA2 | TRUE | Upregulated |
| 0,682947757 | 1,97465E-06 | ACOT7 | FALSE | Upregulated |
| 1,003083763 | 2,28542E-06 | TYROBP | TRUE | Upregulated |
| 0,505686353 | 2,38257E-06 | YWHAZ | FALSE | Upregulated |
| -0,699766572 | 2,38257E-06 | ZNF395 | FALSE | Downregulated |
| 2,314958552 | 2,45244E-06 | DUSP2 | TRUE | Upregulated |
| 0,62117882 | 2,47572E-06 | SLC35F6 | FALSE | Upregulated |
| 1,59684216 | 2,64357E-06 | ACE | TRUE | Upregulated |
| 0,980099003 | 2,64357E-06 | ATP1B3 | FALSE | Upregulated |
| 1,044728786 | 2,77281E-06 | WARS | TRUE | Upregulated |
| -0,783122789 | 2,77399E-06 | L2HGDH | FALSE | Downregulated |
| -1,703862911 | 2,87806E-06 | RANBP3L | TRUE | Downregulated |
| 0,778940765 | 2,89935E-06 | RHOG | FALSE | Upregulated |
| 0,866025594 | 2,96306E-06 | SH3BGRL3 | FALSE | Upregulated |
| 0,895524052 | 3,00403E-06 | RHOC | FALSE | Upregulated |
| 1,2023678 | 3,01758E-06 | CD74 | TRUE | Upregulated |
| 1,507932167 | 3,01758E-06 | FANCD2 | TRUE | Upregulated |
| -0,652619961 | 3,02427E-06 | MTMR4 | FALSE | Downregulated |
| 2,614479943 | 3,04949E-06 | KCNJ5 | TRUE | Upregulated |
| 0,688294306 | 3,09798E-06 | ACTB | FALSE | Upregulated |
| 0,97671625 | 3,12326E-06 | BLVRA | FALSE | Upregulated |
| 0,933952149 | 3,19889E-06 | CASP1 | FALSE | Upregulated |
| 2,490912815 | 3,22181E-06 | CCL19 | TRUE | Upregulated |
| 1,669933709 | 3,22777E-06 | GBP5 | TRUE | Upregulated |
| 0,6606293 | 3,23863E-06 | FAM49B | FALSE | Upregulated |
| 1,402599954 | 3,24736E-06 | CD28 | TRUE | Upregulated |
| 1,159386653 | 3,28097E-06 | HLA-DRA | TRUE | Upregulated |
| 1,1123676 | 3,28673E-06 | ITGA6 | TRUE | Upregulated |
| -0,746414247 | 3,35758E-06 | ACADM | FALSE | Downregulated |
| 0,598260461 | 3,37505E-06 | NINJ1 | FALSE | Upregulated |
| 3,023676544 | 3,50429E-06 | IQGAP3 | TRUE | Upregulated |
| 3,416875461 | 3,55986E-06 | CDC25C | TRUE | Upregulated |
| 1,070158262 | 3,55986E-06 | IRF1 | TRUE | Upregulated |
| -0,674442793 | 3,57957E-06 | ZCCHC9 | FALSE | Downregulated |
| -0,899352149 | 3,74088E-06 | ZKSCAN1 | FALSE | Downregulated |
| 2,284478655 | 3,79372E-06 | EGR2 | TRUE | Upregulated |
| -1,63439327 | 3,80519E-06 | RP11-458D21.1 | TRUE | Downregulated |
| 1,880386695 | 3,82712E-06 | BCL2A1 | TRUE | Upregulated |

| | | | | |
|---|---|---|---|---|
| 1,29096584 | 3,85455E-06 | IGSF6 | TRUE | Upregulated |
| 0,718638959 | 3,85866E-06 | TRIM21 | FALSE | Upregulated |
| 0,369374446 | 3,86434E-06 | RHOA | FALSE | Upregulated |
| -0,795576565 | 3,88414E-06 | TBX3 | FALSE | Downregulated |
| 1,542966857 | 3,90321E-06 | EMR2 | TRUE | Upregulated |
| -1,001235654 | 4,1609E-06 | ACSM3 | TRUE | Downregulated |
| 0,875422687 | 4,17652E-06 | CYBA | FALSE | Upregulated |
| 0,439945921 | 4,30235E-06 | FKBP1A | FALSE | Upregulated |
| 0,494078666 | 4,53937E-06 | CORO1B | FALSE | Upregulated |
| -0,752502741 | 4,62992E-06 | ZNF546 | FALSE | Downregulated |
| 0,718018024 | 4,66625E-06 | ORMDL2 | FALSE | Upregulated |
| 3,887374274 | 4,70663E-06 | RAB3B | TRUE | Upregulated |
| 0,427486254 | 4,78065E-06 | UBE2Z | FALSE | Upregulated |
| 0,796941098 | 4,78566E-06 | HLA-C | FALSE | Upregulated |
| 0,947201726 | 4,7898E-06 | ANKRD13D | FALSE | Upregulated |
| 0,820432417 | 4,82469E-06 | HMGA1 | FALSE | Upregulated |
| 0,679582428 | 4,97255E-06 | HLA-E | FALSE | Upregulated |
| 1,099571218 | 5,02035E-06 | CD53 | TRUE | Upregulated |
| -2,410172126 | 5,0615E-06 | ARNTL | TRUE | Downregulated |
| 1,253789143 | 5,19465E-06 | KRT8 | TRUE | Upregulated |
| 1,380804204 | 5,3996E-06 | SLAMF6 | TRUE | Upregulated |
| 1,322413818 | 5,62396E-06 | IL2RG | TRUE | Upregulated |
| -0,930836621 | 5,95407E-06 | LOC101928499 | FALSE | Downregulated |
| 1,487707588 | 6,18446E-06 | EMR1 | TRUE | Upregulated |
| 0,915445884 | 6,21148E-06 | LCP1 | FALSE | Upregulated |
| 0,6843058 | 6,32183E-06 | ARHGDIB | FALSE | Upregulated |
| 0,337416873 | 6,32183E-06 | CYFIP1 | FALSE | Upregulated |
| 0,984181699 | 6,32183E-06 | HLA-DPB1 | FALSE | Upregulated |
| -0,742622563 | 6,32183E-06 | WDR19 | FALSE | Downregulated |
| 0,949081562 | 6,39728E-06 | CHST2 | FALSE | Upregulated |
| 1,394304361 | 7,29189E-06 | PTPRCAP | TRUE | Upregulated |
| 1,463070417 | 7,50639E-06 | ITGAX | TRUE | Upregulated |
| 0,99659483 | 7,55882E-06 | SPI1 | FALSE | Upregulated |
| 1,7389942 | 7,60717E-06 | MTHFD2 | TRUE | Upregulated |
| 0,861700259 | 7,69393E-06 | FAS | FALSE | Upregulated |
| 1,001292831 | 7,76344E-06 | DOK2 | TRUE | Upregulated |
| -0,31711399 | 7,95441E-06 | FBXO11 | FALSE | Downregulated |
| 0,643790384 | 8,00764E-06 | CXCL16 | FALSE | Upregulated |
| -0,669443527 | 8,0996E-06 | CCDC121 | FALSE | Downregulated |
| 0,570985209 | 8,26272E-06 | ATP6V0D1 | FALSE | Upregulated |
| 0,425253305 | 8,31151E-06 | DBNL | FALSE | Upregulated |
| 0,602887399 | 8,58091E-06 | GM2A | FALSE | Upregulated |
| 0,944078781 | 8,61321E-06 | RHBDF2 | FALSE | Upregulated |
| 2,314413461 | 8,87292E-06 | CCR7 | TRUE | Upregulated |
| 1,169471078 | 8,9E-06 | CORO1A | TRUE | Upregulated |
| 1,358168792 | 8,95666E-06 | CENPH | TRUE | Upregulated |
| 0,803874112 | 9,20615E-06 | COTL1 | FALSE | Upregulated |
| 1,103395995 | 9,24184E-06 | EHD4 | TRUE | Upregulated |
| 0,713426245 | 9,39393E-06 | DYNLT1 | FALSE | Upregulated |
| 1,028632031 | 9,64807E-06 | KLF6 | TRUE | Upregulated |
| 0,808036958 | 1,00533E-05 | LMNB1 | FALSE | Upregulated |

| | | | | |
|---|---|---|---|---|
| 0,926632459 | 1,0229E-05 | GMIP | FALSE | Upregulated |
| 1,082757678 | 1,06579E-05 | STK17B | TRUE | Upregulated |
| 0,932382334 | 1,08283E-05 | CYTH4 | FALSE | Upregulated |
| 0,490134709 | 1,09032E-05 | TAPBP | FALSE | Upregulated |
| 3,113830085 | 1,10021E-05 | LOXL4 | TRUE | Upregulated |
| 1,130613106 | 1,10021E-05 | TUBB2A | TRUE | Upregulated |
| 1,423489957 | 1,10259E-05 | CD2 | TRUE | Upregulated |
| 0,43838715 | 1,11777E-05 | FLII | FALSE | Upregulated |
| 1,398784351 | 1,13077E-05 | IL18BP | TRUE | Upregulated |
| -0,428533532 | 1,13133E-05 | LRPPRC | FALSE | Downregulated |
| -0,766406204 | 1,14612E-05 | LOC101928433 | FALSE | Downregulated |
| 1,094405 | 1,17695E-05 | C1QB | TRUE | Upregulated |
| 1,227133373 | 1,17872E-05 | WDHD1 | TRUE | Upregulated |
| 0,670364245 | 1,1799E-05 | TDRD7 | FALSE | Upregulated |
| 0,962161526 | 1,18317E-05 | LAPTM5 | FALSE | Upregulated |
| 0,676502398 | 1,19035E-05 | TMSB4X | FALSE | Upregulated |
| 2,20466832 | 1,22002E-05 | FCAMR | TRUE | Upregulated |
| 1,077362147 | 1,2474E-05 | UBASH3B | TRUE | Upregulated |
| 4,521062384 | 1,25131E-05 | AKR1B10 | TRUE | Upregulated |
| -0,973398359 | 1,29499E-05 | ACADSB | FALSE | Downregulated |
| 1,78782839 | 1,29499E-05 | CD8A | TRUE | Upregulated |
| -0,8562018 | 1,29499E-05 | SEC61A2 | FALSE | Downregulated |
| 0,999039501 | 1,29631E-05 | PLTP | FALSE | Upregulated |
| 0,630512513 | 1,29666E-05 | CAPNS1 | FALSE | Upregulated |
| 0,585756896 | 1,30583E-05 | LANCL2 | FALSE | Upregulated |
| -1,247307551 | 1,30583E-05 | SLC30A10 | TRUE | Downregulated |
| 0,582209514 | 1,35786E-05 | SKA2 | FALSE | Upregulated |
| 0,888095139 | 1,37089E-05 | YWHAH | FALSE | Upregulated |
| 1,909757955 | 1,38485E-05 | ARHGEF39 | TRUE | Upregulated |
| 1,749136028 | 1,38485E-05 | BCAT1 | TRUE | Upregulated |
| 1,120210557 | 1,42955E-05 | SRGN | TRUE | Upregulated |
| -1,289868181 | 1,44944E-05 | NFIL3 | TRUE | Downregulated |
| 0,53308158 | 1,46083E-05 | ARHGDIA | FALSE | Upregulated |
| 0,634882725 | 1,46684E-05 | IRAK1 | FALSE | Upregulated |
| -0,520640791 | 1,66152E-05 | AUH | FALSE | Downregulated |
| 1,695947774 | 1,67476E-05 | LRCOL1 | TRUE | Upregulated |
| 1,142941956 | 1,68465E-05 | TRIM22 | TRUE | Upregulated |
| 1,726325436 | 1,71803E-05 | CDC25A | TRUE | Upregulated |
| 2,384736269 | 1,77848E-05 | FCGR1A | TRUE | Upregulated |
| 1,814990728 | 1,80806E-05 | GZMB | TRUE | Upregulated |
| -0,607671987 | 1,83298E-05 | CEP120 | FALSE | Downregulated |
| 1,312389202 | 1,83558E-05 | CD37 | TRUE | Upregulated |
| 3,877825883 | 1,862E-05 | LOC100126784 | TRUE | Upregulated |
| 1,504072913 | 1,91477E-05 | PLA2G7 | TRUE | Upregulated |
| -0,37623827 | 1,91523E-05 | ZCCHC7 | FALSE | Downregulated |
| 0,657824646 | 1,96007E-05 | PI4K2A | FALSE | Upregulated |
| -0,774173099 | 1,99273E-05 | AR | FALSE | Downregulated |
| 0,558599805 | 2,00676E-05 | RAN | FALSE | Upregulated |
| 1,282502672 | 2,01551E-05 | CCR5 | TRUE | Upregulated |
| 1,345119035 | 2,06388E-05 | KIAA1524 | TRUE | Upregulated |
| 0,930197927 | 2,06388E-05 | NCKAP1L | FALSE | Upregulated |

| | | | | |
|---|---|---|---|---|
| -0,633188835 | 2,06388E-05 | RBL2 | FALSE | Downregulated |
| 1,112502138 | 2,09675E-05 | RNASE6 | TRUE | Upregulated |
| 0,500335854 | 2,14677E-05 | GNS | FALSE | Upregulated |
| 2,96384013 | 2,16551E-05 | TP73 | TRUE | Upregulated |
| 0,673562978 | 2,21283E-05 | GPC1 | FALSE | Upregulated |
| 0,816306796 | 2,21886E-05 | IKBKG | FALSE | Upregulated |
| 1,984149921 | 2,23014E-05 | ETV7 | TRUE | Upregulated |
| 1,092660803 | 2,34654E-05 | FERMT3 | TRUE | Upregulated |
| -0,834732685 | 2,43764E-05 | GRAMD3 | FALSE | Downregulated |
| 1,322252957 | 2,44912E-05 | SP140 | TRUE | Upregulated |
| 1,366852705 | 2,48355E-05 | C11orf82 | TRUE | Upregulated |
| 0,796088633 | 2,48355E-05 | CARD16 | FALSE | Upregulated |
| 0,526309317 | 2,48355E-05 | HSPA4 | FALSE | Upregulated |
| 0,824548978 | 2,48355E-05 | PYCARD | FALSE | Upregulated |
| 0,676448818 | 2,48419E-05 | ACOT9 | FALSE | Upregulated |
| 1,64843835 | 2,48925E-05 | KIF15 | TRUE | Upregulated |
| 0,862459549 | 2,51267E-05 | GPSM3 | FALSE | Upregulated |
| 1,013767489 | 2,53323E-05 | GBP2 | TRUE | Upregulated |
| 0,428532667 | 2,57466E-05 | PPIA | FALSE | Upregulated |
| 1,275411145 | 2,59924E-05 | FPR3 | TRUE | Upregulated |
| 0,946806488 | 2,62778E-05 | SLC7A7 | FALSE | Upregulated |
| 0,610441001 | 2,65714E-05 | PSMD14 | FALSE | Upregulated |
| 0,911231534 | 2,66015E-05 | SLC1A5 | FALSE | Upregulated |
| -0,507926786 | 2,72404E-05 | PCF11 | FALSE | Downregulated |
| 2,84104123 | 2,75996E-05 | PDCD1 | TRUE | Upregulated |
| 1,074699366 | 2,79158E-05 | MVP | TRUE | Upregulated |
| 0,822870171 | 2,82334E-05 | LST1 | FALSE | Upregulated |
| -1,404531059 | 2,82334E-05 | PPP1R3C | TRUE | Downregulated |
| 1,003902398 | 2,85085E-05 | DRAM1 | TRUE | Upregulated |
| -0,511080494 | 2,98244E-05 | CDADC1 | FALSE | Downregulated |
| 1,419630195 | 2,98274E-05 | TIGIT | TRUE | Upregulated |
| 1,343694692 | 3,09109E-05 | OAS3 | TRUE | Upregulated |
| 1,209904582 | 3,09109E-05 | TCEA2 | TRUE | Upregulated |
| -0,345225926 | 3,13707E-05 | RLF | FALSE | Downregulated |
| 0,286321186 | 3,13738E-05 | XRCC6 | FALSE | Upregulated |
| -0,722080799 | 3,13973E-05 | WDR31 | FALSE | Downregulated |
| 1,560184677 | 3,21733E-05 | RECQL4 | TRUE | Upregulated |
| 0,263275631 | 3,27313E-05 | SZRD1 | FALSE | Upregulated |
| 0,53466261 | 3,30765E-05 | MYO1C | FALSE | Upregulated |
| 1,473808529 | 3,34199E-05 | ARNTL2 | TRUE | Upregulated |
| 1,508498745 | 3,34199E-05 | KDELR3 | TRUE | Upregulated |
| 1,963309825 | 3,34199E-05 | LAMP3 | TRUE | Upregulated |
| -1,16030419 | 3,34952E-05 | AASS | TRUE | Downregulated |
| 0,462854576 | 3,38611E-05 | EIF4A1 | FALSE | Upregulated |
| 1,055179312 | 3,40232E-05 | CYTIP | TRUE | Upregulated |
| -0,671184752 | 3,40232E-05 | ZNF30 | FALSE | Downregulated |
| -0,844214586 | 3,43826E-05 | PTCH1 | FALSE | Downregulated |
| 1,473538218 | 3,44614E-05 | PLAUR | TRUE | Upregulated |
| 0,745127453 | 3,46835E-05 | HSPA8 | FALSE | Upregulated |
| 0,904368023 | 3,53642E-05 | ADPRH | FALSE | Upregulated |
| -0,71475493 | 3,55967E-05 | COBLL1 | FALSE | Downregulated |

| | | | | |
|---|---|---|---|---|
| 0,467311238 | 3,5836E-05 | ATG7 | FALSE | Upregulated |
| -0,691417534 | 3,6278E-05 | PLSCR4 | FALSE | Downregulated |
| 2,566885397 | 3,64356E-05 | TNFRSF12A | TRUE | Upregulated |
| 0,347407937 | 3,6787E-05 | RAB5C | FALSE | Upregulated |
| -0,461215998 | 3,68566E-05 | EIF5 | FALSE | Downregulated |
| 0,668891052 | 3,79791E-05 | PRIM2 | FALSE | Upregulated |
| 0,719446468 | 3,86623E-05 | SLC38A6 | FALSE | Upregulated |
| 2,179292976 | 3,89796E-05 | SIGLEC8 | TRUE | Upregulated |
| 1,03120189 | 3,9072E-05 | FCGR3A | TRUE | Upregulated |
| -0,554297172 | 4,07025E-05 | ZNF615 | FALSE | Downregulated |
| -0,308898701 | 4,16734E-05 | GTPBP10 | FALSE | Downregulated |
| 0,497937736 | 4,16856E-05 | SEPT9 | FALSE | Upregulated |
| 0,982908434 | 4,22526E-05 | PCED1B | FALSE | Upregulated |
| -0,4900819 | 4,24237E-05 | IKZF5 | FALSE | Downregulated |
| -0,464248649 | 4,24237E-05 | ZNF280D | FALSE | Downregulated |
| 2,380518148 | 4,33712E-05 | DERL3 | TRUE | Upregulated |
| -1,662440006 | 4,50643E-05 | C2orf16 | TRUE | Downregulated |
| 2,142299766 | 4,58603E-05 | HAPLN3 | TRUE | Upregulated |
| -1,264790379 | 4,58603E-05 | LOC101929633 | TRUE | Downregulated |
| -0,749348305 | 4,58603E-05 | ZFHX4 | FALSE | Downregulated |
| 0,456802883 | 4,62214E-05 | PFN1 | FALSE | Upregulated |
| 0,692530586 | 4,6232E-05 | GPX1 | FALSE | Upregulated |
| 1,389068636 | 4,6232E-05 | MMP19 | TRUE | Upregulated |
| 0,655832161 | 4,63056E-05 | PPT1 | FALSE | Upregulated |
| 0,394378057 | 4,69239E-05 | ARHGEF18 | FALSE | Upregulated |
| -0,468336756 | 4,69239E-05 | HNRNPDL | FALSE | Downregulated |
| 1,288609071 | 4,69239E-05 | KLHL6 | TRUE | Upregulated |
| 0,6721715 | 4,70596E-05 | INF2 | FALSE | Upregulated |
| 0,724508956 | 4,75425E-05 | DSN1 | FALSE | Upregulated |
| 0,866192173 | 4,88186E-05 | ZMYND15 | FALSE | Upregulated |
| 1,266595875 | 4,9332E-05 | OAS2 | TRUE | Upregulated |
| -1,273600626 | 5,15448E-05 | ARL4D | TRUE | Downregulated |
| 0,689404269 | 5,33706E-05 | ACLY | FALSE | Upregulated |
| 0,962818721 | 5,38564E-05 | IKZF1 | FALSE | Upregulated |
| 2,81228163 | 5,42945E-05 | KIF18B | TRUE | Upregulated |
| 4,060847198 | 5,42945E-05 | LOC102724895 | TRUE | Upregulated |
| 1,613385293 | 5,52268E-05 | IL21R | TRUE | Upregulated |
| -0,657380214 | 5,55325E-05 | MBTD1 | FALSE | Downregulated |
| 1,336437836 | 5,55649E-05 | RHOH | TRUE | Upregulated |
| 0,470349523 | 5,55649E-05 | TSPAN17 | FALSE | Upregulated |
| -0,448292459 | 5,56489E-05 | FOXO4 | FALSE | Downregulated |
| 1,195187839 | 5,62251E-05 | CIITA | TRUE | Upregulated |
| 0,423183264 | 5,63891E-05 | DCTN2 | FALSE | Upregulated |
| 0,597957318 | 5,63891E-05 | GRK6 | FALSE | Upregulated |
| -1,28099767 | 5,7083E-05 | ADRA1A | TRUE | Downregulated |
| 0,599801372 | 5,85572E-05 | GRN | FALSE | Upregulated |
| -0,901545953 | 5,93345E-05 | KIAA1804 | FALSE | Downregulated |
| 1,578238748 | 5,9682E-05 | SGOL1 | TRUE | Upregulated |
| -0,589929209 | 5,98146E-05 | MXI1 | FALSE | Downregulated |
| 1,027262391 | 6,18853E-05 | ITGB2 | TRUE | Upregulated |
| -0,519814805 | 6,22116E-05 | FERMT2 | FALSE | Downregulated |

| | | | | |
|---|---|---|---|---|
| 0,567024617 | 6,22116E-05 | SYNGR2 | FALSE | Upregulated |
| 1,068667715 | 6,23706E-05 | PLEK | TRUE | Upregulated |
| -0,419599197 | 6,24155E-05 | SEPT10 | FALSE | Downregulated |
| 0,586045347 | 6,25275E-05 | ELOVL1 | FALSE | Upregulated |
| 1,400579017 | 6,25646E-05 | CCNE2 | TRUE | Upregulated |
| 0,546556944 | 6,32317E-05 | CDIP1 | FALSE | Upregulated |
| -0,720832264 | 6,32317E-05 | GCDH | FALSE | Downregulated |
| 0,457709356 | 6,42099E-05 | PSMD2 | FALSE | Upregulated |
| -0,628026058 | 6,49781E-05 | SERTAD2 | FALSE | Downregulated |
| 1,562929744 | 6,52115E-05 | SNX20 | TRUE | Upregulated |
| -0,475463229 | 6,55251E-05 | MYO1B | FALSE | Downregulated |
| -0,409021678 | 6,56713E-05 | PTPN4 | FALSE | Downregulated |
| 0,940395887 | 6,6538E-05 | RPS6KA1 | FALSE | Upregulated |
| 0,50045042 | 6,6763E-05 | ITGB5 | FALSE | Upregulated |
| 0,704188281 | 6,69812E-05 | TMBIM1 | FALSE | Upregulated |
| 0,902748574 | 6,77221E-05 | BTK | FALSE | Upregulated |
| 0,434218475 | 7,00793E-05 | SNRPB | FALSE | Upregulated |
| 1,058306721 | 7,02587E-05 | CECR1 | TRUE | Upregulated |
| 0,566354446 | 7,02587E-05 | DDAH2 | FALSE | Upregulated |
| 0,435136108 | 7,02587E-05 | PSMD10 | FALSE | Upregulated |
| 0,729367303 | 7,19059E-05 | CST3 | FALSE | Upregulated |
| 1,991150547 | 7,4291E-05 | HLA-DOB | TRUE | Upregulated |
| -0,452851222 | 7,4291E-05 | ZFP62 | FALSE | Downregulated |
| 1,525789936 | 7,49235E-05 | CD3D | TRUE | Upregulated |
| -0,428335689 | 7,73873E-05 | KRR1 | FALSE | Downregulated |
| -0,897875413 | 7,82697E-05 | KIF27 | FALSE | Downregulated |
| 2,724529706 | 7,89792E-05 | E2F2 | TRUE | Upregulated |
| 0,531367666 | 7,99468E-05 | CTSA | FALSE | Upregulated |
| 0,908308375 | 8,03702E-05 | ZYX | FALSE | Upregulated |
| 0,51416205 | 8,28708E-05 | RQCD1 | FALSE | Upregulated |
| -0,582226725 | 8,30851E-05 | GYS2 | FALSE | Downregulated |
| -0,94394917 | 8,44765E-05 | MEX3A | FALSE | Downregulated |
| 0,441106585 | 8,52893E-05 | LYPLA2 | FALSE | Upregulated |
| -0,82506909 | 8,59672E-05 | GCSHP3 | FALSE | Downregulated |
| 1,98979446 | 8,68398E-05 | DEPDC1B | TRUE | Upregulated |
| -0,59818001 | 8,78154E-05 | ALDH6A1 | FALSE | Downregulated |
| 1,552787708 | 8,84176E-05 | FAT1 | TRUE | Upregulated |
| -0,44282065 | 8,91938E-05 | OARD1 | FALSE | Downregulated |
| -0,236866145 | 8,91938E-05 | ZNF146 | FALSE | Downregulated |
| 0,883910013 | 9,14505E-05 | MSN | FALSE | Upregulated |
| -4,168624292 | 9,16044E-05 | RP11-10L7.1 | TRUE | Downregulated |
| 0,593097174 | 9,49428E-05 | TMEM184B | FALSE | Upregulated |
| -0,317275831 | 9,50357E-05 | ATXN7L3B | FALSE | Downregulated |
| 1,039933269 | 9,52015E-05 | TRIM69 | TRUE | Upregulated |
| 1,646162417 | 9,5204E-05 | BATF | TRUE | Upregulated |
| 1,794878755 | 9,5204E-05 | CD27 | TRUE | Upregulated |
| 1,096469761 | 9,5204E-05 | FANCA | TRUE | Upregulated |
| 0,592373477 | 9,5204E-05 | HIP1R | FALSE | Upregulated |
| 0,963447573 | 9,5204E-05 | RNASEH2A | FALSE | Upregulated |
| 0,52963895 | 9,63476E-05 | DNAJC9 | FALSE | Upregulated |
| -0,580665194 | 9,92205E-05 | A1CF | FALSE | Downregulated |

| | | | | |
|---|---|---|---|---|
| 0,529318517 | 9,92205E-05 | BID | FALSE | Upregulated |
| 1,09031793 | 9,92205E-05 | CSF2RA | TRUE | Upregulated |
| -1,355447484 | 9,92205E-05 | RP11-439L18.3 | TRUE | Downregulated |
| 0,798566614 | 0,000100191 | IRF8 | FALSE | Upregulated |
| 0,759381824 | 0,000100651 | WAS | FALSE | Upregulated |
| 0,930879817 | 0,000101016 | SELPLG | FALSE | Upregulated |
| -0,903311816 | 0,000101402 | BMP4 | FALSE | Downregulated |
| 2,225554672 | 0,000102329 | PLA2G2D | TRUE | Upregulated |
| 3,346800916 | 0,000102717 | TRIM31 | TRUE | Upregulated |
| 1,171570885 | 0,000103212 | ADAP1 | TRUE | Upregulated |
| 1,250187667 | 0,000104505 | LILRB3 | TRUE | Upregulated |
| 0,524677274 | 0,000104505 | POLR3K | FALSE | Upregulated |
| -0,409087198 | 0,000105242 | PDXDC1 | FALSE | Downregulated |
| 0,727908669 | 0,000105242 | TUBB4B | FALSE | Upregulated |
| 2,026574643 | 0,000105568 | UBXN10 | TRUE | Upregulated |
| 1,002228795 | 0,000105911 | C1QC | TRUE | Upregulated |
| -0,645183149 | 0,000106479 | ALAD | FALSE | Downregulated |
| 1,428844648 | 0,000106493 | THBD | TRUE | Upregulated |
| 0,696162806 | 0,000107583 | MYL12B | FALSE | Upregulated |
| 1,037927651 | 0,000107583 | TIMP1 | TRUE | Upregulated |
| 2,943348444 | 0,00010846 | TREM2 | TRUE | Upregulated |
| -0,942118891 | 0,000108867 | GAREM | FALSE | Downregulated |
| 0,355316979 | 0,000110407 | NRAS | FALSE | Upregulated |
| 0,542072596 | 0,00011105 | PTPN6 | FALSE | Upregulated |
| 0,481897836 | 0,000111909 | CYLD | FALSE | Upregulated |
| -1,20484986 | 0,000112556 | CXXC4 | TRUE | Downregulated |
| 0,855977153 | 0,000115173 | EMP3 | FALSE | Upregulated |
| 0,607567463 | 0,000115858 | HNRNPAB | FALSE | Upregulated |
| 0,499765002 | 0,000115858 | MCOLN1 | FALSE | Upregulated |
| 1,030163059 | 0,000116515 | SAMSN1 | TRUE | Upregulated |
| 0,832656299 | 0,000118976 | CTSC | FALSE | Upregulated |
| -0,60677827 | 0,000118976 | MFAP3L | FALSE | Downregulated |
| 0,68022512 | 0,000118976 | RHBDD2 | FALSE | Upregulated |
| 0,959478761 | 0,000119653 | CD86 | FALSE | Upregulated |
| 0,444971373 | 0,000119835 | RALB | FALSE | Upregulated |
| 0,511391153 | 0,000119881 | MRPL17 | FALSE | Upregulated |
| 1,062928146 | 0,00012033 | SLPI | TRUE | Upregulated |
| -0,438110468 | 0,000121599 | BBS4 | FALSE | Downregulated |
| 0,633905709 | 0,000122057 | ITPKC | FALSE | Upregulated |
| 1,113253391 | 0,000122541 | CD247 | TRUE | Upregulated |
| -1,124963768 | 0,000122617 | PITPNM3 | TRUE | Downregulated |
| 1,821622098 | 0,000124069 | LEF1 | TRUE | Upregulated |
| 0,431801428 | 0,000124069 | LRRC42 | FALSE | Upregulated |
| 0,394927494 | 0,000125798 | ETF1 | FALSE | Upregulated |
| -0,71611818 | 0,000125898 | GTF2IRD2B | FALSE | Downregulated |
| 2,729899667 | 0,000125908 | GPRIN1 | TRUE | Upregulated |
| 1,065378001 | 0,000125908 | NEDD9 | TRUE | Upregulated |
| -1,473479775 | 0,000126047 | RP11-12A2.3 | TRUE | Downregulated |
| 0,431956665 | 0,000127789 | CHMP1A | FALSE | Upregulated |
| 1,89605809 | 0,000130058 | CD79A | TRUE | Upregulated |
| 0,477508133 | 0,000130643 | AKIP1 | FALSE | Upregulated |

| | | | | |
|---|---|---|---|---|
| 2,223614922 | 0,000130643 | C16orf59 | TRUE | Upregulated |
| 3,099674162 | 0,000131602 | HS3ST2 | TRUE | Upregulated |
| -0,651121244 | 0,000131909 | ZBTB39 | FALSE | Downregulated |
| -0,598457809 | 0,000134395 | ATF7IP2 | FALSE | Downregulated |
| 0,745668879 | 0,000134395 | PLEKHO1 | FALSE | Upregulated |
| 2,193269763 | 0,00013664 | CDC6 | TRUE | Upregulated |
| 0,603818711 | 0,00013664 | HMGB2 | FALSE | Upregulated |
| -0,400988436 | 0,00013664 | LOC100129034 | FALSE | Downregulated |
| -0,453524256 | 0,000136737 | TSHZ1 | FALSE | Downregulated |
| -0,949641324 | 0,000138037 | CKMT2-AS1 | FALSE | Downregulated |
| 0,861875884 | 0,000138037 | HVCN1 | FALSE | Upregulated |
| 0,369408025 | 0,000139801 | ACTR1A | FALSE | Upregulated |
| 0,523937557 | 0,000139973 | LGMN | FALSE | Upregulated |
| 0,786365804 | 0,000140064 | CD68 | FALSE | Upregulated |
| -1,042818785 | 0,000143443 | LOC102723845 | TRUE | Downregulated |
| -0,601936925 | 0,000145246 | C1orf168 | FALSE | Downregulated |
| 1,637847191 | 0,000145482 | SERPINE1 | TRUE | Upregulated |
| 2,768976251 | 0,000145482 | TROAP | TRUE | Upregulated |
| -0,54329917 | 0,00014771 | AC007246.3 | FALSE | Downregulated |
| 0,379797525 | 0,0001503 | GBA | FALSE | Upregulated |
| 1,275400533 | 0,000151536 | HLA-DQA1 | TRUE | Upregulated |
| 1,441014612 | 0,0001518 | SIGLEC10 | TRUE | Upregulated |
| 1,470911787 | 0,000151844 | CD5 | TRUE | Upregulated |
| 0,887299452 | 0,000151844 | CMKLR1 | FALSE | Upregulated |
| -0,409733908 | 0,00015312 | ZNF12 | FALSE | Downregulated |
| -0,465757812 | 0,000153769 | NR2F2 | FALSE | Downregulated |
| 0,881614684 | 0,000159325 | NFKBIE | FALSE | Upregulated |
| 1,705776675 | 0,000159748 | CD8B | TRUE | Upregulated |
| 2,095652015 | 0,000159748 | LCN2 | TRUE | Upregulated |
| 1,01578556 | 0,00016004 | CENPP | TRUE | Upregulated |
| -0,763134977 | 0,000160792 | CDNF | FALSE | Downregulated |
| -0,448770182 | 0,000160792 | ZNF302 | FALSE | Downregulated |
| 0,40209255 | 0,00016178 | RAB35 | FALSE | Upregulated |
| 1,893811968 | 0,000163954 | CXCR3 | TRUE | Upregulated |
| 1,10163494 | 0,0001644 | GZMA | TRUE | Upregulated |
| 0,975734396 | 0,000167817 | IFI27 | FALSE | Upregulated |
| 0,876263681 | 0,000170229 | PIK3R5 | FALSE | Upregulated |
| 0,703778042 | 0,00017111 | AIF1 | FALSE | Upregulated |
| -1,359484188 | 0,000171218 | CUX2 | TRUE | Downregulated |
| 0,669092183 | 0,000171218 | DCK | FALSE | Upregulated |
| -0,941368055 | 0,000173193 | NIPAL1 | FALSE | Downregulated |
| 3,118266898 | 0,000173554 | GSG2 | TRUE | Upregulated |
| -0,295003146 | 0,00017462 | WAC | FALSE | Downregulated |
| 1,260936313 | 0,000177922 | LSP1 | TRUE | Upregulated |
| 0,564274725 | 0,000177945 | PYGB | FALSE | Upregulated |
| 1,023325215 | 0,000178185 | RGS10 | TRUE | Upregulated |
| 0,474431288 | 0,000179666 | ASNA1 | FALSE | Upregulated |
| -0,412127907 | 0,000180437 | PARP16 | FALSE | Downregulated |
| -0,533321521 | 0,000182226 | BCKDHB | FALSE | Downregulated |
| -2,976305841 | 0,000185906 | GREM1 | TRUE | Downregulated |
| 0,908545312 | 0,000185908 | HLA-DPA1 | FALSE | Upregulated |

| | | | | |
|---|---|---|---|---|
| 0,652657249 | 0,000186166 | TPGS2 | FALSE | Upregulated |
| 0,56785814 | 0,000187058 | DGKZ | FALSE | Upregulated |
| -0,610717425 | 0,000187058 | TMEM170B | FALSE | Downregulated |
| -0,792577618 | 0,000187223 | DNAJC12 | FALSE | Downregulated |
| -0,496021409 | 0,000187868 | SFT2D3 | FALSE | Downregulated |
| 0,552179244 | 0,000187868 | SLC39A3 | FALSE | Upregulated |
| 1,407510908 | 0,000188515 | TPX2 | TRUE | Upregulated |
| 0,662136372 | 0,000189967 | BTN3A3 | FALSE | Upregulated |
| 0,633812392 | 0,000189991 | NCAPG2 | FALSE | Upregulated |
| 0,908567071 | 0,000190761 | PIM2 | FALSE | Upregulated |
| 0,468472966 | 0,000191565 | ATP6V0A1 | FALSE | Upregulated |
| 1,038697457 | 0,000195173 | FCGR2A | TRUE | Upregulated |
| 0,569233576 | 0,000195416 | APEX2 | FALSE | Upregulated |
| 1,036511119 | 0,00019824 | LIMD2 | TRUE | Upregulated |
| -0,47429454 | 0,00019948 | ZNF606 | FALSE | Downregulated |
| 0,641802535 | 0,000208491 | LRP10 | FALSE | Upregulated |
| 1,296757007 | 0,000208491 | MTHFD1L | TRUE | Upregulated |
| -1,125935871 | 0,000208491 | TPRG1 | TRUE | Downregulated |
| 0,581918786 | 0,00020966 | MCM7 | FALSE | Upregulated |
| -0,489318289 | 0,000210111 | SLC46A1 | FALSE | Downregulated |
| 4,955867246 | 0,000213098 | KRT23 | TRUE | Upregulated |
| 2,564322856 | 0,000213098 | NPTX2 | TRUE | Upregulated |
| 1,658445184 | 0,000214392 | LINC00152 | TRUE | Upregulated |
| -0,440934527 | 0,00021667 | ZNF616 | FALSE | Downregulated |
| 0,747539427 | 0,000216987 | PREX1 | FALSE | Upregulated |
| -0,607922216 | 0,000217249 | DMGDH | FALSE | Downregulated |
| -0,497627257 | 0,000217249 | SLC35A3 | FALSE | Downregulated |
| 0,38892609 | 0,000217249 | XPNPEP1 | FALSE | Upregulated |
| 2,727840208 | 0,000220319 | CTLA4 | TRUE | Upregulated |
| 0,657452355 | 0,000222275 | GIPC1 | FALSE | Upregulated |
| 2,587336409 | 0,000222275 | LRRC43 | TRUE | Upregulated |
| -0,54264119 | 0,000223932 | HECTD1 | FALSE | Downregulated |
| 2,244564361 | 0,000226047 | EYS | TRUE | Upregulated |
| 0,85997679 | 0,000226248 | CCNF | FALSE | Upregulated |
| 1,369525077 | 0,000226684 | SEMA4A | TRUE | Upregulated |
| 2,397136154 | 0,000227006 | FCGR1B | TRUE | Upregulated |
| 0,856105077 | 0,000227445 | PILRA | FALSE | Upregulated |
| 0,592371406 | 0,000227785 | PACSIN2 | FALSE | Upregulated |
| 1,336873494 | 0,000227987 | CD7 | TRUE | Upregulated |
| 0,401603558 | 0,000229322 | PGAM1 | FALSE | Upregulated |
| 1,073657249 | 0,000229382 | LRRC25 | TRUE | Upregulated |
| 0,466772623 | 0,000230931 | CD63 | FALSE | Upregulated |
| 2,404504727 | 0,000232122 | CCL18 | TRUE | Upregulated |
| -0,413247098 | 0,000233741 | CEBPZ-AS1 | FALSE | Downregulated |
| 1,195095613 | 0,000233741 | MICAL1 | TRUE | Upregulated |
| -0,481363841 | 0,000234002 | METTL7A | FALSE | Downregulated |
| 3,215187994 | 0,000234153 | LOC102724448 | TRUE | Upregulated |
| 0,912265523 | 0,000234633 | IFIT3 | FALSE | Upregulated |
| 0,642597729 | 0,000235576 | CYTH2 | FALSE | Upregulated |
| 0,577178629 | 0,000236515 | TXNDC9 | FALSE | Upregulated |
| 0,564844625 | 0,000236597 | IFIH1 | FALSE | Upregulated |

| | | | | |
|---|---|---|---|---|
| 0,675717518 | 0,000237737 | ALYREF | FALSE | Upregulated |
| 0,560765804 | 0,000240568 | KIAA0930 | FALSE | Upregulated |
| 0,417836628 | 0,000243651 | ENO1 | FALSE | Upregulated |
| 0,734697288 | 0,000243857 | ZFP36 | FALSE | Upregulated |
| -0,53080894 | 0,000247331 | NMRK1 | FALSE | Downregulated |
| 0,621008166 | 0,000247429 | VRK1 | FALSE | Upregulated |
| 0,980071074 | 0,000250386 | IFI44 | FALSE | Upregulated |
| 1,769280373 | 0,000250386 | IGJ | TRUE | Upregulated |
| -0,834411097 | 0,000250386 | MREG | FALSE | Downregulated |
| -0,453884845 | 0,000250543 | CEP44 | FALSE | Downregulated |
| 0,6796896 | 0,000252385 | ALG1 | FALSE | Upregulated |
| 0,916383551 | 0,000252385 | FAIM | FALSE | Upregulated |
| 1,377271309 | 0,000253687 | ITK | TRUE | Upregulated |
| 1,158648273 | 0,000254765 | MICB | TRUE | Upregulated |
| -0,909241889 | 0,000254997 | LINC01024 | FALSE | Downregulated |
| 0,581083708 | 0,000256257 | ME2 | FALSE | Upregulated |
| -0,629557404 | 0,000256543 | AADAT | FALSE | Downregulated |
| 1,909328256 | 0,000256818 | EGR3 | TRUE | Upregulated |
| 0,44139669 | 0,000258144 | AGFG1 | FALSE | Upregulated |
| 0,73055054 | 0,000258144 | CASP4 | FALSE | Upregulated |
| 1,251415647 | 0,000258144 | F2RL1 | TRUE | Upregulated |
| 0,438984791 | 0,000261984 | SHISA5 | FALSE | Upregulated |
| 0,899160282 | 0,00026357 | C1QA | FALSE | Upregulated |
| 0,893338584 | 0,000265119 | TNFAIP8 | FALSE | Upregulated |
| -0,737586457 | 0,000265674 | MAP2K1 | FALSE | Downregulated |
| -0,764425951 | 0,000265674 | SPEF2 | FALSE | Downregulated |
| -0,562240592 | 0,000266861 | MAF | FALSE | Downregulated |
| 1,957840558 | 0,000267894 | IFI6 | TRUE | Upregulated |
| 0,878667088 | 0,000270723 | CD151 | FALSE | Upregulated |
| 1,365679264 | 0,000272805 | CD69 | TRUE | Upregulated |
| -0,703924954 | 0,000272805 | ZNF432 | FALSE | Downregulated |
| 0,575867627 | 0,000274201 | ANXA11 | FALSE | Upregulated |
| 0,74739025 | 0,000274201 | DNMT1 | FALSE | Upregulated |
| -0,687143396 | 0,000274201 | ZFP1 | FALSE | Downregulated |
| 1,216879737 | 0,000274622 | IKBKE | TRUE | Upregulated |
| -1,237381696 | 0,000274636 | LOC102724711 | TRUE | Downregulated |
| -0,570657513 | 0,000276245 | SKP2 | FALSE | Downregulated |
| 0,868518356 | 0,000276832 | S100A11 | FALSE | Upregulated |
| 1,310516737 | 0,000279799 | CCNE1 | TRUE | Upregulated |
| 1,510943432 | 0,000280259 | TRAT1 | TRUE | Upregulated |
| 0,470209824 | 0,000282053 | MAN2B1 | FALSE | Upregulated |
| 1,341954887 | 0,000283062 | IL18 | TRUE | Upregulated |
| -2,899462391 | 0,000283062 | SLC45A2 | TRUE | Downregulated |
| 0,419954658 | 0,000285447 | SAE1 | FALSE | Upregulated |
| -0,368755158 | 0,000296261 | LINC00657 | FALSE | Downregulated |
| 0,397737642 | 0,000296293 | SPECC1L | FALSE | Upregulated |
| 1,952525336 | 0,000296413 | POU2AF1 | TRUE | Upregulated |
| -0,528003798 | 0,000296469 | DLC1 | FALSE | Downregulated |
| -0,699813482 | 0,000300755 | N4BP2L1 | FALSE | Downregulated |
| 0,408290381 | 0,000300781 | FBXW5 | FALSE | Upregulated |
| 3,251718498 | 0,000300793 | RIBC2 | TRUE | Upregulated |

| | | | | |
|---|---|---|---|---|
| 0,639281053 | 0,000301635 | LIG1 | FALSE | Upregulated |
| 0,476639573 | 0,000303245 | ATP6V1F | FALSE | Upregulated |
| -0,540129708 | 0,000303267 | KLHL24 | FALSE | Downregulated |
| -1,886265657 | 0,000303599 | LOC101929079 | TRUE | Downregulated |
| 0,973911646 | 0,000307683 | FGGY | FALSE | Upregulated |
| 1,004213689 | 0,000309873 | HPSE | TRUE | Upregulated |
| -1,7348076 | 0,000310585 | SCN2A | TRUE | Downregulated |
| 0,659744219 | 0,000311896 | TUBG1 | FALSE | Upregulated |
| 0,655996496 | 0,000312466 | IRF9 | FALSE | Upregulated |
| 0,798795544 | 0,000313912 | LCP2 | FALSE | Upregulated |
| -0,406630907 | 0,000315991 | FAM200B | FALSE | Downregulated |
| -0,523319096 | 0,000320192 | ADAL | FALSE | Downregulated |
| -0,91966307 | 0,000320192 | PPM1K | FALSE | Downregulated |
| 1,414082539 | 0,00032045 | CXCR6 | TRUE | Upregulated |
| -0,376735906 | 0,00032045 | ZSCAN21 | FALSE | Downregulated |
| 0,775798562 | 0,000322171 | CUEDC1 | FALSE | Upregulated |
| 0,961039517 | 0,000323283 | LY96 | FALSE | Upregulated |
| 1,106667719 | 0,000325289 | KRT18 | TRUE | Upregulated |
| 1,351990511 | 0,000328118 | DHRS9 | TRUE | Upregulated |
| 0,681733959 | 0,000328118 | MFSD12 | FALSE | Upregulated |
| 0,616252691 | 0,000328118 | RASSF4 | FALSE | Upregulated |
| 0,310660938 | 0,000329013 | PPP6R1 | FALSE | Upregulated |
| 0,520867816 | 0,000331066 | MAPRE1 | FALSE | Upregulated |
| 1,025266693 | 0,000331724 | CD84 | TRUE | Upregulated |
| 0,79854069 | 0,000331724 | METTL7B | FALSE | Upregulated |
| 0,366607619 | 0,000334466 | C2orf69 | FALSE | Upregulated |
| 0,818485562 | 0,000334466 | HCK | FALSE | Upregulated |
| -0,673242574 | 0,000334466 | SLC25A37 | FALSE | Downregulated |
| 0,498999115 | 0,000335365 | ENTPD6 | FALSE | Upregulated |
| -0,70181591 | 0,000335365 | TIAM1 | FALSE | Downregulated |
| 1,010458016 | 0,00033658 | TNFSF13B | TRUE | Upregulated |
| 1,179925234 | 0,000336758 | PLK4 | TRUE | Upregulated |
| -1,305905365 | 0,000336894 | DCDC5 | TRUE | Downregulated |
| -0,56306193 | 0,000337513 | ALB | FALSE | Downregulated |
| 0,623457807 | 0,00034134 | RAP2B | FALSE | Upregulated |
| -0,917898357 | 0,00034134 | RORC | FALSE | Downregulated |
| -1,119381658 | 0,000345697 | C1orf101 | TRUE | Downregulated |
| 2,130287476 | 0,000345697 | SPATA21 | TRUE | Upregulated |
| 0,464346411 | 0,000346635 | COMMD4 | FALSE | Upregulated |
| -0,540405503 | 0,000346635 | PARD6B | FALSE | Downregulated |
| -0,531010066 | 0,000346775 | ZNF254 | FALSE | Downregulated |
| 0,896212929 | 0,000346809 | TMEM144 | FALSE | Upregulated |
| -0,522951826 | 0,000346809 | ZNF740 | FALSE | Downregulated |
| 0,571923686 | 0,00035031 | DPP3 | FALSE | Upregulated |
| -0,578216553 | 0,000350777 | TTPA | FALSE | Downregulated |
| 1,140658651 | 0,000351058 | PLA2G4C | TRUE | Upregulated |
| 0,729684918 | 0,000355417 | SAMHD1 | FALSE | Upregulated |
| 0,696293851 | 0,000360473 | FBXO6 | FALSE | Upregulated |
| 0,745386884 | 0,000361914 | DTYMK | FALSE | Upregulated |
| 0,37672835 | 0,000361914 | SLC4A2 | FALSE | Upregulated |
| 0,485083067 | 0,000367136 | NMI | FALSE | Upregulated |

| | | | | |
|---|---|---|---|---|
| 3,203049404 | 0,000368037 | CENPM | TRUE | Upregulated |
| -0,412793513 | 0,000368037 | MOCS1 | FALSE | Downregulated |
| 0,954124596 | 0,000371156 | FDXR | FALSE | Upregulated |
| 1,026144033 | 0,000371156 | LPAR5 | TRUE | Upregulated |
| -1,066529657 | 0,000371156 | NDRG1 | TRUE | Downregulated |
| 0,753776539 | 0,000371156 | SERINC2 | FALSE | Upregulated |
| -1,968239758 | 0,000373389 | LOC101927121 | TRUE | Downregulated |
| -0,412075739 | 0,000373404 | ZNF397 | FALSE | Downregulated |
| 0,690607634 | 0,000381217 | GALNT10 | FALSE | Upregulated |
| 0,563393165 | 0,000381217 | GPD2 | FALSE | Upregulated |
| 0,29487704 | 0,000381217 | PPP1CA | FALSE | Upregulated |
| -0,444745453 | 0,000383422 | CTDSP2 | FALSE | Downregulated |
| 0,52074705 | 0,000383841 | MPZL1 | FALSE | Upregulated |
| -1,226971443 | 0,000386846 | RP11-242D8.1 | TRUE | Downregulated |
| 0,617205333 | 0,000388741 | ARL4A | FALSE | Upregulated |
| 2,702515619 | 0,000389087 | CCAT1 | TRUE | Upregulated |
| 0,290893703 | 0,000391913 | RAB1B | FALSE | Upregulated |
| 0,475277688 | 0,000392907 | RNPEPL1 | FALSE | Upregulated |
| 1,75019366 | 0,000394116 | SOX9-AS1 | TRUE | Upregulated |
| -0,4284977 | 0,000395199 | ERBB3 | FALSE | Downregulated |
| -0,527446274 | 0,00039632 | CEP95 | FALSE | Downregulated |
| 1,587311204 | 0,000398036 | FSTL3 | TRUE | Upregulated |
| 0,730571813 | 0,000398036 | LINC00963 | FALSE | Upregulated |
| 0,952049895 | 0,000399399 | PKM | FALSE | Upregulated |
| -0,784984048 | 0,000400316 | ZNF658B | FALSE | Downregulated |
| -1,29386549 | 0,000401621 | RNF152 | TRUE | Downregulated |
| 1,0294937 | 0,000403935 | CCDC109B | TRUE | Upregulated |
| 0,700797746 | 0,000403935 | TGFB1 | FALSE | Upregulated |
| 0,677445348 | 0,000404303 | ATP6V0B | FALSE | Upregulated |
| -0,768947333 | 0,000404303 | CFHR5 | FALSE | Downregulated |
| 1,978757779 | 0,000406912 | C5orf20 | TRUE | Upregulated |
| 0,388439206 | 0,000411097 | ACTR2 | FALSE | Upregulated |
| -0,648396673 | 0,000411948 | MARC2 | FALSE | Downregulated |
| 0,9369546 | 0,00041611 | PTPRC | FALSE | Upregulated |
| 0,432282436 | 0,00041611 | VPS18 | FALSE | Upregulated |
| -0,428616755 | 0,000416778 | ZRANB1 | FALSE | Downregulated |
| -0,573133668 | 0,000418001 | ANGEL1 | FALSE | Downregulated |
| -0,445625167 | 0,000418556 | SOWAHC | FALSE | Downregulated |
| 1,290356124 | 0,000419644 | CSF2RB | TRUE | Upregulated |
| -0,806049048 | 0,000440254 | LNP1 | FALSE | Downregulated |
| 0,909143923 | 0,000440334 | CKAP2 | FALSE | Upregulated |
| 1,02060825 | 0,000440334 | HLA-DOA | TRUE | Upregulated |
| -0,578943039 | 0,000440334 | MPDZ | FALSE | Downregulated |
| 0,59312352 | 0,000440334 | RPSA | FALSE | Upregulated |
| 1,125653702 | 0,000442055 | IKZF3 | TRUE | Upregulated |
| -2,865393968 | 0,000442737 | CRABP2 | TRUE | Downregulated |
| -1,432643253 | 0,000444185 | GPC5 | TRUE | Downregulated |
| -0,29548783 | 0,000444427 | VPS54 | FALSE | Downregulated |
| -0,601955645 | 0,000449465 | PROX1 | FALSE | Downregulated |
| 0,37799769 | 0,000449721 | RAP1A | FALSE | Upregulated |
| 1,190955757 | 0,000449899 | IL12RB1 | TRUE | Upregulated |

| | | | | |
|---|---|---|---|---|
| -1,253745974 | 0,000451538 | LGSN | TRUE | Downregulated |
| 0,711803407 | 0,000453113 | ABR | FALSE | Upregulated |
| -0,307252647 | 0,000453603 | ZNF780A | FALSE | Downregulated |
| 0,362233893 | 0,000457905 | GSR | FALSE | Upregulated |
| -1,171014986 | 0,000459637 | RP11-205M3.3 | TRUE | Downregulated |
| 0,871363668 | 0,000463503 | ARHGAP30 | FALSE | Upregulated |
| -0,441064341 | 0,000465013 | PLK1S1 | FALSE | Downregulated |
| -4,408316651 | 0,000467836 | CDH12 | TRUE | Downregulated |
| 1,005000117 | 0,000470049 | SELM | TRUE | Upregulated |
| 1,084250194 | 0,000475754 | TLR8 | TRUE | Upregulated |
| -0,637676514 | 0,000477199 | ZNF433 | FALSE | Downregulated |
| 0,32071086 | 0,00047776 | CLTA | FALSE | Upregulated |
| -0,485438219 | 0,000477977 | THAP9 | FALSE | Downregulated |
| 0,396250501 | 0,000481239 | NIPA2 | FALSE | Upregulated |
| 2,936787308 | 0,000484839 | SIRPD | TRUE | Upregulated |
| 1,23375515 | 0,000484839 | THEMIS | TRUE | Upregulated |
| -1,543083029 | 0,000485721 | GSTA7P | TRUE | Downregulated |
| 0,844214135 | 0,000488082 | DOCK2 | FALSE | Upregulated |
| -0,989675075 | 0,000488082 | LOC101927809 | FALSE | Downregulated |
| -0,448934599 | 0,000492879 | CYB5D1 | FALSE | Downregulated |
| 2,203818522 | 0,00049564 | DBNDD1 | TRUE | Upregulated |
| 1,664191609 | 0,000501797 | XRCC2 | TRUE | Upregulated |
| 0,464144694 | 0,000503866 | ATP6V0E1 | FALSE | Upregulated |
| -0,813392398 | 0,000505023 | RHOBTB3 | FALSE | Downregulated |
| -0,42355236 | 0,000506469 | ZBTB14 | FALSE | Downregulated |
| -1,503429002 | 0,000507384 | LINC01144 | TRUE | Downregulated |
| 0,776664524 | 0,000508414 | STK17A | FALSE | Upregulated |
| 0,507424255 | 0,000513302 | RCC2 | FALSE | Upregulated |
| 1,539217205 | 0,000516906 | ATP10A | TRUE | Upregulated |
| 1,321839757 | 0,000516906 | CRIP1 | TRUE | Upregulated |
| 0,79939775 | 0,000516906 | DDX60 | FALSE | Upregulated |
| 0,812471112 | 0,000516906 | DEF6 | FALSE | Upregulated |
| -0,41967008 | 0,000516906 | ETAA1 | FALSE | Downregulated |
| 0,882774509 | 0,000516906 | NDE1 | FALSE | Upregulated |
| 0,745016098 | 0,000516906 | RFC4 | FALSE | Upregulated |
| 0,527092709 | 0,000516906 | RNASET2 | FALSE | Upregulated |
| 0,408423067 | 0,000516906 | UBE2A | FALSE | Upregulated |
| 0,742502289 | 0,000517547 | PIK3IP1 | FALSE | Upregulated |
| -0,604438779 | 0,000526245 | SNTB1 | FALSE | Downregulated |
| 0,606586291 | 0,000528988 | HYAL2 | FALSE | Upregulated |
| 2,873353748 | 0,000529451 | HKDC1 | TRUE | Upregulated |
| -1,659124478 | 0,000532198 | RP11-75N4.2 | TRUE | Downregulated |
| 1,230703001 | 0,000533158 | EPSTI1 | TRUE | Upregulated |
| 1,387827537 | 0,000535889 | TRPM2 | TRUE | Upregulated |
| -0,455701905 | 0,000541776 | ZNF33B | FALSE | Downregulated |
| 0,995057091 | 0,00054264 | NUDT1 | FALSE | Upregulated |
| 1,975703852 | 0,000550479 | MTFR2 | TRUE | Upregulated |
| 1,396016203 | 0,000550479 | PTPN7 | TRUE | Upregulated |
| -1,361141119 | 0,000553594 | LINC01093 | TRUE | Downregulated |
| 1,758428481 | 0,000560225 | ZBP1 | TRUE | Upregulated |
| 0,773835534 | 0,000561141 | SLC20A1 | FALSE | Upregulated |

| | | | | |
|---|---|---|---|---|
| 0,407015491 | 0,000566918 | CDC42 | FALSE | Upregulated |
| 0,464883999 | 0,000566918 | PLA2G15 | FALSE | Upregulated |
| 1,04284432 | 0,000569145 | HCST | TRUE | Upregulated |
| 0,980643211 | 0,000569365 | EVI2A | FALSE | Upregulated |
| 0,316608191 | 0,000569365 | TATDN2 | FALSE | Upregulated |
| 0,455006227 | 0,000571369 | AAGAB | FALSE | Upregulated |
| 0,414625848 | 0,000571369 | GNG10 | FALSE | Upregulated |
| 0,536397052 | 0,000571369 | PDZD11 | FALSE | Upregulated |
| 0,442187124 | 0,000572032 | CTSZ | FALSE | Upregulated |
| 0,52696694 | 0,00057207 | NPC2 | FALSE | Upregulated |
| -1,68489521 | 0,000573365 | G6PC | TRUE | Downregulated |
| 0,459324112 | 0,000578721 | S100A16 | FALSE | Upregulated |
| 0,963132843 | 0,000578989 | FBXO2 | FALSE | Upregulated |
| 0,568988052 | 0,000580282 | VPS37C | FALSE | Upregulated |
| 0,705022689 | 0,00058198 | IFITM3 | FALSE | Upregulated |
| -0,555246854 | 0,000583047 | ARVCF | FALSE | Downregulated |
| 0,61349014 | 0,00058474 | UBALD2 | FALSE | Upregulated |
| -0,456169829 | 0,000585015 | EIF4B | FALSE | Downregulated |
| 0,387664185 | 0,000585759 | MPPE1 | FALSE | Upregulated |
| 0,82034484 | 0,000586175 | DYNLL1 | FALSE | Upregulated |
| -0,483547206 | 0,000588649 | KMT2A | FALSE | Downregulated |
| 0,560478567 | 0,000589484 | PPIH | FALSE | Upregulated |
| 0,761357673 | 0,000599755 | LPPR2 | FALSE | Upregulated |
| -0,605625286 | 0,00060283 | ANKRA2 | FALSE | Downregulated |
| 0,666801606 | 0,000604104 | RNF213 | FALSE | Upregulated |
| 1,106318257 | 0,000604169 | POC1A | TRUE | Upregulated |
| 0,297066244 | 0,000604488 | ATP6V1B2 | FALSE | Upregulated |
| 1,596153106 | 0,000606556 | SOCS1 | TRUE | Upregulated |
| 0,712081235 | 0,00060911 | BTN2A2 | FALSE | Upregulated |
| 0,472406754 | 0,000609793 | IFNAR2 | FALSE | Upregulated |
| -1,124070159 | 0,000609793 | ZFP2 | TRUE | Downregulated |
| 1,164919715 | 0,000610802 | CD3E | TRUE | Upregulated |
| 0,417022125 | 0,00062056 | TUBB | FALSE | Upregulated |
| 1,165691592 | 0,000622758 | ZC3H12D | TRUE | Upregulated |
| -0,670610687 | 0,00062402 | KBTBD11 | FALSE | Downregulated |
| 1,270186912 | 0,000626297 | GZMK | TRUE | Upregulated |
| 2,070084222 | 0,000628092 | LOC102723407 | TRUE | Upregulated |
| -0,362267078 | 0,000628852 | FAN1 | FALSE | Downregulated |
| 0,331637998 | 0,000629121 | ACTR3 | FALSE | Upregulated |
| -0,400179004 | 0,000632132 | PATZ1 | FALSE | Downregulated |
| -0,376611297 | 0,000636679 | ZNF181 | FALSE | Downregulated |
| 0,425905431 | 0,000637122 | STX4 | FALSE | Upregulated |
| 0,626130459 | 0,000639931 | DBF4 | FALSE | Upregulated |
| 2,298711991 | 0,000641834 | HAR1A | TRUE | Upregulated |
| 1,005020563 | 0,000642977 | CD96 | TRUE | Upregulated |
| 2,512632545 | 0,000642977 | FAM154B | TRUE | Upregulated |
| 0,62264539 | 0,000642977 | RANGAP1 | FALSE | Upregulated |
| 0,527604625 | 0,000642977 | UNC93B1 | FALSE | Upregulated |
| 1,148854403 | 0,000648841 | MYC | TRUE | Upregulated |
| 1,494946986 | 0,000654531 | RGCC | TRUE | Upregulated |
| -0,517965217 | 0,000662048 | RBM43 | FALSE | Downregulated |

| | | | | |
|---|---|---|---|---|
| -0,573547346 | 0,000665151 | FAM117A | FALSE | Downregulated |
| 0,439991841 | 0,000669033 | COLGALT1 | FALSE | Upregulated |
| 0,679913787 | 0,000671446 | ABI3 | FALSE | Upregulated |
| 0,370694669 | 0,000671446 | MFN2 | FALSE | Upregulated |
| 2,110464815 | 0,000671446 | MZB1 | TRUE | Upregulated |
| -0,481984409 | 0,000675422 | IDH1 | FALSE | Downregulated |
| 0,410395868 | 0,000676876 | ABCF2 | FALSE | Upregulated |
| 1,279003228 | 0,000676876 | HLA-H | TRUE | Upregulated |
| -1,269928799 | 0,000678416 | GNMT | TRUE | Downregulated |
| 1,500858257 | 0,000680358 | GPR171 | TRUE | Upregulated |
| 0,648048684 | 0,000682799 | TCN2 | FALSE | Upregulated |
| 1,377073683 | 0,00068477 | C12orf5 | TRUE | Upregulated |
| -0,720346323 | 0,000689404 | FAM160A1 | FALSE | Downregulated |
| -2,097608233 | 0,000689404 | TRHDE-AS1 | TRUE | Downregulated |
| -0,485577389 | 0,000694054 | ST3GAL1 | FALSE | Downregulated |
| 0,476361054 | 0,000696775 | INTS7 | FALSE | Upregulated |
| 0,465706088 | 0,00070145 | PTMA | FALSE | Upregulated |
| -0,380222576 | 0,000707893 | EIF4A2 | FALSE | Downregulated |
| -1,258528638 | 0,000707893 | LRP4 | TRUE | Downregulated |
| -0,73332338 | 0,000726641 | C7orf63 | FALSE | Downregulated |
| 0,680056246 | 0,000727944 | NIN | FALSE | Upregulated |
| -0,611442201 | 0,000732855 | SLC47A1 | FALSE | Downregulated |
| 1,088258202 | 0,00073444 | SLC7A6 | TRUE | Upregulated |
| 0,986693439 | 0,00073444 | TGFBI | FALSE | Upregulated |
| 0,708390034 | 0,000737547 | NAV2 | FALSE | Upregulated |
| -0,360450197 | 0,00073929 | ARHGAP35 | FALSE | Downregulated |
| 0,50530649 | 0,000742529 | MINA | FALSE | Upregulated |
| 1,208884914 | 0,000751612 | C3orf14 | TRUE | Upregulated |
| -1,737015279 | 0,000755323 | MYH3 | TRUE | Downregulated |
| 0,694377021 | 0,000759452 | UNC119 | FALSE | Upregulated |
| -0,515064563 | 0,000762479 | SBDSP1 | FALSE | Downregulated |
| 2,15317198 | 0,000762726 | KLHDC7B | TRUE | Upregulated |
| 0,534832083 | 0,000764521 | POLE4 | FALSE | Upregulated |
| 0,497230122 | 0,000768123 | CEACAM1 | FALSE | Upregulated |
| -1,353674423 | 0,000775782 | IL1RAP | TRUE | Downregulated |
| 1,681301622 | 0,000783862 | CDCA3 | TRUE | Upregulated |
| 0,533696138 | 0,000783862 | SQSTM1 | FALSE | Upregulated |
| 0,943792167 | 0,000784619 | SYTL3 | FALSE | Upregulated |
| 0,355888478 | 0,000784619 | TRAPPC3 | FALSE | Upregulated |
| 0,391855689 | 0,000789149 | DNASE2 | FALSE | Upregulated |
| -0,550116168 | 0,00078922 | SYNE1 | FALSE | Downregulated |
| -0,830352549 | 0,000790137 | LINC00663 | FALSE | Downregulated |
| 0,471603247 | 0,000790314 | CDC42SE1 | FALSE | Upregulated |
| -0,569819792 | 0,000794183 | DIXDC1 | FALSE | Downregulated |
| 0,313203827 | 0,000794842 | YWHAB | FALSE | Upregulated |
| -0,794793586 | 0,000795717 | CCDC176 | FALSE | Downregulated |
| 0,340032761 | 0,000796793 | PSAP | FALSE | Upregulated |
| 0,477276574 | 0,000799144 | P2RX4 | FALSE | Upregulated |
| -0,385293816 | 0,000802961 | ERMARD | FALSE | Downregulated |
| 0,551915895 | 0,000806137 | STIP1 | FALSE | Upregulated |
| 2,245833676 | 0,000809857 | GDF15 | TRUE | Upregulated |

| | | | | |
|---|---|---|---|---|
| 0,467178984 | 0,000812319 | PSMB5 | FALSE | Upregulated |
| -0,455633207 | 0,000812319 | TTC3 | FALSE | Downregulated |
| -0,371485032 | 0,000814271 | DNAJC24 | FALSE | Downregulated |
| -0,540700061 | 0,000827159 | EHHADH | FALSE | Downregulated |
| -1,531353364 | 0,000828707 | ENHO | TRUE | Downregulated |
| 0,295099201 | 0,000828707 | GRB2 | FALSE | Upregulated |
| -0,743725881 | 0,000837824 | AKAP6 | FALSE | Downregulated |
| -0,889598336 | 0,000838007 | PANK1 | FALSE | Downregulated |
| -1,544644959 | 0,000838007 | RP1-234P15.4 | TRUE | Downregulated |
| 1,316627088 | 0,000839442 | CDCP1 | TRUE | Upregulated |
| 0,919905199 | 0,000840199 | HDX | FALSE | Upregulated |
| -1,546462432 | 0,000840425 | SEC14L3 | TRUE | Downregulated |
| 0,31506683 | 0,000840816 | TBC1D1 | FALSE | Upregulated |
| -0,8117119 | 0,000841192 | PALM3 | FALSE | Downregulated |
| 0,939278683 | 0,000841192 | PRR11 | FALSE | Upregulated |
| 0,504326173 | 0,000845227 | ILK | FALSE | Upregulated |
| 1,400627782 | 0,000847213 | LAMA3 | TRUE | Upregulated |
| -0,565197489 | 0,000854623 | CDC14B | FALSE | Downregulated |
| -1,144278866 | 0,000854623 | PDE8B | TRUE | Downregulated |
| -0,732446093 | 0,000856383 | HNF4A-AS1 | FALSE | Downregulated |
| 1,228094115 | 0,000862249 | MYO1G | TRUE | Upregulated |
| 0,738049618 | 0,000864528 | TNFAIP8L2 | FALSE | Upregulated |
| 0,930271596 | 0,000866235 | NLRC5 | FALSE | Upregulated |
| -0,542988139 | 0,000866997 | SEC14L2 | FALSE | Downregulated |
| 1,220862474 | 0,000868397 | CCL5 | TRUE | Upregulated |
| 0,644187037 | 0,000874227 | TMEM86A | FALSE | Upregulated |
| -0,446190714 | 0,000884081 | WAC-AS1 | FALSE | Downregulated |
| 1,442725711 | 0,000886734 | MDK | TRUE | Upregulated |
| 0,579268977 | 0,000888867 | PARP8 | FALSE | Upregulated |
| 1,490063394 | 0,000902496 | EMILIN2 | TRUE | Upregulated |
| 0,920642757 | 0,000902496 | MORC4 | FALSE | Upregulated |
| 1,118999648 | 0,000902496 | NFE2L3 | TRUE | Upregulated |
| 0,569481731 | 0,000907075 | CTSD | FALSE | Upregulated |
| -0,39496616 | 0,000913919 | USP12 | FALSE | Downregulated |
| 1,177154832 | 0,000915366 | GNA15 | TRUE | Upregulated |
| 0,659140424 | 0,000915366 | SLC36A1 | FALSE | Upregulated |
| 0,434001812 | 0,000919104 | CCT2 | FALSE | Upregulated |
| -0,574073856 | 0,000922263 | CITED2 | FALSE | Downregulated |
| -0,577468894 | 0,000922503 | PALM2 | FALSE | Downregulated |
| 1,336422782 | 0,000925798 | BCL11B | TRUE | Upregulated |
| 0,832442646 | 0,000926728 | SEL1L3 | FALSE | Upregulated |
| 0,384801458 | 0,000928205 | TMEM50A | FALSE | Upregulated |
| -1,091924793 | 0,000928394 | LOC101929761 | TRUE | Downregulated |
| 0,454484996 | 0,000928594 | MYL6 | FALSE | Upregulated |
| 0,864017542 | 0,000930721 | LY86 | FALSE | Upregulated |
| -0,24803615 | 0,000931744 | ATMIN | FALSE | Downregulated |
| -0,679565837 | 0,000932813 | LRP6 | FALSE | Downregulated |
| 1,712212039 | 0,000948777 | SLC1A3 | TRUE | Upregulated |
| 2,618810786 | 0,000948999 | TIFAB | TRUE | Upregulated |
| -0,532015114 | 0,00094982 | EXTL2 | FALSE | Downregulated |
| -0,870142174 | 0,000950321 | GPRASP1 | FALSE | Downregulated |

| | | | | |
|---|---|---|---|---|
| -0,326289352 | 0,000960149 | MAVS | FALSE | Downregulated |
| -1,489543037 | 0,000964208 | EFCAB1 | TRUE | Downregulated |
| 1,047219589 | 0,000964208 | PCED1B-AS1 | TRUE | Upregulated |
| -2,306636063 | 0,000964208 | ST18 | TRUE | Downregulated |
| 1,542769456 | 0,000965697 | NETO2 | TRUE | Upregulated |
| 0,730901358 | 0,000968336 | HN1 | FALSE | Upregulated |
| -0,44964196 | 0,000978098 | ZNF595 | FALSE | Downregulated |
| 0,963376759 | 0,000981364 | GINS3 | FALSE | Upregulated |
| 0,568617925 | 0,000985472 | APOL6 | FALSE | Upregulated |
| 0,399374471 | 0,000990058 | SDCBP | FALSE | Upregulated |
| 4,200890997 | 0,000995723 | ZBED2 | TRUE | Upregulated |
| 0,603708419 | 0,00099683 | JUND | FALSE | Upregulated |
| 0,444919622 | 0,000999422 | PSMA3 | FALSE | Upregulated |
| -0,469001647 | 0,001014748 | PPARA | FALSE | Downregulated |
| -0,461436296 | 0,001030409 | INVS | FALSE | Downregulated |
| 0,443759234 | 0,001037351 | RFWD3 | FALSE | Upregulated |
| -0,507485838 | 0,001044953 | PAIP2B | FALSE | Downregulated |
| 1,164141605 | 0,001047626 | CD300C | TRUE | Upregulated |
| 2,972965528 | 0,00104855 | ALOX15B | TRUE | Upregulated |
| -0,756405229 | 0,001049937 | PTPRD | FALSE | Downregulated |
| -0,426214685 | 0,001060892 | ARHGEF9 | FALSE | Downregulated |
| -0,859071694 | 0,001060892 | LONRF3 | FALSE | Downregulated |
| 0,610430339 | 0,001061141 | PIEZO1 | FALSE | Upregulated |
| -0,418256769 | 0,001063081 | PLG | FALSE | Downregulated |
| 0,856092039 | 0,001063129 | CLEC2D | FALSE | Upregulated |
| 2,305506608 | 0,001065736 | CASP5 | TRUE | Upregulated |
| 0,645448853 | 0,001071748 | STK10 | FALSE | Upregulated |
| -1,483317552 | 0,001074137 | ADAM12 | TRUE | Downregulated |
| -1,183526752 | 0,001074137 | MDN1 | TRUE | Downregulated |
| 1,31688157 | 0,001076809 | SPA17 | TRUE | Upregulated |
| 0,260755711 | 0,001084559 | PSEN1 | FALSE | Upregulated |
| -0,25934639 | 0,001090623 | CUL4B | FALSE | Downregulated |
| 0,357168923 | 0,001092365 | MTOR | FALSE | Upregulated |
| 0,49901525 | 0,001102135 | PSEN2 | FALSE | Upregulated |
| 0,909425236 | 0,00110524 | TRPV2 | FALSE | Upregulated |
| 1,28282147 | 0,001110711 | FASN | TRUE | Upregulated |
| 0,488115796 | 0,001110711 | SYPL1 | FALSE | Upregulated |
| -0,559823422 | 0,00113552 | DST | FALSE | Downregulated |
| -0,59472516 | 0,001140065 | FOXA1 | FALSE | Downregulated |
| -0,674637622 | 0,001140065 | SEMA6A | FALSE | Downregulated |
| 0,447889446 | 0,001154201 | RECQL | FALSE | Upregulated |
| 1,089018708 | 0,001159572 | APOL4 | TRUE | Upregulated |
| 0,316014203 | 0,001159572 | BUB3 | FALSE | Upregulated |
| -0,394192859 | 0,001159572 | LOC100506710 | FALSE | Downregulated |
| 2,138060314 | 0,001159572 | RNASE2 | TRUE | Upregulated |
| -0,540798349 | 0,001159572 | ZNF429 | FALSE | Downregulated |
| 0,764704633 | 0,00116518 | LGALS3BP | FALSE | Upregulated |
| 1,289309872 | 0,00116518 | LILRA6 | TRUE | Upregulated |
| 0,640282152 | 0,00117021 | SPDL1 | FALSE | Upregulated |
| 0,342943655 | 0,001173108 | RAP1GDS1 | FALSE | Upregulated |
| -0,637455033 | 0,001173129 | TMEM25 | FALSE | Downregulated |

| | | | | |
|---|---|---|---|---|
| 1,121748163 | 0,001174176 | LCK | TRUE | Upregulated |
| 0,467847314 | 0,001175534 | CD59 | FALSE | Upregulated |
| -0,524423729 | 0,001180846 | NR1I3 | FALSE | Downregulated |
| -1,665610189 | 0,001184529 | LOC101929552 | TRUE | Downregulated |
| -0,604566512 | 0,001186047 | FLJ31306 | FALSE | Downregulated |
| -0,368883603 | 0,001189761 | CDK7 | FALSE | Downregulated |
| -0,545356929 | 0,001191612 | ZNF518A | FALSE | Downregulated |
| 0,556302008 | 0,001199989 | GMFG | FALSE | Upregulated |
| 0,501353892 | 0,001204378 | HPS1 | FALSE | Upregulated |
| 0,860312194 | 0,001204411 | FAM78A | FALSE | Upregulated |
| 0,668607856 | 0,001204411 | RPL22L1 | FALSE | Upregulated |
| 0,489653339 | 0,001209632 | SMPD1 | FALSE | Upregulated |
| 0,741247886 | 0,001212434 | ITGA4 | FALSE | Upregulated |
| 1,368787122 | 0,001215647 | MAFF | TRUE | Upregulated |
| 1,313287452 | 0,001218944 | HLA-DQB1 | TRUE | Upregulated |
| -0,6306223 | 0,001218944 | IFT172 | FALSE | Downregulated |
| 0,280181032 | 0,001231648 | MAPK1IP1L | FALSE | Upregulated |
| 0,462757129 | 0,001234738 | STAT5A | FALSE | Upregulated |
| 0,339933646 | 0,001242672 | CHFR | FALSE | Upregulated |
| 0,629215409 | 0,00124705 | INO80C | FALSE | Upregulated |
| -0,615109659 | 0,001248714 | CD27-AS1 | FALSE | Downregulated |
| 0,398919617 | 0,001259267 | CAPZA1 | FALSE | Upregulated |
| -0,617654543 | 0,001268681 | SLC17A2 | FALSE | Downregulated |
| 0,777863718 | 0,001271816 | FCER1G | FALSE | Upregulated |
| -0,838400528 | 0,001271816 | LOC101927600 | FALSE | Downregulated |
| -0,691808585 | 0,001276581 | TUBE1 | FALSE | Downregulated |
| -0,78468974 | 0,00128104 | PACSIN3 | FALSE | Downregulated |
| -0,377717231 | 0,00128688 | ZNF37A | FALSE | Downregulated |
| -0,611402667 | 0,001288431 | SMO | FALSE | Downregulated |
| 1,040324335 | 0,001288431 | SPSB1 | TRUE | Upregulated |
| 0,338012198 | 0,001288762 | NECAP2 | FALSE | Upregulated |
| -0,60163051 | 0,001288762 | NOSTRIN | FALSE | Downregulated |
| 0,633670928 | 0,001293999 | NME1 | FALSE | Upregulated |
| 0,688798831 | 0,001295218 | KLF16 | FALSE | Upregulated |
| 0,590821736 | 0,001297619 | DHRS7B | FALSE | Upregulated |
| 1,055628357 | 0,001297752 | AMICA1 | TRUE | Upregulated |
| 1,281735348 | 0,001307796 | ITM2A | TRUE | Upregulated |
| -0,312895383 | 0,001316593 | FAM160B1 | FALSE | Downregulated |
| -0,704324012 | 0,001321517 | GRB7 | FALSE | Downregulated |
| 1,018208646 | 0,001321517 | JUN | TRUE | Upregulated |
| 0,720599836 | 0,001327866 | TTF2 | FALSE | Upregulated |
| -0,660413572 | 0,001332455 | HOMER2 | FALSE | Downregulated |
| -0,379277532 | 0,001336511 | CDKN2AIP | FALSE | Downregulated |
| 1,157898616 | 0,001352263 | LINC00857 | TRUE | Upregulated |
| 0,789088008 | 0,001353084 | LOC81691 | FALSE | Upregulated |
| 0,660685895 | 0,001357573 | LIPA | FALSE | Upregulated |
| 0,866769248 | 0,001362073 | GPR65 | FALSE | Upregulated |
| 0,893318859 | 0,001365558 | SLC38A1 | FALSE | Upregulated |
| 0,534807933 | 0,001371204 | DOLK | FALSE | Upregulated |
| 0,461778318 | 0,001376313 | CSK | FALSE | Upregulated |
| -1,08626005 | 0,001385764 | HERC2P7 | TRUE | Downregulated |

| | | | | |
|---|---|---|---|---|
| 0,390439093 | 0,001392967 | ABHD16A | FALSE | Upregulated |
| 1,613361617 | 0,001394429 | ABCD2 | TRUE | Upregulated |
| -0,640910475 | 0,001397103 | MICU3 | FALSE | Downregulated |
| 1,114632016 | 0,001397103 | SLA2 | TRUE | Upregulated |
| 0,514237878 | 0,001398024 | RHBDD3 | FALSE | Upregulated |
| -0,443886933 | 0,001409347 | ZNF225 | FALSE | Downregulated |
| -0,498685899 | 0,001413816 | TRPM7 | FALSE | Downregulated |
| 0,379011747 | 0,001419891 | PSMC2 | FALSE | Upregulated |
| 0,646788255 | 0,001427174 | ANXA6 | FALSE | Upregulated |
| 0,547165621 | 0,001427174 | LYN | FALSE | Upregulated |
| -2,23989499 | 0,001427174 | NCAM2 | TRUE | Downregulated |
| 0,414137764 | 0,001427174 | TMEM8A | FALSE | Upregulated |
| -0,588366924 | 0,001427174 | ZFP36L1 | FALSE | Downregulated |
| -1,160399803 | 0,001433095 | COL11A2 | TRUE | Downregulated |
| -0,80316374 | 0,001433743 | DNMT3B | FALSE | Downregulated |
| 0,611406762 | 0,001433743 | DTD1 | FALSE | Upregulated |
| 2,298510653 | 0,001436499 | CENPA | TRUE | Upregulated |
| 0,402217533 | 0,001439829 | DESI1 | FALSE | Upregulated |
| 0,543567136 | 0,001439829 | DNAJA1 | FALSE | Upregulated |
| 0,449581246 | 0,001442972 | GNA12 | FALSE | Upregulated |
| 1,525919146 | 0,001444197 | PDGFA | TRUE | Upregulated |
| -0,661092915 | 0,001444197 | SEPT7P2 | FALSE | Downregulated |
| 1,09092858 | 0,001445122 | RP11-290F20.3 | TRUE | Upregulated |
| 0,240486896 | 0,001447925 | GNB1 | FALSE | Upregulated |
| 1,991474492 | 0,001454343 | MYO7B | TRUE | Upregulated |
| 0,308051316 | 0,0014544 | RNH1 | FALSE | Upregulated |
| 0,525423074 | 0,001455777 | STARD3NL | FALSE | Upregulated |
| 0,47530548 | 0,00145689 | NSMCE2 | FALSE | Upregulated |
| -2,953071426 | 0,001459957 | LOC101927170 | TRUE | Downregulated |
| -0,817590549 | 0,001459957 | PLGLA | FALSE | Downregulated |
| 0,367530129 | 0,001483778 | TXNRD1 | FALSE | Upregulated |
| 0,747324175 | 0,00148911 | PLEKHO2 | FALSE | Upregulated |
| 1,495387188 | 0,00148911 | SIGLEC5 | TRUE | Upregulated |
| 0,494538773 | 0,001491519 | PEA15 | FALSE | Upregulated |
| -0,326805469 | 0,001494111 | FRA10AC1 | FALSE | Downregulated |
| 0,268524437 | 0,001501439 | RBBP7 | FALSE | Upregulated |
| -1,38062077 | 0,001501439 | SCG5 | TRUE | Downregulated |
| 0,673667609 | 0,001501439 | WIPF1 | FALSE | Upregulated |
| -0,952121301 | 0,00151904 | ASPA | FALSE | Downregulated |
| 1,168536943 | 0,001519315 | SPOCK2 | TRUE | Upregulated |
| 0,824392651 | 0,001524755 | FGD2 | FALSE | Upregulated |
| 1,987598946 | 0,001524755 | SAPCD2 | TRUE | Upregulated |
| -0,506305912 | 0,001525845 | NAGS | FALSE | Downregulated |
| -0,443693044 | 0,001525845 | ZKSCAN8 | FALSE | Downregulated |
| -0,368630627 | 0,00152841 | NUDT6 | FALSE | Downregulated |
| -0,926186254 | 0,001538331 | GRHL1 | FALSE | Downregulated |
| -0,346981545 | 0,001540125 | GID4 | FALSE | Downregulated |
| 1,569075378 | 0,001540717 | C18orf56 | TRUE | Upregulated |
| -0,944728542 | 0,001548306 | SOX7 | FALSE | Downregulated |
| 0,26135296 | 0,00155879 | NSFL1C | FALSE | Upregulated |
| -0,619868829 | 0,00155879 | TMEM150C | FALSE | Downregulated |

| | | | | |
|---|---|---|---|---|
| -0,465583471 | 0,00155879 | ZNF75D | FALSE | Downregulated |
| 0,771282679 | 0,001565839 | GALNT6 | FALSE | Upregulated |
| -1,304151583 | 0,00156843 | PCK1 | TRUE | Downregulated |
| -1,261694519 | 0,001587374 | CFHR4 | TRUE | Downregulated |
| 0,769726918 | 0,001590289 | ADCY7 | FALSE | Upregulated |
| -0,435733773 | 0,001590289 | LZTFL1 | FALSE | Downregulated |
| 1,907570771 | 0,001592903 | PCDP1 | TRUE | Upregulated |
| -0,534364441 | 0,001594265 | F9 | FALSE | Downregulated |
| 0,471945223 | 0,001594265 | LEPROTL1 | FALSE | Upregulated |
| 0,732248843 | 0,001600464 | KNTC1 | FALSE | Upregulated |
| 0,719143469 | 0,001604778 | FUT4 | FALSE | Upregulated |
| 0,720267863 | 0,001611428 | CASC5 | FALSE | Upregulated |
| 0,368090654 | 0,001611428 | CCT5 | FALSE | Upregulated |
| 0,859367826 | 0,001611428 | SAMD9L | FALSE | Upregulated |
| 0,624093167 | 0,001616615 | C10orf128 | FALSE | Upregulated |
| 1,479712014 | 0,001616615 | CCL21 | TRUE | Upregulated |
| 0,720852832 | 0,001617482 | INPP5D | FALSE | Upregulated |
| -1,486961712 | 0,001620073 | LOC101929143 | TRUE | Downregulated |
| -1,228392197 | 0,001620073 | LOC102724870 | TRUE | Downregulated |
| 0,335150238 | 0,001642711 | PSMA6 | FALSE | Upregulated |
| 1,532328901 | 0,001648823 | BAIAP2L2 | TRUE | Upregulated |
| 0,464483745 | 0,001650936 | TAF10 | FALSE | Upregulated |
| 1,273907853 | 0,001667665 | CD300LF | TRUE | Upregulated |
| 0,699180454 | 0,001675189 | SUSD1 | FALSE | Upregulated |
| -0,83444081 | 0,001691505 | HMGCS2 | FALSE | Downregulated |
| 0,279921784 | 0,001694684 | POLR3C | FALSE | Upregulated |
| 0,535010992 | 0,001701744 | EIF4E3 | FALSE | Upregulated |
| 0,580852976 | 0,001704341 | LAMP2 | FALSE | Upregulated |
| -0,549196866 | 0,001704341 | METAP1D | FALSE | Downregulated |
| 0,343430127 | 0,001704341 | RRAGC | FALSE | Upregulated |
| 0,777465993 | 0,00171538 | SEMA4D | FALSE | Upregulated |
| -0,644494978 | 0,001720027 | CAT | FALSE | Downregulated |
| 0,32308087 | 0,001721871 | TMX1 | FALSE | Upregulated |
| -0,732698825 | 0,001723532 | TMEM220-AS1 | FALSE | Downregulated |
| 1,364359439 | 0,001726042 | BTLA | TRUE | Upregulated |
| 0,441259274 | 0,001727387 | GARS | FALSE | Upregulated |
| -0,771280667 | 0,001737934 | CRY1 | FALSE | Downregulated |
| 0,771338977 | 0,001737934 | FOLR2 | FALSE | Upregulated |
| -2,849697112 | 0,001737934 | KIAA1751 | TRUE | Downregulated |
| 0,347036419 | 0,001737934 | M6PR | FALSE | Upregulated |
| 0,806221256 | 0,001737934 | NEDD4L | FALSE | Upregulated |
| 0,891332641 | 0,001737934 | STX11 | FALSE | Upregulated |
| 0,932133985 | 0,001769096 | SAMD3 | FALSE | Upregulated |
| 0,432020917 | 0,001773478 | SHKBP1 | FALSE | Upregulated |
| -0,725551215 | 0,001778407 | SEMA6D | FALSE | Downregulated |
| 0,598098003 | 0,001785568 | NR2C2AP | FALSE | Upregulated |
| -0,807366396 | 0,001785568 | PMS2CL | FALSE | Downregulated |
| 2,211328948 | 0,001796833 | SCIN | TRUE | Upregulated |
| 0,564480587 | 0,001797606 | NPM3 | FALSE | Upregulated |
| -0,441479162 | 0,001797606 | SS18L1 | FALSE | Downregulated |
| 2,127997068 | 0,001799145 | MIR155HG | TRUE | Upregulated |

| | | | | |
|---|---|---|---|---|
| 0,883709511 | 0,001799866 | DGKA | FALSE | Upregulated |
| -0,553988539 | 0,001800473 | MAOA | FALSE | Downregulated |
| 0,942096015 | 0,001800752 | CD38 | FALSE | Upregulated |
| 0,470362713 | 0,001804971 | FAM50A | FALSE | Upregulated |
| 0,87285274 | 0,001806498 | HPS5 | FALSE | Upregulated |
| -0,33281223 | 0,001812257 | RNF170 | FALSE | Downregulated |
| 0,337874601 | 0,001818635 | PIP4K2C | FALSE | Upregulated |
| 0,891379857 | 0,001829168 | FLVCR2 | FALSE | Upregulated |
| 1,07300161 | 0,001833412 | CD83 | TRUE | Upregulated |
| -0,465472119 | 0,001833412 | ZDHHC17 | FALSE | Downregulated |
| 0,326690773 | 0,001849149 | PTPN1 | FALSE | Upregulated |
| 0,895878054 | 0,001849149 | RUNX3 | FALSE | Upregulated |
| 0,452030492 | 0,001861982 | COQ10B | FALSE | Upregulated |
| 0,811325204 | 0,001868347 | KLF4 | FALSE | Upregulated |
| 0,405615089 | 0,001877832 | HMGN1 | FALSE | Upregulated |
| 0,404427822 | 0,001877832 | TSC22D4 | FALSE | Upregulated |
| 1,395031459 | 0,001885827 | RAD51 | TRUE | Upregulated |
| 0,659812246 | 0,001894325 | HIGD1A | FALSE | Upregulated |
| 0,915607919 | 0,001905093 | MARCH1 | FALSE | Upregulated |
| 0,509845764 | 0,001920444 | LAP3 | FALSE | Upregulated |
| 0,886093269 | 0,001926874 | NFAM1 | FALSE | Upregulated |
| -1,139949182 | 0,001936799 | LOC730101 | TRUE | Downregulated |
| 0,713776881 | 0,001945338 | G6PD | FALSE | Upregulated |
| 0,692403845 | 0,001945338 | MMD | FALSE | Upregulated |
| 0,650958706 | 0,001945338 | POLD1 | FALSE | Upregulated |
| 1,117439259 | 0,001948224 | RHOF | TRUE | Upregulated |
| 0,631187286 | 0,00195451 | SLC38A2 | FALSE | Upregulated |
| -0,699238907 | 0,001974147 | HPR | FALSE | Downregulated |
| -0,449291416 | 0,001985374 | GPATCH11 | FALSE | Downregulated |
| -1,149067123 | 0,001993679 | KCTD21-AS1 | TRUE | Downregulated |
| -0,741947277 | 0,001993679 | ZNF467 | FALSE | Downregulated |
| -0,286573365 | 0,001999776 | CSNK1G3 | FALSE | Downregulated |
| 1,040163984 | 0,002003501 | SLC2A6 | TRUE | Upregulated |
| 0,375110929 | 0,002003652 | ATP13A1 | FALSE | Upregulated |
| 0,545047754 | 0,002003652 | CRLF3 | FALSE | Upregulated |
| 0,632497048 | 0,002003652 | HK1 | FALSE | Upregulated |
| 0,392326694 | 0,002003652 | PGLS | FALSE | Upregulated |
| 0,516013938 | 0,002003652 | TKT | FALSE | Upregulated |
| -0,633419732 | 0,002014364 | LIMCH1 | FALSE | Downregulated |
| 0,511314641 | 0,002031968 | NAGK | FALSE | Upregulated |
| 0,35569017 | 0,002040224 | PSMC3 | FALSE | Upregulated |
| 0,648257586 | 0,002041905 | RHNO1 | FALSE | Upregulated |
| 0,26545228 | 0,002046495 | YWHAG | FALSE | Upregulated |
| -1,323483075 | 0,002050763 | RP11-31F15.2 | TRUE | Downregulated |
| -1,39694403 | 0,002052976 | ASB4 | TRUE | Downregulated |
| 0,406660315 | 0,002052976 | MRPL47 | FALSE | Upregulated |
| 1,264316956 | 0,002053756 | MARCH3 | TRUE | Upregulated |
| 0,991304375 | 0,002056297 | ARRDC2 | FALSE | Upregulated |
| -0,591866976 | 0,002058543 | FAM84B | FALSE | Downregulated |
| -0,354204475 | 0,002058543 | FOXN3 | FALSE | Downregulated |
| 0,705516886 | 0,002059942 | ZNF385A | FALSE | Upregulated |

| | | | | |
|---|---|---|---|---|
| 1,664214391 | 0,002071646 | DNAJC5B | TRUE | Upregulated |
| -0,601323135 | 0,002072036 | C1orf21 | FALSE | Downregulated |
| 1,692397318 | 0,002072036 | CHI3L2 | TRUE | Upregulated |
| 2,424766226 | 0,002072036 | CSPG5 | TRUE | Upregulated |
| 0,755682285 | 0,002072036 | KIAA0513 | FALSE | Upregulated |
| -2,487923715 | 0,002072036 | LOC149684 | TRUE | Downregulated |
| 1,188232663 | 0,002072036 | TESPA1 | TRUE | Upregulated |
| 0,739872603 | 0,002072036 | TMED3 | FALSE | Upregulated |
| 0,731985535 | 0,002072036 | TMEM106A | FALSE | Upregulated |
| -0,970691063 | 0,002072036 | TRNQ | FALSE | Downregulated |
| -0,392235512 | 0,002072036 | ZNF569 | FALSE | Downregulated |
| 0,77274334 | 0,002073474 | ADRBK2 | FALSE | Upregulated |
| 0,287281347 | 0,002073474 | GNAI3 | FALSE | Upregulated |
| 0,353694677 | 0,002109189 | CPSF2 | FALSE | Upregulated |
| 0,545273079 | 0,002115413 | TSPAN4 | FALSE | Upregulated |
| 1,577167347 | 0,002115569 | RP11-701P16.5 | TRUE | Upregulated |
| -0,569465937 | 0,00213234 | IVNS1ABP | FALSE | Downregulated |
| 0,369288426 | 0,00213234 | TMEM87B | FALSE | Upregulated |
| -0,830135644 | 0,002142527 | LINC00999 | FALSE | Downregulated |
| 0,516285893 | 0,002151239 | AGTRAP | FALSE | Upregulated |
| -0,586600222 | 0,00215167 | FAT4 | FALSE | Downregulated |
| 0,927823691 | 0,002158885 | RAMP1 | FALSE | Upregulated |
| -0,351111687 | 0,002185427 | ACAD10 | FALSE | Downregulated |
| 0,692786355 | 0,002187788 | C14orf169 | FALSE | Upregulated |
| -0,945123945 | 0,002193283 | CNGA1 | FALSE | Downregulated |
| 0,70966247 | 0,002193283 | MYO1F | FALSE | Upregulated |
| -0,799474101 | 0,002194567 | INO80B | FALSE | Downregulated |
| 0,463200699 | 0,002201815 | ABCB8 | FALSE | Upregulated |
| 1,432085976 | 0,002209021 | ESCO2 | TRUE | Upregulated |
| 0,419576119 | 0,002231017 | ATP6V1E1 | FALSE | Upregulated |
| -1,787124692 | 0,002233445 | C2orf54 | TRUE | Downregulated |
| 0,313466943 | 0,002233445 | SUB1 | FALSE | Upregulated |
| 0,9358169 | 0,002236595 | LOC102724168 | FALSE | Upregulated |
| 0,398169834 | 0,002239217 | TNFRSF10B | FALSE | Upregulated |
| -0,27161576 | 0,00224638 | ANGEL2 | FALSE | Downregulated |
| 0,503988233 | 0,00224638 | MRPL27 | FALSE | Upregulated |
| -0,269398743 | 0,002255314 | FAM168B | FALSE | Downregulated |
| 0,300863937 | 0,00225583 | SQRDL | FALSE | Upregulated |
| 0,701558676 | 0,002256334 | SYK | FALSE | Upregulated |
| 0,865003408 | 0,002256875 | CD44 | FALSE | Upregulated |
| 1,134752636 | 0,002257464 | RASGRP1 | TRUE | Upregulated |
| 0,410641373 | 0,00226505 | PSMA4 | FALSE | Upregulated |
| -0,447511676 | 0,002266797 | TBC1D8B | FALSE | Downregulated |
| -1,046525499 | 0,002267301 | AC074138.3 | TRUE | Downregulated |
| 2,235926402 | 0,002268064 | FCRL5 | TRUE | Upregulated |
| 1,130752938 | 0,002268064 | LOC100507507 | TRUE | Upregulated |
| 0,609651628 | 0,002268796 | MANEAL | FALSE | Upregulated |
| 1,278370818 | 0,002271901 | LOC101928429 | TRUE | Upregulated |
| 1,297496383 | 0,002273118 | MX1 | TRUE | Upregulated |
| -0,878299555 | 0,002273118 | TTC6 | FALSE | Downregulated |
| 0,446389897 | 0,002284415 | ABHD12 | FALSE | Upregulated |

| | | | | |
|---|---|---|---|---|
| -0,778411689 | 0,002284415 | KDR | FALSE | Downregulated |
| 0,646348569 | 0,002284415 | POLA2 | FALSE | Upregulated |
| -0,479668627 | 0,002290372 | QDPR | FALSE | Downregulated |
| 0,815847449 | 0,002291835 | CHST11 | FALSE | Upregulated |
| -0,342185549 | 0,002291835 | IDS | FALSE | Downregulated |
| 0,341314165 | 0,002298933 | PGK1 | FALSE | Upregulated |
| 0,573684277 | 0,002305216 | TTL | FALSE | Upregulated |
| -0,734039178 | 0,002316653 | HLX | FALSE | Downregulated |
| 0,956138291 | 0,002316653 | LILRA1 | FALSE | Upregulated |
| 0,313383471 | 0,002316653 | NPLOC4 | FALSE | Upregulated |
| 0,409916859 | 0,002316653 | RB1 | FALSE | Upregulated |
| -0,652309367 | 0,002323475 | SKIDA1 | FALSE | Downregulated |
| 0,767277289 | 0,002325205 | BARD1 | FALSE | Upregulated |
| 1,105846222 | 0,002329256 | FAM129A | TRUE | Upregulated |
| -0,488739109 | 0,002339164 | DNAJC27 | FALSE | Downregulated |
| -0,575066093 | 0,002339164 | ZNF248 | FALSE | Downregulated |
| -0,829023831 | 0,002352058 | ENC1 | FALSE | Downregulated |
| -0,335842361 | 0,002352058 | NDUFAF7 | FALSE | Downregulated |
| 0,696023102 | 0,002352058 | SLC25A24 | FALSE | Upregulated |
| -0,482797941 | 0,002362486 | GEMIN4 | FALSE | Downregulated |
| -0,39675498 | 0,00236385 | FAM185A | FALSE | Downregulated |
| -0,280407009 | 0,002375522 | DCAF5 | FALSE | Downregulated |
| -0,551710877 | 0,002375522 | ZHX3 | FALSE | Downregulated |
| 0,559817704 | 0,002384972 | SERPINA7 | FALSE | Upregulated |
| -1,219156979 | 0,002385907 | SLC18A2 | TRUE | Downregulated |
| 0,342610277 | 0,002418304 | TOR1A | FALSE | Upregulated |
| -0,945109979 | 0,002418811 | ADAMTS5 | FALSE | Downregulated |
| -0,53363669 | 0,002423151 | FLT4 | FALSE | Downregulated |
| -0,35598752 | 0,002423151 | MED13L | FALSE | Downregulated |
| 0,227471947 | 0,002436976 | EFTUD2 | FALSE | Upregulated |
| -0,372407903 | 0,002443975 | GRB10 | FALSE | Downregulated |
| -1,143939187 | 0,002472173 | SNORA8 | TRUE | Downregulated |
| -0,420522236 | 0,002473489 | TUBD1 | FALSE | Downregulated |
| 1,27248169 | 0,002480555 | CD6 | TRUE | Upregulated |
| -0,316435679 | 0,002480555 | MKLN1 | FALSE | Downregulated |
| 0,623859664 | 0,002480555 | PLXNA1 | FALSE | Upregulated |
| 0,620201545 | 0,002480555 | TRAF2 | FALSE | Upregulated |
| 0,273610165 | 0,002482894 | RAB9A | FALSE | Upregulated |
| -1,458020021 | 0,002488683 | CTNNA3 | TRUE | Downregulated |
| 0,536359796 | 0,00248995 | ASCC3 | FALSE | Upregulated |
| 0,753321285 | 0,002510191 | PIK3CD | FALSE | Upregulated |
| -1,28026659 | 0,002523902 | COPG2IT1 | TRUE | Downregulated |
| 0,495315915 | 0,00252538 | OSTM1 | FALSE | Upregulated |
| 0,686848415 | 0,00252538 | RARRES3 | FALSE | Upregulated |
| 1,319582865 | 0,002525685 | FAM169A | TRUE | Upregulated |
| 0,708233478 | 0,002528302 | SERTAD1 | FALSE | Upregulated |
| 0,539532811 | 0,002529834 | OPTN | FALSE | Upregulated |
| 0,708080941 | 0,002531145 | CD300A | FALSE | Upregulated |
| 0,578980192 | 0,002531145 | LACC1 | FALSE | Upregulated |
| 0,802122534 | 0,002531145 | PTAFR | FALSE | Upregulated |
| -0,438966167 | 0,00256129 | LMAN1 | FALSE | Downregulated |

| | | | | |
|---|---|---|---|---|
| 0,500125011 | 0,00256129 | TAF9 | FALSE | Upregulated |
| -0,587052792 | 0,002570208 | C2orf72 | FALSE | Downregulated |
| -0,218836055 | 0,002570208 | RBM17 | FALSE | Downregulated |
| 2,04948327 | 0,002570426 | SPC24 | TRUE | Upregulated |
| -0,273122821 | 0,002570426 | UBTF | FALSE | Downregulated |
| -0,957230701 | 0,002582242 | ADCY10 | FALSE | Downregulated |
| 0,532771385 | 0,002582242 | LSM4 | FALSE | Upregulated |
| 0,884292607 | 0,002582242 | RTP4 | FALSE | Upregulated |
| 0,43398056 | 0,002582242 | SLC12A7 | FALSE | Upregulated |
| 0,896926803 | 0,002599621 | SMIM3 | FALSE | Upregulated |
| -0,365426239 | 0,002601539 | NACA | FALSE | Downregulated |
| -0,302636031 | 0,002613012 | RNF146 | FALSE | Downregulated |
| 0,349037192 | 0,002616149 | ARFIP1 | FALSE | Upregulated |
| 3,440449203 | 0,002640149 | LOC101929475 | TRUE | Upregulated |
| 0,753004629 | 0,002669296 | C1orf54 | FALSE | Upregulated |
| -0,55989147 | 0,0026694 | CBS | FALSE | Downregulated |
| -0,369647143 | 0,002676692 | PRKAA1 | FALSE | Downregulated |
| 0,404411234 | 0,002684734 | MAP1LC3B | FALSE | Upregulated |
| -0,332420603 | 0,00269928 | KRIT1 | FALSE | Downregulated |
| -0,46651181 | 0,00269928 | OGDHL | FALSE | Downregulated |
| 0,78523814 | 0,002713513 | BATF2 | FALSE | Upregulated |
| -0,40928556 | 0,002716514 | DCAF11 | FALSE | Downregulated |
| 0,355687073 | 0,002739387 | ATP6AP1 | FALSE | Upregulated |
| -0,459216227 | 0,002741207 | PAH | FALSE | Downregulated |
| -1,351510562 | 0,002741207 | RAD51AP2 | TRUE | Downregulated |
| 0,837614597 | 0,002742507 | AEN | FALSE | Upregulated |
| 3,383623208 | 0,002745517 | CORIN | TRUE | Upregulated |
| -0,342215921 | 0,002745517 | DHFRL1 | FALSE | Downregulated |
| 0,567647962 | 0,002747466 | NUP93 | FALSE | Upregulated |
| 0,395694877 | 0,002753066 | ESYT1 | FALSE | Upregulated |
| 0,537217802 | 0,002774448 | PARP9 | FALSE | Upregulated |
| 0,365611983 | 0,002778067 | CTSB | FALSE | Upregulated |
| 0,470025615 | 0,002780057 | ADPRHL2 | FALSE | Upregulated |
| -0,464251729 | 0,002780057 | RUNDC3B | FALSE | Downregulated |
| 0,503052883 | 0,002782426 | FDX1L | FALSE | Upregulated |
| -0,932658862 | 0,002782426 | MTHFD2L | FALSE | Downregulated |
| 0,865511799 | 0,002792443 | FYB | FALSE | Upregulated |
| -0,410989666 | 0,002804727 | USPL1 | FALSE | Downregulated |
| -0,492998937 | 0,002806603 | OXR1 | FALSE | Downregulated |
| 0,625600723 | 0,002810221 | ARHGAP25 | FALSE | Upregulated |
| -0,756829526 | 0,00281538 | TRIM16L | FALSE | Downregulated |
| 0,260339723 | 0,002824136 | CSNK1D | FALSE | Upregulated |
| -1,932697651 | 0,002834451 | LOC102724329 | TRUE | Downregulated |
| -0,427817239 | 0,002842925 | CDO1 | FALSE | Downregulated |
| 0,935980879 | 0,002845438 | C17orf96 | FALSE | Upregulated |
| -0,42943107 | 0,002851799 | SETD5 | FALSE | Downregulated |
| 4,825823958 | 0,002868045 | PAGE4 | TRUE | Upregulated |
| 0,876490301 | 0,002872622 | AOAH | FALSE | Upregulated |
| -0,333033296 | 0,002892641 | FBXW2 | FALSE | Downregulated |
| -1,420230927 | 0,002892641 | NLRP14 | TRUE | Downregulated |
| -0,457925048 | 0,002892641 | SPEN | FALSE | Downregulated |

| | | | | |
|---|---|---|---|---|
| 0,531636479 | 0,002911451 | BRMS1 | FALSE | Upregulated |
| 0,697220923 | 0,002911451 | PSMC3IP | FALSE | Upregulated |
| 0,710398081 | 0,002912798 | C10orf54 | FALSE | Upregulated |
| 0,921279525 | 0,002926044 | EMB | FALSE | Upregulated |
| 0,679275149 | 0,002929355 | VSIG10 | FALSE | Upregulated |
| 2,732498071 | 0,002934752 | HCN1 | TRUE | Upregulated |
| -0,591938498 | 0,002934752 | ZSCAN16-AS1 | FALSE | Downregulated |
| 0,495217266 | 0,002954425 | SPTAN1 | FALSE | Upregulated |
| -0,41834669 | 0,002962205 | IBA57 | FALSE | Downregulated |
| 0,415793947 | 0,00296864 | TBCB | FALSE | Upregulated |
| -0,558517251 | 0,002969073 | PHLPP1 | FALSE | Downregulated |
| 1,164044114 | 0,002973152 | UCP2 | TRUE | Upregulated |
| 0,949493979 | 0,002974751 | PRF1 | FALSE | Upregulated |
| 0,916442245 | 0,002982883 | FGR | FALSE | Upregulated |
| -0,441869329 | 0,002982883 | PIGV | FALSE | Downregulated |
| 0,629858807 | 0,003006553 | TMEM165 | FALSE | Upregulated |
| 2,489785492 | 0,003018924 | DUSP8 | TRUE | Upregulated |
| 1,606878466 | 0,003029369 | LGALS2 | TRUE | Upregulated |
| 0,476963588 | 0,003046359 | PPP4R1 | FALSE | Upregulated |
| 1,034239579 | 0,00305528 | GNGT2 | TRUE | Upregulated |
| 0,58222061 | 0,003069827 | LMNB2 | FALSE | Upregulated |
| 2,560166322 | 0,003069827 | NRIP3 | TRUE | Upregulated |
| -0,257245773 | 0,003075211 | GPBP1L1 | FALSE | Downregulated |
| -0,779362577 | 0,003076931 | C1QTNF3 | FALSE | Downregulated |
| 0,864888856 | 0,00308038 | ATF3 | FALSE | Upregulated |
| -1,13848482 | 0,003095544 | CELF6 | TRUE | Downregulated |
| 0,981244125 | 0,003111377 | CARD11 | FALSE | Upregulated |
| -1,346956925 | 0,003111377 | HSD17B14 | TRUE | Downregulated |
| -0,283089051 | 0,003140802 | ZNF197 | FALSE | Downregulated |
| -0,316289599 | 0,003155332 | RNF185 | FALSE | Downregulated |
| -0,502339661 | 0,003177841 | ZFP14 | FALSE | Downregulated |
| 0,613446085 | 0,003188186 | ATP8B1 | FALSE | Upregulated |
| -0,762937525 | 0,003188186 | SLC45A3 | FALSE | Downregulated |
| 0,881039549 | 0,003188186 | XRCC3 | FALSE | Upregulated |
| 0,56104548 | 0,003189642 | STAP2 | FALSE | Upregulated |
| 0,666748814 | 0,003199561 | SLC27A4 | FALSE | Upregulated |
| 0,742735146 | 0,00322771 | CD97 | FALSE | Upregulated |
| -1,374241806 | 0,003228005 | RP11-15A1.7 | TRUE | Downregulated |
| 0,786710095 | 0,003228005 | TRAF5 | FALSE | Upregulated |
| 0,753647072 | 0,003235724 | ELF4 | FALSE | Upregulated |
| -0,228552119 | 0,003237546 | MFN1 | FALSE | Downregulated |
| 1,293219256 | 0,003237546 | RLTPR | TRUE | Upregulated |
| -0,60182812 | 0,003243026 | BMI1 | FALSE | Downregulated |
| 2,784282979 | 0,003247673 | CD19 | TRUE | Upregulated |
| 1,029413229 | 0,003247673 | GAS7 | TRUE | Upregulated |
| 1,028935355 | 0,003252083 | RELT | TRUE | Upregulated |
| 0,295207632 | 0,003253423 | ATP6V1G1 | FALSE | Upregulated |
| -0,424905162 | 0,003257934 | ALDH7A1 | FALSE | Downregulated |
| -0,740724034 | 0,003257934 | IGF2 | FALSE | Downregulated |
| 2,979095375 | 0,003257934 | IL8 | TRUE | Upregulated |
| 0,599553508 | 0,003258497 | PPP1R18 | FALSE | Upregulated |

| | | | | |
|---|---|---|---|---|
| -2,765973191 | 0,003263311 | LOC101927032 | TRUE | Downregulated |
| -0,60819743 | 0,003263311 | PLEKHA7 | FALSE | Downregulated |
| 0,419393835 | 0,003263311 | SYS1 | FALSE | Upregulated |
| -0,36467642 | 0,00330394 | EIF4EBP2 | FALSE | Downregulated |
| -0,501303847 | 0,003304467 | NTN4 | FALSE | Downregulated |
| 1,874059676 | 0,003307931 | IGLL1 | TRUE | Upregulated |
| -0,612739495 | 0,003314504 | THSD4 | FALSE | Downregulated |
| 1,054322369 | 0,003321497 | BLM | TRUE | Upregulated |
| 1,513394479 | 0,003330017 | CCL3 | TRUE | Upregulated |
| -0,339978769 | 0,003330017 | PEX12 | FALSE | Downregulated |
| 0,574572784 | 0,003332252 | C12orf49 | FALSE | Upregulated |
| 0,832484859 | 0,003338826 | NIPAL2 | FALSE | Upregulated |
| 0,788279183 | 0,003346543 | CD244 | FALSE | Upregulated |
| -0,65695114 | 0,003346543 | HIC2 | FALSE | Downregulated |
| 0,448222234 | 0,003346543 | WWC3 | FALSE | Upregulated |
| 1,110521025 | 0,003357749 | CDKN1A | TRUE | Upregulated |
| 0,664979474 | 0,003357749 | FAM213B | FALSE | Upregulated |
| 1,103327127 | 0,003357749 | SLC35C1 | TRUE | Upregulated |
| -0,402033808 | 0,003366264 | RBM41 | FALSE | Downregulated |
| 1,910295437 | 0,00337346 | IL4I1 | TRUE | Upregulated |
| 0,578832929 | 0,003379863 | DENND2D | FALSE | Upregulated |
| -0,342338609 | 0,003383075 | AHCTF1 | FALSE | Downregulated |
| 0,588432518 | 0,00338455 | IQGAP1 | FALSE | Upregulated |
| 1,411276458 | 0,003387759 | RAB7B | TRUE | Upregulated |
| 2,075758615 | 0,003387759 | SLC29A4 | TRUE | Upregulated |
| -0,43655189 | 0,003389521 | C4orf29 | FALSE | Downregulated |
| 0,229461033 | 0,003397122 | USP4 | FALSE | Upregulated |
| 0,403663889 | 0,003407012 | SLC38A7 | FALSE | Upregulated |
| 1,732964404 | 0,003408988 | ANKRD22 | TRUE | Upregulated |
| 0,598672203 | 0,003411868 | SRXN1 | FALSE | Upregulated |
| 1,59686033 | 0,003421772 | CRNDE | TRUE | Upregulated |
| 0,882393858 | 0,003421772 | RHOB | FALSE | Upregulated |
| 0,451208207 | 0,003423666 | NACC1 | FALSE | Upregulated |
| 0,730243761 | 0,003432587 | IL15RA | FALSE | Upregulated |
| 3,289831984 | 0,003471866 | STMN2 | TRUE | Upregulated |
| 2,187392072 | 0,003486152 | GTSF1 | TRUE | Upregulated |
| -0,552607602 | 0,003488416 | LIG4 | FALSE | Downregulated |
| 1,568329037 | 0,003495258 | RPS6KL1 | TRUE | Upregulated |
| 0,320396799 | 0,003500152 | CNDP2 | FALSE | Upregulated |
| -0,413541564 | 0,00351635 | ABHD10 | FALSE | Downregulated |
| 0,947867991 | 0,003550887 | CST7 | FALSE | Upregulated |
| 0,49695794 | 0,003567065 | TMEM70 | FALSE | Upregulated |
| 0,789338998 | 0,003568737 | LOC101928154 | FALSE | Upregulated |
| 0,807882628 | 0,003570673 | STBD1 | FALSE | Upregulated |
| 0,970701998 | 0,003573343 | SRGAP1 | FALSE | Upregulated |
| 0,426225662 | 0,003573343 | TRAFD1 | FALSE | Upregulated |
| -0,55858271 | 0,003579471 | AQP11 | FALSE | Downregulated |
| -0,343757487 | 0,003579471 | DHTKD1 | FALSE | Downregulated |
| -2,055613822 | 0,003579471 | MEDAG | TRUE | Downregulated |
| 0,748990801 | 0,003584595 | ODF3B | FALSE | Upregulated |
| 0,867147716 | 0,003587347 | PRKCB | FALSE | Upregulated |

| | | | | |
|---|---|---|---|---|
| 0,405335501 | 0,003593302 | SOCS4 | FALSE | Upregulated |
| 1,007598129 | 0,003602087 | KLRD1 | TRUE | Upregulated |
| 0,822945381 | 0,003626594 | CCR1 | FALSE | Upregulated |
| 0,754992619 | 0,003628722 | NCF2 | FALSE | Upregulated |
| 0,900937071 | 0,003640626 | GAS2L3 | FALSE | Upregulated |
| 3,112034038 | 0,00367134 | LIMS3-LOC440895 | TRUE | Upregulated |
| 0,656750692 | 0,003681192 | KIAA1841 | FALSE | Upregulated |
| 0,433973513 | 0,003699723 | MARS | FALSE | Upregulated |
| -0,468418981 | 0,003714963 | SLC12A6 | FALSE | Downregulated |
| 0,848118891 | 0,003715311 | SEPT1 | FALSE | Upregulated |
| 0,874382985 | 0,003730895 | C5AR1 | FALSE | Upregulated |
| -0,503584269 | 0,003740119 | AKAP9 | FALSE | Downregulated |
| 1,494897869 | 0,003740119 | TRNP1 | TRUE | Upregulated |
| -0,378965533 | 0,003756767 | YOD1 | FALSE | Downregulated |
| -0,513384951 | 0,003763085 | TGFBR3 | FALSE | Downregulated |
| -0,345652959 | 0,00376625 | CLPX | FALSE | Downregulated |
| 0,769211572 | 0,00377791 | APOBEC3C | FALSE | Upregulated |
| 0,336797928 | 0,00377791 | ATP1A1 | FALSE | Upregulated |
| -0,747388959 | 0,00377791 | CTTNBP2 | FALSE | Downregulated |
| 1,752563165 | 0,00377791 | IL2RA | TRUE | Upregulated |
| 0,335428086 | 0,00377791 | RNF19A | FALSE | Upregulated |
| 0,612021973 | 0,00377791 | SLC29A3 | FALSE | Upregulated |
| 1,364525883 | 0,003782074 | FCN1 | TRUE | Upregulated |
| -0,743954523 | 0,003782074 | LRRK2 | FALSE | Downregulated |
| 0,565116656 | 0,003787594 | DCLRE1B | FALSE | Upregulated |
| -2,66083053 | 0,003791081 | LOC101928669 | TRUE | Downregulated |
| 2,711204942 | 0,003808632 | C15orf48 | TRUE | Upregulated |
| 0,599263133 | 0,003808632 | PRKX | FALSE | Upregulated |
| 0,580246496 | 0,003831951 | RBM38 | FALSE | Upregulated |
| 1,486602642 | 0,003836648 | SNAP25-AS1 | TRUE | Upregulated |
| 0,829163454 | 0,003839171 | OAS1 | FALSE | Upregulated |
| 0,935668916 | 0,003841845 | FAIM3 | FALSE | Upregulated |
| -0,587900392 | 0,003846868 | AGL | FALSE | Downregulated |
| -0,66233905 | 0,003847766 | LOC257396 | FALSE | Downregulated |
| -0,2855739 | 0,003850147 | ZNF3 | FALSE | Downregulated |
| 1,089692422 | 0,003853833 | FAM26F | TRUE | Upregulated |
| -0,687917988 | 0,003873018 | ACVR2B | FALSE | Downregulated |
| 0,839196185 | 0,003873188 | RNASE1 | FALSE | Upregulated |
| -0,287098617 | 0,003883682 | PROSC | FALSE | Downregulated |
| 0,881498012 | 0,003887115 | AJUBA | FALSE | Upregulated |
| -0,226984993 | 0,00390854 | RAD17 | FALSE | Downregulated |
| -0,840982905 | 0,003931323 | EXPH5 | FALSE | Downregulated |
| 1,463606826 | 0,003931323 | MPZ | TRUE | Upregulated |
| -0,351367646 | 0,00394385 | EDRF1 | FALSE | Downregulated |
| 0,574606761 | 0,00394385 | LRRC20 | FALSE | Upregulated |
| 0,314635843 | 0,00394385 | SELT | FALSE | Upregulated |
| 0,38794618 | 0,00394385 | SF3B4 | FALSE | Upregulated |
| 1,450677474 | 0,003953451 | OPRL1 | TRUE | Upregulated |
| -0,433926941 | 0,003954092 | RCOR3 | FALSE | Downregulated |
| 0,541634667 | 0,003954205 | ATAD2 | FALSE | Upregulated |
| -0,931290203 | 0,003956611 | BTNL9 | FALSE | Downregulated |

| | | | | |
|---|---|---|---|---|
| 0,350138082 | 0,003967921 | HS1BP3 | FALSE | Upregulated |
| 1,009219815 | 0,003967921 | PTPN22 | TRUE | Upregulated |
| 0,592158026 | 0,003973555 | ZWILCH | FALSE | Upregulated |
| -0,481727941 | 0,003989921 | ALDH5A1 | FALSE | Downregulated |
| -0,498583912 | 0,003989921 | RDH10 | FALSE | Downregulated |
| 0,61485596 | 0,004000053 | PITPNM1 | FALSE | Upregulated |
| 1,091262781 | 0,004005245 | FASLG | TRUE | Upregulated |
| 0,188478299 | 0,004023699 | AP2M1 | FALSE | Upregulated |
| 0,33616339 | 0,00402649 | MLST8 | FALSE | Upregulated |
| 0,278296081 | 0,004033221 | RAC1 | FALSE | Upregulated |
| 0,755809 | 0,00403348 | CEP128 | FALSE | Upregulated |
| -0,232894457 | 0,00403348 | GOPC | FALSE | Downregulated |
| 0,685598087 | 0,004047992 | SMC2 | FALSE | Upregulated |
| 1,148691888 | 0,004049648 | ZNF702P | TRUE | Upregulated |
| -0,849225494 | 0,004056518 | PDE5A | FALSE | Downregulated |
| 1,947133257 | 0,004062819 | FAM19A5 | TRUE | Upregulated |
| -0,44858395 | 0,004079014 | ZNF25 | FALSE | Downregulated |
| 0,306557935 | 0,004081864 | GNA13 | FALSE | Upregulated |
| 0,422010537 | 0,004095848 | CCT6A | FALSE | Upregulated |
| 0,627919116 | 0,004098282 | GMFB | FALSE | Upregulated |
| -0,607278054 | 0,004098282 | SYDE2 | FALSE | Downregulated |
| -2,844640789 | 0,004122176 | THBS4 | TRUE | Downregulated |
| -0,286262626 | 0,004126284 | MRPS14 | FALSE | Downregulated |
| -0,284968593 | 0,00413836 | PPWD1 | FALSE | Downregulated |
| 0,414414126 | 0,004140094 | JDP2 | FALSE | Upregulated |
| -0,422093068 | 0,004140094 | SRSF5 | FALSE | Downregulated |
| 0,342099303 | 0,004141721 | TOMM34 | FALSE | Upregulated |
| 1,196030638 | 0,00415099 | EPHB2 | TRUE | Upregulated |
| -0,29899922 | 0,004154971 | MCCC1 | FALSE | Downregulated |
| -0,518280829 | 0,004173032 | ST13 | FALSE | Downregulated |
| -1,136602958 | 0,004173032 | TUBB1 | TRUE | Downregulated |
| -1,54188828 | 0,004198778 | NPIPB6 | TRUE | Downregulated |
| 0,627319546 | 0,004208317 | TCIRG1 | FALSE | Upregulated |
| -1,368103994 | 0,004211538 | AC097468.4 | TRUE | Downregulated |
| 0,757112573 | 0,004211538 | PLEKHG2 | FALSE | Upregulated |
| 0,490883076 | 0,004211538 | RCC1 | FALSE | Upregulated |
| -0,584990331 | 0,004217391 | SPIN3 | FALSE | Downregulated |
| -1,221599825 | 0,004240243 | NIPSNAP3B | TRUE | Downregulated |
| -0,599003943 | 0,004252256 | HEXIM2 | FALSE | Downregulated |
| 0,27462379 | 0,004281761 | HMGN2 | FALSE | Upregulated |
| -0,463723432 | 0,004304721 | ALDH2 | FALSE | Downregulated |
| -0,547735057 | 0,004324589 | PKD2 | FALSE | Downregulated |
| 0,355593768 | 0,004325761 | SLC39A1 | FALSE | Upregulated |
| 1,16539648 | 0,004325763 | ITPKA | TRUE | Upregulated |
| -2,753935597 | 0,004325763 | RP11-214C8.2 | TRUE | Downregulated |
| 1,22945861 | 0,004325763 | TLR7 | TRUE | Upregulated |
| 0,426168586 | 0,004329179 | FAM21A | FALSE | Upregulated |
| -0,757461241 | 0,004334823 | ZNF737 | FALSE | Downregulated |
| 1,412704617 | 0,004337377 | SIT1 | TRUE | Upregulated |
| -2,582279289 | 0,004339485 | C1QTNF9 | TRUE | Downregulated |
| 0,527510763 | 0,004340239 | GLA | FALSE | Upregulated |

| | | | | |
|---|---|---|---|---|
| -0,529117372 | 0,004360106 | ZNF354C | FALSE | Downregulated |
| 0,420612652 | 0,004360724 | CREB3 | FALSE | Upregulated |
| -0,275419711 | 0,004360724 | PDIK1L | FALSE | Downregulated |
| -0,300464648 | 0,004367929 | STK3 | FALSE | Downregulated |
| 0,839104266 | 0,004398983 | SLC6A6 | FALSE | Upregulated |
| 0,811204147 | 0,00440522 | RASAL3 | FALSE | Upregulated |
| -0,699068838 | 0,004428619 | LIPC | FALSE | Downregulated |
| 0,450038265 | 0,004430653 | WSB2 | FALSE | Upregulated |
| -0,429446928 | 0,004441135 | ACADL | FALSE | Downregulated |
| -1,634915162 | 0,004441135 | C19orf71 | TRUE | Downregulated |
| -0,804906107 | 0,004441135 | SYT1 | FALSE | Downregulated |
| 2,240838827 | 0,004448589 | TNFRSF17 | TRUE | Upregulated |
| -0,718023203 | 0,004457891 | ALDH1L1-AS2 | FALSE | Downregulated |
| 0,275975748 | 0,004478918 | CDC42SE2 | FALSE | Upregulated |
| 0,403492507 | 0,004480271 | MSANTD3 | FALSE | Upregulated |
| -0,404609839 | 0,004480271 | ZNF696 | FALSE | Downregulated |
| -0,59942722 | 0,004487019 | NAPEPLD | FALSE | Downregulated |
| 0,384440841 | 0,004511971 | SPTLC2 | FALSE | Upregulated |
| -0,640186835 | 0,004520465 | SLCO1B1 | FALSE | Downregulated |
| 0,42973209 | 0,004527958 | TMEM140 | FALSE | Upregulated |
| 0,212404674 | 0,004538986 | EXT2 | FALSE | Upregulated |
| -0,629745761 | 0,004538986 | RNF125 | FALSE | Downregulated |
| 1,445879233 | 0,004555987 | HS1BP3-IT1 | TRUE | Upregulated |
| 0,804128992 | 0,004558466 | BTG3 | FALSE | Upregulated |
| 0,320799804 | 0,004570577 | SLBP | FALSE | Upregulated |
| -0,79414548 | 0,004574189 | HCN3 | FALSE | Downregulated |
| -0,295319178 | 0,004613636 | ZNF252P | FALSE | Downregulated |
| -0,538745756 | 0,004632056 | PROX1-AS1 | FALSE | Downregulated |
| 0,842309259 | 0,004632056 | SLC51A | FALSE | Upregulated |
| -0,412136452 | 0,004638973 | ZNF184 | FALSE | Downregulated |
| 0,824472218 | 0,004649992 | KCTD17 | FALSE | Upregulated |
| 0,329529422 | 0,004659071 | CALM3 | FALSE | Upregulated |
| -0,68947221 | 0,004659071 | TPPP2 | FALSE | Downregulated |
| -0,341501189 | 0,004681589 | CELF1 | FALSE | Downregulated |
| 0,342367991 | 0,004687665 | GLRX3 | FALSE | Upregulated |
| 2,409793769 | 0,004689941 | MMP9 | TRUE | Upregulated |
| 3,006962302 | 0,004692085 | CCL20 | TRUE | Upregulated |
| -0,299099659 | 0,004699602 | RPUSD4 | FALSE | Downregulated |
| 0,855438607 | 0,004702944 | ICAM1 | FALSE | Upregulated |
| 0,311131541 | 0,004773819 | ARPC4 | FALSE | Upregulated |
| 0,351516659 | 0,004773819 | CNOT10 | FALSE | Upregulated |
| -0,263341135 | 0,004788999 | BACE1 | FALSE | Downregulated |
| 0,462839425 | 0,00480199 | HAUS8 | FALSE | Upregulated |
| 0,694175219 | 0,004821944 | CHAC2 | FALSE | Upregulated |
| -0,506109151 | 0,004821944 | PTBP2 | FALSE | Downregulated |
| 0,297062187 | 0,004821944 | USP5 | FALSE | Upregulated |
| -0,533024001 | 0,00482317 | PBLD | FALSE | Downregulated |
| 0,492349531 | 0,004842999 | HTATIP2 | FALSE | Upregulated |
| 1,37227753 | 0,004842999 | JAKMIP1 | TRUE | Upregulated |
| -0,543779281 | 0,004842999 | SH3BGRL2 | FALSE | Downregulated |
| 0,733305271 | 0,004849003 | DENND1C | FALSE | Upregulated |

| | | | | |
|---|---|---|---|---|
| 0,349814092 | 0,004858376 | PSMD9 | FALSE | Upregulated |
| -0,342724695 | 0,004864574 | NOP14 | FALSE | Downregulated |
| 0,65322261 | 0,004865518 | TES | FALSE | Upregulated |
| -1,059816756 | 0,004880407 | LOC101927559 | TRUE | Downregulated |
| 0,443019203 | 0,004880407 | UBE2D1 | FALSE | Upregulated |
| 0,784550298 | 0,00490245 | SYT11 | FALSE | Upregulated |
| -0,919255927 | 0,004955973 | CNTFR | FALSE | Downregulated |
| -1,223020092 | 0,004978517 | FAM151A | TRUE | Downregulated |
| 0,731320814 | 0,004980183 | CAPN2 | FALSE | Upregulated |
| 0,789014217 | 0,004982484 | PIK3CG | FALSE | Upregulated |
| -1,25535637 | 0,005013718 | RGPD8 | TRUE | Downregulated |
| -0,350084614 | 0,005013718 | RSAD1 | FALSE | Downregulated |
| 0,320838884 | 0,005023363 | BCAP31 | FALSE | Upregulated |
| 1,033315844 | 0,005023363 | CMPK2 | TRUE | Upregulated |
| 0,269335084 | 0,005024309 | NAPA | FALSE | Upregulated |
| 0,731667721 | 0,005037264 | MYO5A | FALSE | Upregulated |
| -0,469515857 | 0,005037264 | XPNPEP3 | FALSE | Downregulated |
| 1,022300117 | 0,005044463 | FAM222A | TRUE | Upregulated |
| -0,222922129 | 0,005044463 | ZNF644 | FALSE | Downregulated |
| -1,23073892 | 0,005044771 | CYS1 | TRUE | Downregulated |
| 0,522846025 | 0,005055095 | ADAM15 | FALSE | Upregulated |
| 0,29287025 | 0,005065618 | YWHAQ | FALSE | Upregulated |
| 1,141790545 | 0,005068029 | CLDN7 | TRUE | Upregulated |
| -0,641621576 | 0,005068029 | RP5-886K2.3 | FALSE | Downregulated |
| 0,866663964 | 0,005068549 | OSBPL3 | FALSE | Upregulated |
| -0,382717524 | 0,00507088 | C17orf80 | FALSE | Downregulated |
| 1,062428224 | 0,005088885 | LAX1 | TRUE | Upregulated |
| -0,484340944 | 0,005089696 | CFHR2 | FALSE | Downregulated |
| 0,612185997 | 0,005096542 | ALDH3B1 | FALSE | Upregulated |
| 1,700811267 | 0,005096542 | CDCA7 | TRUE | Upregulated |
| -0,371221104 | 0,005096542 | LDHD | FALSE | Downregulated |
| 1,802530619 | 0,005101982 | IFNG | TRUE | Upregulated |
| -0,257107445 | 0,005101982 | TRAPPC8 | FALSE | Downregulated |
| 2,847247428 | 0,005116968 | LOC101059954 | TRUE | Upregulated |
| 3,65412232 | 0,005135005 | ATP6V0D2 | TRUE | Upregulated |
| -1,378087022 | 0,005158312 | CYP3A7 | TRUE | Downregulated |
| 0,401566426 | 0,005158312 | MYO5B | FALSE | Upregulated |
| 0,227322214 | 0,005190639 | KARS | FALSE | Upregulated |
| 0,456935848 | 0,005193477 | BAIAP2L1 | FALSE | Upregulated |
| 0,24820453 | 0,005193477 | CAPZB | FALSE | Upregulated |
| 0,313650822 | 0,005193477 | LAMTOR1 | FALSE | Upregulated |
| -0,330769062 | 0,005193477 | TJP2 | FALSE | Downregulated |
| -0,537975334 | 0,005193696 | MAT1A | FALSE | Downregulated |
| 0,383770211 | 0,005197224 | BLOC1S2 | FALSE | Upregulated |
| 1,096960182 | 0,005197224 | CD3G | TRUE | Upregulated |
| 0,959368023 | 0,005197224 | LILRA5 | FALSE | Upregulated |
| -1,018200964 | 0,005199677 | NDST3 | TRUE | Downregulated |
| 2,45689292 | 0,005225193 | STMND1 | TRUE | Upregulated |
| -0,433020107 | 0,005232496 | ABAT | FALSE | Downregulated |
| -0,459151347 | 0,005233347 | ZNF175 | FALSE | Downregulated |
| 0,501124534 | 0,005244513 | ACER3 | FALSE | Upregulated |

| | | | | |
|---|---|---|---|---|
| -1,082299547 | 0,005269088 | CFHR3 | TRUE | Downregulated |
| -0,408662076 | 0,005269088 | KIAA0907 | FALSE | Downregulated |
| -0,443012351 | 0,005269088 | WDR5B | FALSE | Downregulated |
| 0,804035558 | 0,005297109 | CXorf21 | FALSE | Upregulated |
| 0,532905257 | 0,00530953 | C20orf24 | FALSE | Upregulated |
| 0,640730211 | 0,005352097 | METRNL | FALSE | Upregulated |
| 0,671677761 | 0,005353437 | ANXA5 | FALSE | Upregulated |
| 0,421090795 | 0,005353437 | FAM195B | FALSE | Upregulated |
| 1,033315976 | 0,005412228 | CCND1 | TRUE | Upregulated |
| 0,66541776 | 0,005422935 | RAB31 | FALSE | Upregulated |
| -0,364507657 | 0,005431804 | BBS10 | FALSE | Downregulated |
| 0,503115161 | 0,005431804 | PRKCD | FALSE | Upregulated |
| 1,070789838 | 0,005432592 | GPRIN3 | TRUE | Upregulated |
| 2,524373766 | 0,005467678 | GSDMA | TRUE | Upregulated |
| 0,357283451 | 0,005473272 | DAG1 | FALSE | Upregulated |
| -0,335742559 | 0,005484034 | KLHDC2 | FALSE | Downregulated |
| 0,643976724 | 0,00550119 | E2F3 | FALSE | Upregulated |
| -0,478914803 | 0,00550119 | MCEE | FALSE | Downregulated |
| -0,395480632 | 0,005505706 | MTERFD3 | FALSE | Downregulated |
| 0,51035858 | 0,005511408 | RABEP2 | FALSE | Upregulated |
| 1,225278898 | 0,005517894 | TDGF1 | TRUE | Upregulated |
| -0,463391555 | 0,005519075 | THRB | FALSE | Downregulated |
| -1,627376034 | 0,005519469 | AVPR1A | TRUE | Downregulated |
| 0,302811104 | 0,005538081 | ARPC3 | FALSE | Upregulated |
| 0,458513192 | 0,005538081 | FOCAD | FALSE | Upregulated |
| 1,5624968 | 0,005538621 | MCOLN2 | TRUE | Upregulated |
| 0,600936186 | 0,005591305 | PPA1 | FALSE | Upregulated |
| 0,249850079 | 0,005615901 | ATP6V1C1 | FALSE | Upregulated |
| 0,359750492 | 0,005617355 | CHMP4A | FALSE | Upregulated |
| 1,738530546 | 0,005617355 | PKMYT1 | TRUE | Upregulated |
| 0,88750846 | 0,005617355 | SH2D1A | FALSE | Upregulated |
| -0,348434889 | 0,005617355 | TAF1 | FALSE | Downregulated |
| -0,57365901 | 0,005627829 | ENPP1 | FALSE | Downregulated |
| 0,554386713 | 0,00564859 | PVRL2 | FALSE | Upregulated |
| 2,344542776 | 0,005651083 | CDKN2B-AS1 | TRUE | Upregulated |
| 2,498554307 | 0,005651083 | LOC101927931 | TRUE | Upregulated |
| 0,575220373 | 0,005663754 | GRAMD1A | FALSE | Upregulated |
| -0,382632483 | 0,005674997 | AGTR1 | FALSE | Downregulated |
| -0,242331092 | 0,005696387 | ATPAF1 | FALSE | Downregulated |
| -0,299990021 | 0,005696387 | MED14 | FALSE | Downregulated |
| -0,695321898 | 0,005696387 | ZC3H6 | FALSE | Downregulated |
| 1,010686358 | 0,005697293 | PSTPIP1 | TRUE | Upregulated |
| 2,569375001 | 0,00571222 | SMC1B | TRUE | Upregulated |
| -0,349670343 | 0,005718888 | DNAJC19 | FALSE | Downregulated |
| 1,310898524 | 0,005725693 | HLA-G | TRUE | Upregulated |
| 0,811296513 | 0,005767342 | SLC52A2 | FALSE | Upregulated |
| -0,572364935 | 0,005798726 | SLC35E2B | FALSE | Downregulated |
| -0,61304707 | 0,005817574 | AMY2B | FALSE | Downregulated |
| 0,43118788 | 0,005818776 | PERP | FALSE | Upregulated |
| 0,82352379 | 0,005819671 | FMNL1 | FALSE | Upregulated |
| 0,517798866 | 0,005837637 | TM4SF4 | FALSE | Upregulated |

| | | | | |
|---|---|---|---|---|
| -0,261533103 | 0,005876317 | EIF2A | FALSE | Downregulated |
| 0,494971727 | 0,005876317 | GLRX2 | FALSE | Upregulated |
| 0,994937127 | 0,005876317 | HCG26 | FALSE | Upregulated |
| -2,856988546 | 0,005876317 | SPDYE1 | TRUE | Downregulated |
| -0,455965266 | 0,005876317 | TRAM2-AS1 | FALSE | Downregulated |
| -0,507066855 | 0,005876317 | TRMT10A | FALSE | Downregulated |
| -0,346166346 | 0,005876317 | VAMP4 | FALSE | Downregulated |
| -0,360559865 | 0,005932189 | NUCKS1 | FALSE | Downregulated |
| -0,415641606 | 0,005937233 | GAB1 | FALSE | Downregulated |
| -0,287167853 | 0,005942256 | HERPUD2 | FALSE | Downregulated |
| 0,536297118 | 0,006002176 | SCPEP1 | FALSE | Upregulated |
| 2,283409764 | 0,006008365 | LAIR2 | TRUE | Upregulated |
| -0,399702192 | 0,006025321 | MTX3 | FALSE | Downregulated |
| -0,288937776 | 0,006072894 | LINC00674 | FALSE | Downregulated |
| 0,376639458 | 0,006094446 | UFD1L | FALSE | Upregulated |
| 0,369183911 | 0,006095483 | C1GALT1C1 | FALSE | Upregulated |
| 0,685275472 | 0,006095483 | MFSD6 | FALSE | Upregulated |
| 0,572350517 | 0,006095483 | PVR | FALSE | Upregulated |
| 0,650599864 | 0,006123623 | TXN | FALSE | Upregulated |
| -1,086588939 | 0,006125699 | LOC102724350 | TRUE | Downregulated |
| 0,419430337 | 0,006158439 | LMNA | FALSE | Upregulated |
| 0,483179737 | 0,006172007 | DAPK3 | FALSE | Upregulated |
| 0,406053606 | 0,006199046 | FAM98A | FALSE | Upregulated |
| -0,376156297 | 0,006199046 | ZNF91 | FALSE | Downregulated |
| -0,329758963 | 0,006212959 | CREBBP | FALSE | Downregulated |
| 0,449430513 | 0,00621982 | CLN6 | FALSE | Upregulated |
| 0,707747361 | 0,00621982 | NFATC1 | FALSE | Upregulated |
| 0,255625356 | 0,00621982 | RPA1 | FALSE | Upregulated |
| 0,794716697 | 0,006224927 | PTPRH | FALSE | Upregulated |
| 0,370218461 | 0,006241946 | SMARCA4 | FALSE | Upregulated |
| -0,462193103 | 0,006241946 | ZNF566 | FALSE | Downregulated |
| -0,47576752 | 0,006256246 | LDB2 | FALSE | Downregulated |
| -0,528710983 | 0,00626365 | CPED1 | FALSE | Downregulated |
| -0,518571472 | 0,00626365 | HCFC1R1 | FALSE | Downregulated |
| 1,50427734 | 0,006278858 | CLEC4E | TRUE | Upregulated |
| 1,021635105 | 0,006282117 | JAK3 | TRUE | Upregulated |
| 0,740846409 | 0,006299884 | ARHGAP9 | FALSE | Upregulated |
| -0,513762238 | 0,006326985 | ZNF224 | FALSE | Downregulated |
| 0,635190569 | 0,006333709 | TSSC4 | FALSE | Upregulated |
| -0,867978482 | 0,006334603 | COL21A1 | FALSE | Downregulated |
| -0,972009146 | 0,006375599 | AC004862.6 | FALSE | Downregulated |
| 0,879930072 | 0,006378753 | CAMKK1 | FALSE | Upregulated |
| 0,388266813 | 0,006395035 | PSME1 | FALSE | Upregulated |
| 0,657125912 | 0,00640309 | ARL4C | FALSE | Upregulated |
| 0,790871009 | 0,006415632 | SDF2L1 | FALSE | Upregulated |
| 0,235928359 | 0,006425276 | ATXN10 | FALSE | Upregulated |
| 1,16887335 | 0,006448949 | RARRES1 | TRUE | Upregulated |
| 1,369769317 | 0,006466561 | RPL39L | TRUE | Upregulated |
| 1,165348491 | 0,006471396 | CCL4 | TRUE | Upregulated |
| -0,913818131 | 0,006471396 | LOC102723766 | FALSE | Downregulated |
| 0,370930179 | 0,006471396 | RNF145 | FALSE | Upregulated |

| | | | | |
|---|---|---|---|---|
| 0,588483836 | 0,006471396 | USP18 | FALSE | Upregulated |
| 0,700139572 | 0,006492583 | CSF1R | FALSE | Upregulated |
| -0,442725984 | 0,006497067 | C6 | FALSE | Downregulated |
| 0,346137121 | 0,006497067 | VMP1 | FALSE | Upregulated |
| 1,614649179 | 0,00650128 | PIF1 | TRUE | Upregulated |
| -0,485354549 | 0,00650128 | SLC27A2 | FALSE | Downregulated |
| 0,969930288 | 0,006508439 | ZNF831 | FALSE | Upregulated |
| 0,604789925 | 0,006508599 | ATP13A2 | FALSE | Upregulated |
| 1,531350404 | 0,006538584 | APOBEC3A | TRUE | Upregulated |
| -0,670519304 | 0,006544621 | KLHDC1 | FALSE | Downregulated |
| -1,044075863 | 0,006548534 | LOC102723666 | TRUE | Downregulated |
| 0,814209913 | 0,006548534 | RPS26P11 | FALSE | Upregulated |
| -0,421050822 | 0,006548534 | SETDB2 | FALSE | Downregulated |
| 1,102065189 | 0,006548534 | XCL2 | TRUE | Upregulated |
| 0,341327098 | 0,006567425 | SNX7 | FALSE | Upregulated |
| -0,423716543 | 0,006581906 | PCBD1 | FALSE | Downregulated |
| 0,885011796 | 0,006589582 | NKG7 | FALSE | Upregulated |
| -0,807453564 | 0,00660498 | HSD17B13 | FALSE | Downregulated |
| 0,617801814 | 0,00660498 | HSPB1 | FALSE | Upregulated |
| -0,172746056 | 0,006620237 | DDX42 | FALSE | Downregulated |
| 0,474494203 | 0,006633432 | C7orf50 | FALSE | Upregulated |
| -0,948919263 | 0,006633432 | GSTM4 | FALSE | Downregulated |
| 1,372070582 | 0,006674259 | KLHL29 | TRUE | Upregulated |
| -0,756211379 | 0,006675017 | CSAD | FALSE | Downregulated |
| -0,781489828 | 0,006700186 | LPAL2 | FALSE | Downregulated |
| 0,682427442 | 0,006733198 | MS4A7 | FALSE | Upregulated |
| -2,832233982 | 0,006735632 | C19orf77 | TRUE | Downregulated |
| -0,847531887 | 0,006735632 | CHAD | FALSE | Downregulated |
| 1,350554549 | 0,006735632 | GADD45G | TRUE | Upregulated |
| -1,383179479 | 0,006756003 | FAM9B | TRUE | Downregulated |
| 0,747196571 | 0,006756003 | TSPO | FALSE | Upregulated |
| 0,740946864 | 0,006757091 | TNFRSF10A | FALSE | Upregulated |
| 0,265417118 | 0,006781265 | FIG4 | FALSE | Upregulated |
| 0,756169417 | 0,006803922 | STK39 | FALSE | Upregulated |
| 0,426843381 | 0,006819638 | RBX1 | FALSE | Upregulated |
| 0,645492734 | 0,006821791 | ACP5 | FALSE | Upregulated |
| 0,680306687 | 0,006847337 | DOCK11 | FALSE | Upregulated |
| -1,005277948 | 0,006847337 | MST1L | TRUE | Downregulated |
| -0,503815246 | 0,006857662 | SMIM14 | FALSE | Downregulated |
| 1,641590924 | 0,006859923 | IL1RL1 | TRUE | Upregulated |
| -0,207834901 | 0,006859923 | NFX1 | FALSE | Downregulated |
| -0,451125496 | 0,006859923 | SHPRH | FALSE | Downregulated |
| -0,530613275 | 0,006866917 | CGN | FALSE | Downregulated |
| -0,276127985 | 0,006874205 | ZZZ3 | FALSE | Downregulated |
| -0,668010718 | 0,00689028 | DLEU1 | FALSE | Downregulated |
| 0,490812185 | 0,006901236 | RIPK2 | FALSE | Upregulated |
| 0,353862201 | 0,006924424 | MEA1 | FALSE | Upregulated |
| 0,677844603 | 0,006949775 | MEAF6 | FALSE | Upregulated |
| 0,42798791 | 0,006949775 | SLC25A6 | FALSE | Upregulated |
| 0,624322449 | 0,006973737 | PTGFRN | FALSE | Upregulated |
| 1,344156438 | 0,006992662 | VSTM2L | TRUE | Upregulated |

| | | | | |
|---|---|---|---|---|
| -0,585107384 | 0,006994463 | PPP1R3E | FALSE | Downregulated |
| -0,44399349 | 0,007005726 | SLC41A2 | FALSE | Downregulated |
| -0,693628561 | 0,00703305 | RP11-545I5.3 | FALSE | Downregulated |
| 0,2019413 | 0,007036826 | DHX30 | FALSE | Upregulated |
| 0,305321511 | 0,007064292 | PSMD11 | FALSE | Upregulated |
| 0,497855884 | 0,007064292 | RAB20 | FALSE | Upregulated |
| 0,653543493 | 0,007064517 | SDC3 | FALSE | Upregulated |
| 1,726996508 | 0,007082074 | CENPI | TRUE | Upregulated |
| 0,824936751 | 0,007095837 | CYP11A1 | FALSE | Upregulated |
| 0,524044754 | 0,007096798 | B3GAT3 | FALSE | Upregulated |
| -0,254157661 | 0,007111549 | SECISBP2L | FALSE | Downregulated |
| 0,685219514 | 0,007123199 | IMPDH1 | FALSE | Upregulated |
| 0,40664158 | 0,007124281 | CEP170B | FALSE | Upregulated |
| -0,790347586 | 0,007124281 | MAMLD1 | FALSE | Downregulated |
| -0,366314534 | 0,007138104 | BRWD1 | FALSE | Downregulated |
| -0,582575619 | 0,007139886 | DHRS4-AS1 | FALSE | Downregulated |
| 0,636124425 | 0,007139886 | METTL1 | FALSE | Upregulated |
| -0,662778644 | 0,00716694 | RAD54L2 | FALSE | Downregulated |
| -0,299416162 | 0,007195198 | OMA1 | FALSE | Downregulated |
| -0,508460781 | 0,007208048 | POP7 | FALSE | Downregulated |
| 0,530382311 | 0,007230305 | TRIP6 | FALSE | Upregulated |
| 0,320230037 | 0,007230604 | RPN2 | FALSE | Upregulated |
| -0,370502852 | 0,007295662 | ACVR2A | FALSE | Downregulated |
| 0,348237272 | 0,007299264 | PCSK7 | FALSE | Upregulated |
| 0,320623339 | 0,007299665 | PTGES2 | FALSE | Upregulated |
| 0,492826235 | 0,007332005 | TMEM106C | FALSE | Upregulated |
| -0,656354322 | 0,007340177 | DNAJC25 | FALSE | Downregulated |
| 1,385903391 | 0,007340177 | MNS1 | TRUE | Upregulated |
| 0,581095306 | 0,007340227 | IFI16 | FALSE | Upregulated |
| 0,331383921 | 0,007353684 | DNAJB6 | FALSE | Upregulated |
| 0,297563208 | 0,007359371 | TBC1D22A | FALSE | Upregulated |
| 0,686282656 | 0,007366158 | ABCA7 | FALSE | Upregulated |
| -1,839079085 | 0,007366158 | DNAH12 | TRUE | Downregulated |
| 0,575181047 | 0,007388587 | ITGAL | FALSE | Upregulated |
| -0,372245606 | 0,007388587 | PHLDB2 | FALSE | Downregulated |
| -0,258690539 | 0,007388587 | WDR33 | FALSE | Downregulated |
| -0,993405809 | 0,007422124 | MME | FALSE | Downregulated |
| -0,385967032 | 0,007430574 | BCS1L | FALSE | Downregulated |
| 0,273242604 | 0,007430574 | SNX6 | FALSE | Upregulated |
| 1,293935345 | 0,00743238 | SIRPG | TRUE | Upregulated |
| 0,612171799 | 0,007451741 | RFC3 | FALSE | Upregulated |
| 0,38162239 | 0,00745505 | SAAL1 | FALSE | Upregulated |
| 0,74622859 | 0,007458937 | NCF4 | FALSE | Upregulated |
| -0,636641171 | 0,007465471 | IYD | FALSE | Downregulated |
| 0,328457748 | 0,007465471 | NOP56 | FALSE | Upregulated |
| 0,266377286 | 0,007480358 | SERPINB6 | FALSE | Upregulated |
| 0,523545659 | 0,007484891 | C19orf10 | FALSE | Upregulated |
| -0,398427034 | 0,007485775 | ZNF510 | FALSE | Downregulated |
| -0,748040247 | 0,007502799 | LINC01018 | FALSE | Downregulated |
| 0,47256293 | 0,007502799 | MPHOSPH6 | FALSE | Upregulated |
| 2,415710176 | 0,007502799 | RAB33A | TRUE | Upregulated |

| | | | | |
|---|---|---|---|---|
| -0,987185911 | 0,007505665 | FAM86FP | FALSE | Downregulated |
| 0,735060425 | 0,007505665 | FEN1 | FALSE | Upregulated |
| -1,386106911 | 0,007520924 | LOC102723953 | TRUE | Downregulated |
| -0,367504691 | 0,007520924 | ZBTB10 | FALSE | Downregulated |
| 1,075596551 | 0,007530472 | GPRC5B | TRUE | Upregulated |
| 0,318237344 | 0,007549355 | EMC10 | FALSE | Upregulated |
| -0,818475489 | 0,007564806 | LYVE1 | FALSE | Downregulated |
| -1,663194988 | 0,007616952 | LOC101927635 | TRUE | Downregulated |
| -2,64466704 | 0,007616952 | TNNT2 | TRUE | Downregulated |
| -0,428541231 | 0,007616952 | ZNF445 | FALSE | Downregulated |
| 1,469912826 | 0,00762749 | S100B | TRUE | Upregulated |
| 0,668319737 | 0,007631739 | SAMD9 | FALSE | Upregulated |
| 0,477777074 | 0,007639197 | TRIM14 | FALSE | Upregulated |
| -0,362143704 | 0,007649447 | CLYBL | FALSE | Downregulated |
| 0,838250881 | 0,007653779 | MPV17L2 | FALSE | Upregulated |
| 0,347962898 | 0,007654432 | PARP1 | FALSE | Upregulated |
| -0,380303945 | 0,007666602 | PGM1 | FALSE | Downregulated |
| -0,40533772 | 0,007673726 | ANKRD42 | FALSE | Downregulated |
| -0,304993182 | 0,007673726 | BRD1 | FALSE | Downregulated |
| -1,221532763 | 0,00769413 | AMN | TRUE | Downregulated |
| 0,874013132 | 0,00769413 | C3AR1 | FALSE | Upregulated |
| 0,290887033 | 0,007697972 | PHB | FALSE | Upregulated |
| 0,658612244 | 0,007698437 | AKR1B1 | FALSE | Upregulated |
| 0,735147838 | 0,007698919 | PARVG | FALSE | Upregulated |
| 0,325740793 | 0,007716973 | CD47 | FALSE | Upregulated |
| -0,468572963 | 0,007795617 | ARG1 | FALSE | Downregulated |
| 0,331090165 | 0,007795617 | RPN1 | FALSE | Upregulated |
| -2,341502637 | 0,007831425 | LOC102724208 | TRUE | Downregulated |
| 0,325625368 | 0,007831425 | TMA16 | FALSE | Upregulated |
| 0,3918066 | 0,00783158 | BNIP1 | FALSE | Upregulated |
| -0,412962912 | 0,007843384 | OSGEPL1 | FALSE | Downregulated |
| 0,623573652 | 0,007843384 | TAPSAR1 | FALSE | Upregulated |
| -0,496640642 | 0,007851063 | DNAJC28 | FALSE | Downregulated |
| -0,257855737 | 0,007851063 | PDE8A | FALSE | Downregulated |
| 0,443659303 | 0,007851416 | EML2 | FALSE | Upregulated |
| -0,35065624 | 0,007892513 | DEPDC5 | FALSE | Downregulated |
| -0,730071117 | 0,007895456 | SCN8A | FALSE | Downregulated |
| 0,351597127 | 0,007902355 | MAPK1 | FALSE | Upregulated |
| -0,868605526 | 0,007930896 | SLC16A10 | FALSE | Downregulated |
| -0,618933822 | 0,007941086 | KCNT2 | FALSE | Downregulated |
| 1,09580197 | 0,007953605 | ADORA3 | TRUE | Upregulated |
| -0,609659891 | 0,007954783 | ADHFE1 | FALSE | Downregulated |
| 0,420193485 | 0,007954783 | PNO1 | FALSE | Upregulated |
| 1,097932133 | 0,008001419 | RNF157 | TRUE | Upregulated |
| 0,294983799 | 0,008001419 | SEC22A | FALSE | Upregulated |
| 1,092818611 | 0,008024125 | LOC606724 | TRUE | Upregulated |
| 0,66937192 | 0,008048394 | IKBIP | FALSE | Upregulated |
| -1,250567925 | 0,00807808 | FAT3 | TRUE | Downregulated |
| 0,388193066 | 0,008079361 | CYSTM1 | FALSE | Upregulated |
| 0,52785956 | 0,008079361 | ERI1 | FALSE | Upregulated |
| -0,57187615 | 0,008090188 | ACP6 | FALSE | Downregulated |

| | | | | |
|---|---|---|---|---|
| 0,386430109 | 0,008090188 | PIP4K2A | FALSE | Upregulated |
| 0,656991986 | 0,008090188 | SLC43A2 | FALSE | Upregulated |
| 1,039627128 | 0,008090188 | SPECC1 | TRUE | Upregulated |
| 0,41514848 | 0,008098116 | GTDC1 | FALSE | Upregulated |
| 0,656846021 | 0,008098116 | RCSD1 | FALSE | Upregulated |
| -0,62453218 | 0,008100785 | ANK3 | FALSE | Downregulated |
| -0,409526884 | 0,008100785 | N4BP2 | FALSE | Downregulated |
| -0,345435795 | 0,008122316 | TOM1L1 | FALSE | Downregulated |
| 0,777101422 | 0,008133348 | PTPRO | FALSE | Upregulated |
| -0,909612383 | 0,008148506 | CYP3A4 | FALSE | Downregulated |
| 0,599734378 | 0,008152039 | LHFPL2 | FALSE | Upregulated |
| -0,39468997 | 0,008152039 | MED13 | FALSE | Downregulated |
| -0,361096565 | 0,008162671 | ZNF791 | FALSE | Downregulated |
| 1,697445229 | 0,008172096 | KIAA1644 | TRUE | Upregulated |
| 0,421456246 | 0,0081778 | PHPT1 | FALSE | Upregulated |
| -0,249928595 | 0,008198131 | TRAPPC11 | FALSE | Downregulated |
| -0,349193093 | 0,008200745 | ING1 | FALSE | Downregulated |
| 0,43045646 | 0,008205252 | DPAGT1 | FALSE | Upregulated |
| 0,337639756 | 0,008227878 | TESK1 | FALSE | Upregulated |
| 0,402885298 | 0,008232427 | DTX3L | FALSE | Upregulated |
| 0,705327278 | 0,008233357 | HK3 | FALSE | Upregulated |
| -0,6120584 | 0,008235431 | FAXDC2 | FALSE | Downregulated |
| 0,714297672 | 0,008235431 | SLFN5 | FALSE | Upregulated |
| 0,441747407 | 0,008268096 | DAXX | FALSE | Upregulated |
| 0,80771093 | 0,008268096 | TNFSF8 | FALSE | Upregulated |
| -0,336701909 | 0,008343358 | ECE1 | FALSE | Downregulated |
| -0,434447301 | 0,008352616 | ETFDH | FALSE | Downregulated |
| 0,471230542 | 0,008361128 | TLR3 | FALSE | Upregulated |
| 0,470909319 | 0,008380682 | TMEM185B | FALSE | Upregulated |
| 0,283543374 | 0,00846764 | JMJD8 | FALSE | Upregulated |
| 0,508186665 | 0,00846764 | ZFPM1 | FALSE | Upregulated |
| -0,643898084 | 0,008491743 | ACVR1C | FALSE | Downregulated |
| -0,30075632 | 0,008498384 | SOX13 | FALSE | Downregulated |
| -0,243190874 | 0,008515614 | KIAA1147 | FALSE | Downregulated |
| -0,473453618 | 0,008515614 | MBLAC2 | FALSE | Downregulated |
| 0,522001029 | 0,008516942 | MAP7D1 | FALSE | Upregulated |
| 1,13821879 | 0,008518203 | SH2D2A | TRUE | Upregulated |
| -1,448947639 | 0,008534776 | GOLGA8A | TRUE | Downregulated |
| -0,351883098 | 0,008534776 | MLLT10 | FALSE | Downregulated |
| -0,977396782 | 0,008559171 | AVIL | FALSE | Downregulated |
| -0,505790708 | 0,008573721 | MIR4458HG | FALSE | Downregulated |
| -1,232089151 | 0,008601649 | FAM19A2 | TRUE | Downregulated |
| 0,729111697 | 0,008601649 | PLEKHB2 | FALSE | Upregulated |
| 0,433367609 | 0,008601668 | TRAPPC1 | FALSE | Upregulated |
| -0,316711691 | 0,008608815 | SEPSECS | FALSE | Downregulated |
| -0,269723629 | 0,008618028 | RMND1 | FALSE | Downregulated |
| -0,563836105 | 0,008645041 | L3MBTL3 | FALSE | Downregulated |
| -0,238338373 | 0,008671124 | NDUFA5 | FALSE | Downregulated |
| 0,6022178 | 0,008732604 | P4HA2 | FALSE | Upregulated |
| -0,658105812 | 0,008781373 | CYP2E1 | FALSE | Downregulated |
| 0,855980286 | 0,008781373 | SCML4 | FALSE | Upregulated |

| | | | | |
|---|---|---|---|---|
| 0,25029332 | 0,008791141 | HSBP1 | FALSE | Upregulated |
| -0,250540181 | 0,008791141 | PDS5A | FALSE | Downregulated |
| 1,332950918 | 0,008791141 | SPP1 | TRUE | Upregulated |
| 0,555675436 | 0,00879365 | ZNF579 | FALSE | Upregulated |
| -1,619543755 | 0,008842342 | CBLN4 | TRUE | Downregulated |
| -0,570586685 | 0,0088476 | GLYAT | FALSE | Downregulated |
| 0,483914238 | 0,008859105 | TRADD | FALSE | Upregulated |
| 0,310333443 | 0,008868426 | DRAP1 | FALSE | Upregulated |
| 0,33855083 | 0,008877793 | PCBP1 | FALSE | Upregulated |
| -0,41613004 | 0,00888409 | ZFYVE19 | FALSE | Downregulated |
| 1,831699507 | 0,008905114 | C19orf35 | TRUE | Upregulated |
| -1,765508165 | 0,00890602 | PCOLCE-AS1 | TRUE | Downregulated |
| 0,435141091 | 0,008907089 | PSMG1 | FALSE | Upregulated |
| 2,13507815 | 0,008942473 | ZNF541 | TRUE | Upregulated |
| 0,39652392 | 0,008953826 | SEPN1 | FALSE | Upregulated |
| 0,802843177 | 0,008962511 | P2RY8 | FALSE | Upregulated |
| -0,296912785 | 0,008981268 | IPO5 | FALSE | Downregulated |
| -1,664107911 | 0,008981268 | LOC100996756 | TRUE | Downregulated |
| 0,48999612 | 0,008992044 | SLC25A22 | FALSE | Upregulated |
| 0,414961091 | 0,008997938 | SLC6A12 | FALSE | Upregulated |
| 0,229892834 | 0,009013153 | DNM2 | FALSE | Upregulated |
| 0,684659128 | 0,009020584 | SIGLEC1 | FALSE | Upregulated |
| 0,675040957 | 0,009027101 | ARHGAP27 | FALSE | Upregulated |
| -0,547647137 | 0,009029631 | C11orf65 | FALSE | Downregulated |
| -0,912066455 | 0,009036144 | PDK4 | FALSE | Downregulated |
| 0,441486761 | 0,00903911 | MGRN1 | FALSE | Upregulated |
| -0,361349241 | 0,009050648 | PDCD4 | FALSE | Downregulated |
| -0,70892044 | 0,009057144 | PRLR | FALSE | Downregulated |
| -0,253751127 | 0,009075545 | TRMT1L | FALSE | Downregulated |
| 1,038144216 | 0,009086082 | SLC7A1 | TRUE | Upregulated |
| -0,394026411 | 0,009099711 | C12orf76 | FALSE | Downregulated |
| 0,7863592 | 0,00910368 | DOK3 | FALSE | Upregulated |
| 0,743185832 | 0,00911621 | NEK8 | FALSE | Upregulated |
| 0,464936812 | 0,009118624 | SNX11 | FALSE | Upregulated |
| 0,65862259 | 0,009137047 | ARHGEF3 | FALSE | Upregulated |
| 0,893973142 | 0,009139363 | LTB | FALSE | Upregulated |
| 0,29968271 | 0,009152613 | TRAF7 | FALSE | Upregulated |
| 0,329127922 | 0,009162234 | RNASEK | FALSE | Upregulated |
| 0,556488174 | 0,009162234 | SPAG5 | FALSE | Upregulated |
| -0,840872011 | 0,009180175 | ATP6AP1L | FALSE | Downregulated |
| 0,483274262 | 0,009208264 | CEP41 | FALSE | Upregulated |
| -0,269810494 | 0,009208264 | ZNF22 | FALSE | Downregulated |
| -0,298612301 | 0,009231491 | RPAIN | FALSE | Downregulated |
| -1,276514076 | 0,00923326 | CXCR2 | TRUE | Downregulated |
| 0,669449294 | 0,009262047 | HDAC11 | FALSE | Upregulated |
| 0,417124711 | 0,009262047 | PHLPP2 | FALSE | Upregulated |
| -0,254758975 | 0,009262047 | ZBTB44 | FALSE | Downregulated |
| -0,337765612 | 0,009281899 | LOC100507217 | FALSE | Downregulated |
| -0,341210806 | 0,009309473 | UBR3 | FALSE | Downregulated |
| -0,486297626 | 0,009329193 | SHF | FALSE | Downregulated |
| 0,539903978 | 0,00936949 | FXYD5 | FALSE | Upregulated |

| | | | | |
|---|---|---|---|---|
| -0,524890498 | 0,00936949 | LOC643072 | FALSE | Downregulated |
| -0,706893284 | 0,009376623 | CYP4A11 | FALSE | Downregulated |
| -0,48195313 | 0,009423182 | PUS3 | FALSE | Downregulated |
| 0,396958508 | 0,009435237 | NFATC2 | FALSE | Upregulated |
| 1,020377332 | 0,009461497 | GBP3 | TRUE | Upregulated |
| 0,282890059 | 0,009498085 | CLCN7 | FALSE | Upregulated |
| 0,761893893 | 0,009498085 | GPR34 | FALSE | Upregulated |
| -3,091809506 | 0,009505705 | LOC101926960 | TRUE | Downregulated |
| 0,300363783 | 0,009506386 | EMC1 | FALSE | Upregulated |
| 0,200603741 | 0,009538702 | RTFDC1 | FALSE | Upregulated |
| 2,833673188 | 0,009564744 | MYEOV | TRUE | Upregulated |
| -0,359176684 | 0,009585602 | BRD8 | FALSE | Downregulated |
| 1,066527089 | 0,009609828 | TRIM59 | TRUE | Upregulated |
| -0,425498203 | 0,009617134 | ITIH1 | FALSE | Downregulated |
| -0,426393413 | 0,009626662 | RBM48 | FALSE | Downregulated |
| -0,759293535 | 0,009628457 | BZRAP1 | FALSE | Downregulated |
| -1,026543193 | 0,009690615 | LOC100289098 | TRUE | Downregulated |
| 0,426279189 | 0,009726619 | C12orf4 | FALSE | Upregulated |
| 1,674586471 | 0,009726619 | GLDN | TRUE | Upregulated |
| 0,337462039 | 0,009747855 | HEXB | FALSE | Upregulated |
| 0,789161872 | 0,00975567 | ATP2A3 | FALSE | Upregulated |
| -1,035229846 | 0,009790844 | LOC101927595 | TRUE | Downregulated |
| 0,59745681 | 0,00987195 | ADCY3 | FALSE | Upregulated |
| 0,634522867 | 0,009988327 | ZNF101 | FALSE | Upregulated |
| -1,008041109 | 0,009988676 | PCOLCE2 | TRUE | Downregulated |
| -0,487312474 | 0,010091357 | LARP1B | FALSE | Downregulated |
| -0,365504821 | 0,010107586 | CBX7 | FALSE | Downregulated |
| 0,334563343 | 0,010129466 | CYB5R3 | FALSE | Upregulated |
| 0,782794549 | 0,010129466 | TFEC | FALSE | Upregulated |
| 0,552301058 | 0,010185625 | ADAP2 | FALSE | Upregulated |
| 0,601019813 | 0,010199409 | CMTM3 | FALSE | Upregulated |
| 0,360795423 | 0,010277811 | EIF2B2 | FALSE | Upregulated |
| -0,453829001 | 0,010285626 | DBT | FALSE | Downregulated |
| -0,858571109 | 0,010285626 | LOC102724261 | FALSE | Downregulated |
| 0,536598074 | 0,010304611 | GSTP1 | FALSE | Upregulated |
| 1,611713323 | 0,010329074 | ZNF812 | TRUE | Upregulated |
| 0,91028885 | 0,010358127 | AIM2 | FALSE | Upregulated |
| -0,365603493 | 0,010358127 | PIK3C2G | FALSE | Downregulated |
| -0,604779493 | 0,010364242 | VEGFA | FALSE | Downregulated |
| 0,723220959 | 0,010421015 | CRYM | FALSE | Upregulated |
| -0,459167363 | 0,010421421 | ZNF396 | FALSE | Downregulated |
| -0,751537212 | 0,010438595 | RP11-752G15.6 | FALSE | Downregulated |
| 0,279948856 | 0,010471255 | SNX8 | FALSE | Upregulated |
| 0,940708491 | 0,010483328 | ADM2 | FALSE | Upregulated |
| 0,322576678 | 0,010491151 | HEXA | FALSE | Upregulated |
| -0,67061098 | 0,010491151 | SLC6A16 | FALSE | Downregulated |
| 2,417271671 | 0,010505892 | CILP2 | TRUE | Upregulated |
| -0,476029914 | 0,010522472 | HIBCH | FALSE | Downregulated |
| -0,637463619 | 0,010523496 | ALDOB | FALSE | Downregulated |
| 0,516711861 | 0,010560272 | BYSL | FALSE | Upregulated |
| 0,685436933 | 0,010567033 | DOCK8 | FALSE | Upregulated |

| | | | | |
|---|---|---|---|---|
| 0,623209318 | 0,010578093 | CKLF | FALSE | Upregulated |
| 1,563610892 | 0,010578093 | TNIP3 | TRUE | Upregulated |
| 0,460589667 | 0,010586109 | NME2 | FALSE | Upregulated |
| 0,2995959 | 0,010586109 | PLEKHM2 | FALSE | Upregulated |
| 0,340889084 | 0,010586109 | SH3BGRL | FALSE | Upregulated |
| 0,843586967 | 0,0105979 | PYHIN1 | FALSE | Upregulated |
| 0,257005002 | 0,010621226 | RARS | FALSE | Upregulated |
| 0,89826254 | 0,010655094 | BEST1 | FALSE | Upregulated |
| 0,245074437 | 0,01066231 | DNAJC5 | FALSE | Upregulated |
| -0,518453589 | 0,01066231 | ZNF554 | FALSE | Downregulated |
| 0,931543541 | 0,010716107 | ENKUR | FALSE | Upregulated |
| 0,408602524 | 0,010716107 | HDAC6 | FALSE | Upregulated |
| -0,563692184 | 0,010716107 | ZNF417 | FALSE | Downregulated |
| 0,979974309 | 0,010729485 | EBI3 | FALSE | Upregulated |
| -0,320993105 | 0,010746464 | ZNF451 | FALSE | Downregulated |
| 0,523945346 | 0,010786821 | CD58 | FALSE | Upregulated |
| -1,254805193 | 0,010786821 | LOC102724645 | TRUE | Downregulated |
| -0,601204836 | 0,010801511 | CCT6B | FALSE | Downregulated |
| -0,360451614 | 0,010814678 | MKS1 | FALSE | Downregulated |
| -0,455358904 | 0,010814678 | SIRT1 | FALSE | Downregulated |
| -0,67577497 | 0,010819266 | GLS2 | FALSE | Downregulated |
| -0,581669782 | 0,010819266 | ZNF572 | FALSE | Downregulated |
| -1,445067725 | 0,010849794 | C21orf37 | TRUE | Downregulated |
| -0,480994664 | 0,010850449 | CNST | FALSE | Downregulated |
| 0,538331577 | 0,010850449 | TNFAIP3 | FALSE | Upregulated |
| 0,407660527 | 0,010879793 | GNPDA1 | FALSE | Upregulated |
| -0,237254812 | 0,010879793 | RABGAP1 | FALSE | Downregulated |
| -0,796846832 | 0,010887067 | TRABD2B | FALSE | Downregulated |
| -0,461276402 | 0,010888391 | AOX1 | FALSE | Downregulated |
| 0,847975164 | 0,010888391 | MCTP1 | FALSE | Upregulated |
| 0,252756396 | 0,010931001 | ELAC2 | FALSE | Upregulated |
| -0,386209134 | 0,01097159 | BIRC6 | FALSE | Downregulated |
| -0,596371096 | 0,01097159 | CECR2 | FALSE | Downregulated |
| -0,46385611 | 0,011001862 | BPHL | FALSE | Downregulated |
| 0,320701069 | 0,011018196 | RPA2 | FALSE | Upregulated |
| -0,512228483 | 0,011025315 | C20orf112 | FALSE | Downregulated |
| -0,243789267 | 0,011092134 | DYNLT3 | FALSE | Downregulated |
| 0,476650101 | 0,011105113 | SMCO4 | FALSE | Upregulated |
| 0,448038325 | 0,011124414 | GIMAP2 | FALSE | Upregulated |
| -0,365475716 | 0,011162726 | HLCS | FALSE | Downregulated |
| 0,776365141 | 0,011162726 | LILRA2 | FALSE | Upregulated |
| 0,27975307 | 0,011168831 | CTNNA1 | FALSE | Upregulated |
| -0,422781698 | 0,011170511 | DENND4A | FALSE | Downregulated |
| 1,906996785 | 0,011211624 | LOC100506098 | TRUE | Upregulated |
| 0,410193607 | 0,011214421 | PKN1 | FALSE | Upregulated |
| -0,667659308 | 0,011246106 | CDON | FALSE | Downregulated |
| -0,300516866 | 0,011246106 | NUP88 | FALSE | Downregulated |
| -1,815628876 | 0,011246106 | SLC26A4 | TRUE | Downregulated |
| -0,514208243 | 0,011246106 | SORD | FALSE | Downregulated |
| -0,272106912 | 0,011246106 | TAF4 | FALSE | Downregulated |
| -0,409192257 | 0,011251309 | ERG | FALSE | Downregulated |

| | | | | |
|---|---|---|---|---|
| -0,494437712 | 0,011251309 | MAML3 | FALSE | Downregulated |
| -0,392486543 | 0,011252693 | MAGI3 | FALSE | Downregulated |
| -0,568066518 | 0,01126809 | ZNF527 | FALSE | Downregulated |
| 0,412620886 | 0,011297906 | PARP14 | FALSE | Upregulated |
| -1,235128387 | 0,011303656 | LOC100288974 | TRUE | Downregulated |
| 0,86247761 | 0,011323984 | IL27RA | FALSE | Upregulated |
| 0,750638443 | 0,011334656 | ADA | FALSE | Upregulated |
| 0,633191264 | 0,011334656 | DNAH14 | FALSE | Upregulated |
| -0,37104192 | 0,011334656 | FAM210B | FALSE | Downregulated |
| -0,239831108 | 0,011334656 | INTS10 | FALSE | Downregulated |
| 0,631474536 | 0,011334656 | RHOQ | FALSE | Upregulated |
| -0,783991947 | 0,011334656 | SAMD12 | FALSE | Downregulated |
| -1,301058227 | 0,011334656 | SCN11A | TRUE | Downregulated |
| -0,382418593 | 0,011394932 | ITCH | FALSE | Downregulated |
| -0,602266871 | 0,011419791 | ABCC6P1 | FALSE | Downregulated |
| 0,798976506 | 0,011430408 | CCDC96 | FALSE | Upregulated |
| 0,894827926 | 0,011430408 | KCNAB2 | FALSE | Upregulated |
| -0,846105872 | 0,011467957 | DFFB | FALSE | Downregulated |
| 0,510016367 | 0,011467957 | SAP30 | FALSE | Upregulated |
| 0,333553051 | 0,011467957 | ZNF438 | FALSE | Upregulated |
| 0,436319426 | 0,011495099 | NOP2 | FALSE | Upregulated |
| 0,644220062 | 0,011513933 | HMOX1 | FALSE | Upregulated |
| -0,491750232 | 0,011524927 | C5orf45 | FALSE | Downregulated |
| 0,488127632 | 0,011525065 | KIAA1598 | FALSE | Upregulated |
| -0,282853648 | 0,011525065 | NIPBL | FALSE | Downregulated |
| 0,726594077 | 0,011544223 | EVA1B | FALSE | Upregulated |
| 1,716267728 | 0,011553621 | LIF | TRUE | Upregulated |
| 0,370525314 | 0,01156594 | TNFRSF14 | FALSE | Upregulated |
| 0,297134049 | 0,01156594 | UNC13B | FALSE | Upregulated |
| 0,510792271 | 0,011578263 | C1orf216 | FALSE | Upregulated |
| 0,425877424 | 0,011578263 | CNIH4 | FALSE | Upregulated |
| 0,301999952 | 0,011595364 | RRP36 | FALSE | Upregulated |
| 0,745073855 | 0,011602651 | NLRC4 | FALSE | Upregulated |
| -0,672150743 | 0,011603806 | FAM134B | FALSE | Downregulated |
| -2,129112669 | 0,011677815 | SFRP4 | TRUE | Downregulated |
| 0,681388264 | 0,011678693 | SIGLEC9 | FALSE | Upregulated |
| 0,289229178 | 0,011693989 | YBX1 | FALSE | Upregulated |
| 0,452726555 | 0,01171969 | MIF4GD | FALSE | Upregulated |
| 0,788532302 | 0,011723511 | ELOVL2 | FALSE | Upregulated |
| 0,990574789 | 0,011723511 | HOPX | FALSE | Upregulated |
| -1,142591647 | 0,011725064 | HORMAD2 | TRUE | Downregulated |
| 0,311235825 | 0,011751219 | NUB1 | FALSE | Upregulated |
| 0,629451985 | 0,011751855 | CD33 | FALSE | Upregulated |
| -0,282322183 | 0,011751855 | RHOT1 | FALSE | Downregulated |
| 0,984605332 | 0,011761747 | PNMA2 | FALSE | Upregulated |
| -0,630682129 | 0,011772822 | PTPRB | FALSE | Downregulated |
| -3,098538072 | 0,011822562 | LOC101928430 | TRUE | Downregulated |
| 0,83533478 | 0,011823654 | SLC25A33 | FALSE | Upregulated |
| -0,435723118 | 0,011877567 | PDIA5 | FALSE | Downregulated |
| -0,34425404 | 0,011878513 | KTN1 | FALSE | Downregulated |
| -0,425215334 | 0,011908174 | TNS1 | FALSE | Downregulated |

| | | | | |
|---|---|---|---|---|
| -0,445683196 | 0,011934944 | LINC00094 | FALSE | Downregulated |
| -0,254151166 | 0,011992548 | ABCB7 | FALSE | Downregulated |
| -0,424366983 | 0,012005354 | ZNF503 | FALSE | Downregulated |
| -0,269683643 | 0,012017568 | TMTC4 | FALSE | Downregulated |
| -1,104755935 | 0,012027466 | MANEA-AS1 | TRUE | Downregulated |
| -0,508026765 | 0,012027466 | SPTBN2 | FALSE | Downregulated |
| 0,888495444 | 0,012063751 | LAT | FALSE | Upregulated |
| 1,135443555 | 0,012067956 | P2RY10 | TRUE | Upregulated |
| -0,758653242 | 0,012067956 | THAP7-AS1 | FALSE | Downregulated |
| -0,319860766 | 0,012092079 | AKAP1 | FALSE | Downregulated |
| 0,300891768 | 0,01209238 | RPL36A-HNRNPH2 | FALSE | Upregulated |
| -0,53261468 | 0,012122329 | HAAO | FALSE | Downregulated |
| -0,20778524 | 0,012147302 | CREBL2 | FALSE | Downregulated |
| -0,311968209 | 0,012149238 | ERI2 | FALSE | Downregulated |
| 0,559368363 | 0,012149238 | GLIPR1 | FALSE | Upregulated |
| -0,645981231 | 0,012149238 | SMOC1 | FALSE | Downregulated |
| 0,662650248 | 0,012175209 | THEMIS2 | FALSE | Upregulated |
| -0,26601988 | 0,012182423 | SETD2 | FALSE | Downregulated |
| -0,34486706 | 0,012215234 | RICTOR | FALSE | Downregulated |
| 0,951154274 | 0,012253941 | ENPP7 | FALSE | Upregulated |
| -0,334543208 | 0,012280665 | GOT2 | FALSE | Downregulated |
| -0,604001366 | 0,012280665 | NLGN2 | FALSE | Downregulated |
| 0,359991946 | 0,012280665 | RHEB | FALSE | Upregulated |
| 0,37932387 | 0,012291062 | PNPLA6 | FALSE | Upregulated |
| 0,636704129 | 0,012361323 | TONSL | FALSE | Upregulated |
| -1,029623774 | 0,012372137 | UXT-AS1 | TRUE | Downregulated |
| 1,611167534 | 0,012375193 | CAMK2N2 | TRUE | Upregulated |
| -0,639398198 | 0,012375193 | CCDC7 | FALSE | Downregulated |
| 0,404721501 | 0,01238261 | AGA | FALSE | Upregulated |
| 0,485940807 | 0,01238261 | MOB3A | FALSE | Upregulated |
| -0,961300904 | 0,012429851 | PPP1R1A | FALSE | Downregulated |
| -0,347098201 | 0,012441714 | SCP2 | FALSE | Downregulated |
| -0,303557321 | 0,012470374 | MKL2 | FALSE | Downregulated |
| -0,425426304 | 0,012477345 | KDM7A | FALSE | Downregulated |
| -0,348920349 | 0,012500603 | ASH1L | FALSE | Downregulated |
| 0,485377529 | 0,012509861 | GPR137B | FALSE | Upregulated |
| 0,338570877 | 0,012510397 | TNIP2 | FALSE | Upregulated |
| 0,997161168 | 0,012513876 | CCDC64 | FALSE | Upregulated |
| 0,215164852 | 0,012534081 | DENR | FALSE | Upregulated |
| -0,25956113 | 0,012537408 | FAM168A | FALSE | Downregulated |
| -0,208718883 | 0,012554604 | FXR1 | FALSE | Downregulated |
| -0,520570131 | 0,012584763 | ANO8 | FALSE | Downregulated |
| -0,84748896 | 0,012635707 | TTC18 | FALSE | Downregulated |
| 0,246752331 | 0,012640086 | SYNCRIP | FALSE | Upregulated |
| 0,81704782 | 0,012665395 | MRAS | FALSE | Upregulated |
| -1,001495575 | 0,012684524 | LOC101929691 | TRUE | Downregulated |
| 0,530008892 | 0,012714869 | SP110 | FALSE | Upregulated |
| -0,330877539 | 0,012721186 | ARHGAP29 | FALSE | Downregulated |
| 0,823134982 | 0,012724937 | HSH2D | FALSE | Upregulated |
| 0,740121569 | 0,012724937 | OSMR | FALSE | Upregulated |
| 0,339274966 | 0,012759013 | FAM46A | FALSE | Upregulated |

| | | | | |
|---|---|---|---|---|
| 0,293453432 | 0,012787182 | PPP4C | FALSE | Upregulated |
| -3,462905216 | 0,012791793 | TNMD | TRUE | Downregulated |
| -1,417370364 | 0,012821927 | MCM3AP-AS1 | TRUE | Downregulated |
| 0,993362961 | 0,012874853 | GBP1P1 | FALSE | Upregulated |
| -1,663428045 | 0,012874853 | LOC102723398 | TRUE | Downregulated |
| -1,462276634 | 0,01287668 | ADRB1 | TRUE | Downregulated |
| 2,043625933 | 0,01287668 | LOC101928173 | TRUE | Upregulated |
| -0,528427714 | 0,01287668 | TMEM56 | FALSE | Downregulated |
| -0,351196117 | 0,012917859 | ZFAND1 | FALSE | Downregulated |
| -1,849431878 | 0,012928846 | CYP26A1 | TRUE | Downregulated |
| -0,313213335 | 0,012936386 | IFT88 | FALSE | Downregulated |
| -0,867965642 | 0,0129478 | ENPP3 | FALSE | Downregulated |
| 0,338658241 | 0,012976376 | PARVA | FALSE | Upregulated |
| -0,340562253 | 0,012977696 | RNF168 | FALSE | Downregulated |
| -1,253012382 | 0,013029374 | RP11-996F15.2 | TRUE | Downregulated |
| -0,340328594 | 0,013054833 | HSDL2 | FALSE | Downregulated |
| 1,600601075 | 0,013054833 | NR4A3 | TRUE | Upregulated |
| -0,501391144 | 0,013070503 | PRKAB2 | FALSE | Downregulated |
| -0,51007273 | 0,013086225 | HPRT1 | FALSE | Downregulated |
| 0,739092272 | 0,013100181 | EAF2 | FALSE | Upregulated |
| -0,570162377 | 0,013109643 | FUT10 | FALSE | Downregulated |
| 0,280458526 | 0,0131127 | ANKH | FALSE | Upregulated |
| -0,542016726 | 0,013123539 | BCORL1 | FALSE | Downregulated |
| -0,755205688 | 0,013123539 | BHMT | FALSE | Downregulated |
| 0,351776422 | 0,013123539 | RPL26L1 | FALSE | Upregulated |
| 0,436261685 | 0,013131286 | FBXO25 | FALSE | Upregulated |
| -0,734363704 | 0,013134883 | BCHE | FALSE | Downregulated |
| -0,484419744 | 0,013134883 | F11 | FALSE | Downregulated |
| 0,8655305 | 0,013134883 | INPP4B | FALSE | Upregulated |
| -0,46460619 | 0,013134883 | ZNF558 | FALSE | Downregulated |
| 0,501937561 | 0,013157809 | COQ2 | FALSE | Upregulated |
| 0,623596289 | 0,013167982 | HMHA1 | FALSE | Upregulated |
| 0,37689757 | 0,013186692 | AKIRIN1 | FALSE | Upregulated |
| 0,393036135 | 0,013193121 | CBFB | FALSE | Upregulated |
| 0,982820803 | 0,013193121 | ZAP70 | FALSE | Upregulated |
| 0,88905345 | 0,013209917 | SLC16A6 | FALSE | Upregulated |
| 0,942335835 | 0,013226812 | DUSP1 | FALSE | Upregulated |
| 0,491485371 | 0,013226812 | FTL | FALSE | Upregulated |
| 2,123861981 | 0,013226812 | PHEX | TRUE | Upregulated |
| 1,893651509 | 0,013226812 | RTKN2 | TRUE | Upregulated |
| -0,523755818 | 0,013236581 | GPHN | FALSE | Downregulated |
| -0,690071759 | 0,013238841 | ARHGAP20 | FALSE | Downregulated |
| -1,503868064 | 0,013238841 | CYP1A1 | TRUE | Downregulated |
| 0,223769483 | 0,013238841 | DERL1 | FALSE | Upregulated |
| -1,186928534 | 0,013238841 | DOK6 | TRUE | Downregulated |
| 1,200676348 | 0,013238841 | FBN2 | TRUE | Upregulated |
| 1,381833332 | 0,013238841 | GRIN3A | TRUE | Upregulated |
| 0,278783335 | 0,013238841 | MED15 | FALSE | Upregulated |
| 0,287217291 | 0,013272088 | CAPZA2 | FALSE | Upregulated |
| 0,472278879 | 0,013283059 | PCBP4 | FALSE | Upregulated |
| 0,269743747 | 0,013289214 | CAPN1 | FALSE | Upregulated |

| | | | | |
|---|---|---|---|---|
| 0,241306481 | 0,013291666 | MGAT1 | FALSE | Upregulated |
| 0,57058467 | 0,01330015 | PUSL1 | FALSE | Upregulated |
| -0,252284788 | 0,013302681 | HDLBP | FALSE | Downregulated |
| -0,896272025 | 0,013302681 | WDFY3-AS2 | FALSE | Downregulated |
| 0,344310959 | 0,013356143 | PSMB4 | FALSE | Upregulated |
| -0,618144148 | 0,013382407 | EFCAB7 | FALSE | Downregulated |
| 0,280374822 | 0,013382407 | SNX12 | FALSE | Upregulated |
| -0,375806393 | 0,013415703 | ZNF292 | FALSE | Downregulated |
| 0,674281198 | 0,013421443 | CELF2 | FALSE | Upregulated |
| -0,517264018 | 0,013461048 | ARID4A | FALSE | Downregulated |
| 0,269694897 | 0,013475061 | LLPH | FALSE | Upregulated |
| -0,311986726 | 0,013481025 | BCL2L2 | FALSE | Downregulated |
| -0,341488506 | 0,013481025 | ZNF84 | FALSE | Downregulated |
| 0,733319791 | 0,013482794 | BTG2 | FALSE | Upregulated |
| -0,715693579 | 0,013516767 | TBC1D2 | FALSE | Downregulated |
| 0,438084934 | 0,013520054 | UQCC2 | FALSE | Upregulated |
| 0,655426869 | 0,013527875 | GJA4 | FALSE | Upregulated |
| -0,204349657 | 0,013555214 | PRPF38A | FALSE | Downregulated |
| 0,884065704 | 0,013589322 | LFNG | FALSE | Upregulated |
| 0,627091001 | 0,013589322 | TMEM107 | FALSE | Upregulated |
| -0,706626362 | 0,013595318 | CMBL | FALSE | Downregulated |
| -1,417009645 | 0,013608743 | WDR64 | TRUE | Downregulated |
| 0,908765572 | 0,01365067 | IER3 | FALSE | Upregulated |
| -0,352912871 | 0,01365067 | MORC3 | FALSE | Downregulated |
| 1,497353518 | 0,01365067 | TRIP13 | TRUE | Upregulated |
| 0,372572264 | 0,013683806 | ATL3 | FALSE | Upregulated |
| -0,589023672 | 0,013683806 | SCAND2P | FALSE | Downregulated |
| 0,381114391 | 0,013683806 | SPTBN1 | FALSE | Upregulated |
| 0,244085543 | 0,013700521 | PARN | FALSE | Upregulated |
| -0,237413579 | 0,013727365 | GLTSCR1L | FALSE | Downregulated |
| 0,623252208 | 0,013750899 | SERPINB9 | FALSE | Upregulated |
| 0,673432951 | 0,013770023 | FLT3LG | FALSE | Upregulated |
| -0,468811284 | 0,013770023 | METTL20 | FALSE | Downregulated |
| 0,388314867 | 0,013787288 | TOE1 | FALSE | Upregulated |
| 0,724744579 | 0,013801081 | PFKFB3 | FALSE | Upregulated |
| -1,099242783 | 0,013811626 | LOC100133669 | TRUE | Downregulated |
| 0,466260813 | 0,013813796 | SDR42E1 | FALSE | Upregulated |
| -0,721734191 | 0,013900722 | ZNF334 | FALSE | Downregulated |
| 0,350570291 | 0,013928371 | TMEM9B | FALSE | Upregulated |
| -0,615066039 | 0,013947635 | PNPLA7 | FALSE | Downregulated |
| 1,127216861 | 0,013947635 | UBASH3A | TRUE | Upregulated |
| -1,800400429 | 0,013959465 | LOC101059949 | TRUE | Downregulated |
| -0,491529118 | 0,013959465 | SDPR | FALSE | Downregulated |
| -1,099870715 | 0,013965177 | MBL1P | TRUE | Downregulated |
| 0,420816842 | 0,013965177 | RAB4B | FALSE | Upregulated |
| 0,561247202 | 0,013993195 | CRIP2 | FALSE | Upregulated |
| -1,028424744 | 0,014030229 | FRAS1 | TRUE | Downregulated |
| -0,726908201 | 0,014045714 | PKHD1 | FALSE | Downregulated |
| 0,699565263 | 0,014045714 | STAMBPL1 | FALSE | Upregulated |
| -0,460808061 | 0,014053724 | C11orf54 | FALSE | Downregulated |
| 0,29309965 | 0,014053724 | MTA2 | FALSE | Upregulated |

| | | | | |
|---|---|---|---|---|
| -0,278639296 | 0,014053724 | NADK2 | FALSE | Downregulated |
| 0,355324654 | 0,014053724 | PPAP2A | FALSE | Upregulated |
| 0,734488963 | 0,014053724 | TSPAN1 | FALSE | Upregulated |
| -0,875651514 | 0,014069114 | ARID3C | FALSE | Downregulated |
| 0,803877283 | 0,014069114 | KLRK1 | FALSE | Upregulated |
| 0,951146926 | 0,014077923 | TBC1D10C | FALSE | Upregulated |
| 0,376079028 | 0,014096097 | KLHL18 | FALSE | Upregulated |
| -0,288086373 | 0,014096097 | PPP2R5A | FALSE | Downregulated |
| -2,588240359 | 0,014100011 | FOXH1 | TRUE | Downregulated |
| 0,392162435 | 0,014168567 | SCYL2 | FALSE | Upregulated |
| -0,306626842 | 0,014220943 | KCTD3 | FALSE | Downregulated |
| -0,325762254 | 0,014220943 | LIPT1 | FALSE | Downregulated |
| 0,829721631 | 0,014220943 | LXN | FALSE | Upregulated |
| 2,074839906 | 0,014249595 | CLIC3 | TRUE | Upregulated |
| -0,231810996 | 0,01428913 | SNRNP200 | FALSE | Downregulated |
| -0,296351351 | 0,014339872 | CHD1 | FALSE | Downregulated |
| 0,689626086 | 0,014354038 | MATK | FALSE | Upregulated |
| 0,215997585 | 0,014375635 | NR1H2 | FALSE | Upregulated |
| -0,424584744 | 0,014385432 | CXXC5 | FALSE | Downregulated |
| -0,274274483 | 0,014385432 | MCCC2 | FALSE | Downregulated |
| 0,470608553 | 0,014397671 | NAA40 | FALSE | Upregulated |
| 0,340662692 | 0,01450307 | MAPKAPK3 | FALSE | Upregulated |
| 0,474363828 | 0,014538139 | IFNGR1 | FALSE | Upregulated |
| 1,78053712 | 0,014538139 | MMP7 | TRUE | Upregulated |
| -0,26187915 | 0,014538139 | NDEL1 | FALSE | Downregulated |
| -1,151578477 | 0,014550244 | LOC102546294 | TRUE | Downregulated |
| 0,312169633 | 0,014561451 | BCL7B | FALSE | Upregulated |
| -2,377144993 | 0,014561451 | FAM83C | TRUE | Downregulated |
| 1,160576026 | 0,014600688 | CCL2 | TRUE | Upregulated |
| 0,191896613 | 0,014613721 | ZDHHC3 | FALSE | Upregulated |
| 0,355171151 | 0,014636594 | RRBP1 | FALSE | Upregulated |
| -0,49638259 | 0,014684998 | NR3C2 | FALSE | Downregulated |
| -1,386306258 | 0,014729448 | CALR3 | TRUE | Downregulated |
| -1,485361673 | 0,014729448 | MGAM | TRUE | Downregulated |
| -0,482763406 | 0,014736766 | SLX4IP | FALSE | Downregulated |
| 0,428362685 | 0,014751969 | IQCE | FALSE | Upregulated |
| -0,773562241 | 0,014751969 | SDHAP1 | FALSE | Downregulated |
| 0,519712943 | 0,014751969 | TMCO3 | FALSE | Upregulated |
| 1,358278251 | 0,014834116 | BHLHE41 | TRUE | Upregulated |
| 0,456218481 | 0,014834116 | RAB8B | FALSE | Upregulated |
| 0,534104256 | 0,014834116 | RRAS | FALSE | Upregulated |
| 0,303958273 | 0,014859064 | VAT1 | FALSE | Upregulated |
| 0,310677199 | 0,014904714 | NAA10 | FALSE | Upregulated |
| 0,981163785 | 0,014904714 | OLFML2B | FALSE | Upregulated |
| 0,40761511 | 0,014981645 | PGD | FALSE | Upregulated |
| 0,241744567 | 0,014999947 | WASF2 | FALSE | Upregulated |
| 0,794637317 | 0,015014738 | RTN1 | FALSE | Upregulated |
| -0,30387747 | 0,015065949 | UHRF2 | FALSE | Downregulated |
| -0,276635846 | 0,015095006 | HMGN3 | FALSE | Downregulated |
| 0,401333839 | 0,015095006 | HRAS | FALSE | Upregulated |
| -0,306974501 | 0,015095006 | KLC4 | FALSE | Downregulated |

| | | | | |
|---|---|---|---|---|
| 0,326780964 | 0,015095006 | PIK3CB | FALSE | Upregulated |
| -0,715445685 | 0,01509637 | PNISR | FALSE | Downregulated |
| 1,28465985 | 0,015125646 | HSPA6 | TRUE | Upregulated |
| 0,717292381 | 0,015176222 | CITED4 | FALSE | Upregulated |
| -0,34252745 | 0,015176222 | SPICE1 | FALSE | Downregulated |
| 0,412850495 | 0,015183789 | DTNBP1 | FALSE | Upregulated |
| 0,35380883 | 0,015233633 | C11orf73 | FALSE | Upregulated |
| 0,394199881 | 0,015233633 | PHF5A | FALSE | Upregulated |
| 1,467036231 | 0,015241232 | LOC102723328 | TRUE | Upregulated |
| 0,311910742 | 0,015247588 | GTF3C6 | FALSE | Upregulated |
| 0,951969438 | 0,015287812 | IER5L | FALSE | Upregulated |
| 0,405705209 | 0,015308075 | ASAH1 | FALSE | Upregulated |
| -0,456546376 | 0,015308075 | RBM33 | FALSE | Downregulated |
| 1,254531432 | 0,015311693 | PANDAR | TRUE | Upregulated |
| 0,255316873 | 0,015321741 | POLG | FALSE | Upregulated |
| -0,341599143 | 0,015348033 | C11orf57 | FALSE | Downregulated |
| -0,8074592 | 0,015348033 | C19orf18 | FALSE | Downregulated |
| -0,254374475 | 0,015354196 | WDR89 | FALSE | Downregulated |
| 0,626811181 | 0,015367115 | VAV1 | FALSE | Upregulated |
| 0,277232621 | 0,015370069 | MANBAL | FALSE | Upregulated |
| -0,503675051 | 0,015370069 | ZNF780B | FALSE | Downregulated |
| 0,43399212 | 0,015403492 | TANGO2 | FALSE | Upregulated |
| -0,441694613 | 0,015403492 | ZNF112 | FALSE | Downregulated |
| -0,300269014 | 0,015429269 | ABHD17B | FALSE | Downregulated |
| 1,145231721 | 0,015429269 | LOC100506870 | TRUE | Upregulated |
| -0,312194205 | 0,015451799 | EPM2AIP1 | FALSE | Downregulated |
| 0,809777373 | 0,015523834 | GPX7 | FALSE | Upregulated |
| -0,64324113 | 0,015523834 | ZNF594 | FALSE | Downregulated |
| 0,462962033 | 0,015538636 | HAUS1 | FALSE | Upregulated |
| -0,577162319 | 0,015549103 | AGXT2 | FALSE | Downregulated |
| 0,710751612 | 0,015578565 | ARSJ | FALSE | Upregulated |
| 0,838448888 | 0,015579988 | BAX | FALSE | Upregulated |
| 0,368663696 | 0,015604967 | POLA1 | FALSE | Upregulated |
| -1,288554108 | 0,015606944 | NPIPB9 | TRUE | Downregulated |
| 0,354968137 | 0,015630383 | TMEM199 | FALSE | Upregulated |
| -0,799509232 | 0,0156402 | ZMAT1 | FALSE | Downregulated |
| -0,245642812 | 0,015640403 | FAM8A1 | FALSE | Downregulated |
| -0,304083986 | 0,01565123 | ZNF721 | FALSE | Downregulated |
| 1,118193444 | 0,01572365 | C16orf54 | TRUE | Upregulated |
| -0,69946709 | 0,01572365 | LIPG | FALSE | Downregulated |
| -0,459293595 | 0,015738652 | AMT | FALSE | Downregulated |
| 0,39082074 | 0,015738652 | GPI | FALSE | Upregulated |
| -0,386467601 | 0,015766958 | ACAD11 | FALSE | Downregulated |
| -0,307798701 | 0,015766958 | CCDC126 | FALSE | Downregulated |
| 0,190162423 | 0,015766958 | GGA2 | FALSE | Upregulated |
| -0,438853921 | 0,015766958 | PHYH | FALSE | Downregulated |
| -0,544020103 | 0,015766958 | ZBTB49 | FALSE | Downregulated |
| 0,30356342 | 0,015784146 | TRNAU1AP | FALSE | Upregulated |
| 0,312926377 | 0,015801847 | ABI1 | FALSE | Upregulated |
| -1,48890094 | 0,015805378 | DGKB | TRUE | Downregulated |
| -0,384457739 | 0,015812158 | CALCOCO1 | FALSE | Downregulated |

| | | | | |
|---|---|---|---|---|
| 0,436655512 | 0,015812158 | SEPT6 | FALSE | Upregulated |
| 1,170746619 | 0,015843288 | LOC102724451 | TRUE | Upregulated |
| 0,50609566 | 0,015843288 | PSMC4 | FALSE | Upregulated |
| 2,644106967 | 0,015867713 | C2CD4A | TRUE | Upregulated |
| 0,548901802 | 0,015872768 | BIN2 | FALSE | Upregulated |
| 0,618064994 | 0,015873136 | CYBB | FALSE | Upregulated |
| 0,758627218 | 0,015921376 | PPFIA3 | FALSE | Upregulated |
| -0,324489988 | 0,01595338 | PLEKHA5 | FALSE | Downregulated |
| -0,3186874 | 0,01595338 | ZNF473 | FALSE | Downregulated |
| -1,037758511 | 0,015957787 | LOC101930307 | TRUE | Downregulated |
| -0,395147911 | 0,0159628 | KMT2C | FALSE | Downregulated |
| 0,199104811 | 0,015986339 | PSMC1 | FALSE | Upregulated |
| 0,709610776 | 0,016033087 | ITGB3 | FALSE | Upregulated |
| 0,788117763 | 0,016033087 | MYOF | FALSE | Upregulated |
| 2,264173666 | 0,016035252 | RASGRF1 | TRUE | Upregulated |
| -0,361505716 | 0,016042096 | LIAS | FALSE | Downregulated |
| -0,754164378 | 0,016143634 | GADD45A | FALSE | Downregulated |
| 0,797093111 | 0,016147767 | CD274 | FALSE | Upregulated |
| -0,405551097 | 0,016155069 | THNSL1 | FALSE | Downregulated |
| 1,133876512 | 0,016168338 | TRIM36 | TRUE | Upregulated |
| -0,253929995 | 0,016215609 | NBR1 | FALSE | Downregulated |
| -0,446542778 | 0,016215609 | SNRNP48 | FALSE | Downregulated |
| 0,872378084 | 0,016215609 | TLCD1 | FALSE | Upregulated |
| 0,541282738 | 0,016245993 | RBBP8 | FALSE | Upregulated |
| 0,389182887 | 0,016253627 | DNM1L | FALSE | Upregulated |
| 1,188413685 | 0,016270665 | GRIN2D | TRUE | Upregulated |
| -0,392297172 | 0,016270665 | NAT9 | FALSE | Downregulated |
| -0,459388493 | 0,01630147 | ZNF69 | FALSE | Downregulated |
| 0,428206835 | 0,016333721 | MSH2 | FALSE | Upregulated |
| -1,321296091 | 0,016364881 | PRDM5 | TRUE | Downregulated |
| 0,289005358 | 0,016373963 | ARRB2 | FALSE | Upregulated |
| 0,316670524 | 0,016392798 | NOMO1 | FALSE | Upregulated |
| 0,315255735 | 0,016392798 | STX12 | FALSE | Upregulated |
| 0,998980993 | 0,016400674 | OASL | FALSE | Upregulated |
| -0,394279644 | 0,016412246 | MEIS2 | FALSE | Downregulated |
| 1,528103498 | 0,01643856 | GZMM | TRUE | Upregulated |
| -0,292848513 | 0,01643856 | SGMS1 | FALSE | Downregulated |
| 0,41379988 | 0,016456118 | PTCHD3P1 | FALSE | Upregulated |
| 1,485143218 | 0,016468199 | FZD2 | TRUE | Upregulated |
| 1,016950306 | 0,016468199 | PFKP | TRUE | Upregulated |
| 1,249968088 | 0,016481963 | C1QTNF9B-AS1 | TRUE | Upregulated |
| 0,595840194 | 0,01648481 | ADM | FALSE | Upregulated |
| -0,650236217 | 0,01648481 | RDH16 | FALSE | Downregulated |
| -0,412942395 | 0,016514763 | CREB3L4 | FALSE | Downregulated |
| 0,312414496 | 0,016544375 | UNC45A | FALSE | Upregulated |
| -0,58541152 | 0,016601009 | HECW2 | FALSE | Downregulated |
| -0,484706553 | 0,016612882 | CCDC82 | FALSE | Downregulated |
| 0,29676885 | 0,016612882 | RTCB | FALSE | Upregulated |
| -0,313163438 | 0,016657385 | PKN2 | FALSE | Downregulated |
| -0,461453074 | 0,016680443 | BAHCC1 | FALSE | Downregulated |
| -0,613687599 | 0,016680443 | CREBZF | FALSE | Downregulated |

| | | | | |
|---|---|---|---|---|
| -1,100435697 | 0,016680443 | LOC101929177 | TRUE | Downregulated |
| 0,32388128 | 0,016690903 | HMOX2 | FALSE | Upregulated |
| 0,280966091 | 0,01672056 | TIMM23 | FALSE | Upregulated |
| -0,657481619 | 0,01680824 | RORA | FALSE | Downregulated |
| 1,038691787 | 0,016846826 | SLAMF1 | TRUE | Upregulated |
| 0,644199349 | 0,016895035 | CD180 | FALSE | Upregulated |
| 0,627457664 | 0,016895035 | PAQR8 | FALSE | Upregulated |
| 0,511540963 | 0,016929355 | PSMG3 | FALSE | Upregulated |
| 0,356131576 | 0,016939338 | AHSA1 | FALSE | Upregulated |
| -1,498993141 | 0,016939338 | LGI1 | TRUE | Downregulated |
| 0,437443524 | 0,016939338 | LOC550643 | FALSE | Upregulated |
| 0,715771718 | 0,016959248 | GPR56 | FALSE | Upregulated |
| -0,477107774 | 0,016974841 | HSBP1L1 | FALSE | Downregulated |
| -0,412627162 | 0,017012916 | GOLIM4 | FALSE | Downregulated |
| 0,362838452 | 0,017079564 | MGLL | FALSE | Upregulated |
| 0,286942621 | 0,017091367 | TRAPPC12 | FALSE | Upregulated |
| -0,356429043 | 0,01722895 | CRB3 | FALSE | Downregulated |
| -0,970712081 | 0,017244075 | HNF1A-AS1 | FALSE | Downregulated |
| -0,328421651 | 0,017254075 | SETD6 | FALSE | Downregulated |
| 1,138875413 | 0,017254075 | SYTL1 | TRUE | Upregulated |
| -0,658298719 | 0,017261021 | NR1I2 | FALSE | Downregulated |
| 0,366818799 | 0,017364685 | RPL28 | FALSE | Upregulated |
| -0,18928941 | 0,017378832 | FOPNL | FALSE | Downregulated |
| 0,629932884 | 0,017427507 | IPCEF1 | FALSE | Upregulated |
| -0,734344626 | 0,017451458 | AKR1D1 | FALSE | Downregulated |
| 0,609033383 | 0,017493549 | APOLD1 | FALSE | Upregulated |
| -1,218712751 | 0,017493549 | RP11-793H13.3 | TRUE | Downregulated |
| 0,847060324 | 0,017496329 | RNF24 | FALSE | Upregulated |
| -0,544148719 | 0,01749685 | ZNF608 | FALSE | Downregulated |
| 0,271211037 | 0,017560955 | EMC3 | FALSE | Upregulated |
| -0,303256782 | 0,017635271 | ANKMY2 | FALSE | Downregulated |
| 0,882072073 | 0,017635271 | USP2 | FALSE | Upregulated |
| 1,773313585 | 0,017638246 | PTCRA | TRUE | Upregulated |
| 0,306111141 | 0,017659503 | CIAPIN1 | FALSE | Upregulated |
| 1,566939035 | 0,01766004 | LTA | TRUE | Upregulated |
| -0,743481994 | 0,017706818 | LOC100507577 | FALSE | Downregulated |
| -0,230480808 | 0,01774994 | GXYLT1 | FALSE | Downregulated |
| 0,231422792 | 0,017778889 | GLB1 | FALSE | Upregulated |
| -0,20347447 | 0,017850804 | ZNF24 | FALSE | Downregulated |
| 0,97286891 | 0,017858633 | CCR2 | FALSE | Upregulated |
| 1,584844865 | 0,017898503 | RASD1 | TRUE | Upregulated |
| 0,68900301 | 0,017940069 | TMEM206 | FALSE | Upregulated |
| 0,734054674 | 0,017971511 | GLIPR2 | FALSE | Upregulated |
| 0,428023336 | 0,018000972 | CEP78 | FALSE | Upregulated |
| 0,627513841 | 0,018000972 | RBPMS2 | FALSE | Upregulated |
| -0,321068108 | 0,018000972 | SNX13 | FALSE | Downregulated |
| -0,474627908 | 0,01802583 | PAN2 | FALSE | Downregulated |
| -0,328698421 | 0,018034574 | TADA1 | FALSE | Downregulated |
| -0,422507297 | 0,018090586 | RNF144B | FALSE | Downregulated |
| 0,202269634 | 0,018142226 | WIPI2 | FALSE | Upregulated |
| 0,47105036 | 0,018152611 | HHAT | FALSE | Upregulated |

| | | | | |
|---|---|---|---|---|
| 1,297900308 | 0,018210931 | GNB1L | TRUE | Upregulated |
| -0,265269127 | 0,018230966 | FAM179B | FALSE | Downregulated |
| 1,99978122 | 0,018238234 | C9orf172 | TRUE | Upregulated |
| -2,253049373 | 0,018238234 | HCG14 | TRUE | Downregulated |
| -0,350796608 | 0,018238234 | LETMD1 | FALSE | Downregulated |
| -0,53807882 | 0,018238234 | RBM6 | FALSE | Downregulated |
| -0,263045585 | 0,018238234 | XPC | FALSE | Downregulated |
| -0,535273114 | 0,018238234 | ZNF385B | FALSE | Downregulated |
| 0,661469974 | 0,018265188 | VIM | FALSE | Upregulated |
| 0,835942609 | 0,0183033 | CD5L | FALSE | Upregulated |
| 0,407429204 | 0,0183033 | FUCA2 | FALSE | Upregulated |
| 0,284878688 | 0,0183033 | ITGB1 | FALSE | Upregulated |
| -1,52131975 | 0,0183033 | RUNX2 | TRUE | Downregulated |
| 0,846236777 | 0,0183033 | SLC22A18AS | FALSE | Upregulated |
| -0,537695451 | 0,018379629 | ABCA8 | FALSE | Downregulated |
| -0,2628739 | 0,018431373 | NDUFA10 | FALSE | Downregulated |
| -0,357784995 | 0,018438514 | MON2 | FALSE | Downregulated |
| -0,591541095 | 0,018444717 | NEIL1 | FALSE | Downregulated |
| 0,250395318 | 0,018476912 | PA2G4 | FALSE | Upregulated |
| -0,418494781 | 0,018476912 | PON3 | FALSE | Downregulated |
| 0,316618488 | 0,018525339 | GRWD1 | FALSE | Upregulated |
| 0,286847771 | 0,018529769 | RAB8A | FALSE | Upregulated |
| 0,379141985 | 0,018542904 | GAPDH | FALSE | Upregulated |
| -0,401086434 | 0,018542904 | SMARCA2 | FALSE | Downregulated |
| 0,255919629 | 0,018545022 | VARS | FALSE | Upregulated |
| 0,462031321 | 0,018617765 | PON2 | FALSE | Upregulated |
| 0,275771651 | 0,018621259 | ADAR | FALSE | Upregulated |
| -0,61030324 | 0,018640034 | ST3GAL6 | FALSE | Downregulated |
| -0,525080614 | 0,018667024 | LOC101929291 | FALSE | Downregulated |
| 0,411102897 | 0,018683437 | LRSAM1 | FALSE | Upregulated |
| -0,717555926 | 0,018725247 | NR6A1 | FALSE | Downregulated |
| 0,41512688 | 0,018735138 | SARS2 | FALSE | Upregulated |
| -2,209859822 | 0,018832653 | DNAI1 | TRUE | Downregulated |
| 0,223158358 | 0,018832653 | NDNL2 | FALSE | Upregulated |
| -1,262046901 | 0,01883738 | DLGAP2 | TRUE | Downregulated |
| 0,518151516 | 0,01883738 | EFCAB11 | FALSE | Upregulated |
| 0,759801927 | 0,018862703 | MILR1 | FALSE | Upregulated |
| -0,284532011 | 0,018876311 | PTGR2 | FALSE | Downregulated |
| -1,000881493 | 0,018947134 | ADCY10P1 | TRUE | Downregulated |
| 0,845228305 | 0,018947134 | CLEC12A | FALSE | Upregulated |
| -0,370738053 | 0,018947134 | ZC3H11A | FALSE | Downregulated |
| -0,680884664 | 0,018953017 | DYNC2H1 | FALSE | Downregulated |
| -0,293631435 | 0,018953017 | HIBADH | FALSE | Downregulated |
| 0,805053361 | 0,018953017 | LILRB1 | FALSE | Upregulated |
| -0,1889726 | 0,018953017 | SERINC1 | FALSE | Downregulated |
| -0,484972952 | 0,018953017 | TPPP | FALSE | Downregulated |
| -1,457895618 | 0,018961217 | HRASLS2 | TRUE | Downregulated |
| -1,295979402 | 0,01901184 | GRIP1 | TRUE | Downregulated |
| -1,636692844 | 0,019012056 | CLEC4GP1 | TRUE | Downregulated |
| -0,562925423 | 0,019014752 | VIPR1 | FALSE | Downregulated |
| -0,397277508 | 0,019041815 | ACAT1 | FALSE | Downregulated |

| | | | | |
|---|---|---|---|---|
| -0,35966133 | 0,019041815 | ARL3 | FALSE | Downregulated |
| -0,362142445 | 0,019062713 | POGZ | FALSE | Downregulated |
| -1,19541696 | 0,019089797 | LOC101926980 | TRUE | Downregulated |
| 0,342381232 | 0,019097817 | COX6B1 | FALSE | Upregulated |
| -0,278688826 | 0,019097817 | VHL | FALSE | Downregulated |
| 0,240559558 | 0,01910312 | RALY | FALSE | Upregulated |
| 0,270173695 | 0,019112507 | MAN1B1 | FALSE | Upregulated |
| -0,650427425 | 0,019147976 | PCDH18 | FALSE | Downregulated |
| -0,399839541 | 0,019220955 | CNKSR3 | FALSE | Downregulated |
| 0,491217113 | 0,019220955 | DSEL | FALSE | Upregulated |
| 0,380497804 | 0,01923846 | AGRN | FALSE | Upregulated |
| -0,816679399 | 0,01923866 | KIF26A | FALSE | Downregulated |
| 0,572035261 | 0,019266511 | MLKL | FALSE | Upregulated |
| -0,413767441 | 0,019273891 | MAGI2 | FALSE | Downregulated |
| 0,241517527 | 0,019289848 | STARD3 | FALSE | Upregulated |
| 0,354879057 | 0,019363412 | PSME3 | FALSE | Upregulated |
| -0,237839336 | 0,019372064 | TIAL1 | FALSE | Downregulated |
| 0,457640055 | 0,019373056 | TWF2 | FALSE | Upregulated |
| -0,258963091 | 0,019415897 | CLDN12 | FALSE | Downregulated |
| -1,628143 | 0,019558927 | DLK2 | TRUE | Downregulated |
| 0,261412178 | 0,019558927 | OAZ1 | FALSE | Upregulated |
| 0,457222405 | 0,019558927 | POLB | FALSE | Upregulated |
| 0,284166307 | 0,019558927 | ZBED4 | FALSE | Upregulated |
| -1,272732594 | 0,019586596 | GLUD1P3 | TRUE | Downregulated |
| 0,281731716 | 0,019596648 | GATAD2A | FALSE | Upregulated |
| -0,38013463 | 0,019596648 | PCK2 | FALSE | Downregulated |
| 0,409088812 | 0,019602216 | RRM2B | FALSE | Upregulated |
| -0,215557841 | 0,019624158 | TPP2 | FALSE | Downregulated |
| 0,337894112 | 0,019689775 | ABHD17A | FALSE | Upregulated |
| 1,000475697 | 0,019704323 | SH2B2 | TRUE | Upregulated |
| -0,491952791 | 0,01972689 | HHEX | FALSE | Downregulated |
| -1,247851726 | 0,01973649 | LOC101929443 | TRUE | Downregulated |
| 0,360843423 | 0,019759624 | CLN3 | FALSE | Upregulated |
| -0,52264566 | 0,019767062 | NTHL1 | FALSE | Downregulated |
| 0,829067323 | 0,019770415 | MCHR1 | FALSE | Upregulated |
| 0,248995377 | 0,019772748 | TMEM127 | FALSE | Upregulated |
| -0,306953294 | 0,019783034 | MTHFD1 | FALSE | Downregulated |
| -0,26969138 | 0,019791422 | SEC63 | FALSE | Downregulated |
| 0,216215729 | 0,019799931 | PHC2 | FALSE | Upregulated |
| 0,437793964 | 0,019799931 | SMC4 | FALSE | Upregulated |
| 1,906738063 | 0,019835736 | LOC101060038 | TRUE | Upregulated |
| -0,460976068 | 0,019861277 | SATB1 | FALSE | Downregulated |
| 2,151210931 | 0,019874504 | HTRA4 | TRUE | Upregulated |
| 0,820718771 | 0,019874504 | RSAD2 | FALSE | Upregulated |
| -0,531106209 | 0,019902472 | ZNF354B | FALSE | Downregulated |
| 0,616650074 | 0,019913221 | TMEM201 | FALSE | Upregulated |
| -0,199434715 | 0,019913741 | IARS2 | FALSE | Downregulated |
| 0,322521484 | 0,019913741 | JOSD1 | FALSE | Upregulated |
| -1,071984878 | 0,019913741 | SNX29P2 | TRUE | Downregulated |
| 0,289618117 | 0,019941449 | CAPRIN1 | FALSE | Upregulated |
| -0,780752669 | 0,019941449 | CDHR3 | FALSE | Downregulated |

| | | | | |
|---|---|---|---|---|
| 0,390346114 | 0,019948889 | C1QTNF1 | FALSE | Upregulated |
| 0,515841958 | 0,020014342 | CASZ1 | FALSE | Upregulated |
| 0,269586007 | 0,020014342 | VCP | FALSE | Upregulated |
| -0,634654159 | 0,02002131 | ACOT1 | FALSE | Downregulated |
| -0,347930779 | 0,02002131 | IL1R1 | FALSE | Downregulated |
| 0,741072919 | 0,020122979 | TNFRSF11A | FALSE | Upregulated |
| -0,592817883 | 0,020131932 | CCDC158 | FALSE | Downregulated |
| -0,414193715 | 0,020131932 | MTR | FALSE | Downregulated |
| -0,377798829 | 0,020134818 | MBNL3 | FALSE | Downregulated |
| -0,825196694 | 0,020189098 | RP11-480I12.7 | FALSE | Downregulated |
| -0,409064034 | 0,020284196 | ECM2 | FALSE | Downregulated |
| 0,579814717 | 0,020323384 | KBTBD8 | FALSE | Upregulated |
| 0,616208306 | 0,020345688 | CPVL | FALSE | Upregulated |
| -0,364609778 | 0,020346536 | CHD9 | FALSE | Downregulated |
| 0,651635486 | 0,020346536 | RENBP | FALSE | Upregulated |
| 0,210686677 | 0,020346536 | STIM1 | FALSE | Upregulated |
| -0,440478399 | 0,020346561 | GOLGA4 | FALSE | Downregulated |
| 0,601108924 | 0,020354425 | C1orf162 | FALSE | Upregulated |
| 0,998866698 | 0,020373422 | LY9 | FALSE | Upregulated |
| 0,211196233 | 0,020397639 | ANKFY1 | FALSE | Upregulated |
| -0,440768557 | 0,020422559 | PDK2 | FALSE | Downregulated |
| -0,470583156 | 0,020426368 | TIGD7 | FALSE | Downregulated |
| 0,254462776 | 0,020522629 | GUSB | FALSE | Upregulated |
| -0,310175696 | 0,020567264 | CYB5A | FALSE | Downregulated |
| 0,389607709 | 0,02063402 | MAGED1 | FALSE | Upregulated |
| -0,670800483 | 0,02063402 | SMIM2-AS1 | FALSE | Downregulated |
| -0,241678107 | 0,020649809 | ZNF398 | FALSE | Downregulated |
| -0,364727605 | 0,020651631 | ACOX1 | FALSE | Downregulated |
| -0,766409574 | 0,020671325 | TEN1-CDK3 | FALSE | Downregulated |
| -0,309050021 | 0,020720102 | KIAA2026 | FALSE | Downregulated |
| -0,41495751 | 0,020743531 | SLC27A5 | FALSE | Downregulated |
| -0,724444129 | 0,020802151 | FAM227B | FALSE | Downregulated |
| -0,353151429 | 0,020802151 | ZSCAN26 | FALSE | Downregulated |
| 0,224776761 | 0,020856689 | TSG101 | FALSE | Upregulated |
| -1,322115833 | 0,020884023 | LOC101060212 | TRUE | Downregulated |
| -1,191025116 | 0,020944126 | LINC00346 | TRUE | Downregulated |
| 0,275364272 | 0,020955303 | CC2D1A | FALSE | Upregulated |
| -0,333793233 | 0,020966839 | CBR4 | FALSE | Downregulated |
| -0,335032367 | 0,021050973 | SLC2A2 | FALSE | Downregulated |
| -0,177931496 | 0,02107182 | CSNK2A1 | FALSE | Downregulated |
| -0,2999228 | 0,021075326 | NMD3 | FALSE | Downregulated |
| -0,516673348 | 0,02108919 | CEBPA-AS1 | FALSE | Downregulated |
| -2,047639943 | 0,021092443 | PHOSPHO1 | TRUE | Downregulated |
| -0,351657896 | 0,021110523 | CCS | FALSE | Downregulated |
| 0,375738814 | 0,021174726 | CD40 | FALSE | Upregulated |
| 0,758943803 | 0,021174726 | CEACAM21 | FALSE | Upregulated |
| -0,328372587 | 0,021174726 | ZNF343 | FALSE | Downregulated |
| 0,343240274 | 0,021220438 | CALM2 | FALSE | Upregulated |
| 0,660843263 | 0,021220438 | LPCAT4 | FALSE | Upregulated |
| 1,778882346 | 0,021296192 | SLCO4A1 | TRUE | Upregulated |
| 0,331225673 | 0,021325456 | NADSYN1 | FALSE | Upregulated |

| | | | | |
|---|---|---|---|---|
| -0,856037301 | 0,021358414 | CSRNP3 | FALSE | Downregulated |
| -0,953106848 | 0,021371838 | LOC644794 | FALSE | Downregulated |
| -0,577036337 | 0,021397789 | NUDT13 | FALSE | Downregulated |
| 0,464105652 | 0,02141351 | MAP1LC3A | FALSE | Upregulated |
| -0,540947891 | 0,02141351 | ZBED6 | FALSE | Downregulated |
| 0,264924022 | 0,021436671 | PPP2R1A | FALSE | Upregulated |
| -0,856085289 | 0,021456673 | SH3PXD2A | FALSE | Downregulated |
| 0,771074336 | 0,021472754 | POU2F2 | FALSE | Upregulated |
| 0,352996998 | 0,021480451 | RRP1 | FALSE | Upregulated |
| 0,352178924 | 0,021482159 | CHPF2 | FALSE | Upregulated |
| 0,273025358 | 0,021490203 | CDK5RAP1 | FALSE | Upregulated |
| 2,221402237 | 0,02149629 | PHLDA2 | TRUE | Upregulated |
| 2,123585427 | 0,02149629 | SEPT5-GP1BB | TRUE | Upregulated |
| -0,450000156 | 0,021539188 | ACAA1 | FALSE | Downregulated |
| -0,311414373 | 0,02155456 | KLHL15 | FALSE | Downregulated |
| -0,687150773 | 0,02155456 | LINC00886 | FALSE | Downregulated |
| -0,490187436 | 0,021574917 | HOGA1 | FALSE | Downregulated |
| -0,453386716 | 0,021579638 | TNRC6C | FALSE | Downregulated |
| 0,190633119 | 0,021596678 | PPP1R7 | FALSE | Upregulated |
| 0,600512397 | 0,021642858 | HERC6 | FALSE | Upregulated |
| -0,270900761 | 0,021705732 | CSNK2A2 | FALSE | Downregulated |
| 0,855745466 | 0,02179573 | TAGAP | FALSE | Upregulated |
| 0,688044524 | 0,021807342 | FGD3 | FALSE | Upregulated |
| 0,383813641 | 0,021830855 | ADCK2 | FALSE | Upregulated |
| -0,325864721 | 0,021835054 | CYP4F11 | FALSE | Downregulated |
| -0,451512286 | 0,021842512 | EPB41L4B | FALSE | Downregulated |
| -0,170213346 | 0,0218664 | NUDT21 | FALSE | Downregulated |
| 0,695782491 | 0,021916353 | DSCC1 | FALSE | Upregulated |
| -0,263775918 | 0,022026132 | RAB11FIP2 | FALSE | Downregulated |
| -0,704417055 | 0,022060999 | AC016629.8 | FALSE | Downregulated |
| -2,671854051 | 0,022060999 | WNK4 | TRUE | Downregulated |
| 0,597990594 | 0,022088785 | TNFAIP2 | FALSE | Upregulated |
| -0,474525721 | 0,022167281 | PLA2G16 | FALSE | Downregulated |
| -0,739395841 | 0,022183224 | IL27 | FALSE | Downregulated |
| -0,307824752 | 0,02227656 | ERMAP | FALSE | Downregulated |
| -1,325357309 | 0,02227656 | LOC101927610 | TRUE | Downregulated |
| -0,398972226 | 0,022317192 | HECTD4 | FALSE | Downregulated |
| -0,720554499 | 0,022369069 | GATA6-AS1 | FALSE | Downregulated |
| -0,474679912 | 0,022369069 | TEK | FALSE | Downregulated |
| -0,370522836 | 0,02244254 | ASTE1 | FALSE | Downregulated |
| 0,284462076 | 0,02244254 | DRAM2 | FALSE | Upregulated |
| 0,685658081 | 0,02244254 | KIF21B | FALSE | Upregulated |
| 0,318196916 | 0,022515457 | POMGNT1 | FALSE | Upregulated |
| 0,396722622 | 0,022515457 | RBM45 | FALSE | Upregulated |
| -0,30033685 | 0,022522584 | SIRT3 | FALSE | Downregulated |
| 0,362248194 | 0,022526223 | ACP2 | FALSE | Upregulated |
| 1,179178428 | 0,02253294 | SCD | TRUE | Upregulated |
| 0,356347126 | 0,022584471 | EFHD2 | FALSE | Upregulated |
| 0,236721523 | 0,022589293 | SET | FALSE | Upregulated |
| 0,353551682 | 0,022654003 | EXOC6 | FALSE | Upregulated |
| -0,393752809 | 0,022692282 | BCL7A | FALSE | Downregulated |

| | | | | |
|---|---|---|---|---|
| -1,426123011 | 0,022692282 | LINC01132 | TRUE | Downregulated |
| -1,074280415 | 0,022692282 | RAMP2-AS1 | TRUE | Downregulated |
| -0,786548792 | 0,022692282 | STL | FALSE | Downregulated |
| 0,538683668 | 0,022702236 | RASSF2 | FALSE | Upregulated |
| 0,869719988 | 0,022706666 | SLC2A1 | FALSE | Upregulated |
| 1,162333688 | 0,0227225 | PHLDA3 | TRUE | Upregulated |
| -0,28448123 | 0,022812828 | IMMP2L | FALSE | Downregulated |
| -0,63232736 | 0,022825626 | FSTL1 | FALSE | Downregulated |
| 0,790820627 | 0,022835924 | PLEKHG7 | FALSE | Upregulated |
| -0,390513557 | 0,022883222 | CLOCK | FALSE | Downregulated |
| 0,37301029 | 0,022939369 | RAP2A | FALSE | Upregulated |
| 0,413520175 | 0,022939369 | SUV39H2 | FALSE | Upregulated |
| 0,411034788 | 0,022942604 | TP53BP2 | FALSE | Upregulated |
| 0,379784759 | 0,022965629 | MBNL2 | FALSE | Upregulated |
| -0,455011196 | 0,022965629 | PAXIP1-AS1 | FALSE | Downregulated |
| 0,789592911 | 0,022971074 | GPR137C | FALSE | Upregulated |
| 0,765849032 | 0,022972581 | VSIG4 | FALSE | Upregulated |
| 0,286725213 | 0,023022411 | AGPAT6 | FALSE | Upregulated |
| 0,252435 | 0,02306619 | UBE2N | FALSE | Upregulated |
| -0,407099022 | 0,023109637 | CSPP1 | FALSE | Downregulated |
| 0,760701309 | 0,023132956 | TPM1 | FALSE | Upregulated |
| 0,810904914 | 0,023172272 | TRAIP | FALSE | Upregulated |
| -0,487027741 | 0,023190792 | GNE | FALSE | Downregulated |
| -0,366192011 | 0,023237449 | GNG7 | FALSE | Downregulated |
| 1,072513805 | 0,023270329 | HES4 | TRUE | Upregulated |
| -0,91605195 | 0,023270329 | LOC283922 | FALSE | Downregulated |
| -1,54340446 | 0,023300048 | DDX25 | TRUE | Downregulated |
| 0,434139059 | 0,02333325 | GNG2 | FALSE | Upregulated |
| -0,50900164 | 0,02333325 | ZNF846 | FALSE | Downregulated |
| -0,734579831 | 0,023352132 | EDA | FALSE | Downregulated |
| -0,453921822 | 0,023352132 | PAXBP1 | FALSE | Downregulated |
| 0,682129497 | 0,023573456 | SMOX | FALSE | Upregulated |
| 0,883441555 | 0,023662975 | SIRPB2 | FALSE | Upregulated |
| -0,26716695 | 0,023683263 | SIKE1 | FALSE | Downregulated |
| 0,915992764 | 0,023701363 | LOC154761 | FALSE | Upregulated |
| -0,444791756 | 0,023740271 | FRRS1 | FALSE | Downregulated |
| 0,48382849 | 0,023800052 | PPIL1 | FALSE | Upregulated |
| -0,672258513 | 0,023802134 | PSMD5-AS1 | FALSE | Downregulated |
| -1,963822655 | 0,023813124 | RFPL2 | TRUE | Downregulated |
| -0,384094876 | 0,023814445 | SMARCA1 | FALSE | Downregulated |
| 0,341226771 | 0,02385586 | CNN3 | FALSE | Upregulated |
| -0,214738013 | 0,023864341 | SMAD4 | FALSE | Downregulated |
| 0,8202246 | 0,023869855 | MOB3B | FALSE | Upregulated |
| -0,421106201 | 0,023937889 | NBPF14 | FALSE | Downregulated |
| -1,911191534 | 0,023995708 | DPP10 | TRUE | Downregulated |
| -0,862819542 | 0,024172345 | LOC100287497 | FALSE | Downregulated |
| -0,432337121 | 0,024176326 | BAZ2B | FALSE | Downregulated |
| -1,199929239 | 0,024232404 | CTD-3080P12.3 | TRUE | Downregulated |
| 0,427134722 | 0,024232404 | RHOBTB2 | FALSE | Upregulated |
| 0,810731594 | 0,024238953 | CTSW | FALSE | Upregulated |
| -0,270486746 | 0,024251086 | CEP70 | FALSE | Downregulated |

| | | | | |
|---|---|---|---|---|
| 0,274708175 | 0,024251086 | COPS3 | FALSE | Upregulated |
| -0,347251335 | 0,024258624 | MEST | FALSE | Downregulated |
| 1,968800326 | 0,024285594 | KRT8P41 | TRUE | Upregulated |
| -0,275103096 | 0,024285594 | RSBN1 | FALSE | Downregulated |
| -0,317345953 | 0,024385392 | LRRC58 | FALSE | Downregulated |
| -0,966232351 | 0,024385392 | MOGAT1 | FALSE | Downregulated |
| -0,955044586 | 0,024405376 | C9orf96 | FALSE | Downregulated |
| -0,367400622 | 0,024405376 | CPSF7 | FALSE | Downregulated |
| -0,539422704 | 0,024405376 | LMLN | FALSE | Downregulated |
| 0,405405456 | 0,024405376 | LPGAT1 | FALSE | Upregulated |
| -0,638216011 | 0,024444972 | HGD | FALSE | Downregulated |
| 0,322423749 | 0,024466563 | DOCK9 | FALSE | Upregulated |
| 2,380539781 | 0,024466563 | LOC102724385 | TRUE | Upregulated |
| 0,37335402 | 0,024466563 | SWAP70 | FALSE | Upregulated |
| -0,353824522 | 0,024476162 | PPP3CA | FALSE | Downregulated |
| -0,259940982 | 0,024507379 | MSL2 | FALSE | Downregulated |
| 0,884229696 | 0,024507379 | OSCAR | FALSE | Upregulated |
| -0,533569276 | 0,024507379 | RP11-305E17.6 | FALSE | Downregulated |
| 0,286640644 | 0,024541049 | SLC25A43 | FALSE | Upregulated |
| -1,002546284 | 0,024585869 | LOC100996419 | TRUE | Downregulated |
| 0,82787529 | 0,02463865 | FMN1 | FALSE | Upregulated |
| -0,186971432 | 0,024683694 | PUM1 | FALSE | Downregulated |
| -0,301993445 | 0,024726897 | PMM1 | FALSE | Downregulated |
| 0,924161784 | 0,024739636 | C10orf35 | FALSE | Upregulated |
| -0,469097428 | 0,024739636 | LOC100130744 | FALSE | Downregulated |
| -0,603245446 | 0,024739636 | SDCBP2-AS1 | FALSE | Downregulated |
| -0,952930828 | 0,024739636 | SOAT2 | FALSE | Downregulated |
| 0,279638582 | 0,024799507 | ITFG3 | FALSE | Upregulated |
| 1,516197547 | 0,024800406 | LINC00925 | TRUE | Upregulated |
| -4,703386468 | 0,024800406 | SERPINB2 | TRUE | Downregulated |
| 0,734937032 | 0,024802809 | TRAF3IP3 | FALSE | Upregulated |
| 1,214416408 | 0,024820205 | HCAR3 | TRUE | Upregulated |
| 0,701610043 | 0,024830663 | FCHO1 | FALSE | Upregulated |
| 0,637366875 | 0,024830663 | SLC25A19 | FALSE | Upregulated |
| -0,719229037 | 0,024850085 | ACSM2A | FALSE | Downregulated |
| 0,455095499 | 0,02488998 | RBM3 | FALSE | Upregulated |
| -0,47977685 | 0,024912986 | ADH6 | FALSE | Downregulated |
| 0,472028793 | 0,024912986 | HCLS1 | FALSE | Upregulated |
| -1,09716083 | 0,024912986 | LRRC16B | TRUE | Downregulated |
| 0,511201748 | 0,024912986 | MID1IP1 | FALSE | Upregulated |
| 0,33219728 | 0,024949185 | NOP10 | FALSE | Upregulated |
| 1,841023921 | 0,024953621 | CTSE | TRUE | Upregulated |
| 1,240457454 | 0,024953621 | MTMR11 | TRUE | Upregulated |
| 0,609037381 | 0,024970973 | EPB41L3 | FALSE | Upregulated |
| -0,416497477 | 0,025039897 | RAB40B | FALSE | Downregulated |
| -0,207682663 | 0,025042815 | MIEF1 | FALSE | Downregulated |
| 1,291077012 | 0,025102895 | CD80 | TRUE | Upregulated |
| 0,687623441 | 0,025102895 | ZMAT3 | FALSE | Upregulated |
| 2,2501049 | 0,025111761 | CIDEC | TRUE | Upregulated |
| 0,343475971 | 0,025147797 | MYCBP | FALSE | Upregulated |
| -1,787696446 | 0,025205677 | APLN | TRUE | Downregulated |

| | | | | |
|---|---|---|---|---|
| 0,511814562 | 0,025210723 | ARHGEF5 | FALSE | Upregulated |
| -0,245526347 | 0,025266332 | GID8 | FALSE | Downregulated |
| -0,798176649 | 0,025277537 | PFN2 | FALSE | Downregulated |
| -0,391944646 | 0,02530083 | ZNF551 | FALSE | Downregulated |
| -0,597170436 | 0,025358352 | GPR125 | FALSE | Downregulated |
| 1,45144384 | 0,025358352 | LOC100506411 | TRUE | Upregulated |
| -0,219471453 | 0,025407011 | IMPACT | FALSE | Downregulated |
| -2,691874013 | 0,02544569 | HSPB7 | TRUE | Downregulated |
| 0,982352671 | 0,02545289 | CIT | FALSE | Upregulated |
| -0,331762073 | 0,025476474 | REV1 | FALSE | Downregulated |
| 1,252844101 | 0,02549144 | LOC101930071 | TRUE | Upregulated |
| 0,621781916 | 0,02549144 | NLRC3 | FALSE | Upregulated |
| -0,267003158 | 0,02549144 | PSMD5 | FALSE | Downregulated |
| 1,037098806 | 0,025517013 | GPSM2 | TRUE | Upregulated |
| 0,572400045 | 0,025522021 | ZNF341 | FALSE | Upregulated |
| 0,403539712 | 0,025540757 | GOSR2 | FALSE | Upregulated |
| -0,612298096 | 0,025546857 | ARMCX4 | FALSE | Downregulated |
| 0,361951578 | 0,025546857 | RBM15 | FALSE | Upregulated |
| 0,311488498 | 0,025546857 | ZFYVE27 | FALSE | Upregulated |
| -1,2743398 | 0,025564249 | LOC102724603 | TRUE | Downregulated |
| -0,623399058 | 0,025578079 | STAB2 | FALSE | Downregulated |
| 1,072576987 | 0,025592202 | HRH2 | TRUE | Upregulated |
| -1,425221714 | 0,025595378 | PRIMA1 | TRUE | Downregulated |
| 0,299581903 | 0,025634646 | ALG12 | FALSE | Upregulated |
| 1,98035181 | 0,025634646 | YPEL4 | TRUE | Upregulated |
| -0,481347037 | 0,025646065 | DUSP19 | FALSE | Downregulated |
| -0,328739715 | 0,0256505 | FRMD4B | FALSE | Downregulated |
| 0,27753657 | 0,025662907 | SLC15A4 | FALSE | Upregulated |
| -0,830636768 | 0,025672365 | MIR600HG | FALSE | Downregulated |
| 0,250862832 | 0,025700438 | COMMD9 | FALSE | Upregulated |
| 0,413603901 | 0,025716123 | GYPC | FALSE | Upregulated |
| -0,247966056 | 0,02571838 | N4BP2L2 | FALSE | Downregulated |
| 0,245882427 | 0,025754483 | DENND1A | FALSE | Upregulated |
| -1,593400528 | 0,025834481 | LOC101929658 | TRUE | Downregulated |
| 1,031796595 | 0,025834481 | SAMD11 | TRUE | Upregulated |
| -1,498970633 | 0,025847768 | LOC102723356 | TRUE | Downregulated |
| -0,535503848 | 0,025848565 | FLJ37453 | FALSE | Downregulated |
| -0,398247427 | 0,025848565 | KANSL1L | FALSE | Downregulated |
| 0,303525788 | 0,025848565 | RNF34 | FALSE | Upregulated |
| 0,348853594 | 0,025902206 | ELMO2 | FALSE | Upregulated |
| -0,406388719 | 0,025971847 | ADAMTSL3 | FALSE | Downregulated |
| -1,25377648 | 0,025974904 | SLC28A2 | TRUE | Downregulated |
| 1,018786455 | 0,025991773 | ATP8B3 | TRUE | Upregulated |
| 1,077740132 | 0,025991773 | C1orf51 | TRUE | Upregulated |
| 0,273542148 | 0,025998187 | DENND5A | FALSE | Upregulated |
| -0,588703228 | 0,026007407 | EPB41L4A-AS1 | FALSE | Downregulated |
| -1,52874725 | 0,026008198 | DIO3OS | TRUE | Downregulated |
| 1,007138112 | 0,026014351 | HLA-L | TRUE | Upregulated |
| 0,379138703 | 0,026014351 | SURF2 | FALSE | Upregulated |
| -0,413268857 | 0,026018322 | POLM | FALSE | Downregulated |
| -0,343235294 | 0,026020104 | ZNF654 | FALSE | Downregulated |

| | | | | |
|---|---|---|---|---|
| -0,352339929 | 0,026099809 | HEATR5B | FALSE | Downregulated |
| -1,127801095 | 0,026113078 | LOC101929055 | TRUE | Downregulated |
| -1,090938758 | 0,026228812 | IDO2 | TRUE | Downregulated |
| 0,344627618 | 0,026228812 | ITGB1BP1 | FALSE | Upregulated |
| 0,491607224 | 0,026228812 | RAB3IP | FALSE | Upregulated |
| 0,620588897 | 0,026315023 | SPATA24 | FALSE | Upregulated |
| -1,375076764 | 0,026328559 | CYP2C19 | TRUE | Downregulated |
| 0,502857411 | 0,026406206 | FAM86A | FALSE | Upregulated |
| 0,852635095 | 0,026406206 | PLP2 | FALSE | Upregulated |
| -0,668872694 | 0,026408414 | IL1RL2 | FALSE | Downregulated |
| 0,217960785 | 0,026503001 | PUF60 | FALSE | Upregulated |
| -0,410948929 | 0,026508378 | EYA3 | FALSE | Downregulated |
| -0,688384318 | 0,026508378 | FAM124A | FALSE | Downregulated |
| -0,310844219 | 0,026546044 | ALS2 | FALSE | Downregulated |
| 0,312330193 | 0,026546044 | CD2BP2 | FALSE | Upregulated |
| -0,439489567 | 0,026546044 | ZCWPW1 | FALSE | Downregulated |
| -0,768702151 | 0,026553735 | GOLGA2P5 | FALSE | Downregulated |
| 0,660799579 | 0,026558288 | CHTF18 | FALSE | Upregulated |
| -0,287073327 | 0,026558288 | KIAA1586 | FALSE | Downregulated |
| -0,480532891 | 0,026565915 | AKAP17A | FALSE | Downregulated |
| -1,928870878 | 0,026565915 | MEIOB | TRUE | Downregulated |
| 0,254040834 | 0,026575364 | HCCS | FALSE | Upregulated |
| -1,857821033 | 0,026592164 | CPO | TRUE | Downregulated |
| -0,592230821 | 0,026592164 | HMCN1 | FALSE | Downregulated |
| 0,391786822 | 0,026629879 | HMGN4 | FALSE | Upregulated |
| -0,279450595 | 0,026651419 | TBP | FALSE | Downregulated |
| -0,924362995 | 0,026676833 | CAPN3 | FALSE | Downregulated |
| 0,605885117 | 0,026676833 | GGT1 | FALSE | Upregulated |
| -0,307337372 | 0,026676833 | HSCB | FALSE | Downregulated |
| -0,254635092 | 0,026676833 | MGEA5 | FALSE | Downregulated |
| 0,313108287 | 0,026680505 | MCU | FALSE | Upregulated |
| -1,00821903 | 0,026714175 | SEMA5B | TRUE | Downregulated |
| 0,236109785 | 0,026782331 | MFSD1 | FALSE | Upregulated |
| -0,799370926 | 0,026859099 | LOC101928230 | FALSE | Downregulated |
| -0,69895491 | 0,026867424 | HAO2 | FALSE | Downregulated |
| -0,39572532 | 0,02688622 | SOWAHB | FALSE | Downregulated |
| 1,40781711 | 0,026921684 | LOC101928576 | TRUE | Upregulated |
| -0,628184656 | 0,026934293 | ABHD1 | FALSE | Downregulated |
| 0,692677501 | 0,026985199 | C19orf40 | FALSE | Upregulated |
| -0,25693085 | 0,02701284 | CHRAC1 | FALSE | Downregulated |
| -0,504614239 | 0,027030529 | PSD3 | FALSE | Downregulated |
| 0,873265693 | 0,027030529 | PTGER2 | FALSE | Upregulated |
| 0,694068908 | 0,027030529 | S100A4 | FALSE | Upregulated |
| 0,35127097 | 0,027047617 | ATP6V0C | FALSE | Upregulated |
| -0,521678425 | 0,027064245 | ANKRD24 | FALSE | Downregulated |
| 0,657592527 | 0,027141548 | EVI2B | FALSE | Upregulated |
| -0,528164683 | 0,027148658 | BCL6 | FALSE | Downregulated |
| -0,225748552 | 0,027165042 | HARS2 | FALSE | Downregulated |
| 1,100617892 | 0,027165042 | RAB42 | TRUE | Upregulated |
| -0,451809742 | 0,027177626 | KIAA1217 | FALSE | Downregulated |
| -0,189414324 | 0,027245099 | SENP6 | FALSE | Downregulated |

| | | | | |
|---|---|---|---|---|
| -0,337197816 | 0,027270794 | RRNAD1 | FALSE | Downregulated |
| 0,974140249 | 0,027292717 | FZD7 | FALSE | Upregulated |
| 0,520026793 | 0,027352997 | USB1 | FALSE | Upregulated |
| 0,255252676 | 0,027372627 | LMF2 | FALSE | Upregulated |
| -1,916080762 | 0,027372627 | LOC102724921 | TRUE | Downregulated |
| -0,578962095 | 0,027429748 | ZNF709 | FALSE | Downregulated |
| 0,613082792 | 0,027445655 | MOGAT3 | FALSE | Upregulated |
| 0,854586858 | 0,02747864 | DGCR11 | FALSE | Upregulated |
| -1,805715012 | 0,027488148 | SATL1 | TRUE | Downregulated |
| 0,185041867 | 0,02752364 | EXOC7 | FALSE | Upregulated |
| -0,358464531 | 0,027542601 | BIVM | FALSE | Downregulated |
| -2,353594637 | 0,027548141 | PI16 | TRUE | Downregulated |
| 1,786048804 | 0,027548141 | TSPAN10 | TRUE | Upregulated |
| -0,373291875 | 0,027573244 | TTC30A | FALSE | Downregulated |
| -1,147029967 | 0,027592009 | DRD1 | TRUE | Downregulated |
| -0,316053373 | 0,027615611 | ZEB1 | FALSE | Downregulated |
| -1,199400913 | 0,027705017 | FLJ37201 | TRUE | Downregulated |
| 2,124795077 | 0,027716986 | TFAP2E | TRUE | Upregulated |
| 1,53168964 | 0,027757929 | GRIK4 | TRUE | Upregulated |
| 0,336946787 | 0,027757929 | LNX2 | FALSE | Upregulated |
| 0,280298269 | 0,027757929 | PPP2R4 | FALSE | Upregulated |
| 1,539319731 | 0,027792265 | KEL | TRUE | Upregulated |
| 0,426829921 | 0,027803249 | GOLT1A | FALSE | Upregulated |
| -0,471062806 | 0,027815665 | PCDH1 | FALSE | Downregulated |
| -0,404923984 | 0,027834507 | GPATCH1 | FALSE | Downregulated |
| 0,70509554 | 0,0278613 | IL2RB | FALSE | Upregulated |
| 0,269956538 | 0,027868082 | DYNLRB1 | FALSE | Upregulated |
| -2,095384116 | 0,027903135 | HAO2-IT1 | TRUE | Downregulated |
| 0,518359299 | 0,027983109 | IL10RA | FALSE | Upregulated |
| 0,278258186 | 0,027999045 | MUL1 | FALSE | Upregulated |
| 0,496406354 | 0,028017807 | MBL2 | FALSE | Upregulated |
| 0,41680377 | 0,0280596 | CSF1 | FALSE | Upregulated |
| -0,801485903 | 0,0280596 | LOC100507316 | FALSE | Downregulated |
| 1,304196839 | 0,028061507 | RGS16 | TRUE | Upregulated |
| -0,803036421 | 0,028070019 | NREP | FALSE | Downregulated |
| 2,479369717 | 0,028079597 | DPEP1 | TRUE | Upregulated |
| 0,214510799 | 0,028079597 | SMARCD1 | FALSE | Upregulated |
| 0,263885592 | 0,028079597 | ZFYVE26 | FALSE | Upregulated |
| 0,301595682 | 0,028084502 | RRM1 | FALSE | Upregulated |
| 0,8216051 | 0,028089714 | IGSF23 | FALSE | Upregulated |
| -0,364825342 | 0,028089714 | MUT | FALSE | Downregulated |
| 0,390587711 | 0,028122279 | RPL35 | FALSE | Upregulated |
| -0,541347548 | 0,028173377 | LDLRAD4 | FALSE | Downregulated |
| 0,320393088 | 0,028226377 | IFRD2 | FALSE | Upregulated |
| 0,315251518 | 0,028232204 | CRTC1 | FALSE | Upregulated |
| -2,272661619 | 0,028232204 | LINC01118 | TRUE | Downregulated |
| 0,713768293 | 0,028232204 | SERPINE2 | FALSE | Upregulated |
| -0,415788933 | 0,028232204 | SLC2A11 | FALSE | Downregulated |
| -0,299318709 | 0,028277731 | ESCO1 | FALSE | Downregulated |
| 0,311990942 | 0,028331719 | GLTP | FALSE | Upregulated |
| 0,371611469 | 0,02840378 | POP1 | FALSE | Upregulated |

| | | | | |
|---|---|---|---|---|
| 1,013199313 | 0,028405373 | GPR18 | TRUE | Upregulated |
| 0,259091344 | 0,028405961 | DDRGK1 | FALSE | Upregulated |
| 0,378851535 | 0,028407907 | N6AMT1 | FALSE | Upregulated |
| -0,311829027 | 0,028450344 | IFT122 | FALSE | Downregulated |
| -0,209456203 | 0,028450344 | ZNF268 | FALSE | Downregulated |
| 0,487717428 | 0,028513075 | CORO7 | FALSE | Upregulated |
| -0,319514487 | 0,02861753 | C14orf28 | FALSE | Downregulated |
| 0,238603357 | 0,02861753 | CBX5 | FALSE | Upregulated |
| 0,304343592 | 0,028649494 | IAH1 | FALSE | Upregulated |
| -0,232968673 | 0,028663947 | SOS2 | FALSE | Downregulated |
| 0,408855115 | 0,028663947 | SSBP3 | FALSE | Upregulated |
| -0,362494334 | 0,028725339 | MTMR10 | FALSE | Downregulated |
| 0,387094982 | 0,028745426 | CADM1 | FALSE | Upregulated |
| 0,266406535 | 0,028752381 | SMAD2 | FALSE | Upregulated |
| -0,351266905 | 0,028776932 | ZSWIM8 | FALSE | Downregulated |
| 0,390841244 | 0,028836938 | BOLA3 | FALSE | Upregulated |
| -1,352123896 | 0,028856578 | ADAM22 | TRUE | Downregulated |
| 3,005179493 | 0,02891685 | IRG1 | TRUE | Upregulated |
| 0,7187093 | 0,028936992 | PTGER4 | FALSE | Upregulated |
| 0,283272689 | 0,028952547 | KPNA3 | FALSE | Upregulated |
| -0,32539109 | 0,028987179 | RBM5 | FALSE | Downregulated |
| 0,359554829 | 0,029006219 | ATOX1 | FALSE | Upregulated |
| 0,488246689 | 0,029102341 | BASP1 | FALSE | Upregulated |
| 0,65758758 | 0,029170065 | RCAN3 | FALSE | Upregulated |
| 0,250630608 | 0,029190396 | VBP1 | FALSE | Upregulated |
| -0,508506539 | 0,029283633 | SNX16 | FALSE | Downregulated |
| -0,382971506 | 0,029288878 | TTC33 | FALSE | Downregulated |
| -0,864923457 | 0,029311083 | LINC00641 | FALSE | Downregulated |
| -1,382378221 | 0,029330108 | PCDH19 | TRUE | Downregulated |
| -0,51089019 | 0,029346285 | DNASE1L3 | FALSE | Downregulated |
| -0,291944161 | 0,029354759 | TMEM41B | FALSE | Downregulated |
| -0,336643752 | 0,029380758 | CENPC | FALSE | Downregulated |
| 0,54502847 | 0,029425442 | IFI27L2 | FALSE | Upregulated |
| 0,328927671 | 0,029425442 | OLA1 | FALSE | Upregulated |
| 0,639694027 | 0,029484841 | ALOX5 | FALSE | Upregulated |
| 0,260124475 | 0,029485585 | SUMF2 | FALSE | Upregulated |
| 0,390159906 | 0,029496194 | FRMD8 | FALSE | Upregulated |
| -2,154853943 | 0,029509963 | DKFZp434J0226 | TRUE | Downregulated |
| 0,873950932 | 0,029521634 | TLR10 | FALSE | Upregulated |
| 0,293770126 | 0,029540719 | ICMT | FALSE | Upregulated |
| -1,193787484 | 0,029557772 | EPHB1 | TRUE | Downregulated |
| -0,333019955 | 0,029589477 | CD302 | FALSE | Downregulated |
| 0,27180275 | 0,029591805 | PSMB3 | FALSE | Upregulated |
| 0,434823317 | 0,029616547 | RAPGEF6 | FALSE | Upregulated |
| 0,750272427 | 0,029631266 | LOC115110 | FALSE | Upregulated |
| 1,394567016 | 0,029637951 | C16orf98 | TRUE | Upregulated |
| -0,277217832 | 0,029637951 | DCAF17 | FALSE | Downregulated |
| -0,782111045 | 0,029655265 | LOC101927509 | FALSE | Downregulated |
| 0,468031243 | 0,029691219 | SRM | FALSE | Upregulated |
| -0,991614082 | 0,029755787 | BBOX1 | FALSE | Downregulated |
| -1,293939149 | 0,029816339 | AGBL4 | TRUE | Downregulated |

| | | | | |
|---|---|---|---|---|
| 0,296410286 | 0,029847604 | SLC44A1 | FALSE | Upregulated |
| -0,231863287 | 0,029854004 | EXOSC2 | FALSE | Downregulated |
| -0,400240487 | 0,029903979 | CYP4V2 | FALSE | Downregulated |
| 1,422455832 | 0,029948973 | SHISA3 | TRUE | Upregulated |
| -0,755845931 | 0,029948973 | ZNF530 | FALSE | Downregulated |
| 0,403741649 | 0,030057097 | IFNGR2 | FALSE | Upregulated |
| 0,547399141 | 0,030057742 | SLFN11 | FALSE | Upregulated |
| 0,253054281 | 0,030085835 | LAMP1 | FALSE | Upregulated |
| -0,912239386 | 0,030085835 | SULT1E1 | FALSE | Downregulated |
| 0,501146083 | 0,030168662 | GIMAP1-GIMAP5 | FALSE | Upregulated |
| -0,480748192 | 0,030219928 | KLF12 | FALSE | Downregulated |
| 0,893726348 | 0,030228885 | GIMAP5 | FALSE | Upregulated |
| -0,633245815 | 0,030259608 | UGDH-AS1 | FALSE | Downregulated |
| -0,611936025 | 0,030302857 | LINC00910 | FALSE | Downregulated |
| 0,347946242 | 0,030302857 | RPS12 | FALSE | Upregulated |
| -0,475969169 | 0,030356814 | QPCTL | FALSE | Downregulated |
| 1,476100432 | 0,030411771 | BCAS1 | TRUE | Upregulated |
| 1,786587517 | 0,030451437 | ANKRD36BP2 | TRUE | Upregulated |
| -0,533610261 | 0,030451437 | TSLP | FALSE | Downregulated |
| 0,813156331 | 0,030476666 | DKFZp779M0652 | FALSE | Upregulated |
| -0,39992859 | 0,030476947 | MAGI2-AS3 | FALSE | Downregulated |
| 0,485533863 | 0,030495719 | GMPR | FALSE | Upregulated |
| -1,406656615 | 0,030495719 | LRIT3 | TRUE | Downregulated |
| -0,223840463 | 0,030495719 | TAB3 | FALSE | Downregulated |
| -0,191866957 | 0,030513658 | CAPN7 | FALSE | Downregulated |
| -0,260304023 | 0,030513658 | NR2C2 | FALSE | Downregulated |
| 0,339179782 | 0,030513658 | PLGRKT | FALSE | Upregulated |
| -1,000966071 | 0,030532535 | FLJ13197 | TRUE | Downregulated |
| -2,098111383 | 0,030532535 | RP11-436I24.1 | TRUE | Downregulated |
| -0,561773657 | 0,030550822 | LOC102723701 | FALSE | Downregulated |
| -0,311679393 | 0,030550822 | SLC25A26 | FALSE | Downregulated |
| -0,258671312 | 0,030553418 | CCDC28A | FALSE | Downregulated |
| 0,294395032 | 0,030558079 | ASPSCR1 | FALSE | Upregulated |
| 0,223024523 | 0,030560031 | ENOPH1 | FALSE | Upregulated |
| -0,370937307 | 0,030560031 | TRMT2A | FALSE | Downregulated |
| -0,442704753 | 0,030567061 | APOF | FALSE | Downregulated |
| 0,392174621 | 0,030630452 | TIFA | FALSE | Upregulated |
| 1,143925321 | 0,030630452 | WDR62 | TRUE | Upregulated |
| 3,482935849 | 0,030791567 | CHIT1 | TRUE | Upregulated |
| 0,657300476 | 0,03079936 | EPDR1 | FALSE | Upregulated |
| -0,434438024 | 0,030849448 | HSD17B8 | FALSE | Downregulated |
| -0,189471598 | 0,030929559 | CSDE1 | FALSE | Downregulated |
| -0,330694739 | 0,03094442 | FAM172A | FALSE | Downregulated |
| 0,408587167 | 0,030978535 | GIMAP6 | FALSE | Upregulated |
| 0,268009544 | 0,030994109 | APEH | FALSE | Upregulated |
| 0,232858342 | 0,031014264 | GTF2B | FALSE | Upregulated |
| 0,445152323 | 0,031089461 | OGFRL1 | FALSE | Upregulated |
| -0,248834089 | 0,031108468 | FBXO38 | FALSE | Downregulated |
| -0,286085451 | 0,031153553 | CDKN1B | FALSE | Downregulated |
| 1,79236627 | 0,031153553 | LOC102724907 | TRUE | Upregulated |
| 1,407400103 | 0,031193172 | PCDH20 | TRUE | Upregulated |

| | | | | |
|---|---|---|---|---|
| 0,358152657 | 0,031248796 | DCUN1D5 | FALSE | Upregulated |
| -0,62584348 | 0,031280968 | CDKL3 | FALSE | Downregulated |
| 0,89165573 | 0,03130796 | SIRPB1 | FALSE | Upregulated |
| -0,456077459 | 0,031312326 | GRAMD1C | FALSE | Downregulated |
| -0,224107134 | 0,031312587 | FBXL17 | FALSE | Downregulated |
| 0,214516961 | 0,031335856 | PLAA | FALSE | Upregulated |
| 0,745738735 | 0,031335856 | TMEM156 | FALSE | Upregulated |
| 1,528413565 | 0,031360647 | LOC102724850 | TRUE | Upregulated |
| 1,83115051 | 0,03136819 | GCNT3 | TRUE | Upregulated |
| 0,403853046 | 0,031384946 | PRKCH | FALSE | Upregulated |
| -0,305864354 | 0,031412129 | LYRM5 | FALSE | Downregulated |
| -0,223968909 | 0,031412129 | MAP4K3 | FALSE | Downregulated |
| 0,620132556 | 0,031432092 | FLNA | FALSE | Upregulated |
| -1,030905165 | 0,03143244 | LOC101928816 | TRUE | Downregulated |
| 2,480429305 | 0,031494126 | ARHGAP11B | TRUE | Upregulated |
| 0,252644402 | 0,031494126 | DYNC1LI1 | FALSE | Upregulated |
| -0,610280596 | 0,031494126 | UVSSA | FALSE | Downregulated |
| -0,197230994 | 0,031519722 | PCMTD1 | FALSE | Downregulated |
| 0,757785888 | 0,03153631 | GFI1 | FALSE | Upregulated |
| -0,193727092 | 0,03153631 | SMNDC1 | FALSE | Downregulated |
| -0,906150994 | 0,03154037 | PI4KAP1 | FALSE | Downregulated |
| -0,411123185 | 0,031541894 | TFAP4 | FALSE | Downregulated |
| 0,704734328 | 0,031566268 | TGM2 | FALSE | Upregulated |
| 0,464265133 | 0,031567991 | SH3KBP1 | FALSE | Upregulated |
| -0,743050099 | 0,031621393 | WDR27 | FALSE | Downregulated |
| 0,260259768 | 0,031633104 | EIF2B4 | FALSE | Upregulated |
| 0,346931675 | 0,031633104 | KCTD10 | FALSE | Upregulated |
| 0,366072598 | 0,031637659 | CLEC2B | FALSE | Upregulated |
| 0,937542869 | 0,031637659 | LOC729683 | FALSE | Upregulated |
| 0,618978319 | 0,031637659 | NAT8 | FALSE | Upregulated |
| -0,375406539 | 0,031644832 | RBP5 | FALSE | Downregulated |
| -0,255336415 | 0,031644832 | TAPT1 | FALSE | Downregulated |
| -0,248510362 | 0,031647863 | BCLAF1 | FALSE | Downregulated |
| -0,281883326 | 0,031680496 | KLHDC10 | FALSE | Downregulated |
| 0,457617524 | 0,031680549 | FBXO44 | FALSE | Upregulated |
| -0,451072697 | 0,031707012 | KIAA2018 | FALSE | Downregulated |
| -2,022056564 | 0,031740041 | LOC646903 | TRUE | Downregulated |
| 0,923554819 | 0,031786098 | PLCE1 | FALSE | Upregulated |
| -1,065273065 | 0,031792037 | PLIN1 | TRUE | Downregulated |
| -1,50686857 | 0,031814275 | LRP2 | TRUE | Downregulated |
| -0,260224566 | 0,031849727 | FRYL | FALSE | Downregulated |
| -0,375922109 | 0,031915501 | IRF2BPL | FALSE | Downregulated |
| 0,794029924 | 0,031915501 | LINC00240 | FALSE | Upregulated |
| -1,592056075 | 0,031915501 | LOC100653233 | TRUE | Downregulated |
| 1,035934655 | 0,031915501 | MS4A1 | TRUE | Upregulated |
| 0,657614411 | 0,031915501 | PHLDA1 | FALSE | Upregulated |
| 0,478796281 | 0,031915501 | PPP1R16B | FALSE | Upregulated |
| 0,244114008 | 0,031915501 | RNF10 | FALSE | Upregulated |
| 0,581151209 | 0,031943517 | TRAPPC2P1 | FALSE | Upregulated |
| -1,172795205 | 0,031947793 | ZRANB2-AS1 | TRUE | Downregulated |
| 0,23078275 | 0,031957339 | OTUB1 | FALSE | Upregulated |

| | | | | |
|---|---|---|---|---|
| -0,247080957 | 0,031957339 | WRNIP1 | FALSE | Downregulated |
| -0,291373039 | 0,031962387 | ZNF823 | FALSE | Downregulated |
| -0,293585253 | 0,03199519 | TMEM167B | FALSE | Downregulated |
| 0,496397396 | 0,032011664 | CALR | FALSE | Upregulated |
| -0,478885412 | 0,032011664 | PARP6 | FALSE | Downregulated |
| 0,446329538 | 0,032011664 | PIK3R3 | FALSE | Upregulated |
| -0,385038336 | 0,032020984 | CIRBP | FALSE | Downregulated |
| 0,817218181 | 0,032043631 | VDR | FALSE | Upregulated |
| 0,413703038 | 0,032059414 | ALDH16A1 | FALSE | Upregulated |
| 0,245775524 | 0,032083004 | SENP3 | FALSE | Upregulated |
| 0,410650901 | 0,032124498 | NTMT1 | FALSE | Upregulated |
| 0,340560941 | 0,032147744 | PSMA1 | FALSE | Upregulated |
| -0,417377143 | 0,03214976 | NBR2 | FALSE | Downregulated |
| -0,52840212 | 0,03214976 | SRSF11 | FALSE | Downregulated |
| 0,292826368 | 0,03219576 | SRSF3 | FALSE | Upregulated |
| 0,326279609 | 0,032228645 | CPM | FALSE | Upregulated |
| -0,225741579 | 0,032232371 | SYNJ2BP | FALSE | Downregulated |
| 2,637436848 | 0,032240181 | LPAR3 | TRUE | Upregulated |
| -0,226667012 | 0,032248847 | BMPR2 | FALSE | Downregulated |
| -1,425407775 | 0,032248847 | LINGO4 | TRUE | Downregulated |
| 0,349399303 | 0,032251213 | RFC2 | FALSE | Upregulated |
| -1,002121992 | 0,032251213 | SGK494 | TRUE | Downregulated |
| 0,496645808 | 0,03234724 | FMNL2 | FALSE | Upregulated |
| -1,51637752 | 0,032395283 | KIAA1755 | TRUE | Downregulated |
| 0,238128839 | 0,032395534 | COPS8 | FALSE | Upregulated |
| -0,869740107 | 0,032395534 | PDXDC2P | FALSE | Downregulated |
| 0,466177405 | 0,032395534 | SCLT1 | FALSE | Upregulated |
| -0,642980573 | 0,032395534 | WEE2-AS1 | FALSE | Downregulated |
| 0,337954273 | 0,03244914 | PPM1G | FALSE | Upregulated |
| -0,366376635 | 0,032481034 | IRF6 | FALSE | Downregulated |
| -1,005129047 | 0,032532534 | KCNIP3 | TRUE | Downregulated |
| 0,281107283 | 0,032536705 | WBSCR22 | FALSE | Upregulated |
| -0,523074442 | 0,032607139 | LOC100506730 | FALSE | Downregulated |
| -0,488391143 | 0,032607139 | MTURN | FALSE | Downregulated |
| 0,833339928 | 0,032751832 | LAG3 | FALSE | Upregulated |
| -1,089827286 | 0,032767532 | LOC100134822 | TRUE | Downregulated |
| -0,165826612 | 0,032767532 | PTCD3 | FALSE | Downregulated |
| 1,03035855 | 0,032822476 | NOD2 | TRUE | Upregulated |
| -0,245677343 | 0,032951258 | LIN7C | FALSE | Downregulated |
| -0,787284506 | 0,032967691 | WDR52 | FALSE | Downregulated |
| -0,352115537 | 0,033001425 | ILF3-AS1 | FALSE | Downregulated |
| 0,276557905 | 0,03304205 | UCHL3 | FALSE | Upregulated |
| -0,411395436 | 0,03309505 | NFXL1 | FALSE | Downregulated |
| -0,322543467 | 0,03309505 | RNF43 | FALSE | Downregulated |
| -0,60635892 | 0,033096021 | SLC10A5 | FALSE | Downregulated |
| -0,34100946 | 0,03315826 | ZNF605 | FALSE | Downregulated |
| 0,8214688 | 0,033176893 | LOC643733 | FALSE | Upregulated |
| 0,294916977 | 0,03324084 | LMAN2 | FALSE | Upregulated |
| -0,327841108 | 0,033248083 | KIAA1009 | FALSE | Downregulated |
| -0,620208635 | 0,033248083 | LCA5 | FALSE | Downregulated |
| -0,255036838 | 0,033288249 | KANK1 | FALSE | Downregulated |

| | | | | |
|---|---|---|---|---|
| 0,69456312 | 0,033294849 | LRRC8E | FALSE | Upregulated |
| 0,599121142 | 0,033294849 | TRIM35 | FALSE | Upregulated |
| -0,310892428 | 0,03329607 | ZC3H8 | FALSE | Downregulated |
| 0,816489766 | 0,033316451 | FPR1 | FALSE | Upregulated |
| -0,330195456 | 0,033475893 | TFR2 | FALSE | Downregulated |
| -0,292485187 | 0,033590808 | CCDC59 | FALSE | Downregulated |
| -0,80868407 | 0,033590808 | SLC1A2 | FALSE | Downregulated |
| -1,357202715 | 0,033650991 | TRHDE | TRUE | Downregulated |
| 0,864884326 | 0,033756125 | CD22 | FALSE | Upregulated |
| -0,304402622 | 0,033811854 | ZNF350 | FALSE | Downregulated |
| -0,340753379 | 0,033831873 | ABCA6 | FALSE | Downregulated |
| -0,773584813 | 0,033889561 | C1orf228 | FALSE | Downregulated |
| 0,550245015 | 0,033951519 | LPCAT1 | FALSE | Upregulated |
| 0,499887064 | 0,033993491 | HELZ2 | FALSE | Upregulated |
| 0,526804642 | 0,034021217 | PACS1 | FALSE | Upregulated |
| 0,366012544 | 0,034021217 | SH2B3 | FALSE | Upregulated |
| 0,308421463 | 0,034060271 | NCLN | FALSE | Upregulated |
| -0,480230902 | 0,034167127 | COQ10A | FALSE | Downregulated |
| 0,356488064 | 0,034167127 | P4HB | FALSE | Upregulated |
| -0,232768289 | 0,034170206 | PHF12 | FALSE | Downregulated |
| 0,88288767 | 0,034225669 | INSIG1 | FALSE | Upregulated |
| 0,605009996 | 0,034237012 | PDCD1LG2 | FALSE | Upregulated |
| -2,118214993 | 0,034255831 | BEND6 | TRUE | Downregulated |
| -1,387151316 | 0,034286283 | LOC100287175 | TRUE | Downregulated |
| 0,81641958 | 0,034312026 | PARP15 | FALSE | Upregulated |
| -0,387424473 | 0,034316216 | TEC | FALSE | Downregulated |
| 1,295066199 | 0,034316216 | TTC9 | TRUE | Upregulated |
| 0,37992356 | 0,034391008 | TMEM9 | FALSE | Upregulated |
| -0,599317903 | 0,034449924 | VSNL1 | FALSE | Downregulated |
| -0,388652788 | 0,034464049 | NAAA | FALSE | Downregulated |
| 1,88820319 | 0,034732239 | TAAR3 | TRUE | Upregulated |
| -0,345765101 | 0,034797797 | KMT2D | FALSE | Downregulated |
| -0,425542582 | 0,034799191 | POLR2M | FALSE | Downregulated |
| 0,317487232 | 0,034818055 | RNF166 | FALSE | Upregulated |
| -0,825260365 | 0,034839417 | CATSPERG | FALSE | Downregulated |
| 0,615577916 | 0,034839417 | MRC2 | FALSE | Upregulated |
| 1,082731914 | 0,034919764 | XCL1 | TRUE | Upregulated |
| 0,386861164 | 0,03494642 | GAS2L1 | FALSE | Upregulated |
| -0,447348593 | 0,03494642 | TTC14 | FALSE | Downregulated |
| -0,497086973 | 0,034989905 | SFXN2 | FALSE | Downregulated |
| 0,181353943 | 0,034989905 | VAPA | FALSE | Upregulated |
| -0,863196995 | 0,035032818 | MGC27382 | FALSE | Downregulated |
| 0,580771278 | 0,035170653 | MS4A4A | FALSE | Upregulated |
| 0,502023824 | 0,035170653 | UQCRQ | FALSE | Upregulated |
| -0,426415599 | 0,035207358 | PHYHD1 | FALSE | Downregulated |
| 0,433931822 | 0,035221826 | ABCB1 | FALSE | Upregulated |
| 0,918228115 | 0,035221826 | ATOH8 | FALSE | Upregulated |
| 0,320584942 | 0,035221826 | PAK1 | FALSE | Upregulated |
| 0,455069095 | 0,035241578 | HSPH1 | FALSE | Upregulated |
| -0,381912439 | 0,035241578 | HYLS1 | FALSE | Downregulated |
| -1,050924834 | 0,035241578 | MIR4461 | TRUE | Downregulated |

| | | | | |
|---|---|---|---|---|
| 0,53018286 | 0,035277486 | AK1 | FALSE | Upregulated |
| -0,259610806 | 0,035294479 | DMXL1 | FALSE | Downregulated |
| 0,30215824 | 0,035316554 | B4GALT1 | FALSE | Upregulated |
| -0,509459589 | 0,035369296 | AMDHD1 | FALSE | Downregulated |
| -0,288790035 | 0,035369296 | ZNF275 | FALSE | Downregulated |
| 0,223109627 | 0,035410104 | VAMP3 | FALSE | Upregulated |
| 0,38373324 | 0,035426972 | CYCS | FALSE | Upregulated |
| -0,302085674 | 0,035426972 | RSC1A1 | FALSE | Downregulated |
| -0,564410195 | 0,035426972 | ZNF850 | FALSE | Downregulated |
| 1,940870501 | 0,035448912 | EPHX3 | TRUE | Upregulated |
| -0,397273397 | 0,035492609 | PLA2G12A | FALSE | Downregulated |
| 0,188762057 | 0,035539942 | RNF220 | FALSE | Upregulated |
| 0,48128519 | 0,03557019 | ACYP1 | FALSE | Upregulated |
| -0,758515112 | 0,03557019 | GOLGA8N | FALSE | Downregulated |
| 0,819124922 | 0,03557019 | ZFP69B | FALSE | Upregulated |
| 0,287471975 | 0,035636022 | ARL2BP | FALSE | Upregulated |
| -0,808420247 | 0,035636022 | PLEKHH2 | FALSE | Downregulated |
| 0,36678218 | 0,035636022 | RNF135 | FALSE | Upregulated |
| 0,395087781 | 0,035636022 | RPS19 | FALSE | Upregulated |
| 0,347382298 | 0,035636022 | SNRNP35 | FALSE | Upregulated |
| -0,449377495 | 0,035693627 | KIAA0895 | FALSE | Downregulated |
| 0,180176824 | 0,035706295 | IMMT | FALSE | Upregulated |
| 0,218053595 | 0,035715685 | SCAMP3 | FALSE | Upregulated |
| -0,244523158 | 0,035728022 | DENND4C | FALSE | Downregulated |
| 0,29567311 | 0,035805797 | FAM53B | FALSE | Upregulated |
| -0,22388673 | 0,035870571 | ESYT2 | FALSE | Downregulated |
| 1,062268297 | 0,035871586 | GPR174 | TRUE | Upregulated |
| 0,49432992 | 0,035898018 | FIGNL1 | FALSE | Upregulated |
| -0,338585097 | 0,035898018 | UCK1 | FALSE | Downregulated |
| -0,309425565 | 0,035960806 | CPB2 | FALSE | Downregulated |
| 0,884191289 | 0,035961968 | CCRN4L | FALSE | Upregulated |
| -0,582117987 | 0,035974782 | SLC1A1 | FALSE | Downregulated |
| -0,260569875 | 0,035990732 | C1S | FALSE | Downregulated |
| -0,24911852 | 0,035990732 | ZCCHC8 | FALSE | Downregulated |
| -0,639468508 | 0,036044015 | LOC100506639 | FALSE | Downregulated |
| 0,420129881 | 0,036044015 | MYO19 | FALSE | Upregulated |
| -0,27528653 | 0,036053531 | SUCLG2 | FALSE | Downregulated |
| -0,286756067 | 0,036080462 | CRTC2 | FALSE | Downregulated |
| 0,528647406 | 0,036080462 | DOCK10 | FALSE | Upregulated |
| 0,592841794 | 0,036126681 | SLC2A9 | FALSE | Upregulated |
| -0,33027066 | 0,036126681 | STX18 | FALSE | Downregulated |
| -0,408345802 | 0,036126681 | TSC1 | FALSE | Downregulated |
| -0,354296634 | 0,036126681 | ZNF621 | FALSE | Downregulated |
| -0,70791488 | 0,036153828 | DIRAS3 | FALSE | Downregulated |
| 0,217523211 | 0,036158394 | PSMD13 | FALSE | Upregulated |
| 0,305669785 | 0,036271401 | ADRM1 | FALSE | Upregulated |
| -0,294007248 | 0,036403033 | MAGI1 | FALSE | Downregulated |
| 2,091376417 | 0,036478813 | CCL3L3 | TRUE | Upregulated |
| 0,617235292 | 0,036478813 | FAM110A | FALSE | Upregulated |
| 0,428963999 | 0,036548649 | PLIN3 | FALSE | Upregulated |
| -0,434125083 | 0,036574078 | DMRTA1 | FALSE | Downregulated |

| | | | | |
|---|---|---|---|---|
| 0,253642163 | 0,036575018 | PRMT2 | FALSE | Upregulated |
| -0,766477033 | 0,036593814 | CNDP1 | FALSE | Downregulated |
| 0,381320165 | 0,036599789 | AVPI1 | FALSE | Upregulated |
| -0,653826577 | 0,036599789 | LOC102723665 | FALSE | Downregulated |
| 0,567629737 | 0,036599789 | SLC2A3 | FALSE | Upregulated |
| -0,56656946 | 0,036599789 | ZNF284 | FALSE | Downregulated |
| 0,290817467 | 0,036654109 | TIMELESS | FALSE | Upregulated |
| 0,294697156 | 0,03672793 | PRKD2 | FALSE | Upregulated |
| 0,376498346 | 0,036744323 | OVCA2 | FALSE | Upregulated |
| -0,244302953 | 0,036744323 | VWA8 | FALSE | Downregulated |
| 0,176995883 | 0,036768636 | CCT8 | FALSE | Upregulated |
| -1,475507337 | 0,036768636 | LINC00622 | TRUE | Downregulated |
| 0,455088336 | 0,036787412 | HEG1 | FALSE | Upregulated |
| -0,919282712 | 0,036787412 | SLC4A5 | FALSE | Downregulated |
| -0,172982678 | 0,036840551 | ARFGEF1 | FALSE | Downregulated |
| -0,45278764 | 0,036840735 | KAZN | FALSE | Downregulated |
| 0,328443187 | 0,03687046 | COMMD8 | FALSE | Upregulated |
| 0,329443799 | 0,036886194 | GRK5 | FALSE | Upregulated |
| 0,446401806 | 0,036896922 | NPL | FALSE | Upregulated |
| -0,240654788 | 0,036950959 | PHF2 | FALSE | Downregulated |
| 0,26099025 | 0,036961376 | KIAA1279 | FALSE | Upregulated |
| -0,94872375 | 0,03698207 | LOC101060376 | FALSE | Downregulated |
| -0,661846118 | 0,03701887 | ABCA9 | FALSE | Downregulated |
| -0,457068248 | 0,037065731 | DHX35 | FALSE | Downregulated |
| -1,481822389 | 0,037093293 | ARHGEF38 | TRUE | Downregulated |
| 1,510429043 | 0,037096291 | FCRLA | TRUE | Upregulated |
| 1,223726374 | 0,037110293 | NR1D1 | TRUE | Upregulated |
| -0,936110952 | 0,037110944 | MIR6723 | FALSE | Downregulated |
| -0,278468276 | 0,037196597 | C2orf42 | FALSE | Downregulated |
| -0,918821275 | 0,037238993 | LOC101928370 | FALSE | Downregulated |
| 0,454413854 | 0,037269087 | AKR7A3 | FALSE | Upregulated |
| -0,23609217 | 0,037272801 | ABCD4 | FALSE | Downregulated |
| 0,879038493 | 0,037329223 | CYR61 | FALSE | Upregulated |
| 0,629485585 | 0,037329223 | NCEH1 | FALSE | Upregulated |
| -0,618738744 | 0,037353201 | GBA3 | FALSE | Downregulated |
| 0,541258618 | 0,037353201 | MTFP1 | FALSE | Upregulated |
| -0,378494624 | 0,037353201 | NUAK1 | FALSE | Downregulated |
| -0,299742709 | 0,037353201 | POFUT1 | FALSE | Downregulated |
| -0,694666474 | 0,037357455 | TRIM66 | FALSE | Downregulated |
| -1,197083969 | 0,037376063 | TUBA3FP | TRUE | Downregulated |
| -0,859394684 | 0,037391933 | CYP1A2 | FALSE | Downregulated |
| -0,53398591 | 0,037466048 | DSG1 | FALSE | Downregulated |
| 0,793945531 | 0,037568561 | POLE2 | FALSE | Upregulated |
| -0,317599225 | 0,037568561 | TAB2 | FALSE | Downregulated |
| 0,511993347 | 0,037568561 | TNFRSF11B | FALSE | Upregulated |
| 0,846614172 | 0,037578196 | ANKRD53 | FALSE | Upregulated |
| 0,361918947 | 0,037578196 | SRA1 | FALSE | Upregulated |
| 0,496561906 | 0,037589104 | ACY3 | FALSE | Upregulated |
| 0,296011117 | 0,037589104 | IDH3G | FALSE | Upregulated |
| -0,361378876 | 0,037592093 | MYH10 | FALSE | Downregulated |
| -1,133587758 | 0,03759748 | SELE | TRUE | Downregulated |

| | | | | |
|---|---|---|---|---|
| 1,1980685 | 0,037666871 | DCDC2 | TRUE | Upregulated |
| -1,597488739 | 0,037694878 | MUSTN1 | TRUE | Downregulated |
| -0,27287244 | 0,037736977 | PTPRK | FALSE | Downregulated |
| 1,075865028 | 0,037773692 | FAM115C | TRUE | Upregulated |
| 0,285840245 | 0,037781652 | IPO13 | FALSE | Upregulated |
| -0,756785043 | 0,037794936 | GYLTL1B | FALSE | Downregulated |
| -0,231717559 | 0,037794936 | SPAG16 | FALSE | Downregulated |
| 0,337900653 | 0,037798535 | AP1S1 | FALSE | Upregulated |
| -0,636597395 | 0,037851304 | NEURL2 | FALSE | Downregulated |
| 0,405400684 | 0,037860187 | JAK2 | FALSE | Upregulated |
| -0,701407055 | 0,037860187 | NPR3 | FALSE | Downregulated |
| -0,406824759 | 0,037877465 | ACSM5 | FALSE | Downregulated |
| 0,270246115 | 0,037877465 | CAMSAP1 | FALSE | Upregulated |
| 0,383609691 | 0,037902371 | PPP1R14B | FALSE | Upregulated |
| 1,77855715 | 0,037923937 | LINC00639 | TRUE | Upregulated |
| -0,415560976 | 0,037923937 | TP73-AS1 | FALSE | Downregulated |
| -0,369028022 | 0,037932113 | PON1 | FALSE | Downregulated |
| 0,54621929 | 0,037932113 | RUNX1 | FALSE | Upregulated |
| 0,191035272 | 0,037955482 | KIAA0100 | FALSE | Upregulated |
| 0,451865663 | 0,037955482 | PLBD1 | FALSE | Upregulated |
| 0,607209458 | 0,037955482 | TJP3 | FALSE | Upregulated |
| 0,441745859 | 0,037955482 | TOMM40L | FALSE | Upregulated |
| 0,407827706 | 0,038011807 | MPV17 | FALSE | Upregulated |
| 1,629595085 | 0,038018201 | LOC642131 | TRUE | Upregulated |
| -0,524208366 | 0,038065934 | PITPNA-AS1 | FALSE | Downregulated |
| 0,312223568 | 0,038160785 | RPL18A | FALSE | Upregulated |
| 0,76148921 | 0,038190882 | PLK3 | FALSE | Upregulated |
| 0,215473485 | 0,038190882 | SP100 | FALSE | Upregulated |
| 0,207700359 | 0,038216567 | MCL1 | FALSE | Upregulated |
| 1,184452592 | 0,038218281 | ADD2 | TRUE | Upregulated |
| -0,222338636 | 0,038218281 | USP47 | FALSE | Downregulated |
| -0,657904669 | 0,038251534 | ADRA2B | FALSE | Downregulated |
| -0,751360205 | 0,038251534 | HNRNPU-AS1 | FALSE | Downregulated |
| -0,627029047 | 0,038288613 | ZNF37BP | FALSE | Downregulated |
| -1,344403825 | 0,038289067 | LOC339240 | TRUE | Downregulated |
| 0,331446901 | 0,038289067 | MARCH2 | FALSE | Upregulated |
| 0,389917984 | 0,03833404 | GDA | FALSE | Upregulated |
| 0,807868821 | 0,038375841 | MAP4K1 | FALSE | Upregulated |
| 0,366904109 | 0,038400449 | PDIA3 | FALSE | Upregulated |
| 0,874382395 | 0,038400449 | SLC16A3 | FALSE | Upregulated |
| -0,134699016 | 0,038401115 | MATR3 | FALSE | Downregulated |
| -0,225563835 | 0,038411982 | ZNF480 | FALSE | Downregulated |
| 1,483180841 | 0,03841545 | NRXN2 | TRUE | Upregulated |
| -0,315627118 | 0,038460097 | NUDT16 | FALSE | Downregulated |
| 0,284425429 | 0,038464741 | PREP | FALSE | Upregulated |
| 1,070217125 | 0,038475531 | ASPHD2 | TRUE | Upregulated |
| -1,771987452 | 0,038512363 | LOC101929814 | TRUE | Downregulated |
| 0,262956787 | 0,038545771 | TGFBR1 | FALSE | Upregulated |
| -0,693755951 | 0,038546782 | GPR98 | FALSE | Downregulated |
| -0,567650921 | 0,038546782 | ZNF841 | FALSE | Downregulated |
| 1,315311991 | 0,038564921 | CPNE5 | TRUE | Upregulated |

| | | | | |
|---|---|---|---|---|
| 0,299153995 | 0,038564921 | FIBP | FALSE | Upregulated |
| -1,369181995 | 0,038618565 | DSCAML1 | TRUE | Downregulated |
| -0,617172628 | 0,038618565 | STAG3L3 | FALSE | Downregulated |
| -0,227179522 | 0,038634606 | TYW5 | FALSE | Downregulated |
| -0,440358384 | 0,038660871 | ACOT13 | FALSE | Downregulated |
| -0,276855688 | 0,038666974 | EPB41L5 | FALSE | Downregulated |
| -0,670076882 | 0,038676509 | LSM11 | FALSE | Downregulated |
| -0,384818628 | 0,038715651 | HELZ | FALSE | Downregulated |
| 1,793483723 | 0,038715651 | HIST1H2BN | TRUE | Upregulated |
| -0,590108291 | 0,038796317 | CCT6P3 | FALSE | Downregulated |
| -0,496196915 | 0,038796317 | RECK | FALSE | Downregulated |
| 0,770200188 | 0,038828107 | KCNA3 | FALSE | Upregulated |
| 0,772459447 | 0,038839168 | MANF | FALSE | Upregulated |
| -0,468961961 | 0,038847811 | UBN2 | FALSE | Downregulated |
| -0,825484054 | 0,038875492 | EPB41L1 | FALSE | Downregulated |
| 0,436518448 | 0,038876965 | CCDC107 | FALSE | Upregulated |
| -0,847748262 | 0,038876965 | CYP2B6 | FALSE | Downregulated |
| -0,28104669 | 0,038876965 | FBXO28 | FALSE | Downregulated |
| 0,430175484 | 0,038916729 | HMBS | FALSE | Upregulated |
| 0,414769624 | 0,03893489 | ANKRD44 | FALSE | Upregulated |
| 1,901614803 | 0,038936947 | SAA1 | TRUE | Upregulated |
| -0,287271731 | 0,038975663 | CDK14 | FALSE | Downregulated |
| 1,629386626 | 0,039097309 | OTOA | TRUE | Upregulated |
| 0,353036631 | 0,039139755 | PFDN2 | FALSE | Upregulated |
| -0,212732611 | 0,039139755 | UQCRB | FALSE | Downregulated |
| -0,380711658 | 0,039211545 | GRTP1 | FALSE | Downregulated |
| 0,840256859 | 0,039258419 | AGAP2 | FALSE | Upregulated |
| -0,635892553 | 0,039346182 | EMBP1 | FALSE | Downregulated |
| 0,383161666 | 0,039425712 | ZBTB33 | FALSE | Upregulated |
| -0,564139069 | 0,039543371 | RNPC3 | FALSE | Downregulated |
| 0,456971843 | 0,03956269 | DONSON | FALSE | Upregulated |
| -0,370625688 | 0,039600399 | ZSCAN30 | FALSE | Downregulated |
| 0,489221237 | 0,039649843 | SH2D3C | FALSE | Upregulated |
| 0,228663868 | 0,039744444 | ADRBK1 | FALSE | Upregulated |
| 0,526588314 | 0,039768105 | APOBEC3F | FALSE | Upregulated |
| 0,288158687 | 0,039768105 | CC2D1B | FALSE | Upregulated |
| 0,311982527 | 0,039783349 | MYH9 | FALSE | Upregulated |
| 0,191784215 | 0,039815387 | U2AF2 | FALSE | Upregulated |
| 0,478991592 | 0,039889609 | BAK1 | FALSE | Upregulated |
| -0,466927475 | 0,04003039 | ZNF141 | FALSE | Downregulated |
| -0,249522887 | 0,040060261 | AGO4 | FALSE | Downregulated |
| 0,823867519 | 0,04014514 | LOC102723342 | FALSE | Upregulated |
| -0,615242551 | 0,04014514 | MDM4 | FALSE | Downregulated |
| -0,85647 | 0,04014514 | PDE11A | FALSE | Downregulated |
| 0,437901887 | 0,040156067 | C15orf39 | FALSE | Upregulated |
| 0,7147168 | 0,040181598 | ALOX5AP | FALSE | Upregulated |
| -0,29197369 | 0,040181598 | FBXL3 | FALSE | Downregulated |
| -0,275699408 | 0,040181598 | SNX30 | FALSE | Downregulated |
| 0,160146566 | 0,040229815 | BECN1 | FALSE | Upregulated |
| -0,720098461 | 0,040240458 | LOC102723864 | FALSE | Downregulated |
| -0,308421619 | 0,040247247 | KLHL28 | FALSE | Downregulated |

| | | | | |
|---|---|---|---|---|
| -0,963886308 | 0,040266427 | LOC101928714 | FALSE | Downregulated |
| -0,645694628 | 0,04034536 | LOC100128398 | FALSE | Downregulated |
| -2,445793222 | 0,04034536 | RSPH10B2 | TRUE | Downregulated |
| 0,35335615 | 0,040353475 | C15orf57 | FALSE | Upregulated |
| -0,769380863 | 0,040353475 | VAT1L | FALSE | Downregulated |
| -0,237075227 | 0,040420661 | C1orf115 | FALSE | Downregulated |
| -0,25198797 | 0,040440177 | CXXC1 | FALSE | Downregulated |
| 0,194528618 | 0,040525313 | RNF121 | FALSE | Upregulated |
| 0,917968888 | 0,04055477 | GOLM1 | FALSE | Upregulated |
| 1,002217176 | 0,040559214 | COL8A2 | TRUE | Upregulated |
| -0,679812133 | 0,040623569 | DDT | FALSE | Downregulated |
| -0,971307573 | 0,040722627 | LOC101928826 | FALSE | Downregulated |
| -0,193890938 | 0,040758702 | RPP14 | FALSE | Downregulated |
| -0,328646712 | 0,040761605 | SUGCT | FALSE | Downregulated |
| -0,498514835 | 0,040773707 | PGM5 | FALSE | Downregulated |
| -0,247032835 | 0,040874077 | PIGN | FALSE | Downregulated |
| 0,271649995 | 0,040883135 | TXNDC12 | FALSE | Upregulated |
| -0,23482452 | 0,040918491 | WWP1 | FALSE | Downregulated |
| -0,333631275 | 0,04096227 | C1RL | FALSE | Downregulated |
| 1,043116063 | 0,04096227 | IPO11-LRRC70 | TRUE | Upregulated |
| 0,581985523 | 0,04096227 | TAX1BP3 | FALSE | Upregulated |
| 0,515894903 | 0,04096227 | WRAP53 | FALSE | Upregulated |
| 0,58962822 | 0,040997958 | TUBB6 | FALSE | Upregulated |
| -0,265451201 | 0,041067882 | PAAF1 | FALSE | Downregulated |
| -0,653805672 | 0,04109662 | SAMD5 | FALSE | Downregulated |
| 1,170746232 | 0,041148844 | BCL2L14 | TRUE | Upregulated |
| -1,363490783 | 0,041148844 | DDTL | TRUE | Downregulated |
| 0,372757872 | 0,041265027 | POGLUT1 | FALSE | Upregulated |
| 0,661531385 | 0,041318658 | TMEM229B | FALSE | Upregulated |
| 3,493811836 | 0,041330611 | KRT86 | TRUE | Upregulated |
| -1,947736984 | 0,041403986 | COL19A1 | TRUE | Downregulated |
| 0,537722531 | 0,041447751 | KNSTRN | FALSE | Upregulated |
| 2,192631288 | 0,041447751 | POU5F1B | TRUE | Upregulated |
| -0,335624459 | 0,041479472 | SDC2 | FALSE | Downregulated |
| -1,09494173 | 0,041512765 | STK31 | TRUE | Downregulated |
| -1,164212473 | 0,041523111 | BCRP3 | TRUE | Downregulated |
| -0,2610938 | 0,041538544 | EBAG9 | FALSE | Downregulated |
| 0,624738469 | 0,041538544 | TGFB3 | FALSE | Upregulated |
| 0,372286238 | 0,041546765 | SLC12A4 | FALSE | Upregulated |
| -0,24894318 | 0,041546765 | ZNF770 | FALSE | Downregulated |
| -1,748288874 | 0,041612646 | SLCO1A2 | TRUE | Downregulated |
| -0,273557548 | 0,041665954 | PPP1R13B | FALSE | Downregulated |
| 0,470351154 | 0,04167623 | MAD2L2 | FALSE | Upregulated |
| -2,056647616 | 0,041683591 | NPTXR | TRUE | Downregulated |
| -0,481654056 | 0,041702848 | CRIPAK | FALSE | Downregulated |
| -1,269131153 | 0,041720563 | MAST1 | TRUE | Downregulated |
| 0,188789317 | 0,041720563 | UBE2V2 | FALSE | Upregulated |
| 0,165417709 | 0,041720563 | VDAC1 | FALSE | Upregulated |
| -3,17821129 | 0,041790693 | LOC101929826 | TRUE | Downregulated |
| 1,520483787 | 0,041790693 | TSACC | TRUE | Upregulated |
| 0,620419175 | 0,041804196 | SOD2 | FALSE | Upregulated |

| | | | | |
|---|---|---|---|---|
| 0,669100025 | 0,04186047 | AC009948.5 | FALSE | Upregulated |
| 1,86240831 | 0,04186047 | CCL8 | TRUE | Upregulated |
| -0,548240407 | 0,04186047 | SCAI | FALSE | Downregulated |
| 0,539229059 | 0,041864925 | IER5 | FALSE | Upregulated |
| 0,39233579 | 0,041864925 | MFSD5 | FALSE | Upregulated |
| -0,311271186 | 0,04191964 | LIX1L | FALSE | Downregulated |
| 0,811007272 | 0,041928043 | HULC | FALSE | Upregulated |
| 1,179857389 | 0,041982916 | TUBB2B | TRUE | Upregulated |
| 0,184323378 | 0,042063505 | FAM104A | FALSE | Upregulated |
| 0,647895179 | 0,042103515 | C8orf47 | FALSE | Upregulated |
| 0,220859039 | 0,042103515 | GALNT1 | FALSE | Upregulated |
| -1,116993671 | 0,042103515 | P2RX6 | TRUE | Downregulated |
| 0,275511881 | 0,042103515 | STK24 | FALSE | Upregulated |
| -0,264504677 | 0,042129383 | MSL1 | FALSE | Downregulated |
| 0,428939105 | 0,042132036 | MDM2 | FALSE | Upregulated |
| -0,439355628 | 0,042167693 | LOC101060195 | FALSE | Downregulated |
| -1,61251775 | 0,042230756 | IQUB | TRUE | Downregulated |
| -0,35206168 | 0,042230756 | KIAA1958 | FALSE | Downregulated |
| -0,409495573 | 0,042230756 | USP30 | FALSE | Downregulated |
| 0,296395877 | 0,04227323 | PLCG1 | FALSE | Upregulated |
| 0,277420563 | 0,042379484 | WHSC1 | FALSE | Upregulated |
| -0,63995424 | 0,042416618 | IFITM10 | FALSE | Downregulated |
| -0,387299285 | 0,042416618 | SCML1 | FALSE | Downregulated |
| 1,402434615 | 0,042429452 | TRABD2A | TRUE | Upregulated |
| 0,316893007 | 0,042472598 | TRIAP1 | FALSE | Upregulated |
| 0,392054452 | 0,042567366 | AKNA | FALSE | Upregulated |
| -0,836964971 | 0,042622387 | RAD51-AS1 | FALSE | Downregulated |
| 0,381665734 | 0,042648722 | INCENP | FALSE | Upregulated |
| 0,33208654 | 0,042648722 | PIGU | FALSE | Upregulated |
| 0,353516068 | 0,042727802 | TNS3 | FALSE | Upregulated |
| 0,194243001 | 0,042728264 | AMFR | FALSE | Upregulated |
| -0,283926608 | 0,042728264 | CLUH | FALSE | Downregulated |
| 0,308766702 | 0,042728264 | HIATL1 | FALSE | Upregulated |
| 0,260673544 | 0,042728264 | SDCCAG3 | FALSE | Upregulated |
| -0,194424257 | 0,042757464 | USP25 | FALSE | Downregulated |
| 0,233715393 | 0,042775991 | AIM1 | FALSE | Upregulated |
| -0,622107657 | 0,042812383 | CAPS | FALSE | Downregulated |
| 0,647695893 | 0,042830339 | GALNT16 | FALSE | Upregulated |
| -0,274299731 | 0,042830339 | MTRF1 | FALSE | Downregulated |
| -0,277535547 | 0,042905277 | U2SURP | FALSE | Downregulated |
| 0,595991642 | 0,042924449 | ARHGAP4 | FALSE | Upregulated |
| -0,224269232 | 0,04297844 | PEX19 | FALSE | Downregulated |
| -0,445849061 | 0,043003394 | LOC646719 | FALSE | Downregulated |
| -0,196394597 | 0,043077428 | PCNX | FALSE | Downregulated |
| -0,316277235 | 0,043154984 | HAO1 | FALSE | Downregulated |
| 0,421429813 | 0,043154984 | PPIB | FALSE | Upregulated |
| -0,954532917 | 0,043194146 | MIR198 | FALSE | Downregulated |
| -0,208756614 | 0,04321568 | YTHDC1 | FALSE | Downregulated |
| -1,095568119 | 0,043236486 | LOC102724483 | TRUE | Downregulated |
| 0,231527586 | 0,043271698 | TAF6 | FALSE | Upregulated |
| 0,593644751 | 0,043391301 | SRC | FALSE | Upregulated |

| | | | | |
|---|---|---|---|---|
| -0,913552641 | 0,043398241 | DICER1-AS1 | FALSE | Downregulated |
| 0,301923283 | 0,043455284 | DEGS1 | FALSE | Upregulated |
| -0,251452763 | 0,043474575 | ATG14 | FALSE | Downregulated |
| 0,591013105 | 0,043529983 | PLEKHF1 | FALSE | Upregulated |
| 0,978499595 | 0,043559739 | LRP8 | FALSE | Upregulated |
| -0,893087135 | 0,043559739 | RP11-66A2.2 | FALSE | Downregulated |
| 0,324508845 | 0,043660327 | SLC25A28 | FALSE | Upregulated |
| -0,262280372 | 0,043710461 | DPY19L4 | FALSE | Downregulated |
| 0,698778362 | 0,043802473 | CENPO | FALSE | Upregulated |
| 0,267212863 | 0,043819232 | CSTF3 | FALSE | Upregulated |
| -0,277424678 | 0,043819232 | HKR1 | FALSE | Downregulated |
| -0,369634071 | 0,043866616 | F7 | FALSE | Downregulated |
| -0,379705123 | 0,043866616 | INPP5B | FALSE | Downregulated |
| 0,656672788 | 0,043879464 | FAR2 | FALSE | Upregulated |
| 0,633436822 | 0,043922882 | PDSS1 | FALSE | Upregulated |
| 0,895820568 | 0,043941077 | GBGT1 | FALSE | Upregulated |
| -3,070874054 | 0,044004404 | LOC102723580 | TRUE | Downregulated |
| -0,339259843 | 0,044011908 | RBMS1 | FALSE | Downregulated |
| 1,597453461 | 0,044051531 | ALDH3A1 | TRUE | Upregulated |
| -0,368801886 | 0,04414048 | MTHFS | FALSE | Downregulated |
| -0,450546543 | 0,044151529 | STX1B | FALSE | Downregulated |
| -0,288374078 | 0,044164013 | PCGF6 | FALSE | Downregulated |
| -0,272329178 | 0,04423872 | DCUN1D4 | FALSE | Downregulated |
| 0,237475895 | 0,04424947 | TWF1 | FALSE | Upregulated |
| -0,415165989 | 0,044276572 | RAB28 | FALSE | Downregulated |
| -0,486155082 | 0,044301096 | NPR1 | FALSE | Downregulated |
| 0,774870712 | 0,044321712 | ANKRD29 | FALSE | Upregulated |
| -1,981897834 | 0,044350482 | ESRRB | TRUE | Downregulated |
| -0,586067589 | 0,044381094 | TRPV1 | FALSE | Downregulated |
| 0,619901461 | 0,044389426 | ME1 | FALSE | Upregulated |
| 0,316245473 | 0,044414628 | PELO | FALSE | Upregulated |
| -0,400753038 | 0,044444646 | TGDS | FALSE | Downregulated |
| 0,279330664 | 0,044447046 | SMS | FALSE | Upregulated |
| -0,215208528 | 0,044526564 | ZHX1 | FALSE | Downregulated |
| -0,27118008 | 0,04454451 | SLK | FALSE | Downregulated |
| -0,165137568 | 0,044564815 | OPA1 | FALSE | Downregulated |
| -0,26085661 | 0,044570538 | ROGDI | FALSE | Downregulated |
| 0,273384722 | 0,044575947 | EXOC2 | FALSE | Upregulated |
| -1,959931002 | 0,044586718 | ELMOD1 | TRUE | Downregulated |
| 0,377137309 | 0,044586718 | GOLT1B | FALSE | Upregulated |
| -1,047588992 | 0,044597464 | SYCP3 | TRUE | Downregulated |
| 1,564618341 | 0,044642473 | TICRR | TRUE | Upregulated |
| 0,829621073 | 0,044689809 | ARHGEF16 | FALSE | Upregulated |
| -0,3202474 | 0,044727853 | TSC2 | FALSE | Downregulated |
| 0,468122593 | 0,044743257 | TOR4A | FALSE | Upregulated |
| -0,63338493 | 0,044830895 | CCNB3 | FALSE | Downregulated |
| 1,60622526 | 0,044830895 | FBN3 | TRUE | Upregulated |
| -0,685138922 | 0,044830895 | SOWAHA | FALSE | Downregulated |
| -0,538747933 | 0,044848382 | SLC19A2 | FALSE | Downregulated |
| -0,748643259 | 0,04487112 | AD000090.2 | FALSE | Downregulated |
| -0,181421646 | 0,04492249 | PPIL4 | FALSE | Downregulated |

| | | | | |
|---|---|---|---|---|
| -0,25001664 | 0,044954083 | DIAPH2 | FALSE | Downregulated |
| -2,206248359 | 0,044998591 | FLJ34503 | TRUE | Downregulated |
| 0,477066554 | 0,045009418 | TNFRSF10D | FALSE | Upregulated |
| -0,467714614 | 0,045016347 | STAB1 | FALSE | Downregulated |
| -0,318114836 | 0,045049814 | ZNF26 | FALSE | Downregulated |
| 0,422962708 | 0,045059179 | BTN3A2 | FALSE | Upregulated |
| -0,352837947 | 0,045059179 | LOC102723773 | FALSE | Downregulated |
| -0,252308593 | 0,045059179 | ZBTB11 | FALSE | Downregulated |
| 1,227970489 | 0,045061612 | NRIP2 | TRUE | Upregulated |
| -0,394249941 | 0,04512151 | EPHX2 | FALSE | Downregulated |
| 1,398172301 | 0,04512151 | LINC00892 | TRUE | Upregulated |
| -0,349478715 | 0,045125365 | FAM96A | FALSE | Downregulated |
| 0,39131538 | 0,045125365 | NFKBIA | FALSE | Upregulated |
| 0,268255299 | 0,045171891 | EMC7 | FALSE | Upregulated |
| 0,344120782 | 0,045171891 | SNRNP40 | FALSE | Upregulated |
| 0,41597198 | 0,045171891 | SPAG1 | FALSE | Upregulated |
| 0,23432278 | 0,045171891 | TMX2 | FALSE | Upregulated |
| -0,554497508 | 0,045171891 | ZNF577 | FALSE | Downregulated |
| -0,253000092 | 0,04525045 | KLHL2 | FALSE | Downregulated |
| 0,710633177 | 0,045298962 | CLIP4 | FALSE | Upregulated |
| -0,199023089 | 0,045404233 | BTBD7 | FALSE | Downregulated |
| -0,203469686 | 0,045404233 | USP40 | FALSE | Downregulated |
| 1,891110847 | 0,04546113 | GPR19 | TRUE | Upregulated |
| 0,438922089 | 0,045473269 | HSP90AA1 | FALSE | Upregulated |
| -1,469237625 | 0,045473269 | ZBED3-AS1 | TRUE | Downregulated |
| -0,820175309 | 0,04553723 | LOC100506358 | FALSE | Downregulated |
| -2,028594651 | 0,04553723 | LOC100996813 | TRUE | Downregulated |
| 0,227397935 | 0,045556453 | POP4 | FALSE | Upregulated |
| 0,285413706 | 0,045680015 | SPNS1 | FALSE | Upregulated |
| -1,977768533 | 0,045689614 | C6orf7 | TRUE | Downregulated |
| 0,276021623 | 0,045689614 | DUT | FALSE | Upregulated |
| 0,364372041 | 0,045689614 | GIMAP1 | FALSE | Upregulated |
| -1,077565684 | 0,045689614 | PCLO | TRUE | Downregulated |
| 0,366356195 | 0,045689614 | RND3 | FALSE | Upregulated |
| 0,810234402 | 0,045704119 | BICD1 | FALSE | Upregulated |
| 0,581359305 | 0,045704119 | ELOVL6 | FALSE | Upregulated |
| 2,062880947 | 0,045754052 | GNG4 | TRUE | Upregulated |
| 0,249377579 | 0,045769667 | STAT6 | FALSE | Upregulated |
| -0,307418949 | 0,045786261 | ANKHD1 | FALSE | Downregulated |
| 0,352896495 | 0,045850265 | MRPL54 | FALSE | Upregulated |
| 0,267271876 | 0,045861395 | EIF6 | FALSE | Upregulated |
| -0,98676681 | 0,045862611 | LOC102724889 | FALSE | Downregulated |
| 0,205765848 | 0,045883025 | TRIOBP | FALSE | Upregulated |
| -0,96843998 | 0,046015116 | ATP13A4 | FALSE | Downregulated |
| -0,652081863 | 0,046015116 | ONECUT1 | FALSE | Downregulated |
| 0,996376669 | 0,046080639 | TESC | FALSE | Upregulated |
| -0,45140024 | 0,046088083 | ADCY1 | FALSE | Downregulated |
| -0,283862822 | 0,046102713 | DTWD1 | FALSE | Downregulated |
| -0,604452171 | 0,046128891 | SLC16A1 | FALSE | Downregulated |
| -0,870000498 | 0,046138073 | CHRM2 | FALSE | Downregulated |
| -0,523211308 | 0,046199123 | AMACR | FALSE | Downregulated |

| | | | | |
|---|---|---|---|---|
| 0,279803314 | 0,046199123 | PRDX1 | FALSE | Upregulated |
| 1,11464579 | 0,046426783 | METTL24 | TRUE | Upregulated |
| -2,017640745 | 0,046478164 | FAM9A | TRUE | Downregulated |
| -0,27893364 | 0,046478164 | OCRL | FALSE | Downregulated |
| -0,790317017 | 0,046489532 | JAKMIP2 | FALSE | Downregulated |
| -1,208271194 | 0,046489532 | LOC102724660 | TRUE | Downregulated |
| -0,213490204 | 0,046581056 | MARCH6 | FALSE | Downregulated |
| -0,592657639 | 0,046583338 | NPHP3 | FALSE | Downregulated |
| 0,707416642 | 0,046603769 | ANKRD19P | FALSE | Upregulated |
| 0,40546786 | 0,046603769 | CHD1L | FALSE | Upregulated |
| -1,127189654 | 0,046603769 | EHHADH-AS1 | TRUE | Downregulated |
| -0,269536948 | 0,046603769 | HS2ST1 | FALSE | Downregulated |
| -0,750979885 | 0,046603769 | NPHP3-ACAD11 | FALSE | Downregulated |
| 0,189886331 | 0,046603769 | ZC3H18 | FALSE | Upregulated |
| -0,294953861 | 0,046749535 | TDRP | FALSE | Downregulated |
| -0,252968932 | 0,046769675 | KCTD21 | FALSE | Downregulated |
| 0,954331398 | 0,046771566 | BANK1 | FALSE | Upregulated |
| -0,511009114 | 0,046771566 | MRPS31P5 | FALSE | Downregulated |
| -1,297733185 | 0,046781104 | MAP7D2 | TRUE | Downregulated |
| 0,390183265 | 0,046781104 | SNN | FALSE | Upregulated |
| 0,247505019 | 0,046813695 | GTPBP4 | FALSE | Upregulated |
| 0,462635999 | 0,046844039 | MOSPD1 | FALSE | Upregulated |
| -0,453722629 | 0,0469529 | SERPINA5 | FALSE | Downregulated |
| 1,976487809 | 0,046988445 | NEIL3 | TRUE | Upregulated |
| -0,281595435 | 0,047001159 | KAT8 | FALSE | Downregulated |
| -0,406177234 | 0,047007678 | FAM126B | FALSE | Downregulated |
| 0,467160363 | 0,047012274 | ADAMTS2 | FALSE | Upregulated |
| 0,494729316 | 0,047199244 | SH3BP4 | FALSE | Upregulated |
| -0,33553778 | 0,047219862 | CLCN4 | FALSE | Downregulated |
| -0,386820772 | 0,047342683 | RAB33B | FALSE | Downregulated |
| -0,454769506 | 0,047363239 | ZNF121 | FALSE | Downregulated |
| -0,432997245 | 0,047377599 | DHRS1 | FALSE | Downregulated |
| 2,059298115 | 0,047394824 | AC020571.3 | TRUE | Upregulated |
| -0,498538809 | 0,047398881 | NRN1 | FALSE | Downregulated |
| -0,81416619 | 0,047406434 | C17orf97 | FALSE | Downregulated |
| 0,280108801 | 0,047406434 | RNF19B | FALSE | Upregulated |
| -0,735790939 | 0,047406434 | ST13P4 | FALSE | Downregulated |
| -0,496689913 | 0,047479119 | CCL14 | FALSE | Downregulated |
| 0,291578952 | 0,047479119 | GTF2E1 | FALSE | Upregulated |
| 2,755114003 | 0,047479119 | LINC00313 | TRUE | Upregulated |
| 0,32466692 | 0,047479119 | MAP2K2 | FALSE | Upregulated |
| 0,252729321 | 0,047479119 | PTBP3 | FALSE | Upregulated |
| 0,280748743 | 0,047582807 | KLHL5 | FALSE | Upregulated |
| -0,378382679 | 0,047582807 | RNF217 | FALSE | Downregulated |
| -0,38564648 | 0,047583538 | SAT2 | FALSE | Downregulated |
| -0,762794758 | 0,047583538 | SPTBN4 | FALSE | Downregulated |
| 0,519529341 | 0,047583538 | USP30-AS1 | FALSE | Upregulated |
| 0,745244955 | 0,047592468 | KCNJ2 | FALSE | Upregulated |
| -0,505014601 | 0,047643873 | LOC102606465 | FALSE | Downregulated |
| 1,02642676 | 0,04766268 | FADS2 | TRUE | Upregulated |
| -0,781091536 | 0,047764141 | THRB-AS1 | FALSE | Downregulated |

| | | | | |
|---|---|---|---|---|
| 0,632277577 | 0,047829614 | B3GNT7 | FALSE | Upregulated |
| 0,985728271 | 0,047829614 | CD40LG | FALSE | Upregulated |
| 0,388221672 | 0,047829614 | CERS5 | FALSE | Upregulated |
| 0,24799611 | 0,047829614 | IDH3A | FALSE | Upregulated |
| 0,249863807 | 0,047841619 | PFKL | FALSE | Upregulated |
| -0,976882756 | 0,047841619 | RAD21-AS1 | FALSE | Downregulated |
| -0,3211368 | 0,047841619 | REXO1 | FALSE | Downregulated |
| -1,410368232 | 0,047858198 | PP13 | TRUE | Downregulated |
| -0,324410444 | 0,047893638 | EPN2 | FALSE | Downregulated |
| 0,341701063 | 0,047893638 | GADD45GIP1 | FALSE | Upregulated |
| 0,61055211 | 0,047893638 | SH3BP1 | FALSE | Upregulated |
| 0,542011939 | 0,047896653 | FAM105A | FALSE | Upregulated |
| -0,27263099 | 0,047936642 | RALGAPA1 | FALSE | Downregulated |
| 0,195532001 | 0,047938274 | LRPAP1 | FALSE | Upregulated |
| -0,212905328 | 0,047950803 | STK40 | FALSE | Downregulated |
| -2,479558175 | 0,047951045 | COL18A1-AS2 | TRUE | Downregulated |
| -0,957583064 | 0,047974362 | LOC102723415 | FALSE | Downregulated |
| -0,342643692 | 0,047991732 | ACBD4 | FALSE | Downregulated |
| -0,394457451 | 0,047991732 | C11orf71 | FALSE | Downregulated |
| -0,21790248 | 0,047991732 | HADHA | FALSE | Downregulated |
| 0,653606211 | 0,047991732 | TMEM164 | FALSE | Upregulated |
| -0,274713222 | 0,048049071 | CEP57 | FALSE | Downregulated |
| 0,327999681 | 0,048053853 | TRIM8 | FALSE | Upregulated |
| 1,170508439 | 0,048101675 | FADS1 | TRUE | Upregulated |
| 0,274634121 | 0,048118065 | GTPBP1 | FALSE | Upregulated |
| -0,834191467 | 0,048122397 | NRARP | FALSE | Downregulated |
| 0,278108934 | 0,048157173 | MMGT1 | FALSE | Upregulated |
| -0,191305244 | 0,048240837 | NEK6 | FALSE | Downregulated |
| -0,394328637 | 0,048279923 | SARDH | FALSE | Downregulated |
| 0,539836204 | 0,048298012 | CDCA4 | FALSE | Upregulated |
| 0,311646576 | 0,048306597 | LDLRAP1 | FALSE | Upregulated |
| 0,28678393 | 0,048330122 | ALG9 | FALSE | Upregulated |
| 0,756366383 | 0,048330122 | HK2 | FALSE | Upregulated |
| 0,775517315 | 0,048358175 | SIDT1 | FALSE | Upregulated |
| 1,674939148 | 0,048375726 | F2RL3 | TRUE | Upregulated |
| -0,348714369 | 0,048500183 | F13B | FALSE | Downregulated |
| 0,267942334 | 0,048500183 | POMP | FALSE | Upregulated |
| -0,804661621 | 0,048500183 | PTPRT | FALSE | Downregulated |
| -0,603386754 | 0,048500183 | SLC22A25 | FALSE | Downregulated |
| -0,93009821 | 0,048626183 | SLC25A27 | FALSE | Downregulated |
| -0,701670986 | 0,048633205 | ZNF677 | FALSE | Downregulated |
| 1,443332505 | 0,048658983 | GDNF | TRUE | Upregulated |
| -0,434016531 | 0,048658983 | PHF8 | FALSE | Downregulated |
| 0,250410338 | 0,048668879 | PPP3CC | FALSE | Upregulated |
| -0,85827492 | 0,048692386 | TRNF | FALSE | Downregulated |
| -0,867552418 | 0,048828485 | LOC102724782 | FALSE | Downregulated |
| -0,416194567 | 0,048843093 | UGT2B7 | FALSE | Downregulated |
| -0,411675178 | 0,048944453 | FAM115A | FALSE | Downregulated |
| 0,262754857 | 0,048948161 | CDK2 | FALSE | Upregulated |
| 0,645356414 | 0,048948161 | RHBDF1 | FALSE | Upregulated |
| 0,335424208 | 0,048973872 | CCDC86 | FALSE | Upregulated |

| | | | | |
|---|---|---|---|---|
| 0,327256236 | 0,048983178 | RAD18 | FALSE | Upregulated |
| 0,983093116 | 0,049070708 | RAB39A | FALSE | Upregulated |
| -1,817817465 | 0,049155265 | ANKRD18DP | TRUE | Downregulated |
| -0,387409932 | 0,049173078 | BIVM-ERCC5 | FALSE | Downregulated |
| -0,557313295 | 0,049175809 | C3P1 | FALSE | Downregulated |
| 2,619167078 | 0,049215257 | DNAAF1 | TRUE | Upregulated |
| -0,578151722 | 0,049215257 | TAL1 | FALSE | Downregulated |
| 0,808125232 | 0,049240722 | GPR85 | FALSE | Upregulated |
| 0,229525205 | 0,049269813 | ELK1 | FALSE | Upregulated |
| -0,952189024 | 0,049310083 | AGAP5 | FALSE | Downregulated |
| -0,25532072 | 0,049315029 | MAP3K2 | FALSE | Downregulated |
| 0,282993235 | 0,049323101 | MPDU1 | FALSE | Upregulated |
| -1,258555956 | 0,049474025 | LOC101926935 | TRUE | Downregulated |
| -1,125098118 | 0,049522977 | ANKRD20A9P | TRUE | Downregulated |
| 0,356956272 | 0,049522977 | TMEM104 | FALSE | Upregulated |
| 0,542614006 | 0,049573271 | DDB2 | FALSE | Upregulated |
| 0,829502009 | 0,049603354 | LINC00528 | FALSE | Upregulated |
| 0,470156024 | 0,049603354 | MPEG1 | FALSE | Upregulated |
| 0,645443897 | 0,049736198 | KIF24 | FALSE | Upregulated |
| -0,299457834 | 0,049755237 | RWDD3 | FALSE | Downregulated |
| -0,532367099 | 0,049858813 | HSD11B1L | FALSE | Downregulated |
| 0,3299824 | 0,049880903 | RPLP2 | FALSE | Upregulated |
| 0,281425975 | 0,049924445 | DAD1 | FALSE | Upregulated |
| 2,202827766 | 0,049931435 | SAA2-SAA4 | TRUE | Upregulated |
| -1,298395034 | 0,050002157 | LOC101929625 | FALSE | Downregulated |
| 0,676691053 | 0,050032332 | NR4A2 | FALSE | Upregulated |
| 0,425385952 | 0,050032332 | TLDC1 | FALSE | Upregulated |
| -0,515116545 | 0,050032989 | LINC00478 | FALSE | Downregulated |
| -0,841675427 | 0,050053843 | RASL10B | FALSE | Downregulated |
| -0,2220596 | 0,050053843 | SMARCC2 | FALSE | Downregulated |
| -0,206978848 | 0,050068146 | SDHA | FALSE | Downregulated |
| -0,568242161 | 0,05008389 | ADRA1B | FALSE | Downregulated |
| -2,462873599 | 0,050134228 | LOC101929946 | FALSE | Downregulated |
| -0,534654446 | 0,050159196 | CNTN3 | FALSE | Downregulated |
| 0,311149323 | 0,050160501 | DNAJC10 | FALSE | Upregulated |
| -0,319603276 | 0,050166388 | CFH | FALSE | Downregulated |
| 0,242904194 | 0,050205243 | DNTTIP1 | FALSE | Upregulated |
| 0,649470281 | 0,050222881 | ATAD5 | FALSE | Upregulated |
| 0,320798972 | 0,050252948 | AKIRIN2 | FALSE | Upregulated |
| 0,169552366 | 0,050372072 | ACTR1B | FALSE | Upregulated |
| -0,264289958 | 0,050372072 | ZMYND8 | FALSE | Downregulated |
| 0,563074538 | 0,050373682 | KLF2 | FALSE | Upregulated |
| -1,03430647 | 0,050465514 | WBP2NL | FALSE | Downregulated |
| -0,244965889 | 0,050477154 | MKRN2 | FALSE | Downregulated |
| 0,519630225 | 0,050477154 | PGP | FALSE | Upregulated |
| -0,370896991 | 0,050500248 | FMO3 | FALSE | Downregulated |
| -0,237041077 | 0,050509816 | DERA | FALSE | Downregulated |
| 0,429952197 | 0,050517407 | HM13 | FALSE | Upregulated |
| 1,142327171 | 0,050544604 | RGL4 | FALSE | Upregulated |
| 0,471782509 | 0,050608255 | LRR1 | FALSE | Upregulated |
| 0,325589249 | 0,050608255 | PGAM5 | FALSE | Upregulated |

| | | | | |
|---|---|---|---|---|
| 0,520530245 | 0,050614954 | TUBA1A | FALSE | Upregulated |
| 1,189235991 | 0,050666309 | LOC102723465 | FALSE | Upregulated |
| -1,306216476 | 0,050675071 | LOC101060351 | FALSE | Downregulated |
| 0,308143845 | 0,050734268 | PLD3 | FALSE | Upregulated |
| -0,403120558 | 0,050789947 | ADRB2 | FALSE | Downregulated |
| -3,750199655 | 0,050789947 | LOC102724621 | FALSE | Downregulated |
| 1,130009387 | 0,050851216 | LOC102724608 | FALSE | Upregulated |
| 0,888327664 | 0,050853477 | WISP1 | FALSE | Upregulated |
| -1,172433494 | 0,050866203 | C1orf213 | FALSE | Downregulated |
| -0,39833783 | 0,050866203 | ZNF280C | FALSE | Downregulated |
| -0,410945489 | 0,05109049 | LOC220729 | FALSE | Downregulated |
| 0,337840756 | 0,051125763 | MKL1 | FALSE | Upregulated |
| 0,961398251 | 0,051127569 | CAP2 | FALSE | Upregulated |
| -0,24019819 | 0,051134724 | PURA | FALSE | Downregulated |
| -0,361708165 | 0,051156663 | SHANK3 | FALSE | Downregulated |
| 0,796613861 | 0,051156663 | TOX | FALSE | Upregulated |
| 0,246502495 | 0,051274964 | TXLNA | FALSE | Upregulated |
| -0,27875789 | 0,051369785 | ZNF649 | FALSE | Downregulated |
| 0,41341311 | 0,051442599 | SLC25A45 | FALSE | Upregulated |
| 0,463462209 | 0,051549143 | RPS27L | FALSE | Upregulated |
| 0,219256829 | 0,051549143 | SNX17 | FALSE | Upregulated |
| 0,431591916 | 0,051549143 | ZNF563 | FALSE | Upregulated |
| 0,327850311 | 0,051553497 | UBL7 | FALSE | Upregulated |
| 1,059904926 | 0,051631542 | LOC101929531 | FALSE | Upregulated |
| -0,325204327 | 0,051631542 | WDR81 | FALSE | Downregulated |
| 1,792813748 | 0,051685845 | TUBA4B | FALSE | Upregulated |
| -0,339762819 | 0,051728117 | LINC00938 | FALSE | Downregulated |
| 2,535733272 | 0,051728117 | LOC101927505 | FALSE | Upregulated |
| -0,94067572 | 0,051859863 | LOC100505985 | FALSE | Downregulated |
| 2,130644022 | 0,051949603 | ANTXRLP1 | FALSE | Upregulated |
| 0,361249972 | 0,052206842 | UXS1 | FALSE | Upregulated |
| -0,23098307 | 0,052208888 | JADE3 | FALSE | Downregulated |
| -0,405117787 | 0,05221181 | FLJ10038 | FALSE | Downregulated |
| 1,282104646 | 0,052214881 | FABP4 | FALSE | Upregulated |
| 0,240238144 | 0,052214881 | MRPL4 | FALSE | Upregulated |
| -0,858631886 | 0,052214881 | NPIPB4 | FALSE | Downregulated |
| 0,650131006 | 0,052214881 | NRM | FALSE | Upregulated |
| 0,731090032 | 0,052214881 | RIPPLY1 | FALSE | Upregulated |
| -0,615491082 | 0,052214881 | SRGAP3 | FALSE | Downregulated |
| 0,234783355 | 0,052214881 | VPS33A | FALSE | Upregulated |
| -0,35926179 | 0,052228346 | FAM110B | FALSE | Downregulated |
| -0,577587974 | 0,052228749 | AC007551.3 | FALSE | Downregulated |
| 0,250047696 | 0,05239052 | TMEM123 | FALSE | Upregulated |
| 1,496730727 | 0,052420166 | C19orf84 | FALSE | Upregulated |
| -1,728140198 | 0,052420166 | EDIL3 | FALSE | Downregulated |
| -0,22609914 | 0,052420166 | ELF2 | FALSE | Downregulated |
| -1,48707123 | 0,052420166 | LINC00862 | FALSE | Downregulated |
| 0,536831043 | 0,052568002 | RRS1 | FALSE | Upregulated |
| -0,417732333 | 0,052653297 | C9orf72 | FALSE | Downregulated |
| -0,794453452 | 0,052653297 | RP5-1159O4.2 | FALSE | Downregulated |
| -0,428717552 | 0,052685539 | RP11-983P16.4 | FALSE | Downregulated |

| | | | | |
|---|---|---|---|---|
| -0,383098877 | 0,052695979 | ACAA2 | FALSE | Downregulated |
| 0,297175509 | 0,052695979 | DPH3 | FALSE | Upregulated |
| 0,715278055 | 0,052695979 | PFKFB4 | FALSE | Upregulated |
| 0,495570345 | 0,052695979 | PWWP2B | FALSE | Upregulated |
| -0,711368332 | 0,052695979 | RIC3 | FALSE | Downregulated |
| 0,339568814 | 0,052695979 | RPL36 | FALSE | Upregulated |
| 1,382541849 | 0,052695979 | SRSF12 | FALSE | Upregulated |
| -0,941778904 | 0,052695979 | TNFRSF19 | FALSE | Downregulated |
| 0,527648028 | 0,052732675 | PROCR | FALSE | Upregulated |
| -0,501549296 | 0,052732675 | TMEM8B | FALSE | Downregulated |
| -0,865638295 | 0,052834238 | FAM198A | FALSE | Downregulated |
| -0,965482161 | 0,052834238 | LOC100130992 | FALSE | Downregulated |
| -1,000827139 | 0,053071819 | FAM222A-AS1 | FALSE | Downregulated |
| -0,27580336 | 0,053126774 | KDM1B | FALSE | Downregulated |
| -0,205493114 | 0,053187481 | NFE2L1 | FALSE | Downregulated |
| -0,179270458 | 0,053230884 | SMEK2 | FALSE | Downregulated |
| -0,469604944 | 0,053231803 | ARAP3 | FALSE | Downregulated |
| 1,833937502 | 0,053275395 | LOC100288175 | FALSE | Upregulated |
| 0,211018282 | 0,053343592 | PIN1 | FALSE | Upregulated |
| 0,660659336 | 0,053401425 | ENTPD1 | FALSE | Upregulated |
| -0,361616129 | 0,053591481 | CCNB1IP1 | FALSE | Downregulated |
| -0,225977842 | 0,053748486 | NARS2 | FALSE | Downregulated |
| -0,35958585 | 0,053748486 | NOL9 | FALSE | Downregulated |
| 0,182559431 | 0,053748486 | RRAGA | FALSE | Upregulated |
| -0,34664757 | 0,053832348 | SPSB3 | FALSE | Downregulated |
| 0,304084565 | 0,053846013 | RNF126 | FALSE | Upregulated |
| -0,586575146 | 0,05385371 | SDHAP2 | FALSE | Downregulated |
| 0,258177127 | 0,053886107 | EDF1 | FALSE | Upregulated |
| -1,126766584 | 0,053886107 | LOC102724768 | FALSE | Downregulated |
| 0,951427486 | 0,053886107 | MVD | FALSE | Upregulated |
| -0,294323611 | 0,053904915 | SIAH1 | FALSE | Downregulated |
| 0,445909219 | 0,05395758 | C14orf105 | FALSE | Upregulated |
| 1,597323646 | 0,053965573 | LOC101927070 | FALSE | Upregulated |
| -1,760760822 | 0,053965573 | LOC102723899 | FALSE | Downregulated |
| -0,364268096 | 0,053967125 | ARMC5 | FALSE | Downregulated |
| 1,943189062 | 0,053967125 | LOC102723697 | FALSE | Upregulated |
| 0,364229767 | 0,053967125 | TLR4 | FALSE | Upregulated |
| -0,54906966 | 0,053967125 | ZNF117 | FALSE | Downregulated |
| -0,501235869 | 0,054077052 | MYO16 | FALSE | Downregulated |
| 1,019752503 | 0,054119065 | LOC101928472 | FALSE | Upregulated |
| -0,304109394 | 0,054155702 | UPF2 | FALSE | Downregulated |
| -0,288720407 | 0,054162378 | METTL2B | FALSE | Downregulated |
| 0,319968396 | 0,054162378 | PFDN4 | FALSE | Upregulated |
| 0,33398753 | 0,054393781 | GGCT | FALSE | Upregulated |
| 1,162688612 | 0,054394985 | ZFHX4-AS1 | FALSE | Upregulated |
| -0,20593655 | 0,054460092 | ZC3H7B | FALSE | Downregulated |
| 0,500311865 | 0,054516198 | MFSD10 | FALSE | Upregulated |
| -0,293302055 | 0,054542342 | ALKBH8 | FALSE | Downregulated |
| -0,398383062 | 0,054542342 | C8B | FALSE | Downregulated |
| -1,388537565 | 0,054542342 | CTD-2201I18.1 | FALSE | Downregulated |
| 0,29070028 | 0,054542342 | EFTUD1 | FALSE | Upregulated |

| | | | | |
|---|---|---|---|---|
| 0,3719491 | 0,054542342 | LPXN | FALSE | Upregulated |
| 0,67456632 | 0,054542342 | MX2 | FALSE | Upregulated |
| 0,797533425 | 0,054542342 | PARM1 | FALSE | Upregulated |
| 1,264225345 | 0,054542342 | RAD54L | FALSE | Upregulated |
| -0,288415415 | 0,054542342 | TMEM161B | FALSE | Downregulated |
| 0,274713218 | 0,054565623 | CCT3 | FALSE | Upregulated |
| -1,794336179 | 0,054567032 | AC007879.2 | FALSE | Downregulated |
| 0,435802925 | 0,054567032 | IL16 | FALSE | Upregulated |
| -0,291848937 | 0,054750232 | FZD4 | FALSE | Downregulated |
| -0,35433886 | 0,054783889 | EIF3J-AS1 | FALSE | Downregulated |
| -0,397018287 | 0,054820909 | CLCN5 | FALSE | Downregulated |
| 0,334498042 | 0,054870771 | SLC35B2 | FALSE | Upregulated |
| 0,607546536 | 0,054870771 | SLC35G1 | FALSE | Upregulated |
| -0,258645923 | 0,054913441 | DCLRE1A | FALSE | Downregulated |
| 0,495750284 | 0,054946575 | GSKIP | FALSE | Upregulated |
| 0,35500139 | 0,054963316 | PHTF2 | FALSE | Upregulated |
| -0,492164322 | 0,055098347 | ADH4 | FALSE | Downregulated |
| 0,871678984 | 0,055100719 | LOC102724571 | FALSE | Upregulated |
| 0,471922492 | 0,055175938 | SLC36A4 | FALSE | Upregulated |
| 0,334733585 | 0,055210259 | MBOAT7 | FALSE | Upregulated |
| 1,06605077 | 0,055327285 | NELL2 | FALSE | Upregulated |
| 0,333550164 | 0,055431342 | CYB5R4 | FALSE | Upregulated |
| -2,519238158 | 0,055482777 | AC012309.5 | FALSE | Downregulated |
| 0,458646664 | 0,055482777 | TBC1D7 | FALSE | Upregulated |
| 0,345795051 | 0,055488388 | CYP2U1 | FALSE | Upregulated |
| 0,249878082 | 0,055515778 | CNIH1 | FALSE | Upregulated |
| -0,20443803 | 0,055515778 | CRKL | FALSE | Downregulated |
| 0,290200707 | 0,055515778 | CYB561A3 | FALSE | Upregulated |
| -0,451676223 | 0,055515778 | IMPA2 | FALSE | Downregulated |
| 1,033778809 | 0,055515778 | SEMA7A | FALSE | Upregulated |
| 0,357752279 | 0,055515778 | TXNDC5 | FALSE | Upregulated |
| 0,244936025 | 0,055521283 | ANAPC7 | FALSE | Upregulated |
| -0,372524361 | 0,055566911 | ZSCAN12 | FALSE | Downregulated |
| -0,499832534 | 0,05558629 | SPG20 | FALSE | Downregulated |
| 2,277727499 | 0,055656857 | FAM27B | FALSE | Upregulated |
| -0,238752313 | 0,05566197 | HDGF | FALSE | Downregulated |
| 0,834626219 | 0,05567225 | RASGRF2 | FALSE | Upregulated |
| -0,172036503 | 0,055704282 | DCBLD2 | FALSE | Downregulated |
| 0,324830897 | 0,055728841 | TIMM8B | FALSE | Upregulated |
| -0,193872951 | 0,055846565 | WDR36 | FALSE | Downregulated |
| -0,764010974 | 0,055995225 | HERC2P2 | FALSE | Downregulated |
| 0,253749164 | 0,056030455 | GNB2 | FALSE | Upregulated |
| 0,17596959 | 0,056062489 | UBQLN1 | FALSE | Upregulated |
| 0,316021854 | 0,0561016 | SCOC | FALSE | Upregulated |
| 0,274237293 | 0,056111911 | CHD4 | FALSE | Upregulated |
| 1,982001675 | 0,056152325 | PDZK1IP1 | FALSE | Upregulated |
| 0,637249156 | 0,056168188 | IFITM1 | FALSE | Upregulated |
| -0,563737163 | 0,056177955 | KCNMB3 | FALSE | Downregulated |
| 0,268848235 | 0,05624672 | PDXK | FALSE | Upregulated |
| 0,967641838 | 0,056275565 | IGSF3 | FALSE | Upregulated |
| -0,88282446 | 0,056275565 | LINC00969 | FALSE | Downregulated |

| | | | | |
|---|---|---|---|---|
| -0,738945117 | 0,056283003 | RAPGEFL1 | FALSE | Downregulated |
| 0,753131222 | 0,056304228 | MCTP2 | FALSE | Upregulated |
| -0,305461489 | 0,056351754 | HRSP12 | FALSE | Downregulated |
| -0,369411976 | 0,056417718 | PXMP2 | FALSE | Downregulated |
| 0,532238452 | 0,056423561 | OSBPL11 | FALSE | Upregulated |
| 0,303917791 | 0,056423561 | YARS | FALSE | Upregulated |
| 0,27981719 | 0,056499894 | LSM1 | FALSE | Upregulated |
| 0,868276611 | 0,056525908 | PRKCE | FALSE | Upregulated |
| 0,281905786 | 0,05661582 | C12orf44 | FALSE | Upregulated |
| 0,681399537 | 0,056660833 | ABCB4 | FALSE | Upregulated |
| 0,209654478 | 0,056666649 | CHMP4B | FALSE | Upregulated |
| 0,693690532 | 0,056666649 | WWC1 | FALSE | Upregulated |
| 0,374490906 | 0,056716897 | ZNF544 | FALSE | Upregulated |
| 0,675150873 | 0,05672592 | FBXW9 | FALSE | Upregulated |
| -0,775520632 | 0,056895908 | CCDC183 | FALSE | Downregulated |
| -0,278331086 | 0,056895908 | SLC9B2 | FALSE | Downregulated |
| 0,200257111 | 0,056937218 | SRP14 | FALSE | Upregulated |
| 0,662177353 | 0,056949208 | ADCK1 | FALSE | Upregulated |
| -2,151389973 | 0,056949208 | BDNF | FALSE | Downregulated |
| -1,183115022 | 0,056949208 | LOC102723770 | FALSE | Downregulated |
| -0,252411515 | 0,057009934 | ZNF180 | FALSE | Downregulated |
| 0,214245405 | 0,057024754 | RPTOR | FALSE | Upregulated |
| -0,227321105 | 0,057029258 | ANKRD28 | FALSE | Downregulated |
| -0,579418604 | 0,057033407 | ASPG | FALSE | Downregulated |
| -0,403491223 | 0,057198875 | HFE2 | FALSE | Downregulated |
| 1,617823444 | 0,057253395 | RP11-18H21.1 | FALSE | Upregulated |
| -0,545700045 | 0,057310614 | LOC441155 | FALSE | Downregulated |
| -0,280041248 | 0,057332759 | COPS7B | FALSE | Downregulated |
| 0,321210037 | 0,057332759 | GTPBP2 | FALSE | Upregulated |
| -0,431426676 | 0,057332759 | ZNF772 | FALSE | Downregulated |
| 0,219030214 | 0,057401458 | MXD4 | FALSE | Upregulated |
| 0,653192322 | 0,0574671 | RALGPS1 | FALSE | Upregulated |
| -0,681004236 | 0,057490982 | TMEM44-AS1 | FALSE | Downregulated |
| 0,179540053 | 0,057524329 | KIAA0319L | FALSE | Upregulated |
| -0,226485539 | 0,057524329 | MPP5 | FALSE | Downregulated |
| 0,361886135 | 0,057538852 | CDH1 | FALSE | Upregulated |
| 0,299705672 | 0,057541829 | PDLIM1 | FALSE | Upregulated |
| -0,257612788 | 0,057541829 | THYN1 | FALSE | Downregulated |
| -0,183973519 | 0,05755378 | RAB11B | FALSE | Downregulated |
| -0,57785397 | 0,057599902 | WFDC1 | FALSE | Downregulated |
| -1,32351776 | 0,057691186 | RP5-1050D4.5 | FALSE | Downregulated |
| 0,325100502 | 0,057706567 | MFHAS1 | FALSE | Upregulated |
| -0,367487493 | 0,05771536 | PTMS | FALSE | Downregulated |
| -0,223145884 | 0,057885408 | NDST1 | FALSE | Downregulated |
| -0,269611046 | 0,057914607 | ZNF638 | FALSE | Downregulated |
| -0,475278837 | 0,05792808 | BST1 | FALSE | Downregulated |
| -0,407943582 | 0,05792808 | ZNF251 | FALSE | Downregulated |
| 0,306299491 | 0,057957612 | FUS | FALSE | Upregulated |
| -0,25205902 | 0,057957612 | UNC119B | FALSE | Downregulated |
| -0,223388624 | 0,0579617 | C1orf109 | FALSE | Downregulated |
| 1,119135008 | 0,058018129 | KLRC1 | FALSE | Upregulated |

| | | | | |
|---|---|---|---|---|
| 0,522861284 | 0,058048582 | FGL2 | FALSE | Upregulated |
| -0,884152763 | 0,058079568 | NUTM2B | FALSE | Downregulated |
| 0,264209538 | 0,058239305 | AP2S1 | FALSE | Upregulated |
| 0,562191651 | 0,058263736 | RPP25 | FALSE | Upregulated |
| -0,378516601 | 0,058329155 | IFT46 | FALSE | Downregulated |
| 0,275180344 | 0,058331508 | MDH1 | FALSE | Upregulated |
| 0,5616603 | 0,058384007 | ACRBP | FALSE | Upregulated |
| 0,208408046 | 0,058392251 | AP4E1 | FALSE | Upregulated |
| 0,810494392 | 0,058392251 | AQP4 | FALSE | Upregulated |
| 0,319548661 | 0,058392251 | NAP1L1 | FALSE | Upregulated |
| -0,305660465 | 0,058392251 | POLL | FALSE | Downregulated |
| 1,693259679 | 0,058461645 | C12orf55 | FALSE | Upregulated |
| 0,585366123 | 0,058461645 | SRPX2 | FALSE | Upregulated |
| -0,303912351 | 0,058548932 | IQGAP2 | FALSE | Downregulated |
| -1,072121994 | 0,058548932 | POPDC2 | FALSE | Downregulated |
| -0,855286215 | 0,058555579 | LOC101927331 | FALSE | Downregulated |
| -0,693807617 | 0,058643439 | GOLGA6L9 | FALSE | Downregulated |
| -0,330261477 | 0,058736681 | RMDN2 | FALSE | Downregulated |
| -0,328082784 | 0,058766691 | SERPINF1 | FALSE | Downregulated |
| -0,496845839 | 0,058856719 | ZGPAT | FALSE | Downregulated |
| 0,52851948 | 0,058872648 | NUDT18 | FALSE | Upregulated |
| -0,158940987 | 0,058970081 | ZFYVE9 | FALSE | Downregulated |
| 1,587823287 | 0,059060544 | LOC101927902 | FALSE | Upregulated |
| -0,261449549 | 0,059108162 | ERCC6 | FALSE | Downregulated |
| -1,852723137 | 0,059192927 | UBE2Q2P2 | FALSE | Downregulated |
| 0,244540526 | 0,059241817 | RBM12 | FALSE | Upregulated |
| -0,956993804 | 0,059265404 | PKHD1L1 | FALSE | Downregulated |
| -0,293329464 | 0,059265404 | TTC31 | FALSE | Downregulated |
| -0,56395871 | 0,059265404 | ZNF844 | FALSE | Downregulated |
| 0,274194678 | 0,059315673 | TCEAL1 | FALSE | Upregulated |
| 0,276999033 | 0,059413396 | KRTCAP2 | FALSE | Upregulated |
| 1,096695612 | 0,059420611 | KIAA1211L | FALSE | Upregulated |
| 0,577917772 | 0,059420611 | TLR2 | FALSE | Upregulated |
| -0,441054841 | 0,05946865 | TMCO6 | FALSE | Downregulated |
| 0,657942247 | 0,059487769 | PTGFR | FALSE | Upregulated |
| 0,947588129 | 0,059503643 | ST14 | FALSE | Upregulated |
| -0,85734263 | 0,059523703 | LOC285638 | FALSE | Downregulated |
| -0,266702901 | 0,059574931 | APOB | FALSE | Downregulated |
| 0,428763316 | 0,059612102 | PPM1M | FALSE | Upregulated |
| -0,376047845 | 0,05961531 | KIAA1107 | FALSE | Downregulated |
| 0,232726536 | 0,059692099 | MARK2 | FALSE | Upregulated |
| 0,960330706 | 0,059747176 | LPAR2 | FALSE | Upregulated |
| 1,252354693 | 0,059747176 | PVT1 | FALSE | Upregulated |
| 1,833318692 | 0,059747176 | SFN | FALSE | Upregulated |
| -0,44227135 | 0,059759841 | EGFR | FALSE | Downregulated |
| 0,4860112 | 0,059797047 | SCIMP | FALSE | Upregulated |
| -0,579414874 | 0,059830705 | LOC102724761 | FALSE | Downregulated |
| -0,366130518 | 0,059841831 | GLYCTK | FALSE | Downregulated |
| 0,223554268 | 0,059851215 | SCAMP2 | FALSE | Upregulated |
| -1,112005557 | 0,0598793 | LOC102723576 | FALSE | Downregulated |
| 0,513450514 | 0,059971641 | RGS19 | FALSE | Upregulated |

| | | | | |
|---|---|---|---|---|
| 0,643131812 | 0,060151008 | DAPP1 | FALSE | Upregulated |
| 1,705121732 | 0,060180018 | SLC12A5 | FALSE | Upregulated |
| 0,762439035 | 0,060218554 | CLEC7A | FALSE | Upregulated |
| 0,750360705 | 0,060236833 | CD300E | FALSE | Upregulated |
| -0,491846324 | 0,060272513 | HNF1B | FALSE | Downregulated |
| 1,328156304 | 0,060272513 | LOC101928105 | FALSE | Upregulated |
| -0,299709341 | 0,060376907 | ATG2B | FALSE | Downregulated |
| -0,20991216 | 0,060449888 | PABPC1 | FALSE | Downregulated |
| -1,023990049 | 0,060492143 | RSPH4A | FALSE | Downregulated |
| -0,221689491 | 0,060492472 | AHCYL2 | FALSE | Downregulated |
| -0,915123829 | 0,060492472 | FAM110D | FALSE | Downregulated |
| -0,247831279 | 0,060492472 | RSPH3 | FALSE | Downregulated |
| 1,008264779 | 0,060492472 | SDS | FALSE | Upregulated |
| -0,249796427 | 0,060492472 | UCKL1 | FALSE | Downregulated |
| 0,497581329 | 0,060547984 | SPIN4 | FALSE | Upregulated |
| -0,886367593 | 0,060598975 | LOC101928887 | FALSE | Downregulated |
| 0,300342905 | 0,060666033 | GSDMD | FALSE | Upregulated |
| 0,921743161 | 0,060708396 | MMP24 | FALSE | Upregulated |
| -0,272396932 | 0,060708396 | ZNF507 | FALSE | Downregulated |
| -0,325450787 | 0,060726293 | BOD1L1 | FALSE | Downregulated |
| -0,357184014 | 0,060747272 | INS-IGF2 | FALSE | Downregulated |
| 0,140616411 | 0,060761065 | GRSF1 | FALSE | Upregulated |
| -1,379458431 | 0,060899372 | KIF25 | FALSE | Downregulated |
| -0,250948507 | 0,060947554 | ZNF304 | FALSE | Downregulated |
| 0,27215725 | 0,060977379 | MSH6 | FALSE | Upregulated |
| -0,154635365 | 0,060977379 | TOMM70A | FALSE | Downregulated |
| -1,10420984 | 0,061014713 | GRAPL | FALSE | Downregulated |
| -1,965021547 | 0,061039996 | LOC101927870 | FALSE | Downregulated |
| 1,070790457 | 0,061092207 | ADTRP | FALSE | Upregulated |
| -0,475847811 | 0,061092207 | COX10-AS1 | FALSE | Downregulated |
| 0,704622458 | 0,061120659 | SPSB4 | FALSE | Upregulated |
| -0,509986691 | 0,061120659 | THBS3 | FALSE | Downregulated |
| 0,365556127 | 0,061137165 | APRT | FALSE | Upregulated |
| -0,628716874 | 0,061137165 | SRD5A2 | FALSE | Downregulated |
| -0,278231646 | 0,061139784 | SNX33 | FALSE | Downregulated |
| -0,316087012 | 0,061158662 | COG3 | FALSE | Downregulated |
| -1,109373553 | 0,061161503 | GSTA2 | FALSE | Downregulated |
| -0,306865859 | 0,061262055 | AFMID | FALSE | Downregulated |
| 0,612434052 | 0,061262055 | EIF5A2 | FALSE | Upregulated |
| -0,585602462 | 0,061282771 | S1PR3 | FALSE | Downregulated |
| -0,41574797 | 0,06137701 | LOC101928319 | FALSE | Downregulated |
| -0,412017435 | 0,061385492 | ARSD | FALSE | Downregulated |
| 0,65014478 | 0,06151985 | KIAA1324L | FALSE | Upregulated |
| -0,534505683 | 0,061607907 | HES1 | FALSE | Downregulated |
| -0,190740527 | 0,061655751 | GABPA | FALSE | Downregulated |
| -0,260657418 | 0,061660751 | CCDC174 | FALSE | Downregulated |
| 0,180034142 | 0,06171586 | HNRNPR | FALSE | Upregulated |
| -0,367191592 | 0,061732214 | ADH1B | FALSE | Downregulated |
| -1,119893922 | 0,062002798 | MAP6D1 | FALSE | Downregulated |
| 0,362027001 | 0,062002798 | MYEOV2 | FALSE | Upregulated |
| 0,358939133 | 0,062037708 | BCL2L1 | FALSE | Upregulated |

| | | | | |
|---|---|---|---|---|
| -0,401448043 | 0,062107159 | TPCN2 | FALSE | Downregulated |
| -0,398912659 | 0,062107159 | YBEY | FALSE | Downregulated |
| 0,197066762 | 0,062110913 | PSMD6 | FALSE | Upregulated |
| -0,378040099 | 0,062258325 | LOC101927045 | FALSE | Downregulated |
| 0,492127773 | 0,062353787 | TMEM51 | FALSE | Upregulated |
| -0,347345615 | 0,06235819 | FAM214A | FALSE | Downregulated |
| -0,376358632 | 0,062385285 | MYCBP2 | FALSE | Downregulated |
| 0,182517233 | 0,062385285 | ZNF207 | FALSE | Upregulated |
| -0,663244071 | 0,062494243 | NUDT4P1 | FALSE | Downregulated |
| 0,273991229 | 0,062736635 | ERI3 | FALSE | Upregulated |
| -0,186343637 | 0,062736635 | FUBP3 | FALSE | Downregulated |
| 0,393480037 | 0,062748377 | ZNF593 | FALSE | Upregulated |
| 0,598756099 | 0,062805672 | CXCR2P1 | FALSE | Upregulated |
| -0,754443653 | 0,062823688 | LINC00174 | FALSE | Downregulated |
| -0,38809235 | 0,063034434 | HS3ST3B1 | FALSE | Downregulated |
| 0,173317627 | 0,063086476 | CCNC | FALSE | Upregulated |
| -0,318803274 | 0,063171072 | MAP10 | FALSE | Downregulated |
| -0,526794487 | 0,063185363 | WHAMMP1 | FALSE | Downregulated |
| 0,202689923 | 0,063225818 | PSMB2 | FALSE | Upregulated |
| -0,485208429 | 0,063256251 | RFXAP | FALSE | Downregulated |
| 1,644331064 | 0,063281929 | LOC101927481 | FALSE | Upregulated |
| -0,406453376 | 0,06333904 | SYTL5 | FALSE | Downregulated |
| -0,439439022 | 0,063350572 | DACH1 | FALSE | Downregulated |
| 0,493905164 | 0,06335387 | SLC25A35 | FALSE | Upregulated |
| -1,395280306 | 0,063516601 | XG | FALSE | Downregulated |
| -0,229897047 | 0,063527311 | DHX40 | FALSE | Downregulated |
| 0,241147353 | 0,063595038 | RPP38 | FALSE | Upregulated |
| 0,227537128 | 0,063595038 | TMCO4 | FALSE | Upregulated |
| 0,291637324 | 0,063625801 | FARSA | FALSE | Upregulated |
| -1,00464076 | 0,063644097 | LOC101928143 | FALSE | Downregulated |
| 0,87025267 | 0,063739317 | TFRC | FALSE | Upregulated |
| 1,555234631 | 0,063841598 | RUFY4 | FALSE | Upregulated |
| -2,203538547 | 0,063911464 | LINC00693 | FALSE | Downregulated |
| 0,792463669 | 0,063947215 | NRG1 | FALSE | Upregulated |
| -0,469045283 | 0,063984466 | ZNF587 | FALSE | Downregulated |
| -0,217733686 | 0,064064447 | OTUD4 | FALSE | Downregulated |
| -0,388516595 | 0,064226047 | ARSK | FALSE | Downregulated |
| -0,381890616 | 0,064226047 | PCDH12 | FALSE | Downregulated |
| 0,606661548 | 0,064272043 | VMO1 | FALSE | Upregulated |
| -0,277918553 | 0,064304095 | SNAPC1 | FALSE | Downregulated |
| -1,242716612 | 0,064308011 | LOC101928882 | FALSE | Downregulated |
| 0,379527459 | 0,064314331 | ACTN1 | FALSE | Upregulated |
| -0,488622129 | 0,064314331 | CIDEB | FALSE | Downregulated |
| -1,025636585 | 0,064314331 | KCNH7 | FALSE | Downregulated |
| 0,333834183 | 0,064383928 | TSTA3 | FALSE | Upregulated |
| 0,189773152 | 0,064401762 | PSMD3 | FALSE | Upregulated |
| -0,304005728 | 0,06445722 | NUDT16P1 | FALSE | Downregulated |
| -0,916855392 | 0,064571045 | FAM86HP | FALSE | Downregulated |
| -0,447461489 | 0,064584726 | POLR3GL | FALSE | Downregulated |
| -0,60743015 | 0,064662463 | BCL2L10 | FALSE | Downregulated |
| -0,34865021 | 0,064686507 | GDF2 | FALSE | Downregulated |

| | | | | |
|---|---|---|---|---|
| 0,214409286 | 0,064742647 | GPAA1 | FALSE | Upregulated |
| -0,694312505 | 0,064799398 | CCDC30 | FALSE | Downregulated |
| 0,354484185 | 0,064843191 | RINL | FALSE | Upregulated |
| 0,390983443 | 0,064844173 | ARHGEF2 | FALSE | Upregulated |
| -0,430549119 | 0,064873842 | LOC100132815 | FALSE | Downregulated |
| 0,380780187 | 0,064873842 | RGL1 | FALSE | Upregulated |
| 0,192077037 | 0,06487603 | UTP18 | FALSE | Upregulated |
| -0,323006053 | 0,064910765 | ANKRD16 | FALSE | Downregulated |
| 0,176358755 | 0,064910765 | AREL1 | FALSE | Upregulated |
| 0,256287055 | 0,065074412 | UBA6 | FALSE | Upregulated |
| -0,19128819 | 0,065087261 | TMEM68 | FALSE | Downregulated |
| -0,323756427 | 0,065165095 | NR2C1 | FALSE | Downregulated |
| -0,301419264 | 0,06524386 | AKAP8 | FALSE | Downregulated |
| 0,721436741 | 0,065257999 | EID3 | FALSE | Upregulated |
| 1,078634711 | 0,065302371 | DDIT4 | FALSE | Upregulated |
| 0,936143978 | 0,065382801 | RGAG1 | FALSE | Upregulated |
| 0,250146634 | 0,06542369 | NUDC | FALSE | Upregulated |
| 0,899721443 | 0,065429361 | HIST1H2AE | FALSE | Upregulated |
| -0,47754963 | 0,065440334 | SLC23A2 | FALSE | Downregulated |
| -0,267626201 | 0,065458986 | C1orf56 | FALSE | Downregulated |
| 0,254306989 | 0,065458986 | DYSF | FALSE | Upregulated |
| -0,381762751 | 0,065458986 | GIGYF1 | FALSE | Downregulated |
| 0,451614931 | 0,065458986 | MAFK | FALSE | Upregulated |
| -0,338619422 | 0,065458986 | PDE2A | FALSE | Downregulated |
| 0,296807411 | 0,065458986 | RAD51C | FALSE | Upregulated |
| 0,446029747 | 0,065458986 | TCF7 | FALSE | Upregulated |
| 1,449100936 | 0,065458986 | ZNF683 | FALSE | Upregulated |
| 0,186524084 | 0,065510329 | FKBP15 | FALSE | Upregulated |
| -0,862160233 | 0,065559417 | BMS1P2 | FALSE | Downregulated |
| 0,92318791 | 0,065575836 | GPSM1 | FALSE | Upregulated |
| -0,306453969 | 0,065867382 | TRMT13 | FALSE | Downregulated |
| -0,245903192 | 0,065923614 | HPS4 | FALSE | Downregulated |
| 1,663686473 | 0,065944852 | HLA-DPB2 | FALSE | Upregulated |
| 2,800282546 | 0,066030732 | CEACAM6 | FALSE | Upregulated |
| -0,410080511 | 0,066090719 | ARHGAP32 | FALSE | Downregulated |
| -0,447981995 | 0,066116608 | MORN1 | FALSE | Downregulated |
| 0,260516617 | 0,06612642 | MOV10 | FALSE | Upregulated |
| 0,171423166 | 0,066142379 | USP39 | FALSE | Upregulated |
| -0,274540713 | 0,066142379 | ZCCHC24 | FALSE | Downregulated |
| -0,431839002 | 0,066179177 | C16orf46 | FALSE | Downregulated |
| -0,540602211 | 0,066179177 | EFCAB6 | FALSE | Downregulated |
| 0,180937222 | 0,066189621 | TINF2 | FALSE | Upregulated |
| 1,297923903 | 0,066261183 | KPNA7 | FALSE | Upregulated |
| 0,231870782 | 0,066261183 | RAB1A | FALSE | Upregulated |
| -0,795830442 | 0,066261183 | ZNF239 | FALSE | Downregulated |
| -0,37816375 | 0,066267516 | BTAF1 | FALSE | Downregulated |
| -0,275903963 | 0,066281361 | PCBD2 | FALSE | Downregulated |
| 0,20113372 | 0,066332249 | TOM1 | FALSE | Upregulated |
| 1,524713028 | 0,066354874 | DTNA | FALSE | Upregulated |
| -0,819570029 | 0,066462508 | RP11-677M14.7 | FALSE | Downregulated |
| 0,674014328 | 0,066470899 | APBA2 | FALSE | Upregulated |

| | | | | |
|---|---|---|---|---|
| -0,454134468 | 0,066470899 | LOC100289361 | FALSE | Downregulated |
| 0,208635228 | 0,066470899 | UTP11L | FALSE | Upregulated |
| -0,466254194 | 0,066529643 | ZNF415 | FALSE | Downregulated |
| 0,908197495 | 0,06654581 | LOC101929223 | FALSE | Upregulated |
| 0,376790158 | 0,06654581 | PML | FALSE | Upregulated |
| -0,656011618 | 0,06658226 | LOC441242 | FALSE | Downregulated |
| 0,545814035 | 0,066631924 | IL17RB | FALSE | Upregulated |
| -0,312379861 | 0,066681347 | THUMPD2 | FALSE | Downregulated |
| -0,36124187 | 0,06673652 | IMPA1 | FALSE | Downregulated |
| -0,374812 | 0,06673652 | NXF1 | FALSE | Downregulated |
| -0,693729555 | 0,066749171 | LOC100996724 | FALSE | Downregulated |
| 0,197493258 | 0,066749171 | PSMF1 | FALSE | Upregulated |
| -0,373333045 | 0,066749171 | UACA | FALSE | Downregulated |
| 0,277311762 | 0,066781469 | ATIC | FALSE | Upregulated |
| 1,036736104 | 0,066819024 | CD200 | FALSE | Upregulated |
| -1,362440107 | 0,066822169 | LOC102724402 | FALSE | Downregulated |
| -0,470339645 | 0,066822169 | TPT1-AS1 | FALSE | Downregulated |
| 0,500246316 | 0,06685917 | GIMAP4 | FALSE | Upregulated |
| 0,598888104 | 0,06685917 | TRIM15 | FALSE | Upregulated |
| -0,535394363 | 0,06685917 | ZFP3 | FALSE | Downregulated |
| 0,221513902 | 0,066871815 | TFE3 | FALSE | Upregulated |
| 0,165276997 | 0,066885128 | AP2B1 | FALSE | Upregulated |
| -0,901467436 | 0,067045585 | MN1 | FALSE | Downregulated |
| 0,605948992 | 0,067045707 | CYSLTR1 | FALSE | Upregulated |
| 0,305161011 | 0,067058306 | ANO10 | FALSE | Upregulated |
| -1,092632619 | 0,067131856 | DKFZP434F142 | FALSE | Downregulated |
| -0,841174593 | 0,067131856 | LOC100270804 | FALSE | Downregulated |
| 0,224618086 | 0,067153179 | CCDC94 | FALSE | Upregulated |
| -0,721509583 | 0,067184513 | KANK4 | FALSE | Downregulated |
| 0,425746182 | 0,067184772 | RBL1 | FALSE | Upregulated |
| 0,661089602 | 0,067194373 | BZRAP1-AS1 | FALSE | Upregulated |
| -0,358092047 | 0,067206913 | PHACTR4 | FALSE | Downregulated |
| 0,227182089 | 0,067391863 | GLTPD1 | FALSE | Upregulated |
| 0,892903322 | 0,067560927 | FGFBP2 | FALSE | Upregulated |
| 0,234979631 | 0,067568031 | GSS | FALSE | Upregulated |
| 0,264509649 | 0,067613354 | CCM2 | FALSE | Upregulated |
| 0,260702338 | 0,06763468 | PPIP5K1 | FALSE | Upregulated |
| -0,228884087 | 0,067771519 | HTRA2 | FALSE | Downregulated |
| -0,459010374 | 0,067779354 | TOR3A | FALSE | Downregulated |
| -0,483529137 | 0,067862397 | TBC1D32 | FALSE | Downregulated |
| 0,223553977 | 0,06793331 | GNPTAB | FALSE | Upregulated |
| -0,245901236 | 0,068013908 | LCORL | FALSE | Downregulated |
| -0,267552225 | 0,068060469 | EPM2A | FALSE | Downregulated |
| -0,220186923 | 0,068098679 | AZIN1 | FALSE | Downregulated |
| -0,329519665 | 0,068098679 | ZNF449 | FALSE | Downregulated |
| -0,380782351 | 0,068150635 | TSPAN18 | FALSE | Downregulated |
| 0,437401742 | 0,06816538 | SPTSSA | FALSE | Upregulated |
| -0,197822162 | 0,068172422 | KDM2A | FALSE | Downregulated |
| -0,429348904 | 0,068360074 | NR2F1 | FALSE | Downregulated |
| -0,407664967 | 0,068444823 | NPY1R | FALSE | Downregulated |
| -0,931945897 | 0,068711077 | RP4-756H11.3 | FALSE | Downregulated |

| | | | | |
|---|---|---|---|---|
| 0,537312326 | 0,068711077 | SPN | FALSE | Upregulated |
| 0,345145602 | 0,068766803 | PPAN | FALSE | Upregulated |
| 0,213560272 | 0,068808569 | WWP2 | FALSE | Upregulated |
| 0,482644692 | 0,068847485 | DNAJB11 | FALSE | Upregulated |
| 0,268743812 | 0,06884952 | RFK | FALSE | Upregulated |
| 0,68449291 | 0,068887871 | XYLT1 | FALSE | Upregulated |
| -1,504943019 | 0,069064927 | GPR179 | FALSE | Downregulated |
| -0,512678546 | 0,069068977 | MSANTD2 | FALSE | Downregulated |
| -0,403563728 | 0,069100464 | FAM211B | FALSE | Downregulated |
| -1,415804231 | 0,069100464 | LRP1B | FALSE | Downregulated |
| -0,775550222 | 0,069117534 | LOC101927054 | FALSE | Downregulated |
| -0,726520154 | 0,069181551 | BMS1P5 | FALSE | Downregulated |
| -0,29223507 | 0,069181551 | CLIC4 | FALSE | Downregulated |
| -0,311025721 | 0,069181551 | DMD | FALSE | Downregulated |
| 0,274998204 | 0,069181551 | FAM220A | FALSE | Upregulated |
| -0,434365235 | 0,069181551 | FAM221A | FALSE | Downregulated |
| 0,227736189 | 0,069181551 | MSL3 | FALSE | Upregulated |
| 0,998277932 | 0,069181551 | RP11-503C24.4 | FALSE | Upregulated |
| 0,378816964 | 0,069181551 | SLC9A3R1 | FALSE | Upregulated |
| 0,191894896 | 0,069263432 | UBFD1 | FALSE | Upregulated |
| -0,385924723 | 0,06927406 | AKAP12 | FALSE | Downregulated |
| -0,202186146 | 0,06944424 | STAT5B | FALSE | Downregulated |
| -0,269927548 | 0,069463166 | TBL1X | FALSE | Downregulated |
| -0,898388005 | 0,069521641 | LOC101928434 | FALSE | Downregulated |
| -0,265320054 | 0,069523265 | ZMYM6 | FALSE | Downregulated |
| 0,732434632 | 0,069648282 | ASRGL1 | FALSE | Upregulated |
| -0,334763991 | 0,069648282 | GIPC2 | FALSE | Downregulated |
| -0,628121345 | 0,069952164 | MGC12916 | FALSE | Downregulated |
| -0,314730173 | 0,070044255 | KHDRBS3 | FALSE | Downregulated |
| -0,228473018 | 0,070087404 | DIS3L | FALSE | Downregulated |
| -0,251919868 | 0,070093029 | GTF2H3 | FALSE | Downregulated |
| 0,581599971 | 0,070162079 | PTPRU | FALSE | Upregulated |
| -0,306488755 | 0,070202021 | AZGP1 | FALSE | Downregulated |
| -0,283875633 | 0,070251599 | BDH1 | FALSE | Downregulated |
| -0,804889341 | 0,070251599 | GSTM2 | FALSE | Downregulated |
| -0,34074989 | 0,070251599 | ZNF496 | FALSE | Downregulated |
| 0,219609522 | 0,070333067 | GABARAP | FALSE | Upregulated |
| -0,251381655 | 0,070336961 | SZT2 | FALSE | Downregulated |
| -0,180618809 | 0,070383547 | LSM14B | FALSE | Downregulated |
| -0,413757126 | 0,070383547 | ZNF548 | FALSE | Downregulated |
| -1,122528665 | 0,070399482 | MAP1A | FALSE | Downregulated |
| -0,267184649 | 0,070437651 | KIAA1109 | FALSE | Downregulated |
| -0,237725639 | 0,070449414 | MGA | FALSE | Downregulated |
| -0,28783202 | 0,070474352 | SMIM11 | FALSE | Downregulated |
| 0,199027392 | 0,070478106 | TPD52L2 | FALSE | Upregulated |
| -0,835812967 | 0,070504299 | FLJ44342 | FALSE | Downregulated |
| 0,544182183 | 0,070653105 | ITGAM | FALSE | Upregulated |
| -1,50469223 | 0,070657223 | FLJ42258 | FALSE | Downregulated |
| -0,204152553 | 0,070666685 | ATG4C | FALSE | Downregulated |
| -0,320566809 | 0,070728986 | JRKL | FALSE | Downregulated |
| -0,624440574 | 0,070937982 | ARHGAP10 | FALSE | Downregulated |

| | | | | |
|---|---|---|---|---|
| 0,836220889 | 0,071027416 | P2RY14 | FALSE | Upregulated |
| 2,036530215 | 0,071088111 | CCL25 | FALSE | Upregulated |
| 1,693938829 | 0,071088111 | SAA2 | FALSE | Upregulated |
| 0,828445165 | 0,071205178 | BATF3 | FALSE | Upregulated |
| -0,997957372 | 0,071232173 | LOC339874 | FALSE | Downregulated |
| 0,238893553 | 0,071257601 | COX5A | FALSE | Upregulated |
| 0,188512141 | 0,071274921 | SUPT16H | FALSE | Upregulated |
| -0,20334425 | 0,071297854 | ATL2 | FALSE | Downregulated |
| -0,177134036 | 0,071297854 | MIER3 | FALSE | Downregulated |
| -0,40678121 | 0,071329257 | LINC00667 | FALSE | Downregulated |
| 0,320974833 | 0,071360373 | ORAI1 | FALSE | Upregulated |
| -0,24016362 | 0,071374265 | ZNF7 | FALSE | Downregulated |
| 0,219597705 | 0,07159722 | ARL8A | FALSE | Upregulated |
| -0,7382458 | 0,07159722 | EP400NL | FALSE | Downregulated |
| 0,336187307 | 0,071647536 | CASP8 | FALSE | Upregulated |
| -0,337712228 | 0,071647536 | KMT2E | FALSE | Downregulated |
| 0,349096755 | 0,071647536 | SMYD2 | FALSE | Upregulated |
| 0,566799645 | 0,071669918 | DOK1 | FALSE | Upregulated |
| -1,066187834 | 0,071785634 | CYP3A43 | FALSE | Downregulated |
| 0,619690658 | 0,071785634 | HSPA4L | FALSE | Upregulated |
| 0,176947537 | 0,071820548 | RMDN3 | FALSE | Upregulated |
| -0,128605743 | 0,071857038 | METAP1 | FALSE | Downregulated |
| 0,249291747 | 0,071857038 | MTF1 | FALSE | Upregulated |
| -0,467053202 | 0,071857038 | WDR78 | FALSE | Downregulated |
| 0,581510433 | 0,071913687 | SLC37A1 | FALSE | Upregulated |
| -1,383379094 | 0,071941905 | CYP7A1 | FALSE | Downregulated |
| -0,534523144 | 0,071941905 | HID1 | FALSE | Downregulated |
| -0,790416056 | 0,071941905 | LOC102724493 | FALSE | Downregulated |
| -0,709088736 | 0,071941905 | LOC389834 | FALSE | Downregulated |
| 0,547070723 | 0,072039897 | AGBL3 | FALSE | Upregulated |
| 0,315134115 | 0,072039897 | MICALL1 | FALSE | Upregulated |
| -0,181516896 | 0,072039897 | PRPF31 | FALSE | Downregulated |
| -0,23275834 | 0,072043506 | SFXN1 | FALSE | Downregulated |
| 0,306366161 | 0,072122954 | DTNB | FALSE | Upregulated |
| 0,858286564 | 0,072166891 | PTGDS | FALSE | Upregulated |
| -1,145578881 | 0,072265147 | SPAG8 | FALSE | Downregulated |
| -0,472984384 | 0,07229381 | TSPAN7 | FALSE | Downregulated |
| -0,379400125 | 0,072357117 | PAN3 | FALSE | Downregulated |
| 0,46669179 | 0,072369556 | LOC100505812 | FALSE | Upregulated |
| -0,83173557 | 0,072381184 | LOC101928249 | FALSE | Downregulated |
| 2,538448959 | 0,072381184 | NLRP7 | FALSE | Upregulated |
| 0,406224051 | 0,07238427 | CHPF | FALSE | Upregulated |
| -0,365337298 | 0,07238427 | HOOK1 | FALSE | Downregulated |
| 0,35453073 | 0,07238427 | HSD17B2 | FALSE | Upregulated |
| -1,212463861 | 0,072396613 | ABCA13 | FALSE | Downregulated |
| -0,213371359 | 0,072443706 | EIF3E | FALSE | Downregulated |
| -0,25930072 | 0,072455917 | SRRT | FALSE | Downregulated |
| -0,582423137 | 0,072459452 | SMAD9 | FALSE | Downregulated |
| -0,444695782 | 0,072459452 | ZCCHC6 | FALSE | Downregulated |
| -1,941201321 | 0,072518133 | DENND5B-AS1 | FALSE | Downregulated |
| -0,65996626 | 0,072541297 | BCO2 | FALSE | Downregulated |

| | | | | |
|---|---|---|---|---|
| -0,339298965 | 0,072559116 | AUTS2 | FALSE | Downregulated |
| 0,375015154 | 0,072673965 | C19orf12 | FALSE | Upregulated |
| -0,332514966 | 0,072673965 | DCUN1D2 | FALSE | Downregulated |
| -0,588542059 | 0,072673965 | LOC102723908 | FALSE | Downregulated |
| 0,854663887 | 0,072673965 | SYN1 | FALSE | Upregulated |
| -1,080896233 | 0,072710889 | C11orf85 | FALSE | Downregulated |
| -1,057398746 | 0,072720245 | CYP3A7-CYP3AP1 | FALSE | Downregulated |
| -0,183350563 | 0,072861904 | PDS5B | FALSE | Downregulated |
| -0,568001492 | 0,072886817 | KLF8 | FALSE | Downregulated |
| -0,510627635 | 0,07293922 | ABCC9 | FALSE | Downregulated |
| 0,585093816 | 0,072954411 | HES6 | FALSE | Upregulated |
| -2,225807616 | 0,072954411 | MIR1247 | FALSE | Downregulated |
| 0,250013908 | 0,072954411 | PITPNA | FALSE | Upregulated |
| 0,277826398 | 0,073010344 | ALG5 | FALSE | Upregulated |
| -0,2501046 | 0,073036664 | SDR39U1 | FALSE | Downregulated |
| 0,6423313 | 0,073043067 | LPAR1 | FALSE | Upregulated |
| 0,289397335 | 0,073091989 | KIAA0020 | FALSE | Upregulated |
| -0,498565561 | 0,073103743 | MICALCL | FALSE | Downregulated |
| -0,223570473 | 0,073285101 | COA5 | FALSE | Downregulated |
| -1,160044211 | 0,073285101 | LOC101929112 | FALSE | Downregulated |
| -1,218019115 | 0,073307634 | VIP | FALSE | Downregulated |
| 0,450156134 | 0,073362564 | ZNF710 | FALSE | Upregulated |
| -0,478262744 | 0,073371666 | LINC00324 | FALSE | Downregulated |
| 0,512284459 | 0,073406567 | RAB34 | FALSE | Upregulated |
| 0,223459226 | 0,07352883 | MAZ | FALSE | Upregulated |
| 0,477789484 | 0,07352883 | RBM14-RBM4 | FALSE | Upregulated |
| -0,663092389 | 0,073537339 | LOC102723919 | FALSE | Downregulated |
| 0,347005992 | 0,073588222 | FBXL18 | FALSE | Upregulated |
| -0,656044768 | 0,073614001 | PRR18 | FALSE | Downregulated |
| -0,27482842 | 0,073839583 | RBM12B | FALSE | Downregulated |
| -0,474620996 | 0,073839583 | TRAF3IP2 | FALSE | Downregulated |
| -0,568476052 | 0,073867457 | FLJ38668 | FALSE | Downregulated |
| 0,380707026 | 0,07386872 | PLEKHA8 | FALSE | Upregulated |
| -0,244033319 | 0,073879582 | ORAI3 | FALSE | Downregulated |
| 0,345821881 | 0,074015089 | C18orf32 | FALSE | Upregulated |
| 2,290575183 | 0,074030393 | LOC102724705 | FALSE | Upregulated |
| -0,2481216 | 0,074124944 | TOMM20 | FALSE | Downregulated |
| 0,412617952 | 0,074158908 | QSOX1 | FALSE | Upregulated |
| -0,243641815 | 0,074193559 | ATXN1 | FALSE | Downregulated |
| 0,291395445 | 0,074366173 | ABTB1 | FALSE | Upregulated |
| 0,419665285 | 0,074366173 | AP4M1 | FALSE | Upregulated |
| -0,203725803 | 0,074508382 | ZNF564 | FALSE | Downregulated |
| -0,373925335 | 0,074514458 | PRRG4 | FALSE | Downregulated |
| 0,883675995 | 0,074551211 | LOC102724296 | FALSE | Upregulated |
| -0,266890889 | 0,074637979 | TTC21B | FALSE | Downregulated |
| -0,375040819 | 0,074638845 | DPYD | FALSE | Downregulated |
| -1,811238677 | 0,074638845 | LOC100130285 | FALSE | Downregulated |
| 0,308372382 | 0,074638845 | MOSPD2 | FALSE | Upregulated |
| -0,213804076 | 0,074646568 | C2CD5 | FALSE | Downregulated |
| -0,313049414 | 0,074646568 | COX7C | FALSE | Downregulated |
| -0,650466205 | 0,074646568 | FITM1 | FALSE | Downregulated |

| | | | | |
|---|---|---|---|---|
| -0,4289949 | 0,074679039 | FLJ37035 | FALSE | Downregulated |
| 0,215799125 | 0,074679039 | TOR1B | FALSE | Upregulated |
| 0,928978385 | 0,074682745 | GPR68 | FALSE | Upregulated |
| -0,414057931 | 0,074750441 | ZNF862 | FALSE | Downregulated |
| -0,371644037 | 0,074809665 | AK9 | FALSE | Downregulated |
| -1,536353003 | 0,074810833 | LOC101929555 | FALSE | Downregulated |
| -0,323277544 | 0,074812216 | PAPD7 | FALSE | Downregulated |
| 0,234669278 | 0,07493024 | CYP20A1 | FALSE | Upregulated |
| 0,537614213 | 0,075141097 | KCTD12 | FALSE | Upregulated |
| -0,229291581 | 0,07515886 | MTA1 | FALSE | Downregulated |
| 0,23974835 | 0,075205569 | TPRG1L | FALSE | Upregulated |
| 0,530661027 | 0,075335901 | CD163 | FALSE | Upregulated |
| 1,989372797 | 0,075356316 | LOC644172 | FALSE | Upregulated |
| -1,378411801 | 0,075493318 | ERVFRD-1 | FALSE | Downregulated |
| 0,571386354 | 0,075497225 | SIMC1 | FALSE | Upregulated |
| -0,465179902 | 0,075668034 | WDR90 | FALSE | Downregulated |
| -0,334717448 | 0,075720294 | GPT | FALSE | Downregulated |
| -0,227333269 | 0,075730908 | GPALPP1 | FALSE | Downregulated |
| -0,343047728 | 0,075784359 | ACTR3B | FALSE | Downregulated |
| -0,365352829 | 0,075784359 | FLT1 | FALSE | Downregulated |
| 0,549545012 | 0,075784359 | MBOAT1 | FALSE | Upregulated |
| 0,265022865 | 0,075784359 | SLC35C2 | FALSE | Upregulated |
| 0,247628747 | 0,075846942 | FLOT1 | FALSE | Upregulated |
| -0,509035558 | 0,075846942 | GANC | FALSE | Downregulated |
| 1,097224443 | 0,075846942 | GLI1 | FALSE | Upregulated |
| -2,433955059 | 0,075846942 | LINC01133 | FALSE | Downregulated |
| -0,772935702 | 0,075846942 | LOC101929351 | FALSE | Downregulated |
| -1,109878458 | 0,075846942 | PCP2 | FALSE | Downregulated |
| 0,751561051 | 0,075846942 | TNFSF11 | FALSE | Upregulated |
| 0,368750336 | 0,075846942 | TRPM4 | FALSE | Upregulated |
| -0,344022737 | 0,075915681 | GCKR | FALSE | Downregulated |
| 0,324984125 | 0,075915681 | LINC01146 | FALSE | Upregulated |
| -0,30606459 | 0,07591886 | DET1 | FALSE | Downregulated |
| -0,395362474 | 0,075941472 | CNBP | FALSE | Downregulated |
| -0,203874363 | 0,075995545 | MIB1 | FALSE | Downregulated |
| -0,235691137 | 0,075995545 | PAPD4 | FALSE | Downregulated |
| -0,37656481 | 0,07604937 | SBF2-AS1 | FALSE | Downregulated |
| -1,147857792 | 0,076104456 | ARHGAP28 | FALSE | Downregulated |
| 0,220748684 | 0,076172798 | CREG1 | FALSE | Upregulated |
| -0,530941564 | 0,076172798 | SNHG8 | FALSE | Downregulated |
| -0,249278644 | 0,076202953 | MTFR1 | FALSE | Downregulated |
| 1,556981896 | 0,07621933 | MAL | FALSE | Upregulated |
| 0,205227989 | 0,076359858 | EIF3I | FALSE | Upregulated |
| 1,924523208 | 0,076379262 | DNASE2B | FALSE | Upregulated |
| -0,349434408 | 0,076379262 | XRCC5 | FALSE | Downregulated |
| 0,309504961 | 0,076538392 | FAM177A1 | FALSE | Upregulated |
| -0,271664018 | 0,076830548 | CHD2 | FALSE | Downregulated |
| 0,327488564 | 0,076887254 | CLN8 | FALSE | Upregulated |
| -0,25947723 | 0,076887254 | RPL22 | FALSE | Downregulated |
| -0,381493734 | 0,076912754 | RBBP6 | FALSE | Downregulated |
| 0,256034417 | 0,077075703 | PCGF5 | FALSE | Upregulated |

| | | | | |
|---|---|---|---|---|
| -0,216227667 | 0,077132002 | LARP4 | FALSE | Downregulated |
| 0,510494701 | 0,077249621 | NRROS | FALSE | Upregulated |
| -0,980622324 | 0,077258542 | CACNB2 | FALSE | Downregulated |
| -0,349899925 | 0,077456506 | MAN1A1 | FALSE | Downregulated |
| -0,3624177 | 0,077477772 | UBR4 | FALSE | Downregulated |
| -1,199794127 | 0,07748341 | PPBP | FALSE | Downregulated |
| -0,335560759 | 0,077495113 | C16orf13 | FALSE | Downregulated |
| -0,874582275 | 0,077589242 | LOC102724345 | FALSE | Downregulated |
| 0,42046953 | 0,077589242 | TRAPPC4 | FALSE | Upregulated |
| 0,481473494 | 0,077592185 | FMNL3 | FALSE | Upregulated |
| 0,435845509 | 0,077766983 | SMA5 | FALSE | Upregulated |
| 0,32205956 | 0,077780983 | IFIT5 | FALSE | Upregulated |
| 0,696897502 | 0,077780983 | SREBF1 | FALSE | Upregulated |
| -0,326636432 | 0,077955497 | CYP2C9 | FALSE | Downregulated |
| -0,302302037 | 0,077976483 | HADH | FALSE | Downregulated |
| 0,353504686 | 0,077976483 | RRP7A | FALSE | Upregulated |
| -0,299893638 | 0,077996266 | SMURF2 | FALSE | Downregulated |
| 0,210977632 | 0,078073536 | MRPS7 | FALSE | Upregulated |
| 0,299940207 | 0,078263907 | ATP6V1H | FALSE | Upregulated |
| 0,254916793 | 0,078263907 | SCARB1 | FALSE | Upregulated |
| -0,26022073 | 0,078468547 | RPRD2 | FALSE | Downregulated |
| 0,469417211 | 0,078468547 | UPP1 | FALSE | Upregulated |
| 1,4501044 | 0,07847004 | TMEM132D | FALSE | Upregulated |
| 0,393635194 | 0,07867687 | ARHGAP31 | FALSE | Upregulated |
| -0,194655234 | 0,07867687 | RFC1 | FALSE | Downregulated |
| 0,370331023 | 0,078701704 | VASH1 | FALSE | Upregulated |
| 0,425919123 | 0,078729029 | PLA1A | FALSE | Upregulated |
| -0,488564863 | 0,078788894 | LOC149351 | FALSE | Downregulated |
| -0,11143155 | 0,078809967 | USP7 | FALSE | Downregulated |
| -0,453030437 | 0,078911065 | ACMSD | FALSE | Downregulated |
| 0,344068069 | 0,078911065 | C17orf62 | FALSE | Upregulated |
| -0,162736083 | 0,078911065 | DCAF10 | FALSE | Downregulated |
| -1,620825717 | 0,078930595 | ALOX12 | FALSE | Downregulated |
| 0,301778135 | 0,078930595 | SNX24 | FALSE | Upregulated |
| -0,382273393 | 0,078940212 | FNBP4 | FALSE | Downregulated |
| -0,82570666 | 0,078940212 | HMCN2 | FALSE | Downregulated |
| -0,737197079 | 0,078940212 | LOC101927589 | FALSE | Downregulated |
| 0,145652107 | 0,078940212 | USP22 | FALSE | Upregulated |
| 0,531163865 | 0,078940212 | ZFAND5 | FALSE | Upregulated |
| -1,156602254 | 0,078963926 | LOC100506621 | FALSE | Downregulated |
| -1,054538872 | 0,078978712 | HEATR4 | FALSE | Downregulated |
| 0,324301752 | 0,079013029 | FNBP1 | FALSE | Upregulated |
| -0,37972767 | 0,079013029 | GCAT | FALSE | Downregulated |
| 0,361649912 | 0,079025918 | ZNF267 | FALSE | Upregulated |
| 0,969031377 | 0,079030993 | ULBP3 | FALSE | Upregulated |
| -0,493635067 | 0,07904207 | RP11-328K4.1 | FALSE | Downregulated |
| 0,806509949 | 0,079049558 | C5orf34 | FALSE | Upregulated |
| -0,349611009 | 0,079451273 | EML1 | FALSE | Downregulated |
| 0,626562836 | 0,079462048 | PIGZ | FALSE | Upregulated |
| -0,243312151 | 0,079479062 | ZNF655 | FALSE | Downregulated |
| -1,193360683 | 0,079520735 | MC1R | FALSE | Downregulated |

| | | | | |
|---|---|---|---|---|
| 0,512796406 | 0,079543033 | CSF3R | FALSE | Upregulated |
| 0,189679343 | 0,079569278 | CKAP5 | FALSE | Upregulated |
| -0,25840393 | 0,079569278 | TBC1D12 | FALSE | Downregulated |
| -0,171189833 | 0,079678834 | PHF10 | FALSE | Downregulated |
| -1,542410469 | 0,079678834 | PPFIA4 | FALSE | Downregulated |
| 0,229092052 | 0,079685454 | KLHL22 | FALSE | Upregulated |
| -0,280247013 | 0,079685454 | ZNF865 | FALSE | Downregulated |
| 0,302202296 | 0,079701295 | ATR | FALSE | Upregulated |
| 0,517701983 | 0,079787969 | PDE1B | FALSE | Upregulated |
| -0,264075776 | 0,079932087 | SUN2 | FALSE | Downregulated |
| -1,339058899 | 0,080020335 | CYP4F24P | FALSE | Downregulated |
| -1,671395469 | 0,080020335 | DIO3 | FALSE | Downregulated |
| 0,148451806 | 0,080020335 | MTCH1 | FALSE | Upregulated |
| 0,235536818 | 0,080030047 | HMCES | FALSE | Upregulated |
| 0,265570132 | 0,080069436 | ATP5SL | FALSE | Upregulated |
| -1,117083178 | 0,080202979 | C6orf141 | FALSE | Downregulated |
| -1,752375586 | 0,080212327 | C2CD4C | FALSE | Downregulated |
| -0,48061062 | 0,080212327 | SMIM6 | FALSE | Downregulated |
| 0,755808812 | 0,080392474 | KCNJ10 | FALSE | Upregulated |
| -0,501315496 | 0,080392474 | TTR | FALSE | Downregulated |
| -0,216181343 | 0,080392474 | USP34 | FALSE | Downregulated |
| -0,29330789 | 0,080392474 | USP53 | FALSE | Downregulated |
| -0,538090079 | 0,080464864 | TVP23C | FALSE | Downregulated |
| -0,126696245 | 0,080510228 | API5 | FALSE | Downregulated |
| 0,524804529 | 0,080510228 | CDKN2C | FALSE | Upregulated |
| -0,122363464 | 0,080599885 | NEK9 | FALSE | Downregulated |
| 2,272660415 | 0,080693889 | ACAN | FALSE | Upregulated |
| -1,158564623 | 0,080693889 | NEURL1 | FALSE | Downregulated |
| -0,284284258 | 0,080827727 | BBS9 | FALSE | Downregulated |
| 0,246472062 | 0,080827727 | DEAF1 | FALSE | Upregulated |
| 0,279531857 | 0,080827727 | RC3H2 | FALSE | Upregulated |
| -0,497039833 | 0,080827727 | SCN9A | FALSE | Downregulated |
| 0,219469419 | 0,080905794 | SIAH2 | FALSE | Upregulated |
| -0,507007495 | 0,080981499 | IL6R | FALSE | Downregulated |
| 0,40216557 | 0,080981499 | SRPRB | FALSE | Upregulated |
| -0,19619435 | 0,080981499 | USP48 | FALSE | Downregulated |
| 0,771395486 | 0,080996907 | DAGLA | FALSE | Upregulated |
| 1,579307703 | 0,080998067 | CXCL5 | FALSE | Upregulated |
| 0,338067577 | 0,081039842 | MED27 | FALSE | Upregulated |
| 0,288725083 | 0,08105296 | EMC8 | FALSE | Upregulated |
| 0,478846311 | 0,08105296 | LBH | FALSE | Upregulated |
| 1,38430126 | 0,08105296 | SHC4 | FALSE | Upregulated |
| -1,928975839 | 0,081171299 | LOC102723842 | FALSE | Downregulated |
| -0,174724307 | 0,081171299 | ZNF260 | FALSE | Downregulated |
| 0,400887211 | 0,081172387 | EHBP1L1 | FALSE | Upregulated |
| -0,221006227 | 0,081355902 | DCAF6 | FALSE | Downregulated |
| -0,840145561 | 0,08136414 | LOC100506974 | FALSE | Downregulated |
| 0,911931652 | 0,08136414 | TMCC2 | FALSE | Upregulated |
| 0,570401167 | 0,081371602 | SGK1 | FALSE | Upregulated |
| -0,224135874 | 0,081478711 | BRPF3 | FALSE | Downregulated |
| 0,505899431 | 0,081478711 | PRKRIP1 | FALSE | Upregulated |

| | | | | |
|---|---|---|---|---|
| 0,187440556 | 0,081557515 | HIRA | FALSE | Upregulated |
| -0,239905275 | 0,081557515 | WDR92 | FALSE | Downregulated |
| -0,319942241 | 0,081557515 | ZNF358 | FALSE | Downregulated |
| 0,418288899 | 0,081611575 | AIFM2 | FALSE | Upregulated |
| 0,462094986 | 0,081682429 | TMPRSS2 | FALSE | Upregulated |
| 0,353006625 | 0,081728535 | PDCL3 | FALSE | Upregulated |
| -1,121380786 | 0,081728535 | PHYHIP | FALSE | Downregulated |
| 0,308164523 | 0,081728535 | TMEM251 | FALSE | Upregulated |
| 0,236618742 | 0,081845178 | COPB1 | FALSE | Upregulated |
| -0,296579967 | 0,081845178 | UFL1 | FALSE | Downregulated |
| 0,938818169 | 0,081958657 | CLDN11 | FALSE | Upregulated |
| 0,335221801 | 0,081958657 | RNASEH2B | FALSE | Upregulated |
| -0,361693104 | 0,082041066 | MYSM1 | FALSE | Downregulated |
| 0,607670204 | 0,082088644 | EOMES | FALSE | Upregulated |
| -0,277375674 | 0,082101427 | TNFAIP8L1 | FALSE | Downregulated |
| 0,239346532 | 0,082128231 | SERF2 | FALSE | Upregulated |
| 0,908110563 | 0,08226239 | MLLT11 | FALSE | Upregulated |
| 1,552887752 | 0,082265541 | CXADRP2 | FALSE | Upregulated |
| -1,529945679 | 0,082265541 | GPR112 | FALSE | Downregulated |
| -0,634712633 | 0,082437443 | ZNF204P | FALSE | Downregulated |
| -0,976621808 | 0,082478463 | LOC101929513 | FALSE | Downregulated |
| -1,29521131 | 0,082504213 | LOC100268168 | FALSE | Downregulated |
| -0,244267888 | 0,082621114 | NT5DC1 | FALSE | Downregulated |
| -0,312242763 | 0,082693437 | PRPF39 | FALSE | Downregulated |
| -0,320486979 | 0,08272279 | CBWD2 | FALSE | Downregulated |
| 0,351289817 | 0,082756809 | XRCC4 | FALSE | Upregulated |
| -0,406137749 | 0,082899509 | LOC286437 | FALSE | Downregulated |
| 0,410911545 | 0,082923888 | DAB2IP | FALSE | Upregulated |
| -0,533966938 | 0,082923888 | DUSP10 | FALSE | Downregulated |
| 0,869085378 | 0,082923888 | FOXC1 | FALSE | Upregulated |
| -0,394948275 | 0,082923888 | LBX2-AS1 | FALSE | Downregulated |
| -0,211702691 | 0,082923888 | RPAP1 | FALSE | Downregulated |
| -3,967423732 | 0,082923888 | SFRP2 | FALSE | Downregulated |
| 1,38483681 | 0,082923888 | TMEM217 | FALSE | Upregulated |
| 0,438009857 | 0,082923888 | TRIM11 | FALSE | Upregulated |
| -0,263310595 | 0,082935831 | CYTH1 | FALSE | Downregulated |
| 0,300431872 | 0,082935831 | TOMM5 | FALSE | Upregulated |
| -0,646577285 | 0,082986794 | LOC101928868 | FALSE | Downregulated |
| -0,293062351 | 0,083110104 | C5 | FALSE | Downregulated |
| -0,572971739 | 0,083110104 | CNKSR2 | FALSE | Downregulated |
| 0,83539869 | 0,083110104 | GZMH | FALSE | Upregulated |
| -0,768173389 | 0,083110104 | TENM1 | FALSE | Downregulated |
| 0,296638137 | 0,083160064 | TOR2A | FALSE | Upregulated |
| -0,31193459 | 0,083247165 | PHF7 | FALSE | Downregulated |
| 0,310946295 | 0,083247165 | PRELID1 | FALSE | Upregulated |
| -0,189338074 | 0,083263522 | SLMAP | FALSE | Downregulated |
| -1,114118951 | 0,083325367 | LOC101929561 | FALSE | Downregulated |
| 2,014309924 | 0,083363623 | DCX | FALSE | Upregulated |
| 0,454021955 | 0,083363623 | P2RY13 | FALSE | Upregulated |
| -0,294633436 | 0,083520607 | CTNNBIP1 | FALSE | Downregulated |
| -0,437374593 | 0,083520607 | RAG1 | FALSE | Downregulated |

| | | | | |
|---|---|---|---|---|
| 0,723877075 | 0,083543361 | NRGN | FALSE | Upregulated |
| 0,720949078 | 0,083578006 | GPBAR1 | FALSE | Upregulated |
| -1,018050079 | 0,083623648 | ADARB2 | FALSE | Downregulated |
| -0,572939049 | 0,083675566 | NPIPA2 | FALSE | Downregulated |
| -0,20706694 | 0,083775205 | TNRC6B | FALSE | Downregulated |
| -1,176986134 | 0,083780779 | LINC00671 | FALSE | Downregulated |
| 0,284376641 | 0,083835365 | TBC1D25 | FALSE | Upregulated |
| 0,35364875 | 0,083848214 | RALGDS | FALSE | Upregulated |
| -0,370000672 | 0,0838649 | ABCG8 | FALSE | Downregulated |
| 1,154876286 | 0,083930188 | CLECL1 | FALSE | Upregulated |
| -1,066755942 | 0,083930188 | CXorf66 | FALSE | Downregulated |
| -0,254141708 | 0,083981083 | ERF | FALSE | Downregulated |
| -0,916158195 | 0,083981083 | LOC401127 | FALSE | Downregulated |
| -0,143311085 | 0,083981083 | RSL1D1 | FALSE | Downregulated |
| 1,76937122 | 0,083981083 | SLC45A1 | FALSE | Upregulated |
| -0,219763548 | 0,083981083 | TMEM230 | FALSE | Downregulated |
| 2,42951328 | 0,08408655 | TCN1 | FALSE | Upregulated |
| 0,995681731 | 0,084096553 | FFAR4 | FALSE | Upregulated |
| -0,661286709 | 0,084096553 | STK4-AS1 | FALSE | Downregulated |
| 0,62126261 | 0,084126981 | LOC101927151 | FALSE | Upregulated |
| -0,727540571 | 0,084165794 | LOC101927393 | FALSE | Downregulated |
| 0,210988382 | 0,084215736 | SMARCA5 | FALSE | Upregulated |
| 0,668518622 | 0,084334676 | NOTUM | FALSE | Upregulated |
| 0,294098028 | 0,0845012 | RSU1 | FALSE | Upregulated |
| -0,315935012 | 0,0845012 | ZNF182 | FALSE | Downregulated |
| -0,183499291 | 0,084656805 | SKI | FALSE | Downregulated |
| 0,577249571 | 0,084703825 | RIMS3 | FALSE | Upregulated |
| -0,795959831 | 0,084728949 | LOC101927105 | FALSE | Downregulated |
| 0,199966704 | 0,084733522 | CTDNEP1 | FALSE | Upregulated |
| 0,644273014 | 0,084733522 | KRT222 | FALSE | Upregulated |
| -0,737731888 | 0,084874803 | MEG3 | FALSE | Downregulated |
| 1,775588917 | 0,084874803 | SCG3 | FALSE | Upregulated |
| -0,850163432 | 0,084898119 | TRNS1 | FALSE | Downregulated |
| -0,181935296 | 0,084976505 | RAB2A | FALSE | Downregulated |
| 0,265943292 | 0,085021108 | NUDT16L1 | FALSE | Upregulated |
| -0,169257519 | 0,085065514 | KANSL1 | FALSE | Downregulated |
| -0,725583294 | 0,085065514 | LEKR1 | FALSE | Downregulated |
| 0,83517809 | 0,085135808 | AC011899.9 | FALSE | Upregulated |
| 0,441691539 | 0,085135808 | BAG3 | FALSE | Upregulated |
| 0,321565878 | 0,085204603 | TIMM17A | FALSE | Upregulated |
| 0,32672995 | 0,0852657 | ATAD3A | FALSE | Upregulated |
| -1,193018017 | 0,085266575 | GRIP2 | FALSE | Downregulated |
| 0,25863919 | 0,085309169 | DCAF13 | FALSE | Upregulated |
| -0,723745021 | 0,085309169 | LOC100133331 | FALSE | Downregulated |
| -1,002807885 | 0,08539186 | TREH | FALSE | Downregulated |
| 0,211903538 | 0,085445826 | COX4I1 | FALSE | Upregulated |
| -0,244697179 | 0,085511657 | IRGQ | FALSE | Downregulated |
| -0,719101534 | 0,08560361 | BTBD8 | FALSE | Downregulated |
| 0,741540261 | 0,08560361 | DEFB1 | FALSE | Upregulated |
| -0,550919847 | 0,085649833 | CCDC150 | FALSE | Downregulated |
| 0,236159471 | 0,085649833 | PEF1 | FALSE | Upregulated |

| | | | | |
|---|---|---|---|---|
| 0,743364813 | 0,085649833 | PPP2R2B | FALSE | Upregulated |
| -0,502627078 | 0,085680159 | ASS1 | FALSE | Downregulated |
| 0,624795219 | 0,085838465 | LURAP1L | FALSE | Upregulated |
| -0,259186798 | 0,085838465 | NARG2 | FALSE | Downregulated |
| 0,243061961 | 0,085838465 | PLEKHM1 | FALSE | Upregulated |
| 0,754623194 | 0,085850882 | THBS1 | FALSE | Upregulated |
| -1,375399049 | 0,085851535 | LOC100128276 | FALSE | Downregulated |
| -0,578068651 | 0,085995432 | LOC646976 | FALSE | Downregulated |
| 0,928097932 | 0,086016646 | OXCT2 | FALSE | Upregulated |
| 0,684437323 | 0,086141339 | PLXNA3 | FALSE | Upregulated |
| -0,365029717 | 0,086141339 | ZNF543 | FALSE | Downregulated |
| 0,477451869 | 0,086167357 | FKBP11 | FALSE | Upregulated |
| 0,256952185 | 0,086243255 | ABHD4 | FALSE | Upregulated |
| -1,37496232 | 0,086243255 | SPOCK3 | FALSE | Downregulated |
| 0,327163648 | 0,086490345 | TMEM194A | FALSE | Upregulated |
| -2,19368972 | 0,086634267 | EBF3 | FALSE | Downregulated |
| 0,877680737 | 0,086634267 | EME1 | FALSE | Upregulated |
| -1,885540207 | 0,086710842 | LOC101060264 | FALSE | Downregulated |
| 0,238637168 | 0,086732 | ILF2 | FALSE | Upregulated |
| -0,376045238 | 0,086776004 | C6orf57 | FALSE | Downregulated |
| -0,374191176 | 0,08681237 | RP11-392P7.6 | FALSE | Downregulated |
| -0,317426068 | 0,08684987 | TXNDC16 | FALSE | Downregulated |
| -1,761423474 | 0,086853237 | LOC102724682 | FALSE | Downregulated |
| 0,272283134 | 0,086853237 | PAK1IP1 | FALSE | Upregulated |
| -1,495272797 | 0,086853237 | TLL1 | FALSE | Downregulated |
| -2,055001154 | 0,086953862 | MED15P9 | FALSE | Downregulated |
| -1,136455312 | 0,087094709 | CWH43 | FALSE | Downregulated |
| -0,364296836 | 0,087186709 | USP31 | FALSE | Downregulated |
| -1,550976456 | 0,087244265 | TNR | FALSE | Downregulated |
| 0,992900692 | 0,087277781 | RASEF | FALSE | Upregulated |
| 0,273797435 | 0,087277781 | YIF1A | FALSE | Upregulated |
| -0,585533503 | 0,087312514 | FNIP2 | FALSE | Downregulated |
| 0,208610717 | 0,087364017 | ZDHHC20 | FALSE | Upregulated |
| 0,351714934 | 0,087450643 | FAM69A | FALSE | Upregulated |
| -0,494249517 | 0,087499737 | AC004893.11 | FALSE | Downregulated |
| -0,447019279 | 0,08756951 | ZNF550 | FALSE | Downregulated |
| 0,380177314 | 0,087576095 | DDHD1 | FALSE | Upregulated |
| -0,191962345 | 0,087721014 | C1GALT1 | FALSE | Downregulated |
| -0,324214396 | 0,087875512 | FGD5 | FALSE | Downregulated |
| -0,841329173 | 0,088014347 | ADC | FALSE | Downregulated |
| -0,457082315 | 0,088111567 | ZCWPW2 | FALSE | Downregulated |
| 0,415925588 | 0,088131052 | FUT8 | FALSE | Upregulated |
| 0,482959931 | 0,088136296 | ZNF107 | FALSE | Upregulated |
| -0,495782365 | 0,088193679 | CYP2C8 | FALSE | Downregulated |
| -0,399223392 | 0,088193679 | LOC101928505 | FALSE | Downregulated |
| -0,669443762 | 0,088208458 | C3orf70 | FALSE | Downregulated |
| -0,139944096 | 0,088217817 | UPF1 | FALSE | Downregulated |
| -0,22112704 | 0,088290495 | ORC5 | FALSE | Downregulated |
| -0,366610359 | 0,088352672 | DPYS | FALSE | Downregulated |
| -0,256365507 | 0,088434881 | NEK4 | FALSE | Downregulated |
| -1,755243022 | 0,088434881 | SLC6A4 | FALSE | Downregulated |

| | | | | |
|---|---|---|---|---|
| 0,720341642 | 0,088499159 | IL7R | FALSE | Upregulated |
| 1,780954789 | 0,088499159 | RP11-16E12.2 | FALSE | Upregulated |
| -0,342356018 | 0,088506805 | ZNF680 | FALSE | Downregulated |
| -0,227031875 | 0,088589287 | F2 | FALSE | Downregulated |
| 0,215482121 | 0,088638564 | ERH | FALSE | Upregulated |
| 0,195414887 | 0,088704574 | KCTD9 | FALSE | Upregulated |
| -0,378558186 | 0,088717961 | C2orf88 | FALSE | Downregulated |
| -0,15081132 | 0,088753217 | RAB14 | FALSE | Downregulated |
| 0,221539554 | 0,088772295 | ANAPC2 | FALSE | Upregulated |
| 0,234531977 | 0,088772295 | MRPL22 | FALSE | Upregulated |
| 1,385650602 | 0,088855054 | APOBEC3H | FALSE | Upregulated |
| -0,577389165 | 0,088855054 | C1orf173 | FALSE | Downregulated |
| -0,271187917 | 0,088908981 | CAPN15 | FALSE | Downregulated |
| 1,516279674 | 0,088908981 | ZFP91-CNTF | FALSE | Upregulated |
| 0,388149968 | 0,088996569 | RBP1 | FALSE | Upregulated |
| 0,432440753 | 0,089002041 | S100A10 | FALSE | Upregulated |
| 0,192772106 | 0,089008211 | UBP1 | FALSE | Upregulated |
| 0,241696869 | 0,089041364 | GNG12 | FALSE | Upregulated |
| -0,761709512 | 0,089041364 | LINC01127 | FALSE | Downregulated |
| -1,047051153 | 0,089041364 | SALL4 | FALSE | Downregulated |
| -0,242533664 | 0,089041364 | WASF1 | FALSE | Downregulated |
| -0,446421861 | 0,089102124 | CLEC4G | FALSE | Downregulated |
| 0,233661357 | 0,089123697 | EXOSC9 | FALSE | Upregulated |
| -0,145504643 | 0,089123697 | PUM2 | FALSE | Downregulated |
| -1,176712694 | 0,089138398 | LOC101927645 | FALSE | Downregulated |
| -0,159954092 | 0,089144261 | HMG20A | FALSE | Downregulated |
| -0,228701277 | 0,089406631 | TTC38 | FALSE | Downregulated |
| -0,698734921 | 0,089462045 | GSDMB | FALSE | Downregulated |
| -0,379164524 | 0,089583527 | HNRNPA1L2 | FALSE | Downregulated |
| -1,348001222 | 0,08959411 | COL28A1 | FALSE | Downregulated |
| -0,217656936 | 0,089665253 | ALKBH5 | FALSE | Downregulated |
| -0,426119955 | 0,089709905 | ZNF10 | FALSE | Downregulated |
| 0,977229992 | 0,089745971 | LOC102724141 | FALSE | Upregulated |
| -0,706773548 | 0,089798363 | OAT | FALSE | Downregulated |
| -0,398512392 | 0,089844305 | CLASRP | FALSE | Downregulated |
| 0,330937058 | 0,089881815 | TACC1 | FALSE | Upregulated |
| -0,888403676 | 0,089900159 | TRNL1 | FALSE | Downregulated |
| -0,453396361 | 0,089961341 | GPM6A | FALSE | Downregulated |
| -0,358046217 | 0,090000175 | UBXN2B | FALSE | Downregulated |
| 0,15759815 | 0,090141865 | NUP205 | FALSE | Upregulated |
| -0,235566982 | 0,090189338 | SHPK | FALSE | Downregulated |
| 0,295141148 | 0,090255864 | PGM2 | FALSE | Upregulated |
| -0,405248989 | 0,090293161 | ZSCAN31 | FALSE | Downregulated |
| -0,182072843 | 0,090450918 | C4orf3 | FALSE | Downregulated |
| 0,286207959 | 0,090536861 | MCRS1 | FALSE | Upregulated |
| -0,236873354 | 0,090635889 | ALDH9A1 | FALSE | Downregulated |
| 0,212451303 | 0,090635889 | NHP2L1 | FALSE | Upregulated |
| 0,33638467 | 0,090636328 | INPP4A | FALSE | Upregulated |
| 0,1526817 | 0,090636328 | PPP1R8 | FALSE | Upregulated |
| 0,186490284 | 0,090666762 | RPF1 | FALSE | Upregulated |
| -0,256580425 | 0,090700309 | C21orf33 | FALSE | Downregulated |

| | | | | |
|---|---|---|---|---|
| -0,610046303 | 0,090784801 | UNC5CL | FALSE | Downregulated |
| -0,24559784 | 0,090962146 | RMDN1 | FALSE | Downregulated |
| 0,563943435 | 0,091073289 | PLB1 | FALSE | Upregulated |
| -0,269544999 | 0,091103201 | AASDH | FALSE | Downregulated |
| -1,482821908 | 0,091186964 | LOC101927268 | FALSE | Downregulated |
| 0,533563627 | 0,091188931 | S100A13 | FALSE | Upregulated |
| 0,673359035 | 0,091356743 | C11orf80 | FALSE | Upregulated |
| -0,404044481 | 0,091356743 | C20orf96 | FALSE | Downregulated |
| 0,593961148 | 0,091356743 | FAM83D | FALSE | Upregulated |
| 0,226455383 | 0,091376149 | COMMD5 | FALSE | Upregulated |
| 0,649974859 | 0,09137663 | PAFAH1B3 | FALSE | Upregulated |
| -0,252712587 | 0,09137663 | RIN2 | FALSE | Downregulated |
| -0,229238839 | 0,09137663 | ZADH2 | FALSE | Downregulated |
| -1,783489781 | 0,091387642 | LOC101928233 | FALSE | Downregulated |
| -0,238901459 | 0,091387642 | PDGFC | FALSE | Downregulated |
| -0,223693665 | 0,091405546 | NDFIP2 | FALSE | Downregulated |
| -0,821695548 | 0,091637915 | HTRA3 | FALSE | Downregulated |
| -0,43847665 | 0,091663093 | IKZF4 | FALSE | Downregulated |
| -0,462923403 | 0,091709871 | CDC14A | FALSE | Downregulated |
| 0,255194714 | 0,091731626 | ACOX3 | FALSE | Upregulated |
| 0,182931487 | 0,091731626 | PSMA5 | FALSE | Upregulated |
| -0,722037239 | 0,091818128 | LRRIQ3 | FALSE | Downregulated |
| -0,483311767 | 0,091953585 | LOC729291 | FALSE | Downregulated |
| 0,832115781 | 0,092032547 | FMO2 | FALSE | Upregulated |
| 0,284783231 | 0,092176883 | PPP1R15A | FALSE | Upregulated |
| -0,282066101 | 0,092247458 | SENP7 | FALSE | Downregulated |
| 0,152242568 | 0,09225147 | GAK | FALSE | Upregulated |
| 0,202019513 | 0,092251552 | EIF3K | FALSE | Upregulated |
| -0,294694255 | 0,092260232 | ATP5S | FALSE | Downregulated |
| -0,440927602 | 0,092348739 | LINC01071 | FALSE | Downregulated |
| 0,346314378 | 0,092348739 | LMF1 | FALSE | Upregulated |
| -0,327468119 | 0,092348739 | MTSS1 | FALSE | Downregulated |
| 0,878175915 | 0,092378492 | TMEM45B | FALSE | Upregulated |
| -0,268081224 | 0,092395146 | ZNF195 | FALSE | Downregulated |
| -0,341446136 | 0,092457886 | ANO1 | FALSE | Downregulated |
| 0,376602069 | 0,092457886 | GAS8 | FALSE | Upregulated |
| -0,300695691 | 0,092470906 | IGIP | FALSE | Downregulated |
| -0,516948286 | 0,092470906 | MUC20 | FALSE | Downregulated |
| -0,264172914 | 0,092470906 | SESTD1 | FALSE | Downregulated |
| -0,208165845 | 0,092490654 | CTIF | FALSE | Downregulated |
| -0,530430831 | 0,092511896 | LOC100130093 | FALSE | Downregulated |
| 0,682484917 | 0,092553921 | CCDC110 | FALSE | Upregulated |
| 0,756268575 | 0,092553921 | MEFV | FALSE | Upregulated |
| 0,309966727 | 0,092553921 | MYO1E | FALSE | Upregulated |
| -0,331154051 | 0,092553921 | PHF1 | FALSE | Downregulated |
| -0,245374434 | 0,092553921 | VARS2 | FALSE | Downregulated |
| -0,2570278 | 0,09257389 | NID1 | FALSE | Downregulated |
| 0,249854151 | 0,092610533 | TRMT6 | FALSE | Upregulated |
| 0,525664184 | 0,092696526 | LOC731656 | FALSE | Upregulated |
| 0,25777835 | 0,092696526 | LRCH1 | FALSE | Upregulated |
| 0,390149787 | 0,092696526 | TRPM8 | FALSE | Upregulated |

| | | | | |
|---|---|---|---|---|
| -0,241493021 | 0,092698233 | ZNF623 | FALSE | Downregulated |
| -0,128446763 | 0,092781416 | URI1 | FALSE | Downregulated |
| 0,304144638 | 0,092794854 | CTNNAL1 | FALSE | Upregulated |
| 0,214863478 | 0,093122852 | FASTKD5 | FALSE | Upregulated |
| -1,311833016 | 0,093172944 | RBM44 | FALSE | Downregulated |
| 0,174791453 | 0,093203978 | VDAC3 | FALSE | Upregulated |
| -0,452460014 | 0,093278585 | ENDOG | FALSE | Downregulated |
| 2,133351759 | 0,093381917 | TUBB3 | FALSE | Upregulated |
| 2,007852749 | 0,093520522 | AIM1L | FALSE | Upregulated |
| 0,188556486 | 0,093522368 | TOMM22 | FALSE | Upregulated |
| 0,473617941 | 0,093532205 | SIGLEC7 | FALSE | Upregulated |
| -0,355331953 | 0,09364128 | ZFC3H1 | FALSE | Downregulated |
| 0,329120687 | 0,093643611 | BTN3A1 | FALSE | Upregulated |
| 0,338336858 | 0,093643611 | C15orf41 | FALSE | Upregulated |
| 0,82738477 | 0,093643611 | LYSMD4 | FALSE | Upregulated |
| -1,707262749 | 0,093643611 | RNASE13 | FALSE | Downregulated |
| 0,987729806 | 0,093643611 | TMEM63C | FALSE | Upregulated |
| -0,404814445 | 0,093654356 | HECTD2 | FALSE | Downregulated |
| -0,152069612 | 0,093678118 | ANKRD40 | FALSE | Downregulated |
| -1,921279192 | 0,093678118 | LOC440461 | FALSE | Downregulated |
| -0,303999488 | 0,093718315 | ALG10B | FALSE | Downregulated |
| -0,487674437 | 0,093718315 | XRCC6BP1 | FALSE | Downregulated |
| -0,201839423 | 0,093772192 | PROS1 | FALSE | Downregulated |
| -0,545889918 | 0,093806446 | LINC01000 | FALSE | Downregulated |
| -0,506704715 | 0,093893468 | ZKSCAN7 | FALSE | Downregulated |
| -0,385776615 | 0,093924164 | CLK4 | FALSE | Downregulated |
| -0,608022691 | 0,094028006 | LOC553103 | FALSE | Downregulated |
| -0,415900617 | 0,094046589 | XYLB | FALSE | Downregulated |
| -0,199525752 | 0,094064248 | ANKRD17 | FALSE | Downregulated |
| -0,157030075 | 0,094064248 | GUF1 | FALSE | Downregulated |
| -0,605319495 | 0,094110306 | NPIPA5 | FALSE | Downregulated |
| -0,128457877 | 0,094231156 | C3orf17 | FALSE | Downregulated |
| -0,310742424 | 0,094296249 | ECM1 | FALSE | Downregulated |
| 0,598775234 | 0,094311079 | PER1 | FALSE | Upregulated |
| -0,463847646 | 0,094312474 | DLG4 | FALSE | Downregulated |
| 0,934294404 | 0,094369467 | SLC22A15 | FALSE | Upregulated |
| -2,663363071 | 0,094369845 | DYX1C1 | FALSE | Downregulated |
| 0,671025734 | 0,094369845 | LOC100133445 | FALSE | Upregulated |
| -0,250609222 | 0,094369845 | TFDP2 | FALSE | Downregulated |
| -0,525732035 | 0,094382166 | RP11-178C3.2 | FALSE | Downregulated |
| -0,229133423 | 0,094602436 | LMTK2 | FALSE | Downregulated |
| 0,197938625 | 0,094602436 | MTPN | FALSE | Upregulated |
| 0,492860031 | 0,094612923 | PDE7A | FALSE | Upregulated |
| -0,214454313 | 0,094694247 | AAAS | FALSE | Downregulated |
| 0,39820234 | 0,094694247 | IL1RN | FALSE | Upregulated |
| -0,156470292 | 0,094694247 | PPM1A | FALSE | Downregulated |
| -0,547469958 | 0,094708545 | HERC2P9 | FALSE | Downregulated |
| -0,33405756 | 0,094719497 | PHF3 | FALSE | Downregulated |
| 0,364632924 | 0,094813773 | HMGB3 | FALSE | Upregulated |
| -1,229220957 | 0,094884904 | CCDC64B | FALSE | Downregulated |
| -0,530039072 | 0,094971808 | ALDH1L1 | FALSE | Downregulated |

| | | | | |
|---|---|---|---|---|
| -0,587276208 | 0,094987739 | WNT5B | FALSE | Downregulated |
| -2,245789687 | 0,095030611 | LOC101929462 | FALSE | Downregulated |
| -0,181354338 | 0,095128685 | MAP3K4 | FALSE | Downregulated |
| 1,029261643 | 0,095268493 | LINC01094 | FALSE | Upregulated |
| -0,343397369 | 0,095334755 | TRIB1 | FALSE | Downregulated |
| 0,198933344 | 0,095360744 | NUBP1 | FALSE | Upregulated |
| -3,975616903 | 0,095360744 | TNFRSF6B | FALSE | Downregulated |
| -0,232965834 | 0,095417784 | CPPED1 | FALSE | Downregulated |
| -0,691035289 | 0,095417784 | GLUD1P7 | FALSE | Downregulated |
| 0,20713934 | 0,095635518 | SNIP1 | FALSE | Upregulated |
| 0,452694818 | 0,095679155 | AACS | FALSE | Upregulated |
| -1,305770815 | 0,095730604 | LOC101927842 | FALSE | Downregulated |
| -0,64879076 | 0,095766874 | GNAO1 | FALSE | Downregulated |
| -0,280379913 | 0,095787335 | ANKRD10 | FALSE | Downregulated |
| -0,227513147 | 0,095805019 | FBXO3 | FALSE | Downregulated |
| 1,898806946 | 0,095805019 | LAMP5 | FALSE | Upregulated |
| 0,540523214 | 0,095854936 | DGCR5 | FALSE | Upregulated |
| -0,187321854 | 0,095854936 | FAM193A | FALSE | Downregulated |
| -0,910966932 | 0,095854936 | GNRH1 | FALSE | Downregulated |
| -0,203816168 | 0,095854936 | UBE2O | FALSE | Downregulated |
| -0,364830977 | 0,095854936 | ZNF443 | FALSE | Downregulated |
| -0,583883951 | 0,095987394 | TMTC1 | FALSE | Downregulated |
| -0,398872531 | 0,095989814 | NUTM2A | FALSE | Downregulated |
| 0,254276356 | 0,096013301 | PPME1 | FALSE | Upregulated |
| -0,223426192 | 0,096049245 | AP4S1 | FALSE | Downregulated |
| -0,165225657 | 0,096079491 | CDK13 | FALSE | Downregulated |
| -3,092488478 | 0,096079491 | LOC729222 | FALSE | Downregulated |
| -0,509288453 | 0,096258614 | THUMPD3-AS1 | FALSE | Downregulated |
| 0,215971827 | 0,096280314 | DAZAP1 | FALSE | Upregulated |
| -1,446671024 | 0,096280314 | LOC102723331 | FALSE | Downregulated |
| 0,881469941 | 0,096457129 | TMEM150B | FALSE | Upregulated |
| -0,429232836 | 0,096549223 | LINC00888 | FALSE | Downregulated |
| 0,229153569 | 0,096755608 | DUSP22 | FALSE | Upregulated |
| 0,275722847 | 0,096755608 | TDG | FALSE | Upregulated |
| 0,220702686 | 0,096764674 | MOB1A | FALSE | Upregulated |
| -0,461029184 | 0,096771869 | CASC10 | FALSE | Downregulated |
| -0,206993275 | 0,096771869 | RPRD1A | FALSE | Downregulated |
| -0,206222068 | 0,096881257 | C1orf27 | FALSE | Downregulated |
| -0,424754041 | 0,096943405 | BMPER | FALSE | Downregulated |
| -0,701395906 | 0,096966053 | LOC100506688 | FALSE | Downregulated |
| -0,700883948 | 0,096966053 | TSPAN8 | FALSE | Downregulated |
| -0,582405657 | 0,097094627 | LOC100289495 | FALSE | Downregulated |
| 0,35618573 | 0,097094627 | RAB3IL1 | FALSE | Upregulated |
| 0,296515623 | 0,097121674 | SEC22C | FALSE | Upregulated |
| 0,259990509 | 0,097254062 | LAMTOR5 | FALSE | Upregulated |
| 0,977923092 | 0,097282578 | C11orf21 | FALSE | Upregulated |
| -0,377412391 | 0,097282578 | EBPL | FALSE | Downregulated |
| 0,223050496 | 0,097282578 | MAD1L1 | FALSE | Upregulated |
| -0,429620992 | 0,097282578 | MOGAT2 | FALSE | Downregulated |
| 0,764546757 | 0,097317557 | FPR2 | FALSE | Upregulated |
| -0,830043674 | 0,097317557 | LOC101060581 | FALSE | Downregulated |

| | | | | |
|---|---|---|---|---|
| -0,447003049 | 0,097328957 | RPS6KA6 | FALSE | Downregulated |
| -0,472772838 | 0,097353548 | BTBD16 | FALSE | Downregulated |
| 0,286324913 | 0,097397681 | FAM3C | FALSE | Upregulated |
| -0,22089818 | 0,097397681 | MRPL46 | FALSE | Downregulated |
| 0,219218068 | 0,097397681 | NSMAF | FALSE | Upregulated |
| -0,220032253 | 0,097667801 | KDSR | FALSE | Downregulated |
| -1,324029762 | 0,097687202 | SLC16A12 | FALSE | Downregulated |
| -0,315035062 | 0,097698374 | SLC39A5 | FALSE | Downregulated |
| -0,546849023 | 0,097769114 | PRG4 | FALSE | Downregulated |
| -0,191022976 | 0,097769114 | TTC17 | FALSE | Downregulated |
| -0,393054122 | 0,097781016 | KLHDC9 | FALSE | Downregulated |
| 0,570269714 | 0,097797916 | MARCO | FALSE | Upregulated |
| -0,501597927 | 0,097848133 | ZFYVE28 | FALSE | Downregulated |
| -0,571571619 | 0,097887468 | LOC101928728 | FALSE | Downregulated |
| 1,121566526 | 0,097887468 | MOXD1 | FALSE | Upregulated |
| -0,352296788 | 0,097887867 | ARL16 | FALSE | Downregulated |
| -0,193808148 | 0,097887867 | SIN3A | FALSE | Downregulated |
| 0,372521831 | 0,097998605 | HIST1H2BK | FALSE | Upregulated |
| 0,186286052 | 0,098047108 | AKT1 | FALSE | Upregulated |
| -0,231805692 | 0,098290005 | SOCS5 | FALSE | Downregulated |
| 0,214188656 | 0,098381015 | FKBP3 | FALSE | Upregulated |
| -2,27058145 | 0,098478089 | ACER1 | FALSE | Downregulated |
| -0,739076222 | 0,09862109 | CADM2 | FALSE | Downregulated |
| -0,225833579 | 0,09862109 | LSM6 | FALSE | Downregulated |
| -0,477258469 | 0,098628148 | MACROD2 | FALSE | Downregulated |
| -0,548784588 | 0,098642909 | PCSK4 | FALSE | Downregulated |
| 0,261540279 | 0,09875074 | PIGS | FALSE | Upregulated |
| 0,282341512 | 0,098805502 | SIPA1L3 | FALSE | Upregulated |
| 0,178377869 | 0,09889848 | MGME1 | FALSE | Upregulated |
| -0,252546035 | 0,098923918 | GNAI1 | FALSE | Downregulated |
| 0,372042039 | 0,099102706 | CARD6 | FALSE | Upregulated |
| -0,334914412 | 0,099102706 | MGMT | FALSE | Downregulated |
| 0,221487351 | 0,099102706 | NOC2L | FALSE | Upregulated |
| -0,278384648 | 0,099115826 | PTPRG | FALSE | Downregulated |
| -0,198496458 | 0,09923566 | TSPAN6 | FALSE | Downregulated |
| -0,225213073 | 0,099357851 | LRIF1 | FALSE | Downregulated |
| 0,707412069 | 0,099357851 | PDE6G | FALSE | Upregulated |
| 0,28788082 | 0,099357851 | POLR2L | FALSE | Upregulated |
| -0,95822798 | 0,099402673 | ALPK2 | FALSE | Downregulated |
| -0,262937453 | 0,099475793 | AAMP | FALSE | Downregulated |
| 0,25654342 | 0,099486004 | YEATS2 | FALSE | Upregulated |
| -0,371391537 | 0,099554918 | CCDC84 | FALSE | Downregulated |
| 0,15770993 | 0,099554918 | RANBP3 | FALSE | Upregulated |
| 0,553064702 | 0,099587241 | CALHM2 | FALSE | Upregulated |
| 0,19476061 | 0,099640953 | MBD3 | FALSE | Upregulated |
| 0,24012814 | 0,100023666 | CCDC124 | FALSE | Upregulated |
| -0,343475227 | 0,100134414 | MLPH | FALSE | Downregulated |
| 0,346433592 | 0,100184776 | PECAM1 | FALSE | Upregulated |
| 0,162786234 | 0,100213516 | SARNP | FALSE | Upregulated |
| -0,263013866 | 0,100249613 | GABPB1-AS1 | FALSE | Downregulated |
| 0,812436662 | 0,100249613 | IFI44L | FALSE | Upregulated |

| | | | | |
|---|---|---|---|---|
| -0,649330423 | 0,100249613 | ZNF818P | FALSE | Downregulated |
| -1,096941603 | 0,10044693 | NEK10 | FALSE | Downregulated |
| 1,493012063 | 0,10044693 | TMEM255A | FALSE | Upregulated |
| -0,189003034 | 0,100507709 | ZNF362 | FALSE | Downregulated |
| 0,283545983 | 0,100580217 | AMDHD2 | FALSE | Upregulated |
| -0,431036059 | 0,100580217 | SREK1 | FALSE | Downregulated |
| 0,131445191 | 0,100628853 | NCSTN | FALSE | Upregulated |
| 0,553105676 | 0,100786721 | MGC32805 | FALSE | Upregulated |
| -0,323998293 | 0,100786721 | SEC16B | FALSE | Downregulated |
| -0,251455325 | 0,101018047 | SEMA4G | FALSE | Downregulated |
| -0,379589502 | 0,101020418 | CXorf57 | FALSE | Downregulated |
| -0,329963778 | 0,101116695 | CLK2 | FALSE | Downregulated |
| -0,636582109 | 0,101514088 | PMS2P3 | FALSE | Downregulated |
| 0,444933704 | 0,101660382 | AXL | FALSE | Upregulated |
| -0,271627319 | 0,101660382 | HAGH | FALSE | Downregulated |
| -1,9073663 | 0,101712421 | SAP30L-AS1 | FALSE | Downregulated |
| -0,474628323 | 0,101712421 | STAG3L4 | FALSE | Downregulated |
| -0,420622712 | 0,101780965 | ZNF571 | FALSE | Downregulated |
| 0,672174492 | 0,101781051 | ENPP2 | FALSE | Upregulated |
| -0,439723429 | 0,101782602 | CCDC14 | FALSE | Downregulated |
| 0,301239837 | 0,101782602 | PIGW | FALSE | Upregulated |
| -0,429366278 | 0,101856379 | LCAT | FALSE | Downregulated |
| -0,166301147 | 0,101885385 | SDCCAG8 | FALSE | Downregulated |
| -0,565709357 | 0,101906667 | LPPR1 | FALSE | Downregulated |
| 0,274215757 | 0,101917726 | UGGT1 | FALSE | Upregulated |
| -1,077960208 | 0,101951874 | RP11-452H21.4 | FALSE | Downregulated |
| -1,510171575 | 0,102056319 | CADM3 | FALSE | Downregulated |
| -0,766370191 | 0,102056319 | PSMD6-AS2 | FALSE | Downregulated |
| -0,637626559 | 0,102090784 | PTH1R | FALSE | Downregulated |
| -0,130489053 | 0,102090784 | SETD3 | FALSE | Downregulated |
| -1,969623622 | 0,102186711 | FBXO40 | FALSE | Downregulated |
| -0,719404366 | 0,102186711 | LOC100129534 | FALSE | Downregulated |
| -0,460750089 | 0,102282912 | SNHG3 | FALSE | Downregulated |
| -0,836402285 | 0,102297677 | CCDC144B | FALSE | Downregulated |
| 0,132001116 | 0,102353802 | DCTN1 | FALSE | Upregulated |
| -0,654587204 | 0,102355409 | PMS2P4 | FALSE | Downregulated |
| -0,41233972 | 0,10240692 | ANKAR | FALSE | Downregulated |
| 0,77088217 | 0,10240692 | C9orf9 | FALSE | Upregulated |
| -0,219582792 | 0,102411252 | SENP2 | FALSE | Downregulated |
| 0,548116494 | 0,102415041 | CCDC88B | FALSE | Upregulated |
| -0,295174545 | 0,102415041 | SKIL | FALSE | Downregulated |
| 0,34588295 | 0,102444323 | DHRS7 | FALSE | Upregulated |
| 1,387949764 | 0,10252711 | IFNG-AS1 | FALSE | Upregulated |
| -1,035099729 | 0,10252711 | LOC102724811 | FALSE | Downregulated |
| -0,227705981 | 0,102534257 | MSH3 | FALSE | Downregulated |
| -0,213936098 | 0,10261706 | CTAGE5 | FALSE | Downregulated |
| -0,564429885 | 0,102791692 | DBH-AS1 | FALSE | Downregulated |
| -0,791473437 | 0,10280378 | HAPLN4 | FALSE | Downregulated |
| -0,368415973 | 0,10288682 | CLK1 | FALSE | Downregulated |
| -0,443498256 | 0,10288682 | SPRY4 | FALSE | Downregulated |
| 0,235594035 | 0,103006811 | HIGD2A | FALSE | Upregulated |

| | | | | |
|---|---|---|---|---|
| 0,146366749 | 0,103152913 | SF3A3 | FALSE | Upregulated |
| 0,373105217 | 0,103152913 | SKAP2 | FALSE | Upregulated |
| 0,434282423 | 0,103239247 | TRIB3 | FALSE | Upregulated |
| 0,16544085 | 0,103274236 | CCT4 | FALSE | Upregulated |
| -0,209360887 | 0,103290957 | NAA25 | FALSE | Downregulated |
| 0,36193799 | 0,103305949 | PLBD2 | FALSE | Upregulated |
| -0,453852119 | 0,103415774 | KMT2E-AS1 | FALSE | Downregulated |
| 0,271074955 | 0,103479919 | CDK2AP1 | FALSE | Upregulated |
| -0,277915978 | 0,103580245 | CMTM8 | FALSE | Downregulated |
| -0,122788107 | 0,103580245 | TMEM30A | FALSE | Downregulated |
| -0,505774048 | 0,10359128 | LAD1 | FALSE | Downregulated |
| -1,729403361 | 0,103741223 | LOC400685 | FALSE | Downregulated |
| -0,434170015 | 0,103745936 | FAM76B | FALSE | Downregulated |
| 0,573418755 | 0,103994573 | XAF1 | FALSE | Upregulated |
| -0,441118675 | 0,10409189 | ORC2 | FALSE | Downregulated |
| -0,394853341 | 0,104271763 | GSTZ1 | FALSE | Downregulated |
| -0,22146515 | 0,104402092 | SDHD | FALSE | Downregulated |
| 0,184198173 | 0,10457078 | RBM18 | FALSE | Upregulated |
| -0,244509529 | 0,104627533 | TMEM65 | FALSE | Downregulated |
| -0,408397359 | 0,104725313 | SNED1 | FALSE | Downregulated |
| 2,14397642 | 0,104818201 | CALCA | FALSE | Upregulated |
| 0,240852675 | 0,104818201 | TNFRSF1A | FALSE | Upregulated |
| -0,540885269 | 0,105170702 | AC003665.1 | FALSE | Downregulated |
| 0,181308387 | 0,105170702 | CRK | FALSE | Upregulated |
| -0,670636151 | 0,105170702 | LINC00865 | FALSE | Downregulated |
| 0,637741968 | 0,105170702 | SPIC | FALSE | Upregulated |
| -0,215908134 | 0,105170702 | TAF1D | FALSE | Downregulated |
| -0,515176092 | 0,105170702 | TMEM198B | FALSE | Downregulated |
| 0,336534986 | 0,105175115 | ABCC10 | FALSE | Upregulated |
| 0,192730535 | 0,105175115 | CHCHD3 | FALSE | Upregulated |
| -1,23807567 | 0,105175115 | KLHL7-AS1 | FALSE | Downregulated |
| -0,661214855 | 0,105175115 | LOC102546229 | FALSE | Downregulated |
| 0,199019372 | 0,105175115 | STX7 | FALSE | Upregulated |
| -0,336242316 | 0,105175115 | THAP9-AS1 | FALSE | Downregulated |
| -0,333231468 | 0,105237206 | LIFR | FALSE | Downregulated |
| 0,427524904 | 0,10538989 | ABCG1 | FALSE | Upregulated |
| 0,321099237 | 0,10538989 | EMILIN1 | FALSE | Upregulated |
| -2,510781591 | 0,10538989 | SDC4P | FALSE | Downregulated |
| -0,781770712 | 0,105678942 | TCEAL7 | FALSE | Downregulated |
| -0,316103522 | 0,105686929 | C8A | FALSE | Downregulated |
| -0,240052029 | 0,105686929 | GCSH | FALSE | Downregulated |
| -0,543642168 | 0,105686929 | LINC01089 | FALSE | Downregulated |
| -0,250035484 | 0,105686929 | PCBP2 | FALSE | Downregulated |
| 0,609732613 | 0,105686929 | RIPK3 | FALSE | Upregulated |
| 0,299755814 | 0,105691935 | MEF2BNB | FALSE | Upregulated |
| -0,67460535 | 0,105691935 | RRN3P1 | FALSE | Downregulated |
| -0,464802295 | 0,105691935 | SULT1A1 | FALSE | Downregulated |
| -3,058250418 | 0,105713221 | MIA-RAB4B | FALSE | Downregulated |
| -0,139196076 | 0,105713221 | TBC1D15 | FALSE | Downregulated |
| -0,170668216 | 0,105732271 | SCYL3 | FALSE | Downregulated |
| 0,160618486 | 0,105767283 | EIF2S1 | FALSE | Upregulated |

| | | | | |
|---|---|---|---|---|
| -0,444195961 | 0,105767283 | FAM184A | FALSE | Downregulated |
| 1,702434201 | 0,105767283 | PGAM4 | FALSE | Upregulated |
| -0,33841542 | 0,105767283 | SLC13A5 | FALSE | Downregulated |
| -0,297929076 | 0,105973975 | MFSD9 | FALSE | Downregulated |
| -0,736204031 | 0,105976481 | CPE | FALSE | Downregulated |
| -1,296494439 | 0,105976481 | FAM85A | FALSE | Downregulated |
| 0,263054257 | 0,106000006 | RPLP1 | FALSE | Upregulated |
| -1,955160356 | 0,106091164 | LOC102724328 | FALSE | Downregulated |
| 0,186405952 | 0,106091164 | MLH1 | FALSE | Upregulated |
| 0,469725953 | 0,106102953 | ALDOA | FALSE | Upregulated |
| 0,252343228 | 0,106102953 | TLN1 | FALSE | Upregulated |
| -0,242789011 | 0,106161801 | ESRP2 | FALSE | Downregulated |
| 0,744354482 | 0,106161801 | LOC101928464 | FALSE | Upregulated |
| -0,283625568 | 0,106161801 | PRDX3 | FALSE | Downregulated |
| 0,281996248 | 0,106161801 | SFR1 | FALSE | Upregulated |
| -0,803662667 | 0,106161801 | TRIM73 | FALSE | Downregulated |
| 0,176605876 | 0,106161801 | TRIP4 | FALSE | Upregulated |
| 2,014785274 | 0,106189951 | AC226118.1 | FALSE | Upregulated |
| -0,343236609 | 0,106189951 | KLKB1 | FALSE | Downregulated |
| -0,314687495 | 0,106189951 | WDR91 | FALSE | Downregulated |
| -0,584678152 | 0,106203642 | LINC01061 | FALSE | Downregulated |
| 0,168335659 | 0,106203642 | NFKB1 | FALSE | Upregulated |
| 0,427483711 | 0,106260373 | DOHH | FALSE | Upregulated |
| -0,957861094 | 0,106260373 | LOC100288152 | FALSE | Downregulated |
| 0,281797072 | 0,106409092 | B4GALT2 | FALSE | Upregulated |
| -0,356575287 | 0,106508385 | TMEM39A | FALSE | Downregulated |
| -0,346565364 | 0,106536713 | ONECUT2 | FALSE | Downregulated |
| 0,22952069 | 0,106612624 | CLTB | FALSE | Upregulated |
| 0,300190579 | 0,106628303 | STYXL1 | FALSE | Upregulated |
| 0,309711947 | 0,106629283 | NOMO2 | FALSE | Upregulated |
| -0,360145392 | 0,106637098 | CEP164 | FALSE | Downregulated |
| 0,322012816 | 0,106637098 | DGKQ | FALSE | Upregulated |
| 0,165962399 | 0,106886171 | ADSS | FALSE | Upregulated |
| 0,432135865 | 0,10696324 | FANCG | FALSE | Upregulated |
| 0,772115742 | 0,107104263 | CRTAM | FALSE | Upregulated |
| 0,315932952 | 0,107152692 | MPZL3 | FALSE | Upregulated |
| -1,178109744 | 0,107319707 | INE1 | FALSE | Downregulated |
| -0,28848721 | 0,107356516 | CDK10 | FALSE | Downregulated |
| 0,268814578 | 0,107356516 | POLD4 | FALSE | Upregulated |
| 0,88753552 | 0,107359523 | FAM225A | FALSE | Upregulated |
| -1,718704509 | 0,10746536 | LOC101929950 | FALSE | Downregulated |
| 1,476592382 | 0,10746536 | MLIP-IT1 | FALSE | Upregulated |
| -2,975171046 | 0,107636887 | FAM157A | FALSE | Downregulated |
| -0,395029708 | 0,107636887 | IFRD1 | FALSE | Downregulated |
| 1,369917674 | 0,107636887 | RTEL1-TNFRSF6B | FALSE | Upregulated |
| 1,516796803 | 0,107644406 | FLJ44511 | FALSE | Upregulated |
| -0,185608077 | 0,10788827 | FGFR1OP | FALSE | Downregulated |
| 0,132128905 | 0,10788827 | PDCD6IP | FALSE | Upregulated |
| -2,400676668 | 0,107888374 | LOC101929259 | FALSE | Downregulated |
| -0,221084195 | 0,107929428 | PPP1R10 | FALSE | Downregulated |
| 0,189431037 | 0,107929585 | MRPS23 | FALSE | Upregulated |

| | | | | |
|---|---|---|---|---|
| -0,166818076 | 0,107933038 | HDHD2 | FALSE | Downregulated |
| -0,263224347 | 0,107944215 | APBB2 | FALSE | Downregulated |
| -0,828286102 | 0,108084322 | MTUS2 | FALSE | Downregulated |
| -0,437013922 | 0,108137136 | CORO2A | FALSE | Downregulated |
| 0,182282373 | 0,108178286 | TMOD3 | FALSE | Upregulated |
| -1,246213939 | 0,108369528 | AC018647.3 | FALSE | Downregulated |
| 0,272149939 | 0,108433726 | ATG4A | FALSE | Upregulated |
| -0,594283699 | 0,108467901 | RP11-695J4.2 | FALSE | Downregulated |
| -0,370089199 | 0,108467901 | ZMYND12 | FALSE | Downregulated |
| -0,206404696 | 0,108480649 | BNIP3 | FALSE | Downregulated |
| 0,47698548 | 0,108480649 | VWA1 | FALSE | Upregulated |
| 0,230804268 | 0,108487004 | ADAT1 | FALSE | Upregulated |
| 0,227416722 | 0,108487004 | CDK5RAP2 | FALSE | Upregulated |
| -0,303702926 | 0,108487004 | FBXW7 | FALSE | Downregulated |
| -0,73574474 | 0,108487004 | HCG8 | FALSE | Downregulated |
| -0,736866813 | 0,108487004 | LINC01123 | FALSE | Downregulated |
| -0,821322375 | 0,108487004 | ZNF876P | FALSE | Downregulated |
| 0,578110471 | 0,108522979 | WDFY4 | FALSE | Upregulated |
| -0,755782628 | 0,108567646 | LOC100126582 | FALSE | Downregulated |
| 1,084861708 | 0,108567646 | LOC101928517 | FALSE | Upregulated |
| 0,21557068 | 0,108567646 | ZNF672 | FALSE | Upregulated |
| -0,236970037 | 0,108595301 | TMEM30B | FALSE | Downregulated |
| 0,386202535 | 0,108642232 | ROMO1 | FALSE | Upregulated |
| -0,221805624 | 0,108676107 | BBS2 | FALSE | Downregulated |
| 0,826569672 | 0,108716878 | HRCT1 | FALSE | Upregulated |
| -0,787254684 | 0,108721923 | AGAP4 | FALSE | Downregulated |
| 0,488875416 | 0,108721923 | AP1S2 | FALSE | Upregulated |
| 0,452483317 | 0,108721923 | CFD | FALSE | Upregulated |
| 0,318837892 | 0,108721923 | CNN2 | FALSE | Upregulated |
| -0,310864706 | 0,108736226 | ZCCHC14 | FALSE | Downregulated |
| -0,685330593 | 0,10881557 | C1QTNF7 | FALSE | Downregulated |
| 0,293582516 | 0,108816871 | EIF4G3 | FALSE | Upregulated |
| -0,620382552 | 0,108816871 | LOC101927663 | FALSE | Downregulated |
| 0,264374185 | 0,108816871 | PPAT | FALSE | Upregulated |
| -0,677418528 | 0,108821863 | LAMTOR5-AS1 | FALSE | Downregulated |
| -0,732580056 | 0,108850092 | ZDHHC11B | FALSE | Downregulated |
| 0,130576986 | 0,10886741 | GPR107 | FALSE | Upregulated |
| 0,904105214 | 0,108990663 | LOC100506022 | FALSE | Upregulated |
| 0,754917225 | 0,108990663 | PAPLN | FALSE | Upregulated |
| -1,603984563 | 0,108990663 | RAB9B | FALSE | Downregulated |
| 0,220024202 | 0,109006032 | RETSAT | FALSE | Upregulated |
| 0,302969543 | 0,109017734 | FIZ1 | FALSE | Upregulated |
| -0,216135692 | 0,109059779 | MAP2K5 | FALSE | Downregulated |
| -0,351974752 | 0,109062245 | ZNF19 | FALSE | Downregulated |
| 0,287625603 | 0,109086376 | PIP5K1C | FALSE | Upregulated |
| -0,667943493 | 0,109218953 | LOC101928384 | FALSE | Downregulated |
| -0,215997897 | 0,109218953 | MPI | FALSE | Downregulated |
| 0,556470457 | 0,109218953 | NR1D2 | FALSE | Upregulated |
| -0,323737288 | 0,109218953 | SRD5A1 | FALSE | Downregulated |
| -0,731090289 | 0,109218953 | VN1R1 | FALSE | Downregulated |
| -0,25202774 | 0,109237132 | CFI | FALSE | Downregulated |

| | | | | |
|---|---|---|---|---|
| 0,684321897 | 0,109331361 | GAPT | FALSE | Upregulated |
| 0,388104098 | 0,109331361 | TEAD2 | FALSE | Upregulated |
| 0,142954246 | 0,109340088 | AP2A2 | FALSE | Upregulated |
| 1,983599302 | 0,109340088 | GPA33 | FALSE | Upregulated |
| 0,227927002 | 0,109340088 | SLC38A9 | FALSE | Upregulated |
| -0,399806283 | 0,109358678 | MAP3K5 | FALSE | Downregulated |
| 0,228300441 | 0,109358678 | UBE2M | FALSE | Upregulated |
| 0,39857499 | 0,109378505 | TCEAL3 | FALSE | Upregulated |
| 0,204675483 | 0,109429478 | TRIM38 | FALSE | Upregulated |
| -0,309706449 | 0,109626173 | ZNF34 | FALSE | Downregulated |
| 0,395015043 | 0,109759764 | CLTCL1 | FALSE | Upregulated |
| 1,806564358 | 0,109759764 | GSDMC | FALSE | Upregulated |
| 0,538545476 | 0,109759764 | HEY1 | FALSE | Upregulated |
| 1,511700872 | 0,109759764 | P2RX5 | FALSE | Upregulated |
| -0,478061624 | 0,109759764 | SRRM2 | FALSE | Downregulated |
| 0,416927624 | 0,109759764 | SSX2IP | FALSE | Upregulated |
| -0,364139865 | 0,109759764 | ZBTB48 | FALSE | Downregulated |
| -0,283859946 | 0,109845534 | CYB5D2 | FALSE | Downregulated |
| -0,18848857 | 0,109864065 | ARHGEF7 | FALSE | Downregulated |
| 0,235067254 | 0,109864065 | PRPS2 | FALSE | Upregulated |
| 0,677802461 | 0,109881717 | C11orf63 | FALSE | Upregulated |
| -0,241707439 | 0,10991383 | GPATCH8 | FALSE | Downregulated |
| 0,932235691 | 0,110108833 | ADRA2A | FALSE | Upregulated |
| 0,263613038 | 0,110112819 | PINX1 | FALSE | Upregulated |
| 0,191783892 | 0,110146229 | FNTA | FALSE | Upregulated |
| -0,210777895 | 0,110177581 | PGRMC2 | FALSE | Downregulated |
| -0,710370478 | 0,110177581 | ZNF135 | FALSE | Downregulated |
| 0,235699402 | 0,110180835 | TMEM189 | FALSE | Upregulated |
| -0,315167083 | 0,110453922 | SORL1 | FALSE | Downregulated |
| -0,397207096 | 0,110546372 | DANCR | FALSE | Downregulated |
| -0,925875609 | 0,110546372 | MYH7B | FALSE | Downregulated |
| 0,332229325 | 0,110546372 | SLC2A8 | FALSE | Upregulated |
| 0,373912767 | 0,110546372 | TDRKH | FALSE | Upregulated |
| 0,616994447 | 0,110609853 | SGOL2 | FALSE | Upregulated |
| 0,166281449 | 0,110654431 | COG4 | FALSE | Upregulated |
| 0,406572688 | 0,110672419 | ORAI2 | FALSE | Upregulated |
| -0,316151827 | 0,110782088 | RBFOX2 | FALSE | Downregulated |
| -0,278079567 | 0,110786586 | ZNF691 | FALSE | Downregulated |
| 0,861957755 | 0,11088582 | LGALS4 | FALSE | Upregulated |
| 0,411170947 | 0,110886529 | ANKS6 | FALSE | Upregulated |
| 0,189947709 | 0,11088837 | VPS45 | FALSE | Upregulated |
| 0,85640443 | 0,110938891 | MMS22L | FALSE | Upregulated |
| -0,753937813 | 0,110970244 | MLLT4-AS1 | FALSE | Downregulated |
| -0,232632655 | 0,110972448 | ZNF420 | FALSE | Downregulated |
| 0,297777459 | 0,111029663 | SYNPO | FALSE | Upregulated |
| -0,248066078 | 0,111082094 | AFF4 | FALSE | Downregulated |
| -1,573301901 | 0,111291282 | TMEM178B | FALSE | Downregulated |
| 0,368137381 | 0,111428128 | TMEM234 | FALSE | Upregulated |
| 0,364759969 | 0,111431566 | LRRC16A | FALSE | Upregulated |
| 0,224559117 | 0,111540478 | ERGIC2 | FALSE | Upregulated |
| -0,24733422 | 0,111743488 | NR1H4 | FALSE | Downregulated |

| | | | | |
|---|---|---|---|---|
| 0,384268442 | 0,111743488 | ZDHHC23 | FALSE | Upregulated |
| -0,265142477 | 0,111865979 | ANKZF1 | FALSE | Downregulated |
| -0,185088125 | 0,111865979 | C6orf120 | FALSE | Downregulated |
| 0,120965576 | 0,111899095 | GANAB | FALSE | Upregulated |
| -0,645830697 | 0,111899095 | GPIHBP1 | FALSE | Downregulated |
| -0,173633975 | 0,111899095 | MTIF2 | FALSE | Downregulated |
| -0,256384498 | 0,111899095 | PCTP | FALSE | Downregulated |
| 0,391117822 | 0,111899095 | PPDPF | FALSE | Upregulated |
| -0,689318827 | 0,112028375 | CCNJL | FALSE | Downregulated |
| -0,7539638 | 0,112028375 | GUCA1B | FALSE | Downregulated |
| -0,222464983 | 0,112028375 | PRUNE | FALSE | Downregulated |
| 0,706660725 | 0,112101157 | MFSD2A | FALSE | Upregulated |
| 0,370198892 | 0,112131135 | MARCKS | FALSE | Upregulated |
| -0,354337278 | 0,112198228 | RNR2 | FALSE | Downregulated |
| -1,072558938 | 0,112211121 | LOC102467146 | FALSE | Downregulated |
| -0,805893942 | 0,112211121 | WDR96 | FALSE | Downregulated |
| -0,784033189 | 0,112238405 | NUDT9P1 | FALSE | Downregulated |
| 0,462132867 | 0,112241995 | TMEM243 | FALSE | Upregulated |
| 1,126758424 | 0,112301905 | ADAMTS3 | FALSE | Upregulated |
| 0,462097373 | 0,112301905 | PDE4B | FALSE | Upregulated |
| -0,479168679 | 0,112462315 | SRRM5 | FALSE | Downregulated |
| 0,259530543 | 0,112474937 | CABIN1 | FALSE | Upregulated |
| -0,974215767 | 0,11248981 | LOC728989 | FALSE | Downregulated |
| -0,208270656 | 0,112517387 | BAG4 | FALSE | Downregulated |
| 0,592825914 | 0,112814466 | HEYL | FALSE | Upregulated |
| -0,190160258 | 0,11283247 | ATP13A3 | FALSE | Downregulated |
| -0,214391377 | 0,11283247 | MICU1 | FALSE | Downregulated |
| 0,395115823 | 0,112863367 | NDOR1 | FALSE | Upregulated |
| 0,221700676 | 0,112921514 | NDUFS6 | FALSE | Upregulated |
| -0,205362801 | 0,112993166 | CALM1 | FALSE | Downregulated |
| -0,824562482 | 0,113011365 | LOC643327 | FALSE | Downregulated |
| 4,957424702 | 0,113104917 | RNU1-4 | FALSE | Upregulated |
| 0,429517185 | 0,113374566 | MYL12A | FALSE | Upregulated |
| 0,378200477 | 0,113564441 | PDP1 | FALSE | Upregulated |
| 0,351492065 | 0,11356945 | SCO2 | FALSE | Upregulated |
| 0,281751369 | 0,113574895 | MAGOHB | FALSE | Upregulated |
| 0,19150673 | 0,113583626 | SLC35E3 | FALSE | Upregulated |
| -0,104595317 | 0,113646104 | BAP1 | FALSE | Downregulated |
| 0,190869408 | 0,113647991 | SCAMP4 | FALSE | Upregulated |
| 0,174195257 | 0,113647991 | TFDP1 | FALSE | Upregulated |
| -0,349474275 | 0,113718343 | ZNF584 | FALSE | Downregulated |
| 0,629165689 | 0,11371908 | MFGE8 | FALSE | Upregulated |
| 0,308805849 | 0,11379556 | RAD51D | FALSE | Upregulated |
| 0,43006126 | 0,113808393 | SLC17A3 | FALSE | Upregulated |
| -0,311117304 | 0,113867943 | SH3BP5 | FALSE | Downregulated |
| -0,887127541 | 0,113875292 | LOC102724306 | FALSE | Downregulated |
| -1,020073269 | 0,113901893 | CTB-118N6.3 | FALSE | Downregulated |
| -0,211711393 | 0,114198689 | IBTK | FALSE | Downregulated |
| -0,269847493 | 0,11421404 | SLC20A2 | FALSE | Downregulated |
| 0,441183595 | 0,114293416 | C9orf16 | FALSE | Upregulated |
| -0,889650192 | 0,114293416 | GRHL2 | FALSE | Downregulated |

| | | | | |
|---|---|---|---|---|
| 1,445130125 | 0,114293416 | RP11-27N21.3 | FALSE | Upregulated |
| -0,380208633 | 0,114293416 | SLC24A1 | FALSE | Downregulated |
| -0,161694314 | 0,114361017 | WDR5 | FALSE | Downregulated |
| 0,237864197 | 0,114455355 | NIPAL3 | FALSE | Upregulated |
| -0,253645243 | 0,114490423 | RNF130 | FALSE | Downregulated |
| -1,257885664 | 0,114490423 | SLC4A1 | FALSE | Downregulated |
| -0,386754613 | 0,114490423 | TCP11L2 | FALSE | Downregulated |
| 0,155033343 | 0,114512565 | C10orf76 | FALSE | Upregulated |
| -0,413617187 | 0,114512565 | LOC101927471 | FALSE | Downregulated |
| -0,634411681 | 0,114512565 | LOC101928424 | FALSE | Downregulated |
| 0,580465923 | 0,114512565 | TARP | FALSE | Upregulated |
| -0,184420022 | 0,114512565 | UBR5 | FALSE | Downregulated |
| 0,59516443 | 0,114520549 | FILIP1L | FALSE | Upregulated |
| -0,762509152 | 0,114520549 | YTHDF3-AS1 | FALSE | Downregulated |
| 1,357104297 | 0,114558796 | VSIG2 | FALSE | Upregulated |
| -0,205911971 | 0,114593616 | RBP4 | FALSE | Downregulated |
| -0,375032929 | 0,114616997 | CRHBP | FALSE | Downregulated |
| -0,822079019 | 0,114642955 | LOC102723566 | FALSE | Downregulated |
| 0,122113079 | 0,114660315 | STARD7 | FALSE | Upregulated |
| -0,297689038 | 0,114798487 | RAB30-AS1 | FALSE | Downregulated |
| 0,38646489 | 0,114851247 | RTP3 | FALSE | Upregulated |
| -0,390360115 | 0,114935062 | ANGPTL6 | FALSE | Downregulated |
| -0,471251762 | 0,115066351 | PKI55 | FALSE | Downregulated |
| 0,225711704 | 0,115092155 | C18orf8 | FALSE | Upregulated |
| -0,130703639 | 0,115193602 | ZFYVE20 | FALSE | Downregulated |
| -0,489924667 | 0,115249979 | ZNF540 | FALSE | Downregulated |
| -0,244893622 | 0,115288568 | LZTS3 | FALSE | Downregulated |
| 0,235886412 | 0,115398917 | BRK1 | FALSE | Upregulated |
| -0,427225172 | 0,115406801 | LTB4R | FALSE | Downregulated |
| -1,250629526 | 0,115625271 | CDH4 | FALSE | Downregulated |
| -1,825866161 | 0,115625369 | PROK2 | FALSE | Downregulated |
| -1,912640953 | 0,115629686 | FOXI2 | FALSE | Downregulated |
| 0,841604987 | 0,115748567 | LAMC3 | FALSE | Upregulated |
| -0,487453114 | 0,115748567 | LOC101928707 | FALSE | Downregulated |
| 0,556129928 | 0,115754149 | BBC3 | FALSE | Upregulated |
| 0,328480188 | 0,115754149 | GNB4 | FALSE | Upregulated |
| -1,890010436 | 0,115754149 | LOC101930531 | FALSE | Downregulated |
| 0,232451176 | 0,115754149 | XPR1 | FALSE | Upregulated |
| 1,732599391 | 0,115820171 | RGS17 | FALSE | Upregulated |
| -0,314453817 | 0,115820171 | SEC62 | FALSE | Downregulated |
| -0,227402737 | 0,115845697 | KLHL7 | FALSE | Downregulated |
| -0,745042659 | 0,115845697 | TOB2P1 | FALSE | Downregulated |
| 0,222312322 | 0,116028878 | SLC35A2 | FALSE | Upregulated |
| -1,227031863 | 0,116117166 | CDH26 | FALSE | Downregulated |
| 0,241841302 | 0,116199906 | CASP7 | FALSE | Upregulated |
| 0,286882144 | 0,116216087 | RPS5 | FALSE | Upregulated |
| -0,211897856 | 0,116377257 | HDHD3 | FALSE | Downregulated |
| 0,239653588 | 0,116377257 | MFSD11 | FALSE | Upregulated |
| -0,24317613 | 0,116462879 | VPS13A | FALSE | Downregulated |
| -0,945442781 | 0,116521222 | FAM124B | FALSE | Downregulated |
| -0,428890806 | 0,116630305 | SLC38A4 | FALSE | Downregulated |

| | | | | |
|---|---|---|---|---|
| 0,547841943 | 0,11674167 | C3orf67 | FALSE | Upregulated |
| -1,131971425 | 0,11674167 | MTRNR2L2 | FALSE | Downregulated |
| -0,23743666 | 0,11682783 | GIN1 | FALSE | Downregulated |
| 0,395847521 | 0,116909696 | CTTNBP2NL | FALSE | Upregulated |
| -0,368222069 | 0,116909696 | GEN1 | FALSE | Downregulated |
| 0,234514571 | 0,116909696 | MRPL51 | FALSE | Upregulated |
| -3,865620123 | 0,116938345 | ZNF559-ZNF177 | FALSE | Downregulated |
| -0,278122845 | 0,117103181 | SIK2 | FALSE | Downregulated |
| -1,785156258 | 0,117208687 | CCDC36 | FALSE | Downregulated |
| 0,147048609 | 0,117283506 | KPNA4 | FALSE | Upregulated |
| 1,46887609 | 0,117331432 | STAP1 | FALSE | Upregulated |
| -0,671050641 | 0,117374534 | TRNC | FALSE | Downregulated |
| -0,819960547 | 0,117461442 | LYG1 | FALSE | Downregulated |
| 0,370794225 | 0,117461442 | SMAGP | FALSE | Upregulated |
| -0,89370088 | 0,117583525 | ATAD3C | FALSE | Downregulated |
| -0,255000392 | 0,117583525 | PCYOX1 | FALSE | Downregulated |
| 1,788001216 | 0,117583525 | SPINK1 | FALSE | Upregulated |
| 1,045091596 | 0,117589471 | CMTM1 | FALSE | Upregulated |
| -0,145139393 | 0,117741773 | KCTD18 | FALSE | Downregulated |
| -0,215886878 | 0,117872448 | KLHL20 | FALSE | Downregulated |
| 0,483733187 | 0,118153957 | FCHSD1 | FALSE | Upregulated |
| 1,977290699 | 0,118159261 | TRIM46 | FALSE | Upregulated |
| 0,496561598 | 0,118184045 | CRELD2 | FALSE | Upregulated |
| -0,396069293 | 0,118184045 | LINC00261 | FALSE | Downregulated |
| -0,23968202 | 0,118184045 | PCMTD2 | FALSE | Downregulated |
| 0,220895736 | 0,118184045 | TRMT10C | FALSE | Upregulated |
| -1,993384186 | 0,118191364 | LOC101929396 | FALSE | Downregulated |
| -0,166941136 | 0,118191364 | RYBP | FALSE | Downregulated |
| 0,385324431 | 0,118191364 | TIPIN | FALSE | Upregulated |
| 1,308742794 | 0,118219772 | CHI3L1 | FALSE | Upregulated |
| -1,366121172 | 0,118277824 | LOC101927252 | FALSE | Downregulated |
| -0,793140433 | 0,118500195 | KGFLP1 | FALSE | Downregulated |
| 1,222484746 | 0,118546606 | LOC729083 | FALSE | Upregulated |
| 0,237643965 | 0,118704166 | YKT6 | FALSE | Upregulated |
| -0,212860969 | 0,118706155 | EIF3L | FALSE | Downregulated |
| -0,356183581 | 0,118706155 | MYRIP | FALSE | Downregulated |
| 1,11123186 | 0,118731197 | SEZ6L2 | FALSE | Upregulated |
| 0,342429292 | 0,118731197 | TMEM147 | FALSE | Upregulated |
| 1,540443153 | 0,11877535 | LOC644662 | FALSE | Upregulated |
| -0,246517857 | 0,119057507 | SIRT7 | FALSE | Downregulated |
| -0,399031471 | 0,119156451 | MATN2 | FALSE | Downregulated |
| 0,223684709 | 0,119177603 | ITPK1 | FALSE | Upregulated |
| -0,532696014 | 0,119184018 | RUNX1T1 | FALSE | Downregulated |
| -0,26280719 | 0,119204247 | ANKS4B | FALSE | Downregulated |
| -0,195222081 | 0,119204247 | PLEKHA1 | FALSE | Downregulated |
| -0,432163863 | 0,119214817 | KIAA1407 | FALSE | Downregulated |
| 0,140798736 | 0,119275194 | ARMC8 | FALSE | Upregulated |
| -0,283832408 | 0,119275194 | KDM4B | FALSE | Downregulated |
| -0,714919638 | 0,119275194 | SEMA3B | FALSE | Downregulated |
| -0,121233216 | 0,119278985 | COIL | FALSE | Downregulated |
| 0,336896242 | 0,119278985 | MRPS12 | FALSE | Upregulated |

| | | | | |
|---|---|---|---|---|
| -0,175844458 | 0,119315899 | ARHGAP5 | FALSE | Downregulated |
| 0,237679156 | 0,119315899 | UBA52 | FALSE | Upregulated |
| 0,171629743 | 0,119395595 | ANKRD13A | FALSE | Upregulated |
| 0,22150589 | 0,119464223 | ENTPD4 | FALSE | Upregulated |
| -0,237933015 | 0,11947318 | BLCAP | FALSE | Downregulated |
| -1,439925485 | 0,11947318 | LOC101927458 | FALSE | Downregulated |
| -0,179004806 | 0,11947318 | ST6GAL1 | FALSE | Downregulated |
| -0,259633355 | 0,11947318 | TRAK1 | FALSE | Downregulated |
| 0,41510637 | 0,119625283 | C1orf122 | FALSE | Upregulated |
| 1,011373413 | 0,119645995 | C16orf93 | FALSE | Upregulated |
| 0,30590984 | 0,119709856 | MCM8 | FALSE | Upregulated |
| -0,273813416 | 0,119709856 | TINAGL1 | FALSE | Downregulated |
| -0,248019142 | 0,119709856 | ZBTB5 | FALSE | Downregulated |
| -0,271728932 | 0,119744653 | ZNF557 | FALSE | Downregulated |
| -0,366376752 | 0,1197576 | NUTM2A-AS1 | FALSE | Downregulated |
| -0,299501233 | 0,119816974 | KIAA1143 | FALSE | Downregulated |
| -0,328935408 | 0,119948801 | ZNF493 | FALSE | Downregulated |
| 0,312258696 | 0,120040864 | CBR1 | FALSE | Upregulated |
| 0,198510544 | 0,120079007 | C1QBP | FALSE | Upregulated |
| -0,266830361 | 0,120079007 | GUCD1 | FALSE | Downregulated |
| -1,731066399 | 0,120079007 | LOC100129316 | FALSE | Downregulated |
| 0,158091819 | 0,120093026 | ARF1 | FALSE | Upregulated |
| -0,569609885 | 0,120119795 | LOC101929704 | FALSE | Downregulated |
| 0,591155088 | 0,120394684 | LOC100506100 | FALSE | Upregulated |
| -0,210703154 | 0,120394684 | METTL10 | FALSE | Downregulated |
| -0,226818359 | 0,120394684 | SCAPER | FALSE | Downregulated |
| -0,297515426 | 0,120394684 | ZMYM1 | FALSE | Downregulated |
| 0,340821843 | 0,120420752 | DPH2 | FALSE | Upregulated |
| -2,049810012 | 0,120420752 | PRSS22 | FALSE | Downregulated |
| 0,264498189 | 0,120479001 | APOC2 | FALSE | Upregulated |
| 0,195658076 | 0,120567865 | SNRPG | FALSE | Upregulated |
| 0,196172136 | 0,120567865 | TMEM179B | FALSE | Upregulated |
| 0,308780028 | 0,120594941 | SLC17A5 | FALSE | Upregulated |
| -1,769693093 | 0,120657153 | MFAP5 | FALSE | Downregulated |
| -0,447650554 | 0,120661311 | APLNR | FALSE | Downregulated |
| 1,43202549 | 0,120860457 | HAVCR1 | FALSE | Upregulated |
| -0,164425113 | 0,120931238 | BRD2 | FALSE | Downregulated |
| 0,322768235 | 0,120931238 | CARS | FALSE | Upregulated |
| -0,377140425 | 0,120931238 | CYP39A1 | FALSE | Downregulated |
| -0,611191488 | 0,120931238 | GPR133 | FALSE | Downregulated |
| -0,214014401 | 0,120931238 | KRCC1 | FALSE | Downregulated |
| 0,377667512 | 0,120931238 | LRRC59 | FALSE | Upregulated |
| 0,34914604 | 0,120931238 | PIK3AP1 | FALSE | Upregulated |
| -0,707058334 | 0,120931238 | RHBG | FALSE | Downregulated |
| 0,494255319 | 0,120931238 | SPSB2 | FALSE | Upregulated |
| 1,149952372 | 0,120931238 | TNFRSF4 | FALSE | Upregulated |
| 0,164657058 | 0,121072419 | DAP | FALSE | Upregulated |
| 0,19734301 | 0,121104781 | PSMB1 | FALSE | Upregulated |
| -1,63560199 | 0,121169713 | AL132709.5 | FALSE | Downregulated |
| -1,232473427 | 0,12117184 | LOC101929465 | FALSE | Downregulated |
| -0,608447622 | 0,121257719 | GAS1 | FALSE | Downregulated |

| | | | | |
|---|---|---|---|---|
| -0,336141568 | 0,121327239 | FGFR3 | FALSE | Downregulated |
| -0,403689214 | 0,121327239 | MCF2L | FALSE | Downregulated |
| -0,353199242 | 0,121421369 | LPAR6 | FALSE | Downregulated |
| 1,43846986 | 0,121486381 | DOK7 | FALSE | Upregulated |
| 0,346795166 | 0,121606848 | PSMD1 | FALSE | Upregulated |
| 0,169256206 | 0,121606848 | ZDHHC9 | FALSE | Upregulated |
| 0,542343326 | 0,121877789 | AKAP5 | FALSE | Upregulated |
| -0,242658408 | 0,122114521 | C1R-2 | FALSE | Downregulated |
| 1,529820839 | 0,12213653 | HOXA10 | FALSE | Upregulated |
| -0,148079734 | 0,122282769 | RRN3 | FALSE | Downregulated |
| -0,345052419 | 0,122282769 | ZNF331 | FALSE | Downregulated |
| -0,531229223 | 0,12228895 | ACCS | FALSE | Downregulated |
| -1,620161144 | 0,12228895 | LOC100507530 | FALSE | Downregulated |
| 0,636878961 | 0,122299539 | MSC | FALSE | Upregulated |
| -0,151946827 | 0,122344701 | MAPK14 | FALSE | Downregulated |
| -1,689840423 | 0,122364525 | CASC9 | FALSE | Downregulated |
| -0,49357672 | 0,122364525 | RPL32P3 | FALSE | Downregulated |
| 0,547064959 | 0,122478518 | ABCC4 | FALSE | Upregulated |
| 0,884302271 | 0,122478518 | AUNIP | FALSE | Upregulated |
| 0,218159242 | 0,122532287 | NIP7 | FALSE | Upregulated |
| 0,437418019 | 0,122552157 | RP11-35G9.3 | FALSE | Upregulated |
| 1,26415947 | 0,122571867 | LOC102724908 | FALSE | Upregulated |
| 0,203256009 | 0,122625717 | SLC30A7 | FALSE | Upregulated |
| 1,243969102 | 0,122626292 | FOSL1 | FALSE | Upregulated |
| -0,227558341 | 0,122626292 | ZNF776 | FALSE | Downregulated |
| 0,224584408 | 0,12269674 | CISD1 | FALSE | Upregulated |
| 0,55765328 | 0,12269674 | ZC3H12A | FALSE | Upregulated |
| -0,605787768 | 0,122722538 | ZNRD1-AS1 | FALSE | Downregulated |
| -0,218075633 | 0,122906817 | HMGCL | FALSE | Downregulated |
| -1,223804903 | 0,12292617 | AJAP1 | FALSE | Downregulated |
| 1,389498812 | 0,122945393 | SRD5A3-AS1 | FALSE | Upregulated |
| -1,126462115 | 0,12301547 | CHRDL1 | FALSE | Downregulated |
| -1,195274394 | 0,12301547 | LOC101927192 | FALSE | Downregulated |
| -0,610644712 | 0,123017658 | LINC00950 | FALSE | Downregulated |
| -0,202710646 | 0,123077014 | MED23 | FALSE | Downregulated |
| -1,966568645 | 0,123079652 | LHFPL3-AS2 | FALSE | Downregulated |
| 1,893412089 | 0,123282832 | CABYR | FALSE | Upregulated |
| -1,254747123 | 0,123282832 | DBIL5P | FALSE | Downregulated |
| -0,696708466 | 0,123282832 | LOC100506235 | FALSE | Downregulated |
| -0,409255359 | 0,123282832 | LOC101928061 | FALSE | Downregulated |
| 0,568368982 | 0,123282832 | MBLAC1 | FALSE | Upregulated |
| 0,25226416 | 0,123282832 | RFXANK | FALSE | Upregulated |
| 0,250464662 | 0,123310107 | AP5B1 | FALSE | Upregulated |
| 0,36220853 | 0,123329727 | AZI1 | FALSE | Upregulated |
| -0,446129691 | 0,123329727 | KCNC4 | FALSE | Downregulated |
| -0,710937275 | 0,123329727 | KCNN2 | FALSE | Downregulated |
| 0,506879475 | 0,123399528 | BCL2 | FALSE | Upregulated |
| -1,755466259 | 0,123598419 | MYT1L | FALSE | Downregulated |
| 0,392714289 | 0,123598419 | ODC1 | FALSE | Upregulated |
| -1,457480495 | 0,12360607 | LOC102724539 | FALSE | Downregulated |
| 0,215751581 | 0,123705844 | TMEM62 | FALSE | Upregulated |

| | | | | |
|---|---|---|---|---|
| -0,466180402 | 0,123707365 | ZNF311 | FALSE | Downregulated |
| -1,614077183 | 0,123784184 | ALG1L9P | FALSE | Downregulated |
| -0,183743036 | 0,123784184 | NPEPPS | FALSE | Downregulated |
| -0,225839597 | 0,123890962 | ACBD5 | FALSE | Downregulated |
| 0,400566113 | 0,123931791 | NUDT14 | FALSE | Upregulated |
| -0,583504033 | 0,124014611 | AC144449.1 | FALSE | Downregulated |
| -1,290536975 | 0,124014611 | SLC6A19 | FALSE | Downregulated |
| 0,195599094 | 0,124014611 | UBE2F | FALSE | Upregulated |
| -0,176051572 | 0,124014611 | ZNF766 | FALSE | Downregulated |
| 1,203222747 | 0,124034205 | ALOX15 | FALSE | Upregulated |
| 0,297055694 | 0,124034205 | THOP1 | FALSE | Upregulated |
| -0,309534963 | 0,124127901 | EPHB4 | FALSE | Downregulated |
| -0,211894788 | 0,124132783 | TATDN1 | FALSE | Downregulated |
| 0,244537758 | 0,124142231 | AVEN | FALSE | Upregulated |
| -0,663260044 | 0,124142231 | SLC38A11 | FALSE | Downregulated |
| 0,862560826 | 0,124192604 | HLA-DQB2 | FALSE | Upregulated |
| -0,25282438 | 0,124284459 | HDAC5 | FALSE | Downregulated |
| 0,27022836 | 0,124284459 | YIPF1 | FALSE | Upregulated |
| 1,099702329 | 0,124482799 | CXCL6 | FALSE | Upregulated |
| 0,181224464 | 0,124482799 | HCFC1 | FALSE | Upregulated |
| 0,391743453 | 0,124482799 | HGF | FALSE | Upregulated |
| 1,321362402 | 0,124482799 | LOC102724835 | FALSE | Upregulated |
| -0,337066963 | 0,124482799 | MYCT1 | FALSE | Downregulated |
| -2,691662242 | 0,124482799 | NUP210L | FALSE | Downregulated |
| -0,257577778 | 0,124482799 | PVRL3 | FALSE | Downregulated |
| 0,2935071 | 0,124752103 | TWSG1 | FALSE | Upregulated |
| -0,753248147 | 0,12478997 | LOC100507395 | FALSE | Downregulated |
| -0,251083447 | 0,12478997 | MPHOSPH9 | FALSE | Downregulated |
| -0,34091228 | 0,12478997 | MYO10 | FALSE | Downregulated |
| -2,876703119 | 0,12478997 | RGS7 | FALSE | Downregulated |
| 0,213459808 | 0,12478997 | UFC1 | FALSE | Upregulated |
| -0,241151266 | 0,12484117 | INPP5A | FALSE | Downregulated |
| -0,354960926 | 0,124866101 | ZNF684 | FALSE | Downregulated |
| 0,611881834 | 0,124879708 | CDKN2B | FALSE | Upregulated |
| 0,244661796 | 0,124939084 | LIN7A | FALSE | Upregulated |
| -0,202714934 | 0,124980105 | OBSL1 | FALSE | Downregulated |
| -0,250189771 | 0,125128926 | TRIT1 | FALSE | Downregulated |
| 0,721687588 | 0,125295151 | SGPP2 | FALSE | Upregulated |
| -0,230357217 | 0,125678357 | ANKRD46 | FALSE | Downregulated |
| -0,150472822 | 0,125678357 | PEX13 | FALSE | Downregulated |
| -0,299409784 | 0,125720279 | RALGPS2 | FALSE | Downregulated |
| 2,638371735 | 0,125889715 | FOXD4 | FALSE | Upregulated |
| -0,235403525 | 0,126030147 | RSF1 | FALSE | Downregulated |
| -0,619127832 | 0,126030147 | SH3BP5-AS1 | FALSE | Downregulated |
| -0,481008394 | 0,126038428 | LOC645513 | FALSE | Downregulated |
| -0,377651182 | 0,126090687 | ACACB | FALSE | Downregulated |
| 1,70493548 | 0,126243592 | RP11-475N22.4 | FALSE | Upregulated |
| -0,123939203 | 0,126243592 | UBE4B | FALSE | Downregulated |
| 0,478663649 | 0,126322114 | JUNB | FALSE | Upregulated |
| 0,186812365 | 0,126401328 | ELAVL1 | FALSE | Upregulated |
| -0,338359601 | 0,126427993 | TMEM42 | FALSE | Downregulated |

| | | | | |
|---|---|---|---|---|
| -1,193320916 | 0,126477399 | LOC102723513 | FALSE | Downregulated |
| -0,653222147 | 0,12651643 | C8orf46 | FALSE | Downregulated |
| 0,151263142 | 0,12651643 | SCARB2 | FALSE | Upregulated |
| 1,285688342 | 0,126536166 | C1orf127 | FALSE | Upregulated |
| -0,237466674 | 0,126632115 | PECR | FALSE | Downregulated |
| 1,064709416 | 0,126699385 | TTC16 | FALSE | Upregulated |
| -0,342467181 | 0,127142814 | GABPB2 | FALSE | Downregulated |
| -0,33701204 | 0,127153432 | CPS1 | FALSE | Downregulated |
| -0,175601535 | 0,127181114 | PURB | FALSE | Downregulated |
| 0,172986047 | 0,127256657 | L3MBTL2 | FALSE | Upregulated |
| -0,632687756 | 0,127256657 | NPIPB3 | FALSE | Downregulated |
| -0,363551055 | 0,127425696 | GPR116 | FALSE | Downregulated |
| 1,071591042 | 0,127425696 | MUC1 | FALSE | Upregulated |
| -0,34901018 | 0,127589856 | NRP2 | FALSE | Downregulated |
| -0,522183759 | 0,127677536 | CDKL1 | FALSE | Downregulated |
| -0,264795051 | 0,127677536 | EIF1AX | FALSE | Downregulated |
| -0,391005349 | 0,127696426 | ZNF708 | FALSE | Downregulated |
| -0,773813784 | 0,127726603 | ASB9 | FALSE | Downregulated |
| -0,121125525 | 0,127731443 | GPBP1 | FALSE | Downregulated |
| -0,28206766 | 0,127761491 | SMIM8 | FALSE | Downregulated |
| -1,024801816 | 0,127761491 | VIL1 | FALSE | Downregulated |
| -0,165569897 | 0,127839535 | KIAA1429 | FALSE | Downregulated |
| 0,42092202 | 0,127839535 | SLC9A1 | FALSE | Upregulated |
| -0,475626252 | 0,127920153 | ANGPTL1 | FALSE | Downregulated |
| -0,288298197 | 0,127920153 | INO80D | FALSE | Downregulated |
| -0,904616953 | 0,127920153 | LOC102723647 | FALSE | Downregulated |
| -0,154553801 | 0,128037435 | TMEM248 | FALSE | Downregulated |
| 0,160579471 | 0,128104253 | USP14 | FALSE | Upregulated |
| 0,354954992 | 0,128158165 | ITGAV | FALSE | Upregulated |
| 0,219918284 | 0,128158165 | PSMB6 | FALSE | Upregulated |
| 0,209575127 | 0,12816556 | GUK1 | FALSE | Upregulated |
| -0,429691941 | 0,12816556 | NCMAP | FALSE | Downregulated |
| 0,622554756 | 0,128166216 | H2AFY2 | FALSE | Upregulated |
| 0,648941508 | 0,128166216 | MFSD7 | FALSE | Upregulated |
| 0,55298934 | 0,128189999 | ARL11 | FALSE | Upregulated |
| -0,521819786 | 0,128221355 | HBCBP | FALSE | Downregulated |
| -0,166296598 | 0,128227608 | MYNN | FALSE | Downregulated |
| -0,192049581 | 0,128398231 | GATAD1 | FALSE | Downregulated |
| 0,42594985 | 0,128412575 | NEFH | FALSE | Upregulated |
| 1,244029226 | 0,128471548 | ANXA2P1 | FALSE | Upregulated |
| -0,385462591 | 0,128471548 | C9 | FALSE | Downregulated |
| -0,650277773 | 0,128471548 | LOC101929046 | FALSE | Downregulated |
| -0,160498893 | 0,128471548 | PPP6R3 | FALSE | Downregulated |
| 0,297299767 | 0,128471548 | PRDX4 | FALSE | Upregulated |
| -0,603257622 | 0,128471548 | PREX2 | FALSE | Downregulated |
| -0,756184457 | 0,128471548 | PSD | FALSE | Downregulated |
| -0,403398139 | 0,128471548 | S1PR1 | FALSE | Downregulated |
| -0,839044717 | 0,128514497 | MT1G | FALSE | Downregulated |
| -0,280554451 | 0,128573868 | LOC101927187 | FALSE | Downregulated |
| -0,178505907 | 0,128573868 | PABPC4 | FALSE | Downregulated |
| -0,443751721 | 0,128573868 | PCBP1-AS1 | FALSE | Downregulated |

| | | | | |
|---|---|---|---|---|
| 0,288024496 | 0,128596636 | KIF2A | FALSE | Upregulated |
| 0,283555955 | 0,128596636 | MAP4K2 | FALSE | Upregulated |
| -0,646881107 | 0,128647371 | LOC100507547 | FALSE | Downregulated |
| -0,293017026 | 0,128647371 | ZBTB20 | FALSE | Downregulated |
| 0,298685906 | 0,128701805 | DNAJB4 | FALSE | Upregulated |
| -0,154682088 | 0,128807435 | KIAA0430 | FALSE | Downregulated |
| 0,283460138 | 0,128917119 | MAPK7 | FALSE | Upregulated |
| 0,131642484 | 0,128917119 | NRBP1 | FALSE | Upregulated |
| -0,281122629 | 0,12894813 | NFIA | FALSE | Downregulated |
| -0,283632671 | 0,12894813 | NUBPL | FALSE | Downregulated |
| 0,186923255 | 0,12895344 | GMPS | FALSE | Upregulated |
| -0,134337607 | 0,12895344 | PTK2 | FALSE | Downregulated |
| -0,658313563 | 0,129015372 | F11-AS1 | FALSE | Downregulated |
| 0,187527391 | 0,129074119 | TIMM22 | FALSE | Upregulated |
| -0,478207387 | 0,129180479 | TSNAXIP1 | FALSE | Downregulated |
| -0,206904656 | 0,129222446 | AP1AR | FALSE | Downregulated |
| 0,20014094 | 0,129233906 | TMEM205 | FALSE | Upregulated |
| 0,180370416 | 0,129393751 | EIF2S2 | FALSE | Upregulated |
| -0,491572109 | 0,129393751 | GRK4 | FALSE | Downregulated |
| -1,81257944 | 0,129393751 | RP1L1 | FALSE | Downregulated |
| 0,506926566 | 0,129427337 | BACE2 | FALSE | Upregulated |
| -0,170725738 | 0,129427337 | TOP2B | FALSE | Downregulated |
| -1,080285623 | 0,129504664 | FAM35BP | FALSE | Downregulated |
| -0,287026932 | 0,129617238 | NISCH | FALSE | Downregulated |
| -0,431867211 | 0,129693752 | FAM65C | FALSE | Downregulated |
| 0,195374934 | 0,129693752 | UBAC2 | FALSE | Upregulated |
| -0,3016844 | 0,12969735 | ZFAND4 | FALSE | Downregulated |
| 0,267732255 | 0,129729713 | ERMP1 | FALSE | Upregulated |
| 0,355583888 | 0,129729713 | GLS | FALSE | Upregulated |
| -0,169714529 | 0,129729713 | LRP5 | FALSE | Downregulated |
| 0,177161376 | 0,129729713 | RRP1B | FALSE | Upregulated |
| -0,454566429 | 0,129729713 | ZNF418 | FALSE | Downregulated |
| -0,656216705 | 0,129821721 | RP11-75C9.1 | FALSE | Downregulated |
| -0,648485522 | 0,129832125 | CLEC4M | FALSE | Downregulated |
| -0,38903712 | 0,130011119 | CAV1 | FALSE | Downregulated |
| 0,239997142 | 0,130064618 | MYO1D | FALSE | Upregulated |
| 0,177910882 | 0,130119619 | SCAF1 | FALSE | Upregulated |
| -1,245832886 | 0,130174086 | LOC102723570 | FALSE | Downregulated |
| -0,224127562 | 0,130257252 | SELO | FALSE | Downregulated |
| 0,215450381 | 0,130330963 | HSPBP1 | FALSE | Upregulated |
| 0,299326133 | 0,130371669 | DDX58 | FALSE | Upregulated |
| -0,189295534 | 0,130380617 | DHPS | FALSE | Downregulated |
| 0,487538505 | 0,130409201 | CDYL2 | FALSE | Upregulated |
| -0,786486698 | 0,130489521 | INTS6-AS1 | FALSE | Downregulated |
| -1,478595469 | 0,130570591 | RP11-563J2.2 | FALSE | Downregulated |
| 0,213306283 | 0,130657928 | CIB1 | FALSE | Upregulated |
| -2,628503764 | 0,130679393 | MYBPH | FALSE | Downregulated |
| 0,682050947 | 0,1307046 | P2RY6 | FALSE | Upregulated |
| 0,552523499 | 0,130938745 | FAM83G | FALSE | Upregulated |
| 0,263485938 | 0,131078799 | MSANTD4 | FALSE | Upregulated |
| -0,392221832 | 0,131078799 | TMEM231 | FALSE | Downregulated |

| | | | | |
|---|---|---|---|---|
| -0,537205448 | 0,131272605 | ABI3BP | FALSE | Downregulated |
| 0,860128819 | 0,131272605 | IRF4 | FALSE | Upregulated |
| -2,763461724 | 0,131272605 | LOC100996379 | FALSE | Downregulated |
| 0,359736617 | 0,131290844 | GPRC5C | FALSE | Upregulated |
| -0,23990817 | 0,131290844 | TADA2A | FALSE | Downregulated |
| -0,188422178 | 0,131312904 | KDM4C | FALSE | Downregulated |
| -0,944486144 | 0,131312904 | LOC101929824 | FALSE | Downregulated |
| -0,186272847 | 0,131332432 | SMARCAD1 | FALSE | Downregulated |
| -0,410472274 | 0,131470672 | FGF14-AS2 | FALSE | Downregulated |
| 0,287245942 | 0,131470672 | TMEM138 | FALSE | Upregulated |
| -0,145002302 | 0,13155787 | ATXN7L3 | FALSE | Downregulated |
| -0,487182179 | 0,13156411 | SMA4 | FALSE | Downregulated |
| 0,191440804 | 0,131798291 | U2AF1 | FALSE | Upregulated |
| -0,146314991 | 0,131811851 | IREB2 | FALSE | Downregulated |
| -0,76735091 | 0,131984006 | LOC100506985 | FALSE | Downregulated |
| -0,965989383 | 0,132069145 | ACOT6 | FALSE | Downregulated |
| 1,942794581 | 0,132069145 | SHISA9 | FALSE | Upregulated |
| -0,23617749 | 0,132081133 | GCNT2 | FALSE | Downregulated |
| 0,221153951 | 0,132114077 | VTI1A | FALSE | Upregulated |
| -0,813561669 | 0,132125206 | LOC102724455 | FALSE | Downregulated |
| -0,186039736 | 0,132145191 | DDHD2 | FALSE | Downregulated |
| -0,759113462 | 0,13215857 | AKR1CL1 | FALSE | Downregulated |
| 0,284269187 | 0,13215857 | CSGALNACT2 | FALSE | Upregulated |
| 0,662091251 | 0,13215857 | HOXB7 | FALSE | Upregulated |
| 0,251895069 | 0,13215857 | METTL6 | FALSE | Upregulated |
| 0,162787714 | 0,13215857 | RPE | FALSE | Upregulated |
| -0,190698147 | 0,132180848 | STRN3 | FALSE | Downregulated |
| 0,233581639 | 0,132231261 | DHRSX | FALSE | Upregulated |
| -0,544814017 | 0,132285681 | LOC101927506 | FALSE | Downregulated |
| 1,18196855 | 0,132341126 | HTR7 | FALSE | Upregulated |
| -0,190493802 | 0,132341126 | PHKA2 | FALSE | Downregulated |
| 0,390806302 | 0,132380691 | SPIN2B | FALSE | Upregulated |
| 0,550413452 | 0,132380691 | TRIM6 | FALSE | Upregulated |
| 1,034505135 | 0,132380691 | ULBP1 | FALSE | Upregulated |
| 0,534504729 | 0,132485823 | PLEKHH1 | FALSE | Upregulated |
| -0,306925113 | 0,132544856 | KBTBD3 | FALSE | Downregulated |
| -0,289467288 | 0,132544856 | MET | FALSE | Downregulated |
| -0,239652207 | 0,132697455 | DDX55 | FALSE | Downregulated |
| -0,353821679 | 0,132768157 | CLCN6 | FALSE | Downregulated |
| -0,905656169 | 0,132771006 | EFR3B | FALSE | Downregulated |
| -0,459734609 | 0,132771006 | LOC101927974 | FALSE | Downregulated |
| -0,287323696 | 0,132771006 | ZC3H12C | FALSE | Downregulated |
| -0,189585121 | 0,132816193 | MTF2 | FALSE | Downregulated |
| -0,508687872 | 0,132968976 | NKTR | FALSE | Downregulated |
| -0,43122533 | 0,133183662 | SLC25A23 | FALSE | Downregulated |
| -0,20075974 | 0,133311571 | SRSF4 | FALSE | Downregulated |
| 0,643244901 | 0,133360092 | EVA1C | FALSE | Upregulated |
| 0,264854666 | 0,133416503 | HEATR3 | FALSE | Upregulated |
| -0,541035858 | 0,133416503 | ZNF221 | FALSE | Downregulated |
| 0,626965913 | 0,133417302 | TBX21 | FALSE | Upregulated |
| 0,551951508 | 0,133506436 | GPR132 | FALSE | Upregulated |

| | | | | |
|---|---|---|---|---|
| -0,591150946 | 0,133506436 | LOC102724005 | FALSE | Downregulated |
| -0,832057246 | 0,13367786 | ATM | FALSE | Downregulated |
| -0,215687477 | 0,133688361 | ZCCHC11 | FALSE | Downregulated |
| 0,386213343 | 0,133688696 | TMC6 | FALSE | Upregulated |
| -0,169579464 | 0,13376844 | CGGBP1 | FALSE | Downregulated |
| 0,218402197 | 0,133828907 | DIRC2 | FALSE | Upregulated |
| -0,533001923 | 0,133828907 | ZBTB26 | FALSE | Downregulated |
| 0,146025715 | 0,133837058 | CCT7 | FALSE | Upregulated |
| 1,06079872 | 0,133837058 | PLCXD2 | FALSE | Upregulated |
| 0,538186577 | 0,133861513 | RP11-307C12.11 | FALSE | Upregulated |
| 0,147553611 | 0,133924826 | BSDC1 | FALSE | Upregulated |
| -0,430053171 | 0,133932089 | ZC3H13 | FALSE | Downregulated |
| -0,42961255 | 0,1340842 | LINC00662 | FALSE | Downregulated |
| -0,254607876 | 0,134092816 | NOTCH1 | FALSE | Downregulated |
| -0,16960121 | 0,13420086 | NVL | FALSE | Downregulated |
| -0,259186327 | 0,134416776 | GOLGB1 | FALSE | Downregulated |
| -0,356482078 | 0,134449606 | TRIM52 | FALSE | Downregulated |
| 0,312333809 | 0,134460704 | ZFP69 | FALSE | Upregulated |
| 0,234757612 | 0,134515295 | ETNK1 | FALSE | Upregulated |
| -1,766985357 | 0,134661931 | STON1-GTF2A1L | FALSE | Downregulated |
| -0,918604721 | 0,134665762 | LOC100129722 | FALSE | Downregulated |
| -0,796250753 | 0,134665762 | LOC101927850 | FALSE | Downregulated |
| -0,557036633 | 0,134665762 | LOC101928451 | FALSE | Downregulated |
| 0,669665821 | 0,134665762 | PPM1N | FALSE | Upregulated |
| -0,174149957 | 0,134665762 | TTLL5 | FALSE | Downregulated |
| -0,49042009 | 0,134759993 | GALNT3 | FALSE | Downregulated |
| 0,194075192 | 0,134761489 | KIAA2013 | FALSE | Upregulated |
| -0,421528836 | 0,134891343 | HP | FALSE | Downregulated |
| -0,235633948 | 0,13491773 | BCL9 | FALSE | Downregulated |
| -0,591617782 | 0,13491773 | TAPT1-AS1 | FALSE | Downregulated |
| 1,091221523 | 0,134964769 | CXCR5 | FALSE | Upregulated |
| 1,326332128 | 0,134964769 | TOMM20L | FALSE | Upregulated |
| 0,214850694 | 0,134985203 | SETX | FALSE | Upregulated |
| -0,182777232 | 0,135011048 | KHNYN | FALSE | Downregulated |
| 0,31195327 | 0,135011048 | RPS26 | FALSE | Upregulated |
| -1,027684373 | 0,135015103 | EYA2 | FALSE | Downregulated |
| -0,522749303 | 0,135053627 | RP5-1073O3.7 | FALSE | Downregulated |
| 0,948927731 | 0,135108219 | ITGA2 | FALSE | Upregulated |
| -0,164427113 | 0,135285722 | MRS2 | FALSE | Downregulated |
| -1,368161529 | 0,135430046 | LOC100630923 | FALSE | Downregulated |
| 0,307499891 | 0,135430046 | SPRYD3 | FALSE | Upregulated |
| 0,20875653 | 0,135530339 | SLC25A3 | FALSE | Upregulated |
| 0,404817547 | 0,135575602 | LOC101060179 | FALSE | Upregulated |
| 0,215892242 | 0,135618113 | DNPEP | FALSE | Upregulated |
| 0,698577698 | 0,135627457 | IL12RB2 | FALSE | Upregulated |
| 0,135612436 | 0,135784859 | CDC27 | FALSE | Upregulated |
| 0,12182133 | 0,135795673 | AIDA | FALSE | Upregulated |
| -0,705494445 | 0,135899038 | DNHD1 | FALSE | Downregulated |
| -0,321309842 | 0,13610519 | LOC646762 | FALSE | Downregulated |
| -0,200917652 | 0,136132893 | GNPNAT1 | FALSE | Downregulated |
| -0,655807569 | 0,136271641 | LOC100131564 | FALSE | Downregulated |

| | | | | |
|---|---|---|---|---|
| 0,665230042 | 0,136516672 | HSPA2 | FALSE | Upregulated |
| 0,189661636 | 0,13653313 | BRE | FALSE | Upregulated |
| 0,286610402 | 0,136557953 | TPP1 | FALSE | Upregulated |
| 0,24160177 | 0,136590214 | TRAF3 | FALSE | Upregulated |
| 0,165467333 | 0,136590214 | UBTD2 | FALSE | Upregulated |
| -0,23923146 | 0,136590214 | ZNF330 | FALSE | Downregulated |
| 0,2557867 | 0,136607665 | NOL12 | FALSE | Upregulated |
| -0,179510835 | 0,136607665 | PAPD5 | FALSE | Downregulated |
| 0,259529042 | 0,136755986 | CSTF2 | FALSE | Upregulated |
| 0,169809739 | 0,136789772 | NBN | FALSE | Upregulated |
| -0,320064008 | 0,136800682 | TTC30B | FALSE | Downregulated |
| 0,971154947 | 0,136863856 | SNORD89 | FALSE | Upregulated |
| -0,333441383 | 0,136984471 | CBWD5 | FALSE | Downregulated |
| 0,439008604 | 0,137124171 | WBP5 | FALSE | Upregulated |
| 0,408546653 | 0,137143979 | DSTNP2 | FALSE | Upregulated |
| -1,093913933 | 0,137143979 | EPO | FALSE | Downregulated |
| 1,604700403 | 0,137143979 | LOC101928292 | FALSE | Upregulated |
| -0,257147953 | 0,137298916 | LCOR | FALSE | Downregulated |
| 0,138517533 | 0,137356874 | MTMR14 | FALSE | Upregulated |
| 0,605953779 | 0,137539914 | KLHL25 | FALSE | Upregulated |
| -0,370844606 | 0,13755719 | LOC100507599 | FALSE | Downregulated |
| -0,277339619 | 0,13755719 | MANEA | FALSE | Downregulated |
| -0,318575461 | 0,137703196 | F5 | FALSE | Downregulated |
| -0,450745479 | 0,137703196 | HCG18 | FALSE | Downregulated |
| -0,153981532 | 0,137703196 | MAP4K5 | FALSE | Downregulated |
| -0,176115337 | 0,13774806 | AEBP2 | FALSE | Downregulated |
| -0,229372132 | 0,13779778 | C12orf29 | FALSE | Downregulated |
| -0,501439666 | 0,137827056 | DNM3OS | FALSE | Downregulated |
| 1,35942849 | 0,137853865 | COX4I2 | FALSE | Upregulated |
| 0,549377008 | 0,137888913 | ST8SIA4 | FALSE | Upregulated |
| -0,228681789 | 0,137889606 | RNF139 | FALSE | Downregulated |
| 0,309891527 | 0,137903564 | HAUS7 | FALSE | Upregulated |
| 0,648234111 | 0,137931678 | DAPK2 | FALSE | Upregulated |
| -0,342854248 | 0,137976266 | AGMAT | FALSE | Downregulated |
| 0,153513133 | 0,137976266 | DLST | FALSE | Upregulated |
| -0,367108771 | 0,137976266 | NAALADL2 | FALSE | Downregulated |
| -0,284358319 | 0,137976266 | UBOX5 | FALSE | Downregulated |
| 0,11912575 | 0,138207161 | DDX41 | FALSE | Upregulated |
| -0,181558625 | 0,138245241 | EP300 | FALSE | Downregulated |
| -0,488416441 | 0,138245241 | MYCL | FALSE | Downregulated |
| 1,025753469 | 0,138245241 | POMC | FALSE | Upregulated |
| 1,015394097 | 0,138245241 | RGS9 | FALSE | Upregulated |
| -0,465751337 | 0,138245241 | RP11-96H19.1 | FALSE | Downregulated |
| 0,292552759 | 0,138427883 | TBC1D31 | FALSE | Upregulated |
| 0,390602003 | 0,138449457 | MITF | FALSE | Upregulated |
| 0,263493464 | 0,138449457 | USP38 | FALSE | Upregulated |
| 0,16212366 | 0,138506304 | FBXO33 | FALSE | Upregulated |
| 0,638836305 | 0,138506304 | FBXO41 | FALSE | Upregulated |
| -0,644392517 | 0,138506304 | KLHL4 | FALSE | Downregulated |
| 0,336822916 | 0,138506304 | PAPSS1 | FALSE | Upregulated |
| 0,320254763 | 0,138567321 | CDK18 | FALSE | Upregulated |

| | | | | |
|---|---|---|---|---|
| -0,235991809 | 0,138567321 | TSPAN12 | FALSE | Downregulated |
| -0,166836814 | 0,138583042 | TRIM33 | FALSE | Downregulated |
| -0,832255305 | 0,138604856 | IHH | FALSE | Downregulated |
| -0,283994747 | 0,138662605 | SMLR1 | FALSE | Downregulated |
| -0,189801969 | 0,138662605 | SUPT20H | FALSE | Downregulated |
| -0,284342963 | 0,138762305 | TMEM120A | FALSE | Downregulated |
| -0,158996328 | 0,138831416 | TUG1 | FALSE | Downregulated |
| -0,187681747 | 0,138912548 | CLINT1 | FALSE | Downregulated |
| -2,728156601 | 0,138921331 | LEP | FALSE | Downregulated |
| 0,972869732 | 0,139014001 | EGFR-AS1 | FALSE | Upregulated |
| 0,437573703 | 0,139014001 | PLEKHA4 | FALSE | Upregulated |
| 0,451918581 | 0,139173389 | CD226 | FALSE | Upregulated |
| -0,413632749 | 0,139603936 | AMOTL1 | FALSE | Downregulated |
| -0,968374318 | 0,139667447 | SMC5-AS1 | FALSE | Downregulated |
| -0,621073986 | 0,139684886 | LOC100506060 | FALSE | Downregulated |
| -0,175355622 | 0,139802259 | GATC | FALSE | Downregulated |
| -0,386697322 | 0,139802259 | TRIM52-AS1 | FALSE | Downregulated |
| -1,418158683 | 0,139826712 | LOC100499484-C9ORF174 | FALSE | Downregulated |
| 0,980044711 | 0,139826712 | MEX3B | FALSE | Upregulated |
| 0,444587175 | 0,139918778 | STPG1 | FALSE | Upregulated |
| -0,207178918 | 0,140093966 | DZIP3 | FALSE | Downregulated |
| -0,980796731 | 0,14013131 | SCUBE2 | FALSE | Downregulated |
| 0,565043563 | 0,140295584 | CCDC102B | FALSE | Upregulated |
| -0,261423078 | 0,140326049 | AOC3 | FALSE | Downregulated |
| 0,241777613 | 0,140326049 | FAM89B | FALSE | Upregulated |
| 0,261603633 | 0,140338081 | DPYSL2 | FALSE | Upregulated |
| -0,195471641 | 0,14044085 | RPL5 | FALSE | Downregulated |
| -0,82625013 | 0,140461078 | SV2B | FALSE | Downregulated |
| -0,220750341 | 0,140496231 | GLYATL1 | FALSE | Downregulated |
| 0,373551347 | 0,140501347 | CERS4 | FALSE | Upregulated |
| 0,610583257 | 0,140610043 | CHST9 | FALSE | Upregulated |
| -0,849544534 | 0,140610043 | LRRC3-AS1 | FALSE | Downregulated |
| 0,244393484 | 0,140624907 | EML4 | FALSE | Upregulated |
| 0,334916254 | 0,140624907 | TMEM258 | FALSE | Upregulated |
| -0,337896898 | 0,14064745 | MARCH9 | FALSE | Downregulated |
| 0,534576925 | 0,140675509 | DBNDD2 | FALSE | Upregulated |
| -0,878911111 | 0,140719326 | ROPN1L | FALSE | Downregulated |
| -0,431511491 | 0,140719326 | TAZ | FALSE | Downregulated |
| 0,5955414 | 0,140749323 | CARD9 | FALSE | Upregulated |
| 1,156674356 | 0,140939456 | ESPL1 | FALSE | Upregulated |
| 0,278801845 | 0,140942928 | FRY | FALSE | Upregulated |
| -0,811713763 | 0,140942928 | LOC101928794 | FALSE | Downregulated |
| -0,873363007 | 0,140942928 | LOC102724019 | FALSE | Downregulated |
| -0,136798167 | 0,140990919 | EIF2S3 | FALSE | Downregulated |
| 0,610651363 | 0,140990919 | FNDC5 | FALSE | Upregulated |
| 1,916859904 | 0,140990919 | MAGED4 | FALSE | Upregulated |
| 0,843602537 | 0,140990919 | MEI1 | FALSE | Upregulated |
| -0,506251624 | 0,140990919 | TRNY | FALSE | Downregulated |
| -0,231089421 | 0,141012609 | C16orf80 | FALSE | Downregulated |
| 0,259383611 | 0,141012609 | DDOST | FALSE | Upregulated |

| | | | | |
|---|---|---|---|---|
| -0,588425815 | 0,141012609 | PPP1R32 | FALSE | Downregulated |
| 0,210639911 | 0,141012609 | STXBP2 | FALSE | Upregulated |
| -0,266188688 | 0,141070772 | IFT140 | FALSE | Downregulated |
| -0,15483061 | 0,141172713 | MAGOH | FALSE | Downregulated |
| -0,283991399 | 0,141182977 | LRIG3 | FALSE | Downregulated |
| 0,273597229 | 0,141196795 | NECAB3 | FALSE | Upregulated |
| -2,434125542 | 0,141288882 | CIDEA | FALSE | Downregulated |
| -0,583915584 | 0,141393828 | RP11-761I4.3 | FALSE | Downregulated |
| 0,123054895 | 0,141439638 | DNAJA2 | FALSE | Upregulated |
| -1,264052693 | 0,141533821 | ESPNP | FALSE | Downregulated |
| 0,235036406 | 0,141533821 | STOM | FALSE | Upregulated |
| -0,22818837 | 0,141744665 | ZRANB2 | FALSE | Downregulated |
| 0,267191845 | 0,141844073 | NIPSNAP3A | FALSE | Upregulated |
| -0,383112745 | 0,141940613 | KCND3 | FALSE | Downregulated |
| 0,309036198 | 0,141940613 | PDCD2L | FALSE | Upregulated |
| -0,398397581 | 0,141970649 | LOC285074 | FALSE | Downregulated |
| 0,558471591 | 0,141981168 | ABRACL | FALSE | Upregulated |
| -0,93597135 | 0,141981168 | CYP17A1 | FALSE | Downregulated |
| -0,679544394 | 0,141981168 | KCNJ3 | FALSE | Downregulated |
| 0,338968243 | 0,141981168 | MIER2 | FALSE | Upregulated |
| -1,425796785 | 0,141981168 | SLC26A7 | FALSE | Downregulated |
| 0,179825343 | 0,141989319 | ARF3 | FALSE | Upregulated |
| 0,160050366 | 0,141991904 | YTHDF2 | FALSE | Upregulated |
| 0,40606425 | 0,142055178 | COMTD1 | FALSE | Upregulated |
| -0,386781765 | 0,142055178 | GDPD5 | FALSE | Downregulated |
| 0,394173456 | 0,142123658 | GYG1 | FALSE | Upregulated |
| 0,254489324 | 0,142123658 | ZNF92 | FALSE | Upregulated |
| -0,181368856 | 0,142143801 | KAT6B | FALSE | Downregulated |
| -1,040018645 | 0,142151192 | LOC101928318 | FALSE | Downregulated |
| 0,18775364 | 0,142151192 | SPPL2A | FALSE | Upregulated |
| 0,380085988 | 0,142237708 | HTRA1 | FALSE | Upregulated |
| 0,864852174 | 0,142237708 | LOC101929545 | FALSE | Upregulated |
| 0,430634948 | 0,142237708 | SMPD2 | FALSE | Upregulated |
| -1,383000036 | 0,142237708 | ZNF843 | FALSE | Downregulated |
| -2,028047586 | 0,142256189 | LOC102724694 | FALSE | Downregulated |
| 0,429155716 | 0,142296624 | SFMBT2 | FALSE | Upregulated |
| 0,927464237 | 0,14238364 | HLA-J | FALSE | Upregulated |
| -0,184055881 | 0,142494727 | LEMD3 | FALSE | Downregulated |
| -0,442777645 | 0,142494727 | STOX2 | FALSE | Downregulated |
| -0,544816548 | 0,142582202 | LOC100505902 | FALSE | Downregulated |
| -1,505160878 | 0,142669863 | KCNH2 | FALSE | Downregulated |
| 1,488253395 | 0,142669863 | ZFP57 | FALSE | Upregulated |
| 0,257030561 | 0,142703738 | NTAN1 | FALSE | Upregulated |
| 0,216701844 | 0,14277612 | PDE12 | FALSE | Upregulated |
| -0,279709214 | 0,142855207 | SNHG16 | FALSE | Downregulated |
| 0,309684569 | 0,14298236 | METTL4 | FALSE | Upregulated |
| -0,235869682 | 0,143244841 | ISOC2 | FALSE | Downregulated |
| 0,210058693 | 0,143380672 | PTPMT1 | FALSE | Upregulated |
| -0,29818368 | 0,143385216 | BEND7 | FALSE | Downregulated |
| 0,641917819 | 0,143659834 | DUSP4 | FALSE | Upregulated |
| -0,247222178 | 0,14376278 | GKAP1 | FALSE | Downregulated |

| | | | | |
|---|---|---|---|---|
| -0,141131892 | 0,14376278 | TCEA1 | FALSE | Downregulated |
| -0,526354248 | 0,143772975 | FGF2 | FALSE | Downregulated |
| -0,13939011 | 0,143788129 | UBE4A | FALSE | Downregulated |
| 0,588302797 | 0,143881173 | KLRB1 | FALSE | Upregulated |
| -0,233951165 | 0,144068429 | STX17 | FALSE | Downregulated |
| 1,174214606 | 0,144073499 | FAM155B | FALSE | Upregulated |
| -0,148422898 | 0,144209166 | PPIE | FALSE | Downregulated |
| -0,239099534 | 0,144258494 | FBXO46 | FALSE | Downregulated |
| -0,223923577 | 0,144258494 | PEX7 | FALSE | Downregulated |
| -0,344910505 | 0,144270857 | ATP11C | FALSE | Downregulated |
| 0,526446197 | 0,144270857 | SELL | FALSE | Upregulated |
| -0,414427937 | 0,144365804 | ARHGEF17 | FALSE | Downregulated |
| -0,391751761 | 0,144630206 | SLC9A3R2 | FALSE | Downregulated |
| -0,205782222 | 0,144678546 | DENND5B | FALSE | Downregulated |
| 0,229863196 | 0,144697091 | MAD2L1BP | FALSE | Upregulated |
| -0,35020291 | 0,14476148 | CALML4 | FALSE | Downregulated |
| -0,198638955 | 0,144782003 | BCOR | FALSE | Downregulated |
| 0,29069668 | 0,144782003 | LRRC8A | FALSE | Upregulated |
| -0,859591817 | 0,144927995 | HAP1 | FALSE | Downregulated |
| -0,671304531 | 0,144927995 | MIR210HG | FALSE | Downregulated |
| -0,558142669 | 0,144933478 | CAMK4 | FALSE | Downregulated |
| -0,72796447 | 0,145007966 | LOC100506282 | FALSE | Downregulated |
| 1,02062932 | 0,145007966 | SNORD22 | FALSE | Upregulated |
| -0,650116172 | 0,14504598 | AGBL2 | FALSE | Downregulated |
| -0,208165549 | 0,145208172 | ATXN7 | FALSE | Downregulated |
| -0,5149515 | 0,145208172 | ERVK13-1 | FALSE | Downregulated |
| 0,67185609 | 0,145208172 | MTMR8 | FALSE | Upregulated |
| -0,169264315 | 0,145208172 | ZNF212 | FALSE | Downregulated |
| 0,384064954 | 0,145234034 | KCNQ1 | FALSE | Upregulated |
| 1,165482358 | 0,145276132 | SBSPON | FALSE | Upregulated |
| -0,387546228 | 0,145338032 | C7orf55 | FALSE | Downregulated |
| 0,669520939 | 0,145338032 | DHRS13 | FALSE | Upregulated |
| 0,775556695 | 0,145338032 | FRMPD1 | FALSE | Upregulated |
| -1,831666065 | 0,145338032 | MYCBP2-AS1 | FALSE | Downregulated |
| -0,342794211 | 0,145338032 | UBL7-AS1 | FALSE | Downregulated |
| -0,223646353 | 0,145338032 | ZNF23 | FALSE | Downregulated |
| 0,868310517 | 0,145435641 | LOXL1 | FALSE | Upregulated |
| -1,476152662 | 0,145441186 | LOC101929463 | FALSE | Downregulated |
| -0,337646777 | 0,145606471 | DENND2A | FALSE | Downregulated |
| 0,406569308 | 0,145606471 | PNMA1 | FALSE | Upregulated |
| -0,230386688 | 0,145607992 | DKFZP586I1420 | FALSE | Downregulated |
| -0,713494807 | 0,14564276 | RP5-1057J7.6 | FALSE | Downregulated |
| 0,127771892 | 0,145700256 | GOLGA7 | FALSE | Upregulated |
| 0,181273361 | 0,145842121 | ZC3HAV1 | FALSE | Upregulated |
| 0,275549178 | 0,145873475 | MAP4 | FALSE | Upregulated |
| 0,160469025 | 0,145909086 | RAPGEF1 | FALSE | Upregulated |
| 1,777158482 | 0,14593645 | RNU2-1 | FALSE | Upregulated |
| -0,759419484 | 0,145992714 | LOC102724275 | FALSE | Downregulated |
| 1,558723987 | 0,145992714 | MBOAT4 | FALSE | Upregulated |
| 0,810145125 | 0,145994673 | MIAT | FALSE | Upregulated |
| -0,297317253 | 0,146047293 | UBE2D4 | FALSE | Downregulated |

| | | | | |
|---|---|---|---|---|
| 0,442630624 | 0,146506712 | C5orf56 | FALSE | Upregulated |
| 0,145558844 | 0,146506712 | PPP2CA | FALSE | Upregulated |
| 0,370072191 | 0,146606218 | TMEM102 | FALSE | Upregulated |
| 0,23890385 | 0,146639616 | NDUFAF5 | FALSE | Upregulated |
| 0,316833014 | 0,146647708 | CCDC85B | FALSE | Upregulated |
| 0,962807292 | 0,146647708 | FAM83F | FALSE | Upregulated |
| -0,826905689 | 0,146647708 | LOC102724059 | FALSE | Downregulated |
| 0,241925508 | 0,146647708 | ZBTB38 | FALSE | Upregulated |
| -0,374478727 | 0,146733777 | PRDM11 | FALSE | Downregulated |
| -0,352946071 | 0,146733777 | TIA1 | FALSE | Downregulated |
| -0,217187804 | 0,146733777 | UFM1 | FALSE | Downregulated |
| 0,79804258 | 0,14673602 | ITGB2-AS1 | FALSE | Upregulated |
| 0,143535304 | 0,146746501 | STXBP3 | FALSE | Upregulated |
| -0,871581785 | 0,146863137 | NDNF | FALSE | Downregulated |
| -0,678064814 | 0,14687695 | TBX6 | FALSE | Downregulated |
| 0,314351229 | 0,146920923 | LRMP | FALSE | Upregulated |
| -0,705392155 | 0,146920923 | SYNPO2 | FALSE | Downregulated |
| -0,770138737 | 0,146923463 | AMPH | FALSE | Downregulated |
| -0,207083261 | 0,147049582 | SORBS2 | FALSE | Downregulated |
| -0,507492225 | 0,147074621 | FBXO15 | FALSE | Downregulated |
| -0,693048703 | 0,147243005 | LINC00921 | FALSE | Downregulated |
| -0,262293943 | 0,147243005 | NHLRC2 | FALSE | Downregulated |
| 0,221071901 | 0,147337735 | CRYZ | FALSE | Upregulated |
| -0,751149386 | 0,147337735 | GATA2 | FALSE | Downregulated |
| 1,643240687 | 0,147337735 | PCNA-AS1 | FALSE | Upregulated |
| -0,201296233 | 0,147360749 | TMEM192 | FALSE | Downregulated |
| 1,214870905 | 0,147386311 | APOA4 | FALSE | Upregulated |
| -0,761642388 | 0,147437278 | PDCL3P4 | FALSE | Downregulated |
| 0,238828477 | 0,147482392 | RPS2 | FALSE | Upregulated |
| 0,596525509 | 0,147482392 | TLX1 | FALSE | Upregulated |
| -1,426105748 | 0,147564434 | GSTT2B | FALSE | Downregulated |
| 0,295892267 | 0,147568096 | CLIC2 | FALSE | Upregulated |
| 0,993066867 | 0,147626142 | FAM3B | FALSE | Upregulated |
| -0,660789239 | 0,147626142 | PROM1 | FALSE | Downregulated |
| -0,264748082 | 0,147727218 | PELI1 | FALSE | Downregulated |
| -0,339975823 | 0,147746972 | SMAD6 | FALSE | Downregulated |
| 0,291792285 | 0,148407544 | DSP | FALSE | Upregulated |
| 0,24210708 | 0,148407544 | PARP10 | FALSE | Upregulated |
| -0,223158387 | 0,148453374 | MLLT4 | FALSE | Downregulated |
| -0,151101599 | 0,148468422 | PIK3C2A | FALSE | Downregulated |
| -0,692710707 | 0,148475457 | YBX2 | FALSE | Downregulated |
| -0,232189668 | 0,148616278 | ALG11 | FALSE | Downregulated |
| -0,852914468 | 0,148649737 | CACNA1H | FALSE | Downregulated |
| -0,257775438 | 0,14870628 | MACF1 | FALSE | Downregulated |
| 0,206025411 | 0,148854876 | MTX2 | FALSE | Upregulated |
| 0,41959796 | 0,148881123 | AKR1C3 | FALSE | Upregulated |
| -0,296682151 | 0,148907748 | EI24 | FALSE | Downregulated |
| 0,931350664 | 0,14897254 | AC147651.4 | FALSE | Upregulated |
| -0,126558283 | 0,14897254 | SAR1A | FALSE | Downregulated |
| 0,190564993 | 0,149026988 | NUP188 | FALSE | Upregulated |
| -0,407720449 | 0,149026988 | SIGIRR | FALSE | Downregulated |

| | | | | |
|---|---|---|---|---|
| 0,181097951 | 0,149026988 | VPS25 | FALSE | Upregulated |
| 0,42468286 | 0,149120902 | CEBPB | FALSE | Upregulated |
| -0,302321326 | 0,149120902 | SPRY1 | FALSE | Downregulated |
| -1,847423718 | 0,14915108 | LOC101928926 | FALSE | Downregulated |
| -0,286297944 | 0,14915108 | N6AMT2 | FALSE | Downregulated |
| -0,849314246 | 0,149233415 | RP11-53O19.1 | FALSE | Downregulated |
| -0,489293999 | 0,149276638 | SSTR1 | FALSE | Downregulated |
| 0,320017741 | 0,149282274 | LAT2 | FALSE | Upregulated |
| 0,147692489 | 0,149391643 | ASUN | FALSE | Upregulated |
| 0,165181312 | 0,149391643 | COMMD7 | FALSE | Upregulated |
| 1,590262354 | 0,149391643 | L1CAM | FALSE | Upregulated |
| 0,17074986 | 0,14966598 | FUCA1 | FALSE | Upregulated |
| -0,269006415 | 0,14968916 | MAN2C1 | FALSE | Downregulated |
| 0,806171952 | 0,14968916 | TSC22D3 | FALSE | Upregulated |
| -0,773538468 | 0,149707785 | NSUN7 | FALSE | Downregulated |
| -0,35964573 | 0,149997236 | CCDC122 | FALSE | Downregulated |
| 0,136730137 | 0,149997236 | ELP3 | FALSE | Upregulated |
| 0,723299873 | 0,149997236 | GNLY | FALSE | Upregulated |
| -0,155412245 | 0,149997236 | NOA1 | FALSE | Downregulated |
| 0,185702081 | 0,150070274 | C14orf159 | FALSE | Upregulated |
| 2,219151191 | 0,150534984 | LOC101928722 | FALSE | Upregulated |
| -0,64084004 | 0,150534984 | LRP5L | FALSE | Downregulated |
| 0,176825646 | 0,150583653 | MED8 | FALSE | Upregulated |
| 0,184867984 | 0,150608846 | F11R | FALSE | Upregulated |
| 0,596635995 | 0,150659619 | LUM | FALSE | Upregulated |
| 0,194005689 | 0,150684937 | STX8 | FALSE | Upregulated |
| -0,176241351 | 0,150721425 | ZNF639 | FALSE | Downregulated |
| 0,280068911 | 0,1509185 | ANXA4 | FALSE | Upregulated |
| -0,417048836 | 0,150939016 | TTC36 | FALSE | Downregulated |
| 0,950405839 | 0,151134988 | RGS4 | FALSE | Upregulated |
| -0,390409089 | 0,151147275 | CBFA2T3 | FALSE | Downregulated |
| -0,201296943 | 0,151147275 | SEPHS2 | FALSE | Downregulated |
| -0,474911704 | 0,151195309 | MAN1C1 | FALSE | Downregulated |
| -1,239598188 | 0,151238772 | SLC25A48 | FALSE | Downregulated |
| -1,466903901 | 0,151240096 | LOC101928334 | FALSE | Downregulated |
| 0,209318555 | 0,151324559 | KIF5B | FALSE | Upregulated |
| 1,751342399 | 0,151563375 | MRAP2 | FALSE | Upregulated |
| 0,453194948 | 0,151614981 | TMC8 | FALSE | Upregulated |
| -0,215710571 | 0,151651787 | ARID1A | FALSE | Downregulated |
| -0,185104217 | 0,151710209 | GMCL1 | FALSE | Downregulated |
| -1,046514859 | 0,151710209 | LOC102724569 | FALSE | Downregulated |
| -0,165749386 | 0,151710209 | UBR2 | FALSE | Downregulated |
| -1,30893899 | 0,151727044 | SUGT1P3 | FALSE | Downregulated |
| 0,415680093 | 0,151737172 | DCPS | FALSE | Upregulated |
| 0,358068197 | 0,151750864 | TMEM173 | FALSE | Upregulated |
| -0,193163624 | 0,151794935 | COL4A3BP | FALSE | Downregulated |
| -0,897934852 | 0,15184951 | ADPRHL1 | FALSE | Downregulated |
| 0,344253754 | 0,15184951 | SHB | FALSE | Upregulated |
| -1,04504462 | 0,151913321 | FAM133A | FALSE | Downregulated |
| -0,672341867 | 0,151979387 | CASC2 | FALSE | Downregulated |
| -0,398067152 | 0,152056769 | KLF11 | FALSE | Downregulated |

| | | | | |
|---|---|---|---|---|
| -0,554912571 | 0,152106324 | CYP3A5 | FALSE | Downregulated |
| -0,269388566 | 0,152303204 | CYP4F3 | FALSE | Downregulated |
| -0,686071329 | 0,152303204 | GLIPR1L2 | FALSE | Downregulated |
| 1,498031068 | 0,152360809 | PTGER1 | FALSE | Upregulated |
| 0,38814337 | 0,152371766 | ENTPD7 | FALSE | Upregulated |
| -1,075372263 | 0,152426823 | LOC101060341 | FALSE | Downregulated |
| 1,562077846 | 0,152668384 | SMCO2 | FALSE | Upregulated |
| -0,217097128 | 0,152678785 | SYNJ1 | FALSE | Downregulated |
| -0,388784345 | 0,152728958 | ST6GAL2 | FALSE | Downregulated |
| 0,339042982 | 0,152778966 | ZSWIM3 | FALSE | Upregulated |
| -0,20783979 | 0,152781129 | DGCR8 | FALSE | Downregulated |
| -2,041084172 | 0,153374746 | LOC101929540 | FALSE | Downregulated |
| -0,18436126 | 0,153386838 | APPL2 | FALSE | Downregulated |
| -0,472778522 | 0,153626582 | BMP6 | FALSE | Downregulated |
| 0,85616023 | 0,153637139 | P2RX1 | FALSE | Upregulated |
| 0,290103649 | 0,153732013 | VAC14 | FALSE | Upregulated |
| -2,075549938 | 0,15377309 | MIR6717 | FALSE | Downregulated |
| -1,746269331 | 0,15377309 | RP13-228J13.1 | FALSE | Downregulated |
| -1,344955702 | 0,153837608 | C4orf36 | FALSE | Downregulated |
| 0,298400912 | 0,153919553 | ETHE1 | FALSE | Upregulated |
| -0,93644701 | 0,153919553 | LOC101928895 | FALSE | Downregulated |
| -0,981839723 | 0,153919553 | LOC101929846 | FALSE | Downregulated |
| -0,316099535 | 0,153919553 | ZNF106 | FALSE | Downregulated |
| 0,188293093 | 0,15405157 | HGS | FALSE | Upregulated |
| 0,426102708 | 0,154266966 | LRRC23 | FALSE | Upregulated |
| 0,418403567 | 0,154323278 | FAM65B | FALSE | Upregulated |
| -1,317344201 | 0,154328088 | TRAM1L1 | FALSE | Downregulated |
| -0,217377646 | 0,154563205 | COX7A2L | FALSE | Downregulated |
| -0,586346169 | 0,154563205 | PAN3-AS1 | FALSE | Downregulated |
| 0,380310795 | 0,154563205 | SLC8A1 | FALSE | Upregulated |
| 0,318055708 | 0,154585184 | DNAJC15 | FALSE | Upregulated |
| 0,232807484 | 0,154585184 | EGLN2 | FALSE | Upregulated |
| 0,240056574 | 0,15467432 | COPG2 | FALSE | Upregulated |
| 0,187310448 | 0,15467432 | DGCR2 | FALSE | Upregulated |
| 0,993335024 | 0,15467432 | ENTPD2 | FALSE | Upregulated |
| 0,136353139 | 0,15467432 | SNRPB2 | FALSE | Upregulated |
| 1,059767817 | 0,154681067 | PFN1P2 | FALSE | Upregulated |
| 0,253312716 | 0,15471546 | BOP1 | FALSE | Upregulated |
| 0,246614609 | 0,15471546 | CARM1 | FALSE | Upregulated |
| -0,463994547 | 0,15471546 | LOC101928120 | FALSE | Downregulated |
| 0,409705122 | 0,15471546 | NABP1 | FALSE | Upregulated |
| 0,127926336 | 0,15471546 | RNF14 | FALSE | Upregulated |
| 0,199433478 | 0,15471546 | TUBGCP5 | FALSE | Upregulated |
| -0,174107009 | 0,154743693 | ABL1 | FALSE | Downregulated |
| -0,553918553 | 0,154743693 | PRUNE2 | FALSE | Downregulated |
| -0,177417327 | 0,154841493 | NKIRAS1 | FALSE | Downregulated |
| -0,769796619 | 0,155045177 | TENM3 | FALSE | Downregulated |
| 0,204475437 | 0,155088508 | PSMA7 | FALSE | Upregulated |
| 0,532809736 | 0,155125105 | LRRC31 | FALSE | Upregulated |
| -0,549902459 | 0,155169772 | ELN | FALSE | Downregulated |
| 0,660841788 | 0,155177816 | CLDN2 | FALSE | Upregulated |

| | | | | |
|---|---|---|---|---|
| 1,055536843 | 0,155177816 | LOC102724153 | FALSE | Upregulated |
| -0,178807261 | 0,15526709 | C16orf58 | FALSE | Downregulated |
| 0,316889326 | 0,155336407 | FTSJ1 | FALSE | Upregulated |
| 0,20045461 | 0,155368225 | ATP6AP2 | FALSE | Upregulated |
| 1,655162305 | 0,155368225 | NANOG | FALSE | Upregulated |
| -0,133860638 | 0,155368225 | ZNF322 | FALSE | Downregulated |
| 0,5306981 | 0,155473501 | FAM212B | FALSE | Upregulated |
| 1,434234461 | 0,155577362 | C16orf74 | FALSE | Upregulated |
| 0,763781025 | 0,155577362 | CD24 | FALSE | Upregulated |
| 0,237491342 | 0,155577362 | TP53RK | FALSE | Upregulated |
| -1,582614519 | 0,155577362 | ZAN | FALSE | Downregulated |
| -0,49985318 | 0,155664139 | LRRC4C | FALSE | Downregulated |
| 0,912672926 | 0,155668021 | WIPF3 | FALSE | Upregulated |
| 0,244206658 | 0,155707427 | SIPA1 | FALSE | Upregulated |
| 0,204326556 | 0,155976114 | FUBP1 | FALSE | Upregulated |
| 0,218808904 | 0,15601483 | GDI1 | FALSE | Upregulated |
| -1,457740571 | 0,156032824 | LOC345051 | FALSE | Downregulated |
| -0,184946051 | 0,156123044 | SWT1 | FALSE | Downregulated |
| -1,000542934 | 0,156170189 | RP3-322G13.5 | FALSE | Downregulated |
| -0,155827711 | 0,156170189 | SUV420H1 | FALSE | Downregulated |
| -0,719924308 | 0,156236968 | CREB3L3 | FALSE | Downregulated |
| 0,295024921 | 0,156236968 | TCEB2 | FALSE | Upregulated |
| -0,290707387 | 0,156285195 | NAA16 | FALSE | Downregulated |
| -0,18820187 | 0,156375698 | MAOB | FALSE | Downregulated |
| 0,34346853 | 0,156391824 | EIF2AK2 | FALSE | Upregulated |
| -0,153857696 | 0,156467758 | HELQ | FALSE | Downregulated |
| 0,418507326 | 0,156561396 | UAP1L1 | FALSE | Upregulated |
| 0,312684052 | 0,156600707 | PMM2 | FALSE | Upregulated |
| -0,373703531 | 0,156603404 | ZNF502 | FALSE | Downregulated |
| 0,153152469 | 0,156630666 | EIF4G1 | FALSE | Upregulated |
| 0,644956745 | 0,156630666 | TAF4B | FALSE | Upregulated |
| -0,222718965 | 0,156637946 | GALE | FALSE | Downregulated |
| 0,543597831 | 0,156658472 | PDLIM7 | FALSE | Upregulated |
| 0,168518071 | 0,156705268 | C14orf119 | FALSE | Upregulated |
| -2,131314646 | 0,156904734 | TAS2R31 | FALSE | Downregulated |
| -0,506560746 | 0,156914108 | GRIA3 | FALSE | Downregulated |
| 0,217897824 | 0,156914108 | IFT20 | FALSE | Upregulated |
| 0,397250396 | 0,156914108 | TRAF1 | FALSE | Upregulated |
| -0,111782448 | 0,157310789 | CUL3 | FALSE | Downregulated |
| -0,368992219 | 0,157430215 | MARC1 | FALSE | Downregulated |
| -0,736077809 | 0,157483376 | CHRNA4 | FALSE | Downregulated |
| -0,443118925 | 0,157687079 | LOC101928431 | FALSE | Downregulated |
| -0,447277652 | 0,157719286 | C4A | FALSE | Downregulated |
| -0,711228563 | 0,157788391 | LOC100507520 | FALSE | Downregulated |
| -1,825998577 | 0,157806318 | TMEM240 | FALSE | Downregulated |
| -0,62237726 | 0,157883554 | SEMA6C | FALSE | Downregulated |
| 0,267362744 | 0,15796656 | ARPC5L | FALSE | Upregulated |
| -0,749347289 | 0,158140078 | DPPA4 | FALSE | Downregulated |
| 0,192404137 | 0,158187797 | ATP1B1 | FALSE | Upregulated |
| -0,405916926 | 0,158564631 | AHR | FALSE | Downregulated |
| 0,236163192 | 0,158917742 | CDCA7L | FALSE | Upregulated |

| | | | | |
|---|---|---|---|---|
| 0,19672094 | 0,158917742 | HAT1 | FALSE | Upregulated |
| -0,333273523 | 0,158947629 | ABHD15 | FALSE | Downregulated |
| -0,427570285 | 0,158947629 | RP11-95P2.1 | FALSE | Downregulated |
| 0,136090688 | 0,159173789 | RAP1B | FALSE | Upregulated |
| -0,295603238 | 0,159223858 | USP21 | FALSE | Downregulated |
| 0,156266869 | 0,15942569 | ATRAID | FALSE | Upregulated |
| 0,591483782 | 0,15942569 | SHBG | FALSE | Upregulated |
| 0,617168131 | 0,159737208 | AGAP2-AS1 | FALSE | Upregulated |
| 0,499834389 | 0,159737208 | BEND3 | FALSE | Upregulated |
| -0,284307089 | 0,159737208 | BHLHE40 | FALSE | Downregulated |
| -0,142308414 | 0,159737208 | LRRC47 | FALSE | Downregulated |
| -0,595349134 | 0,159737208 | RP11-372E1.4 | FALSE | Downregulated |
| -0,135728022 | 0,159737208 | SP1 | FALSE | Downregulated |
| -0,252770967 | 0,159737208 | ZNF700 | FALSE | Downregulated |
| -0,187370632 | 0,159755796 | POT1 | FALSE | Downregulated |
| 1,265213708 | 0,159768315 | C12orf39 | FALSE | Upregulated |
| -0,226819066 | 0,159768315 | KDM5B | FALSE | Downregulated |
| -0,273856754 | 0,159768315 | ZNF160 | FALSE | Downregulated |
| -0,421868253 | 0,159826538 | SC5D | FALSE | Downregulated |
| -0,203585017 | 0,15984866 | RBM39 | FALSE | Downregulated |
| 0,137422153 | 0,159969171 | GNA11 | FALSE | Upregulated |
| -0,863507737 | 0,159986508 | DCAF4L1 | FALSE | Downregulated |
| -0,386428362 | 0,159997552 | TBC1D19 | FALSE | Downregulated |
| -1,07934992 | 0,160060513 | ALLC | FALSE | Downregulated |
| 0,219211002 | 0,160060513 | DSE | FALSE | Upregulated |
| -0,284555642 | 0,160162854 | C14orf93 | FALSE | Downregulated |
| -0,225326025 | 0,160162854 | MUM1 | FALSE | Downregulated |
| -0,321181745 | 0,160197874 | LOC730102 | FALSE | Downregulated |
| -0,164202462 | 0,160197874 | MAML1 | FALSE | Downregulated |
| -0,303585383 | 0,160198905 | FBXO4 | FALSE | Downregulated |
| 0,236725055 | 0,160231674 | KLHL21 | FALSE | Upregulated |
| -0,211153657 | 0,160236613 | HLTF | FALSE | Downregulated |
| -1,890359357 | 0,160236613 | LOC102723769 | FALSE | Downregulated |
| 0,390650561 | 0,160398376 | SH3RF3 | FALSE | Upregulated |
| -1,214625868 | 0,160448874 | LOC102724824 | FALSE | Downregulated |
| 0,148051131 | 0,160507669 | YIPF5 | FALSE | Upregulated |
| -0,22470331 | 0,160509864 | CTBP1-AS2 | FALSE | Downregulated |
| 0,163394694 | 0,160530498 | CCDC97 | FALSE | Upregulated |
| -0,16111987 | 0,160541891 | SLC30A5 | FALSE | Downregulated |
| 0,256704565 | 0,160582163 | RELL1 | FALSE | Upregulated |
| -0,548608845 | 0,160681628 | ZNF767 | FALSE | Downregulated |
| 0,503441168 | 0,16069953 | SLC19A1 | FALSE | Upregulated |
| 0,215284366 | 0,160759098 | FAU | FALSE | Upregulated |
| -1,120583687 | 0,160769246 | CYP2G1P | FALSE | Downregulated |
| 0,179385312 | 0,160778964 | IDE | FALSE | Upregulated |
| 0,30642721 | 0,160778964 | KIAA1522 | FALSE | Upregulated |
| 0,247945892 | 0,160778964 | PDIA6 | FALSE | Upregulated |
| 0,923517325 | 0,160778964 | UGT2B17 | FALSE | Upregulated |
| 0,185240607 | 0,16095922 | CCDC22 | FALSE | Upregulated |
| -0,480906752 | 0,160990912 | LOC102723954 | FALSE | Downregulated |
| -0,2306006 | 0,160994826 | GC | FALSE | Downregulated |

| | | | | |
|---|---|---|---|---|
| 0,166994056 | 0,161180038 | NECAP1 | FALSE | Upregulated |
| -0,101201639 | 0,161232132 | PAIP2 | FALSE | Downregulated |
| -0,203992509 | 0,161232132 | ROCK2 | FALSE | Downregulated |
| 0,171455786 | 0,161373191 | NDUFA8 | FALSE | Upregulated |
| -0,694089183 | 0,161526689 | LOC101927402 | FALSE | Downregulated |
| 0,223850179 | 0,161799542 | EIF5A | FALSE | Upregulated |
| 0,194143758 | 0,161848788 | MAP3K3 | FALSE | Upregulated |
| 0,284129241 | 0,162267567 | LIN52 | FALSE | Upregulated |
| -0,538031858 | 0,162276867 | RAB3A | FALSE | Downregulated |
| -0,31738003 | 0,162377137 | ACSM2B | FALSE | Downregulated |
| -0,218499945 | 0,162442611 | NUDT12 | FALSE | Downregulated |
| -0,698663078 | 0,162543223 | SCNN1D | FALSE | Downregulated |
| 0,369891518 | 0,162543223 | ZNF124 | FALSE | Upregulated |
| -0,439867373 | 0,162925767 | JPX | FALSE | Downregulated |
| -0,223632258 | 0,162988089 | ITIH2 | FALSE | Downregulated |
| 0,146445123 | 0,162988089 | PRR13 | FALSE | Upregulated |
| -0,231211377 | 0,162988089 | SPRED2 | FALSE | Downregulated |
| -0,248181499 | 0,162988089 | ZNF552 | FALSE | Downregulated |
| -0,14793817 | 0,163011607 | KLHDC3 | FALSE | Downregulated |
| -0,237727513 | 0,163212898 | AQP9 | FALSE | Downregulated |
| 0,811607929 | 0,163212898 | SPINT1 | FALSE | Upregulated |
| 0,175243112 | 0,163212898 | TCP1 | FALSE | Upregulated |
| 0,271271773 | 0,163429007 | PPIC | FALSE | Upregulated |
| -0,235913058 | 0,163860725 | PMS1 | FALSE | Downregulated |
| 0,490669886 | 0,163923402 | PDE6B | FALSE | Upregulated |
| 0,131411722 | 0,163968327 | PSMD4 | FALSE | Upregulated |
| 0,169146628 | 0,164265771 | VRK2 | FALSE | Upregulated |
| 0,17551047 | 0,16430007 | SNF8 | FALSE | Upregulated |
| -1,024113733 | 0,164345999 | FKBP9L | FALSE | Downregulated |
| 0,231053064 | 0,164345999 | LRRFIP1 | FALSE | Upregulated |
| 1,484436793 | 0,164393103 | ICAM4 | FALSE | Upregulated |
| 0,219604587 | 0,164393103 | JMJD6 | FALSE | Upregulated |
| -0,257426961 | 0,164393103 | LOC93622 | FALSE | Downregulated |
| -3,036868333 | 0,164443431 | KLHL40 | FALSE | Downregulated |
| -0,435987365 | 0,164458415 | WNT5A | FALSE | Downregulated |
| -0,306733981 | 0,164533959 | CD1D | FALSE | Downregulated |
| -0,192979075 | 0,164533959 | TUBGCP4 | FALSE | Downregulated |
| 0,742172165 | 0,164537884 | PLAU | FALSE | Upregulated |
| -0,240360251 | 0,164581433 | SMIM19 | FALSE | Downregulated |
| 0,21845011 | 0,16468275 | MED19 | FALSE | Upregulated |
| -0,675499925 | 0,164692195 | CYP26B1 | FALSE | Downregulated |
| -0,429054816 | 0,164692195 | IMMP1L | FALSE | Downregulated |
| -0,254766532 | 0,164692195 | ZNF697 | FALSE | Downregulated |
| -0,336379123 | 0,164832616 | RAD50 | FALSE | Downregulated |
| 0,419995515 | 0,165010195 | STAT4 | FALSE | Upregulated |
| -0,226977692 | 0,165167778 | WDR59 | FALSE | Downregulated |
| -0,209355642 | 0,165241777 | BOD1 | FALSE | Downregulated |
| -0,283500741 | 0,165358963 | GLCCI1 | FALSE | Downregulated |
| -0,157881406 | 0,165358963 | ZFYVE1 | FALSE | Downregulated |
| 0,267929903 | 0,165386872 | AGPAT1 | FALSE | Upregulated |
| -0,811036751 | 0,165386872 | KIF1A | FALSE | Downregulated |

| | | | | |
|---|---|---|---|---|
| 0,438055587 | 0,165386872 | LIMK2 | FALSE | Upregulated |
| -0,243813674 | 0,165444357 | BHLHB9 | FALSE | Downregulated |
| 1,230721032 | 0,165470929 | CCDC19 | FALSE | Upregulated |
| 0,251302926 | 0,165472359 | SLC41A3 | FALSE | Upregulated |
| -0,163887119 | 0,165479434 | CBLL1 | FALSE | Downregulated |
| 0,195778962 | 0,165933366 | SAYSD1 | FALSE | Upregulated |
| -0,74931677 | 0,165933366 | TTC40 | FALSE | Downregulated |
| 0,611922541 | 0,165958526 | GRAP2 | FALSE | Upregulated |
| -1,147583826 | 0,165958526 | MMP21 | FALSE | Downregulated |
| 0,242088337 | 0,166003659 | DUSP14 | FALSE | Upregulated |
| -1,475729781 | 0,166003659 | KIR2DS4 | FALSE | Downregulated |
| -0,520813407 | 0,166003659 | LOC80154 | FALSE | Downregulated |
| -0,165011468 | 0,16612014 | PEX14 | FALSE | Downregulated |
| -0,258478921 | 0,166132641 | PPP1R26 | FALSE | Downregulated |
| -0,352193249 | 0,166132641 | RBM25 | FALSE | Downregulated |
| 0,144151288 | 0,166225563 | NPTN | FALSE | Upregulated |
| -0,329721363 | 0,166292015 | SLC19A3 | FALSE | Downregulated |
| -0,241429557 | 0,16632924 | ABL2 | FALSE | Downregulated |
| -0,616371941 | 0,166407881 | WNT2 | FALSE | Downregulated |
| 1,814887447 | 0,166433616 | NPAS1 | FALSE | Upregulated |
| -0,131919653 | 0,16647548 | UTP14C | FALSE | Downregulated |
| -0,371878431 | 0,16647548 | ZNF70 | FALSE | Downregulated |
| -0,643643733 | 0,166606701 | LSMEM1 | FALSE | Downregulated |
| -0,400601178 | 0,166606701 | RP11-326F20.5 | FALSE | Downregulated |
| 0,701275167 | 0,166606701 | TMEM171 | FALSE | Upregulated |
| -0,335478037 | 0,166698727 | BAALC | FALSE | Downregulated |
| -1,552040914 | 0,166698727 | LOC101929511 | FALSE | Downregulated |
| 0,303271116 | 0,16679886 | SLC27A3 | FALSE | Upregulated |
| -0,399099003 | 0,166959505 | IRS2 | FALSE | Downregulated |
| -0,177658037 | 0,166999525 | AMZ2 | FALSE | Downregulated |
| -1,520284186 | 0,166999525 | LOC102723840 | FALSE | Downregulated |
| -0,433794912 | 0,167162057 | NPIPA8 | FALSE | Downregulated |
| -0,416748241 | 0,167276012 | USP51 | FALSE | Downregulated |
| 0,286714725 | 0,167343446 | LSM10 | FALSE | Upregulated |
| 0,499230627 | 0,16737065 | FRMD3 | FALSE | Upregulated |
| 0,17093018 | 0,167545 | MTERFD1 | FALSE | Upregulated |
| -0,247989517 | 0,167570479 | NSA2 | FALSE | Downregulated |
| 0,154685268 | 0,167570479 | RNF187 | FALSE | Upregulated |
| -0,560696106 | 0,167570479 | ZNF736 | FALSE | Downregulated |
| 0,20694187 | 0,16771205 | ATG13 | FALSE | Upregulated |
| -0,806518091 | 0,16771205 | CHRNA10 | FALSE | Downregulated |
| 0,151898596 | 0,16771205 | CSNK2B | FALSE | Upregulated |
| -0,376761932 | 0,167736889 | MZF1 | FALSE | Downregulated |
| -0,165748405 | 0,167736889 | SUOX | FALSE | Downregulated |
| -0,322900692 | 0,167736889 | TTBK2 | FALSE | Downregulated |
| -0,691071144 | 0,167737053 | RIBC1 | FALSE | Downregulated |
| 0,191152952 | 0,167785049 | NDUFA6 | FALSE | Upregulated |
| -0,230222677 | 0,167922539 | ANPEP | FALSE | Downregulated |
| 0,421295667 | 0,167923693 | PTPRE | FALSE | Upregulated |
| -2,103504409 | 0,167946089 | LOC102724119 | FALSE | Downregulated |
| 0,176702717 | 0,168176281 | BTBD10 | FALSE | Upregulated |

| | | | | |
|---|---|---|---|---|
| 1,785703383 | 0,168176281 | TMEM40 | FALSE | Upregulated |
| -0,490387044 | 0,168189639 | HSPB9 | FALSE | Downregulated |
| -0,309494704 | 0,168189973 | SCARF1 | FALSE | Downregulated |
| 0,571012156 | 0,168355972 | TMEM100 | FALSE | Upregulated |
| 0,147879483 | 0,168355972 | VPS11 | FALSE | Upregulated |
| -0,146571455 | 0,168360333 | SNX4 | FALSE | Downregulated |
| -0,414085637 | 0,168380925 | DGKE | FALSE | Downregulated |
| 0,345835951 | 0,16845559 | C17orf59 | FALSE | Upregulated |
| -0,210623823 | 0,16845559 | EHMT1 | FALSE | Downregulated |
| -1,18824851 | 0,16845559 | LOC101929195 | FALSE | Downregulated |
| -1,951276751 | 0,168512034 | LHFPL1 | FALSE | Downregulated |
| -0,231742829 | 0,168586632 | SLC16A2 | FALSE | Downregulated |
| -1,11823682 | 0,168665898 | AC002398.5 | FALSE | Downregulated |
| 0,23780253 | 0,168697246 | GOLPH3 | FALSE | Upregulated |
| 0,176153521 | 0,168886721 | DDA1 | FALSE | Upregulated |
| -0,642181492 | 0,168886721 | LINC00939 | FALSE | Downregulated |
| 0,195330811 | 0,168948935 | INTS1 | FALSE | Upregulated |
| -0,322726326 | 0,169050893 | TMEM133 | FALSE | Downregulated |
| -0,830171345 | 0,169125498 | LINC00894 | FALSE | Downregulated |
| -0,711953471 | 0,169252742 | LOC100190986 | FALSE | Downregulated |
| 0,223104302 | 0,169353704 | PSPC1 | FALSE | Upregulated |
| -0,768840081 | 0,169451807 | TSSK6 | FALSE | Downregulated |
| -0,316956127 | 0,169583077 | MOSPD3 | FALSE | Downregulated |
| -0,118351876 | 0,169583077 | RBMX | FALSE | Downregulated |
| -0,231221543 | 0,169583077 | ZNF18 | FALSE | Downregulated |
| 0,526622803 | 0,169640989 | ITLN1 | FALSE | Upregulated |
| 0,541741103 | 0,169753251 | SOGA2 | FALSE | Upregulated |
| 0,760377821 | 0,169760033 | SRRM2-AS1 | FALSE | Upregulated |
| -0,183295424 | 0,169782145 | EPG5 | FALSE | Downregulated |
| -0,200141013 | 0,169785024 | ZDHHC18 | FALSE | Downregulated |
| -0,182276373 | 0,169828532 | POLR1B | FALSE | Downregulated |
| -1,105847731 | 0,170058138 | LOC102724126 | FALSE | Downregulated |
| 0,209914201 | 0,170222401 | NT5C | FALSE | Upregulated |
| -0,526385945 | 0,170301459 | SHH | FALSE | Downregulated |
| 0,235744184 | 0,170404488 | HSP90AB1 | FALSE | Upregulated |
| -0,208359544 | 0,170404488 | PDLIM2 | FALSE | Downregulated |
| 0,122072511 | 0,17042577 | DNAJB12 | FALSE | Upregulated |
| -0,130222044 | 0,17042577 | FEM1B | FALSE | Downregulated |
| -0,180210542 | 0,170517445 | HIPK3 | FALSE | Downregulated |
| -0,257567135 | 0,170587724 | MPLKIP | FALSE | Downregulated |
| 0,868442515 | 0,170587724 | SORT1 | FALSE | Upregulated |
| -0,087972544 | 0,170688391 | C6orf89 | FALSE | Downregulated |
| -1,047048972 | 0,170688391 | PRSS50 | FALSE | Downregulated |
| -0,323756394 | 0,170688391 | RSRC2 | FALSE | Downregulated |
| 0,41537233 | 0,17077551 | CD79B | FALSE | Upregulated |
| -0,801427697 | 0,17077551 | LINC00570 | FALSE | Downregulated |
| 0,282377429 | 0,17084647 | FBXO8 | FALSE | Upregulated |
| -0,213475833 | 0,17084647 | ITGA5 | FALSE | Downregulated |
| -0,168928702 | 0,170876399 | BTBD3 | FALSE | Downregulated |
| 0,226546129 | 0,170876399 | GSTO1 | FALSE | Upregulated |
| -0,575751763 | 0,171248716 | RP11-333E1.1 | FALSE | Downregulated |

| | | | | |
|---|---|---|---|---|
| -0,204216517 | 0,171266375 | LRRC40 | FALSE | Downregulated |
| 0,289431171 | 0,171266375 | PPP1R13L | FALSE | Upregulated |
| -0,127161246 | 0,171288994 | RPL7L1 | FALSE | Downregulated |
| 0,376820326 | 0,171288994 | SLC37A2 | FALSE | Upregulated |
| 0,405542225 | 0,171342707 | PPP1R3D | FALSE | Upregulated |
| 0,146669043 | 0,17139843 | CMTR1 | FALSE | Upregulated |
| -0,441600208 | 0,171476306 | FTCDNL1 | FALSE | Downregulated |
| -0,165697234 | 0,171477134 | GATAD2B | FALSE | Downregulated |
| -0,411280505 | 0,171576432 | MIR22HG | FALSE | Downregulated |
| 0,141724113 | 0,171832111 | PCYT1A | FALSE | Upregulated |
| 0,257627657 | 0,172125934 | HSD3B7 | FALSE | Upregulated |
| -0,245161415 | 0,172125934 | ITIH4 | FALSE | Downregulated |
| 0,270617128 | 0,172155328 | B4GALT5 | FALSE | Upregulated |
| -0,530033063 | 0,172155328 | FGFBP3 | FALSE | Downregulated |
| 0,418428494 | 0,172155328 | RHOD | FALSE | Upregulated |
| 0,164567732 | 0,172155328 | TPI1 | FALSE | Upregulated |
| -0,303138883 | 0,17216111 | TMEM170A | FALSE | Downregulated |
| 0,288935406 | 0,172162314 | ACSS3 | FALSE | Upregulated |
| 0,232680244 | 0,172162314 | SF3B5 | FALSE | Upregulated |
| 0,447263737 | 0,172254448 | CCDC34 | FALSE | Upregulated |
| -1,417825179 | 0,172254448 | LINC00612 | FALSE | Downregulated |
| -0,331780671 | 0,172373711 | PLGLB2 | FALSE | Downregulated |
| 0,139052653 | 0,172415245 | FAM192A | FALSE | Upregulated |
| 1,386141504 | 0,172417109 | LOC102724537 | FALSE | Upregulated |
| -0,630299456 | 0,172417109 | LOC102724655 | FALSE | Downregulated |
| 0,662850323 | 0,17256826 | ALDH1A3 | FALSE | Upregulated |
| 0,38097394 | 0,172593858 | PLCB2 | FALSE | Upregulated |
| -1,67404863 | 0,173067604 | AC007743.1 | FALSE | Downregulated |
| -0,152530969 | 0,173067604 | CCDC93 | FALSE | Downregulated |
| -0,448706241 | 0,173099877 | GPAM | FALSE | Downregulated |
| -0,262769878 | 0,173124728 | METTL21B | FALSE | Downregulated |
| 0,937693079 | 0,173327461 | BMP8B | FALSE | Upregulated |
| 1,328575481 | 0,173362849 | RPH3A | FALSE | Upregulated |
| -0,404990761 | 0,17337258 | KIAA1456 | FALSE | Downregulated |
| -0,271797065 | 0,173453423 | PNPLA2 | FALSE | Downregulated |
| 0,973256084 | 0,173494018 | SLFN12L | FALSE | Upregulated |
| 0,319478056 | 0,173515629 | REEP4 | FALSE | Upregulated |
| -0,158544328 | 0,17375745 | CCDC90B | FALSE | Downregulated |
| 1,442210632 | 0,17375745 | LOC101928373 | FALSE | Upregulated |
| 0,209995563 | 0,173776522 | DHDDS | FALSE | Upregulated |
| -0,152424177 | 0,173776522 | EPC1 | FALSE | Downregulated |
| -0,263394594 | 0,173776522 | PCGF3 | FALSE | Downregulated |
| 0,136804214 | 0,173776522 | VPS35 | FALSE | Upregulated |
| -0,133989183 | 0,173793091 | GDE1 | FALSE | Downregulated |
| 0,2228737 | 0,173888904 | SNTB2 | FALSE | Upregulated |
| -0,187709582 | 0,173896857 | CMTM6 | FALSE | Downregulated |
| -0,223623611 | 0,174048287 | FAM73A | FALSE | Downregulated |
| -0,169123917 | 0,174277719 | SRP9 | FALSE | Downregulated |
| 1,525604245 | 0,174474658 | AP000688.8 | FALSE | Upregulated |
| -0,247395112 | 0,174490917 | BRD4 | FALSE | Downregulated |
| 1,757747768 | 0,174550588 | CHGB | FALSE | Upregulated |

| | | | | |
|---|---|---|---|---|
| -0,605114568 | 0,174729818 | LOC101927258 | FALSE | Downregulated |
| -0,826110162 | 0,174729818 | LOC441454 | FALSE | Downregulated |
| -0,154825032 | 0,174729818 | NSMCE4A | FALSE | Downregulated |
| 0,234175742 | 0,174729818 | ZNF618 | FALSE | Upregulated |
| 1,584524003 | 0,174820657 | CD70 | FALSE | Upregulated |
| 0,19573162 | 0,175197298 | FEZ2 | FALSE | Upregulated |
| -0,187312447 | 0,175332431 | FZD5 | FALSE | Downregulated |
| -0,149254573 | 0,175517865 | MAGEF1 | FALSE | Downregulated |
| 0,231466993 | 0,17570619 | ADCK4 | FALSE | Upregulated |
| 1,432831115 | 0,175720429 | C3orf52 | FALSE | Upregulated |
| -0,273273796 | 0,17576433 | MPHOSPH8 | FALSE | Downregulated |
| -0,274298247 | 0,175888589 | ZNRF3 | FALSE | Downregulated |
| -1,624858426 | 0,176108859 | LOC102723443 | FALSE | Downregulated |
| 0,154866552 | 0,176111002 | XRCC1 | FALSE | Upregulated |
| 0,288922529 | 0,176144607 | COX7B | FALSE | Upregulated |
| -1,141978822 | 0,1761837 | COL6A4P2 | FALSE | Downregulated |
| 0,201729209 | 0,1761837 | ELOF1 | FALSE | Upregulated |
| -0,280326919 | 0,176272283 | ACADS | FALSE | Downregulated |
| -0,2034648 | 0,176479968 | ZNF805 | FALSE | Downregulated |
| 0,217253791 | 0,176555869 | PIAS3 | FALSE | Upregulated |
| 0,270134333 | 0,176567615 | POC5 | FALSE | Upregulated |
| 0,210557346 | 0,176641965 | SMG9 | FALSE | Upregulated |
| -0,241556957 | 0,176656033 | CTBS | FALSE | Downregulated |
| -0,645160783 | 0,176716827 | SLC23A3 | FALSE | Downregulated |
| -0,16187461 | 0,1767445 | LAMTOR3 | FALSE | Downregulated |
| -1,097600616 | 0,1767445 | LOC101928264 | FALSE | Downregulated |
| -0,51617592 | 0,176753533 | UBE2Q2P1 | FALSE | Downregulated |
| 0,179619699 | 0,17693537 | HSPA14 | FALSE | Upregulated |
| 0,118057379 | 0,17695984 | ARL8B | FALSE | Upregulated |
| -1,202352774 | 0,177082262 | GALNT13 | FALSE | Downregulated |
| 0,336966974 | 0,177158034 | CCND2 | FALSE | Upregulated |
| 0,298957813 | 0,177158034 | KMO | FALSE | Upregulated |
| 0,138961405 | 0,177303959 | CCAR2 | FALSE | Upregulated |
| 0,579586996 | 0,177493218 | DGKH | FALSE | Upregulated |
| -0,372210851 | 0,177556232 | LRRC3DN | FALSE | Downregulated |
| 0,485246373 | 0,177556232 | MPPED1 | FALSE | Upregulated |
| 1,134634386 | 0,177556232 | OR2A7 | FALSE | Upregulated |
| -0,215893137 | 0,177556232 | RPL3 | FALSE | Downregulated |
| 0,35185595 | 0,177617423 | LOC101929786 | FALSE | Upregulated |
| 0,225220926 | 0,177763925 | RCCD1 | FALSE | Upregulated |
| -0,187133266 | 0,177820207 | CLASP1 | FALSE | Downregulated |
| 0,35454993 | 0,177849652 | WDR4 | FALSE | Upregulated |
| 0,355642764 | 0,177914284 | EPB41L2 | FALSE | Upregulated |
| 0,276189891 | 0,177914284 | NCAPH2 | FALSE | Upregulated |
| -0,295275494 | 0,177914284 | NHLRC1 | FALSE | Downregulated |
| 0,55547564 | 0,177914284 | NR0B2 | FALSE | Upregulated |
| -0,153897512 | 0,177914284 | SMAD5 | FALSE | Downregulated |
| -0,40211787 | 0,177914284 | ZNF324B | FALSE | Downregulated |
| 0,225796133 | 0,177937722 | C8orf76 | FALSE | Upregulated |
| -0,262522823 | 0,17809557 | SLC4A4 | FALSE | Downregulated |
| -0,332653903 | 0,178137345 | PIGL | FALSE | Downregulated |

| | | | | |
|---|---|---|---|---|
| 0,97174527 | 0,178137679 | C5orf66 | FALSE | Upregulated |
| -0,179291573 | 0,178137679 | CCDC12 | FALSE | Downregulated |
| -0,217214193 | 0,178137679 | SLC25A13 | FALSE | Downregulated |
| -0,631497641 | 0,178145315 | ENO3 | FALSE | Downregulated |
| 0,127860576 | 0,178184064 | CPSF3L | FALSE | Upregulated |
| -0,173783496 | 0,178184064 | PEX5 | FALSE | Downregulated |
| -0,990469111 | 0,178242929 | DHRS2 | FALSE | Downregulated |
| -0,291048734 | 0,178267615 | ZNF587B | FALSE | Downregulated |
| -0,779172622 | 0,17834388 | GSTM3 | FALSE | Downregulated |
| 0,194986331 | 0,178366805 | INPP5K | FALSE | Upregulated |
| -0,675590113 | 0,178385069 | NFATC4 | FALSE | Downregulated |
| 0,166572874 | 0,178408103 | ADAM10 | FALSE | Upregulated |
| -0,212916937 | 0,178408103 | SMAD3 | FALSE | Downregulated |
| 0,444026804 | 0,178483769 | GPX3 | FALSE | Upregulated |
| -0,357780601 | 0,178605489 | APCS | FALSE | Downregulated |
| 0,172798404 | 0,178605489 | C7orf25 | FALSE | Upregulated |
| -0,865051304 | 0,178605489 | LRRN1 | FALSE | Downregulated |
| 0,173710789 | 0,178679531 | UBE2I | FALSE | Upregulated |
| 0,262437919 | 0,178749334 | AAK1 | FALSE | Upregulated |
| 0,210463965 | 0,178841905 | FTSJ3 | FALSE | Upregulated |
| 0,346203818 | 0,178927817 | CTPS1 | FALSE | Upregulated |
| -0,335152334 | 0,179009629 | CES3 | FALSE | Downregulated |
| -0,241399996 | 0,17903002 | SFSWAP | FALSE | Downregulated |
| 0,286777973 | 0,179118561 | ATP9A | FALSE | Upregulated |
| 0,40433534 | 0,179127488 | LOC101928419 | FALSE | Upregulated |
| 0,287201222 | 0,17925545 | CYB561D2 | FALSE | Upregulated |
| -0,557450355 | 0,17925545 | IZUMO4 | FALSE | Downregulated |
| -0,344773358 | 0,179337596 | ANKMY1 | FALSE | Downregulated |
| -0,357847645 | 0,179337596 | LOC100506603 | FALSE | Downregulated |
| 0,097171147 | 0,179373448 | EIF4G2 | FALSE | Upregulated |
| -0,296345539 | 0,179373448 | LOC100505761 | FALSE | Downregulated |
| 0,340304922 | 0,179373448 | PLSCR1 | FALSE | Upregulated |
| 0,185645055 | 0,179639637 | RNASEH1 | FALSE | Upregulated |
| -0,300484653 | 0,180168025 | CCDC66 | FALSE | Downregulated |
| -0,310231401 | 0,180178441 | SMG1 | FALSE | Downregulated |
| -0,214427654 | 0,180486975 | IPP | FALSE | Downregulated |
| -0,821348493 | 0,180977831 | MT1F | FALSE | Downregulated |
| -0,323273899 | 0,181043892 | PKD1P1 | FALSE | Downregulated |
| -0,142973297 | 0,181226615 | DNAJC16 | FALSE | Downregulated |
| 0,42303327 | 0,181226615 | INMT | FALSE | Upregulated |
| -0,256533637 | 0,181228264 | GATA4 | FALSE | Downregulated |
| 0,397974645 | 0,181228264 | PTRH1 | FALSE | Upregulated |
| 0,200090686 | 0,181606794 | ACTR8 | FALSE | Upregulated |
| 1,45317697 | 0,181606794 | IGFBP7-AS1 | FALSE | Upregulated |
| -0,430490457 | 0,181606794 | NOXA1 | FALSE | Downregulated |
| -0,322631142 | 0,181753387 | GREB1L | FALSE | Downregulated |
| -0,397537368 | 0,181753387 | NBPF10 | FALSE | Downregulated |
| -0,276150447 | 0,181855233 | APOH | FALSE | Downregulated |
| -0,233668395 | 0,181995373 | CCNG1 | FALSE | Downregulated |
| 0,75131489 | 0,181995373 | NLRP2 | FALSE | Upregulated |
| -0,256669025 | 0,181995373 | RMND5A | FALSE | Downregulated |

| | | | | |
|---|---|---|---|---|
| 0,135229382 | 0,181995373 | UBA3 | FALSE | Upregulated |
| 0,203559382 | 0,182072586 | SPR | FALSE | Upregulated |
| -0,364032067 | 0,182093547 | FOXA2 | FALSE | Downregulated |
| -0,56064906 | 0,182136269 | PELI2 | FALSE | Downregulated |
| 0,639365516 | 0,182257839 | C19orf57 | FALSE | Upregulated |
| -1,883537698 | 0,182257839 | CXorf30 | FALSE | Downregulated |
| -0,261191633 | 0,182257839 | DCUN1D3 | FALSE | Downregulated |
| 0,218590552 | 0,182325356 | ZNF428 | FALSE | Upregulated |
| 0,232765811 | 0,182365236 | FLAD1 | FALSE | Upregulated |
| -1,683858819 | 0,182365236 | LOC646513 | FALSE | Downregulated |
| -0,566232215 | 0,182365236 | ZNF519 | FALSE | Downregulated |
| 0,422656837 | 0,182396331 | GVINP1 | FALSE | Upregulated |
| 0,228124442 | 0,18246969 | SP2 | FALSE | Upregulated |
| -0,171075198 | 0,182558198 | CASC4 | FALSE | Downregulated |
| 0,890266234 | 0,182558198 | CNR2 | FALSE | Upregulated |
| 1,540219851 | 0,182558198 | FAM159A | FALSE | Upregulated |
| -0,672205797 | 0,182558198 | NYNRIN | FALSE | Downregulated |
| 0,722796304 | 0,182688221 | BEX5 | FALSE | Upregulated |
| -1,245527593 | 0,182884395 | LINC00441 | FALSE | Downregulated |
| 0,225253717 | 0,183048365 | CCHCR1 | FALSE | Upregulated |
| 1,55156397 | 0,183178351 | TNFRSF13C | FALSE | Upregulated |
| 0,198112716 | 0,183225781 | B3GALT6 | FALSE | Upregulated |
| 0,259089459 | 0,183288012 | ABHD14A | FALSE | Upregulated |
| 0,239390668 | 0,183288012 | C8G | FALSE | Upregulated |
| 0,155265968 | 0,183288012 | GTF2E2 | FALSE | Upregulated |
| -0,832746943 | 0,183400137 | NR2F2-AS1 | FALSE | Downregulated |
| -0,817302848 | 0,183566553 | ADH7 | FALSE | Downregulated |
| -0,204234755 | 0,183865548 | HERC2 | FALSE | Downregulated |
| 0,173362834 | 0,183886614 | RUVBL2 | FALSE | Upregulated |
| -0,350328386 | 0,183975972 | FGA | FALSE | Downregulated |
| 0,252015593 | 0,183993908 | STK4 | FALSE | Upregulated |
| -0,239253323 | 0,184064677 | ECSIT | FALSE | Downregulated |
| -0,208679981 | 0,184064677 | GEMIN6 | FALSE | Downregulated |
| 0,240625575 | 0,184064677 | IMPDH2 | FALSE | Upregulated |
| 1,060675026 | 0,184064677 | LOC100129516 | FALSE | Upregulated |
| -0,200988931 | 0,184064677 | SMC3 | FALSE | Downregulated |
| -0,33310818 | 0,184064677 | TAMM41 | FALSE | Downregulated |
| 0,171516871 | 0,184085501 | PSMD12 | FALSE | Upregulated |
| 0,222542695 | 0,184167603 | CALU | FALSE | Upregulated |
| 0,394553436 | 0,184167603 | HIST1H1C | FALSE | Upregulated |
| -0,296628153 | 0,184167603 | TMEM180 | FALSE | Downregulated |
| -0,212282868 | 0,184167603 | ZNF253 | FALSE | Downregulated |
| -0,102509638 | 0,18429195 | MBTPS1 | FALSE | Downregulated |
| -0,2323755 | 0,184308955 | THAP5 | FALSE | Downregulated |
| 0,46949033 | 0,184524904 | CCL15 | FALSE | Upregulated |
| 0,585479024 | 0,18455548 | ADAM28 | FALSE | Upregulated |
| 0,44492375 | 0,184575849 | PALD1 | FALSE | Upregulated |
| 0,429350194 | 0,184584891 | IL15 | FALSE | Upregulated |
| -0,278878287 | 0,1847661 | GATM | FALSE | Downregulated |
| -0,23492723 | 0,1847661 | PRDM10 | FALSE | Downregulated |
| 0,353270625 | 0,184875932 | SIRPA | FALSE | Upregulated |

| | | | | |
|---|---|---|---|---|
| -0,166293753 | 0,185007589 | DHX33 | FALSE | Downregulated |
| -2,053367932 | 0,185098276 | MB | FALSE | Downregulated |
| 0,184845006 | 0,185098276 | SRPK1 | FALSE | Upregulated |
| 0,186964416 | 0,185232643 | EXOSC7 | FALSE | Upregulated |
| -0,162625847 | 0,185232643 | LPHN2 | FALSE | Downregulated |
| 0,212340215 | 0,185237119 | GIT2 | FALSE | Upregulated |
| 0,602552686 | 0,185241091 | ACAP1 | FALSE | Upregulated |
| -0,613883999 | 0,185241091 | C14orf180 | FALSE | Downregulated |
| -1,172007832 | 0,185241091 | LOC101928083 | FALSE | Downregulated |
| -0,187365919 | 0,185241091 | STK11 | FALSE | Downregulated |
| 0,12200883 | 0,18524571 | OTUD5 | FALSE | Upregulated |
| -0,396154973 | 0,185447835 | TMEM67 | FALSE | Downregulated |
| 0,39916543 | 0,185486393 | SEMA3F | FALSE | Upregulated |
| 0,156844486 | 0,185496724 | APLP2 | FALSE | Upregulated |
| -1,708762399 | 0,185496724 | SLC26A9 | FALSE | Downregulated |
| -0,158887159 | 0,185496724 | SLC35A1 | FALSE | Downregulated |
| 0,298686253 | 0,185658267 | TAPBPL | FALSE | Upregulated |
| -0,535450828 | 0,185772168 | DMPK | FALSE | Downregulated |
| -0,685359336 | 0,185772168 | LOC101929715 | FALSE | Downregulated |
| 0,193833408 | 0,185772168 | MRPL15 | FALSE | Upregulated |
| -1,575492443 | 0,185847164 | CCDC178 | FALSE | Downregulated |
| -1,247556791 | 0,185847164 | RP5-1004I9.1 | FALSE | Downregulated |
| -0,214528198 | 0,185903349 | H6PD | FALSE | Downregulated |
| -1,069368309 | 0,185913277 | FAM69C | FALSE | Downregulated |
| -0,15484169 | 0,186038382 | PTER | FALSE | Downregulated |
| 0,557460034 | 0,186056655 | TRG-AS1 | FALSE | Upregulated |
| 0,195866088 | 0,186073949 | RABIF | FALSE | Upregulated |
| -0,157499958 | 0,186110142 | ZNF41 | FALSE | Downregulated |
| -0,295677905 | 0,186150182 | FGFR1 | FALSE | Downregulated |
| 0,303179502 | 0,186301725 | TEF | FALSE | Upregulated |
| -0,132538993 | 0,186357338 | BNIP2 | FALSE | Downregulated |
| -0,169190768 | 0,186404317 | C1orf52 | FALSE | Downregulated |
| 0,343005635 | 0,186404317 | MARVELD1 | FALSE | Upregulated |
| -0,571001116 | 0,186404317 | ZBED3 | FALSE | Downregulated |
| -0,382076113 | 0,186404317 | ZRSR2 | FALSE | Downregulated |
| 0,29822403 | 0,186413488 | ANKRD39 | FALSE | Upregulated |
| 0,142709263 | 0,186413488 | BNIP3L | FALSE | Upregulated |
| -0,25113812 | 0,186413488 | FXN | FALSE | Downregulated |
| -0,242208939 | 0,186413488 | THRA | FALSE | Downregulated |
| -0,310911236 | 0,186499196 | DEPDC7 | FALSE | Downregulated |
| -0,283687777 | 0,186499196 | OSER1-AS1 | FALSE | Downregulated |
| -0,623003363 | 0,186521644 | CLEC1A | FALSE | Downregulated |
| -0,132830856 | 0,186535555 | SETD1A | FALSE | Downregulated |
| 0,185857345 | 0,186549436 | ANKRD52 | FALSE | Upregulated |
| -0,243757591 | 0,18668557 | GRB14 | FALSE | Downregulated |
| -2,126435254 | 0,186731581 | LOC101928349 | FALSE | Downregulated |
| -1,087242408 | 0,186731581 | LOC101929741 | FALSE | Downregulated |
| -0,200651096 | 0,186731581 | POLR1C | FALSE | Downregulated |
| 1,712709862 | 0,186731581 | WNT10A | FALSE | Upregulated |
| 0,198531358 | 0,186772853 | MED16 | FALSE | Upregulated |
| 0,346141763 | 0,186834674 | RAB11FIP5 | FALSE | Upregulated |

| | | | | |
|---|---|---|---|---|
| 0,216235786 | 0,186878247 | RNF216P1 | FALSE | Upregulated |
| -0,229094514 | 0,186883815 | BRWD3 | FALSE | Downregulated |
| 0,779609235 | 0,186883815 | FGF14 | FALSE | Upregulated |
| 0,496564101 | 0,186883815 | HYOU1 | FALSE | Upregulated |
| -0,428315937 | 0,186883815 | LOC100652758 | FALSE | Downregulated |
| -0,423597838 | 0,186883815 | MBOAT2 | FALSE | Downregulated |
| -0,203663146 | 0,186883815 | UBE3A | FALSE | Downregulated |
| -0,255154533 | 0,186883815 | ZNF619 | FALSE | Downregulated |
| -0,543517889 | 0,187262424 | RXFP1 | FALSE | Downregulated |
| -0,218168563 | 0,18738004 | KMT2B | FALSE | Downregulated |
| -0,340494376 | 0,187485936 | ACSL1 | FALSE | Downregulated |
| -1,145781094 | 0,187485936 | CACNA2D3 | FALSE | Downregulated |
| -0,513360808 | 0,187485936 | TMEM232 | FALSE | Downregulated |
| -0,198408907 | 0,187535089 | ILVBL | FALSE | Downregulated |
| 0,422518106 | 0,187535089 | RBP7 | FALSE | Upregulated |
| 0,523067891 | 0,187563951 | LOC102724592 | FALSE | Upregulated |
| -0,434655802 | 0,187849629 | SYNE3 | FALSE | Downregulated |
| -0,131744712 | 0,188173152 | NUP133 | FALSE | Downregulated |
| -1,111616968 | 0,188185883 | C6orf165 | FALSE | Downregulated |
| 0,122672305 | 0,188451281 | NKIRAS2 | FALSE | Upregulated |
| 0,200284226 | 0,188484279 | DNAJB14 | FALSE | Upregulated |
| -0,219417638 | 0,188597802 | DPY19L3 | FALSE | Downregulated |
| 0,236009673 | 0,188744609 | CNOT6 | FALSE | Upregulated |
| 0,582470499 | 0,188746502 | FANCE | FALSE | Upregulated |
| -0,845866183 | 0,188746502 | SEMA3C | FALSE | Downregulated |
| 1,554943099 | 0,188936594 | GPR84 | FALSE | Upregulated |
| -0,283536421 | 0,188987766 | TNK1 | FALSE | Downregulated |
| 0,193147495 | 0,189072126 | NUTF2 | FALSE | Upregulated |
| -0,224657067 | 0,189114525 | CAPRIN2 | FALSE | Downregulated |
| -0,578647112 | 0,189136571 | IRAK1BP1 | FALSE | Downregulated |
| 0,326683897 | 0,189228625 | ITM2C | FALSE | Upregulated |
| 0,216628797 | 0,189283567 | TMX4 | FALSE | Upregulated |
| -1,31035755 | 0,18929129 | MAPK4 | FALSE | Downregulated |
| -0,335435378 | 0,18931289 | TTC26 | FALSE | Downregulated |
| 0,156450924 | 0,18931289 | UQCRFS1 | FALSE | Upregulated |
| 0,248692072 | 0,189448798 | FAM127A | FALSE | Upregulated |
| -0,193506087 | 0,189494061 | FBXO9 | FALSE | Downregulated |
| 0,54134777 | 0,189540671 | LRFN1 | FALSE | Upregulated |
| -0,605294751 | 0,189540671 | ZNF300P1 | FALSE | Downregulated |
| 0,27320199 | 0,189570429 | MIS18BP1 | FALSE | Upregulated |
| 0,14351244 | 0,189692816 | CHMP3 | FALSE | Upregulated |
| 0,136928174 | 0,189692816 | VPS26A | FALSE | Upregulated |
| -0,3813896 | 0,189694676 | TCP10L | FALSE | Downregulated |
| -0,22337105 | 0,189694676 | TYW3 | FALSE | Downregulated |
| -0,181606163 | 0,189885208 | ACAD8 | FALSE | Downregulated |
| 0,892477001 | 0,189905676 | GABRB3 | FALSE | Upregulated |
| -0,482061177 | 0,189905676 | LOC100132111 | FALSE | Downregulated |
| -0,175059118 | 0,189905676 | ZNF592 | FALSE | Downregulated |
| -0,701120553 | 0,189913931 | NPAS3 | FALSE | Downregulated |
| -0,758564419 | 0,189914994 | LRRC24 | FALSE | Downregulated |
| -0,300561733 | 0,189914994 | SLC2A4RG | FALSE | Downregulated |

| | | | | |
|---|---|---|---|---|
| 0,465760308 | 0,189927087 | FLJ32255 | FALSE | Upregulated |
| -0,165419522 | 0,189945271 | MAU2 | FALSE | Downregulated |
| -1,461632067 | 0,189948923 | FAM153A | FALSE | Downregulated |
| -0,218330166 | 0,190284221 | OSBP | FALSE | Downregulated |
| 2,610338731 | 0,190836879 | FGF19 | FALSE | Upregulated |
| -2,338807205 | 0,190836879 | LOC101060569 | FALSE | Downregulated |
| -0,349193828 | 0,19085113 | PNPLA4 | FALSE | Downregulated |
| 0,299209926 | 0,190875883 | SCN1B | FALSE | Upregulated |
| -0,782196502 | 0,191072399 | C8orf44-SGK3 | FALSE | Downregulated |
| -0,516330918 | 0,191081562 | CCT6P1 | FALSE | Downregulated |
| 0,421801264 | 0,191081562 | CEBPD | FALSE | Upregulated |
| -0,504231188 | 0,191081562 | SNHG19 | FALSE | Downregulated |
| -0,162455824 | 0,191193832 | ATG4B | FALSE | Downregulated |
| 0,211552641 | 0,191193832 | GTF3A | FALSE | Upregulated |
| -0,660695582 | 0,191193832 | HS3ST3A1 | FALSE | Downregulated |
| -0,947687405 | 0,191193832 | ITGA8 | FALSE | Downregulated |
| -0,241776229 | 0,191264349 | C11orf95 | FALSE | Downregulated |
| -1,453421203 | 0,191264349 | CNTN5 | FALSE | Downregulated |
| -0,664575925 | 0,191264349 | LOC101928189 | FALSE | Downregulated |
| 0,256336166 | 0,191264349 | MDFIC | FALSE | Upregulated |
| 0,185233712 | 0,191264349 | NHP2 | FALSE | Upregulated |
| 0,767080033 | 0,191264349 | PIP5K1B | FALSE | Upregulated |
| 0,234491455 | 0,191264349 | RP2 | FALSE | Upregulated |
| 0,226766293 | 0,191264349 | RPA3 | FALSE | Upregulated |
| 0,214702219 | 0,191634967 | CHSY1 | FALSE | Upregulated |
| -0,829154153 | 0,191722273 | LOC100288102 | FALSE | Downregulated |
| -0,504334348 | 0,191722273 | ZNF521 | FALSE | Downregulated |
| 1,363781603 | 0,192059999 | LOC101927596 | FALSE | Upregulated |
| -0,788247091 | 0,192059999 | LOC101927880 | FALSE | Downregulated |
| -0,754228528 | 0,192059999 | USH1C | FALSE | Downregulated |
| -0,277042028 | 0,192270434 | THAP6 | FALSE | Downregulated |
| -0,936164343 | 0,192349067 | TAF7L | FALSE | Downregulated |
| 0,312825598 | 0,192349067 | ZNF213 | FALSE | Upregulated |
| -0,351862062 | 0,192417999 | FXYD1 | FALSE | Downregulated |
| -0,164790208 | 0,192889993 | FAM149B1 | FALSE | Downregulated |
| -0,173096295 | 0,19291281 | MRPS9 | FALSE | Downregulated |
| 0,31578942 | 0,192958448 | SFXN3 | FALSE | Upregulated |
| -0,160662951 | 0,193002854 | EFCAB14 | FALSE | Downregulated |
| 0,245072879 | 0,193002854 | TOMM40 | FALSE | Upregulated |
| 0,30271943 | 0,19326596 | FLI1 | FALSE | Upregulated |
| -0,579158741 | 0,193336133 | GNN | FALSE | Downregulated |
| -1,090268161 | 0,193336133 | LOC101927754 | FALSE | Downregulated |
| 0,206181906 | 0,193336133 | ZDHHC12 | FALSE | Upregulated |
| -0,139374868 | 0,193402311 | TMEM242 | FALSE | Downregulated |
| 0,184613247 | 0,193402311 | TRIM37 | FALSE | Upregulated |
| -0,186094896 | 0,193503124 | SLC35D1 | FALSE | Downregulated |
| 0,273459937 | 0,193503124 | ZSWIM7 | FALSE | Upregulated |
| 0,255160163 | 0,193505214 | CTU2 | FALSE | Upregulated |
| 0,288505336 | 0,19357747 | MS4A6A | FALSE | Upregulated |
| -0,189980538 | 0,19358998 | KDM3A | FALSE | Downregulated |
| -1,86605094 | 0,19358998 | LOC285768 | FALSE | Downregulated |

| | | | | |
|---|---|---|---|---|
| -0,315875953 | 0,19358998 | SNHG11 | FALSE | Downregulated |
| -0,241319539 | 0,193607276 | TNRC6A | FALSE | Downregulated |
| -0,571007749 | 0,194030696 | HOTAIRM1 | FALSE | Downregulated |
| -0,362523908 | 0,194080904 | TIGD1 | FALSE | Downregulated |
| -0,262017378 | 0,194293463 | RRAGB | FALSE | Downregulated |
| 0,300285591 | 0,194399436 | PTRHD1 | FALSE | Upregulated |
| -0,236991531 | 0,194819999 | CDC42BPA | FALSE | Downregulated |
| 1,637103973 | 0,194819999 | LOC441455 | FALSE | Upregulated |
| -0,202724557 | 0,194819999 | MRPS24 | FALSE | Downregulated |
| 1,194498792 | 0,194819999 | SLC52A3 | FALSE | Upregulated |
| -0,210939992 | 0,194819999 | TSPAN9 | FALSE | Downregulated |
| 0,196633556 | 0,194819999 | WARS2 | FALSE | Upregulated |
| -0,174158009 | 0,194819999 | WWC2 | FALSE | Downregulated |
| 0,347548208 | 0,194819999 | ZNHIT2 | FALSE | Upregulated |
| -0,963052136 | 0,19520542 | LOC102724474 | FALSE | Downregulated |
| -0,840559668 | 0,195290113 | LOC100996723 | FALSE | Downregulated |
| 0,614392109 | 0,195290113 | PMAIP1 | FALSE | Upregulated |
| 0,243893225 | 0,195301775 | SMG5 | FALSE | Upregulated |
| -0,152504053 | 0,195364329 | WDR70 | FALSE | Downregulated |
| -0,711170831 | 0,195417328 | LOC100506472 | FALSE | Downregulated |
| -1,20545929 | 0,195417328 | NECAB1 | FALSE | Downregulated |
| 0,760430947 | 0,195417328 | VTCN1 | FALSE | Upregulated |
| 0,210061967 | 0,195551614 | SHROOM3 | FALSE | Upregulated |
| 0,603709432 | 0,195567073 | TMEM246 | FALSE | Upregulated |
| 0,2176034 | 0,195614909 | EEF1B2 | FALSE | Upregulated |
| 0,848404569 | 0,195766295 | DDX12P | FALSE | Upregulated |
| -0,382098641 | 0,195766295 | EFHC1 | FALSE | Downregulated |
| 0,5745569 | 0,195766295 | LINC00869 | FALSE | Upregulated |
| -0,883463654 | 0,195766295 | LOC102724192 | FALSE | Downregulated |
| -0,641029813 | 0,195842891 | LOC101928069 | FALSE | Downregulated |
| -0,668808104 | 0,195949971 | TRNI | FALSE | Downregulated |
| -0,138884606 | 0,196015122 | PCID2 | FALSE | Downregulated |
| 0,46442749 | 0,19607578 | VCAM1 | FALSE | Upregulated |
| -0,353573019 | 0,196095677 | UPB1 | FALSE | Downregulated |
| -1,110092419 | 0,19633555 | DCT | FALSE | Downregulated |
| -0,402913763 | 0,19648575 | ETNK2 | FALSE | Downregulated |
| -0,312065148 | 0,19648575 | PRH1-PRR4 | FALSE | Downregulated |
| 1,768328297 | 0,19648575 | SFTA2 | FALSE | Upregulated |
| 0,248018787 | 0,19648575 | UBL5 | FALSE | Upregulated |
| 0,164149804 | 0,196670505 | SKP1 | FALSE | Upregulated |
| 0,23604966 | 0,196670505 | TXNDC17 | FALSE | Upregulated |
| 1,376856414 | 0,196695131 | TTTY14 | FALSE | Upregulated |
| -0,326086179 | 0,196731656 | ZNF214 | FALSE | Downregulated |
| 0,229613965 | 0,196777598 | CHCHD5 | FALSE | Upregulated |
| -0,318788719 | 0,196777598 | MGC27345 | FALSE | Downregulated |
| -0,451014082 | 0,196777598 | PPP1R14A | FALSE | Downregulated |
| 0,250107049 | 0,196819774 | CDC73 | FALSE | Upregulated |
| -0,21332296 | 0,196844301 | CCDC106 | FALSE | Downregulated |
| 1,100291304 | 0,196844301 | NXPH3 | FALSE | Upregulated |
| -0,153890997 | 0,196850859 | GSPT2 | FALSE | Downregulated |
| 0,172928362 | 0,196879263 | MAP3K13 | FALSE | Upregulated |

| | | | | |
|---|---|---|---|---|
| -0,297492998 | 0,196947356 | DISP1 | FALSE | Downregulated |
| -0,519730447 | 0,197371078 | NFYC-AS1 | FALSE | Downregulated |
| 0,246575238 | 0,197457042 | CDR2 | FALSE | Upregulated |
| 0,139658158 | 0,197457042 | NAA20 | FALSE | Upregulated |
| -0,36209988 | 0,197457042 | PLIN5 | FALSE | Downregulated |
| 0,882365695 | 0,197491706 | ICA1L | FALSE | Upregulated |
| -0,568288879 | 0,197532996 | IL17RD | FALSE | Downregulated |
| -0,130091931 | 0,197764891 | COPS2 | FALSE | Downregulated |
| -0,371496522 | 0,197764891 | SGK2 | FALSE | Downregulated |
| 0,319312401 | 0,197959562 | ZXDB | FALSE | Upregulated |
| 0,2303856 | 0,198118172 | FAM118B | FALSE | Upregulated |
| 0,440676649 | 0,198128026 | HYAL3 | FALSE | Upregulated |
| 1,462797313 | 0,198128026 | LINC00900 | FALSE | Upregulated |
| -0,356955696 | 0,198128026 | TUBGCP6 | FALSE | Downregulated |
| -0,131633302 | 0,198287763 | CCNT1 | FALSE | Downregulated |
| 0,914188276 | 0,198287763 | HS3ST1 | FALSE | Upregulated |
| 0,226172611 | 0,198287763 | PPRC1 | FALSE | Upregulated |
| 0,14916214 | 0,198308905 | KIAA0247 | FALSE | Upregulated |
| -0,228772346 | 0,198308905 | OXNAD1 | FALSE | Downregulated |
| -0,130054878 | 0,198391169 | ARMC10 | FALSE | Downregulated |
| -0,683647667 | 0,198391169 | SGCD | FALSE | Downregulated |
| 0,545738506 | 0,198391169 | SLC35G2 | FALSE | Upregulated |
| -0,248046149 | 0,198403684 | AKAP2 | FALSE | Downregulated |
| -0,128489037 | 0,198628319 | TOM1L2 | FALSE | Downregulated |
| 0,378691072 | 0,1987952 | STON2 | FALSE | Upregulated |
| -0,806093112 | 0,198859007 | AC012314.8 | FALSE | Downregulated |
| 0,137349499 | 0,198859007 | FYTTD1 | FALSE | Upregulated |
| -0,545675989 | 0,198859007 | UBAC2-AS1 | FALSE | Downregulated |
| 1,509917735 | 0,198965988 | TCL1A | FALSE | Upregulated |
| 0,18385055 | 0,198971928 | VPS28 | FALSE | Upregulated |
| 0,325965625 | 0,199019061 | SESN1 | FALSE | Upregulated |
| 0,308253075 | 0,199047248 | FLVCR1 | FALSE | Upregulated |
| -1,237084692 | 0,199075365 | LOC100506797 | FALSE | Downregulated |
| 0,19841597 | 0,199133548 | NPC1 | FALSE | Upregulated |
| 0,995073057 | 0,199276802 | C4orf48 | FALSE | Upregulated |
| 0,692091342 | 0,199276802 | LOC101927183 | FALSE | Upregulated |
| -0,185779088 | 0,199311292 | EZH1 | FALSE | Downregulated |
| -0,331329323 | 0,199601012 | ANKS3 | FALSE | Downregulated |
| -0,36506659 | 0,199601012 | KRBA2 | FALSE | Downregulated |
| -0,598648918 | 0,199641052 | ETV2 | FALSE | Downregulated |
| 0,796442601 | 0,19965865 | RP11-65J3.1 | FALSE | Upregulated |
| -0,21191557 | 0,199727943 | AK4 | FALSE | Downregulated |
| -0,173938993 | 0,199727943 | BET1L | FALSE | Downregulated |
| 0,17463708 | 0,199738983 | ZNF562 | FALSE | Upregulated |
| 0,157488221 | 0,199752957 | GNPTG | FALSE | Upregulated |
| -0,525290041 | 0,199754326 | RAB26 | FALSE | Downregulated |
| -0,344453622 | 0,199868443 | DLL4 | FALSE | Downregulated |
| 0,148264819 | 0,199868443 | EFR3A | FALSE | Upregulated |
| -0,490748848 | 0,199868443 | HOXB3 | FALSE | Downregulated |
| -0,268324904 | 0,199868443 | MBD6 | FALSE | Downregulated |
| -0,184204377 | 0,199868443 | RNF123 | FALSE | Downregulated |

| | | | | |
|---|---|---|---|---|
| -1,505158764 | 0,200056795 | PGAM2 | FALSE | Downregulated |
| 0,309910418 | 0,200203631 | BCL2L12 | FALSE | Upregulated |
| 1,4975085 | 0,200203631 | OTOGL | FALSE | Upregulated |
| -0,821768783 | 0,200216896 | PLXNA4 | FALSE | Downregulated |
| -0,151647116 | 0,200216896 | RNF38 | FALSE | Downregulated |
| -0,463879511 | 0,200224134 | CYP2A6 | FALSE | Downregulated |
| -0,11994477 | 0,200244303 | MTAP | FALSE | Downregulated |
| -0,616014241 | 0,200395853 | ANKRD33B | FALSE | Downregulated |
| -0,43214663 | 0,200395853 | ANKRD55 | FALSE | Downregulated |
| -1,17218845 | 0,200395853 | LOC100287934 | FALSE | Downregulated |
| -0,23055817 | 0,200395853 | PTK2B | FALSE | Downregulated |
| -0,544641935 | 0,200395853 | RP11-622A1.2 | FALSE | Downregulated |
| -1,360616877 | 0,20039904 | CRYAB | FALSE | Downregulated |
| -0,563685194 | 0,200416241 | FBLN2 | FALSE | Downregulated |
| -0,668875238 | 0,200416241 | LOC100505650 | FALSE | Downregulated |
| -0,591025999 | 0,200416241 | NGF | FALSE | Downregulated |
| -0,84230597 | 0,20070925 | LOC102723365 | FALSE | Downregulated |
| -0,632387973 | 0,20070925 | LOC102723564 | FALSE | Downregulated |
| -0,772639885 | 0,20070925 | LOC102723648 | FALSE | Downregulated |
| 0,137553704 | 0,200725743 | VPS4A | FALSE | Upregulated |
| 0,316391354 | 0,200734069 | CTR9 | FALSE | Upregulated |
| -0,269390006 | 0,200746459 | BSCL2 | FALSE | Downregulated |
| 0,630220515 | 0,200746459 | COL23A1 | FALSE | Upregulated |
| 0,239719656 | 0,200746459 | DERL2 | FALSE | Upregulated |
| 0,374329391 | 0,200746459 | LINC00959 | FALSE | Upregulated |
| -1,582056773 | 0,200746459 | PAX6 | FALSE | Downregulated |
| -0,162558874 | 0,200897294 | NFIB | FALSE | Downregulated |
| 1,701856272 | 0,200942549 | AC010091.1 | FALSE | Upregulated |
| 0,212290637 | 0,200942549 | NUDT15 | FALSE | Upregulated |
| 0,254188274 | 0,200942549 | SSBP4 | FALSE | Upregulated |
| -0,250479995 | 0,200942549 | TBC1D24 | FALSE | Downregulated |
| -1,026803383 | 0,201161471 | TMEM233 | FALSE | Downregulated |
| 1,485967697 | 0,201189641 | DCAF13P3 | FALSE | Upregulated |
| 1,251833984 | 0,201189641 | RP11-1080G15.1 | FALSE | Upregulated |
| 0,238029662 | 0,201437467 | DNAJC1 | FALSE | Upregulated |
| 1,309779189 | 0,201437467 | LOC101929422 | FALSE | Upregulated |
| 0,197170865 | 0,201437467 | LRRC27 | FALSE | Upregulated |
| -0,276669473 | 0,201620948 | NAT1 | FALSE | Downregulated |
| -1,561842563 | 0,20167356 | LOC101929451 | FALSE | Downregulated |
| 0,595417671 | 0,20167356 | PRKCQ | FALSE | Upregulated |
| 0,636846464 | 0,20167356 | RDH12 | FALSE | Upregulated |
| 0,199269438 | 0,201724931 | SRRD | FALSE | Upregulated |
| -1,924588442 | 0,20173417 | CASQ2 | FALSE | Downregulated |
| 0,228598189 | 0,201800614 | RFT1 | FALSE | Upregulated |
| 0,169498067 | 0,201800614 | VPS16 | FALSE | Upregulated |
| 0,831224453 | 0,202127352 | IL10 | FALSE | Upregulated |
| -0,588922875 | 0,202127352 | LRP2BP | FALSE | Downregulated |
| 0,70130353 | 0,202127352 | SDK1 | FALSE | Upregulated |
| 0,618776259 | 0,202127352 | THBS2 | FALSE | Upregulated |
| 0,626111948 | 0,202308632 | MORC2-AS1 | FALSE | Upregulated |
| -0,994778287 | 0,202344801 | C2orf78 | FALSE | Downregulated |

| | | | | |
|---|---|---|---|---|
| -0,169598719 | 0,202344801 | FBXW4 | FALSE | Downregulated |
| 0,154209807 | 0,202461513 | GTF3C5 | FALSE | Upregulated |
| 0,335286176 | 0,202556462 | SNX10 | FALSE | Upregulated |
| 0,623973556 | 0,20272636 | IL3RA | FALSE | Upregulated |
| 0,418517337 | 0,20272636 | PLIN2 | FALSE | Upregulated |
| 0,395850978 | 0,202769598 | DHCR7 | FALSE | Upregulated |
| -0,752092507 | 0,202769598 | LOC101927284 | FALSE | Downregulated |
| 1,071298307 | 0,202947156 | AC010890.1 | FALSE | Upregulated |
| -0,32675541 | 0,202947156 | G2E3 | FALSE | Downregulated |
| 0,157038101 | 0,202947156 | LUZP6 | FALSE | Upregulated |
| 0,159695545 | 0,202947156 | ZBTB17 | FALSE | Upregulated |
| 0,381287965 | 0,203025319 | DNAJB5 | FALSE | Upregulated |
| -0,233883964 | 0,203093201 | ZNF32 | FALSE | Downregulated |
| -0,618904607 | 0,20316461 | LINC00238 | FALSE | Downregulated |
| -0,714752613 | 0,20316461 | LOC101928068 | FALSE | Downregulated |
| -0,214328167 | 0,203393622 | PHIP | FALSE | Downregulated |
| 0,733231883 | 0,2034116 | PKD2L1 | FALSE | Upregulated |
| 0,237934716 | 0,20400016 | RPL27A | FALSE | Upregulated |
| 0,34818316 | 0,20403669 | LTBP3 | FALSE | Upregulated |
| 1,59855156 | 0,204057954 | MCF2L2 | FALSE | Upregulated |
| 0,345554168 | 0,204081563 | ITPRIP | FALSE | Upregulated |
| 0,403700293 | 0,204081563 | NCS1 | FALSE | Upregulated |
| -0,157141863 | 0,204081563 | NDUFS1 | FALSE | Downregulated |
| 1,041436673 | 0,204134852 | TFF3 | FALSE | Upregulated |
| -0,636217703 | 0,204304971 | LOC101927665 | FALSE | Downregulated |
| -0,178641846 | 0,204381052 | BCKDHA | FALSE | Downregulated |
| -1,544447521 | 0,204426647 | LOC102724930 | FALSE | Downregulated |
| 0,244914372 | 0,204542725 | NT5C3A | FALSE | Upregulated |
| -0,953520028 | 0,204675052 | STARD4-AS1 | FALSE | Downregulated |
| 0,242381923 | 0,204708851 | SLC39A13 | FALSE | Upregulated |
| -0,518716597 | 0,204715891 | AHSA2 | FALSE | Downregulated |
| -1,172368344 | 0,205010817 | GHRL | FALSE | Downregulated |
| 0,273878426 | 0,20501272 | PHF13 | FALSE | Upregulated |
| -0,929010953 | 0,205025459 | HSPB8 | FALSE | Downregulated |
| -0,609725116 | 0,205025459 | UPK3B | FALSE | Downregulated |
| -0,114883964 | 0,205027336 | GTF2A1 | FALSE | Downregulated |
| -1,539308679 | 0,205100851 | CPNE6 | FALSE | Downregulated |
| -0,861914753 | 0,205243879 | KIAA1652 | FALSE | Downregulated |
| -0,09897748 | 0,205243879 | PAPOLA | FALSE | Downregulated |
| 0,14110244 | 0,205243879 | RNF4 | FALSE | Upregulated |
| 0,193346198 | 0,205243879 | TM7SF3 | FALSE | Upregulated |
| -0,124342168 | 0,205243879 | WDR75 | FALSE | Downregulated |
| 0,215870542 | 0,205395142 | FCHO2 | FALSE | Upregulated |
| 0,196392571 | 0,205418071 | ZMPSTE24 | FALSE | Upregulated |
| 0,272933089 | 0,205533176 | BMS1P20 | FALSE | Upregulated |
| 0,397315042 | 0,205533176 | HSPA5 | FALSE | Upregulated |
| -0,759805124 | 0,205594569 | NPM2 | FALSE | Downregulated |
| 0,532412573 | 0,205638466 | DBP | FALSE | Upregulated |
| -0,211262727 | 0,205679335 | LINC00909 | FALSE | Downregulated |
| 0,759215463 | 0,205679335 | LOC388210 | FALSE | Upregulated |
| -0,464094426 | 0,205679335 | RP11-342C23.4 | FALSE | Downregulated |

| | | | | |
|---|---|---|---|---|
| 0,25964168 | 0,205679335 | SLC10A3 | FALSE | Upregulated |
| 0,205714159 | 0,205741741 | NOSIP | FALSE | Upregulated |
| -0,192650654 | 0,205741741 | PRIMPOL | FALSE | Downregulated |
| -0,317087014 | 0,205745369 | BDNF-AS | FALSE | Downregulated |
| -0,31848003 | 0,205745369 | EFNB2 | FALSE | Downregulated |
| 0,181388676 | 0,205745369 | RFC5 | FALSE | Upregulated |
| -0,546986506 | 0,205897083 | CD200R1 | FALSE | Downregulated |
| -0,631219557 | 0,205897083 | FLJ30403 | FALSE | Downregulated |
| -0,19157389 | 0,206319731 | COL18A1 | FALSE | Downregulated |
| -0,450885426 | 0,206319731 | FAM129C | FALSE | Downregulated |
| -0,143751848 | 0,20638159 | MEN1 | FALSE | Downregulated |
| 0,17468831 | 0,20638159 | TIMM50 | FALSE | Upregulated |
| -0,284846381 | 0,206402205 | GYG2 | FALSE | Downregulated |
| 1,57164885 | 0,206402205 | TXLNG2P | FALSE | Upregulated |
| -0,388547866 | 0,206514721 | SYT15 | FALSE | Downregulated |
| -0,428313147 | 0,20660183 | ZNF169 | FALSE | Downregulated |
| -0,222962952 | 0,206622426 | AHNAK | FALSE | Downregulated |
| -0,23987946 | 0,206622426 | CWC22 | FALSE | Downregulated |
| -0,138147058 | 0,206711798 | CIZ1 | FALSE | Downregulated |
| -1,752261401 | 0,207044296 | LOC101928212 | FALSE | Downregulated |
| -0,271695965 | 0,207070569 | CDK11A | FALSE | Downregulated |
| -0,190262942 | 0,207092171 | WDFY3 | FALSE | Downregulated |
| -0,275268598 | 0,207181342 | CEP290 | FALSE | Downregulated |
| 0,202223325 | 0,207181342 | GMNN | FALSE | Upregulated |
| 0,438680343 | 0,207249695 | GPR161 | FALSE | Upregulated |
| 0,146919229 | 0,207249695 | SNAPIN | FALSE | Upregulated |
| 0,620943115 | 0,207249695 | SSR4 | FALSE | Upregulated |
| -0,515819394 | 0,207249695 | TRIL | FALSE | Downregulated |
| -0,547993911 | 0,207270914 | PPP4R4 | FALSE | Downregulated |
| -0,44338371 | 0,207275729 | DNASE1 | FALSE | Downregulated |
| 0,31773218 | 0,207275729 | RASA3 | FALSE | Upregulated |
| -0,209186723 | 0,20731809 | HOOK3 | FALSE | Downregulated |
| -0,227049169 | 0,207350659 | NHSL1 | FALSE | Downregulated |
| -0,171349528 | 0,207350659 | SLC31A1 | FALSE | Downregulated |
| -0,379564233 | 0,207411961 | LOC101929243 | FALSE | Downregulated |
| 0,305120787 | 0,207436825 | LOC102723990 | FALSE | Upregulated |
| 0,181903435 | 0,207539992 | FBXO42 | FALSE | Upregulated |
| -0,21949271 | 0,207731657 | ZBTB41 | FALSE | Downregulated |
| 1,323492476 | 0,207746921 | DPCR1 | FALSE | Upregulated |
| 0,451524249 | 0,207746921 | TRPS1 | FALSE | Upregulated |
| -0,594670112 | 0,207789709 | CPAMD8 | FALSE | Downregulated |
| -0,228673398 | 0,207794798 | SIX5 | FALSE | Downregulated |
| -0,158987149 | 0,207848338 | STAG2 | FALSE | Downregulated |
| 0,308012294 | 0,207955002 | MAP1S | FALSE | Upregulated |
| -0,148380675 | 0,208042139 | THOC1 | FALSE | Downregulated |
| 0,491009184 | 0,208042139 | TMEM54 | FALSE | Upregulated |
| -1,293180935 | 0,20825793 | B3GALT5 | FALSE | Downregulated |
| -0,165342055 | 0,20825793 | SOCS6 | FALSE | Downregulated |
| -0,470892559 | 0,208269286 | C8orf44 | FALSE | Downregulated |
| 0,272319721 | 0,208334663 | SERPINH1 | FALSE | Upregulated |
| 0,184847374 | 0,20855638 | MTCH2 | FALSE | Upregulated |

| | | | | |
|---|---|---|---|---|
| 0,239634273 | 0,20855638 | TTLL12 | FALSE | Upregulated |
| 0,202280392 | 0,208876799 | BRCC3 | FALSE | Upregulated |
| 0,227627125 | 0,209534395 | CXorf38 | FALSE | Upregulated |
| -0,374427383 | 0,209534395 | ENTPD8 | FALSE | Downregulated |
| -0,728263706 | 0,209534395 | FCRLB | FALSE | Downregulated |
| 0,604365876 | 0,209534395 | GTF2H2 | FALSE | Upregulated |
| 0,852392678 | 0,209534395 | MIR4435-1HG | FALSE | Upregulated |
| 0,371040835 | 0,209562345 | IFIT2 | FALSE | Upregulated |
| 0,236834062 | 0,209562345 | RNFT1 | FALSE | Upregulated |
| -0,170652679 | 0,2097618 | TCFL5 | FALSE | Downregulated |
| -0,505680252 | 0,209858666 | MIF | FALSE | Downregulated |
| 0,217903992 | 0,209902014 | INPP1 | FALSE | Upregulated |
| 0,148877634 | 0,209935097 | CCDC127 | FALSE | Upregulated |
| -1,441026094 | 0,210029307 | DOCK3 | FALSE | Downregulated |
| 0,887794956 | 0,210029307 | FBXL16 | FALSE | Upregulated |
| 0,187586293 | 0,210029307 | PTPN23 | FALSE | Upregulated |
| -0,190365002 | 0,210029307 | SOD1 | FALSE | Downregulated |
| 1,397778857 | 0,210029307 | TNFRSF18 | FALSE | Upregulated |
| -0,2000063 | 0,210091609 | KATNAL1 | FALSE | Downregulated |
| -0,190772903 | 0,210179775 | NCBP2-AS2 | FALSE | Downregulated |
| 3,150268456 | 0,210297361 | KRT81 | FALSE | Upregulated |
| -1,420881248 | 0,210320483 | LOC102724901 | FALSE | Downregulated |
| -0,214237595 | 0,210320483 | TMEM57 | FALSE | Downregulated |
| -0,280990888 | 0,21059788 | PPFIBP1 | FALSE | Downregulated |
| 0,299570864 | 0,210628756 | DHRS11 | FALSE | Upregulated |
| -0,14107252 | 0,210662821 | TRIM39 | FALSE | Downregulated |
| -0,386751639 | 0,21083299 | COL25A1 | FALSE | Downregulated |
| 0,171772978 | 0,21083299 | PPP2CB | FALSE | Upregulated |
| 0,234306595 | 0,21083299 | TMEM45A | FALSE | Upregulated |
| 0,591009202 | 0,210939222 | GP1BA | FALSE | Upregulated |
| -0,549277347 | 0,210948086 | LOC150776 | FALSE | Downregulated |
| -0,166359159 | 0,211030823 | ARHGEF12 | FALSE | Downregulated |
| -0,338555399 | 0,211030823 | CDK19 | FALSE | Downregulated |
| -0,865536743 | 0,211030823 | FAM83A-AS1 | FALSE | Downregulated |
| -0,365030201 | 0,211030823 | LOC102724392 | FALSE | Downregulated |
| 0,198805717 | 0,211085129 | FAM103A1 | FALSE | Upregulated |
| 0,244634088 | 0,211131443 | ATRIP | FALSE | Upregulated |
| -0,907099752 | 0,211370953 | BNIPL | FALSE | Downregulated |
| 0,151299429 | 0,211370953 | FEM1C | FALSE | Upregulated |
| 0,173281415 | 0,211370953 | PNKD | FALSE | Upregulated |
| 0,131348131 | 0,211722722 | STAMBP | FALSE | Upregulated |
| -1,554367832 | 0,211723548 | YWHAEP1 | FALSE | Downregulated |
| -1,299052071 | 0,211762688 | NPIPB15 | FALSE | Downregulated |
| -0,293754131 | 0,211956501 | IDNK | FALSE | Downregulated |
| 0,312113693 | 0,212052635 | HLF | FALSE | Upregulated |
| -0,286569204 | 0,212052635 | ZNF565 | FALSE | Downregulated |
| 0,389996849 | 0,212451184 | GALNT18 | FALSE | Upregulated |
| -0,240079929 | 0,21255796 | FAM213A | FALSE | Downregulated |
| -0,235416874 | 0,21255796 | FOXN2 | FALSE | Downregulated |
| -0,567977904 | 0,212564341 | LOC102723804 | FALSE | Downregulated |
| 0,848282401 | 0,21261054 | ARMC4 | FALSE | Upregulated |

| | | | | |
|---|---|---|---|---|
| -2,31103802 | 0,212725056 | MROH7 | FALSE | Downregulated |
| -0,800265501 | 0,21277917 | LOC102724927 | FALSE | Downregulated |
| -0,530933101 | 0,212783727 | AGAP6 | FALSE | Downregulated |
| -0,13392524 | 0,212783727 | KIF3B | FALSE | Downregulated |
| -0,28022823 | 0,212789614 | SPATA5 | FALSE | Downregulated |
| -0,623870662 | 0,212822863 | LOC642236 | FALSE | Downregulated |
| -0,242197125 | 0,213267286 | TET3 | FALSE | Downregulated |
| -0,52744379 | 0,213271909 | GDF7 | FALSE | Downregulated |
| -0,879804624 | 0,213278152 | LOC646471 | FALSE | Downregulated |
| -0,163742325 | 0,213530931 | CYP2J2 | FALSE | Downregulated |
| -1,538373572 | 0,213530931 | LOC101927308 | FALSE | Downregulated |
| -0,18661911 | 0,213530931 | MED28 | FALSE | Downregulated |
| -0,681174413 | 0,213530931 | MORF4L2-AS1 | FALSE | Downregulated |
| -0,167701544 | 0,213530931 | PAPSS2 | FALSE | Downregulated |
| -0,178311007 | 0,213530931 | SHMT2 | FALSE | Downregulated |
| 0,21319964 | 0,213530931 | ZMYND19 | FALSE | Upregulated |
| 0,239920568 | 0,213578445 | MGAT5 | FALSE | Upregulated |
| 0,117901918 | 0,213613717 | DARS | FALSE | Upregulated |
| -0,466187838 | 0,213613717 | LINC00844 | FALSE | Downregulated |
| -0,223789804 | 0,213759462 | HERC3 | FALSE | Downregulated |
| 1,702442235 | 0,213759837 | DMC1 | FALSE | Upregulated |
| 0,217280904 | 0,213759837 | GALNS | FALSE | Upregulated |
| 0,19893505 | 0,213953639 | MRPL43 | FALSE | Upregulated |
| 0,222100008 | 0,214216351 | ABCC3 | FALSE | Upregulated |
| 0,669556322 | 0,214259557 | LOC102724687 | FALSE | Upregulated |
| 0,211904171 | 0,214262493 | CINP | FALSE | Upregulated |
| 0,135759872 | 0,214291346 | GART | FALSE | Upregulated |
| 0,152117654 | 0,214402087 | PDCD10 | FALSE | Upregulated |
| 0,140052461 | 0,214512688 | PYCR2 | FALSE | Upregulated |
| -0,310445201 | 0,214558984 | MIR100HG | FALSE | Downregulated |
| -1,200152798 | 0,214617185 | KCNC2 | FALSE | Downregulated |
| -0,659382707 | 0,214813034 | LOC728554 | FALSE | Downregulated |
| 1,134610663 | 0,21482746 | HSF5 | FALSE | Upregulated |
| 0,335015693 | 0,214966654 | PASK | FALSE | Upregulated |
| -0,256707463 | 0,214966654 | ZNF490 | FALSE | Downregulated |
| -1,520679626 | 0,215010074 | BVES-AS1 | FALSE | Downregulated |
| -0,429944298 | 0,215010074 | OXER1 | FALSE | Downregulated |
| -0,211290799 | 0,215020114 | DVL1 | FALSE | Downregulated |
| 0,199012862 | 0,215150421 | RAB40C | FALSE | Upregulated |
| -0,111784003 | 0,215252984 | BRAP | FALSE | Downregulated |
| -0,254099225 | 0,215316922 | TTC12 | FALSE | Downregulated |
| -0,29106624 | 0,215822509 | C3orf18 | FALSE | Downregulated |
| -2,124914675 | 0,215830163 | REXO1L1P | FALSE | Downregulated |
| -0,220969435 | 0,215919917 | PSD4 | FALSE | Downregulated |
| -0,198381317 | 0,215926807 | LRP1 | FALSE | Downregulated |
| 0,139972263 | 0,21595411 | SPCS1 | FALSE | Upregulated |
| 0,222486213 | 0,215995278 | CKS1B | FALSE | Upregulated |
| 0,24861668 | 0,215995278 | DOK4 | FALSE | Upregulated |
| -1,222985747 | 0,215995278 | WBSCR17 | FALSE | Downregulated |
| -0,944172435 | 0,216106138 | LOC100130987 | FALSE | Downregulated |
| 0,228596773 | 0,216106138 | POLD3 | FALSE | Upregulated |

| | | | | |
|---|---|---|---|---|
| -0,116725267 | 0,216106138 | QKI | FALSE | Downregulated |
| -0,332207717 | 0,216192885 | LINC01184 | FALSE | Downregulated |
| -0,176895926 | 0,216204085 | IMP3 | FALSE | Downregulated |
| -0,253559276 | 0,216362227 | OFD1 | FALSE | Downregulated |
| 0,550762713 | 0,216544417 | LOC101928236 | FALSE | Upregulated |
| -0,145664708 | 0,216632702 | CDC26 | FALSE | Downregulated |
| -0,2169073 | 0,216669784 | SPATA13 | FALSE | Downregulated |
| 0,173376387 | 0,216707778 | POLR2K | FALSE | Upregulated |
| 0,540384706 | 0,216880431 | GSN | FALSE | Upregulated |
| 0,852812108 | 0,216880431 | RPS2P32 | FALSE | Upregulated |
| 0,229527632 | 0,216907135 | PHF23 | FALSE | Upregulated |
| -0,206763023 | 0,216907135 | PNRC1 | FALSE | Downregulated |
| -0,185926871 | 0,216907135 | RERE | FALSE | Downregulated |
| 2,010188731 | 0,216907135 | SAA3P | FALSE | Upregulated |
| -0,19030599 | 0,216907135 | SERPINC1 | FALSE | Downregulated |
| 0,299356604 | 0,216907135 | UQCRBP1 | FALSE | Upregulated |
| 0,280617754 | 0,21696744 | ARHGEF6 | FALSE | Upregulated |
| -0,338412666 | 0,21696744 | BMP10 | FALSE | Downregulated |
| 0,745769396 | 0,21696744 | EFEMP1 | FALSE | Upregulated |
| 0,461051974 | 0,217026552 | NCF1C | FALSE | Upregulated |
| -0,156121184 | 0,217148759 | LOC101928170 | FALSE | Downregulated |
| 0,127222038 | 0,217151054 | HNRNPF | FALSE | Upregulated |
| 0,351150945 | 0,217188733 | LINC01138 | FALSE | Upregulated |
| 0,891023174 | 0,217348318 | C1orf233 | FALSE | Upregulated |
| 0,249825857 | 0,217409538 | SGSH | FALSE | Upregulated |
| -0,198924288 | 0,217409538 | USP8 | FALSE | Downregulated |
| 0,223892712 | 0,217446607 | TEX264 | FALSE | Upregulated |
| 0,375586156 | 0,217500684 | APOBR | FALSE | Upregulated |
| -0,38690988 | 0,217500684 | LINC00341 | FALSE | Downregulated |
| -0,164974686 | 0,217500684 | SRSF8 | FALSE | Downregulated |
| 1,091203145 | 0,217500684 | STYK1 | FALSE | Upregulated |
| 0,139653073 | 0,217534104 | RNF6 | FALSE | Upregulated |
| 0,215695269 | 0,21755434 | ARHGEF1 | FALSE | Upregulated |
| -0,140487247 | 0,217610686 | GTF3C2 | FALSE | Downregulated |
| 0,713896724 | 0,217782694 | TMPRSS9 | FALSE | Upregulated |
| 0,541766263 | 0,217851567 | S100A6 | FALSE | Upregulated |
| -0,195049649 | 0,21793994 | ANAPC16 | FALSE | Downregulated |
| -0,411463419 | 0,21793994 | MAP3K7CL | FALSE | Downregulated |
| -0,842220393 | 0,21793994 | PTPN20B | FALSE | Downregulated |
| -0,297180114 | 0,218134551 | ZNF658 | FALSE | Downregulated |
| 0,151152926 | 0,218251717 | NAE1 | FALSE | Upregulated |
| -0,310654966 | 0,218370144 | LOC389765 | FALSE | Downregulated |
| -0,293452658 | 0,21845568 | ATN1 | FALSE | Downregulated |
| 0,193840993 | 0,218602421 | KCTD2 | FALSE | Upregulated |
| 0,297952474 | 0,218602421 | NKX3-1 | FALSE | Upregulated |
| -0,356132762 | 0,218602421 | PPP1R1C | FALSE | Downregulated |
| -0,096996145 | 0,21889749 | RBM22 | FALSE | Downregulated |
| -0,552484193 | 0,218981378 | LINC00115 | FALSE | Downregulated |
| 0,119145146 | 0,21907267 | PRPF6 | FALSE | Upregulated |
| -0,294953982 | 0,219123473 | AKT3 | FALSE | Downregulated |
| -1,312494096 | 0,219147335 | LOC102723505 | FALSE | Downregulated |

| | | | | |
|---|---|---|---|---|
| -0,220163999 | 0,219152768 | PMS2P5 | FALSE | Downregulated |
| 0,230552128 | 0,219152768 | SPTLC3 | FALSE | Upregulated |
| 0,130750839 | 0,219155949 | C14orf166 | FALSE | Upregulated |
| -0,471089888 | 0,219155949 | C1orf63 | FALSE | Downregulated |
| 1,548036826 | 0,219155949 | CTC-436P18.1 | FALSE | Upregulated |
| 0,32593154 | 0,219155949 | INTS9 | FALSE | Upregulated |
| -0,725098707 | 0,219503286 | LTF | FALSE | Downregulated |
| -0,241779123 | 0,219536355 | C11orf1 | FALSE | Downregulated |
| 0,153189783 | 0,219536355 | DDX56 | FALSE | Upregulated |
| -0,216934055 | 0,219646402 | HACE1 | FALSE | Downregulated |
| -0,656731783 | 0,219646402 | RP11-379B18.5 | FALSE | Downregulated |
| 1,243917512 | 0,219896395 | C5orf64 | FALSE | Upregulated |
| 0,265485283 | 0,219896395 | MAFB | FALSE | Upregulated |
| 0,124168773 | 0,219896395 | MED24 | FALSE | Upregulated |
| -0,339528955 | 0,220024834 | TMEM161B-AS1 | FALSE | Downregulated |
| -0,310040306 | 0,22015315 | ARHGEF15 | FALSE | Downregulated |
| 0,22020776 | 0,22015315 | TEFM | FALSE | Upregulated |
| -0,172434236 | 0,220202244 | SSFA2 | FALSE | Downregulated |
| 0,845883451 | 0,220323519 | BAI1 | FALSE | Upregulated |
| 0,268438975 | 0,220323519 | RPS18 | FALSE | Upregulated |
| 0,372290566 | 0,220406594 | GCNT1 | FALSE | Upregulated |
| -0,934232388 | 0,22074298 | HMGB3P1 | FALSE | Downregulated |
| -0,148114232 | 0,22074298 | NFYA | FALSE | Downregulated |
| -0,176024387 | 0,22074298 | ZNF526 | FALSE | Downregulated |
| -0,153446103 | 0,22075949 | ARID1B | FALSE | Downregulated |
| 0,31048519 | 0,22075949 | CYB561 | FALSE | Upregulated |
| 0,281471852 | 0,22075949 | FABP1 | FALSE | Upregulated |
| 0,142890266 | 0,220810001 | EHMT2 | FALSE | Upregulated |
| 0,787328481 | 0,220971681 | FBXO43 | FALSE | Upregulated |
| -1,6037432 | 0,220971681 | NR2E3 | FALSE | Downregulated |
| -0,210160442 | 0,220973122 | TRIQK | FALSE | Downregulated |
| 0,172844402 | 0,221048542 | SCAMP1 | FALSE | Upregulated |
| -0,207947845 | 0,221110878 | ATAD2B | FALSE | Downregulated |
| -0,1317348 | 0,221143927 | WAPAL | FALSE | Downregulated |
| -0,272906759 | 0,22119252 | RHEBP2 | FALSE | Downregulated |
| -0,506438169 | 0,221338781 | KCNMA1 | FALSE | Downregulated |
| 0,22426676 | 0,221379514 | C9orf69 | FALSE | Upregulated |
| -0,276777136 | 0,221379514 | NUPR1 | FALSE | Downregulated |
| 0,297165473 | 0,221400326 | ABCC1 | FALSE | Upregulated |
| -0,573041077 | 0,221400326 | AOC2 | FALSE | Downregulated |
| 0,475698796 | 0,221400326 | CENPL | FALSE | Upregulated |
| 0,196303323 | 0,221400326 | FAM214B | FALSE | Upregulated |
| -0,14278102 | 0,221540207 | EIF4ENIF1 | FALSE | Downregulated |
| -0,307008106 | 0,221540207 | LOC100129550 | FALSE | Downregulated |
| -0,146476336 | 0,221551835 | SNX25 | FALSE | Downregulated |
| -0,80629011 | 0,221974397 | LOC101926918 | FALSE | Downregulated |
| 0,20057226 | 0,222009878 | PHYKPL | FALSE | Upregulated |
| -0,740524491 | 0,222139783 | TPPP3 | FALSE | Downregulated |
| 1,212853722 | 0,222200969 | KIR2DL4 | FALSE | Upregulated |
| -0,245765769 | 0,222305635 | HPX | FALSE | Downregulated |
| -0,515522712 | 0,222305635 | KCNQ1OT1 | FALSE | Downregulated |

| | | | | |
|---|---|---|---|---|
| 0,831693556 | 0,222305635 | LOC101927886 | FALSE | Upregulated |
| -0,617250223 | 0,222305635 | SPDYE3 | FALSE | Downregulated |
| 0,47430075 | 0,222332493 | B3GALT4 | FALSE | Upregulated |
| -1,848135524 | 0,222332493 | C4orf26 | FALSE | Downregulated |
| -0,440270596 | 0,222332493 | C7orf13 | FALSE | Downregulated |
| 0,100223394 | 0,222332493 | DDB1 | FALSE | Upregulated |
| -0,176469964 | 0,222332493 | ERLIN2 | FALSE | Downregulated |
| 0,158495011 | 0,222332493 | GALM | FALSE | Upregulated |
| -0,422205498 | 0,222332493 | LOC100652824 | FALSE | Downregulated |
| -0,263660111 | 0,222332493 | MPHOSPH10 | FALSE | Downregulated |
| -1,279940197 | 0,222332493 | MSI1 | FALSE | Downregulated |
| -0,503657206 | 0,222332493 | MT1E | FALSE | Downregulated |
| 0,590930277 | 0,222332493 | TLE6 | FALSE | Upregulated |
| 0,143090312 | 0,222332493 | YIPF3 | FALSE | Upregulated |
| -0,584819101 | 0,222354238 | OGFR-AS1 | FALSE | Downregulated |
| -0,257238186 | 0,222446688 | SNAPC5 | FALSE | Downregulated |
| 0,28359223 | 0,222778378 | SLC39A10 | FALSE | Upregulated |
| 1,11336477 | 0,222878626 | CXCL3 | FALSE | Upregulated |
| -0,619959136 | 0,22288075 | MAB21L3 | FALSE | Downregulated |
| 0,17626169 | 0,222913834 | NEU1 | FALSE | Upregulated |
| -0,169134578 | 0,22296767 | VTI1B | FALSE | Downregulated |
| 0,914581782 | 0,223087912 | CTRL | FALSE | Upregulated |
| -0,304981964 | 0,223114046 | CEBPA | FALSE | Downregulated |
| -0,282351193 | 0,223266586 | C1orf35 | FALSE | Downregulated |
| 0,450194105 | 0,223356027 | KCNC3 | FALSE | Upregulated |
| 0,903831471 | 0,223378397 | KCNN4 | FALSE | Upregulated |
| -0,163981177 | 0,223512494 | CHIC2 | FALSE | Downregulated |
| 0,884367071 | 0,223512494 | DNAJC6 | FALSE | Upregulated |
| 0,116491164 | 0,223633172 | NCBP2 | FALSE | Upregulated |
| -0,628934976 | 0,223951775 | RP11-61I13.3 | FALSE | Downregulated |
| -0,191385448 | 0,224001672 | DIEXF | FALSE | Downregulated |
| -0,76106054 | 0,224001672 | RPL23P8 | FALSE | Downregulated |
| 0,173396607 | 0,224027222 | GTF2F2 | FALSE | Upregulated |
| -0,560577137 | 0,224192814 | TTC25 | FALSE | Downregulated |
| -0,457075126 | 0,224225838 | LRAT | FALSE | Downregulated |
| 0,219916705 | 0,224237312 | TMED9 | FALSE | Upregulated |
| 1,238354151 | 0,224288334 | LOC102724827 | FALSE | Upregulated |
| 0,126391189 | 0,224708 | CLPTM1 | FALSE | Upregulated |
| -1,225757936 | 0,224708 | UCHL1 | FALSE | Downregulated |
| 1,02621318 | 0,224873359 | ST8SIA1 | FALSE | Upregulated |
| 0,490327267 | 0,224886302 | RNF150 | FALSE | Upregulated |
| -1,68178261 | 0,225206267 | LOC101929060 | FALSE | Downregulated |
| 0,186407457 | 0,22529517 | PRKACB | FALSE | Upregulated |
| 0,191605266 | 0,225339818 | G6PC3 | FALSE | Upregulated |
| 0,382026346 | 0,225355992 | ZDHHC24 | FALSE | Upregulated |
| 0,294752867 | 0,225465857 | SPIRE2 | FALSE | Upregulated |
| 0,965776411 | 0,225527636 | OR2A20P | FALSE | Upregulated |
| -0,641836133 | 0,225527636 | TDRD6 | FALSE | Downregulated |
| 0,157827239 | 0,225635311 | PRKAG1 | FALSE | Upregulated |
| 0,452059707 | 0,225642957 | PHACTR1 | FALSE | Upregulated |
| -0,434637001 | 0,22574759 | FLJ22763 | FALSE | Downregulated |

| | | | | |
|---|---|---|---|---|
| -1,044232006 | 0,22574759 | LINC00282 | FALSE | Downregulated |
| 0,165171904 | 0,22574759 | OSGIN2 | FALSE | Upregulated |
| -0,218340093 | 0,225916021 | PCCB | FALSE | Downregulated |
| 0,23559803 | 0,225918581 | UQCRH | FALSE | Upregulated |
| -0,68990261 | 0,226155587 | TMEM74B | FALSE | Downregulated |
| 1,267932112 | 0,226158562 | RP11-838N2.4 | FALSE | Upregulated |
| -0,200099097 | 0,226301213 | TESK2 | FALSE | Downregulated |
| -1,793169112 | 0,226867483 | TBC1D3H | FALSE | Downregulated |
| 0,332704147 | 0,226902349 | XXYLT1 | FALSE | Upregulated |
| -0,991436326 | 0,226987918 | CCDC141 | FALSE | Downregulated |
| -1,427995989 | 0,226989946 | HCG17 | FALSE | Downregulated |
| -0,16760866 | 0,226989946 | LARP7 | FALSE | Downregulated |
| -0,303899537 | 0,227199903 | NFAT5 | FALSE | Downregulated |
| 0,404380115 | 0,227278966 | ASTN2 | FALSE | Upregulated |
| -0,235737653 | 0,227278966 | PRDX6 | FALSE | Downregulated |
| -0,344094591 | 0,227448222 | FTCD | FALSE | Downregulated |
| 0,14005651 | 0,227493197 | RELA | FALSE | Upregulated |
| -0,98815205 | 0,22749532 | LOC102724256 | FALSE | Downregulated |
| -2,865844806 | 0,227557107 | TECRL | FALSE | Downregulated |
| -0,239764776 | 0,227616928 | ACSL5 | FALSE | Downregulated |
| -1,180640107 | 0,227622705 | COL24A1 | FALSE | Downregulated |
| -0,19175578 | 0,227622705 | ERCC5 | FALSE | Downregulated |
| -0,281005494 | 0,227622705 | ZNF836 | FALSE | Downregulated |
| 0,189337659 | 0,227661814 | LSR | FALSE | Upregulated |
| -0,243616636 | 0,22771407 | ACOX2 | FALSE | Downregulated |
| 0,943899472 | 0,227854076 | PTGES | FALSE | Upregulated |
| -0,375143232 | 0,227964843 | LOC100506127 | FALSE | Downregulated |
| 0,470561446 | 0,228082081 | SNX22 | FALSE | Upregulated |
| 0,175629867 | 0,228103699 | ACSL3 | FALSE | Upregulated |
| 1,224877486 | 0,228103699 | ARRDC5 | FALSE | Upregulated |
| -0,411649725 | 0,228147925 | PLA2G6 | FALSE | Downregulated |
| -0,186188904 | 0,228206585 | PRR14L | FALSE | Downregulated |
| 0,218128303 | 0,228206585 | SNTA1 | FALSE | Upregulated |
| 0,131974625 | 0,228206585 | THOC7 | FALSE | Upregulated |
| -0,578780082 | 0,22832702 | TMEM178A | FALSE | Downregulated |
| 0,324983665 | 0,228345698 | HS6ST1 | FALSE | Upregulated |
| 0,292306534 | 0,228983179 | ARF4 | FALSE | Upregulated |
| 0,393892718 | 0,228983179 | C19orf38 | FALSE | Upregulated |
| 0,209439664 | 0,228983179 | TRAF4 | FALSE | Upregulated |
| 0,207516814 | 0,229241853 | ULK2 | FALSE | Upregulated |
| 0,233627632 | 0,229282948 | C20orf27 | FALSE | Upregulated |
| -0,193001494 | 0,229282948 | TMEM150A | FALSE | Downregulated |
| 0,199568939 | 0,229441111 | UGCG | FALSE | Upregulated |
| -0,398448261 | 0,229531982 | ZNF891 | FALSE | Downregulated |
| -1,994412855 | 0,22959639 | HMGA2 | FALSE | Downregulated |
| -0,115963984 | 0,229603475 | MARK3 | FALSE | Downregulated |
| -0,615395986 | 0,229603475 | SLC34A1 | FALSE | Downregulated |
| -0,459079699 | 0,229657849 | HUNK | FALSE | Downregulated |
| -0,230374971 | 0,229687784 | FOXJ3 | FALSE | Downregulated |
| -0,730946761 | 0,229723945 | EDAR | FALSE | Downregulated |
| -0,157206218 | 0,229723945 | TCEB3 | FALSE | Downregulated |

| | | | | |
|---|---|---|---|---|
| 0,865852858 | 0,229778984 | LOC102724428 | FALSE | Upregulated |
| -0,144345593 | 0,229787326 | ZXDC | FALSE | Downregulated |
| 0,359166613 | 0,230074349 | ZNF273 | FALSE | Upregulated |
| -0,550371599 | 0,230181799 | FST | FALSE | Downregulated |
| 0,149494753 | 0,230181799 | PIGC | FALSE | Upregulated |
| -0,264694401 | 0,230319248 | DCAF16 | FALSE | Downregulated |
| -1,845207861 | 0,230394598 | LOC101060632 | FALSE | Downregulated |
| -0,205270806 | 0,23081246 | USF2 | FALSE | Downregulated |
| 0,489197443 | 0,230939751 | PCSK5 | FALSE | Upregulated |
| -1,075570437 | 0,231026372 | LOC102723972 | FALSE | Downregulated |
| 0,265005492 | 0,231080667 | FAM117B | FALSE | Upregulated |
| 0,501009677 | 0,231199255 | KLRG1 | FALSE | Upregulated |
| 0,706155172 | 0,231285248 | B3GNT8 | FALSE | Upregulated |
| 0,179182411 | 0,231285248 | C11orf24 | FALSE | Upregulated |
| 0,198749708 | 0,231285248 | NDC1 | FALSE | Upregulated |
| 0,211576172 | 0,231285248 | RBCK1 | FALSE | Upregulated |
| -1,277783011 | 0,231285248 | TMEM108 | FALSE | Downregulated |
| -0,205007934 | 0,231335235 | GOLGA2 | FALSE | Downregulated |
| 0,816878885 | 0,231365055 | TMIGD2 | FALSE | Upregulated |
| 0,197651143 | 0,231499338 | R3HDM4 | FALSE | Upregulated |
| -0,188191264 | 0,231528237 | HNF1A | FALSE | Downregulated |
| 0,149301972 | 0,231528237 | OSER1 | FALSE | Upregulated |
| -0,322457301 | 0,23156532 | RAD52 | FALSE | Downregulated |
| -0,919236481 | 0,231640789 | LINC00202-1 | FALSE | Downregulated |
| 0,233995989 | 0,231654344 | CD3EAP | FALSE | Upregulated |
| -0,563566656 | 0,231661706 | LOC101928913 | FALSE | Downregulated |
| -0,296144517 | 0,231661706 | TRIM2 | FALSE | Downregulated |
| 0,204594546 | 0,231801141 | BAG2 | FALSE | Upregulated |
| 0,210534119 | 0,232012825 | OGFOD3 | FALSE | Upregulated |
| 0,236331552 | 0,232023286 | C4orf32 | FALSE | Upregulated |
| -0,641039994 | 0,232023286 | LOC102723782 | FALSE | Downregulated |
| -0,534644986 | 0,232075135 | LOC101928047 | FALSE | Downregulated |
| 0,225331363 | 0,232212062 | CISD2 | FALSE | Upregulated |
| -0,195387595 | 0,232239122 | CLU | FALSE | Downregulated |
| -0,404666916 | 0,232320837 | KIF17 | FALSE | Downregulated |
| 0,233745128 | 0,232378317 | BLZF1 | FALSE | Upregulated |
| -0,144397714 | 0,232378317 | C16orf70 | FALSE | Downregulated |
| 0,278018315 | 0,232378317 | CD93 | FALSE | Upregulated |
| -1,218125179 | 0,232378317 | FAM41C | FALSE | Downregulated |
| 0,342609293 | 0,232378317 | GPATCH4 | FALSE | Upregulated |
| -0,179168107 | 0,232378317 | HPN | FALSE | Downregulated |
| 0,114967976 | 0,232378317 | RNF40 | FALSE | Upregulated |
| 2,429787881 | 0,232378317 | RNU11 | FALSE | Upregulated |
| -0,738588092 | 0,232378317 | UNC79 | FALSE | Downregulated |
| 0,197275904 | 0,232522847 | DMAP1 | FALSE | Upregulated |
| -0,633167472 | 0,232522847 | LOC101929178 | FALSE | Downregulated |
| 0,855256166 | 0,232522847 | RP11-432I5.1 | FALSE | Upregulated |
| -0,933995027 | 0,232524104 | TCAM1P | FALSE | Downregulated |
| -0,189573449 | 0,232806856 | CDC40 | FALSE | Downregulated |
| 0,42367601 | 0,232806856 | LRRK1 | FALSE | Upregulated |
| -1,286243628 | 0,232834996 | TMEM179 | FALSE | Downregulated |

| | | | | |
|---|---|---|---|---|
| 0,246433467 | 0,232887457 | IPPK | FALSE | Upregulated |
| -0,590430604 | 0,23296066 | LOC613037 | FALSE | Downregulated |
| 0,593625996 | 0,233014603 | BEND4 | FALSE | Upregulated |
| 0,400737907 | 0,233014603 | PLA2G4A | FALSE | Upregulated |
| 0,129763545 | 0,233039205 | CANT1 | FALSE | Upregulated |
| 0,532051085 | 0,233112604 | IL18RAP | FALSE | Upregulated |
| 1,422031807 | 0,233112604 | MTRNR2L1 | FALSE | Upregulated |
| 0,280518327 | 0,233115056 | ERBB2 | FALSE | Upregulated |
| 0,582117034 | 0,233211805 | GADD45B | FALSE | Upregulated |
| 0,184534762 | 0,233211805 | GNG5 | FALSE | Upregulated |
| 0,750559869 | 0,233211805 | IGSF21 | FALSE | Upregulated |
| -1,238512131 | 0,233211805 | RP11-486A14.2 | FALSE | Downregulated |
| -0,88663865 | 0,233211805 | TRPC3 | FALSE | Downregulated |
| -0,150900346 | 0,233331619 | WASL | FALSE | Downregulated |
| -0,215890239 | 0,233513586 | ARGLU1 | FALSE | Downregulated |
| -0,146729842 | 0,233631949 | CRYZL1 | FALSE | Downregulated |
| 0,330086705 | 0,23365147 | GJD3 | FALSE | Upregulated |
| -0,540605214 | 0,233727686 | LOC100128288 | FALSE | Downregulated |
| -0,771682731 | 0,233727686 | LOC102724607 | FALSE | Downregulated |
| -1,03400963 | 0,233727686 | MUC6 | FALSE | Downregulated |
| -0,630367242 | 0,233738399 | LOC101927339 | FALSE | Downregulated |
| -0,926780988 | 0,233755857 | MIR1282 | FALSE | Downregulated |
| 0,684053897 | 0,23416704 | ACSL4 | FALSE | Upregulated |
| -0,754233374 | 0,23416704 | C4orf47 | FALSE | Downregulated |
| 0,233612813 | 0,234272538 | NEDD4 | FALSE | Upregulated |
| -0,757678576 | 0,234273351 | LOC101927894 | FALSE | Downregulated |
| -0,160369303 | 0,234273351 | NDUFV1 | FALSE | Downregulated |
| -0,200025949 | 0,234280783 | ARHGAP42 | FALSE | Downregulated |
| 0,184116219 | 0,234280783 | FTH1 | FALSE | Upregulated |
| 0,130759231 | 0,234280783 | MRPL9 | FALSE | Upregulated |
| 0,175470675 | 0,234318419 | RNF8 | FALSE | Upregulated |
| -0,479331849 | 0,234341878 | LPIN3 | FALSE | Downregulated |
| -0,163851531 | 0,234361492 | SETD7 | FALSE | Downregulated |
| -0,173067992 | 0,234521561 | PTCD2 | FALSE | Downregulated |
| 0,423883715 | 0,234723971 | AP1S3 | FALSE | Upregulated |
| -0,773957866 | 0,234723971 | RP11-290F5.2 | FALSE | Downregulated |
| 0,235251532 | 0,234759429 | USP11 | FALSE | Upregulated |
| -0,892271017 | 0,234819492 | LOC101929571 | FALSE | Downregulated |
| 0,121649125 | 0,234824189 | ARF6 | FALSE | Upregulated |
| 0,373909024 | 0,234824189 | TRPV4 | FALSE | Upregulated |
| 0,329019747 | 0,235017419 | C19orf48 | FALSE | Upregulated |
| 1,349803228 | 0,235017419 | FUT7 | FALSE | Upregulated |
| -0,123638233 | 0,235017419 | FXR2 | FALSE | Downregulated |
| -0,144142774 | 0,235017419 | SPIN1 | FALSE | Downregulated |
| 0,275413535 | 0,235017419 | UBXN8 | FALSE | Upregulated |
| -0,175160306 | 0,235017419 | ZNF346 | FALSE | Downregulated |
| -0,299491497 | 0,235152221 | TTC5 | FALSE | Downregulated |
| 1,105892858 | 0,235159624 | PLEKHN1 | FALSE | Upregulated |
| 0,5679892 | 0,235161551 | RCAN2 | FALSE | Upregulated |
| -0,227692788 | 0,235230147 | NNT-AS1 | FALSE | Downregulated |
| -0,234808213 | 0,235279581 | SGSM2 | FALSE | Downregulated |

| | | | | |
|---|---|---|---|---|
| 1,633943114 | 0,235346871 | LOC102723316 | FALSE | Upregulated |
| -0,192602392 | 0,235346871 | PRPSAP2 | FALSE | Downregulated |
| 0,728183323 | 0,235346871 | SPTSSB | FALSE | Upregulated |
| -0,400384064 | 0,235346871 | ZNF501 | FALSE | Downregulated |
| -0,452507474 | 0,235547485 | GUSBP11 | FALSE | Downregulated |
| 1,349013068 | 0,235642908 | DLG1-AS1 | FALSE | Upregulated |
| 2,029084309 | 0,235642908 | MGC50722 | FALSE | Upregulated |
| -0,794086108 | 0,235791884 | ADAM20P1 | FALSE | Downregulated |
| -0,322453521 | 0,23582292 | C16orf86 | FALSE | Downregulated |
| -1,788562939 | 0,23582292 | KLHL41 | FALSE | Downregulated |
| -0,118352363 | 0,23582292 | VPS52 | FALSE | Downregulated |
| 0,206650398 | 0,235992129 | MORF4L2 | FALSE | Upregulated |
| 0,59203287 | 0,236194907 | C2orf82 | FALSE | Upregulated |
| -0,158408966 | 0,236194907 | R3HDM2 | FALSE | Downregulated |
| -0,265383047 | 0,236389105 | WDR60 | FALSE | Downregulated |
| -0,473796068 | 0,236795878 | ARRDC3 | FALSE | Downregulated |
| 0,318275762 | 0,236795878 | GIMAP7 | FALSE | Upregulated |
| -0,662331529 | 0,236795878 | IGFBP6 | FALSE | Downregulated |
| 0,195076372 | 0,236795878 | IL10RB | FALSE | Upregulated |
| -0,578353779 | 0,236795878 | LINC01004 | FALSE | Downregulated |
| 0,271302901 | 0,236795878 | NOD1 | FALSE | Upregulated |
| -0,593343063 | 0,236795878 | PKNOX2 | FALSE | Downregulated |
| -0,290923299 | 0,236795878 | POLR2J3 | FALSE | Downregulated |
| -0,222612868 | 0,236795878 | QTRT1 | FALSE | Downregulated |
| 1,419580259 | 0,236795878 | VSIG1 | FALSE | Upregulated |
| -0,351455308 | 0,236837582 | AHDC1 | FALSE | Downregulated |
| 0,460407873 | 0,236960379 | RP11-15A1.3 | FALSE | Upregulated |
| -0,405809807 | 0,236964736 | ADCY4 | FALSE | Downregulated |
| 0,468093148 | 0,236964736 | RP11-160O5.1 | FALSE | Upregulated |
| -0,224451743 | 0,236964736 | SLC37A4 | FALSE | Downregulated |
| 0,17517044 | 0,236964736 | TTC4 | FALSE | Upregulated |
| -0,917790213 | 0,236964736 | USP32P1 | FALSE | Downregulated |
| -0,39832174 | 0,237137905 | CCDC73 | FALSE | Downregulated |
| -1,398045027 | 0,237195489 | KIRREL3 | FALSE | Downregulated |
| 0,193630183 | 0,237231287 | LOC100294145 | FALSE | Upregulated |
| 0,203806753 | 0,237231287 | PCIF1 | FALSE | Upregulated |
| 1,154538625 | 0,237231287 | RIMBP3B | FALSE | Upregulated |
| 1,391234687 | 0,237231287 | SLC6A20 | FALSE | Upregulated |
| -0,721696203 | 0,237231287 | VIPR2 | FALSE | Downregulated |
| 0,215826635 | 0,237252152 | H1F0 | FALSE | Upregulated |
| 0,147103097 | 0,237253198 | RCN1 | FALSE | Upregulated |
| 0,333120276 | 0,237338887 | OSGIN1 | FALSE | Upregulated |
| 0,567659097 | 0,237345913 | ST6GALNAC4 | FALSE | Upregulated |
| -0,168429258 | 0,237458425 | UBB | FALSE | Downregulated |
| 0,241993228 | 0,237587783 | HNRNPH2 | FALSE | Upregulated |
| -0,373273202 | 0,23773324 | PEG3 | FALSE | Downregulated |
| -0,219261582 | 0,238065956 | PRPF3 | FALSE | Downregulated |
| -0,760393714 | 0,238116014 | SOCS3 | FALSE | Downregulated |
| 0,122452889 | 0,238152072 | AGPAT3 | FALSE | Upregulated |
| -0,626890144 | 0,238152072 | KIAA1875 | FALSE | Downregulated |
| 0,377050361 | 0,238186115 | PLAC8 | FALSE | Upregulated |

| | | | | |
|---|---|---|---|---|
| 0,91568711 | 0,238195055 | RASL11B | FALSE | Upregulated |
| 0,247963905 | 0,238271478 | SFPQ | FALSE | Upregulated |
| 0,179757952 | 0,238401711 | RBM42 | FALSE | Upregulated |
| -0,247916613 | 0,238451838 | C11orf52 | FALSE | Downregulated |
| 0,139907854 | 0,238484006 | FAM32A | FALSE | Upregulated |
| 0,130391047 | 0,238507966 | SUMO2 | FALSE | Upregulated |
| 0,175298212 | 0,238507966 | SURF4 | FALSE | Upregulated |
| -0,672921735 | 0,238595101 | LOC101928487 | FALSE | Downregulated |
| 0,149228763 | 0,238597828 | ATP6V0A2 | FALSE | Upregulated |
| 0,395155419 | 0,238597828 | C11orf84 | FALSE | Upregulated |
| 0,261709664 | 0,238597828 | FBXO10 | FALSE | Upregulated |
| -0,125815559 | 0,238597828 | SMEK1 | FALSE | Downregulated |
| -0,54543002 | 0,238741469 | LOC101927009 | FALSE | Downregulated |
| 0,732791139 | 0,238839635 | BCAS4 | FALSE | Upregulated |
| -0,851179562 | 0,238839635 | CTC-558O2.1 | FALSE | Downregulated |
| 0,239571348 | 0,238839635 | PPARG | FALSE | Upregulated |
| -0,521231866 | 0,238839635 | QRICH2 | FALSE | Downregulated |
| 0,144429164 | 0,239022547 | PEX26 | FALSE | Upregulated |
| 0,261095188 | 0,239165562 | GBP7 | FALSE | Upregulated |
| -0,705024862 | 0,239199052 | USH2A | FALSE | Downregulated |
| 0,177080735 | 0,23924996 | NFATC2IP | FALSE | Upregulated |
| 0,146360493 | 0,23924996 | SEC24C | FALSE | Upregulated |
| 0,162520551 | 0,23924996 | TGOLN2 | FALSE | Upregulated |
| -0,181789022 | 0,239389253 | ZBED5 | FALSE | Downregulated |
| -0,766513737 | 0,23939656 | LOC101929580 | FALSE | Downregulated |
| -0,239625823 | 0,239410923 | IL33 | FALSE | Downregulated |
| 0,14425907 | 0,239453593 | COPZ1 | FALSE | Upregulated |
| -0,68077891 | 0,239453593 | PLA2G4B | FALSE | Downregulated |
| -0,536365156 | 0,239530736 | PHYHIPL | FALSE | Downregulated |
| 0,215379807 | 0,239609736 | CABLES1 | FALSE | Upregulated |
| 0,132619563 | 0,239717162 | CLTC | FALSE | Upregulated |
| -0,097742157 | 0,239717162 | SLC35E1 | FALSE | Downregulated |
| 0,154854216 | 0,239767573 | COPS4 | FALSE | Upregulated |
| -0,54267954 | 0,239767573 | LOC100506023 | FALSE | Downregulated |
| 0,2584175 | 0,239767573 | LRRC8B | FALSE | Upregulated |
| -0,337054984 | 0,239813323 | SCML2 | FALSE | Downregulated |
| -0,183303954 | 0,239827498 | LYST | FALSE | Downregulated |
| 0,541835458 | 0,239827498 | NAPSB | FALSE | Upregulated |
| 0,496423541 | 0,239827498 | RP1-27K12.2 | FALSE | Upregulated |
| -0,206977079 | 0,23985331 | CCNYL1 | FALSE | Downregulated |
| 0,226174324 | 0,23985331 | FAM127B | FALSE | Upregulated |
| -0,177025657 | 0,239855854 | DLG1 | FALSE | Downregulated |
| -1,032935362 | 0,239856496 | RALYL | FALSE | Downregulated |
| 0,1964982 | 0,239858098 | HSDL1 | FALSE | Upregulated |
| 0,499106514 | 0,239936852 | FAM109B | FALSE | Upregulated |
| -0,254034092 | 0,239941263 | EPHX1 | FALSE | Downregulated |
| -1,060892179 | 0,239941263 | USP44 | FALSE | Downregulated |
| -0,147501701 | 0,239941263 | UTP20 | FALSE | Downregulated |
| -0,129006999 | 0,239973255 | UBE2H | FALSE | Downregulated |
| -0,657453309 | 0,240087135 | CCDC17 | FALSE | Downregulated |
| -0,214876108 | 0,240087135 | WDR73 | FALSE | Downregulated |

| | | | | |
|---|---|---|---|---|
| -0,478839955 | 0,240101227 | PBX1 | FALSE | Downregulated |
| 0,26949798 | 0,240101227 | ZBED6CL | FALSE | Upregulated |
| -1,048685495 | 0,240172015 | ACTG1P4 | FALSE | Downregulated |
| -0,194990611 | 0,240266654 | CDK5RAP3 | FALSE | Downregulated |
| 0,293236599 | 0,240266654 | GDF11 | FALSE | Upregulated |
| -1,098643598 | 0,240266654 | INA | FALSE | Downregulated |
| 1,700188089 | 0,240266654 | LINC00593 | FALSE | Upregulated |
| -0,370485532 | 0,240266654 | NBPF3 | FALSE | Downregulated |
| -0,800664674 | 0,240266654 | PLEKHG5 | FALSE | Downregulated |
| -0,388226636 | 0,240266654 | SH3BGR | FALSE | Downregulated |
| 0,210460632 | 0,240266654 | TMEM41A | FALSE | Upregulated |
| -0,133639289 | 0,240684778 | KIDINS220 | FALSE | Downregulated |
| 0,218681049 | 0,24068978 | CSRNP2 | FALSE | Upregulated |
| 0,176471217 | 0,24068978 | RNF181 | FALSE | Upregulated |
| -0,122226934 | 0,24068978 | TMEM181 | FALSE | Downregulated |
| -0,334502981 | 0,24068978 | ZNF444 | FALSE | Downregulated |
| 0,175079786 | 0,240698141 | CANX | FALSE | Upregulated |
| 1,592647409 | 0,240761827 | LOC102724376 | FALSE | Upregulated |
| -1,436368877 | 0,240761827 | MT1HL1 | FALSE | Downregulated |
| 0,227629887 | 0,240761827 | TTLL4 | FALSE | Upregulated |
| 0,258760568 | 0,240805667 | AKAP7 | FALSE | Upregulated |
| -1,957579511 | 0,240805667 | CTGLF12P | FALSE | Downregulated |
| -0,438762005 | 0,240805667 | LOC101927579 | FALSE | Downregulated |
| -0,941262938 | 0,240805667 | MT1H | FALSE | Downregulated |
| -0,230896444 | 0,240805667 | NFKBIL1 | FALSE | Downregulated |
| -0,251984453 | 0,240805667 | SYNE2 | FALSE | Downregulated |
| 0,163258418 | 0,241104217 | RBM19 | FALSE | Upregulated |
| 0,605362554 | 0,241282234 | LINC00426 | FALSE | Upregulated |
| 0,799101077 | 0,241282234 | LOC100128361 | FALSE | Upregulated |
| -1,74971117 | 0,241282234 | MYOM3 | FALSE | Downregulated |
| -0,344239313 | 0,241282234 | SLC22A9 | FALSE | Downregulated |
| 1,081484405 | 0,241282234 | TKTL1 | FALSE | Upregulated |
| 0,571447759 | 0,241343004 | CCDC28B | FALSE | Upregulated |
| -1,495944373 | 0,241343004 | GPR21 | FALSE | Downregulated |
| -0,887150143 | 0,241343004 | LOC101928829 | FALSE | Downregulated |
| -0,250001183 | 0,241343004 | NSRP1 | FALSE | Downregulated |
| -0,260244964 | 0,241343004 | PDPR | FALSE | Downregulated |
| 0,21124404 | 0,241343004 | PTRH2 | FALSE | Upregulated |
| -1,329796304 | 0,241343004 | RBAK-RBAKDN | FALSE | Downregulated |
| 0,666550517 | 0,241343004 | SIGLEC14 | FALSE | Upregulated |
| -0,409671413 | 0,241343004 | STEAP2 | FALSE | Downregulated |
| -0,147933646 | 0,241343004 | TRAPPC6B | FALSE | Downregulated |
| -0,322752035 | 0,241343004 | ZNF461 | FALSE | Downregulated |
| 0,204979889 | 0,241462969 | ACTN4 | FALSE | Upregulated |
| -0,907299199 | 0,241633441 | C1orf95 | FALSE | Downregulated |
| -0,200190763 | 0,241648387 | CREBRF | FALSE | Downregulated |
| 0,148086017 | 0,241648387 | G3BP1 | FALSE | Upregulated |
| -0,446100905 | 0,24175696 | DNAH1 | FALSE | Downregulated |
| -0,136205232 | 0,241775848 | CTNND1 | FALSE | Downregulated |
| 0,311133299 | 0,241800397 | SLC26A2 | FALSE | Upregulated |
| -0,156323326 | 0,24183163 | MTO1 | FALSE | Downregulated |

| | | | | |
|---|---|---|---|---|
| -0,35562834 | 0,24183163 | SLC17A1 | FALSE | Downregulated |
| -0,123739137 | 0,24183163 | TFAM | FALSE | Downregulated |
| 0,124285564 | 0,24183163 | UBA1 | FALSE | Upregulated |
| -0,845933365 | 0,242091032 | C6orf201 | FALSE | Downregulated |
| 0,258205724 | 0,242091032 | EVI5L | FALSE | Upregulated |
| 0,440274842 | 0,242094821 | FRMD6 | FALSE | Upregulated |
| 0,166340999 | 0,242199895 | NSF | FALSE | Upregulated |
| -0,309601787 | 0,242215473 | FGB | FALSE | Downregulated |
| 0,263085574 | 0,242215473 | IRAK2 | FALSE | Upregulated |
| -0,862218792 | 0,242215473 | LOC100505915 | FALSE | Downregulated |
| 0,46869621 | 0,242215473 | PPP4R1L | FALSE | Upregulated |
| 0,172910126 | 0,242250935 | CNP | FALSE | Upregulated |
| 0,886190553 | 0,242271679 | LINC00880 | FALSE | Upregulated |
| 0,135395335 | 0,242417624 | HTT | FALSE | Upregulated |
| 1,080511829 | 0,24249133 | LYPD1 | FALSE | Upregulated |
| -0,470062076 | 0,242599123 | ANKHD1-EIF4EBP3 | FALSE | Downregulated |
| -0,799239925 | 0,242599123 | FREM2 | FALSE | Downregulated |
| -1,422993186 | 0,242695682 | LOC101928307 | FALSE | Downregulated |
| -0,249543082 | 0,242825863 | FAM13A | FALSE | Downregulated |
| -0,124658229 | 0,242825863 | RAB3GAP2 | FALSE | Downregulated |
| 0,965271601 | 0,242894507 | RP3-395M20.9 | FALSE | Upregulated |
| 0,147926546 | 0,242948371 | NNT | FALSE | Upregulated |
| 0,944967908 | 0,242948371 | VPS9D1-AS1 | FALSE | Upregulated |
| 0,339794682 | 0,243023993 | WDYHV1 | FALSE | Upregulated |
| -0,451431571 | 0,243074323 | P4HA3 | FALSE | Downregulated |
| 0,556486959 | 0,243121746 | PATL2 | FALSE | Upregulated |
| 0,55703097 | 0,243189027 | TNFSF15 | FALSE | Upregulated |
| 0,137237862 | 0,24325476 | EIF3D | FALSE | Upregulated |
| -0,419984691 | 0,243331215 | ZG16 | FALSE | Downregulated |
| -0,606089583 | 0,243350249 | LGR5 | FALSE | Downregulated |
| -0,895783261 | 0,243372003 | S100A1 | FALSE | Downregulated |
| 0,453438312 | 0,243498226 | C18orf54 | FALSE | Upregulated |
| -0,358803026 | 0,243582986 | LOC92249 | FALSE | Downregulated |
| -0,612381035 | 0,243721676 | LOC102724935 | FALSE | Downregulated |
| -0,739306397 | 0,243778123 | RFPL4A | FALSE | Downregulated |
| -0,192850522 | 0,243778123 | SPPL2B | FALSE | Downregulated |
| -0,263025584 | 0,243809142 | AFM | FALSE | Downregulated |
| -0,681567231 | 0,24382531 | BNC2 | FALSE | Downregulated |
| -0,15074174 | 0,24382531 | FMR1 | FALSE | Downregulated |
| -1,026231323 | 0,24382531 | GOLGA8O | FALSE | Downregulated |
| 0,120177611 | 0,243855295 | RAB22A | FALSE | Upregulated |
| -0,454070403 | 0,243928816 | TRAF3IP2-AS1 | FALSE | Downregulated |
| -0,570886082 | 0,243950108 | RP5-1180C10.2 | FALSE | Downregulated |
| 1,244291351 | 0,244055405 | OR2A9P | FALSE | Upregulated |
| 0,691666726 | 0,244064584 | LOC101928399 | FALSE | Upregulated |
| 0,231533501 | 0,244064584 | SBNO2 | FALSE | Upregulated |
| 0,187844126 | 0,244083487 | AIMP2 | FALSE | Upregulated |
| 0,29701436 | 0,244095627 | WLS | FALSE | Upregulated |
| 0,543354105 | 0,244407105 | S100A9 | FALSE | Upregulated |
| -0,26981059 | 0,244429394 | INO80B-WBP1 | FALSE | Downregulated |

| | | | | |
|---|---|---|---|---|
| -0,203817042 | 0,244429394 | PROC | FALSE | Downregulated |
| -0,301849318 | 0,244429394 | TXLNG | FALSE | Downregulated |
| -0,18755113 | 0,244439301 | PPFIBP2 | FALSE | Downregulated |
| -0,297515892 | 0,244439301 | ZNF585B | FALSE | Downregulated |
| -1,005513771 | 0,244468036 | ABO | FALSE | Downregulated |
| 0,367697812 | 0,244468036 | E2F5 | FALSE | Upregulated |
| -1,764271723 | 0,244468036 | FAM221B | FALSE | Downregulated |
| -0,579063656 | 0,244468036 | PLEKHM1P | FALSE | Downregulated |
| -0,206312564 | 0,244561298 | ZNF596 | FALSE | Downregulated |
| -0,252797771 | 0,244569745 | POLG2 | FALSE | Downregulated |
| 0,216661091 | 0,244629913 | ATP5G2 | FALSE | Upregulated |
| 1,272187152 | 0,244629913 | FAM72B | FALSE | Upregulated |
| -1,195363118 | 0,244629913 | LOC101927702 | FALSE | Downregulated |
| -0,343542423 | 0,244629913 | LUC7L2 | FALSE | Downregulated |
| -0,457388902 | 0,244639553 | LRRC37A2 | FALSE | Downregulated |
| -1,09505458 | 0,244921254 | LOC101930129 | FALSE | Downregulated |
| 1,055172542 | 0,245108385 | C14orf64 | FALSE | Upregulated |
| -0,970692201 | 0,245135643 | LOC101559451 | FALSE | Downregulated |
| -0,989912959 | 0,245192823 | SLC5A2 | FALSE | Downregulated |
| -0,214840453 | 0,245198 | MLYCD | FALSE | Downregulated |
| -0,251474911 | 0,24545135 | ZNF81 | FALSE | Downregulated |
| 0,12082696 | 0,245883715 | KIF13B | FALSE | Upregulated |
| -0,246109515 | 0,245905703 | HSD17B6 | FALSE | Downregulated |
| -0,133517448 | 0,245998414 | VPS39 | FALSE | Downregulated |
| -0,379294383 | 0,246078763 | HPGD | FALSE | Downregulated |
| 0,945529759 | 0,246078763 | LINC00996 | FALSE | Upregulated |
| 0,448012743 | 0,246078763 | ZFP82 | FALSE | Upregulated |
| 0,200098867 | 0,246111052 | DCTPP1 | FALSE | Upregulated |
| -1,381481788 | 0,24620494 | COL11A1 | FALSE | Downregulated |
| -0,182188222 | 0,24620494 | FAM120C | FALSE | Downregulated |
| -0,546185351 | 0,246263713 | PVALB | FALSE | Downregulated |
| -0,164109593 | 0,246414797 | GCLC | FALSE | Downregulated |
| 0,201857223 | 0,246485687 | ABCA2 | FALSE | Upregulated |
| -0,364468999 | 0,246485687 | JAM2 | FALSE | Downregulated |
| -0,535432101 | 0,246500489 | GABBR1 | FALSE | Downregulated |
| -0,432981948 | 0,246521824 | RASGRP2 | FALSE | Downregulated |
| -1,030268675 | 0,246521824 | VWA3B | FALSE | Downregulated |
| -0,218842147 | 0,246541244 | APMAP | FALSE | Downregulated |
| 0,188128352 | 0,246541244 | ARHGAP18 | FALSE | Upregulated |
| -0,22267134 | 0,246541244 | BDH2 | FALSE | Downregulated |
| 0,458323238 | 0,246541244 | CR1 | FALSE | Upregulated |
| 0,130820282 | 0,246541244 | DEDD | FALSE | Upregulated |
| -0,524266417 | 0,246541244 | HEXA-AS1 | FALSE | Downregulated |
| 0,42111729 | 0,246541244 | LOC100507642 | FALSE | Upregulated |
| 0,205460159 | 0,246541244 | OSGEP | FALSE | Upregulated |
| 0,338494535 | 0,246541244 | SLC25A51 | FALSE | Upregulated |
| -0,622492963 | 0,246541244 | ZNF137P | FALSE | Downregulated |
| -1,568966607 | 0,246564999 | CKMT2 | FALSE | Downregulated |
| 0,257915393 | 0,246564999 | DPCD | FALSE | Upregulated |
| -0,419120615 | 0,246564999 | LOC101928076 | FALSE | Downregulated |
| 0,783474728 | 0,246564999 | LOC440311 | FALSE | Upregulated |

| | | | | |
|---|---|---|---|---|
| -0,251379883 | 0,246564999 | PAXIP1-AS2 | FALSE | Downregulated |
| -0,163622468 | 0,246564999 | TAF5 | FALSE | Downregulated |
| 0,211304643 | 0,246811105 | APOA1BP | FALSE | Upregulated |
| 0,864333817 | 0,246811105 | CBLN1 | FALSE | Upregulated |
| -0,668995608 | 0,246968677 | CLEC9A | FALSE | Downregulated |
| 1,368254441 | 0,247048404 | FAM132B | FALSE | Upregulated |
| -0,318076307 | 0,247124567 | SPATA33 | FALSE | Downregulated |
| 0,367853885 | 0,247270806 | CDKN2D | FALSE | Upregulated |
| -0,276002327 | 0,247426655 | PHGDH | FALSE | Downregulated |
| -0,200746042 | 0,24747498 | ITPR2 | FALSE | Downregulated |
| 0,193213231 | 0,247563633 | COPE | FALSE | Upregulated |
| 0,279802548 | 0,247593337 | ATP2B4 | FALSE | Upregulated |
| -0,672179484 | 0,247927317 | LOC101926967 | FALSE | Downregulated |
| 1,078343088 | 0,247998682 | DYDC2 | FALSE | Upregulated |
| 0,16115978 | 0,248078571 | EMD | FALSE | Upregulated |
| 0,556471802 | 0,248109115 | ITGA3 | FALSE | Upregulated |
| -0,707591426 | 0,248254567 | RP11-685M7.3 | FALSE | Downregulated |
| 0,311751174 | 0,248363281 | ANXA1 | FALSE | Upregulated |
| -0,975688962 | 0,248384712 | LOC101928803 | FALSE | Downregulated |
| 0,514991972 | 0,248384712 | MAP1B | FALSE | Upregulated |
| -1,036666531 | 0,248433751 | LOC101928123 | FALSE | Downregulated |
| 0,976768723 | 0,248515615 | NKAIN2 | FALSE | Upregulated |
| -0,531115161 | 0,248814935 | LOC102723501 | FALSE | Downregulated |
| -0,45254655 | 0,248841444 | RP5-914P20.5 | FALSE | Downregulated |
| 0,266414795 | 0,248948557 | ALG3 | FALSE | Upregulated |
| -0,191089434 | 0,249036864 | ZNF16 | FALSE | Downregulated |
| 0,806371029 | 0,249097089 | CLEC5A | FALSE | Upregulated |
| -0,255681513 | 0,249097089 | LTBP1 | FALSE | Downregulated |
| 0,485330535 | 0,249201982 | CSTA | FALSE | Upregulated |
| 0,343777949 | 0,249201982 | LPCAT2 | FALSE | Upregulated |
| -1,516509115 | 0,249201982 | SNORA70 | FALSE | Downregulated |
| 0,216057132 | 0,24959836 | RNPEP | FALSE | Upregulated |
| 0,204490776 | 0,249652752 | MRRF | FALSE | Upregulated |
| 0,219184804 | 0,249726805 | SNRPD1 | FALSE | Upregulated |
| -0,293666074 | 0,249745425 | ADK | FALSE | Downregulated |
| -0,705755831 | 0,249753808 | LINC00086 | FALSE | Downregulated |
| 0,156731156 | 0,249846997 | PRMT1 | FALSE | Upregulated |
| -0,178361107 | 0,249854683 | MTMR3 | FALSE | Downregulated |
| -0,298129924 | 0,250562367 | TMEM256 | FALSE | Downregulated |
| -0,316105421 | 0,251025197 | ZNF808 | FALSE | Downregulated |
| 1,63972796 | 0,251124786 | EGOT | FALSE | Upregulated |
| -0,164851425 | 0,251185337 | CCP110 | FALSE | Downregulated |
| -0,403435667 | 0,251185337 | PRKAR2A-AS1 | FALSE | Downregulated |
| -0,132857202 | 0,251313694 | HNRNPD | FALSE | Downregulated |
| 0,16977499 | 0,25137546 | C2orf43 | FALSE | Upregulated |
| 0,173172504 | 0,25139576 | CASK | FALSE | Upregulated |
| 0,252669497 | 0,25139576 | IFT43 | FALSE | Upregulated |
| 0,167332749 | 0,251512002 | TCF3 | FALSE | Upregulated |
| 0,443097682 | 0,251622995 | ARHGAP39 | FALSE | Upregulated |
| 0,239317557 | 0,251724562 | BOK | FALSE | Upregulated |
| 0,215884133 | 0,251724562 | RPS10 | FALSE | Upregulated |

| | | | | |
|---|---|---|---|---|
| -0,122761999 | 0,251724562 | ZDHHC6 | FALSE | Downregulated |
| -0,486768151 | 0,252067679 | PRKG1 | FALSE | Downregulated |
| 0,503379324 | 0,252195196 | MB21D1 | FALSE | Upregulated |
| 0,188647361 | 0,252195196 | MIEN1 | FALSE | Upregulated |
| -0,129289182 | 0,252195196 | RABGGTB | FALSE | Downregulated |
| 0,130517923 | 0,252195196 | SGPL1 | FALSE | Upregulated |
| -0,200623266 | 0,252276034 | ZFYVE16 | FALSE | Downregulated |
| -0,134828154 | 0,252360478 | ELP2 | FALSE | Downregulated |
| 0,393232432 | 0,252508179 | KRBA1 | FALSE | Upregulated |
| 0,124622255 | 0,25258159 | LTA4H | FALSE | Upregulated |
| -0,210854268 | 0,252781525 | C16orf87 | FALSE | Downregulated |
| -0,141936242 | 0,252861592 | KAT7 | FALSE | Downregulated |
| -0,350645428 | 0,25322102 | AGPAT5 | FALSE | Downregulated |
| 1,376048446 | 0,253250209 | LOC102723959 | FALSE | Upregulated |
| 1,013462338 | 0,253502174 | PNMA3 | FALSE | Upregulated |
| -0,251723756 | 0,25355718 | PGBD4 | FALSE | Downregulated |
| -0,269063363 | 0,253594754 | ZNF324 | FALSE | Downregulated |
| -1,289078395 | 0,253735445 | LOC102724230 | FALSE | Downregulated |
| 0,19172965 | 0,253735445 | MKKS | FALSE | Upregulated |
| -0,516706232 | 0,253844685 | LRRC29 | FALSE | Downregulated |
| -0,191768656 | 0,253844685 | PRPSAP1 | FALSE | Downregulated |
| -0,978277428 | 0,253848182 | DPP6 | FALSE | Downregulated |
| 0,131635195 | 0,253859522 | AQR | FALSE | Upregulated |
| -0,346303687 | 0,2539746 | ADAMTS17 | FALSE | Downregulated |
| -0,392753978 | 0,2539746 | PKLR | FALSE | Downregulated |
| -2,106194487 | 0,254141287 | RYR3 | FALSE | Downregulated |
| -0,871170288 | 0,254153794 | CXCR1 | FALSE | Downregulated |
| -1,555977197 | 0,25417816 | LRRC71 | FALSE | Downregulated |
| -0,166483131 | 0,254193178 | VKORC1L1 | FALSE | Downregulated |
| -0,54691855 | 0,254211144 | GOLGA8M | FALSE | Downregulated |
| -0,262913522 | 0,25421925 | ELMOD3 | FALSE | Downregulated |
| -0,856097581 | 0,254237321 | RAB25 | FALSE | Downregulated |
| 0,147814533 | 0,254256388 | ACO2 | FALSE | Upregulated |
| 0,462087881 | 0,254256388 | CYP4F22 | FALSE | Upregulated |
| -1,217723207 | 0,254256388 | WNT7B | FALSE | Downregulated |
| 0,328659924 | 0,254256388 | ZNF35 | FALSE | Upregulated |
| 0,243898168 | 0,25426845 | GCH1 | FALSE | Upregulated |
| -0,360895612 | 0,25426845 | RTTN | FALSE | Downregulated |
| -0,534366978 | 0,254312625 | USP2-AS1 | FALSE | Downregulated |
| 0,891665866 | 0,254325354 | ACHE | FALSE | Upregulated |
| -0,222401831 | 0,254325354 | ANGPTL3 | FALSE | Downregulated |
| -0,240233851 | 0,254325354 | PCED1A | FALSE | Downregulated |
| 1,300807182 | 0,254325354 | RPPH1 | FALSE | Upregulated |
| -0,845942576 | 0,254339969 | RRAD | FALSE | Downregulated |
| -0,299281858 | 0,25452039 | FOXP2 | FALSE | Downregulated |
| 1,046265983 | 0,25452039 | LILRA4 | FALSE | Upregulated |
| -0,615839605 | 0,25452039 | LOC101926956 | FALSE | Downregulated |
| -0,298712558 | 0,25452039 | MRI1 | FALSE | Downregulated |
| -0,141287091 | 0,25452039 | MRPL37 | FALSE | Downregulated |
| 0,234467018 | 0,254607951 | ARRB1 | FALSE | Upregulated |
| -0,487388239 | 0,254674082 | EFCAB13 | FALSE | Downregulated |

| | | | | |
|---|---|---|---|---|
| -0,161877016 | 0,254706067 | LARGE | FALSE | Downregulated |
| -1,186363338 | 0,254706067 | LINC00488 | FALSE | Downregulated |
| 0,133300475 | 0,254706067 | PPIL2 | FALSE | Upregulated |
| -0,155881589 | 0,25477059 | TDRD3 | FALSE | Downregulated |
| 0,128651528 | 0,254835618 | HIF1AN | FALSE | Upregulated |
| -0,265932436 | 0,254894094 | CD36 | FALSE | Downregulated |
| -0,413936095 | 0,254894094 | CDC42EP2 | FALSE | Downregulated |
| 0,28238044 | 0,254894094 | FBXL6 | FALSE | Upregulated |
| -0,199666863 | 0,254894094 | ITPRIPL2 | FALSE | Downregulated |
| -0,832142971 | 0,254894094 | LOC101929065 | FALSE | Downregulated |
| 0,163221813 | 0,254894094 | MAPRE2 | FALSE | Upregulated |
| -0,105308757 | 0,254976683 | NRD1 | FALSE | Downregulated |
| 0,158479921 | 0,254986772 | GAA | FALSE | Upregulated |
| -0,393934805 | 0,25510753 | APBA1 | FALSE | Downregulated |
| -0,226765133 | 0,255275062 | FRAT1 | FALSE | Downregulated |
| -0,799648057 | 0,255275062 | LOC101929506 | FALSE | Downregulated |
| -0,316512753 | 0,255275062 | PEX11A | FALSE | Downregulated |
| 0,19349464 | 0,255276322 | VWA5A | FALSE | Upregulated |
| -0,314408474 | 0,255389833 | C4B | FALSE | Downregulated |
| -0,646468907 | 0,255389833 | NIM1K | FALSE | Downregulated |
| 0,434188533 | 0,255389833 | TBX15 | FALSE | Upregulated |
| 0,427114277 | 0,255474179 | COL4A4 | FALSE | Upregulated |
| -0,192796296 | 0,255474179 | THAP11 | FALSE | Downregulated |
| -0,520219926 | 0,255495109 | LOC101927765 | FALSE | Downregulated |
| -0,174505168 | 0,255503057 | FAM210A | FALSE | Downregulated |
| -0,427218357 | 0,255627303 | GPR182 | FALSE | Downregulated |
| -1,374450411 | 0,255627303 | LOC101930305 | FALSE | Downregulated |
| -0,16170085 | 0,255778855 | CARKD | FALSE | Downregulated |
| -0,498657181 | 0,255868432 | LOC100506083 | FALSE | Downregulated |
| 0,373696546 | 0,255868432 | SLC31A2 | FALSE | Upregulated |
| -0,397515578 | 0,255868432 | ZNF491 | FALSE | Downregulated |
| -0,602268361 | 0,256003198 | KIAA1377 | FALSE | Downregulated |
| -0,379848387 | 0,256229761 | RBM26-AS1 | FALSE | Downregulated |
| -0,163630005 | 0,256292116 | BCDIN3D | FALSE | Downregulated |
| -0,192276883 | 0,256437313 | NFATC3 | FALSE | Downregulated |
| 0,428369472 | 0,256477629 | MYRF | FALSE | Upregulated |
| -0,61173473 | 0,256587721 | CLVS1 | FALSE | Downregulated |
| 1,451999385 | 0,256618306 | TMEM52B | FALSE | Upregulated |
| 0,200166275 | 0,256694437 | LOC102288414 | FALSE | Upregulated |
| 0,174401519 | 0,256742277 | WDR53 | FALSE | Upregulated |
| -0,142704663 | 0,256872081 | HBP1 | FALSE | Downregulated |
| -0,40126264 | 0,256968455 | AC091729.9 | FALSE | Downregulated |
| -0,181321899 | 0,257038749 | C17orf103 | FALSE | Downregulated |
| -0,495821531 | 0,257046455 | AC011747.7 | FALSE | Downregulated |
| -1,14715818 | 0,257046455 | ZNF385D | FALSE | Downregulated |
| 0,61348042 | 0,257074504 | CBR3 | FALSE | Upregulated |
| -0,17133832 | 0,257118742 | ICK | FALSE | Downregulated |
| 0,516098277 | 0,257118742 | PGF | FALSE | Upregulated |
| -0,219659091 | 0,257118742 | POLR2J | FALSE | Downregulated |
| 0,144924001 | 0,257137209 | XPO6 | FALSE | Upregulated |
| -0,378942932 | 0,257268654 | KRTCAP3 | FALSE | Downregulated |

| | | | | |
|---|---|---|---|---|
| 1,061845877 | 0,257269139 | SPNS3 | FALSE | Upregulated |
| -0,489180231 | 0,257389469 | PDZRN4 | FALSE | Downregulated |
| -0,68213538 | 0,257399478 | ESRP1 | FALSE | Downregulated |
| -0,555459828 | 0,257513144 | SLC16A14 | FALSE | Downregulated |
| -0,149474485 | 0,257524717 | SOCS7 | FALSE | Downregulated |
| 0,832309663 | 0,257565096 | CHRNA5 | FALSE | Upregulated |
| -0,188673132 | 0,257580019 | GOLPH3L | FALSE | Downregulated |
| -0,366903735 | 0,257667873 | TCTN2 | FALSE | Downregulated |
| -0,119617378 | 0,257669847 | CLCN3 | FALSE | Downregulated |
| -0,381317291 | 0,257775483 | WASF3 | FALSE | Downregulated |
| 0,434735135 | 0,257779791 | PTPLAD2 | FALSE | Upregulated |
| -0,391760869 | 0,257970562 | FTX | FALSE | Downregulated |
| -0,129392948 | 0,258187595 | SP3 | FALSE | Downregulated |
| 0,22125266 | 0,258327134 | PHF11 | FALSE | Upregulated |
| -0,338601214 | 0,258327134 | PTPN13 | FALSE | Downregulated |
| -0,230196992 | 0,258379068 | ASB3 | FALSE | Downregulated |
| -0,180973973 | 0,258733671 | DDX46 | FALSE | Downregulated |
| -1,1370397 | 0,258874996 | LOC101928344 | FALSE | Downregulated |
| -0,351166785 | 0,25895695 | RP11-513G11.3 | FALSE | Downregulated |
| 0,358078131 | 0,259255585 | PDF | FALSE | Upregulated |
| 0,140201469 | 0,259328772 | UBLCP1 | FALSE | Upregulated |
| -0,549860221 | 0,259467652 | LINC00294 | FALSE | Downregulated |
| 0,20369178 | 0,259548067 | MST4 | FALSE | Upregulated |
| -0,60618009 | 0,259722168 | GLYATL3 | FALSE | Downregulated |
| -0,276525817 | 0,259769352 | B3GNT1 | FALSE | Downregulated |
| -0,306119451 | 0,259769352 | IFNLR1 | FALSE | Downregulated |
| -0,177066005 | 0,260063985 | MDC1 | FALSE | Downregulated |
| 0,162827494 | 0,260063985 | RABGGTA | FALSE | Upregulated |
| 0,763837552 | 0,260081878 | PAK6 | FALSE | Upregulated |
| 0,650908169 | 0,260110203 | LOC102723554 | FALSE | Upregulated |
| -0,136960337 | 0,260110203 | RNF169 | FALSE | Downregulated |
| 0,832862038 | 0,260429665 | AFF2 | FALSE | Upregulated |
| 0,121580884 | 0,260445554 | KDM4A | FALSE | Upregulated |
| -0,793046009 | 0,2604575 | LOC101927838 | FALSE | Downregulated |
| -0,810507816 | 0,260480284 | ADAM23 | FALSE | Downregulated |
| 0,24280734 | 0,26050589 | ZNF513 | FALSE | Upregulated |
| -0,380125473 | 0,260622878 | MRO | FALSE | Downregulated |
| -0,290217749 | 0,260645492 | SYTL2 | FALSE | Downregulated |
| 0,826158834 | 0,2606657 | RERGL | FALSE | Upregulated |
| 0,532992515 | 0,26089059 | GPR82 | FALSE | Upregulated |
| 0,145185896 | 0,261007673 | ATG3 | FALSE | Upregulated |
| -0,43528957 | 0,261055137 | PLAG1 | FALSE | Downregulated |
| 0,695533266 | 0,26112549 | FGF12 | FALSE | Upregulated |
| -0,197817838 | 0,261157806 | TSPAN3 | FALSE | Downregulated |
| -0,13117561 | 0,261247479 | EEF2 | FALSE | Downregulated |
| 0,080468004 | 0,261342152 | IST1 | FALSE | Upregulated |
| 0,420659399 | 0,261342152 | LOXL3 | FALSE | Upregulated |
| -0,271578699 | 0,261362409 | TSPAN33 | FALSE | Downregulated |
| 0,161945577 | 0,261395963 | PRCP | FALSE | Upregulated |
| 0,250510897 | 0,261424827 | LSM7 | FALSE | Upregulated |
| -0,178081251 | 0,261424827 | PTAR1 | FALSE | Downregulated |

| | | | | |
|---|---|---|---|---|
| 0,284278381 | 0,261491778 | RASSF8-AS1 | FALSE | Upregulated |
| -0,317351303 | 0,261559753 | LOC728323 | FALSE | Downregulated |
| 0,084266649 | 0,261569777 | MORF4L1 | FALSE | Upregulated |
| 0,142335503 | 0,261837804 | WDR46 | FALSE | Upregulated |
| 0,190878902 | 0,261837804 | WDR77 | FALSE | Upregulated |
| 0,246645949 | 0,262069301 | EMC9 | FALSE | Upregulated |
| -0,73798112 | 0,262070355 | MAATS1 | FALSE | Downregulated |
| 0,33221081 | 0,262231998 | MIR1181 | FALSE | Upregulated |
| 0,09944104 | 0,262233135 | MRFAP1 | FALSE | Upregulated |
| 1,740469721 | 0,262315023 | KCNQ3 | FALSE | Upregulated |
| 0,916319759 | 0,262315023 | RP11-760N9.1 | FALSE | Upregulated |
| -0,516794363 | 0,262350983 | ZNF503-AS1 | FALSE | Downregulated |
| 0,230404855 | 0,26242324 | PAFAH1B2 | FALSE | Upregulated |
| -0,232830722 | 0,2624348 | BLNK | FALSE | Downregulated |
| 0,859060799 | 0,2624348 | LRFN4 | FALSE | Upregulated |
| -0,296331253 | 0,262536022 | DLGAP1-AS1 | FALSE | Downregulated |
| -0,888062596 | 0,262561663 | LOC102723517 | FALSE | Downregulated |
| 0,14744561 | 0,262561663 | VCPIP1 | FALSE | Upregulated |
| -0,194356505 | 0,262622862 | GPANK1 | FALSE | Downregulated |
| -0,79614722 | 0,262713364 | AC092295.7 | FALSE | Downregulated |
| 0,860538369 | 0,262713364 | FOSB | FALSE | Upregulated |
| 0,237997657 | 0,262713364 | HARS | FALSE | Upregulated |
| 0,546553897 | 0,262713364 | TIMD4 | FALSE | Upregulated |
| -0,406208047 | 0,262713364 | ZNF514 | FALSE | Downregulated |
| -0,288193014 | 0,262753035 | USP49 | FALSE | Downregulated |
| 0,129660726 | 0,262753035 | UTP3 | FALSE | Upregulated |
| 1,217318612 | 0,262837592 | FAM90A1 | FALSE | Upregulated |
| 0,16687606 | 0,263139798 | MEX3C | FALSE | Upregulated |
| 0,648716983 | 0,263153693 | GFPT2 | FALSE | Upregulated |
| 0,267046243 | 0,263362836 | BAZ1A | FALSE | Upregulated |
| -0,221151724 | 0,263362836 | DOPEY1 | FALSE | Downregulated |
| -0,198170614 | 0,263362836 | KCTD6 | FALSE | Downregulated |
| -0,150484269 | 0,263521607 | SPG7 | FALSE | Downregulated |
| -0,335349768 | 0,263663968 | FGG | FALSE | Downregulated |
| 0,300076913 | 0,263780116 | COL6A2 | FALSE | Upregulated |
| -0,720317801 | 0,263817778 | LOC440157 | FALSE | Downregulated |
| -0,700621267 | 0,263951246 | LOC101927330 | FALSE | Downregulated |
| -0,141683504 | 0,263951246 | SAMD4B | FALSE | Downregulated |
| -0,124342528 | 0,263951246 | ZBED1 | FALSE | Downregulated |
| -0,198627413 | 0,263982841 | FAM63A | FALSE | Downregulated |
| -0,880430254 | 0,263982841 | LOC101928774 | FALSE | Downregulated |
| -0,189129789 | 0,264150654 | CHRNB1 | FALSE | Downregulated |
| -0,157352932 | 0,264150654 | CPQ | FALSE | Downregulated |
| -0,164672372 | 0,264156018 | RNF214 | FALSE | Downregulated |
| -0,13184917 | 0,264371556 | BAG1 | FALSE | Downregulated |
| -0,164067467 | 0,264532884 | PEX16 | FALSE | Downregulated |
| -0,141324021 | 0,264660468 | GTF2H5 | FALSE | Downregulated |
| 0,637714237 | 0,264844192 | SBK1 | FALSE | Upregulated |
| 1,8855013 | 0,26513214 | TRNAD17 | FALSE | Upregulated |
| 0,12587989 | 0,265260732 | SNX3 | FALSE | Upregulated |
| -0,376736781 | 0,2652676 | SLC13A3 | FALSE | Downregulated |

| | | | | |
|---|---|---|---|---|
| -0,382194705 | 0,265276916 | ARHGEF37 | FALSE | Downregulated |
| 0,219455743 | 0,265558928 | RDH11 | FALSE | Upregulated |
| -0,303393398 | 0,265650074 | ABCG2 | FALSE | Downregulated |
| 1,416063289 | 0,265854582 | SLC2A14 | FALSE | Upregulated |
| 0,783775009 | 0,266007845 | TREM1 | FALSE | Upregulated |
| 1,547151433 | 0,266024326 | LINC00319 | FALSE | Upregulated |
| 0,334100442 | 0,266112518 | IFIT1 | FALSE | Upregulated |
| -0,244909832 | 0,266123327 | ZBTB8A | FALSE | Downregulated |
| -0,122074649 | 0,266178593 | ASNSD1 | FALSE | Downregulated |
| -0,615358183 | 0,266178593 | COL9A3 | FALSE | Downregulated |
| -0,637647201 | 0,266178593 | KCNAB3 | FALSE | Downregulated |
| -0,386175351 | 0,266178593 | OSBPL6 | FALSE | Downregulated |
| -0,165766386 | 0,266178593 | RPL23 | FALSE | Downregulated |
| -0,167506202 | 0,266178593 | TBC1D17 | FALSE | Downregulated |
| -0,26688835 | 0,266178593 | YPEL1 | FALSE | Downregulated |
| 0,813803143 | 0,266338831 | ATP1A3 | FALSE | Upregulated |
| -0,602744111 | 0,266358926 | NOL4 | FALSE | Downregulated |
| 0,15732706 | 0,266358926 | SF3B14 | FALSE | Upregulated |
| -0,123743967 | 0,26640509 | ZNF777 | FALSE | Downregulated |
| 0,150949601 | 0,266436071 | MRPL28 | FALSE | Upregulated |
| -1,335470254 | 0,266506574 | TTC23L | FALSE | Downregulated |
| -0,503429295 | 0,266673279 | IL17RE | FALSE | Downregulated |
| 0,888827743 | 0,266703447 | CIB2 | FALSE | Upregulated |
| 0,284208741 | 0,266730958 | PAG1 | FALSE | Upregulated |
| -0,328910449 | 0,266730958 | TMEM82 | FALSE | Downregulated |
| -0,437761123 | 0,266733349 | PRICKLE1 | FALSE | Downregulated |
| 0,767335212 | 0,266735959 | LOC102724217 | FALSE | Upregulated |
| 0,215839916 | 0,266962765 | CAV2 | FALSE | Upregulated |
| 0,338940967 | 0,266962765 | IRF7 | FALSE | Upregulated |
| -1,0080112 | 0,266962765 | LOC101929206 | FALSE | Downregulated |
| 0,229031913 | 0,266962765 | SPATA2L | FALSE | Upregulated |
| 0,243531878 | 0,266962765 | TPM4 | FALSE | Upregulated |
| -0,238811405 | 0,266967512 | CAMKMT | FALSE | Downregulated |
| -0,200842365 | 0,266983383 | C1R | FALSE | Downregulated |
| -0,198234188 | 0,267169139 | CHD6 | FALSE | Downregulated |
| 0,204319383 | 0,267169139 | MDH2 | FALSE | Upregulated |
| 0,221701533 | 0,267169139 | PJA1 | FALSE | Upregulated |
| 0,515612879 | 0,267172029 | KCNMB1 | FALSE | Upregulated |
| -0,449183121 | 0,267172029 | LOC100996433 | FALSE | Downregulated |
| -0,308611768 | 0,267447691 | MDP1 | FALSE | Downregulated |
| 1,365210201 | 0,267474212 | SLC44A4 | FALSE | Upregulated |
| -1,14167303 | 0,267507652 | GOLGA8H | FALSE | Downregulated |
| -0,185899306 | 0,267593522 | SWI5 | FALSE | Downregulated |
| 0,315584585 | 0,267723634 | FAM173A | FALSE | Upregulated |
| -0,203665932 | 0,267723634 | MCFD2 | FALSE | Downregulated |
| -0,230041271 | 0,267857513 | STX16 | FALSE | Downregulated |
| -1,193501612 | 0,26797675 | LOC102724281 | FALSE | Downregulated |
| -0,775402753 | 0,267990566 | NDUFA6-AS1 | FALSE | Downregulated |
| 0,343606747 | 0,26801545 | SLC25A15 | FALSE | Upregulated |
| -0,225352101 | 0,268190904 | PIBF1 | FALSE | Downregulated |
| -0,122231285 | 0,268190904 | TARS2 | FALSE | Downregulated |

| | | | | |
|---|---|---|---|---|
| -0,178646537 | 0,268193694 | CTDSPL | FALSE | Downregulated |
| 0,127984233 | 0,268193694 | PPP2R5E | FALSE | Upregulated |
| 1,097369064 | 0,268282169 | LINC00539 | FALSE | Upregulated |
| 0,16635771 | 0,268282408 | AP3B1 | FALSE | Upregulated |
| -0,176228277 | 0,268282408 | ITGA1 | FALSE | Downregulated |
| 0,723967086 | 0,268294613 | B3GNT3 | FALSE | Upregulated |
| 0,12882647 | 0,268294613 | EIF3G | FALSE | Upregulated |
| -0,313433826 | 0,268377199 | CLEC1B | FALSE | Downregulated |
| -0,825884334 | 0,268487908 | PEG10 | FALSE | Downregulated |
| 1,067989464 | 0,268569259 | LOC101928621 | FALSE | Upregulated |
| 0,80663881 | 0,268569259 | TLR9 | FALSE | Upregulated |
| -0,266746241 | 0,268569259 | TNK2 | FALSE | Downregulated |
| -0,164589751 | 0,268774212 | SBNO1 | FALSE | Downregulated |
| 1,39583634 | 0,268774212 | SLC27A6 | FALSE | Upregulated |
| -0,263780984 | 0,268774212 | TMEM176A | FALSE | Downregulated |
| -0,428418535 | 0,26883954 | FAM186B | FALSE | Downregulated |
| -0,133893631 | 0,268862495 | ZMYM2 | FALSE | Downregulated |
| 0,447638502 | 0,269104719 | GRAMD1B | FALSE | Upregulated |
| 0,187931568 | 0,269160527 | FADD | FALSE | Upregulated |
| -0,196575729 | 0,269216853 | TTC28 | FALSE | Downregulated |
| -0,470742844 | 0,269318354 | LOC101927989 | FALSE | Downregulated |
| -0,50854998 | 0,269333465 | PPM1E | FALSE | Downregulated |
| 0,429264468 | 0,269448543 | LDLR | FALSE | Upregulated |
| -0,2718353 | 0,269448543 | MYO9A | FALSE | Downregulated |
| 1,435021866 | 0,269450706 | FRMPD3 | FALSE | Upregulated |
| 1,044150786 | 0,269618473 | HIST1H2BJ | FALSE | Upregulated |
| -0,847054914 | 0,26966795 | GOLGA8I | FALSE | Downregulated |
| 0,711221376 | 0,269848044 | SEPT3 | FALSE | Upregulated |
| 0,117043024 | 0,27003581 | STIM2 | FALSE | Upregulated |
| -0,461851061 | 0,270095234 | LOC100379224 | FALSE | Downregulated |
| 0,160555634 | 0,270161948 | DDX5 | FALSE | Upregulated |
| -0,584983275 | 0,270161948 | DNAH11 | FALSE | Downregulated |
| -1,534200708 | 0,270208525 | GSTM2P1 | FALSE | Downregulated |
| -0,64231433 | 0,270296879 | TTLL3 | FALSE | Downregulated |
| 0,136635097 | 0,270468174 | ASCC2 | FALSE | Upregulated |
| -0,22798333 | 0,27056497 | PPM1H | FALSE | Downregulated |
| -0,284883489 | 0,270586236 | ARL6IP6 | FALSE | Downregulated |
| 0,285032016 | 0,270589398 | C17orf89 | FALSE | Upregulated |
| -0,161286563 | 0,270589398 | MTM1 | FALSE | Downregulated |
| 0,53192509 | 0,270589398 | STMN3 | FALSE | Upregulated |
| 0,368747587 | 0,270756235 | NRTN | FALSE | Upregulated |
| 0,406046311 | 0,271099212 | ADAMTSL5 | FALSE | Upregulated |
| 0,206071579 | 0,271176691 | TIMM17B | FALSE | Upregulated |
| 0,352693399 | 0,271245952 | DLL1 | FALSE | Upregulated |
| -0,525344526 | 0,271245952 | LOC653513 | FALSE | Downregulated |
| -0,365081682 | 0,271363622 | LIPJ | FALSE | Downregulated |
| -0,109817015 | 0,271517221 | ATXN1L | FALSE | Downregulated |
| -0,159281337 | 0,271629219 | KBTBD6 | FALSE | Downregulated |
| 0,816424018 | 0,271633816 | KRT7 | FALSE | Upregulated |
| 0,17569485 | 0,272117754 | AKR7A2 | FALSE | Upregulated |
| 0,515456285 | 0,272284118 | FJX1 | FALSE | Upregulated |

| | | | | |
|---|---|---|---|---|
| 0,116132494 | 0,272465388 | BAG6 | FALSE | Upregulated |
| -0,150965947 | 0,272465388 | LOC101928527 | FALSE | Downregulated |
| -0,117411025 | 0,272465388 | PAFAH2 | FALSE | Downregulated |
| -0,564885502 | 0,272465388 | WDR65 | FALSE | Downregulated |
| -0,327755419 | 0,272570257 | HOXB2 | FALSE | Downregulated |
| 0,322756872 | 0,27274258 | C1orf85 | FALSE | Upregulated |
| -0,253361953 | 0,27274258 | GAS2 | FALSE | Downregulated |
| 1,705375788 | 0,272745493 | LOC102724246 | FALSE | Upregulated |
| 0,180629872 | 0,272745493 | MRPS28 | FALSE | Upregulated |
| 0,224027217 | 0,272811726 | FGD6 | FALSE | Upregulated |
| 0,326486698 | 0,27285709 | SHROOM1 | FALSE | Upregulated |
| 0,156825244 | 0,272979941 | SLC25A39 | FALSE | Upregulated |
| -0,521975739 | 0,273029794 | LOC441124 | FALSE | Downregulated |
| 1,894046066 | 0,273075622 | KRT17 | FALSE | Upregulated |
| -0,187069454 | 0,273075622 | PRDM2 | FALSE | Downregulated |
| -0,700139591 | 0,273407453 | MAGOH2 | FALSE | Downregulated |
| -0,786538714 | 0,273407453 | PKDREJ | FALSE | Downregulated |
| -0,28972101 | 0,273407852 | AFF3 | FALSE | Downregulated |
| -0,571094041 | 0,273412858 | LINC01176 | FALSE | Downregulated |
| -0,175830968 | 0,273451743 | C16orf52 | FALSE | Downregulated |
| -1,545374292 | 0,273451743 | CA3 | FALSE | Downregulated |
| -0,3772995 | 0,273451743 | CCDC152 | FALSE | Downregulated |
| 0,340668425 | 0,273451743 | PRR5L | FALSE | Upregulated |
| 0,153682624 | 0,273451743 | SAMM50 | FALSE | Upregulated |
| -1,227335716 | 0,273521476 | RNF138P1 | FALSE | Downregulated |
| 0,17303578 | 0,273624272 | FARSB | FALSE | Upregulated |
| 0,209964069 | 0,273624272 | PFKFB2 | FALSE | Upregulated |
| -0,388347481 | 0,273747985 | DLG2 | FALSE | Downregulated |
| -1,958369581 | 0,273747985 | LOC100996716 | FALSE | Downregulated |
| 0,17483691 | 0,273747985 | SHQ1 | FALSE | Upregulated |
| -1,464060912 | 0,273747985 | SYT4 | FALSE | Downregulated |
| 0,122373183 | 0,273793062 | C5orf15 | FALSE | Upregulated |
| -0,893322483 | 0,273793062 | LOC102723866 | FALSE | Downregulated |
| 0,220191002 | 0,273793062 | PSMG4 | FALSE | Upregulated |
| -0,470053176 | 0,273793062 | SLC24A3 | FALSE | Downregulated |
| 2,080666709 | 0,273852647 | AGR2 | FALSE | Upregulated |
| -0,282883043 | 0,27387281 | OGT | FALSE | Downregulated |
| -0,673943243 | 0,273878417 | LOC100507460 | FALSE | Downregulated |
| 0,122301554 | 0,274296836 | UEVLD | FALSE | Upregulated |
| -0,493840352 | 0,274457551 | PAX8 | FALSE | Downregulated |
| 0,474516102 | 0,274675658 | FOXD2 | FALSE | Upregulated |
| -0,355367818 | 0,274778871 | C19orf54 | FALSE | Downregulated |
| -0,187385388 | 0,274778871 | CASP8AP2 | FALSE | Downregulated |
| 0,449218929 | 0,274778871 | FDPS | FALSE | Upregulated |
| -0,430919239 | 0,274778871 | KIAA0895L | FALSE | Downregulated |
| -0,162441328 | 0,274778871 | PCCA | FALSE | Downregulated |
| -0,918138481 | 0,274778871 | RHPN1-AS1 | FALSE | Downregulated |
| -0,568332342 | 0,274806596 | SNRK-AS1 | FALSE | Downregulated |
| 0,545111763 | 0,27496784 | TNC | FALSE | Upregulated |
| -0,248728808 | 0,275027373 | TOB2 | FALSE | Downregulated |
| -0,518162567 | 0,275031452 | MMRN1 | FALSE | Downregulated |

| | | | | |
|---|---|---|---|---|
| -0,47005728 | 0,275031452 | WASH5P | FALSE | Downregulated |
| -0,941693929 | 0,275053767 | LAMA1 | FALSE | Downregulated |
| -0,144146156 | 0,275417517 | CCDC43 | FALSE | Downregulated |
| -0,159252716 | 0,275487947 | FAM206A | FALSE | Downregulated |
| 0,149490062 | 0,275511243 | HFE | FALSE | Upregulated |
| 1,344588492 | 0,275516225 | KDM5D | FALSE | Upregulated |
| 0,14235283 | 0,275636145 | CBY1 | FALSE | Upregulated |
| 0,123867515 | 0,275636145 | NUP54 | FALSE | Upregulated |
| -1,340006252 | 0,275650985 | KSR2 | FALSE | Downregulated |
| -0,326092746 | 0,275650985 | POU6F1 | FALSE | Downregulated |
| 0,800836122 | 0,275650985 | TMEM257 | FALSE | Upregulated |
| -0,136645074 | 0,275650985 | ZNF277 | FALSE | Downregulated |
| 0,329356351 | 0,275802186 | GMNC | FALSE | Upregulated |
| -1,268085139 | 0,275886072 | LOC102723917 | FALSE | Downregulated |
| -0,183050244 | 0,275929566 | KIAA1731 | FALSE | Downregulated |
| -0,199587914 | 0,275975821 | TULP3 | FALSE | Downregulated |
| -0,274824688 | 0,276087546 | LOC101928303 | FALSE | Downregulated |
| -0,236008534 | 0,276087546 | RNF180 | FALSE | Downregulated |
| 0,160611016 | 0,276087546 | TYK2 | FALSE | Upregulated |
| -0,777011054 | 0,27610412 | LOC101928182 | FALSE | Downregulated |
| -1,124800689 | 0,276198231 | CLDN22 | FALSE | Downregulated |
| 0,477212052 | 0,276216233 | RPL13P5 | FALSE | Upregulated |
| -0,154832719 | 0,276316333 | TRAP1 | FALSE | Downregulated |
| 0,888918354 | 0,276402539 | ELF5 | FALSE | Upregulated |
| 0,196245552 | 0,276445064 | PTTG1IP | FALSE | Upregulated |
| 0,318529522 | 0,276631464 | ACOT12 | FALSE | Upregulated |
| -0,341898098 | 0,276728553 | PNN | FALSE | Downregulated |
| -0,199039107 | 0,276770797 | CDAN1 | FALSE | Downregulated |
| -0,266077806 | 0,276877991 | TVP23C-CDRT4 | FALSE | Downregulated |
| -1,473468994 | 0,277013483 | SPRN | FALSE | Downregulated |
| 0,204007824 | 0,277013483 | TRAPPC5 | FALSE | Upregulated |
| 0,549823835 | 0,277650341 | TMEM249 | FALSE | Upregulated |
| 0,313247518 | 0,277667522 | ECSCR | FALSE | Upregulated |
| -0,25598555 | 0,277948583 | ZNF470 | FALSE | Downregulated |
| 0,169767186 | 0,278019538 | ACY1 | FALSE | Upregulated |
| 1,376182171 | 0,27820891 | FAM72A | FALSE | Upregulated |
| -1,361120204 | 0,27822062 | ANKRD20A12P | FALSE | Downregulated |
| 1,178418591 | 0,278561875 | LRRIQ1 | FALSE | Upregulated |
| 0,157661073 | 0,278599929 | TFG | FALSE | Upregulated |
| -0,479264592 | 0,278630784 | LOC101929427 | FALSE | Downregulated |
| 0,114146494 | 0,278645359 | TSPYL1 | FALSE | Upregulated |
| -0,760072179 | 0,279055636 | HRC | FALSE | Downregulated |
| -0,200251936 | 0,27905722 | RPS4X | FALSE | Downregulated |
| -0,261111428 | 0,279126029 | SP4 | FALSE | Downregulated |
| 0,474841268 | 0,279154615 | CGREF1 | FALSE | Upregulated |
| 1,021102324 | 0,279237709 | LYPD2 | FALSE | Upregulated |
| 0,749309438 | 0,279300501 | CLIC6 | FALSE | Upregulated |
| 0,168317408 | 0,279394658 | ADCY9 | FALSE | Upregulated |
| -0,225524645 | 0,279394658 | TCEA3 | FALSE | Downregulated |
| -0,253712114 | 0,279406507 | PMEL | FALSE | Downregulated |
| 0,434325886 | 0,279459591 | ZBTB7C | FALSE | Upregulated |

| | | | | |
|---|---|---|---|---|
| -0,390901973 | 0,279583232 | ZNF692 | FALSE | Downregulated |
| -1,141610121 | 0,279648273 | TEKT3 | FALSE | Downregulated |
| 0,708552946 | 0,279691743 | MDGA1 | FALSE | Upregulated |
| -0,70149739 | 0,279691743 | STAC | FALSE | Downregulated |
| 0,276769116 | 0,27982226 | PPAPDC1B | FALSE | Upregulated |
| 1,007713348 | 0,279969934 | OPCML | FALSE | Upregulated |
| 0,313098519 | 0,280072687 | NT5DC2 | FALSE | Upregulated |
| -0,151770866 | 0,280108664 | PDCD11 | FALSE | Downregulated |
| -0,284274261 | 0,280108664 | TRIP10 | FALSE | Downregulated |
| -0,14788401 | 0,28042438 | SF3B1 | FALSE | Downregulated |
| 0,314798544 | 0,280471668 | RPS21 | FALSE | Upregulated |
| -2,11785898 | 0,280471668 | STRA6 | FALSE | Downregulated |
| 0,14071151 | 0,28048333 | REEP3 | FALSE | Upregulated |
| 0,993615288 | 0,280509434 | LIPH | FALSE | Upregulated |
| 0,413275297 | 0,280592845 | MYBL1 | FALSE | Upregulated |
| -0,318265258 | 0,280592845 | NPIPA1 | FALSE | Downregulated |
| -0,658062142 | 0,280652003 | LOC102724766 | FALSE | Downregulated |
| 0,169325515 | 0,280800818 | SNRPA1 | FALSE | Upregulated |
| -0,138780419 | 0,280834523 | PAXIP1 | FALSE | Downregulated |
| 0,299810867 | 0,281048976 | SIGLEC11 | FALSE | Upregulated |
| -0,194416853 | 0,281075915 | RHPN2 | FALSE | Downregulated |
| -0,397945767 | 0,28127734 | GJC3 | FALSE | Downregulated |
| 0,234555533 | 0,281456159 | H2AFJ | FALSE | Upregulated |
| -0,56748663 | 0,281681367 | LRRTM2 | FALSE | Downregulated |
| -0,116936055 | 0,281704505 | MAPK9 | FALSE | Downregulated |
| 0,835416011 | 0,281704505 | RP11-861A13.4 | FALSE | Upregulated |
| 0,156682821 | 0,281813197 | C4orf27 | FALSE | Upregulated |
| -0,601150646 | 0,281813197 | LINC00933 | FALSE | Downregulated |
| -0,26212554 | 0,281874234 | NT5E | FALSE | Downregulated |
| 0,468943962 | 0,281880948 | C7 | FALSE | Upregulated |
| -0,548205774 | 0,281946259 | SPACA6P | FALSE | Downregulated |
| -0,238812069 | 0,28203318 | RAB37 | FALSE | Downregulated |
| 0,207300198 | 0,282237692 | ALG14 | FALSE | Upregulated |
| 0,211733854 | 0,282295559 | SLC15A1 | FALSE | Upregulated |
| -0,89927749 | 0,282297338 | LINC01126 | FALSE | Downregulated |
| 0,25552768 | 0,282535841 | RANGRF | FALSE | Upregulated |
| 0,27802369 | 0,282619787 | LLGL1 | FALSE | Upregulated |
| -0,252181458 | 0,282619787 | LRIG2 | FALSE | Downregulated |
| 0,132138414 | 0,282667292 | FBXO22 | FALSE | Upregulated |
| -0,34715933 | 0,282667292 | ZNF788 | FALSE | Downregulated |
| 0,234329899 | 0,282714558 | IQCG | FALSE | Upregulated |
| 1,241627591 | 0,282714558 | MIR635 | FALSE | Upregulated |
| -0,504219337 | 0,282726696 | BAI3 | FALSE | Downregulated |
| -0,201412648 | 0,282763821 | MIB2 | FALSE | Downregulated |
| -0,292763484 | 0,282763821 | NINL | FALSE | Downregulated |
| -0,281711198 | 0,282784967 | CROT | FALSE | Downregulated |
| 0,25493753 | 0,282784967 | IFI27L1 | FALSE | Upregulated |
| 0,17580359 | 0,282784967 | KIF22 | FALSE | Upregulated |
| 0,219304123 | 0,283214342 | CEP89 | FALSE | Upregulated |
| 0,188127285 | 0,283214342 | GSE1 | FALSE | Upregulated |
| 1,467917752 | 0,283214342 | SIGLEC12 | FALSE | Upregulated |

| | | | | |
|---|---|---|---|---|
| -0,531432937 | 0,283318525 | AC074289.1 | FALSE | Downregulated |
| -0,11677239 | 0,283318525 | GOLGA1 | FALSE | Downregulated |
| -0,142190343 | 0,283318525 | VPS13D | FALSE | Downregulated |
| -0,864552512 | 0,283382329 | LRRC37A11P | FALSE | Downregulated |
| -0,185822398 | 0,283382329 | PHF6 | FALSE | Downregulated |
| 0,448393059 | 0,283382329 | SIPA1L2 | FALSE | Upregulated |
| 0,256260138 | 0,28342964 | USP27X | FALSE | Upregulated |
| -0,1727377 | 0,283676941 | CEP57L1 | FALSE | Downregulated |
| 0,191349652 | 0,283726551 | APAF1 | FALSE | Upregulated |
| -0,155359941 | 0,283726551 | HAUS3 | FALSE | Downregulated |
| 0,145112449 | 0,283726551 | MAEA | FALSE | Upregulated |
| -0,378515923 | 0,283726551 | RP11-3P17.4 | FALSE | Downregulated |
| -0,592535686 | 0,283805199 | RP3-400B16.1 | FALSE | Downregulated |
| -0,392145309 | 0,283841571 | EML5 | FALSE | Downregulated |
| 0,113285048 | 0,283927542 | TSTD2 | FALSE | Upregulated |
| -0,266676428 | 0,284004696 | NAT6 | FALSE | Downregulated |
| -0,341741971 | 0,284015356 | PID1 | FALSE | Downregulated |
| -0,698835147 | 0,284038886 | FLJ38717 | FALSE | Downregulated |
| 0,269939061 | 0,284038886 | STX3 | FALSE | Upregulated |
| 0,278732944 | 0,284172353 | ACTG1 | FALSE | Upregulated |
| 0,159775142 | 0,284172353 | MTMR9 | FALSE | Upregulated |
| 0,143097047 | 0,284172353 | NPM1 | FALSE | Upregulated |
| 2,158848868 | 0,284200091 | ADORA2A | FALSE | Upregulated |
| 0,177570491 | 0,284240258 | TOMM6 | FALSE | Upregulated |
| 0,300229286 | 0,28451105 | LRRC8C | FALSE | Upregulated |
| -0,528961737 | 0,284589788 | FGF23 | FALSE | Downregulated |
| 0,350926769 | 0,28469443 | HHIPL1 | FALSE | Upregulated |
| 0,62388611 | 0,284709773 | EMP1 | FALSE | Upregulated |
| 0,816066841 | 0,284709773 | ROS1 | FALSE | Upregulated |
| 0,965198605 | 0,284993707 | KCNA6 | FALSE | Upregulated |
| -0,27132476 | 0,285040833 | TMX3 | FALSE | Downregulated |
| -0,158096897 | 0,285040833 | ZNF44 | FALSE | Downregulated |
| -0,094922671 | 0,285198981 | ARL14EP | FALSE | Downregulated |
| -1,422964973 | 0,285198981 | LOC100271836 | FALSE | Downregulated |
| 0,197919669 | 0,285198981 | LYPLAL1 | FALSE | Upregulated |
| -0,292838283 | 0,285198981 | SLC7A2 | FALSE | Downregulated |
| 0,136165535 | 0,285198981 | WDFY1 | FALSE | Upregulated |
| -0,565931889 | 0,285267638 | METRN | FALSE | Downregulated |
| 0,199802149 | 0,285327041 | WDR13 | FALSE | Upregulated |
| 0,271523968 | 0,285366656 | PAOX | FALSE | Upregulated |
| 0,149463365 | 0,285442679 | DAGLB | FALSE | Upregulated |
| -0,078536821 | 0,285442679 | RAB5B | FALSE | Downregulated |
| 0,14752195 | 0,285463426 | PMS2P1 | FALSE | Upregulated |
| -0,674785648 | 0,285468999 | LOC101929289 | FALSE | Downregulated |
| -0,26000597 | 0,285619414 | RGAG4 | FALSE | Downregulated |
| -0,328913037 | 0,285706802 | ADAM1A | FALSE | Downregulated |
| -0,332299171 | 0,285706802 | CHN2 | FALSE | Downregulated |
| -0,478921215 | 0,285706802 | LINC01106 | FALSE | Downregulated |
| -0,410952487 | 0,285706802 | PRORSD1P | FALSE | Downregulated |
| -0,226402625 | 0,285771575 | KPNA5 | FALSE | Downregulated |
| -0,36331184 | 0,285771575 | RP11-119F19.2 | FALSE | Downregulated |

| | | | | |
|---|---|---|---|---|
| -0,164462575 | 0,285986457 | BLMH | FALSE | Downregulated |
| -1,543825264 | 0,286106237 | LOC101928668 | FALSE | Downregulated |
| 1,141312054 | 0,286156486 | PRC1-AS1 | FALSE | Upregulated |
| 0,20415832 | 0,286277687 | DIO1 | FALSE | Upregulated |
| 0,10607199 | 0,286304679 | AP3M1 | FALSE | Upregulated |
| -0,652521548 | 0,286498079 | LPA | FALSE | Downregulated |
| -0,402263532 | 0,286498079 | MAPK8IP3 | FALSE | Downregulated |
| -0,411487438 | 0,286566283 | LOC101929130 | FALSE | Downregulated |
| 0,278643598 | 0,286616435 | ANGPTL2 | FALSE | Upregulated |
| 0,66984088 | 0,286616435 | MEIS3 | FALSE | Upregulated |
| -0,539172233 | 0,286637959 | PRSS53 | FALSE | Downregulated |
| -0,144941043 | 0,286661648 | ZNF441 | FALSE | Downregulated |
| -0,574923635 | 0,286857877 | EFS | FALSE | Downregulated |
| 0,136598622 | 0,287047737 | TBCD | FALSE | Upregulated |
| -0,831907668 | 0,287173483 | OGN | FALSE | Downregulated |
| -0,170917086 | 0,28719303 | COX15 | FALSE | Downregulated |
| 0,103148473 | 0,28719303 | RABL3 | FALSE | Upregulated |
| 0,190226592 | 0,2874727 | C14orf2 | FALSE | Upregulated |
| 0,17139656 | 0,287569047 | IARS | FALSE | Upregulated |
| -0,314460788 | 0,287573278 | LOC100507258 | FALSE | Downregulated |
| 0,676834409 | 0,287623761 | COL15A1 | FALSE | Upregulated |
| -0,112476803 | 0,287669959 | CCDC117 | FALSE | Downregulated |
| 1,38908546 | 0,287669959 | PCSK1N | FALSE | Upregulated |
| 0,490775247 | 0,287699256 | LOC101929758 | FALSE | Upregulated |
| 0,237976375 | 0,287708784 | MAP3K14 | FALSE | Upregulated |
| 0,86984877 | 0,287802523 | CTAGE8 | FALSE | Upregulated |
| -0,199869858 | 0,287802523 | POLI | FALSE | Downregulated |
| 0,167820803 | 0,287802523 | POLR2H | FALSE | Upregulated |
| -0,512650021 | 0,287819656 | CRHR1-IT1 | FALSE | Downregulated |
| -0,271810473 | 0,287819656 | FZD8 | FALSE | Downregulated |
| -1,916621156 | 0,287857387 | PLP1 | FALSE | Downregulated |
| -0,342912894 | 0,287865306 | SYNGR1 | FALSE | Downregulated |
| 0,976574494 | 0,288342458 | PDE6C | FALSE | Upregulated |
| -1,410958468 | 0,288431281 | CUZD1 | FALSE | Downregulated |
| -0,147626924 | 0,288922007 | HP1BP3 | FALSE | Downregulated |
| -0,391887174 | 0,288949675 | C14orf37 | FALSE | Downregulated |
| 0,179093115 | 0,288977604 | PPM1D | FALSE | Upregulated |
| -0,298153215 | 0,289273678 | LOC101060181 | FALSE | Downregulated |
| -0,23343406 | 0,289273678 | ZNF485 | FALSE | Downregulated |
| -0,446188256 | 0,289281576 | ERRFI1 | FALSE | Downregulated |
| 0,201239419 | 0,289281576 | UGP2 | FALSE | Upregulated |
| -0,118359937 | 0,289505967 | ZNF629 | FALSE | Downregulated |
| -0,207470684 | 0,289658645 | C4BPA | FALSE | Downregulated |
| 0,093511374 | 0,289658645 | DHX9 | FALSE | Upregulated |
| -0,3395377 | 0,289658645 | FILIP1 | FALSE | Downregulated |
| -0,376540253 | 0,289658645 | GSTA1 | FALSE | Downregulated |
| -0,697966483 | 0,289658645 | LOC101060553 | FALSE | Downregulated |
| 0,31557715 | 0,289658645 | VSIG10L | FALSE | Upregulated |
| -1,133912042 | 0,289774491 | SNORD10 | FALSE | Downregulated |
| 0,205388031 | 0,290163437 | RRP9 | FALSE | Upregulated |
| -0,386854565 | 0,290329149 | CYP4X1 | FALSE | Downregulated |

| | | | | |
|---|---|---|---|---|
| -0,110417065 | 0,290420951 | ARFGAP2 | FALSE | Downregulated |
| 0,093221942 | 0,290547631 | VPS37A | FALSE | Upregulated |
| 0,183824219 | 0,290747336 | PARD3B | FALSE | Upregulated |
| 0,172003358 | 0,290820466 | ARL6IP5 | FALSE | Upregulated |
| 0,181501107 | 0,290837401 | TCF7L2 | FALSE | Upregulated |
| 0,522083699 | 0,290922327 | FCRL3 | FALSE | Upregulated |
| -1,429395725 | 0,291041876 | CSDC2 | FALSE | Downregulated |
| -1,319495784 | 0,291059198 | CPA4 | FALSE | Downregulated |
| 0,163619518 | 0,291059198 | KDELR2 | FALSE | Upregulated |
| -0,136746477 | 0,291065552 | SAP30L | FALSE | Downregulated |
| -0,112032785 | 0,29115773 | SUMO1 | FALSE | Downregulated |
| -0,163986299 | 0,291209155 | FANCF | FALSE | Downregulated |
| -0,818571487 | 0,291282589 | MAMDC2 | FALSE | Downregulated |
| -0,298428645 | 0,29134203 | SEC14L4 | FALSE | Downregulated |
| 0,132627745 | 0,291432401 | TMEM33 | FALSE | Upregulated |
| -0,482241604 | 0,291432401 | ZNF610 | FALSE | Downregulated |
| -0,199162793 | 0,291664661 | ZNF329 | FALSE | Downregulated |
| 1,295237419 | 0,291727188 | CAPN11 | FALSE | Upregulated |
| -0,207610877 | 0,291846575 | EPT1 | FALSE | Downregulated |
| -0,295102669 | 0,291846575 | LOC652276 | FALSE | Downregulated |
| -0,148387549 | 0,291846575 | YES1 | FALSE | Downregulated |
| -1,154985114 | 0,291960588 | LOC100506497 | FALSE | Downregulated |
| -0,290794488 | 0,292140159 | CCNL1 | FALSE | Downregulated |
| -0,136120739 | 0,29216031 | STAU1 | FALSE | Downregulated |
| -0,135416197 | 0,292180858 | FAM120B | FALSE | Downregulated |
| 0,28883198 | 0,29243284 | FAR1 | FALSE | Upregulated |
| 0,354376527 | 0,29243284 | KITLG | FALSE | Upregulated |
| -1,101338506 | 0,29243284 | LOC101928624 | FALSE | Downregulated |
| -0,942706868 | 0,292447605 | LOC101927885 | FALSE | Downregulated |
| -0,255004943 | 0,292590205 | NRBP2 | FALSE | Downregulated |
| -0,470730453 | 0,292702871 | GDPD1 | FALSE | Downregulated |
| 1,250792982 | 0,292702871 | KRTAP5-AS1 | FALSE | Upregulated |
| 0,291827342 | 0,292702871 | MYO5C | FALSE | Upregulated |
| 0,995219666 | 0,292922863 | ANGPTL7 | FALSE | Upregulated |
| 0,162017948 | 0,292922863 | DOLPP1 | FALSE | Upregulated |
| -0,480162063 | 0,292946103 | CTNND2 | FALSE | Downregulated |
| -0,190325712 | 0,292970018 | USP15 | FALSE | Downregulated |
| 0,216645301 | 0,293215619 | APOO | FALSE | Upregulated |
| 0,326352079 | 0,293215619 | FAM46C | FALSE | Upregulated |
| -1,571520071 | 0,293421676 | CABP1 | FALSE | Downregulated |
| 0,198822783 | 0,293434418 | CCDC77 | FALSE | Upregulated |
| -0,444931833 | 0,29364507 | ADAMTSL1 | FALSE | Downregulated |
| 0,135800866 | 0,293679083 | HN1L | FALSE | Upregulated |
| -0,166374118 | 0,293709092 | NT5C3B | FALSE | Downregulated |
| 0,516252984 | 0,293709092 | PLCXD3 | FALSE | Upregulated |
| 0,9220038 | 0,293820862 | HSP90AB4P | FALSE | Upregulated |
| 0,122109836 | 0,29391025 | SF3B3 | FALSE | Upregulated |
| -0,240348897 | 0,293945053 | D2HGDH | FALSE | Downregulated |
| -0,413876522 | 0,293945053 | KLHL3 | FALSE | Downregulated |
| -0,452574969 | 0,293945053 | ZFP37 | FALSE | Downregulated |
| -0,147248223 | 0,293964075 | HDAC3 | FALSE | Downregulated |

| | | | | |
|---|---|---|---|---|
| -0,360466043 | 0,293964075 | LUC7L3 | FALSE | Downregulated |
| 0,332341844 | 0,293964075 | TRIM62 | FALSE | Upregulated |
| 0,198572609 | 0,294008865 | ARAP2 | FALSE | Upregulated |
| -0,707443179 | 0,294008865 | GDPD4 | FALSE | Downregulated |
| 0,261847755 | 0,294008865 | STK11IP | FALSE | Upregulated |
| 0,79389334 | 0,294082713 | AC004019.13 | FALSE | Upregulated |
| -0,840751566 | 0,294140421 | LINC00940 | FALSE | Downregulated |
| 0,439306731 | 0,294140421 | LOXL2 | FALSE | Upregulated |
| -0,189299473 | 0,294140421 | POLR3H | FALSE | Downregulated |
| 1,555516605 | 0,29414906 | FCGR1C | FALSE | Upregulated |
| -0,114880119 | 0,294172879 | C9orf78 | FALSE | Downregulated |
| -0,469647142 | 0,294363205 | KCNJ15 | FALSE | Downregulated |
| -0,467586671 | 0,294482177 | ASPRV1 | FALSE | Downregulated |
| -0,479133094 | 0,294482177 | SNHG18 | FALSE | Downregulated |
| 0,249184856 | 0,294662738 | FGFR2 | FALSE | Upregulated |
| 0,9999493 | 0,294983425 | SCARNA7 | FALSE | Upregulated |
| 0,225069554 | 0,295109801 | C9orf91 | FALSE | Upregulated |
| -0,133757423 | 0,295151603 | SAFB | FALSE | Downregulated |
| -0,585845499 | 0,295234476 | CLDN10 | FALSE | Downregulated |
| -0,668500856 | 0,295356655 | LOC101928055 | FALSE | Downregulated |
| -0,733073452 | 0,295602717 | SUZ12P1 | FALSE | Downregulated |
| 0,566582895 | 0,295745298 | CHKB-AS1 | FALSE | Upregulated |
| 0,161785976 | 0,295745298 | PICK1 | FALSE | Upregulated |
| 0,750327355 | 0,295765396 | RP11-63P12.6 | FALSE | Upregulated |
| -0,179190858 | 0,295933358 | ADI1 | FALSE | Downregulated |
| -0,169239851 | 0,295944155 | NAPB | FALSE | Downregulated |
| 0,160156554 | 0,296072574 | IL18R1 | FALSE | Upregulated |
| 0,158700818 | 0,296183286 | TST | FALSE | Upregulated |
| -0,17468943 | 0,296183286 | ZMYM3 | FALSE | Downregulated |
| -0,157104152 | 0,296354088 | NAA30 | FALSE | Downregulated |
| 0,204687642 | 0,29640762 | ETV6 | FALSE | Upregulated |
| -0,804090203 | 0,296464775 | LOC102724805 | FALSE | Downregulated |
| -0,18064538 | 0,296530948 | COPZ2 | FALSE | Downregulated |
| 0,111688117 | 0,296530948 | UMPS | FALSE | Upregulated |
| -0,134894525 | 0,296607334 | ZER1 | FALSE | Downregulated |
| 0,238565798 | 0,296633687 | ZUFSP | FALSE | Upregulated |
| -0,47149854 | 0,296705989 | LOC146880 | FALSE | Downregulated |
| 0,63241146 | 0,296793384 | B3GNT5 | FALSE | Upregulated |
| 0,319580928 | 0,296793384 | ZDHHC13 | FALSE | Upregulated |
| -1,558550388 | 0,297088784 | EBF2 | FALSE | Downregulated |
| -0,12961554 | 0,297188196 | ATF7 | FALSE | Downregulated |
| -0,257071764 | 0,297329523 | THOC3 | FALSE | Downregulated |
| 0,184518487 | 0,297624532 | TNFAIP1 | FALSE | Upregulated |
| 0,856322472 | 0,297685137 | KCNJ11 | FALSE | Upregulated |
| 0,114299275 | 0,297798532 | DHX15 | FALSE | Upregulated |
| 0,113303618 | 0,297815177 | SDF4 | FALSE | Upregulated |
| -0,223828967 | 0,298144531 | PPT2 | FALSE | Downregulated |
| 0,228873437 | 0,298144531 | TEX30 | FALSE | Upregulated |
| -0,508405747 | 0,298283966 | COL4A5 | FALSE | Downregulated |
| -0,175136687 | 0,298283966 | SFXN5 | FALSE | Downregulated |
| 0,173558272 | 0,298343579 | STX10 | FALSE | Upregulated |

| | | | | |
|---|---|---|---|---|
| 0,121552909 | 0,298374849 | MRPL44 | FALSE | Upregulated |
| 0,165486677 | 0,298395116 | GRINA | FALSE | Upregulated |
| -0,307609169 | 0,298395116 | SEPT4 | FALSE | Downregulated |
| 0,573799382 | 0,298574514 | THY1 | FALSE | Upregulated |
| -0,744487986 | 0,298731245 | ATOH7 | FALSE | Downregulated |
| -0,278598586 | 0,298879329 | FAM69B | FALSE | Downregulated |
| 0,494308123 | 0,299357854 | LINC00861 | FALSE | Upregulated |
| 0,152908501 | 0,299357854 | NEDD1 | FALSE | Upregulated |
| 0,16957044 | 0,299357854 | RAE1 | FALSE | Upregulated |
| -0,199378191 | 0,29961309 | ARHGEF10L | FALSE | Downregulated |
| 1,071595168 | 0,299617083 | LGALS9C | FALSE | Upregulated |
| -0,219221348 | 0,299617083 | ZNF436 | FALSE | Downregulated |
| 0,276394694 | 0,299653035 | CENPQ | FALSE | Upregulated |
| 0,291700995 | 0,299893554 | AS3MT | FALSE | Upregulated |
| -0,195690042 | 0,299893554 | C9orf41 | FALSE | Downregulated |
| -1,085659902 | 0,299893554 | MEG9 | FALSE | Downregulated |
| 0,271647146 | 0,299893554 | PSTPIP2 | FALSE | Upregulated |
| 0,343418753 | 0,30003224 | SPINT2 | FALSE | Upregulated |
| -0,204205911 | 0,300089771 | RECQL5 | FALSE | Downregulated |
| 0,130137477 | 0,30019322 | PARG | FALSE | Upregulated |
| 0,132220278 | 0,30019322 | PI4KA | FALSE | Upregulated |
| -0,315879481 | 0,30019322 | SLC22A1 | FALSE | Downregulated |
| 0,39894435 | 0,300292946 | NCF1 | FALSE | Upregulated |
| 0,167891091 | 0,300378043 | TRAM1 | FALSE | Upregulated |
| -0,531030689 | 0,300451475 | PTK6 | FALSE | Downregulated |
| -0,264489333 | 0,30048657 | RFX3 | FALSE | Downregulated |
| -1,046870675 | 0,3006123 | LOC644554 | FALSE | Downregulated |
| 0,703897577 | 0,300665846 | SPIB | FALSE | Upregulated |
| -0,592904429 | 0,300802528 | LOC100288069 | FALSE | Downregulated |
| -0,85745541 | 0,30091664 | ARSI | FALSE | Downregulated |
| 0,702561739 | 0,30091664 | KCNK13 | FALSE | Upregulated |
| -0,940256137 | 0,30091664 | LOC101929287 | FALSE | Downregulated |
| -0,132804256 | 0,30091664 | TBL1XR1 | FALSE | Downregulated |
| 0,798759047 | 0,30102086 | LOC100287098 | FALSE | Upregulated |
| 0,162473904 | 0,301112047 | COA3 | FALSE | Upregulated |
| 0,170136159 | 0,301112047 | USP46 | FALSE | Upregulated |
| -0,306645239 | 0,301133529 | UPF3A | FALSE | Downregulated |
| -0,783682979 | 0,301231423 | IL20RA | FALSE | Downregulated |
| -0,128094382 | 0,301231423 | PJA2 | FALSE | Downregulated |
| 0,49227149 | 0,301319543 | AEBP1 | FALSE | Upregulated |
| -0,828306905 | 0,301319543 | LOC102723324 | FALSE | Downregulated |
| 0,225094146 | 0,301319543 | RPL31 | FALSE | Upregulated |
| 0,228605097 | 0,301456293 | C9orf142 | FALSE | Upregulated |
| 0,115819333 | 0,301456293 | KIAA0141 | FALSE | Upregulated |
| 0,334866419 | 0,30148571 | CCL15-CCL14 | FALSE | Upregulated |
| 0,093065521 | 0,30148571 | TNPO1 | FALSE | Upregulated |
| -0,247383462 | 0,301489452 | FBXL15 | FALSE | Downregulated |
| 0,158426269 | 0,30153426 | CSE1L | FALSE | Upregulated |
| 0,585094698 | 0,30153426 | LOC101928881 | FALSE | Upregulated |
| 0,17049242 | 0,301606386 | DHRS3 | FALSE | Upregulated |
| -0,166883414 | 0,301637792 | BPTF | FALSE | Downregulated |

| | | | | |
|---|---|---|---|---|
| -0,47813583 | 0,301637792 | CCDC148 | FALSE | Downregulated |
| -0,149023143 | 0,301637792 | KLF3 | FALSE | Downregulated |
| -0,783011758 | 0,301637792 | SOCS2-AS1 | FALSE | Downregulated |
| -0,49211262 | 0,301889052 | PODXL2 | FALSE | Downregulated |
| 0,174964386 | 0,302038313 | LIMA1 | FALSE | Upregulated |
| 0,454286827 | 0,302042416 | CCDC102A | FALSE | Upregulated |
| 0,587835908 | 0,302072321 | FGF7 | FALSE | Upregulated |
| -0,396868435 | 0,302072321 | PRICKLE4 | FALSE | Downregulated |
| -0,097334011 | 0,302110307 | ANKRD13C | FALSE | Downregulated |
| -0,882582351 | 0,302159446 | LOC101926902 | FALSE | Downregulated |
| 0,634064281 | 0,302159446 | SIGLEC17P | FALSE | Upregulated |
| -0,337200786 | 0,302165407 | LCA5L | FALSE | Downregulated |
| 0,283703084 | 0,302165407 | TAF8 | FALSE | Upregulated |
| 0,635718693 | 0,302450928 | PRDM8 | FALSE | Upregulated |
| 0,159634775 | 0,302667428 | RPS16 | FALSE | Upregulated |
| 0,162561729 | 0,302813039 | MEMO1 | FALSE | Upregulated |
| 1,730427567 | 0,302909046 | CCDC177 | FALSE | Upregulated |
| 0,133596725 | 0,303099569 | DRG1 | FALSE | Upregulated |
| 0,114976968 | 0,303115363 | HDAC1 | FALSE | Upregulated |
| -0,155957332 | 0,303115363 | MDM1 | FALSE | Downregulated |
| 0,235018377 | 0,303115363 | NEURL1B | FALSE | Upregulated |
| 0,181913278 | 0,303129135 | OGFR | FALSE | Upregulated |
| 0,20747267 | 0,303229747 | ARID5A | FALSE | Upregulated |
| 0,156513957 | 0,303235691 | KATNB1 | FALSE | Upregulated |
| 0,124845825 | 0,303235691 | MKNK1 | FALSE | Upregulated |
| -0,142490307 | 0,303347804 | C17orf85 | FALSE | Downregulated |
| 0,11287322 | 0,303456715 | TCEAL8 | FALSE | Upregulated |
| 0,204059014 | 0,30347478 | REEP6 | FALSE | Upregulated |
| -0,588977709 | 0,30368999 | LOC101927793 | FALSE | Downregulated |
| -0,156271795 | 0,30368999 | MPP6 | FALSE | Downregulated |
| 0,153928282 | 0,303919288 | ATP11A | FALSE | Upregulated |
| -0,271990194 | 0,304079778 | C2orf74 | FALSE | Downregulated |
| 0,539744769 | 0,304079778 | FAM99B | FALSE | Upregulated |
| 0,365854295 | 0,304162483 | SPIRE1 | FALSE | Upregulated |
| 1,101962682 | 0,304348902 | CNFN | FALSE | Upregulated |
| -1,269844628 | 0,304348902 | CTAGE10P | FALSE | Downregulated |
| 0,242461155 | 0,304348902 | MTSS1L | FALSE | Upregulated |
| 0,135443172 | 0,304348902 | STK38 | FALSE | Upregulated |
| -0,24586664 | 0,304449847 | GPR126 | FALSE | Downregulated |
| -0,223913421 | 0,304474933 | HGFAC | FALSE | Downregulated |
| 0,657074715 | 0,304474933 | SYT13 | FALSE | Upregulated |
| 0,147456632 | 0,304702399 | BTBD6 | FALSE | Upregulated |
| -0,195179075 | 0,304702399 | ESPN | FALSE | Downregulated |
| -0,596814118 | 0,304702399 | LOC101927895 | FALSE | Downregulated |
| 0,136025627 | 0,304702399 | SMARCAL1 | FALSE | Upregulated |
| 0,171353054 | 0,304888437 | TIPARP | FALSE | Upregulated |
| 0,131303555 | 0,304929583 | ATP6V1D | FALSE | Upregulated |
| -0,239187973 | 0,304935705 | NT5DC3 | FALSE | Downregulated |
| -0,18920237 | 0,305216641 | DDX17 | FALSE | Downregulated |
| 0,155816355 | 0,305216641 | GTF2A2 | FALSE | Upregulated |
| 0,120534389 | 0,305216641 | SEP15 | FALSE | Upregulated |

| | | | | |
|---|---|---|---|---|
| 0,157295384 | 0,30530332 | MYO9B | FALSE | Upregulated |
| -0,180355288 | 0,305433612 | TRAM2 | FALSE | Downregulated |
| -0,133431844 | 0,305695625 | RNF113A | FALSE | Downregulated |
| -0,130100156 | 0,305756873 | YIPF4 | FALSE | Downregulated |
| 0,996578213 | 0,305767148 | TIGD3 | FALSE | Upregulated |
| -1,040704545 | 0,30605608 | LOC729224 | FALSE | Downregulated |
| -0,47513353 | 0,306149317 | HOXA5 | FALSE | Downregulated |
| -0,380379525 | 0,306282026 | FAM193B | FALSE | Downregulated |
| -0,214037911 | 0,306298231 | LRRC3 | FALSE | Downregulated |
| -0,210606616 | 0,306308862 | FRAT2 | FALSE | Downregulated |
| -0,174456277 | 0,306323858 | ASXL1 | FALSE | Downregulated |
| 0,13386627 | 0,306323858 | MRPL1 | FALSE | Upregulated |
| 0,306701946 | 0,306394589 | SGCB | FALSE | Upregulated |
| -1,065691432 | 0,306476181 | CCDC144A | FALSE | Downregulated |
| 0,342096116 | 0,30674191 | RASGRP4 | FALSE | Upregulated |
| 0,500180311 | 0,306770523 | SLC4A8 | FALSE | Upregulated |
| -0,380195546 | 0,306812637 | LOC100996492 | FALSE | Downregulated |
| -0,72103874 | 0,306855935 | RPL34-AS1 | FALSE | Downregulated |
| -0,166806006 | 0,306855935 | TRIP11 | FALSE | Downregulated |
| -0,210992152 | 0,306867333 | LEPREL1 | FALSE | Downregulated |
| -0,394563884 | 0,306867333 | LOC653160 | FALSE | Downregulated |
| -0,50251318 | 0,306924482 | FLJ35282 | FALSE | Downregulated |
| -0,381435732 | 0,306924482 | ZNF345 | FALSE | Downregulated |
| -0,409292096 | 0,307241855 | TRNM | FALSE | Downregulated |
| 0,194595582 | 0,307328281 | NEO1 | FALSE | Upregulated |
| 0,181832272 | 0,307348269 | NCKIPSD | FALSE | Upregulated |
| -0,202410425 | 0,307418449 | LYRM9 | FALSE | Downregulated |
| -0,101978298 | 0,307418449 | MARCH8 | FALSE | Downregulated |
| -0,610601596 | 0,307470541 | TEX21P | FALSE | Downregulated |
| -0,364185294 | 0,307490251 | PAXBP1-AS1 | FALSE | Downregulated |
| -0,510151134 | 0,307714397 | CNTNAP3 | FALSE | Downregulated |
| -2,051255813 | 0,307714397 | RNU4-1 | FALSE | Downregulated |
| -1,199693851 | 0,307739165 | LOC101930567 | FALSE | Downregulated |
| -0,283947715 | 0,308062352 | RP9 | FALSE | Downregulated |
| -0,201747115 | 0,308116626 | PC | FALSE | Downregulated |
| 0,264590501 | 0,308143623 | ARMCX6 | FALSE | Upregulated |
| 0,117001545 | 0,308171633 | NSUN4 | FALSE | Upregulated |
| -0,20454908 | 0,308526808 | ZSCAN16 | FALSE | Downregulated |
| -1,081094046 | 0,308558357 | LOC100996808 | FALSE | Downregulated |
| 0,211613292 | 0,308760697 | PRKAB1 | FALSE | Upregulated |
| -0,606466012 | 0,309078445 | C21orf62 | FALSE | Downregulated |
| 0,130212597 | 0,309109444 | DPY30 | FALSE | Upregulated |
| 0,162185914 | 0,309109444 | GNB2L1 | FALSE | Upregulated |
| -0,195197383 | 0,309267758 | TTC28-AS1 | FALSE | Downregulated |
| 0,136877484 | 0,309273447 | SARS | FALSE | Upregulated |
| -0,381117033 | 0,309417895 | RAP2C-AS1 | FALSE | Downregulated |
| -0,153649737 | 0,309435661 | RXRA | FALSE | Downregulated |
| -0,509325974 | 0,30944012 | LINC00342 | FALSE | Downregulated |
| 0,147354876 | 0,309487297 | CCNDBP1 | FALSE | Upregulated |
| 0,203369185 | 0,309487297 | TMEM177 | FALSE | Upregulated |
| -0,24097515 | 0,310046245 | WDR72 | FALSE | Downregulated |

| | | | | |
|---|---|---|---|---|
| 0,158377583 | 0,31007526 | METTL21A | FALSE | Upregulated |
| -1,128738642 | 0,310093166 | LINC00277 | FALSE | Downregulated |
| -0,137661839 | 0,310406776 | NFS1 | FALSE | Downregulated |
| 0,703904064 | 0,310451602 | CDH6 | FALSE | Upregulated |
| 0,136372943 | 0,310451602 | CHCHD2 | FALSE | Upregulated |
| -0,636404738 | 0,310451923 | ASIC3 | FALSE | Downregulated |
| -0,128176872 | 0,310467144 | RBM7 | FALSE | Downregulated |
| -1,206929501 | 0,310512233 | OVCH1 | FALSE | Downregulated |
| -1,572708236 | 0,310596051 | SPIN2A | FALSE | Downregulated |
| -0,126543685 | 0,310689898 | ATXN2 | FALSE | Downregulated |
| -0,192417505 | 0,310689898 | MRPL11 | FALSE | Downregulated |
| 0,146687231 | 0,310689898 | RPA3-AS1 | FALSE | Upregulated |
| 0,105686792 | 0,310719682 | NUP50 | FALSE | Upregulated |
| 0,106994299 | 0,310849339 | ACAD9 | FALSE | Upregulated |
| -0,157520835 | 0,31092883 | TOPORS | FALSE | Downregulated |
| -0,157749029 | 0,310971094 | CCDC101 | FALSE | Downregulated |
| -0,172700253 | 0,310971094 | PSKH1 | FALSE | Downregulated |
| 0,163843904 | 0,310971094 | SEH1L | FALSE | Upregulated |
| 0,147984369 | 0,310998402 | UBE2L3 | FALSE | Upregulated |
| -0,941988973 | 0,311158792 | GTSCR1 | FALSE | Downregulated |
| -0,128370958 | 0,311158792 | LRP11 | FALSE | Downregulated |
| 0,183562773 | 0,311158792 | RPLP0 | FALSE | Upregulated |
| -0,288889206 | 0,311158792 | RWDD2A | FALSE | Downregulated |
| 0,160584425 | 0,311158792 | TAF9B | FALSE | Upregulated |
| 0,692594784 | 0,311508486 | GAL3ST4 | FALSE | Upregulated |
| 0,296784874 | 0,311726554 | UNC13D | FALSE | Upregulated |
| -0,615861642 | 0,311735991 | OLFM2 | FALSE | Downregulated |
| 0,345757733 | 0,311887536 | C1orf198 | FALSE | Upregulated |
| 1,000800161 | 0,311983252 | LMTK3 | FALSE | Upregulated |
| -1,118741326 | 0,311983252 | RP11-230B22.1 | FALSE | Downregulated |
| -0,201333586 | 0,312054984 | NIPSNAP1 | FALSE | Downregulated |
| 0,109288277 | 0,312054984 | PANX1 | FALSE | Upregulated |
| -0,829978651 | 0,312054984 | ZNF676 | FALSE | Downregulated |
| 0,122432074 | 0,312072387 | LARS2 | FALSE | Upregulated |
| -0,546014621 | 0,312234061 | LOC100506124 | FALSE | Downregulated |
| 0,12338877 | 0,312441943 | DCTN6 | FALSE | Upregulated |
| -0,164264785 | 0,312455257 | MYLK | FALSE | Downregulated |
| -0,272908685 | 0,312574751 | HYKK | FALSE | Downregulated |
| 0,12213124 | 0,312665167 | FAM114A2 | FALSE | Upregulated |
| 0,315661074 | 0,312665167 | SDSL | FALSE | Upregulated |
| -0,141716604 | 0,312814622 | XPOT | FALSE | Downregulated |
| 0,403091656 | 0,312858325 | GPR160 | FALSE | Upregulated |
| -0,753578667 | 0,312858325 | MUSK | FALSE | Downregulated |
| 0,175112232 | 0,312858325 | TBL2 | FALSE | Upregulated |
| 0,174114724 | 0,31297032 | MRPL14 | FALSE | Upregulated |
| -1,534022363 | 0,313006595 | GGT8P | FALSE | Downregulated |
| 0,153629252 | 0,313006595 | PDCD6 | FALSE | Upregulated |
| -0,309372169 | 0,31324772 | ARHGEF28 | FALSE | Downregulated |
| 0,153149437 | 0,313262526 | ISOC1 | FALSE | Upregulated |
| -0,155196278 | 0,313262526 | XPA | FALSE | Downregulated |
| 0,143822231 | 0,313369464 | EIF1 | FALSE | Upregulated |

| | | | | |
|---|---|---|---|---|
| 0,893532892 | 0,313483036 | LOC102724476 | FALSE | Upregulated |
| -0,11804563 | 0,313492713 | ING4 | FALSE | Downregulated |
| 0,184772364 | 0,313543454 | NUP85 | FALSE | Upregulated |
| 0,822923333 | 0,313553193 | FAM43B | FALSE | Upregulated |
| 0,214948512 | 0,313664345 | EXO5 | FALSE | Upregulated |
| -0,127438333 | 0,313806456 | ZNF384 | FALSE | Downregulated |
| -0,162342388 | 0,313807388 | HSPA9 | FALSE | Downregulated |
| 0,108959812 | 0,313807388 | RAD1 | FALSE | Upregulated |
| -0,14500306 | 0,313845649 | MCPH1 | FALSE | Downregulated |
| 0,620621437 | 0,313965038 | CNIH3 | FALSE | Upregulated |
| -0,809044478 | 0,314049137 | CES1P2 | FALSE | Downregulated |
| -0,165454078 | 0,314258043 | ECI2 | FALSE | Downregulated |
| -0,329397366 | 0,314274576 | DUSP6 | FALSE | Downregulated |
| -0,733718686 | 0,314305021 | RP11-261C10.3 | FALSE | Downregulated |
| 0,986890842 | 0,314490469 | DKK2 | FALSE | Upregulated |
| -2,142194129 | 0,314527921 | C20orf26 | FALSE | Downregulated |
| -0,356149363 | 0,314527921 | CROCCP2 | FALSE | Downregulated |
| -1,693469818 | 0,314527921 | DIO2 | FALSE | Downregulated |
| 0,415443176 | 0,314527921 | FAM57A | FALSE | Upregulated |
| -0,294299492 | 0,314527921 | HILPDA | FALSE | Downregulated |
| -1,060181813 | 0,314527921 | LOC100505534 | FALSE | Downregulated |
| -0,462275742 | 0,314527921 | LOC101929107 | FALSE | Downregulated |
| 0,158365132 | 0,314655965 | STX6 | FALSE | Upregulated |
| -0,704834806 | 0,314700597 | LOC441666 | FALSE | Downregulated |
| 0,151348812 | 0,314780983 | COA7 | FALSE | Upregulated |
| 0,10638177 | 0,314793379 | CDK9 | FALSE | Upregulated |
| 0,287704887 | 0,314793379 | YIF1B | FALSE | Upregulated |
| -0,210695784 | 0,314799167 | RFX7 | FALSE | Downregulated |
| -0,486795367 | 0,314956872 | ZIC1 | FALSE | Downregulated |
| -0,378758278 | 0,315059046 | FENDRR | FALSE | Downregulated |
| -0,629427145 | 0,315238826 | LOC101928768 | FALSE | Downregulated |
| -0,373038021 | 0,31530556 | KGFLP2 | FALSE | Downregulated |
| 0,940340485 | 0,316100697 | TTTY15 | FALSE | Upregulated |
| -0,652347844 | 0,316192187 | LOC101928810 | FALSE | Downregulated |
| 0,184949248 | 0,316192187 | RGP1 | FALSE | Upregulated |
| -0,363125128 | 0,316322387 | OTUD3 | FALSE | Downregulated |
| -0,573870094 | 0,316379402 | LOC101929385 | FALSE | Downregulated |
| 0,523332483 | 0,316397578 | XK | FALSE | Upregulated |
| -1,263530353 | 0,316402186 | LOC643802 | FALSE | Downregulated |
| 0,19758385 | 0,316619875 | BAD | FALSE | Upregulated |
| -0,203181367 | 0,316979984 | RAB7L1 | FALSE | Downregulated |
| 0,135406693 | 0,317077325 | RFWD2 | FALSE | Upregulated |
| 0,252694681 | 0,317179322 | GGTA1P | FALSE | Upregulated |
| -0,147289609 | 0,317179322 | PRRG1 | FALSE | Downregulated |
| -0,447330327 | 0,317336199 | SYCP2 | FALSE | Downregulated |
| -0,649540106 | 0,317386741 | TMEM182 | FALSE | Downregulated |
| -0,171993603 | 0,317502764 | TFPI | FALSE | Downregulated |
| -0,17391161 | 0,317737591 | BHMT2 | FALSE | Downregulated |
| -0,362821641 | 0,317812177 | ANO3 | FALSE | Downregulated |
| 1,105498893 | 0,317864123 | LOC102723884 | FALSE | Upregulated |
| -0,250359425 | 0,317973093 | DGAT1 | FALSE | Downregulated |

| | | | | |
|---|---|---|---|---|
| 0,156017677 | 0,318063521 | CCDC53 | FALSE | Upregulated |
| -0,440047903 | 0,318063521 | ZNF599 | FALSE | Downregulated |
| 0,209800466 | 0,318092464 | ASAP1 | FALSE | Upregulated |
| -0,633762713 | 0,318092464 | BTBD19 | FALSE | Downregulated |
| -0,168134297 | 0,318092464 | RABL2B | FALSE | Downregulated |
| 0,119160675 | 0,318138788 | ACTR10 | FALSE | Upregulated |
| -0,772226199 | 0,318138788 | C19orf26 | FALSE | Downregulated |
| 0,222846399 | 0,318138788 | DDX28 | FALSE | Upregulated |
| -0,769930565 | 0,318138788 | MTRNR2L6 | FALSE | Downregulated |
| -0,423702184 | 0,318138788 | MYO15B | FALSE | Downregulated |
| -0,093230516 | 0,318155166 | MARCH7 | FALSE | Downregulated |
| -0,517844101 | 0,318167419 | TRIM58 | FALSE | Downregulated |
| -0,306319307 | 0,318226573 | ENGASE | FALSE | Downregulated |
| -0,831652249 | 0,318515778 | BEGAIN | FALSE | Downregulated |
| -0,393420837 | 0,318515778 | MYL6B | FALSE | Downregulated |
| -0,333088781 | 0,318515778 | WHAMMP2 | FALSE | Downregulated |
| -0,156109179 | 0,318567035 | POLR3B | FALSE | Downregulated |
| 0,083185962 | 0,318618993 | C11orf58 | FALSE | Upregulated |
| 0,662596692 | 0,318618993 | GPR55 | FALSE | Upregulated |
| -0,352724779 | 0,318618993 | RHCE | FALSE | Downregulated |
| -0,176283592 | 0,318618993 | SRP54 | FALSE | Downregulated |
| -0,130893854 | 0,318820019 | NOL11 | FALSE | Downregulated |
| 0,159168974 | 0,319118528 | ATP5L | FALSE | Upregulated |
| 0,476911177 | 0,319127206 | KAL1 | FALSE | Upregulated |
| -0,314584903 | 0,319144202 | HHIP | FALSE | Downregulated |
| 0,232865068 | 0,319144202 | PIR | FALSE | Upregulated |
| -0,188613561 | 0,319144202 | SDHAF1 | FALSE | Downregulated |
| 1,123543841 | 0,319324174 | RCVRN | FALSE | Upregulated |
| 0,189048075 | 0,31932744 | TRIM26 | FALSE | Upregulated |
| -0,552405929 | 0,319490461 | C10orf126 | FALSE | Downregulated |
| -0,439703115 | 0,319490461 | CPLX1 | FALSE | Downregulated |
| 0,170035072 | 0,319490461 | TMCC1 | FALSE | Upregulated |
| -1,541302464 | 0,319532829 | LOC102723603 | FALSE | Downregulated |
| -0,148663662 | 0,31956026 | PGPEP1 | FALSE | Downregulated |
| 0,197328726 | 0,320018853 | APBA3 | FALSE | Upregulated |
| -0,224107678 | 0,320126948 | ZNF316 | FALSE | Downregulated |
| -0,123599737 | 0,320260221 | TMEM254 | FALSE | Downregulated |
| -0,227049652 | 0,3202839 | KDM6A | FALSE | Downregulated |
| 0,098454379 | 0,3202839 | RAB10 | FALSE | Upregulated |
| -0,187412885 | 0,320379903 | HSD17B4 | FALSE | Downregulated |
| -0,117969319 | 0,320486455 | TRAF6 | FALSE | Downregulated |
| -0,378543761 | 0,320774324 | ADAMTS7 | FALSE | Downregulated |
| 0,347484522 | 0,32090322 | C11orf48 | FALSE | Upregulated |
| 0,730135029 | 0,32090322 | LYPD5 | FALSE | Upregulated |
| 0,163484096 | 0,320918026 | CEP76 | FALSE | Upregulated |
| 0,440436262 | 0,321002553 | CNTD1 | FALSE | Upregulated |
| -1,691074736 | 0,321002553 | MAFA | FALSE | Downregulated |
| 0,692741824 | 0,321216836 | LOC644090 | FALSE | Upregulated |
| 0,10352463 | 0,321216836 | MBD2 | FALSE | Upregulated |
| 0,13006841 | 0,321338276 | GMPR2 | FALSE | Upregulated |
| 0,178597205 | 0,321533323 | KLHL42 | FALSE | Upregulated |

| | | | | |
|---|---|---|---|---|
| 0,348892932 | 0,321560704 | KLC2 | FALSE | Upregulated |
| 0,202236709 | 0,32170449 | KTI12 | FALSE | Upregulated |
| 0,189785481 | 0,322141332 | CLPB | FALSE | Upregulated |
| 0,340280185 | 0,322240981 | ZNF671 | FALSE | Upregulated |
| -0,54324039 | 0,32251321 | MT1X | FALSE | Downregulated |
| -0,403443278 | 0,32251321 | STAG3 | FALSE | Downregulated |
| -0,760147228 | 0,322613562 | IP6K3 | FALSE | Downregulated |
| 0,759587372 | 0,322802876 | ROR2 | FALSE | Upregulated |
| -0,67221164 | 0,322848406 | LY6G5B | FALSE | Downregulated |
| 0,178406109 | 0,32292148 | RPS10-NUDT3 | FALSE | Upregulated |
| -0,150311745 | 0,323074723 | KAT6A | FALSE | Downregulated |
| -0,113646366 | 0,323243443 | AKT2 | FALSE | Downregulated |
| 0,258229558 | 0,323268186 | FLOT2 | FALSE | Upregulated |
| -0,185613285 | 0,323282598 | ZNF155 | FALSE | Downregulated |
| 1,244242385 | 0,323341022 | RUNDC3A | FALSE | Upregulated |
| -0,479656362 | 0,323347969 | DFNB59 | FALSE | Downregulated |
| -0,262346301 | 0,323441299 | CES1 | FALSE | Downregulated |
| -0,162779838 | 0,323565475 | BAIAP2-AS1 | FALSE | Downregulated |
| -0,172037571 | 0,323639012 | PRR14 | FALSE | Downregulated |
| -0,374811694 | 0,323639012 | ZNF582-AS1 | FALSE | Downregulated |
| -0,547107027 | 0,323834379 | ITGA10 | FALSE | Downregulated |
| -0,163492012 | 0,323834379 | MEF2A | FALSE | Downregulated |
| 0,150340713 | 0,323891267 | ANP32A | FALSE | Upregulated |
| 0,525638504 | 0,32404157 | CBWD6 | FALSE | Upregulated |
| -0,17181707 | 0,32404157 | MAPRE3 | FALSE | Downregulated |
| -0,337892271 | 0,32404157 | NTF3 | FALSE | Downregulated |
| -1,44479325 | 0,32404157 | ZIC4 | FALSE | Downregulated |
| 0,986917615 | 0,324073094 | KRTAP5-6 | FALSE | Upregulated |
| 0,133245668 | 0,324075761 | BCAP29 | FALSE | Upregulated |
| -0,103810113 | 0,324081495 | NUMB | FALSE | Downregulated |
| 1,227797202 | 0,324363838 | LOC102724077 | FALSE | Upregulated |
| -0,915082033 | 0,324365278 | AQP6 | FALSE | Downregulated |
| 0,351807633 | 0,324365278 | DUSP18 | FALSE | Upregulated |
| 0,155446907 | 0,324546938 | GFER | FALSE | Upregulated |
| -0,474437618 | 0,324721824 | GOLGA6A | FALSE | Downregulated |
| 0,260518498 | 0,324721824 | RPUSD1 | FALSE | Upregulated |
| 0,165501707 | 0,324736678 | RPL24 | FALSE | Upregulated |
| 0,182258163 | 0,324769377 | GLTSCR1 | FALSE | Upregulated |
| 0,505708648 | 0,324769377 | PLCL1 | FALSE | Upregulated |
| 0,142774086 | 0,324769377 | SEL1L | FALSE | Upregulated |
| -0,36298681 | 0,324769377 | SH3RF2 | FALSE | Downregulated |
| -0,174545934 | 0,325037787 | CCNT2 | FALSE | Downregulated |
| 0,556248637 | 0,325269285 | CHRDL2 | FALSE | Upregulated |
| 1,25801511 | 0,325269285 | ZFY | FALSE | Upregulated |
| -0,108125628 | 0,325381189 | CAST | FALSE | Downregulated |
| -0,266527064 | 0,325381189 | PGBD5 | FALSE | Downregulated |
| 0,490395818 | 0,325381189 | PLEK2 | FALSE | Upregulated |
| -1,007339003 | 0,325381189 | REM2 | FALSE | Downregulated |
| -0,187283569 | 0,325385351 | HRG | FALSE | Downregulated |
| -1,170230266 | 0,325385351 | LOC101927673 | FALSE | Downregulated |
| -0,237731359 | 0,325385351 | SLC39A14 | FALSE | Downregulated |

| | | | | |
|---|---|---|---|---|
| -0,168585044 | 0,325388371 | SIN3B | FALSE | Downregulated |
| 0,317613628 | 0,325425055 | ZNF517 | FALSE | Upregulated |
| 0,601199644 | 0,325577881 | LOC101927686 | FALSE | Upregulated |
| -0,123532271 | 0,325776416 | ZNF189 | FALSE | Downregulated |
| -0,235231666 | 0,325807836 | CEP63 | FALSE | Downregulated |
| -0,122003438 | 0,325807836 | PPIP5K2 | FALSE | Downregulated |
| -0,223691363 | 0,325807836 | TBCA | FALSE | Downregulated |
| 0,072996519 | 0,325831939 | HNRNPU | FALSE | Upregulated |
| 0,480791711 | 0,326033933 | EMC3-AS1 | FALSE | Upregulated |
| -0,84921593 | 0,326200683 | VWDE | FALSE | Downregulated |
| 0,132691934 | 0,326233796 | CCDC137 | FALSE | Upregulated |
| 0,194793731 | 0,326345019 | VPS29 | FALSE | Upregulated |
| 1,181006383 | 0,326373698 | LOC101928152 | FALSE | Upregulated |
| -0,93222474 | 0,326432537 | PZP | FALSE | Downregulated |
| -0,934384426 | 0,326470055 | LOC101928764 | FALSE | Downregulated |
| 1,053461653 | 0,32651278 | ZG16B | FALSE | Upregulated |
| 0,176890823 | 0,326731084 | KRT10 | FALSE | Upregulated |
| -0,173995384 | 0,326761263 | ECHS1 | FALSE | Downregulated |
| -0,256346204 | 0,326813945 | ZNF529 | FALSE | Downregulated |
| -0,517502509 | 0,327150227 | KCNK1 | FALSE | Downregulated |
| -0,492773798 | 0,327150227 | LOC100287042 | FALSE | Downregulated |
| -0,659561571 | 0,327232455 | LOC101927086 | FALSE | Downregulated |
| 1,093407156 | 0,327271049 | TDRD15 | FALSE | Upregulated |
| -0,175808792 | 0,327322901 | CXADR | FALSE | Downregulated |
| -0,209506271 | 0,327322901 | DNAJC4 | FALSE | Downregulated |
| 0,295749832 | 0,327517022 | ACSL6 | FALSE | Upregulated |
| -0,286811164 | 0,327602855 | MOK | FALSE | Downregulated |
| -0,168309109 | 0,327625312 | CRAMP1L | FALSE | Downregulated |
| -0,204918056 | 0,327654987 | CPEB2 | FALSE | Downregulated |
| -1,553120313 | 0,327654987 | DKK1 | FALSE | Downregulated |
| -0,13365311 | 0,327939318 | LYPLA1 | FALSE | Downregulated |
| -0,283224799 | 0,3279453 | RAD9A | FALSE | Downregulated |
| -0,872395199 | 0,32806542 | LOC101927511 | FALSE | Downregulated |
| -0,183252709 | 0,328073497 | CHD7 | FALSE | Downregulated |
| -0,338224884 | 0,328073497 | LOC102724164 | FALSE | Downregulated |
| 0,262115418 | 0,328196961 | VAV3 | FALSE | Upregulated |
| 0,211483392 | 0,328196961 | YDJC | FALSE | Upregulated |
| 0,152139199 | 0,328243565 | RPS13 | FALSE | Upregulated |
| 1,535573933 | 0,328339869 | C21orf128 | FALSE | Upregulated |
| -0,324613389 | 0,328339869 | RP11-473M20.14 | FALSE | Downregulated |
| 0,191325539 | 0,328339869 | TECPR1 | FALSE | Upregulated |
| 0,092118939 | 0,328339869 | ZDHHC5 | FALSE | Upregulated |
| -0,607037491 | 0,32859285 | PRDM6 | FALSE | Downregulated |
| 0,956261528 | 0,328602599 | LOC102724601 | FALSE | Upregulated |
| 0,225344479 | 0,328672978 | ARMC6 | FALSE | Upregulated |
| -0,257821901 | 0,328728881 | IGF1R | FALSE | Downregulated |
| 0,173852484 | 0,328767197 | RCBTB1 | FALSE | Upregulated |
| 0,27959634 | 0,328794147 | HSPA1B | FALSE | Upregulated |
| -0,457515355 | 0,328797273 | ZDHHC11 | FALSE | Downregulated |
| -0,528704321 | 0,329065115 | FERMT1 | FALSE | Downregulated |
| -0,139465979 | 0,329240527 | TTC19 | FALSE | Downregulated |

| | | | | |
|---|---|---|---|---|
| 0,188211287 | 0,329382531 | AGTPBP1 | FALSE | Upregulated |
| -0,1693696 | 0,329456952 | SERPIND1 | FALSE | Downregulated |
| 0,373821007 | 0,329470678 | C19orf55 | FALSE | Upregulated |
| 0,161436591 | 0,329579148 | CTDP1 | FALSE | Upregulated |
| 0,119098001 | 0,329579148 | PSMB7 | FALSE | Upregulated |
| -0,72742267 | 0,329701493 | ZNF252P-AS1 | FALSE | Downregulated |
| -0,845912407 | 0,329762131 | ESM1 | FALSE | Downregulated |
| 0,559432317 | 0,329762131 | LOC101927069 | FALSE | Upregulated |
| -0,262851035 | 0,329762131 | ZNF276 | FALSE | Downregulated |
| 0,11693338 | 0,329893377 | APTX | FALSE | Upregulated |
| -0,155895602 | 0,330231426 | NUMA1 | FALSE | Downregulated |
| 0,135716443 | 0,330231426 | UTP14A | FALSE | Upregulated |
| 0,312189462 | 0,330260926 | LOC102724967 | FALSE | Upregulated |
| 0,242320029 | 0,330385898 | ETV5 | FALSE | Upregulated |
| -0,614252854 | 0,330502567 | HSD3B2 | FALSE | Downregulated |
| 0,253394051 | 0,330504727 | FBLN7 | FALSE | Upregulated |
| -0,169540959 | 0,330504727 | GS1-251I9.4 | FALSE | Downregulated |
| -0,269318842 | 0,330534001 | KHK | FALSE | Downregulated |
| -0,210519204 | 0,330589446 | LOC100130872 | FALSE | Downregulated |
| -1,444660077 | 0,330609647 | MKRN7P | FALSE | Downregulated |
| 0,088952207 | 0,330693856 | ARPC1A | FALSE | Upregulated |
| -0,964344582 | 0,330693856 | CCDC114 | FALSE | Downregulated |
| -1,58056942 | 0,330693856 | IGSF11 | FALSE | Downregulated |
| 0,208733176 | 0,330693856 | IQCK | FALSE | Upregulated |
| 0,095461739 | 0,330693856 | PSMC6 | FALSE | Upregulated |
| -0,152769432 | 0,330693856 | RAPGEF2 | FALSE | Downregulated |
| 0,088756687 | 0,330693856 | USP10 | FALSE | Upregulated |
| -0,950056869 | 0,330738219 | NPY6R | FALSE | Downregulated |
| -0,12305234 | 0,33077794 | TRIM13 | FALSE | Downregulated |
| 0,28997487 | 0,331099706 | TMEM86B | FALSE | Upregulated |
| -0,21332186 | 0,331135955 | UBXN7 | FALSE | Downregulated |
| 0,239953722 | 0,331143648 | CLDN5 | FALSE | Upregulated |
| -0,581741127 | 0,331155029 | PRRT2 | FALSE | Downregulated |
| -0,576965682 | 0,331244015 | PAQR6 | FALSE | Downregulated |
| 0,151996298 | 0,331244015 | SMIM13 | FALSE | Upregulated |
| -0,103907786 | 0,331394783 | TTC37 | FALSE | Downregulated |
| 0,602797672 | 0,331460569 | IL1B | FALSE | Upregulated |
| -0,104444329 | 0,331531698 | GTF2I | FALSE | Downregulated |
| -0,119314215 | 0,331553542 | CNOT3 | FALSE | Downregulated |
| -0,242186066 | 0,331560477 | RP11-81N13.1 | FALSE | Downregulated |
| -1,289904909 | 0,331859625 | HFM1 | FALSE | Downregulated |
| 0,101098401 | 0,332056993 | PRPF18 | FALSE | Upregulated |
| -0,133103466 | 0,332090948 | CDK17 | FALSE | Downregulated |
| 0,188437455 | 0,332327505 | SLC50A1 | FALSE | Upregulated |
| 0,132922209 | 0,332331755 | CD276 | FALSE | Upregulated |
| -1,173162605 | 0,332331755 | L1TD1 | FALSE | Downregulated |
| -0,095505274 | 0,332331755 | MOAP1 | FALSE | Downregulated |
| -0,47005551 | 0,332373758 | BRCA1 | FALSE | Downregulated |
| -0,177242484 | 0,332412328 | SETMAR | FALSE | Downregulated |
| -0,149572772 | 0,332444973 | MAP7 | FALSE | Downregulated |
| 0,117137108 | 0,332538915 | ECD | FALSE | Upregulated |

| | | | | |
|---|---|---|---|---|
| 0,099659676 | 0,332800705 | PRRC2A | FALSE | Upregulated |
| 0,103115295 | 0,332953055 | IPO9 | FALSE | Upregulated |
| -0,403211092 | 0,333119239 | ATP1B2 | FALSE | Downregulated |
| -0,274130837 | 0,333127057 | FLJ42627 | FALSE | Downregulated |
| -0,938540442 | 0,333128085 | LOC102723997 | FALSE | Downregulated |
| 1,535407974 | 0,333153077 | FIGF | FALSE | Upregulated |
| 0,115380221 | 0,333289539 | ANAPC5 | FALSE | Upregulated |
| 0,208512133 | 0,333289539 | BRI3 | FALSE | Upregulated |
| 0,147341246 | 0,333289539 | MOB3C | FALSE | Upregulated |
| 0,118683444 | 0,333309323 | EXOSC3 | FALSE | Upregulated |
| -0,657422453 | 0,333421131 | CPT1B | FALSE | Downregulated |
| -0,206844857 | 0,333421131 | ORAOV1 | FALSE | Downregulated |
| -0,346014298 | 0,333725074 | PLLP | FALSE | Downregulated |
| 0,123976659 | 0,333824206 | DHX8 | FALSE | Upregulated |
| 0,166959369 | 0,333824206 | LMAN2L | FALSE | Upregulated |
| 0,674330962 | 0,333824206 | MKLN1-AS | FALSE | Upregulated |
| 0,258087744 | 0,333834935 | MMP14 | FALSE | Upregulated |
| 0,527077973 | 0,334139413 | PTK7 | FALSE | Upregulated |
| -0,715560722 | 0,334149405 | ANKLE1 | FALSE | Downregulated |
| -0,104872708 | 0,334297799 | DROSHA | FALSE | Downregulated |
| -1,073245599 | 0,334506945 | SNORA60 | FALSE | Downregulated |
| -1,207262784 | 0,334849172 | HSD3BP4 | FALSE | Downregulated |
| -0,243455445 | 0,334891053 | TRIM16 | FALSE | Downregulated |
| -0,122471409 | 0,334914059 | PPP1R15B | FALSE | Downregulated |
| -0,339349946 | 0,334990171 | CA11 | FALSE | Downregulated |
| -0,921789479 | 0,334990171 | LRRC39 | FALSE | Downregulated |
| 0,112074799 | 0,334990171 | RNF114 | FALSE | Upregulated |
| 0,667748099 | 0,334990171 | RXRG | FALSE | Upregulated |
| -0,097904341 | 0,334990171 | TAF2 | FALSE | Downregulated |
| 0,205748114 | 0,335191366 | EID2 | FALSE | Upregulated |
| 0,243306294 | 0,33554339 | RDH13 | FALSE | Upregulated |
| -0,171096778 | 0,335675363 | THTPA | FALSE | Downregulated |
| 0,159115531 | 0,335977447 | COMMD2 | FALSE | Upregulated |
| 0,594285565 | 0,336015259 | DTHD1 | FALSE | Upregulated |
| -0,586669771 | 0,336015259 | RP11-261N11.8 | FALSE | Downregulated |
| 0,405194676 | 0,336024387 | PLVAP | FALSE | Upregulated |
| -1,417554925 | 0,336287949 | BHLHE40-AS1 | FALSE | Downregulated |
| 0,246369653 | 0,336287949 | C9orf40 | FALSE | Upregulated |
| 0,197661389 | 0,336287949 | CCDC85C | FALSE | Upregulated |
| 0,189109753 | 0,336287949 | DHX57 | FALSE | Upregulated |
| -1,304465257 | 0,336287949 | ERVW-1 | FALSE | Downregulated |
| -0,16954226 | 0,336287949 | FAM126A | FALSE | Downregulated |
| 0,109614715 | 0,336287949 | GPKOW | FALSE | Upregulated |
| -0,488248214 | 0,336287949 | GUSBP4 | FALSE | Downregulated |
| 0,144358845 | 0,336287949 | PPM1F | FALSE | Upregulated |
| 0,305049079 | 0,336287949 | RP6-109B7.3 | FALSE | Upregulated |
| -0,162650904 | 0,336287949 | SYAP1 | FALSE | Downregulated |
| -0,308293858 | 0,336289822 | MLXIPL | FALSE | Downregulated |
| -0,136708164 | 0,336393082 | ZBTB40 | FALSE | Downregulated |
| -0,169537147 | 0,336493243 | SKIV2L | FALSE | Downregulated |
| -0,243816723 | 0,336493243 | TAF1C | FALSE | Downregulated |

| | | | | |
|---|---|---|---|---|
| 1,125630528 | 0,336555836 | LOC102723542 | FALSE | Upregulated |
| 0,197571567 | 0,336555836 | MECR | FALSE | Upregulated |
| -0,242174035 | 0,336575125 | ZNF568 | FALSE | Downregulated |
| 0,666330297 | 0,336727636 | DNAH10 | FALSE | Upregulated |
| -0,312984389 | 0,336727636 | FAM86EP | FALSE | Downregulated |
| -0,555201199 | 0,336853764 | LOC643355 | FALSE | Downregulated |
| 0,130072699 | 0,336880095 | SHARPIN | FALSE | Upregulated |
| -0,175107526 | 0,336984429 | AGK | FALSE | Downregulated |
| -1,347405845 | 0,337206743 | FLNC | FALSE | Downregulated |
| -0,261720516 | 0,337206743 | PLEKHH3 | FALSE | Downregulated |
| -0,560044337 | 0,337397874 | C1orf220 | FALSE | Downregulated |
| 1,066219872 | 0,337397874 | SEMA3B-AS1 | FALSE | Upregulated |
| -0,277386692 | 0,337397874 | TOX2 | FALSE | Downregulated |
| -0,207904807 | 0,337520033 | ZNF484 | FALSE | Downregulated |
| 0,220381944 | 0,337656046 | CCDC57 | FALSE | Upregulated |
| 0,970727683 | 0,337807207 | LOC101928562 | FALSE | Upregulated |
| 0,16876866 | 0,337913359 | DLGAP4 | FALSE | Upregulated |
| 0,20595884 | 0,337913359 | SGTB | FALSE | Upregulated |
| 0,782159487 | 0,337935642 | GGTLC2 | FALSE | Upregulated |
| -0,758560262 | 0,337935642 | LHX6 | FALSE | Downregulated |
| -0,130698489 | 0,338006168 | IPO8 | FALSE | Downregulated |
| 0,14049788 | 0,338021824 | PHF20 | FALSE | Upregulated |
| -0,587093003 | 0,338361945 | FAM84A | FALSE | Downregulated |
| -1,209914301 | 0,338361945 | WDR11-AS1 | FALSE | Downregulated |
| -0,190207022 | 0,338504469 | KDM2B | FALSE | Downregulated |
| -0,339639318 | 0,338504469 | LOC100272217 | FALSE | Downregulated |
| 0,232674719 | 0,338504469 | NXPE3 | FALSE | Upregulated |
| 0,172526675 | 0,338629455 | COQ6 | FALSE | Upregulated |
| -0,215545131 | 0,338802957 | SNAPC4 | FALSE | Downregulated |
| 0,230167361 | 0,338825432 | SLC48A1 | FALSE | Upregulated |
| 0,242626241 | 0,338848806 | MAP3K8 | FALSE | Upregulated |
| 0,373811383 | 0,338868944 | SLC45A4 | FALSE | Upregulated |
| 0,216186476 | 0,338938428 | CBL | FALSE | Upregulated |
| -0,332790616 | 0,338938428 | KAT2A | FALSE | Downregulated |
| -0,169903284 | 0,33915521 | ZNF236 | FALSE | Downregulated |
| -1,28753046 | 0,339205704 | PLN | FALSE | Downregulated |
| 0,134147732 | 0,339280825 | ADSL | FALSE | Upregulated |
| -1,056481586 | 0,339280825 | KCNIP2-AS1 | FALSE | Downregulated |
| -0,289057062 | 0,339280825 | OVGP1 | FALSE | Downregulated |
| -0,084059483 | 0,339280825 | SKIV2L2 | FALSE | Downregulated |
| 0,278500375 | 0,339298129 | LOC642441 | FALSE | Upregulated |
| 0,964458281 | 0,339298129 | RNF219-AS1 | FALSE | Upregulated |
| -0,844478244 | 0,339353996 | CEMP1 | FALSE | Downregulated |
| 0,391571363 | 0,339353996 | ZNF726 | FALSE | Upregulated |
| -0,353181082 | 0,339362522 | COL12A1 | FALSE | Downregulated |
| 0,748241254 | 0,339362522 | KLRC2 | FALSE | Upregulated |
| -0,876793207 | 0,339532485 | FUT3 | FALSE | Downregulated |
| 0,138941935 | 0,339532485 | VAV2 | FALSE | Upregulated |
| 0,082405519 | 0,339641297 | PRCC | FALSE | Upregulated |
| -0,225851779 | 0,339802367 | C9orf3 | FALSE | Downregulated |
| -0,429305962 | 0,339956239 | KIT | FALSE | Downregulated |

| | | | | |
|---|---|---|---|---|
| -0,214178052 | 0,33999925 | RAI14 | FALSE | Downregulated |
| -1,483586149 | 0,34052329 | CSMD3 | FALSE | Downregulated |
| 0,120559303 | 0,340546229 | CBX1 | FALSE | Upregulated |
| 0,345557494 | 0,340547606 | CCDC170 | FALSE | Upregulated |
| -0,152568153 | 0,340547606 | ZNF326 | FALSE | Downregulated |
| -0,215607219 | 0,340750372 | VEGFB | FALSE | Downregulated |
| 0,51675791 | 0,340774081 | DOC2B | FALSE | Upregulated |
| -0,112316976 | 0,340774081 | YTHDF1 | FALSE | Downregulated |
| -0,980175455 | 0,340965915 | TBX1 | FALSE | Downregulated |
| -0,72720396 | 0,341036217 | WDR86 | FALSE | Downregulated |
| 0,191056431 | 0,341043037 | PIGA | FALSE | Upregulated |
| -0,826812979 | 0,341043037 | SULT4A1 | FALSE | Downregulated |
| 0,234098767 | 0,34152337 | CCDC88A | FALSE | Upregulated |
| 0,125111312 | 0,34152337 | DAP3 | FALSE | Upregulated |
| -0,536788108 | 0,34152337 | RP11-143J12.2 | FALSE | Downregulated |
| -0,304558532 | 0,34152337 | VEGFC | FALSE | Downregulated |
| -0,19700829 | 0,341537003 | MTDH | FALSE | Downregulated |
| -0,339121741 | 0,341546076 | IQSEC2 | FALSE | Downregulated |
| -0,266324262 | 0,341571638 | STXBP4 | FALSE | Downregulated |
| -0,206884383 | 0,341656609 | TMEM176B | FALSE | Downregulated |
| -0,470698257 | 0,341656609 | WDR17 | FALSE | Downregulated |
| -0,657066913 | 0,341697639 | MPP3 | FALSE | Downregulated |
| -0,489323779 | 0,341697639 | RP11-314C16.1 | FALSE | Downregulated |
| 0,543926336 | 0,341784431 | LOC101927929 | FALSE | Upregulated |
| 0,693709245 | 0,341814435 | CCL4L1 | FALSE | Upregulated |
| 0,683677009 | 0,341814435 | TNFRSF8 | FALSE | Upregulated |
| -0,079919426 | 0,342222174 | ADNP | FALSE | Downregulated |
| 0,564124379 | 0,342222174 | SPHK1 | FALSE | Upregulated |
| -0,3437743 | 0,342275509 | ATRNL1 | FALSE | Downregulated |
| 0,207913235 | 0,342383006 | KLHL26 | FALSE | Upregulated |
| -0,327892637 | 0,342383006 | WNK3 | FALSE | Downregulated |
| 0,129894594 | 0,342384463 | FRG1 | FALSE | Upregulated |
| -0,560641384 | 0,342384463 | RAB6B | FALSE | Downregulated |
| 0,084067481 | 0,342896288 | AK2 | FALSE | Upregulated |
| -0,087835133 | 0,34290071 | KHSRP | FALSE | Downregulated |
| -0,485672923 | 0,34290071 | LOC101929264 | FALSE | Downregulated |
| 0,171647513 | 0,342921317 | NME6 | FALSE | Upregulated |
| -0,430879491 | 0,343016425 | ACR | FALSE | Downregulated |
| 0,871452161 | 0,343016425 | BDKRB2 | FALSE | Upregulated |
| 0,654080554 | 0,343016425 | HLA-DQA2 | FALSE | Upregulated |
| -1,080650689 | 0,343016425 | KCNC4-AS1 | FALSE | Downregulated |
| 0,326682899 | 0,343016425 | PRSS8 | FALSE | Upregulated |
| 0,171783963 | 0,343016425 | RABGAP1L | FALSE | Upregulated |
| 1,297602819 | 0,343027028 | KRT85 | FALSE | Upregulated |
| -0,600712276 | 0,343030027 | LOC102724450 | FALSE | Downregulated |
| -0,577023819 | 0,343030027 | RP11-101E14.2 | FALSE | Downregulated |
| -0,182780539 | 0,343091357 | IVD | FALSE | Downregulated |
| 0,37553771 | 0,343271664 | ARHGAP44 | FALSE | Upregulated |
| -0,192761455 | 0,343271664 | BCL9L | FALSE | Downregulated |
| -0,237514896 | 0,343312282 | BLOC1S5 | FALSE | Downregulated |
| -0,602304397 | 0,343312282 | SPESP1 | FALSE | Downregulated |

| | | | | |
|---|---|---|---|---|
| 0,382404266 | 0,34349842 | GDPGP1 | FALSE | Upregulated |
| -0,692010024 | 0,343611269 | LOC101928462 | FALSE | Downregulated |
| -0,114924699 | 0,34389283 | DCP1A | FALSE | Downregulated |
| 0,207580368 | 0,343917399 | AMMECR1L | FALSE | Upregulated |
| -0,526434224 | 0,343917399 | CYP2B7P | FALSE | Downregulated |
| -0,132810626 | 0,343917399 | GRPEL2 | FALSE | Downregulated |
| -0,152366505 | 0,343917399 | PLEKHF2 | FALSE | Downregulated |
| -0,126662646 | 0,34398916 | IL13RA1 | FALSE | Downregulated |
| -1,030669932 | 0,344034933 | AXDND1 | FALSE | Downregulated |
| -0,213260072 | 0,344342074 | GLUD1 | FALSE | Downregulated |
| 0,592546595 | 0,344455839 | CCR6 | FALSE | Upregulated |
| 0,197317718 | 0,344459561 | PQLC3 | FALSE | Upregulated |
| 0,191686338 | 0,344485786 | SAC3D1 | FALSE | Upregulated |
| 0,34818539 | 0,344491473 | TNIK | FALSE | Upregulated |
| 0,2986774 | 0,344623523 | BST2 | FALSE | Upregulated |
| -0,178013626 | 0,344660625 | MPST | FALSE | Downregulated |
| 0,241151705 | 0,344660625 | TBXAS1 | FALSE | Upregulated |
| -0,191473381 | 0,344699524 | C8orf82 | FALSE | Downregulated |
| -0,111846324 | 0,344699524 | SLC25A46 | FALSE | Downregulated |
| 0,122601249 | 0,344699524 | SUPT4H1 | FALSE | Upregulated |
| -0,285569742 | 0,344699524 | TIGD5 | FALSE | Downregulated |
| -0,10098896 | 0,344706593 | MBD1 | FALSE | Downregulated |
| 0,218813048 | 0,344781217 | KIAA0556 | FALSE | Upregulated |
| -0,245932154 | 0,344781217 | PRICKLE2 | FALSE | Downregulated |
| 0,456735747 | 0,345133267 | FBXO16 | FALSE | Upregulated |
| 0,592051037 | 0,345133267 | ITPRIPL1 | FALSE | Upregulated |
| 0,142290619 | 0,345138015 | ANTXR2 | FALSE | Upregulated |
| -0,142298485 | 0,34530425 | TMEM161A | FALSE | Downregulated |
| -0,100384915 | 0,345396484 | ADIPOR1 | FALSE | Downregulated |
| -0,801920963 | 0,345396484 | STX19 | FALSE | Downregulated |
| 0,993732877 | 0,345396484 | WFDC2 | FALSE | Upregulated |
| 0,718779296 | 0,345404028 | GPR64 | FALSE | Upregulated |
| -0,273104544 | 0,34564941 | SNHG7 | FALSE | Downregulated |
| 0,292992977 | 0,34564941 | TM4SF5 | FALSE | Upregulated |
| -0,860340563 | 0,345657636 | UCKL1-AS1 | FALSE | Downregulated |
| 0,20369994 | 0,34566275 | DCP2 | FALSE | Upregulated |
| 0,125295035 | 0,346081129 | CCBL2 | FALSE | Upregulated |
| 0,255390398 | 0,346081129 | ITPKB | FALSE | Upregulated |
| 0,187535918 | 0,346166751 | BBS7 | FALSE | Upregulated |
| -0,487048283 | 0,346375073 | COL6A6 | FALSE | Downregulated |
| -0,282796748 | 0,34653224 | SYT9 | FALSE | Downregulated |
| 0,24478764 | 0,346595695 | ALDH1B1 | FALSE | Upregulated |
| -0,15643912 | 0,346810287 | NDUFS4 | FALSE | Downregulated |
| -2,660315129 | 0,346851149 | RNU1-1 | FALSE | Downregulated |
| -0,58497514 | 0,346851149 | SLC3A1 | FALSE | Downregulated |
| -1,943359186 | 0,347083712 | CLVS2 | FALSE | Downregulated |
| -0,807877531 | 0,347104911 | FAM24B | FALSE | Downregulated |
| -0,160986341 | 0,347110639 | DTD2 | FALSE | Downregulated |
| 0,271229632 | 0,34738349 | TTC7A | FALSE | Upregulated |
| -0,282956472 | 0,347623238 | EPOR | FALSE | Downregulated |
| 0,480270652 | 0,347844259 | FICD | FALSE | Upregulated |

| | | | | |
|---|---|---|---|---|
| -0,172561586 | 0,348058615 | AMBP | FALSE | Downregulated |
| -0,207712445 | 0,348166012 | NOTCH2NL | FALSE | Downregulated |
| 0,145497404 | 0,348193735 | ZFP41 | FALSE | Upregulated |
| -0,416132408 | 0,348544906 | LOC100129203 | FALSE | Downregulated |
| -1,119785009 | 0,348812204 | LOC100129138 | FALSE | Downregulated |
| -0,194611242 | 0,348812204 | PHTF1 | FALSE | Downregulated |
| -0,814664964 | 0,34889026 | PLAC8L1 | FALSE | Downregulated |
| -0,262305118 | 0,349244824 | HTR7P1 | FALSE | Downregulated |
| -0,216585822 | 0,349381743 | NEURL4 | FALSE | Downregulated |
| -0,219486258 | 0,349381743 | PXDC1 | FALSE | Downregulated |
| 1,039404872 | 0,349421274 | ELFN2 | FALSE | Upregulated |
| 0,228119896 | 0,349428852 | GNPDA2 | FALSE | Upregulated |
| 0,124945145 | 0,349461863 | HSF1 | FALSE | Upregulated |
| -0,219929188 | 0,349461863 | ZNF518B | FALSE | Downregulated |
| -0,088072852 | 0,349461863 | ZNF664 | FALSE | Downregulated |
| -0,255533826 | 0,349719897 | LOC100506538 | FALSE | Downregulated |
| -0,677049428 | 0,349719897 | PABPC4L | FALSE | Downregulated |
| -0,137004088 | 0,349719897 | SH3RF1 | FALSE | Downregulated |
| 0,119611569 | 0,349887041 | FOXK2 | FALSE | Upregulated |
| 0,260323761 | 0,350278351 | ID2 | FALSE | Upregulated |
| -0,178952902 | 0,350413631 | ZNF717 | FALSE | Downregulated |
| -0,843039329 | 0,350717239 | RP11-57H12.3 | FALSE | Downregulated |
| -0,302656287 | 0,350717239 | WDR66 | FALSE | Downregulated |
| -0,180039624 | 0,350718525 | CNNM2 | FALSE | Downregulated |
| 0,71517188 | 0,35075076 | CYP27C1 | FALSE | Upregulated |
| -0,170118832 | 0,35075076 | REPS2 | FALSE | Downregulated |
| -0,136732211 | 0,35075076 | ZNF768 | FALSE | Downregulated |
| 0,767451683 | 0,350762747 | MGAT3 | FALSE | Upregulated |
| 0,232044201 | 0,351021204 | HARBI1 | FALSE | Upregulated |
| -0,313225731 | 0,351237706 | FAM174B | FALSE | Downregulated |
| -0,221277101 | 0,351237706 | GTPBP3 | FALSE | Downregulated |
| -0,185230558 | 0,351237706 | THAP7 | FALSE | Downregulated |
| -0,240734839 | 0,351237706 | ZDBF2 | FALSE | Downregulated |
| 0,133847923 | 0,351267481 | C2CD3 | FALSE | Upregulated |
| 0,235187317 | 0,351371125 | HSPA1A | FALSE | Upregulated |
| -0,650939193 | 0,351371125 | ZNF638-IT1 | FALSE | Downregulated |
| -0,450105521 | 0,351380777 | EMID1 | FALSE | Downregulated |
| 0,290606874 | 0,351586571 | ZNF185 | FALSE | Upregulated |
| 0,167520955 | 0,351619995 | RAVER1 | FALSE | Upregulated |
| 0,152567981 | 0,351619995 | VPS41 | FALSE | Upregulated |
| -1,368024254 | 0,351643309 | TCAP | FALSE | Downregulated |
| 1,189424103 | 0,35190396 | LOC101060153 | FALSE | Upregulated |
| 0,092375427 | 0,352045509 | UBAP1 | FALSE | Upregulated |
| -0,226589784 | 0,352158639 | GARNL3 | FALSE | Downregulated |
| -0,335736838 | 0,352158639 | LTBP4 | FALSE | Downregulated |
| 0,22983412 | 0,352176931 | BORA | FALSE | Upregulated |
| -0,652225951 | 0,352214486 | MIR6843 | FALSE | Downregulated |
| 0,083287993 | 0,352214486 | UBE3C | FALSE | Upregulated |
| 0,442577072 | 0,352214649 | SLC35F2 | FALSE | Upregulated |
| -1,024045262 | 0,352291397 | LOC101927479 | FALSE | Downregulated |
| 1,046625562 | 0,352466127 | MYO3B | FALSE | Upregulated |

| | | | | |
|---|---|---|---|---|
| -0,200343552 | 0,352466127 | NME4 | FALSE | Downregulated |
| -0,206726949 | 0,352655587 | CAMK2N1 | FALSE | Downregulated |
| 1,619924039 | 0,352655587 | LOC102724971 | FALSE | Upregulated |
| 0,183599533 | 0,352655587 | SNRPD2 | FALSE | Upregulated |
| 0,946316202 | 0,352684412 | SUSD4 | FALSE | Upregulated |
| -0,404070395 | 0,352909867 | NHSL2 | FALSE | Downregulated |
| 1,02726844 | 0,352926211 | GPR141 | FALSE | Upregulated |
| 0,175863999 | 0,352987165 | HPS6 | FALSE | Upregulated |
| -0,433179148 | 0,352987165 | SNCG | FALSE | Downregulated |
| -0,227174237 | 0,352997732 | ARID4B | FALSE | Downregulated |
| -0,553944215 | 0,352997732 | RP11-423H2.3 | FALSE | Downregulated |
| -0,152545306 | 0,353073131 | MRPL33 | FALSE | Downregulated |
| -0,647594853 | 0,353073131 | ZNF727 | FALSE | Downregulated |
| -0,217902105 | 0,353137308 | ATP6V0E2 | FALSE | Downregulated |
| -0,167853822 | 0,353137308 | DDX51 | FALSE | Downregulated |
| 0,422892269 | 0,353137308 | UGT2B11 | FALSE | Upregulated |
| -0,552970249 | 0,353157559 | LOC100129917 | FALSE | Downregulated |
| 0,712732371 | 0,353337004 | CXCL1 | FALSE | Upregulated |
| -0,672385296 | 0,353337004 | FLJ39632 | FALSE | Downregulated |
| -0,280330749 | 0,353337004 | FMO5 | FALSE | Downregulated |
| -1,196997936 | 0,353337004 | LOC100506351 | FALSE | Downregulated |
| -1,317568285 | 0,353337004 | LOC101929375 | FALSE | Downregulated |
| -0,164330915 | 0,353400876 | SYF2 | FALSE | Downregulated |
| -0,158919277 | 0,353541303 | NIT2 | FALSE | Downregulated |
| 0,140052525 | 0,353541303 | NUPL1 | FALSE | Upregulated |
| -0,307148553 | 0,353541303 | RFESD | FALSE | Downregulated |
| -0,825115491 | 0,353541303 | ZNF728 | FALSE | Downregulated |
| -0,178144703 | 0,353589011 | ZNF337 | FALSE | Downregulated |
| 1,168896965 | 0,353676656 | PCSK1 | FALSE | Upregulated |
| 0,476986602 | 0,353676656 | SLC44A3 | FALSE | Upregulated |
| 0,659608187 | 0,35378109 | PMFBP1 | FALSE | Upregulated |
| 0,293211358 | 0,353867946 | LOC101928465 | FALSE | Upregulated |
| -0,42375687 | 0,35416153 | CD248 | FALSE | Downregulated |
| 2,162657894 | 0,354246489 | RP11-513O17.2-2 | FALSE | Upregulated |
| 0,238991896 | 0,354298688 | ST3GAL4 | FALSE | Upregulated |
| -0,146667842 | 0,354307023 | FBXL12 | FALSE | Downregulated |
| 1,006490017 | 0,354412303 | LOC101928955 | FALSE | Upregulated |
| -0,252990582 | 0,354487579 | ZNF266 | FALSE | Downregulated |
| -0,110538167 | 0,35450132 | FAM122A | FALSE | Downregulated |
| -1,334785265 | 0,354516165 | COL6A5 | FALSE | Downregulated |
| 0,172152394 | 0,354516165 | CPNE2 | FALSE | Upregulated |
| -0,099348574 | 0,354516165 | DYRK1A | FALSE | Downregulated |
| 0,259677991 | 0,354516165 | NUDT2 | FALSE | Upregulated |
| -0,638633069 | 0,354528658 | COL9A2 | FALSE | Downregulated |
| 0,830337302 | 0,354587764 | CPNE7 | FALSE | Upregulated |
| -0,819742812 | 0,354587764 | LOC101593348 | FALSE | Downregulated |
| -0,183183229 | 0,354671098 | CCDC125 | FALSE | Downregulated |
| 0,210294855 | 0,354737484 | AAMDC | FALSE | Upregulated |
| -0,116993829 | 0,354737484 | C2orf47 | FALSE | Downregulated |
| -0,295382715 | 0,354737484 | LINC-PINT | FALSE | Downregulated |
| -0,623372183 | 0,354737484 | LOC102724316 | FALSE | Downregulated |

| | | | | |
|---|---|---|---|---|
| -0,292880663 | 0,354737484 | LOC102724873 | FALSE | Downregulated |
| -0,149414957 | 0,354737484 | MAN1A2 | FALSE | Downregulated |
| 0,475090079 | 0,354737484 | RPL23AP64 | FALSE | Upregulated |
| 0,25491309 | 0,354737484 | SEPW1 | FALSE | Upregulated |
| -0,161837987 | 0,355161421 | IRF2BP2 | FALSE | Downregulated |
| -0,091809991 | 0,355771081 | EIF2AK1 | FALSE | Downregulated |
| -0,175017709 | 0,355814268 | CCDC132 | FALSE | Downregulated |
| -1,169536033 | 0,355870828 | LOC100128374 | FALSE | Downregulated |
| -0,146408728 | 0,355870828 | RPS6KA2 | FALSE | Downregulated |
| -0,111689461 | 0,355992472 | C6orf211 | FALSE | Downregulated |
| -0,570977139 | 0,356029521 | FNDC1 | FALSE | Downregulated |
| -0,096477169 | 0,356030284 | OS9 | FALSE | Downregulated |
| -0,148362273 | 0,35606644 | ABCC2 | FALSE | Downregulated |
| 0,566773164 | 0,35606644 | TMEM154 | FALSE | Upregulated |
| -0,142332171 | 0,35606644 | TPCN1 | FALSE | Downregulated |
| -0,113374871 | 0,356482898 | DNMBP | FALSE | Downregulated |
| 0,274472824 | 0,356626567 | KIRREL | FALSE | Upregulated |
| -0,210462635 | 0,356626567 | SELENBP1 | FALSE | Downregulated |
| -0,821444113 | 0,356684236 | FBXW10 | FALSE | Downregulated |
| 0,363379378 | 0,357000632 | PODXL | FALSE | Upregulated |
| 0,852383281 | 0,357038733 | LOC102724049 | FALSE | Upregulated |
| -0,231687568 | 0,357073129 | PDE3B | FALSE | Downregulated |
| -0,429045682 | 0,357289207 | PPP1R9A | FALSE | Downregulated |
| -0,65490111 | 0,357361056 | CTSLP2 | FALSE | Downregulated |
| 0,590541665 | 0,358152429 | SLC9A7P1 | FALSE | Upregulated |
| -0,283226277 | 0,358319847 | EMP2 | FALSE | Downregulated |
| 1,047902193 | 0,358409334 | MDFI | FALSE | Upregulated |
| -0,368826263 | 0,358438087 | INCA1 | FALSE | Downregulated |
| -0,15439798 | 0,358469517 | ING3 | FALSE | Downregulated |
| 0,175442752 | 0,358648094 | DPP7 | FALSE | Upregulated |
| 0,276513387 | 0,358678915 | CCDC113 | FALSE | Upregulated |
| -0,236491829 | 0,358777242 | ZNF662 | FALSE | Downregulated |
| 0,392153198 | 0,358806216 | ARMC9 | FALSE | Upregulated |
| 0,113308853 | 0,358806216 | TBCC | FALSE | Upregulated |
| -0,243542105 | 0,358828087 | FAAH2 | FALSE | Downregulated |
| 0,102655954 | 0,358828087 | SCO1 | FALSE | Upregulated |
| 0,149771977 | 0,358875716 | OPN3 | FALSE | Upregulated |
| -0,411696811 | 0,358875716 | RP1-60O19.1 | FALSE | Downregulated |
| 0,683178074 | 0,358891746 | GPM6B | FALSE | Upregulated |
| -0,969683213 | 0,358891746 | KISS1 | FALSE | Downregulated |
| -0,301178798 | 0,358891746 | LINC00087 | FALSE | Downregulated |
| -1,838893279 | 0,358891746 | PRND | FALSE | Downregulated |
| 0,361656521 | 0,358891746 | PSIMCT-1 | FALSE | Upregulated |
| -0,739276734 | 0,358891746 | RPL21P44 | FALSE | Downregulated |
| 0,980694498 | 0,358953338 | LOC100996557 | FALSE | Upregulated |
| -0,126318891 | 0,359268491 | SRPR | FALSE | Downregulated |
| 0,133284398 | 0,359297112 | HNRNPC | FALSE | Upregulated |
| 0,426173129 | 0,359297112 | RP11-367J11.2 | FALSE | Upregulated |
| -0,153749586 | 0,359481342 | SLC39A8 | FALSE | Downregulated |
| 0,122219315 | 0,359502743 | SNRPE | FALSE | Upregulated |
| -0,646792878 | 0,359845711 | LOC100288911 | FALSE | Downregulated |

| | | | | |
|---|---|---|---|---|
| 0,349112339 | 0,359845711 | MCAM | FALSE | Upregulated |
| -0,138320538 | 0,359845711 | TSR2 | FALSE | Downregulated |
| 0,289995503 | 0,359937907 | TNFSF13 | FALSE | Upregulated |
| -0,13421499 | 0,36011221 | ARID2 | FALSE | Downregulated |
| 0,169373679 | 0,36011221 | NOL6 | FALSE | Upregulated |
| -0,142760048 | 0,36011221 | SSH1 | FALSE | Downregulated |
| 0,380452924 | 0,360260448 | LARP6 | FALSE | Upregulated |
| -0,861451611 | 0,360642727 | LOC102724092 | FALSE | Downregulated |
| -0,273832231 | 0,360882797 | ZBTB3 | FALSE | Downregulated |
| -0,888979463 | 0,36093035 | HEPN1 | FALSE | Downregulated |
| -0,835110418 | 0,36093035 | LRRC15 | FALSE | Downregulated |
| -0,356165159 | 0,36093035 | LRRN3 | FALSE | Downregulated |
| 0,859896622 | 0,36093035 | OLR1 | FALSE | Upregulated |
| 0,157779083 | 0,36093035 | RRP15 | FALSE | Upregulated |
| -0,24570469 | 0,36093035 | SLC2A13 | FALSE | Downregulated |
| -0,205032666 | 0,36093035 | WDR24 | FALSE | Downregulated |
| 0,137750071 | 0,36093035 | ZFP36L2 | FALSE | Upregulated |
| -0,172314444 | 0,36093035 | ZNF782 | FALSE | Downregulated |
| -0,071634413 | 0,361117323 | ZC3H7A | FALSE | Downregulated |
| 0,459475002 | 0,361257937 | LOC101927903 | FALSE | Upregulated |
| -0,099249614 | 0,361402157 | NUP214 | FALSE | Downregulated |
| 0,320522363 | 0,361411972 | DENND6B | FALSE | Upregulated |
| -0,087703353 | 0,361457949 | TRMT5 | FALSE | Downregulated |
| -0,088207462 | 0,36186425 | LSM14A | FALSE | Downregulated |
| 0,145720145 | 0,36188973 | MLF2 | FALSE | Upregulated |
| 0,178884541 | 0,361990549 | LDOC1L | FALSE | Upregulated |
| -0,208263378 | 0,36201611 | PAM | FALSE | Downregulated |
| 0,247353619 | 0,36201611 | STOML1 | FALSE | Upregulated |
| 0,200781712 | 0,362123157 | C8orf37 | FALSE | Upregulated |
| 0,326641383 | 0,362238554 | ARHGEF35 | FALSE | Upregulated |
| -0,205644955 | 0,362362967 | ANP32E | FALSE | Downregulated |
| 1,423627268 | 0,362362967 | EPN3 | FALSE | Upregulated |
| -0,153945027 | 0,362438835 | BMPR1A | FALSE | Downregulated |
| -0,100478849 | 0,362438835 | CNOT2 | FALSE | Downregulated |
| 0,174095138 | 0,362438835 | EHD1 | FALSE | Upregulated |
| -0,347960604 | 0,362438835 | SGCE | FALSE | Downregulated |
| 0,954380102 | 0,362438835 | SPG20OS | FALSE | Upregulated |
| -0,16161407 | 0,362438835 | ZNF202 | FALSE | Downregulated |
| 0,713114342 | 0,362517706 | LOC101927260 | FALSE | Upregulated |
| -0,255052874 | 0,362520275 | CPEB3 | FALSE | Downregulated |
| -0,688785699 | 0,362873466 | GATM-AS1 | FALSE | Downregulated |
| -0,420257293 | 0,362873466 | LOC284023 | FALSE | Downregulated |
| -0,533242361 | 0,362881653 | MAP3K15 | FALSE | Downregulated |
| -1,058593589 | 0,362889787 | LOC101928841 | FALSE | Downregulated |
| -0,148204122 | 0,363115369 | COX19 | FALSE | Downregulated |
| -0,607015107 | 0,363267588 | LINC00266-3 | FALSE | Downregulated |
| -0,734148192 | 0,36345765 | LOC389607 | FALSE | Downregulated |
| 0,171831506 | 0,36345765 | NDUFAF4 | FALSE | Upregulated |
| -0,220395966 | 0,36345765 | TCF7L1 | FALSE | Downregulated |
| -0,197593031 | 0,363684945 | SYT17 | FALSE | Downregulated |
| -0,345660994 | 0,363684945 | TPTE2P5 | FALSE | Downregulated |

| | | | | |
|---|---|---|---|---|
| -0,502263415 | 0,36385365 | JAKMIP3 | FALSE | Downregulated |
| -0,400529746 | 0,363991968 | LOC101928042 | FALSE | Downregulated |
| -0,525208995 | 0,364243121 | GNRHR2 | FALSE | Downregulated |
| 0,641218409 | 0,364274178 | LOC101928070 | FALSE | Upregulated |
| -0,430803781 | 0,364276299 | LOC101929251 | FALSE | Downregulated |
| -0,176232152 | 0,364276299 | TFB1M | FALSE | Downregulated |
| -0,153185914 | 0,364348223 | ALMS1 | FALSE | Downregulated |
| 0,540239978 | 0,364403459 | ID1 | FALSE | Upregulated |
| 0,369610395 | 0,364464804 | CLEC4A | FALSE | Upregulated |
| -0,723602272 | 0,364464804 | LOC101927652 | FALSE | Downregulated |
| 0,223321611 | 0,364490407 | ZBTB45 | FALSE | Upregulated |
| 0,206891811 | 0,364903218 | ENPP4 | FALSE | Upregulated |
| 1,089200817 | 0,364940268 | ASPHD1 | FALSE | Upregulated |
| -0,167211178 | 0,364940268 | FPGS | FALSE | Downregulated |
| -0,453213474 | 0,365074761 | KCNJ14 | FALSE | Downregulated |
| 0,118384333 | 0,365111976 | UBIAD1 | FALSE | Upregulated |
| -0,3602345 | 0,36512182 | BSN | FALSE | Downregulated |
| 0,116035672 | 0,36512182 | URGCP | FALSE | Upregulated |
| -0,25882175 | 0,36520296 | CHCHD10 | FALSE | Downregulated |
| -0,143179809 | 0,36520296 | GGNBP2 | FALSE | Downregulated |
| -0,166979881 | 0,36544157 | APOOL | FALSE | Downregulated |
| -0,182904836 | 0,36544157 | ZNF627 | FALSE | Downregulated |
| -0,198177654 | 0,365475933 | LINC00467 | FALSE | Downregulated |
| -0,672383124 | 0,365475933 | STK32A | FALSE | Downregulated |
| -0,144852211 | 0,365503348 | RAP2C | FALSE | Downregulated |
| -0,605289767 | 0,365508624 | EFHB | FALSE | Downregulated |
| -0,32448193 | 0,365508624 | FOXO1 | FALSE | Downregulated |
| -0,340362237 | 0,365508624 | SLC6A13 | FALSE | Downregulated |
| -0,320704137 | 0,365508624 | SSBP2 | FALSE | Downregulated |
| -0,102674133 | 0,365562665 | INTS8 | FALSE | Downregulated |
| -0,55950846 | 0,365562665 | LINC00265 | FALSE | Downregulated |
| -0,628664168 | 0,365562665 | RSPO2 | FALSE | Downregulated |
| -0,388464726 | 0,365818877 | BMS1P4 | FALSE | Downregulated |
| -0,308313787 | 0,365818877 | NBPF19 | FALSE | Downregulated |
| 0,436783656 | 0,365818877 | SMAD5-AS1 | FALSE | Upregulated |
| 0,297255002 | 0,366035525 | CDC42EP3 | FALSE | Upregulated |
| -0,233723854 | 0,366353245 | TMEM220 | FALSE | Downregulated |
| -1,104762767 | 0,366420813 | DLEU7-AS1 | FALSE | Downregulated |
| 0,213708488 | 0,366498669 | ZKSCAN3 | FALSE | Upregulated |
| 0,845303791 | 0,366660902 | CDKN2A | FALSE | Upregulated |
| -0,089357768 | 0,366692658 | MED4 | FALSE | Downregulated |
| 0,48057989 | 0,366695821 | SH2D1B | FALSE | Upregulated |
| -0,256654613 | 0,36683775 | NLGN4X | FALSE | Downregulated |
| -0,137058875 | 0,366858225 | PEX1 | FALSE | Downregulated |
| 0,143022313 | 0,366915199 | DEDD2 | FALSE | Upregulated |
| 0,250188896 | 0,366915199 | HIVEP3 | FALSE | Upregulated |
| 0,131215776 | 0,366915199 | TMEM11 | FALSE | Upregulated |
| 0,23664562 | 0,366915199 | TMEM214 | FALSE | Upregulated |
| 0,95758784 | 0,366956921 | NCAN | FALSE | Upregulated |
| 0,497017237 | 0,36703389 | ADAT3 | FALSE | Upregulated |
| -0,923727772 | 0,36703389 | BSN-AS2 | FALSE | Downregulated |

| | | | | |
|---|---|---|---|---|
| 0,236846378 | 0,367054753 | GOT1 | FALSE | Upregulated |
| -0,834152761 | 0,367213639 | GS1-259H13.2 | FALSE | Downregulated |
| -0,402767705 | 0,367439821 | FAM13A-AS1 | FALSE | Downregulated |
| 0,618037101 | 0,367491213 | IL10RB-AS1 | FALSE | Upregulated |
| 0,113106687 | 0,367492317 | AUP1 | FALSE | Upregulated |
| -0,802102869 | 0,367492317 | LOC101928558 | FALSE | Downregulated |
| 0,17707534 | 0,367492317 | MINOS1 | FALSE | Upregulated |
| 0,197557365 | 0,367573583 | FUT11 | FALSE | Upregulated |
| -0,110344263 | 0,367573583 | TRUB1 | FALSE | Downregulated |
| -0,163212604 | 0,367636472 | NDUFB7 | FALSE | Downregulated |
| 0,491032035 | 0,367800717 | FAM212A | FALSE | Upregulated |
| -0,501275163 | 0,367913847 | GSTA3 | FALSE | Downregulated |
| -0,131118606 | 0,367913847 | KIAA0195 | FALSE | Downregulated |
| 0,1629483 | 0,367913847 | RPL29 | FALSE | Upregulated |
| 0,154053109 | 0,367927643 | LACE1 | FALSE | Upregulated |
| -0,227499454 | 0,367946504 | ACPL2 | FALSE | Downregulated |
| -0,766502732 | 0,368327261 | OBSCN | FALSE | Downregulated |
| -0,099701773 | 0,368473352 | ARFGEF2 | FALSE | Downregulated |
| 0,16340311 | 0,368473352 | RPL8 | FALSE | Upregulated |
| -0,223803823 | 0,368676107 | RPP21 | FALSE | Downregulated |
| -0,431934552 | 0,368678698 | PRKD1 | FALSE | Downregulated |
| -0,182592202 | 0,368833463 | EEA1 | FALSE | Downregulated |
| 0,127857206 | 0,368865712 | MTMR6 | FALSE | Upregulated |
| 0,170239155 | 0,368956623 | RPS15 | FALSE | Upregulated |
| 0,153221534 | 0,368965341 | AP5M1 | FALSE | Upregulated |
| 0,622213876 | 0,369012387 | LOC102724163 | FALSE | Upregulated |
| -0,154805986 | 0,369073179 | FBXL4 | FALSE | Downregulated |
| -0,495004174 | 0,369073179 | LINC00885 | FALSE | Downregulated |
| -0,804518949 | 0,369073179 | LOC101927135 | FALSE | Downregulated |
| 0,974828176 | 0,369076129 | LOC102724009 | FALSE | Upregulated |
| -0,590017584 | 0,369466626 | PABPC3 | FALSE | Downregulated |
| -1,456849214 | 0,369466626 | SLC35F1 | FALSE | Downregulated |
| 0,219351809 | 0,369487468 | ADAMTSL4 | FALSE | Upregulated |
| 0,144447548 | 0,369556171 | INO80E | FALSE | Upregulated |
| -1,534110473 | 0,369591273 | ANKRD20A3 | FALSE | Downregulated |
| -0,618222885 | 0,369591273 | NOVA1 | FALSE | Downregulated |
| -0,446236005 | 0,369591273 | SYNC | FALSE | Downregulated |
| 0,711216159 | 0,369685093 | B3GAT2 | FALSE | Upregulated |
| 0,84104628 | 0,369685093 | HOXB-AS1 | FALSE | Upregulated |
| 0,497139718 | 0,370059074 | CPZ | FALSE | Upregulated |
| -0,410445589 | 0,370177343 | LOC101927759 | FALSE | Downregulated |
| -0,286892068 | 0,370414845 | ACOT2 | FALSE | Downregulated |
| 0,162991287 | 0,370414845 | SERAC1 | FALSE | Upregulated |
| 0,133632234 | 0,370601051 | TRAPPC13 | FALSE | Upregulated |
| -0,179237581 | 0,370636911 | ZCCHC4 | FALSE | Downregulated |
| 0,284433664 | 0,370660541 | C4orf21 | FALSE | Upregulated |
| 0,112134748 | 0,370776617 | FAM53C | FALSE | Upregulated |
| 0,332317481 | 0,370811091 | C19orf73 | FALSE | Upregulated |
| -0,295737941 | 0,370811091 | USP35 | FALSE | Downregulated |
| -0,302335361 | 0,370856983 | RFX2 | FALSE | Downregulated |
| -0,281195608 | 0,370883504 | SLC6A1 | FALSE | Downregulated |

| | | | | |
|---|---|---|---|---|
| 0,184085888 | 0,371057779 | NGFRAP1 | FALSE | Upregulated |
| 0,155888295 | 0,371057779 | SEC22B | FALSE | Upregulated |
| -0,219748139 | 0,371057779 | SLC22A3 | FALSE | Downregulated |
| -0,188746749 | 0,371069966 | NRIP1 | FALSE | Downregulated |
| 0,110494007 | 0,37115373 | NARS | FALSE | Upregulated |
| -0,134055678 | 0,371207538 | RWDD2B | FALSE | Downregulated |
| -0,383240962 | 0,371207538 | SULT1B1 | FALSE | Downregulated |
| 0,358055644 | 0,371290858 | CDK5R1 | FALSE | Upregulated |
| -0,132870931 | 0,371494406 | MLLT6 | FALSE | Downregulated |
| -0,203701176 | 0,371494406 | SLC30A1 | FALSE | Downregulated |
| -1,275730234 | 0,371815975 | B4GALNT4 | FALSE | Downregulated |
| -0,53784473 | 0,371815975 | LINC00472 | FALSE | Downregulated |
| 0,303220372 | 0,371835295 | TUB | FALSE | Upregulated |
| 0,072776493 | 0,371981603 | AHCYL1 | FALSE | Upregulated |
| 0,37525475 | 0,371981603 | INHBE | FALSE | Upregulated |
| -0,163291983 | 0,371981603 | WWTR1 | FALSE | Downregulated |
| 0,153973772 | 0,372033849 | PMPCA | FALSE | Upregulated |
| 0,179861468 | 0,372148959 | ABCB6 | FALSE | Upregulated |
| 0,236115522 | 0,372148959 | RTN4IP1 | FALSE | Upregulated |
| -0,285108095 | 0,372148959 | SLC25A47 | FALSE | Downregulated |
| -0,26502234 | 0,372263986 | NCKAP5 | FALSE | Downregulated |
| -0,160588399 | 0,372290566 | MRPS18C | FALSE | Downregulated |
| -1,422976731 | 0,372404921 | C3orf20 | FALSE | Downregulated |
| -0,596919464 | 0,372563602 | MDH1B | FALSE | Downregulated |
| 0,20412546 | 0,373036735 | ARSA | FALSE | Upregulated |
| -0,151837423 | 0,373187051 | HADHB | FALSE | Downregulated |
| -0,119881482 | 0,373256513 | APC | FALSE | Downregulated |
| -1,095451532 | 0,373259825 | DOCK9-AS1 | FALSE | Downregulated |
| -0,528025139 | 0,373400338 | LOC101927300 | FALSE | Downregulated |
| -0,248875424 | 0,373420422 | LOC100507412 | FALSE | Downregulated |
| -0,380804155 | 0,373420422 | LTK | FALSE | Downregulated |
| -0,350358125 | 0,373533113 | ADAM8 | FALSE | Downregulated |
| 0,225840925 | 0,373533113 | C19orf82 | FALSE | Upregulated |
| 0,288824453 | 0,373533113 | ZNF28 | FALSE | Upregulated |
| 0,321861914 | 0,373563188 | CSRNP1 | FALSE | Upregulated |
| 0,163174513 | 0,373615811 | POLR1A | FALSE | Upregulated |
| -0,13043828 | 0,373708008 | CNNM3 | FALSE | Downregulated |
| 0,192672196 | 0,373708008 | NAIF1 | FALSE | Upregulated |
| -1,088372551 | 0,373708008 | RBFADN | FALSE | Downregulated |
| 0,604266652 | 0,3739152 | FAM167A | FALSE | Upregulated |
| 1,024345081 | 0,374103718 | LOC100128882 | FALSE | Upregulated |
| 1,082436028 | 0,374103718 | PBX4 | FALSE | Upregulated |
| -0,154309043 | 0,374103718 | RCBTB2 | FALSE | Downregulated |
| 0,302757951 | 0,374103718 | SLFN12 | FALSE | Upregulated |
| 0,155531295 | 0,374103718 | TARS | FALSE | Upregulated |
| -0,236024868 | 0,374312393 | ATXN7L1 | FALSE | Downregulated |
| -0,580163809 | 0,374312393 | CYP2A7 | FALSE | Downregulated |
| 0,202304246 | 0,374312393 | DPM3 | FALSE | Upregulated |
| -0,252706873 | 0,374312393 | GHR | FALSE | Downregulated |
| -0,150476529 | 0,374312393 | MAP2K7 | FALSE | Downregulated |
| 0,116296638 | 0,374312393 | SLC25A5 | FALSE | Upregulated |

| | | | | |
|---|---|---|---|---|
| 0,251450778 | 0,374371228 | SLC25A18 | FALSE | Upregulated |
| 1,104850439 | 0,374400491 | DNAH8 | FALSE | Upregulated |
| -0,094788084 | 0,374519115 | RNF111 | FALSE | Downregulated |
| -0,19439528 | 0,374645436 | PHF21A | FALSE | Downregulated |
| -0,301481591 | 0,374687139 | LOC100133985 | FALSE | Downregulated |
| -0,537331379 | 0,374826098 | RP11-500B12.1 | FALSE | Downregulated |
| -0,184335886 | 0,374969153 | DUS4L | FALSE | Downregulated |
| 0,407940603 | 0,375027627 | KRT19 | FALSE | Upregulated |
| -0,623951611 | 0,375067449 | LOC101927520 | FALSE | Downregulated |
| -0,167430058 | 0,375067449 | MST1 | FALSE | Downregulated |
| 0,198997828 | 0,375207488 | MIS18A | FALSE | Upregulated |
| 1,076776636 | 0,375207488 | P4HA2-AS1 | FALSE | Upregulated |
| -0,163949697 | 0,375632477 | NDRG2 | FALSE | Downregulated |
| -0,14715369 | 0,375757124 | PREPL | FALSE | Downregulated |
| -1,642658735 | 0,375931772 | GABRB2 | FALSE | Downregulated |
| -0,204213504 | 0,376076597 | PLEKHG3 | FALSE | Downregulated |
| 0,149819933 | 0,376093118 | METTL9 | FALSE | Upregulated |
| -0,141831736 | 0,376093118 | OCIAD1 | FALSE | Downregulated |
| 0,097802682 | 0,376111293 | NCBP1 | FALSE | Upregulated |
| -0,344583376 | 0,376111293 | SMCO3 | FALSE | Downregulated |
| -0,418450403 | 0,376376939 | ZNF807 | FALSE | Downregulated |
| -1,085813614 | 0,376382783 | CORO6 | FALSE | Downregulated |
| 0,170355345 | 0,376382783 | EOGT | FALSE | Upregulated |
| -0,221603004 | 0,376587163 | LOC101929145 | FALSE | Downregulated |
| 0,536206026 | 0,37659487 | PLXDC1 | FALSE | Upregulated |
| -0,206184471 | 0,376600123 | CCDC142 | FALSE | Downregulated |
| 0,243154349 | 0,376706213 | PARVB | FALSE | Upregulated |
| -0,776072965 | 0,376854586 | LOC102724156 | FALSE | Downregulated |
| -0,164360111 | 0,376887087 | CDK6 | FALSE | Downregulated |
| 0,890272362 | 0,376887087 | EPHA7 | FALSE | Upregulated |
| -0,280285378 | 0,376887087 | FAM86C2P | FALSE | Downregulated |
| -0,087259903 | 0,376887087 | SCAF8 | FALSE | Downregulated |
| 0,237977205 | 0,376887087 | ZNF786 | FALSE | Upregulated |
| -0,594243851 | 0,377586193 | ATP8B5P | FALSE | Downregulated |
| 0,175366939 | 0,377586193 | BCAR1 | FALSE | Upregulated |
| 0,223116279 | 0,377814986 | FBXO5 | FALSE | Upregulated |
| 0,177944297 | 0,377825993 | ZNF219 | FALSE | Upregulated |
| -0,253889534 | 0,378327597 | UGT2A3 | FALSE | Downregulated |
| 1,254782796 | 0,378529636 | DLK1 | FALSE | Upregulated |
| 0,964598957 | 0,378529636 | MYCN | FALSE | Upregulated |
| -1,046413627 | 0,378724718 | LOC101930452 | FALSE | Downregulated |
| 0,140501365 | 0,378735839 | AARS | FALSE | Upregulated |
| 0,169294777 | 0,378735839 | METTL25 | FALSE | Upregulated |
| 0,273692372 | 0,378860604 | CCDC24 | FALSE | Upregulated |
| -0,288694701 | 0,378860604 | REPS1 | FALSE | Downregulated |
| -0,429018502 | 0,378860604 | TMTC2 | FALSE | Downregulated |
| -0,182480586 | 0,378997268 | PITPNC1 | FALSE | Downregulated |
| 0,187632625 | 0,379093073 | P4HTM | FALSE | Upregulated |
| -0,087832712 | 0,379102479 | AFTPH | FALSE | Downregulated |
| 0,209483894 | 0,379186398 | STX2 | FALSE | Upregulated |
| 0,475072979 | 0,379251041 | COL13A1 | FALSE | Upregulated |

| | | | | |
|---|---|---|---|---|
| 0,116533699 | 0,379251041 | RALBP1 | FALSE | Upregulated |
| 0,239034267 | 0,379251041 | SLCO3A1 | FALSE | Upregulated |
| 0,952246493 | 0,379367728 | LOC101929641 | FALSE | Upregulated |
| 0,701225248 | 0,379367728 | LOC102723722 | FALSE | Upregulated |
| -0,32671802 | 0,379551852 | TSPYL2 | FALSE | Downregulated |
| -0,28294145 | 0,379574574 | HOXA4 | FALSE | Downregulated |
| 0,452892816 | 0,37960832 | TBC1D30 | FALSE | Upregulated |
| -0,381180746 | 0,379636571 | DKFZP434I0714 | FALSE | Downregulated |
| -0,086510115 | 0,379658338 | UBE2R2 | FALSE | Downregulated |
| 0,319702606 | 0,379845621 | SOGA1 | FALSE | Upregulated |
| 0,107526208 | 0,379852648 | MAPKAPK2 | FALSE | Upregulated |
| -0,334277604 | 0,379892537 | AC009299.3 | FALSE | Downregulated |
| 1,075078476 | 0,379892806 | PRR19 | FALSE | Upregulated |
| 0,520545826 | 0,379916472 | MYOM1 | FALSE | Upregulated |
| -0,933945254 | 0,379916472 | NPAP1 | FALSE | Downregulated |
| -0,118978534 | 0,380032329 | FPGT | FALSE | Downregulated |
| 0,468928043 | 0,380105698 | KIAA1211 | FALSE | Upregulated |
| -0,256627434 | 0,380156661 | LOC642361 | FALSE | Downregulated |
| -0,90862899 | 0,380230979 | LOC102723894 | FALSE | Downregulated |
| -0,124775977 | 0,380290672 | ACVR1B | FALSE | Downregulated |
| 0,826541631 | 0,380290672 | HPN-AS1 | FALSE | Upregulated |
| -0,356123211 | 0,380290672 | SSTR2 | FALSE | Downregulated |
| 0,070898036 | 0,38036105 | CNOT7 | FALSE | Upregulated |
| 0,132656354 | 0,380402589 | PIP5K1A | FALSE | Upregulated |
| -1,707656249 | 0,380449621 | COMP | FALSE | Downregulated |
| -0,113025062 | 0,380449621 | LTN1 | FALSE | Downregulated |
| 0,127370242 | 0,380671264 | TCTA | FALSE | Upregulated |
| -0,34557153 | 0,38075928 | TNRC18P1 | FALSE | Downregulated |
| 0,143395569 | 0,380812613 | SEC23A | FALSE | Upregulated |
| -0,219457039 | 0,380825755 | HAUS4 | FALSE | Downregulated |
| -0,165751992 | 0,380839413 | TMEM141 | FALSE | Downregulated |
| -0,135271314 | 0,380996403 | NUDCD1 | FALSE | Downregulated |
| 0,09657723 | 0,381047998 | IL17RA | FALSE | Upregulated |
| 0,13595016 | 0,381188298 | SNRPC | FALSE | Upregulated |
| -0,154933568 | 0,381197014 | BET1 | FALSE | Downregulated |
| -0,114822033 | 0,381197014 | TRIM23 | FALSE | Downregulated |
| -0,161959093 | 0,381296296 | ZDHHC8 | FALSE | Downregulated |
| 0,153687138 | 0,381329131 | A1BG | FALSE | Upregulated |
| 0,189076192 | 0,381329131 | GPR137 | FALSE | Upregulated |
| -0,151486042 | 0,381329131 | KIAA0753 | FALSE | Downregulated |
| 0,142206081 | 0,381329131 | TMEM55B | FALSE | Upregulated |
| -0,134509856 | 0,381329131 | ZC3H10 | FALSE | Downregulated |
| -0,541081108 | 0,381407862 | N4BP2L2-IT2 | FALSE | Downregulated |
| 0,156553127 | 0,381594459 | C12orf45 | FALSE | Upregulated |
| -0,260273311 | 0,381720681 | MAFIP | FALSE | Downregulated |
| -0,144659938 | 0,381894916 | GTF3C4 | FALSE | Downregulated |
| -0,2653997 | 0,381969303 | TMEM14A | FALSE | Downregulated |
| -0,880074241 | 0,382001287 | LRRTM1 | FALSE | Downregulated |
| -0,869851788 | 0,382020282 | LOC101927190 | FALSE | Downregulated |
| -0,199873284 | 0,382020282 | SMPDL3A | FALSE | Downregulated |
| 2,521200698 | 0,382221161 | CKMT1A | FALSE | Upregulated |

| | | | | |
|---|---|---|---|---|
| 0,210095986 | 0,382274557 | SLC9A9 | FALSE | Upregulated |
| 0,11583831 | 0,382322949 | BTBD1 | FALSE | Upregulated |
| 0,146073764 | 0,382322949 | MSRB2 | FALSE | Upregulated |
| -0,209223366 | 0,382358182 | CXCL12 | FALSE | Downregulated |
| -0,159367805 | 0,38240869 | DNAAF2 | FALSE | Downregulated |
| -0,241927282 | 0,382454319 | RP11-290F20.1 | FALSE | Downregulated |
| 0,326454749 | 0,382484619 | B9D1 | FALSE | Upregulated |
| 0,145645198 | 0,382484619 | E2F6 | FALSE | Upregulated |
| -0,349391672 | 0,382484619 | EFCAB4A | FALSE | Downregulated |
| -0,769081318 | 0,382551858 | CDK3 | FALSE | Downregulated |
| 0,130874912 | 0,382562405 | DTWD2 | FALSE | Upregulated |
| -0,157432321 | 0,382722243 | DNAJC17 | FALSE | Downregulated |
| 0,785869625 | 0,382944757 | C1orf167 | FALSE | Upregulated |
| -0,965685625 | 0,383851635 | HCG25 | FALSE | Downregulated |
| -0,662771542 | 0,383851635 | LOC102723641 | FALSE | Downregulated |
| -0,220840151 | 0,383891697 | SCAND3 | FALSE | Downregulated |
| 0,116487053 | 0,383896053 | DKC1 | FALSE | Upregulated |
| -0,355142162 | 0,38394828 | LOC102724832 | FALSE | Downregulated |
| -1,030342929 | 0,384035034 | LOC101929781 | FALSE | Downregulated |
| -0,398318713 | 0,384059717 | INHBB | FALSE | Downregulated |
| -0,199078179 | 0,384079695 | CCDC104 | FALSE | Downregulated |
| -0,219017614 | 0,384079695 | ZNF586 | FALSE | Downregulated |
| 0,397196174 | 0,38412607 | C2orf15 | FALSE | Upregulated |
| 0,102316597 | 0,38412607 | DDX20 | FALSE | Upregulated |
| -0,166672904 | 0,38412607 | ISCA2 | FALSE | Downregulated |
| -0,687542908 | 0,38412607 | LOC100131347 | FALSE | Downregulated |
| -0,56223722 | 0,38412607 | LOC100507656 | FALSE | Downregulated |
| 1,168701233 | 0,38412607 | LOC102723801 | FALSE | Upregulated |
| -0,176968685 | 0,38412607 | PDDC1 | FALSE | Downregulated |
| -1,161147142 | 0,38412607 | RP1 | FALSE | Downregulated |
| -0,073927328 | 0,38412607 | VEZT | FALSE | Downregulated |
| 1,06943519 | 0,38412607 | WDR63 | FALSE | Upregulated |
| 0,281759993 | 0,384226874 | LINC01137 | FALSE | Upregulated |
| -0,384186173 | 0,384226874 | LOC100630918 | FALSE | Downregulated |
| 0,504921005 | 0,384226874 | LOC101928767 | FALSE | Upregulated |
| -0,363934067 | 0,384226874 | LOC102723405 | FALSE | Downregulated |
| -0,158931643 | 0,384247656 | GPX4 | FALSE | Downregulated |
| 0,20293866 | 0,384595106 | SRI | FALSE | Upregulated |
| -0,446119899 | 0,384888904 | PCNXL2 | FALSE | Downregulated |
| -0,869044331 | 0,38494535 | NAPSA | FALSE | Downregulated |
| 1,123397687 | 0,385007962 | HIGD1C | FALSE | Upregulated |
| 0,244913813 | 0,385094587 | C7orf31 | FALSE | Upregulated |
| 0,883669447 | 0,385094587 | HIST1H2BG | FALSE | Upregulated |
| -0,542975639 | 0,385094587 | MT1M | FALSE | Downregulated |
| 1,24124615 | 0,385094587 | ST20-MTHFS | FALSE | Upregulated |
| -0,293009428 | 0,385094587 | TCF21 | FALSE | Downregulated |
| -0,166429299 | 0,385257671 | CTNNBL1 | FALSE | Downregulated |
| 0,089748718 | 0,385375261 | C1orf43 | FALSE | Upregulated |
| -0,405674073 | 0,385375261 | CATSPER3 | FALSE | Downregulated |
| 0,509914491 | 0,385375261 | FTH1P3 | FALSE | Upregulated |
| 0,111638571 | 0,385375261 | MAF1 | FALSE | Upregulated |

| | | | | |
|---|---|---|---|---|
| 0,210665742 | 0,385375261 | MMP15 | FALSE | Upregulated |
| -0,152434153 | 0,385375261 | NDRG3 | FALSE | Downregulated |
| -0,460692271 | 0,385375261 | SMPD3 | FALSE | Downregulated |
| -0,405843194 | 0,385375261 | SPATA41 | FALSE | Downregulated |
| 0,130091178 | 0,385558327 | IER3IP1 | FALSE | Upregulated |
| -0,254159351 | 0,385558327 | PLCXD1 | FALSE | Downregulated |
| -0,329371488 | 0,385558327 | RP5-1024G6.5 | FALSE | Downregulated |
| -0,29707373 | 0,385558327 | SESN3 | FALSE | Downregulated |
| 0,411792698 | 0,385631897 | CHADL | FALSE | Upregulated |
| -0,138658353 | 0,385633843 | KIN | FALSE | Downregulated |
| 0,341056315 | 0,385643156 | FAM167B | FALSE | Upregulated |
| 0,171504587 | 0,385643156 | TMEM187 | FALSE | Upregulated |
| 0,474246734 | 0,385679809 | C17orf51 | FALSE | Upregulated |
| -0,766772751 | 0,385759306 | AC007773.2 | FALSE | Downregulated |
| -1,081393405 | 0,385759306 | CHMP1B2P | FALSE | Downregulated |
| 0,189501298 | 0,385759306 | LOC102723728 | FALSE | Upregulated |
| -0,164503156 | 0,385759306 | TRMT2B | FALSE | Downregulated |
| 0,143728446 | 0,385869223 | LGALS8 | FALSE | Upregulated |
| -0,124584731 | 0,385999573 | UBXN6 | FALSE | Downregulated |
| -0,148378001 | 0,386082534 | PLXNB1 | FALSE | Downregulated |
| 0,443557439 | 0,386292794 | ILDR2 | FALSE | Upregulated |
| -0,227400559 | 0,38662043 | MROH8 | FALSE | Downregulated |
| -0,139085127 | 0,386707374 | INSR | FALSE | Downregulated |
| -0,103382245 | 0,386707374 | TBC1D23 | FALSE | Downregulated |
| -0,335425299 | 0,386707374 | TMEM52 | FALSE | Downregulated |
| -0,929645726 | 0,386884062 | ATP6V1G2-DDX39B | FALSE | Downregulated |
| -0,386938911 | 0,386934733 | ACRC | FALSE | Downregulated |
| 0,291975676 | 0,387045711 | SPATA6L | FALSE | Upregulated |
| -0,463565306 | 0,387319606 | LOC101929140 | FALSE | Downregulated |
| 0,182704502 | 0,387628211 | ANKRD6 | FALSE | Upregulated |
| -0,453942358 | 0,387764499 | LOC100507424 | FALSE | Downregulated |
| -0,101552807 | 0,387764499 | YY1AP1 | FALSE | Downregulated |
| -1,485704209 | 0,387801605 | FAR2P3 | FALSE | Downregulated |
| -0,093310211 | 0,387801605 | TPMT | FALSE | Downregulated |
| 0,121206473 | 0,387971551 | RNF115 | FALSE | Upregulated |
| 1,045059286 | 0,387975868 | IL23R | FALSE | Upregulated |
| -0,147544118 | 0,388055023 | ZNF174 | FALSE | Downregulated |
| 0,169744318 | 0,388131871 | SEC16A | FALSE | Upregulated |
| -0,105595621 | 0,388267476 | TM2D3 | FALSE | Downregulated |
| -0,442786121 | 0,388267543 | MYOM2 | FALSE | Downregulated |
| 0,102288913 | 0,388300852 | SUMO3 | FALSE | Upregulated |
| -0,168662623 | 0,3888287 | ANKS1A | FALSE | Downregulated |
| 0,121740409 | 0,3888287 | TM9SF1 | FALSE | Upregulated |
| -0,134079572 | 0,3888287 | TNKS1BP1 | FALSE | Downregulated |
| 0,243655176 | 0,389419485 | APOC4 | FALSE | Upregulated |
| 0,121411613 | 0,389421983 | RHBDD1 | FALSE | Upregulated |
| 0,127441932 | 0,389448072 | PCMT1 | FALSE | Upregulated |
| -0,298097734 | 0,389474555 | C21orf119 | FALSE | Downregulated |
| -1,377380547 | 0,389474555 | JPH2 | FALSE | Downregulated |
| -0,236493465 | 0,389474555 | SLCO1B3 | FALSE | Downregulated |

| | | | | |
|---|---|---|---|---|
| -0,124913503 | 0,389536501 | TECPR2 | FALSE | Downregulated |
| 0,228481976 | 0,389783451 | SLC22A18 | FALSE | Upregulated |
| 0,200025502 | 0,389872145 | ACAP3 | FALSE | Upregulated |
| -0,267603665 | 0,389872145 | RP1-249H1.4 | FALSE | Downregulated |
| -0,404261973 | 0,390065534 | KCND1 | FALSE | Downregulated |
| 1,174927498 | 0,390081764 | GABBR2 | FALSE | Upregulated |
| -0,367543735 | 0,390097522 | LOC102724833 | FALSE | Downregulated |
| -0,215849515 | 0,390242462 | SLC5A3 | FALSE | Downregulated |
| -0,520761057 | 0,390248412 | GOLGA8B | FALSE | Downregulated |
| -0,594813702 | 0,390248412 | GPR88 | FALSE | Downregulated |
| -0,430124756 | 0,390248412 | NPIPB11 | FALSE | Downregulated |
| 0,286589974 | 0,390248412 | ZNF468 | FALSE | Upregulated |
| -0,70909476 | 0,390547329 | SCXB | FALSE | Downregulated |
| -0,105658125 | 0,390644078 | EPS15 | FALSE | Downregulated |
| 0,154774252 | 0,390795914 | TXNL4A | FALSE | Upregulated |
| 0,144782147 | 0,3908339 | URM1 | FALSE | Upregulated |
| 0,575624787 | 0,39092819 | LOC102724940 | FALSE | Upregulated |
| 0,888351129 | 0,391013557 | CLEC12B | FALSE | Upregulated |
| 0,494527734 | 0,39108879 | LOC100270746 | FALSE | Upregulated |
| 1,142331221 | 0,39108879 | OR2A1 | FALSE | Upregulated |
| 0,152920094 | 0,391124892 | NDUFA3 | FALSE | Upregulated |
| -0,139185704 | 0,391340185 | NAF1 | FALSE | Downregulated |
| -0,225784529 | 0,391340185 | ZNF549 | FALSE | Downregulated |
| -0,180305015 | 0,391394759 | FAM175A | FALSE | Downregulated |
| 0,295479808 | 0,391440712 | GPR183 | FALSE | Upregulated |
| -0,08614856 | 0,391457442 | PPP2R5C | FALSE | Downregulated |
| -0,991619138 | 0,391645587 | SNORD79 | FALSE | Downregulated |
| 0,244483736 | 0,391946474 | CD320 | FALSE | Upregulated |
| 0,361722205 | 0,391946474 | ICOSLG | FALSE | Upregulated |
| 0,227990348 | 0,391965812 | GCA | FALSE | Upregulated |
| -0,983338701 | 0,392187846 | HEMGN | FALSE | Downregulated |
| -0,198521092 | 0,39225924 | HIRIP3 | FALSE | Downregulated |
| -0,171400093 | 0,392518707 | ATRX | FALSE | Downregulated |
| -0,152999088 | 0,392518707 | HSD11B1 | FALSE | Downregulated |
| 0,697160327 | 0,392518707 | PROL1 | FALSE | Upregulated |
| 0,143933247 | 0,392567272 | BPGM | FALSE | Upregulated |
| -0,492135369 | 0,392567272 | CAPN12 | FALSE | Downregulated |
| -0,631120357 | 0,392567272 | FAM27E3 | FALSE | Downregulated |
| 0,56460346 | 0,392638076 | SYCE3 | FALSE | Upregulated |
| -0,102728489 | 0,39276084 | CSRP2BP | FALSE | Downregulated |
| 0,154269505 | 0,392803176 | FAM203A | FALSE | Upregulated |
| 0,292356943 | 0,392876672 | BLOC1S1-RDH5 | FALSE | Upregulated |
| 0,885179266 | 0,393294029 | FAM64A | FALSE | Upregulated |
| -0,423884954 | 0,393455331 | AMHR2 | FALSE | Downregulated |
| -0,142200754 | 0,393455331 | QSER1 | FALSE | Downregulated |
| -0,194188999 | 0,393512699 | TCEANC | FALSE | Downregulated |
| -0,692291389 | 0,393515417 | DNAH7 | FALSE | Downregulated |
| -0,474063137 | 0,39360767 | LOC102724559 | FALSE | Downregulated |
| -0,135285346 | 0,393614756 | APPL1 | FALSE | Downregulated |
| 1,321215396 | 0,39362899 | CDH16 | FALSE | Upregulated |
| -0,260774724 | 0,393677477 | ANXA10 | FALSE | Downregulated |

| | | | | |
|---|---|---|---|---|
| -0,212208383 | 0,393677477 | RHOBTB1 | FALSE | Downregulated |
| -0,307596293 | 0,393688731 | CTF1 | FALSE | Downregulated |
| -0,455809472 | 0,393821851 | LOC102723583 | FALSE | Downregulated |
| -0,097721361 | 0,393950279 | SNX5 | FALSE | Downregulated |
| -0,627472456 | 0,394004327 | SGSM1 | FALSE | Downregulated |
| 0,261320139 | 0,394149388 | PALM | FALSE | Upregulated |
| -0,139150997 | 0,394156424 | TARSL2 | FALSE | Downregulated |
| 0,124193984 | 0,394319598 | DDX52 | FALSE | Upregulated |
| -0,288599825 | 0,394319598 | GALK1 | FALSE | Downregulated |
| -0,932615557 | 0,39434111 | GP5 | FALSE | Downregulated |
| -0,166580427 | 0,394557622 | PCGF1 | FALSE | Downregulated |
| -0,257933707 | 0,39461143 | ZNF497 | FALSE | Downregulated |
| 0,150221698 | 0,394915325 | TARBP2 | FALSE | Upregulated |
| 0,209325914 | 0,394915325 | URB2 | FALSE | Upregulated |
| -0,758099739 | 0,395034719 | LOC100506753 | FALSE | Downregulated |
| 0,203078904 | 0,395256871 | SLX4 | FALSE | Upregulated |
| 0,971474988 | 0,395551735 | OVOL2 | FALSE | Upregulated |
| 0,502271562 | 0,395551735 | SNCAIP | FALSE | Upregulated |
| 0,141845518 | 0,395792445 | TMUB2 | FALSE | Upregulated |
| -0,432491584 | 0,396142561 | LGI4 | FALSE | Downregulated |
| -0,118260995 | 0,396143647 | BIRC2 | FALSE | Downregulated |
| -0,407576104 | 0,396143647 | C7orf55-LUC7L2 | FALSE | Downregulated |
| -0,186571732 | 0,396143647 | DOCK4 | FALSE | Downregulated |
| 0,14140223 | 0,396143647 | GTPBP6 | FALSE | Upregulated |
| -0,176443958 | 0,396555056 | C1orf74 | FALSE | Downregulated |
| -0,1432899 | 0,396612572 | LINC00493 | FALSE | Downregulated |
| 0,148159606 | 0,396639374 | PRKACA | FALSE | Upregulated |
| -0,09754645 | 0,396799844 | PPP2R2D | FALSE | Downregulated |
| -0,411173206 | 0,396925248 | CHRD | FALSE | Downregulated |
| 0,126219051 | 0,396925248 | EXOC6B | FALSE | Upregulated |
| 0,168160177 | 0,396925248 | TCEB1 | FALSE | Upregulated |
| -1,292158134 | 0,396926909 | LOC101927486 | FALSE | Downregulated |
| -1,063379133 | 0,3969476 | LOC101928730 | FALSE | Downregulated |
| 0,182187515 | 0,397289686 | SLC44A2 | FALSE | Upregulated |
| 0,867278957 | 0,397362832 | CCL13 | FALSE | Upregulated |
| -0,191668605 | 0,397362832 | DDX60L | FALSE | Downregulated |
| 0,257674478 | 0,397362832 | DZANK1 | FALSE | Upregulated |
| -0,141460023 | 0,397362832 | GJB1 | FALSE | Downregulated |
| -0,380914045 | 0,397362832 | ISM1 | FALSE | Downregulated |
| -0,12805821 | 0,397362832 | PLOD2 | FALSE | Downregulated |
| -0,34916825 | 0,397362832 | SULF1 | FALSE | Downregulated |
| 0,525514187 | 0,397621017 | TNFRSF10C | FALSE | Upregulated |
| -0,136329407 | 0,397621017 | ZKSCAN2 | FALSE | Downregulated |
| -0,275231912 | 0,397913433 | C17orf67 | FALSE | Downregulated |
| 0,339916526 | 0,397996715 | RGS18 | FALSE | Upregulated |
| 0,230555858 | 0,398027937 | FAM102B | FALSE | Upregulated |
| -0,647151299 | 0,398153506 | GTF2IRD1P1 | FALSE | Downregulated |
| -0,157217079 | 0,398153506 | THEM4 | FALSE | Downregulated |
| 0,188247233 | 0,398453491 | MRPL12 | FALSE | Upregulated |
| -0,245674231 | 0,398453491 | ZNF585A | FALSE | Downregulated |
| -0,396297178 | 0,398570928 | PLEKHB1 | FALSE | Downregulated |

| | | | | |
|---|---|---|---|---|
| -0,505283546 | 0,398860906 | FIGNL2 | FALSE | Downregulated |
| 0,758129129 | 0,398893256 | LINC00605 | FALSE | Upregulated |
| -0,583388978 | 0,398893256 | LOC149134 | FALSE | Downregulated |
| 0,197688447 | 0,398935252 | RASGRP3 | FALSE | Upregulated |
| -0,342719606 | 0,399088972 | RTN4RL1 | FALSE | Downregulated |
| 1,208402361 | 0,399088972 | SEZ6 | FALSE | Upregulated |
| -0,152614115 | 0,399230162 | SLC25A16 | FALSE | Downregulated |
| 0,139945206 | 0,399561948 | LEO1 | FALSE | Upregulated |
| 0,721678025 | 0,399561948 | WISP2 | FALSE | Upregulated |
| -0,164741115 | 0,399563879 | SLTM | FALSE | Downregulated |
| 0,359681562 | 0,399569996 | CCDC151 | FALSE | Upregulated |
| -0,234889916 | 0,399569996 | DCXR | FALSE | Downregulated |
| -0,307131571 | 0,399569996 | DNAJC27-AS1 | FALSE | Downregulated |
| 0,421755372 | 0,399569996 | HNRNPA1P10 | FALSE | Upregulated |
| 0,444215627 | 0,399569996 | PTPLA | FALSE | Upregulated |
| -0,158459625 | 0,39973789 | UTRN | FALSE | Downregulated |
| 0,188261032 | 0,399874596 | TFEB | FALSE | Upregulated |
| -0,217828679 | 0,39993953 | FBXO48 | FALSE | Downregulated |
| -0,460059449 | 0,39993953 | LOC102724814 | FALSE | Downregulated |
| -0,493053189 | 0,39993953 | SEMA3E | FALSE | Downregulated |
| -0,109096142 | 0,400312595 | SBF2 | FALSE | Downregulated |
| 0,907135305 | 0,400422147 | PTCHD4 | FALSE | Upregulated |
| -0,126610873 | 0,400443725 | DSTYK | FALSE | Downregulated |
| -0,431902111 | 0,400443725 | STAG3L5P-PVRIG2P-PILRB | FALSE | Downregulated |
| 0,385513724 | 0,40048206 | BCMO1 | FALSE | Upregulated |
| -0,467802293 | 0,400527742 | ANKRD13B | FALSE | Downregulated |
| 0,108741352 | 0,400527742 | FCHSD2 | FALSE | Upregulated |
| 0,808982208 | 0,400527742 | STC2 | FALSE | Upregulated |
| 0,112532957 | 0,400786993 | C16orf62 | FALSE | Upregulated |
| -0,671475647 | 0,400786993 | CBLN3 | FALSE | Downregulated |
| 1,024113028 | 0,400898791 | C19orf59 | FALSE | Upregulated |
| 0,509945635 | 0,401012801 | GBP6 | FALSE | Upregulated |
| -0,237302777 | 0,401012801 | VCPKMT | FALSE | Downregulated |
| -0,19519238 | 0,401322974 | MMACHC | FALSE | Downregulated |
| -0,179907039 | 0,401335315 | PEX11G | FALSE | Downregulated |
| 0,175356764 | 0,401335315 | SDC1 | FALSE | Upregulated |
| -1,220784265 | 0,40146158 | LOC102724065 | FALSE | Downregulated |
| -0,185806585 | 0,401468638 | ZFX | FALSE | Downregulated |
| -0,139880832 | 0,401527761 | CAMK2D | FALSE | Downregulated |
| 0,411488946 | 0,401527761 | NUDT10 | FALSE | Upregulated |
| -0,158724602 | 0,401634972 | CUTC | FALSE | Downregulated |
| -0,737392164 | 0,401634972 | FLJ20464 | FALSE | Downregulated |
| 0,7079243 | 0,401634972 | RP4-562J12.2 | FALSE | Upregulated |
| -0,262150387 | 0,401720766 | ELMO3 | FALSE | Downregulated |
| -0,172213383 | 0,401738987 | ERO1LB | FALSE | Downregulated |
| 0,19826894 | 0,401788907 | TRIM5 | FALSE | Upregulated |
| -0,806137244 | 0,401809584 | TMPRSS5 | FALSE | Downregulated |
| 0,140762385 | 0,401892298 | MAP3K11 | FALSE | Upregulated |
| 0,155559836 | 0,401892298 | TFPT | FALSE | Upregulated |
| -0,451863144 | 0,401946579 | LINC00638 | FALSE | Downregulated |

| | | | | |
|---|---|---|---|---|
| -0,133404312 | 0,402176608 | GTF3C3 | FALSE | Downregulated |
| 0,169429591 | 0,402176608 | NDUFB2 | FALSE | Upregulated |
| -0,801781261 | 0,402176608 | SIX4 | FALSE | Downregulated |
| -0,086691552 | 0,402264848 | OSBPL2 | FALSE | Downregulated |
| -0,279819252 | 0,40226844 | PLA2R1 | FALSE | Downregulated |
| -1,294119472 | 0,402355039 | CA1 | FALSE | Downregulated |
| 0,16756578 | 0,402426951 | LENG1 | FALSE | Upregulated |
| -0,656411057 | 0,402944218 | FSBP | FALSE | Downregulated |
| 0,434149153 | 0,402944218 | TVP23A | FALSE | Upregulated |
| 0,137455196 | 0,402953639 | SIL1 | FALSE | Upregulated |
| 0,164823802 | 0,40297858 | GPS2 | FALSE | Upregulated |
| -0,23378438 | 0,40297858 | HPS3 | FALSE | Downregulated |
| -0,959203233 | 0,403121556 | SRL | FALSE | Downregulated |
| 0,186473125 | 0,403203432 | PRADC1 | FALSE | Upregulated |
| -0,617823716 | 0,403244138 | LOC152225 | FALSE | Downregulated |
| -0,228392803 | 0,403408148 | UPF3B | FALSE | Downregulated |
| 0,240214008 | 0,403408482 | PRKAR1B | FALSE | Upregulated |
| 0,139559535 | 0,403408482 | TMEM19 | FALSE | Upregulated |
| 0,868012685 | 0,4034995 | LOC102724661 | FALSE | Upregulated |
| 0,166393687 | 0,40365152 | LONRF1 | FALSE | Upregulated |
| -0,133570458 | 0,403686139 | C9orf114 | FALSE | Downregulated |
| -1,087256854 | 0,403686139 | GPR62 | FALSE | Downregulated |
| -0,155134631 | 0,403686139 | PEBP1 | FALSE | Downregulated |
| -0,557980453 | 0,40382253 | TBX10 | FALSE | Downregulated |
| -0,792722819 | 0,403860037 | PADI2 | FALSE | Downregulated |
| -0,687628903 | 0,404005239 | LOC102724314 | FALSE | Downregulated |
| 0,282266282 | 0,404606798 | CLDN3 | FALSE | Upregulated |
| -0,219246589 | 0,404633222 | DMTF1 | FALSE | Downregulated |
| 0,090851982 | 0,404633222 | EXOC4 | FALSE | Upregulated |
| 0,185532018 | 0,404633222 | MZT2B | FALSE | Upregulated |
| 0,116525433 | 0,404689278 | ZNF394 | FALSE | Upregulated |
| -1,004949232 | 0,40484126 | MIR143HG | FALSE | Downregulated |
| 0,135614651 | 0,404850311 | NDUFAB1 | FALSE | Upregulated |
| 0,176462003 | 0,404929787 | STRA13 | FALSE | Upregulated |
| -0,809679779 | 0,40516953 | CBX3P2 | FALSE | Downregulated |
| 0,193841447 | 0,40516953 | SLC26A11 | FALSE | Upregulated |
| 0,206570311 | 0,405437713 | ARHGAP15 | FALSE | Upregulated |
| -0,133648983 | 0,405437713 | CPNE1 | FALSE | Downregulated |
| 0,495951494 | 0,405448292 | PLCH2 | FALSE | Upregulated |
| 0,59703973 | 0,405457681 | FAM81A | FALSE | Upregulated |
| -0,690302911 | 0,40564187 | LOC102724117 | FALSE | Downregulated |
| -0,807979931 | 0,40564187 | NOXO1 | FALSE | Downregulated |
| -0,104914059 | 0,405681671 | HUWE1 | FALSE | Downregulated |
| 0,16005308 | 0,405697344 | PSMG3-AS1 | FALSE | Upregulated |
| -0,764001141 | 0,406307692 | ARHGEF4 | FALSE | Downregulated |
| 0,216226136 | 0,406307692 | TAF13 | FALSE | Upregulated |
| -0,117695718 | 0,406357823 | MPC2 | FALSE | Downregulated |
| 0,14121817 | 0,406357823 | USE1 | FALSE | Upregulated |
| 0,540558358 | 0,406453816 | MTRNR2L9 | FALSE | Upregulated |
| 0,297835131 | 0,406491318 | RP9P | FALSE | Upregulated |
| -0,678538736 | 0,406787066 | LOC102723708 | FALSE | Downregulated |

| | | | | |
|---|---|---|---|---|
| 0,169684054 | 0,406805187 | ZMAT5 | FALSE | Upregulated |
| 0,150867068 | 0,40690489 | FKRP | FALSE | Upregulated |
| 0,579300308 | 0,407109828 | SHC3 | FALSE | Upregulated |
| -0,747379317 | 0,407302988 | DCLK1 | FALSE | Downregulated |
| -0,117956187 | 0,407603868 | ZNF227 | FALSE | Downregulated |
| 0,136332351 | 0,40776665 | DEF8 | FALSE | Upregulated |
| -0,260310005 | 0,40776665 | ZNF425 | FALSE | Downregulated |
| -0,154425316 | 0,407892657 | DYNC2LI1 | FALSE | Downregulated |
| 0,118913654 | 0,408293472 | SGTA | FALSE | Upregulated |
| -0,165834059 | 0,408313982 | RGS14 | FALSE | Downregulated |
| 0,120089753 | 0,408501846 | CHCHD4 | FALSE | Upregulated |
| 0,094988172 | 0,408619868 | TTC27 | FALSE | Upregulated |
| -0,265525381 | 0,408815681 | LOC102723992 | FALSE | Downregulated |
| -0,900391477 | 0,408867602 | AP4B1-AS1 | FALSE | Downregulated |
| 0,186891574 | 0,408867602 | B4GALT7 | FALSE | Upregulated |
| -0,349901751 | 0,408867602 | BEX1 | FALSE | Downregulated |
| 1,015548261 | 0,408867602 | CLEC4D | FALSE | Upregulated |
| -0,642518229 | 0,408867602 | LYPD6B | FALSE | Downregulated |
| -0,093352989 | 0,408867602 | MMS19 | FALSE | Downregulated |
| -0,191955678 | 0,409006169 | DPP4 | FALSE | Downregulated |
| 0,294241879 | 0,409065702 | ACKR2 | FALSE | Upregulated |
| 0,114482484 | 0,409283581 | C19orf43 | FALSE | Upregulated |
| 0,425363209 | 0,409283581 | P2RY12 | FALSE | Upregulated |
| -0,130335985 | 0,409283581 | PIPOX | FALSE | Downregulated |
| 0,253540137 | 0,409283581 | SLC1A4 | FALSE | Upregulated |
| -0,481492613 | 0,409372269 | ADAMTS6 | FALSE | Downregulated |
| 1,096950952 | 0,409482176 | CATSPER1 | FALSE | Upregulated |
| -0,759022251 | 0,409482176 | CCDC39 | FALSE | Downregulated |
| -0,145194003 | 0,409482176 | GLO1 | FALSE | Downregulated |
| 0,824036662 | 0,409482176 | LOC102724197 | FALSE | Upregulated |
| -0,223013385 | 0,409482176 | MSL3P1 | FALSE | Downregulated |
| -0,696503206 | 0,409482176 | SLITRK3 | FALSE | Downregulated |
| 0,542154722 | 0,409551633 | AC010524.2 | FALSE | Upregulated |
| 1,190669904 | 0,409591383 | DPYSL4 | FALSE | Upregulated |
| -0,131837498 | 0,409591383 | PARS2 | FALSE | Downregulated |
| 0,278801569 | 0,409668753 | GPX8 | FALSE | Upregulated |
| 0,612987287 | 0,410055253 | AKR7A2P1 | FALSE | Upregulated |
| 1,080349831 | 0,410055253 | LOC102723996 | FALSE | Upregulated |
| -0,220067009 | 0,410091308 | RBPMS | FALSE | Downregulated |
| 0,733204806 | 0,410148009 | LOC102723610 | FALSE | Upregulated |
| 0,258193564 | 0,41019019 | HENMT1 | FALSE | Upregulated |
| -0,132274771 | 0,41022414 | KIAA1191 | FALSE | Downregulated |
| -0,387457658 | 0,410234849 | CYP4A22 | FALSE | Downregulated |
| 0,327981737 | 0,410252061 | LOC100996740 | FALSE | Upregulated |
| 1,215289587 | 0,410313852 | CAMP | FALSE | Upregulated |
| 0,764726914 | 0,410414196 | MID1-2 | FALSE | Upregulated |
| -0,870333245 | 0,410414196 | PM20D1 | FALSE | Downregulated |
| 0,281011812 | 0,410646005 | SYP | FALSE | Upregulated |
| 0,110977148 | 0,410646005 | UBE2D2 | FALSE | Upregulated |
| 0,107853453 | 0,410840348 | GDAP2 | FALSE | Upregulated |
| 0,160665735 | 0,410840348 | RABAC1 | FALSE | Upregulated |

| | | | | |
|---|---|---|---|---|
| -0,544408377 | 0,410898799 | BRINP1 | FALSE | Downregulated |
| 0,280305259 | 0,411004314 | GAB3 | FALSE | Upregulated |
| 0,46648095 | 0,411127386 | ZSCAN12P1 | FALSE | Upregulated |
| 0,158718134 | 0,411267626 | HNRNPA1 | FALSE | Upregulated |
| -0,722060058 | 0,411267626 | YJEFN3 | FALSE | Downregulated |
| -0,815673219 | 0,411464449 | FAM133DP | FALSE | Downregulated |
| 0,457491064 | 0,411654755 | CREB3L1 | FALSE | Upregulated |
| -0,208664622 | 0,411664754 | C9orf37 | FALSE | Downregulated |
| 1,151068387 | 0,412330667 | LOC102724943 | FALSE | Upregulated |
| 0,983922559 | 0,412428982 | NHLH1 | FALSE | Upregulated |
| -0,222957024 | 0,412583423 | LIME1 | FALSE | Downregulated |
| 1,201047036 | 0,412600328 | SOX8 | FALSE | Upregulated |
| -0,843214903 | 0,412684274 | LOC102724336 | FALSE | Downregulated |
| -0,267928364 | 0,412862467 | NPR2 | FALSE | Downregulated |
| 0,240008771 | 0,412862467 | PSMD7 | FALSE | Upregulated |
| -0,217028049 | 0,413466555 | ZNF559 | FALSE | Downregulated |
| -0,189088668 | 0,413682057 | KLF7 | FALSE | Downregulated |
| -0,128804121 | 0,413797961 | TNKS | FALSE | Downregulated |
| -0,524571305 | 0,413836711 | LOC101928548 | FALSE | Downregulated |
| 0,192877042 | 0,413836711 | TMEM194B | FALSE | Upregulated |
| 0,493775174 | 0,413942118 | LRRC34 | FALSE | Upregulated |
| 0,36196216 | 0,414026321 | C14orf80 | FALSE | Upregulated |
| -0,251690687 | 0,414199796 | KIAA1462 | FALSE | Downregulated |
| -0,967708132 | 0,414211981 | LOC101927896 | FALSE | Downregulated |
| -0,124943264 | 0,414407879 | AGFG2 | FALSE | Downregulated |
| 1,011304597 | 0,414415916 | AC007386.2 | FALSE | Upregulated |
| 1,095563864 | 0,414415916 | AC090044.1 | FALSE | Upregulated |
| -0,798199296 | 0,414415916 | DKFZp667F0711 | FALSE | Downregulated |
| 0,215452232 | 0,414415916 | RAB11FIP4 | FALSE | Upregulated |
| -0,149039675 | 0,414415916 | WHSC1L1 | FALSE | Downregulated |
| -0,472535016 | 0,414467023 | PLGLB1 | FALSE | Downregulated |
| 0,17219679 | 0,414467023 | SPG21 | FALSE | Upregulated |
| 0,107962933 | 0,414639358 | KLC1 | FALSE | Upregulated |
| -0,112391305 | 0,414639358 | PET112 | FALSE | Downregulated |
| 0,632728738 | 0,414686671 | FMO1 | FALSE | Upregulated |
| -0,153811813 | 0,414686671 | PTDSS2 | FALSE | Downregulated |
| 0,273944108 | 0,414686671 | ZNF678 | FALSE | Upregulated |
| -0,107521402 | 0,414776162 | OTUD7B | FALSE | Downregulated |
| -0,941016662 | 0,414797241 | FLJ25363 | FALSE | Downregulated |
| 0,136278306 | 0,414797241 | MGAT4B | FALSE | Upregulated |
| 0,104846153 | 0,414852282 | QRSL1 | FALSE | Upregulated |
| 0,113418803 | 0,414852282 | TMEM209 | FALSE | Upregulated |
| -0,138034712 | 0,414914989 | FBRSL1 | FALSE | Downregulated |
| 0,167237 | 0,415123442 | ATP8B4 | FALSE | Upregulated |
| -0,316707143 | 0,415123442 | LOC113230 | FALSE | Downregulated |
| 0,890497103 | 0,415123442 | LOC400548 | FALSE | Upregulated |
| -0,117140758 | 0,415123442 | ZNF274 | FALSE | Downregulated |
| -0,352017199 | 0,415143271 | PPIL6 | FALSE | Downregulated |
| -0,470950608 | 0,415660749 | TOLLIP-AS1 | FALSE | Downregulated |
| -0,377853089 | 0,415787324 | RP11-279F6.1 | FALSE | Downregulated |
| -0,856678754 | 0,415873432 | LOC102724262 | FALSE | Downregulated |

| | | | | |
|---|---|---|---|---|
| 0,126939192 | 0,416335492 | TP53 | FALSE | Upregulated |
| -0,859590168 | 0,416364267 | ANKRD23 | FALSE | Downregulated |
| 0,483674081 | 0,416364267 | LOC101929612 | FALSE | Upregulated |
| 0,190489447 | 0,416442303 | RPS19BP1 | FALSE | Upregulated |
| -0,706990381 | 0,416568528 | GNRH2 | FALSE | Downregulated |
| -1,238799652 | 0,416643169 | MYL3 | FALSE | Downregulated |
| -0,227525057 | 0,416674564 | MAP3K10 | FALSE | Downregulated |
| 0,27756795 | 0,416741219 | GK | FALSE | Upregulated |
| -0,579950207 | 0,416799108 | IMPG2 | FALSE | Downregulated |
| -0,235241889 | 0,416799108 | NID2 | FALSE | Downregulated |
| 0,283459991 | 0,416899049 | ASCL2 | FALSE | Upregulated |
| 0,519419445 | 0,417057779 | KDM4A-AS1 | FALSE | Upregulated |
| -0,172964735 | 0,417057779 | MUTYH | FALSE | Downregulated |
| -1,00606082 | 0,417197306 | LOC102724339 | FALSE | Downregulated |
| -0,103653194 | 0,417197306 | TCAIM | FALSE | Downregulated |
| 0,166065133 | 0,417460451 | NXT2 | FALSE | Upregulated |
| 0,33096524 | 0,417463874 | LOC102723726 | FALSE | Upregulated |
| 0,416689726 | 0,417556741 | SLC12A8 | FALSE | Upregulated |
| -0,065782377 | 0,417626429 | AP1G1 | FALSE | Downregulated |
| -0,177913637 | 0,417626429 | OGFOD2 | FALSE | Downregulated |
| 0,152662588 | 0,417626429 | SMAD1 | FALSE | Upregulated |
| 0,447597171 | 0,417697649 | SAMD15 | FALSE | Upregulated |
| -0,177979932 | 0,417697649 | WHAMM | FALSE | Downregulated |
| -0,087247506 | 0,417728515 | FAF2 | FALSE | Downregulated |
| 0,509503574 | 0,417728515 | PAPPA | FALSE | Upregulated |
| 0,280342564 | 0,417809935 | RNF207 | FALSE | Upregulated |
| -0,127746551 | 0,417826998 | ASPH | FALSE | Downregulated |
| -0,551477463 | 0,417826998 | LINC00398 | FALSE | Downregulated |
| -0,468023474 | 0,417826998 | MPL | FALSE | Downregulated |
| 0,765474435 | 0,417826998 | NOS2 | FALSE | Upregulated |
| -0,254171863 | 0,417826998 | PAQR7 | FALSE | Downregulated |
| 0,136634496 | 0,417826998 | SEC13 | FALSE | Upregulated |
| 0,225287526 | 0,417826998 | SIRT6 | FALSE | Upregulated |
| -0,151452584 | 0,417826998 | XPO4 | FALSE | Downregulated |
| -0,700672056 | 0,417857833 | LOC101928361 | FALSE | Downregulated |
| -0,972320543 | 0,417934004 | GPR20 | FALSE | Downregulated |
| -0,716918405 | 0,417942516 | GRIK5 | FALSE | Downregulated |
| -0,361729056 | 0,417942516 | NSUN5P1 | FALSE | Downregulated |
| 0,743274163 | 0,41796679 | RP11-182I10.3 | FALSE | Upregulated |
| -0,391046096 | 0,417986276 | MBNL1-AS1 | FALSE | Downregulated |
| 0,307214197 | 0,418057013 | TCTEX1D1 | FALSE | Upregulated |
| -0,80618728 | 0,418216077 | LOC728819 | FALSE | Downregulated |
| 0,209808728 | 0,418222738 | HABP4 | FALSE | Upregulated |
| -0,192386334 | 0,418295116 | ZNF511 | FALSE | Downregulated |
| 0,25317481 | 0,418479886 | EFCAB2 | FALSE | Upregulated |
| -0,198256281 | 0,418479886 | FIGN | FALSE | Downregulated |
| -0,143678881 | 0,418590193 | STRN | FALSE | Downregulated |
| -0,678749992 | 0,418619729 | RET | FALSE | Downregulated |
| 0,219144066 | 0,418735952 | CACNA1D | FALSE | Upregulated |
| 0,152792558 | 0,418735952 | SMIM12 | FALSE | Upregulated |
| -0,713929469 | 0,418777865 | LOC100130539 | FALSE | Downregulated |

| | | | | |
|---|---|---|---|---|
| 1,123538052 | 0,418848774 | SHOX2 | FALSE | Upregulated |
| 0,081370659 | 0,418922262 | ATP6V1A | FALSE | Upregulated |
| -0,451806443 | 0,418922262 | MTMR9LP | FALSE | Downregulated |
| 0,233246345 | 0,419041076 | RGS3 | FALSE | Upregulated |
| 0,297814971 | 0,419041076 | TUSC3 | FALSE | Upregulated |
| -0,562868108 | 0,419204732 | AC007038.7 | FALSE | Downregulated |
| 0,1111139 | 0,419260784 | AP2A1 | FALSE | Upregulated |
| -0,749814151 | 0,419260784 | FAM71F2 | FALSE | Downregulated |
| -0,133437523 | 0,419565044 | FAM133B | FALSE | Downregulated |
| -0,352361032 | 0,419565044 | IGF1 | FALSE | Downregulated |
| -0,652919516 | 0,419565044 | MYO3A | FALSE | Downregulated |
| -0,188310595 | 0,419565044 | SAMD4A | FALSE | Downregulated |
| 0,142242987 | 0,419565044 | TMEM50B | FALSE | Upregulated |
| 0,25171333 | 0,419565044 | VSTM4 | FALSE | Upregulated |
| 0,238616986 | 0,420040756 | FAM60A | FALSE | Upregulated |
| 1,47455363 | 0,420053851 | SNORD3A | FALSE | Upregulated |
| -0,210132188 | 0,420113562 | SPNS2 | FALSE | Downregulated |
| -0,197344616 | 0,420325801 | FOLH1 | FALSE | Downregulated |
| -0,14682394 | 0,42033411 | CIPC | FALSE | Downregulated |
| 0,162783229 | 0,420411997 | ACBD3 | FALSE | Upregulated |
| -0,493807066 | 0,420430261 | LOC101927841 | FALSE | Downregulated |
| 0,161690984 | 0,420430261 | TRIM68 | FALSE | Upregulated |
| 0,468890144 | 0,42045042 | TMEM136 | FALSE | Upregulated |
| -0,169719354 | 0,420611224 | ADH1A | FALSE | Downregulated |
| -0,521194039 | 0,420616992 | SLCO4C1 | FALSE | Downregulated |
| -0,159307028 | 0,420767832 | DGCR6L | FALSE | Downregulated |
| 0,472646364 | 0,420992456 | LINC01171 | FALSE | Upregulated |
| -0,113211942 | 0,420992456 | SMG8 | FALSE | Downregulated |
| 0,62353453 | 0,421120035 | HIST1H2AM | FALSE | Upregulated |
| -0,148209352 | 0,421120035 | RILPL2 | FALSE | Downregulated |
| 0,091947535 | 0,421120035 | SEC11A | FALSE | Upregulated |
| -0,702709664 | 0,421179521 | CALN1 | FALSE | Downregulated |
| -0,13937069 | 0,421363293 | MTRR | FALSE | Downregulated |
| -0,300949694 | 0,421391722 | RIMKLB | FALSE | Downregulated |
| 2,326753882 | 0,421391722 | RNVU1-7 | FALSE | Upregulated |
| 0,186463856 | 0,421472396 | GSTCD | FALSE | Upregulated |
| 0,580795887 | 0,421472396 | LOC101927369 | FALSE | Upregulated |
| 0,720686112 | 0,421472396 | NSG1 | FALSE | Upregulated |
| -0,174151256 | 0,421619809 | ZNF839 | FALSE | Downregulated |
| 0,134832458 | 0,421662003 | PPP2R2A | FALSE | Upregulated |
| 0,13652064 | 0,421727343 | ELF1 | FALSE | Upregulated |
| 0,383502048 | 0,421727343 | ITGA11 | FALSE | Upregulated |
| -0,465088488 | 0,421843783 | RP5-1125A11.1 | FALSE | Downregulated |
| -0,552937942 | 0,421906869 | RP11-637A17.2 | FALSE | Downregulated |
| -0,571260723 | 0,421916299 | AC159540.1 | FALSE | Downregulated |
| 0,252301603 | 0,421916299 | SAA4 | FALSE | Upregulated |
| 0,440831245 | 0,421916299 | SNORA40 | FALSE | Upregulated |
| -0,793266482 | 0,421961948 | SNX29P1 | FALSE | Downregulated |
| -1,14601097 | 0,422217592 | GJB3 | FALSE | Downregulated |
| -0,132042127 | 0,422217592 | KLHL8 | FALSE | Downregulated |
| -0,372607611 | 0,422217592 | LOC102724880 | FALSE | Downregulated |

| | | | | |
|---|---|---|---|---|
| 0,113630196 | 0,422217592 | SH3D19 | FALSE | Upregulated |
| -0,156457137 | 0,422217592 | SNRNP25 | FALSE | Downregulated |
| 0,15977812 | 0,422217592 | UBE2B | FALSE | Upregulated |
| -0,124632654 | 0,422276342 | PTPN18 | FALSE | Downregulated |
| 0,800145284 | 0,422423155 | LOC102724765 | FALSE | Upregulated |
| -0,098344196 | 0,422469015 | CERS2 | FALSE | Downregulated |
| 0,223209639 | 0,422469015 | LRWD1 | FALSE | Upregulated |
| 0,176232218 | 0,422469015 | SMIM4 | FALSE | Upregulated |
| 0,079299347 | 0,422480429 | DHX38 | FALSE | Upregulated |
| -1,145143544 | 0,422480429 | LOC102724349 | FALSE | Downregulated |
| -0,167950788 | 0,422480429 | ZNF583 | FALSE | Downregulated |
| -0,661813653 | 0,422538047 | DPY19L2P3 | FALSE | Downregulated |
| -0,432215954 | 0,422544158 | ZNF667 | FALSE | Downregulated |
| -0,447432167 | 0,422571874 | LOC100288842 | FALSE | Downregulated |
| 0,25867185 | 0,422571874 | MIR497HG | FALSE | Upregulated |
| 0,220379664 | 0,422571874 | MXRA7 | FALSE | Upregulated |
| 0,971549674 | 0,422571874 | OTOF | FALSE | Upregulated |
| -0,360517931 | 0,42266301 | LOC101928403 | FALSE | Downregulated |
| -0,090091655 | 0,422715714 | USP9X | FALSE | Downregulated |
| -0,268076482 | 0,422988238 | LBX2 | FALSE | Downregulated |
| -0,701710844 | 0,423079781 | RCOR2 | FALSE | Downregulated |
| -0,532134226 | 0,423126409 | SLC22A10 | FALSE | Downregulated |
| 0,732138061 | 0,423241984 | MAGI1-IT1 | FALSE | Upregulated |
| -0,179732824 | 0,423523069 | FMO4 | FALSE | Downregulated |
| -1,052350308 | 0,423523069 | IL17D | FALSE | Downregulated |
| -0,518082141 | 0,423523069 | RHPN1 | FALSE | Downregulated |
| 1,503517126 | 0,423523069 | TRNR | FALSE | Upregulated |
| 0,912187244 | 0,423562325 | OSTCP1 | FALSE | Upregulated |
| 0,092784973 | 0,423851748 | GLYR1 | FALSE | Upregulated |
| 0,21522491 | 0,424217133 | FZD6 | FALSE | Upregulated |
| 0,120341388 | 0,424217133 | PIGF | FALSE | Upregulated |
| 0,634251698 | 0,424373818 | LOC388780 | FALSE | Upregulated |
| -0,216564197 | 0,424373818 | RAB3D | FALSE | Downregulated |
| -0,164267923 | 0,424537328 | SERPINA10 | FALSE | Downregulated |
| 0,398857239 | 0,424563486 | CACNB3 | FALSE | Upregulated |
| 0,148592377 | 0,424563486 | RPL12 | FALSE | Upregulated |
| -0,10508836 | 0,424619558 | MRPS10 | FALSE | Downregulated |
| -0,50803165 | 0,424619558 | RP11-434C1.1 | FALSE | Downregulated |
| 0,823661613 | 0,424731407 | KIR3DL2 | FALSE | Upregulated |
| 0,15527983 | 0,42496331 | LAMB2 | FALSE | Upregulated |
| -0,107776518 | 0,425402474 | HINT3 | FALSE | Downregulated |
| 0,317427287 | 0,425480335 | CNTNAP2 | FALSE | Upregulated |
| -0,099018437 | 0,425480335 | MTPAP | FALSE | Downregulated |
| -0,314914787 | 0,425882972 | NEBL | FALSE | Downregulated |
| -0,384891028 | 0,425975234 | RASSF6 | FALSE | Downregulated |
| 0,089782796 | 0,426247565 | LRRC41 | FALSE | Upregulated |
| 0,369298179 | 0,426247565 | METTL12 | FALSE | Upregulated |
| 0,231441143 | 0,426298572 | ACACA | FALSE | Upregulated |
| 0,315932957 | 0,426298572 | KIAA0226L | FALSE | Upregulated |
| -0,207025719 | 0,426298647 | THPO | FALSE | Downregulated |
| -0,270221965 | 0,426318785 | FUZ | FALSE | Downregulated |

| | | | | |
|---|---|---|---|---|
| 0,194935647 | 0,426318785 | PROSER2 | FALSE | Upregulated |
| 0,949009964 | 0,426318785 | RP11-277P12.20 | FALSE | Upregulated |
| 0,322807761 | 0,426507036 | LOC400043 | FALSE | Upregulated |
| -0,244698131 | 0,426529177 | TMPO | FALSE | Downregulated |
| -0,133322505 | 0,426733366 | TOR1AIP1 | FALSE | Downregulated |
| 0,928503253 | 0,426735945 | CNTN2 | FALSE | Upregulated |
| 0,166185468 | 0,426735945 | CTC1 | FALSE | Upregulated |
| -0,147387182 | 0,426735945 | MBD4 | FALSE | Downregulated |
| 0,117996758 | 0,426795197 | MOB4 | FALSE | Upregulated |
| 0,222407072 | 0,426905132 | ABHD11 | FALSE | Upregulated |
| -0,151150034 | 0,426905132 | APOM | FALSE | Downregulated |
| 1,167048834 | 0,426905132 | CCL11 | FALSE | Upregulated |
| 0,110936354 | 0,426905132 | KCTD5 | FALSE | Upregulated |
| 0,095189071 | 0,426905132 | KDELR1 | FALSE | Upregulated |
| -0,664008188 | 0,426905132 | RP11-79E3.3 | FALSE | Downregulated |
| -0,086059133 | 0,426905132 | UBE2G1 | FALSE | Downregulated |
| 0,227817723 | 0,426992144 | FAM86DP | FALSE | Upregulated |
| 0,102464335 | 0,427029224 | SNAP29 | FALSE | Upregulated |
| -0,136996372 | 0,427073853 | CAMK2G | FALSE | Downregulated |
| 0,08963104 | 0,427126477 | ALG13 | FALSE | Upregulated |
| -1,837753257 | 0,427154257 | RP11-24J23.2 | FALSE | Downregulated |
| 1,0393817 | 0,427163583 | ANO4 | FALSE | Upregulated |
| 0,138784304 | 0,427187975 | HSPD1 | FALSE | Upregulated |
| -0,095899547 | 0,427187975 | TATDN3 | FALSE | Downregulated |
| 0,548208125 | 0,427369367 | FKBP1AP1 | FALSE | Upregulated |
| -0,671389163 | 0,427440759 | AOX2P | FALSE | Downregulated |
| -0,172794754 | 0,427440759 | C4orf19 | FALSE | Downregulated |
| -1,140327453 | 0,427440759 | DPRXP4 | FALSE | Downregulated |
| -1,213050274 | 0,427440759 | LOC101927818 | FALSE | Downregulated |
| -0,390050949 | 0,427440759 | PRSS12 | FALSE | Downregulated |
| 0,227640085 | 0,427462969 | KIF3A | FALSE | Upregulated |
| -0,139817649 | 0,4276366 | MKNK2 | FALSE | Downregulated |
| 0,549463987 | 0,4276366 | RP11-475O6.1 | FALSE | Upregulated |
| -0,327157218 | 0,427764307 | COL27A1 | FALSE | Downregulated |
| 0,182214693 | 0,427903559 | AMN1 | FALSE | Upregulated |
| 0,550429362 | 0,427903559 | RP11-466F5.8 | FALSE | Upregulated |
| 0,642019528 | 0,428037516 | LOC101928705 | FALSE | Upregulated |
| -0,15581824 | 0,428402628 | PRODH2 | FALSE | Downregulated |
| -0,220668615 | 0,428448905 | COLEC11 | FALSE | Downregulated |
| -0,304583085 | 0,42866685 | C11orf35 | FALSE | Downregulated |
| 0,215214449 | 0,428797862 | APH1B | FALSE | Upregulated |
| -0,414491767 | 0,428904002 | WNK2 | FALSE | Downregulated |
| -0,288126532 | 0,429030495 | GREB1 | FALSE | Downregulated |
| -0,113581414 | 0,429030495 | MRFAP1L1 | FALSE | Downregulated |
| -0,46392831 | 0,429181826 | GDF6 | FALSE | Downregulated |
| 0,103889144 | 0,429205535 | ILKAP | FALSE | Upregulated |
| 0,113021504 | 0,429297458 | CHUK | FALSE | Upregulated |
| 0,165968098 | 0,429297458 | WDFY2 | FALSE | Upregulated |
| 0,911033559 | 0,42931701 | TMEM151A | FALSE | Upregulated |
| 1,197272102 | 0,429329936 | PNOC | FALSE | Upregulated |
| -0,740069533 | 0,429550091 | TSC22D1-AS1 | FALSE | Downregulated |

| | | | | |
|---|---|---|---|---|
| 0,12230226 | 0,429797526 | NACC2 | FALSE | Upregulated |
| -0,076509829 | 0,430043975 | MRPL16 | FALSE | Downregulated |
| -0,219859073 | 0,430058453 | MYL5 | FALSE | Downregulated |
| -0,593756102 | 0,430127553 | JMJD1C-AS1 | FALSE | Downregulated |
| 0,673044322 | 0,430214601 | SLC38A5 | FALSE | Upregulated |
| -0,506901857 | 0,430237006 | ABCA10 | FALSE | Downregulated |
| -0,214831493 | 0,430244635 | TMEM44 | FALSE | Downregulated |
| 0,182334387 | 0,430261051 | RPS11 | FALSE | Upregulated |
| -0,101800418 | 0,430346326 | METTL16 | FALSE | Downregulated |
| 0,501074211 | 0,430479552 | LINC00672 | FALSE | Upregulated |
| 0,096034667 | 0,430479552 | POLDIP2 | FALSE | Upregulated |
| -0,387190521 | 0,430479552 | RASSF9 | FALSE | Downregulated |
| -0,519420075 | 0,430508255 | LOC101928325 | FALSE | Downregulated |
| -0,25724122 | 0,430586578 | AQP7P1 | FALSE | Downregulated |
| -0,520400559 | 0,430625168 | HEPACAM | FALSE | Downregulated |
| 0,12574975 | 0,43071697 | AXIN1 | FALSE | Upregulated |
| -0,11020886 | 0,430879781 | SORBS1 | FALSE | Downregulated |
| -0,228083985 | 0,430952344 | PEAR1 | FALSE | Downregulated |
| 0,888170305 | 0,43107769 | C1QL1 | FALSE | Upregulated |
| -0,446900794 | 0,43107769 | LOC102724517 | FALSE | Downregulated |
| 0,142279403 | 0,43107769 | UXT | FALSE | Upregulated |
| 0,414653975 | 0,431098205 | GLP2R | FALSE | Upregulated |
| -0,135425577 | 0,431403875 | MEGF8 | FALSE | Downregulated |
| 0,181972773 | 0,431486716 | FEM1A | FALSE | Upregulated |
| 0,082366885 | 0,431536896 | RBBP4 | FALSE | Upregulated |
| 0,905680331 | 0,431564454 | OVCH1-AS1 | FALSE | Upregulated |
| 0,690654526 | 0,431570372 | AC005550.4 | FALSE | Upregulated |
| 0,908624316 | 0,431819528 | DDX11-AS1 | FALSE | Upregulated |
| -0,1843342 | 0,432136013 | PFKFB1 | FALSE | Downregulated |
| -0,141618798 | 0,43217457 | ASB1 | FALSE | Downregulated |
| -0,152255758 | 0,432564643 | STT3A | FALSE | Downregulated |
| 0,53588679 | 0,432564643 | TMC7 | FALSE | Upregulated |
| 0,117692192 | 0,432596113 | SDF2 | FALSE | Upregulated |
| -0,19524596 | 0,432596113 | SPRYD4 | FALSE | Downregulated |
| 0,147126852 | 0,432654798 | RUFY2 | FALSE | Upregulated |
| 0,11010634 | 0,432715863 | MED9 | FALSE | Upregulated |
| -0,19881741 | 0,432715863 | RIC8B | FALSE | Downregulated |
| -0,985483466 | 0,432900141 | TMEM132C | FALSE | Downregulated |
| 0,138792002 | 0,432921281 | C17orf70 | FALSE | Upregulated |
| -0,17564566 | 0,432966345 | SEPP1 | FALSE | Downregulated |
| -0,107695997 | 0,432987966 | ZC3H4 | FALSE | Downregulated |
| -0,112156478 | 0,433015086 | FNIP1 | FALSE | Downregulated |
| -0,101129785 | 0,433209641 | MAPK8 | FALSE | Downregulated |
| -0,146824157 | 0,433354188 | MARCKSL1 | FALSE | Downregulated |
| -0,286402945 | 0,433354188 | PPP1R26-AS1 | FALSE | Downregulated |
| -0,230672279 | 0,433354853 | ARMCX1 | FALSE | Downregulated |
| 0,41809078 | 0,433402437 | LINC00654 | FALSE | Upregulated |
| 0,801072334 | 0,433416808 | LOC101928665 | FALSE | Upregulated |
| 0,17065247 | 0,433424859 | CKAP4 | FALSE | Upregulated |
| -0,104286605 | 0,433505745 | NDFIP1 | FALSE | Downregulated |
| 0,324207495 | 0,433815359 | ETNPPL | FALSE | Upregulated |

| | | | | |
|---|---|---|---|---|
| -0,108531849 | 0,433815359 | SAMD8 | FALSE | Downregulated |
| -1,039257121 | 0,433815359 | SLC5A4 | FALSE | Downregulated |
| 0,453412318 | 0,433815359 | TMEM119 | FALSE | Upregulated |
| 0,367630514 | 0,433957301 | VLDLR | FALSE | Upregulated |
| -0,239764922 | 0,433989913 | FEZ1 | FALSE | Downregulated |
| 1,379885357 | 0,434404218 | UGT8 | FALSE | Upregulated |
| 0,101396336 | 0,43441985 | HNRNPA3 | FALSE | Upregulated |
| 0,12517971 | 0,434420712 | COQ4 | FALSE | Upregulated |
| 0,139417212 | 0,434420712 | LAMTOR4 | FALSE | Upregulated |
| 0,642142987 | 0,434420712 | LINC00571 | FALSE | Upregulated |
| -0,438978272 | 0,434420712 | LOC102724783 | FALSE | Downregulated |
| -0,124139253 | 0,434420712 | LRRC37BP1 | FALSE | Downregulated |
| 0,063233867 | 0,434420712 | NUDCD3 | FALSE | Upregulated |
| 0,151882324 | 0,434437533 | THEM6 | FALSE | Upregulated |
| 0,688644436 | 0,434464795 | PRKY | FALSE | Upregulated |
| 0,175853625 | 0,434464795 | RCN2 | FALSE | Upregulated |
| 0,067005722 | 0,434478555 | KHDRBS1 | FALSE | Upregulated |
| -0,111247557 | 0,434521759 | GOLGA5 | FALSE | Downregulated |
| -0,143567616 | 0,434541424 | ADD3 | FALSE | Downregulated |
| -1,354547316 | 0,434656983 | PTPRZ1 | FALSE | Downregulated |
| 0,134972058 | 0,434887405 | EXOSC1 | FALSE | Upregulated |
| -0,11944031 | 0,434887405 | NRBF2 | FALSE | Downregulated |
| -0,091145754 | 0,435111831 | TMEM106B | FALSE | Downregulated |
| -0,171809365 | 0,435195127 | ABCA1 | FALSE | Downregulated |
| -0,206445541 | 0,435195127 | ATP1A1OS | FALSE | Downregulated |
| -0,576765762 | 0,435195127 | NYAP1 | FALSE | Downregulated |
| 0,164911496 | 0,435258401 | NOP16 | FALSE | Upregulated |
| 0,310016288 | 0,435411941 | EFCAB4B | FALSE | Upregulated |
| 0,183880667 | 0,435458862 | APBB1 | FALSE | Upregulated |
| -0,130079154 | 0,435458862 | ZNF75A | FALSE | Downregulated |
| -0,259116847 | 0,43546997 | C8orf49 | FALSE | Downregulated |
| 0,11173387 | 0,43546997 | NDUFS7 | FALSE | Upregulated |
| -0,158852109 | 0,43546997 | WDR47 | FALSE | Downregulated |
| -0,122046425 | 0,43570028 | SLC25A38 | FALSE | Downregulated |
| -0,274814042 | 0,435781954 | DDX11 | FALSE | Downregulated |
| -0,197347139 | 0,436027654 | ECHDC2 | FALSE | Downregulated |
| -0,115821772 | 0,436111533 | ZZEF1 | FALSE | Downregulated |
| -0,09889636 | 0,43634435 | ADH5 | FALSE | Downregulated |
| 0,645806864 | 0,436393439 | AGAP1-IT1 | FALSE | Upregulated |
| 0,193591345 | 0,436399349 | PKP2 | FALSE | Upregulated |
| 0,226595038 | 0,436453353 | NAB2 | FALSE | Upregulated |
| -0,756710732 | 0,436562951 | FMR1-AS1 | FALSE | Downregulated |
| -0,192989394 | 0,436782541 | ZNF419 | FALSE | Downregulated |
| -0,103039616 | 0,436987277 | ZNF830 | FALSE | Downregulated |
| -0,599570397 | 0,437141474 | LOC102723755 | FALSE | Downregulated |
| 0,761605031 | 0,437263589 | CTHRC1 | FALSE | Upregulated |
| -0,062325255 | 0,437263589 | EIF4H | FALSE | Downregulated |
| 0,101933615 | 0,437263589 | KIAA1430 | FALSE | Upregulated |
| -0,344863614 | 0,437263589 | KLHL13 | FALSE | Downregulated |
| 0,608485756 | 0,437263589 | ZNF826P | FALSE | Upregulated |
| -0,271710424 | 0,437451147 | EPB41L4A | FALSE | Downregulated |

| | | | | |
|---|---|---|---|---|
| -0,168033399 | 0,437451147 | KDM6B | FALSE | Downregulated |
| 0,131527142 | 0,437747348 | SPRTN | FALSE | Upregulated |
| 0,637166482 | 0,437920338 | LOC100996455 | FALSE | Upregulated |
| -0,442571604 | 0,438235782 | ANKRD36B | FALSE | Downregulated |
| -0,571662818 | 0,438380001 | GOLGA6L3 | FALSE | Downregulated |
| 0,23580095 | 0,438474045 | LOC101060398 | FALSE | Upregulated |
| -0,143149907 | 0,438474045 | POLR1E | FALSE | Downregulated |
| 0,423541851 | 0,438551374 | SDCBP2 | FALSE | Upregulated |
| 0,40098206 | 0,4386151 | SLC15A2 | FALSE | Upregulated |
| -0,984239929 | 0,438814333 | FLJ39095 | FALSE | Downregulated |
| -0,738842279 | 0,438862147 | CPB1 | FALSE | Downregulated |
| 0,16056823 | 0,438862147 | TRIB2 | FALSE | Upregulated |
| -0,192666567 | 0,439162211 | ZNF133 | FALSE | Downregulated |
| -0,13058044 | 0,439229308 | MRPL2 | FALSE | Downregulated |
| -0,232371787 | 0,439230717 | ZNF487 | FALSE | Downregulated |
| -0,266316504 | 0,439252195 | RP11-141O11.2 | FALSE | Downregulated |
| -0,119489342 | 0,439268778 | COBL | FALSE | Downregulated |
| -0,955318569 | 0,439268778 | LOC101928482 | FALSE | Downregulated |
| -0,132758096 | 0,439268778 | RNF128 | FALSE | Downregulated |
| -0,190857387 | 0,439297795 | ZC2HC1C | FALSE | Downregulated |
| -0,64133541 | 0,439301064 | LOC102724387 | FALSE | Downregulated |
| 0,250930185 | 0,439506419 | ACSS1 | FALSE | Upregulated |
| -0,125303062 | 0,439506419 | ZBTB2 | FALSE | Downregulated |
| 0,500282327 | 0,439544993 | ARPP21 | FALSE | Upregulated |
| 0,111135099 | 0,439544993 | CDK16 | FALSE | Upregulated |
| -0,574507943 | 0,439544993 | FHL1 | FALSE | Downregulated |
| -0,562462804 | 0,439714442 | FLJ42351 | FALSE | Downregulated |
| 0,110433484 | 0,439779361 | MTERF | FALSE | Upregulated |
| -0,111752157 | 0,439779361 | ZNF318 | FALSE | Downregulated |
| 0,990522314 | 0,439853571 | TBL1Y | FALSE | Upregulated |
| -0,203233627 | 0,439865859 | PTGR1 | FALSE | Downregulated |
| 0,085606017 | 0,439893645 | TRA2B | FALSE | Upregulated |
| -0,605638489 | 0,439991395 | JAKMIP2-AS1 | FALSE | Downregulated |
| 0,107805665 | 0,439991395 | SPATS2L | FALSE | Upregulated |
| 0,142447532 | 0,440041281 | TPRKB | FALSE | Upregulated |
| 0,304929454 | 0,440070375 | DGKG | FALSE | Upregulated |
| -0,124963923 | 0,440132948 | ERLIN1 | FALSE | Downregulated |
| -1,205026849 | 0,440193145 | LOC102724335 | FALSE | Downregulated |
| -1,055362349 | 0,440521857 | KIR2DL3 | FALSE | Downregulated |
| 0,232907063 | 0,440552746 | CBX8 | FALSE | Upregulated |
| 0,603340649 | 0,44057241 | TMEM132A | FALSE | Upregulated |
| -0,290052816 | 0,440579997 | SLC25A34 | FALSE | Downregulated |
| -0,087297817 | 0,440724747 | DSTN | FALSE | Downregulated |
| 0,179815723 | 0,440730688 | SH2D4A | FALSE | Upregulated |
| -0,340448542 | 0,440766601 | MFSD4 | FALSE | Downregulated |
| 0,130542346 | 0,44082898 | PYGL | FALSE | Upregulated |
| 0,234313759 | 0,440923398 | AP1G2 | FALSE | Upregulated |
| -0,292503381 | 0,441002142 | GOLGA6C | FALSE | Downregulated |
| -0,176808409 | 0,441051531 | TMEM143 | FALSE | Downregulated |
| -0,115176916 | 0,441051531 | XIAP | FALSE | Downregulated |
| -0,09419832 | 0,441134624 | BCR | FALSE | Downregulated |

| | | | | |
|---|---|---|---|---|
| 0,533780593 | 0,441135207 | F13A1 | FALSE | Upregulated |
| 0,110704604 | 0,441172799 | TUSC2 | FALSE | Upregulated |
| 0,490956114 | 0,441209996 | TGM1 | FALSE | Upregulated |
| -0,103608478 | 0,441278816 | ARNT | FALSE | Downregulated |
| 0,270272659 | 0,441328096 | CCDC171 | FALSE | Upregulated |
| 0,249040036 | 0,441328096 | HMGN3-AS1 | FALSE | Upregulated |
| 0,61615956 | 0,441328096 | ILDR1 | FALSE | Upregulated |
| 0,511859848 | 0,441328096 | LOC102723901 | FALSE | Upregulated |
| 0,335081005 | 0,441328096 | SERP2 | FALSE | Upregulated |
| 0,137088193 | 0,441405712 | MRPL36 | FALSE | Upregulated |
| -0,387980472 | 0,441444217 | LOC102724770 | FALSE | Downregulated |
| -0,230463074 | 0,441537015 | IGHMBP2 | FALSE | Downregulated |
| -0,833069021 | 0,441537015 | LOC100131496 | FALSE | Downregulated |
| 0,101117493 | 0,441843629 | EPRS | FALSE | Upregulated |
| 0,318146816 | 0,441934785 | NUDT8 | FALSE | Upregulated |
| -0,095615763 | 0,441968798 | AK6 | FALSE | Downregulated |
| -0,27941189 | 0,441968798 | INHBA | FALSE | Downregulated |
| 1,025156834 | 0,441968798 | ZNF695 | FALSE | Upregulated |
| -0,840898513 | 0,441991868 | DSG4 | FALSE | Downregulated |
| -0,27791371 | 0,441991868 | LOC102724795 | FALSE | Downregulated |
| 0,147870358 | 0,441991868 | MRPS17 | FALSE | Upregulated |
| 0,20806887 | 0,442063764 | DYNLL1-AS1 | FALSE | Upregulated |
| -0,673306345 | 0,442151428 | RP11-1134I14.8 | FALSE | Downregulated |
| -0,198742236 | 0,44216392 | ENDOV | FALSE | Downregulated |
| -0,696324115 | 0,44216392 | NOXRED1 | FALSE | Downregulated |
| 0,530158156 | 0,442386876 | ADAMTS15 | FALSE | Upregulated |
| 0,439119236 | 0,442386876 | LOC101927322 | FALSE | Upregulated |
| 0,953234779 | 0,442386876 | RAB40A | FALSE | Upregulated |
| -0,096188653 | 0,442386876 | WBP4 | FALSE | Downregulated |
| 0,106559271 | 0,442460998 | MADD | FALSE | Upregulated |
| 0,183568421 | 0,442617125 | NANS | FALSE | Upregulated |
| 0,325259291 | 0,442654827 | ZNF775 | FALSE | Upregulated |
| -0,360339712 | 0,442710592 | PLAT | FALSE | Downregulated |
| 0,164312649 | 0,442859984 | SLC35B1 | FALSE | Upregulated |
| -0,392145867 | 0,442959639 | LOC101927181 | FALSE | Downregulated |
| -0,081552102 | 0,44321722 | PMPCB | FALSE | Downregulated |
| -0,115125493 | 0,443338869 | COX17 | FALSE | Downregulated |
| -0,116435108 | 0,443338869 | DPH5 | FALSE | Downregulated |
| -0,076018517 | 0,443338869 | SLC30A9 | FALSE | Downregulated |
| 0,238564281 | 0,443387763 | C4orf46 | FALSE | Upregulated |
| -0,11603914 | 0,443387763 | CDH2 | FALSE | Downregulated |
| -0,187179819 | 0,443387763 | DEPTOR | FALSE | Downregulated |
| -0,22999167 | 0,443387763 | HSPA1L | FALSE | Downregulated |
| -0,285568966 | 0,443387763 | MORN4 | FALSE | Downregulated |
| 0,139128672 | 0,443428516 | DNAJC30 | FALSE | Upregulated |
| 0,131821539 | 0,443748153 | LCMT2 | FALSE | Upregulated |
| -0,490517878 | 0,44377234 | CCNT2-AS1 | FALSE | Downregulated |
| 0,186447999 | 0,444128433 | GPCPD1 | FALSE | Upregulated |
| -0,614683417 | 0,444128433 | LOC101929882 | FALSE | Downregulated |
| 0,24429112 | 0,444344024 | TICAM1 | FALSE | Upregulated |
| -0,499394174 | 0,44448959 | GABRE | FALSE | Downregulated |

| | | | | |
|---|---|---|---|---|
| -0,181563094 | 0,44448959 | GBE1 | FALSE | Downregulated |
| -0,108326304 | 0,444519483 | PLCL2 | FALSE | Downregulated |
| -0,416214914 | 0,444539617 | GPRASP2 | FALSE | Downregulated |
| -0,664485272 | 0,444539617 | LOC642648 | FALSE | Downregulated |
| -0,842860489 | 0,444539617 | RSPH10B | FALSE | Downregulated |
| -0,434316669 | 0,444580288 | AGAP9 | FALSE | Downregulated |
| 0,12953188 | 0,44465029 | MRPL13 | FALSE | Upregulated |
| 0,406988337 | 0,444654137 | NOTCH3 | FALSE | Upregulated |
| 0,136043903 | 0,444654137 | TRIM25 | FALSE | Upregulated |
| 0,610403435 | 0,444737665 | LOC100507747 | FALSE | Upregulated |
| -0,177821023 | 0,444737665 | LYNX1 | FALSE | Downregulated |
| -0,10098062 | 0,444737665 | PINK1 | FALSE | Downregulated |
| 0,253851857 | 0,444764079 | MID1 | FALSE | Upregulated |
| 0,221407322 | 0,444834534 | ATP6V1E2 | FALSE | Upregulated |
| 0,606801171 | 0,444834534 | GPR113 | FALSE | Upregulated |
| 0,101963828 | 0,444834534 | UBAP2L | FALSE | Upregulated |
| 0,33542013 | 0,444861828 | SLFN13 | FALSE | Upregulated |
| 0,144482778 | 0,44494875 | FAM207A | FALSE | Upregulated |
| 0,551236709 | 0,445102922 | IQCD | FALSE | Upregulated |
| 0,221383207 | 0,445109716 | CRY2 | FALSE | Upregulated |
| -0,297656271 | 0,445109716 | PCOLCE | FALSE | Downregulated |
| 0,138697385 | 0,445131998 | TRIM32 | FALSE | Upregulated |
| -0,946476322 | 0,445159518 | LOC101928555 | FALSE | Downregulated |
| 0,596659608 | 0,445172354 | ACPP | FALSE | Upregulated |
| -0,10928126 | 0,445172354 | BRMS1L | FALSE | Downregulated |
| 0,296463442 | 0,445172354 | DTX3 | FALSE | Upregulated |
| -0,289035971 | 0,445172354 | NAP1L2 | FALSE | Downregulated |
| 1,262002893 | 0,445172354 | PLEKHS1 | FALSE | Upregulated |
| 0,098939472 | 0,445172354 | STT3B | FALSE | Upregulated |
| 0,229341648 | 0,445205585 | FOXO3 | FALSE | Upregulated |
| 0,204268611 | 0,445205585 | LINC00116 | FALSE | Upregulated |
| 0,144638794 | 0,445205585 | SRGAP2 | FALSE | Upregulated |
| 0,179782864 | 0,445205585 | STAT2 | FALSE | Upregulated |
| -0,795168175 | 0,445218736 | RP11-584P21.2 | FALSE | Downregulated |
| -0,07205239 | 0,445218736 | TFCP2 | FALSE | Downregulated |
| -0,365597909 | 0,445345247 | LOC102724800 | FALSE | Downregulated |
| 0,119036024 | 0,445345247 | PIGX | FALSE | Upregulated |
| 0,833012234 | 0,445864214 | CA12 | FALSE | Upregulated |
| -0,275806304 | 0,445864214 | TMEM255B | FALSE | Downregulated |
| -0,118721654 | 0,445884316 | MTFR1L | FALSE | Downregulated |
| -0,325548111 | 0,44595936 | GGACT | FALSE | Downregulated |
| 0,787997059 | 0,445966935 | FOXS1 | FALSE | Upregulated |
| -0,208061946 | 0,446048196 | ARMCX5-GPRASP2 | FALSE | Downregulated |
| 0,831483008 | 0,446048196 | SPRED3 | FALSE | Upregulated |
| 0,102478677 | 0,446048196 | ZFP64 | FALSE | Upregulated |
| 0,120982659 | 0,446139188 | POLR2G | FALSE | Upregulated |
| 0,097066043 | 0,446270529 | DCTN4 | FALSE | Upregulated |
| 0,075799796 | 0,446270529 | KIAA0196 | FALSE | Upregulated |
| -0,092933255 | 0,446270529 | PRRC2B | FALSE | Downregulated |
| 0,765569773 | 0,446291086 | DARC | FALSE | Upregulated |

| | | | | |
|---|---|---|---|---|
| -0,303152149 | 0,446291086 | EIF1B-AS1 | FALSE | Downregulated |
| 0,190841703 | 0,446322083 | PRIM1 | FALSE | Upregulated |
| 0,172705264 | 0,446380306 | RASSF1 | FALSE | Upregulated |
| 0,229458936 | 0,44647943 | BRIP1 | FALSE | Upregulated |
| 0,110335284 | 0,446512064 | RBM8A | FALSE | Upregulated |
| -0,215093239 | 0,446512064 | RSPO3 | FALSE | Downregulated |
| 0,083068595 | 0,446512565 | TFIP11 | FALSE | Upregulated |
| -0,811400766 | 0,446512565 | TMC5 | FALSE | Downregulated |
| -0,564817201 | 0,446913449 | ADAMTS9-AS2 | FALSE | Downregulated |
| -0,211652763 | 0,446971943 | ADAMTS13 | FALSE | Downregulated |
| -0,602978582 | 0,446971943 | GOLGA6D | FALSE | Downregulated |
| 0,109854244 | 0,447031238 | STRN4 | FALSE | Upregulated |
| 0,847791469 | 0,447035588 | SOGA3 | FALSE | Upregulated |
| -0,112894083 | 0,447169842 | CDC37L1 | FALSE | Downregulated |
| -0,825592203 | 0,447169842 | CRP | FALSE | Downregulated |
| -0,089993979 | 0,447169842 | SCAF11 | FALSE | Downregulated |
| -1,119525016 | 0,447274407 | FAIM2 | FALSE | Downregulated |
| 1,396094807 | 0,447307535 | ADCY8 | FALSE | Upregulated |
| -0,188672537 | 0,447407571 | PTPN9 | FALSE | Downregulated |
| -1,025573437 | 0,447438542 | LOC102723317 | FALSE | Downregulated |
| 0,173438788 | 0,447486795 | FAM217B | FALSE | Upregulated |
| 0,643105506 | 0,447486795 | FRMD7 | FALSE | Upregulated |
| -0,551587897 | 0,447486795 | LOC102724307 | FALSE | Downregulated |
| -0,29973659 | 0,447556778 | HAL | FALSE | Downregulated |
| 0,259643423 | 0,447753324 | ARSB | FALSE | Upregulated |
| 0,244988994 | 0,44775749 | GUSBP1 | FALSE | Upregulated |
| 0,127327619 | 0,44775749 | RNF26 | FALSE | Upregulated |
| -0,491598616 | 0,447761335 | SEC14L5 | FALSE | Downregulated |
| 0,210173455 | 0,447761335 | TMEM139 | FALSE | Upregulated |
| 0,247268463 | 0,447761335 | ZNF628 | FALSE | Upregulated |
| -0,167873518 | 0,447920924 | MAPKAPK5-AS1 | FALSE | Downregulated |
| 0,925126477 | 0,44799375 | AC131097.3 | FALSE | Upregulated |
| -0,076691939 | 0,44799375 | C16orf72 | FALSE | Downregulated |
| -0,215798285 | 0,44799375 | ERN1 | FALSE | Downregulated |
| -2,129814344 | 0,44799375 | GOLGA8J | FALSE | Downregulated |
| -0,221549312 | 0,448126288 | VMAC | FALSE | Downregulated |
| 0,171953979 | 0,448216765 | VIMP | FALSE | Upregulated |
| -0,228447974 | 0,448255554 | KLF13 | FALSE | Downregulated |
| 0,298492637 | 0,448418933 | PLAGL1 | FALSE | Upregulated |
| 0,163404021 | 0,448537273 | DOCK6 | FALSE | Upregulated |
| -0,447776431 | 0,448537273 | GOLGA8R | FALSE | Downregulated |
| 1,303927737 | 0,448629709 | PRR15 | FALSE | Upregulated |
| -0,551876822 | 0,448679805 | COCH | FALSE | Downregulated |
| -1,217906432 | 0,448735176 | BRD7P3 | FALSE | Downregulated |
| 0,514882301 | 0,448735176 | CEACAM4 | FALSE | Upregulated |
| -0,105161785 | 0,448735176 | NCOA2 | FALSE | Downregulated |
| -0,28212859 | 0,448735176 | POLN | FALSE | Downregulated |
| -0,184899205 | 0,448830684 | ITSN1 | FALSE | Downregulated |
| -0,705087693 | 0,448848902 | BPI | FALSE | Downregulated |
| -0,700363547 | 0,448848902 | FRMD6-AS1 | FALSE | Downregulated |
| 0,225023833 | 0,448848902 | LPIN2 | FALSE | Upregulated |

| | | | | |
|---|---|---|---|---|
| -0,213954051 | 0,448848902 | PI4KAP2 | FALSE | Downregulated |
| -0,193666639 | 0,448848902 | PIK3C2B | FALSE | Downregulated |
| 0,135854558 | 0,448848902 | TMEM219 | FALSE | Upregulated |
| 0,290483484 | 0,448879314 | LOC101928377 | FALSE | Upregulated |
| 0,158308178 | 0,448896944 | NSUN5 | FALSE | Upregulated |
| 0,198556324 | 0,448940613 | AAED1 | FALSE | Upregulated |
| 0,135602607 | 0,448940613 | CLIP1 | FALSE | Upregulated |
| 0,138509653 | 0,448940613 | CTSO | FALSE | Upregulated |
| -0,868510978 | 0,448940613 | LDLRAD4-AS1 | FALSE | Downregulated |
| -0,28412538 | 0,44894888 | C8orf48 | FALSE | Downregulated |
| -0,751198526 | 0,449016214 | LOC100996442 | FALSE | Downregulated |
| -0,915572609 | 0,449016214 | TPTE2P1 | FALSE | Downregulated |
| -0,157576086 | 0,449029526 | ZSWIM6 | FALSE | Downregulated |
| -0,234486825 | 0,449030264 | GPER1 | FALSE | Downregulated |
| 0,96859749 | 0,449079772 | LOC102723559 | FALSE | Upregulated |
| -0,687521525 | 0,449121629 | RNF212 | FALSE | Downregulated |
| -0,19308538 | 0,449122615 | C11orf49 | FALSE | Downregulated |
| 0,083608765 | 0,449261787 | AAR2 | FALSE | Upregulated |
| 0,128369627 | 0,449261787 | IP6K1 | FALSE | Upregulated |
| -0,823559328 | 0,449261787 | LOC101929346 | FALSE | Downregulated |
| -0,099119611 | 0,449261787 | MANBA | FALSE | Downregulated |
| 0,201695137 | 0,449261787 | PELI3 | FALSE | Upregulated |
| -1,046282006 | 0,449261787 | PKIA | FALSE | Downregulated |
| 0,102194629 | 0,449261787 | TMEM115 | FALSE | Upregulated |
| -0,195223454 | 0,449261787 | TP53I13 | FALSE | Downregulated |
| -0,373352339 | 0,449454632 | TRO | FALSE | Downregulated |
| -0,289413563 | 0,449463336 | CYFIP2 | FALSE | Downregulated |
| -0,516194944 | 0,449463336 | LOC101926977 | FALSE | Downregulated |
| -0,177106072 | 0,449463336 | MCUR1 | FALSE | Downregulated |
| -0,609725743 | 0,449574708 | SAMD13 | FALSE | Downregulated |
| -0,336658029 | 0,449640604 | CCDC11 | FALSE | Downregulated |
| -0,119130055 | 0,449676271 | DCP1B | FALSE | Downregulated |
| -0,096776899 | 0,449676271 | HNRNPA0 | FALSE | Downregulated |
| 0,170153522 | 0,449676271 | PQLC2 | FALSE | Upregulated |
| 0,119979508 | 0,449677604 | AKTIP | FALSE | Upregulated |
| 0,702481799 | 0,449749238 | FAP | FALSE | Upregulated |
| 0,213148817 | 0,449749238 | IFITM2 | FALSE | Upregulated |
| -0,238577193 | 0,450059516 | FAM35DP | FALSE | Downregulated |
| 0,09982419 | 0,450059516 | SLMO2 | FALSE | Upregulated |
| -0,145110365 | 0,450092821 | GLRX5 | FALSE | Downregulated |
| -0,855326331 | 0,450142223 | ANP32A-IT1 | FALSE | Downregulated |
| 0,10274289 | 0,450280533 | EED | FALSE | Upregulated |
| 0,563367033 | 0,450707551 | TACR2 | FALSE | Upregulated |
| -0,627355884 | 0,450715641 | RBM20 | FALSE | Downregulated |
| -1,070202982 | 0,450744087 | LOC101928062 | FALSE | Downregulated |
| -0,589743913 | 0,450854641 | EMILIN3 | FALSE | Downregulated |
| -0,099012543 | 0,450979419 | YIPF6 | FALSE | Downregulated |
| -1,032218194 | 0,451110943 | FLJ38379 | FALSE | Downregulated |
| 0,101578012 | 0,451206125 | RPAP3 | FALSE | Upregulated |
| -0,897049826 | 0,451206125 | TRNK | FALSE | Downregulated |
| 0,155677963 | 0,451301086 | ABLIM1 | FALSE | Upregulated |

| | | | | |
|---|---|---|---|---|
| -0,186460862 | 0,451448412 | PPP1R12B | FALSE | Downregulated |
| -0,110680115 | 0,451448412 | TMEM218 | FALSE | Downregulated |
| 0,103874738 | 0,451530297 | PIGO | FALSE | Upregulated |
| 0,193272602 | 0,451586319 | AGPAT2 | FALSE | Upregulated |
| -0,135104403 | 0,451586319 | KIF21A | FALSE | Downregulated |
| 0,308038251 | 0,451586319 | NAMPT | FALSE | Upregulated |
| -1,803471164 | 0,451657584 | SLC5A12 | FALSE | Downregulated |
| 1,347521686 | 0,451842893 | FSTL5 | FALSE | Upregulated |
| 0,096094285 | 0,451842893 | KEAP1 | FALSE | Upregulated |
| 0,577588518 | 0,451842893 | SLC25A41 | FALSE | Upregulated |
| 0,135438037 | 0,451842893 | SYNJ2 | FALSE | Upregulated |
| 0,252540411 | 0,451842893 | TRERF1 | FALSE | Upregulated |
| 0,177644459 | 0,451915486 | LSM2 | FALSE | Upregulated |
| 0,100638862 | 0,452034291 | NCAPD2 | FALSE | Upregulated |
| -0,280912733 | 0,452076953 | FAM71E1 | FALSE | Downregulated |
| -0,104426686 | 0,452076953 | FGFRL1 | FALSE | Downregulated |
| -0,103668947 | 0,452076953 | KNG1 | FALSE | Downregulated |
| -0,674851189 | 0,452076953 | LOC286190 | FALSE | Downregulated |
| 0,094360265 | 0,452076953 | NIPA1 | FALSE | Upregulated |
| -0,885704296 | 0,452076953 | NPIPB8 | FALSE | Downregulated |
| -0,550442457 | 0,452076953 | RP5-855F14.1 | FALSE | Downregulated |
| 0,582005952 | 0,452076953 | ZNF804A | FALSE | Upregulated |
| 0,148061462 | 0,452077151 | THOC6 | FALSE | Upregulated |
| -0,113547035 | 0,452927242 | TUBGCP3 | FALSE | Downregulated |
| -0,133251774 | 0,453008072 | RBKS | FALSE | Downregulated |
| -0,744516659 | 0,453042942 | RP11-276H19.1 | FALSE | Downregulated |
| 0,814695843 | 0,453081232 | ZNF280B | FALSE | Upregulated |
| 0,149740303 | 0,453424654 | RNF149 | FALSE | Upregulated |
| 0,518991734 | 0,453456489 | ABCB9 | FALSE | Upregulated |
| -0,131309168 | 0,453644059 | ABCC6 | FALSE | Downregulated |
| -0,174105037 | 0,453644059 | ELL2 | FALSE | Downregulated |
| -0,130872846 | 0,453644059 | OSTC | FALSE | Downregulated |
| -0,239213432 | 0,453644059 | SFRP1 | FALSE | Downregulated |
| 0,677141887 | 0,453717794 | TMEM92 | FALSE | Upregulated |
| -1,262330072 | 0,454061966 | SCHIP1 | FALSE | Downregulated |
| 0,3809786 | 0,454150383 | FAHD2CP | FALSE | Upregulated |
| -0,852091445 | 0,454150383 | LOC101926940 | FALSE | Downregulated |
| 0,55274296 | 0,454150383 | LOC284837 | FALSE | Upregulated |
| 0,102857072 | 0,454150383 | NELFB | FALSE | Upregulated |
| 0,126707963 | 0,454150383 | PKNOX1 | FALSE | Upregulated |
| -0,173299592 | 0,454150383 | SEMA4C | FALSE | Downregulated |
| 0,291464371 | 0,454150383 | TTLL1 | FALSE | Upregulated |
| 0,741972843 | 0,454231647 | TRPC6 | FALSE | Upregulated |
| -0,124316629 | 0,454231647 | ZBTB7B | FALSE | Downregulated |
| -0,142734984 | 0,454302823 | FAM104B | FALSE | Downregulated |
| -0,109026477 | 0,454331237 | COG6 | FALSE | Downregulated |
| -0,179513127 | 0,454331237 | FAM101B | FALSE | Downregulated |
| -0,111975834 | 0,454358297 | RAB11FIP3 | FALSE | Downregulated |
| 0,114825332 | 0,454362151 | B4GALT3 | FALSE | Upregulated |
| 0,223786907 | 0,454362151 | LRRC61 | FALSE | Upregulated |
| 0,185699766 | 0,454561672 | AMOTL2 | FALSE | Upregulated |

| | | | | |
|---|---|---|---|---|
| -0,186187562 | 0,454561672 | SLC33A1 | FALSE | Downregulated |
| -0,706657967 | 0,454665811 | LINC00176 | FALSE | Downregulated |
| 0,087190641 | 0,45477025 | DR1 | FALSE | Upregulated |
| 0,354582838 | 0,45477025 | ZNF681 | FALSE | Upregulated |
| -0,193032717 | 0,45477025 | ZSCAN9 | FALSE | Downregulated |
| -0,179843478 | 0,454927056 | C19orf44 | FALSE | Downregulated |
| -0,984409278 | 0,455291088 | MROH2B | FALSE | Downregulated |
| -0,355765935 | 0,455513597 | LOC101929200 | FALSE | Downregulated |
| 0,947862326 | 0,455744639 | SMKR1 | FALSE | Upregulated |
| -0,137808626 | 0,455847397 | VPS13B | FALSE | Downregulated |
| -0,677467397 | 0,456058438 | ACSM1 | FALSE | Downregulated |
| 0,116715052 | 0,456058438 | ST3GAL2 | FALSE | Upregulated |
| -0,559356055 | 0,45618024 | NEGR1 | FALSE | Downregulated |
| -0,260722883 | 0,4563305 | SH3YL1 | FALSE | Downregulated |
| 0,075034577 | 0,456464076 | CAMTA2 | FALSE | Upregulated |
| 0,792656991 | 0,456728007 | ADAM11 | FALSE | Upregulated |
| -1,34743006 | 0,456739273 | SMPX | FALSE | Downregulated |
| 0,126290799 | 0,456830513 | SLC25A17 | FALSE | Upregulated |
| 0,288289546 | 0,456830513 | TLR5 | FALSE | Upregulated |
| 0,513791274 | 0,456892687 | FFAR2 | FALSE | Upregulated |
| 0,139223822 | 0,456923389 | AGO2 | FALSE | Upregulated |
| 0,19408061 | 0,456923389 | KIF9 | FALSE | Upregulated |
| -0,726945334 | 0,457027242 | CCDC154 | FALSE | Downregulated |
| -0,582619516 | 0,457027242 | CNGA4 | FALSE | Downregulated |
| -1,175776169 | 0,457027242 | DIRAS1 | FALSE | Downregulated |
| -0,100485457 | 0,457308349 | URB1 | FALSE | Downregulated |
| 0,1863873 | 0,457320722 | PRAP1 | FALSE | Upregulated |
| 0,197564426 | 0,457338201 | RPL39 | FALSE | Upregulated |
| -0,113796035 | 0,457498547 | CPNE8 | FALSE | Downregulated |
| 0,367523832 | 0,457638331 | SPDYC | FALSE | Upregulated |
| 0,502845169 | 0,457964858 | RIN1 | FALSE | Upregulated |
| -0,692589699 | 0,458063437 | ADAM33 | FALSE | Downregulated |
| -0,403501527 | 0,458168465 | PRR22 | FALSE | Downregulated |
| -0,19483793 | 0,45821793 | CLEC3B | FALSE | Downregulated |
| -0,08808628 | 0,458230852 | USP28 | FALSE | Downregulated |
| 0,133169592 | 0,458621645 | C6orf47 | FALSE | Upregulated |
| -0,556810427 | 0,458621645 | TRPM3 | FALSE | Downregulated |
| 0,127691867 | 0,459077213 | HNRNPLL | FALSE | Upregulated |
| 0,225170609 | 0,459077213 | RAPGEF4 | FALSE | Upregulated |
| 0,110714042 | 0,459115484 | DBR1 | FALSE | Upregulated |
| -0,335298354 | 0,459217062 | ROR1 | FALSE | Downregulated |
| -0,750324885 | 0,459228414 | RP11-474D1.3 | FALSE | Downregulated |
| 0,280732384 | 0,459228414 | TCHH | FALSE | Upregulated |
| -0,232568342 | 0,459228414 | TM6SF2 | FALSE | Downregulated |
| 0,079226279 | 0,4592285 | ABCF3 | FALSE | Upregulated |
| -0,149989214 | 0,4592285 | FOXRED1 | FALSE | Downregulated |
| 0,199971646 | 0,45929608 | PSIP1 | FALSE | Upregulated |
| 0,867864669 | 0,459360237 | LOC729739 | FALSE | Upregulated |
| 0,540560577 | 0,459500293 | SNAI3 | FALSE | Upregulated |
| -0,202449062 | 0,459935651 | CYB5RL | FALSE | Downregulated |
| 0,797548598 | 0,460011005 | FOXL1 | FALSE | Upregulated |

| | | | | |
|---|---|---|---|---|
| 0,192671708 | 0,460174172 | AMMECR1 | FALSE | Upregulated |
| -0,203900042 | 0,460345664 | SIVA1 | FALSE | Downregulated |
| 0,689715724 | 0,460437166 | LINC01057 | FALSE | Upregulated |
| 0,125834589 | 0,460576339 | GPR108 | FALSE | Upregulated |
| -0,146562616 | 0,460576339 | NAPRT1 | FALSE | Downregulated |
| 0,136039304 | 0,460743805 | COMMD3 | FALSE | Upregulated |
| -0,105711752 | 0,460776624 | THAP1 | FALSE | Downregulated |
| -0,096275465 | 0,460929359 | CUTA | FALSE | Downregulated |
| -0,098417091 | 0,460937031 | FASTKD2 | FALSE | Downregulated |
| 0,232656338 | 0,461156573 | ZNF669 | FALSE | Upregulated |
| 0,236096052 | 0,461251915 | BIN1 | FALSE | Upregulated |
| -0,333395257 | 0,461251915 | DCLK2 | FALSE | Downregulated |
| -0,724086732 | 0,461251915 | RAB3C | FALSE | Downregulated |
| 0,338634299 | 0,461251915 | SDK2 | FALSE | Upregulated |
| -0,242290577 | 0,461297804 | LRRC6 | FALSE | Downregulated |
| -1,314480045 | 0,461507751 | FAM153C | FALSE | Downregulated |
| 1,040665145 | 0,461898953 | OR51E1 | FALSE | Upregulated |
| -0,85262978 | 0,462007763 | AC079145.4 | FALSE | Downregulated |
| 0,071459596 | 0,462007763 | ANXA7 | FALSE | Upregulated |
| 0,262472652 | 0,462100362 | FLJ23867 | FALSE | Upregulated |
| 0,836551232 | 0,462261514 | LINC00877 | FALSE | Upregulated |
| -0,115764857 | 0,462278438 | ALDH4A1 | FALSE | Downregulated |
| -0,303844235 | 0,462278438 | DGAT2 | FALSE | Downregulated |
| -0,557983236 | 0,462278438 | RPL13AP20 | FALSE | Downregulated |
| 0,268642648 | 0,462278438 | UGT3A1 | FALSE | Upregulated |
| 0,203043298 | 0,462305787 | CHST15 | FALSE | Upregulated |
| 0,740198893 | 0,462305787 | FANCB | FALSE | Upregulated |
| -0,178521328 | 0,462305787 | SMTN | FALSE | Downregulated |
| 0,11027317 | 0,462305787 | WDR7 | FALSE | Upregulated |
| 0,177092059 | 0,462416397 | DAO | FALSE | Upregulated |
| -0,509731917 | 0,462416397 | TWIST1 | FALSE | Downregulated |
| -0,107280416 | 0,462496647 | TMEM69 | FALSE | Downregulated |
| -0,084438395 | 0,463260729 | MORC2 | FALSE | Downregulated |
| 0,133688708 | 0,463260729 | MRPL21 | FALSE | Upregulated |
| 0,092491771 | 0,463260729 | PSMC5 | FALSE | Upregulated |
| 0,358824769 | 0,463425554 | GPR37 | FALSE | Upregulated |
| -1,612422158 | 0,463425554 | KBTBD12 | FALSE | Downregulated |
| 0,135851657 | 0,463463132 | C19orf52 | FALSE | Upregulated |
| -1,044357397 | 0,463673493 | NRK | FALSE | Downregulated |
| 0,588335301 | 0,463740221 | LOC102723579 | FALSE | Upregulated |
| -0,1435527 | 0,464242653 | C1orf131 | FALSE | Downregulated |
| 0,173602017 | 0,464242653 | DNMT3A | FALSE | Upregulated |
| -0,831388059 | 0,464242653 | LOC101927733 | FALSE | Downregulated |
| -0,146159687 | 0,46434711 | ZNF2 | FALSE | Downregulated |
| -0,158187497 | 0,464791666 | MOCS2 | FALSE | Downregulated |
| 0,129688245 | 0,4648432 | NDUFA13 | FALSE | Upregulated |
| -0,106002322 | 0,465026082 | ALDH3A2 | FALSE | Downregulated |
| 0,283756691 | 0,465026082 | CDR2L | FALSE | Upregulated |
| -0,244296754 | 0,465046936 | LOC102724905 | FALSE | Downregulated |
| 0,167696403 | 0,465046936 | SLC28A1 | FALSE | Upregulated |
| -0,439566337 | 0,465270826 | RPSAP9 | FALSE | Downregulated |

| | | | | |
|---|---|---|---|---|
| 0,138873967 | 0,465343933 | TSPAN14 | FALSE | Upregulated |
| -0,137007696 | 0,465348817 | NR2F6 | FALSE | Downregulated |
| -0,309078363 | 0,46540147 | LOC401320 | FALSE | Downregulated |
| 0,111650784 | 0,465781237 | TAOK1 | FALSE | Upregulated |
| 0,197073242 | 0,465855728 | ARID5B | FALSE | Upregulated |
| 0,103141064 | 0,465862342 | RNF31 | FALSE | Upregulated |
| -0,087125322 | 0,465862342 | SON | FALSE | Downregulated |
| 0,428051888 | 0,466073632 | FOS | FALSE | Upregulated |
| 0,121389376 | 0,466073632 | INIP | FALSE | Upregulated |
| -0,143565829 | 0,466078956 | KRBOX4 | FALSE | Downregulated |
| 0,146982285 | 0,466696646 | CLSTN1 | FALSE | Upregulated |
| -0,563476274 | 0,46693795 | TFCP2L1 | FALSE | Downregulated |
| -0,244441849 | 0,467383718 | NUMBL | FALSE | Downregulated |
| -0,249706376 | 0,467533781 | FLJ46906 | FALSE | Downregulated |
| 0,200994386 | 0,467597213 | HOMER3 | FALSE | Upregulated |
| -0,097302634 | 0,467607848 | CMPK1 | FALSE | Downregulated |
| 0,164883635 | 0,467880052 | COX10 | FALSE | Upregulated |
| 1,202924387 | 0,467948324 | LOC102724652 | FALSE | Upregulated |
| -0,080757131 | 0,468002909 | ASB7 | FALSE | Downregulated |
| -0,396887079 | 0,468002909 | DNAJA4 | FALSE | Downregulated |
| -0,611521942 | 0,46858638 | TENM4 | FALSE | Downregulated |
| 0,41693598 | 0,468657017 | FBXL19-AS1 | FALSE | Upregulated |
| -0,191901802 | 0,468716643 | IL17RC | FALSE | Downregulated |
| -0,259802549 | 0,468716643 | PDE3A | FALSE | Downregulated |
| 0,080864684 | 0,468766345 | ERCC3 | FALSE | Upregulated |
| 0,17650464 | 0,468766345 | ICAM2 | FALSE | Upregulated |
| 0,122349293 | 0,468766345 | MAN2B2 | FALSE | Upregulated |
| -0,115337204 | 0,468766345 | NUP153 | FALSE | Downregulated |
| -0,148600262 | 0,468766345 | SLC10A7 | FALSE | Downregulated |
| -0,751725576 | 0,468848917 | DYNLRB2 | FALSE | Downregulated |
| -0,621426074 | 0,468848917 | LOC400553 | FALSE | Downregulated |
| 0,817190698 | 0,468948477 | JPH1 | FALSE | Upregulated |
| -0,11922926 | 0,469033809 | C9orf85 | FALSE | Downregulated |
| 0,114987378 | 0,469071054 | SSH3 | FALSE | Upregulated |
| 0,773782119 | 0,469594085 | C2orf81 | FALSE | Upregulated |
| -0,864350464 | 0,469787881 | MMP28 | FALSE | Downregulated |
| 0,166333187 | 0,469824106 | SLC35B4 | FALSE | Upregulated |
| 0,191788074 | 0,469825585 | CFP | FALSE | Upregulated |
| 0,702466328 | 0,470095933 | LOC646127 | FALSE | Upregulated |
| -1,073529122 | 0,4704673 | LOC100132731 | FALSE | Downregulated |
| -0,449268401 | 0,47065874 | BRSK1 | FALSE | Downregulated |
| -0,549098163 | 0,470701335 | FLJ44313 | FALSE | Downregulated |
| -0,196879838 | 0,47113316 | CYP1B1 | FALSE | Downregulated |
| 0,342646326 | 0,471219141 | AMIGO3 | FALSE | Upregulated |
| 0,84000669 | 0,471219141 | AURKAPS1 | FALSE | Upregulated |
| -0,145134121 | 0,471307037 | DDI2 | FALSE | Downregulated |
| -0,155556595 | 0,471307037 | NET1 | FALSE | Downregulated |
| -0,558784713 | 0,471307037 | SPATA25 | FALSE | Downregulated |
| -0,610373954 | 0,471406295 | RP1-257A7.4 | FALSE | Downregulated |
| -0,409059378 | 0,471422467 | EPPK1 | FALSE | Downregulated |
| -0,162452195 | 0,471422467 | FN3K | FALSE | Downregulated |

| | | | | |
|---|---|---|---|---|
| -0,077815131 | 0,471422467 | PRPF8 | FALSE | Downregulated |
| -0,195915727 | 0,471422467 | TOP1P1 | FALSE | Downregulated |
| 0,074399393 | 0,471422467 | TTI1 | FALSE | Upregulated |
| -0,964903631 | 0,471498671 | GJC2 | FALSE | Downregulated |
| -0,087130853 | 0,471498671 | TXNDC15 | FALSE | Downregulated |
| -0,09765809 | 0,471643025 | MTIF3 | FALSE | Downregulated |
| 1,196423742 | 0,471643025 | RP11-82L2.1 | FALSE | Upregulated |
| 0,104318854 | 0,471875622 | C3orf38 | FALSE | Upregulated |
| 0,201493189 | 0,471946474 | ATP2A2 | FALSE | Upregulated |
| 0,241050149 | 0,472311118 | ABCC5 | FALSE | Upregulated |
| -0,112418437 | 0,472311118 | COQ5 | FALSE | Downregulated |
| -0,484287854 | 0,472580667 | FBLN1 | FALSE | Downregulated |
| 0,096267878 | 0,472580667 | SUZ12 | FALSE | Upregulated |
| -0,170606211 | 0,472589887 | MMRN2 | FALSE | Downregulated |
| 0,376015884 | 0,472589887 | ZIK1 | FALSE | Upregulated |
| 0,160844376 | 0,472650828 | GAMT | FALSE | Upregulated |
| 0,414226513 | 0,472660825 | MYZAP | FALSE | Upregulated |
| 0,995483925 | 0,472673623 | DUOX1 | FALSE | Upregulated |
| -0,184472968 | 0,472802626 | ENOSF1 | FALSE | Downregulated |
| 0,58715208 | 0,472904494 | SCARNA9 | FALSE | Upregulated |
| 0,383035794 | 0,472912019 | PA2G4P4 | FALSE | Upregulated |
| -0,254546096 | 0,473036084 | NAP1L3 | FALSE | Downregulated |
| -0,173269627 | 0,473158689 | SERINC5 | FALSE | Downregulated |
| 0,153412578 | 0,473234754 | LYSMD1 | FALSE | Upregulated |
| 0,107511933 | 0,473244195 | GMEB2 | FALSE | Upregulated |
| 0,682944097 | 0,473244195 | LOC101928884 | FALSE | Upregulated |
| -0,197129813 | 0,473465594 | CWC25 | FALSE | Downregulated |
| 0,426905232 | 0,473532828 | QPCT | FALSE | Upregulated |
| -0,098478326 | 0,473646323 | KIAA1468 | FALSE | Downregulated |
| 0,730964493 | 0,473646323 | PGA4 | FALSE | Upregulated |
| -0,107002002 | 0,47376444 | DPM1 | FALSE | Downregulated |
| 0,186693456 | 0,47376444 | TBXA2R | FALSE | Upregulated |
| 0,212497267 | 0,473767708 | NDN | FALSE | Upregulated |
| 0,09512972 | 0,473946082 | ADD1 | FALSE | Upregulated |
| 0,632592556 | 0,473946082 | FCRL1 | FALSE | Upregulated |
| 0,117894666 | 0,473946082 | LATS2 | FALSE | Upregulated |
| -0,178606638 | 0,473985455 | GPATCH2 | FALSE | Downregulated |
| -0,710499713 | 0,474623081 | SH3GL1P1 | FALSE | Downregulated |
| 0,218262672 | 0,474662042 | LAMB3 | FALSE | Upregulated |
| -0,121373563 | 0,474904708 | DICER1 | FALSE | Downregulated |
| -0,905411978 | 0,47490949 | C9orf47 | FALSE | Downregulated |
| 0,120413181 | 0,47490949 | GNAI2 | FALSE | Upregulated |
| -0,17800149 | 0,47490949 | MBIP | FALSE | Downregulated |
| -0,161102341 | 0,47490949 | NEU3 | FALSE | Downregulated |
| -0,581005265 | 0,47490949 | SPATA17 | FALSE | Downregulated |
| -0,096243451 | 0,47490949 | SSU72 | FALSE | Downregulated |
| -0,274533037 | 0,47490949 | ZNF471 | FALSE | Downregulated |
| -0,631750521 | 0,475165358 | LOC100996735 | FALSE | Downregulated |
| -0,158089815 | 0,47541266 | CD14 | FALSE | Downregulated |
| -0,267542685 | 0,47541266 | MECOM | FALSE | Downregulated |
| -0,109126218 | 0,475454132 | IFT74 | FALSE | Downregulated |

| | | | | |
|---|---|---|---|---|
| -0,078213853 | 0,47573825 | KPNA6 | FALSE | Downregulated |
| -0,812614009 | 0,475816815 | SLC22A20 | FALSE | Downregulated |
| -0,796420225 | 0,4758244 | LOC101928428 | FALSE | Downregulated |
| -0,340599635 | 0,4758244 | NPY5R | FALSE | Downregulated |
| 0,138930455 | 0,475946032 | ACTL6A | FALSE | Upregulated |
| -0,442389263 | 0,475946032 | NAPA-AS1 | FALSE | Downregulated |
| -0,084083704 | 0,47606695 | BCCIP | FALSE | Downregulated |
| -0,09431348 | 0,476119729 | EIF2B5 | FALSE | Downregulated |
| 0,730528029 | 0,476169037 | C15orf59 | FALSE | Upregulated |
| -0,692435794 | 0,476169037 | LOC101927734 | FALSE | Downregulated |
| -0,124008275 | 0,476374577 | COQ3 | FALSE | Downregulated |
| -0,715299997 | 0,476374577 | GRIN2A | FALSE | Downregulated |
| 0,116236311 | 0,476374577 | NUP155 | FALSE | Upregulated |
| 0,934993318 | 0,476430176 | LOC102723390 | FALSE | Upregulated |
| -0,445673039 | 0,47657746 | MT1DP | FALSE | Downregulated |
| -0,347768258 | 0,47657746 | PDGFD | FALSE | Downregulated |
| -0,142697933 | 0,476618014 | TPR | FALSE | Downregulated |
| 0,550110255 | 0,476771748 | ARHGAP22 | FALSE | Upregulated |
| 0,125044991 | 0,476903986 | AATF | FALSE | Upregulated |
| 0,108831014 | 0,476908207 | ADO | FALSE | Upregulated |
| -1,022794345 | 0,476964553 | PRH1 | FALSE | Downregulated |
| -0,172367881 | 0,477031867 | AGO3 | FALSE | Downregulated |
| -0,097631419 | 0,477031867 | EIF1AD | FALSE | Downregulated |
| 0,170793785 | 0,477031867 | LINC00843 | FALSE | Upregulated |
| -0,143587009 | 0,477031867 | TLK1 | FALSE | Downregulated |
| -0,371215464 | 0,477243495 | TBC1D3 | FALSE | Downregulated |
| 0,206497258 | 0,477457494 | DGCR14 | FALSE | Upregulated |
| -0,19057757 | 0,477457494 | WHAMMP3 | FALSE | Downregulated |
| 0,317953333 | 0,477519812 | LOC374443 | FALSE | Upregulated |
| 0,094220263 | 0,477522418 | G3BP2 | FALSE | Upregulated |
| 0,140246894 | 0,477522418 | TMEM222 | FALSE | Upregulated |
| 0,18766043 | 0,47782766 | PAQR9 | FALSE | Upregulated |
| 0,132168164 | 0,47782766 | TSEN15 | FALSE | Upregulated |
| -1,022180602 | 0,47782766 | WTAPP1 | FALSE | Downregulated |
| -0,357856474 | 0,477830181 | AGER | FALSE | Downregulated |
| 0,448319977 | 0,477830181 | PLAC9 | FALSE | Upregulated |
| 0,150296675 | 0,477895177 | POMGNT2 | FALSE | Upregulated |
| 0,155466374 | 0,477996853 | RIN3 | FALSE | Upregulated |
| 0,209130115 | 0,478056549 | SAMD1 | FALSE | Upregulated |
| 0,467853994 | 0,478311986 | LOC101929638 | FALSE | Upregulated |
| -0,187150055 | 0,478376826 | NIFK-AS1 | FALSE | Downregulated |
| 0,256274841 | 0,478376826 | PBDC1 | FALSE | Upregulated |
| 1,281776408 | 0,478440405 | LY6K | FALSE | Upregulated |
| -0,33227566 | 0,478473151 | LOC101928762 | FALSE | Downregulated |
| -0,307019854 | 0,4785942 | EDNRA | FALSE | Downregulated |
| 0,096951751 | 0,4786412 | UBE2E1 | FALSE | Upregulated |
| -0,735589126 | 0,478808319 | GK3P | FALSE | Downregulated |
| -0,173661138 | 0,478808319 | KALRN | FALSE | Downregulated |
| -0,107378824 | 0,478808319 | KIF1B | FALSE | Downregulated |
| 0,168175351 | 0,478808319 | NABP2 | FALSE | Upregulated |
| 0,127995393 | 0,478841568 | TLE4 | FALSE | Upregulated |

| | | | | |
|---|---|---|---|---|
| 0,079268803 | 0,479227413 | NGLY1 | FALSE | Upregulated |
| -0,520234007 | 0,479227413 | PDGFRL | FALSE | Downregulated |
| -0,516908397 | 0,479227413 | PRDM16 | FALSE | Downregulated |
| 0,284922436 | 0,479227413 | RNASEK-C17orf49 | FALSE | Upregulated |
| -1,038843608 | 0,479227413 | SLC7A5P2 | FALSE | Downregulated |
| 0,129943143 | 0,479370216 | DCAF15 | FALSE | Upregulated |
| -0,21462525 | 0,479544036 | ENKD1 | FALSE | Downregulated |
| -0,213371159 | 0,479619431 | CCDC15 | FALSE | Downregulated |
| 0,083997495 | 0,479673526 | ANKLE2 | FALSE | Upregulated |
| -0,186355331 | 0,479673526 | CDA | FALSE | Downregulated |
| -0,683798892 | 0,479673526 | FLJ45513 | FALSE | Downregulated |
| 0,526077701 | 0,479673526 | IGFBP1 | FALSE | Upregulated |
| 0,215925642 | 0,479673526 | LRRC4 | FALSE | Upregulated |
| -0,14601913 | 0,479673526 | SIK3 | FALSE | Downregulated |
| -0,102359447 | 0,479989504 | CPD | FALSE | Downregulated |
| 0,99692071 | 0,480153269 | SPATC1 | FALSE | Upregulated |
| -0,197430775 | 0,480294221 | NUDT3 | FALSE | Downregulated |
| 0,670972448 | 0,480302248 | LOC102723652 | FALSE | Upregulated |
| -0,135437882 | 0,48057167 | RAI1 | FALSE | Downregulated |
| 0,420498375 | 0,480834996 | CHEK2 | FALSE | Upregulated |
| 0,18611284 | 0,480844553 | SRD5A3 | FALSE | Upregulated |
| 0,341478206 | 0,480893107 | STK32C | FALSE | Upregulated |
| 0,130172972 | 0,481000543 | ASGR1 | FALSE | Upregulated |
| -1,340728891 | 0,48108964 | SNORA23 | FALSE | Downregulated |
| -0,10325503 | 0,481146256 | DIS3 | FALSE | Downregulated |
| 0,239546994 | 0,481153092 | GATA3 | FALSE | Upregulated |
| 0,298088795 | 0,481265058 | SLCO2A1 | FALSE | Upregulated |
| -0,885941962 | 0,48129501 | SPDYE6 | FALSE | Downregulated |
| 0,138691677 | 0,481368639 | TCTN1 | FALSE | Upregulated |
| -0,102427561 | 0,481506199 | MAL2 | FALSE | Downregulated |
| 0,462472981 | 0,481514273 | LOC100505727 | FALSE | Upregulated |
| -0,391362775 | 0,48152565 | LOC100128494 | FALSE | Downregulated |
| 0,822947039 | 0,481636037 | DKFZp686L13185 | FALSE | Upregulated |
| -0,311123096 | 0,481645861 | LOC100129781 | FALSE | Downregulated |
| -0,401708945 | 0,481746441 | RP1-93I3.1 | FALSE | Downregulated |
| 0,091532219 | 0,481851383 | COPS6 | FALSE | Upregulated |
| -1,627455201 | 0,481980967 | C10orf71 | FALSE | Downregulated |
| -0,144108284 | 0,48216486 | LATS1 | FALSE | Downregulated |
| -0,321030756 | 0,48216486 | MEF2C | FALSE | Downregulated |
| 0,419214544 | 0,482402181 | CD101 | FALSE | Upregulated |
| 0,163092866 | 0,482538009 | USP46-AS1 | FALSE | Upregulated |
| 0,234185795 | 0,482620556 | IRAK3 | FALSE | Upregulated |
| 0,483929396 | 0,482624591 | HTATSF1P2 | FALSE | Upregulated |
| -0,594920872 | 0,482624591 | LOC102723640 | FALSE | Downregulated |
| -0,103025804 | 0,482624591 | NAA35 | FALSE | Downregulated |
| -0,209859419 | 0,482624591 | POLR2J4 | FALSE | Downregulated |
| -0,315850555 | 0,482795598 | LOC101929005 | FALSE | Downregulated |
| -0,087291166 | 0,482812304 | UBE2D3 | FALSE | Downregulated |
| 0,142515039 | 0,482821036 | STEAP3 | FALSE | Upregulated |
| 0,124073331 | 0,482924781 | PTP4A1 | FALSE | Upregulated |
| 0,27466835 | 0,48294293 | LOC101928000 | FALSE | Upregulated |

| | | | | |
|---|---|---|---|---|
| 0,137780786 | 0,483338574 | C19orf24 | FALSE | Upregulated |
| -0,236038051 | 0,483407547 | ST20 | FALSE | Downregulated |
| -0,913364417 | 0,483724711 | EMX2OS | FALSE | Downregulated |
| -0,09973492 | 0,483724711 | MRP63 | FALSE | Downregulated |
| 0,156381399 | 0,48382842 | TP53BP1 | FALSE | Upregulated |
| 0,13161277 | 0,483933372 | HDDC3 | FALSE | Upregulated |
| -0,09176529 | 0,484211479 | COASY | FALSE | Downregulated |
| -0,127082009 | 0,484490644 | CEP350 | FALSE | Downregulated |
| -0,731190724 | 0,484581238 | CTD-2337A12.1 | FALSE | Downregulated |
| -0,274522188 | 0,484616446 | ORM1 | FALSE | Downregulated |
| 0,721952333 | 0,484734687 | ZNF80 | FALSE | Upregulated |
| -0,600519944 | 0,484822092 | LOC102723428 | FALSE | Downregulated |
| -0,095048539 | 0,484822092 | SMAP2 | FALSE | Downregulated |
| -0,099523398 | 0,484882409 | NAA60 | FALSE | Downregulated |
| 0,115626287 | 0,484917561 | ADNP2 | FALSE | Upregulated |
| 0,064412902 | 0,484917561 | FAM120AOS | FALSE | Upregulated |
| 1,058041347 | 0,484917561 | KCNJ13 | FALSE | Upregulated |
| 0,113029135 | 0,484917561 | PDZK1 | FALSE | Upregulated |
| -0,106510595 | 0,484917561 | RXRB | FALSE | Downregulated |
| -0,567328055 | 0,484917561 | ZNF483 | FALSE | Downregulated |
| 0,262442495 | 0,484935202 | BAMBI | FALSE | Upregulated |
| -0,185046864 | 0,484935202 | EGFL7 | FALSE | Downregulated |
| 0,737672166 | 0,484935202 | UPP2 | FALSE | Upregulated |
| -0,794396039 | 0,485327885 | KCNE2 | FALSE | Downregulated |
| 0,185750842 | 0,485338292 | PYROXD1 | FALSE | Upregulated |
| 0,330438033 | 0,48537601 | ABLIM2 | FALSE | Upregulated |
| 0,568858986 | 0,48537601 | FOXP3 | FALSE | Upregulated |
| -0,122415438 | 0,48537601 | LONP2 | FALSE | Downregulated |
| 0,180698235 | 0,48537601 | RRP12 | FALSE | Upregulated |
| -0,820752037 | 0,48537601 | SEC24B-AS1 | FALSE | Downregulated |
| -0,964188213 | 0,485459053 | LOC101927125 | FALSE | Downregulated |
| -0,299065841 | 0,485735354 | COX6C | FALSE | Downregulated |
| 0,119616471 | 0,485794004 | ARHGAP17 | FALSE | Upregulated |
| -0,344833303 | 0,485904144 | LINC01125 | FALSE | Downregulated |
| 0,92431172 | 0,485904144 | PCA3 | FALSE | Upregulated |
| -0,066616245 | 0,485904144 | RAB11A | FALSE | Downregulated |
| -0,224040538 | 0,485904144 | ZBTB7A | FALSE | Downregulated |
| -0,456271218 | 0,485914436 | KCNK5 | FALSE | Downregulated |
| 0,123686983 | 0,485914436 | NCOA4 | FALSE | Upregulated |
| -0,123611186 | 0,485914436 | XRN1 | FALSE | Downregulated |
| -0,278380193 | 0,486186078 | PALM2-AKAP2 | FALSE | Downregulated |
| -0,450004374 | 0,486205377 | RP11-112L6.4 | FALSE | Downregulated |
| 0,451662024 | 0,486410459 | CNNM1 | FALSE | Upregulated |
| -0,194378101 | 0,486675038 | TNFSF10 | FALSE | Downregulated |
| 0,519001503 | 0,486701679 | KLRC4 | FALSE | Upregulated |
| 0,12453331 | 0,486701679 | LEPRE1 | FALSE | Upregulated |
| 0,161074726 | 0,486706484 | GRAP | FALSE | Upregulated |
| 0,113772532 | 0,486722704 | C17orf58 | FALSE | Upregulated |
| 0,2105255 | 0,486816971 | FAM109A | FALSE | Upregulated |
| -0,290496677 | 0,486816971 | ZNF815P | FALSE | Downregulated |
| 0,133367676 | 0,486858436 | NDUFB3 | FALSE | Upregulated |

| | | | | |
|---|---|---|---|---|
| -0,472818169 | 0,487013661 | AC011747.4 | FALSE | Downregulated |
| -0,352300263 | 0,487013661 | ZSWIM5 | FALSE | Downregulated |
| 0,132936124 | 0,48702563 | LINC00997 | FALSE | Upregulated |
| 0,072624265 | 0,487051909 | SPTLC1 | FALSE | Upregulated |
| 0,082066537 | 0,48714845 | BTF3L4 | FALSE | Upregulated |
| 0,092965494 | 0,48714845 | GHITM | FALSE | Upregulated |
| -0,379295607 | 0,48714845 | OSBPL7 | FALSE | Downregulated |
| 0,167433443 | 0,48714845 | THAP8 | FALSE | Upregulated |
| 0,132890323 | 0,487259892 | LZIC | FALSE | Upregulated |
| 0,40169539 | 0,487268128 | CFTR | FALSE | Upregulated |
| 0,212779149 | 0,487268128 | NOL3 | FALSE | Upregulated |
| -0,174512319 | 0,487372671 | DUSP12 | FALSE | Downregulated |
| -0,192898991 | 0,487415328 | FBXO17 | FALSE | Downregulated |
| 0,466095268 | 0,487479942 | AP000688.14 | FALSE | Upregulated |
| -0,118643393 | 0,487479942 | BMP1 | FALSE | Downregulated |
| 0,234270781 | 0,487479942 | CCDC112 | FALSE | Upregulated |
| 0,120565621 | 0,487479942 | COPRS | FALSE | Upregulated |
| -0,150708078 | 0,487479942 | ENPEP | FALSE | Downregulated |
| -0,23207883 | 0,487479942 | LOC102723553 | FALSE | Downregulated |
| -0,157106859 | 0,487479942 | POFUT2 | FALSE | Downregulated |
| -0,692419249 | 0,487601859 | OSR1 | FALSE | Downregulated |
| -0,135373341 | 0,487613581 | ZMYM5 | FALSE | Downregulated |
| -0,160934234 | 0,487674165 | ABHD14B | FALSE | Downregulated |
| 0,726643251 | 0,487682408 | ASB2 | FALSE | Upregulated |
| 0,195678766 | 0,487682408 | TEN1 | FALSE | Upregulated |
| 0,136475049 | 0,487682408 | ZCCHC17 | FALSE | Upregulated |
| -0,513909135 | 0,487714696 | COLEC12 | FALSE | Downregulated |
| -0,402165633 | 0,487837929 | LOC101928588 | FALSE | Downregulated |
| 0,196792782 | 0,487897812 | FAM195A | FALSE | Upregulated |
| 0,174883492 | 0,487942061 | ERAP1 | FALSE | Upregulated |
| 0,506768637 | 0,488139616 | BNC1 | FALSE | Upregulated |
| -0,711998087 | 0,488278509 | LOC101929724 | FALSE | Downregulated |
| -0,132566356 | 0,48847261 | ZNF234 | FALSE | Downregulated |
| -0,951030026 | 0,488683316 | AOC1 | FALSE | Downregulated |
| 0,288250019 | 0,488683316 | DPEP2 | FALSE | Upregulated |
| -1,116395374 | 0,488770258 | LOC102724919 | FALSE | Downregulated |
| -0,116872427 | 0,488770258 | NPRL3 | FALSE | Downregulated |
| -0,169219769 | 0,488834434 | PKD1 | FALSE | Downregulated |
| 0,615851721 | 0,48888059 | CCDC160 | FALSE | Upregulated |
| -0,137091515 | 0,488886809 | LMO7 | FALSE | Downregulated |
| 0,086973947 | 0,489018807 | ARAP1 | FALSE | Upregulated |
| -0,143848559 | 0,489051336 | SGK3 | FALSE | Downregulated |
| 0,110310953 | 0,489098528 | CLEC16A | FALSE | Upregulated |
| -0,133502713 | 0,489098528 | LYRM1 | FALSE | Downregulated |
| -0,352706506 | 0,489335171 | CDHR2 | FALSE | Downregulated |
| 0,970978699 | 0,489335171 | LOC101927775 | FALSE | Upregulated |
| 0,219912995 | 0,489335171 | MXRA5 | FALSE | Upregulated |
| 0,733097655 | 0,489335171 | SLED1 | FALSE | Upregulated |
| -0,377382675 | 0,489335171 | TMEM9B-AS1 | FALSE | Downregulated |
| -0,843909343 | 0,489373164 | FAM74A1 | FALSE | Downregulated |
| -0,360475051 | 0,489373164 | POSTN | FALSE | Downregulated |

| | | | | |
|---|---|---|---|---|
| 0,154611406 | 0,489746439 | EPS8L2 | FALSE | Upregulated |
| 0,472504249 | 0,489746439 | FAM189A1 | FALSE | Upregulated |
| 0,622180253 | 0,489802606 | LOC102724031 | FALSE | Upregulated |
| -0,878608183 | 0,489863932 | TPBGL | FALSE | Downregulated |
| 0,182931576 | 0,489884965 | SLC43A3 | FALSE | Upregulated |
| 0,627524965 | 0,489986029 | LOC101929134 | FALSE | Upregulated |
| 0,091789388 | 0,490337167 | SHFM1 | FALSE | Upregulated |
| -0,123036485 | 0,490436729 | PRKAG2 | FALSE | Downregulated |
| -0,775381835 | 0,490494939 | DLEC1 | FALSE | Downregulated |
| 0,468826376 | 0,490746226 | STK33 | FALSE | Upregulated |
| -0,780851915 | 0,490746226 | ZNF99 | FALSE | Downregulated |
| -0,200110158 | 0,490784112 | CXorf40A | FALSE | Downregulated |
| -0,536915912 | 0,490820432 | LOC102724169 | FALSE | Downregulated |
| 0,076774594 | 0,490827051 | GGPS1 | FALSE | Upregulated |
| -0,882568192 | 0,490827051 | PRRX1 | FALSE | Downregulated |
| -0,207151045 | 0,491095357 | C8orf58 | FALSE | Downregulated |
| -0,374716772 | 0,491462875 | TAS2R20 | FALSE | Downregulated |
| -0,07462537 | 0,491595582 | B4GALT4 | FALSE | Downregulated |
| -1,126719317 | 0,491654361 | EGFL8 | FALSE | Downregulated |
| -0,17265254 | 0,491654361 | TTC7B | FALSE | Downregulated |
| 0,102209506 | 0,491829153 | GORASP1 | FALSE | Upregulated |
| -0,155226648 | 0,492076445 | RCAN1 | FALSE | Downregulated |
| 0,231807409 | 0,492093771 | DCLRE1C | FALSE | Upregulated |
| 0,366592724 | 0,492110668 | LOC100506473 | FALSE | Upregulated |
| 0,581745082 | 0,492110668 | LOC101927792 | FALSE | Upregulated |
| 0,109040596 | 0,492312845 | MRPL18 | FALSE | Upregulated |
| 0,217154899 | 0,492485603 | EXOG | FALSE | Upregulated |
| 0,11907558 | 0,492485603 | LACTB | FALSE | Upregulated |
| 0,149762283 | 0,492925727 | ART4 | FALSE | Upregulated |
| 0,092572532 | 0,492925727 | IGF2R | FALSE | Upregulated |
| -0,42282947 | 0,492925727 | LOC100289230 | FALSE | Downregulated |
| 0,250652841 | 0,49318277 | SOBP | FALSE | Upregulated |
| 0,094284488 | 0,49320065 | ARFRP1 | FALSE | Upregulated |
| -0,282012643 | 0,493292272 | HMGCLL1 | FALSE | Downregulated |
| -0,287050253 | 0,493354037 | LOC284889 | FALSE | Downregulated |
| -0,107305812 | 0,49346832 | AIG1 | FALSE | Downregulated |
| -0,200913264 | 0,49346832 | SLC22A23 | FALSE | Downregulated |
| -0,250504479 | 0,493563069 | SYPL2 | FALSE | Downregulated |
| -0,193142621 | 0,49364295 | PPIF | FALSE | Downregulated |
| 0,206652531 | 0,49378309 | CCZ1 | FALSE | Upregulated |
| -0,067212065 | 0,493899698 | CUL5 | FALSE | Downregulated |
| 0,169039007 | 0,493899698 | NCDN | FALSE | Upregulated |
| -0,536323495 | 0,494197783 | LOC101929619 | FALSE | Downregulated |
| -0,129880488 | 0,494319353 | PQLC1 | FALSE | Downregulated |
| -0,273170925 | 0,494420202 | LOC101930644 | FALSE | Downregulated |
| 0,536594652 | 0,494809793 | LAMC2 | FALSE | Upregulated |
| 0,305095386 | 0,494809793 | LOC101929668 | FALSE | Upregulated |
| -0,244275849 | 0,494852422 | KLLN | FALSE | Downregulated |
| 1,027964899 | 0,495007534 | CDH17 | FALSE | Upregulated |
| 0,092448317 | 0,495007534 | EPS15L1 | FALSE | Upregulated |
| 0,11255228 | 0,495007534 | MCTS1 | FALSE | Upregulated |

| | | | | |
|---|---|---|---|---|
| -0,087615407 | 0,495048614 | PTEN | FALSE | Downregulated |
| 0,339612921 | 0,495361384 | NKD1 | FALSE | Upregulated |
| -0,201902779 | 0,495563493 | MEGF6 | FALSE | Downregulated |
| 0,144855097 | 0,495615069 | FAM208B | FALSE | Upregulated |
| -0,10853086 | 0,495615069 | NAB1 | FALSE | Downregulated |
| -0,106204352 | 0,495615069 | TMF1 | FALSE | Downregulated |
| 0,123943013 | 0,495929716 | ASL | FALSE | Upregulated |
| 0,296196732 | 0,495929716 | ZNF93 | FALSE | Upregulated |
| -0,414928832 | 0,495945197 | LOC200772 | FALSE | Downregulated |
| -0,203303981 | 0,496168065 | MAP2 | FALSE | Downregulated |
| 0,089781261 | 0,496303612 | ARHGEF11 | FALSE | Upregulated |
| -0,904522607 | 0,496303612 | LOC102723696 | FALSE | Downregulated |
| -0,387680712 | 0,496303612 | RP5-1103G7.4 | FALSE | Downregulated |
| 0,08478602 | 0,496370777 | EXOC8 | FALSE | Upregulated |
| -0,121189055 | 0,496370777 | FOXP1 | FALSE | Downregulated |
| -0,528850681 | 0,496370777 | KLHL33 | FALSE | Downregulated |
| -0,896776096 | 0,496370777 | LPPR4 | FALSE | Downregulated |
| -0,239483174 | 0,496370777 | SP5 | FALSE | Downregulated |
| -0,432159931 | 0,496370777 | TPTEP1 | FALSE | Downregulated |
| -0,655413557 | 0,49679088 | FLJ41309 | FALSE | Downregulated |
| 0,149627696 | 0,49679088 | GZF1 | FALSE | Upregulated |
| -0,369732101 | 0,49679088 | MEGF10 | FALSE | Downregulated |
| -0,581026297 | 0,49679088 | SEC14L6 | FALSE | Downregulated |
| 0,137394143 | 0,497214282 | MED17 | FALSE | Upregulated |
| -0,206020318 | 0,497214282 | RP11-317J10.2 | FALSE | Downregulated |
| 0,424588189 | 0,497449877 | LOC389641 | FALSE | Upregulated |
| -0,25749772 | 0,497481382 | CCNL2 | FALSE | Downregulated |
| -0,763924281 | 0,497481382 | DPY19L2 | FALSE | Downregulated |
| 0,472661718 | 0,497562763 | GPX2 | FALSE | Upregulated |
| -0,136857125 | 0,497765652 | METTL15 | FALSE | Downregulated |
| -0,161852627 | 0,49787657 | PRKCA | FALSE | Downregulated |
| 0,594074741 | 0,4978921 | DISC1FP1 | FALSE | Upregulated |
| -0,118762399 | 0,497892919 | CHPT1 | FALSE | Downregulated |
| -0,365001728 | 0,497892919 | CNTN1 | FALSE | Downregulated |
| 0,561113319 | 0,497984181 | HSD3B1 | FALSE | Upregulated |
| -0,33406461 | 0,497984181 | RP6-206I17.2 | FALSE | Downregulated |
| -0,452555964 | 0,497996101 | RP11-611D20.2 | FALSE | Downregulated |
| 0,244038685 | 0,498091242 | USP54 | FALSE | Upregulated |
| -0,332774691 | 0,498165316 | LOC101929072 | FALSE | Downregulated |
| 0,158992685 | 0,49835178 | GLRX | FALSE | Upregulated |
| 0,119790739 | 0,498367333 | FAM92A1 | FALSE | Upregulated |
| 0,093642837 | 0,498590685 | SGSM3 | FALSE | Upregulated |
| 0,664126648 | 0,498658371 | AP000330.8 | FALSE | Upregulated |
| 0,112942874 | 0,498658371 | PACS2 | FALSE | Upregulated |
| -0,173050109 | 0,498821731 | ZNF674 | FALSE | Downregulated |
| 2,771642827 | 0,498914833 | C15orf38-AP3S2 | FALSE | Upregulated |
| -0,323868784 | 0,498914833 | CLUHP3 | FALSE | Downregulated |
| -1,193157378 | 0,498914833 | MATN3 | FALSE | Downregulated |
| -0,150655619 | 0,498914833 | TMEM38B | FALSE | Downregulated |
| -0,129551132 | 0,498990928 | MIS12 | FALSE | Downregulated |
| -0,158017277 | 0,498990928 | ZFAND2A | FALSE | Downregulated |

| | | | | |
|---|---|---|---|---|
| -0,12906238 | 0,499013719 | CRADD | FALSE | Downregulated |
| -0,379163118 | 0,499013719 | LOC644656 | FALSE | Downregulated |
| -0,181995882 | 0,499013719 | ZNF581 | FALSE | Downregulated |
| 0,122372249 | 0,499103733 | CSRP1 | FALSE | Upregulated |
| 1,001415881 | 0,499222769 | NLGN4Y | FALSE | Upregulated |
| 0,218711469 | 0,499222769 | SACS | FALSE | Upregulated |
| 0,456090254 | 0,499222769 | VANGL2 | FALSE | Upregulated |
| 0,523289327 | 0,49949934 | LOC102724040 | FALSE | Upregulated |
| 0,12253174 | 0,49949934 | MXD1 | FALSE | Upregulated |
| 0,099678837 | 0,49949934 | NMT1 | FALSE | Upregulated |
| -0,398669911 | 0,499554656 | CRYGS | FALSE | Downregulated |
| -0,154596732 | 0,49986705 | PPOX | FALSE | Downregulated |
| -0,164630248 | 0,499921612 | LOC101929343 | FALSE | Downregulated |
| -0,634017269 | 0,49993647 | AHNAK2 | FALSE | Downregulated |
| -0,332124646 | 0,499958201 | FREM1 | FALSE | Downregulated |
| 0,108082007 | 0,500162607 | TGFBR2 | FALSE | Upregulated |
| -0,077244751 | 0,500289977 | PHAX | FALSE | Downregulated |
| 0,479027563 | 0,50040818 | C22orf26 | FALSE | Upregulated |
| -0,091278979 | 0,50040818 | HAUS6 | FALSE | Downregulated |
| 0,120208126 | 0,50040818 | JKAMP | FALSE | Upregulated |
| 0,820575809 | 0,500417101 | GAP43 | FALSE | Upregulated |
| 0,102356521 | 0,500418141 | RANBP1 | FALSE | Upregulated |
| -0,326707202 | 0,501087564 | NOVA2 | FALSE | Downregulated |
| -0,167812896 | 0,501114746 | CNPY3 | FALSE | Downregulated |
| -0,51516999 | 0,501442642 | CD300LG | FALSE | Downregulated |
| -0,523906663 | 0,501442642 | LOC101928096 | FALSE | Downregulated |
| 0,282121857 | 0,501442642 | ZNF486 | FALSE | Upregulated |
| -0,779583181 | 0,50155341 | CCDC42B | FALSE | Downregulated |
| -0,205706326 | 0,501555658 | GLIDR | FALSE | Downregulated |
| -1,053720842 | 0,501555658 | LOC101927999 | FALSE | Downregulated |
| -0,384430885 | 0,501561329 | PABPC1L | FALSE | Downregulated |
| -0,218919842 | 0,501561329 | RAPH1 | FALSE | Downregulated |
| 0,153979852 | 0,50159638 | DYNC1I2 | FALSE | Upregulated |
| -0,315752727 | 0,50159638 | LOC100133091 | FALSE | Downregulated |
| 0,634500621 | 0,501676176 | MPO | FALSE | Upregulated |
| 0,150677729 | 0,501814079 | CCSAP | FALSE | Upregulated |
| -0,120652403 | 0,501842977 | KBTBD7 | FALSE | Downregulated |
| 0,32206548 | 0,501842977 | PIK3R6 | FALSE | Upregulated |
| -0,231271969 | 0,501842977 | PRR24 | FALSE | Downregulated |
| 0,637951309 | 0,501842977 | ZPLD1 | FALSE | Upregulated |
| 0,104071907 | 0,5018586 | RNF103 | FALSE | Upregulated |
| -0,141016119 | 0,502015948 | C11orf30 | FALSE | Downregulated |
| 0,117159102 | 0,502122083 | SLIRP | FALSE | Upregulated |
| -0,169613567 | 0,502598963 | RWDD1 | FALSE | Downregulated |
| -0,941349589 | 0,502670034 | MAG | FALSE | Downregulated |
| -0,171528656 | 0,502730232 | TIMP3 | FALSE | Downregulated |
| -0,080943808 | 0,502856137 | UBQLN2 | FALSE | Downregulated |
| -0,884834602 | 0,503118751 | MSH5 | FALSE | Downregulated |
| -0,646609911 | 0,503303882 | NPBWR1 | FALSE | Downregulated |
| -0,948213291 | 0,503303882 | PCAT5 | FALSE | Downregulated |
| 0,083720273 | 0,503429516 | MRPS30 | FALSE | Upregulated |

| | | | | |
|---|---|---|---|---|
| 0,787370479 | 0,503491984 | ATG9B | FALSE | Upregulated |
| 0,150401737 | 0,503491984 | CASP10 | FALSE | Upregulated |
| 0,109810679 | 0,503491984 | SGPP1 | FALSE | Upregulated |
| 0,083860805 | 0,503491984 | VPS72 | FALSE | Upregulated |
| -0,092730292 | 0,503557177 | TMBIM6 | FALSE | Downregulated |
| -0,136743453 | 0,503813776 | FAM229B | FALSE | Downregulated |
| -0,387210542 | 0,503813776 | LOC101928673 | FALSE | Downregulated |
| -0,202022353 | 0,503895591 | IQCC | FALSE | Downregulated |
| -0,377636142 | 0,504076372 | ATHL1 | FALSE | Downregulated |
| 0,130165873 | 0,504105618 | SH3GLB2 | FALSE | Upregulated |
| 0,512460709 | 0,504111537 | LOC339192 | FALSE | Upregulated |
| 0,176712222 | 0,504183302 | PDK3 | FALSE | Upregulated |
| 0,429762819 | 0,504183302 | SLC6A9 | FALSE | Upregulated |
| -0,169818308 | 0,504259118 | EVI5 | FALSE | Downregulated |
| -0,225223373 | 0,504259118 | RP11-60I3.5 | FALSE | Downregulated |
| -0,289721391 | 0,504259118 | RPLP0P2 | FALSE | Downregulated |
| -0,453106988 | 0,504434002 | LOC441204 | FALSE | Downregulated |
| -0,077430472 | 0,504709194 | NUP160 | FALSE | Downregulated |
| -1,031364207 | 0,504841082 | LOC101927249 | FALSE | Downregulated |
| -0,207751452 | 0,504841082 | LRRC37B | FALSE | Downregulated |
| 0,114048744 | 0,504953162 | ARRDC1 | FALSE | Upregulated |
| 0,805699059 | 0,504953162 | RP11-426L16.8 | FALSE | Upregulated |
| -0,091210683 | 0,505618756 | KIAA0947 | FALSE | Downregulated |
| -0,606767742 | 0,505770699 | LOC100653005 | FALSE | Downregulated |
| 0,505417177 | 0,505770699 | SLC22A2 | FALSE | Upregulated |
| 0,604833254 | 0,505850714 | LOC101929280 | FALSE | Upregulated |
| -0,137693123 | 0,50588749 | THOC2 | FALSE | Downregulated |
| 0,215468033 | 0,506096385 | CDC42EP1 | FALSE | Upregulated |
| -0,246240822 | 0,506096385 | HSP90B2P | FALSE | Downregulated |
| -0,072957624 | 0,506096385 | PIAS2 | FALSE | Downregulated |
| -1,055562261 | 0,506096385 | ZNF98 | FALSE | Downregulated |
| 0,555725932 | 0,506100637 | GPR35 | FALSE | Upregulated |
| -0,127653083 | 0,506109982 | CMC4 | FALSE | Downregulated |
| 0,069590661 | 0,506109982 | DDX1 | FALSE | Upregulated |
| -0,317986014 | 0,506109982 | LRRC4B | FALSE | Downregulated |
| -0,409660609 | 0,506171633 | DPH6-AS1 | FALSE | Downregulated |
| 0,092581611 | 0,506171633 | PTPN12 | FALSE | Upregulated |
| -0,136861144 | 0,506171633 | UBE2W | FALSE | Downregulated |
| -0,153606691 | 0,506171633 | ZNF440 | FALSE | Downregulated |
| 0,188300412 | 0,50622402 | AKR1C1 | FALSE | Upregulated |
| 0,16295357 | 0,506415395 | MTMR2 | FALSE | Upregulated |
| 0,165532946 | 0,506511908 | GLTPD2 | FALSE | Upregulated |
| -0,112004518 | 0,506511908 | MRPS26 | FALSE | Downregulated |
| -0,609627455 | 0,506544916 | LINC00598 | FALSE | Downregulated |
| -0,57149285 | 0,506587456 | PF4 | FALSE | Downregulated |
| -0,812348577 | 0,506656222 | SPDYA | FALSE | Downregulated |
| -0,182662774 | 0,506685241 | GSTA4 | FALSE | Downregulated |
| -0,16790869 | 0,506685241 | PRX | FALSE | Downregulated |
| -1,128850796 | 0,507136145 | DGKI | FALSE | Downregulated |
| 0,200690932 | 0,507136145 | LEAP2 | FALSE | Upregulated |
| -0,506919502 | 0,507136145 | TREML2 | FALSE | Downregulated |

| | | | | |
|---|---|---|---|---|
| -0,441399013 | 0,507136145 | UFSP1 | FALSE | Downregulated |
| -0,099006942 | 0,507310744 | NONO | FALSE | Downregulated |
| 0,248274965 | 0,50733095 | APBB3 | FALSE | Upregulated |
| -0,721409627 | 0,507366918 | LOC101928916 | FALSE | Downregulated |
| 0,086123513 | 0,507366918 | STAU2 | FALSE | Upregulated |
| 0,284268804 | 0,507376438 | FZD3 | FALSE | Upregulated |
| 0,627033992 | 0,507376438 | RASD2 | FALSE | Upregulated |
| 0,168534984 | 0,507376438 | SREBF2 | FALSE | Upregulated |
| 0,599754676 | 0,507377219 | DEFB132 | FALSE | Upregulated |
| -0,087063597 | 0,507442273 | CDK12 | FALSE | Downregulated |
| -0,385927386 | 0,507535961 | LINC00471 | FALSE | Downregulated |
| 0,091360211 | 0,507552289 | LRRC8D | FALSE | Upregulated |
| 0,698056468 | 0,507648682 | SCN3A | FALSE | Upregulated |
| -0,41646336 | 0,50765927 | LOC101928947 | FALSE | Downregulated |
| -0,074118067 | 0,507895467 | COPS5 | FALSE | Downregulated |
| 0,056320347 | 0,507895467 | SNX9 | FALSE | Upregulated |
| -0,473920626 | 0,508218211 | LOC101928852 | FALSE | Downregulated |
| -0,365465904 | 0,508218211 | MIR503HG | FALSE | Downregulated |
| 0,282951767 | 0,508218211 | PTPN14 | FALSE | Upregulated |
| 0,790098731 | 0,508218211 | SLC7A4 | FALSE | Upregulated |
| -0,11746291 | 0,5082189 | ASMTL | FALSE | Downregulated |
| 0,270239427 | 0,508262776 | TFPI2 | FALSE | Upregulated |
| 0,220463833 | 0,508274233 | NBEAL2 | FALSE | Upregulated |
| -0,172492391 | 0,508274233 | PNP | FALSE | Downregulated |
| 0,097168931 | 0,508274233 | RBM4 | FALSE | Upregulated |
| -2,538990418 | 0,508406753 | ATP5J2-PTCD1 | FALSE | Downregulated |
| 0,135490538 | 0,508409269 | NUP62 | FALSE | Upregulated |
| 0,884020992 | 0,508409269 | TNFRSF13B | FALSE | Upregulated |
| -0,094910803 | 0,508455319 | ACBD6 | FALSE | Downregulated |
| 0,175359086 | 0,508455319 | GET4 | FALSE | Upregulated |
| -0,144678788 | 0,508455319 | IGFBP3 | FALSE | Downregulated |
| 0,31175801 | 0,508455319 | ITGB4 | FALSE | Upregulated |
| -0,136571149 | 0,508455319 | NUFIP1 | FALSE | Downregulated |
| -0,425013412 | 0,508455319 | SSPO | FALSE | Downregulated |
| -0,245055028 | 0,508671619 | TRNE | FALSE | Downregulated |
| 0,869389699 | 0,508804985 | ADAMTS16 | FALSE | Upregulated |
| 0,118179583 | 0,508804985 | UHRF1BP1 | FALSE | Upregulated |
| -0,341742437 | 0,508830105 | THRSP | FALSE | Downregulated |
| -0,738526734 | 0,508830105 | WWTR1-AS1 | FALSE | Downregulated |
| 0,096772908 | 0,508863214 | HEBP1 | FALSE | Upregulated |
| -0,413584958 | 0,508920304 | PPP1R3G | FALSE | Downregulated |
| -0,125541975 | 0,508926314 | SMYD3 | FALSE | Downregulated |
| 0,089400681 | 0,508980637 | MCMBP | FALSE | Upregulated |
| 0,077568746 | 0,508980637 | TALDO1 | FALSE | Upregulated |
| 0,144996033 | 0,509091178 | MLH3 | FALSE | Upregulated |
| -0,141753788 | 0,509188756 | MAPKBP1 | FALSE | Downregulated |
| 0,245235435 | 0,509207131 | BBS12 | FALSE | Upregulated |
| 0,181626129 | 0,509207131 | DENND3 | FALSE | Upregulated |
| 0,078535982 | 0,509207131 | OSTF1 | FALSE | Upregulated |
| 0,099414136 | 0,509234321 | RIT1 | FALSE | Upregulated |
| -0,061197758 | 0,509234321 | ZNF140 | FALSE | Downregulated |

| | | | | |
|---|---|---|---|---|
| 0,12118157 | 0,509240206 | TBC1D9 | FALSE | Upregulated |
| -0,255903986 | 0,509288634 | CDH19 | FALSE | Downregulated |
| -0,114348852 | 0,509391472 | LLGL2 | FALSE | Downregulated |
| 0,642122181 | 0,509391833 | AC095067.1 | FALSE | Upregulated |
| -0,669673358 | 0,509475805 | MGARP | FALSE | Downregulated |
| -0,684483847 | 0,509478543 | RP11-554F20.1 | FALSE | Downregulated |
| 0,489716898 | 0,509511108 | LOC100130899 | FALSE | Upregulated |
| 0,22213314 | 0,509688058 | C1orf53 | FALSE | Upregulated |
| -0,154499157 | 0,509688058 | SERPINF2 | FALSE | Downregulated |
| -0,39153418 | 0,509734534 | PPIEL | FALSE | Downregulated |
| -1,028199073 | 0,509777048 | LOC101927800 | FALSE | Downregulated |
| -0,596301305 | 0,509777048 | LOC101928238 | FALSE | Downregulated |
| 1,188688166 | 0,509777048 | SNORA14B | FALSE | Upregulated |
| 0,300967686 | 0,509789138 | SYBU | FALSE | Upregulated |
| -0,290397909 | 0,509822419 | DHRS4L1 | FALSE | Downregulated |
| 0,081071049 | 0,509822419 | PIGK | FALSE | Upregulated |
| 0,242325076 | 0,509822419 | RPS10P7 | FALSE | Upregulated |
| 0,416493633 | 0,509932417 | SOX2-OT | FALSE | Upregulated |
| -0,65812138 | 0,51039339 | LINC00890 | FALSE | Downregulated |
| -0,09858912 | 0,510471338 | GEMIN5 | FALSE | Downregulated |
| -1,208549026 | 0,510521474 | GFI1B | FALSE | Downregulated |
| -0,971019227 | 0,510521474 | LOC102724322 | FALSE | Downregulated |
| 0,162331257 | 0,510521474 | NAP1L5 | FALSE | Upregulated |
| 0,210008259 | 0,510521474 | SAMD10 | FALSE | Upregulated |
| -0,132263892 | 0,510521474 | XKR8 | FALSE | Downregulated |
| 0,113823786 | 0,510521474 | ZNF574 | FALSE | Upregulated |
| 0,136656377 | 0,510645435 | L3HYPDH | FALSE | Upregulated |
| -0,177081369 | 0,5106508 | ABCA11P | FALSE | Downregulated |
| -0,283206844 | 0,5106508 | FAM127C | FALSE | Downregulated |
| -1,057695415 | 0,510653023 | NTM | FALSE | Downregulated |
| 0,221230424 | 0,510684056 | LOC100506714 | FALSE | Upregulated |
| -0,238811898 | 0,510684056 | TTBK1 | FALSE | Downregulated |
| 0,190036381 | 0,511174176 | NBPF1 | FALSE | Upregulated |
| -0,472118032 | 0,511307645 | ITLN2 | FALSE | Downregulated |
| -0,7816131 | 0,511402099 | CCDC41-AS1 | FALSE | Downregulated |
| 0,098772315 | 0,511402099 | CDK4 | FALSE | Upregulated |
| -0,139843518 | 0,511402099 | CENPV | FALSE | Downregulated |
| -0,140219199 | 0,511402099 | LGR4 | FALSE | Downregulated |
| 0,261955184 | 0,511402099 | PROZ | FALSE | Upregulated |
| -0,617757587 | 0,511650166 | DEGS2 | FALSE | Downregulated |
| -0,441947032 | 0,511757758 | KCNJ12 | FALSE | Downregulated |
| -0,268704842 | 0,511958327 | LOC102724129 | FALSE | Downregulated |
| 0,09651852 | 0,511958327 | WDR44 | FALSE | Upregulated |
| -0,104008984 | 0,511975066 | HOMEZ | FALSE | Downregulated |
| -0,125164443 | 0,512076568 | SH2B1 | FALSE | Downregulated |
| 0,469087941 | 0,512177974 | SEPT5 | FALSE | Upregulated |
| -0,256435053 | 0,512241187 | EFCC1 | FALSE | Downregulated |
| -0,12393727 | 0,512515491 | PGAP2 | FALSE | Downregulated |
| 0,146922006 | 0,512561348 | ALKBH6 | FALSE | Upregulated |
| 0,088889771 | 0,512561348 | STRIP1 | FALSE | Upregulated |
| -0,101089619 | 0,512591186 | SPIDR | FALSE | Downregulated |

| | | | | |
|---|---|---|---|---|
| -0,147678117 | 0,512738352 | KIAA0355 | FALSE | Downregulated |
| -0,872598186 | 0,512758509 | LOC102724636 | FALSE | Downregulated |
| -0,121992046 | 0,512758509 | PCSK6 | FALSE | Downregulated |
| -0,261150592 | 0,512919984 | SLC25A21-AS1 | FALSE | Downregulated |
| -0,107119361 | 0,513042772 | TFB2M | FALSE | Downregulated |
| 0,06804977 | 0,513048915 | CDV3 | FALSE | Upregulated |
| 0,29380375 | 0,51307152 | LOC101927571 | FALSE | Upregulated |
| -0,101858937 | 0,513361164 | ARL6IP4 | FALSE | Downregulated |
| -1,068224422 | 0,513361164 | PPP1R27 | FALSE | Downregulated |
| -0,133668547 | 0,51343686 | CTSF | FALSE | Downregulated |
| 0,216816177 | 0,51366281 | ATAD3B | FALSE | Upregulated |
| -0,337766517 | 0,51366281 | CAND2 | FALSE | Downregulated |
| -0,510124499 | 0,51366281 | CCER2 | FALSE | Downregulated |
| 0,551905516 | 0,513719342 | TIAM2 | FALSE | Upregulated |
| -0,206980226 | 0,513780985 | CX3CL1 | FALSE | Downregulated |
| -0,487158467 | 0,513931695 | LOC100652901 | FALSE | Downregulated |
| 0,104884335 | 0,513965773 | IKBKB | FALSE | Upregulated |
| -0,170804877 | 0,513965773 | LOC100132352 | FALSE | Downregulated |
| 0,346364632 | 0,514014275 | ACE2 | FALSE | Upregulated |
| -0,165696325 | 0,514119333 | IL1R2 | FALSE | Downregulated |
| -0,456222842 | 0,514236527 | SNTG1 | FALSE | Downregulated |
| 0,217620162 | 0,514834817 | ARL2 | FALSE | Upregulated |
| -0,175425632 | 0,514945632 | OPLAH | FALSE | Downregulated |
| -0,119792723 | 0,515174032 | TMEM186 | FALSE | Downregulated |
| -0,689440969 | 0,515608751 | DPEP3 | FALSE | Downregulated |
| -0,419577701 | 0,515608751 | LGR6 | FALSE | Downregulated |
| 0,356438117 | 0,515608751 | SOWAHD | FALSE | Upregulated |
| -0,142691488 | 0,515608751 | ZNF799 | FALSE | Downregulated |
| 0,368035174 | 0,515631837 | ALDH1L2 | FALSE | Upregulated |
| -0,232089656 | 0,515631837 | C11orf96 | FALSE | Downregulated |
| 0,09633951 | 0,515631837 | GPS1 | FALSE | Upregulated |
| -0,219403927 | 0,515631837 | PDE4DIP | FALSE | Downregulated |
| -0,694142479 | 0,515631837 | RP3-323P13.2 | FALSE | Downregulated |
| 0,097829656 | 0,515735165 | KAT5 | FALSE | Upregulated |
| 0,558701572 | 0,515912613 | LOC645638 | FALSE | Upregulated |
| 0,194067582 | 0,515918524 | C9orf89 | FALSE | Upregulated |
| 0,262376571 | 0,515919921 | FGD1 | FALSE | Upregulated |
| -0,144118347 | 0,515919921 | SLC25A20 | FALSE | Downregulated |
| -0,168292995 | 0,516224085 | UGDH | FALSE | Downregulated |
| -0,1819598 | 0,516289846 | CCBL1 | FALSE | Downregulated |
| 0,832708857 | 0,516289846 | CPS1-IT1 | FALSE | Upregulated |
| 0,494614471 | 0,516289846 | FCER2 | FALSE | Upregulated |
| -0,089926065 | 0,516289846 | TRIM44 | FALSE | Downregulated |
| 0,331970419 | 0,516311134 | LOC102724098 | FALSE | Upregulated |
| -0,772810744 | 0,516547205 | SLC22A24 | FALSE | Downregulated |
| -0,27507268 | 0,516620812 | LOC100507373 | FALSE | Downregulated |
| 0,185262616 | 0,516649891 | CHD3 | FALSE | Upregulated |
| -0,572543914 | 0,516813615 | LTC4S | FALSE | Downregulated |
| -0,151676902 | 0,516813615 | MRPL53 | FALSE | Downregulated |
| -0,38985607 | 0,516912723 | BMS1P6 | FALSE | Downregulated |
| -0,360832328 | 0,51696951 | NS3BP | FALSE | Downregulated |

| | | | | |
|---|---|---|---|---|
| 0,111786218 | 0,516975047 | ZMIZ1 | FALSE | Upregulated |
| 0,120201598 | 0,517234426 | FAM58A | FALSE | Upregulated |
| -0,637619792 | 0,517234426 | LOC101929211 | FALSE | Downregulated |
| -0,177115313 | 0,517234426 | MAP7D3 | FALSE | Downregulated |
| 0,145386364 | 0,517429725 | NDUFV3 | FALSE | Upregulated |
| 0,114046132 | 0,517613879 | MRPS6 | FALSE | Upregulated |
| 0,157315228 | 0,517613879 | SSR3 | FALSE | Upregulated |
| 0,753065655 | 0,517791015 | FAM182A | FALSE | Upregulated |
| 0,716918656 | 0,517854327 | LOC101928699 | FALSE | Upregulated |
| -0,078036155 | 0,517854327 | TADA3 | FALSE | Downregulated |
| -0,085760207 | 0,51798687 | FCF1 | FALSE | Downregulated |
| -0,440313357 | 0,51798687 | SFTPD | FALSE | Downregulated |
| 0,092013504 | 0,518094025 | MICU2 | FALSE | Upregulated |
| -0,136433302 | 0,518094025 | PRRC2C | FALSE | Downregulated |
| -0,641789972 | 0,518094025 | THSD7B | FALSE | Downregulated |
| 0,132232393 | 0,518178416 | ANKEF1 | FALSE | Upregulated |
| -0,151688212 | 0,518178416 | STK35 | FALSE | Downregulated |
| 0,100702683 | 0,518261053 | ERGIC1 | FALSE | Upregulated |
| -0,736930034 | 0,518309812 | LOC102723756 | FALSE | Downregulated |
| 0,766940232 | 0,518458487 | C9orf84 | FALSE | Upregulated |
| 0,100130705 | 0,518458487 | DSC2 | FALSE | Upregulated |
| -0,115059684 | 0,518458487 | FAH | FALSE | Downregulated |
| -0,387989474 | 0,518479166 | CCDC135 | FALSE | Downregulated |
| -0,151074412 | 0,518521865 | MCC | FALSE | Downregulated |
| 1,041704059 | 0,518719058 | CD177 | FALSE | Upregulated |
| -0,073469621 | 0,518862761 | CHD8 | FALSE | Downregulated |
| 0,168428808 | 0,518862761 | LTV1 | FALSE | Upregulated |
| -0,093570515 | 0,518862761 | POM121 | FALSE | Downregulated |
| -0,495248743 | 0,518862761 | ZNF90 | FALSE | Downregulated |
| 0,569827705 | 0,518902728 | ARMC12 | FALSE | Upregulated |
| 0,167876773 | 0,518902728 | GNL2 | FALSE | Upregulated |
| 0,22965232 | 0,518912858 | ITGB3BP | FALSE | Upregulated |
| -0,127619323 | 0,518941683 | SESN2 | FALSE | Downregulated |
| 0,109648507 | 0,519310589 | MRPL48 | FALSE | Upregulated |
| 0,346550572 | 0,519314917 | FHL2 | FALSE | Upregulated |
| -0,12410693 | 0,519314917 | QSOX2 | FALSE | Downregulated |
| 0,122066813 | 0,519314917 | TNIP1 | FALSE | Upregulated |
| -0,095030208 | 0,519374368 | DCAF8 | FALSE | Downregulated |
| -0,236427565 | 0,519374368 | RGS2 | FALSE | Downregulated |
| -0,147843381 | 0,519612194 | ZNF217 | FALSE | Downregulated |
| -1,075782005 | 0,519739044 | LOC102467217 | FALSE | Downregulated |
| 0,22190801 | 0,519824766 | ENAH | FALSE | Upregulated |
| -0,119068696 | 0,519948118 | HNF4G | FALSE | Downregulated |
| 0,530674528 | 0,519998214 | LOC101927681 | FALSE | Upregulated |
| 0,201636285 | 0,519998214 | MAGIX | FALSE | Upregulated |
| -0,204120895 | 0,520017052 | RPS29 | FALSE | Downregulated |
| 0,131812719 | 0,520017052 | RPS9 | FALSE | Upregulated |
| -1,389808483 | 0,520042713 | PIWIL1 | FALSE | Downregulated |
| -0,561096119 | 0,5201323 | PAK7 | FALSE | Downregulated |
| 0,153200716 | 0,520176986 | CCDC130 | FALSE | Upregulated |
| 0,13578231 | 0,520198148 | RPS6 | FALSE | Upregulated |

| | | | | |
|---|---|---|---|---|
| -0,271640596 | 0,520312167 | HTR2B | FALSE | Downregulated |
| 0,098010814 | 0,520312167 | MRPL10 | FALSE | Upregulated |
| 0,094139815 | 0,520312167 | NUPL2 | FALSE | Upregulated |
| -0,350769202 | 0,520333028 | LOC727978 | FALSE | Downregulated |
| 0,609129513 | 0,520688463 | C1orf105 | FALSE | Upregulated |
| 0,081514255 | 0,520688463 | PITPNB | FALSE | Upregulated |
| 0,741806797 | 0,520688463 | SLAMF9 | FALSE | Upregulated |
| -0,999632184 | 0,520756559 | CDC14C | FALSE | Downregulated |
| 0,409182278 | 0,520756559 | LRRC37A6P | FALSE | Upregulated |
| -0,810383397 | 0,520833064 | LOC729080 | FALSE | Downregulated |
| -0,941035401 | 0,520833064 | MURC | FALSE | Downregulated |
| -0,436359518 | 0,520833064 | PCAT6 | FALSE | Downregulated |
| 0,084040264 | 0,520833064 | SNX19 | FALSE | Upregulated |
| 0,308854548 | 0,520833064 | THSD1 | FALSE | Upregulated |
| 0,088033372 | 0,520845711 | EIF4A3 | FALSE | Upregulated |
| -0,12578473 | 0,521166685 | OGG1 | FALSE | Downregulated |
| -0,304894287 | 0,521251376 | C1RL-AS1 | FALSE | Downregulated |
| 0,214930313 | 0,521251376 | EXD3 | FALSE | Upregulated |
| -0,637430053 | 0,521251376 | NAV2-AS4 | FALSE | Downregulated |
| -0,205509256 | 0,521251376 | ZNF814 | FALSE | Downregulated |
| -1,034654482 | 0,521405338 | LMO3 | FALSE | Downregulated |
| 0,333909877 | 0,521573678 | ZNF469 | FALSE | Upregulated |
| -0,638500475 | 0,521581151 | TMC4 | FALSE | Downregulated |
| 0,11729406 | 0,521617467 | IPO4 | FALSE | Upregulated |
| -0,124772617 | 0,521617467 | OCIAD2 | FALSE | Downregulated |
| 0,217049904 | 0,521716929 | FMOD | FALSE | Upregulated |
| 0,074508022 | 0,521763698 | ATP11B | FALSE | Upregulated |
| 0,790938328 | 0,522053645 | FOXN4 | FALSE | Upregulated |
| -0,073459133 | 0,522053645 | RAF1 | FALSE | Downregulated |
| 0,706311571 | 0,522223254 | TRNAE11 | FALSE | Upregulated |
| 0,500387554 | 0,522450769 | LOC100506047 | FALSE | Upregulated |
| 0,5070017 | 0,52246572 | LOC101928178 | FALSE | Upregulated |
| -0,122376305 | 0,52246572 | USP36 | FALSE | Downregulated |
| -0,513151329 | 0,5225731 | ZNF454 | FALSE | Downregulated |
| -0,111714651 | 0,522757897 | STYX | FALSE | Downregulated |
| -0,710882078 | 0,522852638 | IL5RA | FALSE | Downregulated |
| 0,073736401 | 0,522852638 | NDUFA9 | FALSE | Upregulated |
| -0,174739103 | 0,522852638 | PCGF2 | FALSE | Downregulated |
| 0,198378724 | 0,523094598 | EFEMP2 | FALSE | Upregulated |
| 0,174956325 | 0,523259452 | RARA | FALSE | Upregulated |
| 0,749611279 | 0,523365268 | LOC100287072 | FALSE | Upregulated |
| 0,233651984 | 0,523431879 | HSPA13 | FALSE | Upregulated |
| -0,526404197 | 0,523431879 | LOC102723589 | FALSE | Downregulated |
| 0,135908941 | 0,523431879 | SEC61B | FALSE | Upregulated |
| 0,252024183 | 0,523464593 | PCYOX1L | FALSE | Upregulated |
| -1,062228365 | 0,523609196 | LOC100101148 | FALSE | Downregulated |
| -0,717483398 | 0,52375197 | PIFO | FALSE | Downregulated |
| -0,072360482 | 0,523763937 | EAF1 | FALSE | Downregulated |
| 0,698583004 | 0,52384666 | GABRG1 | FALSE | Upregulated |
| -0,099446644 | 0,52384666 | RPUSD3 | FALSE | Downregulated |
| -0,210569146 | 0,523917378 | GREM2 | FALSE | Downregulated |

| | | | | |
|---|---|---|---|---|
| 0,12432608 | 0,524052872 | PTPN11 | FALSE | Upregulated |
| -0,364024667 | 0,524115513 | ZNF789 | FALSE | Downregulated |
| -1,152819168 | 0,524177256 | KRTAP16-1 | FALSE | Downregulated |
| -0,194702101 | 0,524678732 | TRMT10B | FALSE | Downregulated |
| -0,283913058 | 0,524803186 | LOC101927126 | FALSE | Downregulated |
| 0,459978961 | 0,524811075 | NR4A1 | FALSE | Upregulated |
| -0,130435996 | 0,524811075 | NRF1 | FALSE | Downregulated |
| -0,410164255 | 0,525018379 | MYO15A | FALSE | Downregulated |
| 0,16627706 | 0,52502253 | TRIM10 | FALSE | Upregulated |
| -0,197853257 | 0,525101053 | C15orf37 | FALSE | Downregulated |
| 0,482433356 | 0,525101053 | CD1E | FALSE | Upregulated |
| -0,102944769 | 0,52514049 | PI4K2B | FALSE | Downregulated |
| 0,464560382 | 0,525455433 | NPEPL1 | FALSE | Upregulated |
| -0,10505089 | 0,525455433 | TMEM203 | FALSE | Downregulated |
| -0,092786034 | 0,525475031 | COX20 | FALSE | Downregulated |
| 0,16295847 | 0,525501039 | CDPF1 | FALSE | Upregulated |
| -0,05968141 | 0,525501039 | TSNAX | FALSE | Downregulated |
| -0,1929714 | 0,525522893 | CLRN3 | FALSE | Downregulated |
| 0,251830461 | 0,525522893 | GATS | FALSE | Upregulated |
| -0,193891656 | 0,525522893 | NFIC | FALSE | Downregulated |
| -0,394257825 | 0,525522893 | RP11-509J21.1 | FALSE | Downregulated |
| 0,87838823 | 0,525692916 | MUC5B | FALSE | Upregulated |
| 0,080844321 | 0,52579971 | SF3A1 | FALSE | Upregulated |
| -0,254668887 | 0,52579971 | ZNF713 | FALSE | Downregulated |
| -0,333674626 | 0,525856868 | EML6 | FALSE | Downregulated |
| -0,280903464 | 0,525856868 | IQCH | FALSE | Downregulated |
| -0,352425778 | 0,525856868 | KLRAP1 | FALSE | Downregulated |
| 0,278594105 | 0,525856868 | MGC16142 | FALSE | Upregulated |
| 0,118269637 | 0,525856868 | NDUFA11 | FALSE | Upregulated |
| -0,328574594 | 0,525909354 | SEC31B | FALSE | Downregulated |
| -0,141216709 | 0,52637303 | GCC2 | FALSE | Downregulated |
| 0,128140096 | 0,52639586 | ABHD6 | FALSE | Upregulated |
| -0,1208558 | 0,52639586 | GON4L | FALSE | Downregulated |
| 0,105505652 | 0,52639586 | NDUFS3 | FALSE | Upregulated |
| 0,098291647 | 0,526640417 | SCRIB | FALSE | Upregulated |
| -0,135093489 | 0,526682811 | LOC148413 | FALSE | Downregulated |
| 0,082640554 | 0,526718945 | TGS1 | FALSE | Upregulated |
| -0,101536114 | 0,526833505 | BRF2 | FALSE | Downregulated |
| -0,304616383 | 0,526833505 | FAM163B | FALSE | Downregulated |
| -0,618973921 | 0,526833505 | LOC283693 | FALSE | Downregulated |
| 0,332851753 | 0,526844803 | EVC2 | FALSE | Upregulated |
| 0,539931093 | 0,526983962 | SCN7A | FALSE | Upregulated |
| -0,10538479 | 0,527078483 | TYSND1 | FALSE | Downregulated |
| -0,337549342 | 0,527194986 | ST8SIA6 | FALSE | Downregulated |
| 0,181860226 | 0,527352377 | ECHDC3 | FALSE | Upregulated |
| 0,657222148 | 0,527383751 | C10orf107 | FALSE | Upregulated |
| 0,12889481 | 0,527503471 | CACUL1 | FALSE | Upregulated |
| 0,284241183 | 0,527582154 | NLRP3 | FALSE | Upregulated |
| 0,471111119 | 0,527620195 | C1orf204 | FALSE | Upregulated |
| 0,118669228 | 0,527779352 | NDUFA7 | FALSE | Upregulated |
| 0,189496534 | 0,527824928 | TLR1 | FALSE | Upregulated |

| | | | | |
|---|---|---|---|---|
| -0,342340336 | 0,527824928 | ZNF793 | FALSE | Downregulated |
| -0,213438672 | 0,527925261 | MTERFD2 | FALSE | Downregulated |
| 0,086992162 | 0,528096259 | PTPLAD1 | FALSE | Upregulated |
| -0,641122252 | 0,528104231 | FPGT-TNNI3K | FALSE | Downregulated |
| 0,093613252 | 0,528180169 | CD81 | FALSE | Upregulated |
| -0,137275469 | 0,528180726 | GALT | FALSE | Downregulated |
| 0,154936208 | 0,528180726 | SSR2 | FALSE | Upregulated |
| 0,268673824 | 0,528180726 | ULK4 | FALSE | Upregulated |
| -0,315066459 | 0,528188946 | CKB | FALSE | Downregulated |
| -0,084279672 | 0,528188946 | FH | FALSE | Downregulated |
| 0,333256383 | 0,528188946 | IGFBP2 | FALSE | Upregulated |
| -0,068025676 | 0,528259303 | CAMKK2 | FALSE | Downregulated |
| 0,234329562 | 0,52829848 | KPTN | FALSE | Upregulated |
| -0,087593608 | 0,528442997 | ZNF131 | FALSE | Downregulated |
| 0,087605488 | 0,528462311 | PDCL | FALSE | Upregulated |
| -0,125363165 | 0,528601329 | FBXL20 | FALSE | Downregulated |
| -0,230729115 | 0,528654531 | LENG8 | FALSE | Downregulated |
| -0,723289944 | 0,52867473 | GPRC5D | FALSE | Downregulated |
| 0,447430635 | 0,528836742 | BMX | FALSE | Upregulated |
| 0,149017976 | 0,528836742 | TMEM64 | FALSE | Upregulated |
| 0,47438379 | 0,528912289 | RASA4CP | FALSE | Upregulated |
| 0,204411736 | 0,528926058 | AP5S1 | FALSE | Upregulated |
| 0,210952151 | 0,528985884 | MIIP | FALSE | Upregulated |
| 0,209074939 | 0,52914645 | LIN9 | FALSE | Upregulated |
| -0,374541016 | 0,52914645 | SNORA25 | FALSE | Downregulated |
| -0,499842901 | 0,529386238 | LOC101928612 | FALSE | Downregulated |
| 0,268399182 | 0,529386238 | PDE1A | FALSE | Upregulated |
| -0,06952589 | 0,529448128 | RNMT | FALSE | Downregulated |
| -0,640700731 | 0,529544764 | LOC100132356 | FALSE | Downregulated |
| -0,099749878 | 0,529581858 | CEBPG | FALSE | Downregulated |
| -0,218249683 | 0,529953928 | SPRY3 | FALSE | Downregulated |
| -0,189438259 | 0,530046497 | C1orf172 | FALSE | Downregulated |
| -0,707816536 | 0,530046497 | MUC12 | FALSE | Downregulated |
| -0,235238107 | 0,530108225 | LAMA5 | FALSE | Downregulated |
| -0,10217565 | 0,530174604 | ANAPC1 | FALSE | Downregulated |
| -0,152729073 | 0,530174604 | TIMMDC1 | FALSE | Downregulated |
| -0,091418242 | 0,530174604 | USP32 | FALSE | Downregulated |
| -0,501689945 | 0,530190887 | GCK | FALSE | Downregulated |
| 0,382759179 | 0,530409944 | NEURL3 | FALSE | Upregulated |
| -0,453236528 | 0,530455274 | BRINP2 | FALSE | Downregulated |
| 0,047508684 | 0,530546876 | SMU1 | FALSE | Upregulated |
| -0,284803948 | 0,530546876 | ZFHX2 | FALSE | Downregulated |
| 0,297930184 | 0,530594095 | ADAMTS10 | FALSE | Upregulated |
| 0,297753658 | 0,530596185 | C15orf65 | FALSE | Upregulated |
| 0,148720096 | 0,531261702 | CHST13 | FALSE | Upregulated |
| -0,081706907 | 0,53149586 | ZNF143 | FALSE | Downregulated |
| 0,161914718 | 0,531536863 | PPCDC | FALSE | Upregulated |
| 0,546359256 | 0,531570899 | LOC102724554 | FALSE | Upregulated |
| -0,150161859 | 0,531749106 | MAST2 | FALSE | Downregulated |
| -0,432286501 | 0,531800768 | OLFML2A | FALSE | Downregulated |
| -0,171584731 | 0,531854615 | SERP1 | FALSE | Downregulated |

| | | | | |
|---|---|---|---|---|
| 0,259831974 | 0,531926183 | REC8 | FALSE | Upregulated |
| 0,089183179 | 0,531934386 | EPN1 | FALSE | Upregulated |
| -0,10153485 | 0,531987033 | SNX18 | FALSE | Downregulated |
| 0,643375043 | 0,532057671 | FAM135B | FALSE | Upregulated |
| 0,245138876 | 0,532057671 | IGFLR1 | FALSE | Upregulated |
| 0,148451563 | 0,532379407 | HUS1 | FALSE | Upregulated |
| -0,402444693 | 0,532454547 | LOC102724562 | FALSE | Downregulated |
| 0,104298578 | 0,532454547 | SOAT1 | FALSE | Upregulated |
| 0,251272606 | 0,532606113 | FOXO3B | FALSE | Upregulated |
| 0,150620858 | 0,532710561 | FLYWCH1 | FALSE | Upregulated |
| -0,074100208 | 0,532715687 | GNAS | FALSE | Downregulated |
| -0,434317546 | 0,532715687 | LOC100507554 | FALSE | Downregulated |
| 0,233145821 | 0,533200376 | TNFRSF21 | FALSE | Upregulated |
| 0,1047831 | 0,533200376 | XBP1 | FALSE | Upregulated |
| -0,583684836 | 0,533210927 | CCR9 | FALSE | Downregulated |
| -0,129893339 | 0,533266011 | C5orf42 | FALSE | Downregulated |
| 0,169814209 | 0,533266011 | DHX58 | FALSE | Upregulated |
| -0,206700187 | 0,533266011 | KATNAL2 | FALSE | Downregulated |
| -0,160409152 | 0,533266011 | RBM4B | FALSE | Downregulated |
| -0,971266896 | 0,533285399 | ANK1 | FALSE | Downregulated |
| -0,196292262 | 0,533689247 | MERTK | FALSE | Downregulated |
| -0,31063317 | 0,533689247 | TRPC1 | FALSE | Downregulated |
| -0,096660771 | 0,533929664 | ASB8 | FALSE | Downregulated |
| 0,259736189 | 0,533929664 | C15orf52 | FALSE | Upregulated |
| 0,269034158 | 0,53404471 | NFASC | FALSE | Upregulated |
| -0,899347985 | 0,534056095 | MPP4 | FALSE | Downregulated |
| -0,138466445 | 0,534153545 | IL11RA | FALSE | Downregulated |
| -0,145971253 | 0,534255294 | OXLD1 | FALSE | Downregulated |
| -0,177656255 | 0,534364112 | SMC6 | FALSE | Downregulated |
| -0,167346398 | 0,534581227 | IFT80 | FALSE | Downregulated |
| -0,111197572 | 0,534581227 | RALGAPA2 | FALSE | Downregulated |
| 0,184862176 | 0,534621679 | ANG | FALSE | Upregulated |
| 0,296867595 | 0,534621679 | LDLRAD3 | FALSE | Upregulated |
| -1,815526616 | 0,534621679 | MYF6 | FALSE | Downregulated |
| 0,112416588 | 0,534812112 | GHDC | FALSE | Upregulated |
| 0,15618149 | 0,535056215 | IFT52 | FALSE | Upregulated |
| -0,693103135 | 0,535164685 | CATSPERB | FALSE | Downregulated |
| -0,567830926 | 0,535221029 | TPSG1 | FALSE | Downregulated |
| -0,65247167 | 0,535415601 | MIR29C | FALSE | Downregulated |
| -0,97188912 | 0,535415601 | SLC39A12 | FALSE | Downregulated |
| -0,310459161 | 0,535415601 | UBE2QL1 | FALSE | Downregulated |
| -0,284234945 | 0,535654607 | FBN1 | FALSE | Downregulated |
| -0,065848945 | 0,535654607 | PDCD7 | FALSE | Downregulated |
| 0,097771355 | 0,535654607 | SLC43A1 | FALSE | Upregulated |
| -0,194091954 | 0,535796924 | ZNF829 | FALSE | Downregulated |
| 0,612870205 | 0,535866173 | CR2 | FALSE | Upregulated |
| 1,136226891 | 0,535940979 | TNFAIP8L2-SCNM1 | FALSE | Upregulated |
| 0,124382059 | 0,536246728 | IRF3 | FALSE | Upregulated |
| 0,490993426 | 0,536705823 | KRBOX1 | FALSE | Upregulated |
| 0,082280044 | 0,536751174 | BTF3 | FALSE | Upregulated |
| 0,336526529 | 0,536751174 | F3 | FALSE | Upregulated |

| | | | | |
|---|---|---|---|---|
| 0,173157678 | 0,536751174 | GEMIN7 | FALSE | Upregulated |
| -0,163846698 | 0,536751174 | MTHFSD | FALSE | Downregulated |
| -0,066824491 | 0,536751174 | RUFY1 | FALSE | Downregulated |
| -0,442649399 | 0,536768407 | IRX3 | FALSE | Downregulated |
| -0,307184207 | 0,536768407 | SRGAP2C | FALSE | Downregulated |
| -0,11968 | 0,536992143 | SPRY2 | FALSE | Downregulated |
| -0,465488289 | 0,536992143 | TUBB4A | FALSE | Downregulated |
| 0,159389143 | 0,537061027 | HAND2 | FALSE | Upregulated |
| 0,224620347 | 0,537061027 | KLHDC7A | FALSE | Upregulated |
| 0,6391364 | 0,53721449 | AC104820.2 | FALSE | Upregulated |
| -0,105089515 | 0,537367759 | SNHG6 | FALSE | Downregulated |
| -0,112471988 | 0,537564904 | TMEM47 | FALSE | Downregulated |
| 0,099101009 | 0,537696376 | STOML2 | FALSE | Upregulated |
| -0,065667446 | 0,537812143 | NAA50 | FALSE | Downregulated |
| 0,093844772 | 0,537827556 | PSMD8 | FALSE | Upregulated |
| -0,060337049 | 0,537894667 | CSNK1A1 | FALSE | Downregulated |
| -0,630722828 | 0,538085402 | LOC101927910 | FALSE | Downregulated |
| -0,422870446 | 0,538233209 | EBLN2 | FALSE | Downregulated |
| -0,148587471 | 0,538233209 | SURF6 | FALSE | Downregulated |
| -0,15509595 | 0,538233209 | TXNL4B | FALSE | Downregulated |
| 0,280835133 | 0,538233209 | ZNF703 | FALSE | Upregulated |
| 0,105457008 | 0,538687766 | KPNB1 | FALSE | Upregulated |
| 0,080349388 | 0,538771467 | PTGES3 | FALSE | Upregulated |
| 0,110003457 | 0,538771467 | TMED2 | FALSE | Upregulated |
| -0,137030435 | 0,53893079 | ARHGEF40 | FALSE | Downregulated |
| 0,136228887 | 0,538972185 | KIAA0232 | FALSE | Upregulated |
| -0,186172139 | 0,539076849 | LOC100506990 | FALSE | Downregulated |
| -0,075763086 | 0,539101827 | GPN1 | FALSE | Downregulated |
| -0,453402294 | 0,539101827 | MIR564 | FALSE | Downregulated |
| -0,531515822 | 0,539164577 | GPR173 | FALSE | Downregulated |
| 0,058579621 | 0,539377764 | PNRC2 | FALSE | Upregulated |
| 0,082634458 | 0,539377764 | PRKDC | FALSE | Upregulated |
| -0,378415484 | 0,539502809 | FAM78B | FALSE | Downregulated |
| 0,086248835 | 0,539532699 | ATG16L1 | FALSE | Upregulated |
| -0,23576989 | 0,539704325 | CCDC88C | FALSE | Downregulated |
| -0,201789446 | 0,539753398 | MCMDC2 | FALSE | Downregulated |
| -0,122288459 | 0,539768761 | HDAC4 | FALSE | Downregulated |
| 0,080025512 | 0,540195341 | MFAP3 | FALSE | Upregulated |
| 0,076901336 | 0,540523998 | RAD21 | FALSE | Upregulated |
| -0,095488854 | 0,540523998 | ZNF790 | FALSE | Downregulated |
| -0,109397978 | 0,54063303 | RINT1 | FALSE | Downregulated |
| 0,087339969 | 0,54063303 | SUGP1 | FALSE | Upregulated |
| -0,154571852 | 0,540946179 | IFFO2 | FALSE | Downregulated |
| -0,127665241 | 0,540946179 | IRAK4 | FALSE | Downregulated |
| 0,099962644 | 0,540946179 | MID2 | FALSE | Upregulated |
| -1,341613253 | 0,540946179 | TSNAX-DISC1 | FALSE | Downregulated |
| -0,097468909 | 0,540991883 | RPS23 | FALSE | Downregulated |
| 0,133317548 | 0,541034436 | TARBP1 | FALSE | Upregulated |
| 0,179333104 | 0,541097368 | BRI3BP | FALSE | Upregulated |
| -0,11232954 | 0,541097368 | POU2F1 | FALSE | Downregulated |
| 0,225038727 | 0,541097368 | RAMP2 | FALSE | Upregulated |

| | | | | |
|---|---|---|---|---|
| -0,327299753 | 0,541236009 | FBXL22 | FALSE | Downregulated |
| 0,084802171 | 0,541621463 | TAX1BP1 | FALSE | Upregulated |
| -0,072478476 | 0,541986514 | PEX11B | FALSE | Downregulated |
| 0,413406485 | 0,542015955 | LOC100289561 | FALSE | Upregulated |
| -0,611779948 | 0,542104251 | LOC286367 | FALSE | Downregulated |
| -0,085088026 | 0,54225276 | TBC1D13 | FALSE | Downregulated |
| -0,149166306 | 0,542321708 | GYS1 | FALSE | Downregulated |
| 0,134368073 | 0,542321708 | H1FX | FALSE | Upregulated |
| -0,303483979 | 0,542356697 | NSUN5P2 | FALSE | Downregulated |
| 0,254338169 | 0,542390413 | B3GNTL1 | FALSE | Upregulated |
| -0,078895978 | 0,542592684 | CNOT4 | FALSE | Downregulated |
| -1,065360431 | 0,542592684 | FAT2 | FALSE | Downregulated |
| 0,088577187 | 0,542712143 | CHERP | FALSE | Upregulated |
| -0,367091636 | 0,542712143 | EPHA4 | FALSE | Downregulated |
| -0,440311633 | 0,542747232 | CCDC136 | FALSE | Downregulated |
| 0,100477415 | 0,542831657 | PRDX2 | FALSE | Upregulated |
| -0,070863058 | 0,542868491 | MAPKAPK5 | FALSE | Downregulated |
| -0,824563996 | 0,543039357 | C22orf23 | FALSE | Downregulated |
| 0,157112593 | 0,543039357 | DMXL2 | FALSE | Upregulated |
| 0,091207179 | 0,543189729 | FBL | FALSE | Upregulated |
| 0,424092102 | 0,543216224 | FAM166A | FALSE | Upregulated |
| -0,626398642 | 0,543216224 | MCOLN3 | FALSE | Downregulated |
| 0,271346658 | 0,543216224 | PDZRN3 | FALSE | Upregulated |
| -0,078786371 | 0,543216224 | TAB1 | FALSE | Downregulated |
| 0,239393762 | 0,543216224 | TEX9 | FALSE | Upregulated |
| -0,738803389 | 0,543260139 | ATP2C2 | FALSE | Downregulated |
| -1,10448558 | 0,543260139 | HYDIN2 | FALSE | Downregulated |
| -0,155327507 | 0,543260139 | NGFR | FALSE | Downregulated |
| 0,110793415 | 0,543260139 | PARP11 | FALSE | Upregulated |
| -0,328175399 | 0,543317449 | ANGPT2 | FALSE | Downregulated |
| -0,065905082 | 0,54357857 | CIAO1 | FALSE | Downregulated |
| -0,080047256 | 0,543600308 | LARS | FALSE | Downregulated |
| 0,127960585 | 0,543600308 | MARVELD3 | FALSE | Upregulated |
| -0,081433259 | 0,543600308 | NBAS | FALSE | Downregulated |
| -0,697060792 | 0,543606347 | PDLIM3 | FALSE | Downregulated |
| -1,180422733 | 0,54389056 | SHOX | FALSE | Downregulated |
| 0,710573352 | 0,543924521 | LOC102723870 | FALSE | Upregulated |
| 0,127390069 | 0,544020979 | RPL36A | FALSE | Upregulated |
| 0,078466731 | 0,544055597 | DAZAP2 | FALSE | Upregulated |
| 0,282550246 | 0,544196336 | ACSS2 | FALSE | Upregulated |
| -0,552817134 | 0,54421108 | LOC102723958 | FALSE | Downregulated |
| -0,256709958 | 0,54421108 | REP15 | FALSE | Downregulated |
| 0,409289651 | 0,54421108 | WNT4 | FALSE | Upregulated |
| 0,082893115 | 0,544232449 | SUGT1 | FALSE | Upregulated |
| -0,542713079 | 0,5444964 | PANO | FALSE | Downregulated |
| -1,019072838 | 0,5444964 | RNF103-CHMP3 | FALSE | Downregulated |
| 0,08372447 | 0,544707432 | CENPB | FALSE | Upregulated |
| -0,721430829 | 0,544707432 | LINC01105 | FALSE | Downregulated |
| -0,097467822 | 0,544707432 | MRPL24 | FALSE | Downregulated |
| -0,097172183 | 0,545225517 | C12orf65 | FALSE | Downregulated |
| 0,141322944 | 0,545294553 | SEC61A1 | FALSE | Upregulated |

| | | | | |
|---|---|---|---|---|
| -0,064101257 | 0,545294553 | VAMP7 | FALSE | Downregulated |
| -0,270430662 | 0,545294553 | VWCE | FALSE | Downregulated |
| -0,470591001 | 0,545300452 | TYW1B | FALSE | Downregulated |
| 0,694789129 | 0,545319446 | SLITRK2 | FALSE | Upregulated |
| 0,149298627 | 0,545363508 | MED30 | FALSE | Upregulated |
| 0,251107368 | 0,545363508 | USP6NL-IT1 | FALSE | Upregulated |
| -0,304138233 | 0,545399144 | LOC102724872 | FALSE | Downregulated |
| -0,608291031 | 0,545414771 | HLX-AS1 | FALSE | Downregulated |
| -0,130204545 | 0,545468895 | C12orf66 | FALSE | Downregulated |
| -0,08692099 | 0,545468895 | CETN3 | FALSE | Downregulated |
| -0,479940975 | 0,545468895 | NAALADL1 | FALSE | Downregulated |
| 0,158999451 | 0,545468895 | ZEB2 | FALSE | Upregulated |
| -0,085780153 | 0,545726445 | CCNI | FALSE | Downregulated |
| -0,333923848 | 0,545726445 | LOC643770 | FALSE | Downregulated |
| -0,18306485 | 0,545726445 | STARD7-AS1 | FALSE | Downregulated |
| 0,112259931 | 0,545844996 | VPS33B | FALSE | Upregulated |
| -0,526261483 | 0,54595115 | LOC102723927 | FALSE | Downregulated |
| 0,136678012 | 0,546500112 | AMER1 | FALSE | Upregulated |
| -0,160703828 | 0,546500112 | DCHS1 | FALSE | Downregulated |
| 0,088176964 | 0,546500112 | FCGRT | FALSE | Upregulated |
| 0,213923972 | 0,546500112 | PDE4D | FALSE | Upregulated |
| -0,376998946 | 0,546566418 | TP63 | FALSE | Downregulated |
| -0,558487006 | 0,546634468 | HYDIN | FALSE | Downregulated |
| 0,674263153 | 0,546737937 | HIST2H2BC | FALSE | Upregulated |
| -0,263140035 | 0,54673804 | FLRT3 | FALSE | Downregulated |
| -0,112521492 | 0,546770843 | SUMF1 | FALSE | Downregulated |
| -0,148319112 | 0,547063914 | USP13 | FALSE | Downregulated |
| 0,084148096 | 0,547083314 | ARF5 | FALSE | Upregulated |
| -0,145044056 | 0,547083314 | HEMK1 | FALSE | Downregulated |
| -0,184349916 | 0,547107511 | CRIM1 | FALSE | Downregulated |
| -0,125209348 | 0,547122148 | COLEC10 | FALSE | Downregulated |
| -0,163928453 | 0,547122148 | NOTCH4 | FALSE | Downregulated |
| -0,165636322 | 0,547149905 | ADCY5 | FALSE | Downregulated |
| -0,087157514 | 0,547266417 | AARS2 | FALSE | Downregulated |
| 0,17014423 | 0,547266417 | ARRDC4 | FALSE | Upregulated |
| -0,208542473 | 0,547273676 | FOXF1 | FALSE | Downregulated |
| -0,101103426 | 0,547275955 | SIRT5 | FALSE | Downregulated |
| -0,089933447 | 0,547419624 | MON1B | FALSE | Downregulated |
| 0,179841584 | 0,547429353 | CCZ1B | FALSE | Upregulated |
| 0,196446335 | 0,547429353 | PORCN | FALSE | Upregulated |
| 0,132733814 | 0,547429353 | YAP1 | FALSE | Upregulated |
| -0,090040149 | 0,547552174 | ZBTB43 | FALSE | Downregulated |
| 0,10077811 | 0,547659622 | ERCC2 | FALSE | Upregulated |
| 0,086070786 | 0,547659622 | GSTK1 | FALSE | Upregulated |
| -0,087440512 | 0,547659622 | POMT1 | FALSE | Downregulated |
| 0,108332354 | 0,547725151 | IMP4 | FALSE | Upregulated |
| 0,11286752 | 0,547725151 | XPO1 | FALSE | Upregulated |
| 0,113272438 | 0,548013806 | BANP | FALSE | Upregulated |
| 0,07821986 | 0,548013806 | UBAP2 | FALSE | Upregulated |
| 0,262902692 | 0,548216591 | ACKR4 | FALSE | Upregulated |
| 0,104270731 | 0,548345246 | ABT1 | FALSE | Upregulated |

| | | | | |
|---|---|---|---|---|
| 0,080794958 | 0,548345246 | KXD1 | FALSE | Upregulated |
| -0,16936817 | 0,548345246 | PISD | FALSE | Downregulated |
| -0,218172043 | 0,548356171 | IL13RA2 | FALSE | Downregulated |
| -0,120235339 | 0,548488765 | LRRFIP2 | FALSE | Downregulated |
| 0,101134891 | 0,548591411 | RUVBL1 | FALSE | Upregulated |
| -0,133530611 | 0,548591411 | TRPT1 | FALSE | Downregulated |
| 0,632263636 | 0,548673843 | LPL | FALSE | Upregulated |
| 0,135182915 | 0,548673843 | YAF2 | FALSE | Upregulated |
| -0,483836081 | 0,548674212 | KL | FALSE | Downregulated |
| -0,107120283 | 0,548798148 | ZSCAN32 | FALSE | Downregulated |
| 0,433287929 | 0,548902094 | SLMO2-ATP5E | FALSE | Upregulated |
| 0,568883278 | 0,548944548 | CSNK2A3 | FALSE | Upregulated |
| -0,185369578 | 0,549132252 | SMIM10 | FALSE | Downregulated |
| 0,108405025 | 0,549139404 | ENOX2 | FALSE | Upregulated |
| -0,471262339 | 0,549139404 | GP6 | FALSE | Downregulated |
| -1,179433564 | 0,549139404 | LRFN5 | FALSE | Downregulated |
| -0,117574994 | 0,549139404 | NPRL2 | FALSE | Downregulated |
| 0,206873526 | 0,549139404 | TRMT61A | FALSE | Upregulated |
| 0,114678791 | 0,549207503 | ZNF720 | FALSE | Upregulated |
| -0,499498455 | 0,549274419 | GNG3 | FALSE | Downregulated |
| 0,404412951 | 0,549320209 | AGPAT4 | FALSE | Upregulated |
| 0,148567057 | 0,549320209 | DYRK1B | FALSE | Upregulated |
| 0,156476635 | 0,549320209 | NOC4L | FALSE | Upregulated |
| 0,086341679 | 0,549322462 | MED20 | FALSE | Upregulated |
| 0,144824787 | 0,549322462 | POR | FALSE | Upregulated |
| -0,104516208 | 0,549342157 | B3GNT2 | FALSE | Downregulated |
| 0,075769533 | 0,549439652 | RAB3GAP1 | FALSE | Upregulated |
| -0,781872666 | 0,549459312 | SMIM5 | FALSE | Downregulated |
| -0,465558858 | 0,549599572 | TSPEAR-AS1 | FALSE | Downregulated |
| -0,762246627 | 0,549600996 | LINC01010 | FALSE | Downregulated |
| -0,186275016 | 0,549902355 | EHD3 | FALSE | Downregulated |
| 0,077738187 | 0,550197829 | DENND6A | FALSE | Upregulated |
| -0,131944386 | 0,550484124 | SLC10A1 | FALSE | Downregulated |
| 0,369386313 | 0,550531911 | GAS6-AS1 | FALSE | Upregulated |
| 0,524402504 | 0,55065903 | UNC13A | FALSE | Upregulated |
| -0,258068087 | 0,550719796 | SLIT2 | FALSE | Downregulated |
| 0,636630778 | 0,550757004 | VASH2 | FALSE | Upregulated |
| -0,30005428 | 0,550766226 | APCDD1 | FALSE | Downregulated |
| -0,514044801 | 0,550766226 | ENOX1 | FALSE | Downregulated |
| -0,596148098 | 0,550766226 | LOC102724957 | FALSE | Downregulated |
| 0,541814366 | 0,550766226 | RP5-899E9.1 | FALSE | Upregulated |
| -0,136276151 | 0,550881948 | NSUN6 | FALSE | Downregulated |
| -0,610800922 | 0,55097074 | ARL17B | FALSE | Downregulated |
| -0,215518668 | 0,55097074 | RP11-418J17.1 | FALSE | Downregulated |
| 0,74489137 | 0,55097074 | SCTR | FALSE | Upregulated |
| 0,130835514 | 0,551174497 | TIMM10 | FALSE | Upregulated |
| -0,079327017 | 0,551285528 | FAM35A | FALSE | Downregulated |
| 0,209895208 | 0,551285528 | PDIA3P1 | FALSE | Upregulated |
| 0,14331144 | 0,551580066 | DOCK5 | FALSE | Upregulated |
| -0,080355401 | 0,551580066 | UQCRC1 | FALSE | Downregulated |
| -0,185252228 | 0,55161081 | FAM47E-STBD1 | FALSE | Downregulated |

| | | | | |
|---|---|---|---|---|
| -0,145486946 | 0,55161081 | SWSAP1 | FALSE | Downregulated |
| 0,127456005 | 0,551808043 | ABCF1 | FALSE | Upregulated |
| 0,137986802 | 0,551808043 | LYRM4 | FALSE | Upregulated |
| -0,335780421 | 0,552026494 | LOC101928524 | FALSE | Downregulated |
| -0,190633521 | 0,552026494 | LOC102724532 | FALSE | Downregulated |
| -0,765210035 | 0,552485138 | XPNPEP2 | FALSE | Downregulated |
| 0,095749983 | 0,552684973 | PPP1R2 | FALSE | Upregulated |
| -0,113313922 | 0,552684973 | ZNHIT1 | FALSE | Downregulated |
| -0,07983721 | 0,552779968 | C9orf156 | FALSE | Downregulated |
| -0,148073686 | 0,552779968 | IFT81 | FALSE | Downregulated |
| -1,056732456 | 0,552779968 | MIR4426 | FALSE | Downregulated |
| 0,246848364 | 0,552779968 | OLFM1 | FALSE | Upregulated |
| 0,372126251 | 0,552792403 | UQCRHL | FALSE | Upregulated |
| 0,209922902 | 0,55296438 | MXD3 | FALSE | Upregulated |
| -0,095305241 | 0,552979117 | CTSL | FALSE | Downregulated |
| -0,122797531 | 0,552979117 | LRCH4 | FALSE | Downregulated |
| 0,623674924 | 0,553086132 | BIK | FALSE | Upregulated |
| -0,134515131 | 0,553281301 | HYI | FALSE | Downregulated |
| -0,29243306 | 0,553302606 | P4HA1 | FALSE | Downregulated |
| -0,480004996 | 0,553302606 | STAG3L1 | FALSE | Downregulated |
| -0,392277666 | 0,553400811 | GPR158 | FALSE | Downregulated |
| 0,572001461 | 0,55344447 | PTX3 | FALSE | Upregulated |
| 0,148981406 | 0,553493686 | STAT3 | FALSE | Upregulated |
| 0,128292321 | 0,553551944 | RNASEL | FALSE | Upregulated |
| -0,613205385 | 0,553918952 | KIAA1024 | FALSE | Downregulated |
| -0,071829847 | 0,553918952 | RBM10 | FALSE | Downregulated |
| -0,495141884 | 0,554050189 | CSPG4 | FALSE | Downregulated |
| 0,171696347 | 0,554073792 | ASAH2B | FALSE | Upregulated |
| -0,115658477 | 0,554199107 | BOLA1 | FALSE | Downregulated |
| -0,133927935 | 0,554212319 | STARD13 | FALSE | Downregulated |
| -0,084089006 | 0,554586537 | ARV1 | FALSE | Downregulated |
| 0,218913502 | 0,554786303 | NRSN2 | FALSE | Upregulated |
| 0,286481358 | 0,555067674 | LOC100093631 | FALSE | Upregulated |
| -0,373276427 | 0,555385919 | GPR162 | FALSE | Downregulated |
| 0,143934815 | 0,555385919 | ITGA9 | FALSE | Upregulated |
| 0,166664467 | 0,555385919 | KIAA0040 | FALSE | Upregulated |
| -0,07102357 | 0,555385919 | SUPT5H | FALSE | Downregulated |
| -0,612996147 | 0,555391377 | LOC100132813 | FALSE | Downregulated |
| 0,42782152 | 0,55551966 | DUSP9 | FALSE | Upregulated |
| 0,126464475 | 0,555540278 | EXOSC5 | FALSE | Upregulated |
| -0,089419423 | 0,555540278 | PDSS2 | FALSE | Downregulated |
| 0,465261666 | 0,555906496 | LINC00704 | FALSE | Upregulated |
| 0,34706896 | 0,555906496 | SCAMP5 | FALSE | Upregulated |
| -0,1551087 | 0,556080733 | KCTD7 | FALSE | Downregulated |
| -0,881516239 | 0,55656286 | LOC100131395 | FALSE | Downregulated |
| 0,105347565 | 0,55656286 | PES1 | FALSE | Upregulated |
| -0,387574208 | 0,556749594 | ROPN1B | FALSE | Downregulated |
| -0,48737239 | 0,556751794 | LINC00312 | FALSE | Downregulated |
| 0,097218902 | 0,556751794 | NCK1 | FALSE | Upregulated |
| 0,548969888 | 0,556775129 | PRAMENP | FALSE | Upregulated |
| -0,085207274 | 0,556932031 | TRIM41 | FALSE | Downregulated |

| | | | | |
|---|---|---|---|---|
| 0,263548629 | 0,557018138 | RASA4 | FALSE | Upregulated |
| -0,518555879 | 0,557023398 | GPR123 | FALSE | Downregulated |
| -0,296083214 | 0,557023398 | LINC01197 | FALSE | Downregulated |
| 0,470800403 | 0,557023398 | LOC100507336 | FALSE | Upregulated |
| 0,707251383 | 0,557023398 | PCAT1 | FALSE | Upregulated |
| -0,385543808 | 0,55703668 | NDRG4 | FALSE | Downregulated |
| -0,067650014 | 0,557155849 | APEX1 | FALSE | Downregulated |
| -0,106827383 | 0,557291725 | ARMCX5 | FALSE | Downregulated |
| -0,3883719 | 0,557344564 | LINC00852 | FALSE | Downregulated |
| 0,405913501 | 0,557344564 | LRRC1 | FALSE | Upregulated |
| -0,10487065 | 0,557344564 | LYSMD3 | FALSE | Downregulated |
| 0,107898001 | 0,557344564 | SCCPDH | FALSE | Upregulated |
| 0,109610044 | 0,557344564 | UQCR10 | FALSE | Upregulated |
| 0,112671748 | 0,557369808 | RANBP10 | FALSE | Upregulated |
| -0,650062624 | 0,557537478 | LOC101929666 | FALSE | Downregulated |
| -0,195302528 | 0,557537478 | ZNF256 | FALSE | Downregulated |
| -0,310698856 | 0,55756716 | MARK1 | FALSE | Downregulated |
| 0,10540719 | 0,557729255 | DENND1B | FALSE | Upregulated |
| -0,378953693 | 0,557782914 | LOC101928051 | FALSE | Downregulated |
| -0,44021872 | 0,557997239 | SIGLEC15 | FALSE | Downregulated |
| -0,115567126 | 0,55799858 | TASP1 | FALSE | Downregulated |
| 0,155412943 | 0,558205356 | CCNJ | FALSE | Upregulated |
| -0,185172426 | 0,558442152 | KLB | FALSE | Downregulated |
| 0,100783853 | 0,558491096 | UNK | FALSE | Upregulated |
| -0,156012553 | 0,558618645 | CLMN | FALSE | Downregulated |
| -0,100083597 | 0,558618645 | NFRKB | FALSE | Downregulated |
| 0,104075323 | 0,558653033 | UCK2 | FALSE | Upregulated |
| -0,175328286 | 0,558712885 | DAAM2 | FALSE | Downregulated |
| -0,130462882 | 0,558712885 | NOP9 | FALSE | Downregulated |
| -0,591091055 | 0,558849523 | LOC101927879 | FALSE | Downregulated |
| 0,179777596 | 0,55901092 | AP001258.4 | FALSE | Upregulated |
| 0,142390163 | 0,559088958 | RPS27 | FALSE | Upregulated |
| -0,140828001 | 0,55914469 | MEIS1 | FALSE | Downregulated |
| -0,375306616 | 0,559415324 | LOC101927144 | FALSE | Downregulated |
| 0,488618905 | 0,559424964 | TMEM200A | FALSE | Upregulated |
| -0,362577057 | 0,559574432 | DTX2P1-UPK3BP1-PMS2P11 | FALSE | Downregulated |
| 0,081247636 | 0,559684747 | DYNC1H1 | FALSE | Upregulated |
| -0,09802184 | 0,559684747 | SUN1 | FALSE | Downregulated |
| -0,221878511 | 0,559815162 | CX3CR1 | FALSE | Downregulated |
| -1,149852685 | 0,559815162 | LOC102724865 | FALSE | Downregulated |
| -0,754593535 | 0,559815162 | LOC441528 | FALSE | Downregulated |
| -0,101877767 | 0,559815162 | METTL18 | FALSE | Downregulated |
| -0,184733095 | 0,559815162 | PRPF38B | FALSE | Downregulated |
| -1,491073752 | 0,559815162 | RBFOX1 | FALSE | Downregulated |
| 0,098164255 | 0,559815162 | WDR45 | FALSE | Upregulated |
| -0,808152048 | 0,55984335 | ASB11 | FALSE | Downregulated |
| 0,125628874 | 0,55984335 | ULK3 | FALSE | Upregulated |
| 0,17398907 | 0,55984335 | WDR25 | FALSE | Upregulated |
| 0,094603993 | 0,560009557 | CCNH | FALSE | Upregulated |
| 0,685146115 | 0,560009557 | CDH15 | FALSE | Upregulated |

| | | | | |
|---|---|---|---|---|
| 0,203366747 | 0,560009557 | TTC21A | FALSE | Upregulated |
| 0,193942607 | 0,560047781 | ARMC2 | FALSE | Upregulated |
| -0,145755095 | 0,560047781 | CCDC69 | FALSE | Downregulated |
| 0,093884828 | 0,560047781 | LRP3 | FALSE | Upregulated |
| -0,700520093 | 0,560047781 | PRCD | FALSE | Downregulated |
| -0,07447276 | 0,560047781 | SUPV3L1 | FALSE | Downregulated |
| -0,138128235 | 0,560047781 | XDH | FALSE | Downregulated |
| -0,103202024 | 0,560048944 | FARS2 | FALSE | Downregulated |
| 0,433063765 | 0,560048944 | RBMS3-AS3 | FALSE | Upregulated |
| 0,076964729 | 0,560048944 | TSR1 | FALSE | Upregulated |
| -0,143098852 | 0,560081112 | ASAP2 | FALSE | Downregulated |
| -0,121242876 | 0,560081112 | PLAGL2 | FALSE | Downregulated |
| 0,144578508 | 0,560247313 | KCTD13 | FALSE | Upregulated |
| 2,378502781 | 0,560247313 | SIK1 | FALSE | Upregulated |
| -0,481345696 | 0,560376428 | FLJ43681 | FALSE | Downregulated |
| 0,103212322 | 0,560488923 | C1orf50 | FALSE | Upregulated |
| -1,077286257 | 0,560488923 | HAPLN2 | FALSE | Downregulated |
| -0,824101132 | 0,560488923 | LOC100130238 | FALSE | Downregulated |
| -0,515521402 | 0,560488923 | LOC102724026 | FALSE | Downregulated |
| -0,120466545 | 0,560488923 | UBL3 | FALSE | Downregulated |
| -0,072595504 | 0,560488923 | ZNF800 | FALSE | Downregulated |
| -0,383977977 | 0,560782221 | PRKAA2 | FALSE | Downregulated |
| -0,073809158 | 0,561032931 | UBA5 | FALSE | Downregulated |
| 0,553391944 | 0,561133516 | LOC100996634 | FALSE | Upregulated |
| 0,220476841 | 0,561183826 | ALDOC | FALSE | Upregulated |
| -0,138004774 | 0,561264999 | C4BPB | FALSE | Downregulated |
| -0,248687377 | 0,561551798 | TRPC5 | FALSE | Downregulated |
| -0,104712951 | 0,561595864 | CEP104 | FALSE | Downregulated |
| 0,07870673 | 0,561838458 | TXNDC11 | FALSE | Upregulated |
| -0,116750281 | 0,562016872 | EDEM1 | FALSE | Downregulated |
| -0,104890063 | 0,562447666 | NCOA6 | FALSE | Downregulated |
| -0,259346965 | 0,562447666 | TMPPE | FALSE | Downregulated |
| -0,080104754 | 0,562688785 | APPBP2 | FALSE | Downregulated |
| -0,388206853 | 0,562823239 | NEUROD2 | FALSE | Downregulated |
| -0,872370801 | 0,563170753 | MOG | FALSE | Downregulated |
| -0,102362674 | 0,563170753 | PLEC | FALSE | Downregulated |
| -0,092474879 | 0,56334603 | CAAP1 | FALSE | Downregulated |
| -0,081231584 | 0,563471233 | BCL2L13 | FALSE | Downregulated |
| -0,817443392 | 0,563516881 | PTPRVP | FALSE | Downregulated |
| -0,201477233 | 0,563604872 | DHRS4L2 | FALSE | Downregulated |
| -0,135540503 | 0,563604872 | GPLD1 | FALSE | Downregulated |
| 1,05178079 | 0,563604872 | LINC01186 | FALSE | Upregulated |
| 1,919769463 | 0,563604872 | RNU1-28P | FALSE | Upregulated |
| -0,115020234 | 0,563718332 | C16orf45 | FALSE | Downregulated |
| -0,099125627 | 0,563718332 | CACTIN | FALSE | Downregulated |
| 0,166029004 | 0,563718332 | FAM171A1 | FALSE | Upregulated |
| -0,560351172 | 0,563718332 | LOC101928149 | FALSE | Downregulated |
| 0,144203847 | 0,563718332 | MYO6 | FALSE | Upregulated |
| -0,551194978 | 0,563779602 | DNAH5 | FALSE | Downregulated |
| 0,185777061 | 0,56385581 | FAM198B | FALSE | Upregulated |
| 0,112230441 | 0,564230731 | CDKAL1 | FALSE | Upregulated |

| | | | | |
|---|---|---|---|---|
| -0,210569231 | 0,564451802 | BCAT2 | FALSE | Downregulated |
| 0,248149772 | 0,564451802 | C1orf226 | FALSE | Upregulated |
| -0,600536994 | 0,564451802 | EMR3 | FALSE | Downregulated |
| -0,442531877 | 0,564451802 | KLHL23 | FALSE | Downregulated |
| -0,754478277 | 0,564451802 | LIMS3L | FALSE | Downregulated |
| -0,432324185 | 0,564586656 | AC009473.1 | FALSE | Downregulated |
| -0,487572638 | 0,564659019 | LOC100294362 | FALSE | Downregulated |
| 0,243569136 | 0,564659019 | PRRG2 | FALSE | Upregulated |
| 0,178100551 | 0,564659019 | TLR6 | FALSE | Upregulated |
| 0,221495545 | 0,564737938 | COL3A1 | FALSE | Upregulated |
| 0,09228941 | 0,564737938 | NCOR2 | FALSE | Upregulated |
| -0,353624352 | 0,564737938 | SLC34A2 | FALSE | Downregulated |
| 0,289966419 | 0,564864412 | SLC24A4 | FALSE | Upregulated |
| -0,156573828 | 0,565108404 | CASKIN2 | FALSE | Downregulated |
| 0,692212921 | 0,565120515 | PCBP2-OT1 | FALSE | Upregulated |
| -0,095476519 | 0,565128755 | ANO6 | FALSE | Downregulated |
| 0,772402214 | 0,5651941 | CNGB1 | FALSE | Upregulated |
| -0,31480556 | 0,5651941 | GOLGA6B | FALSE | Downregulated |
| -0,309187081 | 0,565215195 | BSPRY | FALSE | Downregulated |
| -0,527463101 | 0,565215195 | C9orf169 | FALSE | Downregulated |
| -0,118138798 | 0,565215195 | CBX6 | FALSE | Downregulated |
| 0,2441672 | 0,565215195 | CRISPLD2 | FALSE | Upregulated |
| -0,800531478 | 0,565215195 | FGF9 | FALSE | Downregulated |
| 0,175679212 | 0,565215195 | FLYWCH2 | FALSE | Upregulated |
| 0,220357598 | 0,565215195 | PLEKHG6 | FALSE | Upregulated |
| -0,251280548 | 0,565215195 | SPATA6 | FALSE | Downregulated |
| 0,080213383 | 0,565215195 | SPTY2D1 | FALSE | Upregulated |
| 0,158724555 | 0,565273245 | EMC6 | FALSE | Upregulated |
| 0,140710603 | 0,565273245 | HIVEP2 | FALSE | Upregulated |
| -0,103223776 | 0,565387075 | ZNF516 | FALSE | Downregulated |
| 0,674775281 | 0,565428574 | ALG1L | FALSE | Upregulated |
| 0,319815099 | 0,565588876 | NBPF25P | FALSE | Upregulated |
| 0,073205072 | 0,565765478 | CD46 | FALSE | Upregulated |
| -0,061112547 | 0,565765478 | DYNC1LI2 | FALSE | Downregulated |
| -0,339956412 | 0,565765478 | NPNT | FALSE | Downregulated |
| -0,346000486 | 0,565765478 | PRRT3-AS1 | FALSE | Downregulated |
| -0,377821346 | 0,565987603 | SLC29A2 | FALSE | Downregulated |
| 0,379656171 | 0,566048691 | NPW | FALSE | Upregulated |
| -0,124910943 | 0,566181399 | FAM98C | FALSE | Downregulated |
| -0,179759769 | 0,566208306 | GPR89A | FALSE | Downregulated |
| 0,691460803 | 0,566257887 | RNF144A-AS1 | FALSE | Upregulated |
| 0,140209088 | 0,566368316 | LOC100862671 | FALSE | Upregulated |
| -0,105790693 | 0,566368316 | TSFM | FALSE | Downregulated |
| -0,742012597 | 0,566685872 | LINC01135 | FALSE | Downregulated |
| -0,622628748 | 0,566932218 | LOC102724028 | FALSE | Downregulated |
| -0,460597127 | 0,566976649 | AC009501.4 | FALSE | Downregulated |
| 0,127248251 | 0,566976649 | LRRC14 | FALSE | Upregulated |
| 0,159218433 | 0,566976649 | VPS53 | FALSE | Upregulated |
| -0,167945962 | 0,566976649 | ZXDA | FALSE | Downregulated |
| -0,116071049 | 0,567130896 | CPOX | FALSE | Downregulated |
| 0,678592491 | 0,567401351 | LOC100288748 | FALSE | Upregulated |

| | | | | |
|---|---|---|---|---|
| 0,100717921 | 0,567401351 | ZSCAN25 | FALSE | Upregulated |
| -0,391575806 | 0,56751179 | PRTG | FALSE | Downregulated |
| -0,105354789 | 0,567683744 | PRPF4B | FALSE | Downregulated |
| -0,266998492 | 0,567743955 | LOC101928498 | FALSE | Downregulated |
| 0,235958866 | 0,5681405 | LOC101929959 | FALSE | Upregulated |
| -0,223173932 | 0,5681405 | RP11-384K6.6 | FALSE | Downregulated |
| -0,206115069 | 0,568216373 | LOC100505942 | FALSE | Downregulated |
| -0,114465043 | 0,568397839 | JMY | FALSE | Downregulated |
| -0,069612326 | 0,568648816 | PRKCI | FALSE | Downregulated |
| 0,097336427 | 0,568663964 | APOE | FALSE | Upregulated |
| 0,243683672 | 0,568663964 | B9D2 | FALSE | Upregulated |
| -0,06898894 | 0,568663964 | DDX47 | FALSE | Downregulated |
| 0,623445215 | 0,568663964 | INPP5J | FALSE | Upregulated |
| 0,152064835 | 0,568663964 | KIF20B | FALSE | Upregulated |
| -0,168237703 | 0,568663964 | LINC00649 | FALSE | Downregulated |
| -0,214597806 | 0,568663964 | MICA | FALSE | Downregulated |
| 0,109985071 | 0,568663964 | MRPS2 | FALSE | Upregulated |
| 0,373518547 | 0,568663964 | NKD2 | FALSE | Upregulated |
| 0,196237363 | 0,568663964 | PSAT1 | FALSE | Upregulated |
| -0,139033156 | 0,568663964 | TF | FALSE | Downregulated |
| -0,186991672 | 0,56915429 | ADH1C | FALSE | Downregulated |
| -0,330960192 | 0,569163922 | SFRP5 | FALSE | Downregulated |
| -0,245356037 | 0,569244964 | ARIH2OS | FALSE | Downregulated |
| -0,094540822 | 0,569244964 | ZFP90 | FALSE | Downregulated |
| -0,175413841 | 0,569339035 | PARD6G | FALSE | Downregulated |
| 0,102207155 | 0,569395127 | ZNF282 | FALSE | Upregulated |
| 0,102000574 | 0,569510002 | AARSD1 | FALSE | Upregulated |
| -0,392699363 | 0,569510002 | CAHM | FALSE | Downregulated |
| -0,543677452 | 0,569521693 | HDC | FALSE | Downregulated |
| 0,329983032 | 0,569547428 | ARHGAP33 | FALSE | Upregulated |
| -0,172256459 | 0,56956759 | APBB1IP | FALSE | Downregulated |
| 0,085466262 | 0,569696315 | SLC9A8 | FALSE | Upregulated |
| -0,47763358 | 0,569906949 | METTL21EP | FALSE | Downregulated |
| -0,258192225 | 0,569906949 | MRGPRF | FALSE | Downregulated |
| -0,1663732 | 0,570064439 | CXCL14 | FALSE | Downregulated |
| -0,128484137 | 0,570064439 | RASAL2 | FALSE | Downregulated |
| 0,083318275 | 0,570086057 | R3HCC1L | FALSE | Upregulated |
| -0,386694682 | 0,570167887 | LOC102723467 | FALSE | Downregulated |
| 0,101291595 | 0,570201802 | LRRC28 | FALSE | Upregulated |
| -0,190871369 | 0,5702601 | AXIN2 | FALSE | Downregulated |
| -0,188018382 | 0,5702601 | C1orf159 | FALSE | Downregulated |
| 0,133889884 | 0,5702601 | DUS3L | FALSE | Upregulated |
| -0,356702351 | 0,5702601 | ITGB8 | FALSE | Downregulated |
| 0,859161606 | 0,5702601 | LINC00891 | FALSE | Upregulated |
| -0,082805071 | 0,5702601 | RPS6KC1 | FALSE | Downregulated |
| -0,272068919 | 0,5702601 | SALL2 | FALSE | Downregulated |
| 0,788043099 | 0,570285853 | LOC101929047 | FALSE | Upregulated |
| 0,871424943 | 0,570350451 | PTPRR | FALSE | Upregulated |
| 0,098161871 | 0,570350451 | RPS6KB2 | FALSE | Upregulated |
| -0,141228298 | 0,570527842 | SUGP2 | FALSE | Downregulated |
| 0,131058198 | 0,570574523 | WDR18 | FALSE | Upregulated |

| | | | | |
|---|---|---|---|---|
| -0,193843276 | 0,570628221 | CFHR1 | FALSE | Downregulated |
| -0,102076416 | 0,570758863 | C2orf49 | FALSE | Downregulated |
| -0,263084501 | 0,570758863 | LOC283177 | FALSE | Downregulated |
| 0,100423025 | 0,570910094 | EEFSEC | FALSE | Upregulated |
| 0,23292515 | 0,571061846 | AMPD3 | FALSE | Upregulated |
| 0,10522047 | 0,571127197 | TMEM134 | FALSE | Upregulated |
| -0,134486226 | 0,571217342 | BDP1 | FALSE | Downregulated |
| 0,068038425 | 0,571334755 | FAM105B | FALSE | Upregulated |
| 1,001772976 | 0,571334755 | LOC102724642 | FALSE | Upregulated |
| 0,449942983 | 0,571624166 | MGC39372 | FALSE | Upregulated |
| 0,085040157 | 0,571865058 | UCHL5 | FALSE | Upregulated |
| 0,106681937 | 0,571976967 | ADIPOR2 | FALSE | Upregulated |
| -0,368023935 | 0,571981319 | DSCAM | FALSE | Downregulated |
| 0,068060762 | 0,572002531 | UBE2J2 | FALSE | Upregulated |
| -0,141228761 | 0,572034466 | LOC100506634 | FALSE | Downregulated |
| -0,132397066 | 0,572034466 | ZNF354A | FALSE | Downregulated |
| 0,530154784 | 0,572042961 | LOC101927789 | FALSE | Upregulated |
| -0,23328331 | 0,57222614 | ARHGEF26 | FALSE | Downregulated |
| -0,835574335 | 0,57222614 | LOC101927594 | FALSE | Downregulated |
| -0,878130417 | 0,57222614 | NXNL2 | FALSE | Downregulated |
| -0,664311849 | 0,57235158 | LOC100287387 | FALSE | Downregulated |
| -0,554660928 | 0,57235158 | LOC102724379 | FALSE | Downregulated |
| 0,07910679 | 0,57235158 | NF2 | FALSE | Upregulated |
| -0,163117261 | 0,57235158 | PER2 | FALSE | Downregulated |
| 0,449163992 | 0,57235158 | RHEBL1 | FALSE | Upregulated |
| -0,396873134 | 0,57235158 | SOCS2 | FALSE | Downregulated |
| -0,232304282 | 0,57235158 | TECTA | FALSE | Downregulated |
| -0,278732963 | 0,57235158 | TTLL7 | FALSE | Downregulated |
| -0,109315819 | 0,57235158 | TVP23B | FALSE | Downregulated |
| 0,559639324 | 0,57235641 | LOC100996273 | FALSE | Upregulated |
| -0,654978848 | 0,572432722 | LOC101926894 | FALSE | Downregulated |
| -0,125826779 | 0,572452338 | NDUFC2 | FALSE | Downregulated |
| -0,14161462 | 0,572570774 | APLF | FALSE | Downregulated |
| 0,158258739 | 0,572668271 | DHX34 | FALSE | Upregulated |
| -0,843740438 | 0,572668271 | DPP10-AS1 | FALSE | Downregulated |
| 0,516372554 | 0,572668271 | LOC101929070 | FALSE | Upregulated |
| 0,130672272 | 0,572763559 | TOP1 | FALSE | Upregulated |
| 0,34706225 | 0,572780843 | LOC100288637 | FALSE | Upregulated |
| 0,185359847 | 0,572780843 | ZNF611 | FALSE | Upregulated |
| -0,29473885 | 0,572822256 | LOC101927287 | FALSE | Downregulated |
| -0,30738191 | 0,573069362 | LOC100233156 | FALSE | Downregulated |
| 0,578076 | 0,573069362 | LOC101927026 | FALSE | Upregulated |
| -0,174998981 | 0,573295386 | CDKN1C | FALSE | Downregulated |
| 0,127278023 | 0,573295386 | SRR | FALSE | Upregulated |
| -0,115323857 | 0,573328331 | HNMT | FALSE | Downregulated |
| -0,156796413 | 0,57337756 | KIAA0754 | FALSE | Downregulated |
| -0,327232395 | 0,573392384 | LOC100996741 | FALSE | Downregulated |
| 0,072032155 | 0,573428697 | PLOD1 | FALSE | Upregulated |
| -0,224190251 | 0,57352047 | RPS6KA5 | FALSE | Downregulated |
| -0,202408887 | 0,573537148 | G0S2 | FALSE | Downregulated |
| -0,399059413 | 0,573552894 | MTL5 | FALSE | Downregulated |

| | | | | |
|---|---|---|---|---|
| 0,077579976 | 0,573552894 | TXNL1 | FALSE | Upregulated |
| 0,475793404 | 0,57359352 | AC058791.1 | FALSE | Upregulated |
| 0,70950515 | 0,57359352 | ADPGK-AS1 | FALSE | Upregulated |
| -0,341080698 | 0,57359352 | ALMS1P | FALSE | Downregulated |
| 0,684457014 | 0,57359352 | APLP1 | FALSE | Upregulated |
| 0,06302434 | 0,57359352 | CDC37 | FALSE | Upregulated |
| 0,108609267 | 0,57359352 | CDIPT | FALSE | Upregulated |
| -0,057355664 | 0,57359352 | CNOT8 | FALSE | Downregulated |
| 0,261193313 | 0,57359352 | DNMBP-AS1 | FALSE | Upregulated |
| 0,465983097 | 0,57359352 | FAM179A | FALSE | Upregulated |
| 0,114696472 | 0,57359352 | GNPAT | FALSE | Upregulated |
| -0,072966596 | 0,57359352 | HBS1L | FALSE | Downregulated |
| 0,33845115 | 0,57359352 | ICA1 | FALSE | Upregulated |
| -0,707205639 | 0,57359352 | LINC00348 | FALSE | Downregulated |
| -0,426124534 | 0,57359352 | LOC101928706 | FALSE | Downregulated |
| -0,998616681 | 0,57359352 | LOC101929038 | FALSE | Downregulated |
| 0,105832833 | 0,57359352 | NDUFS5 | FALSE | Upregulated |
| -0,141960731 | 0,57359352 | PLD1 | FALSE | Downregulated |
| 0,361704381 | 0,57359352 | RP11-408H20.2 | FALSE | Upregulated |
| -0,454843519 | 0,57359352 | RP11-486O12.2 | FALSE | Downregulated |
| -0,097012948 | 0,57359352 | SMC5 | FALSE | Downregulated |
| -0,255274758 | 0,57359352 | SNHG15 | FALSE | Downregulated |
| 0,368961737 | 0,57359352 | SPAG4 | FALSE | Upregulated |
| -0,571726181 | 0,57359352 | TEKT4 | FALSE | Downregulated |
| -0,119281602 | 0,57359352 | THAP2 | FALSE | Downregulated |
| 0,086097151 | 0,57359352 | TROVE2 | FALSE | Upregulated |
| 0,106591387 | 0,57359352 | UNG | FALSE | Upregulated |
| 0,197988605 | 0,57400388 | COL4A2 | FALSE | Upregulated |
| 0,103376358 | 0,57400388 | PXK | FALSE | Upregulated |
| -0,103819063 | 0,574043907 | IFT57 | FALSE | Downregulated |
| -0,094340531 | 0,574043907 | METTL3 | FALSE | Downregulated |
| -0,100894732 | 0,574043907 | RNF138 | FALSE | Downregulated |
| 0,718500784 | 0,574253053 | LOC102723733 | FALSE | Upregulated |
| 0,325022405 | 0,57427627 | C9orf116 | FALSE | Upregulated |
| 0,255981722 | 0,57427627 | CIDECP | FALSE | Upregulated |
| 0,082861549 | 0,574410762 | DPY19L1 | FALSE | Upregulated |
| -0,769976303 | 0,574410762 | LOC102724681 | FALSE | Downregulated |
| -0,087731207 | 0,574572877 | MTMR1 | FALSE | Downregulated |
| -0,115941639 | 0,574831238 | JHDM1D-AS1 | FALSE | Downregulated |
| -0,070698391 | 0,575001514 | VEZF1 | FALSE | Downregulated |
| 0,319495388 | 0,57512849 | ACKR3 | FALSE | Upregulated |
| 0,097333218 | 0,57512849 | DYRK4 | FALSE | Upregulated |
| -0,129595382 | 0,57512849 | FANCL | FALSE | Downregulated |
| -0,114674217 | 0,57512849 | PROSER1 | FALSE | Downregulated |
| 0,386828564 | 0,57512849 | PTP4A3 | FALSE | Upregulated |
| -0,345634325 | 0,57512849 | RP1-232P20.1 | FALSE | Downregulated |
| 0,682484249 | 0,57512849 | TSKS | FALSE | Upregulated |
| -0,147484195 | 0,57512849 | VAMP2 | FALSE | Downregulated |
| -0,710786676 | 0,575193849 | LINC00707 | FALSE | Downregulated |
| 0,389558324 | 0,575196389 | LOC101929115 | FALSE | Upregulated |
| -0,155049779 | 0,575202079 | MPND | FALSE | Downregulated |

| | | | | |
|---|---|---|---|---|
| 0,090129717 | 0,575202079 | TGFBRAP1 | FALSE | Upregulated |
| -0,103571584 | 0,575297478 | HIPK2 | FALSE | Downregulated |
| 0,077377702 | 0,575341757 | EXOSC8 | FALSE | Upregulated |
| 0,184869567 | 0,57537064 | TMEM241 | FALSE | Upregulated |
| -0,082042849 | 0,575409101 | DIDO1 | FALSE | Downregulated |
| -0,35704097 | 0,575409101 | LINC00685 | FALSE | Downregulated |
| 0,390209666 | 0,57553374 | LOC101928806 | FALSE | Upregulated |
| -0,053578912 | 0,57553374 | UFSP2 | FALSE | Downregulated |
| 0,29379747 | 0,575630617 | LOC102724330 | FALSE | Upregulated |
| -0,695136296 | 0,575696723 | KCNA5 | FALSE | Downregulated |
| 0,141167973 | 0,575712309 | SPATS2 | FALSE | Upregulated |
| 0,455390886 | 0,575874926 | AC072062.1 | FALSE | Upregulated |
| 0,067071707 | 0,576028662 | TRIP12 | FALSE | Upregulated |
| -0,105768777 | 0,576256244 | THRAP3 | FALSE | Downregulated |
| -0,1627781 | 0,576278291 | ELFN1 | FALSE | Downregulated |
| -0,637686865 | 0,576646877 | LOC100996643 | FALSE | Downregulated |
| -0,612724867 | 0,576760332 | LOC101929098 | FALSE | Downregulated |
| -0,765134832 | 0,576900608 | DYNAP | FALSE | Downregulated |
| -0,623901087 | 0,576900608 | MED12L | FALSE | Downregulated |
| 0,334860023 | 0,57700499 | NANOS1 | FALSE | Upregulated |
| 0,084424361 | 0,577006474 | SLC35D2 | FALSE | Upregulated |
| -0,17079112 | 0,577227773 | MYL9 | FALSE | Downregulated |
| 0,184709897 | 0,577262732 | PDP2 | FALSE | Upregulated |
| 0,099916827 | 0,577262732 | PRNP | FALSE | Upregulated |
| -0,076201315 | 0,577317913 | DNAJA3 | FALSE | Downregulated |
| -0,132796675 | 0,577481496 | RSBN1L-AS1 | FALSE | Downregulated |
| 0,155625248 | 0,577495138 | ZNF426 | FALSE | Upregulated |
| 0,769242168 | 0,577724276 | CCNI2 | FALSE | Upregulated |
| -0,195524244 | 0,577727555 | LOC388692 | FALSE | Downregulated |
| 0,543739457 | 0,577814243 | CARD14 | FALSE | Upregulated |
| -0,155645572 | 0,577978228 | PRSS23 | FALSE | Downregulated |
| 0,223193209 | 0,578252966 | ANGPT1 | FALSE | Upregulated |
| 0,329698915 | 0,578338721 | ZNF257 | FALSE | Upregulated |
| -0,091217926 | 0,578372516 | TRMT12 | FALSE | Downregulated |
| -0,115271851 | 0,578378672 | ABCB10 | FALSE | Downregulated |
| 0,553185623 | 0,578382232 | LRRN4 | FALSE | Upregulated |
| 0,510490444 | 0,578818401 | LOC100134868 | FALSE | Upregulated |
| 0,467563536 | 0,578843649 | HOXC4 | FALSE | Upregulated |
| -0,35940716 | 0,578843649 | LOC101929670 | FALSE | Downregulated |
| 0,471982886 | 0,578856126 | HAS2 | FALSE | Upregulated |
| -0,370949528 | 0,57887569 | GOLGA6L4 | FALSE | Downregulated |
| 0,733188432 | 0,57887569 | LOC101927811 | FALSE | Upregulated |
| 0,149091105 | 0,57887569 | RPL9 | FALSE | Upregulated |
| 0,649907681 | 0,57887569 | TLDC2 | FALSE | Upregulated |
| -1,115646604 | 0,578878176 | TMOD4 | FALSE | Downregulated |
| -0,111289875 | 0,578921017 | NELFA | FALSE | Downregulated |
| -0,553344765 | 0,578921017 | RP11-333A23.4 | FALSE | Downregulated |
| -0,50128255 | 0,579055569 | CYP2A13 | FALSE | Downregulated |
| -0,698908114 | 0,579068073 | LOC101929295 | FALSE | Downregulated |
| -0,678521223 | 0,579155926 | GIPR | FALSE | Downregulated |
| 0,734477056 | 0,579236983 | GXYLT2 | FALSE | Upregulated |

| | | | | |
|---|---|---|---|---|
| -0,349835065 | 0,579423381 | LOC101927226 | FALSE | Downregulated |
| 0,125139362 | 0,579503738 | EXTL3 | FALSE | Upregulated |
| -0,200929875 | 0,579570796 | CHIC1 | FALSE | Downregulated |
| -0,085880427 | 0,579641675 | CPN2 | FALSE | Downregulated |
| 0,129369504 | 0,579785405 | MGST2 | FALSE | Upregulated |
| 0,411135899 | 0,579903601 | CCDC89 | FALSE | Upregulated |
| -0,576974874 | 0,579903601 | PTPRQ | FALSE | Downregulated |
| 0,116837902 | 0,580266166 | ACTR3C | FALSE | Upregulated |
| -0,416569781 | 0,580266166 | HR | FALSE | Downregulated |
| -0,095127472 | 0,580266166 | WDR6 | FALSE | Downregulated |
| 0,642439729 | 0,580279886 | LOC102723746 | FALSE | Upregulated |
| -0,458100355 | 0,580279886 | MROH2A | FALSE | Downregulated |
| -0,529287166 | 0,580446627 | LOC102724965 | FALSE | Downregulated |
| -0,594860658 | 0,580522012 | LOC100130417 | FALSE | Downregulated |
| -0,751442499 | 0,580561586 | LOC102723750 | FALSE | Downregulated |
| -0,521436142 | 0,580561586 | LOC102723993 | FALSE | Downregulated |
| 0,184925769 | 0,580561586 | ZNF749 | FALSE | Upregulated |
| 0,049131607 | 0,580606893 | SH3GLB1 | FALSE | Upregulated |
| -0,169043439 | 0,580681059 | MLLT3 | FALSE | Downregulated |
| 0,082877351 | 0,580994785 | H3F3AP4 | FALSE | Upregulated |
| -0,540539872 | 0,581120201 | IGF2-AS | FALSE | Downregulated |
| -0,119958895 | 0,581564502 | CPT2 | FALSE | Downregulated |
| 0,460793708 | 0,581977888 | BHLHA15 | FALSE | Upregulated |
| -0,162312828 | 0,582101594 | RARG | FALSE | Downregulated |
| -0,650483242 | 0,582158421 | NRN1L | FALSE | Downregulated |
| -0,182596054 | 0,582210601 | C5orf54 | FALSE | Downregulated |
| 0,457274939 | 0,582247519 | LOC102724615 | FALSE | Upregulated |
| -0,202707318 | 0,582247519 | ZNF431 | FALSE | Downregulated |
| 0,275245266 | 0,582398821 | VNN2 | FALSE | Upregulated |
| -0,201401219 | 0,582593018 | PPP1R3B | FALSE | Downregulated |
| -0,164295907 | 0,582687041 | AHI1 | FALSE | Downregulated |
| 0,079352224 | 0,582687041 | CDC42EP4 | FALSE | Upregulated |
| -0,122550319 | 0,582687041 | PIK3R4 | FALSE | Downregulated |
| 0,545066899 | 0,58269043 | LOC101929426 | FALSE | Upregulated |
| 0,572327408 | 0,582698123 | ADM5 | FALSE | Upregulated |
| 0,180056893 | 0,582752054 | SYNGAP1 | FALSE | Upregulated |
| -0,108887017 | 0,582855483 | MIA3 | FALSE | Downregulated |
| 0,104292797 | 0,582855483 | PPP1R12C | FALSE | Upregulated |
| 0,113521975 | 0,582855483 | PPP1R16A | FALSE | Upregulated |
| 0,094997689 | 0,583124162 | CDK11B | FALSE | Upregulated |
| -0,093626208 | 0,583124162 | RBM26 | FALSE | Downregulated |
| 0,308415551 | 0,583428765 | GALNT12 | FALSE | Upregulated |
| -0,090605732 | 0,583428765 | ZNF747 | FALSE | Downregulated |
| -0,094711623 | 0,583630702 | ATP2B1 | FALSE | Downregulated |
| 0,199367048 | 0,583647736 | PRR26 | FALSE | Upregulated |
| 0,101911577 | 0,584113134 | OTUD6B | FALSE | Upregulated |
| 0,104714157 | 0,584178997 | COMMD6 | FALSE | Upregulated |
| 0,637309032 | 0,584178997 | PRR7 | FALSE | Upregulated |
| -0,45658703 | 0,584397966 | LOC100505570 | FALSE | Downregulated |
| -0,210006665 | 0,584500177 | TLE3 | FALSE | Downregulated |
| 0,696664928 | 0,58451682 | CLDN19 | FALSE | Upregulated |

| | | | | |
|---|---|---|---|---|
| 0,067279706 | 0,584610235 | SRSF9 | FALSE | Upregulated |
| -0,124658148 | 0,584947476 | ZNF778 | FALSE | Downregulated |
| 0,13041166 | 0,585025358 | CASP9 | FALSE | Upregulated |
| -0,102748085 | 0,585058158 | IFFO1 | FALSE | Downregulated |
| 0,079276336 | 0,585066496 | PAWR | FALSE | Upregulated |
| 0,111465991 | 0,585093295 | PLEKHA2 | FALSE | Upregulated |
| 0,064141292 | 0,585097075 | ATP2C1 | FALSE | Upregulated |
| -0,441487709 | 0,585097075 | LOC101928946 | FALSE | Downregulated |
| -0,761071296 | 0,585123087 | EMX2 | FALSE | Downregulated |
| 0,199537968 | 0,585123087 | GAS5 | FALSE | Upregulated |
| -0,599663971 | 0,585123087 | ITPR1-AS1 | FALSE | Downregulated |
| -0,176787367 | 0,585188245 | SLC41A1 | FALSE | Downregulated |
| -0,215650636 | 0,585201825 | RP5-968D22.1 | FALSE | Downregulated |
| 0,146483426 | 0,58528372 | ATAT1 | FALSE | Upregulated |
| -0,238167897 | 0,585332431 | C1orf210 | FALSE | Downregulated |
| 0,141777461 | 0,585332431 | SNRNP27 | FALSE | Upregulated |
| -0,178131805 | 0,585332431 | TAF3 | FALSE | Downregulated |
| -0,155474803 | 0,585389096 | SRRM1 | FALSE | Downregulated |
| 0,1642871 | 0,585400521 | BBS5 | FALSE | Upregulated |
| 0,141195 | 0,585432743 | EIF3J | FALSE | Upregulated |
| -0,382554176 | 0,585432743 | PLXNB3 | FALSE | Downregulated |
| 0,085108902 | 0,585432743 | SHC1 | FALSE | Upregulated |
| 0,11264991 | 0,585432743 | SLC46A3 | FALSE | Upregulated |
| -0,109697967 | 0,585432743 | STRADB | FALSE | Downregulated |
| 0,289188312 | 0,585499547 | NECAB2 | FALSE | Upregulated |
| 0,067602494 | 0,585530448 | WIPF2 | FALSE | Upregulated |
| -0,522365159 | 0,58563233 | CELF4 | FALSE | Downregulated |
| -0,130243723 | 0,58563233 | MINPP1 | FALSE | Downregulated |
| 0,047199003 | 0,58563233 | PWP1 | FALSE | Upregulated |
| 0,119491717 | 0,585657972 | GMEB1 | FALSE | Upregulated |
| 0,081994539 | 0,585657972 | LMBR1 | FALSE | Upregulated |
| -0,302173036 | 0,585657972 | LOC100996457 | FALSE | Downregulated |
| 0,0875024 | 0,585951509 | CYB5B | FALSE | Upregulated |
| -0,229816458 | 0,585951509 | RTEL1 | FALSE | Downregulated |
| 0,09347396 | 0,586085342 | ZBTB4 | FALSE | Upregulated |
| -0,338991412 | 0,586131422 | C2orf40 | FALSE | Downregulated |
| 0,09991062 | 0,586354353 | PDHB | FALSE | Upregulated |
| 0,292108669 | 0,586452213 | EVC | FALSE | Upregulated |
| 0,544239754 | 0,586731336 | FAM19A1 | FALSE | Upregulated |
| -0,1423253 | 0,586786492 | DDC | FALSE | Downregulated |
| -0,07104088 | 0,586786492 | ZCCHC3 | FALSE | Downregulated |
| 0,09417437 | 0,587331354 | DDX49 | FALSE | Upregulated |
| 0,186976143 | 0,587331354 | NCOA7 | FALSE | Upregulated |
| 0,841198726 | 0,587331354 | RP11-148L24.1 | FALSE | Upregulated |
| -0,193507182 | 0,587360709 | ABCA3 | FALSE | Downregulated |
| -0,160117716 | 0,587360709 | FZD1 | FALSE | Downregulated |
| -0,324173701 | 0,587360709 | LINC00884 | FALSE | Downregulated |
| -0,368220883 | 0,587360709 | MLF1 | FALSE | Downregulated |
| 0,13443122 | 0,587360709 | PRAF2 | FALSE | Upregulated |
| -0,094634825 | 0,587360709 | PRMT9 | FALSE | Downregulated |
| 0,505678919 | 0,587360709 | RPL31P11 | FALSE | Upregulated |

| | | | | |
|---|---|---|---|---|
| -0,328783922 | 0,587360709 | SNHG9 | FALSE | Downregulated |
| 0,126239974 | 0,58736073 | MYO7A | FALSE | Upregulated |
| -0,109950743 | 0,587400605 | ARID3B | FALSE | Downregulated |
| 0,255629116 | 0,587400605 | KIF9-AS1 | FALSE | Upregulated |
| 0,153641661 | 0,587400605 | TMEM160 | FALSE | Upregulated |
| 0,294783813 | 0,587431186 | OR7E14P | FALSE | Upregulated |
| 0,085263563 | 0,587431186 | RNF13 | FALSE | Upregulated |
| 0,242548278 | 0,587505281 | FNTB | FALSE | Upregulated |
| -0,12333005 | 0,587759197 | ZNF542 | FALSE | Downregulated |
| 0,127495751 | 0,58783663 | MGST3 | FALSE | Upregulated |
| -0,368130851 | 0,58783663 | MIR17HG | FALSE | Downregulated |
| -0,076847788 | 0,58783663 | PKP4 | FALSE | Downregulated |
| 0,73404085 | 0,58783663 | UNC5A | FALSE | Upregulated |
| -0,144475247 | 0,588225467 | CYP4F2 | FALSE | Downregulated |
| 0,160187513 | 0,588350547 | CDKL5 | FALSE | Upregulated |
| -0,395483985 | 0,58841943 | LOC100507131 | FALSE | Downregulated |
| -0,616039268 | 0,58841943 | M1AP | FALSE | Downregulated |
| -0,05986686 | 0,58841943 | MED29 | FALSE | Downregulated |
| -0,231628597 | 0,58841943 | RAP1GAP | FALSE | Downregulated |
| 0,322631639 | 0,588474206 | TNFRSF25 | FALSE | Upregulated |
| -0,09208953 | 0,58854705 | ELP5 | FALSE | Downregulated |
| -0,081227989 | 0,588766256 | MIPEP | FALSE | Downregulated |
| -0,600820521 | 0,588835107 | RP4-640H8.2 | FALSE | Downregulated |
| 0,076519893 | 0,58888723 | CLASP2 | FALSE | Upregulated |
| -0,224279308 | 0,588922289 | CA14 | FALSE | Downregulated |
| -0,210346556 | 0,588922289 | LOC100506157 | FALSE | Downregulated |
| 0,212368379 | 0,589039108 | AFAP1 | FALSE | Upregulated |
| -0,074055283 | 0,589039108 | STRBP | FALSE | Downregulated |
| 0,054656567 | 0,58909266 | GRIPAP1 | FALSE | Upregulated |
| 0,138686858 | 0,589132677 | PIM3 | FALSE | Upregulated |
| -0,099484708 | 0,589152243 | AGBL5 | FALSE | Downregulated |
| 0,107904733 | 0,589152243 | LOC145783 | FALSE | Upregulated |
| -0,122864386 | 0,589176312 | DECR1 | FALSE | Downregulated |
| 0,069321185 | 0,589208607 | MOB1B | FALSE | Upregulated |
| -0,115733317 | 0,589211693 | ZDHHC21 | FALSE | Downregulated |
| 0,386884287 | 0,589354753 | NMB | FALSE | Upregulated |
| -0,982833472 | 0,589354753 | WNT10B | FALSE | Downregulated |
| -0,378053873 | 0,589385784 | MKX | FALSE | Downregulated |
| -0,07416997 | 0,589385784 | SAP18 | FALSE | Downregulated |
| -0,103240294 | 0,589385784 | ZNF264 | FALSE | Downregulated |
| -0,138850732 | 0,589393328 | RASSF7 | FALSE | Downregulated |
| -0,196953647 | 0,589393328 | RP11-736K20.6 | FALSE | Downregulated |
| -0,525711761 | 0,589531292 | EPB41L4A-AS2 | FALSE | Downregulated |
| -0,19736002 | 0,589542283 | LINC00476 | FALSE | Downregulated |
| 0,151472839 | 0,589784036 | FAM86C1 | FALSE | Upregulated |
| 0,14657566 | 0,589898941 | C19orf70 | FALSE | Upregulated |
| -0,496035412 | 0,589906269 | C5orf27 | FALSE | Downregulated |
| -0,188572255 | 0,589978984 | RAB11B-AS1 | FALSE | Downregulated |
| -0,185269152 | 0,590150447 | FCN2 | FALSE | Downregulated |
| -0,911723128 | 0,590195266 | OLFM4 | FALSE | Downregulated |
| -0,074235765 | 0,590195266 | SORBS3 | FALSE | Downregulated |

| | | | | |
|---|---|---|---|---|
| 0,099839989 | 0,590256865 | BBX | FALSE | Upregulated |
| 0,774416485 | 0,590256865 | FGF18 | FALSE | Upregulated |
| -0,187680816 | 0,590446016 | ADAT2 | FALSE | Downregulated |
| -0,129790346 | 0,590446016 | GAB2 | FALSE | Downregulated |
| 0,178205718 | 0,59064066 | INPP5E | FALSE | Upregulated |
| -0,143436915 | 0,59064066 | TMEM99 | FALSE | Downregulated |
| -0,087160319 | 0,590746153 | AGO1 | FALSE | Downregulated |
| 0,082847926 | 0,590751504 | GPC6 | FALSE | Upregulated |
| 0,141924245 | 0,590751504 | SUPT3H | FALSE | Upregulated |
| 0,092916413 | 0,590803409 | KIAA0586 | FALSE | Upregulated |
| 0,348973897 | 0,591038288 | ARPC4-TTLL3 | FALSE | Upregulated |
| -0,09218442 | 0,59108355 | JUP | FALSE | Downregulated |
| 0,116078653 | 0,591097754 | GIMAP8 | FALSE | Upregulated |
| 0,076318692 | 0,591097754 | HMG20B | FALSE | Upregulated |
| 0,333883804 | 0,591097754 | HOXA1 | FALSE | Upregulated |
| -0,737929603 | 0,591097754 | LGALS8-AS1 | FALSE | Downregulated |
| -0,325886916 | 0,591097754 | PCDH7 | FALSE | Downregulated |
| 0,075399033 | 0,591097754 | POGK | FALSE | Upregulated |
| 0,218455843 | 0,591097754 | SNHG12 | FALSE | Upregulated |
| -0,116369092 | 0,591097754 | TCEANC2 | FALSE | Downregulated |
| -0,144698773 | 0,591268086 | DDR2 | FALSE | Downregulated |
| 0,15459336 | 0,591342601 | GUCY1A3 | FALSE | Upregulated |
| -0,237109445 | 0,591634802 | KIFC2 | FALSE | Downregulated |
| 0,3158166 | 0,591634802 | SOD3 | FALSE | Upregulated |
| -0,17379704 | 0,591978313 | LOC728743 | FALSE | Downregulated |
| 0,165956264 | 0,591978313 | TTC39B | FALSE | Upregulated |
| 0,343219598 | 0,59200202 | COL5A3 | FALSE | Upregulated |
| 0,148321654 | 0,59200202 | DFNA5 | FALSE | Upregulated |
| -0,865237989 | 0,592009282 | CCL17 | FALSE | Downregulated |
| 0,664486042 | 0,592009282 | FCRL2 | FALSE | Upregulated |
| -1,017898956 | 0,592009282 | HNRNPKP3 | FALSE | Downregulated |
| -0,532369318 | 0,592009282 | MROH7-TTC4 | FALSE | Downregulated |
| 0,250873824 | 0,592009282 | MXRA8 | FALSE | Upregulated |
| -0,077536865 | 0,592019893 | CLDN1 | FALSE | Downregulated |
| -0,896180933 | 0,592019893 | CTAGE4 | FALSE | Downregulated |
| -0,335661377 | 0,592083002 | LOC100506036 | FALSE | Downregulated |
| 0,415924871 | 0,59209889 | CDH11 | FALSE | Upregulated |
| -0,481454384 | 0,59209889 | USP12-AS2 | FALSE | Downregulated |
| -0,312211695 | 0,592184726 | MTMR7 | FALSE | Downregulated |
| -0,117746114 | 0,592252704 | TSPYL4 | FALSE | Downregulated |
| 0,086747526 | 0,592252704 | WDR83OS | FALSE | Upregulated |
| -0,295307387 | 0,592290625 | TRNN | FALSE | Downregulated |
| 0,118989611 | 0,592312801 | LIN37 | FALSE | Upregulated |
| -0,492000255 | 0,592413745 | ZNF177 | FALSE | Downregulated |
| 0,070274312 | 0,592748561 | NEDD8 | FALSE | Upregulated |
| -0,133127369 | 0,59279941 | F2R | FALSE | Downregulated |
| -0,349572957 | 0,592854472 | TGFB2 | FALSE | Downregulated |
| -0,054625028 | 0,592854472 | TMEM66 | FALSE | Downregulated |
| -0,405973587 | 0,593053849 | PROB1 | FALSE | Downregulated |
| 0,316117731 | 0,593087707 | PRELID2 | FALSE | Upregulated |
| 0,081789509 | 0,593087707 | TIMM44 | FALSE | Upregulated |

| | | | | |
|---|---|---|---|---|
| -0,137462549 | 0,593127303 | NBEAL1 | FALSE | Downregulated |
| 0,114610504 | 0,593224461 | ALDH1A1 | FALSE | Upregulated |
| -0,167278023 | 0,593451263 | LRIG1 | FALSE | Downregulated |
| -1,174165447 | 0,593451263 | UNC45B | FALSE | Downregulated |
| 0,113606527 | 0,593533889 | DTX2 | FALSE | Upregulated |
| -0,290068307 | 0,593533889 | FLJ36777 | FALSE | Downregulated |
| 0,065677528 | 0,593574782 | CNOT11 | FALSE | Upregulated |
| -0,431239405 | 0,593574782 | HIF1A-AS2 | FALSE | Downregulated |
| -0,236813171 | 0,593574782 | HM13-AS1 | FALSE | Downregulated |
| 0,11181317 | 0,593574782 | LAMC1 | FALSE | Upregulated |
| -0,478793361 | 0,593574782 | LOC102724748 | FALSE | Downregulated |
| 0,462366929 | 0,593574782 | MST1R | FALSE | Upregulated |
| -0,42672067 | 0,593574782 | MTRNR2L8 | FALSE | Downregulated |
| -0,415139922 | 0,593574782 | NACAP1 | FALSE | Downregulated |
| -0,146458889 | 0,593574782 | PIK3R1 | FALSE | Downregulated |
| 0,113152804 | 0,593574782 | SEC14L1 | FALSE | Upregulated |
| 0,059525762 | 0,593574782 | SLC35A4 | FALSE | Upregulated |
| -0,429551684 | 0,59387814 | TEX29 | FALSE | Downregulated |
| 0,081779475 | 0,593981255 | RPL15 | FALSE | Upregulated |
| 0,119779558 | 0,593983006 | CBX3 | FALSE | Upregulated |
| -0,179140614 | 0,593983006 | XRRA1 | FALSE | Downregulated |
| 0,065865528 | 0,594003408 | DDX19A | FALSE | Upregulated |
| -0,067811207 | 0,594003408 | ZNF148 | FALSE | Downregulated |
| -0,079840337 | 0,594020992 | NFYB | FALSE | Downregulated |
| 0,112553946 | 0,594032367 | TMEM39B | FALSE | Upregulated |
| 0,194822031 | 0,594097558 | KLF9 | FALSE | Upregulated |
| 0,087176637 | 0,594216863 | LINC00998 | FALSE | Upregulated |
| 0,116429037 | 0,594252802 | TDP1 | FALSE | Upregulated |
| -0,070959227 | 0,594558061 | C1orf174 | FALSE | Downregulated |
| -0,20748158 | 0,594589581 | LINC01003 | FALSE | Downregulated |
| -0,111596154 | 0,594719891 | REV3L | FALSE | Downregulated |
| 0,509068702 | 0,594928658 | DISP2 | FALSE | Upregulated |
| -0,20119092 | 0,594951406 | DEPDC4 | FALSE | Downregulated |
| -0,094928908 | 0,595053283 | SPOP | FALSE | Downregulated |
| -0,546708936 | 0,595057033 | FDPSP2 | FALSE | Downregulated |
| -0,665206548 | 0,595057033 | SEBOX | FALSE | Downregulated |
| 0,107280638 | 0,595057033 | TMEM60 | FALSE | Upregulated |
| -0,093104172 | 0,595091182 | EML3 | FALSE | Downregulated |
| -0,290430482 | 0,595299535 | CCDC183-AS1 | FALSE | Downregulated |
| -0,223169015 | 0,595421309 | LOC100506713 | FALSE | Downregulated |
| -0,098878607 | 0,595592765 | ANAPC11 | FALSE | Downregulated |
| 0,169446582 | 0,595643933 | C6orf136 | FALSE | Upregulated |
| 0,060708033 | 0,59566365 | OGDH | FALSE | Upregulated |
| -0,120940051 | 0,595747673 | MTUS1 | FALSE | Downregulated |
| 0,540567137 | 0,595855096 | CSMD2 | FALSE | Upregulated |
| 0,101491688 | 0,595855096 | MTMR12 | FALSE | Upregulated |
| -0,072804988 | 0,596157474 | DCUN1D1 | FALSE | Downregulated |
| -0,082627742 | 0,59647408 | NR5A2 | FALSE | Downregulated |
| -0,425325223 | 0,59647408 | PLA2G1B | FALSE | Downregulated |
| -0,100785329 | 0,59647408 | TRIO | FALSE | Downregulated |
| -0,198173219 | 0,59647408 | TTC32 | FALSE | Downregulated |

| | | | | |
|---|---|---|---|---|
| -0,119562912 | 0,59647408 | WDR35 | FALSE | Downregulated |
| 0,077804798 | 0,59647408 | WIZ | FALSE | Upregulated |
| -0,064555731 | 0,596548238 | SPG11 | FALSE | Downregulated |
| 0,495332638 | 0,596571063 | LOC100506860 | FALSE | Upregulated |
| -0,439083554 | 0,596809787 | LOC101927382 | FALSE | Downregulated |
| -0,475467144 | 0,596809787 | MSS51 | FALSE | Downregulated |
| 0,070614551 | 0,596958546 | LSM12 | FALSE | Upregulated |
| -0,061992205 | 0,596961244 | ZKSCAN5 | FALSE | Downregulated |
| -0,764298228 | 0,597016173 | TPTE2P6 | FALSE | Downregulated |
| 0,210261751 | 0,597104242 | FAM216A | FALSE | Upregulated |
| 0,144760035 | 0,597104242 | RPP40 | FALSE | Upregulated |
| -0,364696982 | 0,597155483 | PRKXP1 | FALSE | Downregulated |
| 0,070365639 | 0,597371476 | CHMP1B | FALSE | Upregulated |
| -0,193609432 | 0,597371476 | ETV1 | FALSE | Downregulated |
| 0,067791111 | 0,597371476 | SPPL3 | FALSE | Upregulated |
| -0,183079932 | 0,597371476 | ZBTB21 | FALSE | Downregulated |
| -0,124010585 | 0,597517284 | CYYR1 | FALSE | Downregulated |
| -0,098881656 | 0,597608571 | CLSTN3 | FALSE | Downregulated |
| -0,320356334 | 0,597679723 | KCNQ4 | FALSE | Downregulated |
| -0,308390629 | 0,597978956 | DNAH2 | FALSE | Downregulated |
| -0,357085925 | 0,597978956 | GSTA5 | FALSE | Downregulated |
| -0,587595848 | 0,597978956 | LOC102723643 | FALSE | Downregulated |
| -0,136547891 | 0,597978956 | STK36 | FALSE | Downregulated |
| -0,106510706 | 0,598119552 | KANK2 | FALSE | Downregulated |
| -0,279153877 | 0,598243832 | LOC101929664 | FALSE | Downregulated |
| 0,101467539 | 0,598256074 | LEMD2 | FALSE | Upregulated |
| -0,757830014 | 0,598258022 | CDH3 | FALSE | Downregulated |
| 0,093130006 | 0,598258022 | CFLAR | FALSE | Upregulated |
| -0,269478994 | 0,598258022 | LOC101928657 | FALSE | Downregulated |
| 0,189285197 | 0,598456972 | C6orf123 | FALSE | Upregulated |
| -0,341864636 | 0,598479393 | RP11-475O6.1-2 | FALSE | Downregulated |
| 0,119134645 | 0,59853411 | ZNF259 | FALSE | Upregulated |
| 0,100694853 | 0,598598079 | MRPL52 | FALSE | Upregulated |
| -0,326723265 | 0,598851041 | NLRP12 | FALSE | Downregulated |
| -0,138178569 | 0,598851041 | ZNF570 | FALSE | Downregulated |
| 0,107109261 | 0,598894311 | SLC12A9 | FALSE | Upregulated |
| -0,325336176 | 0,598898304 | LOC101927407 | FALSE | Downregulated |
| -0,490361959 | 0,599175579 | CEL | FALSE | Downregulated |
| -0,228704548 | 0,599227806 | C19orf80 | FALSE | Downregulated |
| -0,13545236 | 0,599476728 | ZNF235 | FALSE | Downregulated |
| 0,099228855 | 0,599880481 | OGFOD1 | FALSE | Upregulated |
| 0,604615523 | 0,600115201 | DNM1P35 | FALSE | Upregulated |
| -0,252565687 | 0,600115201 | LIMD1-AS1 | FALSE | Downregulated |
| -0,355976101 | 0,600115201 | PIPSL | FALSE | Downregulated |
| 0,068044993 | 0,600115201 | SNUPN | FALSE | Upregulated |
| -0,674910259 | 0,600115201 | TMEM72-AS1 | FALSE | Downregulated |
| 0,210816569 | 0,600177895 | BCL3 | FALSE | Upregulated |
| 0,234261583 | 0,600177895 | CDC42BPG | FALSE | Upregulated |
| 0,087176308 | 0,600177895 | HAX1 | FALSE | Upregulated |
| 0,637601149 | 0,600177895 | MTRNR2L7 | FALSE | Upregulated |
| 0,257254332 | 0,600177895 | TCTEX1D2 | FALSE | Upregulated |

| | | | | |
|---|---|---|---|---|
| 0,103115693 | 0,600177895 | XYLT2 | FALSE | Upregulated |
| -0,325412986 | 0,600177895 | ZNF783 | FALSE | Downregulated |
| 0,768443772 | 0,600178589 | HMSD | FALSE | Upregulated |
| -0,071519156 | 0,600364327 | SLC35B3 | FALSE | Downregulated |
| 0,113691494 | 0,600428025 | IQCB1 | FALSE | Upregulated |
| -0,488111844 | 0,600428025 | LOC101927646 | FALSE | Downregulated |
| -0,106418844 | 0,600438261 | PRR3 | FALSE | Downregulated |
| 0,09049904 | 0,600557362 | CARHSP1 | FALSE | Upregulated |
| -0,18929274 | 0,600879109 | IGF2BP2 | FALSE | Downregulated |
| -0,391046627 | 0,600879109 | SAP25 | FALSE | Downregulated |
| 0,148992778 | 0,601051207 | LINC00526 | FALSE | Upregulated |
| -0,139343296 | 0,601185791 | MYLIP | FALSE | Downregulated |
| 0,078399222 | 0,60129397 | COG5 | FALSE | Upregulated |
| 0,096520703 | 0,601339485 | POLR3D | FALSE | Upregulated |
| -0,10011227 | 0,601424424 | PET100 | FALSE | Downregulated |
| -0,191013628 | 0,601424424 | TMEM254-AS1 | FALSE | Downregulated |
| 0,150360912 | 0,601424424 | TMEM27 | FALSE | Upregulated |
| -0,199467884 | 0,601463064 | BCL2L11 | FALSE | Downregulated |
| 0,129057655 | 0,601463064 | RAB11FIP1 | FALSE | Upregulated |
| -0,311650861 | 0,601482462 | ZNF827 | FALSE | Downregulated |
| -0,119368952 | 0,601717205 | L3MBTL4 | FALSE | Downregulated |
| 0,153222884 | 0,601758039 | TUBA4A | FALSE | Upregulated |
| 0,085751081 | 0,601834736 | ATP5H | FALSE | Upregulated |
| -0,592943691 | 0,601834736 | LOC101928044 | FALSE | Downregulated |
| 0,085739043 | 0,601834736 | SIRT2 | FALSE | Upregulated |
| -0,554257256 | 0,601941069 | FOXO6 | FALSE | Downregulated |
| -0,183121123 | 0,601945623 | LOC102724528 | FALSE | Downregulated |
| 0,073961376 | 0,602259067 | E2F4 | FALSE | Upregulated |
| -0,260819086 | 0,602364732 | CCM2L | FALSE | Downregulated |
| 0,411776313 | 0,602364732 | EXTL3-AS1 | FALSE | Upregulated |
| -0,472224224 | 0,602555081 | LOC101929713 | FALSE | Downregulated |
| -0,134659748 | 0,602555081 | NFKBIB | FALSE | Downregulated |
| 0,082247462 | 0,602562923 | ATP5B | FALSE | Upregulated |
| -0,152144512 | 0,602562923 | C5orf55 | FALSE | Downregulated |
| -0,300721636 | 0,602562923 | CCDC65 | FALSE | Downregulated |
| -0,38966839 | 0,602562923 | FAM155A | FALSE | Downregulated |
| 0,232997448 | 0,602819608 | ARG2 | FALSE | Upregulated |
| 0,128575078 | 0,602819608 | LCMT1 | FALSE | Upregulated |
| 0,269690306 | 0,602819608 | MAB21L2 | FALSE | Upregulated |
| -0,080470366 | 0,602819608 | RANBP2 | FALSE | Downregulated |
| 0,098419626 | 0,602820379 | RPL7A | FALSE | Upregulated |
| -0,065984003 | 0,602834008 | BRIX1 | FALSE | Downregulated |
| 0,194407958 | 0,602834008 | ZNF320 | FALSE | Upregulated |
| -0,180779397 | 0,602834008 | ZNF462 | FALSE | Downregulated |
| -0,108828469 | 0,602874571 | NDUFB1 | FALSE | Downregulated |
| 0,175246859 | 0,602960849 | CELSR1 | FALSE | Upregulated |
| -0,096788209 | 0,602960849 | MRPL40 | FALSE | Downregulated |
| -0,182653882 | 0,603023797 | SULT2A1 | FALSE | Downregulated |
| -0,122317579 | 0,603122585 | VANGL1 | FALSE | Downregulated |
| 0,52242333 | 0,603145907 | CIRBP-AS1 | FALSE | Upregulated |
| -0,084936496 | 0,6032729 | FAM219B | FALSE | Downregulated |

| | | | | |
|---|---|---|---|---|
| 0,683832039 | 0,6032729 | KIAA1549L | FALSE | Upregulated |
| -0,087105996 | 0,60360202 | MAN2A1 | FALSE | Downregulated |
| -0,729980474 | 0,603668234 | LOC101929751 | FALSE | Downregulated |
| -0,576403435 | 0,603988385 | LOC102723581 | FALSE | Downregulated |
| 0,059234174 | 0,604005054 | CUX1 | FALSE | Upregulated |
| -0,647983542 | 0,604029737 | FAM95B1 | FALSE | Downregulated |
| -0,059167494 | 0,604029737 | PPCS | FALSE | Downregulated |
| 0,213329975 | 0,604029737 | RAVER2 | FALSE | Upregulated |
| 0,143522082 | 0,604029737 | SKAP1 | FALSE | Upregulated |
| -0,063689946 | 0,604029737 | SUDS3 | FALSE | Downregulated |
| 0,066208049 | 0,604029737 | TDP2 | FALSE | Upregulated |
| 0,527037731 | 0,604117788 | ST3GAL4-AS1 | FALSE | Upregulated |
| 0,067192097 | 0,604327933 | HNRNPL | FALSE | Upregulated |
| -0,197658597 | 0,604391655 | HOXB4 | FALSE | Downregulated |
| -0,691736723 | 0,604538655 | CAMK2A | FALSE | Downregulated |
| -0,596993387 | 0,604592346 | LOC102724280 | FALSE | Downregulated |
| 0,213188878 | 0,604672056 | DBN1 | FALSE | Upregulated |
| 0,485282845 | 0,604672056 | RP11-27K13.3 | FALSE | Upregulated |
| 0,083270422 | 0,604776891 | HACL1 | FALSE | Upregulated |
| 0,095570859 | 0,605053939 | RILP | FALSE | Upregulated |
| -0,105886401 | 0,605053939 | RPL23AP53 | FALSE | Downregulated |
| 0,307551286 | 0,605060048 | LOC100287015 | FALSE | Upregulated |
| 0,119780302 | 0,605140842 | ACVRL1 | FALSE | Upregulated |
| -0,865601862 | 0,605140842 | APC2 | FALSE | Downregulated |
| -0,097736011 | 0,605140842 | B3GALNT2 | FALSE | Downregulated |
| 0,078504109 | 0,605140842 | CSNK1G2 | FALSE | Upregulated |
| 0,302511498 | 0,605140842 | GEM | FALSE | Upregulated |
| 0,128685594 | 0,605140842 | GLMN | FALSE | Upregulated |
| 0,355837513 | 0,605140842 | ID4 | FALSE | Upregulated |
| 0,086513797 | 0,605140842 | INPPL1 | FALSE | Upregulated |
| 0,13353735 | 0,605140842 | IRF5 | FALSE | Upregulated |
| 0,167334891 | 0,605140842 | KIAA1328 | FALSE | Upregulated |
| 0,214831705 | 0,605140842 | PCSK9 | FALSE | Upregulated |
| -1,04651388 | 0,605140842 | POU4F1 | FALSE | Downregulated |
| 0,100455472 | 0,605140842 | STARD10 | FALSE | Upregulated |
| 0,623663928 | 0,605140842 | VAC14-AS1 | FALSE | Upregulated |
| -0,129231614 | 0,605140842 | ZNF589 | FALSE | Downregulated |
| 0,126301302 | 0,605356456 | NUDT7 | FALSE | Upregulated |
| -0,097453376 | 0,60535716 | TOMM7 | FALSE | Downregulated |
| 0,051479289 | 0,60535716 | TUBGCP2 | FALSE | Upregulated |
| -0,124442686 | 0,605683799 | ST3GAL3 | FALSE | Downregulated |
| 0,248416483 | 0,60573567 | TAGLN | FALSE | Upregulated |
| -0,599862964 | 0,606214265 | CTD-2258A20.5 | FALSE | Downregulated |
| 1,013735994 | 0,606214265 | TRNAK16 | FALSE | Upregulated |
| -0,258677582 | 0,606278033 | SPATA1 | FALSE | Downregulated |
| -0,167588497 | 0,60629888 | FAM26E | FALSE | Downregulated |
| 0,110055667 | 0,60629888 | KATNA1 | FALSE | Upregulated |
| 0,294589173 | 0,60629888 | PMEPA1 | FALSE | Upregulated |
| 0,102701465 | 0,60629888 | RNASE4 | FALSE | Upregulated |
| 0,240590049 | 0,60629888 | S1PR5 | FALSE | Upregulated |
| -0,064147815 | 0,60629888 | TLK2 | FALSE | Downregulated |

| | | | | |
|---|---|---|---|---|
| 0,110659362 | 0,60629888 | ZNF576 | FALSE | Upregulated |
| -0,108814006 | 0,606392797 | ALG2 | FALSE | Downregulated |
| -0,105164327 | 0,606392797 | CPSF4 | FALSE | Downregulated |
| -0,141596711 | 0,606392797 | DYRK3 | FALSE | Downregulated |
| 0,535677252 | 0,606392797 | PDX1 | FALSE | Upregulated |
| 0,196677593 | 0,606392797 | TMEM216 | FALSE | Upregulated |
| 0,114053886 | 0,606392797 | TMEM63A | FALSE | Upregulated |
| 0,688096193 | 0,606637713 | LOC101927116 | FALSE | Upregulated |
| -0,282150566 | 0,60665619 | LOC202181 | FALSE | Downregulated |
| 0,080929492 | 0,606746526 | HDAC2 | FALSE | Upregulated |
| -0,146918517 | 0,607044901 | ARHGEF10 | FALSE | Downregulated |
| 0,107449393 | 0,607044901 | DIABLO | FALSE | Upregulated |
| 0,068129621 | 0,607044901 | FAM91A1 | FALSE | Upregulated |
| 0,064223828 | 0,607160093 | TMEM167A | FALSE | Upregulated |
| 0,650029948 | 0,607166708 | PTCD1 | FALSE | Upregulated |
| -0,147299188 | 0,607345958 | DUSP28 | FALSE | Downregulated |
| 0,207541437 | 0,607345958 | FIBIN | FALSE | Upregulated |
| 0,672461248 | 0,607345958 | IGF2BP1 | FALSE | Upregulated |
| -0,478096793 | 0,607345958 | LOC102724420 | FALSE | Downregulated |
| 0,3602944 | 0,607345958 | PCBP3 | FALSE | Upregulated |
| 0,137316368 | 0,607531442 | CADM4 | FALSE | Upregulated |
| -0,768589021 | 0,607531442 | HSFX1 | FALSE | Downregulated |
| -0,155480723 | 0,607776874 | APOC3 | FALSE | Downregulated |
| 0,648575476 | 0,607784334 | OR52N4 | FALSE | Upregulated |
| -0,141877007 | 0,607902198 | DHRS4 | FALSE | Downregulated |
| 0,360505557 | 0,607902198 | PLCH1 | FALSE | Upregulated |
| 0,16572263 | 0,608301997 | TTYH2 | FALSE | Upregulated |
| 0,087031864 | 0,608322164 | COA4 | FALSE | Upregulated |
| 0,253164798 | 0,608322164 | FBLIM1 | FALSE | Upregulated |
| 0,28675515 | 0,608322164 | LOC101928513 | FALSE | Upregulated |
| -0,079100526 | 0,608322164 | YAE1D1 | FALSE | Downregulated |
| 0,08300712 | 0,608385505 | DUSP3 | FALSE | Upregulated |
| 0,177504599 | 0,608725691 | SRPX | FALSE | Upregulated |
| -0,87854626 | 0,608841856 | CALB2 | FALSE | Downregulated |
| 0,069650768 | 0,608993921 | GGA3 | FALSE | Upregulated |
| -0,055040997 | 0,609044554 | GLG1 | FALSE | Downregulated |
| -0,368349365 | 0,609148909 | REREP3 | FALSE | Downregulated |
| 0,111106746 | 0,609167969 | DEK | FALSE | Upregulated |
| -0,09018768 | 0,609211176 | INTS2 | FALSE | Downregulated |
| -0,101809122 | 0,609211176 | MAN2A2 | FALSE | Downregulated |
| -0,596872995 | 0,609211176 | RP11-430B1.2 | FALSE | Downregulated |
| -0,825948944 | 0,609211176 | SCN2B | FALSE | Downregulated |
| -0,39980561 | 0,609211176 | TEX14 | FALSE | Downregulated |
| -0,117485309 | 0,609215637 | IQSEC1 | FALSE | Downregulated |
| 0,687959108 | 0,609215637 | LINC00673 | FALSE | Upregulated |
| -0,081120225 | 0,609215637 | PALB2 | FALSE | Downregulated |
| -0,066549469 | 0,609215637 | SECISBP2 | FALSE | Downregulated |
| -0,353849434 | 0,609298603 | RP11-406A20.4 | FALSE | Downregulated |
| -0,115710965 | 0,609592058 | ZNF232 | FALSE | Downregulated |
| -0,524087157 | 0,609754628 | DNMT3L | FALSE | Downregulated |
| -0,318618447 | 0,609754628 | FAR2P2 | FALSE | Downregulated |

| | | | | |
|---|---|---|---|---|
| 0,15735286 | 0,609754628 | GLUL | FALSE | Upregulated |
| 0,118272275 | 0,609754628 | MLXIP | FALSE | Upregulated |
| 0,24196883 | 0,609754628 | RP11-461O7.1 | FALSE | Upregulated |
| 0,099906103 | 0,609827563 | AP5Z1 | FALSE | Upregulated |
| -0,082531481 | 0,609827563 | CRAT | FALSE | Downregulated |
| 0,746950655 | 0,609827563 | KLHL34 | FALSE | Upregulated |
| -0,411492215 | 0,609827563 | LOC101927206 | FALSE | Downregulated |
| -0,103333095 | 0,609827563 | MRPS36 | FALSE | Downregulated |
| 0,123724393 | 0,609827563 | PSENEN | FALSE | Upregulated |
| -0,112645209 | 0,609827563 | UBA6-AS1 | FALSE | Downregulated |
| -0,161522994 | 0,609878149 | ZNF442 | FALSE | Downregulated |
| -0,229272506 | 0,610102401 | ASPDH | FALSE | Downregulated |
| 0,082296148 | 0,610104638 | CLNS1A | FALSE | Upregulated |
| -0,210042371 | 0,610104638 | ZNF439 | FALSE | Downregulated |
| -0,085874348 | 0,61018309 | POC1B | FALSE | Downregulated |
| -1,068653248 | 0,610240379 | ASB12 | FALSE | Downregulated |
| 0,204965341 | 0,610240379 | PKIB | FALSE | Upregulated |
| 0,088932034 | 0,610321807 | CNNM4 | FALSE | Upregulated |
| 0,073458477 | 0,610321807 | DUSP11 | FALSE | Upregulated |
| -1,043660566 | 0,610321807 | PI3 | FALSE | Downregulated |
| 0,157164699 | 0,610400065 | NDUFC2-KCTD14 | FALSE | Upregulated |
| -0,074364181 | 0,610503151 | COQ7 | FALSE | Downregulated |
| 0,125744824 | 0,610503151 | FAM189B | FALSE | Upregulated |
| -0,930079151 | 0,610515232 | MMP8 | FALSE | Downregulated |
| 0,120460141 | 0,610515232 | SELK | FALSE | Upregulated |
| -0,552639472 | 0,610534336 | LRRC19 | FALSE | Downregulated |
| -0,211318656 | 0,610534336 | NUP62CL | FALSE | Downregulated |
| 0,119902704 | 0,610546567 | FAM114A1 | FALSE | Upregulated |
| -0,155923106 | 0,610573431 | RP5-1033H22.2 | FALSE | Downregulated |
| -0,15307578 | 0,610736879 | LMBR1L | FALSE | Downregulated |
| -0,160752588 | 0,610986487 | GSAP | FALSE | Downregulated |
| 0,375714634 | 0,611088861 | LOC729887 | FALSE | Upregulated |
| 0,378954979 | 0,611245227 | PKD1L2 | FALSE | Upregulated |
| -0,119559113 | 0,611331407 | RELN | FALSE | Downregulated |
| -0,216826362 | 0,611400559 | AFP | FALSE | Downregulated |
| -0,097694247 | 0,611400559 | FAM149A | FALSE | Downregulated |
| 0,087901438 | 0,611400559 | FBXO31 | FALSE | Upregulated |
| -0,668831397 | 0,611400559 | NRAP | FALSE | Downregulated |
| 0,120566684 | 0,611400559 | TTC1 | FALSE | Upregulated |
| -0,06393416 | 0,611470038 | AASDHPPT | FALSE | Downregulated |
| -0,062540285 | 0,611512363 | TADA2B | FALSE | Downregulated |
| -0,091454584 | 0,611590495 | IGFBP4 | FALSE | Downregulated |
| -0,056978026 | 0,611590495 | MAP3K7 | FALSE | Downregulated |
| -0,064801394 | 0,611590495 | NEK7 | FALSE | Downregulated |
| 0,257653594 | 0,611590495 | PRMT6 | FALSE | Upregulated |
| -0,567673129 | 0,611746274 | MYL4 | FALSE | Downregulated |
| 0,07599727 | 0,611813171 | RHOU | FALSE | Upregulated |
| 0,29705965 | 0,611883678 | TNFAIP8L3 | FALSE | Upregulated |
| -0,056986488 | 0,612039379 | EXD2 | FALSE | Downregulated |
| -0,206748443 | 0,612039379 | HLA-F-AS1 | FALSE | Downregulated |
| 0,123518491 | 0,612039379 | RUSC2 | FALSE | Upregulated |

| | | | | |
|---|---|---|---|---|
| 0,054924148 | 0,612039379 | SCYL1 | FALSE | Upregulated |
| 0,302109412 | 0,612039379 | TMEM253 | FALSE | Upregulated |
| 0,10932629 | 0,612164532 | RUNDC1 | FALSE | Upregulated |
| -0,915864716 | 0,612311793 | FLJ20444 | FALSE | Downregulated |
| 0,08458203 | 0,612311793 | SSBP1 | FALSE | Upregulated |
| -0,086170385 | 0,612319115 | ALDH8A1 | FALSE | Downregulated |
| 0,452547799 | 0,612404868 | LOC728026 | FALSE | Upregulated |
| -0,25972232 | 0,612435202 | FAM107A | FALSE | Downregulated |
| 0,350199352 | 0,612435202 | KIAA1324 | FALSE | Upregulated |
| 0,3443706 | 0,612435202 | RNF186 | FALSE | Upregulated |
| -0,121173966 | 0,612435202 | TRAPPC2 | FALSE | Downregulated |
| -0,057455894 | 0,612435202 | UBE2K | FALSE | Downregulated |
| -0,142451062 | 0,612673714 | FHIT | FALSE | Downregulated |
| -0,477656436 | 0,612959413 | VWA2 | FALSE | Downregulated |
| -0,169680457 | 0,613110096 | TMCC1-AS1 | FALSE | Downregulated |
| -0,292302825 | 0,613330627 | C22orf34 | FALSE | Downregulated |
| -0,239883701 | 0,613330627 | LOC100128531 | FALSE | Downregulated |
| 0,266524714 | 0,61347518 | LOC102723389 | FALSE | Upregulated |
| 0,291353695 | 0,613484641 | FRZB | FALSE | Upregulated |
| 0,068219301 | 0,613505485 | PPP1R21 | FALSE | Upregulated |
| -1,051702457 | 0,613529271 | TUSC5 | FALSE | Downregulated |
| -0,168414973 | 0,613529271 | ZNF652 | FALSE | Downregulated |
| 0,181247304 | 0,613551719 | ADAMTSL2 | FALSE | Upregulated |
| 0,225548508 | 0,613551719 | C21orf58 | FALSE | Upregulated |
| 0,179059226 | 0,613551719 | C5orf30 | FALSE | Upregulated |
| 0,724097351 | 0,613551719 | LOC102724593 | FALSE | Upregulated |
| 0,357894213 | 0,613728046 | MAPK8IP2 | FALSE | Upregulated |
| 0,614327599 | 0,613728046 | MPP2 | FALSE | Upregulated |
| 0,091483115 | 0,613728046 | NAA15 | FALSE | Upregulated |
| -0,385096557 | 0,613728046 | NDUFB2-AS1 | FALSE | Downregulated |
| -0,364048462 | 0,613728046 | SH3TC2 | FALSE | Downregulated |
| 0,11109523 | 0,61375325 | MRPL55 | FALSE | Upregulated |
| 0,107891756 | 0,614033021 | ACAP2 | FALSE | Upregulated |
| -0,558090672 | 0,614033021 | MLANA | FALSE | Downregulated |
| 0,102546605 | 0,614033021 | PMVK | FALSE | Upregulated |
| 0,536301351 | 0,614050487 | CCL22 | FALSE | Upregulated |
| -0,506579156 | 0,614052872 | CLUL1 | FALSE | Downregulated |
| -0,440245492 | 0,614052872 | LOC101929887 | FALSE | Downregulated |
| 0,358971297 | 0,614052872 | RAB39B | FALSE | Upregulated |
| 0,13622382 | 0,614278712 | PREB | FALSE | Upregulated |
| 0,05402757 | 0,614278712 | WDR48 | FALSE | Upregulated |
| -0,640933942 | 0,614376467 | HCG23 | FALSE | Downregulated |
| 0,388060222 | 0,614412412 | ANO9 | FALSE | Upregulated |
| 0,080910414 | 0,614414852 | HSPB11 | FALSE | Upregulated |
| -0,093442636 | 0,614469797 | RPL7 | FALSE | Downregulated |
| -0,408922152 | 0,61457923 | RPS15AP10 | FALSE | Downregulated |
| -0,099492805 | 0,614863811 | CBFA2T2 | FALSE | Downregulated |
| -0,122368316 | 0,614912144 | HGSNAT | FALSE | Downregulated |
| -0,077764268 | 0,614912144 | PNPLA8 | FALSE | Downregulated |
| 0,133279689 | 0,614912144 | PTS | FALSE | Upregulated |
| 0,381491296 | 0,615196309 | LOC101927278 | FALSE | Upregulated |

| | | | | |
|---|---|---|---|---|
| -0,649200062 | 0,615196309 | LOC101928369 | FALSE | Downregulated |
| -0,362824743 | 0,615239653 | C3orf35 | FALSE | Downregulated |
| 0,148604386 | 0,615239653 | C3orf62 | FALSE | Upregulated |
| -0,114692595 | 0,615387666 | NCALD | FALSE | Downregulated |
| 0,054887835 | 0,615403216 | FAM175B | FALSE | Upregulated |
| 0,494221987 | 0,615673212 | HERC2P3 | FALSE | Upregulated |
| 0,101379995 | 0,615673212 | MVB12A | FALSE | Upregulated |
| -0,06925601 | 0,615817595 | ATE1 | FALSE | Downregulated |
| 0,495982 | 0,615817595 | TMEM191C | FALSE | Upregulated |
| 0,538262182 | 0,615881613 | LOC101930375 | FALSE | Upregulated |
| -0,100609572 | 0,615881613 | RTN4 | FALSE | Downregulated |
| 0,107456155 | 0,615881613 | SUCO | FALSE | Upregulated |
| -0,211718531 | 0,616083786 | AGPAT9 | FALSE | Downregulated |
| -0,141326427 | 0,616083786 | CA2 | FALSE | Downregulated |
| 0,329994741 | 0,616083786 | CDH24 | FALSE | Upregulated |
| 0,152706742 | 0,616083786 | DNALI1 | FALSE | Upregulated |
| 0,543359097 | 0,616083786 | ENO4 | FALSE | Upregulated |
| 0,141144814 | 0,616083786 | FAHD1 | FALSE | Upregulated |
| -0,539136326 | 0,616083786 | FAM133CP | FALSE | Downregulated |
| -0,986771284 | 0,616083786 | GOLGA8S | FALSE | Downregulated |
| 0,093171501 | 0,616083786 | HEATR5A | FALSE | Upregulated |
| 0,050116333 | 0,616083786 | INO80 | FALSE | Upregulated |
| -0,192342875 | 0,616083786 | LEPR | FALSE | Downregulated |
| -0,192893001 | 0,616083786 | LINC00870 | FALSE | Downregulated |
| -0,666357529 | 0,616083786 | POTEM | FALSE | Downregulated |
| 0,111188255 | 0,616083786 | PPP1R12A | FALSE | Upregulated |
| -0,442874846 | 0,616083786 | RP11-575F12.1 | FALSE | Downregulated |
| 0,088843245 | 0,616083786 | SLC9A6 | FALSE | Upregulated |
| -0,091194057 | 0,616083786 | SNRPA | FALSE | Downregulated |
| 0,109911647 | 0,616083786 | TACC2 | FALSE | Upregulated |
| -0,435357851 | 0,616083786 | TAS2R10 | FALSE | Downregulated |
| 0,082513509 | 0,616083786 | ZDHHC7 | FALSE | Upregulated |
| 0,537374332 | 0,616340372 | HIST2H4B | FALSE | Upregulated |
| 0,113013811 | 0,616350848 | JADE2 | FALSE | Upregulated |
| 0,160429058 | 0,616497396 | ICAM3 | FALSE | Upregulated |
| -0,781222715 | 0,616558466 | GALP | FALSE | Downregulated |
| 0,301257906 | 0,616590279 | BOK-AS1 | FALSE | Upregulated |
| -0,055339434 | 0,616590279 | CLCC1 | FALSE | Downregulated |
| -0,179869632 | 0,616590279 | FRS3 | FALSE | Downregulated |
| 0,140402261 | 0,616590279 | MPC1 | FALSE | Upregulated |
| 0,12360251 | 0,616590279 | NCKAP5L | FALSE | Upregulated |
| -0,122537005 | 0,616590279 | TEAD1 | FALSE | Downregulated |
| -0,065813894 | 0,616590279 | TMEM184C | FALSE | Downregulated |
| -0,43982908 | 0,616639549 | SCARNA12 | FALSE | Downregulated |
| 0,101600637 | 0,616643706 | KIAA1551 | FALSE | Upregulated |
| 0,567798225 | 0,616643706 | MGAT4C | FALSE | Upregulated |
| 0,088651606 | 0,616643706 | PIGT | FALSE | Upregulated |
| -0,507504035 | 0,616643706 | TEX15 | FALSE | Downregulated |
| -0,102567603 | 0,616643706 | YPEL3 | FALSE | Downregulated |
| 0,172844464 | 0,616643706 | ZDHHC1 | FALSE | Upregulated |
| -0,133158453 | 0,616675232 | C15orf61 | FALSE | Downregulated |

| | | | | |
|---|---|---|---|---|
| -0,289599895 | 0,616675232 | CTAGE11P | FALSE | Downregulated |
| -0,067781042 | 0,616675232 | ETV3 | FALSE | Downregulated |
| -0,170426645 | 0,616846788 | C2orf76 | FALSE | Downregulated |
| -1,437038323 | 0,616846788 | LOC102723799 | FALSE | Downregulated |
| -0,066290724 | 0,616846788 | NOL10 | FALSE | Downregulated |
| 0,058823977 | 0,616846788 | RANBP6 | FALSE | Upregulated |
| 0,052441773 | 0,616846788 | STK25 | FALSE | Upregulated |
| 0,079115898 | 0,616846788 | SUCLG1 | FALSE | Upregulated |
| -0,252280305 | 0,616846788 | TMEM55A | FALSE | Downregulated |
| -0,106599648 | 0,616957089 | KLHDC8B | FALSE | Downregulated |
| -1,27331457 | 0,617362778 | TNXA | FALSE | Downregulated |
| -0,385396982 | 0,617427012 | ZNF385C | FALSE | Downregulated |
| -0,986697542 | 0,617686952 | PTHLH | FALSE | Downregulated |
| -0,359671072 | 0,61771094 | ESRRG | FALSE | Downregulated |
| 0,196789158 | 0,618238945 | ME3 | FALSE | Upregulated |
| -0,352005701 | 0,618281537 | LOC102724851 | FALSE | Downregulated |
| 0,095892759 | 0,618281537 | RAB4A | FALSE | Upregulated |
| 0,061809931 | 0,61832097 | SH3GL1 | FALSE | Upregulated |
| 0,138000933 | 0,618514129 | RREB1 | FALSE | Upregulated |
| -0,427350129 | 0,618518179 | RP11-43N5.1 | FALSE | Downregulated |
| -0,116300074 | 0,618614367 | NUDT19 | FALSE | Downregulated |
| 0,077431138 | 0,618733522 | MEPCE | FALSE | Upregulated |
| -0,091386327 | 0,618733522 | TMEM260 | FALSE | Downregulated |
| 0,672116224 | 0,618807889 | ASCL1 | FALSE | Upregulated |
| 0,266886314 | 0,618807889 | GPR143 | FALSE | Upregulated |
| -0,449560802 | 0,618807889 | LGI2 | FALSE | Downregulated |
| 0,352827013 | 0,618807889 | P2RY11 | FALSE | Upregulated |
| -0,584257537 | 0,619029643 | RP11-14D22.5 | FALSE | Downregulated |
| -0,154069208 | 0,619429816 | FDXACB1 | FALSE | Downregulated |
| 0,690933276 | 0,619465148 | TTC24 | FALSE | Upregulated |
| 0,149952293 | 0,619474756 | HIC1 | FALSE | Upregulated |
| -0,518095693 | 0,619474756 | ZRANB2-AS2 | FALSE | Downregulated |
| 0,077181448 | 0,619718673 | BCKDK | FALSE | Upregulated |
| -0,238467597 | 0,619730814 | LINC00961 | FALSE | Downregulated |
| 0,121209115 | 0,619758273 | SLC26A1 | FALSE | Upregulated |
| 0,686893195 | 0,619758273 | TRNH | FALSE | Upregulated |
| -0,078736799 | 0,619777835 | FBRS | FALSE | Downregulated |
| 0,154210741 | 0,619777835 | TELO2 | FALSE | Upregulated |
| -0,1569718 | 0,619787418 | C14orf1 | FALSE | Downregulated |
| -0,065779117 | 0,619787418 | EAPP | FALSE | Downregulated |
| -0,417327347 | 0,619787418 | LOC101927435 | FALSE | Downregulated |
| 0,088507158 | 0,619787418 | TIGD6 | FALSE | Upregulated |
| 0,386258223 | 0,619787418 | UCP3 | FALSE | Upregulated |
| -0,040499317 | 0,619798782 | XPO7 | FALSE | Downregulated |
| 0,635257914 | 0,61988046 | UTY | FALSE | Upregulated |
| -0,062301729 | 0,620242611 | BROX | FALSE | Downregulated |
| -0,223491825 | 0,620242611 | LOC101928101 | FALSE | Downregulated |
| -0,456810312 | 0,620355221 | COL7A1 | FALSE | Downregulated |
| -0,379458259 | 0,620465535 | MESP1 | FALSE | Downregulated |
| 0,094388968 | 0,620527547 | DCTD | FALSE | Upregulated |
| 0,149423465 | 0,620612999 | LILRB5 | FALSE | Upregulated |

| | | | | |
|---|---|---|---|---|
| -0,211655548 | 0,620661574 | ZNF229 | FALSE | Downregulated |
| -0,521200905 | 0,620783903 | LOC101929615 | FALSE | Downregulated |
| 0,102652885 | 0,620783903 | MYD88 | FALSE | Upregulated |
| 0,08086708 | 0,620783903 | RCHY1 | FALSE | Upregulated |
| -0,983727674 | 0,620831759 | PRR5-ARHGAP8 | FALSE | Downregulated |
| -0,105130988 | 0,620895009 | AGMO | FALSE | Downregulated |
| 0,361663436 | 0,620900379 | KIAA1656 | FALSE | Upregulated |
| -0,462055358 | 0,620946054 | STPG2 | FALSE | Downregulated |
| -0,076484418 | 0,621110123 | MYO18A | FALSE | Downregulated |
| -0,062900865 | 0,621125582 | HNRNPUL1 | FALSE | Downregulated |
| 0,25402442 | 0,621170311 | ANO7 | FALSE | Upregulated |
| -0,825977418 | 0,62134389 | SNORA14A | FALSE | Downregulated |
| 0,308949802 | 0,621636419 | F2RL2 | FALSE | Upregulated |
| -0,091956051 | 0,621636419 | FLCN | FALSE | Downregulated |
| 0,122290002 | 0,621636419 | HSD17B12 | FALSE | Upregulated |
| -0,577993395 | 0,621636419 | INSC | FALSE | Downregulated |
| 0,105732396 | 0,621636419 | ITPA | FALSE | Upregulated |
| -0,101303946 | 0,621636419 | PNPT1 | FALSE | Downregulated |
| -0,180597937 | 0,621659319 | DAK | FALSE | Downregulated |
| 0,17986672 | 0,621860667 | ABHD17C | FALSE | Upregulated |
| -0,950338553 | 0,621860667 | UBE2MP1 | FALSE | Downregulated |
| 0,209649398 | 0,621892111 | IQSEC3 | FALSE | Upregulated |
| -0,103090264 | 0,621892111 | KIAA1467 | FALSE | Downregulated |
| -0,077369453 | 0,621892111 | ZCRB1 | FALSE | Downregulated |
| -0,607085635 | 0,621892111 | ZNF571-AS1 | FALSE | Downregulated |
| -0,074131247 | 0,622008349 | ST5 | FALSE | Downregulated |
| -0,086984765 | 0,622403051 | HSF2 | FALSE | Downregulated |
| -0,082335695 | 0,622722535 | C8orf59 | FALSE | Downregulated |
| 0,211709999 | 0,622722535 | GRASP | FALSE | Upregulated |
| 0,454700214 | 0,622759554 | MYOCD | FALSE | Upregulated |
| -0,523288348 | 0,622804341 | LOC101928875 | FALSE | Downregulated |
| -0,329957595 | 0,622804341 | LYPD3 | FALSE | Downregulated |
| 0,19233386 | 0,622804341 | PALLD | FALSE | Upregulated |
| 0,05332692 | 0,622804341 | SART1 | FALSE | Upregulated |
| -0,48700001 | 0,62292443 | ITGA2B | FALSE | Downregulated |
| 0,101580867 | 0,62292443 | MVB12B | FALSE | Upregulated |
| 0,07764557 | 0,622950449 | WDR37 | FALSE | Upregulated |
| -0,133315706 | 0,623360286 | CARF | FALSE | Downregulated |
| 0,262610929 | 0,623360286 | EGLN3 | FALSE | Upregulated |
| 0,215347447 | 0,623360286 | ENO2 | FALSE | Upregulated |
| 0,583144617 | 0,623360286 | KLK10 | FALSE | Upregulated |
| 0,43639209 | 0,623360286 | LRRN2 | FALSE | Upregulated |
| 0,088785613 | 0,623360286 | MNT | FALSE | Upregulated |
| 0,169687233 | 0,623360286 | NEU4 | FALSE | Upregulated |
| -0,666955785 | 0,623360286 | NME9 | FALSE | Downregulated |
| 0,367701897 | 0,623360286 | RNA28S5 | FALSE | Upregulated |
| -0,137607508 | 0,623360286 | SNRNP70 | FALSE | Downregulated |
| -0,141252201 | 0,623448295 | HSPG2 | FALSE | Downregulated |
| 0,306330079 | 0,623448295 | LOC100505564 | FALSE | Upregulated |
| -0,112163723 | 0,623448295 | PGBD2 | FALSE | Downregulated |
| 0,225473978 | 0,623448295 | SLC11A1 | FALSE | Upregulated |

| | | | | |
|---|---|---|---|---|
| -0,081350012 | 0,623621933 | ADCK3 | FALSE | Downregulated |
| 0,335200747 | 0,623621933 | LOC101928054 | FALSE | Upregulated |
| 0,27777644 | 0,623621933 | PINLYP | FALSE | Upregulated |
| 0,092194132 | 0,623621933 | TCOF1 | FALSE | Upregulated |
| 0,122918984 | 0,623673808 | ZNF765 | FALSE | Upregulated |
| -0,073428868 | 0,62390772 | PI4KB | FALSE | Downregulated |
| -0,046457751 | 0,624065028 | BLOC1S6 | FALSE | Downregulated |
| 0,636186101 | 0,624065028 | RP11-58E21.3 | FALSE | Upregulated |
| 0,508497323 | 0,62420351 | LOC100506498 | FALSE | Upregulated |
| 0,128447085 | 0,624299412 | NEK3 | FALSE | Upregulated |
| 0,134104478 | 0,624431424 | CGNL1 | FALSE | Upregulated |
| -0,22315758 | 0,624685723 | MCF2L-AS1 | FALSE | Downregulated |
| -0,267647406 | 0,624850778 | HSF4 | FALSE | Downregulated |
| 0,387518593 | 0,624850778 | LOC440895 | FALSE | Upregulated |
| -0,098449433 | 0,624850778 | MED18 | FALSE | Downregulated |
| 0,666002717 | 0,624996316 | MSTN | FALSE | Upregulated |
| -0,176883081 | 0,625143617 | SOX5 | FALSE | Downregulated |
| 0,101379625 | 0,625182542 | COX6A1 | FALSE | Upregulated |
| -0,520347454 | 0,625182542 | FANK1 | FALSE | Downregulated |
| 0,100169127 | 0,625182542 | HNRNPUL2-BSCL2 | FALSE | Upregulated |
| 0,202682613 | 0,625182542 | SLC12A2 | FALSE | Upregulated |
| 0,234739302 | 0,625222998 | KIAA1683 | FALSE | Upregulated |
| 0,100913692 | 0,625234043 | C14orf142 | FALSE | Upregulated |
| 0,069364808 | 0,6253668 | TIMM10B | FALSE | Upregulated |
| 0,062654778 | 0,625547244 | SMPD4 | FALSE | Upregulated |
| -0,096319567 | 0,625746494 | DMWD | FALSE | Downregulated |
| -0,115904104 | 0,6257587 | SRSF6 | FALSE | Downregulated |
| 0,077747858 | 0,62580709 | CD99 | FALSE | Upregulated |
| -0,080623665 | 0,626067401 | APOPT1 | FALSE | Downregulated |
| -0,084507974 | 0,626067401 | NUCB1 | FALSE | Downregulated |
| 0,101124168 | 0,626067401 | UGT2B4 | FALSE | Upregulated |
| -0,254573418 | 0,626201187 | TDRD10 | FALSE | Downregulated |
| -0,115632882 | 0,626434205 | PGAP1 | FALSE | Downregulated |
| 0,110853631 | 0,627089074 | TRNT1 | FALSE | Upregulated |
| 0,095710442 | 0,627134203 | UBE2J1 | FALSE | Upregulated |
| 0,589147257 | 0,627147205 | KIR3DL1 | FALSE | Upregulated |
| -0,152427516 | 0,627191673 | ZBTB11-AS1 | FALSE | Downregulated |
| 0,335738581 | 0,627203178 | COLCA2 | FALSE | Upregulated |
| 0,117431435 | 0,627203178 | PYCRL | FALSE | Upregulated |
| -0,06368732 | 0,627223132 | RBPJ | FALSE | Downregulated |
| -0,281119033 | 0,627432309 | L3MBTL1 | FALSE | Downregulated |
| 0,242585733 | 0,62775856 | SH2D3A | FALSE | Upregulated |
| 0,14658603 | 0,627931466 | NLE1 | FALSE | Upregulated |
| 0,952325956 | 0,627931466 | TRNAK9 | FALSE | Upregulated |
| -0,504677025 | 0,628020015 | MIR4697HG | FALSE | Downregulated |
| 0,099704256 | 0,628020015 | SH3TC1 | FALSE | Upregulated |
| 0,145213135 | 0,628062098 | SENP8 | FALSE | Upregulated |
| -0,077128009 | 0,628090024 | CCDC58 | FALSE | Downregulated |
| -0,347116659 | 0,628482773 | TIAF1 | FALSE | Downregulated |
| -0,133379473 | 0,628616189 | LOC100287896 | FALSE | Downregulated |
| 0,098892973 | 0,628797007 | RPP30 | FALSE | Upregulated |

| | | | | |
|---|---|---|---|---|
| 0,410914802 | 0,628904339 | CTSV | FALSE | Upregulated |
| 0,164234315 | 0,628904339 | LINC00987 | FALSE | Upregulated |
| 0,112993929 | 0,629045024 | S100PBP | FALSE | Upregulated |
| 0,111713318 | 0,629075046 | FN3KRP | FALSE | Upregulated |
| 0,568628617 | 0,629075046 | LOC101930665 | FALSE | Upregulated |
| 0,347739745 | 0,629075046 | RAB19 | FALSE | Upregulated |
| -0,068353468 | 0,629075046 | YY1 | FALSE | Downregulated |
| -0,106180959 | 0,629075046 | ZNF286A | FALSE | Downregulated |
| -0,390213102 | 0,629203041 | ATXN8OS | FALSE | Downregulated |
| -0,280663234 | 0,629298705 | ST3GAL6-AS1 | FALSE | Downregulated |
| 0,107267759 | 0,629366346 | CEP97 | FALSE | Upregulated |
| 0,10831828 | 0,629366346 | HIF1A | FALSE | Upregulated |
| 0,600652254 | 0,629366346 | TRPC4 | FALSE | Upregulated |
| 0,342607533 | 0,629669232 | STRIP2 | FALSE | Upregulated |
| -0,176101948 | 0,629987525 | GEMIN8P4 | FALSE | Downregulated |
| -1,038149968 | 0,630271848 | MYOZ3 | FALSE | Downregulated |
| -0,129439769 | 0,630620132 | HEXDC | FALSE | Downregulated |
| 0,200547989 | 0,630620132 | IGFBP7 | FALSE | Upregulated |
| -0,227301368 | 0,630620132 | LOC101930114 | FALSE | Downregulated |
| 0,111817659 | 0,630620132 | NPDC1 | FALSE | Upregulated |
| -0,526080906 | 0,630768112 | LOC101928326 | FALSE | Downregulated |
| -0,321590706 | 0,630811863 | LOC102723926 | FALSE | Downregulated |
| -0,302985402 | 0,630811863 | LOC440386 | FALSE | Downregulated |
| 0,583620452 | 0,630998816 | LOC101929609 | FALSE | Upregulated |
| -0,543376259 | 0,630998816 | REG4 | FALSE | Downregulated |
| 0,105868258 | 0,630998816 | ZNF524 | FALSE | Upregulated |
| 0,090572382 | 0,631310955 | ATF6 | FALSE | Upregulated |
| 0,22093859 | 0,631316134 | ROBO3 | FALSE | Upregulated |
| -0,049647764 | 0,631632073 | FTO | FALSE | Downregulated |
| 0,426619606 | 0,631632073 | GAS6-AS2 | FALSE | Upregulated |
| 1,020144063 | 0,631632073 | LOC101927333 | FALSE | Upregulated |
| 0,274500956 | 0,631753053 | SOX4 | FALSE | Upregulated |
| -1,516075073 | 0,631843027 | ADCY2 | FALSE | Downregulated |
| -0,067044821 | 0,631843027 | ADCY6 | FALSE | Downregulated |
| -0,616899772 | 0,631843027 | ADCYAP1R1 | FALSE | Downregulated |
| -0,146884365 | 0,631843027 | AKAP3 | FALSE | Downregulated |
| -0,153417866 | 0,631843027 | ARHGAP24 | FALSE | Downregulated |
| -0,833082361 | 0,631843027 | ASMT | FALSE | Downregulated |
| -0,147771401 | 0,631843027 | FAM180A | FALSE | Downregulated |
| 0,464994693 | 0,631843027 | GPR83 | FALSE | Upregulated |
| 0,10027885 | 0,631843027 | LPCAT3 | FALSE | Upregulated |
| 0,090860372 | 0,631843027 | MRPS34 | FALSE | Upregulated |
| -0,128375599 | 0,631843027 | PANK3 | FALSE | Downregulated |
| 0,43462847 | 0,631843027 | REN | FALSE | Upregulated |
| -0,066575737 | 0,631843027 | SCAF4 | FALSE | Downregulated |
| -0,556984262 | 0,631843027 | SNX31 | FALSE | Downregulated |
| -0,198959173 | 0,631857885 | LOC102723521 | FALSE | Downregulated |
| -0,203537425 | 0,631857885 | SNHG10 | FALSE | Downregulated |
| 0,139582397 | 0,631927892 | STXBP1 | FALSE | Upregulated |
| -0,124055994 | 0,63205206 | SFT2D1 | FALSE | Downregulated |
| -0,64491775 | 0,632332961 | TEX26 | FALSE | Downregulated |

| | | | | |
|---|---|---|---|---|
| 0,714445645 | 0,632355962 | GGT2 | FALSE | Upregulated |
| -0,051602357 | 0,632377461 | H2AFV | FALSE | Downregulated |
| 0,107473005 | 0,632377461 | RILPL1 | FALSE | Upregulated |
| -0,094448687 | 0,632488844 | PHF14 | FALSE | Downregulated |
| -0,093358137 | 0,632507927 | CBX4 | FALSE | Downregulated |
| 0,220671954 | 0,632507927 | CNTNAP1 | FALSE | Upregulated |
| -0,687503637 | 0,632507927 | TSPEAR | FALSE | Downregulated |
| -0,077292675 | 0,632578368 | C22orf29 | FALSE | Downregulated |
| 0,129701995 | 0,632595182 | PHKA1 | FALSE | Upregulated |
| 0,758275406 | 0,632916328 | ELANE | FALSE | Upregulated |
| -0,212539132 | 0,63314562 | TIPARP-AS1 | FALSE | Downregulated |
| 0,105028394 | 0,63340994 | ARL5B | FALSE | Upregulated |
| 0,258775003 | 0,63340994 | KIAA1549 | FALSE | Upregulated |
| 0,090746787 | 0,633436649 | AIP | FALSE | Upregulated |
| -0,062753262 | 0,633436649 | CPNE3 | FALSE | Downregulated |
| 0,133224948 | 0,633436649 | PGBD1 | FALSE | Upregulated |
| -0,574807343 | 0,633436649 | SLFNL1 | FALSE | Downregulated |
| 0,142519483 | 0,633466236 | FES | FALSE | Upregulated |
| 0,288461207 | 0,633686993 | GUSBP5 | FALSE | Upregulated |
| -0,343125335 | 0,633686993 | POU6F2 | FALSE | Downregulated |
| 0,074396187 | 0,633709893 | COMMD1 | FALSE | Upregulated |
| -0,232986337 | 0,633776566 | A2M-AS1 | FALSE | Downregulated |
| -0,091631723 | 0,633776566 | BTRC | FALSE | Downregulated |
| 0,24484405 | 0,633776566 | CLEC10A | FALSE | Upregulated |
| -0,171254356 | 0,633776566 | DLEU2 | FALSE | Downregulated |
| 0,079954479 | 0,633776566 | HEIH | FALSE | Upregulated |
| 0,392764833 | 0,633776566 | LOC407835 | FALSE | Upregulated |
| -0,713436921 | 0,633776566 | LOC644086 | FALSE | Downregulated |
| -0,211835615 | 0,633776566 | SNHG17 | FALSE | Downregulated |
| -0,225416321 | 0,633776566 | TMEM198 | FALSE | Downregulated |
| -0,364512687 | 0,633776566 | TNKS2-AS1 | FALSE | Downregulated |
| 0,153049541 | 0,633776566 | ZNF503-AS2 | FALSE | Upregulated |
| 0,094605437 | 0,633781094 | TBRG4 | FALSE | Upregulated |
| -0,082104382 | 0,633882355 | ELP4 | FALSE | Downregulated |
| 0,092926828 | 0,633952612 | SLC38A3 | FALSE | Upregulated |
| -0,626480373 | 0,634089452 | LOC100132741 | FALSE | Downregulated |
| 0,340151212 | 0,634149327 | LOC101927963 | FALSE | Upregulated |
| -0,279004538 | 0,634381275 | IDH1-AS1 | FALSE | Downregulated |
| 0,261775231 | 0,634406133 | ZNF714 | FALSE | Upregulated |
| -0,087630124 | 0,634478849 | KIAA1715 | FALSE | Downregulated |
| -0,562391361 | 0,635136543 | FAM46B | FALSE | Downregulated |
| -0,086611006 | 0,635211353 | RARRES2 | FALSE | Downregulated |
| -0,247021769 | 0,635340038 | REEP1 | FALSE | Downregulated |
| -0,282261571 | 0,63552727 | LOC100507462 | FALSE | Downregulated |
| -0,109542793 | 0,635787544 | DGKD | FALSE | Downregulated |
| 0,083142114 | 0,63646777 | MRPL50 | FALSE | Upregulated |
| 0,194015086 | 0,63646777 | NCAM1 | FALSE | Upregulated |
| -0,193660998 | 0,63646777 | PDGFB | FALSE | Downregulated |
| -0,556969702 | 0,63646777 | PVRL3-AS1 | FALSE | Downregulated |
| -0,247785008 | 0,63646777 | SLC14A1 | FALSE | Downregulated |
| -0,096616206 | 0,636511389 | C2orf68 | FALSE | Downregulated |

| | | | | |
|---|---|---|---|---|
| -0,706449931 | 0,636562454 | GALNT5 | FALSE | Downregulated |
| -0,072570967 | 0,636562454 | NPAT | FALSE | Downregulated |
| -0,333067975 | 0,636562454 | TMEM105 | FALSE | Downregulated |
| -0,504378085 | 0,636562454 | TRND | FALSE | Downregulated |
| -0,257016255 | 0,636688124 | AC010226.4 | FALSE | Downregulated |
| -0,436720997 | 0,636688124 | LOC102724699 | FALSE | Downregulated |
| -0,587070723 | 0,636769539 | LOC101930332 | FALSE | Downregulated |
| 0,194093518 | 0,63681962 | TTN-AS1 | FALSE | Upregulated |
| -1,034593297 | 0,636896137 | LOC102723630 | FALSE | Downregulated |
| 0,106302646 | 0,637007491 | HEATR2 | FALSE | Upregulated |
| 0,290127907 | 0,637007491 | KCNE4 | FALSE | Upregulated |
| -0,09176979 | 0,637089429 | APP | FALSE | Downregulated |
| -0,090274084 | 0,637195026 | PHLDB3 | FALSE | Downregulated |
| -0,356205608 | 0,637333993 | HCG27 | FALSE | Downregulated |
| 0,069301281 | 0,637534981 | RNGTT | FALSE | Upregulated |
| -0,327473164 | 0,637631723 | DMKN | FALSE | Downregulated |
| -0,109275089 | 0,637644999 | CENPT | FALSE | Downregulated |
| 0,07870673 | 0,638053651 | ACSF3 | FALSE | Upregulated |
| 0,166208322 | 0,638053651 | PVRL1 | FALSE | Upregulated |
| -0,862169216 | 0,638053651 | SCRT2 | FALSE | Downregulated |
| 0,117281994 | 0,638053651 | SNAPC2 | FALSE | Upregulated |
| -0,322059767 | 0,638237771 | AKR1E2 | FALSE | Downregulated |
| 0,075669416 | 0,638237771 | FDX1 | FALSE | Upregulated |
| -0,073891504 | 0,638335799 | CRYL1 | FALSE | Downregulated |
| -0,702682457 | 0,638335799 | FUT2 | FALSE | Downregulated |
| -0,434273837 | 0,638335799 | LOC101927337 | FALSE | Downregulated |
| 0,068564879 | 0,638335799 | NELFCD | FALSE | Upregulated |
| 0,151702193 | 0,638341117 | CHN1 | FALSE | Upregulated |
| -0,064651508 | 0,638586478 | ANKIB1 | FALSE | Downregulated |
| -0,096739461 | 0,638716197 | CTDSPL2 | FALSE | Downregulated |
| -0,15787445 | 0,638716197 | EFNA4 | FALSE | Downregulated |
| -0,117761194 | 0,638831085 | ZNF74 | FALSE | Downregulated |
| 0,049200537 | 0,639075556 | TBK1 | FALSE | Upregulated |
| 0,283716138 | 0,639170902 | MS4A14 | FALSE | Upregulated |
| -0,463933926 | 0,639172425 | CCDC62 | FALSE | Downregulated |
| -0,125471166 | 0,639172425 | FAM102A | FALSE | Downregulated |
| -0,432885605 | 0,639213499 | LOC101928085 | FALSE | Downregulated |
| -0,091026491 | 0,639284523 | ZNF136 | FALSE | Downregulated |
| 0,644352309 | 0,639362151 | HECW1 | FALSE | Upregulated |
| -0,4381831 | 0,639465982 | RP11-401P9.5 | FALSE | Downregulated |
| -0,191455066 | 0,639465982 | TET1 | FALSE | Downregulated |
| 0,071415321 | 0,639465982 | WDR55 | FALSE | Upregulated |
| -0,130366944 | 0,639604451 | RSBN1L | FALSE | Downregulated |
| -0,071554455 | 0,63965423 | EDC4 | FALSE | Downregulated |
| -0,15886191 | 0,63965423 | PNPLA3 | FALSE | Downregulated |
| -0,321876914 | 0,63965423 | ZNF781 | FALSE | Downregulated |
| -0,765567323 | 0,639960896 | LOC102724144 | FALSE | Downregulated |
| 0,205788465 | 0,640016112 | AK7 | FALSE | Upregulated |
| -0,505550542 | 0,640095851 | FHL5 | FALSE | Downregulated |
| -0,224241469 | 0,640095851 | GLT8D2 | FALSE | Downregulated |
| -0,144770062 | 0,640095851 | OSBPL10 | FALSE | Downregulated |

| | | | | |
|---|---|---|---|---|
| -0,062357598 | 0,640095851 | SMIM15 | FALSE | Downregulated |
| -0,10739475 | 0,640202562 | DHRS12 | FALSE | Downregulated |
| 0,216229726 | 0,64030312 | ELOVL2-AS1 | FALSE | Upregulated |
| -0,183423622 | 0,640505015 | SVIL | FALSE | Downregulated |
| 0,078613706 | 0,640885304 | C5orf22 | FALSE | Upregulated |
| -0,088542801 | 0,640939804 | LOC102724364 | FALSE | Downregulated |
| -0,976949321 | 0,640939804 | PMP2 | FALSE | Downregulated |
| -0,416809866 | 0,641028741 | RP11-421P23.1 | FALSE | Downregulated |
| -0,273679982 | 0,641123265 | CYP46A1 | FALSE | Downregulated |
| 0,235365598 | 0,641123265 | SUMO1P3 | FALSE | Upregulated |
| 0,642512857 | 0,641338979 | GRM7 | FALSE | Upregulated |
| -0,662969063 | 0,641462263 | LOC101927720 | FALSE | Downregulated |
| -0,056707908 | 0,641462263 | MRPS35 | FALSE | Downregulated |
| -0,188344599 | 0,64148137 | STARD9 | FALSE | Downregulated |
| 0,062459781 | 0,641497541 | LAPTM4A | FALSE | Upregulated |
| 0,089826219 | 0,641497541 | NAGLU | FALSE | Upregulated |
| 0,087485367 | 0,641497541 | TEP1 | FALSE | Upregulated |
| 0,28177007 | 0,641497541 | TSPAN2 | FALSE | Upregulated |
| 0,149255297 | 0,641832391 | KLHL11 | FALSE | Upregulated |
| 0,149111217 | 0,641915719 | PKDCC | FALSE | Upregulated |
| -0,064759929 | 0,641946769 | TEX2 | FALSE | Downregulated |
| 0,115260802 | 0,642160273 | MFNG | FALSE | Upregulated |
| 0,088345275 | 0,642160273 | TIGD2 | FALSE | Upregulated |
| -0,165530337 | 0,64222523 | RP11-395P17.3 | FALSE | Downregulated |
| 0,119726094 | 0,642433071 | ZNF784 | FALSE | Upregulated |
| -0,0858331 | 0,642485405 | DDX3X | FALSE | Downregulated |
| -1,060757778 | 0,642492851 | CHRNA1 | FALSE | Downregulated |
| 0,052698271 | 0,642492851 | TRAK2 | FALSE | Upregulated |
| 0,07387716 | 0,64261466 | TMEM261 | FALSE | Upregulated |
| -0,601419362 | 0,642814691 | EPGN | FALSE | Downregulated |
| -0,195999026 | 0,642830187 | RGPD1 | FALSE | Downregulated |
| 0,147781489 | 0,642892524 | BAIAP3 | FALSE | Upregulated |
| 0,096080207 | 0,642892524 | FADS3 | FALSE | Upregulated |
| 0,122047325 | 0,642892524 | LNX1 | FALSE | Upregulated |
| -0,070996675 | 0,642892524 | MEF2D | FALSE | Downregulated |
| -0,169980275 | 0,642892524 | RRP7B | FALSE | Downregulated |
| 0,099618482 | 0,642939699 | CPEB4 | FALSE | Upregulated |
| 0,605663327 | 0,642951303 | LOC100128653 | FALSE | Upregulated |
| -0,109938398 | 0,643133889 | CDH5 | FALSE | Downregulated |
| -0,087665202 | 0,643133889 | CHMP4C | FALSE | Downregulated |
| -0,131957487 | 0,643423968 | MEIS3P1 | FALSE | Downregulated |
| -0,081812731 | 0,643430094 | AFF1 | FALSE | Downregulated |
| -0,18276965 | 0,643430094 | AZGP1P1 | FALSE | Downregulated |
| 0,087343316 | 0,643430094 | ERP29 | FALSE | Upregulated |
| 0,654547432 | 0,643468593 | ASAP1-IT2 | FALSE | Upregulated |
| -0,143407766 | 0,643549355 | ADAMTS9 | FALSE | Downregulated |
| 0,152561627 | 0,643549355 | ALG10 | FALSE | Upregulated |
| -0,248861514 | 0,643549355 | ANXA3 | FALSE | Downregulated |
| -0,08862562 | 0,643549355 | C3 | FALSE | Downregulated |
| -0,57581974 | 0,643549355 | CDKL2 | FALSE | Downregulated |
| 0,161105312 | 0,643549355 | EZR | FALSE | Upregulated |

| | | | | |
|---|---|---|---|---|
| 0,061728725 | 0,643549355 | FAM134C | FALSE | Upregulated |
| 0,635825844 | 0,643549355 | FXYD3 | FALSE | Upregulated |
| -0,163549334 | 0,643549355 | ITGAE | FALSE | Downregulated |
| -0,052952596 | 0,643549355 | KDM1A | FALSE | Downregulated |
| 0,431667285 | 0,643549355 | LOC102724037 | FALSE | Upregulated |
| 0,140224562 | 0,643549355 | LYL1 | FALSE | Upregulated |
| 0,195304295 | 0,643549355 | RAD51B | FALSE | Upregulated |
| -0,78462383 | 0,643549355 | RP11-280K24.4 | FALSE | Downregulated |
| 0,111088617 | 0,643549355 | SCAND1 | FALSE | Upregulated |
| -0,152915927 | 0,643549355 | SYNE4 | FALSE | Downregulated |
| 0,080554186 | 0,643549355 | ZCCHC2 | FALSE | Upregulated |
| 0,103659172 | 0,643605368 | C21orf91 | FALSE | Upregulated |
| 0,051880485 | 0,643605368 | GSPT1 | FALSE | Upregulated |
| -0,090072557 | 0,643605368 | RPS28 | FALSE | Downregulated |
| 0,415926685 | 0,643609796 | PSD2 | FALSE | Upregulated |
| 0,093792364 | 0,643675983 | TMA7 | FALSE | Upregulated |
| -0,106350882 | 0,644068519 | DENND2C | FALSE | Downregulated |
| 0,264676661 | 0,644112967 | ANXA13 | FALSE | Upregulated |
| 0,128055899 | 0,644112967 | LOC100506302 | FALSE | Upregulated |
| -0,500898907 | 0,644112967 | LOC101929004 | FALSE | Downregulated |
| 0,372771022 | 0,644112967 | MLC1 | FALSE | Upregulated |
| 0,095825084 | 0,644132016 | ENG | FALSE | Upregulated |
| 0,08284872 | 0,644132016 | SPHK2 | FALSE | Upregulated |
| 0,337884233 | 0,644132016 | ZNF296 | FALSE | Upregulated |
| -0,316476795 | 0,644135785 | SLC22A4 | FALSE | Downregulated |
| 0,216237268 | 0,644186352 | LOC100506922 | FALSE | Upregulated |
| 0,609901497 | 0,644186352 | RIMKLA | FALSE | Upregulated |
| -0,388586735 | 0,644278157 | FCER1A | FALSE | Downregulated |
| 0,189912581 | 0,644540523 | ARHGAP6 | FALSE | Upregulated |
| -0,773788422 | 0,644540523 | CDK15 | FALSE | Downregulated |
| -0,261041108 | 0,644562998 | LRRC55 | FALSE | Downregulated |
| -0,073807975 | 0,644564659 | DOCK7 | FALSE | Downregulated |
| 0,282343177 | 0,644564659 | LRRC46 | FALSE | Upregulated |
| 0,144243722 | 0,644564659 | PDIA4 | FALSE | Upregulated |
| 0,104248436 | 0,644709484 | PDK1 | FALSE | Upregulated |
| 0,241937616 | 0,644716846 | CASS4 | FALSE | Upregulated |
| -0,273887994 | 0,644771817 | LONRF2 | FALSE | Downregulated |
| -0,147608261 | 0,644771817 | MMAB | FALSE | Downregulated |
| -0,108348221 | 0,644808896 | SRSF7 | FALSE | Downregulated |
| 0,053488125 | 0,644819594 | CTNNB1 | FALSE | Upregulated |
| -0,100330984 | 0,644819594 | KDM5C | FALSE | Downregulated |
| 0,08371207 | 0,644824064 | CHMP6 | FALSE | Upregulated |
| 0,367929536 | 0,644824064 | TBX20 | FALSE | Upregulated |
| 0,178781741 | 0,644923261 | MICALL2 | FALSE | Upregulated |
| 0,510481673 | 0,645030195 | CCL24 | FALSE | Upregulated |
| -0,861481997 | 0,645055362 | TRNAK6 | FALSE | Downregulated |
| -0,076507382 | 0,645181139 | CTPS2 | FALSE | Downregulated |
| -0,163739733 | 0,645181139 | TMEM37 | FALSE | Downregulated |
| 0,37325895 | 0,645342346 | PAK3 | FALSE | Upregulated |
| -0,367271324 | 0,645413459 | MEP1B | FALSE | Downregulated |
| -0,285341012 | 0,645425157 | APOC1P1 | FALSE | Downregulated |

| | | | | |
|---|---|---|---|---|
| 0,357925448 | 0,645425157 | LRRTM4 | FALSE | Upregulated |
| 0,141341687 | 0,645854138 | C7orf43 | FALSE | Upregulated |
| -0,074635663 | 0,645854138 | TRRAP | FALSE | Downregulated |
| -0,226378849 | 0,645958071 | KIF12 | FALSE | Downregulated |
| 0,088150281 | 0,645958071 | NUP35 | FALSE | Upregulated |
| -0,131855317 | 0,64598915 | CEP112 | FALSE | Downregulated |
| -0,167364002 | 0,646050289 | HSD11B2 | FALSE | Downregulated |
| 0,446260273 | 0,646103407 | CISH | FALSE | Upregulated |
| 0,343062054 | 0,646103407 | LOC102724751 | FALSE | Upregulated |
| 0,421422806 | 0,646103407 | MFAP2 | FALSE | Upregulated |
| 0,099009945 | 0,646132235 | ABHD8 | FALSE | Upregulated |
| -0,447071635 | 0,646176923 | FAM218A | FALSE | Downregulated |
| -0,281200077 | 0,646241988 | LOC101929356 | FALSE | Downregulated |
| -0,702405125 | 0,646241988 | NRXN1 | FALSE | Downregulated |
| 0,087944058 | 0,646241988 | TULP4 | FALSE | Upregulated |
| -0,090338087 | 0,64625045 | SERGEF | FALSE | Downregulated |
| -0,487146905 | 0,646283345 | RNASE7 | FALSE | Downregulated |
| 0,09770158 | 0,646398968 | NENF | FALSE | Upregulated |
| 0,427260229 | 0,646439404 | KIAA1045 | FALSE | Upregulated |
| 0,8105995 | 0,646439404 | LOC102724951 | FALSE | Upregulated |
| 0,074421187 | 0,646439404 | MRPL35 | FALSE | Upregulated |
| 0,088650795 | 0,646469988 | BCL10 | FALSE | Upregulated |
| -0,258016447 | 0,646469988 | CCDC103 | FALSE | Downregulated |
| 0,514449286 | 0,646469988 | GRIK1-AS2 | FALSE | Upregulated |
| 0,079757943 | 0,646701866 | ICT1 | FALSE | Upregulated |
| 0,395125517 | 0,646772661 | RP11-274B18.2 | FALSE | Upregulated |
| 0,08981283 | 0,646801768 | SMCHD1 | FALSE | Upregulated |
| 0,127928951 | 0,64711368 | PPP1R35 | FALSE | Upregulated |
| -0,110747104 | 0,647121915 | RWDD4 | FALSE | Downregulated |
| -0,141139329 | 0,647121915 | U2AF1L4 | FALSE | Downregulated |
| -0,181917469 | 0,647165885 | RNF165 | FALSE | Downregulated |
| -0,347365582 | 0,647275724 | TRNA | FALSE | Downregulated |
| 0,11729827 | 0,647327138 | TRABD | FALSE | Upregulated |
| 0,145017196 | 0,647338062 | CDK2AP2 | FALSE | Upregulated |
| -0,101790057 | 0,647338062 | PTPLB | FALSE | Downregulated |
| 0,096422286 | 0,647571214 | CHML | FALSE | Upregulated |
| 0,065391753 | 0,647571214 | EGLN1 | FALSE | Upregulated |
| -0,093918562 | 0,647571214 | HERC1 | FALSE | Downregulated |
| -0,122239942 | 0,647571214 | HIST2H2BE | FALSE | Downregulated |
| -0,090602998 | 0,647571214 | INADL | FALSE | Downregulated |
| 0,094167777 | 0,647571214 | NDUFB11 | FALSE | Upregulated |
| -0,129595933 | 0,647571214 | TUBG2 | FALSE | Downregulated |
| 0,889137549 | 0,64758458 | URGCP-MRPS24 | FALSE | Upregulated |
| -0,085579432 | 0,647648395 | FBXW8 | FALSE | Downregulated |
| -0,592369148 | 0,647648395 | RFX8 | FALSE | Downregulated |
| 0,660021753 | 0,647750805 | MORC1 | FALSE | Upregulated |
| 0,200576242 | 0,647750805 | TC2N | FALSE | Upregulated |
| -0,257277785 | 0,6477624 | CAMK2B | FALSE | Downregulated |
| -0,193025746 | 0,6477624 | NPHP1 | FALSE | Downregulated |
| 0,055906369 | 0,6477624 | PRPF19 | FALSE | Upregulated |
| -0,177891736 | 0,647769629 | AQP7P3 | FALSE | Downregulated |

| | | | | |
|---|---|---|---|---|
| 0,301849125 | 0,647769629 | HMGCS1 | FALSE | Upregulated |
| -0,437615262 | 0,647769629 | LOC101927727 | FALSE | Downregulated |
| 0,624063634 | 0,647769629 | SNORA81 | FALSE | Upregulated |
| -0,079266172 | 0,648059918 | CASD1 | FALSE | Downregulated |
| 0,059564732 | 0,648184104 | CHTF8 | FALSE | Upregulated |
| -0,222201574 | 0,648201483 | PGR | FALSE | Downregulated |
| 0,129213121 | 0,648225324 | KLF15 | FALSE | Upregulated |
| 0,414091922 | 0,648388801 | C9orf153 | FALSE | Upregulated |
| -0,100450071 | 0,648388801 | COMT | FALSE | Downregulated |
| 0,462146071 | 0,648799072 | LOC102724248 | FALSE | Upregulated |
| 0,210031004 | 0,64895517 | LOC102724133 | FALSE | Upregulated |
| -0,082850947 | 0,648975327 | HMGXB3 | FALSE | Downregulated |
| 0,377099273 | 0,648975327 | KLRC3 | FALSE | Upregulated |
| 0,150952139 | 0,648975327 | SCNN1A | FALSE | Upregulated |
| -0,304851783 | 0,649287719 | LOC102724017 | FALSE | Downregulated |
| -0,156723336 | 0,649462693 | ARSE | FALSE | Downregulated |
| 0,452703552 | 0,649462693 | FSIP2 | FALSE | Upregulated |
| 0,067313811 | 0,649462693 | SLC25A32 | FALSE | Upregulated |
| 0,091162913 | 0,649603033 | MRTO4 | FALSE | Upregulated |
| 0,078183417 | 0,649603033 | SUV39H1 | FALSE | Upregulated |
| -0,07594344 | 0,649640591 | NUDT5 | FALSE | Downregulated |
| 0,410817956 | 0,649688711 | FKBP5 | FALSE | Upregulated |
| 0,209330287 | 0,649820821 | TPGS1 | FALSE | Upregulated |
| 0,072974238 | 0,650066346 | ATP5C1 | FALSE | Upregulated |
| -0,470885616 | 0,650066346 | LOC392364 | FALSE | Downregulated |
| 0,553587834 | 0,650066346 | SKOR1 | FALSE | Upregulated |
| 0,443601729 | 0,650066346 | SLC14A2 | FALSE | Upregulated |
| 0,118431948 | 0,650066346 | TRIM34 | FALSE | Upregulated |
| -0,188945308 | 0,650132116 | POMK | FALSE | Downregulated |
| -0,653636892 | 0,650197777 | GUSBP3 | FALSE | Downregulated |
| 0,32738611 | 0,650248253 | TICAM2 | FALSE | Upregulated |
| -0,148764403 | 0,650248253 | ZNF620 | FALSE | Downregulated |
| 0,231697344 | 0,650323663 | TSPAN5 | FALSE | Upregulated |
| -0,130485043 | 0,650517481 | PHC1 | FALSE | Downregulated |
| 0,060782578 | 0,650517481 | SNX1 | FALSE | Upregulated |
| 0,624970383 | 0,650594231 | TSSK3 | FALSE | Upregulated |
| 0,075358356 | 0,650719489 | FBXW11 | FALSE | Upregulated |
| -0,481870033 | 0,650811955 | LOC102723988 | FALSE | Downregulated |
| 0,115848066 | 0,651017435 | HAUS5 | FALSE | Upregulated |
| -0,681921747 | 0,651044683 | IGDCC4 | FALSE | Downregulated |
| -0,365431052 | 0,651150227 | LOC102723597 | FALSE | Downregulated |
| -0,246763952 | 0,65125595 | RND2 | FALSE | Downregulated |
| 0,254043064 | 0,651272601 | MTRNR2L3 | FALSE | Upregulated |
| 0,330523859 | 0,651366377 | LIPE-AS1 | FALSE | Upregulated |
| 0,445212365 | 0,651366377 | LOC101930310 | FALSE | Upregulated |
| -0,093870588 | 0,651496475 | FAM45A | FALSE | Downregulated |
| 0,13903799 | 0,651496475 | FNDC4 | FALSE | Upregulated |
| 0,059257079 | 0,651496475 | MINK1 | FALSE | Upregulated |
| 0,063456782 | 0,65152691 | ZCCHC10 | FALSE | Upregulated |
| -0,110867488 | 0,651607075 | PAPOLG | FALSE | Downregulated |
| 0,105499818 | 0,651607075 | RPL10A | FALSE | Upregulated |

| | | | | |
|---|---|---|---|---|
| -0,105736003 | 0,651683594 | FAM162A | FALSE | Downregulated |
| 0,065535015 | 0,651683594 | RNF167 | FALSE | Upregulated |
| -0,368406583 | 0,651900224 | TRPV3 | FALSE | Downregulated |
| -0,135608177 | 0,651925168 | FCN3 | FALSE | Downregulated |
| -0,117298651 | 0,65198574 | AC074286.1 | FALSE | Downregulated |
| -0,392787238 | 0,651990022 | STXBP5-AS1 | FALSE | Downregulated |
| -0,34100644 | 0,652026627 | LOC101928058 | FALSE | Downregulated |
| -0,305188334 | 0,652152563 | GULP1 | FALSE | Downregulated |
| -0,201138027 | 0,652333021 | SHD | FALSE | Downregulated |
| -0,452916949 | 0,652518507 | LOC102724683 | FALSE | Downregulated |
| -0,299841768 | 0,652518507 | TMEM183B | FALSE | Downregulated |
| -0,848991277 | 0,65251902 | EMX1 | FALSE | Downregulated |
| 0,28047857 | 0,652562384 | A1BG-AS1 | FALSE | Upregulated |
| -0,407244433 | 0,652562384 | LOC729162 | FALSE | Downregulated |
| -0,259728807 | 0,652686454 | HAND2-AS1 | FALSE | Downregulated |
| -0,058837212 | 0,65295995 | ZNF317 | FALSE | Downregulated |
| 0,078685732 | 0,653199357 | ATP5G3 | FALSE | Upregulated |
| 0,096832714 | 0,653336883 | FAM111A | FALSE | Upregulated |
| 0,121263658 | 0,653349832 | HSP90B1 | FALSE | Upregulated |
| -0,111350521 | 0,653579635 | ASPN | FALSE | Downregulated |
| -0,230717975 | 0,653579635 | CEACAM19 | FALSE | Downregulated |
| -0,428316439 | 0,653579635 | CLRN1-AS1 | FALSE | Downregulated |
| -0,072355516 | 0,653579635 | TBC1D14 | FALSE | Downregulated |
| 0,521224997 | 0,653579635 | TTLL13 | FALSE | Upregulated |
| -0,132441322 | 0,653650602 | ALKBH2 | FALSE | Downregulated |
| -0,073588234 | 0,653650602 | RNF7 | FALSE | Downregulated |
| 0,098746011 | 0,653675096 | ATG2A | FALSE | Upregulated |
| -0,168037775 | 0,653803588 | CES4A | FALSE | Downregulated |
| 0,057156822 | 0,653803588 | DCBLD1 | FALSE | Upregulated |
| 0,196154283 | 0,653832921 | COL5A1 | FALSE | Upregulated |
| -0,092925218 | 0,653832921 | FAM161B | FALSE | Downregulated |
| 0,201545637 | 0,653832921 | LAMA2 | FALSE | Upregulated |
| 0,71061239 | 0,653832921 | TRNG | FALSE | Upregulated |
| 0,398864307 | 0,653832921 | UNQ6494 | FALSE | Upregulated |
| 0,101374219 | 0,654140602 | ALG6 | FALSE | Upregulated |
| -0,268833371 | 0,654140602 | ESPNL | FALSE | Downregulated |
| -0,308725058 | 0,654493149 | LOC101928111 | FALSE | Downregulated |
| 0,536848142 | 0,654493149 | LOC101928414 | FALSE | Upregulated |
| 0,068228057 | 0,654493149 | TAF12 | FALSE | Upregulated |
| 0,095072472 | 0,654493149 | ZNF555 | FALSE | Upregulated |
| 0,265989866 | 0,654516105 | ZC3HAV1L | FALSE | Upregulated |
| 0,061382496 | 0,654675756 | CHTOP | FALSE | Upregulated |
| -0,12719322 | 0,654737916 | PHC3 | FALSE | Downregulated |
| 0,054026912 | 0,65476129 | CUEDC2 | FALSE | Upregulated |
| -0,235648454 | 0,65481088 | ALOX12P2 | FALSE | Downregulated |
| 0,403007519 | 0,65481088 | SMPDL3B | FALSE | Upregulated |
| -0,406228995 | 0,654823519 | LINC00222 | FALSE | Downregulated |
| -0,131489174 | 0,654830626 | KCNE3 | FALSE | Downregulated |
| -0,376826105 | 0,654857848 | GPR135 | FALSE | Downregulated |
| 0,09991337 | 0,654883599 | FBXL14 | FALSE | Upregulated |
| 0,082641502 | 0,654883599 | TMEM185A | FALSE | Upregulated |

| | | | | |
|---|---|---|---|---|
| -0,061926922 | 0,65495887 | CNPPD1 | FALSE | Downregulated |
| 0,069802679 | 0,65495887 | ELMOD2 | FALSE | Upregulated |
| -0,063118286 | 0,655146365 | NDUFB10 | FALSE | Downregulated |
| 0,272670417 | 0,655240318 | LOC100507054 | FALSE | Upregulated |
| -0,052773766 | 0,655371224 | CRIPT | FALSE | Downregulated |
| 0,07717702 | 0,655395095 | GGCX | FALSE | Upregulated |
| -0,21467144 | 0,655585915 | COLQ | FALSE | Downregulated |
| 0,163307559 | 0,655593855 | C16orf95 | FALSE | Upregulated |
| -0,054303517 | 0,655624089 | SUPT6H | FALSE | Downregulated |
| -0,284931168 | 0,655895975 | LOC100506454 | FALSE | Downregulated |
| 0,132617023 | 0,655958152 | CHRNE | FALSE | Upregulated |
| 0,096088322 | 0,656164227 | ACSF2 | FALSE | Upregulated |
| 0,094042212 | 0,65632765 | CHCHD1 | FALSE | Upregulated |
| 0,331836284 | 0,65632765 | KCNE1 | FALSE | Upregulated |
| 0,32328495 | 0,656361199 | RYR2 | FALSE | Upregulated |
| -0,113417554 | 0,657039497 | ZMYM4 | FALSE | Downregulated |
| 0,144349076 | 0,657324728 | ZNF138 | FALSE | Upregulated |
| -0,42777326 | 0,657623871 | NTN3 | FALSE | Downregulated |
| -0,383768949 | 0,657811151 | C22orf43 | FALSE | Downregulated |
| -0,078522432 | 0,657811151 | TRAPPC9 | FALSE | Downregulated |
| 0,097193658 | 0,657811151 | TTLL11 | FALSE | Upregulated |
| 0,066149 | 0,657974227 | C6orf106 | FALSE | Upregulated |
| 0,093689868 | 0,657974227 | TRMT44 | FALSE | Upregulated |
| -0,089814159 | 0,658248678 | FGFR4 | FALSE | Downregulated |
| -0,106454067 | 0,658254037 | ABCB11 | FALSE | Downregulated |
| -0,056036637 | 0,658254037 | EIF3H | FALSE | Downregulated |
| 0,134578436 | 0,658254037 | PELP1 | FALSE | Upregulated |
| -0,160246766 | 0,658254037 | PLIN4 | FALSE | Downregulated |
| -0,681979561 | 0,658254037 | SLCO5A1 | FALSE | Downregulated |
| 0,188465424 | 0,658466666 | FAM131A | FALSE | Upregulated |
| -0,649614589 | 0,658466666 | LOC100133182 | FALSE | Downregulated |
| 0,103903854 | 0,658594787 | LRRCC1 | FALSE | Upregulated |
| -0,045728182 | 0,65885654 | ATP5A1 | FALSE | Downregulated |
| -0,192149162 | 0,659057318 | TSPYL5 | FALSE | Downregulated |
| 0,541808524 | 0,659061307 | CLEC4C | FALSE | Upregulated |
| 0,279249543 | 0,659061307 | ITGBL1 | FALSE | Upregulated |
| 0,180962593 | 0,659061307 | MTBP | FALSE | Upregulated |
| -0,422038536 | 0,659269535 | LINC00665 | FALSE | Downregulated |
| -0,094854436 | 0,659269535 | ZRANB3 | FALSE | Downregulated |
| -0,451163821 | 0,659424912 | LMCD1-AS1 | FALSE | Downregulated |
| -0,384186008 | 0,659519162 | C9orf139 | FALSE | Downregulated |
| -0,686743997 | 0,659519162 | FSD2 | FALSE | Downregulated |
| -0,810239704 | 0,659519162 | TUBA3D | FALSE | Downregulated |
| -0,208476591 | 0,659539971 | MYLK4 | FALSE | Downregulated |
| -0,229987886 | 0,659539971 | TPI1P2 | FALSE | Downregulated |
| 0,255799625 | 0,659611039 | DNAJC18 | FALSE | Upregulated |
| -0,138126091 | 0,659611039 | GK5 | FALSE | Downregulated |
| 0,391899508 | 0,659611039 | LOC101929381 | FALSE | Upregulated |
| -1,060497622 | 0,659611039 | MYOC | FALSE | Downregulated |
| 0,090597636 | 0,659611039 | RPS25 | FALSE | Upregulated |
| -0,106729852 | 0,659617916 | GUCY1B3 | FALSE | Downregulated |

| | | | | |
|---|---|---|---|---|
| -0,183171716 | 0,659617916 | LOC100129637 | FALSE | Downregulated |
| -0,374251182 | 0,659617916 | LOC102724757 | FALSE | Downregulated |
| 0,093545731 | 0,659617916 | SCFD2 | FALSE | Upregulated |
| 0,233880309 | 0,659617916 | TRIM61 | FALSE | Upregulated |
| 0,16096335 | 0,659648273 | S100A14 | FALSE | Upregulated |
| 0,084254627 | 0,659648273 | WRB | FALSE | Upregulated |
| 0,120383207 | 0,659690617 | TMEM117 | FALSE | Upregulated |
| 0,064858817 | 0,659734964 | SENP1 | FALSE | Upregulated |
| 0,189133781 | 0,659904413 | PHKG1 | FALSE | Upregulated |
| 0,145882741 | 0,660147635 | LHX2 | FALSE | Upregulated |
| 0,297804315 | 0,660147635 | LOC102723422 | FALSE | Upregulated |
| 0,104823157 | 0,660147635 | TSEN34 | FALSE | Upregulated |
| -0,306384089 | 0,660196929 | LOC102724190 | FALSE | Downregulated |
| -0,138277484 | 0,660197735 | FBXO36 | FALSE | Downregulated |
| 0,088868583 | 0,660205169 | GIT1 | FALSE | Upregulated |
| 0,424388884 | 0,660205169 | LOC102724634 | FALSE | Upregulated |
| 0,097884888 | 0,660205169 | RPL32 | FALSE | Upregulated |
| 0,188870343 | 0,660205169 | SYT12 | FALSE | Upregulated |
| 0,192092174 | 0,660205169 | TSTD3 | FALSE | Upregulated |
| 0,782234 | 0,660281419 | LOC650157 | FALSE | Upregulated |
| -0,251868606 | 0,660462567 | LOC730183 | FALSE | Downregulated |
| -0,649748693 | 0,660641186 | ANKRD20A1 | FALSE | Downregulated |
| 0,043049039 | 0,660861291 | TSN | FALSE | Upregulated |
| -0,070616237 | 0,661116518 | SRBD1 | FALSE | Downregulated |
| -0,933143991 | 0,66129438 | DCC | FALSE | Downregulated |
| -0,227156479 | 0,661340945 | DCST2 | FALSE | Downregulated |
| 0,3053191 | 0,661389041 | C6orf132 | FALSE | Upregulated |
| -0,084394324 | 0,661389041 | PDLIM5 | FALSE | Downregulated |
| 0,158650305 | 0,661827628 | HIST1H2AC | FALSE | Upregulated |
| 0,076912893 | 0,661827628 | PFDN5 | FALSE | Upregulated |
| -0,072653674 | 0,661827628 | PIK3CA | FALSE | Downregulated |
| 0,109950332 | 0,661827628 | TUSC1 | FALSE | Upregulated |
| -0,111864108 | 0,661876004 | LOC728769 | FALSE | Downregulated |
| 0,149432307 | 0,661876004 | RARA-AS1 | FALSE | Upregulated |
| 0,584752462 | 0,661948342 | PLSCR3 | FALSE | Upregulated |
| 0,216507107 | 0,661948342 | TUBA8 | FALSE | Upregulated |
| 0,45290366 | 0,661950408 | HIST1H4J | FALSE | Upregulated |
| 0,387153085 | 0,661950408 | LOC646652 | FALSE | Upregulated |
| 0,05928999 | 0,661986205 | UHMK1 | FALSE | Upregulated |
| -0,056070795 | 0,662063411 | COG7 | FALSE | Downregulated |
| 0,068834044 | 0,662063411 | FAM83H | FALSE | Upregulated |
| -0,048283392 | 0,662200308 | DFFA | FALSE | Downregulated |
| -0,062326031 | 0,662200308 | PGGT1B | FALSE | Downregulated |
| -0,065272074 | 0,662329507 | C18orf25 | FALSE | Downregulated |
| -0,057469811 | 0,662346565 | ISG20L2 | FALSE | Downregulated |
| -0,088012703 | 0,6623761 | MAP3K1 | FALSE | Downregulated |
| 0,525190421 | 0,662451037 | LOC101929330 | FALSE | Upregulated |
| -0,103972462 | 0,662765832 | LOC100506746 | FALSE | Downregulated |
| 0,188564285 | 0,662765832 | MRVI1 | FALSE | Upregulated |
| -0,074399663 | 0,662765832 | NOL8 | FALSE | Downregulated |
| 0,109205302 | 0,662765832 | PEMT | FALSE | Upregulated |

| | | | | |
|---|---|---|---|---|
| -0,593488081 | 0,662765832 | RHBDL3 | FALSE | Downregulated |
| 0,243660944 | 0,662765832 | RTN2 | FALSE | Upregulated |
| 0,110671925 | 0,662765832 | THAP3 | FALSE | Upregulated |
| 0,077074652 | 0,662785527 | VPS13C | FALSE | Upregulated |
| -0,443240791 | 0,662834494 | CDRT1 | FALSE | Downregulated |
| 0,047059141 | 0,662834494 | PRKRIR | FALSE | Upregulated |
| 0,052358906 | 0,662834494 | USP1 | FALSE | Upregulated |
| -0,35224609 | 0,663126042 | GNAT2 | FALSE | Downregulated |
| 0,181331076 | 0,663456947 | LOC100129461 | FALSE | Upregulated |
| -0,314536742 | 0,663502099 | CCDC80 | FALSE | Downregulated |
| 0,672929564 | 0,663502099 | HOXA7 | FALSE | Upregulated |
| 0,277093828 | 0,663740514 | ERP27 | FALSE | Upregulated |
| -0,13944652 | 0,663740514 | RGN | FALSE | Downregulated |
| -0,057589915 | 0,663788608 | SNRK | FALSE | Downregulated |
| -0,580625242 | 0,663858412 | ENO1-AS1 | FALSE | Downregulated |
| -0,176728679 | 0,663858412 | INTU | FALSE | Downregulated |
| -0,188765932 | 0,663941374 | ZNF573 | FALSE | Downregulated |
| -0,082330617 | 0,66395741 | ZBTB34 | FALSE | Downregulated |
| 0,514835725 | 0,664029032 | RP11-770J1.3 | FALSE | Upregulated |
| 0,083683637 | 0,6640616 | FIS1 | FALSE | Upregulated |
| 0,338796431 | 0,664132744 | DND1 | FALSE | Upregulated |
| 0,188513245 | 0,664132744 | LOC389906 | FALSE | Upregulated |
| 0,314807854 | 0,664205997 | PKN3 | FALSE | Upregulated |
| 0,781060454 | 0,664286867 | YBX3P1 | FALSE | Upregulated |
| -0,230913545 | 0,664326432 | CCDC168 | FALSE | Downregulated |
| -0,390874933 | 0,66440769 | CTSG | FALSE | Downregulated |
| -0,090951827 | 0,66440769 | ITFG2 | FALSE | Downregulated |
| -0,279769521 | 0,66440769 | LINC01023 | FALSE | Downregulated |
| 0,466234183 | 0,66440769 | LINC01119 | FALSE | Upregulated |
| 1,002572724 | 0,66440769 | LOC102723678 | FALSE | Upregulated |
| -0,172464869 | 0,66440769 | SCARA3 | FALSE | Downregulated |
| -0,07890715 | 0,66440769 | TBC1D4 | FALSE | Downregulated |
| -0,144988044 | 0,664786186 | RPGRIP1L | FALSE | Downregulated |
| 0,094460573 | 0,664865642 | GORASP2 | FALSE | Upregulated |
| 0,052757753 | 0,664967131 | DDX6 | FALSE | Upregulated |
| -0,094872712 | 0,664967131 | TCEAL4 | FALSE | Downregulated |
| 0,115626432 | 0,664967131 | TMEM169 | FALSE | Upregulated |
| 0,209579436 | 0,665043572 | LBP | FALSE | Upregulated |
| 0,655055819 | 0,665043572 | RP11-397C18.2 | FALSE | Upregulated |
| -0,165371183 | 0,665380273 | RND1 | FALSE | Downregulated |
| -0,398824138 | 0,665420753 | C9orf24 | FALSE | Downregulated |
| 0,581492849 | 0,665420753 | CRB2 | FALSE | Upregulated |
| -0,119507456 | 0,665420753 | ZBTB22 | FALSE | Downregulated |
| 0,099516029 | 0,665492518 | SNX29 | FALSE | Upregulated |
| -0,061170987 | 0,665642084 | RSPRY1 | FALSE | Downregulated |
| -0,666082251 | 0,665659764 | MUC16 | FALSE | Downregulated |
| -0,182423875 | 0,66598616 | LOC100506286 | FALSE | Downregulated |
| -0,235806491 | 0,666200723 | EFHD1 | FALSE | Downregulated |
| -0,162227798 | 0,666550286 | LRRC17 | FALSE | Downregulated |
| 0,074489425 | 0,666661421 | CES2 | FALSE | Upregulated |
| 0,350221728 | 0,666661421 | RP11-367N14.2 | FALSE | Upregulated |

| | | | | |
|---|---|---|---|---|
| -0,418564903 | 0,666933663 | SPTB | FALSE | Downregulated |
| -0,104134662 | 0,667015279 | FAM3A | FALSE | Downregulated |
| -0,080161175 | 0,667129487 | SLC8B1 | FALSE | Downregulated |
| -0,095237149 | 0,667161278 | YRDC | FALSE | Downregulated |
| -0,062618709 | 0,667191358 | UTP23 | FALSE | Downregulated |
| -0,106941463 | 0,667208361 | KREMEN1 | FALSE | Downregulated |
| -0,249716918 | 0,667226983 | PILRB | FALSE | Downregulated |
| 0,126909129 | 0,667229694 | SEC23B | FALSE | Upregulated |
| 0,079420916 | 0,667449942 | POLE | FALSE | Upregulated |
| -0,342564792 | 0,668012402 | MYLK3 | FALSE | Downregulated |
| -0,337146506 | 0,668180077 | LSMEM2 | FALSE | Downregulated |
| 0,098428816 | 0,668210625 | APOA2 | FALSE | Upregulated |
| -0,311544699 | 0,668210625 | LOC401286 | FALSE | Downregulated |
| 0,113085502 | 0,668210625 | NSUN3 | FALSE | Upregulated |
| 0,260271727 | 0,668291089 | LOC101928719 | FALSE | Upregulated |
| -0,100543105 | 0,668291089 | TIMM8A | FALSE | Downregulated |
| -0,120672947 | 0,668291089 | ZNF600 | FALSE | Downregulated |
| -0,129340613 | 0,668325539 | OSBPL1A | FALSE | Downregulated |
| 0,164807403 | 0,668325539 | P2RY2 | FALSE | Upregulated |
| 0,273776544 | 0,668326818 | B3GALT2 | FALSE | Upregulated |
| 0,049365551 | 0,668326818 | DNAJC11 | FALSE | Upregulated |
| -0,110118932 | 0,668326818 | DNAJC21 | FALSE | Downregulated |
| -0,621839408 | 0,668326818 | LOC344967 | FALSE | Downregulated |
| -0,098084932 | 0,66839289 | GCGR | FALSE | Downregulated |
| -0,273502734 | 0,668409269 | MAN1B1-AS1 | FALSE | Downregulated |
| 0,058000706 | 0,668409269 | WDR45B | FALSE | Upregulated |
| 0,143644773 | 0,668472684 | RNPS1 | FALSE | Upregulated |
| 0,156834117 | 0,668497305 | THAP10 | FALSE | Upregulated |
| -0,254023192 | 0,668560659 | ADIRF | FALSE | Downregulated |
| 0,055639951 | 0,668560659 | TAOK2 | FALSE | Upregulated |
| 0,13308718 | 0,66863331 | RHOJ | FALSE | Upregulated |
| -0,469886268 | 0,668657053 | ADAMTSL4-AS1 | FALSE | Downregulated |
| 0,084220553 | 0,668657053 | CIC | FALSE | Upregulated |
| -0,062828339 | 0,668682826 | RPS6KB1 | FALSE | Downregulated |
| 0,585865913 | 0,668699464 | AC097500.2 | FALSE | Upregulated |
| -0,277198713 | 0,668699464 | BEND5 | FALSE | Downregulated |
| -0,504095104 | 0,668784742 | AC068831.10 | FALSE | Downregulated |
| -0,376164566 | 0,668784742 | FAM45B | FALSE | Downregulated |
| 0,398223511 | 0,668784742 | PLD4 | FALSE | Upregulated |
| 0,197125479 | 0,668784742 | RAD9B | FALSE | Upregulated |
| -0,089432876 | 0,668784742 | UBL4A | FALSE | Downregulated |
| 0,1132961 | 0,668857535 | ZNF319 | FALSE | Upregulated |
| -0,324263526 | 0,669232539 | HOXA2 | FALSE | Downregulated |
| -0,096156107 | 0,669232539 | SERPINA4 | FALSE | Downregulated |
| -0,874773703 | 0,669507547 | LOC101928552 | FALSE | Downregulated |
| -0,215229806 | 0,669668111 | ITIH5 | FALSE | Downregulated |
| -0,072139752 | 0,669669195 | PRMT3 | FALSE | Downregulated |
| -0,054440542 | 0,669669195 | TEX261 | FALSE | Downregulated |
| 0,372274935 | 0,669801744 | DLGAP3 | FALSE | Upregulated |
| -0,254197025 | 0,669801744 | LINC00337 | FALSE | Downregulated |
| -0,352155719 | 0,669801744 | LOC102723671 | FALSE | Downregulated |

| | | | | |
|---|---|---|---|---|
| 0,047955058 | 0,670050137 | PLRG1 | FALSE | Upregulated |
| -0,232949909 | 0,670050137 | UNC93A | FALSE | Downregulated |
| 0,26636556 | 0,670111541 | OMD | FALSE | Upregulated |
| 0,229226177 | 0,670122861 | SH3D21 | FALSE | Upregulated |
| 0,22586939 | 0,670376176 | CDC42EP5 | FALSE | Upregulated |
| 0,324167196 | 0,670376176 | HIST3H2A | FALSE | Upregulated |
| 0,080609304 | 0,670376176 | LAMTOR2 | FALSE | Upregulated |
| 0,09684534 | 0,670376176 | LINC00984 | FALSE | Upregulated |
| -0,087638767 | 0,670376176 | MRPS18B | FALSE | Downregulated |
| -0,05831273 | 0,670376176 | NCOA1 | FALSE | Downregulated |
| 0,146339883 | 0,670376176 | PLK2 | FALSE | Upregulated |
| 1,499857587 | 0,670376176 | PRAMEF10 | FALSE | Upregulated |
| -0,584703715 | 0,670376176 | RP11-381O7.3 | FALSE | Downregulated |
| -0,076189307 | 0,670376176 | STARD8 | FALSE | Downregulated |
| -0,482541756 | 0,670376176 | WDR93 | FALSE | Downregulated |
| 0,063668223 | 0,670384062 | SMIM7 | FALSE | Upregulated |
| -0,119803087 | 0,670703246 | RNF144A | FALSE | Downregulated |
| -0,396773364 | 0,670766634 | RP11-135J2.4 | FALSE | Downregulated |
| 0,816880167 | 0,670909075 | HSFX2 | FALSE | Upregulated |
| -0,166396229 | 0,670980792 | C15orf62 | FALSE | Downregulated |
| -0,086531768 | 0,670980792 | EMC2 | FALSE | Downregulated |
| 0,098568677 | 0,671218903 | RGS12 | FALSE | Upregulated |
| 0,102116368 | 0,671270648 | CCNK | FALSE | Upregulated |
| 0,103374092 | 0,67127837 | C16orf91 | FALSE | Upregulated |
| -0,09843019 | 0,67127837 | EVL | FALSE | Downregulated |
| -1,146405244 | 0,671350892 | MIR5685 | FALSE | Downregulated |
| -0,531559368 | 0,671361854 | CXCL17 | FALSE | Downregulated |
| -0,503932559 | 0,671361854 | LOC102723627 | FALSE | Downregulated |
| 0,132920577 | 0,671635266 | BRD9 | FALSE | Upregulated |
| -2,325105875 | 0,671663652 | LOC102723360 | FALSE | Downregulated |
| -0,105391722 | 0,671947058 | GCFC2 | FALSE | Downregulated |
| -0,43553014 | 0,671947058 | LOC102724583 | FALSE | Downregulated |
| 0,975941089 | 0,671947058 | LOC730268 | FALSE | Upregulated |
| -0,412260255 | 0,672092107 | H3F3C | FALSE | Downregulated |
| -0,208713901 | 0,672092107 | ZNF880 | FALSE | Downregulated |
| -0,071055248 | 0,672255306 | METTL2A | FALSE | Downregulated |
| 0,121053993 | 0,672255306 | SFT2D2 | FALSE | Upregulated |
| -0,398171546 | 0,672386139 | CACNB4 | FALSE | Downregulated |
| 0,037430552 | 0,672464038 | DDX24 | FALSE | Upregulated |
| -0,315452157 | 0,672464038 | FAM188B | FALSE | Downregulated |
| 0,520509564 | 0,672464038 | KIR2DL1 | FALSE | Upregulated |
| -0,104792067 | 0,672464038 | NCOR1 | FALSE | Downregulated |
| -0,256351145 | 0,672464038 | NUTM2D | FALSE | Downregulated |
| 0,28136566 | 0,672464038 | RUSC1-AS1 | FALSE | Upregulated |
| -0,128433613 | 0,672464038 | TMEM128 | FALSE | Downregulated |
| 0,071666122 | 0,672468816 | FAM96B | FALSE | Upregulated |
| -0,079431705 | 0,672500486 | GIGYF2 | FALSE | Downregulated |
| 0,430131545 | 0,672660866 | RBM12B-AS1 | FALSE | Upregulated |
| -0,543709514 | 0,67270676 | NOS1 | FALSE | Downregulated |
| -0,107856466 | 0,672805749 | RNLS | FALSE | Downregulated |
| 0,178872949 | 0,672815984 | FAM43A | FALSE | Upregulated |

| | | | | |
|---|---|---|---|---|
| 0,071647866 | 0,672843858 | RMI1 | FALSE | Upregulated |
| -0,070668487 | 0,672959591 | TM2D2 | FALSE | Downregulated |
| 0,059001462 | 0,672991548 | ABCE1 | FALSE | Upregulated |
| -0,059774864 | 0,673015177 | TRUB2 | FALSE | Downregulated |
| -0,069795538 | 0,673020902 | EIF2D | FALSE | Downregulated |
| 0,056523481 | 0,673020902 | SF1 | FALSE | Upregulated |
| 0,044389905 | 0,673142271 | ARIH2 | FALSE | Upregulated |
| 0,044196187 | 0,673187196 | VPS36 | FALSE | Upregulated |
| 0,165042597 | 0,673351544 | MGC57346 | FALSE | Upregulated |
| -0,157618942 | 0,673356668 | NLRP1 | FALSE | Downregulated |
| 0,149286149 | 0,673580522 | SLC26A6 | FALSE | Upregulated |
| 0,435573138 | 0,673816208 | PAPPA2 | FALSE | Upregulated |
| -0,272501735 | 0,673910409 | LOC155060 | FALSE | Downregulated |
| 0,492990851 | 0,674582852 | ASB14 | FALSE | Upregulated |
| -0,072006456 | 0,674647611 | TRMT61B | FALSE | Downregulated |
| -0,065794776 | 0,674874112 | OSBPL8 | FALSE | Downregulated |
| -0,070679057 | 0,674950051 | F8A1 | FALSE | Downregulated |
| -0,053041292 | 0,674976174 | VPS26B | FALSE | Downregulated |
| -0,236380501 | 0,675459318 | KIF6 | FALSE | Downregulated |
| -0,410749418 | 0,675495686 | SLC26A8 | FALSE | Downregulated |
| 0,111025581 | 0,675858327 | MICAL3 | FALSE | Upregulated |
| 0,420135023 | 0,676214142 | C1orf106 | FALSE | Upregulated |
| -0,07330521 | 0,676214142 | CRCP | FALSE | Downregulated |
| -0,09112823 | 0,676227638 | CXorf23 | FALSE | Downregulated |
| 0,540648886 | 0,676227638 | PRSS27 | FALSE | Upregulated |
| -0,179497019 | 0,676973034 | DZIP1 | FALSE | Downregulated |
| -0,319196155 | 0,677116357 | SKP1P2 | FALSE | Downregulated |
| 0,054566317 | 0,677305062 | GPATCH3 | FALSE | Upregulated |
| 0,09164993 | 0,677384479 | ALPK1 | FALSE | Upregulated |
| -0,545310237 | 0,677384479 | LOC101928592 | FALSE | Downregulated |
| 0,213176898 | 0,677417778 | TEAD4 | FALSE | Upregulated |
| 0,109872739 | 0,677432611 | DUSP23 | FALSE | Upregulated |
| -0,126239806 | 0,677432611 | FGD4 | FALSE | Downregulated |
| -0,659890377 | 0,677432611 | LOC101929132 | FALSE | Downregulated |
| 0,566939132 | 0,677432611 | LOC101929469 | FALSE | Upregulated |
| 0,065714338 | 0,677432611 | PRKCSH | FALSE | Upregulated |
| 0,09347976 | 0,677529429 | VCL | FALSE | Upregulated |
| 0,067928607 | 0,677592308 | OAZ2 | FALSE | Upregulated |
| 0,117968449 | 0,677939196 | ZNF653 | FALSE | Upregulated |
| -0,333973864 | 0,678058464 | CCDC74A | FALSE | Downregulated |
| -0,071028441 | 0,678058464 | DDX23 | FALSE | Downregulated |
| -0,276196306 | 0,678058464 | LOC101927763 | FALSE | Downregulated |
| -0,348334398 | 0,678058464 | MED4-AS1 | FALSE | Downregulated |
| -0,065575274 | 0,678058464 | PPFIA1 | FALSE | Downregulated |
| -0,103379011 | 0,678068635 | CIR1 | FALSE | Downregulated |
| -0,133730153 | 0,678068635 | RAC3 | FALSE | Downregulated |
| 0,090974193 | 0,678068635 | SFXN4 | FALSE | Upregulated |
| -0,186729931 | 0,678117166 | ACER2 | FALSE | Downregulated |
| 0,074149717 | 0,678117166 | C15orf38 | FALSE | Upregulated |
| -0,083817529 | 0,678117166 | SLC25A4 | FALSE | Downregulated |
| 0,25688712 | 0,678117166 | ZNRF2P1 | FALSE | Upregulated |

| | | | | |
|---|---|---|---|---|
| 0,073027763 | 0,678183499 | PHB2 | FALSE | Upregulated |
| -0,035622285 | 0,678215834 | EIF2B1 | FALSE | Downregulated |
| 0,30535963 | 0,678215834 | PPAPDC1A | FALSE | Upregulated |
| -0,26219207 | 0,678215834 | SERINC4 | FALSE | Downregulated |
| 0,26314981 | 0,67827223 | NLGN3 | FALSE | Upregulated |
| -0,122772687 | 0,67827223 | PIEZO2 | FALSE | Downregulated |
| -0,415789618 | 0,67827223 | SRGAP2D | FALSE | Downregulated |
| -0,053690528 | 0,678317057 | ACP1 | FALSE | Downregulated |
| 0,286614755 | 0,678317057 | CNTD2 | FALSE | Upregulated |
| -1,046175165 | 0,678317057 | KY | FALSE | Downregulated |
| 0,1384255 | 0,678317057 | LAGE3 | FALSE | Upregulated |
| 0,52902035 | 0,678317057 | LOC100653133 | FALSE | Upregulated |
| 0,072291113 | 0,678317057 | POLRMT | FALSE | Upregulated |
| 0,295332816 | 0,678317057 | PPM1J | FALSE | Upregulated |
| 0,065399631 | 0,678317057 | TBCE | FALSE | Upregulated |
| -0,06107032 | 0,678317057 | WBP2 | FALSE | Downregulated |
| 0,104404536 | 0,678317057 | ZFAS1 | FALSE | Upregulated |
| 0,070803 | 0,678514632 | ISCA1 | FALSE | Upregulated |
| -0,064168989 | 0,678514632 | PIKFYVE | FALSE | Downregulated |
| 0,054559748 | 0,678514632 | TCF20 | FALSE | Upregulated |
| 0,509153089 | 0,678687683 | PTH2R | FALSE | Upregulated |
| 0,395384473 | 0,67888034 | CHRM3 | FALSE | Upregulated |
| -0,144645418 | 0,67888034 | KANK3 | FALSE | Downregulated |
| 0,105524894 | 0,67888034 | MIDN | FALSE | Upregulated |
| -0,562550545 | 0,678918318 | CRX | FALSE | Downregulated |
| 0,408491298 | 0,678972911 | DOC2A | FALSE | Upregulated |
| 0,129267058 | 0,678983868 | ECE2 | FALSE | Upregulated |
| 0,441992007 | 0,679120631 | LOC102724584 | FALSE | Upregulated |
| -0,51732933 | 0,6791932 | PEG3-AS1 | FALSE | Downregulated |
| -0,350063616 | 0,679206769 | GUCA2B | FALSE | Downregulated |
| -0,045767464 | 0,679219743 | RABEP1 | FALSE | Downregulated |
| -0,104095889 | 0,679302606 | AMZ2P1 | FALSE | Downregulated |
| 0,080538326 | 0,679311139 | RABL5 | FALSE | Upregulated |
| 0,517337931 | 0,679328009 | AK8 | FALSE | Upregulated |
| -0,110099519 | 0,679328009 | TM4SF18 | FALSE | Downregulated |
| 0,102819131 | 0,679369385 | C19orf25 | FALSE | Upregulated |
| -0,069970662 | 0,679369385 | TSC22D2 | FALSE | Downregulated |
| 0,071852865 | 0,679399528 | RPL18 | FALSE | Upregulated |
| -0,097620249 | 0,679511466 | CAPN5 | FALSE | Downregulated |
| -0,540839846 | 0,679511466 | PRICKLE2-AS1 | FALSE | Downregulated |
| -0,097381191 | 0,679557969 | C10orf25 | FALSE | Downregulated |
| -0,087137498 | 0,679557969 | MIOS | FALSE | Downregulated |
| -0,106338597 | 0,67966985 | C5orf51 | FALSE | Downregulated |
| -0,190413623 | 0,679912145 | MUM1L1 | FALSE | Downregulated |
| 0,080948209 | 0,679932436 | DGUOK | FALSE | Upregulated |
| 0,350260393 | 0,679932436 | LINC00607 | FALSE | Upregulated |
| -0,160905617 | 0,679932436 | TBX2 | FALSE | Downregulated |
| 0,071468982 | 0,679974658 | ST6GALNAC6 | FALSE | Upregulated |
| -0,241795907 | 0,679990315 | NLRP6 | FALSE | Downregulated |
| -0,144384733 | 0,68003935 | HSPA12A | FALSE | Downregulated |
| -0,918342957 | 0,680088969 | DUSP26 | FALSE | Downregulated |

| | | | | |
|---|---|---|---|---|
| 0,107084192 | 0,680215875 | MFAP1 | FALSE | Upregulated |
| 0,20351888 | 0,680224519 | MAP6 | FALSE | Upregulated |
| -0,311798994 | 0,680224519 | RP11-539L10.2 | FALSE | Downregulated |
| -0,051326476 | 0,680226594 | GOSR1 | FALSE | Downregulated |
| -0,408287788 | 0,680226594 | LMX1A | FALSE | Downregulated |
| -0,043545914 | 0,680412284 | SERBP1 | FALSE | Downregulated |
| 0,090781838 | 0,680514781 | CCDC91 | FALSE | Upregulated |
| -0,09802032 | 0,680804917 | RASA2 | FALSE | Downregulated |
| -0,048802636 | 0,680835842 | MED6 | FALSE | Downregulated |
| -0,174933785 | 0,680866271 | LINC00899 | FALSE | Downregulated |
| -0,441265655 | 0,680909459 | ZNF233 | FALSE | Downregulated |
| -0,166946509 | 0,680915838 | NBPF12 | FALSE | Downregulated |
| -0,191208687 | 0,681138502 | LOC101927451 | FALSE | Downregulated |
| 0,48741975 | 0,681138502 | STK32B | FALSE | Upregulated |
| -0,187730062 | 0,681292004 | SLC16A13 | FALSE | Downregulated |
| -0,323725879 | 0,681333334 | CCDC38 | FALSE | Downregulated |
| 0,846263072 | 0,681333334 | RP11-394O9.1 | FALSE | Upregulated |
| -0,043122248 | 0,681409937 | SNW1 | FALSE | Downregulated |
| 0,044196656 | 0,681444886 | CTCF | FALSE | Upregulated |
| 0,603575139 | 0,681444886 | RASA4B | FALSE | Upregulated |
| -0,094333576 | 0,681444886 | SALL1 | FALSE | Downregulated |
| -0,144797911 | 0,681505542 | PIM1 | FALSE | Downregulated |
| -0,080924971 | 0,681753747 | HMGXB4 | FALSE | Downregulated |
| -0,11850206 | 0,681786236 | CCDC18 | FALSE | Downregulated |
| -0,261408177 | 0,682113748 | CELSR3 | FALSE | Downregulated |
| -0,076561304 | 0,682113748 | CENPBD1P1 | FALSE | Downregulated |
| 0,253425545 | 0,682113748 | SNPH | FALSE | Upregulated |
| -0,072633581 | 0,682271548 | JARID2 | FALSE | Downregulated |
| 0,302830806 | 0,682378795 | ADAMTS4 | FALSE | Upregulated |
| -0,347889758 | 0,682421141 | IAPP | FALSE | Downregulated |
| -0,440309762 | 0,682421141 | LOC101928474 | FALSE | Downregulated |
| -0,072136321 | 0,682421141 | PPA2 | FALSE | Downregulated |
| -0,147667152 | 0,682421141 | TMEM79 | FALSE | Downregulated |
| -0,15434467 | 0,682467451 | ZNF837 | FALSE | Downregulated |
| -0,367134249 | 0,683132887 | RGMB-AS1 | FALSE | Downregulated |
| 0,12318149 | 0,683311357 | CAMK1D | FALSE | Upregulated |
| -0,628567031 | 0,683363737 | MSTO2P | FALSE | Downregulated |
| 0,068993587 | 0,683363737 | SEC31A | FALSE | Upregulated |
| 0,498165132 | 0,68358539 | TMLHE-AS1 | FALSE | Upregulated |
| 0,205361435 | 0,683627107 | C21orf67 | FALSE | Upregulated |
| 0,088772831 | 0,683627107 | CCDC61 | FALSE | Upregulated |
| 0,222222063 | 0,683627107 | COL1A1 | FALSE | Upregulated |
| 0,077382339 | 0,683627107 | HINT1 | FALSE | Upregulated |
| 0,298924356 | 0,683627107 | LOC101927011 | FALSE | Upregulated |
| 0,492236954 | 0,683627107 | RN7SL1 | FALSE | Upregulated |
| -0,076135258 | 0,683887503 | FNBP1L | FALSE | Downregulated |
| 0,093386987 | 0,683988611 | FAM219A | FALSE | Upregulated |
| -0,198999437 | 0,684227644 | RNF5P1 | FALSE | Downregulated |
| -0,298050925 | 0,684315639 | HCG16 | FALSE | Downregulated |
| -0,048267639 | 0,684348454 | RAB12 | FALSE | Downregulated |
| 0,301775399 | 0,684519864 | LOC101927377 | FALSE | Upregulated |

| | | | | |
|---|---|---|---|---|
| 0,146614451 | 0,684840745 | CA5A | FALSE | Upregulated |
| 0,063246925 | 0,684840745 | TTC23 | FALSE | Upregulated |
| 0,153600975 | 0,684876826 | EPHA2 | FALSE | Upregulated |
| -0,101430413 | 0,684913367 | ABCG5 | FALSE | Downregulated |
| 0,073522545 | 0,684913367 | ATXN2L | FALSE | Upregulated |
| 0,0529282 | 0,684913367 | BUD31 | FALSE | Upregulated |
| 0,083687471 | 0,684913367 | CASP2 | FALSE | Upregulated |
| 0,074869797 | 0,684913367 | DHX37 | FALSE | Upregulated |
| -0,147722146 | 0,684913367 | LHFP | FALSE | Downregulated |
| -0,101028007 | 0,684913367 | MRPL41 | FALSE | Downregulated |
| 0,087232472 | 0,684913367 | MZT2A | FALSE | Upregulated |
| -0,10870373 | 0,684913367 | PARP2 | FALSE | Downregulated |
| -1,040982777 | 0,684913367 | PTGES3L | FALSE | Downregulated |
| 0,243236093 | 0,684913367 | VIM-AS1 | FALSE | Upregulated |
| 0,424678399 | 0,684963377 | CELP | FALSE | Upregulated |
| -0,091190421 | 0,685020378 | CAD | FALSE | Downregulated |
| -0,175842051 | 0,685334199 | UNC5C | FALSE | Downregulated |
| -0,171139404 | 0,685374218 | LINC00630 | FALSE | Downregulated |
| 0,736396348 | 0,685500847 | ANKRD20A8P | FALSE | Upregulated |
| -0,602077966 | 0,685532191 | ATP1A2 | FALSE | Downregulated |
| 0,220042922 | 0,685532191 | JAG2 | FALSE | Upregulated |
| -0,066803424 | 0,685532191 | NHLRC3 | FALSE | Downregulated |
| -0,078968018 | 0,6858656 | ASB6 | FALSE | Downregulated |
| 0,153519503 | 0,686048073 | MVK | FALSE | Upregulated |
| 0,103903044 | 0,686048073 | SEC11C | FALSE | Upregulated |
| -0,077747936 | 0,686048073 | SLC11A2 | FALSE | Downregulated |
| -0,127719532 | 0,686148949 | PDE10A | FALSE | Downregulated |
| 0,115731532 | 0,686187438 | EIF4E | FALSE | Upregulated |
| 0,116597605 | 0,686239434 | ATP10D | FALSE | Upregulated |
| -0,321820861 | 0,686289481 | GOLGA6L5P | FALSE | Downregulated |
| -0,146993364 | 0,686289481 | ZNF682 | FALSE | Downregulated |
| 0,50816113 | 0,686293536 | ANP32AP1 | FALSE | Upregulated |
| -0,216595689 | 0,686372635 | LOC102724112 | FALSE | Downregulated |
| 0,256630351 | 0,686372635 | LOC441179 | FALSE | Upregulated |
| -0,386392523 | 0,686534719 | FAM66B | FALSE | Downregulated |
| -0,304837642 | 0,686681848 | OR2A1-AS1 | FALSE | Downregulated |
| 0,064023067 | 0,686869963 | DNAJC13 | FALSE | Upregulated |
| -0,109358973 | 0,686869963 | PPP1R11 | FALSE | Downregulated |
| -0,065411217 | 0,686869963 | TRIM24 | FALSE | Downregulated |
| -0,205448398 | 0,687030171 | HAS3 | FALSE | Downregulated |
| 0,080410404 | 0,687030171 | UBA7 | FALSE | Upregulated |
| -0,057257735 | 0,687134438 | SLC39A11 | FALSE | Downregulated |
| 0,068696332 | 0,687134438 | TRA2A | FALSE | Upregulated |
| -0,060216294 | 0,687288054 | KIAA1671 | FALSE | Downregulated |
| -0,273478792 | 0,687288054 | NAGPA-AS1 | FALSE | Downregulated |
| -0,141894444 | 0,687403683 | ZNF852 | FALSE | Downregulated |
| -0,199248257 | 0,687634349 | DNAH6 | FALSE | Downregulated |
| -0,363237204 | 0,687959638 | CLEC4F | FALSE | Downregulated |
| -0,366745185 | 0,687959638 | LOC101928281 | FALSE | Downregulated |
| -0,472726772 | 0,687959638 | LOC102724285 | FALSE | Downregulated |
| -0,071545106 | 0,687959638 | ZNRF2 | FALSE | Downregulated |

| | | | | |
|---|---|---|---|---|
| 0,101000777 | 0,688035176 | EPHA1 | FALSE | Upregulated |
| -0,089945252 | 0,688035176 | ING2 | FALSE | Downregulated |
| -0,530265313 | 0,688035176 | NBPF7 | FALSE | Downregulated |
| 0,049427508 | 0,688035176 | PRKAR2A | FALSE | Upregulated |
| -0,265988263 | 0,688035176 | ZBTB12 | FALSE | Downregulated |
| -0,055696886 | 0,688087662 | SETDB1 | FALSE | Downregulated |
| -0,102342571 | 0,688274494 | NME3 | FALSE | Downregulated |
| 0,152064736 | 0,688406842 | TPM3 | FALSE | Upregulated |
| 0,154500353 | 0,688443349 | ALS2CL | FALSE | Upregulated |
| 0,045420989 | 0,688482486 | ARPP19 | FALSE | Upregulated |
| -0,289591051 | 0,688482486 | CLDND2 | FALSE | Downregulated |
| -0,20370779 | 0,688588699 | FAM151B | FALSE | Downregulated |
| -0,13215335 | 0,688839871 | DNAJC3-AS1 | FALSE | Downregulated |
| -0,09607337 | 0,688839871 | MIA2 | FALSE | Downregulated |
| 0,17228133 | 0,688848712 | TRDMT1 | FALSE | Upregulated |
| 0,052256096 | 0,688895576 | GINM1 | FALSE | Upregulated |
| 0,46865899 | 0,689244295 | RP5-1139I1.1 | FALSE | Upregulated |
| 0,051966563 | 0,689434439 | XPO5 | FALSE | Upregulated |
| -0,130876599 | 0,68946851 | CDH23 | FALSE | Downregulated |
| -0,609700511 | 0,689583257 | LINC00944 | FALSE | Downregulated |
| -0,258118065 | 0,689650185 | VEPH1 | FALSE | Downregulated |
| 0,415190423 | 0,689671897 | PRH2 | FALSE | Upregulated |
| 0,0821974 | 0,689755576 | AIMP1 | FALSE | Upregulated |
| 0,047500298 | 0,689755576 | ATP5F1 | FALSE | Upregulated |
| 0,089186989 | 0,689755576 | CALD1 | FALSE | Upregulated |
| -0,092405445 | 0,689755576 | FRK | FALSE | Downregulated |
| 0,157483395 | 0,689755576 | HIST1H2BC | FALSE | Upregulated |
| -0,061152814 | 0,689755576 | SPAST | FALSE | Downregulated |
| -0,067383413 | 0,689824293 | ATPIF1 | FALSE | Downregulated |
| 0,152637075 | 0,689824293 | SCRN1 | FALSE | Upregulated |
| 0,274213528 | 0,689942526 | LDLRAD2 | FALSE | Upregulated |
| -0,224875025 | 0,690078796 | PODN | FALSE | Downregulated |
| 0,469020799 | 0,690137624 | S100P | FALSE | Upregulated |
| 0,092204994 | 0,690214864 | DEXI | FALSE | Upregulated |
| -0,140207347 | 0,690214864 | SRCIN1 | FALSE | Downregulated |
| -0,184174686 | 0,690214864 | ZNF853 | FALSE | Downregulated |
| -0,080986474 | 0,690754331 | CMAS | FALSE | Downregulated |
| 0,175698987 | 0,690833777 | NFKBID | FALSE | Upregulated |
| -0,074753544 | 0,6909764 | AKAP11 | FALSE | Downregulated |
| 0,323113957 | 0,6909764 | FAM166B | FALSE | Upregulated |
| 0,196589228 | 0,6909764 | FOXD2-AS1 | FALSE | Upregulated |
| -0,044599234 | 0,691125228 | BUD13 | FALSE | Downregulated |
| 0,732778605 | 0,691330593 | TDGF1P3 | FALSE | Upregulated |
| 0,200402393 | 0,691428153 | ELL3 | FALSE | Upregulated |
| 0,061365906 | 0,691552709 | SLC25A44 | FALSE | Upregulated |
| -0,375728865 | 0,691791121 | DLEU2L | FALSE | Downregulated |
| -0,37865023 | 0,691897402 | RP11-298I3.1 | FALSE | Downregulated |
| -0,718359714 | 0,691979493 | CACNA1G | FALSE | Downregulated |
| 0,115975324 | 0,691979493 | ECSCR-2 | FALSE | Upregulated |
| -0,106812093 | 0,691979493 | GCHFR | FALSE | Downregulated |
| -0,381061744 | 0,691979493 | RBM46 | FALSE | Downregulated |

| | | | | |
|---|---|---|---|---|
| -0,050741082 | 0,691979493 | SART3 | FALSE | Downregulated |
| -0,161853605 | 0,691979493 | SHISA4 | FALSE | Downregulated |
| -0,070251118 | 0,692086268 | KAT2B | FALSE | Downregulated |
| 0,279824664 | 0,692086268 | LOC102724159 | FALSE | Upregulated |
| 0,167784914 | 0,692482264 | RTN4RL2 | FALSE | Upregulated |
| -0,244388763 | 0,692681867 | HSD17B3 | FALSE | Downregulated |
| 0,18551472 | 0,692721649 | MSMO1 | FALSE | Upregulated |
| -0,61622795 | 0,69272452 | FAM156A | FALSE | Downregulated |
| 0,273374587 | 0,692838626 | ARSF | FALSE | Upregulated |
| -0,05364711 | 0,692838626 | CYHR1 | FALSE | Downregulated |
| 0,279549791 | 0,692838626 | FAM86B1 | FALSE | Upregulated |
| 0,061710087 | 0,692838626 | MOB2 | FALSE | Upregulated |
| 0,081537399 | 0,692838626 | TTC39C | FALSE | Upregulated |
| 0,07841332 | 0,692965795 | LETM1 | FALSE | Upregulated |
| -0,04920379 | 0,692999929 | DHX32 | FALSE | Downregulated |
| -0,956821896 | 0,693238643 | HOXC8 | FALSE | Downregulated |
| 0,263694954 | 0,693331949 | AC097662.2 | FALSE | Upregulated |
| -0,128980373 | 0,693373751 | ZNF699 | FALSE | Downregulated |
| -0,148422359 | 0,693643927 | TMEM26 | FALSE | Downregulated |
| 0,077600714 | 0,69367168 | TRAPPC2L | FALSE | Upregulated |
| -0,240905757 | 0,693935094 | FSD1L | FALSE | Downregulated |
| -0,496085995 | 0,693935094 | LOC101060400 | FALSE | Downregulated |
| -0,047635521 | 0,693935094 | PPP2R5D | FALSE | Downregulated |
| 0,103286294 | 0,693957824 | CPN1 | FALSE | Upregulated |
| -0,214819951 | 0,693957824 | LRRC56 | FALSE | Downregulated |
| -0,222594949 | 0,694409609 | SLC17A8 | FALSE | Downregulated |
| 0,135465595 | 0,694477293 | LOC102724200 | FALSE | Upregulated |
| -0,184026877 | 0,694490156 | PFKM | FALSE | Downregulated |
| 0,059353661 | 0,694527706 | BTBD2 | FALSE | Upregulated |
| 0,064264676 | 0,694527706 | CALCOCO2 | FALSE | Upregulated |
| -0,202438179 | 0,694585012 | CAPN10-AS1 | FALSE | Downregulated |
| -0,287641357 | 0,694585012 | LOC102723327 | FALSE | Downregulated |
| -0,237742796 | 0,694687406 | RP11-1223D19.1 | FALSE | Downregulated |
| 0,051381027 | 0,694687406 | TUFM | FALSE | Upregulated |
| 0,094686885 | 0,694890989 | SCLY | FALSE | Upregulated |
| 0,337546845 | 0,695018137 | FLJ31104 | FALSE | Upregulated |
| -0,096896709 | 0,695076413 | TENC1 | FALSE | Downregulated |
| -0,16856292 | 0,69550078 | DNA2 | FALSE | Downregulated |
| 0,145268778 | 0,69550078 | ZNF575 | FALSE | Upregulated |
| -0,191985216 | 0,695510767 | ZNF300 | FALSE | Downregulated |
| -0,063168045 | 0,695789794 | TIMM9 | FALSE | Downregulated |
| -0,592832294 | 0,695923857 | SERTAD4 | FALSE | Downregulated |
| -0,104094578 | 0,695923857 | SLC30A4 | FALSE | Downregulated |
| -0,420595151 | 0,695923857 | VENTX | FALSE | Downregulated |
| -0,0878724 | 0,696077003 | CCDC159 | FALSE | Downregulated |
| -0,052047286 | 0,696077003 | DLD | FALSE | Downregulated |
| -0,262590774 | 0,696077003 | EN1 | FALSE | Downregulated |
| 0,050821688 | 0,696124834 | DYM | FALSE | Upregulated |
| 0,081786582 | 0,696453227 | NUDT22 | FALSE | Upregulated |
| 0,068519803 | 0,696525981 | KIF1C | FALSE | Upregulated |
| -0,080791709 | 0,696525981 | RPL4 | FALSE | Downregulated |

| | | | | |
|---|---|---|---|---|
| 0,243924055 | 0,696587024 | HCAR2 | FALSE | Upregulated |
| 0,330984203 | 0,69673273 | LOC101927699 | FALSE | Upregulated |
| -0,062071112 | 0,69673273 | MRPS5 | FALSE | Downregulated |
| -0,216624827 | 0,69673273 | TMEM71 | FALSE | Downregulated |
| 0,475099693 | 0,696736322 | AC104653.1 | FALSE | Upregulated |
| -0,223037111 | 0,696736322 | LOC100133315 | FALSE | Downregulated |
| -0,086633655 | 0,696791171 | TBC1D10A | FALSE | Downregulated |
| -0,066723633 | 0,696839069 | NMT2 | FALSE | Downregulated |
| 0,193774957 | 0,696928222 | PRTFDC1 | FALSE | Upregulated |
| -0,115268251 | 0,696976508 | ENTPD3-AS1 | FALSE | Downregulated |
| 0,330398431 | 0,697072057 | FAM163A | FALSE | Upregulated |
| 0,582243416 | 0,697079039 | SNORA7A | FALSE | Upregulated |
| 0,273254059 | 0,697542816 | ATP6V0E2-AS1 | FALSE | Upregulated |
| 0,052189016 | 0,697548727 | FUNDC1 | FALSE | Upregulated |
| 0,053746749 | 0,697694385 | UBQLN4 | FALSE | Upregulated |
| -0,128411511 | 0,697776998 | C12orf73 | FALSE | Downregulated |
| -0,205932637 | 0,697776998 | EFNA5 | FALSE | Downregulated |
| -0,201481024 | 0,697776998 | PIH1D2 | FALSE | Downregulated |
| -0,056468278 | 0,698005613 | RSL24D1 | FALSE | Downregulated |
| 0,075153694 | 0,698041665 | NADK | FALSE | Upregulated |
| 0,059078099 | 0,698069956 | PCNXL3 | FALSE | Upregulated |
| -0,079197535 | 0,698173532 | TPT1 | FALSE | Downregulated |
| 0,086838944 | 0,698420993 | AHSG | FALSE | Upregulated |
| -0,355362302 | 0,698420993 | MAPT | FALSE | Downregulated |
| -0,106369298 | 0,698448474 | EXOSC6 | FALSE | Downregulated |
| 0,22219513 | 0,698449515 | FCRL6 | FALSE | Upregulated |
| -0,401012776 | 0,698997313 | LOC101929819 | FALSE | Downregulated |
| -0,461625115 | 0,699349508 | C1orf185 | FALSE | Downregulated |
| 0,580563164 | 0,699349508 | LOC102723347 | FALSE | Upregulated |
| 0,083273922 | 0,699349508 | SPATA2 | FALSE | Upregulated |
| -0,057078979 | 0,699349508 | WDR3 | FALSE | Downregulated |
| -0,074807616 | 0,699467129 | F10 | FALSE | Downregulated |
| 0,058035204 | 0,699509533 | UROD | FALSE | Upregulated |
| -0,155355491 | 0,699509533 | ZNF701 | FALSE | Downregulated |
| -0,081353682 | 0,699695898 | ANKRD11 | FALSE | Downregulated |
| 0,145070088 | 0,699859209 | FADS6 | FALSE | Upregulated |
| -0,53471445 | 0,699859209 | GS1-122H1.2 | FALSE | Downregulated |
| 0,051175201 | 0,699859209 | RCOR1 | FALSE | Upregulated |
| 0,064854128 | 0,699927213 | ACOT8 | FALSE | Upregulated |
| 0,501200519 | 0,699927213 | LOC102724426 | FALSE | Upregulated |
| 0,0585896 | 0,699927213 | SDAD1 | FALSE | Upregulated |
| 0,164691028 | 0,699927213 | ZNF607 | FALSE | Upregulated |
| 0,047697524 | 0,700137465 | RIC8A | FALSE | Upregulated |
| 0,167145508 | 0,700262616 | LOC728975 | FALSE | Upregulated |
| -0,04518777 | 0,70027888 | UQCRC2 | FALSE | Downregulated |
| -0,087305241 | 0,700696607 | FKTN | FALSE | Downregulated |
| -0,197410654 | 0,700696607 | KLHL17 | FALSE | Downregulated |
| 0,053694715 | 0,700747226 | C5orf24 | FALSE | Upregulated |
| -0,178612799 | 0,700747226 | GMPPB | FALSE | Downregulated |
| 0,101306747 | 0,700747226 | PNKP | FALSE | Upregulated |
| 0,036015799 | 0,700747226 | QARS | FALSE | Upregulated |

| | | | | |
|---|---|---|---|---|
| 0,337634022 | 0,700881915 | CEACAM3 | FALSE | Upregulated |
| -0,118469426 | 0,700895093 | GLDC | FALSE | Downregulated |
| 0,169406476 | 0,700895093 | IDI1 | FALSE | Upregulated |
| 0,437193866 | 0,700901211 | HORMAD1 | FALSE | Upregulated |
| -0,199453848 | 0,700901211 | LOC102724252 | FALSE | Downregulated |
| -0,088039375 | 0,700914197 | FARP2 | FALSE | Downregulated |
| -0,306908507 | 0,700914197 | P2RX3 | FALSE | Downregulated |
| 0,055495744 | 0,700914197 | PAICS | FALSE | Upregulated |
| 0,198051801 | 0,701117657 | BTC | FALSE | Upregulated |
| 0,07922255 | 0,701202103 | RABGEF1 | FALSE | Upregulated |
| 0,668285221 | 0,701255415 | AMZ1 | FALSE | Upregulated |
| -0,09932761 | 0,701255415 | CNTLN | FALSE | Downregulated |
| -0,523051908 | 0,701255415 | DUSP15 | FALSE | Downregulated |
| -0,300864168 | 0,701255415 | FGF21 | FALSE | Downregulated |
| -0,118359227 | 0,701255415 | JAZF1 | FALSE | Downregulated |
| -0,391715808 | 0,701255415 | LOC102724265 | FALSE | Downregulated |
| 0,073820235 | 0,701255415 | RNF44 | FALSE | Upregulated |
| -0,060632042 | 0,701255415 | SAP30BP | FALSE | Downregulated |
| -0,06960456 | 0,701255415 | SLC37A3 | FALSE | Downregulated |
| -0,089053744 | 0,701255415 | TOP3B | FALSE | Downregulated |
| 0,113347908 | 0,701255415 | ZNF57 | FALSE | Upregulated |
| -0,119908923 | 0,701418044 | ZNF630 | FALSE | Downregulated |
| 0,42253632 | 0,701450484 | CXorf65 | FALSE | Upregulated |
| -0,309444449 | 0,701652149 | KLHL31 | FALSE | Downregulated |
| 0,447544813 | 0,701719346 | RNU6ATAC | FALSE | Upregulated |
| 0,04131073 | 0,701733586 | GNL1 | FALSE | Upregulated |
| -0,198185553 | 0,701821649 | MT2A | FALSE | Downregulated |
| -0,363958523 | 0,701821649 | TENM2 | FALSE | Downregulated |
| -0,099018079 | 0,702095119 | SMAD7 | FALSE | Downregulated |
| 0,112014659 | 0,702154863 | ZNHIT6 | FALSE | Upregulated |
| -0,125464143 | 0,702239415 | AFG3L1P | FALSE | Downregulated |
| 0,56045951 | 0,702239415 | C20orf197 | FALSE | Upregulated |
| -0,082727657 | 0,702239415 | ZNF512B | FALSE | Downregulated |
| -0,349631452 | 0,702346513 | FAM162B | FALSE | Downregulated |
| -0,593957822 | 0,702346513 | PCYT1B | FALSE | Downregulated |
| -0,537421845 | 0,702346513 | TPM2 | FALSE | Downregulated |
| -0,56425933 | 0,702383937 | ESR2 | FALSE | Downregulated |
| -0,129963072 | 0,702400604 | CATSPER2P1 | FALSE | Downregulated |
| 0,0482235 | 0,702564503 | LIMS1 | FALSE | Upregulated |
| 0,342744164 | 0,702564503 | LOC102723558 | FALSE | Upregulated |
| 0,144383845 | 0,702564503 | LOH12CR2 | FALSE | Upregulated |
| 0,071716365 | 0,703014552 | NAGPA | FALSE | Upregulated |
| -0,193206921 | 0,7031181 | NBPF13P | FALSE | Downregulated |
| -1,040274417 | 0,703199155 | TMEM256-PLSCR3 | FALSE | Downregulated |
| 0,347927802 | 0,703398857 | CCR4 | FALSE | Upregulated |
| 0,102843525 | 0,703398857 | GPR146 | FALSE | Upregulated |
| 0,151557877 | 0,70346605 | COL4A1 | FALSE | Upregulated |
| 0,246398193 | 0,703538812 | CUBN | FALSE | Upregulated |
| 0,145596886 | 0,703538812 | RASIP1 | FALSE | Upregulated |
| 0,056638379 | 0,703547038 | ADPGK | FALSE | Upregulated |
| -0,09672705 | 0,703547038 | C10orf118 | FALSE | Downregulated |

| | | | | |
|---|---|---|---|---|
| 0,144952568 | 0,703547038 | FHOD1 | FALSE | Upregulated |
| 0,065447845 | 0,703547038 | OPA3 | FALSE | Upregulated |
| -0,661705674 | 0,703547038 | PKP1 | FALSE | Downregulated |
| 0,074829857 | 0,703547038 | PLXND1 | FALSE | Upregulated |
| 0,234264477 | 0,703549845 | FAM156B | FALSE | Upregulated |
| -0,117196608 | 0,703606912 | RAB13 | FALSE | Downregulated |
| 0,092051114 | 0,703672216 | CARD10 | FALSE | Upregulated |
| -0,062944211 | 0,703672216 | FAM120A | FALSE | Downregulated |
| -0,146238291 | 0,703672216 | ISLR | FALSE | Downregulated |
| -0,296599668 | 0,703672216 | ZBTB16 | FALSE | Downregulated |
| -0,078416155 | 0,703812458 | IFT27 | FALSE | Downregulated |
| -0,061620055 | 0,703812458 | TMEM168 | FALSE | Downregulated |
| -0,243217681 | 0,703843129 | LINC00470 | FALSE | Downregulated |
| 0,082471506 | 0,70388103 | ESRRA | FALSE | Upregulated |
| -0,082223188 | 0,704007562 | SPAG9 | FALSE | Downregulated |
| 0,404515102 | 0,704051344 | LOC102724697 | FALSE | Upregulated |
| -0,255391527 | 0,704064876 | ERAP2 | FALSE | Downregulated |
| -0,579128676 | 0,704134103 | ALAS2 | FALSE | Downregulated |
| -0,171954735 | 0,704134103 | ANTXR1 | FALSE | Downregulated |
| 0,062451132 | 0,704134103 | EIF2B3 | FALSE | Upregulated |
| -0,139320389 | 0,704134103 | SLC25A29 | FALSE | Downregulated |
| 0,089369931 | 0,704134103 | ZNF787 | FALSE | Upregulated |
| -0,24270243 | 0,70426588 | PXDNL | FALSE | Downregulated |
| -0,078792878 | 0,704401862 | ISCU | FALSE | Downregulated |
| 0,104034336 | 0,704451962 | GFOD1 | FALSE | Upregulated |
| -0,117807905 | 0,705225639 | AQP7 | FALSE | Downregulated |
| 0,089356646 | 0,705225639 | BLOC1S3 | FALSE | Upregulated |
| -0,291652757 | 0,705303262 | CARNS1 | FALSE | Downregulated |
| -0,128635756 | 0,705336615 | TLN2 | FALSE | Downregulated |
| -0,6156202 | 0,705557054 | AC105053.3 | FALSE | Downregulated |
| -0,084452846 | 0,705573411 | AKR1C4 | FALSE | Downregulated |
| -0,484196172 | 0,705587069 | SPEG | FALSE | Downregulated |
| -0,176768117 | 0,705587069 | ZNF774 | FALSE | Downregulated |
| 0,09189169 | 0,705717886 | COPG1 | FALSE | Upregulated |
| 0,341966411 | 0,705717886 | IDI2-AS1 | FALSE | Upregulated |
| 0,255767558 | 0,705717886 | SCGB3A1 | FALSE | Upregulated |
| -0,072537528 | 0,705717886 | SREK1IP1 | FALSE | Downregulated |
| 0,22555095 | 0,70576667 | ITGA7 | FALSE | Upregulated |
| 0,598703055 | 0,705797811 | MT1B | FALSE | Upregulated |
| -0,668679002 | 0,705797811 | PTCHD1 | FALSE | Downregulated |
| -0,468923822 | 0,705797811 | SPATA9 | FALSE | Downregulated |
| -0,560956281 | 0,706100397 | SCN3B | FALSE | Downregulated |
| 0,08852082 | 0,706190132 | BZW1 | FALSE | Upregulated |
| -0,176073157 | 0,706602161 | DDX26B | FALSE | Downregulated |
| 0,098239938 | 0,706602161 | EXOSC4 | FALSE | Upregulated |
| 0,114248976 | 0,706602161 | GJB2 | FALSE | Upregulated |
| 0,351765817 | 0,706975395 | EDA2R | FALSE | Upregulated |
| 0,137570532 | 0,707047357 | SLC25A53 | FALSE | Upregulated |
| 0,50080864 | 0,707221224 | MYCBPAP | FALSE | Upregulated |
| 0,098775272 | 0,70737269 | HEY2 | FALSE | Upregulated |
| 0,084079418 | 0,707609627 | DBI | FALSE | Upregulated |

| | | | | |
|---|---|---|---|---|
| 0,100769731 | 0,707666778 | PSTK | FALSE | Upregulated |
| -0,062437744 | 0,707737604 | CHURC1 | FALSE | Downregulated |
| -0,107733072 | 0,707782711 | PFDN1 | FALSE | Downregulated |
| -0,219959732 | 0,707888041 | SCGB1B2P | FALSE | Downregulated |
| 0,0754533 | 0,708134881 | ZNF200 | FALSE | Upregulated |
| -0,614820542 | 0,708292841 | ALS2CR11 | FALSE | Downregulated |
| -0,413712943 | 0,708342425 | LOC102724016 | FALSE | Downregulated |
| 0,096777288 | 0,708396719 | LNPEP | FALSE | Upregulated |
| 0,286363817 | 0,708567511 | LMOD1 | FALSE | Upregulated |
| -0,064950074 | 0,708648618 | MTA3 | FALSE | Downregulated |
| -0,815892489 | 0,708726249 | SNORD55 | FALSE | Downregulated |
| -0,050179437 | 0,708760516 | ARMC1 | FALSE | Downregulated |
| -0,078615098 | 0,708760516 | DENND4B | FALSE | Downregulated |
| 0,293838671 | 0,708760516 | LOC102724791 | FALSE | Upregulated |
| -0,067348908 | 0,708814383 | AHCY | FALSE | Downregulated |
| 0,278014006 | 0,708814383 | ANKRD18A | FALSE | Upregulated |
| -0,369429281 | 0,708814383 | HP09025 | FALSE | Downregulated |
| -0,051762299 | 0,708814383 | METTL13 | FALSE | Downregulated |
| 0,171317586 | 0,708814383 | STEAP4 | FALSE | Upregulated |
| 0,578353292 | 0,709083352 | ZNF724P | FALSE | Upregulated |
| 0,060469077 | 0,709092379 | HEATR6 | FALSE | Upregulated |
| 0,177782137 | 0,709151647 | NKAPP1 | FALSE | Upregulated |
| 0,491492262 | 0,709286114 | C1orf140 | FALSE | Upregulated |
| 0,128430095 | 0,70957292 | RFTN1 | FALSE | Upregulated |
| -0,252219813 | 0,709603039 | COL8A1 | FALSE | Downregulated |
| -0,051039369 | 0,709704657 | PRKD3 | FALSE | Downregulated |
| 0,407233578 | 0,709952162 | HIATL2 | FALSE | Upregulated |
| -0,05930868 | 0,710015374 | MARCH5 | FALSE | Downregulated |
| 0,041170524 | 0,710110827 | EWSR1 | FALSE | Upregulated |
| 0,199938578 | 0,710110827 | LRG1 | FALSE | Upregulated |
| -0,091333629 | 0,710110827 | SAFB2 | FALSE | Downregulated |
| -0,156290746 | 0,710124683 | LOC101929986 | FALSE | Downregulated |
| 0,082705239 | 0,710124683 | UBE2E2 | FALSE | Upregulated |
| -0,328744541 | 0,710197204 | KIAA1614 | FALSE | Downregulated |
| 0,048646903 | 0,710197204 | SH3BP5L | FALSE | Upregulated |
| 0,339634684 | 0,710227553 | IGFBPL1 | FALSE | Upregulated |
| 0,262049463 | 0,710430569 | ACBD7 | FALSE | Upregulated |
| -0,553295077 | 0,7104439 | LINC01091 | FALSE | Downregulated |
| -0,111279744 | 0,710655975 | FBXO27 | FALSE | Downregulated |
| -0,40559816 | 0,710663999 | LINC01016 | FALSE | Downregulated |
| 0,338401462 | 0,710674183 | LOC100507103 | FALSE | Upregulated |
| -0,291488282 | 0,710674183 | SNAP25 | FALSE | Downregulated |
| 0,113660435 | 0,710674183 | SRP14-AS1 | FALSE | Upregulated |
| -0,090732494 | 0,710698867 | LINS | FALSE | Downregulated |
| -0,295413335 | 0,710698867 | LOC102724792 | FALSE | Downregulated |
| -0,504026365 | 0,710907372 | LOC101928261 | FALSE | Downregulated |
| 0,069433599 | 0,711035779 | ADAM17 | FALSE | Upregulated |
| 0,093763668 | 0,711035779 | SERTAD3 | FALSE | Upregulated |
| -0,265351306 | 0,711151852 | C12orf61 | FALSE | Downregulated |
| -0,208833388 | 0,711158372 | LOC101928504 | FALSE | Downregulated |
| -0,55501182 | 0,711158372 | LUZP2 | FALSE | Downregulated |

| | | | | |
|---|---|---|---|---|
| -0,056072491 | 0,711158372 | NFIX | FALSE | Downregulated |
| 0,057854037 | 0,711158372 | ORC3 | FALSE | Upregulated |
| -0,050280191 | 0,711158372 | SMARCB1 | FALSE | Downregulated |
| -0,362522711 | 0,711213489 | RGSL1 | FALSE | Downregulated |
| -0,071536261 | 0,711229045 | USP33 | FALSE | Downregulated |
| -0,065834978 | 0,711271714 | ZNF134 | FALSE | Downregulated |
| -0,146079874 | 0,711286082 | JAM3 | FALSE | Downregulated |
| 0,213652078 | 0,711477696 | PLA2G5 | FALSE | Upregulated |
| 0,066581365 | 0,711547715 | LBR | FALSE | Upregulated |
| -0,351161765 | 0,711547715 | LOC101928635 | FALSE | Downregulated |
| -0,199453136 | 0,711547715 | SDHAP3 | FALSE | Downregulated |
| 0,324568254 | 0,711547715 | ZNF114 | FALSE | Upregulated |
| 0,129806327 | 0,71161477 | EBP | FALSE | Upregulated |
| -0,270553984 | 0,711715078 | ATP6V1G2 | FALSE | Downregulated |
| 0,450057853 | 0,711715078 | LOC101929523 | FALSE | Upregulated |
| 0,289332094 | 0,711765381 | CHAC1 | FALSE | Upregulated |
| 0,044568637 | 0,711773813 | COG2 | FALSE | Upregulated |
| 0,155569211 | 0,711838727 | ZFP92 | FALSE | Upregulated |
| -0,205459653 | 0,711967319 | TOX3 | FALSE | Downregulated |
| 0,397448064 | 0,712042109 | RHCG | FALSE | Upregulated |
| -0,210981029 | 0,712047308 | ARHGEF26-AS1 | FALSE | Downregulated |
| 0,419983686 | 0,712047308 | RGS6 | FALSE | Upregulated |
| 0,08796205 | 0,712407179 | AMOT | FALSE | Upregulated |
| -0,184872249 | 0,71257427 | TSKU | FALSE | Downregulated |
| 0,077741184 | 0,712590454 | B3GALTL | FALSE | Upregulated |
| -0,122693858 | 0,712590454 | FAM47E | FALSE | Downregulated |
| 0,084676516 | 0,712590454 | LMO2 | FALSE | Upregulated |
| 0,351565622 | 0,712632154 | ANKDD1B | FALSE | Upregulated |
| -0,12121047 | 0,712632154 | CEP85L | FALSE | Downregulated |
| 0,381039018 | 0,712651926 | DOK5 | FALSE | Upregulated |
| 0,580060107 | 0,712651926 | FLJ27354 | FALSE | Upregulated |
| 0,201575026 | 0,712651926 | ST6GALNAC2 | FALSE | Upregulated |
| -0,072942889 | 0,712800481 | NOC3L | FALSE | Downregulated |
| -0,083208068 | 0,712800481 | SETD4 | FALSE | Downregulated |
| -0,09494656 | 0,712800481 | SLC9A7 | FALSE | Downregulated |
| -0,036984107 | 0,712894508 | NFYC | FALSE | Downregulated |
| 0,193259414 | 0,712894508 | PCDH17 | FALSE | Upregulated |
| -0,316068717 | 0,713191611 | RBPMS-AS1 | FALSE | Downregulated |
| -0,160803894 | 0,713198857 | LINC01128 | FALSE | Downregulated |
| 0,10331875 | 0,713205547 | MZT1 | FALSE | Upregulated |
| 0,085867898 | 0,713235437 | CRYBG3 | FALSE | Upregulated |
| -0,563429992 | 0,713351393 | FUT5 | FALSE | Downregulated |
| 0,0343323 | 0,713757634 | PAFAH1B1 | FALSE | Upregulated |
| -0,544541187 | 0,713757634 | TMEM191A | FALSE | Downregulated |
| -0,246086614 | 0,713757634 | ZNF790-AS1 | FALSE | Downregulated |
| 0,323268734 | 0,713814667 | LOC101928825 | FALSE | Upregulated |
| -0,041095164 | 0,713944683 | YME1L1 | FALSE | Downregulated |
| -0,102097785 | 0,714001584 | MRPS25 | FALSE | Downregulated |
| -0,639471274 | 0,714001584 | SLC9A2 | FALSE | Downregulated |
| -0,071493422 | 0,714001584 | WRN | FALSE | Downregulated |
| -0,078957051 | 0,714058626 | AFAP1L1 | FALSE | Downregulated |

| | | | | |
|---|---|---|---|---|
| 0,071934446 | 0,714058626 | CMC1 | FALSE | Upregulated |
| -0,585307339 | 0,714058626 | ZNRF3-AS1 | FALSE | Downregulated |
| -0,369546377 | 0,714109042 | POU6F2-AS1 | FALSE | Downregulated |
| 0,122093386 | 0,714151549 | CBWD3 | FALSE | Upregulated |
| 0,108212038 | 0,714151549 | CEP135 | FALSE | Upregulated |
| 0,042078188 | 0,714151549 | SHOC2 | FALSE | Upregulated |
| 0,053972581 | 0,714264517 | HNRNPM | FALSE | Upregulated |
| -0,446090416 | 0,714264517 | TCTE3 | FALSE | Downregulated |
| 0,449862115 | 0,714264517 | ZCCHC18 | FALSE | Upregulated |
| -0,080561769 | 0,714311382 | HSPBAP1 | FALSE | Downregulated |
| -0,06252079 | 0,714665473 | ZNF687 | FALSE | Downregulated |
| -0,265123593 | 0,714802089 | IGSF10 | FALSE | Downregulated |
| -0,056397682 | 0,714835261 | FKBP8 | FALSE | Downregulated |
| -0,066457408 | 0,714884358 | ISY1-RAB43 | FALSE | Downregulated |
| 0,263180703 | 0,714925475 | HSF2BP | FALSE | Upregulated |
| -0,841835644 | 0,715010537 | NPHS1 | FALSE | Downregulated |
| -0,276454249 | 0,715073409 | C2orf27A | FALSE | Downregulated |
| -0,418298792 | 0,715316801 | DNAAF3 | FALSE | Downregulated |
| -0,047470864 | 0,715335132 | HIP1 | FALSE | Downregulated |
| -0,115378236 | 0,715477644 | TTC3P1 | FALSE | Downregulated |
| 0,078929424 | 0,715673489 | KNOP1 | FALSE | Upregulated |
| -0,126317898 | 0,71576984 | ZCCHC16 | FALSE | Downregulated |
| 0,217106274 | 0,715807399 | NAIP | FALSE | Upregulated |
| 0,085356008 | 0,715807399 | ROBO4 | FALSE | Upregulated |
| -0,534759549 | 0,716218593 | AC104777.2 | FALSE | Downregulated |
| -0,528818471 | 0,716218593 | FAM157C | FALSE | Downregulated |
| -0,306738412 | 0,716218593 | LOC102723980 | FALSE | Downregulated |
| -0,244302384 | 0,716218593 | LOC102724852 | FALSE | Downregulated |
| -0,14529859 | 0,716606716 | CACNA2D4 | FALSE | Downregulated |
| -0,709146376 | 0,716606716 | CAPN13 | FALSE | Downregulated |
| 0,391871713 | 0,716606716 | LOC100507195 | FALSE | Upregulated |
| 0,165241337 | 0,716606716 | RP11-15I11.3 | FALSE | Upregulated |
| 0,396633667 | 0,716759217 | FCAR | FALSE | Upregulated |
| 0,61134832 | 0,716759217 | PDE6A | FALSE | Upregulated |
| 0,220787285 | 0,716851581 | ARHGEF19 | FALSE | Upregulated |
| 0,047587343 | 0,716851581 | BPNT1 | FALSE | Upregulated |
| 0,06674721 | 0,716851581 | C17orf75 | FALSE | Upregulated |
| -0,27453255 | 0,716851581 | LOC101929977 | FALSE | Downregulated |
| 0,163989755 | 0,716851581 | LOC102724023 | FALSE | Upregulated |
| -0,600322123 | 0,716851581 | PWRN1 | FALSE | Downregulated |
| -0,110748249 | 0,716851581 | RNF208 | FALSE | Downregulated |
| -0,063123059 | 0,716851581 | SCRN3 | FALSE | Downregulated |
| 0,07930391 | 0,716851581 | SMURF1 | FALSE | Upregulated |
| 0,138005292 | 0,716851581 | STAG3L2 | FALSE | Upregulated |
| 0,098807614 | 0,716851581 | TIMP2 | FALSE | Upregulated |
| 0,059854326 | 0,716851581 | XRN2 | FALSE | Upregulated |
| 0,055224135 | 0,717137501 | CDC16 | FALSE | Upregulated |
| -0,062929021 | 0,717223307 | KLHL36 | FALSE | Downregulated |
| 0,072854902 | 0,717253841 | NDUFAF2 | FALSE | Upregulated |
| -0,760053513 | 0,717253841 | ZNF890P | FALSE | Downregulated |
| -0,348046538 | 0,717318422 | SPDYE5 | FALSE | Downregulated |

| | | | | |
|---|---|---|---|---|
| -0,056045766 | 0,717687888 | EPC2 | FALSE | Downregulated |
| 0,181171533 | 0,717750365 | LOC102723323 | FALSE | Upregulated |
| 0,044405914 | 0,717771687 | GTF2F1 | FALSE | Upregulated |
| 0,154488107 | 0,717993259 | BMP2 | FALSE | Upregulated |
| 0,253539872 | 0,718075934 | SEMA3A | FALSE | Upregulated |
| -0,851193131 | 0,718100811 | SMTNL2 | FALSE | Downregulated |
| 0,106693117 | 0,718326039 | WIPI1 | FALSE | Upregulated |
| -0,64422848 | 0,718743984 | DNAH9 | FALSE | Downregulated |
| -0,117225553 | 0,718743984 | GTF2H2C | FALSE | Downregulated |
| 0,224375926 | 0,718901166 | CDS1 | FALSE | Upregulated |
| -0,057874652 | 0,718901166 | PLEKHM3 | FALSE | Downregulated |
| 0,104984929 | 0,718901166 | VKORC1 | FALSE | Upregulated |
| 0,070469301 | 0,719108784 | GTPBP8 | FALSE | Upregulated |
| 0,119298063 | 0,719242727 | CLPTM1L | FALSE | Upregulated |
| 0,061095411 | 0,719242727 | FYCO1 | FALSE | Upregulated |
| -0,199016888 | 0,719242727 | TMPO-AS1 | FALSE | Downregulated |
| -0,077861422 | 0,719242727 | TSC22D1 | FALSE | Downregulated |
| -0,255249116 | 0,71945763 | ATL1 | FALSE | Downregulated |
| -0,420647676 | 0,71945763 | C21orf88 | FALSE | Downregulated |
| 0,469684895 | 0,71945763 | USP9Y | FALSE | Upregulated |
| -0,090119137 | 0,719546331 | F12 | FALSE | Downregulated |
| 0,039459588 | 0,71958312 | HNRNPK | FALSE | Upregulated |
| 0,508677738 | 0,719625267 | MT1JP | FALSE | Upregulated |
| 0,366045597 | 0,719810973 | RP11-362F19.1 | FALSE | Upregulated |
| -0,064908285 | 0,719810973 | TEAD3 | FALSE | Downregulated |
| 0,134987881 | 0,719891873 | RPH3AL | FALSE | Upregulated |
| 0,32485857 | 0,719891873 | XKR9 | FALSE | Upregulated |
| -0,040211815 | 0,719976756 | CHMP2B | FALSE | Downregulated |
| -0,095445296 | 0,720356121 | SEC61G | FALSE | Downregulated |
| -1,019233159 | 0,720870991 | ANKRD20A2 | FALSE | Downregulated |
| -0,079960602 | 0,720892508 | BTG1 | FALSE | Downregulated |
| 0,134638933 | 0,720915298 | TPK1 | FALSE | Upregulated |
| 0,057051943 | 0,721047488 | ECHDC1 | FALSE | Upregulated |
| 0,158778849 | 0,721047488 | TRIM55 | FALSE | Upregulated |
| 0,235267681 | 0,721177748 | NTRK2 | FALSE | Upregulated |
| 0,976291393 | 0,721177748 | POTEF | FALSE | Upregulated |
| 0,152656384 | 0,721207805 | NXN | FALSE | Upregulated |
| 0,064647611 | 0,721241071 | COA6 | FALSE | Upregulated |
| -0,282787261 | 0,721241071 | SLC26A5 | FALSE | Downregulated |
| -0,777993722 | 0,721241071 | SNORA6 | FALSE | Downregulated |
| -0,095900065 | 0,721454969 | SLC22A7 | FALSE | Downregulated |
| -0,202426851 | 0,721585353 | GLI3 | FALSE | Downregulated |
| -0,050579069 | 0,721585353 | NOL7 | FALSE | Downregulated |
| -0,392617292 | 0,721613317 | C6orf25 | FALSE | Downregulated |
| 0,078667791 | 0,721613317 | PMF1 | FALSE | Upregulated |
| 0,146507062 | 0,721968683 | NUAK2 | FALSE | Upregulated |
| 0,047269729 | 0,722071111 | BRD7 | FALSE | Upregulated |
| -0,093612351 | 0,722112259 | CHORDC1 | FALSE | Downregulated |
| 0,157216791 | 0,722464895 | C1orf112 | FALSE | Upregulated |
| 0,257737832 | 0,722464895 | FAM131B | FALSE | Upregulated |
| -0,363367823 | 0,722464895 | FLJ20712 | FALSE | Downregulated |

| | | | | |
|---|---|---|---|---|
| -0,082821939 | 0,722464895 | MACROD1 | FALSE | Downregulated |
| 0,063049187 | 0,722464895 | MBP | FALSE | Upregulated |
| -0,51741937 | 0,722464895 | TMEM130 | FALSE | Downregulated |
| -0,102405077 | 0,722464895 | TMEM223 | FALSE | Downregulated |
| 0,064586942 | 0,722464895 | ZNRF1 | FALSE | Upregulated |
| -0,575785937 | 0,722471836 | RBM11 | FALSE | Downregulated |
| -0,044555523 | 0,722471836 | WDR11 | FALSE | Downregulated |
| 0,117845856 | 0,722903971 | ERCC6-PGBD3 | FALSE | Upregulated |
| 0,294326924 | 0,722903971 | SPINK5 | FALSE | Upregulated |
| -0,406414547 | 0,722927576 | CACNB1 | FALSE | Downregulated |
| 0,084950184 | 0,722927576 | DPM2 | FALSE | Upregulated |
| -0,215847313 | 0,722927576 | EPHB3 | FALSE | Downregulated |
| 0,479289273 | 0,722927576 | EVPL | FALSE | Upregulated |
| 0,080301323 | 0,722927576 | LGALSL | FALSE | Upregulated |
| -0,142674548 | 0,722927576 | LINC01140 | FALSE | Downregulated |
| 0,214989024 | 0,722927576 | LOC101927934 | FALSE | Upregulated |
| 0,279154178 | 0,722927576 | LOC101928641 | FALSE | Upregulated |
| -0,504208387 | 0,722927576 | LOC101928765 | FALSE | Downregulated |
| 0,478306347 | 0,722927576 | LOC375196 | FALSE | Upregulated |
| -0,082834993 | 0,722927576 | SLC40A1 | FALSE | Downregulated |
| -0,288295152 | 0,722927576 | SLMO1 | FALSE | Downregulated |
| -0,145330396 | 0,722927576 | ZC2HC1A | FALSE | Downregulated |
| 0,837982766 | 0,722927576 | ZNF295-AS1 | FALSE | Upregulated |
| -0,088163374 | 0,722927576 | ZNF688 | FALSE | Downregulated |
| -0,042453637 | 0,722961444 | ATG12 | FALSE | Downregulated |
| -0,09733516 | 0,722961444 | TOB1 | FALSE | Downregulated |
| -0,400373531 | 0,722987197 | LOC645752 | FALSE | Downregulated |
| -0,313104678 | 0,722987197 | MEIG1 | FALSE | Downregulated |
| 0,249782325 | 0,723241951 | LOC102724475 | FALSE | Upregulated |
| -0,393901632 | 0,723597336 | MIR4647 | FALSE | Downregulated |
| 0,063681345 | 0,723859367 | COPB2 | FALSE | Upregulated |
| 0,155581571 | 0,723859367 | FUT6 | FALSE | Upregulated |
| -0,371475647 | 0,723859367 | GFAP | FALSE | Downregulated |
| 0,245882036 | 0,723875669 | LOC101928615 | FALSE | Upregulated |
| 0,071791045 | 0,723985712 | CERK | FALSE | Upregulated |
| -0,346484136 | 0,723985712 | RP5-977B1.7 | FALSE | Downregulated |
| -0,053575528 | 0,723985712 | TNKS2 | FALSE | Downregulated |
| 0,061641828 | 0,724189504 | ATPAF2 | FALSE | Upregulated |
| -0,207748026 | 0,724189504 | BMPR1B | FALSE | Downregulated |
| 0,382858821 | 0,724189504 | LOC102546298 | FALSE | Upregulated |
| 0,048502091 | 0,724189504 | NOLC1 | FALSE | Upregulated |
| -0,372457147 | 0,724219802 | SLC26A3 | FALSE | Downregulated |
| -0,045983429 | 0,724286179 | FECH | FALSE | Downregulated |
| 0,125428557 | 0,724286179 | MPP7 | FALSE | Upregulated |
| -0,077171773 | 0,724286179 | SPAG7 | FALSE | Downregulated |
| -0,068279052 | 0,724317015 | MMP24-AS1 | FALSE | Downregulated |
| 0,632983175 | 0,724422993 | MKRN3 | FALSE | Upregulated |
| 0,389126681 | 0,724511534 | SERPINA12 | FALSE | Upregulated |
| -0,069201134 | 0,724511899 | UBN1 | FALSE | Downregulated |
| 0,066805365 | 0,724580163 | INTS12 | FALSE | Upregulated |
| -0,045118435 | 0,72469922 | ZHX2 | FALSE | Downregulated |

| | | | | |
|---|---|---|---|---|
| -0,172495506 | 0,724947026 | NLRP11 | FALSE | Downregulated |
| -0,129491652 | 0,725341046 | RCN3 | FALSE | Downregulated |
| -0,091797872 | 0,725362658 | CCDC41 | FALSE | Downregulated |
| 0,034035771 | 0,725362658 | KCMF1 | FALSE | Upregulated |
| -0,063951235 | 0,725583015 | NUS1 | FALSE | Downregulated |
| 0,314165489 | 0,725661273 | SNORA61 | FALSE | Upregulated |
| -0,576878397 | 0,725667257 | KIAA2022 | FALSE | Downregulated |
| 0,124618099 | 0,725722865 | CYP2S1 | FALSE | Upregulated |
| 0,057751041 | 0,725722865 | HYAL1 | FALSE | Upregulated |
| -0,47254702 | 0,725983673 | CA8 | FALSE | Downregulated |
| 0,388443096 | 0,725983673 | LOC101928117 | FALSE | Upregulated |
| -0,091737286 | 0,725983673 | UROC1 | FALSE | Downregulated |
| 0,174443917 | 0,726188492 | ARNT2 | FALSE | Upregulated |
| -0,462120683 | 0,726284464 | ZNF878 | FALSE | Downregulated |
| -0,112966483 | 0,726376514 | LRTOMT | FALSE | Downregulated |
| -0,071023343 | 0,72662847 | ELMSAN1 | FALSE | Downregulated |
| 0,431533198 | 0,72662847 | LOC102724176 | FALSE | Upregulated |
| 0,627549816 | 0,726680472 | LOC101928589 | FALSE | Upregulated |
| 0,062642626 | 0,726794836 | SND1 | FALSE | Upregulated |
| -0,056850474 | 0,726845001 | CASC3 | FALSE | Downregulated |
| 0,287625353 | 0,726845001 | IL23A | FALSE | Upregulated |
| 0,051216396 | 0,726845001 | MYH14 | FALSE | Upregulated |
| -0,365190845 | 0,726845001 | ROCK1P1 | FALSE | Downregulated |
| 0,537892244 | 0,726845001 | RPL29P2 | FALSE | Upregulated |
| -0,098291325 | 0,726890773 | JRK | FALSE | Downregulated |
| 0,13284241 | 0,727207381 | MRPS11 | FALSE | Upregulated |
| -0,161725654 | 0,727258503 | EHF | FALSE | Downregulated |
| 0,049689338 | 0,727437492 | STAM | FALSE | Upregulated |
| -0,133212817 | 0,727437492 | ZNF430 | FALSE | Downregulated |
| -0,060757951 | 0,727582683 | BRAF | FALSE | Downregulated |
| 0,099515408 | 0,727585471 | PCAT19 | FALSE | Upregulated |
| 0,064353426 | 0,727710266 | ZNF622 | FALSE | Upregulated |
| 0,078956941 | 0,728038329 | RPS3 | FALSE | Upregulated |
| -0,167205564 | 0,728067199 | CYP21A2 | FALSE | Downregulated |
| -0,197303564 | 0,728067199 | LOC101928274 | FALSE | Downregulated |
| 0,073370841 | 0,728102075 | ACD | FALSE | Upregulated |
| 0,047235553 | 0,728102075 | WDR43 | FALSE | Upregulated |
| -0,193844641 | 0,728199712 | LOC201651 | FALSE | Downregulated |
| -0,075725753 | 0,728199712 | LYAR | FALSE | Downregulated |
| 0,075404673 | 0,728231083 | TGIF2 | FALSE | Upregulated |
| 0,163949533 | 0,728342024 | LSS | FALSE | Upregulated |
| -0,061316133 | 0,728342024 | ZNF561 | FALSE | Downregulated |
| -0,055716701 | 0,728440347 | C1D | FALSE | Downregulated |
| 0,582351347 | 0,728440347 | SERTAD4-AS1 | FALSE | Upregulated |
| -0,284744364 | 0,728554882 | PKD1L1 | FALSE | Downregulated |
| -0,258618323 | 0,728592302 | AGAP11 | FALSE | Downregulated |
| -0,356780322 | 0,728903142 | LOC101929675 | FALSE | Downregulated |
| 0,467859019 | 0,729080688 | LOC440934 | FALSE | Upregulated |
| -0,07429747 | 0,729097613 | KIAA1161 | FALSE | Downregulated |
| 0,097143261 | 0,729097613 | MROH1 | FALSE | Upregulated |
| -0,078830957 | 0,729097613 | POLD2 | FALSE | Downregulated |

| | | | | |
|---|---|---|---|---|
| 0,561382201 | 0,729097613 | TMEM252 | FALSE | Upregulated |
| -0,39036079 | 0,729165848 | C17orf72 | FALSE | Downregulated |
| 0,167525247 | 0,729165848 | GLIS2 | FALSE | Upregulated |
| 0,29714184 | 0,729165848 | GRIK1 | FALSE | Upregulated |
| -0,181491975 | 0,729165848 | RGS7BP | FALSE | Downregulated |
| -0,108916895 | 0,729242333 | C14orf79 | FALSE | Downregulated |
| 0,151879857 | 0,729244789 | BFSP1 | FALSE | Upregulated |
| -0,063551306 | 0,729244789 | MUS81 | FALSE | Downregulated |
| 0,139252747 | 0,729276982 | LOC100996909 | FALSE | Upregulated |
| -0,235156602 | 0,729276982 | MPPED2 | FALSE | Downregulated |
| 0,481569578 | 0,729276982 | TPI1P3 | FALSE | Upregulated |
| 0,24685253 | 0,729315345 | LOC101927178 | FALSE | Upregulated |
| -0,129331695 | 0,729393819 | C14orf132 | FALSE | Downregulated |
| 0,085464185 | 0,729432046 | BANF1 | FALSE | Upregulated |
| -0,083927111 | 0,729432046 | E4F1 | FALSE | Downregulated |
| 0,071450867 | 0,729577041 | AKAP13 | FALSE | Upregulated |
| -0,049446665 | 0,729594113 | SERINC3 | FALSE | Downregulated |
| -0,352349315 | 0,729609965 | CERKL | FALSE | Downregulated |
| -0,08500444 | 0,729626204 | SLC25A30 | FALSE | Downregulated |
| -0,128655626 | 0,729810189 | SEMA6B | FALSE | Downregulated |
| -0,123401544 | 0,729950836 | URAHP | FALSE | Downregulated |
| -0,457034584 | 0,730108364 | FAM209B | FALSE | Downregulated |
| -0,09258313 | 0,730241317 | RITA1 | FALSE | Downregulated |
| -0,314161139 | 0,730301099 | ST8SIA3 | FALSE | Downregulated |
| 0,38495762 | 0,730401197 | LOC102723974 | FALSE | Upregulated |
| -0,251430176 | 0,730653797 | H1FX-AS1 | FALSE | Downregulated |
| -0,048683708 | 0,730653797 | POM121C | FALSE | Downregulated |
| -0,058042084 | 0,730653797 | SNX27 | FALSE | Downregulated |
| 0,051801527 | 0,7308705 | NASP | FALSE | Upregulated |
| 0,050278745 | 0,7308705 | NIF3L1 | FALSE | Upregulated |
| 0,106918502 | 0,7308705 | PPP1R9B | FALSE | Upregulated |
| -0,095028671 | 0,7308705 | RBM28 | FALSE | Downregulated |
| 0,484604604 | 0,7308705 | RNFT2 | FALSE | Upregulated |
| -0,117923478 | 0,7308705 | UNKL | FALSE | Downregulated |
| -0,349131827 | 0,730871068 | LOC102724806 | FALSE | Downregulated |
| 0,105517935 | 0,73096499 | SATB2 | FALSE | Upregulated |
| 0,110091507 | 0,731163136 | SOX12 | FALSE | Upregulated |
| -0,789671501 | 0,731312319 | PAX7 | FALSE | Downregulated |
| 0,116612963 | 0,731331251 | ALPL | FALSE | Upregulated |
| -0,254882042 | 0,731331251 | C15orf43 | FALSE | Downregulated |
| 0,054931008 | 0,73146229 | PIK3R2 | FALSE | Upregulated |
| 0,050710071 | 0,731466447 | CCDC50 | FALSE | Upregulated |
| -0,333140086 | 0,731567987 | LOC100131532 | FALSE | Downregulated |
| 0,680283301 | 0,732066018 | AKR1B15 | FALSE | Upregulated |
| -0,074961564 | 0,732066018 | PPP2R3C | FALSE | Downregulated |
| -0,07903348 | 0,732142668 | CNPY4 | FALSE | Downregulated |
| 0,058783481 | 0,732142668 | SLC25A11 | FALSE | Upregulated |
| 0,691908197 | 0,732219826 | MRPL42P5 | FALSE | Upregulated |
| 1,870977722 | 0,732407005 | RPS4Y1 | FALSE | Upregulated |
| -0,138381772 | 0,732467709 | SHROOM4 | FALSE | Downregulated |
| 0,778336361 | 0,732632253 | PHOSPHO2-KLHL23 | FALSE | Upregulated |

| | | | | |
|---|---|---|---|---|
| 0,158948287 | 0,732723162 | RERG | FALSE | Upregulated |
| -0,116100158 | 0,732833929 | C1QTNF6 | FALSE | Downregulated |
| -0,10517413 | 0,732833929 | LOC100506161 | FALSE | Downregulated |
| -0,076113002 | 0,732833929 | TMEM101 | FALSE | Downregulated |
| -0,090724005 | 0,732971742 | SPARC | FALSE | Downregulated |
| -0,382021894 | 0,733038963 | MS4A4E | FALSE | Downregulated |
| -0,103959673 | 0,733038963 | TREX1 | FALSE | Downregulated |
| 0,452804879 | 0,733207665 | KIAA1199 | FALSE | Upregulated |
| 0,081074202 | 0,733353359 | EVA1A | FALSE | Upregulated |
| -0,339347266 | 0,733353359 | GOLGA6L10 | FALSE | Downregulated |
| -0,884429332 | 0,733353359 | MYO18B | FALSE | Downregulated |
| 0,417669792 | 0,733353359 | S100Z | FALSE | Upregulated |
| 0,086131149 | 0,733353359 | SHE | FALSE | Upregulated |
| -0,185107126 | 0,733353359 | TMED6 | FALSE | Downregulated |
| -1,31263472 | 0,733353359 | TSIX | FALSE | Downregulated |
| -0,035685082 | 0,733353359 | ZMYND11 | FALSE | Downregulated |
| -0,359109616 | 0,733399105 | LOC101929018 | FALSE | Downregulated |
| -0,056659049 | 0,733464651 | SLC38A10 | FALSE | Downregulated |
| 0,249893749 | 0,733473422 | GPAT2 | FALSE | Upregulated |
| -0,040533 | 0,733473422 | HEATR1 | FALSE | Downregulated |
| 0,407769691 | 0,733478201 | XKR6 | FALSE | Upregulated |
| -0,383667836 | 0,733859646 | FLJ45139 | FALSE | Downregulated |
| 0,080971489 | 0,733863992 | JMJD4 | FALSE | Upregulated |
| 0,205535058 | 0,733863992 | STX18-AS1 | FALSE | Upregulated |
| -0,097571104 | 0,733863992 | TAF1A | FALSE | Downregulated |
| -0,052661712 | 0,733958268 | GCC1 | FALSE | Downregulated |
| -0,451403398 | 0,733958268 | TEKT2 | FALSE | Downregulated |
| 0,242342164 | 0,734340239 | FBXO39 | FALSE | Upregulated |
| -0,056447686 | 0,734602457 | ZNF271 | FALSE | Downregulated |
| -0,077392227 | 0,734669727 | FN1 | FALSE | Downregulated |
| -0,118025661 | 0,734669727 | LAMA4 | FALSE | Downregulated |
| 0,091156608 | 0,734669727 | RAB17 | FALSE | Upregulated |
| -0,300193631 | 0,734669727 | SMN2 | FALSE | Downregulated |
| 0,442586729 | 0,734669727 | TRPM1 | FALSE | Upregulated |
| -0,030462186 | 0,734796217 | DDX50 | FALSE | Downregulated |
| -0,40872145 | 0,734796217 | LOC102724012 | FALSE | Downregulated |
| 0,045650078 | 0,734796217 | ORC4 | FALSE | Upregulated |
| -0,258958001 | 0,734796217 | SLC4A10 | FALSE | Downregulated |
| -0,192688089 | 0,734897685 | ELF3 | FALSE | Downregulated |
| -0,226702252 | 0,735053084 | C6orf52 | FALSE | Downregulated |
| 0,670680483 | 0,735053084 | DEFA3 | FALSE | Upregulated |
| 0,181662775 | 0,735053084 | LY6G5C | FALSE | Upregulated |
| 0,089714515 | 0,735053084 | OCLN | FALSE | Upregulated |
| -0,052465335 | 0,735053084 | POLR2D | FALSE | Downregulated |
| 0,092825876 | 0,735053084 | TBC1D2B | FALSE | Upregulated |
| 0,666201973 | 0,735079179 | RNY4 | FALSE | Upregulated |
| 0,475168739 | 0,735219163 | DDX3Y | FALSE | Upregulated |
| 0,356377189 | 0,735256595 | LOC389602 | FALSE | Upregulated |
| 0,502578541 | 0,735643699 | SNORA62 | FALSE | Upregulated |
| -0,038214363 | 0,735684296 | WBP11 | FALSE | Downregulated |
| -0,596704539 | 0,735775417 | LOC101929340 | FALSE | Downregulated |

| | | | | |
|---|---|---|---|---|
| 0,441174528 | 0,735879174 | SLC7A11 | FALSE | Upregulated |
| -0,1622354 | 0,735880549 | AIFM3 | FALSE | Downregulated |
| -0,199996883 | 0,735981771 | GALNT15 | FALSE | Downregulated |
| -0,42430946 | 0,735981771 | KLHL10 | FALSE | Downregulated |
| 0,143039515 | 0,73608423 | DTX1 | FALSE | Upregulated |
| -0,160850279 | 0,73608423 | SLC22A17 | FALSE | Downregulated |
| -0,189773038 | 0,736092029 | CD1C | FALSE | Downregulated |
| -0,072783308 | 0,736092029 | RAB24 | FALSE | Downregulated |
| 0,114866591 | 0,736159786 | ADSSL1 | FALSE | Upregulated |
| -0,135118304 | 0,736277228 | KIAA0825 | FALSE | Downregulated |
| -0,382093619 | 0,736840603 | MORN3 | FALSE | Downregulated |
| -0,288326805 | 0,737132212 | LOC654342 | FALSE | Downregulated |
| -0,297910968 | 0,737147376 | OXTR | FALSE | Downregulated |
| 0,050012357 | 0,737178885 | CLN5 | FALSE | Upregulated |
| -0,277085618 | 0,737185177 | TTYH1 | FALSE | Downregulated |
| -0,16725349 | 0,737417593 | ANK2 | FALSE | Downregulated |
| -0,085318788 | 0,737417593 | CADPS2 | FALSE | Downregulated |
| 0,114392712 | 0,737417593 | WFS1 | FALSE | Upregulated |
| 0,502223989 | 0,737469649 | FAM71D | FALSE | Upregulated |
| 0,077724996 | 0,737469649 | LHPP | FALSE | Upregulated |
| 0,147023691 | 0,737469649 | P2RY1 | FALSE | Upregulated |
| -0,205710825 | 0,737727824 | COL16A1 | FALSE | Downregulated |
| 0,054817716 | 0,73784623 | EPAS1 | FALSE | Upregulated |
| 0,058307677 | 0,73784623 | ZC3HC1 | FALSE | Upregulated |
| -0,141842945 | 0,737896612 | CLDN14 | FALSE | Downregulated |
| -0,069659978 | 0,737896612 | CXorf56 | FALSE | Downregulated |
| 0,269560636 | 0,737896612 | TMEM158 | FALSE | Upregulated |
| -0,061248829 | 0,737914987 | SBDS | FALSE | Downregulated |
| 0,065054219 | 0,737914987 | SETD1B | FALSE | Upregulated |
| -0,410631926 | 0,738108363 | CAPZA3 | FALSE | Downregulated |
| 0,043521996 | 0,738108363 | CLK3 | FALSE | Upregulated |
| -0,094796031 | 0,738137285 | DPH7 | FALSE | Downregulated |
| 1,237869211 | 0,738219966 | GABRA5 | FALSE | Upregulated |
| -0,123572684 | 0,738312102 | ABLIM3 | FALSE | Downregulated |
| -0,140963065 | 0,738312102 | APOA5 | FALSE | Downregulated |
| 0,051663823 | 0,738312102 | KPNA1 | FALSE | Upregulated |
| -0,360947052 | 0,738312102 | LINC00299 | FALSE | Downregulated |
| -0,172767177 | 0,738312102 | LOC595101 | FALSE | Downregulated |
| -0,756283661 | 0,738312102 | OR7D2 | FALSE | Downregulated |
| 0,050160587 | 0,738312102 | TOPBP1 | FALSE | Upregulated |
| 0,049386253 | 0,738542766 | DDX59 | FALSE | Upregulated |
| 0,318456712 | 0,738542766 | PEBP4 | FALSE | Upregulated |
| 0,055183097 | 0,738542766 | PPP1CB | FALSE | Upregulated |
| 0,072780098 | 0,738614926 | DDO | FALSE | Upregulated |
| -0,056988302 | 0,738692898 | LPP | FALSE | Downregulated |
| 0,176880897 | 0,738701623 | EEF1E1 | FALSE | Upregulated |
| 0,437843583 | 0,738701623 | LOC101927259 | FALSE | Upregulated |
| 0,24693692 | 0,738701623 | NCR3 | FALSE | Upregulated |
| -0,139041486 | 0,738701623 | RPL13AP5 | FALSE | Downregulated |
| 0,157080854 | 0,738701623 | SLC16A7 | FALSE | Upregulated |
| -0,053334294 | 0,738844551 | DIMT1 | FALSE | Downregulated |

| | | | | |
|---|---|---|---|---|
| 0,255437712 | 0,739036674 | DACT2 | FALSE | Upregulated |
| 1,613541687 | 0,739152294 | EIF1AY | FALSE | Upregulated |
| 0,171243607 | 0,739291945 | COL1A2 | FALSE | Upregulated |
| 0,402473338 | 0,739447585 | C1orf180 | FALSE | Upregulated |
| -0,046094683 | 0,739447585 | REXO4 | FALSE | Downregulated |
| -0,118648887 | 0,739518709 | CHST3 | FALSE | Downregulated |
| -0,34364465 | 0,739518709 | HRH4 | FALSE | Downregulated |
| 0,05419915 | 0,739518709 | PQBP1 | FALSE | Upregulated |
| 0,05683685 | 0,740143566 | EIF3F | FALSE | Upregulated |
| 0,528474971 | 0,740143566 | LOC102723975 | FALSE | Upregulated |
| -0,075477236 | 0,740177533 | GPN2 | FALSE | Downregulated |
| 0,226986989 | 0,740207077 | CYSLTR2 | FALSE | Upregulated |
| -0,153558954 | 0,740207077 | SAMD14 | FALSE | Downregulated |
| 0,299377099 | 0,740453489 | TMSB4Y | FALSE | Upregulated |
| 0,460142308 | 0,740471466 | LOC102724685 | FALSE | Upregulated |
| -0,055873674 | 0,740471466 | UBAC1 | FALSE | Downregulated |
| 0,081025522 | 0,740514834 | COMMD10 | FALSE | Upregulated |
| 0,28416934 | 0,740545213 | NRCAM | FALSE | Upregulated |
| -0,154513325 | 0,740545213 | THNSL2 | FALSE | Downregulated |
| -0,086717143 | 0,740607694 | ZBTB24 | FALSE | Downregulated |
| -0,046609919 | 0,741080956 | LARP1 | FALSE | Downregulated |
| -0,199571217 | 0,741080956 | LOC101927609 | FALSE | Downregulated |
| -0,226736038 | 0,741392684 | CDHR1 | FALSE | Downregulated |
| -0,300355169 | 0,741436924 | FLJ21408 | FALSE | Downregulated |
| -0,266165269 | 0,741504418 | CCDC153 | FALSE | Downregulated |
| 0,273501738 | 0,741669602 | SHANK1 | FALSE | Upregulated |
| -0,116512933 | 0,741829795 | CD4 | FALSE | Downregulated |
| 0,437631931 | 0,741852717 | CNR1 | FALSE | Upregulated |
| -0,091554501 | 0,741852717 | ISPD | FALSE | Downregulated |
| 0,371733055 | 0,741852717 | LINC00092 | FALSE | Upregulated |
| -0,616246943 | 0,741852717 | MAGED4B | FALSE | Downregulated |
| -0,079769738 | 0,741852717 | OTC | FALSE | Downregulated |
| 0,144060424 | 0,741852744 | TMEM91 | FALSE | Upregulated |
| -0,659223528 | 0,74193791 | SNORA7B | FALSE | Downregulated |
| 0,474272137 | 0,742084277 | COL22A1 | FALSE | Upregulated |
| -0,062123378 | 0,742198623 | RNF5 | FALSE | Downregulated |
| -0,224224906 | 0,74270406 | USP6 | FALSE | Downregulated |
| -0,050255205 | 0,74285339 | CWC15 | FALSE | Downregulated |
| 0,3668134 | 0,74285339 | KC6 | FALSE | Upregulated |
| 0,072569585 | 0,74285339 | KIAA0922 | FALSE | Upregulated |
| -0,181332583 | 0,742924659 | SCD5 | FALSE | Downregulated |
| -0,074964605 | 0,742961202 | MAST3 | FALSE | Downregulated |
| 0,051723991 | 0,742961202 | MRPL42 | FALSE | Upregulated |
| 0,082039551 | 0,742961202 | PRPF40B | FALSE | Upregulated |
| -0,47148368 | 0,742961202 | RAET1E-AS1 | FALSE | Downregulated |
| 0,14195491 | 0,742961202 | TDO2 | FALSE | Upregulated |
| -0,060660036 | 0,74302814 | RTKN | FALSE | Downregulated |
| -0,10543782 | 0,743160611 | ANKRD12 | FALSE | Downregulated |
| -0,66190224 | 0,743160611 | FLJ41200 | FALSE | Downregulated |
| 0,163856716 | 0,743390589 | OXCT1 | FALSE | Upregulated |
| -0,932091286 | 0,743390589 | PERM1 | FALSE | Downregulated |

| | | | | |
|---|---|---|---|---|
| -0,173225138 | 0,743390589 | SCARF2 | FALSE | Downregulated |
| 0,626593384 | 0,743475459 | NPTX1 | FALSE | Upregulated |
| -0,524124056 | 0,743610584 | NXF5 | FALSE | Downregulated |
| 0,064799161 | 0,743613898 | COX16 | FALSE | Upregulated |
| -0,046526544 | 0,744261347 | SLC4A1AP | FALSE | Downregulated |
| -0,21412505 | 0,744332321 | C5orf49 | FALSE | Downregulated |
| -1,333073202 | 0,744354284 | DUSP27 | FALSE | Downregulated |
| -0,065741793 | 0,744383482 | MASP2 | FALSE | Downregulated |
| -0,070986627 | 0,744383482 | NMRAL1 | FALSE | Downregulated |
| 0,058082318 | 0,744383482 | ZNF746 | FALSE | Upregulated |
| -0,067770574 | 0,744457004 | KIAA0226 | FALSE | Downregulated |
| -0,068590631 | 0,744650926 | CYB561D1 | FALSE | Downregulated |
| 0,142209169 | 0,744650926 | ZSWIM4 | FALSE | Upregulated |
| 0,064816031 | 0,74467139 | PRRC1 | FALSE | Upregulated |
| 0,543459044 | 0,744763326 | LOC101060301 | FALSE | Upregulated |
| 0,19505813 | 0,74502657 | SLC2A4 | FALSE | Upregulated |
| -0,067903249 | 0,745049961 | R3HCC1 | FALSE | Downregulated |
| 0,131410203 | 0,745049961 | SOX6 | FALSE | Upregulated |
| -0,049813091 | 0,745049961 | TMEM259 | FALSE | Downregulated |
| -0,509254774 | 0,745375925 | SCRG1 | FALSE | Downregulated |
| -0,061224142 | 0,745522959 | JAGN1 | FALSE | Downregulated |
| 0,065476725 | 0,745657154 | C7orf26 | FALSE | Upregulated |
| 0,087427202 | 0,745657154 | C7orf60 | FALSE | Upregulated |
| 0,167712178 | 0,745657154 | GCSAM | FALSE | Upregulated |
| 0,12071373 | 0,745657154 | LGALS1 | FALSE | Upregulated |
| 0,061690914 | 0,745657154 | MED12 | FALSE | Upregulated |
| 0,181165882 | 0,745657154 | NRG4 | FALSE | Upregulated |
| -0,421665723 | 0,745657154 | VWA8-AS1 | FALSE | Downregulated |
| -0,093281356 | 0,745880744 | CYP8B1 | FALSE | Downregulated |
| 0,371932906 | 0,745975337 | ANKRD20A5P | FALSE | Upregulated |
| 0,347839467 | 0,745975337 | LINC00853 | FALSE | Upregulated |
| -0,34209647 | 0,745975337 | LRRC9 | FALSE | Downregulated |
| -0,184982793 | 0,745975337 | RP11-112H10.4 | FALSE | Downregulated |
| -0,192630015 | 0,745975337 | SLC5A1 | FALSE | Downregulated |
| -0,471639334 | 0,746230295 | SCN5A | FALSE | Downregulated |
| 0,323683501 | 0,746274357 | ZEB2-AS1 | FALSE | Upregulated |
| 0,207581417 | 0,746292071 | MSX1 | FALSE | Upregulated |
| -0,077670663 | 0,746292071 | PPTC7 | FALSE | Downregulated |
| 0,088464369 | 0,746292071 | ZNF879 | FALSE | Upregulated |
| 0,064893063 | 0,746304571 | EIF2AK4 | FALSE | Upregulated |
| 0,054138446 | 0,746401697 | PSMA2 | FALSE | Upregulated |
| -0,101681187 | 0,746401697 | RP11-620J15.3 | FALSE | Downregulated |
| -0,109392227 | 0,746412545 | IDUA | FALSE | Downregulated |
| -0,231494363 | 0,746520942 | LOC283887 | FALSE | Downregulated |
| 0,051135781 | 0,746705343 | CDYL | FALSE | Upregulated |
| 0,377430942 | 0,746705343 | HIST2H2BF | FALSE | Upregulated |
| -0,042101262 | 0,746705343 | PDHX | FALSE | Downregulated |
| -0,457415179 | 0,74673434 | FBXW4P1 | FALSE | Downregulated |
| -1,46433441 | 0,74673434 | RNU1-27P | FALSE | Downregulated |
| -0,09664063 | 0,74673434 | YY2 | FALSE | Downregulated |
| -0,114478831 | 0,746745218 | GLT1D1 | FALSE | Downregulated |

| | | | | |
|---|---|---|---|---|
| 0,058601023 | 0,746795177 | AKR1A1 | FALSE | Upregulated |
| 0,466951753 | 0,746795177 | LOC101927212 | FALSE | Upregulated |
| -0,243354909 | 0,746795177 | LOC102723622 | FALSE | Downregulated |
| -0,578392627 | 0,746829259 | DNM1P41 | FALSE | Downregulated |
| 0,104902897 | 0,746878238 | ROBO1 | FALSE | Upregulated |
| -0,175616963 | 0,746963057 | LOC400927 | FALSE | Downregulated |
| 0,104400872 | 0,746963057 | STS | FALSE | Upregulated |
| 0,04627708 | 0,747143846 | CPSF3 | FALSE | Upregulated |
| -0,236507746 | 0,747143846 | LOC100507472 | FALSE | Downregulated |
| -0,253225381 | 0,747143846 | LOC642846 | FALSE | Downregulated |
| -0,288814643 | 0,747143846 | SGCA | FALSE | Downregulated |
| -0,071477988 | 0,747143846 | SNAP47 | FALSE | Downregulated |
| -0,045723328 | 0,747205275 | NSL1 | FALSE | Downregulated |
| 0,055073033 | 0,747291984 | MLX | FALSE | Upregulated |
| -0,066054893 | 0,747347167 | PLCD1 | FALSE | Downregulated |
| 0,138926629 | 0,747530979 | PER3 | FALSE | Upregulated |
| 0,087049644 | 0,747595118 | ZBTB42 | FALSE | Upregulated |
| 0,141549162 | 0,747648572 | TSPAN15 | FALSE | Upregulated |
| -0,083727906 | 0,747663994 | CCL16 | FALSE | Downregulated |
| 0,057741451 | 0,747674278 | MRPS16 | FALSE | Upregulated |
| 0,059554678 | 0,74785986 | TBPL1 | FALSE | Upregulated |
| 0,231760765 | 0,747964733 | MSH5-SAPCD1 | FALSE | Upregulated |
| -0,098537622 | 0,747964733 | PLXDC2 | FALSE | Downregulated |
| 0,347212918 | 0,748365152 | IGSF22 | FALSE | Upregulated |
| 0,073932981 | 0,748642066 | CAMLG | FALSE | Upregulated |
| 0,07952926 | 0,74885468 | STX5 | FALSE | Upregulated |
| 0,106050353 | 0,74885468 | ZC3H3 | FALSE | Upregulated |
| -0,135456454 | 0,748980195 | CRISPLD1 | FALSE | Downregulated |
| 0,094623734 | 0,749075375 | ZNF675 | FALSE | Upregulated |
| -0,328744742 | 0,749098356 | CLDN4 | FALSE | Downregulated |
| 0,386689681 | 0,749263769 | CYP27B1 | FALSE | Upregulated |
| -0,086847105 | 0,749263769 | ELK3 | FALSE | Downregulated |
| 0,165558134 | 0,749263769 | PIWIL2 | FALSE | Upregulated |
| 0,300080769 | 0,749299448 | ATP5EP2 | FALSE | Upregulated |
| -0,20669637 | 0,749299448 | C1orf116 | FALSE | Downregulated |
| -0,063900281 | 0,749299448 | COA1 | FALSE | Downregulated |
| 0,09960327 | 0,749299448 | FNDC3B | FALSE | Upregulated |
| 0,034866824 | 0,749299448 | IPO7 | FALSE | Upregulated |
| -0,252284092 | 0,749299448 | LOC101927372 | FALSE | Downregulated |
| -0,321093118 | 0,749299448 | LOC645967 | FALSE | Downregulated |
| -0,154414314 | 0,749299448 | LOC728407 | FALSE | Downregulated |
| 0,101852693 | 0,749299448 | MSTO1 | FALSE | Upregulated |
| 0,488315873 | 0,749299448 | PSORS1C3 | FALSE | Upregulated |
| -0,430311322 | 0,749299448 | RYR1 | FALSE | Downregulated |
| 0,405953054 | 0,749299448 | SNHG4 | FALSE | Upregulated |
| -0,094083512 | 0,749299448 | TMEM184A | FALSE | Downregulated |
| -0,106482227 | 0,749461971 | GRAMD4 | FALSE | Downregulated |
| -0,05341444 | 0,749461971 | LRBA | FALSE | Downregulated |
| 0,074705205 | 0,749461971 | TSTD1 | FALSE | Upregulated |
| 0,451576639 | 0,749471351 | LOC101929837 | FALSE | Upregulated |
| 0,042854052 | 0,749481963 | UBA2 | FALSE | Upregulated |

| | | | | |
|---|---|---|---|---|
| 0,521555422 | 0,749743462 | LOC554223 | FALSE | Upregulated |
| -0,480896617 | 0,749743462 | LOC728739 | FALSE | Downregulated |
| 0,297484716 | 0,749743462 | SLC44A5 | FALSE | Upregulated |
| -0,423400748 | 0,749893317 | RASAL1 | FALSE | Downregulated |
| 0,087293454 | 0,749943842 | ALKBH4 | FALSE | Upregulated |
| -0,391677943 | 0,749943842 | FRMD1 | FALSE | Downregulated |
| 0,040220322 | 0,749943842 | TM9SF3 | FALSE | Upregulated |
| 0,059300336 | 0,750046816 | ZNHIT3 | FALSE | Upregulated |
| -0,158467315 | 0,7500645 | HMGN5 | FALSE | Downregulated |
| 0,243393985 | 0,750082036 | RAB38 | FALSE | Upregulated |
| 0,241565298 | 0,750155014 | FAM83H-AS1 | FALSE | Upregulated |
| -0,465943262 | 0,750155014 | LOC102724241 | FALSE | Downregulated |
| -0,059854512 | 0,750192158 | PEX6 | FALSE | Downregulated |
| 0,051751348 | 0,750285035 | GBF1 | FALSE | Upregulated |
| 0,061311088 | 0,750285035 | STK16 | FALSE | Upregulated |
| 0,271929805 | 0,750309735 | ERMN | FALSE | Upregulated |
| -0,041744292 | 0,750419986 | LSM8 | FALSE | Downregulated |
| -0,048420905 | 0,750419986 | NSMCE1 | FALSE | Downregulated |
| 0,350791328 | 0,750505403 | PDZD7 | FALSE | Upregulated |
| 0,098127408 | 0,750611324 | COL6A1 | FALSE | Upregulated |
| 0,206020796 | 0,750611324 | GCNT4 | FALSE | Upregulated |
| -0,15221514 | 0,750611324 | LINC00864 | FALSE | Downregulated |
| 0,05969345 | 0,750611324 | ZDHHC16 | FALSE | Upregulated |
| -0,065089592 | 0,750623219 | MRGBP | FALSE | Downregulated |
| -0,03613706 | 0,75066997 | GCN1L1 | FALSE | Downregulated |
| -0,055538599 | 0,75066997 | OXA1L | FALSE | Downregulated |
| -0,718543615 | 0,750911949 | TMEM212 | FALSE | Downregulated |
| -0,081025882 | 0,750911949 | ZNF79 | FALSE | Downregulated |
| -0,1173302 | 0,751063105 | GTF2IRD1 | FALSE | Downregulated |
| -0,096310678 | 0,751063105 | STK19 | FALSE | Downregulated |
| -0,706946765 | 0,75111036 | CLEC18C | FALSE | Downregulated |
| 0,062783355 | 0,751275066 | GPN3 | FALSE | Upregulated |
| 0,440876993 | 0,751564415 | BARX1 | FALSE | Upregulated |
| -0,45942348 | 0,751657793 | FAM184B | FALSE | Downregulated |
| -0,464503176 | 0,751775082 | LINC00479 | FALSE | Downregulated |
| -0,073085565 | 0,752097607 | NTN1 | FALSE | Downregulated |
| 0,052891669 | 0,752097607 | PTPN21 | FALSE | Upregulated |
| -0,172144264 | 0,752356457 | LOC102724733 | FALSE | Downregulated |
| -0,062290278 | 0,752356457 | NFE2L2 | FALSE | Downregulated |
| 0,101792661 | 0,75239168 | AFAP1L2 | FALSE | Upregulated |
| 0,064514598 | 0,75239168 | JTB | FALSE | Upregulated |
| -0,040373211 | 0,752568653 | CEBPZ | FALSE | Downregulated |
| 0,03660958 | 0,752568653 | PPHLN1 | FALSE | Upregulated |
| -0,121849311 | 0,752570811 | PRELP | FALSE | Downregulated |
| 0,229723205 | 0,752612057 | LINC00920 | FALSE | Upregulated |
| 0,052802381 | 0,752782196 | AZI2 | FALSE | Upregulated |
| -0,053628799 | 0,752901474 | RNF2 | FALSE | Downregulated |
| -0,285348404 | 0,752965066 | RP11-142C4.6 | FALSE | Downregulated |
| -0,052904089 | 0,752965066 | RPL14 | FALSE | Downregulated |
| -0,659246021 | 0,752965066 | SPATA22 | FALSE | Downregulated |
| 0,328065134 | 0,753242772 | SYCE2 | FALSE | Upregulated |

| | | | | |
|---|---|---|---|---|
| -0,76522651 | 0,753327376 | LOC100508046 | FALSE | Downregulated |
| 0,075230659 | 0,753327376 | NDUFAF6 | FALSE | Upregulated |
| -0,071420677 | 0,753379928 | BAZ2A | FALSE | Downregulated |
| 0,501398118 | 0,753379928 | CLCN1 | FALSE | Upregulated |
| 0,144541108 | 0,753379928 | RP11-345L23.1 | FALSE | Upregulated |
| -0,244556986 | 0,753379928 | SSSCA1-AS1 | FALSE | Downregulated |
| 0,151085622 | 0,753458456 | LRRC37A4P | FALSE | Upregulated |
| -0,15099083 | 0,753468093 | CCL28 | FALSE | Downregulated |
| -0,053284235 | 0,753468093 | NOTCH2 | FALSE | Downregulated |
| -0,130524701 | 0,753471133 | FAM228B | FALSE | Downregulated |
| -0,072551448 | 0,753634004 | ATP5J | FALSE | Downregulated |
| -0,067437433 | 0,753645727 | ZNF230 | FALSE | Downregulated |
| -0,059736556 | 0,753707979 | IK | FALSE | Downregulated |
| 0,246125159 | 0,753707979 | STC1 | FALSE | Upregulated |
| 0,038114426 | 0,753736011 | COPA | FALSE | Upregulated |
| 0,207381457 | 0,753909884 | MT1A | FALSE | Upregulated |
| 0,12138161 | 0,753991779 | GATSL3 | FALSE | Upregulated |
| -0,123323243 | 0,754056523 | SULF2 | FALSE | Downregulated |
| -0,06616949 | 0,754102583 | ZNF17 | FALSE | Downregulated |
| -0,130881798 | 0,75417873 | EXOC3L2 | FALSE | Downregulated |
| 0,198616759 | 0,75420924 | LOC100131655 | FALSE | Upregulated |
| -0,08087185 | 0,754223444 | MYPOP | FALSE | Downregulated |
| 0,107680922 | 0,754223909 | LYSMD2 | FALSE | Upregulated |
| -0,078508601 | 0,754292273 | RPRD1B | FALSE | Downregulated |
| -0,733143928 | 0,754302827 | FTH1P18 | FALSE | Downregulated |
| 0,475247596 | 0,754302827 | LINC00506 | FALSE | Upregulated |
| -0,057653544 | 0,754302827 | PIN4 | FALSE | Downregulated |
| 0,054631095 | 0,754302827 | RMND5B | FALSE | Upregulated |
| -0,773106307 | 0,754302827 | TRNAD16 | FALSE | Downregulated |
| -0,073268052 | 0,754373144 | RGMB | FALSE | Downregulated |
| -0,054855342 | 0,754391578 | RBM23 | FALSE | Downregulated |
| -0,430200491 | 0,754391578 | SPTY2D1-AS1 | FALSE | Downregulated |
| -0,045515688 | 0,754455912 | ACIN1 | FALSE | Downregulated |
| -0,036489096 | 0,754658795 | CTTN | FALSE | Downregulated |
| -0,180023981 | 0,754658795 | MIP | FALSE | Downregulated |
| 0,045293962 | 0,754658795 | NHEJ1 | FALSE | Upregulated |
| 0,06362471 | 0,754844101 | MGST1 | FALSE | Upregulated |
| -0,315924904 | 0,754844101 | RNA5-8S5 | FALSE | Downregulated |
| -0,173627218 | 0,754874222 | LOC388906 | FALSE | Downregulated |
| -0,285568497 | 0,754887293 | LOC100287467 | FALSE | Downregulated |
| -0,054302865 | 0,754887293 | PAGR1 | FALSE | Downregulated |
| -0,067348475 | 0,755656217 | CISD3 | FALSE | Downregulated |
| -0,443505552 | 0,755697657 | CAPN8-2 | FALSE | Downregulated |
| 0,032707829 | 0,755697657 | DCAF7 | FALSE | Upregulated |
| -0,130194002 | 0,755697657 | HSD17B7 | FALSE | Downregulated |
| -0,051252906 | 0,755697657 | KDELC2 | FALSE | Downregulated |
| -0,558533881 | 0,755697657 | LOC102724462 | FALSE | Downregulated |
| -0,475199542 | 0,755697657 | LOC388282 | FALSE | Downregulated |
| -0,278187905 | 0,755697657 | LOC441081 | FALSE | Downregulated |
| -0,103890103 | 0,755697657 | MAP3K12 | FALSE | Downregulated |
| -0,039202409 | 0,755697657 | MECP2 | FALSE | Downregulated |

| | | | | |
|---|---|---|---|---|
| 0,182695358 | 0,755697657 | MYEF2 | FALSE | Upregulated |
| -0,039536297 | 0,755697657 | PAK2 | FALSE | Downregulated |
| -0,039642113 | 0,755697657 | SEPT2 | FALSE | Downregulated |
| -0,183885488 | 0,755705153 | LOC727896 | FALSE | Downregulated |
| -0,532595065 | 0,755705153 | RNF157-AS1 | FALSE | Downregulated |
| 0,063171716 | 0,755705153 | RNF25 | FALSE | Upregulated |
| 0,043054791 | 0,755731731 | SYMPK | FALSE | Upregulated |
| 0,461470702 | 0,755917835 | HTR2A-AS1 | FALSE | Upregulated |
| 0,120608048 | 0,756169478 | PCDH9 | FALSE | Upregulated |
| -0,322888812 | 0,756224185 | LOC101928409 | FALSE | Downregulated |
| 0,374831185 | 0,756240335 | LOC102723352 | FALSE | Upregulated |
| -0,083533366 | 0,756510777 | GPR180 | FALSE | Downregulated |
| -0,502131319 | 0,756842891 | ANKRD18B | FALSE | Downregulated |
| -0,16327735 | 0,757027845 | AP000472.2 | FALSE | Downregulated |
| -0,270426258 | 0,757034015 | LOC101929667 | FALSE | Downregulated |
| 0,144950532 | 0,757182193 | MAPK11 | FALSE | Upregulated |
| -0,023628252 | 0,757182193 | MKRN1 | FALSE | Downregulated |
| -0,236964279 | 0,757207632 | BOLA3-AS1 | FALSE | Downregulated |
| -0,070563197 | 0,75727089 | SMDT1 | FALSE | Downregulated |
| 0,247761283 | 0,757346266 | C12orf79 | FALSE | Upregulated |
| -0,066846164 | 0,757346266 | SASH1 | FALSE | Downregulated |
| 0,187176335 | 0,757509295 | SOX9 | FALSE | Upregulated |
| 0,117800569 | 0,757820479 | TBKBP1 | FALSE | Upregulated |
| -0,400668899 | 0,757882377 | LOC101927066 | FALSE | Downregulated |
| -0,167416461 | 0,757882377 | PDZD4 | FALSE | Downregulated |
| 0,206613755 | 0,758000582 | RASGEF1A | FALSE | Upregulated |
| 0,119036963 | 0,758068652 | RAB23 | FALSE | Upregulated |
| 0,18842503 | 0,758068652 | ZNF665 | FALSE | Upregulated |
| -0,264150213 | 0,758098798 | CLMP | FALSE | Downregulated |
| -0,055746326 | 0,758235702 | ANAPC13 | FALSE | Downregulated |
| -0,17519632 | 0,758235702 | GJA5 | FALSE | Downregulated |
| 0,080515753 | 0,758301669 | APIP | FALSE | Upregulated |
| -0,042425207 | 0,758401301 | HERC4 | FALSE | Downregulated |
| -0,425742989 | 0,758539153 | WFIKKN1 | FALSE | Downregulated |
| 0,081461681 | 0,758576656 | COX8A | FALSE | Upregulated |
| -0,483669131 | 0,758576656 | LOC101928082 | FALSE | Downregulated |
| -0,073822874 | 0,758576656 | NUP43 | FALSE | Downregulated |
| 0,096844927 | 0,758576656 | TANC2 | FALSE | Upregulated |
| -0,729737587 | 0,758577508 | AMY2A | FALSE | Downregulated |
| 0,059799125 | 0,758602842 | POLR2E | FALSE | Upregulated |
| 0,351310948 | 0,758613697 | CPB2-AS1 | FALSE | Upregulated |
| -0,186983324 | 0,758631812 | MAP2K6 | FALSE | Downregulated |
| -0,066467747 | 0,758645295 | ESF1 | FALSE | Downregulated |
| -0,08812541 | 0,758645295 | GFRA1 | FALSE | Downregulated |
| -0,209176179 | 0,758645295 | KCNK3 | FALSE | Downregulated |
| 0,271684117 | 0,758645295 | LOC101929482 | FALSE | Upregulated |
| -0,452087974 | 0,758645295 | MYH15 | FALSE | Downregulated |
| 0,320093147 | 0,758645295 | PPFIA2 | FALSE | Upregulated |
| 0,039782263 | 0,758645295 | UBE2E3 | FALSE | Upregulated |
| -0,043559355 | 0,758645295 | ZSCAN29 | FALSE | Downregulated |
| -0,366404949 | 0,758719361 | MDGA2 | FALSE | Downregulated |

| | | | | |
|---|---|---|---|---|
| -0,153926428 | 0,758719361 | PTGES2-AS1 | FALSE | Downregulated |
| -0,055944744 | 0,758828544 | RIF1 | FALSE | Downregulated |
| 0,043297732 | 0,758910916 | METTL14 | FALSE | Upregulated |
| -0,234867076 | 0,758950149 | LOC646626 | FALSE | Downregulated |
| -0,252416421 | 0,759325415 | GDPD3 | FALSE | Downregulated |
| 0,071333111 | 0,759581093 | GEMIN2 | FALSE | Upregulated |
| 0,413257037 | 0,759688396 | PGLYRP1 | FALSE | Upregulated |
| 0,274581873 | 0,759842117 | SLIT1 | FALSE | Upregulated |
| 0,224296498 | 0,759842117 | TACSTD2 | FALSE | Upregulated |
| 0,102892367 | 0,760077833 | WSCD1 | FALSE | Upregulated |
| 0,051038905 | 0,760131855 | DHX29 | FALSE | Upregulated |
| -0,081392029 | 0,760131855 | ST7L | FALSE | Downregulated |
| -0,137958831 | 0,760131855 | ZNF547 | FALSE | Downregulated |
| 0,060876 | 0,76022864 | SGMS2 | FALSE | Upregulated |
| -0,061225605 | 0,760244445 | MFSD8 | FALSE | Downregulated |
| -0,112887696 | 0,760244445 | PPARGC1A | FALSE | Downregulated |
| -0,515275033 | 0,760244445 | RP11-673E1.1 | FALSE | Downregulated |
| -0,198777797 | 0,760246855 | GLRB | FALSE | Downregulated |
| -0,18367535 | 0,760365847 | LOC100509620 | FALSE | Downregulated |
| -0,380238477 | 0,760365847 | OCA2 | FALSE | Downregulated |
| -0,372474383 | 0,760365847 | TMPRSS4 | FALSE | Downregulated |
| -0,047279731 | 0,760446055 | MAPKAP1 | FALSE | Downregulated |
| -0,120635352 | 0,760472446 | ZC3H12B | FALSE | Downregulated |
| -0,350311031 | 0,760641888 | LOC100996761 | FALSE | Downregulated |
| 0,074291291 | 0,760666887 | TLE1 | FALSE | Upregulated |
| -0,095583752 | 0,760673267 | ZNF383 | FALSE | Downregulated |
| 0,402170792 | 0,760678389 | MAB21L1 | FALSE | Upregulated |
| -0,083880464 | 0,760800617 | IER2 | FALSE | Downregulated |
| 0,190003539 | 0,76092712 | EVPLL | FALSE | Upregulated |
| 0,185393427 | 0,76100623 | ASNS | FALSE | Upregulated |
| -0,372141913 | 0,761033461 | LOC101928263 | FALSE | Downregulated |
| -0,273419984 | 0,761042004 | HSPB6 | FALSE | Downregulated |
| -0,105095227 | 0,761042004 | NBPF11 | FALSE | Downregulated |
| 0,251888008 | 0,761109986 | FHDC1 | FALSE | Upregulated |
| 0,07040573 | 0,761109986 | MESDC2 | FALSE | Upregulated |
| 0,077259055 | 0,761350506 | CLDN23 | FALSE | Upregulated |
| 0,493168775 | 0,76170707 | KCNK15 | FALSE | Upregulated |
| -0,102111367 | 0,761881376 | AKR1C6P | FALSE | Downregulated |
| 0,113803221 | 0,761881376 | INPP5F | FALSE | Upregulated |
| 0,315550069 | 0,761881376 | LOC101927279 | FALSE | Upregulated |
| 0,255858242 | 0,76198354 | LOC101929719 | FALSE | Upregulated |
| 0,235909925 | 0,761998378 | ZNF205-AS1 | FALSE | Upregulated |
| 0,187819325 | 0,762128832 | IL34 | FALSE | Upregulated |
| 0,112410914 | 0,762177407 | GALNT4 | FALSE | Upregulated |
| 0,147290469 | 0,762193529 | TXK | FALSE | Upregulated |
| -0,18838364 | 0,762279926 | KCNJ16 | FALSE | Downregulated |
| -0,285857235 | 0,762334177 | LOC101927104 | FALSE | Downregulated |
| 0,066806486 | 0,762471255 | CDC34 | FALSE | Upregulated |
| 0,27027907 | 0,762899647 | GLB1L2 | FALSE | Upregulated |
| 0,06603529 | 0,762899647 | KCTD11 | FALSE | Upregulated |
| -0,042687813 | 0,763050364 | MRPL3 | FALSE | Downregulated |

| | | | | |
|---|---|---|---|---|
| 0,517709632 | 0,76319053 | LOC102724501 | FALSE | Upregulated |
| -2,451941004 | 0,763212078 | LOC102724441 | FALSE | Downregulated |
| 0,052799972 | 0,763212078 | SETD8 | FALSE | Upregulated |
| 0,038024323 | 0,763212078 | ZNF410 | FALSE | Upregulated |
| 0,060558118 | 0,763249776 | THADA | FALSE | Upregulated |
| -0,22701676 | 0,763512059 | IQCA1 | FALSE | Downregulated |
| -0,204889922 | 0,763647796 | MT1L | FALSE | Downregulated |
| 0,025786505 | 0,763752183 | TERF2IP | FALSE | Upregulated |
| -0,234003222 | 0,76386445 | RFPL1S | FALSE | Downregulated |
| 0,065184555 | 0,76386445 | TRANK1 | FALSE | Upregulated |
| -0,115102023 | 0,763909116 | C9orf43 | FALSE | Downregulated |
| 0,269258842 | 0,763909116 | CCDC85A | FALSE | Upregulated |
| 0,106936704 | 0,763937401 | ZNF446 | FALSE | Upregulated |
| 0,176071135 | 0,763956155 | PNMA6A | FALSE | Upregulated |
| -0,110543439 | 0,764031692 | GPT2 | FALSE | Downregulated |
| 0,043028917 | 0,764038101 | BABAM1 | FALSE | Upregulated |
| -0,068554889 | 0,764038101 | SERPINA1 | FALSE | Downregulated |
| -0,097125712 | 0,764052121 | CALCRL | FALSE | Downregulated |
| 0,03597786 | 0,764052121 | CHMP7 | FALSE | Upregulated |
| -0,040263601 | 0,764052121 | EMC4 | FALSE | Downregulated |
| 0,034856702 | 0,764052121 | MAP2K4 | FALSE | Upregulated |
| 0,064854024 | 0,764052121 | RPL13 | FALSE | Upregulated |
| 0,047575054 | 0,764052121 | VRK3 | FALSE | Upregulated |
| -0,082493032 | 0,764052121 | ZNF613 | FALSE | Downregulated |
| -0,174414856 | 0,764071674 | RP4-782L23.1 | FALSE | Downregulated |
| 0,044640406 | 0,764261087 | DNAJC8 | FALSE | Upregulated |
| 0,202767934 | 0,764317081 | MB21D2 | FALSE | Upregulated |
| 0,230365945 | 0,764497749 | ASH1L-AS1 | FALSE | Upregulated |
| -0,063289902 | 0,764497749 | CPSF1 | FALSE | Downregulated |
| -0,04821334 | 0,764497749 | GBA2 | FALSE | Downregulated |
| -0,145157471 | 0,764497749 | GPC4 | FALSE | Downregulated |
| 0,069188609 | 0,764672756 | TAF6L | FALSE | Upregulated |
| -0,132869804 | 0,764672756 | USHBP1 | FALSE | Downregulated |
| -0,082372187 | 0,76475199 | VASN | FALSE | Downregulated |
| -0,06185642 | 0,764795182 | ACN9 | FALSE | Downregulated |
| -0,166164243 | 0,764795182 | ANKRD31 | FALSE | Downregulated |
| -0,30707595 | 0,764795182 | CYP4Z1 | FALSE | Downregulated |
| 0,136449454 | 0,764795182 | KLF5 | FALSE | Upregulated |
| -0,066280447 | 0,764795182 | PEX10 | FALSE | Downregulated |
| -0,083642141 | 0,764798118 | CP | FALSE | Downregulated |
| 0,053519793 | 0,765141099 | WBSCR16 | FALSE | Upregulated |
| 0,347001661 | 0,765725023 | BCL2L15 | FALSE | Upregulated |
| -0,784606183 | 0,765725023 | DAZ4 | FALSE | Downregulated |
| -0,211623148 | 0,765725023 | LPHN3 | FALSE | Downregulated |
| 0,327750747 | 0,765725023 | PRR16 | FALSE | Upregulated |
| 0,060216199 | 0,765725023 | SRFBP1 | FALSE | Upregulated |
| 0,397727735 | 0,765760164 | LOC730257 | FALSE | Upregulated |
| -0,234744555 | 0,765760164 | SCGN | FALSE | Downregulated |
| -0,084254967 | 0,766319064 | GTF2H4 | FALSE | Downregulated |
| -0,02829904 | 0,766387776 | KLHL12 | FALSE | Downregulated |
| 0,070627488 | 0,766560403 | COX7A2 | FALSE | Upregulated |

| | | | | |
|---|---|---|---|---|
| -0,203611848 | 0,766560403 | PAGE5 | FALSE | Downregulated |
| -0,109399519 | 0,766560403 | RRN3P3 | FALSE | Downregulated |
| 0,217643518 | 0,766850106 | YBX3 | FALSE | Upregulated |
| -0,199404733 | 0,76692258 | MEOX2 | FALSE | Downregulated |
| 0,203477404 | 0,76692258 | SPON1 | FALSE | Upregulated |
| -0,077101734 | 0,76692258 | TRIM3 | FALSE | Downregulated |
| -0,110151895 | 0,767069009 | MAPK13 | FALSE | Downregulated |
| -0,054313137 | 0,767069009 | NARFL | FALSE | Downregulated |
| 0,104684911 | 0,767100176 | C6orf1 | FALSE | Upregulated |
| -0,136747677 | 0,767149604 | ZNF154 | FALSE | Downregulated |
| 0,453010501 | 0,767294758 | MEOX1 | FALSE | Upregulated |
| 0,127931249 | 0,767415191 | LIN7B | FALSE | Upregulated |
| 0,169717596 | 0,767698393 | MMP2 | FALSE | Upregulated |
| -0,04800917 | 0,767706191 | BCAS2 | FALSE | Downregulated |
| 0,253159723 | 0,767706191 | EGR1 | FALSE | Upregulated |
| 0,04039774 | 0,767706191 | GTF3C1 | FALSE | Upregulated |
| -0,222465873 | 0,767706191 | KCNB1 | FALSE | Downregulated |
| 0,573118183 | 0,767706191 | LOC101928262 | FALSE | Upregulated |
| 0,225771676 | 0,767706191 | SERF1A | FALSE | Upregulated |
| -0,45809189 | 0,767706191 | SLC10A6 | FALSE | Downregulated |
| -0,081621612 | 0,767706191 | SRP19 | FALSE | Downregulated |
| -0,135855912 | 0,767792817 | ALDH1A2 | FALSE | Downregulated |
| 0,323141945 | 0,767792817 | MMP17 | FALSE | Upregulated |
| 0,028869711 | 0,767940041 | BAG5 | FALSE | Upregulated |
| -0,053301134 | 0,767940041 | FOXJ2 | FALSE | Downregulated |
| -0,088958592 | 0,767940041 | SEMA4B | FALSE | Downregulated |
| -0,331740008 | 0,767940041 | ZNF286B | FALSE | Downregulated |
| -0,423280723 | 0,768035892 | RP11-37B2.1 | FALSE | Downregulated |
| -0,043201783 | 0,768197178 | CRBN | FALSE | Downregulated |
| 0,202233976 | 0,768197178 | CXorf36 | FALSE | Upregulated |
| 0,053322234 | 0,768197178 | RAB30 | FALSE | Upregulated |
| 0,233202156 | 0,768320763 | LOC102724861 | FALSE | Upregulated |
| -0,092165042 | 0,768378133 | LINC00339 | FALSE | Downregulated |
| -0,351630569 | 0,768425154 | GNAT1 | FALSE | Downregulated |
| 0,222030058 | 0,768425154 | SNX15 | FALSE | Upregulated |
| -0,402537255 | 0,768574395 | PABPC1P2 | FALSE | Downregulated |
| 0,04525112 | 0,768574395 | WDR26 | FALSE | Upregulated |
| 0,066532448 | 0,768693085 | UAP1 | FALSE | Upregulated |
| 0,04316364 | 0,768852537 | CPSF6 | FALSE | Upregulated |
| 0,173610827 | 0,768870678 | CTGF | FALSE | Upregulated |
| 0,051036838 | 0,768929363 | CS | FALSE | Upregulated |
| -0,896408466 | 0,769110262 | GSTT2 | FALSE | Downregulated |
| -0,435622485 | 0,769375412 | LOC101929661 | FALSE | Downregulated |
| -0,295118837 | 0,769375412 | LOC102723534 | FALSE | Downregulated |
| -0,418767159 | 0,769375412 | LOC646938 | FALSE | Downregulated |
| -0,184200591 | 0,769375412 | PROM2 | FALSE | Downregulated |
| 0,177781308 | 0,769473005 | LMCD1 | FALSE | Upregulated |
| 0,48808839 | 0,76953349 | PDLIM4 | FALSE | Upregulated |
| 0,056638368 | 0,769834106 | OIP5-AS1 | FALSE | Upregulated |
| 0,086038449 | 0,770038388 | ALKBH1 | FALSE | Upregulated |
| 0,046931637 | 0,770038388 | NDUFA12 | FALSE | Upregulated |

| | | | | |
|---|---|---|---|---|
| 0,123261477 | 0,770096715 | ITPR3 | FALSE | Upregulated |
| -0,234276235 | 0,770178061 | TMEM51-AS1 | FALSE | Downregulated |
| -0,064912166 | 0,770217584 | CCDC25 | FALSE | Downregulated |
| 0,1523571 | 0,770346545 | CLDN9 | FALSE | Upregulated |
| -0,132786524 | 0,770374948 | STON1 | FALSE | Downregulated |
| -0,515974475 | 0,770459121 | CLEC18A | FALSE | Downregulated |
| -0,051279796 | 0,770459121 | METTL17 | FALSE | Downregulated |
| -0,204452386 | 0,770459121 | NACA2 | FALSE | Downregulated |
| -1,014605982 | 0,770459121 | NPTN-IT1 | FALSE | Downregulated |
| -0,071672491 | 0,770629368 | TIRAP | FALSE | Downregulated |
| -0,054312818 | 0,770851371 | EEF1G | FALSE | Downregulated |
| -0,132983899 | 0,770903178 | ATF5 | FALSE | Downregulated |
| -0,029591473 | 0,770903178 | GLE1 | FALSE | Downregulated |
| -0,283794394 | 0,771137005 | LOC100293704 | FALSE | Downregulated |
| 0,072340894 | 0,771158791 | GEMIN8 | FALSE | Upregulated |
| 0,324497523 | 0,771158791 | RP3-510O8.4 | FALSE | Upregulated |
| -0,169320461 | 0,771174788 | FGF1 | FALSE | Downregulated |
| 0,066229453 | 0,771174788 | NDUFS8 | FALSE | Upregulated |
| 0,398408084 | 0,771240947 | MMEL1 | FALSE | Upregulated |
| -0,438656033 | 0,771506037 | LOC102724976 | FALSE | Downregulated |
| -0,102301288 | 0,771531438 | ERV3-1 | FALSE | Downregulated |
| -0,288436064 | 0,771654521 | FRY-AS1 | FALSE | Downregulated |
| -0,326065254 | 0,771720949 | HSPB2 | FALSE | Downregulated |
| 0,068169845 | 0,771998698 | DRG2 | FALSE | Upregulated |
| -0,123072291 | 0,772048258 | TMED8 | FALSE | Downregulated |
| -0,090970219 | 0,772109995 | TP53TG1 | FALSE | Downregulated |
| 0,056680479 | 0,772115584 | ANKRD27 | FALSE | Upregulated |
| 0,053293573 | 0,772115584 | BRF1 | FALSE | Upregulated |
| -0,383368457 | 0,772115584 | GYPE | FALSE | Downregulated |
| -0,375993574 | 0,772300004 | SNORD58C | FALSE | Downregulated |
| 0,403788606 | 0,772365102 | RORB | FALSE | Upregulated |
| -0,403374399 | 0,772365102 | TCP11 | FALSE | Downregulated |
| 0,349573334 | 0,772393005 | RP11-173B14.5 | FALSE | Upregulated |
| 0,253309908 | 0,772406557 | TFAP2A | FALSE | Upregulated |
| -0,168482931 | 0,772477009 | FAM53A | FALSE | Downregulated |
| 0,041227799 | 0,772477009 | FTSJ2 | FALSE | Upregulated |
| -0,310796362 | 0,772477009 | SLC17A7 | FALSE | Downregulated |
| 0,285038851 | 0,77268243 | LOC102724552 | FALSE | Upregulated |
| 0,157317404 | 0,772723541 | A4GALT | FALSE | Upregulated |
| 0,040919163 | 0,772723541 | DUS1L | FALSE | Upregulated |
| -0,409719299 | 0,772723541 | LOC101928967 | FALSE | Downregulated |
| 0,078663531 | 0,772736245 | CRELD1 | FALSE | Upregulated |
| -0,10434076 | 0,773047717 | LOC728290 | FALSE | Downregulated |
| -0,050607466 | 0,773047717 | NICN1 | FALSE | Downregulated |
| 0,053611426 | 0,773047717 | SMYD4 | FALSE | Upregulated |
| 0,154713246 | 0,773047717 | SUSD2 | FALSE | Upregulated |
| -0,082959832 | 0,773047717 | ZFP30 | FALSE | Downregulated |
| 0,040764203 | 0,773119935 | CSTF1 | FALSE | Upregulated |
| -0,048140261 | 0,773119935 | POLR1D | FALSE | Downregulated |
| 0,077340539 | 0,773127473 | IL4R | FALSE | Upregulated |
| 0,067468535 | 0,773211034 | DVL2 | FALSE | Upregulated |

| | | | | |
|---|---|---|---|---|
| -0,377500801 | 0,773223647 | LOC101928911 | FALSE | Downregulated |
| 0,047101148 | 0,773303856 | ZNF706 | FALSE | Upregulated |
| -0,586888768 | 0,773409949 | LOC101929157 | FALSE | Downregulated |
| 0,039168453 | 0,773477689 | RNF41 | FALSE | Upregulated |
| 0,063528785 | 0,773564521 | ENTPD5 | FALSE | Upregulated |
| 0,073341806 | 0,773564521 | RPS14 | FALSE | Upregulated |
| 0,418805212 | 0,773631143 | LINC00540 | FALSE | Upregulated |
| -0,240400405 | 0,773687402 | B3GAT1 | FALSE | Downregulated |
| 0,241478962 | 0,773718242 | SUSD5 | FALSE | Upregulated |
| 0,047776166 | 0,773811363 | CNPY2 | FALSE | Upregulated |
| 1,018150101 | 0,773811363 | SNORD3B-1 | FALSE | Upregulated |
| -0,151731638 | 0,773902351 | ADAM19 | FALSE | Downregulated |
| 0,04588012 | 0,774015773 | SSB | FALSE | Upregulated |
| -0,05346833 | 0,774430242 | DAAM1 | FALSE | Downregulated |
| 0,363887102 | 0,774452425 | B4GALNT1 | FALSE | Upregulated |
| -0,411553699 | 0,774452425 | C21orf15 | FALSE | Downregulated |
| -0,57167313 | 0,774452425 | CRYBB2 | FALSE | Downregulated |
| -0,25332675 | 0,774452425 | FHAD1 | FALSE | Downregulated |
| -0,762519336 | 0,774452425 | GPR156 | FALSE | Downregulated |
| -0,049831959 | 0,774452425 | MED26 | FALSE | Downregulated |
| -0,300033927 | 0,774452425 | MRAP | FALSE | Downregulated |
| -0,075793591 | 0,774452425 | ZNF614 | FALSE | Downregulated |
| 0,050495603 | 0,774464692 | MAX | FALSE | Upregulated |
| -0,45461442 | 0,774580403 | PPAPDC3 | FALSE | Downregulated |
| -0,305083966 | 0,774622522 | LOC101926895 | FALSE | Downregulated |
| 0,287592292 | 0,774666954 | FAM95C | FALSE | Upregulated |
| -0,047855861 | 0,774666954 | TNRC18 | FALSE | Downregulated |
| -0,08737272 | 0,77471518 | FAM161A | FALSE | Downregulated |
| 0,393565412 | 0,77471518 | NCRNA00185 | FALSE | Upregulated |
| 0,095929245 | 0,77471518 | ZSCAN5A | FALSE | Upregulated |
| -0,088751318 | 0,774735981 | ANP32B | FALSE | Downregulated |
| -0,471389821 | 0,774735981 | LMOD3 | FALSE | Downregulated |
| 0,25775097 | 0,774735981 | SCUBE3 | FALSE | Upregulated |
| 0,123992019 | 0,774757853 | MNDA | FALSE | Upregulated |
| -0,035225859 | 0,775083504 | FAM204A | FALSE | Downregulated |
| -0,214618144 | 0,775083504 | MEI4 | FALSE | Downregulated |
| 0,056401003 | 0,775083504 | MRPL19 | FALSE | Upregulated |
| -0,033336241 | 0,775083504 | SS18 | FALSE | Downregulated |
| -0,033009918 | 0,775134925 | CAND1 | FALSE | Downregulated |
| -0,046723777 | 0,775134925 | WDR12 | FALSE | Downregulated |
| -0,036379031 | 0,775211835 | WDR20 | FALSE | Downregulated |
| 0,323008497 | 0,775270941 | FLJ33360 | FALSE | Upregulated |
| 0,105192733 | 0,775274668 | TMEM204 | FALSE | Upregulated |
| 0,155487593 | 0,77532609 | PYCR1 | FALSE | Upregulated |
| 0,081046149 | 0,775493999 | CCDC71L | FALSE | Upregulated |
| 0,057314747 | 0,775783409 | FURIN | FALSE | Upregulated |
| -0,169257542 | 0,775878225 | C5AR2 | FALSE | Downregulated |
| 0,233635822 | 0,775896571 | FBXL2 | FALSE | Upregulated |
| -0,310175372 | 0,775896571 | LOC101929191 | FALSE | Downregulated |
| -0,040527146 | 0,775896571 | NSUN2 | FALSE | Downregulated |
| 0,204912642 | 0,775896571 | ZNF835 | FALSE | Upregulated |

| | | | | |
|---|---|---|---|---|
| 0,070352269 | 0,775928576 | RP11-890B15.2 | FALSE | Upregulated |
| 0,37487687 | 0,776155307 | FA2H | FALSE | Upregulated |
| -0,050865046 | 0,776155307 | SPRYD7 | FALSE | Downregulated |
| 0,189524546 | 0,776155307 | SV2A | FALSE | Upregulated |
| -0,127912301 | 0,776165905 | CES5A | FALSE | Downregulated |
| 0,067911671 | 0,776165905 | MGAT4A | FALSE | Upregulated |
| 0,142848878 | 0,776233995 | ENAM | FALSE | Upregulated |
| 0,093715842 | 0,776376408 | CSRP2 | FALSE | Upregulated |
| -0,228946157 | 0,776508759 | COLGALT2 | FALSE | Downregulated |
| -0,412737438 | 0,776543672 | CES1P1 | FALSE | Downregulated |
| 0,04898199 | 0,776543672 | MAP2K3 | FALSE | Upregulated |
| -0,59219151 | 0,776543672 | PART1 | FALSE | Downregulated |
| 0,048829456 | 0,776543672 | TACO1 | FALSE | Upregulated |
| -0,317082067 | 0,776543672 | ZNF215 | FALSE | Downregulated |
| 0,070393059 | 0,776710817 | ZBTB9 | FALSE | Upregulated |
| 0,102295001 | 0,776812148 | NPC1L1 | FALSE | Upregulated |
| -0,049701898 | 0,776882991 | XAB2 | FALSE | Downregulated |
| 0,064483213 | 0,776917883 | FKBP4 | FALSE | Upregulated |
| -0,419994988 | 0,776979589 | LOC101928838 | FALSE | Downregulated |
| 0,233159859 | 0,77702538 | TRIM29 | FALSE | Upregulated |
| -0,193349063 | 0,777593703 | DNM3 | FALSE | Downregulated |
| 0,05457428 | 0,777629124 | SAV1 | FALSE | Upregulated |
| 0,201644463 | 0,777631959 | LAYN | FALSE | Upregulated |
| -0,078603239 | 0,777663817 | C21orf2 | FALSE | Downregulated |
| 0,440141558 | 0,777738141 | DGCR9 | FALSE | Upregulated |
| 0,114069415 | 0,777738141 | GMDS | FALSE | Upregulated |
| -0,098851234 | 0,777738141 | ORM2 | FALSE | Downregulated |
| -0,065298239 | 0,77795617 | GPATCH2L | FALSE | Downregulated |
| -0,325638303 | 0,778043541 | ACTG2 | FALSE | Downregulated |
| 0,060843175 | 0,778043541 | MASTL | FALSE | Upregulated |
| 0,098330617 | 0,778043541 | UBXN11 | FALSE | Upregulated |
| -0,055369417 | 0,778214304 | ANAPC4 | FALSE | Downregulated |
| 0,486839084 | 0,778214304 | CACNA1I | FALSE | Upregulated |
| -0,10405066 | 0,778214304 | LAPTM4B | FALSE | Downregulated |
| 0,359435508 | 0,778214304 | LOC100130705 | FALSE | Upregulated |
| 0,232986402 | 0,778214304 | LOC102723989 | FALSE | Upregulated |
| -0,055968459 | 0,778214304 | PBX3 | FALSE | Downregulated |
| -0,025707194 | 0,778214304 | POLR2B | FALSE | Downregulated |
| -0,047567024 | 0,778214304 | PTPRF | FALSE | Downregulated |
| -0,415964519 | 0,778214304 | SLC8A3 | FALSE | Downregulated |
| -0,115374268 | 0,778419953 | EFNA2 | FALSE | Downregulated |
| 0,515687932 | 0,778419953 | LOC643387 | FALSE | Upregulated |
| -0,321190938 | 0,778419953 | PF4V1 | FALSE | Downregulated |
| 0,862890994 | 0,778475029 | FBP2 | FALSE | Upregulated |
| -0,049110898 | 0,778475029 | PUS10 | FALSE | Downregulated |
| 0,060174347 | 0,778475029 | RPL38 | FALSE | Upregulated |
| -0,045698427 | 0,778476647 | DIP2B | FALSE | Downregulated |
| -0,235747681 | 0,778476647 | PRKG2 | FALSE | Downregulated |
| 0,042273398 | 0,778493042 | FARP1 | FALSE | Upregulated |
| 0,069983053 | 0,778538405 | TAOK3 | FALSE | Upregulated |
| 0,042128403 | 0,778538405 | TYW1 | FALSE | Upregulated |

| | | | | |
|---|---|---|---|---|
| -0,207233169 | 0,778751007 | TRPM6 | FALSE | Downregulated |
| -0,027777683 | 0,778762228 | KCTD20 | FALSE | Downregulated |
| 0,028235654 | 0,778794785 | RAB21 | FALSE | Upregulated |
| 0,069141379 | 0,778914954 | UQCR11 | FALSE | Upregulated |
| -0,261598995 | 0,778925508 | SNORD104 | FALSE | Downregulated |
| -0,141628229 | 0,778987709 | RP11-115J16.1 | FALSE | Downregulated |
| -0,254224255 | 0,779162742 | RHBDL2 | FALSE | Downregulated |
| -0,058677113 | 0,779162742 | TMTC3 | FALSE | Downregulated |
| -0,282000276 | 0,779173953 | EYA4 | FALSE | Downregulated |
| -0,070984947 | 0,779173953 | SLC25A42 | FALSE | Downregulated |
| 0,063578366 | 0,779173953 | ZSCAN2 | FALSE | Upregulated |
| -0,062244089 | 0,779205347 | FASTK | FALSE | Downregulated |
| 0,490321286 | 0,779205347 | IDI2 | FALSE | Upregulated |
| -0,084010193 | 0,779220577 | RBM47 | FALSE | Downregulated |
| 0,095121743 | 0,779302253 | CEP19 | FALSE | Upregulated |
| 0,063049772 | 0,779399777 | FZR1 | FALSE | Upregulated |
| 0,189027783 | 0,779399777 | LINC01011 | FALSE | Upregulated |
| 0,084082143 | 0,779399777 | PTPRM | FALSE | Upregulated |
| -0,102292833 | 0,779420499 | PDGFRB | FALSE | Downregulated |
| -0,256633827 | 0,77953448 | RIPPLY3 | FALSE | Downregulated |
| -0,034466975 | 0,779638145 | PWWP2A | FALSE | Downregulated |
| 0,329911198 | 0,779638145 | RN7SL2 | FALSE | Upregulated |
| -0,045608972 | 0,779638145 | TOP1MT | FALSE | Downregulated |
| 0,044485228 | 0,779777426 | IGSF8 | FALSE | Upregulated |
| -0,055487204 | 0,779777426 | NUBP2 | FALSE | Downregulated |
| 0,170632195 | 0,779777426 | ZNF667-AS1 | FALSE | Upregulated |
| 0,247422496 | 0,779972812 | FAM223B | FALSE | Upregulated |
| 0,693518267 | 0,779972812 | LOC100131626 | FALSE | Upregulated |
| 0,051776726 | 0,779988954 | BIN3 | FALSE | Upregulated |
| 0,274086452 | 0,779988954 | CLCF1 | FALSE | Upregulated |
| 0,052408176 | 0,779988954 | TTC8 | FALSE | Upregulated |
| 0,374367446 | 0,780009092 | TTLL6 | FALSE | Upregulated |
| 0,047919134 | 0,780021182 | ACO1 | FALSE | Upregulated |
| -0,430402055 | 0,780021182 | GPR110 | FALSE | Downregulated |
| 0,12515146 | 0,780021182 | RAPGEF3 | FALSE | Upregulated |
| 0,712270553 | 0,780021182 | TFF2 | FALSE | Upregulated |
| -0,403892757 | 0,780021182 | UBE2DNL | FALSE | Downregulated |
| -0,303652441 | 0,780077673 | LRGUK | FALSE | Downregulated |
| -0,062023296 | 0,780351271 | ARHGAP19 | FALSE | Downregulated |
| -0,31954952 | 0,780351271 | LOC440354 | FALSE | Downregulated |
| 0,272811419 | 0,780461246 | NAT8B | FALSE | Upregulated |
| 0,20697827 | 0,780472268 | SEMA3G | FALSE | Upregulated |
| -0,043215569 | 0,780576995 | GRPEL1 | FALSE | Downregulated |
| 0,061409053 | 0,780901875 | CAMSAP2 | FALSE | Upregulated |
| 0,071362603 | 0,781268306 | ATP5E | FALSE | Upregulated |
| 0,173745211 | 0,781316607 | DISC1 | FALSE | Upregulated |
| -0,611714081 | 0,781389702 | TFAMP1 | FALSE | Downregulated |
| -0,044219036 | 0,781393654 | GABARAPL2 | FALSE | Downregulated |
| 0,259976849 | 0,781512362 | CAMK1G | FALSE | Upregulated |
| -0,707785052 | 0,781512362 | EGF | FALSE | Downregulated |
| -0,072788968 | 0,781512362 | WBP1L | FALSE | Downregulated |

| | | | | |
|---|---|---|---|---|
| 0,056297355 | 0,781534633 | TPST2 | FALSE | Upregulated |
| 0,287215117 | 0,78157386 | RP11-118B18.1 | FALSE | Upregulated |
| 0,184758807 | 0,78157386 | SLC35E4 | FALSE | Upregulated |
| 0,053010603 | 0,781637879 | C6orf203 | FALSE | Upregulated |
| -0,058966894 | 0,781637879 | CMIP | FALSE | Downregulated |
| -0,097381614 | 0,781706017 | ZNF821 | FALSE | Downregulated |
| -0,129960786 | 0,781795069 | LOC101927943 | FALSE | Downregulated |
| 0,066447937 | 0,781884423 | PEAK1 | FALSE | Upregulated |
| 0,14115139 | 0,781902327 | RELL2 | FALSE | Upregulated |
| -0,221856626 | 0,782089698 | FOXQ1 | FALSE | Downregulated |
| -0,047627406 | 0,78209255 | GALNT11 | FALSE | Downregulated |
| -0,23357837 | 0,782170681 | LOC101928820 | FALSE | Downregulated |
| 0,24466546 | 0,782170681 | ZSCAN23 | FALSE | Upregulated |
| -0,454895441 | 0,782181784 | RAET1G | FALSE | Downregulated |
| -0,305185223 | 0,782181784 | SNORD68 | FALSE | Downregulated |
| 0,0458409 | 0,782228735 | EXOC1 | FALSE | Upregulated |
| -0,260588231 | 0,782304455 | C20orf202 | FALSE | Downregulated |
| -0,242749571 | 0,782304455 | KCNRG | FALSE | Downregulated |
| -0,751244641 | 0,782337021 | PPP1R14C | FALSE | Downregulated |
| -0,066059931 | 0,782422282 | C12orf23 | FALSE | Downregulated |
| -0,059590848 | 0,78270852 | ADARB1 | FALSE | Downregulated |
| -0,07315274 | 0,782769572 | PINK1-AS | FALSE | Downregulated |
| -0,161261425 | 0,782769572 | SCARA5 | FALSE | Downregulated |
| 0,074813763 | 0,782850421 | SFI1 | FALSE | Upregulated |
| 0,070061526 | 0,782850421 | TANC1 | FALSE | Upregulated |
| 0,306495901 | 0,782912948 | KNDC1 | FALSE | Upregulated |
| 0,434849164 | 0,782963533 | FOXJ1 | FALSE | Upregulated |
| -0,079637422 | 0,783000461 | HDAC7 | FALSE | Downregulated |
| -0,067522001 | 0,783180184 | BCAR3 | FALSE | Downregulated |
| -0,34598789 | 0,783180184 | RSPO4 | FALSE | Downregulated |
| 0,175651429 | 0,783219823 | HSD17B1 | FALSE | Upregulated |
| -0,229909411 | 0,783281418 | AC017060.1 | FALSE | Downregulated |
| -0,138868455 | 0,783281418 | ALMS1-IT1 | FALSE | Downregulated |
| -0,042349454 | 0,783281418 | COMMD3-BMI1 | FALSE | Downregulated |
| -0,047605856 | 0,783281418 | PLCB3 | FALSE | Downregulated |
| 0,085980584 | 0,783281418 | ZNF48 | FALSE | Upregulated |
| 0,093650706 | 0,783387688 | PTGS1 | FALSE | Upregulated |
| -0,299250228 | 0,783387688 | TMEFF2 | FALSE | Downregulated |
| -0,095354284 | 0,78368372 | CLCN2 | FALSE | Downregulated |
| -0,101709639 | 0,783714179 | WBP1 | FALSE | Downregulated |
| -0,299176024 | 0,783826223 | LINC00189 | FALSE | Downregulated |
| 1,860484452 | 0,783988849 | EEF1E1-BLOC1S5 | FALSE | Upregulated |
| 0,056792943 | 0,783988849 | FAM174A | FALSE | Upregulated |
| -0,671384925 | 0,783988849 | KCNA7 | FALSE | Downregulated |
| -0,311044187 | 0,783988849 | LOC101928560 | FALSE | Downregulated |
| 0,077979895 | 0,784576265 | ANKRD32 | FALSE | Upregulated |
| 0,033351411 | 0,784576265 | SMG7 | FALSE | Upregulated |
| -0,037292614 | 0,785230108 | FGFR1OP2 | FALSE | Downregulated |
| -0,039740193 | 0,785237798 | ATF2 | FALSE | Downregulated |
| -0,084542532 | 0,785237798 | FAM107B | FALSE | Downregulated |
| 0,170594745 | 0,785237798 | SLC13A2 | FALSE | Upregulated |

| | | | | |
|---|---|---|---|---|
| -0,16025596 | 0,785272641 | CCL23 | FALSE | Downregulated |
| -0,144328159 | 0,785534009 | ATP6V1C2 | FALSE | Downregulated |
| -0,066139174 | 0,785534009 | GLTSCR2 | FALSE | Downregulated |
| -0,073501528 | 0,785561947 | ANXA9 | FALSE | Downregulated |
| 0,05529157 | 0,785561947 | APOL1 | FALSE | Upregulated |
| 0,082109437 | 0,785561947 | ATG10 | FALSE | Upregulated |
| -0,189508409 | 0,785561947 | BVES | FALSE | Downregulated |
| -0,031641886 | 0,785561947 | FAF1 | FALSE | Downregulated |
| -0,374422442 | 0,785561947 | KCNK2 | FALSE | Downregulated |
| -0,191243913 | 0,785561947 | LINC00893 | FALSE | Downregulated |
| 0,072294965 | 0,785561947 | RPL11 | FALSE | Upregulated |
| -0,042724594 | 0,785561947 | TXNRD3 | FALSE | Downregulated |
| 0,032986174 | 0,785561947 | UBE2V1 | FALSE | Upregulated |
| 0,080322235 | 0,785561947 | ZNF283 | FALSE | Upregulated |
| 0,046000132 | 0,785929551 | ALCAM | FALSE | Upregulated |
| -0,176990208 | 0,785929551 | RP11-163N6.2 | FALSE | Downregulated |
| 0,159996475 | 0,786134577 | ANKDD1A | FALSE | Upregulated |
| 0,127893242 | 0,78623729 | RP11-425I13.3 | FALSE | Upregulated |
| 0,199512319 | 0,786289175 | SIGLEC16 | FALSE | Upregulated |
| 0,156566717 | 0,786424042 | SLC16A5 | FALSE | Upregulated |
| -0,051084438 | 0,786424042 | SLC27A1 | FALSE | Downregulated |
| 0,101848645 | 0,786466194 | ARSG | FALSE | Upregulated |
| -0,111956612 | 0,786466194 | CD82 | FALSE | Downregulated |
| 0,392416651 | 0,786499463 | KIAA1257 | FALSE | Upregulated |
| -0,096159234 | 0,786499463 | OPHN1 | FALSE | Downregulated |
| 0,217061554 | 0,786507418 | CAPN6 | FALSE | Upregulated |
| 0,286667656 | 0,786886431 | CNBD2 | FALSE | Upregulated |
| -0,065049831 | 0,786886431 | LOC90784 | FALSE | Downregulated |
| -0,036388576 | 0,786886431 | RBM15B | FALSE | Downregulated |
| -0,107681608 | 0,786886431 | RCE1 | FALSE | Downregulated |
| 0,078219857 | 0,786886431 | TP53INP2 | FALSE | Upregulated |
| -0,226241859 | 0,787153733 | LINC00926 | FALSE | Downregulated |
| 0,253033522 | 0,787153733 | LOC101929796 | FALSE | Upregulated |
| 0,13498415 | 0,787265723 | ZNF404 | FALSE | Upregulated |
| -0,076430068 | 0,787420106 | ECH1 | FALSE | Downregulated |
| 0,060889541 | 0,787491583 | APOA1 | FALSE | Upregulated |
| -0,242750478 | 0,787542053 | GUCY2C | FALSE | Downregulated |
| 0,138727345 | 0,787542053 | LOC100652768 | FALSE | Upregulated |
| -0,06001634 | 0,787542053 | TMEM126A | FALSE | Downregulated |
| -0,164741755 | 0,787551171 | LOC102724229 | FALSE | Downregulated |
| -0,04266221 | 0,787564235 | DDX39B | FALSE | Downregulated |
| -0,061133101 | 0,787685553 | PHKG2 | FALSE | Downregulated |
| 0,071153138 | 0,787713161 | MOCOS | FALSE | Upregulated |
| 0,106316521 | 0,787713161 | RP4-639F20.1 | FALSE | Upregulated |
| 0,188084257 | 0,787717023 | TTC39A | FALSE | Upregulated |
| 0,175774293 | 0,787876183 | FAM86B3P | FALSE | Upregulated |
| 0,040666497 | 0,787922159 | FAM136A | FALSE | Upregulated |
| 0,260180553 | 0,787982065 | LOC101930275 | FALSE | Upregulated |
| 0,255682162 | 0,78811239 | ULBP2 | FALSE | Upregulated |
| 0,239142917 | 0,788113483 | POU5F1 | FALSE | Upregulated |
| -0,204496793 | 0,788113483 | SCUBE1 | FALSE | Downregulated |

| | | | | |
|---|---|---|---|---|
| -0,111005414 | 0,788210591 | EGFLAM | FALSE | Downregulated |
| -0,042635215 | 0,788256574 | SENP5 | FALSE | Downregulated |
| 0,084889981 | 0,788262676 | B4GALT6 | FALSE | Upregulated |
| -0,122716111 | 0,788492981 | ARL17A | FALSE | Downregulated |
| -0,339772576 | 0,788492981 | ART5 | FALSE | Downregulated |
| -0,079035616 | 0,788492981 | BCL7C | FALSE | Downregulated |
| -0,252163967 | 0,788492981 | NEUROD1 | FALSE | Downregulated |
| 0,035326198 | 0,788492981 | RLIM | FALSE | Upregulated |
| -0,069640766 | 0,788492981 | TRAPPC6A | FALSE | Downregulated |
| -0,065448277 | 0,78878393 | CNEP1R1 | FALSE | Downregulated |
| -0,075746142 | 0,78890135 | OIT3 | FALSE | Downregulated |
| 0,066482582 | 0,789067224 | HERPUD1 | FALSE | Upregulated |
| 0,378081423 | 0,789067224 | LOC100506731 | FALSE | Upregulated |
| -0,063403424 | 0,789360002 | THUMPD1 | FALSE | Downregulated |
| -0,056000864 | 0,789774611 | PRMT7 | FALSE | Downregulated |
| 0,071175208 | 0,789941666 | RPL41 | FALSE | Upregulated |
| 0,245206437 | 0,790034712 | LOC101927400 | FALSE | Upregulated |
| 0,057918685 | 0,790034712 | MAPK3 | FALSE | Upregulated |
| -0,227983343 | 0,790153657 | CASP16 | FALSE | Downregulated |
| 0,058940529 | 0,790153657 | NIFK | FALSE | Upregulated |
| 0,352009299 | 0,79032497 | LOC729041 | FALSE | Upregulated |
| -0,160225223 | 0,790631446 | DPT | FALSE | Downregulated |
| -0,061935393 | 0,790834859 | METTL8 | FALSE | Downregulated |
| -0,055641884 | 0,790913907 | PGAP3 | FALSE | Downregulated |
| 0,083957461 | 0,791204761 | CLEC14A | FALSE | Upregulated |
| 0,142414112 | 0,791309137 | SPATA18 | FALSE | Upregulated |
| 0,065832713 | 0,791324558 | MLLT1 | FALSE | Upregulated |
| -0,067467741 | 0,791513257 | AGT | FALSE | Downregulated |
| 0,046551389 | 0,791513257 | CACYBP | FALSE | Upregulated |
| 0,19956595 | 0,791513257 | DDX43 | FALSE | Upregulated |
| -0,227974035 | 0,791606453 | CCDC87 | FALSE | Downregulated |
| -0,115165079 | 0,79160982 | TNN | FALSE | Downregulated |
| 0,279004868 | 0,791794318 | TAS2R14 | FALSE | Upregulated |
| -0,372403208 | 0,791911015 | TMED7-TICAM2 | FALSE | Downregulated |
| -0,031301585 | 0,791932935 | DVL3 | FALSE | Downregulated |
| 0,035117396 | 0,792022625 | TWISTNB | FALSE | Upregulated |
| -0,137258243 | 0,792051581 | IQCH-AS1 | FALSE | Downregulated |
| 0,147334867 | 0,792132007 | B3GALNT1 | FALSE | Upregulated |
| 0,040649015 | 0,792132007 | FBXO45 | FALSE | Upregulated |
| 0,115346798 | 0,792159769 | WASH1 | FALSE | Upregulated |
| 0,04864248 | 0,792289767 | SEPT8 | FALSE | Upregulated |
| -0,38455879 | 0,792681993 | AC073133.1 | FALSE | Downregulated |
| 0,171046558 | 0,792681993 | LOC100129518 | FALSE | Upregulated |
| 0,098552683 | 0,792681993 | OTUB2 | FALSE | Upregulated |
| 0,03872628 | 0,792681993 | PARL | FALSE | Upregulated |
| -0,036921204 | 0,792681993 | SAR1B | FALSE | Downregulated |
| -0,039309813 | 0,792745322 | PPP5C | FALSE | Downregulated |
| 0,058396943 | 0,79307051 | SDC4 | FALSE | Upregulated |
| -0,094142537 | 0,793090875 | FDFT1 | FALSE | Downregulated |
| 0,04124521 | 0,793090875 | HDGFRP2 | FALSE | Upregulated |
| -0,199438496 | 0,793145427 | ZNF578 | FALSE | Downregulated |

| | | | | |
|---|---|---|---|---|
| -0,648705148 | 0,793634031 | HIST2H2AA4 | FALSE | Downregulated |
| -0,039852356 | 0,793634031 | WDR61 | FALSE | Downregulated |
| 0,037505382 | 0,793640276 | CHAMP1 | FALSE | Upregulated |
| 0,453923596 | 0,793640276 | GYG2P1 | FALSE | Upregulated |
| -0,140250224 | 0,793640276 | SLITRK6 | FALSE | Downregulated |
| 1,298845403 | 0,793640276 | TRNAV19 | FALSE | Upregulated |
| 0,087472216 | 0,793817406 | PPP1R3F | FALSE | Upregulated |
| -0,040687396 | 0,793948123 | CUL9 | FALSE | Downregulated |
| 0,114734895 | 0,793948123 | LINGO1 | FALSE | Upregulated |
| 0,11559986 | 0,794467258 | CD160 | FALSE | Upregulated |
| -0,368143066 | 0,794467258 | CR1L | FALSE | Downregulated |
| 0,135210433 | 0,794746648 | SNAI1 | FALSE | Upregulated |
| -0,058393963 | 0,79486448 | MRE11A | FALSE | Downregulated |
| -0,241373404 | 0,795006225 | CPA3 | FALSE | Downregulated |
| -0,149228395 | 0,795006225 | ZNF711 | FALSE | Downregulated |
| -0,046346358 | 0,795129158 | RPL10 | FALSE | Downregulated |
| 0,147072234 | 0,795147044 | NAALAD2 | FALSE | Upregulated |
| 0,079406547 | 0,795296667 | FAM49A | FALSE | Upregulated |
| -0,086441483 | 0,79530263 | ETS2 | FALSE | Downregulated |
| 0,083201989 | 0,795385091 | CBLC | FALSE | Upregulated |
| -0,072477042 | 0,795385091 | DDAH1 | FALSE | Downregulated |
| -0,381284114 | 0,795385091 | LOC102724812 | FALSE | Downregulated |
| -0,092886903 | 0,795385091 | PHLDB1 | FALSE | Downregulated |
| -0,056517143 | 0,795385091 | ROCK1 | FALSE | Downregulated |
| 0,05061767 | 0,795452866 | ETS1 | FALSE | Upregulated |
| 0,096116612 | 0,795452866 | TMEM238 | FALSE | Upregulated |
| -0,087345174 | 0,795452866 | ZNF845 | FALSE | Downregulated |
| -0,246322382 | 0,795646694 | LOC101927469 | FALSE | Downregulated |
| 0,062705427 | 0,795764002 | CARD8 | FALSE | Upregulated |
| 0,638390217 | 0,795764002 | LOC100996517 | FALSE | Upregulated |
| -0,051723792 | 0,795886869 | ABTB2 | FALSE | Downregulated |
| 0,045291201 | 0,795886869 | NR3C1 | FALSE | Upregulated |
| -0,041076037 | 0,79589608 | ZBTB6 | FALSE | Downregulated |
| 0,04926007 | 0,795916855 | UTP15 | FALSE | Upregulated |
| -0,080125252 | 0,796017963 | ZNF211 | FALSE | Downregulated |
| 0,066457972 | 0,796079808 | ARL13B | FALSE | Upregulated |
| -0,319122818 | 0,796079808 | ATP8A2 | FALSE | Downregulated |
| 0,098980414 | 0,796079808 | CAPS2 | FALSE | Upregulated |
| 0,038357157 | 0,796079808 | CECR5 | FALSE | Upregulated |
| 0,341646001 | 0,796079808 | GGTLC1 | FALSE | Upregulated |
| -0,043401552 | 0,796079808 | KIAA1432 | FALSE | Downregulated |
| 0,040702176 | 0,796079808 | METTL5 | FALSE | Upregulated |
| -0,426230866 | 0,796079808 | SNORA55 | FALSE | Downregulated |
| -0,061699204 | 0,796079808 | WWOX | FALSE | Downregulated |
| 0,271987546 | 0,796079808 | ZP3 | FALSE | Upregulated |
| 0,200732341 | 0,796269682 | LOC100996246 | FALSE | Upregulated |
| 0,049602601 | 0,796358693 | TIPRL | FALSE | Upregulated |
| 0,23876074 | 0,796397507 | LOC284865 | FALSE | Upregulated |
| -0,232281512 | 0,796510815 | LOC100507634 | FALSE | Downregulated |
| 0,076747715 | 0,796546401 | CMC2 | FALSE | Upregulated |
| -0,157159955 | 0,796697429 | KCNK17 | FALSE | Downregulated |

| | | | | |
|---|---|---|---|---|
| -0,067975387 | 0,79674284 | ANKRD26 | FALSE | Downregulated |
| -0,087582126 | 0,79674284 | CA13 | FALSE | Downregulated |
| -0,072750918 | 0,79674284 | GATA6 | FALSE | Downregulated |
| -0,040927925 | 0,79674284 | NFU1 | FALSE | Downregulated |
| -0,130114092 | 0,79674284 | SLC25A12 | FALSE | Downregulated |
| 0,037516166 | 0,79686973 | RASA1 | FALSE | Upregulated |
| -0,068431078 | 0,796949139 | MAGEH1 | FALSE | Downregulated |
| 0,042131523 | 0,796949139 | TRMT112 | FALSE | Upregulated |
| -0,036081226 | 0,797446705 | HIAT1 | FALSE | Downregulated |
| 0,061302267 | 0,797446705 | LMBRD2 | FALSE | Upregulated |
| -0,070692376 | 0,797446705 | PPAP2B | FALSE | Downregulated |
| 0,288844823 | 0,797507574 | SNORA33 | FALSE | Upregulated |
| -0,091375355 | 0,797507574 | TSGA10 | FALSE | Downregulated |
| 0,130746728 | 0,797507574 | VNN3 | FALSE | Upregulated |
| 0,191553979 | 0,797517528 | LOC101929265 | FALSE | Upregulated |
| 0,434355109 | 0,797639959 | LINC01181 | FALSE | Upregulated |
| 0,044042349 | 0,797639959 | PLA2G12B | FALSE | Upregulated |
| 0,043181989 | 0,79788197 | CWF19L2 | FALSE | Upregulated |
| -0,242989807 | 0,79788197 | FLJ22184 | FALSE | Downregulated |
| -0,04146521 | 0,79788197 | RRP8 | FALSE | Downregulated |
| -0,040950471 | 0,798029132 | PARD3 | FALSE | Downregulated |
| 0,054802637 | 0,798032386 | COX11 | FALSE | Upregulated |
| -0,35775112 | 0,798147127 | ENDOU | FALSE | Downregulated |
| -0,05822228 | 0,798147127 | FKBP7 | FALSE | Downregulated |
| 0,117289944 | 0,798214927 | PVRIG | FALSE | Upregulated |
| 0,052169033 | 0,798214927 | RBBP9 | FALSE | Upregulated |
| -0,05620894 | 0,798302612 | BBIP1 | FALSE | Downregulated |
| 0,18036888 | 0,79833408 | MMP25 | FALSE | Upregulated |
| -0,063904346 | 0,79833408 | POLK | FALSE | Downregulated |
| 0,544637888 | 0,798376134 | LGALS9B | FALSE | Upregulated |
| 0,146782142 | 0,798376134 | TMEM74 | FALSE | Upregulated |
| 0,057506373 | 0,798395494 | CGRRF1 | FALSE | Upregulated |
| -0,210730519 | 0,79854948 | GOLGA2P7 | FALSE | Downregulated |
| 0,046101782 | 0,798596466 | AURKAIP1 | FALSE | Upregulated |
| -0,230723973 | 0,798596466 | CRTAC1 | FALSE | Downregulated |
| 0,18817739 | 0,798621471 | SNORA67 | FALSE | Upregulated |
| -0,175575376 | 0,798642316 | FAM150B | FALSE | Downregulated |
| -0,347796971 | 0,798642316 | LOC100131257 | FALSE | Downregulated |
| -0,227383662 | 0,798729079 | KIAA1239 | FALSE | Downregulated |
| -0,433169209 | 0,798729079 | LOC102723902 | FALSE | Downregulated |
| -0,077088794 | 0,798729079 | TOPORS-AS1 | FALSE | Downregulated |
| -0,069859204 | 0,798861342 | CEP68 | FALSE | Downregulated |
| 0,218265718 | 0,799082442 | LOC102724970 | FALSE | Upregulated |
| -0,309294849 | 0,799136771 | GPRC5A | FALSE | Downregulated |
| -0,067370926 | 0,799219786 | ZNF414 | FALSE | Downregulated |
| -0,075268659 | 0,799256045 | CDK20 | FALSE | Downregulated |
| -0,056705706 | 0,799256045 | DNAJC2 | FALSE | Downregulated |
| 0,211673526 | 0,799256045 | MS4A2 | FALSE | Upregulated |
| 0,081590276 | 0,799256045 | NOP58 | FALSE | Upregulated |
| 0,283331375 | 0,799256045 | SLC5A10 | FALSE | Upregulated |
| -0,065716155 | 0,799256045 | ZMAT2 | FALSE | Downregulated |

| | | | | |
|---|---|---|---|---|
| 0,053615327 | 0,799396827 | TMEM126B | FALSE | Upregulated |
| 0,043889437 | 0,799605027 | CETN2 | FALSE | Upregulated |
| -0,061278765 | 0,79970507 | C22orf46 | FALSE | Downregulated |
| -0,147248964 | 0,79970507 | NBPF8 | FALSE | Downregulated |
| -0,0973671 | 0,799737114 | SELP | FALSE | Downregulated |
| -0,089950002 | 0,799764577 | GLI4 | FALSE | Downregulated |
| -0,134308233 | 0,799806028 | SPECC1L-ADORA2A | FALSE | Downregulated |
| -0,272436075 | 0,800217712 | RP11-682N22.1 | FALSE | Downregulated |
| 0,043107522 | 0,800250769 | MAPK6 | FALSE | Upregulated |
| -0,286862548 | 0,800250769 | MMP16 | FALSE | Downregulated |
| -0,076894782 | 0,800391458 | PRDM15 | FALSE | Downregulated |
| -0,253676522 | 0,80041147 | LOC439994 | FALSE | Downregulated |
| 0,071610242 | 0,800472006 | CDHR5 | FALSE | Upregulated |
| -0,037925657 | 0,800472006 | DYNLL2 | FALSE | Downregulated |
| 0,125275339 | 0,800472006 | LOC102723983 | FALSE | Upregulated |
| 0,342331908 | 0,800472006 | LOC388499 | FALSE | Upregulated |
| -0,051454103 | 0,800617752 | SRCAP | FALSE | Downregulated |
| 0,042405271 | 0,800653875 | ZW10 | FALSE | Upregulated |
| 0,293754903 | 0,800778849 | SLC4A3 | FALSE | Upregulated |
| 0,102962886 | 0,800825945 | FITM2 | FALSE | Upregulated |
| 0,059950762 | 0,801121894 | SP140L | FALSE | Upregulated |
| 0,054707686 | 0,801121894 | TMEM63B | FALSE | Upregulated |
| -0,054778303 | 0,801121894 | TMPRSS6 | FALSE | Downregulated |
| 0,497711683 | 0,801121894 | XXYLT1-AS2 | FALSE | Upregulated |
| -0,403815688 | 0,801210947 | LOC101928779 | FALSE | Downregulated |
| -0,07911066 | 0,801303051 | ZNF704 | FALSE | Downregulated |
| 0,131536309 | 0,801462389 | WASH2P | FALSE | Upregulated |
| -0,029600635 | 0,801474174 | ARHGAP12 | FALSE | Downregulated |
| 0,052116597 | 0,801474174 | TAF15 | FALSE | Upregulated |
| -0,281636689 | 0,801537184 | CECR6 | FALSE | Downregulated |
| -0,083175534 | 0,801537184 | ITPR1 | FALSE | Downregulated |
| -0,12362617 | 0,801537184 | NPIPB5 | FALSE | Downregulated |
| -0,217026335 | 0,801674999 | LOC399900 | FALSE | Downregulated |
| 0,042986311 | 0,801894983 | IRF2 | FALSE | Upregulated |
| -0,292149918 | 0,801931333 | RPS14P3 | FALSE | Downregulated |
| 0,094687172 | 0,801948083 | PRAM1 | FALSE | Upregulated |
| 0,091296675 | 0,802182381 | ARMCX2 | FALSE | Upregulated |
| 0,051409618 | 0,802182381 | TMLHE | FALSE | Upregulated |
| -0,268732048 | 0,802266632 | UGT2A2 | FALSE | Downregulated |
| -0,446654096 | 0,802398937 | LOC101928045 | FALSE | Downregulated |
| 0,517994827 | 0,802398937 | LOC101928921 | FALSE | Upregulated |
| 0,041852822 | 0,802398937 | PEX2 | FALSE | Upregulated |
| 0,05368466 | 0,802398937 | WEE1 | FALSE | Upregulated |
| -0,326714256 | 0,802743359 | GMCL1P1 | FALSE | Downregulated |
| -0,384416324 | 0,802743359 | MTRNR2L4 | FALSE | Downregulated |
| -0,05071182 | 0,80275937 | TXNRD2 | FALSE | Downregulated |
| -0,040854955 | 0,802976291 | FAM200A | FALSE | Downregulated |
| -0,21610329 | 0,802976291 | HIF3A | FALSE | Downregulated |
| -0,074430289 | 0,80303472 | ABHD5 | FALSE | Downregulated |
| 0,044148573 | 0,803051541 | ABHD3 | FALSE | Upregulated |

| | | | | |
|---|---|---|---|---|
| 0,091975806 | 0,803051541 | ASAP3 | FALSE | Upregulated |
| -0,027623968 | 0,803051541 | ATG5 | FALSE | Downregulated |
| -0,359756266 | 0,803051541 | LOC100188947 | FALSE | Downregulated |
| 0,365296351 | 0,803051541 | SNORA24 | FALSE | Upregulated |
| 0,093563931 | 0,803058443 | TRIM47 | FALSE | Upregulated |
| 0,102208715 | 0,80315496 | ALAS1 | FALSE | Upregulated |
| -0,173102693 | 0,803230685 | MAP1LC3B2 | FALSE | Downregulated |
| 0,061477159 | 0,803230685 | MED22 | FALSE | Upregulated |
| -0,038026394 | 0,803230685 | RAD23A | FALSE | Downregulated |
| -0,077779839 | 0,803230685 | ZNF500 | FALSE | Downregulated |
| 0,046777281 | 0,80324492 | TTC9C | FALSE | Upregulated |
| 0,205716222 | 0,803246318 | SCART1 | FALSE | Upregulated |
| -0,104527934 | 0,803463812 | DAB1 | FALSE | Downregulated |
| 0,055598427 | 0,803537466 | TRMU | FALSE | Upregulated |
| 0,042784022 | 0,80363066 | AMBRA1 | FALSE | Upregulated |
| -0,079583815 | 0,803669145 | HHLA3 | FALSE | Downregulated |
| 0,057330186 | 0,803692042 | EBNA1BP2 | FALSE | Upregulated |
| -0,038933524 | 0,803692042 | SEPHS1 | FALSE | Downregulated |
| 0,39114029 | 0,803721469 | FAXC | FALSE | Upregulated |
| -0,081174765 | 0,803721469 | LIPT2 | FALSE | Downregulated |
| -0,224063582 | 0,803721469 | LOC102724738 | FALSE | Downregulated |
| -0,097097024 | 0,803721469 | SLC2A12 | FALSE | Downregulated |
| 0,893789586 | 0,803811058 | RAD51L3-RFFL | FALSE | Upregulated |
| -0,255817758 | 0,803834369 | PCAT18 | FALSE | Downregulated |
| 0,073101819 | 0,803834369 | WDPCP | FALSE | Upregulated |
| -0,072060241 | 0,803834369 | ZBTB37 | FALSE | Downregulated |
| -0,237319944 | 0,803888696 | LOC101060626 | FALSE | Downregulated |
| -0,108303891 | 0,803888696 | PDE7B | FALSE | Downregulated |
| -0,05690382 | 0,803927713 | RFNG | FALSE | Downregulated |
| -0,086603113 | 0,803994329 | LOC102723779 | FALSE | Downregulated |
| -0,069773698 | 0,804100966 | CCDC120 | FALSE | Downregulated |
| 0,173274537 | 0,804100966 | LOC101928347 | FALSE | Upregulated |
| 0,05984857 | 0,804100966 | ZNF223 | FALSE | Upregulated |
| 0,057752254 | 0,804283306 | C7orf49 | FALSE | Upregulated |
| -0,184155458 | 0,804283306 | UBE2Q2P3 | FALSE | Downregulated |
| -0,111160411 | 0,804344714 | ZFAT | FALSE | Downregulated |
| -0,581307313 | 0,804469409 | AMPD1 | FALSE | Downregulated |
| 0,202839 | 0,804469409 | C17orf49 | FALSE | Upregulated |
| 0,062831372 | 0,804469409 | C5orf28 | FALSE | Upregulated |
| -0,04036069 | 0,804469409 | CDC42BPB | FALSE | Downregulated |
| -0,362165677 | 0,804469409 | LOC101929512 | FALSE | Downregulated |
| -0,220217701 | 0,804469409 | NEXN | FALSE | Downregulated |
| 0,05529755 | 0,804469409 | RPL23AP82 | FALSE | Upregulated |
| 0,065497316 | 0,804469409 | ZNF333 | FALSE | Upregulated |
| -0,039476354 | 0,804520812 | SPOPL | FALSE | Downregulated |
| -0,035018263 | 0,804919641 | PCNP | FALSE | Downregulated |
| 0,074754799 | 0,804919641 | RBMS3 | FALSE | Upregulated |
| -0,044780857 | 0,804919641 | SAT1 | FALSE | Downregulated |
| 0,244554703 | 0,804936503 | ZNF860 | FALSE | Upregulated |
| 0,229179649 | 0,804970723 | SLFNL1-AS1 | FALSE | Upregulated |
| 0,044898141 | 0,804970723 | TOP3A | FALSE | Upregulated |

| | | | | |
|---|---|---|---|---|
| 0,357845816 | 0,804970809 | DYX1C1-CCPG1 | FALSE | Upregulated |
| -0,044030489 | 0,804970809 | EEF1D | FALSE | Downregulated |
| -0,037100569 | 0,804970809 | NDUFS2 | FALSE | Downregulated |
| 0,031434781 | 0,805339899 | TARDBP | FALSE | Upregulated |
| -0,098296291 | 0,805344752 | TMEM17 | FALSE | Downregulated |
| -0,222593832 | 0,80551886 | KCND2 | FALSE | Downregulated |
| 0,078901995 | 0,80551886 | PMS2L2 | FALSE | Upregulated |
| 0,34503728 | 0,80551886 | RP11-809M12.1 | FALSE | Upregulated |
| -0,063279191 | 0,805661418 | SASS6 | FALSE | Downregulated |
| 0,343354003 | 0,805718388 | ARHGEF34P | FALSE | Upregulated |
| 0,031793556 | 0,805718388 | GSK3B | FALSE | Upregulated |
| -0,04963022 | 0,805811368 | USO1 | FALSE | Downregulated |
| 0,143557754 | 0,805882556 | CROCCP3 | FALSE | Upregulated |
| 0,02844175 | 0,805882556 | POLDIP3 | FALSE | Upregulated |
| 0,095466819 | 0,805931492 | ACAT2 | FALSE | Upregulated |
| -0,065131253 | 0,805931492 | C1orf86 | FALSE | Downregulated |
| -0,327884398 | 0,805931492 | FAM150A | FALSE | Downregulated |
| 0,710332919 | 0,806002927 | CTAGE15 | FALSE | Upregulated |
| -0,07458539 | 0,806019651 | CNTN4 | FALSE | Downregulated |
| -0,620621529 | 0,806182368 | HNRNPCP5 | FALSE | Downregulated |
| 0,101226859 | 0,806182368 | ROM1 | FALSE | Upregulated |
| 0,025062715 | 0,806211835 | WDR82 | FALSE | Upregulated |
| -0,081578375 | 0,80628001 | FBP1 | FALSE | Downregulated |
| -0,268829113 | 0,806326132 | CABP7 | FALSE | Downregulated |
| -0,054079748 | 0,806326132 | INSIG2 | FALSE | Downregulated |
| -0,246744236 | 0,806326132 | RP1-1J6.2 | FALSE | Downregulated |
| -0,067070636 | 0,806582235 | NCK2 | FALSE | Downregulated |
| -0,09827698 | 0,806657349 | RGS5 | FALSE | Downregulated |
| -0,071363479 | 0,806770857 | NR2F1-AS1 | FALSE | Downregulated |
| 0,047023041 | 0,806836531 | ZFR | FALSE | Upregulated |
| 0,224847195 | 0,806990259 | LOC100996660 | FALSE | Upregulated |
| -0,062580251 | 0,807021777 | COL5A2 | FALSE | Downregulated |
| 0,300690602 | 0,807295194 | TIMP4 | FALSE | Upregulated |
| 0,06357133 | 0,807753433 | PLD2 | FALSE | Upregulated |
| 0,103918361 | 0,807753433 | TIMM13 | FALSE | Upregulated |
| -0,105417828 | 0,808085235 | FAHD2B | FALSE | Downregulated |
| -0,038641984 | 0,808085235 | TERF1 | FALSE | Downregulated |
| 0,048347201 | 0,808190999 | DALRD3 | FALSE | Upregulated |
| 0,228579511 | 0,808190999 | ELAVL4 | FALSE | Upregulated |
| -0,042300529 | 0,808190999 | NDUFA1 | FALSE | Downregulated |
| -0,03567052 | 0,808363227 | MRPS31 | FALSE | Downregulated |
| 0,06575009 | 0,808476568 | FAM50B | FALSE | Upregulated |
| 0,251496739 | 0,808538209 | CASR | FALSE | Upregulated |
| 0,280924338 | 0,808538209 | CPXM1 | FALSE | Upregulated |
| -0,07887046 | 0,808538209 | SYDE1 | FALSE | Downregulated |
| 0,064641414 | 0,808568189 | GGH | FALSE | Upregulated |
| 0,072632596 | 0,808568189 | RASSF8 | FALSE | Upregulated |
| -0,115548242 | 0,808666786 | ATG16L2 | FALSE | Downregulated |
| -0,034829478 | 0,808666786 | CHMP2A | FALSE | Downregulated |
| 0,135653805 | 0,808666786 | NPHP4 | FALSE | Upregulated |
| -0,365934411 | 0,808666786 | RGMA | FALSE | Downregulated |

| | | | | |
|---|---|---|---|---|
| 0,06033314 | 0,808666786 | RPS24 | FALSE | Upregulated |
| 0,033767292 | 0,808816216 | AES | FALSE | Upregulated |
| 1,1747776 | 0,808816216 | CXCL13 | FALSE | Upregulated |
| 0,044942105 | 0,808816216 | HABP2 | FALSE | Upregulated |
| -0,132102658 | 0,808816216 | LINC00659 | FALSE | Downregulated |
| 0,082015368 | 0,808816216 | LOC102723493 | FALSE | Upregulated |
| -0,059840907 | 0,808816216 | PAQR3 | FALSE | Downregulated |
| 0,035495663 | 0,808816216 | PLEKHA3 | FALSE | Upregulated |
| 0,0318129 | 0,808816216 | RAB18 | FALSE | Upregulated |
| -0,304542476 | 0,808816216 | RP11-286E11.1 | FALSE | Downregulated |
| -0,087982612 | 0,808816216 | STARD5 | FALSE | Downregulated |
| -0,060758364 | 0,808816216 | TMEM116 | FALSE | Downregulated |
| 0,043110956 | 0,808816216 | ZFYVE21 | FALSE | Upregulated |
| -0,105305974 | 0,808816216 | ZNF285 | FALSE | Downregulated |
| 0,06640582 | 0,808984556 | NEIL2 | FALSE | Upregulated |
| 0,108710823 | 0,808984556 | PTGIR | FALSE | Upregulated |
| 0,042130009 | 0,808984556 | TMED5 | FALSE | Upregulated |
| -0,992641855 | 0,809086252 | LOC102724594 | FALSE | Downregulated |
| -0,193810986 | 0,809142223 | NPFF | FALSE | Downregulated |
| 0,202854703 | 0,809392125 | CTBP1-AS | FALSE | Upregulated |
| -0,090408366 | 0,809809372 | HERC5 | FALSE | Downregulated |
| 0,040953061 | 0,809996131 | TCTN3 | FALSE | Upregulated |
| -0,122726431 | 0,810092957 | ACVR2B-AS1 | FALSE | Downregulated |
| -0,327037482 | 0,810137707 | AC093375.1 | FALSE | Downregulated |
| -0,056977199 | 0,81014293 | MALSU1 | FALSE | Downregulated |
| -0,040111025 | 0,810196205 | CMTR2 | FALSE | Downregulated |
| 0,048057484 | 0,810576007 | CCAR1 | FALSE | Upregulated |
| 0,061639588 | 0,810576007 | HSPE1 | FALSE | Upregulated |
| -0,07095011 | 0,810845124 | OTUD1 | FALSE | Downregulated |
| 0,041947385 | 0,810886095 | TMCO1 | FALSE | Upregulated |
| 0,365312391 | 0,810953098 | EDNRB-AS1 | FALSE | Upregulated |
| 0,067624301 | 0,810953098 | UBALD1 | FALSE | Upregulated |
| -0,044804754 | 0,810956193 | ATRN | FALSE | Downregulated |
| 0,041732404 | 0,810956193 | EMG1 | FALSE | Upregulated |
| -0,313216133 | 0,810956193 | RGPD5 | FALSE | Downregulated |
| -0,116415367 | 0,810956193 | SLC5A9 | FALSE | Downregulated |
| 0,07214485 | 0,810956193 | TGIF1 | FALSE | Upregulated |
| -0,056470106 | 0,811129281 | LINC00847 | FALSE | Downregulated |
| -0,040668458 | 0,811129281 | RBMXL1 | FALSE | Downregulated |
| 0,096838749 | 0,811548904 | LOC143666 | FALSE | Upregulated |
| -0,116165652 | 0,811548904 | LPHN1 | FALSE | Downregulated |
| -0,454814713 | 0,811548904 | SSTR5-AS1 | FALSE | Downregulated |
| 0,065898954 | 0,811548904 | USMG5 | FALSE | Upregulated |
| -0,364074626 | 0,811560212 | LRRC63 | FALSE | Downregulated |
| -0,051360457 | 0,811710342 | NIT1 | FALSE | Downregulated |
| -0,430329667 | 0,811822632 | LOC101060026 | FALSE | Downregulated |
| -0,045277868 | 0,811822632 | PPP3R1 | FALSE | Downregulated |
| -0,19364742 | 0,811990823 | GLI2 | FALSE | Downregulated |
| -0,084828997 | 0,812108142 | ABCC6P2 | FALSE | Downregulated |
| -0,197336347 | 0,812108142 | NCR1 | FALSE | Downregulated |
| -0,378728889 | 0,812246778 | CEP170P1 | FALSE | Downregulated |

| | | | | |
|---|---|---|---|---|
| -0,047563428 | 0,812258845 | SMARCD2 | FALSE | Downregulated |
| -0,042046164 | 0,812258845 | ZNF281 | FALSE | Downregulated |
| 0,244570894 | 0,812322083 | APITD1-CORT | FALSE | Upregulated |
| 0,118925306 | 0,812322083 | FBLN5 | FALSE | Upregulated |
| 0,15255284 | 0,812338879 | USP43 | FALSE | Upregulated |
| -0,252988763 | 0,812568922 | GPR97 | FALSE | Downregulated |
| 0,277887302 | 0,812568922 | TCP10L2 | FALSE | Upregulated |
| -0,320667758 | 0,812605811 | CADPS | FALSE | Downregulated |
| -0,047962956 | 0,812605811 | HSD17B10 | FALSE | Downregulated |
| -0,189921322 | 0,812689878 | KIF5C | FALSE | Downregulated |
| -0,042068182 | 0,812689878 | PYURF | FALSE | Downregulated |
| 0,047482056 | 0,812834237 | UBXN2A | FALSE | Upregulated |
| -0,091388954 | 0,812834237 | ZNF785 | FALSE | Downregulated |
| -0,090983101 | 0,81295998 | ATP8B2 | FALSE | Downregulated |
| 0,372789526 | 0,81295998 | LINC00173 | FALSE | Upregulated |
| 0,045885 | 0,81295998 | PIGQ | FALSE | Upregulated |
| -0,259917225 | 0,81295998 | RP11-7F17.5 | FALSE | Downregulated |
| -0,139224574 | 0,813024239 | GNAL | FALSE | Downregulated |
| -0,358817426 | 0,813100165 | HIGD2B | FALSE | Downregulated |
| 0,699806847 | 0,813100165 | LOC100289026 | FALSE | Upregulated |
| -0,190180073 | 0,813100165 | MIR3064 | FALSE | Downregulated |
| 0,163582603 | 0,813100165 | SLC25A21 | FALSE | Upregulated |
| -0,146594891 | 0,813162035 | ANKRD36C | FALSE | Downregulated |
| 0,136930761 | 0,813321718 | CPT1C | FALSE | Upregulated |
| -0,172055483 | 0,813388866 | PFN4 | FALSE | Downregulated |
| 0,155101505 | 0,813478418 | DOPEY2 | FALSE | Upregulated |
| 0,098477117 | 0,813478418 | MGC70870 | FALSE | Upregulated |
| -0,048598584 | 0,813478418 | MRPL32 | FALSE | Downregulated |
| -0,442375562 | 0,813478418 | PLA2G4F | FALSE | Downregulated |
| 0,052263362 | 0,813478418 | SVIP | FALSE | Upregulated |
| 0,298647605 | 0,813756123 | LOC101927764 | FALSE | Upregulated |
| 0,036842877 | 0,813823435 | PXN | FALSE | Upregulated |
| -0,269203369 | 0,814003373 | LOC101927157 | FALSE | Downregulated |
| 0,039985716 | 0,814183152 | SRF | FALSE | Upregulated |
| -0,124478738 | 0,814222398 | HBEGF | FALSE | Downregulated |
| 0,180193838 | 0,81424058 | LOC101928891 | FALSE | Upregulated |
| 0,045128041 | 0,814324082 | VGLL4 | FALSE | Upregulated |
| -0,040954629 | 0,814375479 | DPP8 | FALSE | Downregulated |
| -0,312209954 | 0,814375479 | LOC101927051 | FALSE | Downregulated |
| 0,097596907 | 0,814375479 | NBPF15 | FALSE | Upregulated |
| -0,076219193 | 0,814375479 | PTRF | FALSE | Downregulated |
| 0,236669007 | 0,814375479 | UCA1 | FALSE | Upregulated |
| 0,188702435 | 0,81477357 | HIST2H2AC | FALSE | Upregulated |
| 0,457613547 | 0,815009951 | ATP2A1 | FALSE | Upregulated |
| -0,239939316 | 0,815426278 | PPP1R36 | FALSE | Downregulated |
| 0,044586 | 0,815498003 | CLP1 | FALSE | Upregulated |
| 0,221253968 | 0,815498003 | SPTA1 | FALSE | Upregulated |
| -0,027716834 | 0,815568555 | PANK2 | FALSE | Downregulated |
| 0,145935196 | 0,815588781 | LOX | FALSE | Upregulated |
| 0,172863088 | 0,815696949 | CCDC147 | FALSE | Upregulated |
| 0,568504431 | 0,815740979 | TRNAD8 | FALSE | Upregulated |

| | | | | |
|---|---|---|---|---|
| 0,329432417 | 0,816138481 | BCRP2 | FALSE | Upregulated |
| -0,228871278 | 0,816163029 | LOC101926922 | FALSE | Downregulated |
| -0,15582359 | 0,81631391 | BLK | FALSE | Downregulated |
| 0,425061369 | 0,816518194 | IGSF9B | FALSE | Upregulated |
| -0,164285004 | 0,816518194 | LOC440434 | FALSE | Downregulated |
| -0,035469938 | 0,816518194 | TSPAN31 | FALSE | Downregulated |
| -0,029980628 | 0,816747986 | UBE2G2 | FALSE | Downregulated |
| 0,578067985 | 0,816962529 | LOC101927680 | FALSE | Upregulated |
| -0,407134678 | 0,817174613 | LINC00982 | FALSE | Downregulated |
| -0,03902314 | 0,817189697 | ATP7B | FALSE | Downregulated |
| 0,178474774 | 0,817189697 | NCR3LG1 | FALSE | Upregulated |
| 0,177352733 | 0,81724253 | LOC101926907 | FALSE | Upregulated |
| -0,330951939 | 0,817285018 | PCDH9-AS2 | FALSE | Downregulated |
| -0,06980961 | 0,817450449 | GALNT2 | FALSE | Downregulated |
| 0,164644066 | 0,817450449 | IQCJ-SCHIP1 | FALSE | Upregulated |
| 0,101705757 | 0,817450449 | LDOC1 | FALSE | Upregulated |
| -0,147991923 | 0,817450449 | SYT10 | FALSE | Downregulated |
| -0,059551323 | 0,817450449 | SYVN1 | FALSE | Downregulated |
| 0,158303921 | 0,817480508 | LOC102723602 | FALSE | Upregulated |
| 0,321778512 | 0,817480508 | NDUFAF4P1 | FALSE | Upregulated |
| 0,099775335 | 0,817486723 | HDAC9 | FALSE | Upregulated |
| 0,233915325 | 0,817490761 | CLGN | FALSE | Upregulated |
| -0,064075057 | 0,817490761 | MPZL2 | FALSE | Downregulated |
| 0,034909719 | 0,817490761 | SLC35A5 | FALSE | Upregulated |
| -0,051075705 | 0,817500986 | FAM160A2 | FALSE | Downregulated |
| -0,448348083 | 0,817500986 | HOXD8 | FALSE | Downregulated |
| 0,0712106 | 0,817500986 | WASH3P | FALSE | Upregulated |
| 0,057751974 | 0,817554406 | OLFML3 | FALSE | Upregulated |
| -0,289940245 | 0,817672851 | SOX15 | FALSE | Downregulated |
| -0,056358694 | 0,817692308 | SEPT11 | FALSE | Downregulated |
| -0,047523854 | 0,81826895 | TRMT11 | FALSE | Downregulated |
| -0,170488063 | 0,818407604 | BRICD5 | FALSE | Downregulated |
| -0,040934211 | 0,818407604 | PYGO2 | FALSE | Downregulated |
| -0,080211313 | 0,818407604 | SLC25A36 | FALSE | Downregulated |
| -0,040704631 | 0,818407604 | WIBG | FALSE | Downregulated |
| 0,187733136 | 0,818518897 | PSRC1 | FALSE | Upregulated |
| 0,039780948 | 0,818573222 | ARL1 | FALSE | Upregulated |
| 0,077453987 | 0,818573222 | KRAS | FALSE | Upregulated |
| -0,166045071 | 0,81859801 | BLOC1S4 | FALSE | Downregulated |
| -0,063568243 | 0,81859801 | C15orf40 | FALSE | Downregulated |
| -0,244413093 | 0,81859801 | CLIC5 | FALSE | Downregulated |
| -0,391482788 | 0,81859801 | MADCAM1 | FALSE | Downregulated |
| 0,214092369 | 0,81859801 | NRG2 | FALSE | Upregulated |
| 0,175025396 | 0,81859801 | RP11-579D7.4 | FALSE | Upregulated |
| 0,066269291 | 0,81859801 | SHC2 | FALSE | Upregulated |
| -0,10977029 | 0,81859801 | TLCD2 | FALSE | Downregulated |
| -0,09331656 | 0,81859801 | TPRN | FALSE | Downregulated |
| -0,16482443 | 0,818634829 | RSPH1 | FALSE | Downregulated |
| -0,13305409 | 0,818791926 | PPM1L | FALSE | Downregulated |
| 0,109845927 | 0,818876369 | ABCA4 | FALSE | Upregulated |
| -0,086547787 | 0,818876369 | LOC100132832 | FALSE | Downregulated |

| | | | | |
|---|---|---|---|---|
| -0,166592405 | 0,818876369 | PRSS3 | FALSE | Downregulated |
| 0,099572663 | 0,819170568 | HIST1H2BD | FALSE | Upregulated |
| 0,025651695 | 0,819170568 | ILF3 | FALSE | Upregulated |
| 0,044297974 | 0,819170568 | SLC25A40 | FALSE | Upregulated |
| 0,169426121 | 0,819174963 | GPR114 | FALSE | Upregulated |
| 0,045178481 | 0,819174963 | ZNF76 | FALSE | Upregulated |
| 0,063307479 | 0,81918917 | GPR39 | FALSE | Upregulated |
| 0,21128417 | 0,819458848 | SEPT14 | FALSE | Upregulated |
| -0,035627731 | 0,819526532 | C9orf64 | FALSE | Downregulated |
| -0,029832918 | 0,819526532 | SLAIN2 | FALSE | Downregulated |
| 0,06007762 | 0,81962786 | LDHA | FALSE | Upregulated |
| -0,033756807 | 0,819794518 | ALKBH3 | FALSE | Downregulated |
| -0,173591401 | 0,819794518 | CERS6-AS1 | FALSE | Downregulated |
| -0,018854705 | 0,820045935 | ARIH1 | FALSE | Downregulated |
| 0,026867442 | 0,820045935 | EIF3B | FALSE | Upregulated |
| 0,060834124 | 0,820045935 | FOSL2 | FALSE | Upregulated |
| -0,379739594 | 0,820045935 | SLC7A14 | FALSE | Downregulated |
| 0,163463255 | 0,820045935 | VGLL3 | FALSE | Upregulated |
| 0,08538206 | 0,820159409 | AP3D1 | FALSE | Upregulated |
| 0,032658621 | 0,820226445 | KBTBD4 | FALSE | Upregulated |
| -0,244479211 | 0,820226445 | LOC102724948 | FALSE | Downregulated |
| -0,060911122 | 0,82066195 | HECTD3 | FALSE | Downregulated |
| 0,041587755 | 0,820781824 | MSRB1 | FALSE | Upregulated |
| -0,259684854 | 0,820926267 | LINC00266-1 | FALSE | Downregulated |
| 0,037453249 | 0,820952881 | DARS2 | FALSE | Upregulated |
| -0,077274352 | 0,820952881 | DXO | FALSE | Downregulated |
| 0,124609487 | 0,820972615 | SSC5D | FALSE | Upregulated |
| -0,050251261 | 0,82099269 | APOC1 | FALSE | Downregulated |
| -0,044617478 | 0,821007775 | ARL6IP1 | FALSE | Downregulated |
| 0,050184351 | 0,821007775 | CERS6 | FALSE | Upregulated |
| 0,167677452 | 0,821007775 | CNKSR1 | FALSE | Upregulated |
| -0,09896676 | 0,821007775 | DHODH | FALSE | Downregulated |
| -0,034869437 | 0,821007775 | ELL | FALSE | Downregulated |
| 0,264743902 | 0,821007775 | LOC90246 | FALSE | Upregulated |
| 0,034569784 | 0,821007775 | MFF | FALSE | Upregulated |
| 0,06314519 | 0,82126636 | EIF2AK3 | FALSE | Upregulated |
| -0,047290208 | 0,82126636 | RRAS2 | FALSE | Downregulated |
| 0,025971416 | 0,821472553 | CSTF2T | FALSE | Upregulated |
| -0,254673206 | 0,821472553 | LOC101060494 | FALSE | Downregulated |
| 0,180722537 | 0,821472553 | MACC1 | FALSE | Upregulated |
| 0,067449984 | 0,821472553 | NAT2 | FALSE | Upregulated |
| -0,112460302 | 0,821472553 | USP3-AS1 | FALSE | Downregulated |
| 0,03134758 | 0,82152563 | LANCL1 | FALSE | Upregulated |
| -0,022968301 | 0,821576745 | DNAJC14 | FALSE | Downregulated |
| 0,136340287 | 0,821626328 | COL26A1 | FALSE | Upregulated |
| 0,038338214 | 0,821626328 | EPS8 | FALSE | Upregulated |
| -0,054274983 | 0,821626328 | MAFG | FALSE | Downregulated |
| 0,037564823 | 0,821626328 | UQCC1 | FALSE | Upregulated |
| 0,349508291 | 0,821678813 | DUOX2 | FALSE | Upregulated |
| 0,241714765 | 0,822060679 | ANKRD36BP1 | FALSE | Upregulated |
| 0,036521788 | 0,822060679 | CIRH1A | FALSE | Upregulated |

| | | | | |
|---|---|---|---|---|
| -0,074933807 | 0,822060679 | CTH | FALSE | Downregulated |
| 0,072492312 | 0,822060679 | GLIS3 | FALSE | Upregulated |
| -0,055351749 | 0,822060679 | RFX1 | FALSE | Downregulated |
| -0,249738443 | 0,822060679 | ST7-OT3 | FALSE | Downregulated |
| -0,122058536 | 0,822060679 | SYNM | FALSE | Downregulated |
| -0,095454916 | 0,822290706 | ARID3A | FALSE | Downregulated |
| -0,03632304 | 0,822433578 | SOS1 | FALSE | Downregulated |
| 0,049272178 | 0,82284671 | ERC1 | FALSE | Upregulated |
| 0,123050735 | 0,822977473 | A2M | FALSE | Upregulated |
| -0,070984195 | 0,822977473 | CHKA | FALSE | Downregulated |
| 0,292098287 | 0,822977473 | DNAJC9-AS1 | FALSE | Upregulated |
| -0,063405133 | 0,822977473 | HOOK2 | FALSE | Downregulated |
| 0,148763176 | 0,822977473 | LOC101927057 | FALSE | Upregulated |
| 0,248560177 | 0,822977473 | LOC101929333 | FALSE | Upregulated |
| -0,295638512 | 0,822977473 | LOC646670 | FALSE | Downregulated |
| 0,044262442 | 0,822977473 | RNMTL1 | FALSE | Upregulated |
| 0,288424462 | 0,822977473 | RP11-483E7.1 | FALSE | Upregulated |
| 0,290340146 | 0,822977473 | WBP11P1 | FALSE | Upregulated |
| 0,308948541 | 0,823008468 | LOC101927725 | FALSE | Upregulated |
| -0,06522999 | 0,823356494 | JMJD7 | FALSE | Downregulated |
| 0,030979847 | 0,82355542 | VMA21 | FALSE | Upregulated |
| -0,075063927 | 0,82355542 | ZBTB46 | FALSE | Downregulated |
| 0,10860607 | 0,823710459 | AC022007.5 | FALSE | Upregulated |
| 0,046272557 | 0,823739698 | NLN | FALSE | Upregulated |
| -0,080202826 | 0,823824949 | LOC101927556 | FALSE | Downregulated |
| -0,195294975 | 0,823824949 | LOC101929953 | FALSE | Downregulated |
| 0,05738764 | 0,823824949 | SLC30A6 | FALSE | Upregulated |
| -0,03430514 | 0,82443291 | FBXL5 | FALSE | Downregulated |
| 0,214857716 | 0,82443291 | GOLGA7B | FALSE | Upregulated |
| -0,047690955 | 0,82443291 | GRHPR | FALSE | Downregulated |
| 0,099662695 | 0,82443291 | NKAPL | FALSE | Upregulated |
| 0,146476411 | 0,824435746 | BHLHE22 | FALSE | Upregulated |
| 0,101309967 | 0,824435746 | CHSY3 | FALSE | Upregulated |
| -0,045610618 | 0,824627809 | ABCD3 | FALSE | Downregulated |
| 0,281878916 | 0,824657365 | FGF17 | FALSE | Upregulated |
| -0,055404071 | 0,824657365 | LECT2 | FALSE | Downregulated |
| 0,150482361 | 0,824657365 | LOC102723965 | FALSE | Upregulated |
| 0,101633188 | 0,824734281 | LOC101927160 | FALSE | Upregulated |
| 0,053376845 | 0,824811528 | GCLM | FALSE | Upregulated |
| -0,046421169 | 0,824811528 | PAK4 | FALSE | Downregulated |
| 0,174598753 | 0,824811528 | UBQLNL | FALSE | Upregulated |
| -0,044480268 | 0,825000057 | CRTC3 | FALSE | Downregulated |
| 0,045470645 | 0,825000057 | STK38L | FALSE | Upregulated |
| 0,3720909 | 0,825147905 | RP11-103C16.2 | FALSE | Upregulated |
| -0,146018913 | 0,825454724 | ENTPD1-AS1 | FALSE | Downregulated |
| -0,04986025 | 0,825738017 | PPP1R37 | FALSE | Downregulated |
| -0,124516055 | 0,825976741 | FAM171B | FALSE | Downregulated |
| -0,07026775 | 0,82622627 | LUC7L | FALSE | Downregulated |
| -0,034555711 | 0,82622627 | MMAA | FALSE | Downregulated |
| -0,034420784 | 0,826352697 | AP1M1 | FALSE | Downregulated |
| -0,308505609 | 0,826352697 | CHKB-CPT1B | FALSE | Downregulated |

| | | | | |
|---|---|---|---|---|
| -0,042244108 | 0,826352697 | MOCS3 | FALSE | Downregulated |
| -0,474429656 | 0,826396131 | NEFL | FALSE | Downregulated |
| 0,065137644 | 0,826483106 | FOXRED2 | FALSE | Upregulated |
| 0,074965569 | 0,826483106 | SERPINI1 | FALSE | Upregulated |
| -0,249765032 | 0,826483106 | SULT1C2 | FALSE | Downregulated |
| -0,102396626 | 0,826497719 | NMUR1 | FALSE | Downregulated |
| 0,177866139 | 0,826684062 | ENPP6 | FALSE | Upregulated |
| 0,113606946 | 0,826684062 | GPR176 | FALSE | Upregulated |
| -0,236134965 | 0,826684062 | LPAR4 | FALSE | Downregulated |
| 0,041158552 | 0,826897912 | PSMG2 | FALSE | Upregulated |
| -0,035914653 | 0,827012802 | CNTROB | FALSE | Downregulated |
| 0,024990833 | 0,827012802 | PPP6C | FALSE | Upregulated |
| -0,075004729 | 0,827012802 | ZNF250 | FALSE | Downregulated |
| -0,078888887 | 0,827016753 | NOS3 | FALSE | Downregulated |
| 0,082835124 | 0,827088998 | CBWD1 | FALSE | Upregulated |
| -0,070846071 | 0,827167288 | UBE3D | FALSE | Downregulated |
| -0,204547305 | 0,82738936 | RP11-789C1.1 | FALSE | Downregulated |
| 1,625892672 | 0,827572109 | EEF1A2 | FALSE | Upregulated |
| -0,220144515 | 0,827572109 | LINC00243 | FALSE | Downregulated |
| -0,356772575 | 0,827572109 | RNY3 | FALSE | Downregulated |
| 0,123189443 | 0,827572109 | VWA7 | FALSE | Upregulated |
| -0,054579094 | 0,827845408 | SHROOM2 | FALSE | Downregulated |
| -0,230460265 | 0,82787212 | RP11-81H14.2 | FALSE | Downregulated |
| -0,233941633 | 0,827995393 | KCNG1 | FALSE | Downregulated |
| -0,121922548 | 0,827995393 | LOC100499484 | FALSE | Downregulated |
| -0,562252826 | 0,827995393 | LOC101928510 | FALSE | Downregulated |
| 0,175862708 | 0,827995393 | LOC101928667 | FALSE | Upregulated |
| 0,10484394 | 0,828096876 | ZNF888 | FALSE | Upregulated |
| -0,108575042 | 0,828128823 | DACT1 | FALSE | Downregulated |
| -0,243974233 | 0,828473715 | LOC101930100 | FALSE | Downregulated |
| -0,074848406 | 0,82861778 | PUS7L | FALSE | Downregulated |
| -0,073219114 | 0,828908622 | NEDD8-MDP1 | FALSE | Downregulated |
| 0,098420308 | 0,828934992 | PHACTR3 | FALSE | Upregulated |
| 0,180610672 | 0,828934992 | PRR4 | FALSE | Upregulated |
| -0,092944168 | 0,828971072 | MROH6 | FALSE | Downregulated |
| 0,035292526 | 0,829212059 | SDHAF2 | FALSE | Upregulated |
| 0,037678223 | 0,829212059 | TMED1 | FALSE | Upregulated |
| 0,64985604 | 0,829301243 | TRNAE19 | FALSE | Upregulated |
| -0,057376365 | 0,8293177 | RHOT2 | FALSE | Downregulated |
| 0,165076015 | 0,829369141 | BCYRN1 | FALSE | Upregulated |
| 0,073021377 | 0,829369141 | LOC100506518 | FALSE | Upregulated |
| 0,104149466 | 0,829399695 | ZFP28 | FALSE | Upregulated |
| 0,082088982 | 0,829539653 | AMIGO2 | FALSE | Upregulated |
| -0,572045905 | 0,829539653 | LINC00550 | FALSE | Downregulated |
| 0,040002568 | 0,829695276 | RC3H1 | FALSE | Upregulated |
| 0,188130429 | 0,830059308 | NCF1B | FALSE | Upregulated |
| -0,053223195 | 0,830107694 | FXYD6 | FALSE | Downregulated |
| 0,037006904 | 0,830178464 | FAM13B | FALSE | Upregulated |
| -0,122898968 | 0,830178464 | HESX1 | FALSE | Downregulated |
| -0,041155032 | 0,830178464 | NUDCD2 | FALSE | Downregulated |
| -0,078734636 | 0,830178464 | TPST1 | FALSE | Downregulated |

| | | | | |
|---|---|---|---|---|
| 0,254570922 | 0,830178464 | TSHR | FALSE | Upregulated |
| -0,050921517 | 0,830451085 | ERBB2IP | FALSE | Downregulated |
| -0,041337243 | 0,830451085 | RIOK2 | FALSE | Downregulated |
| -0,053309901 | 0,830520133 | GORAB | FALSE | Downregulated |
| 0,072989705 | 0,830520133 | POMZP3 | FALSE | Upregulated |
| 0,094969279 | 0,830578801 | ADAMTS1 | FALSE | Upregulated |
| 0,170247794 | 0,830578801 | PDE4C | FALSE | Upregulated |
| -0,178602972 | 0,830695098 | GATSL2 | FALSE | Downregulated |
| 0,408828393 | 0,830695098 | LOC102724093 | FALSE | Upregulated |
| 0,168495541 | 0,830695098 | S1PR4 | FALSE | Upregulated |
| -0,167182849 | 0,830845953 | HNRNPA1P33 | FALSE | Downregulated |
| -0,099170102 | 0,830937774 | LRRC48 | FALSE | Downregulated |
| -0,317001605 | 0,830980721 | LOC401463 | FALSE | Downregulated |
| -0,194404401 | 0,831041014 | FGF11 | FALSE | Downregulated |
| 0,0978598 | 0,83110332 | PARD6A | FALSE | Upregulated |
| 0,082748832 | 0,83110332 | RAMP3 | FALSE | Upregulated |
| -0,078500006 | 0,831237676 | CC2D2A | FALSE | Downregulated |
| -0,03639692 | 0,831404757 | ELK4 | FALSE | Downregulated |
| 0,035206524 | 0,831477277 | ERGIC3 | FALSE | Upregulated |
| 0,036741112 | 0,831803931 | SEPT7 | FALSE | Upregulated |
| 0,072080875 | 0,831803931 | SMUG1 | FALSE | Upregulated |
| 0,036599045 | 0,831803931 | ZFPL1 | FALSE | Upregulated |
| 0,256547875 | 0,831915929 | KLC3 | FALSE | Upregulated |
| 0,235044506 | 0,831915929 | LOC100505710 | FALSE | Upregulated |
| 0,038878624 | 0,832125358 | CENPBD1 | FALSE | Upregulated |
| -0,288801516 | 0,832125358 | MEF2C-AS1 | FALSE | Downregulated |
| 0,036970728 | 0,832125358 | VTN | FALSE | Upregulated |
| -0,077554206 | 0,832183108 | CHURC1-FNTB | FALSE | Downregulated |
| -0,129346575 | 0,8326217 | LOC102724535 | FALSE | Downregulated |
| 0,087410067 | 0,8326217 | TMEM81 | FALSE | Upregulated |
| 0,074504028 | 0,832668094 | UBTD1 | FALSE | Upregulated |
| -0,051192526 | 0,832668094 | ZNF707 | FALSE | Downregulated |
| 0,061980561 | 0,833039018 | PAM16 | FALSE | Upregulated |
| -0,054061099 | 0,833123742 | NDUFA2 | FALSE | Downregulated |
| -0,043272379 | 0,833123742 | NKAP | FALSE | Downregulated |
| 0,081509015 | 0,833136843 | EBF4 | FALSE | Upregulated |
| -0,043587272 | 0,833136843 | HNRNPH1 | FALSE | Downregulated |
| -0,566597223 | 0,833136843 | RNU6-36P | FALSE | Downregulated |
| -0,027561405 | 0,833136843 | USP3 | FALSE | Downregulated |
| 0,036385474 | 0,833136843 | ZBTB8OS | FALSE | Upregulated |
| -0,346330945 | 0,833154988 | LOC101928574 | FALSE | Downregulated |
| -0,165915008 | 0,833701571 | MTRNR2L10 | FALSE | Downregulated |
| -0,046689911 | 0,833813559 | PGRMC1 | FALSE | Downregulated |
| -0,042166324 | 0,83381842 | EP400 | FALSE | Downregulated |
| 0,080138674 | 0,83381842 | PMP22 | FALSE | Upregulated |
| 0,24119495 | 0,83381842 | RP11-344B5.2 | FALSE | Upregulated |
| -0,079369037 | 0,833838603 | CROCC | FALSE | Downregulated |
| -0,3065364 | 0,833935662 | BCORP1 | FALSE | Downregulated |
| -0,132066098 | 0,833935662 | C15orf60 | FALSE | Downregulated |
| 0,041454535 | 0,833935662 | LRRC57 | FALSE | Upregulated |
| 0,196808909 | 0,833959096 | EDN2 | FALSE | Upregulated |

| | | | | |
|---|---|---|---|---|
| -0,046739505 | 0,834014153 | C10orf88 | FALSE | Downregulated |
| -0,177287238 | 0,834030463 | LOC102723461 | FALSE | Downregulated |
| -0,2758262 | 0,834310359 | ANP32C | FALSE | Downregulated |
| 0,070127938 | 0,834339084 | LOC647859 | FALSE | Upregulated |
| 0,078421786 | 0,834426087 | CYBRD1 | FALSE | Upregulated |
| -0,210178076 | 0,834426087 | LOC101929647 | FALSE | Downregulated |
| 0,096754844 | 0,834426087 | SUV420H2 | FALSE | Upregulated |
| -0,118011774 | 0,834426087 | TEKT4P2 | FALSE | Downregulated |
| 0,063468545 | 0,834426087 | TSSC1 | FALSE | Upregulated |
| -0,04576313 | 0,834820665 | RTF1 | FALSE | Downregulated |
| 0,047109326 | 0,834883374 | RPS15A | FALSE | Upregulated |
| -0,048076758 | 0,834940892 | DDIT3 | FALSE | Downregulated |
| 0,298602785 | 0,834940892 | FAM83A | FALSE | Upregulated |
| -0,290982763 | 0,834940892 | RP11-353N14.5 | FALSE | Downregulated |
| -0,308453831 | 0,834940892 | TNFAIP6 | FALSE | Downregulated |
| 0,024724992 | 0,834940892 | VTA1 | FALSE | Upregulated |
| 0,115457489 | 0,835049858 | LZTS1 | FALSE | Upregulated |
| -0,026971019 | 0,835153993 | AFG3L2 | FALSE | Downregulated |
| -0,175102003 | 0,835153993 | RP11-503N18.1 | FALSE | Downregulated |
| 0,306171371 | 0,835264642 | LOC102724286 | FALSE | Upregulated |
| -0,046718826 | 0,835264642 | TMEM245 | FALSE | Downregulated |
| 0,039484516 | 0,83545336 | VPS51 | FALSE | Upregulated |
| 0,232142972 | 0,835646065 | CRYBB1 | FALSE | Upregulated |
| 0,278626116 | 0,835646065 | GABRR2 | FALSE | Upregulated |
| 0,041585138 | 0,835646065 | PXMP4 | FALSE | Upregulated |
| 0,163145283 | 0,835747136 | RP11-540A21.2 | FALSE | Upregulated |
| -0,370712953 | 0,836058462 | C9orf129 | FALSE | Downregulated |
| 0,129025163 | 0,836058462 | FCGR3B | FALSE | Upregulated |
| -0,155681211 | 0,836058462 | LOC440300 | FALSE | Downregulated |
| 0,057880333 | 0,836153437 | C19orf68 | FALSE | Upregulated |
| -0,105557714 | 0,836153437 | DNLZ | FALSE | Downregulated |
| -0,139000046 | 0,836153437 | DPYSL3 | FALSE | Downregulated |
| -0,032575429 | 0,836153437 | EID1 | FALSE | Downregulated |
| 0,040014509 | 0,836153437 | EPB41 | FALSE | Upregulated |
| -0,142305572 | 0,836153437 | LOC101928092 | FALSE | Downregulated |
| -0,221921979 | 0,836153437 | NPPA-AS1 | FALSE | Downregulated |
| 0,147333597 | 0,836153437 | PROCA1 | FALSE | Upregulated |
| 0,037793892 | 0,836153437 | SERPINA6 | FALSE | Upregulated |
| -0,048967964 | 0,836153437 | SLC25A10 | FALSE | Downregulated |
| -0,163303124 | 0,836219997 | COL4A3 | FALSE | Downregulated |
| -0,065679605 | 0,836219997 | NBPF9 | FALSE | Downregulated |
| -0,028164764 | 0,836524513 | TBC1D20 | FALSE | Downregulated |
| -0,04356956 | 0,836565288 | SLCO2B1 | FALSE | Downregulated |
| 0,029751225 | 0,83726313 | TMED4 | FALSE | Upregulated |
| -0,025533529 | 0,837412926 | COPS7A | FALSE | Downregulated |
| 0,308763801 | 0,837412926 | TMEM163 | FALSE | Upregulated |
| 0,052024422 | 0,837508896 | EFNB1 | FALSE | Upregulated |
| 0,262101268 | 0,837508896 | TLL2 | FALSE | Upregulated |
| 0,27218174 | 0,837563987 | LOC101928240 | FALSE | Upregulated |
| 0,028273706 | 0,837563987 | RIPK1 | FALSE | Upregulated |
| 0,023715992 | 0,837665816 | CUL1 | FALSE | Upregulated |

| | | | | |
|---|---|---|---|---|
| 0,200955755 | 0,837665816 | LOC101927745 | FALSE | Upregulated |
| -0,455789102 | 0,837717808 | AGAP7 | FALSE | Downregulated |
| -0,079474303 | 0,837717808 | NXT1 | FALSE | Downregulated |
| -0,030213313 | 0,837717808 | PPP4R2 | FALSE | Downregulated |
| -0,263032683 | 0,837717808 | SLC5A11 | FALSE | Downregulated |
| 0,050250085 | 0,837976676 | C10orf32 | FALSE | Upregulated |
| 0,293035105 | 0,837976676 | DGUOK-AS1 | FALSE | Upregulated |
| 0,046590659 | 0,837976676 | FAAH | FALSE | Upregulated |
| -0,244242295 | 0,837976676 | KIR3DX1 | FALSE | Downregulated |
| 0,042773134 | 0,837976676 | MPG | FALSE | Upregulated |
| -0,031238986 | 0,837976676 | UNC50 | FALSE | Downregulated |
| -0,197610646 | 0,8379842 | CNN1 | FALSE | Downregulated |
| -0,168175432 | 0,8379842 | LOC102723824 | FALSE | Downregulated |
| -0,053436101 | 0,8379842 | PABPN1 | FALSE | Downregulated |
| -0,056793416 | 0,838130954 | SHMT1 | FALSE | Downregulated |
| 0,254689514 | 0,838139589 | LOC102724010 | FALSE | Upregulated |
| 0,024727473 | 0,838443603 | STAM2 | FALSE | Upregulated |
| -0,076044391 | 0,83861345 | EPHB6 | FALSE | Downregulated |
| 0,078021218 | 0,838811006 | RAB15 | FALSE | Upregulated |
| 0,040103808 | 0,838896689 | PTPRJ | FALSE | Upregulated |
| -0,063997999 | 0,8390811 | ZNF77 | FALSE | Downregulated |
| -0,120351964 | 0,839083058 | AC159540.1-2 | FALSE | Downregulated |
| 0,05540397 | 0,839083058 | GDAP1 | FALSE | Upregulated |
| -0,079037176 | 0,839083058 | LOC102724250 | FALSE | Downregulated |
| -0,034242343 | 0,839218891 | COX18 | FALSE | Downregulated |
| -1,551869459 | 0,839229673 | GSTM1 | FALSE | Downregulated |
| -0,06498325 | 0,839229673 | LY6E | FALSE | Downregulated |
| -0,074290358 | 0,839601919 | MRC1 | FALSE | Downregulated |
| -0,038372241 | 0,839607971 | FIP1L1 | FALSE | Downregulated |
| -0,146101214 | 0,839607971 | KTN1-AS1 | FALSE | Downregulated |
| 0,315732261 | 0,839612242 | CSTF3-AS1 | FALSE | Upregulated |
| 0,28544262 | 0,839695721 | LOC101926936 | FALSE | Upregulated |
| -0,065162676 | 0,840079997 | C3orf33 | FALSE | Downregulated |
| 0,338994329 | 0,840079997 | LOC101927374 | FALSE | Upregulated |
| -0,128356474 | 0,840079997 | LOC102723616 | FALSE | Downregulated |
| 0,074853848 | 0,840079997 | SLC29A1 | FALSE | Upregulated |
| -0,057304845 | 0,840164707 | ZNF222 | FALSE | Downregulated |
| 0,044156771 | 0,840193904 | WDR83 | FALSE | Upregulated |
| -0,259968799 | 0,840214929 | TRNP | FALSE | Downregulated |
| -0,110239014 | 0,840385542 | FAM118A | FALSE | Downregulated |
| -0,148703005 | 0,840502781 | MLIP | FALSE | Downregulated |
| -0,043597601 | 0,840585117 | KDELC1 | FALSE | Downregulated |
| 0,040702408 | 0,840688191 | ALG8 | FALSE | Upregulated |
| -0,040914499 | 0,840824125 | ADAM9 | FALSE | Downregulated |
| -0,047851932 | 0,840824125 | CDK5 | FALSE | Downregulated |
| 0,147712784 | 0,840824125 | LOC100240735 | FALSE | Upregulated |
| -0,04254335 | 0,840824125 | LRRC45 | FALSE | Downregulated |
| 0,065616234 | 0,840824125 | TUFT1 | FALSE | Upregulated |
| 0,030057608 | 0,840989319 | NUDT9 | FALSE | Upregulated |
| -0,045121191 | 0,840989319 | RPL17 | FALSE | Downregulated |
| -0,033717799 | 0,84100568 | TTI2 | FALSE | Downregulated |

| | | | | |
|---|---|---|---|---|
| 0,028533194 | 0,841089516 | SNX14 | FALSE | Upregulated |
| -0,15008572 | 0,841098442 | LOC101928113 | FALSE | Downregulated |
| -0,037777923 | 0,841382191 | FAM73B | FALSE | Downregulated |
| -0,197228899 | 0,841480045 | LOC101927858 | FALSE | Downregulated |
| 0,036579694 | 0,841535328 | CCDC47 | FALSE | Upregulated |
| 0,228452747 | 0,841604605 | PLEKHG4B | FALSE | Upregulated |
| -0,05220434 | 0,841670699 | TUT1 | FALSE | Downregulated |
| -0,051510925 | 0,841774898 | RAPGEF5 | FALSE | Downregulated |
| -0,030757884 | 0,841903728 | CCNY | FALSE | Downregulated |
| 0,135994452 | 0,841910698 | CXCL2 | FALSE | Upregulated |
| 0,083402026 | 0,841934206 | LOC100505771 | FALSE | Upregulated |
| -0,066208668 | 0,841963862 | BCAM | FALSE | Downregulated |
| -0,193516164 | 0,842020484 | COX7A1 | FALSE | Downregulated |
| 0,067065343 | 0,842020484 | EFNB3 | FALSE | Upregulated |
| 0,107034768 | 0,842032449 | AQP1 | FALSE | Upregulated |
| 0,184158673 | 0,842032449 | MYH11 | FALSE | Upregulated |
| -0,182573289 | 0,84226081 | LOC101927932 | FALSE | Downregulated |
| 0,031249298 | 0,842339167 | ERP44 | FALSE | Upregulated |
| -0,071236868 | 0,842360733 | C12orf60 | FALSE | Downregulated |
| 0,061814488 | 0,842360733 | NSMF | FALSE | Upregulated |
| -0,024441211 | 0,842437265 | CTBP1 | FALSE | Downregulated |
| 0,054006495 | 0,842515963 | RABL6 | FALSE | Upregulated |
| -0,043669527 | 0,842775842 | BACH1 | FALSE | Downregulated |
| -0,366472274 | 0,842775842 | LOC101928976 | FALSE | Downregulated |
| 0,191223507 | 0,842779778 | LOC100505501 | FALSE | Upregulated |
| 0,038379036 | 0,842779778 | SRSF2 | FALSE | Upregulated |
| -0,0280943 | 0,842779778 | WDR41 | FALSE | Downregulated |
| 0,056601621 | 0,842779778 | ZNF100 | FALSE | Upregulated |
| -0,096519331 | 0,842779778 | ZNF287 | FALSE | Downregulated |
| 0,067677344 | 0,842779778 | ZNF506 | FALSE | Upregulated |
| 0,040640338 | 0,842798645 | SDE2 | FALSE | Upregulated |
| 0,035395681 | 0,842872792 | PIGG | FALSE | Upregulated |
| -0,034361201 | 0,843201709 | UGGT2 | FALSE | Downregulated |
| -0,084615823 | 0,843217753 | LENG9 | FALSE | Downregulated |
| 0,094969863 | 0,843239917 | LOC102723863 | FALSE | Upregulated |
| -0,199338638 | 0,843239917 | LRRC69 | FALSE | Downregulated |
| -0,292104606 | 0,843239917 | RAPSN | FALSE | Downregulated |
| 0,066252913 | 0,843239917 | SLC39A4 | FALSE | Upregulated |
| 0,088810941 | 0,843239917 | SUCNR1 | FALSE | Upregulated |
| 0,032261286 | 0,843592206 | NT5C2 | FALSE | Upregulated |
| -0,045925519 | 0,843657639 | GGT7 | FALSE | Downregulated |
| 0,25215133 | 0,843855456 | RP1-69M21.2 | FALSE | Upregulated |
| -0,095329804 | 0,844094702 | CLIP3 | FALSE | Downregulated |
| 0,356679806 | 0,844094702 | LINC00487 | FALSE | Upregulated |
| -0,250043025 | 0,844094702 | LOC101926916 | FALSE | Downregulated |
| 0,104538029 | 0,844094702 | MAGEE1 | FALSE | Upregulated |
| -0,043929358 | 0,844094702 | MSI2 | FALSE | Downregulated |
| 0,150790979 | 0,844094702 | PTCH2 | FALSE | Upregulated |
| -0,229486527 | 0,844094702 | TGM3 | FALSE | Downregulated |
| -0,235467834 | 0,844094702 | TRNS2 | FALSE | Downregulated |
| -0,068023439 | 0,844265216 | PTPRN2 | FALSE | Downregulated |

| | | | | |
|---|---|---|---|---|
| -0,116752257 | 0,844272791 | CLHC1 | FALSE | Downregulated |
| 0,072168632 | 0,844272791 | OSBPL5 | FALSE | Upregulated |
| -0,092135631 | 0,844455178 | LOC101927942 | FALSE | Downregulated |
| 0,1806119 | 0,844455178 | LOXL1-AS1 | FALSE | Upregulated |
| -0,027336657 | 0,844558296 | BFAR | FALSE | Downregulated |
| 0,04585528 | 0,844848136 | HPCAL1 | FALSE | Upregulated |
| 0,218042378 | 0,844848136 | LOC100128108 | FALSE | Upregulated |
| -0,1437861 | 0,844848136 | PRSS35 | FALSE | Downregulated |
| 0,15193994 | 0,844848136 | WNT11 | FALSE | Upregulated |
| 0,047216576 | 0,844848136 | ZMIZ2 | FALSE | Upregulated |
| 0,091354199 | 0,844863815 | NBPF26 | FALSE | Upregulated |
| -0,024764272 | 0,844969886 | ASH2L | FALSE | Downregulated |
| -0,263285288 | 0,844969886 | LOC101927119 | FALSE | Downregulated |
| -0,317801703 | 0,844969886 | LOC101928255 | FALSE | Downregulated |
| -0,078384671 | 0,844969886 | PTN | FALSE | Downregulated |
| 0,115442729 | 0,845180788 | RNF32 | FALSE | Upregulated |
| -0,051125221 | 0,845329928 | FAM20C | FALSE | Downregulated |
| 0,075227607 | 0,845455397 | BCL6B | FALSE | Upregulated |
| 0,183202512 | 0,845458107 | ADD3-AS1 | FALSE | Upregulated |
| -0,063793833 | 0,845521378 | WSB1 | FALSE | Downregulated |
| -0,032818903 | 0,84557658 | DESI2 | FALSE | Downregulated |
| 0,090933062 | 0,845582413 | LOC101929792 | FALSE | Upregulated |
| 0,154451226 | 0,845612398 | SLC28A3 | FALSE | Upregulated |
| 0,202974783 | 0,845790703 | C9orf152 | FALSE | Upregulated |
| -0,084216297 | 0,845966635 | ZNF763 | FALSE | Downregulated |
| -0,04739169 | 0,845981727 | MTHFR | FALSE | Downregulated |
| 0,96034287 | 0,846012091 | SLC6A11 | FALSE | Upregulated |
| 0,083304911 | 0,846012091 | ZNF43 | FALSE | Upregulated |
| -0,040760632 | 0,846049301 | IRF2BP1 | FALSE | Downregulated |
| 0,154383467 | 0,846101373 | SPATA32 | FALSE | Upregulated |
| 0,266866147 | 0,846159314 | INTS4L1 | FALSE | Upregulated |
| -0,093003235 | 0,846323308 | MAP9 | FALSE | Downregulated |
| -0,399732475 | 0,846369563 | MS4A3 | FALSE | Downregulated |
| -0,084292097 | 0,846369563 | ZNF8 | FALSE | Downregulated |
| -0,043705197 | 0,846601195 | SIGMAR1 | FALSE | Downregulated |
| 0,065381337 | 0,846965934 | ASCC1 | FALSE | Upregulated |
| 0,281639417 | 0,847201345 | MIR4442 | FALSE | Upregulated |
| -0,26689327 | 0,847201345 | NME1-NME2 | FALSE | Downregulated |
| 0,245233366 | 0,847201345 | RP11-439L8.4 | FALSE | Upregulated |
| 0,259606158 | 0,847201345 | SOX30 | FALSE | Upregulated |
| -0,045848526 | 0,847208383 | CTNS | FALSE | Downregulated |
| 0,043003 | 0,847208383 | TMEM135 | FALSE | Upregulated |
| 0,236628331 | 0,847219005 | CCDC42 | FALSE | Upregulated |
| 0,041599631 | 0,847369885 | SEC24A | FALSE | Upregulated |
| 0,085870235 | 0,847407094 | KIF3C | FALSE | Upregulated |
| 0,234254173 | 0,847407094 | LOC644936 | FALSE | Upregulated |
| -0,139940956 | 0,847407094 | MIRLET7BHG | FALSE | Downregulated |
| -0,046289436 | 0,847414676 | PHACTR2 | FALSE | Downregulated |
| 0,042941612 | 0,847455623 | ATP5D | FALSE | Upregulated |
| -0,03618773 | 0,847455623 | ATXN3 | FALSE | Downregulated |
| 0,183427463 | 0,847455623 | COL4A6 | FALSE | Upregulated |

| | | | | |
|---|---|---|---|---|
| -0,045239376 | 0,847455623 | ERCC8 | FALSE | Downregulated |
| -0,272392611 | 0,847455623 | LOC101926889 | FALSE | Downregulated |
| -0,038502149 | 0,847455623 | NAT10 | FALSE | Downregulated |
| 1,640016125 | 0,84766567 | RNU2-2P | FALSE | Upregulated |
| -0,072015275 | 0,847685257 | IKZF2 | FALSE | Downregulated |
| 0,205200121 | 0,847745013 | ESYT3 | FALSE | Upregulated |
| 0,17884311 | 0,847745013 | LINC00242 | FALSE | Upregulated |
| -0,033666105 | 0,848064301 | CHCHD7 | FALSE | Downregulated |
| 0,053507054 | 0,848064301 | MRPL23 | FALSE | Upregulated |
| 0,156066409 | 0,84819481 | MFI2-AS1 | FALSE | Upregulated |
| 0,043092834 | 0,848303273 | BTBD9 | FALSE | Upregulated |
| 0,08687166 | 0,848303273 | CABP4 | FALSE | Upregulated |
| -0,039766566 | 0,848303273 | CLUAP1 | FALSE | Downregulated |
| 0,26106028 | 0,848303273 | CRB1 | FALSE | Upregulated |
| -0,037390963 | 0,848303273 | DIP2C | FALSE | Downregulated |
| -0,054231374 | 0,848303273 | FASTKD3 | FALSE | Downregulated |
| -0,127336871 | 0,848303273 | FCGBP | FALSE | Downregulated |
| 0,192829094 | 0,848303273 | HAR1B | FALSE | Upregulated |
| -0,084360612 | 0,848303273 | HELB | FALSE | Downregulated |
| -0,195600592 | 0,848303273 | LOC100506691 | FALSE | Downregulated |
| -0,242245789 | 0,848303273 | LOC102724209 | FALSE | Downregulated |
| -0,160431219 | 0,848303273 | LOC389831 | FALSE | Downregulated |
| 0,102475804 | 0,848303273 | PRKCDBP | FALSE | Upregulated |
| 0,092715783 | 0,848303273 | RAB27B | FALSE | Upregulated |
| -0,054659203 | 0,848303273 | RASGEF1B | FALSE | Downregulated |
| -0,034673874 | 0,848303273 | SLU7 | FALSE | Downregulated |
| 0,258727588 | 0,848303273 | VPREB3 | FALSE | Upregulated |
| -0,041085524 | 0,848303273 | WDR74 | FALSE | Downregulated |
| -0,390304948 | 0,848441813 | CHRM5 | FALSE | Downregulated |
| 0,14301594 | 0,848604718 | LOC100505912 | FALSE | Upregulated |
| 0,137275875 | 0,84881017 | AHCTF1P1 | FALSE | Upregulated |
| 0,158001763 | 0,84881017 | CPXM2 | FALSE | Upregulated |
| -0,023969241 | 0,84881017 | CUL2 | FALSE | Downregulated |
| 0,06432349 | 0,84881017 | IL7 | FALSE | Upregulated |
| 0,346456587 | 0,84881017 | OPRM1 | FALSE | Upregulated |
| 0,286084281 | 0,84881017 | PLA2G2A | FALSE | Upregulated |
| -0,166659503 | 0,84881017 | RP5-1043L13.1-2 | FALSE | Downregulated |
| -0,108881678 | 0,849057698 | FHOD3 | FALSE | Downregulated |
| 0,025692022 | 0,849114684 | YWHAE | FALSE | Upregulated |
| -0,294344436 | 0,849319894 | MAMDC2-AS1 | FALSE | Downregulated |
| 0,090988051 | 0,849340402 | MGC16275 | FALSE | Upregulated |
| 0,064290625 | 0,849340402 | PGM2L1 | FALSE | Upregulated |
| 0,051962864 | 0,849340402 | PUS7 | FALSE | Upregulated |
| 0,05464648 | 0,849427806 | KIF16B | FALSE | Upregulated |
| 0,21891761 | 0,849427806 | ZMYND10 | FALSE | Upregulated |
| -0,347086421 | 0,849457474 | CMA1 | FALSE | Downregulated |
| 0,281113551 | 0,849535299 | C1orf61 | FALSE | Upregulated |
| -0,296212104 | 0,849597774 | LOC150051 | FALSE | Downregulated |
| 0,043404132 | 0,849597774 | NOM1 | FALSE | Upregulated |
| -0,100285064 | 0,849597774 | RP5-1013A22.2 | FALSE | Downregulated |
| 0,037264815 | 0,849649883 | C11orf74 | FALSE | Upregulated |

| | | | | |
|---|---|---|---|---|
| 0,053708201 | 0,849649883 | C6orf48 | FALSE | Upregulated |
| 0,082088004 | 0,849701872 | GPD1L | FALSE | Upregulated |
| 0,031369273 | 0,849745621 | AVL9 | FALSE | Upregulated |
| 0,080858703 | 0,849745621 | SMIM1 | FALSE | Upregulated |
| 0,027306559 | 0,849800444 | CAB39 | FALSE | Upregulated |
| 0,041283118 | 0,849800444 | RPL19 | FALSE | Upregulated |
| 0,036573201 | 0,849800444 | TBC1D8 | FALSE | Upregulated |
| 0,206225268 | 0,849800444 | TDRD1 | FALSE | Upregulated |
| 0,086758341 | 0,849876092 | CILP | FALSE | Upregulated |
| 0,025916229 | 0,849884817 | FAM134A | FALSE | Upregulated |
| -0,108879538 | 0,849884817 | GAS5-AS1 | FALSE | Downregulated |
| -0,032841565 | 0,849884817 | MRPS22 | FALSE | Downregulated |
| -1,607616379 | 0,849884817 | PADI1 | FALSE | Downregulated |
| -0,024909536 | 0,849884817 | PITHD1 | FALSE | Downregulated |
| -0,058798871 | 0,849884817 | PTPDC1 | FALSE | Downregulated |
| 0,14435595 | 0,849884817 | SNORA4 | FALSE | Upregulated |
| -0,032479622 | 0,849884817 | USP19 | FALSE | Downregulated |
| -0,030093979 | 0,849884817 | ZNF45 | FALSE | Downregulated |
| -0,027222214 | 0,849982415 | CREB1 | FALSE | Downregulated |
| -0,433465478 | 0,849982415 | FLJ45974 | FALSE | Downregulated |
| -0,08093697 | 0,849982415 | LOC102724064 | FALSE | Downregulated |
| 0,109828574 | 0,849982415 | LOC283788 | FALSE | Upregulated |
| 0,024027231 | 0,849982415 | SLC39A9 | FALSE | Upregulated |
| -0,316004786 | 0,849982415 | SLC6A2 | FALSE | Downregulated |
| 0,777247516 | 0,849982415 | TRNAG30 | FALSE | Upregulated |
| -0,225517784 | 0,850085559 | FAM169B | FALSE | Downregulated |
| 0,126652012 | 0,850085559 | RPS18P9 | FALSE | Upregulated |
| -0,023530606 | 0,850085559 | SF3B2 | FALSE | Downregulated |
| -0,067835329 | 0,850128898 | CYP4F12 | FALSE | Downregulated |
| -0,039555244 | 0,850175195 | HTATSF1 | FALSE | Downregulated |
| -0,040834577 | 0,850288949 | OAF | FALSE | Downregulated |
| -0,049361537 | 0,850561209 | MALT1 | FALSE | Downregulated |
| 0,053684013 | 0,850599967 | CCDC68 | FALSE | Upregulated |
| 0,121703726 | 0,850599967 | FKBP1B | FALSE | Upregulated |
| -0,106256812 | 0,850599967 | KDM8 | FALSE | Downregulated |
| 0,05198979 | 0,850881296 | BAIAP2 | FALSE | Upregulated |
| -0,155504167 | 0,850881296 | CYP2D7P | FALSE | Downregulated |
| 0,133218906 | 0,850881296 | EBF1 | FALSE | Upregulated |
| -0,23805489 | 0,850896738 | LOC100287808 | FALSE | Downregulated |
| 0,050002252 | 0,850896738 | MED31 | FALSE | Upregulated |
| 0,200941903 | 0,850909179 | DPY19L2P2 | FALSE | Upregulated |
| -0,083537368 | 0,851103788 | LRRC37A3 | FALSE | Downregulated |
| 0,022645301 | 0,851103788 | RAB5A | FALSE | Upregulated |
| -0,136519263 | 0,851337102 | CACNA1C | FALSE | Downregulated |
| -0,241348608 | 0,851435656 | IL1RAPL1 | FALSE | Downregulated |
| 0,048670293 | 0,851467537 | C11orf68 | FALSE | Upregulated |
| -0,065632618 | 0,851575185 | GUCY1A2 | FALSE | Downregulated |
| 0,036510699 | 0,851575185 | NARF | FALSE | Upregulated |
| -0,124247281 | 0,851582101 | BACH2 | FALSE | Downregulated |
| 0,086239266 | 0,851644542 | FGL1 | FALSE | Upregulated |
| -0,026709211 | 0,851703333 | SUCLA2 | FALSE | Downregulated |

| | | | | |
|---|---|---|---|---|
| -0,055361305 | 0,851813171 | ZSCAN18 | FALSE | Downregulated |
| 0,136985034 | 0,852010612 | SLC6A8 | FALSE | Upregulated |
| -0,170751232 | 0,85211484 | FLJ38576 | FALSE | Downregulated |
| -0,340940399 | 0,85213109 | MKRN9P | FALSE | Downregulated |
| -0,098458285 | 0,852136772 | LOC102724481 | FALSE | Downregulated |
| -0,031792449 | 0,852136772 | PPP1CC | FALSE | Downregulated |
| 0,124806468 | 0,852136772 | TRAPPC3L | FALSE | Upregulated |
| -0,111428427 | 0,852136772 | ZSWIM8-AS1 | FALSE | Downregulated |
| -0,201370145 | 0,85214696 | LOC102467080 | FALSE | Downregulated |
| 0,032176883 | 0,85214696 | SRPK2 | FALSE | Upregulated |
| 0,167127661 | 0,85214696 | ST6GALNAC1 | FALSE | Upregulated |
| -0,448536665 | 0,85214696 | UGT2A1 | FALSE | Downregulated |
| -0,035798798 | 0,852237162 | BBS1 | FALSE | Downregulated |
| 0,032451622 | 0,852237162 | C1orf123 | FALSE | Upregulated |
| -0,14542047 | 0,852237162 | SQLE | FALSE | Downregulated |
| -0,146692505 | 0,852237162 | STX16-NPEPL1 | FALSE | Downregulated |
| -0,051638478 | 0,852327919 | RPIA | FALSE | Downregulated |
| -0,182115067 | 0,85257632 | LOC101928725 | FALSE | Downregulated |
| -0,037172963 | 0,85257632 | TXN2 | FALSE | Downregulated |
| -0,04142134 | 0,852874936 | DYRK2 | FALSE | Downregulated |
| 0,18453736 | 0,85304987 | LINC01134 | FALSE | Upregulated |
| -0,19082752 | 0,85304987 | LOC101928676 | FALSE | Downregulated |
| 0,116993093 | 0,85304987 | SLC16A1-AS1 | FALSE | Upregulated |
| 0,125223364 | 0,853070081 | LRRC49 | FALSE | Upregulated |
| -0,09709346 | 0,853070081 | ZNF14 | FALSE | Downregulated |
| 0,097958533 | 0,853094853 | EMCN | FALSE | Upregulated |
| -0,031919141 | 0,853203016 | SBF1 | FALSE | Downregulated |
| -0,16405258 | 0,853572025 | TMPRSS3 | FALSE | Downregulated |
| 0,031881159 | 0,853910842 | ATP5O | FALSE | Upregulated |
| -0,070366792 | 0,854163085 | CNRIP1 | FALSE | Downregulated |
| -0,260688231 | 0,854163085 | RBMXL2 | FALSE | Downregulated |
| 0,082824038 | 0,854279142 | TMEM200B | FALSE | Upregulated |
| 0,08570963 | 0,854389414 | CACHD1 | FALSE | Upregulated |
| -0,270744059 | 0,854416365 | POU2F3 | FALSE | Downregulated |
| 0,090610177 | 0,854416365 | STARD4 | FALSE | Upregulated |
| 0,041279219 | 0,854422189 | DPH1 | FALSE | Upregulated |
| -0,132463679 | 0,854422189 | GDF9 | FALSE | Downregulated |
| -0,035851032 | 0,854422189 | MPRIP | FALSE | Downregulated |
| 0,038155964 | 0,854422189 | PRPF40A | FALSE | Upregulated |
| 0,053196268 | 0,854422189 | RAB27A | FALSE | Upregulated |
| 0,150840791 | 0,854422189 | RNA45S5 | FALSE | Upregulated |
| -0,093860862 | 0,854465528 | CORO2B | FALSE | Downregulated |
| 0,034295915 | 0,854465528 | N4BP1 | FALSE | Upregulated |
| -0,219002442 | 0,854473491 | PTOV1-AS1 | FALSE | Downregulated |
| -0,075574162 | 0,854595872 | LCN12 | FALSE | Downregulated |
| -0,038134423 | 0,854621422 | BLOC1S1 | FALSE | Downregulated |
| -0,031693422 | 0,854621422 | PIGM | FALSE | Downregulated |
| -0,038249699 | 0,854680348 | RPL21 | FALSE | Downregulated |
| 0,029297946 | 0,854680348 | ZNF689 | FALSE | Upregulated |
| 0,059388024 | 0,85468393 | FAM110C | FALSE | Upregulated |
| -0,113978528 | 0,85468393 | GRHL3 | FALSE | Downregulated |

| | | | | |
|---|---|---|---|---|
| 0,268407358 | 0,85468393 | POTEE | FALSE | Upregulated |
| -0,130069493 | 0,855006637 | LOC400863 | FALSE | Downregulated |
| -0,023349381 | 0,855006637 | PRDM4 | FALSE | Downregulated |
| 0,070182814 | 0,855006637 | TTC22 | FALSE | Upregulated |
| -0,062824308 | 0,855266293 | BAAT | FALSE | Downregulated |
| -0,235028948 | 0,855266293 | C10orf90 | FALSE | Downregulated |
| 0,050877624 | 0,855266293 | C11orf83 | FALSE | Upregulated |
| -0,290699933 | 0,855266293 | LINC00675 | FALSE | Downregulated |
| 0,103534109 | 0,855266293 | LOC100499489 | FALSE | Upregulated |
| 0,043084693 | 0,855266293 | ORMDL1 | FALSE | Upregulated |
| 0,231495985 | 0,855266293 | RFPL1 | FALSE | Upregulated |
| 0,087194585 | 0,855271129 | SLC5A6 | FALSE | Upregulated |
| -0,037433027 | 0,855288368 | AMPD2 | FALSE | Downregulated |
| -0,042046369 | 0,855288368 | CNTRL | FALSE | Downregulated |
| -0,022431383 | 0,855288368 | PICALM | FALSE | Downregulated |
| -0,255617886 | 0,855310717 | APOD | FALSE | Downregulated |
| -0,159471858 | 0,855456119 | LOC102723539 | FALSE | Downregulated |
| -0,064380712 | 0,855498129 | LOC102724975 | FALSE | Downregulated |
| 0,023397675 | 0,855732728 | LEPROT | FALSE | Upregulated |
| -0,064708696 | 0,85574859 | ADCK5 | FALSE | Downregulated |
| 0,052849046 | 0,85574859 | DDX19B | FALSE | Upregulated |
| 0,207602371 | 0,85574859 | HERC2P4 | FALSE | Upregulated |
| -0,312785111 | 0,85574859 | LOC102723950 | FALSE | Downregulated |
| -0,034404276 | 0,85574859 | MRPL20 | FALSE | Downregulated |
| -0,030194766 | 0,85574859 | MRPL45 | FALSE | Downregulated |
| -0,117032947 | 0,85574859 | NQO1 | FALSE | Downregulated |
| 0,043758764 | 0,85574859 | PDZD8 | FALSE | Upregulated |
| -0,271463047 | 0,855823773 | RP11-76N22.2 | FALSE | Downregulated |
| 0,085567491 | 0,856090927 | CYP51A1 | FALSE | Upregulated |
| 0,02611951 | 0,856324903 | ATF1 | FALSE | Upregulated |
| -0,114426734 | 0,856324903 | MAMDC4 | FALSE | Downregulated |
| 0,093569788 | 0,85645438 | FAM13C | FALSE | Upregulated |
| 0,058749987 | 0,85645438 | PGM3 | FALSE | Upregulated |
| -0,02597471 | 0,856681171 | CD164 | FALSE | Downregulated |
| -0,133967125 | 0,856681171 | RD3L | FALSE | Downregulated |
| 0,23737277 | 0,85678188 | ADCYAP1 | FALSE | Upregulated |
| 0,166284226 | 0,85678188 | NRXN3 | FALSE | Upregulated |
| 0,295439339 | 0,85678188 | RIMBP2 | FALSE | Upregulated |
| -0,321265756 | 0,856787098 | RP11-197N18.7 | FALSE | Downregulated |
| -0,054698773 | 0,856793722 | GPR89B | FALSE | Downregulated |
| -0,026961825 | 0,8568643 | RNF141 | FALSE | Downregulated |
| -0,164463868 | 0,856976411 | LOC102723981 | FALSE | Downregulated |
| -0,117962436 | 0,856976411 | PRSS36 | FALSE | Downregulated |
| 0,037201824 | 0,856976411 | SMARCE1 | FALSE | Upregulated |
| -0,107434413 | 0,856992522 | LYPD6 | FALSE | Downregulated |
| 1,518877598 | 0,857546103 | LOC102724219 | FALSE | Upregulated |
| -0,091156518 | 0,857723102 | CD34 | FALSE | Downregulated |
| 0,16780226 | 0,857772369 | LAMB2P1 | FALSE | Upregulated |
| -0,090583767 | 0,857772369 | LOC728752 | FALSE | Downregulated |
| -0,039629257 | 0,858016699 | HNF4A | FALSE | Downregulated |
| 0,110543714 | 0,858117041 | C8orf31 | FALSE | Upregulated |

| | | | | |
|---|---|---|---|---|
| -0,031192739 | 0,858117041 | USF1 | FALSE | Downregulated |
| -0,137272905 | 0,858258318 | LOC101927124 | FALSE | Downregulated |
| -0,050486948 | 0,858258318 | MAP4K4 | FALSE | Downregulated |
| 0,026893975 | 0,858373611 | POLR2C | FALSE | Upregulated |
| -0,102942922 | 0,858379233 | BCDIN3D-AS1 | FALSE | Downregulated |
| 0,027454473 | 0,858379233 | COG8 | FALSE | Upregulated |
| 0,156576815 | 0,858379233 | TRPV6 | FALSE | Upregulated |
| -0,094695477 | 0,858416636 | CACNA2D2 | FALSE | Downregulated |
| 0,106564427 | 0,858578045 | KIF19 | FALSE | Upregulated |
| -0,127998288 | 0,858684859 | AATK | FALSE | Downregulated |
| -0,153058988 | 0,858684859 | MOV10L1 | FALSE | Downregulated |
| 0,12182943 | 0,858719411 | TMEM159 | FALSE | Upregulated |
| -0,061291345 | 0,858772826 | PDE4A | FALSE | Downregulated |
| -0,099001765 | 0,85883466 | TOB1-AS1 | FALSE | Downregulated |
| 0,036031046 | 0,859043364 | CACFD1 | FALSE | Upregulated |
| 0,182628584 | 0,859043364 | FAM66C | FALSE | Upregulated |
| -0,321620092 | 0,859043364 | GOLGA8K | FALSE | Downregulated |
| 0,148584347 | 0,859043364 | MIPEPP3 | FALSE | Upregulated |
| 0,036414135 | 0,859090428 | ARFGAP1 | FALSE | Upregulated |
| -0,093195418 | 0,8593652 | LINC00338 | FALSE | Downregulated |
| 0,120854323 | 0,8593652 | TGFBR3L | FALSE | Upregulated |
| -0,145747981 | 0,8593652 | TMEM132B | FALSE | Downregulated |
| -0,202441697 | 0,859372574 | ANKS1B | FALSE | Downregulated |
| -0,270878546 | 0,85943157 | C12orf74 | FALSE | Downregulated |
| 0,220547737 | 0,85943157 | LRRC73 | FALSE | Upregulated |
| -0,042378677 | 0,85943157 | NAA38 | FALSE | Downregulated |
| -0,06382276 | 0,85943157 | PLCB1 | FALSE | Downregulated |
| 0,217076392 | 0,85943157 | SMIM17 | FALSE | Upregulated |
| 0,20740636 | 0,85943157 | XCR1 | FALSE | Upregulated |
| -0,062390735 | 0,859482283 | LOC100289097 | FALSE | Downregulated |
| 0,130680449 | 0,859495907 | KAZALD1 | FALSE | Upregulated |
| 0,040275026 | 0,859567303 | RBM34 | FALSE | Upregulated |
| 0,096393818 | 0,859751477 | NHS | FALSE | Upregulated |
| 0,413416058 | 0,860051526 | LOC100996717 | FALSE | Upregulated |
| -0,06487831 | 0,860051526 | LOC158960 | FALSE | Downregulated |
| -0,042987039 | 0,860059436 | FAM135A | FALSE | Downregulated |
| 0,069745874 | 0,860101015 | FLJ20021 | FALSE | Upregulated |
| -0,030224341 | 0,860296374 | SDHC | FALSE | Downregulated |
| 0,055108362 | 0,860352532 | ELTD1 | FALSE | Upregulated |
| 0,168147993 | 0,860352532 | LSAMP | FALSE | Upregulated |
| 0,121279156 | 0,860352532 | TNXB | FALSE | Upregulated |
| 0,033614768 | 0,860432601 | DSG2 | FALSE | Upregulated |
| -0,053501979 | 0,860499739 | LPP-AS2 | FALSE | Downregulated |
| 0,192047741 | 0,860499739 | NMNAT2 | FALSE | Upregulated |
| -0,039140354 | 0,860499739 | TIE1 | FALSE | Downregulated |
| -0,067140733 | 0,860537343 | ADPRM | FALSE | Downregulated |
| 0,112899516 | 0,860537343 | EDARADD | FALSE | Upregulated |
| 0,110668007 | 0,86074994 | KCNN3 | FALSE | Upregulated |
| -0,155013457 | 0,86074994 | LOC101928153 | FALSE | Downregulated |
| -0,068983572 | 0,860998439 | MGC72080 | FALSE | Downregulated |
| 0,086815088 | 0,861171494 | LOC283070 | FALSE | Upregulated |

| | | | | |
|---|---|---|---|---|
| 0,087744647 | 0,861476861 | SNORA18 | FALSE | Upregulated |
| -0,039848906 | 0,86161767 | DDX31 | FALSE | Downregulated |
| -0,032305623 | 0,861891872 | TTF1 | FALSE | Downregulated |
| -0,264386628 | 0,86189203 | LOC102723775 | FALSE | Downregulated |
| 0,035770619 | 0,86189203 | SSSCA1 | FALSE | Upregulated |
| -0,028458247 | 0,86190531 | GABPB1 | FALSE | Downregulated |
| 0,052335348 | 0,86190531 | ITIH3 | FALSE | Upregulated |
| 0,057010244 | 0,861962287 | CCDC134 | FALSE | Upregulated |
| -0,040159878 | 0,861962287 | ZFHX3 | FALSE | Downregulated |
| 0,033530346 | 0,862112212 | PDHA1 | FALSE | Upregulated |
| -0,064383546 | 0,862151388 | CHKB | FALSE | Downregulated |
| -0,043358756 | 0,862151388 | RPL26 | FALSE | Downregulated |
| 0,091826655 | 0,862374928 | SGK223 | FALSE | Upregulated |
| -0,065722097 | 0,862395652 | CEP72 | FALSE | Downregulated |
| -0,037553557 | 0,862395652 | FAM173B | FALSE | Downregulated |
| 0,186480028 | 0,862395652 | FAM189A2 | FALSE | Upregulated |
| -0,047388196 | 0,862395652 | LRRC2 | FALSE | Downregulated |
| 0,021956714 | 0,862395652 | PHKB | FALSE | Upregulated |
| 0,046017193 | 0,862449935 | EHBP1 | FALSE | Upregulated |
| 0,065038489 | 0,862449935 | STEAP1 | FALSE | Upregulated |
| 0,072678296 | 0,862449935 | ZNF738 | FALSE | Upregulated |
| 0,094318662 | 0,862537618 | SORCS2 | FALSE | Upregulated |
| 0,159592385 | 0,862827366 | C2CD4B | FALSE | Upregulated |
| -0,121485937 | 0,862827366 | CACNA2D1 | FALSE | Downregulated |
| -0,097705267 | 0,862827366 | KLHL32 | FALSE | Downregulated |
| -0,108506403 | 0,862827366 | LOC102724238 | FALSE | Downregulated |
| 0,030059967 | 0,862827366 | MRPS21 | FALSE | Upregulated |
| 0,0327186 | 0,862827366 | SCFD1 | FALSE | Upregulated |
| -0,052926625 | 0,862827366 | TCERG1 | FALSE | Downregulated |
| -0,029432217 | 0,862827366 | VPRBP | FALSE | Downregulated |
| -0,029449411 | 0,863009045 | FBXO7 | FALSE | Downregulated |
| 0,264414114 | 0,863009045 | RALGAPA1P | FALSE | Upregulated |
| 0,021394647 | 0,863009045 | YPEL5 | FALSE | Upregulated |
| 0,232313517 | 0,863082093 | TRIM67 | FALSE | Upregulated |
| 0,047359451 | 0,863150675 | SARM1 | FALSE | Upregulated |
| 0,038950522 | 0,863150675 | SCNM1 | FALSE | Upregulated |
| -0,033002588 | 0,863150675 | ZNF263 | FALSE | Downregulated |
| 0,041956931 | 0,863290536 | PDXP | FALSE | Upregulated |
| -0,022382085 | 0,86337199 | MCM3AP | FALSE | Downregulated |
| 1,16545203 | 0,863430333 | MYLK2 | FALSE | Upregulated |
| -0,060459472 | 0,863430333 | TCF4 | FALSE | Downregulated |
| 0,091475073 | 0,863699418 | LOC101928645 | FALSE | Upregulated |
| -0,055055603 | 0,864143438 | EXOC3L4 | FALSE | Downregulated |
| 0,065128125 | 0,864278634 | P2RX7 | FALSE | Upregulated |
| -0,039187093 | 0,864349976 | USP6NL | FALSE | Downregulated |
| 0,04981431 | 0,864391205 | BAZ1B | FALSE | Upregulated |
| -0,06865937 | 0,864575637 | SLC7A5 | FALSE | Downregulated |
| 0,034974564 | 0,864737829 | EIF3M | FALSE | Upregulated |
| -0,06843223 | 0,865023541 | EIF3C | FALSE | Downregulated |
| -0,057024312 | 0,865023541 | FAM63B | FALSE | Downregulated |
| 0,069096442 | 0,865023541 | GPR157 | FALSE | Upregulated |

| | | | | |
|---|---|---|---|---|
| -0,050894153 | 0,865023541 | GTF2H2B | FALSE | Downregulated |
| 0,102618575 | 0,865023541 | LIPE | FALSE | Upregulated |
| -0,024462486 | 0,865023541 | MRPL38 | FALSE | Downregulated |
| -0,034485633 | 0,865023541 | PIGP | FALSE | Downregulated |
| -0,069036829 | 0,865023541 | SH3PXD2B | FALSE | Downregulated |
| 0,208134425 | 0,865027017 | LOC401010 | FALSE | Upregulated |
| -0,105345607 | 0,865462416 | SGIP1 | FALSE | Downregulated |
| -0,026833031 | 0,865575943 | CASP6 | FALSE | Downregulated |
| -0,019545816 | 0,865629537 | H3F3B | FALSE | Downregulated |
| -0,019808974 | 0,865629537 | SMCR8 | FALSE | Downregulated |
| -0,249051084 | 0,865767639 | LOC100287166 | FALSE | Downregulated |
| -0,125521735 | 0,865767639 | LOC101928958 | FALSE | Downregulated |
| -0,249547211 | 0,865767639 | LOC102724484 | FALSE | Downregulated |
| -0,042525251 | 0,865767639 | RGL2 | FALSE | Downregulated |
| -0,067428235 | 0,865923574 | ARHGAP5-AS1 | FALSE | Downregulated |
| 0,237214374 | 0,865923574 | CCDC155 | FALSE | Upregulated |
| -0,026924349 | 0,865923574 | ITM2B | FALSE | Downregulated |
| 0,144484143 | 0,865923574 | KIF26B | FALSE | Upregulated |
| -0,024098379 | 0,865923574 | PIP4K2B | FALSE | Downregulated |
| 0,051744424 | 0,866044297 | NCL | FALSE | Upregulated |
| 0,045582353 | 0,866083927 | ENDOD1 | FALSE | Upregulated |
| 0,029151193 | 0,866110666 | PPID | FALSE | Upregulated |
| -0,039872105 | 0,866197025 | SSR1 | FALSE | Downregulated |
| 0,077691806 | 0,866440159 | C7orf76 | FALSE | Upregulated |
| 0,046312002 | 0,866572095 | MAT2A | FALSE | Upregulated |
| -0,026615483 | 0,866702876 | PTDSS1 | FALSE | Downregulated |
| -0,038684134 | 0,866702876 | RPL30 | FALSE | Downregulated |
| -0,029474763 | 0,866702876 | ZBTB18 | FALSE | Downregulated |
| 0,044789328 | 0,866833155 | HINFP | FALSE | Upregulated |
| 0,024286239 | 0,866862305 | TM9SF2 | FALSE | Upregulated |
| -0,234033752 | 0,866929688 | SNORD57 | FALSE | Downregulated |
| 0,16942355 | 0,867034855 | LHX9 | FALSE | Upregulated |
| 0,019506166 | 0,867239448 | ZFAND3 | FALSE | Upregulated |
| 0,025873131 | 0,867321084 | ACTR6 | FALSE | Upregulated |
| 0,055500609 | 0,867321084 | FAM89A | FALSE | Upregulated |
| 0,122110005 | 0,867585877 | BACE1-AS | FALSE | Upregulated |
| 0,033002781 | 0,867585877 | MED7 | FALSE | Upregulated |
| 0,02683493 | 0,867677826 | C6orf62 | FALSE | Upregulated |
| 0,218367189 | 0,867677826 | LOC102724732 | FALSE | Upregulated |
| 0,041118784 | 0,867677826 | MORN2 | FALSE | Upregulated |
| 0,149346733 | 0,867677826 | NAT8L | FALSE | Upregulated |
| -0,033686745 | 0,867761665 | MED25 | FALSE | Downregulated |
| -0,019075982 | 0,867939121 | RAD23B | FALSE | Downregulated |
| -0,043754319 | 0,867939121 | ZBED5-AS1 | FALSE | Downregulated |
| 0,023887219 | 0,867955815 | SIDT2 | FALSE | Upregulated |
| -0,104283239 | 0,867955815 | STAG3L5P | FALSE | Downregulated |
| -0,091582937 | 0,868300636 | EDN1 | FALSE | Downregulated |
| -0,105786701 | 0,868300636 | MEF2B | FALSE | Downregulated |
| 0,026904417 | 0,868378412 | FNDC3A | FALSE | Upregulated |
| 0,24197486 | 0,868378412 | LOC284648 | FALSE | Upregulated |
| -0,036300173 | 0,868378412 | LRCH3 | FALSE | Downregulated |

| | | | | |
|---|---|---|---|---|
| 0,30223884 | 0,868378412 | SLC9C2 | FALSE | Upregulated |
| -1,409782418 | 0,868378412 | XIST | FALSE | Downregulated |
| -0,026830954 | 0,868719045 | ESD | FALSE | Downregulated |
| -0,115750691 | 0,868773282 | MST1P2 | FALSE | Downregulated |
| 0,245109505 | 0,868773282 | SLC12A1 | FALSE | Upregulated |
| 0,040586369 | 0,868773282 | TMEM208 | FALSE | Upregulated |
| -0,267682523 | 0,868946272 | RSPO1 | FALSE | Downregulated |
| 0,147382151 | 0,869030188 | ZDHHC15 | FALSE | Upregulated |
| -0,038883388 | 0,869059088 | PTPN3 | FALSE | Downregulated |
| 0,159651751 | 0,869190846 | B4GALNT3 | FALSE | Upregulated |
| -0,069693044 | 0,869271053 | ZNF347 | FALSE | Downregulated |
| -0,229775695 | 0,869313045 | CCDC181 | FALSE | Downregulated |
| 0,083777422 | 0,869313045 | HRH1 | FALSE | Upregulated |
| -0,148518089 | 0,869313045 | LOC102723679 | FALSE | Downregulated |
| -0,072231673 | 0,869313045 | ZNF132 | FALSE | Downregulated |
| 0,037071987 | 0,869484901 | RRAGD | FALSE | Upregulated |
| -0,248755255 | 0,869534065 | RBM5-AS1 | FALSE | Downregulated |
| 0,055941738 | 0,869738175 | RP11-372E1.6 | FALSE | Upregulated |
| -0,069471087 | 0,870007013 | CPT1A | FALSE | Downregulated |
| -0,123240993 | 0,870007013 | FAM211A | FALSE | Downregulated |
| 0,1032123 | 0,870007013 | HPGDS | FALSE | Upregulated |
| 0,238058479 | 0,870007013 | LOC101927986 | FALSE | Upregulated |
| -0,431767812 | 0,870007013 | RMRP | FALSE | Downregulated |
| 0,245149162 | 0,870007013 | RP11-14N7.2 | FALSE | Upregulated |
| -0,201340879 | 0,870007013 | SMG7-AS1 | FALSE | Downregulated |
| -0,069977906 | 0,870224207 | LOC728613 | FALSE | Downregulated |
| -0,037483891 | 0,870224207 | OXSM | FALSE | Downregulated |
| -0,111089722 | 0,870224207 | RGS11 | FALSE | Downregulated |
| 0,028576067 | 0,870224207 | RPF2 | FALSE | Upregulated |
| 0,081869567 | 0,870224207 | SVEP1 | FALSE | Upregulated |
| -0,309836223 | 0,870224207 | ZFR2 | FALSE | Downregulated |
| 0,075202053 | 0,870263257 | PDE9A | FALSE | Upregulated |
| -0,125783881 | 0,870359019 | ANKRD65 | FALSE | Downregulated |
| 0,033217497 | 0,870359019 | ARMCX3 | FALSE | Upregulated |
| -0,033020311 | 0,870359019 | NAPG | FALSE | Downregulated |
| -0,103886453 | 0,870359019 | PPP2R3A | FALSE | Downregulated |
| -0,15508383 | 0,870359019 | SEC1P | FALSE | Downregulated |
| 0,247603892 | 0,870359019 | SERF1B | FALSE | Upregulated |
| 0,08184699 | 0,870393282 | LINC00924 | FALSE | Upregulated |
| 0,14124703 | 0,870393282 | LOC102723672 | FALSE | Upregulated |
| -0,031810809 | 0,870696125 | LOC339803 | FALSE | Downregulated |
| 0,077448609 | 0,870696125 | PTPRS | FALSE | Upregulated |
| -0,031780552 | 0,870696125 | ZNF598 | FALSE | Downregulated |
| -0,062872608 | 0,870827501 | TMOD2 | FALSE | Downregulated |
| 0,043758906 | 0,871190689 | TANGO6 | FALSE | Upregulated |
| 0,163833285 | 0,871267505 | CTCFL | FALSE | Upregulated |
| -0,187701931 | 0,871267505 | NTNG2 | FALSE | Downregulated |
| 0,114814082 | 0,871383216 | CYP21A1P | FALSE | Upregulated |
| -0,034304688 | 0,871503772 | POMT2 | FALSE | Downregulated |
| -0,057778521 | 0,871658842 | ODF2L | FALSE | Downregulated |
| -0,01905821 | 0,871737561 | ARL5A | FALSE | Downregulated |

| | | | | |
|---|---|---|---|---|
| 0,136413179 | 0,871737561 | FLG-AS1 | FALSE | Upregulated |
| -0,121688448 | 0,871737561 | FLVCR1-AS1 | FALSE | Downregulated |
| 0,037719788 | 0,871737561 | OBFC1 | FALSE | Upregulated |
| 0,08918355 | 0,871906361 | LOC102723546 | FALSE | Upregulated |
| 0,020982164 | 0,871906361 | PTBP1 | FALSE | Upregulated |
| -0,040891016 | 0,871957439 | CEP192 | FALSE | Downregulated |
| -0,094660871 | 0,871957439 | CRIP3 | FALSE | Downregulated |
| 0,020529194 | 0,871957439 | HNRNPUL2 | FALSE | Upregulated |
| -0,037499617 | 0,872047942 | CEP170 | FALSE | Downregulated |
| 0,073869465 | 0,872047942 | NOS1AP | FALSE | Upregulated |
| -0,261314655 | 0,872137562 | HRASLS | FALSE | Downregulated |
| 0,035433933 | 0,872153048 | TRIM27 | FALSE | Upregulated |
| -0,063514502 | 0,872454005 | ACTR5 | FALSE | Downregulated |
| -0,069963311 | 0,872493525 | TPBG | FALSE | Downregulated |
| -0,172381213 | 0,872791067 | LOC100505767 | FALSE | Downregulated |
| 0,033923338 | 0,872809779 | ABI2 | FALSE | Upregulated |
| -0,129782561 | 0,87289886 | SEC14L1P1 | FALSE | Downregulated |
| -0,247092439 | 0,872928125 | ZDHHC8P1 | FALSE | Downregulated |
| 0,236525092 | 0,872969827 | ARL14 | FALSE | Upregulated |
| -0,305970223 | 0,873123482 | THEG | FALSE | Downregulated |
| 0,057904319 | 0,873256487 | JOSD2 | FALSE | Upregulated |
| 0,051036093 | 0,873256487 | MAP3K6 | FALSE | Upregulated |
| -0,106674279 | 0,873337651 | LINC00535 | FALSE | Downregulated |
| -0,07451893 | 0,873524659 | CMAHP | FALSE | Downregulated |
| -0,033801737 | 0,873579108 | NCAPD3 | FALSE | Downregulated |
| 0,116262765 | 0,874219165 | LOC554206 | FALSE | Upregulated |
| -0,036303572 | 0,874302576 | EDEM3 | FALSE | Downregulated |
| -0,040128893 | 0,874303137 | C21orf59 | FALSE | Downregulated |
| -0,210645226 | 0,8743875 | BMP8A | FALSE | Downregulated |
| -0,066142756 | 0,8743875 | RDH5 | FALSE | Downregulated |
| -0,029986889 | 0,8743875 | RPL34 | FALSE | Downregulated |
| 1,3552061 | 0,874560675 | TNNT1 | FALSE | Upregulated |
| -0,106819859 | 0,874646127 | LOC646214 | FALSE | Downregulated |
| -0,115195364 | 0,87478191 | LOC102724643 | FALSE | Downregulated |
| 0,020905534 | 0,875136032 | VIPAS39 | FALSE | Upregulated |
| -0,045175577 | 0,875304695 | LOC101927752 | FALSE | Downregulated |
| 0,086534227 | 0,8754399 | FABP3 | FALSE | Upregulated |
| 0,182726382 | 0,875489956 | DNM1P46 | FALSE | Upregulated |
| -0,200507928 | 0,875489956 | HMP19 | FALSE | Downregulated |
| 0,258199811 | 0,875489956 | MIXL1 | FALSE | Upregulated |
| 0,098432007 | 0,875549532 | DKK3 | FALSE | Upregulated |
| 0,038579092 | 0,875563358 | TBC1D16 | FALSE | Upregulated |
| 0,038783911 | 0,8756961 | CBLB | FALSE | Upregulated |
| -0,227503868 | 0,875953865 | CHL1 | FALSE | Downregulated |
| -0,12072539 | 0,875953865 | ZACN | FALSE | Downregulated |
| 0,042991441 | 0,876104656 | TMEM237 | FALSE | Upregulated |
| 0,084774853 | 0,87617755 | FUT1 | FALSE | Upregulated |
| 0,116719537 | 0,876272022 | C17orf53 | FALSE | Upregulated |
| -0,087564638 | 0,876272022 | OSBP2 | FALSE | Downregulated |
| 0,065620951 | 0,876272022 | RNR1 | FALSE | Upregulated |
| 0,089138305 | 0,876307025 | APOBEC3D | FALSE | Upregulated |

| | | | | |
|---|---|---|---|---|
| 0,256198608 | 0,876307025 | C6orf99 | FALSE | Upregulated |
| -0,079329968 | 0,876307025 | MALL | FALSE | Downregulated |
| -0,03524211 | 0,876307025 | MTG2 | FALSE | Downregulated |
| -0,030850334 | 0,876307025 | NEMF | FALSE | Downregulated |
| -0,12926912 | 0,876307025 | NOMO3 | FALSE | Downregulated |
| 0,210068588 | 0,876307025 | PLCD4 | FALSE | Upregulated |
| -0,286556155 | 0,876307025 | THEM5 | FALSE | Downregulated |
| 0,027067705 | 0,876307025 | YIPF2 | FALSE | Upregulated |
| 0,068457576 | 0,876327865 | NUCB2 | FALSE | Upregulated |
| -0,021838312 | 0,876455966 | C8orf33 | FALSE | Downregulated |
| 0,019197022 | 0,876455966 | DIAPH1 | FALSE | Upregulated |
| 0,063184335 | 0,876455966 | NSDHL | FALSE | Upregulated |
| 0,143894941 | 0,876455966 | POM121L9P | FALSE | Upregulated |
| 0,080834849 | 0,876455966 | RPGRIP1 | FALSE | Upregulated |
| 0,042541841 | 0,876468482 | DPH6 | FALSE | Upregulated |
| 0,112072148 | 0,876468482 | GNAZ | FALSE | Upregulated |
| 0,251567633 | 0,876468482 | GNB3 | FALSE | Upregulated |
| -0,314436961 | 0,876468482 | KRT5 | FALSE | Downregulated |
| 0,045693573 | 0,876468482 | SLC39A6 | FALSE | Upregulated |
| 0,085654003 | 0,876574055 | C11orf45 | FALSE | Upregulated |
| 0,08269973 | 0,876574055 | PRNCR1 | FALSE | Upregulated |
| -0,047877882 | 0,876670324 | ABCA5 | FALSE | Downregulated |
| -0,024944859 | 0,87714856 | FRS2 | FALSE | Downregulated |
| -0,165013122 | 0,877184819 | TMEM125 | FALSE | Downregulated |
| 0,1574163 | 0,877290719 | HIST1H3H | FALSE | Upregulated |
| 0,02640221 | 0,877386497 | ZC3H14 | FALSE | Upregulated |
| 0,066541796 | 0,877643159 | ATP8A1 | FALSE | Upregulated |
| -0,0279669 | 0,877643159 | CHP1 | FALSE | Downregulated |
| 0,039598829 | 0,877643159 | CREB3L2 | FALSE | Upregulated |
| 0,055854805 | 0,877643159 | GPR124 | FALSE | Upregulated |
| -0,082095602 | 0,877643159 | HOXB5 | FALSE | Downregulated |
| 0,069037584 | 0,877643159 | LINC01124 | FALSE | Upregulated |
| 0,421415286 | 0,877643159 | LOC102723828 | FALSE | Upregulated |
| -0,121819835 | 0,877643159 | LOC102724187 | FALSE | Downregulated |
| 0,028723363 | 0,877643159 | MTX1 | FALSE | Upregulated |
| -0,031939145 | 0,877643159 | NDUFB4 | FALSE | Downregulated |
| 0,142239957 | 0,877643159 | RHBDL1 | FALSE | Upregulated |
| 0,059420776 | 0,877658932 | RPGR | FALSE | Upregulated |
| -0,074187388 | 0,877731808 | IGF2BP3 | FALSE | Downregulated |
| 0,022408634 | 0,877731808 | LZTR1 | FALSE | Upregulated |
| -0,111113843 | 0,877731808 | SSR4P1 | FALSE | Downregulated |
| -0,046536428 | 0,877830033 | FAM211A-AS1 | FALSE | Downregulated |
| -0,187757759 | 0,877830033 | TPSB2 | FALSE | Downregulated |
| -0,267023994 | 0,877892312 | LOC100505938 | FALSE | Downregulated |
| 0,026925833 | 0,877892312 | RER1 | FALSE | Upregulated |
| -0,037092862 | 0,877892312 | TCHP | FALSE | Downregulated |
| 0,220469996 | 0,877954163 | CCDC108 | FALSE | Upregulated |
| -0,167446478 | 0,878156654 | LOC101928021 | FALSE | Downregulated |
| -0,096801975 | 0,87819112 | GAREML | FALSE | Downregulated |
| -0,221415449 | 0,87819112 | KLHL30 | FALSE | Downregulated |
| 0,226486046 | 0,87819112 | LOC101928323 | FALSE | Upregulated |

| | | | | |
|---|---|---|---|---|
| 0,018080044 | 0,87819112 | PHRF1 | FALSE | Upregulated |
| -0,101375361 | 0,87819112 | RP11-307E17.8 | FALSE | Downregulated |
| -0,038344118 | 0,87819112 | ZNF624 | FALSE | Downregulated |
| 0,14487534 | 0,878239509 | SMOC2 | FALSE | Upregulated |
| 0,040111267 | 0,878257216 | SCRN2 | FALSE | Upregulated |
| -0,022822665 | 0,878440536 | KIFAP3 | FALSE | Downregulated |
| 0,046091208 | 0,878440536 | RP11-85F14.5 | FALSE | Upregulated |
| 0,238791764 | 0,878539463 | SNORA3 | FALSE | Upregulated |
| 0,095211946 | 0,878837847 | USP27X-AS1 | FALSE | Upregulated |
| 0,052704818 | 0,879090097 | LAMB1 | FALSE | Upregulated |
| 0,03765616 | 0,87915458 | CMSS1 | FALSE | Upregulated |
| -0,219544338 | 0,879190817 | LOC100996506 | FALSE | Downregulated |
| 0,215535304 | 0,8792291 | ASIP | FALSE | Upregulated |
| 0,014263702 | 0,879305008 | UBE3B | FALSE | Upregulated |
| -0,142805398 | 0,879314083 | MAP3K14-AS1 | FALSE | Downregulated |
| 0,103421345 | 0,87939994 | LOC102723794 | FALSE | Upregulated |
| -0,080912945 | 0,87939994 | RPL19P12 | FALSE | Downregulated |
| 0,024373437 | 0,87939994 | TMEM5 | FALSE | Upregulated |
| -0,040523275 | 0,879414359 | MON1A | FALSE | Downregulated |
| -0,061272029 | 0,879414359 | ZNF816 | FALSE | Downregulated |
| -0,025229447 | 0,879647727 | PRDX5 | FALSE | Downregulated |
| 0,045695698 | 0,879835037 | LIMD1 | FALSE | Upregulated |
| 0,027059336 | 0,879871114 | SSNA1 | FALSE | Upregulated |
| -0,058651012 | 0,879973348 | C1QTNF5 | FALSE | Downregulated |
| 0,050697917 | 0,879973348 | FAM122C | FALSE | Upregulated |
| -0,039976396 | 0,879973348 | NMNAT3 | FALSE | Downregulated |
| -0,069141099 | 0,879973348 | S1PR2 | FALSE | Downregulated |
| -0,022326108 | 0,879973348 | TMEM14C | FALSE | Downregulated |
| -0,040188852 | 0,880056003 | HDAC10 | FALSE | Downregulated |
| -0,040235784 | 0,880070844 | TSEN2 | FALSE | Downregulated |
| 0,232345599 | 0,880193417 | MSH4 | FALSE | Upregulated |
| -0,141834548 | 0,880193417 | NFE2 | FALSE | Downregulated |
| -0,033514378 | 0,880193417 | SMN1 | FALSE | Downregulated |
| 0,040953596 | 0,88032077 | GALC | FALSE | Upregulated |
| -0,072050992 | 0,880405503 | FANCC | FALSE | Downregulated |
| 0,071369498 | 0,880409195 | NDUFA4L2 | FALSE | Upregulated |
| 0,056830249 | 0,880569092 | FOXA3 | FALSE | Upregulated |
| -0,04835969 | 0,880569092 | PRICKLE3 | FALSE | Downregulated |
| 0,023681955 | 0,880851736 | CDS2 | FALSE | Upregulated |
| 0,073977044 | 0,880851736 | CSGALNACT1 | FALSE | Upregulated |
| -0,036192282 | 0,88090347 | TSHZ2 | FALSE | Downregulated |
| 0,048635902 | 0,880917731 | CENPJ | FALSE | Upregulated |
| -0,017988474 | 0,880974341 | UBE2Q1 | FALSE | Downregulated |
| 0,035299914 | 0,881091466 | HPD | FALSE | Upregulated |
| 0,02234652 | 0,881091466 | MRPL49 | FALSE | Upregulated |
| 0,02782858 | 0,881091466 | NEK1 | FALSE | Upregulated |
| 0,036973382 | 0,881255467 | MARS2 | FALSE | Upregulated |
| -0,031376813 | 0,881255467 | MNAT1 | FALSE | Downregulated |
| -0,296746137 | 0,881459989 | HTR1F | FALSE | Downregulated |
| -0,029185104 | 0,881477505 | ATF4 | FALSE | Downregulated |
| -0,039849226 | 0,881477505 | HEBP2 | FALSE | Downregulated |

| | | | | |
|---|---|---|---|---|
| -0,144673188 | 0,881477505 | SYCE1 | FALSE | Downregulated |
| -0,152706251 | 0,881501348 | LOC101926970 | FALSE | Downregulated |
| 0,042244913 | 0,881503734 | C10orf2 | FALSE | Upregulated |
| 0,214144676 | 0,881503734 | LOC100289455 | FALSE | Upregulated |
| -0,056600867 | 0,881537577 | FAM65A | FALSE | Downregulated |
| -0,040988641 | 0,881824748 | ANAPC15 | FALSE | Downregulated |
| -0,315896339 | 0,881824748 | ATP1A4 | FALSE | Downregulated |
| -0,055604832 | 0,881824748 | GLOD5 | FALSE | Downregulated |
| -0,02360054 | 0,881824748 | LMO4 | FALSE | Downregulated |
| 0,117860834 | 0,881824748 | NAV3 | FALSE | Upregulated |
| 0,09772425 | 0,881824748 | OAZ3 | FALSE | Upregulated |
| 0,025252744 | 0,881860653 | RPL6 | FALSE | Upregulated |
| 0,176253227 | 0,881908449 | C10orf62 | FALSE | Upregulated |
| 0,161036503 | 0,881908449 | DRD4 | FALSE | Upregulated |
| 0,051980912 | 0,881908449 | SPATA20 | FALSE | Upregulated |
| 0,037684136 | 0,88197235 | RBFA | FALSE | Upregulated |
| 0,057102178 | 0,88197235 | SERPINA3 | FALSE | Upregulated |
| 0,062402896 | 0,882021588 | EHD2 | FALSE | Upregulated |
| -0,107367233 | 0,882021588 | FAM83B | FALSE | Downregulated |
| 0,028557925 | 0,882023068 | MOGS | FALSE | Upregulated |
| -0,336322171 | 0,882057831 | RFPL4AL1 | FALSE | Downregulated |
| -0,076271986 | 0,882118404 | NBEA | FALSE | Downregulated |
| 0,093171183 | 0,882466903 | FBXL8 | FALSE | Upregulated |
| 0,159333842 | 0,882475934 | ASIC4 | FALSE | Upregulated |
| 0,035991771 | 0,882475934 | DCTN5 | FALSE | Upregulated |
| 0,041984028 | 0,882545197 | TNFSF12 | FALSE | Upregulated |
| -0,319545329 | 0,882547037 | HPCAL4 | FALSE | Downregulated |
| 0,085153277 | 0,882547037 | KCNAB1 | FALSE | Upregulated |
| -0,170826771 | 0,882547037 | PADI4 | FALSE | Downregulated |
| 0,023159499 | 0,882547037 | TM2D1 | FALSE | Upregulated |
| 0,1729579 | 0,882550698 | SETSIP | FALSE | Upregulated |
| 0,069131924 | 0,882772205 | TM6SF1 | FALSE | Upregulated |
| -0,093109134 | 0,883106524 | CHST1 | FALSE | Downregulated |
| -0,022306854 | 0,883177546 | DNAJC7 | FALSE | Downregulated |
| -0,026948724 | 0,883291573 | ARHGAP21 | FALSE | Downregulated |
| -0,09068778 | 0,883291573 | MSANTD3-TMEFF1 | FALSE | Downregulated |
| -0,129346195 | 0,883293027 | LOC102723340 | FALSE | Downregulated |
| -0,103818015 | 0,883293027 | LTB4R2 | FALSE | Downregulated |
| -0,183380633 | 0,883293027 | NTN5 | FALSE | Downregulated |
| -0,028023032 | 0,883293027 | SSRP1 | FALSE | Downregulated |
| 0,02453998 | 0,883293027 | STRADA | FALSE | Upregulated |
| 0,038412398 | 0,883293027 | TSNARE1 | FALSE | Upregulated |
| -0,058535549 | 0,883293027 | VAMP1 | FALSE | Downregulated |
| 0,108311577 | 0,883398706 | OR7E37P | FALSE | Upregulated |
| -0,19458682 | 0,883398706 | RP11-166O4.5 | FALSE | Downregulated |
| 0,038952729 | 0,883398706 | SLC7A9 | FALSE | Upregulated |
| -0,24148317 | 0,883500599 | LRRC3B | FALSE | Downregulated |
| 0,106859054 | 0,883500599 | NALCN | FALSE | Upregulated |
| 0,100797375 | 0,883500599 | OTUD7A | FALSE | Upregulated |
| 0,037828045 | 0,883669663 | ZBTB47 | FALSE | Upregulated |
| 0,274248413 | 0,883689129 | RP11-138B4.1 | FALSE | Upregulated |

| | | | | |
|---|---|---|---|---|
| -0,038683012 | 0,883692352 | CTBP2 | FALSE | Downregulated |
| -0,020310278 | 0,883707549 | ZNF641 | FALSE | Downregulated |
| 0,103190909 | 0,883896654 | TNF | FALSE | Upregulated |
| 0,025732925 | 0,883947077 | PRKCZ | FALSE | Upregulated |
| -0,045681586 | 0,883947077 | SETBP1 | FALSE | Downregulated |
| 0,027777688 | 0,884107348 | NDUFB8 | FALSE | Upregulated |
| 0,030874292 | 0,884113036 | SF3A2 | FALSE | Upregulated |
| -0,036965331 | 0,884260954 | POLH | FALSE | Downregulated |
| 0,062266793 | 0,884260954 | TYRO3 | FALSE | Upregulated |
| -0,041738245 | 0,88432751 | ZNF335 | FALSE | Downregulated |
| -0,028973339 | 0,884600675 | PCNT | FALSE | Downregulated |
| -0,211854643 | 0,884607874 | LINC00652 | FALSE | Downregulated |
| 0,030743588 | 0,884607874 | PKIG | FALSE | Upregulated |
| -0,035092113 | 0,884607874 | ST3GAL5 | FALSE | Downregulated |
| -0,029249633 | 0,884607874 | TJAP1 | FALSE | Downregulated |
| -0,12829319 | 0,884648359 | SCN4B | FALSE | Downregulated |
| -0,166796832 | 0,884880939 | DIRAS2 | FALSE | Downregulated |
| 0,023919908 | 0,884905793 | ARHGAP1 | FALSE | Upregulated |
| -0,198498474 | 0,884905793 | TWIST2 | FALSE | Downregulated |
| -0,034560901 | 0,884972857 | PP7080 | FALSE | Downregulated |
| -0,201275783 | 0,885047897 | CHST6 | FALSE | Downregulated |
| 0,063772659 | 0,885047897 | GLUD2 | FALSE | Upregulated |
| -0,032448707 | 0,885354572 | RPS17 | FALSE | Downregulated |
| 0,34475213 | 0,885395511 | PPIAL4B | FALSE | Upregulated |
| 0,019089025 | 0,88605189 | RB1CC1 | FALSE | Upregulated |
| 0,047785582 | 0,886068705 | ANKRD9 | FALSE | Upregulated |
| -0,155969727 | 0,886068705 | FER1L4 | FALSE | Downregulated |
| 0,028508225 | 0,886068705 | SPCS2 | FALSE | Upregulated |
| -0,035464983 | 0,886235588 | ZDHHC14 | FALSE | Downregulated |
| -0,08989012 | 0,886359346 | ZNF660 | FALSE | Downregulated |
| -0,072858383 | 0,8863713 | LOC100996255 | FALSE | Downregulated |
| -0,151190532 | 0,886376176 | LOC100996425 | FALSE | Downregulated |
| 0,028067521 | 0,886539039 | AMD1 | FALSE | Upregulated |
| -0,031964928 | 0,886585758 | COQ9 | FALSE | Downregulated |
| -0,084544094 | 0,886949394 | BTN2A3P | FALSE | Downregulated |
| 0,115489851 | 0,886949394 | GFM2 | FALSE | Upregulated |
| 0,082916788 | 0,886949394 | LOC100996664 | FALSE | Upregulated |
| -0,193716568 | 0,886949394 | LOC101929240 | FALSE | Downregulated |
| 0,017939836 | 0,887179277 | DSCR3 | FALSE | Upregulated |
| 0,026226587 | 0,887179277 | PLS1 | FALSE | Upregulated |
| -0,103319963 | 0,887311011 | POLR2J2 | FALSE | Downregulated |
| 0,024851184 | 0,887430008 | NDUFB5 | FALSE | Upregulated |
| -0,081442874 | 0,887683134 | LOC102723897 | FALSE | Downregulated |
| -0,023277086 | 0,88769416 | ARAF | FALSE | Downregulated |
| 0,02853243 | 0,887713836 | MTG1 | FALSE | Upregulated |
| 0,091133177 | 0,887713836 | SRCRB4D | FALSE | Upregulated |
| 0,23537125 | 0,887725048 | CCDC162P | FALSE | Upregulated |
| 1,199186189 | 0,887810203 | ANKRD1 | FALSE | Upregulated |
| -0,131242201 | 0,887810203 | ANXA8 | FALSE | Downregulated |
| -0,042272494 | 0,887810203 | BRD3 | FALSE | Downregulated |
| 0,167340545 | 0,887810203 | KCNK10 | FALSE | Upregulated |

| | | | | |
|---|---|---|---|---|
| -0,040761479 | 0,887810203 | KIAA1919 | FALSE | Downregulated |
| -0,035448178 | 0,887810203 | METAP2 | FALSE | Downregulated |
| -0,022993825 | 0,887810203 | NF1 | FALSE | Downregulated |
| -0,077878668 | 0,887810203 | PDZD2 | FALSE | Downregulated |
| -0,178816799 | 0,887810203 | POM121L8P | FALSE | Downregulated |
| -0,920296056 | 0,887810203 | SLX1B | FALSE | Downregulated |
| 0,025114505 | 0,88784113 | REXO2 | FALSE | Upregulated |
| 0,151632271 | 0,887845544 | TRNW | FALSE | Upregulated |
| -0,056360404 | 0,888014712 | KCTD15 | FALSE | Downregulated |
| 0,019776097 | 0,888028039 | DDX18 | FALSE | Upregulated |
| -0,032499988 | 0,888028039 | ERLEC1 | FALSE | Downregulated |
| 0,029495102 | 0,888028039 | MCM9 | FALSE | Upregulated |
| -0,056162321 | 0,888028039 | RABL2A | FALSE | Downregulated |
| -1,225933608 | 0,888028039 | RNVU1-18 | FALSE | Downregulated |
| 0,180756495 | 0,888028039 | TAS2R5 | FALSE | Upregulated |
| 0,034541612 | 0,888237177 | PDRG1 | FALSE | Upregulated |
| -0,045553606 | 0,888305595 | FCGR2B | FALSE | Downregulated |
| -0,187477829 | 0,888413482 | HAMP | FALSE | Downregulated |
| 0,103239212 | 0,888569112 | PIN4P1 | FALSE | Upregulated |
| -0,405441111 | 0,888742095 | SCARNA10 | FALSE | Downregulated |
| -0,035971418 | 0,888742095 | YLPM1 | FALSE | Downregulated |
| -0,108292711 | 0,889016187 | ZNF625 | FALSE | Downregulated |
| -0,181414159 | 0,88913001 | EEF1DP3 | FALSE | Downregulated |
| -0,58938971 | 0,88913001 | LOC100505841 | FALSE | Downregulated |
| 0,058733854 | 0,88913001 | PDGFRA | FALSE | Upregulated |
| 0,209167598 | 0,88913001 | SLC2A5 | FALSE | Upregulated |
| 0,138930101 | 0,88913001 | TBX18 | FALSE | Upregulated |
| 0,278416892 | 0,88913001 | TPTE2 | FALSE | Upregulated |
| -0,02642774 | 0,889232661 | IGBP1 | FALSE | Downregulated |
| -0,018251618 | 0,889234171 | OXSR1 | FALSE | Downregulated |
| 0,095119305 | 0,889288283 | HCG4 | FALSE | Upregulated |
| -0,073426268 | 0,889330772 | TBX19 | FALSE | Downregulated |
| 0,084257216 | 0,889399204 | LOC101927420 | FALSE | Upregulated |
| 0,026471057 | 0,88953378 | FAM222B | FALSE | Upregulated |
| -0,156086822 | 0,88953378 | RASL12 | FALSE | Downregulated |
| 0,041528285 | 0,889557203 | CTSH | FALSE | Upregulated |
| -0,091567893 | 0,889557203 | TMEM132E | FALSE | Downregulated |
| 0,144484691 | 0,88959166 | AC002116.7 | FALSE | Upregulated |
| 0,046930825 | 0,88959166 | ACTRT3 | FALSE | Upregulated |
| -0,029594466 | 0,88959166 | FOXP4 | FALSE | Downregulated |
| 0,109001029 | 0,88959166 | LOC646278 | FALSE | Upregulated |
| -0,02702601 | 0,88959166 | RPS20 | FALSE | Downregulated |
| 0,019354569 | 0,88959166 | YTHDF3 | FALSE | Upregulated |
| -0,080221244 | 0,889844524 | KIF7 | FALSE | Downregulated |
| -0,134194958 | 0,889844524 | TG | FALSE | Downregulated |
| -0,027068639 | 0,890315909 | BMS1 | FALSE | Downregulated |
| 0,052843899 | 0,890641019 | GPR4 | FALSE | Upregulated |
| -0,028988503 | 0,890880026 | KATNBL1 | FALSE | Downregulated |
| -0,032332392 | 0,890880026 | NELFE | FALSE | Downregulated |
| 0,195777944 | 0,890911829 | GPRIN2 | FALSE | Upregulated |
| -0,178116383 | 0,890911829 | OCLM | FALSE | Downregulated |

| | | | | |
|---|---|---|---|---|
| 0,124254624 | 0,890911829 | PTGDR2 | FALSE | Upregulated |
| 0,09638249 | 0,89115691 | CREB5 | FALSE | Upregulated |
| -0,045773426 | 0,891226736 | LOC100288778 | FALSE | Downregulated |
| 0,033252256 | 0,891276943 | TOR1AIP2 | FALSE | Upregulated |
| 0,155314163 | 0,891419957 | RNA18S5 | FALSE | Upregulated |
| -0,03218195 | 0,891574355 | DCAF4 | FALSE | Downregulated |
| -0,077749566 | 0,891647605 | KDM4D | FALSE | Downregulated |
| 0,035920617 | 0,892189366 | NDUFAF3 | FALSE | Upregulated |
| -0,141788429 | 0,892316236 | LOC101926917 | FALSE | Downregulated |
| -0,02106424 | 0,892316236 | ZBTB1 | FALSE | Downregulated |
| 0,091095825 | 0,892479621 | FHL3 | FALSE | Upregulated |
| 0,020249339 | 0,892479621 | QTRTD1 | FALSE | Upregulated |
| 0,032180603 | 0,892479621 | SNX21 | FALSE | Upregulated |
| -0,057868027 | 0,892479621 | ZNF674-AS1 | FALSE | Downregulated |
| -0,073715512 | 0,892707848 | GJC1 | FALSE | Downregulated |
| -0,017583421 | 0,892747901 | ATAD1 | FALSE | Downregulated |
| -0,023093892 | 0,892747901 | VAPB | FALSE | Downregulated |
| -0,095447525 | 0,892747901 | ZGLP1 | FALSE | Downregulated |
| 0,16359271 | 0,892815537 | POM121L10P | FALSE | Upregulated |
| -0,085747548 | 0,892815537 | SULT1C4 | FALSE | Downregulated |
| 0,042099052 | 0,89294866 | FKBPL | FALSE | Upregulated |
| -0,01875571 | 0,893089036 | KIAA0368 | FALSE | Downregulated |
| -0,203611945 | 0,893089036 | LOC102723735 | FALSE | Downregulated |
| -0,053454386 | 0,893089036 | PPIG | FALSE | Downregulated |
| -0,27500162 | 0,893089036 | PSG4 | FALSE | Downregulated |
| -0,024819947 | 0,893089036 | TOLLIP | FALSE | Downregulated |
| 0,056742635 | 0,893132962 | TSHZ3 | FALSE | Upregulated |
| 0,030209153 | 0,893158953 | TMED7 | FALSE | Upregulated |
| 0,033959606 | 0,8932957 | RBM14 | FALSE | Upregulated |
| 0,029110221 | 0,893408599 | NANP | FALSE | Upregulated |
| -0,179779007 | 0,893547185 | DCD | FALSE | Downregulated |
| 0,034027948 | 0,893643285 | BZW2 | FALSE | Upregulated |
| -0,045866634 | 0,893643285 | RARB | FALSE | Downregulated |
| 0,037101501 | 0,893784399 | PPL | FALSE | Upregulated |
| -0,032324398 | 0,893919612 | ASXL2 | FALSE | Downregulated |
| -0,015904072 | 0,89402397 | TERF2 | FALSE | Downregulated |
| 0,097109296 | 0,894464638 | RP11-844P9.2 | FALSE | Upregulated |
| 0,205577355 | 0,894464638 | SCNN1B | FALSE | Upregulated |
| 0,053546613 | 0,894464638 | ZNF813 | FALSE | Upregulated |
| -0,162301831 | 0,894520919 | GPR63 | FALSE | Downregulated |
| 0,171077599 | 0,894609775 | RBM24 | FALSE | Upregulated |
| -0,054029103 | 0,89467531 | PTGIS | FALSE | Downregulated |
| -0,036972754 | 0,894720715 | C10orf11 | FALSE | Downregulated |
| 0,030409655 | 0,894741603 | PM20D2 | FALSE | Upregulated |
| -0,174653712 | 0,894756748 | RAB6C-AS1 | FALSE | Downregulated |
| -0,068165042 | 0,894771187 | ZNF423 | FALSE | Downregulated |
| 0,04925593 | 0,89478047 | C4orf33 | FALSE | Upregulated |
| 0,044711225 | 0,89478047 | CRYBB2P1 | FALSE | Upregulated |
| 0,022443235 | 0,89478047 | CWF19L1 | FALSE | Upregulated |
| 0,162436244 | 0,89478047 | TSPY26P | FALSE | Upregulated |
| 0,060781671 | 0,89485998 | ABCC11 | FALSE | Upregulated |

| | | | | |
|---|---|---|---|---|
| 0,028531673 | 0,894900772 | ZAK | FALSE | Upregulated |
| -0,026869561 | 0,894940018 | PHF20L1 | FALSE | Downregulated |
| 0,024905112 | 0,894940018 | POLR2F | FALSE | Upregulated |
| -0,018352302 | 0,894960686 | TMEM131 | FALSE | Downregulated |
| 0,025924543 | 0,895025747 | PPAPDC2 | FALSE | Upregulated |
| 0,109287481 | 0,895070555 | CDH13 | FALSE | Upregulated |
| -0,038858438 | 0,895391894 | LUZP1 | FALSE | Downregulated |
| -0,109545331 | 0,895391894 | STX1A | FALSE | Downregulated |
| -0,09672204 | 0,895579512 | ZNF391 | FALSE | Downregulated |
| -0,02433569 | 0,895695515 | NKRF | FALSE | Downregulated |
| 0,023911937 | 0,895701885 | C2 | FALSE | Upregulated |
| -0,035475319 | 0,895742297 | ANAPC10 | FALSE | Downregulated |
| -0,020046536 | 0,895742297 | SAP130 | FALSE | Downregulated |
| 0,077410883 | 0,89588214 | NPAS2 | FALSE | Upregulated |
| -0,225976131 | 0,89592465 | TP53AIP1 | FALSE | Downregulated |
| -0,04281654 | 0,895945852 | PSPH | FALSE | Downregulated |
| 0,262254714 | 0,895967516 | GPR3 | FALSE | Upregulated |
| -0,263713011 | 0,895967516 | TRIM63 | FALSE | Downregulated |
| -0,169791097 | 0,895997436 | LOC100128568 | FALSE | Downregulated |
| -0,228248613 | 0,895997436 | RP11-10A14.5 | FALSE | Downregulated |
| 0,027053653 | 0,895997436 | ZDHHC4 | FALSE | Upregulated |
| 0,033924416 | 0,896105576 | SLC39A7 | FALSE | Upregulated |
| -0,026130419 | 0,896112378 | EEF2K | FALSE | Downregulated |
| -0,02474914 | 0,896212084 | PRR12 | FALSE | Downregulated |
| 0,074940077 | 0,896520132 | LOC102724550 | FALSE | Upregulated |
| -0,12337965 | 0,896621144 | LOC102724436 | FALSE | Downregulated |
| -0,157637824 | 0,896628202 | LOC101927044 | FALSE | Downregulated |
| -0,180922289 | 0,896628202 | LOC102724740 | FALSE | Downregulated |
| -0,052353002 | 0,896628202 | TMEM97 | FALSE | Downregulated |
| 0,018117895 | 0,896650124 | EIF1B | FALSE | Upregulated |
| 0,028577107 | 0,896650124 | FAM21C | FALSE | Upregulated |
| 0,087812121 | 0,896650124 | GAL3ST1 | FALSE | Upregulated |
| -0,028515626 | 0,896650124 | GALK2 | FALSE | Downregulated |
| -0,113633391 | 0,896650124 | SNORA63 | FALSE | Downregulated |
| 0,176154394 | 0,897066914 | ADAMTS14 | FALSE | Upregulated |
| -0,210268644 | 0,897493487 | LOC102723721 | FALSE | Downregulated |
| 0,024141228 | 0,897493487 | SLC7A6OS | FALSE | Upregulated |
| -0,015853888 | 0,897493487 | SNAPC3 | FALSE | Downregulated |
| 0,039434923 | 0,897514032 | AP3M2 | FALSE | Upregulated |
| -0,154161068 | 0,897514032 | NOV | FALSE | Downregulated |
| -0,03548789 | 0,897514032 | RPL13A | FALSE | Downregulated |
| -0,078468217 | 0,897514032 | SEMA3D | FALSE | Downregulated |
| -0,413313011 | 0,897515628 | MYH4 | FALSE | Downregulated |
| -0,066716636 | 0,897515628 | ZNF718 | FALSE | Downregulated |
| 0,094129752 | 0,897801564 | EPCAM | FALSE | Upregulated |
| -0,10349011 | 0,897861944 | PYGO1 | FALSE | Downregulated |
| -0,017641989 | 0,897928716 | CRNKL1 | FALSE | Downregulated |
| -0,167731445 | 0,898311148 | HAGHL | FALSE | Downregulated |
| -0,112385417 | 0,898311148 | SPTBN5 | FALSE | Downregulated |
| 0,197469489 | 0,8983235 | BAI2 | FALSE | Upregulated |
| -0,055132226 | 0,8983235 | DDR1 | FALSE | Downregulated |

| | | | | |
|---|---|---|---|---|
| 0,081226395 | 0,8983235 | FBXO24 | FALSE | Upregulated |
| -0,058877396 | 0,8983235 | GPR75 | FALSE | Downregulated |
| 0,079283933 | 0,8983235 | LOC100288181 | FALSE | Upregulated |
| 0,053490976 | 0,898435308 | TTPAL | FALSE | Upregulated |
| 0,027897859 | 0,898603433 | CFL2 | FALSE | Upregulated |
| -0,024879513 | 0,898603433 | PRKAR1A | FALSE | Downregulated |
| -0,020297301 | 0,898603433 | TMEM43 | FALSE | Downregulated |
| -0,106238704 | 0,898750817 | ASMTL-AS1 | FALSE | Downregulated |
| 0,05479913 | 0,898818508 | TRAF3IP1 | FALSE | Upregulated |
| 0,030841391 | 0,89884777 | CYP7B1 | FALSE | Upregulated |
| 0,026129217 | 0,899053063 | RPL35A | FALSE | Upregulated |
| 0,033958303 | 0,8991076 | FBXO21 | FALSE | Upregulated |
| 0,029307362 | 0,899197384 | INTS5 | FALSE | Upregulated |
| -0,021848049 | 0,899236759 | KRI1 | FALSE | Downregulated |
| -0,212594818 | 0,899236759 | PKP3 | FALSE | Downregulated |
| -0,28239785 | 0,899288873 | CHRNA7 | FALSE | Downregulated |
| -0,028984413 | 0,89937181 | BAHD1 | FALSE | Downregulated |
| -0,026852196 | 0,899566985 | C18orf21 | FALSE | Downregulated |
| 0,131763079 | 0,899669003 | SENCR | FALSE | Upregulated |
| -0,125000882 | 0,899741168 | C6orf164 | FALSE | Downregulated |
| -0,027441882 | 0,899939164 | YEATS4 | FALSE | Downregulated |
| 0,021011193 | 0,899968542 | POLR3A | FALSE | Upregulated |
| 0,093668097 | 0,90002442 | ST7-AS1 | FALSE | Upregulated |
| 0,068905573 | 0,90002442 | VILL | FALSE | Upregulated |
| 0,05793882 | 0,900033706 | GALNT14 | FALSE | Upregulated |
| -0,121263775 | 0,900033706 | S100A12 | FALSE | Downregulated |
| 0,171645131 | 0,900033706 | TPSAB1 | FALSE | Upregulated |
| -0,240243642 | 0,90011058 | C2orf91 | FALSE | Downregulated |
| -0,027067215 | 0,900125699 | ARHGAP26 | FALSE | Downregulated |
| 0,188753098 | 0,900125699 | GRM1 | FALSE | Upregulated |
| 0,202482977 | 0,900125699 | MUC4 | FALSE | Upregulated |
| -0,023611226 | 0,900125699 | PCM1 | FALSE | Downregulated |
| -0,023969617 | 0,900125699 | PDCD5 | FALSE | Downregulated |
| 0,075009703 | 0,900125699 | SPARCL1 | FALSE | Upregulated |
| -0,050347797 | 0,900125699 | TM7SF2 | FALSE | Downregulated |
| 0,076028823 | 0,900132225 | ANXA2R | FALSE | Upregulated |
| 0,049021 | 0,900132225 | CCDC138 | FALSE | Upregulated |
| -0,028244501 | 0,900132225 | ERCC4 | FALSE | Downregulated |
| -0,044796461 | 0,900132225 | FBF1 | FALSE | Downregulated |
| 0,014466161 | 0,900132225 | GTF2H1 | FALSE | Upregulated |
| 0,06993667 | 0,900132225 | LOC101927216 | FALSE | Upregulated |
| 0,029407938 | 0,900132225 | NDUFA4 | FALSE | Upregulated |
| -0,073714539 | 0,900132225 | RP5-1021I20.1 | FALSE | Downregulated |
| 0,133739959 | 0,900434584 | ADORA2B | FALSE | Upregulated |
| 0,01541168 | 0,90053728 | FAM208A | FALSE | Upregulated |
| 0,145975589 | 0,90053728 | NTS | FALSE | Upregulated |
| 0,023866363 | 0,900551288 | UPRT | FALSE | Upregulated |
| -0,031374841 | 0,900637696 | METTL22 | FALSE | Downregulated |
| 0,054554774 | 0,900753714 | SOX18 | FALSE | Upregulated |
| -0,096225677 | 0,900754605 | NEAT1 | FALSE | Downregulated |
| -0,026537706 | 0,900789585 | SACM1L | FALSE | Downregulated |

| | | | | |
|---|---|---|---|---|
| 0,031800976 | 0,90085869 | DNAL4 | FALSE | Upregulated |
| 0,034345348 | 0,90085869 | GNL3 | FALSE | Upregulated |
| -0,14045141 | 0,90085869 | LRRC7 | FALSE | Downregulated |
| -0,164480642 | 0,90085869 | ST7-OT4 | FALSE | Downregulated |
| 0,049595153 | 0,90085869 | ZNF532 | FALSE | Upregulated |
| 0,155729684 | 0,900933289 | KCNJ2-AS1 | FALSE | Upregulated |
| -0,036745306 | 0,900933289 | NGEF | FALSE | Downregulated |
| -0,019846769 | 0,900933289 | PBXIP1 | FALSE | Downregulated |
| 0,030616056 | 0,901003021 | LINC00623 | FALSE | Upregulated |
| -0,061080217 | 0,901003021 | LINC00957 | FALSE | Downregulated |
| -0,022645635 | 0,901003021 | SHANK2 | FALSE | Downregulated |
| -0,028292646 | 0,901003021 | TMEM18 | FALSE | Downregulated |
| 0,164784318 | 0,901315613 | FBXL13 | FALSE | Upregulated |
| 0,027653944 | 0,901315613 | NR1H3 | FALSE | Upregulated |
| 0,016965462 | 0,901315613 | STRAP | FALSE | Upregulated |
| 0,109507894 | 0,901315613 | TMEM121 | FALSE | Upregulated |
| -0,014576707 | 0,901449905 | AP3S1 | FALSE | Downregulated |
| -0,175756728 | 0,901459834 | NCOR1P1 | FALSE | Downregulated |
| -0,025489343 | 0,901459834 | POP5 | FALSE | Downregulated |
| -0,03743857 | 0,90182286 | TM4SF1 | FALSE | Downregulated |
| 0,036082033 | 0,901899959 | CCPG1 | FALSE | Upregulated |
| 0,225851186 | 0,901964998 | SND1-IT1 | FALSE | Upregulated |
| 0,016542537 | 0,902019986 | PLXNB2 | FALSE | Upregulated |
| -0,02206929 | 0,902280253 | ATF7IP | FALSE | Downregulated |
| 0,068169462 | 0,902349282 | PNMAL2 | FALSE | Upregulated |
| 0,081552163 | 0,902349282 | RP5-968D22.3 | FALSE | Upregulated |
| -0,134170171 | 0,902350452 | GDF10 | FALSE | Downregulated |
| -0,032675941 | 0,902502465 | MSRA | FALSE | Downregulated |
| -0,025091952 | 0,902883317 | RPP25L | FALSE | Downregulated |
| -0,213234578 | 0,902891628 | ABCC8 | FALSE | Downregulated |
| -0,043056639 | 0,902891628 | MAPK8IP1 | FALSE | Downregulated |
| -0,019303274 | 0,902891628 | PBRM1 | FALSE | Downregulated |
| -0,026225717 | 0,902891628 | TPD52L1 | FALSE | Downregulated |
| -0,085575027 | 0,902964804 | PNMAL1 | FALSE | Downregulated |
| -0,044179888 | 0,902980402 | REL | FALSE | Downregulated |
| 0,099730705 | 0,902983346 | RP11-473M20.7 | FALSE | Upregulated |
| -0,043942557 | 0,903146663 | MAST4 | FALSE | Downregulated |
| -0,182952545 | 0,903259121 | HIGD1B | FALSE | Downregulated |
| -0,114006528 | 0,903309681 | LOC102724788 | FALSE | Downregulated |
| 0,031575687 | 0,903517703 | SPRED1 | FALSE | Upregulated |
| -0,025882231 | 0,903554004 | C19orf53 | FALSE | Downregulated |
| 1,060218893 | 0,903589654 | HIST2H2AA3 | FALSE | Upregulated |
| 0,068219855 | 0,903589654 | MFI2 | FALSE | Upregulated |
| 0,014739295 | 0,903589654 | TRPC4AP | FALSE | Upregulated |
| -0,328738153 | 0,903609981 | LINC00966 | FALSE | Downregulated |
| 0,042356345 | 0,903841863 | AURKC | FALSE | Upregulated |
| -0,025163858 | 0,904014837 | RPAP2 | FALSE | Downregulated |
| 0,212236774 | 0,904054336 | LOC102724614 | FALSE | Upregulated |
| 0,172007499 | 0,904075381 | HIST1H2AG | FALSE | Upregulated |
| -0,067581871 | 0,904075381 | HSPA12B | FALSE | Downregulated |
| 0,081378923 | 0,904075381 | LOC100506734 | FALSE | Upregulated |

| | | | | |
|---|---|---|---|---|
| -0,082705532 | 0,904140413 | FLT3 | FALSE | Downregulated |
| -0,053870161 | 0,904140413 | ITGB7 | FALSE | Downregulated |
| 0,089017242 | 0,904163161 | C12orf75 | FALSE | Upregulated |
| -0,109501268 | 0,9042309 | CASC1 | FALSE | Downregulated |
| -0,112162042 | 0,9042309 | COX6A2 | FALSE | Downregulated |
| -0,064746433 | 0,9042309 | ZNF165 | FALSE | Downregulated |
| -0,080307819 | 0,904290477 | LOC101928415 | FALSE | Downregulated |
| 0,022999704 | 0,904290477 | PARP3 | FALSE | Upregulated |
| -0,164108132 | 0,904403243 | CALCR | FALSE | Downregulated |
| -0,073395211 | 0,904403243 | LOC101928228 | FALSE | Downregulated |
| 0,225988114 | 0,904403243 | LOC101928466 | FALSE | Upregulated |
| -0,052364694 | 0,904403243 | RBMS2 | FALSE | Downregulated |
| 0,171978698 | 0,904424493 | TMEM31 | FALSE | Upregulated |
| -0,110979471 | 0,904550639 | WT1 | FALSE | Downregulated |
| -0,052544532 | 0,904667281 | AKR7L | FALSE | Downregulated |
| -0,021017451 | 0,904831995 | CCDC92 | FALSE | Downregulated |
| -0,015572774 | 0,90497152 | TAF5L | FALSE | Downregulated |
| 0,026307994 | 0,905202997 | CAMSAP3 | FALSE | Upregulated |
| -0,030653072 | 0,905202997 | DLG5 | FALSE | Downregulated |
| -0,13052537 | 0,905202997 | LOC729732 | FALSE | Downregulated |
| 0,200451253 | 0,905202997 | RGPD2 | FALSE | Upregulated |
| -0,25938643 | 0,905227118 | HES2 | FALSE | Downregulated |
| -0,212815673 | 0,905411909 | CA4 | FALSE | Downregulated |
| -0,058885448 | 0,905411909 | CERCAM | FALSE | Downregulated |
| -0,019771246 | 0,905411909 | EDC3 | FALSE | Downregulated |
| -0,030596221 | 0,905411909 | ERCC6L2 | FALSE | Downregulated |
| 0,031227402 | 0,905411909 | GMDS-AS1 | FALSE | Upregulated |
| -0,119792247 | 0,905411909 | GRIK2 | FALSE | Downregulated |
| 0,018557686 | 0,905411909 | KBTBD2 | FALSE | Upregulated |
| 0,137622918 | 0,905411909 | NACAD | FALSE | Upregulated |
| -0,12273154 | 0,905411909 | NTRK3 | FALSE | Downregulated |
| 0,089565548 | 0,905411909 | PTGS2 | FALSE | Upregulated |
| 0,032134414 | 0,905411909 | SERPINA11 | FALSE | Upregulated |
| 0,024287225 | 0,905411909 | SFMBT1 | FALSE | Upregulated |
| 0,281905705 | 0,905411909 | SYT2 | FALSE | Upregulated |
| 0,019445019 | 0,905456325 | TNPO2 | FALSE | Upregulated |
| 0,087268722 | 0,905521415 | LOC284454 | FALSE | Upregulated |
| 0,103870747 | 0,905719632 | TNFSF14 | FALSE | Upregulated |
| 0,127945774 | 0,905744828 | LINC00839 | FALSE | Upregulated |
| 0,093371048 | 0,905769233 | TMEM200C | FALSE | Upregulated |
| -0,199929623 | 0,905771491 | ITGB6 | FALSE | Downregulated |
| -0,145570049 | 0,905937741 | PWP2 | FALSE | Downregulated |
| 0,018554022 | 0,906047097 | MBNL1 | FALSE | Upregulated |
| 0,049402197 | 0,906047097 | RAB43 | FALSE | Upregulated |
| -0,014498152 | 0,906047097 | TCF12 | FALSE | Downregulated |
| 0,064450719 | 0,906087565 | ADAMTS12 | FALSE | Upregulated |
| 0,01756647 | 0,906087565 | PRPF4 | FALSE | Upregulated |
| 0,039049131 | 0,90619295 | SYTL4 | FALSE | Upregulated |
| 0,018642506 | 0,906218299 | DCAF12 | FALSE | Upregulated |
| -0,059042799 | 0,906313708 | LOC728024 | FALSE | Downregulated |
| -0,049582501 | 0,906388773 | ZSCAN20 | FALSE | Downregulated |

| | | | | |
|---|---|---|---|---|
| -0,046511348 | 0,906419329 | ANKRD35 | FALSE | Downregulated |
| -0,032580065 | 0,906419329 | TPM3P9 | FALSE | Downregulated |
| 0,056348596 | 0,906419329 | UBXN4 | FALSE | Upregulated |
| -0,028114679 | 0,906471815 | ANKRD54 | FALSE | Downregulated |
| 0,06772466 | 0,906471815 | CACNA1A | FALSE | Upregulated |
| -0,412761045 | 0,906471815 | CNTNAP5 | FALSE | Downregulated |
| 0,064340238 | 0,906471815 | CTSK | FALSE | Upregulated |
| -0,039903071 | 0,906471815 | DHFR | FALSE | Downregulated |
| 0,075694974 | 0,906471815 | KLRF1 | FALSE | Upregulated |
| 0,029939436 | 0,906471815 | NMNAT1 | FALSE | Upregulated |
| 0,021006899 | 0,906471815 | RNF20 | FALSE | Upregulated |
| -0,043192348 | 0,906471815 | TGFA | FALSE | Downregulated |
| 0,025522807 | 0,906477556 | FBXO30 | FALSE | Upregulated |
| -0,022398564 | 0,906489712 | NSD1 | FALSE | Downregulated |
| 0,091469855 | 0,906493613 | LOC101929574 | FALSE | Upregulated |
| 0,209400085 | 0,906513557 | LOC102724205 | FALSE | Upregulated |
| 0,09908054 | 0,906584938 | C14orf164 | FALSE | Upregulated |
| 0,133885285 | 0,906610884 | LOC101929918 | FALSE | Upregulated |
| -0,096964043 | 0,906615223 | LOC102060414 | FALSE | Downregulated |
| 0,132979026 | 0,906992643 | SYCP2L | FALSE | Upregulated |
| 0,033086303 | 0,907029363 | ARCN1 | FALSE | Upregulated |
| -0,133427865 | 0,907029363 | C3orf80 | FALSE | Downregulated |
| 0,076894718 | 0,907029363 | ENPP5 | FALSE | Upregulated |
| 0,048945128 | 0,907029363 | EXOC3L1 | FALSE | Upregulated |
| 0,122357134 | 0,907029363 | LOC101927446 | FALSE | Upregulated |
| -0,135934931 | 0,907029363 | REM1 | FALSE | Downregulated |
| 0,091256346 | 0,907171714 | ROBO2 | FALSE | Upregulated |
| 0,024092095 | 0,90726387 | CCDC71 | FALSE | Upregulated |
| -0,118578551 | 0,90726387 | FCGR2C | FALSE | Downregulated |
| -0,125942293 | 0,90726387 | LOC400655 | FALSE | Downregulated |
| -0,024383396 | 0,90726387 | MGAT2 | FALSE | Downregulated |
| 0,049413158 | 0,907667996 | UNC5B | FALSE | Upregulated |
| 0,285863018 | 0,90772993 | TRNAG22 | FALSE | Upregulated |
| -0,03181739 | 0,90772993 | UGT2B15 | FALSE | Downregulated |
| 0,019798532 | 0,90772993 | YARS2 | FALSE | Upregulated |
| 0,177384995 | 0,907735913 | MYRFL | FALSE | Upregulated |
| 0,053272164 | 0,908095836 | FBXO32 | FALSE | Upregulated |
| 0,025919006 | 0,908095836 | FLNB | FALSE | Upregulated |
| 0,087131477 | 0,90812701 | ADORA1 | FALSE | Upregulated |
| -0,014470858 | 0,90812701 | GAPVD1 | FALSE | Downregulated |
| 0,017246475 | 0,90812701 | IKBKAP | FALSE | Upregulated |
| -0,105742055 | 0,90812701 | LINC00965 | FALSE | Downregulated |
| -0,084723594 | 0,90812701 | RANBP17 | FALSE | Downregulated |
| -0,0443912 | 0,90812701 | RP11-1072A3.2 | FALSE | Downregulated |
| -0,024136832 | 0,90812701 | SMYD5 | FALSE | Downregulated |
| -0,064074067 | 0,90812701 | STXBP6 | FALSE | Downregulated |
| -0,031081412 | 0,90812701 | TET2 | FALSE | Downregulated |
| 0,015280448 | 0,90812701 | TK2 | FALSE | Upregulated |
| -0,033400044 | 0,90812701 | ULK1 | FALSE | Downregulated |
| -0,108191775 | 0,90812701 | UTS2B | FALSE | Downregulated |
| 0,021621603 | 0,90812701 | ZNF407 | FALSE | Upregulated |

| | | | | |
|---|---|---|---|---|
| -0,151254729 | 0,908324022 | WTH3DI | FALSE | Downregulated |
| 0,141584412 | 0,908351294 | PDPN | FALSE | Upregulated |
| -0,263098233 | 0,908363067 | KLHL38 | FALSE | Downregulated |
| 0,034416934 | 0,90840153 | IL22RA1 | FALSE | Upregulated |
| -0,04528607 | 0,908418924 | ARHGAP23 | FALSE | Downregulated |
| -0,021258044 | 0,908713418 | ZMYM6NB | FALSE | Downregulated |
| -1,004784623 | 0,908915397 | H19 | FALSE | Downregulated |
| 0,017203809 | 0,908915397 | MRPL39 | FALSE | Upregulated |
| 0,040972015 | 0,908915397 | PLD6 | FALSE | Upregulated |
| 0,047803861 | 0,908915397 | SLC16A4 | FALSE | Upregulated |
| 0,03055313 | 0,908990819 | CFB | FALSE | Upregulated |
| 0,019819053 | 0,909042914 | HIPK1 | FALSE | Upregulated |
| -0,147526492 | 0,909042914 | RNF139-AS1 | FALSE | Downregulated |
| 0,089343541 | 0,909217167 | LOC102724184 | FALSE | Upregulated |
| -0,113524044 | 0,909217167 | SLC13A4 | FALSE | Downregulated |
| -0,029887278 | 0,90947126 | RBBP5 | FALSE | Downregulated |
| 0,042359156 | 0,909538781 | WNT3 | FALSE | Upregulated |
| 0,030735601 | 0,909740255 | PXDN | FALSE | Upregulated |
| 0,01432681 | 0,909812612 | DNTTIP2 | FALSE | Upregulated |
| 0,080958729 | 0,909831675 | CXorf31 | FALSE | Upregulated |
| 0,028251829 | 0,909831675 | DIS3L2 | FALSE | Upregulated |
| 0,026000894 | 0,909831675 | PPP2R5B | FALSE | Upregulated |
| 0,023739811 | 0,909831675 | R3HDM1 | FALSE | Upregulated |
| 0,023113943 | 0,909831675 | SPATA5L1 | FALSE | Upregulated |
| -0,126914683 | 0,909831675 | TEKT5 | FALSE | Downregulated |
| -0,118235824 | 0,909955911 | FBXO22-AS1 | FALSE | Downregulated |
| 0,092852703 | 0,909955911 | GPC3 | FALSE | Upregulated |
| -0,196645599 | 0,909955911 | LOC100505622 | FALSE | Downregulated |
| -0,104721985 | 0,909955911 | LOC102724445 | FALSE | Downregulated |
| 0,372768939 | 0,909955911 | PRODH | FALSE | Upregulated |
| -0,028316434 | 0,909955911 | PRR5 | FALSE | Downregulated |
| -0,042934971 | 0,909955911 | RAD54B | FALSE | Downregulated |
| 0,022074471 | 0,910150831 | SLC3A2 | FALSE | Upregulated |
| 0,023990089 | 0,910447135 | MYBBP1A | FALSE | Upregulated |
| 0,05370414 | 0,910542447 | C17orf100 | FALSE | Upregulated |
| -0,067916355 | 0,910584505 | LOC100506388 | FALSE | Downregulated |
| 0,030400477 | 0,910584505 | MEX3D | FALSE | Upregulated |
| 0,029279326 | 0,910623444 | PLEKHJ1 | FALSE | Upregulated |
| -0,069873489 | 0,910914767 | HNRNPA3P1 | FALSE | Downregulated |
| -0,012016816 | 0,911210993 | PAIP1 | FALSE | Downregulated |
| 0,056969884 | 0,911237769 | ANGPTL4 | FALSE | Upregulated |
| -0,073790096 | 0,911237769 | ELOVL7 | FALSE | Downregulated |
| 0,02278485 | 0,911237769 | H3F3A | FALSE | Upregulated |
| -0,127447418 | 0,911237769 | PPIAL4G | FALSE | Downregulated |
| 0,218156138 | 0,911237769 | SCGB2B3P | FALSE | Upregulated |
| -0,033117692 | 0,911237769 | TMEM80 | FALSE | Downregulated |
| -0,043607429 | 0,911357047 | LRRC37A | FALSE | Downregulated |
| -0,017133333 | 0,911357047 | RANBP9 | FALSE | Downregulated |
| -0,022409313 | 0,911375082 | ORMDL3 | FALSE | Downregulated |
| -0,043217562 | 0,911387764 | SLC17A4 | FALSE | Downregulated |
| -0,022854715 | 0,911414785 | AGAP1 | FALSE | Downregulated |

| | | | | |
|---|---|---|---|---|
| 0,229511231 | 0,91143394 | LOC101928080 | FALSE | Upregulated |
| -0,031080837 | 0,91143394 | SETD9 | FALSE | Downregulated |
| -0,144659644 | 0,911773866 | ARHGAP8 | FALSE | Downregulated |
| 0,038758761 | 0,911773866 | GPR128 | FALSE | Upregulated |
| 0,147599445 | 0,911773866 | KCNQ5 | FALSE | Upregulated |
| 0,049930649 | 0,911773866 | LOC100132891 | FALSE | Upregulated |
| -0,104162248 | 0,911773866 | PTGER3 | FALSE | Downregulated |
| 0,149446117 | 0,911773866 | SLC4A11 | FALSE | Upregulated |
| -0,019408944 | 0,911883368 | CNOT6L | FALSE | Downregulated |
| -0,075072417 | 0,911883368 | DBH | FALSE | Downregulated |
| -0,199674661 | 0,911883368 | LOC101927721 | FALSE | Downregulated |
| 0,016982399 | 0,911883368 | PBX2 | FALSE | Upregulated |
| 0,058445408 | 0,911892614 | GFRA2 | FALSE | Upregulated |
| -0,111596902 | 0,911892614 | LOC102723683 | FALSE | Downregulated |
| -0,155998047 | 0,912095497 | LOC101928659 | FALSE | Downregulated |
| 0,013836433 | 0,912100616 | USP24 | FALSE | Upregulated |
| 0,171277122 | 0,912165898 | CYP4F35P | FALSE | Upregulated |
| 0,024984623 | 0,912174656 | CAMK1 | FALSE | Upregulated |
| -0,026057393 | 0,912174656 | GABARAPL1 | FALSE | Downregulated |
| 0,100248175 | 0,912174656 | HCG4B | FALSE | Upregulated |
| -0,047509773 | 0,912174656 | IGLON5 | FALSE | Downregulated |
| -0,027300041 | 0,912174656 | NUGGC | FALSE | Downregulated |
| 0,025323101 | 0,912174656 | PPP6R2 | FALSE | Upregulated |
| -0,113618524 | 0,912174656 | TECTB | FALSE | Downregulated |
| -0,032274804 | 0,912174656 | TMEM110-MUSTN1 | FALSE | Downregulated |
| 0,104360619 | 0,912408704 | LOC101241902 | FALSE | Upregulated |
| 0,073017925 | 0,912408704 | PAQR5 | FALSE | Upregulated |
| 0,014409655 | 0,912408704 | PTP4A2 | FALSE | Upregulated |
| 0,020982784 | 0,912408704 | TBCK | FALSE | Upregulated |
| 0,084152711 | 0,912492071 | RPL13AP3 | FALSE | Upregulated |
| 0,028246716 | 0,912492071 | TMEM53 | FALSE | Upregulated |
| 0,953756083 | 0,912733768 | GTF2IP1 | FALSE | Upregulated |
| 0,022193821 | 0,912814592 | AGPS | FALSE | Upregulated |
| 0,029275456 | 0,912814592 | FASTKD1 | FALSE | Upregulated |
| -0,065621011 | 0,912814592 | LOC100506258 | FALSE | Downregulated |
| -0,146508637 | 0,912814592 | PI15 | FALSE | Downregulated |
| -0,025023035 | 0,912855253 | COX14 | FALSE | Downregulated |
| -0,046092875 | 0,912976796 | FOLH1B | FALSE | Downregulated |
| -0,059435222 | 0,913061738 | SUSD3 | FALSE | Downregulated |
| -0,033597797 | 0,913200365 | SRSF10 | FALSE | Downregulated |
| 0,028675123 | 0,913200365 | UROS | FALSE | Upregulated |
| -0,069207038 | 0,913361752 | LOC100996763 | FALSE | Downregulated |
| 0,054824971 | 0,913535318 | ACTA2 | FALSE | Upregulated |
| 0,076700467 | 0,913535318 | ATE1-AS1 | FALSE | Upregulated |
| -0,14500238 | 0,913535318 | C1orf229 | FALSE | Downregulated |
| 0,035063923 | 0,913535318 | CHST7 | FALSE | Upregulated |
| 0,02489558 | 0,913535318 | DUS2 | FALSE | Upregulated |
| -0,064727302 | 0,913535318 | LIFR-AS1 | FALSE | Downregulated |
| 0,113731866 | 0,913535318 | LOC101928965 | FALSE | Upregulated |
| 0,166087513 | 0,913535318 | TRNAG20 | FALSE | Upregulated |

| | | | | |
|---|---|---|---|---|
| 0,089850722 | 0,913597295 | ASTN1 | FALSE | Upregulated |
| -0,136688934 | 0,913597295 | C10orf105 | FALSE | Downregulated |
| 0,043956264 | 0,913597295 | COL6A3 | FALSE | Upregulated |
| 0,030357265 | 0,913597295 | GNB5 | FALSE | Upregulated |
| -0,177188602 | 0,913597295 | LOC102724545 | FALSE | Downregulated |
| -0,021212581 | 0,913597295 | MED11 | FALSE | Downregulated |
| 0,096981672 | 0,913597295 | RNF112 | FALSE | Upregulated |
| -0,039365914 | 0,913597295 | TBRG1 | FALSE | Downregulated |
| 0,141108704 | 0,913721768 | LOC102724006 | FALSE | Upregulated |
| 0,037572164 | 0,913868924 | CA5BP1 | FALSE | Upregulated |
| -0,032652769 | 0,913910597 | FSCN1 | FALSE | Downregulated |
| 0,022197055 | 0,914049327 | CDKN2AIPNL | FALSE | Upregulated |
| -0,043267534 | 0,914510511 | GALNT7 | FALSE | Downregulated |
| 0,057971334 | 0,91456208 | RTN4R | FALSE | Upregulated |
| 0,032514306 | 0,914616258 | C19orf47 | FALSE | Upregulated |
| -0,016218265 | 0,914679748 | SNAP23 | FALSE | Downregulated |
| -0,093231046 | 0,914722007 | C1orf192 | FALSE | Downregulated |
| -0,049972912 | 0,914722007 | SLC17A9 | FALSE | Downregulated |
| 0,035677957 | 0,914722709 | SLC7A8 | FALSE | Upregulated |
| 0,021126136 | 0,914765354 | MEGF9 | FALSE | Upregulated |
| 0,264217831 | 0,914772729 | FAM106A | FALSE | Upregulated |
| 0,157323488 | 0,914859464 | CLNK | FALSE | Upregulated |
| 0,035240032 | 0,914864549 | GBAS | FALSE | Upregulated |
| -0,028558315 | 0,914896883 | INHBC | FALSE | Downregulated |
| -0,040503533 | 0,914896883 | OSCP1 | FALSE | Downregulated |
| -0,016300522 | 0,914896883 | RIOK1 | FALSE | Downregulated |
| -0,086263482 | 0,914896883 | RP11-420A23.1 | FALSE | Downregulated |
| -0,04277415 | 0,914896883 | ST6GALNAC3 | FALSE | Downregulated |
| 0,233797485 | 0,914936797 | EPHA10 | FALSE | Upregulated |
| -0,301308867 | 0,915061489 | CACNA1E | FALSE | Downregulated |
| 0,027398007 | 0,915061489 | GNL3L | FALSE | Upregulated |
| -0,034197701 | 0,915159534 | GMPPA | FALSE | Downregulated |
| 0,100624114 | 0,915217844 | AC104134.2 | FALSE | Upregulated |
| 0,078228811 | 0,915217844 | LOC100996760 | FALSE | Upregulated |
| -0,065111853 | 0,915217844 | VPS37D | FALSE | Downregulated |
| 0,126413117 | 0,915294408 | MESTIT1 | FALSE | Upregulated |
| -0,023428478 | 0,91543784 | ATP9B | FALSE | Downregulated |
| 0,063446482 | 0,91543784 | CELSR2 | FALSE | Upregulated |
| 0,020974589 | 0,91543784 | SURF1 | FALSE | Upregulated |
| 0,120362763 | 0,915454059 | PLEKHG4 | FALSE | Upregulated |
| 0,018229618 | 0,915470281 | ZFP91 | FALSE | Upregulated |
| 0,159941408 | 0,915581564 | ACTL10 | FALSE | Upregulated |
| -0,022209938 | 0,915594188 | DOCK1 | FALSE | Downregulated |
| -0,122007272 | 0,915609942 | UCN | FALSE | Downregulated |
| -0,013027406 | 0,915666337 | PDPK1 | FALSE | Downregulated |
| 0,025677786 | 0,915691502 | SPCS3 | FALSE | Upregulated |
| -0,023498203 | 0,915786906 | PET117 | FALSE | Downregulated |
| -0,02644193 | 0,915897714 | FUOM | FALSE | Downregulated |
| -0,082608296 | 0,915928448 | ZNF192P1 | FALSE | Downregulated |
| -0,016052328 | 0,915955202 | TBC1D9B | FALSE | Downregulated |
| 0,023421279 | 0,916145577 | DNAJC22 | FALSE | Upregulated |

| | | | | |
|---|---|---|---|---|
| 0,017786428 | 0,916325596 | CYC1 | FALSE | Upregulated |
| -0,019081771 | 0,916339886 | AKT1S1 | FALSE | Downregulated |
| -0,040541453 | 0,916339886 | EIF5B | FALSE | Downregulated |
| -0,062544378 | 0,916533175 | TRIM45 | FALSE | Downregulated |
| 0,021588658 | 0,916610461 | CYP2R1 | FALSE | Upregulated |
| 0,01989104 | 0,916610461 | FUNDC2 | FALSE | Upregulated |
| 0,018115997 | 0,916610461 | INTS4 | FALSE | Upregulated |
| -0,027132623 | 0,916610461 | WDR34 | FALSE | Downregulated |
| -0,10451193 | 0,916706063 | LOC729737 | FALSE | Downregulated |
| 0,014417852 | 0,916706063 | TMEM109 | FALSE | Upregulated |
| 0,250493494 | 0,916803076 | SRRM3 | FALSE | Upregulated |
| 0,028435971 | 0,916803076 | TRMT1 | FALSE | Upregulated |
| 0,045036789 | 0,916861408 | LOC101928126 | FALSE | Upregulated |
| -0,056933971 | 0,917377297 | KCNMB4 | FALSE | Downregulated |
| -0,024324415 | 0,917445884 | DIP2A | FALSE | Downregulated |
| -0,196968713 | 0,917489275 | FAM106CP | FALSE | Downregulated |
| -0,024894472 | 0,917665255 | NDST2 | FALSE | Downregulated |
| 0,252596016 | 0,917783039 | ART3 | FALSE | Upregulated |
| 0,080161823 | 0,917839491 | LOC102724356 | FALSE | Upregulated |
| 0,024368199 | 0,917874812 | HINT2 | FALSE | Upregulated |
| -0,022430866 | 0,91804257 | EDNRB | FALSE | Downregulated |
| -0,020510867 | 0,91804257 | INTS3 | FALSE | Downregulated |
| 0,024662166 | 0,91804257 | MANSC1 | FALSE | Upregulated |
| -0,089397794 | 0,918235857 | FSD1 | FALSE | Downregulated |
| 0,029451834 | 0,918235857 | ZNF792 | FALSE | Upregulated |
| 0,055875257 | 0,918301805 | FAM86JP | FALSE | Upregulated |
| 0,079851953 | 0,918301805 | ITGAD | FALSE | Upregulated |
| -0,01549407 | 0,918342463 | ASF1A | FALSE | Downregulated |
| -0,054770771 | 0,918342463 | DBF4B | FALSE | Downregulated |
| -0,09393871 | 0,918342463 | GBAP1 | FALSE | Downregulated |
| 0,02354107 | 0,918342463 | RPL37 | FALSE | Upregulated |
| 0,021149312 | 0,918342463 | SMIM20 | FALSE | Upregulated |
| -0,892230094 | 0,918342463 | TRDN | FALSE | Downregulated |
| 0,017486041 | 0,918499473 | ST7 | FALSE | Upregulated |
| 0,028357014 | 0,918549146 | ITSN2 | FALSE | Upregulated |
| 0,214056777 | 0,918549146 | LINC00960 | FALSE | Upregulated |
| -0,077265807 | 0,918549146 | SLITRK4 | FALSE | Downregulated |
| -0,00999523 | 0,918549146 | TNPO3 | FALSE | Downregulated |
| -0,016775759 | 0,918684104 | LMBRD1 | FALSE | Downregulated |
| 0,111492254 | 0,919081629 | LEFTY1 | FALSE | Upregulated |
| 0,12715106 | 0,919081629 | MMP23B | FALSE | Upregulated |
| 0,060876217 | 0,919081629 | SLC16A11 | FALSE | Upregulated |
| 0,170633308 | 0,919227225 | BEND3P3 | FALSE | Upregulated |
| -0,027109998 | 0,919227225 | CABLES2 | FALSE | Downregulated |
| 0,019377364 | 0,919227225 | PIH1D1 | FALSE | Upregulated |
| 0,014906554 | 0,919227225 | RALGAPB | FALSE | Upregulated |
| -0,151840306 | 0,919232741 | RHOV | FALSE | Downregulated |
| 0,063159407 | 0,91927868 | LINC00263 | FALSE | Upregulated |
| 0,017395722 | 0,919386493 | ACVR1 | FALSE | Upregulated |
| 0,132784897 | 0,919386493 | AK5 | FALSE | Upregulated |
| 0,018955427 | 0,919386493 | C2CD2 | FALSE | Upregulated |

| | | | | |
|---|---|---|---|---|
| 0,064563138 | 0,919386493 | CSNK1A1P1 | FALSE | Upregulated |
| -0,076433839 | 0,919386493 | KIAA1244 | FALSE | Downregulated |
| -0,019798119 | 0,919386493 | LDB1 | FALSE | Downregulated |
| 0,128283638 | 0,919386493 | LINC00563 | FALSE | Upregulated |
| 0,027083458 | 0,919386493 | MAP3K9 | FALSE | Upregulated |
| 0,060539108 | 0,919386493 | PDCD4-AS1 | FALSE | Upregulated |
| 0,033251921 | 0,919386493 | RPS3A | FALSE | Upregulated |
| 0,12605289 | 0,919386493 | SLC26A10 | FALSE | Upregulated |
| -0,025032623 | 0,919386493 | UVRAG | FALSE | Downregulated |
| 0,023169391 | 0,919386493 | ZFAND2B | FALSE | Upregulated |
| 0,020188014 | 0,919472623 | ERCC1 | FALSE | Upregulated |
| -0,033699497 | 0,91955931 | CLDN16 | FALSE | Downregulated |
| 0,019159223 | 0,919697023 | PIGB | FALSE | Upregulated |
| 0,023149113 | 0,919753173 | PACRGL | FALSE | Upregulated |
| -0,030930473 | 0,919766495 | ATG4D | FALSE | Downregulated |
| -0,051686414 | 0,919831634 | TMIE | FALSE | Downregulated |
| -0,033293139 | 0,919953879 | CCBE1 | FALSE | Downregulated |
| 0,014848205 | 0,919953879 | PDCD2 | FALSE | Upregulated |
| 0,021144252 | 0,919953879 | RPL37A | FALSE | Upregulated |
| 0,053914269 | 0,919953879 | TSPAN11 | FALSE | Upregulated |
| -0,081342401 | 0,920231499 | LOC100996662 | FALSE | Downregulated |
| -0,080271966 | 0,920398689 | SUGT1P1 | FALSE | Downregulated |
| 0,086715377 | 0,920746703 | CELF5 | FALSE | Upregulated |
| -0,040137404 | 0,920768591 | APOC4-APOC2 | FALSE | Downregulated |
| 0,076159441 | 0,920768591 | CLSTN2 | FALSE | Upregulated |
| -0,016421777 | 0,920768591 | DNASE1L1 | FALSE | Downregulated |
| -0,008941926 | 0,920768591 | HNRNPA2B1 | FALSE | Downregulated |
| -0,024316507 | 0,920768591 | NDUFB6 | FALSE | Downregulated |
| -0,00907347 | 0,920829304 | CUL4A | FALSE | Downregulated |
| 0,028744727 | 0,920829304 | PLEKHG1 | FALSE | Upregulated |
| -0,217253822 | 0,920855226 | PVRL4 | FALSE | Downregulated |
| -0,093866617 | 0,920961811 | ARHGEF33 | FALSE | Downregulated |
| -0,025742356 | 0,920961811 | CEP250 | FALSE | Downregulated |
| 0,022650368 | 0,920961811 | HDAC8 | FALSE | Upregulated |
| 0,017851463 | 0,921015064 | KIAA0391 | FALSE | Upregulated |
| 0,23311285 | 0,921334853 | MMP23A | FALSE | Upregulated |
| -0,087368294 | 0,921375668 | RP11-21L23.3 | FALSE | Downregulated |
| -0,05158511 | 0,921409629 | NES | FALSE | Downregulated |
| 0,023151923 | 0,921409629 | QPRT | FALSE | Upregulated |
| 0,06464305 | 0,921716626 | FGF13 | FALSE | Upregulated |
| 0,061130938 | 0,9217811 | EFNA3 | FALSE | Upregulated |
| -0,117842322 | 0,9217811 | GRID1 | FALSE | Downregulated |
| -0,031451782 | 0,9217811 | ZNF580 | FALSE | Downregulated |
| 0,079065135 | 0,92188276 | MYO1A | FALSE | Upregulated |
| -0,025263917 | 0,922120003 | ZNF205 | FALSE | Downregulated |
| -0,032892296 | 0,92245263 | BMF | FALSE | Downregulated |
| 0,030555808 | 0,922465355 | FAM76A | FALSE | Upregulated |
| 0,026753842 | 0,922475364 | LDHB | FALSE | Upregulated |
| 0,141875527 | 0,922648131 | ANKRD45 | FALSE | Upregulated |
| 0,047419695 | 0,922648131 | BMP5 | FALSE | Upregulated |
| -0,035165432 | 0,922648131 | LINC00574 | FALSE | Downregulated |

| | | | | |
|---|---|---|---|---|
| -0,847568753 | 0,922648131 | RP11-451G4.2 | FALSE | Downregulated |
| 0,12018658 | 0,922648131 | SULT1A3 | FALSE | Upregulated |
| -0,011107829 | 0,922648131 | TM9SF4 | FALSE | Downregulated |
| -0,05892556 | 0,922663776 | C21orf49 | FALSE | Downregulated |
| 0,015653388 | 0,922663776 | LOC100129361 | FALSE | Upregulated |
| 0,052885378 | 0,922926783 | CCDC163P | FALSE | Upregulated |
| -0,166499831 | 0,922950751 | AQPEP | FALSE | Downregulated |
| -0,017304525 | 0,922968661 | CARS2 | FALSE | Downregulated |
| -0,016806743 | 0,923133495 | SRP68 | FALSE | Downregulated |
| 0,034968086 | 0,923159049 | RASL11A | FALSE | Upregulated |
| 0,089699251 | 0,923300115 | LINC01160 | FALSE | Upregulated |
| 0,025318212 | 0,923300115 | RNF215 | FALSE | Upregulated |
| -0,035886843 | 0,923312098 | ZNF668 | FALSE | Downregulated |
| -0,018368523 | 0,923481355 | ACADVL | FALSE | Downregulated |
| 0,017133276 | 0,923481355 | DDX27 | FALSE | Upregulated |
| 0,083146869 | 0,923481355 | LOC101929174 | FALSE | Upregulated |
| -0,018969389 | 0,923481355 | NDUFAF1 | FALSE | Downregulated |
| 0,043603546 | 0,923481355 | SEMA4F | FALSE | Upregulated |
| 0,133330408 | 0,923510238 | CRMP1 | FALSE | Upregulated |
| 0,837770884 | 0,923795487 | MYBPC2 | FALSE | Upregulated |
| -0,076444982 | 0,923802811 | AC093391.2 | FALSE | Downregulated |
| -0,026606961 | 0,923802811 | ANKRD50 | FALSE | Downregulated |
| -0,018946349 | 0,923802811 | PAF1 | FALSE | Downregulated |
| 0,013351706 | 0,923802811 | PARK7 | FALSE | Upregulated |
| -0,029669748 | 0,923802811 | SMAP1 | FALSE | Downregulated |
| -0,247845171 | 0,923802811 | TCEB3B | FALSE | Downregulated |
| -0,188339966 | 0,923802811 | ZNF492 | FALSE | Downregulated |
| 0,110353892 | 0,923984671 | INHBA-AS1 | FALSE | Upregulated |
| -0,020649473 | 0,924244319 | REST | FALSE | Downregulated |
| 0,027565024 | 0,924728023 | DCN | FALSE | Upregulated |
| 0,120560558 | 0,924728023 | LOC100652736 | FALSE | Upregulated |
| -0,1759744 | 0,924728023 | LOC101928477 | FALSE | Downregulated |
| -0,093917805 | 0,924728023 | LOC153684 | FALSE | Downregulated |
| 0,030744876 | 0,924728023 | PGLYRP2 | FALSE | Upregulated |
| -0,120477779 | 0,924728023 | TRNT | FALSE | Downregulated |
| -0,04700541 | 0,92477259 | AKR1C2 | FALSE | Downregulated |
| -0,035545023 | 0,92477259 | LOC102724884 | FALSE | Downregulated |
| -0,014520804 | 0,92477259 | TBC1D5 | FALSE | Downregulated |
| 0,012658653 | 0,92477259 | THUMPD3 | FALSE | Upregulated |
| 0,024869843 | 0,924785172 | CSNK1E | FALSE | Upregulated |
| -0,016368315 | 0,925294923 | CD99L2 | FALSE | Downregulated |
| 0,032395387 | 0,925357686 | FER | FALSE | Upregulated |
| -0,030790348 | 0,925357686 | MFSD3 | FALSE | Downregulated |
| 0,018350193 | 0,925773116 | PLS3 | FALSE | Upregulated |
| -0,012508935 | 0,926146874 | EXOC5 | FALSE | Downregulated |
| 0,16050875 | 0,92616022 | ANO2 | FALSE | Upregulated |
| -0,026195857 | 0,926324718 | OST4 | FALSE | Downregulated |
| 0,027990057 | 0,926344182 | C10orf12 | FALSE | Upregulated |
| 0,140733636 | 0,92639271 | LINC00842 | FALSE | Upregulated |
| -0,102948766 | 0,926798262 | C21orf90 | FALSE | Downregulated |
| 0,134091706 | 0,926953941 | PRSS45 | FALSE | Upregulated |

| | | | | |
|---|---|---|---|---|
| 0,809531927 | 0,927057646 | CASQ1 | FALSE | Upregulated |
| 0,011749839 | 0,927075704 | ITFG1 | FALSE | Upregulated |
| 0,033380224 | 0,927098461 | GFPT1 | FALSE | Upregulated |
| 0,107082203 | 0,927098461 | HSPB2-C11orf52 | FALSE | Upregulated |
| 0,130218104 | 0,927098461 | LOC151174 | FALSE | Upregulated |
| 0,026841591 | 0,927098461 | LSM5 | FALSE | Upregulated |
| -0,045563599 | 0,927098461 | POLR2A | FALSE | Downregulated |
| 0,059082121 | 0,927098461 | WTIP | FALSE | Upregulated |
| 0,017814161 | 0,927152758 | DHX36 | FALSE | Upregulated |
| 0,021534947 | 0,927152758 | PDE6D | FALSE | Upregulated |
| 0,017961479 | 0,927152758 | RFFL | FALSE | Upregulated |
| -0,015138453 | 0,927152758 | UIMC1 | FALSE | Downregulated |
| 0,025271317 | 0,927177071 | LIMS2 | FALSE | Upregulated |
| 0,018774837 | 0,927211288 | SDHB | FALSE | Upregulated |
| -0,067195129 | 0,927318169 | SNHG5 | FALSE | Downregulated |
| 0,186608496 | 0,927391458 | BOLA2B | FALSE | Upregulated |
| -0,139433013 | 0,927391458 | COL10A1 | FALSE | Downregulated |
| 0,032160033 | 0,927391458 | GAN | FALSE | Upregulated |
| -0,130176545 | 0,927391458 | LOC102724384 | FALSE | Downregulated |
| -0,021727157 | 0,927391458 | NME7 | FALSE | Downregulated |
| -0,065764553 | 0,927391458 | NXF3 | FALSE | Downregulated |
| -0,015147883 | 0,927391458 | PATL1 | FALSE | Downregulated |
| 0,033905394 | 0,927391458 | PXN-AS1 | FALSE | Upregulated |
| 0,231848072 | 0,927391458 | RP11-551M15.1 | FALSE | Upregulated |
| -0,021475113 | 0,927391458 | RSRC1 | FALSE | Downregulated |
| -0,023486862 | 0,927517659 | AKAP8L | FALSE | Downregulated |
| 0,01578675 | 0,927517659 | CSNK1G1 | FALSE | Upregulated |
| -0,047578082 | 0,927517659 | KANSL1-AS1 | FALSE | Downregulated |
| -0,016688783 | 0,927517659 | LIG3 | FALSE | Downregulated |
| 0,146372575 | 0,927517659 | LOC101928245 | FALSE | Upregulated |
| -0,058167392 | 0,927517659 | MTCP1 | FALSE | Downregulated |
| 0,013625224 | 0,927517659 | RING1 | FALSE | Upregulated |
| -0,047216757 | 0,927517659 | RP11-98I9.4 | FALSE | Downregulated |
| 0,013871649 | 0,927517659 | SCAP | FALSE | Upregulated |
| -0,02199191 | 0,927597896 | OCEL1 | FALSE | Downregulated |
| 0,028546607 | 0,927603556 | CHCHD6 | FALSE | Upregulated |
| -0,041270775 | 0,927603556 | RPL21P28 | FALSE | Downregulated |
| -0,023618949 | 0,927603556 | TAF1B | FALSE | Downregulated |
| 0,086618613 | 0,927679427 | KCTD14 | FALSE | Upregulated |
| -0,046830098 | 0,927679427 | TNFSF4 | FALSE | Downregulated |
| -0,053045977 | 0,927688328 | FOXN3-AS1 | FALSE | Downregulated |
| -0,018338988 | 0,927732736 | IPO11 | FALSE | Downregulated |
| -0,136060273 | 0,927732736 | LOC101929188 | FALSE | Downregulated |
| 0,08719516 | 0,927732736 | UGT3A2 | FALSE | Upregulated |
| -0,013526355 | 0,927937121 | NUP98 | FALSE | Downregulated |
| 0,049190852 | 0,927950888 | CD163L1 | FALSE | Upregulated |
| -0,016212057 | 0,928179853 | MTTP | FALSE | Downregulated |
| -0,020144603 | 0,928179853 | RPL23A | FALSE | Downregulated |
| 0,025616255 | 0,928179853 | TXNIP | FALSE | Upregulated |
| 0,018492624 | 0,928180563 | CUL7 | FALSE | Upregulated |
| 0,01056552 | 0,928224434 | MRPL30 | FALSE | Upregulated |

| | | | | |
|---|---|---|---|---|
| -0,054847392 | 0,928288412 | LOC101928745 | FALSE | Downregulated |
| -0,02044466 | 0,92836865 | TMEM14B | FALSE | Downregulated |
| 0,015220747 | 0,928462849 | FBXO34 | FALSE | Upregulated |
| 0,14356647 | 0,928462849 | PNLDC1 | FALSE | Upregulated |
| -0,057325283 | 0,928580297 | LOC101928595 | FALSE | Downregulated |
| 0,054172918 | 0,928588498 | CD209 | FALSE | Upregulated |
| 0,689599296 | 0,928717985 | ANKRD2 | FALSE | Upregulated |
| -0,018552939 | 0,928717985 | RPS8 | FALSE | Downregulated |
| 0,182830484 | 0,928733353 | ARGFX | FALSE | Upregulated |
| -0,022229132 | 0,928733353 | DUSP16 | FALSE | Downregulated |
| 0,059982669 | 0,928733353 | EFCAB12 | FALSE | Upregulated |
| 0,014867308 | 0,928733353 | NCOA5 | FALSE | Upregulated |
| -0,14337529 | 0,928733353 | PPT2-EGFL8 | FALSE | Downregulated |
| -0,012903535 | 0,928733353 | REEP5 | FALSE | Downregulated |
| 0,139035565 | 0,928733353 | SNORA29 | FALSE | Upregulated |
| 0,105703955 | 0,929203606 | LOC729652 | FALSE | Upregulated |
| 0,044725983 | 0,929203606 | RNF122 | FALSE | Upregulated |
| -0,050450689 | 0,929392547 | ASAH2 | FALSE | Downregulated |
| 0,134821258 | 0,929392547 | LOC101929566 | FALSE | Upregulated |
| -0,060329082 | 0,929420398 | ALS2CR12 | FALSE | Downregulated |
| -0,054875359 | 0,929529181 | C20orf196 | FALSE | Downregulated |
| 0,015628443 | 0,929619203 | CDK8 | FALSE | Upregulated |
| -0,083572925 | 0,929619203 | EME2 | FALSE | Downregulated |
| 0,012853271 | 0,929661563 | ABHD13 | FALSE | Upregulated |
| 0,013813149 | 0,929661563 | KIAA1033 | FALSE | Upregulated |
| 0,014435406 | 0,929661563 | POLR3F | FALSE | Upregulated |
| 0,011084094 | 0,929724519 | CLDND1 | FALSE | Upregulated |
| -0,016743883 | 0,929759768 | C7orf73 | FALSE | Downregulated |
| 0,028618258 | 0,92984039 | ARMC7 | FALSE | Upregulated |
| -0,067099774 | 0,929873252 | CBR3-AS1 | FALSE | Downregulated |
| -0,027360407 | 0,929882585 | GNG11 | FALSE | Downregulated |
| -0,094673298 | 0,929882585 | PITRM1-AS1 | FALSE | Downregulated |
| 0,036071973 | 0,930120874 | BCL2L2-PABPN1 | FALSE | Upregulated |
| -0,026877442 | 0,930120874 | HDHD1 | FALSE | Downregulated |
| 0,083664218 | 0,930138967 | RASAL2-AS1 | FALSE | Upregulated |
| -0,087467537 | 0,930433352 | DDX11L2 | FALSE | Downregulated |
| -0,080989781 | 0,930433352 | GABRP | FALSE | Downregulated |
| -0,091809208 | 0,930469477 | LOC101409256 | FALSE | Downregulated |
| -0,018096186 | 0,930469477 | MAGT1 | FALSE | Downregulated |
| -0,037484622 | 0,930469477 | PRKAG2-AS1 | FALSE | Downregulated |
| -0,028407444 | 0,930469477 | ZNF582 | FALSE | Downregulated |
| -0,03220856 | 0,930785307 | GIPC3 | FALSE | Downregulated |
| 0,024797856 | 0,930834496 | HMBOX1 | FALSE | Upregulated |
| -0,056719149 | 0,930834496 | IGSF9 | FALSE | Downregulated |
| 0,019688751 | 0,93084041 | THAP4 | FALSE | Upregulated |
| 0,108525029 | 0,931262759 | ABCG4 | FALSE | Upregulated |
| 0,060953774 | 0,931358729 | MINOS1-NBL1 | FALSE | Upregulated |
| 0,012516786 | 0,931415803 | NUP107 | FALSE | Upregulated |
| 0,026455214 | 0,931596774 | KLHDC4 | FALSE | Upregulated |
| -0,044872546 | 0,931728328 | P2RX5-TAX1BP3 | FALSE | Downregulated |
| 0,109353819 | 0,931785737 | CCDC180 | FALSE | Upregulated |

| | | | | |
|---|---|---|---|---|
| 0,014612516 | 0,931785737 | KIF13A | FALSE | Upregulated |
| -0,131537602 | 0,93192474 | IGSF1 | FALSE | Downregulated |
| -0,107478821 | 0,931963802 | RP11-290F20.2 | FALSE | Downregulated |
| 0,075146247 | 0,931963802 | SLX1A-SULT1A3 | FALSE | Upregulated |
| -0,02356235 | 0,931963802 | ZKSCAN4 | FALSE | Downregulated |
| 0,012430021 | 0,93217311 | MED21 | FALSE | Upregulated |
| 0,063368908 | 0,932195981 | BEAN1 | FALSE | Upregulated |
| -0,069841118 | 0,932364883 | SPATC1L | FALSE | Downregulated |
| 0,01760038 | 0,932579591 | TMBIM4 | FALSE | Upregulated |
| -0,012383362 | 0,932953565 | GNAQ | FALSE | Downregulated |
| -0,029094249 | 0,932953565 | POLR3G | FALSE | Downregulated |
| -0,016216095 | 0,933013125 | DDX21 | FALSE | Downregulated |
| 0,093272689 | 0,933022936 | LOC101927512 | FALSE | Upregulated |
| 0,022760523 | 0,933149859 | ELAC1 | FALSE | Upregulated |
| -0,159036078 | 0,93316863 | GAL3ST2 | FALSE | Downregulated |
| 0,087814252 | 0,933283573 | OXT | FALSE | Upregulated |
| -0,031463906 | 0,933283573 | TMCC3 | FALSE | Downregulated |
| -0,149597001 | 0,933283573 | TRIM74 | FALSE | Downregulated |
| -0,16729565 | 0,933330216 | LOC102723446 | FALSE | Downregulated |
| 0,027795927 | 0,933401435 | GAS6 | FALSE | Upregulated |
| 0,056899805 | 0,933473888 | LINC00482 | FALSE | Upregulated |
| 0,655445125 | 0,933683757 | APOBEC2 | FALSE | Upregulated |
| -0,016558364 | 0,933683757 | EDEM2 | FALSE | Downregulated |
| 0,017497296 | 0,933744258 | VPS9D1 | FALSE | Upregulated |
| 0,018246086 | 0,933932374 | CLPP | FALSE | Upregulated |
| 0,01228508 | 0,933932374 | DPF2 | FALSE | Upregulated |
| 0,012428006 | 0,933932374 | NCKAP1 | FALSE | Upregulated |
| -0,025464948 | 0,933932374 | SLC18B1 | FALSE | Downregulated |
| 0,044108404 | 0,934091595 | EPHA3 | FALSE | Upregulated |
| 0,732266517 | 0,934188908 | RPL3L | FALSE | Upregulated |
| -0,008967878 | 0,934196781 | VPS4B | FALSE | Downregulated |
| 0,087031744 | 0,934250982 | CALML6 | FALSE | Upregulated |
| -0,418227336 | 0,934250982 | CRYAA | FALSE | Downregulated |
| -0,013544066 | 0,934250982 | LIN54 | FALSE | Downregulated |
| 0,130167477 | 0,934255706 | CC2D2B | FALSE | Upregulated |
| -0,048019331 | 0,934337717 | LOC100505555 | FALSE | Downregulated |
| 0,675633541 | 0,934400057 | LDB3 | FALSE | Upregulated |
| 0,10110435 | 0,934400057 | SLC52A1 | FALSE | Upregulated |
| 0,072719691 | 0,934423336 | LOC100507670 | FALSE | Upregulated |
| 0,14078767 | 0,934462951 | AACSP1 | FALSE | Upregulated |
| 0,014008896 | 0,934462951 | DCTN3 | FALSE | Upregulated |
| 0,033241927 | 0,934462951 | LOC101929787 | FALSE | Upregulated |
| -0,086291514 | 0,934462951 | LOC102724372 | FALSE | Downregulated |
| -0,040767853 | 0,934462951 | SYT7 | FALSE | Downregulated |
| -0,025105888 | 0,934529129 | CAB39L | FALSE | Downregulated |
| -0,080969767 | 0,934623946 | CATSPER2 | FALSE | Downregulated |
| 0,019026503 | 0,934623946 | CEP85 | FALSE | Upregulated |
| -0,155205921 | 0,934623946 | LOC101929732 | FALSE | Downregulated |
| -0,020686187 | 0,934639923 | SS18L2 | FALSE | Downregulated |
| -0,147772952 | 0,934639923 | TAS2R4 | FALSE | Downregulated |
| -0,062742585 | 0,934639923 | TDRD9 | FALSE | Downregulated |

| | | | | |
|---|---|---|---|---|
| -0,139506585 | 0,934643126 | GABRD | FALSE | Downregulated |
| 0,035314618 | 0,934643126 | LRP12 | FALSE | Upregulated |
| -0,031359236 | 0,934643126 | MRM1 | FALSE | Downregulated |
| 0,080073726 | 0,934692801 | EIF5AL1 | FALSE | Upregulated |
| -0,093136358 | 0,934752411 | LOC101928660 | FALSE | Downregulated |
| 0,113392516 | 0,934757896 | PDE1C | FALSE | Upregulated |
| 0,014473405 | 0,934833155 | PIAS4 | FALSE | Upregulated |
| -0,01222854 | 0,934883344 | PNPO | FALSE | Downregulated |
| -0,086418681 | 0,934883344 | UGT2B28 | FALSE | Downregulated |
| 0,011428982 | 0,935006887 | STAG1 | FALSE | Upregulated |
| 0,076555989 | 0,935059513 | CD300LB | FALSE | Upregulated |
| 0,029705662 | 0,935059513 | ID3 | FALSE | Upregulated |
| 0,118147318 | 0,935059513 | KCNMB2 | FALSE | Upregulated |
| -0,02453885 | 0,935073812 | HAUS2 | FALSE | Downregulated |
| 0,030268978 | 0,935396622 | MAML2 | FALSE | Upregulated |
| -0,074016271 | 0,935432425 | SUMO4 | FALSE | Downregulated |
| -0,066551491 | 0,935646308 | LOC100507487 | FALSE | Downregulated |
| -0,019252866 | 0,935720531 | TBL3 | FALSE | Downregulated |
| 0,017148456 | 0,93603401 | PDAP1 | FALSE | Upregulated |
| -0,046288516 | 0,936056023 | KCNIP2 | FALSE | Downregulated |
| 0,017423539 | 0,936281008 | EARS2 | FALSE | Upregulated |
| -0,029965535 | 0,936281008 | ENTHD2 | FALSE | Downregulated |
| 0,051146796 | 0,936281008 | PRR15L | FALSE | Upregulated |
| 0,023610315 | 0,936417038 | EFNA1 | FALSE | Upregulated |
| -0,040700281 | 0,936934497 | LOC101929747 | FALSE | Downregulated |
| 0,115334773 | 0,936934497 | MAP1LC3C | FALSE | Upregulated |
| -0,101033751 | 0,936935776 | CA9 | FALSE | Downregulated |
| -0,047567816 | 0,936935776 | LOC101927002 | FALSE | Downregulated |
| 0,139916755 | 0,936935776 | ULK4P3 | FALSE | Upregulated |
| 0,026140014 | 0,93705497 | ANO5 | FALSE | Upregulated |
| 0,022437133 | 0,937539798 | ZNF416 | FALSE | Upregulated |
| 0,140977439 | 0,937633544 | AC104777.4 | FALSE | Upregulated |
| -0,030523278 | 0,937703267 | GCOM1 | FALSE | Downregulated |
| 0,092510332 | 0,937703315 | CH25H | FALSE | Upregulated |
| -0,138195363 | 0,937703315 | LOC101928906 | FALSE | Downregulated |
| -0,009859071 | 0,93774232 | JAK1 | FALSE | Downregulated |
| 0,023815873 | 0,937801555 | C12orf43 | FALSE | Upregulated |
| -0,096147753 | 0,938247217 | LOC100272216 | FALSE | Downregulated |
| 0,050056039 | 0,938656971 | SOX17 | FALSE | Upregulated |
| -0,12911246 | 0,938787273 | KCNH1 | FALSE | Downregulated |
| 0,016600595 | 0,938819508 | AIFM1 | FALSE | Upregulated |
| 0,017904257 | 0,938819508 | LACTB2 | FALSE | Upregulated |
| 0,065932222 | 0,938819508 | LOC102723544 | FALSE | Upregulated |
| -0,059243537 | 0,938819508 | NT5M | FALSE | Downregulated |
| 0,054797935 | 0,938819508 | PRRT1 | FALSE | Upregulated |
| -0,059918541 | 0,938822411 | LTBP2 | FALSE | Downregulated |
| -0,008404512 | 0,938822411 | SNRPD3 | FALSE | Downregulated |
| 0,022917941 | 0,93903044 | RUSC1 | FALSE | Upregulated |
| -0,086394074 | 0,93903044 | SCN4A | FALSE | Downregulated |
| -0,025639045 | 0,939046929 | RPL17-C18orf32 | FALSE | Downregulated |
| 0,012763611 | 0,939162991 | ATP5J2 | FALSE | Upregulated |

| | | | | |
|---|---|---|---|---|
| -0,070558896 | 0,939480915 | BTNL8 | FALSE | Downregulated |
| 0,019201236 | 0,939676155 | ALKBH7 | FALSE | Upregulated |
| 0,049484114 | 0,939676155 | MYB | FALSE | Upregulated |
| -0,01626919 | 0,939678052 | DNAJB1 | FALSE | Downregulated |
| -0,116776237 | 0,939770366 | LOC101927345 | FALSE | Downregulated |
| 0,10609546 | 0,939770366 | LOC101928583 | FALSE | Upregulated |
| 0,014104771 | 0,939770925 | NDUFV2 | FALSE | Upregulated |
| 0,094286001 | 0,939812952 | TCEAL2 | FALSE | Upregulated |
| 0,022604048 | 0,939958252 | ARL6 | FALSE | Upregulated |
| 0,02531288 | 0,940053092 | LOC493754 | FALSE | Upregulated |
| -0,061537915 | 0,9404139 | GRM8 | FALSE | Downregulated |
| 0,022108157 | 0,9404139 | LOH12CR1 | FALSE | Upregulated |
| 0,028981419 | 0,9404139 | VNN1 | FALSE | Upregulated |
| 0,020858807 | 0,940447016 | SH3BP2 | FALSE | Upregulated |
| -0,008712346 | 0,940451013 | AGGF1 | FALSE | Downregulated |
| -0,021768872 | 0,940451013 | ARFGAP3 | FALSE | Downregulated |
| -0,014564909 | 0,940451013 | HDDC2 | FALSE | Downregulated |
| -0,041528916 | 0,940451013 | LOC729603 | FALSE | Downregulated |
| -0,023445623 | 0,940451013 | LPIN1 | FALSE | Downregulated |
| 0,016296315 | 0,940451013 | PMS2 | FALSE | Upregulated |
| 0,074313342 | 0,940519986 | LOC101927198 | FALSE | Upregulated |
| -0,068209425 | 0,940621071 | GGT6 | FALSE | Downregulated |
| 0,010877366 | 0,940621071 | LARP4B | FALSE | Upregulated |
| -0,016583324 | 0,940621071 | TTC13 | FALSE | Downregulated |
| 0,426878943 | 0,940707403 | SYNPO2L | FALSE | Upregulated |
| 0,035723009 | 0,940767656 | ARHGEF25 | FALSE | Upregulated |
| -0,014973693 | 0,940767656 | C22orf39 | FALSE | Downregulated |
| -0,133243554 | 0,940767656 | EIF3CL | FALSE | Downregulated |
| -0,0293743 | 0,940767656 | PARK2 | FALSE | Downregulated |
| -0,018678733 | 0,940767656 | RPL36AL | FALSE | Downregulated |
| 0,128322333 | 0,940767656 | SH3GL2 | FALSE | Upregulated |
| 0,020192093 | 0,940767656 | TJP1 | FALSE | Upregulated |
| 0,170728478 | 0,940767656 | TRNAK13 | FALSE | Upregulated |
| -0,070577758 | 0,940921315 | CEACAM22P | FALSE | Downregulated |
| -0,016577485 | 0,940921315 | FRMD4A | FALSE | Downregulated |
| -0,095862265 | 0,940965479 | AC104655.2 | FALSE | Downregulated |
| -0,072191863 | 0,940967912 | LOC101929479 | FALSE | Downregulated |
| 0,016696548 | 0,941003249 | ARFIP2 | FALSE | Upregulated |
| 0,109027596 | 0,941153433 | RP11-661P17.1 | FALSE | Upregulated |
| 0,020047639 | 0,941497261 | WDSUB1 | FALSE | Upregulated |
| 0,02001885 | 0,941655175 | MLEC | FALSE | Upregulated |
| 0,074227656 | 0,941659561 | PARD6G-AS1 | FALSE | Upregulated |
| 0,014419311 | 0,941764993 | SRP72 | FALSE | Upregulated |
| -0,025458902 | 0,941821629 | KSR1 | FALSE | Downregulated |
| 0,014800275 | 0,941821629 | MRPS15 | FALSE | Upregulated |
| 0,241705825 | 0,941821629 | TNFSF12-TNFSF13 | FALSE | Upregulated |
| -0,094337486 | 0,941844711 | RNF175 | FALSE | Downregulated |
| 0,052978998 | 0,942643167 | RAB36 | FALSE | Upregulated |
| -0,016052281 | 0,942697451 | PEX3 | FALSE | Downregulated |
| -0,070392749 | 0,942706307 | TMPRSS13 | FALSE | Downregulated |
| -0,010856915 | 0,942706307 | UBR7 | FALSE | Downregulated |

| | | | | |
|---|---|---|---|---|
| 0,012485857 | 0,942763203 | LSG1 | FALSE | Upregulated |
| -0,059062106 | 0,942884283 | RP11-84C10.4 | FALSE | Downregulated |
| -0,108608381 | 0,943025782 | LOC100507616 | FALSE | Downregulated |
| -0,043832082 | 0,943042523 | VCAN | FALSE | Downregulated |
| -0,013514221 | 0,943183994 | IP6K2 | FALSE | Downregulated |
| 0,049903136 | 0,943641837 | C8orf88 | FALSE | Upregulated |
| -0,050515441 | 0,943641837 | MAPK12 | FALSE | Downregulated |
| -0,097879019 | 0,943641837 | RN7SK | FALSE | Downregulated |
| -0,014473097 | 0,943641837 | RNASEH2C | FALSE | Downregulated |
| -0,013117823 | 0,943641837 | TMEM183A | FALSE | Downregulated |
| 0,02946534 | 0,943686288 | ASB13 | FALSE | Upregulated |
| -0,014576121 | 0,943686288 | FAM178A | FALSE | Downregulated |
| 0,019607809 | 0,943686288 | MICAL2 | FALSE | Upregulated |
| -0,011237234 | 0,943686288 | RTCA | FALSE | Downregulated |
| -0,08913544 | 0,943686288 | TRIM7 | FALSE | Downregulated |
| 0,631388788 | 0,943928552 | BOLA2 | FALSE | Upregulated |
| 0,021524201 | 0,943928552 | CAPN10 | FALSE | Upregulated |
| -0,011661998 | 0,943928552 | KDM3B | FALSE | Downregulated |
| 0,170227526 | 0,943928552 | LINC00485 | FALSE | Upregulated |
| -0,02266372 | 0,943928552 | PPARD | FALSE | Downregulated |
| 0,020627395 | 0,94394246 | PPP2R1B | FALSE | Upregulated |
| -0,036938688 | 0,944014417 | LEPREL2 | FALSE | Downregulated |
| -0,045567268 | 0,944014417 | LOC101928485 | FALSE | Downregulated |
| 0,046529369 | 0,944014417 | SNAI3-AS1 | FALSE | Upregulated |
| -0,09263285 | 0,944014417 | SNORA32 | FALSE | Downregulated |
| 0,014306856 | 0,944014417 | SNRPF | FALSE | Upregulated |
| 0,006922844 | 0,944014417 | SRSF1 | FALSE | Upregulated |
| 0,021690508 | 0,94403189 | CCDC146 | FALSE | Upregulated |
| -0,018188086 | 0,94403189 | FKBP14 | FALSE | Downregulated |
| -0,066714668 | 0,94403189 | PKD1L3 | FALSE | Downregulated |
| -0,016268233 | 0,94403189 | TBCCD1 | FALSE | Downregulated |
| 0,019285032 | 0,944352958 | CRLS1 | FALSE | Upregulated |
| -0,055490053 | 0,944352958 | SLC1A7 | FALSE | Downregulated |
| -0,040053732 | 0,944499702 | LOC646021 | FALSE | Downregulated |
| -0,018693442 | 0,944743264 | MESDC1 | FALSE | Downregulated |
| -0,022625665 | 0,944954792 | KLF10 | FALSE | Downregulated |
| -0,049031763 | 0,944987473 | ALPK3 | FALSE | Downregulated |
| -0,013384109 | 0,945006692 | HNRNPH3 | FALSE | Downregulated |
| -0,13870016 | 0,945329262 | CTNNA2 | FALSE | Downregulated |
| -0,084504626 | 0,945338675 | LOC100288846 | FALSE | Downregulated |
| -0,089833816 | 0,945731795 | PRPH2 | FALSE | Downregulated |
| 0,115456431 | 0,945860617 | SLC7A10 | FALSE | Upregulated |
| -0,027618726 | 0,945899384 | PACRG | FALSE | Downregulated |
| -0,041309506 | 0,946021957 | LOC102723772 | FALSE | Downregulated |
| -0,17192316 | 0,946021957 | MEF2BNB-MEF2B | FALSE | Downregulated |
| 0,017531002 | 0,946132254 | SPATA7 | FALSE | Upregulated |
| -0,013721913 | 0,946415294 | USP37 | FALSE | Downregulated |
| -0,050665842 | 0,946421505 | CHST10 | FALSE | Downregulated |
| 0,016071205 | 0,946421505 | RAB7A | FALSE | Upregulated |
| -0,009637705 | 0,946436035 | APH1A | FALSE | Downregulated |
| 0,045164691 | 0,946436035 | BICC1 | FALSE | Upregulated |

| | | | | |
|---|---|---|---|---|
| 0,109209011 | 0,946436035 | C1QTNF9B | FALSE | Upregulated |
| 0,056902496 | 0,946436035 | C6orf163 | FALSE | Upregulated |
| 0,052274169 | 0,946436035 | LOC101927578 | FALSE | Upregulated |
| -0,019631576 | 0,946436035 | MIEF2 | FALSE | Downregulated |
| 0,604279578 | 0,946436035 | MYH7 | FALSE | Upregulated |
| -0,022016363 | 0,946457709 | SEMA5A | FALSE | Downregulated |
| 0,025758342 | 0,946457709 | SSH2 | FALSE | Upregulated |
| 0,027338877 | 0,946457709 | TLE2 | FALSE | Upregulated |
| -0,030848279 | 0,946593364 | EEPD1 | FALSE | Downregulated |
| -0,016511354 | 0,946671333 | AGXT | FALSE | Downregulated |
| -0,009403606 | 0,946671333 | BICD2 | FALSE | Downregulated |
| -0,050385628 | 0,946671333 | EMR4P | FALSE | Downregulated |
| 0,046850278 | 0,946671333 | LOC101927263 | FALSE | Upregulated |
| 0,018761763 | 0,946671333 | LRFN3 | FALSE | Upregulated |
| 0,029659479 | 0,946750004 | TNRC6C-AS1 | FALSE | Upregulated |
| -0,108260789 | 0,946804762 | AC010127.3 | FALSE | Downregulated |
| 0,09038866 | 0,946804762 | LOC101928801 | FALSE | Upregulated |
| 0,597691405 | 0,946804762 | TNNC2 | FALSE | Upregulated |
| -0,071139146 | 0,947117202 | LINC00907 | FALSE | Downregulated |
| 0,174076467 | 0,947248932 | SLC30A2 | FALSE | Upregulated |
| -0,020041863 | 0,947287093 | CD55 | FALSE | Downregulated |
| 0,025537059 | 0,947287093 | ZDHHC2 | FALSE | Upregulated |
| 0,055120937 | 0,947366047 | PACSIN1 | FALSE | Upregulated |
| -0,014968077 | 0,947366047 | ZNF646 | FALSE | Downregulated |
| 0,113653535 | 0,947408553 | LURAP1 | FALSE | Upregulated |
| -0,010360699 | 0,947427588 | MASP1 | FALSE | Downregulated |
| -0,013808512 | 0,947602395 | C19orf60 | FALSE | Downregulated |
| -0,040467738 | 0,947619031 | DZIP1L | FALSE | Downregulated |
| -0,059474373 | 0,947642382 | GPR153 | FALSE | Downregulated |
| -0,09377896 | 0,947645998 | LOC158434 | FALSE | Downregulated |
| 0,015660744 | 0,947645998 | MARK4 | FALSE | Upregulated |
| 0,0129649 | 0,947645998 | NTPCR | FALSE | Upregulated |
| 0,020359945 | 0,947645998 | RABEPK | FALSE | Upregulated |
| -0,049005954 | 0,947712783 | LOC728392 | FALSE | Downregulated |
| -0,034600116 | 0,947712783 | SPAG5-AS1 | FALSE | Downregulated |
| 0,109879827 | 0,948235081 | MTRNR2L5 | FALSE | Upregulated |
| 0,012369661 | 0,948235081 | NAP1L4 | FALSE | Upregulated |
| 0,13692156 | 0,948245015 | LOC101059936 | FALSE | Upregulated |
| 0,069635507 | 0,948364219 | FXYD7 | FALSE | Upregulated |
| 0,079711288 | 0,948364219 | MMP11 | FALSE | Upregulated |
| 0,091215316 | 0,948421398 | CFL1P1 | FALSE | Upregulated |
| -0,050430432 | 0,948421398 | HOXA3 | FALSE | Downregulated |
| -0,086631821 | 0,948421398 | LOC644249 | FALSE | Downregulated |
| -0,01064445 | 0,948421398 | TRIM28 | FALSE | Downregulated |
| 0,01433889 | 0,948421398 | VPS37B | FALSE | Upregulated |
| 0,010845966 | 0,94843982 | ATF6B | FALSE | Upregulated |
| -0,012688278 | 0,94843982 | C2orf44 | FALSE | Downregulated |
| 0,045506659 | 0,948449976 | BTBD11 | FALSE | Upregulated |
| -0,082830247 | 0,948449976 | TXLNB | FALSE | Downregulated |
| -0,044469013 | 0,948487216 | XAGE3 | FALSE | Downregulated |
| 0,066736883 | 0,948851378 | LOC100996695 | FALSE | Upregulated |

| | | | | |
|---|---|---|---|---|
| -0,054405269 | 0,948851378 | LOC102724312 | FALSE | Downregulated |
| -0,010262617 | 0,948851378 | MAGED2 | FALSE | Downregulated |
| 0,013963027 | 0,948879144 | ECI1 | FALSE | Upregulated |
| -0,013056744 | 0,948879144 | MRPS27 | FALSE | Downregulated |
| 0,013772727 | 0,948879144 | NDUFB9 | FALSE | Upregulated |
| -0,014436948 | 0,948879144 | SLC25A1 | FALSE | Downregulated |
| 0,022856684 | 0,948879144 | TCF25 | FALSE | Upregulated |
| 0,527092871 | 0,948916911 | ACTN2 | FALSE | Upregulated |
| 0,055438939 | 0,948916911 | LOC728763 | FALSE | Upregulated |
| 0,108169142 | 0,948931648 | ERBB4 | FALSE | Upregulated |
| -0,008570076 | 0,948931648 | RARS2 | FALSE | Downregulated |
| 0,016582673 | 0,948953397 | GFOD2 | FALSE | Upregulated |
| -0,132837983 | 0,948975636 | CD1B | FALSE | Downregulated |
| 0,028656089 | 0,948975636 | CLDN15 | FALSE | Upregulated |
| 0,020644653 | 0,948975636 | GPD1 | FALSE | Upregulated |
| -0,029953121 | 0,948978794 | CCDC40 | FALSE | Downregulated |
| -0,019871029 | 0,948978794 | CREM | FALSE | Downregulated |
| 0,02114687 | 0,948978794 | FRG1B | FALSE | Upregulated |
| 0,021582444 | 0,948978794 | PAMR1 | FALSE | Upregulated |
| -0,037925794 | 0,948978794 | PRRT3 | FALSE | Downregulated |
| -0,238404414 | 0,949008467 | SMTNL1 | FALSE | Downregulated |
| 0,093860321 | 0,949018047 | KCNH3 | FALSE | Upregulated |
| -0,053065372 | 0,949082468 | C1QTNF2 | FALSE | Downregulated |
| -0,017326843 | 0,949082468 | PLXNA2 | FALSE | Downregulated |
| 0,189728278 | 0,949103946 | PADI6 | FALSE | Upregulated |
| -0,059163812 | 0,949157061 | AP1M2 | FALSE | Downregulated |
| 0,126018877 | 0,949157061 | LOC101927143 | FALSE | Upregulated |
| -0,045949831 | 0,949157061 | LOC648987 | FALSE | Downregulated |
| 0,022684166 | 0,949157061 | SLC25A5-AS1 | FALSE | Upregulated |
| 0,152310518 | 0,949157061 | SNORA44 | FALSE | Upregulated |
| -0,084174133 | 0,949284403 | LOC100506901 | FALSE | Downregulated |
| -0,017846313 | 0,949474614 | C12orf57 | FALSE | Downregulated |
| -0,009642807 | 0,949495314 | CDC5L | FALSE | Downregulated |
| -0,039640671 | 0,949495314 | TSPAN13 | FALSE | Downregulated |
| -0,091350949 | 0,949525865 | LOC101929494 | FALSE | Downregulated |
| -0,036148351 | 0,949734196 | LOC100129148 | FALSE | Downregulated |
| -0,008763266 | 0,94988512 | CASP3 | FALSE | Downregulated |
| 0,07601105 | 0,94988512 | FAM83E | FALSE | Upregulated |
| -0,046219209 | 0,94988512 | LOC102723780 | FALSE | Downregulated |
| -0,014284767 | 0,94988512 | NUP37 | FALSE | Downregulated |
| 0,010527045 | 0,950095977 | FGD5-AS1 | FALSE | Upregulated |
| -0,060209457 | 0,95040423 | SEZ6L | FALSE | Downregulated |
| -0,008348672 | 0,950677491 | FBXO18 | FALSE | Downregulated |
| 0,552771291 | 0,950722589 | CKM | FALSE | Upregulated |
| -0,01001903 | 0,950722589 | SIPA1L1 | FALSE | Downregulated |
| -0,010590553 | 0,950910224 | DCAKD | FALSE | Downregulated |
| -0,140940286 | 0,951021772 | ATP12A | FALSE | Downregulated |
| 0,014523766 | 0,951021772 | EIF4E2 | FALSE | Upregulated |
| -0,08107145 | 0,951021772 | FOLR1 | FALSE | Downregulated |
| 0,083085989 | 0,951021772 | HMGN2P46 | FALSE | Upregulated |
| 0,015026869 | 0,951021772 | NOP14-AS1 | FALSE | Upregulated |

| | | | | |
|---|---|---|---|---|
| -0,015916768 | 0,951021772 | RUFY3 | FALSE | Downregulated |
| -0,025428273 | 0,951171992 | AMIGO1 | FALSE | Downregulated |
| -0,085667355 | 0,951171992 | KRT80 | FALSE | Downregulated |
| 0,05592715 | 0,951171992 | LETM2 | FALSE | Upregulated |
| 0,045943954 | 0,951171992 | LOC101927060 | FALSE | Upregulated |
| -0,009892026 | 0,951171992 | LONP1 | FALSE | Downregulated |
| 0,015676286 | 0,951171992 | PEPD | FALSE | Upregulated |
| 0,012462998 | 0,951203093 | HECA | FALSE | Upregulated |
| 0,075857074 | 0,951290328 | C19orf33 | FALSE | Upregulated |
| -0,018964282 | 0,951639314 | CCNG2 | FALSE | Downregulated |
| 0,402106523 | 0,951639314 | MYH6 | FALSE | Upregulated |
| -0,109632393 | 0,951997233 | RAB6C | FALSE | Downregulated |
| -0,123586292 | 0,952107468 | ABCC13 | FALSE | Downregulated |
| 0,016443058 | 0,952107468 | CA5B | FALSE | Upregulated |
| -0,044402757 | 0,952107468 | CCDC13 | FALSE | Downregulated |
| 0,023070612 | 0,952107468 | CYP2D6 | FALSE | Upregulated |
| 0,020191643 | 0,952107468 | FAM129B | FALSE | Upregulated |
| -0,06427192 | 0,952107468 | ISLR2 | FALSE | Downregulated |
| -0,199074282 | 0,952107468 | TRNAG19 | FALSE | Downregulated |
| -0,036991249 | 0,952163508 | ANKRD34A | FALSE | Downregulated |
| 0,02597113 | 0,952390844 | PANX2 | FALSE | Upregulated |
| 0,014858301 | 0,952390844 | TRIM65 | FALSE | Upregulated |
| -0,006985434 | 0,952390844 | VWA9 | FALSE | Downregulated |
| -0,072798226 | 0,952410667 | TSPAN32 | FALSE | Downregulated |
| -0,019420199 | 0,952771635 | OLFML1 | FALSE | Downregulated |
| 0,086817124 | 0,952771635 | RP11-219B17.1 | FALSE | Upregulated |
| 0,009338327 | 0,952771635 | ZC3H15 | FALSE | Upregulated |
| -0,010101085 | 0,953057369 | COG1 | FALSE | Downregulated |
| -0,113873801 | 0,953057369 | PIR-FIGF | FALSE | Downregulated |
| -0,020347899 | 0,953057369 | SPP2 | FALSE | Downregulated |
| -0,015277682 | 0,953188928 | ATP7A | FALSE | Downregulated |
| 0,016248612 | 0,953188928 | CYP2C18 | FALSE | Upregulated |
| -0,069216938 | 0,953188928 | LOC101928102 | FALSE | Downregulated |
| 0,01415588 | 0,953188928 | MTRF1L | FALSE | Upregulated |
| -0,013492444 | 0,953188928 | SCMH1 | FALSE | Downregulated |
| -0,051464007 | 0,953306312 | TRNV | FALSE | Downregulated |
| -0,012544733 | 0,953397994 | AKAP10 | FALSE | Downregulated |
| 0,040808943 | 0,953468729 | DLG5-AS1 | FALSE | Upregulated |
| 0,04120857 | 0,953468729 | WBSCR27 | FALSE | Upregulated |
| 0,01326219 | 0,954073834 | ARL15 | FALSE | Upregulated |
| 0,02366053 | 0,954073834 | ZNF670 | FALSE | Upregulated |
| 0,025923614 | 0,954085762 | ZNF366 | FALSE | Upregulated |
| 0,044596028 | 0,954088953 | LRRC66 | FALSE | Upregulated |
| -0,019426414 | 0,954142344 | NRP1 | FALSE | Downregulated |
| 0,075813639 | 0,954167418 | C16orf89 | FALSE | Upregulated |
| 0,025274534 | 0,954420681 | CYB5R2 | FALSE | Upregulated |
| 0,021418579 | 0,954602214 | ADORA2A-AS1 | FALSE | Upregulated |
| 0,017113258 | 0,954602214 | C2CD2L | FALSE | Upregulated |
| -0,513772767 | 0,954602214 | TNNT3 | FALSE | Downregulated |
| -0,070660339 | 0,954638393 | LOC102724676 | FALSE | Downregulated |
| 0,016541607 | 0,954935048 | PITPNM2 | FALSE | Upregulated |

| | | | | |
|---|---|---|---|---|
| 0,024072292 | 0,955207005 | CLEC11A | FALSE | Upregulated |
| -0,01536006 | 0,955280605 | DTX4 | FALSE | Downregulated |
| -0,012162835 | 0,955280605 | MTFMT | FALSE | Downregulated |
| -0,103190657 | 0,955352617 | DKK4 | FALSE | Downregulated |
| -0,012807716 | 0,955364685 | POLR2I | FALSE | Downregulated |
| 0,015292098 | 0,955364685 | TBC1D22B | FALSE | Upregulated |
| 0,012939451 | 0,955383951 | DNAL1 | FALSE | Upregulated |
| -0,036572576 | 0,955478815 | C6orf226 | FALSE | Downregulated |
| 0,010268355 | 0,955478815 | CHM | FALSE | Upregulated |
| -0,015176204 | 0,955478815 | DHX16 | FALSE | Downregulated |
| -0,017220213 | 0,955478815 | ESAM | FALSE | Downregulated |
| -0,079759074 | 0,955478815 | RGPD3 | FALSE | Downregulated |
| 0,158371629 | 0,955495221 | KLRC4-KLRK1 | FALSE | Upregulated |
| 0,106404573 | 0,955539954 | LOC101927012 | FALSE | Upregulated |
| 0,070974573 | 0,955539954 | RP11-426A6.5 | FALSE | Upregulated |
| -0,112813905 | 0,955555289 | GABARAPL3 | FALSE | Downregulated |
| 0,06975411 | 0,955555289 | KCNH8 | FALSE | Upregulated |
| -0,063921249 | 0,955555289 | KIAA0408 | FALSE | Downregulated |
| -0,007875583 | 0,955773199 | CDC23 | FALSE | Downregulated |
| 0,049947264 | 0,955804079 | CASKIN1 | FALSE | Upregulated |
| 0,011400225 | 0,955855267 | IFNAR1 | FALSE | Upregulated |
| -0,058195046 | 0,956048119 | FSIP1 | FALSE | Downregulated |
| 0,038309957 | 0,956433669 | PAX5 | FALSE | Upregulated |
| 0,013228314 | 0,956433669 | PCYT2 | FALSE | Upregulated |
| -0,075708435 | 0,956575754 | DKFZP434K028 | FALSE | Downregulated |
| -0,02155432 | 0,956675853 | PPP2R3B | FALSE | Downregulated |
| -0,097127994 | 0,956688569 | BLOC1S5-TXNDC5 | FALSE | Downregulated |
| -0,01491734 | 0,956732153 | F8 | FALSE | Downregulated |
| 0,034337552 | 0,956732153 | ZNF382 | FALSE | Upregulated |
| -0,009859872 | 0,956732153 | ZNF512 | FALSE | Downregulated |
| 0,031973566 | 0,956746127 | CTC-338M12.4 | FALSE | Upregulated |
| 0,017482295 | 0,956746127 | DAPK1 | FALSE | Upregulated |
| 0,014070286 | 0,956746127 | DLAT | FALSE | Upregulated |
| 0,016413936 | 0,956789573 | BGN | FALSE | Upregulated |
| -0,02175626 | 0,956859756 | C5orf63 | FALSE | Downregulated |
| 0,008081698 | 0,956859756 | PPP3CB | FALSE | Upregulated |
| 0,01400548 | 0,956859756 | TMEM175 | FALSE | Upregulated |
| 0,071163427 | 0,956880583 | DDIT4L | FALSE | Upregulated |
| 0,022615957 | 0,956880583 | ZNF83 | FALSE | Upregulated |
| 0,081424636 | 0,956942363 | CASC15 | FALSE | Upregulated |
| -0,016406098 | 0,956942363 | MITD1 | FALSE | Downregulated |
| -0,014652459 | 0,957149063 | TP53INP1 | FALSE | Downregulated |
| -0,045064363 | 0,957202703 | ATP5L2 | FALSE | Downregulated |
| -0,485663082 | 0,957202703 | CSRP3 | FALSE | Downregulated |
| 0,065390318 | 0,957202703 | ERVMER34-1 | FALSE | Upregulated |
| 0,010021464 | 0,957202703 | INTS6 | FALSE | Upregulated |
| 0,095235834 | 0,957202703 | MAK | FALSE | Upregulated |
| 0,053124143 | 0,957202703 | MUC3A | FALSE | Upregulated |
| 0,487558172 | 0,957202703 | MYPN | FALSE | Upregulated |
| -0,02842349 | 0,957202703 | ZNF71 | FALSE | Downregulated |
| -0,040776116 | 0,957202703 | ZNF833P | FALSE | Downregulated |

| | | | | |
|---|---|---|---|---|
| -0,044150418 | 0,957225533 | RP11-557H15.4 | FALSE | Downregulated |
| -0,029903544 | 0,957369635 | ATXN7L2 | FALSE | Downregulated |
| 0,482435091 | 0,957369635 | LOC102724843 | FALSE | Upregulated |
| -0,076708838 | 0,95740325 | TCTE1 | FALSE | Downregulated |
| 0,007346181 | 0,957435434 | POLR3E | FALSE | Upregulated |
| 0,027508736 | 0,95746771 | WDR54 | FALSE | Upregulated |
| -0,052576808 | 0,957533203 | LOC101927546 | FALSE | Downregulated |
| 0,019653309 | 0,957533203 | SLC2A10 | FALSE | Upregulated |
| -0,050474288 | 0,95756031 | DCLK3 | FALSE | Downregulated |
| -0,010549928 | 0,95756031 | MAT2B | FALSE | Downregulated |
| 0,009000644 | 0,95756031 | MED10 | FALSE | Upregulated |
| -0,012654817 | 0,95756031 | METTL23 | FALSE | Downregulated |
| 0,009736699 | 0,957612078 | PTOV1 | FALSE | Upregulated |
| -0,007853175 | 0,957826565 | MED1 | FALSE | Downregulated |
| 0,016582289 | 0,958120662 | ANKRD37 | FALSE | Upregulated |
| 0,069965876 | 0,958223436 | SIGLEC6 | FALSE | Upregulated |
| -0,079067218 | 0,958365297 | RRH | FALSE | Downregulated |
| -0,030461436 | 0,95845048 | LY75 | FALSE | Downregulated |
| -0,015364251 | 0,958483309 | DAB2 | FALSE | Downregulated |
| -0,033951094 | 0,958490943 | CCDC157 | FALSE | Downregulated |
| 0,0133864 | 0,958619498 | CMTM4 | FALSE | Upregulated |
| 0,020894268 | 0,958619498 | HEPH | FALSE | Upregulated |
| -0,014821466 | 0,958619498 | WRAP73 | FALSE | Downregulated |
| -0,04116329 | 0,958722439 | COLCA1 | FALSE | Downregulated |
| -0,010727734 | 0,958722439 | FAHD2A | FALSE | Downregulated |
| -0,061454386 | 0,958722439 | FAM228A | FALSE | Downregulated |
| 0,033784132 | 0,958722439 | TAT | FALSE | Upregulated |
| -0,054649142 | 0,958873317 | SIM1 | FALSE | Downregulated |
| 0,076825289 | 0,958939944 | LOXHD1 | FALSE | Upregulated |
| 0,049873867 | 0,958939944 | TMED10P1 | FALSE | Upregulated |
| 0,015032769 | 0,959058164 | ZNF408 | FALSE | Upregulated |
| 0,087096992 | 0,959322405 | DSC3 | FALSE | Upregulated |
| -0,05683946 | 0,959351938 | CTAGE7P | FALSE | Downregulated |
| 0,098272007 | 0,959351938 | GALNTL6 | FALSE | Upregulated |
| -0,033081594 | 0,959369606 | KCNK6 | FALSE | Downregulated |
| 0,012420978 | 0,959369606 | MCAT | FALSE | Upregulated |
| -0,013879799 | 0,959460257 | BTD | FALSE | Downregulated |
| -0,015295335 | 0,959460257 | SLC25A14 | FALSE | Downregulated |
| -0,015959548 | 0,959703873 | IDH2 | FALSE | Downregulated |
| 0,02130008 | 0,959907977 | RAI2 | FALSE | Upregulated |
| -0,013203309 | 0,959907977 | SLC23A1 | FALSE | Downregulated |
| 0,00928431 | 0,959996468 | UBXN1 | FALSE | Upregulated |
| 0,452663792 | 0,960077749 | MYBPC1 | FALSE | Upregulated |
| 0,006844896 | 0,9600981 | CNOT1 | FALSE | Upregulated |
| -0,041048855 | 0,960160043 | SCARNA17 | FALSE | Downregulated |
| 0,013935753 | 0,960413477 | SLC22A5 | FALSE | Upregulated |
| 0,013662104 | 0,960439543 | JADE1 | FALSE | Upregulated |
| -0,013331661 | 0,960439543 | ZNRD1 | FALSE | Downregulated |
| -0,083281885 | 0,96067964 | C3orf83 | FALSE | Downregulated |
| -0,014563789 | 0,96067964 | CD2AP | FALSE | Downregulated |
| -0,029211147 | 0,960714265 | LINC00883 | FALSE | Downregulated |

| | | | | |
|---|---|---|---|---|
| 0,075735161 | 0,960728894 | A2MP1 | FALSE | Upregulated |
| -0,138880732 | 0,960766174 | RGPD6 | FALSE | Downregulated |
| -0,007585761 | 0,961087264 | EXOSC10 | FALSE | Downregulated |
| -0,042422422 | 0,961142584 | HSD17B7P2 | FALSE | Downregulated |
| 0,016470017 | 0,961314597 | FKBP2 | FALSE | Upregulated |
| -0,063522523 | 0,961314597 | JPH3 | FALSE | Downregulated |
| 0,022657157 | 0,961314597 | MSRB3 | FALSE | Upregulated |
| 0,010420872 | 0,961314597 | PRMT5 | FALSE | Upregulated |
| -0,008465529 | 0,961360033 | SLC35F5 | FALSE | Downregulated |
| -0,017320351 | 0,961463845 | NAV1 | FALSE | Downregulated |
| -0,029660609 | 0,961617173 | AIF1L | FALSE | Downregulated |
| -0,012950396 | 0,96171854 | CHST14 | FALSE | Downregulated |
| -0,012961626 | 0,961812121 | GAR1 | FALSE | Downregulated |
| -0,078948154 | 0,96181675 | LOC101927254 | FALSE | Downregulated |
| 0,035333041 | 0,961859822 | GUSBP2 | FALSE | Upregulated |
| 0,047885727 | 0,961859822 | LOC101926994 | FALSE | Upregulated |
| -0,012825324 | 0,961859822 | PLEKHA6 | FALSE | Downregulated |
| -0,0148244 | 0,961902438 | CCDC23 | FALSE | Downregulated |
| -0,012233294 | 0,961902438 | HEXIM1 | FALSE | Downregulated |
| -0,194647523 | 0,961902438 | LOC101060389 | FALSE | Downregulated |
| -0,022932925 | 0,961902438 | ZNF773 | FALSE | Downregulated |
| -0,039831961 | 0,962378533 | FUT8-AS1 | FALSE | Downregulated |
| 0,094860102 | 0,962378533 | LOC100507387 | FALSE | Upregulated |
| 0,014024018 | 0,962378533 | PPIL3 | FALSE | Upregulated |
| -0,03801737 | 0,962643344 | LOC101929374 | FALSE | Downregulated |
| -0,023062865 | 0,962643344 | PRKCQ-AS1 | FALSE | Downregulated |
| -0,023398993 | 0,962818163 | CETP | FALSE | Downregulated |
| -0,018813811 | 0,962818163 | DLG3 | FALSE | Downregulated |
| -0,009355594 | 0,962818163 | NAGA | FALSE | Downregulated |
| 0,028547549 | 0,962818163 | PCP4L1 | FALSE | Upregulated |
| 0,424664862 | 0,962955869 | LMOD2 | FALSE | Upregulated |
| -0,425241706 | 0,962955869 | XIRP2 | FALSE | Downregulated |
| 0,044047587 | 0,963147332 | MATN1-AS1 | FALSE | Upregulated |
| -0,00704915 | 0,963242256 | WNK1 | FALSE | Downregulated |
| 0,010940216 | 0,963300452 | FOXK1 | FALSE | Upregulated |
| -0,012403676 | 0,96360298 | ZSWIM1 | FALSE | Downregulated |
| 0,018193426 | 0,963616416 | KIFC3 | FALSE | Upregulated |
| -0,010051769 | 0,963822769 | C11orf31 | FALSE | Downregulated |
| 0,027458472 | 0,963822769 | C16orf96 | FALSE | Upregulated |
| -0,102150849 | 0,963822769 | JSRP1 | FALSE | Downregulated |
| 0,01287379 | 0,963822769 | LEPREL4 | FALSE | Upregulated |
| -0,058534093 | 0,963822769 | LOC102724737 | FALSE | Downregulated |
| 0,041310186 | 0,963822769 | LOC90834 | FALSE | Upregulated |
| 0,006982458 | 0,963822769 | MIER1 | FALSE | Upregulated |
| 0,016945293 | 0,963822769 | TMEM98 | FALSE | Upregulated |
| 0,010275409 | 0,963822769 | TRAPPC10 | FALSE | Upregulated |
| 0,057162331 | 0,963955919 | AC005594.3 | FALSE | Upregulated |
| 0,035398663 | 0,964074959 | LINC00310 | FALSE | Upregulated |
| 0,050596063 | 0,96416839 | CACTIN-AS1 | FALSE | Upregulated |
| 0,03142889 | 0,96416839 | LOC101928158 | FALSE | Upregulated |
| -0,061171172 | 0,964199839 | KLHL14 | FALSE | Downregulated |

| | | | | |
|---|---|---|---|---|
| -0,09794728 | 0,964199839 | LOC100505498 | FALSE | Downregulated |
| -0,075748576 | 0,964199839 | MSANTD1 | FALSE | Downregulated |
| -0,00822751 | 0,964199839 | UHRF1BP1L | FALSE | Downregulated |
| 0,01169104 | 0,964199839 | ZSCAN22 | FALSE | Upregulated |
| 0,017120504 | 0,964246765 | FBXL7 | FALSE | Upregulated |
| 0,046900353 | 0,964270853 | LOC102724541 | FALSE | Upregulated |
| -0,011357752 | 0,964333324 | MBD5 | FALSE | Downregulated |
| 0,010033768 | 0,964352537 | MMADHC | FALSE | Upregulated |
| -0,086279361 | 0,964352537 | SLC22A11 | FALSE | Downregulated |
| 0,007924941 | 0,964500486 | ASGR2 | FALSE | Upregulated |
| -0,016253884 | 0,964500486 | C8orf4 | FALSE | Downregulated |
| 0,025586049 | 0,96458278 | NRG3 | FALSE | Upregulated |
| 0,006798493 | 0,96458278 | RNF11 | FALSE | Upregulated |
| 0,008450232 | 0,964615612 | PIGH | FALSE | Upregulated |
| 0,013406449 | 0,964615612 | PLCG2 | FALSE | Upregulated |
| -0,017135186 | 0,964639619 | ASB16-AS1 | FALSE | Downregulated |
| 0,012170026 | 0,964639619 | POLE3 | FALSE | Upregulated |
| 0,01188656 | 0,964702557 | CWC27 | FALSE | Upregulated |
| -0,050623669 | 0,964754947 | PSPN | FALSE | Downregulated |
| 0,011395809 | 0,964776698 | GSK3A | FALSE | Upregulated |
| 0,011048735 | 0,964782595 | ATP5I | FALSE | Upregulated |
| 0,055004822 | 0,964782595 | N4BP3 | FALSE | Upregulated |
| 0,010370299 | 0,964962578 | AP3S2 | FALSE | Upregulated |
| 0,011138557 | 0,964962578 | NGRN | FALSE | Upregulated |
| -0,07164375 | 0,964962578 | SSUH2 | FALSE | Downregulated |
| -0,072856939 | 0,965031214 | LOC102724560 | FALSE | Downregulated |
| 0,012652566 | 0,965038173 | LZTS2 | FALSE | Upregulated |
| 0,009360084 | 0,965126436 | ZNF226 | FALSE | Upregulated |
| -0,009645986 | 0,965238651 | SMG6 | FALSE | Downregulated |
| 0,010212435 | 0,965243308 | DOT1L | FALSE | Upregulated |
| -0,022662508 | 0,965390787 | LINC01006 | FALSE | Downregulated |
| 0,06174557 | 0,965487521 | LOC100129215 | FALSE | Upregulated |
| 0,073039917 | 0,965508581 | UNC80 | FALSE | Upregulated |
| 0,015505842 | 0,965809914 | GGT5 | FALSE | Upregulated |
| -0,010107859 | 0,966113774 | DUSP7 | FALSE | Downregulated |
| 0,010477001 | 0,966113774 | RAB6A | FALSE | Upregulated |
| -0,024677654 | 0,966129258 | LOC102724105 | FALSE | Downregulated |
| 0,010705784 | 0,966186514 | PUS1 | FALSE | Upregulated |
| 0,032339249 | 0,966279719 | C1orf186 | FALSE | Upregulated |
| 0,044821439 | 0,966279719 | LOC100996746 | FALSE | Upregulated |
| 0,038899824 | 0,966279719 | PLCB4 | FALSE | Upregulated |
| 0,097540196 | 0,966379988 | LOC100505874 | FALSE | Upregulated |
| 0,014323399 | 0,966379988 | SYNRG | FALSE | Upregulated |
| -0,020306834 | 0,966523301 | ZNF528 | FALSE | Downregulated |
| -0,033344591 | 0,966563287 | LOC284581 | FALSE | Downregulated |
| -0,032774765 | 0,966606895 | LRCH2 | FALSE | Downregulated |
| -0,022482322 | 0,966606895 | WWC2-AS2 | FALSE | Downregulated |
| -0,092067976 | 0,966974108 | TRNAG4 | FALSE | Downregulated |
| 0,026553058 | 0,967036529 | RNF39 | FALSE | Upregulated |
| -0,00822027 | 0,967102423 | AGAP3 | FALSE | Downregulated |
| -0,032516761 | 0,967102423 | BEX2 | FALSE | Downregulated |

| | | | | |
|---|---|---|---|---|
| -0,008821228 | 0,967102423 | C12orf10 | FALSE | Downregulated |
| -0,03923846 | 0,967102423 | PPAP2C | FALSE | Downregulated |
| -0,009680409 | 0,967102423 | RBMX2 | FALSE | Downregulated |
| -0,009964491 | 0,967102423 | TPRA1 | FALSE | Downregulated |
| 0,048528278 | 0,967121065 | LOC101929074 | FALSE | Upregulated |
| 0,014349112 | 0,967121065 | PALMD | FALSE | Upregulated |
| 0,046436056 | 0,967121065 | PPP5D1 | FALSE | Upregulated |
| 0,02375318 | 0,967370403 | SEPSECS-AS1 | FALSE | Upregulated |
| 0,02896934 | 0,967400337 | KCNS3 | FALSE | Upregulated |
| 0,012083264 | 0,96751683 | CHID1 | FALSE | Upregulated |
| 0,004565358 | 0,96751683 | PTPRA | FALSE | Upregulated |
| 0,011658214 | 0,96751683 | SEC24B | FALSE | Upregulated |
| 0,010258306 | 0,967674006 | ING5 | FALSE | Upregulated |
| -0,009805128 | 0,967674088 | BRAT1 | FALSE | Downregulated |
| 0,019275709 | 0,967674088 | HCG11 | FALSE | Upregulated |
| 0,011510034 | 0,967674088 | NUP210 | FALSE | Upregulated |
| -0,009442717 | 0,96772717 | CCDC115 | FALSE | Downregulated |
| -0,009544184 | 0,967900027 | USP42 | FALSE | Downregulated |
| 0,058453704 | 0,968016562 | CBLN2 | FALSE | Upregulated |
| -0,041232938 | 0,968016562 | ELOVL4 | FALSE | Downregulated |
| 0,080179065 | 0,968243643 | HOXD-AS1 | FALSE | Upregulated |
| 0,065451415 | 0,968243643 | LOC102723709 | FALSE | Upregulated |
| -0,013710353 | 0,968243643 | UGT2B10 | FALSE | Downregulated |
| 0,008630346 | 0,968417475 | TAF11 | FALSE | Upregulated |
| -0,083333687 | 0,968469852 | ANGPT4 | FALSE | Downregulated |
| -0,330338596 | 0,968469852 | ATP1B4 | FALSE | Downregulated |
| -0,006456742 | 0,968469852 | ERAL1 | FALSE | Downregulated |
| -0,035981414 | 0,968469852 | RP11-356I2.4 | FALSE | Downregulated |
| -0,013653366 | 0,968469852 | ZBTB25 | FALSE | Downregulated |
| -0,034085898 | 0,968507014 | RGPD4 | FALSE | Downregulated |
| -0,362561845 | 0,968507014 | SLN | FALSE | Downregulated |
| -0,014606488 | 0,968687059 | RGL3 | FALSE | Downregulated |
| -0,083130657 | 0,968722661 | PPP1R2P3 | FALSE | Downregulated |
| -0,008326804 | 0,968722661 | SIAE | FALSE | Downregulated |
| 0,024426979 | 0,968722661 | SLCO1B7 | FALSE | Upregulated |
| 0,007061445 | 0,968842103 | OSBPL9 | FALSE | Upregulated |
| 0,052451315 | 0,96893435 | SLC9A5 | FALSE | Upregulated |
| 0,011531631 | 0,96893435 | STXBP5 | FALSE | Upregulated |
| -0,04932311 | 0,96894887 | MAEL | FALSE | Downregulated |
| 0,019995451 | 0,969224736 | LOC101927340 | FALSE | Upregulated |
| -0,008635675 | 0,969320972 | SERF2-C15ORF63 | FALSE | Downregulated |
| -0,008178248 | 0,969373078 | IPMK | FALSE | Downregulated |
| -0,034834702 | 0,969490133 | LOC101928584 | FALSE | Downregulated |
| -0,005982486 | 0,969562838 | DNAJB2 | FALSE | Downregulated |
| -0,018903371 | 0,969622961 | IGFALS | FALSE | Downregulated |
| -0,006225354 | 0,969622961 | PRPS1 | FALSE | Downregulated |
| -0,024228535 | 0,969650962 | MFAP4 | FALSE | Downregulated |
| 0,014297009 | 0,969682051 | FETUB | FALSE | Upregulated |
| 0,038541821 | 0,969869992 | TIGD4 | FALSE | Upregulated |
| 0,011943211 | 0,969928394 | BEX4 | FALSE | Upregulated |
| -0,063989396 | 0,969928394 | FAM196B | FALSE | Downregulated |

| | | | | |
|---|---|---|---|---|
| 0,016638179 | 0,969941975 | PMF1-BGLAP | FALSE | Upregulated |
| -0,140653174 | 0,969941975 | RP4-669L17.10-2 | FALSE | Downregulated |
| 0,009963505 | 0,969946555 | ZNF567 | FALSE | Upregulated |
| -0,007395302 | 0,970021505 | AK3 | FALSE | Downregulated |
| 0,061564324 | 0,970021505 | CPEB1 | FALSE | Upregulated |
| -0,007892878 | 0,970021505 | ETFA | FALSE | Downregulated |
| -0,026003431 | 0,970021505 | LOC101928830 | FALSE | Downregulated |
| -0,006100272 | 0,970021505 | MRPS18A | FALSE | Downregulated |
| 0,014275647 | 0,970021505 | RDX | FALSE | Upregulated |
| -0,008136529 | 0,970021505 | TEX10 | FALSE | Downregulated |
| -0,016630376 | 0,970021505 | ZNF460 | FALSE | Downregulated |
| -0,034321227 | 0,970144385 | ITGB1BP2 | FALSE | Downregulated |
| 0,015199731 | 0,97018022 | ZFPM2 | FALSE | Upregulated |
| 0,061383578 | 0,970577297 | PDZD3 | FALSE | Upregulated |
| -0,041296554 | 0,970577297 | SMG1P1 | FALSE | Downregulated |
| -0,016430137 | 0,970577297 | TP53I11 | FALSE | Downregulated |
| -0,008211561 | 0,970583722 | RYK | FALSE | Downregulated |
| 0,125638897 | 0,971279135 | HCCAT5 | FALSE | Upregulated |
| 0,004644448 | 0,971479684 | RDH14 | FALSE | Upregulated |
| -0,008199159 | 0,971540215 | COX5B | FALSE | Downregulated |
| 0,007140738 | 0,971540215 | GPR155 | FALSE | Upregulated |
| -0,076139693 | 0,971540215 | LOC285556 | FALSE | Downregulated |
| 0,017521914 | 0,971540215 | SRGAP2B | FALSE | Upregulated |
| -0,014089341 | 0,971540215 | UST | FALSE | Downregulated |
| 0,028235558 | 0,971590958 | NNMT | FALSE | Upregulated |
| -0,009907298 | 0,971595075 | CCSER2 | FALSE | Downregulated |
| 0,028844785 | 0,971595075 | SSPN | FALSE | Upregulated |
| -0,007590349 | 0,971595075 | TMEM129 | FALSE | Downregulated |
| -0,029185539 | 0,971595075 | ZMIZ1-AS1 | FALSE | Downregulated |
| 0,008866944 | 0,971630264 | EEF1A1 | FALSE | Upregulated |
| -0,048801549 | 0,971636698 | LINC00881 | FALSE | Downregulated |
| 0,059224423 | 0,971817059 | LOC102724135 | FALSE | Upregulated |
| 0,00654285 | 0,971817059 | MBTPS2 | FALSE | Upregulated |
| -0,306697188 | 0,971817059 | TTN | FALSE | Downregulated |
| 0,018012846 | 0,971817059 | ZDHHC19 | FALSE | Upregulated |
| -0,010076113 | 0,971817059 | ZYG11B | FALSE | Downregulated |
| -0,006485141 | 0,971928451 | BCAS3 | FALSE | Downregulated |
| -0,022636396 | 0,972138999 | C14orf182 | FALSE | Downregulated |
| 0,006766416 | 0,972138999 | FAM188A | FALSE | Upregulated |
| -0,013255299 | 0,972138999 | FKBP10 | FALSE | Downregulated |
| -0,125003353 | 0,972138999 | LOC100288570 | FALSE | Downregulated |
| -0,007847145 | 0,972138999 | MARVELD2 | FALSE | Downregulated |
| 0,006644315 | 0,972138999 | YTHDC2 | FALSE | Upregulated |
| 0,036654306 | 0,972138999 | ZNF321P | FALSE | Upregulated |
| 0,008288213 | 0,972199024 | ODF2 | FALSE | Upregulated |
| -0,003753213 | 0,972202595 | PIK3C3 | FALSE | Downregulated |
| -0,030670563 | 0,972288627 | RPL13AP6 | FALSE | Downregulated |
| 0,044740873 | 0,972335906 | FLJ38723 | FALSE | Upregulated |
| -0,014944205 | 0,972335906 | SNAI2 | FALSE | Downregulated |
| -0,006822888 | 0,972372915 | KYNU | FALSE | Downregulated |
| -0,03858481 | 0,97241285 | LOC284412 | FALSE | Downregulated |

| | | | | |
|---|---|---|---|---|
| 0,287489996 | 0,972450581 | SLX1A | FALSE | Upregulated |
| -0,034731552 | 0,972485962 | CYP1B1-AS1 | FALSE | Downregulated |
| 0,01176273 | 0,972488582 | FBXL19 | FALSE | Upregulated |
| 0,032432295 | 0,972803487 | DLEU7 | FALSE | Upregulated |
| -0,309835439 | 0,972803487 | RNU1-2 | FALSE | Downregulated |
| -0,017343726 | 0,97300508 | THSD7A | FALSE | Downregulated |
| -0,008196587 | 0,973061135 | APITD1 | FALSE | Downregulated |
| -0,038716733 | 0,973061135 | LINC00624 | FALSE | Downregulated |
| 0,016766361 | 0,973070183 | PHOSPHO2 | FALSE | Upregulated |
| -0,048560751 | 0,973070183 | PTPRG-AS1 | FALSE | Downregulated |
| 0,015923922 | 0,973070183 | TMOD1 | FALSE | Upregulated |
| 0,005817859 | 0,973070183 | UTP6 | FALSE | Upregulated |
| 0,036475147 | 0,973238037 | FAM227A | FALSE | Upregulated |
| 0,008669641 | 0,973245602 | GNA14 | FALSE | Upregulated |
| 0,007509783 | 0,973593883 | TMEM110 | FALSE | Upregulated |
| -0,016979375 | 0,973753777 | NINJ2 | FALSE | Downregulated |
| 0,298962575 | 0,974018194 | ACTA1 | FALSE | Upregulated |
| -0,008266969 | 0,974018194 | ACYP2 | FALSE | Downregulated |
| -0,033184111 | 0,974018194 | C16orf71 | FALSE | Downregulated |
| 0,03348305 | 0,974018194 | CCSER1 | FALSE | Upregulated |
| 0,022599892 | 0,974018194 | CHST4 | FALSE | Upregulated |
| 0,035763059 | 0,974018194 | FAM201A | FALSE | Upregulated |
| -0,013597424 | 0,974018194 | IGFBP5 | FALSE | Downregulated |
| 0,028993781 | 0,974018194 | LOC101928291 | FALSE | Upregulated |
| -0,043523499 | 0,974018194 | LOC101928512 | FALSE | Downregulated |
| 0,016201116 | 0,974018194 | NAT14 | FALSE | Upregulated |
| 0,02424546 | 0,974018194 | PAX8-AS1 | FALSE | Upregulated |
| 0,035749082 | 0,974018194 | RRN3P2 | FALSE | Upregulated |
| 0,004540517 | 0,974018194 | SUPT7L | FALSE | Upregulated |
| 0,04200257 | 0,974217014 | LINC00525 | FALSE | Upregulated |
| -0,01925712 | 0,974243852 | SLC16A9 | FALSE | Downregulated |
| -0,045073425 | 0,974523479 | CDHR4 | FALSE | Downregulated |
| 0,050189772 | 0,974631398 | PODNL1 | FALSE | Upregulated |
| 0,010031619 | 0,974660363 | LSM3 | FALSE | Upregulated |
| -0,007029637 | 0,974692819 | LTBR | FALSE | Downregulated |
| 0,006578503 | 0,974993609 | CYB5R1 | FALSE | Upregulated |
| 0,035421944 | 0,974993609 | HSPA7 | FALSE | Upregulated |
| -0,039586212 | 0,974993609 | LOC102723820 | FALSE | Downregulated |
| 0,009450671 | 0,974993609 | MIPOL1 | FALSE | Upregulated |
| 0,088497336 | 0,975079312 | HHATL | FALSE | Upregulated |
| 0,010826226 | 0,975143931 | SERPING1 | FALSE | Upregulated |
| 0,012893039 | 0,975174986 | ATP2B2 | FALSE | Upregulated |
| -0,060414206 | 0,975224406 | NBL1 | FALSE | Downregulated |
| 0,050754431 | 0,97522487 | APOL5 | FALSE | Upregulated |
| 0,004596777 | 0,97522487 | CDC123 | FALSE | Upregulated |
| -0,00845274 | 0,97522487 | DECR2 | FALSE | Downregulated |
| 0,020088927 | 0,97522487 | LOC101926931 | FALSE | Upregulated |
| 0,035350197 | 0,97522487 | LOC101929687 | FALSE | Upregulated |
| -0,053423186 | 0,97522487 | LOC102724081 | FALSE | Downregulated |
| 0,023341884 | 0,97522487 | ZNF208 | FALSE | Upregulated |
| 0,005460655 | 0,97522487 | ZNF609 | FALSE | Upregulated |

| | | | | |
|---|---|---|---|---|
| -0,043173246 | 0,97553468 | ADRA2C | FALSE | Downregulated |
| -0,007502289 | 0,97553468 | GOLGA3 | FALSE | Downregulated |
| -0,009238158 | 0,97553468 | ZNF771 | FALSE | Downregulated |
| 0,281929147 | 0,975599561 | MYL2 | FALSE | Upregulated |
| 0,041390969 | 0,975601024 | HS6ST2 | FALSE | Upregulated |
| -0,008840653 | 0,975601331 | DDX10 | FALSE | Downregulated |
| 0,033729169 | 0,975603548 | MYLK-AS1 | FALSE | Upregulated |
| -0,050921372 | 0,975792723 | RP4-794H19.4 | FALSE | Downregulated |
| 0,032221064 | 0,975893098 | LOC102723311 | FALSE | Upregulated |
| 0,01358504 | 0,975893098 | RAP1GAP2 | FALSE | Upregulated |
| -0,006323412 | 0,975976122 | RPS27A | FALSE | Downregulated |
| 0,055336371 | 0,97618356 | LOC100506114 | FALSE | Upregulated |
| -0,015634282 | 0,97618356 | RFTN2 | FALSE | Downregulated |
| -0,004301599 | 0,97641371 | MAK16 | FALSE | Downregulated |
| -0,02122492 | 0,976531869 | LOC100506675 | FALSE | Downregulated |
| -0,009837371 | 0,976531869 | TBCEL | FALSE | Downregulated |
| 0,008373161 | 0,976649728 | MYADM | FALSE | Upregulated |
| -0,271842309 | 0,976703255 | ACTC1 | FALSE | Downregulated |
| 0,005087257 | 0,976821404 | FYN | FALSE | Upregulated |
| 0,018458072 | 0,976845045 | LOC102724061 | FALSE | Upregulated |
| 0,011949541 | 0,976845045 | NQO2 | FALSE | Upregulated |
| -0,042237816 | 0,976999926 | ZNF20 | FALSE | Downregulated |
| -0,025304305 | 0,976999926 | ZNF883 | FALSE | Downregulated |
| 0,004029596 | 0,977102535 | THOC5 | FALSE | Upregulated |
| 0,019541337 | 0,977363751 | PTGDR | FALSE | Upregulated |
| 0,004660015 | 0,977532349 | STUB1 | FALSE | Upregulated |
| 0,011962355 | 0,977607391 | KCTD1 | FALSE | Upregulated |
| -0,004381841 | 0,977978668 | SEC23IP | FALSE | Downregulated |
| -0,043651149 | 0,977978668 | TUBBP5 | FALSE | Downregulated |
| 0,008501751 | 0,978138181 | EIF4EBP1 | FALSE | Upregulated |
| -0,008720126 | 0,978138181 | SMC1A | FALSE | Downregulated |
| 0,014080707 | 0,978138181 | ZNF525 | FALSE | Upregulated |
| -0,038387953 | 0,978442123 | PKD2L2 | FALSE | Downregulated |
| -0,032301804 | 0,978484881 | FLJ22447 | FALSE | Downregulated |
| -0,015823676 | 0,978509691 | UBAP1L | FALSE | Downregulated |
| -0,027305768 | 0,978549432 | LOC101928862 | FALSE | Downregulated |
| -0,010312149 | 0,978666057 | LRRC32 | FALSE | Downregulated |
| -0,029778892 | 0,978816315 | USP32P2 | FALSE | Downregulated |
| 0,050804485 | 0,978872736 | SPOCK1 | FALSE | Upregulated |
| -0,006924244 | 0,978878936 | HSD17B11 | FALSE | Downregulated |
| 0,249969787 | 0,979232143 | SMYD1 | FALSE | Upregulated |
| -0,020101503 | 0,979284905 | SNCA | FALSE | Downregulated |
| 0,010255686 | 0,979386987 | FANCM | FALSE | Upregulated |
| 0,003753072 | 0,979386987 | GLOD4 | FALSE | Upregulated |
| 0,017360666 | 0,979386987 | RP11-132A1.4 | FALSE | Upregulated |
| -0,006982086 | 0,979592313 | BRPF1 | FALSE | Downregulated |
| -0,005065112 | 0,979592313 | CTDSP1 | FALSE | Downregulated |
| -0,050467299 | 0,979592313 | FLJ16779 | FALSE | Downregulated |
| 0,01537386 | 0,979592313 | MRPL45P2 | FALSE | Upregulated |
| -0,024200358 | 0,979592313 | RP5-1074L1.1 | FALSE | Downregulated |
| -0,012187503 | 0,979700151 | COL14A1 | FALSE | Downregulated |

| | | | | |
|---|---|---|---|---|
| -0,030148672 | 0,979912035 | ERI3-IT1 | FALSE | Downregulated |
| -0,049928875 | 0,979938851 | SPDYE2 | FALSE | Downregulated |
| 0,028744325 | 0,98000615 | DCHS2 | FALSE | Upregulated |
| 0,005715535 | 0,98000615 | FKBP9 | FALSE | Upregulated |
| -0,022292287 | 0,98000615 | RASSF1-AS1 | FALSE | Downregulated |
| -0,00392017 | 0,98000615 | RTN3 | FALSE | Downregulated |
| 0,004814594 | 0,98000615 | TIMM21 | FALSE | Upregulated |
| 0,037574868 | 0,980018563 | CD207 | FALSE | Upregulated |
| 0,011987522 | 0,980018563 | FAM99A | FALSE | Upregulated |
| 0,037202338 | 0,980018563 | GDNF-AS1 | FALSE | Upregulated |
| -0,050006318 | 0,980018563 | LOC101926969 | FALSE | Downregulated |
| 0,021813856 | 0,980018563 | LOC102724507 | FALSE | Upregulated |
| -0,021121882 | 0,980018563 | NME5 | FALSE | Downregulated |
| -0,00567799 | 0,980040421 | DHCR24 | FALSE | Downregulated |
| 0,047384104 | 0,980040421 | TAF1L | FALSE | Upregulated |
| 0,210237064 | 0,980040421 | TRIM54 | FALSE | Upregulated |
| 0,008136992 | 0,980048053 | CCDC167 | FALSE | Upregulated |
| 0,006579383 | 0,980048053 | NLRX1 | FALSE | Upregulated |
| 0,041368172 | 0,980067958 | RPSAP52 | FALSE | Upregulated |
| 0,004254505 | 0,980084903 | TMED10 | FALSE | Upregulated |
| 0,027635958 | 0,980089976 | HNRNPCL1 | FALSE | Upregulated |
| 0,193264981 | 0,980089976 | LOC102724828 | FALSE | Upregulated |
| 0,029207534 | 0,980089976 | PLD5 | FALSE | Upregulated |
| -0,004319583 | 0,980179234 | NUFIP2 | FALSE | Downregulated |
| 0,009674706 | 0,980179234 | PIDD | FALSE | Upregulated |
| 0,003486719 | 0,980237065 | CCDC6 | FALSE | Upregulated |
| 0,005769462 | 0,980237065 | CHDH | FALSE | Upregulated |
| 0,00775748 | 0,980237065 | DNAJC3 | FALSE | Upregulated |
| 0,016668345 | 0,98061765 | CNTNAP3B | FALSE | Upregulated |
| -0,025552392 | 0,98061765 | FLJ44635 | FALSE | Downregulated |
| 0,006080956 | 0,98096152 | RPUSD2 | FALSE | Upregulated |
| 0,006258445 | 0,981172137 | RPL27 | FALSE | Upregulated |
| -0,003301029 | 0,981172137 | WDTC1 | FALSE | Downregulated |
| 0,036011461 | 0,981272843 | LOC102724024 | FALSE | Upregulated |
| 0,003770705 | 0,981272843 | PSME4 | FALSE | Upregulated |
| 0,0050693 | 0,981377567 | CFDP1 | FALSE | Upregulated |
| 0,02743706 | 0,981377567 | DIRC3 | FALSE | Upregulated |
| 0,01822121 | 0,981377567 | MALAT1 | FALSE | Upregulated |
| 0,003610237 | 0,981377567 | TMEM59 | FALSE | Upregulated |
| -0,006739748 | 0,981377567 | TRIM56 | FALSE | Downregulated |
| -0,003915929 | 0,981377567 | VDAC2 | FALSE | Downregulated |
| -0,006774665 | 0,981377567 | ZNF626 | FALSE | Downregulated |
| 0,003002856 | 0,981519327 | KANSL3 | FALSE | Upregulated |
| 0,003424907 | 0,981699542 | QRICH1 | FALSE | Upregulated |
| -0,009126394 | 0,981958064 | TMEM56-RWDD3 | FALSE | Downregulated |
| -0,048247001 | 0,981963956 | PYGM | FALSE | Downregulated |
| 0,015234208 | 0,982157671 | HOMER1 | FALSE | Upregulated |
| -0,015035528 | 0,982157671 | LOC100996573 | FALSE | Downregulated |
| -0,011038299 | 0,982157671 | MAPK10 | FALSE | Downregulated |
| 0,212577629 | 0,982157671 | MYOZ2 | FALSE | Upregulated |
| 0,003529658 | 0,982157671 | REPIN1 | FALSE | Upregulated |

| | | | | |
|---|---|---|---|---|
| 0,004924695 | 0,982157671 | RNF219 | FALSE | Upregulated |
| 0,037572061 | 0,982157671 | SLC35F3 | FALSE | Upregulated |
| -0,019673175 | 0,982157671 | SYCE1L | FALSE | Downregulated |
| 0,02930861 | 0,982157671 | SYN3 | FALSE | Upregulated |
| -0,010563781 | 0,982157671 | TMEM120B | FALSE | Downregulated |
| 0,004250757 | 0,982157671 | WTAP | FALSE | Upregulated |
| 0,010975349 | 0,982171904 | TGFB1I1 | FALSE | Upregulated |
| 0,056844375 | 0,982275267 | ZNF625-ZNF20 | FALSE | Upregulated |
| 0,061599245 | 0,982432753 | RP11-707A18.1 | FALSE | Upregulated |
| -0,020491423 | 0,982631151 | ZNF648 | FALSE | Downregulated |
| -0,038342985 | 0,98275678 | ANXA8L1 | FALSE | Downregulated |
| -0,035696643 | 0,98275678 | USP50 | FALSE | Downregulated |
| -0,046432245 | 0,982792193 | C11orf70 | FALSE | Downregulated |
| -0,008846962 | 0,982804013 | RCL1 | FALSE | Downregulated |
| 0,024684042 | 0,982853402 | CRYBB3 | FALSE | Upregulated |
| -0,009888412 | 0,982999976 | DFNB31 | FALSE | Downregulated |
| -0,008268309 | 0,983089479 | CD9 | FALSE | Downregulated |
| 0,006793487 | 0,983089479 | CYP27A1 | FALSE | Upregulated |
| 0,014124856 | 0,983089479 | SMARCD3 | FALSE | Upregulated |
| -0,035685055 | 0,983157489 | COL17A1 | FALSE | Downregulated |
| -0,007425062 | 0,983186044 | TCP11L1 | FALSE | Downregulated |
| 0,034117779 | 0,983220584 | FLJ45825 | FALSE | Upregulated |
| -0,006951809 | 0,983227441 | CCDC149 | FALSE | Downregulated |
| -0,032587657 | 0,983227441 | IL12A | FALSE | Downregulated |
| -0,007202866 | 0,983227441 | LOC101927371 | FALSE | Downregulated |
| -0,025731432 | 0,983227441 | MCF2 | FALSE | Downregulated |
| -0,005860482 | 0,983388262 | GLB1L | FALSE | Downregulated |
| 0,003521415 | 0,983388262 | PITRM1 | FALSE | Upregulated |
| 0,183421338 | 0,983400217 | TNNI1 | FALSE | Upregulated |
| -0,00943345 | 0,983433906 | CTU1 | FALSE | Downregulated |
| -0,019476867 | 0,983439489 | RASL10A | FALSE | Downregulated |
| -0,003903404 | 0,983439489 | RPS6KA3 | FALSE | Downregulated |
| 0,028934989 | 0,983439489 | WNT2B | FALSE | Upregulated |
| 0,015084724 | 0,983439489 | ZYG11A | FALSE | Upregulated |
| -0,003308854 | 0,9835596 | ATG9A | FALSE | Downregulated |
| 0,194080954 | 0,9835596 | IGFN1 | FALSE | Upregulated |
| -0,016392538 | 0,9835596 | LOC100506014 | FALSE | Downregulated |
| -0,026144133 | 0,9835596 | LOC285389 | FALSE | Downregulated |
| 0,004766036 | 0,9835596 | PTPN2 | FALSE | Upregulated |
| 0,00669728 | 0,9835596 | SEC24D | FALSE | Upregulated |
| 0,004397223 | 0,9835596 | TECR | FALSE | Upregulated |
| -0,004041476 | 0,9835596 | VPS8 | FALSE | Downregulated |
| 0,038460419 | 0,983740073 | FIRRE | FALSE | Upregulated |
| -0,011284022 | 0,983740753 | ZNF85 | FALSE | Downregulated |
| 0,021655015 | 0,983822703 | CMYA5 | FALSE | Upregulated |
| -0,010187954 | 0,984058796 | DPF3 | FALSE | Downregulated |
| -0,034769838 | 0,984058796 | RPL23AP32 | FALSE | Downregulated |
| 0,016648178 | 0,984061815 | CBX2 | FALSE | Upregulated |
| -0,004198284 | 0,984061815 | ELMO1 | FALSE | Downregulated |
| -0,009706184 | 0,984061815 | HOXB6 | FALSE | Downregulated |
| -0,023844035 | 0,984061815 | LOC101929533 | FALSE | Downregulated |

| | | | | |
|---|---|---|---|---|
| 0,022419489 | 0,984061815 | LOC102724611 | FALSE | Upregulated |
| -0,01679381 | 0,984061815 | STOX1 | FALSE | Downregulated |
| -0,012034353 | 0,984225885 | TMEM88 | FALSE | Downregulated |
| 0,04515969 | 0,984272587 | SH3GL1P2 | FALSE | Upregulated |
| -0,18309241 | 0,984333138 | MYH1 | FALSE | Downregulated |
| 0,004318633 | 0,984336797 | PPM1B | FALSE | Upregulated |
| 0,01700852 | 0,984348083 | DOCK9-AS2 | FALSE | Upregulated |
| 0,046160703 | 0,984419688 | GUCY2EP | FALSE | Upregulated |
| 0,012943647 | 0,984524138 | IL1RAPL2 | FALSE | Upregulated |
| -0,003912098 | 0,984524138 | PANK4 | FALSE | Downregulated |
| -0,020328041 | 0,984524138 | STAC3 | FALSE | Downregulated |
| -0,012870508 | 0,984524138 | VWF | FALSE | Downregulated |
| 0,004573751 | 0,984563361 | ERICH1 | FALSE | Upregulated |
| 0,00663408 | 0,984563361 | GLCE | FALSE | Upregulated |
| -0,01317571 | 0,984615635 | ARL10 | FALSE | Downregulated |
| -0,020882431 | 0,984615635 | LOC102724235 | FALSE | Downregulated |
| 0,002871594 | 0,984615635 | NGDN | FALSE | Upregulated |
| 0,129455224 | 0,984615635 | TNNI2 | FALSE | Upregulated |
| -0,178532426 | 0,984615635 | XIRP1 | FALSE | Downregulated |
| 0,176806618 | 0,984792252 | CACNA1S | FALSE | Upregulated |
| 0,010011758 | 0,984814953 | HDGFRP3 | FALSE | Upregulated |
| -0,01274484 | 0,984817817 | SERHL2 | FALSE | Downregulated |
| -0,01781849 | 0,985030271 | CCDC81 | FALSE | Downregulated |
| -0,005660809 | 0,985030271 | RASSF5 | FALSE | Downregulated |
| -0,002522764 | 0,985064886 | FAM199X | FALSE | Downregulated |
| 0,016905579 | 0,985179605 | HIST1H3E | FALSE | Upregulated |
| -0,028269206 | 0,985179605 | LRRC2-AS1 | FALSE | Downregulated |
| -0,005246297 | 0,985179605 | TSR3 | FALSE | Downregulated |
| -0,023478334 | 0,985309025 | STAR | FALSE | Downregulated |
| -0,007176372 | 0,985859048 | CHST12 | FALSE | Downregulated |
| 0,003865326 | 0,985943508 | CAMTA1 | FALSE | Upregulated |
| 0,015492526 | 0,986075711 | ACOT11 | FALSE | Upregulated |
| -0,014954318 | 0,986075711 | ETV4 | FALSE | Downregulated |
| -0,007710996 | 0,986075711 | NRL | FALSE | Downregulated |
| -0,005736803 | 0,986075711 | TBC1D10B | FALSE | Downregulated |
| -0,00442464 | 0,986075711 | TSEN54 | FALSE | Downregulated |
| 0,004327835 | 0,986114452 | NDUFC1 | FALSE | Upregulated |
| -0,009619466 | 0,986119951 | EID2B | FALSE | Downregulated |
| 0,031295266 | 0,986119951 | LINC00311 | FALSE | Upregulated |
| -0,021110787 | 0,986218672 | C1orf177 | FALSE | Downregulated |
| 0,011507318 | 0,986218672 | CCDC8 | FALSE | Upregulated |
| -0,008940553 | 0,986218672 | JAG1 | FALSE | Downregulated |
| -0,012628504 | 0,986271836 | S100A8 | FALSE | Downregulated |
| 0,015681702 | 0,986362817 | LOC644961 | FALSE | Upregulated |
| -0,045895199 | 0,986381424 | HCN4 | FALSE | Downregulated |
| 0,004886125 | 0,986586334 | C20orf194 | FALSE | Upregulated |
| -0,005991507 | 0,986595742 | UBE2Q2 | FALSE | Downregulated |
| 0,027718555 | 0,986744123 | C1orf145 | FALSE | Upregulated |
| 0,002535472 | 0,986744123 | CCDC9 | FALSE | Upregulated |
| 0,01640663 | 0,986744123 | DYNC1I1 | FALSE | Upregulated |
| -0,00398984 | 0,986744123 | GLT8D1 | FALSE | Downregulated |

| | | | | |
|---|---|---|---|---|
| -0,003430689 | 0,986744123 | RPS7 | FALSE | Downregulated |
| -0,002815332 | 0,986812646 | PRKRA | FALSE | Downregulated |
| -0,005497548 | 0,98686283 | ALDH18A1 | FALSE | Downregulated |
| -0,01636777 | 0,98686283 | FAM229A | FALSE | Downregulated |
| 0,003564586 | 0,98686283 | FAM98B | FALSE | Upregulated |
| 0,002253698 | 0,98686283 | KDM5A | FALSE | Upregulated |
| -0,010413745 | 0,98686283 | LOC101928192 | FALSE | Downregulated |
| 0,002101673 | 0,98686283 | LYRM2 | FALSE | Upregulated |
| -0,018921894 | 0,98686283 | MAMSTR | FALSE | Downregulated |
| -0,008551287 | 0,98686283 | NEK11 | FALSE | Downregulated |
| 0,003199339 | 0,986894239 | RNF216 | FALSE | Upregulated |
| 0,005272425 | 0,987272747 | LOC642852 | FALSE | Upregulated |
| -0,00296167 | 0,987272747 | RAB2B | FALSE | Downregulated |
| 0,003383029 | 0,987348104 | LAS1L | FALSE | Upregulated |
| 0,025384312 | 0,987375136 | ZHX1-C8ORF76 | FALSE | Upregulated |
| 0,004261483 | 0,987450146 | SLAIN1 | FALSE | Upregulated |
| -0,018771383 | 0,987495207 | CACNG8 | FALSE | Downregulated |
| -0,033840447 | 0,987495207 | LOC101928195 | FALSE | Downregulated |
| -0,010850392 | 0,987495207 | PABPC5 | FALSE | Downregulated |
| -0,011387914 | 0,987495207 | PYROXD2 | FALSE | Downregulated |
| 0,023039647 | 0,987495207 | RNY5 | FALSE | Upregulated |
| -0,002852123 | 0,987495207 | TMEM87A | FALSE | Downregulated |
| -0,003734792 | 0,987495207 | ZNF764 | FALSE | Downregulated |
| -0,042198973 | 0,987805521 | CRISP3 | FALSE | Downregulated |
| 0,015535485 | 0,987981939 | LOC101927837 | FALSE | Upregulated |
| 0,00369009 | 0,987981939 | TPD52 | FALSE | Upregulated |
| 0,138014172 | 0,988028607 | DES | FALSE | Upregulated |
| 0,009182711 | 0,988126164 | DACT3 | FALSE | Upregulated |
| 0,015059107 | 0,988126164 | LOC102723928 | FALSE | Upregulated |
| 0,033856004 | 0,988393828 | AC096559.1 | FALSE | Upregulated |
| 0,004568756 | 0,988393828 | ACOT4 | FALSE | Upregulated |
| -0,013101529 | 0,988393828 | EPS8L1 | FALSE | Downregulated |
| -0,019407122 | 0,988759469 | C9orf163 | FALSE | Downregulated |
| 0,001905715 | 0,988825131 | EXOC3 | FALSE | Upregulated |
| -0,015728149 | 0,989018226 | LOC102723582 | FALSE | Downregulated |
| -0,02480473 | 0,989118238 | CHRFAM7A | FALSE | Downregulated |
| 0,004992362 | 0,989420215 | SNHG1 | FALSE | Upregulated |
| 0,003378117 | 0,989437127 | USP16 | FALSE | Upregulated |
| 0,006074945 | 0,989565751 | CEP152 | FALSE | Upregulated |
| 0,002810792 | 0,989608327 | CCDC51 | FALSE | Upregulated |
| 0,008208022 | 0,989608327 | RP11-696N14.1 | FALSE | Upregulated |
| 0,016050219 | 0,989638541 | C9orf66 | FALSE | Upregulated |
| -0,002392115 | 0,989638541 | GFM1 | FALSE | Downregulated |
| -0,023756889 | 0,989638541 | HCN2 | FALSE | Downregulated |
| -0,004082645 | 0,989638541 | HIVEP1 | FALSE | Downregulated |
| -0,002500145 | 0,989638541 | IL6ST | FALSE | Downregulated |
| 0,00286183 | 0,989638541 | KCNJ8 | FALSE | Upregulated |
| -0,01663636 | 0,989638541 | LINC01021 | FALSE | Downregulated |
| 0,016476827 | 0,989638541 | LOC101927354 | FALSE | Upregulated |
| 0,017205802 | 0,989638541 | LOC101928059 | FALSE | Upregulated |
| 0,011595349 | 0,989638541 | PDZD9 | FALSE | Upregulated |

| | | | | |
|---|---|---|---|---|
| 0,003523002 | 0,989638541 | PGS1 | FALSE | Upregulated |
| 0,009753489 | 0,989638541 | POF1B | FALSE | Upregulated |
| 0,005960377 | 0,989638541 | PRG2 | FALSE | Upregulated |
| -0,013064295 | 0,989638541 | STRC | FALSE | Downregulated |
| 0,003151367 | 0,989638541 | USP20 | FALSE | Upregulated |
| -0,111752316 | 0,98983489 | MYOZ1 | FALSE | Downregulated |
| 0,011454457 | 0,989949222 | PIWIL4 | FALSE | Upregulated |
| -0,003880186 | 0,990054971 | AADAC | FALSE | Downregulated |
| -0,001656635 | 0,990128055 | CHMP5 | FALSE | Downregulated |
| 0,003625369 | 0,990128055 | LCLAT1 | FALSE | Upregulated |
| -0,002152593 | 0,990128055 | ZFAND6 | FALSE | Downregulated |
| -0,003470225 | 0,990199054 | FAM20A | FALSE | Downregulated |
| -0,004096413 | 0,990508778 | LOC102724002 | FALSE | Downregulated |
| -0,014409858 | 0,990536539 | OSGEPL1-AS1 | FALSE | Downregulated |
| -0,005758409 | 0,990597213 | SLC35E2 | FALSE | Downregulated |
| -0,018490003 | 0,990597213 | TRIM9 | FALSE | Downregulated |
| 0,013790548 | 0,990709954 | TRNL2 | FALSE | Upregulated |
| -0,001724149 | 0,990840291 | RIOK3 | FALSE | Downregulated |
| 0,003320203 | 0,990886905 | ATP5G1 | FALSE | Upregulated |
| 0,004443223 | 0,990932172 | TMEM38A | FALSE | Upregulated |
| -0,008748847 | 0,991153204 | DPY19L1P1 | FALSE | Downregulated |
| 0,013807539 | 0,991304365 | PHKA2-AS1 | FALSE | Upregulated |
| -0,002339308 | 0,991402686 | CRTAP | FALSE | Downregulated |
| 0,002427113 | 0,991839953 | BTN2A1 | FALSE | Upregulated |
| 0,019102177 | 0,991850897 | NEB | FALSE | Upregulated |
| 0,009580359 | 0,991850897 | WDR86-AS1 | FALSE | Upregulated |
| -0,004455033 | 0,991924816 | RPL23AP7 | FALSE | Downregulated |
| -0,002862518 | 0,99196198 | AQP3 | FALSE | Downregulated |
| 0,016949315 | 0,99196198 | LOC101926943 | FALSE | Upregulated |
| -0,002051811 | 0,99196198 | NCOA3 | FALSE | Downregulated |
| 0,003573643 | 0,99196198 | SULT1A2 | FALSE | Upregulated |
| -0,001246811 | 0,992248856 | FAM20B | FALSE | Downregulated |
| -0,00137801 | 0,992248856 | IDH3B | FALSE | Downregulated |
| 0,015303858 | 0,992248856 | LOC101928692 | FALSE | Upregulated |
| -0,004446106 | 0,992248856 | SIRT4 | FALSE | Downregulated |
| -0,007613797 | 0,992248856 | SLIT3 | FALSE | Downregulated |
| 0,015318166 | 0,992296749 | KCNIP1 | FALSE | Upregulated |
| -0,01158366 | 0,992296749 | RP11-399K21.14 | FALSE | Downregulated |
| -0,093053142 | 0,992381453 | MYOT | FALSE | Downregulated |
| 0,002340172 | 0,992609168 | FAM122B | FALSE | Upregulated |
| 0,007638808 | 0,99267499 | BCL11A | FALSE | Upregulated |
| -0,004696348 | 0,99267499 | C10orf10 | FALSE | Downregulated |
| -0,002676384 | 0,99267499 | DUSP5 | FALSE | Downregulated |
| 0,011622781 | 0,99267499 | EFCAB5 | FALSE | Upregulated |
| 0,001362513 | 0,99267499 | HMGB1 | FALSE | Upregulated |
| 0,017646809 | 0,99267499 | LINC00260 | FALSE | Upregulated |
| 0,016735158 | 0,99267499 | LOC641746 | FALSE | Upregulated |
| -0,086472532 | 0,99267499 | MYH2 | FALSE | Downregulated |
| 0,001810365 | 0,99267499 | PARP4 | FALSE | Upregulated |
| -0,002801989 | 0,99267499 | RBM27 | FALSE | Downregulated |
| -0,007172615 | 0,99267499 | RP11-434D9.1 | FALSE | Downregulated |

| | | | | |
|---|---|---|---|---|
| -0,00200963 | 0,99267499 | SNX2 | FALSE | Downregulated |
| 0,007719569 | 0,99267499 | TMEM236 | FALSE | Upregulated |
| 0,003799114 | 0,99267499 | ZEB1-AS1 | FALSE | Upregulated |
| 0,001385377 | 0,992688857 | NLK | FALSE | Upregulated |
| -0,008740376 | 0,992948853 | GSTM5 | FALSE | Downregulated |
| 0,014872047 | 0,992948853 | LDHC | FALSE | Upregulated |
| 0,002433017 | 0,992948853 | MRPS33 | FALSE | Upregulated |
| 0,008843825 | 0,992954979 | C18orf42 | FALSE | Upregulated |
| 0,00570223 | 0,993189791 | NUDT17 | FALSE | Upregulated |
| 0,002056988 | 0,993357728 | RALA | FALSE | Upregulated |
| 0,002630436 | 0,993542894 | ESR1 | FALSE | Upregulated |
| -0,017919604 | 0,993542894 | HIST2H4A | FALSE | Downregulated |
| -0,00123491 | 0,993542894 | UBR1 | FALSE | Downregulated |
| -0,00186367 | 0,993771882 | ELP6 | FALSE | Downregulated |
| 0,003037822 | 0,99381313 | C17orf107 | FALSE | Upregulated |
| -0,00358027 | 0,99381313 | GNG12-AS1 | FALSE | Downregulated |
| -0,00376852 | 0,99381313 | MPV17L | FALSE | Downregulated |
| -0,002044557 | 0,993917294 | GGA1 | FALSE | Downregulated |
| -0,001819133 | 0,994150684 | PFAS | FALSE | Downregulated |
| -0,001586218 | 0,994205855 | ISY1 | FALSE | Downregulated |
| 0,004975129 | 0,994208859 | GATA5 | FALSE | Upregulated |
| 0,002539179 | 0,994208859 | GTF2IRD2 | FALSE | Upregulated |
| -0,071741533 | 0,994208859 | LOC101928874 | FALSE | Downregulated |
| 0,012716268 | 0,994208859 | TACR1 | FALSE | Upregulated |
| -0,005718156 | 0,994272228 | LRRC70 | FALSE | Downregulated |
| -0,003208286 | 0,994272228 | RNASEH1-AS1 | FALSE | Downregulated |
| 0,003374638 | 0,994272228 | SLC25A25 | FALSE | Upregulated |
| 0,010821539 | 0,994380337 | LINC01152 | FALSE | Upregulated |
| -0,067594496 | 0,994425131 | HYPK | FALSE | Downregulated |
| -0,003438231 | 0,994425131 | LOC100996497 | FALSE | Downregulated |
| 0,004098189 | 0,994579987 | HMGCR | FALSE | Upregulated |
| 0,002105374 | 0,99458214 | EIF3A | FALSE | Upregulated |
| 0,001347459 | 0,99458214 | ERO1L | FALSE | Upregulated |
| 0,002332049 | 0,99458214 | PPARGC1B | FALSE | Upregulated |
| -0,002126335 | 0,99458214 | ZC4H2 | FALSE | Downregulated |
| -0,001354555 | 0,994601359 | ETFB | FALSE | Downregulated |
| -0,001522073 | 0,994755691 | PFDN6 | FALSE | Downregulated |
| 0,002717019 | 0,994755691 | RIPK4 | FALSE | Upregulated |
| -0,001273312 | 0,994765565 | SUFU | FALSE | Downregulated |
| 0,001820747 | 0,994765565 | THG1L | FALSE | Upregulated |
| 0,001416736 | 0,994794183 | MRPL34 | FALSE | Upregulated |
| 0,00944007 | 0,995032163 | PRSS16 | FALSE | Upregulated |
| -0,008811366 | 0,995295364 | CALML3 | FALSE | Downregulated |
| -0,001464169 | 0,995356108 | ENY2 | FALSE | Downregulated |
| 0,002168018 | 0,995476241 | B3GNT9 | FALSE | Upregulated |
| -0,003471835 | 0,995554426 | RP11-346I3.4 | FALSE | Downregulated |
| -0,003491558 | 0,995562935 | SLC9B1 | FALSE | Downregulated |
| 0,006485355 | 0,995765837 | ASIC1 | FALSE | Upregulated |
| -0,007880541 | 0,995765837 | IL20RB | FALSE | Downregulated |
| 0,003150391 | 0,99576792 | RPSAP58 | FALSE | Upregulated |
| -0,000913086 | 0,99590284 | ANKRD49 | FALSE | Downregulated |

| | | | | |
|---|---|---|---|---|
| 0,002065818 | 0,996043838 | ERICH2 | FALSE | Upregulated |
| -0,01064827 | 0,996043838 | LOC101060363 | FALSE | Downregulated |
| 0,002575149 | 0,996043838 | RP11-521B24.3 | FALSE | Upregulated |
| -0,001457248 | 0,996043838 | SMARCC1 | FALSE | Downregulated |
| 0,048786647 | 0,996260963 | ACTN3 | FALSE | Upregulated |
| 0,006570376 | 0,996260963 | HOXB-AS3 | FALSE | Upregulated |
| 0,001301697 | 0,996277678 | BLVRB | FALSE | Upregulated |
| 0,00133539 | 0,996440313 | ABHD2 | FALSE | Upregulated |
| 0,001458764 | 0,996440313 | C3orf58 | FALSE | Upregulated |
| 0,000866456 | 0,996440313 | ENSA | FALSE | Upregulated |
| -0,003896317 | 0,996440313 | MGP | FALSE | Downregulated |
| -0,002502951 | 0,996610268 | CD109 | FALSE | Downregulated |
| 0,001230062 | 0,996701049 | AP4B1 | FALSE | Upregulated |
| -0,004388579 | 0,996701049 | BOC | FALSE | Downregulated |
| -0,003897971 | 0,996701049 | CCDC173 | FALSE | Downregulated |
| -0,001570661 | 0,996701049 | CLIP2 | FALSE | Downregulated |
| 0,000679959 | 0,996701049 | KLHL9 | FALSE | Upregulated |
| 0,008139649 | 0,996701049 | OMP | FALSE | Upregulated |
| -0,000859119 | 0,996701049 | TRIM4 | FALSE | Downregulated |
| 0,002189854 | 0,996712785 | ISYNA1 | FALSE | Upregulated |
| 0,000901324 | 0,996760749 | BMP2K | FALSE | Upregulated |
| 0,003747741 | 0,996996651 | HOXA-AS2 | FALSE | Upregulated |
| -0,002360839 | 0,996996651 | NBPF20 | FALSE | Downregulated |
| -0,004067182 | 0,997051783 | EPHA1-AS1 | FALSE | Downregulated |
| 0,001611742 | 0,997163062 | DNAJB9 | FALSE | Upregulated |
| 0,006359366 | 0,997315799 | LOC102723693 | FALSE | Upregulated |
| -0,000973115 | 0,997404276 | DNPH1 | FALSE | Downregulated |
| 0,001125403 | 0,99769817 | FUK | FALSE | Upregulated |
| 0,001267335 | 0,99769817 | ZNF597 | FALSE | Upregulated |
| 0,001295059 | 0,997975508 | GJA1 | FALSE | Upregulated |
| -0,001451524 | 0,997975508 | RSG1 | FALSE | Downregulated |
| 0,004953165 | 0,998025523 | CHODL | FALSE | Upregulated |
| 0,000862483 | 0,998140869 | TMUB1 | FALSE | Upregulated |
| -0,001620904 | 0,998290323 | HHIP-AS1 | FALSE | Downregulated |
| 0,000813789 | 0,998450212 | NRDE2 | FALSE | Upregulated |
| -0,002857126 | 0,998450212 | TCF15 | FALSE | Downregulated |
| 0,001529684 | 0,998697917 | PLCD3 | FALSE | Upregulated |
| -0,001024191 | 0,998769505 | EIF4EBP3 | FALSE | Downregulated |
| 0,005501018 | 0,998863378 | GUCY1B2 | FALSE | Upregulated |
| -0,000798543 | 0,998863378 | PIAS1 | FALSE | Downregulated |
| -0,003115399 | 0,998891281 | DLGAP1 | FALSE | Downregulated |
| 0,000568957 | 0,999175538 | C19orf66 | FALSE | Upregulated |
| 0,000640407 | 0,999175538 | CCDC3 | FALSE | Upregulated |
| 0,000794924 | 0,999175538 | CYGB | FALSE | Upregulated |
| -0,000564904 | 0,999175538 | FAM160B2 | FALSE | Downregulated |
| -0,001358967 | 0,999175538 | GSTO2 | FALSE | Downregulated |
| 0,000433016 | 0,999175538 | HCFC2 | FALSE | Upregulated |
| 0,016960117 | 0,999175538 | MYL1 | FALSE | Upregulated |
| -0,000572513 | 0,999175538 | PRKAR2B | FALSE | Downregulated |
| 0,002794316 | 0,999175538 | WASH7P | FALSE | Upregulated |
| 0,000504043 | 0,999175538 | ZNF761 | FALSE | Upregulated |

| | | | | |
|---|---|---|---|---|
| 0,001190425 | 0,999205406 | ANKRD36 | FALSE | Upregulated |
| 0,000425352 | 0,999205406 | CEPT1 | FALSE | Upregulated |
| 0,000508296 | 0,999205406 | CYTH3 | FALSE | Upregulated |
| 0,000564327 | 0,999205406 | EXT1 | FALSE | Upregulated |
| -0,000329673 | 0,999205406 | GDI2 | FALSE | Downregulated |
| -0,002313739 | 0,999205406 | LOC100506082 | FALSE | Downregulated |
| -0,001989271 | 0,999205406 | MAFG-AS1 | FALSE | Downregulated |
| 0,014179881 | 0,999205406 | MYLPF | FALSE | Upregulated |
| -0,002393587 | 0,999205406 | OOEP | FALSE | Downregulated |
| -0,00033398 | 0,999205406 | PIGY | FALSE | Downregulated |
| -0,000937902 | 0,999274723 | LOC101928235 | FALSE | Downregulated |
| 0,000619842 | 0,999345036 | CD99P1 | FALSE | Upregulated |
| 0,000416919 | 0,999345036 | CXorf40B | FALSE | Upregulated |
| -0,000221131 | 0,999345036 | IMPAD1 | FALSE | Downregulated |
| -0,000207889 | 0,999345036 | TANK | FALSE | Downregulated |
| 0,000457566 | 0,99940023 | FLRT2 | FALSE | Upregulated |
| -0,000679815 | 0,99940023 | FXYD2 | FALSE | Downregulated |
| 0,001962494 | 0,99940023 | HIST1H3D | FALSE | Upregulated |
| 0,000203579 | 0,99940023 | KANSL2 | FALSE | Upregulated |
| 0,000603306 | 0,99940023 | LINC00936 | FALSE | Upregulated |
| -0,001183528 | 0,99940023 | NOX4 | FALSE | Downregulated |
| 0,000295794 | 0,99940023 | PLEKHA8P1 | FALSE | Upregulated |
| 0,000107434 | 0,999459432 | DDX54 | FALSE | Upregulated |
| 0,000128219 | 0,999459432 | NOB1 | FALSE | Upregulated |
| -0,006992371 | 0,999459432 | TNNC1 | FALSE | Downregulated |
| 0,000188245 | 0,999459432 | ZNF142 | FALSE | Upregulated |
| -0,000632532 | 0,999826768 | CACNA1F | FALSE | Downregulated |
| -0,00059347 | 0,999826768 | CTAGE1 | FALSE | Downregulated |
| -0,000256815 | 0,999826768 | GHRLOS | FALSE | Downregulated |
| 0,000102259 | 0,999826768 | IWS1 | FALSE | Upregulated |
| 0,000421059 | 0,999826768 | LOC102724231 | FALSE | Upregulated |
| 0,000133769 | 0,999826768 | SLC4A7 | FALSE | Upregulated |
| -6,4577E-05 | 0,999890556 | DPP9 | FALSE | Downregulated |
| -0,000416661 | 0,999890556 | TPSD1 | FALSE | Downregulated |
| 4,42698E-05 | 0,999890556 | USP45 | FALSE | Upregulated |
| 8,86994E-05 | 0,999931914 | AHRR | FALSE | Upregulated |
| 0 | 1 | C3orf56 | FALSE | Upregulated |
| -1,00353E-05 | 1 | DMTN | FALSE | Downregulated |
| -1,021157667 | | A4GNT | | Downregulated |
| 1,974650338 | | ABCA12 | | Upregulated |
| -1,100166986 | | ABCA17P | | Downregulated |
| -0,419626994 | | ABCB5 | FALSE | Downregulated |
| -0,033409254 | | ABCC12 | FALSE | Downregulated |
| 0,068258212 | | ABHD12B | FALSE | Upregulated |
| -1,863209224 | | ABHD16B | | Downregulated |
| -0,117671621 | | AC002480.3 | FALSE | Downregulated |
| 0,66209391 | | AC003102.3 | FALSE | Upregulated |
| -1,525127849 | | AC005022.1 | | Downregulated |
| 0,247502558 | | AC006145.4 | FALSE | Upregulated |
| -1,50958891 | | AC006272.1 | | Downregulated |
| -0,834851255 | | AC007405.4 | FALSE | Downregulated |

| | | | |
|---|---|---|---|
| 1,199337593 | AC007750.5 | | Upregulated |
| -1,355529656 | AC007966.1 | | Downregulated |
| -0,787734456 | AC073283.4 | FALSE | Downregulated |
| -0,570284604 | AC079610.2 | FALSE | Downregulated |
| -1,216823946 | AC087859.1 | | Downregulated |
| 0,052806983 | AC098823.3 | FALSE | Upregulated |
| 0,019839916 | AC104024.1 | FALSE | Upregulated |
| -0,108165722 | AC104654.2 | FALSE | Downregulated |
| 2,163289499 | ACOXL | | Upregulated |
| 0,729517367 | ADAD2 | FALSE | Upregulated |
| -0,689482252 | ADAM20 | FALSE | Downregulated |
| 0,125730522 | ADAM21 | FALSE | Upregulated |
| 0,595902199 | ADAM32 | FALSE | Upregulated |
| 0,396878843 | ADAMTS8 | FALSE | Upregulated |
| -0,036676756 | ADGB | FALSE | Downregulated |
| -0,033154322 | ADIPOQ-AS1 | FALSE | Downregulated |
| 0,101700757 | AFAP1-AS1 | FALSE | Upregulated |
| 0,44734113 | AGPAT4-IT1 | FALSE | Upregulated |
| -0,68250621 | AIPL1 | FALSE | Downregulated |
| -1,081615355 | AL022476.2 | | Downregulated |
| 0,574606146 | AL109763.2 | FALSE | Upregulated |
| -1,065327952 | ALK | | Downregulated |
| 1,017423465 | AMH | | Upregulated |
| -0,556493391 | ANKK1 | FALSE | Downregulated |
| -1,591796287 | ANKRD30A | | Downregulated |
| -0,48632092 | ANKRD30B | FALSE | Downregulated |
| 0,254088313 | ANKRD30BP3 | FALSE | Upregulated |
| -0,817868817 | ANKRD44-IT1 | FALSE | Downregulated |
| 0,35121202 | ANKRD61 | FALSE | Upregulated |
| -0,198680553 | ANKRD62 | FALSE | Downregulated |
| 0,770073823 | APOOP5 | FALSE | Upregulated |
| 0,560192537 | AQP8 | FALSE | Upregulated |
| -0,632060056 | ARC | FALSE | Downregulated |
| 1,65824724 | AREG | | Upregulated |
| -1,350615498 | ARHGAP40 | | Downregulated |
| 1,213950965 | ARL9 | | Upregulated |
| 0,252404406 | ARRDC3-AS1 | FALSE | Upregulated |
| 0,21206137 | ARTN | FALSE | Upregulated |
| -1,3793518 | ARX | | Downregulated |
| -0,342900389 | ASB16 | FALSE | Downregulated |
| 0,93143676 | ASB9P1 | FALSE | Upregulated |
| 0,20970374 | ATCAY | FALSE | Upregulated |
| 1,192858545 | ATP10B | | Upregulated |
| -0,355856284 | ATP2B3 | FALSE | Downregulated |
| -0,332780422 | ATP6V1B1 | FALSE | Downregulated |
| -0,627534911 | AZU1 | FALSE | Downregulated |
| -1,824040295 | B3GALT1 | | Downregulated |
| 0,116160465 | BARX2 | FALSE | Upregulated |
| 0,404267047 | BEND2 | FALSE | Upregulated |
| -0,588097588 | BEST4 | FALSE | Downregulated |
| -0,011316506 | BHLHE23 | FALSE | Downregulated |

| | | | |
|---|---|---|---|
| 0,718581127 | BMP3 | FALSE | Upregulated |
| 0,030059109 | BPIFB2 | FALSE | Upregulated |
| 1,043843809 | BRE-AS1 | | Upregulated |
| 0,273246245 | BREA2 | FALSE | Upregulated |
| 0,531442372 | BRSK2 | FALSE | Upregulated |
| 0,490803281 | BTNL3 | FALSE | Upregulated |
| -0,388570603 | C10orf111 | FALSE | Downregulated |
| -0,515392286 | C10orf67 | FALSE | Downregulated |
| -1,389066519 | C10orf85 | | Downregulated |
| -2,177248935 | C10orf95 | | Downregulated |
| 1,095355407 | C12orf42 | | Upregulated |
| -0,703674048 | C12orf68 | FALSE | Downregulated |
| -2,130118094 | C15orf26 | | Downregulated |
| -0,875172951 | C16orf3 | FALSE | Downregulated |
| 0,016529224 | C17orf104 | FALSE | Upregulated |
| 1,016185232 | C17orf66 | | Upregulated |
| 0,735582637 | C18orf61 | FALSE | Upregulated |
| 1,212223002 | C19orf45 | | Upregulated |
| 0,707337176 | C19orf83 | FALSE | Upregulated |
| 0,398703165 | C1orf110 | FALSE | Upregulated |
| 1,412478245 | C1orf111 | | Upregulated |
| -0,479841609 | C1orf189 | FALSE | Downregulated |
| -0,244010219 | C1orf200 | FALSE | Downregulated |
| -0,452001665 | C1QL2 | FALSE | Downregulated |
| -1,309444804 | C1QL3 | | Downregulated |
| 0,809848769 | C1QTNF1-AS1 | FALSE | Upregulated |
| -0,657157165 | C1QTNF4 | FALSE | Downregulated |
| -1,505951353 | C20orf166-AS1 | | Downregulated |
| -1,16035493 | C21orf140 | | Downregulated |
| -0,042050829 | C22orf31 | FALSE | Downregulated |
| 0,551727323 | C3orf27 | FALSE | Upregulated |
| 0,442181119 | C3orf36 | FALSE | Upregulated |
| 0,408902244 | C4orf50 | FALSE | Upregulated |
| 2,499809607 | C4orf6 | | Upregulated |
| -0,194177438 | C5orf60 | FALSE | Downregulated |
| -0,261808861 | C6orf222 | FALSE | Downregulated |
| -0,937377286 | C6orf223 | FALSE | Downregulated |
| -0,761412929 | C9orf170 | FALSE | Downregulated |
| -2,028449547 | C9orf173 | | Downregulated |
| -0,49999691 | CACNA1B | FALSE | Downregulated |
| -0,782516549 | CACNA1C-AS1 | FALSE | Downregulated |
| 0,668795111 | CACNG3 | FALSE | Upregulated |
| -0,127794855 | CACNG4 | FALSE | Downregulated |
| -0,706127592 | CALB1 | FALSE | Downregulated |
| 0,207663742 | CAPN14 | FALSE | Upregulated |
| 0,277183851 | CAPN8 | FALSE | Upregulated |
| -2,56006685 | CCDC116 | | Downregulated |
| -1,226091312 | CCDC175 | | Downregulated |
| -0,25895933 | CCDC74B | FALSE | Downregulated |
| -0,808237224 | CCDC78 | FALSE | Downregulated |
| 0,348091835 | CCDC79 | FALSE | Upregulated |

| | | | |
|---|---|---|---|
| 0,729742385 | CCR10 | FALSE | Upregulated |
| 1,741908772 | CCR8 | | Upregulated |
| 0,011103554 | CD1A | FALSE | Upregulated |
| -0,430614714 | CD81-AS1 | FALSE | Downregulated |
| 0,996561102 | CDC42P3 | FALSE | Upregulated |
| -0,009979074 | CDCP2 | FALSE | Downregulated |
| -0,302855827 | CDH22 | FALSE | Downregulated |
| 0,439094794 | CEACAM20 | FALSE | Upregulated |
| -0,654761661 | CEACAM8 | FALSE | Downregulated |
| -0,072771502 | CECR5-AS1 | FALSE | Downregulated |
| 0,945321157 | CECR7 | FALSE | Upregulated |
| -0,476204468 | CERS1 | FALSE | Downregulated |
| -0,212810597 | CETN4P | FALSE | Downregulated |
| 0,250851193 | CHD5 | FALSE | Upregulated |
| 0,636455896 | CHIA | FALSE | Upregulated |
| 0,190639561 | CHRM3-AS2 | FALSE | Upregulated |
| 0,622836993 | CHRNA2 | FALSE | Upregulated |
| -0,471253808 | CHRNB2 | FALSE | Downregulated |
| -0,445948982 | CHST5 | FALSE | Downregulated |
| -0,04275263 | CHST8 | FALSE | Downregulated |
| 0,720805025 | CITED1 | FALSE | Upregulated |
| 1,179996007 | CKMT1B | | Upregulated |
| -1,140048101 | CLCNKA | | Downregulated |
| 0,968155265 | CLEC17A | FALSE | Upregulated |
| -0,654966021 | CLEC19A | FALSE | Downregulated |
| 0,893138629 | CLK2P | FALSE | Upregulated |
| -1,23308319 | CMTM2 | | Downregulated |
| 0,507619008 | CNIH2 | FALSE | Upregulated |
| 0,144014843 | CNTNAP4 | FALSE | Upregulated |
| 0,316512454 | COL2A1 | FALSE | Upregulated |
| 0,732473544 | CORT | FALSE | Upregulated |
| 0,550331866 | CPA1 | FALSE | Upregulated |
| -0,553287951 | CPEB2-AS1 | FALSE | Downregulated |
| -0,025751687 | CPLX4 | FALSE | Downregulated |
| 1,40459361 | CRLF1 | | Upregulated |
| -0,943168693 | CRYBA4 | FALSE | Downregulated |
| 1,047057177 | CRYM-AS1 | | Upregulated |
| -0,195289824 | CSAG3 | FALSE | Downregulated |
| 0,322534612 | CSMD1 | FALSE | Upregulated |
| 0,374830123 | CSNK1A1L | FALSE | Upregulated |
| 0,786997842 | CTD-2127H9.1 | FALSE | Upregulated |
| 0,245612517 | CTXN1 | FALSE | Upregulated |
| -0,067677451 | CXADRP3 | FALSE | Downregulated |
| -1,089125754 | CXorf22 | | Downregulated |
| -0,796481925 | CXorf58 | FALSE | Downregulated |
| 0,053603948 | CXorf64 | FALSE | Upregulated |
| -0,164029286 | CXXC1P1 | FALSE | Downregulated |
| 0,700756215 | CYP26C1 | FALSE | Upregulated |
| -0,722057931 | CYP4F8 | FALSE | Downregulated |
| -1,215562386 | CYTL1 | | Downregulated |
| -0,539841333 | DCST1 | FALSE | Downregulated |

| | | | |
|---|---|---|---|
| -0,627814987 | DGCR10 | FALSE | Downregulated |
| 1,327297383 | DHDH | | Upregulated |
| -0,945997093 | DKFZP434A062 | FALSE | Downregulated |
| -0,449084131 | DLGAP1-AS3 | FALSE | Downregulated |
| 0,975692649 | DLX4 | FALSE | Upregulated |
| -1,67975783 | DMBX1 | | Downregulated |
| -0,141685114 | DMRT2 | FALSE | Downregulated |
| -0,31168236 | DNAH17 | FALSE | Downregulated |
| 1,180031779 | DNAH3 | | Upregulated |
| -0,684523429 | DNAJB13 | FALSE | Downregulated |
| -0,651880637 | DNAJB7 | FALSE | Downregulated |
| -1,310776156 | DNASE1L2 | | Downregulated |
| -0,759860826 | DNER | FALSE | Downregulated |
| 0,379286011 | DNM1P50 | FALSE | Upregulated |
| -1,185534793 | DOC2GP | | Downregulated |
| 0,513006811 | DPH3P1 | FALSE | Upregulated |
| -1,066263276 | DPYSL5 | | Downregulated |
| -0,217051416 | DQX1 | FALSE | Downregulated |
| -0,423541549 | DRC1 | FALSE | Downregulated |
| 0,185915672 | DSC1 | FALSE | Upregulated |
| 0,898245257 | DSCR9 | FALSE | Upregulated |
| -0,738691111 | DUOXA1 | FALSE | Downregulated |
| 0,290356457 | ECEL1 | FALSE | Upregulated |
| -0,382645534 | ECT2L | FALSE | Downregulated |
| -0,392076839 | EFCAB10 | FALSE | Downregulated |
| 1,265275476 | EFHC2 | | Upregulated |
| -1,318894946 | EGFEM1P | | Downregulated |
| 1,143613264 | EGFL6 | | Upregulated |
| -0,555844527 | EIF3IP1 | FALSE | Downregulated |
| -0,121596556 | ELAVL3 | FALSE | Downregulated |
| 0,973919343 | ELOVL3 | FALSE | Upregulated |
| -0,365402376 | ENTPD3 | FALSE | Downregulated |
| -2,592630838 | EPB42 | | Downregulated |
| 0,091968691 | EPHA6 | FALSE | Upregulated |
| 0,030870146 | EPHA8 | FALSE | Upregulated |
| 1,063219353 | EPS8L3 | | Upregulated |
| 0,063974711 | ERC2 | FALSE | Upregulated |
| 1,605568388 | EREG | | Upregulated |
| -1,761266784 | ESRG | | Downregulated |
| 1,822221389 | FABP5P3 | | Upregulated |
| -0,347411807 | FABP6 | FALSE | Downregulated |
| -0,275212175 | FAM132A | FALSE | Downregulated |
| -0,081148976 | FAM154A | FALSE | Downregulated |
| -1,940958402 | FAM170A | | Downregulated |
| 0,123263504 | FAM171A2 | FALSE | Upregulated |
| -0,242895424 | FAM177B | FALSE | Downregulated |
| 1,181679919 | FAM178B | | Upregulated |
| 0,438810503 | FAM183B | FALSE | Upregulated |
| -0,758822817 | FAM186A | FALSE | Downregulated |
| -0,767054263 | FAM194A | FALSE | Downregulated |
| -1,099450437 | FAM209A | | Downregulated |

| | | | |
|---|---|---|---|
| -0,175788535 | FAM212B-AS1 | FALSE | Downregulated |
| 0,569926173 | FAM226A | FALSE | Upregulated |
| 0,372218844 | FAM226B | FALSE | Upregulated |
| 1,104390936 | FAM27C | | Upregulated |
| 0,753260008 | FAM3D | FALSE | Upregulated |
| 2,073877727 | FAM57B | | Upregulated |
| -0,190188347 | FAM66D | FALSE | Downregulated |
| 0,597003142 | FAM71F1 | FALSE | Upregulated |
| -0,698455941 | FAM86KP | FALSE | Downregulated |
| -0,221874934 | FAM92A1P2 | FALSE | Downregulated |
| 0,018913889 | FAM95A | FALSE | Upregulated |
| -0,101556091 | FBLL1 | FALSE | Downregulated |
| -1,129928341 | FBXL21 | | Downregulated |
| -0,423165776 | FCRL4 | FALSE | Downregulated |
| 0,224286272 | FER1L6 | FALSE | Upregulated |
| -0,287698492 | FGD5P1 | FALSE | Downregulated |
| 0,114827427 | FGF10 | FALSE | Upregulated |
| -0,367922516 | FGF13-AS1 | FALSE | Downregulated |
| 0,154011996 | FGF20 | FALSE | Upregulated |
| -0,475925389 | FLG2 | FALSE | Downregulated |
| -2,318389363 | FLJ30375 | | Downregulated |
| -1,466209085 | FLJ30679 | | Downregulated |
| -1,466854743 | FLJ31356 | | Downregulated |
| 0,462036176 | FLJ31945 | FALSE | Upregulated |
| 1,694383903 | FLJ35934 | | Upregulated |
| -0,826069572 | FLJ36000 | FALSE | Downregulated |
| -0,907903748 | FLJ38122 | FALSE | Downregulated |
| -0,95428749 | FLJ41481 | FALSE | Downregulated |
| 0,074206091 | FLJ42393 | FALSE | Upregulated |
| -0,91440883 | FLJ43315 | FALSE | Downregulated |
| -0,127728018 | FLJ45079 | FALSE | Downregulated |
| -1,027073537 | FLJ45872 | | Downregulated |
| 1,219197439 | FLRT1 | | Upregulated |
| -1,332003054 | FMN2 | | Downregulated |
| -0,929398407 | FMO6P | FALSE | Downregulated |
| -0,013890097 | FMR1NB | FALSE | Downregulated |
| 0,154391595 | FNDC8 | FALSE | Upregulated |
| -0,62988557 | FOXD1 | FALSE | Downregulated |
| 1,932694995 | FOXD4L1 | | Upregulated |
| 0,938021572 | FOXF2 | FALSE | Upregulated |
| -1,150751531 | FRMD5 | | Downregulated |
| -1,547644079 | FRMPD4 | | Downregulated |
| -0,738133824 | FSCN2 | FALSE | Downregulated |
| 2,791297202 | FSTL4 | | Upregulated |
| -0,399388661 | FUT9 | FALSE | Downregulated |
| -1,431584797 | FZD10-AS1 | | Downregulated |
| 0,126228137 | GABRA2 | FALSE | Upregulated |
| -0,68293776 | GABRA4 | FALSE | Downregulated |
| 0,057445207 | GAD1 | FALSE | Upregulated |
| -0,346336119 | GALNT8 | FALSE | Downregulated |
| -0,697399753 | GALNT9 | FALSE | Downregulated |

| | | | |
|---|---|---|---|
| -0,665977955 | GATA3-AS1 | FALSE | Downregulated |
| -0,597843234 | GCSAML | FALSE | Downregulated |
| -0,346348709 | GGN | FALSE | Downregulated |
| -0,406851214 | GJA3 | FALSE | Downregulated |
| 0,736381151 | GLB1L3 | FALSE | Upregulated |
| 0,985637066 | GLIPR1L1 | FALSE | Upregulated |
| 0,072111872 | GLIS1 | FALSE | Upregulated |
| -0,076434955 | GLRA1 | FALSE | Downregulated |
| 0,276570942 | GNAS-AS1 | FALSE | Upregulated |
| -0,630812051 | GP2 | FALSE | Downregulated |
| -0,094746266 | GPC2 | FALSE | Downregulated |
| -0,882022824 | GPR1 | FALSE | Downregulated |
| -1,245753581 | GPR1-AS | | Downregulated |
| 0,363424385 | GPR15 | FALSE | Upregulated |
| 0,460928047 | GPR17 | FALSE | Upregulated |
| 1,294848049 | GPR37L1 | | Upregulated |
| -0,345505325 | GRIA1 | FALSE | Downregulated |
| 0,54125594 | GRID2 | FALSE | Upregulated |
| -0,077553116 | GRID2IP | FALSE | Downregulated |
| -0,085781777 | GRIK1-AS1 | FALSE | Downregulated |
| 0,213907289 | GRIK3 | FALSE | Upregulated |
| -0,449677669 | GRIN2B | FALSE | Downregulated |
| 0,473083423 | GRIN2C | FALSE | Upregulated |
| -1,400992123 | GRIN3B | | Downregulated |
| -0,58337768 | GRK5-IT1 | FALSE | Downregulated |
| -0,543529816 | GRM2 | FALSE | Downregulated |
| -3,060090631 | GRM3 | | Downregulated |
| 0,910143036 | GRM4 | FALSE | Upregulated |
| 0,185530956 | GRM6 | FALSE | Upregulated |
| -0,130636723 | GRPR | FALSE | Downregulated |
| -0,157315685 | GSG1 | FALSE | Downregulated |
| -1,481085366 | GSG1L | | Downregulated |
| -1,151597661 | GSN-AS1 | | Downregulated |
| 0,47566392 | GTSE1-AS1 | FALSE | Upregulated |
| 1,037288642 | GUCA2A | | Upregulated |
| -1,308774484 | GYPA | | Downregulated |
| -0,582461823 | H2BFXP | FALSE | Downregulated |
| -0,354659634 | HAPLN1 | FALSE | Downregulated |
| -0,354901727 | HAS2-AS1 | FALSE | Downregulated |
| 0,57628587 | HCAR1 | FALSE | Upregulated |
| 0,851424736 | HERC2P10 | FALSE | Upregulated |
| 0,920273621 | HHIPL2 | FALSE | Upregulated |
| 0,275090466 | HHLA2 | FALSE | Upregulated |
| 0,142793003 | HIPK4 | FALSE | Upregulated |
| -1,14782538 | HIST1H1E | | Downregulated |
| 0,100946204 | HIST1H2AK | FALSE | Upregulated |
| -0,453158269 | HIST1H2BO | FALSE | Downregulated |
| 0,203150583 | HIST1H4C | FALSE | Upregulated |
| 1,637816968 | HIST1H4H | | Upregulated |
| -0,621197085 | HIST2H3D | FALSE | Downregulated |
| 1,195713929 | HIST4H4 | | Upregulated |

| | | | |
|---|---|---|---|
| -0,799188382 | HRASLS5 | FALSE | Downregulated |
| -1,108290161 | HS6ST3 | | Downregulated |
| -1,534469352 | HTR2A | | Downregulated |
| -0,782672562 | HTR4 | FALSE | Downregulated |
| -0,598221175 | ICAM5 | FALSE | Downregulated |
| -0,134050309 | IFIT1B | FALSE | Downregulated |
| -1,021775959 | IFITM4P | | Downregulated |
| 1,562542507 | IFNL1 | | Upregulated |
| -0,026591498 | IGBP1P1 | FALSE | Downregulated |
| -1,596600238 | IGLL3P | | Downregulated |
| 0,012359023 | IGSF5 | FALSE | Upregulated |
| 0,063419768 | IL12B | FALSE | Upregulated |
| -0,400199689 | IL21R-AS1 | FALSE | Downregulated |
| -0,080258708 | IL24 | FALSE | Downregulated |
| -0,05516671 | IL31RA | FALSE | Downregulated |
| -0,210196306 | IL36G | FALSE | Downregulated |
| -0,316217889 | IL9R | FALSE | Downregulated |
| -0,607881867 | INE2 | FALSE | Downregulated |
| 1,474177012 | INGX | | Upregulated |
| -0,841268161 | INHA | FALSE | Downregulated |
| -1,445865655 | INTS4L2 | | Downregulated |
| 0,590242802 | IPO9-AS1 | FALSE | Upregulated |
| 0,171365493 | ISL2 | FALSE | Upregulated |
| 0,149730274 | JAZF1-AS1 | FALSE | Upregulated |
| -0,8328995 | JPH4 | FALSE | Downregulated |
| 0,365437849 | KATNBL1P6 | FALSE | Upregulated |
| 1,655439895 | KCNA2 | | Upregulated |
| -0,485419437 | KCNE1L | FALSE | Downregulated |
| 0,271197641 | KCNF1 | FALSE | Upregulated |
| 0,435707694 | KCNH4 | FALSE | Upregulated |
| -1,480802962 | KCNH6 | | Downregulated |
| -0,282365415 | KCNIP4 | FALSE | Downregulated |
| -0,31185716 | KCNJ1 | FALSE | Downregulated |
| 1,574463564 | KCNJ4 | | Upregulated |
| 0,274779736 | KCNJ9 | FALSE | Upregulated |
| 0,372404427 | KCNK7 | FALSE | Upregulated |
| -0,822300275 | KCNQ2 | FALSE | Downregulated |
| -0,514132479 | KCNS1 | FALSE | Downregulated |
| 0,270996281 | KCNT1 | FALSE | Upregulated |
| -0,369744138 | KCP | FALSE | Downregulated |
| -1,037587143 | KCTD16 | | Downregulated |
| 0,173500661 | KHDRBS2 | FALSE | Upregulated |
| -0,296420533 | KIAA0319 | FALSE | Downregulated |
| -0,909761653 | KIF28P | FALSE | Downregulated |
| -0,318241157 | KIF5A | FALSE | Downregulated |
| -1,121270599 | KLF14 | | Downregulated |
| -0,839081589 | KLHDC8A | FALSE | Downregulated |
| 0,422833472 | KLHL35 | FALSE | Upregulated |
| 0,778232405 | KLK11 | FALSE | Upregulated |
| 0,586817058 | KLK14 | FALSE | Upregulated |
| -1,315130538 | KRT1 | | Downregulated |

| | | | |
|---|---|---|---|
| 1,311725993 | KRT15 | | Upregulated |
| 0,092586275 | KRT18P55 | FALSE | Upregulated |
| -0,120017147 | KRT27 | FALSE | Downregulated |
| 1,10618393 | KRT36 | | Upregulated |
| -0,898241337 | KRT42P | FALSE | Downregulated |
| -0,759072326 | KRT72 | FALSE | Downregulated |
| -1,571150095 | KRTAP5-2 | | Downregulated |
| 0,597799269 | KRTAP5-8 | FALSE | Upregulated |
| -0,928764356 | KRTAP5-9 | FALSE | Downregulated |
| -0,312967596 | LAMB4 | FALSE | Downregulated |
| 0,611186777 | LANCL3 | FALSE | Upregulated |
| -1,050929695 | LCE2D | | Downregulated |
| -0,447374986 | LCN10 | FALSE | Downregulated |
| -0,415391526 | LCTL | FALSE | Downregulated |
| 0,654641882 | LDLRAD1 | FALSE | Upregulated |
| -0,671990925 | LEMD1-AS1 | FALSE | Downregulated |
| -0,909455751 | LGALS12 | FALSE | Downregulated |
| 1,852617206 | LGALS17A | | Upregulated |
| 0,665615569 | LGI3 | FALSE | Upregulated |
| -1,357723374 | LHFPL5 | | Downregulated |
| -0,09653609 | LINC00032 | FALSE | Downregulated |
| 0,668551432 | LINC00167 | FALSE | Upregulated |
| -1,92842587 | LINC00202-2 | | Downregulated |
| -1,176030213 | LINC00271 | | Downregulated |
| 0,629124972 | LINC00278 | FALSE | Upregulated |
| -1,506712215 | LINC00298 | | Downregulated |
| -1,554278215 | LINC00327 | | Downregulated |
| -0,314918931 | LINC00330 | FALSE | Downregulated |
| 0,017522352 | LINC00461 | FALSE | Upregulated |
| -0,608319805 | LINC00494 | FALSE | Downregulated |
| 0,397200277 | LINC00515 | FALSE | Upregulated |
| -0,79736984 | LINC00629 | FALSE | Downregulated |
| 0,771605891 | LINC00643 | FALSE | Upregulated |
| -0,88287887 | LINC00664 | FALSE | Downregulated |
| 1,205124185 | LINC00854 | | Upregulated |
| -0,276570201 | LINC00887 | FALSE | Downregulated |
| 0,189636742 | LINC00889 | FALSE | Upregulated |
| -0,679461531 | LINC00896 | FALSE | Downregulated |
| 0,496039818 | LINC00908 | FALSE | Upregulated |
| -0,068006555 | LINC00923 | FALSE | Downregulated |
| -0,267848744 | LINC00941 | FALSE | Downregulated |
| -1,43508536 | LINC00942 | | Downregulated |
| 1,403230463 | LINC00954 | | Upregulated |
| 0,125431243 | LINC00958 | FALSE | Upregulated |
| -0,789143643 | LINC00964 | FALSE | Downregulated |
| 0,547297724 | LINC00968 | FALSE | Upregulated |
| 1,952628973 | LINC00977 | | Upregulated |
| -0,069982904 | LINC00989 | FALSE | Downregulated |
| 0,776706313 | LINC01013 | FALSE | Upregulated |
| -1,318290594 | LINC01087 | | Downregulated |
| 0,79916941 | LINC01108 | FALSE | Upregulated |

| | | | |
|---|---|---|---|
| -0,882820471 | LINC01122 | FALSE | Downregulated |
| -0,770074641 | LINC01139 | FALSE | Downregulated |
| 0,492920327 | LINC01141 | FALSE | Upregulated |
| 0,551364593 | LINC01150 | FALSE | Upregulated |
| -0,177593997 | LINGO2 | FALSE | Downregulated |
| 2,156532844 | LIPN | | Upregulated |
| -0,422460412 | LOC100127974 | FALSE | Downregulated |
| -0,540402969 | LOC100128164 | FALSE | Downregulated |
| -0,153420645 | LOC100128573 | FALSE | Downregulated |
| -0,882252508 | LOC100128593 | FALSE | Downregulated |
| -0,24050655 | LOC100129473 | FALSE | Downregulated |
| -1,279795462 | LOC100129617 | | Downregulated |
| 0,822632137 | LOC100130075 | FALSE | Upregulated |
| 1,645878405 | LOC100130298 | | Upregulated |
| -1,034371025 | LOC100130520 | | Downregulated |
| -1,268437077 | LOC100130876 | | Downregulated |
| -1,08817165 | LOC100131289 | | Downregulated |
| -2,496906061 | LOC100131315 | | Downregulated |
| -1,479639776 | LOC100131825 | | Downregulated |
| -0,904003801 | LOC100132354 | FALSE | Downregulated |
| -0,190171336 | LOC100134040 | FALSE | Downregulated |
| -1,700586844 | LOC100134368 | | Downregulated |
| -0,542892313 | LOC100288162 | FALSE | Downregulated |
| 0,011624835 | LOC100289092 | FALSE | Upregulated |
| 0,184928348 | LOC100289333 | FALSE | Upregulated |
| 0,236051148 | LOC100289511 | FALSE | Upregulated |
| 1,342657022 | LOC100335030 | | Upregulated |
| 0,402275258 | LOC100422556 | FALSE | Upregulated |
| -0,114486482 | LOC100505625 | FALSE | Downregulated |
| -0,39987912 | LOC100505685 | FALSE | Downregulated |
| 0,736187162 | LOC100505774 | FALSE | Upregulated |
| -2,288905133 | LOC100506253 | | Downregulated |
| 0,525081178 | LOC100506504 | FALSE | Upregulated |
| 2,422425637 | LOC100506526 | | Upregulated |
| -0,567740285 | LOC100506859 | FALSE | Downregulated |
| -0,088017291 | LOC100507033 | FALSE | Downregulated |
| 0,294040228 | LOC100507156 | FALSE | Upregulated |
| -1,856097669 | LOC100507346 | | Downregulated |
| -0,467970306 | LOC100507403 | FALSE | Downregulated |
| -0,156220523 | LOC100616530 | FALSE | Downregulated |
| -0,749762614 | LOC100652930 | FALSE | Downregulated |
| -1,380481435 | LOC100653515 | | Downregulated |
| 0,365681754 | LOC100996369 | FALSE | Upregulated |
| -0,392068322 | LOC100996405 | FALSE | Downregulated |
| -0,549028576 | LOC100996549 | FALSE | Downregulated |
| -0,389354823 | LOC100996696 | FALSE | Downregulated |
| -0,908659582 | LOC100996873 | FALSE | Downregulated |
| -0,598204136 | LOC101059906 | FALSE | Downregulated |
| -0,262312304 | LOC101059976 | FALSE | Downregulated |
| -0,499089789 | LOC101926887 | FALSE | Downregulated |
| -0,074971616 | LOC101926896 | FALSE | Downregulated |

| | | | |
|---|---|---|---|
| 0,555609506 | LOC101926997 | FALSE | Upregulated |
| -0,761697874 | LOC101927031 | FALSE | Downregulated |
| 0,061385912 | LOC101927085 | FALSE | Upregulated |
| -0,534682879 | LOC101927131 | FALSE | Downregulated |
| -0,512782027 | LOC101927185 | FALSE | Downregulated |
| -0,23687454 | LOC101927262 | FALSE | Downregulated |
| 0,041336929 | LOC101927326 | FALSE | Upregulated |
| -0,82078847 | LOC101927366 | FALSE | Downregulated |
| 0,425805234 | LOC101927375 | FALSE | Upregulated |
| -0,289268168 | LOC101927437 | FALSE | Downregulated |
| -1,361301288 | LOC101927568 | | Downregulated |
| -2,448575436 | LOC101927582 | | Downregulated |
| 0,286631564 | LOC101927617 | FALSE | Upregulated |
| -2,005445746 | LOC101927632 | | Downregulated |
| 0,362913664 | LOC101927685 | FALSE | Upregulated |
| 1,374702617 | LOC101927696 | | Upregulated |
| -0,455962575 | LOC101927697 | FALSE | Downregulated |
| -1,500176452 | LOC101927705 | | Downregulated |
| 1,385419497 | LOC101927757 | | Upregulated |
| -0,819103587 | LOC101927762 | FALSE | Downregulated |
| 0,39197463 | LOC101927799 | FALSE | Upregulated |
| 0,304797215 | LOC101927803 | FALSE | Upregulated |
| 0,183618011 | LOC101927814 | FALSE | Upregulated |
| 1,084741793 | LOC101927848 | | Upregulated |
| -1,562102797 | LOC101927854 | | Downregulated |
| -0,494284704 | LOC101927875 | FALSE | Downregulated |
| 0,69792002 | LOC101927898 | FALSE | Upregulated |
| -0,344411466 | LOC101927954 | FALSE | Downregulated |
| -1,923919816 | LOC101927971 | | Downregulated |
| -1,026693466 | LOC101928011 | | Downregulated |
| -0,288372954 | LOC101928027 | FALSE | Downregulated |
| 0,443938482 | LOC101928039 | FALSE | Upregulated |
| -1,586162605 | LOC101928066 | | Downregulated |
| -0,03142 | LOC101928109 | FALSE | Downregulated |
| 0,050942303 | LOC101928139 | FALSE | Upregulated |
| -0,496264007 | LOC101928171 | FALSE | Downregulated |
| 0,42019711 | LOC101928188 | FALSE | Upregulated |
| -1,425018871 | LOC101928231 | | Downregulated |
| 0,124084812 | LOC101928279 | FALSE | Upregulated |
| 0,308820104 | LOC101928284 | FALSE | Upregulated |
| 0,25951336 | LOC101928317 | FALSE | Upregulated |
| -0,228421888 | LOC101928343 | FALSE | Downregulated |
| -1,601075045 | LOC101928355 | | Downregulated |
| 0,07150668 | LOC101928360 | FALSE | Upregulated |
| 0,79978398 | LOC101928402 | FALSE | Upregulated |
| -0,216024919 | LOC101928407 | FALSE | Downregulated |
| 0,211213214 | LOC101928439 | FALSE | Upregulated |
| -1,630019525 | LOC101928445 | | Downregulated |
| -0,635782872 | LOC101928484 | FALSE | Downregulated |
| -0,798089995 | LOC101928514 | FALSE | Downregulated |
| -1,408225334 | LOC101928530 | | Downregulated |

| | | | |
|---|---|---|---|
| 0,072749563 | LOC101928628 | FALSE | Upregulated |
| -1,9525794 | LOC101928632 | | Downregulated |
| -1,708524689 | LOC101928671 | | Downregulated |
| -0,425773202 | LOC101928690 | FALSE | Downregulated |
| 1,664623487 | LOC101928716 | | Upregulated |
| 0,212945139 | LOC101928717 | FALSE | Upregulated |
| 0,091826427 | LOC101928733 | FALSE | Upregulated |
| -1,403911198 | LOC101928738 | | Downregulated |
| -0,808068222 | LOC101928814 | FALSE | Downregulated |
| -0,963516167 | LOC101928824 | FALSE | Downregulated |
| -0,864462458 | LOC101928867 | FALSE | Downregulated |
| -0,421562261 | LOC101928910 | FALSE | Downregulated |
| -0,196543511 | LOC101928961 | FALSE | Downregulated |
| -1,462271215 | LOC101928963 | | Downregulated |
| -2,139255574 | LOC101928981 | | Downregulated |
| 0,546174665 | LOC101928993 | FALSE | Upregulated |
| -3,161472701 | LOC101929008 | | Downregulated |
| -0,797879541 | LOC101929017 | FALSE | Downregulated |
| -0,975155376 | LOC101929035 | FALSE | Downregulated |
| -1,704830922 | LOC101929051 | | Downregulated |
| -0,121812314 | LOC101929054 | FALSE | Downregulated |
| -1,0295938 | LOC101929057 | | Downregulated |
| -0,061881687 | LOC101929078 | FALSE | Downregulated |
| -0,779976357 | LOC101929085 | FALSE | Downregulated |
| 1,308719307 | LOC101929125 | | Upregulated |
| -0,135569092 | LOC101929149 | FALSE | Downregulated |
| -1,147350179 | LOC101929151 | | Downregulated |
| -0,233422198 | LOC101929159 | FALSE | Downregulated |
| -0,612773124 | LOC101929165 | FALSE | Downregulated |
| 1,237261913 | LOC101929173 | | Upregulated |
| -0,149303144 | LOC101929192 | FALSE | Downregulated |
| -0,023069498 | LOC101929216 | FALSE | Downregulated |
| -0,138630635 | LOC101929227 | FALSE | Downregulated |
| -0,4763918 | LOC101929241 | FALSE | Downregulated |
| -0,370074404 | LOC101929248 | FALSE | Downregulated |
| -1,958976011 | LOC101929270 | | Downregulated |
| 1,222239201 | LOC101929272 | | Upregulated |
| -0,122004688 | LOC101929336 | FALSE | Downregulated |
| -1,309643919 | LOC101929354 | | Downregulated |
| -0,030795101 | LOC101929409 | FALSE | Downregulated |
| -1,819893965 | LOC101929414 | | Downregulated |
| -1,558122722 | LOC101929431 | | Downregulated |
| -1,144428903 | LOC101929434 | | Downregulated |
| 0,248700085 | LOC101929447 | FALSE | Upregulated |
| 0,285229231 | LOC101929490 | FALSE | Upregulated |
| 0,460325826 | LOC101929491 | FALSE | Upregulated |
| -0,175098653 | LOC101929500 | FALSE | Downregulated |
| 0,322872148 | LOC101929521 | FALSE | Upregulated |
| -0,087939671 | LOC101929536 | FALSE | Downregulated |
| 0,578457694 | LOC101929560 | FALSE | Upregulated |
| 0,520944761 | LOC101929587 | FALSE | Upregulated |

| | | | |
|---|---|---|---|
| -1,085111372 | LOC101929634 | | Downregulated |
| 0,117824341 | LOC101929692 | FALSE | Upregulated |
| 0,068031717 | LOC101929714 | FALSE | Upregulated |
| -0,603421392 | LOC101929726 | FALSE | Downregulated |
| 1,603537277 | LOC101929790 | | Upregulated |
| -1,388118017 | LOC101929866 | | Downregulated |
| -0,790502335 | LOC101929937 | FALSE | Downregulated |
| 0,388952859 | LOC101930026 | FALSE | Upregulated |
| -1,598427351 | LOC101930137 | | Downregulated |
| 0,551295967 | LOC101930370 | FALSE | Upregulated |
| -0,6693338 | LOC101930580 | FALSE | Downregulated |
| -1,766567115 | LOC102467073 | | Downregulated |
| 0,185906645 | LOC102467147 | FALSE | Upregulated |
| -0,414044303 | LOC102467214 | FALSE | Downregulated |
| -0,719952808 | LOC102723305 | FALSE | Downregulated |
| 0,044658968 | LOC102723310 | FALSE | Upregulated |
| -0,697407514 | LOC102723320 | FALSE | Downregulated |
| -2,000897468 | LOC102723353 | | Downregulated |
| -0,913147814 | LOC102723354 | FALSE | Downregulated |
| -0,118557326 | LOC102723361 | FALSE | Downregulated |
| -0,186124309 | LOC102723363 | FALSE | Downregulated |
| -1,212113268 | LOC102723366 | | Downregulated |
| 0,885662685 | LOC102723371 | FALSE | Upregulated |
| -2,056589171 | LOC102723382 | | Downregulated |
| -0,651229823 | LOC102723399 | FALSE | Downregulated |
| 0,363946651 | LOC102723408 | FALSE | Upregulated |
| 0,361650143 | LOC102723419 | FALSE | Upregulated |
| -0,906325007 | LOC102723420 | FALSE | Downregulated |
| -0,741498152 | LOC102723437 | FALSE | Downregulated |
| -0,172170659 | LOC102723458 | FALSE | Downregulated |
| 0,150730638 | LOC102723475 | FALSE | Upregulated |
| -0,447476669 | LOC102723489 | FALSE | Downregulated |
| 0,649532963 | LOC102723507 | FALSE | Upregulated |
| 0,567059016 | LOC102723510 | FALSE | Upregulated |
| 0,49930147 | LOC102723543 | FALSE | Upregulated |
| -0,43752298 | LOC102723545 | FALSE | Downregulated |
| -0,659587675 | LOC102723561 | FALSE | Downregulated |
| -1,403808023 | LOC102723595 | | Downregulated |
| 0,56059528 | LOC102723619 | FALSE | Upregulated |
| -0,639390558 | LOC102723654 | FALSE | Downregulated |
| -1,02638125 | LOC102723656 | | Downregulated |
| -0,788282632 | LOC102723659 | FALSE | Downregulated |
| 0,728086935 | LOC102723682 | FALSE | Upregulated |
| -1,023731035 | LOC102723694 | | Downregulated |
| -0,212527293 | LOC102723714 | FALSE | Downregulated |
| 0,717951716 | LOC102723724 | FALSE | Upregulated |
| -0,256486494 | LOC102723729 | FALSE | Downregulated |
| -0,581664974 | LOC102723730 | FALSE | Downregulated |
| -1,591816229 | LOC102723754 | | Downregulated |
| 1,556069309 | LOC102723761 | | Upregulated |
| -0,017061964 | LOC102723811 | FALSE | Downregulated |

| | | | |
|---|---|---|---|
| -0,471199001 | LOC102723814 | FALSE | Downregulated |
| 0,37013563 | LOC102723837 | FALSE | Upregulated |
| 1,108268576 | LOC102723838 | | Upregulated |
| -0,429115912 | LOC102723867 | FALSE | Downregulated |
| 0,257353398 | LOC102723877 | FALSE | Upregulated |
| -0,019548556 | LOC102723891 | FALSE | Downregulated |
| 0,33530248 | LOC102723913 | FALSE | Upregulated |
| 0,249858508 | LOC102723922 | FALSE | Upregulated |
| 0,35889871 | LOC102723923 | FALSE | Upregulated |
| -1,584831594 | LOC102723943 | | Downregulated |
| -2,102880832 | LOC102724003 | | Downregulated |
| 0,195034777 | LOC102724004 | FALSE | Upregulated |
| -0,994212153 | LOC102724057 | FALSE | Downregulated |
| 0,785281996 | LOC102724069 | FALSE | Upregulated |
| -1,279700858 | LOC102724075 | | Downregulated |
| -0,071604686 | LOC102724106 | FALSE | Downregulated |
| -0,177928358 | LOC102724108 | FALSE | Downregulated |
| -1,423259242 | LOC102724113 | | Downregulated |
| -2,454814519 | LOC102724122 | | Downregulated |
| -0,087170065 | LOC102724143 | FALSE | Downregulated |
| 0,267910394 | LOC102724149 | FALSE | Upregulated |
| -1,680716937 | LOC102724177 | | Downregulated |
| -0,320375487 | LOC102724193 | FALSE | Downregulated |
| -0,190374228 | LOC102724196 | FALSE | Downregulated |
| -0,521000866 | LOC102724259 | FALSE | Downregulated |
| 0,667019961 | LOC102724271 | FALSE | Upregulated |
| -1,713334474 | LOC102724273 | | Downregulated |
| -0,594014489 | LOC102724292 | FALSE | Downregulated |
| -0,273062823 | LOC102724342 | FALSE | Downregulated |
| -0,023115443 | LOC102724348 | FALSE | Downregulated |
| 0,11084586 | LOC102724351 | FALSE | Upregulated |
| -0,955744382 | LOC102724377 | FALSE | Downregulated |
| 1,380525869 | LOC102724378 | | Upregulated |
| 0,953532879 | LOC102724393 | FALSE | Upregulated |
| 0,137780552 | LOC102724400 | FALSE | Upregulated |
| -0,404608599 | LOC102724423 | FALSE | Downregulated |
| 2,499094592 | LOC102724424 | | Upregulated |
| 0,764324521 | LOC102724425 | FALSE | Upregulated |
| -0,325893092 | LOC102724440 | FALSE | Downregulated |
| -0,594197552 | LOC102724449 | FALSE | Downregulated |
| -2,096864423 | LOC102724480 | | Downregulated |
| 0,690865637 | LOC102724488 | FALSE | Upregulated |
| -1,392825139 | LOC102724508 | | Downregulated |
| 0,091348934 | LOC102724518 | FALSE | Upregulated |
| -0,216565412 | LOC102724536 | FALSE | Downregulated |
| -0,063338148 | LOC102724543 | FALSE | Downregulated |
| 0,72347771 | LOC102724689 | FALSE | Upregulated |
| -0,600190408 | LOC102724696 | FALSE | Downregulated |
| 0,58031764 | LOC102724703 | FALSE | Upregulated |
| -1,713531695 | LOC102724720 | | Downregulated |
| -0,795153638 | LOC102724731 | FALSE | Downregulated |

| | | | |
|---|---|---|---|
| 0,354240234 | LOC102724758 | FALSE | Upregulated |
| -1,781672423 | LOC102724759 | | Downregulated |
| 0,650315593 | LOC102724797 | FALSE | Upregulated |
| -0,610498621 | LOC102724810 | FALSE | Downregulated |
| 0,654917456 | LOC102724826 | FALSE | Upregulated |
| -0,609089153 | LOC102724830 | FALSE | Downregulated |
| 0,934460472 | LOC102724848 | FALSE | Upregulated |
| -2,689827964 | LOC102724874 | | Downregulated |
| 0,52384335 | LOC102724875 | FALSE | Upregulated |
| -1,361116079 | LOC102724883 | | Downregulated |
| 0,426257337 | LOC102724898 | FALSE | Upregulated |
| 0,106384109 | LOC102724914 | FALSE | Upregulated |
| -0,462128676 | LOC102724944 | FALSE | Downregulated |
| -1,340200556 | LOC102724956 | | Downregulated |
| 0,062859683 | LOC151760 | FALSE | Upregulated |
| -1,135841572 | LOC256880 | | Downregulated |
| -0,050647059 | LOC257358 | FALSE | Downregulated |
| -0,534814066 | LOC283683 | FALSE | Downregulated |
| -0,967559769 | LOC283731 | FALSE | Downregulated |
| 1,338743923 | LOC284009 | | Upregulated |
| -0,73953225 | LOC284344 | FALSE | Downregulated |
| -0,311364487 | LOC284600 | FALSE | Downregulated |
| -0,087844222 | LOC285043 | FALSE | Downregulated |
| -3,210238616 | LOC285095 | | Downregulated |
| 0,716010261 | LOC285819 | FALSE | Upregulated |
| 0,231996693 | LOC286178 | FALSE | Upregulated |
| 0,996607567 | LOC341056 | FALSE | Upregulated |
| -0,531269015 | LOC388242 | FALSE | Downregulated |
| -0,472938774 | LOC390956 | FALSE | Downregulated |
| -0,184374834 | LOC391722 | FALSE | Downregulated |
| 1,67638531 | LOC399815 | | Upregulated |
| 0,009342889 | LOC400499 | FALSE | Upregulated |
| -1,06670453 | LOC400891 | | Downregulated |
| -2,211571374 | LOC401098 | | Downregulated |
| -1,332290426 | LOC440292 | | Downregulated |
| 0,736096303 | LOC441956 | FALSE | Upregulated |
| -0,162902378 | LOC494127 | FALSE | Downregulated |
| 0,280496827 | LOC494141 | FALSE | Upregulated |
| -1,524159942 | LOC613038 | | Downregulated |
| -1,93352357 | LOC642249 | | Downregulated |
| -0,410220245 | LOC642934 | FALSE | Downregulated |
| -0,862847329 | LOC643406 | FALSE | Downregulated |
| -0,842304585 | LOC643711 | FALSE | Downregulated |
| -0,469111294 | LOC644919 | FALSE | Downregulated |
| -0,179716574 | LOC645485 | FALSE | Downregulated |
| 0,20033565 | LOC646588 | FALSE | Upregulated |
| -0,244585139 | LOC650368 | FALSE | Downregulated |
| 1,552703681 | LOC653786 | | Upregulated |
| 0,565240876 | LOC727710 | FALSE | Upregulated |
| 1,066553688 | LOC728637 | | Upregulated |
| 0,221801729 | LOC728715 | FALSE | Upregulated |

| | | | |
|---|---|---|---|
| -0,510495194 | LOC729468 | FALSE | Downregulated |
| -0,561274736 | LOC729506 | FALSE | Downregulated |
| -1,393399317 | LOC729658 | | Downregulated |
| -0,950675946 | LOC729770 | FALSE | Downregulated |
| 0,110347091 | LOC730338 | FALSE | Upregulated |
| -1,798485259 | LOC731157 | | Downregulated |
| -0,401046026 | LOC91450 | FALSE | Downregulated |
| -0,322051722 | LRRC10B | FALSE | Downregulated |
| -0,367284411 | LRRC36 | FALSE | Downregulated |
| -0,372042496 | LRRC3C | FALSE | Downregulated |
| -1,157013612 | LRRN4CL | | Downregulated |
| 0,483473853 | LUCAT1 | FALSE | Upregulated |
| 0,163730353 | LY86-AS1 | FALSE | Upregulated |
| -0,014646684 | MAGEL2 | FALSE | Downregulated |
| -1,629097545 | MALRD1 | | Downregulated |
| -0,91335415 | MAP2K4P1 | FALSE | Downregulated |
| 0,078584308 | MAPK15 | FALSE | Upregulated |
| 0,06989461 | MARCH4 | FALSE | Upregulated |
| 1,512531397 | MCCD1 | | Upregulated |
| -0,414657948 | MEIS1-AS2 | FALSE | Downregulated |
| -0,559507467 | MEIS1-AS3 | FALSE | Downregulated |
| 1,173988679 | MEP1A | | Upregulated |
| 1,695317996 | MESP2 | | Upregulated |
| -1,085695824 | MFSD2B | | Downregulated |
| 0,671108734 | MFSD6L | FALSE | Upregulated |
| 0,26427542 | MGAT5B | FALSE | Upregulated |
| -0,200773777 | MIMT1 | FALSE | Downregulated |
| -0,637311672 | MINOS1P1 | FALSE | Downregulated |
| -0,607361355 | MIR1184-1 | FALSE | Downregulated |
| 0,50731627 | MIR142 | FALSE | Upregulated |
| 0,909485106 | MIR181A2HG | FALSE | Upregulated |
| 0,33756619 | MIR2861 | FALSE | Upregulated |
| 0,249886296 | MIR3936 | FALSE | Upregulated |
| 0,717058452 | MIR4767 | FALSE | Upregulated |
| -1,686698454 | MIR5010 | | Downregulated |
| -1,568768135 | MIR6772 | | Downregulated |
| 0,312505248 | MLK7-AS1 | FALSE | Upregulated |
| 1,270362504 | MMP1 | | Upregulated |
| -0,463835742 | MOBP | FALSE | Downregulated |
| 0,002246145 | MRPL23-AS1 | FALSE | Upregulated |
| -0,159756105 | MSLN | FALSE | Downregulated |
| 1,361292338 | MSMB | | Upregulated |
| -0,562003997 | MSMP | FALSE | Downregulated |
| -1,045922384 | MSX2 | | Downregulated |
| 0,954549233 | MSX2P1 | FALSE | Upregulated |
| -0,579708429 | MT1IP | FALSE | Downregulated |
| -1,01988571 | MUC2 | | Downregulated |
| -0,028289389 | MUC5AC | FALSE | Downregulated |
| 0,374107965 | MYBPC3 | FALSE | Upregulated |
| 1,762436532 | MYCNOS | | Upregulated |
| -0,147045658 | MYT1 | FALSE | Downregulated |

| | | | |
|---|---|---|---|
| -0,267544291 | NALCN-AS1 | FALSE | Downregulated |
| -1,542265424 | NAP1L6 | | Downregulated |
| -1,727141522 | NBEAP1 | | Downregulated |
| -0,540852774 | NCAM1-AS1 | FALSE | Downregulated |
| 1,452443787 | NCCRP1 | | Upregulated |
| 0,480083692 | NCRUPAR | FALSE | Upregulated |
| -0,959537704 | NEFM | FALSE | Downregulated |
| -1,707364473 | NEK5 | | Downregulated |
| -0,191509202 | NETO1 | FALSE | Downregulated |
| 0,432454787 | NEXN-AS1 | FALSE | Upregulated |
| 0,319885608 | NHLRC4 | FALSE | Upregulated |
| -0,877011128 | NKPD1 | FALSE | Downregulated |
| -0,386930548 | NKX2-3 | FALSE | Downregulated |
| 1,125091337 | NLGN1 | | Upregulated |
| -0,31915599 | NLRP8 | FALSE | Downregulated |
| 0,247617022 | NLRP9 | FALSE | Upregulated |
| -0,075345169 | NME8 | FALSE | Downregulated |
| -0,538064653 | NNAT | FALSE | Downregulated |
| 0,552015324 | NOG | FALSE | Upregulated |
| -0,38583691 | NOX5 | FALSE | Downregulated |
| -0,149144648 | NPB | FALSE | Downregulated |
| 0,313953808 | NPSR1-AS1 | FALSE | Upregulated |
| 0,840672149 | NR2E1 | FALSE | Upregulated |
| -1,578052472 | NT5C1B-RDH14 | | Downregulated |
| -1,416554223 | NTNG1 | | Downregulated |
| 0,155433251 | NTRK1 | FALSE | Upregulated |
| -0,382071596 | NTSR1 | FALSE | Downregulated |
| -1,835273112 | NUDT11 | | Downregulated |
| -1,190350775 | NUTM2G | | Downregulated |
| -0,147373864 | NWD1 | FALSE | Downregulated |
| -1,023146454 | NXPH1 | | Downregulated |
| -0,629749829 | NXPH2 | FALSE | Downregulated |
| 0,091597581 | NXPH4 | FALSE | Upregulated |
| 1,79822468 | NYAP2 | | Upregulated |
| 1,544435343 | ODF3L1 | | Upregulated |
| 2,127721556 | ODF3L2 | | Upregulated |
| 0,18807324 | OLAH | FALSE | Upregulated |
| 0,273525102 | OPN1SW | FALSE | Upregulated |
| 0,672071106 | OPRK1 | FALSE | Upregulated |
| 1,308731299 | OR10R2 | | Upregulated |
| 0,519031686 | OR2C1 | FALSE | Upregulated |
| 1,329038174 | OR51B5 | | Upregulated |
| -0,404456293 | OR6W1P | FALSE | Downregulated |
| 1,392679105 | OSM | | Upregulated |
| -0,175295644 | OSR2 | FALSE | Downregulated |
| -0,279713662 | OSTCP2 | FALSE | Downregulated |
| -0,487169402 | OTUD6A | FALSE | Downregulated |
| 0,930086045 | OTX1 | FALSE | Upregulated |
| -1,593882887 | OVOL1 | | Downregulated |
| 0,464294112 | P2RX2 | FALSE | Upregulated |
| -0,314265791 | PAPL | FALSE | Downregulated |

| | | | |
|---|---|---|---|
| -0,813519385 | PAPOLB | FALSE | Downregulated |
| -0,742916047 | PAPPA-AS1 | FALSE | Downregulated |
| -0,486327213 | PAX2 | FALSE | Downregulated |
| -0,348064552 | PCAT14 | FALSE | Downregulated |
| -0,596514917 | PCDH10 | FALSE | Downregulated |
| -0,13580506 | PCDH15 | FALSE | Downregulated |
| 0,025271229 | PCDH8 | FALSE | Upregulated |
| -0,241493034 | PCSK2 | FALSE | Downregulated |
| -0,141253493 | PDILT | FALSE | Downregulated |
| -1,336723349 | PGM5-AS1 | | Downregulated |
| -0,000315224 | PGM5P2 | FALSE | Downregulated |
| -0,99832602 | PLCZ1 | FALSE | Downregulated |
| 0,292726121 | PLSCR2 | FALSE | Upregulated |
| 1,720463585 | PNMA5 | | Upregulated |
| 1,14625167 | PNMT | | Upregulated |
| 0,240298982 | POU3F1 | FALSE | Upregulated |
| 0,655590547 | POU3F2 | FALSE | Upregulated |
| -0,626786493 | POU6F2-AS2 | FALSE | Downregulated |
| -0,894997254 | PPEF1 | FALSE | Downregulated |
| -0,82360118 | PPIAP30 | FALSE | Downregulated |
| 0,980857828 | PPP2R2C | FALSE | Upregulated |
| -0,640971406 | PRO2852 | FALSE | Downregulated |
| -1,343241305 | PROK1 | | Downregulated |
| -0,004097671 | PROKR2 | FALSE | Downregulated |
| -0,812800137 | PROSER2-AS1 | FALSE | Downregulated |
| 0,490300981 | PROX2 | FALSE | Upregulated |
| -0,120154342 | PRPH | FALSE | Downregulated |
| -0,360531043 | PRRT4 | FALSE | Downregulated |
| 0,953644415 | PRSS21 | FALSE | Upregulated |
| -0,755836744 | PRSS30P | FALSE | Downregulated |
| -0,333572185 | PRSS42 | FALSE | Downregulated |
| 0,176981587 | PTCHD2 | FALSE | Upregulated |
| -0,772865586 | PTCHD3 | FALSE | Downregulated |
| 0,2727966 | PTGER4P2-CDK2AP2P2 | FALSE | Upregulated |
| -0,796878121 | PURG | FALSE | Downregulated |
| -1,3679685 | PXT1 | | Downregulated |
| 0,167453664 | PYY2 | FALSE | Upregulated |
| 0,139771822 | QRFPR | FALSE | Upregulated |
| 0,196288363 | RAB44 | FALSE | Upregulated |
| -0,252057531 | RADIL | FALSE | Downregulated |
| 0,625939875 | RAET1K | FALSE | Upregulated |
| -1,249743323 | RASGEF1C | | Downregulated |
| 0,991939222 | RASSF10 | FALSE | Upregulated |
| 0,286398206 | RBBP8NL | FALSE | Upregulated |
| -0,273703153 | RBP2 | FALSE | Downregulated |
| 0,253563841 | RCAN3AS | FALSE | Upregulated |
| -0,153479329 | RD3 | FALSE | Downregulated |
| 1,772354117 | RDM1 | | Upregulated |
| -0,747172484 | REEP2 | FALSE | Downregulated |
| -0,359314851 | RFPL3S | FALSE | Downregulated |
| -2,267130729 | RGPD4-AS1 | | Downregulated |

| | | | |
|---|---|---|---|
| 0,519202718 | RGS13 | FALSE | Upregulated |
| -1,215438169 | RGS22 | | Downregulated |
| -0,669513403 | RGS9BP | FALSE | Downregulated |
| -0,993201712 | RHD | FALSE | Downregulated |
| -0,414814707 | RHOXF1 | FALSE | Downregulated |
| 1,249456802 | RIMS1 | | Upregulated |
| 0,416177583 | RIMS2 | FALSE | Upregulated |
| 1,874321369 | RLN1 | | Upregulated |
| 0,510365247 | RLN2 | FALSE | Upregulated |
| -0,862951948 | RNA5S1 | FALSE | Downregulated |
| 0,225391966 | RNA5S10 | FALSE | Upregulated |
| -0,789329181 | RNA5S11 | FALSE | Downregulated |
| | RNA5S12 | | |
| | RNA5S13 | | |
| | RNA5S14 | | |
| | RNA5S15 | | |
| | RNA5S16 | | |
| -0,557336974 | RNA5S17 | FALSE | Downregulated |
| 0 | RNA5S2 | FALSE | Upregulated |
| 0,072730206 | RNA5S3 | FALSE | Upregulated |
| | RNA5S4 | | |
| 0 | RNA5S5 | FALSE | Upregulated |
| | RNA5S6 | | |
| | RNA5S7 | | |
| | RNA5S8 | | |
| 0,969965875 | RNF148 | FALSE | Upregulated |
| 0,174746558 | RNF17 | FALSE | Upregulated |
| -0,139931409 | RNF182 | FALSE | Downregulated |
| -0,985499626 | RNU6-26P | FALSE | Downregulated |
| 0,541249859 | RNU6-31P | FALSE | Upregulated |
| -0,514324926 | RNU6-46P | FALSE | Downregulated |
| 0,409694884 | RNY1 | FALSE | Upregulated |
| -1,068770075 | RP1-90L6.2 | | Downregulated |
| -0,540152248 | RP11-1057B8.2 | FALSE | Downregulated |
| -0,467584832 | RP11-106M7.1 | FALSE | Downregulated |
| -1,283638864 | RP11-1079K10.5 | | Downregulated |
| -0,688308055 | RP11-133O22.6 | FALSE | Downregulated |
| 0,231522 | RP11-134P9.3 | FALSE | Upregulated |
| 0,190234289 | RP11-143M1.2 | FALSE | Upregulated |
| 0,721537795 | RP11-154D17.1 | FALSE | Upregulated |
| -0,547802376 | RP11-158M2.5 | FALSE | Downregulated |
| -0,036649839 | RP11-170N11.1 | FALSE | Downregulated |
| -1,270937808 | RP11-175K6.1 | | Downregulated |
| -0,649302193 | RP11-17A4.2 | FALSE | Downregulated |
| 1,623119 | RP11-193H5.1 | | Upregulated |
| 0,693001499 | RP11-219G17.4 | FALSE | Upregulated |
| 1,204970949 | RP11-227H15.4 | | Upregulated |
| -1,011268428 | RP11-231I16.1 | | Downregulated |
| -1,718981708 | RP11-260E18.1 | | Downregulated |
| -1,817735793 | RP11-264C15.2 | | Downregulated |
| -0,263683949 | RP11-26J3.1 | FALSE | Downregulated |

| | | | |
|---|---|---|---|
| 0,036690295 | RP11-294O2.2 | FALSE | Upregulated |
| -1,144959523 | RP11-319G6.1 | | Downregulated |
| 0,358996162 | RP11-320G24.1 | FALSE | Upregulated |
| -1,216333134 | RP11-341A22.2 | | Downregulated |
| -0,563678998 | RP11-365O16.6 | FALSE | Downregulated |
| 0,97464747 | RP11-374P20.4 | FALSE | Upregulated |
| -0,094666298 | RP11-379F4.4 | FALSE | Downregulated |
| -0,635486257 | RP11-383C5.3 | FALSE | Downregulated |
| -1,021624116 | RP11-430H10.1 | | Downregulated |
| 0,757540893 | RP11-445H22.3 | FALSE | Upregulated |
| -0,57213665 | RP11-446H18.5 | FALSE | Downregulated |
| -0,484592389 | RP11-449D8.1 | FALSE | Downregulated |
| -2,253726035 | RP11-492A10.1 | | Downregulated |
| 1,146312609 | RP11-508P1.2 | | Upregulated |
| -0,843329923 | RP11-534N16.1 | FALSE | Downregulated |
| -0,717151407 | RP11-536C5.7 | FALSE | Downregulated |
| 1,477510963 | RP11-538P18.2 | | Upregulated |
| -0,119742458 | RP11-542B15.1 | FALSE | Downregulated |
| -0,917808184 | RP11-544M22.1 | FALSE | Downregulated |
| -0,434800438 | RP11-554A11.6 | FALSE | Downregulated |
| 0,298015818 | RP11-576I22.2 | FALSE | Upregulated |
| 0,685535851 | RP11-588G21.2 | FALSE | Upregulated |
| -0,118272882 | RP11-593F23.1 | FALSE | Downregulated |
| 0,996379433 | RP11-710F7.3 | FALSE | Upregulated |
| -0,284490021 | RP11-723O4.2 | FALSE | Downregulated |
| 1,050166108 | RP11-725G5.2 | | Upregulated |
| 0,659535804 | RP11-770E5.1 | FALSE | Upregulated |
| 0,358806146 | RP11-829H16.3 | FALSE | Upregulated |
| -0,03277924 | RP11-840I19.3 | FALSE | Downregulated |
| 0,130349879 | RP11-864J10.4 | FALSE | Upregulated |
| -1,837758425 | RP11-885B4.2 | | Downregulated |
| -1,111533594 | RP11-887P2.5 | | Downregulated |
| 1,54351861 | RP11-973H7.4 | | Upregulated |
| 0,154226217 | RP3-399L15.3 | FALSE | Upregulated |
| 0,72760288 | RP3-400N23.6 | FALSE | Upregulated |
| -1,231797332 | RP3-476K8.3 | | Downregulated |
| 0,866516695 | RP3-522P13.3 | FALSE | Upregulated |
| -0,268206944 | RP4-565E6.1 | FALSE | Downregulated |
| 0,436937239 | RP4-724E16.2 | FALSE | Upregulated |
| -1,547901551 | RP5-1142A6.2 | | Downregulated |
| -0,63965757 | RPA4 | FALSE | Downregulated |
| -0,492445522 | RPEL1 | FALSE | Downregulated |
| -0,590708624 | RPS4Y2 | FALSE | Downregulated |
| 0,499562635 | SAG | FALSE | Upregulated |
| -1,706444225 | SCG2 | | Downregulated |
| -1,434740286 | SCGB2B2 | | Downregulated |
| -0,594303774 | SCHLAP1 | FALSE | Downregulated |
| 1,107398211 | SCT | | Upregulated |
| 0,568208567 | SERPINA9 | FALSE | Upregulated |
| 0,829154495 | SFTPB | FALSE | Upregulated |
| -2,342237992 | SGCZ | | Downregulated |

| | | | |
|---|---|---|---|
| 0,132474576 | SH2D5 | FALSE | Upregulated |
| 0,251579939 | SH2D6 | FALSE | Upregulated |
| 0,032604972 | SH3RF3-AS1 | FALSE | Upregulated |
| 0,909656439 | SHISA2 | FALSE | Upregulated |
| -1,078130947 | SIM2 | | Downregulated |
| -0,713390009 | SIX3 | FALSE | Downregulated |
| -0,573600561 | SLC10A4 | FALSE | Downregulated |
| -0,033810095 | SLC12A3 | FALSE | Downregulated |
| -0,666058597 | SLC13A1 | FALSE | Downregulated |
| 0,379726799 | SLC18A1 | FALSE | Upregulated |
| -0,011178164 | SLC22A14 | FALSE | Downregulated |
| -2,377681575 | SLC24A2 | | Downregulated |
| 0,596424289 | SLC2A1-AS1 | FALSE | Upregulated |
| -0,827712539 | SLC39A2 | FALSE | Downregulated |
| 0,147876337 | SLC46A2 | FALSE | Upregulated |
| -0,35994229 | SLC5A5 | FALSE | Downregulated |
| 0,155683563 | SLC6A7 | FALSE | Upregulated |
| 0,140820017 | SLC8A2 | FALSE | Upregulated |
| -1,135657229 | SLCO1C1 | | Downregulated |
| -0,573937438 | SLITRK5 | FALSE | Downregulated |
| 0,426905892 | SMARCA5-AS1 | FALSE | Upregulated |
| 0,608204259 | SMIM2 | FALSE | Upregulated |
| 1,719670396 | SMIM22 | | Upregulated |
| -0,413483781 | SNORA31 | FALSE | Downregulated |
| 0,155088097 | SNORA41 | FALSE | Upregulated |
| 0 | SNORA59A | FALSE | Upregulated |
| -0,725618312 | SNORA59B | FALSE | Downregulated |
| 0,736652815 | SNORA65 | FALSE | Upregulated |
| -0,328729683 | SNORD103A | FALSE | Downregulated |
| -0,454023033 | SNORD103B | FALSE | Downregulated |
| 0,743956214 | SNORD16 | FALSE | Upregulated |
| -0,268220673 | SNORD17 | FALSE | Downregulated |
| 0,068584867 | SNORD35B | FALSE | Upregulated |
| 1,167707978 | SNORD3B-2 | | Upregulated |
| -1,592487067 | SNORD3C | | Downregulated |
| 0,327882992 | SNORD45B | FALSE | Upregulated |
| -0,509993352 | SNORD4A | FALSE | Downregulated |
| -0,316859847 | SNORD54 | FALSE | Downregulated |
| -1,077126005 | SNORD56 | | Downregulated |
| -0,900329292 | SNORD58A | FALSE | Downregulated |
| 0,45920107 | SNORD6 | FALSE | Upregulated |
| 0,387362415 | SNORD62A | FALSE | Upregulated |
| 1,23472573 | SNORD62B | | Upregulated |
| 0,933737889 | SNORD74 | FALSE | Upregulated |
| -0,041550272 | SNORD76 | FALSE | Downregulated |
| -1,199518746 | SORCS1 | | Downregulated |
| -0,60156209 | SOX11 | FALSE | Downregulated |
| 0,305424539 | SOX2 | FALSE | Upregulated |
| 0,632976698 | SP6 | FALSE | Upregulated |
| -0,673020757 | SPAG17 | FALSE | Downregulated |
| -0,634814276 | SPATA31D1 | FALSE | Downregulated |

| | | | |
|---|---|---|---|
| -1,268029853 | SPEF1 | | Downregulated |
| -0,541882911 | SPOCD1 | FALSE | Downregulated |
| -0,114522265 | SRD5A1P1 | FALSE | Downregulated |
| 0,369718423 | SRGAP2-AS1 | FALSE | Upregulated |
| -0,264942753 | ST7-AS2 | FALSE | Downregulated |
| -2,508737345 | ST8SIA2 | | Downregulated |
| -1,731163359 | ST8SIA5 | | Downregulated |
| -0,6201207 | STAC2 | FALSE | Downregulated |
| -0,103862828 | STEAP1B | FALSE | Downregulated |
| 0,480770946 | SULT2B1 | FALSE | Upregulated |
| 0,183918878 | SUMO1P1 | FALSE | Upregulated |
| -0,670137 | SV2C | FALSE | Downregulated |
| -0,707457809 | SVILP1 | FALSE | Downregulated |
| -0,551515535 | SYCP1 | FALSE | Downregulated |
| 0,001096181 | SYN2 | FALSE | Upregulated |
| -1,048739252 | SYNDIG1L | | Downregulated |
| 0,421399331 | SYNGR3 | FALSE | Upregulated |
| 0,706838256 | SYNGR4 | FALSE | Upregulated |
| -0,630381728 | SYT6 | FALSE | Downregulated |
| -0,733863265 | TAS1R1 | FALSE | Downregulated |
| 0,123871692 | TAS1R3 | FALSE | Upregulated |
| 1,856691422 | TAS2R60 | | Upregulated |
| 0,739661182 | TBC1D27 | FALSE | Upregulated |
| -0,791862742 | TBC1D29 | FALSE | Downregulated |
| -1,815670699 | TBC1D3P1-DHX40P1 | | Downregulated |
| -0,192439046 | TCERG1L | FALSE | Downregulated |
| 0,002128402 | TCTEX1D4 | FALSE | Upregulated |
| -0,19793236 | TEPP | FALSE | Downregulated |
| 1,334517378 | TEX11 | | Upregulated |
| -0,025748452 | TEX12 | FALSE | Downregulated |
| 0,183526727 | TEX22 | FALSE | Upregulated |
| 0,53465492 | TFAP2A-AS1 | FALSE | Upregulated |
| 0,053666344 | TFAP2C | FALSE | Upregulated |
| 0,387234503 | TH | FALSE | Upregulated |
| 0,012758106 | TINCR | FALSE | Upregulated |
| -0,390165906 | TM4SF19-AS1 | FALSE | Downregulated |
| -2,141715216 | TMC1 | | Downregulated |
| -1,489524869 | TMC3 | | Downregulated |
| -0,261905248 | TMEM213 | FALSE | Downregulated |
| 0,148144892 | TMEM221 | FALSE | Upregulated |
| 0,380660818 | TMEM59L | FALSE | Upregulated |
| 0,369372204 | TMEM72 | FALSE | Upregulated |
| 0 | TMPRSS7 | FALSE | Upregulated |
| -0,084419156 | TMSB15B | FALSE | Downregulated |
| 2,730671375 | TNFSF18 | | Upregulated |
| 1,664006626 | TNFSF9 | | Upregulated |
| 1,669576687 | TNS4 | | Upregulated |
| 0,095363142 | TPH1 | FALSE | Upregulated |
| -1,097534374 | TREML1 | | Downregulated |
| -0,81330929 | TRIM17 | FALSE | Downregulated |
| -0,091261109 | TRIM40 | FALSE | Downregulated |

| | | | |
|---|---|---|---|
| -0,280791881 | TRIM50 | FALSE | Downregulated |
| -3,100230926 | TRNAD14 | | Downregulated |
| -0,959538661 | TRNAD15 | FALSE | Downregulated |
| -0,279585374 | TRNAG15 | FALSE | Downregulated |
| -0,090445922 | TRNAG21 | FALSE | Downregulated |
| -0,088033562 | TRNAG23 | FALSE | Downregulated |
| -0,715286499 | TRNAG27 | FALSE | Downregulated |
| -0,512900185 | TRNAH1 | FALSE | Downregulated |
| 0,099240906 | TRNAH10 | FALSE | Upregulated |
| -0,682212456 | TRNAH2 | FALSE | Downregulated |
| -0,848315115 | TRNAH3 | FALSE | Downregulated |
| -0,111961767 | TRNAH4 | FALSE | Downregulated |
| -0,273865599 | TRNAH5 | FALSE | Downregulated |
| 1,503015245 | TRNAH6 | | Upregulated |
| -0,320624416 | TRNAH7 | FALSE | Downregulated |
| -0,256921569 | TRNAH9 | FALSE | Downregulated |
| -1,524635417 | TRNAK1 | | Downregulated |
| 0,074149489 | TRNAK14 | FALSE | Upregulated |
| -0,039186191 | TRNAK2 | FALSE | Downregulated |
| -1,459243061 | TRNAK20 | | Downregulated |
| -0,912315506 | TRNAK22 | FALSE | Downregulated |
| 0,009724644 | TRNAK26 | FALSE | Upregulated |
| -1,811887974 | TRNAK3 | | Downregulated |
| -0,612021328 | TRNAK33 | FALSE | Downregulated |
| -1,110190195 | TRNAK34 | | Downregulated |
| 0 | TRNAV-CAC | FALSE | Upregulated |
| -0,508258414 | TRNAV11 | FALSE | Downregulated |
| 0 | TRNAV2 | FALSE | Upregulated |
| -0,942444084 | TRNAV21 | FALSE | Downregulated |
| 0 | TRNAV22 | FALSE | Upregulated |
| 0 | TRNAV25 | FALSE | Upregulated |
| 0 | TRNAV29 | FALSE | Upregulated |
| -0,798192588 | TRNAV4 | FALSE | Downregulated |
| 0 | TRNAV6 | FALSE | Upregulated |
| 0 | TRNAV9 | FALSE | Upregulated |
| 1,20560458 | TRPA1 | | Upregulated |
| 0,810782196 | TRPC2 | FALSE | Upregulated |
| 0,396405815 | TRPC5OS | FALSE | Upregulated |
| 0,137261384 | TSPO2 | FALSE | Upregulated |
| -1,643950812 | TSSK4 | | Downregulated |
| 0,344949256 | TTLL2 | FALSE | Upregulated |
| 0,885009411 | TTLL9 | FALSE | Upregulated |
| 1,263491304 | TTTY10 | | Upregulated |
| -0,420156803 | TUBA3E | FALSE | Downregulated |
| -0,13395686 | TUBB8 | FALSE | Downregulated |
| -1,388522831 | TULP2 | | Downregulated |
| 1,107711333 | TXNDC2 | | Upregulated |
| -0,427844917 | UNQ9370 | FALSE | Downregulated |
| -0,08154522 | UPK1A | FALSE | Downregulated |
| 0,133419721 | UPK1B | FALSE | Upregulated |
| -0,382450915 | VAX2 | FALSE | Downregulated |

| | | | |
|---|---|---|---|
| 0,610873084 | VENTXP7 | FALSE | Upregulated |
| 0,219303272 | VGLL1 | FALSE | Upregulated |
| -0,795505783 | VIT | FALSE | Downregulated |
| 2,44404878 | VSTM1 | | Upregulated |
| 0,285472498 | VWA3A | FALSE | Upregulated |
| 0,597328575 | VWA5B2 | FALSE | Upregulated |
| 0,903701337 | WDR16 | FALSE | Upregulated |
| -0,977263961 | WDR88 | FALSE | Downregulated |
| 0,607503228 | WEE2 | FALSE | Upregulated |
| -1,359516927 | WFDC3 | | Downregulated |
| -0,317684847 | WFIKKN2 | FALSE | Downregulated |
| -0,739491016 | WNT7A | FALSE | Downregulated |
| -1,09183314 | WNT9A | | Downregulated |
| 0,547042939 | WNT9B | FALSE | Upregulated |
| 0,868076553 | WSCD2 | FALSE | Upregulated |
| -1,973414506 | WT1-AS | | Downregulated |
| 0,189231955 | XKRX | FALSE | Upregulated |
| -1,245616443 | XRCC6P5 | | Downregulated |
| -1,659186387 | ZBTB20-AS1 | | Downregulated |
| 2,027826264 | ZBTB32 | | Upregulated |
| 0,125069504 | ZBTB8B | FALSE | Upregulated |
| 1,499806788 | ZCCHC12 | | Upregulated |
| -2,284080249 | ZIM2 | | Downregulated |
| -1,568474217 | ZNF32-AS1 | | Downregulated |
| 0,306902931 | ZNF32-AS2 | FALSE | Upregulated |
| 0,579760326 | ZNF365 | FALSE | Upregulated |
| -1,497801893 | ZNF474 | | Downregulated |
| -1,704651939 | ZNF488 | | Downregulated |
| 0,064331904 | ZNF664-FAM101A | FALSE | Upregulated |
| 0,102772543 | ZNF705E | FALSE | Upregulated |
| -1,2900165 | ZNF716 | | Downregulated |
| -0,169355496 | ZNF730 | FALSE | Downregulated |
| -0,296924617 | ZNF732 | FALSE | Downregulated |
| -0,769520142 | ZPBP | FALSE | Downregulated |

<u>subTCMR vs. NHR</u>

| log2FoldChange | padj | gene_names | significance | direction |
|---|---|---|---|---|
| 1,902852785 | 0,00053959 | UBD | TRUE | Upregulated |
| 2,76054727 | 0,006677259 | ADAMDEC1 | TRUE | Upregulated |
| 1,85718399 | 0,006677259 | CCR7 | TRUE | Upregulated |
| 1,956210746 | 0,006677259 | CDC20 | TRUE | Upregulated |
| 4,844399926 | 0,006677259 | CHIT1 | TRUE | Upregulated |
| 2,457120813 | 0,006677259 | FAM154B | TRUE | Upregulated |

| | | | | |
|---|---|---|---|---|
| 0,386269997 | 0,006677259 | INTS1 | FALSE | Upregulated |
| 3,902705097 | 0,006677259 | KRTAP16-1 | TRUE | Upregulated |
| 1,692013155 | 0,006677259 | LGALS2 | TRUE | Upregulated |
| 1,909801456 | 0,006677259 | LOC102723407 | TRUE | Upregulated |
| 1,330445974 | 0,006677259 | NAIP | TRUE | Upregulated |
| 0,669788019 | 0,006677259 | POLD1 | FALSE | Upregulated |
| 3,568917671 | 0,006677259 | STMN2 | TRUE | Upregulated |
| 1,408758003 | 0,007007649 | A1BG-AS1 | TRUE | Upregulated |
| 0,341606181 | 0,007007649 | CHERP | FALSE | Upregulated |
| -0,309368239 | 0,007007649 | FRA10AC1 | FALSE | Downregulated |
| -0,232843019 | 0,008396404 | ZNF644 | FALSE | Downregulated |
| -0,352213872 | 0,010590082 | NACA | FALSE | Downregulated |
| -1,573922806 | 0,013342768 | HAP1 | TRUE | Downregulated |
| -0,370378518 | 0,016771249 | LIPT1 | FALSE | Downregulated |
| 0,654654113 | 0,017901051 | CHST2 | FALSE | Upregulated |
| 0,299795672 | 0,021741652 | CLCN7 | FALSE | Upregulated |
| -0,277816897 | 0,022674717 | BMPR2 | FALSE | Downregulated |
| 1,173884938 | 0,022674717 | LRCOL1 | TRUE | Upregulated |
| 2,319602266 | 0,022674717 | RN7SL2 | TRUE | Upregulated |
| 0,346576564 | 0,022674717 | SEPT9 | FALSE | Upregulated |
| 1,539129611 | 0,022862833 | CCL19 | TRUE | Upregulated |
| 0,42860142 | 0,022862833 | GPI | FALSE | Upregulated |
| 1,700738544 | 0,022862833 | UBE2C | TRUE | Upregulated |
| 0,282308094 | 0,024011758 | FLII | FALSE | Upregulated |
| 1,803114211 | 0,024011758 | LOC102723722 | TRUE | Upregulated |
| -0,295534739 | 0,024011758 | MTIF3 | FALSE | Downregulated |

| | | | | |
|---|---|---|---|---|
| 1,242940887 | 0,024011758 | RGCC | TRUE | Upregulated |
| -2,554258268 | 0,024011758 | RP11-10L7.1 | TRUE | Downregulated |
| -2,63362431 | 0,024011758 | THBS4 | TRUE | Downregulated |
| 2,337244638 | 0,024537914 | MUC13 | TRUE | Upregulated |
| 0,710362309 | 0,024621861 | ZNF628 | FALSE | Upregulated |
| -0,389040064 | 0,025032271 | AC007246.3 | FALSE | Downregulated |
| 7,16892858 | 0,025032271 | GABRA5 | TRUE | Upregulated |
| -0,334706465 | 0,025032271 | KRCC1 | FALSE | Downregulated |
| -0,164517121 | 0,025032271 | MATR3 | FALSE | Downregulated |
| 1,591700986 | 0,026968463 | APOBEC3B | TRUE | Upregulated |
| -2,459763756 | 0,026968463 | MYL4 | TRUE | Downregulated |
| 0,905992488 | 0,030206888 | KCNAB2 | FALSE | Upregulated |
| 1,849215398 | 0,033048378 | MMP7 | TRUE | Upregulated |
| -0,295621479 | 0,033488059 | NFU1 | FALSE | Downregulated |
| 0,232983563 | 0,033495218 | AP1B1 | FALSE | Upregulated |
| 0,517332024 | 0,033495218 | BYSL | FALSE | Upregulated |
| 0,225387427 | 0,033495218 | CSNK1D | FALSE | Upregulated |
| -0,314601702 | 0,033495218 | IFT88 | FALSE | Downregulated |
| -0,327529657 | 0,033495218 | LOC100507217 | FALSE | Downregulated |
| 2,682631575 | 0,033495218 | MTRNR2L1 | TRUE | Upregulated |
| 0,525507675 | 0,033495218 | RBM38 | FALSE | Upregulated |
| 2,173411106 | 0,033495218 | RP11-14D22.5 | TRUE | Upregulated |
| -2,387598854 | 0,033495218 | RP11-436I24.1 | TRUE | Downregulated |
| 1,616631539 | 0,033495218 | SPATA21 | TRUE | Upregulated |
| 0,344012152 | 0,033532065 | MED16 | FALSE | Upregulated |
| -1,191765775 | 0,033532065 | S100A8 | TRUE | Downregulated |

| | | | | |
|---|---|---|---|---|
| -0,935288953 | 0,033532065 | SNORA8 | FALSE | Downregulated |
| 0,258116402 | 0,035274191 | ARHGEF18 | FALSE | Upregulated |
| -0,657186583 | 0,035301515 | TCP10L | FALSE | Downregulated |
| 1,189632654 | 0,036726146 | HLA-G | TRUE | Upregulated |
| 0,301697561 | 0,037421255 | ATG7 | FALSE | Upregulated |
| -0,401899556 | 0,037421255 | THUMPD2 | FALSE | Downregulated |
| -0,500171963 | 0,037421255 | ZSCAN16-AS1 | FALSE | Downregulated |
| 1,720471388 | 0,03809132 | LOC101926970 | TRUE | Upregulated |
| 0,253325701 | 0,03809132 | STARD3 | FALSE | Upregulated |
| -0,340325551 | 0,03809132 | WAC-AS1 | FALSE | Downregulated |
| 0,753124752 | 0,039686546 | CD247 | FALSE | Upregulated |
| 3,268662413 | 0,040427105 | CAPN13 | TRUE | Upregulated |
| -0,233974655 | 0,040427105 | DYNLT3 | FALSE | Downregulated |
| -0,222160368 | 0,040427105 | FAM204A | FALSE | Downregulated |
| -0,197037631 | 0,040446157 | SNW1 | FALSE | Downregulated |
| 1,222895821 | 0,041169679 | COL5A3 | TRUE | Upregulated |
| -0,878688032 | 0,041169679 | PRUNE2 | FALSE | Downregulated |
| 2,092195838 | 0,041169679 | TNFRSF17 | TRUE | Upregulated |
| 0,851546906 | 0,041169679 | TP53I3 | FALSE | Upregulated |
| 1,035131022 | 0,041702487 | RRM2 | TRUE | Upregulated |
| 0,37160561 | 0,041702487 | ZNF200 | FALSE | Upregulated |
| -0,274017154 | 0,042192654 | ARV1 | FALSE | Downregulated |
| -0,255863744 | 0,042192654 | CDC26 | FALSE | Downregulated |
| -0,20720189 | 0,042192654 | CHMP5 | FALSE | Downregulated |
| -0,426413799 | 0,042192654 | F7 | FALSE | Downregulated |
| 1,35352407 | 0,042192654 | FAM111B | TRUE | Upregulated |

| | | | | |
|---|---|---|---|---|
| 0,293194421 | 0,042192654 | HGS | FALSE | Upregulated |
| -0,339077826 | 0,042192654 | METTL18 | FALSE | Downregulated |
| 1,509882772 | 0,042192654 | PLA2G2D | TRUE | Upregulated |
| 2,492869332 | 0,042192654 | SCN5A | TRUE | Upregulated |
| -0,309546259 | 0,043956545 | HNRNPH3 | FALSE | Downregulated |
| -0,913443975 | 0,04542291 | LOC102546229 | FALSE | Downregulated |
| -1,148559263 | 0,04542291 | MTUS2 | TRUE | Downregulated |
| 1,053691146 | 0,04542291 | NCAPG | TRUE | Upregulated |
| 0,335420384 | 0,04542291 | RNPEPL1 | FALSE | Upregulated |
| 0,534477091 | 0,04542291 | ZSWIM3 | FALSE | Upregulated |
| -0,21365916 | 0,04680692 | PTP4A2 | FALSE | Downregulated |
| -0,30979416 | 0,04680692 | TMEM41B | FALSE | Downregulated |
| 1,272505683 | 0,049571567 | ZNF578 | TRUE | Upregulated |
| 1,558232363 | 0,049588708 | E2F1 | TRUE | Upregulated |
| 2,8533973 | 0,049678543 | CACNA1I | TRUE | Upregulated |
| -0,275106455 | 0,049678543 | CEBPZ-AS1 | FALSE | Downregulated |
| -0,390663495 | 0,049678543 | COX7C | FALSE | Downregulated |
| -1,968483417 | 0,049678543 | MEIOB | TRUE | Downregulated |
| 0,321640417 | 0,049678543 | MYO1C | FALSE | Upregulated |
| 0,404433199 | 0,049678543 | PLOD3 | FALSE | Upregulated |
| -0,487966124 | 0,049678543 | SEC61A2 | FALSE | Downregulated |
| 0,218660054 | 0,051196409 | BAG6 | FALSE | Upregulated |
| -0,204861006 | 0,051196409 | PPIL4 | FALSE | Downregulated |
| 3,493822883 | 0,051196409 | TFAMP1 | FALSE | Upregulated |
| 0,373303396 | 0,051243059 | HPS1 | FALSE | Upregulated |
| 0,68253553 | 0,051243059 | LIMD2 | FALSE | Upregulated |

| | | | | |
|---|---|---|---|---|
| 0,30142382 | 0,051243059 | PCBP1 | FALSE | Upregulated |
| -0,198025194 | 0,051243059 | RABEP1 | FALSE | Downregulated |
| -0,323973128 | 0,051243059 | RPL22 | FALSE | Downregulated |
| 0,885139461 | 0,051553912 | SPOCK2 | FALSE | Upregulated |
| -0,163894677 | 0,051734639 | PAPOLA | FALSE | Downregulated |
| 1,219977728 | 0,051873461 | SOX9-AS1 | FALSE | Upregulated |
| 0,286979405 | 0,054104282 | ANAPC2 | FALSE | Upregulated |
| -1,225444313 | 0,054104282 | MYH3 | FALSE | Downregulated |
| 2,130956467 | 0,054104282 | RN7SL1 | FALSE | Upregulated |
| -1,935235416 | 0,054104282 | SFRP4 | FALSE | Downregulated |
| 0,281120526 | 0,05433884 | PFKL | FALSE | Upregulated |
| 0,303237667 | 0,054580646 | FBXO42 | FALSE | Upregulated |
| -0,431540614 | 0,05509659 | LINC00467 | FALSE | Downregulated |
| 0,546090157 | 0,056888742 | SLC36A4 | FALSE | Upregulated |
| -0,586002492 | 0,058743678 | RPS29 | FALSE | Downregulated |
| -0,403216934 | 0,060587339 | PLSCR4 | FALSE | Downregulated |
| 0,888487245 | 0,06242955 | CD7 | FALSE | Upregulated |
| 1,224665277 | 0,06242955 | CXCL10 | FALSE | Upregulated |
| -0,866067136 | 0,06242955 | DNM1 | FALSE | Downregulated |
| 0,263989563 | 0,06242955 | EMC1 | FALSE | Upregulated |
| -1,019398234 | 0,06242955 | HPGDS | FALSE | Downregulated |
| -0,33584409 | 0,06242955 | LIX1L | FALSE | Downregulated |
| 0,798922149 | 0,06242955 | LSP1 | FALSE | Upregulated |
| -0,436064875 | 0,06242955 | MIR4458HG | FALSE | Downregulated |
| -0,245540832 | 0,06242955 | SPAG16 | FALSE | Downregulated |
| 1,812275756 | 0,06242955 | VSIG2 | FALSE | Upregulated |

| | | | | |
|---|---|---|---|---|
| 0,77862628 | 0,062563181 | ANKRD19P | FALSE | Upregulated |
| 1,733236762 | 0,062563181 | MIR155HG | FALSE | Upregulated |
| 0,933079656 | 0,062563181 | TYMS | FALSE | Upregulated |
| 2,370211789 | 0,0631509 | CCNI2 | FALSE | Upregulated |
| -0,429201263 | 0,0631509 | HCFC1R1 | FALSE | Downregulated |
| 0,609606473 | 0,0631509 | ITGB2 | FALSE | Upregulated |
| 1,36441346 | 0,0631509 | MYBL2 | FALSE | Upregulated |
| 0,666630033 | 0,0631509 | PRDM1 | FALSE | Upregulated |
| 3,456608884 | 0,0631509 | RP11-394O9.1 | FALSE | Upregulated |
| -0,247839641 | 0,0631509 | SEPT10 | FALSE | Downregulated |
| 1,007514548 | 0,0631509 | TFRC | FALSE | Upregulated |
| 0,462533193 | 0,0631509 | ZNF319 | FALSE | Upregulated |
| -0,26318625 | 0,063252934 | SSB | FALSE | Downregulated |
| 1,366235457 | 0,065004543 | CDC45 | FALSE | Upregulated |
| -0,254130007 | 0,065004543 | CHRAC1 | FALSE | Downregulated |
| -0,30657034 | 0,065004543 | COA6 | FALSE | Downregulated |
| 0,263906074 | 0,065004543 | FBXW5 | FALSE | Upregulated |
| 0,242903367 | 0,065004543 | JMJD8 | FALSE | Upregulated |
| -1,068642915 | 0,065004543 | LOC100130992 | FALSE | Downregulated |
| 1,123041432 | 0,065015214 | IGJ | FALSE | Upregulated |
| -1,793915429 | 0,065015214 | PADI2 | FALSE | Downregulated |
| 0,26209143 | 0,065015214 | UNC13B | FALSE | Upregulated |
| 0,267102392 | 0,066198609 | GPS1 | FALSE | Upregulated |
| 0,182603414 | 0,067313093 | PHRF1 | FALSE | Upregulated |
| 0,21411159 | 0,068716714 | DNAJC5 | FALSE | Upregulated |
| -0,174845639 | 0,069113226 | FOPNL | FALSE | Downregulated |

| | | | | |
|---|---|---|---|---|
| 1,137429125 | 0,069162951 | LEF1 | FALSE | Upregulated |
| 1,935751881 | 0,069162951 | MMP9 | FALSE | Upregulated |
| 0,323428215 | 0,069599947 | ENTPD6 | FALSE | Upregulated |
| 2,124961525 | 0,069621045 | TTLL6 | FALSE | Upregulated |
| -0,876340656 | 0,071487917 | LOC102724870 | FALSE | Downregulated |
| -1,536168346 | 0,071487917 | MEDAG | FALSE | Downregulated |
| 0,350558307 | 0,071487917 | SIPA1 | FALSE | Upregulated |
| -0,219964061 | 0,071487917 | UQCRB | FALSE | Downregulated |
| -0,190954098 | 0,071627471 | ARPP19 | FALSE | Downregulated |
| 0,209130716 | 0,071627471 | TINF2 | FALSE | Upregulated |
| 0,373446991 | 0,072246856 | RHBDD3 | FALSE | Upregulated |
| -0,334578655 | 0,079557834 | MRPS18C | FALSE | Downregulated |
| 1,723666225 | 0,079728217 | GTSF1 | FALSE | Upregulated |
| -0,565544875 | 0,08109258 | SMIM6 | FALSE | Downregulated |
| -0,286178886 | 0,081097346 | NUCKS1 | FALSE | Downregulated |
| -0,310912109 | 0,081278104 | PEX7 | FALSE | Downregulated |
| 0,41977229 | 0,081903567 | ALYREF | FALSE | Upregulated |
| 2,004288728 | 0,081903567 | CD207 | FALSE | Upregulated |
| 2,892002577 | 0,082066 | MIR4426 | FALSE | Upregulated |
| 0,728815836 | 0,082738567 | CD28 | FALSE | Upregulated |
| 0,255817332 | 0,08291283 | CC2D1A | FALSE | Upregulated |
| 1,160375512 | 0,083941861 | CD79A | FALSE | Upregulated |
| 0,39147081 | 0,083941861 | GAS2L1 | FALSE | Upregulated |
| 0,386623998 | 0,083941861 | GPX1 | FALSE | Upregulated |
| -0,217417327 | 0,083941861 | SRP14 | FALSE | Downregulated |
| 2,961187553 | 0,084750539 | KCNQ3 | FALSE | Upregulated |

| | | | | |
|---|---|---|---|---|
| 1,107147863 | 0,084934498 | LDLRAD2 | FALSE | Upregulated |
| -0,20713192 | 0,085561303 | BTBD7 | FALSE | Downregulated |
| -0,187032043 | 0,085561303 | BUD13 | FALSE | Downregulated |
| 0,380182368 | 0,085561303 | SARS2 | FALSE | Upregulated |
| 0,365635622 | 0,0862565 | ATG2A | FALSE | Upregulated |
| 0,329191684 | 0,087025858 | FAM203A | FALSE | Upregulated |
| 1,175670607 | 0,087105409 | CXCR3 | FALSE | Upregulated |
| -2,65965944 | 0,08842361 | CA3 | FALSE | Downregulated |
| 1,198091746 | 0,089261238 | CENPK | FALSE | Upregulated |
| 0,311085053 | 0,089452673 | CDIP1 | FALSE | Upregulated |
| 0,377175091 | 0,089452673 | CTSD | FALSE | Upregulated |
| -0,177136484 | 0,089452673 | FXR1 | FALSE | Downregulated |
| 2,0622572 | 0,089761427 | EVPL | FALSE | Upregulated |
| 0,275004079 | 0,09027332 | RPS6KA4 | FALSE | Upregulated |
| -0,224027429 | 0,090501595 | DNAJC7 | FALSE | Downregulated |
| 0,283234073 | 0,090640435 | PARS2 | FALSE | Upregulated |
| 0,485870879 | 0,090769222 | CYTH4 | FALSE | Upregulated |
| 2,760472212 | 0,090860295 | SCRT2 | FALSE | Upregulated |
| -0,34674063 | 0,09105094 | ATF7IP2 | FALSE | Downregulated |
| -0,211310206 | 0,09105094 | GPN1 | FALSE | Downregulated |
| 0,745466734 | 0,09105094 | HLA-DQA1 | FALSE | Upregulated |
| 2,366751984 | 0,09105094 | LOC102724895 | FALSE | Upregulated |
| 0,438306212 | 0,09105094 | MANEAL | FALSE | Upregulated |
| 0,226646614 | 0,091572706 | USP5 | FALSE | Upregulated |
| -0,528614839 | 0,093956362 | SPG20 | FALSE | Downregulated |
| 1,445535147 | 0,094591343 | SCN7A | FALSE | Upregulated |

| | | | | |
|---|---|---|---|---|
| -0,433976849 | 0,095052073 | CCDC23 | FALSE | Downregulated |
| 0,242619038 | 0,095075614 | ACO2 | FALSE | Upregulated |
| 0,551748195 | 0,095075614 | ARHGAP27 | FALSE | Upregulated |
| 0,33601466 | 0,095075614 | PYGB | FALSE | Upregulated |
| -0,172439996 | 0,095769238 | WAC | FALSE | Downregulated |
| 0,226894292 | 0,096062568 | VARS | FALSE | Upregulated |
| 1,943028184 | 0,097552454 | CDKN2B-AS1 | FALSE | Upregulated |
| 0,184779656 | 0,097552454 | WDTC1 | FALSE | Upregulated |
| 0,234783292 | 0,097892613 | AP1M1 | FALSE | Upregulated |
| 0,389157646 | 0,097892613 | CNN2 | FALSE | Upregulated |
| 0,337594311 | 0,097892613 | DGKZ | FALSE | Upregulated |
| 1,2263874 | 0,097892613 | LCN2 | FALSE | Upregulated |
| 0,516835421 | 0,097892613 | LTBP3 | FALSE | Upregulated |
| 1,050634861 | 0,098399064 | CXCL9 | FALSE | Upregulated |
| 0,1844732 | 0,098399064 | DNM2 | FALSE | Upregulated |
| -0,37406414 | 0,098399064 | FLT4 | FALSE | Downregulated |
| -0,738384934 | 0,098399064 | HM13-AS1 | FALSE | Downregulated |
| 1,302192609 | 0,098399064 | MEX3B | FALSE | Upregulated |
| -0,342308197 | 0,098399064 | NSA2 | FALSE | Downregulated |
| 0,369974611 | 0,098399064 | USP11 | FALSE | Upregulated |
| -0,251369765 | 0,098399064 | ZNF441 | FALSE | Downregulated |
| 0,235589777 | 0,099699973 | TBCD | FALSE | Upregulated |
| 0,815117184 | 0,100689082 | RP11-696N14.1 | FALSE | Upregulated |
| -0,195881718 | 0,100689082 | TMEM248 | FALSE | Downregulated |
| 1,457924497 | 0,100809597 | FAM19A5 | FALSE | Upregulated |
| 1,77372497 | 0,100809597 | LOC101927481 | FALSE | Upregulated |

| | | | | |
|---|---|---|---|---|
| -0,451056171 | 0,100809597 | PFDN1 | FALSE | Downregulated |
| 0,336633316 | 0,100809597 | PHF5A | FALSE | Upregulated |
| -0,208875518 | 0,100809597 | PPWD1 | FALSE | Downregulated |
| -0,250579478 | 0,100809597 | SGMS1 | FALSE | Downregulated |
| -0,221148454 | 0,100809597 | THOC1 | FALSE | Downregulated |
| -0,285212075 | 0,101563903 | FBXL3 | FALSE | Downregulated |
| 0,285630731 | 0,101817839 | ATP6V0D1 | FALSE | Upregulated |
| 1,869551879 | 0,103687537 | CRMP1 | FALSE | Upregulated |
| 0,49523109 | 0,103687537 | G6PD | FALSE | Upregulated |
| -1,498132277 | 0,103687537 | LOC102723324 | FALSE | Downregulated |
| 0,826768133 | 0,103687537 | SEMA4A | FALSE | Upregulated |
| 0,459311987 | 0,103687537 | TCIRG1 | FALSE | Upregulated |
| -0,218532522 | 0,103687537 | ZNF480 | FALSE | Downregulated |
| 1,882340147 | 0,104192436 | SLC12A5 | FALSE | Upregulated |
| 0,634523942 | 0,105631745 | ATP2A3 | FALSE | Upregulated |
| 1,043980222 | 0,105631745 | GRIN2D | FALSE | Upregulated |
| -0,265865974 | 0,105631745 | HOMEZ | FALSE | Downregulated |
| -0,21439043 | 0,105631745 | WDR89 | FALSE | Downregulated |
| 0,267646451 | 0,105879887 | ACSF3 | FALSE | Upregulated |
| 0,175747051 | 0,105943924 | GAK | FALSE | Upregulated |
| 0,705069606 | 0,105943924 | MICAL1 | FALSE | Upregulated |
| -0,20864283 | 0,105943924 | UBE2D3 | FALSE | Downregulated |
| -1,101977003 | 0,106035957 | C21orf37 | FALSE | Downregulated |
| -2,344699979 | 0,106035957 | CDH12 | FALSE | Downregulated |
| 0,147736542 | 0,106035957 | GRSF1 | FALSE | Upregulated |
| 0,310493137 | 0,106035957 | PNPLA6 | FALSE | Upregulated |

| | | | | |
|---|---|---|---|---|
| -0,368346835 | 0,106035957 | TP53INP1 | FALSE | Downregulated |
| -1,294812557 | 0,106035957 | TRHDE-AS1 | FALSE | Downregulated |
| -0,2632144 | 0,106803541 | LSM6 | FALSE | Downregulated |
| 0,232168295 | 0,107052366 | LAMP1 | FALSE | Upregulated |
| 0,472425783 | 0,10719021 | SLC27A4 | FALSE | Upregulated |
| -1,564328091 | 0,107745181 | STPG2 | FALSE | Downregulated |
| -0,470671096 | 0,108523941 | TMEM160 | FALSE | Downregulated |
| -0,272483468 | 0,10967253 | CCDC59 | FALSE | Downregulated |
| 0,369342855 | 0,10967253 | PRIM2 | FALSE | Upregulated |
| -0,333552441 | 0,109725926 | NDUFB1 | FALSE | Downregulated |
| 0,325422278 | 0,110902977 | HIP1R | FALSE | Upregulated |
| -0,211837819 | 0,111158443 | FUNDC1 | FALSE | Downregulated |
| 1,249618887 | 0,111158443 | MELK | FALSE | Upregulated |
| 0,712819112 | 0,11224381 | ITLN1 | FALSE | Upregulated |
| -0,448666216 | 0,11224381 | PLGLB2 | FALSE | Downregulated |
| -0,667424534 | 0,11224381 | TMTC1 | FALSE | Downregulated |
| 1,865425332 | 0,112319831 | RP11-27N21.3 | FALSE | Upregulated |
| -0,38715272 | 0,112601587 | DDX60L | FALSE | Downregulated |
| 0,154750248 | 0,113049099 | DHX30 | FALSE | Upregulated |
| -0,349752292 | 0,114237921 | AKAP2 | FALSE | Downregulated |
| -0,288728691 | 0,114237921 | COQ3 | FALSE | Downregulated |
| -0,206030875 | 0,114237921 | EXOSC10 | FALSE | Downregulated |
| 0,927430765 | 0,114237921 | FAM99B | FALSE | Upregulated |
| 0,240813563 | 0,114237921 | GLTPD1 | FALSE | Upregulated |
| 0,678747981 | 0,114237921 | GPX7 | FALSE | Upregulated |
| 0,784626149 | 0,114237921 | HLA-H | FALSE | Upregulated |

| | | | | |
|---|---|---|---|---|
| 1,264918781 | 0,114237921 | IL2RA | FALSE | Upregulated |
| 0,715605306 | 0,114237921 | ITGAX | FALSE | Upregulated |
| -0,257144448 | 0,114237921 | KIAA1586 | FALSE | Downregulated |
| -1,115953342 | 0,114237921 | LINC00086 | FALSE | Downregulated |
| 1,292418062 | 0,114237921 | MOXD1 | FALSE | Upregulated |
| -0,203075126 | 0,114237921 | MRPS14 | FALSE | Downregulated |
| 0,241014318 | 0,114237921 | NDUFS7 | FALSE | Upregulated |
| -0,233432462 | 0,114237921 | PARD3 | FALSE | Downregulated |
| -0,171370368 | 0,114237921 | PPP2R5C | FALSE | Downregulated |
| -0,231966795 | 0,114237921 | TERF1 | FALSE | Downregulated |
| 0,391186889 | 0,114597336 | ALG1 | FALSE | Upregulated |
| 1,031679926 | 0,114597336 | BLOC1S4 | FALSE | Upregulated |
| -0,507732433 | 0,114597336 | LINC00478 | FALSE | Downregulated |
| -0,384020396 | 0,114597336 | NOSTRIN | FALSE | Downregulated |
| 1,504520689 | 0,114597336 | PBK | FALSE | Upregulated |
| -0,462473298 | 0,114597336 | POLR3GL | FALSE | Downregulated |
| 0,313530505 | 0,115196863 | ABCB8 | FALSE | Upregulated |
| 0,578485125 | 0,115196863 | ITGAM | FALSE | Upregulated |
| -0,22726369 | 0,115196863 | KRR1 | FALSE | Downregulated |
| 2,216452789 | 0,115196863 | LINC00506 | FALSE | Upregulated |
| 1,605035068 | 0,115196863 | LOC100272216 | FALSE | Upregulated |
| -1,164316922 | 0,115196863 | MGAM | FALSE | Downregulated |
| -0,256767597 | 0,115196863 | ZMYM6NB | FALSE | Downregulated |
| -1,081335828 | 0,11600267 | CALR3 | FALSE | Downregulated |
| 0,984664904 | 0,11600267 | MALAT1 | FALSE | Upregulated |
| -0,290875587 | 0,11600267 | SERF2-C15ORF63 | FALSE | Downregulated |

| | | | | |
|---|---|---|---|---|
| 0,233188866 | 0,118484232 | IGSF8 | FALSE | Upregulated |
| 0,362258022 | 0,119031584 | SCRN2 | FALSE | Upregulated |
| 0,215385716 | 0,119192034 | ADRBK1 | FALSE | Upregulated |
| 0,960987315 | 0,119192034 | GPR171 | FALSE | Upregulated |
| 0,543661391 | 0,119192034 | NUDT18 | FALSE | Upregulated |
| -0,244783983 | 0,119192034 | SEPT7 | FALSE | Downregulated |
| -0,233416555 | 0,119433081 | AMZ2 | FALSE | Downregulated |
| 0,153325777 | 0,119433081 | EFTUD2 | FALSE | Upregulated |
| 0,925106372 | 0,119433081 | FCGBP | FALSE | Upregulated |
| 0,217069183 | 0,119433081 | RPTOR | FALSE | Upregulated |
| 0,7958498 | 0,119724165 | CD22 | FALSE | Upregulated |
| -0,578310446 | 0,119724165 | FAM174B | FALSE | Downregulated |
| -2,323989208 | 0,119724165 | HSPB7 | FALSE | Downregulated |
| -0,270465721 | 0,119724165 | PIN4 | FALSE | Downregulated |
| -1,066797236 | 0,119724165 | PRDM5 | FALSE | Downregulated |
| 0,860766513 | 0,119816179 | IL21R | FALSE | Upregulated |
| -1,561264325 | 0,120000017 | CACNA2D3 | FALSE | Downregulated |
| 0,453839854 | 0,120000017 | GMIP | FALSE | Upregulated |
| -0,787363689 | 0,120000017 | IRAK1BP1 | FALSE | Downregulated |
| 2,443276795 | 0,120000017 | NPAS1 | FALSE | Upregulated |
| 0,236025074 | 0,120000017 | RPL36A-HNRNPH2 | FALSE | Upregulated |
| -0,222254711 | 0,123342565 | NDUFAF7 | FALSE | Downregulated |
| 0,40194052 | 0,123468459 | ZNF879 | FALSE | Upregulated |
| 0,180333311 | 0,12417101 | GRB2 | FALSE | Upregulated |
| 0,715179652 | 0,125007192 | IKBKE | FALSE | Upregulated |
| 0,348093542 | 0,125229858 | VPS37C | FALSE | Upregulated |

| | | | | |
|---|---|---|---|---|
| 0,649308534 | 0,125481744 | ADAP1 | FALSE | Upregulated |
| -0,497756318 | 0,125592733 | ZCWPW2 | FALSE | Downregulated |
| 0,246914827 | 0,126223739 | ERGIC1 | FALSE | Upregulated |
| -0,232943178 | 0,126223739 | ERMARD | FALSE | Downregulated |
| 0,723102689 | 0,127829434 | TIGIT | FALSE | Upregulated |
| -1,116711745 | 0,128382941 | ATP5L2 | FALSE | Downregulated |
| -3,187645279 | 0,129734676 | MYBPH | FALSE | Downregulated |
| 0,262918423 | 0,130303126 | DNPEP | FALSE | Upregulated |
| 2,236924109 | 0,130303126 | MYEOV | FALSE | Upregulated |
| 0,378535841 | 0,130303126 | ZNF408 | FALSE | Upregulated |
| -0,334923932 | 0,130895498 | MPLKIP | FALSE | Downregulated |
| 0,542707334 | 0,131512316 | PLEKHB2 | FALSE | Upregulated |
| 0,307031558 | 0,132604859 | ABCD1 | FALSE | Upregulated |
| -0,235009884 | 0,132604859 | DIMT1 | FALSE | Downregulated |
| 0,304240297 | 0,13296269 | BSG | FALSE | Upregulated |
| 0,80109091 | 0,13296269 | CLHC1 | FALSE | Upregulated |
| 1,323110443 | 0,13296269 | KIAA1644 | FALSE | Upregulated |
| -0,176552134 | 0,13296269 | NDUFA5 | FALSE | Downregulated |
| -0,295008522 | 0,13296269 | SMIM11 | FALSE | Downregulated |
| -0,20815648 | 0,13296269 | TMTC4 | FALSE | Downregulated |
| 2,030477497 | 0,133691008 | IL8 | FALSE | Upregulated |
| 0,934215005 | 0,135528322 | ATP10A | FALSE | Upregulated |
| 0,247858236 | 0,135891407 | TPRG1L | FALSE | Upregulated |
| 0,301825672 | 0,13592157 | SMPD1 | FALSE | Upregulated |
| 0,303554446 | 0,136838909 | POLR1A | FALSE | Upregulated |
| -0,231479989 | 0,13818175 | MRFAP1L1 | FALSE | Downregulated |

| | | | | |
|---|---|---|---|---|
| 0,502695724 | 0,138952417 | PIM2 | FALSE | Upregulated |
| -0,824819859 | 0,139488953 | NIPSNAP3B | FALSE | Downregulated |
| 1,487825264 | 0,139856372 | LOC642131 | FALSE | Upregulated |
| -0,379444494 | 0,14044505 | TBCA | FALSE | Downregulated |
| 0,896852512 | 0,141499051 | IL32 | FALSE | Upregulated |
| 0,336757538 | 0,141499051 | INF2 | FALSE | Upregulated |
| -0,230376563 | 0,141499051 | THAP1 | FALSE | Downregulated |
| 0,301754703 | 0,142199251 | TNFRSF1B | FALSE | Upregulated |
| -0,971709383 | 0,142748191 | STK31 | FALSE | Downregulated |
| 0,917449481 | 0,143671492 | BUB1B | FALSE | Upregulated |
| 0,917922937 | 0,143671492 | FCN1 | FALSE | Upregulated |
| 2,143258175 | 0,143697981 | LAMP5 | FALSE | Upregulated |
| 0,493662578 | 0,143939601 | ITGA6 | FALSE | Upregulated |
| -0,70968338 | 0,143979237 | VNN3 | FALSE | Downregulated |
| -0,199226494 | 0,143999309 | USP47 | FALSE | Downregulated |
| -0,617916325 | 0,14406516 | FAM129C | FALSE | Downregulated |
| 1,65358381 | 0,145642478 | LYPD2 | FALSE | Upregulated |
| 0,768587339 | 0,145642478 | PARM1 | FALSE | Upregulated |
| 1,978688996 | 0,145642478 | RPL23AP32 | FALSE | Upregulated |
| 0,491586583 | 0,145642478 | SASH3 | FALSE | Upregulated |
| -0,29804477 | 0,145642478 | TMEM18 | FALSE | Downregulated |
| -0,270511947 | 0,145642478 | TMEM65 | FALSE | Downregulated |
| -0,380658412 | 0,145642478 | TSPAN18 | FALSE | Downregulated |
| -0,234809043 | 0,145642478 | ZCRB1 | FALSE | Downregulated |
| 1,524568912 | 0,14646442 | NXPH3 | FALSE | Upregulated |
| -0,486794615 | 0,146896202 | IGF2 | FALSE | Downregulated |

| | | | | |
|---|---|---|---|---|
| 0,251195168 | 0,149875506 | ICMT | FALSE | Upregulated |
| 0,279153721 | 0,149875506 | MYH9 | FALSE | Upregulated |
| 0,441217239 | 0,149875506 | PIGR | FALSE | Upregulated |
| -0,389937435 | 0,149875506 | RWDD1 | FALSE | Downregulated |
| 0,212820509 | 0,149875506 | URB1 | FALSE | Upregulated |
| -0,169253132 | 0,149881387 | SMEK2 | FALSE | Downregulated |
| -0,293013714 | 0,149881387 | TYW3 | FALSE | Downregulated |
| 0,690977044 | 0,149881387 | ZWINT | FALSE | Upregulated |
| 1,574521014 | 0,15058054 | CCL22 | FALSE | Upregulated |
| 0,74444836 | 0,15089151 | CD300C | FALSE | Upregulated |
| -0,49741786 | 0,15089151 | SLC45A3 | FALSE | Downregulated |
| -1,046559524 | 0,15089151 | SQLE | FALSE | Downregulated |
| 2,870221316 | 0,150986078 | DCC | FALSE | Upregulated |
| 0,255157574 | 0,150986078 | ESYT1 | FALSE | Upregulated |
| -0,7969753 | 0,150986078 | LOC102724655 | FALSE | Downregulated |
| 0,268557639 | 0,150986078 | PLCG1 | FALSE | Upregulated |
| 1,929949302 | 0,151623215 | IGFBP7-AS1 | FALSE | Upregulated |
| 0,225306842 | 0,151741522 | ARFIP1 | FALSE | Upregulated |
| -0,226860903 | 0,151741522 | CALM1 | FALSE | Downregulated |
| 0,260103732 | 0,151741522 | CCM2 | FALSE | Upregulated |
| 0,766988311 | 0,151741522 | CCNA2 | FALSE | Upregulated |
| 0,581822783 | 0,151741522 | CD96 | FALSE | Upregulated |
| -0,545223274 | 0,151741522 | CHIC1 | FALSE | Downregulated |
| 0,417299969 | 0,151741522 | CORO7 | FALSE | Upregulated |
| 2,418813029 | 0,151741522 | DNAAF1 | FALSE | Upregulated |
| 0,554950208 | 0,151741522 | ECT2 | FALSE | Upregulated |

| | | | | |
|---|---|---|---|---|
| -0,186094745 | 0,151741522 | EMC4 | FALSE | Downregulated |
| 0,914597849 | 0,151741522 | KIF20A | FALSE | Upregulated |
| 0,963540246 | 0,151741522 | LOC389641 | FALSE | Upregulated |
| 0,754346878 | 0,151741522 | MFSD7 | FALSE | Upregulated |
| -0,226848688 | 0,151741522 | MRPS9 | FALSE | Downregulated |
| -0,268536528 | 0,151741522 | NDUFS4 | FALSE | Downregulated |
| 1,005056104 | 0,151741522 | PANDAR | FALSE | Upregulated |
| 1,882586079 | 0,151741522 | PDZK1IP1 | FALSE | Upregulated |
| 0,305071707 | 0,151741522 | PKN1 | FALSE | Upregulated |
| 0,363459835 | 0,151741522 | POR | FALSE | Upregulated |
| 0,638254488 | 0,151741522 | PTPRCAP | FALSE | Upregulated |
| -0,279068149 | 0,151741522 | SGMS2 | FALSE | Downregulated |
| 0,457352519 | 0,151741522 | SMA5 | FALSE | Upregulated |
| 0,319509475 | 0,151741522 | SPTAN1 | FALSE | Upregulated |
| -0,193586773 | 0,151741522 | TMEM14C | FALSE | Downregulated |
| -0,172492063 | 0,151741522 | TPMT | FALSE | Downregulated |
| 0,148188825 | 0,151741522 | VWA9 | FALSE | Upregulated |
| 0,822265278 | 0,151741522 | ZIK1 | FALSE | Upregulated |
| -0,386497857 | 0,152303264 | ACOT13 | FALSE | Downregulated |
| 0,930824216 | 0,152303264 | CD27 | FALSE | Upregulated |
| 0,593802688 | 0,152303264 | EMB | FALSE | Upregulated |
| -0,275911398 | 0,152303264 | IFT57 | FALSE | Downregulated |
| 0,252887203 | 0,152303264 | IPO13 | FALSE | Upregulated |
| 0,908842153 | 0,152303264 | LRRC46 | FALSE | Upregulated |
| -0,299701476 | 0,152303264 | LRRFIP2 | FALSE | Downregulated |
| -0,22358466 | 0,152303264 | R3HDM2 | FALSE | Downregulated |

| | | | | |
|---|---|---|---|---|
| 0,709049217 | 0,152303264 | TOP2A | FALSE | Upregulated |
| -0,261050632 | 0,152518025 | C1orf56 | FALSE | Downregulated |
| 1,052145097 | 0,152518025 | DTL | FALSE | Upregulated |
| 0,713762961 | 0,154078335 | THEMIS | FALSE | Upregulated |
| -0,330364468 | 0,154216952 | C11orf1 | FALSE | Downregulated |
| 2,768812303 | 0,154216952 | CEACAM6 | FALSE | Upregulated |
| -0,277258757 | 0,154216952 | PTPN2 | FALSE | Downregulated |
| 0,566384242 | 0,154598001 | FMNL1 | FALSE | Upregulated |
| 0,318499229 | 0,154598001 | TECPR1 | FALSE | Upregulated |
| 0,254090762 | 0,155246454 | SMIM13 | FALSE | Upregulated |
| 0,241868973 | 0,155773631 | NACC2 | FALSE | Upregulated |
| 0,744838618 | 0,156180601 | PNMA2 | FALSE | Upregulated |
| 0,284599713 | 0,156747137 | AP5B1 | FALSE | Upregulated |
| 0,522066937 | 0,157509139 | FLNA | FALSE | Upregulated |
| 0,274023422 | 0,157509139 | FOXP4 | FALSE | Upregulated |
| -0,1561089 | 0,157509139 | SENP6 | FALSE | Downregulated |
| -0,241747511 | 0,158651137 | APOPT1 | FALSE | Downregulated |
| 0,278204141 | 0,159867436 | NANP | FALSE | Upregulated |
| 0,813096879 | 0,159916802 | THBS2 | FALSE | Upregulated |
| -1,124560055 | 0,160596086 | DLK2 | FALSE | Downregulated |
| 1,189263998 | 0,160596086 | LOC100505570 | FALSE | Upregulated |
| -0,541435853 | 0,160616077 | PCDH9 | FALSE | Downregulated |
| 0,796198883 | 0,161885061 | FOXM1 | FALSE | Upregulated |
| 0,359709767 | 0,161885061 | NCKAP5L | FALSE | Upregulated |
| -0,347549394 | 0,161885061 | PHF3 | FALSE | Downregulated |
| 2,502713417 | 0,161885061 | TCN1 | FALSE | Upregulated |

| | | | | |
|---|---|---|---|---|
| -1,206098798 | 0,161885061 | TFCP2L1 | FALSE | Downregulated |
| 0,688712088 | 0,161885061 | ZNF831 | FALSE | Upregulated |
| 3,157533894 | 0,161988577 | PAGE4 | FALSE | Upregulated |
| -0,761177499 | 0,161988577 | SOAT2 | FALSE | Downregulated |
| 0,201578697 | 0,161988577 | TMEM43 | FALSE | Upregulated |
| 0,506972053 | 0,162013943 | CEP128 | FALSE | Upregulated |
| 0,78897798 | 0,162107367 | PTPN7 | FALSE | Upregulated |
| -0,551098095 | 0,162107367 | RGS2 | FALSE | Downregulated |
| -0,307405667 | 0,162243182 | NGRN | FALSE | Downregulated |
| -1,346753463 | 0,162327572 | LOC101929796 | FALSE | Downregulated |
| 0,300726774 | 0,162327572 | THEM6 | FALSE | Upregulated |
| 1,19246187 | 0,162779076 | MCM10 | FALSE | Upregulated |
| 0,51984967 | 0,162779076 | MCM5 | FALSE | Upregulated |
| -0,25112699 | 0,162779076 | OCIAD1 | FALSE | Downregulated |
| 0,223998033 | 0,162779076 | USP19 | FALSE | Upregulated |
| 1,174995408 | 0,163076267 | DERL3 | FALSE | Upregulated |
| 0,348453169 | 0,163076267 | PQLC2 | FALSE | Upregulated |
| -0,491151561 | 0,163549063 | C1QTNF3 | FALSE | Downregulated |
| 0,238779875 | 0,16362332 | CYB5R3 | FALSE | Upregulated |
| -1,395122172 | 0,16362332 | LOC102723570 | FALSE | Downregulated |
| 1,765194867 | 0,164254883 | FAM182A | FALSE | Upregulated |
| 0,254401307 | 0,16443691 | MCOLN1 | FALSE | Upregulated |
| -0,269382447 | 0,165406821 | ALKBH8 | FALSE | Downregulated |
| 0,381689311 | 0,165406821 | DSEL | FALSE | Upregulated |
| -0,176903914 | 0,165406821 | MAP4K5 | FALSE | Downregulated |
| -0,166456886 | 0,165406821 | MBD2 | FALSE | Downregulated |

| | | | | |
|---|---|---|---|---|
| 1,271294975 | 0,165406821 | MND1 | FALSE | Upregulated |
| 1,793622902 | 0,165406821 | NRIP3 | FALSE | Upregulated |
| 1,182956822 | 0,165547861 | E2F8 | FALSE | Upregulated |
| -0,198323637 | 0,165716835 | C9orf156 | FALSE | Downregulated |
| -0,150950944 | 0,165716835 | CREBL2 | FALSE | Downregulated |
| 1,22824846 | 0,165716835 | DEPDC1 | FALSE | Upregulated |
| -0,167410029 | 0,166018392 | PEX13 | FALSE | Downregulated |
| -0,155329226 | 0,166322502 | QKI | FALSE | Downregulated |
| 2,572872458 | 0,167493279 | KRT23 | FALSE | Upregulated |
| -0,195202389 | 0,167493279 | MRPS31 | FALSE | Downregulated |
| -0,574406464 | 0,168182904 | CNGA1 | FALSE | Downregulated |
| -0,726515596 | 0,168692467 | UXT-AS1 | FALSE | Downregulated |
| 0,2211941 | 0,16936396 | APEH | FALSE | Upregulated |
| -0,200345802 | 0,169544469 | DNAJC8 | FALSE | Downregulated |
| -0,63404908 | 0,169647347 | IGF2BP3 | FALSE | Downregulated |
| -0,857596122 | 0,169647347 | LINC00885 | FALSE | Downregulated |
| 0,356987196 | 0,169647347 | LMF1 | FALSE | Upregulated |
| -0,368303089 | 0,16980334 | LOC643072 | FALSE | Downregulated |
| 0,634439437 | 0,170076157 | CD2 | FALSE | Upregulated |
| 0,307807162 | 0,170259568 | GPN2 | FALSE | Upregulated |
| -0,503572 | 0,170988377 | APOC4-APOC2 | FALSE | Downregulated |
| -0,193421241 | 0,170988377 | CCDC90B | FALSE | Downregulated |
| 0,53472104 | 0,170988377 | DGKA | FALSE | Upregulated |
| -1,659659407 | 0,170988377 | UCHL1 | FALSE | Downregulated |
| 0,28284803 | 0,171092243 | ABCA2 | FALSE | Upregulated |
| 0,816551298 | 0,171092243 | CAPG | FALSE | Upregulated |

| | | | | |
|---|---|---|---|---|
| 0,61791184 | 0,171699578 | KLHL6 | FALSE | Upregulated |
| -0,654012187 | 0,171699578 | RP11-458D21.1 | FALSE | Downregulated |
| -0,91865312 | 0,171713445 | LINC00685 | FALSE | Downregulated |
| 0,190958277 | 0,171713445 | POLG | FALSE | Upregulated |
| -0,382658342 | 0,17305409 | CNBP | FALSE | Downregulated |
| -0,35139199 | 0,174649296 | CCDC104 | FALSE | Downregulated |
| -2,09389542 | 0,174649296 | EBF3 | FALSE | Downregulated |
| 1,210373109 | 0,174649296 | EYS | FALSE | Upregulated |
| -0,229356984 | 0,174649296 | HSPB11 | FALSE | Downregulated |
| 1,186774022 | 0,174649296 | IGLL1 | FALSE | Upregulated |
| 1,136316402 | 0,174649296 | KCNJ5 | FALSE | Upregulated |
| 0,778811923 | 0,174649296 | TRPM2 | FALSE | Upregulated |
| 0,289914712 | 0,174931935 | ABCC3 | FALSE | Upregulated |
| 1,11608096 | 0,174931935 | GABRB3 | FALSE | Upregulated |
| -0,307161772 | 0,174931935 | LOC102723773 | FALSE | Downregulated |
| -0,277965797 | 0,174931935 | PHF14 | FALSE | Downregulated |
| -1,508030219 | 0,174931935 | RP11-214C8.2 | FALSE | Downregulated |
| -0,114731623 | 0,174931935 | ZNF146 | FALSE | Downregulated |
| -0,202008684 | 0,175210795 | C1orf115 | FALSE | Downregulated |
| 0,41272788 | 0,175361334 | ANKRD13D | FALSE | Upregulated |
| 2,213343158 | 0,175361334 | CHGB | FALSE | Upregulated |
| -0,66582694 | 0,175361334 | DKFZP434I0714 | FALSE | Downregulated |
| 0,400577656 | 0,175361334 | FAM86C1 | FALSE | Upregulated |
| 0,706460157 | 0,175361334 | GPRIN3 | FALSE | Upregulated |
| 0,250472231 | 0,175361334 | GRINA | FALSE | Upregulated |
| 0,236563452 | 0,175361334 | H2AFY | FALSE | Upregulated |

| | | | | |
|---|---|---|---|---|
| 0,899297631 | 0,175361334 | LAMC3 | FALSE | Upregulated |
| -0,437286651 | 0,175361334 | NRN1 | FALSE | Downregulated |
| -0,451295157 | 0,175361334 | PALM3 | FALSE | Downregulated |
| 0,85602818 | 0,175361334 | PTK7 | FALSE | Upregulated |
| 1,744766271 | 0,175361334 | RHOV | FALSE | Upregulated |
| -0,232075767 | 0,175361334 | SEPHS2 | FALSE | Downregulated |
| 1,668583216 | 0,175361334 | SOGA3 | FALSE | Upregulated |
| -0,248356937 | 0,175538575 | SNX2 | FALSE | Downregulated |
| -0,391139427 | 0,176458501 | CLEC3B | FALSE | Downregulated |
| 0,198048402 | 0,176458501 | CTDNEP1 | FALSE | Upregulated |
| -0,655216075 | 0,176458501 | PTH1R | FALSE | Downregulated |
| -1,006539056 | 0,176458501 | SCUBE1 | FALSE | Downregulated |
| 1,626710971 | 0,176458501 | TIFAB | FALSE | Upregulated |
| -0,25406702 | 0,176458501 | TUBD1 | FALSE | Downregulated |
| -0,27973855 | 0,176458501 | ZCCHC9 | FALSE | Downregulated |
| -0,181238385 | 0,176458501 | ZNF33A | FALSE | Downregulated |
| -0,240898948 | 0,177364845 | FIP1L1 | FALSE | Downregulated |
| 2,206738837 | 0,177455681 | DLK1 | FALSE | Upregulated |
| 0,923928994 | 0,177755618 | IDO1 | FALSE | Upregulated |
| -0,94021129 | 0,178080012 | MPPED2 | FALSE | Downregulated |
| -0,226148641 | 0,178080012 | VAMP4 | FALSE | Downregulated |
| -0,21478348 | 0,179824398 | CCDC92 | FALSE | Downregulated |
| -1,457313384 | 0,179824398 | LOC102724265 | FALSE | Downregulated |
| -1,79572875 | 0,179824398 | PI16 | FALSE | Downregulated |
| 0,279466746 | 0,179824398 | TRAF4 | FALSE | Upregulated |
| 0,254411318 | 0,180139618 | NADSYN1 | FALSE | Upregulated |

| | | | | |
|---|---|---|---|---|
| 0,283953489 | 0,180806611 | NOL6 | FALSE | Upregulated |
| 0,284631286 | 0,181020502 | HDAC6 | FALSE | Upregulated |
| 1,126766756 | 0,181020502 | ICOS | FALSE | Upregulated |
| -0,288401924 | 0,182233678 | NIFK | FALSE | Downregulated |
| -0,256979486 | 0,182233678 | SNAPC1 | FALSE | Downregulated |
| -0,246374836 | 0,183070455 | LYRM5 | FALSE | Downregulated |
| -0,225356912 | 0,183124582 | DPH5 | FALSE | Downregulated |
| 0,695829854 | 0,183725385 | LAX1 | FALSE | Upregulated |
| 0,20633937 | 0,183904657 | PCNXL3 | FALSE | Upregulated |
| 0,266316133 | 0,183904657 | SLCO2B1 | FALSE | Upregulated |
| 0,161494165 | 0,184195911 | KIAA0319L | FALSE | Upregulated |
| -0,799645035 | 0,184233941 | LOC101928887 | FALSE | Downregulated |
| 0,435127728 | 0,184494344 | TMEM102 | FALSE | Upregulated |
| 1,072539731 | 0,185413182 | POLQ | FALSE | Upregulated |
| -0,187638059 | 0,186131931 | NVL | FALSE | Downregulated |
| 0,223104389 | 0,186299266 | SCRIB | FALSE | Upregulated |
| -0,708501016 | 0,187416657 | LOC102724800 | FALSE | Downregulated |
| 0,197040415 | 0,187416657 | SNX8 | FALSE | Upregulated |
| 0,753983127 | 0,187416657 | TMEM246 | FALSE | Upregulated |
| 0,638949467 | 0,187445764 | LOC102724592 | FALSE | Upregulated |
| -0,224855655 | 0,187569787 | AP4S1 | FALSE | Downregulated |
| -0,277560109 | 0,187569787 | LACTB2 | FALSE | Downregulated |
| 0,294878517 | 0,187569787 | SEC16A | FALSE | Upregulated |
| -0,281950543 | 0,187569787 | SETMAR | FALSE | Downregulated |
| 1,021865167 | 0,187569787 | SIGLEC8 | FALSE | Upregulated |
| -0,333660209 | 0,187569787 | UBE2D4 | FALSE | Downregulated |

| | | | | |
|---|---|---|---|---|
| -0,391244197 | 0,187776078 | CALML4 | FALSE | Downregulated |
| 0,317177662 | 0,188226943 | DAPK3 | FALSE | Upregulated |
| 0,461606831 | 0,188226943 | HLA-F | FALSE | Upregulated |
| 0,235561628 | 0,188226943 | SLC35A2 | FALSE | Upregulated |
| -0,297682136 | 0,189932287 | TCEAL4 | FALSE | Downregulated |
| -1,875390187 | 0,190166342 | MFAP5 | FALSE | Downregulated |
| 0,222407703 | 0,19057541 | C22orf29 | FALSE | Upregulated |
| 1,89091625 | 0,191318051 | AC020571.3 | FALSE | Upregulated |
| 0,37811364 | 0,191363515 | SLC15A3 | FALSE | Upregulated |
| -0,124993626 | 0,191433169 | KLHL12 | FALSE | Downregulated |
| -0,370127815 | 0,191485715 | MAP2K1 | FALSE | Downregulated |
| 0,427033293 | 0,19202481 | ZYX | FALSE | Upregulated |
| 0,771367991 | 0,1929056 | CD6 | FALSE | Upregulated |
| 0,908528773 | 0,1929056 | NUF2 | FALSE | Upregulated |
| 0,524961942 | 0,1929056 | RUNX3 | FALSE | Upregulated |
| 1,93686848 | 0,193232996 | CAPN11 | FALSE | Upregulated |
| 0,519478103 | 0,193232996 | RNASE1 | FALSE | Upregulated |
| 0,174227946 | 0,193481193 | EIF4G1 | FALSE | Upregulated |
| -0,84148409 | 0,194176963 | AC097468.4 | FALSE | Downregulated |
| 0,396157597 | 0,194257014 | ALDH3B1 | FALSE | Upregulated |
| 0,279550691 | 0,194257014 | DPH1 | FALSE | Upregulated |
| 0,581434652 | 0,194257014 | FAIM3 | FALSE | Upregulated |
| 0,918128476 | 0,194257014 | HLA-DQB2 | FALSE | Upregulated |
| -1,839264383 | 0,194380119 | PROK2 | FALSE | Downregulated |
| -0,197664747 | 0,194380119 | TRAPPC6B | FALSE | Downregulated |
| -0,291550564 | 0,195094162 | RPL27 | FALSE | Downregulated |

| | | | | |
|---|---|---|---|---|
| -0,476969157 | 0,196306333 | ZBTB21 | FALSE | Downregulated |
| -0,228051579 | 0,196817462 | PLK1S1 | FALSE | Downregulated |
| 0,852603242 | 0,19687583 | COL8A2 | FALSE | Upregulated |
| 0,262766837 | 0,197381643 | PIGS | FALSE | Upregulated |
| -0,616942948 | 0,197381643 | RP11-242D8.1 | FALSE | Downregulated |
| 0,618729181 | 0,197391609 | CD3E | FALSE | Upregulated |
| 0,415207706 | 0,197493706 | MYO1F | FALSE | Upregulated |
| 0,58418459 | 0,200048082 | TCF19 | FALSE | Upregulated |
| 0,500300661 | 0,200772032 | SLC38A1 | FALSE | Upregulated |
| -1,619982939 | 0,201453061 | DKFZp434J0226 | FALSE | Downregulated |
| 0,246700755 | 0,201453061 | FIBP | FALSE | Upregulated |
| -0,14367126 | 0,201453061 | HNRNPK | FALSE | Downregulated |
| 0,680977358 | 0,201453061 | KRT19 | FALSE | Upregulated |
| -0,316702703 | 0,201453061 | LRRCC1 | FALSE | Downregulated |
| -0,326585638 | 0,201453061 | RAB33B | FALSE | Downregulated |
| 0,490220902 | 0,201453061 | RNASE6 | FALSE | Upregulated |
| -0,221463805 | 0,201453061 | RNF7 | FALSE | Downregulated |
| -0,212730101 | 0,201453061 | RPL5 | FALSE | Downregulated |
| -0,208711438 | 0,201453061 | SAP30BP | FALSE | Downregulated |
| 0,698745181 | 0,201453061 | SIGLEC10 | FALSE | Upregulated |
| 0,851266707 | 0,201453061 | SYTL1 | FALSE | Upregulated |
| -0,427779899 | 0,201731614 | MEX3A | FALSE | Downregulated |
| 0,927711548 | 0,202470615 | CXCL11 | FALSE | Upregulated |
| 1,558106978 | 0,202470615 | RNF219-AS1 | FALSE | Upregulated |
| -0,4181294 | 0,202760414 | LINC01184 | FALSE | Downregulated |
| -0,187133001 | 0,202790398 | SNAP29 | FALSE | Downregulated |

| | | | | |
|---|---|---|---|---|
| -0,15997635 | 0,202790398 | UBTF | FALSE | Downregulated |
| 0,521777969 | 0,202911961 | AOAH | FALSE | Upregulated |
| 0,239096937 | 0,202911961 | FTH1 | FALSE | Upregulated |
| 0,231399875 | 0,202911961 | INPP5K | FALSE | Upregulated |
| 0,400406091 | 0,202911961 | MMP14 | FALSE | Upregulated |
| -0,195330411 | 0,202911961 | UTP6 | FALSE | Downregulated |
| -0,313809936 | 0,203161197 | LEPREL1 | FALSE | Downregulated |
| 1,169919995 | 0,204398401 | IL4I1 | FALSE | Upregulated |
| -0,211870016 | 0,204565284 | DDX27 | FALSE | Downregulated |
| 0,637458215 | 0,205003749 | SPSB4 | FALSE | Upregulated |
| 0,485527923 | 0,205130634 | APOBEC3G | FALSE | Upregulated |
| 0,195218881 | 0,205130634 | UBC | FALSE | Upregulated |
| -0,198410737 | 0,205753814 | AZIN1 | FALSE | Downregulated |
| 0,437197472 | 0,205753814 | IKZF1 | FALSE | Upregulated |
| 1,151178206 | 0,206659811 | CXCL6 | FALSE | Upregulated |
| -0,20112418 | 0,207096938 | C1orf27 | FALSE | Downregulated |
| -0,272269814 | 0,208497044 | CCDC121 | FALSE | Downregulated |
| 1,2014288 | 0,208497044 | CCL18 | FALSE | Upregulated |
| 0,439918493 | 0,208497044 | CYP2D6 | FALSE | Upregulated |
| -0,881863435 | 0,208497044 | HMGCS1 | FALSE | Downregulated |
| -0,564742786 | 0,208497044 | HSPB9 | FALSE | Downregulated |
| 0,65059192 | 0,208497044 | PSTPIP1 | FALSE | Upregulated |
| 1,163898399 | 0,208497044 | PVT1 | FALSE | Upregulated |
| 0,747224961 | 0,208497044 | SNX20 | FALSE | Upregulated |
| -0,187805871 | 0,208497044 | TXNL1 | FALSE | Downregulated |
| -0,614322979 | 0,209667032 | AC004862.6 | FALSE | Downregulated |

| | | | | |
|---|---|---|---|---|
| -0,309380874 | 0,209667032 | C16orf13 | FALSE | Downregulated |
| -0,156380037 | 0,209667032 | COPS5 | FALSE | Downregulated |
| 0,858187737 | 0,209667032 | FTH1P3 | FALSE | Upregulated |
| -0,235702934 | 0,209667032 | SMC3 | FALSE | Downregulated |
| -0,162422514 | 0,209667032 | WDR20 | FALSE | Downregulated |
| -0,595725899 | 0,21003583 | AK7 | FALSE | Downregulated |
| -0,880236578 | 0,21003583 | ATM | FALSE | Downregulated |
| -0,333724152 | 0,21003583 | SLC25A37 | FALSE | Downregulated |
| 0,24697666 | 0,21003583 | SMG9 | FALSE | Upregulated |
| 0,60187969 | 0,210332162 | FBXW9 | FALSE | Upregulated |
| 0,802729931 | 0,211508928 | BTLA | FALSE | Upregulated |
| 0,530062775 | 0,212253202 | AJUBA | FALSE | Upregulated |
| -0,441844115 | 0,212253202 | EPB41L4A-AS1 | FALSE | Downregulated |
| -0,357670321 | 0,212253202 | LIMCH1 | FALSE | Downregulated |
| -0,272138142 | 0,212253202 | PAIP2B | FALSE | Downregulated |
| 0,375348473 | 0,212253202 | PITPNM1 | FALSE | Upregulated |
| -0,487185961 | 0,212341378 | IGF2BP2 | FALSE | Downregulated |
| -0,19797476 | 0,212512324 | CETN3 | FALSE | Downregulated |
| 0,277387469 | 0,212676625 | ATP6V1H | FALSE | Upregulated |
| -0,415433971 | 0,212676625 | CCDC176 | FALSE | Downregulated |
| -1,921528703 | 0,212676625 | FBXO40 | FALSE | Downregulated |
| -0,191401088 | 0,212676625 | OMA1 | FALSE | Downregulated |
| -0,284005108 | 0,212676625 | SMARCA1 | FALSE | Downregulated |
| 0,314795804 | 0,212676625 | TPGS2 | FALSE | Upregulated |
| 0,48424218 | 0,212806724 | CORO1A | FALSE | Upregulated |
| 0,40267297 | 0,212806724 | INPP5D | FALSE | Upregulated |

| | | | | |
|---|---|---|---|---|
| 0,263279364 | 0,213028534 | DCAF15 | FALSE | Upregulated |
| -0,217353938 | 0,213153628 | UBE2L3 | FALSE | Downregulated |
| 0,272795406 | 0,213228799 | BTBD9 | FALSE | Upregulated |
| -0,160296516 | 0,213228799 | DSTN | FALSE | Downregulated |
| -0,373769532 | 0,213228799 | GCSHP3 | FALSE | Downregulated |
| 3,286313331 | 0,213228799 | PADI6 | FALSE | Upregulated |
| 0,228598154 | 0,213228799 | SND1 | FALSE | Upregulated |
| 0,171317608 | 0,213228799 | TAOK2 | FALSE | Upregulated |
| -0,388507576 | 0,213228799 | TNFSF10 | FALSE | Downregulated |
| -0,2603175 | 0,213228799 | USF2 | FALSE | Downregulated |
| 0,213641547 | 0,213559621 | ATP13A1 | FALSE | Upregulated |
| -0,224386109 | 0,213559621 | RPL23 | FALSE | Downregulated |
| 0,246813201 | 0,214666335 | SETD1B | FALSE | Upregulated |
| 0,875450467 | 0,215639586 | CLDN11 | FALSE | Upregulated |
| -0,40822857 | 0,2165314 | BMP4 | FALSE | Downregulated |
| 0,462043886 | 0,2165314 | CLEC2D | FALSE | Upregulated |
| 0,316632016 | 0,2165314 | FBXL18 | FALSE | Upregulated |
| 1,3547013 | 0,2165314 | HAR1A | FALSE | Upregulated |
| -0,525883762 | 0,2165314 | HSD17B7 | FALSE | Downregulated |
| 0,35151831 | 0,2165314 | LAT2 | FALSE | Upregulated |
| 0,663763049 | 0,2165314 | LOC102723342 | FALSE | Upregulated |
| 0,191310252 | 0,2165314 | MTOR | FALSE | Upregulated |
| 0,243777205 | 0,2165314 | POLR2E | FALSE | Upregulated |
| 0,145240429 | 0,2165314 | RAB1B | FALSE | Upregulated |
| -0,102048039 | 0,2165314 | USP7 | FALSE | Downregulated |
| 1,350426395 | 0,216608398 | FCRL5 | FALSE | Upregulated |

| | | | | |
|---|---|---|---|---|
| 0,947119476 | 0,217038259 | AUNIP | FALSE | Upregulated |
| -0,217527451 | 0,217038259 | PCM1 | FALSE | Downregulated |
| -0,843979149 | 0,21723175 | DLG5-AS1 | FALSE | Downregulated |
| -0,222069479 | 0,21723175 | POMP | FALSE | Downregulated |
| 0,737091877 | 0,217329635 | LY9 | FALSE | Upregulated |
| 0,685777266 | 0,217463409 | NPHP4 | FALSE | Upregulated |
| 0,320999351 | 0,217899504 | ZNF544 | FALSE | Upregulated |
| 2,775930142 | 0,218284747 | RP11-551M15.1 | FALSE | Upregulated |
| 0,226583028 | 0,219028048 | PREP | FALSE | Upregulated |
| 0,320419368 | 0,219028048 | RHOT2 | FALSE | Upregulated |
| 0,19137985 | 0,219028048 | TMEM115 | FALSE | Upregulated |
| -0,230033764 | 0,219028048 | ZNF638 | FALSE | Downregulated |
| -0,121681385 | 0,219493998 | URI1 | FALSE | Downregulated |
| -0,393059569 | 0,219620971 | ACSM3 | FALSE | Downregulated |
| 0,148386977 | 0,219620971 | ELP3 | FALSE | Upregulated |
| -0,367299693 | 0,219620971 | SKAP1 | FALSE | Downregulated |
| -0,282730127 | 0,219620971 | UBE2W | FALSE | Downregulated |
| 0,635413901 | 0,219646994 | CXCR4 | FALSE | Upregulated |
| -1,004485458 | 0,219646994 | ITLN2 | FALSE | Downregulated |
| 1,501113851 | 0,219646994 | LMTK3 | FALSE | Upregulated |
| 0,747147962 | 0,219646994 | PAPLN | FALSE | Upregulated |
| -0,16656847 | 0,219646994 | REEP5 | FALSE | Downregulated |
| -0,309578094 | 0,219646994 | SNRNP48 | FALSE | Downregulated |
| 0,790954057 | 0,219737679 | RLTPR | FALSE | Upregulated |
| -0,287135814 | 0,220593787 | ZNF429 | FALSE | Downregulated |
| -0,316674372 | 0,221136849 | ATP5E | FALSE | Downregulated |

| | | | | |
|---|---|---|---|---|
| 0,577013979 | 0,221560975 | LYZ | FALSE | Upregulated |
| 0,400378224 | 0,22234571 | ADPRH | FALSE | Upregulated |
| 0,980931098 | 0,22234571 | CYP2D7P | FALSE | Upregulated |
| 0,817622674 | 0,22234571 | GPSM1 | FALSE | Upregulated |
| 1,157191194 | 0,22234571 | GZMM | FALSE | Upregulated |
| -0,161497043 | 0,22234571 | MAPK9 | FALSE | Downregulated |
| -0,212494322 | 0,22234571 | TMCO1 | FALSE | Downregulated |
| 0,899093256 | 0,222444288 | SPINT1 | FALSE | Upregulated |
| -0,780858965 | 0,222444288 | TRNS1 | FALSE | Downregulated |
| -1,024820539 | 0,223039011 | COLEC12 | FALSE | Downregulated |
| -0,777844991 | 0,223039011 | HS3ST3A1 | FALSE | Downregulated |
| -0,257916346 | 0,223039011 | TMEM57 | FALSE | Downregulated |
| 0,371487713 | 0,223039011 | ZNF579 | FALSE | Upregulated |
| -0,111294101 | 0,223553362 | PAIP2 | FALSE | Downregulated |
| 1,549405798 | 0,223679715 | C12orf55 | FALSE | Upregulated |
| -1,84561267 | 0,223679715 | LOC101930531 | FALSE | Downregulated |
| 0,269455188 | 0,22380435 | AGRN | FALSE | Upregulated |
| 0,140990412 | 0,22380435 | CAMTA2 | FALSE | Upregulated |
| 0,80342559 | 0,22380435 | CCL21 | FALSE | Upregulated |
| 0,499746388 | 0,22380435 | CCNB1 | FALSE | Upregulated |
| 0,529492383 | 0,22380435 | CIITA | FALSE | Upregulated |
| 0,257346485 | 0,22380435 | FAM89B | FALSE | Upregulated |
| 0,622101616 | 0,22380435 | HMMR | FALSE | Upregulated |
| 0,208202701 | 0,22380435 | LASP1 | FALSE | Upregulated |
| -1,67585431 | 0,22380435 | LOC101929781 | FALSE | Downregulated |
| 0,799131265 | 0,22380435 | MMP24 | FALSE | Upregulated |

| | | | | |
|---|---|---|---|---|
| 0,242129433 | 0,22380435 | NAGPA | FALSE | Upregulated |
| -0,213940479 | 0,22380435 | PCF11 | FALSE | Downregulated |
| -0,191339334 | 0,22380435 | RPAIN | FALSE | Downregulated |
| 1,407477741 | 0,22380435 | RUFY4 | FALSE | Upregulated |
| -1,067456637 | 0,22380435 | RUNX2 | FALSE | Downregulated |
| 0,769768202 | 0,22380435 | SAMD11 | FALSE | Upregulated |
| -0,181172061 | 0,22380435 | SAP18 | FALSE | Downregulated |
| 0,345767727 | 0,22380435 | SLC25A45 | FALSE | Upregulated |
| -0,414149173 | 0,22380435 | SLC9A3R2 | FALSE | Downregulated |
| -0,41618176 | 0,22380435 | SMIM10 | FALSE | Downregulated |
| 0,286156083 | 0,22380435 | TKT | FALSE | Upregulated |
| -0,216073237 | 0,22380435 | TSR2 | FALSE | Downregulated |
| -0,156616026 | 0,224848312 | MIER3 | FALSE | Downregulated |
| 0,665493436 | 0,224848312 | OSCAR | FALSE | Upregulated |
| -0,196911599 | 0,225975729 | FRG1 | FALSE | Downregulated |
| 0,642593133 | 0,225975729 | RHOF | FALSE | Upregulated |
| 0,567936166 | 0,226125337 | CARD11 | FALSE | Upregulated |
| 2,104078154 | 0,226125337 | LINC00319 | FALSE | Upregulated |
| 0,303566422 | 0,226125337 | NPDC1 | FALSE | Upregulated |
| 0,165608109 | 0,226125337 | TUFM | FALSE | Upregulated |
| 0,471206903 | 0,226125337 | ZNF575 | FALSE | Upregulated |
| -0,802801437 | 0,226477063 | CXCR2 | FALSE | Downregulated |
| 0,393590082 | 0,226477063 | ITGA4 | FALSE | Upregulated |
| 0,452873402 | 0,226477063 | MVP | FALSE | Upregulated |
| -0,158862536 | 0,226700739 | EIF2A | FALSE | Downregulated |
| -2,232256011 | 0,226700739 | KLHL41 | FALSE | Downregulated |

| | | | | |
|---|---|---|---|---|
| 0,570499665 | 0,226700739 | LOC115110 | FALSE | Upregulated |
| 0,222531091 | 0,226700739 | POLRMT | FALSE | Upregulated |
| 0,246890061 | 0,226700739 | THADA | FALSE | Upregulated |
| 0,428561668 | 0,226700739 | UNC13D | FALSE | Upregulated |
| -0,152215156 | 0,227179932 | APEX1 | FALSE | Downregulated |
| 0,234924382 | 0,227179932 | DHDDS | FALSE | Upregulated |
| -0,380969184 | 0,227179932 | KCNE3 | FALSE | Downregulated |
| 0,904403713 | 0,229542306 | ABCD2 | FALSE | Upregulated |
| 1,510886927 | 0,229542306 | RIMBP3B | FALSE | Upregulated |
| 0,7863197 | 0,229542306 | SHCBP1 | FALSE | Upregulated |
| 0,739147888 | 0,229542306 | TVP23A | FALSE | Upregulated |
| 0,180100001 | 0,229718112 | CAPN1 | FALSE | Upregulated |
| 0,280732996 | 0,229718112 | MAST3 | FALSE | Upregulated |
| 0,169624107 | 0,22979733 | HIRA | FALSE | Upregulated |
| -0,272234945 | 0,22979733 | MAP10 | FALSE | Downregulated |
| -0,53653868 | 0,22987537 | NR1D2 | FALSE | Downregulated |
| 0,487466002 | 0,22987537 | PTPRH | FALSE | Upregulated |
| -0,474733031 | 0,22987537 | RP11-141O11.2 | FALSE | Downregulated |
| -0,185881472 | 0,230117746 | BCLAF1 | FALSE | Downregulated |
| 1,218785187 | 0,230117746 | DISC1FP1 | FALSE | Upregulated |
| -0,186142962 | 0,230117746 | HMGN3 | FALSE | Downregulated |
| 0,909170289 | 0,230117746 | LOC101929545 | FALSE | Upregulated |
| 0,470846021 | 0,230117746 | MRC2 | FALSE | Upregulated |
| -0,293000264 | 0,230117746 | NET1 | FALSE | Downregulated |
| 0,564420885 | 0,230117746 | PSIMCT-1 | FALSE | Upregulated |
| 0,608044265 | 0,230117746 | RHOH | FALSE | Upregulated |

| | | | | |
|---|---|---|---|---|
| -0,151162536 | 0,230117746 | RNF111 | FALSE | Downregulated |
| -0,289841453 | 0,230117746 | RPL26 | FALSE | Downregulated |
| -0,24401851 | 0,230117746 | SESTD1 | FALSE | Downregulated |
| 0,267524897 | 0,230117746 | SLC12A9 | FALSE | Upregulated |
| 0,345234588 | 0,230230262 | ZFP36 | FALSE | Upregulated |
| 2,91037854 | 0,230778109 | LINC00966 | FALSE | Upregulated |
| -0,33431833 | 0,230778109 | NUPR1 | FALSE | Downregulated |
| -0,158697159 | 0,230778109 | UBE2E3 | FALSE | Downregulated |
| -1,219580034 | 0,230827517 | MUSTN1 | FALSE | Downregulated |
| -0,250314891 | 0,230827517 | SYF2 | FALSE | Downregulated |
| 0,917780087 | 0,231035004 | HLA-DOB | FALSE | Upregulated |
| 0,302847329 | 0,231035004 | TSPAN4 | FALSE | Upregulated |
| -0,187115826 | 0,231439098 | ESD | FALSE | Downregulated |
| 0,72344604 | 0,231861368 | FZD7 | FALSE | Upregulated |
| 0,406904909 | 0,231861368 | PIK3R5 | FALSE | Upregulated |
| -1,070379816 | 0,231861368 | PTGER3 | FALSE | Downregulated |
| -0,171149209 | 0,23186332 | LAPTM4A | FALSE | Downregulated |
| 0,517178391 | 0,23230584 | IL2RB | FALSE | Upregulated |
| -0,155482631 | 0,232712945 | CNOT2 | FALSE | Downregulated |
| -0,569588544 | 0,233142731 | BFSP1 | FALSE | Downregulated |
| -1,342313245 | 0,233142731 | GREM1 | FALSE | Downregulated |
| -0,748685412 | 0,233142731 | HTRA3 | FALSE | Downregulated |
| -0,135060197 | 0,233142731 | NRD1 | FALSE | Downregulated |
| -0,199333192 | 0,233142731 | PPID | FALSE | Downregulated |
| -0,228376464 | 0,233142731 | RBM41 | FALSE | Downregulated |
| -0,24042656 | 0,233142731 | ZMYM3 | FALSE | Downregulated |

| | | | | |
|---|---|---|---|---|
| 0,415389865 | 0,233142731 | ZNF101 | FALSE | Upregulated |
| -0,209078451 | 0,233642976 | DNAJC19 | FALSE | Downregulated |
| -0,486041312 | 0,233642976 | RFX2 | FALSE | Downregulated |
| -0,341115015 | 0,233642976 | RNR2 | FALSE | Downregulated |
| -2,294873259 | 0,23410612 | MB | FALSE | Downregulated |
| -1,564138005 | 0,23410612 | TNNT2 | FALSE | Downregulated |
| 0,172842181 | 0,23410612 | YIPF3 | FALSE | Upregulated |
| 0,198043498 | 0,234349307 | PRKCSH | FALSE | Upregulated |
| -0,163553534 | 0,234451502 | UBA2 | FALSE | Downregulated |
| -0,265652384 | 0,234816632 | TULP3 | FALSE | Downregulated |
| -0,210729542 | 0,234842987 | KLHL20 | FALSE | Downregulated |
| 0,258081275 | 0,235106421 | ARHGEF1 | FALSE | Upregulated |
| 0,288434368 | 0,235106421 | ITPKC | FALSE | Upregulated |
| -0,172464602 | 0,235106421 | LSM12 | FALSE | Downregulated |
| 1,83714532 | 0,235106421 | RNU2-1 | FALSE | Upregulated |
| -0,263950577 | 0,235312009 | ARGLU1 | FALSE | Downregulated |
| 0,659751875 | 0,235312009 | BIRC5 | FALSE | Upregulated |
| -0,286439006 | 0,235312009 | C22orf46 | FALSE | Downregulated |
| 0,154573044 | 0,235312009 | CCAR2 | FALSE | Upregulated |
| -0,142285386 | 0,235312009 | LYRM7 | FALSE | Downregulated |
| 0,270608093 | 0,235312009 | MAGED1 | FALSE | Upregulated |
| -0,792387025 | 0,235312009 | MPL | FALSE | Downregulated |
| 0,720069243 | 0,235312009 | SLAMF8 | FALSE | Upregulated |
| 0,4101257 | 0,235312009 | TRIB3 | FALSE | Upregulated |
| -0,475573586 | 0,235312009 | WEE2-AS1 | FALSE | Downregulated |
| 0,394708812 | 0,235312009 | ZNF710 | FALSE | Upregulated |

| | | | | |
|---|---|---|---|---|
| 1,217851276 | 0,236400858 | CASR | FALSE | Upregulated |
| 0,943304984 | 0,236400858 | DNAJC5B | FALSE | Upregulated |
| 0,27662091 | 0,236400858 | GPC1 | FALSE | Upregulated |
| 0,220382317 | 0,236400858 | MAN2B1 | FALSE | Upregulated |
| 0,297336218 | 0,236400858 | PPAN | FALSE | Upregulated |
| 0,19212284 | 0,236400858 | RNF19A | FALSE | Upregulated |
| -0,221977438 | 0,236400858 | TYSND1 | FALSE | Downregulated |
| 0,684148915 | 0,23649275 | CD5 | FALSE | Upregulated |
| 0,729765303 | 0,237790968 | TLR7 | FALSE | Upregulated |
| -0,141234555 | 0,238066832 | CLDND1 | FALSE | Downregulated |
| -0,222758121 | 0,238066832 | DAAM1 | FALSE | Downregulated |
| -0,281669132 | 0,238066832 | PTPLB | FALSE | Downregulated |
| -1,080921115 | 0,238066832 | S100A12 | FALSE | Downregulated |
| 0,230349107 | 0,238066832 | SLC38A7 | FALSE | Upregulated |
| 0,500276806 | 0,238133422 | DOK1 | FALSE | Upregulated |
| -0,901506706 | 0,238133422 | MCM3AP-AS1 | FALSE | Downregulated |
| -0,127959335 | 0,238356487 | CSNK1A1 | FALSE | Downregulated |
| 0,799451491 | 0,238356487 | FRMPD1 | FALSE | Upregulated |
| 0,731126207 | 0,238356487 | GBGT1 | FALSE | Upregulated |
| -0,75322986 | 0,238356487 | RP11-15A1.7 | FALSE | Downregulated |
| -0,678699685 | 0,238356487 | S100A9 | FALSE | Downregulated |
| 0,625475284 | 0,238437854 | CENPU | FALSE | Upregulated |
| -0,218230629 | 0,238460531 | CSE1L | FALSE | Downregulated |
| -0,151771107 | 0,238460531 | PHAX | FALSE | Downregulated |
| 0,180180444 | 0,238460531 | RNF10 | FALSE | Upregulated |
| 0,654884081 | 0,238460531 | RP11-367J11.2 | FALSE | Upregulated |

| | | | | |
|---|---|---|---|---|
| 1,538855304 | 0,238460531 | SEPT5-GP1BB | FALSE | Upregulated |
| -0,128096215 | 0,23871821 | ARL14EP | FALSE | Downregulated |
| 0,767064526 | 0,23871821 | C16orf54 | FALSE | Upregulated |
| 0,297810552 | 0,23871821 | CARD10 | FALSE | Upregulated |
| 0,483677615 | 0,23871821 | CXorf21 | FALSE | Upregulated |
| -0,169295512 | 0,23871821 | EPS15 | FALSE | Downregulated |
| -0,990818475 | 0,23871821 | PTPN20B | FALSE | Downregulated |
| -0,731916033 | 0,23871821 | TRIM58 | FALSE | Downregulated |
| -0,166489496 | 0,23871821 | ZZZ3 | FALSE | Downregulated |
| 1,199376401 | 0,23873458 | LEFTY1 | FALSE | Upregulated |
| 0,171309979 | 0,238814662 | YWHAQ | FALSE | Upregulated |
| 0,253531553 | 0,239227094 | BAHD1 | FALSE | Upregulated |
| -0,451524739 | 0,239227094 | C1orf53 | FALSE | Downregulated |
| 1,132726776 | 0,239227094 | CTCFL | FALSE | Upregulated |
| 0,599414935 | 0,239227094 | LOC100652768 | FALSE | Upregulated |
| 0,273985996 | 0,239227094 | MBOAT7 | FALSE | Upregulated |
| 0,159111575 | 0,239227094 | PSMD3 | FALSE | Upregulated |
| 2,01350956 | 0,239227094 | SHISA9 | FALSE | Upregulated |
| 0,710684813 | 0,239227094 | ST6GALNAC4 | FALSE | Upregulated |
| 0,448069849 | 0,240602876 | FAM78A | FALSE | Upregulated |
| -0,239197662 | 0,240835226 | GPATCH11 | FALSE | Downregulated |
| 0,228920152 | 0,240835226 | MAP3K11 | FALSE | Upregulated |
| -0,325212928 | 0,240835226 | MITD1 | FALSE | Downregulated |
| 0,627193837 | 0,240835226 | MYO1G | FALSE | Upregulated |
| 0,242607823 | 0,240835226 | PAK4 | FALSE | Upregulated |
| 0,140483466 | 0,240835226 | PPP1CA | FALSE | Upregulated |

| | | | | |
|---|---|---|---|---|
| -0,640996563 | 0,242061994 | LOC101927589 | FALSE | Downregulated |
| -0,553986233 | 0,242301618 | HERC2P7 | FALSE | Downregulated |
| -0,140132736 | 0,242551681 | DNTTIP2 | FALSE | Downregulated |
| -1,688719038 | 0,243115532 | C19orf77 | FALSE | Downregulated |
| 0,123980294 | 0,243115532 | EXT2 | FALSE | Upregulated |
| 0,447489366 | 0,243115532 | ZNF813 | FALSE | Upregulated |
| 0,36549055 | 0,243312189 | CTSC | FALSE | Upregulated |
| -0,339398707 | 0,243312189 | DUSP19 | FALSE | Downregulated |
| -0,119063487 | 0,243312189 | GPBP1 | FALSE | Downregulated |
| 0,375302522 | 0,243312189 | HLA-DPB1 | FALSE | Upregulated |
| 0,812176542 | 0,243312189 | LGALS4 | FALSE | Upregulated |
| 1,798023981 | 0,243312189 | MORC1 | FALSE | Upregulated |
| 1,334458436 | 0,245520062 | AC104655.2 | FALSE | Upregulated |
| 0,216763386 | 0,245520062 | JUP | FALSE | Upregulated |
| -0,218439408 | 0,245520062 | KTN1 | FALSE | Downregulated |
| 0,237060645 | 0,245520062 | PHYKPL | FALSE | Upregulated |
| -0,302564517 | 0,245520062 | RPL13A | FALSE | Downregulated |
| 0,994185638 | 0,245520062 | TRPV6 | FALSE | Upregulated |
| -0,173797998 | 0,245747275 | DCUN1D1 | FALSE | Downregulated |
| 0,283169113 | 0,245747275 | H1FX | FALSE | Upregulated |
| 0,485629837 | 0,245756557 | FAM110A | FALSE | Upregulated |
| -0,537682016 | 0,245756557 | LOC102723766 | FALSE | Downregulated |
| 0,327738313 | 0,245756557 | NR2C2AP | FALSE | Upregulated |
| 1,086166828 | 0,245756557 | SCARNA9 | FALSE | Upregulated |
| -0,171842479 | 0,245756557 | SERINC3 | FALSE | Downregulated |
| 1,550183998 | 0,245921213 | RBM5-AS1 | FALSE | Upregulated |

| | | | | |
|---|---|---|---|---|
| -0,278358076 | 0,245950121 | METAP2 | FALSE | Downregulated |
| 1,189583122 | 0,246068736 | FAM95C | FALSE | Upregulated |
| 0,709997358 | 0,246068736 | HLA-L | FALSE | Upregulated |
| 0,224624537 | 0,246068736 | PLEC | FALSE | Upregulated |
| -0,215698052 | 0,246068736 | RBM26 | FALSE | Downregulated |
| 0,177866506 | 0,246068736 | SEC24C | FALSE | Upregulated |
| 0,296415231 | 0,246068736 | SGSH | FALSE | Upregulated |
| -1,525423954 | 0,246068736 | SPDYE1 | FALSE | Downregulated |
| 0,396394089 | 0,246242018 | DEF6 | FALSE | Upregulated |
| 0,868195 | 0,246287218 | EFEMP1 | FALSE | Upregulated |
| 0,169124101 | 0,246287218 | GBA | FALSE | Upregulated |
| -0,154973177 | 0,247453709 | BCCIP | FALSE | Downregulated |
| -0,252277075 | 0,247545287 | RUNDC3B | FALSE | Downregulated |
| 0,218606868 | 0,247683488 | CC2D1B | FALSE | Upregulated |
| -0,414901286 | 0,248413948 | NPIPA1 | FALSE | Downregulated |
| -0,14603746 | 0,248525258 | COPS2 | FALSE | Downregulated |
| -0,223993907 | 0,248525258 | ZNF595 | FALSE | Downregulated |
| 1,382036678 | 0,249013145 | HKDC1 | FALSE | Upregulated |
| 0,142124004 | 0,249949826 | ABCF3 | FALSE | Upregulated |
| 1,899850411 | 0,250148612 | DPEP1 | FALSE | Upregulated |
| 0,476110015 | 0,250238968 | FITM2 | FALSE | Upregulated |
| 0,203957174 | 0,250238968 | HMCES | FALSE | Upregulated |
| -0,396984593 | 0,250238968 | LOC100289097 | FALSE | Downregulated |
| -0,121902537 | 0,250238968 | UFSP2 | FALSE | Downregulated |
| -0,196782571 | 0,250238968 | YES1 | FALSE | Downregulated |
| 0,402030921 | 0,250732292 | ADCY7 | FALSE | Upregulated |

| | | | | |
|---|---|---|---|---|
| -0,187960846 | 0,250732292 | EIF5 | FALSE | Downregulated |
| -0,299075982 | 0,250732292 | KPNA5 | FALSE | Downregulated |
| -0,297526754 | 0,250954192 | DNAJC28 | FALSE | Downregulated |
| -0,219883939 | 0,251461276 | ZNF326 | FALSE | Downregulated |
| -1,063181504 | 0,251790157 | DNAH12 | FALSE | Downregulated |
| 0,19383497 | 0,251790157 | EPAS1 | FALSE | Upregulated |
| 0,208281187 | 0,252533803 | CLEC16A | FALSE | Upregulated |
| 0,344064516 | 0,252533803 | IRF8 | FALSE | Upregulated |
| -0,970271883 | 0,25270761 | LOC102723356 | FALSE | Downregulated |
| -0,198179298 | 0,25270761 | PCNXL4 | FALSE | Downregulated |
| 0,443274221 | 0,25270761 | PHF19 | FALSE | Upregulated |
| -0,364398839 | 0,25270761 | SMC6 | FALSE | Downregulated |
| -0,216704624 | 0,25270761 | SNRPF | FALSE | Downregulated |
| -0,359698222 | 0,25270761 | TTC5 | FALSE | Downregulated |
| 0,218546524 | 0,25270761 | ZBTB7B | FALSE | Upregulated |
| 0,37148943 | 0,252880625 | LPPR2 | FALSE | Upregulated |
| 0,58064698 | 0,253007571 | SPON2 | FALSE | Upregulated |
| -0,247749956 | 0,253173859 | CMC4 | FALSE | Downregulated |
| 0,185898278 | 0,253173859 | CORO1B | FALSE | Upregulated |
| -0,222408868 | 0,253173859 | COX7A2L | FALSE | Downregulated |
| -0,378118032 | 0,253173859 | LOC100507412 | FALSE | Downregulated |
| 3,310714843 | 0,254256307 | CTAGE15 | FALSE | Upregulated |
| -0,241717854 | 0,254270785 | ACADL | FALSE | Downregulated |
| 0,162785895 | 0,254270785 | PSMF1 | FALSE | Upregulated |
| 0,497583471 | 0,254270785 | SAMD3 | FALSE | Upregulated |
| 0,406182832 | 0,254270785 | SH3RF3 | FALSE | Upregulated |

| | | | | |
|---|---|---|---|---|
| -0,449979337 | 0,254969266 | LOC102723845 | FALSE | Downregulated |
| 0,579141106 | 0,255412384 | PLK3 | FALSE | Upregulated |
| 0,17338848 | 0,255417158 | TRIM28 | FALSE | Upregulated |
| 0,409059977 | 0,25546703 | TMEM86B | FALSE | Upregulated |
| -0,218639257 | 0,25582868 | LAS1L | FALSE | Downregulated |
| -0,287146614 | 0,255910564 | IL33 | FALSE | Downregulated |
| 0,462764565 | 0,255910564 | TMC8 | FALSE | Upregulated |
| -0,146919801 | 0,255967891 | HMGB1 | FALSE | Downregulated |
| 0,13490921 | 0,255967891 | MOAP1 | FALSE | Upregulated |
| -0,385715387 | 0,256200756 | TMEM98 | FALSE | Downregulated |
| 0,502754867 | 0,256590283 | CALHM2 | FALSE | Upregulated |
| -0,734486409 | 0,256846543 | TRNI | FALSE | Downregulated |
| -0,202052439 | 0,257645386 | AUH | FALSE | Downregulated |
| -0,198800939 | 0,258125952 | ARL6IP4 | FALSE | Downregulated |
| 0,955236127 | 0,258125952 | DNAH10 | FALSE | Upregulated |
| -0,300496903 | 0,258125952 | GAB2 | FALSE | Downregulated |
| 0,582285209 | 0,258125952 | MMP19 | FALSE | Upregulated |
| -0,859749781 | 0,258125952 | TPPP3 | FALSE | Downregulated |
| 0,238725817 | 0,258125952 | YIF1A | FALSE | Upregulated |
| -0,270086158 | 0,258671666 | SMARCC1 | FALSE | Downregulated |
| 0,173798625 | 0,258832202 | URGCP | FALSE | Upregulated |
| -0,194176646 | 0,259297993 | ZBTB8OS | FALSE | Downregulated |
| 0,403458229 | 0,259443516 | PIK3CD | FALSE | Upregulated |
| 0,287059143 | 0,260038674 | PRKAB1 | FALSE | Upregulated |
| -0,340251918 | 0,260984143 | TMEM223 | FALSE | Downregulated |
| -0,163455524 | 0,261314976 | CHMP2A | FALSE | Downregulated |

| | | | | |
|---|---|---|---|---|
| 0,214703827 | 0,261314976 | MRPS26 | FALSE | Upregulated |
| 0,238857836 | 0,261787436 | ATP9B | FALSE | Upregulated |
| 0,38577877 | 0,261787436 | HLA-B | FALSE | Upregulated |
| -0,197862808 | 0,261787436 | IFT74 | FALSE | Downregulated |
| -0,97210125 | 0,261787436 | PDGFRL | FALSE | Downregulated |
| -0,121292102 | 0,261807457 | ATMIN | FALSE | Downregulated |
| 0,241753282 | 0,261807457 | CAPNS1 | FALSE | Upregulated |
| -1,728348935 | 0,261807457 | CPNE6 | FALSE | Downregulated |
| 0,674974651 | 0,261807457 | GRHL3 | FALSE | Upregulated |
| -0,703405939 | 0,261807457 | KCNIP3 | FALSE | Downregulated |
| -0,739138277 | 0,261807457 | KGFLP1 | FALSE | Downregulated |
| -0,320236593 | 0,261807457 | NGFR | FALSE | Downregulated |
| 0,570749939 | 0,261807457 | PRC1 | FALSE | Upregulated |
| -0,129313067 | 0,261807457 | PTK2 | FALSE | Downregulated |
| 0,238569848 | 0,261807457 | RPL28 | FALSE | Upregulated |
| -0,266555237 | 0,261807457 | SNRPD1 | FALSE | Downregulated |
| 0,219569234 | 0,261807457 | SPNS1 | FALSE | Upregulated |
| -0,308780685 | 0,261807457 | USP13 | FALSE | Downregulated |
| -1,386025059 | 0,262701046 | ANKRD23 | FALSE | Downregulated |
| 0,170050351 | 0,262701046 | CCDC97 | FALSE | Upregulated |
| -0,232081204 | 0,262701046 | HTATSF1 | FALSE | Downregulated |
| 0,15895579 | 0,262701046 | UBE2Z | FALSE | Upregulated |
| 0,322040074 | 0,262919699 | INO80C | FALSE | Upregulated |
| -0,186494259 | 0,262919699 | MTO1 | FALSE | Downregulated |
| -0,484234795 | 0,262919699 | SAMD5 | FALSE | Downregulated |
| 1,272593006 | 0,263126488 | CCAT1 | FALSE | Upregulated |

| | | | | |
|---|---|---|---|---|
| 0,197178431 | 0,263126488 | LARGE | FALSE | Upregulated |
| 0,252224103 | 0,263732984 | PHPT1 | FALSE | Upregulated |
| -0,187239306 | 0,264182075 | DCAF6 | FALSE | Downregulated |
| 0,179490593 | 0,264400478 | SUGP1 | FALSE | Upregulated |
| 0,176822679 | 0,264964574 | ATG16L1 | FALSE | Upregulated |
| -0,231390398 | 0,264964574 | CRB3 | FALSE | Downregulated |
| -0,182322662 | 0,265662934 | ACAD10 | FALSE | Downregulated |
| -0,15291517 | 0,266576607 | HDAC1 | FALSE | Downregulated |
| 0,326509827 | 0,26706953 | C19orf47 | FALSE | Upregulated |
| -0,305954032 | 0,26706953 | HIRIP3 | FALSE | Downregulated |
| 0,235885958 | 0,26706953 | TRIAP1 | FALSE | Upregulated |
| 0,359770968 | 0,267586525 | CHRNE | FALSE | Upregulated |
| -0,353643108 | 0,267586525 | EFNB2 | FALSE | Downregulated |
| 0,19177307 | 0,267586525 | ITFG3 | FALSE | Upregulated |
| 3,49972057 | 0,267586525 | PRODH | FALSE | Upregulated |
| -0,182553521 | 0,268019747 | ACBD6 | FALSE | Downregulated |
| -0,503373911 | 0,268019747 | C19orf18 | FALSE | Downregulated |
| 0,240517764 | 0,268019747 | CLN3 | FALSE | Upregulated |
| 0,2861622 | 0,268019747 | CYTH2 | FALSE | Upregulated |
| 0,178491568 | 0,268019747 | EPN1 | FALSE | Upregulated |
| 0,347705418 | 0,268019747 | FBXL6 | FALSE | Upregulated |
| -0,224669987 | 0,268019747 | ING3 | FALSE | Downregulated |
| -0,244303051 | 0,268019747 | ISCU | FALSE | Downregulated |
| -0,210547954 | 0,268019747 | MED28 | FALSE | Downregulated |
| -0,128243557 | 0,268019747 | PRPF38A | FALSE | Downregulated |
| -0,243957541 | 0,268019747 | RABGEF1 | FALSE | Downregulated |

| | | | | |
|---|---|---|---|---|
| -0,16232852 | 0,268019747 | TOP2B | FALSE | Downregulated |
| 0,265277782 | 0,268019747 | VOPP1 | FALSE | Upregulated |
| 0,285033649 | 0,268019747 | WDR18 | FALSE | Upregulated |
| -0,211148701 | 0,268419248 | RNF168 | FALSE | Downregulated |
| 0,225042434 | 0,269148806 | ATP5SL | FALSE | Upregulated |
| 0,625833634 | 0,269252686 | CRIP1 | FALSE | Upregulated |
| 0,473266142 | 0,269252686 | EEPD1 | FALSE | Upregulated |
| 1,532232405 | 0,269252686 | GPR141 | FALSE | Upregulated |
| 0,133311046 | 0,269252686 | NKIRAS2 | FALSE | Upregulated |
| -0,19908263 | 0,269387401 | FBXO3 | FALSE | Downregulated |
| 0,192628927 | 0,269571438 | SAMM50 | FALSE | Upregulated |
| 0,330524013 | 0,270774805 | CSTB | FALSE | Upregulated |
| 0,526076288 | 0,270973368 | WDFY4 | FALSE | Upregulated |
| 1,65569296 | 0,271578567 | CD19 | FALSE | Upregulated |
| 1,090283699 | 0,271651129 | CENPI | FALSE | Upregulated |
| 0,284764365 | 0,271669076 | BCL2L1 | FALSE | Upregulated |
| 0,264509097 | 0,271669076 | PFKFB2 | FALSE | Upregulated |
| 0,491292883 | 0,271825759 | CYFIP2 | FALSE | Upregulated |
| 0,556009586 | 0,271825759 | F2RL1 | FALSE | Upregulated |
| 0,386098368 | 0,271825759 | TAP1 | FALSE | Upregulated |
| -0,194871491 | 0,27206899 | CETN2 | FALSE | Downregulated |
| -0,191288854 | 0,27206899 | EBAG9 | FALSE | Downregulated |
| -0,221836732 | 0,27206899 | GNAI1 | FALSE | Downregulated |
| -0,174828044 | 0,27206899 | HINT3 | FALSE | Downregulated |
| 0,226985641 | 0,27206899 | LEPRE1 | FALSE | Upregulated |
| 1,309273526 | 0,27206899 | LOC102724197 | FALSE | Upregulated |

| | | | | |
|---|---|---|---|---|
| 0,912069489 | 0,27206899 | LOC389831 | FALSE | Upregulated |
| 0,28070245 | 0,27206899 | RHOG | FALSE | Upregulated |
| -1,154361129 | 0,27206899 | SPOCK3 | FALSE | Downregulated |
| 0,109850829 | 0,272243004 | QARS | FALSE | Upregulated |
| 0,510182435 | 0,273366845 | CD37 | FALSE | Upregulated |
| 0,331960913 | 0,273366845 | CTSS | FALSE | Upregulated |
| -0,16076307 | 0,273366845 | NSMCE4A | FALSE | Downregulated |
| 0,849419693 | 0,273366845 | SIGLEC17P | FALSE | Upregulated |
| -0,184336797 | 0,274610179 | HDGF | FALSE | Downregulated |
| -0,363297789 | 0,27510771 | AGXT2 | FALSE | Downregulated |
| -0,355090481 | 0,27510771 | CCDC122 | FALSE | Downregulated |
| 0,138759157 | 0,27510771 | USP39 | FALSE | Upregulated |
| -0,930096875 | 0,275356112 | CYP2C19 | FALSE | Downregulated |
| -0,129795701 | 0,275356112 | IARS2 | FALSE | Downregulated |
| -0,627434798 | 0,275356112 | LINC01093 | FALSE | Downregulated |
| -0,279683987 | 0,275356112 | RAB40B | FALSE | Downregulated |
| -0,209536163 | 0,275356112 | RPS27A | FALSE | Downregulated |
| 0,809927629 | 0,275356112 | ZNF835 | FALSE | Upregulated |
| -1,31315606 | 0,275842023 | FAM169B | FALSE | Downregulated |
| -0,611899395 | 0,275842023 | RP11-12A2.3 | FALSE | Downregulated |
| -0,207456731 | 0,275943523 | GGNBP2 | FALSE | Downregulated |
| -0,274905114 | 0,275943523 | GRTP1 | FALSE | Downregulated |
| -0,531554925 | 0,276441694 | LOC102724832 | FALSE | Downregulated |
| 0,907247591 | 0,277582676 | EGR3 | FALSE | Upregulated |
| 0,611770183 | 0,277582676 | ITK | FALSE | Upregulated |
| 0,927785555 | 0,277582676 | RMI2 | FALSE | Upregulated |

| | | | | |
|---|---|---|---|---|
| 0,208734863 | 0,277631729 | NFRKB | FALSE | Upregulated |
| 0,842042733 | 0,277631729 | UBXN10 | FALSE | Upregulated |
| 0,553484211 | 0,277816352 | CENPE | FALSE | Upregulated |
| 1,669667384 | 0,277816352 | POU5F1B | FALSE | Upregulated |
| -0,880794989 | 0,277926983 | SOCS3 | FALSE | Downregulated |
| 0,119244019 | 0,278489993 | RTFDC1 | FALSE | Upregulated |
| 0,205340043 | 0,278968939 | CORO1C | FALSE | Upregulated |
| 0,148778101 | 0,278968939 | GLYR1 | FALSE | Upregulated |
| -0,122244787 | 0,278968939 | KHSRP | FALSE | Downregulated |
| -0,250158145 | 0,278968939 | LYAR | FALSE | Downregulated |
| 1,526271807 | 0,278968939 | MYO3B | FALSE | Upregulated |
| -0,262736426 | 0,278968939 | NFIA | FALSE | Downregulated |
| -0,287475667 | 0,278968939 | RWDD4 | FALSE | Downregulated |
| 0,419182306 | 0,279046158 | CYTIP | FALSE | Upregulated |
| 1,340514806 | 0,279046158 | LOC101927902 | FALSE | Upregulated |
| 0,179331407 | 0,279046158 | RNF31 | FALSE | Upregulated |
| 0,210230337 | 0,279046158 | SMYD4 | FALSE | Upregulated |
| 0,157370091 | 0,279218485 | SF3A1 | FALSE | Upregulated |
| -0,609933468 | 0,279522968 | CACNA1A | FALSE | Downregulated |
| 0,213051594 | 0,279522968 | NINJ1 | FALSE | Upregulated |
| 0,363172164 | 0,279522968 | TSSC4 | FALSE | Upregulated |
| 0,173321151 | 0,279917284 | ARF3 | FALSE | Upregulated |
| 0,421273011 | 0,279917284 | ARHGAP9 | FALSE | Upregulated |
| 0,133400961 | 0,279917284 | CPSF3L | FALSE | Upregulated |
| 1,002765001 | 0,279917284 | DBNDD1 | FALSE | Upregulated |
| -0,134250674 | 0,279917284 | DCAF10 | FALSE | Downregulated |

| | | | | |
|---|---|---|---|---|
| -2,552637028 | 0,279917284 | FAM157A | FALSE | Downregulated |
| 0,340075476 | 0,279917284 | FBXL19 | FALSE | Upregulated |
| -0,153170903 | 0,279917284 | FGFR1OP2 | FALSE | Downregulated |
| -0,462459594 | 0,279917284 | IGLON5 | FALSE | Downregulated |
| 0,300217967 | 0,279917284 | LINC00843 | FALSE | Upregulated |
| 0,182833338 | 0,279917284 | LYPLA2 | FALSE | Upregulated |
| -0,218938517 | 0,279917284 | MTERFD3 | FALSE | Downregulated |
| 0,999510673 | 0,279917284 | MZB1 | FALSE | Upregulated |
| -0,438004411 | 0,279917284 | OTUB2 | FALSE | Downregulated |
| 1,223954857 | 0,279917284 | PAPPA2 | FALSE | Upregulated |
| 0,285783575 | 0,279917284 | RHBDD2 | FALSE | Upregulated |
| -0,286012489 | 0,279917284 | SFPQ | FALSE | Downregulated |
| 0,282500596 | 0,279917284 | SYVN1 | FALSE | Upregulated |
| 0,527772355 | 0,279917284 | ZNF486 | FALSE | Upregulated |
| 0,216635222 | 0,279917284 | ZNF574 | FALSE | Upregulated |
| -0,263725965 | 0,280426541 | ANGPTL3 | FALSE | Downregulated |
| 0,330571175 | 0,280426541 | HIGD1A | FALSE | Upregulated |
| -0,111538302 | 0,280426541 | NUDT21 | FALSE | Downregulated |
| 0,154025091 | 0,280426541 | SIDT2 | FALSE | Upregulated |
| 0,813987005 | 0,280426541 | TMEM200C | FALSE | Upregulated |
| 0,238298739 | 0,280738726 | ACOT7 | FALSE | Upregulated |
| -0,303242689 | 0,280738726 | ARL16 | FALSE | Downregulated |
| 0,200581324 | 0,280738726 | ATP6V0A1 | FALSE | Upregulated |
| -0,196313784 | 0,280738726 | CDADC1 | FALSE | Downregulated |
| -0,176981805 | 0,280738726 | CDK7 | FALSE | Downregulated |
| 0,140383623 | 0,280738726 | CDK9 | FALSE | Upregulated |

| | | | | |
|---|---|---|---|---|
| -0,184175996 | 0,280738726 | CHIC2 | FALSE | Downregulated |
| -0,1945072 | 0,280738726 | CTSL | FALSE | Downregulated |
| -0,197344527 | 0,280738726 | EPB41L5 | FALSE | Downregulated |
| 1,915315104 | 0,280738726 | FAT2 | FALSE | Upregulated |
| 0,626628922 | 0,280738726 | FBXO41 | FALSE | Upregulated |
| 0,756808365 | 0,280738726 | GAS6-AS1 | FALSE | Upregulated |
| -0,308790797 | 0,280738726 | GYG2 | FALSE | Downregulated |
| -0,876261488 | 0,280738726 | HSD17B7P2 | FALSE | Downregulated |
| -1,561157984 | 0,280738726 | LINC01010 | FALSE | Downregulated |
| 0,545185511 | 0,280738726 | MOB3B | FALSE | Upregulated |
| 0,773879675 | 0,280738726 | NETO2 | FALSE | Upregulated |
| 0,186024594 | 0,280738726 | OBSL1 | FALSE | Upregulated |
| -1,044626163 | 0,280738726 | OGN | FALSE | Downregulated |
| -0,145363786 | 0,280738726 | PDS5A | FALSE | Downregulated |
| 0,292708976 | 0,280738726 | PIEZO1 | FALSE | Upregulated |
| 1,078299475 | 0,280738726 | RP11-861A13.4 | FALSE | Upregulated |
| -0,822442848 | 0,280738726 | SLCO4C1 | FALSE | Downregulated |
| 0,172767896 | 0,280738726 | SNX12 | FALSE | Upregulated |
| 0,294153362 | 0,280738726 | STAP2 | FALSE | Upregulated |
| -0,31788247 | 0,280738726 | SYTL5 | FALSE | Downregulated |
| 0,198198867 | 0,280738726 | TESK1 | FALSE | Upregulated |
| 1,172783313 | 0,280882484 | KCNK10 | FALSE | Upregulated |
| -0,150174759 | 0,281093319 | C5orf15 | FALSE | Downregulated |
| -0,27930943 | 0,281108529 | BLNK | FALSE | Downregulated |
| 0,231488779 | 0,281108529 | CEP170B | FALSE | Upregulated |
| 0,2044134 | 0,281108529 | COLGALT1 | FALSE | Upregulated |

| | | | | |
|---|---|---|---|---|
| 1,477910573 | 0,281108529 | LRRIQ1 | FALSE | Upregulated |
| -0,239207169 | 0,282076019 | EIF4E2 | FALSE | Downregulated |
| -0,568096117 | 0,2824702 | LOC101928424 | FALSE | Downregulated |
| 0,207940315 | 0,282853206 | ATG13 | FALSE | Upregulated |
| -0,315890361 | 0,283454353 | EXOSC6 | FALSE | Downregulated |
| 0,214118161 | 0,283454353 | GTPBP6 | FALSE | Upregulated |
| -0,59136667 | 0,283454353 | TUBB1 | FALSE | Downregulated |
| 0,748886542 | 0,28355382 | C1orf186 | FALSE | Upregulated |
| -0,223252874 | 0,28355382 | DTD2 | FALSE | Downregulated |
| -0,909380759 | 0,28355382 | HRASLS2 | FALSE | Downregulated |
| 0,418921997 | 0,28355382 | MID1 | FALSE | Upregulated |
| -0,249785973 | 0,28355382 | SLC41A2 | FALSE | Downregulated |
| 0,585455676 | 0,284075064 | KDELR3 | FALSE | Upregulated |
| -0,959887893 | 0,284296899 | AC092295.7 | FALSE | Downregulated |
| -0,16867408 | 0,284296899 | EFCAB14 | FALSE | Downregulated |
| -0,212606794 | 0,284840479 | IK | FALSE | Downregulated |
| -0,200138096 | 0,285036419 | NUDCD1 | FALSE | Downregulated |
| 0,201374934 | 0,285183607 | ARL2BP | FALSE | Upregulated |
| -0,188875501 | 0,285570531 | PRKAA1 | FALSE | Downregulated |
| 0,871682967 | 0,285790665 | SLMO2-ATP5E | FALSE | Upregulated |
| 0,390129752 | 0,286096811 | CSF1R | FALSE | Upregulated |
| 0,188454773 | 0,286096811 | EMD | FALSE | Upregulated |
| -0,726483026 | 0,286096811 | INTS6-AS1 | FALSE | Downregulated |
| 0,764157427 | 0,286096811 | MMS22L | FALSE | Upregulated |
| -0,339445171 | 0,286096811 | RPL17-C18orf32 | FALSE | Downregulated |
| -0,280445251 | 0,286096811 | SEC62 | FALSE | Downregulated |

| | | | | |
|---|---|---|---|---|
| 0,363466077 | 0,286096811 | SLC7A7 | FALSE | Upregulated |
| -0,180333536 | 0,286246003 | BCAS2 | FALSE | Downregulated |
| -0,16736041 | 0,286246003 | HNRNPM | FALSE | Downregulated |
| -0,181249884 | 0,28786606 | HTRA2 | FALSE | Downregulated |
| 0,850371544 | 0,28786606 | SKA3 | FALSE | Upregulated |
| -0,236732591 | 0,287982693 | DCTPP1 | FALSE | Downregulated |
| 0,247397348 | 0,287982693 | DPAGT1 | FALSE | Upregulated |
| 0,478317069 | 0,287982693 | MCM6 | FALSE | Upregulated |
| -0,410357812 | 0,287982693 | PORCN | FALSE | Downregulated |
| -0,254135688 | 0,287982693 | RPL36AL | FALSE | Downregulated |
| -0,226909437 | 0,287982693 | RPS28 | FALSE | Downregulated |
| 0,194366247 | 0,287982693 | ZNF687 | FALSE | Upregulated |
| 1,239199677 | 0,288010594 | CTSE | FALSE | Upregulated |
| 0,299575451 | 0,28864665 | TCN2 | FALSE | Upregulated |
| 0,842768644 | 0,288703906 | ADRA2A | FALSE | Upregulated |
| -0,592027644 | 0,288703906 | C17orf97 | FALSE | Downregulated |
| 0,761527928 | 0,289111177 | HJURP | FALSE | Upregulated |
| 0,723615841 | 0,289111177 | SNPH | FALSE | Upregulated |
| 0,235943102 | 0,292382712 | GRN | FALSE | Upregulated |
| -0,167956114 | 0,292642889 | ELF2 | FALSE | Downregulated |
| 1,492332266 | 0,29276614 | C3orf52 | FALSE | Upregulated |
| -0,193733298 | 0,29276614 | CCNH | FALSE | Downregulated |
| -0,254145292 | 0,29276614 | NAA38 | FALSE | Downregulated |
| -1,088135353 | 0,29276614 | SLC4A1 | FALSE | Downregulated |
| 0,546174745 | 0,29277096 | KCNA3 | FALSE | Upregulated |
| 0,352974441 | 0,292910535 | NDOR1 | FALSE | Upregulated |

| | | | | |
|---|---|---|---|---|
| 0,992406807 | 0,293328998 | SHISA3 | FALSE | Upregulated |
| -0,783761358 | 0,293328998 | TEX21P | FALSE | Downregulated |
| -1,210551956 | 0,293627229 | LINC00890 | FALSE | Downregulated |
| 0,151995559 | 0,293627229 | LZTR1 | FALSE | Upregulated |
| 0,173614365 | 0,293627229 | TOR1A | FALSE | Upregulated |
| -0,187913036 | 0,293795769 | CHURC1 | FALSE | Downregulated |
| 0,201926991 | 0,294685576 | CLUH | FALSE | Upregulated |
| 0,85367638 | 0,294685576 | FFAR4 | FALSE | Upregulated |
| -0,185669793 | 0,294685576 | HNRNPC | FALSE | Downregulated |
| 0,237469532 | 0,294685576 | PTTG1IP | FALSE | Upregulated |
| -0,551266 | 0,295265045 | MT1E | FALSE | Downregulated |
| 0,398136079 | 0,295492147 | ZNF749 | FALSE | Upregulated |
| 0,387429178 | 0,295494824 | CD53 | FALSE | Upregulated |
| 0,31713802 | 0,295494824 | PRKX | FALSE | Upregulated |
| 0,550947136 | 0,295643399 | PAQR4 | FALSE | Upregulated |
| 0,518251798 | 0,295643399 | PIGZ | FALSE | Upregulated |
| 0,661747098 | 0,295643399 | PLK1 | FALSE | Upregulated |
| -0,19507464 | 0,295970123 | SSBP1 | FALSE | Downregulated |
| -0,286518228 | 0,296059019 | AR | FALSE | Downregulated |
| 0,443253307 | 0,296726475 | CHTF18 | FALSE | Upregulated |
| -0,203558848 | 0,296726475 | RICTOR | FALSE | Downregulated |
| -0,209164986 | 0,297886067 | SPOP | FALSE | Downregulated |
| 0,556854454 | 0,29880647 | ADAMTS12 | FALSE | Upregulated |
| -0,387673197 | 0,29880647 | FDFT1 | FALSE | Downregulated |
| -0,12156251 | 0,298911458 | CUL4B | FALSE | Downregulated |
| 0,182896874 | 0,299214393 | ZBTB18 | FALSE | Upregulated |

| | | | | |
|---|---|---|---|---|
| 1,261789208 | 0,299350495 | SCARNA7 | FALSE | Upregulated |
| -0,306861429 | 0,299908541 | C12orf43 | FALSE | Downregulated |
| 0,212025811 | 0,299908541 | SLC35C2 | FALSE | Upregulated |
| -1,868135452 | 0,299908541 | TCAP | FALSE | Downregulated |
| -0,348881234 | 0,299908541 | ZCCHC6 | FALSE | Downregulated |
| 0,416748797 | 0,300280802 | TBKBP1 | FALSE | Upregulated |
| 1,507033994 | 0,300480401 | CILP2 | FALSE | Upregulated |
| -1,291607189 | 0,300480401 | COL19A1 | FALSE | Downregulated |
| -0,170083639 | 0,300480401 | MAPKAP1 | FALSE | Downregulated |
| -0,986338865 | 0,300535264 | IP6K3 | FALSE | Downregulated |
| 2,537828939 | 0,300535264 | KRT86 | FALSE | Upregulated |
| 0,664732043 | 0,301000924 | C9orf9 | FALSE | Upregulated |
| -0,227303885 | 0,301000924 | HRSP12 | FALSE | Downregulated |
| -0,141133763 | 0,301000924 | SMEK1 | FALSE | Downregulated |
| 0,445781473 | 0,301538875 | GPR56 | FALSE | Upregulated |
| -0,274287428 | 0,301538875 | HIBCH | FALSE | Downregulated |
| 0,182081223 | 0,301757382 | CCDC22 | FALSE | Upregulated |
| -0,222624224 | 0,301757382 | PET100 | FALSE | Downregulated |
| 1,672137496 | 0,301757382 | SLC51B | FALSE | Upregulated |
| 0,279551543 | 0,302024681 | NTMT1 | FALSE | Upregulated |
| -0,131341134 | 0,302024681 | UBE2V1 | FALSE | Downregulated |
| -0,134415463 | 0,302024681 | ZNF780A | FALSE | Downregulated |
| 0,405064914 | 0,302149021 | AURKA | FALSE | Upregulated |
| -0,224525627 | 0,302149021 | NDUFAF2 | FALSE | Downregulated |
| 0,192538819 | 0,302149021 | STX4 | FALSE | Upregulated |
| 0,282463833 | 0,302733711 | ADAM15 | FALSE | Upregulated |

| | | | | |
|---|---|---|---|---|
| 1,055759553 | 0,302733711 | TNFRSF9 | FALSE | Upregulated |
| -0,365920662 | 0,302904656 | PLD6 | FALSE | Downregulated |
| -0,265469612 | 0,303017935 | GPATCH1 | FALSE | Downregulated |
| 1,200482306 | 0,303017935 | PDX1 | FALSE | Upregulated |
| 1,404416399 | 0,304107943 | YPEL4 | FALSE | Upregulated |
| -0,215082955 | 0,304202212 | C19orf53 | FALSE | Downregulated |
| 0,127032398 | 0,304202212 | DFFA | FALSE | Upregulated |
| 0,232996111 | 0,304521823 | PPP2R5B | FALSE | Upregulated |
| 0,293646591 | 0,304521823 | SLC26A11 | FALSE | Upregulated |
| 0,248374035 | 0,305037789 | SEC22C | FALSE | Upregulated |
| 1,868692435 | 0,30539699 | CACNA1G | FALSE | Upregulated |
| -0,12798087 | 0,30539699 | DCBLD2 | FALSE | Downregulated |
| -0,3364942 | 0,30539699 | FAM171A1 | FALSE | Downregulated |
| 0,259089452 | 0,30539699 | IQCE | FALSE | Upregulated |
| 1,427828865 | 0,30539699 | NEIL3 | FALSE | Upregulated |
| -0,16284277 | 0,30539699 | NIF3L1 | FALSE | Downregulated |
| 0,699191125 | 0,30539699 | UBASH3A | FALSE | Upregulated |
| 0,433022889 | 0,30539699 | ZNF320 | FALSE | Upregulated |
| -0,128865484 | 0,30539699 | ZNF322 | FALSE | Downregulated |
| -0,298058231 | 0,305795346 | CD1D | FALSE | Downregulated |
| 2,04088472 | 0,305795346 | NPTX1 | FALSE | Upregulated |
| 0,219847374 | 0,305795346 | PIAS3 | FALSE | Upregulated |
| 0,168157831 | 0,307485234 | PGK1 | FALSE | Upregulated |
| -0,158391918 | 0,307616319 | ATG4C | FALSE | Downregulated |
| -2,121865967 | 0,307616319 | HMGA2 | FALSE | Downregulated |
| 0,181322768 | 0,307616319 | NUP188 | FALSE | Upregulated |

| | | | | |
|---|---|---|---|---|
| 0,22664636 | 0,307993029 | LMNA | FALSE | Upregulated |
| 0,575999109 | 0,308007257 | GZMK | FALSE | Upregulated |
| -0,292046429 | 0,308091632 | C1orf21 | FALSE | Downregulated |
| -0,308456818 | 0,308091632 | CDNF | FALSE | Downregulated |
| 0,634694592 | 0,308091632 | GINS2 | FALSE | Upregulated |
| 0,551404251 | 0,308091632 | MKI67 | FALSE | Upregulated |
| -0,155206223 | 0,308091632 | NCOA1 | FALSE | Downregulated |
| 0,179362885 | 0,308091632 | PTOV1 | FALSE | Upregulated |
| 0,323102559 | 0,308091632 | SERINC2 | FALSE | Upregulated |
| 0,159938563 | 0,308091632 | SNX17 | FALSE | Upregulated |
| 0,148141647 | 0,308344366 | GTF3C1 | FALSE | Upregulated |
| -0,738352921 | 0,310698669 | C2orf40 | FALSE | Downregulated |
| -0,141333927 | 0,310698669 | CDC123 | FALSE | Downregulated |
| 0,428359852 | 0,310698669 | EZH2 | FALSE | Upregulated |
| 0,753069338 | 0,310698669 | ITGB2-AS1 | FALSE | Upregulated |
| 0,132761628 | 0,310698669 | PPP6R1 | FALSE | Upregulated |
| -0,210236627 | 0,311168471 | ABHD10 | FALSE | Downregulated |
| -0,139974518 | 0,311168471 | SDCCAG8 | FALSE | Downregulated |
| -0,358861964 | 0,311203696 | DNAJC25 | FALSE | Downregulated |
| 1,456439777 | 0,311203696 | GSG2 | FALSE | Upregulated |
| -0,905278858 | 0,311203696 | TRHDE | FALSE | Downregulated |
| 0,201442471 | 0,311369654 | POMGNT1 | FALSE | Upregulated |
| 0,422790517 | 0,311719344 | NIPAL2 | FALSE | Upregulated |
| 0,250939953 | 0,311719344 | ULK3 | FALSE | Upregulated |
| 0,231224192 | 0,311731111 | ABTB1 | FALSE | Upregulated |
| -0,123830882 | 0,311731111 | CHMP2B | FALSE | Downregulated |

| | | | | |
|---|---|---|---|---|
| 0,489577261 | 0,311731111 | COL4A4 | FALSE | Upregulated |
| 1,106439476 | 0,311913589 | GRIK4 | FALSE | Upregulated |
| 0,15846757 | 0,311913589 | SORBS3 | FALSE | Upregulated |
| 0,163437438 | 0,312540418 | AIM1 | FALSE | Upregulated |
| 0,830016974 | 0,312540418 | ANKRD18A | FALSE | Upregulated |
| -0,132846625 | 0,312540418 | BNIP2 | FALSE | Downregulated |
| -0,179246521 | 0,312540418 | CDK4 | FALSE | Downregulated |
| 0,142481961 | 0,312540418 | CLK3 | FALSE | Upregulated |
| -0,815335626 | 0,312540418 | FAM124B | FALSE | Downregulated |
| -2,115477664 | 0,312540418 | FLJ20444 | FALSE | Downregulated |
| -0,172945002 | 0,312540418 | H3F3AP4 | FALSE | Downregulated |
| -0,680568352 | 0,312540418 | LOC100506675 | FALSE | Downregulated |
| -0,811639246 | 0,312540418 | LOC441454 | FALSE | Downregulated |
| -0,23160515 | 0,312540418 | MRRF | FALSE | Downregulated |
| 0,14151464 | 0,312540418 | NRAS | FALSE | Upregulated |
| 0,160166178 | 0,312540418 | SCAF1 | FALSE | Upregulated |
| -0,196111956 | 0,312540418 | SMS | FALSE | Downregulated |
| 0,213012575 | 0,312540418 | TLN1 | FALSE | Upregulated |
| -0,437818575 | 0,312540418 | TMEM255B | FALSE | Downregulated |
| -0,277173525 | 0,312540418 | ZNF484 | FALSE | Downregulated |
| -0,633697737 | 0,312937926 | FGF23 | FALSE | Downregulated |
| 0,315535863 | 0,312937926 | FKBPL | FALSE | Upregulated |
| -0,616348999 | 0,312937926 | MANEA-AS1 | FALSE | Downregulated |
| -0,282341798 | 0,312937926 | PKD2 | FALSE | Downregulated |
| -0,190819527 | 0,313436457 | FUNDC2 | FALSE | Downregulated |
| -0,217211866 | 0,313887985 | TFPI | FALSE | Downregulated |

| | | | | |
|---|---|---|---|---|
| 1,417344709 | 0,313887985 | TRPC4 | FALSE | Upregulated |
| -0,512528002 | 0,314558836 | ARID3C | FALSE | Downregulated |
| -0,247309202 | 0,314558836 | DEK | FALSE | Downregulated |
| 0,735594533 | 0,314558836 | PHLDA3 | FALSE | Upregulated |
| 0,640965565 | 0,314558836 | PTK6 | FALSE | Upregulated |
| -1,01934986 | 0,314558836 | RET | FALSE | Downregulated |
| 0,49694883 | 0,314558836 | SLC7A6 | FALSE | Upregulated |
| 0,235488508 | 0,315018409 | TMEM184B | FALSE | Upregulated |
| -0,385347305 | 0,315018409 | XRRA1 | FALSE | Downregulated |
| -0,158453541 | 0,315344913 | RNF170 | FALSE | Downregulated |
| 0,410364842 | 0,315376403 | RENBP | FALSE | Upregulated |
| -0,151200543 | 0,315376403 | SMAD5 | FALSE | Downregulated |
| -0,190695651 | 0,315846588 | C5orf22 | FALSE | Downregulated |
| -0,463155784 | 0,315846588 | CDKL1 | FALSE | Downregulated |
| -1,108493126 | 0,315846588 | CRABP2 | FALSE | Downregulated |
| 0,170931463 | 0,315846588 | ERGIC3 | FALSE | Upregulated |
| -0,224469977 | 0,315846588 | GPX4 | FALSE | Downregulated |
| -0,178663778 | 0,315846588 | HIBADH | FALSE | Downregulated |
| 0,350299772 | 0,315846588 | KNTC1 | FALSE | Upregulated |
| 0,546149862 | 0,315846588 | PARPBP | FALSE | Upregulated |
| -0,253850851 | 0,315846588 | PON3 | FALSE | Downregulated |
| 0,582659739 | 0,315846588 | SLAMF7 | FALSE | Upregulated |
| 0,516439503 | 0,315846588 | TMEM198 | FALSE | Upregulated |
| -0,26618403 | 0,315846588 | TSR3 | FALSE | Downregulated |
| 0,147741682 | 0,315846588 | WDR1 | FALSE | Upregulated |
| -0,188033428 | 0,315846588 | ZNF569 | FALSE | Downregulated |

| | | | | |
|---|---|---|---|---|
| -0,288710097 | 0,316125582 | RAB28 | FALSE | Downregulated |
| -0,163700001 | 0,316125582 | RPAP3 | FALSE | Downregulated |
| 0,328336181 | 0,316328685 | FBLN7 | FALSE | Upregulated |
| 0,916778265 | 0,316328685 | GABRP | FALSE | Upregulated |
| 1,181299299 | 0,316328685 | LOC102724835 | FALSE | Upregulated |
| 0,218255816 | 0,316328685 | PCSK6 | FALSE | Upregulated |
| -0,286879442 | 0,316328685 | SDPR | FALSE | Downregulated |
| -0,150906031 | 0,316328685 | WBP4 | FALSE | Downregulated |
| 0,491050782 | 0,316367123 | RP11-307C12.11 | FALSE | Upregulated |
| -0,234361625 | 0,316534575 | MUT | FALSE | Downregulated |
| -0,257058748 | 0,316534575 | SIRT1 | FALSE | Downregulated |
| -0,328661551 | 0,316534575 | SNX16 | FALSE | Downregulated |
| -0,1857972 | 0,316534575 | STK11 | FALSE | Downregulated |
| 2,242774507 | 0,316534575 | SYT2 | FALSE | Upregulated |
| 0,21812406 | 0,316588128 | CAMSAP3 | FALSE | Upregulated |
| -0,777971379 | 0,316588128 | RP11-112L6.4 | FALSE | Downregulated |
| -1,055400019 | 0,317565207 | LOC102724208 | FALSE | Downregulated |
| 0,866045387 | 0,317795088 | SCART1 | FALSE | Upregulated |
| -0,161216713 | 0,318320799 | TIMM10B | FALSE | Downregulated |
| -0,15361373 | 0,318320799 | USP48 | FALSE | Downregulated |
| -0,29441217 | 0,318597964 | LSM3 | FALSE | Downregulated |
| -0,170006567 | 0,318601221 | COX20 | FALSE | Downregulated |
| -0,27015257 | 0,318601221 | GMDS-AS1 | FALSE | Downregulated |
| 0,558817814 | 0,318601221 | HELLS | FALSE | Upregulated |
| -0,466337974 | 0,318601221 | LOC101929633 | FALSE | Downregulated |
| -0,158414003 | 0,318603285 | SUOX | FALSE | Downregulated |

| | | | | |
|---|---|---|---|---|
| 1,286625647 | 0,318842224 | MAL | FALSE | Upregulated |
| 0,3771793 | 0,31912105 | CPVL | FALSE | Upregulated |
| 1,343737308 | 0,319185109 | TLDC2 | FALSE | Upregulated |
| -0,671118227 | 0,319431355 | CREB3L3 | FALSE | Downregulated |
| 0,187974506 | 0,319431355 | RABGGTA | FALSE | Upregulated |
| -0,168386985 | 0,319431355 | TCEB3 | FALSE | Downregulated |
| -0,192258014 | 0,319431355 | ZNF346 | FALSE | Downregulated |
| 0,247406407 | 0,319585777 | PDXP | FALSE | Upregulated |
| 0,119388924 | 0,319585777 | STK25 | FALSE | Upregulated |
| -0,347082528 | 0,319674462 | BST1 | FALSE | Downregulated |
| 0,16043707 | 0,319674462 | DHX8 | FALSE | Upregulated |
| 0,559295318 | 0,319775067 | GPNMB | FALSE | Upregulated |
| -0,176807925 | 0,319775067 | YAE1D1 | FALSE | Downregulated |
| 0,133444688 | 0,320613701 | ERCC3 | FALSE | Upregulated |
| -0,271644588 | 0,320613701 | IVNS1ABP | FALSE | Downregulated |
| -0,241063988 | 0,320613701 | OXR1 | FALSE | Downregulated |
| 0,90267408 | 0,320613701 | PCDP1 | FALSE | Upregulated |
| -0,35697659 | 0,320613701 | RP11-317J10.2 | FALSE | Downregulated |
| -0,3063552 | 0,320613701 | SYDE2 | FALSE | Downregulated |
| 0,166272373 | 0,320613701 | ZBTB17 | FALSE | Upregulated |
| 1,147018395 | 0,321078863 | RTEL1-TNFRSF6B | FALSE | Upregulated |
| -0,257102076 | 0,321735047 | KIAA1143 | FALSE | Downregulated |
| -0,136621629 | 0,32255545 | YWHAE | FALSE | Downregulated |
| 1,130997796 | 0,322621486 | LOC100505710 | FALSE | Upregulated |
| -0,536313154 | 0,322621486 | MEF2C | FALSE | Downregulated |
| 1,372737226 | 0,324071101 | RP11-353N14.5 | FALSE | Upregulated |

| | | | | |
|---|---|---|---|---|
| 0,218033617 | 0,324399866 | PLD3 | FALSE | Upregulated |
| -0,216583213 | 0,324533484 | ARL3 | FALSE | Downregulated |
| 0,517353045 | 0,324533484 | FANCD2 | FALSE | Upregulated |
| 0,55023447 | 0,324533484 | ITGB4 | FALSE | Upregulated |
| 1,089790573 | 0,324533484 | LOXL4 | FALSE | Upregulated |
| 0,271333422 | 0,324533484 | RANGAP1 | FALSE | Upregulated |
| -0,130459474 | 0,324533484 | TPP2 | FALSE | Downregulated |
| -0,182791084 | 0,325321492 | WIBG | FALSE | Downregulated |
| 0,802222184 | 0,325538091 | GFM2 | FALSE | Upregulated |
| 0,60577218 | 0,325838691 | GINS4 | FALSE | Upregulated |
| -0,180888554 | 0,326053451 | NAP1L4 | FALSE | Downregulated |
| 0,295583156 | 0,326212553 | TGFB1 | FALSE | Upregulated |
| 0,186122621 | 0,326319198 | ARPC5 | FALSE | Upregulated |
| 0,347529446 | 0,326779051 | ANKS6 | FALSE | Upregulated |
| -0,19929243 | 0,326779051 | C11orf57 | FALSE | Downregulated |
| 0,379407105 | 0,326779051 | NFATC1 | FALSE | Upregulated |
| -1,472378934 | 0,326779051 | NRAP | FALSE | Downregulated |
| 0,363950241 | 0,326779051 | TTYH3 | FALSE | Upregulated |
| -0,133158213 | 0,326779051 | ZCCHC7 | FALSE | Downregulated |
| -0,438607015 | 0,326875399 | SAMD12 | FALSE | Downregulated |
| 0,159503018 | 0,327316543 | GPAA1 | FALSE | Upregulated |
| 0,374845188 | 0,327326799 | ABCA7 | FALSE | Upregulated |
| 0,786906422 | 0,327326799 | AP000688.14 | FALSE | Upregulated |
| -0,192329352 | 0,327326799 | CDK14 | FALSE | Downregulated |
| 0,171392778 | 0,327326799 | CHMP1A | FALSE | Upregulated |
| 0,873203214 | 0,327326799 | CLIC6 | FALSE | Upregulated |

| | | | | |
|---|---|---|---|---|
| 0,862254045 | 0,327326799 | GAL3ST4 | FALSE | Upregulated |
| 0,177616697 | 0,327326799 | GMEB2 | FALSE | Upregulated |
| 0,315100491 | 0,327326799 | GPRC5C | FALSE | Upregulated |
| -0,269183251 | 0,327326799 | HSD17B6 | FALSE | Downregulated |
| 0,446726593 | 0,327326799 | LAIR1 | FALSE | Upregulated |
| 0,478906939 | 0,327326799 | LOC389906 | FALSE | Upregulated |
| 0,216911405 | 0,327326799 | MARS | FALSE | Upregulated |
| -0,137325024 | 0,327326799 | PRR13 | FALSE | Downregulated |
| 0,252891045 | 0,327326799 | ROBO4 | FALSE | Upregulated |
| -0,447452956 | 0,327326799 | RP5-968D22.1 | FALSE | Downregulated |
| -0,207720807 | 0,327326799 | SREK1IP1 | FALSE | Downregulated |
| -0,322426135 | 0,327916447 | ADK | FALSE | Downregulated |
| 0,231908739 | 0,327916447 | APEX2 | FALSE | Upregulated |
| -0,177424806 | 0,327916447 | CCDC58 | FALSE | Downregulated |
| 0,240047673 | 0,327916447 | CCDC61 | FALSE | Upregulated |
| 0,219772487 | 0,327916447 | DMWD | FALSE | Upregulated |
| 0,141442602 | 0,327916447 | ELAC2 | FALSE | Upregulated |
| 0,969235644 | 0,327916447 | ENTPD2 | FALSE | Upregulated |
| 0,260290564 | 0,327916447 | EPS8L2 | FALSE | Upregulated |
| 0,148127678 | 0,327916447 | FAM91A1 | FALSE | Upregulated |
| 0,25054635 | 0,327916447 | FBXL14 | FALSE | Upregulated |
| -0,363894165 | 0,327916447 | FEZ1 | FALSE | Downregulated |
| 0,17921015 | 0,327916447 | HEXA | FALSE | Upregulated |
| 0,311025903 | 0,327916447 | IL16 | FALSE | Upregulated |
| 0,231841672 | 0,327916447 | JHDM1D-AS1 | FALSE | Upregulated |
| 0,409436921 | 0,327916447 | LILRB4 | FALSE | Upregulated |

| | | | | |
|---|---|---|---|---|
| -0,778368479 | 0,327916447 | LOC101927880 | FALSE | Downregulated |
| -0,955891194 | 0,327916447 | LOC102724402 | FALSE | Downregulated |
| 0,482222815 | 0,327916447 | LOXL3 | FALSE | Upregulated |
| -0,267689868 | 0,327916447 | MINPP1 | FALSE | Downregulated |
| 0,144113437 | 0,327916447 | PLOD1 | FALSE | Upregulated |
| -0,23666547 | 0,327916447 | TRAM2-AS1 | FALSE | Downregulated |
| 0,421384686 | 0,327916447 | TRPV2 | FALSE | Upregulated |
| 0,322889984 | 0,327916447 | ZDHHC23 | FALSE | Upregulated |
| -0,372953517 | 0,328333357 | PDP2 | FALSE | Downregulated |
| 0,355607783 | 0,328333357 | RPS6KA1 | FALSE | Upregulated |
| -0,830704587 | 0,328333357 | TRNV | FALSE | Downregulated |
| 0,11182589 | 0,328333357 | USP22 | FALSE | Upregulated |
| 0,168744519 | 0,328449314 | GMCL1 | FALSE | Upregulated |
| 0,742858094 | 0,328535407 | HS1BP3-IT1 | FALSE | Upregulated |
| 0,561424228 | 0,328724825 | FASLG | FALSE | Upregulated |
| 1,139879903 | 0,328724825 | FBN3 | FALSE | Upregulated |
| -0,138399887 | 0,328809658 | MAGOH | FALSE | Downregulated |
| 0,223962891 | 0,328809658 | RNASET2 | FALSE | Upregulated |
| 1,010069211 | 0,328914768 | LIF | FALSE | Upregulated |
| 0,23533306 | 0,329089661 | PP7080 | FALSE | Upregulated |
| -0,357422935 | 0,329089661 | ZNF829 | FALSE | Downregulated |
| 1,802245872 | 0,329275283 | LOC101929475 | FALSE | Upregulated |
| 1,229772386 | 0,329275283 | RP11-18H21.1 | FALSE | Upregulated |
| 0,284906299 | 0,329516887 | KIAA0556 | FALSE | Upregulated |
| 0,852888979 | 0,329516887 | KIF18A | FALSE | Upregulated |
| 0,188183772 | 0,329516887 | MAPK1 | FALSE | Upregulated |

| | | | | |
|---|---|---|---|---|
| -0,133508083 | 0,329516887 | SLAIN2 | FALSE | Downregulated |
| -0,317702764 | 0,329516887 | ZNF527 | FALSE | Downregulated |
| 0,169560348 | 0,329617055 | BCAS3 | FALSE | Upregulated |
| 0,62405187 | 0,329617055 | CD8A | FALSE | Upregulated |
| -0,175892752 | 0,329617055 | IRF2 | FALSE | Downregulated |
| 0,474004395 | 0,330151057 | IKZF3 | FALSE | Upregulated |
| 0,69799076 | 0,330920286 | ANLN | FALSE | Upregulated |
| 0,303249652 | 0,3310119 | ZBTB45 | FALSE | Upregulated |
| 0,483880587 | 0,331238807 | ABCB4 | FALSE | Upregulated |
| 0,185230569 | 0,331238807 | ARHGDIA | FALSE | Upregulated |
| 0,151259826 | 0,331238807 | ETV3 | FALSE | Upregulated |
| 0,2174577 | 0,331238807 | FAM122B | FALSE | Upregulated |
| 0,89961584 | 0,331238807 | SEZ6L | FALSE | Upregulated |
| -0,258569562 | 0,331238807 | ST13 | FALSE | Downregulated |
| 1,153551047 | 0,331238807 | XCR1 | FALSE | Upregulated |
| 0,424950777 | 0,331392064 | FANCI | FALSE | Upregulated |
| 0,359262871 | 0,331392064 | MAGIX | FALSE | Upregulated |
| -0,336357797 | 0,331718128 | CAV1 | FALSE | Downregulated |
| 0,378166874 | 0,331718128 | DOCK8 | FALSE | Upregulated |
| -0,882905628 | 0,331942712 | LOC152225 | FALSE | Downregulated |
| 0,213970187 | 0,332235199 | AKIRIN1 | FALSE | Upregulated |
| -0,340395061 | 0,332235199 | CCL14 | FALSE | Downregulated |
| 0,303471684 | 0,332235199 | CHPF | FALSE | Upregulated |
| -1,925722812 | 0,332235199 | LOC102723580 | FALSE | Downregulated |
| 0,593969073 | 0,332235199 | LOC102724535 | FALSE | Upregulated |
| -0,288421806 | 0,332235199 | MBIP | FALSE | Downregulated |

| | | | | |
|---|---|---|---|---|
| 3,693289811 | 0,332235199 | RNU1-28P | FALSE | Upregulated |
| -0,153247201 | 0,332235199 | SHFM1 | FALSE | Downregulated |
| 0,93072034 | 0,332341498 | BMP8B | FALSE | Upregulated |
| -0,637460394 | 0,332341498 | DNAJA4 | FALSE | Downregulated |
| 1,152267304 | 0,332341498 | KCNH8 | FALSE | Upregulated |
| 0,18872892 | 0,332341498 | MOGS | FALSE | Upregulated |
| 0,83366085 | 0,332953402 | DEPDC1B | FALSE | Upregulated |
| 1,000793469 | 0,333185222 | ARMC12 | FALSE | Upregulated |
| -0,396062923 | 0,333185222 | BCL2L2-PABPN1 | FALSE | Downregulated |
| 1,222179841 | 0,333185222 | CCDC19 | FALSE | Upregulated |
| 0,352433444 | 0,333185222 | CMTM7 | FALSE | Upregulated |
| -0,39436558 | 0,333185222 | COL25A1 | FALSE | Downregulated |
| 0,976618617 | 0,333185222 | OIP5 | FALSE | Upregulated |
| -0,230550331 | 0,333185222 | PPP2R3C | FALSE | Downregulated |
| 0,300196582 | 0,333185222 | TRAF2 | FALSE | Upregulated |
| 0,262594162 | 0,333212291 | CBR1 | FALSE | Upregulated |
| 0,404498924 | 0,333212291 | DLL1 | FALSE | Upregulated |
| 0,607132971 | 0,333212291 | ITGA3 | FALSE | Upregulated |
| 0,16298815 | 0,333212291 | MADD | FALSE | Upregulated |
| 0,275067655 | 0,333646993 | ARSA | FALSE | Upregulated |
| -0,196825508 | 0,333646993 | HSPA9 | FALSE | Downregulated |
| -0,396058752 | 0,333646993 | PEG3 | FALSE | Downregulated |
| 0,812492081 | 0,333646993 | POU2AF1 | FALSE | Upregulated |
| -0,690172591 | 0,333646993 | SNCA | FALSE | Downregulated |
| 0,318568003 | 0,333646993 | SYTL4 | FALSE | Upregulated |
| -1,852773413 | 0,333853979 | CA1 | FALSE | Downregulated |

| | | | | |
|---|---|---|---|---|
| -0,61919628 | 0,333973293 | MYB | FALSE | Downregulated |
| 0,243911329 | 0,333973293 | TMEM63A | FALSE | Upregulated |
| 1,140344651 | 0,334285468 | TP73 | FALSE | Upregulated |
| 0,159698186 | 0,334674859 | ELK1 | FALSE | Upregulated |
| -0,207318414 | 0,334674859 | STX17 | FALSE | Downregulated |
| 0,27533045 | 0,334747421 | ANKRD44 | FALSE | Upregulated |
| 0,489594743 | 0,334963212 | APOL4 | FALSE | Upregulated |
| 0,718060703 | 0,334963212 | ARHGEF39 | FALSE | Upregulated |
| 0,732286845 | 0,334963212 | BUB1 | FALSE | Upregulated |
| 0,218023505 | 0,334963212 | C19orf52 | FALSE | Upregulated |
| 1,560813095 | 0,334963212 | CCL20 | FALSE | Upregulated |
| 0,512115564 | 0,334963212 | CDCA8 | FALSE | Upregulated |
| 0,549833224 | 0,334963212 | GBP3 | FALSE | Upregulated |
| -0,129417809 | 0,334963212 | GGPS1 | FALSE | Downregulated |
| 0,424184529 | 0,334963212 | HLA-DOA | FALSE | Upregulated |
| 0,609047445 | 0,334963212 | KIFC1 | FALSE | Upregulated |
| 1,025215948 | 0,334963212 | MGAT3 | FALSE | Upregulated |
| -0,53780819 | 0,334963212 | NPIPB3 | FALSE | Downregulated |
| -0,799736416 | 0,334963212 | NR1D1 | FALSE | Downregulated |
| 1,368372558 | 0,334963212 | OR2A9P | FALSE | Upregulated |
| 0,605443384 | 0,334963212 | PNMA6A | FALSE | Upregulated |
| -0,309551914 | 0,334963212 | RAD50 | FALSE | Downregulated |
| -0,565625621 | 0,334963212 | RASSF6 | FALSE | Downregulated |
| 1,227971091 | 0,334963212 | RP11-430B1.2 | FALSE | Upregulated |
| -0,232907063 | 0,334963212 | SERPINF1 | FALSE | Downregulated |
| -0,193191173 | 0,334963212 | SFT2D3 | FALSE | Downregulated |

| | | | | |
|---|---|---|---|---|
| 0,62190535 | 0,334963212 | SH2B2 | FALSE | Upregulated |
| -0,25668294 | 0,334963212 | TCEA3 | FALSE | Downregulated |
| -1,524459291 | 0,334963212 | TNFAIP6 | FALSE | Downregulated |
| 0,746369414 | 0,334963212 | VTCN1 | FALSE | Upregulated |
| 0,104068868 | 0,334963212 | WDR82 | FALSE | Upregulated |
| -0,589177668 | 0,334963212 | ZIC1 | FALSE | Downregulated |
| 0,177812752 | 0,33596279 | RNASEK | FALSE | Upregulated |
| 1,163422956 | 0,33596279 | RTKN2 | FALSE | Upregulated |
| 0,387379861 | 0,33596279 | TRAF5 | FALSE | Upregulated |
| 0,427444913 | 0,336267129 | CDYL2 | FALSE | Upregulated |
| 0,171629879 | 0,336410628 | MPP1 | FALSE | Upregulated |
| -0,284932415 | 0,336410628 | PIR | FALSE | Downregulated |
| 0,257687268 | 0,336410628 | PRKCD | FALSE | Upregulated |
| 0,566937001 | 0,336873498 | CD69 | FALSE | Upregulated |
| 0,10297982 | 0,336907531 | DDB1 | FALSE | Upregulated |
| 0,450782349 | 0,33714642 | RHBDF1 | FALSE | Upregulated |
| 0,806417014 | 0,337651873 | CASKIN1 | FALSE | Upregulated |
| 0,535353718 | 0,337651873 | CHST9 | FALSE | Upregulated |
| 0,125441679 | 0,337651873 | COPA | FALSE | Upregulated |
| -0,677378913 | 0,337651873 | MSX1 | FALSE | Downregulated |
| 0,713897051 | 0,337651873 | SDK1 | FALSE | Upregulated |
| 0,216547087 | 0,338324546 | ZNF764 | FALSE | Upregulated |
| 0,174626853 | 0,338446893 | AMBRA1 | FALSE | Upregulated |
| -0,828907983 | 0,338446893 | CBLN4 | FALSE | Downregulated |
| 0,141017485 | 0,338446893 | SMPD4 | FALSE | Upregulated |
| 0,337875306 | 0,338859451 | NAB2 | FALSE | Upregulated |

| | | | | |
|---|---|---|---|---|
| 0,158496469 | 0,338859451 | PGAM1 | FALSE | Upregulated |
| 0,652042225 | 0,339248915 | LOC100131655 | FALSE | Upregulated |
| 0,403865368 | 0,339309839 | RASIP1 | FALSE | Upregulated |
| 0,17712144 | 0,339309839 | ZNF672 | FALSE | Upregulated |
| 0,188620726 | 0,340056246 | MOV10 | FALSE | Upregulated |
| 0,161067626 | 0,340056246 | TRAF7 | FALSE | Upregulated |
| 0,147282347 | 0,34011597 | AKT1 | FALSE | Upregulated |
| 0,167328594 | 0,340315253 | TRAPPC12 | FALSE | Upregulated |
| 0,201641742 | 0,340526346 | ARFGAP1 | FALSE | Upregulated |
| 0,390114951 | 0,340526346 | CECR1 | FALSE | Upregulated |
| -0,178611449 | 0,340835889 | PYURF | FALSE | Downregulated |
| 0,422131977 | 0,341238297 | TYMP | FALSE | Upregulated |
| 0,2289257 | 0,341305073 | APOA1 | FALSE | Upregulated |
| 2,595115647 | 0,341305073 | CACNA1E | FALSE | Upregulated |
| -0,50483419 | 0,341305073 | COL11A2 | FALSE | Downregulated |
| 1,530291372 | 0,341631076 | AKR1B10 | FALSE | Upregulated |
| -1,312242422 | 0,341631076 | FOLR1 | FALSE | Downregulated |
| 0,859343019 | 0,341631076 | LOC440311 | FALSE | Upregulated |
| -0,485988682 | 0,341725587 | COX6C | FALSE | Downregulated |
| 1,121435439 | 0,3418124 | E2F2 | FALSE | Upregulated |
| -0,170220683 | 0,3418124 | MRPS22 | FALSE | Downregulated |
| 0,293863037 | 0,342758341 | GRAMD1A | FALSE | Upregulated |
| 2,457418436 | 0,342758341 | LOC101928510 | FALSE | Upregulated |
| 0,216217623 | 0,342871104 | ACP2 | FALSE | Upregulated |
| -0,273017528 | 0,342871104 | AK9 | FALSE | Downregulated |
| 0,281328789 | 0,342871104 | ALKBH4 | FALSE | Upregulated |

| | | | | |
|---|---|---|---|---|
| -0,152174695 | 0,342871104 | ARPC3 | FALSE | Downregulated |
| 0,725884185 | 0,342871104 | AURKB | FALSE | Upregulated |
| 0,160424334 | 0,342871104 | BTBD2 | FALSE | Upregulated |
| -0,147162985 | 0,342871104 | C1GALT1 | FALSE | Downregulated |
| -0,241407063 | 0,342871104 | CFHR2 | FALSE | Downregulated |
| 0,401050086 | 0,342871104 | CKS2 | FALSE | Upregulated |
| -0,341766086 | 0,342871104 | CLEC4G | FALSE | Downregulated |
| 0,199663328 | 0,342871104 | CLIC1 | FALSE | Upregulated |
| -0,129979195 | 0,342871104 | DCAF5 | FALSE | Downregulated |
| -0,180784538 | 0,342871104 | DDX23 | FALSE | Downregulated |
| -0,133276574 | 0,342871104 | DDX47 | FALSE | Downregulated |
| 0,924103627 | 0,342871104 | FZD2 | FALSE | Upregulated |
| 0,351109046 | 0,342871104 | KIAA0513 | FALSE | Upregulated |
| 0,722221758 | 0,342871104 | LOC102723554 | FALSE | Upregulated |
| -0,51798632 | 0,342871104 | LOC730101 | FALSE | Downregulated |
| 0,492517482 | 0,342871104 | LRRC37A4P | FALSE | Upregulated |
| -0,642781455 | 0,342871104 | METRN | FALSE | Downregulated |
| -0,43619083 | 0,342871104 | MTHFD2L | FALSE | Downregulated |
| 0,154582116 | 0,342871104 | MXD4 | FALSE | Upregulated |
| 0,661789584 | 0,342871104 | MYZAP | FALSE | Upregulated |
| -0,134341137 | 0,342871104 | NLK | FALSE | Downregulated |
| 0,843400459 | 0,342871104 | NRIP2 | FALSE | Upregulated |
| 0,386844317 | 0,342871104 | PARVG | FALSE | Upregulated |
| 0,152370373 | 0,342871104 | PEX26 | FALSE | Upregulated |
| 0,116778266 | 0,342871104 | POLDIP3 | FALSE | Upregulated |
| 0,759065077 | 0,342871104 | RASEF | FALSE | Upregulated |

| | | | | |
|---|---|---|---|---|
| 0,884988711 | 0,342871104 | RGS9 | FALSE | Upregulated |
| 0,172295272 | 0,342871104 | RPN1 | FALSE | Upregulated |
| -0,621990901 | 0,342871104 | SNAI3-AS1 | FALSE | Downregulated |
| -1,338937651 | 0,342871104 | SRL | FALSE | Downregulated |
| -0,187564491 | 0,342871104 | TOPORS | FALSE | Downregulated |
| -0,156700445 | 0,342871104 | VBP1 | FALSE | Downregulated |
| 0,190089947 | 0,343122039 | AAGAB | FALSE | Upregulated |
| -0,194773316 | 0,343122039 | GIN1 | FALSE | Downregulated |
| -1,942933587 | 0,343122039 | LOC285556 | FALSE | Downregulated |
| 0,321909415 | 0,343318973 | HLA-DMB | FALSE | Upregulated |
| -0,263870923 | 0,343457911 | ANAPC15 | FALSE | Downregulated |
| -0,547142452 | 0,343457911 | CLEC4M | FALSE | Downregulated |
| -0,25295912 | 0,343457911 | SH3BP5 | FALSE | Downregulated |
| 0,872011315 | 0,343605587 | PMFBP1 | FALSE | Upregulated |
| 0,208662534 | 0,344276652 | C7orf26 | FALSE | Upregulated |
| 0,466905305 | 0,344290379 | AMICA1 | FALSE | Upregulated |
| 0,204214798 | 0,344290379 | IRF2BP1 | FALSE | Upregulated |
| 0,176078165 | 0,344290379 | MAD1L1 | FALSE | Upregulated |
| 0,371826053 | 0,344557714 | LOC102723553 | FALSE | Upregulated |
| 0,13323303 | 0,344557714 | TATDN2 | FALSE | Upregulated |
| 0,665378159 | 0,344809402 | ZNF208 | FALSE | Upregulated |
| -0,253279827 | 0,345101103 | WHAMM | FALSE | Downregulated |
| 0,161227771 | 0,345508621 | AVL9 | FALSE | Upregulated |
| -0,224412951 | 0,345508621 | CCDC25 | FALSE | Downregulated |
| 0,560546818 | 0,345508621 | GAPT | FALSE | Upregulated |
| 0,722717778 | 0,345508621 | TMCC2 | FALSE | Upregulated |

| | | | | |
|---|---|---|---|---|
| 0,229415215 | 0,345884837 | CLPB | FALSE | Upregulated |
| -0,140499076 | 0,346000443 | PDCD2 | FALSE | Downregulated |
| 0,729093179 | 0,346264793 | ASF1B | FALSE | Upregulated |
| -0,747817553 | 0,346264793 | BCRP3 | FALSE | Downregulated |
| 0,153054719 | 0,346264793 | CNDP2 | FALSE | Upregulated |
| -0,141469391 | 0,346264793 | HNRNPD | FALSE | Downregulated |
| 0,177334405 | 0,346536446 | B4GALT3 | FALSE | Upregulated |
| 0,941631366 | 0,346536446 | VWA2 | FALSE | Upregulated |
| 0,25234258 | 0,34671042 | BRMS1 | FALSE | Upregulated |
| 0,412213335 | 0,34671042 | RCAN3 | FALSE | Upregulated |
| -0,180231691 | 0,347130023 | C8orf59 | FALSE | Downregulated |
| 1,575997786 | 0,347130023 | DUOX1 | FALSE | Upregulated |
| 0,381326498 | 0,347130023 | FERMT3 | FALSE | Upregulated |
| 0,678664148 | 0,347130023 | LOC101927183 | FALSE | Upregulated |
| -0,62408412 | 0,347130023 | LOC101927665 | FALSE | Downregulated |
| 1,752200502 | 0,347130023 | LPAR3 | FALSE | Upregulated |
| 0,965411716 | 0,347130023 | LTA | FALSE | Upregulated |
| 0,766515868 | 0,347130023 | METTL24 | FALSE | Upregulated |
| 0,149126317 | 0,347130023 | MLH1 | FALSE | Upregulated |
| 0,213583507 | 0,347130023 | NIT1 | FALSE | Upregulated |
| 0,125622622 | 0,347130023 | RRAGA | FALSE | Upregulated |
| 0,322903686 | 0,347130023 | SLC1A5 | FALSE | Upregulated |
| -0,129810121 | 0,347130023 | ZNF410 | FALSE | Downregulated |
| 0,1386294 | 0,34733937 | BUB3 | FALSE | Upregulated |
| -0,124631945 | 0,34774957 | ANGEL2 | FALSE | Downregulated |
| 0,137728331 | 0,34774957 | CLTC | FALSE | Upregulated |

| | | | | |
|---|---|---|---|---|
| 1,230336219 | 0,34774957 | LINC00639 | FALSE | Upregulated |
| -0,398321464 | 0,34774957 | RGS5 | FALSE | Downregulated |
| -0,371725146 | 0,34774957 | STAG3L4 | FALSE | Downregulated |
| 0,231360391 | 0,34774957 | UBE2E2 | FALSE | Upregulated |
| 0,761498472 | 0,34832183 | NELL2 | FALSE | Upregulated |
| -0,177272089 | 0,348423206 | F8A1 | FALSE | Downregulated |
| 0,238578165 | 0,348423206 | PIGW | FALSE | Upregulated |
| -0,228792259 | 0,348423206 | TMEM101 | FALSE | Downregulated |
| -0,214804688 | 0,349590522 | RPL4 | FALSE | Downregulated |
| 0,817218948 | 0,349590522 | TTC9 | FALSE | Upregulated |
| 0,882482346 | 0,349757549 | EXO1 | FALSE | Upregulated |
| 0,145622135 | 0,349757549 | KEAP1 | FALSE | Upregulated |
| 0,161438594 | 0,349877177 | CDK5RAP1 | FALSE | Upregulated |
| -0,150559125 | 0,350374878 | MRPS10 | FALSE | Downregulated |
| -0,258006585 | 0,350430505 | ZFP36L1 | FALSE | Downregulated |
| 0,576944652 | 0,350494899 | CCDC183-AS1 | FALSE | Upregulated |
| 1,112958112 | 0,350538811 | CTLA4 | FALSE | Upregulated |
| 0,461710868 | 0,350538811 | SAMD14 | FALSE | Upregulated |
| -1,171882902 | 0,350538811 | STAR | FALSE | Downregulated |
| 0,200718647 | 0,350538811 | TNFRSF14 | FALSE | Upregulated |
| -1,046003569 | 0,35058235 | AHNAK2 | FALSE | Downregulated |
| 0,300224197 | 0,35058235 | ATP1A1OS | FALSE | Upregulated |
| 0,636878395 | 0,35058235 | P2RY10 | FALSE | Upregulated |
| 1,206394774 | 0,352409472 | LOC101928621 | FALSE | Upregulated |
| 0,221789905 | 0,353289498 | TBL3 | FALSE | Upregulated |
| 1,00617398 | 0,353597575 | LOC101927789 | FALSE | Upregulated |

| | | | | |
|---|---|---|---|---|
| 0,467642493 | 0,353907495 | ENO2 | FALSE | Upregulated |
| 0,592723197 | 0,353907495 | PATL2 | FALSE | Upregulated |
| 0,471716416 | 0,353907495 | SLCO2A1 | FALSE | Upregulated |
| 0,444755836 | 0,353928463 | PTGER4 | FALSE | Upregulated |
| -0,192006066 | 0,35457022 | STYX | FALSE | Downregulated |
| 0,58272278 | 0,354610309 | DAGLA | FALSE | Upregulated |
| 0,410983709 | 0,354610309 | SEPT1 | FALSE | Upregulated |
| 0,140103532 | 0,354610309 | SF1 | FALSE | Upregulated |
| -0,771723191 | 0,354610309 | SYCE1L | FALSE | Downregulated |
| -0,169335701 | 0,354610309 | ZNF271 | FALSE | Downregulated |
| -0,286644474 | 0,354726017 | KLHDC4 | FALSE | Downregulated |
| 1,227289385 | 0,355393065 | ADAM11 | FALSE | Upregulated |
| 0,471605317 | 0,355393065 | LOC400043 | FALSE | Upregulated |
| -0,188248268 | 0,355393065 | MRPL48 | FALSE | Downregulated |
| -0,582466102 | 0,355393065 | SCARA5 | FALSE | Downregulated |
| 0,270668297 | 0,355393065 | TRIP6 | FALSE | Upregulated |
| 0,320957628 | 0,355393065 | TTF2 | FALSE | Upregulated |
| 0,60895773 | 0,355393065 | WISP1 | FALSE | Upregulated |
| -0,589082178 | 0,355871666 | FLJ36777 | FALSE | Downregulated |
| -0,329339407 | 0,356013262 | COX10-AS1 | FALSE | Downregulated |
| 0,42927276 | 0,356016765 | B3GNT7 | FALSE | Upregulated |
| 0,178873114 | 0,356016765 | DEPDC5 | FALSE | Upregulated |
| 1,410628441 | 0,356016765 | PGAM4 | FALSE | Upregulated |
| 0,164462185 | 0,356016765 | PTGES2 | FALSE | Upregulated |
| -0,399969082 | 0,356068546 | SH3BGR | FALSE | Downregulated |
| 0,83780344 | 0,356161732 | C5orf20 | FALSE | Upregulated |

| | | | | |
|---|---|---|---|---|
| 1,164211811 | 0,356161732 | MED12L | FALSE | Upregulated |
| -0,197734545 | 0,356161732 | SMARCE1 | FALSE | Downregulated |
| -0,131177997 | 0,356161732 | SUDS3 | FALSE | Downregulated |
| 0,277658203 | 0,356249833 | PROSER2 | FALSE | Upregulated |
| -0,093758106 | 0,356844191 | CNOT7 | FALSE | Downregulated |
| -0,162549684 | 0,356844191 | UHRF1BP1L | FALSE | Downregulated |
| -0,253870786 | 0,356844191 | ZMAT2 | FALSE | Downregulated |
| -0,127697147 | 0,357043351 | BACE1 | FALSE | Downregulated |
| 0,41396756 | 0,357043351 | DGCR5 | FALSE | Upregulated |
| -0,127667365 | 0,357043351 | GSK3B | FALSE | Downregulated |
| 1,432519946 | 0,357043351 | IL23R | FALSE | Upregulated |
| 0,505502143 | 0,357043351 | MAD2L1 | FALSE | Upregulated |
| 0,281676837 | 0,357043351 | NOD1 | FALSE | Upregulated |
| -0,194102885 | 0,357043351 | ZNF696 | FALSE | Downregulated |
| -0,142239048 | 0,357499266 | ARHGAP5 | FALSE | Downregulated |
| 0,183193134 | 0,357931452 | PLEKHM1 | FALSE | Upregulated |
| -0,280129786 | 0,357931452 | PPP1R11 | FALSE | Downregulated |
| -0,228972703 | 0,357931452 | RGS14 | FALSE | Downregulated |
| 0,693467383 | 0,357931452 | VSTM2L | FALSE | Upregulated |
| -0,357086795 | 0,357931452 | ZCCHC16 | FALSE | Downregulated |
| 0,165275128 | 0,358062241 | ZBED4 | FALSE | Upregulated |
| -0,204996423 | 0,358098079 | MAPRE3 | FALSE | Downregulated |
| 0,532671751 | 0,358239535 | C16orf96 | FALSE | Upregulated |
| 1,180535294 | 0,358239535 | CXCL5 | FALSE | Upregulated |
| 0,460428038 | 0,358239535 | LXN | FALSE | Upregulated |
| -0,723559394 | 0,358239535 | NEUROD2 | FALSE | Downregulated |

| | | | | |
|---|---|---|---|---|
| -0,111024843 | 0,358239535 | TBK1 | FALSE | Downregulated |
| -0,229733106 | 0,358239535 | THAP2 | FALSE | Downregulated |
| -0,216064454 | 0,358239535 | TRPM7 | FALSE | Downregulated |
| 0,249033038 | 0,358239535 | ZBTB9 | FALSE | Upregulated |
| 1,167656412 | 0,358867686 | HLA-DPB2 | FALSE | Upregulated |
| -0,264199854 | 0,358867686 | TIMMDC1 | FALSE | Downregulated |
| 0,546985455 | 0,359050014 | CDK1 | FALSE | Upregulated |
| 0,789938786 | 0,359115565 | ADD2 | FALSE | Upregulated |
| 0,501202961 | 0,359115565 | FAM57A | FALSE | Upregulated |
| -0,271767851 | 0,359115565 | RASGEF1B | FALSE | Downregulated |
| 0,230787142 | 0,359115565 | RNFT1 | FALSE | Upregulated |
| 0,896761767 | 0,359882025 | FBXL16 | FALSE | Upregulated |
| -0,242529604 | 0,359882025 | MBLAC2 | FALSE | Downregulated |
| 1,246565013 | 0,360404735 | HIST2H2BC | FALSE | Upregulated |
| -0,216057744 | 0,360450154 | MAF | FALSE | Downregulated |
| 0,328418692 | 0,360469275 | ARL4C | FALSE | Upregulated |
| 1,055613413 | 0,360469275 | FAM155B | FALSE | Upregulated |
| -0,225667293 | 0,360545333 | TTC28-AS1 | FALSE | Downregulated |
| -0,180554616 | 0,360545333 | XPA | FALSE | Downregulated |
| -0,120424947 | 0,360575419 | PCMTD1 | FALSE | Downregulated |
| 0,345896666 | 0,360749719 | H2AFX | FALSE | Upregulated |
| 0,430526727 | 0,360749719 | KRT18 | FALSE | Upregulated |
| -0,129803167 | 0,360926988 | PIK3C2A | FALSE | Downregulated |
| 0,698599212 | 0,360926988 | TTK | FALSE | Upregulated |
| 0,182622937 | 0,361609591 | PARVA | FALSE | Upregulated |
| 0,257660297 | 0,361609591 | WDR24 | FALSE | Upregulated |

| | | | | |
|---|---|---|---|---|
| -0,375381978 | 0,36172363 | CDKL3 | FALSE | Downregulated |
| -0,426424694 | 0,361928821 | ATP6AP1L | FALSE | Downregulated |
| 0,309175623 | 0,361928821 | CCND2 | FALSE | Upregulated |
| 0,724677604 | 0,361928821 | DLGAP5 | FALSE | Upregulated |
| 0,366434728 | 0,361928821 | HAVCR2 | FALSE | Upregulated |
| 1,335069051 | 0,361928821 | LINC00842 | FALSE | Upregulated |
| -0,181913806 | 0,361928821 | PRDX1 | FALSE | Downregulated |
| 0,22850271 | 0,361928821 | RUNDC1 | FALSE | Upregulated |
| 0,536395701 | 0,361928821 | SPECC1 | FALSE | Upregulated |
| 0,418893975 | 0,362130838 | CYP11A1 | FALSE | Upregulated |
| 0,283995044 | 0,362283544 | C15orf39 | FALSE | Upregulated |
| -0,337427193 | 0,362479562 | KLB | FALSE | Downregulated |
| -0,139074509 | 0,362838811 | NEDD8 | FALSE | Downregulated |
| 0,203745321 | 0,363036816 | MAPRE1 | FALSE | Upregulated |
| -0,171339155 | 0,363372026 | NMD3 | FALSE | Downregulated |
| -0,17521071 | 0,363372026 | PDCD5 | FALSE | Downregulated |
| 0,368779571 | 0,363372026 | RP11-395P17.3 | FALSE | Upregulated |
| -0,230724882 | 0,363372026 | TMEM208 | FALSE | Downregulated |
| 1,116550516 | 0,363475174 | CTHRC1 | FALSE | Upregulated |
| -0,170880895 | 0,363475174 | RPS23 | FALSE | Downregulated |
| -0,122743705 | 0,363801159 | ADIPOR1 | FALSE | Downregulated |
| -0,139948044 | 0,363801159 | EXOSC2 | FALSE | Downregulated |
| 0,167548102 | 0,363801159 | GNS | FALSE | Upregulated |
| -0,134801587 | 0,363801159 | HELQ | FALSE | Downregulated |
| 0,337781765 | 0,363801159 | HMHA1 | FALSE | Upregulated |
| -1,1567651 | 0,363801159 | LOC646903 | FALSE | Downregulated |

| | | | | |
|---|---|---|---|---|
| 0,674063638 | 0,363801159 | LPAR2 | FALSE | Upregulated |
| 0,281044896 | 0,363801159 | MROH1 | FALSE | Upregulated |
| -0,259392872 | 0,363801159 | PAXIP1-AS2 | FALSE | Downregulated |
| 0,121923589 | 0,363801159 | PTBP1 | FALSE | Upregulated |
| -0,177018736 | 0,363801159 | RPS7 | FALSE | Downregulated |
| -0,174549617 | 0,363801159 | SLK | FALSE | Downregulated |
| -0,200657034 | 0,363801159 | THRAP3 | FALSE | Downregulated |
| 0,738456018 | 0,363801793 | ZBP1 | FALSE | Upregulated |
| -0,679466576 | 0,363944938 | BRSK1 | FALSE | Downregulated |
| 0,205261576 | 0,363944938 | CABIN1 | FALSE | Upregulated |
| -0,776075124 | 0,363944938 | DGKB | FALSE | Downregulated |
| 1,577572113 | 0,363944938 | LIMS3-LOC440895 | FALSE | Upregulated |
| -0,829989995 | 0,363944938 | LOC100506621 | FALSE | Downregulated |
| 1,349287564 | 0,363944938 | LOC101927931 | FALSE | Upregulated |
| 0,386430026 | 0,363944938 | MATK | FALSE | Upregulated |
| -1,536985173 | 0,363944938 | PKIA | FALSE | Downregulated |
| 0,171472625 | 0,363944938 | RETSAT | FALSE | Upregulated |
| -0,565685125 | 0,363944938 | RND2 | FALSE | Downregulated |
| -0,187442087 | 0,363944938 | ZEB1 | FALSE | Downregulated |
| -0,480991495 | 0,364573336 | ATP1B2 | FALSE | Downregulated |
| -0,178278032 | 0,364573336 | CCDC174 | FALSE | Downregulated |
| -0,999977806 | 0,364573336 | LOC101928264 | FALSE | Downregulated |
| 1,062538884 | 0,364573336 | PDCD1 | FALSE | Upregulated |
| -0,266187722 | 0,364573336 | SRI | FALSE | Downregulated |
| 1,130450604 | 0,364573336 | TSPAN10 | FALSE | Upregulated |
| -0,510712772 | 0,36477731 | JAKMIP2 | FALSE | Downregulated |

| | | | | |
|---|---|---|---|---|
| -0,186689444 | 0,36525058 | C11orf74 | FALSE | Downregulated |
| -0,161701728 | 0,36525058 | C17orf85 | FALSE | Downregulated |
| 0,425848468 | 0,36525058 | CCDC88B | FALSE | Upregulated |
| 0,368546691 | 0,36525058 | CD74 | FALSE | Upregulated |
| -0,172462615 | 0,36525058 | EEF1D | FALSE | Downregulated |
| 0,176565648 | 0,36525058 | ISOC1 | FALSE | Upregulated |
| 0,2190879 | 0,36525058 | ZNF512B | FALSE | Upregulated |
| 0,564375256 | 0,365509149 | LINC00528 | FALSE | Upregulated |
| 0,301064501 | 0,365916732 | ZNF500 | FALSE | Upregulated |
| 0,754708536 | 0,365938041 | ADTRP | FALSE | Upregulated |
| 0,449644195 | 0,365938041 | ARHGAP11A | FALSE | Upregulated |
| -0,408996801 | 0,365938041 | ARHGEF35 | FALSE | Downregulated |
| -0,200956585 | 0,365938041 | COMMD6 | FALSE | Downregulated |
| -0,176693826 | 0,365938041 | CRTC2 | FALSE | Downregulated |
| 0,631418573 | 0,365938041 | DOPEY2 | FALSE | Upregulated |
| 1,574259821 | 0,365938041 | GABBR2 | FALSE | Upregulated |
| 0,211190972 | 0,365938041 | GAPDH | FALSE | Upregulated |
| -0,186722591 | 0,365938041 | GDI2 | FALSE | Downregulated |
| 0,64400548 | 0,365938041 | HES4 | FALSE | Upregulated |
| 1,884477849 | 0,365938041 | LINC00313 | FALSE | Upregulated |
| -0,315958503 | 0,365938041 | LOC100506730 | FALSE | Downregulated |
| -0,597071677 | 0,365938041 | LOC642236 | FALSE | Downregulated |
| -0,521766822 | 0,365938041 | MT2A | FALSE | Downregulated |
| 1,031354616 | 0,365938041 | OR2A7 | FALSE | Upregulated |
| 0,152522949 | 0,365938041 | POLR3A | FALSE | Upregulated |
| 0,490697222 | 0,365938041 | RELT | FALSE | Upregulated |

| | | | | |
|---|---|---|---|---|
| -0,213406282 | 0,365938041 | SASH1 | FALSE | Downregulated |
| 0,408996332 | 0,365938041 | SELM | FALSE | Upregulated |
| -0,272239543 | 0,365938041 | SLC39A6 | FALSE | Downregulated |
| -0,278958597 | 0,365938041 | UGDH | FALSE | Downregulated |
| 0,198013616 | 0,366267202 | A1BG | FALSE | Upregulated |
| -0,203399813 | 0,366267202 | CLIC4 | FALSE | Downregulated |
| 0,792263665 | 0,366267202 | ITGA2 | FALSE | Upregulated |
| 0,660648033 | 0,366267202 | KIF4A | FALSE | Upregulated |
| -0,144606785 | 0,366267202 | N4BP2L2 | FALSE | Downregulated |
| -0,109001603 | 0,366267202 | PSMC1 | FALSE | Downregulated |
| 0,257766197 | 0,366267202 | SARM1 | FALSE | Upregulated |
| 0,166361573 | 0,36673208 | C2CD3 | FALSE | Upregulated |
| 1,218960884 | 0,36673208 | DOK7 | FALSE | Upregulated |
| 0,741741762 | 0,36673208 | EGR2 | FALSE | Upregulated |
| -1,197227218 | 0,36673208 | LOC101927044 | FALSE | Downregulated |
| -1,465987353 | 0,36673208 | RGMA | FALSE | Downregulated |
| 0,508110995 | 0,36673208 | TK1 | FALSE | Upregulated |
| 0,199209521 | 0,367004987 | PPP1R12C | FALSE | Upregulated |
| -0,155261254 | 0,367825986 | MED20 | FALSE | Downregulated |
| 0,377743821 | 0,368492635 | E2F5 | FALSE | Upregulated |
| 0,450273851 | 0,368492635 | KCNC3 | FALSE | Upregulated |
| 0,134309158 | 0,368492635 | RNF4 | FALSE | Upregulated |
| -0,118036668 | 0,368492635 | UBE2K | FALSE | Downregulated |
| -0,151317881 | 0,368595616 | C2CD5 | FALSE | Downregulated |
| 0,257057201 | 0,368963095 | CCDC130 | FALSE | Upregulated |
| -0,208883458 | 0,368963095 | HDAC8 | FALSE | Downregulated |

| | | | | |
|---|---|---|---|---|
| 0,257375132 | 0,368963095 | RASSF3 | FALSE | Upregulated |
| 0,112732675 | 0,368963095 | SART1 | FALSE | Upregulated |
| 0,322221641 | 0,369294688 | FLYWCH2 | FALSE | Upregulated |
| 0,348380087 | 0,369597985 | CAPN2 | FALSE | Upregulated |
| -0,131924128 | 0,369597985 | CHMP3 | FALSE | Downregulated |
| 0,592548019 | 0,369597985 | FAM169A | FALSE | Upregulated |
| -0,727197597 | 0,369597985 | NDNF | FALSE | Downregulated |
| -0,156670806 | 0,369597985 | OARD1 | FALSE | Downregulated |
| -0,189275998 | 0,369730725 | CPB2 | FALSE | Downregulated |
| 0,377479244 | 0,370127682 | ARSB | FALSE | Upregulated |
| -0,291879296 | 0,370127682 | DNAJC12 | FALSE | Downregulated |
| -0,953203888 | 0,370127682 | LOC101928281 | FALSE | Downregulated |
| -0,489858609 | 0,370127682 | LOC102723666 | FALSE | Downregulated |
| -0,18769267 | 0,370127682 | PIK3C2G | FALSE | Downregulated |
| 0,323800407 | 0,370127682 | PLTP | FALSE | Upregulated |
| -0,103625603 | 0,370127682 | RAB11A | FALSE | Downregulated |
| 0,231660907 | 0,370127682 | TMEM9 | FALSE | Upregulated |
| 0,146055497 | 0,370127682 | TRAPPC3 | FALSE | Upregulated |
| 0,330169552 | 0,370127682 | ZSCAN5A | FALSE | Upregulated |
| -0,216856003 | 0,370347509 | SPAG7 | FALSE | Downregulated |
| 0,937834687 | 0,370543522 | COL7A1 | FALSE | Upregulated |
| -0,301152788 | 0,370543522 | TAMM41 | FALSE | Downregulated |
| 0,763035934 | 0,371539523 | GLI2 | FALSE | Upregulated |
| -1,143424927 | 0,371539523 | IL5RA | FALSE | Downregulated |
| 0,555571387 | 0,371539523 | NCAPH | FALSE | Upregulated |
| 0,352936345 | 0,371777987 | HLA-DRA | FALSE | Upregulated |

| | | | | |
|---|---|---|---|---|
| -0,097586049 | 0,371777987 | RBM17 | FALSE | Downregulated |
| 0,134856692 | 0,371963465 | ENOPH1 | FALSE | Upregulated |
| -0,217803684 | 0,372316148 | C9orf41 | FALSE | Downregulated |
| -0,232895743 | 0,372316148 | FBXW7 | FALSE | Downregulated |
| 0,247860475 | 0,372379173 | LRFN3 | FALSE | Upregulated |
| -0,171107477 | 0,372379173 | NUDT5 | FALSE | Downregulated |
| 0,239390775 | 0,372379173 | TIRAP | FALSE | Upregulated |
| -0,457387225 | 0,372510867 | P4HA3 | FALSE | Downregulated |
| -0,255142301 | 0,372510867 | ZCWPW1 | FALSE | Downregulated |
| 0,178808331 | 0,372547194 | FBXO31 | FALSE | Upregulated |
| 0,310197145 | 0,372547194 | HLA-A | FALSE | Upregulated |
| 0,750129603 | 0,372547194 | LOC101928291 | FALSE | Upregulated |
| -0,277360153 | 0,372547194 | MOSPD3 | FALSE | Downregulated |
| 0,219975561 | 0,372564689 | CLN6 | FALSE | Upregulated |
| 0,510152288 | 0,372564689 | RASGRP1 | FALSE | Upregulated |
| 0,169877626 | 0,37275945 | ZFP36L2 | FALSE | Upregulated |
| -0,537800401 | 0,372854858 | GPC5 | FALSE | Downregulated |
| -0,244092051 | 0,372878145 | C19orf66 | FALSE | Downregulated |
| -0,26015507 | 0,372878145 | CHORDC1 | FALSE | Downregulated |
| -1,591706628 | 0,372878145 | FLNC | FALSE | Downregulated |
| -2,602355986 | 0,372878145 | KLHL40 | FALSE | Downregulated |
| 0,211919151 | 0,372878145 | RGP1 | FALSE | Upregulated |
| 0,12245363 | 0,37300222 | C10orf76 | FALSE | Upregulated |
| -0,288963643 | 0,373473766 | RBM47 | FALSE | Downregulated |
| -0,147042929 | 0,373894684 | ATP13A3 | FALSE | Downregulated |
| 0,655476534 | 0,373894684 | CDHR1 | FALSE | Upregulated |

| | | | | |
|---|---|---|---|---|
| 1,300997158 | 0,373894684 | LOC101926943 | FALSE | Upregulated |
| 0,236251868 | 0,373894684 | MR1 | FALSE | Upregulated |
| -0,173674136 | 0,374286626 | FAM185A | FALSE | Downregulated |
| 1,410860198 | 0,37437378 | DPP10-AS1 | FALSE | Upregulated |
| -0,72720553 | 0,37437378 | KCNJ12 | FALSE | Downregulated |
| -0,320156124 | 0,374464107 | CHCHD10 | FALSE | Downregulated |
| -0,420645237 | 0,374464107 | NAP1L2 | FALSE | Downregulated |
| 1,215011186 | 0,374464107 | NCAN | FALSE | Upregulated |
| 0,157235374 | 0,374464107 | NUPL2 | FALSE | Upregulated |
| -0,176140366 | 0,374727816 | APPL1 | FALSE | Downregulated |
| -1,781579873 | 0,374747058 | CASQ2 | FALSE | Downregulated |
| 1,375172051 | 0,374747058 | SLC45A1 | FALSE | Upregulated |
| -0,22628452 | 0,374762566 | ZFP14 | FALSE | Downregulated |
| -0,298209514 | 0,374825736 | ANP32B | FALSE | Downregulated |
| 1,37477979 | 0,374825736 | CCL3L3 | FALSE | Upregulated |
| 1,277351692 | 0,374825736 | GPRIN2 | FALSE | Upregulated |
| 0,317673083 | 0,374825736 | LOC100506518 | FALSE | Upregulated |
| 0,433606229 | 0,374825736 | LOC101927160 | FALSE | Upregulated |
| 0,159148393 | 0,374825736 | PLEKHM3 | FALSE | Upregulated |
| -0,225784574 | 0,374825736 | PON1 | FALSE | Downregulated |
| -0,117548465 | 0,374825736 | PPCS | FALSE | Downregulated |
| 0,335951908 | 0,374825736 | PXN-AS1 | FALSE | Upregulated |
| 0,187579413 | 0,374825736 | SOCS4 | FALSE | Upregulated |
| -0,140091308 | 0,374825736 | STK3 | FALSE | Downregulated |
| -0,652820655 | 0,374825736 | TRAPPC3L | FALSE | Downregulated |
| -0,186447326 | 0,374825736 | ZC3H8 | FALSE | Downregulated |

| | | | | |
|---|---|---|---|---|
| -0,135990028 | 0,374825736 | ZNF22 | FALSE | Downregulated |
| 0,941661437 | 0,375050545 | LOC101927792 | FALSE | Upregulated |
| 0,205655301 | 0,375050545 | UGGT1 | FALSE | Upregulated |
| 0,861570264 | 0,375868764 | SLIT1 | FALSE | Upregulated |
| 0,119347196 | 0,376372343 | TFIP11 | FALSE | Upregulated |
| 0,171325255 | 0,376603444 | NOMO1 | FALSE | Upregulated |
| -0,410316622 | 0,376603444 | SHISA4 | FALSE | Downregulated |
| -0,242560872 | 0,376780266 | PABPN1 | FALSE | Downregulated |
| 0,636959816 | 0,377164113 | GUSBP2 | FALSE | Upregulated |
| -0,253575433 | 0,377164113 | UACA | FALSE | Downregulated |
| 0,332575187 | 0,377164113 | WFS1 | FALSE | Upregulated |
| 0,596428328 | 0,377837457 | NYNRIN | FALSE | Upregulated |
| 0,385326443 | 0,378155632 | PRKAG2-AS1 | FALSE | Upregulated |
| 0,328605738 | 0,378629394 | LAMB3 | FALSE | Upregulated |
| -0,35976509 | 0,378629394 | LOC102724250 | FALSE | Downregulated |
| -0,227069882 | 0,378629394 | MRPS33 | FALSE | Downregulated |
| -0,143185904 | 0,378870711 | PINK1 | FALSE | Downregulated |
| -0,102303677 | 0,378870711 | RSL1D1 | FALSE | Downregulated |
| -0,357180565 | 0,379244651 | CYP2C8 | FALSE | Downregulated |
| 0,213570277 | 0,379244651 | RBM15 | FALSE | Upregulated |
| -0,123617471 | 0,379255354 | CSNK1G3 | FALSE | Downregulated |
| 0,305929233 | 0,379470584 | ADCY3 | FALSE | Upregulated |
| 0,172268848 | 0,379470584 | AKTIP | FALSE | Upregulated |
| 0,177243598 | 0,379470584 | BRF1 | FALSE | Upregulated |
| -0,157540992 | 0,379470584 | CREBBP | FALSE | Downregulated |
| 0,211945154 | 0,379470584 | DDOST | FALSE | Upregulated |

| | | | | |
|---|---|---|---|---|
| 0,430245371 | 0,379470584 | ENTPD1 | FALSE | Upregulated |
| 0,282251117 | 0,379470584 | ETV5 | FALSE | Upregulated |
| 0,569739578 | 0,379470584 | F3 | FALSE | Upregulated |
| 0,920652692 | 0,379470584 | FAM227A | FALSE | Upregulated |
| 0,541303438 | 0,379470584 | FAT1 | FALSE | Upregulated |
| -0,167441376 | 0,379470584 | GTF2F2 | FALSE | Downregulated |
| 0,965663921 | 0,379470584 | IQGAP3 | FALSE | Upregulated |
| -0,172930187 | 0,379470584 | ISY1 | FALSE | Downregulated |
| 0,281419605 | 0,379470584 | ITGAL | FALSE | Upregulated |
| 1,210558189 | 0,379470584 | KSR2 | FALSE | Upregulated |
| -0,130673963 | 0,379470584 | LANCL1 | FALSE | Downregulated |
| -0,685587554 | 0,379470584 | LOC101927284 | FALSE | Downregulated |
| -0,440857669 | 0,379470584 | LOC653160 | FALSE | Downregulated |
| -1,073139722 | 0,379470584 | LRRC39 | FALSE | Downregulated |
| -0,232296045 | 0,379470584 | MOCS2 | FALSE | Downregulated |
| -0,135507644 | 0,379470584 | MRPL49 | FALSE | Downregulated |
| -0,24018931 | 0,379470584 | NME4 | FALSE | Downregulated |
| 0,385703974 | 0,379470584 | PLIN2 | FALSE | Upregulated |
| -0,200274963 | 0,379470584 | PPP3CA | FALSE | Downregulated |
| -0,138954148 | 0,379470584 | PTER | FALSE | Downregulated |
| 0,147771945 | 0,379470584 | PXN | FALSE | Upregulated |
| -0,229683636 | 0,379470584 | RAB30-AS1 | FALSE | Downregulated |
| 0,790745395 | 0,379470584 | SPIB | FALSE | Upregulated |
| 1,339945395 | 0,379470584 | TFAP2E | FALSE | Upregulated |
| -0,218451138 | 0,379470584 | THOC2 | FALSE | Downregulated |
| 0,241999646 | 0,379470584 | TMBIM1 | FALSE | Upregulated |

| | | | | |
|---|---|---|---|---|
| 0,818825751 | 0,379470584 | TMIGD2 | FALSE | Upregulated |
| -0,161783504 | 0,379470584 | TSHZ1 | FALSE | Downregulated |
| 0,259152776 | 0,379470584 | WDR60 | FALSE | Upregulated |
| -0,166214059 | 0,379470584 | ZADH2 | FALSE | Downregulated |
| -0,138435224 | 0,379470584 | ZNF143 | FALSE | Downregulated |
| 0,215220809 | 0,379470584 | ZNF329 | FALSE | Upregulated |
| 1,207048559 | 0,380020375 | LOC100131257 | FALSE | Upregulated |
| 0,568507164 | 0,380141834 | MTRNR2L3 | FALSE | Upregulated |
| -0,156503675 | 0,380141834 | RBM8A | FALSE | Downregulated |
| 0,128627397 | 0,380141834 | VPS11 | FALSE | Upregulated |
| 0,123979176 | 0,380411974 | CLTA | FALSE | Upregulated |
| 0,319539652 | 0,380411974 | DOK2 | FALSE | Upregulated |
| -1,0917822 | 0,38053177 | FAM83C | FALSE | Downregulated |
| -0,20951391 | 0,380632645 | GNPAT | FALSE | Downregulated |
| -0,534682069 | 0,380632645 | LRP2BP | FALSE | Downregulated |
| -0,164146921 | 0,380632645 | SLC25A43 | FALSE | Downregulated |
| 0,20684303 | 0,380722921 | DDX39A | FALSE | Upregulated |
| -0,199406239 | 0,380722921 | RPL19 | FALSE | Downregulated |
| 0,198091922 | 0,380722921 | VASP | FALSE | Upregulated |
| -0,20648522 | 0,381344709 | APOC1 | FALSE | Downregulated |
| 0,344461056 | 0,381344709 | ODF3B | FALSE | Upregulated |
| 0,770182467 | 0,381345597 | SHC3 | FALSE | Upregulated |
| 0,230228616 | 0,381398915 | SLC12A4 | FALSE | Upregulated |
| -0,265804393 | 0,381398915 | SNRNP27 | FALSE | Downregulated |
| -0,129273059 | 0,381890079 | YTHDC1 | FALSE | Downregulated |
| 0,32670941 | 0,382210613 | DOCK2 | FALSE | Upregulated |

| | | | | |
|---|---|---|---|---|
| -0,659252554 | 0,382210613 | USP6 | FALSE | Downregulated |
| -0,391140967 | 0,382216744 | CD82 | FALSE | Downregulated |
| 0,322315686 | 0,382216744 | ITPRIP | FALSE | Upregulated |
| 0,366238033 | 0,382283238 | SPN | FALSE | Upregulated |
| 0,149606907 | 0,382583418 | AP2A1 | FALSE | Upregulated |
| 0,139796613 | 0,382583418 | ARHGEF11 | FALSE | Upregulated |
| -0,13036495 | 0,382583418 | PROSC | FALSE | Downregulated |
| 0,246407797 | 0,382767361 | DGKQ | FALSE | Upregulated |
| -0,160815782 | 0,383218168 | VTI1B | FALSE | Downregulated |
| 0,284924735 | 0,383279864 | TMEM234 | FALSE | Upregulated |
| 0,342780883 | 0,383311672 | CEP72 | FALSE | Upregulated |
| 0,856041597 | 0,383311672 | LOC102723311 | FALSE | Upregulated |
| -0,150406893 | 0,383452739 | CASC4 | FALSE | Downregulated |
| 0,838117202 | 0,383452739 | CPNE5 | FALSE | Upregulated |
| -0,516375762 | 0,383452739 | CYP1A2 | FALSE | Downregulated |
| -0,763387018 | 0,383452739 | FFAR2 | FALSE | Downregulated |
| -0,287643043 | 0,383452739 | FKBP2 | FALSE | Downregulated |
| -0,506401757 | 0,383452739 | HBEGF | FALSE | Downregulated |
| -0,471065565 | 0,383452739 | IGF1 | FALSE | Downregulated |
| -0,257035279 | 0,383452739 | MAP4K4 | FALSE | Downregulated |
| 0,456062262 | 0,383452739 | NFE2L3 | FALSE | Upregulated |
| 0,144792476 | 0,383452739 | OSER1 | FALSE | Upregulated |
| -0,26258664 | 0,383452739 | PLA2G16 | FALSE | Downregulated |
| -0,388856942 | 0,383452739 | RP11-119F19.2 | FALSE | Downregulated |
| -0,671322571 | 0,383452739 | RUSC1-AS1 | FALSE | Downregulated |
| 0,332116621 | 0,383452739 | SEMA4D | FALSE | Upregulated |

| | | | | |
|---|---|---|---|---|
| 0,128690688 | 0,383452739 | SH3BP5L | FALSE | Upregulated |
| 0,367240876 | 0,383452739 | SPATA24 | FALSE | Upregulated |
| 1,145604328 | 0,383452739 | TKTL1 | FALSE | Upregulated |
| -0,17156975 | 0,384485064 | ALDH8A1 | FALSE | Downregulated |
| -0,332824891 | 0,384485064 | ANXA10 | FALSE | Downregulated |
| 0,447912201 | 0,384485064 | BAX | FALSE | Upregulated |
| -0,15125555 | 0,384485064 | BCAP29 | FALSE | Downregulated |
| 0,45996702 | 0,384485064 | CDC42BPG | FALSE | Upregulated |
| 1,050030019 | 0,384485064 | CDKN2A | FALSE | Upregulated |
| 0,383042105 | 0,384485064 | EVI2A | FALSE | Upregulated |
| 0,438619366 | 0,384485064 | FAM83G | FALSE | Upregulated |
| -0,176089733 | 0,384485064 | FBXL12 | FALSE | Downregulated |
| -0,169046351 | 0,384485064 | HMGCL | FALSE | Downregulated |
| -1,18226903 | 0,384485064 | LOC100996808 | FALSE | Downregulated |
| 0,228127655 | 0,384485064 | LRRC14 | FALSE | Upregulated |
| 0,689072623 | 0,384485064 | MSH5-SAPCD1 | FALSE | Upregulated |
| -0,147140368 | 0,384485064 | NOL11 | FALSE | Downregulated |
| -0,238911056 | 0,384485064 | PTMS | FALSE | Downregulated |
| 0,132617732 | 0,384485064 | SYMPK | FALSE | Upregulated |
| 0,328297219 | 0,384539604 | ARHGEF3 | FALSE | Upregulated |
| 0,324833359 | 0,384539604 | MAFIP | FALSE | Upregulated |
| -0,613223805 | 0,384539604 | MIR4461 | FALSE | Downregulated |
| 1,846329009 | 0,384539604 | ZBED2 | FALSE | Upregulated |
| -0,451426953 | 0,384875399 | GSTM4 | FALSE | Downregulated |
| 0,184889066 | 0,385093592 | CCDC71 | FALSE | Upregulated |
| -0,249870405 | 0,385093592 | KLF7 | FALSE | Downregulated |

| | | | | |
|---|---|---|---|---|
| 0,566405438 | 0,385093592 | RARRES1 | FALSE | Upregulated |
| -0,221359251 | 0,385242483 | POLR2J | FALSE | Downregulated |
| -0,122958473 | 0,385410966 | C14orf166 | FALSE | Downregulated |
| -0,266202372 | 0,385410966 | CBWD5 | FALSE | Downregulated |
| 0,249109681 | 0,385410966 | COTL1 | FALSE | Upregulated |
| 0,761623976 | 0,385410966 | KIF26B | FALSE | Upregulated |
| -0,151397559 | 0,385410966 | OSBPL9 | FALSE | Downregulated |
| 1,090931595 | 0,385410966 | SCN3A | FALSE | Upregulated |
| 0,387309389 | 0,385410966 | SLC6A6 | FALSE | Upregulated |
| -0,193045071 | 0,385410966 | TAF15 | FALSE | Downregulated |
| -0,257005858 | 0,385410966 | TMEM150C | FALSE | Downregulated |
| -0,17606928 | 0,385410966 | ZNF420 | FALSE | Downregulated |
| -0,153049242 | 0,385482125 | NUDT6 | FALSE | Downregulated |
| 0,250778521 | 0,38605081 | FLYWCH1 | FALSE | Upregulated |
| 0,293012201 | 0,38605081 | RASA3 | FALSE | Upregulated |
| -0,172722836 | 0,38605081 | RPS20 | FALSE | Downregulated |
| 0,349976731 | 0,386434164 | APOBEC3C | FALSE | Upregulated |
| -0,211353637 | 0,386434164 | DDIT3 | FALSE | Downregulated |
| 0,558827429 | 0,386434164 | EPHB2 | FALSE | Upregulated |
| 0,204537247 | 0,386434164 | GRK6 | FALSE | Upregulated |
| -0,209016052 | 0,386471665 | ALB | FALSE | Downregulated |
| 0,987934545 | 0,386471665 | C1orf106 | FALSE | Upregulated |
| 0,217262428 | 0,386471665 | CYB5R4 | FALSE | Upregulated |
| -0,216898646 | 0,386471665 | DBI | FALSE | Downregulated |
| 0,244530116 | 0,386471665 | DSN1 | FALSE | Upregulated |
| -0,195855623 | 0,386471665 | DTX3L | FALSE | Downregulated |

| | | | | |
|---|---|---|---|---|
| 0,538411598 | 0,386471665 | EGLN3 | FALSE | Upregulated |
| 0,161151876 | 0,386471665 | LIG3 | FALSE | Upregulated |
| -1,163874209 | 0,386471665 | LINC00488 | FALSE | Downregulated |
| 0,57071682 | 0,386471665 | LOC101928429 | FALSE | Upregulated |
| -1,158190321 | 0,386471665 | MAPK4 | FALSE | Downregulated |
| 0,354409987 | 0,386471665 | NLRC3 | FALSE | Upregulated |
| 0,333340802 | 0,386471665 | OSBPL5 | FALSE | Upregulated |
| 0,366280762 | 0,386471665 | PLEKHF1 | FALSE | Upregulated |
| -0,181759007 | 0,386471665 | PRNP | FALSE | Downregulated |
| -0,139923797 | 0,386471665 | PSMA6 | FALSE | Downregulated |
| 0,582512581 | 0,386471665 | PTGDS | FALSE | Upregulated |
| -0,26163333 | 0,386471665 | RBBP6 | FALSE | Downregulated |
| -0,198715391 | 0,386471665 | SETDB2 | FALSE | Downregulated |
| -0,42443133 | 0,386471665 | SMCO3 | FALSE | Downregulated |
| 0,361735356 | 0,386471665 | TRIM22 | FALSE | Upregulated |
| -0,507370123 | 0,386471665 | TSPAN8 | FALSE | Downregulated |
| -0,156749103 | 0,386471665 | UBXN1 | FALSE | Downregulated |
| -0,129712169 | 0,386471665 | VMA21 | FALSE | Downregulated |
| 0,246581069 | 0,386513162 | KLHL26 | FALSE | Upregulated |
| 0,482154236 | 0,386513162 | LRRC4B | FALSE | Upregulated |
| 1,02049962 | 0,38662794 | HPN-AS1 | FALSE | Upregulated |
| 0,240298776 | 0,387517512 | SERPINA7 | FALSE | Upregulated |
| 0,229798845 | 0,387584071 | NOMO2 | FALSE | Upregulated |
| 0,303984558 | 0,38777807 | COL6A2 | FALSE | Upregulated |
| -0,205692483 | 0,38777807 | ZNF445 | FALSE | Downregulated |
| -0,552661797 | 0,388465129 | C14orf182 | FALSE | Downregulated |

| | | | | |
|---|---|---|---|---|
| 0,270816224 | 0,388465129 | PREX1 | FALSE | Upregulated |
| 1,041560888 | 0,388537538 | CPNE7 | FALSE | Upregulated |
| 1,046879294 | 0,388537538 | E2F7 | FALSE | Upregulated |
| -0,655885565 | 0,388537538 | EN1 | FALSE | Downregulated |
| 0,503017123 | 0,388537538 | FABP5 | FALSE | Upregulated |
| -1,105317352 | 0,388537538 | KCNC2 | FALSE | Downregulated |
| -0,675942341 | 0,388537538 | LOC102723602 | FALSE | Downregulated |
| -0,180087855 | 0,388537538 | MRPL13 | FALSE | Downregulated |
| -0,284326463 | 0,388537538 | MTURN | FALSE | Downregulated |
| 1,074711264 | 0,388537538 | SLC26A10 | FALSE | Upregulated |
| 0,222298773 | 0,388537538 | SLC39A13 | FALSE | Upregulated |
| -0,169077158 | 0,388537538 | U2SURP | FALSE | Downregulated |
| -0,227394869 | 0,388688197 | CITED2 | FALSE | Downregulated |
| 0,849952731 | 0,388802847 | DNAJC6 | FALSE | Upregulated |
| -0,135944507 | 0,388802847 | FAM114A2 | FALSE | Downregulated |
| -0,13176627 | 0,388802847 | FAM168A | FALSE | Downregulated |
| -0,163527151 | 0,388802847 | LRIF1 | FALSE | Downregulated |
| 0,463625058 | 0,388802847 | MCM2 | FALSE | Upregulated |
| 0,315165117 | 0,388802847 | MCM4 | FALSE | Upregulated |
| -0,346999425 | 0,388802847 | MTERFD2 | FALSE | Downregulated |
| -0,122380053 | 0,388802847 | PRPF31 | FALSE | Downregulated |
| -0,533975135 | 0,388937919 | BRCA1 | FALSE | Downregulated |
| 0,205012761 | 0,389089047 | ENG | FALSE | Upregulated |
| 0,900402128 | 0,389089047 | GDF15 | FALSE | Upregulated |
| -0,527073234 | 0,389089047 | LOC101927287 | FALSE | Downregulated |
| 1,417767428 | 0,389089047 | MTRNR2L5 | FALSE | Upregulated |

| | | | | |
|---|---|---|---|---|
| -0,790149093 | 0,389089047 | NEGR1 | FALSE | Downregulated |
| -0,99384941 | 0,389089047 | PHOSPHO1 | FALSE | Downregulated |
| -0,174109688 | 0,389089047 | RMDN1 | FALSE | Downregulated |
| 0,316716836 | 0,389089047 | TTC7A | FALSE | Upregulated |
| 0,137966458 | 0,389089047 | UQCRFS1 | FALSE | Upregulated |
| 0,172171755 | 0,389089047 | VPS51 | FALSE | Upregulated |
| -0,188835229 | 0,389089047 | ZAK | FALSE | Downregulated |
| -0,217513888 | 0,389175702 | ATP5J | FALSE | Downregulated |
| 0,457660749 | 0,389175702 | CLEC12A | FALSE | Upregulated |
| 0,909734688 | 0,389175702 | LINC00925 | FALSE | Upregulated |
| 0,337012534 | 0,389175702 | STK17B | FALSE | Upregulated |
| -0,141487959 | 0,389260405 | NAE1 | FALSE | Downregulated |
| 0,300278332 | 0,389260405 | SYK | FALSE | Upregulated |
| -0,216525713 | 0,389260405 | TMEM170B | FALSE | Downregulated |
| 0,745993569 | 0,389401224 | LOC729887 | FALSE | Upregulated |
| -0,308928474 | 0,389401224 | RP9 | FALSE | Downregulated |
| 0,185388606 | 0,389641969 | RILP | FALSE | Upregulated |
| 0,495864438 | 0,389998825 | C15orf65 | FALSE | Upregulated |
| 0,414112119 | 0,390070621 | LRRC8E | FALSE | Upregulated |
| -0,25073172 | 0,39043488 | ZMYM4 | FALSE | Downregulated |
| 0,972071721 | 0,390455849 | DOC2A | FALSE | Upregulated |
| -0,251424665 | 0,390455849 | GDAP1 | FALSE | Downregulated |
| 0,973307114 | 0,390455849 | TFF3 | FALSE | Upregulated |
| 0,140555757 | 0,390730576 | NECAP1 | FALSE | Upregulated |
| 0,397835699 | 0,390730576 | ZNF285 | FALSE | Upregulated |
| 0,124835882 | 0,390998317 | COG4 | FALSE | Upregulated |

| | | | | |
|---|---|---|---|---|
| -0,300597908 | 0,390998317 | PTPRD | FALSE | Downregulated |
| 0,157888501 | 0,390998317 | VAT1 | FALSE | Upregulated |
| -0,142856694 | 0,391262276 | ARF5 | FALSE | Downregulated |
| -0,586248435 | 0,391262276 | HSD17B14 | FALSE | Downregulated |
| 0,856351852 | 0,391262276 | LOC101928576 | FALSE | Upregulated |
| -0,299050632 | 0,391544015 | RP11-290F20.1 | FALSE | Downregulated |
| 0,086357381 | 0,392087729 | NUDCD3 | FALSE | Upregulated |
| -1,268056884 | 0,392529411 | BMP8A | FALSE | Downregulated |
| -0,35609484 | 0,393075762 | ACAT2 | FALSE | Downregulated |
| 0,503893619 | 0,393075762 | APCDD1 | FALSE | Upregulated |
| -0,250674398 | 0,393075762 | C2orf72 | FALSE | Downregulated |
| 0,16700171 | 0,393075762 | CD63 | FALSE | Upregulated |
| 0,244674551 | 0,393075762 | CHID1 | FALSE | Upregulated |
| -0,157797515 | 0,393075762 | COX17 | FALSE | Downregulated |
| 0,138144956 | 0,393075762 | DENND1A | FALSE | Upregulated |
| -0,191028349 | 0,393075762 | FAM21C | FALSE | Downregulated |
| 0,26418539 | 0,393075762 | FBXO44 | FALSE | Upregulated |
| -0,501544758 | 0,393075762 | HOTAIRM1 | FALSE | Downregulated |
| 0,260645533 | 0,393075762 | IKBKG | FALSE | Upregulated |
| 0,185009561 | 0,393075762 | KBTBD7 | FALSE | Upregulated |
| -0,32831763 | 0,393075762 | KMT2E-AS1 | FALSE | Downregulated |
| 0,330092414 | 0,393075762 | LBH | FALSE | Upregulated |
| 0,285786214 | 0,393075762 | LINC00526 | FALSE | Upregulated |
| 0,390108786 | 0,393075762 | LPAR5 | FALSE | Upregulated |
| 1,047176401 | 0,393075762 | NKAIN2 | FALSE | Upregulated |
| 0,889842546 | 0,393075762 | NRXN2 | FALSE | Upregulated |

| | | | | |
|---|---|---|---|---|
| 0,391783141 | 0,393075762 | NUDT1 | FALSE | Upregulated |
| -0,156592219 | 0,393075762 | RIT1 | FALSE | Downregulated |
| -1,002782802 | 0,393075762 | RP11-333A23.4 | FALSE | Downregulated |
| 0,179002766 | 0,393075762 | ZSCAN25 | FALSE | Upregulated |
| 0,260484269 | 0,393079787 | ERAP1 | FALSE | Upregulated |
| 0,347269891 | 0,393079787 | LOC101928154 | FALSE | Upregulated |
| -0,140015509 | 0,393358125 | ZCCHC10 | FALSE | Downregulated |
| 0,236970665 | 0,393525741 | DUS3L | FALSE | Upregulated |
| 0,300491392 | 0,393525741 | E2F3 | FALSE | Upregulated |
| -0,367652514 | 0,394072215 | TTC6 | FALSE | Downregulated |
| 0,283323988 | 0,39427804 | ARHGEF5 | FALSE | Upregulated |
| -0,222946107 | 0,39427804 | DST | FALSE | Downregulated |
| 0,267159209 | 0,394424975 | ZBTB48 | FALSE | Upregulated |
| -0,251763113 | 0,394467294 | ZNF771 | FALSE | Downregulated |
| 0,227688164 | 0,394823081 | ATAD3A | FALSE | Upregulated |
| -0,139937911 | 0,394823081 | FBXL5 | FALSE | Downregulated |
| 0,206799553 | 0,395581201 | CXorf38 | FALSE | Upregulated |
| -0,260508609 | 0,395789426 | WDR72 | FALSE | Downregulated |
| -1,207794972 | 0,39605549 | CRYAB | FALSE | Downregulated |
| -0,255808802 | 0,396178266 | SPATA5 | FALSE | Downregulated |
| -0,222815962 | 0,396429624 | CBX3 | FALSE | Downregulated |
| -0,120787802 | 0,396429624 | KIAA0368 | FALSE | Downregulated |
| -0,193940552 | 0,396429624 | ZNF254 | FALSE | Downregulated |
| 0,115192346 | 0,396593517 | BSDC1 | FALSE | Upregulated |
| -0,328165529 | 0,396593517 | CASC10 | FALSE | Downregulated |
| -0,271402175 | 0,396593517 | TMEM133 | FALSE | Downregulated |

| | | | | |
|---|---|---|---|---|
| 0,145561414 | 0,396930168 | BCAP31 | FALSE | Upregulated |
| -0,239743769 | 0,396930168 | CIR1 | FALSE | Downregulated |
| -0,292576098 | 0,396930168 | COLEC11 | FALSE | Downregulated |
| -0,257079519 | 0,396930168 | MYCT1 | FALSE | Downregulated |
| -0,093023646 | 0,396930168 | PAIP1 | FALSE | Downregulated |
| 0,153402525 | 0,396930168 | RALB | FALSE | Upregulated |
| 0,289602804 | 0,396933746 | LOC100506161 | FALSE | Upregulated |
| 1,038774365 | 0,397537526 | ANKRD20A5P | FALSE | Upregulated |
| 1,922859967 | 0,397537526 | TRNAD17 | FALSE | Upregulated |
| 0,599118349 | 0,397772019 | BANK1 | FALSE | Upregulated |
| 0,252586135 | 0,397772019 | VASH1 | FALSE | Upregulated |
| -0,204260813 | 0,398456328 | CTNNBL1 | FALSE | Downregulated |
| 0,530829532 | 0,398456328 | EVC | FALSE | Upregulated |
| 0,608824901 | 0,398456328 | LOC101928092 | FALSE | Upregulated |
| 0,332416107 | 0,398456328 | PCED1B | FALSE | Upregulated |
| 0,198969235 | 0,398668555 | NELFA | FALSE | Upregulated |
| -0,267596297 | 0,398668555 | NPY1R | FALSE | Downregulated |
| -0,104921228 | 0,398867138 | UBE2Q1 | FALSE | Downregulated |
| -0,206140693 | 0,399240716 | C16orf87 | FALSE | Downregulated |
| 0,255359199 | 0,399304726 | DCLRE1B | FALSE | Upregulated |
| -0,312020159 | 0,399687667 | CCDC15 | FALSE | Downregulated |
| -0,824745672 | 0,399719215 | DIO3OS | FALSE | Downregulated |
| -0,150259276 | 0,399719215 | RSBN1 | FALSE | Downregulated |
| -0,833246803 | 0,399795774 | ST18 | FALSE | Downregulated |
| 1,952418565 | 0,400221688 | KRT17 | FALSE | Upregulated |
| 0,522950423 | 0,400221688 | LOC201651 | FALSE | Upregulated |

| | | | | |
|---|---|---|---|---|
| -0,623198622 | 0,400517047 | CCDC135 | FALSE | Downregulated |
| -0,181951398 | 0,400517047 | HINT1 | FALSE | Downregulated |
| -0,436491941 | 0,400517047 | PDE8B | FALSE | Downregulated |
| 0,21435669 | 0,400517047 | PEPD | FALSE | Upregulated |
| -1,100378855 | 0,400517047 | TRNT | FALSE | Downregulated |
| -0,122874873 | 0,400517047 | ZBTB44 | FALSE | Downregulated |
| 0,165403092 | 0,400930384 | SSH3 | FALSE | Upregulated |
| 0,125863918 | 0,401447643 | PIP4K2B | FALSE | Upregulated |
| 1,360456403 | 0,401921723 | LINC00900 | FALSE | Upregulated |
| 0,111763782 | 0,401921723 | RAB5C | FALSE | Upregulated |
| -0,560496423 | 0,401921723 | SYNPO2 | FALSE | Downregulated |
| 0,39529035 | 0,401921723 | TRIM15 | FALSE | Upregulated |
| -0,210212403 | 0,401921723 | TTPA | FALSE | Downregulated |
| 0,38749707 | 0,403120692 | VAMP5 | FALSE | Upregulated |
| 0,352558135 | 0,403187849 | TNFRSF10A | FALSE | Upregulated |
| 0,620412222 | 0,403272766 | EMR4P | FALSE | Upregulated |
| -0,511821371 | 0,403407291 | LINC01144 | FALSE | Downregulated |
| 0,15018923 | 0,403564471 | TM9SF1 | FALSE | Upregulated |
| 0,14120663 | 0,404175156 | SGSM3 | FALSE | Upregulated |
| -0,160453735 | 0,404748053 | HAT1 | FALSE | Downregulated |
| -0,146436529 | 0,404748053 | RPRD1A | FALSE | Downregulated |
| -0,840569628 | 0,404794376 | SYCE1 | FALSE | Downregulated |
| -0,314120169 | 0,405020333 | CIDEB | FALSE | Downregulated |
| 0,225610919 | 0,405020333 | LRP10 | FALSE | Upregulated |
| -0,338670145 | 0,40534722 | ANKRD35 | FALSE | Downregulated |
| -0,11018475 | 0,40534722 | ATPAF1 | FALSE | Downregulated |

| | | | | |
|---|---|---|---|---|
| 0,794844228 | 0,40534722 | FLJ43681 | FALSE | Upregulated |
| 0,456291507 | 0,40534722 | PTPN22 | FALSE | Upregulated |
| 0,153766099 | 0,40534722 | SHC1 | FALSE | Upregulated |
| 0,141634178 | 0,405365717 | TMEM259 | FALSE | Upregulated |
| -0,140419792 | 0,405613569 | GTF2H5 | FALSE | Downregulated |
| 0,43527471 | 0,405613569 | SNX22 | FALSE | Upregulated |
| -0,215477078 | 0,407162656 | NTN4 | FALSE | Downregulated |
| 0,648114309 | 0,407192184 | C18orf56 | FALSE | Upregulated |
| -0,245851908 | 0,407192184 | DNAJC21 | FALSE | Downregulated |
| 0,250527697 | 0,407192184 | FBXO10 | FALSE | Upregulated |
| 0,212579992 | 0,407192184 | FUCA2 | FALSE | Upregulated |
| 0,220362484 | 0,407192184 | GOSR2 | FALSE | Upregulated |
| -0,232207281 | 0,407192184 | PHF21A | FALSE | Downregulated |
| -0,468904191 | 0,407459609 | DCDC5 | FALSE | Downregulated |
| 0,175717458 | 0,407700883 | PIGQ | FALSE | Upregulated |
| 0,19695775 | 0,408044357 | ADARB1 | FALSE | Upregulated |
| 0,207996911 | 0,408044357 | ADCK2 | FALSE | Upregulated |
| -0,609118049 | 0,408044357 | CHEK2 | FALSE | Downregulated |
| 0,16947642 | 0,408044357 | COL18A1 | FALSE | Upregulated |
| 0,212291788 | 0,408044357 | COMT | FALSE | Upregulated |
| 0,159581832 | 0,408044357 | GNB2 | FALSE | Upregulated |
| -0,238181734 | 0,408044357 | KIAA1107 | FALSE | Downregulated |
| -0,518799566 | 0,408044357 | LOC102723501 | FALSE | Downregulated |
| -0,10117141 | 0,408044357 | MARCH7 | FALSE | Downregulated |
| -0,277521245 | 0,408044357 | MGC27345 | FALSE | Downregulated |
| 0,148472843 | 0,408044357 | MTA2 | FALSE | Upregulated |

| | | | | |
|---|---|---|---|---|
| -0,192040271 | 0,408044357 | MXI1 | FALSE | Downregulated |
| 0,163772794 | 0,408044357 | NCK1 | FALSE | Upregulated |
| -0,184785758 | 0,408044357 | NDUFS5 | FALSE | Downregulated |
| -1,039415078 | 0,408729769 | ZP3 | FALSE | Downregulated |
| -0,314149098 | 0,408732248 | ENDOG | FALSE | Downregulated |
| 0,300704036 | 0,408732248 | INTS9 | FALSE | Upregulated |
| -0,138724062 | 0,408732248 | PAPD5 | FALSE | Downregulated |
| -0,114906482 | 0,408732248 | TARDBP | FALSE | Downregulated |
| -0,249734986 | 0,408732248 | TMEM25 | FALSE | Downregulated |
| 0,076802624 | 0,409218076 | BAP1 | FALSE | Upregulated |
| -0,271340953 | 0,409298313 | EIF4E | FALSE | Downregulated |
| -0,222244571 | 0,409298313 | TGFBR3 | FALSE | Downregulated |
| 0,29071708 | 0,409334353 | LAPTM5 | FALSE | Upregulated |
| -0,148225769 | 0,409334353 | RPS6KA3 | FALSE | Downregulated |
| -0,163715862 | 0,409362779 | ANP32A | FALSE | Downregulated |
| -0,181406303 | 0,409362779 | BCL9 | FALSE | Downregulated |
| 0,4006028 | 0,410644653 | ZFP92 | FALSE | Upregulated |
| 1,14971245 | 0,411700965 | KRT8P41 | FALSE | Upregulated |
| -0,49301342 | 0,411700965 | PAX8 | FALSE | Downregulated |
| 0,127545802 | 0,41182518 | DBNL | FALSE | Upregulated |
| 0,114958653 | 0,411955149 | AMFR | FALSE | Upregulated |
| 0,182377483 | 0,411955149 | SF3A2 | FALSE | Upregulated |
| -0,799294207 | 0,412004469 | FAM133A | FALSE | Downregulated |
| -1,434186272 | 0,412004469 | PLN | FALSE | Downregulated |
| -0,157823815 | 0,412240061 | AIG1 | FALSE | Downregulated |
| -0,403987834 | 0,412594115 | SGCE | FALSE | Downregulated |

| | | | | |
|---|---|---|---|---|
| -0,120524198 | 0,412735754 | BTF3L4 | FALSE | Downregulated |
| 0,20208627 | 0,412735754 | PPP1R16A | FALSE | Upregulated |
| -0,191123036 | 0,412735754 | VWA5A | FALSE | Downregulated |
| 0,382635478 | 0,412899498 | TUSC3 | FALSE | Upregulated |
| 0,851012926 | 0,413079575 | GLDN | FALSE | Upregulated |
| -0,328599748 | 0,413090722 | DSTNP2 | FALSE | Downregulated |
| 0,356909576 | 0,413090722 | GPR157 | FALSE | Upregulated |
| -0,229254806 | 0,413325467 | ZNF433 | FALSE | Downregulated |
| 0,50832717 | 0,413440927 | TLR10 | FALSE | Upregulated |
| -0,305687059 | 0,413567465 | C14orf1 | FALSE | Downregulated |
| 0,822568505 | 0,413567465 | CAMK2N2 | FALSE | Upregulated |
| -0,180517862 | 0,413567465 | ELP6 | FALSE | Downregulated |
| 0,445816263 | 0,413567465 | FBLN5 | FALSE | Upregulated |
| 0,498153574 | 0,413567465 | PTTG1 | FALSE | Upregulated |
| 0,162465335 | 0,414120008 | INPPL1 | FALSE | Upregulated |
| 0,342140278 | 0,414356846 | SECTM1 | FALSE | Upregulated |
| 0,729530629 | 0,414463061 | TRIM29 | FALSE | Upregulated |
| -0,223508597 | 0,414958364 | BPHL | FALSE | Downregulated |
| -0,127993576 | 0,414958364 | C6orf211 | FALSE | Downregulated |
| 0,129522133 | 0,414958364 | CHMP1B | FALSE | Upregulated |
| 0,369968802 | 0,414958364 | FANCA | FALSE | Upregulated |
| 0,199485323 | 0,414958364 | GNPDA1 | FALSE | Upregulated |
| -0,613974052 | 0,414958364 | IGFBP6 | FALSE | Downregulated |
| 0,334557057 | 0,414958364 | LINC00959 | FALSE | Upregulated |
| -0,087796788 | 0,414958364 | RBM22 | FALSE | Downregulated |
| -0,117443525 | 0,414958364 | ZNF260 | FALSE | Downregulated |

| | | | | |
|---|---|---|---|---|
| 0,972659924 | 0,414961439 | LOC389602 | FALSE | Upregulated |
| -0,186053761 | 0,414965275 | PIGV | FALSE | Downregulated |
| 0,13732046 | 0,415195036 | DCTN2 | FALSE | Upregulated |
| 0,294137405 | 0,415195036 | HIC1 | FALSE | Upregulated |
| 0,11337027 | 0,415195036 | PHC2 | FALSE | Upregulated |
| 0,880967093 | 0,415195036 | SPDYE5 | FALSE | Upregulated |
| 0,543803226 | 0,415195036 | TBC1D30 | FALSE | Upregulated |
| -0,445298537 | 0,415195036 | ZNF774 | FALSE | Downregulated |
| 0,38413241 | 0,415354714 | LRRC25 | FALSE | Upregulated |
| -0,145523813 | 0,415702738 | COPS4 | FALSE | Downregulated |
| 0,27294716 | 0,415702738 | RAB11FIP4 | FALSE | Upregulated |
| 0,983749413 | 0,416032657 | FAM133CP | FALSE | Upregulated |
| 0,092953616 | 0,416032657 | NCSTN | FALSE | Upregulated |
| 0,310173122 | 0,416032657 | SELPLG | FALSE | Upregulated |
| 0,183076213 | 0,416032657 | TMLHE | FALSE | Upregulated |
| 0,993825407 | 0,416241819 | C19orf33 | FALSE | Upregulated |
| -0,416510687 | 0,416241819 | UNC5CL | FALSE | Downregulated |
| -1,039641405 | 0,416619461 | LOC102723997 | FALSE | Downregulated |
| 0,217771873 | 0,416736195 | RFX1 | FALSE | Upregulated |
| 0,249324657 | 0,416810918 | SLC30A4 | FALSE | Upregulated |
| -0,179667676 | 0,417100258 | C18orf21 | FALSE | Downregulated |
| 0,224443675 | 0,41711913 | FBXO21 | FALSE | Upregulated |
| 0,487750069 | 0,417312739 | TPX2 | FALSE | Upregulated |
| -0,348126812 | 0,417546663 | SEMA6B | FALSE | Downregulated |
| -0,312275576 | 0,417563195 | FAM47E | FALSE | Downregulated |
| -0,604534821 | 0,417563195 | TRNL1 | FALSE | Downregulated |

| | | | | |
|---|---|---|---|---|
| -0,29686439 | 0,417563195 | ZNF415 | FALSE | Downregulated |
| 0,267571894 | 0,417616355 | WIPF1 | FALSE | Upregulated |
| 2,090189928 | 0,417707138 | AGR2 | FALSE | Upregulated |
| -1,316723866 | 0,418342156 | SLC5A4 | FALSE | Downregulated |
| -0,139566646 | 0,418463466 | SEC63 | FALSE | Downregulated |
| 0,08160272 | 0,418849184 | AP2M1 | FALSE | Upregulated |
| 0,170931979 | 0,418849184 | C12orf10 | FALSE | Upregulated |
| -0,756541281 | 0,418849184 | HRC | FALSE | Downregulated |
| -0,530957217 | 0,418849184 | IL1RAPL2 | FALSE | Downregulated |
| -0,351356097 | 0,418849184 | LOC101928707 | FALSE | Downregulated |
| -0,190394165 | 0,418849184 | SUGCT | FALSE | Downregulated |
| 0,185013245 | 0,418849184 | TARBP2 | FALSE | Upregulated |
| 0,185714979 | 0,418849184 | ZNF646 | FALSE | Upregulated |
| 0,263661794 | 0,419034856 | CCND3 | FALSE | Upregulated |
| -0,807871548 | 0,419034856 | PEG10 | FALSE | Downregulated |
| -0,173174522 | 0,419034856 | TBL1X | FALSE | Downregulated |
| 0,340159885 | 0,419034856 | TRMT61A | FALSE | Upregulated |
| -0,159065997 | 0,419718298 | SETD8 | FALSE | Downregulated |
| 0,23490055 | 0,419785674 | B4GALT7 | FALSE | Upregulated |
| 0,118651927 | 0,419785674 | GLB1 | FALSE | Upregulated |
| -0,269932914 | 0,419785674 | LOC101928505 | FALSE | Downregulated |
| -0,552612886 | 0,419785674 | LOC102723583 | FALSE | Downregulated |
| 0,21605987 | 0,419785674 | MAP4K2 | FALSE | Upregulated |
| -1,557743465 | 0,419785674 | MYL3 | FALSE | Downregulated |
| -0,660254468 | 0,419785674 | NPIPB6 | FALSE | Downregulated |
| -0,235389575 | 0,419785674 | OST4 | FALSE | Downregulated |

| | | | | |
|---|---|---|---|---|
| -0,413829359 | 0,419785674 | POMK | FALSE | Downregulated |
| 0,487154029 | 0,419785674 | TBC1D10C | FALSE | Upregulated |
| -0,121189653 | 0,420178037 | ACP1 | FALSE | Downregulated |
| -0,163573029 | 0,420178037 | CEP57 | FALSE | Downregulated |
| 0,159552256 | 0,420178037 | PRR12 | FALSE | Upregulated |
| -0,245895371 | 0,420178037 | PXDC1 | FALSE | Downregulated |
| -0,260567226 | 0,420747013 | EBPL | FALSE | Downregulated |
| 0,205350174 | 0,420747013 | NDUFB6 | FALSE | Upregulated |
| 0,202367141 | 0,421167824 | AMDHD2 | FALSE | Upregulated |
| -0,109566545 | 0,421965058 | ATAD1 | FALSE | Downregulated |
| -0,17176642 | 0,421965058 | C6orf203 | FALSE | Downregulated |
| -0,511006941 | 0,421965058 | CADM2 | FALSE | Downregulated |
| 0,171928278 | 0,421965058 | CUL7 | FALSE | Upregulated |
| 0,88026831 | 0,421965058 | FER1L4 | FALSE | Upregulated |
| -1,063309849 | 0,421965058 | FOXH1 | FALSE | Downregulated |
| -0,406496029 | 0,421965058 | IDI1 | FALSE | Downregulated |
| 0,790580973 | 0,421965058 | LOC100128361 | FALSE | Upregulated |
| -0,585644349 | 0,421965058 | LOC401127 | FALSE | Downregulated |
| 0,217319712 | 0,421965058 | MAPK7 | FALSE | Upregulated |
| -0,204947631 | 0,421965058 | MINOS1 | FALSE | Downregulated |
| 0,191035598 | 0,421965058 | NCKIPSD | FALSE | Upregulated |
| -0,303447729 | 0,421965058 | P2RY13 | FALSE | Downregulated |
| 0,159319943 | 0,421965058 | STX12 | FALSE | Upregulated |
| 0,37523629 | 0,421965058 | TCEA2 | FALSE | Upregulated |
| -0,125932641 | 0,422448624 | DAZAP2 | FALSE | Downregulated |
| 0,15054444 | 0,422529154 | CFL1 | FALSE | Upregulated |

| | | | | |
|---|---|---|---|---|
| -0,15311387 | 0,422529154 | CMTM6 | FALSE | Downregulated |
| 0,181049329 | 0,422529154 | CSK | FALSE | Upregulated |
| -0,932753067 | 0,422529154 | FAM66B | FALSE | Downregulated |
| 0,589088644 | 0,422529154 | FGFBP2 | FALSE | Upregulated |
| -0,164532016 | 0,422529154 | IL18R1 | FALSE | Downregulated |
| -0,143101558 | 0,422529154 | MRPL42 | FALSE | Downregulated |
| 0,136188581 | 0,422529154 | PI4KA | FALSE | Upregulated |
| 1,474613965 | 0,422529154 | RBM11 | FALSE | Upregulated |
| -0,662987701 | 0,422529154 | RP11-685M7.3 | FALSE | Downregulated |
| -0,178522612 | 0,422529154 | RWDD3 | FALSE | Downregulated |
| -0,98248212 | 0,422529154 | SLC17A7 | FALSE | Downregulated |
| 0,171138832 | 0,422529154 | SPATA5L1 | FALSE | Upregulated |
| 0,118375009 | 0,422529154 | VPS26B | FALSE | Upregulated |
| 0,516031929 | 0,422529154 | XCL2 | FALSE | Upregulated |
| -0,238734769 | 0,422529154 | ZNF493 | FALSE | Downregulated |
| -0,883314548 | 0,422650879 | OBSCN | FALSE | Downregulated |
| -0,246205642 | 0,422888189 | ITSN1 | FALSE | Downregulated |
| -0,155930019 | 0,423173076 | CACYBP | FALSE | Downregulated |
| 0,258500944 | 0,423309607 | ABR | FALSE | Upregulated |
| -0,366910493 | 0,42333796 | AQP7P3 | FALSE | Downregulated |
| 1,134079839 | 0,42333796 | B4GALNT1 | FALSE | Upregulated |
| -0,165680266 | 0,42333796 | FERMT2 | FALSE | Downregulated |
| -0,280220812 | 0,42333796 | TMEM256 | FALSE | Downregulated |
| -0,818064462 | 0,424069144 | CXCR1 | FALSE | Downregulated |
| -0,153397309 | 0,424170916 | HACL1 | FALSE | Downregulated |
| 0,123442687 | 0,424232277 | SGPL1 | FALSE | Upregulated |

| | | | | |
|---|---|---|---|---|
| 0,181042233 | 0,424824165 | STAT5A | FALSE | Upregulated |
| -0,147990293 | 0,425131429 | COMMD1 | FALSE | Downregulated |
| -0,371024877 | 0,425201818 | ASPA | FALSE | Downregulated |
| -0,240296901 | 0,425201818 | FBXO4 | FALSE | Downregulated |
| 0,130835229 | 0,425201818 | GSR | FALSE | Upregulated |
| -0,283539978 | 0,425201818 | JAZF1 | FALSE | Downregulated |
| -0,118889209 | 0,425201818 | TWISTNB | FALSE | Downregulated |
| 0,279553542 | 0,425285025 | TNFRSF10D | FALSE | Upregulated |
| 0,164695483 | 0,425466371 | DEAF1 | FALSE | Upregulated |
| -0,182335937 | 0,425466371 | NFATC3 | FALSE | Downregulated |
| -0,191681003 | 0,425757188 | IFT27 | FALSE | Downregulated |
| 0,151005248 | 0,425757188 | WDR37 | FALSE | Upregulated |
| 0,147925534 | 0,425850513 | SBF1 | FALSE | Upregulated |
| 0,129245261 | 0,426212025 | COPS6 | FALSE | Upregulated |
| -0,53271136 | 0,42636451 | LRRC55 | FALSE | Downregulated |
| -0,771169102 | 0,426537042 | GRIP2 | FALSE | Downregulated |
| -0,177372613 | 0,426537042 | MNAT1 | FALSE | Downregulated |
| 0,16589052 | 0,426537042 | SCARB1 | FALSE | Upregulated |
| -0,254854663 | 0,426718796 | TBC1D10B | FALSE | Downregulated |
| -0,227669956 | 0,426719828 | BEND7 | FALSE | Downregulated |
| 1,002791744 | 0,426719828 | LOC100652736 | FALSE | Upregulated |
| -0,501820192 | 0,427450825 | IL1RAP | FALSE | Downregulated |
| -1,263153172 | 0,427450825 | LOC102723443 | FALSE | Downregulated |
| 0,156887523 | 0,427450825 | UNC45A | FALSE | Upregulated |
| -0,269592947 | 0,427565597 | C16orf46 | FALSE | Downregulated |
| -0,208515736 | 0,427565597 | MANEA | FALSE | Downregulated |

| | | | | |
|---|---|---|---|---|
| 0,447776122 | 0,428099856 | ACE | FALSE | Upregulated |
| 0,511709296 | 0,428099856 | BCL11B | FALSE | Upregulated |
| -0,388282136 | 0,428099856 | C14orf37 | FALSE | Downregulated |
| 0,712008823 | 0,428099856 | DUSP2 | FALSE | Upregulated |
| -0,185135623 | 0,428099856 | RBMX2 | FALSE | Downregulated |
| 0,535119426 | 0,428099856 | SSR4 | FALSE | Upregulated |
| 0,166758712 | 0,428099856 | STARD8 | FALSE | Upregulated |
| 0,116758147 | 0,428099856 | YTHDF3 | FALSE | Upregulated |
| -0,195942355 | 0,428099856 | ZNF557 | FALSE | Downregulated |
| -0,83315078 | 0,428341309 | KIF25 | FALSE | Downregulated |
| -0,381788549 | 0,428459386 | C8orf48 | FALSE | Downregulated |
| 0,395178995 | 0,428459386 | TTLL1 | FALSE | Upregulated |
| -0,739538686 | 0,428648768 | KLHL31 | FALSE | Downregulated |
| 0,453185119 | 0,428874674 | KLRB1 | FALSE | Upregulated |
| 0,272996973 | 0,428874674 | LRRC59 | FALSE | Upregulated |
| -0,15086759 | 0,428874674 | MAP3K2 | FALSE | Downregulated |
| -0,758666966 | 0,428874674 | RASL10A | FALSE | Downregulated |
| 0,384322913 | 0,429149702 | LRRK1 | FALSE | Upregulated |
| -0,808756425 | 0,429638129 | MTRNR2L2 | FALSE | Downregulated |
| -0,168826886 | 0,429861566 | HSF2 | FALSE | Downregulated |
| -0,520390149 | 0,429919078 | ANGPT2 | FALSE | Downregulated |
| 0,198200193 | 0,429919078 | MCM7 | FALSE | Upregulated |
| -0,16981024 | 0,430135434 | GJB1 | FALSE | Downregulated |
| 0,518601145 | 0,430135434 | ZNF702P | FALSE | Upregulated |
| -0,158489108 | 0,430479481 | RPL34 | FALSE | Downregulated |
| 0,199348007 | 0,431140391 | HPCAL1 | FALSE | Upregulated |

| | | | | |
|---|---|---|---|---|
| 0,315580541 | 0,431140391 | LOC102724200 | FALSE | Upregulated |
| -1,19937389 | 0,43159711 | RYR1 | FALSE | Downregulated |
| 0,581202338 | 0,431855655 | GCNT4 | FALSE | Upregulated |
| -0,092110507 | 0,432322154 | ZNF140 | FALSE | Downregulated |
| -0,676560879 | 0,432527379 | POU6F2 | FALSE | Downregulated |
| 0,120986912 | 0,432670195 | UTP18 | FALSE | Upregulated |
| 0,154810362 | 0,432670195 | XAB2 | FALSE | Upregulated |
| 1,174025828 | 0,432756628 | ATG9B | FALSE | Upregulated |
| 0,474373664 | 0,432756628 | BICD1 | FALSE | Upregulated |
| -0,145848888 | 0,432756628 | C3orf38 | FALSE | Downregulated |
| -0,156916365 | 0,432756628 | C7orf73 | FALSE | Downregulated |
| 0,332958048 | 0,432756628 | CD244 | FALSE | Upregulated |
| -0,232765874 | 0,432756628 | CD27-AS1 | FALSE | Downregulated |
| 0,309035965 | 0,432756628 | CD97 | FALSE | Upregulated |
| 0,670243879 | 0,432756628 | CDKN3 | FALSE | Upregulated |
| 0,320572333 | 0,432756628 | GGT1 | FALSE | Upregulated |
| -0,187622115 | 0,432756628 | GKAP1 | FALSE | Downregulated |
| -0,365106593 | 0,432756628 | IFITM10 | FALSE | Downregulated |
| -0,15853033 | 0,432756628 | ING1 | FALSE | Downregulated |
| 0,442528648 | 0,432756628 | LFNG | FALSE | Upregulated |
| 0,278324085 | 0,432756628 | LIMK1 | FALSE | Upregulated |
| 0,87963763 | 0,432756628 | LINC00892 | FALSE | Upregulated |
| 0,103057678 | 0,432756628 | RHOA | FALSE | Upregulated |
| 0,395521087 | 0,432756628 | RTN1 | FALSE | Upregulated |
| 0,629547041 | 0,432756628 | SLC1A7 | FALSE | Upregulated |
| -0,453120138 | 0,432756628 | SLC24A3 | FALSE | Downregulated |

| | | | | |
|---|---|---|---|---|
| 0,261967181 | 0,432756628 | SLC29A3 | FALSE | Upregulated |
| 0,355834394 | 0,432756628 | TNFSF13B | FALSE | Upregulated |
| 1,090075125 | 0,432756628 | TRIM67 | FALSE | Upregulated |
| -0,106664371 | 0,432756628 | WDR75 | FALSE | Downregulated |
| -0,202301218 | 0,432756628 | XPNPEP3 | FALSE | Downregulated |
| 0,118464335 | 0,433471842 | DAP | FALSE | Upregulated |
| -0,357663141 | 0,433801426 | EMBP1 | FALSE | Downregulated |
| 0,300156938 | 0,434127516 | AKR1B1 | FALSE | Upregulated |
| 0,444124996 | 0,434127516 | MTHFD1L | FALSE | Upregulated |
| 0,11630877 | 0,434127516 | NAPA | FALSE | Upregulated |
| 0,541049346 | 0,434127516 | SGPP2 | FALSE | Upregulated |
| 0,251194734 | 0,434230794 | ZSCAN18 | FALSE | Upregulated |
| 0,679841016 | 0,434432255 | OPRL1 | FALSE | Upregulated |
| -0,08754492 | 0,434676292 | COIL | FALSE | Downregulated |
| 0,225526421 | 0,434712742 | GIPC1 | FALSE | Upregulated |
| 0,627269429 | 0,434712742 | SRRM2-AS1 | FALSE | Upregulated |
| 0,166806239 | 0,434948921 | BMP1 | FALSE | Upregulated |
| 0,379987781 | 0,434948921 | TECTA | FALSE | Upregulated |
| -0,317000144 | 0,435724883 | EIF5B | FALSE | Downregulated |
| 0,748386111 | 0,435725191 | NCF1B | FALSE | Upregulated |
| 0,159665011 | 0,435848292 | ASNA1 | FALSE | Upregulated |
| 0,1725322 | 0,435848292 | IMP4 | FALSE | Upregulated |
| -0,639808691 | 0,435848292 | TUBA3FP | FALSE | Downregulated |
| -0,585555161 | 0,436520265 | C5orf49 | FALSE | Downregulated |
| 0,27684389 | 0,436520265 | RHBDF2 | FALSE | Upregulated |
| 0,154661971 | 0,436520265 | VPS18 | FALSE | Upregulated |

| | | | | |
|---|---|---|---|---|
| 0,940150839 | 0,436733326 | DIRAS2 | FALSE | Upregulated |
| -0,173124415 | 0,437403928 | ABHD17A | FALSE | Downregulated |
| 0,407270295 | 0,437403928 | ALOX5AP | FALSE | Upregulated |
| 0,222940135 | 0,437403928 | APOL2 | FALSE | Upregulated |
| -0,126342576 | 0,437403928 | BABAM1 | FALSE | Downregulated |
| -0,221493461 | 0,437403928 | C11orf54 | FALSE | Downregulated |
| 0,352629202 | 0,437403928 | CPT1A | FALSE | Upregulated |
| 1,240733188 | 0,437403928 | CRB2 | FALSE | Upregulated |
| 0,490142511 | 0,437403928 | CYR61 | FALSE | Upregulated |
| -0,284663202 | 0,437403928 | DNALI1 | FALSE | Downregulated |
| 0,996178789 | 0,437403928 | ERCC6L | FALSE | Upregulated |
| 0,559593447 | 0,437403928 | GPR18 | FALSE | Upregulated |
| -0,179673749 | 0,437403928 | HAO1 | FALSE | Downregulated |
| 0,261667603 | 0,437403928 | HARBI1 | FALSE | Upregulated |
| 0,157883564 | 0,437403928 | ITGB5 | FALSE | Upregulated |
| 1,018081708 | 0,437403928 | KLC3 | FALSE | Upregulated |
| 1,301758624 | 0,437403928 | LOC100126784 | FALSE | Upregulated |
| -0,446972217 | 0,437403928 | LOC102724133 | FALSE | Downregulated |
| -0,358002524 | 0,437403928 | LUC7L3 | FALSE | Downregulated |
| 0,188280219 | 0,437403928 | OGFOD3 | FALSE | Upregulated |
| 0,171152184 | 0,437403928 | PLA2G15 | FALSE | Upregulated |
| 0,17493225 | 0,437403928 | PMS2 | FALSE | Upregulated |
| 0,172599795 | 0,437403928 | PPME1 | FALSE | Upregulated |
| 0,394948035 | 0,437403928 | PRICKLE4 | FALSE | Upregulated |
| -0,301030416 | 0,437403928 | RPS6KA6 | FALSE | Downregulated |
| 0,174062586 | 0,437403928 | SAAL1 | FALSE | Upregulated |

| | | | | |
|---|---|---|---|---|
| -0,538220667 | 0,437403928 | SERPINE1 | FALSE | Downregulated |
| 0,088501037 | 0,437403928 | SLC35E1 | FALSE | Upregulated |
| -0,124136573 | 0,437403928 | SNAP23 | FALSE | Downregulated |
| -0,139844989 | 0,437403928 | STRN3 | FALSE | Downregulated |
| -0,20006629 | 0,437403928 | SUCO | FALSE | Downregulated |
| 0,643943038 | 0,437403928 | TACSTD2 | FALSE | Upregulated |
| -0,182818403 | 0,437403928 | TRAM2 | FALSE | Downregulated |
| -0,13450349 | 0,437403928 | TSPAN6 | FALSE | Downregulated |
| -0,277129645 | 0,437403928 | TTC26 | FALSE | Downregulated |
| 0,23441042 | 0,437403928 | TUBB4B | FALSE | Upregulated |
| 0,231539017 | 0,437403928 | TWF2 | FALSE | Upregulated |
| 0,155758869 | 0,437403928 | TYK2 | FALSE | Upregulated |
| 0,682946254 | 0,437648183 | RPS6KL1 | FALSE | Upregulated |
| -0,240156488 | 0,437690279 | AKAP12 | FALSE | Downregulated |
| -0,153585243 | 0,437690279 | BCDIN3D | FALSE | Downregulated |
| -0,355502054 | 0,437690279 | SOBP | FALSE | Downregulated |
| 0,19618253 | 0,437690279 | TOR2A | FALSE | Upregulated |
| 0,309027464 | 0,437690279 | ZNF341 | FALSE | Upregulated |
| 0,257060546 | 0,437903018 | UNC119 | FALSE | Upregulated |
| 0,258517966 | 0,437947833 | BTN2A2 | FALSE | Upregulated |
| 0,433403878 | 0,437947833 | SLFN13 | FALSE | Upregulated |
| -0,116901588 | 0,438068007 | THOC7 | FALSE | Downregulated |
| 0,30470991 | 0,438138064 | MFSD10 | FALSE | Upregulated |
| -2,508128851 | 0,438138064 | TECRL | FALSE | Downregulated |
| 0,227550851 | 0,43818167 | ALDH16A1 | FALSE | Upregulated |
| -0,271230627 | 0,438255723 | DNASE1L3 | FALSE | Downregulated |

| | | | | |
|---|---|---|---|---|
| 0,508260864 | 0,438255723 | ST8SIA6 | FALSE | Upregulated |
| -0,782275658 | 0,438520184 | LOC102724738 | FALSE | Downregulated |
| 0,269180686 | 0,438520184 | MOSPD1 | FALSE | Upregulated |
| 0,341061693 | 0,438529777 | ARHGAP4 | FALSE | Upregulated |
| 0,14285586 | 0,438529777 | MARCH5 | FALSE | Upregulated |
| -0,185738853 | 0,438750738 | MARCKSL1 | FALSE | Downregulated |
| 0,452625274 | 0,439246583 | ADAMTS10 | FALSE | Upregulated |
| -0,22022152 | 0,439246583 | ANAPC10 | FALSE | Downregulated |
| -0,273206366 | 0,439246583 | ANGPTL6 | FALSE | Downregulated |
| -1,966419134 | 0,439246583 | COMP | FALSE | Downregulated |
| -0,394553245 | 0,439246583 | CYP51A1 | FALSE | Downregulated |
| -0,205029975 | 0,439246583 | EEA1 | FALSE | Downregulated |
| 0,500439003 | 0,439246583 | FANCE | FALSE | Upregulated |
| 0,574564752 | 0,439246583 | FOXC1 | FALSE | Upregulated |
| 0,146818462 | 0,439246583 | GAA | FALSE | Upregulated |
| -0,652719216 | 0,439246583 | GRM8 | FALSE | Downregulated |
| 0,221059904 | 0,439246583 | HLA-C | FALSE | Upregulated |
| 0,470358805 | 0,439246583 | KIF14 | FALSE | Upregulated |
| 0,914685513 | 0,439246583 | LOC100996273 | FALSE | Upregulated |
| -0,348162354 | 0,439246583 | LOC102724761 | FALSE | Downregulated |
| 0,223527061 | 0,439246583 | LPXN | FALSE | Upregulated |
| 0,206909716 | 0,439246583 | MAP4 | FALSE | Upregulated |
| -0,153614521 | 0,439246583 | MAX | FALSE | Downregulated |
| 0,312734619 | 0,439246583 | NEDD4L | FALSE | Upregulated |
| 0,124875969 | 0,439246583 | NIPA1 | FALSE | Upregulated |
| -0,351309235 | 0,439246583 | PBDC1 | FALSE | Downregulated |

| | | | | |
|---|---|---|---|---|
| -0,398576513 | 0,439246583 | PHACTR3 | FALSE | Downregulated |
| 0,341701174 | 0,439246583 | PIK3CG | FALSE | Upregulated |
| 0,326723683 | 0,439246583 | PTPRC | FALSE | Upregulated |
| -0,218848471 | 0,439246583 | RBFOX2 | FALSE | Downregulated |
| 0,534883827 | 0,439246583 | SLAMF1 | FALSE | Upregulated |
| 0,23270553 | 0,439246583 | SNAPC2 | FALSE | Upregulated |
| -0,119676244 | 0,439246583 | SUV420H1 | FALSE | Downregulated |
| -0,146970713 | 0,439246583 | ZNF302 | FALSE | Downregulated |
| -0,204840769 | 0,43929552 | F9 | FALSE | Downregulated |
| -0,258839149 | 0,43929552 | SNHG11 | FALSE | Downregulated |
| -0,908876688 | 0,439514233 | AC002116.7 | FALSE | Downregulated |
| 0,12641368 | 0,439514233 | ARPC2 | FALSE | Upregulated |
| 0,465287157 | 0,439646288 | AMIGO3 | FALSE | Upregulated |
| 1,166528695 | 0,439646288 | APLP1 | FALSE | Upregulated |
| 0,315112675 | 0,439646288 | DDB2 | FALSE | Upregulated |
| 0,434779275 | 0,439646288 | FMN1 | FALSE | Upregulated |
| -0,630110056 | 0,439646288 | LINC00852 | FALSE | Downregulated |
| -0,150790395 | 0,439646288 | NRBF2 | FALSE | Downregulated |
| 1,810174307 | 0,439646288 | SNORD3A | FALSE | Upregulated |
| 0,145009908 | 0,439646288 | TNRC18 | FALSE | Upregulated |
| 0,334343012 | 0,439646288 | TRPV4 | FALSE | Upregulated |
| 0,249088732 | 0,439646288 | URB2 | FALSE | Upregulated |
| -0,335677243 | 0,439646288 | ZFYVE28 | FALSE | Downregulated |
| 0,330324824 | 0,439646288 | ZNF816 | FALSE | Upregulated |
| -0,172953283 | 0,440252729 | ALDH7A1 | FALSE | Downregulated |
| 0,501415512 | 0,440252729 | GPR82 | FALSE | Upregulated |

| | | | | |
|---|---|---|---|---|
| 0,516844417 | 0,440785017 | FBXL8 | FALSE | Upregulated |
| 0,388005069 | 0,440802947 | SH2D1A | FALSE | Upregulated |
| -0,395208673 | 0,441462605 | AFP | FALSE | Downregulated |
| -1,396279613 | 0,441462605 | MURC | FALSE | Downregulated |
| 0,129369711 | 0,441474986 | RAP1GDS1 | FALSE | Upregulated |
| -0,245732907 | 0,441745715 | GABPB2 | FALSE | Downregulated |
| -0,182963332 | 0,441976743 | DNAAF2 | FALSE | Downregulated |
| -0,104397212 | 0,442150408 | COPS7A | FALSE | Downregulated |
| 0,355622387 | 0,442150408 | MICALL2 | FALSE | Upregulated |
| 0,20291302 | 0,442605701 | HSD3B7 | FALSE | Upregulated |
| 0,753850359 | 0,442605701 | NAT8L | FALSE | Upregulated |
| 0,194080538 | 0,442605701 | PSMG3-AS1 | FALSE | Upregulated |
| 0,180003272 | 0,442605701 | SEPN1 | FALSE | Upregulated |
| 0,276337543 | 0,4428382 | ITPKB | FALSE | Upregulated |
| 0,290218349 | 0,4428382 | TMEM106A | FALSE | Upregulated |
| 0,43192238 | 0,442867084 | CIDECP | FALSE | Upregulated |
| -0,257282638 | 0,442867084 | CXorf57 | FALSE | Downregulated |
| -0,135712616 | 0,442867084 | FMR1 | FALSE | Downregulated |
| -0,77153389 | 0,442867084 | KL | FALSE | Downregulated |
| 0,194131892 | 0,442867084 | WDR13 | FALSE | Upregulated |
| -0,2059751 | 0,442867084 | WDR19 | FALSE | Downregulated |
| -0,525170671 | 0,443278101 | PPP2R3A | FALSE | Downregulated |
| 0,21317624 | 0,44345825 | CTU2 | FALSE | Upregulated |
| -0,219946922 | 0,44346628 | ZNF490 | FALSE | Downregulated |
| 0,914549568 | 0,444437576 | ANKS1B | FALSE | Upregulated |
| -0,195332623 | 0,444437576 | ARVCF | FALSE | Downregulated |

| | | | | |
|---|---|---|---|---|
| -0,118408911 | 0,444437576 | BRIX1 | FALSE | Downregulated |
| -0,149313812 | 0,444437576 | CAAP1 | FALSE | Downregulated |
| -0,830257842 | 0,444437576 | CTSV | FALSE | Downregulated |
| -0,28562571 | 0,444437576 | DLL4 | FALSE | Downregulated |
| -0,563711942 | 0,444437576 | PCDH7 | FALSE | Downregulated |
| 0,209537889 | 0,444437576 | SNX29 | FALSE | Upregulated |
| -0,184336957 | 0,444667249 | AIMP1 | FALSE | Downregulated |
| 1,018886854 | 0,444667249 | ANGPTL7 | FALSE | Upregulated |
| -0,234453118 | 0,444667249 | CCDC82 | FALSE | Downregulated |
| 0,489720729 | 0,444667249 | CD1C | FALSE | Upregulated |
| 2,143719214 | 0,444667249 | LINC00550 | FALSE | Upregulated |
| -0,412922273 | 0,444667249 | LOC101928230 | FALSE | Downregulated |
| 0,161076458 | 0,444667249 | MMGT1 | FALSE | Upregulated |
| -0,188541274 | 0,444667249 | MTRF1L | FALSE | Downregulated |
| -0,397875229 | 0,444667249 | NAMPT | FALSE | Downregulated |
| 0,162223397 | 0,444667249 | NUMA1 | FALSE | Upregulated |
| -0,599045353 | 0,444667249 | PAGE5 | FALSE | Downregulated |
| -0,188607151 | 0,444667249 | RBM34 | FALSE | Downregulated |
| -0,138931923 | 0,444667249 | RFWD2 | FALSE | Downregulated |
| -0,102726194 | 0,444667249 | RLF | FALSE | Downregulated |
| 0,202631049 | 0,444667249 | RNF135 | FALSE | Upregulated |
| -0,697067892 | 0,444667249 | RP11-452H21.4 | FALSE | Downregulated |
| 0,462225136 | 0,444667249 | SUSD2 | FALSE | Upregulated |
| 0,273297823 | 0,444667249 | TAGLN2 | FALSE | Upregulated |
| 0,304525216 | 0,444667249 | TRAF1 | FALSE | Upregulated |
| 0,16204013 | 0,444667249 | TTC9C | FALSE | Upregulated |

| | | | | |
|---|---|---|---|---|
| -0,140536945 | 0,444667249 | WDR44 | FALSE | Downregulated |
| 0,50417092 | 0,444667249 | WNK2 | FALSE | Upregulated |
| 0,131236757 | 0,444667249 | XPNPEP1 | FALSE | Upregulated |
| -0,141430349 | 0,445299408 | CDKN2AIP | FALSE | Downregulated |
| 0,388826214 | 0,445299408 | TRDMT1 | FALSE | Upregulated |
| 0,126285941 | 0,445711482 | C2orf69 | FALSE | Upregulated |
| 0,166143267 | 0,445711482 | CHMP6 | FALSE | Upregulated |
| 2,029367468 | 0,445711482 | LOC100508046 | FALSE | Upregulated |
| -0,549585819 | 0,445711482 | LOC102724559 | FALSE | Downregulated |
| -0,546260261 | 0,445711482 | LOC102724889 | FALSE | Downregulated |
| -0,288455982 | 0,445711482 | LRIG1 | FALSE | Downregulated |
| 0,101838111 | 0,445711482 | MTMR14 | FALSE | Upregulated |
| 0,268702994 | 0,445711482 | PPDPF | FALSE | Upregulated |
| -0,331307285 | 0,445711482 | ZKSCAN7 | FALSE | Downregulated |
| -0,241641167 | 0,445808629 | ANO8 | FALSE | Downregulated |
| 0,275614706 | 0,445808629 | C14orf169 | FALSE | Upregulated |
| -0,165744269 | 0,445808629 | CDC40 | FALSE | Downregulated |
| 0,482719244 | 0,445808629 | CLDN7 | FALSE | Upregulated |
| 0,128412337 | 0,445808629 | COPS8 | FALSE | Upregulated |
| -0,423085252 | 0,445808629 | CSRNP3 | FALSE | Downregulated |
| 0,181150992 | 0,445808629 | CXCL16 | FALSE | Upregulated |
| -0,199661344 | 0,445808629 | GOLIM4 | FALSE | Downregulated |
| 0,270239515 | 0,445808629 | IL10RA | FALSE | Upregulated |
| 0,291201715 | 0,445808629 | KCNQ1 | FALSE | Upregulated |
| 0,186665276 | 0,445808629 | KIAA0930 | FALSE | Upregulated |
| 0,57247976 | 0,445808629 | LOC100288069 | FALSE | Upregulated |

| | | | | |
|---|---|---|---|---|
| 0,230019077 | 0,445808629 | MYO19 | FALSE | Upregulated |
| 0,24164893 | 0,445808629 | PLEKHO1 | FALSE | Upregulated |
| -0,143283218 | 0,445808629 | RPF2 | FALSE | Downregulated |
| 0,233710456 | 0,445808629 | SH3BGRL3 | FALSE | Upregulated |
| 0,165901325 | 0,445808629 | SNX33 | FALSE | Upregulated |
| 0,642572538 | 0,445808629 | TIAF1 | FALSE | Upregulated |
| 0,377479299 | 0,445808629 | TRAF3IP3 | FALSE | Upregulated |
| 0,324081068 | 0,445808629 | ZNF28 | FALSE | Upregulated |
| 0,339586641 | 0,445862403 | SULF2 | FALSE | Upregulated |
| 0,200186562 | 0,446315 | BRPF1 | FALSE | Upregulated |
| 0,979972173 | 0,446315 | SLC4A3 | FALSE | Upregulated |
| 0,132457704 | 0,446403946 | NELFCD | FALSE | Upregulated |
| 0,262376298 | 0,446525226 | HSPG2 | FALSE | Upregulated |
| 0,596228121 | 0,446525226 | UHRF1 | FALSE | Upregulated |
| -0,140694809 | 0,446623827 | DIAPH2 | FALSE | Downregulated |
| 0,275984159 | 0,446623827 | EMP3 | FALSE | Upregulated |
| 0,219301406 | 0,446623827 | RCN2 | FALSE | Upregulated |
| -0,189950731 | 0,446623827 | RPS4X | FALSE | Downregulated |
| 0,247345974 | 0,446623827 | SCO2 | FALSE | Upregulated |
| 0,470540712 | 0,446623827 | TESPA1 | FALSE | Upregulated |
| -0,210694263 | 0,446623827 | UQCR11 | FALSE | Downregulated |
| 0,525650544 | 0,446623827 | ZNF665 | FALSE | Upregulated |
| -0,536803232 | 0,446932477 | FAM110D | FALSE | Downregulated |
| -0,109846064 | 0,446932477 | INTS10 | FALSE | Downregulated |
| 0,829972064 | 0,446932477 | LOC100293704 | FALSE | Upregulated |
| -0,245952647 | 0,446936351 | CCDC84 | FALSE | Downregulated |

| | | | | |
|---|---|---|---|---|
| 0,159473792 | 0,446936351 | RBM42 | FALSE | Upregulated |
| 0,499806867 | 0,446992498 | LOC102724330 | FALSE | Upregulated |
| -0,212769069 | 0,446992498 | RFX7 | FALSE | Downregulated |
| 0,482910921 | 0,447161171 | NAPSB | FALSE | Upregulated |
| -0,326953382 | 0,447233249 | SNAI2 | FALSE | Downregulated |
| -0,118649201 | 0,447839874 | SUPV3L1 | FALSE | Downregulated |
| -0,105812136 | 0,447994689 | CCDC6 | FALSE | Downregulated |
| -1,07391261 | 0,448769829 | HAMP | FALSE | Downregulated |
| 0,213166705 | 0,448769829 | P4HTM | FALSE | Upregulated |
| -0,238964848 | 0,44882684 | DDX19B | FALSE | Downregulated |
| -0,239352457 | 0,44903511 | FABP1 | FALSE | Downregulated |
| -0,368131392 | 0,44903511 | TRNY | FALSE | Downregulated |
| -0,201516057 | 0,44903511 | ZNF546 | FALSE | Downregulated |
| 0,483548337 | 0,449153886 | AEBP1 | FALSE | Upregulated |
| -0,148075931 | 0,449153886 | ARHGAP21 | FALSE | Downregulated |
| 0,21126376 | 0,449153886 | E4F1 | FALSE | Upregulated |
| -0,161755113 | 0,449153886 | PDLIM2 | FALSE | Downregulated |
| -0,576915949 | 0,449363734 | LINC00346 | FALSE | Downregulated |
| -0,139010152 | 0,449363734 | MED13L | FALSE | Downregulated |
| 0,216118205 | 0,449864423 | EID2 | FALSE | Upregulated |
| -0,173291249 | 0,450715241 | PSPC1 | FALSE | Downregulated |
| -0,291090308 | 0,45191656 | LRRC3DN | FALSE | Downregulated |
| 0,354679686 | 0,451937761 | KPNA2 | FALSE | Upregulated |
| -0,152266113 | 0,451937761 | SAT1 | FALSE | Downregulated |
| -0,175232847 | 0,45258155 | ENY2 | FALSE | Downregulated |
| 0,325419834 | 0,45258155 | FDXR | FALSE | Upregulated |

| | | | | |
|---|---|---|---|---|
| -0,160541894 | 0,45258155 | GCSH | FALSE | Downregulated |
| 0,626985457 | 0,453475707 | HSPA6 | FALSE | Upregulated |
| 0,294508279 | 0,45394092 | ADSSL1 | FALSE | Upregulated |
| 0,547384384 | 0,454212085 | CFTR | FALSE | Upregulated |
| 0,127152646 | 0,454341893 | CTNNA1 | FALSE | Upregulated |
| -0,148757881 | 0,454341893 | PIK3CA | FALSE | Downregulated |
| -0,222760964 | 0,454591826 | SETD9 | FALSE | Downregulated |
| 0,314914002 | 0,454873861 | HMGN3-AS1 | FALSE | Upregulated |
| -0,182171161 | 0,454873861 | RPL37 | FALSE | Downregulated |
| -0,181190188 | 0,454893248 | EMC2 | FALSE | Downregulated |
| -0,185452907 | 0,45513223 | ACTL6A | FALSE | Downregulated |
| -0,230550756 | 0,45513223 | APLF | FALSE | Downregulated |
| 0,202068872 | 0,45513223 | HMGN4 | FALSE | Upregulated |
| 0,194919739 | 0,45513223 | SLC41A3 | FALSE | Upregulated |
| -0,302525627 | 0,45513223 | TPST1 | FALSE | Downregulated |
| -0,24088003 | 0,45513223 | ZKSCAN1 | FALSE | Downregulated |
| -0,949577131 | 0,455156314 | AC007879.2 | FALSE | Downregulated |
| 0,107793311 | 0,455156314 | CLPTM1 | FALSE | Upregulated |
| -0,296083751 | 0,455156314 | FAM184A | FALSE | Downregulated |
| -0,220517588 | 0,455156314 | GLRX | FALSE | Downregulated |
| -0,159369985 | 0,455156314 | HLTF | FALSE | Downregulated |
| 0,443628512 | 0,455156314 | KLHL17 | FALSE | Upregulated |
| 0,617666185 | 0,455423296 | AC011747.4 | FALSE | Upregulated |
| -0,346690033 | 0,455423296 | MAP3K7CL | FALSE | Downregulated |
| -0,761217963 | 0,455423296 | RP5-1043L13.1-2 | FALSE | Downregulated |
| -0,147691318 | 0,455423296 | RWDD2B | FALSE | Downregulated |

| | | | | |
|---|---|---|---|---|
| -0,593595035 | 0,455633442 | GRIP1 | FALSE | Downregulated |
| -0,727388286 | 0,455633442 | KLHL33 | FALSE | Downregulated |
| -0,122237362 | 0,455633442 | LINC00674 | FALSE | Downregulated |
| -0,475587422 | 0,455633442 | RASSF9 | FALSE | Downregulated |
| -0,123520489 | 0,455633442 | TCAIM | FALSE | Downregulated |
| -0,166362201 | 0,455671162 | TMEM161B | FALSE | Downregulated |
| 0,473033133 | 0,456278312 | CD3D | FALSE | Upregulated |
| 0,215241169 | 0,456278312 | IPPK | FALSE | Upregulated |
| -0,198010693 | 0,456278312 | SERPINA10 | FALSE | Downregulated |
| 0,396340842 | 0,45655104 | ST8SIA4 | FALSE | Upregulated |
| 1,08606788 | 0,4568058 | LOC101928240 | FALSE | Upregulated |
| 0,164692403 | 0,457097974 | ZNF76 | FALSE | Upregulated |
| -0,25324315 | 0,45710685 | CEBPA-AS1 | FALSE | Downregulated |
| 0,103630618 | 0,45710685 | DERL1 | FALSE | Upregulated |
| -0,108264299 | 0,45710685 | DROSHA | FALSE | Downregulated |
| 0,258520359 | 0,45710685 | FAM129B | FALSE | Upregulated |
| 0,17915903 | 0,45710685 | FOXK1 | FALSE | Upregulated |
| -0,172776754 | 0,45710685 | GABPB1-AS1 | FALSE | Downregulated |
| -0,112476278 | 0,45710685 | INTS8 | FALSE | Downregulated |
| -0,313903555 | 0,45710685 | LRRC6 | FALSE | Downregulated |
| 0,267056878 | 0,45710685 | MMD | FALSE | Upregulated |
| 0,088604864 | 0,45710685 | PDCD6IP | FALSE | Upregulated |
| -0,108373667 | 0,45710685 | PPIE | FALSE | Downregulated |
| 0,102750605 | 0,45710685 | PRRC2A | FALSE | Upregulated |
| -0,248161457 | 0,45710685 | RSRC2 | FALSE | Downregulated |
| 0,270018164 | 0,45710685 | SH2D3C | FALSE | Upregulated |

| | | | | |
|---|---|---|---|---|
| 0,846346232 | 0,45710685 | TNFRSF4 | FALSE | Upregulated |
| 1,382112037 | 0,45710685 | WNT10A | FALSE | Upregulated |
| 0,592183842 | 0,45710685 | XCL1 | FALSE | Upregulated |
| 0,334487649 | 0,45710685 | ZDHHC24 | FALSE | Upregulated |
| -0,169279814 | 0,457182264 | ANAPC11 | FALSE | Downregulated |
| 0,199315231 | 0,457182264 | BAD | FALSE | Upregulated |
| -0,179632178 | 0,457182264 | CAMK1 | FALSE | Downregulated |
| -0,216365788 | 0,457182264 | DDX10 | FALSE | Downregulated |
| 0,156183941 | 0,457182264 | KDELR2 | FALSE | Upregulated |
| 0,61890412 | 0,457182264 | LOC100505727 | FALSE | Upregulated |
| 0,777235697 | 0,457182264 | LOC102723331 | FALSE | Upregulated |
| 0,309136516 | 0,457182264 | NDE1 | FALSE | Upregulated |
| -0,152499716 | 0,457182264 | SSRP1 | FALSE | Downregulated |
| -0,205209833 | 0,457182264 | ZMYM1 | FALSE | Downregulated |
| 0,764470513 | 0,457224549 | MUC1 | FALSE | Upregulated |
| 0,574938699 | 0,457601736 | CLSPN | FALSE | Upregulated |
| -0,446489886 | 0,457672436 | RP11-279F6.1 | FALSE | Downregulated |
| 0,135327341 | 0,457755742 | ATP1A1 | FALSE | Upregulated |
| 0,259442434 | 0,457755742 | GAS6 | FALSE | Upregulated |
| -0,554672989 | 0,457755742 | LOC100129518 | FALSE | Downregulated |
| -0,509756736 | 0,457755742 | LOC727896 | FALSE | Downregulated |
| -0,25727217 | 0,457755742 | NR2F1 | FALSE | Downregulated |
| -0,181013363 | 0,457755742 | SBDSP1 | FALSE | Downregulated |
| 0,371023262 | 0,457755742 | TMEM164 | FALSE | Upregulated |
| -0,170536514 | 0,457755742 | ZNF330 | FALSE | Downregulated |
| 0,173762832 | 0,457944768 | NUDT15 | FALSE | Upregulated |

| | | | | |
|---|---|---|---|---|
| 0,221174752 | 0,458150659 | SDR42E1 | FALSE | Upregulated |
| -0,225014304 | 0,458682989 | CNTLN | FALSE | Downregulated |
| 0,466626575 | 0,458682989 | DZIP1L | FALSE | Upregulated |
| 1,291881562 | 0,458682989 | HCN2 | FALSE | Upregulated |
| 0,297038064 | 0,458682989 | IER5 | FALSE | Upregulated |
| 0,800572618 | 0,458682989 | IGFBPL1 | FALSE | Upregulated |
| -0,130260015 | 0,458682989 | IPO5 | FALSE | Downregulated |
| 0,899938056 | 0,458682989 | NSG1 | FALSE | Upregulated |
| -0,13197435 | 0,458682989 | REEP3 | FALSE | Downregulated |
| -0,343729201 | 0,458742577 | SOX7 | FALSE | Downregulated |
| 0,163386546 | 0,459025072 | NCLN | FALSE | Upregulated |
| -0,2819061 | 0,45911124 | ADH4 | FALSE | Downregulated |
| 0,146466686 | 0,459570381 | CS | FALSE | Upregulated |
| -0,748813603 | 0,459570381 | PP13 | FALSE | Downregulated |
| -0,254324755 | 0,459570381 | SEMA4B | FALSE | Downregulated |
| 0,722397414 | 0,459570381 | ST6GALNAC1 | FALSE | Upregulated |
| 0,128967106 | 0,459693993 | C1orf109 | FALSE | Upregulated |
| 0,130309315 | 0,459693993 | SENP3 | FALSE | Upregulated |
| -0,121294626 | 0,460025969 | ANKRD49 | FALSE | Downregulated |
| 0,134703592 | 0,460025969 | CALM3 | FALSE | Upregulated |
| 0,127200416 | 0,460025969 | EIF4A3 | FALSE | Upregulated |
| 0,292958573 | 0,460025969 | GLI4 | FALSE | Upregulated |
| 0,401080068 | 0,460025969 | ISG20 | FALSE | Upregulated |
| 0,102183131 | 0,460025969 | NUP205 | FALSE | Upregulated |
| -1,279583866 | 0,460025969 | SLC8A3 | FALSE | Downregulated |
| -0,23048946 | 0,460268127 | CD55 | FALSE | Downregulated |

| | | | | |
|---|---|---|---|---|
| -0,508126686 | 0,46066729 | LOC100509620 | FALSE | Downregulated |
| -0,52633995 | 0,460826524 | FGF13 | FALSE | Downregulated |
| 0,172783492 | 0,461345896 | BCL10 | FALSE | Upregulated |
| -0,122525785 | 0,461345896 | DCTN6 | FALSE | Downregulated |
| -0,282620721 | 0,461345896 | GRAMD4 | FALSE | Downregulated |
| 0,206588271 | 0,461483562 | LSM4 | FALSE | Upregulated |
| -0,143299044 | 0,461483562 | TMEM11 | FALSE | Downregulated |
| -0,58630828 | 0,461487988 | LOC101927895 | FALSE | Downregulated |
| 0,125698846 | 0,462007849 | GTF3C5 | FALSE | Upregulated |
| -0,453554451 | 0,462007849 | LOC101927595 | FALSE | Downregulated |
| 0,486440579 | 0,46288624 | ARHGEF26-AS1 | FALSE | Upregulated |
| -1,047086301 | 0,463321121 | GNAT1 | FALSE | Downregulated |
| -0,419788623 | 0,463321121 | SGOL2 | FALSE | Downregulated |
| -0,298873365 | 0,463321121 | ZRSR2 | FALSE | Downregulated |
| 1,190593152 | 0,463338429 | AP000688.8 | FALSE | Upregulated |
| 1,027929118 | 0,46336337 | C2orf81 | FALSE | Upregulated |
| 0,147490216 | 0,463463927 | SLC25A3 | FALSE | Upregulated |
| -0,082952716 | 0,463491082 | C3orf17 | FALSE | Downregulated |
| 0,303537828 | 0,463491082 | C5orf63 | FALSE | Upregulated |
| 0,256262113 | 0,463491082 | KLF16 | FALSE | Upregulated |
| 1,095547139 | 0,463491082 | PCBP2-OT1 | FALSE | Upregulated |
| 0,617329484 | 0,463491082 | SNAP25-AS1 | FALSE | Upregulated |
| -0,122034681 | 0,463491082 | WDR70 | FALSE | Downregulated |
| 0,449012419 | 0,46359197 | LOC100507507 | FALSE | Upregulated |
| -0,142567669 | 0,464057833 | NDUFV2 | FALSE | Downregulated |
| -0,999402661 | 0,464328443 | DDIT4L | FALSE | Downregulated |

| | | | | |
|---|---|---|---|---|
| 1,502588961 | 0,464328443 | GPR3 | FALSE | Upregulated |
| 0,231548567 | 0,464328443 | MICAL3 | FALSE | Upregulated |
| -0,157478701 | 0,464328443 | PDCD4 | FALSE | Downregulated |
| 0,227801913 | 0,464328443 | RFX5 | FALSE | Upregulated |
| 0,214045911 | 0,464328443 | SMCO4 | FALSE | Upregulated |
| -0,175112808 | 0,464328443 | TMTC3 | FALSE | Downregulated |
| 0,681778229 | 0,464439649 | DDX12P | FALSE | Upregulated |
| 0,219556515 | 0,464463959 | ZNF513 | FALSE | Upregulated |
| 0,272624243 | 0,464554429 | WDR25 | FALSE | Upregulated |
| 0,263194698 | 0,465882057 | TEN1 | FALSE | Upregulated |
| 0,208140309 | 0,46610378 | GEMIN8 | FALSE | Upregulated |
| -0,175675553 | 0,466278786 | NDUFB7 | FALSE | Downregulated |
| 0,128108295 | 0,466278786 | VPS45 | FALSE | Upregulated |
| -0,463047594 | 0,46636979 | LOC101927765 | FALSE | Downregulated |
| 0,288201319 | 0,46636979 | SLC43A2 | FALSE | Upregulated |
| -0,177192809 | 0,466379164 | A1CF | FALSE | Downregulated |
| 0,336430526 | 0,466379164 | BTG2 | FALSE | Upregulated |
| -0,178433488 | 0,466379164 | CCNYL1 | FALSE | Downregulated |
| -0,159373537 | 0,466379164 | FAM120C | FALSE | Downregulated |
| -0,155398078 | 0,466379164 | SNHG6 | FALSE | Downregulated |
| -0,272284416 | 0,466379164 | SRSF11 | FALSE | Downregulated |
| 1,126939125 | 0,466379164 | TUBBP5 | FALSE | Upregulated |
| -0,095054246 | 0,466379164 | UBE2R2 | FALSE | Downregulated |
| -0,122349089 | 0,466379164 | ZXDC | FALSE | Downregulated |
| 0,255417426 | 0,46652471 | FBXO27 | FALSE | Upregulated |
| -0,215580618 | 0,46652471 | SLC35B4 | FALSE | Downregulated |

| | | | | |
|---|---|---|---|---|
| 0,226766153 | 0,466697704 | MAP7D1 | FALSE | Upregulated |
| 0,919461879 | 0,467148879 | CR2 | FALSE | Upregulated |
| 0,291325838 | 0,467148879 | TNFAIP2 | FALSE | Upregulated |
| 0,422408353 | 0,467564165 | PAFAH1B3 | FALSE | Upregulated |
| 0,311819949 | 0,467806667 | PTAFR | FALSE | Upregulated |
| 0,151196863 | 0,467867649 | MRPL15 | FALSE | Upregulated |
| 0,212623158 | 0,46793997 | AKNA | FALSE | Upregulated |
| -0,337976173 | 0,46793997 | FAM124A | FALSE | Downregulated |
| 0,554102843 | 0,46793997 | GINS1 | FALSE | Upregulated |
| 0,393884739 | 0,46793997 | NFASC | FALSE | Upregulated |
| -0,265891183 | 0,468760848 | DCXR | FALSE | Downregulated |
| -0,15599759 | 0,468858701 | MRPL50 | FALSE | Downregulated |
| -0,382252118 | 0,468858701 | POLR2A | FALSE | Downregulated |
| 0,959564147 | 0,468969541 | SERPINA12 | FALSE | Upregulated |
| 0,312603629 | 0,468969541 | VIM | FALSE | Upregulated |
| 0,462296179 | 0,469132845 | C11orf63 | FALSE | Upregulated |
| 0,400893271 | 0,469132845 | CCL28 | FALSE | Upregulated |
| 0,20738596 | 0,469132845 | FAM160B2 | FALSE | Upregulated |
| -0,220514926 | 0,469132845 | FLT1 | FALSE | Downregulated |
| -0,090883848 | 0,469132845 | GRIPAP1 | FALSE | Downregulated |
| 0,285752715 | 0,469132845 | KLHL11 | FALSE | Upregulated |
| -0,190952546 | 0,469132845 | PELI1 | FALSE | Downregulated |
| -0,14821697 | 0,469132845 | SAYSD1 | FALSE | Downregulated |
| 0,100791768 | 0,469132845 | VPS52 | FALSE | Upregulated |
| 0,172995998 | 0,469132845 | YARS | FALSE | Upregulated |
| 0,184193472 | 0,469976016 | MVB12B | FALSE | Upregulated |

| | | | | |
|---|---|---|---|---|
| -0,150302824 | 0,470057945 | ESCO1 | FALSE | Downregulated |
| 0,529803762 | 0,470151383 | ENTPD1-AS1 | FALSE | Upregulated |
| 0,89951337 | 0,470151383 | LINC00853 | FALSE | Upregulated |
| 0,1267174 | 0,470291211 | HMG20B | FALSE | Upregulated |
| -0,951415076 | 0,470291211 | RPS14P3 | FALSE | Downregulated |
| -0,096863435 | 0,470291211 | SS18 | FALSE | Downregulated |
| -0,248884047 | 0,470291211 | TRIM52 | FALSE | Downregulated |
| 0,472228664 | 0,470434798 | SLC16A3 | FALSE | Upregulated |
| -0,216559955 | 0,47046624 | C10orf11 | FALSE | Downregulated |
| 0,289630972 | 0,470841427 | ZNF668 | FALSE | Upregulated |
| -0,279832715 | 0,471610026 | NFKBIZ | FALSE | Downregulated |
| -0,658045908 | 0,471610026 | SEMA3C | FALSE | Downregulated |
| -0,15431676 | 0,471721442 | CFDP1 | FALSE | Downregulated |
| 0,248434279 | 0,471721442 | GPSM3 | FALSE | Upregulated |
| -0,219237565 | 0,471721442 | MCUR1 | FALSE | Downregulated |
| 0,256884176 | 0,471721442 | MORN1 | FALSE | Upregulated |
| -0,195098392 | 0,471721442 | RUFY3 | FALSE | Downregulated |
| -0,27906028 | 0,471721442 | TMEM161B-AS1 | FALSE | Downregulated |
| 0,167575501 | 0,47194645 | B3GALNT2 | FALSE | Upregulated |
| 0,122644545 | 0,47194645 | DYNLL2 | FALSE | Upregulated |
| 0,288980862 | 0,47194645 | HCP5 | FALSE | Upregulated |
| 0,407949817 | 0,47194645 | LAT | FALSE | Upregulated |
| -0,138987392 | 0,47194645 | TECPR2 | FALSE | Downregulated |
| 0,164673902 | 0,471952305 | GNA12 | FALSE | Upregulated |
| -0,17399668 | 0,471952305 | ITPR2 | FALSE | Downregulated |
| 0,323037904 | 0,471952305 | KCTD12 | FALSE | Upregulated |

| | | | | |
|---|---|---|---|---|
| 0,173842932 | 0,471952305 | SYPL1 | FALSE | Upregulated |
| -0,227025381 | 0,472014466 | SBF2-AS1 | FALSE | Downregulated |
| 0,167572305 | 0,472109806 | CHDH | FALSE | Upregulated |
| -0,495273564 | 0,472109806 | LOC727978 | FALSE | Downregulated |
| -0,171061644 | 0,472109806 | MPHOSPH9 | FALSE | Downregulated |
| -0,437384459 | 0,472109806 | RANBP3L | FALSE | Downregulated |
| -0,634116769 | 0,472109806 | TMEM182 | FALSE | Downregulated |
| -0,097380105 | 0,472227236 | MARK3 | FALSE | Downregulated |
| -0,081503032 | 0,472227236 | VEZT | FALSE | Downregulated |
| -1,052755103 | 0,472470286 | FAM46B | FALSE | Downregulated |
| 0,214454553 | 0,472520588 | GOLT1A | FALSE | Upregulated |
| -0,155673929 | 0,472520588 | NAF1 | FALSE | Downregulated |
| -0,753921154 | 0,472520588 | RASD1 | FALSE | Downregulated |
| 0,336286979 | 0,473048337 | FGD3 | FALSE | Upregulated |
| 0,691504103 | 0,473277573 | CCNB2 | FALSE | Upregulated |
| -0,385818933 | 0,473277573 | FITM1 | FALSE | Downregulated |
| 0,576526313 | 0,473277573 | GTSE1 | FALSE | Upregulated |
| -0,862612323 | 0,473277573 | LOC101928083 | FALSE | Downregulated |
| -0,800276574 | 0,473277573 | LOC149684 | FALSE | Downregulated |
| -0,875178616 | 0,473277573 | LRRC15 | FALSE | Downregulated |
| 0,685303251 | 0,473277573 | MKLN1-AS | FALSE | Upregulated |
| 0,188271426 | 0,473277573 | PHF23 | FALSE | Upregulated |
| 0,126670132 | 0,473277573 | PIP4K2C | FALSE | Upregulated |
| -1,602349366 | 0,473277573 | SSTR5-AS1 | FALSE | Downregulated |
| 0,125899339 | 0,473277573 | TBC1D13 | FALSE | Upregulated |
| 0,190937525 | 0,473277573 | ZNF576 | FALSE | Upregulated |

| | | | | |
|---|---|---|---|---|
| -0,138342627 | 0,473295484 | ERO1L | FALSE | Downregulated |
| 0,609442424 | 0,473295484 | LINC00920 | FALSE | Upregulated |
| 1,003114855 | 0,473295484 | LOC102724907 | FALSE | Upregulated |
| 0,241497129 | 0,473713472 | CASKIN2 | FALSE | Upregulated |
| 0,862294216 | 0,473914464 | C19orf35 | FALSE | Upregulated |
| -0,197504599 | 0,473914464 | SLC10A1 | FALSE | Downregulated |
| 0,452550975 | 0,473914464 | SLC24A4 | FALSE | Upregulated |
| 0,187454605 | 0,47405954 | SNRNP40 | FALSE | Upregulated |
| 0,184417987 | 0,474424543 | ESRRA | FALSE | Upregulated |
| -1,886031956 | 0,474424543 | MYOZ3 | FALSE | Downregulated |
| 0,335856979 | 0,474563558 | ACKR2 | FALSE | Upregulated |
| -1,231369628 | 0,474668666 | ADAMTS16 | FALSE | Downregulated |
| -0,260562066 | 0,474668666 | G2E3 | FALSE | Downregulated |
| 1,20965372 | 0,474668666 | RIBC2 | FALSE | Upregulated |
| -0,231467977 | 0,474668666 | RP11-392P7.6 | FALSE | Downregulated |
| -0,131565987 | 0,474668666 | SDHAF2 | FALSE | Downregulated |
| 0,148712499 | 0,474800787 | ALG12 | FALSE | Upregulated |
| 0,105984556 | 0,474800787 | ARF1 | FALSE | Upregulated |
| -0,145669354 | 0,474800787 | C4orf27 | FALSE | Downregulated |
| 0,255635503 | 0,474800787 | CUEDC1 | FALSE | Upregulated |
| -1,014130871 | 0,474800787 | GSTT2B | FALSE | Downregulated |
| -0,201653174 | 0,474800787 | HPX | FALSE | Downregulated |
| 0,347862248 | 0,474800787 | KLC2 | FALSE | Upregulated |
| -0,681270197 | 0,474800787 | LOC102724192 | FALSE | Downregulated |
| -0,306699658 | 0,474800787 | LOC92249 | FALSE | Downregulated |
| 1,249261689 | 0,474800787 | MUC5B | FALSE | Upregulated |

| | | | | |
|---|---|---|---|---|
| 0,243064267 | 0,474800787 | NAV2 | FALSE | Upregulated |
| -0,200605686 | 0,474800787 | PALM2 | FALSE | Downregulated |
| -0,175037632 | 0,474800787 | PEBP1 | FALSE | Downregulated |
| 0,845723559 | 0,474800787 | PTPRQ | FALSE | Upregulated |
| -0,120068241 | 0,474800787 | RNF38 | FALSE | Downregulated |
| -0,615756039 | 0,474800787 | SIGLEC16 | FALSE | Downregulated |
| 0,149409355 | 0,474800787 | TNFRSF10B | FALSE | Upregulated |
| 0,14795214 | 0,474800787 | TPCN1 | FALSE | Upregulated |
| -0,190829709 | 0,475104378 | ALG10B | FALSE | Downregulated |
| 0,308495097 | 0,475104378 | CA5A | FALSE | Upregulated |
| 1,063396288 | 0,475277226 | MTRNR2L7 | FALSE | Upregulated |
| -0,781032664 | 0,475277226 | PF4 | FALSE | Downregulated |
| 0,152682114 | 0,475348707 | ZKSCAN2 | FALSE | Upregulated |
| 0,167649812 | 0,475582205 | ASB1 | FALSE | Upregulated |
| -0,227603052 | 0,475582205 | CCDC66 | FALSE | Downregulated |
| -0,133992317 | 0,475582205 | CHD1 | FALSE | Downregulated |
| -1,005702341 | 0,475582205 | CRP | FALSE | Downregulated |
| 0,196113235 | 0,475582205 | DNAL4 | FALSE | Upregulated |
| 0,126411299 | 0,475582205 | EMC3 | FALSE | Upregulated |
| 0,167384964 | 0,475582205 | FAM98A | FALSE | Upregulated |
| -0,381097266 | 0,475582205 | HMGCLL1 | FALSE | Downregulated |
| -1,774643618 | 0,475582205 | LOC101060569 | FALSE | Downregulated |
| 0,326370912 | 0,475582205 | NEK8 | FALSE | Upregulated |
| -0,132323942 | 0,475582205 | RRP8 | FALSE | Downregulated |
| -0,262145299 | 0,475582205 | RSPO3 | FALSE | Downregulated |
| -0,386211081 | 0,475582205 | STEAP4 | FALSE | Downregulated |

| | | | | |
|---|---|---|---|---|
| -0,3646977 | 0,475582205 | VMO1 | FALSE | Downregulated |
| 0,263599105 | 0,475582205 | ZMYND15 | FALSE | Upregulated |
| 0,482176061 | 0,475582205 | ZNF793 | FALSE | Upregulated |
| 0,411722315 | 0,4756438 | NOTUM | FALSE | Upregulated |
| 0,512014031 | 0,475985936 | SH2D1B | FALSE | Upregulated |
| 0,776755808 | 0,476167642 | ROS1 | FALSE | Upregulated |
| -0,745385025 | 0,476341609 | NFE2 | FALSE | Downregulated |
| 0,393798281 | 0,476430521 | CR1 | FALSE | Upregulated |
| -0,140440382 | 0,476609277 | CCDC126 | FALSE | Downregulated |
| 0,675261266 | 0,476609277 | DND1 | FALSE | Upregulated |
| -0,733994354 | 0,476609277 | FAM35BP | FALSE | Downregulated |
| 0,473812672 | 0,476609277 | IGFBP2 | FALSE | Upregulated |
| 0,646383895 | 0,476609277 | RIN1 | FALSE | Upregulated |
| -0,188110393 | 0,476609277 | SPTLC3 | FALSE | Downregulated |
| 0,226824745 | 0,476646045 | ARHGEF2 | FALSE | Upregulated |
| -0,158572292 | 0,476792783 | C14orf28 | FALSE | Downregulated |
| -0,314186143 | 0,476792783 | NSDHL | FALSE | Downregulated |
| 0,177440344 | 0,476792783 | TOE1 | FALSE | Upregulated |
| 0,102107945 | 0,477032674 | MED24 | FALSE | Upregulated |
| -0,123471996 | 0,477208996 | ERP44 | FALSE | Downregulated |
| -0,294605552 | 0,477208996 | FAM134B | FALSE | Downregulated |
| 0,126321732 | 0,477521659 | SEPHS1 | FALSE | Upregulated |
| -0,121687781 | 0,47754517 | ACTR6 | FALSE | Downregulated |
| -0,843468461 | 0,47754517 | BPI | FALSE | Downregulated |
| -0,345439763 | 0,477643831 | C1orf101 | FALSE | Downregulated |
| -0,236453833 | 0,477643831 | RP11-983P16.4 | FALSE | Downregulated |

| | | | | |
|---|---|---|---|---|
| -0,173386911 | 0,477687675 | PAH | FALSE | Downregulated |
| 0,149604736 | 0,477722097 | ASPSCR1 | FALSE | Upregulated |
| -0,157044184 | 0,477837825 | ZNF615 | FALSE | Downregulated |
| -0,872078445 | 0,477874498 | DEGS2 | FALSE | Downregulated |
| -0,130843664 | 0,478031605 | AGO4 | FALSE | Downregulated |
| 0,740541198 | 0,478031605 | C5orf66 | FALSE | Upregulated |
| -0,179744357 | 0,478031605 | HNMT | FALSE | Downregulated |
| -0,505748575 | 0,478031605 | LOC101928826 | FALSE | Downregulated |
| -0,590738092 | 0,478031605 | POPDC2 | FALSE | Downregulated |
| -0,179183099 | 0,478031605 | PRDM10 | FALSE | Downregulated |
| 0,321538456 | 0,478031605 | RGS10 | FALSE | Upregulated |
| -0,117140858 | 0,478031605 | SETD2 | FALSE | Downregulated |
| -0,240296321 | 0,478031605 | SLC43A3 | FALSE | Downregulated |
| 0,341005855 | 0,478031605 | SLC52A2 | FALSE | Upregulated |
| -0,18427753 | 0,478031605 | TFB1M | FALSE | Downregulated |
| -0,159005393 | 0,478031605 | ZKSCAN8 | FALSE | Downregulated |
| 0,273546561 | 0,478213216 | C10orf54 | FALSE | Upregulated |
| 0,195148534 | 0,478213216 | CLN8 | FALSE | Upregulated |
| 0,248021327 | 0,478213216 | NUTM2A | FALSE | Upregulated |
| -0,249264836 | 0,478213216 | RBM6 | FALSE | Downregulated |
| -0,506656707 | 0,478406472 | LINC00638 | FALSE | Downregulated |
| -0,501028211 | 0,47858572 | NUDT9P1 | FALSE | Downregulated |
| -2,349978333 | 0,479259267 | SFRP2 | FALSE | Downregulated |
| 1,030356155 | 0,479259267 | TMEM132C | FALSE | Upregulated |
| -0,103026861 | 0,479259267 | USP25 | FALSE | Downregulated |
| -0,167438741 | 0,479377116 | BHMT2 | FALSE | Downregulated |

| | | | | |
|---|---|---|---|---|
| 0,426144356 | 0,479631025 | MAP4K1 | FALSE | Upregulated |
| -0,515693766 | 0,479797124 | LRRTM2 | FALSE | Downregulated |
| 0,47082447 | 0,479797124 | MTHFD2 | FALSE | Upregulated |
| -0,263598073 | 0,479797124 | NLGN2 | FALSE | Downregulated |
| -0,137309993 | 0,479797124 | PDCD11 | FALSE | Downregulated |
| -0,43194426 | 0,479797124 | SYNM | FALSE | Downregulated |
| -0,505708848 | 0,479797124 | ZNF876P | FALSE | Downregulated |
| -0,258880802 | 0,479968173 | CCT6B | FALSE | Downregulated |
| 0,777829335 | 0,479968173 | CXCR5 | FALSE | Upregulated |
| -0,166730507 | 0,479968173 | NMRK1 | FALSE | Downregulated |
| 0,265858315 | 0,479968173 | RAB11FIP5 | FALSE | Upregulated |
| -0,213685332 | 0,480008312 | C19orf25 | FALSE | Downregulated |
| -0,2552962 | 0,480008312 | CROT | FALSE | Downregulated |
| 0,151315207 | 0,480255181 | BBS1 | FALSE | Upregulated |
| -0,857983851 | 0,480421134 | LOC101927260 | FALSE | Downregulated |
| -0,0956941 | 0,48057219 | PANK2 | FALSE | Downregulated |
| 0,170893868 | 0,48057219 | PSMB10 | FALSE | Upregulated |
| 0,114330536 | 0,480594147 | ANAPC5 | FALSE | Upregulated |
| -0,277020786 | 0,480594147 | CLEC1B | FALSE | Downregulated |
| 0,110248261 | 0,480594147 | DCBLD1 | FALSE | Upregulated |
| 0,173100097 | 0,480594147 | ILK | FALSE | Upregulated |
| 0,171336576 | 0,480595466 | RCBTB1 | FALSE | Upregulated |
| 0,157684554 | 0,480595466 | RECQL | FALSE | Upregulated |
| 0,153745244 | 0,480660805 | S100A16 | FALSE | Upregulated |
| -0,144541693 | 0,480697075 | EXOSC9 | FALSE | Downregulated |
| 0,422854788 | 0,480885903 | LINC00857 | FALSE | Upregulated |

| | | | | |
|---|---|---|---|---|
| -0,125343778 | 0,480885903 | SRP9 | FALSE | Downregulated |
| 0,214653421 | 0,481514207 | NAA40 | FALSE | Upregulated |
| -0,558046747 | 0,481530649 | LOC101928487 | FALSE | Downregulated |
| -0,097913349 | 0,481530649 | MAP3K7 | FALSE | Downregulated |
| -0,145893543 | 0,481594208 | NTPCR | FALSE | Downregulated |
| 0,128793475 | 0,482279059 | ARL8A | FALSE | Upregulated |
| -0,160863368 | 0,482331159 | CDO1 | FALSE | Downregulated |
| -0,140198437 | 0,482335822 | UBB | FALSE | Downregulated |
| 0,467254938 | 0,482564167 | CDCP1 | FALSE | Upregulated |
| 0,298235389 | 0,482564167 | HK3 | FALSE | Upregulated |
| -0,564293885 | 0,482564167 | MT1G | FALSE | Downregulated |
| -0,363125902 | 0,482781107 | ABI3BP | FALSE | Downregulated |
| 0,169877711 | 0,482781107 | DOT1L | FALSE | Upregulated |
| 0,973249719 | 0,482781107 | HSF5 | FALSE | Upregulated |
| -0,293608686 | 0,482781107 | KCNC4 | FALSE | Downregulated |
| 0,433266565 | 0,482781107 | LOC100133445 | FALSE | Upregulated |
| 0,565681788 | 0,482781107 | PROCA1 | FALSE | Upregulated |
| 1,082866791 | 0,482781107 | RAB3B | FALSE | Upregulated |
| 0,332565328 | 0,482781107 | TGFBI | FALSE | Upregulated |
| -0,792988785 | 0,482781107 | XG | FALSE | Downregulated |
| -0,091228324 | 0,4827902 | LSM14A | FALSE | Downregulated |
| 0,134205919 | 0,482819608 | ATP6AP1 | FALSE | Upregulated |
| -2,08335579 | 0,482819608 | C10orf71 | FALSE | Downregulated |
| -0,180049839 | 0,482819608 | SFSWAP | FALSE | Downregulated |
| -0,130872067 | 0,482819608 | SUPT20H | FALSE | Downregulated |
| 1,374152503 | 0,483316738 | TUBB3 | FALSE | Upregulated |

| | | | | |
|---|---|---|---|---|
| 0,180681485 | 0,483692321 | GGA1 | FALSE | Upregulated |
| -0,189594067 | 0,484296014 | ACADM | FALSE | Downregulated |
| -0,149892377 | 0,484531615 | ACBD5 | FALSE | Downregulated |
| 0,575798702 | 0,484531615 | BACE1-AS | FALSE | Upregulated |
| 1,098035379 | 0,484531615 | CLNK | FALSE | Upregulated |
| -0,26624395 | 0,484531615 | GHR | FALSE | Downregulated |
| -0,283508146 | 0,484531615 | KIAA1407 | FALSE | Downregulated |
| 0,117027131 | 0,484531615 | LARS2 | FALSE | Upregulated |
| -0,132407096 | 0,484531615 | NAA35 | FALSE | Downregulated |
| -0,299020185 | 0,484531615 | RAB15 | FALSE | Downregulated |
| 0,158002732 | 0,484531615 | VPS9D1 | FALSE | Upregulated |
| -0,225071081 | 0,484573249 | FOLH1 | FALSE | Downregulated |
| 0,19372071 | 0,484683642 | RBFA | FALSE | Upregulated |
| 0,448167503 | 0,484805452 | GPBAR1 | FALSE | Upregulated |
| -0,162277214 | 0,484805452 | PRKACA | FALSE | Downregulated |
| 0,137775715 | 0,484888093 | ACAD8 | FALSE | Upregulated |
| -0,136190738 | 0,484888093 | ALCAM | FALSE | Downregulated |
| 0,234806599 | 0,484888093 | ARHGAP31 | FALSE | Upregulated |
| 1,036465203 | 0,484888093 | C9orf172 | FALSE | Upregulated |
| 0,385485782 | 0,484888093 | GMPPB | FALSE | Upregulated |
| 0,202881169 | 0,484888093 | INCENP | FALSE | Upregulated |
| 0,13065806 | 0,484888093 | MAZ | FALSE | Upregulated |
| 0,30589085 | 0,484888093 | RAB34 | FALSE | Upregulated |
| -0,223910573 | 0,484888093 | TEK | FALSE | Downregulated |
| -0,256218349 | 0,484997177 | KCNT2 | FALSE | Downregulated |
| -0,218859493 | 0,484997177 | LOC646762 | FALSE | Downregulated |

| | | | | |
|---|---|---|---|---|
| 0,157332131 | 0,485171538 | ZNF282 | FALSE | Upregulated |
| -0,160711606 | 0,48559934 | IDH1 | FALSE | Downregulated |
| 0,278170746 | 0,485945376 | HLA-DPA1 | FALSE | Upregulated |
| -0,1916742 | 0,485945376 | LDB2 | FALSE | Downregulated |
| 0,125075009 | 0,486437425 | LMF2 | FALSE | Upregulated |
| -0,255088046 | 0,486437425 | NOP58 | FALSE | Downregulated |
| 0,398139135 | 0,486485815 | LCK | FALSE | Upregulated |
| -0,48024643 | 0,486558191 | FAM151A | FALSE | Downregulated |
| 0,187878555 | 0,486611574 | DLAT | FALSE | Upregulated |
| 0,697422964 | 0,486611574 | FLJ22184 | FALSE | Upregulated |
| -0,482806816 | 0,486865146 | EFCAB1 | FALSE | Downregulated |
| 0,143683751 | 0,486936614 | ATP11A | FALSE | Upregulated |
| -0,242189172 | 0,486936614 | MAP7D3 | FALSE | Downregulated |
| 0,140487062 | 0,486936614 | TFG | FALSE | Upregulated |
| 0,623195183 | 0,487130658 | FAM188B | FALSE | Upregulated |
| 0,382540335 | 0,487466123 | ANGPTL4 | FALSE | Upregulated |
| 0,124322898 | 0,487466123 | RAB35 | FALSE | Upregulated |
| -0,124024537 | 0,487764506 | PTPN12 | FALSE | Downregulated |
| -0,18137838 | 0,487965093 | NOP14-AS1 | FALSE | Downregulated |
| 0,578837869 | 0,487965093 | TMEM154 | FALSE | Upregulated |
| 0,131312505 | 0,487965093 | TOP1MT | FALSE | Upregulated |
| 0,309574136 | 0,488434897 | CRYM | FALSE | Upregulated |
| 0,149441735 | 0,488434897 | SRSF2 | FALSE | Upregulated |
| 0,717161477 | 0,488805047 | LOC101926994 | FALSE | Upregulated |
| -0,247294435 | 0,489064739 | ANO5 | FALSE | Downregulated |
| -0,17078989 | 0,489064739 | CDK2AP1 | FALSE | Downregulated |

| | | | | |
|---|---|---|---|---|
| -0,222125617 | 0,489064739 | HFE2 | FALSE | Downregulated |
| 0,636749984 | 0,489064739 | ORC1 | FALSE | Upregulated |
| -0,17153758 | 0,489076731 | XPO4 | FALSE | Downregulated |
| -0,103977617 | 0,489323069 | ALG13 | FALSE | Downregulated |
| -0,462789312 | 0,489488141 | GJA5 | FALSE | Downregulated |
| -0,161293448 | 0,489747888 | TADA2A | FALSE | Downregulated |
| -0,735006491 | 0,489886603 | CHRDL1 | FALSE | Downregulated |
| -0,289516029 | 0,490127645 | INO80B | FALSE | Downregulated |
| 0,477716422 | 0,490127645 | SGOL1 | FALSE | Upregulated |
| 0,71582561 | 0,4902617 | FAM223B | FALSE | Upregulated |
| -0,167947074 | 0,490485599 | INADL | FALSE | Downregulated |
| 0,147288034 | 0,490485599 | ZNF134 | FALSE | Upregulated |
| -0,178521251 | 0,490560475 | AOC3 | FALSE | Downregulated |
| 0,125318237 | 0,490560475 | COG1 | FALSE | Upregulated |
| 0,776815264 | 0,490560475 | GPR35 | FALSE | Upregulated |
| -0,134178327 | 0,490560475 | TCFL5 | FALSE | Downregulated |
| 0,38905952 | 0,490695261 | GFI1 | FALSE | Upregulated |
| 0,374389343 | 0,490695261 | LILRB3 | FALSE | Upregulated |
| -0,131786607 | 0,490695261 | MRPS5 | FALSE | Downregulated |
| -0,146226926 | 0,490695261 | PLS3 | FALSE | Downregulated |
| 0,131241693 | 0,490695261 | RAB1A | FALSE | Upregulated |
| -0,491271344 | 0,490695261 | RGPD8 | FALSE | Downregulated |
| 0,556391599 | 0,490695261 | SIGLEC14 | FALSE | Upregulated |
| -0,435381739 | 0,490695261 | SNCG | FALSE | Downregulated |
| -0,316107482 | 0,490695261 | TP53I11 | FALSE | Downregulated |
| 0,688045481 | 0,49071629 | LOC339192 | FALSE | Upregulated |

| | | | | |
|---|---|---|---|---|
| 0,277683238 | 0,491397929 | AXL | FALSE | Upregulated |
| 0,121149991 | 0,491863465 | CHFR | FALSE | Upregulated |
| -0,125034022 | 0,491863465 | DDHD2 | FALSE | Downregulated |
| 0,168637128 | 0,491863465 | ELMO2 | FALSE | Upregulated |
| 0,628169816 | 0,491863465 | F13A1 | FALSE | Upregulated |
| -0,575227483 | 0,491863465 | GPR158 | FALSE | Downregulated |
| 0,148395506 | 0,491863465 | ILVBL | FALSE | Upregulated |
| -0,124894722 | 0,491863465 | JADE3 | FALSE | Downregulated |
| -0,152976992 | 0,491863465 | LGALS8 | FALSE | Downregulated |
| -0,554239327 | 0,491863465 | LOC101927841 | FALSE | Downregulated |
| 0,624319702 | 0,491863465 | LOC728763 | FALSE | Upregulated |
| -0,111079402 | 0,491863465 | MOB1B | FALSE | Downregulated |
| 0,162821714 | 0,491863465 | NAGLU | FALSE | Upregulated |
| -0,170668093 | 0,491863465 | NUP37 | FALSE | Downregulated |
| 0,113418352 | 0,491863465 | PSMC5 | FALSE | Upregulated |
| 0,668852575 | 0,491863465 | RGS4 | FALSE | Upregulated |
| -0,179655095 | 0,491863465 | RPL23AP82 | FALSE | Downregulated |
| 0,26641506 | 0,491863465 | TELO2 | FALSE | Upregulated |
| 1,036573999 | 0,491863465 | TRIM31 | FALSE | Upregulated |
| -0,410928458 | 0,491863465 | UBAP1L | FALSE | Downregulated |
| 0,196122618 | 0,492514552 | DOK4 | FALSE | Upregulated |
| 0,144843295 | 0,492514552 | GTF2A2 | FALSE | Upregulated |
| -0,149140633 | 0,492514552 | KDM1B | FALSE | Downregulated |
| -0,769263593 | 0,492514552 | LOC101927192 | FALSE | Downregulated |
| -0,840839842 | 0,492514552 | LOC101929814 | FALSE | Downregulated |
| 0,220384967 | 0,492514552 | NME1 | FALSE | Upregulated |

| | | | | |
|---|---|---|---|---|
| 0,193956024 | 0,492514552 | PPIC | FALSE | Upregulated |
| -0,109261856 | 0,492514552 | RAD21 | FALSE | Downregulated |
| -0,119960653 | 0,492514552 | RBM7 | FALSE | Downregulated |
| -0,101613997 | 0,492514552 | SARNP | FALSE | Downregulated |
| 0,406363829 | 0,492514552 | SREBF1 | FALSE | Upregulated |
| 0,313150869 | 0,492514552 | SYT11 | FALSE | Upregulated |
| 0,222766921 | 0,492617676 | CDHR5 | FALSE | Upregulated |
| -0,14345747 | 0,492669118 | PSMA7 | FALSE | Downregulated |
| -0,207388492 | 0,493456465 | FLJ31306 | FALSE | Downregulated |
| -0,387162328 | 0,493456465 | MSMO1 | FALSE | Downregulated |
| -0,207109017 | 0,493456465 | PSENEN | FALSE | Downregulated |
| -0,266331694 | 0,493456465 | PSMD7 | FALSE | Downregulated |
| -0,118881937 | 0,493723141 | SBF2 | FALSE | Downregulated |
| -0,521382013 | 0,493953968 | GLYATL3 | FALSE | Downregulated |
| 0,939616861 | 0,493953968 | SIGLEC6 | FALSE | Upregulated |
| -0,272661496 | 0,494562146 | GPM6A | FALSE | Downregulated |
| 0,073566562 | 0,494562146 | SZRD1 | FALSE | Upregulated |
| -0,209549337 | 0,494577294 | MPHOSPH10 | FALSE | Downregulated |
| 0,515774436 | 0,494638319 | MS4A1 | FALSE | Upregulated |
| -0,42434957 | 0,494706946 | CD248 | FALSE | Downregulated |
| 1,190260609 | 0,495804255 | ANXA8L1 | FALSE | Upregulated |
| -0,185151226 | 0,495804255 | CWC22 | FALSE | Downregulated |
| 0,131415443 | 0,495804255 | DGCR2 | FALSE | Upregulated |
| -0,104009961 | 0,495804255 | IMPACT | FALSE | Downregulated |
| 0,142586942 | 0,495804255 | KANSL2 | FALSE | Upregulated |
| 0,376417631 | 0,495804255 | LAMA2 | FALSE | Upregulated |

| | | | | |
|---|---|---|---|---|
| 0,164819071 | 0,495804255 | WDR77 | FALSE | Upregulated |
| 0,330471649 | 0,495804255 | ZNF8 | FALSE | Upregulated |
| -0,17385209 | 0,496045642 | AZGP1 | FALSE | Downregulated |
| 0,127626377 | 0,496045642 | CPSF2 | FALSE | Upregulated |
| 0,184137449 | 0,496045642 | FOCAD | FALSE | Upregulated |
| -0,23545455 | 0,496045642 | GLYAT | FALSE | Downregulated |
| 0,715914263 | 0,496045642 | MYO7B | FALSE | Upregulated |
| 0,194663343 | 0,496045642 | NCAPG2 | FALSE | Upregulated |
| 0,10394217 | 0,496045642 | NDNL2 | FALSE | Upregulated |
| 0,146472911 | 0,496045642 | PDXK | FALSE | Upregulated |
| -0,478990241 | 0,496045642 | RAMP2-AS1 | FALSE | Downregulated |
| 0,168436102 | 0,496045642 | RAVER1 | FALSE | Upregulated |
| 0,205897953 | 0,496045642 | RRP7A | FALSE | Upregulated |
| 0,228588073 | 0,496045642 | SLC36A1 | FALSE | Upregulated |
| 0,633806905 | 0,496062185 | STC1 | FALSE | Upregulated |
| 0,100248121 | 0,496084905 | ZC3H18 | FALSE | Upregulated |
| -0,226848245 | 0,496089673 | CAT | FALSE | Downregulated |
| 0,191654963 | 0,496089673 | EVA1A | FALSE | Upregulated |
| 0,416693795 | 0,496089673 | GNA15 | FALSE | Upregulated |
| 0,093526191 | 0,496089673 | KCTD18 | FALSE | Upregulated |
| -0,170291218 | 0,496089673 | KCTD6 | FALSE | Downregulated |
| -0,131634497 | 0,496089673 | TBCE | FALSE | Downregulated |
| -0,805399733 | 0,496118615 | LINC00598 | FALSE | Downregulated |
| -0,107912709 | 0,497030836 | ZNF3 | FALSE | Downregulated |
| 0,131479721 | 0,497087425 | BCKDK | FALSE | Upregulated |
| 0,155409999 | 0,497087425 | CTDP1 | FALSE | Upregulated |

| | | | | |
|---|---|---|---|---|
| 0,133477074 | 0,497087425 | PPP2CB | FALSE | Upregulated |
| -0,129630262 | 0,497087425 | RAB8A | FALSE | Downregulated |
| -0,742141988 | 0,497087425 | RHBDL2 | FALSE | Downregulated |
| 0,176260212 | 0,497087425 | TEX264 | FALSE | Upregulated |
| -0,318416256 | 0,497213068 | AASS | FALSE | Downregulated |
| 0,138431417 | 0,497213068 | FAM219B | FALSE | Upregulated |
| -0,451396734 | 0,497255835 | MMRN1 | FALSE | Downregulated |
| 0,683095711 | 0,497284927 | LOC101929882 | FALSE | Upregulated |
| 0,893236706 | 0,497284927 | LOC102724584 | FALSE | Upregulated |
| 0,14462439 | 0,497284927 | MYO9B | FALSE | Upregulated |
| -0,158103214 | 0,497284927 | PRPF40A | FALSE | Downregulated |
| 0,28243129 | 0,497504303 | FAM35DP | FALSE | Upregulated |
| 0,372415025 | 0,497846495 | LTB | FALSE | Upregulated |
| 0,574164472 | 0,497846495 | PLXNB3 | FALSE | Upregulated |
| 0,426913415 | 0,497911814 | DGCR11 | FALSE | Upregulated |
| -0,33686635 | 0,497911814 | DLG2 | FALSE | Downregulated |
| -0,145340833 | 0,497911814 | PPA2 | FALSE | Downregulated |
| 0,256617458 | 0,497911814 | SLC25A24 | FALSE | Upregulated |
| -0,147058467 | 0,497944189 | TBC1D8 | FALSE | Downregulated |
| 0,349149611 | 0,497944189 | ZFAT | FALSE | Upregulated |
| -0,110464492 | 0,498288919 | RBPJ | FALSE | Downregulated |
| 0,763470642 | 0,498288919 | TTC16 | FALSE | Upregulated |
| 0,762829463 | 0,498352038 | ACHE | FALSE | Upregulated |
| -0,342088658 | 0,498352038 | TRNQ | FALSE | Downregulated |
| 1,334165096 | 0,498465721 | OPRM1 | FALSE | Upregulated |
| -0,156154002 | 0,499082601 | RBM5 | FALSE | Downregulated |

| | | | | |
|---|---|---|---|---|
| -0,169184471 | 0,499143347 | LOC102288414 | FALSE | Downregulated |
| 0,63103412 | 0,499147223 | HLA-J | FALSE | Upregulated |
| -0,122870381 | 0,499147223 | KAT6B | FALSE | Downregulated |
| -0,958467956 | 0,499147223 | RBM24 | FALSE | Downregulated |
| -0,105356682 | 0,499147223 | TLK2 | FALSE | Downregulated |
| -0,201140477 | 0,499147223 | TMEM176B | FALSE | Downregulated |
| -0,154198148 | 0,499224987 | COMMD2 | FALSE | Downregulated |
| -0,232942336 | 0,499224987 | NIFK-AS1 | FALSE | Downregulated |
| -0,104201816 | 0,499628672 | TADA2B | FALSE | Downregulated |
| 0,090755838 | 0,500096815 | ACTR1B | FALSE | Upregulated |
| -0,238434201 | 0,500096815 | C7orf63 | FALSE | Downregulated |
| -0,118526332 | 0,500096815 | CCDC50 | FALSE | Downregulated |
| 0,402651517 | 0,500096815 | CSTA | FALSE | Upregulated |
| 0,646913798 | 0,500096815 | CTBP1-AS | FALSE | Upregulated |
| 0,507701443 | 0,500096815 | DOC2B | FALSE | Upregulated |
| -0,156011753 | 0,500096815 | KMT2A | FALSE | Downregulated |
| -0,436631042 | 0,500096815 | PI4KAP1 | FALSE | Downregulated |
| 0,549432111 | 0,500096815 | TMEM150B | FALSE | Upregulated |
| 0,156103414 | 0,500096815 | USP46 | FALSE | Upregulated |
| -0,145884349 | 0,500375176 | AHCY | FALSE | Downregulated |
| 0,095507017 | 0,500883972 | DNAJC11 | FALSE | Upregulated |
| -0,195408995 | 0,501328776 | C8A | FALSE | Downregulated |
| -0,166166492 | 0,501328776 | GYS2 | FALSE | Downregulated |
| -0,173290103 | 0,501328776 | LMO7 | FALSE | Downregulated |
| 0,182656799 | 0,501328776 | SRA1 | FALSE | Upregulated |
| -0,113838162 | 0,50197082 | NDEL1 | FALSE | Downregulated |

| | | | | |
|---|---|---|---|---|
| 0,236051446 | 0,502166488 | ZNF213 | FALSE | Upregulated |
| 0,6636263 | 0,502175476 | CD1E | FALSE | Upregulated |
| 0,913869053 | 0,502175476 | F2RL3 | FALSE | Upregulated |
| -0,228383073 | 0,502284284 | AGMAT | FALSE | Downregulated |
| -0,142321825 | 0,502284284 | ATXN3 | FALSE | Downregulated |
| -0,133702227 | 0,502284284 | ETAA1 | FALSE | Downregulated |
| 1,504896338 | 0,502284284 | FIGF | FALSE | Upregulated |
| 0,359956466 | 0,502284284 | KLF5 | FALSE | Upregulated |
| 1,179699925 | 0,502284284 | LOC102723996 | FALSE | Upregulated |
| -0,792288254 | 0,502284284 | LOC102724256 | FALSE | Downregulated |
| 0,667343717 | 0,502284284 | SPATA25 | FALSE | Upregulated |
| -0,732941727 | 0,502284284 | TACR2 | FALSE | Downregulated |
| -0,115792413 | 0,502284284 | TTLL5 | FALSE | Downregulated |
| -0,278302823 | 0,502284284 | UPP1 | FALSE | Downregulated |
| 0,519357934 | 0,502332375 | CD40LG | FALSE | Upregulated |
| 0,115917362 | 0,502332375 | CECR5 | FALSE | Upregulated |
| 0,120610874 | 0,502332375 | G3BP1 | FALSE | Upregulated |
| 0,174340815 | 0,502332375 | GLTSCR1 | FALSE | Upregulated |
| -0,137143027 | 0,502332375 | RNF2 | FALSE | Downregulated |
| 0,541595549 | 0,502332375 | SCD | FALSE | Upregulated |
| 0,101909578 | 0,502332375 | SDF4 | FALSE | Upregulated |
| 0,149442786 | 0,502332375 | SPHK2 | FALSE | Upregulated |
| 0,127278358 | 0,502383483 | ZFYVE26 | FALSE | Upregulated |
| 0,332391577 | 0,502588329 | CENPF | FALSE | Upregulated |
| -0,299979805 | 0,502871516 | ELOVL6 | FALSE | Downregulated |
| -0,149866122 | 0,502871516 | LEO1 | FALSE | Downregulated |

| | | | | |
|---|---|---|---|---|
| -0,344403605 | 0,502871516 | LPPR1 | FALSE | Downregulated |
| 0,248862062 | 0,502871516 | NCKAP1L | FALSE | Upregulated |
| 0,094365575 | 0,502871516 | SLC35A4 | FALSE | Upregulated |
| 0,276817802 | 0,503027848 | KIFC3 | FALSE | Upregulated |
| -0,125834004 | 0,503152592 | BIRC2 | FALSE | Downregulated |
| 0,417407943 | 0,503152592 | CGREF1 | FALSE | Upregulated |
| -0,753437193 | 0,503152592 | GPR123 | FALSE | Downregulated |
| -0,248652819 | 0,503152592 | RAB13 | FALSE | Downregulated |
| -0,308424407 | 0,503152592 | TCTN2 | FALSE | Downregulated |
| -0,219194853 | 0,503152592 | TM4SF18 | FALSE | Downregulated |
| 0,170294494 | 0,503224394 | AP1S1 | FALSE | Upregulated |
| -0,231462405 | 0,503293981 | PARP6 | FALSE | Downregulated |
| 0,35368827 | 0,503293981 | SLAMF6 | FALSE | Upregulated |
| 0,387918496 | 0,503364942 | GLIS2 | FALSE | Upregulated |
| 0,580089342 | 0,503512068 | ALPK2 | FALSE | Upregulated |
| -0,219833477 | 0,503603661 | EVI5 | FALSE | Downregulated |
| 0,41122122 | 0,503603661 | SHBG | FALSE | Upregulated |
| -0,752965337 | 0,503603661 | SIM1 | FALSE | Downregulated |
| 0,289566065 | 0,503635627 | IRF7 | FALSE | Upregulated |
| 0,252706682 | 0,503831918 | ZNF385A | FALSE | Upregulated |
| -0,727733344 | 0,504047598 | CMYA5 | FALSE | Downregulated |
| 0,269990557 | 0,504047598 | NFKB2 | FALSE | Upregulated |
| -0,607260991 | 0,504300347 | EFHB | FALSE | Downregulated |
| -0,092148266 | 0,504300347 | PSMD4 | FALSE | Downregulated |
| -0,179874933 | 0,504300347 | SKIL | FALSE | Downregulated |
| -0,159306151 | 0,504536474 | NDRG3 | FALSE | Downregulated |

| | | | | |
|---|---|---|---|---|
| -1,01853752 | 0,504892737 | IL1RAPL1 | FALSE | Downregulated |
| 0,356809663 | 0,504892737 | RALGPS1 | FALSE | Upregulated |
| -0,368177432 | 0,505167946 | SSTR2 | FALSE | Downregulated |
| 0,444886593 | 0,505610617 | SH3D21 | FALSE | Upregulated |
| -0,199833265 | 0,505610617 | UGT2B15 | FALSE | Downregulated |
| -0,120184528 | 0,505670043 | WASL | FALSE | Downregulated |
| -0,091080304 | 0,505826444 | FAF1 | FALSE | Downregulated |
| -0,11026722 | 0,505880547 | REXO4 | FALSE | Downregulated |
| -0,404033403 | 0,506106037 | ISM1 | FALSE | Downregulated |
| 0,146618733 | 0,506106037 | MAP1LC3B | FALSE | Upregulated |
| -0,084910994 | 0,506106037 | RAD17 | FALSE | Downregulated |
| 0,208612972 | 0,506106037 | RAPGEF6 | FALSE | Upregulated |
| -0,219996254 | 0,506809869 | CEP63 | FALSE | Downregulated |
| -0,388211682 | 0,506809869 | GNAO1 | FALSE | Downregulated |
| -0,205073958 | 0,506809869 | PDCL3 | FALSE | Downregulated |
| 0,282086122 | 0,506809869 | RBPMS2 | FALSE | Upregulated |
| 0,440955819 | 0,506924103 | ITM2A | FALSE | Upregulated |
| 0,514007047 | 0,507306665 | THY1 | FALSE | Upregulated |
| -0,121780749 | 0,507450324 | NFIB | FALSE | Downregulated |
| -0,222453995 | 0,507915647 | C2orf88 | FALSE | Downregulated |
| 0,186409728 | 0,507915647 | FRY | FALSE | Upregulated |
| -0,209148447 | 0,507977792 | PRAF2 | FALSE | Downregulated |
| 0,222929346 | 0,508196055 | CDPF1 | FALSE | Upregulated |
| 0,299979129 | 0,508196055 | CLEC11A | FALSE | Upregulated |
| -0,7650711 | 0,508214954 | AC002398.5 | FALSE | Downregulated |
| 0,257507078 | 0,508214954 | ANXA5 | FALSE | Upregulated |

| | | | | |
|---|---|---|---|---|
| 0,206877416 | 0,508214954 | BAIAP2 | FALSE | Upregulated |
| -0,239018195 | 0,508214954 | CFHR5 | FALSE | Downregulated |
| -0,681097249 | 0,508214954 | CTC-558O2.1 | FALSE | Downregulated |
| 0,237466977 | 0,508214954 | DAB2IP | FALSE | Upregulated |
| 0,157077201 | 0,508214954 | DPH3 | FALSE | Upregulated |
| -0,518594039 | 0,508214954 | FAM78B | FALSE | Downregulated |
| -0,70457127 | 0,508214954 | KIAA1755 | FALSE | Downregulated |
| -1,11402175 | 0,508214954 | LOC101928430 | FALSE | Downregulated |
| 1,138543534 | 0,508214954 | MAG | FALSE | Upregulated |
| 0,150872358 | 0,508214954 | MNT | FALSE | Upregulated |
| 1,082656791 | 0,508214954 | MTRNR2L4 | FALSE | Upregulated |
| -0,154762701 | 0,508214954 | NKAP | FALSE | Downregulated |
| -0,112897914 | 0,508214954 | NOL7 | FALSE | Downregulated |
| 0,191770096 | 0,508214954 | PARP8 | FALSE | Upregulated |
| 0,145259916 | 0,508214954 | PICK1 | FALSE | Upregulated |
| 0,153354071 | 0,508214954 | POLR3K | FALSE | Upregulated |
| -0,195077122 | 0,508214954 | RCAN1 | FALSE | Downregulated |
| -0,349158025 | 0,508214954 | RP11-342C23.4 | FALSE | Downregulated |
| -0,103688717 | 0,508214954 | SART3 | FALSE | Downregulated |
| 0,27269337 | 0,508214954 | TAP2 | FALSE | Upregulated |
| -0,210511609 | 0,508214954 | UBXN2B | FALSE | Downregulated |
| 0,11943867 | 0,508214954 | WWP2 | FALSE | Upregulated |
| -0,901863527 | 0,508529067 | BEND6 | FALSE | Downregulated |
| 0,168463791 | 0,508529067 | SH3GLB2 | FALSE | Upregulated |
| -0,167940551 | 0,508529067 | THAP5 | FALSE | Downregulated |
| -0,371857047 | 0,509158563 | RPLP0P2 | FALSE | Downregulated |

| | | | | |
|---|---|---|---|---|
| -0,330722735 | 0,509280753 | ACSM2A | FALSE | Downregulated |
| 1,262657549 | 0,509280753 | AIM1L | FALSE | Upregulated |
| 0,145295976 | 0,509280753 | CCDC124 | FALSE | Upregulated |
| 0,156378126 | 0,509280753 | CD59 | FALSE | Upregulated |
| -0,108215784 | 0,509280753 | CDC5L | FALSE | Downregulated |
| -0,120218002 | 0,509280753 | CLDN1 | FALSE | Downregulated |
| 0,250338821 | 0,509280753 | CMTM3 | FALSE | Upregulated |
| -0,138553199 | 0,509280753 | CPPED1 | FALSE | Downregulated |
| -0,114754328 | 0,509280753 | CPSF6 | FALSE | Downregulated |
| 0,128137921 | 0,509280753 | ELMOD2 | FALSE | Upregulated |
| 0,3526622 | 0,509280753 | EOMES | FALSE | Upregulated |
| 0,651937353 | 0,509280753 | FAM83F | FALSE | Upregulated |
| -0,11880934 | 0,509280753 | FBXO38 | FALSE | Downregulated |
| -0,473436097 | 0,509280753 | FLJ30403 | FALSE | Downregulated |
| 0,6151648 | 0,509280753 | GNB1L | FALSE | Upregulated |
| -0,102530768 | 0,509280753 | HDHD2 | FALSE | Downregulated |
| 0,403080566 | 0,509280753 | LUM | FALSE | Upregulated |
| -0,170276475 | 0,509280753 | PCCB | FALSE | Downregulated |
| -0,140664025 | 0,509280753 | PLEKHA5 | FALSE | Downregulated |
| -0,670325092 | 0,509280753 | PPBP | FALSE | Downregulated |
| -0,103044281 | 0,509280753 | PRKCI | FALSE | Downregulated |
| 0,319283289 | 0,509280753 | SRPX2 | FALSE | Upregulated |
| -0,861666042 | 0,509280753 | TMPRSS5 | FALSE | Downregulated |
| 0,182884061 | 0,509280753 | ZNF219 | FALSE | Upregulated |
| -0,283741043 | 0,509823594 | C9orf16 | FALSE | Downregulated |
| 0,41086858 | 0,509917995 | FAM109B | FALSE | Upregulated |

| | | | | |
|---|---|---|---|---|
| -0,589021995 | 0,510053756 | ATP8B5P | FALSE | Downregulated |
| 0,20922819 | 0,510053756 | GLMN | FALSE | Upregulated |
| 0,827121239 | 0,510053756 | TAS2R14 | FALSE | Upregulated |
| -0,236825164 | 0,510883704 | CYP1B1 | FALSE | Downregulated |
| 0,40126813 | 0,510883704 | ENPP7 | FALSE | Upregulated |
| 0,169104651 | 0,510883704 | MST1 | FALSE | Upregulated |
| -0,155579888 | 0,511000653 | NUDT16 | FALSE | Downregulated |
| 0,894189982 | 0,511035905 | IFNG-AS1 | FALSE | Upregulated |
| 0,124171581 | 0,511035905 | PLEKHM2 | FALSE | Upregulated |
| 0,45925347 | 0,511316687 | C5AR2 | FALSE | Upregulated |
| -0,154199695 | 0,511316687 | RBM12B | FALSE | Downregulated |
| 0,86369883 | 0,511316687 | SLC5A10 | FALSE | Upregulated |
| 1,306055945 | 0,511316687 | ST20-MTHFS | FALSE | Upregulated |
| -0,124687142 | 0,511348415 | AHCTF1 | FALSE | Downregulated |
| 0,327867973 | 0,511348415 | BACE2 | FALSE | Upregulated |
| -0,096938932 | 0,511348415 | EXOC5 | FALSE | Downregulated |
| 0,670308166 | 0,511348415 | RPL21P44 | FALSE | Upregulated |
| -0,076424034 | 0,511479089 | CUL3 | FALSE | Downregulated |
| 0,303084658 | 0,511673386 | MSR1 | FALSE | Upregulated |
| -0,242598838 | 0,512095684 | ARL6IP6 | FALSE | Downregulated |
| -0,137730438 | 0,512104649 | PARP3 | FALSE | Downregulated |
| 0,492491114 | 0,512392383 | FSTL3 | FALSE | Upregulated |
| -0,152547598 | 0,512392383 | PDAP1 | FALSE | Downregulated |
| 0,695084981 | 0,512392383 | ZNF804A | FALSE | Upregulated |
| -0,109953141 | 0,512403971 | ACTR10 | FALSE | Downregulated |
| 0,451465011 | 0,512403971 | FAHD2CP | FALSE | Upregulated |

| | | | | |
|---|---|---|---|---|
| 0,489273923 | 0,512403971 | FAM156B | FALSE | Upregulated |
| -0,228968357 | 0,512403971 | FBXO5 | FALSE | Downregulated |
| 0,637277614 | 0,512403971 | GTF2IRD1P1 | FALSE | Upregulated |
| -0,100819595 | 0,512403971 | GXYLT1 | FALSE | Downregulated |
| -0,167919196 | 0,512403971 | MBNL3 | FALSE | Downregulated |
| -0,140007741 | 0,512403971 | MPP6 | FALSE | Downregulated |
| -2,034517541 | 0,512403971 | MYO18B | FALSE | Downregulated |
| 0,213182137 | 0,512403971 | PMM2 | FALSE | Upregulated |
| 0,399167062 | 0,512929132 | PLXNA3 | FALSE | Upregulated |
| -0,120440203 | 0,512955433 | ATG3 | FALSE | Downregulated |
| 0,248433134 | 0,512955433 | MAFK | FALSE | Upregulated |
| -0,244668248 | 0,512955433 | MAP3K5 | FALSE | Downregulated |
| 0,797988606 | 0,513258182 | LOC100129516 | FALSE | Upregulated |
| -0,275952996 | 0,513258182 | LUC7L2 | FALSE | Downregulated |
| 0,506458092 | 0,513258182 | ST14 | FALSE | Upregulated |
| -0,16415365 | 0,513923195 | PCBD1 | FALSE | Downregulated |
| -0,928331882 | 0,514107469 | FAM41C | FALSE | Downregulated |
| -0,446409703 | 0,514107469 | POSTN | FALSE | Downregulated |
| -0,154820329 | 0,514243464 | DDX55 | FALSE | Downregulated |
| 0,463620742 | 0,514243464 | LOC101927903 | FALSE | Upregulated |
| 1,149787244 | 0,514243464 | SLC6A20 | FALSE | Upregulated |
| 0,310939193 | 0,514310034 | NFAM1 | FALSE | Upregulated |
| 0,465482614 | 0,514511769 | ITPKA | FALSE | Upregulated |
| -0,102707673 | 0,514511769 | PDIK1L | FALSE | Downregulated |
| -1,68409855 | 0,514511769 | PI3 | FALSE | Downregulated |
| 0,291856667 | 0,514655852 | COL4A2 | FALSE | Upregulated |

| | | | | |
|---|---|---|---|---|
| -0,394317441 | 0,514655852 | CUX2 | FALSE | Downregulated |
| 0,280780638 | 0,514655852 | DENND1C | FALSE | Upregulated |
| 0,12124172 | 0,514655852 | LEMD3 | FALSE | Upregulated |
| 0,196649142 | 0,514655852 | RAD51D | FALSE | Upregulated |
| -0,299919782 | 0,514655852 | RFESD | FALSE | Downregulated |
| -0,778578425 | 0,514655852 | TRNW | FALSE | Downregulated |
| 0,670303549 | 0,515066165 | ZNF812 | FALSE | Upregulated |
| 0,319612018 | 0,515383807 | DENND6B | FALSE | Upregulated |
| 0,129024315 | 0,515383807 | SLC15A4 | FALSE | Upregulated |
| -0,189662738 | 0,515403894 | ASB3 | FALSE | Downregulated |
| -0,397381799 | 0,515403894 | DBP | FALSE | Downregulated |
| -0,463472517 | 0,515403894 | FAM19A2 | FALSE | Downregulated |
| 0,205359671 | 0,515403894 | NUP93 | FALSE | Upregulated |
| 0,134016749 | 0,515403894 | VMP1 | FALSE | Upregulated |
| -0,751071596 | 0,515403894 | YJEFN3 | FALSE | Downregulated |
| 0,325020335 | 0,515530266 | ITPR3 | FALSE | Upregulated |
| -0,350121375 | 0,515723889 | CD274 | FALSE | Downregulated |
| 0,14133593 | 0,515810709 | ZNF263 | FALSE | Upregulated |
| 0,723245415 | 0,515935511 | GOLGA7B | FALSE | Upregulated |
| 0,323818358 | 0,515935511 | MGC57346 | FALSE | Upregulated |
| -0,105724439 | 0,515935511 | ORC4 | FALSE | Downregulated |
| 0,627836585 | 0,515935511 | RGL4 | FALSE | Upregulated |
| 0,324784983 | 0,515946092 | AFAP1 | FALSE | Upregulated |
| -0,117777258 | 0,515946092 | EIF3E | FALSE | Downregulated |
| -0,207011122 | 0,515946092 | SERINC5 | FALSE | Downregulated |
| -0,12943496 | 0,516044401 | LARP7 | FALSE | Downregulated |

| | | | | |
|---|---|---|---|---|
| 0,289125268 | 0,516044401 | LOC101928465 | FALSE | Upregulated |
| 0,291836417 | 0,516044401 | PDE7A | FALSE | Upregulated |
| -0,192909192 | 0,516044401 | TRMT10A | FALSE | Downregulated |
| -0,897433022 | 0,516221449 | CACNB1 | FALSE | Downregulated |
| -0,165268597 | 0,516221449 | CDK6 | FALSE | Downregulated |
| -0,220707133 | 0,516221449 | SEPT7P2 | FALSE | Downregulated |
| 0,215366272 | 0,516352966 | RHOC | FALSE | Upregulated |
| 0,371928243 | 0,516706735 | KLRD1 | FALSE | Upregulated |
| 0,166091913 | 0,516706735 | SLC6A12 | FALSE | Upregulated |
| 0,18603079 | 0,516888561 | AVPI1 | FALSE | Upregulated |
| -0,178370186 | 0,516888561 | DMGDH | FALSE | Downregulated |
| 0,225963812 | 0,517118414 | NOTCH4 | FALSE | Upregulated |
| 0,16939253 | 0,517297678 | LAMB2 | FALSE | Upregulated |
| 0,14793744 | 0,517299062 | IARS | FALSE | Upregulated |
| -0,134309018 | 0,51734218 | ATP5H | FALSE | Downregulated |
| 0,123537276 | 0,51734218 | CCDC137 | FALSE | Upregulated |
| -0,097798388 | 0,51734218 | MAPKAPK5 | FALSE | Downregulated |
| 0,187320139 | 0,51734218 | MED22 | FALSE | Upregulated |
| 0,575805753 | 0,51734218 | MTRNR2L10 | FALSE | Upregulated |
| 0,453283395 | 0,51734218 | WDR65 | FALSE | Upregulated |
| 0,144974446 | 0,517415426 | FAM53B | FALSE | Upregulated |
| 0,79904572 | 0,517415426 | LINC00996 | FALSE | Upregulated |
| -0,671137472 | 0,51808079 | LOC100268168 | FALSE | Downregulated |
| 0,51257672 | 0,51808079 | LOC100505564 | FALSE | Upregulated |
| 0,148189919 | 0,51808079 | RFWD3 | FALSE | Upregulated |
| 0,12984007 | 0,51808079 | RXRB | FALSE | Upregulated |

| | | | | |
|---|---|---|---|---|
| -0,105863952 | 0,51808079 | SP3 | FALSE | Downregulated |
| -0,104873868 | 0,51808079 | TAB3 | FALSE | Downregulated |
| -0,25366105 | 0,51825224 | MLLT3 | FALSE | Downregulated |
| -0,209009108 | 0,518327696 | AFM | FALSE | Downregulated |
| 0,107887323 | 0,518327696 | HTT | FALSE | Upregulated |
| 0,741854303 | 0,518327696 | LOC102724611 | FALSE | Upregulated |
| -0,866363957 | 0,518327696 | LOC102724908 | FALSE | Downregulated |
| -0,147629182 | 0,518327696 | SLIRP | FALSE | Downregulated |
| 0,652626428 | 0,518327696 | SNORD89 | FALSE | Upregulated |
| 0,354757587 | 0,519117883 | PLXNC1 | FALSE | Upregulated |
| -0,206549175 | 0,519160774 | PRRG4 | FALSE | Downregulated |
| -0,287866438 | 0,519160774 | RP11-60I3.5 | FALSE | Downregulated |
| -0,086051541 | 0,519214645 | ARL5A | FALSE | Downregulated |
| 0,208539944 | 0,519214645 | MFSD12 | FALSE | Upregulated |
| -0,508865952 | 0,519214645 | MORF4L2-AS1 | FALSE | Downregulated |
| 0,174603125 | 0,519214645 | OSTM1 | FALSE | Upregulated |
| 0,564609934 | 0,519229862 | CNNM1 | FALSE | Upregulated |
| -0,134863563 | 0,519370234 | PTPN18 | FALSE | Downregulated |
| -0,361475842 | 0,520400207 | PFN2 | FALSE | Downregulated |
| 0,170331276 | 0,520447555 | EHD1 | FALSE | Upregulated |
| -0,288606555 | 0,520516317 | LOC101929787 | FALSE | Downregulated |
| 0,473196356 | 0,520516317 | SDCBP2 | FALSE | Upregulated |
| -0,164804207 | 0,520550281 | TRMT13 | FALSE | Downregulated |
| 0,2880948 | 0,520579841 | CAPS2 | FALSE | Upregulated |
| -0,115193601 | 0,520587136 | WWP1 | FALSE | Downregulated |
| 0,272464128 | 0,520639142 | ACSL6 | FALSE | Upregulated |

| | | | | |
|---|---|---|---|---|
| -0,611604837 | 0,520639142 | AVPR1A | FALSE | Downregulated |
| 0,399468301 | 0,520639142 | CD72 | FALSE | Upregulated |
| -0,186008761 | 0,520639142 | CYP4V2 | FALSE | Downregulated |
| 0,113794847 | 0,520639142 | MYH14 | FALSE | Upregulated |
| -0,14240038 | 0,520639142 | PPP3R1 | FALSE | Downregulated |
| 0,092029669 | 0,520639142 | PRPF8 | FALSE | Upregulated |
| -0,13777552 | 0,520639142 | RALGAPA1 | FALSE | Downregulated |
| 0,980283106 | 0,520639142 | RASAL1 | FALSE | Upregulated |
| 0,465046006 | 0,520639142 | TRAT1 | FALSE | Upregulated |
| -0,123451113 | 0,520771572 | MPC2 | FALSE | Downregulated |
| 0,349949855 | 0,52087271 | CST7 | FALSE | Upregulated |
| 0,657579458 | 0,52087271 | EME2 | FALSE | Upregulated |
| 0,124752492 | 0,52087271 | KIAA2013 | FALSE | Upregulated |
| 0,53425198 | 0,52087271 | RAB7B | FALSE | Upregulated |
| 0,944800457 | 0,521116721 | FAM150A | FALSE | Upregulated |
| -0,284087507 | 0,521409396 | IL6R | FALSE | Downregulated |
| 0,133840118 | 0,521409396 | NHP2 | FALSE | Upregulated |
| 0,250637196 | 0,521613578 | METTL1 | FALSE | Upregulated |
| -0,231630612 | 0,521613578 | NRP1 | FALSE | Downregulated |
| -0,176308958 | 0,521613578 | WDR47 | FALSE | Downregulated |
| -0,301235133 | 0,522021015 | IL13RA2 | FALSE | Downregulated |
| -0,344154931 | 0,522021015 | LOC100506286 | FALSE | Downregulated |
| 0,39018375 | 0,522021015 | TAGLN | FALSE | Upregulated |
| -0,129719865 | 0,522021015 | TRAP1 | FALSE | Downregulated |
| -0,18136376 | 0,522036543 | GIPC2 | FALSE | Downregulated |
| 0,127848465 | 0,522036543 | GLRX3 | FALSE | Upregulated |

| | | | | |
|---|---|---|---|---|
| -0,2443482 | 0,522036543 | LNP1 | FALSE | Downregulated |
| 0,165294974 | 0,522036543 | MASTL | FALSE | Upregulated |
| -0,434474338 | 0,522036543 | NBPF8 | FALSE | Downregulated |
| 0,328025775 | 0,522036543 | OSBPL3 | FALSE | Upregulated |
| -0,141889928 | 0,522036543 | PCGF5 | FALSE | Downregulated |
| 0,441265304 | 0,522036543 | RECQL4 | FALSE | Upregulated |
| -0,144927311 | 0,522036543 | RNF219 | FALSE | Downregulated |
| -0,184245159 | 0,522036543 | SLAIN1 | FALSE | Downregulated |
| -0,215556776 | 0,52203868 | LOC101060195 | FALSE | Downregulated |
| 0,157900735 | 0,52203868 | RCC2 | FALSE | Upregulated |
| 0,578763099 | 0,52203868 | SSSCA1-AS1 | FALSE | Upregulated |
| -0,180178145 | 0,522566563 | AQP3 | FALSE | Downregulated |
| -0,279962222 | 0,522566563 | CBWD1 | FALSE | Downregulated |
| -0,187233958 | 0,522566563 | JADE1 | FALSE | Downregulated |
| -0,490873124 | 0,522566563 | LOC102724645 | FALSE | Downregulated |
| -0,2449231 | 0,522566563 | ZNF708 | FALSE | Downregulated |
| 0,804651955 | 0,522888281 | DISP2 | FALSE | Upregulated |
| 0,288783114 | 0,522888281 | GPR65 | FALSE | Upregulated |
| -0,400498082 | 0,522888281 | LOC100506060 | FALSE | Downregulated |
| -0,302942584 | 0,522888281 | RP11-178C3.2 | FALSE | Downregulated |
| -0,162418781 | 0,523234133 | APOOL | FALSE | Downregulated |
| -0,251752502 | 0,523234133 | C7orf55 | FALSE | Downregulated |
| -0,130802062 | 0,523234133 | DUSP3 | FALSE | Downregulated |
| 0,427411895 | 0,523234133 | DUSP4 | FALSE | Upregulated |
| -0,083515171 | 0,523234133 | H3F3B | FALSE | Downregulated |
| -0,163534931 | 0,523234133 | KANK2 | FALSE | Downregulated |

| | | | | |
|---|---|---|---|---|
| 0,529179015 | 0,523234133 | LAMP3 | FALSE | Upregulated |
| 1,012457328 | 0,523234133 | LOC100288175 | FALSE | Upregulated |
| 0,410140307 | 0,523234133 | MAP1B | FALSE | Upregulated |
| -0,117939947 | 0,523234133 | PARP16 | FALSE | Downregulated |
| -0,944719747 | 0,523234133 | PDLIM3 | FALSE | Downregulated |
| 0,152343942 | 0,523234133 | PTPN6 | FALSE | Upregulated |
| -0,098914378 | 0,523393647 | GLTSCR1L | FALSE | Downregulated |
| -0,222546459 | 0,523393647 | SULT1A2 | FALSE | Downregulated |
| -0,148499338 | 0,523505891 | METTL2B | FALSE | Downregulated |
| 0,978868175 | 0,52375235 | SLITRK2 | FALSE | Upregulated |
| 0,155365344 | 0,523916637 | FARSA | FALSE | Upregulated |
| -0,221166328 | 0,523916637 | NCL | FALSE | Downregulated |
| -0,213616914 | 0,523919966 | C1orf35 | FALSE | Downregulated |
| 0,561979888 | 0,523919966 | LOC102724187 | FALSE | Upregulated |
| -0,195705415 | 0,523919966 | NSRP1 | FALSE | Downregulated |
| 0,242931863 | 0,524622254 | NLE1 | FALSE | Upregulated |
| -0,14393365 | 0,524622254 | UBE3A | FALSE | Downregulated |
| -0,393121822 | 0,524657324 | FPR1 | FALSE | Downregulated |
| -0,139442004 | 0,524799173 | ICT1 | FALSE | Downregulated |
| -0,100073563 | 0,524799173 | PDE8A | FALSE | Downregulated |
| -0,365530742 | 0,524845847 | EDNRA | FALSE | Downregulated |
| 0,217479541 | 0,524845847 | LIPA | FALSE | Upregulated |
| -0,137594683 | 0,524845847 | SNX13 | FALSE | Downregulated |
| -1,631517345 | 0,524845847 | TMOD4 | FALSE | Downregulated |
| -0,346614954 | 0,524857441 | LOC553103 | FALSE | Downregulated |
| 0,631589491 | 0,524857441 | SPC25 | FALSE | Upregulated |

| | | | | |
|---|---|---|---|---|
| 0,630498371 | 0,524977938 | LOC102723340 | FALSE | Upregulated |
| 0,712883271 | 0,525139852 | LINC00571 | FALSE | Upregulated |
| -0,373004153 | 0,525283932 | LOC102724833 | FALSE | Downregulated |
| 0,177411511 | 0,525512069 | ABHD2 | FALSE | Upregulated |
| 0,105624876 | 0,525512069 | ARAP1 | FALSE | Upregulated |
| 0,414140179 | 0,525512069 | PCP4L1 | FALSE | Upregulated |
| 0,254190686 | 0,525512069 | SIGLEC11 | FALSE | Upregulated |
| -0,161933264 | 0,525512069 | TTC28 | FALSE | Downregulated |
| -0,102025787 | 0,525512069 | USP28 | FALSE | Downregulated |
| 0,128733272 | 0,525512069 | ZNF790 | FALSE | Upregulated |
| -0,334242098 | 0,525757569 | TPTE2P5 | FALSE | Downregulated |
| 0,384100677 | 0,526198861 | CCND1 | FALSE | Upregulated |
| -0,298550031 | 0,526198861 | HPGD | FALSE | Downregulated |
| 0,136117605 | 0,526198861 | PITPNA | FALSE | Upregulated |
| 0,283626019 | 0,526198861 | PPP1R3D | FALSE | Upregulated |
| 0,988511531 | 0,526422908 | LOC101929950 | FALSE | Upregulated |
| 0,084617026 | 0,526422908 | TERF2 | FALSE | Upregulated |
| -0,932297897 | 0,526590946 | LOC102724682 | FALSE | Downregulated |
| 0,249106141 | 0,526590946 | SPATA20 | FALSE | Upregulated |
| 0,621216033 | 0,526740736 | CDH6 | FALSE | Upregulated |
| 0,357568304 | 0,526793322 | FAM212B | FALSE | Upregulated |
| 0,225708071 | 0,526793322 | PPP1R16B | FALSE | Upregulated |
| -0,1596003 | 0,527055297 | AKAP8 | FALSE | Downregulated |
| 0,298706768 | 0,527055297 | FBXO2 | FALSE | Upregulated |
| -0,146751115 | 0,527055297 | INTS12 | FALSE | Downregulated |
| 0,564713659 | 0,527055297 | MEI1 | FALSE | Upregulated |

| | | | | |
|---|---|---|---|---|
| -0,139496152 | 0,527091315 | MRPS15 | FALSE | Downregulated |
| 0,239152714 | 0,527091315 | UBE3D | FALSE | Upregulated |
| 0,63017931 | 0,527135701 | KCNK13 | FALSE | Upregulated |
| -0,863331716 | 0,527504519 | NPY6R | FALSE | Downregulated |
| 0,502794052 | 0,527504519 | TMEM45B | FALSE | Upregulated |
| -0,130474057 | 0,527515798 | EPM2AIP1 | FALSE | Downregulated |
| 0,623394861 | 0,527515798 | LOC101930071 | FALSE | Upregulated |
| 0,134903422 | 0,527515798 | MSRB1 | FALSE | Upregulated |
| 0,360169676 | 0,527515798 | MYC | FALSE | Upregulated |
| 0,2659068 | 0,527515798 | RACGAP1 | FALSE | Upregulated |
| 0,371834328 | 0,527607958 | PLVAP | FALSE | Upregulated |
| 0,169775103 | 0,527758089 | SCFD2 | FALSE | Upregulated |
| -0,067472738 | 0,527758089 | SRSF1 | FALSE | Downregulated |
| 0,541987027 | 0,527758089 | ZACN | FALSE | Upregulated |
| 0,130914772 | 0,527859569 | MAP3K3 | FALSE | Upregulated |
| -0,091211641 | 0,527881577 | CHMP7 | FALSE | Downregulated |
| -0,258639059 | 0,527881577 | LINC01024 | FALSE | Downregulated |
| 0,106863494 | 0,527881577 | PSAP | FALSE | Upregulated |
| -0,154111951 | 0,527881577 | ZNF292 | FALSE | Downregulated |
| -0,102010759 | 0,527923598 | BRD7 | FALSE | Downregulated |
| -0,445198654 | 0,527923598 | GADD45B | FALSE | Downregulated |
| 0,37460083 | 0,52827797 | LINC00338 | FALSE | Upregulated |
| -0,370708989 | 0,52827797 | NBPF20 | FALSE | Downregulated |
| -0,143262441 | 0,52827797 | TTC3 | FALSE | Downregulated |
| 0,175797073 | 0,528321506 | ABCB6 | FALSE | Upregulated |
| 0,185353848 | 0,528321506 | FGD6 | FALSE | Upregulated |

| | | | | |
|---|---|---|---|---|
| 0,353698884 | 0,528321506 | PHACTR1 | FALSE | Upregulated |
| -0,271229829 | 0,528321506 | SLC25A23 | FALSE | Downregulated |
| 0,153428005 | 0,528321506 | THAP11 | FALSE | Upregulated |
| 0,125203548 | 0,528415988 | CAPRIN1 | FALSE | Upregulated |
| 0,096603707 | 0,528415988 | NCBP1 | FALSE | Upregulated |
| -0,096596647 | 0,528415988 | NUP160 | FALSE | Downregulated |
| 0,17024207 | 0,528415988 | TUBA1B | FALSE | Upregulated |
| 1,034072708 | 0,529300168 | MDFI | FALSE | Upregulated |
| 0,830811908 | 0,529340805 | AP000330.8 | FALSE | Upregulated |
| 0,156798149 | 0,529340805 | RNF126 | FALSE | Upregulated |
| 0,141466513 | 0,529347591 | ZNHIT3 | FALSE | Upregulated |
| 0,356285159 | 0,529440279 | SLC16A13 | FALSE | Upregulated |
| 0,129351797 | 0,529489413 | ADO | FALSE | Upregulated |
| -0,224802169 | 0,529489413 | LOC286437 | FALSE | Downregulated |
| 0,325106373 | 0,52950266 | FGR | FALSE | Upregulated |
| 0,528583352 | 0,52950266 | RP11-611D20.2 | FALSE | Upregulated |
| 0,727384154 | 0,529572893 | SLC26A8 | FALSE | Upregulated |
| -0,102816031 | 0,530070202 | MED21 | FALSE | Downregulated |
| -0,115296959 | 0,530267401 | EIF1AD | FALSE | Downregulated |
| 0,347668944 | 0,530649761 | NR4A2 | FALSE | Upregulated |
| 0,189841961 | 0,530649761 | SEC61A1 | FALSE | Upregulated |
| 0,296864577 | 0,530649761 | TSHZ3 | FALSE | Upregulated |
| -0,740049739 | 0,530769646 | FLJ42258 | FALSE | Downregulated |
| -0,123044874 | 0,530769646 | NONO | FALSE | Downregulated |
| -0,401930389 | 0,530769646 | NRARP | FALSE | Downregulated |
| -0,219054898 | 0,530769646 | PDE3B | FALSE | Downregulated |

| | | | | |
|---|---|---|---|---|
| 0,181035266 | 0,530769646 | PPTC7 | FALSE | Upregulated |
| 0,174153265 | 0,530769646 | TMEM175 | FALSE | Upregulated |
| 0,086553057 | 0,53082066 | SCYL1 | FALSE | Upregulated |
| 0,240688777 | 0,530897237 | MARVELD1 | FALSE | Upregulated |
| -0,582570486 | 0,53118437 | AGBL4 | FALSE | Downregulated |
| -0,268802193 | 0,53118437 | GCOM1 | FALSE | Downregulated |
| 0,181810498 | 0,53118437 | RASAL2 | FALSE | Upregulated |
| -0,250464329 | 0,531240511 | ARMCX1 | FALSE | Downregulated |
| -0,373889823 | 0,531240511 | HMGCR | FALSE | Downregulated |
| 0,112112123 | 0,531240511 | MFF | FALSE | Upregulated |
| 0,809647713 | 0,531240511 | NPTX2 | FALSE | Upregulated |
| 0,431717756 | 0,53129734 | CROCCP3 | FALSE | Upregulated |
| 0,082015142 | 0,531299787 | MTCH1 | FALSE | Upregulated |
| -0,184255534 | 0,531338113 | MLLT1 | FALSE | Downregulated |
| 0,318314711 | 0,531338113 | TNFSF8 | FALSE | Upregulated |
| -0,121307769 | 0,531338113 | ZNF280D | FALSE | Downregulated |
| -0,49637186 | 0,531424803 | ADAMTS6 | FALSE | Downregulated |
| -0,111365332 | 0,531424803 | CBX1 | FALSE | Downregulated |
| 0,487518639 | 0,531424803 | CHEK1 | FALSE | Upregulated |
| 0,231873588 | 0,531424803 | CYB561 | FALSE | Upregulated |
| -0,143165913 | 0,531424803 | DIEXF | FALSE | Downregulated |
| 0,161191084 | 0,531424803 | EMC8 | FALSE | Upregulated |
| 0,316444643 | 0,531424803 | GINS3 | FALSE | Upregulated |
| -0,136503819 | 0,531424803 | ICK | FALSE | Downregulated |
| 0,825208441 | 0,531424803 | LOC102724248 | FALSE | Upregulated |
| -1,102939678 | 0,531424803 | LOC102724681 | FALSE | Downregulated |

| | | | | |
|---|---|---|---|---|
| 0,372380011 | 0,531424803 | NLRP3 | FALSE | Upregulated |
| 0,819493819 | 0,531424803 | NOS2 | FALSE | Upregulated |
| 0,181885019 | 0,531424803 | RASSF4 | FALSE | Upregulated |
| -0,354479117 | 0,531424803 | SNHG19 | FALSE | Downregulated |
| 0,286093715 | 0,531424803 | TUBB6 | FALSE | Upregulated |
| -0,100197044 | 0,531424803 | UBQLN2 | FALSE | Downregulated |
| 0,386026668 | 0,531424803 | WASH2P | FALSE | Upregulated |
| 0,158255399 | 0,531424803 | ZMYND19 | FALSE | Upregulated |
| -0,821447762 | 0,531453725 | GPR112 | FALSE | Downregulated |
| -1,053265183 | 0,531453725 | LOC101928967 | FALSE | Downregulated |
| 0,099228393 | 0,531541123 | SH3GL1 | FALSE | Upregulated |
| -0,739795824 | 0,531743264 | ELAVL4 | FALSE | Downregulated |
| -0,148052477 | 0,531743264 | MRPL40 | FALSE | Downregulated |
| -0,30439771 | 0,531764151 | FLJ38668 | FALSE | Downregulated |
| -0,105812523 | 0,531764151 | NOL10 | FALSE | Downregulated |
| -0,276286323 | 0,531764151 | PCDH18 | FALSE | Downregulated |
| 1,040596675 | 0,531957588 | SMC1B | FALSE | Upregulated |
| 0,422541175 | 0,5322827 | ACAP1 | FALSE | Upregulated |
| 0,213915303 | 0,5322827 | AKR7A3 | FALSE | Upregulated |
| 0,152841806 | 0,5322827 | ASGR1 | FALSE | Upregulated |
| -0,391880527 | 0,5322827 | ASPRV1 | FALSE | Downregulated |
| 0,286422098 | 0,5322827 | CD163 | FALSE | Upregulated |
| 0,405427563 | 0,5322827 | CLEC10A | FALSE | Upregulated |
| 0,12508147 | 0,5322827 | CTDSP1 | FALSE | Upregulated |
| 0,117295777 | 0,5322827 | DHPS | FALSE | Upregulated |
| -0,279483793 | 0,5322827 | DHRS4L2 | FALSE | Downregulated |

| | | | | |
|---|---|---|---|---|
| -0,131720092 | 0,5322827 | DZIP3 | FALSE | Downregulated |
| 0,263165245 | 0,5322827 | ELMO3 | FALSE | Upregulated |
| -0,630264142 | 0,5322827 | FAM27E3 | FALSE | Downregulated |
| -0,800673534 | 0,5322827 | FLJ20712 | FALSE | Downregulated |
| 0,263371781 | 0,5322827 | IMPDH1 | FALSE | Upregulated |
| 0,870555849 | 0,5322827 | JPH3 | FALSE | Upregulated |
| 0,203324849 | 0,5322827 | KIAA1522 | FALSE | Upregulated |
| 0,331784538 | 0,5322827 | LGALS3 | FALSE | Upregulated |
| -0,366836951 | 0,5322827 | LOC100129781 | FALSE | Downregulated |
| 0,87417711 | 0,5322827 | LOC101928117 | FALSE | Upregulated |
| -0,501293479 | 0,5322827 | NIM1K | FALSE | Downregulated |
| -0,202664014 | 0,5322827 | ONECUT2 | FALSE | Downregulated |
| 0,208241601 | 0,5322827 | PCNA | FALSE | Upregulated |
| 0,18193812 | 0,5322827 | PPP1R14B | FALSE | Upregulated |
| 0,115299254 | 0,5322827 | PPP2R1A | FALSE | Upregulated |
| 0,354892025 | 0,5322827 | RELB | FALSE | Upregulated |
| -0,132685716 | 0,5322827 | SCRN3 | FALSE | Downregulated |
| -0,12569021 | 0,5322827 | SRF | FALSE | Downregulated |
| 0,220597364 | 0,5322827 | SRPRB | FALSE | Upregulated |
| -0,098670454 | 0,5322827 | ZNF197 | FALSE | Downregulated |
| -0,111632579 | 0,5322827 | ZNF384 | FALSE | Downregulated |
| -0,227798169 | 0,5322827 | ZXDB | FALSE | Downregulated |
| -0,137419883 | 0,532394525 | KIN | FALSE | Downregulated |
| -0,140153878 | 0,532394525 | NARF | FALSE | Downregulated |
| 0,112773454 | 0,532394525 | PKP4 | FALSE | Upregulated |
| -0,125333314 | 0,532394525 | TRIM24 | FALSE | Downregulated |

| | | | | |
|---|---|---|---|---|
| -0,142996819 | 0,532394525 | TXN2 | FALSE | Downregulated |
| 0,167935095 | 0,532394525 | ZSCAN2 | FALSE | Upregulated |
| 0,604748971 | 0,532869063 | PLEKHG5 | FALSE | Upregulated |
| 0,874418689 | 0,533030479 | CCL24 | FALSE | Upregulated |
| -0,249315967 | 0,533030479 | MDP1 | FALSE | Downregulated |
| 0,973984017 | 0,533288392 | LOC101928955 | FALSE | Upregulated |
| -0,184168935 | 0,533335818 | COBLL1 | FALSE | Downregulated |
| -0,136624097 | 0,533335818 | ELMO1 | FALSE | Downregulated |
| 0,939111239 | 0,533384345 | POTEM | FALSE | Upregulated |
| -0,333209205 | 0,533400816 | NUP62CL | FALSE | Downregulated |
| -0,160984168 | 0,534383788 | ALDH6A1 | FALSE | Downregulated |
| -0,084008993 | 0,534383788 | ANKRD13C | FALSE | Downregulated |
| 0,324831863 | 0,534383788 | ANXA2 | FALSE | Upregulated |
| -0,164841452 | 0,534383788 | CCDC132 | FALSE | Downregulated |
| -0,740337256 | 0,534383788 | DBIL5P | FALSE | Downregulated |
| 0,671635608 | 0,534383788 | DHRS2 | FALSE | Upregulated |
| -0,138219622 | 0,534383788 | ISY1-RAB43 | FALSE | Downregulated |
| -0,4764431 | 0,534383788 | LOC100506985 | FALSE | Downregulated |
| -1,003710934 | 0,534383788 | LOC101928841 | FALSE | Downregulated |
| 0,531977268 | 0,534383788 | LOC102724768 | FALSE | Upregulated |
| 0,367598777 | 0,535414346 | EMR2 | FALSE | Upregulated |
| -0,121034067 | 0,535464352 | ZNF37A | FALSE | Downregulated |
| 0,266498353 | 0,535594056 | FCGR3A | FALSE | Upregulated |
| 0,243804691 | 0,535594056 | RGL3 | FALSE | Upregulated |
| 0,747623864 | 0,535594056 | SRSF12 | FALSE | Upregulated |
| -0,204850559 | 0,535594056 | ZNF106 | FALSE | Downregulated |

| | | | | |
|---|---|---|---|---|
| 0,26157618 | 0,535606717 | FMNL3 | FALSE | Upregulated |
| -0,156197742 | 0,535773302 | CXADR | FALSE | Downregulated |
| 0,230732583 | 0,535990438 | CRIP2 | FALSE | Upregulated |
| -0,280080524 | 0,535998528 | ENAH | FALSE | Downregulated |
| 0,273009284 | 0,536248245 | ALDOA | FALSE | Upregulated |
| -0,150743993 | 0,536248245 | CEP120 | FALSE | Downregulated |
| 0,116987378 | 0,536248245 | CTIF | FALSE | Upregulated |
| -0,248852384 | 0,536248245 | DLEU1 | FALSE | Downregulated |
| 0,364770697 | 0,536248245 | GFRA2 | FALSE | Upregulated |
| -0,323912708 | 0,536248245 | HLA-F-AS1 | FALSE | Downregulated |
| -0,930894709 | 0,536248245 | LOC102723352 | FALSE | Downregulated |
| -0,679280297 | 0,536248245 | MEIG1 | FALSE | Downregulated |
| 0,14403752 | 0,536248245 | MFSD11 | FALSE | Upregulated |
| 0,248036269 | 0,536248245 | NFKBIE | FALSE | Upregulated |
| 0,218621526 | 0,536248245 | OPLAH | FALSE | Upregulated |
| 0,296400905 | 0,536248245 | RASAL3 | FALSE | Upregulated |
| -0,244113864 | 0,536248245 | TMEM231 | FALSE | Downregulated |
| 0,162994147 | 0,536248245 | UNC93B1 | FALSE | Upregulated |
| 0,220249224 | 0,536339862 | PGBD1 | FALSE | Upregulated |
| -0,345496681 | 0,536339862 | RP11-480I12.7 | FALSE | Downregulated |
| 0,155818455 | 0,536798242 | CXorf56 | FALSE | Upregulated |
| -0,299773007 | 0,536798242 | SYPL2 | FALSE | Downregulated |
| 0,40051222 | 0,536798242 | ZAP70 | FALSE | Upregulated |
| -0,119342482 | 0,536901712 | BBS4 | FALSE | Downregulated |
| -0,25742692 | 0,536901712 | GJA1 | FALSE | Downregulated |
| -0,908898606 | 0,536901712 | LOC101927170 | FALSE | Downregulated |

| | | | | |
|---|---|---|---|---|
| -0,230610582 | 0,536901712 | NUDT3 | FALSE | Downregulated |
| -0,150589137 | 0,536901712 | ZSCAN26 | FALSE | Downregulated |
| 0,211177424 | 0,536938764 | RIC8B | FALSE | Upregulated |
| 0,088824722 | 0,536938764 | UBQLN1 | FALSE | Upregulated |
| -0,101462724 | 0,537048344 | DLD | FALSE | Downregulated |
| -0,405026969 | 0,537048344 | SUMO1P3 | FALSE | Downregulated |
| 0,135428954 | 0,537048344 | UBE2M | FALSE | Upregulated |
| 0,35517874 | 0,537137598 | GPC4 | FALSE | Upregulated |
| 0,875701279 | 0,537137598 | LILRA4 | FALSE | Upregulated |
| -0,21159572 | 0,537137598 | LTV1 | FALSE | Downregulated |
| -0,148533251 | 0,537141576 | GOLPH3L | FALSE | Downregulated |
| -0,32863914 | 0,53736915 | GRIA3 | FALSE | Downregulated |
| 0,264876901 | 0,537430975 | ELF4 | FALSE | Upregulated |
| 0,484053241 | 0,537487367 | CKAP2L | FALSE | Upregulated |
| -1,500751201 | 0,537487367 | SMPX | FALSE | Downregulated |
| 0,300616155 | 0,537487367 | SOX18 | FALSE | Upregulated |
| -0,112450169 | 0,537487367 | TMEM5 | FALSE | Downregulated |
| -0,604284765 | 0,537487367 | TNKS2-AS1 | FALSE | Downregulated |
| 0,252492418 | 0,537990839 | RAC2 | FALSE | Upregulated |
| -0,117567045 | 0,538069666 | CEP70 | FALSE | Downregulated |
| 0,56533607 | 0,538069666 | IGSF21 | FALSE | Upregulated |
| -0,126980756 | 0,538069666 | METTL10 | FALSE | Downregulated |
| 0,297211047 | 0,538069666 | PRSS8 | FALSE | Upregulated |
| -0,088315291 | 0,538618997 | GPBP1L1 | FALSE | Downregulated |
| 0,90798523 | 0,538665861 | LOC101929240 | FALSE | Upregulated |
| -0,173344988 | 0,538840911 | MAGI2 | FALSE | Downregulated |

| | | | | |
|---|---|---|---|---|
| -0,102767409 | 0,539094002 | RMND1 | FALSE | Downregulated |
| -0,154159855 | 0,539166608 | KDELC1 | FALSE | Downregulated |
| 0,139944267 | 0,539314062 | BRK1 | FALSE | Upregulated |
| 0,41070258 | 0,539314062 | CCR2 | FALSE | Upregulated |
| -0,175030243 | 0,539314062 | JMJD1C | FALSE | Downregulated |
| 0,152669036 | 0,539314062 | LDLRAP1 | FALSE | Upregulated |
| -0,514603347 | 0,539314062 | LOC101928472 | FALSE | Downregulated |
| -0,541115817 | 0,539314062 | LOC102724450 | FALSE | Downregulated |
| -0,335984868 | 0,539314062 | LRAT | FALSE | Downregulated |
| -0,284161108 | 0,539314062 | NRL | FALSE | Downregulated |
| 0,124686082 | 0,539314062 | RBM19 | FALSE | Upregulated |
| -0,584363445 | 0,539314062 | RNA28S5 | FALSE | Downregulated |
| -0,108663041 | 0,539314062 | SPIN1 | FALSE | Downregulated |
| 0,266267796 | 0,539314062 | URAHP | FALSE | Upregulated |
| -0,127246476 | 0,539424204 | EXOC6B | FALSE | Downregulated |
| 0,201043616 | 0,539795361 | TMEM110-MUSTN1 | FALSE | Upregulated |
| 0,526055287 | 0,53982804 | PIPSL | FALSE | Upregulated |
| 0,681698446 | 0,53982804 | TRABD2A | FALSE | Upregulated |
| 0,177249462 | 0,53982804 | WDR35 | FALSE | Upregulated |
| 0,2027336 | 0,539878357 | KIF16B | FALSE | Upregulated |
| 0,217400678 | 0,539878357 | YWHAH | FALSE | Upregulated |
| -0,376916097 | 0,540144853 | LRP4 | FALSE | Downregulated |
| -0,157887609 | 0,540252274 | APITD1 | FALSE | Downregulated |
| 0,183377301 | 0,540295223 | GAMT | FALSE | Upregulated |
| 0,659604953 | 0,540700297 | TNFRSF12A | FALSE | Upregulated |
| -0,099799251 | 0,540700297 | TRIOBP | FALSE | Downregulated |

| | | | | |
|---|---|---|---|---|
| 0,187050793 | 0,540716038 | FMO4 | FALSE | Upregulated |
| -0,238454651 | 0,540772082 | CPEB3 | FALSE | Downregulated |
| -0,099844838 | 0,540772082 | GNAQ | FALSE | Downregulated |
| 0,295086707 | 0,541685308 | P2RY2 | FALSE | Upregulated |
| 0,171800059 | 0,541941975 | ZNF627 | FALSE | Upregulated |
| 1,184279298 | 0,542493029 | CCL17 | FALSE | Upregulated |
| -0,258343956 | 0,542493029 | PPP1R1C | FALSE | Downregulated |
| 0,097628098 | 0,542493029 | TTC27 | FALSE | Upregulated |
| -0,61398477 | 0,542527999 | FHL1 | FALSE | Downregulated |
| -0,285131551 | 0,542858183 | NPIPA8 | FALSE | Downregulated |
| 0,16543694 | 0,542858183 | PI4K2A | FALSE | Upregulated |
| -0,629755113 | 0,542892606 | LETM2 | FALSE | Downregulated |
| -0,146831438 | 0,542892606 | SIRT7 | FALSE | Downregulated |
| 0,275331065 | 0,542982891 | MOGAT3 | FALSE | Upregulated |
| 0,259329135 | 0,543404383 | TONSL | FALSE | Upregulated |
| 0,901806607 | 0,543664427 | ATP2C2 | FALSE | Upregulated |
| 0,284773047 | 0,543664427 | TJP3 | FALSE | Upregulated |
| -0,107945122 | 0,543670036 | NFYA | FALSE | Downregulated |
| -0,285913185 | 0,543670036 | SH3PXD2B | FALSE | Downregulated |
| -0,398063371 | 0,543981164 | HEXA-AS1 | FALSE | Downregulated |
| -0,104068459 | 0,544164323 | RFC1 | FALSE | Downregulated |
| -0,304742313 | 0,544164323 | SMAD9 | FALSE | Downregulated |
| 0,14600154 | 0,544431444 | PMPCA | FALSE | Upregulated |
| 0,347086931 | 0,54449732 | MICB | FALSE | Upregulated |
| 0,509035233 | 0,544570177 | ACSL4 | FALSE | Upregulated |
| -0,452496207 | 0,544570177 | LINC01176 | FALSE | Downregulated |

| | | | | |
|---|---|---|---|---|
| 0,146882997 | 0,544607872 | CYSTM1 | FALSE | Upregulated |
| 0,332316322 | 0,544607872 | RAPGEF3 | FALSE | Upregulated |
| 0,50159649 | 0,544818883 | SPON1 | FALSE | Upregulated |
| 0,116815701 | 0,545531527 | SRPR | FALSE | Upregulated |
| -0,321181816 | 0,545634851 | C10orf10 | FALSE | Downregulated |
| 0,101571387 | 0,545634851 | GTF3C2 | FALSE | Upregulated |
| 0,472862108 | 0,545634851 | LOC440386 | FALSE | Upregulated |
| 0,118917921 | 0,545634851 | MARK2 | FALSE | Upregulated |
| 0,136799969 | 0,545634851 | PRKD2 | FALSE | Upregulated |
| 0,449341133 | 0,545634851 | RPL23AP64 | FALSE | Upregulated |
| -0,256305301 | 0,545634851 | SEPT4 | FALSE | Downregulated |
| 0,487595725 | 0,545809223 | HRH2 | FALSE | Upregulated |
| 0,160708789 | 0,545809223 | RPUSD2 | FALSE | Upregulated |
| 0,747641283 | 0,545903971 | HTR7 | FALSE | Upregulated |
| 0,165239213 | 0,545903971 | NUP62 | FALSE | Upregulated |
| 0,159084877 | 0,545903971 | RFC2 | FALSE | Upregulated |
| -0,111452079 | 0,546038104 | ATP5J2 | FALSE | Downregulated |
| -0,143854068 | 0,546143767 | GCNT2 | FALSE | Downregulated |
| 0,878620241 | 0,546143767 | HS3ST2 | FALSE | Upregulated |
| 0,095106382 | 0,546143767 | SPG11 | FALSE | Upregulated |
| 1,835910512 | 0,546143767 | TFF2 | FALSE | Upregulated |
| -0,121280874 | 0,546568 | ORC5 | FALSE | Downregulated |
| -0,262218593 | 0,546568 | PPM1K | FALSE | Downregulated |
| 0,293010499 | 0,546659199 | GPR34 | FALSE | Upregulated |
| -0,316558494 | 0,546659199 | NCAM1 | FALSE | Downregulated |
| 0,429716374 | 0,547102 | C8orf31 | FALSE | Upregulated |

| | | | | |
|---|---|---|---|---|
| 0,431424337 | 0,547102 | HMCN2 | FALSE | Upregulated |
| 0,172050559 | 0,547268499 | DOLK | FALSE | Upregulated |
| -0,255951378 | 0,547268499 | UPF3A | FALSE | Downregulated |
| 0,286182427 | 0,547484016 | GVINP1 | FALSE | Upregulated |
| -0,303401776 | 0,547484016 | LYVE1 | FALSE | Downregulated |
| 0,199416345 | 0,547569157 | ABCG8 | FALSE | Upregulated |
| -0,429875797 | 0,547569157 | AC074289.1 | FALSE | Downregulated |
| 0,201946709 | 0,547569157 | CCDC120 | FALSE | Upregulated |
| 0,509896738 | 0,547569157 | KLHL29 | FALSE | Upregulated |
| -0,813742105 | 0,547569157 | OSR1 | FALSE | Downregulated |
| -0,138702366 | 0,547569157 | RNF5 | FALSE | Downregulated |
| -0,248977501 | 0,547569157 | ZNF138 | FALSE | Downregulated |
| 0,226037229 | 0,547698944 | ANKAR | FALSE | Upregulated |
| 0,119008114 | 0,547698944 | DNTTIP1 | FALSE | Upregulated |
| -0,10843099 | 0,547698944 | KCTD9 | FALSE | Downregulated |
| 0,402711536 | 0,547698944 | KIF23 | FALSE | Upregulated |
| -0,67287976 | 0,547698944 | LOC101927652 | FALSE | Downregulated |
| 0,123858205 | 0,547698944 | NOTCH2 | FALSE | Upregulated |
| -0,214203738 | 0,547698944 | RP11-81N13.1 | FALSE | Downregulated |
| -0,137679986 | 0,547698944 | RSC1A1 | FALSE | Downregulated |
| -0,091964127 | 0,547698944 | SUMO1 | FALSE | Downregulated |
| -0,374958349 | 0,547698944 | UBAC2-AS1 | FALSE | Downregulated |
| -0,135815601 | 0,547698944 | ZBTB10 | FALSE | Downregulated |
| 0,479682405 | 0,547698944 | ZSCAN12P1 | FALSE | Upregulated |
| -0,312412676 | 0,548189047 | FBXO32 | FALSE | Downregulated |
| 0,436410498 | 0,548189047 | LOC100506473 | FALSE | Upregulated |

| | | | | |
|---|---|---|---|---|
| -0,408625935 | 0,548189047 | LOC101927009 | FALSE | Downregulated |
| 0,402533582 | 0,548189047 | MIR210HG | FALSE | Upregulated |
| -0,105349371 | 0,548189047 | ST6GAL1 | FALSE | Downregulated |
| -0,111197578 | 0,548189047 | UBE2E1 | FALSE | Downregulated |
| 0,32225332 | 0,548189047 | ZNF607 | FALSE | Upregulated |
| 0,277247704 | 0,548338542 | ADAMTSL2 | FALSE | Upregulated |
| -0,204880217 | 0,548338542 | HAUS2 | FALSE | Downregulated |
| -0,300949598 | 0,548357952 | ARID3A | FALSE | Downregulated |
| 0,266829567 | 0,548357952 | CCDC24 | FALSE | Upregulated |
| -0,127107769 | 0,548357952 | CCDC47 | FALSE | Downregulated |
| 0,236907536 | 0,548357952 | DGCR14 | FALSE | Upregulated |
| -0,338945136 | 0,548357952 | HOXB3 | FALSE | Downregulated |
| 0,176720843 | 0,548357952 | MECR | FALSE | Upregulated |
| -0,14122134 | 0,548357952 | NUDT12 | FALSE | Downregulated |
| 0,390548752 | 0,548357952 | PDZD4 | FALSE | Upregulated |
| -0,155684368 | 0,548357952 | SMIM19 | FALSE | Downregulated |
| 0,217574162 | 0,548357952 | SPIRE2 | FALSE | Upregulated |
| -0,106394668 | 0,548357952 | TEX2 | FALSE | Downregulated |
| -0,193148408 | 0,548357952 | ZFP1 | FALSE | Downregulated |
| 0,40003591 | 0,548794981 | ADAM28 | FALSE | Upregulated |
| 0,73393738 | 0,548794981 | CLDN4 | FALSE | Upregulated |
| -0,309561962 | 0,548794981 | COL21A1 | FALSE | Downregulated |
| 0,769431922 | 0,548794981 | GPR83 | FALSE | Upregulated |
| 0,626387683 | 0,548794981 | PCDH20 | FALSE | Upregulated |
| 0,257667403 | 0,548794981 | ZNF425 | FALSE | Upregulated |
| 1,742952178 | 0,549133512 | LOC730268 | FALSE | Upregulated |

| | | | | |
|---|---|---|---|---|
| -0,163383185 | 0,549285176 | CNNM2 | FALSE | Downregulated |
| 0,08317722 | 0,549338206 | GCN1L1 | FALSE | Upregulated |
| -0,152892068 | 0,549345912 | AP5Z1 | FALSE | Downregulated |
| 0,716772478 | 0,549611728 | LOC102724850 | FALSE | Upregulated |
| 0,317082219 | 0,549668328 | FPR3 | FALSE | Upregulated |
| -0,522882379 | 0,549678808 | NT5M | FALSE | Downregulated |
| -0,163243705 | 0,550124993 | PFDN2 | FALSE | Downregulated |
| -0,205842434 | 0,550124993 | ZNF674 | FALSE | Downregulated |
| 0,526772908 | 0,550382622 | ZNF192P1 | FALSE | Upregulated |
| -0,134370851 | 0,551136453 | C17orf58 | FALSE | Downregulated |
| -0,421928102 | 0,551136453 | LGI4 | FALSE | Downregulated |
| 0,196052248 | 0,551136453 | TRABD | FALSE | Upregulated |
| 0,279484667 | 0,551136453 | ZNF468 | FALSE | Upregulated |
| -0,232884494 | 0,551416328 | ACADSB | FALSE | Downregulated |
| -0,244362963 | 0,551416328 | IGHMBP2 | FALSE | Downregulated |
| 0,643226673 | 0,551416328 | KRT7 | FALSE | Upregulated |
| 0,566596115 | 0,551416328 | LOC644961 | FALSE | Upregulated |
| -0,146367099 | 0,551416328 | PRPSAP1 | FALSE | Downregulated |
| 0,145727064 | 0,551416328 | SEC22B | FALSE | Upregulated |
| -0,320703362 | 0,551416328 | SLC16A10 | FALSE | Downregulated |
| -0,080957758 | 0,551416328 | SLC30A9 | FALSE | Downregulated |
| -0,10751847 | 0,551416328 | TM2D1 | FALSE | Downregulated |
| -0,110482864 | 0,551585155 | NDST1 | FALSE | Downregulated |
| 0,760488074 | 0,551699363 | FCRLA | FALSE | Upregulated |
| -0,125891522 | 0,551699363 | KYNU | FALSE | Downregulated |
| 0,592685136 | 0,551699363 | MCOLN2 | FALSE | Upregulated |

| | | | | |
|---|---|---|---|---|
| 0,304449484 | 0,551699363 | NBPF12 | FALSE | Upregulated |
| 0,24025494 | 0,551764718 | TTC39B | FALSE | Upregulated |
| -0,172609426 | 0,552073515 | C21orf59 | FALSE | Downregulated |
| -0,153509927 | 0,552073515 | LZIC | FALSE | Downregulated |
| -0,100090745 | 0,552126258 | ISG20L2 | FALSE | Downregulated |
| 0,184145938 | 0,552126258 | MZT2B | FALSE | Upregulated |
| 0,298566879 | 0,552282099 | KCTD17 | FALSE | Upregulated |
| -0,616411487 | 0,552827322 | TREML2 | FALSE | Downregulated |
| 0,199704385 | 0,552827322 | ZKSCAN3 | FALSE | Upregulated |
| 0,170616162 | 0,553022457 | RNASEL | FALSE | Upregulated |
| 1,305074653 | 0,553340835 | EGOT | FALSE | Upregulated |
| -0,168813865 | 0,553340835 | EPT1 | FALSE | Downregulated |
| -0,816009228 | 0,553340835 | HSPB2 | FALSE | Downregulated |
| -0,11601255 | 0,553340835 | KCTD5 | FALSE | Downregulated |
| 0,551937075 | 0,553340835 | MEIS3 | FALSE | Upregulated |
| -0,298065388 | 0,553340835 | NUCB2 | FALSE | Downregulated |
| 0,322351082 | 0,553340835 | SUV420H2 | FALSE | Upregulated |
| -0,174653088 | 0,553340835 | ZNF624 | FALSE | Downregulated |
| 0,297710964 | 0,553414952 | CSF2RA | FALSE | Upregulated |
| -0,14613403 | 0,553661058 | AAMP | FALSE | Downregulated |
| -0,112827982 | 0,554128449 | DYNC1LI1 | FALSE | Downregulated |
| -0,114484833 | 0,554238536 | GFM1 | FALSE | Downregulated |
| -0,158180997 | 0,554266279 | CPSF4 | FALSE | Downregulated |
| -0,082577282 | 0,554348133 | TCF12 | FALSE | Downregulated |
| 0,234136198 | 0,554378899 | SPI1 | FALSE | Upregulated |
| -0,415958008 | 0,554635202 | KCNJ3 | FALSE | Downregulated |

| | | | | |
|---|---|---|---|---|
| 0,155589419 | 0,554635202 | PCYT2 | FALSE | Upregulated |
| -0,134572304 | 0,554696854 | UFM1 | FALSE | Downregulated |
| 1,049220679 | 0,554827211 | GUSBP3 | FALSE | Upregulated |
| -0,141328908 | 0,554827211 | OSTC | FALSE | Downregulated |
| -0,172166954 | 0,554861406 | CWC27 | FALSE | Downregulated |
| 0,288042512 | 0,554861406 | HHIPL1 | FALSE | Upregulated |
| 0,271071088 | 0,554861406 | TMEM243 | FALSE | Upregulated |
| -0,638934652 | 0,555064885 | ITGA8 | FALSE | Downregulated |
| 0,156225489 | 0,555064885 | PSEN2 | FALSE | Upregulated |
| -0,105020287 | 0,555231503 | PTPN4 | FALSE | Downregulated |
| 0,145088876 | 0,555368828 | AIP | FALSE | Upregulated |
| -0,655705699 | 0,556110757 | RBPMS-AS1 | FALSE | Downregulated |
| 0,187979489 | 0,556441025 | LSM10 | FALSE | Upregulated |
| 0,326220713 | 0,556972745 | PYHIN1 | FALSE | Upregulated |
| 0,999407729 | 0,557198342 | LOC101927345 | FALSE | Upregulated |
| -0,364196679 | 0,55732029 | CMPK2 | FALSE | Downregulated |
| -0,45163319 | 0,55732029 | ENHO | FALSE | Downregulated |
| -0,180138266 | 0,55732029 | HEBP2 | FALSE | Downregulated |
| 0,683514571 | 0,55732029 | ISLR2 | FALSE | Upregulated |
| 0,320293817 | 0,55732029 | LOC101928377 | FALSE | Upregulated |
| -0,087117022 | 0,55732029 | OS9 | FALSE | Downregulated |
| 0,317648059 | 0,55732029 | TLR8 | FALSE | Upregulated |
| -0,22703506 | 0,55732029 | XYLB | FALSE | Downregulated |
| 0,37809248 | 0,557974796 | THBD | FALSE | Upregulated |
| -0,111618865 | 0,55798594 | FAM149B1 | FALSE | Downregulated |
| 0,1544956 | 0,55798594 | KCTD10 | FALSE | Upregulated |

| | | | | |
|---|---|---|---|---|
| -0,128057023 | 0,55798594 | NOL8 | FALSE | Downregulated |
| -0,135829789 | 0,55811889 | B3GNT2 | FALSE | Downregulated |
| -0,201206675 | 0,55811889 | C6orf57 | FALSE | Downregulated |
| -0,249878765 | 0,55811889 | LOC102724532 | FALSE | Downregulated |
| -0,404242017 | 0,55811889 | TMTC2 | FALSE | Downregulated |
| -0,162094396 | 0,558236483 | CLEC2B | FALSE | Downregulated |
| 0,501577709 | 0,55855122 | AC097662.2 | FALSE | Upregulated |
| 0,608728108 | 0,55855122 | FAM166B | FALSE | Upregulated |
| -0,664082649 | 0,55855122 | LINC00862 | FALSE | Downregulated |
| -0,333326394 | 0,55855122 | LOC101927340 | FALSE | Downregulated |
| 0,516805724 | 0,55855122 | LOC102724356 | FALSE | Upregulated |
| -0,756845292 | 0,55855122 | LRRC9 | FALSE | Downregulated |
| -0,255509246 | 0,55855122 | PNPLA3 | FALSE | Downregulated |
| -0,10492712 | 0,55855122 | TMEM254 | FALSE | Downregulated |
| 0,379790581 | 0,559001535 | DCST2 | FALSE | Upregulated |
| 0,528486463 | 0,55901531 | LOC100289561 | FALSE | Upregulated |
| 0,116506985 | 0,55901531 | PIK3R2 | FALSE | Upregulated |
| 0,190276501 | 0,55901531 | RALGDS | FALSE | Upregulated |
| -0,253552731 | 0,55901531 | TIGD5 | FALSE | Downregulated |
| -0,280050203 | 0,55901531 | TVP23C | FALSE | Downregulated |
| 0,106762778 | 0,559714565 | IGF2R | FALSE | Upregulated |
| -0,187631654 | 0,559716011 | AQP11 | FALSE | Downregulated |
| -0,143041406 | 0,559716011 | CTBP1-AS2 | FALSE | Downregulated |
| 0,466849297 | 0,559716011 | FAM86B3P | FALSE | Upregulated |
| 0,491331855 | 0,559716011 | LOC101927069 | FALSE | Upregulated |
| 0,101766435 | 0,559716011 | METTL13 | FALSE | Upregulated |

| | | | | |
|---|---|---|---|---|
| -0,140199248 | 0,559716011 | PDHB | FALSE | Downregulated |
| 0,157309169 | 0,559716011 | PPP1R37 | FALSE | Upregulated |
| 0,163553808 | 0,559716011 | TNS3 | FALSE | Upregulated |
| -0,313120813 | 0,559976402 | RP5-1024G6.5 | FALSE | Downregulated |
| 1,020770061 | 0,560123659 | AC010127.3 | FALSE | Upregulated |
| -1,830479346 | 0,560123659 | EGF | FALSE | Downregulated |
| 0,413569663 | 0,560123659 | PLCE1 | FALSE | Upregulated |
| 0,39563443 | 0,560176196 | ARHGEF16 | FALSE | Upregulated |
| 0,116041971 | 0,560176196 | CHUK | FALSE | Upregulated |
| 0,134563596 | 0,560176196 | IDH3G | FALSE | Upregulated |
| -1,172996189 | 0,560176196 | RP11-10A14.5 | FALSE | Downregulated |
| 0,143625122 | 0,560176196 | SIN3B | FALSE | Upregulated |
| -0,233573524 | 0,560176196 | TRIM52-AS1 | FALSE | Downregulated |
| -0,192777987 | 0,560290786 | APOF | FALSE | Downregulated |
| -0,139688817 | 0,560290786 | CAMK2G | FALSE | Downregulated |
| 0,949216713 | 0,560290786 | CASC15 | FALSE | Upregulated |
| 0,205072391 | 0,560290786 | CCDC88A | FALSE | Upregulated |
| -1,116424573 | 0,560290786 | CEP170P1 | FALSE | Downregulated |
| 0,666749635 | 0,560290786 | ILDR1 | FALSE | Upregulated |
| -0,479870075 | 0,560290786 | KCNE4 | FALSE | Downregulated |
| 0,32042817 | 0,560290786 | KIF24 | FALSE | Upregulated |
| 0,807542342 | 0,56037683 | APOBEC3H | FALSE | Upregulated |
| 0,669537173 | 0,56037683 | CDH4 | FALSE | Upregulated |
| -0,168464788 | 0,56037683 | CNPY4 | FALSE | Downregulated |
| -0,166875252 | 0,56037683 | ETV6 | FALSE | Downregulated |
| 0,171615847 | 0,56037683 | GIMAP2 | FALSE | Upregulated |

| | | | | |
|---|---|---|---|---|
| -0,180355585 | 0,56037683 | GOLGA4 | FALSE | Downregulated |
| 0,909389334 | 0,56037683 | LOC101930665 | FALSE | Upregulated |
| -0,138534075 | 0,56037683 | LTBR | FALSE | Downregulated |
| 0,197344639 | 0,56037683 | MDM2 | FALSE | Upregulated |
| -0,161340786 | 0,56037683 | NHLRC2 | FALSE | Downregulated |
| -0,429867228 | 0,56037683 | NUTM2B | FALSE | Downregulated |
| -0,164352357 | 0,56037683 | SGK3 | FALSE | Downregulated |
| 0,170186967 | 0,56037683 | SGSM2 | FALSE | Upregulated |
| 0,169101888 | 0,560422567 | FBXO25 | FALSE | Upregulated |
| 0,12514587 | 0,560422567 | FBXW11 | FALSE | Upregulated |
| -0,270728278 | 0,560422567 | HCG18 | FALSE | Downregulated |
| -0,252477742 | 0,560557867 | GLOD5 | FALSE | Downregulated |
| -0,823174698 | 0,560557867 | LRRC19 | FALSE | Downregulated |
| -0,103987487 | 0,560611126 | ABCE1 | FALSE | Downregulated |
| 0,083349134 | 0,560611126 | RNF40 | FALSE | Upregulated |
| 0,0863155 | 0,560942327 | KIAA0100 | FALSE | Upregulated |
| -0,151863343 | 0,560942327 | MTG2 | FALSE | Downregulated |
| -0,29362808 | 0,560942327 | MYCL | FALSE | Downregulated |
| -0,250192241 | 0,561366776 | CTTNBP2 | FALSE | Downregulated |
| -0,292454724 | 0,561485208 | EFNA2 | FALSE | Downregulated |
| 0,322761463 | 0,561507714 | ADAMTSL5 | FALSE | Upregulated |
| 0,344557493 | 0,561507714 | CD83 | FALSE | Upregulated |
| -0,214234377 | 0,561507714 | CHST7 | FALSE | Downregulated |
| 0,144077171 | 0,561507714 | CLPP | FALSE | Upregulated |
| 0,153264985 | 0,561507714 | DUS2 | FALSE | Upregulated |
| 0,107294133 | 0,561507714 | GNPTG | FALSE | Upregulated |

| | | | | |
|---|---|---|---|---|
| 0,109697612 | 0,561507714 | MRPS7 | FALSE | Upregulated |
| 0,20365391 | 0,561507714 | NOC4L | FALSE | Upregulated |
| 0,176931571 | 0,561507714 | PLXNA2 | FALSE | Upregulated |
| 0,33051213 | 0,561507714 | POC1A | FALSE | Upregulated |
| -0,813667766 | 0,561507714 | RP3-323P13.2 | FALSE | Downregulated |
| 0,923789081 | 0,561507714 | VPREB3 | FALSE | Upregulated |
| 0,115949221 | 0,561649465 | FBL | FALSE | Upregulated |
| -0,757683784 | 0,561649465 | LOC101929195 | FALSE | Downregulated |
| -0,128769725 | 0,561649465 | MLLT10 | FALSE | Downregulated |
| 0,117147071 | 0,561649465 | PEX2 | FALSE | Upregulated |
| 0,224918288 | 0,56218248 | CDKL5 | FALSE | Upregulated |
| -0,095149582 | 0,56218248 | CIZ1 | FALSE | Downregulated |
| -0,885339647 | 0,56218248 | LOC101927733 | FALSE | Downregulated |
| -0,525763901 | 0,56218248 | RP11-406A20.4 | FALSE | Downregulated |
| -0,139958816 | 0,56218248 | SDE2 | FALSE | Downregulated |
| -0,314309151 | 0,56218248 | ZMAT1 | FALSE | Downregulated |
| 0,276860872 | 0,562310235 | AEN | FALSE | Upregulated |
| 0,113560188 | 0,562310235 | EIF4A1 | FALSE | Upregulated |
| 0,259258578 | 0,562310235 | SLA | FALSE | Upregulated |
| 0,09532818 | 0,562310235 | WASF2 | FALSE | Upregulated |
| 0,149420091 | 0,562316343 | QSOX2 | FALSE | Upregulated |
| 0,750076296 | 0,562407379 | HSP90AB4P | FALSE | Upregulated |
| -0,479797194 | 0,562407379 | TENM3 | FALSE | Downregulated |
| -0,264536392 | 0,562407379 | VSNL1 | FALSE | Downregulated |
| 0,247132959 | 0,563281373 | RIPK4 | FALSE | Upregulated |
| -0,439365583 | 0,563358418 | AGAP4 | FALSE | Downregulated |

| | | | | |
|---|---|---|---|---|
| -0,172127069 | 0,563358418 | FGD5 | FALSE | Downregulated |
| -0,229916995 | 0,563358418 | MARC1 | FALSE | Downregulated |
| -0,139333552 | 0,563358418 | XRN1 | FALSE | Downregulated |
| 0,205771326 | 0,563358418 | ZNF563 | FALSE | Upregulated |
| 0,864260863 | 0,563547049 | CSPG5 | FALSE | Upregulated |
| -0,143963776 | 0,563547049 | NOC3L | FALSE | Downregulated |
| 0,617909803 | 0,563547049 | RP11-367N14.2 | FALSE | Upregulated |
| 0,657259113 | 0,563712559 | AC004019.13 | FALSE | Upregulated |
| 0,136257476 | 0,563724579 | BLCAP | FALSE | Upregulated |
| -0,130551497 | 0,563724579 | CCDC51 | FALSE | Downregulated |
| -0,173097273 | 0,563765461 | HELZ | FALSE | Downregulated |
| 0,332204653 | 0,564091992 | ARL11 | FALSE | Upregulated |
| 0,899765281 | 0,564091992 | CDH15 | FALSE | Upregulated |
| -0,137784388 | 0,564091992 | CDKN2AIPNL | FALSE | Downregulated |
| 0,261129168 | 0,564091992 | EXD3 | FALSE | Upregulated |
| 0,165924771 | 0,564091992 | GCLM | FALSE | Upregulated |
| 0,306076154 | 0,564091992 | MPV17L2 | FALSE | Upregulated |
| 0,127885178 | 0,564091992 | MTF1 | FALSE | Upregulated |
| -0,133689413 | 0,564091992 | PIGY | FALSE | Downregulated |
| 0,177867837 | 0,564091992 | SLC30A6 | FALSE | Upregulated |
| 0,416671547 | 0,564091992 | STMN3 | FALSE | Upregulated |
| 0,134469051 | 0,564091992 | ZFYVE27 | FALSE | Upregulated |
| -0,300105011 | 0,564259182 | HIST1H2BD | FALSE | Downregulated |
| 0,132679951 | 0,564259182 | RB1 | FALSE | Upregulated |
| -0,150179222 | 0,564259182 | RPL38 | FALSE | Downregulated |
| -0,326930665 | 0,564259182 | ZNF204P | FALSE | Downregulated |

| | | | | |
|---|---|---|---|---|
| -0,609798879 | 0,564474792 | AC009501.4 | FALSE | Downregulated |
| -0,097588657 | 0,564474792 | NCKAP1 | FALSE | Downregulated |
| -0,291625626 | 0,564888324 | SVIL | FALSE | Downregulated |
| 0,366803572 | 0,56513906 | C7 | FALSE | Upregulated |
| -0,091653736 | 0,56513906 | PARK7 | FALSE | Downregulated |
| 0,549959643 | 0,56513906 | RAB38 | FALSE | Upregulated |
| -0,154929369 | 0,565493261 | DNAJC2 | FALSE | Downregulated |
| 0,10857068 | 0,565493261 | FBXO45 | FALSE | Upregulated |
| -0,107256991 | 0,56589188 | CASP6 | FALSE | Downregulated |
| -0,152573084 | 0,56589188 | OGG1 | FALSE | Downregulated |
| 0,279939483 | 0,56589188 | PFKFB3 | FALSE | Upregulated |
| 0,152436896 | 0,56589188 | TMEM245 | FALSE | Upregulated |
| 0,390898036 | 0,56589188 | VN1R1 | FALSE | Upregulated |
| 0,164300083 | 0,565933987 | ACOT9 | FALSE | Upregulated |
| -0,116547492 | 0,565933987 | CELF1 | FALSE | Downregulated |
| 0,146779215 | 0,565933987 | DNAL1 | FALSE | Upregulated |
| 0,109481019 | 0,565933987 | MAN1B1 | FALSE | Upregulated |
| -0,168889411 | 0,565933987 | MTHFS | FALSE | Downregulated |
| 1,01112729 | 0,565933987 | PBX4 | FALSE | Upregulated |
| -0,302253303 | 0,565933987 | PDE7B | FALSE | Downregulated |
| -0,327165968 | 0,565933987 | PLAG1 | FALSE | Downregulated |
| -0,453261604 | 0,565933987 | RPS18P9 | FALSE | Downregulated |
| 0,160773799 | 0,565933987 | SURF2 | FALSE | Upregulated |
| -0,187850317 | 0,565933987 | ZNF280C | FALSE | Downregulated |
| -0,125147135 | 0,566236332 | RNF214 | FALSE | Downregulated |
| -0,125549897 | 0,566428479 | PTCD2 | FALSE | Downregulated |

| | | | | |
|---|---|---|---|---|
| -0,333112937 | 0,566554826 | BCO2 | FALSE | Downregulated |
| -0,327988753 | 0,566554826 | LOC80154 | FALSE | Downregulated |
| -0,379480414 | 0,566708515 | CYP2B6 | FALSE | Downregulated |
| -0,450152911 | 0,566708515 | DOK6 | FALSE | Downregulated |
| 0,184147521 | 0,566708515 | MAT2A | FALSE | Upregulated |
| -0,154243552 | 0,566708515 | PRR3 | FALSE | Downregulated |
| 0,933726889 | 0,566708515 | TTTY14 | FALSE | Upregulated |
| -0,518534678 | 0,566733827 | C4orf47 | FALSE | Downregulated |
| -0,776857709 | 0,566733827 | INA | FALSE | Downregulated |
| 0,107778964 | 0,566733827 | NNT | FALSE | Upregulated |
| -0,419764105 | 0,566733827 | SGIP1 | FALSE | Downregulated |
| 0,281607657 | 0,566733827 | ZNF347 | FALSE | Upregulated |
| 0,194951466 | 0,566984408 | RAB3IL1 | FALSE | Upregulated |
| -0,151958001 | 0,56710275 | USP16 | FALSE | Downregulated |
| -0,155658252 | 0,567304433 | MRE11A | FALSE | Downregulated |
| -0,119324034 | 0,567304433 | MYO1B | FALSE | Downregulated |
| 0,165261737 | 0,567304433 | TRIB2 | FALSE | Upregulated |
| 0,261019613 | 0,567632482 | EBF4 | FALSE | Upregulated |
| 0,498343069 | 0,567750707 | BTBD19 | FALSE | Upregulated |
| 0,737753055 | 0,567750707 | CASP5 | FALSE | Upregulated |
| -0,255677571 | 0,567750707 | GPRASP1 | FALSE | Downregulated |
| 0,302092135 | 0,567750707 | KIF3C | FALSE | Upregulated |
| 0,589898296 | 0,567750707 | LOC102723328 | FALSE | Upregulated |
| 0,108395433 | 0,567750707 | M6PR | FALSE | Upregulated |
| -0,136581317 | 0,567750707 | PCBD2 | FALSE | Downregulated |
| 0,14619667 | 0,567750707 | PGAP2 | FALSE | Upregulated |

| | | | | |
|---|---|---|---|---|
| 0,107878584 | 0,567750707 | WDR46 | FALSE | Upregulated |
| 0,148193065 | 0,567750707 | XYLT2 | FALSE | Upregulated |
| 0,158283238 | 0,567930597 | CYP7B1 | FALSE | Upregulated |
| -0,129119683 | 0,567930597 | HSCB | FALSE | Downregulated |
| 0,149688685 | 0,56841807 | AGO2 | FALSE | Upregulated |
| -0,115304295 | 0,56841807 | CCDC12 | FALSE | Downregulated |
| 0,13853594 | 0,56841807 | FLCN | FALSE | Upregulated |
| 0,411259284 | 0,56841807 | LOC100288181 | FALSE | Upregulated |
| 0,439374157 | 0,56841807 | LOC100506734 | FALSE | Upregulated |
| -0,888003187 | 0,56841807 | LOC101060264 | FALSE | Downregulated |
| -0,589032684 | 0,56841807 | LOC102467146 | FALSE | Downregulated |
| -0,450893253 | 0,568465934 | ALPK3 | FALSE | Downregulated |
| -0,258127222 | 0,568465934 | C1QTNF5 | FALSE | Downregulated |
| -0,125362505 | 0,568465934 | KAT6A | FALSE | Downregulated |
| -0,108152689 | 0,568465934 | STAG2 | FALSE | Downregulated |
| 0,198768902 | 0,568714137 | ADAMTSL4 | FALSE | Upregulated |
| -0,134890139 | 0,568714137 | MORC3 | FALSE | Downregulated |
| 0,147910872 | 0,568714137 | THAP4 | FALSE | Upregulated |
| -0,144044362 | 0,568714137 | UPF2 | FALSE | Downregulated |
| 0,629954374 | 0,568763841 | HS3ST1 | FALSE | Upregulated |
| -0,109870492 | 0,568763841 | SLBP | FALSE | Downregulated |
| 0,100983088 | 0,568797141 | DCTN4 | FALSE | Upregulated |
| 0,180049634 | 0,568797141 | DHRS7B | FALSE | Upregulated |
| -0,095648374 | 0,568797141 | MIER1 | FALSE | Downregulated |
| 0,409677333 | 0,568797141 | P2RY6 | FALSE | Upregulated |
| 0,140279583 | 0,568797141 | PARP11 | FALSE | Upregulated |

| | | | | |
|---|---|---|---|---|
| 0,136800403 | 0,568797141 | PPAP2A | FALSE | Upregulated |
| 0,28566334 | 0,568797141 | SLC51A | FALSE | Upregulated |
| 0,116327013 | 0,568797141 | SUFU | FALSE | Upregulated |
| 0,271342819 | 0,568797141 | ZNF517 | FALSE | Upregulated |
| 0,113532117 | 0,568947346 | ERH | FALSE | Upregulated |
| -0,24957085 | 0,569039239 | PLGLA | FALSE | Downregulated |
| -0,096750797 | 0,569106141 | MTIF2 | FALSE | Downregulated |
| -2,881306139 | 0,569115589 | DUSP27 | FALSE | Downregulated |
| 0,98886276 | 0,569228398 | C15orf48 | FALSE | Upregulated |
| 0,11653831 | 0,569228398 | CHCHD4 | FALSE | Upregulated |
| -0,279281751 | 0,569228398 | EXPH5 | FALSE | Downregulated |
| -0,380011564 | 0,569228398 | HNF1A-AS1 | FALSE | Downregulated |
| 0,271320259 | 0,569228398 | HPS5 | FALSE | Upregulated |
| -0,449919435 | 0,569228398 | LOC101926956 | FALSE | Downregulated |
| -0,31266482 | 0,569228398 | NBPF26 | FALSE | Downregulated |
| -0,23309375 | 0,569228398 | NFIC | FALSE | Downregulated |
| 0,318732382 | 0,569228398 | PRRG2 | FALSE | Upregulated |
| 0,095467605 | 0,569228398 | RARS | FALSE | Upregulated |
| 0,249641778 | 0,569228398 | SIGLEC1 | FALSE | Upregulated |
| -0,136577724 | 0,569228398 | SMAD3 | FALSE | Downregulated |
| 0,58609666 | 0,569228398 | TMED10P1 | FALSE | Upregulated |
| -0,066978337 | 0,569228398 | TMEM30A | FALSE | Downregulated |
| -0,114267627 | 0,569228398 | TMEM50A | FALSE | Downregulated |
| 0,180042487 | 0,569228398 | TPM3P9 | FALSE | Upregulated |
| 0,156786162 | 0,569228398 | TRMT44 | FALSE | Upregulated |
| -0,075544566 | 0,569228398 | USP10 | FALSE | Downregulated |

| | | | | |
|---|---|---|---|---|
| -0,122832094 | 0,569228398 | ZFYVE21 | FALSE | Downregulated |
| -0,183533137 | 0,56923846 | F11 | FALSE | Downregulated |
| -0,179278506 | 0,569712212 | GCDH | FALSE | Downregulated |
| 0,997362337 | 0,569712212 | SNORA81 | FALSE | Upregulated |
| 0,125964357 | 0,569730242 | PDCD6 | FALSE | Upregulated |
| -0,22749647 | 0,570094977 | BMPER | FALSE | Downregulated |
| -0,097391505 | 0,570224608 | SMARCB1 | FALSE | Downregulated |
| 0,275202431 | 0,570839768 | C21orf119 | FALSE | Upregulated |
| 0,580312606 | 0,570839768 | LOC102724217 | FALSE | Upregulated |
| 0,353272544 | 0,570839768 | TAGAP | FALSE | Upregulated |
| -0,135372588 | 0,571235854 | PSME1 | FALSE | Downregulated |
| 0,380005354 | 0,571235854 | ZNF681 | FALSE | Upregulated |
| 0,119861892 | 0,571531639 | BCKDHA | FALSE | Upregulated |
| -0,161251162 | 0,571660591 | CTNNAL1 | FALSE | Downregulated |
| -0,406390506 | 0,571660591 | KCNK6 | FALSE | Downregulated |
| -0,252232412 | 0,57185565 | TTC36 | FALSE | Downregulated |
| 0,435820435 | 0,571929537 | IL34 | FALSE | Upregulated |
| -0,422568186 | 0,571987073 | LOC101928816 | FALSE | Downregulated |
| 0,246885734 | 0,571987073 | TMEM107 | FALSE | Upregulated |
| -0,338741545 | 0,572300825 | LOC101928101 | FALSE | Downregulated |
| -0,16452181 | 0,572300825 | METTL22 | FALSE | Downregulated |
| -0,487470944 | 0,572357489 | BVES | FALSE | Downregulated |
| -0,143548494 | 0,572357489 | NDUFA7 | FALSE | Downregulated |
| -0,17291915 | 0,572357489 | PLA2G12A | FALSE | Downregulated |
| -0,193990813 | 0,572405704 | GCHFR | FALSE | Downregulated |
| 0,934232162 | 0,572423436 | C1QL1 | FALSE | Upregulated |

| | | | | |
|---|---|---|---|---|
| 0,174161529 | 0,572423436 | SMYD2 | FALSE | Upregulated |
| 0,18681307 | 0,57255316 | ACAP3 | FALSE | Upregulated |
| 0,141765959 | 0,572561358 | ZNF142 | FALSE | Upregulated |
| -0,351550087 | 0,572571545 | RTN4R | FALSE | Downregulated |
| -0,131525898 | 0,572571545 | ZFR | FALSE | Downregulated |
| -0,176685948 | 0,572571545 | ZNF688 | FALSE | Downregulated |
| -0,524765262 | 0,572729493 | LOC102724329 | FALSE | Downregulated |
| -0,267633635 | 0,572854437 | AC007551.3 | FALSE | Downregulated |
| 0,447432911 | 0,573179189 | C1orf116 | FALSE | Upregulated |
| 0,267404608 | 0,573281326 | CD48 | FALSE | Upregulated |
| -0,171904246 | 0,573281326 | NDUFA2 | FALSE | Downregulated |
| 0,363291784 | 0,573415277 | CARD9 | FALSE | Upregulated |
| -0,804337153 | 0,573415277 | TXLNB | FALSE | Downregulated |
| -0,219550718 | 0,573496613 | BCL6 | FALSE | Downregulated |
| -0,491679603 | 0,573496613 | CACNB2 | FALSE | Downregulated |
| -0,060650229 | 0,573496613 | HNRNPU | FALSE | Downregulated |
| 0,327418557 | 0,573496613 | LRRC31 | FALSE | Upregulated |
| -0,270519795 | 0,573496613 | TBC1D2 | FALSE | Downregulated |
| -0,202919228 | 0,573496613 | TCERG1 | FALSE | Downregulated |
| 0,199693561 | 0,573496613 | TNK2 | FALSE | Upregulated |
| -0,127414715 | 0,573496613 | WASF1 | FALSE | Downregulated |
| 0,166886794 | 0,573598365 | C7orf50 | FALSE | Upregulated |
| 0,087615769 | 0,573629011 | FXR2 | FALSE | Upregulated |
| 0,491150871 | 0,573629011 | TMPRSS9 | FALSE | Upregulated |
| -0,276297451 | 0,573917406 | BCHE | FALSE | Downregulated |
| 0,110765179 | 0,573917406 | SOCS7 | FALSE | Upregulated |

| | | | | |
|---|---|---|---|---|
| -0,081479848 | 0,573938472 | PPHLN1 | FALSE | Downregulated |
| -0,143872666 | 0,573977112 | WHSC1L1 | FALSE | Downregulated |
| -0,226654319 | 0,574003673 | ABCG2 | FALSE | Downregulated |
| 0,251673874 | 0,574003673 | IPCEF1 | FALSE | Upregulated |
| -0,173351973 | 0,574003673 | SCML1 | FALSE | Downregulated |
| 0,105721155 | 0,57435207 | ABL1 | FALSE | Upregulated |
| -0,208144056 | 0,57435207 | CRHBP | FALSE | Downregulated |
| -0,150960144 | 0,57435207 | MRPS17 | FALSE | Downregulated |
| 0,782148781 | 0,574504293 | C14orf64 | FALSE | Upregulated |
| 1,021322684 | 0,574504293 | COL22A1 | FALSE | Upregulated |
| 0,434645899 | 0,574504293 | FLT3 | FALSE | Upregulated |
| 0,086629223 | 0,574504293 | MBD1 | FALSE | Upregulated |
| 0,164848092 | 0,574504293 | N6AMT1 | FALSE | Upregulated |
| 0,129816519 | 0,574504293 | PRPS2 | FALSE | Upregulated |
| -0,145743314 | 0,574504293 | R3HCC1 | FALSE | Downregulated |
| -0,147306595 | 0,574504293 | RBM43 | FALSE | Downregulated |
| 0,129101924 | 0,574504293 | TRAPPC9 | FALSE | Upregulated |
| -0,163833111 | 0,574504293 | ZNF567 | FALSE | Downregulated |
| -0,191361721 | 0,574785417 | HIVEP1 | FALSE | Downregulated |
| -0,248767984 | 0,574785417 | SFRP1 | FALSE | Downregulated |
| 0,154003868 | 0,574913946 | CENPN | FALSE | Upregulated |
| -0,266466431 | 0,574913946 | CMBL | FALSE | Downregulated |
| 0,147716595 | 0,574913946 | FAM63A | FALSE | Upregulated |
| 1,027990534 | 0,574913946 | FXYD3 | FALSE | Upregulated |
| 0,323706474 | 0,574913946 | NUDT8 | FALSE | Upregulated |
| 0,228482529 | 0,574913946 | ZNF808 | FALSE | Upregulated |

| | | | | |
|---|---|---|---|---|
| -0,164762701 | 0,574934977 | BBS7 | FALSE | Downregulated |
| -0,19211545 | 0,574934977 | PARP2 | FALSE | Downregulated |
| -0,097005273 | 0,574934977 | ZHX1 | FALSE | Downregulated |
| -0,354836742 | 0,575238504 | DNAH6 | FALSE | Downregulated |
| 0,919633632 | 0,575238504 | ELFN2 | FALSE | Upregulated |
| 0,618142835 | 0,575238504 | LOC153684 | FALSE | Upregulated |
| -0,104143583 | 0,575238504 | MCCC2 | FALSE | Downregulated |
| -0,125475197 | 0,575238504 | MRPL20 | FALSE | Downregulated |
| -0,10210164 | 0,575238504 | NDUFS2 | FALSE | Downregulated |
| -0,36519504 | 0,575238504 | PIWIL2 | FALSE | Downregulated |
| 0,243354007 | 0,575238504 | SEL1L3 | FALSE | Upregulated |
| -0,131255297 | 0,575238504 | SLC35A3 | FALSE | Downregulated |
| 0,369105566 | 0,575379922 | A2M | FALSE | Upregulated |
| -0,200077761 | 0,575379922 | ATN1 | FALSE | Downregulated |
| -0,184657939 | 0,575379922 | C5orf51 | FALSE | Downregulated |
| -0,145240114 | 0,575379922 | CAPRIN2 | FALSE | Downregulated |
| 0,13920459 | 0,575379922 | CPM | FALSE | Upregulated |
| 0,089616312 | 0,575379922 | DLST | FALSE | Upregulated |
| 0,467419541 | 0,575379922 | EME1 | FALSE | Upregulated |
| -0,216976166 | 0,575379922 | FAXDC2 | FALSE | Downregulated |
| -0,197800422 | 0,575379922 | IDNK | FALSE | Downregulated |
| 0,088423827 | 0,575379922 | IL17RA | FALSE | Upregulated |
| 0,578345291 | 0,575379922 | IQCD | FALSE | Upregulated |
| 0,502388946 | 0,575379922 | IRF4 | FALSE | Upregulated |
| 0,935721354 | 0,575379922 | KIR2DL1 | FALSE | Upregulated |
| 0,086859174 | 0,575379922 | LRRC47 | FALSE | Upregulated |

| | | | | |
|---|---|---|---|---|
| 0,230085892 | 0,575379922 | MAST4 | FALSE | Upregulated |
| 0,099203781 | 0,575379922 | OTUB1 | FALSE | Upregulated |
| 0,144855339 | 0,575379922 | PARP10 | FALSE | Upregulated |
| 0,178814342 | 0,575379922 | RABEP2 | FALSE | Upregulated |
| -0,193611791 | 0,575379922 | RBM28 | FALSE | Downregulated |
| -0,156582633 | 0,575379922 | RMDN2 | FALSE | Downregulated |
| 0,082746025 | 0,575379922 | RNF220 | FALSE | Upregulated |
| -1,034090539 | 0,575379922 | RSPH10B2 | FALSE | Downregulated |
| -0,127549727 | 0,575379922 | SBDS | FALSE | Downregulated |
| -0,138414389 | 0,575379922 | TAB2 | FALSE | Downregulated |
| 0,110646085 | 0,575379922 | TMEM209 | FALSE | Upregulated |
| -0,127480955 | 0,575379922 | ZNRF2 | FALSE | Downregulated |
| 0,150729284 | 0,575390333 | CDAN1 | FALSE | Upregulated |
| -0,198407667 | 0,575390333 | SEMA6A | FALSE | Downregulated |
| -1,062140559 | 0,575639918 | NEB | FALSE | Downregulated |
| 0,157313194 | 0,575873727 | HEXIM1 | FALSE | Upregulated |
| -0,142961751 | 0,576225777 | STX18 | FALSE | Downregulated |
| -0,147835629 | 0,57634704 | PTPRG | FALSE | Downregulated |
| 0,754593878 | 0,576648862 | LRRC43 | FALSE | Upregulated |
| 1,014961115 | 0,576648862 | PRR19 | FALSE | Upregulated |
| -0,399392131 | 0,576989498 | GNAL | FALSE | Downregulated |
| -0,335176962 | 0,577254645 | LOC100996433 | FALSE | Downregulated |
| 0,127680856 | 0,577336271 | ADNP2 | FALSE | Upregulated |
| 0,209044922 | 0,577336271 | ARHGEF10 | FALSE | Upregulated |
| -0,175127845 | 0,577336271 | ARL13B | FALSE | Downregulated |
| 0,278113578 | 0,577336271 | C1orf198 | FALSE | Upregulated |

| | | | | |
|---|---|---|---|---|
| 0,57230991 | 0,577336271 | CDH11 | FALSE | Upregulated |
| 0,119980928 | 0,577336271 | COPB1 | FALSE | Upregulated |
| -0,115529373 | 0,577336271 | ERI2 | FALSE | Downregulated |
| -0,103449377 | 0,577336271 | EXOSC8 | FALSE | Downregulated |
| 0,116016612 | 0,577336271 | KIAA0195 | FALSE | Upregulated |
| -0,104968596 | 0,577336271 | NASP | FALSE | Downregulated |
| 0,094184673 | 0,577336271 | NUP54 | FALSE | Upregulated |
| 0,282306129 | 0,577336271 | PRKCB | FALSE | Upregulated |
| 0,121586279 | 0,577336271 | PRUNE | FALSE | Upregulated |
| 0,147848132 | 0,577336271 | TBC1D25 | FALSE | Upregulated |
| -0,07404885 | 0,577336271 | TFCP2 | FALSE | Downregulated |
| 0,231112182 | 0,577336271 | TLR6 | FALSE | Upregulated |
| 0,280712819 | 0,577336271 | TMEM206 | FALSE | Upregulated |
| 0,205595711 | 0,577336271 | VPS53 | FALSE | Upregulated |
| 0,359427621 | 0,577336271 | WDR76 | FALSE | Upregulated |
| 0,13765592 | 0,577336271 | YEATS2 | FALSE | Upregulated |
| 0,393014141 | 0,577433545 | C12orf5 | FALSE | Upregulated |
| -0,122641063 | 0,577557403 | CYB5A | FALSE | Downregulated |
| 0,426486867 | 0,577672008 | KIAA1211 | FALSE | Upregulated |
| -0,252487193 | 0,577709961 | ADAM1A | FALSE | Downregulated |
| 0,386347076 | 0,577721431 | AQP4 | FALSE | Upregulated |
| 0,608672096 | 0,577721431 | MTRNR2L6 | FALSE | Upregulated |
| 0,122546385 | 0,577721431 | TP53 | FALSE | Upregulated |
| -0,372585328 | 0,578359247 | LINC00115 | FALSE | Downregulated |
| -1,205251734 | 0,578359247 | LOC101929540 | FALSE | Downregulated |
| -0,535927472 | 0,578359247 | SPAG8 | FALSE | Downregulated |

| | | | | |
|---|---|---|---|---|
| 0,409150359 | 0,578411991 | GALNT15 | FALSE | Upregulated |
| -0,139231033 | 0,57845177 | RNF130 | FALSE | Downregulated |
| -0,172048459 | 0,578938853 | BANF1 | FALSE | Downregulated |
| -0,634414767 | 0,578938853 | C2CD4B | FALSE | Downregulated |
| -0,215289461 | 0,578938853 | CXCL14 | FALSE | Downregulated |
| -0,692452988 | 0,578938853 | EYA4 | FALSE | Downregulated |
| 0,146149345 | 0,578938853 | INPP1 | FALSE | Upregulated |
| 0,151258008 | 0,578938853 | KCTD11 | FALSE | Upregulated |
| -0,505196561 | 0,578938853 | LOC101927121 | FALSE | Downregulated |
| 0,277224816 | 0,578938853 | ME1 | FALSE | Upregulated |
| -0,160971097 | 0,578938853 | NIPSNAP1 | FALSE | Downregulated |
| 0,172468332 | 0,578938853 | NUP210 | FALSE | Upregulated |
| -0,383665982 | 0,578938853 | STK32C | FALSE | Downregulated |
| -0,113204591 | 0,578938853 | TMEM69 | FALSE | Downregulated |
| 0,23748632 | 0,578938853 | ZNF503-AS2 | FALSE | Upregulated |
| -0,106020324 | 0,579015655 | DNAJC24 | FALSE | Downregulated |
| -0,142385188 | 0,579180416 | RALA | FALSE | Downregulated |
| 0,1241488 | 0,579208631 | HSPBP1 | FALSE | Upregulated |
| -0,151841139 | 0,579817609 | ECSIT | FALSE | Downregulated |
| 0,273724377 | 0,579817609 | GALNT3 | FALSE | Upregulated |
| -0,114333304 | 0,579817609 | IL6ST | FALSE | Downregulated |
| -0,276776264 | 0,579817609 | LOC100128398 | FALSE | Downregulated |
| 0,252429903 | 0,579817609 | PLEKHG2 | FALSE | Upregulated |
| 0,16422176 | 0,579817609 | TMEM104 | FALSE | Upregulated |
| 0,871775851 | 0,579817609 | WFDC2 | FALSE | Upregulated |
| 0,243573388 | 0,579817609 | ZNF611 | FALSE | Upregulated |

| | | | | |
|---|---|---|---|---|
| -0,135233615 | 0,579967234 | CLU | FALSE | Downregulated |
| 0,100731456 | 0,579974612 | DCAF12 | FALSE | Upregulated |
| -0,152307055 | 0,579974612 | RBM48 | FALSE | Downregulated |
| 0,549251076 | 0,579974612 | RP11-475O6.1 | FALSE | Upregulated |
| 0,231182022 | 0,580155685 | CDH23 | FALSE | Upregulated |
| 0,135166495 | 0,580155685 | EXOSC1 | FALSE | Upregulated |
| 0,137781868 | 0,580155685 | FIS1 | FALSE | Upregulated |
| 0,76909769 | 0,580155685 | OLR1 | FALSE | Upregulated |
| 0,234407393 | 0,580414534 | AP1G2 | FALSE | Upregulated |
| -0,2239905 | 0,580414534 | APCS | FALSE | Downregulated |
| 0,126193797 | 0,580414534 | ATP2B1 | FALSE | Upregulated |
| 0,112378801 | 0,580414534 | ATP7B | FALSE | Upregulated |
| -0,137942114 | 0,580414534 | BDH1 | FALSE | Downregulated |
| 0,272275443 | 0,580414534 | CITED4 | FALSE | Upregulated |
| -0,208112839 | 0,580414534 | EGFR | FALSE | Downregulated |
| 0,147504744 | 0,580414534 | FAM160A2 | FALSE | Upregulated |
| 0,312842514 | 0,580414534 | FAM186B | FALSE | Upregulated |
| -0,215452441 | 0,580414534 | HHLA3 | FALSE | Downregulated |
| 0,849582843 | 0,580414534 | HIST1H2BN | FALSE | Upregulated |
| 0,461705517 | 0,580414534 | IPO11-LRRC70 | FALSE | Upregulated |
| 0,973937496 | 0,580414534 | LDHC | FALSE | Upregulated |
| -0,198386513 | 0,580414534 | LINC00667 | FALSE | Downregulated |
| -0,398975613 | 0,580414534 | LOC100270804 | FALSE | Downregulated |
| -1,121101619 | 0,580414534 | LOC102724694 | FALSE | Downregulated |
| 0,097825785 | 0,580414534 | NPLOC4 | FALSE | Upregulated |
| 1,089331886 | 0,580414534 | PCSK1N | FALSE | Upregulated |

| | | | | |
|---|---|---|---|---|
| -0,265975118 | 0,580414534 | PMF1-BGLAP | FALSE | Downregulated |
| -0,135137999 | 0,580414534 | RAB32 | FALSE | Downregulated |
| -0,612891909 | 0,580414534 | RP11-43N5.1 | FALSE | Downregulated |
| 0,334082196 | 0,580414534 | SERPINB8 | FALSE | Upregulated |
| 0,936001064 | 0,580414534 | SLC26A9 | FALSE | Upregulated |
| 0,224556162 | 0,580414534 | SMC2 | FALSE | Upregulated |
| 0,104318137 | 0,580414534 | TNPO2 | FALSE | Upregulated |
| 0,308270734 | 0,580414534 | ZNF229 | FALSE | Upregulated |
| 0,759372026 | 0,580656389 | TROAP | FALSE | Upregulated |
| -0,334075238 | 0,580687862 | LOC101927559 | FALSE | Downregulated |
| -0,317206929 | 0,580800124 | AD000090.2 | FALSE | Downregulated |
| -0,131247245 | 0,581033904 | CBR4 | FALSE | Downregulated |
| -0,514763279 | 0,581033904 | CCDC80 | FALSE | Downregulated |
| -0,185006141 | 0,581033904 | FLJ10038 | FALSE | Downregulated |
| -0,296853884 | 0,581033904 | LOC101928076 | FALSE | Downregulated |
| -0,646540087 | 0,581033904 | LOC102723428 | FALSE | Downregulated |
| 0,222406473 | 0,581033904 | TAF8 | FALSE | Upregulated |
| -0,201390992 | 0,581033904 | UBL7-AS1 | FALSE | Downregulated |
| 0,257047031 | 0,581109539 | AMIGO1 | FALSE | Upregulated |
| -0,181243638 | 0,581176201 | F2R | FALSE | Downregulated |
| 0,111958643 | 0,58124945 | MRPL28 | FALSE | Upregulated |
| 0,508225836 | 0,581518844 | B3GNT8 | FALSE | Upregulated |
| -0,345170338 | 0,581714223 | RNF5P1 | FALSE | Downregulated |
| 0,555880625 | 0,581923772 | CCDC87 | FALSE | Upregulated |
| 0,370130388 | 0,582089929 | EVA1C | FALSE | Upregulated |
| 0,15018071 | 0,582089929 | TCTN1 | FALSE | Upregulated |

| | | | | |
|---|---|---|---|---|
| -0,402396855 | 0,582288853 | KIF6 | FALSE | Downregulated |
| 1,139605627 | 0,582429385 | AC010091.1 | FALSE | Upregulated |
| 0,204572026 | 0,582429385 | BCL2L12 | FALSE | Upregulated |
| 0,174813039 | 0,582429385 | SLC25A22 | FALSE | Upregulated |
| -0,174833199 | 0,582460312 | CBS | FALSE | Downregulated |
| -0,241271337 | 0,582460312 | SLC22A9 | FALSE | Downregulated |
| -0,147200214 | 0,582540167 | EDEM1 | FALSE | Downregulated |
| 0,607069166 | 0,582540167 | LOC102723928 | FALSE | Upregulated |
| -0,573799599 | 0,58264008 | CWH43 | FALSE | Downregulated |
| -0,078490747 | 0,58264008 | EIF2AK1 | FALSE | Downregulated |
| 0,193397089 | 0,58264008 | ERBB2 | FALSE | Upregulated |
| 0,115307907 | 0,58264008 | HS1BP3 | FALSE | Upregulated |
| 0,122455197 | 0,58264008 | MTMR9 | FALSE | Upregulated |
| 0,8443827 | 0,58264008 | PTCHD4 | FALSE | Upregulated |
| -0,12874576 | 0,58264008 | TIPRL | FALSE | Downregulated |
| 0,219321651 | 0,58264008 | TTC3P1 | FALSE | Upregulated |
| -0,177970632 | 0,58264008 | ZFHX4 | FALSE | Downregulated |
| 0,165346539 | 0,58264008 | ZNF34 | FALSE | Upregulated |
| -0,94554095 | 0,582698434 | CA8 | FALSE | Downregulated |
| -1,568470672 | 0,583025545 | AMPD1 | FALSE | Downregulated |
| 0,098213964 | 0,583025545 | CHTOP | FALSE | Upregulated |
| -0,166620207 | 0,583025545 | EYA3 | FALSE | Downregulated |
| 0,152587288 | 0,583025545 | FZR1 | FALSE | Upregulated |
| -0,253864465 | 0,583025545 | LGALS1 | FALSE | Downregulated |
| 0,1377962 | 0,583025545 | LIN7A | FALSE | Upregulated |
| -0,397103881 | 0,583025545 | LINC00294 | FALSE | Downregulated |

| | | | | |
|---|---|---|---|---|
| -0,179191538 | 0,583025545 | SLC9A7 | FALSE | Downregulated |
| -0,155341472 | 0,583025545 | TMEM45A | FALSE | Downregulated |
| 0,148314898 | 0,583025545 | USP6NL | FALSE | Upregulated |
| 0,418924385 | 0,583049974 | BATF3 | FALSE | Upregulated |
| 0,164985304 | 0,583218394 | TMUB1 | FALSE | Upregulated |
| 0,117619322 | 0,583350392 | PRCP | FALSE | Upregulated |
| -1,338742366 | 0,583418825 | FAR2P3 | FALSE | Downregulated |
| -0,678479538 | 0,583418825 | HYDIN | FALSE | Downregulated |
| -0,215900654 | 0,583418825 | LOC100289361 | FALSE | Downregulated |
| -0,277567003 | 0,583418825 | LOC102724795 | FALSE | Downregulated |
| 0,12720548 | 0,583418825 | NPRL3 | FALSE | Upregulated |
| -0,143065618 | 0,583418825 | ZNF605 | FALSE | Downregulated |
| -0,188602214 | 0,583486666 | GPATCH2 | FALSE | Downregulated |
| -0,127873343 | 0,583486666 | LYST | FALSE | Downregulated |
| 0,127730547 | 0,583486666 | TNFRSF1A | FALSE | Upregulated |
| -0,284387945 | 0,583779845 | IQSEC2 | FALSE | Downregulated |
| 0,22247331 | 0,58390223 | HVCN1 | FALSE | Upregulated |
| 0,431917947 | 0,58390223 | MAP3K15 | FALSE | Upregulated |
| 0,181027398 | 0,58390223 | PML | FALSE | Upregulated |
| 0,268932365 | 0,584137402 | FLVCR2 | FALSE | Upregulated |
| -0,357809735 | 0,584137402 | LOC100506688 | FALSE | Downregulated |
| 0,140109448 | 0,584167229 | CERK | FALSE | Upregulated |
| -0,266526152 | 0,584167229 | SESN3 | FALSE | Downregulated |
| 0,271095269 | 0,584229409 | BTG3 | FALSE | Upregulated |
| -0,5651768 | 0,584408241 | LOC101929506 | FALSE | Downregulated |
| -0,185179844 | 0,584408241 | PPFIBP1 | FALSE | Downregulated |

| | | | | |
|---|---|---|---|---|
| 0,362462829 | 0,584672016 | IER5L | FALSE | Upregulated |
| -0,133566228 | 0,584672016 | MED19 | FALSE | Downregulated |
| 0,13779905 | 0,584698905 | LRRC45 | FALSE | Upregulated |
| 0,186412548 | 0,584762281 | LRRC20 | FALSE | Upregulated |
| -0,270742634 | 0,584762281 | PDE5A | FALSE | Downregulated |
| 0,167488889 | 0,584762281 | SNX11 | FALSE | Upregulated |
| 0,300660005 | 0,584807967 | LILRA1 | FALSE | Upregulated |
| -0,11596166 | 0,584907505 | CLNS1A | FALSE | Downregulated |
| -0,150306031 | 0,584907505 | KNOP1 | FALSE | Downregulated |
| 0,289683 | 0,584907505 | LOC102724168 | FALSE | Upregulated |
| 0,600424393 | 0,584946063 | FAM179A | FALSE | Upregulated |
| -0,78825268 | 0,584946063 | LOC100506797 | FALSE | Downregulated |
| -0,164938322 | 0,584946063 | RPS24 | FALSE | Downregulated |
| 0,085060682 | 0,585251256 | RANBP6 | FALSE | Upregulated |
| 0,266461938 | 0,585629287 | STAT1 | FALSE | Upregulated |
| -0,647735909 | 0,58600254 | AJAP1 | FALSE | Downregulated |
| -0,23565388 | 0,586095193 | FAM26E | FALSE | Downregulated |
| -0,160239248 | 0,586095193 | HIF1A | FALSE | Downregulated |
| 0,128779279 | 0,586610882 | ALG9 | FALSE | Upregulated |
| -0,237922835 | 0,586660773 | RFTN1 | FALSE | Downregulated |
| 0,160289609 | 0,586730059 | BZW2 | FALSE | Upregulated |
| 0,232787844 | 0,586730059 | C1QTNF6 | FALSE | Upregulated |
| -0,401773152 | 0,586730059 | CYP26B1 | FALSE | Downregulated |
| 0,074837748 | 0,586730059 | DNAJB12 | FALSE | Upregulated |
| 0,458834554 | 0,586730059 | FGF7 | FALSE | Upregulated |
| -0,146845079 | 0,586730059 | HDAC4 | FALSE | Downregulated |

| | | | | |
|---|---|---|---|---|
| 0,166362069 | 0,586730059 | INS-IGF2 | FALSE | Upregulated |
| -0,213796217 | 0,586730059 | LMLN | FALSE | Downregulated |
| -0,547783368 | 0,586730059 | LOC100996756 | FALSE | Downregulated |
| -0,394371123 | 0,586730059 | LOC101927331 | FALSE | Downregulated |
| -0,155869637 | 0,586730059 | MPDZ | FALSE | Downregulated |
| 0,156550115 | 0,586730059 | MRPL54 | FALSE | Upregulated |
| -0,095479576 | 0,586730059 | NBR1 | FALSE | Downregulated |
| 0,331468828 | 0,586730059 | PBX1 | FALSE | Upregulated |
| 0,098761407 | 0,586730059 | PRKDC | FALSE | Upregulated |
| -0,438998537 | 0,586730059 | RP4-756H11.3 | FALSE | Downregulated |
| 0,092131757 | 0,586730059 | SEC23IP | FALSE | Upregulated |
| 0,088863211 | 0,586730059 | SMARCD1 | FALSE | Upregulated |
| 0,196874409 | 0,586730059 | THOC3 | FALSE | Upregulated |
| -0,35988601 | 0,586730059 | TRNC | FALSE | Downregulated |
| 0,335738057 | 0,586730059 | XYLT1 | FALSE | Upregulated |
| -0,129135649 | 0,586730059 | ZNF226 | FALSE | Downregulated |
| -0,16582468 | 0,586751486 | INPP5B | FALSE | Downregulated |
| -0,106063964 | 0,586959372 | DHFRL1 | FALSE | Downregulated |
| -0,124512903 | 0,586959372 | FAM210A | FALSE | Downregulated |
| 0,111594095 | 0,586959372 | FAM83H | FALSE | Upregulated |
| -0,99323641 | 0,586959372 | IL17D | FALSE | Downregulated |
| -0,196088402 | 0,586959372 | LBX2-AS1 | FALSE | Downregulated |
| -0,725822412 | 0,586959372 | LOC101928102 | FALSE | Downregulated |
| -0,634107546 | 0,586959372 | NEUROD1 | FALSE | Downregulated |
| -0,093200162 | 0,586959372 | SECISBP2 | FALSE | Downregulated |
| 0,209985788 | 0,586959372 | TEAD2 | FALSE | Upregulated |

| | | | | |
|---|---|---|---|---|
| 0,103424372 | 0,586959372 | TMX2 | FALSE | Upregulated |
| 0,133228693 | 0,587324757 | CDC42SE1 | FALSE | Upregulated |
| 0,152838655 | 0,58746764 | BTG1 | FALSE | Upregulated |
| 0,57607929 | 0,58746764 | C18orf42 | FALSE | Upregulated |
| -0,711839373 | 0,58746764 | CCDC39 | FALSE | Downregulated |
| 0,078950196 | 0,58746764 | DVL3 | FALSE | Upregulated |
| -0,759039058 | 0,58746764 | GALNT13 | FALSE | Downregulated |
| 0,193081292 | 0,58746764 | HM13 | FALSE | Upregulated |
| -0,124047065 | 0,58746764 | METTL7A | FALSE | Downregulated |
| 0,388812369 | 0,58746764 | PFKP | FALSE | Upregulated |
| 0,338986695 | 0,58746764 | SLA2 | FALSE | Upregulated |
| 0,237064944 | 0,58746764 | SYNGAP1 | FALSE | Upregulated |
| 0,156946794 | 0,58746764 | TTLL12 | FALSE | Upregulated |
| 0,103104752 | 0,58746764 | WDR26 | FALSE | Upregulated |
| 0,092112979 | 0,587482112 | SQRDL | FALSE | Upregulated |
| 0,275854324 | 0,587482112 | ZNF43 | FALSE | Upregulated |
| 0,256917034 | 0,587602467 | EIF3C | FALSE | Upregulated |
| -0,43783663 | 0,58782362 | CXCL2 | FALSE | Downregulated |
| -0,095276448 | 0,58782362 | METTL14 | FALSE | Downregulated |
| 0,257447437 | 0,58782362 | MXD3 | FALSE | Upregulated |
| 1,123546711 | 0,587831329 | C9orf129 | FALSE | Upregulated |
| 0,210920532 | 0,587831329 | ZNF48 | FALSE | Upregulated |
| 0,196967121 | 0,588094378 | ITGAV | FALSE | Upregulated |
| 0,297108348 | 0,588208661 | DTX3 | FALSE | Upregulated |
| -0,381256412 | 0,588208661 | RP11-31F15.2 | FALSE | Downregulated |
| 0,162217854 | 0,588437494 | SBNO2 | FALSE | Upregulated |

| | | | | |
|---|---|---|---|---|
| -0,951578083 | 0,588606211 | EMX2OS | FALSE | Downregulated |
| -0,602776638 | 0,588634095 | FKBP9L | FALSE | Downregulated |
| -0,129177656 | 0,588634095 | SMIM20 | FALSE | Downregulated |
| -1,642744043 | 0,588871437 | KCNA7 | FALSE | Downregulated |
| 0,074035322 | 0,588871437 | PUM1 | FALSE | Upregulated |
| -0,348055668 | 0,588949949 | FABP3 | FALSE | Downregulated |
| -0,197533707 | 0,588956957 | B3GNT1 | FALSE | Downregulated |
| 0,413683409 | 0,588956957 | BEND4 | FALSE | Upregulated |
| 0,551684228 | 0,588956957 | CRNDE | FALSE | Upregulated |
| -0,157535136 | 0,588956957 | FAM117A | FALSE | Downregulated |
| 0,527279939 | 0,588956957 | KIAA1656 | FALSE | Upregulated |
| 0,187200752 | 0,588956957 | MMP15 | FALSE | Upregulated |
| -0,262811462 | 0,588956957 | RP11-96H19.1 | FALSE | Downregulated |
| 0,095151957 | 0,588956957 | TBC1D9B | FALSE | Upregulated |
| -0,073042409 | 0,588956957 | TMEM66 | FALSE | Downregulated |
| -0,108810762 | 0,588956957 | ZFP62 | FALSE | Downregulated |
| 0,209766664 | 0,589098921 | LCP2 | FALSE | Upregulated |
| -0,149127521 | 0,589098921 | MACF1 | FALSE | Downregulated |
| 0,614339715 | 0,589098921 | ROR2 | FALSE | Upregulated |
| 0,495522708 | 0,589098921 | SIT1 | FALSE | Upregulated |
| 0,728531406 | 0,589098921 | ZNF233 | FALSE | Upregulated |
| -0,197374919 | 0,589197576 | DYRK3 | FALSE | Downregulated |
| 0,136597545 | 0,589197576 | FAM149A | FALSE | Upregulated |
| -0,309873797 | 0,589197576 | GYLTL1B | FALSE | Downregulated |
| -0,218169213 | 0,589197576 | LDLRAD4 | FALSE | Downregulated |
| 0,13792466 | 0,589197576 | SEC24A | FALSE | Upregulated |

| | | | | |
|---|---|---|---|---|
| -0,169446004 | 0,589197576 | SLC47A1 | FALSE | Downregulated |
| -0,114401405 | 0,589197576 | SPIDR | FALSE | Downregulated |
| 0,101386903 | 0,589313062 | ATF6B | FALSE | Upregulated |
| 0,826240764 | 0,589313062 | CDC25C | FALSE | Upregulated |
| -0,431122027 | 0,589313062 | PKHD1L1 | FALSE | Downregulated |
| -0,316846154 | 0,589313062 | RP5-914P20.5 | FALSE | Downregulated |
| -0,465518832 | 0,589315093 | MT1DP | FALSE | Downregulated |
| 0,188372021 | 0,589382134 | ANXA6 | FALSE | Upregulated |
| 0,690628207 | 0,589382134 | LOC102724948 | FALSE | Upregulated |
| -0,10791431 | 0,589462796 | DCAF8 | FALSE | Downregulated |
| -0,150826493 | 0,589462796 | DNPH1 | FALSE | Downregulated |
| -0,497640666 | 0,589462796 | LOC101927635 | FALSE | Downregulated |
| -0,131140304 | 0,589462796 | RPL37A | FALSE | Downregulated |
| -0,153624747 | 0,589940432 | PHACTR2 | FALSE | Downregulated |
| 0,186536387 | 0,589970948 | HCLS1 | FALSE | Upregulated |
| -0,103248357 | 0,589970948 | PDCL | FALSE | Downregulated |
| 0,255594528 | 0,589970948 | SAMSN1 | FALSE | Upregulated |
| 0,676778863 | 0,590160135 | FAM43B | FALSE | Upregulated |
| 0,950232365 | 0,590209614 | FUT2 | FALSE | Upregulated |
| 0,143976889 | 0,590493055 | SLC20A2 | FALSE | Upregulated |
| -0,119325889 | 0,590553883 | SPRYD7 | FALSE | Downregulated |
| -0,138814293 | 0,5906929 | ERICH1 | FALSE | Downregulated |
| 0,323833447 | 0,5906929 | SLC2A6 | FALSE | Upregulated |
| -0,380281794 | 0,5906929 | TSPAN13 | FALSE | Downregulated |
| -0,276185934 | 0,590985399 | CTF1 | FALSE | Downregulated |
| -0,099724547 | 0,591091466 | MRPL37 | FALSE | Downregulated |

| | | | | |
|---|---|---|---|---|
| -0,139227715 | 0,591298825 | CEP95 | FALSE | Downregulated |
| 0,280210064 | 0,591298825 | RAMP1 | FALSE | Upregulated |
| -0,96175905 | 0,591407274 | TPM2 | FALSE | Downregulated |
| 0,124515138 | 0,591431966 | MPRIP | FALSE | Upregulated |
| 0,352702285 | 0,59179619 | IL18 | FALSE | Upregulated |
| -0,439253183 | 0,59179619 | PRRT3-AS1 | FALSE | Downregulated |
| -0,373721875 | 0,591821898 | LOC102724493 | FALSE | Downregulated |
| 0,378071197 | 0,591821898 | NUDT17 | FALSE | Upregulated |
| 0,549448117 | 0,592163977 | FREM2 | FALSE | Upregulated |
| -0,194864553 | 0,592304055 | FAM76A | FALSE | Downregulated |
| 0,282576066 | 0,592404691 | ME3 | FALSE | Upregulated |
| -0,629128237 | 0,592404691 | RBM20 | FALSE | Downregulated |
| 0,163956678 | 0,592404691 | RNF215 | FALSE | Upregulated |
| 0,130634901 | 0,592436304 | LANCL2 | FALSE | Upregulated |
| 0,22232421 | 0,59289881 | MAP3K12 | FALSE | Upregulated |
| -0,8249855 | 0,593609132 | DPY19L2 | FALSE | Downregulated |
| 0,266243856 | 0,593609132 | EVA1B | FALSE | Upregulated |
| 0,163817288 | 0,593836842 | PNPLA2 | FALSE | Upregulated |
| -0,102960422 | 0,593908759 | DHX33 | FALSE | Downregulated |
| 0,394866908 | 0,593921267 | ADC | FALSE | Upregulated |
| 0,477600803 | 0,593921267 | C17orf53 | FALSE | Upregulated |
| 0,11909455 | 0,593921267 | DYSF | FALSE | Upregulated |
| -0,094951722 | 0,593921267 | ECD | FALSE | Downregulated |
| -0,185293851 | 0,593921267 | IWS1 | FALSE | Downregulated |
| -0,223652951 | 0,593921267 | LIPC | FALSE | Downregulated |
| 0,23975342 | 0,593921267 | LOC81691 | FALSE | Upregulated |

| | | | | |
|---|---|---|---|---|
| -0,161890165 | 0,593921267 | MAGI2-AS3 | FALSE | Downregulated |
| -0,128568806 | 0,593921267 | NMI | FALSE | Downregulated |
| -0,142335233 | 0,593921267 | RAB7A | FALSE | Downregulated |
| 0,174866796 | 0,593921267 | SLC30A1 | FALSE | Upregulated |
| -0,093536076 | 0,594632649 | CNOT4 | FALSE | Downregulated |
| 0,824803519 | 0,594735364 | LOC102724661 | FALSE | Upregulated |
| -0,137519417 | 0,594833105 | APBB2 | FALSE | Downregulated |
| 0,093690559 | 0,594833105 | DUS1L | FALSE | Upregulated |
| -0,165202925 | 0,595113813 | LOC101928433 | FALSE | Downregulated |
| -0,121444068 | 0,595156809 | APOE | FALSE | Downregulated |
| 0,166996855 | 0,595156809 | BOK | FALSE | Upregulated |
| -0,07233187 | 0,595216462 | CUL5 | FALSE | Downregulated |
| -0,18248111 | 0,595216462 | NFKBIB | FALSE | Downregulated |
| -0,110185062 | 0,595578829 | CTAGE5 | FALSE | Downregulated |
| 0,112468921 | 0,595578829 | CYP20A1 | FALSE | Upregulated |
| -0,128809298 | 0,595578829 | ZRANB2 | FALSE | Downregulated |
| 0,110803233 | 0,595678358 | USF1 | FALSE | Upregulated |
| 0,33777835 | 0,595971702 | RGS1 | FALSE | Upregulated |
| -0,274146159 | 0,596197059 | DAB1 | FALSE | Downregulated |
| 0,413164683 | 0,596209175 | GEM | FALSE | Upregulated |
| -0,288040007 | 0,596425263 | SRD5A2 | FALSE | Downregulated |
| -0,278521752 | 0,59649657 | DNAJC27-AS1 | FALSE | Downregulated |
| 0,10132274 | 0,59649657 | NDUFA8 | FALSE | Upregulated |
| 0,157405934 | 0,59649657 | TMEM177 | FALSE | Upregulated |
| -0,108272176 | 0,596543015 | NAA25 | FALSE | Downregulated |
| -0,70535042 | 0,59726583 | CCDC160 | FALSE | Downregulated |

| | | | | |
|---|---|---|---|---|
| -0,321025779 | 0,59726583 | INHBE | FALSE | Downregulated |
| -0,099376984 | 0,597352982 | ZNF277 | FALSE | Downregulated |
| -0,169574984 | 0,597402819 | EI24 | FALSE | Downregulated |
| 0,1119927 | 0,597402819 | MOB4 | FALSE | Upregulated |
| -0,587485529 | 0,597453686 | INE1 | FALSE | Downregulated |
| 0,082894334 | 0,597461316 | EXOC4 | FALSE | Upregulated |
| -0,076123375 | 0,597461316 | MED4 | FALSE | Downregulated |
| -0,16990675 | 0,597502356 | BIVM-ERCC5 | FALSE | Downregulated |
| 0,381152882 | 0,597502356 | EVC2 | FALSE | Upregulated |
| 0,500297698 | 0,597724946 | ESRRG | FALSE | Upregulated |
| -0,093623672 | 0,597919249 | MRPL3 | FALSE | Downregulated |
| -0,08498684 | 0,597966809 | TRAPPC11 | FALSE | Downregulated |
| 0,218492686 | 0,598052288 | C9orf89 | FALSE | Upregulated |
| -0,388420903 | 0,598052288 | GOLGA6B | FALSE | Downregulated |
| 0,814414138 | 0,598052288 | LOC102724740 | FALSE | Upregulated |
| -0,112815586 | 0,598052288 | MRPL35 | FALSE | Downregulated |
| -0,734710915 | 0,598135824 | DNAI1 | FALSE | Downregulated |
| 0,090801834 | 0,598345409 | DDX20 | FALSE | Upregulated |
| 0,391005829 | 0,598526441 | CIT | FALSE | Upregulated |
| 0,098253713 | 0,598888457 | ASCC2 | FALSE | Upregulated |
| -0,12732636 | 0,598888457 | NAA15 | FALSE | Downregulated |
| 0,229976751 | 0,599079888 | GALNT6 | FALSE | Upregulated |
| -0,125534574 | 0,599079888 | KIAA1109 | FALSE | Downregulated |
| -0,200581923 | 0,599079888 | SLC24A1 | FALSE | Downregulated |
| -0,379454651 | 0,59920456 | SYNC | FALSE | Downregulated |
| 0,128269359 | 0,599310165 | ALG8 | FALSE | Upregulated |

| | | | | |
|---|---|---|---|---|
| 0,103105308 | 0,599310165 | YWHAZ | FALSE | Upregulated |
| -0,096821606 | 0,59943964 | CBLL1 | FALSE | Downregulated |
| 0,43662068 | 0,59943964 | NOD2 | FALSE | Upregulated |
| 0,189917087 | 0,59943964 | SAMHD1 | FALSE | Upregulated |
| -0,285141753 | 0,599724522 | C1orf173 | FALSE | Downregulated |
| -0,170029103 | 0,599724522 | MARC2 | FALSE | Downregulated |
| -0,109726969 | 0,599724522 | NOP14 | FALSE | Downregulated |
| -0,095501555 | 0,599724522 | TAF4 | FALSE | Downregulated |
| -0,150684467 | 0,599749245 | ITPA | FALSE | Downregulated |
| 0,69575608 | 0,600113633 | CHI3L1 | FALSE | Upregulated |
| -0,161413284 | 0,600138043 | BTD | FALSE | Downregulated |
| 0,381788198 | 0,600247229 | ZNF469 | FALSE | Upregulated |
| 0,612521188 | 0,600302 | LOC286190 | FALSE | Upregulated |
| 0,350261244 | 0,600303353 | GSN | FALSE | Upregulated |
| 0,104888194 | 0,600303353 | IDE | FALSE | Upregulated |
| 0,091143108 | 0,600303353 | MINK1 | FALSE | Upregulated |
| -0,133986471 | 0,600585204 | NUP35 | FALSE | Downregulated |
| 0,870951565 | 0,600952635 | ALOX15B | FALSE | Upregulated |
| -0,16780046 | 0,60105755 | NAA16 | FALSE | Downregulated |
| -0,250154753 | 0,60105755 | SRRM5 | FALSE | Downregulated |
| 0,244529998 | 0,601078091 | FMOD | FALSE | Upregulated |
| 0,080000308 | 0,601210129 | STIM1 | FALSE | Upregulated |
| 0,133837774 | 0,601345834 | ABHD12 | FALSE | Upregulated |
| 0,654639015 | 0,601347476 | OR2A20P | FALSE | Upregulated |
| -0,725066126 | 0,601347476 | PPFIA4 | FALSE | Downregulated |
| -0,143752056 | 0,601347476 | ZCCHC17 | FALSE | Downregulated |

| | | | | |
|---|---|---|---|---|
| -0,184323718 | 0,601494948 | ECH1 | FALSE | Downregulated |
| 0,308365185 | 0,602037368 | ACTA2 | FALSE | Upregulated |
| -0,101156773 | 0,602037368 | FBXW2 | FALSE | Downregulated |
| -1,609721945 | 0,602037368 | KBTBD12 | FALSE | Downregulated |
| -0,602861344 | 0,602037368 | LOC101927469 | FALSE | Downregulated |
| -0,11517749 | 0,602123242 | SLC35D1 | FALSE | Downregulated |
| 0,194913624 | 0,602405187 | BASP1 | FALSE | Upregulated |
| -0,477752976 | 0,602405187 | LOC102724019 | FALSE | Downregulated |
| 0,204109537 | 0,602537035 | SESN1 | FALSE | Upregulated |
| -0,155629794 | 0,602785844 | TXNDC16 | FALSE | Downregulated |
| 0,140113986 | 0,602819992 | USP20 | FALSE | Upregulated |
| 0,398933894 | 0,602836832 | PMEPA1 | FALSE | Upregulated |
| -0,269304784 | 0,60286029 | CCNB3 | FALSE | Downregulated |
| -0,618648917 | 0,60286029 | DPY19L2P3 | FALSE | Downregulated |
| 0,290171989 | 0,60286029 | KLRK1 | FALSE | Upregulated |
| -0,444361013 | 0,60286029 | LOC101927226 | FALSE | Downregulated |
| 0,100510436 | 0,60286029 | MED15 | FALSE | Upregulated |
| -0,107798405 | 0,60286029 | RANBP2 | FALSE | Downregulated |
| -0,133267212 | 0,60286029 | TULP4 | FALSE | Downregulated |
| 0,142679955 | 0,603008188 | MVB12A | FALSE | Upregulated |
| -0,173611723 | 0,603112992 | AMZ2P1 | FALSE | Downregulated |
| -0,143990854 | 0,603133838 | NME6 | FALSE | Downregulated |
| -0,090672517 | 0,603257526 | MYNN | FALSE | Downregulated |
| -0,111250902 | 0,603533964 | BCL2L2 | FALSE | Downregulated |
| 0,105902142 | 0,603533964 | BRE | FALSE | Upregulated |
| 0,201419569 | 0,603533964 | CCBL1 | FALSE | Upregulated |

| | | | | |
|---|---|---|---|---|
| 0,182170846 | 0,603533964 | CYBA | FALSE | Upregulated |
| 0,105618133 | 0,603533964 | ITGB1 | FALSE | Upregulated |
| 0,254841918 | 0,603533964 | PDE1B | FALSE | Upregulated |
| -0,113932408 | 0,603533964 | PPP1R10 | FALSE | Downregulated |
| -0,11832003 | 0,603533964 | PRKAR1A | FALSE | Downregulated |
| 0,201688679 | 0,603533964 | TRAPPC4 | FALSE | Upregulated |
| 0,109564626 | 0,603533964 | ZNF562 | FALSE | Upregulated |
| 0,309233339 | 0,604636151 | ABCA4 | FALSE | Upregulated |
| -0,500250694 | 0,604636151 | ACBD7 | FALSE | Downregulated |
| 0,176132911 | 0,604636151 | B3GAT3 | FALSE | Upregulated |
| 0,680028587 | 0,604636151 | C9orf66 | FALSE | Upregulated |
| 0,814529676 | 0,604636151 | CXADRP2 | FALSE | Upregulated |
| -0,148007617 | 0,604636151 | EBNA1BP2 | FALSE | Downregulated |
| -0,121427415 | 0,604636151 | EIF4B | FALSE | Downregulated |
| -0,758583677 | 0,604636151 | ELMOD1 | FALSE | Downregulated |
| -0,233161127 | 0,604636151 | EMP2 | FALSE | Downregulated |
| 0,320477282 | 0,604636151 | EMR1 | FALSE | Upregulated |
| 0,178349909 | 0,604636151 | FAM117B | FALSE | Upregulated |
| -0,151672436 | 0,604636151 | FOXJ3 | FALSE | Downregulated |
| 0,174031444 | 0,604636151 | FTL | FALSE | Upregulated |
| -0,186184118 | 0,604636151 | GNL2 | FALSE | Downregulated |
| -0,213899366 | 0,604636151 | LINC00888 | FALSE | Downregulated |
| 0,702203929 | 0,604636151 | LOC101928624 | FALSE | Upregulated |
| -0,788228771 | 0,604636151 | LOC102724697 | FALSE | Downregulated |
| -0,211390518 | 0,604636151 | LOC647859 | FALSE | Downregulated |
| 0,146025876 | 0,604636151 | LRCH4 | FALSE | Upregulated |

| | | | | |
|---|---|---|---|---|
| 0,092406481 | 0,604636151 | MAPKAPK2 | FALSE | Upregulated |
| -0,210384375 | 0,604636151 | MATN2 | FALSE | Downregulated |
| 0,062085828 | 0,604636151 | MBTPS1 | FALSE | Upregulated |
| 0,209530803 | 0,604636151 | MSN | FALSE | Upregulated |
| 0,136756784 | 0,604636151 | OGFR | FALSE | Upregulated |
| 0,257147264 | 0,604636151 | PDGFRA | FALSE | Upregulated |
| 0,202959579 | 0,604636151 | PGM3 | FALSE | Upregulated |
| 0,174254159 | 0,604636151 | PHF13 | FALSE | Upregulated |
| -0,088996898 | 0,604636151 | PIGK | FALSE | Downregulated |
| -0,075835063 | 0,604636151 | RAB5A | FALSE | Downregulated |
| 0,919124031 | 0,604636151 | RASGRF1 | FALSE | Upregulated |
| -0,202460621 | 0,604636151 | RP5-886K2.3 | FALSE | Downregulated |
| 0,276479242 | 0,604636151 | SLC37A1 | FALSE | Upregulated |
| -0,13684651 | 0,604636151 | SMN1 | FALSE | Downregulated |
| -0,155931348 | 0,604636151 | SNHG16 | FALSE | Downregulated |
| 0,146563186 | 0,604636151 | SPATA2 | FALSE | Upregulated |
| -0,596920843 | 0,604636151 | STAC3 | FALSE | Downregulated |
| -0,224422292 | 0,604636151 | TGFA | FALSE | Downregulated |
| -0,113901182 | 0,604636151 | TM2D2 | FALSE | Downregulated |
| -0,072663837 | 0,604636151 | ZFAND3 | FALSE | Downregulated |
| 0,90018181 | 0,605233409 | CNR1 | FALSE | Upregulated |
| 0,091400422 | 0,605558154 | FKBP1A | FALSE | Upregulated |
| -0,128988175 | 0,605558154 | PBX3 | FALSE | Downregulated |
| 0,195928722 | 0,605558154 | POLA2 | FALSE | Upregulated |
| 0,177528777 | 0,605585029 | KIAA1919 | FALSE | Upregulated |
| -0,10917611 | 0,605710625 | MAPK6 | FALSE | Downregulated |

| | | | | |
|---|---|---|---|---|
| 0,124958885 | 0,605731226 | CTSA | FALSE | Upregulated |
| -0,509221825 | 0,605731226 | LOC101928891 | FALSE | Downregulated |
| -0,246342891 | 0,605731226 | TAL1 | FALSE | Downregulated |
| 0,403297473 | 0,605747688 | TMEM249 | FALSE | Upregulated |
| -0,636502658 | 0,605800046 | LOC101928361 | FALSE | Downregulated |
| 0,071200935 | 0,605800046 | MRPL30 | FALSE | Upregulated |
| -0,136030339 | 0,60582469 | ARL15 | FALSE | Downregulated |
| -0,661597538 | 0,60582469 | FAM218A | FALSE | Downregulated |
| 0,127392743 | 0,60582469 | GARS | FALSE | Upregulated |
| -0,189403051 | 0,60582469 | NAT2 | FALSE | Downregulated |
| -0,18509083 | 0,60582604 | MRPS25 | FALSE | Downregulated |
| -0,605831217 | 0,606323096 | C1orf95 | FALSE | Downregulated |
| 0,16883529 | 0,606323096 | GLA | FALSE | Upregulated |
| -0,716006443 | 0,606323096 | LOC102724957 | FALSE | Downregulated |
| 0,1330118 | 0,606397714 | CREB3 | FALSE | Upregulated |
| -0,249995325 | 0,606397714 | FUZ | FALSE | Downregulated |
| 0,310426822 | 0,606397714 | GPR132 | FALSE | Upregulated |
| -0,401171749 | 0,606397714 | GSTA7P | FALSE | Downregulated |
| 0,309023101 | 0,606397714 | LOC100996255 | FALSE | Upregulated |
| -0,139734188 | 0,606397714 | USP15 | FALSE | Downregulated |
| 0,419469785 | 0,606397714 | WBSCR27 | FALSE | Upregulated |
| 0,607442253 | 0,606408066 | LOC102724156 | FALSE | Upregulated |
| 0,173078903 | 0,606624635 | C19orf10 | FALSE | Upregulated |
| 0,302078819 | 0,606624635 | HSH2D | FALSE | Upregulated |
| 0,142702397 | 0,606624635 | SIPA1L3 | FALSE | Upregulated |
| -0,109001788 | 0,606624635 | SRSF4 | FALSE | Downregulated |

| | | | | |
|---|---|---|---|---|
| -0,155022291 | 0,607226703 | RNF144B | FALSE | Downregulated |
| 0,299569694 | 0,607308789 | APOL3 | FALSE | Upregulated |
| 0,138464982 | 0,607308789 | RAI1 | FALSE | Upregulated |
| -0,322468528 | 0,607568699 | DMPK | FALSE | Downregulated |
| -1,060634855 | 0,607568699 | FSD2 | FALSE | Downregulated |
| 0,225573339 | 0,607568699 | MTFP1 | FALSE | Upregulated |
| -0,418336558 | 0,607568699 | NPAS3 | FALSE | Downregulated |
| 0,088863028 | 0,607568699 | RPE | FALSE | Upregulated |
| 0,137621861 | 0,607584292 | ZSWIM8 | FALSE | Upregulated |
| 0,142445341 | 0,607611777 | MPST | FALSE | Upregulated |
| -0,131394104 | 0,607893876 | PEX3 | FALSE | Downregulated |
| -0,54153076 | 0,608084664 | LOC339240 | FALSE | Downregulated |
| 0,728460765 | 0,608084664 | TMEM132D | FALSE | Upregulated |
| 0,273760077 | 0,608379181 | LILRA2 | FALSE | Upregulated |
| 0,150183055 | 0,608379181 | TBC1D16 | FALSE | Upregulated |
| 0,210282537 | 0,608735331 | C17orf107 | FALSE | Upregulated |
| -0,122425397 | 0,60891389 | SNRPA1 | FALSE | Downregulated |
| -0,236876361 | 0,608958198 | EZR | FALSE | Downregulated |
| -0,16041597 | 0,609225958 | GYPC | FALSE | Downregulated |
| 0,106620181 | 0,609264567 | IDH3A | FALSE | Upregulated |
| -0,101779409 | 0,609374799 | CSNK2A2 | FALSE | Downregulated |
| 0,810504319 | 0,609374799 | PRSS16 | FALSE | Upregulated |
| -1,087738872 | 0,609374799 | SPOCK1 | FALSE | Downregulated |
| -0,268089273 | 0,609769374 | CA11 | FALSE | Downregulated |
| -0,245009032 | 0,609769374 | IFI44 | FALSE | Downregulated |
| 0,124457433 | 0,609769374 | IP6K1 | FALSE | Upregulated |

| | | | | |
|---|---|---|---|---|
| 0,663665987 | 0,609769374 | LINC00441 | FALSE | Upregulated |
| -0,079974447 | 0,609769374 | PRPF18 | FALSE | Downregulated |
| -0,107500428 | 0,609769374 | USP12 | FALSE | Downregulated |
| 0,200508383 | 0,609769374 | WLS | FALSE | Upregulated |
| 0,311569316 | 0,609909414 | CENPW | FALSE | Upregulated |
| 0,303251672 | 0,609909414 | EHF | FALSE | Upregulated |
| 0,128429559 | 0,609909414 | F10 | FALSE | Upregulated |
| 0,087650069 | 0,609909414 | GINM1 | FALSE | Upregulated |
| -0,22444863 | 0,609909414 | GPATCH4 | FALSE | Downregulated |
| 0,12292997 | 0,609909414 | HNF1A | FALSE | Upregulated |
| -0,107022312 | 0,609909414 | HP1BP3 | FALSE | Downregulated |
| -0,245341523 | 0,609909414 | LCA5 | FALSE | Downregulated |
| 0,486229857 | 0,609909414 | LOC101929580 | FALSE | Upregulated |
| 0,459550778 | 0,609909414 | LOC102724451 | FALSE | Upregulated |
| -0,135400838 | 0,609909414 | N4BP2 | FALSE | Downregulated |
| 0,134979551 | 0,609909414 | NFATC2 | FALSE | Upregulated |
| -0,279759937 | 0,609909414 | PER3 | FALSE | Downregulated |
| 0,163210466 | 0,609909414 | POC5 | FALSE | Upregulated |
| 0,288500417 | 0,609909414 | PRF1 | FALSE | Upregulated |
| -0,933083904 | 0,609909414 | RAPSN | FALSE | Downregulated |
| -0,138239634 | 0,609909414 | ROCK1 | FALSE | Downregulated |
| 0,242541989 | 0,609909414 | SLC25A29 | FALSE | Upregulated |
| -0,502945125 | 0,609909414 | SLC4A10 | FALSE | Downregulated |
| 0,115586127 | 0,609909414 | SZT2 | FALSE | Upregulated |
| 0,578655023 | 0,609909414 | TG | FALSE | Upregulated |
| -0,378468986 | 0,609909414 | TIMD4 | FALSE | Downregulated |

| | | | | |
|---|---|---|---|---|
| -0,279635359 | 0,609909414 | TMEM200B | FALSE | Downregulated |
| 0,247460187 | 0,609909414 | TSPO | FALSE | Upregulated |
| 0,083114724 | 0,609909414 | UBA1 | FALSE | Upregulated |
| -0,087489659 | 0,609909414 | YY1AP1 | FALSE | Downregulated |
| -0,198617705 | 0,609909414 | ZNF529 | FALSE | Downregulated |
| 0,241867967 | 0,609975111 | CHST11 | FALSE | Upregulated |
| 0,147409974 | 0,609975111 | MICALL1 | FALSE | Upregulated |
| 0,207385362 | 0,60997788 | CXorf40A | FALSE | Upregulated |
| -0,094224689 | 0,610246006 | ACIN1 | FALSE | Downregulated |
| 0,331203348 | 0,610246006 | ANKRD29 | FALSE | Upregulated |
| 0,544889151 | 0,610246006 | AP1M2 | FALSE | Upregulated |
| 0,250135192 | 0,610246006 | C16orf95 | FALSE | Upregulated |
| 0,130181736 | 0,610246006 | C17orf70 | FALSE | Upregulated |
| 0,271162312 | 0,610246006 | CSRNP1 | FALSE | Upregulated |
| -0,219377888 | 0,610246006 | CYP2E1 | FALSE | Downregulated |
| 0,168890507 | 0,610246006 | ETHE1 | FALSE | Upregulated |
| 0,88028098 | 0,610246006 | HCN1 | FALSE | Upregulated |
| 0,257329649 | 0,610246006 | ITGB3 | FALSE | Upregulated |
| -0,290865929 | 0,610246006 | KIF26A | FALSE | Downregulated |
| 0,257284512 | 0,610246006 | KRT8 | FALSE | Upregulated |
| -0,287164229 | 0,610246006 | LOC100289495 | FALSE | Downregulated |
| 0,549876517 | 0,610246006 | LOC101928070 | FALSE | Upregulated |
| -0,100800968 | 0,610246006 | NCOA5 | FALSE | Downregulated |
| 0,295854099 | 0,610246006 | PODXL | FALSE | Upregulated |
| 0,203816698 | 0,610246006 | TBC1D7 | FALSE | Upregulated |
| 0,230854764 | 0,610246006 | THEMIS2 | FALSE | Upregulated |

| | | | | |
|---|---|---|---|---|
| 0,212263121 | 0,610246006 | TRIM11 | FALSE | Upregulated |
| 0,563759734 | 0,610246006 | UTS2B | FALSE | Upregulated |
| 0,85193581 | 0,610246006 | ZG16B | FALSE | Upregulated |
| -0,205047537 | 0,610396435 | C9 | FALSE | Downregulated |
| 0,18154868 | 0,610768077 | DISP1 | FALSE | Upregulated |
| 0,442761551 | 0,610768077 | RHPN1 | FALSE | Upregulated |
| 0,253821985 | 0,610794544 | SPINT2 | FALSE | Upregulated |
| -0,109154208 | 0,61096438 | MGA | FALSE | Downregulated |
| -0,176387152 | 0,61124762 | KBTBD11 | FALSE | Downregulated |
| 0,194371608 | 0,61124762 | MAP1S | FALSE | Upregulated |
| 0,199393794 | 0,611297845 | COL6A1 | FALSE | Upregulated |
| -0,278180823 | 0,611297845 | EFCC1 | FALSE | Downregulated |
| 0,234612321 | 0,611297845 | FGD2 | FALSE | Upregulated |
| -0,27954856 | 0,611297845 | LOC101928728 | FALSE | Downregulated |
| -0,090636366 | 0,611297845 | SOS2 | FALSE | Downregulated |
| 0,254847005 | 0,611297845 | ZNF674-AS1 | FALSE | Upregulated |
| 0,289508771 | 0,611408023 | PARD6A | FALSE | Upregulated |
| -0,231673234 | 0,611408023 | RP11-328K4.1 | FALSE | Downregulated |
| 0,166204924 | 0,611465321 | RHOBTB2 | FALSE | Upregulated |
| -0,09478137 | 0,611465321 | ZNF564 | FALSE | Downregulated |
| 0,841660664 | 0,61175397 | CLEC4C | FALSE | Upregulated |
| -0,226486621 | 0,61175397 | LRRK2 | FALSE | Downregulated |
| -0,172696826 | 0,611816596 | TNFSF12 | FALSE | Downregulated |
| 0,123146679 | 0,612084871 | NT5C | FALSE | Upregulated |
| -0,235824435 | 0,612087901 | ACSS1 | FALSE | Downregulated |
| 0,126762128 | 0,612087901 | SYS1 | FALSE | Upregulated |

| | | | | |
|---|---|---|---|---|
| -0,264939455 | 0,612641234 | AKR1D1 | FALSE | Downregulated |
| -0,117068974 | 0,612641234 | CRTAP | FALSE | Downregulated |
| -0,147265826 | 0,612641234 | ERBB2IP | FALSE | Downregulated |
| -0,200918328 | 0,612641234 | FIGNL1 | FALSE | Downregulated |
| -0,210533885 | 0,612641234 | LOC100506603 | FALSE | Downregulated |
| 0,603078194 | 0,612641234 | LOC100996455 | FALSE | Upregulated |
| -0,155224278 | 0,612641234 | MDFIC | FALSE | Downregulated |
| 0,10757517 | 0,612641234 | NOP56 | FALSE | Upregulated |
| -0,064175625 | 0,612641234 | SNRPD3 | FALSE | Downregulated |
| -0,222943424 | 0,612715924 | CEP19 | FALSE | Downregulated |
| -0,714107792 | 0,612715924 | PZP | FALSE | Downregulated |
| -0,378903968 | 0,612913162 | ADAM12 | FALSE | Downregulated |
| 0,428143453 | 0,612913162 | B3GALT2 | FALSE | Upregulated |
| 0,067666864 | 0,612952194 | DHX9 | FALSE | Upregulated |
| -0,13322238 | 0,612952194 | DNAJC17 | FALSE | Downregulated |
| 0,72283153 | 0,612952194 | FLJ33360 | FALSE | Upregulated |
| 0,464496085 | 0,613216381 | PAPPA | FALSE | Upregulated |
| 0,09919409 | 0,613558992 | ACTR2 | FALSE | Upregulated |
| -0,163224138 | 0,613558992 | LOC100130872 | FALSE | Downregulated |
| 0,973337762 | 0,613708601 | PCYT1B | FALSE | Upregulated |
| 0,273947393 | 0,613777046 | ABCC11 | FALSE | Upregulated |
| -0,352955916 | 0,613838677 | TEAD4 | FALSE | Downregulated |
| -0,105358376 | 0,613924027 | ADSL | FALSE | Downregulated |
| -0,136846687 | 0,613924027 | DYNC2LI1 | FALSE | Downregulated |
| -0,35186527 | 0,613924027 | ZNF239 | FALSE | Downregulated |
| -0,430418429 | 0,614032325 | CACNA2D1 | FALSE | Downregulated |

| | | | | |
|---|---|---|---|---|
| 0,352242992 | 0,61427586 | GPRC5B | FALSE | Upregulated |
| -0,518036751 | 0,614504469 | C6orf141 | FALSE | Downregulated |
| -0,087490765 | 0,614504469 | WDR41 | FALSE | Downregulated |
| 0,163225544 | 0,61454473 | NCAPH2 | FALSE | Upregulated |
| -0,09601343 | 0,61482849 | LOC100129361 | FALSE | Downregulated |
| 0,814665558 | 0,61516468 | GABRR2 | FALSE | Upregulated |
| 0,28394837 | 0,61516468 | HSPA4L | FALSE | Upregulated |
| -0,252314307 | 0,61516468 | PNISR | FALSE | Downregulated |
| 0,217849303 | 0,615307773 | CD151 | FALSE | Upregulated |
| 0,342553093 | 0,615307773 | FOXD2 | FALSE | Upregulated |
| 0,361330518 | 0,615307773 | HOXB7 | FALSE | Upregulated |
| 0,463226472 | 0,615307773 | LOC101929574 | FALSE | Upregulated |
| 0,067068616 | 0,615307773 | UPF1 | FALSE | Upregulated |
| -0,145234503 | 0,615445741 | LINC00938 | FALSE | Downregulated |
| 0,179685805 | 0,615445741 | PPP1R18 | FALSE | Upregulated |
| -0,124981067 | 0,615776821 | INVS | FALSE | Downregulated |
| 0,128018355 | 0,61620762 | ASL | FALSE | Upregulated |
| -0,718784492 | 0,61620762 | LOC101930129 | FALSE | Downregulated |
| -0,086874174 | 0,61620762 | MAGEF1 | FALSE | Downregulated |
| -0,540461262 | 0,61620762 | MYLK3 | FALSE | Downregulated |
| -0,174117957 | 0,61620762 | NR3C2 | FALSE | Downregulated |
| 0,452725431 | 0,61620762 | PRKAA2 | FALSE | Upregulated |
| 0,209325229 | 0,61620762 | RGS3 | FALSE | Upregulated |
| 0,180229642 | 0,61620762 | WAS | FALSE | Upregulated |
| 0,174680474 | 0,616342413 | PVRL2 | FALSE | Upregulated |
| -2,026082517 | 0,616500177 | MYF6 | FALSE | Downregulated |

| | | | | |
|---|---|---|---|---|
| 0,286979122 | 0,616686046 | AKAP5 | FALSE | Upregulated |
| -0,182830763 | 0,616686046 | BAZ1A | FALSE | Downregulated |
| 0,280653182 | 0,616686046 | CDR2L | FALSE | Upregulated |
| 0,696231147 | 0,616686046 | EFCAB5 | FALSE | Upregulated |
| -0,172466878 | 0,616686046 | IFT81 | FALSE | Downregulated |
| 0,23855569 | 0,616686046 | PRKRIP1 | FALSE | Upregulated |
| 0,103670791 | 0,617446297 | CDK16 | FALSE | Upregulated |
| 0,310601957 | 0,617446297 | GDPGP1 | FALSE | Upregulated |
| -0,132991624 | 0,617446297 | MAGI3 | FALSE | Downregulated |
| 0,144039154 | 0,617446297 | NEO1 | FALSE | Upregulated |
| -0,106696422 | 0,617446297 | RANBP1 | FALSE | Downregulated |
| 0,137634201 | 0,617446297 | SLC25A6 | FALSE | Upregulated |
| 0,331285379 | 0,617446297 | SORCS2 | FALSE | Upregulated |
| 0,905994198 | 0,617849688 | WTAPP1 | FALSE | Upregulated |
| -0,19912623 | 0,618054059 | PLSCR1 | FALSE | Downregulated |
| -0,149791296 | 0,618054059 | WWOX | FALSE | Downregulated |
| -0,158674311 | 0,618341749 | OFD1 | FALSE | Downregulated |
| 0,115882647 | 0,618341749 | PIK3CB | FALSE | Upregulated |
| -0,163605708 | 0,618638622 | CLIC2 | FALSE | Downregulated |
| -0,14918565 | 0,619314397 | THUMPD1 | FALSE | Downregulated |
| -0,225895818 | 0,619463845 | PAQR7 | FALSE | Downregulated |
| 0,685351932 | 0,619492297 | C1orf105 | FALSE | Upregulated |
| -0,091566719 | 0,619492297 | RTCA | FALSE | Downregulated |
| 0,217584567 | 0,61963868 | ADCK5 | FALSE | Upregulated |
| -0,14083005 | 0,61963868 | DET1 | FALSE | Downregulated |
| 0,330658329 | 0,61963868 | KIAA0101 | FALSE | Upregulated |

| | | | | |
|---|---|---|---|---|
| 0,290780861 | 0,61963868 | KIAA0226L | FALSE | Upregulated |
| -0,506599834 | 0,61963868 | LOC101929977 | FALSE | Downregulated |
| -0,121809164 | 0,61963868 | MRPS24 | FALSE | Downregulated |
| -0,089951284 | 0,61963868 | PTEN | FALSE | Downregulated |
| -0,09924517 | 0,61963868 | SDAD1 | FALSE | Downregulated |
| 0,303393476 | 0,61963868 | ZFP82 | FALSE | Upregulated |
| 0,716203225 | 0,61963868 | ZNF99 | FALSE | Upregulated |
| -0,169691657 | 0,619708049 | HSBP1L1 | FALSE | Downregulated |
| 0,182148713 | 0,619839617 | HLA-DMA | FALSE | Upregulated |
| 0,156809646 | 0,619995092 | GOLT1B | FALSE | Upregulated |
| -0,166552449 | 0,620179328 | SH2D4A | FALSE | Downregulated |
| 0,522121242 | 0,620226661 | CASC1 | FALSE | Upregulated |
| -0,294904646 | 0,620226661 | GRASP | FALSE | Downregulated |
| 0,481910982 | 0,620339322 | PPAPDC1A | FALSE | Upregulated |
| 0,499865697 | 0,620859957 | BCAS4 | FALSE | Upregulated |
| -0,150673575 | 0,620859957 | DCUN1D2 | FALSE | Downregulated |
| -0,193281566 | 0,620859957 | IL1RN | FALSE | Downregulated |
| 0,712495454 | 0,620859957 | LIPH | FALSE | Upregulated |
| -0,087768685 | 0,620859957 | PSMA5 | FALSE | Downregulated |
| -0,225106355 | 0,620859957 | SLC29A1 | FALSE | Downregulated |
| 0,26149423 | 0,620859957 | ZNF132 | FALSE | Upregulated |
| 0,200989835 | 0,620976664 | CLTCL1 | FALSE | Upregulated |
| 0,424175457 | 0,620976664 | LOC101926967 | FALSE | Upregulated |
| 0,194027006 | 0,620976664 | PPAPDC1B | FALSE | Upregulated |
| 0,520661147 | 0,621139652 | GNAT2 | FALSE | Upregulated |
| 0,290714132 | 0,621155527 | DBNDD2 | FALSE | Upregulated |

| | | | | |
|---|---|---|---|---|
| -0,652376819 | 0,621687123 | AQP6 | FALSE | Downregulated |
| -0,776238753 | 0,621687123 | LINC00277 | FALSE | Downregulated |
| -0,10078747 | 0,621687123 | PEX5 | FALSE | Downregulated |
| -0,119438111 | 0,621687123 | PHLDB2 | FALSE | Downregulated |
| 0,260878705 | 0,621695379 | APBB3 | FALSE | Upregulated |
| 0,494335425 | 0,621695379 | C11orf21 | FALSE | Upregulated |
| -0,172141488 | 0,621695379 | CYP27A1 | FALSE | Downregulated |
| -0,59320941 | 0,621695379 | LINC00940 | FALSE | Downregulated |
| -0,110198154 | 0,621695379 | MRPL24 | FALSE | Downregulated |
| -0,087529218 | 0,621695379 | NSUN4 | FALSE | Downregulated |
| 0,203148671 | 0,621695379 | ODC1 | FALSE | Upregulated |
| 0,480897695 | 0,621695379 | PIP5K1B | FALSE | Upregulated |
| -0,13601885 | 0,621695379 | RPL17 | FALSE | Downregulated |
| 0,404938713 | 0,621741859 | CDCA5 | FALSE | Upregulated |
| 0,161515114 | 0,621741859 | HINFP | FALSE | Upregulated |
| -0,102933264 | 0,621741859 | RNF115 | FALSE | Downregulated |
| -0,184459713 | 0,621831317 | FAM160A1 | FALSE | Downregulated |
| 0,199783966 | 0,621912723 | B4GALT6 | FALSE | Upregulated |
| -0,601699625 | 0,621912723 | BEGAIN | FALSE | Downregulated |
| -0,062084672 | 0,621912723 | C11orf58 | FALSE | Downregulated |
| 0,485497766 | 0,621912723 | CNIH3 | FALSE | Upregulated |
| 0,536773935 | 0,621912723 | DLEU7 | FALSE | Upregulated |
| 0,75489564 | 0,621912723 | GCNT3 | FALSE | Upregulated |
| -0,179419768 | 0,621912723 | GPHN | FALSE | Downregulated |
| -0,103715498 | 0,621912723 | HNRNPDL | FALSE | Downregulated |
| 0,564506152 | 0,621912723 | LHX6 | FALSE | Upregulated |

| | | | | |
|---|---|---|---|---|
| -0,607230459 | 0,621912723 | LOC102724541 | FALSE | Downregulated |
| -0,494232158 | 0,621912723 | OR2A1-AS1 | FALSE | Downregulated |
| 0,553171132 | 0,621912723 | SCARNA12 | FALSE | Upregulated |
| 0,128419091 | 0,621912723 | XPO1 | FALSE | Upregulated |
| 0,257330415 | 0,621912723 | ZMAT3 | FALSE | Upregulated |
| -0,087536595 | 0,621983338 | CCDC93 | FALSE | Downregulated |
| -0,08158378 | 0,621983338 | CCT4 | FALSE | Downregulated |
| 0,125157328 | 0,621983338 | NUBP2 | FALSE | Upregulated |
| -0,290418799 | 0,621983338 | REP15 | FALSE | Downregulated |
| -0,279606035 | 0,621983338 | STL | FALSE | Downregulated |
| -0,191202506 | 0,621983338 | ZNF675 | FALSE | Downregulated |
| -0,229101674 | 0,622012908 | RDH16 | FALSE | Downregulated |
| -0,610813807 | 0,622883661 | MT1H | FALSE | Downregulated |
| 0,283722324 | 0,62354936 | C3AR1 | FALSE | Upregulated |
| 0,166762569 | 0,62354936 | DBF4 | FALSE | Upregulated |
| 0,148358565 | 0,62354936 | LRSAM1 | FALSE | Upregulated |
| -0,267725447 | 0,62354936 | POLN | FALSE | Downregulated |
| 0,120592507 | 0,62354936 | RFK | FALSE | Upregulated |
| 0,087041456 | 0,62354936 | SCAMP3 | FALSE | Upregulated |
| -0,122506023 | 0,623623307 | BPGM | FALSE | Downregulated |
| -0,203903218 | 0,623623307 | CCZ1B | FALSE | Downregulated |
| -0,568502135 | 0,623623307 | EIF5AL1 | FALSE | Downregulated |
| -0,193323438 | 0,623623307 | FAM107B | FALSE | Downregulated |
| 1,88023615 | 0,623623307 | HCCAT5 | FALSE | Upregulated |
| 0,627152844 | 0,623623307 | IGF2-AS | FALSE | Upregulated |
| -0,125026298 | 0,623623307 | ITIH2 | FALSE | Downregulated |

| | | | | |
|---|---|---|---|---|
| 0,697035157 | 0,623623307 | LOC102724676 | FALSE | Upregulated |
| 0,687907152 | 0,623623307 | MORN3 | FALSE | Upregulated |
| -0,717422911 | 0,623623307 | PDZD7 | FALSE | Downregulated |
| 0,12221086 | 0,623623307 | SHQ1 | FALSE | Upregulated |
| -0,129645363 | 0,623623307 | ZNHIT1 | FALSE | Downregulated |
| -0,18553307 | 0,624031895 | SETBP1 | FALSE | Downregulated |
| -0,467041493 | 0,624100902 | CAHM | FALSE | Downregulated |
| 0,281500482 | 0,624100902 | LOC101929792 | FALSE | Upregulated |
| -0,193544679 | 0,624100902 | PAMR1 | FALSE | Downregulated |
| -0,254578399 | 0,624100902 | S1PR3 | FALSE | Downregulated |
| -0,339327854 | 0,624100902 | SLC18A2 | FALSE | Downregulated |
| -0,147777831 | 0,624100902 | ZSWIM6 | FALSE | Downregulated |
| -0,101868455 | 0,624191188 | ALDH3A2 | FALSE | Downregulated |
| 0,166600159 | 0,624191188 | C9orf142 | FALSE | Upregulated |
| 0,144485544 | 0,624191188 | DTX2 | FALSE | Upregulated |
| -0,281297923 | 0,624191188 | DZIP1 | FALSE | Downregulated |
| 0,210177587 | 0,624191188 | ENKD1 | FALSE | Upregulated |
| -0,099725097 | 0,624191188 | EPC2 | FALSE | Downregulated |
| -0,429050503 | 0,624191188 | FAM212A | FALSE | Downregulated |
| 0,267980005 | 0,624191188 | IL2RG | FALSE | Upregulated |
| 0,340199119 | 0,624191188 | IL7R | FALSE | Upregulated |
| 0,497033524 | 0,624191188 | KIAA1211L | FALSE | Upregulated |
| -0,181119383 | 0,624191188 | LOC100132352 | FALSE | Downregulated |
| -0,198167879 | 0,624191188 | MYL5 | FALSE | Downregulated |
| 1,071617383 | 0,624191188 | PKP1 | FALSE | Upregulated |
| 0,178843962 | 0,624191188 | SCN1B | FALSE | Upregulated |

| | | | | |
|---|---|---|---|---|
| 0,175851173 | 0,624191188 | SPDL1 | FALSE | Upregulated |
| 0,163048557 | 0,624191188 | SPRYD3 | FALSE | Upregulated |
| 0,313907984 | 0,624191188 | TLCD1 | FALSE | Upregulated |
| 0,444303227 | 0,624191188 | UFSP1 | FALSE | Upregulated |
| -0,079535954 | 0,624191188 | ZNF268 | FALSE | Downregulated |
| 0,166221817 | 0,624191188 | ZNF316 | FALSE | Upregulated |
| 0,106255013 | 0,624450151 | CNNM3 | FALSE | Upregulated |
| 0,547032032 | 0,624477998 | LAMC2 | FALSE | Upregulated |
| -0,12007051 | 0,624477998 | NAT10 | FALSE | Downregulated |
| -0,190206531 | 0,624477998 | PSIP1 | FALSE | Downregulated |
| 0,099965875 | 0,624477998 | VPS33A | FALSE | Upregulated |
| 0,207523154 | 0,624477998 | ZNF250 | FALSE | Upregulated |
| 0,129367715 | 0,624627893 | ARFIP2 | FALSE | Upregulated |
| -0,121215487 | 0,624627893 | C2orf49 | FALSE | Downregulated |
| -0,332982769 | 0,624627893 | OAT | FALSE | Downregulated |
| 0,426125206 | 0,624627893 | PPP1R3G | FALSE | Upregulated |
| -0,131971054 | 0,624627893 | RPL12 | FALSE | Downregulated |
| 0,105571912 | 0,624627893 | SLC38A10 | FALSE | Upregulated |
| 0,154763139 | 0,624627893 | SRSF7 | FALSE | Upregulated |
| 0,233556095 | 0,624627893 | ZDHHC1 | FALSE | Upregulated |
| -2,094098858 | 0,624719648 | ADCY2 | FALSE | Downregulated |
| -0,839821024 | 0,624719648 | APOD | FALSE | Downregulated |
| -0,510928234 | 0,624719648 | CYP1A1 | FALSE | Downregulated |
| 0,156412943 | 0,624719648 | FASTKD1 | FALSE | Upregulated |
| -0,239448798 | 0,624719648 | GALNT18 | FALSE | Downregulated |
| -0,824579736 | 0,624719648 | HEMGN | FALSE | Downregulated |

| | | | | |
|---|---|---|---|---|
| 0,120590541 | 0,624719648 | IFT20 | FALSE | Upregulated |
| 0,539785506 | 0,624719648 | LOC100294362 | FALSE | Upregulated |
| -0,168842595 | 0,624719648 | LOC101929343 | FALSE | Downregulated |
| 0,107981433 | 0,624719648 | NCOR2 | FALSE | Upregulated |
| 0,131419544 | 0,624719648 | PCIF1 | FALSE | Upregulated |
| 0,130873664 | 0,624719648 | PELO | FALSE | Upregulated |
| 0,187140428 | 0,624719648 | PPM1M | FALSE | Upregulated |
| -0,484075658 | 0,624719648 | RYR2 | FALSE | Downregulated |
| -0,449540682 | 0,624719648 | SLC25A21 | FALSE | Downregulated |
| 0,617052813 | 0,624719648 | WISP2 | FALSE | Upregulated |
| 0,931009861 | 0,624719648 | ZFY | FALSE | Upregulated |
| 0,101159867 | 0,624719648 | ZNF394 | FALSE | Upregulated |
| -0,689684993 | 0,624881714 | DIO3 | FALSE | Downregulated |
| 0,133477897 | 0,624881714 | GIT1 | FALSE | Upregulated |
| -0,295348794 | 0,624881714 | SULF1 | FALSE | Downregulated |
| -0,136523124 | 0,624881714 | UFL1 | FALSE | Downregulated |
| 0,409119176 | 0,625182972 | SLC34A2 | FALSE | Upregulated |
| -0,396226658 | 0,625188808 | FMO2 | FALSE | Downregulated |
| -0,098403343 | 0,625188808 | FPGT | FALSE | Downregulated |
| 0,86054414 | 0,625188808 | RORB | FALSE | Upregulated |
| -0,535804131 | 0,625188808 | SLMO1 | FALSE | Downregulated |
| 0,136727784 | 0,625188808 | TRMT2B | FALSE | Upregulated |
| -0,135345494 | 0,625218389 | CMTM8 | FALSE | Downregulated |
| -0,346128812 | 0,625282613 | LOC102724782 | FALSE | Downregulated |
| -0,440334228 | 0,625417177 | SLITRK4 | FALSE | Downregulated |
| -0,162378038 | 0,62548771 | NUDT4 | FALSE | Downregulated |

| | | | | |
|---|---|---|---|---|
| 0,129909249 | 0,62548771 | PRMT7 | FALSE | Upregulated |
| 0,223270579 | 0,625495168 | SLC25A35 | FALSE | Upregulated |
| 0,157825859 | 0,625505278 | BRI3 | FALSE | Upregulated |
| -0,181297046 | 0,625532748 | ABCA5 | FALSE | Downregulated |
| -0,712052182 | 0,625532748 | FOXO6 | FALSE | Downregulated |
| -0,102813103 | 0,625532748 | WWC2 | FALSE | Downregulated |
| 0,323951334 | 0,625700137 | FNDC5 | FALSE | Upregulated |
| 0,141317754 | 0,625761359 | CDK10 | FALSE | Upregulated |
| -0,581192947 | 0,625858245 | AC018647.3 | FALSE | Downregulated |
| 0,155583957 | 0,626179587 | COPG1 | FALSE | Upregulated |
| 0,352024364 | 0,626316476 | AGAP2-AS1 | FALSE | Upregulated |
| -0,464979238 | 0,626316476 | C19orf71 | FALSE | Downregulated |
| -0,076363097 | 0,626316476 | FAF2 | FALSE | Downregulated |
| 0,177869694 | 0,626316476 | GSTP1 | FALSE | Upregulated |
| 0,176357842 | 0,626316476 | NIN | FALSE | Upregulated |
| -0,143443426 | 0,626316476 | SNRPD2 | FALSE | Downregulated |
| -0,275262204 | 0,626540603 | SOWAHA | FALSE | Downregulated |
| 0,106332371 | 0,627167216 | DSC2 | FALSE | Upregulated |
| -0,115937141 | 0,627650586 | KPNB1 | FALSE | Downregulated |
| -0,097266288 | 0,627650586 | NKIRAS1 | FALSE | Downregulated |
| -0,076003278 | 0,627702929 | GDE1 | FALSE | Downregulated |
| -0,655088234 | 0,627703359 | ITGA2B | FALSE | Downregulated |
| 0,133464095 | 0,627898634 | ACTB | FALSE | Upregulated |
| 0,150541837 | 0,627898634 | ART4 | FALSE | Upregulated |
| 0,746591458 | 0,627898634 | EEF1DP3 | FALSE | Upregulated |
| 0,202826089 | 0,627898634 | FBXO36 | FALSE | Upregulated |

| | | | | |
|---|---|---|---|---|
| 0,15478517 | 0,627898634 | HYAL2 | FALSE | Upregulated |
| 0,274666688 | 0,627898634 | JUN | FALSE | Upregulated |
| -0,335816666 | 0,627898634 | KCND1 | FALSE | Downregulated |
| -0,241389004 | 0,627898634 | LOC101928419 | FALSE | Downregulated |
| -0,073699388 | 0,627898634 | MED29 | FALSE | Downregulated |
| 0,498830925 | 0,627898634 | MIPEPP3 | FALSE | Upregulated |
| 0,125991845 | 0,627898634 | MPG | FALSE | Upregulated |
| -0,230823902 | 0,627898634 | PLEKHA4 | FALSE | Downregulated |
| 0,109357233 | 0,627898634 | RNF123 | FALSE | Upregulated |
| 0,148783486 | 0,627898634 | TARBP1 | FALSE | Upregulated |
| 0,108487071 | 0,627898634 | TGFBRAP1 | FALSE | Upregulated |
| 0,250787447 | 0,627898634 | TMEM81 | FALSE | Upregulated |
| -0,105929419 | 0,627937265 | CDH2 | FALSE | Downregulated |
| 0,306337604 | 0,62798547 | RIPK3 | FALSE | Upregulated |
| -0,12562933 | 0,62819916 | CD302 | FALSE | Downregulated |
| -0,121536326 | 0,628282597 | AATF | FALSE | Downregulated |
| -0,226754301 | 0,628282597 | ADPRM | FALSE | Downregulated |
| -0,228945796 | 0,628282597 | AGPAT5 | FALSE | Downregulated |
| 0,652674072 | 0,628282597 | ALOX15 | FALSE | Upregulated |
| -0,067604812 | 0,628282597 | ARFGEF1 | FALSE | Downregulated |
| 0,318165275 | 0,628282597 | ARNTL2 | FALSE | Upregulated |
| -0,090155574 | 0,628282597 | ATL2 | FALSE | Downregulated |
| -0,182972315 | 0,628282597 | ATP11C | FALSE | Downregulated |
| 0,258175947 | 0,628282597 | BCL2 | FALSE | Upregulated |
| -0,132168442 | 0,628282597 | BRWD3 | FALSE | Downregulated |
| 0,580028338 | 0,628282597 | C16orf59 | FALSE | Upregulated |

| | | | | |
|---|---|---|---|---|
| 0,658665019 | 0,628282597 | C1orf177 | FALSE | Upregulated |
| 0,205995745 | 0,628282597 | CDK2AP2 | FALSE | Upregulated |
| 0,072967537 | 0,628282597 | CIAO1 | FALSE | Upregulated |
| 0,779115848 | 0,628282597 | CTAGE1 | FALSE | Upregulated |
| -0,26917483 | 0,628282597 | DDT | FALSE | Downregulated |
| -0,289296202 | 0,628282597 | ENPP3 | FALSE | Downregulated |
| 0,245101395 | 0,628282597 | EXOC3L1 | FALSE | Upregulated |
| -0,11640798 | 0,628282597 | FAM133B | FALSE | Downregulated |
| 0,65362547 | 0,628282597 | FAP | FALSE | Upregulated |
| -0,278120327 | 0,628282597 | FGF2 | FALSE | Downregulated |
| 0,101383701 | 0,628282597 | GBF1 | FALSE | Upregulated |
| 0,191402183 | 0,628282597 | GEMIN7 | FALSE | Upregulated |
| 0,090244039 | 0,628282597 | GGA3 | FALSE | Upregulated |
| -0,159592989 | 0,628282597 | HGSNAT | FALSE | Downregulated |
| -0,153202561 | 0,628282597 | HNRNPH2 | FALSE | Downregulated |
| 0,082818434 | 0,628282597 | KIAA0141 | FALSE | Upregulated |
| 0,14213051 | 0,628282597 | LDOC1L | FALSE | Upregulated |
| -0,874901457 | 0,628282597 | LIMS3L | FALSE | Downregulated |
| -0,240520703 | 0,628282597 | LIPG | FALSE | Downregulated |
| 0,577370666 | 0,628282597 | LOC100996746 | FALSE | Upregulated |
| -0,354059315 | 0,628282597 | LOC102723389 | FALSE | Downregulated |
| 0,120512338 | 0,628282597 | LRP1 | FALSE | Upregulated |
| 0,237369937 | 0,628282597 | LY96 | FALSE | Upregulated |
| 0,302377771 | 0,628282597 | MBLAC1 | FALSE | Upregulated |
| -0,255502677 | 0,628282597 | MGC70870 | FALSE | Downregulated |
| -0,233695425 | 0,628282597 | MGC72080 | FALSE | Downregulated |

| | | | | |
|---|---|---|---|---|
| 0,141960877 | 0,628282597 | MKL1 | FALSE | Upregulated |
| 0,142258299 | 0,628282597 | MRPL27 | FALSE | Upregulated |
| 1,194105186 | 0,628282597 | NLRP7 | FALSE | Upregulated |
| -0,094989413 | 0,628282597 | PATL1 | FALSE | Downregulated |
| -0,097009667 | 0,628282597 | PET112 | FALSE | Downregulated |
| 0,12467573 | 0,628282597 | POMT2 | FALSE | Upregulated |
| 0,12620122 | 0,628282597 | PPM1D | FALSE | Upregulated |
| 0,127898187 | 0,628282597 | RANBP10 | FALSE | Upregulated |
| 0,147317364 | 0,628282597 | RCC1 | FALSE | Upregulated |
| 0,109869302 | 0,628282597 | RFC5 | FALSE | Upregulated |
| -0,08886902 | 0,628282597 | RIOK1 | FALSE | Downregulated |
| 0,223038706 | 0,628282597 | RNASEH2A | FALSE | Upregulated |
| -0,242689652 | 0,628282597 | RP11-473M20.14 | FALSE | Downregulated |
| 0,42112663 | 0,628282597 | RP11-844P9.2 | FALSE | Upregulated |
| 0,451144014 | 0,628282597 | RPL39L | FALSE | Upregulated |
| -0,156458907 | 0,628282597 | RPS14 | FALSE | Downregulated |
| 0,253150566 | 0,628282597 | RPS26P11 | FALSE | Upregulated |
| 0,228519135 | 0,628282597 | SDSL | FALSE | Upregulated |
| -0,147423746 | 0,628282597 | SEPT11 | FALSE | Downregulated |
| 0,174912604 | 0,628282597 | SHC2 | FALSE | Upregulated |
| -0,088864226 | 0,628282597 | SLC4A1AP | FALSE | Downregulated |
| -0,078324254 | 0,628282597 | SMAD4 | FALSE | Downregulated |
| 0,429404717 | 0,628282597 | SPP1 | FALSE | Upregulated |
| 0,159453314 | 0,628282597 | ST3GAL3 | FALSE | Upregulated |
| -0,204889788 | 0,628282597 | STARD7-AS1 | FALSE | Downregulated |
| -0,097470505 | 0,628282597 | TAPT1 | FALSE | Downregulated |

| | | | | |
|---|---|---|---|---|
| 0,123063661 | 0,628282597 | TIPARP | FALSE | Upregulated |
| -0,15367817 | 0,628282597 | TMEM116 | FALSE | Downregulated |
| 0,111373069 | 0,628282597 | TMEM62 | FALSE | Upregulated |
| 0,08786076 | 0,628282597 | TOM1 | FALSE | Upregulated |
| 0,070024383 | 0,628282597 | TRPC4AP | FALSE | Upregulated |
| 0,11528534 | 0,628282597 | TSC22D4 | FALSE | Upregulated |
| 0,235457851 | 0,628282597 | UAP1L1 | FALSE | Upregulated |
| 0,119475487 | 0,628282597 | UBA52 | FALSE | Upregulated |
| -0,361134444 | 0,628282597 | UPK3B | FALSE | Downregulated |
| 0,100976388 | 0,628282597 | UQCC1 | FALSE | Upregulated |
| 0,194534493 | 0,628282597 | UQCRQ | FALSE | Upregulated |
| -0,126214345 | 0,628282597 | ZNF195 | FALSE | Downregulated |
| -0,129679836 | 0,628282597 | ZNF236 | FALSE | Downregulated |
| -0,145062662 | 0,628282597 | ZNF518A | FALSE | Downregulated |
| 0,252179882 | 0,628282597 | ZNF671 | FALSE | Upregulated |
| -0,260562575 | 0,628343267 | DNM3OS | FALSE | Downregulated |
| -0,111758021 | 0,628343267 | FOXJ2 | FALSE | Downregulated |
| -0,226837723 | 0,628343267 | TTR | FALSE | Downregulated |
| 0,577453482 | 0,628393058 | LSAMP | FALSE | Upregulated |
| -0,134812233 | 0,628393058 | NAT9 | FALSE | Downregulated |
| -0,125057463 | 0,628557901 | MXD1 | FALSE | Downregulated |
| 0,218058375 | 0,628557901 | POU6F1 | FALSE | Upregulated |
| 0,160884963 | 0,628571573 | KIAA1598 | FALSE | Upregulated |
| -0,23677262 | 0,628571573 | SOX5 | FALSE | Downregulated |
| -0,110233583 | 0,628675888 | GTF2H3 | FALSE | Downregulated |
| -0,241802328 | 0,628675888 | TPM3 | FALSE | Downregulated |

| | | | | |
|---|---|---|---|---|
| -0,983866749 | 0,628741854 | PRH1 | FALSE | Downregulated |
| -0,224857721 | 0,628768463 | SLC1A1 | FALSE | Downregulated |
| 0,085698344 | 0,628897172 | CPNE3 | FALSE | Upregulated |
| -0,140176314 | 0,628897172 | CXorf23 | FALSE | Downregulated |
| -0,974534723 | 0,628956839 | SERTAD4 | FALSE | Downregulated |
| 0,259865844 | 0,629061372 | SPIRE1 | FALSE | Upregulated |
| -0,171871316 | 0,62939614 | FNBP4 | FALSE | Downregulated |
| 0,286014762 | 0,62939614 | SRGAP1 | FALSE | Upregulated |
| 0,110720149 | 0,62939614 | SURF4 | FALSE | Upregulated |
| 0,740499474 | 0,629618067 | AXDND1 | FALSE | Upregulated |
| 0,316853665 | 0,629618067 | CCL5 | FALSE | Upregulated |
| -0,418078169 | 0,629618067 | DPY19L1P1 | FALSE | Downregulated |
| 0,219808801 | 0,629618067 | FAIM | FALSE | Upregulated |
| 0,115873464 | 0,629618067 | INIP | FALSE | Upregulated |
| 0,130993117 | 0,629618067 | KRT10 | FALSE | Upregulated |
| -0,166369932 | 0,629618067 | LSM5 | FALSE | Downregulated |
| -0,258611249 | 0,629618067 | PRG4 | FALSE | Downregulated |
| 0,55186641 | 0,629618067 | RERGL | FALSE | Upregulated |
| 0,11520374 | 0,629618067 | SPTLC2 | FALSE | Upregulated |
| -0,123391402 | 0,629618067 | ST3GAL1 | FALSE | Downregulated |
| 0,186333356 | 0,629867854 | STK11IP | FALSE | Upregulated |
| 0,148754784 | 0,630006335 | AAK1 | FALSE | Upregulated |
| 0,116583087 | 0,630189215 | PEX6 | FALSE | Upregulated |
| 0,208988989 | 0,630189215 | PTGS1 | FALSE | Upregulated |
| 0,301767521 | 0,630310513 | EPHA3 | FALSE | Upregulated |
| 0,105160368 | 0,63039134 | RPP38 | FALSE | Upregulated |

| | | | | |
|---|---|---|---|---|
| 0,739061168 | 0,63039134 | STAP1 | FALSE | Upregulated |
| -0,160724997 | 0,630392256 | ANP32E | FALSE | Downregulated |
| 0,422588097 | 0,630392256 | BEAN1 | FALSE | Upregulated |
| -0,221249998 | 0,630392256 | FGG | FALSE | Downregulated |
| -0,387284885 | 0,630392256 | LOC100505985 | FALSE | Downregulated |
| -0,17377101 | 0,630788558 | ENPP1 | FALSE | Downregulated |
| 0,158788299 | 0,630788558 | PAPOLG | FALSE | Upregulated |
| -0,127980225 | 0,630788558 | SLC16A2 | FALSE | Downregulated |
| 0,415140003 | 0,630974459 | CHST10 | FALSE | Upregulated |
| -0,191920225 | 0,630974459 | LOC257396 | FALSE | Downregulated |
| -0,172829231 | 0,630974459 | TM4SF1 | FALSE | Downregulated |
| -0,620869613 | 0,630974952 | ESM1 | FALSE | Downregulated |
| 0,421458011 | 0,630974952 | LGR6 | FALSE | Upregulated |
| 0,114546312 | 0,630974952 | SF3B4 | FALSE | Upregulated |
| -0,587691113 | 0,631378509 | CEACAM3 | FALSE | Downregulated |
| 0,101291302 | 0,631378509 | GEMIN5 | FALSE | Upregulated |
| -1,003687451 | 0,631378509 | LMOD3 | FALSE | Downregulated |
| 0,310638768 | 0,631378509 | ROR1 | FALSE | Upregulated |
| -0,401880609 | 0,631378509 | TOLLIP-AS1 | FALSE | Downregulated |
| -0,090077671 | 0,631378509 | TTC17 | FALSE | Downregulated |
| 0,445280051 | 0,631511097 | OXCT2 | FALSE | Upregulated |
| -0,051316028 | 0,63154027 | HNRNPA2B1 | FALSE | Downregulated |
| -0,091886411 | 0,631701651 | TSPAN31 | FALSE | Downregulated |
| -0,665087166 | 0,632208939 | GTSCR1 | FALSE | Downregulated |
| 0,07087817 | 0,632208939 | NSFL1C | FALSE | Upregulated |
| 0,118492197 | 0,632208939 | OIP5-AS1 | FALSE | Upregulated |

| | | | | |
|---|---|---|---|---|
| -0,251808464 | 0,632208939 | TLE3 | FALSE | Downregulated |
| 0,180345824 | 0,632208939 | TMEM165 | FALSE | Upregulated |
| -0,124201824 | 0,632236555 | EDNRB | FALSE | Downregulated |
| -0,088245145 | 0,632236555 | FAM35A | FALSE | Downregulated |
| -0,151103187 | 0,632497464 | ANKRA2 | FALSE | Downregulated |
| -0,491849058 | 0,632497464 | PHYHIP | FALSE | Downregulated |
| -0,09666104 | 0,632497464 | SOS1 | FALSE | Downregulated |
| -0,103512244 | 0,632603729 | KDSR | FALSE | Downregulated |
| -0,12146612 | 0,632603729 | TMEM47 | FALSE | Downregulated |
| 0,246060695 | 0,632894975 | ALAS1 | FALSE | Upregulated |
| 0,119535609 | 0,632894975 | FAM103A1 | FALSE | Upregulated |
| 0,411563726 | 0,632894975 | TMEM136 | FALSE | Upregulated |
| -0,113420062 | 0,633032171 | C22orf39 | FALSE | Downregulated |
| 0,081103351 | 0,633108569 | VPS4A | FALSE | Upregulated |
| -0,077953579 | 0,63349417 | CLCN3 | FALSE | Downregulated |
| 0,154465641 | 0,63349417 | FAM3A | FALSE | Upregulated |
| 0,146958649 | 0,63349417 | LIN37 | FALSE | Upregulated |
| -0,15545295 | 0,63349417 | SAT2 | FALSE | Downregulated |
| 0,135937519 | 0,63355234 | DPP7 | FALSE | Upregulated |
| -0,094908722 | 0,63355234 | LOC101928170 | FALSE | Downregulated |
| -0,335902395 | 0,63355234 | LOC101929351 | FALSE | Downregulated |
| 0,132544554 | 0,63355234 | NUDT22 | FALSE | Upregulated |
| 0,123580062 | 0,63355234 | PLXND1 | FALSE | Upregulated |
| 0,097994815 | 0,63355234 | PPP4C | FALSE | Upregulated |
| 0,101318528 | 0,633745096 | BET1L | FALSE | Upregulated |
| -0,221659094 | 0,633848079 | SLC6A1 | FALSE | Downregulated |

| | | | | |
|---|---|---|---|---|
| -0,252353877 | 0,633927566 | ANKRD55 | FALSE | Downregulated |
| 0,131080919 | 0,633927566 | APAF1 | FALSE | Upregulated |
| 0,13594623 | 0,633927566 | ATR | FALSE | Upregulated |
| 0,080718287 | 0,633927566 | CAP1 | FALSE | Upregulated |
| 0,126097963 | 0,633927566 | CHPF2 | FALSE | Upregulated |
| -0,385674043 | 0,633927566 | DNAH11 | FALSE | Downregulated |
| 0,138255826 | 0,633927566 | GADD45GIP1 | FALSE | Upregulated |
| 0,329317396 | 0,633927566 | LINC00659 | FALSE | Upregulated |
| -0,378371813 | 0,633927566 | LOC101927609 | FALSE | Downregulated |
| 0,455058584 | 0,633927566 | LOC102724475 | FALSE | Upregulated |
| 0,288384884 | 0,633927566 | MAPK11 | FALSE | Upregulated |
| 0,086383043 | 0,633927566 | MASP1 | FALSE | Upregulated |
| 0,112210482 | 0,633927566 | MEGF9 | FALSE | Upregulated |
| 0,123558605 | 0,633927566 | NDC1 | FALSE | Upregulated |
| 0,205700671 | 0,633927566 | SLFN11 | FALSE | Upregulated |
| 0,107443811 | 0,633927566 | THYN1 | FALSE | Upregulated |
| -0,484668159 | 0,633927566 | TRNA | FALSE | Downregulated |
| 0,328872634 | 0,6349043 | HOXA5 | FALSE | Upregulated |
| -0,175352336 | 0,634987356 | LOC646719 | FALSE | Downregulated |
| -0,294529199 | 0,634987356 | MUM1L1 | FALSE | Downregulated |
| -0,405110095 | 0,635216121 | COL9A3 | FALSE | Downregulated |
| -0,563290839 | 0,635271129 | ENOX1 | FALSE | Downregulated |
| -0,144760094 | 0,635344067 | MTR | FALSE | Downregulated |
| 0,184586338 | 0,635383921 | ALDH1B1 | FALSE | Upregulated |
| 0,13510645 | 0,635383921 | ITIH4 | FALSE | Upregulated |
| 0,090208614 | 0,635391344 | ADCY6 | FALSE | Upregulated |

| | | | | |
|---|---|---|---|---|
| -0,337963685 | 0,635391344 | AGBL2 | FALSE | Downregulated |
| 0,561319723 | 0,635391344 | C20orf202 | FALSE | Upregulated |
| -0,335565408 | 0,635391344 | CAPN3 | FALSE | Downregulated |
| -0,141461896 | 0,635391344 | CCDC159 | FALSE | Downregulated |
| 0,120254309 | 0,635391344 | CEP76 | FALSE | Upregulated |
| 0,095870774 | 0,635391344 | COX4I1 | FALSE | Upregulated |
| 0,113165888 | 0,635391344 | DDX49 | FALSE | Upregulated |
| -0,09613759 | 0,635391344 | DERA | FALSE | Downregulated |
| 0,09791328 | 0,635391344 | EDC4 | FALSE | Upregulated |
| 0,184802064 | 0,635391344 | FAM86A | FALSE | Upregulated |
| 0,156394447 | 0,635391344 | FUT11 | FALSE | Upregulated |
| 0,770566865 | 0,635391344 | HAVCR1 | FALSE | Upregulated |
| -0,46335269 | 0,635391344 | HEATR4 | FALSE | Downregulated |
| 0,102892481 | 0,635391344 | KLC4 | FALSE | Upregulated |
| 0,365329075 | 0,635391344 | LOC101928189 | FALSE | Upregulated |
| -0,714681382 | 0,635391344 | LOC102723643 | FALSE | Downregulated |
| 0,461988721 | 0,635391344 | LOC102724312 | FALSE | Upregulated |
| 0,113870316 | 0,635391344 | LOC339803 | FALSE | Upregulated |
| 0,295465769 | 0,635391344 | LPIN3 | FALSE | Upregulated |
| 0,178723004 | 0,635391344 | LRRC16A | FALSE | Upregulated |
| -0,50903408 | 0,635391344 | MESP1 | FALSE | Downregulated |
| -0,087441299 | 0,635391344 | NBN | FALSE | Downregulated |
| 0,201576665 | 0,635391344 | POLR2J4 | FALSE | Upregulated |
| -0,08594864 | 0,635391344 | QTRTD1 | FALSE | Downregulated |
| -0,210919223 | 0,635391344 | RP11-545I5.3 | FALSE | Downregulated |
| 0,437703818 | 0,635391344 | SEMA7A | FALSE | Upregulated |

| | | | | |
|---|---|---|---|---|
| 0,166487499 | 0,635391344 | SFI1 | FALSE | Upregulated |
| -0,050808072 | 0,635391344 | SMU1 | FALSE | Downregulated |
| -0,11245134 | 0,635391344 | SOD1 | FALSE | Downregulated |
| 0,097652698 | 0,635391344 | SPPL2A | FALSE | Upregulated |
| -0,090597967 | 0,635391344 | SUGT1 | FALSE | Downregulated |
| 0,085582639 | 0,635391344 | TAB1 | FALSE | Upregulated |
| -0,368065151 | 0,635391344 | TCEAL7 | FALSE | Downregulated |
| -0,136491947 | 0,635391344 | TLK1 | FALSE | Downregulated |
| -0,452750509 | 0,635391344 | TREH | FALSE | Downregulated |
| -0,345165765 | 0,635391344 | TRNF | FALSE | Downregulated |
| 0,492124153 | 0,635391344 | TTYH1 | FALSE | Upregulated |
| 0,160009297 | 0,635391344 | TXNIP | FALSE | Upregulated |
| 0,060020103 | 0,635391344 | XRCC6 | FALSE | Upregulated |
| -0,216599286 | 0,635391344 | ZBTB7A | FALSE | Downregulated |
| -0,154060115 | 0,635391344 | ZFX | FALSE | Downregulated |
| -0,089154117 | 0,635391344 | ZSCAN21 | FALSE | Downregulated |
| 0,265073148 | 0,635441212 | FAR2 | FALSE | Upregulated |
| -0,13952631 | 0,635441212 | GSK3A | FALSE | Downregulated |
| -0,098827417 | 0,635441212 | HIPK3 | FALSE | Downregulated |
| -0,129622565 | 0,635441212 | RBL2 | FALSE | Downregulated |
| 0,13052072 | 0,635795027 | GOLGA3 | FALSE | Upregulated |
| 0,151422977 | 0,635795027 | STX5 | FALSE | Upregulated |
| -0,103622128 | 0,635887217 | NDUFA1 | FALSE | Downregulated |
| -0,157377573 | 0,635887217 | SYNE2 | FALSE | Downregulated |
| 0,593755492 | 0,636025601 | ANKDD1B | FALSE | Upregulated |
| -0,07846499 | 0,636025601 | APH1A | FALSE | Downregulated |

| | | | | |
|---|---|---|---|---|
| -0,10063601 | 0,636025601 | BAG4 | FALSE | Downregulated |
| -0,145605413 | 0,636025601 | ENPEP | FALSE | Downregulated |
| 0,459412869 | 0,636025601 | FAM131B | FALSE | Upregulated |
| -0,08406731 | 0,636025601 | HERC4 | FALSE | Downregulated |
| 0,161562707 | 0,636025601 | LOC100862671 | FALSE | Upregulated |
| 0,114581874 | 0,636025601 | MDH1 | FALSE | Upregulated |
| 0,104602073 | 0,636025601 | NAA10 | FALSE | Upregulated |
| -0,073733385 | 0,636025601 | PIAS2 | FALSE | Downregulated |
| 0,139407823 | 0,636025601 | PIP5K1C | FALSE | Upregulated |
| 0,472932279 | 0,636025601 | RP3-322G13.5 | FALSE | Upregulated |
| 0,106931115 | 0,636025601 | TUBGCP5 | FALSE | Upregulated |
| 0,218577325 | 0,636025601 | ZNF324B | FALSE | Upregulated |
| -0,52899004 | 0,636183207 | DDTL | FALSE | Downregulated |
| -0,540789755 | 0,636183207 | GUCA2B | FALSE | Downregulated |
| -0,417032393 | 0,636183207 | LOC102724306 | FALSE | Downregulated |
| -0,127653383 | 0,636183207 | ZC3H11A | FALSE | Downregulated |
| -0,084185582 | 0,636233422 | EPC1 | FALSE | Downregulated |
| -0,135504157 | 0,636233422 | EXTL2 | FALSE | Downregulated |
| 0,149605987 | 0,636233422 | RAP2B | FALSE | Upregulated |
| -0,13152268 | 0,636240475 | QDPR | FALSE | Downregulated |
| -0,157104604 | 0,636279964 | IFT46 | FALSE | Downregulated |
| -0,106563062 | 0,63628367 | PEX1 | FALSE | Downregulated |
| -0,32249359 | 0,636737985 | LOC101927393 | FALSE | Downregulated |
| -0,556312137 | 0,636997873 | ALLC | FALSE | Downregulated |
| 0,094359785 | 0,637085862 | NELFB | FALSE | Upregulated |
| -0,131266969 | 0,637292961 | LCOR | FALSE | Downregulated |

| | | | | |
|---|---|---|---|---|
| -0,127501867 | 0,63737134 | PARD6B | FALSE | Downregulated |
| -0,218569962 | 0,63737134 | ZNF737 | FALSE | Downregulated |
| 0,103300326 | 0,6380704 | KLHL22 | FALSE | Upregulated |
| -0,63542824 | 0,6380704 | LOC102723972 | FALSE | Downregulated |
| -0,355115114 | 0,6380704 | MX1 | FALSE | Downregulated |
| 0,112046183 | 0,638344263 | PACS2 | FALSE | Upregulated |
| 0,14767558 | 0,638568151 | ACLY | FALSE | Upregulated |
| -0,328401205 | 0,638654702 | LOC101929072 | FALSE | Downregulated |
| -0,16133416 | 0,638654702 | RNLS | FALSE | Downregulated |
| 0,126158085 | 0,638730771 | LEPROTL1 | FALSE | Upregulated |
| 0,142417272 | 0,638730771 | PTCHD3P1 | FALSE | Upregulated |
| 0,337943488 | 0,638748466 | CCRN4L | FALSE | Upregulated |
| -0,146523567 | 0,638748466 | MTUS1 | FALSE | Downregulated |
| 0,220579962 | 0,638748466 | ZNF852 | FALSE | Upregulated |
| -0,388741784 | 0,638773357 | MT1X | FALSE | Downregulated |
| 0,43602324 | 0,638773357 | ZNF625 | FALSE | Upregulated |
| -0,358651811 | 0,638799657 | LINC00238 | FALSE | Downregulated |
| -0,589021931 | 0,639120788 | AC007773.2 | FALSE | Downregulated |
| 0,271804566 | 0,639120788 | CD52 | FALSE | Upregulated |
| -0,098723397 | 0,639120788 | CLPX | FALSE | Downregulated |
| -0,194789345 | 0,639120788 | IYD | FALSE | Downregulated |
| 0,146882135 | 0,639120788 | KTI12 | FALSE | Upregulated |
| -0,45525988 | 0,639120788 | MT1F | FALSE | Downregulated |
| -0,07962788 | 0,639120788 | ZNF641 | FALSE | Downregulated |
| -0,476135393 | 0,639120788 | ZNF883 | FALSE | Downregulated |
| -0,395269742 | 0,639176593 | DPPA4 | FALSE | Downregulated |

| | | | | |
|---|---|---|---|---|
| -0,188793094 | 0,639176593 | TXLNG | FALSE | Downregulated |
| -0,555158472 | 0,63927678 | LRRC37A11P | FALSE | Downregulated |
| 0,151482429 | 0,639810866 | BTN3A3 | FALSE | Upregulated |
| -0,246582203 | 0,639810866 | CCDC150 | FALSE | Downregulated |
| -1,454460551 | 0,640391721 | CHRNA1 | FALSE | Downregulated |
| -0,143800021 | 0,640391721 | METTL21B | FALSE | Downregulated |
| 0,934992608 | 0,640422012 | ANTXRLP1 | FALSE | Upregulated |
| -0,76220477 | 0,640422012 | DCD | FALSE | Downregulated |
| 0,679621948 | 0,640422012 | DLEC1 | FALSE | Upregulated |
| -0,132035166 | 0,640422012 | HERC3 | FALSE | Downregulated |
| -0,619586642 | 0,640422012 | KIAA0408 | FALSE | Downregulated |
| -0,122607009 | 0,640422012 | PCTP | FALSE | Downregulated |
| -0,064686305 | 0,640422012 | SERINC1 | FALSE | Downregulated |
| 0,211769077 | 0,640422012 | SLC9A1 | FALSE | Upregulated |
| 0,5906898 | 0,640422012 | UCA1 | FALSE | Upregulated |
| -0,076817137 | 0,640543783 | PRKAR2A | FALSE | Downregulated |
| -0,187467572 | 0,640543783 | UPF3B | FALSE | Downregulated |
| 0,155470434 | 0,64101805 | ATAD2 | FALSE | Upregulated |
| -0,212811361 | 0,64101805 | CATSPER2P1 | FALSE | Downregulated |
| 0,356930982 | 0,64101805 | CHST1 | FALSE | Upregulated |
| 0,368428542 | 0,64101805 | GATA2 | FALSE | Upregulated |
| 0,166185084 | 0,64101805 | IQGAP1 | FALSE | Upregulated |
| -0,119424898 | 0,64101805 | SLC25A13 | FALSE | Downregulated |
| -0,084703495 | 0,641204174 | TM2D3 | FALSE | Downregulated |
| 0,155285257 | 0,641221238 | TMEM143 | FALSE | Upregulated |
| -0,108036013 | 0,641253057 | FUBP1 | FALSE | Downregulated |

| | | | | |
|---|---|---|---|---|
| -0,162088218 | 0,641253057 | MFAP1 | FALSE | Downregulated |
| -0,542084595 | 0,641270809 | CSPG4 | FALSE | Downregulated |
| -0,191950157 | 0,641270809 | FGB | FALSE | Downregulated |
| -0,236407999 | 0,641421023 | MVK | FALSE | Downregulated |
| 0,130340961 | 0,641421023 | TRIM8 | FALSE | Upregulated |
| -0,491330841 | 0,642164276 | TUBB4A | FALSE | Downregulated |
| 1,082266855 | 0,642188065 | AMZ1 | FALSE | Upregulated |
| 0,199178406 | 0,642303224 | AK1 | FALSE | Upregulated |
| 0,111109618 | 0,642303224 | DEDD2 | FALSE | Upregulated |
| 0,172376401 | 0,642303224 | HEG1 | FALSE | Upregulated |
| -0,649971112 | 0,642303224 | LOC286367 | FALSE | Downregulated |
| 0,164283231 | 0,642303224 | NGEF | FALSE | Upregulated |
| -0,586137829 | 0,642303224 | NOX4 | FALSE | Downregulated |
| -0,169871884 | 0,642303224 | SLC39A14 | FALSE | Downregulated |
| 0,095983624 | 0,642303224 | SMG8 | FALSE | Upregulated |
| -0,11576635 | 0,642303224 | THAP9 | FALSE | Downregulated |
| -0,122501288 | 0,642303224 | TNFAIP8L1 | FALSE | Downregulated |
| 0,108163957 | 0,642303224 | TTC4 | FALSE | Upregulated |
| -0,110547223 | 0,642303224 | WDFY3 | FALSE | Downregulated |
| -0,157024464 | 0,642521449 | SURF6 | FALSE | Downregulated |
| -0,158753734 | 0,642616219 | PAXIP1-AS1 | FALSE | Downregulated |
| -0,443387233 | 0,642870127 | LOC100507656 | FALSE | Downregulated |
| -0,105385884 | 0,642870866 | MRPL18 | FALSE | Downregulated |
| -0,24841441 | 0,643416866 | ADCY4 | FALSE | Downregulated |
| 0,438271939 | 0,643416866 | WNT4 | FALSE | Upregulated |
| 0,525462367 | 0,643535945 | AC104134.2 | FALSE | Upregulated |

| | | | | |
|---|---|---|---|---|
| -0,069443444 | 0,643535945 | ANKRD40 | FALSE | Downregulated |
| -0,266034432 | 0,643535945 | CILP | FALSE | Downregulated |
| -0,108293515 | 0,643535945 | FBXO9 | FALSE | Downregulated |
| -0,215491815 | 0,643535945 | LOC101927600 | FALSE | Downregulated |
| -0,129968237 | 0,643535945 | NDUFA4 | FALSE | Downregulated |
| -0,590444129 | 0,643535945 | SMG1P1 | FALSE | Downregulated |
| 0,135162322 | 0,643535945 | UBE2B | FALSE | Upregulated |
| -0,429078877 | 0,643686447 | ATAD3C | FALSE | Downregulated |
| 0,282912066 | 0,643686447 | CD5L | FALSE | Upregulated |
| 0,533280808 | 0,643686447 | CMTM1 | FALSE | Upregulated |
| -0,940474487 | 0,643686447 | CSDC2 | FALSE | Downregulated |
| 0,193758492 | 0,643686447 | CTH | FALSE | Upregulated |
| 0,598424282 | 0,643686447 | LOC100505767 | FALSE | Upregulated |
| -0,402761866 | 0,643686447 | LOC101928794 | FALSE | Downregulated |
| 0,226534425 | 0,643686447 | NBPF3 | FALSE | Upregulated |
| 0,07647389 | 0,643686447 | RIC8A | FALSE | Upregulated |
| -0,299990147 | 0,643686447 | ZNF736 | FALSE | Downregulated |
| 0,568673272 | 0,643781164 | IFNG | FALSE | Upregulated |
| -0,413378278 | 0,643781164 | RP11-423H2.3 | FALSE | Downregulated |
| -0,191046586 | 0,643978886 | AQP7 | FALSE | Downregulated |
| 0,110013798 | 0,643978886 | WDR6 | FALSE | Upregulated |
| -0,695486472 | 0,643998255 | DPEP3 | FALSE | Downregulated |
| -0,186590739 | 0,644041135 | ZBED6 | FALSE | Downregulated |
| -0,469960886 | 0,644222165 | LINC01132 | FALSE | Downregulated |
| -0,101867214 | 0,644248171 | ZRANB1 | FALSE | Downregulated |
| 0,125782054 | 0,644405279 | MORF4L2 | FALSE | Upregulated |

-0,232320472 0,64444433 LOC101927809 FALSE Downregulated

-0,289759945 0,644575763 ANKHD1-EIF4EBP3 FALSE Downregulated

0,100061958 0,644575763 ATP5B FALSE Upregulated

0,382556002 0,644575763 CCL2 FALSE Upregulated

-0,162431618 0,644575763 EFNA1 FALSE Downregulated

0,126138896 0,644575763 ELOVL1 FALSE Upregulated

-0,957009191 0,644575763 LOC285768 FALSE Downregulated

0,178144116 0,644575763 SERP1 FALSE Upregulated

0,186624185 0,644575763 TACC3 FALSE Upregulated

-0,118985523 0,644575763 TSPAN12 FALSE Downregulated

0,144566052 0,644575763 ZNF787 FALSE Upregulated

-0,084160382 0,644699628 TRIM33 FALSE Downregulated

-0,137627853 0,64477324 ALKBH7 FALSE Downregulated

0,210393289 0,644812778 CHST12 FALSE Upregulated

0,20325113 0,644854983 TMPRSS2 FALSE Upregulated

0,102427804 0,644969899 AGFG1 FALSE Upregulated

-0,088049026 0,645010799 MIB1 FALSE Downregulated

0,358706266 0,645010799 NBPF25P FALSE Upregulated

-0,17369746 0,645047742 AKT3 FALSE Downregulated

-0,442142993 0,645047742 LOC101928325 FALSE Downregulated

-0,246716262 0,645047742 PPP1R26-AS1 FALSE Downregulated

-0,083628041 0,64509387 AK6 FALSE Downregulated

-0,256641462 0,645350201 TNN FALSE Downregulated

-0,358598131 0,645576742 BMPR1B FALSE Downregulated

-0,313438497 0,645673384 LOC101928384 FALSE Downregulated

| | | | | |
|---|---|---|---|---|
| 0,235252289 | 0,645692251 | ALOX5 | FALSE | Upregulated |
| -0,152099007 | 0,645809912 | LARP1B | FALSE | Downregulated |
| 0,224289274 | 0,645809912 | ZNF273 | FALSE | Upregulated |
| 0,153749265 | 0,646133027 | CEP41 | FALSE | Upregulated |
| -0,20826423 | 0,646214118 | AMOTL1 | FALSE | Downregulated |
| -0,23736719 | 0,646366532 | TTBK1 | FALSE | Downregulated |
| -0,08528995 | 0,646436058 | LINC00657 | FALSE | Downregulated |
| -0,315964907 | 0,646589112 | LOC102724350 | FALSE | Downregulated |
| 0,124357618 | 0,646589112 | TPRKB | FALSE | Upregulated |
| -0,32945967 | 0,646703827 | FST | FALSE | Downregulated |
| 0,080481311 | 0,647024927 | REPIN1 | FALSE | Upregulated |
| 0,363556383 | 0,647127113 | ARL10 | FALSE | Upregulated |
| 0,3903228 | 0,647127113 | CD8B | FALSE | Upregulated |
| -0,328444737 | 0,647127113 | LOC100499489 | FALSE | Downregulated |
| -0,298392483 | 0,647127113 | PLEKHH2 | FALSE | Downregulated |
| 0,775679877 | 0,647127113 | SPRED3 | FALSE | Upregulated |
| 0,190128768 | 0,647127113 | TES | FALSE | Upregulated |
| 0,119634076 | 0,64714555 | EIF2AK4 | FALSE | Upregulated |
| 0,299961615 | 0,64714555 | FAM129A | FALSE | Upregulated |
| -0,223929905 | 0,64714555 | LHFP | FALSE | Downregulated |
| -0,316500569 | 0,64714555 | LRRIQ3 | FALSE | Downregulated |
| 0,148162066 | 0,647210462 | CRLF3 | FALSE | Upregulated |
| -0,072112174 | 0,647433387 | RARS2 | FALSE | Downregulated |
| -0,218454966 | 0,647577072 | CYP4A11 | FALSE | Downregulated |
| -0,305578662 | 0,647624826 | KCNMB4 | FALSE | Downregulated |
| -0,234996049 | 0,647702729 | LINC00886 | FALSE | Downregulated |

| | | | | |
|---|---|---|---|---|
| -0,244535825 | 0,647777768 | GREB1 | FALSE | Downregulated |
| -0,366684438 | 0,64789668 | ADIRF | FALSE | Downregulated |
| -0,134992611 | 0,64789668 | SERPINA1 | FALSE | Downregulated |
| 0,513943389 | 0,648015986 | NRCAM | FALSE | Upregulated |
| 0,315499754 | 0,648183256 | CTNND2 | FALSE | Upregulated |
| -0,113589456 | 0,648183256 | TMBIM4 | FALSE | Downregulated |
| 0,09078749 | 0,648183256 | UBIAD1 | FALSE | Upregulated |
| 0,686824317 | 0,648240904 | ANP32A-IT1 | FALSE | Upregulated |
| 0,13116646 | 0,648240904 | P4HB | FALSE | Upregulated |
| 0,643128102 | 0,648240904 | PNMA3 | FALSE | Upregulated |
| -0,082199115 | 0,648300925 | CREB1 | FALSE | Downregulated |
| 0,118244591 | 0,648606853 | MYO1D | FALSE | Upregulated |
| 0,411888284 | 0,648707229 | IQCJ-SCHIP1 | FALSE | Upregulated |
| -0,510580136 | 0,648707229 | SUMO4 | FALSE | Downregulated |
| 0,173826999 | 0,648889927 | RAB3IP | FALSE | Upregulated |
| 0,080074311 | 0,648889927 | RNH1 | FALSE | Upregulated |
| -0,614981775 | 0,649132186 | LOC102724965 | FALSE | Downregulated |
| -0,080392232 | 0,649132186 | NUP107 | FALSE | Downregulated |
| 0,081981614 | 0,649234508 | ACTR1A | FALSE | Upregulated |
| 0,45213041 | 0,64928869 | DIAPH3 | FALSE | Upregulated |
| -0,647658739 | 0,64933543 | TEKT4 | FALSE | Downregulated |
| -0,105607661 | 0,649586347 | TNKS1BP1 | FALSE | Downregulated |
| 0,350023072 | 0,649620612 | FHL2 | FALSE | Upregulated |
| 0,140940436 | 0,64995392 | SALL1 | FALSE | Upregulated |
| -0,264517082 | 0,650034901 | RNASEK-C17orf49 | FALSE | Downregulated |
| -0,208739814 | 0,650067383 | LOC100507258 | FALSE | Downregulated |

| | | | | |
|---|---|---|---|---|
| -0,246962232 | 0,650067383 | MROH6 | FALSE | Downregulated |
| -0,100771008 | 0,650067383 | SENP2 | FALSE | Downregulated |
| 0,222127985 | 0,650067383 | STOX2 | FALSE | Upregulated |
| -0,060799632 | 0,650146391 | AIDA | FALSE | Downregulated |
| 1,184552067 | 0,650178845 | HSFX2 | FALSE | Upregulated |
| -0,089563053 | 0,650433754 | SAFB | FALSE | Downregulated |
| -0,437013969 | 0,650464789 | TBX10 | FALSE | Downregulated |
| 0,660000137 | 0,650478164 | PRH2 | FALSE | Upregulated |
| 0,142239554 | 0,650480227 | SLC39A7 | FALSE | Upregulated |
| -0,280283344 | 0,650480227 | TRIM6 | FALSE | Downregulated |
| 0,212661027 | 0,650480227 | ZNF669 | FALSE | Upregulated |
| -0,12466598 | 0,650564518 | ITPRIPL2 | FALSE | Downregulated |
| 0,121940335 | 0,651476084 | PLCD1 | FALSE | Upregulated |
| 0,345635687 | 0,651611196 | TAF4B | FALSE | Upregulated |
| -0,12641843 | 0,651733927 | MRPS18B | FALSE | Downregulated |
| -0,16325914 | 0,651908987 | ZNF100 | FALSE | Downregulated |
| -0,101889261 | 0,65231456 | ZNF180 | FALSE | Downregulated |
| -0,526495043 | 0,652315391 | HSPB8 | FALSE | Downregulated |
| -0,156757016 | 0,652315391 | SLC17A2 | FALSE | Downregulated |
| -0,15370658 | 0,652568913 | RBM27 | FALSE | Downregulated |
| 0,119655585 | 0,652568913 | TTC13 | FALSE | Upregulated |
| 0,107566139 | 0,652601009 | DSG2 | FALSE | Upregulated |
| 0,14442237 | 0,652601009 | FIZ1 | FALSE | Upregulated |
| 0,09241566 | 0,652601009 | TOR1B | FALSE | Upregulated |
| -0,073671211 | 0,652653602 | HNRNPR | FALSE | Downregulated |
| 0,549132795 | 0,652979587 | C16orf98 | FALSE | Upregulated |

| | | | | |
|---|---|---|---|---|
| -0,836926945 | 0,652979587 | CAMK2A | FALSE | Downregulated |
| 0,096158414 | 0,652979587 | STAT6 | FALSE | Upregulated |
| 0,082360674 | 0,652979587 | TMEM184C | FALSE | Upregulated |
| 0,103217024 | 0,653087897 | TMEM161A | FALSE | Upregulated |
| 0,136553489 | 0,653224066 | CAMLG | FALSE | Upregulated |
| 0,080961465 | 0,653576781 | RALY | FALSE | Upregulated |
| -0,123572601 | 0,653576781 | USP42 | FALSE | Downregulated |
| -0,192280399 | 0,653835316 | HS6ST1 | FALSE | Downregulated |
| 1,108572467 | 0,654085055 | ANKRD20A8P | FALSE | Upregulated |
| 0,081372991 | 0,654085055 | ATP6V1G1 | FALSE | Upregulated |
| 0,429742961 | 0,654085055 | FCAMR | FALSE | Upregulated |
| -0,094011737 | 0,654085055 | NDUFA12 | FALSE | Downregulated |
| -0,111404368 | 0,654085055 | TMEM189 | FALSE | Downregulated |
| -0,083297505 | 0,654102635 | BAG1 | FALSE | Downregulated |
| -0,093333521 | 0,654102635 | CCDC43 | FALSE | Downregulated |
| 0,130024929 | 0,654102635 | CHCHD5 | FALSE | Upregulated |
| 0,462875344 | 0,654102635 | FKBP1AP1 | FALSE | Upregulated |
| 0,168760304 | 0,654102635 | FLI1 | FALSE | Upregulated |
| -1,605754797 | 0,654102635 | RBFOX1 | FALSE | Downregulated |
| 0,127925691 | 0,654102635 | YPEL3 | FALSE | Upregulated |
| 0,740799048 | 0,654210021 | SLC4A11 | FALSE | Upregulated |
| 0,2674197 | 0,654637749 | C15orf52 | FALSE | Upregulated |
| -0,177587213 | 0,654637749 | C19orf82 | FALSE | Downregulated |
| 0,506582841 | 0,654637749 | FGF11 | FALSE | Upregulated |
| 0,879539361 | 0,654637749 | FGF18 | FALSE | Upregulated |
| 0,146741337 | 0,654637749 | GLRX2 | FALSE | Upregulated |

| | | | | |
|---|---|---|---|---|
| -0,909464411 | 0,654637749 | LOC102724593 | FALSE | Downregulated |
| -0,167859185 | 0,654637749 | LRP6 | FALSE | Downregulated |
| -0,077422116 | 0,654637749 | NGDN | FALSE | Downregulated |
| 0,114573588 | 0,654637749 | PCBP2 | FALSE | Upregulated |
| 0,480004277 | 0,654637749 | RP11-466F5.8 | FALSE | Upregulated |
| -0,204184588 | 0,654637749 | TAF3 | FALSE | Downregulated |
| 0,173515238 | 0,654637749 | TGIF1 | FALSE | Upregulated |
| 0,199758643 | 0,654637749 | TRAF3IP2 | FALSE | Upregulated |
| -0,097266411 | 0,654637749 | TROVE2 | FALSE | Downregulated |
| 0,868587984 | 0,654637749 | UNC5A | FALSE | Upregulated |
| -0,114959372 | 0,654637749 | USP8 | FALSE | Downregulated |
| -0,087930004 | 0,654779792 | FAM179B | FALSE | Downregulated |
| 0,201371157 | 0,655254433 | WRAP53 | FALSE | Upregulated |
| 0,29784697 | 0,65534064 | GDF7 | FALSE | Upregulated |
| 0,299222789 | 0,65534064 | NRG3 | FALSE | Upregulated |
| 0,607100765 | 0,655545279 | LOC101927910 | FALSE | Upregulated |
| 0,106631275 | 0,656058175 | AXIN1 | FALSE | Upregulated |
| 0,113038543 | 0,656217386 | AARSD1 | FALSE | Upregulated |
| -0,10939776 | 0,656217386 | PRIMPOL | FALSE | Downregulated |
| 0,297221384 | 0,656217386 | ZC3H12D | FALSE | Upregulated |
| 0,384292299 | 0,656388111 | BMS1P6 | FALSE | Upregulated |
| 0,41399847 | 0,656754666 | ADORA1 | FALSE | Upregulated |
| 0,22207616 | 0,657125917 | ABCA3 | FALSE | Upregulated |
| -0,093835381 | 0,657188734 | COA5 | FALSE | Downregulated |
| 0,159263469 | 0,657194811 | RNF213 | FALSE | Upregulated |
| -0,559328127 | 0,657224055 | LOC101927337 | FALSE | Downregulated |

| | | | | |
|---|---|---|---|---|
| -0,237376785 | 0,657317412 | CCDC34 | FALSE | Downregulated |
| -0,221040368 | 0,657317412 | CSAD | FALSE | Downregulated |
| 0,12160214 | 0,657317412 | GON4L | FALSE | Upregulated |
| -0,074517732 | 0,657317412 | ZNF207 | FALSE | Downregulated |
| -0,398057244 | 0,657524956 | LOC102723953 | FALSE | Downregulated |
| -0,070121256 | 0,657524956 | SP1 | FALSE | Downregulated |
| 0,123056457 | 0,657674799 | RFXANK | FALSE | Upregulated |
| -0,091781877 | 0,657674799 | SPATS2L | FALSE | Downregulated |
| -0,738203279 | 0,657674799 | TRNAE11 | FALSE | Downregulated |
| -0,128271914 | 0,658309125 | PROSER1 | FALSE | Downregulated |
| 0,09862429 | 0,658309125 | TMEM87B | FALSE | Upregulated |
| 0,092884035 | 0,658518398 | PSMD12 | FALSE | Upregulated |
| 0,164629152 | 0,65855093 | ISPD | FALSE | Upregulated |
| 0,278527361 | 0,65855093 | LOC101927151 | FALSE | Upregulated |
| -0,127856449 | 0,65855093 | MIOS | FALSE | Downregulated |
| -0,662357751 | 0,65856743 | EMR3 | FALSE | Downregulated |
| 0,27262503 | 0,658618814 | CLIP4 | FALSE | Upregulated |
| -0,251643086 | 0,658628738 | MNDA | FALSE | Downregulated |
| -0,426018324 | 0,658628738 | MYH7B | FALSE | Downregulated |
| 0,326650934 | 0,65898617 | CD209 | FALSE | Upregulated |
| 0,11395537 | 0,65898617 | PSMB5 | FALSE | Upregulated |
| -0,260172837 | 0,65898617 | XAF1 | FALSE | Downregulated |
| 0,129005316 | 0,659060555 | TGIF2 | FALSE | Upregulated |
| 0,081818567 | 0,659060555 | UBTD2 | FALSE | Upregulated |
| 0,14833921 | 0,659258432 | TRMT1 | FALSE | Upregulated |
| -0,160022125 | 0,659258432 | VASN | FALSE | Downregulated |

| | | | | |
|---|---|---|---|---|
| 0,387520718 | 0,659258432 | VEPH1 | FALSE | Upregulated |
| -0,865797353 | 0,659457749 | ASB11 | FALSE | Downregulated |
| 0,170532233 | 0,659457749 | ZNF548 | FALSE | Upregulated |
| -0,403331571 | 0,659554053 | AHCTF1P1 | FALSE | Downregulated |
| 0,414013518 | 0,659554053 | LAYN | FALSE | Upregulated |
| -0,498382389 | 0,659554053 | RPL13AP20 | FALSE | Downregulated |
| -0,302126442 | 0,659775766 | CELF6 | FALSE | Downregulated |
| 0,496849391 | 0,659775766 | LOC284837 | FALSE | Upregulated |
| 0,351641468 | 0,660037445 | C15orf60 | FALSE | Upregulated |
| 0,126731806 | 0,660037445 | COG3 | FALSE | Upregulated |
| -0,173871958 | 0,660037445 | DFNA5 | FALSE | Downregulated |
| -0,18526044 | 0,660037445 | DLGAP1-AS1 | FALSE | Downregulated |
| -0,132301117 | 0,660037445 | FAM96A | FALSE | Downregulated |
| 0,199914434 | 0,660037445 | FOLR2 | FALSE | Upregulated |
| 0,110938255 | 0,660037445 | GTF3C4 | FALSE | Upregulated |
| -0,364072812 | 0,660037445 | GZMH | FALSE | Downregulated |
| 0,192291345 | 0,660037445 | HLX | FALSE | Upregulated |
| -0,248002329 | 0,660037445 | IL27 | FALSE | Downregulated |
| 0,38797067 | 0,660037445 | ITGB8 | FALSE | Upregulated |
| -0,62955499 | 0,660037445 | LOC101927734 | FALSE | Downregulated |
| -0,62561948 | 0,660037445 | LOC645638 | FALSE | Downregulated |
| 0,122558273 | 0,660037445 | MAP3K1 | FALSE | Upregulated |
| -0,106657359 | 0,660037445 | MED11 | FALSE | Downregulated |
| 0,242303273 | 0,660037445 | MORN4 | FALSE | Upregulated |
| -0,080344195 | 0,660037445 | MRS2 | FALSE | Downregulated |
| 0,157427885 | 0,660037445 | NEURL1B | FALSE | Upregulated |

| | | | | |
|---|---|---|---|---|
| 0,979838581 | 0,660037445 | NTM | FALSE | Upregulated |
| -0,19422267 | 0,660037445 | PEX11A | FALSE | Downregulated |
| 0,072293509 | 0,660037445 | PLXNB2 | FALSE | Upregulated |
| -0,157505417 | 0,660037445 | PRKAB2 | FALSE | Downregulated |
| -0,065750552 | 0,660037445 | RAB14 | FALSE | Downregulated |
| 0,222363952 | 0,660037445 | RPH3AL | FALSE | Upregulated |
| -0,131912292 | 0,660037445 | RPL36A | FALSE | Downregulated |
| 0,623410698 | 0,660037445 | SKA1 | FALSE | Upregulated |
| 0,216012089 | 0,660037445 | TMEM201 | FALSE | Upregulated |
| -0,534832948 | 0,660037445 | ZBED3-AS1 | FALSE | Downregulated |
| -0,128185206 | 0,660037445 | ZNF782 | FALSE | Downregulated |
| 0,200017395 | 0,660187164 | SLC37A2 | FALSE | Upregulated |
| 0,203721017 | 0,660243195 | ALG10 | FALSE | Upregulated |
| -0,144472491 | 0,660243195 | ATP5G1 | FALSE | Downregulated |
| -0,341952414 | 0,660243195 | LOC100133669 | FALSE | Downregulated |
| -0,155448334 | 0,660243195 | NME3 | FALSE | Downregulated |
| 0,43123235 | 0,660391127 | WNT11 | FALSE | Upregulated |
| -0,793957567 | 0,660593793 | CHL1 | FALSE | Downregulated |
| -0,090554628 | 0,660593793 | KPNA1 | FALSE | Downregulated |
| 0,083860908 | 0,660593793 | MKNK1 | FALSE | Upregulated |
| 0,150245602 | 0,660593793 | TCEB2 | FALSE | Upregulated |
| -0,09591558 | 0,660727145 | PSMD8 | FALSE | Downregulated |
| 0,174409939 | 0,66080654 | GMFB | FALSE | Upregulated |
| -0,1543401 | 0,661072423 | CCDC41 | FALSE | Downregulated |
| -0,116321818 | 0,661072423 | MIEN1 | FALSE | Downregulated |
| -0,093193653 | 0,661072423 | ZC3H14 | FALSE | Downregulated |

| | | | | |
|---|---|---|---|---|
| -0,066745971 | 0,661346134 | ATXN7L3B | FALSE | Downregulated |
| 0,093602987 | 0,661346134 | C14orf159 | FALSE | Upregulated |
| -0,560148421 | 0,661346134 | CYP2A13 | FALSE | Downregulated |
| 0,181666177 | 0,661346134 | ECHDC3 | FALSE | Upregulated |
| 0,389814236 | 0,661346134 | FBN2 | FALSE | Upregulated |
| 0,141829942 | 0,661346134 | FN3KRP | FALSE | Upregulated |
| -0,100087951 | 0,661346134 | PLOD2 | FALSE | Downregulated |
| 0,11634587 | 0,661346134 | RRP15 | FALSE | Upregulated |
| 0,228000328 | 0,661346134 | SDHAP2 | FALSE | Upregulated |
| -0,059686397 | 0,661346134 | SKIV2L2 | FALSE | Downregulated |
| 0,134063721 | 0,661398708 | EML2 | FALSE | Upregulated |
| -0,075957706 | 0,661398708 | RLIM | FALSE | Downregulated |
| -0,053779537 | 0,661621336 | API5 | FALSE | Downregulated |
| 0,215730187 | 0,661621336 | C4orf21 | FALSE | Upregulated |
| -0,33424835 | 0,661621336 | CRHR1-IT1 | FALSE | Downregulated |
| -0,141157589 | 0,661621336 | GLB1L | FALSE | Downregulated |
| 0,313195096 | 0,661621336 | KIF7 | FALSE | Upregulated |
| -0,566719813 | 0,661621336 | LOC100128276 | FALSE | Downregulated |
| -0,56250654 | 0,661621336 | LOC101929741 | FALSE | Downregulated |
| 0,582438363 | 0,661621336 | PLD4 | FALSE | Upregulated |
| -0,101251045 | 0,661621336 | POLR2F | FALSE | Downregulated |
| 0,610775017 | 0,661621336 | RP3-395M20.9 | FALSE | Upregulated |
| 0,168537558 | 0,661621336 | TVP23C-CDRT4 | FALSE | Upregulated |
| 0,331609885 | 0,661758595 | CXCR6 | FALSE | Upregulated |
| 0,173071502 | 0,662016541 | SHB | FALSE | Upregulated |
| 0,614191848 | 0,662179283 | SCIN | FALSE | Upregulated |

| | | | | |
|---|---|---|---|---|
| 0,230605726 | 0,662187486 | FYB | FALSE | Upregulated |
| 0,195291005 | 0,662187486 | RCSD1 | FALSE | Upregulated |
| 0,214474932 | 0,662187486 | SLC25A15 | FALSE | Upregulated |
| 0,613114084 | 0,662187486 | ST8SIA1 | FALSE | Upregulated |
| 0,201063264 | 0,662598831 | TNFAIP8 | FALSE | Upregulated |
| 0,247390376 | 0,662860722 | DTX1 | FALSE | Upregulated |
| 0,199867695 | 0,662860722 | FAM173A | FALSE | Upregulated |
| -0,330584423 | 0,662860722 | LOC101930307 | FALSE | Downregulated |
| 0,509580181 | 0,662860722 | MROH2A | FALSE | Upregulated |
| -0,095205465 | 0,662860722 | PQBP1 | FALSE | Downregulated |
| 0,100501265 | 0,663286204 | LRP3 | FALSE | Upregulated |
| 0,347627703 | 0,663286204 | SRCRB4D | FALSE | Upregulated |
| -0,099738611 | 0,663363705 | KLHDC10 | FALSE | Downregulated |
| 0,084232417 | 0,663430478 | TXNDC11 | FALSE | Upregulated |
| -0,815057335 | 0,663461343 | CR1L | FALSE | Downregulated |
| -0,311126543 | 0,663473768 | CTC-338M12.4 | FALSE | Downregulated |
| -0,098321391 | 0,663473768 | FZD5 | FALSE | Downregulated |
| -0,20689766 | 0,663473768 | NQO2 | FALSE | Downregulated |
| -0,293367466 | 0,663473768 | PAN3-AS1 | FALSE | Downregulated |
| -0,079494649 | 0,663473768 | SET | FALSE | Downregulated |
| -0,096431015 | 0,663736945 | HDAC2 | FALSE | Downregulated |
| 0,10221183 | 0,663736945 | PARP1 | FALSE | Upregulated |
| 0,150803474 | 0,663736945 | ZNF333 | FALSE | Upregulated |
| 0,17501922 | 0,663858563 | GDF11 | FALSE | Upregulated |
| -0,105406143 | 0,663858563 | KAT2B | FALSE | Downregulated |
| -0,137721452 | 0,663858563 | RAB7L1 | FALSE | Downregulated |

| | | | | |
|---|---|---|---|---|
| -0,117063155 | 0,66414313 | LNX2 | FALSE | Downregulated |
| 0,22412881 | 0,664356627 | STAG3L3 | FALSE | Upregulated |
| -0,10287763 | 0,664407059 | ATXN7 | FALSE | Downregulated |
| -0,211335559 | 0,664407059 | LINC00999 | FALSE | Downregulated |
| 0,156401772 | 0,665340271 | RITA1 | FALSE | Upregulated |
| -0,153806421 | 0,665657723 | GBP7 | FALSE | Downregulated |
| -0,077084007 | 0,665730043 | AASDHPPT | FALSE | Downregulated |
| -0,886587763 | 0,665789795 | LINC00173 | FALSE | Downregulated |
| -0,972824513 | 0,665877195 | ANK1 | FALSE | Downregulated |
| 0,429146891 | 0,665877195 | C3orf35 | FALSE | Upregulated |
| -0,214729102 | 0,665877195 | C8orf49 | FALSE | Downregulated |
| -0,072226944 | 0,665877195 | CRIPT | FALSE | Downregulated |
| -0,145795661 | 0,665877195 | GLCCI1 | FALSE | Downregulated |
| -0,113269278 | 0,665877195 | HCFC2 | FALSE | Downregulated |
| 0,693689995 | 0,665877195 | MMP28 | FALSE | Upregulated |
| -0,109828738 | 0,665877195 | MRPL33 | FALSE | Downregulated |
| -0,126337285 | 0,665877195 | MYD88 | FALSE | Downregulated |
| 0,084800639 | 0,665877195 | NECAP2 | FALSE | Upregulated |
| -0,441266862 | 0,665877195 | REREP3 | FALSE | Downregulated |
| -0,121270089 | 0,665877195 | RPS17 | FALSE | Downregulated |
| 0,45756519 | 0,665877195 | TMC7 | FALSE | Upregulated |
| 0,079198386 | 0,665877195 | ZNF317 | FALSE | Upregulated |
| -0,19734365 | 0,665877195 | ZNF444 | FALSE | Downregulated |
| 0,083516907 | 0,665955575 | AP4E1 | FALSE | Upregulated |
| -0,408521091 | 0,665955575 | LRRC3-AS1 | FALSE | Downregulated |
| -0,299074452 | 0,665955575 | PRKCDBP | FALSE | Downregulated |

| | | | | |
|---|---|---|---|---|
| 0,77983329 | 0,665955575 | PRSS27 | FALSE | Upregulated |
| -0,14448012 | 0,665955575 | TTC1 | FALSE | Downregulated |
| 0,694008077 | 0,665990064 | CYP27B1 | FALSE | Upregulated |
| -0,259336392 | 0,666039283 | GPR182 | FALSE | Downregulated |
| -0,072330077 | 0,666039283 | TMEM181 | FALSE | Downregulated |
| 0,664082165 | 0,666039283 | ZFP91-CNTF | FALSE | Upregulated |
| -0,121869978 | 0,666120924 | CFH | FALSE | Downregulated |
| 0,438098369 | 0,666120924 | LOXL1 | FALSE | Upregulated |
| -0,189830271 | 0,666120924 | PDE4A | FALSE | Downregulated |
| -0,705657547 | 0,666329188 | SNORD10 | FALSE | Downregulated |
| 0,45558138 | 0,666643343 | GATSL2 | FALSE | Upregulated |
| -0,120341555 | 0,666775511 | KLHL24 | FALSE | Downregulated |
| 0,168185448 | 0,666775511 | LCP1 | FALSE | Upregulated |
| -0,111538852 | 0,666775511 | NCOA6 | FALSE | Downregulated |
| 0,101462394 | 0,666866751 | IKBKB | FALSE | Upregulated |
| 0,188610342 | 0,666892115 | BTK | FALSE | Upregulated |
| 0,103640505 | 0,666892115 | C11orf24 | FALSE | Upregulated |
| 0,524090334 | 0,666892115 | CTSG | FALSE | Upregulated |
| 0,114993096 | 0,666892115 | DMAP1 | FALSE | Upregulated |
| -0,169807445 | 0,666892115 | EMC6 | FALSE | Downregulated |
| -0,094575152 | 0,666892115 | GRPEL2 | FALSE | Downregulated |
| -0,285300178 | 0,666892115 | HMGN5 | FALSE | Downregulated |
| -0,091629427 | 0,666892115 | LYPLA1 | FALSE | Downregulated |
| 0,109074301 | 0,666892115 | MRPL17 | FALSE | Upregulated |
| 0,453701631 | 0,666892115 | NCR3 | FALSE | Upregulated |
| 0,126477193 | 0,666892115 | PEX10 | FALSE | Upregulated |

| | | | | |
|---|---|---|---|---|
| -0,270049821 | 0,666892115 | RSAD2 | FALSE | Downregulated |
| -0,529642183 | 0,666892115 | S100A1 | FALSE | Downregulated |
| 0,185575987 | 0,666892115 | WARS | FALSE | Upregulated |
| 0,350279365 | 0,66696865 | RDH12 | FALSE | Upregulated |
| -0,106984102 | 0,667667825 | GNAI2 | FALSE | Downregulated |
| 0,31978357 | 0,668029911 | VWA7 | FALSE | Upregulated |
| 0,653628645 | 0,668166572 | PGA4 | FALSE | Upregulated |
| 0,138016085 | 0,66817898 | ARHGEF40 | FALSE | Upregulated |
| 0,151861553 | 0,66817898 | DDX28 | FALSE | Upregulated |
| 0,308521644 | 0,66817898 | KLHL32 | FALSE | Upregulated |
| 0,498536057 | 0,66817898 | LOC102724428 | FALSE | Upregulated |
| 0,45681341 | 0,66817898 | LOC90834 | FALSE | Upregulated |
| -0,210545657 | 0,66817898 | PSAT1 | FALSE | Downregulated |
| -0,145767018 | 0,66817898 | SRD5A1 | FALSE | Downregulated |
| -0,061086535 | 0,66817898 | TOMM70A | FALSE | Downregulated |
| 0,096954859 | 0,66817898 | TSPAN17 | FALSE | Upregulated |
| 0,638499808 | 0,668355472 | AC005594.3 | FALSE | Upregulated |
| -0,122650551 | 0,668355472 | BIVM | FALSE | Downregulated |
| -0,088012607 | 0,668355472 | DKC1 | FALSE | Downregulated |
| 0,185438877 | 0,668355472 | FUT4 | FALSE | Upregulated |
| -0,74584983 | 0,668355472 | LOC101930452 | FALSE | Downregulated |
| 0,197332715 | 0,668355472 | NUSAP1 | FALSE | Upregulated |
| 0,06976159 | 0,668355472 | OGDH | FALSE | Upregulated |
| -0,204409961 | 0,668355472 | OSCP1 | FALSE | Downregulated |
| 0,195705134 | 0,668355472 | PWWP2B | FALSE | Upregulated |
| -0,179557988 | 0,668355472 | SLCO1B3 | FALSE | Downregulated |

| | | | | |
|---|---|---|---|---|
| 0,270665404 | 0,668355472 | SVEP1 | FALSE | Upregulated |
| 0,115371157 | 0,668355472 | TBC1D9 | FALSE | Upregulated |
| 0,789620158 | 0,668355472 | TDRD15 | FALSE | Upregulated |
| 0,14277546 | 0,668355472 | TSEN34 | FALSE | Upregulated |
| -0,427223455 | 0,668380248 | GBP6 | FALSE | Downregulated |
| 0,211648866 | 0,668380248 | HID1 | FALSE | Upregulated |
| 0,230369495 | 0,668380248 | HPSE | FALSE | Upregulated |
| 0,144883614 | 0,668380248 | SPATA2L | FALSE | Upregulated |
| 0,690481196 | 0,668628073 | KIR3DL2 | FALSE | Upregulated |
| -0,106248601 | 0,668628073 | KLHL8 | FALSE | Downregulated |
| -0,422898634 | 0,668628073 | LOC102724603 | FALSE | Downregulated |
| -0,375573337 | 0,668628073 | ZRANB2-AS1 | FALSE | Downregulated |
| 0,732090939 | 0,668648167 | PTH2R | FALSE | Upregulated |
| 0,145295008 | 0,668721558 | RINL | FALSE | Upregulated |
| 0,092401461 | 0,668793874 | MAEA | FALSE | Upregulated |
| 0,065447893 | 0,668793874 | RPL7L1 | FALSE | Upregulated |
| -0,15930525 | 0,668862431 | INO80B-WBP1 | FALSE | Downregulated |
| 0,079121684 | 0,668862431 | MGME1 | FALSE | Upregulated |
| 0,252843261 | 0,668867164 | ATAD5 | FALSE | Upregulated |
| -0,127982056 | 0,668867164 | COLEC10 | FALSE | Downregulated |
| -0,099106904 | 0,668867164 | TRNAU1AP | FALSE | Downregulated |
| 0,250677667 | 0,668989481 | MYOF | FALSE | Upregulated |
| 0,083976155 | 0,669392002 | FBXO22 | FALSE | Upregulated |
| -0,066988193 | 0,669666524 | CLCC1 | FALSE | Downregulated |
| -0,126346115 | 0,669666524 | DCAF4 | FALSE | Downregulated |
| 0,120601126 | 0,669807316 | IRAK1 | FALSE | Upregulated |

| | | | | |
|---|---|---|---|---|
| -0,342457898 | 0,669825045 | COL6A6 | FALSE | Downregulated |
| -0,113503227 | 0,669838437 | GPN3 | FALSE | Downregulated |
| 0,148030044 | 0,669838437 | SLC26A1 | FALSE | Upregulated |
| -0,263028969 | 0,669838437 | TMEM44-AS1 | FALSE | Downregulated |
| -0,360675042 | 0,669851938 | FERMT1 | FALSE | Downregulated |
| 0,149418235 | 0,669851938 | RAB11FIP1 | FALSE | Upregulated |
| 0,131175852 | 0,670004726 | PPP1R26 | FALSE | Upregulated |
| -0,171545824 | 0,670004726 | PTPDC1 | FALSE | Downregulated |
| 0,170649091 | 0,670523544 | CASZ1 | FALSE | Upregulated |
| -0,233074163 | 0,670523544 | EGFLAM | FALSE | Downregulated |
| 0,488344036 | 0,670523544 | EPHA1-AS1 | FALSE | Upregulated |
| 0,765914804 | 0,670523544 | FAM72B | FALSE | Upregulated |
| -0,193139832 | 0,670523544 | PRPF38B | FALSE | Downregulated |
| -0,344470031 | 0,670523544 | SGK494 | FALSE | Downregulated |
| 0,136278344 | 0,670527072 | GNL3L | FALSE | Upregulated |
| 0,341198529 | 0,670545173 | LOC100128494 | FALSE | Upregulated |
| 0,186439852 | 0,670545173 | ORAI2 | FALSE | Upregulated |
| -0,074743797 | 0,670545173 | SNX4 | FALSE | Downregulated |
| 0,221031773 | 0,670545173 | STAG3L2 | FALSE | Upregulated |
| 0,092656685 | 0,670604638 | TRIM4 | FALSE | Upregulated |
| 0,95737264 | 0,670604638 | TRNG | FALSE | Upregulated |
| 0,091323861 | 0,670604638 | TXNRD1 | FALSE | Upregulated |
| -0,145091812 | 0,670609056 | TSC1 | FALSE | Downregulated |
| -0,159263244 | 0,671100318 | ARID4A | FALSE | Downregulated |
| -0,822436686 | 0,671100318 | ASCL1 | FALSE | Downregulated |
| -0,174229337 | 0,671100318 | GSTZ1 | FALSE | Downregulated |

| | | | | |
|---|---|---|---|---|
| -0,09345462 | 0,671100318 | KLHDC2 | FALSE | Downregulated |
| -0,303850833 | 0,671100318 | KLHL4 | FALSE | Downregulated |
| 1,127546285 | 0,671100318 | LOC102724865 | FALSE | Upregulated |
| -0,09094051 | 0,671100318 | PRMT2 | FALSE | Downregulated |
| -0,128750622 | 0,671100318 | SMARCA2 | FALSE | Downregulated |
| -0,361797449 | 0,671247601 | LOC646214 | FALSE | Downregulated |
| 0,15084654 | 0,671292237 | FAM50B | FALSE | Upregulated |
| 1,162244387 | 0,671510628 | LOC102724971 | FALSE | Upregulated |
| 0,665752848 | 0,671510628 | LOC401010 | FALSE | Upregulated |
| 0,108696807 | 0,671510628 | TIGD6 | FALSE | Upregulated |
| 0,176078639 | 0,671880332 | PLA1A | FALSE | Upregulated |
| -0,121827068 | 0,672089371 | ASTE1 | FALSE | Downregulated |
| 0,22154295 | 0,672089371 | CD44 | FALSE | Upregulated |
| -0,164484397 | 0,672089371 | SPEF2 | FALSE | Downregulated |
| -0,117314701 | 0,672282823 | FAM58A | FALSE | Downregulated |
| 0,106695794 | 0,672282823 | HERC2 | FALSE | Upregulated |
| -0,408059509 | 0,672282823 | RP11-53O19.1 | FALSE | Downregulated |
| -0,123151226 | 0,672282823 | RSRC1 | FALSE | Downregulated |
| -0,158623413 | 0,672282823 | SPRYD4 | FALSE | Downregulated |
| -0,111934682 | 0,672282823 | SSSCA1 | FALSE | Downregulated |
| 0,549606324 | 0,672642576 | PITRM1-AS1 | FALSE | Upregulated |
| 0,274665534 | 0,672642576 | RP11-98I9.4 | FALSE | Upregulated |
| 0,405190769 | 0,672642576 | RPL23P8 | FALSE | Upregulated |
| -0,087108995 | 0,672642576 | ZNF181 | FALSE | Downregulated |
| 0,262794366 | 0,672649431 | BIRC3 | FALSE | Upregulated |
| -0,798973309 | 0,672727465 | PGAM2 | FALSE | Downregulated |

| | | | | |
|---|---|---|---|---|
| 0,169968534 | 0,672794735 | PGLYRP2 | FALSE | Upregulated |
| 0,234094174 | 0,672824214 | PTPRU | FALSE | Upregulated |
| -0,077374772 | 0,673259811 | C4orf3 | FALSE | Downregulated |
| 0,383706961 | 0,673259811 | ZBTB12 | FALSE | Upregulated |
| 0,184428183 | 0,67335047 | MAPK8IP1 | FALSE | Upregulated |
| -0,104573893 | 0,673552635 | AFF1 | FALSE | Downregulated |
| -0,134738002 | 0,673552635 | ARG1 | FALSE | Downregulated |
| 0,126543626 | 0,673552635 | BOP1 | FALSE | Upregulated |
| 0,190524824 | 0,673552635 | FAM60A | FALSE | Upregulated |
| -0,207614786 | 0,673552635 | HERC5 | FALSE | Downregulated |
| 0,168440151 | 0,673552635 | KIAA0754 | FALSE | Upregulated |
| -0,087331605 | 0,673552635 | KIAA1671 | FALSE | Downregulated |
| 0,132193462 | 0,673552635 | NOP2 | FALSE | Upregulated |
| -0,087015822 | 0,673694759 | DRG1 | FALSE | Downregulated |
| 0,104754467 | 0,673728085 | NDUFAB1 | FALSE | Upregulated |
| 0,451248116 | 0,673808002 | LINC00926 | FALSE | Upregulated |
| -0,162152864 | 0,673808207 | MYRIP | FALSE | Downregulated |
| -0,269187423 | 0,673819342 | STAG3 | FALSE | Downregulated |
| 0,639753361 | 0,673857203 | GPRIN1 | FALSE | Upregulated |
| 0,209195285 | 0,673857203 | NPR2 | FALSE | Upregulated |
| -0,096090092 | 0,673857203 | RNF103 | FALSE | Downregulated |
| 0,078093556 | 0,673857203 | SF3B3 | FALSE | Upregulated |
| -0,163834712 | 0,673857203 | ZNF684 | FALSE | Downregulated |
| -0,092982185 | 0,674286896 | LOC101928527 | FALSE | Downregulated |
| 0,271211818 | 0,674437656 | DACT1 | FALSE | Upregulated |
| 0,642966873 | 0,674703575 | ANKRD36BP2 | FALSE | Upregulated |

| | | | | |
|---|---|---|---|---|
| 0,065766102 | 0,674703575 | AP2B1 | FALSE | Upregulated |
| -0,085613905 | 0,674703575 | ARID1B | FALSE | Downregulated |
| -0,116349842 | 0,674703575 | ATG2B | FALSE | Downregulated |
| 0,258521456 | 0,674703575 | C19orf55 | FALSE | Upregulated |
| -0,334905613 | 0,674703575 | CCDC103 | FALSE | Downregulated |
| 0,237799128 | 0,674703575 | HDAC9 | FALSE | Upregulated |
| 0,269706023 | 0,674703575 | LINC00864 | FALSE | Upregulated |
| 0,130730385 | 0,674703575 | MAN2C1 | FALSE | Upregulated |
| -0,203982316 | 0,674703575 | MCMDC2 | FALSE | Downregulated |
| 0,166546375 | 0,674703575 | PPA1 | FALSE | Upregulated |
| 0,168278449 | 0,674703575 | PRSS23 | FALSE | Upregulated |
| -0,295607706 | 0,674703575 | SLC38A11 | FALSE | Downregulated |
| -0,113957935 | 0,674703575 | TMEM186 | FALSE | Downregulated |
| 0,709809121 | 0,674703575 | TPSAB1 | FALSE | Upregulated |
| -0,064543926 | 0,674703575 | VTA1 | FALSE | Downregulated |
| -0,080013196 | 0,674703575 | ZC3H15 | FALSE | Downregulated |
| -0,305791241 | 0,674703575 | ZNF610 | FALSE | Downregulated |
| -0,124747087 | 0,674703575 | ZNF621 | FALSE | Downregulated |
| -0,133446087 | 0,674947812 | LINC00094 | FALSE | Downregulated |
| -0,208251305 | 0,675149592 | RP11-1072A3.2 | FALSE | Downregulated |
| -0,434328304 | 0,675208045 | LOC101929824 | FALSE | Downregulated |
| 0,091840969 | 0,675465303 | PSMD2 | FALSE | Upregulated |
| 0,272156671 | 0,675465303 | RAD9B | FALSE | Upregulated |
| 0,155910366 | 0,675892722 | TMEM80 | FALSE | Upregulated |
| -0,146896532 | 0,676513439 | RAI14 | FALSE | Downregulated |
| 0,292279638 | 0,67658474 | CDKN1A | FALSE | Upregulated |

| | | | | |
|---|---|---|---|---|
| 0,536420345 | 0,67658474 | LOC101928105 | FALSE | Upregulated |
| 0,491040156 | 0,67658474 | PAK6 | FALSE | Upregulated |
| 0,271259758 | 0,67658474 | RASA4 | FALSE | Upregulated |
| 0,109646051 | 0,67658474 | SMYD5 | FALSE | Upregulated |
| 0,112317459 | 0,67658474 | TRIM25 | FALSE | Upregulated |
| 0,12943063 | 0,67659177 | PPT1 | FALSE | Upregulated |
| -0,237442827 | 0,676615962 | BCL2L10 | FALSE | Downregulated |
| -0,78677304 | 0,676615962 | CORO6 | FALSE | Downregulated |
| -0,0659849 | 0,676615962 | FEM1B | FALSE | Downregulated |
| 0,451718048 | 0,676689456 | HAPLN3 | FALSE | Upregulated |
| -0,179436239 | 0,676949732 | ABCC6P1 | FALSE | Downregulated |
| 0,506416834 | 0,676949732 | C6orf164 | FALSE | Upregulated |
| -0,079251695 | 0,676949732 | CSTF1 | FALSE | Downregulated |
| -0,105659069 | 0,676949732 | DICER1 | FALSE | Downregulated |
| 0,111320346 | 0,676949732 | GHDC | FALSE | Upregulated |
| 0,182633036 | 0,676949732 | KIF3A | FALSE | Upregulated |
| 0,530692946 | 0,676949732 | LOC729162 | FALSE | Upregulated |
| 0,098918406 | 0,676949732 | NUPL1 | FALSE | Upregulated |
| -0,111587154 | 0,676949732 | PXK | FALSE | Downregulated |
| -0,544724761 | 0,676949732 | SPTA1 | FALSE | Downregulated |
| 0,425888956 | 0,676949732 | STK33 | FALSE | Upregulated |
| -0,166874308 | 0,677022004 | RPL39 | FALSE | Downregulated |
| -0,10816794 | 0,677100365 | FTSJ3 | FALSE | Downregulated |
| -0,50256966 | 0,677100365 | HSPB6 | FALSE | Downregulated |
| 0,578113413 | 0,677100365 | LOC100996634 | FALSE | Upregulated |
| 0,541707587 | 0,677100365 | LOC101928149 | FALSE | Upregulated |

| | | | | |
|---|---|---|---|---|
| -0,092725638 | 0,677339807 | CPN2 | FALSE | Downregulated |
| 0,166782724 | 0,677339807 | EFCAB11 | FALSE | Upregulated |
| -0,086791207 | 0,677339807 | SMARCAD1 | FALSE | Downregulated |
| -0,066296638 | 0,677339807 | YPEL5 | FALSE | Downregulated |
| 0,281990735 | 0,677339807 | ZNF14 | FALSE | Upregulated |
| -0,238197118 | 0,677414811 | ANO3 | FALSE | Downregulated |
| -0,769698003 | 0,677414811 | LOC730257 | FALSE | Downregulated |
| 0,594490433 | 0,677844723 | ALDH3A1 | FALSE | Upregulated |
| -0,13655691 | 0,678149994 | OLFML3 | FALSE | Downregulated |
| -0,187751326 | 0,678286963 | MMAB | FALSE | Downregulated |
| 1,209013707 | 0,6782937 | MMP23A | FALSE | Upregulated |
| -0,104418603 | 0,678455137 | MUS81 | FALSE | Downregulated |
| -0,446757931 | 0,678455137 | NEURL1 | FALSE | Downregulated |
| 0,304942972 | 0,678455137 | PHYHIPL | FALSE | Upregulated |
| -0,220145115 | 0,678455137 | RP11-418J17.1 | FALSE | Downregulated |
| 0,099413451 | 0,678455137 | ZC3HC1 | FALSE | Upregulated |
| 0,700482534 | 0,678576346 | FAM209B | FALSE | Upregulated |
| -0,165038102 | 0,678576346 | TRPM8 | FALSE | Downregulated |
| 0,151573761 | 0,678806831 | TMEM194B | FALSE | Upregulated |
| 0,310111767 | 0,678942091 | GRAP2 | FALSE | Upregulated |
| -0,622400137 | 0,678942091 | SNX29P1 | FALSE | Downregulated |
| -0,442415193 | 0,6790002 | GPR88 | FALSE | Downregulated |
| 0,13350533 | 0,6790002 | RGL2 | FALSE | Upregulated |
| 0,165067804 | 0,679054445 | CFP | FALSE | Upregulated |
| 0,449104682 | 0,679060486 | HLA-DQA2 | FALSE | Upregulated |
| 0,325374326 | 0,679060486 | SLC12A8 | FALSE | Upregulated |

| | | | | |
|---|---|---|---|---|
| 0,099287105 | 0,679181879 | PCSK7 | FALSE | Upregulated |
| 0,290434231 | 0,679243435 | SLC2A1 | FALSE | Upregulated |
| -0,089472287 | 0,679362427 | PURA | FALSE | Downregulated |
| -0,754867311 | 0,679487135 | ADCYAP1R1 | FALSE | Downregulated |
| -0,056323519 | 0,679487135 | BLOC1S6 | FALSE | Downregulated |
| 0,092745171 | 0,679487135 | TOMM34 | FALSE | Upregulated |
| -0,54372281 | 0,679487135 | UNQ6494 | FALSE | Downregulated |
| 0,919267777 | 0,679626908 | ARHGAP11B | FALSE | Upregulated |
| 0,200512585 | 0,679628027 | C1QB | FALSE | Upregulated |
| -0,09082243 | 0,679628027 | C1S | FALSE | Downregulated |
| 0,621449681 | 0,679628027 | CENPA | FALSE | Upregulated |
| -0,429831083 | 0,679628027 | LINC00398 | FALSE | Downregulated |
| 0,120579104 | 0,679628027 | PPRC1 | FALSE | Upregulated |
| 0,3932401 | 0,679628027 | SBK1 | FALSE | Upregulated |
| -0,182568101 | 0,679686988 | MYL9 | FALSE | Downregulated |
| -0,120532745 | 0,679948379 | C1R | FALSE | Downregulated |
| 0,165355302 | 0,679948379 | HIC2 | FALSE | Upregulated |
| 0,142653674 | 0,679948379 | LAMP2 | FALSE | Upregulated |
| -0,236881559 | 0,679948379 | TPRG1 | FALSE | Downregulated |
| 0,068662934 | 0,679990364 | NUP50 | FALSE | Upregulated |
| -0,171668997 | 0,680212904 | ZNF658B | FALSE | Downregulated |
| 0,262954928 | 0,680238558 | BRCA2 | FALSE | Upregulated |
| 0,242429223 | 0,680238558 | LOC102723521 | FALSE | Upregulated |
| 0,177993201 | 0,680238558 | LYL1 | FALSE | Upregulated |
| 0,13010905 | 0,680238558 | MRPL55 | FALSE | Upregulated |
| 0,097538193 | 0,680238558 | PIP5K1A | FALSE | Upregulated |

| | | | | |
|---|---|---|---|---|
| -0,682107025 | 0,680238558 | TEKT3 | FALSE | Downregulated |
| -0,113474274 | 0,680238558 | TRAPPC2L | FALSE | Downregulated |
| -0,586795397 | 0,680238642 | RFPL2 | FALSE | Downregulated |
| 0,203974441 | 0,680347409 | TLE2 | FALSE | Upregulated |
| 0,088871523 | 0,680584146 | KRI1 | FALSE | Upregulated |
| -0,104848439 | 0,680584146 | MDC1 | FALSE | Downregulated |
| -0,11373638 | 0,680584146 | PRR14 | FALSE | Downregulated |
| 0,20454143 | 0,680584146 | TNFSF13 | FALSE | Upregulated |
| -0,172022533 | 0,68088255 | NAALADL2 | FALSE | Downregulated |
| -0,168938403 | 0,680936531 | MIR100HG | FALSE | Downregulated |
| -0,187687031 | 0,680936531 | ODF2L | FALSE | Downregulated |
| -0,073991724 | 0,680936531 | VDAC3 | FALSE | Downregulated |
| -0,337099069 | 0,681109234 | FPR2 | FALSE | Downregulated |
| 0,393190931 | 0,681109234 | LRRC34 | FALSE | Upregulated |
| -0,346422501 | 0,681109234 | RAD21-AS1 | FALSE | Downregulated |
| 0,151378087 | 0,681149865 | DPCD | FALSE | Upregulated |
| 0,133467838 | 0,68150821 | KIF2A | FALSE | Upregulated |
| 0,243407318 | 0,681578645 | CYSLTR1 | FALSE | Upregulated |
| 0,231272091 | 0,681631926 | EXOC3L2 | FALSE | Upregulated |
| -0,129404636 | 0,681790599 | CAV2 | FALSE | Downregulated |
| 0,127423307 | 0,681824404 | TRIM68 | FALSE | Upregulated |
| 0,581475753 | 0,681824404 | VENTX | FALSE | Upregulated |
| 0,207594214 | 0,681824404 | ZNF83 | FALSE | Upregulated |
| -0,505446172 | 0,681951161 | CYP4F24P | FALSE | Downregulated |
| -0,083663362 | 0,681951161 | ENSA | FALSE | Downregulated |
| 0,30254417 | 0,681951161 | ZNF300P1 | FALSE | Upregulated |

| | | | | |
|---|---|---|---|---|
| -0,080811193 | 0,682128949 | PPIP5K2 | FALSE | Downregulated |
| -0,08925184 | 0,682174193 | APPL2 | FALSE | Downregulated |
| 0,446038375 | 0,682174193 | C22orf26 | FALSE | Upregulated |
| 0,18315279 | 0,682174193 | CD320 | FALSE | Upregulated |
| -0,323664291 | 0,682174193 | LOC100126582 | FALSE | Downregulated |
| 0,364161895 | 0,682174193 | LOC100506870 | FALSE | Upregulated |
| 0,645134165 | 0,682174193 | LYPD1 | FALSE | Upregulated |
| -0,111537876 | 0,682174193 | MRPL32 | FALSE | Downregulated |
| -0,276393965 | 0,682174193 | RXFP1 | FALSE | Downregulated |
| -0,122715818 | 0,682174193 | TMEM126A | FALSE | Downregulated |
| 0,362652845 | 0,682174193 | TMEM171 | FALSE | Upregulated |
| 0,089274606 | 0,682174193 | TMEM179B | FALSE | Upregulated |
| 0,669256174 | 0,682174193 | ZNF80 | FALSE | Upregulated |
| -0,194746618 | 0,682195856 | AACS | FALSE | Downregulated |
| 0,097336645 | 0,682195856 | ALDH4A1 | FALSE | Upregulated |
| 0,06951961 | 0,682195856 | ATP2C1 | FALSE | Upregulated |
| -0,140332148 | 0,682195856 | BAZ2B | FALSE | Downregulated |
| 0,35625539 | 0,682195856 | CPZ | FALSE | Upregulated |
| 0,225723091 | 0,682195856 | CX3CR1 | FALSE | Upregulated |
| -0,169012298 | 0,682195856 | CYP39A1 | FALSE | Downregulated |
| -1,274972394 | 0,682195856 | DUSP26 | FALSE | Downregulated |
| 0,188509369 | 0,682195856 | EFEMP2 | FALSE | Upregulated |
| 0,265563575 | 0,682195856 | GPR137C | FALSE | Upregulated |
| 0,22087987 | 0,682195856 | NCEH1 | FALSE | Upregulated |
| -0,31758512 | 0,682195856 | NPIPB4 | FALSE | Downregulated |
| 0,07669021 | 0,682195856 | NUP98 | FALSE | Upregulated |

| | | | | |
|---|---|---|---|---|
| -0,105395031 | 0,682195856 | OPN3 | FALSE | Downregulated |
| -0,662507331 | 0,682195856 | RAB9B | FALSE | Downregulated |
| 0,089971034 | 0,682195856 | RHBDD1 | FALSE | Upregulated |
| 0,071065263 | 0,682195856 | RPA1 | FALSE | Upregulated |
| -0,147460946 | 0,682195856 | SMAD7 | FALSE | Downregulated |
| 0,293460193 | 0,682195856 | SPATA18 | FALSE | Upregulated |
| 0,158047833 | 0,682195856 | SRM | FALSE | Upregulated |
| 0,09917097 | 0,682195856 | TOP3A | FALSE | Upregulated |
| -0,236039773 | 0,682195856 | TSPYL2 | FALSE | Downregulated |
| 0,510264481 | 0,682195856 | ZNF321P | FALSE | Upregulated |
| 0,20941623 | 0,682195856 | ZNF367 | FALSE | Upregulated |
| 0,216927881 | 0,682195856 | ZNF837 | FALSE | Upregulated |
| -0,111416583 | 0,682210026 | TPST2 | FALSE | Downregulated |
| -0,12387908 | 0,682932376 | MANSC1 | FALSE | Downregulated |
| -0,164622818 | 0,683125181 | TBXA2R | FALSE | Downregulated |
| 0,122984077 | 0,683275204 | HPS6 | FALSE | Upregulated |
| 0,307109886 | 0,683388207 | BTN2A3P | FALSE | Upregulated |
| -0,445337841 | 0,683543983 | AC009473.1 | FALSE | Downregulated |
| -0,532299347 | 0,683543983 | FAM69C | FALSE | Downregulated |
| -0,121982861 | 0,683543983 | TIMM8B | FALSE | Downregulated |
| -0,110234856 | 0,683627697 | RBKS | FALSE | Downregulated |
| -0,101198841 | 0,683944615 | ATP5G3 | FALSE | Downregulated |
| 0,089542485 | 0,683944615 | EMC10 | FALSE | Upregulated |
| -0,382892846 | 0,683944615 | LOC101929513 | FALSE | Downregulated |
| 0,33959512 | 0,683944615 | P2RY14 | FALSE | Upregulated |
| -0,223100091 | 0,683944615 | SLC13A3 | FALSE | Downregulated |

| | | | | |
|---|---|---|---|---|
| 0,242409664 | 0,683944615 | TTN-AS1 | FALSE | Upregulated |
| -0,245831484 | 0,684037301 | LOC102724711 | FALSE | Downregulated |
| 0,201993385 | 0,6840823 | C1QC | FALSE | Upregulated |
| 0,184114331 | 0,68411065 | CKLF | FALSE | Upregulated |
| -0,430384602 | 0,684311689 | ACOT6 | FALSE | Downregulated |
| 0,506678949 | 0,684311689 | SEZ6L2 | FALSE | Upregulated |
| -0,277325309 | 0,684434311 | ZNF726 | FALSE | Downregulated |
| 0,091235677 | 0,684468782 | CDC42 | FALSE | Upregulated |
| 0,179288584 | 0,684481195 | ZNF587 | FALSE | Upregulated |
| 0,174072764 | 0,684637606 | RASSF2 | FALSE | Upregulated |
| -0,110563245 | 0,684750571 | BCKDHB | FALSE | Downregulated |
| 0,157917171 | 0,684773926 | IFITM3 | FALSE | Upregulated |
| 0,207846073 | 0,684773926 | ZNF785 | FALSE | Upregulated |
| -1,175710241 | 0,684850874 | KLHL38 | FALSE | Downregulated |
| -0,259936253 | 0,684850874 | PDK4 | FALSE | Downregulated |
| -0,109703406 | 0,684850874 | TMEM110 | FALSE | Downregulated |
| -0,220957058 | 0,684884328 | KRTCAP3 | FALSE | Downregulated |
| -0,100956057 | 0,684935171 | BMS1 | FALSE | Downregulated |
| -1,465647438 | 0,684935171 | SERPINB2 | FALSE | Downregulated |
| -0,178447307 | 0,684959045 | APOC4 | FALSE | Downregulated |
| -0,084866314 | 0,685155804 | YBX1 | FALSE | Downregulated |
| -0,136112046 | 0,68531483 | DNAJB4 | FALSE | Downregulated |
| -0,378063363 | 0,68531483 | FAM9B | FALSE | Downregulated |
| 0,103557954 | 0,68531483 | IPMK | FALSE | Upregulated |
| -0,048234491 | 0,68531483 | RAB5B | FALSE | Downregulated |
| 0,102095313 | 0,685370438 | ADAT1 | FALSE | Upregulated |

| | | | | |
|---|---|---|---|---|
| 0,61850297 | 0,685370438 | POM121L10P | FALSE | Upregulated |
| 0,125692767 | 0,685370438 | TBC1D10A | FALSE | Upregulated |
| -0,103844144 | 0,685370438 | TBC1D12 | FALSE | Downregulated |
| -0,138878763 | 0,685370438 | XRCC5 | FALSE | Downregulated |
| -0,341997584 | 0,685446439 | MN1 | FALSE | Downregulated |
| -0,254762678 | 0,685663068 | MYL6B | FALSE | Downregulated |
| -0,114175285 | 0,685663068 | SCMH1 | FALSE | Downregulated |
| -0,174591163 | 0,685882727 | LOC100506538 | FALSE | Downregulated |
| -0,154668486 | 0,685882727 | VAMP2 | FALSE | Downregulated |
| 0,148345898 | 0,68597686 | HEY2 | FALSE | Upregulated |
| 0,105905358 | 0,68597686 | PCMTD2 | FALSE | Upregulated |
| 0,130126921 | 0,686091215 | CTC1 | FALSE | Upregulated |
| 0,333855726 | 0,686091215 | KIF2C | FALSE | Upregulated |
| 0,501852517 | 0,686091215 | SNORD22 | FALSE | Upregulated |
| -0,080879449 | 0,686091215 | UBR5 | FALSE | Downregulated |
| -0,105281416 | 0,686224669 | ANAPC16 | FALSE | Downregulated |
| 0,237654441 | 0,686224669 | B9D1 | FALSE | Upregulated |
| -0,165235204 | 0,686224669 | BAALC | FALSE | Downregulated |
| -0,16630811 | 0,686224669 | C20orf96 | FALSE | Downregulated |
| -0,23801882 | 0,686224669 | FBXO15 | FALSE | Downregulated |
| 0,108141699 | 0,686224669 | GIT2 | FALSE | Upregulated |
| 0,099364611 | 0,686224669 | GRWD1 | FALSE | Upregulated |
| -0,129188411 | 0,686224669 | IFIT5 | FALSE | Downregulated |
| -0,129326762 | 0,686224669 | KANSL1L | FALSE | Downregulated |
| -0,377046569 | 0,686224669 | LOC101928143 | FALSE | Downregulated |
| -0,086740643 | 0,686224669 | PSMD5 | FALSE | Downregulated |

| | | | | |
|---|---|---|---|---|
| -0,184063872 | 0,686224669 | RDX | FALSE | Downregulated |
| 0,235333103 | 0,686224669 | TNIK | FALSE | Upregulated |
| -0,108170721 | 0,686224669 | USP33 | FALSE | Downregulated |
| 0,16194705 | 0,686224669 | ZNF581 | FALSE | Upregulated |
| -0,101716413 | 0,686224669 | ZNF84 | FALSE | Downregulated |
| 0,607675403 | 0,686258476 | LINC00605 | FALSE | Upregulated |
| 0,11843017 | 0,686356493 | FLAD1 | FALSE | Upregulated |
| 0,077540498 | 0,686372333 | RPF1 | FALSE | Upregulated |
| -0,855589307 | 0,686646985 | ALAS2 | FALSE | Downregulated |
| -0,116443124 | 0,686646985 | C14orf2 | FALSE | Downregulated |
| -0,100772549 | 0,686646985 | CLYBL | FALSE | Downregulated |
| 0,086314054 | 0,686646985 | CYC1 | FALSE | Upregulated |
| 0,222802212 | 0,686646985 | INHBA | FALSE | Upregulated |
| 0,431782956 | 0,686646985 | PIWIL4 | FALSE | Upregulated |
| -0,097213615 | 0,686646985 | PLG | FALSE | Downregulated |
| -0,088080907 | 0,686646985 | RBAK | FALSE | Downregulated |
| -0,059950396 | 0,686646985 | SERBP1 | FALSE | Downregulated |
| 0,176117704 | 0,686646985 | SIRT6 | FALSE | Upregulated |
| -0,0938269 | 0,686646985 | SNX27 | FALSE | Downregulated |
| -0,217197825 | 0,686646985 | ZC3H12B | FALSE | Downregulated |
| -0,096838991 | 0,686887762 | EZH1 | FALSE | Downregulated |
| 0,230655465 | 0,687077699 | C11orf48 | FALSE | Upregulated |
| -0,091991473 | 0,687077699 | CALCOCO2 | FALSE | Downregulated |
| 0,189473208 | 0,687077699 | CX3CL1 | FALSE | Upregulated |
| -0,078227927 | 0,687077699 | PSMB7 | FALSE | Downregulated |
| -0,623129334 | 0,687484051 | ITPR1-AS1 | FALSE | Downregulated |

| | | | | |
|---|---|---|---|---|
| -0,219554658 | 0,687484051 | LOC102723665 | FALSE | Downregulated |
| 0,36892621 | 0,687484051 | LRP8 | FALSE | Upregulated |
| 0,116615986 | 0,687484051 | PTDSS2 | FALSE | Upregulated |
| -0,348920859 | 0,687484051 | RP11-379B18.5 | FALSE | Downregulated |
| -0,263079413 | 0,687484051 | ST13P4 | FALSE | Downregulated |
| -0,273771919 | 0,687550737 | KCTD21-AS1 | FALSE | Downregulated |
| 0,111954287 | 0,687567356 | FAM21A | FALSE | Upregulated |
| 0,471220483 | 0,687567356 | FCER1A | FALSE | Upregulated |
| -0,104126753 | 0,687567356 | IGFBP4 | FALSE | Downregulated |
| -0,099306459 | 0,687567356 | IRGQ | FALSE | Downregulated |
| -0,458351885 | 0,687567356 | LOC102724791 | FALSE | Downregulated |
| -0,157782728 | 0,687567356 | MTHFSD | FALSE | Downregulated |
| -0,075070359 | 0,687567356 | SUCLA2 | FALSE | Downregulated |
| 0,668532112 | 0,687567356 | TPTE2P1 | FALSE | Upregulated |
| 0,288499661 | 0,687576555 | ZNF807 | FALSE | Upregulated |
| 0,122419689 | 0,687816781 | C9orf69 | FALSE | Upregulated |
| -0,313201075 | 0,687816781 | LOC285638 | FALSE | Downregulated |
| 0,09801107 | 0,687816781 | NIP7 | FALSE | Upregulated |
| 0,164652993 | 0,687876529 | AZI1 | FALSE | Upregulated |
| -0,565622987 | 0,687876529 | LOC100506753 | FALSE | Downregulated |
| 0,093631447 | 0,687876529 | MAP2K3 | FALSE | Upregulated |
| -0,089128679 | 0,687876529 | PDSS2 | FALSE | Downregulated |
| -0,156029351 | 0,687876529 | SRSF10 | FALSE | Downregulated |
| -0,071911501 | 0,687876529 | TMEM242 | FALSE | Downregulated |
| -0,200454905 | 0,687956759 | AQP7P1 | FALSE | Downregulated |
| 0,067468773 | 0,688058849 | ARF6 | FALSE | Upregulated |

| | | | | |
|---|---|---|---|---|
| -0,836834829 | 0,688058849 | ATP1A2 | FALSE | Downregulated |
| -0,415234112 | 0,688058849 | ATP6V0E2-AS1 | FALSE | Downregulated |
| -0,055544948 | 0,688058849 | GLE1 | FALSE | Downregulated |
| 0,273363198 | 0,688058849 | GNGT2 | FALSE | Upregulated |
| 0,225024514 | 0,688058849 | KPTN | FALSE | Upregulated |
| -0,087927156 | 0,688058849 | MAL2 | FALSE | Downregulated |
| -0,119546974 | 0,688058849 | SNRNP35 | FALSE | Downregulated |
| 0,270101097 | 0,688058849 | SOGA2 | FALSE | Upregulated |
| 0,243032776 | 0,688123087 | ARRDC2 | FALSE | Upregulated |
| 0,179628895 | 0,688123087 | PLEKHO2 | FALSE | Upregulated |
| -0,093543713 | 0,688179226 | CYB5B | FALSE | Downregulated |
| 0,109405401 | 0,688179226 | IPO4 | FALSE | Upregulated |
| 0,372177808 | 0,688179226 | LOC102724852 | FALSE | Upregulated |
| 0,126705218 | 0,688838135 | DIS3L2 | FALSE | Upregulated |
| 0,075710173 | 0,688838135 | FBXO33 | FALSE | Upregulated |
| -0,103614866 | 0,689009951 | SLC25A26 | FALSE | Downregulated |
| 0,185355871 | 0,689098302 | GPR124 | FALSE | Upregulated |
| 0,508203562 | 0,689098302 | KLRC2 | FALSE | Upregulated |
| -0,140561712 | 0,689098302 | RNF180 | FALSE | Downregulated |
| 0,100552843 | 0,689098302 | SMARCA4 | FALSE | Upregulated |
| 0,140466832 | 0,689183851 | USP21 | FALSE | Upregulated |
| 0,236098996 | 0,68920536 | C11orf35 | FALSE | Upregulated |
| 0,231917019 | 0,689255732 | ZNF773 | FALSE | Upregulated |
| 0,29825797 | 0,689265232 | NKD1 | FALSE | Upregulated |
| -0,092861746 | 0,689304809 | ANKMY2 | FALSE | Downregulated |
| -0,087282522 | 0,689316602 | CPD | FALSE | Downregulated |

| | | | | |
|---|---|---|---|---|
| -0,062151124 | 0,689316602 | DYNC1LI2 | FALSE | Downregulated |
| -0,193499181 | 0,689419838 | PPP2R3B | FALSE | Downregulated |
| 0,297059111 | 0,689449246 | CHAF1B | FALSE | Upregulated |
| -0,066237853 | 0,689449246 | CTNNB1 | FALSE | Downregulated |
| 0,094790176 | 0,689855443 | CIAPIN1 | FALSE | Upregulated |
| -0,087504913 | 0,689855443 | MIPEP | FALSE | Downregulated |
| 0,242417093 | 0,689855443 | P2RY1 | FALSE | Upregulated |
| -0,188904927 | 0,689936273 | ZDHHC2 | FALSE | Downregulated |
| 0,076823118 | 0,690739271 | PPIL2 | FALSE | Upregulated |
| 0,09536551 | 0,690770406 | AGAP3 | FALSE | Upregulated |
| -0,171582641 | 0,690770406 | KIAA1804 | FALSE | Downregulated |
| 0,907013691 | 0,690770406 | MAGED4 | FALSE | Upregulated |
| 0,379634144 | 0,690770406 | MIAT | FALSE | Upregulated |
| -0,08294075 | 0,690770406 | NSMCE1 | FALSE | Downregulated |
| -0,493234003 | 0,690770406 | PRSS50 | FALSE | Downregulated |
| -0,117798158 | 0,690770406 | RAB6A | FALSE | Downregulated |
| 0,086861795 | 0,690770406 | SH3D19 | FALSE | Upregulated |
| -0,123900637 | 0,690770406 | SMDT1 | FALSE | Downregulated |
| -0,07392668 | 0,690770406 | SNX14 | FALSE | Downregulated |
| 0,090899051 | 0,690770406 | VTN | FALSE | Upregulated |
| 0,161488497 | 0,690926504 | PPP1R35 | FALSE | Upregulated |
| -0,13937915 | 0,691057164 | MED31 | FALSE | Downregulated |
| -0,049945646 | 0,691061739 | DDX50 | FALSE | Downregulated |
| -0,270027282 | 0,691281032 | EPSTI1 | FALSE | Downregulated |
| -0,125242891 | 0,691290895 | MESDC2 | FALSE | Downregulated |
| 0,288836767 | 0,691290895 | MSC | FALSE | Upregulated |

| | | | | |
|---|---|---|---|---|
| -0,145450425 | 0,691290895 | TTC30B | FALSE | Downregulated |
| 0,075249419 | 0,691290895 | ZC3H7B | FALSE | Upregulated |
| 0,074777788 | 0,691290895 | ZDHHC9 | FALSE | Upregulated |
| 0,819596118 | 0,691340354 | B4GALNT4 | FALSE | Upregulated |
| 0,171178159 | 0,691340354 | BCAM | FALSE | Upregulated |
| 0,262866192 | 0,691340354 | BZRAP1-AS1 | FALSE | Upregulated |
| 0,22745605 | 0,691340354 | C5AR1 | FALSE | Upregulated |
| 0,151296319 | 0,691340354 | CCDC146 | FALSE | Upregulated |
| -0,138487744 | 0,691340354 | CLDN23 | FALSE | Downregulated |
| 0,25955837 | 0,691340354 | CTSW | FALSE | Upregulated |
| -0,147507183 | 0,691340354 | DDAH1 | FALSE | Downregulated |
| 0,356288082 | 0,691340354 | DLGAP2 | FALSE | Upregulated |
| 0,341080003 | 0,691340354 | ESPNL | FALSE | Upregulated |
| 0,289374401 | 0,691340354 | F11-AS1 | FALSE | Upregulated |
| -0,09156867 | 0,691340354 | FAM120A | FALSE | Downregulated |
| -0,404547356 | 0,691340354 | FBLN1 | FALSE | Downregulated |
| 0,112380442 | 0,691340354 | FLNB | FALSE | Upregulated |
| -0,152365227 | 0,691340354 | FSCN1 | FALSE | Downregulated |
| 0,087695611 | 0,691340354 | GATAD2A | FALSE | Upregulated |
| 0,577160946 | 0,691340354 | HAO2-IT1 | FALSE | Upregulated |
| 0,116519346 | 0,691340354 | HLA-E | FALSE | Upregulated |
| -0,149533383 | 0,691340354 | HMGB3 | FALSE | Downregulated |
| -0,198179399 | 0,691340354 | HMGCS2 | FALSE | Downregulated |
| -0,433024575 | 0,691340354 | HR | FALSE | Downregulated |
| -0,129339861 | 0,691340354 | IGFBP3 | FALSE | Downregulated |
| -0,236825809 | 0,691340354 | IGFBP7 | FALSE | Downregulated |

| | | | | |
|---|---|---|---|---|
| -0,453686272 | 0,691340354 | ITGB1BP2 | FALSE | Downregulated |
| 0,165241133 | 0,691340354 | ITM2C | FALSE | Upregulated |
| 0,692488738 | 0,691340354 | KCNIP1 | FALSE | Upregulated |
| -0,082462043 | 0,691340354 | KRIT1 | FALSE | Downregulated |
| 0,196890756 | 0,691340354 | LILRB2 | FALSE | Upregulated |
| -0,457982394 | 0,691340354 | LINC00671 | FALSE | Downregulated |
| -0,128127323 | 0,691340354 | LINC01146 | FALSE | Downregulated |
| -0,477440308 | 0,691340354 | LINC01160 | FALSE | Downregulated |
| 0,314849054 | 0,691340354 | LOC100233156 | FALSE | Upregulated |
| -0,27530941 | 0,691340354 | LOC101928403 | FALSE | Downregulated |
| -0,319579338 | 0,691340354 | LOC102723564 | FALSE | Downregulated |
| -0,099488783 | 0,691340354 | LRRC40 | FALSE | Downregulated |
| -0,424406696 | 0,691340354 | MAP1A | FALSE | Downregulated |
| 0,36531866 | 0,691340354 | MAP1LC3B2 | FALSE | Upregulated |
| -0,194510621 | 0,691340354 | MSANTD2 | FALSE | Downregulated |
| 0,091276303 | 0,691340354 | MUL1 | FALSE | Upregulated |
| -0,969213391 | 0,691340354 | MYOM3 | FALSE | Downregulated |
| -0,122556598 | 0,691340354 | NR2C1 | FALSE | Downregulated |
| -0,125362904 | 0,691340354 | PAPD7 | FALSE | Downregulated |
| 0,087114776 | 0,691340354 | PARP4 | FALSE | Upregulated |
| -0,126411774 | 0,691340354 | PGS1 | FALSE | Downregulated |
| -0,318288691 | 0,691340354 | PSMD6-AS2 | FALSE | Downregulated |
| 0,109814754 | 0,691340354 | RAB4A | FALSE | Upregulated |
| 0,265628889 | 0,691340354 | RNF24 | FALSE | Upregulated |
| -0,381019543 | 0,691340354 | ROPN1B | FALSE | Downregulated |
| 0,27424683 | 0,691340354 | RP11-346I3.4 | FALSE | Upregulated |

| | | | | |
|---|---|---|---|---|
| -0,419123955 | 0,691340354 | RP11-579D7.4 | FALSE | Downregulated |
| -0,104822932 | 0,691340354 | RPAP2 | FALSE | Downregulated |
| 0,196167588 | 0,691340354 | RRS1 | FALSE | Upregulated |
| -0,070195173 | 0,691340354 | SCO1 | FALSE | Downregulated |
| 0,132927825 | 0,691340354 | SLC44A2 | FALSE | Upregulated |
| 0,124486278 | 0,691340354 | SMG5 | FALSE | Upregulated |
| 0,078291678 | 0,691340354 | SNAPIN | FALSE | Upregulated |
| 0,341154639 | 0,691340354 | SOWAHD | FALSE | Upregulated |
| -0,289440151 | 0,691340354 | SPATA41 | FALSE | Downregulated |
| -0,544592063 | 0,691340354 | SPTB | FALSE | Downregulated |
| -0,157530396 | 0,691340354 | TBCEL | FALSE | Downregulated |
| 0,207089933 | 0,691340354 | TPK1 | FALSE | Upregulated |
| 0,264107217 | 0,691340354 | TXK | FALSE | Upregulated |
| -0,242871732 | 0,691340354 | UNC5C | FALSE | Downregulated |
| 0,078626248 | 0,691340354 | UQCRC1 | FALSE | Upregulated |
| 0,179606206 | 0,691340354 | ZC3H3 | FALSE | Upregulated |
| -0,134832032 | 0,691340354 | ZFAND4 | FALSE | Downregulated |
| -0,081967989 | 0,691340354 | ZNF131 | FALSE | Downregulated |
| -0,093290021 | 0,691340354 | ZNF281 | FALSE | Downregulated |
| -0,128065476 | 0,691409395 | AKAP9 | FALSE | Downregulated |
| 0,18080528 | 0,691580111 | AKR1C6P | FALSE | Upregulated |
| -0,094748547 | 0,691580111 | SLC7A6OS | FALSE | Downregulated |
| -0,085876582 | 0,691580111 | SNRPG | FALSE | Downregulated |
| -0,085961017 | 0,691580111 | ZNF770 | FALSE | Downregulated |
| -0,180366187 | 0,691659757 | ALDOB | FALSE | Downregulated |
| -0,526661599 | 0,691659757 | C5orf27 | FALSE | Downregulated |

| | | | | |
|---|---|---|---|---|
| 0,226771877 | 0,691659757 | CCDC109B | FALSE | Upregulated |
| 0,10258752 | 0,691659757 | COX15 | FALSE | Upregulated |
| 0,140792176 | 0,691659757 | FEM1A | FALSE | Upregulated |
| -0,204887813 | 0,691659757 | FILIP1 | FALSE | Downregulated |
| 0,221916177 | 0,691659757 | HIST1H2BC | FALSE | Upregulated |
| -0,476208845 | 0,691659757 | KCNRG | FALSE | Downregulated |
| 0,144042288 | 0,691659757 | MAP3K14 | FALSE | Upregulated |
| 0,22359626 | 0,691659757 | RP11-521B24.3 | FALSE | Upregulated |
| -0,10802014 | 0,691659757 | TESK2 | FALSE | Downregulated |
| -0,172109621 | 0,691659757 | TMEM220-AS1 | FALSE | Downregulated |
| -0,879701906 | 0,691685698 | MT1B | FALSE | Downregulated |
| -0,246638106 | 0,691685698 | NPIPA5 | FALSE | Downregulated |
| -0,106588595 | 0,691685698 | ZNF184 | FALSE | Downregulated |
| 0,258042277 | 0,69210299 | MRGPRF | FALSE | Upregulated |
| 0,468174868 | 0,692161353 | LOC102724040 | FALSE | Upregulated |
| -0,122211231 | 0,692161353 | TCHP | FALSE | Downregulated |
| -0,113099459 | 0,692368157 | RTF1 | FALSE | Downregulated |
| 0,175794424 | 0,692463978 | ZNF506 | FALSE | Upregulated |
| -0,121845476 | 0,692529291 | NUFIP1 | FALSE | Downregulated |
| -0,225413193 | 0,692582887 | RPL13AP5 | FALSE | Downregulated |
| 0,188833008 | 0,692734657 | C1orf85 | FALSE | Upregulated |
| 0,489450077 | 0,692734657 | CLEC4F | FALSE | Upregulated |
| -0,154001615 | 0,692734657 | DDR2 | FALSE | Downregulated |
| 0,683873486 | 0,692734657 | LOC101928583 | FALSE | Upregulated |
| 0,751458718 | 0,692746359 | CENPM | FALSE | Upregulated |
| -0,180461885 | 0,692746359 | PTPRB | FALSE | Downregulated |

| | | | | |
|---|---|---|---|---|
| -0,075940295 | 0,692746359 | UHMK1 | FALSE | Downregulated |
| 0,141763366 | 0,693038022 | AAMDC | FALSE | Upregulated |
| 0,103493247 | 0,693038022 | ABHD4 | FALSE | Upregulated |
| 0,08869785 | 0,693038022 | HFE | FALSE | Upregulated |
| 0,161637981 | 0,693038022 | LOC102723990 | FALSE | Upregulated |
| -0,194712982 | 0,693038022 | NEFH | FALSE | Downregulated |
| 0,319729272 | 0,693038022 | PSD | FALSE | Upregulated |
| -0,134527429 | 0,693038022 | PXMP2 | FALSE | Downregulated |
| -0,343870178 | 0,693038022 | SCN2A | FALSE | Downregulated |
| -0,149089319 | 0,693038022 | TTC14 | FALSE | Downregulated |
| 0,156400862 | 0,693083392 | ADAP2 | FALSE | Upregulated |
| 0,119177599 | 0,693083392 | ANKRD54 | FALSE | Upregulated |
| -0,081597071 | 0,693083392 | ATG4B | FALSE | Downregulated |
| -0,223131077 | 0,693083392 | CCDC88C | FALSE | Downregulated |
| 0,297377704 | 0,693083392 | CELSR2 | FALSE | Upregulated |
| -0,302958541 | 0,693083392 | ETV2 | FALSE | Downregulated |
| 0,223066078 | 0,693083392 | FAM71E1 | FALSE | Upregulated |
| 0,149492842 | 0,693083392 | FAS | FALSE | Upregulated |
| -0,225745502 | 0,693083392 | GSTA1 | FALSE | Downregulated |
| 0,669089724 | 0,693083392 | LAIR2 | FALSE | Upregulated |
| 0,278185192 | 0,693083392 | LINC00174 | FALSE | Upregulated |
| 1,219384892 | 0,693083392 | LOC101927333 | FALSE | Upregulated |
| -0,082800097 | 0,693083392 | MPP5 | FALSE | Downregulated |
| 0,115689195 | 0,693083392 | PACRGL | FALSE | Upregulated |
| 0,606451764 | 0,693083392 | RNF138P1 | FALSE | Upregulated |
| -0,246757211 | 0,693083392 | RP11-695J4.2 | FALSE | Downregulated |

| | | | | |
|---|---|---|---|---|
| -0,11061786 | 0,693083392 | RPL23A | FALSE | Downregulated |
| 0,416874544 | 0,693083392 | S1PR4 | FALSE | Upregulated |
| -0,174299747 | 0,693083392 | SLC7A2 | FALSE | Downregulated |
| -0,128084305 | 0,693083392 | TBC1D24 | FALSE | Downregulated |
| -0,198368824 | 0,693083392 | TM7SF2 | FALSE | Downregulated |
| 0,13963961 | 0,693083392 | TTL | FALSE | Upregulated |
| 0,224529726 | 0,693083392 | WDHD1 | FALSE | Upregulated |
| -0,195514804 | 0,693083392 | WHAMMP1 | FALSE | Downregulated |
| -0,308487844 | 0,693083392 | ZDHHC11B | FALSE | Downregulated |
| 0,532282027 | 0,693443992 | ESPL1 | FALSE | Upregulated |
| 0,112783013 | 0,693535464 | SPRY2 | FALSE | Upregulated |
| 0,355610911 | 0,693656221 | FAM83B | FALSE | Upregulated |
| 0,060730188 | 0,693656221 | H2AFV | FALSE | Upregulated |
| -0,397449733 | 0,693656221 | RNF186 | FALSE | Downregulated |
| 0,414210935 | 0,693676512 | LOC388210 | FALSE | Upregulated |
| -0,11402133 | 0,69378687 | C1RL | FALSE | Downregulated |
| -0,151040143 | 0,694115101 | ADH6 | FALSE | Downregulated |
| -0,36328559 | 0,694143767 | DPH6-AS1 | FALSE | Downregulated |
| -0,152233292 | 0,694190596 | HOMER2 | FALSE | Downregulated |
| 0,111725261 | 0,69430237 | GRK5 | FALSE | Upregulated |
| 0,330309801 | 0,69430237 | LOC100507520 | FALSE | Upregulated |
| 0,589708656 | 0,69430237 | LOC102724336 | FALSE | Upregulated |
| 0,147397031 | 0,69430237 | ZNF565 | FALSE | Upregulated |
| 0,119647363 | 0,694674425 | ACYP2 | FALSE | Upregulated |
| 0,152482101 | 0,694674425 | YPEL1 | FALSE | Upregulated |
| -1,12222202 | 0,694840636 | LOC101928921 | FALSE | Downregulated |

| | | | | |
|---|---|---|---|---|
| -0,099898344 | 0,694994884 | TARSL2 | FALSE | Downregulated |
| -0,131110113 | 0,695016519 | C4orf19 | FALSE | Downregulated |
| -0,072433416 | 0,695343995 | RABGGTB | FALSE | Downregulated |
| 0,73902959 | 0,695382411 | POTEE | FALSE | Upregulated |
| 0,334260204 | 0,695586052 | C17orf51 | FALSE | Upregulated |
| 0,338124717 | 0,695816554 | EMILIN2 | FALSE | Upregulated |
| 0,102811334 | 0,696203606 | UCK2 | FALSE | Upregulated |
| -0,525332268 | 0,696242502 | CYP7A1 | FALSE | Downregulated |
| 0,403492274 | 0,696242502 | TICAM2 | FALSE | Upregulated |
| -0,071696325 | 0,696518058 | CCDC9 | FALSE | Downregulated |
| 0,148332414 | 0,696518058 | FUK | FALSE | Upregulated |
| 0,423090614 | 0,696518058 | ZNF296 | FALSE | Upregulated |
| 0,122192897 | 0,696551284 | NUGGC | FALSE | Upregulated |
| 0,221814406 | 0,69667944 | CXCR2P1 | FALSE | Upregulated |
| -0,312698849 | 0,69667944 | LOC101926931 | FALSE | Downregulated |
| 0,139202131 | 0,69667944 | OSER1-AS1 | FALSE | Upregulated |
| 0,114346175 | 0,697071902 | DENND4B | FALSE | Upregulated |
| -0,077186482 | 0,697071902 | GID8 | FALSE | Downregulated |
| 0,322452507 | 0,697071902 | KCNN3 | FALSE | Upregulated |
| -0,175623075 | 0,697071902 | NEIL1 | FALSE | Downregulated |
| 0,168236139 | 0,697071902 | PGBD5 | FALSE | Upregulated |
| 0,114393989 | 0,697071902 | SCOC | FALSE | Upregulated |
| 0,189048327 | 0,697071902 | SLC4A7 | FALSE | Upregulated |
| 0,094621381 | 0,697071902 | STK24 | FALSE | Upregulated |
| -0,086572066 | 0,697071902 | ZNF318 | FALSE | Downregulated |
| 0,166563954 | 0,697071902 | ZNF383 | FALSE | Upregulated |

| | | | | |
|---|---|---|---|---|
| -0,068512637 | 0,697527696 | GNAS | FALSE | Downregulated |
| -0,238991245 | 0,697527696 | LOC101927506 | FALSE | Downregulated |
| -0,279094654 | 0,698108395 | CLDN9 | FALSE | Downregulated |
| 0,357478144 | 0,698132496 | ORC6 | FALSE | Upregulated |
| -0,312084431 | 0,698205199 | MIR6723 | FALSE | Downregulated |
| -0,348396728 | 0,698314269 | GNRHR2 | FALSE | Downregulated |
| -0,10430833 | 0,698362053 | CHD2 | FALSE | Downregulated |
| 0,059143378 | 0,698531269 | EXOC7 | FALSE | Upregulated |
| -0,135634146 | 0,698760468 | ABHD14B | FALSE | Downregulated |
| 0,145581855 | 0,698760468 | ARL6 | FALSE | Upregulated |
| -0,086197961 | 0,698760468 | ATG14 | FALSE | Downregulated |
| 0,052205085 | 0,698760468 | BAG5 | FALSE | Upregulated |
| -0,087104329 | 0,698760468 | BRPF3 | FALSE | Downregulated |
| -0,400504731 | 0,698760468 | C15orf43 | FALSE | Downregulated |
| 0,096693537 | 0,698760468 | EDEM2 | FALSE | Upregulated |
| 0,427185542 | 0,698760468 | FCGR1A | FALSE | Upregulated |
| 0,276393785 | 0,698760468 | GBP5 | FALSE | Upregulated |
| 0,257887088 | 0,698760468 | GRAMD1B | FALSE | Upregulated |
| -0,149307833 | 0,698760468 | HSP90B1 | FALSE | Downregulated |
| 0,139507477 | 0,698760468 | ITGA9 | FALSE | Upregulated |
| 0,333840865 | 0,698760468 | LINC00482 | FALSE | Upregulated |
| 0,870064868 | 0,698760468 | LINC00891 | FALSE | Upregulated |
| 0,154877777 | 0,698760468 | LINC00963 | FALSE | Upregulated |
| -0,73728402 | 0,698760468 | LOC100132741 | FALSE | Downregulated |
| 0,168183653 | 0,698760468 | LRWD1 | FALSE | Upregulated |
| 0,273049069 | 0,698760468 | MCTP2 | FALSE | Upregulated |

| | | | | |
|---|---|---|---|---|
| -0,340749216 | 0,698760468 | MVD | FALSE | Downregulated |
| 0,622078858 | 0,698760468 | OPCML | FALSE | Upregulated |
| -0,139992209 | 0,698760468 | PAXBP1 | FALSE | Downregulated |
| -0,081693582 | 0,698760468 | PAXIP1 | FALSE | Downregulated |
| 0,49961045 | 0,698760468 | PLCXD2 | FALSE | Upregulated |
| 0,30888614 | 0,698760468 | PLCXD3 | FALSE | Upregulated |
| -0,142815093 | 0,698760468 | PLD1 | FALSE | Downregulated |
| -0,110748236 | 0,698760468 | PQLC1 | FALSE | Downregulated |
| -0,593161814 | 0,698760468 | RBM12B-AS1 | FALSE | Downregulated |
| 0,108980755 | 0,698760468 | RC3H2 | FALSE | Upregulated |
| 0,166797608 | 0,698760468 | RPUSD1 | FALSE | Upregulated |
| -0,169339764 | 0,698760468 | SEMA6D | FALSE | Downregulated |
| 0,266733339 | 0,698760468 | SGK223 | FALSE | Upregulated |
| 0,362126263 | 0,698760468 | SIRPG | FALSE | Upregulated |
| -0,160761739 | 0,698760468 | SRRM1 | FALSE | Downregulated |
| -0,292367276 | 0,698760468 | TPI1P2 | FALSE | Downregulated |
| -0,221686304 | 0,698760468 | ZC2HC1A | FALSE | Downregulated |
| -0,156846716 | 0,698760468 | ZNF141 | FALSE | Downregulated |
| -0,157562178 | 0,699024131 | WHAMMP3 | FALSE | Downregulated |
| 0,117550311 | 0,699079106 | ANKRD16 | FALSE | Upregulated |
| -0,528175181 | 0,699079106 | COL28A1 | FALSE | Downregulated |
| -0,172130401 | 0,699079106 | HECW2 | FALSE | Downregulated |
| 0,141529714 | 0,699079106 | IFT172 | FALSE | Upregulated |
| 0,25248273 | 0,699079106 | LINC00924 | FALSE | Upregulated |
| -0,387878234 | 0,699079106 | LOC102724660 | FALSE | Downregulated |
| -0,103588964 | 0,699079106 | NR1H4 | FALSE | Downregulated |

| | | | | |
|---|---|---|---|---|
| -0,832171934 | 0,699079106 | RIMBP2 | FALSE | Downregulated |
| -0,094893997 | 0,699079106 | RMI1 | FALSE | Downregulated |
| -0,062917593 | 0,699079106 | SMNDC1 | FALSE | Downregulated |
| -0,138191538 | 0,699079106 | TNRC6C | FALSE | Downregulated |
| -0,115621433 | 0,699268679 | BLVRB | FALSE | Downregulated |
| 0,118523661 | 0,699268679 | C1QTNF1 | FALSE | Upregulated |
| -0,07757234 | 0,699268679 | C2orf47 | FALSE | Downregulated |
| 0,10130945 | 0,699268679 | HMGXB3 | FALSE | Upregulated |
| 0,112693558 | 0,699268679 | NACC1 | FALSE | Upregulated |
| -0,16229885 | 0,699268679 | PLXDC2 | FALSE | Downregulated |
| 0,201290369 | 0,699274878 | PRR5L | FALSE | Upregulated |
| -0,066068752 | 0,699274878 | RAB22A | FALSE | Downregulated |
| -0,352076858 | 0,699307627 | LRRC49 | FALSE | Downregulated |
| -0,132046886 | 0,69951955 | CCNK | FALSE | Downregulated |
| -0,130646234 | 0,699790664 | DPP9 | FALSE | Downregulated |
| -0,948395247 | 0,699790664 | JPH2 | FALSE | Downregulated |
| 0,061386308 | 0,700125883 | RABL3 | FALSE | Upregulated |
| 0,111576503 | 0,700183458 | CDCA7L | FALSE | Upregulated |
| -0,381720482 | 0,700183458 | SOX2-OT | FALSE | Downregulated |
| -0,118020668 | 0,700538523 | DENND4A | FALSE | Downregulated |
| -0,156478181 | 0,700538523 | KLF15 | FALSE | Downregulated |
| -0,617054993 | 0,700538523 | LOC644554 | FALSE | Downregulated |
| -0,116919748 | 0,700538523 | TINAGL1 | FALSE | Downregulated |
| -0,094927615 | 0,700637497 | ARHGEF9 | FALSE | Downregulated |
| -0,058019363 | 0,700637497 | BRAP | FALSE | Downregulated |
| 0,451315963 | 0,700637497 | CEACAM22P | FALSE | Upregulated |

| | | | | |
|---|---|---|---|---|
| 0,151241356 | 0,700637497 | HAUS4 | FALSE | Upregulated |
| 0,09017903 | 0,700637497 | SLC43A1 | FALSE | Upregulated |
| -0,134497162 | 0,700637497 | SNAPC5 | FALSE | Downregulated |
| -0,092840751 | 0,700637497 | TGFBR2 | FALSE | Downregulated |
| 0,058297038 | 0,700637497 | ZDHHC5 | FALSE | Upregulated |
| -0,083144034 | 0,700695506 | MRP63 | FALSE | Downregulated |
| 0,251053257 | 0,700758992 | CCT6P1 | FALSE | Upregulated |
| 0,156262243 | 0,701407354 | QSOX1 | FALSE | Upregulated |
| -0,241993567 | 0,701479233 | LOC100505902 | FALSE | Downregulated |
| -0,404790674 | 0,70154554 | LOC102724316 | FALSE | Downregulated |
| 0,069820978 | 0,701615418 | WIPF2 | FALSE | Upregulated |
| 0,351007502 | 0,701708749 | CD24 | FALSE | Upregulated |
| 0,486152943 | 0,701708749 | ZNF114 | FALSE | Upregulated |
| 0,090292633 | 0,70237095 | ABHD16A | FALSE | Upregulated |
| 0,131748654 | 0,702561035 | ABCB11 | FALSE | Upregulated |
| 0,565203171 | 0,702561035 | C19orf84 | FALSE | Upregulated |
| 0,071162164 | 0,702561035 | COG7 | FALSE | Upregulated |
| -0,474016791 | 0,702561035 | GUCY2C | FALSE | Downregulated |
| 0,293846339 | 0,702561035 | HOPX | FALSE | Upregulated |
| 0,319541765 | 0,702561035 | IL12RB2 | FALSE | Upregulated |
| -0,170948101 | 0,702561035 | IL7 | FALSE | Downregulated |
| -0,072090364 | 0,702561035 | KIDINS220 | FALSE | Downregulated |
| 0,110655097 | 0,702561035 | KLHL21 | FALSE | Upregulated |
| -0,431021445 | 0,702561035 | LOC101928882 | FALSE | Downregulated |
| 0,207641027 | 0,702561035 | NEDD9 | FALSE | Upregulated |
| -1,419354622 | 0,702561035 | PPP1R14C | FALSE | Downregulated |

| | | | | |
|---|---|---|---|---|
| 0,114963347 | 0,702561035 | RAP2A | FALSE | Upregulated |
| 0,542096334 | 0,702561035 | RASL12 | FALSE | Upregulated |
| -0,095725566 | 0,702561035 | RIF1 | FALSE | Downregulated |
| -0,53150259 | 0,702561035 | RNA5-8S5 | FALSE | Downregulated |
| 0,089818318 | 0,702561035 | RPA2 | FALSE | Upregulated |
| 0,153864287 | 0,702561035 | RREB1 | FALSE | Upregulated |
| 0,235196981 | 0,702561035 | RTEL1 | FALSE | Upregulated |
| -0,053079897 | 0,702561035 | SETD3 | FALSE | Downregulated |
| -0,113201497 | 0,702561035 | WDR5B | FALSE | Downregulated |
| 0,086525333 | 0,702620356 | EPG5 | FALSE | Upregulated |
| 0,672783265 | 0,702754213 | CHMP1B2P | FALSE | Upregulated |
| -0,227171146 | 0,702754213 | ORM1 | FALSE | Downregulated |
| 0,231244471 | 0,702754213 | PALM2-AKAP2 | FALSE | Upregulated |
| 0,297546166 | 0,702754213 | RNF157 | FALSE | Upregulated |
| 0,360463193 | 0,702754213 | RP11-500B12.1 | FALSE | Upregulated |
| 0,183933269 | 0,702781933 | APOLD1 | FALSE | Upregulated |
| -0,274673269 | 0,702781933 | RP11-75C9.1 | FALSE | Downregulated |
| -0,096828168 | 0,702815033 | C9orf85 | FALSE | Downregulated |
| -0,087330242 | 0,702815033 | CDC37L1 | FALSE | Downregulated |
| -0,19094778 | 0,702815033 | RARA-AS1 | FALSE | Downregulated |
| 0,095609927 | 0,703310555 | BLMH | FALSE | Upregulated |
| 0,25902981 | 0,703310555 | KCNJ2 | FALSE | Upregulated |
| -0,087113268 | 0,703310555 | PGRMC2 | FALSE | Downregulated |
| 0,198031468 | 0,703310555 | RAI2 | FALSE | Upregulated |
| -0,208312642 | 0,703310555 | SLC22A25 | FALSE | Downregulated |
| -0,154268057 | 0,703310555 | ZNF846 | FALSE | Downregulated |

| | | | | |
|---|---|---|---|---|
| 0,101396804 | 0,703476627 | KCTD2 | FALSE | Upregulated |
| 0,522035696 | 0,703476627 | MYOCD | FALSE | Upregulated |
| -0,127337786 | 0,703476627 | POLE3 | FALSE | Downregulated |
| 0,382132348 | 0,703476627 | SLC22A15 | FALSE | Upregulated |
| -0,288572118 | 0,703476627 | SLCO1B7 | FALSE | Downregulated |
| 0,069546209 | 0,703476627 | SYNCRIP | FALSE | Upregulated |
| -0,105883624 | 0,703476627 | TMX4 | FALSE | Downregulated |
| 0,143607448 | 0,703476627 | ZNF593 | FALSE | Upregulated |
| 0,093881461 | 0,703634367 | CTSZ | FALSE | Upregulated |
| 0,560019887 | 0,703634367 | LOC102723652 | FALSE | Upregulated |
| -0,102015782 | 0,703634367 | SS18L1 | FALSE | Downregulated |
| 0,083357541 | 0,703690489 | PLCL2 | FALSE | Upregulated |
| -0,095642711 | 0,703700113 | APOB | FALSE | Downregulated |
| -0,124033089 | 0,703700113 | INO80D | FALSE | Downregulated |
| -0,080838488 | 0,704568371 | ANKRD28 | FALSE | Downregulated |
| -0,15216942 | 0,704625506 | ACP6 | FALSE | Downregulated |
| 0,149415706 | 0,704625506 | TNFAIP3 | FALSE | Upregulated |
| 0,091958519 | 0,704625506 | TSC22D2 | FALSE | Upregulated |
| -0,501583667 | 0,704934194 | CACNB4 | FALSE | Downregulated |
| 0,092171037 | 0,704940858 | SHPK | FALSE | Upregulated |
| 0,088318927 | 0,705038695 | PPP2R4 | FALSE | Upregulated |
| -0,328147434 | 0,705340022 | C22orf34 | FALSE | Downregulated |
| 0,164670566 | 0,705340022 | PTGFRN | FALSE | Upregulated |
| -1,145500257 | 0,705340022 | RYR3 | FALSE | Downregulated |
| -0,190407426 | 0,705340022 | TMEM238 | FALSE | Downregulated |
| -1,295655618 | 0,705727333 | HOXC8 | FALSE | Downregulated |

| | | | | |
|---|---|---|---|---|
| -0,159962158 | 0,705878927 | ARMCX5-GPRASP2 | FALSE | Downregulated |
| -0,084595462 | 0,705878927 | DYNLRB1 | FALSE | Downregulated |
| 0,350555803 | 0,705878927 | PLAU | FALSE | Upregulated |
| -0,144805697 | 0,705878927 | TMEM42 | FALSE | Downregulated |
| -0,185454111 | 0,705940642 | FAM65A | FALSE | Downregulated |
| 0,165557478 | 0,705940642 | KSR1 | FALSE | Upregulated |
| -0,259331046 | 0,705940642 | ZNF818P | FALSE | Downregulated |
| 0,288131136 | 0,706064835 | CD3G | FALSE | Upregulated |
| -0,184743741 | 0,706064835 | CD4 | FALSE | Downregulated |
| -0,224819751 | 0,706064835 | CD99P1 | FALSE | Downregulated |
| -0,135516952 | 0,706064835 | EIF3A | FALSE | Downregulated |
| -0,14346862 | 0,706064835 | EIF3J | FALSE | Downregulated |
| 0,137628622 | 0,706064835 | XRCC4 | FALSE | Upregulated |
| 0,275765795 | 0,706103576 | ABLIM2 | FALSE | Upregulated |
| -0,52827634 | 0,706103576 | HP09025 | FALSE | Downregulated |
| -0,315133713 | 0,706103576 | KLRAP1 | FALSE | Downregulated |
| 0,218529478 | 0,706103576 | TGFB1I1 | FALSE | Upregulated |
| -0,110897245 | 0,706103576 | ZNF25 | FALSE | Downregulated |
| 0,191292228 | 0,706103576 | ZNF462 | FALSE | Upregulated |
| 0,178690468 | 0,70613029 | TMEM56-RWDD3 | FALSE | Upregulated |
| -0,54389642 | 0,706530046 | ACTG1P4 | FALSE | Downregulated |
| -0,145740496 | 0,706530046 | CCDC167 | FALSE | Downregulated |
| 0,239682092 | 0,706530046 | HYOU1 | FALSE | Upregulated |
| 0,102362722 | 0,706530046 | JKAMP | FALSE | Upregulated |
| -0,465338648 | 0,706530046 | METTL21EP | FALSE | Downregulated |

| | | | | |
|---|---|---|---|---|
| -0,123309271 | 0,706530046 | NGFRAP1 | FALSE | Downregulated |
| -0,27561423 | 0,706781488 | BLM | FALSE | Downregulated |
| 0,190696104 | 0,706781488 | CDCA4 | FALSE | Upregulated |
| -0,232804156 | 0,706838567 | LOC102724261 | FALSE | Downregulated |
| 0,503631915 | 0,706838567 | UGT2B28 | FALSE | Upregulated |
| -0,231597249 | 0,706957458 | CHSY3 | FALSE | Downregulated |
| -0,113666987 | 0,706957458 | DNAJC27 | FALSE | Downregulated |
| 0,162734957 | 0,707003784 | P4HA2 | FALSE | Upregulated |
| -0,15414693 | 0,707038251 | TAF13 | FALSE | Downregulated |
| -0,151699333 | 0,707046686 | CLK4 | FALSE | Downregulated |
| 0,386717277 | 0,707291297 | COLCA2 | FALSE | Upregulated |
| 0,122224661 | 0,707291297 | ORAI1 | FALSE | Upregulated |
| -0,094960812 | 0,70752066 | CPNE1 | FALSE | Downregulated |
| -0,077726387 | 0,707526668 | NDFIP1 | FALSE | Downregulated |
| -0,060730423 | 0,707756474 | CTTN | FALSE | Downregulated |
| -0,342904935 | 0,707756474 | HCG4 | FALSE | Downregulated |
| -0,105840774 | 0,707901409 | PRDM2 | FALSE | Downregulated |
| 0,283331033 | 0,707934163 | AOC2 | FALSE | Upregulated |
| -0,222137813 | 0,708176997 | HBCBP | FALSE | Downregulated |
| -0,102486469 | 0,708228874 | HIPK2 | FALSE | Downregulated |
| 0,094824814 | 0,708228874 | ZFP41 | FALSE | Upregulated |
| -0,299856034 | 0,708369989 | KCNN2 | FALSE | Downregulated |
| 0,118848114 | 0,708417564 | AHSG | FALSE | Upregulated |
| -0,110752272 | 0,708417564 | CYB561D1 | FALSE | Downregulated |
| -0,574533234 | 0,708417564 | HIST1H4J | FALSE | Downregulated |
| -0,074972192 | 0,708417564 | PEX19 | FALSE | Downregulated |

| | | | | |
|---|---|---|---|---|
| -0,09490379 | 0,708417564 | SETD6 | FALSE | Downregulated |
| 0,493728157 | 0,708484689 | AC058791.1 | FALSE | Upregulated |
| 0,185415125 | 0,708484689 | AXIN2 | FALSE | Upregulated |
| -0,104641356 | 0,708484689 | CCS | FALSE | Downregulated |
| 0,140417717 | 0,708484689 | CDK18 | FALSE | Upregulated |
| 0,068266737 | 0,708484689 | CNOT11 | FALSE | Upregulated |
| -0,105846647 | 0,708484689 | DLC1 | FALSE | Downregulated |
| 0,208396653 | 0,708484689 | KIF17 | FALSE | Upregulated |
| -0,406934135 | 0,708484689 | LOC102723622 | FALSE | Downregulated |
| -0,158375085 | 0,708484689 | LY6E | FALSE | Downregulated |
| -0,184969845 | 0,708484689 | RAPH1 | FALSE | Downregulated |
| -0,162400581 | 0,708484689 | RBM25 | FALSE | Downregulated |
| 0,063859731 | 0,708484689 | SETD1A | FALSE | Upregulated |
| 0,396081883 | 0,708484689 | TUBB2B | FALSE | Upregulated |
| 0,155717385 | 0,708484689 | WIPI1 | FALSE | Upregulated |
| -0,083618019 | 0,708484689 | YARS2 | FALSE | Downregulated |
| 0,096447938 | 0,708498591 | UNK | FALSE | Upregulated |
| 0,404905618 | 0,708674104 | SCGN | FALSE | Upregulated |
| 0,162581508 | 0,708674104 | TCF7 | FALSE | Upregulated |
| -0,459384892 | 0,708745059 | GOLGA6D | FALSE | Downregulated |
| 0,210061092 | 0,708745059 | ROM1 | FALSE | Upregulated |
| -0,108970606 | 0,708772701 | TSPAN3 | FALSE | Downregulated |
| -0,182237359 | 0,70900608 | GPR125 | FALSE | Downregulated |
| 0,115035216 | 0,709190855 | GTPBP2 | FALSE | Upregulated |
| 0,166890599 | 0,709849106 | HTRA1 | FALSE | Upregulated |
| 0,095835031 | 0,709849106 | RTKN | FALSE | Upregulated |

| | | | | |
|---|---|---|---|---|
| -0,171635391 | 0,70989902 | MRPS31P5 | FALSE | Downregulated |
| -0,369561874 | 0,70989902 | MT1M | FALSE | Downregulated |
| 0,101237045 | 0,70989902 | PLXNB1 | FALSE | Upregulated |
| 0,587929886 | 0,70989902 | STYK1 | FALSE | Upregulated |
| 0,196441557 | 0,70989902 | TRERF1 | FALSE | Upregulated |
| 0,086460823 | 0,710024551 | ANAPC7 | FALSE | Upregulated |
| 0,134954664 | 0,710024551 | C20orf24 | FALSE | Upregulated |
| 0,458821793 | 0,710024551 | CDCA7 | FALSE | Upregulated |
| -0,742272408 | 0,710024551 | CES1P1 | FALSE | Downregulated |
| -0,160652313 | 0,710024551 | HPR | FALSE | Downregulated |
| 0,270184481 | 0,710024551 | INHBB | FALSE | Upregulated |
| -0,203508369 | 0,710024551 | NAP1L3 | FALSE | Downregulated |
| 0,189377463 | 0,710024551 | NBEAL2 | FALSE | Upregulated |
| -0,474715595 | 0,710024551 | SNORD104 | FALSE | Downregulated |
| -0,116080285 | 0,710024551 | UGP2 | FALSE | Downregulated |
| 0,046952042 | 0,710024551 | ZC3H7A | FALSE | Upregulated |
| 0,176549226 | 0,710024551 | ZNF620 | FALSE | Upregulated |
| -0,128249261 | 0,710201663 | DDX58 | FALSE | Downregulated |
| -0,151884781 | 0,710386626 | ADCY1 | FALSE | Downregulated |
| 0,079202435 | 0,710386626 | ARAF | FALSE | Upregulated |
| -0,427853189 | 0,710386626 | ENPP6 | FALSE | Downregulated |
| 0,153618537 | 0,710386626 | GMPPA | FALSE | Upregulated |
| -0,351424623 | 0,710386626 | LOC102724483 | FALSE | Downregulated |
| -0,500180987 | 0,710386626 | NCAM2 | FALSE | Downregulated |
| -0,216498123 | 0,710386626 | RPL21P28 | FALSE | Downregulated |
| 0,403585462 | 0,71043162 | NDUFB2-AS1 | FALSE | Upregulated |

| | | | | |
|---|---|---|---|---|
| 0,072003117 | 0,71043162 | VAMP3 | FALSE | Upregulated |
| 0,242278818 | 0,710685037 | LOC401320 | FALSE | Upregulated |
| -0,124007601 | 0,710744034 | NAAA | FALSE | Downregulated |
| 0,059058377 | 0,710765521 | DDX1 | FALSE | Upregulated |
| 0,387785517 | 0,710765521 | GPAT2 | FALSE | Upregulated |
| -0,502526399 | 0,710765521 | LOC100130539 | FALSE | Downregulated |
| -0,31007802 | 0,710765521 | LOC100507395 | FALSE | Downregulated |
| -0,298426891 | 0,710765521 | MLLT4-AS1 | FALSE | Downregulated |
| 0,116331591 | 0,710765521 | QTRT1 | FALSE | Upregulated |
| 0,934851219 | 0,710765521 | SCGB2B3P | FALSE | Upregulated |
| -0,28626441 | 0,710765521 | SIRPB1 | FALSE | Downregulated |
| 0,143481796 | 0,710765521 | SLC33A1 | FALSE | Upregulated |
| 0,239394361 | 0,710765521 | TMEM232 | FALSE | Upregulated |
| -0,073034992 | 0,710765521 | TRUB1 | FALSE | Downregulated |
| -0,121089999 | 0,710769357 | LMO2 | FALSE | Downregulated |
| -0,39067039 | 0,71080067 | SERF1A | FALSE | Downregulated |
| -0,132222395 | 0,710988208 | FAM13A | FALSE | Downregulated |
| 0,51734086 | 0,710988208 | ZNF843 | FALSE | Upregulated |
| -0,586950346 | 0,71105551 | LOC100630923 | FALSE | Downregulated |
| 0,096411251 | 0,71105551 | SPR | FALSE | Upregulated |
| -0,103875043 | 0,711114885 | WDR59 | FALSE | Downregulated |
| 0,136927692 | 0,711137961 | CYB561D2 | FALSE | Upregulated |
| -0,184514315 | 0,711137961 | TICAM1 | FALSE | Downregulated |
| -0,137719195 | 0,711252027 | PDCD2L | FALSE | Downregulated |
| -0,279544463 | 0,711252027 | RP6-206I17.2 | FALSE | Downregulated |
| 0,306179842 | 0,711252027 | TNC | FALSE | Upregulated |

| | | | | |
|---|---|---|---|---|
| -0,104721336 | 0,711320186 | ADAL | FALSE | Downregulated |
| -0,081878271 | 0,711320186 | CREG1 | FALSE | Downregulated |
| -0,204038142 | 0,711320186 | IL1RL2 | FALSE | Downregulated |
| 0,334992777 | 0,711320186 | LINC00152 | FALSE | Upregulated |
| -0,537539013 | 0,711320186 | PRAMENP | FALSE | Downregulated |
| 0,136301906 | 0,711320186 | RPS19BP1 | FALSE | Upregulated |
| -0,177505903 | 0,711320186 | SSH2 | FALSE | Downregulated |
| -0,221149968 | 0,711341999 | SH3RF2 | FALSE | Downregulated |
| -0,541970111 | 0,711353403 | USP44 | FALSE | Downregulated |
| 0,10419442 | 0,711404633 | C6orf47 | FALSE | Upregulated |
| 0,582632174 | 0,711412832 | PF4V1 | FALSE | Upregulated |
| 0,077929629 | 0,711500537 | ANKH | FALSE | Upregulated |
| 0,406741669 | 0,711500537 | CDT1 | FALSE | Upregulated |
| -0,07917177 | 0,711500537 | PSMD11 | FALSE | Downregulated |
| 0,440978663 | 0,71160235 | LOC101927932 | FALSE | Upregulated |
| 0,099512135 | 0,711609951 | ACOX3 | FALSE | Upregulated |
| -0,139370932 | 0,711609951 | ATP6V0E2 | FALSE | Downregulated |
| -0,114862843 | 0,711609951 | BRD4 | FALSE | Downregulated |
| 0,59585728 | 0,711609951 | C21orf88 | FALSE | Upregulated |
| 0,078620244 | 0,711609951 | CUL9 | FALSE | Upregulated |
| 0,074922778 | 0,711609951 | FAM53C | FALSE | Upregulated |
| -0,329986457 | 0,711609951 | ICA1 | FALSE | Downregulated |
| 0,244998778 | 0,711609951 | INPP4B | FALSE | Upregulated |
| -0,558086302 | 0,711609951 | KIAA1024 | FALSE | Downregulated |
| 0,640910023 | 0,711609951 | LINC00539 | FALSE | Upregulated |
| 0,467247304 | 0,711609951 | LOC100506411 | FALSE | Upregulated |

| | | | | |
|---|---|---|---|---|
| 0,076508374 | 0,711609951 | SLC4A2 | FALSE | Upregulated |
| 0,226294257 | 0,711609951 | TLR5 | FALSE | Upregulated |
| -0,139561649 | 0,711621613 | SORD | FALSE | Downregulated |
| 0,106000735 | 0,711621613 | VPS13B | FALSE | Upregulated |
| 0,102920237 | 0,711663368 | LOC100294145 | FALSE | Upregulated |
| 0,076370657 | 0,712001105 | MCMBP | FALSE | Upregulated |
| -0,091288927 | 0,712109129 | SOCS5 | FALSE | Downregulated |
| 0,14718058 | 0,712243008 | RRP12 | FALSE | Upregulated |
| 0,440916732 | 0,712413321 | RHBDL1 | FALSE | Upregulated |
| 0,32235461 | 0,712470584 | FSD1L | FALSE | Upregulated |
| 0,314751336 | 0,712470584 | LOC101928673 | FALSE | Upregulated |
| 0,133488882 | 0,712470584 | MAST2 | FALSE | Upregulated |
| -0,135370381 | 0,712470584 | NABP2 | FALSE | Downregulated |
| -0,102952569 | 0,712470584 | RCBTB2 | FALSE | Downregulated |
| 0,117722041 | 0,712470584 | UVRAG | FALSE | Upregulated |
| 0,063951502 | 0,712570451 | CCNT1 | FALSE | Upregulated |
| 0,205782326 | 0,712570451 | EPDR1 | FALSE | Upregulated |
| 0,180117125 | 0,712570451 | PNMA1 | FALSE | Upregulated |
| 0,30205234 | 0,712570451 | PRKCE | FALSE | Upregulated |
| 0,256583901 | 0,712570451 | RNF150 | FALSE | Upregulated |
| -0,075372936 | 0,712842012 | ADD1 | FALSE | Downregulated |
| -0,124876297 | 0,712842012 | ZCCHC14 | FALSE | Downregulated |
| 0,592839282 | 0,712973949 | C9orf163 | FALSE | Upregulated |
| 0,243282727 | 0,713177927 | MCF2L-AS1 | FALSE | Upregulated |
| 0,437765084 | 0,713177927 | SLC22A2 | FALSE | Upregulated |
| -0,131883907 | 0,713177927 | TMEM64 | FALSE | Downregulated |

| | | | | |
|---|---|---|---|---|
| -0,080085758 | 0,713177927 | TXNRD3 | FALSE | Downregulated |
| -0,180346497 | 0,713298865 | ZNF594 | FALSE | Downregulated |
| 0,167585893 | 0,713331983 | FAM86DP | FALSE | Upregulated |
| 0,142382393 | 0,713501932 | FTSJ1 | FALSE | Upregulated |
| -0,072166031 | 0,713758176 | PTGES3 | FALSE | Downregulated |
| -0,076736552 | 0,714019162 | NIPBL | FALSE | Downregulated |
| -0,251310944 | 0,714019162 | PREX2 | FALSE | Downregulated |
| 0,143856197 | 0,714041065 | PLEKHA7 | FALSE | Upregulated |
| 0,111456287 | 0,714237208 | AGA | FALSE | Upregulated |
| -0,094789625 | 0,714543164 | C17orf80 | FALSE | Downregulated |
| -0,110164655 | 0,714543164 | CARM1 | FALSE | Downregulated |
| -0,36356099 | 0,714724811 | PRSS3 | FALSE | Downregulated |
| 0,072885233 | 0,714792498 | POM121C | FALSE | Upregulated |
| -0,091273358 | 0,714792498 | RBM39 | FALSE | Downregulated |
| -0,378576292 | 0,714792498 | RP11-503C24.4 | FALSE | Downregulated |
| -0,107927627 | 0,714792498 | RTN4 | FALSE | Downregulated |
| 0,188986663 | 0,714792498 | TUBB2A | FALSE | Upregulated |
| 1,043111836 | 0,715056256 | LRFN5 | FALSE | Upregulated |
| 0,25745395 | 0,715169315 | ASPM | FALSE | Upregulated |
| 0,075339102 | 0,715169315 | CCDC127 | FALSE | Upregulated |
| -0,1093249 | 0,715169315 | SIGMAR1 | FALSE | Downregulated |
| 0,062002288 | 0,71530571 | ANKFY1 | FALSE | Upregulated |
| 0,26609889 | 0,71530571 | LY75 | FALSE | Upregulated |
| -0,083265632 | 0,71530571 | PNPLA8 | FALSE | Downregulated |
| 0,100898788 | 0,71530571 | RAB40C | FALSE | Upregulated |
| 0,228599305 | 0,71530571 | RAD51B | FALSE | Upregulated |

| | | | | |
|---|---|---|---|---|
| -0,471533072 | 0,71530571 | SEC1P | FALSE | Downregulated |
| -0,07093873 | 0,715552854 | ANKRD13A | FALSE | Downregulated |
| 0,088185341 | 0,715552854 | COBL | FALSE | Upregulated |
| 0,265317536 | 0,715552854 | FAM163B | FALSE | Upregulated |
| -0,155986645 | 0,715552854 | PALMD | FALSE | Downregulated |
| 0,122274708 | 0,715552854 | SLC35B1 | FALSE | Upregulated |
| -0,096134732 | 0,715552854 | ZMIZ1 | FALSE | Downregulated |
| -0,280817044 | 0,715582334 | EFNA5 | FALSE | Downregulated |
| 0,264140359 | 0,715582334 | LOC100507547 | FALSE | Upregulated |
| -0,214176103 | 0,715582334 | NPIPA2 | FALSE | Downregulated |
| -0,507973764 | 0,715582334 | RP11-298I3.1 | FALSE | Downregulated |
| 0,257943406 | 0,715630391 | ADM2 | FALSE | Upregulated |
| 0,259977716 | 0,715630391 | AQP1 | FALSE | Upregulated |
| 0,102828108 | 0,715630391 | ASB6 | FALSE | Upregulated |
| 0,116247445 | 0,715630391 | BCAR1 | FALSE | Upregulated |
| -0,806701825 | 0,715630391 | BLOC1S5-TXNDC5 | FALSE | Downregulated |
| 0,231490581 | 0,715630391 | CAMKK1 | FALSE | Upregulated |
| 0,138702584 | 0,715630391 | CNPY3 | FALSE | Upregulated |
| 0,289578722 | 0,715630391 | EML6 | FALSE | Upregulated |
| -0,151169405 | 0,715630391 | FBXO48 | FALSE | Downregulated |
| 0,160002491 | 0,715630391 | FES | FALSE | Upregulated |
| 0,083691675 | 0,715630391 | HSPA14 | FALSE | Upregulated |
| 0,113568812 | 0,715630391 | IVD | FALSE | Upregulated |
| 0,490127633 | 0,715630391 | LOC100507103 | FALSE | Upregulated |
| -0,371203161 | 0,715630391 | LOC101060212 | FALSE | Downregulated |
| 0,748331943 | 0,715630391 | LOC102724385 | FALSE | Upregulated |

| | | | | |
|---|---|---|---|---|
| -0,13400019 | 0,715630391 | MMRN2 | FALSE | Downregulated |
| 0,120136467 | 0,715630391 | NISCH | FALSE | Upregulated |
| -0,329830022 | 0,715630391 | NSUN7 | FALSE | Downregulated |
| 0,12576752 | 0,715630391 | OGFOD2 | FALSE | Upregulated |
| 0,081407427 | 0,715630391 | PIKFYVE | FALSE | Upregulated |
| -0,089998178 | 0,715630391 | PMM1 | FALSE | Downregulated |
| -0,108390086 | 0,715630391 | POLR3H | FALSE | Downregulated |
| 0,105876546 | 0,715630391 | RPL29 | FALSE | Upregulated |
| 0,083064854 | 0,715630391 | SGTA | FALSE | Upregulated |
| 0,093899138 | 0,715630391 | VARS2 | FALSE | Upregulated |
| -0,0912856 | 0,715630391 | WHSC1 | FALSE | Downregulated |
| 0,176008372 | 0,715630391 | ZBTB3 | FALSE | Upregulated |
| 0,212650211 | 0,715630391 | ZNF491 | FALSE | Upregulated |
| -0,348100404 | 0,715701089 | MGP | FALSE | Downregulated |
| -0,100919629 | 0,715749697 | PKIG | FALSE | Downregulated |
| -0,194128121 | 0,716058785 | DLEU2 | FALSE | Downregulated |
| -0,104043372 | 0,716122085 | PHIP | FALSE | Downregulated |
| -0,348270403 | 0,716159508 | LOC100996760 | FALSE | Downregulated |
| -0,102387079 | 0,716177993 | ZNF428 | FALSE | Downregulated |
| 0,112968981 | 0,71664017 | ANXA11 | FALSE | Upregulated |
| 0,089424821 | 0,716762233 | ACADVL | FALSE | Upregulated |
| -0,218215745 | 0,716762233 | ACCS | FALSE | Downregulated |
| -0,246159589 | 0,716762233 | IQCH | FALSE | Downregulated |
| 0,350438476 | 0,716827905 | ANKRD36 | FALSE | Upregulated |
| -0,066889381 | 0,716827905 | KLHL9 | FALSE | Downregulated |
| -0,23517092 | 0,717006972 | CYP3A4 | FALSE | Downregulated |

| | | | | |
|---|---|---|---|---|
| -0,070270952 | 0,717006972 | MRPL44 | FALSE | Downregulated |
| 0,069306191 | 0,717006972 | RCOR1 | FALSE | Upregulated |
| -0,422133654 | 0,717158507 | ABCA13 | FALSE | Downregulated |
| 0,182672787 | 0,717158507 | EHD4 | FALSE | Upregulated |
| -0,275292353 | 0,717158507 | MEG3 | FALSE | Downregulated |
| 0,121065636 | 0,717158507 | SLC4A4 | FALSE | Upregulated |
| -0,215234159 | 0,717287965 | GOLGA6C | FALSE | Downregulated |
| 0,179298363 | 0,717435168 | ZNF35 | FALSE | Upregulated |
| 0,082334258 | 0,717531252 | MTERF | FALSE | Upregulated |
| -0,405865437 | 0,717862045 | WDR86-AS1 | FALSE | Downregulated |
| 0,345676068 | 0,718255478 | CXorf36 | FALSE | Upregulated |
| 0,102923081 | 0,718266465 | EGLN2 | FALSE | Upregulated |
| -0,187982348 | 0,718266465 | TMEM37 | FALSE | Downregulated |
| -0,192317478 | 0,718279982 | GATA3 | FALSE | Downregulated |
| 0,100830437 | 0,718786183 | CNN3 | FALSE | Upregulated |
| 0,089762001 | 0,719030706 | CIB1 | FALSE | Upregulated |
| 0,124830082 | 0,719030706 | FANCL | FALSE | Upregulated |
| 0,164743022 | 0,719030706 | FAR1 | FALSE | Upregulated |
| -0,394064807 | 0,719030706 | KCNE1 | FALSE | Downregulated |
| 0,514172842 | 0,719030706 | LRRC7 | FALSE | Upregulated |
| -0,330462409 | 0,719030706 | NLRP12 | FALSE | Downregulated |
| 0,406070072 | 0,719030706 | SLC26A4 | FALSE | Upregulated |
| 0,097528782 | 0,719030706 | STK16 | FALSE | Upregulated |
| 0,139503202 | 0,719030706 | TMSB10 | FALSE | Upregulated |
| -0,099584384 | 0,719030706 | TOMM6 | FALSE | Downregulated |
| 0,284705707 | 0,719030706 | YTHDF3-AS1 | FALSE | Upregulated |

| | | | | |
|---|---|---|---|---|
| 0,113588935 | 0,719261716 | CLSTN1 | FALSE | Upregulated |
| 0,086170041 | 0,719261716 | NOC2L | FALSE | Upregulated |
| -0,334510822 | 0,719702895 | LOC101926980 | FALSE | Downregulated |
| -0,278962762 | 0,719705104 | VCAN | FALSE | Downregulated |
| 0,86817792 | 0,719759712 | GALNTL6 | FALSE | Upregulated |
| -0,164039116 | 0,719759712 | HTR7P1 | FALSE | Downregulated |
| -0,131706596 | 0,719777485 | CDK11A | FALSE | Downregulated |
| -0,167805132 | 0,719777485 | CDKN1C | FALSE | Downregulated |
| -0,372482547 | 0,719777485 | CNTD2 | FALSE | Downregulated |
| 0,180945833 | 0,719777485 | GREM2 | FALSE | Upregulated |
| -0,223287039 | 0,719777485 | LOC101929959 | FALSE | Downregulated |
| -0,078070772 | 0,719777485 | METTL5 | FALSE | Downregulated |
| 0,648502307 | 0,719777485 | NPTXR | FALSE | Upregulated |
| 0,614269524 | 0,719777485 | RP1L1 | FALSE | Upregulated |
| 0,088281055 | 0,719793053 | ATP6AP2 | FALSE | Upregulated |
| 0,201119507 | 0,719793053 | OSMR | FALSE | Upregulated |
| -0,30704979 | 0,719793053 | RIBC1 | FALSE | Downregulated |
| 0,178391106 | 0,719844254 | CDC42EP1 | FALSE | Upregulated |
| 0,3090924 | 0,719877717 | COL16A1 | FALSE | Upregulated |
| -0,326932355 | 0,719877717 | OAZ3 | FALSE | Downregulated |
| 0,386721218 | 0,719954617 | LOC101928806 | FALSE | Upregulated |
| 0,249731582 | 0,719956407 | LOC101927989 | FALSE | Upregulated |
| -0,196614381 | 0,719956407 | MBOAT2 | FALSE | Downregulated |
| 0,274871171 | 0,720163134 | DGKH | FALSE | Upregulated |
| 0,09895138 | 0,720163134 | MCM9 | FALSE | Upregulated |
| 0,164017572 | 0,720163134 | TMC6 | FALSE | Upregulated |

| | | | | |
|---|---|---|---|---|
| -0,058088839 | 0,720163134 | UBAP1 | FALSE | Downregulated |
| 0,806341128 | 0,720177076 | CABYR | FALSE | Upregulated |
| -0,611703586 | 0,720177076 | INSC | FALSE | Downregulated |
| -0,072734889 | 0,720177076 | KIAA1033 | FALSE | Downregulated |
| 0,565923795 | 0,720177076 | KIR3DX1 | FALSE | Upregulated |
| 0,564717144 | 0,720177076 | LOC101060038 | FALSE | Upregulated |
| 0,294874858 | 0,720177076 | LOC101929715 | FALSE | Upregulated |
| 0,069577731 | 0,720177076 | MEN1 | FALSE | Upregulated |
| -0,134160813 | 0,720177076 | OCLN | FALSE | Downregulated |
| -0,092107443 | 0,720177076 | PAF1 | FALSE | Downregulated |
| -0,715100526 | 0,720177076 | PCDH9-AS2 | FALSE | Downregulated |
| 0,137235777 | 0,720177076 | PLEKHA8 | FALSE | Upregulated |
| 0,116668747 | 0,720177076 | ZNF286A | FALSE | Upregulated |
| 0,11430315 | 0,720283485 | NIPSNAP3A | FALSE | Upregulated |
| 0,077207979 | 0,7206767 | ZFP64 | FALSE | Upregulated |
| -0,37450328 | 0,7207493 | OLFML2A | FALSE | Downregulated |
| -0,112155408 | 0,7207493 | PRPF3 | FALSE | Downregulated |
| 0,189082035 | 0,7207493 | USP35 | FALSE | Upregulated |
| 0,166412591 | 0,720835604 | DOCK10 | FALSE | Upregulated |
| 0,303960762 | 0,720835604 | L3MBTL1 | FALSE | Upregulated |
| -0,250592812 | 0,720835604 | NKAPP1 | FALSE | Downregulated |
| -0,392146499 | 0,720835604 | RP11-539L10.2 | FALSE | Downregulated |
| -0,109735193 | 0,720999042 | KMT2C | FALSE | Downregulated |
| -0,800864877 | 0,721275888 | LINC01133 | FALSE | Downregulated |
| 0,130353628 | 0,721320243 | IFNGR1 | FALSE | Upregulated |
| -0,072529473 | 0,721387247 | AUP1 | FALSE | Downregulated |

| | | | | |
|---|---|---|---|---|
| -0,17953577 | 0,721723364 | LOC728290 | FALSE | Downregulated |
| 0,191471043 | 0,721941971 | C6orf123 | FALSE | Upregulated |
| -0,068521327 | 0,721941971 | FBXL17 | FALSE | Downregulated |
| -0,200222805 | 0,721941971 | GMDS | FALSE | Downregulated |
| 0,182330506 | 0,721941971 | HIST1H1C | FALSE | Upregulated |
| -0,084002162 | 0,721941971 | KIF1B | FALSE | Downregulated |
| -0,182039498 | 0,721941971 | LOC729291 | FALSE | Downregulated |
| 0,068536916 | 0,721941971 | MRPL38 | FALSE | Upregulated |
| 0,110466102 | 0,721941971 | MSANTD4 | FALSE | Upregulated |
| 0,746462517 | 0,721941971 | MUC16 | FALSE | Upregulated |
| 0,132268255 | 0,721941971 | SERPINH1 | FALSE | Upregulated |
| 0,258487382 | 0,721941971 | SLC35C1 | FALSE | Upregulated |
| 0,063353896 | 0,721941971 | STRAP | FALSE | Upregulated |
| -0,108676845 | 0,721941971 | TMEM60 | FALSE | Downregulated |
| 0,136453431 | 0,721941971 | ZBTB42 | FALSE | Upregulated |
| -0,102917723 | 0,721941971 | ZDHHC17 | FALSE | Downregulated |
| 0,545958893 | 0,722004156 | LRRN4 | FALSE | Upregulated |
| 0,139813262 | 0,722004156 | ZNF331 | FALSE | Upregulated |
| 0,227822473 | 0,722005887 | NRM | FALSE | Upregulated |
| -0,078207024 | 0,722067368 | EIF4A2 | FALSE | Downregulated |
| 0,13533546 | 0,722067368 | HAUS5 | FALSE | Upregulated |
| 0,125429457 | 0,722067368 | MON1A | FALSE | Upregulated |
| 0,139012312 | 0,722067368 | TUBA1C | FALSE | Upregulated |
| -0,366716534 | 0,722233926 | C11orf85 | FALSE | Downregulated |
| -0,175123505 | 0,722537168 | SIGIRR | FALSE | Downregulated |
| -0,06945504 | 0,722560557 | SNX6 | FALSE | Downregulated |

| | | | | |
|---|---|---|---|---|
| 0,07967665 | 0,722560557 | UBE2I | FALSE | Upregulated |
| 0,243550058 | 0,722768064 | C11orf82 | FALSE | Upregulated |
| 0,110670641 | 0,722768064 | CHCHD1 | FALSE | Upregulated |
| 0,555560203 | 0,722768064 | LOC100506498 | FALSE | Upregulated |
| 0,203851495 | 0,722768064 | PLA2G6 | FALSE | Upregulated |
| -0,096286578 | 0,722768064 | RNMTL1 | FALSE | Downregulated |
| 0,173735871 | 0,722910523 | ACOT12 | FALSE | Upregulated |
| 0,110717065 | 0,722910523 | B4GALT2 | FALSE | Upregulated |
| -0,103609432 | 0,722910523 | C12orf45 | FALSE | Downregulated |
| 0,104231775 | 0,722910523 | C8orf76 | FALSE | Upregulated |
| 0,172462384 | 0,722910523 | DOHH | FALSE | Upregulated |
| 0,238734865 | 0,722910523 | GPR160 | FALSE | Upregulated |
| 0,092245684 | 0,722910523 | INO80E | FALSE | Upregulated |
| 0,119659034 | 0,722910523 | LYSMD1 | FALSE | Upregulated |
| -0,097858397 | 0,722910523 | LZTS3 | FALSE | Downregulated |
| -0,098465217 | 0,722910523 | MTX3 | FALSE | Downregulated |
| 0,091320906 | 0,722910523 | NUP155 | FALSE | Upregulated |
| 0,265222182 | 0,722910523 | PLAUR | FALSE | Upregulated |
| 0,182030484 | 0,722910523 | RRN3P3 | FALSE | Upregulated |
| -0,153763728 | 0,722910523 | SIVA1 | FALSE | Downregulated |
| 0,184748761 | 0,722910523 | ST6GALNAC3 | FALSE | Upregulated |
| 0,099326667 | 0,722910523 | STX10 | FALSE | Upregulated |
| -0,095587608 | 0,722910523 | TAF11 | FALSE | Downregulated |
| 0,492418703 | 0,722910523 | UNC13A | FALSE | Upregulated |
| -0,151899931 | 0,723216263 | IMPA2 | FALSE | Downregulated |
| -0,461333005 | 0,723216263 | RP11-142C4.6 | FALSE | Downregulated |

| | | | | |
|---|---|---|---|---|
| 0,098517766 | 0,723246837 | FAM214B | FALSE | Upregulated |
| 0,784848052 | 0,723390817 | AACSP1 | FALSE | Upregulated |
| -0,385763236 | 0,723390817 | ADAM22 | FALSE | Downregulated |
| 0,053710547 | 0,723390817 | ARL8B | FALSE | Upregulated |
| -0,179986587 | 0,723390817 | ASAP3 | FALSE | Downregulated |
| -0,204064357 | 0,723390817 | BMP6 | FALSE | Downregulated |
| 0,471290697 | 0,723390817 | C9orf169 | FALSE | Upregulated |
| 0,238663311 | 0,723390817 | CDK5R1 | FALSE | Upregulated |
| 0,17395679 | 0,723390817 | FANCG | FALSE | Upregulated |
| 0,165013302 | 0,723390817 | FBF1 | FALSE | Upregulated |
| -0,156312868 | 0,723390817 | FCN3 | FALSE | Downregulated |
| 0,171057175 | 0,723390817 | FTCD | FALSE | Upregulated |
| 0,068316644 | 0,723390817 | GPATCH3 | FALSE | Upregulated |
| -0,318598046 | 0,723390817 | GSTA3 | FALSE | Downregulated |
| -0,173872221 | 0,723390817 | HERC6 | FALSE | Downregulated |
| -0,271866494 | 0,723390817 | INSIG1 | FALSE | Downregulated |
| -0,582290237 | 0,723390817 | LOC101928428 | FALSE | Downregulated |
| 0,204843142 | 0,723390817 | LOC102724064 | FALSE | Upregulated |
| 0,338172506 | 0,723390817 | LTF | FALSE | Upregulated |
| -0,125890928 | 0,723390817 | MAN1A1 | FALSE | Downregulated |
| -0,397247135 | 0,723390817 | MAP7D2 | FALSE | Downregulated |
| 0,060332848 | 0,723390817 | MAPK1IP1L | FALSE | Upregulated |
| -0,126340037 | 0,723390817 | METTL20 | FALSE | Downregulated |
| -0,107603483 | 0,723390817 | OXNAD1 | FALSE | Downregulated |
| -0,517011672 | 0,723390817 | RALYL | FALSE | Downregulated |
| -0,097347037 | 0,723390817 | REV1 | FALSE | Downregulated |

| | | | | |
|---|---|---|---|---|
| -0,472326038 | 0,723390817 | RHPN1-AS1 | FALSE | Downregulated |
| 0,514388444 | 0,723390817 | RP11-760N9.1 | FALSE | Upregulated |
| 0,229412722 | 0,723390817 | TC2N | FALSE | Upregulated |
| -0,689402143 | 0,723390817 | TMED7-TICAM2 | FALSE | Downregulated |
| 0,439281599 | 0,723390817 | TMEM257 | FALSE | Upregulated |
| 0,076508296 | 0,723390817 | TNKS2 | FALSE | Upregulated |
| -0,580537984 | 0,723390817 | TSHR | FALSE | Downregulated |
| 0,102821358 | 0,723390817 | TXNL4A | FALSE | Upregulated |
| -0,101168936 | 0,723390817 | ZNF654 | FALSE | Downregulated |
| 0,185592289 | 0,72341567 | SYT15 | FALSE | Upregulated |
| 0,237420117 | 0,723558522 | HHIP-AS1 | FALSE | Upregulated |
| -0,142259507 | 0,723558522 | NRP2 | FALSE | Downregulated |
| -0,054088629 | 0,723558522 | PUM2 | FALSE | Downregulated |
| -0,544055191 | 0,723558522 | SLC6A4 | FALSE | Downregulated |
| 0,201080897 | 0,723777692 | LEPR | FALSE | Upregulated |
| 0,151031133 | 0,723777692 | PELI3 | FALSE | Upregulated |
| 0,094703138 | 0,723890541 | LSM1 | FALSE | Upregulated |
| -0,528482039 | 0,723890541 | REM1 | FALSE | Downregulated |
| -1,218706149 | 0,724204151 | ADORA2A | FALSE | Downregulated |
| 0,579502551 | 0,724204151 | ATP8A2 | FALSE | Upregulated |
| 0,236120183 | 0,724204151 | FZD3 | FALSE | Upregulated |
| 0,210214662 | 0,724204151 | HOXB6 | FALSE | Upregulated |
| 0,098777794 | 0,724302783 | ALG5 | FALSE | Upregulated |
| 0,070446061 | 0,724302783 | GALNT1 | FALSE | Upregulated |
| -0,140503954 | 0,724302783 | HSPH1 | FALSE | Downregulated |
| -0,053936916 | 0,724302783 | TBC1D15 | FALSE | Downregulated |

| | | | | |
|---|---|---|---|---|
| 0,148133608 | 0,724302783 | TOPORS-AS1 | FALSE | Upregulated |
| 0,12091421 | 0,724302783 | ZBTB33 | FALSE | Upregulated |
| 0,191881833 | 0,724356747 | ABHD17C | FALSE | Upregulated |
| 0,113434087 | 0,724356747 | C7orf49 | FALSE | Upregulated |
| -0,3723501 | 0,724356747 | DDX43 | FALSE | Downregulated |
| 0,337217523 | 0,724356747 | GDF9 | FALSE | Upregulated |
| 0,121757554 | 0,724356747 | TLR4 | FALSE | Upregulated |
| 0,161006087 | 0,724368138 | FBP1 | FALSE | Upregulated |
| 0,629252382 | 0,724368138 | LOC101927119 | FALSE | Upregulated |
| -0,092743741 | 0,724382383 | UFD1L | FALSE | Downregulated |
| -0,142327426 | 0,724405225 | CYP8B1 | FALSE | Downregulated |
| -0,097261223 | 0,724405225 | FAM210B | FALSE | Downregulated |
| 0,067994161 | 0,724405225 | FIG4 | FALSE | Upregulated |
| 0,282648977 | 0,724405225 | HNRNPA1P10 | FALSE | Upregulated |
| 0,331921292 | 0,724405225 | IGSF3 | FALSE | Upregulated |
| -0,269096269 | 0,724405225 | LOC644794 | FALSE | Downregulated |
| -0,12620995 | 0,724405225 | NBR2 | FALSE | Downregulated |
| -0,595364722 | 0,724405225 | SAA2 | FALSE | Downregulated |
| -0,086025654 | 0,724405225 | SLC25A11 | FALSE | Downregulated |
| 0,334017253 | 0,724405225 | SNHG5 | FALSE | Upregulated |
| 0,380308738 | 0,724405225 | SYT13 | FALSE | Upregulated |
| 0,067706129 | 0,724405225 | TM9SF2 | FALSE | Upregulated |
| -0,171988358 | 0,724405225 | TMEM14A | FALSE | Downregulated |
| 0,111751465 | 0,724509883 | TSHZ2 | FALSE | Upregulated |
| 0,186753222 | 0,724509883 | TSNAXIP1 | FALSE | Upregulated |
| -0,135497916 | 0,724509883 | ZNF518B | FALSE | Downregulated |

| | | | | |
|---|---|---|---|---|
| 0,102085999 | 0,724606777 | DVL1 | FALSE | Upregulated |
| -0,579656536 | 0,724660472 | IL20RB | FALSE | Downregulated |
| -0,114941548 | 0,724660472 | ZNF449 | FALSE | Downregulated |
| 0,160697633 | 0,724934904 | ANXA1 | FALSE | Upregulated |
| 0,310107579 | 0,724934904 | ARHGAP33 | FALSE | Upregulated |
| 0,133752147 | 0,724934904 | ATP6V0B | FALSE | Upregulated |
| -0,259296145 | 0,724934904 | BTBD8 | FALSE | Downregulated |
| 0,506558492 | 0,724934904 | DRD4 | FALSE | Upregulated |
| 0,347711183 | 0,724934904 | MNS1 | FALSE | Upregulated |
| 0,100988088 | 0,724934904 | TBL2 | FALSE | Upregulated |
| -0,050773041 | 0,724934904 | TSN | FALSE | Downregulated |
| 0,129550985 | 0,725206135 | COX7B | FALSE | Upregulated |
| 0,138897969 | 0,725281817 | TOMM40L | FALSE | Upregulated |
| 0,438033024 | 0,725359222 | C22orf43 | FALSE | Upregulated |
| -0,0548964 | 0,725359222 | CCT8 | FALSE | Downregulated |
| 0,132887968 | 0,725418364 | PKD1 | FALSE | Upregulated |
| -0,143774182 | 0,72573773 | CDA | FALSE | Downregulated |
| 0,12621837 | 0,725759275 | NPM3 | FALSE | Upregulated |
| -0,167272864 | 0,725877519 | OSGIN1 | FALSE | Downregulated |
| -0,095633029 | 0,725879862 | C1orf50 | FALSE | Downregulated |
| 0,184893768 | 0,725879862 | FEN1 | FALSE | Upregulated |
| -0,093614283 | 0,725879862 | TFDP2 | FALSE | Downregulated |
| -0,183059978 | 0,725894845 | LINC-PINT | FALSE | Downregulated |
| 0,270108489 | 0,725989673 | NRGN | FALSE | Upregulated |
| -0,263753087 | 0,726088026 | C1QTNF7 | FALSE | Downregulated |
| -0,123407835 | 0,726165009 | LOC102723728 | FALSE | Downregulated |

| | | | | |
|---|---|---|---|---|
| -0,354561353 | 0,726286594 | USH2A | FALSE | Downregulated |
| -0,261644995 | 0,726504774 | LDLRAD3 | FALSE | Downregulated |
| 0,560294115 | 0,726773837 | FAM135B | FALSE | Upregulated |
| -0,488924351 | 0,726968804 | AGAP1-IT1 | FALSE | Downregulated |
| -0,124115357 | 0,726968804 | PGAP1 | FALSE | Downregulated |
| 0,144069025 | 0,726978791 | CDK20 | FALSE | Upregulated |
| -0,155176405 | 0,727060342 | ANKRD9 | FALSE | Downregulated |
| 0,169351374 | 0,727120155 | LOC101060398 | FALSE | Upregulated |
| -0,076277349 | 0,727170789 | APC | FALSE | Downregulated |
| 0,214468713 | 0,727170789 | ARHGEF25 | FALSE | Upregulated |
| 0,319455558 | 0,727170789 | ATP8B3 | FALSE | Upregulated |
| 0,084435899 | 0,727170789 | BCL7B | FALSE | Upregulated |
| -0,097626293 | 0,727170789 | BLOC1S1 | FALSE | Downregulated |
| -0,133500721 | 0,727170789 | FAM115A | FALSE | Downregulated |
| 0,136060517 | 0,727170789 | GALNT10 | FALSE | Upregulated |
| -0,116286424 | 0,727170789 | GALT | FALSE | Downregulated |
| 0,129430052 | 0,727170789 | HMBOX1 | FALSE | Upregulated |
| -0,174653167 | 0,727170789 | LOC102724392 | FALSE | Downregulated |
| 0,091386748 | 0,727170789 | PDE12 | FALSE | Upregulated |
| 0,110292723 | 0,727170789 | UAP1 | FALSE | Upregulated |
| -0,095678346 | 0,727235625 | DNAJB1 | FALSE | Downregulated |
| 0,115347511 | 0,72745193 | TMEM70 | FALSE | Upregulated |
| -0,115538374 | 0,727468473 | GATA4 | FALSE | Downregulated |
| 0,129709346 | 0,727517864 | ACTN1 | FALSE | Upregulated |
| 0,394683739 | 0,727517864 | B3GNT3 | FALSE | Upregulated |
| 0,198489166 | 0,727517864 | C19orf38 | FALSE | Upregulated |

| | | | | |
|---|---|---|---|---|
| -0,206095496 | 0,727517864 | LOC102723908 | FALSE | Downregulated |
| 0,208413105 | 0,727517864 | NFIL3 | FALSE | Upregulated |
| -0,086056493 | 0,727517864 | PPP1R13B | FALSE | Downregulated |
| 0,146182703 | 0,727517864 | TMEM220 | FALSE | Upregulated |
| 0,10430533 | 0,727517864 | TMEM63B | FALSE | Upregulated |
| 0,136464212 | 0,727517864 | TOB1 | FALSE | Upregulated |
| 0,561626469 | 0,727517864 | TREM2 | FALSE | Upregulated |
| 0,122731677 | 0,727517864 | ZBED5-AS1 | FALSE | Upregulated |
| 0,246355892 | 0,72762701 | LOXL2 | FALSE | Upregulated |
| 0,093431226 | 0,727674602 | AGAP1 | FALSE | Upregulated |
| 0,103231537 | 0,727674602 | BOLA1 | FALSE | Upregulated |
| 0,112641578 | 0,727674602 | FN1 | FALSE | Upregulated |
| 0,119703682 | 0,727674602 | UROS | FALSE | Upregulated |
| 0,24813679 | 0,727692716 | CTSK | FALSE | Upregulated |
| -0,083291498 | 0,727692716 | IKZF5 | FALSE | Downregulated |
| 0,134674798 | 0,727692716 | RUSC1 | FALSE | Upregulated |
| 0,130418007 | 0,727692716 | ST7L | FALSE | Upregulated |
| -0,121067453 | 0,727776651 | PPARG | FALSE | Downregulated |
| -0,113453678 | 0,727776651 | YRDC | FALSE | Downregulated |
| -0,09920578 | 0,727859176 | ADI1 | FALSE | Downregulated |
| 0,2499965 | 0,727859176 | DUSP1 | FALSE | Upregulated |
| -0,374350711 | 0,72791136 | CCDC173 | FALSE | Downregulated |
| 0,272393111 | 0,72791136 | TNFSF11 | FALSE | Upregulated |
| 0,446288202 | 0,727929789 | TMEM200A | FALSE | Upregulated |
| 0,081249235 | 0,727957547 | ZBTB40 | FALSE | Upregulated |
| -0,080446682 | 0,728050328 | GPALPP1 | FALSE | Downregulated |

| | | | | |
|---|---|---|---|---|
| 0,076627399 | 0,728368337 | PIGO | FALSE | Upregulated |
| 0,302371593 | 0,728449597 | ANKRD13B | FALSE | Upregulated |
| -0,143992009 | 0,728449597 | CLCN6 | FALSE | Downregulated |
| -0,10786182 | 0,728449597 | CNTRL | FALSE | Downregulated |
| 0,09900231 | 0,728449597 | KMT2B | FALSE | Upregulated |
| 0,321598723 | 0,728449597 | LOC100506454 | FALSE | Upregulated |
| 0,588586844 | 0,728449597 | LOC100506497 | FALSE | Upregulated |
| -0,252926245 | 0,728449597 | LOC102724105 | FALSE | Downregulated |
| -0,28688554 | 0,728449597 | MYOM2 | FALSE | Downregulated |
| -0,085515756 | 0,728449597 | OXA1L | FALSE | Downregulated |
| 0,263264014 | 0,728449597 | PCDH17 | FALSE | Upregulated |
| 0,094406425 | 0,728449597 | PXMP4 | FALSE | Upregulated |
| 0,210264942 | 0,728449597 | SUCNR1 | FALSE | Upregulated |
| 0,156764094 | 0,728449597 | TAPSAR1 | FALSE | Upregulated |
| -0,688197544 | 0,728449597 | TRPM1 | FALSE | Downregulated |
| 0,305811945 | 0,728644762 | FHL3 | FALSE | Upregulated |
| -0,565429839 | 0,728732414 | MESTIT1 | FALSE | Downregulated |
| 0,087994187 | 0,728745301 | MRPS16 | FALSE | Upregulated |
| -0,116038558 | 0,728755677 | PLEKHA6 | FALSE | Downregulated |
| 0,518590053 | 0,728755677 | PTX3 | FALSE | Upregulated |
| 0,126395652 | 0,728755677 | SLC38A6 | FALSE | Upregulated |
| 0,521141131 | 0,729063139 | CH25H | FALSE | Upregulated |
| -0,51011377 | 0,729063139 | MUC6 | FALSE | Downregulated |
| -0,123216571 | 0,729107785 | SMIM14 | FALSE | Downregulated |
| -0,379888618 | 0,72917542 | MTL5 | FALSE | Downregulated |
| -0,151422916 | 0,729249132 | SPP2 | FALSE | Downregulated |

| | | | | |
|---|---|---|---|---|
| -0,075412148 | 0,729269669 | IDS | FALSE | Downregulated |
| -0,279020568 | 0,729269669 | NPIPB11 | FALSE | Downregulated |
| 0,400334405 | 0,729288017 | CCDC89 | FALSE | Upregulated |
| 0,104645791 | 0,729339257 | ADCK4 | FALSE | Upregulated |
| -0,095633853 | 0,729339257 | AIFM1 | FALSE | Downregulated |
| 0,236544927 | 0,729339257 | APBA2 | FALSE | Upregulated |
| 0,090897634 | 0,729339257 | DIP2C | FALSE | Upregulated |
| 0,202793744 | 0,729339257 | FAM228B | FALSE | Upregulated |
| -0,146765702 | 0,729339257 | GSTA4 | FALSE | Downregulated |
| -0,122074899 | 0,729339257 | HSPE1 | FALSE | Downregulated |
| -0,304868272 | 0,729339257 | IFI44L | FALSE | Downregulated |
| 0,21898171 | 0,729339257 | JAG1 | FALSE | Upregulated |
| 0,697082619 | 0,729339257 | LOC102723542 | FALSE | Upregulated |
| -0,180151133 | 0,729339257 | MIR22HG | FALSE | Downregulated |
| -0,187659045 | 0,729339257 | MRPS11 | FALSE | Downregulated |
| 0,745845891 | 0,729339257 | RASA4B | FALSE | Upregulated |
| -0,18118873 | 0,729339257 | RNASEH1-AS1 | FALSE | Downregulated |
| 0,078198393 | 0,729339257 | RUVBL2 | FALSE | Upregulated |
| -0,102771723 | 0,729339257 | SPEN | FALSE | Downregulated |
| 0,087847946 | 0,729339257 | STXBP2 | FALSE | Upregulated |
| -0,303401792 | 0,729339257 | TNFRSF19 | FALSE | Downregulated |
| -0,241726736 | 0,729339257 | TPGS1 | FALSE | Downregulated |
| 0,091606699 | 0,729442706 | RAN | FALSE | Upregulated |
| -0,161490794 | 0,729723454 | POMZP3 | FALSE | Downregulated |
| 0,494906818 | 0,729735877 | TMEM92 | FALSE | Upregulated |
| 0,063982074 | 0,730071089 | HMGN2 | FALSE | Upregulated |

| | | | | |
|---|---|---|---|---|
| -0,094487249 | 0,730176843 | KATNAL1 | FALSE | Downregulated |
| 0,435871848 | 0,730176843 | TMEM132A | FALSE | Upregulated |
| -0,122182406 | 0,730570357 | DYNC1I2 | FALSE | Downregulated |
| -0,312622995 | 0,730679735 | LOC101929055 | FALSE | Downregulated |
| -0,280629261 | 0,730690462 | CASC2 | FALSE | Downregulated |
| -0,17153658 | 0,730841068 | LOC149351 | FALSE | Downregulated |
| -0,19686501 | 0,730966412 | HSP90B2P | FALSE | Downregulated |
| 0,089662831 | 0,730968459 | TECR | FALSE | Upregulated |
| -0,283971205 | 0,731074656 | TRNN | FALSE | Downregulated |
| 0,366467811 | 0,731339578 | PACSIN1 | FALSE | Upregulated |
| 0,575407127 | 0,731339578 | SOX15 | FALSE | Upregulated |
| -0,141748856 | 0,731361877 | CMC2 | FALSE | Downregulated |
| 0,090570544 | 0,731361877 | SPICE1 | FALSE | Upregulated |
| -0,170762077 | 0,731560076 | HP | FALSE | Downregulated |
| -0,170324158 | 0,731560076 | KDR | FALSE | Downregulated |
| -0,382953809 | 0,731560076 | LOC100996723 | FALSE | Downregulated |
| 0,088492495 | 0,731560076 | RERE | FALSE | Upregulated |
| 0,111208282 | 0,731560076 | TMEM2 | FALSE | Upregulated |
| -0,068278752 | 0,731560076 | TRIP4 | FALSE | Downregulated |
| -0,15370272 | 0,732017191 | AMDHD1 | FALSE | Downregulated |
| 0,236735286 | 0,732416792 | ZNF775 | FALSE | Upregulated |
| 0,244995141 | 0,732466651 | ISG15 | FALSE | Upregulated |
| -0,111879391 | 0,732466651 | MBTD1 | FALSE | Downregulated |
| 0,112283076 | 0,732605892 | CPT2 | FALSE | Upregulated |
| -0,129214472 | 0,732628314 | ZNF267 | FALSE | Downregulated |
| -0,080694574 | 0,732686739 | EDRF1 | FALSE | Downregulated |

| | | | | |
|---|---|---|---|---|
| -0,093070173 | 0,732725835 | ELF1 | FALSE | Downregulated |
| 0,066990786 | 0,732725835 | PYCR2 | FALSE | Upregulated |
| -0,090020696 | 0,732746905 | ZNF805 | FALSE | Downregulated |
| 0,377320184 | 0,732927505 | DDX25 | FALSE | Upregulated |
| -0,086786334 | 0,732927505 | MID2 | FALSE | Downregulated |
| 0,12564321 | 0,732927505 | SARDH | FALSE | Upregulated |
| -0,143773351 | 0,732927505 | TUBE1 | FALSE | Downregulated |
| -0,059297041 | 0,732927505 | UBE2G1 | FALSE | Downregulated |
| 0,643768299 | 0,733074634 | MPP2 | FALSE | Upregulated |
| -0,120045906 | 0,733314297 | SSBP4 | FALSE | Downregulated |
| -0,492953434 | 0,733571252 | DCLK1 | FALSE | Downregulated |
| -0,480900371 | 0,733571252 | LRP1B | FALSE | Downregulated |
| 0,701015333 | 0,733620066 | COL6A5 | FALSE | Upregulated |
| 0,10407702 | 0,733620066 | OSGEP | FALSE | Upregulated |
| 0,279712327 | 0,733620066 | RCAN2 | FALSE | Upregulated |
| 0,116332085 | 0,733678897 | TMEM138 | FALSE | Upregulated |
| -0,103545524 | 0,733683231 | APOM | FALSE | Downregulated |
| 0,603417527 | 0,733683231 | MYCN | FALSE | Upregulated |
| 0,407624718 | 0,733840213 | ATXN8OS | FALSE | Upregulated |
| 0,140613758 | 0,734030537 | LOC642852 | FALSE | Upregulated |
| -0,167111331 | 0,734089686 | ORM2 | FALSE | Downregulated |
| 0,151559937 | 0,734089686 | TOR4A | FALSE | Upregulated |
| 0,195886065 | 0,734128639 | LAMA5 | FALSE | Upregulated |
| 0,073620118 | 0,734332206 | ADAM10 | FALSE | Upregulated |
| 0,215454834 | 0,734527648 | LOC100506713 | FALSE | Upregulated |
| 0,831317175 | 0,734533347 | PIR-FIGF | FALSE | Upregulated |

| | | | | |
|---|---|---|---|---|
| 0,115698836 | 0,734566808 | JADE2 | FALSE | Upregulated |
| -0,357647175 | 0,734566808 | LOC100288152 | FALSE | Downregulated |
| -0,218185758 | 0,734601935 | LOC100507642 | FALSE | Downregulated |
| -0,181324829 | 0,734601935 | REPS1 | FALSE | Downregulated |
| 0,256260386 | 0,734614744 | CCRL2 | FALSE | Upregulated |
| 0,337868237 | 0,734614744 | FAM211A | FALSE | Upregulated |
| -0,082975339 | 0,734614744 | SDHD | FALSE | Downregulated |
| -0,08109719 | 0,734707801 | ZNF721 | FALSE | Downregulated |
| 0,385670308 | 0,734928918 | RP5-855F14.1 | FALSE | Upregulated |
| 0,117914636 | 0,734942757 | TMEM185B | FALSE | Upregulated |
| 0,165031743 | 0,735213692 | LPCAT1 | FALSE | Upregulated |
| -0,13730557 | 0,7355649 | AP3M2 | FALSE | Downregulated |
| 0,174196204 | 0,735586415 | ADAMTS17 | FALSE | Upregulated |
| 0,740656754 | 0,735586415 | ANO4 | FALSE | Upregulated |
| -0,051575308 | 0,735586415 | ARPC1A | FALSE | Downregulated |
| -0,113443812 | 0,735586415 | CYP2U1 | FALSE | Downregulated |
| -0,120665515 | 0,735586415 | DOCK4 | FALSE | Downregulated |
| 0,571825389 | 0,735586415 | ENO4 | FALSE | Upregulated |
| 0,07035923 | 0,735586415 | ETF1 | FALSE | Upregulated |
| 0,143730765 | 0,735586415 | FHIT | FALSE | Upregulated |
| -0,051686541 | 0,735586415 | GTPBP10 | FALSE | Downregulated |
| 0,157014093 | 0,735586415 | NEDD8-MDP1 | FALSE | Upregulated |
| -0,17570553 | 0,735586415 | PIM1 | FALSE | Downregulated |
| -0,089172872 | 0,735586415 | PSMA4 | FALSE | Downregulated |
| 0,096025138 | 0,735586415 | TIGD2 | FALSE | Upregulated |
| 0,07854986 | 0,735586415 | VAMP8 | FALSE | Upregulated |

| | | | | |
|---|---|---|---|---|
| -0,115428947 | 0,735586415 | YLPM1 | FALSE | Downregulated |
| 0,178498596 | 0,735762626 | LY86 | FALSE | Upregulated |
| 0,138873198 | 0,735762626 | RPL22L1 | FALSE | Upregulated |
| -0,618002216 | 0,735897248 | CCL8 | FALSE | Downregulated |
| 0,411419877 | 0,735897248 | LOC101929953 | FALSE | Upregulated |
| 0,096450517 | 0,735952284 | NCAPD3 | FALSE | Upregulated |
| -0,110632307 | 0,736350028 | GIMAP8 | FALSE | Downregulated |
| -0,771732908 | 0,736350028 | LOC100131395 | FALSE | Downregulated |
| 0,152013443 | 0,736350028 | PLEKHH3 | FALSE | Upregulated |
| -0,22125831 | 0,736350028 | REC8 | FALSE | Downregulated |
| -0,072792928 | 0,736350028 | WDR3 | FALSE | Downregulated |
| 0,545679235 | 0,736355273 | C10orf105 | FALSE | Upregulated |
| 0,212019723 | 0,736355273 | C17orf96 | FALSE | Upregulated |
| 0,359512306 | 0,736355273 | C1QTNF9B-AS1 | FALSE | Upregulated |
| -0,137059879 | 0,736355273 | DHX35 | FALSE | Downregulated |
| -0,52610688 | 0,736355273 | LOC101930100 | FALSE | Downregulated |
| 0,575556643 | 0,736355273 | LRRC73 | FALSE | Upregulated |
| -0,124523658 | 0,736355273 | MEIS1 | FALSE | Downregulated |
| -0,087841264 | 0,736355273 | NDUFAF1 | FALSE | Downregulated |
| -1,378505471 | 0,736355273 | PERM1 | FALSE | Downregulated |
| -0,112218571 | 0,736355273 | PRR5 | FALSE | Downregulated |
| -0,134904836 | 0,736355273 | PSMD1 | FALSE | Downregulated |
| -0,170816853 | 0,736355273 | STARD5 | FALSE | Downregulated |
| -0,066871239 | 0,736355273 | UTP3 | FALSE | Downregulated |
| 0,075658923 | 0,736355273 | VPS25 | FALSE | Upregulated |
| 0,103679427 | 0,736357049 | PSD4 | FALSE | Upregulated |

| | | | | |
|---|---|---|---|---|
| -0,104938349 | 0,736779371 | NHSL1 | FALSE | Downregulated |
| -0,208524191 | 0,736995958 | CXXC4 | FALSE | Downregulated |
| 0,332576603 | 0,736995958 | HOXA3 | FALSE | Upregulated |
| 0,27631736 | 0,736995958 | SLC16A5 | FALSE | Upregulated |
| -0,145988369 | 0,736995958 | YY2 | FALSE | Downregulated |
| 0,105434188 | 0,737055787 | SIX5 | FALSE | Upregulated |
| 0,128081973 | 0,737104841 | C11orf83 | FALSE | Upregulated |
| -0,179177731 | 0,737104841 | PNN | FALSE | Downregulated |
| -0,059125724 | 0,737104841 | PWWP2A | FALSE | Downregulated |
| 0,250173655 | 0,737359483 | LOC101929200 | FALSE | Upregulated |
| 0,721230929 | 0,737528124 | TNFRSF13B | FALSE | Upregulated |
| 0,227682777 | 0,737939267 | ARHGAP39 | FALSE | Upregulated |
| 0,104475104 | 0,737939267 | CPNE2 | FALSE | Upregulated |
| -0,094583979 | 0,737939267 | EFTUD1 | FALSE | Downregulated |
| -0,114215817 | 0,738065777 | CRYBG3 | FALSE | Downregulated |
| 0,059229458 | 0,738065777 | MCM3AP | FALSE | Upregulated |
| 0,092700287 | 0,738065777 | SLC35F6 | FALSE | Upregulated |
| 0,094384678 | 0,738142509 | HEATR5A | FALSE | Upregulated |
| 0,095066319 | 0,738142509 | SLC12A7 | FALSE | Upregulated |
| 0,103413053 | 0,738214671 | KIAA1161 | FALSE | Upregulated |
| -0,170386979 | 0,738214671 | RP11-326F20.5 | FALSE | Downregulated |
| 0,068534276 | 0,738214671 | SETDB1 | FALSE | Upregulated |
| -0,199178991 | 0,738214671 | TIMM13 | FALSE | Downregulated |
| 0,307282596 | 0,738214671 | TSSK6 | FALSE | Upregulated |
| -0,230288642 | 0,738297964 | LOC101929747 | FALSE | Downregulated |
| 0,189375226 | 0,738297964 | RBP7 | FALSE | Upregulated |

| | | | | |
|---|---|---|---|---|
| 0,111649997 | 0,738297964 | RRM2B | FALSE | Upregulated |
| 0,288928428 | 0,738297964 | SOX9 | FALSE | Upregulated |
| 0,335186298 | 0,738297964 | VGLL3 | FALSE | Upregulated |
| -0,159305066 | 0,738346815 | ALDH1L1-AS2 | FALSE | Downregulated |
| 0,35369846 | 0,738471656 | RP11-557H15.4 | FALSE | Upregulated |
| -0,169382948 | 0,738494821 | AP5S1 | FALSE | Downregulated |
| -0,143382474 | 0,738494821 | ZNF559 | FALSE | Downregulated |
| -0,450269241 | 0,738494821 | ZNF676 | FALSE | Downregulated |
| 0,164178802 | 0,738780886 | ENTHD2 | FALSE | Upregulated |
| -0,127777746 | 0,738780886 | KLKB1 | FALSE | Downregulated |
| -0,051164694 | 0,738780886 | OPA1 | FALSE | Downregulated |
| -0,14505396 | 0,738780886 | PISD | FALSE | Downregulated |
| 0,206813682 | 0,738780886 | ZNF439 | FALSE | Upregulated |
| 0,234447389 | 0,739049343 | BMS1P4 | FALSE | Upregulated |
| 0,247660908 | 0,739361982 | FUT1 | FALSE | Upregulated |
| -0,319874032 | 0,739361982 | LOC101929385 | FALSE | Downregulated |
| 0,082036448 | 0,739563754 | NSF | FALSE | Upregulated |
| -0,187058634 | 0,739696576 | LRRC17 | FALSE | Downregulated |
| -0,124701451 | 0,73975833 | HSD17B12 | FALSE | Downregulated |
| 0,108317782 | 0,739840226 | LONRF1 | FALSE | Upregulated |
| -0,226248418 | 0,739871571 | ASPDH | FALSE | Downregulated |
| 0,455117284 | 0,739871571 | LHX9 | FALSE | Upregulated |
| -0,479723382 | 0,739871571 | LOC400655 | FALSE | Downregulated |
| 0,065564018 | 0,739920169 | CSRP2BP | FALSE | Upregulated |
| -0,360895651 | 0,739920169 | CYP3A43 | FALSE | Downregulated |
| -0,135026689 | 0,739920169 | KAZN | FALSE | Downregulated |

| | | | | |
|---|---|---|---|---|
| 0,357209915 | 0,739920169 | MTMR11 | FALSE | Upregulated |
| 0,502460443 | 0,739920169 | NMNAT2 | FALSE | Upregulated |
| 0,082513515 | 0,739920169 | PUS10 | FALSE | Upregulated |
| -0,082046226 | 0,739920169 | SLU7 | FALSE | Downregulated |
| -0,189917356 | 0,739920169 | THAP7-AS1 | FALSE | Downregulated |
| 0,124119904 | 0,739920169 | TRIB1 | FALSE | Upregulated |
| -0,579593712 | 0,739924704 | LOC644936 | FALSE | Downregulated |
| 0,214179027 | 0,740357168 | ABCC4 | FALSE | Upregulated |
| 0,131973766 | 0,740357168 | C1orf216 | FALSE | Upregulated |
| -0,167846365 | 0,740357168 | CNTN3 | FALSE | Downregulated |
| 0,104099892 | 0,740357168 | DUSP14 | FALSE | Upregulated |
| -0,151152098 | 0,74038457 | KIF27 | FALSE | Downregulated |
| -0,384543342 | 0,740400352 | CHAC1 | FALSE | Downregulated |
| 0,125851594 | 0,740400352 | TUSC1 | FALSE | Upregulated |
| -0,105340678 | 0,740464656 | ALDH2 | FALSE | Downregulated |
| 0,473772301 | 0,740464656 | C9orf153 | FALSE | Upregulated |
| -0,060455234 | 0,740464656 | CUEDC2 | FALSE | Downregulated |
| 0,217535439 | 0,740464656 | LOC284889 | FALSE | Upregulated |
| 0,091909338 | 0,740464656 | LONP2 | FALSE | Upregulated |
| 0,336239182 | 0,740464656 | LTBP2 | FALSE | Upregulated |
| 0,170832124 | 0,740464656 | MIR1181 | FALSE | Upregulated |
| -0,142367122 | 0,740585981 | FRG1B | FALSE | Downregulated |
| 0,207368189 | 0,740615682 | BTNL9 | FALSE | Upregulated |
| -0,106971121 | 0,740615682 | FKBP4 | FALSE | Downregulated |
| -0,220305271 | 0,740893255 | TLCD2 | FALSE | Downregulated |
| -0,594892258 | 0,740963445 | KCNQ5 | FALSE | Downregulated |

| | | | | |
|---|---|---|---|---|
| 0,263875406 | 0,740996786 | OLFML2B | FALSE | Upregulated |
| 0,535645901 | 0,740996786 | TMEM108 | FALSE | Upregulated |
| -0,125622575 | 0,741067937 | ASAP2 | FALSE | Downregulated |
| -0,055799611 | 0,741067937 | CTBP1 | FALSE | Downregulated |
| -0,2742479 | 0,741067937 | FAM198A | FALSE | Downregulated |
| 0,074628602 | 0,741067937 | KIF13A | FALSE | Upregulated |
| -0,113953981 | 0,741067937 | KMT2E | FALSE | Downregulated |
| -0,095113427 | 0,741067937 | LIMA1 | FALSE | Downregulated |
| 0,497948482 | 0,741067937 | LOC102724634 | FALSE | Upregulated |
| -0,267916509 | 0,741067937 | TENM1 | FALSE | Downregulated |
| -0,118706961 | 0,741067937 | TET2 | FALSE | Downregulated |
| -0,516229347 | 0,741067937 | TRAM1L1 | FALSE | Downregulated |
| 0,352631724 | 0,74144459 | NAV3 | FALSE | Upregulated |
| 0,086321031 | 0,741588558 | NFATC2IP | FALSE | Upregulated |
| 0,061164109 | 0,741700552 | FUBP3 | FALSE | Upregulated |
| -0,470501509 | 0,741700552 | HCG14 | FALSE | Downregulated |
| -0,577174558 | 0,741700552 | LOC101928669 | FALSE | Downregulated |
| 0,087769793 | 0,741723482 | ACTR8 | FALSE | Upregulated |
| -0,404805718 | 0,741723482 | ANKLE1 | FALSE | Downregulated |
| -0,086647338 | 0,741723482 | PFDN5 | FALSE | Downregulated |
| -0,111467038 | 0,741723482 | RDH11 | FALSE | Downregulated |
| -0,329171849 | 0,741723482 | RP11-475O6.1-2 | FALSE | Downregulated |
| -0,172331674 | 0,741755601 | ABCC9 | FALSE | Downregulated |
| 0,09123271 | 0,741755601 | ARL6IP5 | FALSE | Upregulated |
| 0,292245294 | 0,741755601 | ATHL1 | FALSE | Upregulated |
| -0,048568466 | 0,741755601 | CSNK2A1 | FALSE | Downregulated |

| | | | | |
|---|---|---|---|---|
| 0,317110406 | 0,741755601 | CYP3A7 | FALSE | Upregulated |
| -0,104629501 | 0,741755601 | DIABLO | FALSE | Downregulated |
| 0,097765202 | 0,741755601 | DNAJC10 | FALSE | Upregulated |
| -0,081360739 | 0,741755601 | EMG1 | FALSE | Downregulated |
| 0,099079883 | 0,741755601 | FKRP | FALSE | Upregulated |
| 0,570506999 | 0,741755601 | FOXS1 | FALSE | Upregulated |
| -0,097604581 | 0,741755601 | HECTD1 | FALSE | Downregulated |
| 0,091545711 | 0,741755601 | IFRD2 | FALSE | Upregulated |
| 0,095286888 | 0,741755601 | LAMTOR4 | FALSE | Upregulated |
| -0,281586092 | 0,741755601 | LOC613037 | FALSE | Downregulated |
| -0,11466596 | 0,741755601 | OTC | FALSE | Downregulated |
| 0,096357901 | 0,741755601 | POLR2I | FALSE | Upregulated |
| 0,700709115 | 0,741755601 | PPT2-EGFL8 | FALSE | Upregulated |
| 0,254012622 | 0,741755601 | PRMT6 | FALSE | Upregulated |
| -0,224365808 | 0,741755601 | RIC3 | FALSE | Downregulated |
| -0,070556547 | 0,741755601 | RSL24D1 | FALSE | Downregulated |
| 0,110139748 | 0,741755601 | STK4 | FALSE | Upregulated |
| -0,175579205 | 0,741755601 | SULT2A1 | FALSE | Downregulated |
| 0,062135811 | 0,741755601 | TBC1D20 | FALSE | Upregulated |
| -0,070762615 | 0,741755601 | ZNF830 | FALSE | Downregulated |
| 0,127434995 | 0,741768045 | ARMC6 | FALSE | Upregulated |
| 0,472622164 | 0,741768045 | LINC00839 | FALSE | Upregulated |
| -0,25090185 | 0,741768045 | LINC01106 | FALSE | Downregulated |
| -0,253583172 | 0,741768045 | LOC101929107 | FALSE | Downregulated |
| 0,538627451 | 0,741768045 | LOC90246 | FALSE | Upregulated |
| 0,069036346 | 0,741768045 | LONP1 | FALSE | Upregulated |

| | | | | |
|---|---|---|---|---|
| -0,116888606 | 0,741768045 | RNASEH2B | FALSE | Downregulated |
| 0,114920541 | 0,741768045 | STARD13 | FALSE | Upregulated |
| -0,428598167 | 0,741768045 | STXBP5-AS1 | FALSE | Downregulated |
| 0,165178941 | 0,741768045 | VNN1 | FALSE | Upregulated |
| -1,009906073 | 0,741834372 | RFPL4AL1 | FALSE | Downregulated |
| 0,105233852 | 0,741834372 | TXNDC9 | FALSE | Upregulated |
| 0,237932427 | 0,741875294 | HK2 | FALSE | Upregulated |
| 0,063135472 | 0,741901955 | MED1 | FALSE | Upregulated |
| -0,090693982 | 0,741901955 | NARG2 | FALSE | Downregulated |
| 0,067142155 | 0,741901955 | NARS | FALSE | Upregulated |
| 0,088142241 | 0,741901955 | PSME2 | FALSE | Upregulated |
| 0,172563582 | 0,741961091 | CEBPB | FALSE | Upregulated |
| 0,271905146 | 0,741961091 | CLDN2 | FALSE | Upregulated |
| 0,166984501 | 0,741961091 | FOXA3 | FALSE | Upregulated |
| -0,145530306 | 0,741961091 | IGF1R | FALSE | Downregulated |
| 0,119248158 | 0,741961091 | NRDE2 | FALSE | Upregulated |
| -0,345105583 | 0,741961091 | RPL13AP3 | FALSE | Downregulated |
| -0,281792833 | 0,741961091 | SCG5 | FALSE | Downregulated |
| -0,079809075 | 0,741961091 | SSNA1 | FALSE | Downregulated |
| -0,675872657 | 0,741961091 | TRNAG20 | FALSE | Downregulated |
| 0,249663256 | 0,742069441 | CCDC148 | FALSE | Upregulated |
| -0,13191941 | 0,742069441 | CD36 | FALSE | Downregulated |
| 0,133341594 | 0,742069441 | CDC25B | FALSE | Upregulated |
| -0,269895344 | 0,742069441 | CPE | FALSE | Downregulated |
| 0,169740016 | 0,742069441 | CYP2S1 | FALSE | Upregulated |
| 0,187797234 | 0,742069441 | HSPA13 | FALSE | Upregulated |

| | | | | |
|---|---|---|---|---|
| 0,240954291 | 0,742069441 | LOC100506358 | FALSE | Upregulated |
| 0,484568665 | 0,742069441 | LOC102467080 | FALSE | Upregulated |
| -0,161816504 | 0,742069441 | LOC642441 | FALSE | Downregulated |
| 0,31519418 | 0,742069441 | LOC728392 | FALSE | Upregulated |
| 0,192869819 | 0,742069441 | MARCH1 | FALSE | Upregulated |
| -0,103457363 | 0,742069441 | MST4 | FALSE | Downregulated |
| -0,124261772 | 0,742069441 | PAM | FALSE | Downregulated |
| 0,477676068 | 0,742069441 | RP11-399K21.14 | FALSE | Upregulated |
| 0,078089864 | 0,742069441 | RTCB | FALSE | Upregulated |
| 0,142172377 | 0,742069441 | SERPINB1 | FALSE | Upregulated |
| -0,112795399 | 0,742069441 | TMEM120A | FALSE | Downregulated |
| -0,083381289 | 0,742069441 | TTF1 | FALSE | Downregulated |
| 0,07649093 | 0,742069441 | TUSC2 | FALSE | Upregulated |
| 0,260677859 | 0,742116289 | MARK1 | FALSE | Upregulated |
| -0,163723559 | 0,742418207 | RNPS1 | FALSE | Downregulated |
| -0,140204935 | 0,742452068 | GPR116 | FALSE | Downregulated |
| -0,189122062 | 0,742487781 | GPR183 | FALSE | Downregulated |
| -0,094561078 | 0,742627397 | KIF21A | FALSE | Downregulated |
| -0,080048574 | 0,742627397 | MCPH1 | FALSE | Downregulated |
| 0,101763712 | 0,742627397 | RPP30 | FALSE | Upregulated |
| 0,104398225 | 0,743216233 | IMPDH2 | FALSE | Upregulated |
| 0,118246106 | 0,743227806 | C18orf32 | FALSE | Upregulated |
| 0,425253678 | 0,743227806 | CBLN3 | FALSE | Upregulated |
| -0,234960323 | 0,743227806 | KCNMA1 | FALSE | Downregulated |
| 0,061710393 | 0,743227806 | YIPF5 | FALSE | Upregulated |
| -0,051325272 | 0,743228283 | AP3S1 | FALSE | Downregulated |

| | | | | |
|---|---|---|---|---|
| 0,085612688 | 0,743376964 | ZNF623 | FALSE | Upregulated |
| -0,079166184 | 0,743559897 | ASB8 | FALSE | Downregulated |
| -0,387296467 | 0,743559897 | C14orf164 | FALSE | Downregulated |
| 0,157643699 | 0,743559897 | LINC00574 | FALSE | Upregulated |
| 0,09406591 | 0,743559897 | PSKH1 | FALSE | Upregulated |
| 0,077672936 | 0,743667852 | DAZAP1 | FALSE | Upregulated |
| -0,399679716 | 0,744096941 | AC147651.4 | FALSE | Downregulated |
| -0,303314166 | 0,744335566 | BOK-AS1 | FALSE | Downregulated |
| -0,239725037 | 0,744335566 | RP5-1013A22.2 | FALSE | Downregulated |
| 0,246483665 | 0,744774055 | AP000472.2 | FALSE | Upregulated |
| -0,040470818 | 0,744774055 | DDX42 | FALSE | Downregulated |
| -0,133771638 | 0,744774055 | HSP90AA1 | FALSE | Downregulated |
| -0,165729334 | 0,744774055 | SGCB | FALSE | Downregulated |
| 0,138187091 | 0,744774055 | USP18 | FALSE | Upregulated |
| -0,072975886 | 0,744805965 | DCAKD | FALSE | Downregulated |
| -0,109444267 | 0,745156159 | FAM135A | FALSE | Downregulated |
| -0,167893412 | 0,745156159 | PTPN13 | FALSE | Downregulated |
| -1,090601566 | 0,745156159 | UNC45B | FALSE | Downregulated |
| -0,091235919 | 0,745202691 | GIGYF2 | FALSE | Downregulated |
| 0,377024225 | 0,745317768 | ABCB9 | FALSE | Upregulated |
| 0,227110051 | 0,745317768 | SLC22A18AS | FALSE | Upregulated |
| 0,139944846 | 0,745317768 | SYNRG | FALSE | Upregulated |
| -0,100289692 | 0,745393244 | RRAGD | FALSE | Downregulated |
| 0,092512633 | 0,745482626 | EIF5A | FALSE | Upregulated |
| 0,156929252 | 0,745573252 | MYO16 | FALSE | Upregulated |
| 0,173932906 | 0,745573252 | ZDHHC13 | FALSE | Upregulated |

| | | | | |
|---|---|---|---|---|
| 0,09232463 | 0,745611235 | CALM2 | FALSE | Upregulated |
| 0,189433901 | 0,745611235 | PTPRO | FALSE | Upregulated |
| -0,122731649 | 0,745611235 | SATB1 | FALSE | Downregulated |
| -0,239523244 | 0,745611235 | SPIC | FALSE | Downregulated |
| 0,041662464 | 0,745611235 | UBE3B | FALSE | Upregulated |
| -0,063469957 | 0,745685377 | ABCB7 | FALSE | Downregulated |
| -0,068963838 | 0,745685377 | MED14 | FALSE | Downregulated |
| -0,330520489 | 0,74572163 | LOC101928881 | FALSE | Downregulated |
| -0,627209932 | 0,74572163 | PDE1C | FALSE | Downregulated |
| 0,351349141 | 0,74572163 | VANGL2 | FALSE | Upregulated |
| -0,061076408 | 0,74572163 | WDR33 | FALSE | Downregulated |
| 0,127511128 | 0,74572163 | ZNF784 | FALSE | Upregulated |
| -0,103401648 | 0,745749436 | DDO | FALSE | Downregulated |
| -0,101410564 | 0,745820096 | PNPT1 | FALSE | Downregulated |
| -0,14896043 | 0,745848512 | LRR1 | FALSE | Downregulated |
| -0,093702678 | 0,745900887 | CLIP1 | FALSE | Downregulated |
| 0,137469952 | 0,745900887 | KLF10 | FALSE | Upregulated |
| 0,075681273 | 0,745900887 | RRP36 | FALSE | Upregulated |
| 0,371477298 | 0,745900887 | SIGLEC15 | FALSE | Upregulated |
| 0,134237694 | 0,746048217 | AGPAT2 | FALSE | Upregulated |
| -0,073767551 | 0,746048217 | CLINT1 | FALSE | Downregulated |
| -0,154829833 | 0,746048217 | PARD6G | FALSE | Downregulated |
| 0,164853909 | 0,746109951 | C5orf54 | FALSE | Upregulated |
| 0,068792209 | 0,746109951 | COG8 | FALSE | Upregulated |
| -0,116501652 | 0,746109951 | NEU3 | FALSE | Downregulated |
| 0,089794389 | 0,746109951 | POLE | FALSE | Upregulated |

| | | | | |
|---|---|---|---|---|
| -0,185184122 | 0,746109951 | ZSCAN20 | FALSE | Downregulated |
| 0,306972503 | 0,746315234 | ITGBL1 | FALSE | Upregulated |
| -0,610154769 | 0,746479049 | SPEG | FALSE | Downregulated |
| 0,071224925 | 0,746524537 | ARFRP1 | FALSE | Upregulated |
| -0,212203472 | 0,746524537 | PRORSD1P | FALSE | Downregulated |
| 0,077952169 | 0,746524537 | RNF13 | FALSE | Upregulated |
| 0,084675557 | 0,746577859 | PHB2 | FALSE | Upregulated |
| -0,676388299 | 0,746662598 | EMX2 | FALSE | Downregulated |
| 0,260054175 | 0,746676336 | ANXA3 | FALSE | Upregulated |
| -0,086299902 | 0,746676336 | CCT3 | FALSE | Downregulated |
| -0,262100594 | 0,746676336 | GOLGA6A | FALSE | Downregulated |
| -0,258023056 | 0,746676336 | LOC283788 | FALSE | Downregulated |
| -0,101264275 | 0,746833362 | ACD | FALSE | Downregulated |
| 0,10383116 | 0,746833362 | DGCR6L | FALSE | Upregulated |
| 0,297671245 | 0,746833362 | LOC101928719 | FALSE | Upregulated |
| -0,299812785 | 0,746833362 | LOC102723566 | FALSE | Downregulated |
| -0,292917311 | 0,746833362 | MTMR9LP | FALSE | Downregulated |
| 0,053499306 | 0,746833362 | WIPI2 | FALSE | Upregulated |
| 0,103511447 | 0,746906967 | USP38 | FALSE | Upregulated |
| 0,181707818 | 0,747142924 | EPHA2 | FALSE | Upregulated |
| 0,560671952 | 0,74726288 | DUSP8 | FALSE | Upregulated |
| 0,124880265 | 0,747359212 | HS3ST3B1 | FALSE | Upregulated |
| -0,244718894 | 0,747359212 | LOC101927759 | FALSE | Downregulated |
| -0,110551506 | 0,747360971 | PCGF3 | FALSE | Downregulated |
| -0,381340704 | 0,747382047 | GOLGA6L5P | FALSE | Downregulated |
| -0,346803905 | 0,747382047 | MKX | FALSE | Downregulated |

| | | | | |
|---|---|---|---|---|
| 0,572553819 | 0,747382047 | PHEX | FALSE | Upregulated |
| -0,133226636 | 0,747382047 | PHGDH | FALSE | Downregulated |
| 0,389266001 | 0,747382047 | RASA4CP | FALSE | Upregulated |
| 0,34793261 | 0,747489581 | MPP3 | FALSE | Upregulated |
| 0,186325596 | 0,747489581 | TNRC6C-AS1 | FALSE | Upregulated |
| 0,093396329 | 0,747532686 | CCT6A | FALSE | Upregulated |
| -0,470487904 | 0,747532686 | HRH4 | FALSE | Downregulated |
| 0,071382924 | 0,747532686 | MED8 | FALSE | Upregulated |
| -0,826230952 | 0,747532686 | PPP1R27 | FALSE | Downregulated |
| 0,176280871 | 0,747532686 | PTPRE | FALSE | Upregulated |
| -0,062986771 | 0,747532686 | SKI | FALSE | Downregulated |
| -0,252421162 | 0,747941635 | LINC01197 | FALSE | Downregulated |
| 0,157078298 | 0,747982612 | DNAH14 | FALSE | Upregulated |
| -0,370005327 | 0,747982612 | LOC101929561 | FALSE | Downregulated |
| 0,682057248 | 0,747982612 | LOC102724560 | FALSE | Upregulated |
| 0,527603562 | 0,747982612 | LOC645752 | FALSE | Upregulated |
| 0,132787185 | 0,747982612 | MCM3 | FALSE | Upregulated |
| -0,131412791 | 0,747982612 | MYLIP | FALSE | Downregulated |
| 0,103690993 | 0,747982612 | PUS1 | FALSE | Upregulated |
| 0,657094515 | 0,747982612 | SNORA24 | FALSE | Upregulated |
| -0,138860412 | 0,747982612 | ZDBF2 | FALSE | Downregulated |
| 0,921608223 | 0,748257102 | DEFA3 | FALSE | Upregulated |
| 0,07942745 | 0,748279708 | CCNDBP1 | FALSE | Upregulated |
| -0,128252639 | 0,748279708 | TPCN2 | FALSE | Downregulated |
| -0,222960283 | 0,748313153 | ARRDC3 | FALSE | Downregulated |
| 0,186274052 | 0,748313153 | CD84 | FALSE | Upregulated |

| | | | | |
|---|---|---|---|---|
| 0,170238051 | 0,748313153 | CELF2 | FALSE | Upregulated |
| 0,072794464 | 0,748313153 | KANK1 | FALSE | Upregulated |
| 0,068218922 | 0,748313153 | TRUB2 | FALSE | Upregulated |
| 0,2350014 | 0,748349366 | ARNT2 | FALSE | Upregulated |
| -0,082262941 | 0,748349366 | CENPBD1 | FALSE | Downregulated |
| 0,09267931 | 0,748349366 | EARS2 | FALSE | Upregulated |
| -0,122223451 | 0,748349366 | GNB4 | FALSE | Downregulated |
| 0,183678053 | 0,748416841 | ALS2CL | FALSE | Upregulated |
| -0,232866586 | 0,748416841 | ANK2 | FALSE | Downregulated |
| 0,172799651 | 0,748733391 | ABCC5 | FALSE | Upregulated |
| 0,254807362 | 0,748733391 | LY6G5C | FALSE | Upregulated |
| -0,170301052 | 0,748733391 | TMEM204 | FALSE | Downregulated |
| 0,243432494 | 0,748744138 | CCM2L | FALSE | Upregulated |
| 0,067565336 | 0,748744138 | DNAJA3 | FALSE | Upregulated |
| -0,101434821 | 0,748744138 | PDZD11 | FALSE | Downregulated |
| 0,057375941 | 0,748744138 | POLR3E | FALSE | Upregulated |
| 0,056226563 | 0,748744138 | UBE2V2 | FALSE | Upregulated |
| 0,503888684 | 0,748870616 | BDKRB2 | FALSE | Upregulated |
| 0,103300226 | 0,748870616 | EOGT | FALSE | Upregulated |
| -0,082260728 | 0,748870616 | ZNF823 | FALSE | Downregulated |
| 0,108468855 | 0,749065022 | EXOSC5 | FALSE | Upregulated |
| -0,115090018 | 0,749065022 | MYADM | FALSE | Downregulated |
| 0,094690815 | 0,749065022 | SLC38A3 | FALSE | Upregulated |
| -0,131423951 | 0,749065022 | TMEM56 | FALSE | Downregulated |
| -0,076408888 | 0,749166648 | CHM | FALSE | Downregulated |
| -0,246010189 | 0,749166648 | LOC100133331 | FALSE | Downregulated |

| | | | | |
|---|---|---|---|---|
| 0,151863458 | 0,749166648 | SDC3 | FALSE | Upregulated |
| -0,09200622 | 0,749208659 | CHMP4C | FALSE | Downregulated |
| -0,110696521 | 0,749208659 | KIAA0232 | FALSE | Downregulated |
| 0,084036418 | 0,749208659 | ZBTB2 | FALSE | Upregulated |
| -0,067241678 | 0,749232902 | HNRNPA3 | FALSE | Downregulated |
| -0,209613359 | 0,749232902 | LINC00844 | FALSE | Downregulated |
| 0,968978684 | 0,749232902 | SNORA44 | FALSE | Upregulated |
| -0,135314786 | 0,749288171 | ACPL2 | FALSE | Downregulated |
| -0,157144064 | 0,749288171 | ZNF709 | FALSE | Downregulated |
| 0,220585312 | 0,749288171 | ZNF71 | FALSE | Upregulated |
| 0,288155966 | 0,749381963 | NUTM2D | FALSE | Upregulated |
| -0,112360536 | 0,749381963 | YAF2 | FALSE | Downregulated |
| 0,078309312 | 0,749493778 | ZNF768 | FALSE | Upregulated |
| 0,187546873 | 0,749683986 | TNFRSF21 | FALSE | Upregulated |
| 0,09181113 | 0,749712184 | NOSIP | FALSE | Upregulated |
| 0,133593277 | 0,749712184 | SKAP2 | FALSE | Upregulated |
| -0,10371872 | 0,749712437 | HMGB2 | FALSE | Downregulated |
| -0,118665515 | 0,75008581 | PBLD | FALSE | Downregulated |
| -0,146384311 | 0,750116284 | FAM89A | FALSE | Downregulated |
| -0,310058408 | 0,750116284 | MEOX2 | FALSE | Downregulated |
| 0,128627394 | 0,750116284 | ZNF836 | FALSE | Upregulated |
| -0,088571624 | 0,750117833 | ARMCX3 | FALSE | Downregulated |
| 0,149308702 | 0,750117833 | C7orf43 | FALSE | Upregulated |
| -0,124724079 | 0,750117833 | ESR1 | FALSE | Downregulated |
| -0,386154343 | 0,750117833 | JMJD1C-AS1 | FALSE | Downregulated |
| -0,490420136 | 0,750117833 | LOC100287934 | FALSE | Downregulated |

| | | | | |
|---|---|---|---|---|
| 0,470199632 | 0,750117833 | LOC101929287 | FALSE | Upregulated |
| -0,30145765 | 0,750117833 | LOC648987 | FALSE | Downregulated |
| 0,564914575 | 0,750117833 | SPATA9 | FALSE | Upregulated |
| 0,139454307 | 0,750117833 | ZNF426 | FALSE | Upregulated |
| 0,287196042 | 0,750117833 | ZNF827 | FALSE | Upregulated |
| -0,225525199 | 0,750327068 | C3orf70 | FALSE | Downregulated |
| 0,452828709 | 0,750327068 | FAM66C | FALSE | Upregulated |
| 0,558906656 | 0,750327068 | FBXL13 | FALSE | Upregulated |
| -0,106964669 | 0,750327068 | FXN | FALSE | Downregulated |
| 0,289977429 | 0,750327068 | NPNT | FALSE | Upregulated |
| 0,173470761 | 0,750327068 | PKM | FALSE | Upregulated |
| 0,048781687 | 0,750327068 | ZDHHC3 | FALSE | Upregulated |
| 0,07690819 | 0,750327068 | ZZEF1 | FALSE | Upregulated |
| 0,152390589 | 0,750332326 | PILRA | FALSE | Upregulated |
| -0,410880421 | 0,75051644 | LOC100506047 | FALSE | Downregulated |
| 0,571580792 | 0,750533755 | C1orf180 | FALSE | Upregulated |
| -0,060423436 | 0,750702368 | WDR36 | FALSE | Downregulated |
| 0,078938167 | 0,75094486 | CAPZA1 | FALSE | Upregulated |
| 0,188839102 | 0,75094486 | COL3A1 | FALSE | Upregulated |
| -0,047554127 | 0,75094486 | PNRC2 | FALSE | Downregulated |
| 0,881528036 | 0,750995541 | CTGLF12P | FALSE | Upregulated |
| -0,090889553 | 0,750995541 | SIAH1 | FALSE | Downregulated |
| 0,310595695 | 0,751092031 | CUBN | FALSE | Upregulated |
| -0,089896717 | 0,751211111 | FOXP1 | FALSE | Downregulated |
| -0,169364411 | 0,751265036 | LIN9 | FALSE | Downregulated |
| -0,526544619 | 0,751341094 | TRNS2 | FALSE | Downregulated |

| | | | | |
|---|---|---|---|---|
| 0,200802735 | 0,751341094 | USHBP1 | FALSE | Upregulated |
| 0,210686822 | 0,751798583 | CENPH | FALSE | Upregulated |
| 0,523762067 | 0,751798583 | ZNF541 | FALSE | Upregulated |
| -0,067484418 | 0,751916737 | CLDN12 | FALSE | Downregulated |
| -1,805591469 | 0,751916737 | LOC100505841 | FALSE | Downregulated |
| -0,08674546 | 0,752044035 | RNF26 | FALSE | Downregulated |
| -0,088313837 | 0,752145825 | RSF1 | FALSE | Downregulated |
| 0,110868941 | 0,75222443 | ANXA4 | FALSE | Upregulated |
| -0,087849676 | 0,75222443 | DDX46 | FALSE | Downregulated |
| 0,375960033 | 0,75222443 | EMILIN3 | FALSE | Upregulated |
| -0,077487317 | 0,75222443 | ERBB3 | FALSE | Downregulated |
| 0,119732051 | 0,75222443 | GNB5 | FALSE | Upregulated |
| 0,26339079 | 0,75222443 | RASGRF2 | FALSE | Upregulated |
| 0,162721456 | 0,75222443 | SHROOM1 | FALSE | Upregulated |
| 0,246960695 | 0,75222443 | SRGAP2C | FALSE | Upregulated |
| -0,223114191 | 0,75222443 | TMIE | FALSE | Downregulated |
| -0,111104588 | 0,75222443 | UXS1 | FALSE | Downregulated |
| 0,064335174 | 0,752279853 | CYHR1 | FALSE | Upregulated |
| -0,08971188 | 0,752290412 | AFF4 | FALSE | Downregulated |
| 0,457366488 | 0,752290412 | CHRM3 | FALSE | Upregulated |
| -0,290083314 | 0,752290412 | SEC14L3 | FALSE | Downregulated |
| -0,138265559 | 0,752290412 | SERPING1 | FALSE | Downregulated |
| -0,120169335 | 0,752290412 | SMO | FALSE | Downregulated |
| -0,063425679 | 0,752290412 | ZNF398 | FALSE | Downregulated |
| 0,143540928 | 0,752335624 | ACY3 | FALSE | Upregulated |
| -0,290347738 | 0,752335624 | ASB9 | FALSE | Downregulated |

| | | | | |
|---|---|---|---|---|
| 0,093019212 | 0,752335624 | CHSY1 | FALSE | Upregulated |
| 0,343358972 | 0,752402788 | KIAA1324 | FALSE | Upregulated |
| -0,09120108 | 0,752542553 | GLO1 | FALSE | Downregulated |
| -0,323192126 | 0,752707129 | LOC101928767 | FALSE | Downregulated |
| 0,074875447 | 0,752844885 | YIPF2 | FALSE | Upregulated |
| -0,122681407 | 0,752889676 | NCK2 | FALSE | Downregulated |
| 0,096478214 | 0,752889676 | TDG | FALSE | Upregulated |
| 0,233251914 | 0,753070829 | BMS1P5 | FALSE | Upregulated |
| -0,154861394 | 0,753070829 | CCDC14 | FALSE | Downregulated |
| 0,424170457 | 0,753070829 | LOC102723558 | FALSE | Upregulated |
| -0,143173101 | 0,753070829 | RECK | FALSE | Downregulated |
| -0,229231169 | 0,753153129 | ACSS2 | FALSE | Downregulated |
| 0,156455531 | 0,753153129 | CYP4F12 | FALSE | Upregulated |
| -0,151667715 | 0,753153129 | PTPRN2 | FALSE | Downregulated |
| 0,535394654 | 0,753236074 | C10orf107 | FALSE | Upregulated |
| 0,16056981 | 0,753446448 | CCNF | FALSE | Upregulated |
| 0,229422371 | 0,753446448 | IL12RB1 | FALSE | Upregulated |
| -0,113132019 | 0,753604018 | RNF217 | FALSE | Downregulated |
| 0,526970581 | 0,753810562 | CXCL3 | FALSE | Upregulated |
| -0,51328867 | 0,753810562 | LOC101927032 | FALSE | Downregulated |
| 0,403911985 | 0,753810562 | PAK7 | FALSE | Upregulated |
| 0,233811284 | 0,753810562 | TMEM100 | FALSE | Upregulated |
| 0,078975253 | 0,753823192 | AK3 | FALSE | Upregulated |
| 0,38754219 | 0,753823192 | LOC101929719 | FALSE | Upregulated |
| -0,086165876 | 0,753823192 | SLC9B2 | FALSE | Downregulated |
| -0,10901095 | 0,753823192 | SMLR1 | FALSE | Downregulated |

| | | | | |
|---|---|---|---|---|
| -0,567793281 | 0,753842938 | CCL13 | FALSE | Downregulated |
| 0,10395011 | 0,753842938 | CSRNP2 | FALSE | Upregulated |
| 0,161661952 | 0,753842938 | FAM65C | FALSE | Upregulated |
| -0,044773292 | 0,753842938 | KCTD20 | FALSE | Downregulated |
| -0,432718376 | 0,753842938 | LOC102723866 | FALSE | Downregulated |
| -0,108118425 | 0,753842938 | NFKBIL1 | FALSE | Downregulated |
| -0,09887989 | 0,753842938 | PMS2P5 | FALSE | Downregulated |
| 0,179603167 | 0,753842938 | RPP25 | FALSE | Upregulated |
| 0,714901689 | 0,753954511 | SNORA55 | FALSE | Upregulated |
| -0,145520364 | 0,754421333 | TCF25 | FALSE | Downregulated |
| 0,45223384 | 0,754483438 | LOC101929206 | FALSE | Upregulated |
| 0,083716961 | 0,754483438 | MRPL34 | FALSE | Upregulated |
| -0,154893469 | 0,754483438 | TBC1D19 | FALSE | Downregulated |
| 0,410693201 | 0,754504526 | CATSPER2 | FALSE | Upregulated |
| -0,212299531 | 0,754741442 | RAB3A | FALSE | Downregulated |
| 0,112879815 | 0,754852295 | ARL4A | FALSE | Upregulated |
| -0,218259519 | 0,754852295 | DDX26B | FALSE | Downregulated |
| -0,074146232 | 0,754852295 | INTS4 | FALSE | Downregulated |
| -0,081997106 | 0,754852295 | LDHD | FALSE | Downregulated |
| 0,128492387 | 0,754852295 | MARCH9 | FALSE | Upregulated |
| -0,14995221 | 0,754862637 | LINC01071 | FALSE | Downregulated |
| 0,070358301 | 0,754862637 | SCAMP2 | FALSE | Upregulated |
| -0,108083475 | 0,754862637 | STYXL1 | FALSE | Downregulated |
| -0,295451722 | 0,754862637 | SYCP3 | FALSE | Downregulated |
| -0,472575917 | 0,754862637 | VWA3B | FALSE | Downregulated |
| 0,137788857 | 0,754862637 | ZNF211 | FALSE | Upregulated |

| | | | | |
|---|---|---|---|---|
| 0,11370375 | 0,754862637 | ZNF217 | FALSE | Upregulated |
| -0,104141269 | 0,754991327 | RBP5 | FALSE | Downregulated |
| -0,080645736 | 0,754991327 | RXRA | FALSE | Downregulated |
| 0,074463149 | 0,754991327 | SAE1 | FALSE | Upregulated |
| 0,688582561 | 0,754991327 | SH3GL2 | FALSE | Upregulated |
| -0,122696674 | 0,754991327 | STAT3 | FALSE | Downregulated |
| 0,129558252 | 0,75504977 | ATG4D | FALSE | Upregulated |
| -0,098567465 | 0,75504977 | DDX17 | FALSE | Downregulated |
| 0,20361974 | 0,75504977 | FLJ32255 | FALSE | Upregulated |
| -0,098212852 | 0,75504977 | HNRNPUL2-BSCL2 | FALSE | Downregulated |
| -0,555664128 | 0,75504977 | LOC101928263 | FALSE | Downregulated |
| 0,117207583 | 0,75504977 | MFSD5 | FALSE | Upregulated |
| 0,280900599 | 0,75504977 | SFRP5 | FALSE | Upregulated |
| -0,448154949 | 0,75504977 | SLC25A48 | FALSE | Downregulated |
| 0,207172337 | 0,75504977 | SP140 | FALSE | Upregulated |
| 0,05718148 | 0,755127914 | POLR3C | FALSE | Upregulated |
| -0,240634969 | 0,755392696 | ASXL3 | FALSE | Downregulated |
| -0,147468404 | 0,755696295 | ADAMTS9 | FALSE | Downregulated |
| 0,453500037 | 0,755696295 | ADORA2B | FALSE | Upregulated |
| -0,436443295 | 0,755696295 | CALN1 | FALSE | Downregulated |
| -0,058422145 | 0,755696295 | FAM105B | FALSE | Downregulated |
| -0,339976092 | 0,755696295 | KDM4A-AS1 | FALSE | Downregulated |
| 0,073771299 | 0,755696295 | PPP1CC | FALSE | Upregulated |
| 0,35889441 | 0,755696295 | SEPT5 | FALSE | Upregulated |
| -0,224707527 | 0,755696295 | SLC25A12 | FALSE | Downregulated |
| 0,088181571 | 0,755696295 | URM1 | FALSE | Upregulated |

| | | | | |
|---|---|---|---|---|
| -0,279157773 | 0,755798455 | LOC101929143 | FALSE | Downregulated |
| 0,208015991 | 0,755813753 | LOC102724023 | FALSE | Upregulated |
| 0,076014767 | 0,755868397 | MRPL10 | FALSE | Upregulated |
| 0,103901474 | 0,755868397 | POLD2 | FALSE | Upregulated |
| -0,167853673 | 0,755868397 | SC5D | FALSE | Downregulated |
| -0,406132694 | 0,755868397 | SHANK1 | FALSE | Downregulated |
| -0,108286737 | 0,755868397 | TMEM53 | FALSE | Downregulated |
| 0,258261518 | 0,755868397 | VPS37D | FALSE | Upregulated |
| 0,252134383 | 0,755896947 | CASS4 | FALSE | Upregulated |
| 0,065708275 | 0,755896947 | NBAS | FALSE | Upregulated |
| -0,173459579 | 0,756019861 | G0S2 | FALSE | Downregulated |
| 0,532418022 | 0,756019861 | GIPR | FALSE | Upregulated |
| -0,47260774 | 0,756019861 | KCNG1 | FALSE | Downregulated |
| -0,455490573 | 0,75616739 | SULT4A1 | FALSE | Downregulated |
| -0,356182491 | 0,756372218 | HOXA2 | FALSE | Downregulated |
| -0,203685117 | 0,756495767 | FGD1 | FALSE | Downregulated |
| 0,07156604 | 0,756647566 | VCPIP1 | FALSE | Upregulated |
| 0,123937753 | 0,756818847 | STK36 | FALSE | Upregulated |
| -0,191392074 | 0,756949206 | EDA | FALSE | Downregulated |
| -0,565463959 | 0,757048996 | ART5 | FALSE | Downregulated |
| -0,082132738 | 0,757048996 | RNF181 | FALSE | Downregulated |
| -0,081574186 | 0,757130848 | ANAPC13 | FALSE | Downregulated |
| -0,087060427 | 0,757130848 | CTDSPL | FALSE | Downregulated |
| -0,161157824 | 0,757130848 | WDYHV1 | FALSE | Downregulated |
| 0,053680159 | 0,757232622 | AP3M1 | FALSE | Upregulated |
| -0,092415446 | 0,757232622 | GAB1 | FALSE | Downregulated |

| | | | | |
|---|---|---|---|---|
| 0,244359044 | 0,757232622 | LOC154761 | FALSE | Upregulated |
| -0,150667715 | 0,757232622 | LOC642361 | FALSE | Downregulated |
| 0,36983178 | 0,757232622 | MS4A2 | FALSE | Upregulated |
| -0,05703005 | 0,757232622 | NDUFA9 | FALSE | Downregulated |
| -0,19718683 | 0,757232622 | RP5-1073O3.7 | FALSE | Downregulated |
| 0,092642897 | 0,757232622 | SKIV2L | FALSE | Upregulated |
| 0,150627098 | 0,757232622 | SPIN2B | FALSE | Upregulated |
| 0,142132308 | 0,757232622 | TMEM128 | FALSE | Upregulated |
| -0,634319861 | 0,757245026 | PRRX1 | FALSE | Downregulated |
| -1,888094917 | 0,757295822 | ATP5J2-PTCD1 | FALSE | Downregulated |
| 0,230280034 | 0,757295822 | IQCH-AS1 | FALSE | Upregulated |
| 0,202631857 | 0,757370543 | EFCAB4B | FALSE | Upregulated |
| 0,226747373 | 0,757370543 | FAM222A | FALSE | Upregulated |
| -0,105805538 | 0,757370543 | FARP2 | FALSE | Downregulated |
| 0,093719064 | 0,757370543 | FCHO2 | FALSE | Upregulated |
| -0,504773184 | 0,757370543 | PRR16 | FALSE | Downregulated |
| 0,392970457 | 0,757370543 | TRIP13 | FALSE | Upregulated |
| 0,18732282 | 0,757707982 | GZMA | FALSE | Upregulated |
| 0,943057591 | 0,757718094 | AGAP7 | FALSE | Upregulated |
| -0,53950013 | 0,757718094 | ALG1L9P | FALSE | Downregulated |
| 0,262216372 | 0,757718094 | CD300LF | FALSE | Upregulated |
| -0,113952923 | 0,757718094 | CNST | FALSE | Downregulated |
| 0,098770555 | 0,757718094 | DCUN1D5 | FALSE | Upregulated |
| 0,074583594 | 0,757718094 | KAT5 | FALSE | Upregulated |
| 0,116481045 | 0,757718094 | KREMEN1 | FALSE | Upregulated |
| -0,067035209 | 0,757718094 | PDXDC1 | FALSE | Downregulated |

| | | | | |
|---|---|---|---|---|
| 0,460589065 | 0,757718094 | RP11-118B18.1 | FALSE | Upregulated |
| -0,157066319 | 0,757718094 | SLC1A4 | FALSE | Downregulated |
| 0,109398883 | 0,757718094 | TANGO2 | FALSE | Upregulated |
| 0,084588887 | 0,757718094 | VGLL4 | FALSE | Upregulated |
| 0,121660957 | 0,757718094 | ZNF792 | FALSE | Upregulated |
| 0,160700454 | 0,757718094 | ZNF841 | FALSE | Upregulated |
| 0,150649189 | 0,757718094 | ZNHIT2 | FALSE | Upregulated |
| 0,078909451 | 0,757729115 | DENND5B | FALSE | Upregulated |
| 0,118694029 | 0,757729115 | MMACHC | FALSE | Upregulated |
| 0,373620974 | 0,757729115 | RNF112 | FALSE | Upregulated |
| 0,134577253 | 0,757729115 | SFXN3 | FALSE | Upregulated |
| 0,563042582 | 0,757729115 | SRD5A3-AS1 | FALSE | Upregulated |
| 0,115968626 | 0,757729115 | WRAP73 | FALSE | Upregulated |
| 0,226001092 | 0,75788488 | CACNA2D2 | FALSE | Upregulated |
| 0,24621023 | 0,75788488 | CCDC64 | FALSE | Upregulated |
| 0,120177108 | 0,75788488 | DHRS7 | FALSE | Upregulated |
| -0,090523477 | 0,75788488 | RRNAD1 | FALSE | Downregulated |
| -0,290758816 | 0,75788488 | SLIT3 | FALSE | Downregulated |
| 0,180603288 | 0,757942786 | KIAA0825 | FALSE | Upregulated |
| -0,484179209 | 0,757987039 | C4orf36 | FALSE | Downregulated |
| -0,520897323 | 0,757987039 | ERVMER34-1 | FALSE | Downregulated |
| -0,091665137 | 0,757987039 | PAK1IP1 | FALSE | Downregulated |
| 0,082243302 | 0,758382497 | CYB5R1 | FALSE | Upregulated |
| -0,088722244 | 0,758382497 | DKFZP586I1420 | FALSE | Downregulated |
| -0,481957782 | 0,758382497 | ESYT3 | FALSE | Downregulated |
| -0,082341858 | 0,758382497 | FAM46A | FALSE | Downregulated |

| | | | | |
|---|---|---|---|---|
| -0,091208313 | 0,758382497 | GS1-251I9.4 | FALSE | Downregulated |
| -0,554686234 | 0,758382497 | KLHL14 | FALSE | Downregulated |
| -0,158135515 | 0,758382497 | LAGE3 | FALSE | Downregulated |
| 0,131409891 | 0,758382497 | LOC101927371 | FALSE | Upregulated |
| -0,066530791 | 0,758382497 | MAF1 | FALSE | Downregulated |
| -0,057448489 | 0,758382497 | STIM2 | FALSE | Downregulated |
| 0,258808116 | 0,758382497 | VWF | FALSE | Upregulated |
| -0,127176003 | 0,758382497 | ZEB2 | FALSE | Downregulated |
| -0,503779905 | 0,758393921 | C6orf25 | FALSE | Downregulated |
| -0,980884825 | 0,758393921 | LOC729222 | FALSE | Downregulated |
| -0,330357986 | 0,758393921 | NLRP2 | FALSE | Downregulated |
| -0,125606312 | 0,758393921 | RBM4B | FALSE | Downregulated |
| -0,296926787 | 0,758523828 | CREB5 | FALSE | Downregulated |
| 0,233618675 | 0,758523828 | FAM118A | FALSE | Upregulated |
| 0,107538889 | 0,758523828 | GORAB | FALSE | Upregulated |
| -0,221661208 | 0,758523828 | MYLK4 | FALSE | Downregulated |
| -0,094959715 | 0,758523828 | NDUFB3 | FALSE | Downregulated |
| -0,097552684 | 0,758523828 | PFDN4 | FALSE | Downregulated |
| -0,070718361 | 0,758523828 | R3HCC1L | FALSE | Downregulated |
| -0,096152007 | 0,758523828 | SDC2 | FALSE | Downregulated |
| 0,310457131 | 0,758523828 | SDS | FALSE | Upregulated |
| -0,066944764 | 0,758523828 | SLMO2 | FALSE | Downregulated |
| -0,067897856 | 0,758523828 | TBL1XR1 | FALSE | Downregulated |
| 0,16697719 | 0,758523828 | TMEM79 | FALSE | Upregulated |
| 0,110757101 | 0,758859341 | ACVRL1 | FALSE | Upregulated |
| -0,063961917 | 0,758859341 | CHMP4B | FALSE | Downregulated |

| | | | | |
|---|---|---|---|---|
| 0,115009057 | 0,758859341 | LZTS2 | FALSE | Upregulated |
| -0,101532295 | 0,759059199 | DNAJC1 | FALSE | Downregulated |
| 0,292141733 | 0,759059199 | KCNAB3 | FALSE | Upregulated |
| -0,16860685 | 0,759059199 | LOC102724905 | FALSE | Downregulated |
| 0,05788731 | 0,759059199 | PCYT1A | FALSE | Upregulated |
| 0,137698376 | 0,759129298 | ALKBH2 | FALSE | Upregulated |
| 0,058201787 | 0,759129298 | DHX15 | FALSE | Upregulated |
| -0,159929065 | 0,759129298 | DLG3 | FALSE | Downregulated |
| 0,217013468 | 0,759129298 | GAS7 | FALSE | Upregulated |
| -0,425280028 | 0,759129298 | LOC101927104 | FALSE | Downregulated |
| -0,37055831 | 0,759129298 | SGSM1 | FALSE | Downregulated |
| -0,091885421 | 0,759129298 | SMCHD1 | FALSE | Downregulated |
| 0,238550503 | 0,759129298 | TLX1 | FALSE | Upregulated |
| -0,114212152 | 0,759207686 | C12orf57 | FALSE | Downregulated |
| -0,051326317 | 0,759207686 | PSMC6 | FALSE | Downregulated |
| 0,082352637 | 0,759207686 | RAP2C | FALSE | Upregulated |
| 0,172921316 | 0,759207686 | STK39 | FALSE | Upregulated |
| 0,299190929 | 0,759250678 | LOC101927060 | FALSE | Upregulated |
| 0,387227558 | 0,759511683 | FAM3B | FALSE | Upregulated |
| -0,227802569 | 0,759521769 | RP1-27K12.2 | FALSE | Downregulated |
| -0,158925042 | 0,759555683 | EXOG | FALSE | Downregulated |
| -0,163239275 | 0,759586378 | HSPA12A | FALSE | Downregulated |
| -0,174674178 | 0,759679933 | GNG12-AS1 | FALSE | Downregulated |
| -0,223149722 | 0,759679933 | LOC100287497 | FALSE | Downregulated |
| 0,356661768 | 0,759679933 | P2RX1 | FALSE | Upregulated |
| 0,103564357 | 0,759679933 | PDRG1 | FALSE | Upregulated |

-0,097158427 0,759679933 RSU1 FALSE Downregulated

-0,107740874 0,759812 AADAT FALSE Downregulated

0,215071331 0,759850403 LOC101927126 FALSE Upregulated

0,112952357 0,759850403 TENC1 FALSE Upregulated

0,105841488 0,760389042 TTLL4 FALSE Upregulated

-0,12980684 0,760407703 BLOC1S5 FALSE Downregulated

-0,070505499 0,760532497 CHCHD2 FALSE Downregulated

-0,059735218 0,760712062 TUG1 FALSE Downregulated

-0,546321565 0,760775859 ATP6V1G2-DDX39B FALSE Downregulated

-0,083194504 0,760775859 ERCC1 FALSE Downregulated

-0,084460473 0,760775859 MED7 FALSE Downregulated

0,468472442 0,760775859 NACAD FALSE Upregulated

0,187226275 0,760775859 SLC25A34 FALSE Upregulated

-0,121975223 0,760775859 SRP19 FALSE Downregulated

-0,117852978 0,760775859 TNK1 FALSE Downregulated

0,121954469 0,760775859 ZNF235 FALSE Upregulated

0,1958301 0,760775859 ZNF525 FALSE Upregulated

0,302140546 0,760775859 ZNF833P FALSE Upregulated

-0,19210417 0,760892695 IFITM1 FALSE Downregulated

0,197149758 0,761056682 MRAS FALSE Upregulated

-0,088546958 0,761056682 TCOF1 FALSE Downregulated

0,387758924 0,761289004 LOC102724436 FALSE Upregulated

0,184520133 0,761375864 BAMBI FALSE Upregulated

0,269495523 0,761375864 EPHB3 FALSE Upregulated

0,105940369 0,761375864 FRAT1 FALSE Upregulated

| | | | | |
|---|---|---|---|---|
| 0,126250892 | 0,761375864 | HK1 | FALSE | Upregulated |
| -0,533288056 | 0,761375864 | LOC100507530 | FALSE | Downregulated |
| -0,162345214 | 0,761375864 | LOC101929243 | FALSE | Downregulated |
| 0,152785732 | 0,761375864 | MFSD6 | FALSE | Upregulated |
| -0,249421498 | 0,761375864 | PODXL2 | FALSE | Downregulated |
| -0,108695505 | 0,761606814 | FAM162A | FALSE | Downregulated |
| -0,25558602 | 0,761606814 | LOC101928504 | FALSE | Downregulated |
| -0,052810548 | 0,761756086 | TAF2 | FALSE | Downregulated |
| 0,537045443 | 0,761756086 | TAS2R5 | FALSE | Upregulated |
| -0,068134354 | 0,761756086 | UIMC1 | FALSE | Downregulated |
| 0,192977061 | 0,762214189 | C19orf40 | FALSE | Upregulated |
| -0,474276908 | 0,762214189 | FDPSP2 | FALSE | Downregulated |
| -0,081510016 | 0,762214189 | MAGI1 | FALSE | Downregulated |
| 0,091791799 | 0,762214189 | PGAP3 | FALSE | Upregulated |
| 0,449711494 | 0,762214189 | ZRANB2-AS2 | FALSE | Upregulated |
| 0,092750521 | 0,762272354 | FAM195B | FALSE | Upregulated |
| -0,145211691 | 0,762375758 | CDK19 | FALSE | Downregulated |
| -0,135400596 | 0,762375758 | CENPJ | FALSE | Downregulated |
| 0,144002608 | 0,762375758 | ECE2 | FALSE | Upregulated |
| -0,796187325 | 0,762375758 | EGFL8 | FALSE | Downregulated |
| -0,201397141 | 0,762375758 | FAM167B | FALSE | Downregulated |
| -0,085529567 | 0,762375758 | FNBP1L | FALSE | Downregulated |
| 0,136076197 | 0,762375758 | FZD6 | FALSE | Upregulated |
| -0,190784945 | 0,762375758 | INTU | FALSE | Downregulated |
| 0,521257929 | 0,762375758 | LOC101926916 | FALSE | Upregulated |
| 0,273689536 | 0,762375758 | LOC101928415 | FALSE | Upregulated |

| | | | | |
|---|---|---|---|---|
| 0,236985722 | 0,762375758 | LOC101929251 | FALSE | Upregulated |
| -0,119547382 | 0,762375758 | LOH12CR1 | FALSE | Downregulated |
| 0,175821783 | 0,762375758 | LPCAT4 | FALSE | Upregulated |
| 0,058607485 | 0,762375758 | NFKB1 | FALSE | Upregulated |
| 0,084743366 | 0,762375758 | POP5 | FALSE | Upregulated |
| 0,434458437 | 0,762375758 | PSD2 | FALSE | Upregulated |
| -0,077232719 | 0,762375758 | RPL10 | FALSE | Downregulated |
| -0,123131543 | 0,762375758 | SREBF2 | FALSE | Downregulated |
| -0,088893803 | 0,762375758 | STRN | FALSE | Downregulated |
| 0,089161944 | 0,762375758 | SUMF1 | FALSE | Upregulated |
| 0,093280758 | 0,762375758 | TBCCD1 | FALSE | Upregulated |
| -0,060038116 | 0,762375758 | TDP2 | FALSE | Downregulated |
| 0,072608026 | 0,762375758 | UTP14A | FALSE | Upregulated |
| -0,27062077 | 0,762384787 | LOC101929140 | FALSE | Downregulated |
| -0,086462883 | 0,762384787 | MRPL19 | FALSE | Downregulated |
| -0,255693429 | 0,762464647 | CLEC1A | FALSE | Downregulated |
| 0,066024274 | 0,762464647 | DYNC1H1 | FALSE | Upregulated |
| 0,228150332 | 0,762464647 | HSPA12B | FALSE | Upregulated |
| 0,074891513 | 0,762464647 | PCMT1 | FALSE | Upregulated |
| -0,061046232 | 0,762464647 | RAPGEF1 | FALSE | Downregulated |
| 0,932448859 | 0,762464647 | SNORA6 | FALSE | Upregulated |
| -0,220289026 | 0,762570096 | ERRFI1 | FALSE | Downregulated |
| 0,088238932 | 0,762570096 | INTS7 | FALSE | Upregulated |
| 0,256703423 | 0,762570096 | LINC00969 | FALSE | Upregulated |
| -0,085134141 | 0,762570096 | MSRB2 | FALSE | Downregulated |
| 0,121375511 | 0,762570096 | NT5DC3 | FALSE | Upregulated |

| | | | | |
|---|---|---|---|---|
| -0,224722658 | 0,762570096 | PRKG1 | FALSE | Downregulated |
| 0,093082818 | 0,762570096 | TP53RK | FALSE | Upregulated |
| 0,118699804 | 0,7626531 | DAO | FALSE | Upregulated |
| 0,09412541 | 0,762860711 | GALNS | FALSE | Upregulated |
| 0,042348683 | 0,762860711 | GANAB | FALSE | Upregulated |
| -0,220763222 | 0,762860711 | KLHL13 | FALSE | Downregulated |
| 0,160373039 | 0,762860711 | LOC102724873 | FALSE | Upregulated |
| 0,200646766 | 0,762860711 | MCTP1 | FALSE | Upregulated |
| 0,117234335 | 0,762860711 | ZNF416 | FALSE | Upregulated |
| 0,162581562 | 0,762989198 | RP11-35G9.3 | FALSE | Upregulated |
| -0,083066482 | 0,763065277 | COX19 | FALSE | Downregulated |
| 0,172380479 | 0,763065277 | TMEM17 | FALSE | Upregulated |
| 0,455573913 | 0,763261711 | CABP7 | FALSE | Upregulated |
| -0,079283329 | 0,763261711 | CYB5D1 | FALSE | Downregulated |
| 0,0879476 | 0,763261711 | FAM220A | FALSE | Upregulated |
| -0,121055136 | 0,763261711 | UGT2B7 | FALSE | Downregulated |
| 0,071383697 | 0,763334055 | MRPL4 | FALSE | Upregulated |
| 0,303993014 | 0,763334055 | NAGPA-AS1 | FALSE | Upregulated |
| 0,126288888 | 0,763334055 | TMCO3 | FALSE | Upregulated |
| 0,125698939 | 0,763334055 | ZWILCH | FALSE | Upregulated |
| -0,043682379 | 0,764037509 | PTPRA | FALSE | Downregulated |
| -0,504839811 | 0,764136086 | CSMD2 | FALSE | Downregulated |
| 0,232924834 | 0,764156029 | CHRM2 | FALSE | Upregulated |
| 0,086382717 | 0,764156029 | ETNK1 | FALSE | Upregulated |
| 0,3332532 | 0,764156029 | HSF2BP | FALSE | Upregulated |
| -0,273951864 | 0,764156029 | LOC100996741 | FALSE | Downregulated |

| | | | | |
|---|---|---|---|---|
| -0,061573372 | 0,764156029 | SMIM15 | FALSE | Downregulated |
| 0,170330613 | 0,764156029 | STIL | FALSE | Upregulated |
| 0,121749241 | 0,764156029 | TRIM34 | FALSE | Upregulated |
| -0,071343991 | 0,764156029 | TTI2 | FALSE | Downregulated |
| 0,063456379 | 0,765138758 | CPSF3 | FALSE | Upregulated |
| 0,655026257 | 0,765138758 | KRTAP5-AS1 | FALSE | Upregulated |
| -0,242568198 | 0,76521365 | GPR85 | FALSE | Downregulated |
| -0,100775439 | 0,76521365 | PRPF39 | FALSE | Downregulated |
| 1,159892356 | 0,765228445 | SCARNA10 | FALSE | Upregulated |
| 0,183695554 | 0,76536585 | C8orf47 | FALSE | Upregulated |
| 0,150569365 | 0,76536585 | SLC17A3 | FALSE | Upregulated |
| -0,157302898 | 0,76536585 | ZC3H13 | FALSE | Downregulated |
| 0,235324433 | 0,765385362 | LZTS1 | FALSE | Upregulated |
| -0,054840126 | 0,765395667 | AGPAT3 | FALSE | Downregulated |
| 0,086536174 | 0,765395667 | C11orf73 | FALSE | Upregulated |
| -0,06070413 | 0,765395667 | CRKL | FALSE | Downregulated |
| 0,138038799 | 0,765395667 | CRYBB2P1 | FALSE | Upregulated |
| -0,117414567 | 0,765395667 | F5 | FALSE | Downregulated |
| -0,043272121 | 0,765395667 | KCMF1 | FALSE | Downregulated |
| 0,083638794 | 0,765395667 | SEC13 | FALSE | Upregulated |
| 0,070679785 | 0,765395667 | SIAH2 | FALSE | Upregulated |
| -0,128059361 | 0,765395667 | SMAD6 | FALSE | Downregulated |
| -0,31611177 | 0,765395667 | TDRD9 | FALSE | Downregulated |
| 0,163657977 | 0,765395667 | ZNF311 | FALSE | Upregulated |
| 0,135521601 | 0,765496694 | APH1B | FALSE | Upregulated |
| -0,455820986 | 0,765496694 | ARHGEF4 | FALSE | Downregulated |

| | | | | |
|---|---|---|---|---|
| 0,175076002 | 0,765496694 | ARSJ | FALSE | Upregulated |
| -0,46828063 | 0,765496694 | CERKL | FALSE | Downregulated |
| 0,329930025 | 0,765496694 | DDIT4 | FALSE | Upregulated |
| -0,10852249 | 0,765496694 | INHBC | FALSE | Downregulated |
| 0,205254467 | 0,765496694 | KIFC2 | FALSE | Upregulated |
| -0,197582616 | 0,765496694 | LOC100132111 | FALSE | Downregulated |
| -0,24852893 | 0,765496694 | LOC101927181 | FALSE | Downregulated |
| -0,442141924 | 0,765496694 | LOC101928965 | FALSE | Downregulated |
| 0,066770446 | 0,765496694 | MFN2 | FALSE | Upregulated |
| -0,370613196 | 0,765496694 | PRKG2 | FALSE | Downregulated |
| 0,263539707 | 0,765496694 | PRR22 | FALSE | Upregulated |
| 0,57161406 | 0,765496694 | SOX30 | FALSE | Upregulated |
| -0,205820826 | 0,765496694 | TIPARP-AS1 | FALSE | Downregulated |
| 0,19720428 | 0,765496694 | ZDHHC19 | FALSE | Upregulated |
| -0,34104741 | 0,765634304 | PRIMA1 | FALSE | Downregulated |
| -0,560546808 | 0,76567943 | ANP32C | FALSE | Downregulated |
| -0,378198952 | 0,76567943 | STAG3L1 | FALSE | Downregulated |
| 0,105995277 | 0,765730571 | SEPT6 | FALSE | Upregulated |
| 0,087205112 | 0,765764325 | AHSA1 | FALSE | Upregulated |
| -0,209825453 | 0,765764325 | CNDP1 | FALSE | Downregulated |
| 0,223617006 | 0,765764325 | FAM86JP | FALSE | Upregulated |
| -0,078727071 | 0,765764325 | SCFD1 | FALSE | Downregulated |
| 0,368296997 | 0,766211496 | TMPRSS3 | FALSE | Upregulated |
| -0,093288388 | 0,766389942 | TASP1 | FALSE | Downregulated |
| 0,872175283 | 0,766397304 | AKR1B15 | FALSE | Upregulated |
| -0,483086946 | 0,766397304 | CCDC154 | FALSE | Downregulated |

| | | | | |
|---|---|---|---|---|
| 0,117897199 | 0,766397304 | CCDC57 | FALSE | Upregulated |
| -0,137176755 | 0,766397304 | FZD1 | FALSE | Downregulated |
| -0,090803785 | 0,766397304 | GPANK1 | FALSE | Downregulated |
| -0,374313453 | 0,766397304 | HIF1A-AS2 | FALSE | Downregulated |
| -0,647665042 | 0,766397304 | HIGD2B | FALSE | Downregulated |
| -0,195968284 | 0,766397304 | LOC101927579 | FALSE | Downregulated |
| 0,363702333 | 0,766397304 | LOC101929571 | FALSE | Upregulated |
| -0,07450989 | 0,766397304 | NHLRC3 | FALSE | Downregulated |
| 0,104107991 | 0,766397304 | POLH | FALSE | Upregulated |
| 0,133846907 | 0,766397304 | SEC23B | FALSE | Upregulated |
| 0,037471314 | 0,766397304 | TERF2IP | FALSE | Upregulated |
| 0,232876394 | 0,766397304 | TRG-AS1 | FALSE | Upregulated |
| -0,086135058 | 0,766397304 | UHRF1BP1 | FALSE | Downregulated |
| 0,107506096 | 0,766527846 | RPS19 | FALSE | Upregulated |
| -0,083155111 | 0,766599837 | BPTF | FALSE | Downregulated |
| 0,104599216 | 0,766599837 | GLTSCR2 | FALSE | Upregulated |
| -0,353760334 | 0,766599837 | SCUBE2 | FALSE | Downregulated |
| 0,181233149 | 0,766614204 | C11orf84 | FALSE | Upregulated |
| 0,082084126 | 0,766614204 | DEF8 | FALSE | Upregulated |
| -0,084069544 | 0,766614204 | IBA57 | FALSE | Downregulated |
| 0,048377164 | 0,766614204 | KANSL3 | FALSE | Upregulated |
| -0,253205292 | 0,766614204 | LOC101927420 | FALSE | Downregulated |
| 0,112047325 | 0,766614204 | PLD2 | FALSE | Upregulated |
| 0,109267015 | 0,766614204 | SLC25A42 | FALSE | Upregulated |
| -0,136639221 | 0,766753818 | OSBPL1A | FALSE | Downregulated |
| 0,159669788 | 0,76705829 | ABHD1 | FALSE | Upregulated |

| | | | | |
|---|---|---|---|---|
| 0,118676434 | 0,76710244 | ABCC10 | FALSE | Upregulated |
| 0,086378618 | 0,76710244 | ATF6 | FALSE | Upregulated |
| 0,227187559 | 0,76710244 | CATSPER3 | FALSE | Upregulated |
| -0,434001168 | 0,76710244 | GNRH2 | FALSE | Downregulated |
| -0,38080079 | 0,76710244 | LOC101929658 | FALSE | Downregulated |
| 0,56826762 | 0,76710244 | LOC151174 | FALSE | Upregulated |
| -0,333349284 | 0,76710244 | NCR3LG1 | FALSE | Downregulated |
| -0,12257693 | 0,76710244 | PANK3 | FALSE | Downregulated |
| 0,084487203 | 0,76710244 | RHEB | FALSE | Upregulated |
| 0,077710411 | 0,76710244 | TBCK | FALSE | Upregulated |
| -0,24372671 | 0,767108933 | ADORA3 | FALSE | Downregulated |
| 0,111603592 | 0,767108933 | C16orf91 | FALSE | Upregulated |
| 0,387432597 | 0,767108933 | C16orf93 | FALSE | Upregulated |
| -0,213305247 | 0,767108933 | GAS1 | FALSE | Downregulated |
| 0,071749757 | 0,767108933 | KIAA1432 | FALSE | Upregulated |
| -0,276994427 | 0,767108933 | LOC100190986 | FALSE | Downregulated |
| -0,360571139 | 0,767108933 | LOC101928462 | FALSE | Downregulated |
| 0,31836797 | 0,767108933 | LOC646278 | FALSE | Upregulated |
| -0,074375669 | 0,767108933 | NDUFA6 | FALSE | Downregulated |
| -0,094806103 | 0,767108933 | NMRAL1 | FALSE | Downregulated |
| 0,244808079 | 0,767108933 | PPM1L | FALSE | Upregulated |
| -0,223965231 | 0,767108933 | PPP1R1A | FALSE | Downregulated |
| 0,338770293 | 0,767108933 | RAB39B | FALSE | Upregulated |
| -0,051149084 | 0,767108933 | RUFY1 | FALSE | Downregulated |
| 0,229888381 | 0,767108933 | SIRPB2 | FALSE | Upregulated |
| -0,155457813 | 0,767108933 | STEAP1 | FALSE | Downregulated |

| | | | | |
|---|---|---|---|---|
| -0,056089725 | 0,767108933 | ZDHHC6 | FALSE | Downregulated |
| 0,227470352 | 0,76718958 | CSF2RB | FALSE | Upregulated |
| -0,204322408 | 0,76718958 | FAM127C | FALSE | Downregulated |
| -0,092232156 | 0,76718958 | OCIAD2 | FALSE | Downregulated |
| -0,368088795 | 0,767394687 | CLEC4GP1 | FALSE | Downregulated |
| -0,237448598 | 0,767473569 | FHOD3 | FALSE | Downregulated |
| 0,642611296 | 0,767473569 | HIGD1C | FALSE | Upregulated |
| -0,093885135 | 0,767966293 | RPS15A | FALSE | Downregulated |
| -0,15567421 | 0,768013562 | IFIT1 | FALSE | Downregulated |
| 0,364840612 | 0,768013562 | KLRC4 | FALSE | Upregulated |
| 0,290349372 | 0,768013562 | PDGFA | FALSE | Upregulated |
| -0,084697679 | 0,768013562 | PTP4A1 | FALSE | Downregulated |
| 0,06651962 | 0,768187853 | ADPGK | FALSE | Upregulated |
| 0,386829175 | 0,768187853 | CCL4L1 | FALSE | Upregulated |
| -0,394575678 | 0,768187853 | FAM45B | FALSE | Downregulated |
| -0,077251362 | 0,768187853 | FEZ2 | FALSE | Downregulated |
| 0,056417815 | 0,768187853 | KDM4A | FALSE | Upregulated |
| 0,076491406 | 0,768187853 | MRPL22 | FALSE | Upregulated |
| -0,145968215 | 0,768187853 | PACSIN3 | FALSE | Downregulated |
| -0,117092831 | 0,768187853 | SEC61G | FALSE | Downregulated |
| 0,201862577 | 0,768187853 | SHD | FALSE | Upregulated |
| -0,079048864 | 0,768187853 | SOAT1 | FALSE | Downregulated |
| 0,138198333 | 0,768187853 | SYDE1 | FALSE | Upregulated |
| 0,117734841 | 0,76823648 | C12orf49 | FALSE | Upregulated |
| -0,423321727 | 0,768306075 | FLJ20464 | FALSE | Downregulated |
| 0,056711475 | 0,768494324 | ADSS | FALSE | Upregulated |

| | | | | |
|---|---|---|---|---|
| -0,075407047 | 0,768696069 | LCORL | FALSE | Downregulated |
| -0,229336559 | 0,768696069 | P4HA1 | FALSE | Downregulated |
| -0,196915087 | 0,768696069 | RAP1GAP | FALSE | Downregulated |
| 0,394326803 | 0,768696069 | RASAL2-AS1 | FALSE | Upregulated |
| -0,173798284 | 0,768696069 | RBM26-AS1 | FALSE | Downregulated |
| -0,305700907 | 0,768696069 | RP11-101E14.2 | FALSE | Downregulated |
| -0,072608153 | 0,768696069 | RPL14 | FALSE | Downregulated |
| -0,082900559 | 0,768696069 | RPS8 | FALSE | Downregulated |
| -0,729650222 | 0,768696069 | SIGLEC12 | FALSE | Downregulated |
| -0,219980433 | 0,768696069 | TSPAN11 | FALSE | Downregulated |
| -0,120897996 | 0,768696069 | UBXN8 | FALSE | Downregulated |
| 0,169158352 | 0,768935881 | C5orf56 | FALSE | Upregulated |
| 0,084756519 | 0,769037914 | ECI1 | FALSE | Upregulated |
| -0,137010599 | 0,769037914 | FAM69B | FALSE | Downregulated |
| -0,209762654 | 0,769064955 | NES | FALSE | Downregulated |
| -0,425654778 | 0,769194237 | NAT8B | FALSE | Downregulated |
| -0,494358649 | 0,769436492 | CTD-2337A12.1 | FALSE | Downregulated |
| 0,449598577 | 0,769507977 | CDRT1 | FALSE | Upregulated |
| 0,061201848 | 0,769509027 | CRK | FALSE | Upregulated |
| 0,077968961 | 0,769509027 | GGCX | FALSE | Upregulated |
| 0,161660278 | 0,769509027 | HGD | FALSE | Upregulated |
| 0,324034298 | 0,769576788 | BOLA3-AS1 | FALSE | Upregulated |
| 0,100937556 | 0,769592267 | ATP8B4 | FALSE | Upregulated |
| 0,069891874 | 0,769592267 | CSNK1G1 | FALSE | Upregulated |
| 0,071257677 | 0,769592267 | EPS8 | FALSE | Upregulated |
| -0,073467763 | 0,769592267 | GCLC | FALSE | Downregulated |

| | | | | |
|---|---|---|---|---|
| -0,132493652 | 0,769592267 | GIMAP1-GIMAP5 | FALSE | Downregulated |
| 0,074080911 | 0,769592267 | PSMD9 | FALSE | Upregulated |
| 0,066820279 | 0,769592267 | SLC35E3 | FALSE | Upregulated |
| 0,325603334 | 0,769948448 | DCDC2 | FALSE | Upregulated |
| 0,089039755 | 0,769948448 | DNAJC30 | FALSE | Upregulated |
| -0,067550302 | 0,769948448 | GABPB1 | FALSE | Downregulated |
| 0,922558923 | 0,769948448 | MGC50722 | FALSE | Upregulated |
| 0,106725923 | 0,769948448 | MYO7A | FALSE | Upregulated |
| 0,102367682 | 0,769948448 | NT5C3A | FALSE | Upregulated |
| 0,068066505 | 0,769948448 | SUMF2 | FALSE | Upregulated |
| 0,061569086 | 0,769948448 | TMEM127 | FALSE | Upregulated |
| 0,107307933 | 0,769948448 | TSEN2 | FALSE | Upregulated |
| -0,237400698 | 0,770008889 | FOXN3-AS1 | FALSE | Downregulated |
| -0,070126754 | 0,770188745 | C1D | FALSE | Downregulated |
| -0,056626239 | 0,770209049 | TMEM106B | FALSE | Downregulated |
| 0,104842946 | 0,770312136 | FRMD8 | FALSE | Upregulated |
| 0,170087998 | 0,77033116 | SCRN1 | FALSE | Upregulated |
| -0,107677282 | 0,770340831 | BHLHE40 | FALSE | Downregulated |
| 0,109697047 | 0,770345394 | AMMECR1L | FALSE | Upregulated |
| -0,378367833 | 0,770345394 | LOC101927382 | FALSE | Downregulated |
| 0,321392771 | 0,770345394 | LRRC1 | FALSE | Upregulated |
| -0,072689661 | 0,770345394 | STX8 | FALSE | Downregulated |
| 0,282106005 | 0,770413734 | OMD | FALSE | Upregulated |
| -0,487457584 | 0,770421704 | LOC102546298 | FALSE | Downregulated |
| -0,28780712 | 0,770421704 | RSPH4A | FALSE | Downregulated |
| 0,486373225 | 0,770573096 | SLC22A20 | FALSE | Upregulated |

| | | | | |
|---|---|---|---|---|
| 0,099755514 | 0,770612538 | MALSU1 | FALSE | Upregulated |
| 0,235398976 | 0,77063131 | AGAP2 | FALSE | Upregulated |
| -0,405316344 | 0,770655805 | LOC388780 | FALSE | Downregulated |
| -0,134156384 | 0,770655902 | LOC100288778 | FALSE | Downregulated |
| -0,129570053 | 0,770789987 | AMY2B | FALSE | Downregulated |
| -0,04896576 | 0,770789987 | SF3A3 | FALSE | Downregulated |
| 0,196759602 | 0,770789987 | TBX15 | FALSE | Upregulated |
| -0,175684916 | 0,771236606 | LOC100272217 | FALSE | Downregulated |
| 0,077279846 | 0,771283451 | MBP | FALSE | Upregulated |
| 0,089718796 | 0,771283451 | NR2F6 | FALSE | Upregulated |
| 0,061251359 | 0,77163591 | XRCC1 | FALSE | Upregulated |
| 0,466221765 | 0,77184642 | NTS | FALSE | Upregulated |
| 0,520530503 | 0,771990803 | GMCL1P1 | FALSE | Upregulated |
| -0,091149732 | 0,771991639 | TMA7 | FALSE | Downregulated |
| -0,324415174 | 0,77205647 | HNRNPA1P33 | FALSE | Downregulated |
| -0,610388255 | 0,772154336 | DYX1C1-CCPG1 | FALSE | Downregulated |
| -0,330344536 | 0,772154336 | FADS1 | FALSE | Downregulated |
| 0,116258827 | 0,772154336 | GPR39 | FALSE | Upregulated |
| 0,291780439 | 0,772154336 | LYPD3 | FALSE | Upregulated |
| -0,340658558 | 0,772169321 | HOXA-AS2 | FALSE | Downregulated |
| 0,097186478 | 0,772216978 | NLRX1 | FALSE | Upregulated |
| -0,172518382 | 0,772225844 | MICA | FALSE | Downregulated |
| 0,183795046 | 0,772257632 | PKIB | FALSE | Upregulated |
| -0,216931413 | 0,772579306 | ENAM | FALSE | Downregulated |
| 0,107393375 | 0,772579306 | LIG1 | FALSE | Upregulated |
| 0,27273165 | 0,772579306 | LOC101929356 | FALSE | Upregulated |

| | | | | |
|---|---|---|---|---|
| 0,127015811 | 0,772579306 | NSMF | FALSE | Upregulated |
| 0,107544854 | 0,772579306 | SHF | FALSE | Upregulated |
| 0,337282694 | 0,772579306 | SLC26A5 | FALSE | Upregulated |
| -0,172560136 | 0,772588888 | HSD17B13 | FALSE | Downregulated |
| -0,149257964 | 0,772588888 | LHX2 | FALSE | Downregulated |
| -0,106540047 | 0,772588888 | TBX3 | FALSE | Downregulated |
| 0,222889416 | 0,772830683 | LOC100093631 | FALSE | Upregulated |
| -0,072860099 | 0,772830683 | MOB1A | FALSE | Downregulated |
| -0,131323894 | 0,772830683 | PAG1 | FALSE | Downregulated |
| 0,205105206 | 0,772830683 | PDLIM7 | FALSE | Upregulated |
| -0,142817134 | 0,772830683 | PUS7L | FALSE | Downregulated |
| -0,059710901 | 0,772830683 | SLMAP | FALSE | Downregulated |
| 0,068661513 | 0,772830683 | TAPBP | FALSE | Upregulated |
| -0,12797506 | 0,772830683 | TDRKH | FALSE | Downregulated |
| 0,072048104 | 0,772830683 | TMA16 | FALSE | Upregulated |
| 1,930121265 | 0,773362854 | C15orf38-AP3S2 | FALSE | Upregulated |
| -0,102986863 | 0,773362854 | L2HGDH | FALSE | Downregulated |
| -0,078229547 | 0,773362854 | PDHA1 | FALSE | Downregulated |
| 0,089333396 | 0,773362854 | PLGRKT | FALSE | Upregulated |
| 0,10633141 | 0,773362854 | TUT1 | FALSE | Upregulated |
| -0,109904419 | 0,773378863 | CAMK2N1 | FALSE | Downregulated |
| -0,092485263 | 0,773378863 | ERG | FALSE | Downregulated |
| 0,579675012 | 0,773378863 | NCRNA00185 | FALSE | Upregulated |
| -0,235934364 | 0,773585858 | SULT1E1 | FALSE | Downregulated |
| -0,301671117 | 0,773675576 | CHRNA10 | FALSE | Downregulated |
| -0,149216249 | 0,773675576 | SLC19A2 | FALSE | Downregulated |

| | | | | |
|---|---|---|---|---|
| 0,144587536 | 0,773685539 | C19orf48 | FALSE | Upregulated |
| 0,39197931 | 0,773734804 | LOC102724314 | FALSE | Upregulated |
| -0,064909401 | 0,773879963 | RALBP1 | FALSE | Downregulated |
| 0,094536007 | 0,773879963 | UBL7 | FALSE | Upregulated |
| -0,047250212 | 0,774192669 | GOPC | FALSE | Downregulated |
| 0,072979736 | 0,774192669 | MLLT6 | FALSE | Upregulated |
| 0,133550152 | 0,774192669 | USB1 | FALSE | Upregulated |
| 0,467371086 | 0,774192669 | WDR93 | FALSE | Upregulated |
| 0,078089925 | 0,774202157 | GTPBP1 | FALSE | Upregulated |
| -0,059734896 | 0,77431925 | FAM134C | FALSE | Downregulated |
| 0,050343435 | 0,774553957 | RBBP4 | FALSE | Upregulated |
| 0,064997249 | 0,77460893 | TBC1D22A | FALSE | Upregulated |
| 0,233014372 | 0,774890222 | DHRS9 | FALSE | Upregulated |
| 0,472789448 | 0,774890222 | HOXB-AS1 | FALSE | Upregulated |
| 0,416013437 | 0,774890222 | MYLK-AS1 | FALSE | Upregulated |
| -0,217800539 | 0,774890222 | RP11-333E1.1 | FALSE | Downregulated |
| -0,116510678 | 0,774890222 | TIAM1 | FALSE | Downregulated |
| -0,101654646 | 0,775187872 | DECR2 | FALSE | Downregulated |
| -0,075484607 | 0,775187872 | IGBP1 | FALSE | Downregulated |
| -0,104999947 | 0,775187872 | PARP9 | FALSE | Downregulated |
| 0,226543453 | 0,775187872 | ZNF503-AS1 | FALSE | Upregulated |
| 0,068011622 | 0,77522136 | F11R | FALSE | Upregulated |
| -0,083851124 | 0,775223429 | ERP29 | FALSE | Downregulated |
| 0,079584644 | 0,775223429 | PSMB4 | FALSE | Upregulated |
| -0,337839246 | 0,775440682 | PCDH19 | FALSE | Downregulated |
| -0,122137857 | 0,775440682 | SLC5A3 | FALSE | Downregulated |

| | | | | |
|---|---|---|---|---|
| -0,094593894 | 0,77548764 | PTPN3 | FALSE | Downregulated |
| 0,190674227 | 0,775491942 | NKG7 | FALSE | Upregulated |
| 0,062735517 | 0,775531245 | RRP1B | FALSE | Upregulated |
| -0,180880521 | 0,775689316 | CLDN14 | FALSE | Downregulated |
| -0,057390432 | 0,775689316 | NOA1 | FALSE | Downregulated |
| 0,418388845 | 0,77570031 | HSPA7 | FALSE | Upregulated |
| 0,291387715 | 0,775700717 | GRHL2 | FALSE | Upregulated |
| -0,222487785 | 0,775700717 | LAMTOR5-AS1 | FALSE | Downregulated |
| -0,055626962 | 0,775700717 | OSTF1 | FALSE | Downregulated |
| -0,09747077 | 0,775700717 | PHF7 | FALSE | Downregulated |
| -0,068512441 | 0,775781893 | ARHGEF12 | FALSE | Downregulated |
| 0,119388493 | 0,775781893 | CD93 | FALSE | Upregulated |
| 0,108397454 | 0,775781893 | GALC | FALSE | Upregulated |
| 0,133816862 | 0,775781893 | LLGL1 | FALSE | Upregulated |
| 0,216621845 | 0,775781893 | LOC374443 | FALSE | Upregulated |
| -0,326450161 | 0,775781893 | TWIST1 | FALSE | Downregulated |
| 0,10156861 | 0,775890888 | ACSL5 | FALSE | Upregulated |
| 0,096314025 | 0,775890888 | ACTR3C | FALSE | Upregulated |
| 0,38889056 | 0,775890888 | C1orf233 | FALSE | Upregulated |
| -0,068070722 | 0,775890888 | C1QBP | FALSE | Downregulated |
| -0,054956935 | 0,775890888 | C9orf78 | FALSE | Downregulated |
| -0,083704424 | 0,775890888 | INPP5A | FALSE | Downregulated |
| 0,149906209 | 0,775890888 | LINC01138 | FALSE | Upregulated |
| 0,403035458 | 0,775890888 | MST1R | FALSE | Upregulated |
| -0,480147821 | 0,775890888 | PLEKHG4B | FALSE | Downregulated |
| 0,076337034 | 0,775890888 | ST6GALNAC6 | FALSE | Upregulated |

| | | | | |
|---|---|---|---|---|
| 0,070403412 | 0,775890888 | UGGT2 | FALSE | Upregulated |
| -0,240412198 | 0,775890888 | UNC93A | FALSE | Downregulated |
| 0,103528612 | 0,776135182 | F8 | FALSE | Upregulated |
| 0,472854507 | 0,776135182 | LOC102724737 | FALSE | Upregulated |
| 0,091991352 | 0,776135182 | RBX1 | FALSE | Upregulated |
| 0,092411544 | 0,776135763 | DIP2A | FALSE | Upregulated |
| 0,276084742 | 0,776135763 | SAMD15 | FALSE | Upregulated |
| 0,077289212 | 0,776464341 | ATRN | FALSE | Upregulated |
| 0,081304331 | 0,776464341 | ITGA5 | FALSE | Upregulated |
| 0,137643894 | 0,776464341 | MEGF6 | FALSE | Upregulated |
| -0,111759511 | 0,776464341 | NAT1 | FALSE | Downregulated |
| -0,087261548 | 0,776464341 | ZNF26 | FALSE | Downregulated |
| 0,38028147 | 0,776491382 | TENM4 | FALSE | Upregulated |
| -0,128839985 | 0,776582282 | DUSP28 | FALSE | Downregulated |
| -0,126361876 | 0,776582282 | ELTD1 | FALSE | Downregulated |
| 0,374389628 | 0,776582282 | LOC102724231 | FALSE | Upregulated |
| 0,126191607 | 0,776582282 | PCGF2 | FALSE | Upregulated |
| 0,172444487 | 0,776599895 | C15orf62 | FALSE | Upregulated |
| -0,117998481 | 0,776599895 | C9orf72 | FALSE | Downregulated |
| 0,085876359 | 0,776599895 | MGLL | FALSE | Upregulated |
| -0,068850719 | 0,776599895 | RPL15 | FALSE | Downregulated |
| -0,457581664 | 0,776599895 | SLCO4A1 | FALSE | Downregulated |
| 0,124415923 | 0,776599895 | UQCRBP1 | FALSE | Upregulated |
| -0,4241924 | 0,776599895 | VWDE | FALSE | Downregulated |
| -0,067138604 | 0,776600972 | IPO8 | FALSE | Downregulated |
| 0,053255112 | 0,776600972 | KIF13B | FALSE | Upregulated |

| | | | | |
|---|---|---|---|---|
| -0,109554198 | 0,776600972 | LIFR | FALSE | Downregulated |
| -0,138957924 | 0,776600972 | LRRC32 | FALSE | Downregulated |
| -0,218618449 | 0,776821152 | CCDC151 | FALSE | Downregulated |
| -0,054060295 | 0,776821152 | SECISBP2L | FALSE | Downregulated |
| -0,087234038 | 0,776821152 | SLC25A16 | FALSE | Downregulated |
| -0,337590206 | 0,77687446 | FOXQ1 | FALSE | Downregulated |
| 0,142978596 | 0,776894818 | ICAM3 | FALSE | Upregulated |
| 0,184899617 | 0,776894818 | LINGO1 | FALSE | Upregulated |
| -0,332418039 | 0,776894818 | LOC101928058 | FALSE | Downregulated |
| 0,185991603 | 0,776894818 | PCSK9 | FALSE | Upregulated |
| -0,084112554 | 0,776894818 | POU2F1 | FALSE | Downregulated |
| -0,161509337 | 0,776894818 | PTN | FALSE | Downregulated |
| -0,07745705 | 0,776894818 | SCAPER | FALSE | Downregulated |
| -0,187131738 | 0,776894818 | TMEM52 | FALSE | Downregulated |
| 0,175929022 | 0,776894818 | TSPAN1 | FALSE | Upregulated |
| -0,085515248 | 0,777049207 | ATL3 | FALSE | Downregulated |
| -0,066190517 | 0,777049207 | DCTN3 | FALSE | Downregulated |
| 0,094281448 | 0,777049207 | DDI2 | FALSE | Upregulated |
| 0,080820193 | 0,777049207 | ERI3 | FALSE | Upregulated |
| 0,493882655 | 0,777049207 | LINC00092 | FALSE | Upregulated |
| 0,438472121 | 0,777049207 | LOC100128108 | FALSE | Upregulated |
| -0,410022248 | 0,777049207 | LOC100505501 | FALSE | Downregulated |
| 0,054389831 | 0,777049207 | LRPAP1 | FALSE | Upregulated |
| -0,07868852 | 0,777049207 | POLR1C | FALSE | Downregulated |
| -0,118140719 | 0,777049207 | SLC2A4RG | FALSE | Downregulated |
| 0,270714385 | 0,777049207 | SNORA40 | FALSE | Upregulated |

| | | | | |
|---|---|---|---|---|
| 0,113700116 | 0,777049207 | ZNF335 | FALSE | Upregulated |
| 0,3743506 | 0,777117221 | KLRC3 | FALSE | Upregulated |
| -0,085965378 | 0,777117221 | RNF216P1 | FALSE | Downregulated |
| 0,349133889 | 0,777117221 | UCP3 | FALSE | Upregulated |
| 0,192654753 | 0,77724758 | NCF1C | FALSE | Upregulated |
| -0,102566875 | 0,777401183 | GZF1 | FALSE | Downregulated |
| -0,385060637 | 0,777401183 | RHEBL1 | FALSE | Downregulated |
| -0,225946618 | 0,777401183 | VNN2 | FALSE | Downregulated |
| 0,098526329 | 0,777484159 | DRG2 | FALSE | Upregulated |
| 0,370774002 | 0,777484159 | RP1-257A7.4 | FALSE | Upregulated |
| 0,28520804 | 0,777507983 | MIRLET7BHG | FALSE | Upregulated |
| 0,091222644 | 0,777616903 | WDR81 | FALSE | Upregulated |
| 0,357691374 | 0,777780714 | CLMP | FALSE | Upregulated |
| 0,06811564 | 0,778068629 | MOCS1 | FALSE | Upregulated |
| 0,41463895 | 0,778232754 | DEFB132 | FALSE | Upregulated |
| 0,126547593 | 0,778232754 | DTYMK | FALSE | Upregulated |
| -0,202136975 | 0,778232754 | NPIPB5 | FALSE | Downregulated |
| 0,180498918 | 0,778232754 | TEX9 | FALSE | Upregulated |
| 0,275311487 | 0,778232754 | TNXB | FALSE | Upregulated |
| -0,093480121 | 0,778232754 | TTC30A | FALSE | Downregulated |
| 0,111115216 | 0,778232754 | UROC1 | FALSE | Upregulated |
| 0,069988682 | 0,778232754 | ZNF616 | FALSE | Upregulated |
| 0,104942166 | 0,778308315 | CABLES2 | FALSE | Upregulated |
| 0,119757006 | 0,778308315 | PYCARD | FALSE | Upregulated |
| -0,093735174 | 0,778308315 | RABAC1 | FALSE | Downregulated |
| 0,152276202 | 0,778356219 | MS4A4A | FALSE | Upregulated |

| | | | | |
|---|---|---|---|---|
| 0,087248414 | 0,778379433 | GRHPR | FALSE | Upregulated |
| -0,06527557 | 0,778579194 | COQ7 | FALSE | Downregulated |
| 0,120104735 | 0,778731637 | NXPE3 | FALSE | Upregulated |
| -0,210824959 | 0,778731637 | RPL19P12 | FALSE | Downregulated |
| -0,120836371 | 0,778837989 | PLBD1 | FALSE | Downregulated |
| -0,256045268 | 0,778837989 | PMAIP1 | FALSE | Downregulated |
| -0,111123368 | 0,778837989 | USP46-AS1 | FALSE | Downregulated |
| 0,128754941 | 0,779045372 | CYB5RL | FALSE | Upregulated |
| 0,092067714 | 0,779045372 | TSC22D1 | FALSE | Upregulated |
| -0,080829079 | 0,779231157 | GEMIN6 | FALSE | Downregulated |
| 0,186742293 | 0,779439885 | AC009948.5 | FALSE | Upregulated |
| -0,101810458 | 0,779648243 | CCSER2 | FALSE | Downregulated |
| 0,084524768 | 0,77968189 | COQ10B | FALSE | Upregulated |
| 0,990163968 | 0,77968189 | CRISP3 | FALSE | Upregulated |
| 0,190322719 | 0,779699515 | BSN | FALSE | Upregulated |
| 0,452062856 | 0,779699515 | CNBD2 | FALSE | Upregulated |
| 0,140910317 | 0,779699515 | HSPA1L | FALSE | Upregulated |
| -0,29823169 | 0,779699515 | LOC101927610 | FALSE | Downregulated |
| 0,083138297 | 0,779699515 | PIP4K2A | FALSE | Upregulated |
| -0,210021094 | 0,779699515 | S1PR5 | FALSE | Downregulated |
| -0,244260713 | 0,779786079 | CNTNAP3 | FALSE | Downregulated |
| 0,202535545 | 0,779904484 | PDZRN3 | FALSE | Upregulated |
| -0,074320008 | 0,779930484 | ALDH9A1 | FALSE | Downregulated |
| -0,078248603 | 0,779930484 | ASH1L | FALSE | Downregulated |
| 0,144838091 | 0,779930484 | DRAM1 | FALSE | Upregulated |
| -0,118145726 | 0,779930484 | FBXL15 | FALSE | Downregulated |

| | | | | |
|---|---|---|---|---|
| 0,124570814 | 0,779930484 | GRB7 | FALSE | Upregulated |
| -0,197755983 | 0,779930484 | LOC283070 | FALSE | Downregulated |
| -0,325418399 | 0,779930484 | LRIT3 | FALSE | Downregulated |
| 0,44521436 | 0,779930484 | SYCP2L | FALSE | Upregulated |
| -0,053044086 | 0,779930484 | USP9X | FALSE | Downregulated |
| -0,098978641 | 0,780486125 | ZNF396 | FALSE | Downregulated |
| 0,099674794 | 0,780558009 | ZNFX1 | FALSE | Upregulated |
| 0,096272048 | 0,780772118 | HAUS8 | FALSE | Upregulated |
| -0,073740679 | 0,780772118 | SOWAHC | FALSE | Downregulated |
| 0,25516761 | 0,780772234 | ENPP5 | FALSE | Upregulated |
| 0,249056221 | 0,780772234 | ILDR2 | FALSE | Upregulated |
| 0,285412219 | 0,780772234 | NALCN | FALSE | Upregulated |
| 0,098043729 | 0,780772234 | NECAB3 | FALSE | Upregulated |
| -0,194670373 | 0,780772234 | SNHG17 | FALSE | Downregulated |
| 0,110296452 | 0,780872999 | HAUS7 | FALSE | Upregulated |
| -0,06995539 | 0,780872999 | KIAA0391 | FALSE | Downregulated |
| -0,139471798 | 0,781575054 | LINC00663 | FALSE | Downregulated |
| 0,480068277 | 0,781575054 | LOC101927070 | FALSE | Upregulated |
| -0,117843376 | 0,781575054 | LOC730102 | FALSE | Downregulated |
| 0,06039842 | 0,781575054 | PLEKHA3 | FALSE | Upregulated |
| -0,282750843 | 0,781594327 | LOC101927300 | FALSE | Downregulated |
| 0,295484798 | 0,781772285 | GFPT2 | FALSE | Upregulated |
| 0,083729508 | 0,781901783 | CCDC106 | FALSE | Upregulated |
| -0,070386524 | 0,781901783 | GNG10 | FALSE | Downregulated |
| 0,071746637 | 0,781955139 | EIF3F | FALSE | Upregulated |
| -0,053277516 | 0,782153194 | CDC23 | FALSE | Downregulated |

| | | | | |
|---|---|---|---|---|
| -0,09535982 | 0,782250153 | PRELID1 | FALSE | Downregulated |
| 0,085531791 | 0,78257406 | ELMSAN1 | FALSE | Upregulated |
| 0,125599714 | 0,78259606 | PIK3IP1 | FALSE | Upregulated |
| 0,315667103 | 0,782649869 | PCBP3 | FALSE | Upregulated |
| 0,117087569 | 0,782649869 | SRXN1 | FALSE | Upregulated |
| 0,174024586 | 0,782649869 | TRABD2B | FALSE | Upregulated |
| 0,081228652 | 0,782727669 | ATIC | FALSE | Upregulated |
| -0,152615901 | 0,782727669 | CNRIP1 | FALSE | Downregulated |
| -0,243989181 | 0,782727669 | CTGF | FALSE | Downregulated |
| 0,097902703 | 0,782727669 | DIXDC1 | FALSE | Upregulated |
| 0,768265702 | 0,782727669 | LOC102724951 | FALSE | Upregulated |
| -0,06257757 | 0,782727669 | POGK | FALSE | Downregulated |
| -0,238570843 | 0,782727669 | PPM1N | FALSE | Downregulated |
| -0,324960502 | 0,782727669 | RNA45S5 | FALSE | Downregulated |
| 0,388903951 | 0,782727669 | SLC44A5 | FALSE | Upregulated |
| 0,045156538 | 0,782727669 | USP4 | FALSE | Upregulated |
| -0,196034834 | 0,782727669 | ZNF287 | FALSE | Downregulated |
| -0,260683063 | 0,782771261 | AKR1CL1 | FALSE | Downregulated |
| -0,164840986 | 0,782771261 | CDKN2D | FALSE | Downregulated |
| -0,056333714 | 0,782771261 | FAM8A1 | FALSE | Downregulated |
| -0,357103298 | 0,782771261 | FHDC1 | FALSE | Downregulated |
| -0,453784097 | 0,782771261 | GRIN2A | FALSE | Downregulated |
| -0,239713131 | 0,782771261 | LINC00263 | FALSE | Downregulated |
| 0,058219177 | 0,782771261 | LRP5 | FALSE | Upregulated |
| -0,050976808 | 0,782771261 | RAB12 | FALSE | Downregulated |
| 0,109967221 | 0,782771261 | RPS18 | FALSE | Upregulated |

| | | | | |
|---|---|---|---|---|
| -0,07422553 | 0,78289552 | PIPOX | FALSE | Downregulated |
| -0,248842282 | 0,782897124 | BBOX1 | FALSE | Downregulated |
| 0,332478148 | 0,783023183 | LOC646471 | FALSE | Upregulated |
| -0,162237046 | 0,783023183 | WHAMMP2 | FALSE | Downregulated |
| -0,114175998 | 0,783042844 | GAS2 | FALSE | Downregulated |
| 0,119204968 | 0,783253342 | DCHS1 | FALSE | Upregulated |
| 0,081557319 | 0,783253342 | GM2A | FALSE | Upregulated |
| -0,404856563 | 0,783253342 | HNRNPCL1 | FALSE | Downregulated |
| 0,499562463 | 0,783253342 | LOC102724765 | FALSE | Upregulated |
| 0,077263439 | 0,783253342 | MTMR3 | FALSE | Upregulated |
| -0,148861707 | 0,783253342 | NABP1 | FALSE | Downregulated |
| 0,227730558 | 0,783253342 | PRKD1 | FALSE | Upregulated |
| -0,061079006 | 0,783253342 | SLC35A1 | FALSE | Downregulated |
| -0,108342092 | 0,783253342 | THAP9-AS1 | FALSE | Downregulated |
| -0,121968286 | 0,783253342 | TRIM16 | FALSE | Downregulated |
| 0,313044115 | 0,783526166 | CDCA3 | FALSE | Upregulated |
| 0,453682385 | 0,783613557 | AP4B1-AS1 | FALSE | Upregulated |
| 0,127922551 | 0,783613557 | STXBP1 | FALSE | Upregulated |
| -0,07834604 | 0,783613557 | ZNF225 | FALSE | Downregulated |
| 0,112002047 | 0,783635236 | D2HGDH | FALSE | Upregulated |
| 0,318579566 | 0,783985612 | FAM83H-AS1 | FALSE | Upregulated |
| 0,146133767 | 0,783998579 | EIF4EBP3 | FALSE | Upregulated |
| -0,59975218 | 0,783998579 | IGSF1 | FALSE | Downregulated |
| -0,064720393 | 0,784068718 | KDM4C | FALSE | Downregulated |
| -0,151510373 | 0,784410825 | CCDC138 | FALSE | Downregulated |
| 0,344608416 | 0,784410825 | CRTAC1 | FALSE | Upregulated |

| | | | | |
|---|---|---|---|---|
| -0,06931896 | 0,784410825 | DCAF17 | FALSE | Downregulated |
| -0,080008786 | 0,784410825 | DHCR24 | FALSE | Downregulated |
| -0,399515359 | 0,784410825 | DLEU2L | FALSE | Downregulated |
| 0,276151803 | 0,784410825 | LOC100289230 | FALSE | Upregulated |
| -0,18133573 | 0,784410825 | PRKCQ-AS1 | FALSE | Downregulated |
| 0,077204773 | 0,784410825 | RNF145 | FALSE | Upregulated |
| -0,114805595 | 0,784410825 | SPIN3 | FALSE | Downregulated |
| -0,067336206 | 0,784436276 | ALKBH5 | FALSE | Downregulated |
| 0,259056855 | 0,784560037 | CBWD6 | FALSE | Upregulated |
| -0,100587002 | 0,784560037 | PTBP2 | FALSE | Downregulated |
| 0,103422888 | 0,784560037 | SLC17A5 | FALSE | Upregulated |
| -0,051337751 | 0,784664598 | VPS35 | FALSE | Downregulated |
| -0,062598248 | 0,784664598 | WRNIP1 | FALSE | Downregulated |
| -0,651152984 | 0,784735894 | MEOX1 | FALSE | Downregulated |
| -0,105036523 | 0,784751276 | TANGO6 | FALSE | Downregulated |
| 0,059734644 | 0,785030458 | COASY | FALSE | Upregulated |
| -0,408663105 | 0,785030458 | SENCR | FALSE | Downregulated |
| -0,0577464 | 0,785084521 | CTNND1 | FALSE | Downregulated |
| 0,298418984 | 0,785170492 | H1FX-AS1 | FALSE | Upregulated |
| 0,084315712 | 0,785170492 | PDLIM1 | FALSE | Upregulated |
| 0,260628136 | 0,78534796 | LOC102723794 | FALSE | Upregulated |
| -0,171154616 | 0,78534796 | PLA2G4A | FALSE | Downregulated |
| -0,19379717 | 0,78534796 | ZNF521 | FALSE | Downregulated |
| 0,072132947 | 0,785472273 | ADCK3 | FALSE | Upregulated |
| 0,61782815 | 0,785472273 | DUOX2 | FALSE | Upregulated |
| 0,318669446 | 0,785585609 | LOC101928895 | FALSE | Upregulated |

| | | | | |
|---|---|---|---|---|
| 0,275346988 | 0,785585609 | LTB4R2 | FALSE | Upregulated |
| -0,126557937 | 0,785597915 | ZNF585B | FALSE | Downregulated |
| 0,117939187 | 0,78561565 | NRBP2 | FALSE | Upregulated |
| -0,102301281 | 0,786069783 | CCSAP | FALSE | Downregulated |
| 0,134575501 | 0,786192819 | ACP5 | FALSE | Upregulated |
| 0,080047169 | 0,786192819 | MTFMT | FALSE | Upregulated |
| 0,08602149 | 0,786192819 | RPL8 | FALSE | Upregulated |
| 0,157551277 | 0,786212799 | IL17RB | FALSE | Upregulated |
| 0,550087923 | 0,786212799 | PTCD1 | FALSE | Upregulated |
| -0,151378164 | 0,786257321 | ZNF117 | FALSE | Downregulated |
| 0,096299767 | 0,786257321 | ZNF707 | FALSE | Upregulated |
| 0,196380421 | 0,786420921 | ELN | FALSE | Upregulated |
| 0,081566412 | 0,786511813 | KIAA0753 | FALSE | Upregulated |
| 0,144533711 | 0,786529971 | ARHGEF17 | FALSE | Upregulated |
| -0,281373519 | 0,786529971 | EBF1 | FALSE | Downregulated |
| 0,098549589 | 0,786529971 | ERCC6L2 | FALSE | Upregulated |
| -0,476411874 | 0,786529971 | FCAR | FALSE | Downregulated |
| 0,155956586 | 0,786529971 | KLF2 | FALSE | Upregulated |
| 0,385960187 | 0,786529971 | LOC102723683 | FALSE | Upregulated |
| 0,255618943 | 0,786529971 | PAQR5 | FALSE | Upregulated |
| -0,65156949 | 0,786529971 | RSPO1 | FALSE | Downregulated |
| 0,167827463 | 0,786529971 | SH3BP1 | FALSE | Upregulated |
| 0,056756771 | 0,786529971 | SNRK | FALSE | Upregulated |
| -0,13021881 | 0,786529971 | VEGFA | FALSE | Downregulated |
| -0,056848136 | 0,786529971 | VPS72 | FALSE | Downregulated |
| 0,106297668 | 0,786529971 | ZNF414 | FALSE | Upregulated |

| | | | | |
|---|---|---|---|---|
| -0,054333091 | 0,78658812 | PCID2 | FALSE | Downregulated |
| -0,103928918 | 0,78658812 | PPOX | FALSE | Downregulated |
| -0,056117085 | 0,78658812 | PSMD6 | FALSE | Downregulated |
| -0,232926423 | 0,78658812 | SMARCD3 | FALSE | Downregulated |
| -0,200098068 | 0,78658812 | SNORA18 | FALSE | Downregulated |
| -0,077208771 | 0,786639832 | CYP4F11 | FALSE | Downregulated |
| 0,062772827 | 0,786648862 | CCNI | FALSE | Upregulated |
| -0,108967547 | 0,786648862 | CGN | FALSE | Downregulated |
| 0,076591706 | 0,786648862 | MAN1A2 | FALSE | Upregulated |
| 0,107260612 | 0,786648862 | PGBD4 | FALSE | Upregulated |
| 0,25145523 | 0,786648862 | PROM2 | FALSE | Upregulated |
| -0,162859633 | 0,786648862 | TRAF3IP1 | FALSE | Downregulated |
| 0,065089566 | 0,786648862 | VRK3 | FALSE | Upregulated |
| 0,112145533 | 0,786766585 | ERI1 | FALSE | Upregulated |
| 0,250875824 | 0,786837859 | LOC100507462 | FALSE | Upregulated |
| 0,105065119 | 0,786837859 | ZNF79 | FALSE | Upregulated |
| -0,143277698 | 0,786888392 | CDC14A | FALSE | Downregulated |
| -0,077084634 | 0,78702287 | AGPS | FALSE | Downregulated |
| 0,041433614 | 0,78702287 | DDX41 | FALSE | Upregulated |
| -0,690757535 | 0,78702287 | DMC1 | FALSE | Downregulated |
| 0,544923304 | 0,78702287 | GSDMA | FALSE | Upregulated |
| -0,73648788 | 0,78702287 | PYGM | FALSE | Downregulated |
| -0,052142464 | 0,78702287 | SEPT2 | FALSE | Downregulated |
| -0,067595553 | 0,787067489 | MTA1 | FALSE | Downregulated |
| -0,04783122 | 0,787067734 | CAPN7 | FALSE | Downregulated |
| -0,128332323 | 0,787205185 | ARAP3 | FALSE | Downregulated |

| | | | | |
|---|---|---|---|---|
| -0,079022798 | 0,787256552 | ZSCAN32 | FALSE | Downregulated |
| -0,082210684 | 0,787409108 | P2RX4 | FALSE | Downregulated |
| -0,086650272 | 0,787619627 | SENP7 | FALSE | Downregulated |
| 0,321249603 | 0,787661888 | FAM189A1 | FALSE | Upregulated |
| -0,178771644 | 0,787712644 | CD160 | FALSE | Downregulated |
| -0,097578262 | 0,787737967 | FRK | FALSE | Downregulated |
| 0,624848892 | 0,787750699 | TCL1A | FALSE | Upregulated |
| 0,078789748 | 0,787750699 | ZNF516 | FALSE | Upregulated |
| 0,085407398 | 0,787967298 | ACOX1 | FALSE | Upregulated |
| 0,118423693 | 0,787967298 | DNMT1 | FALSE | Upregulated |
| 0,604427103 | 0,787972165 | LOC102723390 | FALSE | Upregulated |
| -0,106830693 | 0,788422649 | APIP | FALSE | Downregulated |
| -0,301132045 | 0,788422649 | BCYRN1 | FALSE | Downregulated |
| 0,069741649 | 0,788422649 | CRAT | FALSE | Upregulated |
| 0,441529746 | 0,788422649 | FPGT-TNNI3K | FALSE | Upregulated |
| 0,081574313 | 0,788422649 | LACE1 | FALSE | Upregulated |
| -0,060523319 | 0,788422649 | LOC100129034 | FALSE | Downregulated |
| 0,423525168 | 0,788422649 | LOC100132356 | FALSE | Upregulated |
| -0,13563768 | 0,788422649 | LOC101928061 | FALSE | Downregulated |
| 0,241816807 | 0,788422649 | LOC101928595 | FALSE | Upregulated |
| 0,50281882 | 0,788422649 | MGAT4C | FALSE | Upregulated |
| -0,077320291 | 0,788422649 | MKS1 | FALSE | Downregulated |
| -0,129195852 | 0,788422649 | MREG | FALSE | Downregulated |
| 0,073517679 | 0,788422649 | ZNF746 | FALSE | Upregulated |
| -0,347236884 | 0,788770454 | ADAM23 | FALSE | Downregulated |
| -0,059619714 | 0,788770454 | ATE1 | FALSE | Downregulated |

| | | | | |
|---|---|---|---|---|
| -0,558568046 | 0,788770454 | CTAGE10P | FALSE | Downregulated |
| -0,145015386 | 0,788770454 | IRS2 | FALSE | Downregulated |
| 1,027906739 | 0,788770454 | KLRC4-KLRK1 | FALSE | Upregulated |
| -0,465259995 | 0,788770814 | CCDC114 | FALSE | Downregulated |
| -0,831646551 | 0,788770814 | LOC100505874 | FALSE | Downregulated |
| -0,170523883 | 0,788770814 | LOC100652824 | FALSE | Downregulated |
| -0,100264028 | 0,788770814 | NBPF14 | FALSE | Downregulated |
| 0,186933112 | 0,789143524 | LOC100507373 | FALSE | Upregulated |
| 0,090121193 | 0,789253044 | AGXT | FALSE | Upregulated |
| 0,073903727 | 0,789298928 | AP2S1 | FALSE | Upregulated |
| -0,105881283 | 0,789298928 | CCL16 | FALSE | Downregulated |
| -0,058111123 | 0,789298928 | NSL1 | FALSE | Downregulated |
| -0,115624606 | 0,789298928 | RHOBTB1 | FALSE | Downregulated |
| 0,258067499 | 0,789298928 | SLC13A2 | FALSE | Upregulated |
| -0,125560647 | 0,789298928 | TREX1 | FALSE | Downregulated |
| -0,10977474 | 0,78936323 | CCPG1 | FALSE | Downregulated |
| 0,190317162 | 0,789593488 | ARHGAP44 | FALSE | Upregulated |
| -0,131665103 | 0,789593488 | GIMAP7 | FALSE | Downregulated |
| -0,335981791 | 0,789593488 | NACAP1 | FALSE | Downregulated |
| 0,310540916 | 0,789593488 | RP11-540A21.2 | FALSE | Upregulated |
| 0,224061071 | 0,789612748 | DAPK2 | FALSE | Upregulated |
| 0,364643232 | 0,789612748 | KLHL7-AS1 | FALSE | Upregulated |
| -0,45629002 | 0,789612748 | LINC00266-1 | FALSE | Downregulated |
| -0,069580806 | 0,789612748 | MLX | FALSE | Downregulated |
| 0,436442229 | 0,789612748 | MMP16 | FALSE | Upregulated |
| 0,243925751 | 0,789612748 | NS3BP | FALSE | Upregulated |

| | | | | |
|---|---|---|---|---|
| 0,83824929 | 0,789612748 | PRR5-ARHGAP8 | FALSE | Upregulated |
| -0,926912635 | 0,789612748 | TRNAG22 | FALSE | Downregulated |
| 0,096696565 | 0,78997446 | PEAK1 | FALSE | Upregulated |
| -0,086666013 | 0,78997446 | SMAD1 | FALSE | Downregulated |
| 0,098774862 | 0,790040401 | ANKRD6 | FALSE | Upregulated |
| 0,443816571 | 0,790040401 | RP3-510O8.4 | FALSE | Upregulated |
| 0,421830652 | 0,790413569 | LOC101929533 | FALSE | Upregulated |
| 0,105611059 | 0,790494863 | ACSS3 | FALSE | Upregulated |
| -0,080539613 | 0,790494863 | COX16 | FALSE | Downregulated |
| -0,107317697 | 0,790494863 | COX8A | FALSE | Downregulated |
| 0,066650534 | 0,790494863 | FASTKD5 | FALSE | Upregulated |
| -0,284365007 | 0,790494863 | LOC101929443 | FALSE | Downregulated |
| 0,099763942 | 0,790494863 | MIF4GD | FALSE | Upregulated |
| -0,06964449 | 0,790494863 | PTPRK | FALSE | Downregulated |
| 0,324447057 | 0,790522639 | ADAMTS9-AS2 | FALSE | Upregulated |
| 0,127242333 | 0,790831515 | ZNF786 | FALSE | Upregulated |
| 0,151389728 | 0,790923228 | ARSG | FALSE | Upregulated |
| -0,053588077 | 0,790923228 | PPP6R3 | FALSE | Downregulated |
| 0,096698907 | 0,791087629 | HEATR2 | FALSE | Upregulated |
| 0,33757346 | 0,791087629 | LOC101928153 | FALSE | Upregulated |
| -0,063719067 | 0,791087629 | SORBS1 | FALSE | Downregulated |
| -0,080284122 | 0,791091735 | HNRNPH1 | FALSE | Downregulated |
| -0,175772444 | 0,791091735 | LRRC4C | FALSE | Downregulated |
| 0,18298037 | 0,791225638 | CEACAM21 | FALSE | Upregulated |
| 0,32717221 | 0,791225638 | SLX1A-SULT1A3 | FALSE | Upregulated |
| -0,105967869 | 0,791225638 | SMPDL3A | FALSE | Downregulated |

| | | | | |
|---|---|---|---|---|
| -0,306818891 | 0,791225638 | SNTG1 | FALSE | Downregulated |
| -0,061816921 | 0,791225638 | TNRC6B | FALSE | Downregulated |
| -0,094756375 | 0,791225638 | TSTD1 | FALSE | Downregulated |
| 0,097306923 | 0,79126532 | DHRS12 | FALSE | Upregulated |
| -0,099940795 | 0,791403865 | ABCA1 | FALSE | Downregulated |
| 0,168611288 | 0,791637364 | AC003665.1 | FALSE | Upregulated |
| 0,139479449 | 0,791637364 | CBX8 | FALSE | Upregulated |
| -0,156314239 | 0,791894923 | SLC16A4 | FALSE | Downregulated |
| 0,048008378 | 0,791894923 | UBE4A | FALSE | Upregulated |
| 0,074559826 | 0,791894923 | ZNF598 | FALSE | Upregulated |
| 0,098590146 | 0,79219955 | PQLC3 | FALSE | Upregulated |
| 0,050301744 | 0,79219955 | TRA2B | FALSE | Upregulated |
| 0,274474827 | 0,792322904 | RANBP17 | FALSE | Upregulated |
| -0,119601281 | 0,792596451 | COQ10A | FALSE | Downregulated |
| 0,07367325 | 0,792596451 | PTPN23 | FALSE | Upregulated |
| 0,079812769 | 0,792596451 | RNF44 | FALSE | Upregulated |
| 0,219868288 | 0,792596451 | SH2D3A | FALSE | Upregulated |
| 0,557091215 | 0,792596451 | STMND1 | FALSE | Upregulated |
| -0,122156395 | 0,792771798 | TOP1P1 | FALSE | Downregulated |
| 0,121662777 | 0,792851838 | ARID5B | FALSE | Upregulated |
| -0,237357901 | 0,792851838 | CAMK2B | FALSE | Downregulated |
| 0,290862285 | 0,792851838 | EDAR | FALSE | Upregulated |
| 0,299491529 | 0,792851838 | MTRNR2L9 | FALSE | Upregulated |
| 0,095336571 | 0,792851838 | NEDD4 | FALSE | Upregulated |
| -0,560035869 | 0,792851838 | RP11-381O7.3 | FALSE | Downregulated |
| 0,427575627 | 0,792851838 | SLC26A3 | FALSE | Upregulated |

| | | | | |
|---|---|---|---|---|
| 0,226271478 | 0,792915683 | CDHR2 | FALSE | Upregulated |
| -0,148174278 | 0,792915683 | XRCC6BP1 | FALSE | Downregulated |
| -0,30984071 | 0,793114505 | C1orf213 | FALSE | Downregulated |
| -0,349973046 | 0,793114505 | NEXN | FALSE | Downregulated |
| -0,101148696 | 0,793116561 | RBM33 | FALSE | Downregulated |
| -0,096433511 | 0,793354589 | DUS4L | FALSE | Downregulated |
| 0,083168457 | 0,793354589 | FAM173B | FALSE | Upregulated |
| 0,090910694 | 0,793354589 | HRAS | FALSE | Upregulated |
| 0,263577267 | 0,793354589 | TMEM159 | FALSE | Upregulated |
| 0,068155039 | 0,793383187 | DPY19L4 | FALSE | Upregulated |
| -0,146884303 | 0,793383187 | FCN2 | FALSE | Downregulated |
| 0,495248345 | 0,793383187 | P2RX5 | FALSE | Upregulated |
| 0,434368419 | 0,793383187 | ZPLD1 | FALSE | Upregulated |
| 0,112945602 | 0,793535811 | SEC24D | FALSE | Upregulated |
| 0,215199033 | 0,793809429 | TMEM88 | FALSE | Upregulated |
| -0,128568476 | 0,793930066 | APOC3 | FALSE | Downregulated |
| 0,389438156 | 0,793930066 | GDNF | FALSE | Upregulated |
| 0,128339301 | 0,793930066 | LILRB5 | FALSE | Upregulated |
| 0,078513326 | 0,793930066 | PRPSAP2 | FALSE | Upregulated |
| -0,062788497 | 0,793930066 | SENP1 | FALSE | Downregulated |
| 0,077896834 | 0,794045309 | DENND1B | FALSE | Upregulated |
| -0,226646014 | 0,794075605 | CAND2 | FALSE | Downregulated |
| -0,078154499 | 0,794075605 | R3HDM1 | FALSE | Downregulated |
| 0,653240931 | 0,794075605 | ULK4P3 | FALSE | Upregulated |
| 0,515736911 | 0,794118257 | FAM74A1 | FALSE | Upregulated |
| 0,259089655 | 0,7947231 | GDF6 | FALSE | Upregulated |

| | | | | |
|---|---|---|---|---|
| -0,054621894 | 0,794761957 | KIAA1429 | FALSE | Downregulated |
| -0,173431116 | 0,794761957 | ZG16 | FALSE | Downregulated |
| -0,361473158 | 0,795117765 | STARD4-AS1 | FALSE | Downregulated |
| 0,362800571 | 0,795139371 | RAD54L | FALSE | Upregulated |
| 0,452222377 | 0,795345198 | KC6 | FALSE | Upregulated |
| 0,174668218 | 0,795345198 | MARCO | FALSE | Upregulated |
| 0,064881871 | 0,795486144 | FBXO7 | FALSE | Upregulated |
| 0,113583156 | 0,795486144 | JMJD7 | FALSE | Upregulated |
| -0,152000447 | 0,795486144 | KCNMB3 | FALSE | Downregulated |
| 0,055010084 | 0,795516096 | HNRNPL | FALSE | Upregulated |
| -0,325237796 | 0,795516096 | RPL34-AS1 | FALSE | Downregulated |
| -0,105596376 | 0,795826396 | BMS1P20 | FALSE | Downregulated |
| 0,272140684 | 0,795826396 | CBR3 | FALSE | Upregulated |
| -0,335248118 | 0,795832041 | LINC00622 | FALSE | Downregulated |
| -0,052163212 | 0,795832041 | ZHX2 | FALSE | Downregulated |
| 0,162684115 | 0,795903667 | SYTL3 | FALSE | Upregulated |
| -0,329106959 | 0,796056618 | FLJ39632 | FALSE | Downregulated |
| -0,09248792 | 0,796098084 | NNT-AS1 | FALSE | Downregulated |
| -0,063649677 | 0,796143959 | U2AF1 | FALSE | Downregulated |
| -0,329564541 | 0,796336974 | GOLGA6L3 | FALSE | Downregulated |
| 0,054195069 | 0,796791973 | ACTR3 | FALSE | Upregulated |
| 0,073933035 | 0,796791973 | ADCY9 | FALSE | Upregulated |
| -0,082141443 | 0,796791973 | ATAD2B | FALSE | Downregulated |
| 0,066944176 | 0,796791973 | CTSB | FALSE | Upregulated |
| -0,069327478 | 0,796791973 | EDF1 | FALSE | Downregulated |
| 0,04573209 | 0,796791973 | EIF3B | FALSE | Upregulated |

| | | | | |
|---|---|---|---|---|
| 0,492819645 | 0,796791973 | FRMD1 | FALSE | Upregulated |
| 0,094902 | 0,796791973 | GCGR | FALSE | Upregulated |
| 0,150223023 | 0,796791973 | INPP5F | FALSE | Upregulated |
| 0,51185774 | 0,796791973 | LINC01152 | FALSE | Upregulated |
| -0,443464958 | 0,796791973 | LOC100996442 | FALSE | Downregulated |
| -0,385760383 | 0,796791973 | LOC101928635 | FALSE | Downregulated |
| -0,409617806 | 0,796791973 | LOC101929555 | FALSE | Downregulated |
| 0,588994944 | 0,796791973 | RALGAPA1P | FALSE | Upregulated |
| -0,581520267 | 0,796791973 | RP13-228J13.1 | FALSE | Downregulated |
| -0,241299063 | 0,796791973 | RP5-1057J7.6 | FALSE | Downregulated |
| -0,124050034 | 0,796791973 | SLC2A13 | FALSE | Downregulated |
| 0,115255279 | 0,796791973 | ZBTB22 | FALSE | Upregulated |
| -0,043616427 | 0,796791973 | ZFYVE9 | FALSE | Downregulated |
| 0,658265504 | 0,797026054 | HOXA7 | FALSE | Upregulated |
| 0,627060279 | 0,797026054 | LOC100271836 | FALSE | Upregulated |
| 0,165304387 | 0,797026054 | PDSS1 | FALSE | Upregulated |
| -0,400749696 | 0,797026054 | REN | FALSE | Downregulated |
| -0,12454906 | 0,797026054 | TRIP10 | FALSE | Downregulated |
| -0,329862114 | 0,797026054 | WDR86 | FALSE | Downregulated |
| 0,067722047 | 0,797253713 | ABCF2 | FALSE | Upregulated |
| 0,610389107 | 0,797253713 | ANKRD20A1 | FALSE | Upregulated |
| 0,069218585 | 0,797253713 | ANKRD52 | FALSE | Upregulated |
| -0,085454468 | 0,797253713 | ATG4A | FALSE | Downregulated |
| 0,277518108 | 0,797253713 | CTD-3080P12.3 | FALSE | Upregulated |
| 0,452135465 | 0,797253713 | DARC | FALSE | Upregulated |
| -0,258339378 | 0,797253713 | EPHA4 | FALSE | Downregulated |

| | | | | |
|---|---|---|---|---|
| -0,081585154 | 0,797253713 | LRRFIP1 | FALSE | Downregulated |
| 0,057648689 | 0,797253713 | MAML1 | FALSE | Upregulated |
| -0,08218249 | 0,797253713 | MRGBP | FALSE | Downregulated |
| -0,128789728 | 0,797253713 | NINL | FALSE | Downregulated |
| 0,057098341 | 0,797253713 | PIN1 | FALSE | Upregulated |
| -0,210007927 | 0,797253713 | SDK2 | FALSE | Downregulated |
| -0,052915139 | 0,797253713 | SON | FALSE | Downregulated |
| -0,134274495 | 0,797253713 | TSPAN7 | FALSE | Downregulated |
| -0,054217994 | 0,797253713 | UBA5 | FALSE | Downregulated |
| 0,470209006 | 0,797603025 | LOC102724006 | FALSE | Upregulated |
| -0,044449457 | 0,797612581 | GNL1 | FALSE | Downregulated |
| -0,077443848 | 0,797612581 | MAP2K7 | FALSE | Downregulated |
| -0,099316115 | 0,797612581 | SAFB2 | FALSE | Downregulated |
| -0,155938411 | 0,797651956 | NOXA1 | FALSE | Downregulated |
| -0,079003501 | 0,797651956 | RPL21 | FALSE | Downregulated |
| -0,082434922 | 0,797832717 | IQGAP2 | FALSE | Downregulated |
| 0,16943192 | 0,797832717 | KRBA1 | FALSE | Upregulated |
| 0,074860618 | 0,797865028 | ANKRD46 | FALSE | Upregulated |
| 0,048587611 | 0,797865028 | MIEF1 | FALSE | Upregulated |
| -0,207672606 | 0,797865028 | RP11-372E1.4 | FALSE | Downregulated |
| 0,102392985 | 0,797865028 | TUBG1 | FALSE | Upregulated |
| -0,068725576 | 0,797865028 | ZNF304 | FALSE | Downregulated |
| 0,16457432 | 0,797876758 | CROCCP2 | FALSE | Upregulated |
| 0,065715581 | 0,797876758 | DPM1 | FALSE | Upregulated |
| -0,058282716 | 0,797876758 | EIF2B5 | FALSE | Downregulated |
| -0,363679498 | 0,797876758 | GPR173 | FALSE | Downregulated |

| | | | | |
|---|---|---|---|---|
| -0,325395024 | 0,797876758 | KLHL23 | FALSE | Downregulated |
| 0,22981101 | 0,797876758 | LOC101929758 | FALSE | Upregulated |
| -0,250635529 | 0,797876758 | LRRC16B | FALSE | Downregulated |
| -0,070839087 | 0,797876758 | MTCH2 | FALSE | Downregulated |
| -0,119607736 | 0,797876758 | RFX3 | FALSE | Downregulated |
| 0,08498641 | 0,797876758 | RHPN2 | FALSE | Upregulated |
| 0,185785134 | 0,797876758 | RP11-345L23.1 | FALSE | Upregulated |
| -0,042390199 | 0,797876758 | TNPO1 | FALSE | Downregulated |
| 0,114394255 | 0,797876758 | ZSCAN9 | FALSE | Upregulated |
| -0,06373477 | 0,797911236 | CDK17 | FALSE | Downregulated |
| -0,076863723 | 0,797911236 | KIAA1715 | FALSE | Downregulated |
| 0,384864978 | 0,797911236 | LOC101593348 | FALSE | Upregulated |
| -0,310642764 | 0,797911236 | S100B | FALSE | Downregulated |
| -0,064214159 | 0,797911236 | SRSF8 | FALSE | Downregulated |
| -0,193264723 | 0,798036415 | LOC728752 | FALSE | Downregulated |
| 0,08677263 | 0,798036415 | ZNF761 | FALSE | Upregulated |
| 0,490733621 | 0,798089071 | KRTAP5-6 | FALSE | Upregulated |
| 0,085867883 | 0,798089071 | PNO1 | FALSE | Upregulated |
| 0,262494465 | 0,798457831 | DTX2P1-UPK3BP1-PMS2P11 | FALSE | Upregulated |
| -0,223700314 | 0,798457831 | THRSP | FALSE | Downregulated |
| -0,143607743 | 0,798457831 | TMCC1-AS1 | FALSE | Downregulated |
| 0,225188728 | 0,798457831 | ZSWIM8-AS1 | FALSE | Upregulated |
| 0,162287667 | 0,798476787 | CCDC171 | FALSE | Upregulated |
| -0,145268423 | 0,798476787 | FSTL1 | FALSE | Downregulated |
| -0,056245476 | 0,798476787 | FTSJ2 | FALSE | Downregulated |

| | | | | |
|---|---|---|---|---|
| 0,121209772 | 0,798476787 | GUCY1A3 | FALSE | Upregulated |
| 0,097070382 | 0,79851843 | TBC1D22B | FALSE | Upregulated |
| -0,267632796 | 0,798937549 | ZNF790-AS1 | FALSE | Downregulated |
| -0,196022585 | 0,799122984 | ADAMTSL1 | FALSE | Downregulated |
| 0,075769946 | 0,799122984 | ADRM1 | FALSE | Upregulated |
| 0,049103583 | 0,799122984 | AREL1 | FALSE | Upregulated |
| 0,086693656 | 0,799122984 | C14orf142 | FALSE | Upregulated |
| 0,135565156 | 0,799122984 | DOCK11 | FALSE | Upregulated |
| 0,085020928 | 0,799122984 | GTDC1 | FALSE | Upregulated |
| 0,205793447 | 0,799122984 | HAS3 | FALSE | Upregulated |
| -0,11505662 | 0,799122984 | KLF12 | FALSE | Downregulated |
| 0,543990443 | 0,799122984 | LOC100507616 | FALSE | Upregulated |
| 0,265370649 | 0,799122984 | LOC101929223 | FALSE | Upregulated |
| -0,07956844 | 0,799122984 | MED13 | FALSE | Downregulated |
| 0,043889519 | 0,799122984 | PPP6C | FALSE | Upregulated |
| -0,086698152 | 0,799122984 | SKP2 | FALSE | Downregulated |
| 0,064044319 | 0,799122984 | SLC35D2 | FALSE | Upregulated |
| -0,301870449 | 0,799122984 | STRIP2 | FALSE | Downregulated |
| -0,395148399 | 0,799166461 | ESPNP | FALSE | Downregulated |
| -0,24943229 | 0,799166461 | LOC102723782 | FALSE | Downregulated |
| 0,101298915 | 0,799166461 | RASSF1 | FALSE | Upregulated |
| -0,413254917 | 0,799236654 | GS1-259H13.2 | FALSE | Downregulated |
| 0,069820701 | 0,799286281 | NUP153 | FALSE | Upregulated |
| 0,082888997 | 0,79930279 | CPSF1 | FALSE | Upregulated |
| -0,074489523 | 0,799357949 | IPO11 | FALSE | Downregulated |
| 0,044651289 | 0,799357949 | VAPA | FALSE | Upregulated |

| | | | | |
|---|---|---|---|---|
| 0,15768069 | 0,799460045 | CENPP | FALSE | Upregulated |
| 0,444663301 | 0,799460045 | LOC100506098 | FALSE | Upregulated |
| 0,227497995 | 0,799460045 | LOC100996573 | FALSE | Upregulated |
| 0,102698154 | 0,799460045 | MYPOP | FALSE | Upregulated |
| 0,105378326 | 0,799460045 | NEURL4 | FALSE | Upregulated |
| 0,112885523 | 0,799460045 | PLXNA1 | FALSE | Upregulated |
| 0,051249729 | 0,799460045 | PUF60 | FALSE | Upregulated |
| 0,583893563 | 0,799460045 | TRNH | FALSE | Upregulated |
| -0,082906379 | 0,799460045 | WWTR1 | FALSE | Downregulated |
| -0,096488317 | 0,799460045 | ZBTB47 | FALSE | Downregulated |
| -0,051645831 | 0,799656864 | PDS5B | FALSE | Downregulated |
| -0,144985271 | 0,79969685 | PPARGC1A | FALSE | Downregulated |
| -0,235640533 | 0,799834318 | FCGR3B | FALSE | Downregulated |
| 0,510023236 | 0,799834318 | LOC729080 | FALSE | Upregulated |
| -0,432974743 | 0,799834318 | LRRTM1 | FALSE | Downregulated |
| -0,125018171 | 0,799834318 | NBPF1 | FALSE | Downregulated |
| 0,147639563 | 0,799834318 | TAX1BP3 | FALSE | Upregulated |
| -0,24456723 | 0,799846058 | FAM86HP | FALSE | Downregulated |
| 0,086876635 | 0,799846058 | TMEM222 | FALSE | Upregulated |
| -0,070005277 | 0,799846058 | YOD1 | FALSE | Downregulated |
| -0,206484577 | 0,800050403 | ACR | FALSE | Downregulated |
| 0,120559363 | 0,800050403 | ATP1B3 | FALSE | Upregulated |
| -0,052164572 | 0,800050403 | BNIP3L | FALSE | Downregulated |
| 0,086157762 | 0,800050403 | CLCN4 | FALSE | Upregulated |
| -0,082875083 | 0,800050403 | CMIP | FALSE | Downregulated |
| 0,07285741 | 0,800050403 | FAHD2A | FALSE | Upregulated |

| | | | | |
|---|---|---|---|---|
| -0,10183697 | 0,800050403 | H2AFJ | FALSE | Downregulated |
| 0,166520836 | 0,800050403 | HYAL3 | FALSE | Upregulated |
| 0,186666591 | 0,800050403 | LEPREL2 | FALSE | Upregulated |
| -0,296073511 | 0,800050403 | LOC102724607 | FALSE | Downregulated |
| 0,514821815 | 0,800050403 | LOC644662 | FALSE | Upregulated |
| 0,09856024 | 0,800050403 | MPZL3 | FALSE | Upregulated |
| 0,213931344 | 0,800050403 | PIK3R6 | FALSE | Upregulated |
| 0,295944275 | 0,800050403 | PPM1J | FALSE | Upregulated |
| -0,973212629 | 0,800050403 | RMRP | FALSE | Downregulated |
| -0,096580732 | 0,800050403 | RRAGB | FALSE | Downregulated |
| -0,738445767 | 0,800050403 | SCHIP1 | FALSE | Downregulated |
| -0,406501293 | 0,800050403 | TSC22D1-AS1 | FALSE | Downregulated |
| 0,129418615 | 0,800050403 | ZBTB11-AS1 | FALSE | Upregulated |
| -0,17861616 | 0,800284914 | SCML4 | FALSE | Downregulated |
| -0,131134302 | 0,800348553 | ERICH2 | FALSE | Downregulated |
| 0,24523081 | 0,800485478 | ALDH1L2 | FALSE | Upregulated |
| 0,060873328 | 0,800485478 | ARHGAP1 | FALSE | Upregulated |
| 0,075702546 | 0,800485478 | DHX37 | FALSE | Upregulated |
| -0,117976276 | 0,800485478 | EIF2AK2 | FALSE | Downregulated |
| 0,145138232 | 0,800485478 | FOXF1 | FALSE | Upregulated |
| 0,079621171 | 0,800485478 | GALK2 | FALSE | Upregulated |
| 0,233664051 | 0,800485478 | GAREML | FALSE | Upregulated |
| -0,126484737 | 0,800485478 | HIVEP3 | FALSE | Downregulated |
| -0,100587452 | 0,800485478 | MGMT | FALSE | Downregulated |
| -0,4116443 | 0,800485478 | MRAP | FALSE | Downregulated |
| 0,41007419 | 0,800485478 | NPBWR1 | FALSE | Upregulated |

| | | | | |
|---|---|---|---|---|
| 0,050443246 | 0,800485478 | PRPF19 | FALSE | Upregulated |
| -0,058753766 | 0,800485478 | PTDSS1 | FALSE | Downregulated |
| -0,073990391 | 0,800485478 | PTPRJ | FALSE | Downregulated |
| -0,06864088 | 0,800485478 | RBM23 | FALSE | Downregulated |
| 0,186420105 | 0,800485478 | RP11-425I13.3 | FALSE | Upregulated |
| 0,660737347 | 0,800485478 | SLC22A11 | FALSE | Upregulated |
| 0,088951066 | 0,800485478 | SLC25A10 | FALSE | Upregulated |
| 0,047562392 | 0,800485478 | SLC39A9 | FALSE | Upregulated |
| -0,097251526 | 0,800485478 | TP53BP1 | FALSE | Downregulated |
| -0,094016279 | 0,800485478 | TRPT1 | FALSE | Downregulated |
| 0,142514298 | 0,800485478 | UBASH3B | FALSE | Upregulated |
| 0,329361487 | 0,800485478 | VIL1 | FALSE | Upregulated |
| 0,052037099 | 0,800485478 | ZNF629 | FALSE | Upregulated |
| 0,147048616 | 0,800756501 | COL6A3 | FALSE | Upregulated |
| -0,054279188 | 0,800756501 | HBS1L | FALSE | Downregulated |
| -0,070323339 | 0,800756501 | KIF1C | FALSE | Downregulated |
| -0,264743505 | 0,800756501 | LOC339874 | FALSE | Downregulated |
| 0,127867396 | 0,800756501 | PACS1 | FALSE | Upregulated |
| -0,100630176 | 0,800756501 | PROX1-AS1 | FALSE | Downregulated |
| 0,092405859 | 0,800756501 | RBM45 | FALSE | Upregulated |
| -1,008353592 | 0,800756501 | RNU6-36P | FALSE | Downregulated |
| 0,066825103 | 0,800862199 | NSMAF | FALSE | Upregulated |
| 0,531180872 | 0,800869316 | CNFN | FALSE | Upregulated |
| -0,271143753 | 0,80092554 | COL8A1 | FALSE | Downregulated |
| -0,095544326 | 0,800928666 | ARMC5 | FALSE | Downregulated |
| 0,054132885 | 0,800928666 | ATRAID | FALSE | Upregulated |

| | | | | |
|---|---|---|---|---|
| 0,262298258 | 0,800928666 | BEX5 | FALSE | Upregulated |
| 0,141816148 | 0,800928666 | CROCC | FALSE | Upregulated |
| -0,123814277 | 0,800928666 | SNED1 | FALSE | Downregulated |
| -0,082704757 | 0,80093943 | NUDT16P1 | FALSE | Downregulated |
| 0,23848681 | 0,80096236 | CTAGE11P | FALSE | Upregulated |
| 0,233783966 | 0,801154318 | KLRF1 | FALSE | Upregulated |
| -0,074975541 | 0,80127102 | TRIP11 | FALSE | Downregulated |
| 0,075701169 | 0,801746192 | ASMTL | FALSE | Upregulated |
| 0,117827533 | 0,801746192 | GLIPR1 | FALSE | Upregulated |
| -0,168786916 | 0,801979835 | ADCY10 | FALSE | Downregulated |
| -0,066426977 | 0,801979835 | AP1AR | FALSE | Downregulated |
| -0,144527366 | 0,801979835 | C17orf67 | FALSE | Downregulated |
| -0,475218369 | 0,801979835 | C1orf127 | FALSE | Downregulated |
| 0,087521109 | 0,801979835 | CPEB4 | FALSE | Upregulated |
| -0,046271741 | 0,801979835 | DSCR3 | FALSE | Downregulated |
| -0,061312243 | 0,801979835 | FOXO4 | FALSE | Downregulated |
| 0,10969283 | 0,801979835 | GTF2H4 | FALSE | Upregulated |
| 0,188435655 | 0,801979835 | KANSL1-AS1 | FALSE | Upregulated |
| 0,070807221 | 0,801979835 | KIAA1191 | FALSE | Upregulated |
| 0,273817505 | 0,801979835 | LOC102724766 | FALSE | Upregulated |
| 0,074291403 | 0,801979835 | MYL6 | FALSE | Upregulated |
| 0,215006105 | 0,801979835 | NRG1 | FALSE | Upregulated |
| 0,084085436 | 0,801979835 | SESN2 | FALSE | Upregulated |
| -0,045856142 | 0,801979835 | SHOC2 | FALSE | Downregulated |
| -0,10655293 | 0,801979835 | SPAG1 | FALSE | Downregulated |
| 0,104340219 | 0,801979835 | ULK1 | FALSE | Upregulated |

| | | | | |
|---|---|---|---|---|
| -0,316751081 | 0,80239251 | LOC102723461 | FALSE | Downregulated |
| 0,218423962 | 0,802459326 | TMPO-AS1 | FALSE | Upregulated |
| 0,051124795 | 0,802569459 | SRSF9 | FALSE | Upregulated |
| 0,057577372 | 0,802588684 | PHB | FALSE | Upregulated |
| -0,085319568 | 0,802640826 | NAGS | FALSE | Downregulated |
| -0,14844936 | 0,802745716 | RP6-109B7.3 | FALSE | Downregulated |
| 0,212998123 | 0,802930538 | FAM171B | FALSE | Upregulated |
| 0,656121442 | 0,802978914 | CSMD3 | FALSE | Upregulated |
| -0,078877208 | 0,803025063 | AASDH | FALSE | Downregulated |
| -0,08773408 | 0,803025063 | FAM111A | FALSE | Downregulated |
| -0,288310307 | 0,803025063 | LOC101926935 | FALSE | Downregulated |
| -0,086134074 | 0,803025063 | METTL15 | FALSE | Downregulated |
| -0,091614559 | 0,803025063 | MORN2 | FALSE | Downregulated |
| -0,060614019 | 0,803025063 | NT5C2 | FALSE | Downregulated |
| -0,256004707 | 0,803025063 | RP1-232P20.1 | FALSE | Downregulated |
| -0,073641581 | 0,803025063 | TBC1D4 | FALSE | Downregulated |
| 0,119660862 | 0,803054761 | CCBE1 | FALSE | Upregulated |
| 0,057386686 | 0,803054761 | FEM1C | FALSE | Upregulated |
| 0,110002615 | 0,803054761 | SLC18B1 | FALSE | Upregulated |
| 0,148731002 | 0,803054761 | SLC25A53 | FALSE | Upregulated |
| -0,290259168 | 0,803054761 | TMEM105 | FALSE | Downregulated |
| 0,098537589 | 0,803054761 | XDH | FALSE | Upregulated |
| 0,075565602 | 0,803064839 | ANAPC4 | FALSE | Upregulated |
| -0,897297394 | 0,803064839 | ART3 | FALSE | Downregulated |
| 0,10490578 | 0,803064839 | C6orf48 | FALSE | Upregulated |
| 0,221253816 | 0,803064839 | DICER1-AS1 | FALSE | Upregulated |

| | | | | |
|---|---|---|---|---|
| -0,188518992 | 0,803064839 | ICOSLG | FALSE | Downregulated |
| -0,202759529 | 0,803064839 | LOC100506083 | FALSE | Downregulated |
| 0,080345503 | 0,803064839 | MPZL1 | FALSE | Upregulated |
| 0,094684762 | 0,803064839 | RPL10A | FALSE | Upregulated |
| -0,067229056 | 0,803064839 | TRAPPC13 | FALSE | Downregulated |
| 0,060971007 | 0,803064839 | WIZ | FALSE | Upregulated |
| 0,219310286 | 0,803189428 | JAG2 | FALSE | Upregulated |
| -0,073840535 | 0,803189428 | QSER1 | FALSE | Downregulated |
| -0,246956795 | 0,803253566 | CRYGS | FALSE | Downregulated |
| -0,38240287 | 0,803306771 | CEMP1 | FALSE | Downregulated |
| -0,111273343 | 0,803306771 | SMAP1 | FALSE | Downregulated |
| 0,082291828 | 0,803869572 | PERP | FALSE | Upregulated |
| 0,132100544 | 0,803954661 | BRIP1 | FALSE | Upregulated |
| -0,125949561 | 0,804192471 | BRD9 | FALSE | Downregulated |
| -0,069698876 | 0,804192471 | METTL3 | FALSE | Downregulated |
| -0,347366037 | 0,804192471 | NRXN3 | FALSE | Downregulated |
| -0,060067907 | 0,804498186 | IL13RA1 | FALSE | Downregulated |
| -0,116213235 | 0,804498186 | ZNF354B | FALSE | Downregulated |
| 0,054465932 | 0,804633829 | SCAP | FALSE | Upregulated |
| -0,210856653 | 0,804863248 | LOC100129534 | FALSE | Downregulated |
| -0,139225064 | 0,804963196 | BTBD16 | FALSE | Downregulated |
| -0,234773615 | 0,804963196 | CCDC28B | FALSE | Downregulated |
| -0,344331394 | 0,804963196 | EGFR-AS1 | FALSE | Downregulated |
| -0,084649445 | 0,804963196 | FAM161B | FALSE | Downregulated |
| 0,436019687 | 0,804963196 | KIF18B | FALSE | Upregulated |
| -0,276963305 | 0,804963196 | RP11-434C1.1 | FALSE | Downregulated |

| | | | | |
|---|---|---|---|---|
| -0,494267696 | 0,805014542 | KIR2DS4 | FALSE | Downregulated |
| -0,047315199 | 0,805078789 | ATP5F1 | FALSE | Downregulated |
| 0,103713097 | 0,805236691 | DTD1 | FALSE | Upregulated |
| -0,071638659 | 0,805263792 | EEF2K | FALSE | Downregulated |
| 1,208362653 | 0,805263792 | LOC100288570 | FALSE | Upregulated |
| 0,048489539 | 0,805263792 | UBE2G2 | FALSE | Upregulated |
| -0,061316478 | 0,805293257 | RPL6 | FALSE | Downregulated |
| -0,121563862 | 0,805366587 | C3orf62 | FALSE | Downregulated |
| -0,318437443 | 0,805366587 | GDPD3 | FALSE | Downregulated |
| 0,043182948 | 0,805366587 | RNF14 | FALSE | Upregulated |
| 0,123622211 | 0,805366587 | STOML1 | FALSE | Upregulated |
| 0,105940309 | 0,80570242 | AC074286.1 | FALSE | Upregulated |
| -0,086092942 | 0,80570242 | CHML | FALSE | Downregulated |
| -0,3209088 | 0,80570242 | DSCAML1 | FALSE | Downregulated |
| 0,059661483 | 0,80570242 | EDC3 | FALSE | Upregulated |
| -0,056572845 | 0,80570242 | G3BP2 | FALSE | Downregulated |
| 0,101487384 | 0,80570242 | GPR137B | FALSE | Upregulated |
| 0,751357608 | 0,80570242 | IRG1 | FALSE | Upregulated |
| -0,172682121 | 0,80570242 | LOC441242 | FALSE | Downregulated |
| -0,061632449 | 0,80570242 | PSMB1 | FALSE | Downregulated |
| 0,043661916 | 0,80570242 | RDH14 | FALSE | Upregulated |
| -0,088409743 | 0,80570242 | TMEM187 | FALSE | Downregulated |
| 0,083649891 | 0,80570242 | UBL4A | FALSE | Upregulated |
| -0,071954736 | 0,80570242 | ZNF622 | FALSE | Downregulated |
| 0,129235663 | 0,806014936 | ADM | FALSE | Upregulated |
| 0,092808142 | 0,806014936 | ZNF496 | FALSE | Upregulated |

| | | | | |
|---|---|---|---|---|
| -0,080801183 | 0,80602883 | ABCA6 | FALSE | Downregulated |
| -0,099676442 | 0,80602883 | C11orf71 | FALSE | Downregulated |
| 0,054822219 | 0,80602883 | C1orf174 | FALSE | Upregulated |
| -0,181581654 | 0,80602883 | C1orf228 | FALSE | Downregulated |
| 0,082471613 | 0,80602883 | CBFB | FALSE | Upregulated |
| -0,066437265 | 0,80602883 | CEP44 | FALSE | Downregulated |
| 0,078394779 | 0,80602883 | COPZ2 | FALSE | Upregulated |
| -0,112356771 | 0,80602883 | DHRS4-AS1 | FALSE | Downregulated |
| -0,058084269 | 0,80602883 | DPY30 | FALSE | Downregulated |
| -0,355482526 | 0,80602883 | EDN2 | FALSE | Downregulated |
| 0,171993138 | 0,80602883 | FUT6 | FALSE | Upregulated |
| -0,097403688 | 0,80602883 | GUCY1B3 | FALSE | Downregulated |
| 0,287987891 | 0,80602883 | LOC102060414 | FALSE | Upregulated |
| -0,402845971 | 0,80602883 | LOC102724031 | FALSE | Downregulated |
| -0,074432847 | 0,80602883 | MEGF8 | FALSE | Downregulated |
| -0,307861321 | 0,80602883 | MIR564 | FALSE | Downregulated |
| 0,086037894 | 0,80602883 | NRF1 | FALSE | Upregulated |
| 0,150902058 | 0,80602883 | NTF3 | FALSE | Upregulated |
| 0,14469912 | 0,80602883 | PLIN4 | FALSE | Upregulated |
| 0,061030556 | 0,80602883 | PSMC3 | FALSE | Upregulated |
| 0,3027958 | 0,80602883 | RGS16 | FALSE | Upregulated |
| -0,057706647 | 0,80602883 | RHOT1 | FALSE | Downregulated |
| 0,080815637 | 0,80602883 | RNASE4 | FALSE | Upregulated |
| 0,125943489 | 0,80602883 | SAMD1 | FALSE | Upregulated |
| -0,089530925 | 0,80602883 | SP140L | FALSE | Downregulated |
| 0,047220726 | 0,80602883 | SUPT7L | FALSE | Upregulated |

| | | | | |
|---|---|---|---|---|
| 0,122529521 | 0,80602883 | UBE2L6 | FALSE | Upregulated |
| 0,184291547 | 0,80602883 | VWCE | FALSE | Upregulated |
| 0,078507566 | 0,80602883 | WDR74 | FALSE | Upregulated |
| -0,210395969 | 0,80602883 | WNT2 | FALSE | Downregulated |
| -0,097094458 | 0,80611706 | APOH | FALSE | Downregulated |
| 0,084119729 | 0,806172823 | C19orf24 | FALSE | Upregulated |
| 0,068518437 | 0,806172823 | C2orf44 | FALSE | Upregulated |
| -0,61735035 | 0,806172823 | CHRFAM7A | FALSE | Downregulated |
| 0,20048757 | 0,806172823 | ELL3 | FALSE | Upregulated |
| -0,056486625 | 0,806172823 | GHITM | FALSE | Downregulated |
| -0,650737268 | 0,806172823 | LOC101927818 | FALSE | Downregulated |
| 0,094172276 | 0,806172823 | MSRA | FALSE | Upregulated |
| 0,057177871 | 0,806172823 | RAD23A | FALSE | Upregulated |
| -0,181746653 | 0,806172823 | SLC5A9 | FALSE | Downregulated |
| -0,057177975 | 0,806172823 | SUZ12 | FALSE | Downregulated |
| -0,111629237 | 0,806292636 | RSBN1L | FALSE | Downregulated |
| 0,074292222 | 0,806313285 | RINT1 | FALSE | Upregulated |
| -0,029370836 | 0,806391791 | MKRN1 | FALSE | Downregulated |
| -0,221925181 | 0,806391791 | PODN | FALSE | Downregulated |
| 0,069882976 | 0,806470202 | CCT2 | FALSE | Upregulated |
| -0,090986359 | 0,806470202 | EIF3J-AS1 | FALSE | Downregulated |
| 0,127586796 | 0,806470202 | GFPT1 | FALSE | Upregulated |
| -0,048777235 | 0,806470202 | MRPS35 | FALSE | Downregulated |
| 0,147571068 | 0,806470202 | RAB43 | FALSE | Upregulated |
| -0,371463316 | 0,806470202 | TENM2 | FALSE | Downregulated |
| 0,09322268 | 0,806470202 | TIMM8A | FALSE | Upregulated |

| | | | | |
|---|---|---|---|---|
| 0,061973995 | 0,806470202 | UBAC2 | FALSE | Upregulated |
| -0,075531833 | 0,806470202 | ZNF174 | FALSE | Downregulated |
| -0,21067974 | 0,806723562 | LRP5L | FALSE | Downregulated |
| -0,0961118 | 0,806723562 | PHACTR4 | FALSE | Downregulated |
| -0,121687108 | 0,806869824 | ANK3 | FALSE | Downregulated |
| -0,050604891 | 0,806902813 | RAC1 | FALSE | Downregulated |
| 0,240183562 | 0,807138858 | LINC00884 | FALSE | Upregulated |
| 0,228552725 | 0,807297554 | SLC4A8 | FALSE | Upregulated |
| -1,0280766 | 0,807297554 | TNXA | FALSE | Downregulated |
| 0,364388254 | 0,807509132 | SMPDL3B | FALSE | Upregulated |
| 0,220338696 | 0,807543702 | CACNB3 | FALSE | Upregulated |
| 0,052353221 | 0,807543702 | DPF2 | FALSE | Upregulated |
| -0,072683256 | 0,807543702 | GPATCH8 | FALSE | Downregulated |
| -0,182377955 | 0,807543702 | KRT222 | FALSE | Downregulated |
| 0,09511685 | 0,807543702 | SERPINA11 | FALSE | Upregulated |
| 0,215513281 | 0,807612181 | NECAB2 | FALSE | Upregulated |
| -0,08336355 | 0,807649105 | BHLHB9 | FALSE | Downregulated |
| 0,2562942 | 0,807649105 | LOC101929612 | FALSE | Upregulated |
| 0,083354391 | 0,807649105 | SH3TC1 | FALSE | Upregulated |
| -0,080177868 | 0,807689137 | TMEM14B | FALSE | Downregulated |
| 0,088152924 | 0,807726936 | DHX57 | FALSE | Upregulated |
| 0,510565041 | 0,807726936 | NBPF7 | FALSE | Upregulated |
| 0,079871131 | 0,807726936 | PSMA1 | FALSE | Upregulated |
| 0,37193416 | 0,807737394 | GAS6-AS2 | FALSE | Upregulated |
| -0,083288973 | 0,807737394 | HNRNPA1 | FALSE | Downregulated |
| 0,074659223 | 0,807737394 | MYO5B | FALSE | Upregulated |

| | | | | |
|---|---|---|---|---|
| 0,070067441 | 0,807866651 | KAT8 | FALSE | Upregulated |
| -0,076904921 | 0,807866651 | YEATS4 | FALSE | Downregulated |
| -0,305513255 | 0,808135874 | LOC100652901 | FALSE | Downregulated |
| -0,212597765 | 0,808210304 | COPG2IT1 | FALSE | Downregulated |
| 0,109773506 | 0,808210304 | CYTH3 | FALSE | Upregulated |
| -0,09133237 | 0,808210304 | FAM214A | FALSE | Downregulated |
| 0,440214945 | 0,808210304 | KCNE2 | FALSE | Upregulated |
| 0,086102338 | 0,808210304 | MBD5 | FALSE | Upregulated |
| 0,297955188 | 0,808210304 | PDZD9 | FALSE | Upregulated |
| -0,079305773 | 0,808210304 | USP36 | FALSE | Downregulated |
| -0,137577527 | 0,808486033 | PARK2 | FALSE | Downregulated |
| -0,073732666 | 0,808486033 | RALGAPA2 | FALSE | Downregulated |
| 0,319883202 | 0,808486033 | RPS15AP10 | FALSE | Upregulated |
| 0,084582492 | 0,80850259 | SMURF1 | FALSE | Upregulated |
| -0,045593546 | 0,808601104 | B4GALT4 | FALSE | Downregulated |
| -0,070442635 | 0,808601104 | EPB41 | FALSE | Downregulated |
| -0,251262901 | 0,808601104 | FAR2P2 | FALSE | Downregulated |
| -0,05527079 | 0,808601104 | FGFR1OP | FALSE | Downregulated |
| 0,112408445 | 0,808601104 | KLHDC9 | FALSE | Upregulated |
| -0,500880777 | 0,808601104 | LOC101930567 | FALSE | Downregulated |
| 0,089707697 | 0,808601104 | MACROD1 | FALSE | Upregulated |
| -0,05674429 | 0,808601104 | PGGT1B | FALSE | Downregulated |
| -0,54224846 | 0,808601104 | PLA2G2A | FALSE | Downregulated |
| 0,075834175 | 0,808601104 | TRAPPC10 | FALSE | Upregulated |
| 0,065426994 | 0,808625873 | CNOT10 | FALSE | Upregulated |
| 0,153267715 | 0,808625873 | KIF21B | FALSE | Upregulated |

| | | | | |
|---|---|---|---|---|
| 0,214542498 | 0,808625873 | LINC00861 | FALSE | Upregulated |
| -0,087101303 | 0,808625873 | MARVELD3 | FALSE | Downregulated |
| -0,077695805 | 0,808625873 | TRAF3 | FALSE | Downregulated |
| -0,06462115 | 0,808638592 | DBR1 | FALSE | Downregulated |
| 0,068893476 | 0,808638592 | GTF3C3 | FALSE | Upregulated |
| -0,055887459 | 0,808638592 | LLPH | FALSE | Downregulated |
| 0,765543621 | 0,808638592 | LOC101928722 | FALSE | Upregulated |
| 0,259943761 | 0,808638592 | LOC102724643 | FALSE | Upregulated |
| -0,054521827 | 0,808638592 | MKLN1 | FALSE | Downregulated |
| -0,110832867 | 0,808638592 | ZNF658 | FALSE | Downregulated |
| 0,85611824 | 0,808683252 | RGS7 | FALSE | Upregulated |
| 0,30865234 | 0,808709374 | C12orf79 | FALSE | Upregulated |
| 0,052127552 | 0,808709374 | CAB39 | FALSE | Upregulated |
| 0,309140585 | 0,808709374 | CDC6 | FALSE | Upregulated |
| -0,075834804 | 0,808709374 | COA1 | FALSE | Downregulated |
| -0,293864499 | 0,808709374 | CTB-118N6.3 | FALSE | Downregulated |
| 0,074527423 | 0,808709374 | HSPD1 | FALSE | Upregulated |
| 0,259390762 | 0,808709374 | KCNK3 | FALSE | Upregulated |
| 0,415942245 | 0,808709374 | LOC101928474 | FALSE | Upregulated |
| -0,274779355 | 0,808709374 | LPA | FALSE | Downregulated |
| -0,057411324 | 0,808709374 | MGEA5 | FALSE | Downregulated |
| 0,057053992 | 0,808709374 | NF2 | FALSE | Upregulated |
| -0,059202044 | 0,808709374 | SLC25A32 | FALSE | Downregulated |
| -0,387282962 | 0,808709374 | UCKL1-AS1 | FALSE | Downregulated |
| 0,088102596 | 0,808709374 | VCL | FALSE | Upregulated |
| 0,412156368 | 0,808724393 | ESRRB | FALSE | Upregulated |

| | | | | |
|---|---|---|---|---|
| 0,05675032 | 0,808820625 | PARG | FALSE | Upregulated |
| 0,075425061 | 0,808820625 | PRRC1 | FALSE | Upregulated |
| -0,15792199 | 0,808820625 | RP11-384K6.6 | FALSE | Downregulated |
| -0,073014705 | 0,808820625 | TDRP | FALSE | Downregulated |
| 0,472369295 | 0,808820625 | TPBGL | FALSE | Upregulated |
| 0,224877886 | 0,809160406 | MTCP1 | FALSE | Upregulated |
| 0,284493148 | 0,809160406 | P2RY11 | FALSE | Upregulated |
| -0,085839874 | 0,809286962 | DIO1 | FALSE | Downregulated |
| -0,284906672 | 0,809286962 | FBXO39 | FALSE | Downregulated |
| -0,172344453 | 0,809286962 | GOLGA2P5 | FALSE | Downregulated |
| 0,114328025 | 0,809286962 | HNF4A-AS1 | FALSE | Upregulated |
| 0,068675546 | 0,809286962 | WDR45 | FALSE | Upregulated |
| -0,120847547 | 0,809499099 | HYKK | FALSE | Downregulated |
| 0,070364233 | 0,809499099 | ZNRF1 | FALSE | Upregulated |
| -0,128943522 | 0,809531727 | AMACR | FALSE | Downregulated |
| 0,131914417 | 0,809531727 | LOC102724884 | FALSE | Upregulated |
| -0,088699808 | 0,809531727 | UQCRH | FALSE | Downregulated |
| -0,387772745 | 0,809571953 | NPPA-AS1 | FALSE | Downregulated |
| -0,152000984 | 0,809607308 | ASCL2 | FALSE | Downregulated |
| -0,05758621 | 0,809607308 | FAM120B | FALSE | Downregulated |
| -0,070382379 | 0,809607308 | GLYATL1 | FALSE | Downregulated |
| -0,083248003 | 0,809607308 | HSD17B4 | FALSE | Downregulated |
| 0,08547401 | 0,809607308 | LAP3 | FALSE | Upregulated |
| 0,148255241 | 0,809607308 | LIPJ | FALSE | Upregulated |
| 0,398712448 | 0,809607308 | MPO | FALSE | Upregulated |
| -0,063512205 | 0,809607308 | NDFIP2 | FALSE | Downregulated |

| | | | | |
|---|---|---|---|---|
| 0,100303368 | 0,809607308 | PARP12 | FALSE | Upregulated |
| -0,352792637 | 0,809607308 | PROL1 | FALSE | Downregulated |
| 0,059322269 | 0,809705618 | CBY1 | FALSE | Upregulated |
| 0,133303988 | 0,809705618 | MRM1 | FALSE | Upregulated |
| -0,581707852 | 0,809705618 | SNORA7A | FALSE | Downregulated |
| -0,177096313 | 0,809705618 | STARD4 | FALSE | Downregulated |
| -0,097512301 | 0,809705618 | ZNF248 | FALSE | Downregulated |
| 0,094166236 | 0,809955862 | MAP3K9 | FALSE | Upregulated |
| -0,087482114 | 0,809977908 | PDIA5 | FALSE | Downregulated |
| 0,107172122 | 0,810021661 | ARHGAP15 | FALSE | Upregulated |
| -0,068332143 | 0,810021661 | HABP2 | FALSE | Downregulated |
| -0,048334516 | 0,810021661 | MAK16 | FALSE | Downregulated |
| -0,072129518 | 0,810021661 | NAPG | FALSE | Downregulated |
| -0,158657157 | 0,810021661 | SSTR1 | FALSE | Downregulated |
| 0,193231477 | 0,810021661 | STXBP6 | FALSE | Upregulated |
| 0,187482729 | 0,810025866 | IER3 | FALSE | Upregulated |
| 0,342580999 | 0,810230496 | SLFNL1-AS1 | FALSE | Upregulated |
| 0,082909215 | 0,810330704 | TRMU | FALSE | Upregulated |
| 0,038540292 | 0,810359974 | DCTN1 | FALSE | Upregulated |
| 0,342826224 | 0,810359974 | LOC101928178 | FALSE | Upregulated |
| -0,076014171 | 0,810359974 | MTMR4 | FALSE | Downregulated |
| -0,107128026 | 0,810359974 | PRIM1 | FALSE | Downregulated |
| -0,098181412 | 0,810359974 | RPS27 | FALSE | Downregulated |
| 0,107545143 | 0,810492164 | ATP8B1 | FALSE | Upregulated |
| -0,152959258 | 0,810545317 | ATP8A1 | FALSE | Downregulated |
| 0,548565716 | 0,810545317 | KDM5D | FALSE | Upregulated |

| | | | | |
|---|---|---|---|---|
| -0,19774166 | 0,810545317 | RAD51-AS1 | FALSE | Downregulated |
| 0,139502961 | 0,810545317 | SLC2A9 | FALSE | Upregulated |
| -0,191807215 | 0,810608857 | LOC101928645 | FALSE | Downregulated |
| -0,056439842 | 0,810608857 | UBLCP1 | FALSE | Downregulated |
| -0,211435104 | 0,810688782 | C14orf180 | FALSE | Downregulated |
| -0,088389623 | 0,810688782 | C8orf82 | FALSE | Downregulated |
| -0,061793332 | 0,810688782 | CASC3 | FALSE | Downregulated |
| -0,096709498 | 0,810688782 | CASP9 | FALSE | Downregulated |
| -0,32856726 | 0,810688782 | CTD-2201I18.1 | FALSE | Downregulated |
| 0,257622076 | 0,810688782 | ERAP2 | FALSE | Upregulated |
| 0,085537261 | 0,810688782 | KATNA1 | FALSE | Upregulated |
| -0,917314491 | 0,810688782 | LOC102723678 | FALSE | Downregulated |
| -0,126754714 | 0,810688782 | LOC285074 | FALSE | Downregulated |
| 0,083019087 | 0,810688782 | MBNL2 | FALSE | Upregulated |
| 0,059980608 | 0,810688782 | SMARCAL1 | FALSE | Upregulated |
| -0,126471598 | 0,810688782 | TBRG1 | FALSE | Downregulated |
| -0,075650178 | 0,810688782 | ZNF75D | FALSE | Downregulated |
| 0,257566066 | 0,810853965 | PLCH2 | FALSE | Upregulated |
| 0,380522065 | 0,810910559 | ACSM1 | FALSE | Upregulated |
| 0,116991673 | 0,810910559 | BAK1 | FALSE | Upregulated |
| 0,189725749 | 0,810910559 | C14orf80 | FALSE | Upregulated |
| -0,100581435 | 0,810910559 | C19orf12 | FALSE | Downregulated |
| -0,249687893 | 0,810910559 | LOC101927793 | FALSE | Downregulated |
| -0,056139554 | 0,810910559 | NPEPPS | FALSE | Downregulated |
| 0,181587155 | 0,810910559 | TMEM54 | FALSE | Upregulated |
| -0,051436253 | 0,810910559 | UBR7 | FALSE | Downregulated |

| | | | | |
|---|---|---|---|---|
| 0,090724294 | 0,810974205 | ABHD14A | FALSE | Upregulated |
| -0,075910355 | 0,810974205 | C12orf76 | FALSE | Downregulated |
| -0,100155632 | 0,810974205 | CCDC69 | FALSE | Downregulated |
| -0,05167388 | 0,810974205 | CERS2 | FALSE | Downregulated |
| 0,170217173 | 0,810974205 | CRISPLD2 | FALSE | Upregulated |
| 0,168312306 | 0,810974205 | DAPP1 | FALSE | Upregulated |
| 0,044165239 | 0,810974205 | DDX54 | FALSE | Upregulated |
| -0,176289065 | 0,810974205 | GOLGA8N | FALSE | Downregulated |
| -0,046711386 | 0,810974205 | IPO9 | FALSE | Downregulated |
| -0,080296506 | 0,810974205 | KIAA1551 | FALSE | Downregulated |
| -0,317476634 | 0,810974205 | LOC102723671 | FALSE | Downregulated |
| 0,22518108 | 0,810974205 | MAMDC4 | FALSE | Upregulated |
| 0,320578409 | 0,810974205 | MATN1-AS1 | FALSE | Upregulated |
| 0,049109921 | 0,810974205 | MCL1 | FALSE | Upregulated |
| 0,312273213 | 0,810974205 | MEP1B | FALSE | Upregulated |
| 0,065883682 | 0,810974205 | PI4K2B | FALSE | Upregulated |
| 0,055922091 | 0,810974205 | PRKRA | FALSE | Upregulated |
| -0,302734353 | 0,810974205 | PROB1 | FALSE | Downregulated |
| -0,302037137 | 0,810974205 | SKP1P2 | FALSE | Downregulated |
| -0,124038834 | 0,810974205 | SREK1 | FALSE | Downregulated |
| -0,077382924 | 0,810974205 | SRP54 | FALSE | Downregulated |
| -0,404565124 | 0,810974205 | TMC4 | FALSE | Downregulated |
| 0,091762386 | 0,810974205 | TMEM194A | FALSE | Upregulated |
| -0,12117649 | 0,810974205 | U2AF1L4 | FALSE | Downregulated |
| 0,33410652 | 0,810974205 | USP12-AS2 | FALSE | Upregulated |
| -0,237493799 | 0,810974205 | WDR96 | FALSE | Downregulated |

| | | | | |
|---|---|---|---|---|
| -0,093692083 | 0,810974205 | WDSUB1 | FALSE | Downregulated |
| -0,082710627 | 0,810974205 | XKR8 | FALSE | Downregulated |
| 0,165953117 | 0,811190792 | RIMS3 | FALSE | Upregulated |
| -0,28443257 | 0,811197206 | LOC102723327 | FALSE | Downregulated |
| 0,427537263 | 0,811219227 | C10orf62 | FALSE | Upregulated |
| 0,193843684 | 0,811219227 | FAM26F | FALSE | Upregulated |
| 0,172187876 | 0,811219227 | IL27RA | FALSE | Upregulated |
| 0,078626195 | 0,811219227 | TBRG4 | FALSE | Upregulated |
| 0,46569088 | 0,811373542 | KRT80 | FALSE | Upregulated |
| -0,09217412 | 0,811489592 | MIA2 | FALSE | Downregulated |
| 0,436649505 | 0,81158545 | ADRA2C | FALSE | Upregulated |
| 0,08119355 | 0,81158545 | ATOX1 | FALSE | Upregulated |
| 0,192107548 | 0,81158545 | KIF9-AS1 | FALSE | Upregulated |
| 0,067110419 | 0,81158545 | PTPMT1 | FALSE | Upregulated |
| -0,068702271 | 0,81158545 | SMC5 | FALSE | Downregulated |
| 0,121561131 | 0,81158545 | STXBP4 | FALSE | Upregulated |
| 0,088905053 | 0,81158545 | TEC | FALSE | Upregulated |
| 0,077393415 | 0,81158545 | TRIM27 | FALSE | Upregulated |
| 0,339657978 | 0,812104596 | LOC101927837 | FALSE | Upregulated |
| 0,137585005 | 0,812172313 | FAM46C | FALSE | Upregulated |
| 0,328014814 | 0,812172313 | LOC101927699 | FALSE | Upregulated |
| -0,083612047 | 0,812172313 | ME2 | FALSE | Downregulated |
| 0,078515595 | 0,812172313 | PINX1 | FALSE | Upregulated |
| 0,061481232 | 0,812223124 | MTMR6 | FALSE | Upregulated |
| -0,115927747 | 0,812409238 | ADAMTS13 | FALSE | Downregulated |
| 0,1656205 | 0,812409238 | AGPAT9 | FALSE | Upregulated |

| | | | | |
|---|---|---|---|---|
| 0,08910148 | 0,812409238 | AGTRAP | FALSE | Upregulated |
| 0,133837519 | 0,812409238 | ARSE | FALSE | Upregulated |
| 0,476637983 | 0,812409238 | CLIC3 | FALSE | Upregulated |
| 0,146078469 | 0,812409238 | DFNB31 | FALSE | Upregulated |
| 0,087281858 | 0,812409238 | DVL2 | FALSE | Upregulated |
| 0,059417854 | 0,812409238 | ENO1 | FALSE | Upregulated |
| 0,083070577 | 0,812409238 | ERMP1 | FALSE | Upregulated |
| -0,06477257 | 0,812409238 | FAM206A | FALSE | Downregulated |
| -0,265667104 | 0,812409238 | FLVCR1-AS1 | FALSE | Downregulated |
| 0,370601678 | 0,812409238 | FRY-AS1 | FALSE | Upregulated |
| -0,048108491 | 0,812409238 | GOLGA1 | FALSE | Downregulated |
| -0,284352638 | 0,812409238 | GSTA2 | FALSE | Downregulated |
| 0,073233034 | 0,812409238 | KIF22 | FALSE | Upregulated |
| -0,094125135 | 0,812409238 | LCLAT1 | FALSE | Downregulated |
| -0,443368236 | 0,812409238 | LINC00525 | FALSE | Downregulated |
| -0,118106269 | 0,812409238 | LINC00649 | FALSE | Downregulated |
| 0,157677965 | 0,812409238 | LINC01124 | FALSE | Upregulated |
| 0,209779241 | 0,812409238 | LOC101928228 | FALSE | Upregulated |
| 0,392707799 | 0,812409238 | LOC102723610 | FALSE | Upregulated |
| 0,422292233 | 0,812409238 | MAEL | FALSE | Upregulated |
| -0,521065548 | 0,812409238 | MT1HL1 | FALSE | Downregulated |
| -0,071048673 | 0,812409238 | NEK4 | FALSE | Downregulated |
| 0,143775064 | 0,812409238 | NLRP1 | FALSE | Upregulated |
| -0,109049412 | 0,812409238 | PCAT19 | FALSE | Downregulated |
| 0,059333139 | 0,812409238 | PFN1 | FALSE | Upregulated |
| -0,103312245 | 0,812409238 | PSTK | FALSE | Downregulated |

| | | | | |
|---|---|---|---|---|
| -0,216748407 | 0,812409238 | RP11-75N4.2 | FALSE | Downregulated |
| 0,072182404 | 0,812409238 | RRBP1 | FALSE | Upregulated |
| -0,047995075 | 0,812409238 | SEC11A | FALSE | Downregulated |
| -0,2296383 | 0,812409238 | SLC16A1-AS1 | FALSE | Downregulated |
| -0,086139705 | 0,812409238 | TACC2 | FALSE | Downregulated |
| -0,056321065 | 0,812409238 | TMBIM6 | FALSE | Downregulated |
| -1,108210406 | 0,812409238 | TRNAG19 | FALSE | Downregulated |
| -0,079515547 | 0,812409238 | USP53 | FALSE | Downregulated |
| -0,490168031 | 0,812409238 | XKR6 | FALSE | Downregulated |
| -0,079632519 | 0,812409238 | ZFYVE19 | FALSE | Downregulated |
| -0,097248104 | 0,812409238 | ZNF485 | FALSE | Downregulated |
| 0,057346363 | 0,812409238 | ZNF609 | FALSE | Upregulated |
| 0,16968414 | 0,812742246 | DNAH1 | FALSE | Upregulated |
| -0,19570477 | 0,812826284 | RP5-1159O4.2 | FALSE | Downregulated |
| -0,105350464 | 0,812874972 | HDHD1 | FALSE | Downregulated |
| 0,290361287 | 0,813134253 | LOC102724307 | FALSE | Upregulated |
| -0,126828906 | 0,813134253 | LRRC4 | FALSE | Downregulated |
| -0,082424647 | 0,813217694 | LAMC1 | FALSE | Downregulated |
| -0,059346047 | 0,813458873 | COG5 | FALSE | Downregulated |
| -0,519674392 | 0,813463521 | ANP32AP1 | FALSE | Downregulated |
| 0,05248799 | 0,813463521 | EIF3D | FALSE | Upregulated |
| -0,171739874 | 0,813463521 | LDLR | FALSE | Downregulated |
| 0,188494763 | 0,813463521 | PLA2G7 | FALSE | Upregulated |
| -0,061746685 | 0,813463521 | TTC19 | FALSE | Downregulated |
| 0,167670372 | 0,813463521 | ZNF573 | FALSE | Upregulated |
| -0,084461515 | 0,813624792 | ABHD8 | FALSE | Downregulated |

| | | | | |
|---|---|---|---|---|
| -0,089508044 | 0,813624792 | AUTS2 | FALSE | Downregulated |
| 0,359902554 | 0,813953328 | CTAGE8 | FALSE | Upregulated |
| -0,075251253 | 0,814099003 | UXT | FALSE | Downregulated |
| 0,069179098 | 0,81413889 | CD2BP2 | FALSE | Upregulated |
| -0,064171272 | 0,81413889 | CSTF3 | FALSE | Downregulated |
| 0,038997077 | 0,81413889 | DCAF7 | FALSE | Upregulated |
| 0,060422249 | 0,81413889 | DENND5A | FALSE | Upregulated |
| -0,064635741 | 0,81413889 | EMC7 | FALSE | Downregulated |
| -0,074271571 | 0,81413889 | FAM172A | FALSE | Downregulated |
| 0,238687083 | 0,81413889 | HCAR2 | FALSE | Upregulated |
| 0,145298901 | 0,81413889 | KLF8 | FALSE | Upregulated |
| 0,181644363 | 0,81413889 | LOC102723983 | FALSE | Upregulated |
| -0,070449365 | 0,81413889 | PREPL | FALSE | Downregulated |
| -0,056003325 | 0,81413889 | UTP20 | FALSE | Downregulated |
| 0,038132325 | 0,814182861 | GPR107 | FALSE | Upregulated |
| 0,081058259 | 0,814182861 | NDUFAF4 | FALSE | Upregulated |
| 0,05611353 | 0,814182861 | RNF167 | FALSE | Upregulated |
| 0,336956994 | 0,814182861 | ST8SIA3 | FALSE | Upregulated |
| -0,322349063 | 0,814263102 | LOC654342 | FALSE | Downregulated |
| -0,142960914 | 0,814526455 | LOC100505942 | FALSE | Downregulated |
| 0,337821427 | 0,814526455 | LOC102723517 | FALSE | Upregulated |
| 0,24835282 | 0,814548706 | FADS2 | FALSE | Upregulated |
| 0,076682169 | 0,814548706 | POLA1 | FALSE | Upregulated |
| 0,084310451 | 0,814627819 | DCTN5 | FALSE | Upregulated |
| -0,055516525 | 0,814663424 | CHCHD3 | FALSE | Downregulated |
| 0,08799035 | 0,814663424 | WDFY2 | FALSE | Upregulated |

| | | | | |
|---|---|---|---|---|
| 0,088444791 | 0,814752009 | ZNF799 | FALSE | Upregulated |
| -0,11211929 | 0,814935754 | CWC25 | FALSE | Downregulated |
| 0,272666351 | 0,814935754 | LOC101927086 | FALSE | Upregulated |
| 0,162540303 | 0,814935754 | MCAM | FALSE | Upregulated |
| 0,047323848 | 0,814935754 | RAB18 | FALSE | Upregulated |
| 0,27260217 | 0,814935754 | SNORA4 | FALSE | Upregulated |
| 0,065472761 | 0,814968465 | PGLS | FALSE | Upregulated |
| -0,114154509 | 0,814972017 | ADHFE1 | FALSE | Downregulated |
| 0,15174563 | 0,814972017 | ASTN2 | FALSE | Upregulated |
| -0,074802282 | 0,814972017 | GC | FALSE | Downregulated |
| -0,277173114 | 0,814972017 | IQCA1 | FALSE | Downregulated |
| 0,368347122 | 0,814972017 | KEL | FALSE | Upregulated |
| 0,086962061 | 0,814972017 | NSUN5 | FALSE | Upregulated |
| 0,296514029 | 0,814972017 | TEX14 | FALSE | Upregulated |
| -0,054532256 | 0,814972017 | XPC | FALSE | Downregulated |
| -0,058214145 | 0,815039994 | DPY19L1 | FALSE | Downregulated |
| 0,201969395 | 0,815039994 | LOC100996664 | FALSE | Upregulated |
| -0,100955601 | 0,815039994 | PRKAR2B | FALSE | Downregulated |
| -0,1524785 | 0,815039994 | SCARA3 | FALSE | Downregulated |
| -0,064069493 | 0,815311095 | PPP1R2 | FALSE | Downregulated |
| -0,079946871 | 0,815535145 | RPL30 | FALSE | Downregulated |
| 0,179126622 | 0,815812146 | CYP4F22 | FALSE | Upregulated |
| 0,476916608 | 0,815812146 | DPCR1 | FALSE | Upregulated |
| 0,122516641 | 0,815812146 | FOXA2 | FALSE | Upregulated |
| 0,100770858 | 0,815812146 | IER2 | FALSE | Upregulated |
| 0,707300489 | 0,815812146 | LINC00960 | FALSE | Upregulated |

| | | | | |
|---|---|---|---|---|
| -0,295246362 | 0,815846014 | LOC101928706 | FALSE | Downregulated |
| 0,334910962 | 0,815846014 | NR4A3 | FALSE | Upregulated |
| 0,168201906 | 0,815846014 | PCED1B-AS1 | FALSE | Upregulated |
| -0,071791899 | 0,81585393 | AMPD2 | FALSE | Downregulated |
| -0,158962285 | 0,81585393 | ANGPT1 | FALSE | Downregulated |
| 0,504286783 | 0,81585393 | BAI2 | FALSE | Upregulated |
| -0,057637975 | 0,81585393 | C19orf43 | FALSE | Downregulated |
| 0,249174535 | 0,81585393 | CD101 | FALSE | Upregulated |
| 0,059049915 | 0,81585393 | CD81 | FALSE | Upregulated |
| 0,246250066 | 0,81585393 | FAM150B | FALSE | Upregulated |
| -0,109081899 | 0,81585393 | GGTA1P | FALSE | Downregulated |
| -0,423935113 | 0,81585393 | GPR63 | FALSE | Downregulated |
| 0,065355528 | 0,81585393 | HEIH | FALSE | Upregulated |
| -0,072705825 | 0,81585393 | IL1R1 | FALSE | Downregulated |
| -0,098038434 | 0,81585393 | KDM6A | FALSE | Downregulated |
| 0,188838274 | 0,81585393 | KIF12 | FALSE | Upregulated |
| 0,088111849 | 0,81585393 | MLH3 | FALSE | Upregulated |
| -0,101736349 | 0,81585393 | NCDN | FALSE | Downregulated |
| -0,088058678 | 0,81585393 | NDUFAF6 | FALSE | Downregulated |
| 0,234784794 | 0,81585393 | OTUD7A | FALSE | Upregulated |
| -0,108217876 | 0,81585393 | POLR2J3 | FALSE | Downregulated |
| -0,113511692 | 0,81585393 | PPIF | FALSE | Downregulated |
| -0,108154603 | 0,81585393 | RPP40 | FALSE | Downregulated |
| 0,094864081 | 0,81585393 | TFEB | FALSE | Upregulated |
| 0,134036245 | 0,81585393 | TRAPPC2P1 | FALSE | Upregulated |
| 0,14671536 | 0,81585393 | TUB | FALSE | Upregulated |

| | | | | |
|---|---|---|---|---|
| 0,255232359 | 0,815864985 | LOC102724687 | FALSE | Upregulated |
| 0,19581262 | 0,815870919 | METTL12 | FALSE | Upregulated |
| -0,20293043 | 0,815870919 | NAALAD2 | FALSE | Downregulated |
| 0,053934305 | 0,81588187 | GNA13 | FALSE | Upregulated |
| -0,170338517 | 0,81588187 | LOC101928868 | FALSE | Downregulated |
| 0,10753156 | 0,81588187 | NAT6 | FALSE | Upregulated |
| 0,053692887 | 0,81588187 | UNC50 | FALSE | Upregulated |
| -0,087058173 | 0,81588187 | ZNF524 | FALSE | Downregulated |
| 0,071145098 | 0,815998586 | VPS13A | FALSE | Upregulated |
| 0,052258452 | 0,816184778 | FAM32A | FALSE | Upregulated |
| -0,127094516 | 0,816420428 | RCL1 | FALSE | Downregulated |
| -0,065196708 | 0,816516948 | ERGIC2 | FALSE | Downregulated |
| 0,457436532 | 0,816516948 | FOXL1 | FALSE | Upregulated |
| -0,04487138 | 0,816516948 | GLOD4 | FALSE | Downregulated |
| -0,047339919 | 0,816516948 | RAD1 | FALSE | Downregulated |
| -0,068525605 | 0,816622838 | ZNF507 | FALSE | Downregulated |
| 0,161811682 | 0,816655902 | AMPD3 | FALSE | Upregulated |
| 0,070466054 | 0,816655902 | CRTC1 | FALSE | Upregulated |
| 0,101254968 | 0,816655902 | DNAJC15 | FALSE | Upregulated |
| -0,404378372 | 0,816655902 | LOC100130285 | FALSE | Downregulated |
| -0,098519175 | 0,816655902 | NHLRC1 | FALSE | Downregulated |
| -0,257985855 | 0,816655902 | NPIPB9 | FALSE | Downregulated |
| -0,135506451 | 0,816655902 | RAB11B-AS1 | FALSE | Downregulated |
| 0,086600911 | 0,816655902 | SDC1 | FALSE | Upregulated |
| -0,356060781 | 0,816655902 | SLC29A4 | FALSE | Downregulated |
| -0,062568843 | 0,816655902 | TAF5 | FALSE | Downregulated |

| | | | | |
|---|---|---|---|---|
| -0,093438754 | 0,816655902 | UBXN7 | FALSE | Downregulated |
| 0,068230263 | 0,816655902 | WARS2 | FALSE | Upregulated |
| -0,148903805 | 0,816655902 | ZNF501 | FALSE | Downregulated |
| -0,110895208 | 0,816655902 | ZNF572 | FALSE | Downregulated |
| 0,108128749 | 0,816774541 | ABCC1 | FALSE | Upregulated |
| -0,502447233 | 0,816774541 | ANKRD20A12P | FALSE | Downregulated |
| 0,093334581 | 0,816774541 | ANO1 | FALSE | Upregulated |
| -0,410500807 | 0,816774541 | C1orf167 | FALSE | Downregulated |
| -0,113196062 | 0,816774541 | C2orf74 | FALSE | Downregulated |
| 0,093014473 | 0,816774541 | CADPS2 | FALSE | Upregulated |
| -0,073349938 | 0,816774541 | CFL2 | FALSE | Downregulated |
| 0,186765137 | 0,816774541 | EP400NL | FALSE | Upregulated |
| 0,096071283 | 0,816774541 | FAM189B | FALSE | Upregulated |
| -0,157214513 | 0,816774541 | FLJ22763 | FALSE | Downregulated |
| -0,089631885 | 0,816774541 | FMO3 | FALSE | Downregulated |
| -0,196005296 | 0,816774541 | GNN | FALSE | Downregulated |
| -1,134764545 | 0,816774541 | GOLGA8J | FALSE | Downregulated |
| 0,053137486 | 0,816774541 | GUSB | FALSE | Upregulated |
| 0,049813994 | 0,816774541 | HIF1AN | FALSE | Upregulated |
| 0,056526045 | 0,816774541 | HSF1 | FALSE | Upregulated |
| -0,088172191 | 0,816774541 | IQSEC1 | FALSE | Downregulated |
| -0,134810345 | 0,816774541 | ITGAE | FALSE | Downregulated |
| 0,066900441 | 0,816774541 | JARID2 | FALSE | Upregulated |
| -0,100371993 | 0,816774541 | KIAA2018 | FALSE | Downregulated |
| -0,183748721 | 0,816774541 | LOC100128531 | FALSE | Downregulated |
| -0,190213227 | 0,816774541 | LOC101929046 | FALSE | Downregulated |

| | | | | |
|---|---|---|---|---|
| -0,179067281 | 0,816774541 | LOC101929668 | FALSE | Downregulated |
| -0,468399987 | 0,816774541 | LOC102724901 | FALSE | Downregulated |
| 0,18000519 | 0,816774541 | LOC283177 | FALSE | Upregulated |
| -0,065486448 | 0,816774541 | LRRC37BP1 | FALSE | Downregulated |
| 0,092410534 | 0,816774541 | MED27 | FALSE | Upregulated |
| -0,0709642 | 0,816774541 | MRPL43 | FALSE | Downregulated |
| 0,239922474 | 0,816774541 | MST1P2 | FALSE | Upregulated |
| -0,089537339 | 0,816774541 | N6AMT2 | FALSE | Downregulated |
| 0,386113627 | 0,816774541 | NAPSA | FALSE | Upregulated |
| -0,064559669 | 0,816774541 | PIGX | FALSE | Downregulated |
| -0,140158189 | 0,816774541 | PLAC8 | FALSE | Downregulated |
| 0,143350643 | 0,816774541 | RAP1GAP2 | FALSE | Upregulated |
| -0,076543136 | 0,816774541 | REPS2 | FALSE | Downregulated |
| 0,201066021 | 0,816774541 | SLC7A1 | FALSE | Upregulated |
| 0,227712576 | 0,816774541 | SOD3 | FALSE | Upregulated |
| -0,082759466 | 0,816774541 | ST3GAL5 | FALSE | Downregulated |
| 0,063395568 | 0,816774541 | STOML2 | FALSE | Upregulated |
| -0,094539921 | 0,816774541 | TCEANC | FALSE | Downregulated |
| 0,296938752 | 0,816774541 | TMEM158 | FALSE | Upregulated |
| -0,074879079 | 0,816774541 | TNIP1 | FALSE | Downregulated |
| -0,165899879 | 0,816774541 | TSPYL5 | FALSE | Downregulated |
| 0,126181421 | 0,816774541 | TTYH2 | FALSE | Upregulated |
| 0,077685725 | 0,816774541 | ZFAND2B | FALSE | Upregulated |
| 0,124819457 | 0,816774541 | ZNF497 | FALSE | Upregulated |
| 0,14974199 | 0,816774541 | ZNF763 | FALSE | Upregulated |
| -0,179061981 | 0,817066801 | THRB-AS1 | FALSE | Downregulated |

| | | | | |
|---|---|---|---|---|
| -0,282831557 | 0,817410112 | C2orf27A | FALSE | Downregulated |
| 0,464354306 | 0,817410112 | RPH3A | FALSE | Upregulated |
| -0,363469839 | 0,817469941 | RGSL1 | FALSE | Downregulated |
| -0,100016459 | 0,81762816 | BAZ1B | FALSE | Downregulated |
| -0,121317948 | 0,817698377 | GPR89A | FALSE | Downregulated |
| 0,341737165 | 0,817698377 | LOC102723579 | FALSE | Upregulated |
| -0,097526029 | 0,817840847 | KIAA1217 | FALSE | Downregulated |
| 0,211130928 | 0,818003467 | STAG3L5P-PVRIG2P-PILRB | FALSE | Upregulated |
| 0,067374259 | 0,818189082 | ACY1 | FALSE | Upregulated |
| 0,146405193 | 0,818226036 | NBPF15 | FALSE | Upregulated |
| 0,08411715 | 0,818275104 | ZNF182 | FALSE | Upregulated |
| 0,097530435 | 0,818464491 | NEIL2 | FALSE | Upregulated |
| -0,057030726 | 0,818530635 | FAM200B | FALSE | Downregulated |
| 0,540079646 | 0,818530635 | SNORA62 | FALSE | Upregulated |
| -0,153295745 | 0,818810156 | MAPK10 | FALSE | Downregulated |
| -0,106198228 | 0,818891096 | PAQR9 | FALSE | Downregulated |
| 0,362337665 | 0,819173779 | SPC24 | FALSE | Upregulated |
| 0,059999195 | 0,819259764 | PMS2P1 | FALSE | Upregulated |
| -0,058703275 | 0,819353825 | SWT1 | FALSE | Downregulated |
| -0,212445906 | 0,819467189 | CNTNAP3B | FALSE | Downregulated |
| -0,041976108 | 0,819467189 | RAB10 | FALSE | Downregulated |
| 0,064497043 | 0,819596292 | MTX2 | FALSE | Upregulated |
| -0,058348928 | 0,81967663 | CSNK1G2 | FALSE | Downregulated |
| -0,077290226 | 0,820101898 | AGK | FALSE | Downregulated |
| -0,070950438 | 0,820195755 | PRPF4B | FALSE | Downregulated |

| | | | | |
|---|---|---|---|---|
| -0,167241091 | 0,820266212 | COL27A1 | FALSE | Downregulated |
| -0,063831867 | 0,820266212 | FAM96B | FALSE | Downregulated |
| -0,118660816 | 0,820266212 | FXYD1 | FALSE | Downregulated |
| 0,589392745 | 0,820266212 | LOC554223 | FALSE | Upregulated |
| -0,214657335 | 0,820266212 | LOC643770 | FALSE | Downregulated |
| -0,053152198 | 0,820266212 | PAICS | FALSE | Downregulated |
| 0,095199054 | 0,820266212 | RASSF5 | FALSE | Upregulated |
| 0,081186124 | 0,820266212 | SEC14L1 | FALSE | Upregulated |
| 0,505404374 | 0,820266212 | SFN | FALSE | Upregulated |
| -0,048617422 | 0,820266212 | SNX3 | FALSE | Downregulated |
| 0,130309293 | 0,820266212 | SRCIN1 | FALSE | Upregulated |
| 0,07989814 | 0,820266212 | TMSB4X | FALSE | Upregulated |
| -0,095506243 | 0,820471174 | IL4R | FALSE | Downregulated |
| -0,086635422 | 0,820471174 | PHTF1 | FALSE | Downregulated |
| 0,121681107 | 0,820471174 | PIDD | FALSE | Upregulated |
| 0,21091877 | 0,820471174 | PRR15L | FALSE | Upregulated |
| 0,08233181 | 0,820471174 | RPS3 | FALSE | Upregulated |
| -0,105473256 | 0,820471174 | THPO | FALSE | Downregulated |
| 0,264401453 | 0,820523954 | SCARNA17 | FALSE | Upregulated |
| -0,305107122 | 0,820774633 | CCER2 | FALSE | Downregulated |
| 0,097782256 | 0,820774633 | DENND2D | FALSE | Upregulated |
| 0,068614264 | 0,820774633 | EEFSEC | FALSE | Upregulated |
| 0,194099639 | 0,820774633 | LOC102724733 | FALSE | Upregulated |
| -0,086953298 | 0,820774633 | PSMG4 | FALSE | Downregulated |
| -0,094407312 | 0,820774633 | SH3BGRL2 | FALSE | Downregulated |
| -0,124133504 | 0,820838058 | GIPC3 | FALSE | Downregulated |

| | | | | |
|---|---|---|---|---|
| -0,07983571 | 0,820838058 | MFAP3L | FALSE | Downregulated |
| -0,266584228 | 0,820838058 | PCAT6 | FALSE | Downregulated |
| 0,247319124 | 0,82142258 | FCRLB | FALSE | Upregulated |
| 0,140683474 | 0,82142258 | NLRC5 | FALSE | Upregulated |
| 0,218626711 | 0,82142258 | SH2D2A | FALSE | Upregulated |
| 0,095671652 | 0,82142258 | THAP3 | FALSE | Upregulated |
| 0,364862845 | 0,82142258 | VPS9D1-AS1 | FALSE | Upregulated |
| -0,119434057 | 0,82142258 | WDR78 | FALSE | Downregulated |
| 0,053445627 | 0,82142258 | YY1 | FALSE | Upregulated |
| -0,065886063 | 0,821458004 | ZNF33B | FALSE | Downregulated |
| 0,208928294 | 0,821530277 | FJX1 | FALSE | Upregulated |
| -0,057189739 | 0,821530277 | IMPAD1 | FALSE | Downregulated |
| 0,063855767 | 0,821530277 | KDM3A | FALSE | Upregulated |
| -0,175631232 | 0,821530277 | NPHP3-ACAD11 | FALSE | Downregulated |
| 0,260843486 | 0,821530277 | PKD2L1 | FALSE | Upregulated |
| 0,089467931 | 0,821530277 | POFUT2 | FALSE | Upregulated |
| -0,177948496 | 0,821530277 | SIPA1L2 | FALSE | Downregulated |
| 0,08322801 | 0,821530277 | SPSB3 | FALSE | Upregulated |
| 0,298816162 | 0,821534519 | GSTM5 | FALSE | Upregulated |
| 0,063361863 | 0,821579891 | KLHL36 | FALSE | Upregulated |
| 0,239134307 | 0,821579891 | LOC155060 | FALSE | Upregulated |
| -0,113560226 | 0,821579891 | PELP1 | FALSE | Downregulated |
| -0,069464918 | 0,821579891 | TMEM150A | FALSE | Downregulated |
| -0,103792845 | 0,821599631 | PDK3 | FALSE | Downregulated |
| -0,12333214 | 0,821599631 | ST3GAL6 | FALSE | Downregulated |
| 0,359995028 | 0,821634615 | LOC101929134 | FALSE | Upregulated |

| | | | | |
|---|---|---|---|---|
| -0,172908613 | 0,821969775 | DHRS4L1 | FALSE | Downregulated |
| -0,077157988 | 0,821969775 | ESF1 | FALSE | Downregulated |
| -0,112462616 | 0,821969775 | SLC20A1 | FALSE | Downregulated |
| 0,123802049 | 0,822016725 | C1QA | FALSE | Upregulated |
| 0,273532543 | 0,822016725 | C1QTNF2 | FALSE | Upregulated |
| 0,04010496 | 0,822016725 | DHX38 | FALSE | Upregulated |
| 0,211872261 | 0,822016725 | DPT | FALSE | Upregulated |
| 0,053801495 | 0,822016725 | KLC1 | FALSE | Upregulated |
| 0,075411826 | 0,822016725 | SF3B5 | FALSE | Upregulated |
| 0,079047323 | 0,822016725 | SQSTM1 | FALSE | Upregulated |
| -0,331256073 | 0,822115924 | CHRNA5 | FALSE | Downregulated |
| 0,081186433 | 0,822115924 | CISD2 | FALSE | Upregulated |
| 0,130958518 | 0,822115924 | FAM65B | FALSE | Upregulated |
| 0,235285663 | 0,822115924 | FBXL19-AS1 | FALSE | Upregulated |
| 0,90646045 | 0,822115924 | FGF19 | FALSE | Upregulated |
| 0,100531851 | 0,822115924 | GARNL3 | FALSE | Upregulated |
| 0,055053991 | 0,822115924 | GSTK1 | FALSE | Upregulated |
| -0,058968793 | 0,822115924 | IMMP2L | FALSE | Downregulated |
| 0,242732463 | 0,822115924 | LINC01011 | FALSE | Upregulated |
| 0,341724221 | 0,822115924 | LRRN2 | FALSE | Upregulated |
| 0,214251573 | 0,822115924 | NFATC4 | FALSE | Upregulated |
| 0,062535204 | 0,822115924 | NMT2 | FALSE | Upregulated |
| -0,071055326 | 0,822115924 | OLA1 | FALSE | Downregulated |
| 0,093902391 | 0,822115924 | RAB37 | FALSE | Upregulated |
| 0,06423475 | 0,822115924 | TBC1D17 | FALSE | Upregulated |
| 0,135281666 | 0,822115924 | TIMP1 | FALSE | Upregulated |

| | | | | |
|---|---|---|---|---|
| -0,063722938 | 0,822115924 | ZFP90 | FALSE | Downregulated |
| -0,074043281 | 0,822115924 | ZNF18 | FALSE | Downregulated |
| 0,136750302 | 0,822220317 | CCL15-CCL14 | FALSE | Upregulated |
| -0,09517911 | 0,822220317 | MIPOL1 | FALSE | Downregulated |
| 0,058041171 | 0,822220317 | ZNF706 | FALSE | Upregulated |
| 0,097090208 | 0,822441687 | CCDC68 | FALSE | Upregulated |
| -0,08104534 | 0,822579925 | BCL7A | FALSE | Downregulated |
| 0,079676783 | 0,822743193 | ASAH1 | FALSE | Upregulated |
| -0,141333033 | 0,822743193 | CAPS | FALSE | Downregulated |
| -0,042669147 | 0,822743193 | CDC37 | FALSE | Downregulated |
| 0,037917809 | 0,822743193 | FAM20B | FALSE | Upregulated |
| -0,043368549 | 0,822743193 | KPNA6 | FALSE | Downregulated |
| 0,333861151 | 0,822743193 | LOC101927745 | FALSE | Upregulated |
| 0,06758963 | 0,822743193 | SLC25A40 | FALSE | Upregulated |
| 0,105197363 | 0,822743193 | SNRNP70 | FALSE | Upregulated |
| 0,156068893 | 0,822743193 | TRPS1 | FALSE | Upregulated |
| -0,409534352 | 0,822837059 | LOC101928326 | FALSE | Downregulated |
| -0,066330192 | 0,822837059 | PCGF6 | FALSE | Downregulated |
| -0,189629883 | 0,822837059 | PDXDC2P | FALSE | Downregulated |
| 0,084789612 | 0,822973776 | APBA3 | FALSE | Upregulated |
| 0,064399338 | 0,822973776 | FAM73B | FALSE | Upregulated |
| 0,15658578 | 0,822973776 | FOXO3B | FALSE | Upregulated |
| -0,076810194 | 0,822973776 | INSIG2 | FALSE | Downregulated |
| -0,22475953 | 0,822973776 | LOC100505650 | FALSE | Downregulated |
| 0,635878086 | 0,822973776 | LOC102723603 | FALSE | Upregulated |
| 0,418128793 | 0,822973776 | PRPH2 | FALSE | Upregulated |

| | | | | |
|---|---|---|---|---|
| -0,071640346 | 0,823098151 | MKKS | FALSE | Downregulated |
| 0,460459194 | 0,823098151 | WBP11P1 | FALSE | Upregulated |
| -0,092692741 | 0,823098151 | ZFAS1 | FALSE | Downregulated |
| 0,245895265 | 0,823121123 | LOC102723926 | FALSE | Upregulated |
| -0,122799456 | 0,823510303 | CLCN2 | FALSE | Downregulated |
| -0,292062106 | 0,823510303 | RP5-1050D4.5 | FALSE | Downregulated |
| -0,156995557 | 0,823688057 | ARHGAP10 | FALSE | Downregulated |
| -0,041203673 | 0,823688057 | SPTLC1 | FALSE | Downregulated |
| 0,030117326 | 0,82381981 | CUL4A | FALSE | Upregulated |
| 0,153827804 | 0,82381981 | GATS | FALSE | Upregulated |
| -0,078423196 | 0,823842916 | PRDX3 | FALSE | Downregulated |
| 0,288370671 | 0,823862946 | LOC100130899 | FALSE | Upregulated |
| -0,257432674 | 0,824145069 | MAP6D1 | FALSE | Downregulated |
| 0,042573599 | 0,824203104 | EXD2 | FALSE | Upregulated |
| 0,208427847 | 0,824203104 | LAMA3 | FALSE | Upregulated |
| -0,13771808 | 0,824203104 | LOC100506639 | FALSE | Downregulated |
| -0,07787549 | 0,824203104 | MDH2 | FALSE | Downregulated |
| 0,059635217 | 0,824203104 | NCOA3 | FALSE | Upregulated |
| -0,244189741 | 0,824203104 | NNMT | FALSE | Downregulated |
| 0,145454717 | 0,824203104 | PCSK4 | FALSE | Upregulated |
| 0,085876003 | 0,824203104 | SHROOM2 | FALSE | Upregulated |
| 0,123385197 | 0,824732267 | NT5DC2 | FALSE | Upregulated |
| 0,158872106 | 0,824732267 | STON1 | FALSE | Upregulated |
| 0,136326189 | 0,824920952 | CD79B | FALSE | Upregulated |
| -0,154296728 | 0,824920952 | GJC3 | FALSE | Downregulated |
| -0,133731437 | 0,824920952 | LINC01018 | FALSE | Downregulated |

| | | | | |
|---|---|---|---|---|
| 0,130575969 | 0,824920952 | ZNF418 | FALSE | Upregulated |
| 0,16193364 | 0,825102945 | IQSEC3 | FALSE | Upregulated |
| 0,069634108 | 0,825102945 | RPL18A | FALSE | Upregulated |
| -0,045602899 | 0,825131275 | GNAI3 | FALSE | Downregulated |
| -0,055152136 | 0,825131895 | SMARCA5 | FALSE | Downregulated |
| 0,049139944 | 0,825174215 | AQR | FALSE | Upregulated |
| -0,080119477 | 0,825174215 | C5orf42 | FALSE | Downregulated |
| -0,072338641 | 0,825174215 | FADD | FALSE | Downregulated |
| -0,118207159 | 0,825174215 | FCER1G | FALSE | Downregulated |
| -0,096573638 | 0,825174215 | FGFR1 | FALSE | Downregulated |
| -0,032552112 | 0,825174215 | METAP1 | FALSE | Downregulated |
| 0,115538332 | 0,825179984 | C7orf31 | FALSE | Upregulated |
| -0,05670072 | 0,825179984 | DRAP1 | FALSE | Downregulated |
| -0,187915319 | 0,825179984 | S100A6 | FALSE | Downregulated |
| -0,107717488 | 0,825217156 | HECTD2 | FALSE | Downregulated |
| -0,048655876 | 0,825217156 | SAR1B | FALSE | Downregulated |
| -0,227109003 | 0,825250421 | CHRNA4 | FALSE | Downregulated |
| -0,363693229 | 0,825250421 | SCN4A | FALSE | Downregulated |
| -0,136531421 | 0,825258255 | C19orf54 | FALSE | Downregulated |
| 0,258634794 | 0,825258255 | CCDC147 | FALSE | Upregulated |
| 0,050706894 | 0,825258255 | L3MBTL2 | FALSE | Upregulated |
| -0,068228657 | 0,825458083 | CYTH1 | FALSE | Downregulated |
| -0,373347565 | 0,825458083 | RP11-182I10.3 | FALSE | Downregulated |
| -0,215715511 | 0,825458083 | SLC25A27 | FALSE | Downregulated |
| -0,075195929 | 0,825458083 | ZNF17 | FALSE | Downregulated |
| -0,093051785 | 0,825463361 | AIF1 | FALSE | Downregulated |

| | | | | |
|---|---|---|---|---|
| 0,268751953 | 0,825463361 | GPC3 | FALSE | Upregulated |
| -0,211772217 | 0,825463361 | LOC101928235 | FALSE | Downregulated |
| 0,327717297 | 0,825463361 | LOC101928725 | FALSE | Upregulated |
| 0,578135507 | 0,825463361 | SPDYE2 | FALSE | Upregulated |
| 0,059994137 | 0,825463361 | TRRAP | FALSE | Upregulated |
| 0,380210835 | 0,825524943 | LINC01021 | FALSE | Upregulated |
| 0,223713913 | 0,825529826 | AGAP9 | FALSE | Upregulated |
| -0,074953735 | 0,825678661 | AMBP | FALSE | Downregulated |
| -0,049776514 | 0,825678661 | ELP2 | FALSE | Downregulated |
| -0,067854704 | 0,825678661 | PIGP | FALSE | Downregulated |
| 0,046925699 | 0,825678661 | ZNF777 | FALSE | Upregulated |
| -0,14143071 | 0,825727077 | LRP12 | FALSE | Downregulated |
| -0,035961684 | 0,825900809 | FBXO11 | FALSE | Downregulated |
| -0,089841477 | 0,825939426 | KBTBD3 | FALSE | Downregulated |
| -0,066754518 | 0,826255436 | ITGA1 | FALSE | Downregulated |
| 0,13015275 | 0,826295284 | TRIM47 | FALSE | Upregulated |
| 0,080694033 | 0,826356756 | TTLL11 | FALSE | Upregulated |
| 0,345692307 | 0,826378926 | DCT | FALSE | Upregulated |
| -0,203840919 | 0,826378926 | FGF1 | FALSE | Downregulated |
| -0,180312498 | 0,826378926 | LOC101929691 | FALSE | Downregulated |
| 0,071157842 | 0,826378926 | PNRC1 | FALSE | Upregulated |
| 0,10794496 | 0,826436518 | ATP13A2 | FALSE | Upregulated |
| -0,043750785 | 0,826436518 | COG2 | FALSE | Downregulated |
| 0,131802089 | 0,826436518 | LBX2 | FALSE | Upregulated |
| 0,116849638 | 0,826445852 | CMKLR1 | FALSE | Upregulated |
| 0,087661937 | 0,826445852 | MICAL2 | FALSE | Upregulated |

| | | | | |
|---|---|---|---|---|
| 0,078981599 | 0,826559287 | PHKG2 | FALSE | Upregulated |
| -0,212535315 | 0,826701684 | SCGB1B2P | FALSE | Downregulated |
| 0,172166568 | 0,826742859 | LOC100630918 | FALSE | Upregulated |
| -0,199145171 | 0,827022857 | ANKRD31 | FALSE | Downregulated |
| 0,108605941 | 0,827022857 | AP4M1 | FALSE | Upregulated |
| -0,076974333 | 0,827022857 | SLC46A3 | FALSE | Downregulated |
| -0,081640265 | 0,827022857 | SMIM8 | FALSE | Downregulated |
| -0,063554651 | 0,827086944 | AKR1A1 | FALSE | Downregulated |
| 0,206180228 | 0,827086944 | ANKDD1A | FALSE | Upregulated |
| 0,108461959 | 0,827086944 | CACNA1D | FALSE | Upregulated |
| 0,064609988 | 0,827086944 | COMMD4 | FALSE | Upregulated |
| 0,208265788 | 0,827086944 | FCRL6 | FALSE | Upregulated |
| 0,301422599 | 0,827086944 | FLJ38576 | FALSE | Upregulated |
| -0,149232564 | 0,827086944 | GALNT16 | FALSE | Downregulated |
| -0,050089089 | 0,827086944 | GTF2B | FALSE | Downregulated |
| 0,081032717 | 0,827086944 | IQCB1 | FALSE | Upregulated |
| 0,341475314 | 0,827086944 | LOC101927446 | FALSE | Upregulated |
| 0,583550425 | 0,827086944 | LOC102723902 | FALSE | Upregulated |
| -0,067348183 | 0,827086944 | MRPS6 | FALSE | Downregulated |
| 0,061776244 | 0,827086944 | PEF1 | FALSE | Upregulated |
| -0,065946229 | 0,827086944 | POFUT1 | FALSE | Downregulated |
| -0,124981123 | 0,827086944 | RNPC3 | FALSE | Downregulated |
| 0,385614346 | 0,827086944 | RP11-584P21.2 | FALSE | Upregulated |
| 0,232276557 | 0,827086944 | RPSAP9 | FALSE | Upregulated |
| 0,087780905 | 0,827086944 | SLC22A5 | FALSE | Upregulated |
| -0,164891612 | 0,827086944 | TRNM | FALSE | Downregulated |

| | | | | |
|---|---|---|---|---|
| 0,409396991 | 0,827086944 | TRPC6 | FALSE | Upregulated |
| -0,053702464 | 0,827106862 | CWC15 | FALSE | Downregulated |
| -0,126849603 | 0,827106862 | MPP7 | FALSE | Downregulated |
| 0,247596062 | 0,827167206 | LINC00310 | FALSE | Upregulated |
| 0,058379296 | 0,827193303 | PTBP3 | FALSE | Upregulated |
| 0,470744813 | 0,827222473 | DKFZp686L13185 | FALSE | Upregulated |
| 0,131089528 | 0,827378118 | ATP8B2 | FALSE | Upregulated |
| 0,11039328 | 0,827378118 | PTRF | FALSE | Upregulated |
| 0,146662833 | 0,827485979 | ELOVL2 | FALSE | Upregulated |
| 0,255124692 | 0,827485979 | ID4 | FALSE | Upregulated |
| -0,357548593 | 0,827485979 | POU6F2-AS1 | FALSE | Downregulated |
| -0,312976835 | 0,827485979 | TFAP2A | FALSE | Downregulated |
| 0,396991317 | 0,827572464 | C15orf59 | FALSE | Upregulated |
| -0,41627495 | 0,827572464 | C16orf89 | FALSE | Downregulated |
| 0,073794121 | 0,827572464 | FGFR4 | FALSE | Upregulated |
| -0,330673949 | 0,827572464 | FLJ31104 | FALSE | Downregulated |
| 0,040667199 | 0,827572464 | GAPVD1 | FALSE | Upregulated |
| -0,09072858 | 0,827572464 | HECTD3 | FALSE | Downregulated |
| -0,131904992 | 0,827572464 | HIST1H2AC | FALSE | Downregulated |
| 0,068646254 | 0,827572464 | JMJD6 | FALSE | Upregulated |
| -0,13932989 | 0,827572464 | LOC100130093 | FALSE | Downregulated |
| -0,225894305 | 0,827572464 | PDCL3P4 | FALSE | Downregulated |
| -0,393701694 | 0,827572464 | TGM3 | FALSE | Downregulated |
| -0,050263772 | 0,827572464 | ZNF766 | FALSE | Downregulated |
| -0,136546153 | 0,82823994 | CES4A | FALSE | Downregulated |
| 0,078393463 | 0,82834564 | ACBD4 | FALSE | Upregulated |

| | | | | |
|---|---|---|---|---|
| -0,106226039 | 0,82834564 | ATXN7L1 | FALSE | Downregulated |
| -0,049491462 | 0,82834564 | EIF4ENIF1 | FALSE | Downregulated |
| 0,092343592 | 0,82834564 | EMC9 | FALSE | Upregulated |
| 0,079712498 | 0,82834564 | FOXN2 | FALSE | Upregulated |
| 0,087105342 | 0,82834564 | MALT1 | FALSE | Upregulated |
| -0,158317038 | 0,82834564 | RP11-439L18.3 | FALSE | Downregulated |
| -0,075898358 | 0,82834564 | RPS15 | FALSE | Downregulated |
| 0,248946025 | 0,828514977 | ARSF | FALSE | Upregulated |
| -0,098305571 | 0,828514977 | HNRNPA1L2 | FALSE | Downregulated |
| 0,179670617 | 0,828788289 | COL1A2 | FALSE | Upregulated |
| -0,486926725 | 0,828866259 | KLHL30 | FALSE | Downregulated |
| -0,043339457 | 0,829181098 | DYRK1A | FALSE | Downregulated |
| 0,123953935 | 0,829181098 | FGL2 | FALSE | Upregulated |
| 0,055250878 | 0,829181098 | MPPE1 | FALSE | Upregulated |
| -0,068869327 | 0,829181098 | NIT2 | FALSE | Downregulated |
| -0,163920261 | 0,829677716 | GOLGA6L9 | FALSE | Downregulated |
| -0,138941 | 0,829677716 | WNK3 | FALSE | Downregulated |
| 0,161043124 | 0,829793901 | ACER2 | FALSE | Upregulated |
| -0,11233584 | 0,829793901 | ACVR2B | FALSE | Downregulated |
| 0,246148323 | 0,829793901 | C12orf61 | FALSE | Upregulated |
| -0,110338039 | 0,829793901 | C14orf79 | FALSE | Downregulated |
| 0,093744814 | 0,829793901 | C19orf44 | FALSE | Upregulated |
| 0,063020448 | 0,829793901 | CASK | FALSE | Upregulated |
| 0,055882002 | 0,829793901 | COX18 | FALSE | Upregulated |
| -0,038465142 | 0,829793901 | CUL1 | FALSE | Downregulated |
| -0,070647262 | 0,829793901 | DDAH2 | FALSE | Downregulated |

| | | | | |
|---|---|---|---|---|
| -0,075346887 | 0,829793901 | EIF1AX | FALSE | Downregulated |
| -0,043491263 | 0,829793901 | EIF2S1 | FALSE | Downregulated |
| 0,098631888 | 0,829793901 | ENOSF1 | FALSE | Upregulated |
| 0,071397352 | 0,829793901 | FAM174A | FALSE | Upregulated |
| -0,10541046 | 0,829793901 | FAM221A | FALSE | Downregulated |
| 0,359590027 | 0,829793901 | FLJ22447 | FALSE | Upregulated |
| -0,064772632 | 0,829793901 | GNG5 | FALSE | Downregulated |
| 0,037164369 | 0,829793901 | GTF2H1 | FALSE | Upregulated |
| -0,097322277 | 0,829793901 | HIST2H2BE | FALSE | Downregulated |
| -0,07033386 | 0,829793901 | ITCH | FALSE | Downregulated |
| -0,175452051 | 0,829793901 | KANK4 | FALSE | Downregulated |
| 1,072373866 | 0,829793901 | KRT81 | FALSE | Upregulated |
| -0,15115425 | 0,829793901 | LOC100129637 | FALSE | Downregulated |
| -0,268429129 | 0,829793901 | LRRN1 | FALSE | Downregulated |
| 0,080223347 | 0,829793901 | ORMDL2 | FALSE | Upregulated |
| -0,086346353 | 0,829793901 | PHF1 | FALSE | Downregulated |
| 0,13156509 | 0,829793901 | PLEK | FALSE | Upregulated |
| 0,481244268 | 0,829793901 | PLSCR3 | FALSE | Upregulated |
| -0,101531971 | 0,829793901 | RASSF8 | FALSE | Downregulated |
| 0,069053076 | 0,829793901 | RBBP9 | FALSE | Upregulated |
| -0,407249516 | 0,829793901 | RP11-426A6.5 | FALSE | Downregulated |
| 0,089719152 | 0,829793901 | SERTAD3 | FALSE | Upregulated |
| 0,053765153 | 0,829793901 | SKP1 | FALSE | Upregulated |
| -0,070454522 | 0,829793901 | SNTB2 | FALSE | Downregulated |
| 0,205841309 | 0,829793901 | SOX17 | FALSE | Upregulated |
| -0,085518833 | 0,829793901 | TJP1 | FALSE | Downregulated |

| | | | | |
|---|---|---|---|---|
| -0,05600314 | 0,829793901 | TMCO4 | FALSE | Downregulated |
| -0,057921729 | 0,829793901 | TMEM123 | FALSE | Downregulated |
| 0,405005009 | 0,829793901 | TRNP | FALSE | Upregulated |
| 0,053107389 | 0,829793901 | UBE2D2 | FALSE | Upregulated |
| -0,06114435 | 0,829793901 | UBR3 | FALSE | Downregulated |
| 0,068480733 | 0,829793901 | UBXN2A | FALSE | Upregulated |
| 0,316315454 | 0,829793901 | ULBP1 | FALSE | Upregulated |
| -0,153462057 | 0,829793901 | ZNF677 | FALSE | Downregulated |
| -0,31387833 | 0,830039917 | SCXB | FALSE | Downregulated |
| -0,308381701 | 0,830071588 | C2orf78 | FALSE | Downregulated |
| -0,180349106 | 0,830071588 | EDN1 | FALSE | Downregulated |
| 0,126264283 | 0,830071588 | GANC | FALSE | Upregulated |
| 0,036820165 | 0,830071588 | GGA2 | FALSE | Upregulated |
| -0,066803057 | 0,830071588 | LINC00493 | FALSE | Downregulated |
| 0,259823239 | 0,830071588 | LOC101927377 | FALSE | Upregulated |
| -0,08277153 | 0,830071588 | PROX1 | FALSE | Downregulated |
| 0,152542405 | 0,830071588 | PTGIR | FALSE | Upregulated |
| -0,412547445 | 0,830071588 | TIGD3 | FALSE | Downregulated |
| 0,124331754 | 0,830126269 | FAM105A | FALSE | Upregulated |
| 0,233086778 | 0,830126269 | LOC102724059 | FALSE | Upregulated |
| 0,272063434 | 0,830126269 | SNAP25 | FALSE | Upregulated |
| -0,092670356 | 0,830144285 | RPIA | FALSE | Downregulated |
| 0,150459164 | 0,830159188 | CCR5 | FALSE | Upregulated |
| 0,151299483 | 0,830234408 | ZNF530 | FALSE | Upregulated |
| -0,072071257 | 0,830235347 | OSGEPL1 | FALSE | Downregulated |
| 0,048076551 | 0,830235347 | TRAF6 | FALSE | Upregulated |

| | | | | |
|---|---|---|---|---|
| -0,112554508 | 0,830419969 | ACACA | FALSE | Downregulated |
| 0,0486716 | 0,830419969 | APTX | FALSE | Upregulated |
| 0,095278196 | 0,830419969 | ARHGEF6 | FALSE | Upregulated |
| -0,112571364 | 0,830419969 | BAAT | FALSE | Downregulated |
| -0,082663244 | 0,830419969 | CEP250 | FALSE | Downregulated |
| -0,094109184 | 0,830419969 | DEPTOR | FALSE | Downregulated |
| 0,184352081 | 0,830419969 | EEF1E1 | FALSE | Upregulated |
| -0,083109502 | 0,830419969 | FAM175A | FALSE | Downregulated |
| -0,107273623 | 0,830419969 | FNDC3B | FALSE | Downregulated |
| -0,139298713 | 0,830419969 | GPAM | FALSE | Downregulated |
| -0,220016779 | 0,830419969 | GPSM2 | FALSE | Downregulated |
| 0,088328564 | 0,830419969 | HGFAC | FALSE | Upregulated |
| 0,062160128 | 0,830419969 | HPN | FALSE | Upregulated |
| -0,094343916 | 0,830419969 | HPRT1 | FALSE | Downregulated |
| 0,076981821 | 0,830419969 | HSPBAP1 | FALSE | Upregulated |
| 0,104346542 | 0,830419969 | LINC00116 | FALSE | Upregulated |
| -0,24189722 | 0,830419969 | LINC00570 | FALSE | Downregulated |
| -0,068858255 | 0,830419969 | LMAN1 | FALSE | Downregulated |
| 0,399439206 | 0,830419969 | LOC102723974 | FALSE | Upregulated |
| 0,140396609 | 0,830419969 | LSM11 | FALSE | Upregulated |
| -0,134984031 | 0,830419969 | MAMLD1 | FALSE | Downregulated |
| 0,064864424 | 0,830419969 | MAPKAPK3 | FALSE | Upregulated |
| 0,172448176 | 0,830419969 | MDN1 | FALSE | Upregulated |
| 0,172939857 | 0,830419969 | MFGE8 | FALSE | Upregulated |
| -0,069294552 | 0,830419969 | MRPL36 | FALSE | Downregulated |
| -0,075304499 | 0,830419969 | NTAN1 | FALSE | Downregulated |

| | | | | |
|---|---|---|---|---|
| -0,081599876 | 0,830419969 | PIGA | FALSE | Downregulated |
| -0,116748931 | 0,830419969 | PKDCC | FALSE | Downregulated |
| 0,127766512 | 0,830419969 | PKI55 | FALSE | Upregulated |
| 0,083103492 | 0,830419969 | PLEKHJ1 | FALSE | Upregulated |
| -0,235894098 | 0,830419969 | RAB6B | FALSE | Downregulated |
| -0,067821949 | 0,830419969 | SETD5 | FALSE | Downregulated |
| -0,06652738 | 0,830419969 | SLC39A8 | FALSE | Downregulated |
| -0,070360812 | 0,830419969 | SPRED2 | FALSE | Downregulated |
| -0,100483547 | 0,830419969 | TAF1A | FALSE | Downregulated |
| 0,083086068 | 0,830419969 | TRNT1 | FALSE | Upregulated |
| -0,223716018 | 0,830419969 | TSPAN2 | FALSE | Downregulated |
| 0,130198003 | 0,830419969 | TTC21A | FALSE | Upregulated |
| -0,142063034 | 0,830419969 | ZNF701 | FALSE | Downregulated |
| 0,107962827 | 0,830419969 | ZXDA | FALSE | Upregulated |
| -0,297436447 | 0,830592903 | LOC101928612 | FALSE | Downregulated |
| 0,108311148 | 0,830737473 | STK17A | FALSE | Upregulated |
| -0,080602396 | 0,830764798 | ALAD | FALSE | Downregulated |
| -0,059210396 | 0,830764798 | GTF3C6 | FALSE | Downregulated |
| -0,122497972 | 0,830781693 | CELSR1 | FALSE | Downregulated |
| -0,072116566 | 0,830828671 | AQP9 | FALSE | Downregulated |
| 0,154562706 | 0,830850076 | FRMD3 | FALSE | Upregulated |
| 0,110310533 | 0,830850076 | VCPKMT | FALSE | Upregulated |
| 0,350234596 | 0,830855092 | KIAA1045 | FALSE | Upregulated |
| -0,12005896 | 0,831053751 | LAMA4 | FALSE | Downregulated |
| -0,191971289 | 0,831163223 | USP2-AS1 | FALSE | Downregulated |
| 0,297925727 | 0,83123556 | LOC101927435 | FALSE | Upregulated |

| | | | | |
|---|---|---|---|---|
| -0,24038195 | 0,831342783 | PRSS35 | FALSE | Downregulated |
| -0,04519858 | 0,831342783 | RTN3 | FALSE | Downregulated |
| 0,059560731 | 0,831391422 | TXNDC12 | FALSE | Upregulated |
| 0,188887607 | 0,831393277 | IL18RAP | FALSE | Upregulated |
| -0,195619093 | 0,831393277 | LOC100996419 | FALSE | Downregulated |
| -0,131996346 | 0,831393277 | TCF21 | FALSE | Downregulated |
| -0,04522713 | 0,831415425 | ANKLE2 | FALSE | Downregulated |
| 0,05381732 | 0,83151264 | SENP5 | FALSE | Upregulated |
| -0,249577542 | 0,831616615 | GRAPL | FALSE | Downregulated |
| -0,099756163 | 0,831673745 | CASP1 | FALSE | Downregulated |
| 0,386170758 | 0,831673745 | PDE6C | FALSE | Upregulated |
| 0,093726051 | 0,83168141 | LMNB2 | FALSE | Upregulated |
| 0,076291846 | 0,831687144 | ALG2 | FALSE | Upregulated |
| 0,097552722 | 0,831687144 | CLMN | FALSE | Upregulated |
| 0,476636677 | 0,831687144 | COL10A1 | FALSE | Upregulated |
| 0,126430751 | 0,831687144 | GALNT4 | FALSE | Upregulated |
| 0,089772751 | 0,831687144 | HMGA1 | FALSE | Upregulated |
| -0,536077215 | 0,831687144 | IGDCC4 | FALSE | Downregulated |
| -0,523990655 | 0,831687144 | LOC101929047 | FALSE | Downregulated |
| -0,391220375 | 0,831687144 | MEG9 | FALSE | Downregulated |
| 0,218494465 | 0,831687144 | PRRT1 | FALSE | Upregulated |
| -0,074381 | 0,831687144 | STT3A | FALSE | Downregulated |
| 0,062568325 | 0,831961426 | RPS6KA2 | FALSE | Upregulated |
| -0,281075816 | 0,832522666 | MOV10L1 | FALSE | Downregulated |
| 0,067007966 | 0,832522666 | SACM1L | FALSE | Upregulated |
| -0,059900878 | 0,832522666 | SNX30 | FALSE | Downregulated |

| | | | | |
|---|---|---|---|---|
| 0,04393587 | 0,832522666 | TMEM131 | FALSE | Upregulated |
| 0,090057808 | 0,832522666 | ZNF74 | FALSE | Upregulated |
| 0,111236471 | 0,832549113 | BBS5 | FALSE | Upregulated |
| 0,134528966 | 0,832603636 | SDF2L1 | FALSE | Upregulated |
| -0,379776443 | 0,832615417 | LOC102724016 | FALSE | Downregulated |
| 0,064989077 | 0,832615417 | PRMT5 | FALSE | Upregulated |
| 0,065411627 | 0,8326837 | FKBP9 | FALSE | Upregulated |
| -0,055843403 | 0,8326837 | RBP4 | FALSE | Downregulated |
| -0,064304382 | 0,832709914 | TBPL1 | FALSE | Downregulated |
| -0,09434411 | 0,832777975 | FIGN | FALSE | Downregulated |
| 0,227827067 | 0,832828984 | CXorf31 | FALSE | Upregulated |
| -0,340979385 | 0,832893844 | LOC101927727 | FALSE | Downregulated |
| 0,080980335 | 0,832946883 | POMGNT2 | FALSE | Upregulated |
| -0,043806283 | 0,832948816 | RAB3GAP2 | FALSE | Downregulated |
| -0,468296163 | 0,832949812 | HIGD1B | FALSE | Downregulated |
| -0,156589237 | 0,832980916 | DGAT2 | FALSE | Downregulated |
| -0,385354148 | 0,83304705 | RP11-474D1.3 | FALSE | Downregulated |
| -0,354652283 | 0,833281248 | APOA4 | FALSE | Downregulated |
| 0,23520131 | 0,833281248 | CACNA1C | FALSE | Upregulated |
| 0,329098005 | 0,833281248 | FRMD7 | FALSE | Upregulated |
| -0,209010253 | 0,833281248 | GSTO2 | FALSE | Downregulated |
| 0,125989527 | 0,833578208 | GTF2IRD1 | FALSE | Upregulated |
| -0,126107666 | 0,833630002 | ABLIM3 | FALSE | Downregulated |
| 0,107566299 | 0,833630002 | C17orf59 | FALSE | Upregulated |
| -0,290885893 | 0,833630002 | LOC101928946 | FALSE | Downregulated |
| 0,276676573 | 0,833630002 | LOC101929846 | FALSE | Upregulated |

| | | | | |
|---|---|---|---|---|
| 0,060533837 | 0,833630002 | TNKS | FALSE | Upregulated |
| -0,452299543 | 0,833701224 | ADAMTS14 | FALSE | Downregulated |
| -0,063704277 | 0,833930736 | PLEKHF2 | FALSE | Downregulated |
| 0,247282584 | 0,834049025 | CDCA2 | FALSE | Upregulated |
| -0,274643788 | 0,834049025 | FLG-AS1 | FALSE | Downregulated |
| -0,07221361 | 0,834049025 | FPGS | FALSE | Downregulated |
| 0,259674289 | 0,834193669 | LOC100507131 | FALSE | Upregulated |
| -0,078078687 | 0,834210002 | ABCF1 | FALSE | Downregulated |
| 0,068208517 | 0,834210841 | ADIPOR2 | FALSE | Upregulated |
| 0,381643114 | 0,834210841 | ANXA2P1 | FALSE | Upregulated |
| 0,112327468 | 0,834210841 | IQCC | FALSE | Upregulated |
| 0,202569634 | 0,834210841 | LOC101928464 | FALSE | Upregulated |
| -0,049489783 | 0,834210841 | PBRM1 | FALSE | Downregulated |
| 0,304277592 | 0,834210841 | RPS2P32 | FALSE | Upregulated |
| -0,104969097 | 0,834210841 | SLC7A8 | FALSE | Downregulated |
| -0,156702815 | 0,834243846 | RTN4RL1 | FALSE | Downregulated |
| 0,337020786 | 0,834283046 | RP11-563J2.2 | FALSE | Upregulated |
| 0,093201957 | 0,834283046 | STK10 | FALSE | Upregulated |
| -0,366458879 | 0,834331446 | SLC14A2 | FALSE | Downregulated |
| -0,082325868 | 0,8343823 | ACOX2 | FALSE | Downregulated |
| 0,325975239 | 0,8343823 | B3GAT2 | FALSE | Upregulated |
| 0,226796125 | 0,8343823 | LRRC70 | FALSE | Upregulated |
| -0,06096456 | 0,8343823 | SH3BGRL | FALSE | Downregulated |
| 0,045631594 | 0,8343823 | UEVLD | FALSE | Upregulated |
| -0,162115394 | 0,8343823 | ZNF853 | FALSE | Downregulated |
| 0,057838253 | 0,834413111 | SLC25A39 | FALSE | Upregulated |

| | | | | |
|---|---|---|---|---|
| -0,065261088 | 0,834475256 | DDX3X | FALSE | Downregulated |
| 0,12841586 | 0,834475256 | GBP1 | FALSE | Upregulated |
| -0,073285092 | 0,834475256 | GEMIN4 | FALSE | Downregulated |
| -0,316029759 | 0,834475256 | MMP21 | FALSE | Downregulated |
| -0,07924149 | 0,834475256 | PPP1R12A | FALSE | Downregulated |
| 0,352071603 | 0,834475256 | SBSPON | FALSE | Upregulated |
| 0,136561379 | 0,834475256 | TMEM120B | FALSE | Upregulated |
| -0,071777211 | 0,834475256 | TMEM135 | FALSE | Downregulated |
| -0,185463579 | 0,834475256 | TSPAN5 | FALSE | Downregulated |
| 0,111813669 | 0,834542103 | C4B | FALSE | Upregulated |
| 0,058323758 | 0,834542103 | FYCO1 | FALSE | Upregulated |
| 0,26503373 | 0,834542103 | MAPK8IP2 | FALSE | Upregulated |
| 0,06963452 | 0,834542103 | NR1H3 | FALSE | Upregulated |
| -0,21275148 | 0,834542103 | TNFAIP8L3 | FALSE | Downregulated |
| -0,124538188 | 0,834551372 | CHN2 | FALSE | Downregulated |
| 0,113157633 | 0,834578016 | PPP1R3F | FALSE | Upregulated |
| 0,073878112 | 0,83467244 | FAM3C | FALSE | Upregulated |
| 0,189622854 | 0,83485851 | SEC31B | FALSE | Upregulated |
| -0,080026747 | 0,834886368 | EIF4E3 | FALSE | Downregulated |
| -0,106046654 | 0,834987882 | CLDN16 | FALSE | Downregulated |
| 0,067992153 | 0,834987882 | R3HDM4 | FALSE | Upregulated |
| 0,078837125 | 0,834987882 | ZNF358 | FALSE | Upregulated |
| -0,096593351 | 0,835136246 | ARHGAP32 | FALSE | Downregulated |
| -0,040811855 | 0,835136246 | CUX1 | FALSE | Downregulated |
| -0,067653671 | 0,835136246 | ZNF510 | FALSE | Downregulated |
| -0,193590268 | 0,835166646 | CYS1 | FALSE | Downregulated |

| | | | | |
|---|---|---|---|---|
| -0,121681179 | 0,835193021 | APLNR | FALSE | Downregulated |
| 0,0654954 | 0,835193021 | COX6B1 | FALSE | Upregulated |
| 0,317066744 | 0,835193021 | FAM85A | FALSE | Upregulated |
| 0,078937946 | 0,835193021 | LMBRD2 | FALSE | Upregulated |
| 0,109564233 | 0,835193021 | MCF2L | FALSE | Upregulated |
| 0,048367242 | 0,835193021 | PRPS1 | FALSE | Upregulated |
| 0,073387136 | 0,835193021 | RAB24 | FALSE | Upregulated |
| 0,046012822 | 0,835193021 | RALGAPB | FALSE | Upregulated |
| -0,098182984 | 0,835193021 | SCAND3 | FALSE | Downregulated |
| 0,05498804 | 0,835193021 | ZNF592 | FALSE | Upregulated |
| -0,081829733 | 0,8353033 | AOX1 | FALSE | Downregulated |
| -0,196039649 | 0,835471339 | CCL23 | FALSE | Downregulated |
| -0,159788819 | 0,835471339 | LOC101927943 | FALSE | Downregulated |
| 0,060318962 | 0,835471339 | MOB3C | FALSE | Upregulated |
| -0,07439198 | 0,835471339 | TOMM5 | FALSE | Downregulated |
| 0,215160576 | 0,835780181 | HSD17B1 | FALSE | Upregulated |
| -0,545034616 | 0,835865268 | EIF3CL | FALSE | Downregulated |
| -0,050188848 | 0,835865268 | FASTKD2 | FALSE | Downregulated |
| -0,051854408 | 0,835865268 | KNG1 | FALSE | Downregulated |
| -0,18811596 | 0,835865268 | MOGAT1 | FALSE | Downregulated |
| 0,102141455 | 0,835865268 | PDP1 | FALSE | Upregulated |
| 0,089603644 | 0,835865268 | PECAM1 | FALSE | Upregulated |
| -0,068072935 | 0,835865268 | VPS33B | FALSE | Downregulated |
| 0,154061899 | 0,835865268 | ZC3H12A | FALSE | Upregulated |
| 0,834566859 | 0,835868381 | RNU11 | FALSE | Upregulated |
| -0,176226987 | 0,836002198 | LOC102723323 | FALSE | Downregulated |

| | | | | |
|---|---|---|---|---|
| -0,480816046 | 0,836002198 | UBE2DNL | FALSE | Downregulated |
| 0,130498912 | 0,836032627 | BCAT2 | FALSE | Upregulated |
| 0,169556982 | 0,836032627 | HCG26 | FALSE | Upregulated |
| -0,208138357 | 0,836064173 | LOC100288842 | FALSE | Downregulated |
| 0,227720032 | 0,836393633 | MYOM1 | FALSE | Upregulated |
| -0,089289885 | 0,836401945 | AMN1 | FALSE | Downregulated |
| 0,073166713 | 0,836401945 | FAAH | FALSE | Upregulated |
| 0,201531018 | 0,836409524 | CCL4 | FALSE | Upregulated |
| -0,223251942 | 0,836409524 | LOC100134822 | FALSE | Downregulated |
| -0,116063966 | 0,836409524 | SCML2 | FALSE | Downregulated |
| 0,060758878 | 0,836409524 | SLC38A9 | FALSE | Upregulated |
| -0,215893388 | 0,836531803 | ITGA10 | FALSE | Downregulated |
| -0,110260724 | 0,83665439 | ACVR1C | FALSE | Downregulated |
| 0,390635001 | 0,83665439 | ADM5 | FALSE | Upregulated |
| 0,1270744 | 0,83665439 | PDIA3P1 | FALSE | Upregulated |
| 0,115390204 | 0,83665439 | PLCB2 | FALSE | Upregulated |
| -0,061896144 | 0,83665439 | SRRT | FALSE | Downregulated |
| 0,104475829 | 0,83665439 | TMEM173 | FALSE | Upregulated |
| 0,115532456 | 0,836722082 | FAM198B | FALSE | Upregulated |
| 0,185310069 | 0,836722082 | MAB21L2 | FALSE | Upregulated |
| 0,496248268 | 0,836722082 | NRXN1 | FALSE | Upregulated |
| 0,073555059 | 0,836919494 | ABHD6 | FALSE | Upregulated |
| 0,056763509 | 0,836919494 | ACO1 | FALSE | Upregulated |
| -0,063857883 | 0,836919494 | ARL6IP1 | FALSE | Downregulated |
| 0,0686122 | 0,836919494 | CBX4 | FALSE | Upregulated |
| 0,250817125 | 0,836919494 | CYP3A7-CYP3AP1 | FALSE | Upregulated |

| | | | | |
|---|---|---|---|---|
| -0,086847076 | 0,836919494 | FRRS1 | FALSE | Downregulated |
| -0,052817266 | 0,836919494 | GATAD1 | FALSE | Downregulated |
| 0,197629941 | 0,836919494 | GP1BA | FALSE | Upregulated |
| -0,07472906 | 0,836919494 | NENF | FALSE | Downregulated |
| -0,05060366 | 0,836919494 | OTUD4 | FALSE | Downregulated |
| -0,051316237 | 0,836919494 | SAP30L | FALSE | Downregulated |
| -0,112498537 | 0,836919494 | SERPINI1 | FALSE | Downregulated |
| -0,080228723 | 0,836919494 | ZNF259 | FALSE | Downregulated |
| -0,327262748 | 0,837123536 | DPP6 | FALSE | Downregulated |
| -0,108906081 | 0,837123536 | GK5 | FALSE | Downregulated |
| -0,052067796 | 0,837123536 | PRDX5 | FALSE | Downregulated |
| -0,364952613 | 0,837123536 | RNASE7 | FALSE | Downregulated |
| -0,065797913 | 0,837123536 | SCCPDH | FALSE | Downregulated |
| 0,108197573 | 0,837123536 | TNFRSF11B | FALSE | Upregulated |
| -0,244589726 | 0,837123536 | TRPM6 | FALSE | Downregulated |
| 0,075394796 | 0,837258168 | FOXRED1 | FALSE | Upregulated |
| -0,047260082 | 0,837258168 | HERPUD2 | FALSE | Downregulated |
| -0,2750476 | 0,837258168 | LGI1 | FALSE | Downregulated |
| 0,084452148 | 0,837258168 | RAB20 | FALSE | Upregulated |
| -0,036388077 | 0,837258168 | SAR1A | FALSE | Downregulated |
| -0,045720238 | 0,837258168 | SDHA | FALSE | Downregulated |
| 0,087276145 | 0,837258168 | SNAPC4 | FALSE | Upregulated |
| 0,102684779 | 0,837281557 | CHST15 | FALSE | Upregulated |
| 0,113603859 | 0,837320787 | LRRC37B | FALSE | Upregulated |
| -0,098010726 | 0,837320787 | SEMA5A | FALSE | Downregulated |
| -0,357886554 | 0,837365351 | AC005550.4 | FALSE | Downregulated |

| | | | | |
|---|---|---|---|---|
| -0,074577874 | 0,837365351 | CGRRF1 | FALSE | Downregulated |
| 0,202892357 | 0,837375245 | DNAJC18 | FALSE | Upregulated |
| 0,285967758 | 0,837375245 | SPINK5 | FALSE | Upregulated |
| 0,175661508 | 0,837552184 | SLC35G2 | FALSE | Upregulated |
| -0,073572492 | 0,837704839 | RP2 | FALSE | Downregulated |
| -0,124076036 | 0,837704839 | VEGFC | FALSE | Downregulated |
| 0,18476669 | 0,83781258 | ANKRD34A | FALSE | Upregulated |
| -0,111884501 | 0,83781258 | GLIDR | FALSE | Downregulated |
| -0,068042654 | 0,83781258 | HMGXB4 | FALSE | Downregulated |
| 0,161620764 | 0,83781258 | LARP6 | FALSE | Upregulated |
| 0,321290583 | 0,83781258 | PPP1R36 | FALSE | Upregulated |
| 0,123579116 | 0,83781258 | SRGN | FALSE | Upregulated |
| -0,035139156 | 0,83781258 | ZMYND11 | FALSE | Downregulated |
| -0,057809677 | 0,837825431 | ACOT8 | FALSE | Downregulated |
| 0,147136917 | 0,837838383 | FAM43A | FALSE | Upregulated |
| -0,044058103 | 0,837838383 | USP40 | FALSE | Downregulated |
| 0,095724208 | 0,838074503 | GNPDA2 | FALSE | Upregulated |
| 0,102477913 | 0,838322537 | ACTG1 | FALSE | Upregulated |
| 0,055587483 | 0,838322537 | AGPAT6 | FALSE | Upregulated |
| -0,250311129 | 0,838322537 | LINC00266-3 | FALSE | Downregulated |
| -0,056727135 | 0,838322537 | TAOK1 | FALSE | Downregulated |
| -0,071764928 | 0,838384837 | SCNM1 | FALSE | Downregulated |
| 0,174281542 | 0,838397769 | ASAH2 | FALSE | Upregulated |
| 0,040955164 | 0,838397769 | ASB7 | FALSE | Upregulated |
| 0,08310721 | 0,838397769 | C11orf68 | FALSE | Upregulated |
| 0,135627451 | 0,838397769 | CCDC152 | FALSE | Upregulated |

| | | | | |
|---|---|---|---|---|
| -0,049505748 | 0,838397769 | CENPB | FALSE | Downregulated |
| 0,151724901 | 0,838397769 | CENPO | FALSE | Upregulated |
| -0,182197896 | 0,838397769 | CFHR4 | FALSE | Downregulated |
| -0,340440945 | 0,838397769 | CLIC5 | FALSE | Downregulated |
| -0,071003341 | 0,838397769 | CPSF7 | FALSE | Downregulated |
| 0,302007564 | 0,838397769 | CYP2G1P | FALSE | Upregulated |
| 0,055948026 | 0,838397769 | EIF2B3 | FALSE | Upregulated |
| -0,050665897 | 0,838397769 | EP300 | FALSE | Downregulated |
| 0,308406487 | 0,838397769 | ERVFRD-1 | FALSE | Upregulated |
| -0,061016619 | 0,838397769 | ETFA | FALSE | Downregulated |
| 0,100811133 | 0,838397769 | EXOC3L4 | FALSE | Upregulated |
| 0,056362268 | 0,838397769 | FAM188A | FALSE | Upregulated |
| 0,093706095 | 0,838397769 | FAM211A-AS1 | FALSE | Upregulated |
| -0,073123272 | 0,838397769 | FASTK | FALSE | Downregulated |
| -0,192896024 | 0,838397769 | FBXL22 | FALSE | Downregulated |
| -0,156796052 | 0,838397769 | FGFBP3 | FALSE | Downregulated |
| 0,173555228 | 0,838397769 | GDPD1 | FALSE | Upregulated |
| -0,079504325 | 0,838397769 | GMFG | FALSE | Downregulated |
| -0,235376077 | 0,838397769 | GOLGA6L4 | FALSE | Downregulated |
| 0,232500743 | 0,838397769 | GPR68 | FALSE | Upregulated |
| 0,064343247 | 0,838397769 | HNRNPLL | FALSE | Upregulated |
| -0,136379403 | 0,838397769 | IL15 | FALSE | Downregulated |
| -0,075372595 | 0,838397769 | KDM7A | FALSE | Downregulated |
| 0,083149628 | 0,838397769 | LEPREL4 | FALSE | Upregulated |
| -0,368627236 | 0,838397769 | LOC100132813 | FALSE | Downregulated |
| 0,312756439 | 0,838397769 | LOC100996660 | FALSE | Upregulated |

| | | | | |
|---|---|---|---|---|
| -0,323179728 | 0,838397769 | LOC101927646 | FALSE | Downregulated |
| 0,184581988 | 0,838397769 | LOC101928192 | FALSE | Upregulated |
| 0,359406902 | 0,838397769 | LOC101928560 | FALSE | Upregulated |
| 0,202028531 | 0,838397769 | LOC101929552 | FALSE | Upregulated |
| 0,132735434 | 0,838397769 | LOC102724164 | FALSE | Upregulated |
| 0,227793997 | 0,838397769 | LOC102724296 | FALSE | Upregulated |
| -0,266033165 | 0,838397769 | LOC102724751 | FALSE | Downregulated |
| 0,239541874 | 0,838397769 | LRRC37A6P | FALSE | Upregulated |
| -0,045387337 | 0,838397769 | MAVS | FALSE | Downregulated |
| -0,439947933 | 0,838397769 | MEF2C-AS1 | FALSE | Downregulated |
| -0,056105975 | 0,838397769 | MICU1 | FALSE | Downregulated |
| -0,314000113 | 0,838397769 | MSS51 | FALSE | Downregulated |
| 0,08316046 | 0,838397769 | NAIF1 | FALSE | Upregulated |
| 0,118266534 | 0,838397769 | NCF2 | FALSE | Upregulated |
| 0,052863518 | 0,838397769 | NHP2L1 | FALSE | Upregulated |
| 0,057303019 | 0,838397769 | NKRF | FALSE | Upregulated |
| -0,057971839 | 0,838397769 | ORAI3 | FALSE | Downregulated |
| -0,494929793 | 0,838397769 | PHLDA2 | FALSE | Downregulated |
| -0,409528905 | 0,838397769 | PIFO | FALSE | Downregulated |
| -0,240226092 | 0,838397769 | PLAC9 | FALSE | Downregulated |
| -0,120959512 | 0,838397769 | PRLR | FALSE | Downregulated |
| 0,096032003 | 0,838397769 | PSMG3 | FALSE | Upregulated |
| -0,307379661 | 0,838397769 | RBM44 | FALSE | Downregulated |
| 0,410175795 | 0,838397769 | RFPL1 | FALSE | Upregulated |
| 0,081723316 | 0,838397769 | RP11-890B15.2 | FALSE | Upregulated |
| 0,063328923 | 0,838397769 | SAV1 | FALSE | Upregulated |

| | | | | |
|---|---|---|---|---|
| 0,046844387 | 0,838397769 | SLC30A5 | FALSE | Upregulated |
| 0,075484061 | 0,838397769 | SLC35B2 | FALSE | Upregulated |
| -0,113271087 | 0,838397769 | SLC38A4 | FALSE | Downregulated |
| -0,151149394 | 0,838397769 | SLC5A6 | FALSE | Downregulated |
| 0,140098406 | 0,838397769 | SMIM3 | FALSE | Upregulated |
| 0,197034829 | 0,838397769 | SNRK-AS1 | FALSE | Upregulated |
| 0,154827293 | 0,838397769 | THNSL2 | FALSE | Upregulated |
| 0,090678939 | 0,838397769 | TMEM86A | FALSE | Upregulated |
| 0,037054276 | 0,838397769 | TRMT5 | FALSE | Upregulated |
| 0,177345146 | 0,838397769 | UBE2QL1 | FALSE | Upregulated |
| 0,12455627 | 0,838397769 | ZNF682 | FALSE | Upregulated |
| 0,085621885 | 0,838467281 | RPS11 | FALSE | Upregulated |
| -0,060407345 | 0,838777183 | HSDL2 | FALSE | Downregulated |
| 0,069255098 | 0,839025588 | ACTN4 | FALSE | Upregulated |
| -0,104038699 | 0,839025588 | AFF3 | FALSE | Downregulated |
| 0,347003476 | 0,839025588 | C3orf80 | FALSE | Upregulated |
| 0,112684566 | 0,839025588 | CYBB | FALSE | Upregulated |
| -0,077459764 | 0,839025588 | ECM1 | FALSE | Downregulated |
| 0,323544601 | 0,839025588 | FAM201A | FALSE | Upregulated |
| -0,097578282 | 0,839025588 | NTHL1 | FALSE | Downregulated |
| 0,127755155 | 0,839055051 | INMT | FALSE | Upregulated |
| 0,057562079 | 0,839055051 | NAB1 | FALSE | Upregulated |
| -0,062034125 | 0,839086527 | H3F3A | FALSE | Downregulated |
| -0,398384595 | 0,839086527 | HORMAD1 | FALSE | Downregulated |
| 0,064033221 | 0,839086527 | SHKBP1 | FALSE | Upregulated |
| 0,09687165 | 0,839086527 | TMEM184A | FALSE | Upregulated |

| | | | | |
|---|---|---|---|---|
| -0,073850449 | 0,839152569 | ACAT1 | FALSE | Downregulated |
| 0,409745116 | 0,839152569 | CA12 | FALSE | Upregulated |
| 0,165506611 | 0,839152569 | CD163L1 | FALSE | Upregulated |
| 0,100187825 | 0,839152569 | CLIP2 | FALSE | Upregulated |
| -0,052297954 | 0,839152569 | FRYL | FALSE | Downregulated |
| -0,169367945 | 0,839152569 | GJC1 | FALSE | Downregulated |
| 0,113522175 | 0,839152569 | KNSTRN | FALSE | Upregulated |
| 0,060308536 | 0,839152569 | LATS2 | FALSE | Upregulated |
| -0,047615791 | 0,839152569 | MBNL1 | FALSE | Downregulated |
| -0,187074049 | 0,839152569 | PCK1 | FALSE | Downregulated |
| 0,049114313 | 0,839152569 | POP4 | FALSE | Upregulated |
| 0,147392628 | 0,839152569 | PVRIG | FALSE | Upregulated |
| -0,137402104 | 0,839152569 | S100A4 | FALSE | Downregulated |
| 0,302673437 | 0,839152569 | SLFN12L | FALSE | Upregulated |
| 0,200129931 | 0,839152569 | TMEM132E | FALSE | Upregulated |
| -0,611558862 | 0,839152569 | TRIM63 | FALSE | Downregulated |
| -0,142066809 | 0,839176518 | ZNF514 | FALSE | Downregulated |
| 0,053435765 | 0,839239095 | ACVR1B | FALSE | Upregulated |
| 0,687765647 | 0,839239095 | ADCY8 | FALSE | Upregulated |
| -0,09269876 | 0,839239095 | ARID4B | FALSE | Downregulated |
| -0,092085483 | 0,839239095 | CEP164 | FALSE | Downregulated |
| 0,055845328 | 0,839239095 | COX5A | FALSE | Upregulated |
| -0,279746105 | 0,839239095 | LOC102724474 | FALSE | Downregulated |
| -0,079517855 | 0,839239095 | PHYH | FALSE | Downregulated |
| 0,081303312 | 0,839239095 | TACC1 | FALSE | Upregulated |
| -0,058946041 | 0,839239095 | UBA6 | FALSE | Downregulated |

| | | | | |
|---|---|---|---|---|
| 0,416632113 | 0,839239095 | ZNF20 | FALSE | Upregulated |
| 0,049648813 | 0,839243731 | ARPC4 | FALSE | Upregulated |
| -0,232091294 | 0,839243731 | RSPH1 | FALSE | Downregulated |
| -0,07857898 | 0,839302491 | ADAMTSL3 | FALSE | Downregulated |
| -0,066473032 | 0,839302491 | APP | FALSE | Downregulated |
| -0,271878121 | 0,839302491 | ARHGEF38 | FALSE | Downregulated |
| -0,265766729 | 0,839302491 | C1orf192 | FALSE | Downregulated |
| -0,326875564 | 0,839302491 | C6orf165 | FALSE | Downregulated |
| -0,164777548 | 0,839302491 | EMCN | FALSE | Downregulated |
| 0,33632257 | 0,839302491 | FAM189A2 | FALSE | Upregulated |
| 0,3636899 | 0,839302491 | FAM19A1 | FALSE | Upregulated |
| 0,187950774 | 0,839302491 | FLJ35282 | FALSE | Upregulated |
| 0,153295743 | 0,839302491 | FLRT3 | FALSE | Upregulated |
| 0,118151935 | 0,839302491 | GJA4 | FALSE | Upregulated |
| -0,036221664 | 0,839302491 | GTF2A1 | FALSE | Downregulated |
| 0,148164024 | 0,839302491 | HUNK | FALSE | Upregulated |
| -0,0495618 | 0,839302491 | LIN7C | FALSE | Downregulated |
| 0,052381343 | 0,839302491 | LMBR1 | FALSE | Upregulated |
| -0,123938282 | 0,839302491 | LOC101927974 | FALSE | Downregulated |
| 0,31119982 | 0,839302491 | LOC102724554 | FALSE | Upregulated |
| -0,058717699 | 0,839302491 | MAP7 | FALSE | Downregulated |
| -0,058168365 | 0,839302491 | MTA3 | FALSE | Downregulated |
| -0,257385841 | 0,839302491 | NOVA1 | FALSE | Downregulated |
| 0,1209228 | 0,839302491 | PANK1 | FALSE | Upregulated |
| 0,264264078 | 0,839302491 | PKD1L1 | FALSE | Upregulated |
| 0,078761018 | 0,839302491 | POGLUT1 | FALSE | Upregulated |

| | | | | |
|---|---|---|---|---|
| -0,057461521 | 0,839302491 | RMND5B | FALSE | Downregulated |
| 0,2938587 | 0,839302491 | RP11-789C1.1 | FALSE | Upregulated |
| 0,127839398 | 0,839302491 | RSG1 | FALSE | Upregulated |
| -0,192253404 | 0,839302491 | RTN2 | FALSE | Downregulated |
| 0,083536323 | 0,839302491 | SLC10A3 | FALSE | Upregulated |
| -0,045012978 | 0,839302491 | SLC25A46 | FALSE | Downregulated |
| -0,046651003 | 0,839302491 | STUB1 | FALSE | Downregulated |
| 0,146394519 | 0,839302491 | TFPI2 | FALSE | Upregulated |
| -0,079996041 | 0,839302491 | TMEM38B | FALSE | Downregulated |
| -0,093007519 | 0,839302491 | TMEM99 | FALSE | Downregulated |
| 0,038995682 | 0,839302491 | TRAK2 | FALSE | Upregulated |
| -0,042871303 | 0,839302491 | TRMT1L | FALSE | Downregulated |
| -0,100382724 | 0,839302491 | USP49 | FALSE | Downregulated |
| -0,056687739 | 0,839302491 | WBSCR22 | FALSE | Downregulated |
| -0,117595586 | 0,839420907 | ARMCX4 | FALSE | Downregulated |
| -0,079642388 | 0,839420907 | CAPN5 | FALSE | Downregulated |
| 0,17754492 | 0,839420907 | GIMAP5 | FALSE | Upregulated |
| -0,121847008 | 0,839608046 | ACTR5 | FALSE | Downregulated |
| 0,321354831 | 0,839608046 | LOC100287098 | FALSE | Upregulated |
| 0,311885392 | 0,839608046 | LOC101926917 | FALSE | Upregulated |
| -0,0982252 | 0,839608046 | MEIS3P1 | FALSE | Downregulated |
| 0,349776333 | 0,839608046 | MMP17 | FALSE | Upregulated |
| -0,106403672 | 0,839608046 | NUDT2 | FALSE | Downregulated |
| 0,758447225 | 0,839608046 | PHOSPHO2-KLHL23 | FALSE | Upregulated |
| -0,052431139 | 0,839608046 | PTGR2 | FALSE | Downregulated |
| 0,506101388 | 0,839608046 | RGPD2 | FALSE | Upregulated |

| | | | | |
|---|---|---|---|---|
| -0,231138197 | 0,839608046 | ROPN1L | FALSE | Downregulated |
| 0,101548278 | 0,839608046 | SUSD1 | FALSE | Upregulated |
| 0,060721535 | 0,839608046 | TMEM129 | FALSE | Upregulated |
| -0,087966126 | 0,839608046 | ZNF554 | FALSE | Downregulated |
| 0,061634301 | 0,83967135 | ABT1 | FALSE | Upregulated |
| -0,083628915 | 0,839749172 | ITSN2 | FALSE | Downregulated |
| 0,249005418 | 0,839782854 | ACOT11 | FALSE | Upregulated |
| 0,578040346 | 0,839782854 | CALCA | FALSE | Upregulated |
| -0,062859255 | 0,839782854 | HLCS | FALSE | Downregulated |
| -0,055312728 | 0,839782854 | KLF3 | FALSE | Downregulated |
| 0,183666339 | 0,839782854 | OGFR-AS1 | FALSE | Upregulated |
| -0,048240445 | 0,839782854 | SIN3A | FALSE | Downregulated |
| 0,131310959 | 0,839793024 | RASGRP4 | FALSE | Upregulated |
| 0,043586155 | 0,84001666 | GPKOW | FALSE | Upregulated |
| -0,334066923 | 0,840703537 | LOC101927885 | FALSE | Downregulated |
| 0,236719009 | 0,840703537 | RP11-84C10.4 | FALSE | Upregulated |
| -0,065189802 | 0,84089896 | ABI2 | FALSE | Downregulated |
| 0,208071284 | 0,840953868 | BHLHE22 | FALSE | Upregulated |
| 0,478158886 | 0,840953868 | CUZD1 | FALSE | Upregulated |
| 0,139673835 | 0,840953868 | GPR161 | FALSE | Upregulated |
| 0,141498603 | 0,840953868 | PCOLCE | FALSE | Upregulated |
| 0,241147587 | 0,840953868 | PTCH2 | FALSE | Upregulated |
| 0,357319223 | 0,840953868 | RP11-344B5.2 | FALSE | Upregulated |
| 0,252422215 | 0,840953868 | STX1A | FALSE | Upregulated |
| -0,078027708 | 0,841118644 | EVL | FALSE | Downregulated |
| -0,137688933 | 0,841209766 | ONECUT1 | FALSE | Downregulated |

| | | | | |
|---|---|---|---|---|
| 0,047151351 | 0,841209766 | PPIA | FALSE | Upregulated |
| 0,211480165 | 0,841209766 | RP11-434D9.1 | FALSE | Upregulated |
| -0,103889469 | 0,841414622 | PRDM11 | FALSE | Downregulated |
| -0,07816967 | 0,841575447 | DECR1 | FALSE | Downregulated |
| -0,095647686 | 0,841575447 | OLFML1 | FALSE | Downregulated |
| -0,241637367 | 0,841882257 | ADH7 | FALSE | Downregulated |
| 0,325747596 | 0,841882257 | DKFZp667F0711 | FALSE | Upregulated |
| -0,19099816 | 0,841882257 | EFNA3 | FALSE | Downregulated |
| 0,336122993 | 0,841882257 | FCRL1 | FALSE | Upregulated |
| 0,248411222 | 0,841882257 | LOC100507554 | FALSE | Upregulated |
| 0,290067222 | 0,841882257 | LOC101241902 | FALSE | Upregulated |
| -0,13540922 | 0,841882257 | NDRG1 | FALSE | Downregulated |
| -0,133933902 | 0,841882257 | OSBPL6 | FALSE | Downregulated |
| 0,450647356 | 0,841882257 | OTOF | FALSE | Upregulated |
| 0,144281198 | 0,841882257 | POU2F2 | FALSE | Upregulated |
| 0,107000028 | 0,841882257 | SLC2A10 | FALSE | Upregulated |
| 0,242096634 | 0,841882257 | SOCS2 | FALSE | Upregulated |
| 0,093782685 | 0,841882257 | SPAG5 | FALSE | Upregulated |
| 0,071710256 | 0,841882257 | TEFM | FALSE | Upregulated |
| 0,101968737 | 0,841882257 | VSIG10 | FALSE | Upregulated |
| -0,112565125 | 0,841882257 | WNT3 | FALSE | Downregulated |
| 0,081826806 | 0,841882257 | ZNF440 | FALSE | Upregulated |
| -0,084802308 | 0,841993207 | HIVEP2 | FALSE | Downregulated |
| 0,105042407 | 0,841993207 | SLC25A18 | FALSE | Upregulated |
| 0,115090702 | 0,842240587 | ZNF256 | FALSE | Upregulated |
| 0,15664601 | 0,842357954 | MCHR1 | FALSE | Upregulated |

| | | | | |
|---|---|---|---|---|
| 0,184043142 | 0,842369488 | TOB2P1 | FALSE | Upregulated |
| -0,256504357 | 0,842704024 | PSRC1 | FALSE | Downregulated |
| -0,075846794 | 0,842724788 | PRDX6 | FALSE | Downregulated |
| 0,117695405 | 0,842784487 | BATF2 | FALSE | Upregulated |
| -0,142140521 | 0,842784487 | DIRAS3 | FALSE | Downregulated |
| -0,153859211 | 0,842784487 | LOC101927451 | FALSE | Downregulated |
| 0,196265231 | 0,842784487 | MARCH3 | FALSE | Upregulated |
| 0,18015782 | 0,842784487 | OASL | FALSE | Upregulated |
| 0,075238129 | 0,842784487 | REEP6 | FALSE | Upregulated |
| -0,347286475 | 0,842784487 | SUGT1P3 | FALSE | Downregulated |
| 0,105390865 | 0,843406573 | LOC100506990 | FALSE | Upregulated |
| 0,180667303 | 0,843406573 | ZNF711 | FALSE | Upregulated |
| 0,043775642 | 0,843681894 | CSNK2B | FALSE | Upregulated |
| 0,190209688 | 0,843822223 | OSBPL7 | FALSE | Upregulated |
| -0,068724619 | 0,844145217 | PM20D2 | FALSE | Downregulated |
| -0,143387572 | 0,844314368 | DFFB | FALSE | Downregulated |
| 0,104035714 | 0,844314368 | KCND3 | FALSE | Upregulated |
| 0,084167238 | 0,844314368 | MAFB | FALSE | Upregulated |
| 0,115584414 | 0,844314368 | PAQR8 | FALSE | Upregulated |
| 0,405765076 | 0,844314368 | TLL2 | FALSE | Upregulated |
| -0,063379273 | 0,844314368 | TSFM | FALSE | Downregulated |
| 0,078534778 | 0,844314368 | WDR31 | FALSE | Upregulated |
| 0,092882833 | 0,844420893 | PPARGC1B | FALSE | Upregulated |
| 0,077926714 | 0,844472242 | JMJD4 | FALSE | Upregulated |
| -0,104667414 | 0,844492337 | RHOBTB3 | FALSE | Downregulated |
| 0,172849637 | 0,844554196 | PDGFD | FALSE | Upregulated |

| | | | | |
|---|---|---|---|---|
| -0,103405007 | 0,844573989 | DDX60 | FALSE | Downregulated |
| 0,109250785 | 0,844573989 | MEAF6 | FALSE | Upregulated |
| -0,067595919 | 0,844573989 | MIA3 | FALSE | Downregulated |
| 0,324538401 | 0,844573989 | TLL1 | FALSE | Upregulated |
| 0,066809619 | 0,844653232 | BAG2 | FALSE | Upregulated |
| 0,04987641 | 0,844653232 | FGFRL1 | FALSE | Upregulated |
| 0,263362783 | 0,844653232 | IGFBP1 | FALSE | Upregulated |
| -0,458474477 | 0,844653232 | LUZP2 | FALSE | Downregulated |
| -0,036876194 | 0,844653232 | MTAP | FALSE | Downregulated |
| 0,113172807 | 0,844653232 | NCMAP | FALSE | Upregulated |
| 0,042306507 | 0,844653232 | RIPK1 | FALSE | Upregulated |
| -0,252251981 | 0,844653232 | RSPO2 | FALSE | Downregulated |
| -0,224368559 | 0,844653232 | SLC28A2 | FALSE | Downregulated |
| 0,077477511 | 0,844653232 | TXNDC5 | FALSE | Upregulated |
| -0,139192145 | 0,844653232 | ZNF345 | FALSE | Downregulated |
| -0,061123401 | 0,844724283 | CBX7 | FALSE | Downregulated |
| 0,226325542 | 0,844724283 | EHHADH-AS1 | FALSE | Upregulated |
| -0,060621798 | 0,844724283 | LSR | FALSE | Downregulated |
| 0,060430579 | 0,844724283 | POLR2H | FALSE | Upregulated |
| 0,239157188 | 0,845188031 | SH3TC2 | FALSE | Upregulated |
| -0,075885607 | 0,845302595 | CEP97 | FALSE | Downregulated |
| 0,470519745 | 0,845302595 | L1CAM | FALSE | Upregulated |
| -0,081087322 | 0,845302595 | MEX3D | FALSE | Downregulated |
| -0,088886669 | 0,845302595 | MYO10 | FALSE | Downregulated |
| -0,072952954 | 0,845502589 | SIK3 | FALSE | Downregulated |
| -0,106762299 | 0,845545298 | AFG3L1P | FALSE | Downregulated |

| | | | | |
|---|---|---|---|---|
| -0,147416488 | 0,845545298 | ANTXR1 | FALSE | Downregulated |
| -0,05594807 | 0,845545298 | ANTXR2 | FALSE | Downregulated |
| 0,09338504 | 0,845545298 | ARF4 | FALSE | Upregulated |
| 0,285623058 | 0,845545298 | BHLHA15 | FALSE | Upregulated |
| -0,101701967 | 0,845545298 | CASC5 | FALSE | Downregulated |
| 0,167766336 | 0,845545298 | CCDC102A | FALSE | Upregulated |
| 0,068759772 | 0,845545298 | CEP85 | FALSE | Upregulated |
| -0,052052181 | 0,845545298 | COG6 | FALSE | Downregulated |
| -0,053261882 | 0,845545298 | COL4A3BP | FALSE | Downregulated |
| 0,430152436 | 0,845545298 | DDX11-AS1 | FALSE | Upregulated |
| -0,059625396 | 0,845545298 | DNAJB14 | FALSE | Downregulated |
| 0,168725661 | 0,845545298 | EBI3 | FALSE | Upregulated |
| 0,091212328 | 0,845545298 | HIST1H2BK | FALSE | Upregulated |
| 0,054835226 | 0,845545298 | IER3IP1 | FALSE | Upregulated |
| -0,058527309 | 0,845545298 | INSR | FALSE | Downregulated |
| -0,445412859 | 0,845545298 | KIAA1199 | FALSE | Downregulated |
| -0,067716815 | 0,845545298 | KLHL42 | FALSE | Downregulated |
| -0,324678846 | 0,845545298 | LOC101926922 | FALSE | Downregulated |
| -0,312186115 | 0,845545298 | LOC101928238 | FALSE | Downregulated |
| -0,729380954 | 0,845545298 | LOC102723630 | FALSE | Downregulated |
| -0,086717173 | 0,845545298 | LOC220729 | FALSE | Downregulated |
| -0,06776456 | 0,845545298 | LYRM1 | FALSE | Downregulated |
| 0,249221586 | 0,845545298 | MIR3064 | FALSE | Upregulated |
| -0,050666571 | 0,845545298 | NUP88 | FALSE | Downregulated |
| -0,071214273 | 0,845545298 | OCEL1 | FALSE | Downregulated |
| -0,087058329 | 0,845545298 | OGFRL1 | FALSE | Downregulated |

| | | | | |
|---|---|---|---|---|
| -0,050242428 | 0,845545298 | PIGN | FALSE | Downregulated |
| 0,118082007 | 0,845545298 | SFMBT2 | FALSE | Upregulated |
| 0,171317655 | 0,845545298 | SIDT1 | FALSE | Upregulated |
| -0,35548597 | 0,845545298 | SLCO1A2 | FALSE | Downregulated |
| 0,356980935 | 0,845545298 | STC2 | FALSE | Upregulated |
| -0,079538041 | 0,845545298 | TFAP4 | FALSE | Downregulated |
| -0,124003174 | 0,845545298 | TMEM26 | FALSE | Downregulated |
| 0,162522215 | 0,845545298 | TSKU | FALSE | Upregulated |
| -0,042156451 | 0,845545298 | UBE2H | FALSE | Downregulated |
| -0,12550793 | 0,845545298 | ZNF431 | FALSE | Downregulated |
| 0,055517245 | 0,845545298 | ZNF526 | FALSE | Upregulated |
| 0,067121629 | 0,845782196 | MRTO4 | FALSE | Upregulated |
| 0,391227844 | 0,845944825 | FAM153A | FALSE | Upregulated |
| -0,067652239 | 0,845944825 | PPP6R2 | FALSE | Downregulated |
| 0,127800945 | 0,846105753 | SMA4 | FALSE | Upregulated |
| -0,178776602 | 0,846107854 | TRIM45 | FALSE | Downregulated |
| -0,045677776 | 0,846111243 | TGS1 | FALSE | Downregulated |
| -0,075548085 | 0,84622639 | APOA2 | FALSE | Downregulated |
| 0,301074985 | 0,846505183 | KCNN4 | FALSE | Upregulated |
| 0,10471986 | 0,846505183 | LOC101060181 | FALSE | Upregulated |
| -0,051574631 | 0,846505183 | OSBPL8 | FALSE | Downregulated |
| -0,05015013 | 0,846577048 | FOXN3 | FALSE | Downregulated |
| -0,062458312 | 0,846577048 | HSD17B10 | FALSE | Downregulated |
| 0,369282731 | 0,846577048 | KCNJ2-AS1 | FALSE | Upregulated |
| 0,059898168 | 0,846577048 | MGAT4B | FALSE | Upregulated |
| 0,135766555 | 0,846577048 | SRGAP2B | FALSE | Upregulated |

| | | | | |
|---|---|---|---|---|
| -0,04793841 | 0,846661338 | LRRC8D | FALSE | Downregulated |
| -0,051964412 | 0,846663276 | GPC6 | FALSE | Downregulated |
| 0,056485246 | 0,846803694 | MEA1 | FALSE | Upregulated |
| 0,055391803 | 0,846868327 | UFC1 | FALSE | Upregulated |
| 0,057949429 | 0,846921538 | AARS | FALSE | Upregulated |
| -0,065469474 | 0,846921538 | ACN9 | FALSE | Downregulated |
| -0,047437064 | 0,846921538 | EGLN1 | FALSE | Downregulated |
| -0,226393479 | 0,846921538 | LOC101927578 | FALSE | Downregulated |
| 0,089329388 | 0,846921538 | NPL | FALSE | Upregulated |
| -0,087789332 | 0,846921538 | PPP1R12B | FALSE | Downregulated |
| 0,062788415 | 0,846921538 | SHROOM3 | FALSE | Upregulated |
| 0,398501124 | 0,847002216 | ALG1L | FALSE | Upregulated |
| -0,10731414 | 0,847002216 | ZNF124 | FALSE | Downregulated |
| -0,090774066 | 0,847007149 | SP4 | FALSE | Downregulated |
| -0,10515078 | 0,847294963 | BMP10 | FALSE | Downregulated |
| 0,067642858 | 0,847294963 | C2orf68 | FALSE | Upregulated |
| -0,308610492 | 0,847294963 | CALML6 | FALSE | Downregulated |
| -0,05750263 | 0,847294963 | CRCP | FALSE | Downregulated |
| -0,080210752 | 0,847294963 | EXO5 | FALSE | Downregulated |
| -0,045068056 | 0,847294963 | FAM193A | FALSE | Downregulated |
| 0,124176075 | 0,847294963 | FHOD1 | FALSE | Upregulated |
| 0,274404896 | 0,847294963 | GNG3 | FALSE | Upregulated |
| -0,070801102 | 0,847294963 | GPS2 | FALSE | Downregulated |
| 0,308532335 | 0,847294963 | HDC | FALSE | Upregulated |
| 0,158380323 | 0,847294963 | HOXB5 | FALSE | Upregulated |
| -0,189910962 | 0,847294963 | LOC100506974 | FALSE | Downregulated |

| | | | | |
|---|---|---|---|---|
| -0,383409935 | 0,847294963 | LOC101926894 | FALSE | Downregulated |
| 0,201118934 | 0,847294963 | LOC101927263 | FALSE | Upregulated |
| 0,272068154 | 0,847294963 | LOC102724788 | FALSE | Upregulated |
| -0,369478451 | 0,847294963 | LOC345051 | FALSE | Downregulated |
| -0,243430376 | 0,847294963 | MDH1B | FALSE | Downregulated |
| 0,224429022 | 0,847294963 | NEAT1 | FALSE | Upregulated |
| 0,241048669 | 0,847294963 | PLCH1 | FALSE | Upregulated |
| -0,204925665 | 0,847294963 | PPIEL | FALSE | Downregulated |
| -0,060055174 | 0,847294963 | RIN2 | FALSE | Downregulated |
| 0,117326378 | 0,847294963 | SIGLEC9 | FALSE | Upregulated |
| -0,041321855 | 0,847294963 | SNX5 | FALSE | Downregulated |
| 0,073879502 | 0,847294963 | SPATA7 | FALSE | Upregulated |
| 0,153900232 | 0,847294963 | SPTBN4 | FALSE | Upregulated |
| 0,030866122 | 0,847294963 | STARD7 | FALSE | Upregulated |
| 0,07436741 | 0,847294963 | TDP1 | FALSE | Upregulated |
| -0,386299401 | 0,847294963 | WTH3DI | FALSE | Downregulated |
| 0,306819661 | 0,847296448 | CCR9 | FALSE | Upregulated |
| 0,060224601 | 0,847296448 | MED12 | FALSE | Upregulated |
| 0,063273298 | 0,847296448 | UQCR10 | FALSE | Upregulated |
| 0,115696766 | 0,847721771 | SLFN5 | FALSE | Upregulated |
| 0,10875107 | 0,847792672 | TMCC3 | FALSE | Upregulated |
| -0,035801939 | 0,84806779 | C16orf72 | FALSE | Downregulated |
| 0,040497323 | 0,84806779 | DHX32 | FALSE | Upregulated |
| -0,071589355 | 0,84806779 | ERCC8 | FALSE | Downregulated |
| -0,08948114 | 0,84806779 | HAAO | FALSE | Downregulated |
| -0,130879277 | 0,84806779 | HAO2 | FALSE | Downregulated |

| | | | | |
|---|---|---|---|---|
| -0,028235567 | 0,84806779 | NEK9 | FALSE | Downregulated |
| -0,108235605 | 0,848075905 | ZNF850 | FALSE | Downregulated |
| 0,034701087 | 0,848159751 | GOLGA7 | FALSE | Upregulated |
| 0,296995922 | 0,848159751 | LOC101927681 | FALSE | Upregulated |
| 0,035859137 | 0,848159751 | YME1L1 | FALSE | Upregulated |
| -0,067838744 | 0,848176695 | CISD3 | FALSE | Downregulated |
| -0,069685669 | 0,848176695 | TPR | FALSE | Downregulated |
| -0,078911078 | 0,848219772 | ZNF354C | FALSE | Downregulated |
| 0,164795263 | 0,848348283 | MFI2 | FALSE | Upregulated |
| 0,091957837 | 0,848643155 | GALNT2 | FALSE | Upregulated |
| 0,115528345 | 0,848664012 | ARL2 | FALSE | Upregulated |
| -0,204535853 | 0,848664012 | RP11-163N6.2 | FALSE | Downregulated |
| -0,065226815 | 0,848818372 | ZBTB38 | FALSE | Downregulated |
| 0,054886424 | 0,848818372 | ZNF44 | FALSE | Upregulated |
| 0,102660693 | 0,848818799 | C1orf172 | FALSE | Upregulated |
| 0,255938242 | 0,848826179 | CCDC153 | FALSE | Upregulated |
| 0,075356355 | 0,848826179 | ZSWIM1 | FALSE | Upregulated |
| -0,038831931 | 0,848831021 | ARFGAP2 | FALSE | Downregulated |
| -0,305772576 | 0,848831021 | DCHS2 | FALSE | Downregulated |
| 0,081146171 | 0,848831021 | HUS1 | FALSE | Upregulated |
| 0,052322835 | 0,848831021 | PTPRF | FALSE | Upregulated |
| 0,040802314 | 0,848831021 | UBE2J2 | FALSE | Upregulated |
| 0,228787844 | 0,848917212 | ASPHD2 | FALSE | Upregulated |
| 0,141710311 | 0,848923166 | CCDC96 | FALSE | Upregulated |
| 0,178001441 | 0,848923166 | FBXO16 | FALSE | Upregulated |
| -0,115814788 | 0,848929212 | HHIP | FALSE | Downregulated |

| | | | | |
|---|---|---|---|---|
| 0,07022968 | 0,848929212 | RNF149 | FALSE | Upregulated |
| -0,057864234 | 0,848929212 | TADA1 | FALSE | Downregulated |
| 0,204571592 | 0,849002774 | ZGLP1 | FALSE | Upregulated |
| 0,100056651 | 0,849073216 | CTTNBP2NL | FALSE | Upregulated |
| -0,239565713 | 0,849102352 | LOC101060553 | FALSE | Downregulated |
| -0,054757844 | 0,849295522 | METTL17 | FALSE | Downregulated |
| -0,050907144 | 0,849295522 | MKL2 | FALSE | Downregulated |
| -0,091846706 | 0,849295522 | SLC19A3 | FALSE | Downregulated |
| 0,12574282 | 0,849400146 | GATA6-AS1 | FALSE | Upregulated |
| -0,103141526 | 0,849478131 | ZNF585A | FALSE | Downregulated |
| 0,09503286 | 0,849633718 | LOC100507599 | FALSE | Upregulated |
| -0,03757487 | 0,849633718 | TCEA1 | FALSE | Downregulated |
| 0,266122529 | 0,849661011 | FCER2 | FALSE | Upregulated |
| -0,212559914 | 0,849661011 | LINC00472 | FALSE | Downregulated |
| 0,238322726 | 0,849661011 | RGPD4 | FALSE | Upregulated |
| -0,109604058 | 0,849829754 | PMS2CL | FALSE | Downregulated |
| 1,377768232 | 0,849829754 | SIK1 | FALSE | Upregulated |
| -0,075169885 | 0,849916989 | GPT | FALSE | Downregulated |
| 0,100778988 | 0,849916989 | HCK | FALSE | Upregulated |
| 0,195400502 | 0,850099871 | BATF | FALSE | Upregulated |
| 0,077728952 | 0,850099871 | C14orf93 | FALSE | Upregulated |
| 0,225662421 | 0,850099871 | CBX2 | FALSE | Upregulated |
| 0,082596178 | 0,850099871 | ENPP4 | FALSE | Upregulated |
| -0,112584689 | 0,850099871 | ENTPD8 | FALSE | Downregulated |
| 0,035506468 | 0,850099871 | GLG1 | FALSE | Upregulated |
| -0,075721019 | 0,850099871 | KDM6B | FALSE | Downregulated |

| | | | | |
|---|---|---|---|---|
| -0,100325198 | 0,850099871 | SLC39A4 | FALSE | Downregulated |
| -0,05918234 | 0,850099871 | TJAP1 | FALSE | Downregulated |
| 0,216553013 | 0,850099871 | WDR64 | FALSE | Upregulated |
| 0,135400649 | 0,850099871 | ZNF713 | FALSE | Upregulated |
| -0,095990848 | 0,85017602 | AKAP6 | FALSE | Downregulated |
| -0,098497521 | 0,850217885 | IFITM2 | FALSE | Downregulated |
| -0,831511863 | 0,850330653 | SLC5A12 | FALSE | Downregulated |
| -0,050084657 | 0,850330653 | UCHL5 | FALSE | Downregulated |
| 0,041263126 | 0,850531578 | C8orf33 | FALSE | Upregulated |
| 0,247195523 | 0,850531578 | CAPN6 | FALSE | Upregulated |
| -0,252508476 | 0,850531578 | CELF5 | FALSE | Downregulated |
| -0,136825901 | 0,850531578 | FENDRR | FALSE | Downregulated |
| 0,065285581 | 0,850531578 | HDDC3 | FALSE | Upregulated |
| 0,126839884 | 0,850531578 | IGSF6 | FALSE | Upregulated |
| 0,267326821 | 0,850531578 | MGC39372 | FALSE | Upregulated |
| 0,136636342 | 0,850531578 | RAVER2 | FALSE | Upregulated |
| 0,064215079 | 0,850531578 | RRP1 | FALSE | Upregulated |
| -0,079252636 | 0,850531578 | SASS6 | FALSE | Downregulated |
| 0,073609706 | 0,850531578 | TIMM10 | FALSE | Upregulated |
| 0,096805046 | 0,850531578 | TMEM139 | FALSE | Upregulated |
| -0,18110484 | 0,850539464 | SLC17A8 | FALSE | Downregulated |
| 0,057671259 | 0,850541615 | B3GALT6 | FALSE | Upregulated |
| -0,080797606 | 0,850541615 | C5orf45 | FALSE | Downregulated |
| -0,046005122 | 0,850541615 | NOB1 | FALSE | Downregulated |
| 0,074261307 | 0,850541615 | NOP9 | FALSE | Upregulated |
| -0,221795081 | 0,851025348 | NPM2 | FALSE | Downregulated |

| | | | | |
|---|---|---|---|---|
| -0,319211019 | 0,851070387 | TEKT5 | FALSE | Downregulated |
| 0,321754626 | 0,851194597 | TDRD1 | FALSE | Upregulated |
| -0,347025302 | 0,851219101 | MS4A4E | FALSE | Downregulated |
| -0,075894209 | 0,851219101 | TLR3 | FALSE | Downregulated |
| 0,065599836 | 0,851249129 | COQ6 | FALSE | Upregulated |
| -0,386584277 | 0,851407804 | GABRG1 | FALSE | Downregulated |
| 0,036515914 | 0,851407804 | UTP14C | FALSE | Upregulated |
| 0,154847802 | 0,851496401 | SYBU | FALSE | Upregulated |
| -0,267597829 | 0,851561871 | HCG4B | FALSE | Downregulated |
| 0,313256892 | 0,851561871 | LOC102724037 | FALSE | Upregulated |
| 0,062479059 | 0,851561871 | MON2 | FALSE | Upregulated |
| -0,051616339 | 0,851561871 | REXO2 | FALSE | Downregulated |
| 0,078329503 | 0,851561871 | VAC14 | FALSE | Upregulated |
| 0,185681528 | 0,851584073 | NOTCH3 | FALSE | Upregulated |
| 0,668505705 | 0,851662097 | CLEC18C | FALSE | Upregulated |
| 0,075175804 | 0,851662097 | DGKD | FALSE | Upregulated |
| -0,059985613 | 0,851662097 | DSE | FALSE | Downregulated |
| 0,210421802 | 0,851662097 | FAM115C | FALSE | Upregulated |
| -0,081280604 | 0,851662097 | FAT4 | FALSE | Downregulated |
| 0,064493999 | 0,851662097 | GLT8D1 | FALSE | Upregulated |
| -0,087646844 | 0,851662097 | GNE | FALSE | Downregulated |
| 0,234286496 | 0,851662097 | LOC100996662 | FALSE | Upregulated |
| -0,35443429 | 0,851662097 | LPL | FALSE | Downregulated |
| 0,112553425 | 0,851662097 | PCBP1-AS1 | FALSE | Upregulated |
| -0,158804415 | 0,851662097 | PFKFB4 | FALSE | Downregulated |
| -0,038569422 | 0,851662097 | RB1CC1 | FALSE | Downregulated |

| | | | | |
|---|---|---|---|---|
| -0,042121455 | 0,851662097 | SP100 | FALSE | Downregulated |
| 0,059647184 | 0,851662097 | TMUB2 | FALSE | Upregulated |
| -0,136541342 | 0,851662097 | UBXN4 | FALSE | Downregulated |
| -0,058751509 | 0,851662097 | VPS41 | FALSE | Downregulated |
| -0,086712968 | 0,851662097 | ZNF470 | FALSE | Downregulated |
| -0,05531807 | 0,851746809 | CNP | FALSE | Downregulated |
| 0,052244063 | 0,851746809 | DAGLB | FALSE | Upregulated |
| 0,108264132 | 0,851746809 | PHLDB1 | FALSE | Upregulated |
| 0,060658499 | 0,851746809 | SMG6 | FALSE | Upregulated |
| 0,089791368 | 0,851819815 | WDPCP | FALSE | Upregulated |
| 0,26621071 | 0,851910175 | RP11-432I5.1 | FALSE | Upregulated |
| -0,150437183 | 0,851938853 | EID2B | FALSE | Downregulated |
| -0,368526365 | 0,851938853 | LOC101928344 | FALSE | Downregulated |
| -0,064324294 | 0,851972276 | GSE1 | FALSE | Downregulated |
| 0,268342736 | 0,851972276 | POM121L9P | FALSE | Upregulated |
| 0,058187798 | 0,851972276 | SFMBT1 | FALSE | Upregulated |
| -0,113822579 | 0,851972276 | WBP5 | FALSE | Downregulated |
| 0,223259821 | 0,851972276 | WDR62 | FALSE | Upregulated |
| 0,100394754 | 0,851972276 | ZBTB46 | FALSE | Upregulated |
| -0,123971462 | 0,852118884 | MPV17L | FALSE | Downregulated |
| 0,203799813 | 0,852235865 | GUSBP5 | FALSE | Upregulated |
| 0,134726871 | 0,852235865 | ZBTB26 | FALSE | Upregulated |
| -0,060418837 | 0,852235865 | ZNF253 | FALSE | Downregulated |
| -0,353922724 | 0,85225837 | HOXB-AS3 | FALSE | Downregulated |
| 0,033350719 | 0,852281737 | RAD23B | FALSE | Upregulated |
| 0,0888906 | 0,852281737 | RGAG4 | FALSE | Upregulated |

| | | | | |
|---|---|---|---|---|
| -0,159468858 | 0,852281737 | RP1-60O19.1 | FALSE | Downregulated |
| -0,05582284 | 0,852380929 | KIF5B | FALSE | Downregulated |
| 0,387473172 | 0,852380929 | LOC101928173 | FALSE | Upregulated |
| 0,058163862 | 0,852380929 | TOR1AIP1 | FALSE | Upregulated |
| -0,463439085 | 0,852405527 | C1QTNF9B | FALSE | Downregulated |
| 0,398769886 | 0,852690695 | ZCCHC18 | FALSE | Upregulated |
| 0,066124825 | 0,853092162 | HERC1 | FALSE | Upregulated |
| -0,057581553 | 0,853157761 | DCAF11 | FALSE | Downregulated |
| -0,138050279 | 0,853157761 | EIF1B-AS1 | FALSE | Downregulated |
| 0,153599303 | 0,853157761 | RP11-115J16.1 | FALSE | Upregulated |
| 0,073194907 | 0,853409239 | TIMM17A | FALSE | Upregulated |
| -0,056956045 | 0,853481122 | ABCC2 | FALSE | Downregulated |
| 0,058635846 | 0,853481122 | AKR7A2 | FALSE | Upregulated |
| 0,179216616 | 0,853481122 | ANKRD33B | FALSE | Upregulated |
| 0,183815655 | 0,853481122 | ANXA13 | FALSE | Upregulated |
| -0,06952092 | 0,853481122 | BAZ2A | FALSE | Downregulated |
| -0,080918189 | 0,853481122 | FAM84B | FALSE | Downregulated |
| 0,40663649 | 0,853481122 | FLJ44511 | FALSE | Upregulated |
| 0,099291508 | 0,853481122 | GBP4 | FALSE | Upregulated |
| 0,077902746 | 0,853481122 | IQCK | FALSE | Upregulated |
| -0,334533282 | 0,853481122 | LOC101060494 | FALSE | Downregulated |
| -0,182964204 | 0,853481122 | LOC102546294 | FALSE | Downregulated |
| -0,227195692 | 0,853481122 | LOC102724507 | FALSE | Downregulated |
| 0,065127487 | 0,853481122 | NDRG2 | FALSE | Upregulated |
| -0,116375797 | 0,853481122 | RTTN | FALSE | Downregulated |
| 0,380857506 | 0,853481122 | SLC52A3 | FALSE | Upregulated |

| | | | | |
|---|---|---|---|---|
| -0,078529866 | 0,853481122 | STK35 | FALSE | Downregulated |
| -0,106863094 | 0,853481122 | SYNE4 | FALSE | Downregulated |
| -0,114068602 | 0,853481122 | TMEM97 | FALSE | Downregulated |
| 0,04866332 | 0,853481122 | VAPB | FALSE | Upregulated |
| 0,08196553 | 0,853481122 | ZFP69 | FALSE | Upregulated |
| -0,207534142 | 0,853817637 | FIGNL2 | FALSE | Downregulated |
| 0,207637405 | 0,853817637 | LOC101928947 | FALSE | Upregulated |
| -0,277659908 | 0,853817637 | MIR1282 | FALSE | Downregulated |
| 0,366426954 | 0,854025521 | FANCB | FALSE | Upregulated |
| -0,045418282 | 0,854334484 | AEBP2 | FALSE | Downregulated |
| 0,157985186 | 0,854403803 | C3orf14 | FALSE | Upregulated |
| 0,066054647 | 0,854816966 | FBXL20 | FALSE | Upregulated |
| 0,095220443 | 0,854816966 | SATB2 | FALSE | Upregulated |
| 0,22695302 | 0,85482468 | ETV4 | FALSE | Upregulated |
| 0,165008136 | 0,854877967 | LOC100505555 | FALSE | Upregulated |
| -0,142127416 | 0,855341184 | USP6NL-IT1 | FALSE | Downregulated |
| 0,302650497 | 0,855362807 | LOC101929070 | FALSE | Upregulated |
| -0,209101456 | 0,855362807 | PAQR6 | FALSE | Downregulated |
| -0,049817291 | 0,855362807 | PPFIA1 | FALSE | Downregulated |
| 0,056070989 | 0,855499659 | KATNB1 | FALSE | Upregulated |
| 0,069350318 | 0,855499659 | RPL36 | FALSE | Upregulated |
| 0,108738718 | 0,855743459 | EFHC1 | FALSE | Upregulated |
| -0,093655935 | 0,855743459 | FLJ37035 | FALSE | Downregulated |
| -0,202659228 | 0,855911031 | F2RL2 | FALSE | Downregulated |
| -0,357001436 | 0,855911031 | TMC5 | FALSE | Downregulated |
| -0,066916613 | 0,855950746 | DMD | FALSE | Downregulated |

| | | | | |
|---|---|---|---|---|
| -0,053605927 | 0,856022847 | ACSL3 | FALSE | Downregulated |
| -0,059944094 | 0,856022847 | FZD4 | FALSE | Downregulated |
| 0,211639575 | 0,856022847 | TSC22D3 | FALSE | Upregulated |
| -0,031399903 | 0,856032956 | AK2 | FALSE | Downregulated |
| 0,054493577 | 0,856032956 | C2orf43 | FALSE | Upregulated |
| -0,079815383 | 0,856032956 | CPED1 | FALSE | Downregulated |
| -0,125972347 | 0,856032956 | FAM227B | FALSE | Downregulated |
| 0,224723354 | 0,856032956 | IGSF10 | FALSE | Upregulated |
| -0,126820979 | 0,856032956 | LOC100507577 | FALSE | Downregulated |
| -0,235018548 | 0,856032956 | LOC101926907 | FALSE | Downregulated |
| 0,118044364 | 0,856058352 | SRC | FALSE | Upregulated |
| -0,098647407 | 0,856121741 | ITPR1 | FALSE | Downregulated |
| -0,250831867 | 0,856121741 | ZNF385C | FALSE | Downregulated |
| 0,490355703 | 0,856230105 | KLHL34 | FALSE | Upregulated |
| -0,063750678 | 0,856327281 | CRADD | FALSE | Downregulated |
| 0,34405782 | 0,856343542 | ACTG2 | FALSE | Upregulated |
| 0,325548037 | 0,856343542 | DTNA | FALSE | Upregulated |
| -0,06797145 | 0,856343542 | KIAA1467 | FALSE | Downregulated |
| -0,03961858 | 0,856343542 | KIF3B | FALSE | Downregulated |
| -0,086372265 | 0,856343542 | LOC101928319 | FALSE | Downregulated |
| 0,090203132 | 0,856343542 | TUBG2 | FALSE | Upregulated |
| -0,043408175 | 0,856343542 | VDAC2 | FALSE | Downregulated |
| 0,06225477 | 0,856402039 | MYCBP | FALSE | Upregulated |
| -0,044604953 | 0,856496242 | CCNY | FALSE | Downregulated |
| 0,290073252 | 0,856583784 | AFF2 | FALSE | Upregulated |
| -0,073336426 | 0,856583784 | SLC27A2 | FALSE | Downregulated |

| | | | | |
|---|---|---|---|---|
| -0,062789967 | 0,85690534 | ECHS1 | FALSE | Downregulated |
| -0,060768091 | 0,857130256 | CEP350 | FALSE | Downregulated |
| -0,054709285 | 0,857130256 | POLR2G | FALSE | Downregulated |
| -0,050337875 | 0,857130256 | TMEM218 | FALSE | Downregulated |
| -0,091902453 | 0,85735903 | DACH1 | FALSE | Downregulated |
| 0,048904569 | 0,857601929 | DARS2 | FALSE | Upregulated |
| -0,054372646 | 0,857601929 | POC1B | FALSE | Downregulated |
| 0,367623978 | 0,857624182 | LOC102723627 | FALSE | Upregulated |
| 0,042204212 | 0,857681112 | ATG9A | FALSE | Upregulated |
| 0,168009554 | 0,857681112 | GAS5-AS1 | FALSE | Upregulated |
| 0,415726066 | 0,857683397 | CCDC155 | FALSE | Upregulated |
| 0,210610912 | 0,857683397 | SNCAIP | FALSE | Upregulated |
| 0,157593956 | 0,85789506 | C21orf67 | FALSE | Upregulated |
| -0,101713822 | 0,85789506 | CCDC158 | FALSE | Downregulated |
| -0,062876441 | 0,85789506 | EFHD2 | FALSE | Downregulated |
| -0,1327905 | 0,85789506 | FGL1 | FALSE | Downregulated |
| 0,10455279 | 0,85789506 | KIRREL | FALSE | Upregulated |
| -0,349327642 | 0,85789506 | MSI1 | FALSE | Downregulated |
| -0,037213915 | 0,85789506 | TK2 | FALSE | Downregulated |
| -0,077459818 | 0,85789506 | ZFAND2A | FALSE | Downregulated |
| -0,086327351 | 0,858192443 | PHF8 | FALSE | Downregulated |
| -0,071849699 | 0,858303262 | BOLA3 | FALSE | Downregulated |
| 0,063150468 | 0,858303262 | FUS | FALSE | Upregulated |
| -0,056643519 | 0,858303262 | INTS2 | FALSE | Downregulated |
| -0,080381695 | 0,858303262 | RGL1 | FALSE | Downregulated |
| 0,069234042 | 0,858303262 | TAF9 | FALSE | Upregulated |

| | | | | |
|---|---|---|---|---|
| -0,364296609 | 0,858352854 | CNTN5 | FALSE | Downregulated |
| 0,116371444 | 0,858352854 | FLT3LG | FALSE | Upregulated |
| -0,041118773 | 0,858355168 | AES | FALSE | Downregulated |
| -0,037275772 | 0,858355168 | ARMC10 | FALSE | Downregulated |
| -0,140402946 | 0,858355168 | C1orf210 | FALSE | Downregulated |
| -0,079083719 | 0,858355168 | ZBTB8A | FALSE | Downregulated |
| 0,138487987 | 0,858435648 | AKR1C2 | FALSE | Upregulated |
| 0,131277282 | 0,858939581 | EIF5A2 | FALSE | Upregulated |
| -0,116266869 | 0,858939581 | RAB23 | FALSE | Downregulated |
| -0,381535542 | 0,858939581 | RSPO4 | FALSE | Downregulated |
| -0,215805134 | 0,858939581 | VIPR2 | FALSE | Downregulated |
| -0,064591681 | 0,858939581 | ZNF395 | FALSE | Downregulated |
| -0,067426105 | 0,859243227 | ANKEF1 | FALSE | Downregulated |
| 0,174480185 | 0,859243227 | CCDC8 | FALSE | Upregulated |
| -0,040490149 | 0,859243227 | FAM160B1 | FALSE | Downregulated |
| 0,117924671 | 0,859243227 | PRR26 | FALSE | Upregulated |
| -0,063468638 | 0,859243227 | RABGAP1L | FALSE | Downregulated |
| 0,402091375 | 0,859243227 | TUBA4B | FALSE | Upregulated |
| 0,252325299 | 0,859264792 | C1orf204 | FALSE | Upregulated |
| -0,04717702 | 0,859264792 | FKBP8 | FALSE | Downregulated |
| 0,124348058 | 0,859264792 | GLUD2 | FALSE | Upregulated |
| 0,070958688 | 0,859264792 | GMEB1 | FALSE | Upregulated |
| 0,039418226 | 0,859264792 | LTA4H | FALSE | Upregulated |
| -0,056591086 | 0,859264792 | PHF20L1 | FALSE | Downregulated |
| 0,100659846 | 0,859264792 | REL | FALSE | Upregulated |
| 0,041412029 | 0,859264792 | RNF146 | FALSE | Upregulated |

| | | | | |
|---|---|---|---|---|
| -0,071464627 | 0,859264792 | YAP1 | FALSE | Downregulated |
| 0,090714821 | 0,859276348 | MAP3K6 | FALSE | Upregulated |
| -0,065469581 | 0,859318097 | MTMR10 | FALSE | Downregulated |
| -0,044459439 | 0,859333435 | ARHGAP35 | FALSE | Downregulated |
| 0,130541825 | 0,859333435 | CDHR3 | FALSE | Upregulated |
| -0,08170286 | 0,859333435 | COQ2 | FALSE | Downregulated |
| 1,387220433 | 0,859333435 | CRYAA | FALSE | Upregulated |
| -0,324683241 | 0,859333435 | DNMT3L | FALSE | Downregulated |
| -0,117341387 | 0,859333435 | DUSP10 | FALSE | Downregulated |
| -0,151281012 | 0,859333435 | ELOVL2-AS1 | FALSE | Downregulated |
| 0,080103604 | 0,859333435 | EXOSC4 | FALSE | Upregulated |
| 0,072425846 | 0,859333435 | EXTL3 | FALSE | Upregulated |
| 0,06398622 | 0,859333435 | FAM207A | FALSE | Upregulated |
| 0,077915279 | 0,859333435 | FOSL2 | FALSE | Upregulated |
| 0,217559996 | 0,859333435 | GABRE | FALSE | Upregulated |
| -0,124361484 | 0,859333435 | GPD1L | FALSE | Downregulated |
| 0,242446644 | 0,859333435 | GPR153 | FALSE | Upregulated |
| -0,044838175 | 0,859333435 | HAUS6 | FALSE | Downregulated |
| -0,037491125 | 0,859333435 | HNRNPF | FALSE | Downregulated |
| -0,231370375 | 0,859333435 | ITPRIPL1 | FALSE | Downregulated |
| 0,03381654 | 0,859333435 | KIAA0196 | FALSE | Upregulated |
| -0,036794319 | 0,859333435 | LEPROT | FALSE | Downregulated |
| 0,169754966 | 0,859333435 | LINC00869 | FALSE | Upregulated |
| -0,201505794 | 0,859333435 | LINC01171 | FALSE | Downregulated |
| 0,205056294 | 0,859333435 | LOC101927144 | FALSE | Upregulated |
| 0,227920826 | 0,859333435 | LOC101929638 | FALSE | Upregulated |

| | | | | |
|---|---|---|---|---|
| -0,332884734 | 0,859333435 | LOC101929724 | FALSE | Downregulated |
| -0,192830146 | 0,859333435 | LOC643327 | FALSE | Downregulated |
| -0,277999879 | 0,859333435 | LOC729737 | FALSE | Downregulated |
| 0,070014914 | 0,859333435 | MAFG | FALSE | Upregulated |
| 0,306640925 | 0,859333435 | MCF2 | FALSE | Upregulated |
| -0,082449232 | 0,859333435 | MICU3 | FALSE | Downregulated |
| 0,08076724 | 0,859333435 | MPZL2 | FALSE | Upregulated |
| 0,103098497 | 0,859333435 | NBPF11 | FALSE | Upregulated |
| 0,150310079 | 0,859333435 | NSUN5P1 | FALSE | Upregulated |
| 0,028624296 | 0,859333435 | PAFAH1B1 | FALSE | Upregulated |
| 0,071140272 | 0,859333435 | PDK1 | FALSE | Upregulated |
| -0,094853827 | 0,859333435 | PGM5 | FALSE | Downregulated |
| 1,103717266 | 0,859333435 | PRAMEF10 | FALSE | Upregulated |
| -0,055956657 | 0,859333435 | PRKACB | FALSE | Downregulated |
| 0,11293018 | 0,859333435 | PTGIS | FALSE | Upregulated |
| -0,046304848 | 0,859333435 | SNRPE | FALSE | Downregulated |
| 0,234087648 | 0,859333435 | SORT1 | FALSE | Upregulated |
| 0,180527786 | 0,859333435 | SOX4 | FALSE | Upregulated |
| 0,112234177 | 0,859333435 | STAT4 | FALSE | Upregulated |
| 0,086438365 | 0,859333435 | STX1B | FALSE | Upregulated |
| 0,314637715 | 0,859333435 | TMEM217 | FALSE | Upregulated |
| 0,037624897 | 0,859333435 | VIPAS39 | FALSE | Upregulated |
| -0,089191231 | 0,859333435 | ZBTB49 | FALSE | Downregulated |
| -0,116615772 | 0,859333435 | ZNF334 | FALSE | Downregulated |
| -0,068284235 | 0,859365245 | ATRX | FALSE | Downregulated |
| -0,036414009 | 0,859365245 | CNOT1 | FALSE | Downregulated |

| | | | | |
|---|---|---|---|---|
| -0,288769665 | 0,859422855 | FOSB | FALSE | Downregulated |
| 0,169421382 | 0,859425353 | KCNJ14 | FALSE | Upregulated |
| -0,055661288 | 0,859575183 | ACVR2A | FALSE | Downregulated |
| -0,219853924 | 0,859575183 | BRICD5 | FALSE | Downregulated |
| -0,084242345 | 0,859575183 | PIK3AP1 | FALSE | Downregulated |
| -0,155502823 | 0,859575183 | PTPRT | FALSE | Downregulated |
| 0,048139462 | 0,859575183 | SLC9A8 | FALSE | Upregulated |
| -0,141047841 | 0,859780266 | HNRNPU-AS1 | FALSE | Downregulated |
| 0,0880335 | 0,859858469 | GLS | FALSE | Upregulated |
| 0,065898824 | 0,860317825 | ABCB10 | FALSE | Upregulated |
| 0,07792156 | 0,860317825 | C1orf86 | FALSE | Upregulated |
| -0,119935189 | 0,860317825 | CCDC73 | FALSE | Downregulated |
| -0,129588712 | 0,860317825 | FNIP2 | FALSE | Downregulated |
| -0,074384192 | 0,860317825 | PUS3 | FALSE | Downregulated |
| -0,039500354 | 0,860317825 | TEX261 | FALSE | Downregulated |
| 0,16556968 | 0,860317825 | WDR17 | FALSE | Upregulated |
| 0,053835101 | 0,860318941 | COQ4 | FALSE | Upregulated |
| 0,235064516 | 0,860318941 | CPXM2 | FALSE | Upregulated |
| -0,286221997 | 0,860318941 | LINC00907 | FALSE | Downregulated |
| 0,314609056 | 0,860318941 | LOC101928660 | FALSE | Upregulated |
| -0,328169667 | 0,860318941 | LOC729224 | FALSE | Downregulated |
| -0,383130088 | 0,860318941 | PLCD4 | FALSE | Downregulated |
| 0,0508138 | 0,860318941 | RNASEH1 | FALSE | Upregulated |
| -0,281361165 | 0,860318941 | RP11-63P12.6 | FALSE | Downregulated |
| 0,080554429 | 0,860360099 | EIF2AK3 | FALSE | Upregulated |
| 0,064349286 | 0,860854084 | ANO10 | FALSE | Upregulated |

| | | | | |
|---|---|---|---|---|
| 0,188275181 | 0,861275209 | AC011899.9 | FALSE | Upregulated |
| -0,110868327 | 0,861275209 | APOBR | FALSE | Downregulated |
| 0,056717367 | 0,861275209 | ETS1 | FALSE | Upregulated |
| 0,046845881 | 0,861275209 | MTF2 | FALSE | Upregulated |
| -0,05796643 | 0,861275209 | RAE1 | FALSE | Downregulated |
| 0,147678423 | 0,861275209 | RP11-160O5.1 | FALSE | Upregulated |
| 0,053203312 | 0,861275209 | ZNF275 | FALSE | Upregulated |
| 0,086306721 | 0,86128339 | LGALS9 | FALSE | Upregulated |
| -0,073330631 | 0,86128339 | MLEC | FALSE | Downregulated |
| 0,161283981 | 0,86128339 | PTGER2 | FALSE | Upregulated |
| 0,176664737 | 0,861624076 | KIF15 | FALSE | Upregulated |
| -0,178089189 | 0,861624076 | MEGF10 | FALSE | Downregulated |
| 0,129000522 | 0,861624076 | NAT14 | FALSE | Upregulated |
| -0,302917303 | 0,861624076 | NECAB1 | FALSE | Downregulated |
| -0,098911443 | 0,861652035 | RFXAP | FALSE | Downregulated |
| -0,090666047 | 0,861689079 | ENTPD3-AS1 | FALSE | Downregulated |
| -0,207787171 | 0,861689079 | OR7E37P | FALSE | Downregulated |
| -0,102270854 | 0,861689079 | SLC41A1 | FALSE | Downregulated |
| -0,064201854 | 0,861689079 | UBE2D1 | FALSE | Downregulated |
| 0,114970405 | 0,861873255 | GAS5 | FALSE | Upregulated |
| -0,262820265 | 0,861873255 | KRBOX1 | FALSE | Downregulated |
| -0,055886641 | 0,861885912 | ZMPSTE24 | FALSE | Downregulated |
| 0,118062953 | 0,861918965 | MBOAT1 | FALSE | Upregulated |
| -0,077373712 | 0,861918965 | PUS7 | FALSE | Downregulated |
| -0,128913576 | 0,861918965 | RTN4RL2 | FALSE | Downregulated |
| 0,104723807 | 0,86199307 | CD86 | FALSE | Upregulated |

| | | | | |
|---|---|---|---|---|
| -0,067183818 | 0,86199307 | JRKL | FALSE | Downregulated |
| -0,063221143 | 0,86199307 | SERAC1 | FALSE | Downregulated |
| -0,089190456 | 0,86199307 | STX3 | FALSE | Downregulated |
| -0,267564856 | 0,86199307 | TCF15 | FALSE | Downregulated |
| -0,066549476 | 0,862137605 | DTNB | FALSE | Downregulated |
| -0,18298766 | 0,862203969 | KCNJ16 | FALSE | Downregulated |
| -0,167323231 | 0,862203969 | LINC01127 | FALSE | Downregulated |
| -0,076541141 | 0,862246553 | ACAA1 | FALSE | Downregulated |
| -0,328382039 | 0,862246553 | EDIL3 | FALSE | Downregulated |
| -0,097015716 | 0,862337983 | ACMSD | FALSE | Downregulated |
| -0,129724416 | 0,862337983 | DPF3 | FALSE | Downregulated |
| -0,142765688 | 0,862337983 | VAT1L | FALSE | Downregulated |
| -0,106603468 | 0,862356281 | ZNF446 | FALSE | Downregulated |
| -0,052211751 | 0,862412147 | NEDD1 | FALSE | Downregulated |
| -0,245934494 | 0,862613432 | CCDC81 | FALSE | Downregulated |
| -0,312216821 | 0,862613432 | LOC101929819 | FALSE | Downregulated |
| -0,188639112 | 0,862721535 | C1orf51 | FALSE | Downregulated |
| -0,035471348 | 0,862721535 | CUL2 | FALSE | Downregulated |
| 0,171702087 | 0,862721535 | LOC101927105 | FALSE | Upregulated |
| 0,082120587 | 0,862721535 | NSUN3 | FALSE | Upregulated |
| -0,115317655 | 0,862721535 | PSMD5-AS1 | FALSE | Downregulated |
| -0,105604379 | 0,862721535 | SLC17A4 | FALSE | Downregulated |
| 0,313636894 | 0,862721535 | SYCE2 | FALSE | Upregulated |
| 0,181684585 | 0,862765685 | CRTAM | FALSE | Upregulated |
| -0,066578197 | 0,862788134 | C11orf95 | FALSE | Downregulated |
| -0,116573866 | 0,862788134 | FTCDNL1 | FALSE | Downregulated |

| | | | | |
|---|---|---|---|---|
| -0,342397839 | 0,862788134 | LOC102724028 | FALSE | Downregulated |
| -0,333328989 | 0,862788134 | LOC392364 | FALSE | Downregulated |
| -0,05499948 | 0,862788134 | MEMO1 | FALSE | Downregulated |
| 0,087480621 | 0,862907706 | MBL2 | FALSE | Upregulated |
| 0,041526023 | 0,862907706 | PSMB2 | FALSE | Upregulated |
| 0,105817631 | 0,863009187 | EPB41L3 | FALSE | Upregulated |
| -0,038759541 | 0,863009187 | GABPA | FALSE | Downregulated |
| 0,457348941 | 0,863009187 | MSTO2P | FALSE | Upregulated |
| -0,071925393 | 0,863009187 | RALGPS2 | FALSE | Downregulated |
| 0,153476928 | 0,863051237 | ADAM19 | FALSE | Upregulated |
| 0,102418046 | 0,863051237 | ARHGAP30 | FALSE | Upregulated |
| 0,132761971 | 0,863051237 | C3orf67 | FALSE | Upregulated |
| -0,192472502 | 0,863051237 | CAPN12 | FALSE | Downregulated |
| -0,104230796 | 0,863051237 | CC2D2A | FALSE | Downregulated |
| 0,18977815 | 0,863051237 | CELSR3 | FALSE | Upregulated |
| -0,51334865 | 0,863051237 | CLEC18A | FALSE | Downregulated |
| 0,147141849 | 0,863051237 | COLQ | FALSE | Upregulated |
| 0,099295173 | 0,863051237 | CRY1 | FALSE | Upregulated |
| -0,027009782 | 0,863051237 | DDX24 | FALSE | Downregulated |
| -0,049462613 | 0,863051237 | DOCK7 | FALSE | Downregulated |
| -0,052085735 | 0,863051237 | ILF2 | FALSE | Downregulated |
| -0,588575786 | 0,863051237 | JSRP1 | FALSE | Downregulated |
| -0,06843457 | 0,863051237 | LGR4 | FALSE | Downregulated |
| -0,074864528 | 0,863051237 | MCC | FALSE | Downregulated |
| -0,073011465 | 0,863051237 | SORL1 | FALSE | Downregulated |
| -0,035491409 | 0,863051237 | TAF5L | FALSE | Downregulated |

| | | | | |
|---|---|---|---|---|
| 0,394350946 | 0,863051237 | TRIM9 | FALSE | Upregulated |
| -0,041697052 | 0,863051237 | ZNF252P | FALSE | Downregulated |
| 0,078237073 | 0,863686805 | ARPC1B | FALSE | Upregulated |
| -0,115004893 | 0,863686805 | ASPG | FALSE | Downregulated |
| -0,112198188 | 0,863686805 | DYNC2H1 | FALSE | Downregulated |
| 0,044663123 | 0,863686805 | EIF3K | FALSE | Upregulated |
| -0,086016574 | 0,863686805 | IL17RC | FALSE | Downregulated |
| 0,109382181 | 0,863686805 | LOC100506714 | FALSE | Upregulated |
| 0,095305067 | 0,863686805 | PSMB9 | FALSE | Upregulated |
| -0,189531664 | 0,863686805 | SNHG9 | FALSE | Downregulated |
| -0,055149643 | 0,863686805 | USPL1 | FALSE | Downregulated |
| 0,062213946 | 0,863686805 | WWC3 | FALSE | Upregulated |
| 0,080586178 | 0,863686805 | ZNF584 | FALSE | Upregulated |
| -0,044187008 | 0,863817374 | ANKRD17 | FALSE | Downregulated |
| -0,203401585 | 0,863817374 | NACA2 | FALSE | Downregulated |
| 0,067391873 | 0,863832196 | TCEANC2 | FALSE | Upregulated |
| -0,058613329 | 0,863911231 | APOL1 | FALSE | Downregulated |
| -0,068754706 | 0,863914008 | CBWD2 | FALSE | Downregulated |
| 0,211920046 | 0,863914008 | SLC6A9 | FALSE | Upregulated |
| 0,276836937 | 0,863914008 | TECTB | FALSE | Upregulated |
| -0,439265024 | 0,863914008 | TMEM191A | FALSE | Downregulated |
| -0,095400887 | 0,863914008 | ZNF630 | FALSE | Downregulated |
| 0,048210066 | 0,863971871 | MEPCE | FALSE | Upregulated |
| 0,051924789 | 0,864274249 | ASPH | FALSE | Upregulated |
| 0,274606826 | 0,864274249 | BCAS1 | FALSE | Upregulated |
| 0,063103682 | 0,864274249 | CTNNBIP1 | FALSE | Upregulated |

| | | | | |
|---|---|---|---|---|
| -0,036712797 | 0,864274249 | FAM192A | FALSE | Downregulated |
| -0,290407809 | 0,864274249 | FXYD7 | FALSE | Downregulated |
| -0,075040256 | 0,864274249 | GATM | FALSE | Downregulated |
| 0,261262681 | 0,864274249 | LYPD5 | FALSE | Upregulated |
| 0,082731883 | 0,864274249 | PVR | FALSE | Upregulated |
| 0,042699252 | 0,864436381 | PTPN1 | FALSE | Upregulated |
| -0,262374647 | 0,864453844 | COL4A6 | FALSE | Downregulated |
| -0,053132403 | 0,864647413 | ELP4 | FALSE | Downregulated |
| -0,045664216 | 0,864647413 | POLR2D | FALSE | Downregulated |
| 0,156337227 | 0,864647413 | TRPC1 | FALSE | Upregulated |
| 0,042512663 | 0,864655264 | RNF169 | FALSE | Upregulated |
| -0,253024007 | 0,864660433 | CNGA4 | FALSE | Downregulated |
| 0,063481665 | 0,864660433 | ENTPD5 | FALSE | Upregulated |
| -0,151786304 | 0,864660433 | EPB41L1 | FALSE | Downregulated |
| -0,367125775 | 0,864660433 | JPH1 | FALSE | Downregulated |
| 0,10403621 | 0,864660433 | MYO5A | FALSE | Upregulated |
| -0,048199513 | 0,864660433 | NR5A2 | FALSE | Downregulated |
| -0,235205053 | 0,864660433 | NYAP1 | FALSE | Downregulated |
| 0,038895776 | 0,864660433 | PARN | FALSE | Upregulated |
| 0,127317038 | 0,864660433 | PDE9A | FALSE | Upregulated |
| 0,047838926 | 0,864660433 | PSMC2 | FALSE | Upregulated |
| 0,082911333 | 0,864660433 | PTRHD1 | FALSE | Upregulated |
| 0,091181353 | 0,864660433 | SRP14-AS1 | FALSE | Upregulated |
| 0,130159121 | 0,864660433 | SYT12 | FALSE | Upregulated |
| -0,050259954 | 0,864660433 | TMEM192 | FALSE | Downregulated |
| -0,17628958 | 0,864660433 | WTIP | FALSE | Downregulated |

| | | | | |
|---|---|---|---|---|
| -0,04396851 | 0,864660433 | ZFAND6 | FALSE | Downregulated |
| -0,165152859 | 0,864722506 | GSTM2 | FALSE | Downregulated |
| -0,036897508 | 0,864808016 | CASP3 | FALSE | Downregulated |
| 0,070076625 | 0,864808016 | MUTYH | FALSE | Upregulated |
| -0,088179778 | 0,864808016 | PSPH | FALSE | Downregulated |
| 0,417665161 | 0,864808016 | TMEM130 | FALSE | Upregulated |
| -0,033611887 | 0,865136344 | EIF2S3 | FALSE | Downregulated |
| 0,277193784 | 0,865136344 | IDI2-AS1 | FALSE | Upregulated |
| 0,150904966 | 0,865136344 | LAG3 | FALSE | Upregulated |
| 0,092479828 | 0,865136344 | NAV1 | FALSE | Upregulated |
| -0,045882654 | 0,865136344 | NR2C2 | FALSE | Downregulated |
| 0,064481929 | 0,865136344 | SNAP47 | FALSE | Upregulated |
| 0,048545617 | 0,865136344 | SUCLG1 | FALSE | Upregulated |
| -0,036916459 | 0,865136344 | SUPT6H | FALSE | Downregulated |
| 0,272616817 | 0,865136344 | TIAM2 | FALSE | Upregulated |
| -0,175312999 | 0,865136344 | ZNF667-AS1 | FALSE | Downregulated |
| 0,165465476 | 0,86520628 | UBE2T | FALSE | Upregulated |
| 0,250766319 | 0,86544614 | ERMN | FALSE | Upregulated |
| 0,08136199 | 0,86544614 | SLC38A2 | FALSE | Upregulated |
| -0,099697945 | 0,86544614 | YPEL2 | FALSE | Downregulated |
| -0,189610677 | 0,865582276 | LOC101927850 | FALSE | Downregulated |
| -0,081115696 | 0,866254303 | PTS | FALSE | Downregulated |
| 0,056165771 | 0,866254303 | USE1 | FALSE | Upregulated |
| -0,159626547 | 0,866296991 | CCDC168 | FALSE | Downregulated |
| -0,181185259 | 0,866296991 | DBH | FALSE | Downregulated |
| 0,445720328 | 0,866296991 | DSC3 | FALSE | Upregulated |

| | | | | |
|---|---|---|---|---|
| -0,16554215 | 0,866296991 | ELF3 | FALSE | Downregulated |
| 0,149366628 | 0,866296991 | MAGEE1 | FALSE | Upregulated |
| -0,124729808 | 0,866296991 | S1PR2 | FALSE | Downregulated |
| -0,325672718 | 0,866296991 | ST7-OT3 | FALSE | Downregulated |
| -0,087157743 | 0,866296991 | ZNF582 | FALSE | Downregulated |
| -0,048310097 | 0,866306279 | KCTD3 | FALSE | Downregulated |
| 0,051219706 | 0,866306279 | NAGA | FALSE | Upregulated |
| -0,268679847 | 0,866438823 | CBLN1 | FALSE | Downregulated |
| -0,526678556 | 0,866438823 | LOC101928589 | FALSE | Downregulated |
| 0,203072988 | 0,866438823 | MLIP | FALSE | Upregulated |
| -0,246954517 | 0,866438823 | NAALADL1 | FALSE | Downregulated |
| 0,046265448 | 0,866438823 | NIPA2 | FALSE | Upregulated |
| 0,117046315 | 0,866438823 | NLRC4 | FALSE | Upregulated |
| 0,073344484 | 0,866438823 | PPT2 | FALSE | Upregulated |
| 0,063042405 | 0,866438823 | RPLP2 | FALSE | Upregulated |
| 0,104265639 | 0,866438823 | SPIN4 | FALSE | Upregulated |
| 0,052358801 | 0,866438823 | ZNF438 | FALSE | Upregulated |
| 0,078296483 | 0,866557102 | ELK3 | FALSE | Upregulated |
| 0,206219939 | 0,866557102 | NKD2 | FALSE | Upregulated |
| 0,054208082 | 0,866565199 | ZNF343 | FALSE | Upregulated |
| 0,132661951 | 0,866589464 | C19orf80 | FALSE | Upregulated |
| 0,323028762 | 0,866589464 | HIATL2 | FALSE | Upregulated |
| 0,05883338 | 0,866589464 | HSDL1 | FALSE | Upregulated |
| -0,308621559 | 0,866589464 | LOC102724092 | FALSE | Downregulated |
| -0,173608429 | 0,866589464 | PILRB | FALSE | Downregulated |
| 0,275748376 | 0,866589464 | STX19 | FALSE | Upregulated |

| | | | | |
|---|---|---|---|---|
| -0,104364189 | 0,866718722 | LINC00910 | FALSE | Downregulated |
| -0,057086677 | 0,867060912 | CLUAP1 | FALSE | Downregulated |
| -0,093861812 | 0,867603545 | LOC101929786 | FALSE | Downregulated |
| 0,035779005 | 0,867603545 | THUMPD3 | FALSE | Upregulated |
| 0,100293159 | 0,86760526 | ADRBK2 | FALSE | Upregulated |
| -0,140790775 | 0,86760526 | USP3-AS1 | FALSE | Downregulated |
| -0,495429183 | 0,867647882 | LINC00982 | FALSE | Downregulated |
| 0,084950625 | 0,867729738 | ZNF283 | FALSE | Upregulated |
| 0,217926516 | 0,867785187 | CLEC4E | FALSE | Upregulated |
| 0,131146831 | 0,867785187 | COL5A1 | FALSE | Upregulated |
| -0,061986334 | 0,867785187 | GOLGB1 | FALSE | Downregulated |
| -0,10595947 | 0,867785187 | HOXA4 | FALSE | Downregulated |
| -0,050084142 | 0,867785187 | LINC00998 | FALSE | Downregulated |
| -0,249095187 | 0,867785187 | LOC101927369 | FALSE | Downregulated |
| -0,19906967 | 0,867785187 | LRCH2 | FALSE | Downregulated |
| -0,073304609 | 0,867785187 | MRPL53 | FALSE | Downregulated |
| -0,58828251 | 0,867785187 | PLP1 | FALSE | Downregulated |
| -0,086981393 | 0,867785187 | PRICKLE3 | FALSE | Downregulated |
| 0,04879047 | 0,867785187 | RSPH3 | FALSE | Upregulated |
| 0,075659535 | 0,867785187 | SELENBP1 | FALSE | Upregulated |
| -0,046113053 | 0,867785187 | TAF12 | FALSE | Downregulated |
| 0,233658751 | 0,867785187 | UGT2B17 | FALSE | Upregulated |
| -0,058801174 | 0,867785187 | ZNF136 | FALSE | Downregulated |
| -0,325099137 | 0,867785187 | ZNF385D | FALSE | Downregulated |
| 0,044202783 | 0,867875038 | ATP6V0A2 | FALSE | Upregulated |
| -0,068811001 | 0,867875038 | TPP1 | FALSE | Downregulated |

| | | | | |
|---|---|---|---|---|
| 0,064468184 | 0,867875038 | ZNF555 | FALSE | Upregulated |
| 0,068267645 | 0,867958357 | DEXI | FALSE | Upregulated |
| 0,033467746 | 0,867958357 | FAM175B | FALSE | Upregulated |
| 0,042962861 | 0,867958357 | KIAA0947 | FALSE | Upregulated |
| -0,092609704 | 0,867958357 | LIPT2 | FALSE | Downregulated |
| 0,316479736 | 0,867958357 | MAGI1-IT1 | FALSE | Upregulated |
| 0,089685545 | 0,867958357 | RARRES3 | FALSE | Upregulated |
| -0,050203563 | 0,867958357 | ZMYND8 | FALSE | Downregulated |
| 0,256489761 | 0,867958357 | ZSCAN23 | FALSE | Upregulated |
| -0,055668088 | 0,868105764 | CENPBD1P1 | FALSE | Downregulated |
| 0,315522764 | 0,868105764 | LOC729652 | FALSE | Upregulated |
| 0,089850684 | 0,868105764 | MAP3K10 | FALSE | Upregulated |
| 0,042833439 | 0,868105764 | NUFIP2 | FALSE | Upregulated |
| -0,035143145 | 0,868105764 | OXSR1 | FALSE | Downregulated |
| 0,04674862 | 0,868105764 | PAPSS2 | FALSE | Upregulated |
| 0,049645946 | 0,868105764 | SHMT2 | FALSE | Upregulated |
| 0,09471094 | 0,868105764 | ZNF461 | FALSE | Upregulated |
| 0,038965521 | 0,868347546 | CD46 | FALSE | Upregulated |
| -0,072123899 | 0,868347546 | LOC100506746 | FALSE | Downregulated |
| 0,078562282 | 0,868347546 | MRPS12 | FALSE | Upregulated |
| -0,128713506 | 0,868347546 | TNRC18P1 | FALSE | Downregulated |
| -0,225526904 | 0,868388279 | BCL2L14 | FALSE | Downregulated |
| 0,101202864 | 0,868388279 | C4orf46 | FALSE | Upregulated |
| -0,203648364 | 0,868388279 | EPHB1 | FALSE | Downregulated |
| 0,092872064 | 0,868388279 | GLUL | FALSE | Upregulated |
| 0,287842954 | 0,868388279 | LOC102724806 | FALSE | Upregulated |

| | | | | |
|---|---|---|---|---|
| -0,248268795 | 0,868388279 | LOC407835 | FALSE | Downregulated |
| -0,063967982 | 0,868388279 | STEAP3 | FALSE | Downregulated |
| 0,044855149 | 0,868388279 | ZBTB6 | FALSE | Upregulated |
| 0,085482338 | 0,868422694 | RDH13 | FALSE | Upregulated |
| 0,206380198 | 0,868422694 | SLC1A3 | FALSE | Upregulated |
| 0,077098184 | 0,868526261 | DYRK1B | FALSE | Upregulated |
| -0,067807411 | 0,868526261 | ZBTB39 | FALSE | Downregulated |
| 0,109999682 | 0,868542432 | GK | FALSE | Upregulated |
| 0,067287459 | 0,868542432 | RPSA | FALSE | Upregulated |
| -0,060133899 | 0,868577356 | ABAT | FALSE | Downregulated |
| 0,22063922 | 0,868738631 | BRINP2 | FALSE | Upregulated |
| -0,17071786 | 0,868738631 | FAT3 | FALSE | Downregulated |
| -0,047693816 | 0,868738631 | INTS6 | FALSE | Downregulated |
| -0,050679878 | 0,868738631 | ZC3H10 | FALSE | Downregulated |
| -0,066576415 | 0,868769851 | RAB4B | FALSE | Downregulated |
| -0,40954481 | 0,868895196 | BARX1 | FALSE | Downregulated |
| 0,032188286 | 0,868906438 | LYRM2 | FALSE | Upregulated |
| 0,0332257 | 0,868906438 | STAM2 | FALSE | Upregulated |
| 0,038278035 | 0,868913727 | BPNT1 | FALSE | Upregulated |
| -0,094904083 | 0,869097112 | TCF4 | FALSE | Downregulated |
| -0,103599644 | 0,869097112 | ZNF107 | FALSE | Downregulated |
| 0,054739049 | 0,869195521 | C12orf44 | FALSE | Upregulated |
| 0,200583986 | 0,869236603 | LOC101927330 | FALSE | Upregulated |
| -0,0467534 | 0,869263054 | KLHL2 | FALSE | Downregulated |
| -0,047121441 | 0,869621974 | CWF19L2 | FALSE | Downregulated |
| 0,05595298 | 0,869621974 | DALRD3 | FALSE | Upregulated |

| | | | | |
|---|---|---|---|---|
| 0,250535783 | 0,869621974 | KIAA1614 | FALSE | Upregulated |
| -0,062539418 | 0,869644089 | AFAP1L1 | FALSE | Downregulated |
| 0,157484359 | 0,869644089 | CCNE1 | FALSE | Upregulated |
| 0,047117788 | 0,869644089 | CD47 | FALSE | Upregulated |
| 0,039287355 | 0,869644089 | FAM122A | FALSE | Upregulated |
| -0,166788055 | 0,869644089 | GUCA1B | FALSE | Downregulated |
| 0,312287096 | 0,869644089 | KNDC1 | FALSE | Upregulated |
| -0,056578122 | 0,869644089 | LAMTOR5 | FALSE | Downregulated |
| -0,148721535 | 0,869644089 | LOC101927216 | FALSE | Downregulated |
| -0,057129856 | 0,869644089 | MASP2 | FALSE | Downregulated |
| 0,0652741 | 0,869644089 | METTL8 | FALSE | Upregulated |
| -0,030890266 | 0,869644089 | NRBP1 | FALSE | Downregulated |
| 0,037835526 | 0,869644089 | RBM15B | FALSE | Upregulated |
| -0,219336052 | 0,869644089 | SUGT1P1 | FALSE | Downregulated |
| -0,043239693 | 0,869644089 | UTP23 | FALSE | Downregulated |
| -0,036605361 | 0,869644089 | WDR5 | FALSE | Downregulated |
| 0,076170751 | 0,869644089 | ZNF765 | FALSE | Upregulated |
| 0,201898021 | 0,869705731 | BCL11A | FALSE | Upregulated |
| -0,048744518 | 0,869705731 | PIGF | FALSE | Downregulated |
| 0,260592319 | 0,869761191 | LAMB2P1 | FALSE | Upregulated |
| -0,208183829 | 0,869761191 | LOC100505912 | FALSE | Downregulated |
| 0,06613627 | 0,869761191 | TSTA3 | FALSE | Upregulated |
| 0,237590995 | 0,869912302 | ICA1L | FALSE | Upregulated |
| 0,097531634 | 0,869912302 | KLF11 | FALSE | Upregulated |
| -0,124287829 | 0,870149891 | MGC32805 | FALSE | Downregulated |
| -0,047046162 | 0,870149891 | OSGIN2 | FALSE | Downregulated |

| | | | | |
|---|---|---|---|---|
| -0,190408518 | 0,870213621 | DRD1 | FALSE | Downregulated |
| -0,257434979 | 0,870213621 | KCTD14 | FALSE | Downregulated |
| -0,068699532 | 0,870213621 | L3MBTL4 | FALSE | Downregulated |
| -0,185885239 | 0,870213621 | LOC101927057 | FALSE | Downregulated |
| -0,050800708 | 0,870213621 | PDGFC | FALSE | Downregulated |
| 0,056585736 | 0,870213621 | PHLDB3 | FALSE | Upregulated |
| 0,192678342 | 0,870213621 | SIGLEC5 | FALSE | Upregulated |
| 0,074597417 | 0,870213621 | ZNF432 | FALSE | Upregulated |
| -0,104844747 | 0,870228838 | DNAJB5 | FALSE | Downregulated |
| 0,036275122 | 0,870228838 | VPS26A | FALSE | Upregulated |
| -0,055141695 | 0,870244889 | ABCC6 | FALSE | Downregulated |
| -0,362773941 | 0,870244889 | ADCYAP1 | FALSE | Downregulated |
| 0,050704585 | 0,870244889 | AP3B1 | FALSE | Upregulated |
| 0,055572515 | 0,870244889 | AP5M1 | FALSE | Upregulated |
| -0,074760285 | 0,870244889 | C10orf2 | FALSE | Downregulated |
| -0,147579933 | 0,870244889 | C9orf96 | FALSE | Downregulated |
| 0,033974842 | 0,870244889 | CCNC | FALSE | Upregulated |
| -0,255898022 | 0,870244889 | CD300LB | FALSE | Downregulated |
| 0,212749884 | 0,870244889 | DOCK9-AS2 | FALSE | Upregulated |
| -0,244111206 | 0,870244889 | EXTL3-AS1 | FALSE | Downregulated |
| 0,126796171 | 0,870244889 | FAM83D | FALSE | Upregulated |
| -0,103723619 | 0,870244889 | HES1 | FALSE | Downregulated |
| 0,082030009 | 0,870244889 | IFT80 | FALSE | Upregulated |
| 0,17375451 | 0,870244889 | IL3RA | FALSE | Upregulated |
| 0,120497076 | 0,870244889 | ISYNA1 | FALSE | Upregulated |
| -0,195528344 | 0,870244889 | KCNH7 | FALSE | Downregulated |

| | | | | |
|---|---|---|---|---|
| -0,079626758 | 0,870244889 | KCTD7 | FALSE | Downregulated |
| 0,427057135 | 0,870244889 | LOC100133182 | FALSE | Upregulated |
| -0,301613481 | 0,870244889 | LOC101929074 | FALSE | Downregulated |
| 0,266186133 | 0,870244889 | LOC400553 | FALSE | Upregulated |
| -0,131768058 | 0,870244889 | LOC728024 | FALSE | Downregulated |
| -0,159070291 | 0,870244889 | LRRC56 | FALSE | Downregulated |
| -0,095456356 | 0,870244889 | LTB4R | FALSE | Downregulated |
| 0,048367413 | 0,870244889 | MAPRE2 | FALSE | Upregulated |
| -0,059797621 | 0,870244889 | MED18 | FALSE | Downregulated |
| -0,054606147 | 0,870244889 | MEF2A | FALSE | Downregulated |
| 0,634279287 | 0,870244889 | MEF2BNB-MEF2B | FALSE | Upregulated |
| 0,039722676 | 0,870244889 | MFSD1 | FALSE | Upregulated |
| -0,05577928 | 0,870244889 | MKNK2 | FALSE | Downregulated |
| 0,062937633 | 0,870244889 | NDST2 | FALSE | Upregulated |
| -0,078149033 | 0,870244889 | NEK3 | FALSE | Downregulated |
| 0,035499195 | 0,870244889 | NUP214 | FALSE | Upregulated |
| -0,047717074 | 0,870244889 | PAAF1 | FALSE | Downregulated |
| -0,073232158 | 0,870244889 | PEMT | FALSE | Downregulated |
| 0,319631413 | 0,870244889 | PWP2 | FALSE | Upregulated |
| 0,076292788 | 0,870244889 | RAB17 | FALSE | Upregulated |
| 0,099907294 | 0,870244889 | RAD54B | FALSE | Upregulated |
| -0,148470571 | 0,870244889 | SLITRK6 | FALSE | Downregulated |
| -0,068494055 | 0,870244889 | SRR | FALSE | Downregulated |
| -0,107308638 | 0,870244889 | SYT1 | FALSE | Downregulated |
| 0,149105707 | 0,870244889 | TBX21 | FALSE | Upregulated |
| -0,047287204 | 0,870244889 | TMEM261 | FALSE | Downregulated |

| | | | | |
|---|---|---|---|---|
| -0,058640813 | 0,870244889 | TPT1 | FALSE | Downregulated |
| 0,422968621 | 0,870244889 | TXLNG2P | FALSE | Upregulated |
| 0,039869145 | 0,870244889 | UBAP2 | FALSE | Upregulated |
| 0,084473955 | 0,870244889 | WASH3P | FALSE | Upregulated |
| -0,064041583 | 0,870244889 | ZNF566 | FALSE | Downregulated |
| -0,459010003 | 0,870244889 | ZNF695 | FALSE | Downregulated |
| -0,049801951 | 0,870279456 | CHMP4A | FALSE | Downregulated |
| 0,06241779 | 0,870294736 | MZT2A | FALSE | Upregulated |
| 0,23620272 | 0,870442393 | IL1RL1 | FALSE | Upregulated |
| -0,180844976 | 0,870442393 | KCNQ4 | FALSE | Downregulated |
| 0,145381041 | 0,870442393 | PRTFDC1 | FALSE | Upregulated |
| -0,044438395 | 0,870442393 | SPAST | FALSE | Downregulated |
| -0,178073833 | 0,870528757 | ARHGEF19 | FALSE | Downregulated |
| -0,063155661 | 0,870528757 | GGCT | FALSE | Downregulated |
| 0,138970905 | 0,870528757 | LOC100506922 | FALSE | Upregulated |
| -0,134738118 | 0,870528757 | NREP | FALSE | Downregulated |
| -0,025436872 | 0,870528757 | POLR2B | FALSE | Downregulated |
| 0,071650176 | 0,870528757 | PRADC1 | FALSE | Upregulated |
| -0,046192868 | 0,870528757 | PRMT1 | FALSE | Downregulated |
| -0,193235947 | 0,870528757 | PRRT2 | FALSE | Downregulated |
| 0,269585046 | 0,870528757 | RP5-977B1.7 | FALSE | Upregulated |
| -0,065015028 | 0,870528757 | SEC61B | FALSE | Downregulated |
| -0,179185685 | 0,870579186 | MS4A14 | FALSE | Downregulated |
| 0,095682196 | 0,870640853 | MS4A7 | FALSE | Upregulated |
| -0,147047895 | 0,870656445 | CD200R1 | FALSE | Downregulated |
| 0,069141505 | 0,870656445 | CFB | FALSE | Upregulated |

| | | | | |
|---|---|---|---|---|
| 0,141779646 | 0,870656445 | LOC100133091 | FALSE | Upregulated |
| 0,059715362 | 0,870656445 | RBCK1 | FALSE | Upregulated |
| -0,050747439 | 0,870656445 | SLC3A2 | FALSE | Downregulated |
| -0,101683347 | 0,870656445 | VSIG10L | FALSE | Downregulated |
| -0,108717249 | 0,870656445 | WFDC1 | FALSE | Downregulated |
| 0,082791919 | 0,870656445 | ZNF653 | FALSE | Upregulated |
| 0,418556683 | 0,870678984 | MSTN | FALSE | Upregulated |
| 0,068445194 | 0,870739582 | AMER1 | FALSE | Upregulated |
| -0,094450087 | 0,870739582 | APOBEC3F | FALSE | Downregulated |
| 0,073300675 | 0,870739582 | C15orf41 | FALSE | Upregulated |
| 0,101968006 | 0,870739582 | CKAP2 | FALSE | Upregulated |
| -0,204700394 | 0,870739582 | FAM86B1 | FALSE | Downregulated |
| 0,071722453 | 0,870739582 | GCH1 | FALSE | Upregulated |
| 0,062243633 | 0,870739582 | HNRNPAB | FALSE | Upregulated |
| 0,109590279 | 0,870739582 | ISLR | FALSE | Upregulated |
| -0,031057519 | 0,870739582 | KARS | FALSE | Downregulated |
| -0,201351694 | 0,870739582 | MLF1 | FALSE | Downregulated |
| -0,028784264 | 0,870739582 | NFYC | FALSE | Downregulated |
| -0,053467321 | 0,870739582 | PTMA | FALSE | Downregulated |
| -0,147884967 | 0,870739582 | QRICH2 | FALSE | Downregulated |
| 0,266521243 | 0,870739582 | TLR9 | FALSE | Upregulated |
| 0,340113616 | 0,870739582 | TMPRSS4 | FALSE | Upregulated |
| 0,081793975 | 0,870739582 | ZNF600 | FALSE | Upregulated |
| 0,058011281 | 0,870739582 | ZNF720 | FALSE | Upregulated |
| 0,187949528 | 0,870819723 | RAB39A | FALSE | Upregulated |
| 0,707144177 | 0,871014977 | GPR156 | FALSE | Upregulated |

| | | | | |
|---|---|---|---|---|
| 0,149371369 | 0,871056115 | THSD1 | FALSE | Upregulated |
| -0,134217153 | 0,871290173 | AC022007.5 | FALSE | Downregulated |
| 0,071149156 | 0,871290173 | BAHCC1 | FALSE | Upregulated |
| 0,06858776 | 0,871290173 | COMMD10 | FALSE | Upregulated |
| -0,136119093 | 0,871290173 | DNLZ | FALSE | Downregulated |
| -0,075642638 | 0,871290173 | DPYS | FALSE | Downregulated |
| 0,063767102 | 0,871290173 | DUSP16 | FALSE | Upregulated |
| -0,048017451 | 0,871290173 | ECE1 | FALSE | Downregulated |
| 0,12562718 | 0,871290173 | EFCAB4A | FALSE | Upregulated |
| -0,052525669 | 0,871290173 | FARSB | FALSE | Downregulated |
| 0,251086831 | 0,871290173 | FLJ44635 | FALSE | Upregulated |
| 0,056470686 | 0,871290173 | FRMD4A | FALSE | Upregulated |
| -0,640986793 | 0,871290173 | FTH1P18 | FALSE | Downregulated |
| 0,035728805 | 0,871290173 | GOSR1 | FALSE | Upregulated |
| 0,190926517 | 0,871290173 | GPR174 | FALSE | Upregulated |
| 0,07195649 | 0,871290173 | GRAP | FALSE | Upregulated |
| -0,111301738 | 0,871290173 | GRHL1 | FALSE | Downregulated |
| 0,201838916 | 0,871290173 | HCAR3 | FALSE | Upregulated |
| -0,067854751 | 0,871290173 | HYI | FALSE | Downregulated |
| 0,112032529 | 0,871290173 | KAT2A | FALSE | Upregulated |
| 0,20630043 | 0,871290173 | KCNB1 | FALSE | Upregulated |
| -0,055316073 | 0,871290173 | KDM5B | FALSE | Downregulated |
| 0,070525028 | 0,871290173 | LIMS2 | FALSE | Upregulated |
| -0,150817911 | 0,871290173 | LOC101927258 | FALSE | Downregulated |
| 0,317645547 | 0,871290173 | LOC102723756 | FALSE | Upregulated |
| 0,04631815 | 0,871290173 | LRBA | FALSE | Upregulated |

| | | | | |
|---|---|---|---|---|
| -0,182923561 | 0,871290173 | MAN1B1-AS1 | FALSE | Downregulated |
| -0,049890157 | 0,871290173 | MDM1 | FALSE | Downregulated |
| -0,066827738 | 0,871290173 | METAP1D | FALSE | Downregulated |
| -0,119725686 | 0,871290173 | MGC12916 | FALSE | Downregulated |
| 0,130310433 | 0,871290173 | MGC16275 | FALSE | Upregulated |
| -0,070588972 | 0,871290173 | MYEOV2 | FALSE | Downregulated |
| 0,14545601 | 0,871290173 | NHS | FALSE | Upregulated |
| 0,03540548 | 0,871290173 | PAFAH2 | FALSE | Upregulated |
| -0,108661591 | 0,871290173 | PANX2 | FALSE | Downregulated |
| -0,214159096 | 0,871290173 | PCP2 | FALSE | Downregulated |
| -0,04141232 | 0,871290173 | PDCD10 | FALSE | Downregulated |
| -0,129812635 | 0,871290173 | PRR18 | FALSE | Downregulated |
| -0,153554445 | 0,871290173 | PRRT3 | FALSE | Downregulated |
| -0,025839333 | 0,871290173 | PWP1 | FALSE | Downregulated |
| 0,093189289 | 0,871290173 | RHOQ | FALSE | Upregulated |
| 0,06632901 | 0,871290173 | RND3 | FALSE | Upregulated |
| -0,053491254 | 0,871290173 | RNF139 | FALSE | Downregulated |
| 0,109303675 | 0,871290173 | RNF207 | FALSE | Upregulated |
| 0,174534335 | 0,871290173 | RP11-307E17.8 | FALSE | Upregulated |
| 0,047733677 | 0,871290173 | RPA3-AS1 | FALSE | Upregulated |
| 0,072184019 | 0,871290173 | RPS26 | FALSE | Upregulated |
| -0,072076797 | 0,871290173 | SPRY1 | FALSE | Downregulated |
| 0,209227014 | 0,871290173 | SPTSSB | FALSE | Upregulated |
| -0,121652043 | 0,871290173 | TEN1-CDK3 | FALSE | Downregulated |
| -0,062162176 | 0,871290173 | THRA | FALSE | Downregulated |
| -0,047681123 | 0,871290173 | TOLLIP | FALSE | Downregulated |

| | | | | |
|---|---|---|---|---|
| -0,057622297 | 0,871290173 | TSEN15 | FALSE | Downregulated |
| -0,088215949 | 0,871290173 | VAV3 | FALSE | Downregulated |
| 0,035285305 | 0,871290173 | WDR11 | FALSE | Upregulated |
| -0,046338459 | 0,871290173 | YTHDC2 | FALSE | Downregulated |
| 0,04980352 | 0,871290173 | ZDHHC18 | FALSE | Upregulated |
| -0,05963546 | 0,871290173 | ZNF596 | FALSE | Downregulated |
| 0,086266808 | 0,87150446 | CAMK1D | FALSE | Upregulated |
| 0,534210162 | 0,87150446 | CIDEA | FALSE | Upregulated |
| -0,137363005 | 0,87150446 | FBN1 | FALSE | Downregulated |
| -0,054950516 | 0,871731551 | UBN1 | FALSE | Downregulated |
| -0,275781796 | 0,871975468 | LOC729083 | FALSE | Downregulated |
| -0,062858843 | 0,871980418 | AKR1C4 | FALSE | Downregulated |
| -0,179508611 | 0,871980418 | AMPH | FALSE | Downregulated |
| -0,063952035 | 0,871980418 | CXXC5 | FALSE | Downregulated |
| 0,238925658 | 0,871980418 | GCK | FALSE | Upregulated |
| 0,143279997 | 0,871980418 | JAK3 | FALSE | Upregulated |
| -0,261032513 | 0,871980418 | LOC102724026 | FALSE | Downregulated |
| 0,234614263 | 0,871980418 | LYPD6B | FALSE | Upregulated |
| -0,056420682 | 0,871980418 | MRPL52 | FALSE | Downregulated |
| -0,047770387 | 0,871980418 | NR2F2 | FALSE | Downregulated |
| -0,091285074 | 0,871980418 | PDE10A | FALSE | Downregulated |
| 0,105564457 | 0,871980418 | PDF | FALSE | Upregulated |
| -0,067599682 | 0,871980418 | POLK | FALSE | Downregulated |
| -0,048585918 | 0,871980418 | TRMT12 | FALSE | Downregulated |
| 0,08482924 | 0,87199005 | ACACB | FALSE | Upregulated |
| 0,050103183 | 0,87199005 | AKIP1 | FALSE | Upregulated |

| | | | | |
|---|---|---|---|---|
| 0,064756689 | 0,87199005 | ATP7A | FALSE | Upregulated |
| -0,236675341 | 0,87199005 | CCDC64B | FALSE | Downregulated |
| -0,064661599 | 0,87199005 | CDC14B | FALSE | Downregulated |
| 0,090813236 | 0,87199005 | CRIM1 | FALSE | Upregulated |
| -0,048947672 | 0,87199005 | DCP1B | FALSE | Downregulated |
| 0,114900183 | 0,87199005 | EAF2 | FALSE | Upregulated |
| 0,024403943 | 0,87199005 | EIF4G2 | FALSE | Upregulated |
| -0,083385378 | 0,87199005 | FAM161A | FALSE | Downregulated |
| 0,070009322 | 0,87199005 | FBXO8 | FALSE | Upregulated |
| 0,039844922 | 0,87199005 | FCF1 | FALSE | Upregulated |
| 0,081017033 | 0,87199005 | GREB1L | FALSE | Upregulated |
| -0,831133487 | 0,87199005 | GSTT2 | FALSE | Downregulated |
| 0,093836508 | 0,87199005 | HSD11B1L | FALSE | Upregulated |
| 0,368419493 | 0,87199005 | INTS4L1 | FALSE | Upregulated |
| 0,296559173 | 0,87199005 | LOC101928884 | FALSE | Upregulated |
| 0,15382588 | 0,87199005 | LOC101929664 | FALSE | Upregulated |
| 0,147900185 | 0,87199005 | LOC102724872 | FALSE | Upregulated |
| 0,145590962 | 0,87199005 | LOC150776 | FALSE | Upregulated |
| -0,061979431 | 0,87199005 | MAGOHB | FALSE | Downregulated |
| 0,048046387 | 0,87199005 | MTFR1L | FALSE | Upregulated |
| 0,048160683 | 0,87199005 | MTHFD1 | FALSE | Upregulated |
| -0,064353632 | 0,87199005 | MYO1E | FALSE | Downregulated |
| -0,070561367 | 0,87199005 | NCOR1 | FALSE | Downregulated |
| 0,079655245 | 0,87199005 | OTUD1 | FALSE | Upregulated |
| -0,076429773 | 0,87199005 | PRX | FALSE | Downregulated |
| 0,109318518 | 0,87199005 | RND1 | FALSE | Upregulated |

| | | | | |
|---|---|---|---|---|
| 0,037434896 | 0,87199005 | RNF41 | FALSE | Upregulated |
| 0,120327885 | 0,87199005 | SLC12A2 | FALSE | Upregulated |
| 0,047117519 | 0,87199005 | TMF1 | FALSE | Upregulated |
| -0,063450262 | 0,87199005 | TTC33 | FALSE | Downregulated |
| 0,042653762 | 0,87199005 | ZC3H4 | FALSE | Upregulated |
| 0,052480635 | 0,87199005 | ZNF23 | FALSE | Upregulated |
| -0,093156826 | 0,87199005 | ZNF699 | FALSE | Downregulated |
| 0,290434686 | 0,872025815 | TSACC | FALSE | Upregulated |
| -0,188618134 | 0,872238274 | SOCS1 | FALSE | Downregulated |
| -0,279803561 | 0,872423899 | KCNJ11 | FALSE | Downregulated |
| -0,174429706 | 0,872423899 | RP5-968D22.3 | FALSE | Downregulated |
| -0,03559334 | 0,872423899 | STAG1 | FALSE | Downregulated |
| -0,247288667 | 0,872593149 | CYP1B1-AS1 | FALSE | Downregulated |
| -0,067443552 | 0,872593149 | PAFAH1B2 | FALSE | Downregulated |
| 0,084099523 | 0,872641951 | ADAMTS2 | FALSE | Upregulated |
| 0,066818251 | 0,872641951 | PEX11G | FALSE | Upregulated |
| -0,129805206 | 0,872897627 | SLC16A7 | FALSE | Downregulated |
| -0,042490704 | 0,872950336 | SLC35F5 | FALSE | Downregulated |
| 0,19281405 | 0,873211438 | LOC101927929 | FALSE | Upregulated |
| 0,067379834 | 0,873283286 | CDH1 | FALSE | Upregulated |
| 0,032854011 | 0,873283286 | HNRNPUL2 | FALSE | Upregulated |
| -0,674954561 | 0,873283286 | MYOC | FALSE | Downregulated |
| 0,070973071 | 0,873283286 | PCBP4 | FALSE | Upregulated |
| -0,428399496 | 0,873283286 | PPAPDC3 | FALSE | Downregulated |
| -0,263676042 | 0,873283286 | RRN3P2 | FALSE | Downregulated |
| -0,054609719 | 0,873283286 | TMEM50B | FALSE | Downregulated |

| | | | | |
|---|---|---|---|---|
| -0,065617622 | 0,873469549 | FAM110B | FALSE | Downregulated |
| 0,331138143 | 0,873469549 | WNT2B | FALSE | Upregulated |
| 0,044358205 | 0,87355199 | FBXW4 | FALSE | Upregulated |
| 0,037901115 | 0,87355199 | RNF187 | FALSE | Upregulated |
| 0,073111592 | 0,873659384 | LSM2 | FALSE | Upregulated |
| 0,103379069 | 0,873705306 | FGGY | FALSE | Upregulated |
| 0,042240215 | 0,873705306 | PAWR | FALSE | Upregulated |
| -0,21192558 | 0,873705306 | RASSF1-AS1 | FALSE | Downregulated |
| 0,082773193 | 0,873705306 | STAB1 | FALSE | Upregulated |
| 0,068371816 | 0,873896925 | EMILIN1 | FALSE | Upregulated |
| -0,265960535 | 0,873896925 | GRIK5 | FALSE | Downregulated |
| 0,042590175 | 0,873896925 | LARP4 | FALSE | Upregulated |
| -0,166581531 | 0,873896925 | SLC34A1 | FALSE | Downregulated |
| -0,049013419 | 0,873896925 | SLC44A1 | FALSE | Downregulated |
| 0,039601458 | 0,873896925 | STT3B | FALSE | Upregulated |
| -0,060501391 | 0,873896925 | ZNF175 | FALSE | Downregulated |
| -0,810774738 | 0,873896925 | ZNF559-ZNF177 | FALSE | Downregulated |
| -0,193651548 | 0,874104512 | LOC102723422 | FALSE | Downregulated |
| -0,095017394 | 0,874124243 | CKMT2-AS1 | FALSE | Downregulated |
| -0,043153903 | 0,874124243 | NPM1 | FALSE | Downregulated |
| -2,194358632 | 0,874191642 | TRDN | FALSE | Downregulated |
| -0,075115828 | 0,874191642 | UBALD1 | FALSE | Downregulated |
| 0,03892484 | 0,874242426 | STAT5B | FALSE | Upregulated |
| 0,067774636 | 0,874253561 | CA5B | FALSE | Upregulated |
| 0,045664765 | 0,874253561 | LOC100506710 | FALSE | Upregulated |
| -0,07874367 | 0,874253561 | RANGRF | FALSE | Downregulated |

| | | | | |
|---|---|---|---|---|
| -0,184193371 | 0,874253561 | TP63 | FALSE | Downregulated |
| 0,073310963 | 0,874282997 | PLEKHG1 | FALSE | Upregulated |
| -0,036132449 | 0,874618369 | BRD2 | FALSE | Downregulated |
| -0,075756756 | 0,874823567 | BEX4 | FALSE | Downregulated |
| 0,042227807 | 0,874968045 | C6orf106 | FALSE | Upregulated |
| -0,168784918 | 0,874968045 | KIAA1875 | FALSE | Downregulated |
| -0,37705492 | 0,874968045 | LOC102724484 | FALSE | Downregulated |
| -0,231632974 | 0,874968045 | LOC102724569 | FALSE | Downregulated |
| -0,144774686 | 0,874968045 | SSC5D | FALSE | Downregulated |
| 0,298247818 | 0,875064833 | LINC01057 | FALSE | Upregulated |
| 0,332255995 | 0,875076852 | SLC22A24 | FALSE | Upregulated |
| -0,136015385 | 0,875086028 | ZNF703 | FALSE | Downregulated |
| 0,059644001 | 0,875183345 | ISCA2 | FALSE | Upregulated |
| -0,146277605 | 0,875231335 | IL17RE | FALSE | Downregulated |
| -0,076958571 | 0,875310387 | FAM211B | FALSE | Downregulated |
| -0,102490331 | 0,875429146 | JAM2 | FALSE | Downregulated |
| -0,351720807 | 0,87585248 | ZAN | FALSE | Downregulated |
| -0,105628833 | 0,8759707 | LOC100133985 | FALSE | Downregulated |
| 0,381454042 | 0,8759707 | RPPH1 | FALSE | Upregulated |
| -0,121252415 | 0,8759707 | TMEM91 | FALSE | Downregulated |
| -0,082175712 | 0,875991482 | RPS3A | FALSE | Downregulated |
| -0,146638563 | 0,876078881 | APOBEC3D | FALSE | Downregulated |
| -0,043739328 | 0,876078881 | BNIP3 | FALSE | Downregulated |
| -0,055282733 | 0,876078881 | CASP8AP2 | FALSE | Downregulated |
| 0,032785579 | 0,876078881 | FAM168B | FALSE | Upregulated |
| 0,147359839 | 0,876078881 | FASN | FALSE | Upregulated |

| | | | | |
|---|---|---|---|---|
| 0,113888327 | 0,876078881 | GLIPR2 | FALSE | Upregulated |
| 0,074048811 | 0,876078881 | LRMP | FALSE | Upregulated |
| 0,187001461 | 0,876078881 | MB21D2 | FALSE | Upregulated |
| 0,061249593 | 0,876078881 | METTL25 | FALSE | Upregulated |
| 0,058983599 | 0,876078881 | NOM1 | FALSE | Upregulated |
| -0,192533672 | 0,876078881 | RD3L | FALSE | Downregulated |
| -0,15222427 | 0,876078881 | RNF32 | FALSE | Downregulated |
| 0,068049204 | 0,876078881 | TBC1D31 | FALSE | Upregulated |
| 0,05013798 | 0,876078881 | TM7SF3 | FALSE | Upregulated |
| 0,073978971 | 0,876078881 | TPPP | FALSE | Upregulated |
| -0,110029237 | 0,876078881 | TYRO3 | FALSE | Downregulated |
| -0,049606818 | 0,876097852 | IFNAR1 | FALSE | Downregulated |
| -0,425596477 | 0,876331291 | HSFX1 | FALSE | Downregulated |
| 0,259938251 | 0,876583114 | CCDC141 | FALSE | Upregulated |
| -0,036223661 | 0,876616165 | HNRNPUL1 | FALSE | Downregulated |
| 0,122615053 | 0,876616165 | INCA1 | FALSE | Upregulated |
| -0,040408708 | 0,876616165 | TAX1BP1 | FALSE | Downregulated |
| 0,216495057 | 0,876616165 | TMEM63C | FALSE | Upregulated |
| 0,128192789 | 0,876698571 | HEYL | FALSE | Upregulated |
| 0,165242392 | 0,876698571 | PINLYP | FALSE | Upregulated |
| 0,120188803 | 0,876698571 | PPFIA3 | FALSE | Upregulated |
| 0,081293741 | 0,876698571 | TMEM214 | FALSE | Upregulated |
| 0,067830644 | 0,876733613 | EPHB4 | FALSE | Upregulated |
| 0,145489264 | 0,876814591 | CKB | FALSE | Upregulated |
| -0,055917726 | 0,876814591 | KIAA0020 | FALSE | Downregulated |
| -0,22430145 | 0,876814591 | LINC01094 | FALSE | Downregulated |

| | | | | |
|---|---|---|---|---|
| 0,208804992 | 0,876814591 | LOC101927372 | FALSE | Upregulated |
| -0,248950178 | 0,876814591 | SLC13A4 | FALSE | Downregulated |
| 0,049145994 | 0,876814591 | SUV39H1 | FALSE | Upregulated |
| -0,051980046 | 0,876814591 | TMED2 | FALSE | Downregulated |
| -0,041419372 | 0,876852014 | NFIX | FALSE | Downregulated |
| 0,157371596 | 0,877045221 | GOLM1 | FALSE | Upregulated |
| 0,256823461 | 0,87712175 | LRFN4 | FALSE | Upregulated |
| -0,399582994 | 0,87712175 | RP1 | FALSE | Downregulated |
| 0,378871516 | 0,877184595 | ERVW-1 | FALSE | Upregulated |
| -0,074876867 | 0,877220902 | KIAA0895 | FALSE | Downregulated |
| -0,135031614 | 0,877220902 | LBP | FALSE | Downregulated |
| -0,141402007 | 0,877424795 | MGC27382 | FALSE | Downregulated |
| -0,261735589 | 0,877424795 | ZNF177 | FALSE | Downregulated |
| -0,081216045 | 0,877427022 | CALR | FALSE | Downregulated |
| 0,035590604 | 0,877427022 | CANT1 | FALSE | Upregulated |
| -0,035709351 | 0,877427022 | CDC42SE2 | FALSE | Downregulated |
| -0,055405775 | 0,877427022 | CDIPT | FALSE | Downregulated |
| -0,433334375 | 0,877427022 | CKMT2 | FALSE | Downregulated |
| 0,073855818 | 0,877427022 | CPS1 | FALSE | Upregulated |
| 0,053090225 | 0,877427022 | CSRP1 | FALSE | Upregulated |
| 0,043435875 | 0,877427022 | FAM200A | FALSE | Upregulated |
| 0,280683414 | 0,877427022 | FSIP2 | FALSE | Upregulated |
| -0,068265534 | 0,877427022 | GNL3 | FALSE | Downregulated |
| -0,078734849 | 0,877427022 | HABP4 | FALSE | Downregulated |
| 0,04934393 | 0,877427022 | HECA | FALSE | Upregulated |
| -0,063306584 | 0,877427022 | HYLS1 | FALSE | Downregulated |

| | | | | |
|---|---|---|---|---|
| 0,061075926 | 0,877427022 | KDM5C | FALSE | Upregulated |
| -0,119816164 | 0,877427022 | LOC101927942 | FALSE | Downregulated |
| -0,108911116 | 0,877427022 | LOC101928126 | FALSE | Downregulated |
| 0,122958499 | 0,877427022 | LOC102723863 | FALSE | Upregulated |
| -0,10162581 | 0,877427022 | LRRC23 | FALSE | Downregulated |
| -0,238110602 | 0,877427022 | MAP3K14-AS1 | FALSE | Downregulated |
| 0,040385173 | 0,877427022 | MBD3 | FALSE | Upregulated |
| -0,078864055 | 0,877427022 | MIER2 | FALSE | Downregulated |
| -0,093916427 | 0,877427022 | NIPAL1 | FALSE | Downregulated |
| 0,134466231 | 0,877427022 | PCSK5 | FALSE | Upregulated |
| -0,064993703 | 0,877427022 | PIBF1 | FALSE | Downregulated |
| -0,245159043 | 0,877427022 | POMC | FALSE | Downregulated |
| -0,050982396 | 0,877427022 | PPM1B | FALSE | Downregulated |
| 0,026823621 | 0,877427022 | RAB21 | FALSE | Upregulated |
| -0,063602219 | 0,877427022 | SLC27A5 | FALSE | Downregulated |
| 0,189680079 | 0,877427022 | SLC6A8 | FALSE | Upregulated |
| -0,207240212 | 0,877427022 | SMC5-AS1 | FALSE | Downregulated |
| -0,053711271 | 0,877427022 | SRSF5 | FALSE | Downregulated |
| 0,070523524 | 0,877427022 | STAT2 | FALSE | Upregulated |
| -0,050327116 | 0,877427022 | TAF9B | FALSE | Downregulated |
| -0,155986962 | 0,877427022 | TAT | FALSE | Downregulated |
| 0,070481438 | 0,877427022 | VEGFB | FALSE | Upregulated |
| 0,056869064 | 0,877614054 | ACAD11 | FALSE | Upregulated |
| 0,061178242 | 0,877614054 | ALG14 | FALSE | Upregulated |
| 0,032084099 | 0,877614054 | ATXN7L3 | FALSE | Upregulated |
| 0,053398197 | 0,877614054 | C3 | FALSE | Upregulated |

| | | | | |
|---|---|---|---|---|
| 0,031303713 | 0,877614054 | CAMKK2 | FALSE | Upregulated |
| -0,043832877 | 0,877614054 | CDK8 | FALSE | Downregulated |
| -0,042235161 | 0,877614054 | CIRH1A | FALSE | Downregulated |
| -0,056492568 | 0,877614054 | COX14 | FALSE | Downregulated |
| 0,06155539 | 0,877614054 | DTNBP1 | FALSE | Upregulated |
| 0,030001808 | 0,877614054 | EWSR1 | FALSE | Upregulated |
| 0,273348548 | 0,877614054 | FLJ45513 | FALSE | Upregulated |
| 0,074344843 | 0,877614054 | HHEX | FALSE | Upregulated |
| 0,083708155 | 0,877614054 | HOMER3 | FALSE | Upregulated |
| 0,069171107 | 0,877614054 | HOOK1 | FALSE | Upregulated |
| 0,029995079 | 0,877614054 | IMMT | FALSE | Upregulated |
| -0,235052608 | 0,877614054 | KCND2 | FALSE | Downregulated |
| 0,135108273 | 0,877614054 | LIMD1-AS1 | FALSE | Upregulated |
| -0,314505354 | 0,877614054 | LOC101928044 | FALSE | Downregulated |
| -0,115516567 | 0,877614054 | LOC101929986 | FALSE | Downregulated |
| -0,035466542 | 0,877614054 | MRPL9 | FALSE | Downregulated |
| 0,084714317 | 0,877614054 | MSTO1 | FALSE | Upregulated |
| -0,067961408 | 0,877614054 | OXLD1 | FALSE | Downregulated |
| 0,038837648 | 0,877614054 | PARL | FALSE | Upregulated |
| -0,033244528 | 0,877614054 | PHF10 | FALSE | Downregulated |
| -0,065962716 | 0,877614054 | PPM1H | FALSE | Downregulated |
| 0,043051084 | 0,877614054 | RBM4 | FALSE | Upregulated |
| 0,040851646 | 0,877614054 | RNF113A | FALSE | Upregulated |
| -0,226424886 | 0,877614054 | RP11-261C10.3 | FALSE | Downregulated |
| -0,060186927 | 0,877614054 | RPS10 | FALSE | Downregulated |
| 0,068208215 | 0,877614054 | SERPINF2 | FALSE | Upregulated |

| | | | | |
|---|---|---|---|---|
| 0,287403056 | 0,877614054 | SHC4 | FALSE | Upregulated |
| 0,154434536 | 0,877614054 | SLC16A14 | FALSE | Upregulated |
| 0,039265258 | 0,877614054 | SLC35A5 | FALSE | Upregulated |
| -0,114568375 | 0,877614054 | TCHH | FALSE | Downregulated |
| 0,144245055 | 0,877614054 | TOX | FALSE | Upregulated |
| 0,119125383 | 0,877614054 | TTC18 | FALSE | Upregulated |
| -0,083152475 | 0,877614054 | TUBA4A | FALSE | Downregulated |
| 0,033557429 | 0,877614054 | USP1 | FALSE | Upregulated |
| 0,092772722 | 0,877614054 | UST | FALSE | Upregulated |
| 0,141774516 | 0,877614054 | ZFP69B | FALSE | Upregulated |
| -0,136696107 | 0,877614054 | ZNF519 | FALSE | Downregulated |
| -0,040588224 | 0,877618399 | BTBD3 | FALSE | Downregulated |
| 0,089313787 | 0,87779591 | ABCG1 | FALSE | Upregulated |
| 0,064041637 | 0,87779591 | CASP8 | FALSE | Upregulated |
| -0,081589825 | 0,87779591 | SLCO1B1 | FALSE | Downregulated |
| 0,044243029 | 0,877912766 | UBE2A | FALSE | Upregulated |
| 0,091434602 | 0,877912766 | UBE2S | FALSE | Upregulated |
| 0,13318822 | 0,877953933 | BEST1 | FALSE | Upregulated |
| 0,092516677 | 0,877953933 | C1orf54 | FALSE | Upregulated |
| 0,238932305 | 0,877953933 | CYP27C1 | FALSE | Upregulated |
| -0,195480614 | 0,877953933 | FGF14 | FALSE | Downregulated |
| -0,200320819 | 0,877953933 | RFPL4A | FALSE | Downregulated |
| 0,056150091 | 0,878223414 | ZBTB41 | FALSE | Upregulated |
| 0,430206769 | 0,878449499 | NLGN4Y | FALSE | Upregulated |
| 0,059301632 | 0,878449499 | TXNDC17 | FALSE | Upregulated |
| 0,244942408 | 0,8784847 | B4GALNT3 | FALSE | Upregulated |

| | | | | |
|---|---|---|---|---|
| 0,155440165 | 0,8784847 | DKK3 | FALSE | Upregulated |
| -0,080786508 | 0,8784847 | ECSCR-2 | FALSE | Downregulated |
| -0,047611009 | 0,8784847 | ENTPD4 | FALSE | Downregulated |
| -0,085254355 | 0,8784847 | FGA | FALSE | Downregulated |
| 0,049346238 | 0,8784847 | GFER | FALSE | Upregulated |
| 0,041043643 | 0,8784847 | GMPS | FALSE | Upregulated |
| -0,049588979 | 0,8784847 | NT5C3B | FALSE | Downregulated |
| -0,036739044 | 0,8784847 | PCNP | FALSE | Downregulated |
| -0,037141806 | 0,8784847 | PRKD3 | FALSE | Downregulated |
| -0,03880333 | 0,8784847 | QRSL1 | FALSE | Downregulated |
| 0,049297226 | 0,8784847 | RABIF | FALSE | Upregulated |
| -0,315301906 | 0,8784847 | RNF175 | FALSE | Downregulated |
| -0,093939343 | 0,8784847 | RP1-249H1.4 | FALSE | Downregulated |
| 0,099776145 | 0,8784847 | SEMA3F | FALSE | Upregulated |
| -0,033990553 | 0,8784847 | TALDO1 | FALSE | Downregulated |
| 0,107387837 | 0,87851632 | COL4A1 | FALSE | Upregulated |
| -0,13266047 | 0,878941434 | IZUMO4 | FALSE | Downregulated |
| 0,159524133 | 0,87906402 | CCDC65 | FALSE | Upregulated |
| -0,030966385 | 0,87906402 | GSPT1 | FALSE | Downregulated |
| 0,285686164 | 0,87906402 | LOC101927190 | FALSE | Upregulated |
| -0,076143728 | 0,87906402 | LOC101929145 | FALSE | Downregulated |
| 0,208109321 | 0,87906402 | LOC102724141 | FALSE | Upregulated |
| 0,103553959 | 0,87906402 | LOH12CR2 | FALSE | Upregulated |
| -0,161201222 | 0,87906402 | PLCL1 | FALSE | Downregulated |
| 0,10858661 | 0,87906402 | SIMC1 | FALSE | Upregulated |
| -0,098200006 | 0,87906402 | SLC10A5 | FALSE | Downregulated |

| | | | | |
|---|---|---|---|---|
| -0,032948221 | 0,87906402 | TRAPPC8 | FALSE | Downregulated |
| 0,128149832 | 0,87906402 | ZBTB7C | FALSE | Upregulated |
| 0,184266054 | 0,87908104 | LOC102723647 | FALSE | Upregulated |
| 0,031719621 | 0,879082274 | RNMT | FALSE | Upregulated |
| -0,189879011 | 0,879146648 | LOC101928548 | FALSE | Downregulated |
| -0,055636488 | 0,879146648 | RAD51C | FALSE | Downregulated |
| 0,113745469 | 0,879146648 | SLC17A9 | FALSE | Upregulated |
| -0,492909516 | 0,879218767 | LOC101928195 | FALSE | Downregulated |
| 0,240355883 | 0,879218767 | LOC102724387 | FALSE | Upregulated |
| 0,085164691 | 0,879218767 | PGM2L1 | FALSE | Upregulated |
| 0,04366836 | 0,879218767 | TGOLN2 | FALSE | Upregulated |
| 0,066874618 | 0,879218767 | TRADD | FALSE | Upregulated |
| -0,043373362 | 0,879218767 | XPOT | FALSE | Downregulated |
| -0,154807584 | 0,879399377 | FLJ44342 | FALSE | Downregulated |
| -0,185454116 | 0,879399377 | GULP1 | FALSE | Downregulated |
| 0,206856202 | 0,879399377 | KLRC1 | FALSE | Upregulated |
| 0,080960708 | 0,879399377 | RASSF8-AS1 | FALSE | Upregulated |
| 0,041141111 | 0,879399377 | SNF8 | FALSE | Upregulated |
| 0,226278924 | 0,879455599 | LOC100507634 | FALSE | Upregulated |
| -0,095962242 | 0,879455599 | SLC17A1 | FALSE | Downregulated |
| -0,039356788 | 0,879455599 | YIPF4 | FALSE | Downregulated |
| -0,05084167 | 0,879497418 | DNAJC9 | FALSE | Downregulated |
| -0,1633589 | 0,879497418 | LOC102724571 | FALSE | Downregulated |
| 0,049824914 | 0,87953174 | PYGL | FALSE | Upregulated |
| -0,068042337 | 0,879586763 | RABL6 | FALSE | Downregulated |
| -0,051127477 | 0,879608413 | DOCK1 | FALSE | Downregulated |

| | | | | |
|---|---|---|---|---|
| -0,041257156 | 0,879621914 | C18orf25 | FALSE | Downregulated |
| 0,062763069 | 0,879683583 | BZW1 | FALSE | Upregulated |
| -0,176938988 | 0,879683583 | CYP21A1P | FALSE | Downregulated |
| -0,04685527 | 0,879683583 | DHX36 | FALSE | Downregulated |
| -0,051429177 | 0,879683583 | KLHL28 | FALSE | Downregulated |
| 0,331278457 | 0,879683583 | LINC00348 | FALSE | Upregulated |
| -0,108110272 | 0,879683583 | PKLR | FALSE | Downregulated |
| -0,109241556 | 0,879683583 | PRKAR2A-AS1 | FALSE | Downregulated |
| 0,132258529 | 0,879683583 | PRNCR1 | FALSE | Upregulated |
| -0,066303309 | 0,879683583 | SCAND1 | FALSE | Downregulated |
| -0,04268723 | 0,879683583 | WDR12 | FALSE | Downregulated |
| -0,065514907 | 0,879711807 | FN3K | FALSE | Downregulated |
| -0,175485601 | 0,879711807 | LOC102723780 | FALSE | Downregulated |
| -0,488697791 | 0,879711807 | LOC102724093 | FALSE | Downregulated |
| 0,099942694 | 0,879725129 | LINC01137 | FALSE | Upregulated |
| -0,247318405 | 0,879725129 | LOC102724748 | FALSE | Downregulated |
| 0,068383104 | 0,879725129 | MAGEH1 | FALSE | Upregulated |
| -0,054042913 | 0,879725129 | OAF | FALSE | Downregulated |
| 0,097462957 | 0,879948444 | RCN3 | FALSE | Upregulated |
| 0,237923271 | 0,879951912 | ACPP | FALSE | Upregulated |
| -0,296056925 | 0,879951912 | CLDN22 | FALSE | Downregulated |
| -0,065065273 | 0,879951912 | DUSP5 | FALSE | Downregulated |
| -0,036419106 | 0,879951912 | KDM2A | FALSE | Downregulated |
| -0,039806018 | 0,879951912 | LMBRD1 | FALSE | Downregulated |
| -0,070817881 | 0,879951912 | N4BP2L1 | FALSE | Downregulated |
| -0,046818141 | 0,879951912 | NT5DC1 | FALSE | Downregulated |

| | | | | |
|---|---|---|---|---|
| -0,248122116 | 0,879951912 | PFN1P2 | FALSE | Downregulated |
| -0,098818238 | 0,879951912 | RHOD | FALSE | Downregulated |
| -0,314573723 | 0,879951912 | SAA1 | FALSE | Downregulated |
| 0,038972794 | 0,879951912 | TYW1 | FALSE | Upregulated |
| -0,038850106 | 0,879956743 | CMPK1 | FALSE | Downregulated |
| -0,038102657 | 0,879956743 | CUTA | FALSE | Downregulated |
| 0,045521082 | 0,879956743 | EXOSC7 | FALSE | Upregulated |
| -0,181483506 | 0,879956743 | LINC00893 | FALSE | Downregulated |
| -0,127267717 | 0,879966143 | AIM2 | FALSE | Downregulated |
| 0,135433275 | 0,880030875 | H2AFY2 | FALSE | Upregulated |
| 0,04167479 | 0,880030875 | NFS1 | FALSE | Upregulated |
| -0,054228914 | 0,880030875 | PET117 | FALSE | Downregulated |
| -0,030734944 | 0,880235887 | ATXN10 | FALSE | Downregulated |
| 0,111382339 | 0,880235887 | FLRT2 | FALSE | Upregulated |
| 0,039173361 | 0,880235887 | LPHN2 | FALSE | Upregulated |
| 0,079620584 | 0,880298661 | BIN2 | FALSE | Upregulated |
| -0,05397696 | 0,880298661 | BNIP1 | FALSE | Downregulated |
| -0,043629247 | 0,880298661 | PSMA2 | FALSE | Downregulated |
| -0,068434857 | 0,880298661 | ZC4H2 | FALSE | Downregulated |
| 0,058047159 | 0,880438269 | CSTF2 | FALSE | Upregulated |
| 0,129956019 | 0,880438269 | LOC101930114 | FALSE | Upregulated |
| -0,372047562 | 0,880438269 | LPPR4 | FALSE | Downregulated |
| 0,048435542 | 0,880509271 | EML3 | FALSE | Upregulated |
| 0,390029993 | 0,880511642 | CNGB1 | FALSE | Upregulated |
| 0,293132921 | 0,880511642 | IGSF22 | FALSE | Upregulated |
| -0,24566056 | 0,880511642 | LOC101927252 | FALSE | Downregulated |

| | | | | |
|---|---|---|---|---|
| 0,263471287 | 0,880511642 | LOC101928021 | FALSE | Upregulated |
| -0,040039536 | 0,880511642 | MED10 | FALSE | Downregulated |
| 0,12980973 | 0,880511642 | MGC16142 | FALSE | Upregulated |
| -0,092185837 | 0,880511642 | SAA4 | FALSE | Downregulated |
| -0,25192539 | 0,880511642 | SEPT14 | FALSE | Downregulated |
| -0,252098669 | 0,880519813 | LOC100996735 | FALSE | Downregulated |
| 0,051713054 | 0,880563673 | ABCD3 | FALSE | Upregulated |
| -0,077274403 | 0,880661667 | ADCY5 | FALSE | Downregulated |
| -0,071445006 | 0,880761535 | C9orf3 | FALSE | Downregulated |
| 0,068807954 | 0,880761535 | CBL | FALSE | Upregulated |
| 0,28966972 | 0,880761535 | DNM1P46 | FALSE | Upregulated |
| -0,649459496 | 0,880761535 | HHATL | FALSE | Downregulated |
| 1,04646048 | 0,880761535 | RNU1-4 | FALSE | Upregulated |
| -0,055506361 | 0,880761535 | SMIM12 | FALSE | Downregulated |
| 0,117509485 | 0,880761535 | ZNF547 | FALSE | Upregulated |
| 0,07402875 | 0,880795211 | JRK | FALSE | Upregulated |
| 0,064769546 | 0,880838818 | BTN3A1 | FALSE | Upregulated |
| 0,218869381 | 0,880858175 | BTNL8 | FALSE | Upregulated |
| 0,08813692 | 0,880858175 | CYGB | FALSE | Upregulated |
| -0,038854284 | 0,880858175 | RCN1 | FALSE | Downregulated |
| 0,064786598 | 0,880858175 | ZNF436 | FALSE | Upregulated |
| -0,208307743 | 0,880904096 | CCDC38 | FALSE | Downregulated |
| 0,107160104 | 0,880904096 | ITGB7 | FALSE | Upregulated |
| -0,135377427 | 0,880904096 | LEKR1 | FALSE | Downregulated |
| -0,091650431 | 0,880904096 | LOC101060179 | FALSE | Downregulated |
| -0,118526781 | 0,880904096 | MAP9 | FALSE | Downregulated |

| | | | | |
|---|---|---|---|---|
| -0,038813807 | 0,880904096 | PTPLAD1 | FALSE | Downregulated |
| -0,091204269 | 0,880904096 | SNHG3 | FALSE | Downregulated |
| -0,049026674 | 0,8811231 | JAGN1 | FALSE | Downregulated |
| -0,02838149 | 0,881173887 | CNOT8 | FALSE | Downregulated |
| -0,162230539 | 0,881173887 | LOC102724517 | FALSE | Downregulated |
| -0,146964501 | 0,881173887 | LOC388906 | FALSE | Downregulated |
| 0,129077771 | 0,881173887 | SLC1A2 | FALSE | Upregulated |
| 0,059926982 | 0,881173887 | ZDHHC14 | FALSE | Upregulated |
| -0,122170102 | 0,881279198 | CCDC30 | FALSE | Downregulated |
| 0,05459481 | 0,881279198 | RPS12 | FALSE | Upregulated |
| 0,049156722 | 0,881466462 | ARHGAP17 | FALSE | Upregulated |
| -0,067207726 | 0,881636835 | MYCBP2 | FALSE | Downregulated |
| -0,068498058 | 0,881645501 | CEP290 | FALSE | Downregulated |
| -0,058994509 | 0,881645501 | CREB3L4 | FALSE | Downregulated |
| -0,057983725 | 0,881645501 | FAM229B | FALSE | Downregulated |
| 0,028244643 | 0,881645501 | FTO | FALSE | Upregulated |
| -0,303929719 | 0,881645501 | LOC100287387 | FALSE | Downregulated |
| -0,187573732 | 0,881645501 | LOC102723467 | FALSE | Downregulated |
| -0,078566357 | 0,881645501 | PAOX | FALSE | Downregulated |
| 0,061657866 | 0,881645501 | SPG21 | FALSE | Upregulated |
| 0,0614162 | 0,881645501 | TOP3B | FALSE | Upregulated |
| 0,047769222 | 0,881654262 | IAH1 | FALSE | Upregulated |
| 0,146640347 | 0,881654262 | KAL1 | FALSE | Upregulated |
| -0,19257039 | 0,881654262 | MAST1 | FALSE | Downregulated |
| 0,068857178 | 0,881654262 | SSR2 | FALSE | Upregulated |
| -0,128201963 | 0,881654262 | ZNF165 | FALSE | Downregulated |

| | | | | |
|---|---|---|---|---|
| 0,069603629 | 0,881765634 | C2CD2L | FALSE | Upregulated |
| 0,058381045 | 0,881765634 | CCDC77 | FALSE | Upregulated |
| 0,050164165 | 0,881765634 | DHRS3 | FALSE | Upregulated |
| 0,062896298 | 0,881765634 | EFNB1 | FALSE | Upregulated |
| -0,299813402 | 0,881765634 | PM20D1 | FALSE | Downregulated |
| 0,069122242 | 0,881765634 | RBM3 | FALSE | Upregulated |
| 0,040351196 | 0,881765634 | SNRPB | FALSE | Upregulated |
| -0,231859238 | 0,881765634 | TREM1 | FALSE | Downregulated |
| 0,056699129 | 0,881765634 | WDR73 | FALSE | Upregulated |
| 0,236524257 | 0,881765634 | XKR9 | FALSE | Upregulated |
| -0,061369878 | 0,881765634 | ZNF223 | FALSE | Downregulated |
| -0,065042128 | 0,881834811 | C5orf28 | FALSE | Downregulated |
| -0,060970266 | 0,881834811 | CARD8 | FALSE | Downregulated |
| -0,03543826 | 0,881881268 | NFE2L1 | FALSE | Downregulated |
| 0,062034204 | 0,881887065 | NOL9 | FALSE | Upregulated |
| 0,100246863 | 0,881971168 | CDKN2C | FALSE | Upregulated |
| 0,068121473 | 0,88203354 | LCMT1 | FALSE | Upregulated |
| -0,114335232 | 0,882118529 | OAS2 | FALSE | Downregulated |
| 0,499916969 | 0,882118529 | TRNAG4 | FALSE | Upregulated |
| 0,053897003 | 0,882172998 | BACH1 | FALSE | Upregulated |
| 0,116170282 | 0,882172998 | C1orf112 | FALSE | Upregulated |
| -0,355518331 | 0,882172998 | LOC100287072 | FALSE | Downregulated |
| -0,035878799 | 0,882172998 | MRPL39 | FALSE | Downregulated |
| 0,116397517 | 0,882172998 | MYRF | FALSE | Upregulated |
| -0,105819028 | 0,882549396 | AZGP1P1 | FALSE | Downregulated |
| 0,117081547 | 0,882549396 | LOC102723864 | FALSE | Upregulated |

| | | | | |
|---|---|---|---|---|
| 0,044281877 | 0,882603226 | AGFG2 | FALSE | Upregulated |
| -0,072741216 | 0,882603226 | ANKRD37 | FALSE | Downregulated |
| -0,038348234 | 0,882603226 | FAM136A | FALSE | Downregulated |
| 0,046441865 | 0,882603226 | GNG12 | FALSE | Upregulated |
| 0,173546395 | 0,882603226 | HOXA1 | FALSE | Upregulated |
| 0,238460112 | 0,882603226 | HSD3B1 | FALSE | Upregulated |
| 0,088060769 | 0,882603226 | IKBIP | FALSE | Upregulated |
| 0,243683383 | 0,882603226 | LOC102724420 | FALSE | Upregulated |
| 0,048472041 | 0,882603226 | MRPL2 | FALSE | Upregulated |
| -0,276806144 | 0,882603226 | PADI4 | FALSE | Downregulated |
| 0,081787035 | 0,882603226 | PUSL1 | FALSE | Upregulated |
| -0,08546551 | 0,882603226 | RGN | FALSE | Downregulated |
| 0,263071638 | 0,882603226 | RP11-57H12.3 | FALSE | Upregulated |
| 0,037086328 | 0,882603226 | ZMYM2 | FALSE | Upregulated |
| -0,194818623 | 0,882693455 | NR2F2-AS1 | FALSE | Downregulated |
| -0,043673949 | 0,882847636 | AKAP1 | FALSE | Downregulated |
| 0,038066649 | 0,882847636 | ARHGEF7 | FALSE | Upregulated |
| 0,044569625 | 0,882847636 | BBS2 | FALSE | Upregulated |
| 0,163240152 | 0,882847636 | COL13A1 | FALSE | Upregulated |
| 0,087226333 | 0,882847636 | CORO2A | FALSE | Upregulated |
| -0,182021428 | 0,882847636 | FAM84A | FALSE | Downregulated |
| -0,050650914 | 0,882847636 | H6PD | FALSE | Downregulated |
| 0,214574633 | 0,882847636 | HIF3A | FALSE | Upregulated |
| 0,100894575 | 0,882847636 | KLHDC7A | FALSE | Upregulated |
| 0,292805572 | 0,882847636 | LOC102724281 | FALSE | Upregulated |
| 0,139906482 | 0,882847636 | MB21D1 | FALSE | Upregulated |

| | | | | |
|---|---|---|---|---|
| -0,077445034 | 0,882847636 | MROH8 | FALSE | Downregulated |
| 0,114562082 | 0,882847636 | OLFM1 | FALSE | Upregulated |
| -0,083566969 | 0,882847636 | PDIA4 | FALSE | Downregulated |
| -0,04411319 | 0,882847636 | PIGG | FALSE | Downregulated |
| 0,124550497 | 0,882847636 | ROBO3 | FALSE | Upregulated |
| 0,261291141 | 0,882847636 | RPL31P11 | FALSE | Upregulated |
| -0,092593721 | 0,882847636 | SIGLEC7 | FALSE | Downregulated |
| -0,045013962 | 0,882870221 | DCUN1D4 | FALSE | Downregulated |
| -0,029161317 | 0,883043651 | ATP6V1A | FALSE | Downregulated |
| 0,061089917 | 0,883043651 | NUP43 | FALSE | Upregulated |
| 0,078303063 | 0,883093081 | BLVRA | FALSE | Upregulated |
| 0,039739631 | 0,883093081 | CAPZA2 | FALSE | Upregulated |
| -0,044581471 | 0,883093081 | DTWD2 | FALSE | Downregulated |
| -0,045521052 | 0,883093081 | FLOT1 | FALSE | Downregulated |
| 0,073475272 | 0,883093081 | GNG11 | FALSE | Upregulated |
| -0,05784829 | 0,883093081 | IGIP | FALSE | Downregulated |
| 0,09183294 | 0,883093081 | LONRF3 | FALSE | Upregulated |
| 0,068496373 | 0,883093081 | MAP1LC3A | FALSE | Upregulated |
| -0,083104732 | 0,883093081 | PNPLA4 | FALSE | Downregulated |
| 0,08853571 | 0,883093081 | RBM14-RBM4 | FALSE | Upregulated |
| -0,120863536 | 0,883093081 | RPSAP58 | FALSE | Downregulated |
| -0,320243618 | 0,883093081 | S100P | FALSE | Downregulated |
| 0,110807842 | 0,883093081 | SNHG12 | FALSE | Upregulated |
| -0,041253885 | 0,883093081 | TIMM21 | FALSE | Downregulated |
| 0,09871071 | 0,883093081 | TRIM69 | FALSE | Upregulated |
| -0,084381118 | 0,883093081 | TRMT10B | FALSE | Downregulated |

| | | | | |
|---|---|---|---|---|
| -0,070164931 | 0,883093081 | ZNF443 | FALSE | Downregulated |
| 0,161162209 | 0,88309775 | ALS2CR12 | FALSE | Upregulated |
| 0,178700906 | 0,88309775 | LOC554206 | FALSE | Upregulated |
| 0,094186449 | 0,88309775 | TRIM35 | FALSE | Upregulated |
| 0,063901834 | 0,883226204 | DCP2 | FALSE | Upregulated |
| 0,098046218 | 0,883226204 | PRELP | FALSE | Upregulated |
| 0,076827842 | 0,883266336 | GOT1 | FALSE | Upregulated |
| -0,12426113 | 0,883266336 | ZNRD1-AS1 | FALSE | Downregulated |
| -0,359447107 | 0,883524659 | GAP43 | FALSE | Downregulated |
| -0,127192214 | 0,883524659 | NHSL2 | FALSE | Downregulated |
| -0,062262006 | 0,883560014 | B4GALT5 | FALSE | Downregulated |
| -0,093503348 | 0,883560014 | PPIG | FALSE | Downregulated |
| -0,18055038 | 0,883560014 | SELE | FALSE | Downregulated |
| 0,08679662 | 0,883582014 | GAREM | FALSE | Upregulated |
| -0,038750867 | 0,883604847 | PROS1 | FALSE | Downregulated |
| 0,244690894 | 0,883759666 | CCR4 | FALSE | Upregulated |
| 0,136751843 | 0,883759666 | DACT3 | FALSE | Upregulated |
| -0,060921119 | 0,883759666 | GAR1 | FALSE | Downregulated |
| 0,209443679 | 0,883759666 | GP6 | FALSE | Upregulated |
| 0,111302606 | 0,883759666 | HES6 | FALSE | Upregulated |
| 0,178645705 | 0,883759666 | KIAA1244 | FALSE | Upregulated |
| -0,142153186 | 0,883759666 | LIFR-AS1 | FALSE | Downregulated |
| -0,044122139 | 0,883759666 | MED26 | FALSE | Downregulated |
| -0,065568475 | 0,883759666 | MPV17 | FALSE | Downregulated |
| -0,095626045 | 0,883759666 | PID1 | FALSE | Downregulated |
| -0,072084757 | 0,883759666 | PNP | FALSE | Downregulated |

| | | | | |
|---|---|---|---|---|
| 0,139109674 | 0,883759666 | RASL10B | FALSE | Upregulated |
| -0,100686354 | 0,883759666 | RCE1 | FALSE | Downregulated |
| -0,091116242 | 0,883759666 | RGS19 | FALSE | Downregulated |
| 0,122643852 | 0,883759666 | RP11-290F20.3 | FALSE | Upregulated |
| -0,049823512 | 0,883759666 | RPL3 | FALSE | Downregulated |
| 0,040964061 | 0,883759666 | RPN2 | FALSE | Upregulated |
| -0,241663385 | 0,883759666 | SLITRK3 | FALSE | Downregulated |
| 0,341015187 | 0,883759666 | SMIM5 | FALSE | Upregulated |
| 0,053819879 | 0,883759666 | SPATA13 | FALSE | Upregulated |
| 0,138748491 | 0,883759666 | TDRD10 | FALSE | Upregulated |
| -0,057029619 | 0,883759666 | THNSL1 | FALSE | Downregulated |
| -0,064552333 | 0,883759666 | TM4SF4 | FALSE | Downregulated |
| 0,733703453 | 0,883759666 | TRNAE19 | FALSE | Upregulated |
| -0,078112729 | 0,883759666 | ZBTB25 | FALSE | Downregulated |
| -0,077834852 | 0,883759666 | ZNF704 | FALSE | Downregulated |
| 0,140033502 | 0,883820165 | EMID1 | FALSE | Upregulated |
| -0,071421657 | 0,883934814 | CCDC71L | FALSE | Downregulated |
| -0,07487532 | 0,883934814 | RNF144A | FALSE | Downregulated |
| 0,142174297 | 0,883940398 | HLA-DQB1 | FALSE | Upregulated |
| 0,084817589 | 0,883940398 | TMEM38A | FALSE | Upregulated |
| -0,172923313 | 0,884030372 | NAPA-AS1 | FALSE | Downregulated |
| -0,612319655 | 0,884030372 | TRNAD8 | FALSE | Downregulated |
| 0,049236003 | 0,884053356 | DHRSX | FALSE | Upregulated |
| -0,036742161 | 0,884078748 | MAPK8 | FALSE | Downregulated |
| -0,052552103 | 0,884081857 | BBS9 | FALSE | Downregulated |
| 0,030253697 | 0,884151922 | HMG20A | FALSE | Upregulated |

| | | | | |
|---|---|---|---|---|
| 0,289766201 | 0,884392971 | AC104820.2 | FALSE | Upregulated |
| -0,049012739 | 0,884392971 | ADAM9 | FALSE | Downregulated |
| -0,289407065 | 0,884392971 | B3GALT5 | FALSE | Downregulated |
| -0,040645048 | 0,884392971 | BRMS1L | FALSE | Downregulated |
| 0,086969417 | 0,884392971 | C1orf162 | FALSE | Upregulated |
| 0,062276113 | 0,884392971 | CASP10 | FALSE | Upregulated |
| -0,073009826 | 0,884392971 | ENDOV | FALSE | Downregulated |
| -0,180029156 | 0,884392971 | ESRP1 | FALSE | Downregulated |
| 0,117018205 | 0,884392971 | ETNPPL | FALSE | Upregulated |
| -0,042404827 | 0,884392971 | GNPNAT1 | FALSE | Downregulated |
| 0,055563688 | 0,884392971 | HPD | FALSE | Upregulated |
| -0,2035048 | 0,884392971 | LOC102724861 | FALSE | Downregulated |
| 0,216259311 | 0,884392971 | LOC102724970 | FALSE | Upregulated |
| 0,162536482 | 0,884392971 | SERINC4 | FALSE | Upregulated |
| 0,143549029 | 0,884392971 | SPA17 | FALSE | Upregulated |
| 0,101492921 | 0,884392971 | SYNE3 | FALSE | Upregulated |
| -0,121400569 | 0,884392971 | ZNF718 | FALSE | Downregulated |
| 0,06267187 | 0,884516673 | PRKCH | FALSE | Upregulated |
| -0,357325822 | 0,884607735 | RAET1E-AS1 | FALSE | Downregulated |
| -0,075750015 | 0,884607735 | ZNF385B | FALSE | Downregulated |
| 0,072629295 | 0,884841444 | GPR89B | FALSE | Upregulated |
| 0,070723239 | 0,884890945 | CASP4 | FALSE | Upregulated |
| -0,137282177 | 0,884890945 | LRG1 | FALSE | Downregulated |
| 0,158846001 | 0,884890945 | MTMR8 | FALSE | Upregulated |
| -0,126851339 | 0,884890945 | SH3BP5-AS1 | FALSE | Downregulated |
| 0,150858919 | 0,884990652 | ANKRD36C | FALSE | Upregulated |

| | | | | |
|---|---|---|---|---|
| -0,079466766 | 0,884990652 | CBLC | FALSE | Downregulated |
| 0,213759132 | 0,884990652 | CNR2 | FALSE | Upregulated |
| -0,510984612 | 0,884990652 | GOLGA8S | FALSE | Downregulated |
| 0,037129555 | 0,884990652 | LTN1 | FALSE | Upregulated |
| -0,042346178 | 0,884990652 | MRPL46 | FALSE | Downregulated |
| 0,029258196 | 0,884990652 | NCBP2 | FALSE | Upregulated |
| 0,083837481 | 0,884990652 | PASK | FALSE | Upregulated |
| -0,142188922 | 0,884990652 | PVALB | FALSE | Downregulated |
| 0,120369544 | 0,884990652 | SNHG15 | FALSE | Upregulated |
| 0,133919191 | 0,884990652 | TMEM55A | FALSE | Upregulated |
| 0,048180632 | 0,884990652 | ZDHHC4 | FALSE | Upregulated |
| -0,099587057 | 0,885004081 | RP11-736K20.6 | FALSE | Downregulated |
| -0,082732024 | 0,8850477 | CLRN3 | FALSE | Downregulated |
| 0,128663254 | 0,8850477 | KLRG1 | FALSE | Upregulated |
| -0,083948169 | 0,8850477 | LRRC8C | FALSE | Downregulated |
| -0,029578177 | 0,8850477 | MARCH8 | FALSE | Downregulated |
| -0,051326636 | 0,8850477 | NDUFA3 | FALSE | Downregulated |
| -0,041049602 | 0,8850477 | NDUFB5 | FALSE | Downregulated |
| 0,04472427 | 0,8850477 | NDUFB8 | FALSE | Upregulated |
| -0,042930557 | 0,8850477 | RPUSD3 | FALSE | Downregulated |
| -0,042478487 | 0,8850477 | RSAD1 | FALSE | Downregulated |
| -0,099497045 | 0,8850477 | SOD2 | FALSE | Downregulated |
| 0,187286556 | 0,8850477 | SPATC1L | FALSE | Upregulated |
| 0,031656245 | 0,8850477 | USP3 | FALSE | Upregulated |
| -0,187523194 | 0,885055852 | IRX3 | FALSE | Downregulated |
| -0,173517323 | 0,885084999 | PLA2G4B | FALSE | Downregulated |

| | | | | |
|---|---|---|---|---|
| 0,025776328 | 0,885317328 | FAM120AOS | FALSE | Upregulated |
| -0,121795987 | 0,885317328 | LOC102724005 | FALSE | Downregulated |
| -0,090296391 | 0,885317328 | LPCAT2 | FALSE | Downregulated |
| 0,531988569 | 0,885458834 | TRNR | FALSE | Upregulated |
| 0,201623106 | 0,885467631 | COCH | FALSE | Upregulated |
| 0,137798879 | 0,885492742 | CHRD | FALSE | Upregulated |
| -0,046522988 | 0,885492742 | COA3 | FALSE | Downregulated |
| 0,06775383 | 0,885492742 | LOC100505761 | FALSE | Upregulated |
| 0,102225078 | 0,885492742 | NRTN | FALSE | Upregulated |
| 0,268129968 | 0,885593965 | AC095067.1 | FALSE | Upregulated |
| 0,128077598 | 0,885593965 | CDC7 | FALSE | Upregulated |
| 0,070252006 | 0,885593965 | PCDH1 | FALSE | Upregulated |
| -0,075385087 | 0,885593965 | RNF125 | FALSE | Downregulated |
| 0,221007165 | 0,885714764 | PIF1 | FALSE | Upregulated |
| 0,042803553 | 0,885714764 | TUBGCP4 | FALSE | Upregulated |
| 0,082202821 | 0,885714764 | WDR90 | FALSE | Upregulated |
| 0,058144439 | 0,885752569 | CYP4F3 | FALSE | Upregulated |
| 0,237195261 | 0,885752569 | LRRC69 | FALSE | Upregulated |
| -0,159630807 | 0,885752569 | RP11-143J12.2 | FALSE | Downregulated |
| -0,211522106 | 0,885810997 | EGR1 | FALSE | Downregulated |
| -0,076596733 | 0,885810997 | LIME1 | FALSE | Downregulated |
| -0,275792312 | 0,885810997 | LINC00299 | FALSE | Downregulated |
| 0,17029466 | 0,885810997 | NEURL3 | FALSE | Upregulated |
| 0,047744923 | 0,885810997 | SLC25A17 | FALSE | Upregulated |
| -0,038713388 | 0,885810997 | ZC3HAV1 | FALSE | Downregulated |
| -0,06513365 | 0,885883228 | CARS | FALSE | Downregulated |

| | | | | |
|---|---|---|---|---|
| 0,072880001 | 0,885883228 | SRD5A3 | FALSE | Upregulated |
| -0,059993903 | 0,885883228 | ZNF112 | FALSE | Downregulated |
| -0,06207009 | 0,886023142 | ALKBH6 | FALSE | Downregulated |
| -0,08365236 | 0,886023142 | CA13 | FALSE | Downregulated |
| 0,126148349 | 0,886023142 | CDKN2B | FALSE | Upregulated |
| 0,064192367 | 0,886023142 | FAM217B | FALSE | Upregulated |
| 0,06883973 | 0,886023142 | IFT52 | FALSE | Upregulated |
| -0,278866229 | 0,886023142 | LOC100130417 | FALSE | Downregulated |
| -0,128617761 | 0,886023142 | LOC100289098 | FALSE | Downregulated |
| 0,156276926 | 0,886023142 | LYSMD4 | FALSE | Upregulated |
| -0,046942526 | 0,886023142 | MCU | FALSE | Downregulated |
| -0,094926953 | 0,886023142 | SLC2A12 | FALSE | Downregulated |
| 0,601367371 | 0,886023142 | TRNAD16 | FALSE | Upregulated |
| -0,076294724 | 0,886023142 | ZNF214 | FALSE | Downregulated |
| 0,05318399 | 0,886023142 | ZNF618 | FALSE | Upregulated |
| 0,045516489 | 0,886315742 | AMD1 | FALSE | Upregulated |
| -0,048431331 | 0,886315742 | CCAR1 | FALSE | Downregulated |
| 0,170061081 | 0,886315742 | CCNT2-AS1 | FALSE | Upregulated |
| -0,069058934 | 0,886315742 | CTSH | FALSE | Downregulated |
| -0,029185829 | 0,886315742 | EXOC3 | FALSE | Downregulated |
| -0,051349516 | 0,886315742 | EXOC6 | FALSE | Downregulated |
| 0,059600137 | 0,886315742 | LITAF | FALSE | Upregulated |
| 0,040045038 | 0,886315742 | LRRC42 | FALSE | Upregulated |
| -0,053152556 | 0,886315742 | MRPS36 | FALSE | Downregulated |
| 0,062586786 | 0,886315742 | NFXL1 | FALSE | Upregulated |
| -0,04821097 | 0,886315742 | PECR | FALSE | Downregulated |

| | | | | |
|---|---|---|---|---|
| -0,048058825 | 0,886315742 | PFDN6 | FALSE | Downregulated |
| -0,035722281 | 0,886315742 | PHF12 | FALSE | Downregulated |
| -0,120915811 | 0,886315742 | PPIL6 | FALSE | Downregulated |
| 0,158873196 | 0,886315742 | RGS11 | FALSE | Upregulated |
| 0,045589607 | 0,88651193 | BRWD1 | FALSE | Upregulated |
| -0,028295927 | 0,88651193 | CDC27 | FALSE | Downregulated |
| 0,052702585 | 0,88651193 | PGRMC1 | FALSE | Upregulated |
| 0,068990713 | 0,886597821 | ZNF133 | FALSE | Upregulated |
| 0,18050626 | 0,886629443 | PRKXP1 | FALSE | Upregulated |
| -0,032675427 | 0,88698574 | DR1 | FALSE | Downregulated |
| 0,049140341 | 0,887182453 | FURIN | FALSE | Upregulated |
| -0,062217526 | 0,887214149 | EHBP1 | FALSE | Downregulated |
| -0,084247457 | 0,887214149 | MYL12A | FALSE | Downregulated |
| 0,030794635 | 0,887216474 | ACAD9 | FALSE | Upregulated |
| -0,201675785 | 0,887216474 | LOC102724608 | FALSE | Downregulated |
| -0,043905442 | 0,887216474 | ZNF606 | FALSE | Downregulated |
| 0,043437193 | 0,887295177 | CDK2 | FALSE | Upregulated |
| -0,219930053 | 0,887295177 | FHAD1 | FALSE | Downregulated |
| -0,259516792 | 0,887295177 | FMR1-AS1 | FALSE | Downregulated |
| -0,165537746 | 0,887295177 | GNRH1 | FALSE | Downregulated |
| -0,091229746 | 0,887295177 | IRAK3 | FALSE | Downregulated |
| -0,363907442 | 0,887295177 | KIAA1257 | FALSE | Downregulated |
| 0,046867385 | 0,887295177 | KRTCAP2 | FALSE | Upregulated |
| 0,381323147 | 0,887295177 | LINC00675 | FALSE | Upregulated |
| 0,140634508 | 0,887295177 | LOC101928274 | FALSE | Upregulated |
| -0,172640531 | 0,887295177 | LOC102724017 | FALSE | Downregulated |

| | | | | |
|---|---|---|---|---|
| 0,339109686 | 0,887295177 | LOC441528 | FALSE | Upregulated |
| -0,117243521 | 0,887295177 | LSS | FALSE | Downregulated |
| -0,158775064 | 0,887295177 | MAPK12 | FALSE | Downregulated |
| -0,074790871 | 0,887295177 | NFAT5 | FALSE | Downregulated |
| 0,154460822 | 0,887295177 | PABPC1L | FALSE | Upregulated |
| -0,495493309 | 0,887295177 | RNY4 | FALSE | Downregulated |
| 0,046147766 | 0,887295177 | SEC23A | FALSE | Upregulated |
| -0,125493153 | 0,887295177 | SERP2 | FALSE | Downregulated |
| 0,12384361 | 0,887295177 | SUSD3 | FALSE | Upregulated |
| 0,192527452 | 0,887295177 | TMEM132B | FALSE | Upregulated |
| 0,083350387 | 0,887295177 | YIF1B | FALSE | Upregulated |
| 0,033378978 | 0,887295177 | ZNF148 | FALSE | Upregulated |
| 0,085710514 | 0,887295177 | ZNF814 | FALSE | Upregulated |
| -0,152032101 | 0,887346069 | LOC102723415 | FALSE | Downregulated |
| -0,048718956 | 0,887361645 | CFI | FALSE | Downregulated |
| -0,035414918 | 0,887361645 | RPUSD4 | FALSE | Downregulated |
| 0,082283054 | 0,887379394 | TBC1D32 | FALSE | Upregulated |
| -0,043151352 | 0,887514669 | WDR92 | FALSE | Downregulated |
| 0,102195035 | 0,887693819 | CCNL2 | FALSE | Upregulated |
| 0,281682511 | 0,887693819 | HIST2H4B | FALSE | Upregulated |
| -0,109459247 | 0,88777858 | C17orf100 | FALSE | Downregulated |
| 0,101879352 | 0,887865523 | TFEC | FALSE | Upregulated |
| 0,09472239 | 0,887975284 | ZFAND5 | FALSE | Upregulated |
| -0,087551272 | 0,887987188 | AHDC1 | FALSE | Downregulated |
| 0,124709157 | 0,887987188 | LOC100506100 | FALSE | Upregulated |
| -0,048145961 | 0,887987188 | MMP24-AS1 | FALSE | Downregulated |

| | | | | |
|---|---|---|---|---|
| -0,049061631 | 0,888028453 | ANKHD1 | FALSE | Downregulated |
| -0,160353314 | 0,888028453 | BLK | FALSE | Downregulated |
| 0,058548705 | 0,888028453 | CYCS | FALSE | Upregulated |
| 0,069964898 | 0,888028453 | GAS8 | FALSE | Upregulated |
| 0,192410833 | 0,888028453 | GRIN3A | FALSE | Upregulated |
| 0,13934477 | 0,888028453 | RHBG | FALSE | Upregulated |
| -0,209189073 | 0,888028453 | TRPV3 | FALSE | Downregulated |
| -0,091555883 | 0,888053113 | ADRA1B | FALSE | Downregulated |
| 0,24765038 | 0,888113114 | APITD1-CORT | FALSE | Upregulated |
| -0,039668445 | 0,888156729 | VPRBP | FALSE | Downregulated |
| -0,055267396 | 0,888209468 | GRB14 | FALSE | Downregulated |
| -0,0374491 | 0,888209468 | RAB11FIP2 | FALSE | Downregulated |
| 0,211229689 | 0,888384401 | CLEC5A | FALSE | Upregulated |
| 0,101207766 | 0,888602375 | TMOD1 | FALSE | Upregulated |
| -0,183401187 | 0,888820993 | ATP6V1G2 | FALSE | Downregulated |
| 0,25522508 | 0,888820993 | NAV2-AS4 | FALSE | Upregulated |
| -0,048902201 | 0,888820993 | NUDT16L1 | FALSE | Downregulated |
| 0,143669023 | 0,888820993 | PKNOX2 | FALSE | Upregulated |
| -0,069925356 | 0,888820993 | PPCDC | FALSE | Downregulated |
| 0,131122272 | 0,888820993 | SNHG18 | FALSE | Upregulated |
| 0,049164035 | 0,888820993 | VPS37B | FALSE | Upregulated |
| 0,153056956 | 0,888829911 | LOC101927934 | FALSE | Upregulated |
| -0,154730777 | 0,888839553 | LOC101927339 | FALSE | Downregulated |
| -0,029937843 | 0,888855815 | TOM1L2 | FALSE | Downregulated |
| -0,181849294 | 0,888875084 | AKR1E2 | FALSE | Downregulated |
| 0,03436299 | 0,888888679 | KIFAP3 | FALSE | Upregulated |

| | | | | |
|---|---|---|---|---|
| -0,070209164 | 0,888905913 | DEPDC7 | FALSE | Downregulated |
| -0,124397276 | 0,88911124 | CCDC163P | FALSE | Downregulated |
| 0,033857799 | 0,889171046 | EXOC8 | FALSE | Upregulated |
| 0,267882631 | 0,889171046 | N4BP3 | FALSE | Upregulated |
| -0,051517676 | 0,889236358 | AFMID | FALSE | Downregulated |
| 0,046730478 | 0,889245431 | NIPAL3 | FALSE | Upregulated |
| 0,156431431 | 0,889260918 | LOC728554 | FALSE | Upregulated |
| -0,067653511 | 0,889383753 | GRAMD1C | FALSE | Downregulated |
| 0,075601628 | 0,889383753 | MXRA7 | FALSE | Upregulated |
| 0,072118122 | 0,889383753 | ZNF862 | FALSE | Upregulated |
| -0,056896653 | 0,88941939 | ECM2 | FALSE | Downregulated |
| -0,043221902 | 0,88941939 | SBNO1 | FALSE | Downregulated |
| -0,212476019 | 0,889450062 | ARMC4 | FALSE | Downregulated |
| 0,093750625 | 0,889450062 | GEMIN8P4 | FALSE | Upregulated |
| -0,141659266 | 0,889450062 | LOC102724098 | FALSE | Downregulated |
| -0,145094564 | 0,889450062 | MIR503HG | FALSE | Downregulated |
| -0,048723151 | 0,889450062 | ZBTB5 | FALSE | Downregulated |
| -0,046876115 | 0,889494297 | PGM1 | FALSE | Downregulated |
| -0,074052859 | 0,889583428 | CEP135 | FALSE | Downregulated |
| -0,107156722 | 0,889583428 | ZNF582-AS1 | FALSE | Downregulated |
| 0,060872553 | 0,889869601 | GPLD1 | FALSE | Upregulated |
| 0,043392075 | 0,889869601 | HSPA4 | FALSE | Upregulated |
| 0,235317045 | 0,889869601 | PEBP4 | FALSE | Upregulated |
| -0,097442454 | 0,889949716 | AC016629.8 | FALSE | Downregulated |
| 0,042209709 | 0,889949716 | CDKN1B | FALSE | Upregulated |
| -0,047734015 | 0,889949716 | FBXW8 | FALSE | Downregulated |

| | | | | |
|---|---|---|---|---|
| -0,091483753 | 0,889949716 | GALNT7 | FALSE | Downregulated |
| 0,064467366 | 0,889949716 | GSTCD | FALSE | Upregulated |
| -0,046512656 | 0,889949716 | KCNJ8 | FALSE | Downregulated |
| 0,118086948 | 0,889949716 | LOC100379224 | FALSE | Upregulated |
| 0,038488021 | 0,889949716 | RHOU | FALSE | Upregulated |
| 0,254524727 | 0,889949716 | RP11-682N22.1 | FALSE | Upregulated |
| -0,263444451 | 0,889949716 | UCN | FALSE | Downregulated |
| 0,307467614 | 0,890057672 | LOC158434 | FALSE | Upregulated |
| 0,081656552 | 0,890141913 | TUBGCP6 | FALSE | Upregulated |
| 0,049960845 | 0,890172457 | PHF6 | FALSE | Upregulated |
| 0,03248405 | 0,890560435 | AKT2 | FALSE | Upregulated |
| -0,191116825 | 0,890560435 | CARNS1 | FALSE | Downregulated |
| 0,364606763 | 0,890560435 | DNASE2B | FALSE | Upregulated |
| 0,061816251 | 0,890560435 | EPHX2 | FALSE | Upregulated |
| 0,127761609 | 0,890560435 | FBLIM1 | FALSE | Upregulated |
| -0,057126965 | 0,890560435 | FNBP1 | FALSE | Downregulated |
| -0,240358839 | 0,890560435 | LOC100131532 | FALSE | Downregulated |
| -0,220151871 | 0,890560435 | LOC101928774 | FALSE | Downregulated |
| -0,030543715 | 0,890560435 | MECP2 | FALSE | Downregulated |
| -0,04120169 | 0,890560435 | POLR2K | FALSE | Downregulated |
| -0,042044733 | 0,890560435 | SIRT3 | FALSE | Downregulated |
| -0,073749339 | 0,890560435 | STK19 | FALSE | Downregulated |
| -0,125582054 | 0,890560435 | SYCP2 | FALSE | Downregulated |
| 0,115268541 | 0,890560435 | TNFSF4 | FALSE | Upregulated |
| 0,207534676 | 0,890560435 | TNIP3 | FALSE | Upregulated |
| 0,054872581 | 0,890560435 | TRIM65 | FALSE | Upregulated |

| | | | | |
|---|---|---|---|---|
| -0,133372843 | 0,89059834 | LOC102724061 | FALSE | Downregulated |
| 0,123954994 | 0,890686247 | MAP6 | FALSE | Upregulated |
| 0,067842864 | 0,890724011 | PLEKHA8P1 | FALSE | Upregulated |
| -0,156267518 | 0,890735831 | C10orf126 | FALSE | Downregulated |
| 0,255097663 | 0,890735831 | MIR4697HG | FALSE | Upregulated |
| 0,13189696 | 0,890735831 | TMED6 | FALSE | Upregulated |
| -0,08187357 | 0,890735831 | ZGPAT | FALSE | Downregulated |
| -0,041043345 | 0,890925979 | SDF2 | FALSE | Downregulated |
| 0,197801713 | 0,890969391 | AC017060.1 | FALSE | Upregulated |
| 0,034881101 | 0,890969391 | APLP2 | FALSE | Upregulated |
| -0,064826924 | 0,890969391 | CEP68 | FALSE | Downregulated |
| 0,1859655 | 0,890969391 | LOC101927011 | FALSE | Upregulated |
| -0,148422797 | 0,890969391 | NQO1 | FALSE | Downregulated |
| -0,035564595 | 0,890969391 | TANK | FALSE | Downregulated |
| -0,034016659 | 0,890969391 | ZFYVE1 | FALSE | Downregulated |
| -0,053551875 | 0,891086133 | C1orf168 | FALSE | Downregulated |
| 0,236166537 | 0,891086133 | CACTIN-AS1 | FALSE | Upregulated |
| -0,052725828 | 0,891086133 | CAMSAP2 | FALSE | Downregulated |
| -0,08730678 | 0,891086133 | CD180 | FALSE | Downregulated |
| 0,245846861 | 0,891086133 | CYP4Z1 | FALSE | Upregulated |
| 0,186830642 | 0,891086133 | SAP25 | FALSE | Upregulated |
| 0,272688708 | 0,891086133 | TPSB2 | FALSE | Upregulated |
| 0,063815234 | 0,891099169 | MRPL23 | FALSE | Upregulated |
| -0,064693826 | 0,891099169 | USMG5 | FALSE | Downregulated |
| 0,144016284 | 0,89121288 | FICD | FALSE | Upregulated |
| -0,120485894 | 0,891297907 | MME | FALSE | Downregulated |

| | | | | |
|---|---|---|---|---|
| 0,054395216 | 0,891505248 | SEC14L2 | FALSE | Upregulated |
| -0,050318669 | 0,891539926 | ANKZF1 | FALSE | Downregulated |
| -0,036395165 | 0,891539926 | ATP6V1D | FALSE | Downregulated |
| -0,038659417 | 0,891539926 | DIP2B | FALSE | Downregulated |
| 0,02150365 | 0,891539926 | EIF2B1 | FALSE | Upregulated |
| -0,151236893 | 0,891539926 | JAKMIP3 | FALSE | Downregulated |
| 0,096454515 | 0,891539926 | KITLG | FALSE | Upregulated |
| -0,362964259 | 0,891539926 | SCN3B | FALSE | Downregulated |
| 0,071489404 | 0,891539926 | TBXAS1 | FALSE | Upregulated |
| 0,05658543 | 0,891539926 | THG1L | FALSE | Upregulated |
| -0,0282306 | 0,891539926 | VPS54 | FALSE | Downregulated |
| 0,062980759 | 0,891713694 | HSD17B8 | FALSE | Upregulated |
| 0,04069411 | 0,891740701 | TMED1 | FALSE | Upregulated |
| -0,036197895 | 0,891740701 | TRIM44 | FALSE | Downregulated |
| 0,050382595 | 0,891740701 | WEE1 | FALSE | Upregulated |
| 0,143847109 | 0,892136887 | BICC1 | FALSE | Upregulated |
| 0,229673024 | 0,892136887 | CEL | FALSE | Upregulated |
| 0,085906678 | 0,892136887 | KATNAL2 | FALSE | Upregulated |
| -0,081371027 | 0,892136887 | LINC00324 | FALSE | Downregulated |
| -0,204790821 | 0,892136887 | LINGO4 | FALSE | Downregulated |
| -0,053457024 | 0,892136887 | MRPL11 | FALSE | Downregulated |
| 0,214918271 | 0,892182333 | LOC100507336 | FALSE | Upregulated |
| 0,23825402 | 0,892215687 | TMEM125 | FALSE | Upregulated |
| -0,040384381 | 0,892274399 | AKT1S1 | FALSE | Downregulated |
| 0,130566915 | 0,892274399 | ZNF135 | FALSE | Upregulated |
| -0,036503545 | 0,892291049 | ZNF639 | FALSE | Downregulated |

| | | | | |
|---|---|---|---|---|
| -0,026073185 | 0,892339056 | ZNF664 | FALSE | Downregulated |
| -0,048545217 | 0,89252842 | NDUFA11 | FALSE | Downregulated |
| 0,161526704 | 0,892656504 | LOC284454 | FALSE | Upregulated |
| 0,240851086 | 0,892730499 | HLX-AS1 | FALSE | Upregulated |
| 0,046515187 | 0,892730499 | SRRD | FALSE | Upregulated |
| -0,042182825 | 0,892762652 | ZNF655 | FALSE | Downregulated |
| 0,075291223 | 0,892803802 | ACSL1 | FALSE | Upregulated |
| -0,105441979 | 0,893121467 | ADAMTS7 | FALSE | Downregulated |
| -0,253872905 | 0,893121467 | CBX3P2 | FALSE | Downregulated |
| -0,270399095 | 0,893121467 | LOC440354 | FALSE | Downregulated |
| 0,051087993 | 0,893121467 | MARK4 | FALSE | Upregulated |
| 0,039950563 | 0,893121467 | RAB11FIP3 | FALSE | Upregulated |
| 0,033018689 | 0,893121467 | SEP15 | FALSE | Upregulated |
| 0,191219896 | 0,893121467 | SPTBN5 | FALSE | Upregulated |
| 0,08411584 | 0,893200033 | PALM | FALSE | Upregulated |
| -0,134549548 | 0,893200033 | PROM1 | FALSE | Downregulated |
| -0,044769036 | 0,893356001 | BLOC1S2 | FALSE | Downregulated |
| -0,115699742 | 0,893629646 | LILRA5 | FALSE | Downregulated |
| -0,041896302 | 0,893673946 | PEX16 | FALSE | Downregulated |
| -0,296371423 | 0,893791152 | C12orf39 | FALSE | Downregulated |
| 0,074447498 | 0,893791152 | NPR1 | FALSE | Upregulated |
| 0,057784072 | 0,893791152 | RIN3 | FALSE | Upregulated |
| 0,212769457 | 0,893791152 | TIGD4 | FALSE | Upregulated |
| -0,036587988 | 0,89409891 | BRD1 | FALSE | Downregulated |
| -0,220491013 | 0,89409891 | COX7A1 | FALSE | Downregulated |
| -0,119938209 | 0,89409891 | GPIHBP1 | FALSE | Downregulated |

| | | | | |
|---|---|---|---|---|
| 0,065997318 | 0,89409891 | PMEL | FALSE | Upregulated |
| -0,155763222 | 0,89409891 | RP11-996F15.2 | FALSE | Downregulated |
| -0,080724209 | 0,89409891 | SAMD9 | FALSE | Downregulated |
| -0,036094913 | 0,89409891 | WBP2 | FALSE | Downregulated |
| -0,034767362 | 0,894141948 | C6orf120 | FALSE | Downregulated |
| -0,176425038 | 0,894183573 | PYGO1 | FALSE | Downregulated |
| -0,092929817 | 0,894201864 | EPB41L4A | FALSE | Downregulated |
| 0,24120806 | 0,894201864 | ERI3-IT1 | FALSE | Upregulated |
| 0,045536156 | 0,894201864 | TMEM9B | FALSE | Upregulated |
| -0,05851208 | 0,894207824 | TOP1 | FALSE | Downregulated |
| -0,169516795 | 0,894263265 | LOC441204 | FALSE | Downregulated |
| -0,128269214 | 0,894390847 | KCNAB1 | FALSE | Downregulated |
| -0,309587231 | 0,894401458 | LOC101928307 | FALSE | Downregulated |
| -0,034562162 | 0,894401458 | SCAF4 | FALSE | Downregulated |
| 0,042677019 | 0,894433981 | TMEM203 | FALSE | Upregulated |
| 0,039346299 | 0,894586989 | RBMXL1 | FALSE | Upregulated |
| -0,047386378 | 0,894828454 | AK4 | FALSE | Downregulated |
| -0,087926766 | 0,894828454 | AMIGO2 | FALSE | Downregulated |
| -0,046240978 | 0,894828454 | ARMCX5 | FALSE | Downregulated |
| -0,03799862 | 0,894828454 | BTBD10 | FALSE | Downregulated |
| -0,106108653 | 0,894828454 | C8orf44 | FALSE | Downregulated |
| -0,060190885 | 0,894828454 | C9orf91 | FALSE | Downregulated |
| -0,267977824 | 0,894828454 | CADM3 | FALSE | Downregulated |
| -0,183741406 | 0,894828454 | CCDC136 | FALSE | Downregulated |
| -0,037061939 | 0,894828454 | CCDC28A | FALSE | Downregulated |
| 0,034186358 | 0,894828454 | CNPPD1 | FALSE | Upregulated |

| | | | | |
|---|---|---|---|---|
| 0,038886564 | 0,894828454 | CNPY2 | FALSE | Upregulated |
| -0,048484198 | 0,894828454 | COMMD3 | FALSE | Downregulated |
| -0,15540271 | 0,894828454 | CTNNA3 | FALSE | Downregulated |
| -0,136795571 | 0,894828454 | DHRS13 | FALSE | Downregulated |
| 0,037886925 | 0,894828454 | DHTKD1 | FALSE | Upregulated |
| -0,030345276 | 0,894828454 | EIF1B | FALSE | Downregulated |
| 0,143685858 | 0,894828454 | ELOVL7 | FALSE | Upregulated |
| 0,139663481 | 0,894828454 | FCRL3 | FALSE | Upregulated |
| -0,122680257 | 0,894828454 | FOXD2-AS1 | FALSE | Downregulated |
| -0,057309729 | 0,894828454 | GDF2 | FALSE | Downregulated |
| 0,080013897 | 0,894828454 | GJD3 | FALSE | Upregulated |
| 0,070283409 | 0,894828454 | GMPR | FALSE | Upregulated |
| 0,231474699 | 0,894828454 | H3F3C | FALSE | Upregulated |
| -0,077371644 | 0,894828454 | IKZF4 | FALSE | Downregulated |
| 0,041802758 | 0,894828454 | ISCA1 | FALSE | Upregulated |
| -0,029428961 | 0,894828454 | KANSL1 | FALSE | Downregulated |
| -0,337292012 | 0,894828454 | KCNK2 | FALSE | Downregulated |
| -0,04680131 | 0,894828454 | KIAA0907 | FALSE | Downregulated |
| 0,182213909 | 0,894828454 | LIPE-AS1 | FALSE | Upregulated |
| -0,128758657 | 0,894828454 | LOC101928657 | FALSE | Downregulated |
| -0,261696695 | 0,894828454 | LOC101928875 | FALSE | Downregulated |
| -0,088368654 | 0,894828454 | LOC388692 | FALSE | Downregulated |
| -0,44010344 | 0,894828454 | MAGED4B | FALSE | Downregulated |
| -0,044065401 | 0,894828454 | MAN2B2 | FALSE | Downregulated |
| 0,061826387 | 0,894828454 | NXF1 | FALSE | Upregulated |
| -0,167820756 | 0,894828454 | OLFM2 | FALSE | Downregulated |

| | | | | |
|---|---|---|---|---|
| 0,109727424 | 0,894828454 | PDE1A | FALSE | Upregulated |
| -0,191512673 | 0,894828454 | PRR4 | FALSE | Downregulated |
| 0,110393196 | 0,894828454 | PTPN14 | FALSE | Upregulated |
| 0,042341086 | 0,894828454 | RFFL | FALSE | Upregulated |
| -1,894972976 | 0,894828454 | RP11-451G4.2 | FALSE | Downregulated |
| 0,087850523 | 0,894828454 | RP11-513G11.3 | FALSE | Upregulated |
| 0,076836775 | 0,894828454 | SIRPA | FALSE | Upregulated |
| -0,173306401 | 0,894828454 | SPESP1 | FALSE | Downregulated |
| -0,03739617 | 0,894828454 | ST5 | FALSE | Downregulated |
| 0,037251757 | 0,894828454 | TIMM23 | FALSE | Upregulated |
| 0,031026412 | 0,894828454 | TMEM167A | FALSE | Upregulated |
| 0,061275634 | 0,894828454 | TMEM237 | FALSE | Upregulated |
| -0,686761145 | 0,894828454 | TNFRSF6B | FALSE | Downregulated |
| -0,037710002 | 0,894828454 | TRMT112 | FALSE | Downregulated |
| 0,04333214 | 0,894828454 | VPS13C | FALSE | Upregulated |
| 0,036252522 | 0,894828454 | XPO6 | FALSE | Upregulated |
| -0,042222174 | 0,894828454 | ZCCHC11 | FALSE | Downregulated |
| 0,038162892 | 0,894828454 | ZNF227 | FALSE | Upregulated |
| -0,168522951 | 0,894828454 | ZNF781 | FALSE | Downregulated |
| -0,107584842 | 0,894828454 | ZNF815P | FALSE | Downregulated |
| -0,038819222 | 0,894828454 | ZW10 | FALSE | Downregulated |
| 0,137699549 | 0,895091634 | BCAT1 | FALSE | Upregulated |
| -0,139297392 | 0,895091634 | BMS1P2 | FALSE | Downregulated |
| 0,054439395 | 0,895091634 | CADM1 | FALSE | Upregulated |
| 0,040337038 | 0,895091634 | CAMSAP1 | FALSE | Upregulated |
| 0,062973481 | 0,895091634 | CPN1 | FALSE | Upregulated |

| | | | | |
|---|---|---|---|---|
| 0,2263493 | 0,895091634 | CTRL | FALSE | Upregulated |
| 0,173039801 | 0,895091634 | DTHD1 | FALSE | Upregulated |
| -0,211163556 | 0,895091634 | FAM163A | FALSE | Downregulated |
| -0,072315228 | 0,895091634 | GJB2 | FALSE | Downregulated |
| -0,04719876 | 0,895091634 | KIAA1009 | FALSE | Downregulated |
| 0,075461434 | 0,895091634 | METRNL | FALSE | Upregulated |
| -0,040932007 | 0,895091634 | NEK1 | FALSE | Downregulated |
| -0,033440775 | 0,895091634 | NUDT9 | FALSE | Downregulated |
| -0,191571543 | 0,895091634 | PABPC4L | FALSE | Downregulated |
| -0,041477809 | 0,895091634 | PANK4 | FALSE | Downregulated |
| -0,292578207 | 0,895091634 | PI15 | FALSE | Downregulated |
| 0,116405784 | 0,895091634 | RIPPLY1 | FALSE | Upregulated |
| 0,096902452 | 0,895091634 | SLC35G1 | FALSE | Upregulated |
| 0,078432868 | 0,895091634 | ST6GAL2 | FALSE | Upregulated |
| 0,047168826 | 0,895091634 | SYNGR2 | FALSE | Upregulated |
| 1,214603905 | 0,895091634 | SYNPO2L | FALSE | Upregulated |
| 0,222268098 | 0,895091634 | USP32P2 | FALSE | Upregulated |
| -0,031458264 | 0,895091634 | ZNF800 | FALSE | Downregulated |
| 0,219725297 | 0,895119594 | ATP1A3 | FALSE | Upregulated |
| -0,262079696 | 0,895119594 | KCNH2 | FALSE | Downregulated |
| -0,270279617 | 0,895119594 | LOC102723894 | FALSE | Downregulated |
| 0,182550956 | 0,895119594 | NMB | FALSE | Upregulated |
| 0,043497875 | 0,895119594 | PRR14L | FALSE | Upregulated |
| 0,033416202 | 0,895119594 | TMX1 | FALSE | Upregulated |
| 0,129040205 | 0,895402116 | KCNJ10 | FALSE | Upregulated |
| -0,050927449 | 0,895517169 | C1orf131 | FALSE | Downregulated |

| | | | | |
|---|---|---|---|---|
| 0,027083778 | 0,895517169 | MAP2K4 | FALSE | Upregulated |
| 0,067360892 | 0,895710825 | ARHGAP25 | FALSE | Upregulated |
| -0,157957999 | 0,895710825 | ESCO2 | FALSE | Downregulated |
| -0,104099413 | 0,895767356 | CACHD1 | FALSE | Downregulated |
| -0,037544636 | 0,895878912 | DMXL1 | FALSE | Downregulated |
| -0,521456439 | 0,895878912 | LOC100996717 | FALSE | Downregulated |
| -0,035290098 | 0,895878912 | NADK2 | FALSE | Downregulated |
| 0,03499532 | 0,895878912 | SLC25A5 | FALSE | Upregulated |
| 0,261556415 | 0,895923154 | LOC101928059 | FALSE | Upregulated |
| 0,370328373 | 0,896065258 | PSORS1C3 | FALSE | Upregulated |
| 0,051716959 | 0,896088371 | GGT7 | FALSE | Upregulated |
| -0,057292299 | 0,896088371 | MTSS1 | FALSE | Downregulated |
| 0,423963228 | 0,896198493 | MAFA | FALSE | Upregulated |
| -0,029187413 | 0,896232812 | CCDC117 | FALSE | Downregulated |
| -0,306074469 | 0,896232812 | LOC101928414 | FALSE | Downregulated |
| -0,081370493 | 0,896232812 | TM6SF2 | FALSE | Downregulated |
| -0,102137669 | 0,896322718 | CA14 | FALSE | Downregulated |
| -0,115228044 | 0,896322718 | CATSPERG | FALSE | Downregulated |
| 0,032224263 | 0,896322718 | CKAP5 | FALSE | Upregulated |
| -0,043868444 | 0,896322718 | ESRP2 | FALSE | Downregulated |
| -0,276917592 | 0,896322718 | LOC100996643 | FALSE | Downregulated |
| -0,104273697 | 0,896322718 | LRRC48 | FALSE | Downregulated |
| 0,032520238 | 0,896322718 | SNX1 | FALSE | Upregulated |
| 0,472887806 | 0,896322718 | URGCP-MRPS24 | FALSE | Upregulated |
| -0,045759448 | 0,896322718 | USO1 | FALSE | Downregulated |
| 0,059135099 | 0,896322718 | ZKSCAN4 | FALSE | Upregulated |

| | | | | |
|---|---|---|---|---|
| -0,051640187 | 0,896322718 | ZNF337 | FALSE | Downregulated |
| -0,034001529 | 0,896534082 | ATF7 | FALSE | Downregulated |
| -0,149396282 | 0,896677076 | IDO2 | FALSE | Downregulated |
| -0,033699399 | 0,896677076 | ZNF362 | FALSE | Downregulated |
| 0,083182235 | 0,896761499 | CHST3 | FALSE | Upregulated |
| -0,061897251 | 0,896787313 | SLC9A3R1 | FALSE | Downregulated |
| -0,207416901 | 0,896788312 | AC072062.1 | FALSE | Downregulated |
| -0,071031653 | 0,896788312 | CHKA | FALSE | Downregulated |
| 0,039159107 | 0,896788312 | HDHD3 | FALSE | Upregulated |
| -0,076540979 | 0,896788312 | LCAT | FALSE | Downregulated |
| -0,033709913 | 0,896842231 | CRBN | FALSE | Downregulated |
| -0,02988895 | 0,896864309 | PEX11B | FALSE | Downregulated |
| 0,032105843 | 0,896864309 | SNUPN | FALSE | Upregulated |
| -0,057182386 | 0,896864309 | ZNF626 | FALSE | Downregulated |
| -0,035233242 | 0,896897633 | COMMD3-BMI1 | FALSE | Downregulated |
| 0,182040921 | 0,896897633 | HCG27 | FALSE | Upregulated |
| -0,033872729 | 0,896897633 | RNF185 | FALSE | Downregulated |
| -0,072405741 | 0,896897633 | RNF208 | FALSE | Downregulated |
| -0,064913669 | 0,896897633 | ZNF324 | FALSE | Downregulated |
| 0,061934073 | 0,896897633 | ZNF511 | FALSE | Upregulated |
| -0,077590217 | 0,896932616 | C1orf122 | FALSE | Downregulated |
| -0,053370303 | 0,896932616 | CMTM4 | FALSE | Downregulated |
| 0,070569553 | 0,896932616 | FMNL2 | FALSE | Upregulated |
| 0,042526373 | 0,896932616 | INTS3 | FALSE | Upregulated |
| -0,291514113 | 0,896932616 | KCNC4-AS1 | FALSE | Downregulated |
| -0,192387787 | 0,896932616 | LOC101928820 | FALSE | Downregulated |

| | | | | |
|---|---|---|---|---|
| -0,209140241 | 0,896932616 | MAMDC2 | FALSE | Downregulated |
| -0,054712121 | 0,896932616 | NUAK1 | FALSE | Downregulated |
| 0,036382667 | 0,896932616 | PRKAG1 | FALSE | Upregulated |
| 0,050735945 | 0,896932616 | RFT1 | FALSE | Upregulated |
| 0,159696344 | 0,896932616 | SEC14L5 | FALSE | Upregulated |
| -0,127660769 | 0,896932616 | SLC4A5 | FALSE | Downregulated |
| 0,056766164 | 0,896932616 | SSBP3 | FALSE | Upregulated |
| -0,041196354 | 0,896932616 | WBSCR16 | FALSE | Downregulated |
| 0,028187385 | 0,89702044 | U2AF2 | FALSE | Upregulated |
| 0,188038145 | 0,89710339 | P2RX3 | FALSE | Upregulated |
| 0,391684421 | 0,897193413 | TMEM252 | FALSE | Upregulated |
| 0,198170576 | 0,897267801 | LOC101928318 | FALSE | Upregulated |
| 0,085315831 | 0,898143301 | PVRL1 | FALSE | Upregulated |
| 0,215530765 | 0,898282684 | ATP5EP2 | FALSE | Upregulated |
| -0,041957302 | 0,898282684 | CASD1 | FALSE | Downregulated |
| 0,04199888 | 0,898282684 | CCDC101 | FALSE | Upregulated |
| 0,041806176 | 0,898282684 | COPB2 | FALSE | Upregulated |
| 0,22476872 | 0,898282684 | CTAGE7P | FALSE | Upregulated |
| 0,026959868 | 0,898282684 | IDH3B | FALSE | Upregulated |
| -0,157034252 | 0,898282684 | IHH | FALSE | Downregulated |
| 0,051414126 | 0,898282684 | TRAPPC5 | FALSE | Upregulated |
| -0,115372459 | 0,898298169 | LOC101928042 | FALSE | Downregulated |
| -0,079618968 | 0,898352699 | COMTD1 | FALSE | Downregulated |
| 0,031696208 | 0,898352699 | RNF6 | FALSE | Upregulated |
| -0,145983557 | 0,898352699 | TDRD6 | FALSE | Downregulated |
| -0,069430873 | 0,898376206 | ZSCAN31 | FALSE | Downregulated |

| | | | | |
|---|---|---|---|---|
| 0,031922304 | 0,898617253 | EEF2 | FALSE | Upregulated |
| 0,245515792 | 0,898617253 | RASD2 | FALSE | Upregulated |
| -0,053719402 | 0,898638065 | SLC15A1 | FALSE | Downregulated |
| -0,095586119 | 0,898733059 | PAXBP1-AS1 | FALSE | Downregulated |
| -0,038141132 | 0,898740935 | DHX29 | FALSE | Downregulated |
| 0,122211812 | 0,898740935 | KDM4D | FALSE | Upregulated |
| 0,048551224 | 0,898746326 | CALCOCO1 | FALSE | Upregulated |
| 0,056000732 | 0,898746326 | CKAP4 | FALSE | Upregulated |
| 0,128624632 | 0,898752596 | BCDIN3D-AS1 | FALSE | Upregulated |
| 0,077408503 | 0,898752596 | NOL3 | FALSE | Upregulated |
| 0,059308064 | 0,898752596 | SEMA4C | FALSE | Upregulated |
| -0,195074827 | 0,898752596 | TRPC3 | FALSE | Downregulated |
| -0,070888132 | 0,899383224 | B3GNT9 | FALSE | Downregulated |
| 0,046197435 | 0,89953412 | TRAFD1 | FALSE | Upregulated |
| -0,08030497 | 0,899553001 | CHKB | FALSE | Downregulated |
| 0,043781965 | 0,899553001 | CHPT1 | FALSE | Upregulated |
| -0,746007842 | 0,899553001 | RP4-669L17.10-2 | FALSE | Downregulated |
| 0,041571276 | 0,899728609 | RPS16 | FALSE | Upregulated |
| -0,055451083 | 0,899912916 | DTX4 | FALSE | Downregulated |
| 0,085647084 | 0,899912916 | MIR497HG | FALSE | Upregulated |
| 0,069094615 | 0,899937172 | KIAA0040 | FALSE | Upregulated |
| 0,187225914 | 0,899937172 | LOC102723755 | FALSE | Upregulated |
| 0,211253448 | 0,899937172 | LOC102723927 | FALSE | Upregulated |
| -0,10220896 | 0,899937172 | LRRN3 | FALSE | Downregulated |
| 0,038399696 | 0,899937172 | OAZ2 | FALSE | Upregulated |
| -0,291383312 | 0,899937172 | RBAK-RBAKDN | FALSE | Downregulated |

| | | | | |
|---|---|---|---|---|
| -0,089785095 | 0,899937172 | SLC31A2 | FALSE | Downregulated |
| -0,203044191 | 0,899937172 | TBX20 | FALSE | Downregulated |
| -0,090862696 | 0,899937172 | USP54 | FALSE | Downregulated |
| -0,062682294 | 0,899937172 | YBEY | FALSE | Downregulated |
| 0,05616304 | 0,900091506 | ARRB1 | FALSE | Upregulated |
| -0,103416463 | 0,900541137 | NINJ2 | FALSE | Downregulated |
| -0,088086496 | 0,900556624 | GBA3 | FALSE | Downregulated |
| -0,057874378 | 0,900556624 | RSBN1L-AS1 | FALSE | Downregulated |
| 0,276708409 | 0,900556624 | SLED1 | FALSE | Upregulated |
| 0,04959706 | 0,90066946 | HINT2 | FALSE | Upregulated |
| 0,225660923 | 0,900815802 | LOC283693 | FALSE | Upregulated |
| -0,027029567 | 0,900834034 | GUF1 | FALSE | Downregulated |
| -0,277468082 | 0,900834034 | HIST2H2BF | FALSE | Downregulated |
| -0,18271379 | 0,900834034 | LOC441666 | FALSE | Downregulated |
| -1,825067171 | 0,900834034 | MYH2 | FALSE | Downregulated |
| -0,028293376 | 0,900834034 | NGLY1 | FALSE | Downregulated |
| -0,286325605 | 0,900834034 | PEG3-AS1 | FALSE | Downregulated |
| -0,054714334 | 0,900834034 | PGBD2 | FALSE | Downregulated |
| -0,034227092 | 0,900834034 | SUPT4H1 | FALSE | Downregulated |
| -0,102382096 | 0,900936009 | BMP5 | FALSE | Downregulated |
| 0,046394825 | 0,900936009 | CALU | FALSE | Upregulated |
| 0,071038791 | 0,900936009 | CHAF1A | FALSE | Upregulated |
| 0,299505153 | 0,900936009 | LOC100653133 | FALSE | Upregulated |
| 0,120951913 | 0,900936009 | NOVA2 | FALSE | Upregulated |
| -0,098010948 | 0,900936009 | NXN | FALSE | Downregulated |
| -0,115230182 | 0,900936009 | VILL | FALSE | Downregulated |

| | | | | |
|---|---|---|---|---|
| -0,053803658 | 0,900947926 | AKAP8L | FALSE | Downregulated |
| 0,139590039 | 0,900947926 | LGR5 | FALSE | Upregulated |
| 0,048749374 | 0,900947926 | TRANK1 | FALSE | Upregulated |
| 0,249029538 | 0,900948162 | MIR4647 | FALSE | Upregulated |
| -0,091163459 | 0,900948162 | NRSN2 | FALSE | Downregulated |
| -0,042473711 | 0,900948162 | PSMA3 | FALSE | Downregulated |
| -0,043424452 | 0,900948162 | RNF128 | FALSE | Downregulated |
| 0,055121761 | 0,900948162 | VRK1 | FALSE | Upregulated |
| 0,037114229 | 0,901251421 | CRYL1 | FALSE | Upregulated |
| 0,061639048 | 0,901251421 | ZNF222 | FALSE | Upregulated |
| -0,162058246 | 0,901278109 | LOC101928113 | FALSE | Downregulated |
| -0,038299262 | 0,901734986 | SSFA2 | FALSE | Downregulated |
| -0,122899626 | 0,901800191 | C7orf55-LUC7L2 | FALSE | Downregulated |
| 0,055746421 | 0,901800191 | CEP89 | FALSE | Upregulated |
| 0,044783892 | 0,901800191 | CYB561A3 | FALSE | Upregulated |
| -0,060545128 | 0,901800191 | DCN | FALSE | Downregulated |
| -0,051925238 | 0,901800191 | EIF4G3 | FALSE | Downregulated |
| 0,03712129 | 0,901800191 | EXOC1 | FALSE | Upregulated |
| 0,184091293 | 0,901800191 | GPR55 | FALSE | Upregulated |
| 0,367682275 | 0,901800191 | LOC102724144 | FALSE | Upregulated |
| -0,064945432 | 0,901800191 | NT5E | FALSE | Downregulated |
| 0,07476872 | 0,901800191 | NUDT14 | FALSE | Upregulated |
| 0,026244158 | 0,901800191 | PDCD7 | FALSE | Upregulated |
| 0,045503358 | 0,901800191 | SLC25A1 | FALSE | Upregulated |
| -0,196591905 | 0,901800191 | SPATA17 | FALSE | Downregulated |
| 0,048134123 | 0,901800191 | SRGAP2 | FALSE | Upregulated |

| | | | | |
|---|---|---|---|---|
| -0,052097077 | 0,901814153 | C4BPA | FALSE | Downregulated |
| -0,047870502 | 0,901814153 | CHD6 | FALSE | Downregulated |
| -0,078427454 | 0,901814153 | CHN1 | FALSE | Downregulated |
| -0,040007588 | 0,901814153 | NAA30 | FALSE | Downregulated |
| -0,09441555 | 0,901814153 | TMED8 | FALSE | Downregulated |
| 0,059641641 | 0,901836471 | SUGP2 | FALSE | Upregulated |
| 0,033164089 | 0,901851803 | COPZ1 | FALSE | Upregulated |
| 0,036088813 | 0,901851803 | DESI2 | FALSE | Upregulated |
| 0,037074548 | 0,901851803 | GORASP1 | FALSE | Upregulated |
| 0,046223003 | 0,901851803 | HDAC5 | FALSE | Upregulated |
| 0,204376501 | 0,901851803 | LOC102723640 | FALSE | Upregulated |
| -0,055507428 | 0,901851803 | RASA2 | FALSE | Downregulated |
| 0,157577932 | 0,901875602 | JAKMIP1 | FALSE | Upregulated |
| 0,069553848 | 0,901886436 | CES1 | FALSE | Upregulated |
| 0,142422871 | 0,901886436 | GAL3ST1 | FALSE | Upregulated |
| -0,045726188 | 0,901886436 | ZNF865 | FALSE | Downregulated |
| -0,035847799 | 0,902077233 | DDA1 | FALSE | Downregulated |
| -0,128421263 | 0,902228908 | ZNF382 | FALSE | Downregulated |
| 0,193289318 | 0,902288608 | ZFHX4-AS1 | FALSE | Upregulated |
| 0,164629419 | 0,902549445 | AGPAT4 | FALSE | Upregulated |
| -0,073702205 | 0,902549445 | AURKC | FALSE | Downregulated |
| 0,108458662 | 0,902549445 | C21orf58 | FALSE | Upregulated |
| -0,11852604 | 0,902549445 | IL17RD | FALSE | Downregulated |
| -0,046733647 | 0,902549445 | SMYD3 | FALSE | Downregulated |
| -0,045422848 | 0,902549445 | UNG | FALSE | Downregulated |
| 0,052845404 | 0,902549445 | ZNF583 | FALSE | Upregulated |

| | | | | |
|---|---|---|---|---|
| -0,043414018 | 0,902572797 | CASP7 | FALSE | Downregulated |
| 0,06067803 | 0,902572797 | PNKP | FALSE | Upregulated |
| -0,093552753 | 0,902572797 | SGK1 | FALSE | Downregulated |
| -0,062737338 | 0,902572797 | TEF | FALSE | Downregulated |
| -0,055459303 | 0,902572797 | TTC12 | FALSE | Downregulated |
| 0,041408638 | 0,902572797 | VTI1A | FALSE | Upregulated |
| -0,032406463 | 0,902673145 | EXOSC3 | FALSE | Downregulated |
| -0,246376529 | 0,902673145 | FAM83E | FALSE | Downregulated |
| 0,080717431 | 0,902722944 | SULT1A1 | FALSE | Upregulated |
| 0,027570581 | 0,902724915 | AFG3L2 | FALSE | Upregulated |
| 0,068430305 | 0,902724915 | AIFM2 | FALSE | Upregulated |
| 0,031376188 | 0,902724915 | ASF1A | FALSE | Upregulated |
| 0,034956688 | 0,902724915 | ASGR2 | FALSE | Upregulated |
| -0,044575798 | 0,902724915 | ASXL1 | FALSE | Downregulated |
| -0,100437546 | 0,902724915 | ATF5 | FALSE | Downregulated |
| 0,058457888 | 0,902724915 | ATP9A | FALSE | Upregulated |
| -0,04910767 | 0,902724915 | C6 | FALSE | Downregulated |
| -0,058316236 | 0,902724915 | CD2AP | FALSE | Downregulated |
| -0,074500723 | 0,902724915 | CEP85L | FALSE | Downregulated |
| 0,048672593 | 0,902724915 | CHD7 | FALSE | Upregulated |
| 0,024273798 | 0,902724915 | CYFIP1 | FALSE | Upregulated |
| -0,05262486 | 0,902724915 | CYP2C9 | FALSE | Downregulated |
| -0,041227641 | 0,902724915 | DTWD1 | FALSE | Downregulated |
| -0,069756873 | 0,902724915 | FAM102B | FALSE | Downregulated |
| -0,055520587 | 0,902724915 | FAM114A1 | FALSE | Downregulated |
| -0,182714197 | 0,902724915 | LOC100507670 | FALSE | Downregulated |

| | | | | |
|---|---|---|---|---|
| -0,11939114 | 0,902724915 | LOC101928047 | FALSE | Downregulated |
| 0,18674281 | 0,902724915 | LOC102723539 | FALSE | Upregulated |
| 0,128413074 | 0,902724915 | LOC202181 | FALSE | Upregulated |
| -0,210594527 | 0,902724915 | MAPT | FALSE | Downregulated |
| 0,239985491 | 0,902724915 | MCOLN3 | FALSE | Upregulated |
| -0,056514097 | 0,902724915 | MYL12B | FALSE | Downregulated |
| 0,060886985 | 0,902724915 | NAP1L5 | FALSE | Upregulated |
| 0,047075563 | 0,902724915 | NOTCH1 | FALSE | Upregulated |
| 0,043853323 | 0,902724915 | POLR3D | FALSE | Upregulated |
| 0,030941557 | 0,902724915 | RING1 | FALSE | Upregulated |
| -0,040142517 | 0,902724915 | ROCK2 | FALSE | Downregulated |
| 0,037478043 | 0,902724915 | SDCCAG3 | FALSE | Upregulated |
| -0,058758858 | 0,902724915 | SEC11C | FALSE | Downregulated |
| 0,057523313 | 0,902724915 | SNN | FALSE | Upregulated |
| 0,051054566 | 0,902724915 | STIP1 | FALSE | Upregulated |
| -0,058252091 | 0,902724915 | STXBP5 | FALSE | Downregulated |
| 0,056507568 | 0,902724915 | VIMP | FALSE | Upregulated |
| 0,036369974 | 0,902724915 | VPS16 | FALSE | Upregulated |
| 0,080638716 | 0,90274719 | ATAD3B | FALSE | Upregulated |
| 0,188873835 | 0,90274719 | HIST3H2A | FALSE | Upregulated |
| -0,188928727 | 0,90274719 | LOC101927206 | FALSE | Downregulated |
| -0,055593169 | 0,90274719 | LYRM4 | FALSE | Downregulated |
| 0,044222836 | 0,90274719 | MAD2L1BP | FALSE | Upregulated |
| -0,045486538 | 0,90274719 | POGZ | FALSE | Downregulated |
| 0,106096119 | 0,90274719 | TAPT1-AS1 | FALSE | Upregulated |
| -0,05792209 | 0,90274719 | TP73-AS1 | FALSE | Downregulated |

| | | | | |
|---|---|---|---|---|
| 0,172660855 | 0,902867256 | LY6G5B | FALSE | Upregulated |
| -0,039288635 | 0,902917845 | ANAPC1 | FALSE | Downregulated |
| -0,041416176 | 0,902917845 | ERF | FALSE | Downregulated |
| -0,024221141 | 0,902917845 | MRPL16 | FALSE | Downregulated |
| 0,056367489 | 0,902917845 | NFKBIA | FALSE | Upregulated |
| 0,033620661 | 0,902917845 | WDFY1 | FALSE | Upregulated |
| 0,095601697 | 0,90297371 | EML5 | FALSE | Upregulated |
| -0,089172448 | 0,902980073 | WASF3 | FALSE | Downregulated |
| 0,033628947 | 0,903037858 | C6orf62 | FALSE | Upregulated |
| 0,089010612 | 0,903039431 | ZNF738 | FALSE | Upregulated |
| 0,194285641 | 0,903120644 | C9orf139 | FALSE | Upregulated |
| -0,219197253 | 0,90312958 | LMX1A | FALSE | Downregulated |
| 0,121940321 | 0,90312958 | LOC102724112 | FALSE | Upregulated |
| -0,038685249 | 0,903403755 | GPR155 | FALSE | Downregulated |
| -0,258759161 | 0,903403755 | PTCRA | FALSE | Downregulated |
| 0,055361895 | 0,903403755 | PXDN | FALSE | Upregulated |
| -0,041236775 | 0,903403755 | SIL1 | FALSE | Downregulated |
| 0,059949414 | 0,903403755 | SLC27A3 | FALSE | Upregulated |
| 0,054394972 | 0,90346226 | LRRC3 | FALSE | Upregulated |
| 0,33503375 | 0,903506225 | SLC2A14 | FALSE | Upregulated |
| -0,142878501 | 0,903678873 | SGCD | FALSE | Downregulated |
| 0,025868298 | 0,903824902 | PLRG1 | FALSE | Upregulated |
| 0,079334385 | 0,903866718 | ARHGEF28 | FALSE | Upregulated |
| 0,035480126 | 0,903993573 | ACVR1 | FALSE | Upregulated |
| -0,144948219 | 0,903993573 | FOS | FALSE | Downregulated |
| 0,130540759 | 0,903993573 | SLC5A1 | FALSE | Upregulated |

| | | | | |
|---|---|---|---|---|
| 0,038149372 | 0,904198621 | C12orf65 | FALSE | Upregulated |
| 0,038102639 | 0,904198621 | IMP3 | FALSE | Upregulated |
| -0,232756235 | 0,904198621 | LOC101928764 | FALSE | Downregulated |
| -0,051143975 | 0,904198621 | MTHFR | FALSE | Downregulated |
| 0,182262888 | 0,904234951 | ANO9 | FALSE | Upregulated |
| 0,086986074 | 0,904234951 | DDX11 | FALSE | Upregulated |
| -0,056411458 | 0,904234951 | MRPL41 | FALSE | Downregulated |
| -0,102738544 | 0,904234951 | UBE2Q2P1 | FALSE | Downregulated |
| 0,112354814 | 0,904290069 | A2M-AS1 | FALSE | Upregulated |
| 0,127088875 | 0,904290069 | C20orf196 | FALSE | Upregulated |
| -0,170029422 | 0,904290069 | DACT2 | FALSE | Downregulated |
| -0,116295359 | 0,904290069 | GCSAM | FALSE | Downregulated |
| 0,329533824 | 0,904290069 | LOC644172 | FALSE | Upregulated |
| -0,060101948 | 0,904290069 | PIM3 | FALSE | Downregulated |
| 0,048698468 | 0,904290069 | PPP4R1 | FALSE | Upregulated |
| 0,096878762 | 0,904336049 | AP1S3 | FALSE | Upregulated |
| -0,050374372 | 0,904336049 | F13B | FALSE | Downregulated |
| -0,140522556 | 0,904336049 | LOC100506472 | FALSE | Downregulated |
| 0,065882941 | 0,904336049 | PSD3 | FALSE | Upregulated |
| 0,029854724 | 0,904336049 | RNF114 | FALSE | Upregulated |
| -0,100968622 | 0,904336049 | RP11-205M3.3 | FALSE | Downregulated |
| 0,074762717 | 0,904336049 | SERPINB9 | FALSE | Upregulated |
| 0,113642333 | 0,904336049 | SLC9B1 | FALSE | Upregulated |
| 0,024467817 | 0,904336049 | SMCR8 | FALSE | Upregulated |
| -0,027902464 | 0,904344265 | CRNKL1 | FALSE | Downregulated |
| -0,068956111 | 0,904344265 | GLDC | FALSE | Downregulated |

| | | | | |
|---|---|---|---|---|
| -0,06191266 | 0,904344265 | HOGA1 | FALSE | Downregulated |
| -0,1484566 | 0,904344265 | LOC100288637 | FALSE | Downregulated |
| 0,25928874 | 0,904344265 | LOC102724601 | FALSE | Upregulated |
| 0,232685652 | 0,9043544 | ANKRD36BP1 | FALSE | Upregulated |
| 0,058279435 | 0,9043544 | CCDC142 | FALSE | Upregulated |
| -0,054478523 | 0,9043544 | FAM20C | FALSE | Downregulated |
| -0,036915836 | 0,9043544 | FDX1 | FALSE | Downregulated |
| 0,060336281 | 0,9043544 | GTPBP3 | FALSE | Upregulated |
| 0,059308983 | 0,9043544 | LOC100130744 | FALSE | Upregulated |
| -0,876493748 | 0,9043544 | LOC102724594 | FALSE | Downregulated |
| -0,127293544 | 0,9043544 | LYPD6 | FALSE | Downregulated |
| -0,049179426 | 0,9043544 | MOSPD2 | FALSE | Downregulated |
| -0,099092632 | 0,9043544 | NMUR1 | FALSE | Downregulated |
| -0,066022664 | 0,9043544 | PHC1 | FALSE | Downregulated |
| -0,187805609 | 0,9043544 | PKD1L3 | FALSE | Downregulated |
| 0,06519091 | 0,9043544 | PRDM15 | FALSE | Upregulated |
| -0,026483194 | 0,9043544 | SMG7 | FALSE | Downregulated |
| 0,184237337 | 0,9043544 | SNAI3 | FALSE | Upregulated |
| 0,066027287 | 0,904371259 | CCDC3 | FALSE | Upregulated |
| 0,238856573 | 0,904371259 | CIRBP-AS1 | FALSE | Upregulated |
| -0,068300583 | 0,904371259 | DHFR | FALSE | Downregulated |
| 0,03131316 | 0,904371259 | EHMT2 | FALSE | Upregulated |
| -0,073381614 | 0,904371259 | FAM76B | FALSE | Downregulated |
| 0,089223545 | 0,904371259 | FTX | FALSE | Upregulated |
| 0,176059839 | 0,904371259 | NR4A1 | FALSE | Upregulated |
| 0,128412889 | 0,904371259 | P2RY12 | FALSE | Upregulated |

| | | | | |
|---|---|---|---|---|
| 0,029754733 | 0,904371259 | PSME4 | FALSE | Upregulated |
| 0,043260521 | 0,904371259 | SLC11A2 | FALSE | Upregulated |
| -0,060726319 | 0,904371259 | SLX4 | FALSE | Downregulated |
| 0,278305045 | 0,904371259 | TMLHE-AS1 | FALSE | Upregulated |
| -0,042962346 | 0,904541476 | NFE2L2 | FALSE | Downregulated |
| 0,03622688 | 0,9045904 | TMEM168 | FALSE | Upregulated |
| -0,050672752 | 0,904677717 | CBLB | FALSE | Downregulated |
| -0,041648954 | 0,904677717 | LACTB | FALSE | Downregulated |
| -0,09149931 | 0,904677717 | LINC01089 | FALSE | Downregulated |
| -0,213836797 | 0,904677717 | LOC102724372 | FALSE | Downregulated |
| -0,058827204 | 0,904677717 | LRRC8B | FALSE | Downregulated |
| -0,055148904 | 0,904761935 | ADH1B | FALSE | Downregulated |
| -0,132734004 | 0,90487828 | GLP2R | FALSE | Downregulated |
| -0,049288617 | 0,904940113 | ANGEL1 | FALSE | Downregulated |
| -0,023049795 | 0,905293019 | MRFAP1 | FALSE | Downregulated |
| 0,207984847 | 0,905293019 | NOXRED1 | FALSE | Upregulated |
| 0,079724408 | 0,905293019 | SLC25A47 | FALSE | Upregulated |
| 0,036660288 | 0,905346618 | ATP1B1 | FALSE | Upregulated |
| 0,070813534 | 0,905346618 | SLC26A2 | FALSE | Upregulated |
| -0,287651736 | 0,905351108 | NOS1 | FALSE | Downregulated |
| 0,079806916 | 0,905464202 | LOC100506127 | FALSE | Upregulated |
| 0,042156728 | 0,905665207 | CACFD1 | FALSE | Upregulated |
| -0,130794029 | 0,905665207 | LOC102724229 | FALSE | Downregulated |
| -0,108987953 | 0,905665207 | MST1L | FALSE | Downregulated |
| 0,032351579 | 0,905665207 | OTUD7B | FALSE | Upregulated |
| 0,114328916 | 0,905665207 | SCARF2 | FALSE | Upregulated |

| | | | | |
|---|---|---|---|---|
| 0,044988755 | 0,905885698 | FAM49B | FALSE | Upregulated |
| -0,036414601 | 0,906090215 | ATP5C1 | FALSE | Downregulated |
| -0,049617276 | 0,906090215 | CALD1 | FALSE | Downregulated |
| -0,039609684 | 0,906090215 | CARHSP1 | FALSE | Downregulated |
| -0,036125057 | 0,906090215 | CHCHD7 | FALSE | Downregulated |
| -0,382950336 | 0,906090215 | CTAGE4 | FALSE | Downregulated |
| 0,062083014 | 0,906090215 | FOXRED2 | FALSE | Upregulated |
| 0,053854918 | 0,906090215 | GPR137 | FALSE | Upregulated |
| -0,081598161 | 0,906090215 | HCN3 | FALSE | Downregulated |
| -0,09081636 | 0,906090215 | LOC102723954 | FALSE | Downregulated |
| 0,044003778 | 0,906090215 | MTMR12 | FALSE | Upregulated |
| 0,13424191 | 0,906090215 | MTMR7 | FALSE | Upregulated |
| 0,114463741 | 0,906090215 | NDC80 | FALSE | Upregulated |
| 0,127632576 | 0,906090215 | PLEKHM1P | FALSE | Upregulated |
| 0,047242492 | 0,906090215 | UGT2B4 | FALSE | Upregulated |
| -0,095566817 | 0,906090215 | ZSWIM4 | FALSE | Downregulated |
| -0,221672477 | 0,906325098 | ASIC4 | FALSE | Downregulated |
| 0,069474518 | 0,906382492 | C8orf4 | FALSE | Upregulated |
| -0,077797551 | 0,906382492 | EPHB6 | FALSE | Downregulated |
| -0,038746686 | 0,906382492 | KLHL15 | FALSE | Downregulated |
| -0,057464202 | 0,906382492 | LTBP1 | FALSE | Downregulated |
| 0,170404418 | 0,906382492 | RAB19 | FALSE | Upregulated |
| -0,033508819 | 0,906506732 | MTERFD1 | FALSE | Downregulated |
| -0,199423382 | 0,906506732 | PARD6G-AS1 | FALSE | Downregulated |
| 0,027076635 | 0,906506732 | TSPYL1 | FALSE | Upregulated |
| 0,091124922 | 0,906617438 | CDH19 | FALSE | Upregulated |

| | | | | |
|---|---|---|---|---|
| -0,192590988 | 0,906659934 | LOC101927546 | FALSE | Downregulated |
| 0,043191061 | 0,906659934 | SWI5 | FALSE | Upregulated |
| -0,049929424 | 0,907050338 | VPS29 | FALSE | Downregulated |
| 0,066756163 | 0,907093146 | ATP2A2 | FALSE | Upregulated |
| 0,134519068 | 0,907237785 | EPPK1 | FALSE | Upregulated |
| 0,057336896 | 0,907280534 | ANXA9 | FALSE | Upregulated |
| -0,036903304 | 0,907280534 | LPP | FALSE | Downregulated |
| 0,04347688 | 0,907304683 | BTRC | FALSE | Upregulated |
| 0,049159531 | 0,907304683 | CNIH4 | FALSE | Upregulated |
| -0,113101136 | 0,907304683 | LOC102723546 | FALSE | Downregulated |
| 0,049941942 | 0,907326933 | CAD | FALSE | Upregulated |
| 0,182912206 | 0,907333505 | WT1 | FALSE | Upregulated |
| 0,037570709 | 0,907393747 | ZDHHC7 | FALSE | Upregulated |
| 0,052389763 | 0,907674634 | AMOT | FALSE | Upregulated |
| 0,226952935 | 0,907674634 | GFAP | FALSE | Upregulated |
| 0,207939503 | 0,907674634 | LOC101928705 | FALSE | Upregulated |
| -0,034275856 | 0,907674634 | ORC3 | FALSE | Downregulated |
| 0,071484037 | 0,907674634 | PNPLA7 | FALSE | Upregulated |
| -0,030480946 | 0,907674634 | SAP130 | FALSE | Downregulated |
| 0,045191711 | 0,907674634 | SLC25A28 | FALSE | Upregulated |
| 0,039428472 | 0,907710577 | C1GALT1C1 | FALSE | Upregulated |
| 0,076937214 | 0,907926411 | CYBRD1 | FALSE | Upregulated |
| 0,090503388 | 0,907926411 | DNASE1 | FALSE | Upregulated |
| -0,104655586 | 0,907926411 | FDPS | FALSE | Downregulated |
| -0,194517344 | 0,907926411 | FLJ21408 | FALSE | Downregulated |
| 0,171995114 | 0,907926411 | FLJ42351 | FALSE | Upregulated |

| | | | | |
|---|---|---|---|---|
| 0,133450668 | 0,907926411 | GZMB | FALSE | Upregulated |
| 0,077814248 | 0,907926411 | KIAA1456 | FALSE | Upregulated |
| -0,040568551 | 0,907926411 | PPP2R2A | FALSE | Downregulated |
| -0,056664856 | 0,907926411 | SLC2A8 | FALSE | Downregulated |
| -0,075182018 | 0,907956809 | KBTBD8 | FALSE | Downregulated |
| -0,06738154 | 0,908018181 | FUT8 | FALSE | Downregulated |
| -0,037318337 | 0,908124125 | ATXN1 | FALSE | Downregulated |
| 0,097322905 | 0,908124125 | GUSBP11 | FALSE | Upregulated |
| -0,134598896 | 0,908124125 | LOC101928158 | FALSE | Downregulated |
| -1,726048618 | 0,908124125 | MYL1 | FALSE | Downregulated |
| 0,042195346 | 0,908124125 | NDUFA13 | FALSE | Upregulated |
| -0,166988595 | 0,908124125 | RP11-637A17.2 | FALSE | Downregulated |
| -0,041081525 | 0,908126634 | PES1 | FALSE | Downregulated |
| -0,035713135 | 0,908129748 | EIF1 | FALSE | Downregulated |
| -0,200630466 | 0,908129748 | LOC102724117 | FALSE | Downregulated |
| 0,128513683 | 0,908129748 | LOC644656 | FALSE | Upregulated |
| -0,113550064 | 0,908129748 | NPY5R | FALSE | Downregulated |
| -0,108499979 | 0,908269865 | ALMS1-IT1 | FALSE | Downregulated |
| 0,212855433 | 0,908269865 | CIB2 | FALSE | Upregulated |
| -0,22951717 | 0,908269865 | HSPB2-C11orf52 | FALSE | Downregulated |
| 0,060277796 | 0,908269865 | LIMD1 | FALSE | Upregulated |
| -0,045561189 | 0,908438351 | FAM104B | FALSE | Downregulated |
| -0,118596867 | 0,908487269 | PGF | FALSE | Downregulated |
| -0,155383119 | 0,908488155 | FAM225A | FALSE | Downregulated |
| 0,136971777 | 0,908501625 | LOC643355 | FALSE | Upregulated |
| 0,062588 | 0,908501625 | REEP4 | FALSE | Upregulated |

| | | | | |
|---|---|---|---|---|
| 0,098197802 | 0,908501625 | TMEM156 | FALSE | Upregulated |
| -0,178964527 | 0,90853651 | ZDHHC15 | FALSE | Downregulated |
| -0,045643441 | 0,908682925 | RPL13 | FALSE | Downregulated |
| 0,076735354 | 0,908752739 | KANK3 | FALSE | Upregulated |
| 0,065487379 | 0,908865711 | RTP3 | FALSE | Upregulated |
| -0,227098564 | 0,908907465 | DOK5 | FALSE | Downregulated |
| 0,033665261 | 0,908907465 | PI4KB | FALSE | Upregulated |
| -0,054632981 | 0,908907465 | ZSWIM7 | FALSE | Downregulated |
| 0,047017349 | 0,909019609 | POLR2L | FALSE | Upregulated |
| -0,281669077 | 0,909024847 | AC093375.1 | FALSE | Downregulated |
| 0,057894288 | 0,909024847 | CARD16 | FALSE | Upregulated |
| -0,060405862 | 0,909024847 | CLK1 | FALSE | Downregulated |
| -0,043437972 | 0,909024847 | EEF1B2 | FALSE | Downregulated |
| -0,290335949 | 0,909024847 | LOC388499 | FALSE | Downregulated |
| 0,04497533 | 0,909024847 | SLC39A3 | FALSE | Upregulated |
| 0,086288043 | 0,909024847 | ZNF169 | FALSE | Upregulated |
| 0,084197576 | 0,909024847 | ZNF460 | FALSE | Upregulated |
| -0,207408614 | 0,909212859 | COL6A4P2 | FALSE | Downregulated |
| 0,06293556 | 0,909212859 | FLVCR1 | FALSE | Upregulated |
| -0,033277866 | 0,909212859 | GALM | FALSE | Downregulated |
| 0,612001272 | 0,909212859 | LOC100289026 | FALSE | Upregulated |
| 0,082064501 | 0,909212859 | LOC102724967 | FALSE | Upregulated |
| -0,04131586 | 0,909212859 | METTL9 | FALSE | Downregulated |
| 0,261058557 | 0,909212859 | MLIP-IT1 | FALSE | Upregulated |
| 0,211558532 | 0,909212859 | NTN3 | FALSE | Upregulated |
| 0,045135646 | 0,909212859 | OGDHL | FALSE | Upregulated |

| | | | | |
|---|---|---|---|---|
| 0,082217722 | 0,909212859 | STPG1 | FALSE | Upregulated |
| -0,073829265 | 0,909310227 | DLG4 | FALSE | Downregulated |
| -0,044254274 | 0,909310227 | TRAK1 | FALSE | Downregulated |
| -0,053552027 | 0,909428125 | COX10 | FALSE | Downregulated |
| -0,081475678 | 0,909460676 | GALK1 | FALSE | Downregulated |
| 0,118945577 | 0,909460676 | HSPA2 | FALSE | Upregulated |
| 0,018255001 | 0,909460676 | IST1 | FALSE | Upregulated |
| -0,123465548 | 0,909460676 | MBL1P | FALSE | Downregulated |
| -0,103269975 | 0,909460676 | MIF | FALSE | Downregulated |
| 0,044750273 | 0,909460676 | NPRL2 | FALSE | Upregulated |
| -0,035021253 | 0,909460676 | NUB1 | FALSE | Downregulated |
| -0,045681854 | 0,909460676 | PCK2 | FALSE | Downregulated |
| 0,077976084 | 0,909460676 | PDE4B | FALSE | Upregulated |
| 0,202980424 | 0,909460676 | RASL11B | FALSE | Upregulated |
| 0,048472183 | 0,909460676 | RPS6 | FALSE | Upregulated |
| 0,12605052 | 0,909460676 | TMEM9B-AS1 | FALSE | Upregulated |
| 0,028867099 | 0,909460676 | TRIM39 | FALSE | Upregulated |
| -0,043483791 | 0,909460676 | UBA7 | FALSE | Downregulated |
| -0,099498295 | 0,909460676 | WDFY3-AS2 | FALSE | Downregulated |
| -0,035456443 | 0,909460676 | WDR7 | FALSE | Downregulated |
| -0,04890141 | 0,909460676 | ZCCHC4 | FALSE | Downregulated |
| -0,047611693 | 0,909460676 | ZRANB3 | FALSE | Downregulated |
| 0,021519673 | 0,909578527 | PTCD3 | FALSE | Upregulated |
| -0,05245805 | 0,909690306 | MGST2 | FALSE | Downregulated |
| -0,112660652 | 0,909690306 | VLDLR | FALSE | Downregulated |
| 0,263518281 | 0,909804967 | LOC646670 | FALSE | Upregulated |

| | | | | |
|---|---|---|---|---|
| 0,093343314 | 0,909804967 | PHOSPHO2 | FALSE | Upregulated |
| 0,106797803 | 0,909938764 | SALL2 | FALSE | Upregulated |
| -0,029115788 | 0,909938764 | SYNJ2BP | FALSE | Downregulated |
| 0,08957914 | 0,909938764 | ZNF528 | FALSE | Upregulated |
| -0,045385623 | 0,909969773 | ILF3-AS1 | FALSE | Downregulated |
| -0,744196641 | 0,910170118 | SNORD3B-1 | FALSE | Downregulated |
| 0,048260926 | 0,910176788 | POP1 | FALSE | Upregulated |
| -0,091398012 | 0,910323271 | AC009299.3 | FALSE | Downregulated |
| -0,126379441 | 0,910323271 | ALDH1A3 | FALSE | Downregulated |
| 0,065334567 | 0,910323271 | CHD3 | FALSE | Upregulated |
| 0,037093639 | 0,910323271 | CYLD | FALSE | Upregulated |
| 0,065991763 | 0,910323271 | FAM213B | FALSE | Upregulated |
| -0,070194815 | 0,910323271 | FAM47E-STBD1 | FALSE | Downregulated |
| 0,039741131 | 0,910323271 | GLTP | FALSE | Upregulated |
| -0,174496761 | 0,910323271 | LOC102723465 | FALSE | Downregulated |
| 0,076091008 | 0,910323271 | LOC102723992 | FALSE | Upregulated |
| 0,11803795 | 0,910323271 | LOC102724770 | FALSE | Upregulated |
| 0,108769651 | 0,910323271 | LOC102724880 | FALSE | Upregulated |
| -0,089444352 | 0,910323271 | LOC646976 | FALSE | Downregulated |
| 0,034387792 | 0,910323271 | MAOB | FALSE | Upregulated |
| -0,095705938 | 0,910323271 | NCF1 | FALSE | Downregulated |
| -0,038201187 | 0,910323271 | OCRL | FALSE | Downregulated |
| -0,046929467 | 0,910323271 | PMF1 | FALSE | Downregulated |
| 0,029144246 | 0,910323271 | PPP2R5E | FALSE | Upregulated |
| -0,078778412 | 0,910323271 | PRR24 | FALSE | Downregulated |
| -0,192267249 | 0,910323271 | SEC14L6 | FALSE | Downregulated |

| | | | | |
|---|---|---|---|---|
| -1,694194942 | 0,910323271 | SLN | FALSE | Downregulated |
| -0,078983398 | 0,910323271 | SRRM2 | FALSE | Downregulated |
| 0,082347227 | 0,910323271 | STAMBPL1 | FALSE | Upregulated |
| -0,096016141 | 0,910323271 | TSTD3 | FALSE | Downregulated |
| 0,071433384 | 0,910323271 | ZC3H6 | FALSE | Upregulated |
| -0,082196297 | 0,910355955 | CEP152 | FALSE | Downregulated |
| -0,497549512 | 0,910355955 | CLVS2 | FALSE | Downregulated |
| -0,035171555 | 0,910355955 | IBTK | FALSE | Downregulated |
| 0,067376385 | 0,910355955 | ID2 | FALSE | Upregulated |
| 0,083874146 | 0,910355955 | LOC100505771 | FALSE | Upregulated |
| -0,042543901 | 0,910355955 | TFR2 | FALSE | Downregulated |
| 0,046865651 | 0,910355955 | THOC6 | FALSE | Upregulated |
| -0,093171536 | 0,910355955 | WNT5B | FALSE | Downregulated |
| -0,023909613 | 0,910355955 | ZNF24 | FALSE | Downregulated |
| -0,039495749 | 0,910587194 | CAMTA1 | FALSE | Downregulated |
| 0,065310785 | 0,910587194 | HN1 | FALSE | Upregulated |
| -0,122296103 | 0,910587194 | LINC00342 | FALSE | Downregulated |
| 0,245847385 | 0,91081937 | CCDC42 | FALSE | Upregulated |
| -0,036086757 | 0,91081937 | CCP110 | FALSE | Downregulated |
| -0,149314259 | 0,910858992 | LOC101929079 | FALSE | Downregulated |
| -0,26381678 | 0,911147645 | GDNF-AS1 | FALSE | Downregulated |
| 0,159670378 | 0,911147645 | SEC14L1P1 | FALSE | Upregulated |
| 0,019826124 | 0,911374684 | SH3GLB1 | FALSE | Upregulated |
| -0,034802718 | 0,911468679 | DDX52 | FALSE | Downregulated |
| 0,041961638 | 0,911468679 | RNF166 | FALSE | Upregulated |
| -0,06816451 | 0,911572584 | AFAP1L2 | FALSE | Downregulated |

| | | | | |
|---|---|---|---|---|
| 0,038245823 | 0,911572584 | CACTIN | FALSE | Upregulated |
| 0,078552513 | 0,911572584 | DHCR7 | FALSE | Upregulated |
| 0,0609833 | 0,911572584 | DONSON | FALSE | Upregulated |
| -0,03412895 | 0,911572584 | ELK4 | FALSE | Downregulated |
| 0,058163266 | 0,911572584 | FAM102A | FALSE | Upregulated |
| -0,181182593 | 0,911572584 | HMGB3P1 | FALSE | Downregulated |
| -0,041839695 | 0,911572584 | HOOK3 | FALSE | Downregulated |
| 0,067917018 | 0,911572584 | MAP2 | FALSE | Upregulated |
| 0,115136587 | 0,911572584 | SLC16A11 | FALSE | Upregulated |
| 0,142722625 | 0,911572584 | SSPO | FALSE | Upregulated |
| -0,041719551 | 0,911572584 | TARS | FALSE | Downregulated |
| 0,037315196 | 0,911572584 | TST | FALSE | Upregulated |
| 0,028086467 | 0,91169783 | ABHD13 | FALSE | Upregulated |
| 0,064342807 | 0,91169783 | ANKMY1 | FALSE | Upregulated |
| -0,088106683 | 0,91169783 | BCL3 | FALSE | Downregulated |
| -0,278618958 | 0,91169783 | CCDC108 | FALSE | Downregulated |
| 0,036302581 | 0,91169783 | DAG1 | FALSE | Upregulated |
| -0,060941837 | 0,91169783 | EHBP1L1 | FALSE | Downregulated |
| -0,063112902 | 0,91169783 | FLOT2 | FALSE | Downregulated |
| -0,075273496 | 0,91169783 | HEPH | FALSE | Downregulated |
| -0,111943806 | 0,91169783 | LOC100506023 | FALSE | Downregulated |
| 0,19887439 | 0,91169783 | LOC101559451 | FALSE | Upregulated |
| 0,171879183 | 0,91169783 | LOC101928096 | FALSE | Upregulated |
| 0,166005335 | 0,91169783 | LOC101928584 | FALSE | Upregulated |
| 0,158496702 | 0,91169783 | LOC102723770 | FALSE | Upregulated |
| -0,059185947 | 0,91169783 | LPAR6 | FALSE | Downregulated |

| | | | | |
|---|---|---|---|---|
| -0,075513978 | 0,91169783 | NEU4 | FALSE | Downregulated |
| 0,029511099 | 0,91169783 | PBX2 | FALSE | Upregulated |
| 0,045086727 | 0,91169783 | PGAM5 | FALSE | Upregulated |
| 0,093193858 | 0,91169783 | PLA2G4C | FALSE | Upregulated |
| -0,057878531 | 0,91169783 | PSMC4 | FALSE | Downregulated |
| 0,04545763 | 0,91169783 | RAD18 | FALSE | Upregulated |
| 0,035005778 | 0,91169783 | SEL1L | FALSE | Upregulated |
| 0,089434976 | 0,91169783 | SEMA4F | FALSE | Upregulated |
| -0,039943091 | 0,91169783 | SKA2 | FALSE | Downregulated |
| -0,287800944 | 0,91169783 | STON1-GTF2A1L | FALSE | Downregulated |
| 0,031153953 | 0,91169783 | STRIP1 | FALSE | Upregulated |
| 0,048630918 | 0,91169783 | SYNPO | FALSE | Upregulated |
| 0,046866897 | 0,91169783 | TCEB1 | FALSE | Upregulated |
| 0,060147106 | 0,91175462 | CD68 | FALSE | Upregulated |
| 0,225247519 | 0,911906199 | RNASE13 | FALSE | Upregulated |
| -0,039798457 | 0,912205646 | BAIAP2-AS1 | FALSE | Downregulated |
| -0,057166431 | 0,912205646 | CP | FALSE | Downregulated |
| 0,058305394 | 0,912205646 | CYP4F2 | FALSE | Upregulated |
| -0,051650435 | 0,912205646 | DSP | FALSE | Downregulated |
| -0,11245342 | 0,912205646 | FXYD2 | FALSE | Downregulated |
| -0,106335097 | 0,912205646 | LINC00641 | FALSE | Downregulated |
| 0,211288096 | 0,912205646 | LMCD1-AS1 | FALSE | Upregulated |
| -0,162350628 | 0,912205646 | LOC100288911 | FALSE | Downregulated |
| 0,172348836 | 0,912205646 | LOC102724805 | FALSE | Upregulated |
| 0,043911344 | 0,912205646 | MAP2K2 | FALSE | Upregulated |
| -0,149623293 | 0,912205646 | PFN4 | FALSE | Downregulated |

| | | | | |
|---|---|---|---|---|
| 0,144936732 | 0,912205646 | PTGS2 | FALSE | Upregulated |
| 0,297359386 | 0,912205646 | TBL1Y | FALSE | Upregulated |
| 0,059208226 | 0,912205646 | TLDC1 | FALSE | Upregulated |
| -0,051975332 | 0,912283544 | APRT | FALSE | Downregulated |
| 0,181158776 | 0,912283544 | ARHGAP22 | FALSE | Upregulated |
| -0,027000551 | 0,912283544 | DDX18 | FALSE | Downregulated |
| 0,031816316 | 0,912283544 | HDGFRP2 | FALSE | Upregulated |
| -0,055885007 | 0,912283544 | KMO | FALSE | Downregulated |
| 0,032494066 | 0,912283544 | POM121 | FALSE | Upregulated |
| 0,186397895 | 0,912283544 | TAS2R10 | FALSE | Upregulated |
| -0,233060173 | 0,912283544 | TNR | FALSE | Downregulated |
| -0,051407798 | 0,91230979 | CYYR1 | FALSE | Downregulated |
| 0,112746105 | 0,91230979 | DEFB1 | FALSE | Upregulated |
| -0,066211498 | 0,91230979 | KHK | FALSE | Downregulated |
| 0,073269853 | 0,912395109 | DZANK1 | FALSE | Upregulated |
| 0,043154609 | 0,912440262 | KLHL18 | FALSE | Upregulated |
| 0,036117092 | 0,912449501 | BRAF | FALSE | Upregulated |
| 0,224287926 | 0,912449501 | HEPN1 | FALSE | Upregulated |
| -0,026622666 | 0,912449501 | LIMS1 | FALSE | Downregulated |
| 0,043626493 | 0,912449501 | METTL6 | FALSE | Upregulated |
| -0,026468042 | 0,912449501 | NPTN | FALSE | Downregulated |
| 0,115005354 | 0,912449501 | PDCD4-AS1 | FALSE | Upregulated |
| -0,369403861 | 0,912449501 | SPIN2A | FALSE | Downregulated |
| 0,122713513 | 0,912449501 | TNFSF15 | FALSE | Upregulated |
| 0,041862327 | 0,912587862 | TRIT1 | FALSE | Upregulated |
| -0,075903898 | 0,912748429 | ARHGAP20 | FALSE | Downregulated |

| | | | | |
|---|---|---|---|---|
| 0,092908181 | 0,912748429 | ARIH2OS | FALSE | Upregulated |
| 0,062486948 | 0,912748429 | CES3 | FALSE | Upregulated |
| 0,071907996 | 0,912748429 | CHURC1-FNTB | FALSE | Upregulated |
| 0,031626557 | 0,912748429 | EED | FALSE | Upregulated |
| -0,034034911 | 0,912748429 | FAM13B | FALSE | Downregulated |
| 0,039950636 | 0,912748429 | HADHB | FALSE | Upregulated |
| 0,055984296 | 0,912748429 | HMBS | FALSE | Upregulated |
| 0,146944543 | 0,912748429 | LOC101929374 | FALSE | Upregulated |
| -0,174002868 | 0,912748429 | LOC101929619 | FALSE | Downregulated |
| -0,03389745 | 0,912748429 | MTTP | FALSE | Downregulated |
| 0,02443853 | 0,912748429 | NR1H2 | FALSE | Upregulated |
| 0,042802293 | 0,912748429 | ODF2 | FALSE | Upregulated |
| -0,044735039 | 0,912748429 | PARP14 | FALSE | Downregulated |
| -0,112021596 | 0,912748429 | PPM1E | FALSE | Downregulated |
| -0,038555205 | 0,912748429 | RPRD2 | FALSE | Downregulated |
| 0,058035206 | 0,912748429 | SH3KBP1 | FALSE | Upregulated |
| 0,114339024 | 0,912748429 | THBS1 | FALSE | Upregulated |
| 0,227477083 | 0,912789556 | MID1-2 | FALSE | Upregulated |
| -0,203448533 | 0,912823843 | C9orf152 | FALSE | Downregulated |
| 0,037911998 | 0,912829275 | CHD4 | FALSE | Upregulated |
| 0,043470373 | 0,912829275 | ERC1 | FALSE | Upregulated |
| -0,117817774 | 0,912829275 | LOC100287042 | FALSE | Downregulated |
| -0,040659052 | 0,912829275 | MRPS34 | FALSE | Downregulated |
| 0,032464847 | 0,912829275 | NFYB | FALSE | Upregulated |
| 0,045594209 | 0,912829275 | ZNF32 | FALSE | Upregulated |
| 0,059603611 | 0,912960986 | DMTN | FALSE | Upregulated |

| | | | | |
|---|---|---|---|---|
| -0,04231308 | 0,91297038 | TRIQK | FALSE | Downregulated |
| -0,326559743 | 0,913017386 | FAXC | FALSE | Downregulated |
| -0,241787823 | 0,913313895 | LOC100499484-C9ORF174 | FALSE | Downregulated |
| 0,062806777 | 0,913344861 | ANKRD24 | FALSE | Upregulated |
| 0,160226391 | 0,913344861 | C2orf54 | FALSE | Upregulated |
| 0,049070366 | 0,913344861 | DENND2C | FALSE | Upregulated |
| -0,105698904 | 0,913344861 | HULC | FALSE | Downregulated |
| -0,026543826 | 0,913344861 | PDHX | FALSE | Downregulated |
| 0,029206693 | 0,913344861 | POLR2C | FALSE | Upregulated |
| 0,044066149 | 0,913344861 | WSB2 | FALSE | Upregulated |
| 0,035760393 | 0,91354182 | HS2ST1 | FALSE | Upregulated |
| -0,070323867 | 0,91355503 | C12orf60 | FALSE | Downregulated |
| 0,065324964 | 0,91355503 | CALCRL | FALSE | Upregulated |
| 0,068203826 | 0,91355503 | CLPTM1L | FALSE | Upregulated |
| -0,101954446 | 0,91355503 | DCLK2 | FALSE | Downregulated |
| -0,046441257 | 0,91355503 | GPATCH2L | FALSE | Downregulated |
| -0,051526689 | 0,91355503 | LDHB | FALSE | Downregulated |
| -0,054227334 | 0,91355503 | LOC728769 | FALSE | Downregulated |
| 0,04541784 | 0,91355503 | PGD | FALSE | Upregulated |
| 0,036116358 | 0,91355503 | PRDX2 | FALSE | Upregulated |
| 0,906642074 | 0,91355503 | RNU1-27P | FALSE | Upregulated |
| -0,025651543 | 0,91355503 | SNRNP200 | FALSE | Downregulated |
| 0,031148962 | 0,91355503 | SOX13 | FALSE | Upregulated |
| -0,021232815 | 0,91355503 | ZFYVE20 | FALSE | Downregulated |
| -0,060225204 | 0,91355503 | ZNF121 | FALSE | Downregulated |

| | | | | |
|---|---|---|---|---|
| -0,040412791 | 0,913598355 | LIAS | FALSE | Downregulated |
| 0,525928495 | 0,913620737 | DAZ4 | FALSE | Upregulated |
| 0,047242649 | 0,913837106 | AP4B1 | FALSE | Upregulated |
| -0,031523192 | 0,914101027 | TRIM41 | FALSE | Downregulated |
| -0,197132144 | 0,914119326 | CELF4 | FALSE | Downregulated |
| -0,119768719 | 0,914119326 | LOC101927402 | FALSE | Downregulated |
| 0,150924155 | 0,914119326 | LOC102724851 | FALSE | Upregulated |
| -0,027855652 | 0,914119326 | PNPO | FALSE | Downregulated |
| 0,114810814 | 0,9141387 | ANKRD53 | FALSE | Upregulated |
| 0,137839005 | 0,9141387 | KTN1-AS1 | FALSE | Upregulated |
| 0,20103036 | 0,914215095 | LOC645967 | FALSE | Upregulated |
| 0,062862099 | 0,914215095 | MPEG1 | FALSE | Upregulated |
| -0,054894596 | 0,914215095 | SLC22A7 | FALSE | Downregulated |
| 0,041428955 | 0,914460962 | LEMD2 | FALSE | Upregulated |
| -0,016289655 | 0,914635193 | ARIH1 | FALSE | Downregulated |
| 0,06011156 | 0,914635193 | CNTN4 | FALSE | Upregulated |
| 0,098437827 | 0,914635193 | CORO2B | FALSE | Upregulated |
| 0,037022751 | 0,914635193 | DOLPP1 | FALSE | Upregulated |
| 0,104465103 | 0,914635193 | LOC646021 | FALSE | Upregulated |
| -0,136259623 | 0,914635193 | MT1L | FALSE | Downregulated |
| 0,0275248 | 0,914635193 | PRRC2B | FALSE | Upregulated |
| 0,038071674 | 0,914635193 | SLC9A6 | FALSE | Upregulated |
| -0,131525359 | 0,914635193 | TGFBR3L | FALSE | Downregulated |
| -0,04039311 | 0,914635193 | TMEM134 | FALSE | Downregulated |
| 0,221776703 | 0,914635193 | ZMYND10 | FALSE | Upregulated |
| -0,058416777 | 0,914635193 | ZMYND12 | FALSE | Downregulated |

| | | | | |
|---|---|---|---|---|
| 0,024893287 | 0,914698919 | RPP14 | FALSE | Upregulated |
| -0,046817961 | 0,914757719 | PDDC1 | FALSE | Downregulated |
| -0,044853875 | 0,914757719 | RPS9 | FALSE | Downregulated |
| -0,229751133 | 0,914757719 | SYN3 | FALSE | Downregulated |
| 0,234142792 | 0,91499281 | ASB2 | FALSE | Upregulated |
| -0,065532723 | 0,91499281 | RP5-1033H22.2 | FALSE | Downregulated |
| 0,131584562 | 0,915081728 | NANOS1 | FALSE | Upregulated |
| -0,101895089 | 0,915081728 | ZFP37 | FALSE | Downregulated |
| -0,057899491 | 0,915182354 | CGNL1 | FALSE | Downregulated |
| 0,112217766 | 0,915182354 | CHADL | FALSE | Upregulated |
| -0,04856064 | 0,915182354 | GCC2 | FALSE | Downregulated |
| 0,074378271 | 0,915182354 | MAPK13 | FALSE | Upregulated |
| 0,060422008 | 0,915182354 | SMAGP | FALSE | Upregulated |
| -0,060415435 | 0,915182354 | ZNF549 | FALSE | Downregulated |
| -0,037774195 | 0,915256091 | EHMT1 | FALSE | Downregulated |
| 0,024448119 | 0,915256091 | FBXO18 | FALSE | Upregulated |
| 0,02580902 | 0,91540182 | TMEM109 | FALSE | Upregulated |
| 0,037913521 | 0,915437035 | ATP6V1E1 | FALSE | Upregulated |
| 0,091555944 | 0,915500708 | ALDH1A2 | FALSE | Upregulated |
| 0,056509651 | 0,915500708 | CCNG2 | FALSE | Upregulated |
| 0,06111822 | 0,915574539 | GCA | FALSE | Upregulated |
| 0,062429879 | 0,915654192 | ASAH2B | FALSE | Upregulated |
| 0,069113006 | 0,915654192 | CD33 | FALSE | Upregulated |
| 0,048846539 | 0,915654192 | CPEB2 | FALSE | Upregulated |
| -0,068080743 | 0,915654192 | DNAJB11 | FALSE | Downregulated |
| 0,053047466 | 0,915654192 | EIF4EBP1 | FALSE | Upregulated |

| | | | | |
|---|---|---|---|---|
| 0,179206064 | 0,915654192 | LOC102723708 | FALSE | Upregulated |
| -0,032471959 | 0,915654192 | NPAT | FALSE | Downregulated |
| -0,035169479 | 0,915689928 | OPA3 | FALSE | Downregulated |
| -0,06670158 | 0,915689928 | PLCXD1 | FALSE | Downregulated |
| 0,08461081 | 0,915752834 | FAHD2B | FALSE | Upregulated |
| -0,246947338 | 0,915752834 | FGF17 | FALSE | Downregulated |
| -0,070691192 | 0,915752834 | GLT1D1 | FALSE | Downregulated |
| -0,056141132 | 0,915752834 | IDH2 | FALSE | Downregulated |
| 0,16486063 | 0,915752834 | LOC399900 | FALSE | Upregulated |
| 0,187914821 | 0,915752834 | LOC646652 | FALSE | Upregulated |
| 0,046459715 | 0,915752834 | RECQL5 | FALSE | Upregulated |
| -0,030799564 | 0,915752834 | SCAMP4 | FALSE | Downregulated |
| 0,03532351 | 0,916059876 | ANO6 | FALSE | Upregulated |
| 0,035911815 | 0,916059876 | DGCR8 | FALSE | Upregulated |
| 0,307295236 | 0,916059876 | FAM72A | FALSE | Upregulated |
| 0,114121685 | 0,916169344 | PCNXL2 | FALSE | Upregulated |
| 0,097308172 | 0,916322247 | CENPL | FALSE | Upregulated |
| -0,032617886 | 0,916322247 | DNAJC13 | FALSE | Downregulated |
| -0,061712324 | 0,916322247 | PSTPIP2 | FALSE | Downregulated |
| 0,159670979 | 0,916322247 | ZNF648 | FALSE | Upregulated |
| -0,055208251 | 0,916664621 | TMEM39A | FALSE | Downregulated |
| -0,037296761 | 0,916787901 | AGTR1 | FALSE | Downregulated |
| -0,045368887 | 0,916787901 | CCDC91 | FALSE | Downregulated |
| 0,271211682 | 0,916787901 | MIR1247 | FALSE | Upregulated |
| 0,029059073 | 0,916787901 | SIPA1L1 | FALSE | Upregulated |
| 0,117057461 | 0,916826274 | CCDC157 | FALSE | Upregulated |

| | | | | |
|---|---|---|---|---|
| 0,109880331 | 0,916826274 | CRIP3 | FALSE | Upregulated |
| 0,172043671 | 0,916826274 | IQUB | FALSE | Upregulated |
| -0,130999359 | 0,916826274 | MT1A | FALSE | Downregulated |
| -0,039254115 | 0,916826274 | PPARA | FALSE | Downregulated |
| 0,042349389 | 0,916989008 | AKIRIN2 | FALSE | Upregulated |
| -0,080455438 | 0,916989008 | DEPDC4 | FALSE | Downregulated |
| -0,343601291 | 0,916989008 | LOC101060301 | FALSE | Downregulated |
| -0,150067027 | 0,916989008 | LOC101928810 | FALSE | Downregulated |
| -0,046541586 | 0,916989008 | PHF11 | FALSE | Downregulated |
| 0,163026267 | 0,916989008 | SCN4B | FALSE | Upregulated |
| -0,066574687 | 0,916989008 | SNHG7 | FALSE | Downregulated |
| 0,347286404 | 0,917207889 | LOC101060026 | FALSE | Upregulated |
| 0,266458851 | 0,917207889 | LOC101928730 | FALSE | Upregulated |
| -0,272238127 | 0,917356103 | HIST1H2AG | FALSE | Downregulated |
| 0,028003207 | 0,917356103 | KBTBD4 | FALSE | Upregulated |
| 0,0263493 | 0,91739532 | BROX | FALSE | Upregulated |
| 0,047361725 | 0,91739532 | DCUN1D3 | FALSE | Upregulated |
| -0,049184709 | 0,917444355 | CTBP2 | FALSE | Downregulated |
| -0,32334128 | 0,917444355 | LOC100506114 | FALSE | Downregulated |
| 0,079575866 | 0,917444355 | TBX2 | FALSE | Upregulated |
| -0,022957864 | 0,917452276 | DENR | FALSE | Downregulated |
| 0,04975374 | 0,917452276 | PDK2 | FALSE | Upregulated |
| -0,036112476 | 0,917545774 | CISD1 | FALSE | Downregulated |
| -0,022569823 | 0,917545774 | FAM208A | FALSE | Downregulated |
| -0,156942223 | 0,917545774 | ZBTB16 | FALSE | Downregulated |
| -0,15142723 | 0,91757316 | ASMTL-AS1 | FALSE | Downregulated |

| | | | | |
|---|---|---|---|---|
| -0,026255881 | 0,91757316 | BFAR | FALSE | Downregulated |
| -0,230123417 | 0,91757316 | CA9 | FALSE | Downregulated |
| -0,063051705 | 0,91757316 | CENPQ | FALSE | Downregulated |
| -0,043850252 | 0,91757316 | ETFDH | FALSE | Downregulated |
| -0,154287864 | 0,91757316 | MAATS1 | FALSE | Downregulated |
| -0,053200638 | 0,91757316 | PAN3 | FALSE | Downregulated |
| -0,045561505 | 0,91757316 | PDIA3 | FALSE | Downregulated |
| 0,234378033 | 0,91757316 | RNF139-AS1 | FALSE | Upregulated |
| 0,036756793 | 0,91757316 | RNF19B | FALSE | Upregulated |
| 0,165625763 | 0,91757316 | RP11-486O12.2 | FALSE | Upregulated |
| 0,032641171 | 0,91757316 | SETD7 | FALSE | Upregulated |
| 0,101112381 | 0,91757316 | SNAI1 | FALSE | Upregulated |
| -0,037900155 | 0,91757316 | SNRPA | FALSE | Downregulated |
| 0,122565601 | 0,91757316 | VIM-AS1 | FALSE | Upregulated |
| -0,154644004 | 0,917652937 | ADAM20P1 | FALSE | Downregulated |
| 0,031708483 | 0,917652937 | CCT5 | FALSE | Upregulated |
| 0,051257537 | 0,917652937 | CERS5 | FALSE | Upregulated |
| -0,364031783 | 0,917652937 | CTNNA2 | FALSE | Downregulated |
| -0,053441109 | 0,917652937 | FGFR3 | FALSE | Downregulated |
| -0,062248246 | 0,917652937 | HPS3 | FALSE | Downregulated |
| -1,309231599 | 0,917652937 | LOC102724828 | FALSE | Downregulated |
| 0,087731784 | 0,917652937 | MAPK8IP3 | FALSE | Upregulated |
| 0,219011528 | 0,917652937 | MMP11 | FALSE | Upregulated |
| 0,0818297 | 0,917652937 | PALD1 | FALSE | Upregulated |
| 0,027820263 | 0,917652937 | PPP4R2 | FALSE | Upregulated |
| 0,058140881 | 0,917652937 | RBMS3 | FALSE | Upregulated |

| | | | | |
|---|---|---|---|---|
| -0,032441546 | 0,917652937 | RNF20 | FALSE | Downregulated |
| -0,116951399 | 0,917652937 | SPACA6P | FALSE | Downregulated |
| -0,044523311 | 0,917652937 | TIMM17B | FALSE | Downregulated |
| -0,02078528 | 0,917652937 | VPS37A | FALSE | Downregulated |
| -0,0924918 | 0,917652937 | WWC1 | FALSE | Downregulated |
| -0,039687333 | 0,91772358 | TLE4 | FALSE | Downregulated |
| -0,039850246 | 0,917864631 | MEST | FALSE | Downregulated |
| -0,030790691 | 0,917864631 | SPOPL | FALSE | Downregulated |
| 0,040980181 | 0,917868243 | HAGH | FALSE | Upregulated |
| 0,058538003 | 0,917868243 | LOC100132815 | FALSE | Upregulated |
| -0,224767673 | 0,917868243 | TCEAL2 | FALSE | Downregulated |
| 0,027380921 | 0,917868243 | TFDP1 | FALSE | Upregulated |
| 0,056061678 | 0,917868243 | ZFP30 | FALSE | Upregulated |
| -0,091646054 | 0,917898975 | DPEP2 | FALSE | Downregulated |
| 0,08877949 | 0,917898975 | FAM131A | FALSE | Upregulated |
| 0,085315243 | 0,917898975 | FAM193B | FALSE | Upregulated |
| 0,028487615 | 0,917898975 | KXD1 | FALSE | Upregulated |
| 0,174616033 | 0,917926572 | CXCL1 | FALSE | Upregulated |
| -0,035524021 | 0,917926572 | RAB30 | FALSE | Downregulated |
| -0,043889737 | 0,917926572 | USP45 | FALSE | Downregulated |
| 0,183793645 | 0,917951113 | FCGR2C | FALSE | Upregulated |
| -0,074111078 | 0,917971316 | STBD1 | FALSE | Downregulated |
| -0,064296686 | 0,917971316 | ZNF70 | FALSE | Downregulated |
| -0,247664458 | 0,917996997 | LOC101928801 | FALSE | Downregulated |
| 0,055435546 | 0,918031484 | ANKRD32 | FALSE | Upregulated |
| 0,206101159 | 0,918031484 | RNA18S5 | FALSE | Upregulated |

| | | | | |
|---|---|---|---|---|
| -0,124686009 | 0,918031484 | RP1-93I3.1 | FALSE | Downregulated |
| -0,032213549 | 0,918031484 | SRPK2 | FALSE | Downregulated |
| -0,09407853 | 0,918031484 | WDR52 | FALSE | Downregulated |
| -0,047066284 | 0,918031484 | ZBTB20 | FALSE | Downregulated |
| 0,042393872 | 0,918405513 | EEF1A1 | FALSE | Upregulated |
| -0,037827814 | 0,918405513 | FRMD4B | FALSE | Downregulated |
| 0,043153423 | 0,918405513 | NAPRT1 | FALSE | Upregulated |
| -0,097818527 | 0,918412115 | LOC101928498 | FALSE | Downregulated |
| -0,186661725 | 0,918429449 | LGI2 | FALSE | Downregulated |
| 0,137004763 | 0,918445354 | CBR3-AS1 | FALSE | Upregulated |
| 0,033824637 | 0,91845122 | ABI1 | FALSE | Upregulated |
| 0,033048286 | 0,918476901 | MPI | FALSE | Upregulated |
| -0,034979433 | 0,91850338 | RNF8 | FALSE | Downregulated |
| 0,035173205 | 0,918554912 | DUT | FALSE | Upregulated |
| -0,150819764 | 0,918554912 | SPATA32 | FALSE | Downregulated |
| -0,037405404 | 0,918645635 | COPRS | FALSE | Downregulated |
| 0,038629933 | 0,9186911 | SPPL2B | FALSE | Upregulated |
| -0,085665533 | 0,918955726 | LOC101930644 | FALSE | Downregulated |
| 0,063344922 | 0,918955726 | NDUFC2-KCTD14 | FALSE | Upregulated |
| 0,110785048 | 0,918955726 | RP11-112H10.4 | FALSE | Upregulated |
| 0,286871818 | 0,919042297 | HECW1 | FALSE | Upregulated |
| -0,125177293 | 0,919113948 | C9orf116 | FALSE | Downregulated |
| -0,112689263 | 0,919218887 | MSANTD3-TMEFF1 | FALSE | Downregulated |
| 0,1461918 | 0,919224472 | BHLHE41 | FALSE | Upregulated |
| 0,143084025 | 0,919224472 | CERS6-AS1 | FALSE | Upregulated |

| | | | | |
|---|---|---|---|---|
| 0,072657711 | 0,919224472 | KCTD15 | FALSE | Upregulated |
| -0,283888263 | 0,919224472 | SSUH2 | FALSE | Downregulated |
| 0,04831957 | 0,919264815 | DNAJC4 | FALSE | Upregulated |
| 0,029662717 | 0,919264815 | HN1L | FALSE | Upregulated |
| -0,059999603 | 0,919264815 | LHFPL2 | FALSE | Downregulated |
| -0,034510153 | 0,919264815 | SNRPC | FALSE | Downregulated |
| 0,154008632 | 0,919271552 | RP11-65J3.1 | FALSE | Upregulated |
| -0,070527951 | 0,919518675 | FAM86C2P | FALSE | Downregulated |
| -0,029137925 | 0,91975323 | HBP1 | FALSE | Downregulated |
| -0,022293239 | 0,919765366 | ARHGAP12 | FALSE | Downregulated |
| -0,025473365 | 0,919765366 | FECH | FALSE | Downregulated |
| 0,076659303 | 0,919765366 | LENG8 | FALSE | Upregulated |
| -0,091992156 | 0,919765366 | LOC102723405 | FALSE | Downregulated |
| -0,04588656 | 0,919765366 | LPGAT1 | FALSE | Downregulated |
| -0,028429494 | 0,919765366 | MANBA | FALSE | Downregulated |
| -0,195604093 | 0,919765366 | PCAT18 | FALSE | Downregulated |
| 0,033684215 | 0,919765366 | PKN2 | FALSE | Upregulated |
| 0,041525401 | 0,919765366 | PMVK | FALSE | Upregulated |
| 0,036020529 | 0,919765366 | PRMT9 | FALSE | Upregulated |
| -0,058997909 | 0,919765366 | RTN4IP1 | FALSE | Downregulated |
| 0,14269287 | 0,919765366 | SEMA3G | FALSE | Upregulated |
| 0,171618728 | 0,919765366 | SMN2 | FALSE | Upregulated |
| -0,040020757 | 0,919765366 | TSC2 | FALSE | Downregulated |
| 0,067198346 | 0,919765366 | ZEB1-AS1 | FALSE | Upregulated |
| 0,041596559 | 0,919765366 | ZNF230 | FALSE | Upregulated |
| 0,156106329 | 0,919765366 | ZNF90 | FALSE | Upregulated |

| | | | | |
|---|---|---|---|---|
| 0,102496033 | 0,919790799 | CYP4A22 | FALSE | Upregulated |
| -0,024966499 | 0,919790799 | HSBP1 | FALSE | Downregulated |
| -0,104660389 | 0,919790799 | MBNL1-AS1 | FALSE | Downregulated |
| 0,028113937 | 0,919790799 | METTL16 | FALSE | Upregulated |
| -0,065934621 | 0,919790799 | MOK | FALSE | Downregulated |
| 0,037704731 | 0,919790799 | RPL7 | FALSE | Upregulated |
| 0,074405398 | 0,919790799 | TMEM254-AS1 | FALSE | Upregulated |
| -0,129477273 | 0,91989992 | ADAT3 | FALSE | Downregulated |
| -0,307571377 | 0,91989992 | FAM156A | FALSE | Downregulated |
| 0,024529211 | 0,91989992 | KIAA0430 | FALSE | Upregulated |
| 0,053077825 | 0,91989992 | TBC1D2B | FALSE | Upregulated |
| -0,051419278 | 0,919966923 | CLCN5 | FALSE | Downregulated |
| 0,301788655 | 0,920075359 | LOC101926960 | FALSE | Upregulated |
| -0,062627944 | 0,920249209 | SLC25A5-AS1 | FALSE | Downregulated |
| -0,11699641 | 0,920252634 | ST3GAL6-AS1 | FALSE | Downregulated |
| 0,084219674 | 0,920567413 | GAS2L3 | FALSE | Upregulated |
| 0,076537681 | 0,920578215 | CRELD2 | FALSE | Upregulated |
| -0,102311244 | 0,920590553 | FBLN2 | FALSE | Downregulated |
| 0,18128104 | 0,920590553 | LOC101929211 | FALSE | Upregulated |
| -0,190255259 | 0,920678739 | RP1-1J6.2 | FALSE | Downregulated |
| -0,226687906 | 0,920678739 | SLC52A1 | FALSE | Downregulated |
| -0,052466275 | 0,920678739 | STX2 | FALSE | Downregulated |
| -0,082363605 | 0,920683069 | LOC101929761 | FALSE | Downregulated |
| -0,124646545 | 0,920789701 | PAX8-AS1 | FALSE | Downregulated |
| 0,149130805 | 0,920822243 | FAM155A | FALSE | Upregulated |
| -0,044929479 | 0,920822243 | MARS2 | FALSE | Downregulated |

| | | | | |
|---|---|---|---|---|
| 0,026865812 | 0,920853366 | RELA | FALSE | Upregulated |
| -0,043487319 | 0,921083353 | ALDH1A1 | FALSE | Downregulated |
| -0,06512393 | 0,921083353 | AP001258.4 | FALSE | Downregulated |
| -0,06472677 | 0,921083353 | FANCM | FALSE | Downregulated |
| 0,158608613 | 0,921083353 | FBXO43 | FALSE | Upregulated |
| -0,061981642 | 0,921083353 | IFNLR1 | FALSE | Downregulated |
| 0,108089197 | 0,921083353 | LILRA6 | FALSE | Upregulated |
| -0,053657086 | 0,921083353 | TMEM169 | FALSE | Downregulated |
| -0,437908683 | 0,921083353 | UGT2A1 | FALSE | Downregulated |
| 0,040537655 | 0,921083353 | ZDHHC8 | FALSE | Upregulated |
| 0,262653623 | 0,921346083 | LOC101928261 | FALSE | Upregulated |
| 0,075985186 | 0,921346083 | PRG2 | FALSE | Upregulated |
| -0,216723404 | 0,921371829 | ANKRD18DP | FALSE | Downregulated |
| -0,238164446 | 0,921371829 | ASB14 | FALSE | Downregulated |
| -0,033058634 | 0,921371829 | ATPIF1 | FALSE | Downregulated |
| -0,082822814 | 0,921371829 | C7orf13 | FALSE | Downregulated |
| -0,081697359 | 0,921371829 | LDOC1 | FALSE | Downregulated |
| 0,107855896 | 0,921371829 | LINC00939 | FALSE | Upregulated |
| -0,186279776 | 0,921371829 | LOC728026 | FALSE | Downregulated |
| -0,026559999 | 0,921371829 | MARCH6 | FALSE | Downregulated |
| 0,033928638 | 0,921371829 | MAT2B | FALSE | Upregulated |
| -0,074106445 | 0,921371829 | PPP1R3B | FALSE | Downregulated |
| 0,058081952 | 0,921371829 | RFC4 | FALSE | Upregulated |
| -0,034742682 | 0,921371829 | RUVBL1 | FALSE | Downregulated |
| -0,028239553 | 0,921371829 | TPI1 | FALSE | Downregulated |
| -0,221303692 | 0,921371829 | UBE2Q2P2 | FALSE | Downregulated |

| | | | | |
|---|---|---|---|---|
| 0,075728055 | 0,921371829 | ZNF891 | FALSE | Upregulated |
| 0,111917894 | 0,921372099 | COL26A1 | FALSE | Upregulated |
| 0,031268946 | 0,921372099 | UBXN6 | FALSE | Upregulated |
| -0,159437949 | 0,921441248 | CD80 | FALSE | Downregulated |
| -0,02548321 | 0,921441248 | PPP3CB | FALSE | Downregulated |
| -0,196740692 | 0,921462893 | ELF5 | FALSE | Downregulated |
| 0,082241047 | 0,921581445 | LIN7B | FALSE | Upregulated |
| -0,04910264 | 0,921765528 | DPH6 | FALSE | Downregulated |
| 0,083861261 | 0,921765528 | LOC101929704 | FALSE | Upregulated |
| -0,115878808 | 0,921765528 | NLRP6 | FALSE | Downregulated |
| -0,063842629 | 0,921765528 | RAD9A | FALSE | Downregulated |
| -0,068579434 | 0,921765528 | TM4SF5 | FALSE | Downregulated |
| 0,04811153 | 0,921765528 | TRAPPC2 | FALSE | Upregulated |
| 0,041431875 | 0,922124385 | COX6A1 | FALSE | Upregulated |
| 0,179375268 | 0,922176002 | LOC101929647 | FALSE | Upregulated |
| 0,039178108 | 0,922204189 | POLR1E | FALSE | Upregulated |
| 0,158822335 | 0,922246139 | ZNF252P-AS1 | FALSE | Upregulated |
| -0,033277259 | 0,922346135 | HEXB | FALSE | Downregulated |
| -0,085350898 | 0,922346135 | SELL | FALSE | Downregulated |
| 0,045418465 | 0,922380835 | TP53BP2 | FALSE | Upregulated |
| 0,032555995 | 0,922458435 | CPNE8 | FALSE | Upregulated |
| -0,065263336 | 0,922501018 | LOC102723779 | FALSE | Downregulated |
| -0,041003405 | 0,922667914 | B3GALTL | FALSE | Downregulated |
| -0,128558926 | 0,922667914 | CEP55 | FALSE | Downregulated |
| -0,123164584 | 0,922667914 | GLRB | FALSE | Downregulated |
| -0,142378961 | 0,922667914 | LOC101927686 | FALSE | Downregulated |

| | | | | |
|---|---|---|---|---|
| -0,036281168 | 0,922667914 | RNF138 | FALSE | Downregulated |
| 0,212666501 | 0,922795937 | LOC102724476 | FALSE | Upregulated |
| -0,033286742 | 0,922982522 | DDX21 | FALSE | Downregulated |
| 0,02585081 | 0,923040677 | BCR | FALSE | Upregulated |
| 0,128674611 | 0,923040677 | DSCAM | FALSE | Upregulated |
| -0,060716619 | 0,923040677 | RP11-372E1.6 | FALSE | Downregulated |
| 0,045526555 | 0,923040677 | SLC25A20 | FALSE | Upregulated |
| -0,03550558 | 0,923040677 | SYNJ1 | FALSE | Downregulated |
| -0,07473131 | 0,923040677 | TPRN | FALSE | Downregulated |
| 0,01958713 | 0,923040677 | UBE3C | FALSE | Upregulated |
| -0,207569413 | 0,923232811 | MGARP | FALSE | Downregulated |
| 0,023161386 | 0,923232811 | NUMB | FALSE | Upregulated |
| -0,046617021 | 0,923232811 | SUV39H2 | FALSE | Downregulated |
| -0,02998839 | 0,923312104 | ZFP91 | FALSE | Downregulated |
| 0,034965765 | 0,923377168 | TMEM30B | FALSE | Upregulated |
| 0,029740636 | 0,92339671 | FBXO34 | FALSE | Upregulated |
| -0,127729956 | 0,92339671 | LINC01023 | FALSE | Downregulated |
| -0,053869397 | 0,92339671 | QPCTL | FALSE | Downregulated |
| 0,133421184 | 0,92339671 | XRCC2 | FALSE | Upregulated |
| -0,119317586 | 0,923447822 | LINC00471 | FALSE | Downregulated |
| 0,020209688 | 0,923495993 | ATP5A1 | FALSE | Upregulated |
| -0,492991955 | 0,923495993 | PTGES3L | FALSE | Downregulated |
| -0,034958957 | 0,923639856 | PPIP5K1 | FALSE | Downregulated |
| 0,044105144 | 0,923639856 | SOWAHB | FALSE | Upregulated |
| -0,073273592 | 0,923722113 | SH3YL1 | FALSE | Downregulated |
| 0,366212018 | 0,923827371 | CRYBB2 | FALSE | Upregulated |

| | | | | |
|---|---|---|---|---|
| 0,047602063 | 0,923827371 | HARS | FALSE | Upregulated |
| -0,120379803 | 0,923943937 | CCDC144B | FALSE | Downregulated |
| -0,38322232 | 0,923943937 | PTHLH | FALSE | Downregulated |
| -0,046832961 | 0,924025974 | COL5A2 | FALSE | Downregulated |
| 0,273634242 | 0,924025974 | CPS1-IT1 | FALSE | Upregulated |
| -0,040069063 | 0,924025974 | DYRK2 | FALSE | Downregulated |
| -0,039186239 | 0,924098647 | DDX51 | FALSE | Downregulated |
| 0,027765645 | 0,924164833 | APPBP2 | FALSE | Upregulated |
| 0,029049471 | 0,924164833 | C16orf58 | FALSE | Upregulated |
| 0,032655313 | 0,924164833 | DESI1 | FALSE | Upregulated |
| -0,035740415 | 0,924164833 | EIF2B2 | FALSE | Downregulated |
| 0,031768119 | 0,924164833 | MAP2K5 | FALSE | Upregulated |
| -0,035061133 | 0,924164833 | MRPL51 | FALSE | Downregulated |
| 0,042091723 | 0,924164833 | POLE4 | FALSE | Upregulated |
| -0,299739255 | 0,924164833 | SLCO5A1 | FALSE | Downregulated |
| -0,330794843 | 0,924164833 | TBC1D3H | FALSE | Downregulated |
| -0,180451837 | 0,924164833 | TBX18 | FALSE | Downregulated |
| 0,037202864 | 0,924164833 | TOMM7 | FALSE | Upregulated |
| -0,024800512 | 0,924164833 | USP14 | FALSE | Downregulated |
| -0,051535477 | 0,924164833 | ZNF780B | FALSE | Downregulated |
| 0,087682066 | 0,924164833 | ZNF888 | FALSE | Upregulated |
| -0,035236571 | 0,924179284 | CNNM4 | FALSE | Downregulated |
| 0,082767132 | 0,924218166 | SNHG10 | FALSE | Upregulated |
| 0,038979718 | 0,924335205 | B2M | FALSE | Upregulated |
| 0,122227342 | 0,924335205 | SEMA3E | FALSE | Upregulated |
| 0,067168361 | 0,924475101 | AKR1C3 | FALSE | Upregulated |

| | | | | |
|---|---|---|---|---|
| 0,114557273 | 0,924475101 | C19orf57 | FALSE | Upregulated |
| -0,086138324 | 0,924475101 | HDGFRP3 | FALSE | Downregulated |
| 0,21046026 | 0,924475101 | HERC2P4 | FALSE | Upregulated |
| -0,070568571 | 0,924475101 | LINC00341 | FALSE | Downregulated |
| 0,167819064 | 0,924475101 | LOC100507747 | FALSE | Upregulated |
| -0,056829221 | 0,924475101 | PIEZO2 | FALSE | Downregulated |
| 0,055819432 | 0,924475101 | PIGL | FALSE | Upregulated |
| 0,01855256 | 0,924475101 | VPS4B | FALSE | Upregulated |
| -0,071504982 | 0,924480152 | AP1S2 | FALSE | Downregulated |
| 0,143906615 | 0,924480152 | ASTN1 | FALSE | Upregulated |
| -0,092185519 | 0,924480152 | AVIL | FALSE | Downregulated |
| -0,034917074 | 0,924480152 | CLP1 | FALSE | Downregulated |
| 0,030060147 | 0,924480152 | COPS3 | FALSE | Upregulated |
| -0,146192686 | 0,924480152 | CYP17A1 | FALSE | Downregulated |
| -0,032360244 | 0,924480152 | DCLRE1A | FALSE | Downregulated |
| -0,029997214 | 0,924480152 | DIS3L | FALSE | Downregulated |
| 0,057301012 | 0,924480152 | FAM110C | FALSE | Upregulated |
| -0,035459004 | 0,924480152 | FBRSL1 | FALSE | Downregulated |
| -0,032958791 | 0,924480152 | FBXO28 | FALSE | Downregulated |
| 0,130968052 | 0,924480152 | GHRLOS | FALSE | Upregulated |
| 0,046565753 | 0,924480152 | GLYCTK | FALSE | Upregulated |
| 1,500062519 | 0,924480152 | H19 | FALSE | Upregulated |
| 0,104700343 | 0,924480152 | LINC00936 | FALSE | Upregulated |
| -0,063252035 | 0,924480152 | NCKAP5 | FALSE | Downregulated |
| 0,043237145 | 0,924480152 | NXT2 | FALSE | Upregulated |
| -0,04872225 | 0,924480152 | SAMD4A | FALSE | Downregulated |

| | | | | |
|---|---|---|---|---|
| -0,036435442 | 0,924480152 | TMEM185A | FALSE | Downregulated |
| 0,030641475 | 0,924480152 | TMEM205 | FALSE | Upregulated |
| -0,145866848 | 0,924480152 | UBQLNL | FALSE | Downregulated |
| -0,065121833 | 0,924480152 | ZNF652 | FALSE | Downregulated |
| 0,111962331 | 0,924480152 | ZNF667 | FALSE | Upregulated |
| 0,042811695 | 0,924480152 | ZSCAN22 | FALSE | Upregulated |
| -0,060080327 | 0,924609395 | EFNB3 | FALSE | Downregulated |
| -0,040085913 | 0,924655297 | TMEM141 | FALSE | Downregulated |
| -0,139018418 | 0,924683058 | AGAP11 | FALSE | Downregulated |
| 0,171360303 | 0,924683058 | AOX2P | FALSE | Upregulated |
| -0,027763665 | 0,924683058 | BCL2L13 | FALSE | Downregulated |
| 0,031298922 | 0,924683058 | EIF4EBP2 | FALSE | Upregulated |
| 0,094047952 | 0,924683058 | FAM13A-AS1 | FALSE | Upregulated |
| -0,102358911 | 0,924683058 | GBP1P1 | FALSE | Downregulated |
| -0,062077635 | 0,924683058 | HNF1B | FALSE | Downregulated |
| -0,164488782 | 0,924683058 | LOC102724615 | FALSE | Downregulated |
| -0,023431635 | 0,924683058 | PCNX | FALSE | Downregulated |
| -0,031281924 | 0,924683058 | PIGH | FALSE | Downregulated |
| -0,026597073 | 0,924683058 | PITRM1 | FALSE | Downregulated |
| -0,055808187 | 0,924683058 | PRICKLE2 | FALSE | Downregulated |
| 0,079522928 | 0,924683058 | SERPINE2 | FALSE | Upregulated |
| -0,031065296 | 0,924683058 | TAF1D | FALSE | Downregulated |
| -0,032511343 | 0,924683058 | TRA2A | FALSE | Downregulated |
| -0,050300081 | 0,924683058 | TXNL4B | FALSE | Downregulated |
| -0,068365733 | 0,924683058 | UVSSA | FALSE | Downregulated |
| 0,112588734 | 0,924683921 | RP11-793H13.3 | FALSE | Upregulated |

| | | | | |
|---|---|---|---|---|
| 0,098760091 | 0,92474913 | LOC102723897 | FALSE | Upregulated |
| -0,028112845 | 0,92474913 | RPS6KB1 | FALSE | Downregulated |
| -0,040487739 | 0,924974808 | CDC34 | FALSE | Downregulated |
| 0,04970617 | 0,924974808 | SMTN | FALSE | Upregulated |
| 0,030123276 | 0,924995828 | POLR1B | FALSE | Upregulated |
| 0,02410774 | 0,925138131 | KLHDC3 | FALSE | Upregulated |
| 0,147823288 | 0,925138131 | LOC101929065 | FALSE | Upregulated |
| 0,023691548 | 0,925138131 | MORC2 | FALSE | Upregulated |
| -0,043740778 | 0,925149431 | ARID5A | FALSE | Downregulated |
| 0,056638233 | 0,925149431 | IFRD1 | FALSE | Upregulated |
| -0,051045902 | 0,925149431 | LNX1 | FALSE | Downregulated |
| 0,051279848 | 0,925149431 | PKP2 | FALSE | Upregulated |
| 0,060836443 | 0,925238698 | ATP6V1E2 | FALSE | Upregulated |
| -1,041558508 | 0,925238698 | LOC102723360 | FALSE | Downregulated |
| -0,04722557 | 0,925238698 | PHLPP1 | FALSE | Downregulated |
| -0,06807112 | 0,925238698 | USP51 | FALSE | Downregulated |
| 0,053136622 | 0,925314714 | C19orf70 | FALSE | Upregulated |
| -0,141548419 | 0,925314714 | COL9A2 | FALSE | Downregulated |
| -0,661661677 | 0,925314714 | LOC101060389 | FALSE | Downregulated |
| 0,035112871 | 0,925314714 | MAGT1 | FALSE | Upregulated |
| -0,075151991 | 0,925314714 | RPS10P7 | FALSE | Downregulated |
| 0,056527663 | 0,925314714 | SPTSSA | FALSE | Upregulated |
| 0,040671889 | 0,925314714 | ZNF700 | FALSE | Upregulated |
| 0,02897933 | 0,925336925 | SARS | FALSE | Upregulated |
| 0,117262349 | 0,925336925 | TNFRSF25 | FALSE | Upregulated |
| -0,126049673 | 0,925361133 | LOC101929670 | FALSE | Downregulated |

| | | | | |
|---|---|---|---|---|
| 0,186567164 | 0,925361831 | DNAH5 | FALSE | Upregulated |
| 0,071451209 | 0,925361831 | FOLH1B | FALSE | Upregulated |
| -0,047514136 | 0,925412066 | ELL2 | FALSE | Downregulated |
| -0,11000744 | 0,925412066 | LOC730183 | FALSE | Downregulated |
| 0,094848002 | 0,925412066 | PNMAL2 | FALSE | Upregulated |
| 0,050924099 | 0,925412066 | ZNF570 | FALSE | Upregulated |
| 0,061413211 | 0,92551071 | LOC102723701 | FALSE | Upregulated |
| -0,048385756 | 0,92551071 | MYSM1 | FALSE | Downregulated |
| 0,080718392 | 0,925521752 | C7orf76 | FALSE | Upregulated |
| 0,098329661 | 0,925521752 | FAM107A | FALSE | Upregulated |
| 0,17434637 | 0,925521752 | FSIP1 | FALSE | Upregulated |
| -0,030271059 | 0,925521752 | GSS | FALSE | Downregulated |
| 0,029214428 | 0,925521752 | HEBP1 | FALSE | Upregulated |
| -0,066700862 | 0,925521752 | IFI27 | FALSE | Downregulated |
| 0,131440352 | 0,925521752 | ITGAD | FALSE | Upregulated |
| 0,135272048 | 0,925521752 | LOC100129722 | FALSE | Upregulated |
| -0,060726438 | 0,925521752 | LPIN2 | FALSE | Downregulated |
| 0,044828148 | 0,925521752 | NSUN6 | FALSE | Upregulated |
| -0,171665924 | 0,925521752 | PANO | FALSE | Downregulated |
| 0,050396597 | 0,925521752 | PIK3R3 | FALSE | Upregulated |
| 0,032760418 | 0,925521752 | SETX | FALSE | Upregulated |
| -0,044966508 | 0,925521752 | TAOK3 | FALSE | Downregulated |
| -0,063082427 | 0,925521752 | ZNF284 | FALSE | Downregulated |
| 0,053137326 | 0,925521752 | ZNF772 | FALSE | Upregulated |
| 0,056241212 | 0,925533947 | C17orf89 | FALSE | Upregulated |
| 0,056559844 | 0,925533947 | C3orf33 | FALSE | Upregulated |

| | | | | |
|---|---|---|---|---|
| 0,051763946 | 0,925533947 | LST1 | FALSE | Upregulated |
| -0,063952156 | 0,925533947 | MICALCL | FALSE | Downregulated |
| 0,029391453 | 0,925533947 | MON1B | FALSE | Upregulated |
| 0,125600097 | 0,925533947 | SLC35E4 | FALSE | Upregulated |
| 0,022960903 | 0,925533947 | TM9SF3 | FALSE | Upregulated |
| -0,063818337 | 0,925586546 | GIMAP4 | FALSE | Downregulated |
| 0,024111763 | 0,925586546 | NAA20 | FALSE | Upregulated |
| -0,089642444 | 0,925586546 | RP11-761I4.3 | FALSE | Downregulated |
| -0,026461713 | 0,925586546 | ZNF212 | FALSE | Downregulated |
| -0,044341288 | 0,925593971 | DPM2 | FALSE | Downregulated |
| 0,119546755 | 0,925593971 | SCN11A | FALSE | Upregulated |
| -0,219031462 | 0,925593971 | TIMP4 | FALSE | Downregulated |
| -0,039135195 | 0,925616263 | CUTC | FALSE | Downregulated |
| -0,034938838 | 0,925616263 | SLC2A2 | FALSE | Downregulated |
| 0,152837881 | 0,925678862 | PPAP2C | FALSE | Upregulated |
| -0,162485452 | 0,925781735 | LINC00202-1 | FALSE | Downregulated |
| -0,076125977 | 0,925886654 | HCG11 | FALSE | Downregulated |
| 0,135754202 | 0,925886654 | SNX15 | FALSE | Upregulated |
| 0,123694662 | 0,925944692 | C12orf75 | FALSE | Upregulated |
| -0,053875574 | 0,925944692 | RHNO1 | FALSE | Downregulated |
| 0,165450493 | 0,925952812 | APLN | FALSE | Upregulated |
| -0,167890009 | 0,925952812 | ARSI | FALSE | Downregulated |
| 0,095264542 | 0,925952812 | ASRGL1 | FALSE | Upregulated |
| -0,033917199 | 0,925952812 | ATP5L | FALSE | Downregulated |
| -0,071839508 | 0,925952812 | LOC100652758 | FALSE | Downregulated |
| 0,055574514 | 0,925952812 | MSL3P1 | FALSE | Upregulated |

| | | | | |
|---|---|---|---|---|
| 0,035816989 | 0,925952812 | MYBBP1A | FALSE | Upregulated |
| -0,151770222 | 0,925952812 | SEPT3 | FALSE | Downregulated |
| 0,15017755 | 0,925952812 | ZNF454 | FALSE | Upregulated |
| -0,064252749 | 0,926062276 | PGP | FALSE | Downregulated |
| 0,259378735 | 0,926122127 | CLDN19 | FALSE | Upregulated |
| -0,093003933 | 0,926363741 | KLHL25 | FALSE | Downregulated |
| -0,03227404 | 0,926363741 | TRMT61B | FALSE | Downregulated |
| -0,241805708 | 0,926731359 | CASC9 | FALSE | Downregulated |
| 0,063775624 | 0,926740803 | EFCAB2 | FALSE | Upregulated |
| 0,098571926 | 0,926740803 | LOC102723804 | FALSE | Upregulated |
| 0,029305206 | 0,926740803 | SF3B1 | FALSE | Upregulated |
| 0,110113697 | 0,926740803 | TDGF1 | FALSE | Upregulated |
| -0,025891751 | 0,926740803 | TMOD3 | FALSE | Downregulated |
| 0,118928395 | 0,926777661 | GOLGA8R | FALSE | Upregulated |
| -0,08386918 | 0,926777661 | ULK4 | FALSE | Downregulated |
| 0,01832583 | 0,92692941 | PRCC | FALSE | Upregulated |
| 0,080899803 | 0,92692941 | SPSB1 | FALSE | Upregulated |
| -0,054598561 | 0,926953284 | ZNF608 | FALSE | Downregulated |
| -0,087117444 | 0,927044595 | CD34 | FALSE | Downregulated |
| -0,043000407 | 0,927044595 | ZC3H12C | FALSE | Downregulated |
| 0,048522136 | 0,927249191 | C15orf40 | FALSE | Upregulated |
| -0,055134829 | 0,927249191 | FAAH2 | FALSE | Downregulated |
| 0,030668653 | 0,927249191 | GALNT11 | FALSE | Upregulated |
| -0,073677332 | 0,927249191 | LOC100506157 | FALSE | Downregulated |
| -0,083116196 | 0,927249191 | LOC101927509 | FALSE | Downregulated |
| -0,140041163 | 0,927249191 | MYO1A | FALSE | Downregulated |

| | | | | |
|---|---|---|---|---|
| 0,075758772 | 0,927249191 | OTUD3 | FALSE | Upregulated |
| 0,042043504 | 0,927249191 | RPL35 | FALSE | Upregulated |
| -0,181447512 | 0,92737314 | LOC101928676 | FALSE | Downregulated |
| 0,031160852 | 0,92737314 | RCHY1 | FALSE | Upregulated |
| 0,053237143 | 0,927493473 | PTPRM | FALSE | Upregulated |
| 0,062214618 | 0,927696456 | RUNX1 | FALSE | Upregulated |
| -0,035574881 | 0,927757573 | SFXN5 | FALSE | Downregulated |
| 0,230094013 | 0,927817688 | BCORP1 | FALSE | Upregulated |
| 0,038947746 | 0,927817688 | SVIP | FALSE | Upregulated |
| -0,037083265 | 0,927817688 | TMEM126B | FALSE | Downregulated |
| 0,061220138 | 0,927874951 | ALPL | FALSE | Upregulated |
| 0,031848686 | 0,927874951 | ATPAF2 | FALSE | Upregulated |
| -0,161785281 | 0,927874951 | C16orf71 | FALSE | Downregulated |
| 0,060298341 | 0,927874951 | ENTPD7 | FALSE | Upregulated |
| 0,076770918 | 0,927874951 | FCHO1 | FALSE | Upregulated |
| -0,035294211 | 0,927874951 | LETMD1 | FALSE | Downregulated |
| -0,155376971 | 0,927874951 | MUSK | FALSE | Downregulated |
| -0,038980549 | 0,927874951 | SDC4 | FALSE | Downregulated |
| 0,168921126 | 0,928112519 | LOC101926902 | FALSE | Upregulated |
| 0,132263718 | 0,928112519 | LOC101928055 | FALSE | Upregulated |
| 0,050994278 | 0,928171509 | HDAC7 | FALSE | Upregulated |
| 0,174884131 | 0,928171509 | KPNA7 | FALSE | Upregulated |
| 0,157464367 | 0,928171509 | TMPRSS13 | FALSE | Upregulated |
| 0,035773787 | 0,928171509 | TTC31 | FALSE | Upregulated |
| 0,047354758 | 0,928524791 | AKAP7 | FALSE | Upregulated |
| -0,031672499 | 0,928524791 | BBS10 | FALSE | Downregulated |

| | | | | |
|---|---|---|---|---|
| -0,051195465 | 0,928524791 | C9orf37 | FALSE | Downregulated |
| 0,09341477 | 0,928524791 | KCNJ15 | FALSE | Upregulated |
| 0,051687356 | 0,928524791 | RBP1 | FALSE | Upregulated |
| -0,030020766 | 0,928524791 | ROGDI | FALSE | Downregulated |
| -0,096677416 | 0,928524791 | SEMA6C | FALSE | Downregulated |
| -0,067174319 | 0,928524791 | SPRY3 | FALSE | Downregulated |
| 0,100961461 | 0,928551839 | RP11-132A1.4 | FALSE | Upregulated |
| -0,23153414 | 0,928616208 | CBLN2 | FALSE | Downregulated |
| 0,254258226 | 0,928616208 | LOC100506731 | FALSE | Upregulated |
| 0,067539264 | 0,928616208 | NRROS | FALSE | Upregulated |
| -0,056897186 | 0,928669656 | BRI3BP | FALSE | Downregulated |
| -0,069068769 | 0,928669656 | CCR1 | FALSE | Downregulated |
| 0,026558605 | 0,928669656 | GDAP2 | FALSE | Upregulated |
| -0,035582911 | 0,928669656 | KIAA1731 | FALSE | Downregulated |
| -0,199203452 | 0,928669656 | LINC00612 | FALSE | Downregulated |
| -0,169938532 | 0,928669656 | LOC101929713 | FALSE | Downregulated |
| 0,02320816 | 0,928669656 | MGAT1 | FALSE | Upregulated |
| 0,038905761 | 0,928669656 | NELFE | FALSE | Upregulated |
| 0,046929661 | 0,928669656 | PLEKHG3 | FALSE | Upregulated |
| -0,255727018 | 0,928669656 | RP11-14N7.2 | FALSE | Downregulated |
| -0,030923928 | 0,928669656 | SEC31A | FALSE | Downregulated |
| -0,02566316 | 0,928669656 | UBP1 | FALSE | Downregulated |
| -0,041958713 | 0,928669656 | ZSCAN16 | FALSE | Downregulated |
| 0,126461583 | 0,928887308 | ATL1 | FALSE | Upregulated |
| -0,138403049 | 0,928887308 | LOC101926918 | FALSE | Downregulated |
| 0,04232998 | 0,928932525 | C20orf27 | FALSE | Upregulated |

| | | | | |
|---|---|---|---|---|
| 0,047181194 | 0,928932525 | SCPEP1 | FALSE | Upregulated |
| 0,174215273 | 0,928939759 | CRYBB3 | FALSE | Upregulated |
| -0,022152582 | 0,928939759 | RAB11B | FALSE | Downregulated |
| -0,041631582 | 0,929085158 | CIRBP | FALSE | Downregulated |
| -0,204405541 | 0,929085158 | TCP10L2 | FALSE | Downregulated |
| 0,039078062 | 0,929253938 | ITIH1 | FALSE | Upregulated |
| -0,077011437 | 0,929373229 | NKAPL | FALSE | Downregulated |
| 0,040416506 | 0,929496545 | CDK5 | FALSE | Upregulated |
| 0,027118768 | 0,929496545 | SEC22A | FALSE | Upregulated |
| 0,086784462 | 0,929669347 | SLC11A1 | FALSE | Upregulated |
| 0,034148965 | 0,929800035 | IFNAR2 | FALSE | Upregulated |
| -0,026412486 | 0,929800035 | MRPS23 | FALSE | Downregulated |
| -0,08527074 | 0,929800035 | P2RX5-TAX1BP3 | FALSE | Downregulated |
| 0,064253846 | 0,930134514 | HENMT1 | FALSE | Upregulated |
| -0,070166742 | 0,930134514 | STX11 | FALSE | Downregulated |
| -0,031476395 | 0,930134514 | TFB2M | FALSE | Downregulated |
| -0,141986823 | 0,930134514 | TNFSF14 | FALSE | Downregulated |
| -0,054984979 | 0,93049895 | FGD4 | FALSE | Downregulated |
| 0,023709041 | 0,930649679 | VEZF1 | FALSE | Upregulated |
| 0,047624188 | 0,930717597 | PPIB | FALSE | Upregulated |
| -0,069593411 | 0,930749307 | GRK4 | FALSE | Downregulated |
| 0,179813753 | 0,930751644 | C6orf7 | FALSE | Upregulated |
| 0,061563081 | 0,930751644 | HMCN1 | FALSE | Upregulated |
| 1,409504514 | 0,930751644 | LOC102724219 | FALSE | Upregulated |
| 0,040306929 | 0,930751644 | NDUFB2 | FALSE | Upregulated |
| -0,088703768 | 0,930751644 | NR0B2 | FALSE | Downregulated |

| | | | | |
|---|---|---|---|---|
| 0,076293824 | 0,930751644 | PCYOX1L | FALSE | Upregulated |
| 0,393132126 | 0,930751644 | PRND | FALSE | Upregulated |
| -0,036872415 | 0,930751644 | REXO1 | FALSE | Downregulated |
| 0,035427933 | 0,930751644 | SERPIND1 | FALSE | Upregulated |
| 0,196459459 | 0,930751644 | ST3GAL4-AS1 | FALSE | Upregulated |
| -0,05398456 | 0,930751644 | TYROBP | FALSE | Downregulated |
| -0,025938179 | 0,93083677 | ABCD4 | FALSE | Downregulated |
| 0,123847156 | 0,930867517 | NPW | FALSE | Upregulated |
| 0,029753477 | 0,930935047 | CLASP1 | FALSE | Upregulated |
| 0,06118985 | 0,930935047 | KIF11 | FALSE | Upregulated |
| -0,043029843 | 0,930935047 | RAPGEF5 | FALSE | Downregulated |
| 0,035291417 | 0,930935047 | TRIO | FALSE | Upregulated |
| -0,043464472 | 0,930935047 | ZYG11B | FALSE | Downregulated |
| 0,081046401 | 0,931029192 | LOC102723919 | FALSE | Upregulated |
| -0,070013956 | 0,931029192 | NR6A1 | FALSE | Downregulated |
| 0,063054261 | 0,931029192 | OSBPL11 | FALSE | Upregulated |
| 0,050069207 | 0,931029192 | PTCH1 | FALSE | Upregulated |
| -0,048271362 | 0,931092567 | CCDC85B | FALSE | Downregulated |
| 0,119409027 | 0,931092567 | LOC101928051 | FALSE | Upregulated |
| -0,032945836 | 0,931092567 | SLC27A1 | FALSE | Downregulated |
| 0,022468392 | 0,931092567 | STAMBP | FALSE | Upregulated |
| -0,042238448 | 0,931092567 | TAF1B | FALSE | Downregulated |
| -0,044689302 | 0,931092567 | YDJC | FALSE | Downregulated |
| -0,068311387 | 0,931110715 | NBPF19 | FALSE | Downregulated |
| -0,126860221 | 0,931114111 | HSD3B2 | FALSE | Downregulated |
| 0,078784838 | 0,931160116 | ADCK1 | FALSE | Upregulated |

| | | | | |
|---|---|---|---|---|
| -0,108992466 | 0,931160116 | AGAP5 | FALSE | Downregulated |
| 0,150377843 | 0,931160116 | ANXA8 | FALSE | Upregulated |
| -0,064380302 | 0,931160116 | CCT6P3 | FALSE | Downregulated |
| 0,073363019 | 0,931160116 | CD38 | FALSE | Upregulated |
| 0,133762687 | 0,931160116 | CDH13 | FALSE | Upregulated |
| -0,037442625 | 0,931160116 | CERS6 | FALSE | Downregulated |
| -0,038485315 | 0,931160116 | FAM118B | FALSE | Downregulated |
| -0,199113599 | 0,931160116 | FLJ38723 | FALSE | Downregulated |
| -0,028909181 | 0,931160116 | HDAC3 | FALSE | Downregulated |
| -0,180720942 | 0,931160116 | HERC2P3 | FALSE | Downregulated |
| -0,018495258 | 0,931160116 | INO80 | FALSE | Downregulated |
| -0,088208523 | 0,931160116 | LINC00865 | FALSE | Downregulated |
| 0,080603632 | 0,931160116 | LOC101928451 | FALSE | Upregulated |
| 0,239165402 | 0,931160116 | LOC102723735 | FALSE | Upregulated |
| 0,116548893 | 0,931160116 | LOC102724455 | FALSE | Upregulated |
| -0,020401744 | 0,931160116 | PRPF6 | FALSE | Downregulated |
| 0,153156209 | 0,931160116 | RIPPLY3 | FALSE | Upregulated |
| 0,098669284 | 0,931160116 | RNF39 | FALSE | Upregulated |
| 0,130546506 | 0,931160116 | RP11-420A23.1 | FALSE | Upregulated |
| 0,089217691 | 0,931160116 | SEPSECS-AS1 | FALSE | Upregulated |
| -0,05566138 | 0,931160116 | SEPW1 | FALSE | Downregulated |
| -0,033494149 | 0,931160116 | SPRTN | FALSE | Downregulated |
| 0,027454631 | 0,931160116 | ST7 | FALSE | Upregulated |
| 0,074257681 | 0,931160116 | TET1 | FALSE | Upregulated |
| -0,196526647 | 0,931160116 | TRIM7 | FALSE | Downregulated |
| -0,040597641 | 0,931160116 | UBL5 | FALSE | Downregulated |

| | | | | |
|---|---|---|---|---|
| 0,048110803 | 0,931160116 | ZNF251 | FALSE | Upregulated |
| -0,03803091 | 0,931160116 | ZNF92 | FALSE | Downregulated |
| 0,03163379 | 0,931179956 | PSMG2 | FALSE | Upregulated |
| -0,049289827 | 0,931256149 | CARF | FALSE | Downregulated |
| 0,027800523 | 0,931256149 | HIPK1 | FALSE | Upregulated |
| -0,105272412 | 0,931256149 | MDK | FALSE | Downregulated |
| 0,099090038 | 0,931256149 | MIP | FALSE | Upregulated |
| -0,18561965 | 0,931256149 | RP11-276H19.1 | FALSE | Downregulated |
| 0,026503151 | 0,931256149 | RRAGC | FALSE | Upregulated |
| 0,02809299 | 0,931256149 | TTC23 | FALSE | Upregulated |
| -0,101096295 | 0,931256149 | YBX2 | FALSE | Downregulated |
| -0,032497731 | 0,931256149 | ZNF75A | FALSE | Downregulated |
| 0,096712836 | 0,931356436 | ANO7 | FALSE | Upregulated |
| -0,157656825 | 0,931865756 | RGMB-AS1 | FALSE | Downregulated |
| -0,114928104 | 0,931865756 | TMEM119 | FALSE | Downregulated |
| 0,182004804 | 0,931877064 | GOLGA8O | FALSE | Upregulated |
| -0,09799963 | 0,931944909 | SPDYC | FALSE | Downregulated |
| 0,168166569 | 0,931950409 | LINC00880 | FALSE | Upregulated |
| -0,035313248 | 0,931950409 | RPLP1 | FALSE | Downregulated |
| -0,072240131 | 0,931966279 | OXER1 | FALSE | Downregulated |
| -0,142900606 | 0,93198172 | LOC102724163 | FALSE | Downregulated |
| -0,111078563 | 0,931986823 | CYP2B7P | FALSE | Downregulated |
| 0,028964546 | 0,932016885 | PNKD | FALSE | Upregulated |
| -0,026583835 | 0,932029033 | CDC42EP4 | FALSE | Downregulated |
| 0,085167848 | 0,932029033 | FAM13C | FALSE | Upregulated |
| -0,048230901 | 0,932029033 | HAND2 | FALSE | Downregulated |

| | | | | |
|---|---|---|---|---|
| -0,124802605 | 0,932090985 | LOC102724190 | FALSE | Downregulated |
| -0,040089072 | 0,932110456 | ERO1LB | FALSE | Downregulated |
| 0,155680448 | 0,932265561 | CPA3 | FALSE | Upregulated |
| -0,042964023 | 0,93234937 | AGT | FALSE | Downregulated |
| 0,109571094 | 0,93234937 | SCAMP5 | FALSE | Upregulated |
| 0,037571651 | 0,93234937 | ZNF691 | FALSE | Upregulated |
| 0,032240356 | 0,932388635 | ARRDC1 | FALSE | Upregulated |
| -0,047620425 | 0,932388635 | BMI1 | FALSE | Downregulated |
| 0,043183808 | 0,932388635 | C11orf52 | FALSE | Upregulated |
| -0,092711148 | 0,932388635 | C2orf15 | FALSE | Downregulated |
| -0,057708365 | 0,932388635 | CBWD3 | FALSE | Downregulated |
| 0,040983611 | 0,932388635 | CHST14 | FALSE | Upregulated |
| -0,177259171 | 0,932388635 | CLUL1 | FALSE | Downregulated |
| -0,027194882 | 0,932388635 | CWF19L1 | FALSE | Downregulated |
| 0,105187477 | 0,932388635 | DNAH2 | FALSE | Upregulated |
| -0,035347034 | 0,932388635 | DNAJC22 | FALSE | Downregulated |
| 0,027890794 | 0,932388635 | DNASE2 | FALSE | Upregulated |
| -0,071107529 | 0,932388635 | DOK3 | FALSE | Downregulated |
| 0,03217608 | 0,932388635 | EPM2A | FALSE | Upregulated |
| -0,020309105 | 0,932388635 | FAM199X | FALSE | Downregulated |
| 0,169351775 | 0,932388635 | FAM71F2 | FALSE | Upregulated |
| -0,027065467 | 0,932388635 | GNPTAB | FALSE | Downregulated |
| -0,073249782 | 0,932388635 | IGFLR1 | FALSE | Downregulated |
| -0,266406517 | 0,932388635 | ITGB6 | FALSE | Downregulated |
| 0,188353881 | 0,932388635 | LINC01016 | FALSE | Upregulated |
| -0,080695522 | 0,932388635 | LOC100507316 | FALSE | Downregulated |

| | | | | |
|---|---|---|---|---|
| 0,169328064 | 0,932388635 | LOC102723534 | FALSE | Upregulated |
| -0,053585496 | 0,932388635 | MRI1 | FALSE | Downregulated |
| 0,033733605 | 0,932388635 | MRPS28 | FALSE | Upregulated |
| -1,376165143 | 0,932388635 | MYH7 | FALSE | Downregulated |
| 0,033530479 | 0,932388635 | NCOA4 | FALSE | Upregulated |
| -0,067884957 | 0,932388635 | PDE3A | FALSE | Downregulated |
| -0,073446358 | 0,932388635 | PDGFB | FALSE | Downregulated |
| -0,151261629 | 0,932388635 | POU5F1 | FALSE | Downregulated |
| 0,055732862 | 0,932388635 | RAB31 | FALSE | Upregulated |
| 0,166185432 | 0,932388635 | RBM46 | FALSE | Upregulated |
| 0,15852372 | 0,932388635 | RP11-401P9.5 | FALSE | Upregulated |
| -0,161662359 | 0,932388635 | RP11-575F12.1 | FALSE | Downregulated |
| 0,127631486 | 0,932388635 | SALL4 | FALSE | Upregulated |
| 0,019561109 | 0,932388635 | SNAPC3 | FALSE | Upregulated |
| -0,108827738 | 0,932388635 | SNORA25 | FALSE | Downregulated |
| 0,026519795 | 0,932388635 | SPECC1L | FALSE | Upregulated |
| -0,026171098 | 0,932388635 | STAU1 | FALSE | Downregulated |
| 0,068073358 | 0,932388635 | SYP | FALSE | Upregulated |
| -1,37767061 | 0,932388635 | TNNT3 | FALSE | Downregulated |
| 0,023342946 | 0,932388635 | UBFD1 | FALSE | Upregulated |
| -0,033924658 | 0,932388635 | WRN | FALSE | Downregulated |
| -0,041555858 | 0,932629332 | ACTR3B | FALSE | Downregulated |
| 0,069110417 | 0,932629332 | AGBL3 | FALSE | Upregulated |
| 0,145827999 | 0,932629332 | AKR7A2P1 | FALSE | Upregulated |
| 0,037229457 | 0,932629332 | ARHGDIB | FALSE | Upregulated |
| 0,041438926 | 0,932629332 | ATRIP | FALSE | Upregulated |

| | | | | |
|---|---|---|---|---|
| 0,040000143 | 0,932629332 | C21orf91 | FALSE | Upregulated |
| 0,041621628 | 0,932629332 | CEP78 | FALSE | Upregulated |
| 0,061157226 | 0,932629332 | DCPS | FALSE | Upregulated |
| 0,057558879 | 0,932629332 | FGF14-AS2 | FALSE | Upregulated |
| 0,046299978 | 0,932629332 | GPCPD1 | FALSE | Upregulated |
| 0,142778544 | 0,932629332 | GPR135 | FALSE | Upregulated |
| -0,120130836 | 0,932629332 | GSTM3 | FALSE | Downregulated |
| -0,116628187 | 0,932629332 | ID1 | FALSE | Downregulated |
| 0,042865009 | 0,932629332 | IMPA1 | FALSE | Upregulated |
| 0,079609541 | 0,932629332 | KIAA1524 | FALSE | Upregulated |
| 0,06564355 | 0,932629332 | KLF9 | FALSE | Upregulated |
| 0,074556356 | 0,932629332 | LINC00899 | FALSE | Upregulated |
| 0,118827775 | 0,932629332 | LOC100507460 | FALSE | Upregulated |
| 0,08057147 | 0,932629332 | LOC101927663 | FALSE | Upregulated |
| -0,021800061 | 0,932629332 | LSM14B | FALSE | Downregulated |
| 0,027791212 | 0,932629332 | MAP3K13 | FALSE | Upregulated |
| 0,045854619 | 0,932629332 | METTL4 | FALSE | Upregulated |
| -0,065489765 | 0,932629332 | NPHP3 | FALSE | Downregulated |
| -0,052230657 | 0,932629332 | PER2 | FALSE | Downregulated |
| -0,029401769 | 0,932629332 | PLCB3 | FALSE | Downregulated |
| -0,039069711 | 0,932629332 | PRRC2C | FALSE | Downregulated |
| -0,043905692 | 0,932629332 | PYCRL | FALSE | Downregulated |
| -0,047195166 | 0,932629332 | RABEPK | FALSE | Downregulated |
| -0,079842671 | 0,932629332 | RERG | FALSE | Downregulated |
| 0,091974907 | 0,932629332 | RP5-1021I20.1 | FALSE | Upregulated |
| -0,064452604 | 0,932629332 | SIRT4 | FALSE | Downregulated |

| | | | | |
|---|---|---|---|---|
| 0,795837935 | 0,932629332 | SLC6A11 | FALSE | Upregulated |
| -0,03185038 | 0,932629332 | ZDHHC16 | FALSE | Downregulated |
| -0,042281233 | 0,932825645 | RPRD1B | FALSE | Downregulated |
| 0,05561989 | 0,932840221 | FLJ42627 | FALSE | Upregulated |
| 0,111787283 | 0,932873989 | ZNRF2P1 | FALSE | Upregulated |
| -0,058281158 | 0,932912714 | C4orf33 | FALSE | Downregulated |
| 0,095045608 | 0,933367268 | AC159540.1-2 | FALSE | Upregulated |
| -1,362901356 | 0,933367268 | ACTC1 | FALSE | Downregulated |
| -0,037624486 | 0,933367268 | ANKS1A | FALSE | Downregulated |
| -0,025358023 | 0,933367268 | ARNT | FALSE | Downregulated |
| 0,038684905 | 0,933367268 | C15orf57 | FALSE | Upregulated |
| 0,030055802 | 0,933367268 | CDK5RAP2 | FALSE | Upregulated |
| 0,058962377 | 0,933367268 | DNAJB9 | FALSE | Upregulated |
| 1,356977535 | 0,933367268 | LOC102724843 | FALSE | Upregulated |
| -0,188764932 | 0,933367268 | MAFG-AS1 | FALSE | Downregulated |
| 0,100637626 | 0,933367268 | MYEF2 | FALSE | Upregulated |
| -0,041402268 | 0,933367268 | NDUFAF3 | FALSE | Downregulated |
| 0,156070711 | 0,933367268 | NRG2 | FALSE | Upregulated |
| 0,099009141 | 0,933367268 | PLA2G5 | FALSE | Upregulated |
| 0,02231996 | 0,933367268 | SCAF11 | FALSE | Upregulated |
| 0,092981979 | 0,933367268 | SCD5 | FALSE | Upregulated |
| -0,088785903 | 0,933367268 | SPATA1 | FALSE | Downregulated |
| 0,05099657 | 0,9334516 | FUT10 | FALSE | Upregulated |
| -0,210449303 | 0,933479824 | BDNF | FALSE | Downregulated |
| 0,163900728 | 0,933479824 | CLCF1 | FALSE | Upregulated |
| -0,033450563 | 0,933479824 | COX5B | FALSE | Downregulated |

| | | | | |
|---|---|---|---|---|
| -0,054847066 | 0,933479824 | CSRP2 | FALSE | Downregulated |
| -0,047154591 | 0,933479824 | GRAMD3 | FALSE | Downregulated |
| 0,145318838 | 0,933479824 | LINC00965 | FALSE | Upregulated |
| -0,030927057 | 0,933479824 | NARFL | FALSE | Downregulated |
| -0,037898394 | 0,933479824 | POLD3 | FALSE | Downregulated |
| 0,107582911 | 0,933479824 | PYROXD2 | FALSE | Upregulated |
| 0,107148233 | 0,933479824 | RP5-1125A11.1 | FALSE | Upregulated |
| 0,023022004 | 0,933479824 | SPCS1 | FALSE | Upregulated |
| 0,089368804 | 0,933572294 | LOC643733 | FALSE | Upregulated |
| 0,147536912 | 0,933572294 | OXTR | FALSE | Upregulated |
| -0,069056915 | 0,933629897 | ABCA9 | FALSE | Downregulated |
| 0,037915376 | 0,933629897 | ACBD3 | FALSE | Upregulated |
| -0,06106847 | 0,933629897 | ARHGAP23 | FALSE | Downregulated |
| -0,101908918 | 0,933629897 | ASB4 | FALSE | Downregulated |
| 0,056048499 | 0,933629897 | BAIAP3 | FALSE | Upregulated |
| -0,212392911 | 0,933629897 | CDKL2 | FALSE | Downregulated |
| 0,02921194 | 0,933629897 | COMMD5 | FALSE | Upregulated |
| 0,075120588 | 0,933629897 | DGKG | FALSE | Upregulated |
| -0,059300184 | 0,933629897 | EBP | FALSE | Downregulated |
| 0,025155307 | 0,933629897 | EID1 | FALSE | Upregulated |
| 0,028657191 | 0,933629897 | EIF2D | FALSE | Upregulated |
| -0,244967527 | 0,933629897 | FAM157C | FALSE | Downregulated |
| 0,133743489 | 0,933629897 | FSD1 | FALSE | Upregulated |
| -0,149220219 | 0,933629897 | GPM6B | FALSE | Downregulated |
| 0,060826045 | 0,933629897 | GPT2 | FALSE | Upregulated |
| -0,033956995 | 0,933629897 | GSTO1 | FALSE | Downregulated |

| | | | | |
|---|---|---|---|---|
| 0,018178029 | 0,933629897 | IPO7 | FALSE | Upregulated |
| 0,21483876 | 0,933629897 | KLK10 | FALSE | Upregulated |
| -0,05318723 | 0,933629897 | LINC00261 | FALSE | Downregulated |
| 0,186767577 | 0,933629897 | LOC100507195 | FALSE | Upregulated |
| 0,138885226 | 0,933629897 | LOC101927278 | FALSE | Upregulated |
| 0,076673068 | 0,933629897 | MYBL1 | FALSE | Upregulated |
| -0,039671316 | 0,933629897 | NDUFC2 | FALSE | Downregulated |
| -0,032214868 | 0,933629897 | NID1 | FALSE | Downregulated |
| -0,131371224 | 0,933629897 | NPFF | FALSE | Downregulated |
| -0,047444744 | 0,933629897 | PPIL1 | FALSE | Downregulated |
| 0,038995401 | 0,933629897 | RGMB | FALSE | Upregulated |
| 0,022330511 | 0,933629897 | RYBP | FALSE | Upregulated |
| 0,061621125 | 0,933629897 | SCN8A | FALSE | Upregulated |
| -0,121944333 | 0,933629897 | SEMA3A | FALSE | Downregulated |
| 0,038307146 | 0,933629897 | SETD4 | FALSE | Upregulated |
| -0,190375802 | 0,933629897 | SULT1A3 | FALSE | Downregulated |
| -0,03944377 | 0,933629897 | TET3 | FALSE | Downregulated |
| 0,034634734 | 0,933629897 | TMEM140 | FALSE | Upregulated |
| -0,048414987 | 0,933629897 | USP27X | FALSE | Downregulated |
| 0,022861234 | 0,933629897 | YTHDF1 | FALSE | Upregulated |
| -0,028342085 | 0,933629897 | ZBTB14 | FALSE | Downregulated |
| -0,06546896 | 0,933629897 | ZNF37BP | FALSE | Downregulated |
| 0,056017648 | 0,933629897 | ZNF487 | FALSE | Upregulated |
| -0,097175492 | 0,933736831 | LOC100133315 | FALSE | Downregulated |
| -0,056874634 | 0,933737084 | HMOX1 | FALSE | Downregulated |
| 0,029652329 | 0,933737084 | TRMT10C | FALSE | Upregulated |

| | | | | |
|---|---|---|---|---|
| -0,027234513 | 0,933749809 | ZBTB11 | FALSE | Downregulated |
| -0,168781615 | 0,933784862 | LOC101929675 | FALSE | Downregulated |
| -0,120724516 | 0,933881884 | B3GNT5 | FALSE | Downregulated |
| 0,105097185 | 0,933881884 | HSD17B3 | FALSE | Upregulated |
| -0,268621368 | 0,933881884 | LOC100505938 | FALSE | Downregulated |
| -0,146884531 | 0,933991736 | LOC100131347 | FALSE | Downregulated |
| -0,038149222 | 0,934009612 | C11orf30 | FALSE | Downregulated |
| 0,020607636 | 0,934009612 | CHD8 | FALSE | Upregulated |
| 0,044672249 | 0,934009612 | CRELD1 | FALSE | Upregulated |
| 0,116251967 | 0,934009612 | LOC101928958 | FALSE | Upregulated |
| 0,151809877 | 0,934009612 | SNORA61 | FALSE | Upregulated |
| 0,219081726 | 0,934009612 | USP9Y | FALSE | Upregulated |
| 0,024257469 | 0,934017665 | ATXN2 | FALSE | Upregulated |
| 0,050002871 | 0,934017665 | BDNF-AS | FALSE | Upregulated |
| -0,047306092 | 0,934017665 | NR2F1-AS1 | FALSE | Downregulated |
| -0,167199911 | 0,934017665 | SLC38A5 | FALSE | Downregulated |
| 0,022600048 | 0,934017665 | TCF20 | FALSE | Upregulated |
| 0,057216331 | 0,934017665 | TMPO | FALSE | Upregulated |
| 0,272065974 | 0,934017665 | TRIM74 | FALSE | Upregulated |
| -0,031291162 | 0,934125507 | RPL35A | FALSE | Downregulated |
| -0,073719962 | 0,934165419 | RASGRP2 | FALSE | Downregulated |
| -0,041818281 | 0,934184788 | AMT | FALSE | Downregulated |
| 0,165133686 | 0,934184788 | LOC101929687 | FALSE | Upregulated |
| 0,079891126 | 0,934184788 | NBEA | FALSE | Upregulated |
| -0,050580388 | 0,934184788 | ZNF57 | FALSE | Downregulated |
| -0,029509238 | 0,934220296 | HKR1 | FALSE | Downregulated |

| | | | | |
|---|---|---|---|---|
| 0,030382741 | 0,934220296 | KLHL5 | FALSE | Upregulated |
| 0,039415998 | 0,934303823 | FAM219A | FALSE | Upregulated |
| 0,160669738 | 0,934303823 | LOC102723958 | FALSE | Upregulated |
| -0,084578205 | 0,934303823 | PLCD3 | FALSE | Downregulated |
| 0,193884319 | 0,934303823 | RP11-290F20.2 | FALSE | Upregulated |
| -0,059587837 | 0,934303823 | RPGR | FALSE | Downregulated |
| 0,116540708 | 0,934303823 | SCGB3A1 | FALSE | Upregulated |
| -0,034212709 | 0,934303823 | THTPA | FALSE | Downregulated |
| 0,040202932 | 0,934333362 | ASCC3 | FALSE | Upregulated |
| -0,03538281 | 0,934333362 | C5 | FALSE | Downregulated |
| -0,033208711 | 0,934333362 | CAPN15 | FALSE | Downregulated |
| 0,065091464 | 0,934333362 | FCHSD1 | FALSE | Upregulated |
| -0,064190199 | 0,934333362 | MDM4 | FALSE | Downregulated |
| -0,06872607 | 0,934333362 | PLEKHG6 | FALSE | Downregulated |
| 0,079078001 | 0,934333362 | RP11-15A1.3 | FALSE | Upregulated |
| -0,054764369 | 0,934333362 | UPB1 | FALSE | Downregulated |
| -0,078480608 | 0,934449684 | GGACT | FALSE | Downregulated |
| 0,02507854 | 0,934449684 | KHNYN | FALSE | Upregulated |
| 0,020905933 | 0,934449684 | UMPS | FALSE | Upregulated |
| 0,224020716 | 0,934449684 | ZFP57 | FALSE | Upregulated |
| -0,099401464 | 0,935026939 | MAP2K6 | FALSE | Downregulated |
| -0,048044481 | 0,935061368 | ALDH18A1 | FALSE | Downregulated |
| 0,096130813 | 0,935153179 | EMC3-AS1 | FALSE | Upregulated |
| -0,059115354 | 0,935230383 | ACOT1 | FALSE | Downregulated |
| 0,046898033 | 0,935230383 | ATAT1 | FALSE | Upregulated |
| 0,058506541 | 0,935230383 | BIN1 | FALSE | Upregulated |

| | | | | |
|---|---|---|---|---|
| -0,041885135 | 0,935230383 | BSCL2 | FALSE | Downregulated |
| 0,05417389 | 0,935230383 | C8orf58 | FALSE | Upregulated |
| -0,026175626 | 0,935230383 | CD276 | FALSE | Downregulated |
| 0,106807792 | 0,935230383 | CLDN10 | FALSE | Upregulated |
| 0,791829412 | 0,935230383 | EIF1AY | FALSE | Upregulated |
| 0,093713627 | 0,935230383 | FAM53A | FALSE | Upregulated |
| 0,032780645 | 0,935230383 | IFFO1 | FALSE | Upregulated |
| -0,032478033 | 0,935230383 | LCMT2 | FALSE | Downregulated |
| -0,139774849 | 0,935230383 | LOC102723980 | FALSE | Downregulated |
| -0,040396365 | 0,935230383 | MPHOSPH8 | FALSE | Downregulated |
| -0,072638515 | 0,935230383 | PLAGL1 | FALSE | Downregulated |
| 0,020063389 | 0,935230383 | RANBP3 | FALSE | Upregulated |
| 0,032571901 | 0,935230383 | RPL7A | FALSE | Upregulated |
| 0,04522269 | 0,935230383 | SCARF1 | FALSE | Upregulated |
| -0,062405932 | 0,935230383 | SMIM2-AS1 | FALSE | Downregulated |
| 0,028816893 | 0,935230383 | SNX18 | FALSE | Upregulated |
| 0,203473957 | 0,935230383 | STK32B | FALSE | Upregulated |
| -0,023847001 | 0,935230383 | STRBP | FALSE | Downregulated |
| -0,052662491 | 0,935230383 | TOR3A | FALSE | Downregulated |
| -0,118675457 | 0,935230383 | YBX3 | FALSE | Downregulated |
| -0,029597263 | 0,935324172 | C9orf114 | FALSE | Downregulated |
| 0,047516893 | 0,935324172 | DHRS4 | FALSE | Upregulated |
| -0,119858007 | 0,935324172 | LINC00894 | FALSE | Downregulated |
| 0,090475679 | 0,935324172 | LOC100506388 | FALSE | Upregulated |
| 0,025133258 | 0,935324172 | MAU2 | FALSE | Upregulated |
| 0,099232083 | 0,935413 | BCL2A1 | FALSE | Upregulated |

| | | | | |
|---|---|---|---|---|
| 0,027722317 | 0,935413 | CARKD | FALSE | Upregulated |
| 0,094243427 | 0,935413 | LOC606724 | FALSE | Upregulated |
| 0,027858504 | 0,935413 | PHF20 | FALSE | Upregulated |
| -0,115011852 | 0,935413 | SSPN | FALSE | Downregulated |
| 0,042155894 | 0,935413 | THAP6 | FALSE | Upregulated |
| 0,047377386 | 0,935616929 | EGFL7 | FALSE | Upregulated |
| 0,141225457 | 0,93565916 | SAMD13 | FALSE | Upregulated |
| -0,023508002 | 0,935993907 | FH | FALSE | Downregulated |
| -0,055186927 | 0,936003501 | CCDC7 | FALSE | Downregulated |
| -0,185410822 | 0,936003501 | LOC101929098 | FALSE | Downregulated |
| -0,098228441 | 0,936003501 | LOC102724238 | FALSE | Downregulated |
| -0,038193632 | 0,936003501 | MGAT5 | FALSE | Downregulated |
| -0,026323424 | 0,936003501 | PATZ1 | FALSE | Downregulated |
| -0,122854337 | 0,936003501 | RP11-290F5.2 | FALSE | Downregulated |
| -0,020930328 | 0,936003501 | SUPT5H | FALSE | Downregulated |
| 0,128826553 | 0,936003501 | VIP | FALSE | Upregulated |
| 0,101663852 | 0,936101277 | GNLY | FALSE | Upregulated |
| 0,021574 | 0,936101277 | SCYL3 | FALSE | Upregulated |
| 0,037129515 | 0,936101277 | ZMIZ2 | FALSE | Upregulated |
| -0,047027282 | 0,936295932 | ABHD5 | FALSE | Downregulated |
| 0,073314186 | 0,936334785 | FRMD6 | FALSE | Upregulated |
| 0,055788207 | 0,936334785 | KIAA1328 | FALSE | Upregulated |
| -0,072557662 | 0,936334785 | NDUFA4L2 | FALSE | Downregulated |
| 0,050518615 | 0,936413874 | BMF | FALSE | Upregulated |
| -0,098434175 | 0,936413874 | LOC101928830 | FALSE | Downregulated |
| -0,062793231 | 0,936522977 | THAP10 | FALSE | Downregulated |

| | | | | |
|---|---|---|---|---|
| -0,031634206 | 0,93658433 | DEGS1 | FALSE | Downregulated |
| -0,15295307 | 0,93658433 | FAM162B | FALSE | Downregulated |
| -0,308368385 | 0,93658433 | LOC101059936 | FALSE | Downregulated |
| 0,094460483 | 0,93658433 | LOC101929177 | FALSE | Upregulated |
| 0,019896675 | 0,936684725 | ATP6V1C1 | FALSE | Upregulated |
| -0,044425115 | 0,936684725 | GPR146 | FALSE | Downregulated |
| 0,096462144 | 0,936684725 | MIR198 | FALSE | Upregulated |
| 0,053311666 | 0,936684725 | MITF | FALSE | Upregulated |
| -0,038960654 | 0,936684725 | NRIP1 | FALSE | Downregulated |
| 0,020227478 | 0,936684725 | PITHD1 | FALSE | Upregulated |
| -0,019294504 | 0,936684725 | RRN3 | FALSE | Downregulated |
| -0,147703967 | 0,936684725 | SATL1 | FALSE | Downregulated |
| 0,095909234 | 0,936684725 | SLC44A3 | FALSE | Upregulated |
| -0,040213845 | 0,936684725 | SMC4 | FALSE | Downregulated |
| -0,023077047 | 0,936684725 | UBR2 | FALSE | Downregulated |
| 0,024125309 | 0,936684725 | WDR61 | FALSE | Upregulated |
| -0,021610054 | 0,936684725 | WNK1 | FALSE | Downregulated |
| 0,043174058 | 0,936938744 | BGN | FALSE | Upregulated |
| -0,029884668 | 0,936938744 | BRD8 | FALSE | Downregulated |
| -0,019782642 | 0,936938744 | MAPK14 | FALSE | Downregulated |
| -0,035644646 | 0,936938744 | SERTAD2 | FALSE | Downregulated |
| 0,075881334 | 0,936938744 | STK4-AS1 | FALSE | Upregulated |
| 0,093393265 | 0,937307322 | LINC01004 | FALSE | Upregulated |
| -0,116003839 | 0,937307322 | LOC102723576 | FALSE | Downregulated |
| 0,055916173 | 0,937307322 | MRC1 | FALSE | Upregulated |
| 0,144407777 | 0,937307322 | RP11-79E3.3 | FALSE | Upregulated |

| | | | | |
|---|---|---|---|---|
| -0,086140017 | 0,937307322 | UCP2 | FALSE | Downregulated |
| -0,019995512 | 0,937651898 | RAF1 | FALSE | Downregulated |
| 0,108010382 | 0,937687718 | AATK | FALSE | Upregulated |
| 0,058995667 | 0,937687718 | BCL6B | FALSE | Upregulated |
| 0,040895142 | 0,937687718 | HSPA8 | FALSE | Upregulated |
| -0,155562413 | 0,937687718 | LINC00665 | FALSE | Downregulated |
| 0,045872874 | 0,937687718 | PHKA1 | FALSE | Upregulated |
| -0,036236968 | 0,937687718 | TRIM21 | FALSE | Downregulated |
| 0,027653836 | 0,937717371 | DDX5 | FALSE | Upregulated |
| -0,095685018 | 0,937717371 | HORMAD2 | FALSE | Downregulated |
| -0,155507457 | 0,937717371 | MFAP2 | FALSE | Downregulated |
| 0,044419793 | 0,937717371 | OIT3 | FALSE | Upregulated |
| 0,078305671 | 0,937717371 | ZNF93 | FALSE | Upregulated |
| -0,189985721 | 0,937799602 | ARHGAP8 | FALSE | Downregulated |
| -0,050372466 | 0,937799602 | PI4KAP2 | FALSE | Downregulated |
| -0,292802426 | 0,937813384 | LOC102724652 | FALSE | Downregulated |
| 0,049123834 | 0,937813384 | RPS27L | FALSE | Upregulated |
| -0,041591972 | 0,937823109 | ARFGAP3 | FALSE | Downregulated |
| 0,079429318 | 0,937823109 | ARMC9 | FALSE | Upregulated |
| 0,050757255 | 0,937823109 | ASCC1 | FALSE | Upregulated |
| -0,021933125 | 0,937823109 | ATF1 | FALSE | Downregulated |
| -0,028030271 | 0,937823109 | AURKAIP1 | FALSE | Downregulated |
| 0,032123682 | 0,937823109 | BET1 | FALSE | Upregulated |
| -0,132987729 | 0,937823109 | BOC | FALSE | Downregulated |
| -0,02259152 | 0,937823109 | C5orf24 | FALSE | Downregulated |
| -0,029967801 | 0,937823109 | CAMK2D | FALSE | Downregulated |

| | | | | |
|---|---|---|---|---|
| 0,072945148 | 0,937823109 | CLIP3 | FALSE | Upregulated |
| 0,035339333 | 0,937823109 | CTSF | FALSE | Upregulated |
| -0,037634379 | 0,937823109 | DBT | FALSE | Downregulated |
| 0,048873648 | 0,937823109 | DYNLL1-AS1 | FALSE | Upregulated |
| 0,213013812 | 0,937823109 | FLJ39095 | FALSE | Upregulated |
| -0,112310012 | 0,937823109 | GNAZ | FALSE | Downregulated |
| 0,029194484 | 0,937823109 | HIGD2A | FALSE | Upregulated |
| 0,114974905 | 0,937823109 | LOC100507472 | FALSE | Upregulated |
| -0,079845481 | 0,937823109 | LOC102724481 | FALSE | Downregulated |
| 0,097496764 | 0,937823109 | LOC102724783 | FALSE | Upregulated |
| 0,02222725 | 0,937823109 | MTPAP | FALSE | Upregulated |
| -0,039676753 | 0,937823109 | ORAOV1 | FALSE | Downregulated |
| 0,130767509 | 0,937823109 | PKDREJ | FALSE | Upregulated |
| -0,240757178 | 0,937823109 | PNLDC1 | FALSE | Downregulated |
| 0,026258936 | 0,937823109 | PRKCZ | FALSE | Upregulated |
| -0,035830107 | 0,937823109 | PSMG1 | FALSE | Downregulated |
| 0,099342031 | 0,937823109 | PXDNL | FALSE | Upregulated |
| -0,086803879 | 0,937823109 | PYCR1 | FALSE | Downregulated |
| 0,065439262 | 0,937823109 | RHOB | FALSE | Upregulated |
| -0,036815586 | 0,937823109 | RPL32 | FALSE | Downregulated |
| -0,051128008 | 0,937823109 | S100A10 | FALSE | Downregulated |
| -0,019553345 | 0,937823109 | SCARB2 | FALSE | Downregulated |
| -0,044446371 | 0,937823109 | SLC48A1 | FALSE | Downregulated |
| 0,021569585 | 0,937823109 | SUMO3 | FALSE | Upregulated |
| -0,035823689 | 0,937823109 | THRB | FALSE | Downregulated |
| 0,114237774 | 0,937823109 | TMEM236 | FALSE | Upregulated |

| | | | | |
|---|---|---|---|---|
| -0,033781527 | 0,937823109 | TPD52 | FALSE | Downregulated |
| 0,035533553 | 0,937823109 | TRMT2A | FALSE | Upregulated |
| 0,031525038 | 0,937823109 | TSPAN9 | FALSE | Upregulated |
| -0,189618906 | 0,938086762 | M1AP | FALSE | Downregulated |
| -0,085981003 | 0,938183411 | TTC25 | FALSE | Downregulated |
| -1,275961904 | 0,938231951 | CSRP3 | FALSE | Downregulated |
| 0,023611009 | 0,938231951 | SMAP2 | FALSE | Upregulated |
| -0,029313881 | 0,938231951 | ZNF91 | FALSE | Downregulated |
| -0,113100424 | 0,938265487 | COLCA1 | FALSE | Downregulated |
| -0,073097544 | 0,938265487 | LINC01006 | FALSE | Downregulated |
| -0,046661512 | 0,938265487 | PHC3 | FALSE | Downregulated |
| 0,034907409 | 0,938435639 | HRG | FALSE | Upregulated |
| -0,024448296 | 0,938435639 | NCOA2 | FALSE | Downregulated |
| -0,023410178 | 0,938435639 | OAZ1 | FALSE | Downregulated |
| 0,032919444 | 0,938435639 | TNFAIP1 | FALSE | Upregulated |
| 0,026669324 | 0,938435639 | UBAC1 | FALSE | Upregulated |
| 1,130416595 | 0,938835433 | EEF1A2 | FALSE | Upregulated |
| 0,268568665 | 0,938836036 | GOLGA8K | FALSE | Upregulated |
| 0,041066254 | 0,938836036 | GTF2IRD2B | FALSE | Upregulated |
| -0,030968947 | 0,938836036 | GTF3A | FALSE | Downregulated |
| -0,128956782 | 0,938836036 | IL10RB-AS1 | FALSE | Downregulated |
| -0,023917048 | 0,938836036 | KIAA1468 | FALSE | Downregulated |
| 0,060829886 | 0,938836036 | LINC00476 | FALSE | Upregulated |
| -0,111670944 | 0,938836036 | LPHN3 | FALSE | Downregulated |
| -0,05722294 | 0,938836036 | NCOA7 | FALSE | Downregulated |
| 0,037913313 | 0,938836036 | NME2 | FALSE | Upregulated |

| | | | | |
|---|---|---|---|---|
| 0,037320827 | 0,938836036 | NOP16 | FALSE | Upregulated |
| 0,021429901 | 0,938836036 | PSMD13 | FALSE | Upregulated |
| 0,045958535 | 0,938836036 | PYROXD1 | FALSE | Upregulated |
| 0,861724823 | 0,938836036 | RPS4Y1 | FALSE | Upregulated |
| 0,059930978 | 0,938836036 | SLC7A5 | FALSE | Upregulated |
| 0,024956669 | 0,938836036 | SNIP1 | FALSE | Upregulated |
| 0,070479571 | 0,938836036 | TRIM55 | FALSE | Upregulated |
| -0,018801367 | 0,939205764 | PHKB | FALSE | Downregulated |
| 0,204238815 | 0,939205764 | SLAMF9 | FALSE | Upregulated |
| -1,256302885 | 0,939303558 | MYBPC1 | FALSE | Downregulated |
| 0,115023403 | 0,939303558 | NEK2 | FALSE | Upregulated |
| 0,036757606 | 0,939303558 | NMNAT1 | FALSE | Upregulated |
| -0,083428685 | 0,939539718 | LOC102724550 | FALSE | Downregulated |
| 0,067343114 | 0,939542574 | SLC6A13 | FALSE | Upregulated |
| 0,110299879 | 0,9395946 | LOC102723648 | FALSE | Upregulated |
| 0,029798349 | 0,939673393 | ADAM17 | FALSE | Upregulated |
| -0,027109144 | 0,939673393 | C2CD2 | FALSE | Downregulated |
| 0,18094261 | 0,939673393 | CATSPERB | FALSE | Upregulated |
| 0,055594211 | 0,939673393 | IFI30 | FALSE | Upregulated |
| 0,05577988 | 0,939673393 | IFIT2 | FALSE | Upregulated |
| -0,130293669 | 0,939673393 | LOC440895 | FALSE | Downregulated |
| 0,064399125 | 0,939673393 | MSRB3 | FALSE | Upregulated |
| 0,02739648 | 0,939673393 | NDUFS6 | FALSE | Upregulated |
| 0,101488081 | 0,939673393 | PLGLB1 | FALSE | Upregulated |
| -0,023009311 | 0,939673393 | PPP1R15B | FALSE | Downregulated |
| 0,026052308 | 0,939673393 | PTPN21 | FALSE | Upregulated |

| | | | | |
|---|---|---|---|---|
| -0,037700026 | 0,939673393 | TLE1 | FALSE | Downregulated |
| 0,032343941 | 0,939673393 | TMED7 | FALSE | Upregulated |
| 0,023193374 | 0,939673393 | TMEM33 | FALSE | Upregulated |
| -0,051226653 | 0,939673393 | UGT2A3 | FALSE | Downregulated |
| -0,149555658 | 0,939766802 | C9orf24 | FALSE | Downregulated |
| 0,088558714 | 0,939834291 | ATOH8 | FALSE | Upregulated |
| 0,178826633 | 0,939926424 | POM121L8P | FALSE | Upregulated |
| -0,031725046 | 0,939949479 | CTSO | FALSE | Downregulated |
| 0,122822041 | 0,939951508 | ADAMTS15 | FALSE | Upregulated |
| 0,17970961 | 0,939951508 | BCL2L15 | FALSE | Upregulated |
| 0,085605424 | 0,939951508 | CCDC110 | FALSE | Upregulated |
| -0,115095164 | 0,939951508 | CLDND2 | FALSE | Downregulated |
| 0,142815208 | 0,939951508 | DDX11L2 | FALSE | Upregulated |
| 0,020353946 | 0,939951508 | DNAJC16 | FALSE | Upregulated |
| 0,110081703 | 0,939951508 | FNDC1 | FALSE | Upregulated |
| -0,15962218 | 0,939951508 | LINC00176 | FALSE | Downregulated |
| -0,146591542 | 0,939951508 | LINC00222 | FALSE | Downregulated |
| -0,116682361 | 0,939951508 | LOC101927178 | FALSE | Downregulated |
| -1,250110568 | 0,939951508 | LOC101928874 | FALSE | Downregulated |
| -0,16838628 | 0,939951508 | NTNG2 | FALSE | Downregulated |
| 0,064974891 | 0,939951508 | PPP1R14A | FALSE | Upregulated |
| -0,044270232 | 0,939951508 | RPL9 | FALSE | Downregulated |
| -0,042953249 | 0,939951508 | SLC9A9 | FALSE | Downregulated |
| 0,036092828 | 0,940036375 | ZMAT5 | FALSE | Upregulated |
| -0,095592256 | 0,940223719 | RP11-1223D19.1 | FALSE | Downregulated |
| 0,040360076 | 0,940410688 | UBR4 | FALSE | Upregulated |

| | | | | |
|---|---|---|---|---|
| 0,072876519 | 0,940651246 | ABRACL | FALSE | Upregulated |
| -0,039632462 | 0,940651246 | GCFC2 | FALSE | Downregulated |
| 0,044656471 | 0,940651246 | GET4 | FALSE | Upregulated |
| 0,080403588 | 0,940651246 | ITIH5 | FALSE | Upregulated |
| 0,030470712 | 0,940651246 | KATNBL1 | FALSE | Upregulated |
| 0,026469864 | 0,940651246 | LOC100506548 | FALSE | Upregulated |
| 0,033796201 | 0,940651246 | PHLPP2 | FALSE | Upregulated |
| 0,035602006 | 0,940651246 | TAF6L | FALSE | Upregulated |
| 0,029271798 | 0,940651246 | TMEM19 | FALSE | Upregulated |
| 0,035438488 | 0,940651246 | TMEM251 | FALSE | Upregulated |
| -0,441271831 | 0,940697579 | ANKRD20A2 | FALSE | Downregulated |
| 0,03629082 | 0,940697579 | CSGALNACT2 | FALSE | Upregulated |
| 0,066546373 | 0,940697579 | XRCC3 | FALSE | Upregulated |
| 0,052907655 | 0,940793463 | SMOC1 | FALSE | Upregulated |
| 0,04847696 | 0,940888339 | ABHD11 | FALSE | Upregulated |
| -0,023313965 | 0,940888339 | AHCYL2 | FALSE | Downregulated |
| -0,029581996 | 0,940888339 | C11orf31 | FALSE | Downregulated |
| -0,031047135 | 0,940888339 | CHRNB1 | FALSE | Downregulated |
| 0,020180499 | 0,940888339 | CMTR1 | FALSE | Upregulated |
| -0,15840907 | 0,940888339 | CSNK2A3 | FALSE | Downregulated |
| -0,037764931 | 0,940888339 | CYP2C18 | FALSE | Downregulated |
| -0,026215516 | 0,940888339 | DSTYK | FALSE | Downregulated |
| 0,029574338 | 0,940888339 | EIF3M | FALSE | Upregulated |
| 0,025899146 | 0,940888339 | F2 | FALSE | Upregulated |
| 0,040854355 | 0,940888339 | IFT43 | FALSE | Upregulated |
| -0,041526105 | 0,940888339 | KALRN | FALSE | Downregulated |

| | | | | |
|---|---|---|---|---|
| 0,087900903 | 0,940888339 | KCNMB1 | FALSE | Upregulated |
| 0,033920146 | 0,940888339 | KIAA0922 | FALSE | Upregulated |
| -0,059693993 | 0,940888339 | LCN12 | FALSE | Downregulated |
| 0,082521748 | 0,940888339 | LOC101927054 | FALSE | Upregulated |
| -0,069481268 | 0,940888339 | LOC101928000 | FALSE | Downregulated |
| 0,056004469 | 0,940888339 | LOC441155 | FALSE | Upregulated |
| 0,079202867 | 0,940888339 | LOC595101 | FALSE | Upregulated |
| -0,068276364 | 0,940888339 | LOC728975 | FALSE | Downregulated |
| -0,104511707 | 0,940888339 | LRRC24 | FALSE | Downregulated |
| 0,044865891 | 0,940888339 | MPND | FALSE | Upregulated |
| -0,029741651 | 0,940888339 | NDUFB4 | FALSE | Downregulated |
| 0,026691887 | 0,940888339 | NEU1 | FALSE | Upregulated |
| -0,027130579 | 0,940888339 | PDZK1 | FALSE | Downregulated |
| 0,031628181 | 0,940888339 | POLL | FALSE | Upregulated |
| -0,031258958 | 0,940888339 | RCOR3 | FALSE | Downregulated |
| 0,054917412 | 0,940888339 | RORC | FALSE | Upregulated |
| 0,135417549 | 0,940888339 | SGCA | FALSE | Upregulated |
| 0,109609539 | 0,940888339 | SLC3A1 | FALSE | Upregulated |
| 0,168454865 | 0,940888339 | SNORA33 | FALSE | Upregulated |
| -0,058692964 | 0,940888339 | SPATA6L | FALSE | Downregulated |
| -0,218935249 | 0,940888339 | SPINK1 | FALSE | Downregulated |
| 0,030428375 | 0,940888339 | SRCAP | FALSE | Upregulated |
| 0,057572236 | 0,940888339 | SYNGR1 | FALSE | Upregulated |
| 0,042588013 | 0,940888339 | TIMP3 | FALSE | Upregulated |
| 0,045705145 | 0,940888339 | TMCO6 | FALSE | Upregulated |
| 0,066233119 | 0,940888339 | TMEM229B | FALSE | Upregulated |

| | | | | |
|---|---|---|---|---|
| -0,050514568 | 0,940888339 | TMEM8B | FALSE | Downregulated |
| 0,032239824 | 0,940888339 | TMPRSS6 | FALSE | Upregulated |
| -0,02255078 | 0,940888339 | TRIM13 | FALSE | Downregulated |
| -0,020381113 | 0,940888339 | TXNDC15 | FALSE | Downregulated |
| -0,019052904 | 0,940888339 | UBA3 | FALSE | Downregulated |
| -0,047130244 | 0,940888339 | UBE2Q2 | FALSE | Downregulated |
| 0,076080415 | 0,940888339 | USP2 | FALSE | Upregulated |
| -0,016596606 | 0,940888339 | VPS36 | FALSE | Downregulated |
| 0,032078515 | 0,940888339 | ZFYVE16 | FALSE | Upregulated |
| 0,095589754 | 0,940888339 | ZNF137P | FALSE | Upregulated |
| -0,03349635 | 0,940888339 | ZNF155 | FALSE | Downregulated |
| 0,049841424 | 0,940890538 | MLKL | FALSE | Upregulated |
| 0,037958255 | 0,940890538 | TMEM106C | FALSE | Upregulated |
| -0,139514047 | 0,940890538 | UGT3A2 | FALSE | Downregulated |
| 0,077154609 | 0,940890538 | ZNF599 | FALSE | Upregulated |
| 0,061995346 | 0,94094295 | LOC100996909 | FALSE | Upregulated |
| 0,034346848 | 0,940978811 | APMAP | FALSE | Upregulated |
| -0,094297306 | 0,940990318 | C2orf82 | FALSE | Downregulated |
| 0,034422819 | 0,940990318 | DAXX | FALSE | Upregulated |
| -0,074995564 | 0,940990318 | MFSD4 | FALSE | Downregulated |
| 0,024629982 | 0,94099119 | STRN4 | FALSE | Upregulated |
| -0,023376661 | 0,94099119 | ZNF689 | FALSE | Downregulated |
| -0,0474875 | 0,941081481 | FOXP2 | FALSE | Downregulated |
| 0,018418059 | 0,941396456 | CCT7 | FALSE | Upregulated |
| -0,184242662 | 0,941512544 | CCDC42B | FALSE | Downregulated |
| -0,022334946 | 0,941512544 | FCHSD2 | FALSE | Downregulated |

| | | | | |
|---|---|---|---|---|
| 0,021759878 | 0,941512544 | PURB | FALSE | Upregulated |
| -0,027559046 | 0,941658934 | ATF4 | FALSE | Downregulated |
| 0,072754422 | 0,941658934 | CCDC102B | FALSE | Upregulated |
| 0,077747862 | 0,941658934 | HERC2P2 | FALSE | Upregulated |
| 0,051408311 | 0,941658934 | LOC100505812 | FALSE | Upregulated |
| -0,1560991 | 0,941658934 | LOC101927279 | FALSE | Downregulated |
| -0,13087095 | 0,941658934 | LOC102723679 | FALSE | Downregulated |
| 0,13434534 | 0,941658934 | MYO3A | FALSE | Upregulated |
| 0,049678825 | 0,94190019 | CD9 | FALSE | Upregulated |
| -0,029055205 | 0,94206751 | BID | FALSE | Downregulated |
| 0,098406485 | 0,94206751 | CCDC17 | FALSE | Upregulated |
| -0,095694568 | 0,94206751 | CDS1 | FALSE | Downregulated |
| 0,126507032 | 0,94206751 | DUSP9 | FALSE | Upregulated |
| 0,0754908 | 0,94206751 | GPR176 | FALSE | Upregulated |
| 0,164622154 | 0,94206751 | INHBA-AS1 | FALSE | Upregulated |
| 0,086724899 | 0,94206751 | KIF19 | FALSE | Upregulated |
| 0,037174498 | 0,94206751 | LOC550643 | FALSE | Upregulated |
| -0,036019331 | 0,94206751 | MEF2BNB | FALSE | Downregulated |
| 0,021996198 | 0,94206751 | PABPC4 | FALSE | Upregulated |
| 0,019586623 | 0,94206751 | RBM10 | FALSE | Upregulated |
| 0,202843595 | 0,94206751 | RIMKLA | FALSE | Upregulated |
| 0,030780739 | 0,94206751 | RPS2 | FALSE | Upregulated |
| -0,021803088 | 0,94206751 | SLC35B3 | FALSE | Downregulated |
| -0,024157077 | 0,94206751 | SRP68 | FALSE | Downregulated |
| 0,018638768 | 0,94206751 | STXBP3 | FALSE | Upregulated |
| 0,017135927 | 0,94206751 | WDR48 | FALSE | Upregulated |

| | | | | |
|---|---|---|---|---|
| -0,21103123 | 0,94206751 | ZHX1-C8ORF76 | FALSE | Downregulated |
| -0,045335548 | 0,942080944 | ABCA8 | FALSE | Downregulated |
| -0,055779572 | 0,942080944 | MLXIPL | FALSE | Downregulated |
| -0,039113234 | 0,942080944 | ZNF81 | FALSE | Downregulated |
| 0,092393773 | 0,942087093 | PABPC5 | FALSE | Upregulated |
| -0,03718879 | 0,942087093 | TSNARE1 | FALSE | Downregulated |
| 0,086591684 | 0,942103486 | LIPE | FALSE | Upregulated |
| 0,196296609 | 0,942103486 | NCOR1P1 | FALSE | Upregulated |
| -0,030562325 | 0,942103486 | RNF25 | FALSE | Downregulated |
| 0,158484111 | 0,942103486 | TEX15 | FALSE | Upregulated |
| 0,083632918 | 0,942223346 | LINC00921 | FALSE | Upregulated |
| -0,028333398 | 0,942247351 | FAU | FALSE | Downregulated |
| 0,145965559 | 0,94236339 | LINC00243 | FALSE | Upregulated |
| -0,025940978 | 0,94236339 | LMTK2 | FALSE | Downregulated |
| 0,028591808 | 0,94236339 | TTC21B | FALSE | Upregulated |
| 0,039506639 | 0,94236339 | ZNF589 | FALSE | Upregulated |
| -0,020863488 | 0,942541303 | STK40 | FALSE | Downregulated |
| 0,051463395 | 0,942551879 | ABCC6P2 | FALSE | Upregulated |
| -0,062913638 | 0,942551879 | ADAMTS5 | FALSE | Downregulated |
| -0,023100004 | 0,942551879 | ADAR | FALSE | Downregulated |
| -0,045315176 | 0,942551879 | ARHGEF15 | FALSE | Downregulated |
| -0,059996376 | 0,942551879 | ATF3 | FALSE | Downregulated |
| 0,026368065 | 0,942551879 | BMP2K | FALSE | Upregulated |
| 0,077540311 | 0,942551879 | CD300E | FALSE | Upregulated |
| 0,078368291 | 0,942551879 | CLEC7A | FALSE | Upregulated |
| -0,024951946 | 0,942551879 | CNIH1 | FALSE | Downregulated |

| | | | | |
|---|---|---|---|---|
| -0,041197069 | 0,942551879 | DUSP12 | FALSE | Downregulated |
| 0,023690389 | 0,942551879 | EIF2B4 | FALSE | Upregulated |
| 0,028275052 | 0,942551879 | FAM98B | FALSE | Upregulated |
| -0,045074708 | 0,942551879 | FCGR2B | FALSE | Downregulated |
| -0,111201095 | 0,942551879 | FLJ38717 | FALSE | Downregulated |
| 0,075584345 | 0,942551879 | GLUD1P7 | FALSE | Upregulated |
| 0,028916121 | 0,942551879 | IPP | FALSE | Upregulated |
| -0,022388656 | 0,942551879 | ITM2B | FALSE | Downregulated |
| -0,188718892 | 0,942551879 | KCNA5 | FALSE | Downregulated |
| 0,177498667 | 0,942551879 | KIAA1751 | FALSE | Upregulated |
| 0,033659061 | 0,942551879 | KLHDC8B | FALSE | Upregulated |
| -0,107263373 | 0,942551879 | LOC102724184 | FALSE | Downregulated |
| -0,08433972 | 0,942551879 | LOC653513 | FALSE | Downregulated |
| -0,02778223 | 0,942551879 | MSH6 | FALSE | Downregulated |
| 0,022498753 | 0,942551879 | MTPN | FALSE | Upregulated |
| -0,041130671 | 0,942551879 | MTSS1L | FALSE | Downregulated |
| -1,193002323 | 0,942551879 | MYH1 | FALSE | Downregulated |
| 0,020225976 | 0,942551879 | POLDIP2 | FALSE | Upregulated |
| -0,069718118 | 0,942551879 | PRICKLE1 | FALSE | Downregulated |
| 0,198783068 | 0,942551879 | RP11-838N2.4 | FALSE | Upregulated |
| 0,023200266 | 0,942551879 | SAMD8 | FALSE | Upregulated |
| -0,073141559 | 0,942551879 | SH3PXD2A | FALSE | Downregulated |
| -0,902742566 | 0,942551879 | SLX1B | FALSE | Downregulated |
| -0,02384179 | 0,942551879 | SMAD2 | FALSE | Downregulated |
| -0,021871317 | 0,942551879 | SMARCC2 | FALSE | Downregulated |
| -0,060494065 | 0,942551879 | SSBP2 | FALSE | Downregulated |

| | | | | |
|---|---|---|---|---|
| -0,021988809 | 0,942551879 | SSU72 | FALSE | Downregulated |
| 0,055013133 | 0,942551879 | ST20 | FALSE | Upregulated |
| 0,026606592 | 0,942551879 | TMEM167B | FALSE | Upregulated |
| -0,10297035 | 0,942551879 | TMEM183B | FALSE | Downregulated |
| -0,307121943 | 0,942551879 | TRNAK13 | FALSE | Downregulated |
| -0,067712873 | 0,942551879 | TRPC5 | FALSE | Downregulated |
| 0,021273395 | 0,942551879 | TYW5 | FALSE | Upregulated |
| 0,119688957 | 0,942551879 | WIPF3 | FALSE | Upregulated |
| -0,033000879 | 0,942551879 | ZNF2 | FALSE | Downregulated |
| 0,032827312 | 0,942551879 | ZNF717 | FALSE | Upregulated |
| 0,165814363 | 0,942569314 | LOC101928916 | FALSE | Upregulated |
| -0,03126575 | 0,942607941 | ATP5D | FALSE | Downregulated |
| -0,176169665 | 0,942607941 | EPHA7 | FALSE | Downregulated |
| 0,028225976 | 0,942607941 | GDI1 | FALSE | Upregulated |
| 0,095210277 | 0,942607941 | LOC100287015 | FALSE | Upregulated |
| 0,15448663 | 0,942607941 | LOC102724379 | FALSE | Upregulated |
| 0,051595504 | 0,942607941 | TSGA10 | FALSE | Upregulated |
| -0,023899593 | 0,942723948 | MSL3 | FALSE | Downregulated |
| 0,031518668 | 0,942723948 | SCYL2 | FALSE | Upregulated |
| 0,102560623 | 0,942958339 | DKFZP434F142 | FALSE | Upregulated |
| 0,020958091 | 0,942958339 | KIAA0247 | FALSE | Upregulated |
| -0,094982223 | 0,94319877 | APOC1P1 | FALSE | Downregulated |
| 0,031817935 | 0,94319877 | C10orf88 | FALSE | Upregulated |
| 0,095892597 | 0,94319877 | C6orf52 | FALSE | Upregulated |
| 0,034618102 | 0,94319877 | DOPEY1 | FALSE | Upregulated |
| 0,017956235 | 0,94319877 | GNA11 | FALSE | Upregulated |

| | | | | |
|---|---|---|---|---|
| -0,079805526 | 0,94319877 | LINC01125 | FALSE | Downregulated |
| 0,109887083 | 0,94319877 | LOC102723365 | FALSE | Upregulated |
| 0,077483982 | 0,94319877 | PLEKHB1 | FALSE | Upregulated |
| -0,035677173 | 0,94319877 | SNTB1 | FALSE | Downregulated |
| -0,03583168 | 0,94319877 | WDR91 | FALSE | Downregulated |
| -0,035426446 | 0,94319877 | ZNF542 | FALSE | Downregulated |
| -0,038691302 | 0,943292654 | BTAF1 | FALSE | Downregulated |
| -0,123062004 | 0,943417966 | LOC101927645 | FALSE | Downregulated |
| -0,06180347 | 0,943443427 | HELB | FALSE | Downregulated |
| 0,137590538 | 0,943443427 | HOXC4 | FALSE | Upregulated |
| 0,021766961 | 0,943443427 | KAT7 | FALSE | Upregulated |
| -0,058873833 | 0,943443427 | MRO | FALSE | Downregulated |
| -0,019626066 | 0,943443427 | RAB2A | FALSE | Downregulated |
| 0,201876092 | 0,943443427 | RP11-76N22.2 | FALSE | Upregulated |
| 0,092723515 | 0,943443427 | SYN1 | FALSE | Upregulated |
| 0,06698609 | 0,943443427 | ZNF154 | FALSE | Upregulated |
| -0,117667311 | 0,943443427 | ZNF205-AS1 | FALSE | Downregulated |
| -0,01828929 | 0,943469255 | GTF2I | FALSE | Downregulated |
| 0,11320479 | 0,943469255 | LOC149134 | FALSE | Upregulated |
| -0,054443898 | 0,943469255 | RABL2A | FALSE | Downregulated |
| -0,162324803 | 0,943469255 | TICRR | FALSE | Downregulated |
| -0,090514907 | 0,943957536 | LOC100288974 | FALSE | Downregulated |
| -0,148181711 | 0,944073023 | GPR97 | FALSE | Downregulated |
| -0,017896805 | 0,944073023 | PRDM4 | FALSE | Downregulated |
| -0,206590219 | 0,944073023 | SLC2A5 | FALSE | Downregulated |
| 0,024306083 | 0,9441767 | AZI2 | FALSE | Upregulated |

| | | | | |
|---|---|---|---|---|
| 0,033812343 | 0,9441767 | C12orf66 | FALSE | Upregulated |
| -0,036677502 | 0,9441767 | C17orf62 | FALSE | Downregulated |
| 0,023977265 | 0,9441767 | PIAS4 | FALSE | Upregulated |
| 0,049638028 | 0,9441767 | PLIN5 | FALSE | Upregulated |
| 0,1542442 | 0,9441767 | TMEM191C | FALSE | Upregulated |
| 0,106024143 | 0,944209608 | DCAF4L1 | FALSE | Upregulated |
| -0,029350457 | 0,944467319 | ARHGAP42 | FALSE | Downregulated |
| 0,075771007 | 0,944467319 | CCNJL | FALSE | Upregulated |
| 0,023181339 | 0,944467319 | CXXC1 | FALSE | Upregulated |
| -0,040383778 | 0,944467319 | DMXL2 | FALSE | Downregulated |
| -0,092087739 | 0,944467319 | LINC00535 | FALSE | Downregulated |
| -0,090384293 | 0,944467319 | LOC102724935 | FALSE | Downregulated |
| -0,028673277 | 0,944467319 | NADK | FALSE | Downregulated |
| 0,113417628 | 0,944467319 | PKD1L2 | FALSE | Upregulated |
| -0,170996848 | 0,944467319 | SMIM17 | FALSE | Downregulated |
| 0,098476548 | 0,944467319 | SV2B | FALSE | Upregulated |
| -0,031375247 | 0,944467319 | TMEM41A | FALSE | Downregulated |
| 0,049310756 | 0,944467319 | UBTD1 | FALSE | Upregulated |
| 0,277824706 | 0,944508915 | ANKRD20A3 | FALSE | Upregulated |
| 0,073089182 | 0,944508915 | ATXN7L2 | FALSE | Upregulated |
| 0,06453761 | 0,944508915 | BEND3 | FALSE | Upregulated |
| -0,030572921 | 0,944508915 | C16orf45 | FALSE | Downregulated |
| 0,117141265 | 0,944508915 | C6orf201 | FALSE | Upregulated |
| -0,046928077 | 0,944508915 | CA5BP1 | FALSE | Downregulated |
| 0,025590244 | 0,944508915 | CANX | FALSE | Upregulated |
| 0,119204825 | 0,944508915 | CCDC74A | FALSE | Upregulated |

| | | | | |
|---|---|---|---|---|
| -0,023236603 | 0,944508915 | CD99L2 | FALSE | Downregulated |
| -0,018239763 | 0,944508915 | CEBPZ | FALSE | Downregulated |
| -0,032283707 | 0,944508915 | CLOCK | FALSE | Downregulated |
| -0,025511835 | 0,944508915 | COA7 | FALSE | Downregulated |
| 0,061068759 | 0,944508915 | COL12A1 | FALSE | Upregulated |
| -0,135235685 | 0,944508915 | CPO | FALSE | Downregulated |
| -0,035834351 | 0,944508915 | CRLS1 | FALSE | Downregulated |
| 0,11783852 | 0,944508915 | CTSLP2 | FALSE | Upregulated |
| -0,105465165 | 0,944508915 | EFR3B | FALSE | Downregulated |
| 0,074033775 | 0,944508915 | ENPP2 | FALSE | Upregulated |
| 0,140420905 | 0,944508915 | EPB41L4A-AS2 | FALSE | Upregulated |
| 0,028869094 | 0,944508915 | FAM73A | FALSE | Upregulated |
| 0,151508351 | 0,944508915 | FOSL1 | FALSE | Upregulated |
| -0,12131202 | 0,944508915 | GLI1 | FALSE | Downregulated |
| -0,030660747 | 0,944508915 | GNG7 | FALSE | Downregulated |
| 0,087847793 | 0,944508915 | HAPLN4 | FALSE | Upregulated |
| 0,03166181 | 0,944508915 | HSD17B11 | FALSE | Upregulated |
| -0,030237559 | 0,944508915 | HSP90AB1 | FALSE | Downregulated |
| 0,02154757 | 0,944508915 | KDM3B | FALSE | Upregulated |
| -0,017767553 | 0,944508915 | KIAA1147 | FALSE | Downregulated |
| 0,026105478 | 0,944508915 | LMAN2 | FALSE | Upregulated |
| 0,033130391 | 0,944508915 | LOC101927187 | FALSE | Upregulated |
| -0,096912344 | 0,944508915 | LOC101927322 | FALSE | Downregulated |
| -0,126390884 | 0,944508915 | LOC101928829 | FALSE | Downregulated |
| -0,147438761 | 0,944508915 | LOC728819 | FALSE | Downregulated |
| -0,04341711 | 0,944508915 | MAD2L2 | FALSE | Downregulated |

| | | | | |
|---|---|---|---|---|
| 0,142174872 | 0,944508915 | MAMSTR | FALSE | Upregulated |
| 0,042831238 | 0,944508915 | MAP3K8 | FALSE | Upregulated |
| -0,037568955 | 0,944508915 | MIDN | FALSE | Downregulated |
| -0,396512214 | 0,944508915 | MIR5685 | FALSE | Downregulated |
| -0,10102945 | 0,944508915 | MMP25 | FALSE | Downregulated |
| 0,242604584 | 0,944508915 | NBL1 | FALSE | Upregulated |
| -0,026665154 | 0,944508915 | NEMF | FALSE | Downregulated |
| -0,042552824 | 0,944508915 | NKX3-1 | FALSE | Downregulated |
| -0,019707929 | 0,944508915 | RAB3GAP1 | FALSE | Downregulated |
| 0,046252555 | 0,944508915 | RAD52 | FALSE | Upregulated |
| 0,046035 | 0,944508915 | RARB | FALSE | Upregulated |
| -0,164881115 | 0,944508915 | RHCG | FALSE | Downregulated |
| 0,147454905 | 0,944508915 | RP11-362F19.1 | FALSE | Upregulated |
| -0,02587532 | 0,944508915 | RRM1 | FALSE | Downregulated |
| -0,039702503 | 0,944508915 | SAP30 | FALSE | Downregulated |
| -0,023164014 | 0,944508915 | SEPSECS | FALSE | Downregulated |
| 0,025956988 | 0,944508915 | SIRT5 | FALSE | Upregulated |
| 0,127839804 | 0,944508915 | SMOC2 | FALSE | Upregulated |
| 0,086359554 | 0,944508915 | SNX29P2 | FALSE | Upregulated |
| 0,029250245 | 0,944508915 | STARD10 | FALSE | Upregulated |
| 0,108305656 | 0,944508915 | STX16-NPEPL1 | FALSE | Upregulated |
| -0,025787608 | 0,944508915 | SUN1 | FALSE | Downregulated |
| -0,037618201 | 0,944508915 | SWSAP1 | FALSE | Downregulated |
| -0,11859929 | 0,944508915 | TAF7L | FALSE | Downregulated |
| 0,043311642 | 0,944508915 | TIA1 | FALSE | Upregulated |
| -0,027029943 | 0,944508915 | TMEM55B | FALSE | Downregulated |

| | | | | |
|---|---|---|---|---|
| -1,154766791 | 0,944508915 | TNNC2 | FALSE | Downregulated |
| 0,057880793 | 0,944508915 | TRNE | FALSE | Upregulated |
| -0,029595526 | 0,944508915 | UBE2J1 | FALSE | Downregulated |
| 0,025013482 | 0,944508915 | ZNF407 | FALSE | Upregulated |
| -0,037872995 | 0,944508915 | ZNF614 | FALSE | Downregulated |
| -0,043107999 | 0,944508915 | ZNF77 | FALSE | Downregulated |
| 0,137406751 | 0,944660205 | PLAC8L1 | FALSE | Upregulated |
| -0,029839825 | 0,944696214 | DUSP7 | FALSE | Downregulated |
| -0,061705746 | 0,944696214 | IGFALS | FALSE | Downregulated |
| -0,10789854 | 0,944696214 | LOC101928852 | FALSE | Downregulated |
| 0,03123582 | 0,944696214 | LPCAT3 | FALSE | Upregulated |
| -0,069402073 | 0,944696214 | MANF | FALSE | Downregulated |
| -0,10164327 | 0,944696214 | SLC22A4 | FALSE | Downregulated |
| 0,025197397 | 0,944696214 | SORBS2 | FALSE | Upregulated |
| -0,022078517 | 0,944696214 | ZCCHC8 | FALSE | Downregulated |
| 0,025764269 | 0,944746948 | RPS13 | FALSE | Upregulated |
| 0,05219442 | 0,944758122 | KCTD1 | FALSE | Upregulated |
| -0,208033271 | 0,944758122 | LOC102723709 | FALSE | Downregulated |
| -0,098950645 | 0,944758122 | MYO15A | FALSE | Downregulated |
| -0,05976867 | 0,944758122 | P2RY8 | FALSE | Downregulated |
| 0,03096846 | 0,944758122 | PDIA6 | FALSE | Upregulated |
| -0,01996904 | 0,944776018 | EFR3A | FALSE | Downregulated |
| -0,158768028 | 0,944776018 | FAM133DP | FALSE | Downregulated |
| -0,029920335 | 0,944776018 | ZNF234 | FALSE | Downregulated |
| 0,017536331 | 0,944847787 | EAF1 | FALSE | Upregulated |
| -0,030726967 | 0,944847787 | FAM45A | FALSE | Downregulated |

| | | | | |
|---|---|---|---|---|
| 0,036825982 | 0,944847787 | HAUS1 | FALSE | Upregulated |
| -0,186951697 | 0,944847787 | HSD3BP4 | FALSE | Downregulated |
| -0,150602287 | 0,944847787 | SIX4 | FALSE | Downregulated |
| -0,048205178 | 0,944913617 | HILPDA | FALSE | Downregulated |
| 0,101643359 | 0,944913617 | LOC440300 | FALSE | Upregulated |
| -0,038409929 | 0,944913617 | SMC1A | FALSE | Downregulated |
| 0,047363446 | 0,944913617 | TMED3 | FALSE | Upregulated |
| -0,05880333 | 0,944913617 | TPBG | FALSE | Downregulated |
| -0,098424307 | 0,944913617 | USH1C | FALSE | Downregulated |
| 0,067476438 | 0,944913617 | ZNF423 | FALSE | Upregulated |
| 0,027756793 | 0,944993213 | ARHGAP18 | FALSE | Upregulated |
| 0,055572326 | 0,944993213 | FCGR2A | FALSE | Upregulated |
| -0,028739137 | 0,944993213 | JTB | FALSE | Downregulated |
| -0,057931557 | 0,944993213 | LOC100996497 | FALSE | Downregulated |
| 0,099750725 | 0,944993213 | LOC101928111 | FALSE | Upregulated |
| 0,068844338 | 0,944993213 | NDST3 | FALSE | Upregulated |
| 0,020787304 | 0,944993213 | NDUFB10 | FALSE | Upregulated |
| -0,037901017 | 0,944993213 | PPAP2B | FALSE | Downregulated |
| 0,066844541 | 0,944993213 | RNF122 | FALSE | Upregulated |
| 0,07277537 | 0,944993213 | RP11-622A1.2 | FALSE | Upregulated |
| 0,062962095 | 0,944993213 | SLC19A1 | FALSE | Upregulated |
| 0,041427299 | 0,944993213 | SPNS2 | FALSE | Upregulated |
| -0,142383688 | 0,945011494 | TRNL2 | FALSE | Downregulated |
| -0,02033215 | 0,945019284 | ZBTB1 | FALSE | Downregulated |
| -0,039077014 | 0,94505009 | DDC | FALSE | Downregulated |
| 0,045651921 | 0,94505009 | PEAR1 | FALSE | Upregulated |

| | | | | |
|---|---|---|---|---|
| -0,047923494 | 0,945061382 | CREBZF | FALSE | Downregulated |
| 0,166986208 | 0,945061382 | CTD-2258A20.5 | FALSE | Upregulated |
| -0,041367257 | 0,945061382 | PTGR1 | FALSE | Downregulated |
| -0,036389057 | 0,945081024 | C10orf118 | FALSE | Downregulated |
| 0,164384672 | 0,945081024 | CCDC180 | FALSE | Upregulated |
| 0,016129122 | 0,945081024 | DARS | FALSE | Upregulated |
| -0,024613749 | 0,945081024 | DUSP22 | FALSE | Downregulated |
| -0,067690324 | 0,945081024 | FAM86FP | FALSE | Downregulated |
| 0,046755343 | 0,945081024 | GAN | FALSE | Upregulated |
| -0,026896225 | 0,945081024 | GPR108 | FALSE | Downregulated |
| -0,085312417 | 0,945081024 | GPRASP2 | FALSE | Downregulated |
| -0,072795744 | 0,945081024 | KIAA1683 | FALSE | Downregulated |
| -0,117367808 | 0,945081024 | LOC102723989 | FALSE | Downregulated |
| -0,034804929 | 0,945081024 | LRRC8A | FALSE | Downregulated |
| -0,023241672 | 0,945081024 | PPP1CB | FALSE | Downregulated |
| 0,138365592 | 0,945081024 | PRKY | FALSE | Upregulated |
| 0,078869596 | 0,945081024 | RP1-234P15.4 | FALSE | Upregulated |
| -0,041954263 | 0,945081024 | ZNHIT6 | FALSE | Downregulated |
| 0,043692237 | 0,945178665 | ANKS3 | FALSE | Upregulated |
| 0,056403305 | 0,945178665 | BCL2L11 | FALSE | Upregulated |
| 0,030640614 | 0,945178665 | CTBS | FALSE | Upregulated |
| -0,08258005 | 0,945178665 | FLJ13197 | FALSE | Downregulated |
| 0,047214592 | 0,945178665 | GSKIP | FALSE | Upregulated |
| -0,076339732 | 0,945178665 | LOC100131564 | FALSE | Downregulated |
| 0,050659286 | 0,945178665 | MACROD2 | FALSE | Upregulated |
| 0,113029918 | 0,945178665 | MC1R | FALSE | Upregulated |

| | | | | |
|---|---|---|---|---|
| -0,030729334 | 0,945178665 | MCAT | FALSE | Downregulated |
| -0,022881429 | 0,945178665 | MOB2 | FALSE | Downregulated |
| -0,024885623 | 0,945178665 | PIGB | FALSE | Downregulated |
| 0,033677345 | 0,945178665 | PJA1 | FALSE | Upregulated |
| 0,100737349 | 0,945178665 | PRTG | FALSE | Upregulated |
| 0,031252248 | 0,945178665 | TRAPPC1 | FALSE | Upregulated |
| 0,05236577 | 0,945334678 | CSF3R | FALSE | Upregulated |
| 0,167254699 | 0,945334678 | CXorf65 | FALSE | Upregulated |
| 0,054656362 | 0,945334678 | EVI2B | FALSE | Upregulated |
| -0,024707978 | 0,945334678 | HDDC2 | FALSE | Downregulated |
| -0,036999784 | 0,945334678 | LMNB1 | FALSE | Downregulated |
| 0,162547543 | 0,945334678 | LOC101929463 | FALSE | Upregulated |
| 0,072712584 | 0,945334678 | LRFN1 | FALSE | Upregulated |
| 0,09314081 | 0,945334678 | RGAG1 | FALSE | Upregulated |
| 0,064218126 | 0,945334678 | RGS18 | FALSE | Upregulated |
| 0,037409757 | 0,945334678 | TRIM5 | FALSE | Upregulated |
| -0,022359155 | 0,945551955 | FNIP1 | FALSE | Downregulated |
| 0,022990245 | 0,945551955 | MRPS21 | FALSE | Upregulated |
| 0,030589421 | 0,945551955 | SWAP70 | FALSE | Upregulated |
| -0,042024944 | 0,945564378 | HLF | FALSE | Downregulated |
| 0,105671202 | 0,945564378 | HTATSF1P2 | FALSE | Upregulated |
| 0,105720741 | 0,945564378 | LOC100507487 | FALSE | Upregulated |
| 0,062663585 | 0,945564378 | MRVI1 | FALSE | Upregulated |
| 0,117947546 | 0,945572558 | PAK3 | FALSE | Upregulated |
| -0,030309263 | 0,945648044 | LOC145783 | FALSE | Downregulated |
| -0,027605574 | 0,945685609 | STK38L | FALSE | Downregulated |

| | | | | |
|---|---|---|---|---|
| -0,021730426 | 0,945716672 | AARS2 | FALSE | Downregulated |
| 0,191002738 | 0,945716672 | ASIP | FALSE | Upregulated |
| 0,025685942 | 0,945716672 | C15orf38 | FALSE | Upregulated |
| 0,091301681 | 0,945716672 | CCDC13 | FALSE | Upregulated |
| 0,039794347 | 0,945716672 | CCNJ | FALSE | Upregulated |
| 0,123912738 | 0,945716672 | DNAH7 | FALSE | Upregulated |
| 0,083413111 | 0,945716672 | LOC101928513 | FALSE | Upregulated |
| 0,086187279 | 0,945716672 | LOC102724275 | FALSE | Upregulated |
| 0,013202737 | 0,945716672 | PIK3C3 | FALSE | Upregulated |
| -0,056719 | 0,945716672 | PITPNM3 | FALSE | Downregulated |
| 0,017543394 | 0,945716672 | QRICH1 | FALSE | Upregulated |
| 0,023831449 | 0,945716672 | SCAMP1 | FALSE | Upregulated |
| 0,035139442 | 0,945716672 | SGTB | FALSE | Upregulated |
| 0,194694184 | 0,945716672 | SLC12A1 | FALSE | Upregulated |
| 0,094693247 | 0,945716672 | SSR4P1 | FALSE | Upregulated |
| -0,071816617 | 0,945716672 | TUBA8 | FALSE | Downregulated |
| 0,073025332 | 0,945716672 | UGT2B11 | FALSE | Upregulated |
| 0,094012154 | 0,945716672 | UQCRHL | FALSE | Upregulated |
| -0,179559142 | 0,945716672 | WNK4 | FALSE | Downregulated |
| 0,040271578 | 0,945737258 | AMMECR1 | FALSE | Upregulated |
| -0,017675358 | 0,945798033 | CAST | FALSE | Downregulated |
| -0,199863361 | 0,945798033 | MROH2B | FALSE | Downregulated |
| 0,212345759 | 0,945798407 | LURAP1 | FALSE | Upregulated |
| 0,082678588 | 0,945798407 | LYG1 | FALSE | Upregulated |
| -0,034461187 | 0,945815779 | ACER3 | FALSE | Downregulated |
| 0,053085666 | 0,945815779 | DAK | FALSE | Upregulated |

| | | | | |
|---|---|---|---|---|
| 0,093435777 | 0,945815779 | ERP27 | FALSE | Upregulated |
| 0,039948035 | 0,945815779 | GPD1 | FALSE | Upregulated |
| 0,031240732 | 0,945815779 | GPD2 | FALSE | Upregulated |
| -0,036383523 | 0,94581934 | DNMT3A | FALSE | Downregulated |
| -0,029589731 | 0,94581934 | GLRX5 | FALSE | Downregulated |
| -0,066017327 | 0,94581934 | PFKM | FALSE | Downregulated |
| -0,050249899 | 0,94581934 | TMEM144 | FALSE | Downregulated |
| 0,012510815 | 0,94581934 | TNPO3 | FALSE | Upregulated |
| 0,02218824 | 0,94587564 | ECHDC1 | FALSE | Upregulated |
| -0,104015557 | 0,94587564 | PRDM8 | FALSE | Downregulated |
| 0,072917851 | 0,94587564 | TOB1-AS1 | FALSE | Upregulated |
| -0,064646454 | 0,945901065 | TARP | FALSE | Downregulated |
| 0,037984312 | 0,945947656 | TMEM147 | FALSE | Upregulated |
| 0,031193739 | 0,945947656 | TMEM39B | FALSE | Upregulated |
| 0,138932118 | 0,946000651 | DPP10 | FALSE | Upregulated |
| 0,131763286 | 0,946143503 | EDA2R | FALSE | Upregulated |
| -0,024071709 | 0,946143503 | SLC25A38 | FALSE | Downregulated |
| 0,032400424 | 0,946161951 | BLZF1 | FALSE | Upregulated |
| -0,047990237 | 0,946161951 | C1orf159 | FALSE | Downregulated |
| -0,051970068 | 0,946161951 | HSD11B2 | FALSE | Downregulated |
| 0,132558877 | 0,946161951 | KLHDC7B | FALSE | Upregulated |
| -0,038860005 | 0,946161951 | LPIN1 | FALSE | Downregulated |
| -0,040847812 | 0,946161951 | RARA | FALSE | Downregulated |
| -0,035486778 | 0,946172557 | GGH | FALSE | Downregulated |
| 0,106910309 | 0,946172557 | LOC642846 | FALSE | Upregulated |
| 0,021657808 | 0,946172557 | SHARPIN | FALSE | Upregulated |

| | | | | |
|---|---|---|---|---|
| 0,030535194 | 0,946172557 | SSR1 | FALSE | Upregulated |
| 0,066059595 | 0,946325081 | HRH1 | FALSE | Upregulated |
| 0,118847389 | 0,946504043 | ARHGEF33 | FALSE | Upregulated |
| 0,023111821 | 0,946504043 | BCOR | FALSE | Upregulated |
| -0,065161052 | 0,946533051 | TMPPE | FALSE | Downregulated |
| 0,031516936 | 0,946539309 | LINC00847 | FALSE | Upregulated |
| 0,074476815 | 0,946539309 | LOC101928370 | FALSE | Upregulated |
| -0,048734002 | 0,946562912 | LINC00987 | FALSE | Downregulated |
| 0,146049594 | 0,946562912 | LOC101929191 | FALSE | Upregulated |
| -0,029654694 | 0,946562912 | PLEKHA2 | FALSE | Downregulated |
| 0,051358902 | 0,946562912 | SELP | FALSE | Upregulated |
| 0,03346713 | 0,946594126 | GDA | FALSE | Upregulated |
| 0,047548502 | 0,946594126 | RPGRIP1L | FALSE | Upregulated |
| -0,113731522 | 0,946594126 | TYW1B | FALSE | Downregulated |
| 0,069315794 | 0,946594126 | WASH5P | FALSE | Upregulated |
| -0,066177077 | 0,946638763 | ACKR4 | FALSE | Downregulated |
| 0,032176468 | 0,946638763 | OVCA2 | FALSE | Upregulated |
| -0,067029595 | 0,947100104 | CCDC11 | FALSE | Downregulated |
| -0,172984906 | 0,947100104 | RP11-439L8.4 | FALSE | Downregulated |
| -0,01558591 | 0,947162259 | ARMC8 | FALSE | Downregulated |
| -0,108272517 | 0,947162259 | LSMEM2 | FALSE | Downregulated |
| -0,02749612 | 0,947162259 | MGST1 | FALSE | Downregulated |
| -0,038577277 | 0,947162259 | PIAS1 | FALSE | Downregulated |
| 0,019264333 | 0,947193473 | ADH5 | FALSE | Upregulated |
| -0,037296201 | 0,947284407 | RAB27A | FALSE | Downregulated |
| -0,022248916 | 0,947313521 | ABHD17B | FALSE | Downregulated |

| | | | | |
|---|---|---|---|---|
| 0,126975513 | 0,947412324 | GPR113 | FALSE | Upregulated |
| 0,027294954 | 0,947412324 | IRF2BP2 | FALSE | Upregulated |
| -0,121816803 | 0,947412324 | SLC25A41 | FALSE | Downregulated |
| 0,025367812 | 0,947412324 | WDR53 | FALSE | Upregulated |
| -0,033624557 | 0,947430203 | ZHX3 | FALSE | Downregulated |
| 0,020454383 | 0,947438672 | FRS2 | FALSE | Upregulated |
| -0,10883849 | 0,947438672 | GOLGA2P7 | FALSE | Downregulated |
| -0,046029997 | 0,947438672 | ORC2 | FALSE | Downregulated |
| -0,049432681 | 0,947438672 | PMP22 | FALSE | Downregulated |
| -0,102814018 | 0,947471662 | ADPRHL1 | FALSE | Downregulated |
| -0,016172327 | 0,947471662 | ATG12 | FALSE | Downregulated |
| -0,033899582 | 0,947471662 | BDP1 | FALSE | Downregulated |
| 0,013679605 | 0,947471662 | BECN1 | FALSE | Upregulated |
| -0,025681707 | 0,947471662 | C12orf29 | FALSE | Downregulated |
| 0,064394254 | 0,947471662 | C18orf54 | FALSE | Upregulated |
| -0,049248921 | 0,947471662 | C3P1 | FALSE | Downregulated |
| 0,052650585 | 0,947471662 | C4A | FALSE | Upregulated |
| 0,098832318 | 0,947471662 | C6orf163 | FALSE | Upregulated |
| 0,010652485 | 0,947471662 | C6orf89 | FALSE | Upregulated |
| -0,097361578 | 0,947471662 | C8orf44-SGK3 | FALSE | Downregulated |
| -0,040063902 | 0,947471662 | CCDC18 | FALSE | Downregulated |
| 0,172999581 | 0,947471662 | CCDC181 | FALSE | Upregulated |
| -0,019340136 | 0,947471662 | CDK12 | FALSE | Downregulated |
| 0,06406949 | 0,947471662 | CLEC4A | FALSE | Upregulated |
| 0,069970837 | 0,947471662 | CLUHP3 | FALSE | Upregulated |
| 0,02827865 | 0,947471662 | COPG2 | FALSE | Upregulated |

| | | | | |
|---|---|---|---|---|
| 0,041992356 | 0,947471662 | CTPS1 | FALSE | Upregulated |
| 0,049990008 | 0,947471662 | DCLRE1C | FALSE | Upregulated |
| -0,114855818 | 0,947471662 | FABP4 | FALSE | Downregulated |
| -0,06022233 | 0,947471662 | FANCC | FALSE | Downregulated |
| 0,020481611 | 0,947471662 | FNDC3A | FALSE | Upregulated |
| -0,067291469 | 0,947471662 | GLT8D2 | FALSE | Downregulated |
| -0,089658965 | 0,947471662 | GPR162 | FALSE | Downregulated |
| -0,019145184 | 0,947471662 | HADHA | FALSE | Downregulated |
| 0,055536895 | 0,947471662 | ICAM1 | FALSE | Upregulated |
| -0,058124273 | 0,947471662 | KGFLP2 | FALSE | Downregulated |
| 0,196284508 | 0,947471662 | KIR2DL3 | FALSE | Upregulated |
| -0,045983294 | 0,947471662 | KRBA2 | FALSE | Downregulated |
| -0,038533496 | 0,947471662 | L3MBTL3 | FALSE | Downregulated |
| 0,019758776 | 0,947471662 | LMO4 | FALSE | Upregulated |
| -0,040361434 | 0,947471662 | LOC100129550 | FALSE | Downregulated |
| -0,074839098 | 0,947471662 | LOC100506258 | FALSE | Downregulated |
| -0,119089072 | 0,947471662 | LOC101060341 | FALSE | Downregulated |
| 0,123800757 | 0,947471662 | LOC101060626 | FALSE | Upregulated |
| 0,09683044 | 0,947471662 | LOC102724159 | FALSE | Upregulated |
| -0,019075847 | 0,947471662 | MRPS18A | FALSE | Downregulated |
| 0,024390883 | 0,947471662 | MRPS27 | FALSE | Upregulated |
| -0,029038607 | 0,947471662 | OXSM | FALSE | Downregulated |
| 0,01719178 | 0,947471662 | PANX1 | FALSE | Upregulated |
| -0,029943297 | 0,947471662 | PCGF1 | FALSE | Downregulated |
| 0,084156488 | 0,947471662 | PRSS36 | FALSE | Upregulated |
| -0,09047661 | 0,947471662 | RAB36 | FALSE | Downregulated |

| | | | | |
|---|---|---|---|---|
| 0,019471573 | 0,947471662 | RBM18 | FALSE | Upregulated |
| 0,074841522 | 0,947471662 | RP11-66A2.2 | FALSE | Upregulated |
| 0,052221544 | 0,947471662 | RPL32P3 | FALSE | Upregulated |
| 0,022770048 | 0,947471662 | SHISA5 | FALSE | Upregulated |
| 0,02842598 | 0,947471662 | SP2 | FALSE | Upregulated |
| -0,024638844 | 0,947471662 | SPCS2 | FALSE | Downregulated |
| -0,095072953 | 0,947471662 | SPHK1 | FALSE | Downregulated |
| 0,055038326 | 0,947471662 | SPSB2 | FALSE | Upregulated |
| 0,035404141 | 0,947471662 | SSR3 | FALSE | Upregulated |
| 0,022696028 | 0,947471662 | TACO1 | FALSE | Upregulated |
| -0,017401357 | 0,947471662 | TCEAL8 | FALSE | Downregulated |
| -0,022499732 | 0,947471662 | TCTN3 | FALSE | Downregulated |
| -0,034950848 | 0,947471662 | TEX30 | FALSE | Downregulated |
| 0,053210524 | 0,947471662 | TLR2 | FALSE | Upregulated |
| -0,039635985 | 0,947471662 | TRIM2 | FALSE | Downregulated |
| -0,084348864 | 0,947471662 | TTC40 | FALSE | Downregulated |
| 0,046545384 | 0,947471662 | VAV1 | FALSE | Upregulated |
| 0,024779011 | 0,947471662 | ZBED5 | FALSE | Upregulated |
| -0,026491193 | 0,947471662 | ZNF16 | FALSE | Downregulated |
| -0,083920405 | 0,947471662 | ZNF789 | FALSE | Downregulated |
| 0,072507852 | 0,947533921 | ITGA11 | FALSE | Upregulated |
| -0,039139135 | 0,947539767 | BORA | FALSE | Downregulated |
| -0,050298754 | 0,947616625 | ETNK2 | FALSE | Downregulated |
| 0,028853256 | 0,947643614 | FADS3 | FALSE | Upregulated |
| 0,043107956 | 0,947643614 | LOC101927471 | FALSE | Upregulated |
| -0,053098343 | 0,947643614 | PRR11 | FALSE | Downregulated |

| | | | | |
|---|---|---|---|---|
| 0,178934405 | 0,947643614 | UTY | FALSE | Upregulated |
| 0,082570523 | 0,947643614 | XK | FALSE | Upregulated |
| -0,027622849 | 0,947835862 | COMMD8 | FALSE | Downregulated |
| 0,021177678 | 0,947835862 | GBA2 | FALSE | Upregulated |
| 0,025402225 | 0,947875006 | MED25 | FALSE | Upregulated |
| -0,058323769 | 0,947904903 | ARL4D | FALSE | Downregulated |
| -0,027132426 | 0,947904903 | CREBRF | FALSE | Downregulated |
| 0,144199433 | 0,947904903 | FANK1 | FALSE | Upregulated |
| -0,019521501 | 0,948268068 | NDUFA10 | FALSE | Downregulated |
| 0,023038762 | 0,948268068 | PGPEP1 | FALSE | Upregulated |
| 0,047323474 | 0,948268068 | ZNF577 | FALSE | Upregulated |
| -0,031264981 | 0,948482753 | MAPKBP1 | FALSE | Downregulated |
| -0,098713067 | 0,948517057 | AC093391.2 | FALSE | Downregulated |
| -0,025782708 | 0,948949734 | NOP10 | FALSE | Downregulated |
| 0,073270076 | 0,94897424 | GATA5 | FALSE | Upregulated |
| -0,029105771 | 0,94897424 | KHDRBS3 | FALSE | Downregulated |
| 0,051237704 | 0,94897424 | LAD1 | FALSE | Upregulated |
| 0,097831196 | 0,94897424 | LOC102723901 | FALSE | Upregulated |
| -0,020608668 | 0,94897424 | NAA60 | FALSE | Downregulated |
| -0,023208122 | 0,94897424 | PCCA | FALSE | Downregulated |
| -0,108667491 | 0,94911049 | ATOH7 | FALSE | Downregulated |
| -0,029610983 | 0,94911049 | C12orf4 | FALSE | Downregulated |
| 0,025126259 | 0,94911049 | C17orf103 | FALSE | Upregulated |
| -0,041582113 | 0,94911049 | ERN1 | FALSE | Downregulated |
| -0,042415368 | 0,94911049 | FAM122C | FALSE | Downregulated |
| 0,036931324 | 0,94911049 | GFOD1 | FALSE | Upregulated |

| | | | | |
|---|---|---|---|---|
| -0,042314376 | 0,94911049 | LAMB1 | FALSE | Downregulated |
| 0,089567254 | 0,94911049 | LOC100129917 | FALSE | Upregulated |
| 0,060987484 | 0,94911049 | LPAR1 | FALSE | Upregulated |
| 0,042593846 | 0,94911049 | NLGN4X | FALSE | Upregulated |
| 0,113720493 | 0,94911049 | ZNF483 | FALSE | Upregulated |
| 0,032812339 | 0,949140077 | MCM8 | FALSE | Upregulated |
| 0,033578366 | 0,949149801 | ANKRD50 | FALSE | Upregulated |
| -0,022161101 | 0,949149801 | ARID2 | FALSE | Downregulated |
| 0,026552628 | 0,949149801 | BIRC6 | FALSE | Upregulated |
| -0,038708427 | 0,949149801 | FAHD1 | FALSE | Downregulated |
| -0,063991203 | 0,949149801 | PPP1R32 | FALSE | Downregulated |
| 0,043287099 | 0,949149801 | RAC3 | FALSE | Upregulated |
| 0,015365569 | 0,949149801 | RBBP7 | FALSE | Upregulated |
| -0,047713446 | 0,949149801 | RORA | FALSE | Downregulated |
| -0,147049269 | 0,949150278 | TSPAN32 | FALSE | Downregulated |
| 0,092693583 | 0,949150278 | UNC79 | FALSE | Upregulated |
| 0,051691489 | 0,94923939 | DDR1 | FALSE | Upregulated |
| -0,120953148 | 0,949321753 | LOC101927400 | FALSE | Downregulated |
| 0,078719684 | 0,949321753 | LOC102724345 | FALSE | Upregulated |
| -0,04435273 | 0,949321753 | MERTK | FALSE | Downregulated |
| 0,046943659 | 0,949321753 | PDGFRB | FALSE | Upregulated |
| -0,041591848 | 0,949321753 | SNX10 | FALSE | Downregulated |
| 0,061160862 | 0,949321753 | TRAF3IP2-AS1 | FALSE | Upregulated |
| 0,015421557 | 0,949429026 | YWHAG | FALSE | Upregulated |
| -0,021329827 | 0,949563159 | ATP5O | FALSE | Downregulated |
| 0,1233041 | 0,949563159 | FBXO22-AS1 | FALSE | Upregulated |

| | | | | |
|---|---|---|---|---|
| -0,122489427 | 0,949563159 | LINC01126 | FALSE | Downregulated |
| 0,032379331 | 0,949563159 | LINS | FALSE | Upregulated |
| -0,127845136 | 0,949563159 | LOC101927754 | FALSE | Downregulated |
| -0,08629115 | 0,949563159 | LOC200772 | FALSE | Downregulated |
| 0,03076595 | 0,949563159 | PITPNC1 | FALSE | Upregulated |
| 0,020447785 | 0,949563159 | POMT1 | FALSE | Upregulated |
| 0,028390486 | 0,949563159 | ZFHX3 | FALSE | Upregulated |
| 0,064207735 | 0,949588804 | MYO15B | FALSE | Upregulated |
| 0,029250717 | 0,949824927 | AGTPBP1 | FALSE | Upregulated |
| 0,035878972 | 0,949824927 | AMOTL2 | FALSE | Upregulated |
| 0,048310124 | 0,949824927 | CCDC112 | FALSE | Upregulated |
| 0,03471333 | 0,949824927 | DNAJC3 | FALSE | Upregulated |
| 0,033817256 | 0,949824927 | FAM69A | FALSE | Upregulated |
| 0,043377225 | 0,949824927 | FOXO3 | FALSE | Upregulated |
| 0,019001795 | 0,949824927 | LAMTOR3 | FALSE | Upregulated |
| 0,049674684 | 0,949824927 | MORC4 | FALSE | Upregulated |
| -0,185268907 | 0,949824927 | MSH5 | FALSE | Downregulated |
| -0,029445136 | 0,949824927 | NAGK | FALSE | Downregulated |
| 0,020864074 | 0,949824927 | NMT1 | FALSE | Upregulated |
| 0,022444301 | 0,949824927 | PAGR1 | FALSE | Upregulated |
| 0,021783766 | 0,949824927 | PALB2 | FALSE | Upregulated |
| -0,049097214 | 0,949824927 | PDCD1LG2 | FALSE | Downregulated |
| -0,110699992 | 0,949824927 | PLA2G1B | FALSE | Downregulated |
| 0,04462405 | 0,949824927 | RPS21 | FALSE | Upregulated |
| -0,05491523 | 0,949824927 | RPS6KA5 | FALSE | Downregulated |
| 0,025440872 | 0,949824927 | TEP1 | FALSE | Upregulated |

| | | | | |
|---|---|---|---|---|
| 0,028526623 | 0,949824927 | UBL3 | FALSE | Upregulated |
| -0,019596254 | 0,949824927 | VRK2 | FALSE | Downregulated |
| -0,124523025 | 0,949851127 | GPR179 | FALSE | Downregulated |
| -0,031163918 | 0,949934963 | STRA13 | FALSE | Downregulated |
| 0,10530665 | 0,94999921 | ANKRD22 | FALSE | Upregulated |
| -0,045338434 | 0,94999921 | EFNA4 | FALSE | Downregulated |
| -0,065989768 | 0,94999921 | LOC100129203 | FALSE | Downregulated |
| 0,096203885 | 0,94999921 | LOC401286 | FALSE | Upregulated |
| -0,025112279 | 0,950113964 | BMPR1A | FALSE | Downregulated |
| -0,026628267 | 0,950113964 | PPM1G | FALSE | Downregulated |
| -0,026863797 | 0,950114403 | KIAA0226 | FALSE | Downregulated |
| -0,0311282 | 0,950114403 | RBBP5 | FALSE | Downregulated |
| -0,031562539 | 0,950181946 | KIAA0355 | FALSE | Downregulated |
| 0,026266051 | 0,950322253 | ANPEP | FALSE | Upregulated |
| -0,020732674 | 0,950322253 | DIS3 | FALSE | Downregulated |
| 0,072079767 | 0,950322253 | LOC441124 | FALSE | Upregulated |
| 0,024285207 | 0,950322253 | LZTFL1 | FALSE | Upregulated |
| -0,02185216 | 0,950322253 | TTC38 | FALSE | Downregulated |
| -0,035574631 | 0,950322253 | ZC2HC1C | FALSE | Downregulated |
| 0,055533801 | 0,95035651 | LINC00630 | FALSE | Upregulated |
| 0,099639314 | 0,95035651 | MACC1 | FALSE | Upregulated |
| 0,028324487 | 0,950383697 | CACUL1 | FALSE | Upregulated |
| -0,225561255 | 0,950383697 | FCGR1C | FALSE | Downregulated |
| 0,049621102 | 0,950383697 | SLC25A25 | FALSE | Upregulated |
| 0,034485244 | 0,950383697 | TRPM4 | FALSE | Upregulated |
| 0,034581519 | 0,950446245 | MTMR2 | FALSE | Upregulated |

| | | | | |
|---|---|---|---|---|
| 0,040649535 | 0,950634095 | ACTRT3 | FALSE | Upregulated |
| 0,094819716 | 0,950634095 | HCG16 | FALSE | Upregulated |
| 0,026760113 | 0,950634095 | MAN2A2 | FALSE | Upregulated |
| -0,017189549 | 0,950634095 | NEK7 | FALSE | Downregulated |
| -0,038581174 | 0,950634095 | ROMO1 | FALSE | Downregulated |
| 0,034752623 | 0,950634095 | SPATS2 | FALSE | Upregulated |
| 0,019803334 | 0,950634095 | STX7 | FALSE | Upregulated |
| -0,021852297 | 0,950634095 | SUCLG2 | FALSE | Downregulated |
| 0,035176621 | 0,950634095 | TAPBPL | FALSE | Upregulated |
| -0,041386129 | 0,950738694 | GDPD5 | FALSE | Downregulated |
| 0,131270094 | 0,950738694 | PLD5 | FALSE | Upregulated |
| 0,07045581 | 0,950784466 | B3GALNT1 | FALSE | Upregulated |
| -0,02953922 | 0,950784466 | CMSS1 | FALSE | Downregulated |
| -0,042454969 | 0,950784466 | KLF6 | FALSE | Downregulated |
| -0,033315372 | 0,950784466 | PINK1-AS | FALSE | Downregulated |
| -0,072107913 | 0,950784466 | SEMA5B | FALSE | Downregulated |
| -0,038651667 | 0,950784466 | SLC22A18 | FALSE | Downregulated |
| 0,048962411 | 0,950789899 | LCA5L | FALSE | Upregulated |
| -0,057188804 | 0,950789899 | PHKG1 | FALSE | Downregulated |
| 0,039774573 | 0,950789899 | ZNF276 | FALSE | Upregulated |
| 0,200271139 | 0,950789899 | ZNRF3-AS1 | FALSE | Upregulated |
| 0,09506185 | 0,950794001 | LOC102724811 | FALSE | Upregulated |
| -0,027075396 | 0,950794001 | UTRN | FALSE | Downregulated |
| -0,024378786 | 0,950804021 | LRRC27 | FALSE | Downregulated |
| -0,019251518 | 0,95099104 | BTBD1 | FALSE | Downregulated |
| 0,019889446 | 0,95099104 | FCGRT | FALSE | Upregulated |

| | | | | |
|---|---|---|---|---|
| 0,068008897 | 0,95099104 | FREM1 | FALSE | Upregulated |
| -0,051343534 | 0,95099104 | LINC00870 | FALSE | Downregulated |
| 0,113891474 | 0,95099104 | LOC102724126 | FALSE | Upregulated |
| 0,078195141 | 0,95099104 | LOC102724814 | FALSE | Upregulated |
| 0,032866183 | 0,95099104 | LYNX1 | FALSE | Upregulated |
| -0,079341712 | 0,95099104 | PRELID2 | FALSE | Downregulated |
| -0,027852791 | 0,95099104 | SPAG9 | FALSE | Downregulated |
| 0,026834234 | 0,95099104 | TIE1 | FALSE | Upregulated |
| 0,103561392 | 0,95099104 | TMEM121 | FALSE | Upregulated |
| 0,025395636 | 0,951085509 | DCTD | FALSE | Upregulated |
| -0,075477346 | 0,951185492 | TAS2R20 | FALSE | Downregulated |
| -0,090835219 | 0,951361854 | CNKSR1 | FALSE | Downregulated |
| 0,027937753 | 0,951361854 | MLYCD | FALSE | Upregulated |
| -0,140057912 | 0,951519628 | DYNLRB2 | FALSE | Downregulated |
| 0,077582636 | 0,951519628 | EFS | FALSE | Upregulated |
| -1,015130975 | 0,951519628 | MYL2 | FALSE | Downregulated |
| -0,028115164 | 0,951566337 | ATP5G2 | FALSE | Downregulated |
| -0,043932133 | 0,951573305 | ADH1C | FALSE | Downregulated |
| 0,100038799 | 0,951718627 | EPS8L1 | FALSE | Upregulated |
| -0,054773163 | 0,951718627 | GPR98 | FALSE | Downregulated |
| 0,122363756 | 0,951718627 | LOC100653005 | FALSE | Upregulated |
| -0,135045392 | 0,951718627 | LRGUK | FALSE | Downregulated |
| -0,020454654 | 0,951718627 | MYO18A | FALSE | Downregulated |
| -0,013949318 | 0,951718627 | PSEN1 | FALSE | Downregulated |
| -0,195754685 | 0,951718627 | SPRN | FALSE | Downregulated |
| -0,077921531 | 0,951745012 | DISC1 | FALSE | Downregulated |

| | | | | |
|---|---|---|---|---|
| 0,034217268 | 0,951745012 | LUZP1 | FALSE | Upregulated |
| 0,066179295 | 0,95186343 | C11orf80 | FALSE | Upregulated |
| -0,031861682 | 0,951917742 | ACSM5 | FALSE | Downregulated |
| -0,836795334 | 0,951917742 | EEF1E1-BLOC1S5 | FALSE | Downregulated |
| -0,023866869 | 0,951917742 | ERCC5 | FALSE | Downregulated |
| -0,022330352 | 0,951917742 | ERCC6 | FALSE | Downregulated |
| -0,028802996 | 0,951917742 | LGALSL | FALSE | Downregulated |
| -0,029820835 | 0,951917742 | LINC00984 | FALSE | Downregulated |
| 0,022128412 | 0,951917742 | MAN2A1 | FALSE | Upregulated |
| -0,027089732 | 0,951917742 | NUP85 | FALSE | Downregulated |
| -0,028089274 | 0,951917742 | ORMDL1 | FALSE | Downregulated |
| 0,094679274 | 0,951917742 | RP11-473M20.7 | FALSE | Upregulated |
| -0,153428231 | 0,951917742 | RSPH10B | FALSE | Downregulated |
| -0,021194319 | 0,951917742 | SH3RF1 | FALSE | Downregulated |
| 0,022247747 | 0,951917742 | SRP72 | FALSE | Upregulated |
| -0,017102709 | 0,951917742 | TATDN3 | FALSE | Downregulated |
| -0,054551787 | 0,951917742 | VCAM1 | FALSE | Downregulated |
| -0,059585317 | 0,951953669 | BEX1 | FALSE | Downregulated |
| -0,01654241 | 0,951953669 | LARP4B | FALSE | Downregulated |
| -0,294600237 | 0,951953669 | SMTNL2 | FALSE | Downregulated |
| -0,046899578 | 0,951984872 | C14orf132 | FALSE | Downregulated |
| 0,05034074 | 0,952056942 | ALDOC | FALSE | Upregulated |
| -0,045032453 | 0,952056942 | AP3D1 | FALSE | Downregulated |
| -0,01573164 | 0,952056942 | ARFGEF2 | FALSE | Downregulated |
| 0,030586784 | 0,952056942 | ASAP1 | FALSE | Upregulated |
| 0,029182027 | 0,952056942 | C12orf23 | FALSE | Upregulated |

| | | | | |
|---|---|---|---|---|
| -0,041921644 | 0,952056942 | C15orf37 | FALSE | Downregulated |
| 0,175195775 | 0,952056942 | C21orf15 | FALSE | Upregulated |
| -0,030169728 | 0,952056942 | CD14 | FALSE | Downregulated |
| -0,023609535 | 0,952056942 | CEP57L1 | FALSE | Downregulated |
| -0,022209874 | 0,952056942 | CES2 | FALSE | Downregulated |
| 0,10328852 | 0,952056942 | CHI3L2 | FALSE | Upregulated |
| -0,038509987 | 0,952056942 | DANCR | FALSE | Downregulated |
| -0,077441314 | 0,952056942 | EFCAB12 | FALSE | Downregulated |
| 0,039999437 | 0,952056942 | EPB41L2 | FALSE | Upregulated |
| 0,125876696 | 0,952056942 | FLJ41309 | FALSE | Upregulated |
| -0,024322013 | 0,952056942 | GTF2E1 | FALSE | Downregulated |
| 0,02663931 | 0,952056942 | GUCD1 | FALSE | Upregulated |
| -0,067679916 | 0,952056942 | HESX1 | FALSE | Downregulated |
| 0,057983892 | 0,952056942 | KIAA0895L | FALSE | Upregulated |
| 0,074351338 | 0,952056942 | LOC100506282 | FALSE | Upregulated |
| 0,127739596 | 0,952056942 | LOC102724921 | FALSE | Upregulated |
| -0,121807696 | 0,952056942 | LOC439994 | FALSE | Downregulated |
| 0,092321202 | 0,952056942 | LOC440157 | FALSE | Upregulated |
| -0,023827455 | 0,952056942 | NDUFB9 | FALSE | Downregulated |
| 0,164342093 | 0,952056942 | OVCH1 | FALSE | Upregulated |
| 0,06369867 | 0,952056942 | PARP15 | FALSE | Upregulated |
| 0,055443051 | 0,952056942 | PTPRS | FALSE | Upregulated |
| 0,028960275 | 0,952056942 | RBM14 | FALSE | Upregulated |
| -0,027047122 | 0,952056942 | RBMS1 | FALSE | Downregulated |
| 0,111588007 | 0,952056942 | RCOR2 | FALSE | Upregulated |
| -0,997307085 | 0,952056942 | RNVU1-18 | FALSE | Downregulated |

| | | | | |
|---|---|---|---|---|
| 0,057074928 | 0,952056942 | RP11-15I11.3 | FALSE | Upregulated |
| -0,107923172 | 0,952056942 | RRAD | FALSE | Downregulated |
| -0,052465267 | 0,952056942 | RUNX1T1 | FALSE | Downregulated |
| 0,067842489 | 0,952056942 | SDHAP3 | FALSE | Upregulated |
| -0,036512934 | 0,952056942 | SERPINA5 | FALSE | Downregulated |
| 0,034508406 | 0,952056942 | TCF7L1 | FALSE | Upregulated |
| -0,117783535 | 0,952056942 | TPSG1 | FALSE | Downregulated |
| -0,03030999 | 0,952056942 | TRIM56 | FALSE | Downregulated |
| -0,033451536 | 0,952056942 | USP31 | FALSE | Downregulated |
| 0,059699017 | 0,952056942 | ZFHX2 | FALSE | Upregulated |
| -0,029666732 | 0,952056942 | ZNF205 | FALSE | Downregulated |
| -0,014059433 | 0,952087363 | VAMP7 | FALSE | Downregulated |
| 0,013016997 | 0,952172906 | AGGF1 | FALSE | Upregulated |
| 0,187062901 | 0,95227693 | NANOG | FALSE | Upregulated |
| 0,158387751 | 0,952282153 | RGPD3 | FALSE | Upregulated |
| -0,018202656 | 0,952399909 | EIF2S2 | FALSE | Downregulated |
| 0,02237505 | 0,952399909 | ERCC2 | FALSE | Upregulated |
| -0,017414824 | 0,952399909 | LARS | FALSE | Downregulated |
| -0,022462239 | 0,952399909 | LBR | FALSE | Downregulated |
| 0,039899714 | 0,952399909 | LGALS3BP | FALSE | Upregulated |
| 0,143194515 | 0,952399909 | MMP23B | FALSE | Upregulated |
| 0,032626593 | 0,952399909 | PAN2 | FALSE | Upregulated |
| -0,020381457 | 0,952399909 | TIMM9 | FALSE | Downregulated |
| -0,019885767 | 0,952399909 | ZNF274 | FALSE | Downregulated |
| -0,02394271 | 0,952452212 | ABTB2 | FALSE | Downregulated |
| 0,041276656 | 0,952452212 | ID3 | FALSE | Upregulated |

| | | | | |
|---|---|---|---|---|
| 0,022689853 | 0,952452212 | RIOK2 | FALSE | Upregulated |
| -0,016088644 | 0,952526041 | EIF3H | FALSE | Downregulated |
| 0,018788472 | 0,952526041 | ELAVL1 | FALSE | Upregulated |
| 0,023660044 | 0,952526041 | G6PC3 | FALSE | Upregulated |
| -0,072206431 | 0,952526041 | LOC100996457 | FALSE | Downregulated |
| 0,050292504 | 0,952526041 | PALLD | FALSE | Upregulated |
| -0,027283986 | 0,952544311 | ASXL2 | FALSE | Downregulated |
| 0,03502804 | 0,952544311 | C11orf49 | FALSE | Upregulated |
| 0,029358025 | 0,952544311 | CREB3L2 | FALSE | Upregulated |
| -0,028873107 | 0,952544311 | DLG5 | FALSE | Downregulated |
| -0,120055547 | 0,952544311 | FAM24B | FALSE | Downregulated |
| -0,105409785 | 0,952544311 | LOC100505915 | FALSE | Downregulated |
| 0,023211857 | 0,952544311 | SEH1L | FALSE | Upregulated |
| 0,030195771 | 0,952544311 | SLC25A30 | FALSE | Upregulated |
| -0,132816893 | 0,952544311 | TRND | FALSE | Downregulated |
| -0,015666423 | 0,952581895 | ASUN | FALSE | Downregulated |
| -0,10638477 | 0,952581895 | C19orf26 | FALSE | Downregulated |
| -0,043125026 | 0,952581895 | ECSCR | FALSE | Downregulated |
| 0,084319445 | 0,952581895 | EMP1 | FALSE | Upregulated |
| 0,03458014 | 0,952581895 | ITIH3 | FALSE | Upregulated |
| -0,022355165 | 0,952581895 | NUS1 | FALSE | Downregulated |
| -0,012901276 | 0,952581895 | RBMX | FALSE | Downregulated |
| 0,043737219 | 0,952581895 | SLC2A3 | FALSE | Upregulated |
| -0,034495394 | 0,952789414 | C3orf18 | FALSE | Downregulated |
| 0,028140447 | 0,952789414 | CENPV | FALSE | Upregulated |
| -0,136112084 | 0,952789414 | GK3P | FALSE | Downregulated |

| | | | | |
|---|---|---|---|---|
| -0,033370439 | 0,952789414 | ICAM2 | FALSE | Downregulated |
| -0,014225317 | 0,952789414 | ITFG1 | FALSE | Downregulated |
| -0,103064579 | 0,952789414 | LOC101060351 | FALSE | Downregulated |
| 0,104160406 | 0,952789414 | LOC102724445 | FALSE | Upregulated |
| -0,132902598 | 0,952789414 | LOC102724539 | FALSE | Downregulated |
| -0,111075463 | 0,952789414 | LTC4S | FALSE | Downregulated |
| -0,085075755 | 0,952789414 | N4BP2L2-IT2 | FALSE | Downregulated |
| -0,021206781 | 0,952789414 | NICN1 | FALSE | Downregulated |
| -0,08240442 | 0,952789414 | PCLO | FALSE | Downregulated |
| -0,01897431 | 0,952789414 | RPS6KC1 | FALSE | Downregulated |
| -0,026285807 | 0,952789414 | SMURF2 | FALSE | Downregulated |
| -0,044394712 | 0,952789414 | STAB2 | FALSE | Downregulated |
| 0,016104054 | 0,952789414 | ZBED1 | FALSE | Upregulated |
| 0,035826709 | 0,952789414 | ZNF580 | FALSE | Upregulated |
| 0,14286743 | 0,952853123 | SUSD4 | FALSE | Upregulated |
| 0,081463718 | 0,952884231 | EPCAM | FALSE | Upregulated |
| -0,032610987 | 0,953095711 | LOC101927752 | FALSE | Downregulated |
| -0,01977119 | 0,953095711 | RER1 | FALSE | Downregulated |
| 0,02032257 | 0,953095711 | SDCBP | FALSE | Upregulated |
| 0,024054804 | 0,953122767 | ALG11 | FALSE | Upregulated |
| -0,018214076 | 0,953148405 | CLASP2 | FALSE | Downregulated |
| 0,06468219 | 0,953148405 | TRO | FALSE | Upregulated |
| -0,023700702 | 0,953165497 | BBX | FALSE | Downregulated |
| -0,0945926 | 0,953165497 | KIAA1652 | FALSE | Downregulated |
| 0,028614651 | 0,953165497 | LYN | FALSE | Upregulated |
| 0,048589647 | 0,953165497 | TGFB3 | FALSE | Upregulated |

| | | | | |
|---|---|---|---|---|
| -0,02239936 | 0,953284082 | ATP6V0E1 | FALSE | Downregulated |
| -0,018399815 | 0,953284082 | C16orf62 | FALSE | Downregulated |
| -0,118406976 | 0,953284082 | RNASE2 | FALSE | Downregulated |
| 0,050419451 | 0,953284082 | SDHAP1 | FALSE | Upregulated |
| 0,023216926 | 0,953284082 | SEMA4G | FALSE | Upregulated |
| 0,019042335 | 0,953284082 | TIMM22 | FALSE | Upregulated |
| 0,041277326 | 0,953284082 | USP30-AS1 | FALSE | Upregulated |
| 0,044381367 | 0,953284082 | ZNF670 | FALSE | Upregulated |
| -0,112271907 | 0,953501274 | HAS2 | FALSE | Downregulated |
| 0,052258263 | 0,953501274 | LOC100996492 | FALSE | Upregulated |
| -0,049321786 | 0,953501274 | SRGAP3 | FALSE | Downregulated |
| 0,143283199 | 0,953501274 | TMEM255A | FALSE | Upregulated |
| -0,01375745 | 0,953610154 | CSDE1 | FALSE | Downregulated |
| -0,026690588 | 0,953610154 | FKBP7 | FALSE | Downregulated |
| 0,034098313 | 0,953610154 | PREB | FALSE | Upregulated |
| -0,07133825 | 0,95383885 | C6orf226 | FALSE | Downregulated |
| -0,200487468 | 0,95383885 | LOC101927800 | FALSE | Downregulated |
| 0,305157525 | 0,95383885 | LOC102723828 | FALSE | Upregulated |
| -0,089964326 | 0,95383885 | PKN3 | FALSE | Downregulated |
| -0,036470475 | 0,95383885 | RAB3D | FALSE | Downregulated |
| -0,014869466 | 0,953870462 | PPP2R5D | FALSE | Downregulated |
| 0,048455629 | 0,953870462 | UGT3A1 | FALSE | Upregulated |
| -0,037126308 | 0,954274942 | CLEC14A | FALSE | Downregulated |
| -0,049609625 | 0,954274942 | GADD45A | FALSE | Downregulated |
| -0,054737641 | 0,95450786 | AC144449.1 | FALSE | Downregulated |
| -0,111933904 | 0,954620614 | FKBP5 | FALSE | Downregulated |

| | | | | |
|---|---|---|---|---|
| -0,05466549 | 0,954772922 | CSGALNACT1 | FALSE | Downregulated |
| 0,106032877 | 0,955020155 | TNFRSF8 | FALSE | Upregulated |
| -0,032956518 | 0,955020155 | ZBED6CL | FALSE | Downregulated |
| 0,019751381 | 0,955064645 | ARL1 | FALSE | Upregulated |
| 0,139825252 | 0,955064645 | BIK | FALSE | Upregulated |
| -0,022102308 | 0,955064645 | CFLAR | FALSE | Downregulated |
| -0,017954978 | 0,955064645 | DENND4C | FALSE | Downregulated |
| -0,02054024 | 0,955064645 | ERLIN2 | FALSE | Downregulated |
| 0,032541506 | 0,955064645 | EXT1 | FALSE | Upregulated |
| 0,179971903 | 0,955064645 | HRASLS | FALSE | Upregulated |
| 0,045801065 | 0,955064645 | ITGB3BP | FALSE | Upregulated |
| -0,013947918 | 0,955064645 | KPNA4 | FALSE | Downregulated |
| 0,097344853 | 0,955064645 | LOC102724792 | FALSE | Upregulated |
| 0,134771643 | 0,955064645 | NPAP1 | FALSE | Upregulated |
| 0,031372294 | 0,955064645 | PLCG2 | FALSE | Upregulated |
| 0,023851733 | 0,955064645 | PSME3 | FALSE | Upregulated |
| -0,128165684 | 0,955064645 | RAB3C | FALSE | Downregulated |
| -0,090912103 | 0,955064645 | RP11-701P16.5 | FALSE | Downregulated |
| 0,030831175 | 0,955064645 | RPL41 | FALSE | Upregulated |
| -0,019215449 | 0,955064645 | SDHC | FALSE | Downregulated |
| 0,043064956 | 0,955064645 | SRPX | FALSE | Upregulated |
| 0,01965688 | 0,955064645 | TMEM183A | FALSE | Upregulated |
| 0,035432873 | 0,955064645 | TMX3 | FALSE | Upregulated |
| -0,024250042 | 0,955064645 | TVP23B | FALSE | Downregulated |
| -0,085634593 | 0,955064645 | ZMIZ1-AS1 | FALSE | Downregulated |
| -0,023800258 | 0,955071579 | CENPC | FALSE | Downregulated |

| | | | | |
|---|---|---|---|---|
| -0,034092826 | 0,955071579 | CXorf40B | FALSE | Downregulated |
| -0,037705477 | 0,955071579 | FNDC4 | FALSE | Downregulated |
| -0,023780535 | 0,955071579 | GTPBP8 | FALSE | Downregulated |
| -0,048337999 | 0,955071579 | LINC01000 | FALSE | Downregulated |
| -0,027188146 | 0,955071579 | MFSD9 | FALSE | Downregulated |
| 0,067952908 | 0,955071579 | SLC35F2 | FALSE | Upregulated |
| -0,02728428 | 0,955271937 | GFOD2 | FALSE | Downregulated |
| 0,081692017 | 0,955271937 | LOC284581 | FALSE | Upregulated |
| 0,041646894 | 0,955271937 | OPHN1 | FALSE | Upregulated |
| -0,020047056 | 0,955315942 | KPNA3 | FALSE | Downregulated |
| -0,041722012 | 0,955473811 | CFD | FALSE | Downregulated |
| -0,042128652 | 0,955473811 | GPR4 | FALSE | Downregulated |
| -0,024976314 | 0,955571975 | AHNAK | FALSE | Downregulated |
| -0,037814832 | 0,955571975 | AKAP3 | FALSE | Downregulated |
| -0,032999341 | 0,955571975 | ARSD | FALSE | Downregulated |
| 0,027587208 | 0,955571975 | C1orf74 | FALSE | Upregulated |
| 0,065291214 | 0,955571975 | CPAMD8 | FALSE | Upregulated |
| 0,019339256 | 0,955571975 | DRAM2 | FALSE | Upregulated |
| -0,021467928 | 0,955571975 | EEF1G | FALSE | Downregulated |
| 0,024223178 | 0,955571975 | ERCC4 | FALSE | Upregulated |
| 0,015228273 | 0,955571975 | FAM134A | FALSE | Upregulated |
| -0,068097679 | 0,955571975 | GTF2H2 | FALSE | Downregulated |
| -0,026907872 | 0,955571975 | ING2 | FALSE | Downregulated |
| -0,072231376 | 0,955571975 | KIAA1377 | FALSE | Downregulated |
| -0,073164845 | 0,955571975 | LOC101060581 | FALSE | Downregulated |
| -0,022855491 | 0,955571975 | LRCH1 | FALSE | Downregulated |

| | | | | |
|---|---|---|---|---|
| 0,01861639 | 0,955571975 | MEF2D | FALSE | Upregulated |
| 0,033891224 | 0,955571975 | PPARD | FALSE | Upregulated |
| -0,017092772 | 0,955571975 | PPP5C | FALSE | Downregulated |
| -0,068511505 | 0,955571975 | SERHL2 | FALSE | Downregulated |
| -0,029689042 | 0,955571975 | ZNF680 | FALSE | Downregulated |
| 0,018022573 | 0,95565454 | MANBAL | FALSE | Upregulated |
| 0,01922365 | 0,955676967 | PIGM | FALSE | Upregulated |
| -0,022839627 | 0,955901157 | MSANTD3 | FALSE | Downregulated |
| 0,028898883 | 0,955901157 | SLC7A9 | FALSE | Upregulated |
| -0,097674261 | 0,955901157 | SUSD5 | FALSE | Downregulated |
| -0,134346479 | 0,955949542 | TBX1 | FALSE | Downregulated |
| -0,025833503 | 0,955974739 | AGMO | FALSE | Downregulated |
| -0,064210269 | 0,955974739 | C2orf16 | FALSE | Downregulated |
| 0,050646882 | 0,955974739 | DUSP18 | FALSE | Upregulated |
| -0,031267192 | 0,955974739 | FASTKD3 | FALSE | Downregulated |
| 0,043509689 | 0,955974739 | IMMP1L | FALSE | Upregulated |
| 0,027671534 | 0,955974739 | LRIG3 | FALSE | Upregulated |
| -0,028219439 | 0,955974739 | MLXIP | FALSE | Downregulated |
| -0,077122807 | 0,955974739 | P2RX6 | FALSE | Downregulated |
| 0,016605271 | 0,955974739 | PABPC1 | FALSE | Upregulated |
| 0,028013212 | 0,955974739 | PAQR3 | FALSE | Upregulated |
| 0,031054929 | 0,955974739 | RASGRP3 | FALSE | Upregulated |
| -0,019494641 | 0,955974739 | SHANK2 | FALSE | Downregulated |
| 0,058500048 | 0,955974739 | SLC16A6 | FALSE | Upregulated |
| -0,022992033 | 0,955974739 | STOM | FALSE | Downregulated |
| 0,019596085 | 0,955974739 | TUBB | FALSE | Upregulated |

| | | | | |
|---|---|---|---|---|
| 0,030482692 | 0,955974739 | ZFPM1 | FALSE | Upregulated |
| 0,032511394 | 0,956068226 | ABI3 | FALSE | Upregulated |
| 0,019599152 | 0,956068226 | ZNF7 | FALSE | Upregulated |
| 0,022451983 | 0,956126453 | MTRR | FALSE | Upregulated |
| 0,02258276 | 0,956126453 | PRKAG2 | FALSE | Upregulated |
| -0,035151408 | 0,956126453 | RP11-305E17.6 | FALSE | Downregulated |
| 0,045893643 | 0,956126453 | SLFN12 | FALSE | Upregulated |
| -0,020358104 | 0,956126453 | ST3GAL2 | FALSE | Downregulated |
| 0,0509607 | 0,956162569 | TSPAN15 | FALSE | Upregulated |
| 0,083548116 | 0,956205644 | AC012314.8 | FALSE | Upregulated |
| 0,029573007 | 0,956205644 | ARCN1 | FALSE | Upregulated |
| -0,827981313 | 0,956205644 | ATP1B4 | FALSE | Downregulated |
| -0,161235969 | 0,956205644 | BCRP2 | FALSE | Downregulated |
| 0,031810411 | 0,956205644 | C10orf12 | FALSE | Upregulated |
| 0,022339089 | 0,956205644 | C1R-2 | FALSE | Upregulated |
| 0,190743353 | 0,956205644 | C3orf20 | FALSE | Upregulated |
| -0,029232558 | 0,956205644 | CAMKMT | FALSE | Downregulated |
| -0,018132857 | 0,956205644 | CDC42BPB | FALSE | Downregulated |
| -0,11941185 | 0,956205644 | CLRN1-AS1 | FALSE | Downregulated |
| 0,019190899 | 0,956205644 | CPQ | FALSE | Upregulated |
| -0,127450737 | 0,956205644 | CRYBB1 | FALSE | Downregulated |
| -0,029693718 | 0,956205644 | CST3 | FALSE | Downregulated |
| 0,025961328 | 0,956205644 | CTNS | FALSE | Upregulated |
| 0,536552645 | 0,956205644 | CXCL13 | FALSE | Upregulated |
| 0,017310072 | 0,956205644 | DAP3 | FALSE | Upregulated |
| -0,015634394 | 0,956205644 | DEDD | FALSE | Downregulated |

| | | | | |
|---|---|---|---|---|
| -0,071007435 | 0,956205644 | EDARADD | FALSE | Downregulated |
| -0,015843525 | 0,956205644 | EIF3G | FALSE | Downregulated |
| -0,017365752 | 0,956205644 | ELL | FALSE | Downregulated |
| 0,039247837 | 0,956205644 | EPOR | FALSE | Upregulated |
| -0,121956937 | 0,956205644 | FAM228A | FALSE | Downregulated |
| -0,034299622 | 0,956205644 | FAM49A | FALSE | Downregulated |
| 0,026783423 | 0,956205644 | FDX1L | FALSE | Upregulated |
| 0,085331101 | 0,956205644 | FGF12 | FALSE | Upregulated |
| 0,043832697 | 0,956205644 | GAB3 | FALSE | Upregulated |
| 0,025668128 | 0,956205644 | GORASP2 | FALSE | Upregulated |
| 0,018761555 | 0,956205644 | HYAL1 | FALSE | Upregulated |
| 0,029047405 | 0,956205644 | INPP4A | FALSE | Upregulated |
| -0,025596121 | 0,956205644 | IRF6 | FALSE | Downregulated |
| 0,09768969 | 0,956205644 | LOC101928862 | FALSE | Upregulated |
| -0,045278459 | 0,956205644 | LOC143666 | FALSE | Downregulated |
| 0,0778843 | 0,956205644 | LOC440434 | FALSE | Upregulated |
| 0,100592768 | 0,956205644 | LOC441081 | FALSE | Upregulated |
| 0,024610657 | 0,956205644 | MAPK3 | FALSE | Upregulated |
| -0,012032778 | 0,956205644 | MFN1 | FALSE | Downregulated |
| -0,677797901 | 0,956205644 | MYH6 | FALSE | Downregulated |
| -0,911650985 | 0,956205644 | MYOT | FALSE | Downregulated |
| 0,156786136 | 0,956205644 | OVCH1-AS1 | FALSE | Upregulated |
| -0,023133569 | 0,956205644 | PCYOX1 | FALSE | Downregulated |
| -0,058561661 | 0,956205644 | PGR | FALSE | Downregulated |
| -0,021468117 | 0,956205644 | PIGT | FALSE | Downregulated |
| -0,037034128 | 0,956205644 | PITPNA-AS1 | FALSE | Downregulated |

| | | | | |
|---|---|---|---|---|
| -0,028545709 | 0,956205644 | RAB8B | FALSE | Downregulated |
| 0,034196598 | 0,956205644 | RBPMS | FALSE | Upregulated |
| 0,059531733 | 0,956205644 | RGS7BP | FALSE | Upregulated |
| 0,02693873 | 0,956205644 | RMND5A | FALSE | Upregulated |
| -0,028296472 | 0,956205644 | SCLY | FALSE | Downregulated |
| -0,033035163 | 0,956205644 | SKIDA1 | FALSE | Downregulated |
| -0,026011512 | 0,956205644 | SLC10A7 | FALSE | Downregulated |
| -0,040520618 | 0,956205644 | SLC22A1 | FALSE | Downregulated |
| 0,013664284 | 0,956205644 | SNRPB2 | FALSE | Upregulated |
| 0,047612722 | 0,956205644 | STEAP2 | FALSE | Upregulated |
| -0,056228485 | 0,956205644 | SULT1B1 | FALSE | Downregulated |
| -0,026164381 | 0,956205644 | THAP7 | FALSE | Downregulated |
| 0,043356022 | 0,956205644 | THUMPD3-AS1 | FALSE | Upregulated |
| 0,012184657 | 0,956205644 | TM9SF4 | FALSE | Upregulated |
| 0,123084134 | 0,956205644 | TMEFF2 | FALSE | Upregulated |
| -0,021977776 | 0,956205644 | TMEM260 | FALSE | Downregulated |
| 0,089455112 | 0,956205644 | TMEM51-AS1 | FALSE | Upregulated |
| -0,064739967 | 0,956205644 | TOX3 | FALSE | Downregulated |
| -0,016888918 | 0,956205644 | UBAP2L | FALSE | Downregulated |
| -0,248819491 | 0,956205644 | ZNF625-ZNF20 | FALSE | Downregulated |
| -0,159823199 | 0,956333678 | MYRFL | FALSE | Downregulated |
| 0,036509213 | 0,956371616 | CRY2 | FALSE | Upregulated |
| -0,025818554 | 0,956371616 | LENG1 | FALSE | Downregulated |
| 0,139152532 | 0,956371616 | LINC00881 | FALSE | Upregulated |
| 0,052570538 | 0,956371616 | ZFP28 | FALSE | Upregulated |
| -0,140812494 | 0,956494414 | CLEC12B | FALSE | Downregulated |

| | | | | |
|---|---|---|---|---|
| 0,024803998 | 0,956651267 | SNTA1 | FALSE | Upregulated |
| 0,016536858 | 0,956723979 | PBXIP1 | FALSE | Upregulated |
| -0,021832504 | 0,956727034 | BTN2A1 | FALSE | Downregulated |
| -0,10379799 | 0,956727034 | CNN1 | FALSE | Downregulated |
| -0,03226288 | 0,956727034 | DENND2A | FALSE | Downregulated |
| 0,028685519 | 0,956880758 | MIS18BP1 | FALSE | Upregulated |
| 0,110920375 | 0,956956488 | CYP26A1 | FALSE | Upregulated |
| 0,024430832 | 0,956956488 | PTRH2 | FALSE | Upregulated |
| 0,064667902 | 0,956956488 | SMAD5-AS1 | FALSE | Upregulated |
| 0,092300776 | 0,957005784 | B3GAT1 | FALSE | Upregulated |
| -0,07947853 | 0,957005784 | ST7-AS1 | FALSE | Downregulated |
| 0,050152398 | 0,957064563 | ADAM8 | FALSE | Upregulated |
| 0,03173916 | 0,957064563 | AKAP17A | FALSE | Upregulated |
| -0,014683482 | 0,957064563 | DDX19A | FALSE | Downregulated |
| -0,899981511 | 0,957064563 | LOC102724441 | FALSE | Downregulated |
| 0,022642873 | 0,957064563 | PFAS | FALSE | Upregulated |
| 0,014071979 | 0,957064563 | RAB9A | FALSE | Upregulated |
| -0,033486396 | 0,957064563 | TCP11L1 | FALSE | Downregulated |
| -0,077348931 | 0,957064563 | TGFB2 | FALSE | Downregulated |
| -0,021384484 | 0,957064563 | ZCCHC24 | FALSE | Downregulated |
| -0,029922533 | 0,957064563 | ZFC3H1 | FALSE | Downregulated |
| -0,04350153 | 0,957101765 | SLPI | FALSE | Downregulated |
| -0,02874794 | 0,95712327 | ANKRD26 | FALSE | Downregulated |
| -0,07074284 | 0,95712327 | BTBD11 | FALSE | Downregulated |
| 0,037864598 | 0,95712327 | C12orf73 | FALSE | Upregulated |
| 0,023406615 | 0,95712327 | CCHCR1 | FALSE | Upregulated |

| | | | | |
|---|---|---|---|---|
| -0,031222997 | 0,95712327 | CREM | FALSE | Downregulated |
| -0,047759668 | 0,957138181 | GPR75 | FALSE | Downregulated |
| 0,021669251 | 0,957242591 | CRTC3 | FALSE | Upregulated |
| -0,037139696 | 0,957242591 | ZNF442 | FALSE | Downregulated |
| 0,020187037 | 0,957267951 | MCTS1 | FALSE | Upregulated |
| 0,014058126 | 0,957334539 | TSTD2 | FALSE | Upregulated |
| -0,076772317 | 0,957372599 | C21orf62 | FALSE | Downregulated |
| 0,120996333 | 0,957507348 | LOC102723820 | FALSE | Upregulated |
| 0,028325769 | 0,9576291 | IRF9 | FALSE | Upregulated |
| 0,027057007 | 0,9576291 | PRDX4 | FALSE | Upregulated |
| 0,029358036 | 0,957707395 | ADRB2 | FALSE | Upregulated |
| -0,015835564 | 0,957707395 | BTF3 | FALSE | Downregulated |
| 0,026612484 | 0,957707395 | C4orf32 | FALSE | Upregulated |
| 0,082420465 | 0,957707395 | CYP2A7 | FALSE | Upregulated |
| 0,02102704 | 0,957707395 | FARS2 | FALSE | Upregulated |
| -0,047791145 | 0,957707395 | FIBIN | FALSE | Downregulated |
| -0,01575985 | 0,957707395 | HUWE1 | FALSE | Downregulated |
| -0,086037211 | 0,957707395 | KIF5C | FALSE | Downregulated |
| 0,126690739 | 0,957707395 | LOC101928512 | FALSE | Upregulated |
| 0,049619938 | 0,957707395 | LOC101929130 | FALSE | Upregulated |
| 0,063505589 | 0,957707395 | LOC101929264 | FALSE | Upregulated |
| 0,164125692 | 0,957707395 | LOC388282 | FALSE | Upregulated |
| -0,068801407 | 0,957707395 | LONRF2 | FALSE | Downregulated |
| 0,020639314 | 0,957707395 | METTL21A | FALSE | Upregulated |
| 0,045629549 | 0,957707395 | RP9P | FALSE | Upregulated |
| -0,038366285 | 0,957707395 | SDCBP2-AS1 | FALSE | Downregulated |

| | | | | |
|---|---|---|---|---|
| -0,013120498 | 0,957707395 | SF3B2 | FALSE | Downregulated |
| 0,03038739 | 0,957707395 | TIMP2 | FALSE | Upregulated |
| 0,057358015 | 0,957713047 | CAPN10-AS1 | FALSE | Upregulated |
| 0,061683962 | 0,957713047 | COL1A1 | FALSE | Upregulated |
| -0,048336297 | 0,957713047 | NPR3 | FALSE | Downregulated |
| 0,071156102 | 0,957713047 | POF1B | FALSE | Upregulated |
| -0,019155524 | 0,957713047 | TRIM37 | FALSE | Downregulated |
| -0,114855151 | 0,957713047 | UGT2A2 | FALSE | Downregulated |
| 0,047142642 | 0,957713047 | ZFP2 | FALSE | Upregulated |
| 0,025384364 | 0,957753512 | ARL5B | FALSE | Upregulated |
| -0,018899544 | 0,957753512 | CEBPG | FALSE | Downregulated |
| -0,047877956 | 0,957806894 | C19orf73 | FALSE | Downregulated |
| 0,09862423 | 0,957806894 | FMO1 | FALSE | Upregulated |
| -0,019812277 | 0,957836307 | DLG1 | FALSE | Downregulated |
| -0,015348301 | 0,957836307 | EAPP | FALSE | Downregulated |
| -0,016126298 | 0,957836307 | FUCA1 | FALSE | Downregulated |
| 0,133127462 | 0,957836307 | INPP5J | FALSE | Upregulated |
| -0,06053663 | 0,957836307 | LOC102723772 | FALSE | Downregulated |
| -0,018585373 | 0,957836307 | PAPD4 | FALSE | Downregulated |
| -0,053900055 | 0,957836307 | RPGRIP1 | FALSE | Downregulated |
| -0,024305016 | 0,957836307 | SDHAF1 | FALSE | Downregulated |
| -0,033519105 | 0,957836307 | TCP11L2 | FALSE | Downregulated |
| 0,017699625 | 0,957836307 | TMEM230 | FALSE | Upregulated |
| 0,023266568 | 0,957836307 | TPRA1 | FALSE | Upregulated |
| -0,027392866 | 0,958043517 | CSF1 | FALSE | Downregulated |
| 0,028237504 | 0,958043517 | EPHA1 | FALSE | Upregulated |

| | | | | |
|---|---|---|---|---|
| 0,026044751 | 0,958043517 | FXYD6 | FALSE | Upregulated |
| 0,056136635 | 0,958043517 | RELL2 | FALSE | Upregulated |
| 0,018190971 | 0,958043517 | UHRF2 | FALSE | Upregulated |
| -0,028443981 | 0,958223771 | GLUD1 | FALSE | Downregulated |
| 0,072568817 | 0,958241408 | C1orf220 | FALSE | Upregulated |
| 0,027524537 | 0,958241408 | DCK | FALSE | Upregulated |
| -0,021060777 | 0,958241408 | FAH | FALSE | Downregulated |
| -0,089069177 | 0,958241408 | NDUFA6-AS1 | FALSE | Downregulated |
| 0,088463816 | 0,958241408 | PLCB4 | FALSE | Upregulated |
| 0,028374645 | 0,958241408 | PON2 | FALSE | Upregulated |
| 0,028641939 | 0,958241408 | PPP2R1B | FALSE | Upregulated |
| -0,035943313 | 0,958311923 | DXO | FALSE | Downregulated |
| -0,053315067 | 0,958311923 | RAPGEFL1 | FALSE | Downregulated |
| 0,044599957 | 0,958311923 | TMEM216 | FALSE | Upregulated |
| 0,034320671 | 0,95832069 | AHI1 | FALSE | Upregulated |
| 0,02202257 | 0,95832069 | ANKRD42 | FALSE | Upregulated |
| 0,012875743 | 0,95832069 | ATXN1L | FALSE | Upregulated |
| 0,025297987 | 0,95832069 | CDR2 | FALSE | Upregulated |
| 0,025306514 | 0,95832069 | FAM98C | FALSE | Upregulated |
| -0,102421273 | 0,95832069 | GGTLC2 | FALSE | Downregulated |
| 0,016597628 | 0,95832069 | GMPR2 | FALSE | Upregulated |
| 0,033258148 | 0,95832069 | HSPB1 | FALSE | Upregulated |
| 0,0246793 | 0,95832069 | LINC00623 | FALSE | Upregulated |
| -0,044490421 | 0,95832069 | LINC01140 | FALSE | Downregulated |
| -0,107017596 | 0,95832069 | LOC101927458 | FALSE | Downregulated |
| 0,106703017 | 0,95832069 | LOC102723582 | FALSE | Upregulated |

| | | | | |
|---|---|---|---|---|
| -0,030764126 | 0,95832069 | PAM16 | FALSE | Downregulated |
| 0,022378116 | 0,95832069 | PARD3B | FALSE | Upregulated |
| -0,024333449 | 0,95832069 | SHPRH | FALSE | Downregulated |
| -0,050379361 | 0,95832069 | SLIT2 | FALSE | Downregulated |
| 0,022347342 | 0,95832069 | TRIM32 | FALSE | Upregulated |
| 0,018618311 | 0,95832069 | UCHL3 | FALSE | Upregulated |
| 0,021991638 | 0,95832069 | ZNF740 | FALSE | Upregulated |
| 0,117903746 | 0,958363846 | GRID1 | FALSE | Upregulated |
| -0,053006723 | 0,958402569 | GPR133 | FALSE | Downregulated |
| -0,057751692 | 0,958425799 | LOC102724252 | FALSE | Downregulated |
| 0,021064287 | 0,958430858 | AGBL5 | FALSE | Upregulated |
| 0,091374578 | 0,958430858 | PTGDR2 | FALSE | Upregulated |
| -0,034492928 | 0,958480567 | DAAM2 | FALSE | Downregulated |
| -0,028000032 | 0,958480567 | LOC101927045 | FALSE | Downregulated |
| -0,049154087 | 0,958480567 | NUDT4P1 | FALSE | Downregulated |
| -0,016633506 | 0,958655008 | CDC16 | FALSE | Downregulated |
| -0,028174943 | 0,958655008 | MOB3A | FALSE | Downregulated |
| -0,140749184 | 0,958655008 | NPIPB8 | FALSE | Downregulated |
| -0,074993125 | 0,958655008 | RP11-261N11.8 | FALSE | Downregulated |
| 0,020028731 | 0,958655008 | STX6 | FALSE | Upregulated |
| -0,04179607 | 0,95865616 | BLOC1S1-RDH5 | FALSE | Downregulated |
| 0,047626996 | 0,95865616 | MILR1 | FALSE | Upregulated |
| 0,061912552 | 0,95865616 | RP5-1180C10.2 | FALSE | Upregulated |
| 0,087318655 | 0,95865616 | STK32A | FALSE | Upregulated |
| 0,025305594 | 0,958683692 | CDC73 | FALSE | Upregulated |
| -0,017625465 | 0,958683841 | KDELC2 | FALSE | Downregulated |

| | | | | |
|---|---|---|---|---|
| 0,029758261 | 0,958683841 | LRIG2 | FALSE | Upregulated |
| -0,018140626 | 0,958683841 | MSL1 | FALSE | Downregulated |
| -0,082876794 | 0,958683841 | NOMO3 | FALSE | Downregulated |
| -0,03109637 | 0,958683841 | PIK3R1 | FALSE | Downregulated |
| -0,020486054 | 0,958683841 | PKNOX1 | FALSE | Downregulated |
| 0,074668322 | 0,95870893 | CYSLTR2 | FALSE | Upregulated |
| -0,017961946 | 0,958730911 | PLA2G12B | FALSE | Downregulated |
| 0,026359201 | 0,958730911 | S100PBP | FALSE | Upregulated |
| -0,040240916 | 0,958730911 | SAMD9L | FALSE | Downregulated |
| 0,013580981 | 0,958730911 | TFAM | FALSE | Upregulated |
| 0,128708333 | 0,958763427 | LOC644249 | FALSE | Upregulated |
| -0,025910236 | 0,958845073 | DOCK6 | FALSE | Downregulated |
| 0,013531723 | 0,958864686 | ATP11B | FALSE | Upregulated |
| -0,072035277 | 0,958864686 | CLEC9A | FALSE | Downregulated |
| -0,018462691 | 0,958864686 | KBTBD6 | FALSE | Downregulated |
| -0,025211524 | 0,958864686 | PIK3R4 | FALSE | Downregulated |
| 0,074945173 | 0,958864686 | PLXDC1 | FALSE | Upregulated |
| -0,017067156 | 0,958864686 | ZNF397 | FALSE | Downregulated |
| 0,028245027 | 0,958987109 | AGO3 | FALSE | Upregulated |
| -0,023830847 | 0,958987109 | ARAP2 | FALSE | Downregulated |
| -0,017183229 | 0,958987109 | ARRB2 | FALSE | Downregulated |
| -0,012088574 | 0,958987109 | CAND1 | FALSE | Downregulated |
| 0,019067536 | 0,958987109 | COA4 | FALSE | Upregulated |
| 0,145267048 | 0,958987109 | FAM83A | FALSE | Upregulated |
| 0,015865181 | 0,958987109 | FOXK2 | FALSE | Upregulated |
| 0,05566745 | 0,958987109 | LINC00883 | FALSE | Upregulated |

| | | | | |
|---|---|---|---|---|
| -0,082281673 | 0,958987109 | LMOD1 | FALSE | Downregulated |
| -0,097911049 | 0,958987109 | LOC102724699 | FALSE | Downregulated |
| 0,01953915 | 0,958987109 | NUDCD2 | FALSE | Upregulated |
| 0,047910009 | 0,958987109 | PER1 | FALSE | Upregulated |
| 0,027816587 | 0,958987109 | PFKFB1 | FALSE | Upregulated |
| -0,136409983 | 0,958987109 | PRICKLE2-AS1 | FALSE | Downregulated |
| 0,038433423 | 0,958987109 | RPL23AP7 | FALSE | Upregulated |
| 0,080372158 | 0,958987109 | SLC9A7P1 | FALSE | Upregulated |
| 0,024503871 | 0,958987109 | TOMM40 | FALSE | Upregulated |
| -0,034631733 | 0,958987109 | TPPP2 | FALSE | Downregulated |
| 0,023448783 | 0,958987109 | TRIM26 | FALSE | Upregulated |
| -0,024793049 | 0,958987109 | TSPYL4 | FALSE | Downregulated |
| 0,026555821 | 0,958987109 | UBALD2 | FALSE | Upregulated |
| 0,12313487 | 0,958987109 | UPP2 | FALSE | Upregulated |
| -0,853610599 | 0,958987109 | XIRP2 | FALSE | Downregulated |
| -0,021473543 | 0,958987109 | ZNF264 | FALSE | Downregulated |
| 0,088287247 | 0,959031568 | IL10 | FALSE | Upregulated |
| -0,020644897 | 0,959160141 | RPL24 | FALSE | Downregulated |
| 0,017692103 | 0,959353104 | FBRS | FALSE | Upregulated |
| -0,107180931 | 0,959353104 | HAR1B | FALSE | Downregulated |
| -0,125377285 | 0,959353104 | KCNIP2-AS1 | FALSE | Downregulated |
| 0,102528359 | 0,959353104 | LOC101409256 | FALSE | Upregulated |
| 0,02545529 | 0,959353104 | PHTF2 | FALSE | Upregulated |
| 0,015228724 | 0,959353104 | TBCC | FALSE | Upregulated |
| -0,02351986 | 0,959353104 | ZNF503 | FALSE | Downregulated |
| 0,084220763 | 0,959409734 | ARHGAP28 | FALSE | Upregulated |

| | | | | |
|---|---|---|---|---|
| 0,022338721 | 0,959409734 | EPN2 | FALSE | Upregulated |
| -0,039242381 | 0,959409734 | VWA1 | FALSE | Downregulated |
| -0,034524583 | 0,959421831 | KLHDC1 | FALSE | Downregulated |
| -0,027168192 | 0,959502713 | EPHX1 | FALSE | Downregulated |
| -0,028825564 | 0,959502713 | GCAT | FALSE | Downregulated |
| 0,095279271 | 0,959502713 | LOC101059949 | FALSE | Upregulated |
| 0,018092506 | 0,959502713 | SNX7 | FALSE | Upregulated |
| 0,027728836 | 0,959502713 | ZNF587B | FALSE | Upregulated |
| 0,035505522 | 0,959531856 | FAM109A | FALSE | Upregulated |
| -0,026520173 | 0,959552473 | ACAA2 | FALSE | Downregulated |
| -0,052828097 | 0,959552473 | ACRC | FALSE | Downregulated |
| -0,028646862 | 0,959552473 | BTN3A2 | FALSE | Downregulated |
| 0,02909407 | 0,959552473 | C11orf65 | FALSE | Upregulated |
| 0,0223586 | 0,959552473 | CASP2 | FALSE | Upregulated |
| 0,019320691 | 0,959552473 | DAD1 | FALSE | Upregulated |
| 0,075781262 | 0,959552473 | EBLN2 | FALSE | Upregulated |
| -0,180471239 | 0,959552473 | IDI2 | FALSE | Downregulated |
| 0,023137767 | 0,959552473 | ING5 | FALSE | Upregulated |
| -0,06818553 | 0,959552473 | LINC00933 | FALSE | Downregulated |
| -0,077563769 | 0,959552473 | LOC101928054 | FALSE | Downregulated |
| 0,139961798 | 0,959552473 | LOC101928292 | FALSE | Upregulated |
| 0,111134589 | 0,959552473 | LOC101929918 | FALSE | Upregulated |
| 0,041401301 | 0,959552473 | LOC731656 | FALSE | Upregulated |
| -0,014387609 | 0,959552473 | MAP3K4 | FALSE | Downregulated |
| 0,019835642 | 0,959552473 | MLLT4 | FALSE | Upregulated |
| -0,151348809 | 0,959552473 | MT1JP | FALSE | Downregulated |

| | | | | |
|---|---|---|---|---|
| 0,054235583 | 0,959552473 | PDZD2 | FALSE | Upregulated |
| 0,025215659 | 0,959552473 | POLM | FALSE | Upregulated |
| -0,020684426 | 0,959552473 | RNF43 | FALSE | Downregulated |
| -0,063541178 | 0,959552473 | SCNN1D | FALSE | Downregulated |
| 0,019323069 | 0,959552473 | SERPINC1 | FALSE | Upregulated |
| 0,023559991 | 0,959552473 | SLC50A1 | FALSE | Upregulated |
| -0,079196788 | 0,959552473 | SYCE3 | FALSE | Downregulated |
| 0,055899894 | 0,959552473 | TEKT4P2 | FALSE | Upregulated |
| 0,059378774 | 0,959552473 | TMEM178A | FALSE | Upregulated |
| 0,013694516 | 0,959552473 | TMEM59 | FALSE | Upregulated |
| 0,123700199 | 0,959552473 | VASH2 | FALSE | Upregulated |
| 0,046273484 | 0,959552473 | VSIG4 | FALSE | Upregulated |
| 0,064448206 | 0,959552473 | XAGE3 | FALSE | Upregulated |
| 0,032746584 | 0,959552473 | ZNF266 | FALSE | Upregulated |
| -0,027154777 | 0,959694405 | DPP4 | FALSE | Downregulated |
| -0,83369983 | 0,959694405 | GTF2IP1 | FALSE | Downregulated |
| -0,031451176 | 0,959694405 | LOC389765 | FALSE | Downregulated |
| 0,047826919 | 0,959694405 | SPATA6 | FALSE | Upregulated |
| -0,016622966 | 0,959694405 | TJP2 | FALSE | Downregulated |
| -0,099807389 | 0,959694405 | TMEM233 | FALSE | Downregulated |
| -0,01654751 | 0,959703889 | CCDC94 | FALSE | Downregulated |
| 0,118933587 | 0,959748212 | PDPN | FALSE | Upregulated |
| -0,128606952 | 0,959878664 | RP11-166O4.5 | FALSE | Downregulated |
| -0,053599747 | 0,959964495 | AC011747.7 | FALSE | Downregulated |
| -0,028040737 | 0,959964495 | ACOT4 | FALSE | Downregulated |
| -0,050162283 | 0,959964495 | AIFM3 | FALSE | Downregulated |

| | | | | |
|---|---|---|---|---|
| -0,024097432 | 0,959964495 | ARPC5L | FALSE | Downregulated |
| -0,042919693 | 0,959964495 | ATRNL1 | FALSE | Downregulated |
| -0,014883829 | 0,959964495 | C16orf70 | FALSE | Downregulated |
| -0,82889129 | 0,959964495 | CKM | FALSE | Downregulated |
| -0,022305395 | 0,959964495 | CPOX | FALSE | Downregulated |
| -0,049763994 | 0,959964495 | CYP21A2 | FALSE | Downregulated |
| 0,027237362 | 0,959964495 | DCAF16 | FALSE | Upregulated |
| 0,024236954 | 0,959964495 | DNAJA1 | FALSE | Upregulated |
| 0,032800709 | 0,959964495 | HGF | FALSE | Upregulated |
| 0,021898048 | 0,959964495 | JMY | FALSE | Upregulated |
| -0,026395003 | 0,959964495 | LIG4 | FALSE | Downregulated |
| -0,112715636 | 0,959964495 | LINC00189 | FALSE | Downregulated |
| -0,048977414 | 0,959964495 | MALL | FALSE | Downregulated |
| -0,021526768 | 0,959964495 | MIS12 | FALSE | Downregulated |
| -0,011814526 | 0,959964495 | NAA50 | FALSE | Downregulated |
| -0,011986573 | 0,959964495 | PICALM | FALSE | Downregulated |
| 0,022923308 | 0,959964495 | PLAGL2 | FALSE | Upregulated |
| -0,023994784 | 0,959964495 | SERPINA4 | FALSE | Downregulated |
| -0,022787076 | 0,959964495 | SLTM | FALSE | Downregulated |
| 0,013894547 | 0,959964495 | XPO5 | FALSE | Upregulated |
| -0,018939969 | 0,959964495 | ZNF776 | FALSE | Downregulated |
| -0,033121123 | 0,959997261 | SH3BP4 | FALSE | Downregulated |
| -0,019737741 | 0,960133904 | AKAP11 | FALSE | Downregulated |
| 0,038908319 | 0,960133904 | C9orf43 | FALSE | Upregulated |
| -0,054180595 | 0,960207047 | C8orf46 | FALSE | Downregulated |
| -0,033650891 | 0,960207047 | FDXACB1 | FALSE | Downregulated |

| | | | | |
|---|---|---|---|---|
| -0,080365781 | 0,960207047 | LOC101929531 | FALSE | Downregulated |
| -0,018040029 | 0,960207047 | PIH1D1 | FALSE | Downregulated |
| -0,027804223 | 0,960207047 | RASSF7 | FALSE | Downregulated |
| -0,01625289 | 0,960207047 | SOCS6 | FALSE | Downregulated |
| 0,013242836 | 0,960207047 | YWHAB | FALSE | Upregulated |
| 0,014128652 | 0,960248198 | TMED4 | FALSE | Upregulated |
| 0,067421426 | 0,960294876 | BEND5 | FALSE | Upregulated |
| 0,087052861 | 0,960294876 | DCLK3 | FALSE | Upregulated |
| 0,015451702 | 0,960381902 | FYN | FALSE | Upregulated |
| 0,098631703 | 0,960381902 | LINC01134 | FALSE | Upregulated |
| -0,034328326 | 0,960381902 | LOC102724975 | FALSE | Downregulated |
| -0,021963721 | 0,960381902 | MCRS1 | FALSE | Downregulated |
| 0,049241902 | 0,960381902 | PMS2P3 | FALSE | Upregulated |
| -0,022264097 | 0,960410443 | NME7 | FALSE | Downregulated |
| -0,023425265 | 0,960414822 | ABLIM1 | FALSE | Downregulated |
| -0,040614636 | 0,960414822 | HOXB4 | FALSE | Downregulated |
| -0,088708812 | 0,960414822 | LINC00607 | FALSE | Downregulated |
| 0,057599392 | 0,960414822 | LINC01123 | FALSE | Upregulated |
| 0,085921071 | 0,96046582 | LOC102723641 | FALSE | Upregulated |
| 0,031665803 | 0,96060867 | GLIS3 | FALSE | Upregulated |
| 0,122151894 | 0,96060867 | LOC101927051 | FALSE | Upregulated |
| -0,069260574 | 0,96060867 | LOC101927763 | FALSE | Downregulated |
| 0,047449137 | 0,96060867 | LOC102723726 | FALSE | Upregulated |
| -0,029498092 | 0,960628256 | ZNF568 | FALSE | Downregulated |
| 0,023292225 | 0,960736194 | NTN1 | FALSE | Upregulated |
| -0,02732332 | 0,960785581 | PLIN3 | FALSE | Downregulated |

| | | | | |
|---|---|---|---|---|
| -0,014321068 | 0,960785581 | ZSCAN29 | FALSE | Downregulated |
| 0,009723432 | 0,960806942 | KHDRBS1 | FALSE | Upregulated |
| 0,060656676 | 0,960806942 | LOC101928434 | FALSE | Upregulated |
| -0,034418446 | 0,960806942 | SPRY4 | FALSE | Downregulated |
| 0,053777157 | 0,960806942 | TCTEX1D2 | FALSE | Upregulated |
| 0,031504166 | 0,960978349 | FLJ37453 | FALSE | Upregulated |
| -0,029559226 | 0,96105273 | ABCA11P | FALSE | Downregulated |
| -0,025452736 | 0,96105273 | ACADS | FALSE | Downregulated |
| -0,017492767 | 0,96105273 | C2orf42 | FALSE | Downregulated |
| 0,089080036 | 0,96105273 | CES1P2 | FALSE | Upregulated |
| -0,044985834 | 0,96105273 | CNTNAP2 | FALSE | Downregulated |
| -0,042432397 | 0,96105273 | DHODH | FALSE | Downregulated |
| 0,063652859 | 0,96105273 | EVPLL | FALSE | Upregulated |
| 0,093629785 | 0,96105273 | LOC101927858 | FALSE | Upregulated |
| 0,066618726 | 0,96105273 | NLGN3 | FALSE | Upregulated |
| -0,044158768 | 0,96105273 | RGPD1 | FALSE | Downregulated |
| -0,028405279 | 0,96105273 | TMEM258 | FALSE | Downregulated |
| -0,021479404 | 0,961066672 | ABL2 | FALSE | Downregulated |
| 0,038797476 | 0,961066672 | C5orf30 | FALSE | Upregulated |
| -0,153709935 | 0,961066672 | ERBB4 | FALSE | Downregulated |
| -0,021877679 | 0,961066672 | MCFD2 | FALSE | Downregulated |
| -0,017632024 | 0,961066672 | NSD1 | FALSE | Downregulated |
| -0,04035197 | 0,961066672 | SOX6 | FALSE | Downregulated |
| -0,095444702 | 0,961073752 | CAMK1G | FALSE | Downregulated |
| -0,021531004 | 0,961073752 | NDUFB11 | FALSE | Downregulated |
| 0,020262058 | 0,9614351 | FBXO46 | FALSE | Upregulated |

| | | | | |
|---|---|---|---|---|
| 0,043989522 | 0,9614351 | ZNF221 | FALSE | Upregulated |
| 0,037598714 | 0,961669227 | PLA2R1 | FALSE | Upregulated |
| -0,011282525 | 0,961679499 | CDV3 | FALSE | Downregulated |
| 0,085357063 | 0,961679499 | FAM81A | FALSE | Upregulated |
| -0,050287492 | 0,961679499 | KCNQ1OT1 | FALSE | Downregulated |
| -0,024993506 | 0,961679499 | SH3BP2 | FALSE | Downregulated |
| -0,112271206 | 0,961792478 | KISS1 | FALSE | Downregulated |
| 0,033174436 | 0,961905003 | EHD3 | FALSE | Upregulated |
| -0,024217555 | 0,961905003 | HERPUD1 | FALSE | Downregulated |
| 0,075885994 | 0,961905003 | LOC101928399 | FALSE | Upregulated |
| -0,029812204 | 0,961977297 | RBBP8 | FALSE | Downregulated |
| -0,050436705 | 0,962014918 | DNMBP-AS1 | FALSE | Downregulated |
| -0,035122248 | 0,962014918 | GLS2 | FALSE | Downregulated |
| 0,01126138 | 0,962014918 | WBP11 | FALSE | Upregulated |
| 0,023308219 | 0,962014918 | ZNF619 | FALSE | Upregulated |
| -0,035871314 | 0,96217142 | NBPF10 | FALSE | Downregulated |
| -0,034170208 | 0,962179264 | GPER1 | FALSE | Downregulated |
| 0,014748504 | 0,962179264 | ILKAP | FALSE | Upregulated |
| -0,07382082 | 0,962179264 | PABPC3 | FALSE | Downregulated |
| -0,021496618 | 0,962236389 | TMED9 | FALSE | Downregulated |
| -0,12188258 | 0,962236389 | TMEM179 | FALSE | Downregulated |
| -0,030884123 | 0,962349761 | FAM195A | FALSE | Downregulated |
| -0,024834885 | 0,962349761 | PRPF40B | FALSE | Downregulated |
| 0,018346532 | 0,962349761 | RQCD1 | FALSE | Upregulated |
| 0,018676843 | 0,962532181 | RPP25L | FALSE | Upregulated |
| -0,048464221 | 0,962615732 | LOC101929005 | FALSE | Downregulated |

| | | | | |
|---|---|---|---|---|
| 0,019485257 | 0,962727659 | JDP2 | FALSE | Upregulated |
| -0,034106963 | 0,962727659 | KIAA1462 | FALSE | Downregulated |
| 0,044338154 | 0,962727659 | PLEKHH1 | FALSE | Upregulated |
| -0,032022996 | 0,962766819 | BAG3 | FALSE | Downregulated |
| -0,024486501 | 0,962766819 | GIMAP6 | FALSE | Downregulated |
| -0,06882983 | 0,962823061 | KAZALD1 | FALSE | Downregulated |
| 0,036562124 | 0,962834605 | RAMP3 | FALSE | Upregulated |
| -0,086403775 | 0,96306204 | DYNC1I1 | FALSE | Downregulated |
| -0,060145877 | 0,963159266 | C5orf34 | FALSE | Downregulated |
| -0,090069813 | 0,963159266 | FCGR1B | FALSE | Downregulated |
| -0,052670748 | 0,963159266 | LOC100128288 | FALSE | Downregulated |
| -0,04870789 | 0,963159266 | LOC101927571 | FALSE | Downregulated |
| -0,21786395 | 0,963159266 | MROH7 | FALSE | Downregulated |
| 0,095449254 | 0,963159266 | PTGES | FALSE | Upregulated |
| -0,011067346 | 0,963159266 | SCAF8 | FALSE | Downregulated |
| 0,016727288 | 0,963159266 | UNC119B | FALSE | Upregulated |
| 0,019288605 | 0,963159266 | ZBTB34 | FALSE | Upregulated |
| -0,0653028 | 0,963372164 | LOC101928068 | FALSE | Downregulated |
| -0,019503288 | 0,963382789 | HIATL1 | FALSE | Downregulated |
| -0,099476482 | 0,963382789 | SLC6A19 | FALSE | Downregulated |
| -0,716479827 | 0,963453977 | ACTN2 | FALSE | Downregulated |
| 0,038850545 | 0,96350241 | ALDH1L1 | FALSE | Upregulated |
| -0,013973757 | 0,96350241 | ERAL1 | FALSE | Downregulated |
| -0,016598679 | 0,96350241 | HPS4 | FALSE | Downregulated |
| -0,059031186 | 0,96350241 | LOC102723616 | FALSE | Downregulated |
| -0,030547877 | 0,96350241 | RFC3 | FALSE | Downregulated |

| | | | | |
|---|---|---|---|---|
| 0,021827325 | 0,96350241 | RNPEP | FALSE | Upregulated |
| -0,025935618 | 0,963528895 | ARSK | FALSE | Downregulated |
| -0,014239734 | 0,963528895 | FAN1 | FALSE | Downregulated |
| 0,036842327 | 0,963528895 | SP5 | FALSE | Upregulated |
| 0,041478319 | 0,963528895 | TCTEX1D1 | FALSE | Upregulated |
| 0,28932958 | 0,963528895 | TNFSF12-TNFSF13 | FALSE | Upregulated |
| 0,018574369 | 0,96369286 | AIMP2 | FALSE | Upregulated |
| 0,045826581 | 0,96369286 | ARG2 | FALSE | Upregulated |
| 0,058067488 | 0,96369286 | ASNS | FALSE | Upregulated |
| -0,021359725 | 0,96369286 | BAIAP2L1 | FALSE | Downregulated |
| 0,022572372 | 0,96369286 | BCL9L | FALSE | Upregulated |
| -0,029332633 | 0,96369286 | C9orf40 | FALSE | Downregulated |
| 0,025394683 | 0,96369286 | CHD1L | FALSE | Upregulated |
| -0,021438318 | 0,96369286 | CYB5D2 | FALSE | Downregulated |
| -0,009521795 | 0,96369286 | DNAJC14 | FALSE | Downregulated |
| -0,15322259 | 0,96369286 | GSTM2P1 | FALSE | Downregulated |
| -0,014174227 | 0,96369286 | HCFC1 | FALSE | Downregulated |
| -0,013795224 | 0,96369286 | HNRNPA0 | FALSE | Downregulated |
| -0,031838331 | 0,96369286 | HSPA1B | FALSE | Downregulated |
| -0,066902166 | 0,96369286 | LINC00265 | FALSE | Downregulated |
| -0,06573119 | 0,96369286 | LOC100506014 | FALSE | Downregulated |
| 0,131674799 | 0,96369286 | LOC100506082 | FALSE | Upregulated |
| -0,060171241 | 0,96369286 | LOC101929178 | FALSE | Downregulated |
| 0,05124215 | 0,96369286 | LRRC29 | FALSE | Upregulated |
| 0,024861556 | 0,96369286 | MAML3 | FALSE | Upregulated |
| -0,020145767 | 0,96369286 | MINA | FALSE | Downregulated |

| | | | | |
|---|---|---|---|---|
| 0,01679078 | 0,96369286 | MTM1 | FALSE | Upregulated |
| 0,037942494 | 0,96369286 | NCS1 | FALSE | Upregulated |
| -0,053405527 | 0,96369286 | NRG4 | FALSE | Downregulated |
| 0,046378144 | 0,96369286 | PLK4 | FALSE | Upregulated |
| 0,019783609 | 0,96369286 | RCCD1 | FALSE | Upregulated |
| 0,022076361 | 0,96369286 | RELL1 | FALSE | Upregulated |
| 0,027337156 | 0,96369286 | RPP21 | FALSE | Upregulated |
| -0,014090592 | 0,96369286 | RSPRY1 | FALSE | Downregulated |
| -0,135383981 | 0,96369286 | SCTR | FALSE | Downregulated |
| 0,032173711 | 0,96369286 | SEC14L4 | FALSE | Upregulated |
| -0,019838897 | 0,96369286 | SERGEF | FALSE | Downregulated |
| 0,01583724 | 0,96369286 | SLC31A1 | FALSE | Upregulated |
| 0,09016593 | 0,96369286 | SLC5A2 | FALSE | Upregulated |
| 0,017172033 | 0,96369286 | SRSF3 | FALSE | Upregulated |
| 0,013791872 | 0,96369286 | STAU2 | FALSE | Upregulated |
| 0,03025006 | 0,96369286 | TANC2 | FALSE | Upregulated |
| 0,064388016 | 0,96369286 | TESC | FALSE | Upregulated |
| -0,221106969 | 0,96369286 | TSNAX-DISC1 | FALSE | Downregulated |
| 0,017567689 | 0,96369286 | USP37 | FALSE | Upregulated |
| -0,027003719 | 0,963696047 | ECHDC2 | FALSE | Downregulated |
| -0,041373899 | 0,963696047 | TRIM66 | FALSE | Downregulated |
| -0,07604296 | 0,963712748 | AC159540.1 | FALSE | Downregulated |
| 0,015453539 | 0,96375438 | GTPBP4 | FALSE | Upregulated |
| -0,087341925 | 0,96382316 | RP11-421P23.1 | FALSE | Downregulated |
| -0,074970913 | 0,963848197 | LOC101927520 | FALSE | Downregulated |
| -0,064170916 | 0,963880006 | ANKRD20A9P | FALSE | Downregulated |

| | | | | |
|---|---|---|---|---|
| -0,042360139 | 0,963880006 | LOC102724129 | FALSE | Downregulated |
| 0,021337345 | 0,963880006 | SPCS3 | FALSE | Upregulated |
| -0,108300002 | 0,963899991 | C2CD4C | FALSE | Downregulated |
| 0,060579909 | 0,963899991 | CAP2 | FALSE | Upregulated |
| -0,041646652 | 0,963899991 | CYP2A6 | FALSE | Downregulated |
| -0,110337082 | 0,963899991 | DLGAP1 | FALSE | Downregulated |
| 0,030141459 | 0,963899991 | GGT5 | FALSE | Upregulated |
| 0,02177599 | 0,963899991 | HECTD4 | FALSE | Upregulated |
| -0,021430081 | 0,963899991 | LATS1 | FALSE | Downregulated |
| 0,023142365 | 0,963899991 | MESDC1 | FALSE | Upregulated |
| -0,033705263 | 0,963899991 | PLK2 | FALSE | Downregulated |
| 0,023797891 | 0,963899991 | TANC1 | FALSE | Upregulated |
| 0,038302509 | 0,963905817 | FAM99A | FALSE | Upregulated |
| 0,050665207 | 0,963905817 | KDM8 | FALSE | Upregulated |
| 0,029943315 | 0,963905817 | LMBR1L | FALSE | Upregulated |
| 0,072693086 | 0,963905817 | LOC646626 | FALSE | Upregulated |
| -0,01432787 | 0,963905817 | MMAA | FALSE | Downregulated |
| 0,016593122 | 0,963905817 | MSH3 | FALSE | Upregulated |
| -0,041486788 | 0,963905817 | TNFRSF11A | FALSE | Downregulated |
| 0,037941312 | 0,963956789 | PHLDA1 | FALSE | Upregulated |
| 0,298413078 | 0,963956789 | RNU1-1 | FALSE | Upregulated |
| -0,748635275 | 0,963978769 | ACTA1 | FALSE | Downregulated |
| 0,02138982 | 0,963978769 | ALDH5A1 | FALSE | Upregulated |
| 0,02998159 | 0,963978769 | ANG | FALSE | Upregulated |
| -0,019825192 | 0,963978769 | ANKS4B | FALSE | Downregulated |
| -0,056083529 | 0,963978769 | ATP13A4 | FALSE | Downregulated |

| | | | | |
|---|---|---|---|---|
| -0,036456491 | 0,963978769 | C11orf96 | FALSE | Downregulated |
| -0,01465338 | 0,963978769 | C1orf52 | FALSE | Downregulated |
| -0,068265307 | 0,963978769 | CACNA1H | FALSE | Downregulated |
| 0,026286728 | 0,963978769 | CCDC107 | FALSE | Upregulated |
| 0,015316484 | 0,963978769 | CMTR2 | FALSE | Upregulated |
| -0,027466567 | 0,963978769 | DGAT1 | FALSE | Downregulated |
| 0,070528326 | 0,963978769 | FAM166A | FALSE | Upregulated |
| 0,0284942 | 0,963978769 | FBXO17 | FALSE | Upregulated |
| -0,03750505 | 0,963978769 | JAM3 | FALSE | Downregulated |
| -0,0514857 | 0,963978769 | LOC100506235 | FALSE | Downregulated |
| -0,094051306 | 0,963978769 | LOC101928558 | FALSE | Downregulated |
| -0,175757943 | 0,963978769 | LOC101929826 | FALSE | Downregulated |
| -0,113222719 | 0,963978769 | LOC101930375 | FALSE | Downregulated |
| 0,014972755 | 0,963978769 | MED9 | FALSE | Upregulated |
| -0,030867693 | 0,963978769 | MYO5C | FALSE | Downregulated |
| -0,059986057 | 0,963978769 | NOL4 | FALSE | Downregulated |
| 0,099056347 | 0,963978769 | PLEKHG4 | FALSE | Upregulated |
| 0,015477521 | 0,963978769 | POT1 | FALSE | Upregulated |
| -0,026102194 | 0,963978769 | PTPN9 | FALSE | Downregulated |
| -0,119638136 | 0,963978769 | RGPD5 | FALSE | Downregulated |
| 0,036460349 | 0,963978769 | RP4-639F20.1 | FALSE | Upregulated |
| 0,098963396 | 0,963978769 | RP4-640H8.2 | FALSE | Upregulated |
| 0,014120832 | 0,963978769 | SAMD4B | FALSE | Upregulated |
| -0,056270139 | 0,963978769 | SMPD3 | FALSE | Downregulated |
| -0,109573892 | 0,963978769 | SNORA32 | FALSE | Downregulated |
| -0,020715751 | 0,963978769 | SNX24 | FALSE | Downregulated |

| | | | | |
|---|---|---|---|---|
| -0,012591451 | 0,963978769 | TMEM68 | FALSE | Downregulated |
| 0,074815535 | 0,963991098 | CD300LG | FALSE | Upregulated |
| 0,02465277 | 0,963991098 | LACC1 | FALSE | Upregulated |
| -0,04410945 | 0,963991098 | LPHN1 | FALSE | Downregulated |
| -0,037826373 | 0,963991098 | SHROOM4 | FALSE | Downregulated |
| 0,069439594 | 0,963991098 | SNORA67 | FALSE | Upregulated |
| 0,030035291 | 0,963991098 | STS | FALSE | Upregulated |
| 0,018711235 | 0,963991098 | TMEM219 | FALSE | Upregulated |
| -0,019703288 | 0,964078386 | ANKRD10 | FALSE | Downregulated |
| -0,023375886 | 0,964078386 | FOXA1 | FALSE | Downregulated |
| 0,024200112 | 0,964078386 | IQCG | FALSE | Upregulated |
| 0,14419395 | 0,964078386 | LOC101927594 | FALSE | Upregulated |
| -0,110571069 | 0,964078386 | LOC102723505 | FALSE | Downregulated |
| 0,019827851 | 0,964078386 | METTL23 | FALSE | Upregulated |
| 0,023698539 | 0,964078386 | NOTCH2NL | FALSE | Upregulated |
| 0,014580577 | 0,964078386 | USP34 | FALSE | Upregulated |
| -0,013980165 | 0,964099324 | RAP1A | FALSE | Downregulated |
| -0,020428181 | 0,964206633 | DPYSL2 | FALSE | Downregulated |
| 0,073296798 | 0,964206633 | LOC102724940 | FALSE | Upregulated |
| 0,0132676 | 0,964206633 | MAGED2 | FALSE | Upregulated |
| 0,023003309 | 0,964206633 | SMIM4 | FALSE | Upregulated |
| -0,086254526 | 0,964293301 | ELOVL4 | FALSE | Downregulated |
| -0,038573669 | 0,964293301 | LOC100132891 | FALSE | Downregulated |
| 0,0340163 | 0,964297123 | ASS1 | FALSE | Upregulated |
| -0,015245058 | 0,964297123 | CARS2 | FALSE | Downregulated |
| 0,033333574 | 0,964297123 | CFHR1 | FALSE | Upregulated |

| | | | | |
|---|---|---|---|---|
| 0,017641907 | 0,964297123 | ECI2 | FALSE | Upregulated |
| -0,016647413 | 0,964297123 | ERLIN1 | FALSE | Downregulated |
| 0,01821822 | 0,964297123 | FBXL4 | FALSE | Upregulated |
| -0,027379681 | 0,964297123 | GLCE | FALSE | Downregulated |
| 0,021902785 | 0,964297123 | HTATIP2 | FALSE | Upregulated |
| 0,087370036 | 0,964297123 | LOC102724235 | FALSE | Upregulated |
| 0,040999729 | 0,964297123 | NEBL | FALSE | Upregulated |
| 0,012763443 | 0,964297123 | NOLC1 | FALSE | Upregulated |
| -0,023854517 | 0,964297123 | PITPNM2 | FALSE | Downregulated |
| -0,015180776 | 0,964297123 | PSMB3 | FALSE | Downregulated |
| 0,023976253 | 0,964297123 | RPL11 | FALSE | Upregulated |
| -0,071860088 | 0,964297123 | RPL13AP6 | FALSE | Downregulated |
| -0,015086985 | 0,964297123 | TDRD3 | FALSE | Downregulated |
| -0,022264716 | 0,964297123 | TRIM14 | FALSE | Downregulated |
| 0,035361532 | 0,964297123 | ZNF430 | FALSE | Upregulated |
| 0,052903848 | 0,964297123 | ZNF660 | FALSE | Upregulated |
| -0,073590011 | 0,964297123 | ZNF727 | FALSE | Downregulated |
| 0,0659354 | 0,964346608 | ABCA10 | FALSE | Upregulated |
| -0,03292123 | 0,964348507 | AC004893.11 | FALSE | Downregulated |
| 0,027755884 | 0,964348507 | ARRDC4 | FALSE | Upregulated |
| 0,013866757 | 0,964348507 | C9orf64 | FALSE | Upregulated |
| -0,028987158 | 0,964348507 | CCDC134 | FALSE | Downregulated |
| -0,019104402 | 0,964348507 | COX11 | FALSE | Downregulated |
| 0,018451928 | 0,964348507 | FBXO30 | FALSE | Upregulated |
| 0,043654468 | 0,964348507 | FNTB | FALSE | Upregulated |
| -0,012308037 | 0,964348507 | KDELR1 | FALSE | Downregulated |

| | | | | |
|---|---|---|---|---|
| -0,064677991 | 0,964348507 | NME5 | FALSE | Downregulated |
| -0,012979067 | 0,964348507 | RANBP9 | FALSE | Downregulated |
| -0,020719797 | 0,964348507 | SH2B3 | FALSE | Downregulated |
| -0,020727458 | 0,964348507 | SYNE1 | FALSE | Downregulated |
| 0,018750972 | 0,964443998 | ADD3 | FALSE | Upregulated |
| -0,040994377 | 0,964443998 | ARL17A | FALSE | Downregulated |
| 0,109298018 | 0,964443998 | CPB2-AS1 | FALSE | Upregulated |
| -0,049132404 | 0,964443998 | PPP1R9A | FALSE | Downregulated |
| -0,015252696 | 0,964526784 | HEATR6 | FALSE | Downregulated |
| -0,018507842 | 0,964580474 | LETM1 | FALSE | Downregulated |
| 0,026785436 | 0,964580474 | VKORC1 | FALSE | Upregulated |
| 0,037271024 | 0,964622121 | ADRA2B | FALSE | Upregulated |
| -0,02663101 | 0,964622121 | DHRS11 | FALSE | Downregulated |
| 0,009463374 | 0,964622121 | GNB1 | FALSE | Upregulated |
| 0,011494585 | 0,964622121 | PAK2 | FALSE | Upregulated |
| -0,016389304 | 0,964622121 | PSMB6 | FALSE | Downregulated |
| -0,017571737 | 0,964622121 | SLC8B1 | FALSE | Downregulated |
| 0,075283596 | 0,964622121 | TSPEAR-AS1 | FALSE | Upregulated |
| 0,020445018 | 0,96467998 | ACAP2 | FALSE | Upregulated |
| -0,037856038 | 0,964712098 | SLC35E2 | FALSE | Downregulated |
| -0,016007743 | 0,964793727 | ARHGAP29 | FALSE | Downregulated |
| -0,020160542 | 0,964793727 | FAM177A1 | FALSE | Downregulated |
| -0,085022716 | 0,964793727 | RP11-274B18.2 | FALSE | Downregulated |
| 0,077514916 | 0,964793727 | TEX29 | FALSE | Upregulated |
| 0,028635888 | 0,964842508 | FAM63B | FALSE | Upregulated |
| 0,056330296 | 0,964842508 | LOC101928588 | FALSE | Upregulated |

| | | | | |
|---|---|---|---|---|
| 0,016725991 | 0,964842508 | SCP2 | FALSE | Upregulated |
| 0,039260842 | 0,964842527 | BMP2 | FALSE | Upregulated |
| 0,083347815 | 0,964842527 | LOC100996695 | FALSE | Upregulated |
| 0,04657772 | 0,964842527 | LOC283922 | FALSE | Upregulated |
| -0,019603209 | 0,964842527 | NAP1L1 | FALSE | Downregulated |
| 0,014114357 | 0,964842527 | PLEKHA1 | FALSE | Upregulated |
| 0,029447061 | 0,964842527 | TLR1 | FALSE | Upregulated |
| -0,104261382 | 0,964842527 | WNT7B | FALSE | Downregulated |
| 0,025153987 | 0,964978216 | FGFR2 | FALSE | Upregulated |
| 0,184155607 | 0,964978216 | FSTL5 | FALSE | Upregulated |
| -0,018856201 | 0,964978216 | HNF4A | FALSE | Downregulated |
| -0,026805008 | 0,964978216 | KIF9 | FALSE | Downregulated |
| 0,016295114 | 0,964978216 | KLHL7 | FALSE | Upregulated |
| -0,031085228 | 0,965077139 | STMN1 | FALSE | Downregulated |
| -0,033037299 | 0,965284253 | ADAT2 | FALSE | Downregulated |
| -0,037701555 | 0,965284253 | CHAD | FALSE | Downregulated |
| 0,064855493 | 0,965284253 | ROBO2 | FALSE | Upregulated |
| -0,014202997 | 0,965394325 | CYP2J2 | FALSE | Downregulated |
| 0,010746472 | 0,965394325 | FAM104A | FALSE | Upregulated |
| -0,031077341 | 0,965430398 | SLC6A16 | FALSE | Downregulated |
| -0,713503277 | 0,965430398 | XIST | FALSE | Downregulated |
| -0,031407535 | 0,965569686 | AHR | FALSE | Downregulated |
| 0,02021612 | 0,965569686 | ALPK1 | FALSE | Upregulated |
| 0,069857736 | 0,965569686 | APOBEC3A | FALSE | Upregulated |
| -0,015416783 | 0,965569686 | CTPS2 | FALSE | Downregulated |
| -0,03265521 | 0,965569686 | DGKE | FALSE | Downregulated |

| | | | | |
|---|---|---|---|---|
| 0,021740814 | 0,965569686 | H2AFZ | FALSE | Upregulated |
| -0,10538616 | 0,965569686 | HIST1H2BJ | FALSE | Downregulated |
| 0,024264675 | 0,965569686 | HOOK2 | FALSE | Upregulated |
| -0,029886863 | 0,965569686 | KLF13 | FALSE | Downregulated |
| -0,042823658 | 0,965569686 | LOC100996724 | FALSE | Downregulated |
| 0,092063666 | 0,965569686 | LOC101927512 | FALSE | Upregulated |
| -0,260864315 | 0,965569686 | LOC102723799 | FALSE | Downregulated |
| 0,045573159 | 0,965569686 | MRPL45P2 | FALSE | Upregulated |
| -0,037380816 | 0,965569686 | NKTR | FALSE | Downregulated |
| -0,012285165 | 0,965569686 | PRPF4 | FALSE | Downregulated |
| -0,053170364 | 0,965569686 | PTGDR | FALSE | Downregulated |
| -0,020861768 | 0,965569686 | RDH10 | FALSE | Downregulated |
| -0,021144741 | 0,965569686 | RILPL1 | FALSE | Downregulated |
| 0,04070482 | 0,965569686 | SHH | FALSE | Upregulated |
| 0,01715692 | 0,965569686 | TEX10 | FALSE | Upregulated |
| 0,015443745 | 0,965569686 | VKORC1L1 | FALSE | Upregulated |
| -0,022157394 | 0,965666899 | IFI35 | FALSE | Downregulated |
| 0,05587329 | 0,965746663 | LOC729683 | FALSE | Upregulated |
| 0,08391701 | 0,965934467 | FRMD6-AS1 | FALSE | Upregulated |
| 0,308874652 | 0,965934467 | RAD51L3-RFFL | FALSE | Upregulated |
| 0,121502486 | 0,965934467 | XPNPEP2 | FALSE | Upregulated |
| -0,073389877 | 0,966062469 | FLJ44313 | FALSE | Downregulated |
| 0,017142615 | 0,966113208 | C16orf80 | FALSE | Upregulated |
| 0,045337902 | 0,966113208 | ENKUR | FALSE | Upregulated |
| 0,033876434 | 0,966113208 | JPX | FALSE | Upregulated |
| -0,125843183 | 0,966113208 | LOC102724241 | FALSE | Downregulated |

| | | | | |
|---|---|---|---|---|
| -0,017848752 | 0,966113208 | TCF7L2 | FALSE | Downregulated |
| -0,022799563 | 0,966113208 | TF | FALSE | Downregulated |
| 0,079189847 | 0,966160166 | CCDC85A | FALSE | Upregulated |
| 0,049556337 | 0,96616128 | LOC101928913 | FALSE | Upregulated |
| 0,077563241 | 0,96616128 | LOC102723589 | FALSE | Upregulated |
| -0,021192311 | 0,966180878 | TWSG1 | FALSE | Downregulated |
| -0,019908947 | 0,966231846 | KDM4B | FALSE | Downregulated |
| 0,059872358 | 0,966310424 | RASGEF1A | FALSE | Upregulated |
| 0,035711248 | 0,966310424 | RTP4 | FALSE | Upregulated |
| 0,05373652 | 0,966337772 | LINC00337 | FALSE | Upregulated |
| -0,019235182 | 0,966430595 | CNOT6 | FALSE | Downregulated |
| 0,160937818 | 0,966430595 | HIST2H4A | FALSE | Upregulated |
| 0,031626733 | 0,966505379 | ACOT2 | FALSE | Upregulated |
| 0,125006825 | 0,966505379 | APOL5 | FALSE | Upregulated |
| 0,019084245 | 0,966505379 | ATP5I | FALSE | Upregulated |
| 0,040213246 | 0,966505379 | B9D2 | FALSE | Upregulated |
| 0,046149844 | 0,966505379 | BCMO1 | FALSE | Upregulated |
| -0,027829347 | 0,966505379 | C16orf86 | FALSE | Downregulated |
| -0,034226754 | 0,966505379 | CEBPD | FALSE | Downregulated |
| 0,011884075 | 0,966505379 | DNAJB2 | FALSE | Upregulated |
| 0,018691386 | 0,966505379 | DNM1L | FALSE | Upregulated |
| 0,048524193 | 0,966505379 | HCG8 | FALSE | Upregulated |
| 0,039409447 | 0,966505379 | KIAA1324L | FALSE | Upregulated |
| -0,032363882 | 0,966505379 | LINC00087 | FALSE | Downregulated |
| -0,017440403 | 0,966505379 | LMAN2L | FALSE | Downregulated |
| -0,048142484 | 0,966505379 | MFAP4 | FALSE | Downregulated |

| | | | | |
|---|---|---|---|---|
| 0,024922876 | 0,966505379 | MZT1 | FALSE | Upregulated |
| 0,026198384 | 0,966505379 | NBPF9 | FALSE | Upregulated |
| -0,0724688 | 0,966505379 | NEK10 | FALSE | Downregulated |
| -0,018150838 | 0,966505379 | NSMCE2 | FALSE | Downregulated |
| 0,095307659 | 0,966505379 | OTOA | FALSE | Upregulated |
| 0,012608535 | 0,966505379 | PJA2 | FALSE | Upregulated |
| 0,015716813 | 0,966505379 | PPAPDC2 | FALSE | Upregulated |
| 0,018063532 | 0,966505379 | PSMD14 | FALSE | Upregulated |
| 0,701359619 | 0,966505379 | RNU2-2P | FALSE | Upregulated |
| 0,019950415 | 0,966505379 | STARD3NL | FALSE | Upregulated |
| -0,057789367 | 0,966505379 | SV2A | FALSE | Downregulated |
| 0,014597081 | 0,966505379 | TBC1D14 | FALSE | Upregulated |
| 0,011732914 | 0,966505379 | TIAL1 | FALSE | Upregulated |
| -0,026321663 | 0,966505379 | TMEM27 | FALSE | Downregulated |
| -0,06203257 | 0,966505379 | TNFRSF10C | FALSE | Downregulated |
| -0,01539932 | 0,966505379 | UPRT | FALSE | Downregulated |
| -0,095142198 | 0,966505379 | WBSCR17 | FALSE | Downregulated |
| 0,029040041 | 0,966505379 | XXYLT1 | FALSE | Upregulated |
| -0,018940345 | 0,966561826 | CKS1B | FALSE | Downregulated |
| -0,062555716 | 0,966597852 | ARPP21 | FALSE | Downregulated |
| 0,014182833 | 0,966614474 | KCTD21 | FALSE | Upregulated |
| -0,041328633 | 0,966616472 | SLC22A17 | FALSE | Downregulated |
| 0,019483029 | 0,96662069 | SFXN4 | FALSE | Upregulated |
| -0,038158552 | 0,966658739 | CES5A | FALSE | Downregulated |
| 0,035099399 | 0,966658739 | MAN1C1 | FALSE | Upregulated |
| -0,037645793 | 0,966658739 | MTBP | FALSE | Downregulated |

| | | | | |
|---|---|---|---|---|
| -0,027165699 | 0,966658739 | PARVB | FALSE | Downregulated |
| -0,012995019 | 0,966658739 | ZNF12 | FALSE | Downregulated |
| 0,012090544 | 0,966781296 | CHAMP1 | FALSE | Upregulated |
| 0,013285082 | 0,966919959 | MICU2 | FALSE | Upregulated |
| -0,017127462 | 0,966919959 | TRMT11 | FALSE | Downregulated |
| 0,020660894 | 0,966922112 | CNEP1R1 | FALSE | Upregulated |
| 0,046323865 | 0,966922112 | PDE11A | FALSE | Upregulated |
| -0,031786929 | 0,966922112 | RP11-752G15.6 | FALSE | Downregulated |
| 0,012394253 | 0,966922112 | SNX25 | FALSE | Upregulated |
| 0,014598158 | 0,966922112 | SRPK1 | FALSE | Upregulated |
| 0,013591911 | 0,966950427 | C7orf25 | FALSE | Upregulated |
| 0,015389716 | 0,966950427 | ELP5 | FALSE | Upregulated |
| -0,050737427 | 0,966950427 | FBXO24 | FALSE | Downregulated |
| -0,074500931 | 0,966950427 | JAKMIP2-AS1 | FALSE | Downregulated |
| 0,031341557 | 0,966950427 | KLF4 | FALSE | Upregulated |
| -0,085437381 | 0,966950427 | LOC101928409 | FALSE | Downregulated |
| 0,042242414 | 0,966950427 | RAB26 | FALSE | Upregulated |
| -0,011729087 | 0,96700443 | HIAT1 | FALSE | Downregulated |
| 0,205752482 | 0,96700443 | HIST2H2AA4 | FALSE | Upregulated |
| 0,029234489 | 0,96700443 | IFI27L2 | FALSE | Upregulated |
| -0,012110628 | 0,96700443 | RNF141 | FALSE | Downregulated |
| -0,030119043 | 0,96700443 | SAMD10 | FALSE | Downregulated |
| 0,024141619 | 0,96700443 | THSD4 | FALSE | Upregulated |
| 0,010657841 | 0,967020146 | AAR2 | FALSE | Upregulated |
| 0,021668781 | 0,967163451 | ENDOD1 | FALSE | Upregulated |
| 0,028297164 | 0,967439188 | SCIMP | FALSE | Upregulated |

| | | | | |
|---|---|---|---|---|
| -0,032399745 | 0,9676039 | CLDN3 | FALSE | Downregulated |
| -0,017284845 | 0,9676039 | FAM126A | FALSE | Downregulated |
| -0,015493474 | 0,9676039 | FANCF | FALSE | Downregulated |
| 0,01843246 | 0,9676039 | PTK2B | FALSE | Upregulated |
| 0,085365281 | 0,967718972 | LPAR4 | FALSE | Upregulated |
| 0,013886316 | 0,967989382 | ATF7IP | FALSE | Upregulated |
| 0,057609241 | 0,967989382 | ETV7 | FALSE | Upregulated |
| 0,012875894 | 0,967989382 | GRPEL1 | FALSE | Upregulated |
| 0,027246679 | 0,968158241 | CCNL1 | FALSE | Upregulated |
| -0,05986697 | 0,968158241 | KIF1A | FALSE | Downregulated |
| 0,021789934 | 0,968158241 | POLG2 | FALSE | Upregulated |
| 0,023084037 | 0,968273022 | TTC7B | FALSE | Upregulated |
| -0,041943853 | 0,968460978 | ZNF300 | FALSE | Downregulated |
| 0,024292106 | 0,968487759 | ABHD15 | FALSE | Upregulated |
| 0,039151178 | 0,968487759 | LRRC37A2 | FALSE | Upregulated |
| 0,015561741 | 0,968487759 | PCNT | FALSE | Upregulated |
| -0,026812351 | 0,968487759 | SENP8 | FALSE | Downregulated |
| -0,012993735 | 0,968487759 | SIKE1 | FALSE | Downregulated |
| -0,021561878 | 0,968487759 | WBP1L | FALSE | Downregulated |
| 0,047224623 | 0,96850685 | AC010226.4 | FALSE | Upregulated |
| -0,058212867 | 0,96850685 | ADARB2 | FALSE | Downregulated |
| 0,008110178 | 0,96850685 | ADNP | FALSE | Upregulated |
| -0,016178521 | 0,96850685 | ALMS1 | FALSE | Downregulated |
| 0,052046838 | 0,96850685 | BACH2 | FALSE | Upregulated |
| 0,031899659 | 0,96850685 | CD226 | FALSE | Upregulated |
| -0,016824094 | 0,96850685 | CIC | FALSE | Downregulated |

| | | | | |
|---|---|---|---|---|
| 0,113428069 | 0,96850685 | FOXN4 | FALSE | Upregulated |
| 0,034186703 | 0,96850685 | FOXO1 | FALSE | Upregulated |
| -0,012894578 | 0,96850685 | GCC1 | FALSE | Downregulated |
| 0,02776952 | 0,96850685 | IRF1 | FALSE | Upregulated |
| 0,021940518 | 0,96850685 | LNPEP | FALSE | Upregulated |
| -0,068953149 | 0,96850685 | LOC102723672 | FALSE | Downregulated |
| 0,027724129 | 0,96850685 | LRRC61 | FALSE | Upregulated |
| 0,014857855 | 0,96850685 | MEX3C | FALSE | Upregulated |
| -0,105845914 | 0,96850685 | MYT1L | FALSE | Downregulated |
| -0,020333954 | 0,96850685 | OPTN | FALSE | Downregulated |
| 0,022171184 | 0,96850685 | RHEBP2 | FALSE | Upregulated |
| -0,012860325 | 0,96850685 | RNGTT | FALSE | Downregulated |
| 0,026884881 | 0,96850685 | ROBO1 | FALSE | Upregulated |
| -0,027788612 | 0,96850685 | S100A11 | FALSE | Downregulated |
| 0,036678953 | 0,96850685 | SLC25A33 | FALSE | Upregulated |
| -0,045162931 | 0,96850685 | SPARCL1 | FALSE | Downregulated |
| 0,023542761 | 0,96850685 | SUPT3H | FALSE | Upregulated |
| -0,016939573 | 0,96850685 | TAF10 | FALSE | Downregulated |
| 0,012102935 | 0,96850685 | TMED10 | FALSE | Upregulated |
| -0,483760196 | 0,96850685 | TNNI2 | FALSE | Downregulated |
| -0,021550528 | 0,96850685 | TRIM3 | FALSE | Downregulated |
| 0,038415977 | 0,96850685 | WDR54 | FALSE | Upregulated |
| 0,011239042 | 0,96850685 | YTHDF2 | FALSE | Upregulated |
| 0,029948051 | 0,96850685 | ZFPM2 | FALSE | Upregulated |
| -0,02242976 | 0,96850685 | ZUFSP | FALSE | Downregulated |
| -0,050079019 | 0,968735873 | IDH1-AS1 | FALSE | Downregulated |

| | | | | |
|---|---|---|---|---|
| -0,032315915 | 0,968735873 | JUNB | FALSE | Downregulated |
| 0,010787163 | 0,968804135 | PLAA | FALSE | Upregulated |
| 0,032662623 | 0,968804135 | RAMP2 | FALSE | Upregulated |
| -0,013147262 | 0,96881658 | SPG7 | FALSE | Downregulated |
| 0,056167447 | 0,968888772 | ARPC4-TTLL3 | FALSE | Upregulated |
| -0,079232107 | 0,968888772 | ASIC1 | FALSE | Downregulated |
| -0,031832817 | 0,968888772 | BZRAP1 | FALSE | Downregulated |
| 0,022674111 | 0,968888772 | DPH2 | FALSE | Upregulated |
| -0,021602728 | 0,968888772 | FAM126B | FALSE | Downregulated |
| -0,026409396 | 0,968888772 | FAM180A | FALSE | Downregulated |
| 0,024561559 | 0,968888772 | FZD8 | FALSE | Upregulated |
| -0,046474221 | 0,968888772 | GOLGA8M | FALSE | Downregulated |
| -0,042392206 | 0,968888772 | GSDMB | FALSE | Downregulated |
| 0,028991419 | 0,968888772 | INPP5E | FALSE | Upregulated |
| 0,061608807 | 0,968888772 | LOC101929112 | FALSE | Upregulated |
| -0,031667225 | 0,968888772 | LOC645513 | FALSE | Downregulated |
| -0,656874223 | 0,968888772 | MYLPF | FALSE | Downregulated |
| -0,039281435 | 0,968888772 | PLEKHG7 | FALSE | Downregulated |
| -0,040587683 | 0,968888772 | PMS2P4 | FALSE | Downregulated |
| -0,038724737 | 0,968888772 | PPP1R3C | FALSE | Downregulated |
| 0,014108838 | 0,968888772 | PYGO2 | FALSE | Upregulated |
| 0,015559882 | 0,968888772 | RARRES2 | FALSE | Upregulated |
| 0,065723891 | 0,968888772 | RP11-21L23.3 | FALSE | Upregulated |
| 0,06616737 | 0,968888772 | SUZ12P1 | FALSE | Upregulated |
| -0,020423796 | 0,968888772 | SYT17 | FALSE | Downregulated |
| -0,041524788 | 0,968888772 | ZNF880 | FALSE | Downregulated |

| | | | | |
|---|---|---|---|---|
| 0,022742202 | 0,968975196 | CAB39L | FALSE | Upregulated |
| -0,647880776 | 0,968975196 | DES | FALSE | Downregulated |
| -0,014375305 | 0,968975196 | ERMAP | FALSE | Downregulated |
| -0,050128457 | 0,968975196 | GALNT12 | FALSE | Downregulated |
| -0,020606844 | 0,968975196 | LDHA | FALSE | Downregulated |
| 0,009136694 | 0,968975196 | PRKRIR | FALSE | Upregulated |
| 0,012740822 | 0,968975196 | VHL | FALSE | Upregulated |
| -0,015136125 | 0,968975196 | ZMYM6 | FALSE | Downregulated |
| 0,044934131 | 0,968975196 | ZSWIM5 | FALSE | Upregulated |
| -0,01286725 | 0,969086109 | DHX40 | FALSE | Downregulated |
| 0,047878222 | 0,969086109 | LOC100507424 | FALSE | Upregulated |
| -0,019549719 | 0,969176518 | THOP1 | FALSE | Downregulated |
| -0,017031876 | 0,969199788 | TTC39C | FALSE | Downregulated |
| -0,01300084 | 0,96944255 | NDUFV1 | FALSE | Downregulated |
| -0,03690352 | 0,96944255 | NFKBID | FALSE | Downregulated |
| 0,017515896 | 0,969466423 | GABARAPL1 | FALSE | Upregulated |
| 0,01249513 | 0,969504314 | DDRGK1 | FALSE | Upregulated |
| 0,015046502 | 0,969540209 | DYRK4 | FALSE | Upregulated |
| -0,113754598 | 0,969540209 | RAB40A | FALSE | Downregulated |
| 0,025020915 | 0,969540209 | SLCO3A1 | FALSE | Upregulated |
| -0,029957255 | 0,969540209 | WDR66 | FALSE | Downregulated |
| 0,105278017 | 0,969540671 | KIR3DL1 | FALSE | Upregulated |
| -0,077229382 | 0,969540671 | LOC102723899 | FALSE | Downregulated |
| -0,047600513 | 0,969540671 | PRSS53 | FALSE | Downregulated |
| 0,020503342 | 0,969540671 | TRAPPC6A | FALSE | Upregulated |
| -0,023020619 | 0,969540671 | TTBK2 | FALSE | Downregulated |

| | | | | |
|---|---|---|---|---|
| -0,094143862 | 0,96955374 | LOC101928233 | FALSE | Downregulated |
| 0,015734891 | 0,969648829 | MARVELD2 | FALSE | Upregulated |
| -0,008178701 | 0,969831294 | TSNAX | FALSE | Downregulated |
| -0,060815606 | 0,9699142 | AC010524.2 | FALSE | Downregulated |
| 0,062471799 | 0,96992185 | GDPD4 | FALSE | Upregulated |
| 0,017135731 | 0,96992185 | PEA15 | FALSE | Upregulated |
| -0,085965822 | 0,970073669 | PPIAL4G | FALSE | Downregulated |
| -0,05285092 | 0,97017971 | LOC441179 | FALSE | Downregulated |
| 0,014938344 | 0,97017971 | MTG1 | FALSE | Upregulated |
| 0,010944291 | 0,97017971 | PITPNB | FALSE | Upregulated |
| -0,02904728 | 0,97030761 | PDE4DIP | FALSE | Downregulated |
| 0,021343443 | 0,97035509 | C19orf68 | FALSE | Upregulated |
| -0,05703566 | 0,970385376 | LOC101928768 | FALSE | Downregulated |
| -0,043302459 | 0,970666184 | ALOX12P2 | FALSE | Downregulated |
| 0,024725302 | 0,970666184 | RARG | FALSE | Upregulated |
| 0,060967811 | 0,970666184 | RFPL1S | FALSE | Upregulated |
| 0,015738294 | 0,970687867 | MFSD8 | FALSE | Upregulated |
| -0,636252939 | 0,970700053 | SMYD1 | FALSE | Downregulated |
| 0,080403749 | 0,970821582 | C21orf90 | FALSE | Upregulated |
| 0,025852193 | 0,9709096 | GTF2H2C | FALSE | Upregulated |
| -0,635503795 | 0,9709096 | MYPN | FALSE | Downregulated |
| -0,015909877 | 0,9709096 | RRAS2 | FALSE | Downregulated |
| 0,021061756 | 0,9709096 | THAP8 | FALSE | Upregulated |
| -0,012046163 | 0,970996201 | TWF1 | FALSE | Downregulated |
| -0,015713776 | 0,971240237 | COPE | FALSE | Downregulated |
| -0,026116093 | 0,971240237 | DSG1 | FALSE | Downregulated |

| | | | | |
|---|---|---|---|---|
| -0,026025148 | 0,971240237 | GSAP | FALSE | Downregulated |
| -0,084933585 | 0,971240237 | LOC101927354 | FALSE | Downregulated |
| -0,034576234 | 0,971258398 | TM6SF1 | FALSE | Downregulated |
| 0,099224295 | 0,971279694 | AK5 | FALSE | Upregulated |
| 0,031272435 | 0,971279694 | AKR7L | FALSE | Upregulated |
| 0,016041469 | 0,971279694 | CCNT2 | FALSE | Upregulated |
| -0,010969272 | 0,971279694 | CGGBP1 | FALSE | Downregulated |
| 0,011817425 | 0,971279694 | FGD5-AS1 | FALSE | Upregulated |
| -0,04538348 | 0,971279694 | GPR143 | FALSE | Downregulated |
| 0,026988628 | 0,971279694 | LINC00662 | FALSE | Upregulated |
| -0,0343905 | 0,971279694 | LINC01128 | FALSE | Downregulated |
| -0,101665385 | 0,971279694 | LOC102724732 | FALSE | Downregulated |
| 0,071615692 | 0,971279694 | LOC642648 | FALSE | Upregulated |
| 0,085574465 | 0,971279694 | MIR29C | FALSE | Upregulated |
| -0,037323577 | 0,971279694 | MIR600HG | FALSE | Downregulated |
| -0,068033457 | 0,971279694 | MLC1 | FALSE | Downregulated |
| 0,01575245 | 0,971279694 | MRPL21 | FALSE | Upregulated |
| -0,073266324 | 0,971279694 | PKMYT1 | FALSE | Downregulated |
| 0,010648548 | 0,971279694 | SPPL3 | FALSE | Upregulated |
| 0,02034985 | 0,971279694 | TPM4 | FALSE | Upregulated |
| -0,016274572 | 0,971279694 | ULK2 | FALSE | Downregulated |
| -0,020706393 | 0,971372481 | AADAC | FALSE | Downregulated |
| 0,086039571 | 0,971459397 | SETSIP | FALSE | Upregulated |
| -0,065645038 | 0,97146274 | CASP16 | FALSE | Downregulated |
| -0,093691342 | 0,971506233 | GOLGA8H | FALSE | Downregulated |
| 0,012083926 | 0,971506233 | PPP2R5A | FALSE | Upregulated |

| | | | | |
|---|---|---|---|---|
| 0,072371418 | 0,97169039 | ADAMTS3 | FALSE | Upregulated |
| 0,009708399 | 0,97169039 | BICD2 | FALSE | Upregulated |
| 0,014152039 | 0,97169039 | BIN3 | FALSE | Upregulated |
| -0,016764521 | 0,97169039 | C8G | FALSE | Downregulated |
| 0,021527448 | 0,97169039 | CCDC149 | FALSE | Upregulated |
| 0,015827144 | 0,97169039 | CEACAM1 | FALSE | Upregulated |
| -0,01594156 | 0,97169039 | COQ9 | FALSE | Downregulated |
| 0,014421912 | 0,97169039 | CRYZ | FALSE | Upregulated |
| -0,011319631 | 0,97169039 | CRYZL1 | FALSE | Downregulated |
| -0,029755427 | 0,97169039 | CTU1 | FALSE | Downregulated |
| 0,033760127 | 0,97169039 | DNA2 | FALSE | Upregulated |
| -0,020882093 | 0,97169039 | DUSP23 | FALSE | Downregulated |
| 0,068967292 | 0,97169039 | EPO | FALSE | Upregulated |
| 0,072998838 | 0,97169039 | LOC100653233 | FALSE | Upregulated |
| -0,040937707 | 0,97169039 | LOC284023 | FALSE | Downregulated |
| -0,01692103 | 0,97169039 | MFHAS1 | FALSE | Downregulated |
| 0,033902963 | 0,97169039 | MPPED1 | FALSE | Upregulated |
| -0,085213564 | 0,97169039 | NOV | FALSE | Downregulated |
| 0,008917445 | 0,97169039 | PPP1R8 | FALSE | Upregulated |
| 0,023436961 | 0,97169039 | RBL1 | FALSE | Upregulated |
| -0,079578397 | 0,97169039 | ROCK1P1 | FALSE | Downregulated |
| -0,101942086 | 0,97169039 | RP11-483E7.1 | FALSE | Downregulated |
| -0,014471011 | 0,97169039 | SDHB | FALSE | Downregulated |
| -0,013467606 | 0,97169039 | SERF2 | FALSE | Downregulated |
| 0,058494956 | 0,97169039 | SLC28A3 | FALSE | Upregulated |
| -0,020881711 | 0,97169039 | TMEM180 | FALSE | Downregulated |

| | | | | |
|---|---|---|---|---|
| 0,024543646 | 0,97169039 | TXN | FALSE | Upregulated |
| 0,014055534 | 0,97169039 | ZBTB4 | FALSE | Upregulated |
| 0,01745788 | 0,97169039 | ZNF551 | FALSE | Upregulated |
| 0,013541578 | 0,97169039 | ZNF561 | FALSE | Upregulated |
| 0,032577012 | 0,97169039 | ZNF692 | FALSE | Upregulated |
| 0,058286824 | 0,97177019 | CD200 | FALSE | Upregulated |
| 0,012376677 | 0,97177019 | RAB2B | FALSE | Upregulated |
| -0,612295908 | 0,971906961 | MYOZ2 | FALSE | Downregulated |
| 0,038628463 | 0,972007134 | EFCAB13 | FALSE | Upregulated |
| 0,075484892 | 0,972007134 | GOLGA6L10 | FALSE | Upregulated |
| -0,018759125 | 0,972007134 | RELN | FALSE | Downregulated |
| 0,033258041 | 0,972175426 | HAL | FALSE | Upregulated |
| 0,035094725 | 0,972175426 | KLHL3 | FALSE | Upregulated |
| 0,010611603 | 0,972175426 | NSUN2 | FALSE | Upregulated |
| 0,016252097 | 0,972211544 | ARID3B | FALSE | Upregulated |
| -0,014309599 | 0,972211544 | C16orf52 | FALSE | Downregulated |
| 0,022636255 | 0,972211544 | CEBPA | FALSE | Upregulated |
| 0,012004344 | 0,972211544 | CNTROB | FALSE | Upregulated |
| -0,022927454 | 0,972211544 | CRIPAK | FALSE | Downregulated |
| 0,011317506 | 0,972211544 | E2F4 | FALSE | Upregulated |
| -0,042848734 | 0,972211544 | ENO3 | FALSE | Downregulated |
| 0,024951723 | 0,972211544 | FKBP10 | FALSE | Upregulated |
| -0,055698833 | 0,972211544 | GOLGA8A | FALSE | Downregulated |
| 0,033247255 | 0,972211544 | HCST | FALSE | Upregulated |
| -0,030996269 | 0,972211544 | LINC01003 | FALSE | Downregulated |
| 0,023099716 | 0,972211544 | LOC493754 | FALSE | Upregulated |

| | | | | |
|---|---|---|---|---|
| 0,067437076 | 0,972211544 | LOXL1-AS1 | FALSE | Upregulated |
| -0,019647113 | 0,972211544 | MLPH | FALSE | Downregulated |
| 0,017484853 | 0,972211544 | NUBPL | FALSE | Upregulated |
| -0,028987909 | 0,972211544 | PDE4D | FALSE | Downregulated |
| 0,048964869 | 0,972211544 | PNMAL1 | FALSE | Upregulated |
| 0,043277613 | 0,972211544 | PRKCQ | FALSE | Upregulated |
| 0,013936117 | 0,972211544 | SIAE | FALSE | Upregulated |
| 0,010673127 | 0,972211544 | SNX19 | FALSE | Upregulated |
| -0,043120294 | 0,972211544 | TBC1D3 | FALSE | Downregulated |
| 0,055199326 | 0,972211544 | TGM1 | FALSE | Upregulated |
| 0,009705378 | 0,972211544 | TSG101 | FALSE | Upregulated |
| -0,013673605 | 0,972303795 | MRPL47 | FALSE | Downregulated |
| -0,014525667 | 0,972325312 | CYP2R1 | FALSE | Downregulated |
| 0,016981563 | 0,972517541 | CCDC125 | FALSE | Upregulated |
| -0,056343694 | 0,972517541 | CXorf66 | FALSE | Downregulated |
| -0,04044592 | 0,972517541 | EID3 | FALSE | Downregulated |
| 0,025714764 | 0,972517541 | JOSD2 | FALSE | Upregulated |
| -0,012798682 | 0,972517541 | PLS1 | FALSE | Downregulated |
| -0,016756183 | 0,972517541 | TOR1AIP2 | FALSE | Downregulated |
| -0,015930143 | 0,972536089 | BRAT1 | FALSE | Downregulated |
| -0,017038014 | 0,972635781 | BDH2 | FALSE | Downregulated |
| -0,025735545 | 0,972635781 | LOC101927556 | FALSE | Downregulated |
| 0,046350044 | 0,972635781 | PAX5 | FALSE | Upregulated |
| 0,025828392 | 0,972635781 | SCNN1A | FALSE | Upregulated |
| 0,014028362 | 0,972635781 | TCEAL1 | FALSE | Upregulated |
| 0,013354133 | 0,972635781 | TMEM8A | FALSE | Upregulated |

| | | | | |
|---|---|---|---|---|
| 0,019340948 | 0,972689725 | MIS18A | FALSE | Upregulated |
| -0,013335406 | 0,972689725 | NUTF2 | FALSE | Downregulated |
| 0,037656929 | 0,972689725 | RNF152 | FALSE | Upregulated |
| 0,098689632 | 0,972726409 | SPDYE6 | FALSE | Upregulated |
| -0,018643413 | 0,972793849 | EML1 | FALSE | Downregulated |
| 0,024337821 | 0,972854518 | NDN | FALSE | Upregulated |
| -0,017804537 | 0,972866101 | BCAR3 | FALSE | Downregulated |
| -0,023164653 | 0,972866101 | CERS4 | FALSE | Downregulated |
| -0,019861726 | 0,972866101 | DMRTA1 | FALSE | Downregulated |
| 0,052475813 | 0,972866101 | MPZ | FALSE | Upregulated |
| 0,009651028 | 0,972866101 | PPP2R2D | FALSE | Upregulated |
| -0,027197686 | 0,972880614 | SLC26A6 | FALSE | Downregulated |
| -0,026342594 | 0,972908328 | SERPINA3 | FALSE | Downregulated |
| 0,039938065 | 0,972989668 | BAI3 | FALSE | Upregulated |
| 0,012286211 | 0,972989668 | C2 | FALSE | Upregulated |
| 0,014765412 | 0,972989668 | HNF4G | FALSE | Upregulated |
| 0,012084956 | 0,972989668 | KIAA1279 | FALSE | Upregulated |
| 0,024220593 | 0,972989668 | S1PR1 | FALSE | Upregulated |
| -0,034990552 | 0,972989668 | ZNF767 | FALSE | Downregulated |
| -0,040618556 | 0,973006519 | PRSS12 | FALSE | Downregulated |
| -0,021106277 | 0,973233062 | LOC100506302 | FALSE | Downregulated |
| -0,017342099 | 0,973344704 | AGPAT1 | FALSE | Downregulated |
| -0,033327174 | 0,973344704 | B3GNTL1 | FALSE | Downregulated |
| -0,009631937 | 0,973344704 | HIP1 | FALSE | Downregulated |
| 0,011172825 | 0,973422038 | DNASE1L1 | FALSE | Upregulated |
| 0,087656488 | 0,973422038 | PLEKHN1 | FALSE | Upregulated |

| | | | | |
|---|---|---|---|---|
| -0,019198006 | 0,973422038 | PPP1R13L | FALSE | Downregulated |
| 0,017403524 | 0,973551402 | ZNF160 | FALSE | Upregulated |
| -0,020560693 | 0,973563572 | C15orf61 | FALSE | Downregulated |
| -0,015466302 | 0,973563572 | CCNG1 | FALSE | Downregulated |
| 0,025191745 | 0,973563572 | CHAC2 | FALSE | Upregulated |
| 0,010998796 | 0,973563572 | HCCS | FALSE | Upregulated |
| -0,016528284 | 0,973563572 | IRF2BPL | FALSE | Downregulated |
| -0,021764198 | 0,973563572 | MID1IP1 | FALSE | Downregulated |
| -0,017452222 | 0,973563572 | MSH2 | FALSE | Downregulated |
| 0,043914628 | 0,973563572 | PA2G4P4 | FALSE | Upregulated |
| -0,013234781 | 0,973563572 | POLR3B | FALSE | Downregulated |
| 0,029561972 | 0,973563572 | SLC30A10 | FALSE | Upregulated |
| 0,012674682 | 0,973563572 | ZNF473 | FALSE | Upregulated |
| -0,007843542 | 0,973582684 | ILF3 | FALSE | Downregulated |
| -0,049667475 | 0,973582684 | PIN4P1 | FALSE | Downregulated |
| -0,016234259 | 0,973697621 | IRAK4 | FALSE | Downregulated |
| 0,012917111 | 0,97385156 | CLTB | FALSE | Upregulated |
| 0,016284058 | 0,97385156 | MYH10 | FALSE | Upregulated |
| 0,043763215 | 0,973950713 | CHST4 | FALSE | Upregulated |
| 0,068577432 | 0,973950713 | OXT | FALSE | Upregulated |
| 0,075861355 | 0,973950713 | PPFIA2 | FALSE | Upregulated |
| 0,037114441 | 0,973950713 | SPECC1L-ADORA2A | FALSE | Upregulated |
| -0,030572693 | 0,973969941 | ADAMTS1 | FALSE | Downregulated |
| -0,037520637 | 0,974044402 | VDR | FALSE | Downregulated |
| 0,013634613 | 0,974125782 | LRRC28 | FALSE | Upregulated |
| 0,073241904 | 0,97425574 | ABO | FALSE | Upregulated |

| | | | | |
|---|---|---|---|---|
| -0,008412229 | 0,974340909 | C1orf43 | FALSE | Downregulated |
| -0,025682832 | 0,974340909 | GUSBP1 | FALSE | Downregulated |
| 0,052626891 | 0,974340909 | HEPACAM | FALSE | Upregulated |
| 0,010812919 | 0,974340909 | NF1 | FALSE | Upregulated |
| -0,023808259 | 0,974340909 | SERTAD1 | FALSE | Downregulated |
| 0,011890123 | 0,974340909 | XIAP | FALSE | Upregulated |
| -0,025406637 | 0,974392127 | ENGASE | FALSE | Downregulated |
| 0,023596116 | 0,974392127 | SLC8A1 | FALSE | Upregulated |
| 0,049770225 | 0,974477408 | BAIAP2L2 | FALSE | Upregulated |
| -0,055676747 | 0,974477408 | LOC102723398 | FALSE | Downregulated |
| -0,012435163 | 0,974477408 | MTX1 | FALSE | Downregulated |
| -0,07415685 | 0,974609427 | LOC102724683 | FALSE | Downregulated |
| 0,036351335 | 0,974650932 | AGAP6 | FALSE | Upregulated |
| 0,032033561 | 0,974650932 | SYT7 | FALSE | Upregulated |
| 0,060008087 | 0,974841511 | LOC644090 | FALSE | Upregulated |
| 0,016789747 | 0,974841511 | UQCC2 | FALSE | Upregulated |
| 0,557346796 | 0,974885644 | ANKRD1 | FALSE | Upregulated |
| -0,064122061 | 0,974885644 | MTFR2 | FALSE | Downregulated |
| -0,049914128 | 0,974885644 | SPAG4 | FALSE | Downregulated |
| -0,018427725 | 0,974915943 | PPIL3 | FALSE | Downregulated |
| 0,01008225 | 0,974915943 | PPP1R21 | FALSE | Upregulated |
| 0,015717789 | 0,974915943 | SNX21 | FALSE | Upregulated |
| 0,009988638 | 0,974915943 | UBQLN4 | FALSE | Upregulated |
| 0,024739943 | 0,974915943 | VSTM4 | FALSE | Upregulated |
| 0,009579821 | 0,974962689 | ARMC1 | FALSE | Upregulated |
| -0,020306128 | 0,974962689 | ATP2B4 | FALSE | Downregulated |

| | | | | |
|---|---|---|---|---|
| -0,014078897 | 0,974962689 | AVEN | FALSE | Downregulated |
| 0,061829112 | 0,974962689 | CDH26 | FALSE | Upregulated |
| -0,019016962 | 0,974962689 | GLTPD2 | FALSE | Downregulated |
| 0,042902584 | 0,974962689 | LOX | FALSE | Upregulated |
| 0,030174856 | 0,974962689 | TDO2 | FALSE | Upregulated |
| -0,018068927 | 0,974962689 | TGDS | FALSE | Downregulated |
| 0,011239421 | 0,974962689 | TMEM87A | FALSE | Upregulated |
| -0,012052595 | 0,974962689 | WDR83OS | FALSE | Downregulated |
| -0,012930733 | 0,974978631 | JOSD1 | FALSE | Downregulated |
| -0,013086409 | 0,974978631 | MPDU1 | FALSE | Downregulated |
| -0,010483953 | 0,974978631 | TRIM23 | FALSE | Downregulated |
| 0,051311578 | 0,975093032 | LOC101926977 | FALSE | Upregulated |
| 0,066745674 | 0,975093032 | LOC101930275 | FALSE | Upregulated |
| 0,023207714 | 0,975093032 | RWDD2A | FALSE | Upregulated |
| -0,030865369 | 0,975098516 | AC074138.3 | FALSE | Downregulated |
| -0,05864466 | 0,975098516 | LOC102724169 | FALSE | Downregulated |
| -0,017634801 | 0,975098516 | MBD6 | FALSE | Downregulated |
| 0,011751407 | 0,975098516 | TIMM50 | FALSE | Upregulated |
| -0,029318964 | 0,975099944 | APOA5 | FALSE | Downregulated |
| -0,015996499 | 0,975099944 | KDM2B | FALSE | Downregulated |
| -0,039309942 | 0,975099944 | NUDT10 | FALSE | Downregulated |
| -0,108976764 | 0,975136393 | CCDC162P | FALSE | Downregulated |
| -0,093547663 | 0,975136393 | CHKB-CPT1B | FALSE | Downregulated |
| -0,125920621 | 0,975136393 | DYX1C1 | FALSE | Downregulated |
| -0,026730927 | 0,975136393 | FLJ20021 | FALSE | Downregulated |
| -0,031672655 | 0,975136393 | HTR2B | FALSE | Downregulated |

| | | | | |
|---|---|---|---|---|
| -0,01221341 | 0,975136393 | PRMT3 | FALSE | Downregulated |
| -0,017975653 | 0,975147104 | SLX4IP | FALSE | Downregulated |
| 0,026076325 | 0,975147104 | TRPV1 | FALSE | Upregulated |
| -0,012749519 | 0,975149049 | SLC46A1 | FALSE | Downregulated |
| -0,024823384 | 0,975242366 | ARMCX2 | FALSE | Downregulated |
| -0,014340606 | 0,975242366 | FAM50A | FALSE | Downregulated |
| 0,01131 | 0,975242366 | TXLNA | FALSE | Upregulated |
| -0,020670076 | 0,975276507 | CTR9 | FALSE | Downregulated |
| 0,020393211 | 0,97528455 | CA2 | FALSE | Upregulated |
| 0,00939486 | 0,97528455 | CHTF8 | FALSE | Upregulated |
| 0,016586032 | 0,97528455 | MPHOSPH6 | FALSE | Upregulated |
| 0,010713585 | 0,97528455 | ZDHHC20 | FALSE | Upregulated |
| 0,01682408 | 0,975376045 | APOO | FALSE | Upregulated |
| -0,025050687 | 0,975376045 | BST2 | FALSE | Downregulated |
| 0,059894098 | 0,975376045 | CLECL1 | FALSE | Upregulated |
| 0,010289098 | 0,975376045 | COMMD9 | FALSE | Upregulated |
| 0,007617726 | 0,975376045 | CTCF | FALSE | Upregulated |
| 0,023150703 | 0,975376045 | IDUA | FALSE | Upregulated |
| 0,038044616 | 0,975376045 | LOC101928236 | FALSE | Upregulated |
| -0,022297614 | 0,975376045 | OSBPL10 | FALSE | Downregulated |
| 0,071312033 | 0,975376045 | RP11-7F17.5 | FALSE | Upregulated |
| 0,008560671 | 0,975376045 | ZKSCAN5 | FALSE | Upregulated |
| 0,008136384 | 0,975660358 | RAP1B | FALSE | Upregulated |
| 0,064655682 | 0,97574623 | NTRK3 | FALSE | Upregulated |
| -0,084646532 | 0,975779392 | AC068831.10 | FALSE | Downregulated |
| 0,010794504 | 0,975779392 | SLC25A44 | FALSE | Upregulated |

| | | | | |
|---|---|---|---|---|
| 0,00738141 | 0,975855153 | ATG5 | FALSE | Upregulated |
| -0,012932971 | 0,975855153 | TRMT6 | FALSE | Downregulated |
| 0,015757652 | 0,975967587 | FAM208B | FALSE | Upregulated |
| -0,051949651 | 0,975967587 | FUT8-AS1 | FALSE | Downregulated |
| 0,015142808 | 0,975967587 | IFT140 | FALSE | Upregulated |
| 0,010376759 | 0,975967587 | LAMTOR1 | FALSE | Upregulated |
| -0,019742531 | 0,975967587 | LOC100287896 | FALSE | Downregulated |
| -0,109510625 | 0,975967587 | LOC102723975 | FALSE | Downregulated |
| -0,081405659 | 0,975967587 | OCLM | FALSE | Downregulated |
| 0,019492644 | 0,975967587 | TP53I13 | FALSE | Upregulated |
| -0,011665138 | 0,976135945 | COQ5 | FALSE | Downregulated |
| 0,01723475 | 0,976135945 | RPL31 | FALSE | Upregulated |
| -0,046933762 | 0,976154637 | NDRG4 | FALSE | Downregulated |
| 0,032216208 | 0,976154637 | WWC2-AS2 | FALSE | Upregulated |
| -0,027232147 | 0,97628396 | BHMT | FALSE | Downregulated |
| -0,035493874 | 0,97628396 | CNTD1 | FALSE | Downregulated |
| -0,020860951 | 0,97628396 | HELZ2 | FALSE | Downregulated |
| -0,065668912 | 0,97628396 | HIST1H3H | FALSE | Downregulated |
| -0,011822741 | 0,97628396 | MTRF1 | FALSE | Downregulated |
| -0,045658519 | 0,97628396 | QPCT | FALSE | Downregulated |
| -0,010907171 | 0,97628396 | TIMM44 | FALSE | Downregulated |
| -0,015568124 | 0,976289793 | FAM127B | FALSE | Downregulated |
| -0,015683516 | 0,976289793 | FKBP14 | FALSE | Downregulated |
| 0,025880165 | 0,976289793 | GPX8 | FALSE | Upregulated |
| 0,031989021 | 0,976289793 | SPAG5-AS1 | FALSE | Upregulated |
| 0,009603928 | 0,976667331 | ANKIB1 | FALSE | Upregulated |

| | | | | |
|---|---|---|---|---|
| 0,028784104 | 0,976667331 | CCL15 | FALSE | Upregulated |
| -0,017191613 | 0,976667331 | CHST13 | FALSE | Downregulated |
| -0,13388623 | 0,976667331 | NUP210L | FALSE | Downregulated |
| -0,010346423 | 0,976667331 | RPAP1 | FALSE | Downregulated |
| 0,015923562 | 0,976667331 | SHANK3 | FALSE | Upregulated |
| -0,014091092 | 0,976667331 | THEM4 | FALSE | Downregulated |
| -0,017679207 | 0,976667331 | TIGD7 | FALSE | Downregulated |
| 0,038418673 | 0,976824626 | C10orf35 | FALSE | Upregulated |
| -0,061414792 | 0,97684539 | USP32P1 | FALSE | Downregulated |
| 0,038657931 | 0,977006078 | MEFV | FALSE | Upregulated |
| -0,012096192 | 0,977118678 | ZNF791 | FALSE | Downregulated |
| 0,036241525 | 0,977195326 | AGER | FALSE | Upregulated |
| -0,049577421 | 0,977195326 | LOC100288102 | FALSE | Downregulated |
| -0,06168595 | 0,977195326 | LOC101927842 | FALSE | Downregulated |
| -0,017480844 | 0,977195326 | MIEF2 | FALSE | Downregulated |
| 0,051488591 | 0,977195326 | MIR6843 | FALSE | Upregulated |
| -0,008073086 | 0,977195326 | NUP133 | FALSE | Downregulated |
| -0,012609167 | 0,977195326 | YKT6 | FALSE | Downregulated |
| -0,018931321 | 0,977195326 | ZNF586 | FALSE | Downregulated |
| 0,012116244 | 0,97720164 | ELOF1 | FALSE | Upregulated |
| -0,048561008 | 0,977295036 | LOC101927894 | FALSE | Downregulated |
| -0,011258599 | 0,977340469 | AGO1 | FALSE | Downregulated |
| 0,037408294 | 0,97740451 | GLI3 | FALSE | Upregulated |
| -0,024171409 | 0,977440782 | GCNT1 | FALSE | Downregulated |
| 0,074373801 | 0,977440782 | LOC100996761 | FALSE | Upregulated |
| 0,019549376 | 0,977440782 | LOC101928303 | FALSE | Upregulated |

| | | | | |
|---|---|---|---|---|
| -0,027542881 | 0,977440782 | STARD9 | FALSE | Downregulated |
| 0,01109405 | 0,977440782 | TMED5 | FALSE | Upregulated |
| -0,030693055 | 0,977476922 | CAMK4 | FALSE | Downregulated |
| 0,01336571 | 0,977476922 | DLGAP4 | FALSE | Upregulated |
| 0,038897212 | 0,977612471 | CSNK1A1P1 | FALSE | Upregulated |
| 0,016508704 | 0,977612471 | POP7 | FALSE | Upregulated |
| -0,015590029 | 0,977612471 | SPRED1 | FALSE | Downregulated |
| 0,007419603 | 0,977692628 | AFTPH | FALSE | Upregulated |
| 0,028072397 | 0,977692628 | ARHGAP6 | FALSE | Upregulated |
| 0,00785968 | 0,977692628 | ATP6V1B2 | FALSE | Upregulated |
| -0,063193776 | 0,977692628 | LINC00282 | FALSE | Downregulated |
| 0,021136531 | 0,977692628 | MOGAT2 | FALSE | Upregulated |
| 0,016901504 | 0,977692628 | PRKCA | FALSE | Upregulated |
| 0,008165553 | 0,977692628 | RNF121 | FALSE | Upregulated |
| 0,023054508 | 0,977692628 | SACS | FALSE | Upregulated |
| 0,017653698 | 0,977692628 | SFT2D1 | FALSE | Upregulated |
| 0,009495136 | 0,977692628 | UTP11L | FALSE | Upregulated |
| -0,040798964 | 0,977692628 | ZC3HAV1L | FALSE | Downregulated |
| -0,012628751 | 0,977789317 | BRCC3 | FALSE | Downregulated |
| -0,020943757 | 0,977789317 | LRTOMT | FALSE | Downregulated |
| 0,012787093 | 0,977851213 | ARHGAP26 | FALSE | Upregulated |
| 0,014177128 | 0,977851213 | ATP5S | FALSE | Upregulated |
| -0,014735953 | 0,977851213 | L3HYPDH | FALSE | Downregulated |
| -0,021241655 | 0,977851213 | NUDT13 | FALSE | Downregulated |
| 0,0257829 | 0,977851213 | RBMS2 | FALSE | Upregulated |
| 0,036076855 | 0,977884896 | CDC42EP5 | FALSE | Upregulated |

| | | | | |
|---|---|---|---|---|
| -0,035332217 | 0,977884896 | GABBR1 | FALSE | Downregulated |
| -0,08519845 | 0,977884896 | GPR62 | FALSE | Downregulated |
| -0,013675152 | 0,977884896 | RPS25 | FALSE | Downregulated |
| -0,009298259 | 0,977884896 | STAM | FALSE | Downregulated |
| -0,011995481 | 0,977884896 | UTP15 | FALSE | Downregulated |
| 0,097955888 | 0,977902042 | C2CD4A | FALSE | Upregulated |
| -0,056428297 | 0,977902042 | FAM229A | FALSE | Downregulated |
| 0,058196182 | 0,977902042 | LOC100287467 | FALSE | Upregulated |
| -0,046966434 | 0,977902042 | LOC102724927 | FALSE | Downregulated |
| 0,03780289 | 0,977902042 | MAFF | FALSE | Upregulated |
| 0,015868444 | 0,977902042 | MGST3 | FALSE | Upregulated |
| -0,01257498 | 0,977902042 | MMADHC | FALSE | Downregulated |
| 0,022247264 | 0,977902042 | MXRA5 | FALSE | Upregulated |
| -0,024176517 | 0,977902042 | NAT8 | FALSE | Downregulated |
| -0,028256371 | 0,977902042 | NPHP1 | FALSE | Downregulated |
| -0,077249847 | 0,977902042 | OCA2 | FALSE | Downregulated |
| 0,016758141 | 0,977902042 | PHYHD1 | FALSE | Upregulated |
| -0,113515363 | 0,977902042 | RNF103-CHMP3 | FALSE | Downregulated |
| -0,024356774 | 0,977902042 | RRP7B | FALSE | Downregulated |
| 0,020670391 | 0,977902042 | SOX12 | FALSE | Upregulated |
| 0,037984565 | 0,977902042 | TRIM59 | FALSE | Upregulated |
| 0,021239872 | 0,977902042 | WDR4 | FALSE | Upregulated |
| -0,013484432 | 0,977902042 | WDR83 | FALSE | Downregulated |
| 0,026999777 | 0,977902042 | ZNF471 | FALSE | Upregulated |
| 0,025198401 | 0,977961137 | ACRBP | FALSE | Upregulated |
| 0,015121689 | 0,977961137 | APOL6 | FALSE | Upregulated |

| | | | | |
|---|---|---|---|---|
| -0,018930415 | 0,977961137 | C14orf105 | FALSE | Downregulated |
| 0,066380957 | 0,977961137 | CACNG8 | FALSE | Upregulated |
| 0,018116624 | 0,977961137 | CADM4 | FALSE | Upregulated |
| -0,02532613 | 0,977961137 | COL14A1 | FALSE | Downregulated |
| 0,036967419 | 0,977961137 | DNHD1 | FALSE | Upregulated |
| -0,014661922 | 0,977961137 | DYNLT1 | FALSE | Downregulated |
| 0,01097938 | 0,977961137 | EIF6 | FALSE | Upregulated |
| 0,023907354 | 0,977961137 | HSPA5 | FALSE | Upregulated |
| 0,017157007 | 0,977961137 | IRAK2 | FALSE | Upregulated |
| -0,074049839 | 0,977961137 | KCNA6 | FALSE | Downregulated |
| -0,013733842 | 0,977961137 | MEIS2 | FALSE | Downregulated |
| -0,015859581 | 0,977961137 | SEC24B | FALSE | Downregulated |
| 0,022045691 | 0,977961137 | SGK2 | FALSE | Upregulated |
| -0,011645082 | 0,977968122 | ZNF649 | FALSE | Downregulated |
| 0,011539888 | 0,978006442 | MLF2 | FALSE | Upregulated |
| -0,016409744 | 0,978006442 | POLB | FALSE | Downregulated |
| -0,081762029 | 0,97809381 | GGTLC1 | FALSE | Downregulated |
| 0,012276884 | 0,97809381 | GMNN | FALSE | Upregulated |
| 0,066669698 | 0,97809381 | OSGEPL1-AS1 | FALSE | Upregulated |
| 0,009742463 | 0,978136579 | STK38 | FALSE | Upregulated |
| -0,009461631 | 0,978310247 | NARS2 | FALSE | Downregulated |
| 0,054950488 | 0,978358131 | KIAA1239 | FALSE | Upregulated |
| 0,016764011 | 0,978420021 | ABCB1 | FALSE | Upregulated |
| -0,012647038 | 0,978559088 | DPY19L3 | FALSE | Downregulated |
| 0,02388397 | 0,978559088 | NCF4 | FALSE | Upregulated |
| -0,014217056 | 0,978559088 | PDZD8 | FALSE | Downregulated |

| | | | | |
|---|---|---|---|---|
| 0,013798518 | 0,978559088 | POLI | FALSE | Upregulated |
| -0,012727652 | 0,978670921 | PSMB8 | FALSE | Downregulated |
| -0,491874061 | 0,978689899 | RNU1-2 | FALSE | Downregulated |
| -0,062889699 | 0,9787453 | ADAM33 | FALSE | Downregulated |
| -0,049981115 | 0,978766101 | COLGALT2 | FALSE | Downregulated |
| -0,018909061 | 0,978766101 | TMEM44 | FALSE | Downregulated |
| 0,08307291 | 0,978766101 | YWHAEP1 | FALSE | Upregulated |
| 0,008776246 | 0,978802444 | TBC1D5 | FALSE | Upregulated |
| 0,009549579 | 0,978802444 | UBE2O | FALSE | Upregulated |
| 0,030482569 | 0,978879885 | AHSA2 | FALSE | Upregulated |
| -0,009678134 | 0,978879885 | GABARAPL2 | FALSE | Downregulated |
| -0,013429489 | 0,978879885 | HADH | FALSE | Downregulated |
| 0,028439414 | 0,978879885 | RAB27B | FALSE | Upregulated |
| 0,010232005 | 0,978879885 | TGFBR1 | FALSE | Upregulated |
| -0,034776183 | 0,978879885 | TMEM74 | FALSE | Downregulated |
| -0,020653071 | 0,978890829 | GYG1 | FALSE | Downregulated |
| 0,033789997 | 0,978890829 | LOC400927 | FALSE | Upregulated |
| 0,02530245 | 0,978928393 | SMIM1 | FALSE | Upregulated |
| 0,04125414 | 0,979056288 | TMEM74B | FALSE | Upregulated |
| -0,00689593 | 0,979067074 | AP2A2 | FALSE | Downregulated |
| 0,012930587 | 0,979067074 | APOC2 | FALSE | Upregulated |
| -0,0166082 | 0,979067074 | C7orf60 | FALSE | Downregulated |
| 0,072919743 | 0,979067074 | CCDC144A | FALSE | Upregulated |
| 0,022333957 | 0,979067074 | EFCAB6 | FALSE | Upregulated |
| -0,022885891 | 0,979067074 | FAM86EP | FALSE | Downregulated |
| 0,016730164 | 0,979067074 | GATA6 | FALSE | Upregulated |

| | | | | |
|---|---|---|---|---|
| -0,066226801 | 0,979067074 | GHRL | FALSE | Downregulated |
| -0,056152278 | 0,979067074 | GLB1L2 | FALSE | Downregulated |
| 0,008400769 | 0,979067074 | IKBKAP | FALSE | Upregulated |
| 0,023717656 | 0,979067074 | LOC101928120 | FALSE | Upregulated |
| -0,049035847 | 0,979067074 | LOC101929174 | FALSE | Downregulated |
| -0,023762322 | 0,979067074 | OAS1 | FALSE | Downregulated |
| -0,027966849 | 0,979067074 | OAS3 | FALSE | Downregulated |
| 0,047329156 | 0,979067074 | PLXNA4 | FALSE | Upregulated |
| 0,020458473 | 0,979067074 | RASL11A | FALSE | Upregulated |
| 0,022036696 | 0,979067074 | TMEM241 | FALSE | Upregulated |
| -0,051694538 | 0,979448289 | COX6A2 | FALSE | Downregulated |
| 0,015051894 | 0,979448289 | VANGL1 | FALSE | Upregulated |
| -0,030733451 | 0,979528536 | DKFZp779M0652 | FALSE | Downregulated |
| 0,011516844 | 0,979528536 | LYSMD3 | FALSE | Upregulated |
| -0,011425115 | 0,979528536 | NPC1 | FALSE | Downregulated |
| 0,018130823 | 0,979528536 | UGT2B10 | FALSE | Upregulated |
| -0,014157604 | 0,979616856 | PDE2A | FALSE | Downregulated |
| 0,016849763 | 0,979673808 | FXYD5 | FALSE | Upregulated |
| 0,007106066 | 0,979674017 | PPM1A | FALSE | Upregulated |
| -0,011127793 | 0,979732256 | FAM178A | FALSE | Downregulated |
| 0,059895494 | 0,97975336 | LOC101927511 | FALSE | Upregulated |
| 0,011420377 | 0,97975336 | RPL26L1 | FALSE | Upregulated |
| -0,01413174 | 0,979916396 | ADH1A | FALSE | Downregulated |
| -0,040555106 | 0,979916396 | GADD45G | FALSE | Downregulated |
| -0,01266166 | 0,979916396 | RUFY2 | FALSE | Downregulated |
| 0,062472328 | 0,979916396 | ZNF683 | FALSE | Upregulated |

| | | | | |
|---|---|---|---|---|
| 0,05015991 | 0,980000136 | BNC1 | FALSE | Upregulated |
| 0,017014145 | 0,980000136 | DAPK1 | FALSE | Upregulated |
| -0,010422558 | 0,980000136 | METTL2A | FALSE | Downregulated |
| -0,028751423 | 0,980000136 | NFYC-AS1 | FALSE | Downregulated |
| 0,080729802 | 0,980010906 | SLC7A11 | FALSE | Upregulated |
| 0,008961785 | 0,980123625 | EPS15L1 | FALSE | Upregulated |
| -0,045552393 | 0,980162193 | ADAMTS4 | FALSE | Downregulated |
| 0,010678583 | 0,980190486 | HMOX2 | FALSE | Upregulated |
| 0,03069925 | 0,980400593 | RP11-461O7.1 | FALSE | Upregulated |
| 0,020580249 | 0,980448209 | THBS3 | FALSE | Upregulated |
| 0,009493742 | 0,980450435 | AAAS | FALSE | Upregulated |
| -0,012173681 | 0,980450435 | AP3S2 | FALSE | Downregulated |
| 0,010975421 | 0,980450435 | ATP6V1F | FALSE | Upregulated |
| 0,109996243 | 0,980450435 | BOLA2B | FALSE | Upregulated |
| -0,009024124 | 0,980450435 | CCBL2 | FALSE | Downregulated |
| -0,014418355 | 0,980450435 | CDH5 | FALSE | Downregulated |
| -0,039711424 | 0,980450435 | DNM3 | FALSE | Downregulated |
| 0,046894891 | 0,980450435 | FGF21 | FALSE | Upregulated |
| -0,017881454 | 0,980450435 | GBAS | FALSE | Downregulated |
| -0,039664809 | 0,980450435 | GLUD1P3 | FALSE | Downregulated |
| 0,012624813 | 0,980450435 | IFIH1 | FALSE | Upregulated |
| -0,065804212 | 0,980450435 | LOC102724262 | FALSE | Downregulated |
| -0,066889003 | 0,980450435 | LOC646127 | FALSE | Downregulated |
| -0,020335757 | 0,980450435 | LYSMD2 | FALSE | Downregulated |
| -0,046581684 | 0,980450435 | NPEPL1 | FALSE | Downregulated |
| 0,031314314 | 0,980450435 | NSUN5P2 | FALSE | Upregulated |

| | | | | |
|---|---|---|---|---|
| -0,046614178 | 0,980450435 | PRDM16 | FALSE | Downregulated |
| -0,011398176 | 0,980450435 | RYK | FALSE | Downregulated |
| 0,017935964 | 0,980450435 | SFXN2 | FALSE | Upregulated |
| -0,008600305 | 0,980450435 | SLC39A11 | FALSE | Downregulated |
| 0,007704198 | 0,980450435 | TADA3 | FALSE | Upregulated |
| 0,01575803 | 0,980450435 | TOB2 | FALSE | Upregulated |
| -0,013401977 | 0,980450435 | ZDHHC21 | FALSE | Downregulated |
| 0,028455443 | 0,980450435 | ZNF85 | FALSE | Upregulated |
| 0,013701113 | 0,980450987 | COX7A2 | FALSE | Upregulated |
| 0,009747503 | 0,980451824 | DNAJB6 | FALSE | Upregulated |
| -0,012468093 | 0,980451824 | HACE1 | FALSE | Downregulated |
| -0,033403934 | 0,980451824 | HSF4 | FALSE | Downregulated |
| -0,467119957 | 0,980451824 | LMOD2 | FALSE | Downregulated |
| -0,057330392 | 0,980451824 | ZNF728 | FALSE | Downregulated |
| -0,012471557 | 0,980466168 | CTDSPL2 | FALSE | Downregulated |
| -0,053589754 | 0,980466168 | LOC101928517 | FALSE | Downregulated |
| 0,038565504 | 0,980466168 | NTRK2 | FALSE | Upregulated |
| -0,01291767 | 0,980466168 | PGM2 | FALSE | Downregulated |
| -0,02228187 | 0,980466168 | WNT5A | FALSE | Downregulated |
| -0,012417126 | 0,980466168 | ZMYM5 | FALSE | Downregulated |
| 0,03754094 | 0,980537539 | C8orf88 | FALSE | Upregulated |
| -0,013401001 | 0,980537539 | LHPP | FALSE | Downregulated |
| -0,029453739 | 0,980537539 | PPP4R4 | FALSE | Downregulated |
| 0,009689823 | 0,980537539 | RBM12 | FALSE | Upregulated |
| -0,019043884 | 0,980667188 | LOC102723493 | FALSE | Downregulated |
| -0,011424023 | 0,980695916 | CDK5RAP3 | FALSE | Downregulated |

| | | | | |
|---|---|---|---|---|
| -0,011844 | 0,980695916 | MRPS2 | FALSE | Downregulated |
| 0,033951252 | 0,980695916 | STAG3L5P | FALSE | Upregulated |
| -0,009013368 | 0,980695916 | TFE3 | FALSE | Downregulated |
| -0,058924199 | 0,980799786 | CCDC62 | FALSE | Downregulated |
| -0,02213495 | 0,980799786 | FADS6 | FALSE | Downregulated |
| -0,01020789 | 0,980868745 | GOT2 | FALSE | Downregulated |
| 0,011808552 | 0,980870804 | CHD9 | FALSE | Upregulated |
| 0,009988552 | 0,980870804 | CTDSP2 | FALSE | Upregulated |
| 0,008451647 | 0,980870804 | EIF3I | FALSE | Upregulated |
| -0,011854758 | 0,980870804 | MBD4 | FALSE | Downregulated |
| 0,010194022 | 0,980870804 | MTFR1 | FALSE | Upregulated |
| 0,012217097 | 0,980870804 | NDUFC1 | FALSE | Upregulated |
| 0,008028642 | 0,980870804 | PA2G4 | FALSE | Upregulated |
| 0,02678433 | 0,980870804 | SLC45A4 | FALSE | Upregulated |
| -0,018689286 | 0,980870804 | SPATA33 | FALSE | Downregulated |
| 0,05151846 | 0,980870804 | TMSB4Y | FALSE | Upregulated |
| 0,062436661 | 0,980870804 | TTTY15 | FALSE | Upregulated |
| 0,008201003 | 0,980943587 | KBTBD2 | FALSE | Upregulated |
| -0,021103137 | 0,980943587 | ZFP3 | FALSE | Downregulated |
| 0,016144782 | 0,981016509 | PAPSS1 | FALSE | Upregulated |
| 0,080704522 | 0,981097058 | SERF1B | FALSE | Upregulated |
| -0,008085685 | 0,981197331 | CNOT3 | FALSE | Downregulated |
| 0,041935618 | 0,981197331 | GPR114 | FALSE | Upregulated |
| -0,05203992 | 0,981197331 | HIST1H2AM | FALSE | Downregulated |
| 0,034268276 | 0,981197331 | MAB21L3 | FALSE | Upregulated |
| 0,006811862 | 0,98132296 | ASH2L | FALSE | Upregulated |

| | | | | |
|---|---|---|---|---|
| -0,01519442 | 0,98132296 | SPTBN2 | FALSE | Downregulated |
| 0,016650616 | 0,98132296 | ZNF662 | FALSE | Upregulated |
| -0,006761215 | 0,981394089 | HEATR1 | FALSE | Downregulated |
| 0,055619331 | 0,981394089 | LOC100134868 | FALSE | Upregulated |
| 0,03391365 | 0,981394089 | PTPLA | FALSE | Upregulated |
| -0,034950122 | 0,981443677 | SYT10 | FALSE | Downregulated |
| -0,006971716 | 0,981530246 | PPP2CA | FALSE | Downregulated |
| 0,010564358 | 0,981530246 | PTAR1 | FALSE | Upregulated |
| -0,02325895 | 0,981530246 | RP11-3P17.4 | FALSE | Downregulated |
| 0,012376825 | 0,981580239 | TSEN54 | FALSE | Upregulated |
| -0,007502239 | 0,981677356 | BUD31 | FALSE | Downregulated |
| 0,028433216 | 0,981677356 | POLE2 | FALSE | Upregulated |
| -0,056552977 | 0,981828041 | C4orf48 | FALSE | Downregulated |
| 0,014842838 | 0,981828041 | GPR180 | FALSE | Upregulated |
| -0,404077588 | 0,981828041 | GSTM1 | FALSE | Downregulated |
| -0,048350026 | 0,981828041 | LOC101927886 | FALSE | Downregulated |
| 0,031460383 | 0,981828041 | MMP2 | FALSE | Upregulated |
| -0,0465661 | 0,981828041 | RXRG | FALSE | Downregulated |
| 0,021101537 | 0,981828041 | S100A14 | FALSE | Upregulated |
| -0,019273454 | 0,982007051 | NOS3 | FALSE | Downregulated |
| 0,082278399 | 0,982014606 | AOC1 | FALSE | Upregulated |
| 0,01870314 | 0,982014606 | ASB16-AS1 | FALSE | Upregulated |
| 0,01331748 | 0,982014606 | DHX16 | FALSE | Upregulated |
| -0,007207686 | 0,982014606 | FKBP15 | FALSE | Downregulated |
| 0,019899309 | 0,982014606 | GMNC | FALSE | Upregulated |
| -0,009685214 | 0,982014606 | HAX1 | FALSE | Downregulated |

| | | | | |
|---|---|---|---|---|
| 0,009904917 | 0,982014606 | LDB1 | FALSE | Upregulated |
| -0,047563874 | 0,982014606 | LINC00312 | FALSE | Downregulated |
| -0,065688647 | 0,982014606 | LINC00563 | FALSE | Downregulated |
| 0,039803409 | 0,982014606 | LOC100506022 | FALSE | Upregulated |
| 0,052124353 | 0,982014606 | LOC100506691 | FALSE | Upregulated |
| -0,044109358 | 0,982014606 | LOC102723544 | FALSE | Downregulated |
| 0,008617142 | 0,982014606 | PHKA2 | FALSE | Upregulated |
| -0,030783917 | 0,982014606 | SEMA3B | FALSE | Downregulated |
| 0,018959554 | 0,982014606 | TLN2 | FALSE | Upregulated |
| 0,017727756 | 0,982014606 | TSLP | FALSE | Upregulated |
| 0,017909799 | 0,982014606 | VMAC | FALSE | Upregulated |
| -0,017344799 | 0,982014606 | ZNF10 | FALSE | Downregulated |
| 0,011279991 | 0,982014606 | ZNF202 | FALSE | Upregulated |
| -0,014051995 | 0,982233111 | ASPN | FALSE | Downregulated |
| -0,01842648 | 0,982233111 | ERCC6-PGBD3 | FALSE | Downregulated |
| 0,055695054 | 0,982233111 | LOC101926895 | FALSE | Upregulated |
| 0,009996938 | 0,982233111 | RNASEH2C | FALSE | Upregulated |
| 0,069367128 | 0,982416 | FAM95B1 | FALSE | Upregulated |
| 0,007210535 | 0,982416 | GART | FALSE | Upregulated |
| 0,023400091 | 0,982416 | RAP2C-AS1 | FALSE | Upregulated |
| 0,007140588 | 0,982416 | TBC1D1 | FALSE | Upregulated |
| -0,017865668 | 0,982416 | ZNF550 | FALSE | Downregulated |
| -0,009928257 | 0,982416579 | ALS2 | FALSE | Downregulated |
| 0,049331087 | 0,982416579 | LOC101929887 | FALSE | Upregulated |
| 0,062895468 | 0,982416579 | MDGA2 | FALSE | Upregulated |
| 0,011470905 | 0,982416579 | RPA3 | FALSE | Upregulated |

| | | | | |
|---|---|---|---|---|
| -0,015160893 | 0,982416579 | SLC22A3 | FALSE | Downregulated |
| 0,033915028 | 0,982416579 | STX18-AS1 | FALSE | Upregulated |
| -0,020055126 | 0,982486432 | PRAM1 | FALSE | Downregulated |
| -0,012821233 | 0,982486432 | SAC3D1 | FALSE | Downregulated |
| 0,01146199 | 0,982486432 | TFPT | FALSE | Upregulated |
| -0,058975757 | 0,982532184 | LOC101927157 | FALSE | Downregulated |
| -0,015779797 | 0,98254656 | LOC101929291 | FALSE | Downregulated |
| -0,01569 | 0,982620401 | AAED1 | FALSE | Downregulated |
| 0,015655549 | 0,982620401 | ARMC7 | FALSE | Upregulated |
| -0,012467907 | 0,982620401 | C10orf32 | FALSE | Downregulated |
| -0,029093096 | 0,982620401 | C21orf49 | FALSE | Downregulated |
| -0,006276905 | 0,982620401 | CAPZB | FALSE | Downregulated |
| -0,011119532 | 0,982620401 | CEPT1 | FALSE | Downregulated |
| -0,057198982 | 0,982620401 | COL24A1 | FALSE | Downregulated |
| 0,038850738 | 0,982620401 | COL4A3 | FALSE | Upregulated |
| 0,006108391 | 0,982620401 | DIAPH1 | FALSE | Upregulated |
| 0,049559783 | 0,982620401 | DLGAP3 | FALSE | Upregulated |
| -0,01530953 | 0,982620401 | DMTF1 | FALSE | Downregulated |
| -0,01333746 | 0,982620401 | DOCK5 | FALSE | Downregulated |
| -0,011802513 | 0,982620401 | EHHADH | FALSE | Downregulated |
| 0,043156365 | 0,982620401 | FAM167A | FALSE | Upregulated |
| -0,042568765 | 0,982620401 | FAM83A-AS1 | FALSE | Downregulated |
| -0,016957909 | 0,982620401 | FETUB | FALSE | Downregulated |
| -0,015457923 | 0,982620401 | GEN1 | FALSE | Downregulated |
| 0,012300602 | 0,982620401 | GIMAP1 | FALSE | Upregulated |
| -0,03576892 | 0,982620401 | GOLGA8B | FALSE | Downregulated |

| | | | | |
|---|---|---|---|---|
| 0,016766166 | 0,982620401 | GPR128 | FALSE | Upregulated |
| 0,014751701 | 0,982620401 | HEXIM2 | FALSE | Upregulated |
| -0,022828017 | 0,982620401 | HEY1 | FALSE | Downregulated |
| -0,043876342 | 0,982620401 | HIST1H3E | FALSE | Downregulated |
| 0,015377433 | 0,982620401 | IFI16 | FALSE | Upregulated |
| -0,013981505 | 0,982620401 | KCTD13 | FALSE | Downregulated |
| -0,014651749 | 0,982620401 | KIF20B | FALSE | Downregulated |
| 0,053162911 | 0,982620401 | LINC00624 | FALSE | Upregulated |
| -0,02447191 | 0,982620401 | LOC100996740 | FALSE | Downregulated |
| 0,031875402 | 0,982620401 | LOC101928714 | FALSE | Upregulated |
| -0,029340136 | 0,982620401 | LOC101928745 | FALSE | Downregulated |
| -0,038167083 | 0,982620401 | LOC101929265 | FALSE | Downregulated |
| -0,038035022 | 0,982620401 | LOC101929289 | FALSE | Downregulated |
| 0,046165641 | 0,982620401 | LOC101929479 | FALSE | Upregulated |
| 0,053222676 | 0,982620401 | LOC102724583 | FALSE | Upregulated |
| 0,048435231 | 0,982620401 | LOC284865 | FALSE | Upregulated |
| -0,022670795 | 0,982620401 | LRRC37A3 | FALSE | Downregulated |
| -0,00664664 | 0,982620401 | LRRC41 | FALSE | Downregulated |
| -0,007372951 | 0,982620401 | MCCC1 | FALSE | Downregulated |
| -0,006713723 | 0,982620401 | MMS19 | FALSE | Downregulated |
| -0,043467034 | 0,982620401 | MTRNR2L8 | FALSE | Downregulated |
| -0,017378243 | 0,982620401 | MZF1 | FALSE | Downregulated |
| 0,00967163 | 0,982620401 | NDUFS3 | FALSE | Upregulated |
| 0,011687368 | 0,982620401 | NOL12 | FALSE | Upregulated |
| 0,013808507 | 0,982620401 | PCDH12 | FALSE | Upregulated |
| -0,024608891 | 0,982620401 | PDE6B | FALSE | Downregulated |

| | | | | |
|---|---|---|---|---|
| 0,011199536 | 0,982620401 | POLD4 | FALSE | Upregulated |
| 0,035720758 | 0,982620401 | POLR2J2 | FALSE | Upregulated |
| 0,012215544 | 0,982620401 | PPIH | FALSE | Upregulated |
| 0,022539899 | 0,982620401 | PROZ | FALSE | Upregulated |
| -0,007420974 | 0,982620401 | RASA1 | FALSE | Downregulated |
| 0,016848138 | 0,982620401 | RHOJ | FALSE | Upregulated |
| 0,00693658 | 0,982620401 | RNF11 | FALSE | Upregulated |
| 0,021954184 | 0,982620401 | RNF165 | FALSE | Upregulated |
| -0,042768091 | 0,982620401 | RP11-356I2.4 | FALSE | Downregulated |
| 0,020728297 | 0,982620401 | S100A13 | FALSE | Upregulated |
| 0,008495922 | 0,982620401 | SFXN1 | FALSE | Upregulated |
| 0,017913113 | 0,982620401 | SLC25A36 | FALSE | Upregulated |
| 0,013496033 | 0,982620401 | SLC28A1 | FALSE | Upregulated |
| -0,382127304 | 0,982620401 | SLX1A | FALSE | Downregulated |
| 0,016357315 | 0,982620401 | SNHG1 | FALSE | Upregulated |
| 0,010585009 | 0,982620401 | SRFBP1 | FALSE | Upregulated |
| 0,009198979 | 0,982620401 | SSH1 | FALSE | Upregulated |
| -0,056040409 | 0,982620401 | THSD7B | FALSE | Downregulated |
| -0,104141377 | 0,982620401 | TNFAIP8L2-SCNM1 | FALSE | Downregulated |
| -0,007623878 | 0,982620401 | TOMM22 | FALSE | Downregulated |
| -0,016210797 | 0,982620401 | TP53TG1 | FALSE | Downregulated |
| 0,007452286 | 0,982620401 | TPD52L2 | FALSE | Upregulated |
| -0,033403814 | 0,982620401 | TRIM36 | FALSE | Downregulated |
| 0,043475754 | 0,982620401 | TRPM3 | FALSE | Upregulated |
| -0,037169131 | 0,982620401 | UBE2Q2P3 | FALSE | Downregulated |

| | | | | |
|---|---|---|---|---|
| 0,025571942 | 0,982620401 | ZNF404 | FALSE | Upregulated |
| -0,016503552 | 0,982620401 | ZNF597 | FALSE | Downregulated |
| 0,008692088 | 0,982710443 | ABHD3 | FALSE | Upregulated |
| -0,015560896 | 0,982710443 | AKR1C1 | FALSE | Downregulated |
| -0,019729459 | 0,982710443 | ANGPTL1 | FALSE | Downregulated |
| 0,013512182 | 0,982710443 | APBB1 | FALSE | Upregulated |
| -0,010242245 | 0,982710443 | C4orf29 | FALSE | Downregulated |
| 0,012225065 | 0,982710443 | CENPT | FALSE | Upregulated |
| 0,015066197 | 0,982710443 | CEP112 | FALSE | Upregulated |
| 0,07116264 | 0,982710443 | DDX3Y | FALSE | Upregulated |
| -0,028472164 | 0,982710443 | EFHD1 | FALSE | Downregulated |
| 0,032959484 | 0,982710443 | FAM222A-AS1 | FALSE | Upregulated |
| -0,018424325 | 0,982710443 | IGFBP5 | FALSE | Downregulated |
| -0,009091773 | 0,982710443 | ITPK1 | FALSE | Downregulated |
| -0,007595163 | 0,982710443 | LIN54 | FALSE | Downregulated |
| -0,034320452 | 0,982710443 | LOC101928347 | FALSE | Downregulated |
| -0,043711188 | 0,982710443 | LOC101929465 | FALSE | Downregulated |
| 0,087706789 | 0,982710443 | LOC102724376 | FALSE | Upregulated |
| 0,007664261 | 0,982710443 | LUZP6 | FALSE | Upregulated |
| -0,0069063 | 0,982710443 | MAP4K3 | FALSE | Downregulated |
| -0,005833647 | 0,982710443 | PDPK1 | FALSE | Downregulated |
| 0,05286533 | 0,982710443 | RP11-27K13.3 | FALSE | Upregulated |
| 0,018193485 | 0,982710443 | SCN9A | FALSE | Upregulated |
| -0,009348971 | 0,982710443 | SELO | FALSE | Downregulated |
| -0,01256673 | 0,982710443 | STX16 | FALSE | Downregulated |
| 0,017284596 | 0,982710443 | TRIM16L | FALSE | Upregulated |

| | | | | |
|---|---|---|---|---|
| 0,00720304 | 0,982710443 | WDR45B | FALSE | Upregulated |
| -0,012194571 | 0,982710443 | ZNF232 | FALSE | Downregulated |
| 0,056434033 | 0,982710443 | ZNF286B | FALSE | Upregulated |
| -0,011370204 | 0,982710443 | ZNF30 | FALSE | Downregulated |
| 0,051721423 | 0,982788619 | LOC101929018 | FALSE | Upregulated |
| 0,022481725 | 0,982891891 | ADRA1A | FALSE | Upregulated |
| 0,044006578 | 0,982891891 | IL20RA | FALSE | Upregulated |
| -0,014719876 | 0,982891891 | PRH1-PRR4 | FALSE | Downregulated |
| 0,028868625 | 0,982891891 | RP11-314C16.1 | FALSE | Upregulated |
| 0,017435898 | 0,982891891 | ZNF821 | FALSE | Upregulated |
| 0,035618018 | 0,982903621 | ASIC3 | FALSE | Upregulated |
| 0,015319316 | 0,982903621 | LUC7L | FALSE | Upregulated |
| 0,010956497 | 0,982917761 | ARHGAP19 | FALSE | Upregulated |
| -0,040634651 | 0,982917761 | BNIPL | FALSE | Downregulated |
| -0,02090024 | 0,982917761 | CCDC170 | FALSE | Downregulated |
| 0,036727813 | 0,982917761 | CCR6 | FALSE | Upregulated |
| 0,01077238 | 0,982917761 | DDX31 | FALSE | Upregulated |
| 0,02084546 | 0,982917761 | DSCC1 | FALSE | Upregulated |
| -0,006480829 | 0,982917761 | FYTTD1 | FALSE | Downregulated |
| -0,010616258 | 0,982917761 | INTS5 | FALSE | Downregulated |
| -0,008436379 | 0,982917761 | IP6K2 | FALSE | Downregulated |
| -0,005649393 | 0,982917761 | JAK1 | FALSE | Downregulated |
| 0,016260065 | 0,982917761 | LEAP2 | FALSE | Upregulated |
| 0,016926283 | 0,982917761 | PLCB1 | FALSE | Upregulated |
| -0,05160193 | 0,982917761 | PPP5D1 | FALSE | Downregulated |
| -0,022551302 | 0,982917761 | RFTN2 | FALSE | Downregulated |

| | | | | |
|---|---|---|---|---|
| -0,048125612 | 0,982917761 | RP11-135J2.4 | FALSE | Downregulated |
| 0,059682962 | 0,982917761 | SLC5A11 | FALSE | Upregulated |
| -0,054536677 | 0,982917761 | SULT1C2 | FALSE | Downregulated |
| 0,033956816 | 0,982917761 | TTLL3 | FALSE | Upregulated |
| -0,013648394 | 0,982917761 | ZNF419 | FALSE | Downregulated |
| 0,007905879 | 0,982918029 | WTAP | FALSE | Upregulated |
| 0,030834287 | 0,983189577 | ANKRD36B | FALSE | Upregulated |
| 0,025068251 | 0,983189577 | C1RL-AS1 | FALSE | Upregulated |
| -0,005077796 | 0,983189577 | DNAJA2 | FALSE | Downregulated |
| 0,015928775 | 0,983189577 | DNMT3B | FALSE | Upregulated |
| -0,016622863 | 0,983189577 | FBXL7 | FALSE | Downregulated |
| 0,043744058 | 0,983189577 | GOLGA8I | FALSE | Upregulated |
| 0,012103063 | 0,983189577 | HDAC10 | FALSE | Upregulated |
| 0,034914296 | 0,983189577 | KCNK5 | FALSE | Upregulated |
| -0,111911328 | 0,983189577 | LOC102724621 | FALSE | Downregulated |
| 0,026140172 | 0,983189577 | OSBP2 | FALSE | Upregulated |
| -0,008011954 | 0,983189577 | POLR1D | FALSE | Downregulated |
| -0,016282595 | 0,983189577 | PSMC3IP | FALSE | Downregulated |
| 0,04061911 | 0,983189577 | RBMS3-AS3 | FALSE | Upregulated |
| 0,018188849 | 0,983189577 | UGDH-AS1 | FALSE | Upregulated |
| -0,005819515 | 0,983189577 | UQCRC2 | FALSE | Downregulated |
| 0,030430673 | 0,983189577 | ZYG11A | FALSE | Upregulated |
| 0,016892849 | 0,983358279 | CCDC113 | FALSE | Upregulated |
| -0,015527774 | 0,983376123 | KIAA1841 | FALSE | Downregulated |
| -0,390738577 | 0,983495705 | CACNA1S | FALSE | Downregulated |
| -0,009277872 | 0,983528316 | LRRC57 | FALSE | Downregulated |

| | | | | |
|---|---|---|---|---|
| -0,028019859 | 0,983528316 | PLEK2 | FALSE | Downregulated |
| 0,011909656 | 0,983528316 | SLC2A11 | FALSE | Upregulated |
| -0,01157005 | 0,983528316 | TIFA | FALSE | Downregulated |
| -0,016527067 | 0,983528316 | TMEM51 | FALSE | Downregulated |
| -0,021984127 | 0,98362343 | C1orf226 | FALSE | Downregulated |
| 0,063826211 | 0,98362343 | LOC101926889 | FALSE | Upregulated |
| 0,015116732 | 0,98362343 | RP11-620J15.3 | FALSE | Upregulated |
| 0,008701773 | 0,98362343 | ZCCHC2 | FALSE | Upregulated |
| 0,012573518 | 0,983645441 | DAB2 | FALSE | Upregulated |
| -0,058249292 | 0,983660208 | CALML3 | FALSE | Downregulated |
| -0,050722623 | 0,983660208 | DPY19L2P2 | FALSE | Downregulated |
| -0,009838151 | 0,983660208 | EML4 | FALSE | Downregulated |
| 0,006601899 | 0,983660208 | MRPS30 | FALSE | Upregulated |
| -0,030345778 | 0,983703216 | CREB3L1 | FALSE | Downregulated |
| 0,008556508 | 0,983789343 | HMGN1 | FALSE | Upregulated |
| -0,006218519 | 0,98387695 | ASNSD1 | FALSE | Downregulated |
| 0,01758869 | 0,984057351 | LOC102724528 | FALSE | Upregulated |
| -0,044055676 | 0,98408145 | SAPCD2 | FALSE | Downregulated |
| -0,034527864 | 0,984215667 | IL1B | FALSE | Downregulated |
| -0,023247349 | 0,984215667 | TBX19 | FALSE | Downregulated |
| -0,060740773 | 0,984215667 | TRNK | FALSE | Downregulated |
| -0,011478055 | 0,984228754 | ZNF839 | FALSE | Downregulated |
| -0,026643472 | 0,984231728 | COL4A5 | FALSE | Downregulated |
| -0,008853334 | 0,984231728 | ZFAND1 | FALSE | Downregulated |
| -0,379202701 | 0,984235512 | CASQ1 | FALSE | Downregulated |
| 0,013517141 | 0,984248108 | GTF2IRD2 | FALSE | Upregulated |

| | | | | |
|---|---|---|---|---|
| -0,0126004 | 0,984252431 | EVI5L | FALSE | Downregulated |
| -0,023825373 | 0,984264011 | ACVR2B-AS1 | FALSE | Downregulated |
| -0,009668408 | 0,984264011 | GSDMD | FALSE | Downregulated |
| -0,009248919 | 0,984264011 | TPD52L1 | FALSE | Downregulated |
| 0,007621319 | 0,984356367 | FKBP3 | FALSE | Upregulated |
| -0,022797207 | 0,984356367 | LTK | FALSE | Downregulated |
| 0,013675137 | 0,984356367 | NUTM2A-AS1 | FALSE | Upregulated |
| -0,02349302 | 0,984356367 | OXCT1 | FALSE | Downregulated |
| 0,012992342 | 0,984356367 | PRAP1 | FALSE | Upregulated |
| 0,008542482 | 0,984356367 | SURF1 | FALSE | Upregulated |
| -0,007943684 | 0,984356367 | TAF1 | FALSE | Downregulated |
| 0,037047318 | 0,984356367 | ZNF638-IT1 | FALSE | Upregulated |
| -0,063464324 | 0,984387032 | LOC101928369 | FALSE | Downregulated |
| 0,017635042 | 0,984426427 | FRS3 | FALSE | Upregulated |
| -0,048081164 | 0,984426427 | LOC100506860 | FALSE | Downregulated |
| 0,009936778 | 0,984466718 | ERLEC1 | FALSE | Upregulated |
| -0,010343282 | 0,984466718 | MED17 | FALSE | Downregulated |
| -0,050988094 | 0,984474413 | C17orf72 | FALSE | Downregulated |
| 0,007916966 | 0,984751533 | GUK1 | FALSE | Upregulated |
| 0,019104417 | 0,984751533 | LTBP4 | FALSE | Upregulated |
| -0,04490715 | 0,984753003 | LOC101928803 | FALSE | Downregulated |
| -0,016927194 | 0,984753003 | PACRG | FALSE | Downregulated |
| -0,03270566 | 0,984794796 | LRRC66 | FALSE | Downregulated |
| 0,060513322 | 0,984798934 | C9orf84 | FALSE | Upregulated |
| -0,046259627 | 0,984798934 | LOC101927135 | FALSE | Downregulated |
| -0,009754846 | 0,98492264 | EDEM3 | FALSE | Downregulated |

| | | | | |
|---|---|---|---|---|
| -0,047640754 | 0,98492264 | LOC102724824 | FALSE | Downregulated |
| -0,016387134 | 0,98492264 | LRRC37A | FALSE | Downregulated |
| 0,005159032 | 0,98492264 | OTUD5 | FALSE | Upregulated |
| -0,013357143 | 0,98492264 | RRAS | FALSE | Downregulated |
| -0,011589768 | 0,98492264 | SHE | FALSE | Downregulated |
| 0,010120787 | 0,98492264 | SIK2 | FALSE | Upregulated |
| 0,009960614 | 0,98492264 | SNRNP25 | FALSE | Upregulated |
| 0,004245166 | 0,98492264 | SNX9 | FALSE | Upregulated |
| -0,054547293 | 0,98492264 | TMEM178B | FALSE | Downregulated |
| 0,00907823 | 0,98492264 | WRB | FALSE | Upregulated |
| -0,013295168 | 0,984929656 | ATP10D | FALSE | Downregulated |
| 0,008277558 | 0,984929656 | CCDC53 | FALSE | Upregulated |
| 0,010946059 | 0,984929656 | CSNK1E | FALSE | Upregulated |
| 0,006946002 | 0,984929656 | DUSP11 | FALSE | Upregulated |
| 0,035301691 | 0,984929656 | IMPG2 | FALSE | Upregulated |
| 0,011571249 | 0,984929656 | LIN52 | FALSE | Upregulated |
| 0,036055717 | 0,984929656 | LOC101928667 | FALSE | Upregulated |
| 0,061110576 | 0,984929656 | LOC102724636 | FALSE | Upregulated |
| -0,009255966 | 0,984929656 | LRCH3 | FALSE | Downregulated |
| 0,006629435 | 0,984929656 | PEX14 | FALSE | Upregulated |
| 0,022843088 | 0,984929656 | PLP2 | FALSE | Upregulated |
| 0,047904542 | 0,984929656 | RP5-899E9.1 | FALSE | Upregulated |
| 0,011760387 | 0,984929656 | SLC13A5 | FALSE | Upregulated |
| 0,010527965 | 0,984929656 | SLC40A1 | FALSE | Upregulated |
| 0,036495117 | 0,984929656 | SNORA63 | FALSE | Upregulated |
| 0,040072484 | 0,984929656 | TCAM1P | FALSE | Upregulated |

| | | | | |
|---|---|---|---|---|
| -0,359781397 | 0,984929656 | TNNC1 | FALSE | Downregulated |
| -0,004807215 | 0,984929656 | TUBGCP2 | FALSE | Downregulated |
| 0,015417413 | 0,984929656 | WSCD1 | FALSE | Upregulated |
| 0,006231701 | 0,984929656 | ZCCHC3 | FALSE | Upregulated |
| -0,016382211 | 0,984929656 | ZNF185 | FALSE | Downregulated |
| -0,02326713 | 0,984944135 | CCNE2 | FALSE | Downregulated |
| -0,032536559 | 0,984944135 | SFTPD | FALSE | Downregulated |
| -0,014793077 | 0,984944135 | SLC23A2 | FALSE | Downregulated |
| 0,034915741 | 0,985278463 | MFI2-AS1 | FALSE | Upregulated |
| 0,018260772 | 0,985367133 | APBA1 | FALSE | Upregulated |
| -0,02708693 | 0,985367133 | CDC25A | FALSE | Downregulated |
| 0,150314199 | 0,985367133 | NPTN-IT1 | FALSE | Upregulated |
| -0,022551138 | 0,985388735 | PELI2 | FALSE | Downregulated |
| -0,006598404 | 0,985459297 | DDX59 | FALSE | Downregulated |
| -0,040554925 | 0,985671988 | CCSER1 | FALSE | Downregulated |
| 0,026995539 | 0,985742369 | AMN | FALSE | Upregulated |
| 0,032072386 | 0,985792639 | C6orf132 | FALSE | Upregulated |
| -0,013228797 | 0,985792639 | DENND3 | FALSE | Downregulated |
| -0,037844322 | 0,985792639 | LINC00704 | FALSE | Downregulated |
| 0,034503538 | 0,985792639 | LOC101927963 | FALSE | Upregulated |
| -0,012372832 | 0,985792639 | SMG1 | FALSE | Downregulated |
| -0,005181845 | 0,985801155 | PMPCB | FALSE | Downregulated |
| -0,040017452 | 0,985986403 | SCUBE3 | FALSE | Downregulated |
| -0,007884763 | 0,985986403 | SEPT8 | FALSE | Downregulated |
| 0,014391817 | 0,986067895 | RAD54L2 | FALSE | Upregulated |
| -0,008318937 | 0,986469163 | AKAP10 | FALSE | Downregulated |

| | | | | |
|---|---|---|---|---|
| -0,058536052 | 0,986629692 | ARHGEF34P | FALSE | Downregulated |
| -0,010466586 | 0,986629692 | MAOA | FALSE | Downregulated |
| 0,009166025 | 0,986785212 | ANKRD11 | FALSE | Upregulated |
| -0,021797416 | 0,986785212 | CPLX1 | FALSE | Downregulated |
| -0,01107721 | 0,986785212 | EPB41L4B | FALSE | Downregulated |
| -0,00690773 | 0,986785212 | MBTPS2 | FALSE | Downregulated |
| 0,010226835 | 0,986785212 | MTDH | FALSE | Upregulated |
| -0,005438483 | 0,986785212 | RIOK3 | FALSE | Downregulated |
| 0,016372611 | 0,986785212 | ZNF540 | FALSE | Upregulated |
| -0,030184422 | 0,986853873 | CLSTN2 | FALSE | Downregulated |
| 0,014380762 | 0,986853873 | DNAJC3-AS1 | FALSE | Upregulated |
| -0,007948714 | 0,986875033 | C17orf75 | FALSE | Downregulated |
| 0,00929616 | 0,986875033 | CEP170 | FALSE | Upregulated |
| -0,011929705 | 0,986875033 | HEXDC | FALSE | Downregulated |
| 0,036674342 | 0,986875033 | IAPP | FALSE | Upregulated |
| -0,030062077 | 0,986875033 | LOC283887 | FALSE | Downregulated |
| 0,011399336 | 0,986875033 | MS4A6A | FALSE | Upregulated |
| 0,007080442 | 0,986875033 | PPP3CC | FALSE | Upregulated |
| -0,016574083 | 0,986875033 | RDH5 | FALSE | Downregulated |
| -0,014655223 | 0,986875033 | SYT9 | FALSE | Downregulated |
| 0,015982533 | 0,986890813 | ARHGAP5-AS1 | FALSE | Upregulated |
| 0,008415633 | 0,986890813 | ISOC2 | FALSE | Upregulated |
| -0,023785214 | 0,987091691 | ST6GALNAC2 | FALSE | Downregulated |
| 0,009998608 | 0,987107129 | CCDC85C | FALSE | Upregulated |
| 0,022570725 | 0,987107129 | NGF | FALSE | Upregulated |
| -0,011337471 | 0,987107129 | SLC25A14 | FALSE | Downregulated |

| | | | | |
|---|---|---|---|---|
| -0,0084679 | 0,987107129 | SYNJ2 | FALSE | Downregulated |
| 0,313050522 | 0,987107129 | TNNI1 | FALSE | Upregulated |
| 0,0090224 | 0,987126743 | PIGU | FALSE | Upregulated |
| -0,037350705 | 0,987150389 | FOXP3 | FALSE | Downregulated |
| 0,03021199 | 0,987250202 | ANKRD65 | FALSE | Upregulated |
| -0,031657728 | 0,987250202 | COL15A1 | FALSE | Downregulated |
| -0,009568374 | 0,987250202 | FAM127A | FALSE | Downregulated |
| -0,03410313 | 0,987250202 | FBXL2 | FALSE | Downregulated |
| 0,010056722 | 0,987250202 | IFNGR2 | FALSE | Upregulated |
| 0,007227814 | 0,987250202 | KIAA2026 | FALSE | Upregulated |
| 0,031990854 | 0,987250202 | LOC100287175 | FALSE | Upregulated |
| 0,027686798 | 0,987250202 | LOC100507054 | FALSE | Upregulated |
| 0,015906371 | 0,987250202 | LOC101928431 | FALSE | Upregulated |
| -0,034730568 | 0,987250202 | LOC102723597 | FALSE | Downregulated |
| 0,054315799 | 0,987250202 | LOXHD1 | FALSE | Upregulated |
| 0,00682445 | 0,987250202 | MED23 | FALSE | Upregulated |
| -0,006876528 | 0,987250202 | MTMR1 | FALSE | Downregulated |
| -0,015968718 | 0,987250202 | MUC20 | FALSE | Downregulated |
| -0,015316034 | 0,987250202 | NXT1 | FALSE | Downregulated |
| 0,004861801 | 0,987250202 | OSBPL2 | FALSE | Upregulated |
| -0,00816651 | 0,987250202 | PAK1 | FALSE | Downregulated |
| -0,010738069 | 0,987250202 | POLR2M | FALSE | Downregulated |
| 0,027448425 | 0,987250202 | RP11-509J21.1 | FALSE | Upregulated |
| -0,012881757 | 0,987250202 | TCEAL3 | FALSE | Downregulated |
| -0,007452587 | 0,987250202 | TIMELESS | FALSE | Downregulated |
| -0,014296842 | 0,987250202 | UBXN11 | FALSE | Downregulated |

| | | | | |
|---|---|---|---|---|
| -0,010372584 | 0,987250202 | ZNF558 | FALSE | Downregulated |
| -0,040346535 | 0,987250202 | ZNF860 | FALSE | Downregulated |
| 0,015499804 | 0,987253708 | PTRH1 | FALSE | Upregulated |
| -0,052382781 | 0,987279546 | AC104653.1 | FALSE | Downregulated |
| -0,018684443 | 0,987279546 | PTGFR | FALSE | Downregulated |
| 0,005067861 | 0,987279546 | TTC37 | FALSE | Upregulated |
| 0,010153001 | 0,987639973 | NMNAT3 | FALSE | Upregulated |
| 0,006903371 | 0,987667539 | NR3C1 | FALSE | Upregulated |
| -0,008855218 | 0,987667539 | ZNF552 | FALSE | Downregulated |
| -0,02703849 | 0,987713209 | TNF | FALSE | Downregulated |
| -0,006098482 | 0,987713209 | TOPBP1 | FALSE | Downregulated |
| -0,005703832 | 0,987845357 | ALKBH3 | FALSE | Downregulated |
| 0,017918179 | 0,987845357 | BBC3 | FALSE | Upregulated |
| 0,016856885 | 0,987845357 | CCDC40 | FALSE | Upregulated |
| 0,044916088 | 0,987845357 | DENND5B-AS1 | FALSE | Upregulated |
| 0,008783047 | 0,987845357 | ESPN | FALSE | Upregulated |
| 0,007061755 | 0,987845357 | EXOC2 | FALSE | Upregulated |
| -0,011590964 | 0,987845357 | HSPA1A | FALSE | Downregulated |
| 0,019559848 | 0,987845357 | IGSF23 | FALSE | Upregulated |
| 0,015721069 | 0,987845357 | LIMK2 | FALSE | Upregulated |
| 0,01797083 | 0,987845357 | MX2 | FALSE | Upregulated |
| 0,007949115 | 0,987845357 | OTUD6B | FALSE | Upregulated |
| -0,106499669 | 0,987845357 | RP11-513O17.2-2 | FALSE | Downregulated |
| 0,040419971 | 0,987845357 | RP4-562J12.2 | FALSE | Upregulated |
| -0,009473943 | 0,987845357 | SLC39A5 | FALSE | Downregulated |
| 0,132751881 | 0,987845357 | SMTNL1 | FALSE | Upregulated |

| | | | | |
|---|---|---|---|---|
| -0,005290262 | 0,987845357 | SUMO2 | FALSE | Downregulated |
| 0,022139352 | 0,987845357 | TBX6 | FALSE | Upregulated |
| 0,004906041 | 0,987845357 | THOC5 | FALSE | Upregulated |
| -0,019424868 | 0,987845357 | TRIL | FALSE | Downregulated |
| -0,01560909 | 0,987845357 | TTC32 | FALSE | Downregulated |
| -0,004008891 | 0,987845357 | UBE4B | FALSE | Downregulated |
| 0,004564342 | 0,987845357 | USP24 | FALSE | Upregulated |
| 0,004180017 | 0,987845357 | VDAC1 | FALSE | Upregulated |
| 0,02686076 | 0,987845357 | ZNF391 | FALSE | Upregulated |
| -0,01454271 | 0,987845357 | ZNF532 | FALSE | Downregulated |
| 0,008896513 | 0,987845357 | ZNF697 | FALSE | Upregulated |
| -0,022538392 | 0,987874071 | GUSBP4 | FALSE | Downregulated |
| -0,01072307 | 0,987896793 | CXCL12 | FALSE | Downregulated |
| 0,008960606 | 0,987915928 | GNA14 | FALSE | Upregulated |
| 0,021963699 | 0,988068312 | ANXA2R | FALSE | Upregulated |
| -0,008569751 | 0,988068312 | BOD1L1 | FALSE | Downregulated |
| 0,010337209 | 0,988068312 | C20orf112 | FALSE | Upregulated |
| 0,038942318 | 0,988068312 | CISH | FALSE | Upregulated |
| -0,005465175 | 0,988068312 | DNMBP | FALSE | Downregulated |
| 0,010868685 | 0,988068312 | FBXO6 | FALSE | Upregulated |
| 0,00902593 | 0,988068312 | FUOM | FALSE | Upregulated |
| 0,021098432 | 0,988068312 | GPR37 | FALSE | Upregulated |
| -0,010031316 | 0,988068312 | HEMK1 | FALSE | Downregulated |
| 0,00937392 | 0,988068312 | IL11RA | FALSE | Upregulated |
| -0,004641223 | 0,988068312 | KDM5A | FALSE | Downregulated |
| -0,025240464 | 0,988068312 | LOC100506036 | FALSE | Downregulated |

| | | | | |
|---|---|---|---|---|
| -0,010257346 | 0,988068312 | LOC100506634 | FALSE | Downregulated |
| -0,017968889 | 0,988068312 | LURAP1L | FALSE | Downregulated |
| -0,057576185 | 0,988068312 | SAA2-SAA4 | FALSE | Downregulated |
| 0,013504071 | 0,988068312 | SCAI | FALSE | Upregulated |
| -0,006945214 | 0,988068312 | SGPP1 | FALSE | Downregulated |
| -0,023443245 | 0,988068312 | SLC2A4 | FALSE | Downregulated |
| -0,006358259 | 0,988068312 | SLC30A7 | FALSE | Downregulated |
| -0,009741384 | 0,988068312 | SRSF6 | FALSE | Downregulated |
| 0,010529493 | 0,988068312 | TMEM170A | FALSE | Upregulated |
| 0,011361754 | 0,988068312 | ZNF417 | FALSE | Upregulated |
| -0,009712278 | 0,988140405 | C4BPB | FALSE | Downregulated |
| 0,28683066 | 0,988144468 | LDB3 | FALSE | Upregulated |
| 0,008580338 | 0,988201109 | ACSF2 | FALSE | Upregulated |
| -0,01072514 | 0,988201109 | PLBD2 | FALSE | Downregulated |
| 0,037122041 | 0,98839229 | C1QTNF9 | FALSE | Upregulated |
| -0,012312418 | 0,98839229 | ETS2 | FALSE | Downregulated |
| -0,018802194 | 0,98839229 | NLRP11 | FALSE | Downregulated |
| -0,28749495 | 0,98839229 | TTN | FALSE | Downregulated |
| -0,035021668 | 0,988638323 | GRIK2 | FALSE | Downregulated |
| 0,006093727 | 0,988661131 | CNOT6L | FALSE | Upregulated |
| -0,02153242 | 0,988661131 | RAD51AP2 | FALSE | Downregulated |
| 0,006719177 | 0,988661131 | RC3H1 | FALSE | Upregulated |
| 0,071010897 | 0,988661131 | TRNAK6 | FALSE | Upregulated |
| 0,009101616 | 0,988722107 | ZNF778 | FALSE | Upregulated |
| -0,020178141 | 0,988785906 | AIF1L | FALSE | Downregulated |
| -0,023810071 | 0,988785906 | ALMS1P | FALSE | Downregulated |

| | | | | |
|---|---|---|---|---|
| -0,00902652 | 0,988785906 | BLOC1S3 | FALSE | Downregulated |
| 0,00679656 | 0,988785906 | CDK11B | FALSE | Upregulated |
| 0,005012145 | 0,988785906 | DYM | FALSE | Upregulated |
| -0,009420504 | 0,988785906 | F12 | FALSE | Downregulated |
| -0,013891192 | 0,988785906 | FLJ46906 | FALSE | Downregulated |
| -0,012978773 | 0,988785906 | IFIT3 | FALSE | Downregulated |
| -0,010326539 | 0,988785906 | IL22RA1 | FALSE | Downregulated |
| -0,009470867 | 0,988785906 | JAK2 | FALSE | Downregulated |
| 0,023245934 | 0,988785906 | KCNS3 | FALSE | Upregulated |
| 0,010613206 | 0,988785906 | NAPEPLD | FALSE | Upregulated |
| 0,013497236 | 0,988785906 | TMOD2 | FALSE | Upregulated |
| 0,006972866 | 0,988785906 | TTC8 | FALSE | Upregulated |
| 0,02336946 | 0,988785906 | USP43 | FALSE | Upregulated |
| 0,009927637 | 0,988785906 | ZNF543 | FALSE | Upregulated |
| -0,006850022 | 0,98880142 | SLC37A3 | FALSE | Downregulated |
| -0,022180036 | 0,988811495 | MEF2B | FALSE | Downregulated |
| -0,011622477 | 0,988811495 | PMS2L2 | FALSE | Downregulated |
| -0,034486652 | 0,988811495 | RP11-1134I14.8 | FALSE | Downregulated |
| 0,006790715 | 0,988868098 | RPS6KB2 | FALSE | Upregulated |
| 0,006744721 | 0,988884236 | DIRC2 | FALSE | Upregulated |
| -0,045164521 | 0,988977845 | RN7SK | FALSE | Downregulated |
| 0,003278701 | 0,989032432 | MORF4L1 | FALSE | Upregulated |
| -0,009244151 | 0,989189404 | ZNF224 | FALSE | Downregulated |
| -0,02748745 | 0,989220894 | ARNTL | FALSE | Downregulated |
| -0,007929749 | 0,989220894 | CD40 | FALSE | Downregulated |
| -0,031109183 | 0,989220894 | GATM-AS1 | FALSE | Downregulated |

| | | | | |
|---|---|---|---|---|
| -0,054918842 | 0,989220894 | GXYLT2 | FALSE | Downregulated |
| 0,028502036 | 0,989220894 | MDGA1 | FALSE | Upregulated |
| 0,038038846 | 0,989336827 | CECR6 | FALSE | Upregulated |
| 0,006099 | 0,989336827 | DPP8 | FALSE | Upregulated |
| 0,030570383 | 0,989336827 | IL23A | FALSE | Upregulated |
| -0,006623347 | 0,989336827 | KIAA0586 | FALSE | Downregulated |
| -0,004456151 | 0,989336827 | MED6 | FALSE | Downregulated |
| 0,019673831 | 0,989336827 | NPAS2 | FALSE | Upregulated |
| -0,008042653 | 0,989336827 | NR1I3 | FALSE | Downregulated |
| -0,006857686 | 0,989336827 | TRAM1 | FALSE | Downregulated |
| -0,007953171 | 0,989336827 | YIPF1 | FALSE | Downregulated |
| -0,02753318 | 0,989388973 | LOC101929115 | FALSE | Downregulated |
| 0,246406064 | 0,989667142 | ANKRD2 | FALSE | Upregulated |
| -0,023741889 | 0,989667142 | AOC4P | FALSE | Downregulated |
| -0,014250838 | 0,989667142 | ARHGEF37 | FALSE | Downregulated |
| 0,016204717 | 0,989667142 | ATG16L2 | FALSE | Upregulated |
| 0,010104623 | 0,989667142 | BCORL1 | FALSE | Upregulated |
| -0,027295712 | 0,989667142 | BMX | FALSE | Downregulated |
| -0,018380963 | 0,989667142 | C11orf45 | FALSE | Downregulated |
| -0,028512653 | 0,989667142 | C17orf49 | FALSE | Downregulated |
| -0,005747533 | 0,989667142 | C1orf123 | FALSE | Downregulated |
| 0,025307648 | 0,989667142 | CCL3 | FALSE | Upregulated |
| 0,005915582 | 0,989667142 | CD99 | FALSE | Upregulated |
| -0,01758503 | 0,989667142 | DBF4B | FALSE | Downregulated |
| 0,005465918 | 0,989667142 | DDX39B | FALSE | Upregulated |
| 0,009500217 | 0,989667142 | ELMOD3 | FALSE | Upregulated |

| | | | | |
|---|---|---|---|---|
| 0,008388056 | 0,989667142 | GEMIN2 | FALSE | Upregulated |
| -0,007152786 | 0,989667142 | HSD11B1 | FALSE | Downregulated |
| -0,02264539 | 0,989667142 | LINC00470 | FALSE | Downregulated |
| -0,034435387 | 0,989667142 | LOC101928123 | FALSE | Downregulated |
| 0,029515255 | 0,989667142 | LOC102723513 | FALSE | Upregulated |
| 0,024178491 | 0,989667142 | LOC102724562 | FALSE | Upregulated |
| 0,010289627 | 0,989667142 | LPP-AS2 | FALSE | Upregulated |
| -0,007297053 | 0,989667142 | MARCH2 | FALSE | Downregulated |
| -0,014366661 | 0,989667142 | MECOM | FALSE | Downregulated |
| -0,023990907 | 0,989667142 | MLLT11 | FALSE | Downregulated |
| 0,005605862 | 0,989667142 | MLST8 | FALSE | Upregulated |
| -0,006415512 | 0,989667142 | MOCS3 | FALSE | Downregulated |
| -0,036007824 | 0,989667142 | MROH7-TTC4 | FALSE | Downregulated |
| 0,00644656 | 0,989667142 | NUCB1 | FALSE | Upregulated |
| -0,026714692 | 0,989667142 | PDE4C | FALSE | Downregulated |
| 0,00515535 | 0,989667142 | PHF2 | FALSE | Upregulated |
| 0,011053593 | 0,989667142 | POLR3G | FALSE | Upregulated |
| 0,022902016 | 0,989667142 | RAD51 | FALSE | Upregulated |
| -0,007599494 | 0,989667142 | RPL23AP53 | FALSE | Downregulated |
| -0,011070712 | 0,989667142 | SLC22A23 | FALSE | Downregulated |
| -0,007317757 | 0,989667142 | SLC25A4 | FALSE | Downregulated |
| -0,007307537 | 0,989667142 | STRADB | FALSE | Downregulated |
| 0,006816944 | 0,989667142 | TBCB | FALSE | Upregulated |
| 0,015657007 | 0,989667142 | TPM1 | FALSE | Upregulated |
| 0,011165412 | 0,989667142 | TPT1-AS1 | FALSE | Upregulated |
| 0,013125664 | 0,989667142 | TRIM62 | FALSE | Upregulated |

| | | | | |
|---|---|---|---|---|
| -0,003020433 | 0,989667142 | XPO7 | FALSE | Downregulated |
| -0,011209932 | 0,989667142 | ZNF467 | FALSE | Downregulated |
| -0,006849669 | 0,989794643 | OGFOD1 | FALSE | Downregulated |
| 0,01789697 | 0,989891844 | CFHR3 | FALSE | Upregulated |
| 0,033628886 | 0,989891844 | LOC100996425 | FALSE | Upregulated |
| 0,00543918 | 0,989891844 | WDR55 | FALSE | Upregulated |
| -0,009309887 | 0,990051361 | ESAM | FALSE | Downregulated |
| 0,032753025 | 0,990051361 | FBXW10 | FALSE | Upregulated |
| -0,034670918 | 0,990051361 | LOC101929280 | FALSE | Downregulated |
| -0,007044167 | 0,990051361 | PMS1 | FALSE | Downregulated |
| 0,005629539 | 0,990051361 | TCTA | FALSE | Upregulated |
| -0,017574064 | 0,990310197 | PCOLCE2 | FALSE | Downregulated |
| 0,014166042 | 0,990465329 | CRISPLD1 | FALSE | Upregulated |
| 0,007990188 | 0,990465329 | MET | FALSE | Upregulated |
| 0,024896119 | 0,990465329 | SLC22A10 | FALSE | Upregulated |
| -0,267986896 | 0,990465329 | TNNT1 | FALSE | Downregulated |
| -0,011904943 | 0,990513149 | PROCR | FALSE | Downregulated |
| 0,017164085 | 0,990580863 | LOC101928762 | FALSE | Upregulated |
| -0,011609107 | 0,990607463 | C6orf1 | FALSE | Downregulated |
| -0,007243997 | 0,990607463 | CBX6 | FALSE | Downregulated |
| 0,030097026 | 0,990607463 | GRIK1 | FALSE | Upregulated |
| 0,0315178 | 0,990607463 | LOC101929381 | FALSE | Upregulated |
| 0,004709855 | 0,990607463 | MFAP3 | FALSE | Upregulated |
| 0,009397489 | 0,990607463 | PIK3C2B | FALSE | Upregulated |
| 0,024742316 | 0,990607463 | STAC | FALSE | Upregulated |
| 0,010433885 | 0,990607463 | TNFAIP8L2 | FALSE | Upregulated |

| | | | | |
|---|---|---|---|---|
| 0,005349394 | 0,990607463 | TRIM38 | FALSE | Upregulated |
| 0,005588362 | 0,990683165 | SLC39A1 | FALSE | Upregulated |
| 0,041087105 | 0,990721847 | NPIPB15 | FALSE | Upregulated |
| -0,009897919 | 0,990721847 | SFT2D2 | FALSE | Downregulated |
| -0,007575731 | 0,990845958 | FAM213A | FALSE | Downregulated |
| -0,005186936 | 0,990845958 | GOLGA5 | FALSE | Downregulated |
| 0,010609524 | 0,990845958 | SMUG1 | FALSE | Upregulated |
| -0,005991646 | 0,990845958 | TEAD3 | FALSE | Downregulated |
| -0,029235159 | 0,990847008 | LRP2 | FALSE | Downregulated |
| -0,005269472 | 0,990847008 | MKRN2 | FALSE | Downregulated |
| 0,017518571 | 0,990896936 | A4GALT | FALSE | Upregulated |
| -0,006305468 | 0,990896936 | E2F6 | FALSE | Downregulated |
| 0,019424764 | 0,990896936 | IGSF9 | FALSE | Upregulated |
| 0,004556735 | 0,990896936 | SUB1 | FALSE | Upregulated |
| 0,008116271 | 0,990957822 | C20orf194 | FALSE | Upregulated |
| -0,011686922 | 0,991137778 | LAPTM4B | FALSE | Downregulated |
| 0,01833023 | 0,991148113 | ACE2 | FALSE | Upregulated |
| -0,009599947 | 0,991148113 | ATG10 | FALSE | Downregulated |
| -0,009232767 | 0,991148113 | CHCHD6 | FALSE | Downregulated |
| 0,004432047 | 0,991148113 | ESYT2 | FALSE | Upregulated |
| -0,013538793 | 0,991148113 | FAM216A | FALSE | Downregulated |
| 0,038128111 | 0,991148113 | FLJ34503 | FALSE | Upregulated |
| 0,006164165 | 0,991148113 | GALE | FALSE | Upregulated |
| 0,041070921 | 0,991148113 | PABPC1P2 | FALSE | Upregulated |
| 0,079230037 | 0,991148113 | RGPD6 | FALSE | Upregulated |
| 0,00825651 | 0,991148113 | RUSC2 | FALSE | Upregulated |

| | | | | |
|---|---|---|---|---|
| 0,004780125 | 0,991148113 | SPTY2D1 | FALSE | Upregulated |
| 0,009386677 | 0,991148113 | TAF1C | FALSE | Upregulated |
| -0,258102221 | 0,991148113 | XIRP1 | FALSE | Downregulated |
| 0,005910984 | 0,991148113 | XPR1 | FALSE | Upregulated |
| 0,024903511 | 0,991152266 | LOC102723824 | FALSE | Upregulated |
| -0,010010903 | 0,991183704 | VIPR1 | FALSE | Downregulated |
| -0,009648645 | 0,991242023 | ARMCX6 | FALSE | Downregulated |
| -0,038240784 | 0,991242023 | HS6ST2 | FALSE | Downregulated |
| -0,016550843 | 0,991256753 | AMHR2 | FALSE | Downregulated |
| 0,002714312 | 0,991256753 | AP1G1 | FALSE | Upregulated |
| 0,007036746 | 0,991256753 | ARHGEF10L | FALSE | Upregulated |
| 0,00326768 | 0,991256753 | ARIH2 | FALSE | Upregulated |
| 0,011209691 | 0,991256753 | ASB13 | FALSE | Upregulated |
| -0,00438656 | 0,991256753 | ATF2 | FALSE | Downregulated |
| -0,009094786 | 0,991256753 | BCL7C | FALSE | Downregulated |
| 0,016073387 | 0,991256753 | BTC | FALSE | Upregulated |
| 0,024005203 | 0,991256753 | CEACAM4 | FALSE | Upregulated |
| 0,007172479 | 0,991256753 | CEP192 | FALSE | Upregulated |
| 0,006091275 | 0,991256753 | CLSTN3 | FALSE | Upregulated |
| 0,006201629 | 0,991256753 | CMAS | FALSE | Upregulated |
| 0,014786037 | 0,991256753 | CNTNAP1 | FALSE | Upregulated |
| 0,00850074 | 0,991256753 | DHRS1 | FALSE | Upregulated |
| 0,005879494 | 0,991256753 | DOCK9 | FALSE | Upregulated |
| 0,008786551 | 0,991256753 | DPH7 | FALSE | Upregulated |
| -0,010350024 | 0,991256753 | ERV3-1 | FALSE | Downregulated |
| 0,016270379 | 0,991256753 | FAM151B | FALSE | Upregulated |

| | | | | |
|---|---|---|---|---|
| -0,005506884 | 0,991256753 | FAM222B | FALSE | Downregulated |
| 0,034654021 | 0,991256753 | FLJ45139 | FALSE | Upregulated |
| -0,047532054 | 0,991256753 | GGT2 | FALSE | Downregulated |
| 0,059111099 | 0,991256753 | GUCY1B2 | FALSE | Upregulated |
| 0,004113713 | 0,991256753 | HARS2 | FALSE | Upregulated |
| 0,242214936 | 0,991256753 | IGFN1 | FALSE | Upregulated |
| 0,018635784 | 0,991256753 | KCNK17 | FALSE | Upregulated |
| -0,004367465 | 0,991256753 | KIAA1430 | FALSE | Downregulated |
| -0,005808657 | 0,991256753 | LAMTOR2 | FALSE | Downregulated |
| -0,019927507 | 0,991256753 | LOC101927407 | FALSE | Downregulated |
| 0,025398361 | 0,991256753 | LOC101927838 | FALSE | Upregulated |
| -0,028346005 | 0,991256753 | LOC101928085 | FALSE | Downregulated |
| 0,016167698 | 0,991256753 | LOC101929427 | FALSE | Upregulated |
| -0,042319937 | 0,991256753 | LOC101929837 | FALSE | Downregulated |
| -0,034008139 | 0,991256753 | LOC285389 | FALSE | Downregulated |
| 0,01059858 | 0,991256753 | LOC728743 | FALSE | Upregulated |
| -0,007316934 | 0,991256753 | LOC90784 | FALSE | Downregulated |
| -0,011696077 | 0,991256753 | LPAL2 | FALSE | Downregulated |
| 0,006810497 | 0,991256753 | LYPLAL1 | FALSE | Upregulated |
| 0,006724849 | 0,991256753 | NDUFS8 | FALSE | Upregulated |
| -0,003110366 | 0,991256753 | NFX1 | FALSE | Downregulated |
| 0,012904006 | 0,991256753 | NOS1AP | FALSE | Upregulated |
| -0,006779836 | 0,991256753 | OBFC1 | FALSE | Downregulated |
| 0,017425876 | 0,991256753 | OR7E14P | FALSE | Upregulated |
| 0,0063374 | 0,991256753 | PPAT | FALSE | Upregulated |
| 0,014981845 | 0,991256753 | PPP4R1L | FALSE | Upregulated |

| | | | | |
|---|---|---|---|---|
| -0,006654635 | 0,991256753 | RFNG | FALSE | Downregulated |
| -0,024998612 | 0,991256753 | RP11-503N18.1 | FALSE | Downregulated |
| -0,008609385 | 0,991256753 | SCLT1 | FALSE | Downregulated |
| 0,021713303 | 0,991256753 | STOX1 | FALSE | Upregulated |
| -0,005887812 | 0,991256753 | SYAP1 | FALSE | Downregulated |
| -0,041340115 | 0,991256753 | TEX26 | FALSE | Downregulated |
| -0,008792501 | 0,991256753 | TSPAN33 | FALSE | Downregulated |
| -0,010575614 | 0,991256753 | TUBA1A | FALSE | Downregulated |
| 0,004629069 | 0,991256753 | VWA8 | FALSE | Upregulated |
| 0,007971771 | 0,991256753 | ZBTB24 | FALSE | Upregulated |
| 0,007197088 | 0,991256753 | ZNRD1 | FALSE | Upregulated |
| -0,007444024 | 0,991347723 | HSD17B2 | FALSE | Downregulated |
| 0,015951518 | 0,991353539 | ADCY10P1 | FALSE | Upregulated |
| 0,00712393 | 0,991353539 | C10orf25 | FALSE | Upregulated |
| 0,012706241 | 0,991353539 | CABP4 | FALSE | Upregulated |
| -0,0293674 | 0,991353539 | CDK3 | FALSE | Downregulated |
| 0,017840635 | 0,991353539 | CYP46A1 | FALSE | Upregulated |
| -0,008233564 | 0,991353539 | DDHD1 | FALSE | Downregulated |
| -0,008573953 | 0,991353539 | DHX58 | FALSE | Downregulated |
| -0,010693443 | 0,991353539 | ENC1 | FALSE | Downregulated |
| -0,032271499 | 0,991353539 | FHL5 | FALSE | Downregulated |
| -0,011377289 | 0,991353539 | HDX | FALSE | Downregulated |
| -0,007899892 | 0,991353539 | HHAT | FALSE | Downregulated |
| -0,021671973 | 0,991353539 | IFI6 | FALSE | Downregulated |
| -0,035395133 | 0,991353539 | LOC102724285 | FALSE | Downregulated |
| 0,029535936 | 0,991353539 | LOC729732 | FALSE | Upregulated |

| | | | | |
|---|---|---|---|---|
| -0,004098225 | 0,991353539 | LRPPRC | FALSE | Downregulated |
| 0,00436238 | 0,991353539 | MSL2 | FALSE | Upregulated |
| -0,009427218 | 0,991353539 | NID2 | FALSE | Downregulated |
| 0,006084784 | 0,991353539 | PTPN11 | FALSE | Upregulated |
| -0,236832071 | 0,991353539 | RPL3L | FALSE | Downregulated |
| -0,007749879 | 0,991353539 | SPARC | FALSE | Downregulated |
| 0,017184907 | 0,991353539 | SULT1C4 | FALSE | Upregulated |
| 0,009742467 | 0,991353539 | TIGD1 | FALSE | Upregulated |
| 0,019834394 | 0,991353539 | TPTEP1 | FALSE | Upregulated |
| 0,009118098 | 0,991353539 | WSB1 | FALSE | Upregulated |
| -0,013156399 | 0,991371103 | CERCAM | FALSE | Downregulated |
| -0,016507649 | 0,991371103 | CPT1C | FALSE | Downregulated |
| 0,004180796 | 0,991395761 | NHEJ1 | FALSE | Upregulated |
| 0,020411529 | 0,991395761 | TRNP1 | FALSE | Upregulated |
| -0,027934163 | 0,991444279 | GDF10 | FALSE | Downregulated |
| -0,036753785 | 0,991445537 | WASH7P | FALSE | Downregulated |
| -0,034885905 | 0,991498396 | COX4I2 | FALSE | Downregulated |
| 0,004568891 | 0,99156143 | ZBTB43 | FALSE | Upregulated |
| 0,007996235 | 0,991819396 | ALKBH1 | FALSE | Upregulated |
| 0,019321101 | 0,991819396 | BNC2 | FALSE | Upregulated |
| -0,020342594 | 0,991819396 | BRINP1 | FALSE | Downregulated |
| -0,004111788 | 0,991819396 | C14orf119 | FALSE | Downregulated |
| 0,007476746 | 0,991819396 | C21orf2 | FALSE | Upregulated |
| -0,010884065 | 0,991819396 | CYB5R2 | FALSE | Downregulated |
| -0,033819281 | 0,991819396 | DYDC2 | FALSE | Downregulated |
| -0,008979231 | 0,991819396 | EFCAB7 | FALSE | Downregulated |

| | | | | |
|---|---|---|---|---|
| -0,018695236 | 0,991819396 | G6PC | FALSE | Downregulated |
| 0,022694558 | 0,991819396 | GGT6 | FALSE | Upregulated |
| 0,009425022 | 0,991819396 | IL15RA | FALSE | Upregulated |
| 0,012490056 | 0,991819396 | IL18BP | FALSE | Upregulated |
| 0,007567351 | 0,991819396 | IRF5 | FALSE | Upregulated |
| 0,019173914 | 0,991819396 | LOC102724175 | FALSE | Upregulated |
| -0,006373822 | 0,991819396 | LOC148413 | FALSE | Downregulated |
| 0,026866521 | 0,991819396 | LOC284412 | FALSE | Upregulated |
| -0,020558411 | 0,991819396 | MEI4 | FALSE | Downregulated |
| 0,007082528 | 0,991819396 | MOCOS | FALSE | Upregulated |
| -0,01383709 | 0,991819396 | MXRA8 | FALSE | Downregulated |
| -0,004854354 | 0,991819396 | NUDC | FALSE | Downregulated |
| -0,022027355 | 0,991819396 | PCOLCE-AS1 | FALSE | Downregulated |
| 0,027647726 | 0,991819396 | SLC16A12 | FALSE | Upregulated |
| -0,011499188 | 0,991819396 | SMOX | FALSE | Downregulated |
| -0,004615369 | 0,991819396 | TBP | FALSE | Downregulated |
| 0,013809588 | 0,991819396 | TRAIP | FALSE | Upregulated |
| -0,006352433 | 0,991819396 | UBA6-AS1 | FALSE | Downregulated |
| 0,00656225 | 0,991819396 | UBOX5 | FALSE | Upregulated |
| -0,00417702 | 0,991819396 | UROD | FALSE | Downregulated |
| -0,004572948 | 0,991819396 | XRN2 | FALSE | Downregulated |
| 0,004070932 | 0,991819396 | ZNF41 | FALSE | Upregulated |
| -0,010724208 | 0,991819396 | ZNF788 | FALSE | Downregulated |
| 0,028353641 | 0,991864838 | CPB1 | FALSE | Upregulated |
| -0,08204947 | 0,991864838 | FBP2 | FALSE | Downregulated |
| 0,00893549 | 0,991864838 | ZNF845 | FALSE | Upregulated |

| | | | | |
|---|---|---|---|---|
| -0,053787106 | 0,991880498 | PVRL4 | FALSE | Downregulated |
| -0,023854352 | 0,991880498 | SOCS2-AS1 | FALSE | Downregulated |
| -0,01405362 | 0,991880498 | ZBED3 | FALSE | Downregulated |
| -0,00906549 | 0,992137341 | C6orf136 | FALSE | Downregulated |
| -0,010059 | 0,992137341 | CDC42EP3 | FALSE | Downregulated |
| -0,017634794 | 0,992137341 | CHRDL2 | FALSE | Downregulated |
| -0,026513194 | 0,992137341 | DIRC3 | FALSE | Downregulated |
| -0,00546014 | 0,992137341 | GOLGA2 | FALSE | Downregulated |
| 0,007534497 | 0,992137341 | GTF2H2B | FALSE | Upregulated |
| 0,007054477 | 0,992137341 | NANS | FALSE | Upregulated |
| 0,004155817 | 0,992137341 | PSMD10 | FALSE | Upregulated |
| 0,010131679 | 0,992137341 | SNHG8 | FALSE | Upregulated |
| -0,006151654 | 0,992137341 | SS18L2 | FALSE | Downregulated |
| -0,00910814 | 0,992137341 | UNKL | FALSE | Downregulated |
| -0,009642731 | 0,992137341 | WBP1 | FALSE | Downregulated |
| -0,015424382 | 0,992165615 | CNTN1 | FALSE | Downregulated |
| 0,003276236 | 0,992215509 | GTF2F1 | FALSE | Upregulated |
| -0,187049809 | 0,992281215 | APOBEC2 | FALSE | Downregulated |
| 0,017180771 | 0,992281215 | ATE1-AS1 | FALSE | Upregulated |
| 0,005602751 | 0,992281215 | BBIP1 | FALSE | Upregulated |
| 0,012406005 | 0,992281215 | FILIP1L | FALSE | Upregulated |
| -0,012846434 | 0,992281215 | LGSN | FALSE | Downregulated |
| -0,010885301 | 0,992281215 | PLLP | FALSE | Downregulated |
| -0,00541659 | 0,992281215 | SH2B1 | FALSE | Downregulated |
| 0,011212028 | 0,992281215 | WASH1 | FALSE | Upregulated |
| -0,019540151 | 0,992302445 | AC007038.7 | FALSE | Downregulated |

| | | | | |
|---|---|---|---|---|
| 0,009856942 | 0,992302445 | AC091729.9 | FALSE | Upregulated |
| -0,005543492 | 0,992302445 | ADPRHL2 | FALSE | Downregulated |
| -0,00223876 | 0,992302445 | AHCYL1 | FALSE | Downregulated |
| -0,005652551 | 0,992302445 | ALG6 | FALSE | Downregulated |
| 0,007690991 | 0,992302445 | ANGPTL2 | FALSE | Upregulated |
| -0,004509251 | 0,992302445 | ARID1A | FALSE | Downregulated |
| 0,009135188 | 0,992302445 | ATP2B2 | FALSE | Upregulated |
| -0,008549163 | 0,992302445 | BARD1 | FALSE | Downregulated |
| -0,006262863 | 0,992302445 | C3orf58 | FALSE | Downregulated |
| 0,005376391 | 0,992302445 | CABLES1 | FALSE | Upregulated |
| 0,008112641 | 0,992302445 | CD300A | FALSE | Upregulated |
| -0,004794088 | 0,992302445 | CEP104 | FALSE | Downregulated |
| -0,024185247 | 0,992302445 | CLGN | FALSE | Downregulated |
| -0,010467435 | 0,992302445 | DBH-AS1 | FALSE | Downregulated |
| 0,010372135 | 0,992302445 | DBN1 | FALSE | Upregulated |
| 0,00339595 | 0,992302445 | DCP1A | FALSE | Upregulated |
| -0,003647059 | 0,992302445 | DDX56 | FALSE | Downregulated |
| -0,003729593 | 0,992302445 | DIDO1 | FALSE | Downregulated |
| -0,009692706 | 0,992302445 | EHD2 | FALSE | Downregulated |
| -0,00692247 | 0,992302445 | FAM101B | FALSE | Downregulated |
| 0,024815091 | 0,992302445 | FUT3 | FALSE | Upregulated |
| 0,003912871 | 0,992302445 | GABARAP | FALSE | Upregulated |
| 0,009645919 | 0,992302445 | GATSL3 | FALSE | Upregulated |
| -0,018852192 | 0,992302445 | GPX2 | FALSE | Downregulated |
| -0,005453114 | 0,992302445 | HEATR3 | FALSE | Downregulated |
| -0,005265581 | 0,992302445 | KIAA1958 | FALSE | Downregulated |

| | | | | |
|---|---|---|---|---|
| -0,009627272 | 0,992302445 | KLLN | FALSE | Downregulated |
| 0,00958758 | 0,992302445 | LENG9 | FALSE | Upregulated |
| 0,0072829 | 0,992302445 | LINC00339 | FALSE | Upregulated |
| 0,004894523 | 0,992302445 | LINC00909 | FALSE | Upregulated |
| -0,019903834 | 0,992302445 | LOC100996246 | FALSE | Downregulated |
| 0,041442754 | 0,992302445 | LOC101927999 | FALSE | Upregulated |
| 0,013546913 | 0,992302445 | LOC101928069 | FALSE | Upregulated |
| 0,014740024 | 0,992302445 | LOC101928485 | FALSE | Upregulated |
| -0,007254421 | 0,992302445 | LOC101928499 | FALSE | Downregulated |
| -0,015121172 | 0,992302445 | LOC101928524 | FALSE | Downregulated |
| -0,007985347 | 0,992302445 | LOC728323 | FALSE | Downregulated |
| -0,020441376 | 0,992302445 | LRRTM4 | FALSE | Downregulated |
| -0,008694563 | 0,992302445 | MAML2 | FALSE | Downregulated |
| 0,004876169 | 0,992302445 | MYLK | FALSE | Upregulated |
| 0,005074232 | 0,992302445 | NDUFAF5 | FALSE | Upregulated |
| -0,033539122 | 0,992302445 | NME1-NME2 | FALSE | Downregulated |
| 0,009008556 | 0,992302445 | NR1I2 | FALSE | Upregulated |
| 0,008784731 | 0,992302445 | NUMBL | FALSE | Upregulated |
| 0,01782202 | 0,992302445 | PRDM6 | FALSE | Upregulated |
| -0,040997912 | 0,992302445 | RAB6C | FALSE | Downregulated |
| 0,004339824 | 0,992302445 | RAPGEF2 | FALSE | Upregulated |
| -0,010399595 | 0,992302445 | RHCE | FALSE | Downregulated |
| -0,005240338 | 0,992302445 | RILPL2 | FALSE | Downregulated |
| 0,009861895 | 0,992302445 | RIMKLB | FALSE | Upregulated |
| 0,023241821 | 0,992302445 | RNF212 | FALSE | Upregulated |
| -0,004227571 | 0,992302445 | RNF216 | FALSE | Downregulated |

| | | | | |
|---|---|---|---|---|
| 0,014508098 | 0,992302445 | RP11-61I13.3 | FALSE | Upregulated |
| 0,015831165 | 0,992302445 | RP3-400B16.1 | FALSE | Upregulated |
| 0,013514131 | 0,992302445 | RP4-782L23.1 | FALSE | Upregulated |
| -0,020707205 | 0,992302445 | RP5-1074L1.1 | FALSE | Downregulated |
| -0,012327269 | 0,992302445 | RRN3P1 | FALSE | Downregulated |
| 0,0058863 | 0,992302445 | SEC16B | FALSE | Upregulated |
| -0,006019965 | 0,992302445 | SELK | FALSE | Downregulated |
| -0,004165414 | 0,992302445 | SF3B14 | FALSE | Downregulated |
| 0,00579063 | 0,992302445 | SLC23A1 | FALSE | Upregulated |
| 0,006547621 | 0,992302445 | SLC35E2B | FALSE | Upregulated |
| 0,00432977 | 0,992302445 | SRBD1 | FALSE | Upregulated |
| -0,025518101 | 0,992302445 | SRGAP2D | FALSE | Downregulated |
| 0,007191684 | 0,992302445 | TRIM10 | FALSE | Upregulated |
| -0,170503956 | 0,992302445 | TRIM54 | FALSE | Downregulated |
| 0,00940784 | 0,992302445 | TTC22 | FALSE | Upregulated |
| -0,017131385 | 0,992302445 | TTC39A | FALSE | Downregulated |
| -0,004366469 | 0,992302445 | TUBGCP3 | FALSE | Downregulated |
| -0,003172947 | 0,992302445 | WAPAL | FALSE | Downregulated |
| -0,015016348 | 0,992302445 | WBP2NL | FALSE | Downregulated |
| 0,003440407 | 0,992302445 | WDR43 | FALSE | Upregulated |
| 0,004356792 | 0,992302445 | ZNF451 | FALSE | Upregulated |
| 0,008302173 | 0,992302445 | ZNF502 | FALSE | Upregulated |
| -0,00588555 | 0,992302445 | ZSCAN12 | FALSE | Downregulated |
| -0,005669344 | 0,992302445 | ZSCAN30 | FALSE | Downregulated |
| -0,007213066 | 0,992308317 | C5orf55 | FALSE | Downregulated |
| 0,003184833 | 0,992308317 | CD164 | FALSE | Upregulated |

| | | | | |
|---|---|---|---|---|
| 0,004330465 | 0,992308317 | IFT122 | FALSE | Upregulated |
| -0,022625748 | 0,992308317 | LOC102723993 | FALSE | Downregulated |
| -0,006799498 | 0,992308317 | MARCKS | FALSE | Downregulated |
| 0,002069623 | 0,9923894 | EIF4H | FALSE | Upregulated |
| -0,004685979 | 0,9923894 | MRPL14 | FALSE | Downregulated |
| -0,010351835 | 0,9923894 | WDR27 | FALSE | Downregulated |
| 0,027901192 | 0,992453761 | CAPZA3 | FALSE | Upregulated |
| -0,015570889 | 0,992497489 | HRCT1 | FALSE | Downregulated |
| -0,003823268 | 0,992670585 | ETFB | FALSE | Downregulated |
| -0,18006819 | 0,99268086 | ACTN3 | FALSE | Downregulated |
| -0,02053876 | 0,99268086 | ADD3-AS1 | FALSE | Downregulated |
| 0,006245104 | 0,99268086 | ANKRD39 | FALSE | Upregulated |
| -0,012403496 | 0,99268086 | DFNB59 | FALSE | Downregulated |
| -0,016269051 | 0,99268086 | DMKN | FALSE | Downregulated |
| 0,008612014 | 0,99268086 | DUSP6 | FALSE | Upregulated |
| 0,007335958 | 0,99268086 | DYNLL1 | FALSE | Upregulated |
| 0,018593106 | 0,99268086 | LOC101929625 | FALSE | Upregulated |
| 0,007810941 | 0,99268086 | LOC158960 | FALSE | Upregulated |
| -0,0033017 | 0,99268086 | LRP11 | FALSE | Downregulated |
| -0,00599436 | 0,99268086 | LSM7 | FALSE | Downregulated |
| -0,005845651 | 0,99268086 | MFNG | FALSE | Downregulated |
| -0,004985269 | 0,99268086 | MIB2 | FALSE | Downregulated |
| -0,005403032 | 0,99268086 | NCALD | FALSE | Downregulated |
| 0,003347798 | 0,99268086 | NDUFS1 | FALSE | Upregulated |
| -0,005678767 | 0,99268086 | PCED1A | FALSE | Downregulated |
| 0,022841619 | 0,99268086 | SH3GL1P1 | FALSE | Upregulated |

| | | | | |
|---|---|---|---|---|
| -0,00297379 | 0,99268086 | SUPT16H | FALSE | Downregulated |
| 0,008159746 | 0,99268086 | TAZ | FALSE | Upregulated |
| 0,008567028 | 0,99268086 | TTPAL | FALSE | Upregulated |
| -0,007039773 | 0,99268086 | ZNF571 | FALSE | Downregulated |
| 0,005579507 | 0,992682617 | CLK2 | FALSE | Upregulated |
| 0,003913382 | 0,992721246 | N4BP1 | FALSE | Upregulated |
| 0,032134838 | 0,992752792 | LOC102724135 | FALSE | Upregulated |
| -0,004311563 | 0,992804019 | AKAP13 | FALSE | Downregulated |
| 0,007890623 | 0,992804019 | C2orf76 | FALSE | Upregulated |
| -0,005966246 | 0,992804019 | CAPN10 | FALSE | Downregulated |
| -0,004811932 | 0,992804019 | CCDC86 | FALSE | Downregulated |
| 0,004650875 | 0,992804019 | CDC42BPA | FALSE | Upregulated |
| 0,009251653 | 0,992804019 | CDC42EP2 | FALSE | Upregulated |
| 0,003327751 | 0,992804019 | CDYL | FALSE | Upregulated |
| -0,004503585 | 0,992804019 | CINP | FALSE | Downregulated |
| -0,017653711 | 0,992804019 | GBAP1 | FALSE | Downregulated |
| -0,012227169 | 0,992804019 | HNRNPA3P1 | FALSE | Downregulated |
| -0,012768163 | 0,992804019 | KCNK1 | FALSE | Downregulated |
| 0,00447643 | 0,992804019 | KMT2D | FALSE | Upregulated |
| 0,004072067 | 0,992804019 | LGMN | FALSE | Upregulated |
| -0,013624915 | 0,992804019 | LINC00426 | FALSE | Downregulated |
| -0,010455723 | 0,992804019 | LINC00950 | FALSE | Downregulated |
| -0,011251341 | 0,992804019 | LOC100499484 | FALSE | Downregulated |
| -0,011735213 | 0,992804019 | LOC101927002 | FALSE | Downregulated |
| -0,004183221 | 0,992804019 | LOC102724364 | FALSE | Downregulated |
| 0,011030394 | 0,992804019 | LOC146880 | FALSE | Upregulated |

| | | | | |
|---|---|---|---|---|
| -0,014231115 | 0,992804019 | MINOS1-NBL1 | FALSE | Downregulated |
| 0,014852275 | 0,992804019 | MIR17HG | FALSE | Upregulated |
| -0,161273088 | 0,992804019 | MYOZ1 | FALSE | Downregulated |
| -0,00850676 | 0,992804019 | PKHD1 | FALSE | Downregulated |
| 0,006847506 | 0,992804019 | PRKAR1B | FALSE | Upregulated |
| 0,01060787 | 0,992804019 | PTGES2-AS1 | FALSE | Upregulated |
| 0,006678922 | 0,992804019 | SCAND2P | FALSE | Upregulated |
| -0,011540052 | 0,992804019 | SLC14A1 | FALSE | Downregulated |
| -0,024282381 | 0,992804019 | SLC26A7 | FALSE | Downregulated |
| -0,007804278 | 0,992804019 | SMPD2 | FALSE | Downregulated |
| 0,012129785 | 0,992804019 | SPDYE3 | FALSE | Upregulated |
| 0,003976936 | 0,992804019 | SUN2 | FALSE | Upregulated |
| 0,013485748 | 0,992804019 | TMEM253 | FALSE | Upregulated |
| 0,005683791 | 0,992804019 | ZNF19 | FALSE | Upregulated |
| 0,013981336 | 0,992804019 | ZNF257 | FALSE | Upregulated |
| 0,005419355 | 0,992804019 | ZNF354A | FALSE | Upregulated |
| 0,003529615 | 0,992804019 | ZNF512 | FALSE | Upregulated |
| -0,013697325 | 0,992804019 | ZNF783 | FALSE | Downregulated |
| 0,164880038 | 0,992830246 | HYPK | FALSE | Upregulated |
| -0,003230636 | 0,992830246 | VPS13D | FALSE | Downregulated |
| -0,012057418 | 0,992976623 | LOC101060376 | FALSE | Downregulated |
| -0,004850449 | 0,992976623 | MGAT4A | FALSE | Downregulated |
| 0,013766273 | 0,992995075 | LOC400863 | FALSE | Upregulated |
| 0,016729447 | 0,993079461 | NCR1 | FALSE | Upregulated |
| 0,0051474 | 0,993261436 | JUND | FALSE | Upregulated |
| 0,008036378 | 0,993362384 | CACNA2D4 | FALSE | Upregulated |

| | | | | |
|---|---|---|---|---|
| 0,010575941 | 0,993362384 | CEACAM19 | FALSE | Upregulated |
| 0,003744303 | 0,993362384 | COPS7B | FALSE | Upregulated |
| -0,006348575 | 0,993362384 | FER | FALSE | Downregulated |
| 0,013252446 | 0,993362384 | FLJ37201 | FALSE | Upregulated |
| 0,006594302 | 0,993362384 | GUCY1A2 | FALSE | Upregulated |
| 0,004700905 | 0,993362384 | IRS1 | FALSE | Upregulated |
| -0,017131927 | 0,993362384 | LOC101929667 | FALSE | Downregulated |
| -0,017283577 | 0,993362384 | LOC102723981 | FALSE | Downregulated |
| 0,003828517 | 0,993362384 | MGAT2 | FALSE | Upregulated |
| -0,01247123 | 0,993362384 | MORC2-AS1 | FALSE | Downregulated |
| -0,005324079 | 0,993362384 | PDPR | FALSE | Downregulated |
| 0,011657302 | 0,993362384 | SLC15A2 | FALSE | Upregulated |
| -0,010216132 | 0,993390862 | ATP6V1C2 | FALSE | Downregulated |
| 0,003871285 | 0,99358446 | IL10RB | FALSE | Upregulated |
| -0,027298135 | 0,993640556 | IGF2BP1 | FALSE | Downregulated |
| -0,005856332 | 0,993812817 | ACYP1 | FALSE | Downregulated |
| 0,00801198 | 0,993812817 | ADA | FALSE | Upregulated |
| 0,004509225 | 0,993812817 | CD3EAP | FALSE | Upregulated |
| 0,003687704 | 0,993812817 | EP400 | FALSE | Upregulated |
| -0,002804327 | 0,993812817 | GATC | FALSE | Downregulated |
| -0,006348664 | 0,993812817 | GBP2 | FALSE | Downregulated |
| -0,005427012 | 0,993812817 | GFRA1 | FALSE | Downregulated |
| 0,007555272 | 0,993812817 | GPX3 | FALSE | Upregulated |
| -0,002672572 | 0,993812817 | GTF2E2 | FALSE | Downregulated |
| 0,004987791 | 0,993812817 | GYS1 | FALSE | Upregulated |
| 0,006659533 | 0,993812817 | IKZF2 | FALSE | Upregulated |

| | | | | |
|---|---|---|---|---|
| -0,017495442 | 0,993812817 | LOC102724153 | FALSE | Downregulated |
| 0,012270171 | 0,993812817 | NXF3 | FALSE | Upregulated |
| 0,003808513 | 0,993812817 | OSBP | FALSE | Upregulated |
| 0,009632003 | 0,993812817 | PDZRN4 | FALSE | Upregulated |
| 0,003565456 | 0,993812817 | PPFIBP2 | FALSE | Upregulated |
| -0,002421568 | 0,993812817 | RMDN3 | FALSE | Downregulated |
| -0,003403571 | 0,993812817 | RNF34 | FALSE | Downregulated |
| 0,005329039 | 0,993812817 | RP11-85F14.5 | FALSE | Upregulated |
| 0,00248275 | 0,993812817 | SERPINB6 | FALSE | Upregulated |
| -0,005331891 | 0,993812817 | SLC39A10 | FALSE | Downregulated |
| -0,003574716 | 0,993812817 | SMARCD2 | FALSE | Downregulated |
| -0,004156795 | 0,993812817 | TDRD7 | FALSE | Downregulated |
| 0,008168226 | 0,993812817 | TGM2 | FALSE | Upregulated |
| -0,00373095 | 0,993812817 | TMEM199 | FALSE | Downregulated |
| 0,010874854 | 0,993812817 | TMEM71 | FALSE | Upregulated |
| -0,002827961 | 0,993812817 | ZER1 | FALSE | Downregulated |
| -0,004589305 | 0,993812817 | ZNRF3 | FALSE | Downregulated |
| 0,004851474 | 0,993887158 | ALG3 | FALSE | Upregulated |
| 0,00346121 | 0,993887158 | BCS1L | FALSE | Upregulated |
| 0,00811894 | 0,993887158 | CD109 | FALSE | Upregulated |
| 0,006115421 | 0,993887158 | CDON | FALSE | Upregulated |
| -0,025150257 | 0,993887158 | CIDEC | FALSE | Downregulated |
| 0,014186913 | 0,993887158 | CPT1B | FALSE | Upregulated |
| 0,003493697 | 0,993887158 | ENOX2 | FALSE | Upregulated |
| -0,007108495 | 0,993887158 | FLJ23867 | FALSE | Downregulated |
| -0,011768074 | 0,993887158 | HIST1H2AE | FALSE | Downregulated |

| | | | | |
|---|---|---|---|---|
| -0,017947944 | 0,993887158 | LAMA1 | FALSE | Downregulated |
| -0,01331932 | 0,993887158 | LOC100240735 | FALSE | Downregulated |
| 0,016631966 | 0,993887158 | LOC101928825 | FALSE | Upregulated |
| 0,005244209 | 0,993887158 | MPC1 | FALSE | Upregulated |
| 0,004386929 | 0,993887158 | NDUFV3 | FALSE | Upregulated |
| 0,002716924 | 0,993887158 | NUBP1 | FALSE | Upregulated |
| -0,004131851 | 0,993887158 | PC | FALSE | Downregulated |
| -0,003781108 | 0,993887158 | PPP1R15A | FALSE | Downregulated |
| 0,006292881 | 0,993887158 | SLC25A51 | FALSE | Upregulated |
| -0,00631241 | 0,993887158 | STON2 | FALSE | Downregulated |
| 0,002736713 | 0,993887158 | TAF6 | FALSE | Upregulated |
| -0,005014826 | 0,993887158 | TP53INP2 | FALSE | Downregulated |
| -0,010087171 | 0,993887158 | TRIM61 | FALSE | Downregulated |
| -0,002684461 | 0,993887158 | VCP | FALSE | Downregulated |
| 0,00339776 | 0,993887158 | ZNF350 | FALSE | Upregulated |
| -0,007245378 | 0,993887158 | ZNF366 | FALSE | Downregulated |
| 0,006741629 | 0,993887158 | ZNF678 | FALSE | Upregulated |
| 0,010668746 | 0,993907403 | ITGA7 | FALSE | Upregulated |
| 0,005148969 | 0,99397258 | ADORA2A-AS1 | FALSE | Upregulated |
| 0,001589674 | 0,99397258 | ANXA7 | FALSE | Upregulated |
| 0,005384302 | 0,99397258 | ARMC2 | FALSE | Upregulated |
| 0,003401352 | 0,99397258 | ATP6V0C | FALSE | Upregulated |
| 0,002723026 | 0,99397258 | ATXN2L | FALSE | Upregulated |
| -0,00301015 | 0,99397258 | B4GALT1 | FALSE | Downregulated |
| -0,006065871 | 0,99397258 | BBS12 | FALSE | Downregulated |
| -0,008681215 | 0,99397258 | BSPRY | FALSE | Downregulated |

| | | | | |
|---|---|---|---|---|
| 0,002919441 | 0,99397258 | C19orf60 | FALSE | Upregulated |
| -0,002803629 | 0,99397258 | C21orf33 | FALSE | Downregulated |
| 0,005735179 | 0,99397258 | CCZ1 | FALSE | Upregulated |
| -0,010311559 | 0,99397258 | CDH24 | FALSE | Downregulated |
| -0,001876048 | 0,99397258 | CDK13 | FALSE | Downregulated |
| 0,003594725 | 0,99397258 | CNKSR3 | FALSE | Upregulated |
| 0,00757779 | 0,99397258 | CNTFR | FALSE | Upregulated |
| -0,002455978 | 0,99397258 | COMMD7 | FALSE | Downregulated |
| -0,026865379 | 0,99397258 | CORIN | FALSE | Downregulated |
| -0,003321883 | 0,99397258 | DGUOK | FALSE | Downregulated |
| -0,002818773 | 0,99397258 | EIF3L | FALSE | Downregulated |
| 0,004466649 | 0,99397258 | ELFN1 | FALSE | Upregulated |
| -0,003305011 | 0,99397258 | FAM92A1 | FALSE | Downregulated |
| -0,005980009 | 0,99397258 | FKBP11 | FALSE | Downregulated |
| 0,005516844 | 0,99397258 | FMO5 | FALSE | Upregulated |
| -0,004147786 | 0,99397258 | FRAT2 | FALSE | Downregulated |
| 0,015056017 | 0,99397258 | FSBP | FALSE | Upregulated |
| 0,00216662 | 0,99397258 | GID4 | FALSE | Upregulated |
| -0,004161678 | 0,99397258 | GIGYF1 | FALSE | Downregulated |
| 0,013057854 | 0,99397258 | GPR64 | FALSE | Upregulated |
| -0,002584983 | 0,99397258 | GRB10 | FALSE | Downregulated |
| 0,002714505 | 0,99397258 | HAUS3 | FALSE | Upregulated |
| -0,005377615 | 0,99397258 | HDAC11 | FALSE | Downregulated |
| 0,001944538 | 0,99397258 | HDLBP | FALSE | Upregulated |
| 0,003406766 | 0,99397258 | HEATR5B | FALSE | Upregulated |
| -0,009130032 | 0,99397258 | HOMER1 | FALSE | Downregulated |

| | | | | |
|---|---|---|---|---|
| 0,003916107 | 0,99397258 | IFFO2 | FALSE | Upregulated |
| -0,004560029 | 0,99397258 | IL1R2 | FALSE | Downregulated |
| -0,001744423 | 0,99397258 | IREB2 | FALSE | Downregulated |
| 0,003357327 | 0,99397258 | ITGB1BP1 | FALSE | Upregulated |
| -0,001820498 | 0,99397258 | KDM1A | FALSE | Downregulated |
| -0,009618903 | 0,99397258 | KIAA1549 | FALSE | Downregulated |
| -0,005448242 | 0,99397258 | KRAS | FALSE | Downregulated |
| -0,011540112 | 0,99397258 | LOC100270746 | FALSE | Downregulated |
| 0,005118709 | 0,99397258 | LOC102606465 | FALSE | Upregulated |
| -0,004913378 | 0,99397258 | LOC102724002 | FALSE | Downregulated |
| 0,019270198 | 0,99397258 | LOC102724930 | FALSE | Upregulated |
| -0,003862733 | 0,99397258 | LRRC2 | FALSE | Downregulated |
| 0,002573197 | 0,99397258 | LSG1 | FALSE | Upregulated |
| -0,001965734 | 0,99397258 | LSM8 | FALSE | Downregulated |
| -0,012215667 | 0,99397258 | MAGOH2 | FALSE | Downregulated |
| 0,008698829 | 0,99397258 | MFSD2A | FALSE | Upregulated |
| 0,014329968 | 0,99397258 | MIR4435-1HG | FALSE | Upregulated |
| -0,004141427 | 0,99397258 | MRPL12 | FALSE | Downregulated |
| 0,0147696 | 0,99397258 | MUC3A | FALSE | Upregulated |
| 0,003883339 | 0,99397258 | NBEAL1 | FALSE | Upregulated |
| 0,00269272 | 0,99397258 | NCBP2-AS2 | FALSE | Upregulated |
| -0,007320383 | 0,99397258 | NEK11 | FALSE | Downregulated |
| -0,005787556 | 0,99397258 | NEURL2 | FALSE | Downregulated |
| -0,003203187 | 0,99397258 | NPC2 | FALSE | Downregulated |
| 0,002657423 | 0,99397258 | ORMDL3 | FALSE | Upregulated |
| 0,006068786 | 0,99397258 | P2RX7 | FALSE | Upregulated |

| | | | | |
|---|---|---|---|---|
| -0,003219948 | 0,99397258 | PACSIN2 | FALSE | Downregulated |
| 0,004342907 | 0,99397258 | PKD1P1 | FALSE | Upregulated |
| 0,010709872 | 0,99397258 | PLIN1 | FALSE | Upregulated |
| -0,004017157 | 0,99397258 | PPL | FALSE | Downregulated |
| -0,004197707 | 0,99397258 | PPP1R3E | FALSE | Downregulated |
| 0,008894457 | 0,99397258 | PPP2R2B | FALSE | Upregulated |
| -0,010650971 | 0,99397258 | RAB42 | FALSE | Downregulated |
| -0,002966399 | 0,99397258 | REST | FALSE | Downregulated |
| -0,0038537 | 0,99397258 | RGS12 | FALSE | Downregulated |
| -0,017817677 | 0,99397258 | RP11-173B14.5 | FALSE | Downregulated |
| -0,011283044 | 0,99397258 | RP11-408H20.2 | FALSE | Downregulated |
| 0,00361859 | 0,99397258 | RRP9 | FALSE | Upregulated |
| -0,00902059 | 0,99397258 | SEMA3D | FALSE | Downregulated |
| -0,003539023 | 0,99397258 | SFR1 | FALSE | Downregulated |
| -0,006814639 | 0,99397258 | SLC16A1 | FALSE | Downregulated |
| -0,008554285 | 0,99397258 | SLC23A3 | FALSE | Downregulated |
| -0,003061281 | 0,99397258 | SPTBN1 | FALSE | Downregulated |
| 0,013240794 | 0,99397258 | STRC | FALSE | Upregulated |
| 0,004819197 | 0,99397258 | SYTL2 | FALSE | Upregulated |
| -0,002270737 | 0,99397258 | TBC1D23 | FALSE | Downregulated |
| -0,002620299 | 0,99397258 | TCF3 | FALSE | Downregulated |
| 0,003134326 | 0,99397258 | TMCC1 | FALSE | Upregulated |
| -0,006359028 | 0,99397258 | TMEM198B | FALSE | Downregulated |
| 0,009902182 | 0,99397258 | TRIM73 | FALSE | Upregulated |
| 0,002117994 | 0,99397258 | TRIP12 | FALSE | Upregulated |
| 0,075864799 | 0,99397258 | TRNAV19 | FALSE | Upregulated |

| | | | | |
|---|---|---|---|---|
| 0,00176805 | 0,99397258 | TTI1 | FALSE | Upregulated |
| -0,002280986 | 0,99397258 | UBE2N | FALSE | Downregulated |
| -0,006634614 | 0,99397258 | UNC5B | FALSE | Downregulated |
| -0,00946294 | 0,99397258 | USP27X-AS1 | FALSE | Downregulated |
| 0,004528267 | 0,99397258 | ZBTB37 | FALSE | Upregulated |
| -0,017172161 | 0,99397258 | ZNF215 | FALSE | Downregulated |
| -0,004515539 | 0,99397258 | ZNF613 | FALSE | Downregulated |
| -0,009913683 | 0,99397258 | ZNF714 | FALSE | Downregulated |
| -0,002397155 | 0,994076645 | CHP1 | FALSE | Downregulated |
| 0,007968351 | 0,994103802 | LSMEM1 | FALSE | Upregulated |
| 0,113569219 | 0,994103802 | PADI1 | FALSE | Upregulated |
| -0,011287928 | 0,994142727 | LOC101928182 | FALSE | Downregulated |
| 0,002677023 | 0,994142727 | NAPB | FALSE | Upregulated |
| 0,003646698 | 0,994158776 | DPM3 | FALSE | Upregulated |
| -0,002386254 | 0,994265558 | PRRG1 | FALSE | Downregulated |
| -0,002540465 | 0,994432744 | BOD1 | FALSE | Downregulated |
| 0,008596533 | 0,994432744 | LOC100506124 | FALSE | Upregulated |
| -0,004108388 | 0,994638127 | MFSD3 | FALSE | Downregulated |
| -0,015363751 | 0,99471252 | CPXM1 | FALSE | Downregulated |
| -0,002914672 | 0,994757075 | APOA1BP | FALSE | Downregulated |
| 0,003012328 | 0,994757075 | DERL2 | FALSE | Upregulated |
| 0,014894482 | 0,994757075 | DNAAF3 | FALSE | Upregulated |
| 0,005719251 | 0,994757075 | GALNT14 | FALSE | Upregulated |
| -0,006164206 | 0,994757075 | LINC00957 | FALSE | Downregulated |
| 0,00569946 | 0,994757075 | LOC113230 | FALSE | Upregulated |
| 0,003534137 | 0,994757075 | MYO6 | FALSE | Upregulated |

| | | | | |
|---|---|---|---|---|
| 0,006761301 | 0,994757075 | NBPF13P | FALSE | Upregulated |
| -0,002635681 | 0,994757075 | NLN | FALSE | Downregulated |
| -0,002279526 | 0,994757075 | TATDN1 | FALSE | Downregulated |
| -0,002991681 | 0,994757075 | TNS1 | FALSE | Downregulated |
| -0,002662626 | 0,994792249 | BANP | FALSE | Downregulated |
| -0,016352192 | 0,994792249 | LOC101928562 | FALSE | Downregulated |
| 0,002937324 | 0,994792249 | RPL27A | FALSE | Upregulated |
| -0,003398904 | 0,994910664 | SHMT1 | FALSE | Downregulated |
| 0,01112544 | 0,994935052 | BAI1 | FALSE | Upregulated |
| -0,003514678 | 0,994935052 | C10orf128 | FALSE | Downregulated |
| -0,007535626 | 0,994935052 | HAND2-AS1 | FALSE | Downregulated |
| -0,003283711 | 0,994935052 | NUDT7 | FALSE | Downregulated |
| 0,001916968 | 0,994935052 | PIGC | FALSE | Upregulated |
| -0,003188377 | 0,994935052 | RIPK2 | FALSE | Downregulated |
| 0,003268994 | 0,994935052 | TEAD1 | FALSE | Upregulated |
| -0,002486588 | 0,994935052 | TXNRD2 | FALSE | Downregulated |
| 0,005024067 | 0,995013369 | CLDN15 | FALSE | Upregulated |
| 0,008295179 | 0,995013369 | LOC102723965 | FALSE | Upregulated |
| -0,001564745 | 0,995013369 | NEK6 | FALSE | Downregulated |
| 0,001776124 | 0,995049546 | CBX5 | FALSE | Upregulated |
| 0,003043857 | 0,995108689 | MAPKAPK5-AS1 | FALSE | Upregulated |
| -0,005565702 | 0,995165612 | LILRB1 | FALSE | Downregulated |
| -0,002116438 | 0,995248565 | SIRT2 | FALSE | Downregulated |
| 0,006605219 | 0,995534806 | RPL13P5 | FALSE | Upregulated |
| 0,002904721 | 0,995536821 | ABCG5 | FALSE | Upregulated |
| 0,007549456 | 0,995536821 | AHRR | FALSE | Upregulated |

| | | | | |
|---|---|---|---|---|
| -0,003548976 | 0,995536821 | APBB1IP | FALSE | Downregulated |
| 0,003704404 | 0,995536821 | ARHGAP24 | FALSE | Upregulated |
| 0,011760763 | 0,995536821 | ARL17B | FALSE | Upregulated |
| -0,005343339 | 0,995536821 | B3GALT4 | FALSE | Downregulated |
| -0,001993345 | 0,995536821 | C18orf8 | FALSE | Downregulated |
| 0,003137489 | 0,995536821 | C8B | FALSE | Upregulated |
| 0,011994107 | 0,995536821 | CARD14 | FALSE | Upregulated |
| -0,003346168 | 0,995536821 | CARD6 | FALSE | Downregulated |
| 0,005872494 | 0,995536821 | CCDC183 | FALSE | Upregulated |
| 0,002858654 | 0,995536821 | CCNB1IP1 | FALSE | Upregulated |
| 0,006635603 | 0,995536821 | CLVS1 | FALSE | Upregulated |
| -0,001586107 | 0,995536821 | DENND6A | FALSE | Downregulated |
| 0,013031344 | 0,995536821 | DKK2 | FALSE | Upregulated |
| -0,002586084 | 0,995536821 | DPP3 | FALSE | Downregulated |
| 0,006946217 | 0,995536821 | FKBP1B | FALSE | Upregulated |
| -0,016065518 | 0,995536821 | GJB3 | FALSE | Downregulated |
| -0,002667808 | 0,995536821 | LECT2 | FALSE | Downregulated |
| 0,006571528 | 0,995536821 | LINC00654 | FALSE | Upregulated |
| -0,005194228 | 0,995536821 | LINC01061 | FALSE | Downregulated |
| -0,009521677 | 0,995536821 | LOC100130987 | FALSE | Downregulated |
| -0,01604018 | 0,995536821 | LOC646938 | FALSE | Downregulated |
| -0,004731682 | 0,995536821 | LOC728613 | FALSE | Downregulated |
| -0,005520162 | 0,995536821 | LOC729603 | FALSE | Downregulated |
| 0,003210381 | 0,995536821 | MED30 | FALSE | Upregulated |
| 0,011251294 | 0,995536821 | NOXO1 | FALSE | Upregulated |
| -0,002377368 | 0,995536821 | PRODH2 | FALSE | Downregulated |

| | | | | |
|---|---|---|---|---|
| -0,014635634 | 0,995536821 | PRR7 | FALSE | Downregulated |
| 0,00234375 | 0,995536821 | QPRT | FALSE | Upregulated |
| -0,003395657 | 0,995536821 | RAG1 | FALSE | Downregulated |
| -0,006449024 | 0,995536821 | REEP1 | FALSE | Downregulated |
| -0,002650249 | 0,995536821 | SLC37A4 | FALSE | Downregulated |
| -0,011594559 | 0,995536821 | SNORD68 | FALSE | Downregulated |
| -0,00455556 | 0,995536821 | SOGA1 | FALSE | Downregulated |
| -0,004927727 | 0,995536821 | THSD7A | FALSE | Downregulated |
| 0,003263227 | 0,995536821 | TMEM176A | FALSE | Upregulated |
| -0,001963721 | 0,995536821 | TNIP2 | FALSE | Downregulated |
| 0,001874233 | 0,995536821 | VAV2 | FALSE | Upregulated |
| 0,004573365 | 0,995672093 | CMAHP | FALSE | Upregulated |
| 0,00257493 | 0,995672093 | ELAC1 | FALSE | Upregulated |
| -0,002640285 | 0,995672093 | GNG2 | FALSE | Downregulated |
| 0,005546551 | 0,995672093 | KCNIP2 | FALSE | Upregulated |
| -0,002225099 | 0,995672093 | LINC00997 | FALSE | Downregulated |
| 0,003834422 | 0,995672093 | MIIP | FALSE | Upregulated |
| 0,002547273 | 0,995672093 | NUDT19 | FALSE | Upregulated |
| 0,007154742 | 0,995693187 | BEX2 | FALSE | Upregulated |
| 0,007174481 | 0,995796387 | SLC29A2 | FALSE | Upregulated |
| 0,010547809 | 0,995801508 | LOC101927879 | FALSE | Upregulated |
| -0,0807085 | 0,995840401 | HIST2H2AA3 | FALSE | Downregulated |
| -0,00211517 | 0,995840401 | RPS10-NUDT3 | FALSE | Downregulated |
| -0,001525138 | 0,995913572 | ZNF45 | FALSE | Downregulated |
| -0,002105365 | 0,995964821 | GOLPH3 | FALSE | Downregulated |
| 0,012776593 | 0,995973894 | LOC101927066 | FALSE | Upregulated |

| | | | | |
|---|---|---|---|---|
| 0,0043218 | 0,996088465 | LOC100129461 | FALSE | Upregulated |
| 0,005071685 | 0,996111261 | SLC16A9 | FALSE | Upregulated |
| -0,012153266 | 0,996137471 | GPR20 | FALSE | Downregulated |
| 0,0019004 | 0,99614595 | RABL2B | FALSE | Upregulated |
| -0,002540216 | 0,996210162 | AGL | FALSE | Downregulated |
| -0,00194163 | 0,996210162 | CBFA2T2 | FALSE | Downregulated |
| -0,010222507 | 0,996210162 | CELP | FALSE | Downregulated |
| -0,002769056 | 0,996210162 | CLASRP | FALSE | Downregulated |
| -0,001876647 | 0,996210162 | CMC1 | FALSE | Downregulated |
| 0,006516952 | 0,996210162 | GSTA5 | FALSE | Upregulated |
| 0,002184019 | 0,996210162 | LYRM9 | FALSE | Upregulated |
| 0,00807006 | 0,996210162 | MED4-AS1 | FALSE | Upregulated |
| 0,002054356 | 0,996210162 | PDE6D | FALSE | Upregulated |
| -0,001444207 | 0,996210162 | POLR3F | FALSE | Downregulated |
| 0,008056299 | 0,996210162 | RAB25 | FALSE | Upregulated |
| 0,001366988 | 0,996210162 | ZNF189 | FALSE | Upregulated |
| -0,008482078 | 0,996210162 | ZNF826P | FALSE | Downregulated |
| 0,005342861 | 0,996277177 | LOC100996763 | FALSE | Upregulated |
| -0,001335978 | 0,996288581 | CDS2 | FALSE | Downregulated |
| 0,007072944 | 0,996560888 | LOC101928641 | FALSE | Upregulated |
| 0,004090059 | 0,996560888 | PTPLAD2 | FALSE | Upregulated |
| -0,009094974 | 0,996560888 | RP11-81H14.2 | FALSE | Downregulated |
| 0,003081449 | 0,996560888 | TMEM82 | FALSE | Upregulated |
| 0,006716039 | 0,996628203 | ASH1L-AS1 | FALSE | Upregulated |
| -0,003786007 | 0,996628203 | SLC25A21-AS1 | FALSE | Downregulated |
| 0,002811092 | 0,996628203 | SSX2IP | FALSE | Upregulated |

| | | | | |
|---|---|---|---|---|
| 0,001356054 | 0,996628203 | USP32 | FALSE | Upregulated |
| 0,002820562 | 0,996740042 | CBFA2T3 | FALSE | Upregulated |
| -0,007552307 | 0,996740042 | LOC389607 | FALSE | Downregulated |
| 0,00440919 | 0,996740042 | PLAT | FALSE | Upregulated |
| 0,004753851 | 0,996840558 | PDE6G | FALSE | Upregulated |
| -0,001998504 | 0,996937502 | FAM20A | FALSE | Downregulated |
| 0,001442261 | 0,996937502 | ZNF747 | FALSE | Upregulated |
| 0,006860237 | 0,996951932 | EYA2 | FALSE | Upregulated |
| -0,001720172 | 0,996951932 | ITFG2 | FALSE | Downregulated |
| -0,001752267 | 0,996951932 | KRBOX4 | FALSE | Downregulated |
| -0,006081336 | 0,996951932 | LOC728989 | FALSE | Downregulated |
| 0,001688912 | 0,996951932 | MSI2 | FALSE | Upregulated |
| 0,001232911 | 0,996951932 | NCAPD2 | FALSE | Upregulated |
| 0,00494918 | 0,996951932 | RP5-1103G7.4 | FALSE | Upregulated |
| -0,001435331 | 0,996951932 | SDR39U1 | FALSE | Downregulated |
| -0,00206294 | 0,996951932 | USP30 | FALSE | Downregulated |
| -0,003940747 | 0,996955298 | LOC389834 | FALSE | Downregulated |
| -0,001628657 | 0,996964281 | CDKAL1 | FALSE | Downregulated |
| 0,003396831 | 0,996964281 | ERVK13-1 | FALSE | Upregulated |
| 0,009726588 | 0,996964281 | LDLRAD4-AS1 | FALSE | Upregulated |
| -0,007214845 | 0,996964281 | LOC101927198 | FALSE | Downregulated |
| 0,001550737 | 0,996964281 | RABL5 | FALSE | Upregulated |
| -0,001668831 | 0,996964281 | SLC12A6 | FALSE | Downregulated |
| 0,009033055 | 0,996964281 | SLC9A5 | FALSE | Upregulated |
| 0,005568678 | 0,996999717 | LINC00672 | FALSE | Upregulated |
| -0,001714724 | 0,996999717 | MGRN1 | FALSE | Downregulated |

| | | | | |
|---|---|---|---|---|
| -0,002057059 | 0,997052272 | BRD3 | FALSE | Downregulated |
| 0,002180103 | 0,997052272 | IFI27L1 | FALSE | Upregulated |
| -0,002900885 | 0,997052272 | RNR1 | FALSE | Downregulated |
| 0,002798041 | 0,997143873 | ZNF844 | FALSE | Upregulated |
| -0,003009737 | 0,997184687 | CNKSR2 | FALSE | Downregulated |
| -0,003701087 | 0,997184687 | KIT | FALSE | Downregulated |
| 0,001089481 | 0,997579745 | MRPL45 | FALSE | Upregulated |
| 0,004080923 | 0,997579745 | RAD51AP1 | FALSE | Upregulated |
| -0,001191961 | 0,997579745 | SERPINA6 | FALSE | Downregulated |
| -0,001995404 | 0,997579745 | TSSC1 | FALSE | Downregulated |
| -0,00258647 | 0,997660539 | ETV1 | FALSE | Downregulated |
| -0,011378171 | 0,997660539 | LOC101928477 | FALSE | Downregulated |
| 0,053774454 | 0,997660539 | MYBPC2 | FALSE | Upregulated |
| -0,002137634 | 0,997660539 | TIPIN | FALSE | Downregulated |
| 0,001444103 | 0,997661883 | IRF3 | FALSE | Upregulated |
| 0,000722523 | 0,997690289 | PPP1R7 | FALSE | Upregulated |
| 0,001313348 | 0,997690289 | UGCG | FALSE | Upregulated |
| 0,001962669 | 0,997749098 | CECR2 | FALSE | Upregulated |
| 0,001781201 | 0,997749098 | GPR126 | FALSE | Upregulated |
| -0,003299454 | 0,997749098 | LOC100129148 | FALSE | Downregulated |
| -0,002327467 | 0,997749098 | LOC100132832 | FALSE | Downregulated |
| 0,004735921 | 0,997749098 | LOC101927124 | FALSE | Upregulated |
| 0,009910274 | 0,997749098 | NRK | FALSE | Upregulated |
| 0,008475094 | 0,997749098 | PRSS45 | FALSE | Upregulated |
| 0,001116021 | 0,997749098 | TOM1L1 | FALSE | Upregulated |
| 0,001920771 | 0,997806721 | AS3MT | FALSE | Upregulated |

| | | | | |
|---|---|---|---|---|
| 0,002701495 | 0,997806721 | C1orf63 | FALSE | Upregulated |
| -0,001224499 | 0,997806721 | CIPC | FALSE | Downregulated |
| 0,001180415 | 0,997806721 | CRAMP1L | FALSE | Upregulated |
| -0,002627779 | 0,997806721 | CYP4X1 | FALSE | Downregulated |
| -0,005087478 | 0,997806721 | LOC102724757 | FALSE | Downregulated |
| 0,001060427 | 0,997806721 | PPM1F | FALSE | Upregulated |
| -0,001218848 | 0,997806721 | PROC | FALSE | Downregulated |
| 0,001330367 | 0,997806721 | REV3L | FALSE | Upregulated |
| 0,001039466 | 0,997806721 | RPL18 | FALSE | Upregulated |
| 0,000901802 | 0,997806721 | SELT | FALSE | Upregulated |
| -0,000759832 | 0,997806721 | TARS2 | FALSE | Downregulated |
| -0,00100931 | 0,997806721 | VPS8 | FALSE | Downregulated |
| -0,001560056 | 0,997860921 | ACSM2B | FALSE | Downregulated |
| 0,001311598 | 0,997860921 | CSPP1 | FALSE | Upregulated |
| -0,001688854 | 0,997860921 | DHX34 | FALSE | Downregulated |
| -0,002708766 | 0,997860921 | LOC728407 | FALSE | Downregulated |
| 0,001659222 | 0,997860921 | MYO9A | FALSE | Upregulated |
| -0,001366533 | 0,997865624 | GCKR | FALSE | Downregulated |
| -0,00362955 | 0,997951549 | CHKB-AS1 | FALSE | Downregulated |
| -0,000818041 | 0,997951549 | FNTA | FALSE | Downregulated |
| 0,005337654 | 0,997951549 | LOC102724552 | FALSE | Upregulated |
| -0,002882158 | 0,997951549 | PIH1D2 | FALSE | Downregulated |
| -0,001624103 | 0,997951549 | ST3GAL4 | FALSE | Downregulated |
| 0,005807857 | 0,99799648 | BSN-AS2 | FALSE | Upregulated |
| 0,002982472 | 0,998057297 | ACKR3 | FALSE | Upregulated |
| 0,003093351 | 0,998057297 | FRZB | FALSE | Upregulated |

| | | | | |
|---|---|---|---|---|
| -0,000739555 | 0,998057297 | GSPT2 | FALSE | Downregulated |
| -0,001082888 | 0,998057297 | H1F0 | FALSE | Downregulated |
| 0,000951594 | 0,998057297 | LRRC58 | FALSE | Upregulated |
| -0,001746484 | 0,998057297 | OVGP1 | FALSE | Downregulated |
| -0,001074323 | 0,998057297 | PVRL3 | FALSE | Downregulated |
| -0,006863417 | 0,998057297 | SNORA70 | FALSE | Downregulated |
| -0,000772051 | 0,998057297 | STRADA | FALSE | Downregulated |
| -0,001094709 | 0,998057297 | TNRC6A | FALSE | Downregulated |
| -0,004869884 | 0,998057297 | ULBP2 | FALSE | Downregulated |
| 0,002065688 | 0,998126421 | ARHGEF26 | FALSE | Upregulated |
| -0,000708026 | 0,998126421 | CLN5 | FALSE | Downregulated |
| 0,001100925 | 0,998126421 | FKTN | FALSE | Upregulated |
| 0,000777773 | 0,998166145 | ZFPL1 | FALSE | Upregulated |
| 0,000855063 | 0,998213025 | LLGL2 | FALSE | Upregulated |
| -0,002723938 | 0,998213025 | LMCD1 | FALSE | Downregulated |
| -0,001068424 | 0,998213025 | MCEE | FALSE | Downregulated |
| 0,001479707 | 0,99821869 | LOC652276 | FALSE | Upregulated |
| -0,001404661 | 0,998313079 | ANKRD12 | FALSE | Downregulated |
| -0,002292635 | 0,998313079 | TTLL7 | FALSE | Downregulated |
| -0,002018214 | 0,998503149 | CYP3A5 | FALSE | Downregulated |
| 0,003323408 | 0,998581325 | HIST2H2AC | FALSE | Upregulated |
| -0,004587943 | 0,998581325 | LOC101927026 | FALSE | Downregulated |
| 0,000657716 | 0,998581325 | UBE2F | FALSE | Upregulated |
| -0,000824096 | 0,998642601 | CCDC115 | FALSE | Downregulated |
| -0,000695967 | 0,998705737 | DCAF13 | FALSE | Downregulated |
| -0,000947413 | 0,998705737 | DPYD | FALSE | Downregulated |

| | | | | |
|---|---|---|---|---|
| 0,000674941 | 0,998705737 | ELOVL5 | FALSE | Upregulated |
| -0,001972008 | 0,998705737 | FRAS1 | FALSE | Downregulated |
| 0,000483973 | 0,998705737 | ING4 | FALSE | Upregulated |
| 0,02418333 | 0,998705737 | MYLK2 | FALSE | Upregulated |
| 0,000730745 | 0,998705737 | PDLIM5 | FALSE | Upregulated |
| 0,001545778 | 0,998705737 | PLB1 | FALSE | Upregulated |
| -0,001201793 | 0,998705737 | PPP1R9B | FALSE | Downregulated |
| 0,001182101 | 0,998705737 | RAPGEF4 | FALSE | Upregulated |
| 0,00360229 | 0,998705737 | SLC45A2 | FALSE | Upregulated |
| -0,00059942 | 0,998705737 | TCP1 | FALSE | Downregulated |
| -0,002770703 | 0,998705737 | ULBP3 | FALSE | Downregulated |
| 0,001311367 | 0,998705737 | VAMP1 | FALSE | Upregulated |
| 0,000517592 | 0,998751306 | SMIM7 | FALSE | Upregulated |
| 0,000660265 | 0,998751306 | TBC1D8B | FALSE | Upregulated |
| -0,012590814 | 0,998751306 | TSIX | FALSE | Downregulated |
| -0,000696503 | 0,998751306 | UCK1 | FALSE | Downregulated |
| -0,000500972 | 0,998751306 | YIPF6 | FALSE | Downregulated |
| -0,001789228 | 0,998751306 | ZDHHC11 | FALSE | Downregulated |
| 0,001373897 | 0,9988881 | CETP | FALSE | Upregulated |
| 0,000390602 | 0,9988881 | RABGAP1 | FALSE | Upregulated |
| -0,000429787 | 0,998928161 | EPRS | FALSE | Downregulated |
| 0,002659586 | 0,998928161 | LINC00242 | FALSE | Upregulated |
| 0,000767358 | 0,998928161 | MAT1A | FALSE | Upregulated |
| 0,001008525 | 0,998928161 | TOX2 | FALSE | Upregulated |
| -0,000518292 | 0,998928161 | UCKL1 | FALSE | Downregulated |
| -0,00069218 | 0,999156587 | ZNF69 | FALSE | Downregulated |

| | | | | |
|---|---|---|---|---|
| -0,001000215 | 0,99919914 | RP11-95P2.1 | FALSE | Downregulated |
| -0,002823691 | 0,999224851 | LOC101927268 | FALSE | Downregulated |
| 0,004821322 | 0,999242435 | ATP2A1 | FALSE | Upregulated |
| -0,001602555 | 0,999242435 | COL23A1 | FALSE | Downregulated |
| -0,000766475 | 0,999242435 | OGT | FALSE | Downregulated |
| -0,000492624 | 0,999242435 | ZDHHC12 | FALSE | Downregulated |
| -0,001193758 | 0,99939729 | LINC00240 | FALSE | Downregulated |
| -0,002036898 | 0,999407537 | MYH11 | FALSE | Downregulated |
| 0,000325804 | 0,999407537 | TSR1 | FALSE | Upregulated |
| -0,001211698 | 0,999431669 | RP11-677M14.7 | FALSE | Downregulated |
| 0,003830286 | 0,999431669 | TNMD | FALSE | Upregulated |
| -0,000243703 | 0,999547077 | DDX6 | FALSE | Downregulated |
| -0,000809209 | 0,999547077 | HERC2P9 | FALSE | Downregulated |
| -0,001718845 | 0,999547077 | LOC101929482 | FALSE | Downregulated |
| 0,00040739 | 0,999547077 | MUM1 | FALSE | Upregulated |
| 0,000681903 | 0,999547077 | NPC1L1 | FALSE | Upregulated |
| 0,000412115 | 0,999547077 | TSPAN14 | FALSE | Upregulated |
| 0,000643535 | 0,999547077 | TUFT1 | FALSE | Upregulated |
| 0,001306099 | 0,999665127 | ADRB1 | FALSE | Upregulated |
| -0,000317948 | 0,999695105 | ANKRD27 | FALSE | Downregulated |
| 0,000243493 | 0,999695105 | BTBD6 | FALSE | Upregulated |
| 0,00042277 | 0,999695105 | CD58 | FALSE | Upregulated |
| -0,000246532 | 0,999695105 | FARP1 | FALSE | Downregulated |
| -0,000252628 | 0,999695105 | GATAD2B | FALSE | Downregulated |
| 0,00090148 | 0,999695105 | GNMT | FALSE | Upregulated |
| 0,000359679 | 0,999695105 | LOC93622 | FALSE | Upregulated |

| | | | | |
|---|---|---|---|---|
| -0,000913562 | 0,999695105 | NLRP14 | FALSE | Downregulated |
| -0,000325504 | 0,999695105 | RPS5 | FALSE | Downregulated |
| 0,000428705 | 0,999695105 | SP110 | FALSE | Upregulated |
| -0,000460304 | 0,999695105 | TMEM117 | FALSE | Downregulated |
| 0,001890214 | 0,999758114 | ASPHD1 | FALSE | Upregulated |
| -0,010472364 | 0,999758114 | BOLA2 | FALSE | Downregulated |
| -0,000257666 | 0,999758114 | GBE1 | FALSE | Downregulated |
| -0,000314417 | 0,999758114 | HOXB2 | FALSE | Downregulated |
| -0,000154603 | 0,999758114 | LARP1 | FALSE | Downregulated |
| -0,001130839 | 0,999758114 | LGALS9C | FALSE | Downregulated |
| 0,000524584 | 0,999758114 | LINC00961 | FALSE | Upregulated |
| 0,000680725 | 0,999758114 | LOC101928249 | FALSE | Upregulated |
| -0,00025156 | 0,999758114 | METTL7B | FALSE | Downregulated |
| -0,000132004 | 0,999758114 | PEX12 | FALSE | Downregulated |
| -0,001311892 | 0,999758114 | PHKA2-AS1 | FALSE | Downregulated |
| -0,000799002 | 0,999758114 | PSPN | FALSE | Downregulated |
| 0,001131136 | 0,999758114 | RGS6 | FALSE | Upregulated |
| -0,000191936 | 0,999758114 | RPLP0 | FALSE | Downregulated |
| 0,000226994 | 0,999758114 | SEPP1 | FALSE | Upregulated |
| 0,000343465 | 0,999758114 | SLC25A19 | FALSE | Upregulated |
| 0,000126786 | 0,999758114 | TOMM20 | FALSE | Upregulated |
| 0,000100132 | 0,999758114 | UBR1 | FALSE | Upregulated |
| 0,000173724 | 0,999758114 | VPS39 | FALSE | Upregulated |
| -0,00013011 | 0,999758114 | XBP1 | FALSE | Downregulated |
| -0,000156362 | 0,999780738 | C8orf37 | FALSE | Downregulated |
| 7,11336E-05 | 0,999780738 | CSTF2T | FALSE | Upregulated |

0,000459381 0,999780738 PTP4A3 FALSE Upregulated

-0,000386692 0,999819205 LOC101928615 FALSE Downregulated

-0,000139131 0,999884836 UBN2 FALSE Downregulated

-0,000204263 0,999923222 GLIPR1L2 FALSE Downregulated

-6,38015E-05 0,999923222 GNB2L1 FALSE Downregulated

-4,40509E-05 0,999923222 MRPL1 FALSE Downregulated

0,00077861 0,999923222 TMEM256-PLSCR3 FALSE Upregulated

0,000119288 0,999923222 TMEM67 FALSE Upregulated

3,56381E-05 0,999947042 BRF2 FALSE Upregulated

-5,81869E-05 0,999947042 CLDN5 FALSE Downregulated

-3,98932E-05 0,999947042 WDR34 FALSE Downregulated

-5,04275E-05 0,999967435 NUAK2 FALSE Downregulated

7,1311E-06 0,999981589 DPYSL3 FALSE Upregulated

1,91242E-05 0,999981589 TLE6 FALSE Upregulated

-3,08337E-06 0,999981589 VPS28 FALSE Downregulated

0,263981551 A2MP1 FALSE Upregulated

0,157368803 A4GNT FALSE Upregulated

0,644822764 ABCA12 FALSE Upregulated

-1,12590781 ABCA17P Downregulated

-0,461319731 ABCB5 FALSE Downregulated

-0,691540001 ABCC12 FALSE Downregulated

1,372942551 ABCC13 Upregulated

0,646587041 ABCC8 FALSE Upregulated

-0,426639002 ABCG4 FALSE Downregulated

0,435981333 ABHD12B FALSE Upregulated

-0,934305996 ABHD16B FALSE Downregulated

| | | | |
|---|---|---|---|
| 0,250175492 | AC002480.3 | FALSE | Upregulated |
| 1,184800323 | AC003102.3 | | Upregulated |
| -0,634846795 | AC005022.1 | FALSE | Downregulated |
| 0,007899179 | AC006145.4 | FALSE | Upregulated |
| -0,900123323 | AC006272.1 | FALSE | Downregulated |
| 0,536215288 | AC007386.2 | FALSE | Upregulated |
| -0,270548019 | AC007405.4 | FALSE | Downregulated |
| -0,413369956 | AC007743.1 | FALSE | Downregulated |
| -0,112408717 | AC007750.5 | FALSE | Downregulated |
| -0,501378634 | AC007966.1 | FALSE | Downregulated |
| 0,6658346 | AC010890.1 | FALSE | Upregulated |
| -1,276054372 | AC012309.5 | | Downregulated |
| -0,496884103 | AC073133.1 | FALSE | Downregulated |
| -0,05051798 | AC073283.4 | FALSE | Downregulated |
| 0,415308903 | AC079145.4 | FALSE | Upregulated |
| 0,350408505 | AC079610.2 | FALSE | Upregulated |
| -1,310783522 | AC087859.1 | | Downregulated |
| 0,300608376 | AC090044.1 | FALSE | Upregulated |
| 1,39043883 | AC096559.1 | | Upregulated |
| -0,0159006 | AC097500.2 | FALSE | Downregulated |
| 0,143797647 | AC098823.3 | FALSE | Upregulated |
| -0,127443193 | AC104024.1 | FALSE | Downregulated |
| 0,50782265 | AC104654.2 | FALSE | Upregulated |
| -0,745258436 | AC104777.2 | FALSE | Downregulated |
| 0,5921689 | AC104777.4 | FALSE | Upregulated |
| -0,185486752 | AC105053.3 | FALSE | Downregulated |

| | | | |
|---|---|---|---|
| 1,009296569 | AC131097.3 | | Upregulated |
| -0,368448818 | AC226118.1 | FALSE | Downregulated |
| 0,338263669 | ACAN | FALSE | Upregulated |
| 0,502641801 | ACER1 | FALSE | Upregulated |
| 0,93350053 | ACOXL | FALSE | Upregulated |
| 0,188240816 | ACTL10 | FALSE | Upregulated |
| 0,78280531 | ADAD2 | FALSE | Upregulated |
| -0,016832481 | ADAM20 | FALSE | Downregulated |
| -0,563826835 | ADAM21 | FALSE | Downregulated |
| 1,03331579 | ADAM32 | | Upregulated |
| 0,46769197 | ADAMTS8 | FALSE | Upregulated |
| 0,503981796 | ADAMTSL4-AS1 | FALSE | Upregulated |
| 0,885007024 | ADGB | FALSE | Upregulated |
| 0,529870302 | ADIPOQ-AS1 | FALSE | Upregulated |
| -0,106009779 | ADPGK-AS1 | FALSE | Downregulated |
| 0,288632605 | AFAP1-AS1 | FALSE | Upregulated |
| -0,153730914 | AGPAT4-IT1 | FALSE | Downregulated |
| -0,574034609 | AIPL1 | FALSE | Downregulated |
| 0,716611358 | AK8 | FALSE | Upregulated |
| -0,55724855 | AL022476.2 | FALSE | Downregulated |
| 0,630372587 | AL109763.2 | FALSE | Upregulated |
| 0,404684804 | AL132709.5 | FALSE | Upregulated |
| -0,551322225 | ALK | FALSE | Downregulated |
| -0,738532879 | ALOX12 | FALSE | Downregulated |
| 0,083863116 | ALS2CR11 | FALSE | Upregulated |
| 0,505559976 | AMH | FALSE | Upregulated |

| | | | |
|---|---|---|---|
| -1,116411642 | AMY2A | | Downregulated |
| 1,347481961 | ANGPT4 | | Upregulated |
| 0,025489334 | ANKK1 | FALSE | Upregulated |
| -0,497402669 | ANKRD18B | FALSE | Downregulated |
| -1,051427869 | ANKRD30A | | Downregulated |
| -0,494048395 | ANKRD30B | FALSE | Downregulated |
| -0,49961853 | ANKRD30BP3 | FALSE | Downregulated |
| 1,311477125 | ANKRD44-IT1 | | Upregulated |
| 0,328277111 | ANKRD45 | FALSE | Upregulated |
| 0,351210614 | ANKRD61 | FALSE | Upregulated |
| -0,64974451 | ANKRD62 | FALSE | Downregulated |
| -0,311105576 | ANO2 | FALSE | Downregulated |
| 1,343012353 | APC2 | | Upregulated |
| 0,215990413 | APOOP5 | FALSE | Upregulated |
| -0,323201797 | AQP8 | FALSE | Downregulated |
| -0,442224669 | AQPEP | FALSE | Downregulated |
| -0,46673215 | ARC | FALSE | Downregulated |
| 0,064774293 | AREG | FALSE | Upregulated |
| 1,463004786 | ARGFX | | Upregulated |
| 0,03871669 | ARHGAP40 | FALSE | Upregulated |
| 0,274347971 | ARL14 | FALSE | Upregulated |
| 1,053409132 | ARL9 | | Upregulated |
| 1,425477856 | ARRDC3-AS1 | | Upregulated |
| 0,736527753 | ARRDC5 | FALSE | Upregulated |
| 0,749232833 | ARTN | FALSE | Upregulated |
| -0,143446783 | ARX | FALSE | Downregulated |

| | | | |
|---|---|---|---|
| 1,384898563 | ASAP1-IT2 | | Upregulated |
| 0,168999349 | ASB12 | FALSE | Upregulated |
| -0,815375586 | ASB16 | FALSE | Downregulated |
| -0,765516455 | ASB9P1 | FALSE | Downregulated |
| -0,648252125 | ASMT | FALSE | Downregulated |
| 0,359630406 | ATCAY | FALSE | Upregulated |
| 1,415924014 | ATP10B | | Upregulated |
| 0,501858524 | ATP12A | FALSE | Upregulated |
| 1,968258007 | ATP1A4 | | Upregulated |
| 0,598019195 | ATP2B3 | FALSE | Upregulated |
| 2,334391747 | ATP6V0D2 | | Upregulated |
| -0,520260266 | ATP6V1B1 | FALSE | Downregulated |
| 0,431581317 | AURKAPS1 | FALSE | Upregulated |
| 1,16172935 | AZU1 | | Upregulated |
| -0,133797646 | B3GALT1 | FALSE | Downregulated |
| 0,296350717 | BARX2 | FALSE | Upregulated |
| -0,073718955 | BEND2 | FALSE | Downregulated |
| 0,913088202 | BEND3P3 | FALSE | Upregulated |
| 0,626915436 | BEST4 | FALSE | Upregulated |
| 1,064236706 | BHLHE23 | | Upregulated |
| 0,100998505 | BHLHE40-AS1 | FALSE | Upregulated |
| 0,506720145 | BMP3 | FALSE | Upregulated |
| 1,243914649 | BPIFB2 | | Upregulated |
| 0,331714106 | BRD7P3 | FALSE | Upregulated |
| 1,170703885 | BRE-AS1 | | Upregulated |
| -0,545006064 | BREA2 | FALSE | Downregulated |

| | | | |
|---|---|---|---|
| 0,801706311 | BRSK2 | FALSE | Upregulated |
| -1,413453752 | BTNL3 | | Downregulated |
| -1,053019317 | BVES-AS1 | | Downregulated |
| -0,499691409 | C10orf111 | FALSE | Downregulated |
| -0,729933968 | C10orf67 | FALSE | Downregulated |
| -0,63117494 | C10orf85 | FALSE | Downregulated |
| -0,784455676 | C10orf90 | FALSE | Downregulated |
| -0,185993857 | C10orf95 | FALSE | Downregulated |
| 1,055024607 | C11orf70 | | Upregulated |
| 1,43598537 | C12orf42 | | Upregulated |
| -0,073962581 | C12orf68 | FALSE | Downregulated |
| -0,165515562 | C12orf74 | FALSE | Downregulated |
| -0,830359377 | C15orf26 | FALSE | Downregulated |
| -0,031974314 | C16orf3 | FALSE | Downregulated |
| 0,104531131 | C16orf74 | FALSE | Upregulated |
| 0,618267915 | C17orf104 | FALSE | Upregulated |
| 0,471429276 | C17orf66 | FALSE | Upregulated |
| 1,100196328 | C18orf61 | | Upregulated |
| 0,975804949 | C19orf45 | FALSE | Upregulated |
| -1,384680156 | C19orf59 | | Downregulated |
| -0,157613725 | C19orf83 | FALSE | Downregulated |
| -0,09294099 | C1orf110 | FALSE | Downregulated |
| -0,112096781 | C1orf111 | FALSE | Downregulated |
| 0,080105271 | C1orf140 | FALSE | Upregulated |
| 0,109407199 | C1orf145 | FALSE | Upregulated |
| -0,430315604 | C1orf185 | FALSE | Downregulated |

| | | | |
|---|---|---|---|
| 0,087944707 | C1orf189 | FALSE | Upregulated |
| 0,1169153 | C1orf200 | FALSE | Upregulated |
| -0,143252982 | C1orf229 | FALSE | Downregulated |
| -0,544547046 | C1orf61 | FALSE | Downregulated |
| -0,938686847 | C1QL2 | FALSE | Downregulated |
| 0,122742978 | C1QL3 | FALSE | Upregulated |
| -0,060458335 | C1QTNF1-AS1 | FALSE | Downregulated |
| 0,867247836 | C1QTNF4 | FALSE | Upregulated |
| 0,08359686 | C20orf166-AS1 | FALSE | Upregulated |
| 1,059914031 | C20orf197 | | Upregulated |
| -1,597898936 | C20orf26 | | Downregulated |
| 0,186716228 | C21orf128 | FALSE | Upregulated |
| -0,328346564 | C21orf140 | FALSE | Downregulated |
| 0,346200367 | C22orf23 | FALSE | Upregulated |
| 0,024385583 | C22orf31 | FALSE | Upregulated |
| 0,595142332 | C2orf91 | FALSE | Upregulated |
| -0,134511523 | C3orf27 | FALSE | Downregulated |
| -0,385073938 | C3orf36 | FALSE | Downregulated |
| 1,484294907 | C3orf56 | | Upregulated |
| -0,093376213 | C3orf83 | FALSE | Downregulated |
| -1,287540735 | C4orf26 | | Downregulated |
| -0,095745901 | C4orf50 | FALSE | Downregulated |
| 0,487714183 | C4orf6 | FALSE | Upregulated |
| 0,489595426 | C5orf60 | FALSE | Upregulated |
| 0,156718752 | C5orf64 | FALSE | Upregulated |
| -0,477966261 | C6orf222 | FALSE | Downregulated |

| | | | |
|---|---|---|---|
| -0,346360852 | C6orf223 | FALSE | Downregulated |
| 0,015177511 | C6orf99 | FALSE | Upregulated |
| -1,034468265 | C9orf170 | | Downregulated |
| 0,405356381 | C9orf173 | FALSE | Upregulated |
| -0,05810795 | C9orf47 | FALSE | Downregulated |
| 0,181794741 | CA4 | FALSE | Upregulated |
| 0,663853865 | CABP1 | FALSE | Upregulated |
| 0,254286283 | CACNA1B | FALSE | Upregulated |
| 0,632996992 | CACNA1C-AS1 | FALSE | Upregulated |
| 0,93621319 | CACNA1F | FALSE | Upregulated |
| 1,509833342 | CACNG3 | | Upregulated |
| -0,052903444 | CACNG4 | FALSE | Downregulated |
| -0,70081853 | CADPS | FALSE | Downregulated |
| 1,618217453 | CALB1 | | Upregulated |
| 0,076410792 | CALB2 | FALSE | Upregulated |
| 0,046066015 | CALCR | FALSE | Upregulated |
| 0,382503301 | CAMP | FALSE | Upregulated |
| 0,795908336 | CAPN14 | FALSE | Upregulated |
| -0,450876002 | CAPN8 | FALSE | Downregulated |
| 0,174855602 | CAPN8-2 | FALSE | Upregulated |
| 0,550064091 | CATSPER1 | FALSE | Upregulated |
| 1,002145049 | CC2D2B | | Upregulated |
| -1,057710242 | CCDC116 | | Downregulated |
| -0,794821873 | CCDC175 | FALSE | Downregulated |
| 0,601294352 | CCDC177 | FALSE | Upregulated |
| 0,159106904 | CCDC178 | FALSE | Upregulated |

| | | | |
|---|---|---|---|
| -0,194607589 | CCDC36 | FALSE | Downregulated |
| -0,883900022 | CCDC41-AS1 | FALSE | Downregulated |
| 0,05389853 | CCDC74B | FALSE | Upregulated |
| -1,105193087 | CCDC78 | | Downregulated |
| -0,440528483 | CCDC79 | FALSE | Downregulated |
| -0,876317473 | CCL11 | FALSE | Downregulated |
| 1,536636209 | CCL25 | | Upregulated |
| -0,677395722 | CCR10 | FALSE | Downregulated |
| 1,757484211 | CCR8 | | Upregulated |
| 0,123518688 | CD177 | FALSE | Upregulated |
| 0,454156219 | CD1A | FALSE | Upregulated |
| 1,304322568 | CD1B | | Upregulated |
| -0,573657964 | CD70 | FALSE | Downregulated |
| -0,71391736 | CD81-AS1 | FALSE | Downregulated |
| 0,991772062 | CDC14C | FALSE | Upregulated |
| 0,655550712 | CDC42P3 | FALSE | Upregulated |
| 0,159223462 | CDCP2 | FALSE | Upregulated |
| 1,567856693 | CDH16 | | Upregulated |
| 0,832319876 | CDH17 | FALSE | Upregulated |
| -0,905460687 | CDH22 | FALSE | Downregulated |
| 0,343243421 | CDH3 | FALSE | Upregulated |
| 0,213949566 | CDHR4 | FALSE | Upregulated |
| 0,195531597 | CDK15 | FALSE | Upregulated |
| 0,524546411 | CEACAM20 | FALSE | Upregulated |
| -0,111236613 | CEACAM8 | FALSE | Downregulated |
| 0,722419514 | CECR5-AS1 | FALSE | Upregulated |

| | | | |
|---|---|---|---|
| 0,269679846 | CECR7 | FALSE | Upregulated |
| -0,37040852 | CERS1 | FALSE | Downregulated |
| 0,345203051 | CETN4P | FALSE | Upregulated |
| 0,34113053 | CFL1P1 | FALSE | Upregulated |
| -0,086833297 | CHD5 | FALSE | Downregulated |
| -1,145528693 | CHIA | | Downregulated |
| -0,67725708 | CHODL | FALSE | Downregulated |
| 1,112482508 | CHRM3-AS2 | | Upregulated |
| 1,173502186 | CHRM5 | | Upregulated |
| -0,105685661 | CHRNA2 | FALSE | Downregulated |
| -0,867296506 | CHRNA7 | FALSE | Downregulated |
| 0,521433999 | CHRNB2 | FALSE | Upregulated |
| 0,34488245 | CHST5 | FALSE | Upregulated |
| 0,46229539 | CHST6 | FALSE | Upregulated |
| 0,116293975 | CHST8 | FALSE | Upregulated |
| 0,772427878 | CITED1 | FALSE | Upregulated |
| 0,995407579 | CKMT1A | FALSE | Upregulated |
| -0,092546052 | CKMT1B | FALSE | Downregulated |
| 0,507706092 | CLCN1 | FALSE | Upregulated |
| -1,285858716 | CLCNKA | | Downregulated |
| 0,345953717 | CLEC17A | FALSE | Upregulated |
| -0,674990647 | CLEC19A | FALSE | Downregulated |
| -0,392224125 | CLEC4D | FALSE | Downregulated |
| 0,120914167 | CLK2P | FALSE | Upregulated |
| -0,311112481 | CMA1 | FALSE | Downregulated |
| -1,421166823 | CMTM2 | | Downregulated |

| | | | |
|---|---|---|---|
| -0,385856423 | CNIH2 | FALSE | Downregulated |
| 0,365941821 | CNTN2 | FALSE | Upregulated |
| 0,063684003 | CNTNAP4 | FALSE | Upregulated |
| 1,298414649 | CNTNAP5 | | Upregulated |
| -0,412743117 | COL11A1 | FALSE | Downregulated |
| 0,078646807 | COL17A1 | FALSE | Upregulated |
| -0,125551741 | COL18A1-AS2 | FALSE | Downregulated |
| -0,033965229 | COL2A1 | FALSE | Downregulated |
| 1,499548992 | CORT | | Upregulated |
| 1,097179439 | CPA1 | | Upregulated |
| -0,208749063 | CPA4 | FALSE | Downregulated |
| -0,1346281 | CPEB1 | FALSE | Downregulated |
| 0,008598448 | CPEB2-AS1 | FALSE | Upregulated |
| 0,213311361 | CPLX4 | FALSE | Upregulated |
| 0,92836921 | CRB1 | FALSE | Upregulated |
| 0,040597778 | CRLF1 | FALSE | Upregulated |
| -0,830105055 | CRX | FALSE | Downregulated |
| -0,754728706 | CRYBA4 | FALSE | Downregulated |
| 0,484757288 | CRYM-AS1 | FALSE | Upregulated |
| 0,228487379 | CSAG3 | FALSE | Upregulated |
| 0,080883203 | CSMD1 | FALSE | Upregulated |
| 0,395663876 | CSNK1A1L | FALSE | Upregulated |
| 0,570854876 | CSTF3-AS1 | FALSE | Upregulated |
| 1,407844126 | CTC-436P18.1 | | Upregulated |
| 0,319504476 | CTD-2127H9.1 | FALSE | Upregulated |
| -0,210721804 | CTXN1 | FALSE | Downregulated |

| | | | |
|---|---|---|---|
| -0,001208572 | CXADRP3 | FALSE | Downregulated |
| -0,513253683 | CXCL17 | FALSE | Downregulated |
| -0,461806126 | CXorf22 | FALSE | Downregulated |
| -0,275384942 | CXorf30 | FALSE | Downregulated |
| 0,097184361 | CXorf58 | FALSE | Upregulated |
| 0,170870859 | CXorf64 | FALSE | Upregulated |
| 0,533372328 | CXXC1P1 | FALSE | Upregulated |
| 0,603594147 | CYP26C1 | FALSE | Upregulated |
| -0,160135409 | CYP4F35P | FALSE | Downregulated |
| 0,41477803 | CYP4F8 | FALSE | Upregulated |
| -1,167964726 | CYTL1 | | Downregulated |
| 1,313681469 | DCAF13P3 | | Upregulated |
| 0,40768698 | DCST1 | FALSE | Upregulated |
| -0,147207478 | DCX | FALSE | Downregulated |
| 0,355289328 | DGCR10 | FALSE | Upregulated |
| 1,630464055 | DGCR9 | | Upregulated |
| 0,415656944 | DGKI | FALSE | Upregulated |
| 0,328970392 | DGUOK-AS1 | FALSE | Upregulated |
| -0,059211742 | DHDH | FALSE | Downregulated |
| -0,277519169 | DIO2 | FALSE | Downregulated |
| -1,006058663 | DIRAS1 | | Downregulated |
| 0,314514536 | DKFZP434A062 | FALSE | Upregulated |
| 0,364745969 | DKFZP434K028 | FALSE | Upregulated |
| 0,518275051 | DKK1 | FALSE | Upregulated |
| -0,088066761 | DKK4 | FALSE | Downregulated |
| -0,295803257 | DLEU7-AS1 | FALSE | Downregulated |

| | | | |
|---|---|---|---|
| 1,150711264 | DLG1-AS1 | | Upregulated |
| -0,383405783 | DLGAP1-AS3 | FALSE | Downregulated |
| 0,593891666 | DLX4 | FALSE | Upregulated |
| -1,092111091 | DMBX1 | | Downregulated |
| -0,35150008 | DMRT2 | FALSE | Downregulated |
| 0,412458608 | DNAH17 | FALSE | Upregulated |
| 0,456719951 | DNAH3 | FALSE | Upregulated |
| 0,762517049 | DNAH8 | FALSE | Upregulated |
| -0,402973884 | DNAH9 | FALSE | Downregulated |
| 0,715474468 | DNAJB13 | FALSE | Upregulated |
| -0,944916874 | DNAJB7 | FALSE | Downregulated |
| -0,463003096 | DNAJC9-AS1 | FALSE | Downregulated |
| 0,804763857 | DNASE1L2 | FALSE | Upregulated |
| 0,471544536 | DNER | FALSE | Upregulated |
| 0,140942653 | DNM1P35 | FALSE | Upregulated |
| 1,20848149 | DNM1P41 | | Upregulated |
| 1,006194909 | DNM1P50 | | Upregulated |
| -1,13753936 | DOC2GP | | Downregulated |
| 0,014719973 | DOCK3 | FALSE | Upregulated |
| -0,292429772 | DOCK9-AS1 | FALSE | Downregulated |
| 0,674005524 | DPH3P1 | FALSE | Upregulated |
| 0,739973449 | DPRXP4 | FALSE | Upregulated |
| 0,036612953 | DPYSL4 | FALSE | Upregulated |
| -0,541843623 | DPYSL5 | FALSE | Downregulated |
| 1,123273048 | DQX1 | | Upregulated |
| -0,013018026 | DRC1 | FALSE | Downregulated |

| | | | |
|---|---|---|---|
| 1,697412317 | DSC1 | | Upregulated |
| 0,335214867 | DSCR9 | FALSE | Upregulated |
| 0,047484487 | DSG4 | FALSE | Upregulated |
| -0,684842008 | DUOXA1 | FALSE | Downregulated |
| -0,434452249 | DUSP15 | FALSE | Downregulated |
| 0,266113686 | DYNAP | FALSE | Upregulated |
| -2,581341037 | EBF2 | | Downregulated |
| 1,337973195 | ECEL1 | | Upregulated |
| 1,06587656 | ECT2L | | Upregulated |
| -0,508048591 | EDNRB-AS1 | FALSE | Downregulated |
| -0,6402275 | EFCAB10 | FALSE | Downregulated |
| 1,283779499 | EFHC2 | | Upregulated |
| -0,942678132 | EGFEM1P | FALSE | Downregulated |
| 0,131419968 | EGFL6 | FALSE | Upregulated |
| -0,283668507 | EIF3IP1 | FALSE | Downregulated |
| 0,982684808 | ELANE | FALSE | Upregulated |
| -0,208701708 | ELAVL3 | FALSE | Downregulated |
| 1,13419891 | ELOVL3 | | Upregulated |
| 0,218759532 | EMX1 | FALSE | Upregulated |
| -0,697822716 | ENDOU | FALSE | Downregulated |
| -0,247842711 | ENO1-AS1 | FALSE | Downregulated |
| 0,392684124 | ENTPD3 | FALSE | Upregulated |
| -1,863068761 | EPB42 | | Downregulated |
| -0,012866852 | EPGN | FALSE | Downregulated |
| 0,681558452 | EPHA10 | FALSE | Upregulated |
| 1,050582663 | EPHA6 | | Upregulated |

| | | | |
|---|---|---|---|
| 1,376504567 | EPHA8 | | Upregulated |
| 0,953460413 | EPHX3 | FALSE | Upregulated |
| 1,259247436 | EPN3 | | Upregulated |
| 1,555276853 | EPS8L3 | | Upregulated |
| 0,462207129 | ERC2 | FALSE | Upregulated |
| 0,633498503 | EREG | FALSE | Upregulated |
| 0,125490078 | ESR2 | FALSE | Upregulated |
| -1,424471794 | ESRG | | Downregulated |
| 0,320660268 | FA2H | FALSE | Upregulated |
| 0,637012506 | FABP5P3 | FALSE | Upregulated |
| -0,428439808 | FABP6 | FALSE | Downregulated |
| 0,372886641 | FAIM2 | FALSE | Upregulated |
| 0,132414772 | FAM106A | FALSE | Upregulated |
| 0,245888989 | FAM106CP | FALSE | Upregulated |
| -0,6262612 | FAM132A | FALSE | Downregulated |
| 0,600611737 | FAM132B | FALSE | Upregulated |
| -0,781201239 | FAM153C | FALSE | Downregulated |
| -0,004078106 | FAM154A | FALSE | Downregulated |
| 0,632159602 | FAM159A | FALSE | Upregulated |
| -0,306493601 | FAM170A | FALSE | Downregulated |
| 0,63038699 | FAM171A2 | FALSE | Upregulated |
| 0,290998765 | FAM177B | FALSE | Upregulated |
| 0,630754837 | FAM178B | FALSE | Upregulated |
| 0,425718175 | FAM183B | FALSE | Upregulated |
| -0,490761707 | FAM184B | FALSE | Downregulated |
| -0,533361807 | FAM186A | FALSE | Downregulated |

| | | | |
|---|---|---|---|
| 0,166636958 | FAM194A | FALSE | Upregulated |
| 1,278939308 | FAM196B | | Upregulated |
| -1,172766827 | FAM209A | | Downregulated |
| 0,281061005 | FAM212B-AS1 | FALSE | Upregulated |
| -0,529400302 | FAM221B | FALSE | Downregulated |
| 0,725179362 | FAM226A | FALSE | Upregulated |
| -0,244214927 | FAM226B | FALSE | Downregulated |
| 0,656390856 | FAM27B | FALSE | Upregulated |
| 1,213432603 | FAM27C | | Upregulated |
| -0,301946135 | FAM3D | FALSE | Downregulated |
| 0,39379707 | FAM57B | FALSE | Upregulated |
| 0,820407801 | FAM64A | FALSE | Upregulated |
| -0,901106975 | FAM66D | FALSE | Downregulated |
| 1,297572475 | FAM71D | | Upregulated |
| 0,2782842 | FAM71F1 | FALSE | Upregulated |
| -0,930746202 | FAM86KP | FALSE | Downregulated |
| 0,377172684 | FAM90A1 | FALSE | Upregulated |
| 0,622660535 | FAM92A1P2 | FALSE | Upregulated |
| -0,508071409 | FAM95A | FALSE | Downregulated |
| -2,243208643 | FAM9A | | Downregulated |
| 0,581186575 | FBLL1 | FALSE | Upregulated |
| 0,78590076 | FBXL21 | FALSE | Upregulated |
| -0,172700688 | FBXW4P1 | FALSE | Downregulated |
| 0,973434434 | FCRL2 | FALSE | Upregulated |
| 0,402309745 | FCRL4 | FALSE | Upregulated |
| 0,610282272 | FER1L6 | FALSE | Upregulated |

| | | | |
|---|---|---|---|
| 0,502128407 | FGD5P1 | FALSE | Upregulated |
| -0,105766637 | FGF10 | FALSE | Downregulated |
| 0,960803814 | FGF13-AS1 | FALSE | Upregulated |
| 0,833947213 | FGF20 | FALSE | Upregulated |
| -0,135127021 | FGF9 | FALSE | Downregulated |
| 0,209889556 | FIRRE | FALSE | Upregulated |
| 0,101565649 | FLG2 | FALSE | Upregulated |
| -0,335595027 | FLJ16779 | FALSE | Downregulated |
| 0,330452061 | FLJ25363 | FALSE | Upregulated |
| 1,153455708 | FLJ27354 | | Upregulated |
| -0,21010451 | FLJ30375 | FALSE | Downregulated |
| -0,599828662 | FLJ30679 | FALSE | Downregulated |
| -0,788968066 | FLJ31356 | FALSE | Downregulated |
| -0,374609374 | FLJ31945 | FALSE | Downregulated |
| 0,90624499 | FLJ35934 | FALSE | Upregulated |
| -0,008882851 | FLJ36000 | FALSE | Downregulated |
| -0,155939564 | FLJ38122 | FALSE | Downregulated |
| 0,600308204 | FLJ38379 | FALSE | Upregulated |
| 0,643413821 | FLJ41200 | FALSE | Upregulated |
| 0,47439712 | FLJ41481 | FALSE | Upregulated |
| 0,494860927 | FLJ42393 | FALSE | Upregulated |
| 0,229112864 | FLJ43315 | FALSE | Upregulated |
| -0,405806279 | FLJ45079 | FALSE | Downregulated |
| 0,186022735 | FLJ45825 | FALSE | Upregulated |
| 0,739640204 | FLJ45872 | FALSE | Upregulated |
| 0,166257401 | FLJ45974 | FALSE | Upregulated |

| | | | |
|---|---|---|---|
| -0,22847051 | FLRT1 | FALSE | Downregulated |
| 0,502238991 | FMN2 | FALSE | Upregulated |
| -0,404076838 | FMO6P | FALSE | Downregulated |
| 0,416276835 | FMR1NB | FALSE | Upregulated |
| 0,536054208 | FNDC8 | FALSE | Upregulated |
| -0,617043349 | FOXD1 | FALSE | Downregulated |
| 0,387728593 | FOXD4 | FALSE | Upregulated |
| 0,05465006 | FOXD4L1 | FALSE | Upregulated |
| 0,56467799 | FOXF2 | FALSE | Upregulated |
| -1,01796533 | FOXI2 | | Downregulated |
| 0,526567717 | FOXJ1 | FALSE | Upregulated |
| -0,381103135 | FRMD5 | FALSE | Downregulated |
| 1,670376149 | FRMPD3 | | Upregulated |
| -0,351306445 | FRMPD4 | FALSE | Downregulated |
| -1,298997809 | FSCN2 | | Downregulated |
| 0,752919538 | FSTL4 | FALSE | Upregulated |
| 0,254830975 | FUT5 | FALSE | Upregulated |
| 0,586641016 | FUT7 | FALSE | Upregulated |
| 0,403464545 | FUT9 | FALSE | Upregulated |
| -0,812741379 | FZD10-AS1 | FALSE | Downregulated |
| 0,268377099 | GABARAPL3 | FALSE | Upregulated |
| 0,014150401 | GABRA2 | FALSE | Upregulated |
| -0,631245167 | GABRA4 | FALSE | Downregulated |
| 0,393202259 | GABRB2 | FALSE | Upregulated |
| 0,008312293 | GABRD | FALSE | Upregulated |
| 0,808819515 | GAD1 | FALSE | Upregulated |

| | | | |
|---|---|---|---|
| -0,138467935 | GAL3ST2 | FALSE | Downregulated |
| -0,095952027 | GALNT5 | FALSE | Downregulated |
| 1,220260945 | GALNT8 | | Upregulated |
| -1,306795469 | GALNT9 | | Downregulated |
| -1,71088379 | GALP | | Downregulated |
| -0,502953985 | GATA3-AS1 | FALSE | Downregulated |
| 0,69864292 | GCSAML | FALSE | Upregulated |
| 2,055012535 | GFI1B | | Upregulated |
| 0,434746479 | GGN | FALSE | Upregulated |
| -0,870717862 | GGT8P | FALSE | Downregulated |
| 0,227339724 | GJA3 | FALSE | Upregulated |
| 0,096236122 | GJC2 | FALSE | Upregulated |
| 0,364281492 | GLB1L3 | FALSE | Upregulated |
| 0,522257431 | GLIPR1L1 | FALSE | Upregulated |
| 0,034107721 | GLIS1 | FALSE | Upregulated |
| 0,462632848 | GLRA1 | FALSE | Upregulated |
| -1,289082705 | GNAS-AS1 | | Downregulated |
| -0,25568179 | GNB3 | FALSE | Downregulated |
| 1,25659221 | GNG4 | | Upregulated |
| 1,026727639 | GP2 | | Upregulated |
| -0,114946157 | GP5 | FALSE | Downregulated |
| 0,366531964 | GPA33 | FALSE | Upregulated |
| 0,591743979 | GPC2 | FALSE | Upregulated |
| -0,946931891 | GPR1 | FALSE | Downregulated |
| -0,375655337 | GPR1-AS | FALSE | Downregulated |
| 0,037863527 | GPR110 | FALSE | Upregulated |

| | | | |
|---|---|---|---|
| -0,078174071 | GPR15 | FALSE | Downregulated |
| 0,701531051 | GPR17 | FALSE | Upregulated |
| -0,224542391 | GPR19 | FALSE | Downregulated |
| -0,926632735 | GPR21 | FALSE | Downregulated |
| -0,301534502 | GPR37L1 | FALSE | Downregulated |
| 0,905931678 | GPR84 | FALSE | Upregulated |
| -0,657391197 | GPRC5A | FALSE | Downregulated |
| -0,008515297 | GPRC5D | FALSE | Downregulated |
| -0,039422906 | GRIA1 | FALSE | Downregulated |
| -0,681632234 | GRID2 | FALSE | Downregulated |
| 1,557575679 | GRID2IP | | Upregulated |
| -0,258104935 | GRIK1-AS1 | FALSE | Downregulated |
| -0,00210525 | GRIK1-AS2 | FALSE | Downregulated |
| 0,20272318 | GRIK3 | FALSE | Upregulated |
| 0,675769113 | GRIN2B | FALSE | Upregulated |
| 0,722971351 | GRIN2C | FALSE | Upregulated |
| -1,099995808 | GRIN3B | | Downregulated |
| -0,003446627 | GRK5-IT1 | FALSE | Downregulated |
| -0,481797705 | GRM1 | FALSE | Downregulated |
| 0,414750402 | GRM2 | FALSE | Upregulated |
| -0,477350923 | GRM3 | FALSE | Downregulated |
| 0,577824747 | GRM4 | FALSE | Upregulated |
| -0,039570543 | GRM6 | FALSE | Downregulated |
| 0,909856067 | GRM7 | FALSE | Upregulated |
| -0,795228395 | GRPR | FALSE | Downregulated |
| 0,635176011 | GS1-122H1.2 | FALSE | Upregulated |

| | | | |
|---|---|---|---|
| -0,024680403 | GSDMC | FALSE | Downregulated |
| 0,140064335 | GSG1 | FALSE | Upregulated |
| -0,685109671 | GSG1L | FALSE | Downregulated |
| -1,725016098 | GSN-AS1 | | Downregulated |
| 0,201455243 | GTSE1-AS1 | FALSE | Upregulated |
| 0,657541483 | GUCA2A | FALSE | Upregulated |
| -0,775366136 | GUCY2EP | FALSE | Downregulated |
| 0,855358744 | GYG2P1 | FALSE | Upregulated |
| -0,035668981 | GYPA | FALSE | Downregulated |
| -0,315487866 | GYPE | FALSE | Downregulated |
| -0,264292919 | H2BFXP | FALSE | Downregulated |
| -0,06356798 | HAGHL | FALSE | Downregulated |
| -1,280506019 | HAPLN1 | | Downregulated |
| 1,845658291 | HAPLN2 | | Upregulated |
| -0,416943344 | HAS2-AS1 | FALSE | Downregulated |
| -0,69519063 | HCAR1 | FALSE | Downregulated |
| -0,278153575 | HCG17 | FALSE | Downregulated |
| -1,321712724 | HCG23 | | Downregulated |
| -0,079360854 | HCG25 | FALSE | Downregulated |
| 1,353004607 | HCN4 | | Upregulated |
| 0,921353444 | HERC2P10 | FALSE | Upregulated |
| -0,02103084 | HES2 | FALSE | Downregulated |
| -0,270424741 | HFM1 | FALSE | Downregulated |
| 0,962612927 | HHIPL2 | FALSE | Upregulated |
| 0,900420235 | HHLA2 | FALSE | Upregulated |
| -0,307863781 | HIPK4 | FALSE | Downregulated |

| | | | |
|---|---|---|---|
| -0,466582078 | HIST1H1E | FALSE | Downregulated |
| 0,248186847 | HIST1H2AK | FALSE | Upregulated |
| 0,106883162 | HIST1H2BG | FALSE | Upregulated |
| -0,859788927 | HIST1H2BO | FALSE | Downregulated |
| 0,330173131 | HIST1H3D | FALSE | Upregulated |
| 1,462528444 | HIST1H4C | | Upregulated |
| 0,748121511 | HIST1H4H | FALSE | Upregulated |
| -0,454734091 | HIST2H3D | FALSE | Downregulated |
| 0,975333716 | HIST4H4 | FALSE | Upregulated |
| 0,688442055 | HMGN2P46 | FALSE | Upregulated |
| -0,397076762 | HMP19 | FALSE | Downregulated |
| -0,16657853 | HMSD | FALSE | Downregulated |
| 0,550765981 | HNRNPCP5 | FALSE | Upregulated |
| -0,318562094 | HNRNPKP3 | FALSE | Downregulated |
| 0,693409198 | HOXA10 | FALSE | Upregulated |
| 1,044167206 | HOXD-AS1 | | Upregulated |
| -0,520298228 | HOXD8 | FALSE | Downregulated |
| 0,840013708 | HPCAL4 | FALSE | Upregulated |
| -0,152893626 | HRASLS5 | FALSE | Downregulated |
| -0,593519947 | HS6ST3 | FALSE | Downregulated |
| 1,096373077 | HTR1F | | Upregulated |
| -0,629792938 | HTR2A | FALSE | Downregulated |
| 0,351312207 | HTR2A-AS1 | FALSE | Upregulated |
| 0,299161438 | HTR4 | FALSE | Upregulated |
| 0,782333337 | HTRA4 | FALSE | Upregulated |
| -1,201684267 | HYDIN2 | | Downregulated |

| | | | |
|---|---|---|---|
| 0,617716687 | ICAM4 | FALSE | Upregulated |
| -0,160395971 | ICAM5 | FALSE | Downregulated |
| -0,85270777 | IFIT1B | FALSE | Downregulated |
| -0,279117644 | IFITM4P | FALSE | Downregulated |
| 0,605788033 | IFNL1 | FALSE | Upregulated |
| -0,49582388 | IGBP1P1 | FALSE | Downregulated |
| -0,007279862 | IGLL3P | FALSE | Downregulated |
| -0,263775862 | IGSF11 | FALSE | Downregulated |
| -0,380610627 | IGSF5 | FALSE | Downregulated |
| 1,338431738 | IGSF9B | | Upregulated |
| -0,029215191 | IL12A | FALSE | Downregulated |
| -0,519451185 | IL12B | FALSE | Downregulated |
| 0,217548709 | IL21R-AS1 | FALSE | Upregulated |
| -0,118417052 | IL24 | FALSE | Downregulated |
| -0,283882866 | IL31RA | FALSE | Downregulated |
| 0,812932916 | IL36G | FALSE | Upregulated |
| -0,004909579 | IL9R | FALSE | Downregulated |
| -0,542358943 | INE2 | FALSE | Downregulated |
| -0,261877954 | INGX | FALSE | Downregulated |
| -0,898138049 | INHA | FALSE | Downregulated |
| -0,182435168 | INTS4L2 | FALSE | Downregulated |
| 0,123555327 | IPO9-AS1 | FALSE | Upregulated |
| 1,174000419 | ISL2 | | Upregulated |
| 0,781459339 | JAZF1-AS1 | FALSE | Upregulated |
| 1,16615341 | JPH4 | | Upregulated |
| 0,560506138 | KATNBL1P6 | FALSE | Upregulated |

| | | | |
|---|---|---|---|
| -0,171937026 | KCNA2 | FALSE | Downregulated |
| 0,350779027 | KCNE1L | FALSE | Upregulated |
| 0,09611745 | KCNF1 | FALSE | Upregulated |
| -1,255834554 | KCNH1 | | Downregulated |
| 0,77535573 | KCNH3 | FALSE | Upregulated |
| -0,106068047 | KCNH4 | FALSE | Downregulated |
| -0,720729704 | KCNH6 | FALSE | Downregulated |
| -0,439622455 | KCNIP4 | FALSE | Downregulated |
| 0,404106101 | KCNJ1 | FALSE | Upregulated |
| 1,065228279 | KCNJ13 | | Upregulated |
| 1,081713733 | KCNJ4 | | Upregulated |
| -0,456110967 | KCNJ9 | FALSE | Downregulated |
| -1,371591982 | KCNK15 | | Downregulated |
| 0,288113629 | KCNK7 | FALSE | Upregulated |
| 0,628026703 | KCNMB2 | FALSE | Upregulated |
| -0,039242251 | KCNQ2 | FALSE | Downregulated |
| 0,073904833 | KCNS1 | FALSE | Upregulated |
| 1,316831402 | KCNT1 | | Upregulated |
| -0,233887695 | KCP | FALSE | Downregulated |
| 0,647769435 | KCTD16 | FALSE | Upregulated |
| -1,064356337 | KHDRBS2 | | Downregulated |
| 0,602332158 | KIAA0319 | FALSE | Upregulated |
| 0,871475754 | KIAA1549L | FALSE | Upregulated |
| 0,355216104 | KIAA2022 | FALSE | Upregulated |
| -0,410842873 | KIF28P | FALSE | Downregulated |
| -0,378503465 | KIF5A | FALSE | Downregulated |

| | | | |
|---|---|---|---|
| 0,017893253 | KIR2DL4 | FALSE | Upregulated |
| -1,031881313 | KIRREL3 | | Downregulated |
| -0,365171707 | KLF14 | FALSE | Downregulated |
| -0,940937539 | KLHDC8A | FALSE | Downregulated |
| -0,449596838 | KLHL10 | FALSE | Downregulated |
| 0,650306079 | KLHL35 | FALSE | Upregulated |
| 1,264550236 | KLK11 | | Upregulated |
| 1,339051907 | KLK14 | | Upregulated |
| -0,152715546 | KRT1 | FALSE | Downregulated |
| 0,000398681 | KRT15 | FALSE | Upregulated |
| 0,276701117 | KRT18P55 | FALSE | Upregulated |
| -0,308091164 | KRT27 | FALSE | Downregulated |
| 0,800607716 | KRT36 | FALSE | Upregulated |
| -0,118668164 | KRT42P | FALSE | Downregulated |
| 0,210147791 | KRT5 | FALSE | Upregulated |
| 0,135998966 | KRT72 | FALSE | Upregulated |
| 0,889955205 | KRT85 | FALSE | Upregulated |
| -0,236773729 | KRTAP5-2 | FALSE | Downregulated |
| 0,364467158 | KRTAP5-8 | FALSE | Upregulated |
| 0,451297084 | KRTAP5-9 | FALSE | Upregulated |
| 0,193812541 | KY | FALSE | Upregulated |
| -1,299577466 | L1TD1 | | Downregulated |
| 0,9948055 | LAMB4 | FALSE | Upregulated |
| 0,303491317 | LANCL3 | FALSE | Upregulated |
| 0,50426836 | LCE2D | FALSE | Upregulated |
| 0,228539507 | LCN10 | FALSE | Upregulated |

| | | | |
|---|---|---|---|
| 0,622336163 | LCTL | FALSE | Upregulated |
| 0,892394185 | LDLRAD1 | FALSE | Upregulated |
| 0,544656189 | LEMD1-AS1 | FALSE | Upregulated |
| -3,060210274 | LEP | | Downregulated |
| 0,369648387 | LGALS12 | FALSE | Upregulated |
| -0,435213265 | LGALS17A | FALSE | Downregulated |
| -0,591085275 | LGALS8-AS1 | FALSE | Downregulated |
| 0,54392381 | LGALS9B | FALSE | Upregulated |
| 0,720867003 | LGI3 | FALSE | Upregulated |
| -0,703319816 | LHFPL1 | FALSE | Downregulated |
| -0,54890144 | LHFPL3-AS2 | FALSE | Downregulated |
| -1,384331183 | LHFPL5 | | Downregulated |
| -0,273432853 | LINC00032 | FALSE | Downregulated |
| 1,059601828 | LINC00167 | | Upregulated |
| -1,973869874 | LINC00202-2 | | Downregulated |
| 0,645535161 | LINC00260 | FALSE | Upregulated |
| -0,16486526 | LINC00271 | FALSE | Downregulated |
| 0,011040658 | LINC00278 | FALSE | Upregulated |
| -0,309597783 | LINC00298 | FALSE | Downregulated |
| -0,864912639 | LINC00311 | FALSE | Downregulated |
| -1,163631867 | LINC00327 | | Downregulated |
| 0,360644789 | LINC00330 | FALSE | Upregulated |
| 0,042836937 | LINC00461 | FALSE | Upregulated |
| -0,052018087 | LINC00479 | FALSE | Downregulated |
| 0,442360436 | LINC00485 | FALSE | Upregulated |
| 0,172253005 | LINC00487 | FALSE | Upregulated |

| | | | |
|---|---|---|---|
| 0,334639677 | LINC00494 | FALSE | Upregulated |
| -0,625554938 | LINC00515 | FALSE | Downregulated |
| 0,98831019 | LINC00540 | FALSE | Upregulated |
| 0,754431032 | LINC00593 | FALSE | Upregulated |
| -0,388569899 | LINC00629 | FALSE | Downregulated |
| -0,41911236 | LINC00643 | FALSE | Downregulated |
| 0,567343065 | LINC00652 | FALSE | Upregulated |
| 0,042270277 | LINC00664 | FALSE | Upregulated |
| 1,185561218 | LINC00673 | | Upregulated |
| -1,55933443 | LINC00693 | | Downregulated |
| 0,060450531 | LINC00707 | FALSE | Upregulated |
| 0,197530047 | LINC00854 | FALSE | Upregulated |
| -0,123396288 | LINC00877 | FALSE | Downregulated |
| 0,890176485 | LINC00887 | FALSE | Upregulated |
| 0,443308778 | LINC00889 | FALSE | Upregulated |
| -0,185756575 | LINC00896 | FALSE | Downregulated |
| -0,89668428 | LINC00908 | FALSE | Downregulated |
| -0,248515653 | LINC00923 | FALSE | Downregulated |
| 0,609973845 | LINC00941 | FALSE | Upregulated |
| -0,303315007 | LINC00942 | FALSE | Downregulated |
| -1,296505717 | LINC00944 | | Downregulated |
| 0,981046119 | LINC00954 | FALSE | Upregulated |
| -0,622525746 | LINC00958 | FALSE | Downregulated |
| -0,548022694 | LINC00964 | FALSE | Downregulated |
| 0,540478263 | LINC00968 | FALSE | Upregulated |
| 0,575962451 | LINC00977 | FALSE | Upregulated |

| | | | |
|---|---|---|---|
| 0,072698406 | LINC00989 | FALSE | Upregulated |
| 0,951516729 | LINC01013 | FALSE | Upregulated |
| -0,709907527 | LINC01087 | FALSE | Downregulated |
| -0,085511141 | LINC01091 | FALSE | Downregulated |
| -0,244866461 | LINC01105 | FALSE | Downregulated |
| 1,041160727 | LINC01108 | | Upregulated |
| -0,384087821 | LINC01118 | FALSE | Downregulated |
| 0,023781918 | LINC01119 | FALSE | Upregulated |
| -0,274899664 | LINC01122 | FALSE | Downregulated |
| 0,325995093 | LINC01135 | FALSE | Upregulated |
| -1,386948023 | LINC01139 | | Downregulated |
| 0,271259301 | LINC01141 | FALSE | Upregulated |
| 0,878952691 | LINC01150 | FALSE | Upregulated |
| -0,356993126 | LINC01181 | FALSE | Downregulated |
| 1,192550505 | LINC01186 | | Upregulated |
| 0,566303924 | LINGO2 | FALSE | Upregulated |
| 1,282311416 | LIPN | | Upregulated |
| 0,177796256 | LMO3 | FALSE | Upregulated |
| -0,179450796 | LOC100101148 | FALSE | Downregulated |
| -0,630190776 | LOC100127974 | FALSE | Downregulated |
| -0,202029832 | LOC100128164 | FALSE | Downregulated |
| -0,854271873 | LOC100128374 | FALSE | Downregulated |
| -0,3550369 | LOC100128568 | FALSE | Downregulated |
| 0,059966824 | LOC100128573 | FALSE | Upregulated |
| -0,494396566 | LOC100128593 | FALSE | Downregulated |
| 0,583932355 | LOC100128653 | FALSE | Upregulated |

| | | | |
|---|---|---|---|
| -0,515642003 | LOC100128882 | FALSE | Downregulated |
| -0,065511972 | LOC100129138 | FALSE | Downregulated |
| 0,559106998 | LOC100129215 | FALSE | Upregulated |
| -0,248543137 | LOC100129316 | FALSE | Downregulated |
| 0,788849747 | LOC100129473 | FALSE | Upregulated |
| -1,478618278 | LOC100129617 | | Downregulated |
| 0,118550646 | LOC100130075 | FALSE | Upregulated |
| -0,897470196 | LOC100130238 | FALSE | Downregulated |
| 0,488319381 | LOC100130298 | FALSE | Upregulated |
| -0,077041125 | LOC100130520 | FALSE | Downregulated |
| 0,353009548 | LOC100130705 | FALSE | Upregulated |
| 0,932611558 | LOC100130876 | FALSE | Upregulated |
| -0,845795184 | LOC100131289 | FALSE | Downregulated |
| -0,88697176 | LOC100131315 | FALSE | Downregulated |
| -0,341939207 | LOC100131496 | FALSE | Downregulated |
| -1,209231394 | LOC100131626 | | Downregulated |
| -0,329921987 | LOC100131825 | FALSE | Downregulated |
| -0,851468618 | LOC100132354 | FALSE | Downregulated |
| -0,634091382 | LOC100132731 | FALSE | Downregulated |
| -0,226619118 | LOC100134040 | FALSE | Downregulated |
| -1,098264015 | LOC100134368 | | Downregulated |
| -0,986270388 | LOC100188947 | FALSE | Downregulated |
| -1,269601328 | LOC100287166 | | Downregulated |
| -0,265081623 | LOC100287808 | FALSE | Downregulated |
| 0,712673992 | LOC100288162 | FALSE | Upregulated |
| -0,588307677 | LOC100288748 | FALSE | Downregulated |

| | | | |
|---|---|---|---|
| -0,230186579 | LOC100288846 | FALSE | Downregulated |
| -0,662041416 | LOC100289092 | FALSE | Downregulated |
| 0,1060787 | LOC100289333 | FALSE | Upregulated |
| 0,385673119 | LOC100289455 | FALSE | Upregulated |
| 0,470696233 | LOC100289511 | FALSE | Upregulated |
| 0,606358784 | LOC100335030 | FALSE | Upregulated |
| -0,323591358 | LOC100422556 | FALSE | Downregulated |
| 2,046839819 | LOC100505498 | | Upregulated |
| -0,740868263 | LOC100505534 | FALSE | Downregulated |
| 0,371383713 | LOC100505622 | FALSE | Upregulated |
| -1,097394038 | LOC100505625 | | Downregulated |
| 0,640779012 | LOC100505685 | FALSE | Upregulated |
| 0,188420811 | LOC100505774 | FALSE | Upregulated |
| -1,383133364 | LOC100506253 | | Downregulated |
| -0,429117405 | LOC100506351 | FALSE | Downregulated |
| 0,5334752 | LOC100506504 | FALSE | Upregulated |
| 0,13987293 | LOC100506526 | FALSE | Upregulated |
| 0,157771174 | LOC100506859 | FALSE | Upregulated |
| -0,490279349 | LOC100506901 | FALSE | Downregulated |
| 0,882353292 | LOC100507033 | FALSE | Upregulated |
| -0,17883875 | LOC100507156 | FALSE | Downregulated |
| 0,163209652 | LOC100507346 | FALSE | Upregulated |
| 0,513528729 | LOC100507387 | FALSE | Upregulated |
| -0,739455005 | LOC100507403 | FALSE | Downregulated |
| 0,253457508 | LOC100616530 | FALSE | Upregulated |
| -0,810734699 | LOC100652930 | FALSE | Downregulated |

| | | | |
|---|---|---|---|
| -1,119349927 | LOC100653515 | | Downregulated |
| 1,045345703 | LOC100996369 | | Upregulated |
| -0,51400014 | LOC100996379 | FALSE | Downregulated |
| 0,339593562 | LOC100996405 | FALSE | Upregulated |
| -0,319543203 | LOC100996506 | FALSE | Downregulated |
| 0,951994688 | LOC100996517 | FALSE | Upregulated |
| -0,956240481 | LOC100996549 | FALSE | Downregulated |
| -0,315048613 | LOC100996557 | FALSE | Downregulated |
| -0,124228877 | LOC100996696 | FALSE | Downregulated |
| -1,114267801 | LOC100996716 | | Downregulated |
| -1,053799067 | LOC100996813 | | Downregulated |
| -0,430266091 | LOC100996873 | FALSE | Downregulated |
| 0,341637092 | LOC101059906 | FALSE | Upregulated |
| 1,90502531 | LOC101059954 | | Upregulated |
| -0,70168141 | LOC101059976 | FALSE | Downregulated |
| 1,184995303 | LOC101060153 | | Upregulated |
| 0,610289648 | LOC101060363 | FALSE | Upregulated |
| -0,227738805 | LOC101060400 | FALSE | Downregulated |
| -0,602289815 | LOC101060632 | FALSE | Downregulated |
| -1,166682447 | LOC101926887 | | Downregulated |
| -0,31657226 | LOC101926896 | FALSE | Downregulated |
| -0,697730932 | LOC101926936 | FALSE | Downregulated |
| -0,499658327 | LOC101926940 | FALSE | Downregulated |
| -0,952480242 | LOC101926969 | FALSE | Downregulated |
| -0,219430442 | LOC101926997 | FALSE | Downregulated |
| -0,226076511 | LOC101927012 | FALSE | Downregulated |

| | | | |
|---|---|---|---|
| 1,348966272 | LOC101927031 | | Upregulated |
| 0,670949825 | LOC101927085 | FALSE | Upregulated |
| -0,291067501 | LOC101927116 | FALSE | Downregulated |
| -0,629319974 | LOC101927125 | FALSE | Downregulated |
| 0,404351477 | LOC101927131 | FALSE | Upregulated |
| 1,496053794 | LOC101927143 | | Upregulated |
| -0,268143576 | LOC101927185 | FALSE | Downregulated |
| 0,26270738 | LOC101927212 | FALSE | Upregulated |
| 0,426577574 | LOC101927249 | FALSE | Upregulated |
| 1,072527287 | LOC101927254 | | Upregulated |
| -0,639737662 | LOC101927259 | FALSE | Downregulated |
| -0,215402031 | LOC101927262 | FALSE | Downregulated |
| -1,323110715 | LOC101927308 | | Downregulated |
| -0,917045406 | LOC101927326 | FALSE | Downregulated |
| -0,359341758 | LOC101927366 | FALSE | Downregulated |
| 0,80528335 | LOC101927374 | FALSE | Upregulated |
| -0,271210885 | LOC101927375 | FALSE | Downregulated |
| -0,471062668 | LOC101927437 | FALSE | Downregulated |
| -0,725518822 | LOC101927479 | FALSE | Downregulated |
| 0,62133287 | LOC101927486 | FALSE | Upregulated |
| 1,390478919 | LOC101927505 | | Upregulated |
| -1,469408999 | LOC101927568 | | Downregulated |
| -1,097975386 | LOC101927582 | | Downregulated |
| 0,03921428 | LOC101927596 | FALSE | Upregulated |
| 0,400031185 | LOC101927617 | FALSE | Upregulated |
| -0,834120561 | LOC101927632 | FALSE | Downregulated |

| | | | |
|---|---|---|---|
| -0,135509497 | LOC101927673 | FALSE | Downregulated |
| -0,183976285 | LOC101927680 | FALSE | Downregulated |
| 0,645507018 | LOC101927685 | FALSE | Upregulated |
| 0,594796633 | LOC101927696 | FALSE | Upregulated |
| -1,317885479 | LOC101927697 | | Downregulated |
| -0,462117698 | LOC101927702 | FALSE | Downregulated |
| -0,670713902 | LOC101927705 | FALSE | Downregulated |
| -0,560394528 | LOC101927720 | FALSE | Downregulated |
| 0,353053725 | LOC101927721 | FALSE | Upregulated |
| 0,0474354 | LOC101927725 | FALSE | Upregulated |
| -0,02828514 | LOC101927757 | FALSE | Downregulated |
| -0,392237027 | LOC101927762 | FALSE | Downregulated |
| -0,212274382 | LOC101927764 | FALSE | Downregulated |
| 1,319211507 | LOC101927775 | | Upregulated |
| 0,538133282 | LOC101927799 | FALSE | Upregulated |
| 0,142376882 | LOC101927803 | FALSE | Upregulated |
| 0,054474814 | LOC101927811 | FALSE | Upregulated |
| -0,415251172 | LOC101927814 | FALSE | Downregulated |
| -0,25039306 | LOC101927848 | FALSE | Downregulated |
| -1,267577059 | LOC101927854 | | Downregulated |
| -0,870542335 | LOC101927870 | FALSE | Downregulated |
| 0,34114764 | LOC101927875 | FALSE | Upregulated |
| -0,382502871 | LOC101927896 | FALSE | Downregulated |
| 0,400926981 | LOC101927898 | FALSE | Upregulated |
| 0,290062685 | LOC101927954 | FALSE | Upregulated |
| -0,856026202 | LOC101927971 | FALSE | Downregulated |

| | | | |
|---|---|---|---|
| -0,117479931 | LOC101927986 | FALSE | Downregulated |
| -0,164265705 | LOC101928011 | FALSE | Downregulated |
| 0,635973947 | LOC101928027 | FALSE | Upregulated |
| -0,169381236 | LOC101928039 | FALSE | Downregulated |
| 0,093558653 | LOC101928045 | FALSE | Upregulated |
| 0,287541464 | LOC101928062 | FALSE | Upregulated |
| -1,376046372 | LOC101928066 | | Downregulated |
| 0,656144144 | LOC101928080 | FALSE | Upregulated |
| 0,026117719 | LOC101928082 | FALSE | Upregulated |
| 1,002178347 | LOC101928109 | | Upregulated |
| 0,147849598 | LOC101928139 | FALSE | Upregulated |
| 0,019719446 | LOC101928152 | FALSE | Upregulated |
| -0,288446168 | LOC101928171 | FALSE | Downregulated |
| -0,31008063 | LOC101928188 | FALSE | Downregulated |
| -1,74999081 | LOC101928212 | | Downregulated |
| -1,208984342 | LOC101928231 | | Downregulated |
| -0,382858263 | LOC101928245 | FALSE | Downregulated |
| 0,69236843 | LOC101928255 | FALSE | Upregulated |
| 1,129271706 | LOC101928262 | | Upregulated |
| -0,011643077 | LOC101928279 | FALSE | Downregulated |
| -0,408676559 | LOC101928284 | FALSE | Downregulated |
| -0,65537435 | LOC101928317 | FALSE | Downregulated |
| 1,196154074 | LOC101928323 | | Upregulated |
| -0,385012836 | LOC101928334 | FALSE | Downregulated |
| 0,317892849 | LOC101928343 | FALSE | Upregulated |
| -0,119707728 | LOC101928349 | FALSE | Downregulated |

| | | | |
|---|---|---|---|
| 0,263125767 | LOC101928355 | FALSE | Upregulated |
| 0,280559898 | LOC101928360 | FALSE | Upregulated |
| -0,487267336 | LOC101928373 | FALSE | Downregulated |
| 0,143319259 | LOC101928402 | FALSE | Upregulated |
| 0,723548466 | LOC101928407 | FALSE | Upregulated |
| -0,088962223 | LOC101928439 | FALSE | Downregulated |
| -0,005854835 | LOC101928445 | FALSE | Downregulated |
| 1,796570089 | LOC101928466 | | Upregulated |
| 0,231347668 | LOC101928482 | FALSE | Upregulated |
| -0,057560109 | LOC101928484 | FALSE | Downregulated |
| -0,582110493 | LOC101928514 | FALSE | Downregulated |
| 0,735504226 | LOC101928530 | FALSE | Upregulated |
| 0,709817298 | LOC101928552 | FALSE | Upregulated |
| 0,383761493 | LOC101928555 | FALSE | Upregulated |
| 1,212508195 | LOC101928574 | | Upregulated |
| 0,71552298 | LOC101928592 | FALSE | Upregulated |
| -0,080335403 | LOC101928628 | FALSE | Downregulated |
| -1,204897518 | LOC101928632 | | Downregulated |
| 0,532491967 | LOC101928659 | FALSE | Upregulated |
| -0,09743078 | LOC101928665 | FALSE | Downregulated |
| 0,499383561 | LOC101928668 | FALSE | Upregulated |
| -0,178647757 | LOC101928671 | FALSE | Downregulated |
| 0,013446129 | LOC101928690 | FALSE | Upregulated |
| 0,006810888 | LOC101928692 | FALSE | Upregulated |
| -0,098856551 | LOC101928699 | FALSE | Downregulated |
| 0,130185413 | LOC101928716 | FALSE | Upregulated |

| | | | |
|---|---|---|---|
| 1,109189818 | LOC101928717 | | Upregulated |
| 0,460543423 | LOC101928733 | FALSE | Upregulated |
| -1,310110703 | LOC101928738 | | Downregulated |
| -0,28292978 | LOC101928765 | FALSE | Downregulated |
| -0,269267861 | LOC101928779 | FALSE | Downregulated |
| -1,313415899 | LOC101928814 | | Downregulated |
| -0,326024902 | LOC101928824 | FALSE | Downregulated |
| -0,002359353 | LOC101928838 | FALSE | Downregulated |
| 0,89575391 | LOC101928867 | FALSE | Upregulated |
| 0,420820978 | LOC101928906 | FALSE | Upregulated |
| 0,355800089 | LOC101928910 | FALSE | Upregulated |
| -0,07283028 | LOC101928911 | FALSE | Downregulated |
| -0,06300356 | LOC101928926 | FALSE | Downregulated |
| -0,192426255 | LOC101928961 | FALSE | Downregulated |
| -0,854849802 | LOC101928963 | FALSE | Downregulated |
| -0,873257688 | LOC101928976 | FALSE | Downregulated |
| -0,974459177 | LOC101928981 | FALSE | Downregulated |
| -0,306324624 | LOC101928993 | FALSE | Downregulated |
| 0,303120799 | LOC101929004 | FALSE | Upregulated |
| -0,827381993 | LOC101929008 | FALSE | Downregulated |
| -0,728643595 | LOC101929017 | FALSE | Downregulated |
| 0,699980818 | LOC101929035 | FALSE | Upregulated |
| 0,623742158 | LOC101929038 | FALSE | Upregulated |
| 0,04226871 | LOC101929051 | FALSE | Upregulated |
| -0,072088521 | LOC101929054 | FALSE | Downregulated |
| 0,016097428 | LOC101929057 | FALSE | Upregulated |

| | | | |
|---|---|---|---|
| -1,038726462 | LOC101929060 | | Downregulated |
| -0,545476168 | LOC101929078 | FALSE | Downregulated |
| -0,682138632 | LOC101929085 | FALSE | Downregulated |
| -0,634635584 | LOC101929125 | FALSE | Downregulated |
| 0,049271788 | LOC101929132 | FALSE | Upregulated |
| 0,049187678 | LOC101929149 | FALSE | Upregulated |
| -0,295988782 | LOC101929151 | FALSE | Downregulated |
| 0,608384952 | LOC101929157 | FALSE | Upregulated |
| 0,206623042 | LOC101929159 | FALSE | Upregulated |
| 1,0945315 | LOC101929165 | | Upregulated |
| 0,610946753 | LOC101929173 | FALSE | Upregulated |
| 0,170495669 | LOC101929188 | FALSE | Upregulated |
| 0,910122903 | LOC101929192 | FALSE | Upregulated |
| -0,056868158 | LOC101929216 | FALSE | Downregulated |
| 0,819961025 | LOC101929227 | FALSE | Upregulated |
| 0,163874081 | LOC101929241 | FALSE | Upregulated |
| 0,35164584 | LOC101929248 | FALSE | Upregulated |
| -0,643337779 | LOC101929259 | FALSE | Downregulated |
| -0,607544547 | LOC101929270 | FALSE | Downregulated |
| 0,556361044 | LOC101929272 | FALSE | Upregulated |
| -0,445675583 | LOC101929295 | FALSE | Downregulated |
| -0,156302213 | LOC101929330 | FALSE | Downregulated |
| -0,023557093 | LOC101929333 | FALSE | Downregulated |
| -0,172300375 | LOC101929336 | FALSE | Downregulated |
| 1,146191756 | LOC101929340 | | Upregulated |
| -0,655861986 | LOC101929346 | FALSE | Downregulated |

| | | | |
|---|---|---|---|
| 0,295128687 | LOC101929354 | FALSE | Upregulated |
| -0,16640911 | LOC101929375 | FALSE | Downregulated |
| -0,869387477 | LOC101929396 | FALSE | Downregulated |
| 0,136426015 | LOC101929409 | FALSE | Upregulated |
| -0,82665373 | LOC101929414 | FALSE | Downregulated |
| -0,242664642 | LOC101929422 | FALSE | Downregulated |
| 0,190114247 | LOC101929426 | FALSE | Upregulated |
| 0,13156158 | LOC101929431 | FALSE | Upregulated |
| -0,347727983 | LOC101929434 | FALSE | Downregulated |
| -0,588403873 | LOC101929447 | FALSE | Downregulated |
| -0,610584598 | LOC101929451 | FALSE | Downregulated |
| -1,376972272 | LOC101929462 | | Downregulated |
| 0,833096248 | LOC101929469 | FALSE | Upregulated |
| -0,202669464 | LOC101929490 | FALSE | Downregulated |
| 0,877007784 | LOC101929491 | FALSE | Upregulated |
| 0,971497469 | LOC101929494 | FALSE | Upregulated |
| 0,357717587 | LOC101929500 | FALSE | Upregulated |
| -0,286204792 | LOC101929511 | FALSE | Downregulated |
| 0,592085487 | LOC101929512 | FALSE | Upregulated |
| -0,164955021 | LOC101929521 | FALSE | Downregulated |
| 0,024844881 | LOC101929523 | FALSE | Upregulated |
| 0,201148104 | LOC101929536 | FALSE | Upregulated |
| 0,757616042 | LOC101929560 | FALSE | Upregulated |
| 0,206711414 | LOC101929566 | FALSE | Upregulated |
| 1,175141497 | LOC101929587 | | Upregulated |
| 0,374068698 | LOC101929609 | FALSE | Upregulated |

| | | | |
|---|---|---|---|
| -0,177128458 | LOC101929615 | FALSE | Downregulated |
| -0,207517329 | LOC101929634 | FALSE | Downregulated |
| 0,021184604 | LOC101929641 | FALSE | Upregulated |
| 0,099056536 | LOC101929661 | FALSE | Upregulated |
| -0,14641877 | LOC101929666 | FALSE | Downregulated |
| -0,204273997 | LOC101929692 | FALSE | Downregulated |
| 0,891260307 | LOC101929714 | FALSE | Upregulated |
| -1,312234374 | LOC101929726 | | Downregulated |
| -0,025588949 | LOC101929732 | FALSE | Downregulated |
| 0,893441976 | LOC101929751 | FALSE | Upregulated |
| 0,496738045 | LOC101929790 | FALSE | Upregulated |
| -0,839873947 | LOC101929866 | FALSE | Downregulated |
| -0,028529209 | LOC101929937 | FALSE | Downregulated |
| -0,749066154 | LOC101929946 | FALSE | Downregulated |
| -0,794700324 | LOC101930026 | FALSE | Downregulated |
| 0,275938732 | LOC101930137 | FALSE | Upregulated |
| 0,226964975 | LOC101930305 | FALSE | Upregulated |
| -0,719103142 | LOC101930310 | FALSE | Downregulated |
| -0,495785835 | LOC101930332 | FALSE | Downregulated |
| 0,273526164 | LOC101930370 | FALSE | Upregulated |
| 0,785843181 | LOC101930580 | FALSE | Upregulated |
| -0,784485035 | LOC102467073 | FALSE | Downregulated |
| 0,788791982 | LOC102467147 | FALSE | Upregulated |
| -0,545137169 | LOC102467214 | FALSE | Downregulated |
| 0,123407444 | LOC102467217 | FALSE | Upregulated |
| 0,596082268 | LOC102723305 | FALSE | Upregulated |

| | | | |
|---|---|---|---|
| 0,481607159 | LOC102723310 | FALSE | Upregulated |
| -0,515913389 | LOC102723316 | FALSE | Downregulated |
| -0,267338546 | LOC102723317 | FALSE | Downregulated |
| 0,425946569 | LOC102723320 | FALSE | Upregulated |
| -0,413657064 | LOC102723347 | FALSE | Downregulated |
| -0,212922469 | LOC102723353 | FALSE | Downregulated |
| -0,582973842 | LOC102723354 | FALSE | Downregulated |
| -0,680835435 | LOC102723361 | FALSE | Downregulated |
| 0,041205316 | LOC102723363 | FALSE | Upregulated |
| -1,165806759 | LOC102723366 | | Downregulated |
| 1,587158334 | LOC102723371 | | Upregulated |
| -0,219357811 | LOC102723382 | FALSE | Downregulated |
| 0,76490912 | LOC102723399 | FALSE | Upregulated |
| 0,851389602 | LOC102723408 | FALSE | Upregulated |
| 0,448149557 | LOC102723419 | FALSE | Upregulated |
| -0,761763564 | LOC102723420 | FALSE | Downregulated |
| 0,084869835 | LOC102723437 | FALSE | Upregulated |
| 0,042261554 | LOC102723446 | FALSE | Upregulated |
| 0,24181392 | LOC102723458 | FALSE | Upregulated |
| -0,544497026 | LOC102723475 | FALSE | Downregulated |
| 0,63300843 | LOC102723489 | FALSE | Upregulated |
| 0,35864989 | LOC102723507 | FALSE | Upregulated |
| 1,044393228 | LOC102723510 | | Upregulated |
| 0,197404257 | LOC102723543 | FALSE | Upregulated |
| 0,205385542 | LOC102723545 | FALSE | Upregulated |
| 1,452592406 | LOC102723559 | | Upregulated |

| | | | |
|---|---|---|---|
| -0,818168308 | LOC102723561 | FALSE | Downregulated |
| -0,353738811 | LOC102723581 | FALSE | Downregulated |
| -1,164885975 | LOC102723595 | | Downregulated |
| 1,22200981 | LOC102723619 | | Upregulated |
| -0,140576099 | LOC102723654 | FALSE | Downregulated |
| -1,010452759 | LOC102723656 | | Downregulated |
| 0,572797024 | LOC102723659 | FALSE | Upregulated |
| 1,229347049 | LOC102723682 | | Upregulated |
| 0,332232203 | LOC102723693 | FALSE | Upregulated |
| -1,363283703 | LOC102723694 | | Downregulated |
| 0,507658828 | LOC102723696 | FALSE | Upregulated |
| 0,84316497 | LOC102723697 | FALSE | Upregulated |
| 0,485686531 | LOC102723714 | FALSE | Upregulated |
| -0,611691431 | LOC102723721 | FALSE | Downregulated |
| 0,996579046 | LOC102723724 | FALSE | Upregulated |
| -0,629084104 | LOC102723729 | FALSE | Downregulated |
| -0,393793142 | LOC102723730 | FALSE | Downregulated |
| 0,397301217 | LOC102723733 | FALSE | Upregulated |
| 0,843845458 | LOC102723746 | FALSE | Upregulated |
| 0,464645129 | LOC102723750 | FALSE | Upregulated |
| -0,895944183 | LOC102723754 | FALSE | Downregulated |
| 0,446339519 | LOC102723761 | FALSE | Upregulated |
| 0,425063126 | LOC102723769 | FALSE | Upregulated |
| -0,327320724 | LOC102723775 | FALSE | Downregulated |
| 0,672612969 | LOC102723801 | FALSE | Upregulated |
| -0,265718504 | LOC102723811 | FALSE | Downregulated |

| | | | |
|---|---|---|---|
| 0,368209881 | LOC102723814 | FALSE | Upregulated |
| -0,157817811 | LOC102723837 | FALSE | Downregulated |
| -1,330937 | LOC102723838 | | Downregulated |
| -0,161942802 | LOC102723840 | FALSE | Downregulated |
| -0,834931612 | LOC102723842 | FALSE | Downregulated |
| 0,211404957 | LOC102723867 | FALSE | Upregulated |
| 0,033040573 | LOC102723870 | FALSE | Upregulated |
| -0,000289407 | LOC102723877 | FALSE | Downregulated |
| 0,969176039 | LOC102723884 | FALSE | Upregulated |
| 0,011038753 | LOC102723891 | FALSE | Upregulated |
| 0,009613243 | LOC102723913 | FALSE | Upregulated |
| -0,268835324 | LOC102723917 | FALSE | Downregulated |
| -0,330204104 | LOC102723922 | FALSE | Downregulated |
| 0,027839886 | LOC102723923 | FALSE | Upregulated |
| -1,044001771 | LOC102723943 | | Downregulated |
| -1,762976212 | LOC102723950 | | Downregulated |
| 0,822756876 | LOC102723959 | FALSE | Upregulated |
| -0,447403011 | LOC102723988 | FALSE | Downregulated |
| -1,929870447 | LOC102724003 | | Downregulated |
| -0,479797467 | LOC102724004 | FALSE | Downregulated |
| 0,722692983 | LOC102724009 | FALSE | Upregulated |
| 0,955735279 | LOC102724010 | FALSE | Upregulated |
| 0,006314384 | LOC102724012 | FALSE | Upregulated |
| -0,085970385 | LOC102724024 | FALSE | Downregulated |
| 0,099282274 | LOC102724049 | FALSE | Upregulated |
| -0,438499285 | LOC102724057 | FALSE | Downregulated |

| | | | |
|---|---|---|---|
| -0,08765791 | LOC102724065 | FALSE | Downregulated |
| 0,224338009 | LOC102724069 | FALSE | Upregulated |
| 0,081392312 | LOC102724075 | FALSE | Upregulated |
| 0,573708704 | LOC102724077 | FALSE | Upregulated |
| 0,940666238 | LOC102724081 | FALSE | Upregulated |
| 0,160489984 | LOC102724106 | FALSE | Upregulated |
| 0,616361305 | LOC102724108 | FALSE | Upregulated |
| 0,026825591 | LOC102724113 | FALSE | Upregulated |
| 0,372887015 | LOC102724119 | FALSE | Upregulated |
| -1,369674402 | LOC102724122 | | Downregulated |
| -1,157874573 | LOC102724143 | | Downregulated |
| 0,237417944 | LOC102724149 | FALSE | Upregulated |
| 0,344946131 | LOC102724176 | FALSE | Upregulated |
| -0,456490054 | LOC102724177 | FALSE | Downregulated |
| 0,433877296 | LOC102724193 | FALSE | Upregulated |
| 0,1638725 | LOC102724196 | FALSE | Upregulated |
| -1,297232654 | LOC102724205 | | Downregulated |
| 0,124156336 | LOC102724209 | FALSE | Upregulated |
| -0,631759455 | LOC102724230 | FALSE | Downregulated |
| -1,231030449 | LOC102724246 | | Downregulated |
| 0,348575417 | LOC102724259 | FALSE | Upregulated |
| -0,909645494 | LOC102724271 | FALSE | Downregulated |
| -0,505700399 | LOC102724273 | FALSE | Downregulated |
| -0,036170673 | LOC102724280 | FALSE | Downregulated |
| 0,606437257 | LOC102724286 | FALSE | Upregulated |
| -0,834921922 | LOC102724292 | FALSE | Downregulated |

| | | | |
|---|---|---|---|
| -0,549546788 | LOC102724322 | FALSE | Downregulated |
| -0,255147205 | LOC102724328 | FALSE | Downregulated |
| 0,550746501 | LOC102724335 | FALSE | Upregulated |
| 0,403622163 | LOC102724339 | FALSE | Upregulated |
| 0,704601036 | LOC102724342 | FALSE | Upregulated |
| -0,464472714 | LOC102724348 | FALSE | Downregulated |
| -0,652467065 | LOC102724349 | FALSE | Downregulated |
| 0,786158631 | LOC102724351 | FALSE | Upregulated |
| -0,109316951 | LOC102724377 | FALSE | Downregulated |
| 0,278947122 | LOC102724378 | FALSE | Upregulated |
| 0,190146473 | LOC102724384 | FALSE | Upregulated |
| 0,457723864 | LOC102724393 | FALSE | Upregulated |
| -0,420149037 | LOC102724400 | FALSE | Downregulated |
| 0,709595487 | LOC102724423 | FALSE | Upregulated |
| 1,154697762 | LOC102724424 | | Upregulated |
| 0,518757362 | LOC102724425 | FALSE | Upregulated |
| 0,692689876 | LOC102724426 | FALSE | Upregulated |
| -0,765061144 | LOC102724440 | FALSE | Downregulated |
| 0,490309905 | LOC102724448 | FALSE | Upregulated |
| 1,021576294 | LOC102724449 | | Upregulated |
| 0,61707484 | LOC102724462 | FALSE | Upregulated |
| -1,336043736 | LOC102724480 | | Downregulated |
| -1,233827573 | LOC102724488 | | Downregulated |
| -0,193950281 | LOC102724501 | FALSE | Downregulated |
| -0,371694737 | LOC102724508 | FALSE | Downregulated |
| -0,160890111 | LOC102724518 | FALSE | Downregulated |

| | | | |
|---|---|---|---|
| -0,84698688 | LOC102724536 | FALSE | Downregulated |
| -0,082301757 | LOC102724537 | FALSE | Downregulated |
| -0,044511337 | LOC102724543 | FALSE | Downregulated |
| 0,148182989 | LOC102724545 | FALSE | Upregulated |
| 1,157434553 | LOC102724614 | | Upregulated |
| 1,527057125 | LOC102724642 | | Upregulated |
| -0,571638397 | LOC102724685 | FALSE | Downregulated |
| -1,300440793 | LOC102724689 | | Downregulated |
| 0,119698361 | LOC102724696 | FALSE | Upregulated |
| -0,303683024 | LOC102724703 | FALSE | Downregulated |
| 1,114085677 | LOC102724705 | | Upregulated |
| -1,20456572 | LOC102724720 | | Downregulated |
| -0,948257174 | LOC102724731 | FALSE | Downregulated |
| 0,628387513 | LOC102724758 | FALSE | Upregulated |
| 0,111122988 | LOC102724759 | FALSE | Upregulated |
| 1,172246424 | LOC102724797 | | Upregulated |
| 0,439070543 | LOC102724810 | FALSE | Upregulated |
| -0,383191464 | LOC102724812 | FALSE | Downregulated |
| 1,068264132 | LOC102724826 | | Upregulated |
| 1,129684884 | LOC102724827 | | Upregulated |
| 0,662517372 | LOC102724830 | FALSE | Upregulated |
| 0,404904943 | LOC102724848 | FALSE | Upregulated |
| -0,882241992 | LOC102724874 | FALSE | Downregulated |
| 1,035167285 | LOC102724875 | | Upregulated |
| 0,357741613 | LOC102724883 | FALSE | Upregulated |
| -0,59163251 | LOC102724898 | FALSE | Downregulated |

| | | | |
|---|---|---|---|
| -0,777610623 | LOC102724914 | FALSE | Downregulated |
| -0,542088457 | LOC102724919 | FALSE | Downregulated |
| 0,241796521 | LOC102724943 | FALSE | Upregulated |
| 1,045904147 | LOC102724944 | | Upregulated |
| 0,273557354 | LOC102724956 | FALSE | Upregulated |
| 1,281492247 | LOC102724976 | | Upregulated |
| 0,123849603 | LOC150051 | FALSE | Upregulated |
| -0,006367872 | LOC151760 | FALSE | Downregulated |
| -0,186988386 | LOC256880 | FALSE | Downregulated |
| 0,435437885 | LOC257358 | FALSE | Upregulated |
| -1,618585822 | LOC283683 | | Downregulated |
| -1,202211935 | LOC283731 | | Downregulated |
| 1,01718112 | LOC284009 | | Upregulated |
| -2,055499369 | LOC284344 | | Downregulated |
| 0,730279554 | LOC284600 | FALSE | Upregulated |
| 0,452243533 | LOC284648 | FALSE | Upregulated |
| 0,443701832 | LOC285043 | FALSE | Upregulated |
| -2,244481479 | LOC285095 | | Downregulated |
| -0,004055103 | LOC285819 | FALSE | Downregulated |
| -0,804812864 | LOC286178 | FALSE | Downregulated |
| 0,887565466 | LOC341056 | FALSE | Upregulated |
| 0,137462026 | LOC344967 | FALSE | Upregulated |
| -0,193233966 | LOC375196 | FALSE | Downregulated |
| 1,048235174 | LOC388242 | | Upregulated |
| 0,235863775 | LOC390956 | FALSE | Upregulated |
| 0,14985946 | LOC391722 | FALSE | Upregulated |

| | | | |
|---|---|---|---|
| 0,680895314 | LOC399815 | FALSE | Upregulated |
| 0,399618819 | LOC400499 | FALSE | Upregulated |
| 1,084845134 | LOC400548 | | Upregulated |
| -0,236958268 | LOC400685 | FALSE | Downregulated |
| 0,313077166 | LOC400891 | FALSE | Upregulated |
| -1,219807531 | LOC401098 | | Downregulated |
| 0,465235603 | LOC401463 | FALSE | Upregulated |
| -0,347017024 | LOC440292 | FALSE | Downregulated |
| -0,31062459 | LOC440461 | FALSE | Downregulated |
| 0,568623916 | LOC440934 | FALSE | Upregulated |
| 2,174712858 | LOC441455 | | Upregulated |
| 0,405419304 | LOC441956 | FALSE | Upregulated |
| -0,405002536 | LOC494127 | FALSE | Downregulated |
| 1,261596549 | LOC494141 | | Upregulated |
| -0,323455627 | LOC613038 | FALSE | Downregulated |
| 0,477263115 | LOC641746 | FALSE | Upregulated |
| -0,93651536 | LOC642249 | FALSE | Downregulated |
| 0,099773772 | LOC642934 | FALSE | Upregulated |
| -0,225421902 | LOC643387 | FALSE | Downregulated |
| 0,606169443 | LOC643406 | FALSE | Upregulated |
| 0,842571946 | LOC643711 | FALSE | Upregulated |
| -0,660482504 | LOC643802 | FALSE | Downregulated |
| -0,401618689 | LOC644086 | FALSE | Downregulated |
| 0,803670945 | LOC644919 | FALSE | Upregulated |
| -0,136063858 | LOC645485 | FALSE | Downregulated |
| -0,208093127 | LOC646513 | FALSE | Downregulated |

| | | | |
|---|---|---|---|
| 0,065685773 | LOC646588 | FALSE | Upregulated |
| 0,109244335 | LOC650157 | FALSE | Upregulated |
| 0,553842825 | LOC650368 | FALSE | Upregulated |
| 0,600993823 | LOC653786 | FALSE | Upregulated |
| 1,019143788 | LOC727710 | | Upregulated |
| 0,637655987 | LOC728637 | FALSE | Upregulated |
| -0,188897715 | LOC728715 | FALSE | Downregulated |
| -0,683950387 | LOC728739 | FALSE | Downregulated |
| 0,090305551 | LOC729041 | FALSE | Upregulated |
| -0,958765874 | LOC729468 | FALSE | Downregulated |
| 0,915945663 | LOC729506 | FALSE | Upregulated |
| -0,984455592 | LOC729658 | FALSE | Downregulated |
| 0,368436347 | LOC729739 | FALSE | Upregulated |
| 0,063380081 | LOC729770 | FALSE | Upregulated |
| 0,481436401 | LOC730338 | FALSE | Upregulated |
| 0,222474475 | LOC731157 | FALSE | Upregulated |
| 0,586427131 | LOC91450 | FALSE | Upregulated |
| 0,348337941 | LRRC10B | FALSE | Upregulated |
| 1,174207992 | LRRC2-AS1 | | Upregulated |
| 0,124106302 | LRRC36 | FALSE | Upregulated |
| -0,22828628 | LRRC3B | FALSE | Downregulated |
| 0,384903489 | LRRC3C | FALSE | Upregulated |
| -0,742207228 | LRRC63 | FALSE | Downregulated |
| 0,391557882 | LRRC71 | FALSE | Upregulated |
| -1,217769503 | LRRN4CL | | Downregulated |
| 0,767237491 | LUCAT1 | FALSE | Upregulated |

| | | | |
|---|---|---|---|
| 1,328796839 | LY6K | | Upregulated |
| 0,308965943 | LY86-AS1 | FALSE | Upregulated |
| -0,719437369 | MAB21L1 | FALSE | Downregulated |
| 1,361516644 | MADCAM1 | | Upregulated |
| 0,801010643 | MAGEL2 | FALSE | Upregulated |
| -0,164704347 | MAK | FALSE | Downregulated |
| 0,210032278 | MALRD1 | FALSE | Upregulated |
| -0,095846558 | MAMDC2-AS1 | FALSE | Downregulated |
| 0,030099162 | MAP1LC3C | FALSE | Upregulated |
| -1,051429091 | MAP2K4P1 | | Downregulated |
| 0,351951575 | MAPK15 | FALSE | Upregulated |
| -0,32179977 | MARCH4 | FALSE | Downregulated |
| 0,237129064 | MATN3 | FALSE | Upregulated |
| 0,879145161 | MBOAT4 | FALSE | Upregulated |
| 1,364342416 | MCCD1 | | Upregulated |
| 1,182981508 | MCF2L2 | | Upregulated |
| -0,738985263 | MED15P9 | FALSE | Downregulated |
| -0,229091053 | MEIS1-AS2 | FALSE | Downregulated |
| 1,853308957 | MEIS1-AS3 | | Upregulated |
| 0,838748685 | MEP1A | FALSE | Upregulated |
| 1,19509371 | MESP2 | | Upregulated |
| -0,384430136 | MFSD2B | FALSE | Downregulated |
| 0,975379567 | MFSD6L | FALSE | Upregulated |
| 0,067615481 | MGAT5B | FALSE | Upregulated |
| 0,608344947 | MIA-RAB4B | FALSE | Upregulated |
| -0,692556306 | MIMT1 | FALSE | Downregulated |

| | | | |
|---|---|---|---|
| -0,216724235 | MINOS1P1 | FALSE | Downregulated |
| -0,702600845 | MIR1184-1 | FALSE | Downregulated |
| -0,370570691 | MIR142 | FALSE | Downregulated |
| 0,492032962 | MIR143HG | FALSE | Upregulated |
| 0,53373528 | MIR181A2HG | FALSE | Upregulated |
| 0,808398405 | MIR2861 | FALSE | Upregulated |
| -0,127475313 | MIR3936 | FALSE | Downregulated |
| 0,220068075 | MIR4442 | FALSE | Upregulated |
| 0,431058806 | MIR4767 | FALSE | Upregulated |
| -1,259160383 | MIR5010 | | Downregulated |
| 1,309945595 | MIR635 | | Upregulated |
| -0,369448005 | MIR6717 | FALSE | Downregulated |
| 0,264195978 | MIR6772 | FALSE | Upregulated |
| 1,068433613 | MIXL1 | | Upregulated |
| 0,177533021 | MKRN3 | FALSE | Upregulated |
| -0,738505485 | MKRN7P | FALSE | Downregulated |
| -0,070623986 | MKRN9P | FALSE | Downregulated |
| -1,098245537 | MLANA | | Downregulated |
| 0,514941407 | MLK7-AS1 | FALSE | Upregulated |
| 0,900585126 | MMEL1 | FALSE | Upregulated |
| -0,028450298 | MMP1 | FALSE | Downregulated |
| 0,182073901 | MMP8 | FALSE | Upregulated |
| 0,353285613 | MOBP | FALSE | Upregulated |
| 0,80516866 | MOG | FALSE | Upregulated |
| 0,689482888 | MPP4 | FALSE | Upregulated |
| 0,757596529 | MRAP2 | FALSE | Upregulated |

| | | | |
|---|---|---|---|
| 1,086076565 | MRPL23-AS1 | | Upregulated |
| 0,295886365 | MRPL42P5 | FALSE | Upregulated |
| 0,657829972 | MS4A3 | FALSE | Upregulated |
| 0,23977842 | MSANTD1 | FALSE | Upregulated |
| 0,73119196 | MSH4 | FALSE | Upregulated |
| -0,169045119 | MSLN | FALSE | Downregulated |
| 1,417759055 | MSMB | | Upregulated |
| 0,834506131 | MSMP | FALSE | Upregulated |
| -0,454362834 | MSX2 | FALSE | Downregulated |
| 1,156902153 | MSX2P1 | | Upregulated |
| 0,52017399 | MT1IP | FALSE | Upregulated |
| -0,708115178 | MUC12 | FALSE | Downregulated |
| 0,223339019 | MUC2 | FALSE | Upregulated |
| 1,356054977 | MUC4 | | Upregulated |
| 1,729022805 | MUC5AC | | Upregulated |
| 0,537815032 | MYBPC3 | FALSE | Upregulated |
| -0,725914959 | MYCBP2-AS1 | FALSE | Downregulated |
| 0,88610969 | MYCBPAP | FALSE | Upregulated |
| 1,07013281 | MYCNOS | | Upregulated |
| 0,320420134 | MYH15 | FALSE | Upregulated |
| -1,007093061 | MYH4 | | Downregulated |
| -0,197262808 | MYT1 | FALSE | Downregulated |
| 0,136692158 | NALCN-AS1 | FALSE | Upregulated |
| -0,507426834 | NAP1L6 | FALSE | Downregulated |
| -0,967960942 | NBEAP1 | FALSE | Downregulated |
| -0,566542815 | NCAM1-AS1 | FALSE | Downregulated |

| | | | |
|---|---|---|---|
| 1,098889994 | NCCRP1 | | Upregulated |
| 0,322970034 | NCRUPAR | FALSE | Upregulated |
| 0,793678777 | NDUFAF4P1 | FALSE | Upregulated |
| 1,181905958 | NEFL | Upregulated | |
| -0,210527257 | NEFM | FALSE | Downregulated |
| -0,243327465 | NEK5 | FALSE | Downregulated |
| 0,053561592 | NETO1 | FALSE | Upregulated |
| 0,773759519 | NEXN-AS1 | FALSE | Upregulated |
| -0,508396391 | NHLH1 | FALSE | Downregulated |
| 0,042160183 | NHLRC4 | FALSE | Upregulated |
| 0,44377702 | NKPD1 | FALSE | Upregulated |
| 0,521988552 | NKX2-3 | FALSE | Upregulated |
| 0,085298087 | NLGN1 | FALSE | Upregulated |
| 0,590202169 | NLRP8 | FALSE | Upregulated |
| 0,588985194 | NLRP9 | FALSE | Upregulated |
| -0,230053217 | NME8 | FALSE | Downregulated |
| -0,297954857 | NME9 | FALSE | Downregulated |
| -0,087175395 | NNAT | FALSE | Downregulated |
| 0,624013625 | NOG | FALSE | Upregulated |
| -0,230375647 | NOX5 | FALSE | Downregulated |
| -0,876324118 | NPB | FALSE | Downregulated |
| 1,560575152 | NPHS1 | | Upregulated |
| 0,733187934 | NPSR1-AS1 | FALSE | Upregulated |
| -0,022121016 | NR2E1 | FALSE | Downregulated |
| 0,262692986 | NR2E3 | FALSE | Upregulated |
| -0,831758871 | NRN1L | FALSE | Downregulated |

| | | | |
|---|---|---|---|
| -0,775192369 | NT5C1B-RDH14 | FALSE | Downregulated |
| 0,011733642 | NTN5 | FALSE | Upregulated |
| 0,161916597 | NTNG1 | FALSE | Upregulated |
| -0,121689539 | NTRK1 | FALSE | Downregulated |
| -0,035281955 | NTSR1 | FALSE | Downregulated |
| -1,446251879 | NUDT11 | | Downregulated |
| -0,551463297 | NUTM2G | FALSE | Downregulated |
| 0,875881802 | NWD1 | FALSE | Upregulated |
| 1,165390532 | NXF5 | | Upregulated |
| 0,214939423 | NXNL2 | FALSE | Upregulated |
| -1,655280711 | NXPH1 | | Downregulated |
| 0,059243048 | NXPH2 | FALSE | Upregulated |
| 1,086641224 | NXPH4 | | Upregulated |
| 0,905099308 | NYAP2 | FALSE | Upregulated |
| -0,155019367 | ODF3L1 | FALSE | Downregulated |
| 1,100437795 | ODF3L2 | | Upregulated |
| -0,972632566 | OLAH | FALSE | Downregulated |
| -1,205433025 | OLFM4 | | Downregulated |
| 0,532174448 | OMP | FALSE | Upregulated |
| -0,706021878 | OOEP | FALSE | Downregulated |
| 0,485163351 | OPN1SW | FALSE | Upregulated |
| 1,113466683 | OPRK1 | | Upregulated |
| 0,287730313 | OR10R2 | FALSE | Upregulated |
| -0,413508029 | OR2A1 | FALSE | Downregulated |
| -0,278218664 | OR2C1 | FALSE | Downregulated |
| -0,077823419 | OR51B5 | FALSE | Downregulated |

| | | | |
|---|---|---|---|
| 0,458252248 | OR51E1 | FALSE | Upregulated |
| 0,889055646 | OR52N4 | FALSE | Upregulated |
| -0,13350014 | OR6W1P | FALSE | Downregulated |
| 1,163603881 | OR7D2 | | Upregulated |
| 0,540395635 | OSM | FALSE | Upregulated |
| -0,069041721 | OSR2 | FALSE | Downregulated |
| 0,517811559 | OSTCP1 | FALSE | Upregulated |
| -0,058620116 | OSTCP2 | FALSE | Downregulated |
| 1,191544193 | OTOGL | | Upregulated |
| 0,321557059 | OTUD6A | FALSE | Upregulated |
| 0,359848998 | OTX1 | FALSE | Upregulated |
| 0,062783576 | OVOL1 | FALSE | Upregulated |
| 0,546834762 | OVOL2 | FALSE | Upregulated |
| -0,750071795 | P2RX2 | FALSE | Downregulated |
| 0,430218264 | P4HA2-AS1 | FALSE | Upregulated |
| -0,669784243 | PAPL | FALSE | Downregulated |
| -0,263059082 | PAPOLB | FALSE | Downregulated |
| 0,352286039 | PAPPA-AS1 | FALSE | Upregulated |
| 1,043033747 | PART1 | | Upregulated |
| 0,545130793 | PAX2 | FALSE | Upregulated |
| -0,233958938 | PAX6 | FALSE | Downregulated |
| -1,668449863 | PAX7 | | Downregulated |
| -1,576111957 | PCA3 | | Downregulated |
| -0,926881151 | PCAT1 | FALSE | Downregulated |
| 0,292503858 | PCAT14 | FALSE | Upregulated |
| -0,135837982 | PCAT5 | FALSE | Downregulated |

| | | | |
|---|---|---|---|
| 0,748049259 | PCDH10 | FALSE | Upregulated |
| -0,524213104 | PCDH15 | FALSE | Downregulated |
| -0,308880652 | PCDH8 | FALSE | Downregulated |
| 0,87576448 | PCNA-AS1 | FALSE | Upregulated |
| 1,711931577 | PCSK1 | | Upregulated |
| 1,442177306 | PCSK2 | | Upregulated |
| 1,02476708 | PDE6A | | Upregulated |
| 0,46773382 | PDILT | FALSE | Upregulated |
| -0,215612806 | PDLIM4 | FALSE | Downregulated |
| 0,721766839 | PDZD3 | FALSE | Upregulated |
| -1,434021881 | PGLYRP1 | | Downregulated |
| 0,32406015 | PGM5-AS1 | FALSE | Upregulated |
| 1,111673047 | PGM5P2 | | Upregulated |
| 0,796305631 | PIWIL1 | FALSE | Upregulated |
| 0,019451984 | PKD2L2 | FALSE | Upregulated |
| 0,158797914 | PKP3 | FALSE | Upregulated |
| 0,315293846 | PLA2G4F | FALSE | Upregulated |
| 0,357754039 | PLCZ1 | FALSE | Upregulated |
| 1,533868035 | PLEKHS1 | | Upregulated |
| 0,370870374 | PLSCR2 | FALSE | Upregulated |
| 0,157885906 | PMP2 | FALSE | Upregulated |
| 0,755904782 | PNMA5 | FALSE | Upregulated |
| 0,314156279 | PNMT | FALSE | Upregulated |
| 1,624082096 | PNOC | | Upregulated |
| 1,007926246 | PODNL1 | | Upregulated |
| 2,164883641 | POTEF | | Upregulated |

| | | | |
|---|---|---|---|
| 0,984203402 | POU2F3 | FALSE | Upregulated |
| 0,686981838 | POU3F1 | FALSE | Upregulated |
| -0,602233588 | POU3F2 | FALSE | Downregulated |
| 0,14194142 | POU4F1 | FALSE | Upregulated |
| 0,143733331 | POU6F2-AS2 | FALSE | Upregulated |
| -0,398276476 | PPEF1 | FALSE | Downregulated |
| -0,595671649 | PPIAL4B | FALSE | Downregulated |
| 0,033208771 | PPIAP30 | FALSE | Upregulated |
| 0,153111127 | PPP1R2P3 | FALSE | Upregulated |
| -0,000275723 | PPP2R2C | FALSE | Downregulated |
| 1,151068608 | PRC1-AS1 | | Upregulated |
| 0,388345158 | PRCD | FALSE | Upregulated |
| -0,072346293 | PRO2852 | FALSE | Downregulated |
| 0,3633449 | PROK1 | FALSE | Upregulated |
| 1,172715386 | PROKR2 | | Upregulated |
| 0,059688664 | PROSER2-AS1 | FALSE | Upregulated |
| -0,41524016 | PROX2 | FALSE | Downregulated |
| 1,191896606 | PRPH | | Upregulated |
| 1,01769824 | PRR15 | | Upregulated |
| 0,404496559 | PRRT4 | FALSE | Upregulated |
| 0,370766803 | PRSS21 | FALSE | Upregulated |
| 0,335632598 | PRSS22 | FALSE | Upregulated |
| 0,005177387 | PRSS30P | FALSE | Upregulated |
| 0,915208484 | PRSS42 | FALSE | Upregulated |
| -0,647666197 | PSG4 | FALSE | Downregulated |
| 0,052676022 | PTCHD1 | FALSE | Upregulated |

| | | | |
|---|---|---|---|
| -0,620838342 | PTCHD2 | FALSE | Downregulated |
| -1,455565289 | PTCHD3 | | Downregulated |
| 1,277600774 | PTGER1 | | Upregulated |
| -0,493850675 | PTGER4P2-CDK2AP2P2 | FALSE | Downregulated |
| -0,071435786 | PTOV1-AS1 | FALSE | Downregulated |
| 0,305374485 | PTPRG-AS1 | FALSE | Upregulated |
| 2,072238817 | PTPRR | | Upregulated |
| 0,277091085 | PTPRVP | FALSE | Upregulated |
| 1,533765298 | PTPRZ1 | | Upregulated |
| 0,298211618 | PURG | FALSE | Upregulated |
| -0,013286506 | PVRL3-AS1 | FALSE | Downregulated |
| 1,238397582 | PWRN1 | | Upregulated |
| -1,008825706 | PXT1 | | Downregulated |
| 0,349605065 | PYY2 | FALSE | Upregulated |
| 1,361101713 | QRFPR | | Upregulated |
| 1,358321626 | RAB33A | | Upregulated |
| 0,637971479 | RAB44 | FALSE | Upregulated |
| -0,004726261 | RAB6C-AS1 | FALSE | Downregulated |
| -0,381088374 | RADIL | FALSE | Downregulated |
| -0,396331252 | RAET1G | FALSE | Downregulated |
| 0,553087947 | RAET1K | FALSE | Upregulated |
| -0,255017858 | RASGEF1C | FALSE | Downregulated |
| 0,181137682 | RASSF10 | FALSE | Upregulated |
| 0,689509966 | RBBP8NL | FALSE | Upregulated |
| -0,846692498 | RBFADN | FALSE | Downregulated |
| -0,229732307 | RBMXL2 | FALSE | Downregulated |

| | | | |
|---|---|---|---|
| 0,555255264 | RBP2 | FALSE | Upregulated |
| 1,056184748 | RCAN3AS | | Upregulated |
| 0,038135875 | RCVRN | FALSE | Upregulated |
| 0,251189468 | RD3 | FALSE | Upregulated |
| -0,052189565 | RDM1 | FALSE | Downregulated |
| -0,160593448 | REEP2 | FALSE | Downregulated |
| -0,768531124 | REG4 | FALSE | Downregulated |
| 0,114961976 | REM2 | FALSE | Upregulated |
| -1,695676804 | REXO1L1P | | Downregulated |
| -0,346955721 | RFPL3S | FALSE | Downregulated |
| 0,141234736 | RFX8 | FALSE | Upregulated |
| -1,319858153 | RGPD4-AS1 | | Downregulated |
| -0,073365463 | RGS13 | FALSE | Downregulated |
| 0,976816474 | RGS17 | FALSE | Upregulated |
| 0,413352896 | RGS22 | FALSE | Upregulated |
| -0,985609662 | RGS9BP | FALSE | Downregulated |
| -0,496882457 | RHBDL3 | FALSE | Downregulated |
| -0,750862619 | RHD | FALSE | Downregulated |
| -0,256566753 | RHOXF1 | FALSE | Downregulated |
| -0,27139567 | RIMS1 | FALSE | Downregulated |
| -0,511238258 | RIMS2 | FALSE | Downregulated |
| 0,634435365 | RLN1 | FALSE | Upregulated |
| -0,052447301 | RLN2 | FALSE | Downregulated |
| 0,122481092 | RNA5S1 | FALSE | Upregulated |
| 0 | RNA5S10 | FALSE | Upregulated |
| -0,781026209 | RNA5S11 | FALSE | Downregulated |

| | | | |
|---|---|---|---|
| | RNA5S12 | | |
| | RNA5S13 | | |
| | RNA5S14 | | |
| | RNA5S15 | | |
| | RNA5S16 | | |
| -0,140113742 | RNA5S17 | FALSE | Downregulated |
| -0,184353968 | RNA5S2 | FALSE | Downregulated |
| 0 | RNA5S3 | FALSE | Upregulated |
| | RNA5S4 | | |
| 0,163225105 | RNA5S5 | FALSE | Upregulated |
| | RNA5S6 | | |
| | RNA5S7 | | |
| | RNA5S8 | | |
| -0,960807477 | RNF144A-AS1 | FALSE | Downregulated |
| 1,05497975 | RNF148 | | Upregulated |
| -0,259588964 | RNF157-AS1 | FALSE | Downregulated |
| 0,863914038 | RNF17 | FALSE | Upregulated |
| 1,086638013 | RNF182 | | Upregulated |
| 0,055596036 | RNFT2 | FALSE | Upregulated |
| 0,114513421 | RNU4-1 | FALSE | Upregulated |
| 0,459407072 | RNU6-26P | FALSE | Upregulated |
| 0,634690433 | RNU6-31P | FALSE | Upregulated |
| -0,317342321 | RNU6-46P | FALSE | Downregulated |
| -0,793334894 | RNU6ATAC | FALSE | Downregulated |
| 0,263004011 | RNVU1-7 | FALSE | Upregulated |
| -0,252119393 | RNY1 | FALSE | Downregulated |

| | | | |
|---|---|---|---|
| 0,745740375 | RNY3 | FALSE | Upregulated |
| -0,61170525 | RNY5 | FALSE | Downregulated |
| -0,571388472 | RP1-69M21.2 | FALSE | Downregulated |
| -1,675129993 | RP1-90L6.2 | | Downregulated |
| 0,690346094 | RP11-103C16.2 | FALSE | Upregulated |
| 0,386511798 | RP11-1057B8.2 | FALSE | Upregulated |
| 0,535079417 | RP11-106M7.1 | FALSE | Upregulated |
| -1,233998768 | RP11-1079K10.5 | | Downregulated |
| 0,117745618 | RP11-1080G15.1 | FALSE | Upregulated |
| 0,010289281 | RP11-133O22.6 | FALSE | Upregulated |
| -0,386582976 | RP11-134P9.3 | FALSE | Downregulated |
| -0,505635379 | RP11-138B4.1 | FALSE | Downregulated |
| 0,673139063 | RP11-143M1.2 | FALSE | Upregulated |
| -0,092482059 | RP11-148L24.1 | FALSE | Downregulated |
| 1,351625671 | RP11-154D17.1 | | Upregulated |
| -0,39872137 | RP11-158M2.5 | FALSE | Downregulated |
| 1,392780618 | RP11-16E12.2 | | Upregulated |
| 0,740052252 | RP11-170N11.1 | FALSE | Upregulated |
| 0,427513745 | RP11-175K6.1 | FALSE | Upregulated |
| -0,474758902 | RP11-17A4.2 | FALSE | Downregulated |
| 1,002585107 | RP11-193H5.1 | | Upregulated |
| 0,542525431 | RP11-197N18.7 | FALSE | Upregulated |
| -0,532189976 | RP11-219B17.1 | FALSE | Downregulated |
| 0,294192237 | RP11-219G17.4 | FALSE | Upregulated |
| 0,268883936 | RP11-227H15.4 | FALSE | Upregulated |
| 0,265646989 | RP11-230B22.1 | FALSE | Upregulated |

| | | | |
|---|---|---|---|
| -1,222072715 | RP11-231I16.1 | | Downregulated |
| 1,013345422 | RP11-24J23.2 | | Upregulated |
| -1,373219983 | RP11-260E18.1 | | Downregulated |
| -0,659098432 | RP11-264C15.2 | FALSE | Downregulated |
| -0,073694233 | RP11-26J3.1 | FALSE | Downregulated |
| 0,426645546 | RP11-277P12.20 | FALSE | Upregulated |
| 0,741340998 | RP11-280K24.4 | FALSE | Upregulated |
| 0,658549631 | RP11-286E11.1 | FALSE | Upregulated |
| 0,673972825 | RP11-294O2.2 | FALSE | Upregulated |
| 0,802687196 | RP11-319G6.1 | FALSE | Upregulated |
| -0,744151654 | RP11-320G24.1 | FALSE | Downregulated |
| -0,83124925 | RP11-341A22.2 | FALSE | Downregulated |
| 0,301590942 | RP11-365O16.6 | FALSE | Upregulated |
| -0,386424161 | RP11-374P20.4 | FALSE | Downregulated |
| 0,824389029 | RP11-379F4.4 | FALSE | Upregulated |
| -0,469595602 | RP11-37B2.1 | FALSE | Downregulated |
| -0,075953356 | RP11-383C5.3 | FALSE | Downregulated |
| 0,165900711 | RP11-397C18.2 | FALSE | Upregulated |
| -0,157596652 | RP11-426L16.8 | FALSE | Downregulated |
| 0,057494319 | RP11-430H10.1 | FALSE | Upregulated |
| 0,451482004 | RP11-445H22.3 | FALSE | Upregulated |
| 0,611774904 | RP11-446H18.5 | FALSE | Upregulated |
| -0,447811166 | RP11-449D8.1 | FALSE | Downregulated |
| 0,440529023 | RP11-475N22.4 | FALSE | Upregulated |
| -0,864171703 | RP11-486A14.2 | FALSE | Downregulated |
| -0,645741424 | RP11-492A10.1 | FALSE | Downregulated |

| | | | |
|---|---|---|---|
| 0,476754119 | RP11-508P1.2 | FALSE | Upregulated |
| -0,333696466 | RP11-534N16.1 | FALSE | Downregulated |
| -0,204056063 | RP11-536C5.7 | FALSE | Downregulated |
| 0,565934259 | RP11-538P18.2 | FALSE | Upregulated |
| 1,146465101 | RP11-542B15.1 | | Upregulated |
| -0,470512572 | RP11-544M22.1 | FALSE | Downregulated |
| -0,951495755 | RP11-554A11.6 | FALSE | Downregulated |
| -0,86050018 | RP11-554F20.1 | FALSE | Downregulated |
| -0,540503802 | RP11-576I22.2 | FALSE | Downregulated |
| 0,037992175 | RP11-588G21.2 | FALSE | Upregulated |
| 0,702103378 | RP11-58E21.3 | FALSE | Upregulated |
| 0,679349508 | RP11-593F23.1 | FALSE | Upregulated |
| -0,598759928 | RP11-661P17.1 | FALSE | Downregulated |
| 0,500744242 | RP11-673E1.1 | FALSE | Upregulated |
| -1,058318074 | RP11-707A18.1 | | Downregulated |
| -0,061685846 | RP11-710F7.3 | FALSE | Downregulated |
| -0,090909218 | RP11-723O4.2 | FALSE | Downregulated |
| 1,223070392 | RP11-725G5.2 | | Upregulated |
| 0,497864024 | RP11-770E5.1 | FALSE | Upregulated |
| -0,3533687 | RP11-770J1.3 | FALSE | Downregulated |
| 1,23456897 | RP11-809M12.1 | | Upregulated |
| -0,220021245 | RP11-829H16.3 | FALSE | Downregulated |
| -0,059601012 | RP11-82L2.1 | FALSE | Downregulated |
| 0,016268158 | RP11-840I19.3 | FALSE | Upregulated |
| -0,291946823 | RP11-864J10.4 | FALSE | Downregulated |
| -1,322282532 | RP11-885B4.2 | | Downregulated |

| | | | |
|---|---|---|---|
| -0,785497326 | RP11-887P2.5 | FALSE | Downregulated |
| 0,344722767 | RP11-973H7.4 | FALSE | Upregulated |
| 0,825093217 | RP3-399L15.3 | FALSE | Upregulated |
| -0,634224777 | RP3-400N23.6 | FALSE | Downregulated |
| -1,241453585 | RP3-476K8.3 | | Downregulated |
| -0,489497671 | RP3-522P13.3 | FALSE | Downregulated |
| -0,67976284 | RP4-565E6.1 | FALSE | Downregulated |
| 0,868051796 | RP4-724E16.2 | FALSE | Upregulated |
| -1,797506208 | RP4-794H19.4 | | Downregulated |
| 0,179943973 | RP5-1004I9.1 | FALSE | Upregulated |
| -0,526406906 | RP5-1139I1.1 | FALSE | Downregulated |
| -0,56264645 | RP5-1142A6.2 | FALSE | Downregulated |
| 1,390898783 | RPA4 | | Upregulated |
| 0,269270362 | RPEL1 | FALSE | Upregulated |
| 1,06692972 | RPL29P2 | | Upregulated |
| 0,540666894 | RPS4Y2 | FALSE | Upregulated |
| -0,573517885 | RPSAP52 | FALSE | Downregulated |
| 0,587428791 | RRH | FALSE | Upregulated |
| 0,442831319 | RUNDC3A | FALSE | Upregulated |
| -0,182512396 | S100Z | FALSE | Downregulated |
| 0,595620751 | SAA3P | FALSE | Upregulated |
| 0,326219042 | SAG | FALSE | Upregulated |
| -0,227724155 | SAP30L-AS1 | FALSE | Downregulated |
| -0,66618997 | SCG2 | FALSE | Downregulated |
| 1,233242068 | SCG3 | | Upregulated |
| 0,12749869 | SCGB2B2 | FALSE | Upregulated |

| | | | |
|---|---|---|---|
| -0,409623535 | SCHLAP1 | FALSE | Downregulated |
| -1,204256433 | SCN2B | | Downregulated |
| -0,046234588 | SCNN1B | FALSE | Downregulated |
| -0,071781128 | SCRG1 | FALSE | Downregulated |
| -0,203234044 | SCT | FALSE | Downregulated |
| -0,184841504 | SDC4P | FALSE | Downregulated |
| 0,045929502 | SEBOX | FALSE | Upregulated |
| 0,113849626 | SEC24B-AS1 | FALSE | Upregulated |
| 0,209421059 | SEMA3B-AS1 | FALSE | Upregulated |
| 1,148135236 | SERPINA9 | | Upregulated |
| 0,234635575 | SERTAD4-AS1 | FALSE | Upregulated |
| -0,059823486 | SEZ6 | FALSE | Downregulated |
| 1,44924722 | SFTA2 | | Upregulated |
| -0,386527609 | SFTPB | FALSE | Downregulated |
| 0,281714418 | SGCZ | FALSE | Upregulated |
| -0,339303066 | SH2D5 | FALSE | Downregulated |
| 0,451615283 | SH2D6 | FALSE | Upregulated |
| -0,498770294 | SH3GL1P2 | FALSE | Downregulated |
| 0,110761033 | SH3RF3-AS1 | FALSE | Upregulated |
| -0,563768537 | SHISA2 | FALSE | Downregulated |
| 0,645980957 | SHOX | FALSE | Upregulated |
| 0,932828305 | SHOX2 | FALSE | Upregulated |
| -0,393578705 | SIM2 | FALSE | Downregulated |
| 1,331288683 | SIRPD | | Upregulated |
| -0,796800815 | SIX3 | FALSE | Downregulated |
| -0,227712567 | SKOR1 | FALSE | Downregulated |

| | | | |
|---|---|---|---|
| -0,353285602 | SLC10A4 | FALSE | Downregulated |
| 0,03877803 | SLC10A6 | FALSE | Upregulated |
| 0,535433108 | SLC12A3 | FALSE | Upregulated |
| 0,52098264 | SLC13A1 | FALSE | Upregulated |
| 1,133025992 | SLC18A1 | | Upregulated |
| 0,244418029 | SLC22A14 | FALSE | Upregulated |
| -0,943413999 | SLC24A2 | FALSE | Downregulated |
| 0,016766716 | SLC27A6 | FALSE | Upregulated |
| 0,352535808 | SLC2A1-AS1 | FALSE | Upregulated |
| 0,895302908 | SLC30A2 | FALSE | Upregulated |
| 0,46915807 | SLC35F1 | FALSE | Upregulated |
| 1,157316393 | SLC35F3 | | Upregulated |
| -0,347624404 | SLC39A12 | FALSE | Downregulated |
| -1,0134973 | SLC39A2 | | Downregulated |
| 2,056229326 | SLC44A4 | | Upregulated |
| -0,585283998 | SLC46A2 | FALSE | Downregulated |
| 0,268861224 | SLC5A5 | FALSE | Upregulated |
| -0,245250903 | SLC6A2 | FALSE | Downregulated |
| 0,822982858 | SLC6A7 | FALSE | Upregulated |
| 0,61213292 | SLC7A10 | FALSE | Upregulated |
| -0,06827862 | SLC7A14 | FALSE | Downregulated |
| -0,030534586 | SLC7A4 | FALSE | Downregulated |
| -0,914106832 | SLC7A5P2 | FALSE | Downregulated |
| 0,233204543 | SLC8A2 | FALSE | Upregulated |
| -1,198622392 | SLC9A2 | | Downregulated |
| 0,657182453 | SLC9C2 | FALSE | Upregulated |

| | | | |
|---|---|---|---|
| -0,570281849 | SLCO1C1 | FALSE | Downregulated |
| 0,126040391 | SLFNL1 | FALSE | Upregulated |
| -1,174284353 | SLITRK5 | | Downregulated |
| 1,315635298 | SMARCA5-AS1 | | Upregulated |
| 0,561600483 | SMCO2 | FALSE | Upregulated |
| 0,39997285 | SMG7-AS1 | FALSE | Upregulated |
| 1,081273879 | SMIM2 | | Upregulated |
| 0,661112175 | SMIM22 | FALSE | Upregulated |
| 0,810285544 | SMKR1 | FALSE | Upregulated |
| 1,165714201 | SND1-IT1 | | Upregulated |
| 0,561974341 | SNHG4 | FALSE | Upregulated |
| 0,542804436 | SNORA14A | FALSE | Upregulated |
| 0,657102214 | SNORA14B | FALSE | Upregulated |
| -0,052646084 | SNORA23 | FALSE | Downregulated |
| -0,156484608 | SNORA29 | FALSE | Downregulated |
| 0,869836922 | SNORA3 | FALSE | Upregulated |
| -0,574841535 | SNORA31 | FALSE | Downregulated |
| 0,660191647 | SNORA41 | FALSE | Upregulated |
| 0,564721539 | SNORA59A | FALSE | Upregulated |
| 0,799511956 | SNORA59B | FALSE | Upregulated |
| -0,463041689 | SNORA60 | FALSE | Downregulated |
| 1,487203876 | SNORA65 | | Upregulated |
| -1,544356163 | SNORA7B | | Downregulated |
| -0,446687077 | SNORD103A | FALSE | Downregulated |
| -0,180218651 | SNORD103B | FALSE | Downregulated |
| 0,745342025 | SNORD16 | FALSE | Upregulated |

| | | | |
|---|---|---|---|
| 0,297299242 | SNORD17 | FALSE | Upregulated |
| -0,158207811 | SNORD35B | FALSE | Downregulated |
| 0,264201675 | SNORD3B-2 | FALSE | Upregulated |
| -1,007876147 | SNORD3C | | Downregulated |
| -0,288266444 | SNORD45B | FALSE | Downregulated |
| -0,182565149 | SNORD4A | FALSE | Downregulated |
| -0,27632255 | SNORD54 | FALSE | Downregulated |
| 1,628365037 | SNORD55 | | Upregulated |
| -0,110087072 | SNORD56 | FALSE | Downregulated |
| -0,199048793 | SNORD57 | FALSE | Downregulated |
| -0,746578525 | SNORD58A | FALSE | Downregulated |
| -0,617706695 | SNORD58C | FALSE | Downregulated |
| -0,399299434 | SNORD6 | FALSE | Downregulated |
| 0,487851392 | SNORD62A | FALSE | Upregulated |
| -1,631418026 | SNORD62B | | Downregulated |
| 1,245340555 | SNORD74 | | Upregulated |
| -1,203702312 | SNORD76 | | Downregulated |
| -0,361081614 | SNORD79 | FALSE | Downregulated |
| -0,629209009 | SNX31 | FALSE | Downregulated |
| -0,827916989 | SORCS1 | FALSE | Downregulated |
| 0,02369203 | SOX11 | FALSE | Upregulated |
| 0,712626685 | SOX2 | FALSE | Upregulated |
| 0,713140166 | SOX8 | FALSE | Upregulated |
| 0,712063243 | SP6 | FALSE | Upregulated |
| -0,490581886 | SPAG17 | FALSE | Downregulated |
| -0,322331982 | SPATA22 | FALSE | Downregulated |

| | | | |
|---|---|---|---|
| 1,504357352 | SPATA31D1 | | Upregulated |
| 1,205440108 | SPATC1 | | Upregulated |
| -0,079339412 | SPDYA | FALSE | Downregulated |
| 0,521449467 | SPEF1 | FALSE | Upregulated |
| -0,646923941 | SPG20OS | FALSE | Downregulated |
| 0,587941428 | SPNS3 | FALSE | Upregulated |
| 0,032139086 | SPOCD1 | FALSE | Upregulated |
| -0,281589328 | SPTY2D1-AS1 | FALSE | Downregulated |
| 0,408262947 | SRD5A1P1 | FALSE | Upregulated |
| 1,355617986 | SRGAP2-AS1 | | Upregulated |
| -1,16188592 | SRRM3 | | Downregulated |
| 1,043937798 | ST7-AS2 | | Upregulated |
| -0,091131206 | ST7-OT4 | FALSE | Downregulated |
| -1,237469493 | ST8SIA2 | | Downregulated |
| 0,31263987 | ST8SIA5 | FALSE | Upregulated |
| -0,088530717 | STAC2 | FALSE | Downregulated |
| -0,528803838 | STEAP1B | FALSE | Downregulated |
| -2,006070863 | STRA6 | | Downregulated |
| 0,798347475 | SULT2B1 | FALSE | Upregulated |
| 0,633538332 | SUMO1P1 | FALSE | Upregulated |
| -0,055847615 | SV2C | FALSE | Downregulated |
| -1,271950309 | SVILP1 | | Downregulated |
| 0,127882561 | SYCP1 | FALSE | Upregulated |
| 0,872110446 | SYN2 | FALSE | Upregulated |
| 0,878029422 | SYNDIG1L | FALSE | Upregulated |
| 0,320249528 | SYNGR3 | FALSE | Upregulated |

| | | | |
|---|---|---|---|
| -0,096232543 | SYNGR4 | FALSE | Downregulated |
| -0,253578148 | SYT4 | FALSE | Downregulated |
| 0,445149153 | SYT6 | FALSE | Upregulated |
| 0,520158316 | TAAR3 | FALSE | Upregulated |
| -0,22450876 | TACR1 | FALSE | Downregulated |
| -0,371485263 | TAF1L | FALSE | Downregulated |
| -0,287752937 | TAS1R1 | FALSE | Downregulated |
| 0,331899746 | TAS1R3 | FALSE | Upregulated |
| 0,459333278 | TAS2R31 | FALSE | Upregulated |
| 0,213200928 | TAS2R4 | FALSE | Upregulated |
| 0,743545122 | TAS2R60 | FALSE | Upregulated |
| 0,539573544 | TBC1D27 | FALSE | Upregulated |
| -0,022885187 | TBC1D29 | FALSE | Downregulated |
| 0,704427372 | TBC1D3P1-DHX40P1 | FALSE | Upregulated |
| 1,66278785 | TCEB3B | | Upregulated |
| 0,328315191 | TCERG1L | FALSE | Upregulated |
| -0,602593061 | TCP11 | FALSE | Downregulated |
| -0,250281634 | TCTE1 | FALSE | Downregulated |
| -0,155028759 | TCTE3 | FALSE | Downregulated |
| 0,795727919 | TCTEX1D4 | FALSE | Upregulated |
| -0,884902711 | TDGF1P3 | FALSE | Downregulated |
| -0,504886537 | TEKT2 | FALSE | Downregulated |
| 0,901396871 | TEPP | FALSE | Upregulated |
| 0,210746529 | TEX11 | FALSE | Upregulated |
| 0,064475633 | TEX12 | FALSE | Upregulated |
| 0,30532236 | TEX22 | FALSE | Upregulated |

| | | | |
|---|---|---|---|
| -0,310622373 | TFAP2A-AS1 | FALSE | Downregulated |
| 0,568757642 | TFAP2C | FALSE | Upregulated |
| 0,572777092 | TH | FALSE | Upregulated |
| -0,622738685 | THEG | FALSE | Downregulated |
| 1,126245938 | THEM5 | | Upregulated |
| 0,523866569 | TINCR | FALSE | Upregulated |
| 0,725477664 | TM4SF19-AS1 | FALSE | Upregulated |
| -1,123073184 | TMC1 | | Downregulated |
| -0,186708515 | TMC3 | FALSE | Downregulated |
| 1,053299332 | TMEM151A | | Upregulated |
| 1,305300926 | TMEM163 | | Upregulated |
| 0,707253658 | TMEM212 | FALSE | Upregulated |
| 0,649203785 | TMEM213 | FALSE | Upregulated |
| 1,515646239 | TMEM221 | | Upregulated |
| -0,353460394 | TMEM240 | FALSE | Downregulated |
| 0,617913525 | TMEM31 | FALSE | Upregulated |
| -0,955481533 | TMEM40 | FALSE | Downregulated |
| 1,087623211 | TMEM52B | | Upregulated |
| -0,577369208 | TMEM59L | FALSE | Downregulated |
| -0,089779405 | TMEM72 | FALSE | Downregulated |
| -0,440230901 | TMEM72-AS1 | FALSE | Downregulated |
| -0,084549366 | TMPRSS7 | FALSE | Downregulated |
| 0,682452585 | TMSB15B | FALSE | Upregulated |
| 0,490500804 | TNFRSF13C | FALSE | Upregulated |
| 1,101825782 | TNFRSF18 | | Upregulated |
| -0,587255684 | TNFSF18 | FALSE | Downregulated |

| | | | |
|---|---|---|---|
| -0,170368411 | TNFSF9 | FALSE | Downregulated |
| 0,303220315 | TNS4 | FALSE | Upregulated |
| -0,454606003 | TOMM20L | FALSE | Downregulated |
| 0,925140882 | TP53AIP1 | FALSE | Upregulated |
| -0,079510134 | TPH1 | FALSE | Downregulated |
| 0,397909647 | TPI1P3 | FALSE | Upregulated |
| 0,805898155 | TPSD1 | FALSE | Upregulated |
| 1,106627774 | TPTE2 | | Upregulated |
| 0,082261292 | TPTE2P6 | FALSE | Upregulated |
| -0,680990728 | TREML1 | FALSE | Downregulated |
| -0,047377128 | TRIM17 | FALSE | Downregulated |
| 0,010334736 | TRIM40 | FALSE | Upregulated |
| 0,411848358 | TRIM46 | FALSE | Upregulated |
| 0,583832601 | TRIM50 | FALSE | Upregulated |
| -2,28019844 | TRNAD14 | | Downregulated |
| 0,944676047 | TRNAD15 | FALSE | Upregulated |
| -0,369784192 | TRNAG15 | FALSE | Downregulated |
| -0,070680966 | TRNAG21 | FALSE | Downregulated |
| 0,064192863 | TRNAG23 | FALSE | Upregulated |
| -0,15087473 | TRNAG27 | FALSE | Downregulated |
| 0,390889736 | TRNAG30 | FALSE | Upregulated |
| 0,805352324 | TRNAH1 | FALSE | Upregulated |
| 0,185338331 | TRNAH10 | FALSE | Upregulated |
| 0,477426081 | TRNAH2 | FALSE | Upregulated |
| -0,916651946 | TRNAH3 | FALSE | Downregulated |
| 0,854813507 | TRNAH4 | FALSE | Upregulated |

| | | | |
|---|---|---|---|
| -0,010115346 | TRNAH5 | FALSE | Downregulated |
| 0,527788774 | TRNAH6 | FALSE | Upregulated |
| -0,059573731 | TRNAH7 | FALSE | Downregulated |
| -0,254451633 | TRNAH9 | FALSE | Downregulated |
| -0,687014303 | TRNAK1 | FALSE | Downregulated |
| -0,192090215 | TRNAK14 | FALSE | Downregulated |
| 0,672355025 | TRNAK16 | FALSE | Upregulated |
| -0,120967483 | TRNAK2 | FALSE | Downregulated |
| -0,378780781 | TRNAK20 | FALSE | Downregulated |
| -0,362550826 | TRNAK22 | FALSE | Downregulated |
| -0,402692765 | TRNAK26 | FALSE | Downregulated |
| -0,032650271 | TRNAK3 | FALSE | Downregulated |
| -0,959765018 | TRNAK33 | FALSE | Downregulated |
| -0,930362664 | TRNAK34 | FALSE | Downregulated |
| 0,406584683 | TRNAK9 | FALSE | Upregulated |
| 0 | TRNAV-CAC | FALSE | Upregulated |
| 0,392528479 | TRNAV11 | FALSE | Upregulated |
| 0 | TRNAV2 | FALSE | Upregulated |
| 0,173747006 | TRNAV21 | FALSE | Upregulated |
| 0 | TRNAV22 | FALSE | Upregulated |
| 0 | TRNAV25 | FALSE | Upregulated |
| -0,17694106 | TRNAV29 | FALSE | Downregulated |
| -0,81175215 | TRNAV4 | FALSE | Downregulated |
| -0,097280695 | TRNAV6 | FALSE | Downregulated |
| 0 | TRNAV9 | FALSE | Upregulated |
| 1,135718124 | TRPA1 | | Upregulated |

| | | | |
|---|---|---|---|
| 1,111302453 | TRPC2 | | Upregulated |
| -0,103115988 | TRPC5OS | FALSE | Downregulated |
| 0,537365187 | TSKS | FALSE | Upregulated |
| 0,01271953 | TSPEAR | FALSE | Upregulated |
| -0,186290225 | TSPO2 | FALSE | Downregulated |
| -0,104461101 | TSPY26P | FALSE | Downregulated |
| 0,600652296 | TSSK3 | FALSE | Upregulated |
| -1,42924322 | TSSK4 | | Downregulated |
| 1,027775183 | TTC23L | | Upregulated |
| -0,672298461 | TTC24 | FALSE | Downregulated |
| -0,35527751 | TTLL13 | FALSE | Downregulated |
| 0,589883603 | TTLL2 | FALSE | Upregulated |
| 1,286941665 | TTLL9 | | Upregulated |
| -0,023201949 | TTTY10 | FALSE | Downregulated |
| 0,187308227 | TUBA3D | FALSE | Upregulated |
| -1,736630397 | TUBA3E | | Downregulated |
| -0,290416795 | TUBB8 | FALSE | Downregulated |
| -0,891529873 | TULP2 | FALSE | Downregulated |
| -1,864810677 | TUSC5 | | Downregulated |
| 0,821191297 | TWIST2 | FALSE | Upregulated |
| -0,715952002 | TXNDC2 | FALSE | Downregulated |
| 0,67328439 | UBE2MP1 | FALSE | Upregulated |
| 1,811374582 | UGT8 | | Upregulated |
| 0,579910628 | UNC80 | FALSE | Upregulated |
| 0,083354319 | UNQ9370 | FALSE | Upregulated |
| 0,319048435 | UPK1A | FALSE | Upregulated |

| | | | |
|---|---|---|---|
| -0,01504622 | UPK1B | FALSE | Downregulated |
| -0,238647687 | USP50 | FALSE | Downregulated |
| -0,482257226 | VAC14-AS1 | FALSE | Downregulated |
| 0,036673006 | VAX2 | FALSE | Upregulated |
| -0,049625537 | VENTXP7 | FALSE | Downregulated |
| 1,429810472 | VGLL1 | | Upregulated |
| -1,695844754 | VIT | | Downregulated |
| 1,282149745 | VSIG1 | | Upregulated |
| 1,117204588 | VSTM1 | | Upregulated |
| 1,173980754 | VWA3A | | Upregulated |
| 0,147360919 | VWA5B2 | FALSE | Upregulated |
| 0,588892305 | VWA8-AS1 | FALSE | Upregulated |
| 0,20246811 | WDR11-AS1 | FALSE | Upregulated |
| 0,747089344 | WDR16 | FALSE | Upregulated |
| 0,118883102 | WDR63 | FALSE | Upregulated |
| -1,437578081 | WDR88 | | Downregulated |
| 1,343332187 | WEE2 | | Upregulated |
| -0,280457133 | WFDC3 | FALSE | Downregulated |
| -0,443786818 | WFIKKN1 | FALSE | Downregulated |
| 0,907952931 | WFIKKN2 | FALSE | Upregulated |
| 0,230011001 | WNT10B | FALSE | Upregulated |
| -1,03527894 | WNT7A | | Downregulated |
| -0,67831567 | WNT9A | FALSE | Downregulated |
| -0,842078877 | WNT9B | FALSE | Downregulated |
| -0,299839012 | WSCD2 | FALSE | Downregulated |
| -1,338994619 | WT1-AS | | Downregulated |

| | | | |
|---|---|---|---|
| -0,150794921 | WWTR1-AS1 | FALSE | Downregulated |
| -0,251317098 | XKRX | FALSE | Downregulated |
| -0,431885621 | XRCC6P5 | FALSE | Downregulated |
| 1,059884796 | XXYLT1-AS2 | | Upregulated |
| 0,331958895 | YBX3P1 | FALSE | Upregulated |
| -1,488568074 | ZBTB20-AS1 | | Downregulated |
| 0,231260517 | ZBTB32 | FALSE | Upregulated |
| 0,6213867 | ZBTB8B | FALSE | Upregulated |
| 2,412902922 | ZCCHC12 | | Upregulated |
| 0,156893965 | ZDHHC8P1 | FALSE | Upregulated |
| -0,273480907 | ZEB2-AS1 | FALSE | Downregulated |
| 0,040048239 | ZFR2 | FALSE | Upregulated |
| -0,624403367 | ZIC4 | FALSE | Downregulated |
| -1,1310052 | ZIM2 | | Downregulated |
| 0,690213491 | ZNF280B | FALSE | Upregulated |
| 0,191507223 | ZNF295-AS1 | FALSE | Upregulated |
| 0,29298008 | ZNF32-AS1 | FALSE | Upregulated |
| -0,14974638 | ZNF32-AS2 | FALSE | Downregulated |
| 0,344483707 | ZNF365 | FALSE | Upregulated |
| -1,103444425 | ZNF474 | | Downregulated |
| -0,189863106 | ZNF488 | FALSE | Downregulated |
| 0,702767675 | ZNF492 | FALSE | Upregulated |
| -0,386349182 | ZNF571-AS1 | FALSE | Downregulated |
| -0,325728729 | ZNF664-FAM101A | FALSE | Downregulated |
| 0,728896207 | ZNF705E | FALSE | Upregulated |
| -0,53596721 | ZNF716 | FALSE | Downregulated |

| | | | |
|---|---|---|---|
| -0,128289825 | ZNF724P | FALSE | Downregulated |
| 0,673857538 | ZNF730 | FALSE | Upregulated |
| 0,253401313 | ZNF732 | FALSE | Upregulated |
| -0,673471194 | ZNF878 | FALSE | Downregulated |
| -1,798888804 | ZNF890P | | Downregulated |
| -1,111657935 | ZNF98 | | Downregulated |
| 0,014229914 | ZPBP | FALSE | Upregulated |

**Suppl. Table 2: Differentially expressed genes of the discovery cohort**

Adjustment of p-value by false discovery rate.

**Suppl. Table 3: Pathway analysis within the discovery cohort**

**cABMR upregulated**

| q-value | Pathway name | Pathway ID | | q-value | pathway | external_id |
|---|---|---|---|---|---|---|
| 1.275380 | response to chemokine | GO:1990868 | | 2.7355 | Extracellular matrix organization | R-HSA-1474244 |
| 2.646931 | chemokine-mediated signaling pathway | GO:0070098 | | 5.2426 | Chemokine receptors bind chemokines | R-HSA-380108 |
| 4.411598 | generation of neurons | GO:0048699 | | 4.2824 | Viral protein interaction with cytokine and cytokine receptor - Homo sapiens (human) | path:hsa04061 |
| 7.474346 | positive regulation of cellular component movement | GO:0051272 | | 2.1045 | ECM-receptor interaction - Homo sapiens (human) | path:hsa04512 |
| 1.580162 | regulation of ion transport | GO:0043269 | | 2.1045 | Cytokine-cytokine receptor interaction - Homo sapiens (human) | path:hsa04060 |
| 4.006603 | cellular response to cytokine stimulus | GO:0071345 | | 2.4715 | Collagen formation | R-HSA-1474290 |
| 1.920425 | neutrophil migration | GO:1990266 | | 4.4162 | Assembly of collagen fibrils and other multimeric structures | R-HSA-2022090 |
| 1.970920 | positive regulation of cell-cell adhesion | GO:0022409 | | 9.7189 | Hematopoietic cell lineage - Homo sapiens (human) | path:hsa04640 |
| 2.201478 | positive regulation of cell differentiation | GO:0045597 | | 4.3660 | Amoebiasis - Homo sapiens (human) | path:hsa05146 |
| 5.395637 | regulation of leukocyte cell-cell adhesion | GO:1903037 | | 5.7129 | NF-kappa B signaling pathway - Homo sapiens (human) | path:hsa04064 |
| 1.566200 | neutrophil chemotaxis | GO:0030593 | | 9.2249 | Integrin cell surface interactions | R-HSA-216083 |
| 1.693270 | regulation of transmembrane transport | GO:0034762 | | 9.4584 | Focal adhesion - Homo sapiens (human) | path:hsa04510 |
| 1.693270 | neuron projection development | GO:0031175 | | 1.7446 | Peptide ligand-binding receptors | R-HSA-375276 |
| 1.693270 | ion transmembrane transport | GO:0034220 | | 4.0590 | Collagen chain trimerization | R-HSA-8948216 |
| 1.693270 | regulation of lymphocyte activation | GO:0051249 | | 5.4651 | Signal Transduction | R-HSA-162582 |
| 4.284858 | cation transport | GO:0006812 | | 5.4667 | Cell adhesion molecules - Homo sapiens (human) | path:hsa04514 |
| 4.695359 | hemopoiesis | GO:0030097 | | 5.6045 | Nervous system development | R-HSA-9675108 |
| 4.695359 | ERK1 and ERK2 cascade | GO:0070371 | | 0.0001 | Laminin interactions | R-HSA-3000157 |
| 5.865580 | ion homeostasis | GO:0050801 | | 0.0001 | Axon guidance | R-HSA-422475 |
| 6.907301 | positive regulation of cell communication | GO:0010647 | | 0.0001 | Class A/1 (Rhodopsin-like receptors) | R-HSA-373076 |
| 9.358787 | cell morphogenesis involved in differentiation | GO:0000904 | | 0.0001 | Developmental Biology | R-HSA-1266738 |
| 9.958293 | regulation of nervous system development | GO:0051960 | | 0.0001 | Rheumatoid arthritis - Homo sapiens (human) | path:hsa05323 |
| 1.010780 | anterograde trans-synaptic signaling | GO:0098916 | | 0.0001 | Small cell lung cancer - Homo sapiens (human) | path:hsa05222 |
| 1.092958 | regulation of leukocyte differentiation | GO:1902105 | | 0.0001 | Chemokine signaling pathway - Homo sapiens (human) | path:hsa04062 |
| 1.306154 | positive regulation of nervous system development | GO:0051962 | | 0.0005 | Human papillomavirus infection - Homo sapiens (human) | path:hsa05165 |
| 1.306154 | cellular cation homeostasis | GO:0030003 | | 0.0005 | Non-integrin membrane-ECM interactions | R-HSA-3000171 |
| 1.681516 | regulation of cell development | GO:0060284 | | 0.0005 | TNF signaling pathway - Homo sapiens (human) | path:hsa04668 |
| 4.010459 | positive regulation of cell proliferation | GO:0008284 | | 0.0006 | Degradation of the extracellular matrix | R-HSA-1474228 |
| 4.320829 | positive regulation of leukocyte activation | GO:0002696 | | 0.0006 | Collagen biosynthesis and modifying enzymes | R-HSA-1650814 |
| 5.454800 | calcium ion transmembrane import into cytosol | GO:0097553 | | 0.0006 | GPCR ligand binding | R-HSA-500792 |
| 6.217301 | regulation of intracellular signal transduction | GO:1902531 | | 0.0006 | Unwinding of DNA | R-HSA-176974 |
| 6.648382 | positive regulation of response to external stimulus | GO:0032103 | | 0.0007 | O-linked glycosylation | R-HSA-5173105 |
| 8.473623 | regulation of kidney development | GO:0090183 | | 0.0007 | G alpha (i) signalling events | R-HSA-418594 |
| 8.691489 | positive regulation of cell activation | GO:0050867 | | 0.0007 | NCAM1 interactions | R-HSA-419037 |
| 0.000106 | positive regulation of kidney development | GO:0090184 | | 0.0012 | AGE-RAGE signaling pathway in diabetic complications - Homo sapiens (human) | path:hsa04933 |
| 0.000108 | gliogenesis | GO:0042063 | | 0.0012 | O-glycosylation of TSR domain-containing proteins | R-HSA-5173214 |
| 0.000143 | kidney epithelium development | GO:0072073 | | 0.0012 | Signaling by GPCR | R-HSA-372790 |
| 0.000198 | positive regulation of hemopoiesis | GO:1903708 | | 0.0012 | Pathways in cancer - Homo sapiens (human) | path:hsa05200 |
| 0.000201 | calcium-mediated signaling | GO:0019722 | | 0.0014 | ECM proteoglycans | R-HSA-3000178 |
| 0.000204 | regulation of cell projection organization | GO:0031344 | | 0.0014 | Toxoplasmosis - Homo sapiens (human) | path:hsa05145 |
| 0.000255 | positive regulation of signal transduction | GO:0009967 | | 0.0021 | TNFR2 non-canonical NF-kB pathway | R-HSA-5668541 |
| 0.000373 | regulation of programmed cell death | GO:0043067 | | 0.0022 | Arrhythmogenic right ventricular cardiomyopathy - Homo sapiens (human) | path:hsa05412 |
| 0.000506 | renal vesicle morphogenesis | GO:0072077 | | 0.0023 | Axon guidance - Homo sapiens (human) | path:hsa04360 |
| 0.000509 | regulation of phosphorus metabolic process | GO:0051174 | | 0.0026 | PI3K-Akt signaling pathway - Homo sapiens (human) | path:hsa04151 |
| 0.000656 | neuroinflammatory response | GO:0150076 | | 0.0027 | Human T-cell leukemia virus 1 infection - Homo sapiens (human) | path:hsa05166 |
| 0.000656 | regulation of glutamate receptor signaling pathway | GO:1900449 | | 0.0028 | Cell junction organization | R-HSA-446728 |
| 0.000766 | regulation of neurological system process | GO:0031644 | | 0.0031 | Hypertrophic cardiomyopathy - Homo sapiens (human) | path:hsa05410 |
| 0.000869 | ureteric bud development | GO:0001657 | | 0.0034 | Primary immunodeficiency - Homo sapiens (human) | path:hsa05340 |
| 0.000890 | negative regulation of cell communication | GO:0010648 | | 0.0043 | Cell cycle - Homo sapiens (human) | path:hsa04110 |
| 0.000890 | positive regulation of leukocyte migration | GO:0002687 | | 0.0054 | Dilated cardiomyopathy - Homo sapiens (human) | path:hsa05414 |
| 0.000994 | response to interferon-gamma | GO:0034341 | | 0.0075 | GPCR downstream signalling | R-HSA-388396 |
| 0.001040 | acid secretion | GO:0046717 | | 0.0077 | NCAM signaling for neurite out-growth | R-HSA-375165 |
| 0.001102 | nephron epithelium morphogenesis | GO:0072088 | | 0.0095 | Calcium signaling pathway - Homo sapiens (human) | path:hsa04020 |
| 0.001227 | phosphorylation | GO:0016310 | | 0.0102 | Regulation of commissural axon pathfinding by SLIT and ROBO | R-HSA-428542 |
| 0.001230 | regulation of cell-matrix adhesion | GO:0001952 | | 0.0106 | Elastic fibre formation | R-HSA-1566948 |
| 0.001367 | myeloid leukocyte differentiation | GO:0002573 | | 0.0131 | Mitotic G1 phase and G1/S transition | R-HSA-453279 |
| 0.001438 | mesonephros development | GO:0001823 | | 0.0137 | IL-17 signaling pathway - Homo sapiens (human) | path:hsa04657 |
| 0.001524 | negative regulation of cell proliferation | GO:0008285 | | 0.0142 | GnRH secretion - Homo sapiens (human) | path:hsa04929 |
| 0.001742 | neuron projection extension involved in neuron projection gu | GO:1902284 | | 0.0145 | Activation of ATR in response to replication stress | R-HSA-176187 |
| 0.002073 | negative regulation of cell differentiation | GO:0045596 | | 0.0145 | Interaction between L1 and Ankyrins | R-HSA-445095 |
| 0.002616 | metanephros morphogenesis | GO:0003338 | | 0.0145 | Activation of Matrix Metalloproteinases | R-HSA-1592389 |
| 0.002654 | negative regulation of signal transduction | GO:0009968 | | 0.0145 | TNFs bind their physiological receptors | R-HSA-5669034 |
| 0.002744 | dendritic cell differentiation | GO:0097028 | | 0.0151 | Type I hemidesmosome assembly | R-HSA-446107 |
| 0.002905 | regulation of cell morphogenesis | GO:0022604 | | 0.0174 | Molecules associated with elastic fibres | R-HSA-2129379 |
| 0.003111 | regulation of secretion | GO:0051046 | | 0.0209 | MAPK signaling pathway - Homo sapiens (human) | path:hsa04010 |
| 0.003243 | sensory perception of pain | GO:0019233 | | 0.0225 | Crosslinking of collagen fibrils | R-HSA-2243919 |
| 0.003243 | positive regulation of cell-substrate adhesion | GO:0010811 | | 0.0225 | Caspase activation via Dependence Receptors in the absence of ligand | R-HSA-418889 |
| 0.003496 | glycoprotein biosynthetic process | GO:0009101 | | 0.0225 | Cross-presentation of soluble exogenous antigens (endosomes) | R-HSA-1236978 |
| 0.004058 | G1/S transition of mitotic cell cycle | GO:0000082 | | 0.0251 | L1CAM interactions | R-HSA-373760 |
| 0.004058 | regulation of GTPase activity | GO:0043087 | | 0.0251 | Bladder cancer - Homo sapiens (human) | path:hsa05219 |
| 0.004859 | branching involved in ureteric bud morphogenesis | GO:0001658 | | 0.0253 | Cell Cycle Checkpoints | R-HSA-69620 |
| 0.004951 | metanephros development | GO:0001656 | | 0.0253 | Estrogen signaling pathway - Homo sapiens (human) | path:hsa04915 |
| 0.005328 | regulation of leukocyte proliferation | GO:0070663 | | 0.0253 | G1/S-Specific Transcription | R-HSA-69205 |
| 0.005394 | T cell chemotaxis | GO:0010818 | | 0.0253 | Potassium Channels | R-HSA-1296071 |
| 0.005688 | regulation of antigen receptor-mediated signaling pathway | GO:0050854 | | 0.0253 | Protein digestion and absorption - Homo sapiens (human) | path:hsa04974 |
| 0.005830 | eosinophil chemotaxis | GO:0048245 | | 0.0253 | Th1 and Th2 cell differentiation - Homo sapiens (human) | path:hsa04658 |
| 0.006163 | negative regulation of cellular component organization | GO:0051129 | | 0.0262 | Activation of the pre-replicative complex | R-HSA-68962 |
| 0.006163 | negative regulation of cellular component movement | GO:0051271 | | 0.0339 | Orc1 removal from chromatin | R-HSA-68949 |
| 0.006564 | smooth muscle contraction | GO:0006939 | | 0.0353 | Amplification of signal from unattached kinetochores via a MAD2 inhibitory signal | R-HSA-141444 |
| 0.007204 | mitotic spindle assembly checkpoint | GO:0007094 | | 0.0353 | Amplification of signal from the kinetochores | R-HSA-141424 |
| 0.007204 | pulmonary valve development | GO:0003177 | | 0.0362 | Signaling by PDGF | R-HSA-186797 |
| 0.007204 | positive regulation of cell adhesion mediated by integrin | GO:0033630 | | 0.0385 | Scavenging by Class A Receptors | R-HSA-3000480 |
| 0.007204 | response to tumor necrosis factor | GO:0034612 | | 0.0385 | Other semaphorin interactions | R-HSA-416700 |
| 0.007410 | regulation of response to wounding | GO:1903034 | | 0.0385 | Condensation of Prometaphase Chromosomes | R-HSA-2514853 |
| 0.007880 | response to calcium ion | GO:0051592 | | 0.0385 | Resolution of Sister Chromatid Cohesion | R-HSA-2500257 |
| 0.008098 | regulation of cellular protein metabolic process | GO:0032268 | | 0.0392 | African trypanosomiasis - Homo sapiens (human) | path:hsa05143 |
| 0.009046 | cell-substrate junction assembly | GO:0007044 | | 0.0392 | G1/S Transition | R-HSA-69206 |
| 0.009323 | epithelial cell differentiation involved in kidney development | GO:0035850 | | 0.0439 | Breast cancer - Homo sapiens (human) | path:hsa05224 |
| 0.009594 | regulation of transforming growth factor beta production | GO:0071634 | | 0.0439 | RHO GTPases Activate Formins | R-HSA-5663220 |
| 0.010394 | anion transport | GO:0006820 | | 0.0439 | DNA Replication Pre-Initiation | R-HSA-69002 |
| 0.010394 | astrocyte differentiation | GO:0048708 | | 0.0439 | EML4 and NUDC in mitotic spindle formation | R-HSA-9648025 |
| 0.010673 | epidermis morphogenesis | GO:0048730 | | 0.0439 | Syndecan interactions | R-HSA-3000170 |
| 0.010709 | aminoglycan biosynthetic process | GO:0006023 | | 0.0439 | Assembly of the pre-replicative complex | R-HSA-68867 |
| 0.010709 | regulation of receptor binding | GO:1900120 | | 0.0446 | Legionellosis - Homo sapiens (human) | path:hsa05134 |
| 0.010891 | regulation of ion transmembrane transporter activity | GO:0032412 | | 0.0446 | Cell-Cell communication | R-HSA-1500931 |
| 0.011352 | hepatocyte differentiation | GO:0070365 | | 0.0446 | Intestinal immune network for IgA production - Homo sapiens (human) | path:hsa04672 |
| 0.011352 | negative regulation of cell division | GO:0051782 | | 0.0502 | Neurofascin interactions | R-HSA-447043 |
| 0.012319 | T-helper 1 type immune response | GO:0042088 | | 0.0502 | Mitotic Spindle Checkpoint | R-HSA-69618 |
| 0.012394 | peptide secretion | GO:0002790 | | 0.0559 | DNA Replication | R-HSA-69306 |
| 0.012453 | negative regulation of cell death | GO:0060548 | | 0.0559 | T cell receptor signaling pathway - Homo sapiens (human) | path:hsa04660 |
| 0.012453 | fever generation | GO:0001660 | | 0.0559 | C-type lectin receptor signaling pathway - Homo sapiens (human) | path:hsa04625 |
| 0.012453 | cellular response to amino acid stimulus | GO:0071230 | | 0.0589 | TP53 Regulates Transcription of Genes Involved in G1 Cell Cycle Arrest | R-HSA-6804116 |
| 0.012453 | eosinophil migration | GO:0072677 | | 0.0589 | Wnt signaling pathway - Homo sapiens (human) | path:hsa04310 |
| 0.013082 | regulation of antigen processing and presentation of peptide ( | GO:0002580 | | 0.0596 | Proteoglycans in cancer - Homo sapiens (human) | path:hsa05205 |
| 0.013370 | T cell mediated immunity | GO:0002456 | | 0.0596 | Malaria - Homo sapiens (human) | path:hsa05144 |
| 0.013448 | regulation of embryonic development | GO:0045995 | | 0.0596 | Phase 0 - rapid depolarisation | R-HSA-5576892 |
| 0.013475 | neuronal action potential | GO:0019228 | | 0.0626 | Cushing syndrome - Homo sapiens (human) | path:hsa04934 |
| 0.013475 | regulation of inflammatory response | GO:0050727 | | 0.0689 | RUNX3 regulates WNT signaling | R-HSA-8951430 |
| 0.013567 | pseudopodium organization | GO:0031268 | | 0.0689 | Netrin mediated repulsion signals | R-HSA-418886 |
| 0.013567 | vascular wound healing | GO:0061042 | | 0.0689 | Hippo signaling pathway - Homo sapiens (human) | path:hsa04390 |
| 0.013655 | regulation of morphogenesis of an epithelium | GO:1905330 | | 0.0698 | DNA strand elongation | R-HSA-69190 |
| 0.013988 | regulation of vasculature development | GO:1901342 | | 0.0698 | Glycosphingolipid biosynthesis - globo and isoglobo series - Homo sapiens (human) | path:hsa00603 |
| 0.014217 | cellular response to growth factor stimulus | GO:0071363 | | 0.0717 | Graft-versus-host disease - Homo sapiens (human) | path:hsa05332 |
| 0.014866 | regulation of leukocyte chemotaxis | GO:0002688 | | 0.0717 | RHOF GTPase cycle | R-HSA-9035034 |
| 0.015206 | regulation of cellular extravasation | GO:0002691 | | 0.0734 | Adrenaline signalling through Alpha-2 adrenergic receptor | R-HSA-392023 |
| 0.015269 | regulation of protein metabolic process | GO:0051246 | | | | |
| 0.015761 | cardiac muscle cell contraction | GO:0086003 | | | | |
| 0.015761 | regulation of lymphocyte migration | GO:2000401 | | | | |
| 0.015882 | positive regulation of tolerance induction | GO:0002645 | | | | |
| 0.015882 | regulation of dendritic cell antigen processing and presentati | GO:0002604 | | | | |
| 0.016238 | chordate embryonic development | GO:0043009 | | | | |
| 0.017109 | membrane depolarization during action potential | GO:0086010 | | | | |
| 0.019049 | negative regulation of nervous system development | GO:0051961 | | | | |
| 0.019955 | aortic valve development | GO:0003176 | | | | |
| 0.020537 | chronic inflammatory response | GO:0002544 | | | | |
| 0.020744 | metanephric tubule formation | GO:0072174 | | | | |
| 0.021136 | regulation of T cell tolerance induction | GO:0002664 | | | | |
| 0.021136 | establishment of T cell polarity | GO:0001768 | | | | |
| 0.021136 | natural killer cell chemotaxis | GO:0035747 | | | | |
| 0.021136 | negative regulation of vasculature development | GO:1901343 | | | | |
| 0.021274 | regulation of beta-catenin-TCF complex assembly | GO:1904863 | | | | |
| 0.021709 | positive regulation of cytokine production | GO:0001819 | | | | |
| 0.021709 | negative regulation of response to external stimulus | GO:0032102 | | | | |
| 0.021709 | negative regulation of cell-cell adhesion | GO:0022408 | | | | |
| 0.022405 | neural crest cell differentiation | GO:0014033 | | | | |
| 0.022405 | embryonic epithelial tube formation | GO:0001838 | | | | |
| 0.023906 | myeloid dendritic cell differentiation | GO:0043011 | | | | |
| 0.024096 | oligodendrocyte differentiation | GO:0048709 | | | | |
| 0.024096 | regulation of extent of cell growth | GO:0061387 | | | | |
| 0.024559 | visual system development | GO:0150063 | | | | |
| 0.025053 | eye morphogenesis | GO:0048592 | | | | |
| 0.027419 | negative regulation of chromosome segregation | GO:0051985 | | | | |
| 0.027419 | regulation of mononuclear cell migration | GO:0071675 | | | | |
| 0.028565 | calcium ion import into cytosol | GO:1902656 | | | | |
| 0.029476 | central nervous system neuron development | GO:0021954 | | | | |
| 0.029586 | membrane raft distribution | GO:0031580 | | | | |
| 0.029586 | anterior/posterior axon guidance | GO:0033564 | | | | |
| 0.029586 | positive regulation of fever generation | GO:0031622 | | | | |
| 0.029586 | negative regulation of cytokine production | GO:0001818 | | | | |
| 0.029601 | positive regulation of cell death | GO:0010942 | | | | |
| 0.029601 | signal transduction involved in DNA integrity checkpoint | GO:0072401 | | | | |
| 0.029601 | signal transduction involved in DNA damage checkpoint | GO:0072422 | | | | |
| 0.029856 | negative regulation of cell-substrate adhesion | GO:0010812 | | | | |
| 0.029856 | DNA replication initiation | GO:0006270 | | | | |
| 0.029944 | negative regulation of hydrolase activity | GO:0051346 | | | | |
| 0.030035 | skin epidermis development | GO:0098773 | | | | |
| 0.030035 | regulation of animal organ morphogenesis | GO:2000027 | | | | |
| 0.031926 | fatty acid derivative transport | GO:1901571 | | | | |
| 0.032004 | poly-N-acetyllactosamine metabolic process | GO:0030309 | | | | |
| 0.032618 | positive regulation of transferase activity | GO:0051347 | | | | |
| 0.032618 | regulation of blood circulation | GO:1903522 | | | | |
| 0.032618 | regulation of actin filament-based movement | GO:1903115 | | | | |
| 0.032618 | eye development | GO:0001654 | | | | |
| 0.033294 | actin polymerization or depolymerization | GO:0008154 | | | | |
| 0.033956 | cellular response to metal ion | GO:0071248 | | | | |
| 0.035920 | cellular response to alkaloid | GO:0071312 | | | | |
| 0.035920 | regulation of epithelial cell differentiation | GO:0030856 | | | | |
| 0.035920 | response to alkaloid | GO:0043279 | | | | |
| 0.035920 | regulation of neutrophil migration | GO:1902622 | | | | |
| 0.036261 | stress-activated MAPK cascade | GO:0051403 | | | | |
| 0.037714 | regulation of smooth muscle cell proliferation | GO:0048660 | | | | |
| 0.038447 | establishment of planar polarity of embryonic epithelium | GO:0042249 | | | | |
| 0.038447 | macrophage cytokine production | GO:0010934 | | | | |
| 0.040544 | positive regulation of immune effector process | GO:0002699 | | | | |
| 0.041489 | negative regulation of response to wounding | GO:1903035 | | | | |
| 0.041569 | negative regulation of blood vessel morphogenesis | GO:2000181 | | | | |
| 0.041649 | negative regulation of cell activation | GO:0050866 | | | | |
| 0.042132 | organic anion transport | GO:0015711 | | | | |
| 0.042532 | leukocyte apoptotic process | GO:0071887 | | | | |
| 0.042532 | negative regulation of developmental growth | GO:0048640 | | | | |
| 0.043364 | negative regulation of leukocyte migration | GO:0002686 | | | | |
| 0.043364 | positive regulation of leukocyte mediated cytotoxicity | GO:0001912 | | | | |
| 0.043364 | regulation of actin binding | GO:1904616 | | | | |
| 0.043364 | negative regulation of transcription involved in G1/S transitic | GO:0071930 | | | | |
| 0.043364 | regulation of eosinophil degranulation | GO:0043309 | | | | |
| 0.043364 | chemorepulsion involved in embryonic olfactory bulb interne | GO:0021834 | | | | |
| 0.043394 | negative regulation of cell growth | GO:0030308 | | | | |
| 0.044824 | homophilic cell adhesion via plasma membrane adhesion mo | GO:0007156 | | | | |
| 0.044981 | regulation of membrane invagination | GO:1905153 | | | | |
| 0.046963 | neural crest cell migration | GO:0001755 | | | | |
| 0.050187 | endothelium development | GO:0003158 | | | | |
| 0.050187 | positive regulation of antigen processing and presentation | GO:0002579 | | | | |
| 0.050187 | negative regulation of antigen processing and presentation | GO:0002578 | | | | |
| 0.050187 | positive regulation of isomerase activity | GO:0010912 | | | | |

#### cABMR downregulated

| q-value | Pathway name | Pathway ID | | q-value | pathway | external_id |
|---|---|---|---|---|---|---|
| 1.46742728627e-08 | striated muscle contrac | GO:0006941 | | 1.24810951124e-05 | Striated Muscle Contraction | R-HSA-390522 |
| 1.84580502769e-05 | heart contraction | GO:0060047 | | 0.00115279815982 | Muscle contraction | R-HSA-397014 |
| 7.60445938487e-05 | positive regulation of n | GO:0051962 | | 0.0122557917854 | Cardiac muscle contraction - Homo sapiens (human) | path:hsa04260 |
| 0.000126892461586 | regulation of blood circ | GO:1903522 | | 0.0162178060674 | Acetylcholine inhibits contraction of outer hair cells | R-HSA-9667769 |
| 0.000245927647039 | actin-myosin filament s | GO:0033275 | | 0.0255244225315 | Fatty acid metabolism | R-HSA-8978868 |
| 0.00147157148441 | striated muscle cell dev | GO:0055002 | | 0.0255244225315 | Synthesis of epoxy (EET) and dihydroxyeicosatrienoic acids (DHET) | R-HSA-2142670 |
| 0.00158325091342 | regulation of nervous s | GO:0051960 | | 0.0255244225315 | Synthesis of (16-20)-hydroxyeicosatetraenoic acids (HETE) | R-HSA-2142816 |
| 0.00297117625647 | sarcomere organization | GO:0045214 | | 0.0255244225315 | Ca2+ activated K+ channels | R-HSA-1296052 |
| 0.0057611469702 | regulation of cell develo | GO:0060284 | | 0.0255244225315 | Erythrocytes take up oxygen and release carbon dioxide | R-HSA-1247673 |
| 0.0057611469702 | striated muscle cell diff | GO:0051146 | | 0.0376620868706 | Metabolism of lipids | R-HSA-556833 |
| 0.00803155100891 | positive regulation of c | GO:0051130 | | 0.0376620868706 | Reversible hydration of carbon dioxide | R-HSA-1475029 |
| 0.00803155100891 | positive regulation of c | GO:0045597 | | 0.0376620868706 | Erythrocytes take up carbon dioxide and release oxygen | R-HSA-1237044 |
| 0.00993965752525 | positive regulation of s | GO:0051965 | | 0.0376620868706 | O2/CO2 exchange in erythrocytes | R-HSA-1480926 |
| 0.0200072170783 | generation of neurons | GO:0048699 | | 0.046239462302 | Potassium Channels | R-HSA-1296071 |
| 0.0263213232855 | G2 DNA damage checkp | GO:0031572 | | 0.046239462302 | Metabolism | R-HSA-1430728 |
| 0.0319122602578 | regulation of synapse o | GO:0050807 | | 0.0524859028965 | Nitrogen metabolism - Homo sapiens (human) | path:hsa00910 |
| 0.0319122602578 | regulation of postsynap | GO:0099150 | | 0.0681695196718 | Receptor-type tyrosine-protein phosphatases | R-HSA-388844 |
| 0.0319122602578 | regulation of postsynap | GO:0150052 | | 0.0708687277869 | Xenobiotics | R-HSA-211981 |
| 0.0319122602578 | lipid droplet formation | GO:0140042 | | | | |
| 0.039858659738 | regulation of excitatory | GO:1904889 | | | | |
| 0.039858659738 | regulation of muscle sy | GO:0090257 | | | | |
| 0.039858659738 | positive regulation of c | GO:0030307 | | | | |
| 0.039858659738 | ribonucleotide metabo | GO:0009259 | | | | |
| 0.0424714222866 | fatty acid biosynthetic | GO:0006633 | | | | |
| 0.0438049961342 | purine nucleotide meta | GO:0006163 | | | | |
| 0.0438049961342 | terpenoid biosynthetic | GO:0016114 | | | | |
| 0.0470497378576 | regulation of cell proje | GO:0031344 | | | | |
| 0.0499817587862 | amyloid-beta metaboli | GO:0050435 | | | | |
| 0.0499817587862 | striated muscle tissue c | GO:0014706 | | | | |
| 0.0503994339216 | cardiac muscle tissue m | GO:0055008 | | | | |
| 0.0503994339216 | nucleotide metabolic p | GO:0009117 | | | | |
| 0.0503994339216 | cell morphogenesis inv | GO:0000904 | | | | |
| 0.0609496425618 | cardiac muscle cell con | GO:0086003 | | | | |
| 0.0609496425618 | cellular response to ret | GO:0071300 | | | | |
| 0.0609496425618 | regulation of lipid bios | GO:0046890 | | | | |
| 0.0609496425618 | postsynaptic specializa | GO:0098698 | | | | |
| 0.0609496425618 | neuron projection deve | GO:0031175 | | | | |
| 0.0619133661072 | terpenoid metabolic pr | GO:0006721 | | | | |
| 0.0619133661072 | carboxylic acid biosynt | GO:0046394 | | | | |
| 0.0664696142355 | cardiac muscle cell dev | GO:0055013 | | | | |

#### clinTCMR upregulated

| q-value | Pathway nam | Pathway ID | | q-value | pathway | external_id |
|---|---|---|---|---|---|---|
| 1.44700856178e-23 | cellular respo | GO:0071345 | | 1.02081160447e-25 | Cell Cycle | R-HSA-1640170 |
| 4.6217721049e-22 | response to in | GO:0034341 | | 2.07987830736e-24 | Cell Cycle, Mitotic | R-HSA-69278 |
| 4.6217721049e-22 | nuclear divisi | GO:0000280 | | 2.2180507734e-24 | Cell Cycle Checkpoints | R-HSA-69620 |
| 1.11572035177e-21 | regulation of l | GO:1903037 | | 7.83687297536e-20 | Cell cycle - Homo sapiens (human) | path:hsa04110 |
| 4.54432635023e-21 | regulation of l | GO:0051249 | | 5.28917257083e-18 | Immune System | R-HSA-168256 |
| 8.09037142645e-21 | positive regul | GO:0022409 | | 8.53513070558e-16 | Resolution of Sister Chromatid Cohesion | R-HSA-2500257 |
| 7.49315761371e-20 | hemopoiesis | GO:0030097 | | 3.82927535556e-15 | G1/S Transition | R-HSA-69206 |
| 3.07171975633e-18 | positive regul | GO:0050867 | | 1.55649496598e-14 | Mitotic G1 phase and G1/S transition | R-HSA-453279 |
| 3.10631532346e-18 | positive regul | GO:0002696 | | 1.14432240478e-13 | Separation of Sister Chromatids | R-HSA-2467813 |
| 1.4987247952e-16 | regulation of l | GO:1902105 | | 1.18616935007e-13 | Amplification of signal from unattached kinetochores via a MAD2 inhibitory signal | R-HSA-141444 |
| 1.76953248005e-16 | positive regul | GO:0050778 | | 1.18616935007e-13 | Amplification of signal from the kinetochores | R-HSA-141424 |
| 8.48334305377e-15 | regulation of c | GO:0010564 | | 1.38944690518e-13 | Mitotic Spindle Checkpoint | R-HSA-69618 |
| 2.24754647778e-14 | DNA replicati | GO:0006260 | | 1.88640215328e-13 | Cytokine Signaling in Immune system | R-HSA-1280215 |
| 5.32629194452e-14 | positive regul | GO:0001819 | | 1.88640215328e-13 | Mitotic Anaphase | R-HSA-68882 |
| 1.20696333095e-13 | neutrophil mi | GO:1990266 | | 2.08048130289e-13 | Mitotic Metaphase and Anaphase | R-HSA-2555396 |
| 2.45182519777e-13 | G1/S transitio | GO:0000082 | | 2.26334666653e-13 | Human T-cell leukemia virus 1 infection - Homo sapiens (human) | path:hsa05166 |
| 4.41226238653e-13 | regulation of l | GO:0070663 | | 2.26334666653e-13 | EML4 and NUDC in mitotic spindle formation | R-HSA-9648025 |
| 4.41226238653e-13 | regulation of r | GO:0007346 | | 3.92745991229e-13 | Epstein-Barr virus infection - Homo sapiens (human) | path:hsa05169 |
| 6.85178886773e-13 | neutrophil ch | GO:0030593 | | 6.56498532858e-13 | Interferon gamma signaling | R-HSA-877300 |
| 2.18548919242e-12 | chemokine-m | GO:0070098 | | 2.82704542481e-12 | Mitotic Prometaphase | R-HSA-68877 |
| 3.4201940285e-12 | interferon-gar | GO:0060333 | | 4.39083040641e-12 | S Phase | R-HSA-69242 |
| 1.67227693017e-11 | response to ch | GO:1990868 | | 4.68158930942e-12 | RHO GTPases Activate Formins | R-HSA-5663220 |
| 3.23260755501e-11 | negative regul | GO:0050866 | | 2.23573467346e-11 | Interferon Signaling | R-HSA-913531 |
| 6.94353172536e-11 | positive regul | GO:1903708 | | 3.09862351391e-11 | Chemokine receptors bind chemokines | R-HSA-380108 |
| 2.05247969563e-10 | regulation of c | GO:0031347 | | 3.18248768183e-11 | Antigen processing and presentation - Homo sapiens (human) | path:hsa04612 |
| 3.61335608464e-10 | DNA replicati | GO:0006270 | | 3.91367540013e-11 | Graft-versus-host disease - Homo sapiens (human) | path:hsa05332 |
| 5.28328859021e-10 | negative regul | GO:0002695 | | 5.7417183929e-11 | Type I diabetes mellitus - Homo sapiens (human) | path:hsa04940 |
| 5.75979423449e-10 | regulation of i | GO:0032649 | | 5.7417183929e-11 | Immunoregulatory interactions between a Lymphoid and a non-Lymphoid cell | R-HSA-198933 |
| 7.6466039927e-10 | positive regul | GO:0002699 | | 5.7417183929e-11 | Activation of ATR in response to replication stress | R-HSA-176187 |
| 1.47097038751e-09 | positive regul | GO:0008284 | | 5.7417183929e-11 | Allograft rejection - Homo sapiens (human) | path:hsa05330 |
| 1.56782374782e-09 | mitotic spind | GO:0007094 | | 5.7417183929e-11 | DNA Replication | R-HSA-69306 |
| 1.96558267491e-09 | negative regul | GO:0045786 | | 6.32980238548e-11 | G1/S-Specific Transcription | R-HSA-69205 |
| 6.33571254845e-09 | negative regul | GO:0051985 | | 7.33906278836e-11 | Viral protein interaction with cytokine and cytokine receptor - Homo sapiens (human) | path:hsa04061 |
| 1.0502210138e-08 | phosphorylat | GO:0016310 | | 8.59563708089e-11 | Cytokine-cytokine receptor interaction - Homo sapiens (human) | path:hsa04060 |
| 1.24180943535e-08 | positive regul | GO:0031349 | | 3.15618949001e-10 | Adaptive Immune System | R-HSA-1280218 |
| 1.44633972992e-08 | positive regul | GO:0045787 | | 7.70243575052e-10 | Unwinding of DNA | R-HSA-176974 |
| 1.44633972992e-08 | negative regul | GO:0001818 | | 7.95090472099e-10 | Primary immunodeficiency - Homo sapiens (human) | path:hsa05340 |
| 1.69348397544e-08 | positive regul | GO:0032103 | | 1.05492571561e-09 | Synthesis of DNA | R-HSA-69239 |
| 2.17662647899e-08 | regulation of a | GO:0002819 | | 2.30177724221e-09 | Hematopoietic cell lineage - Homo sapiens (human) | path:hsa04640 |
| 2.33143605343e-08 | G2/M transiti | GO:0000086 | | 2.66384127492e-09 | G2/M Checkpoints | R-HSA-69481 |
| 4.41700643549e-08 | centromere c | GO:0034508 | | 4.63057574011e-09 | Rheumatoid arthritis - Homo sapiens (human) | path:hsa05323 |
| 4.41700643549e-08 | antigen proce | GO:0019886 | | 8.17180282073e-09 | Cell adhesion molecules - Homo sapiens (human) | path:hsa04514 |
| 4.41700643549e-08 | dendritic cell | GO:0097028 | | 1.1435773096e-08 | Autoimmune thyroid disease - Homo sapiens (human) | path:hsa05320 |
| 5.59274953924e-08 | negative regul | GO:0022408 | | 1.64539517434e-08 | M Phase | R-HSA-68886 |
| 6.55366252126e-08 | T-helper 1 typ | GO:0042088 | | 2.55862300451e-08 | Switching of origins to a post-replicative state | R-HSA-69052 |
| 6.63594263235e-08 | regulation of i | GO:0032655 | | 2.72711911607e-08 | Polo-like kinase mediated events | R-HSA-156711 |
| 6.63594263235e-08 | leukocyte apo | GO:0071887 | | 2.74824421345e-08 | Activation of the pre-replicative complex | R-HSA-68962 |
| 7.21408785562e-08 | regulation of v | GO:0050792 | | 5.69031586488e-08 | Phagosome - Homo sapiens (human) | path:hsa04145 |
| 9.77863187402e-08 | regulation of p | GO:0051174 | | 6.95404763887e-08 | Viral myocarditis - Homo sapiens (human) | path:hsa05416 |
| 1.37503121528e-07 | negative regul | GO:0048525 | | 9.79315322388e-08 | APC/C-mediated degradation of cell cycle proteins | R-HSA-174143 |
| 1.4740946887e-07 | regulation of l | GO:0002688 | | 9.79315322388e-08 | Regulation of mitotic cell cycle | R-HSA-453276 |
| 1.88410890756e-07 | regulation of i | GO:1902531 | | 1.12550046594e-07 | DNA Replication Pre-Initiation | R-HSA-69002 |
| 2.19065974615e-07 | positive regul | GO:0001912 | | 1.2058754448e-07 | Cyclin A/B1/B2 associated events during G2/M transition | R-HSA-69273 |
| 2.32010366786e-07 | regulation of l | GO:0002703 | | 1.65651034498e-07 | Th1 and Th2 cell differentiation - Homo sapiens (human) | path:hsa04658 |
| 3.01115557753e-07 | positive regul | GO:0002687 | | 1.87004452533e-07 | G0 and Early G1 | R-HSA-1538133 |
| 3.33427541204e-07 | regulation of T | GO:0002664 | | 2.17222472302e-07 | Intestinal immune network for IgA production - Homo sapiens (human) | path:hsa04672 |
| 3.45154348676e-07 | regulation of c | GO:0033043 | | 2.78247649843e-07 | Assembly of the pre-replicative complex | R-HSA-68867 |
| 5.75717438621e-07 | myeloid leukc | GO:0002573 | | 2.78247649843e-07 | Orc1 removal from chromatin | R-HSA-68949 |
| 6.56566776545e-07 | type I interfe | GO:0060337 | | 3.05271507099e-07 | Activation of APC/C and APC/C:Cdc20 mediated degradation of mitotic proteins | R-HSA-176814 |
| 6.59379450334e-07 | regulation of p | GO:0043067 | | 3.10762976453e-07 | Th17 cell differentiation - Homo sapiens (human) | path:hsa04659 |
| 6.71422069371e-07 | regulation of i | GO:0045088 | | 3.1615982693e-07 | Chemokine signaling pathway - Homo sapiens (human) | path:hsa04062 |
| 7.29597370875e-07 | DNA recombir | GO:0006310 | | 3.17608620977e-07 | Signal Transduction | R-HSA-162582 |
| 7.29597370875e-07 | signal transdu | GO:0072401 | | 5.6707365788e-07 | TP53 Regulates Transcription of Cell Cycle Genes | R-HSA-6791312 |
| 7.29597370875e-07 | signal transdu | GO:0072422 | | 9.68755246053e-07 | Interferon alpha/beta signaling | R-HSA-909733 |
| 9.93711925185e-07 | DNA conform | GO:0071103 | | 9.94155634173e-07 | Measles - Homo sapiens (human) | path:hsa05162 |
| 9.96003606802e-07 | positive regul | GO:0002579 | | 1.6391062777e-06 | Inflammatory bowel disease - Homo sapiens (human) | path:hsa05321 |
| 1.06550396243e-06 | positive regul | GO:0045597 | | 1.95260673532e-06 | RHO GTPase Effectors | R-HSA-195258 |
| 1.20378874276e-06 | response to tu | GO:0034612 | | 2.15441551809e-06 | Human immunodeficiency virus 1 infection - Homo sapiens (human) | path:hsa05170 |
| 1.23225017497e-06 | negative regul | GO:0050777 | | 2.26986595313e-06 | APC/C:Cdc20 mediated degradation of mitotic proteins | R-HSA-176409 |
| 1.35675456288e-06 | response to ty | GO:0034340 | | 2.26986595313e-06 | DNA strand elongation | R-HSA-69190 |
| 1.52127992804e-06 | regulation of i | GO:0032653 | | 3.66034388274e-06 | TP53 Regulates Transcription of Genes Involved in G1 Cell Cycle Arrest | R-HSA-6804116 |
| 1.52127992804e-06 | mitotic DNA c | GO:0044773 | | 4.63047497162e-06 | Mitotic G2-G2/M phases | R-HSA-453274 |
| 1.60781450182e-06 | regulation of c | GO:0032268 | | 4.63489818319e-06 | Peptide ligand-binding receptors | R-HSA-375276 |
| 1.70393374125e-06 | regulation of c | GO:1904029 | | 5.08798873501e-06 | Kinesins | R-HSA-983189 |
| 2.56372431981e-06 | calcium ion tr | GO:0097553 | | 6.31539823855e-06 | Cellular senescence - Homo sapiens (human) | path:hsa04218 |
| 2.61406894022e-06 | DNA repair | GO:0006281 | | 6.48438631496e-06 | p53 signaling pathway - Homo sapiens (human) | path:hsa04115 |
| 2.65277451945e-06 | regulation of i | GO:0050727 | | 6.48438631496e-06 | Translocation of ZAP-70 to Immunological synapse | R-HSA-202430 |
| 2.71440386697e-06 | chromatin rer | GO:0031055 | | 1.24212573303e-05 | Costimulation by the CD28 family | R-HSA-388841 |
| 2.75400807205e-06 | T cell mediate | GO:0002456 | | 1.24212573303e-05 | The role of GTSE1 in G2/M progression after G2 checkpoint | R-HSA-8852276 |
| 3.34764983977e-06 | positive regul | GO:0051347 | | 1.43422037367e-05 | G2/M Transition | R-HSA-69275 |
| 3.60753673141e-06 | ERK1 and ERK | GO:0070371 | | 1.52724889787e-05 | Tuberculosis - Homo sapiens (human) | path:hsa05152 |
| 3.74498851232e-06 | regulation of p | GO:0051246 | | 1.60186738265e-05 | Phosphorylation of CD3 and TCR zeta chains | R-HSA-202427 |
| 4.05056390755e-06 | antigen recep | GO:0050851 | | 1.62588105205e-05 | Chagas disease - Homo sapiens (human) | path:hsa05142 |
| 4.13527754237e-06 | regulation of c | GO:0002718 | | 1.87964005291e-05 | Antigen processing-Cross presentation | R-HSA-1236975 |
| 4.50037200743e-06 | negative regul | GO:0002698 | | 2.07687900616e-05 | PD-1 signaling | R-HSA-389948 |
| 4.6300946483e-06 | antigen proce | GO:0019885 | | 2.64519873578e-05 | Staphylococcus aureus infection - Homo sapiens (human) | path:hsa05150 |
| 5.82300247937e-06 | regulation of r | GO:1902622 | | 3.6552889282e-05 | Deposition of new CENPA-containing nucleosomes at the centromere | R-HSA-606279 |
| 5.92174226131e-06 | G1 DNA dama | GO:0044783 | | 3.6552889282e-05 | Nucleosome assembly | R-HSA-774815 |
| 5.92951539391e-06 | protein-DNA c | GO:0065004 | | 3.83184010471e-05 | Endosomal/Vacuolar pathway | R-HSA-1236977 |
| 6.04048810665e-06 | positive regul | GO:0051272 | | 3.95167027392e-05 | Class A/1 (Rhodopsin-like receptors) | R-HSA-373076 |
| 9.96939589237e-06 | DNA replicati | GO:0034724 | | 5.18888111653e-05 | Leishmaniasis - Homo sapiens (human) | path:hsa05140 |
| 9.96939589237e-06 | response to in | GO:0070670 | | 6.5133283952e-05 | Osteoclast differentiation - Homo sapiens (human) | path:hsa04380 |
| 1.00836220977e-05 | regulation of i | GO:0032675 | | 7.08243388704e-05 | Regulation of APC/C activators between G1/S and early anaphase | R-HSA-176408 |
| 1.18706925438e-05 | T cell chemota | GO:0010818 | | 8.68654234789e-05 | T cell receptor signaling pathway - Homo sapiens (human) | path:hsa04660 |
| 1.25191139111e-05 | DNA damage r | GO:0030330 | | 8.68654234789e-05 | G2/M DNA replication checkpoint | R-HSA-69478 |
| 1.51939130524e-05 | eosinophil ch | GO:0048245 | | 8.82876375486e-05 | Influenza A - Homo sapiens (human) | path:hsa05164 |
| 2.00227389899e-05 | regulation of T | GO:0001914 | | 8.82876375486e-05 | Signaling by Rho GTPases, Miro GTPases and RHOBTB3 | R-HSA-9716542 |
| 3.09533217108e-05 | negative regul | GO:0001911 | | 9.82255869418e-05 | Signaling by Rho GTPases | R-HSA-194315 |
| 3.75946006058e-05 | regulation of a | GO:0050854 | | 0.000101906924707 | APC:Cdc20 mediated degradation of cell cycle proteins prior to satisfation of the cell cycle checkpoint | R-HSA-179419 |
| 4.1878253211e-05 | regulation of p | GO:0002700 | | 0.000130529001084 | Interleukin-2 family signaling | R-HSA-451927 |
| 5.11271326016e-05 | negative regul | GO:0051782 | | 0.000133725884594 | TNFR2 non-canonical NF-kB pathway | R-HSA-5668541 |
| 5.7055202925e-05 | eosinophil mi | GO:0072677 | | 0.000183247256991 | Antigen Presentation: Folding, assembly and peptide loading of class I MHC | R-HSA-983170 |
| 6.25670138096e-05 | viral life cycle | GO:0019058 | | 0.000192875004788 | Generation of second messenger molecules | R-HSA-202433 |
| 6.25670138096e-05 | response to in | GO:0070555 | | 0.000194672242675 | Toxoplasmosis - Homo sapiens (human) | path:hsa05145 |
| 6.76913795153e-05 | regulation of r | GO:0042269 | | 0.000228539575597 | Phosphorylation of Emi1 | R-HSA-176417 |
| 8.88609261878e-05 | G2 DNA dama | GO:0031572 | | 0.000287443159122 | G alpha (i) signalling events | R-HSA-418594 |
| 9.67759174155e-05 | positive regul | GO:0002645 | | 0.00029756516604 | Natural killer cell mediated cytotoxicity - Homo sapiens (human) | path:hsa04650 |
| 9.67759174155e-05 | regulation of c | GO:0002604 | | 0.000326865836359 | Toll-like receptor signaling pathway - Homo sapiens (human) | path:hsa04620 |
| 9.9225771645e-05 | positive regul | GO:0051130 | | 0.000378929382438 | Condensation of Prometaphase Chromosomes | R-HSA-2514853 |
| 0.000102673506529 | regulation of i | GO:0032663 | | 0.000394077096749 | Resolution of D-loop Structures through Synthesis-Dependent Strand Annealing (SDSA) | R-HSA-5693554 |
| 0.000102708315382 | viral genome r | GO:0019079 | | 0.000433108740976 | Human cytomegalovirus infection - Homo sapiens (human) | path:hsa05163 |
| 0.000102708315382 | positive regul | GO:0009967 | | 0.000478054037647 | COPI-dependent Golgi-to-ER retrograde traffic | R-HSA-6811434 |
| 0.000102708315382 | regulation of C | GO:2000348 | | 0.00048031422462 | Activation of NIMA Kinases NEK9, NEK6, NEK7 | R-HSA-2980767 |
| 0.000102708315382 | regulation of T | GO:2001188 | | 0.000606037229529 | MHC class II antigen presentation | R-HSA-2132295 |
| 0.000102708315382 | antigen proce | GO:0002476 | | 0.000606037229529 | Cdc20:Phospho-APC/C mediated degradation of Cyclin A | R-HSA-174184 |
| 0.000102708315382 | regulation of c | GO:0032642 | | 0.000606037229529 | Cyclin E associated events during G1/S transition | R-HSA-69202 |
| 0.00011291468807 | histone excha | GO:0043486 | | 0.000606037229529 | Cyclin A:Cdk2-associated events at S phase entry | R-HSA-69656 |
| 0.000147813893445 | natural killer c | GO:0035747 | | 0.000606037229529 | Pathways in cancer - Homo sapiens (human) | path:hsa05200 |
| 0.000157088373253 | regulation of i | GO:0032652 | | 0.000684622349284 | TCR signaling | R-HSA-202403 |
| 0.000206501710664 | positive regul | GO:0010647 | | 0.000689491657406 | G1/S DNA Damage Checkpoints | R-HSA-69615 |
| 0.000243533927953 | regulation of C | GO:0043087 | | 0.000742481782615 | APC/C:Cdh1 mediated degradation of Cdc20 and other APC/C:Cdh1 targeted proteins in late mitosis/early G1 | R-HSA-174178 |
| 0.000259390324005 | neuroinflamm | GO:0150076 | | 0.000758156704333 | Viral carcinogenesis - Homo sapiens (human) | path:hsa05203 |
| 0.000273298417281 | pattern recog | GO:0002221 | | 0.000914130202358 | Innate Immune System | R-HSA-168249 |
| 0.000293023396539 | regulation of i | GO:0032673 | | 0.00093696958742 | Downstream TCR signaling | R-HSA-202424 |
| 0.000308758519732 | antigen proce | GO:0042590 | | 0.00096017378375 | PD-L1 expression and PD-1 checkpoint pathway in cancer - Homo sapiens (human) | path:hsa05235 |
| 0.000331331161368 | negative regul | GO:0060548 | | 0.00107255785598 | Homologous DNA Pairing and Strand Exchange | R-HSA-5693579 |
| 0.000356102173737 | response to X- | GO:0010165 | | 0.0010898769859 | Hepatitis B - Homo sapiens (human) | path:hsa05161 |
| 0.000406214092773 | interleukin-21 | GO:0038114 | | 0.00124798742473 | GPCR ligand binding | R-HSA-500792 |
| 0.000406214092773 | response to in | GO:0098756 | | 0.00128028153925 | Small cell lung cancer - Homo sapiens (human) | path:hsa05222 |
| 0.000406214687882 | negative regul | GO:0031348 | | 0.00131127819243 | Resolution of D-loop Structures through Holliday Junction Intermediates | R-HSA-5693568 |
| 0.00042608661061 | establishment | GO:0040001 | | 0.00135738495529 | NOD-like receptor signaling pathway - Homo sapiens (human) | path:hsa04621 |
| 0.000473560439213 | cellular cation | GO:0030003 | | 0.00139103541071 | E2F-enabled inhibition of pre-replication complex formation | R-HSA-113507 |
| 0.000528327743386 | regulation of r | GO:0045637 | | 0.00150559479063 | Transcription of E2F targets under negative control by DREAM complex | R-HSA-1362277 |
| 0.000543839462162 | neutrophil m | GO:0002446 | | 0.00187260282821 | Resolution of D-Loop Structures | R-HSA-5693537 |
| 0.00068086211587 | positive regul | GO:0010912 | | 0.00265307985115 | Oocyte meiosis - Homo sapiens (human) | path:hsa04114 |
| 0.000738183358975 | regulation of r | GO:0070507 | | 0.00265783216404 | Progesterone-mediated oocyte maturation - Homo sapiens (human) | path:hsa04914 |
| 0.000738183358975 | negative regul | GO:1903707 | | 0.00266450041628 | TP53 Regulates Transcription of Genes Involved in G2 Cell Cycle Arrest | R-HSA-6804114 |
| 0.00081699258902 | regulation of s | GO:0060142 | | 0.00266450041628 | SCF(Skp2)-mediated degradation of p27/p21 | R-HSA-187577 |
| 0.000986928157104 | regulation of i | GO:0032677 | | 0.00266450041628 | APC-Cdc20 mediated degradation of Nek2A | R-HSA-179409 |
| 0.00106635475079 | mitotic DNA r | GO:0033314 | | 0.00322852866704 | Transcription of E2F targets under negative control by p107 (RBL1) and p130 (RBL2) in complex with HDAC1 | R-HSA-1362300 |
| 0.00108433321721 | regulation of f | GO:0048145 | | 0.00343335129786 | Other semaphorin interactions | R-HSA-416700 |
| 0.00109581301921 | regulation of l | GO:2000401 | | 0.00343707508883 | Chromosome Maintenance | R-HSA-73886 |
| 0.00109581301921 | regulation of l | GO:0051052 | | 0.00351087860416 | Presynaptic phase of homologous DNA pairing and strand exchange | R-HSA-5693616 |
| 0.00147763543116 | negative regul | GO:0050858 | | 0.00365387033243 | Systemic lupus erythematosus - Homo sapiens (human) | path:hsa05322 |
| 0.00147763543116 | DNA strand el | GO:0022616 | | 0.00381193005929 | CDK-mediated phosphorylation and removal of Cdc6 | R-HSA-69017 |
| 0.00166836073901 | nucleosome a | GO:0006334 | | 0.00381193005929 | TNFs bind their physiological receptors | R-HSA-5669034 |
| 0.00167728604573 | regulation of a | GO:0002580 | | 0.00438394633967 | Transcriptional Regulation by TP53 | R-HSA-3700989 |
| 0.00212439300773 | liver regenera | GO:0097421 | | 0.00444148222119 | Interleukin-2 signaling | R-HSA-9020558 |
| 0.00217922025938 | positive regul | GO:0051345 | | 0.00451823598588 | Asthma - Homo sapiens (human) | path:hsa05310 |
| 0.00273256345301 | positive regul | GO:0010942 | | 0.00457620339517 | Bladder cancer - Homo sapiens (human) | path:hsa05219 |
| 0.0027495672821 | calcium-medi | GO:0019722 | | 0.0048178854461 | Hemostasis | R-HSA-109582 |
| 0.00280922355902 | negative regul | GO:0008285 | | 0.00519345745242 | Fanconi anemia pathway - Homo sapiens (human) | path:hsa03460 |
| 0.00314572875543 | regulation of a | GO:0002583 | | 0.00519345745242 | Phosphorylation of the APC/C | R-HSA-176412 |
| 0.00319712825525 | ATP-depender | GO:0043044 | | 0.00584325065886 | HDR through Homologous Recombination (HRR) | R-HSA-5685942 |
| 0.00341361132003 | establishment | GO:0051294 | | 0.00593940227264 | Chk1/Chk2(Cds1) mediated inactivation of Cyclin B:Cdk1 complex | R-HSA-75035 |
| 0.00386675087607 | regulation of t | GO:0032479 | | 0.00630335866772 | E2F mediated regulation of DNA replication | R-HSA-113510 |
| 0.00393190014409 | regulation of r | GO:1903034 | | 0.00630335866772 | Inactivation of APC/C via direct inhibition of the APC/C complex | R-HSA-141430 |
| 0.00442549541398 | regulation of t | GO:1903555 | | 0.00630335866772 | Inhibition of the proteolytic activity of APC/C required for the onset of anaphase by mitotic spindle checkpo | R-HSA-141405 |
| 0.00442792517384 | negative regul | GO:0051129 | | 0.00655653814624 | Kaposi sarcoma-associated herpesvirus infection - Homo sapiens (human) | path:hsa05167 |
| 0.00449196559723 | regulation of r | GO:0071675 | | 0.00712362396848 | Factors involved in megakaryocyte development and platelet production | R-HSA-983231 |
| 0.00462752000935 | response to U | GO:0009411 | | 0.0074327485163 | JAK-STAT signaling pathway - Homo sapiens (human) | path:hsa04630 |
| 0.00473390735665 | positive regul | GO:0031325 | | 0.0076201050957 | TP53 regulates transcription of several additional cell death genes whose specific roles in p53-dependent apo | R-HSA-6803205 |
| 0.00488313788911 | lateral attach | GO:0099607 | | 0.0076201050957 | Golgi Cisternae Pericentriolar Stack Reorganization | R-HSA-162658 |
| 0.00488313788911 | regulation of C | GO:0032832 | | 0.0076201050957 | CDT1 association with the CDC6:ORC:origin complex | R-HSA-68827 |
| 0.00488313788911 | interleukin-10 | GO:0140105 | | 0.00821307414804 | Golgi-to-ER retrograde transport | R-HSA-8856688 |
| 0.00492277372904 | regulation of l | GO:0002911 | | 0.00941221973575 | Transcriptional Regulation by E2F6 | R-HSA-8953750 |
| 0.00492277372904 | interleukin-4- | GO:0035771 | | 0.00941544113445 | Assembly of collagen fibrils and other multimeric structures | R-HSA-2022090 |
| 0.00504875184211 | nuclear envel | GO:0051081 | | 0.00959244611742 | NF-kappa B signaling pathway - Homo sapiens (human) | path:hsa04064 |
| 0.00504875184211 | regulation of g | GO:0032645 | | 0.0096781248591 | Signaling by GPCR | R-HSA-372790 |
| 0.00506573648347 | negative regul | GO:0032102 | | 0.00972020242416 | Interleukin-4 and Interleukin-13 signaling | R-HSA-6785807 |
| 0.00543145147844 | positive regul | GO:0051173 | | 0.0106120836771 | DNA replication - Homo sapiens (human) | path:hsa03030 |
| 0.0056622112241 | regulation of T | GO:0045622 | | 0.0106152690623 | APC/C:Cdc20 mediated degradation of Cyclin B | R-HSA-174048 |
| 0.00627312989754 | cellular prote | GO:0006464 | | 0.0106170972566 | Neutrophil degranulation | R-HSA-6798695 |
| 0.00639167662515 | positive regul | GO:0010604 | | 0.0111631255588 | Signaling by Interleukins | R-HSA-449147 |
| 0.00659636804429 | response to ga | GO:0010332 | | 0.0113201402171 | Pancreatic cancer - Homo sapiens (human) | path:hsa05212 |
| 0.00689552762295 | Ras protein sig | GO:0007265 | | 0.0119110798128 | Antiviral mechanism by IFN-stimulated genes | R-HSA-1169410 |
| 0.00755990822937 | ion homeosta | GO:0050801 | | 0.0120276873165 | RUNX3 regulates WNT signaling | R-HSA-8951430 |
| 0.00788727828513 | regulation of i | GO:0002861 | | 0.0120276873165 | CDC6 association with the ORC:origin complex | R-HSA-68689 |
| 0.00788727828513 | exit from mito | GO:0010458 | | 0.0133934980322 | Hepatitis C - Homo sapiens (human) | path:hsa05160 |
| 0.00837614228015 | positive regul | GO:0051781 | | 0.0144619146956 | Interleukin receptor SHC signaling | R-HSA-912526 |
| 0.00970108523425 | DNA biosynth | GO:0071897 | | 0.0149688254067 | TNF receptor superfamily (TNFSF) members mediating non-canonical NF-kB pathway | R-HSA-5676594 |
| 0.00970108523425 | negative regul | GO:1903035 | | 0.015467098305 | B cell receptor signaling pathway - Homo sapiens (human) | path:hsa04662 |
| 0.0102170003348 | negative regul | GO:0002578 | | 0.0167753352661 | The role of Nef in HIV-1 replication and disease pathogenesis | R-HSA-164952 |
| 0.0102170003348 | interleukin-9- | GO:0038113 | | 0.0168626171697 | TNF signaling pathway - Homo sapiens (human) | path:hsa04668 |
| 0.010494383596 | regulation of t | GO:0002828 | | 0.0168626171697 | Interleukin-9 signaling | R-HSA-8985947 |
| 0.0105432260509 | cortical cytos | GO:0030865 | | 0.0179692560255 | Phosphorylation of proteins involved in the G2/M transition by Cyclin A:Cdc2 complexes | R-HSA-170145 |
| 0.012185028532 | epithelial cell | GO:1904019 | | 0.0179692560255 | Deregulated CDK5 triggers multiple neurodegenerative pathways in Alzheimer,s disease models | R-HSA-8862803 |
| 0.0123196973173 | macrophage r | GO:0035691 | | 0.0179692560255 | Neurodegenerative Diseases | R-HSA-8863678 |
| 0.0123196973173 | negative regul | GO:0071930 | | 0.0218420254004 | p53-Dependent G1 DNA Damage Response | R-HSA-69563 |
| 0.0123196973173 | regulation of p | GO:2000361 | | 0.0218420254004 | p53-Dependent G1/S DNA damage checkpoint | R-HSA-69580 |
| 0.0123196973173 | regulation of g | GO:0071661 | | 0.022667013675 | RUNX1 and FOXP3 control the development of regulatory T lymphocytes (Tregs) | R-HSA-8877330 |
| 0.0123196973173 | negative regul | GO:2000524 | | 0.0235541610096 | Signaling by SCF-KIT | R-HSA-1433557 |
| 0.0134823876561 | regulation of c | GO:1901976 | | 0.0276720700847 | C-type lectin receptor signaling pathway - Homo sapiens (human) | path:hsa04625 |
| 0.0135135578945 | response to in | GO:0071104 | | 0.0298114778937 | Dissolution of Fibrin Clot | R-HSA-75205 |
| 0.0135273368956 | regulation of g | GO:1903018 | | 0.0303482540752 | Homology Directed Repair | R-HSA-5693538 |
| 0.0135273368956 | regulation of l | GO:0043300 | | 0.0307553679514 | Nef-mediates down modulation of cell surface receptors by recruiting them to clathrin adapters | R-HSA-164938 |
| 0.0135273368956 | regulation of i | GO:0032674 | | 0.0317189133243 | GPCR downstream signalling | R-HSA-388396 |
| 0.0162073554806 | positive regul | GO:0033630 | | 0.0328321833416 | Phosphorylation of proteins involved in G1/S transition by active Cyclin E:Cdk2 complexes | R-HSA-69200 |
| 0.0164966964408 | entry into hos | GO:0044409 | | 0.0328321833416 | Interleukin-18 signaling | R-HSA-9012546 |
| 0.0164966964408 | entry into cell | GO:0051806 | | 0.0357315066468 | Diseases of programmed cell death | R-HSA-9645723 |
| 0.0175074973764 | regulation of c | GO:0033632 | | 0.0370897153082 | Human papillomavirus infection - Homo sapiens (human) | path:hsa05165 |
| 0.0183631928685 | regulation of a | GO:0032956 | | 0.0370897153082 | CD28 dependent Vav1 pathway | R-HSA-389359 |
| 0.0190906128256 | positive regul | GO:0051984 | | | | |
| 0.0202671321549 | intrinsic apop | GO:0008630 | | | | |
| 0.0218123976627 | regulation of r | GO:1900120 | | | | |
| 0.0218123976627 | regulation of p | GO:0032069 | | | | |
| 0.0218123976627 | B cell mediate | GO:0019724 | | | | |
| 0.0218123976627 | negative regul | GO:0051348 | | | | |
| 0.0218123976627 | establishment | GO:0001768 | | | | |
| 0.0218123976627 | interleukin-2- | GO:0038110 | | | | |
| 0.0218382646407 | cellular respo | GO:0072757 | | | | |
| 0.0218382646407 | microglial cel | GO:0002282 | | | | |
| 0.0231028815748 | regulation of c | GO:0001959 | | | | |
| 0.0252100765493 | regulation of l | GO:0090183 | | | | |
| 0.0257538068086 | regulation of r | GO:1905521 | | | | |
| 0.0273521515857 | regulation of u | GO:0051438 | | | | |
| 0.0277312756525 | regulation of r | GO:1901214 | | | | |
| 0.0281056653381 | integrin activa | GO:0033622 | | | | |
| 0.029436663271 | regulation of r | GO:0043030 | | | | |
| 0.029436663271 | negative regul | GO:0009968 | | | | |
| 0.0311639550021 | negative regul | GO:0034122 | | | | |
| 0.0323950118079 | response to in | GO:0070669 | | | | |
| 0.0323950118079 | deoxyribonuc | GO:0009263 | | | | |
| 0.0326046856119 | regulation of e | GO:0030856 | | | | |
| 0.032877751627 | tumor necros | GO:0033209 | | | | |
| 0.032877751627 | response to ca | GO:1901563 | | | | |
| 0.032877751627 | regulation of r | GO:0030885 | | | | |
| 0.032877751627 | strand invasio | GO:0042148 | | | | |
| 0.032877751627 | T cell recepto | GO:0033153 | | | | |
| 0.032877751627 | intrinsic apop | GO:0072332 | | | | |
| 0.0331504210747 | positive regul | GO:0060760 | | | | |

#### clinTCMR downregulated

| q-value | Pathway name | Pathway ID | | q-value | pathway | external_id |
|---|---|---|---|---|---|---|
| 0.013005514 | terpenoid me | GO:0006721 | | 0.0416183130504 | Xenobiotics | R-HSA-211981 |
| 0.017965699 | positive regula | GO:0003099 | | 0.0416183130504 | Cytochrome P450 - arranged by substrate type | R-HSA-211897 |
| 0.017965699 | receptor inter | GO:0031623 | | 0.043704648867 | Synthesis of epoxy (EET) and dihydroxyeicosatrienoic acids (DHET) | R-HSA-2142670 |
| 0.035935276 | regulation of r | GO:0002090 | | 0.043704648867 | Adrenoceptors | R-HSA-390696 |
| 0.035935276 | adenylate cyc | GO:0071880 | | 0.043704648867 | Synthesis of (16-20)-hydroxyeicosatetraenoic acids (HETE) | R-HSA-2142816 |
| 0.035935276 | positive regula | GO:0001996 | | 0.0648019943333 | Amine ligand-binding receptors | R-HSA-375280 |
| 0.035935276 | striated musc | GO:0014706 | | 0.0648019943333 | Phospholipase D signaling pathway - Homo sapiens (human) | path:hsa04072 |
| 0.035935276 | heart contrac | GO:0060047 | | 0.0675257625006 | LGI-ADAM interactions | R-HSA-5682910 |
| 0.037205891 | regulation of l | GO:1903522 | | 0.0684107402393 | Cocaine addiction - Homo sapiens (human) | path:hsa05030 |
| 0.037205891 | protein locali | GO:1903539 | | 0.0684727404739 | Phase I - Functionalization of compounds | R-HSA-211945 |
| 0.037205891 | lipid hydroxyl | GO:0002933 | | 0.0838921982904 | Muscle contraction | R-HSA-397014 |
| 0.037205891 | embryonic lin | GO:0030326 | | 0.0996222323838 | Neuroactive ligand-receptor interaction - Homo sapiens (human) | path:hsa04080 |
| 0.048396877 | regulation of t | GO:0035296 | | 0.10171975073 | Phase 0 - rapid depolarisation | R-HSA-5576892 |
| 0.048396877 | regulation of l | GO:0050880 | | | | |
| 0.048396877 | cellular gluca | GO:0006073 | | | | |
| 0.048396877 | glucan metab | GO:0044042 | | | | |
| 0.052588294 | striated musc | GO:0051146 | | | | |
| 0.056468517 | positive regula | GO:0003084 | | | | |
| 0.056468517 | regulation of r | GO:0031644 | | | | |
| 0.071912878 | regulation of s | GO:0003044 | | | | |
| 0.083213855 | negative regul | GO:0003085 | | | | |
| 0.083213855 | hepatocyte di | GO:0070365 | | | | |
| 0.089771562 | heart field spe | GO:0003128 | | | | |
| 0.092463645 | regulation of s | GO:2001014 | | | | |
| 0.092463645 | response to vi | GO:0033189 | | | | |
| 0.095719497 | cardiac muscl | GO:0055008 | | | | |
| 0.096773677 | positive regula | GO:0051965 | | | | |

#### subTCMR upregulated

| q-value | Pathway name | Pathway ID | | q-value | pathway | external_id |
|---|---|---|---|---|---|---|
| 7.699705570 | dendritic cell | GO:0097028 | | 0.00618439117185 | Conversion from APC/C:Cdc20 to APC/C:Cdh1 in late anaphase | R-HSA-176407 |
| 0.003167739 | positive regula | GO:0002579 | | 0.00618439117185 | Inactivation of APC/C via direct inhibition of the APC/C complex | R-HSA-141430 |
| 0.003167739 | regulation of i | GO:0032655 | | 0.00618439117185 | Inhibition of the proteolytic activity of APC/C required for the onset of anaphase by mitotic spindle checkpo | R-HSA-141405 |
| 0.003167739 | regulation of c | GO:0002604 | | 0.00618439117185 | APC/C:Cdc20 mediated degradation of Securin | R-HSA-174154 |
| 0.003167739 | establishment | GO:0001768 | | 0.00618439117185 | APC/C:Cdc20 mediated degradation of Cyclin B | R-HSA-174048 |
| 0.003722666 | regulation of p | GO:0043067 | | 0.00618439117185 | APC-Cdc20 mediated degradation of Nek2A | R-HSA-179409 |
| 0.009937072 | regulation of c | GO:0010564 | | 0.00618439117185 | Cdc20:Phospho-APC/C mediated degradation of Cyclin A | R-HSA-174184 |
| 0.009937072 | regulation of i | GO:0002861 | | 0.00618439117185 | APC:Cdc20 mediated degradation of cell cycle proteins prior to satisfation of the cell cycle checkpoint | R-HSA-179419 |
| 0.009937072 | exit from mitc | GO:0010458 | | 0.00618439117185 | APC/C:Cdh1 mediated degradation of Cdc20 and other APC/C:Cdh1 targeted proteins in late mitosis/early G1 | R-HSA-174178 |
| 0.012682835 | regulation of l | GO:1903037 | | 0.00618439117185 | APC/C:Cdc20 mediated degradation of mitotic proteins | R-HSA-176409 |
| 0.018213374 | regulation of r | GO:1902622 | | 0.00618439117185 | Activation of APC/C and APC/C:Cdc20 mediated degradation of mitotic proteins | R-HSA-176814 |
| 0.018213374 | regulation of g | GO:1903018 | | 0.00712382235667 | Regulation of APC/C activators between G1/S and early anaphase | R-HSA-176408 |
| 0.018213374 | negative regul | GO:0022408 | | 0.00804189974273 | RND1 GTPase cycle | R-HSA-9696273 |
| 0.018213374 | negative regul | GO:0060548 | | 0.00804189974273 | APC/C-mediated degradation of cell cycle proteins | R-HSA-174143 |
| 0.019260146 | positive regula | GO:0051347 | | 0.00804189974273 | Regulation of mitotic cell cycle | R-HSA-453276 |
| 0.019260146 | regulation of u | GO:0051438 | | 0.0148176240291 | Chemokine receptors bind chemokines | R-HSA-380108 |
| 0.019260146 | response to in | GO:0034341 | | 0.0153418859511 | Human T-cell leukemia virus 1 infection - Homo sapiens (human) | path:hsa05166 |
| 0.019260146 | myeloid leukc | GO:0002573 | | 0.0253907516152 | Cell Cycle, Mitotic | R-HSA-69278 |
| 0.019260146 | nuclear divisic | GO:0000280 | | 0.0262954762582 | Collagen formation | R-HSA-1474290 |
| 0.021926119 | positive regula | GO:0001819 | | 0.0272205976168 | Extracellular matrix organization | R-HSA-1474244 |
| 0.027205945 | regulation of l | GO:0070663 | | | | |
| 0.033584183 | regulation of l | GO:0051249 | | | | |
| 0.036144267 | positive regula | GO:0022409 | | | | |
| 0.036144267 | chemokine-m | GO:0070098 | | | | |
| 0.036144267 | hemopoiesis | GO:0030097 | | | | |
| 0.036144267 | positive regula | GO:0051130 | | | | |
| 0.036144267 | response to ch | GO:1990868 | | | | |
| 0.036144267 | leukocyte apo | GO:0071887 | | | | |
| 0.036144267 | regulation of i | GO:0032652 | | | | |
| 0.036144267 | neutrophil ch | GO:0030593 | | | | |
| 0.036144267 | positive regula | GO:0050778 | | | | |
| 0.036144267 | response to tu | GO:0034612 | | | | |
| 0.036144267 | negative regul | GO:0045786 | | | | |

#### subTCMR downregulated

| q-value | Pathway name | Pathway ID |
|---|---|---|
| 0.003756328 | neutrophil ch | GO:0030593 |
| 0.003756328 | neutrophil mi | GO:1990266 |
| 0.016023216 | positive regula | GO:0032103 |
| 0.039590551 | regulation of s | GO:0051046 |

**Suppl. Table 4: Unique differentially expressed genes**
cABMR upregulated

| gene name | log2fold change | padj |
|---|---|---|
| PLOD3 | 1,02880407 | 7,40951E-12 |
| FLNA | 1,6317091 | 4,34351E-11 |
| CFTR | 2,56282409 | 1,10167E-09 |
| MMP14 | 1,24253869 | 1,32729E-09 |
| CXCL6 | 3,55217543 | 1,44719E-09 |
| PAPLN | 2,30990727 | 2,21808E-09 |
| KRT19 | 2,03488675 | 2,96993E-09 |
| CLIC6 | 3,14608598 | 4,61834E-09 |
| COL4A2 | 1,43061686 | 8,16685E-09 |
| VTCN1 | 2,70031649 | 8,32979E-09 |
| BACE2 | 1,57633617 | 9,35002E-09 |
| THBS2 | 2,1790276 | 2,16129E-08 |
| LAMC2 | 3,12526582 | 2,7787E-08 |
| FTH1P3 | 2,42175413 | 3,65955E-08 |
| MTRNR2L8 | 3,06667593 | 3,99767E-08 |
| EFEMP1 | 2,64369218 | 4,14407E-08 |
| SLC12A2 | 1,57940533 | 4,14407E-08 |
| ADRA2A | 2,58621974 | 6,15533E-08 |
| ITGA3 | 2,04262344 | 7,2481E-08 |
| CLDN11 | 2,40443531 | 9,15553E-08 |
| CDH6 | 2,85102344 | 1,0331E-07 |
| RGS4 | 2,86194925 | 1,12572E-07 |
| PMEPA1 | 2,09626818 | 1,17097E-07 |
| ITGB8 | 2,27762763 | 1,2321E-07 |
| SPINT1 | 2,48555881 | 1,30775E-07 |
| RHBDF2 | 1,06574828 | 1,33846E-07 |
| MOXD1 | 2,93781329 | 1,71634E-07 |
| PTK7 | 2,13469537 | 2,20399E-07 |
| FLJ44635 | 2,98668606 | 2,40494E-07 |
| SLC25A24 | 1,07393754 | 2,50943E-07 |
| KIRREL | 1,16540021 | 2,56718E-07 |
| FAM133CP | 3,50601986 | 2,83479E-07 |
| LAMC3 | 2,26357045 | 3,07012E-07 |
| COL4A1 | 1,36736863 | 3,21127E-07 |
| WISP1 | 1,97936823 | 3,21127E-07 |
| CLEC2D | 1,22271988 | 3,25463E-07 |
| MRC2 | 1,27525494 | 3,82347E-07 |
| ATP2A3 | 1,37048508 | 3,95807E-07 |
| ARNT2 | 1,65465794 | 4,09126E-07 |
| LXN | 1,50286637 | 4,47423E-07 |
| ANXA3 | 1,82444201 | 4,84118E-07 |
| PODXL | 1,50682244 | 4,98298E-07 |
| GPR56 | 1,307493 | 5,85467E-07 |
| ITGB4 | 1,65623936 | 6,07194E-07 |
| WNK2 | 1,86706092 | 6,97634E-07 |

**cABMR downregulated**

| gene name | log2fold change | padj |
|---|---|---|
| ACOT13 | -1,0339213 | 1,8504E-08 |
| ZSCAN16-AS1 | -1,0099268 | 2,7664E-08 |
| SMIM6 | -1,2901073 | 6,7369E-08 |
| AQP7P3 | -1,4487478 | 9,1555E-08 |
| ATM | -2,283682 | 5,8547E-07 |
| CHCHD10 | -1,0229407 | 2,5423E-06 |
| EPB41L4A-AS1 | -1,0708142 | 3,7203E-06 |
| SLC9A3R2 | -1,0185579 | 4,0002E-06 |
| DKFZP434I0714 | -1,5061009 | 6,504E-06 |
| PTH1R | -1,4549927 | 8,1461E-06 |
| RPS29 | -1,0123515 | 9,5665E-06 |
| FITM1 | -1,3547668 | 1,3638E-05 |
| DHRS4L2 | -1,0660973 | 1,5962E-05 |
| LOC102724655 | -1,6198801 | 1,8286E-05 |
| TCP10L | -1,0114306 | 1,9355E-05 |
| COX6C | -1,3190488 | 2,516E-05 |
| AQP7P1 | -1,0145102 | 3,2207E-05 |
| SLCO4C1 | -2,0039156 | 3,5429E-05 |
| MYB | -1,5491519 | 0,00010772 |
| AC004862.6 | -1,2410272 | 0,00011014 |
| C14orf37 | -1,1325489 | 0,00012241 |
| ASPDH | -1,1959075 | 0,00015313 |
| HM13-AS1 | -1,2076247 | 0,00017688 |
| C1orf173 | -1,0890138 | 0,00018763 |
| CNGA1 | -1,0502723 | 0,00019969 |
| EMBP1 | -1,0020902 | 0,00019975 |
| LOC101929781 | -3,7397835 | 0,00021523 |
| LOC102724832 | -1,1428225 | 0,00024773 |
| HAO2 | -1,0056348 | 0,00028629 |
| RP11-423H2.3 | -1,7387241 | 0,00029112 |
| NEUROD2 | -1,815604 | 0,0003088 |
| IGF1 | -1,182198 | 0,00031315 |
| LOC100509620 | -1,4245046 | 0,00038421 |
| FGF23 | -1,4031726 | 0,00048534 |
| METRN | -1,4662193 | 0,00050083 |
| RND2 | -1,331744 | 0,00050281 |
| LOC101928384 | -1,2129626 | 0,00056736 |
| MPL | -1,5020018 | 0,0007778 |
| BRCA1 | -1,2610385 | 0,00087431 |
| HSPB9 | -1,0136028 | 0,00089685 |
| LOC101928230 | -1,0576306 | 0,00092364 |
| LOC102723782 | -1,4722794 | 0,00118725 |
| FBXO40 | -3,3120842 | 0,00139643 |
| CA1 | -4,0581878 | 0,00152059 |
| ARID3C | -1,0149793 | 0,00156684 |

**clinTCMR upregulated**

| gene name | log2fold change | padj |
|---|---|---|
| BIRC5 | 2,92840489 | 5,78565E-19 |
| MKI67 | 2,86448938 | 7,91385E-19 |
| HMMR | 2,68370876 | 3,29788E-18 |
| CDK1 | 2,64706258 | 2,0257E-15 |
| NUSAP1 | 1,71979787 | 4,85211E-15 |
| CCNB1 | 1,9664764 | 5,40764E-15 |
| PTTG1 | 2,54110015 | 1,04363E-14 |
| PRC1 | 2,37560699 | 1,46334E-14 |
| CENPF | 2,01795702 | 3,1056E-14 |
| TK1 | 2,42311328 | 1,44913E-13 |
| TUBA1B | 1,05901111 | 5,87302E-13 |
| NDC80 | 2,32985683 | 9,60612E-13 |
| AURKA | 1,63108124 | 1,57267E-12 |
| TACC3 | 1,38838226 | 2,05012E-12 |
| KIF23 | 2,37866906 | 2,41517E-12 |
| STAT1 | 1,78627004 | 4,18686E-12 |
| HLA-F | 1,58906853 | 6,12565E-12 |
| UBE2T | 2,39102821 | 1,3795E-11 |
| CENPU | 2,26190271 | 1,98841E-11 |
| KIF2C | 2,46346748 | 2,64046E-11 |
| IFI30 | 1,55968251 | 3,62393E-11 |
| MCM4 | 1,42531282 | 6,75031E-11 |
| HLA-A | 1,36772809 | 6,78273E-11 |
| ORC6 | 2,71410529 | 6,78273E-11 |
| PIGR | 1,33109084 | 1,20323E-10 |
| GBP1 | 1,58993419 | 2,49122E-10 |
| KPNA2 | 1,69411643 | 3,96494E-10 |
| GTSE1 | 2,56316201 | 3,97165E-10 |
| CDCA8 | 1,86087302 | 4,63818E-10 |
| CKAP2L | 2,50958768 | 5,52909E-10 |
| MAD2L1 | 1,99517743 | 5,52909E-10 |
| KIF4A | 2,55873272 | 7,93007E-10 |
| CDCA2 | 2,71400885 | 8,48899E-10 |
| PAQR4 | 1,95465046 | 8,48904E-10 |
| RAD51AP1 | 2,34075262 | 1,13391E-09 |
| CENPE | 1,89224033 | 1,21741E-09 |
| HLA-B | 1,33845002 | 1,28043E-09 |
| SKA3 | 2,81035242 | 1,32053E-09 |
| FANCI | 1,58109395 | 1,63261E-09 |
| LGALS9 | 1,14967923 | 2,34858E-09 |
| KIAA0101 | 1,91245777 | 3,00318E-09 |
| TMSB10 | 1,08695508 | 3,80111E-09 |
| PCNA | 1,08127547 | 4,49925E-09 |
| CENPW | 1,75131493 | 5,18504E-09 |
| PSMB9 | 1,26219426 | 5,45497E-09 |

**clinTCMR downregulated**

| gene name | log2fold change | padj |
|---|---|---|
| NFKBIZ | -1,2808978 | 1,34166E-08 |
| ASXL3 | -1,9961655 | 1,38239E-07 |
| YPEL2 | -1,1889795 | 4,70271E-07 |
| LOC102724175 | -2,4586822 | 5,98809E-07 |
| AOC4P | -2,371319 | 1,03141E-06 |
| RANBP3L | -1,7038629 | 2,87806E-06 |
| ARNTL | -2,4101721 | 5,0615E-06 |
| SLC30A10 | -1,2473076 | 1,30583E-05 |
| NFIL3 | -1,2898682 | 1,44944E-05 |
| PPP1R3C | -1,4045311 | 2,82334E-05 |
| AASS | -1,1603042 | 3,34952E-05 |
| C2orf16 | -1,66244 | 4,50643E-05 |
| ARL4D | -1,2736006 | 5,15448E-05 |
| ADRA1A | -1,2809977 | 5,7083E-05 |
| RP11-439L18.3 | -1,3554475 | 9,92205E-05 |
| CXXC4 | -1,2048499 | 0,000112556 |
| PITPNM3 | -1,1249638 | 0,000122617 |
| GREM1 | -2,9763058 | 0,000185906 |
| TPRG1 | -1,1259359 | 0,000208491 |
| LOC102724711 | -1,2373817 | 0,000274636 |
| SLC45A2 | -2,8994624 | 0,000283062 |
| LOC101929079 | -1,8862657 | 0,000303599 |
| SCN2A | -1,7348076 | 0,000310585 |
| C1orf101 | -1,1193817 | 0,000345697 |
| NDRG1 | -1,0665297 | 0,000371156 |
| LOC101927121 | -1,9682398 | 0,000373389 |
| RNF152 | -1,2938655 | 0,000401621 |
| CRABP2 | -2,865394 | 0,000442737 |
| LGSN | -1,253746 | 0,000451538 |
| RP11-205M3.3 | -1,171015 | 0,000459637 |
| CDH12 | -4,4083167 | 0,000467836 |
| GSTA7P | -1,543083 | 0,000485721 |
| LINC01144 | -1,503429 | 0,000507384 |
| G6PC | -1,6848952 | 0,000573365 |
| ZFP2 | -1,1240702 | 0,000609793 |
| LRP4 | -1,2585286 | 0,000707893 |
| RP1-234P15.4 | -1,544645 | 0,000838007 |
| SEC14L3 | -1,5464624 | 0,000840425 |
| PDE8B | -1,1442789 | 0,000854623 |
| LOC101929761 | -1,0919248 | 0,000928394 |
| ST18 | -2,3066361 | 0,000964208 |
| ADAM12 | -1,4833176 | 0,001074137 |
| MDN1 | -1,1835268 | 0,001074137 |
| LOC101929552 | -1,6656102 | 0,001184529 |
| HERC2P7 | -1,08626 | 0,001385764 |

#### subTCMR upregulated

| gene name | log2fold change | padj |
|---|---|---|
| GABRA5 | 7,16892858 | 0,02503227 |
| COL5A3 | 1,22289582 | 0,04116968 |

#### subTCMR downregulated

| gene name | log2fold change | padj |
|---|---|---|
| S100A8 | -1,1917658 | 0,03353207 |

#### Suppl. table 5: Unique pathways of rejection phenotypes

#### cABMR upregulated

| q-value | Pathway name | Pathway ID | | q-value | Pathway Name | Pathway ID |
|---|---|---|---|---|---|---|
| 6.391643732 | generation of neurons | GO:0048699 | | 4.49936080192e-17 | Extracellular matrix organization | R-HSA-1474244 |
| 3.573964542 | regulation of ion transp | GO:0043269 | | 3.43269960376e-10 | ECM-receptor interaction - Homo sapiens (human) | path:hsa04512 |
| 1.444600475 | cell morphogenesis inv | GO:0000904 | | 1.16710095112e-06 | Amoebiasis - Homo sapiens (human) | path:hsa05146 |
| 3.357734210 | neuron projection deve | GO:0031175 | | 1.16710095112e-06 | Collagen formation | R-HSA-1474290 |
| 5.107898978 | anterograde trans-syna | GO:0098916 | | 1.91491076318e-06 | Nervous system development | R-HSA-9675108 |
| 7.539803052 | positive regulation of c | GO:0051272 | | 2.91153518707e-06 | Axon guidance | R-HSA-422475 |
| 7.539803052 | regulation of transmem | GO:0034762 | | 5.03963516316e-06 | Collagen chain trimerization | R-HSA-8948216 |
| 1.009954376 | ion transmembrane tra | GO:0034220 | | 5.68640082099e-06 | Developmental Biology | R-HSA-1266738 |
| 2.130425360 | regulation of cell devel | GO:0060284 | | 9.76211530619e-06 | Integrin cell surface interactions | R-HSA-216083 |
| 5.992460355 | regulation of nervous s | GO:0051960 | | 1.13493958538e-05 | Focal adhesion - Homo sapiens (human) | path:hsa04510 |
| 8.896620098 | kidney epithelium deve | GO:0072073 | | 6.07510821316e-05 | Assembly of collagen fibrils and other multimeric structures | R-HSA-2022090 |
| 0.000171535 | positive regulation of c | GO:0045597 | | 6.07510821316e-05 | Collagen biosynthesis and modifying enzymes | R-HSA-1650814 |
| 0.000370052 | cation transport | GO:0006812 | | 7.50141668881e-05 | O-linked glycosylation | R-HSA-5173105 |
| 0.000496884 | gliogenesis | GO:0042063 | | 0.000159286762823 | Human papillomavirus infection - Homo sapiens (human) | path:hsa05165 |
| 0.000686635 | ureteric bud developm | GO:0001657 | | 0.000222086458078 | AGE-RAGE signaling pathway in diabetic complications - Homo sapiens (human) | path:hsa04933 |
| 0.000778245 | regulation of secretion | GO:0051046 | | 0.000276379155677 | Hypertrophic cardiomyopathy - Homo sapiens (human) | path:hsa05410 |
| 0.001067713 | peptide secretion | GO:0002790 | | 0.000276379155677 | NCAM1 interactions | R-HSA-419037 |
| 0.001067713 | mesonephros developm | GO:0001823 | | 0.000415475639831 | O-glycosylation of TSR domain-containing proteins | R-HSA-5173214 |
| 0.001073522 | visual system developm | GO:0150063 | | 0.000443703866154 | Dilated cardiomyopathy - Homo sapiens (human) | path:hsa05414 |
| 0.001073522 | acid secretion | GO:0046717 | | 0.000502142089952 | Laminin interactions | R-HSA-3000157 |
| 0.001210435 | regulation of cell-matri | GO:0001952 | | 0.000600069514559 | Axon guidance - Homo sapiens (human) | path:hsa04360 |
| 0.001210435 | positive regulation of n | GO:0051962 | | 0.000891038079112 | Arrhythmogenic right ventricular cardiomyopathy - Homo sapiens (human) | path:hsa05412 |
| 0.001474144 | smooth muscle contrac | GO:0006939 | | 0.000920207136594 | PI3K-Akt signaling pathway - Homo sapiens (human) | path:hsa04151 |
| 0.001474144 | neuron projection exte | GO:1902284 | | 0.00107749060582 | Small cell lung cancer - Homo sapiens (human) | path:hsa05222 |
| 0.001685476 | eye development | GO:0001654 | | 0.00107749060582 | ECM proteoglycans | R-HSA-3000178 |
| 0.001959775 | regulation of transform | GO:0071634 | | 0.00165041565183 | NCAM signaling for neurite out-growth | R-HSA-375165 |
| 0.001959775 | aminoglycan biosynthe | GO:0006023 | | 0.00245046084566 | Molecules associated with elastic fibres | R-HSA-2129379 |
| 0.002056240 | anion transport | GO:0006820 | | 0.00275713948757 | Regulation of commissural axon pathfinding by SLIT and ROBO | R-HSA-428542 |
| 0.002120781 | pulmonary valve devel | GO:0003177 | | 0.00275713948757 | Cell junction organization | R-HSA-446728 |
| 0.002139556 | regulation of cell proje | GO:0031344 | | 0.00275713948757 | NF-kappa B signaling pathway - Homo sapiens (human) | path:hsa04064 |
| 0.002574062 | epidermis morphogene | GO:0048730 | | 0.00275713948757 | Potassium Channels | R-HSA-1296071 |
| 0.002581462 | glycoprotein biosynthe | GO:0009101 | | 0.00275713948757 | Protein digestion and absorption - Homo sapiens (human) | path:hsa04974 |
| 0.003823403 | negative regulation of c | GO:0051271 | | 0.00298341344721 | Non-integrin membrane-ECM interactions | R-HSA-3000171 |
| 0.004196545 | ion homeostasis | GO:0050801 | | 0.00545117349238 | Elastic fibre formation | R-HSA-1566948 |
| 0.004199291 | nephron epithelium m | GO:0072088 | | 0.00575103513193 | Calcium signaling pathway - Homo sapiens (human) | path:hsa04020 |
| 0.004247037 | negative regulation of c | GO:0045596 | | 0.00633207369626 | Caspase activation via Dependence Receptors in the absence of ligand | R-HSA-418889 |
| 0.004820463 | response to chemokine | GO:1990868 | | 0.00823986319004 | L1CAM interactions | R-HSA-373760 |
| 0.005549641 | cell-substrate junction | GO:0007044 | | 0.00854983856129 | Chemokine receptors bind chemokines | R-HSA-380108 |
| 0.005549641 | aortic valve developme | GO:0003176 | | 0.00854983856129 | Degradation of the extracellular matrix | R-HSA-1474228 |
| 0.005601823 | negative regulation of r | GO:0051961 | | 0.00917962323084 | Scavenging by Class A Receptors | R-HSA-3000480 |
| 0.005998822 | fever generation | GO:0001660 | | 0.00995227992475 | Interaction between L1 and Ankyrins | R-HSA-445095 |
| 0.006227096 | negative regulation of c | GO:0010812 | | 0.00995227992475 | GnRH secretion - Homo sapiens (human) | path:hsa04929 |
| 0.006227096 | cellular response to am | GO:0071230 | | 0.0162700590177 | Pathways in cancer - Homo sapiens (human) | path:hsa05200 |
| 0.006602150 | renal vesicle morphoge | GO:0072077 | | 0.0206268571334 | Viral protein interaction with cytokine and cytokine receptor - Homo sapiens (human) | path:hsa04061 |
| 0.007017688 | negative regulation of c | GO:0010648 | | 0.021511209595 | Insulin secretion - Homo sapiens (human) | path:hsa04911 |
| 0.007625417 | homophilic cell adhesi | GO:0007156 | | 0.0233880203489 | Neurofascin interactions | R-HSA-447043 |
| 0.007625417 | positive regulation of k | GO:0090184 | | 0.0270868690599 | Glycosphingolipid biosynthesis - globo and isoglobo series - Homo sapiens (human) | path:hsa00603 |
| 0.007625417 | cellular response to gro | GO:0071363 | | 0.0274189467688 | Neuronal System | R-HSA-112316 |
| 0.007625417 | regulation of neurologi | GO:0031644 | | 0.0277668327927 | Proteoglycans in cancer - Homo sapiens (human) | path:hsa05205 |
| 0.007952605 | oligodendrocyte differe | GO:0048709 | | 0.0290623545895 | Parathyroid hormone synthesis, secretion and action - Homo sapiens (human) | path:hsa04928 |
| 0.008347351 | branching involved in u | GO:0001658 | | 0.0309409625344 | Formation of Fibrin Clot (Clotting Cascade) | R-HSA-140877 |
| 0.008347351 | neuroinflammatory res | GO:0150076 | | 0.0313629305606 | Netrin mediated repulsion signals | R-HSA-418886 |
| 0.009202443 | regulation of kidney de | GO:0090183 | | 0.0313629305606 | Muscle contraction | R-HSA-397014 |
| 0.009932622 | eye morphogenesis | GO:0048592 | | 0.0313629305606 | Cell-Cell communication | R-HSA-1500931 |
| 0.011065704 | metanephros morphog | GO:0003338 | | 0.0323235361606 | Cell-cell junction organization | R-HSA-421270 |
| 0.011065704 | negative regulation of c | GO:0008285 | | 0.0354152521796 | Glycosphingolipid biosynthesis - lacto and neolacto series - Homo sapiens (human) | path:hsa00601 |
| 0.011116001 | regulation of blood circ | GO:1903522 | | 0.0378869265355 | O-linked glycosylation of mucins | R-HSA-913709 |
| 0.011281243 | central nervous system | GO:0021954 | | 0.040455871754 | Relaxin signaling pathway - Homo sapiens (human) | path:hsa04926 |
| 0.011281243 | regulation of cell morp | GO:0022604 | | 0.0431782276601 | Hematopoietic cell lineage - Homo sapiens (human) | path:hsa04640 |
| 0.011281243 | astrocyte differentiatio | GO:0048708 | | 0.0431782276601 | Signaling by PDGF | R-HSA-186797 |
| 0.011679533 | metanephric tubule for | GO:0072174 | | 0.0431782276601 | Adrenaline signalling through Alpha-2 adrenergic receptor | R-HSA-392023 |
| 0.011840949 | skin epidermis develop | GO:0098773 | | 0.0431782276601 | Cell adhesion molecules - Homo sapiens (human) | path:hsa04514 |
| 0.012478991 | chordate embryonic de | GO:0043009 | | 0.0435045454814 | Activation of Matrix Metalloproteinases | R-HSA-1592389 |
| 0.012478991 | negative regulation of c | GO:0030308 | | 0.0517521218859 | GP1b-IX-V activation signalling | R-HSA-430116 |
| 0.012478991 | regulation of ion transm | GO:0032412 | | 0.0517521218859 | Crosslinking of collagen fibrils | R-HSA-2243919 |
| 0.013135236 | neuronal action potent | GO:0019228 | | 0.0517521218859 | Voltage gated Potassium channels | R-HSA-1296072 |
| 0.015012746 | photoreceptor cell diff | GO:0046530 | | 0.057969409861 | Syndecan interactions | R-HSA-3000170 |
| 0.015012746 | regulation of cellular ex | GO:0002691 | | 0.057969409861 | Cushing syndrome - Homo sapiens (human) | path:hsa04934 |
| 0.015131638 | poly-N-acetyllactosami | GO:0030309 | | 0.0603429619954 | Cardiac conduction | R-HSA-5576891 |
| 0.015293248 | regulation of beta-cate | GO:1904863 | | 0.0625629798257 | Hippo signaling pathway - Homo sapiens (human) | path:hsa04390 |
| 0.016176554 | chemokine-mediated s | GO:0070098 | | 0.0752217324081 | cAMP signaling pathway - Homo sapiens (human) | path:hsa04024 |
| 0.016176554 | membrane depolarizat | GO:0086010 | | 0.0767530822503 | Phase 0 - rapid depolarisation | R-HSA-5576892 |
| 0.016176554 | positive regulation of c | GO:0010811 | | 0.07875621606 | Cytokine-cytokine receptor interaction - Homo sapiens (human) | path:hsa04060 |
| 0.016176554 | anterior/posterior axo | GO:0033564 | | 0.0792713038148 | Presynaptic depolarization and calcium channel opening | R-HSA-112308 |
| 0.016176554 | positive regulation of f | GO:0031622 | | | | |
| 0.017706699 | organic anion transpor | GO:0015711 | | | | |
| 0.017706699 | neural crest cell differe | GO:0014033 | | | | |
| 0.020781268 | Wnt signaling pathway | GO:0016055 | | | | |
| 0.021490934 | cardiac muscle cell con | GO:0086003 | | | | |
| 0.021490934 | cellular response to ret | GO:0071300 | | | | |
| 0.021661102 | negative regulation of c | GO:0061037 | | | | |
| 0.022614945 | regulation of vasculatu | GO:1901342 | | | | |
| 0.022614945 | fatty acid derivative tra | GO:1901571 | | | | |
| 0.022614945 | regulation of glutamate | GO:1900449 | | | | |
| 0.022704136 | positive regulation of c | GO:0010647 | | | | |
| 0.022704136 | response to calcium ion | GO:0051592 | | | | |
| 0.022987610 | ERK1 and ERK2 cascade | GO:0070371 | | | | |
| 0.023150167 | adherens junction asse | GO:0034333 | | | | |
| 0.023307326 | epithelial to mesenchy | GO:0001837 | | | | |
| 0.024676844 | regulation of embryoni | GO:0045995 | | | | |
| 0.024780180 | vascular wound healing | GO:0061042 | | | | |
| 0.024780180 | cellular cation homeos | GO:0030003 | | | | |
| 0.024780180 | metanephros developm | GO:0001656 | | | | |
| 0.024780180 | axon extension | GO:0048675 | | | | |
| 0.024890854 | regulation of peptide tr | GO:0090087 | | | | |
| 0.025550913 | embryonic epithelial tu | GO:0001838 | | | | |
| 0.025550913 | positive regulation of c | GO:0008284 | | | | |
| 0.025550913 | regulation of animal or | GO:2000027 | | | | |
| 0.025550913 | response to alkaloid | GO:0043279 | | | | |
| 0.025550913 | sensory perception of p | GO:0019233 | | | | |
| 0.026586567 | regulation of Wnt signa | GO:0030111 | | | | |
| 0.027008036 | regulation of actin filan | GO:1903115 | | | | |
| 0.027539028 | negative regulation of s | GO:0009968 | | | | |
| 0.029849889 | regulation of actin binc | GO:1904616 | | | | |
| 0.029849889 | chemorepulsion involv | GO:0021834 | | | | |
| 0.032161458 | peptide hormone secre | GO:0030072 | | | | |
| 0.034076886 | regulation of extent of | GO:0061387 | | | | |
| 0.034945633 | calcium-mediated sign | GO:0019722 | | | | |
| 0.034973864 | positive regulation of le | GO:0002687 | | | | |
| 0.034973864 | transmembrane recept | GO:0007178 | | | | |
| 0.036797256 | glycosaminoglycan me | GO:0030203 | | | | |
| 0.037964212 | glomerular visceral epi | GO:0072112 | | | | |
| 0.038875379 | regulation of tube diam | GO:0035296 | | | | |
| 0.038875379 | regulation of blood ves | GO:0050880 | | | | |
| 0.039064153 | cellular response to alk | GO:0071312 | | | | |
| 0.041026297 | protein O-linked glycos | GO:0006493 | | | | |
| 0.042665137 | negative regulation of v | GO:1901343 | | | | |
| 0.042862310 | sphingolipid biosynthe | GO:0030148 | | | | |
| 0.042862310 | central nervous system | GO:0021953 | | | | |
| 0.042862310 | regulation of morphog | GO:1905330 | | | | |
| 0.044181747 | female gonad developm | GO:0008585 | | | | |
| 0.046039769 | ear development | GO:0043583 | | | | |
| 0.046039769 | mechanoreceptor diffe | GO:0042490 | | | | |
| 0.046039769 | negative regulation of l | GO:0045833 | | | | |
| 0.046039769 | glycolipid metabolic pr | GO:0006664 | | | | |
| 0.047426156 | epithelial cell differenti | GO:0035850 | | | | |
| 0.047426156 | cellular response to mc | GO:0071315 | | | | |
| 0.047426156 | glycine import across p | GO:1903804 | | | | |
| 0.047426156 | astrocyte fate commitm | GO:0060018 | | | | |
| 0.051223076 | collagen-activated tyro | GO:0038063 | | | | |
| 0.051391693 | regulation of cell-subst | GO:0090109 | | | | |
| 0.052590423 | regulation of epithelial | GO:0030856 | | | | |
| 0.053729318 | regulation of muscle sy | GO:0090257 | | | | |
| 0.054241495 | protein localization to | GO:0034394 | | | | |
| 0.054241495 | neural crest cell develo | GO:0014032 | | | | |
| 0.054927781 | apoptotic process invo | GO:1902742 | | | | |
| 0.057008171 | organic acid transport | GO:0015849 | | | | |
| 0.058997005 | aorta development | GO:0035904 | | | | |

#### cABMR downregulated

| q-value | Pathway nam | Pathway ID | | q-value | Pathway Name | Pathway ID |
|---|---|---|---|---|---|---|
| 1.561392894 | striated musc | GO:0006941 | | 0.0264612160857 | Acetylcholine inhibits contraction of outer hair cells | R-HSA-9667769 |
| 0.000548214 | positive regul | GO:0051962 | | 0.0264612160857 | Striated Muscle Contraction | R-HSA-390522 |
| 0.002937431 | heart contrac | GO:0060047 | | 0.0264612160857 | Ca2+ activated K+ channels | R-HSA-1296052 |
| 0.003476793 | regulation of | GO:0051960 | | 0.0264612160857 | Erythrocytes take up oxygen and release carbon dioxide | R-HSA-1247673 |
| 0.003960488 | regulation of | GO:0060284 | | 0.0264612160857 | Reversible hydration of carbon dioxide | R-HSA-1475029 |
| 0.005453322 | positive regul | GO:0045597 | | 0.0264612160857 | Muscle contraction | R-HSA-397014 |
| 0.007236527 | positive regul | GO:0051130 | | 0.0264612160857 | Potassium Channels | R-HSA-1296071 |
| 0.010885675 | regulation of | GO:1903522 | | 0.0264612160857 | Erythrocytes take up carbon dioxide and release oxygen | R-HSA-1237044 |
| 0.013607050 | striated musc | GO:0055002 | | 0.0264612160857 | O2/CO2 exchange in erythrocytes | R-HSA-1480926 |
| 0.013607050 | G2 DNA dama | GO:0031572 | | 0.0410009144298 | Nitrogen metabolism - Homo sapiens (human) | path:hsa00910 |
| 0.024968505 | lipid droplet f | GO:0140042 | | 0.0626488661276 | Recruitment and ATM-mediated phosphorylation of repair and signaling proteins at DNA double strand brea | R-HSA-5693565 |
| 0.053453346 | cardiac muscl | GO:0086003 | | 0.0626488661276 | DNA Double Strand Break Response | R-HSA-5693606 |
| 0.053453346 | cellular respo | GO:0071300 | | 0.0626488661276 | Cholesterol biosynthesis | R-HSA-191273 |
| 0.053453346 | generation of | GO:0048699 | | 0.0626488661276 | Activation of gene expression by SREBF (SREBP) | R-HSA-2426168 |
| 0.053453346 | cardiac muscl | GO:0055013 | | 0.0626488661276 | Resolution of D-loop Structures through Synthesis-Dependent Strand Annealing (SDSA) | R-HSA-5693554 |
| 0.053453346 | regulation of | GO:0090257 | | 0.0626488661276 | Activation of G protein gated Potassium channels | R-HSA-1296041 |
| 0.053453346 | cellular respo | GO:0071396 | | 0.0626488661276 | G protein gated Potassium channels | R-HSA-1296059 |
| 0.053453346 | striated musc | GO:0014706 | | 0.0626488661276 | Inhibition of voltage gated Ca2+ channels via Gbeta/gamma subunits | R-HSA-997272 |
| 0.053453346 | cardiac muscl | GO:0086001 | | 0.0626488661276 | Cardiac muscle contraction - Homo sapiens (human) | path:hsa04260 |
| 0.053453346 | ribonucleotid | GO:0009259 | | | | |
| 0.053453346 | regulation of | GO:0031344 | | | | |
| 0.053453346 | pyridine-cont | GO:0072525 | | | | |
| 0.053453346 | regulation of | GO:1902930 | | | | |
| 0.057463987 | purine nucleo | GO:0006163 | | | | |
| 0.057463987 | DNA double-s | GO:0000729 | | | | |
| 0.057705814 | nucleotide m | GO:0009117 | | | | |
| 0.059036739 | striated musc | GO:0051146 | | | | |
| 0.059208158 | disruption of | GO:0051818 | | | | |
| 0.059208158 | regulation of | GO:0022604 | | | | |
| 0.065811192 | chemokine-m | GO:0070098 | | | | |
| 0.066717387 | regulation of | GO:2001233 | | | | |

#### clinTCMR upregulated

| q-value | Pathway Name | Pathway ID |
|---|---|---|
| 3.93247437637e-15 | response to interferon-gamma | GO:0034341 |
| 5.63938166694e-15 | nuclear division | GO:0000280 |
| 1.75446454509e-11 | cellular response to cytokine stimulus | GO:0071345 |
| 3.13603055428e-10 | regulation of mitotic cell cycle | GO:0007346 |
| 3.13603055428e-10 | regulation of cell cycle process | GO:0010564 |
| 3.13603055428e-10 | negative regulation of cell activation | GO:0050866 |
| 3.13603055428e-10 | positive regulation of immune response | GO:0050778 |
| 7.30362786868e-10 | interferon-gamma-mediated signaling pathway | GO:0060333 |
| 2.7831561662e-09 | negative regulation of leukocyte activation | GO:0002695 |
| 5.04174017034e-09 | regulation of leukocyte differentiation | GO:1902105 |
| 8.78927715353e-09 | regulation of leukocyte proliferation | GO:0070663 |
| 1.0046458658e-08 | regulation of leukocyte cell-cell adhesion | GO:1903037 |
| 1.36249834311e-08 | positive regulation of cell cycle | GO:0045787 |
| 1.71153747323e-08 | antigen processing and presentation of exogenous peptide antigen via MHC class II | GO:0019886 |
| 2.35209724921e-08 | positive regulation of cell activation | GO:0050867 |
| 2.5713157548e-08 | regulation of lymphocyte activation | GO:0051249 |
| 4.34175705325e-08 | regulation of adaptive immune response | GO:0002819 |
| 4.72590776822e-08 | positive regulation of leukocyte activation | GO:0002696 |
| 4.94698692915e-08 | hemopoiesis | GO:0030097 |
| 5.87765231631e-08 | positive regulation of cell-cell adhesion | GO:0022409 |
| 5.87765231631e-08 | positive regulation of cytokine production | GO:0001819 |
| 2.88986976626e-07 | regulation of interleukin-10 production | GO:0032653 |
| 3.06687610006e-07 | positive regulation of immune effector process | GO:0002699 |
| 3.98838921471e-07 | DNA replication | GO:0006260 |
| 5.35347918106e-07 | G2/M transition of mitotic cell cycle | GO:0000086 |
| 6.57618582875e-07 | regulation of leukocyte mediated immunity | GO:0002703 |
| 1.3629420469e-06 | G1/S transition of mitotic cell cycle | GO:0000082 |
| 1.40643965371e-06 | negative regulation of immune effector process | GO:0002698 |
| 1.40643965371e-06 | regulation of defense response | GO:0031347 |
| 2.36346349194e-06 | negative regulation of immune response | GO:0050777 |
| 2.91172761308e-06 | positive regulation of leukocyte mediated cytotoxicity | GO:0001912 |
| 4.04755290414e-06 | positive regulation of cell proliferation | GO:0008284 |
| 4.04755290414e-06 | type I interferon signaling pathway | GO:0060337 |
| 5.85011479582e-06 | DNA repair | GO:0006281 |
| 7.20465257026e-06 | response to type I interferon | GO:0034340 |
| 7.41660951787e-06 | response to X-ray | GO:0010165 |
| 7.41660951787e-06 | mitotic DNA damage checkpoint | GO:0044773 |
| 7.41660951787e-06 | negative regulation of cytokine production | GO:0001818 |
| 7.9110837734e-06 | regulation of cyclin-dependent protein kinase activity | GO:1904029 |
| 8.30917117971e-06 | positive regulation of hemopoiesis | GO:1903708 |
| 8.30917117971e-06 | neutrophil chemotaxis | GO:0030593 |
| 1.02684621681e-05 | negative regulation of leukocyte mediated cytotoxicity | GO:0001911 |
| 1.02684621681e-05 | negative regulation of cell-cell adhesion | GO:0022408 |
| 1.02684621681e-05 | regulation of interferon-gamma production | GO:0032649 |
| 1.02684621681e-05 | regulation of T cell activation via T cell receptor contact with antigen bound to MHC molecule on antigen pre | GO:2001188 |
| 2.07704937641e-05 | regulation of T cell mediated cytotoxicity | GO:0001914 |
| 2.23182701579e-05 | positive regulation of leukocyte migration | GO:0002687 |
| 2.86288424052e-05 | DNA replication initiation | GO:0006270 |
| 3.02870832446e-05 | regulation of viral process | GO:0050792 |
| 3.40772256314e-05 | regulation of interleukin-2 production | GO:0032663 |
| 3.81302454785e-05 | regulation of interleukin-12 production | GO:0032655 |
| 4.15522761912e-05 | neutrophil migration | GO:1990266 |
| 4.36954428149e-05 | regulation of innate immune response | GO:0045088 |
| 4.48914056633e-05 | regulation of interleukin-1 production | GO:0032652 |
| 4.86808436746e-05 | regulation of natural killer cell mediated cytotoxicity | GO:0042269 |
| 5.74556910541e-05 | G1 DNA damage checkpoint | GO:0044783 |
| 6.01977288821e-05 | T cell mediated immunity | GO:0002456 |
| 6.0626280674e-05 | regulation of cytokine production involved in immune response | GO:0002718 |
| 6.3277734208e-05 | negative regulation of viral process | GO:0048525 |
| 6.85503030632e-05 | positive regulation of defense response | GO:0031349 |
| 7.42269673969e-05 | negative regulation of cell cycle | GO:0045786 |
| 9.37241529239e-05 | establishment of mitotic spindle localization | GO:0040001 |
| 0.000131669873197 | response to interleukin-4 | GO:0070670 |
| 0.000161208391901 | regulation of leukocyte chemotaxis | GO:0002688 |
| 0.000238918417824 | centromere complex assembly | GO:0034508 |
| 0.000291794690305 | regulation of protein metabolic process | GO:0051246 |
| 0.000317006723148 | T-helper 1 type immune response | GO:0042088 |
| 0.000341046502118 | DNA damage response, signal transduction by p53 class mediator | GO:0030330 |
| 0.000341046502118 | regulation of interleukin-6 production | GO:0032675 |
| 0.000412881822035 | regulation of cellular protein metabolic process | GO:0032268 |
| 0.000448992028701 | DNA conformation change | GO:0071103 |
| 0.000547355123984 | regulation of CD40 signaling pathway | GO:2000348 |
| 0.000785239737424 | pattern recognition receptor signaling pathway | GO:0002221 |
| 0.000793133690897 | liver regeneration | GO:0097421 |
| 0.000793133690897 | negative regulation of chromosome segregation | GO:0051985 |
| 0.000841927853705 | antigen processing and presentation of endogenous peptide antigen via MHC class I | GO:0019885 |
| 0.000885299651844 | interleukin-4-mediated signaling pathway | GO:0035771 |
| 0.000885842742527 | positive regulation of cell differentiation | GO:0045597 |
| 0.000885842742527 | signal transduction involved in DNA integrity checkpoint | GO:0072401 |
| 0.000885842742527 | signal transduction involved in DNA damage checkpoint | GO:0072422 |
| 0.000968010428963 | negative regulation of defense response | GO:0031348 |
| 0.00113926941786 | establishment of spindle orientation | GO:0051294 |
| 0.00118966671955 | regulation of phosphorus metabolic process | GO:0051174 |
| 0.0012143398231 | regulation of microtubule cytoskeleton organization | GO:0070507 |
| 0.00123915847521 | leukocyte apoptotic process | GO:0071887 |
| 0.00123915847521 | interleukin-21-mediated signaling pathway | GO:0038114 |
| 0.00123915847521 | response to interleukin-21 | GO:0098756 |
| 0.00149360629329 | interleukin-10-mediated signaling pathway | GO:0140105 |
| 0.00154406087105 | antigen processing and presentation of exogenous peptide antigen via MHC class I | GO:0042590 |
| 0.00156498602333 | mitotic spindle assembly checkpoint | GO:0007094 |
| 0.00173739460047 | positive regulation of antigen processing and presentation | GO:0002579 |
| 0.00173739460047 | interleukin-9-mediated signaling pathway | GO:0038113 |
| 0.00189353881594 | DNA recombination | GO:0006310 |
| 0.00193167673722 | response to interleukin-1 | GO:0070555 |
| 0.00195592369857 | regulation of chemokine production | GO:0032642 |
| 0.00215253405742 | viral life cycle | GO:0019058 |
| 0.00227939879668 | regulation of nuclease activity | GO:0032069 |
| 0.00227984767684 | phosphorylation | GO:0016310 |
| 0.00227984767684 | response to interleukin-9 | GO:0071104 |
| 0.00256802234378 | regulation of neutrophil migration | GO:1902622 |
| 0.00256802234378 | positive regulation of response to external stimulus | GO:0032103 |
| 0.00273792376935 | regulation of production of molecular mediator of immune response | GO:0002700 |
| 0.00276834935434 | neuroinflammatory response | GO:0150076 |
| 0.00308634606159 | regulation of mast cell activation | GO:0033003 |
| 0.00329931625549 | protein-DNA complex assembly | GO:0065004 |
| 0.00351714676788 | negative regulation of hemopoiesis | GO:1903707 |
| 0.00359059408529 | regulation of prostaglandin-E synthase activity | GO:2000361 |
| 0.00359059408529 | negative regulation of T cell costimulation | GO:2000524 |
| 0.00359059408529 | regulation of organelle organization | GO:0033043 |
| 0.00359059408529 | regulation of leukocyte degranulation | GO:0043300 |
| 0.00364230733605 | regulation of T cell tolerance induction | GO:0002664 |
| 0.00420345205117 | viral genome replication | GO:0019079 |
| 0.0042192803841 | chromatin remodeling at centromere | GO:0031055 |
| 0.0042192803841 | dendritic cell differentiation | GO:0097028 |
| 0.00470858274037 | negative regulation of antigen receptor-mediated signaling pathway | GO:0050858 |
| 0.00470858274037 | DNA strand elongation | GO:0022616 |
| 0.00470858274037 | positive regulation of blood coagulation | GO:0030194 |
| 0.00532128851032 | positive regulation of coagulation | GO:0050820 |
| 0.00537615508748 | positive regulation of cellular component movement | GO:0051272 |
| 0.00591322074673 | regulation of interleukin-4 production | GO:0032673 |
| 0.00591322074673 | response to tumor necrosis factor | GO:0034612 |
| 0.00620127336384 | response to gamma radiation | GO:0010332 |
| 0.00622898050924 | cellular response to camptothecin | GO:0072757 |
| 0.00622898050924 | microglial cell activation involved in immune response | GO:0002282 |
| 0.00643186270996 | regulation of cell cycle checkpoint | GO:1901976 |
| 0.0070407655747 | regulation of macrophage activation | GO:0043030 |
| 0.0070407655747 | DNA replication-independent nucleosome organization | GO:0034724 |
| 0.0088160704485 | G2 DNA damage checkpoint | GO:0031572 |
| 0.00940613216591 | regulation of cellular protein localization | GO:1903827 |
| 0.009410026976 | histone exchange | GO:0043486 |
| 0.0095791886366 | antigen receptor-mediated signaling pathway | GO:0050851 |
| 0.0095791886366 | response to camptothecin | GO:1901563 |
| 0.0095791886366 | strand invasion | GO:0042148 |
| 0.00970496475991 | regulation of myeloid cell differentiation | GO:0045637 |
| 0.00970496475991 | positive regulation of cell division | GO:0051781 |
| 0.00970770779021 | negative regulation of cell death | GO:0060548 |
| 0.00970770779021 | neutrophil mediated immunity | GO:0002446 |
| 0.00977384846101 | positive regulation of transferase activity | GO:0051347 |
| 0.0107165765433 | regulation of programmed cell death | GO:0043067 |
| 0.010954330774 | regulation of T-helper cell differentiation | GO:0045622 |
| 0.0129149543883 | regulation of inflammatory response | GO:0050727 |
| 0.0133193193559 | antigen processing and presentation of endogenous peptide antigen via MHC class Ib | GO:0002476 |
| 0.0133558012239 | eosinophil chemotaxis | GO:0048245 |
| 0.0166128055382 | negative regulation of cell proliferation | GO:0008285 |
| 0.0177471796708 | myeloid leukocyte differentiation | GO:0002573 |
| 0.0179494072512 | regulation of lymphocyte anergy | GO:0002911 |
| 0.0217187433762 | telomere organization | GO:0032200 |
| 0.0217187433762 | negative regulation of response to external stimulus | GO:0032102 |
| 0.0217187433762 | eosinophil migration | GO:0072677 |
| 0.0217187433762 | intrinsic apoptotic signaling pathway by p53 class mediator | GO:0072332 |
| 0.0227514189635 | microglial cell migration | GO:1904124 |
| 0.0227514189635 | regulation of interleukin-18 production | GO:0032661 |
| 0.0237769481532 | integrin activation | GO:0033622 |
| 0.024682548232 | regulation of interleukin-8 production | GO:0032677 |
| 0.024682548232 | positive regulation of response to wounding | GO:1903036 |
| 0.024682548232 | regulation of mononuclear cell migration | GO:0071675 |
| 0.0257307053501 | regulation of fibroblast proliferation | GO:0048145 |
| 0.0257307053501 | regulation of syncytium formation by plasma membrane fusion | GO:0060142 |
| 0.0258427818907 | DNA-dependent DNA replication maintenance of fidelity | GO:0045005 |
| 0.0258427818907 | mast cell mediated immunity | GO:0002448 |
| 0.0271642509132 | regulation of tumor necrosis factor superfamily cytokine production | GO:1903555 |
| 0.0271642509132 | positive regulation of isomerase activity | GO:0010912 |
| 0.0277391692967 | T cell chemotaxis | GO:0010818 |
| 0.0287990667807 | nucleosome assembly | GO:0006334 |
| 0.030694436228 | regulation of epithelial cell differentiation | GO:0030856 |

| q-value | Pathway Name | Pathway ID |
|---|---|---|
| 4.82769898433e-14 | Cell Cycle, Mitotic | R-HSA-69278 |
| 4.82769898433e-14 | Cell Cycle | R-HSA-1640170 |
| 1.54624933424e-12 | Cell Cycle Checkpoints | R-HSA-69620 |
| 3.83338687284e-12 | Cell cycle - Homo sapiens (human) | path:hsa04110 |
| 3.83338687284e-12 | Immune System | R-HSA-168256 |
| 3.2556442652e-11 | Interferon gamma signaling | R-HSA-877300 |
| 6.24129255209e-11 | Interferon Signaling | R-HSA-913531 |
| 7.02326041974e-10 | Cytokine Signaling in Immune system | R-HSA-1280215 |
| 1.22784648842e-08 | G1/S Transition | R-HSA-69206 |
| 1.28043672888e-08 | Immunoregulatory interactions between a Lymphoid and a non-Lymphoid cell | R-HSA-198933 |
| 1.60166662048e-08 | Graft-versus-host disease - Homo sapiens (human) | path:hsa05332 |
| 1.60166662048e-08 | Resolution of Sister Chromatid Cohesion | R-HSA-2500257 |
| 1.74123876438e-08 | Epstein-Barr virus infection - Homo sapiens (human) | path:hsa05169 |
| 5.0445713799e-08 | Mitotic Anaphase | R-HSA-68882 |
| 5.0445713799e-08 | Allograft rejection - Homo sapiens (human) | path:hsa05330 |
| 5.0445713799e-08 | S Phase | R-HSA-69242 |
| 5.0445713799e-08 | Mitotic G1 phase and G1/S transition | R-HSA-453279 |
| 5.0445713799e-08 | Mitotic Metaphase and Anaphase | R-HSA-2555396 |
| 9.48405956932e-08 | Antigen processing and presentation - Homo sapiens (human) | path:hsa04612 |
| 1.06582634213e-07 | Autoimmune thyroid disease - Homo sapiens (human) | path:hsa05320 |
| 1.28952621082e-07 | Separation of Sister Chromatids | R-HSA-2467813 |
| 1.87328865061e-07 | Type I diabetes mellitus - Homo sapiens (human) | path:hsa04940 |
| 3.80228435983e-07 | Viral myocarditis - Homo sapiens (human) | path:hsa05416 |
| 4.98896042004e-07 | Adaptive Immune System | R-HSA-1280218 |
| 8.91906579746e-07 | Phagosome - Homo sapiens (human) | path:hsa04145 |
| 8.91906579746e-07 | Amplification of signal from unattached kinetochores via a MAD2 inhibitory signal | R-HSA-141444 |
| 8.91906579746e-07 | Amplification of signal from the kinetochores | R-HSA-141424 |
| 9.22719777333e-07 | Mitotic Spindle Checkpoint | R-HSA-69618 |
| 1.36410820236e-06 | EML4 and NUDC in mitotic spindle formation | R-HSA-9648025 |
| 2.51170090949e-06 | Interferon alpha/beta signaling | R-HSA-909733 |
| 4.04157913128e-06 | Mitotic Prometaphase | R-HSA-68877 |
| 5.26603936483e-06 | Human T-cell leukemia virus 1 infection - Homo sapiens (human) | path:hsa05166 |
| 8.93383170133e-06 | G1/S-Specific Transcription | R-HSA-69205 |
| 1.34487864297e-05 | RHO GTPases Activate Formins | R-HSA-5663220 |
| 2.27231997158e-05 | p53 signaling pathway - Homo sapiens (human) | path:hsa04115 |
| 2.32966842434e-05 | Kinesins | R-HSA-983189 |
| 2.32966842434e-05 | Natural killer cell mediated cytotoxicity - Homo sapiens (human) | path:hsa04650 |
| 4.45030319925e-05 | G2/M Transition | R-HSA-69275 |
| 4.98421558554e-05 | Mitotic G2-G2/M phases | R-HSA-453274 |
| 4.98421558554e-05 | G2/M Checkpoints | R-HSA-69481 |
| 5.10868375822e-05 | DNA Replication | R-HSA-69306 |
| 5.6074442096e-05 | Human immunodeficiency virus 1 infection - Homo sapiens (human) | path:hsa05170 |
| 6.64140775675e-05 | Cyclin A/B1/B2 associated events during G2/M transition | R-HSA-69273 |
| 7.31332690653e-05 | TP53 Regulates Transcription of Cell Cycle Genes | R-HSA-6791312 |
| 7.56274892298e-05 | Activation of ATR in response to replication stress | R-HSA-176187 |
| 7.56274892298e-05 | M Phase | R-HSA-68886 |
| 7.7001719348e-05 | Cell adhesion molecules - Homo sapiens (human) | path:hsa04514 |
| 7.7001719348e-05 | G0 and Early G1 | R-HSA-1538133 |
| 0.000110604331489 | Polo-like kinase mediated events | R-HSA-156711 |
| 0.000117524498993 | Interleukin-2 family signaling | R-HSA-451927 |
| 0.000125520238215 | Cellular senescence - Homo sapiens (human) | path:hsa04218 |
| 0.000125520238215 | Rheumatoid arthritis - Homo sapiens (human) | path:hsa05323 |
| 0.000125520238215 | Th1 and Th2 cell differentiation - Homo sapiens (human) | path:hsa04658 |
| 0.000149244459233 | Staphylococcus aureus infection - Homo sapiens (human) | path:hsa05150 |
| 0.000164311042027 | Synthesis of DNA | R-HSA-69239 |
| 0.000165720375758 | Cyclin A:Cdk2-associated events at S phase entry | R-HSA-69656 |
| 0.00017352522492 | Measles - Homo sapiens (human) | path:hsa05162 |
| 0.000177472365925 | TP53 Regulates Transcription of Genes Involved in G2 Cell Cycle Arrest | R-HSA-6804114 |
| 0.000293815619567 | Intestinal immune network for IgA production - Homo sapiens (human) | path:hsa04672 |
| 0.000293815619567 | Switching of origins to a post-replicative state | R-HSA-69052 |
| 0.000329800285557 | Inflammatory bowel disease - Homo sapiens (human) | path:hsa05321 |
| 0.000365417519903 | The role of GTSE1 in G2/M progression after G2 checkpoint | R-HSA-8852276 |
| 0.000391343030156 | RHO GTPase Effectors | R-HSA-195258 |
| 0.000391343030156 | Th17 cell differentiation - Homo sapiens (human) | path:hsa04659 |
| 0.000429717053127 | Tuberculosis - Homo sapiens (human) | path:hsa05152 |
| 0.000526843005962 | Hemostasis | R-HSA-109582 |
| 0.000571057681894 | G2/M DNA replication checkpoint | R-HSA-69478 |
| 0.000585453980511 | Antiviral mechanism by IFN-stimulated genes | R-HSA-1169410 |
| 0.000639965994975 | Fanconi anemia pathway - Homo sapiens (human) | path:hsa03460 |
| 0.000679796682511 | Endosomal/Vacuolar pathway | R-HSA-1236977 |
| 0.000857077516555 | Leishmaniasis - Homo sapiens (human) | path:hsa05140 |
| 0.000857077516555 | Pancreatic cancer - Homo sapiens (human) | path:hsa05212 |
| 0.000857077516555 | Antigen processing-Cross presentation | R-HSA-1236975 |
| 0.000963634077174 | Factors involved in megakaryocyte development and platelet production | R-HSA-983231 |
| 0.00111885720301 | Resolution of D-loop Structures through Synthesis-Dependent Strand Annealing (SDSA) | R-HSA-5693554 |
| 0.00114625114112 | MHC class II antigen presentation | R-HSA-2132295 |
| 0.00121079618242 | Chagas disease - Homo sapiens (human) | path:hsa05142 |
| 0.00128884428117 | Translocation of ZAP-70 to Immunological synapse | R-HSA-202430 |
| 0.00133022187 | COPI-dependent Golgi-to-ER retrograde traffic | R-HSA-6811434 |
| 0.00133022187 | APC/C-mediated degradation of cell cycle proteins | R-HSA-174143 |
| 0.00133022187 | Regulation of mitotic cell cycle | R-HSA-453276 |
| 0.00144781567247 | Cyclin E associated events during G1/S transition | R-HSA-69202 |
| 0.00144781567247 | Oocyte meiosis - Homo sapiens (human) | path:hsa04114 |
| 0.00197186746342 | Phosphorylation of CD3 and TCR zeta chains | R-HSA-202427 |
| 0.00221721050579 | SCF(Skp2)-mediated degradation of p27/p21 | R-HSA-187577 |
| 0.0022205751991 | PD-1 signaling | R-HSA-389948 |
| 0.0022205751991 | APC/C:Cdc20 mediated degradation of mitotic proteins | R-HSA-176409 |
| 0.00248967599055 | Activation of APC/C and APC/C:Cdc20 mediated degradation of mitotic proteins | R-HSA-176814 |
| 0.00248967599055 | Resolution of D-loop Structures through Holliday Junction Intermediates | R-HSA-5693568 |
| 0.00248967599055 | Activation of the pre-replicative complex | R-HSA-68962 |
| 0.00317921540336 | Assembly of the pre-replicative complex | R-HSA-68867 |
| 0.00322466940545 | Resolution of D-Loop Structures | R-HSA-5693537 |
| 0.00322466940545 | E2F-enabled inhibition of pre-replication complex formation | R-HSA-113507 |
| 0.00322466940545 | Interleukin-9 signaling | R-HSA-8985947 |
| 0.00370421454369 | G1/S DNA Damage Checkpoints | R-HSA-69615 |
| 0.00397964819145 | Primary immunodeficiency - Homo sapiens (human) | path:hsa05340 |
| 0.00397964819145 | DNA Replication Pre-Initiation | R-HSA-69002 |
| 0.00415108268472 | Costimulation by the CD28 family | R-HSA-388841 |
| 0.00415108268472 | Progesterone-mediated oocyte maturation - Homo sapiens (human) | path:hsa04914 |
| 0.00471729410027 | Osteoclast differentiation - Homo sapiens (human) | path:hsa04380 |
| 0.00590600860909 | Antigen Presentation: Folding, assembly and peptide loading of class I MHC | R-HSA-983170 |
| 0.00668799366701 | Systemic lupus erythematosus - Homo sapiens (human) | path:hsa05322 |
| 0.00668799366701 | Generation of second messenger molecules | R-HSA-202433 |
| 0.00668799366701 | Homologous DNA Pairing and Strand Exchange | R-HSA-5693579 |
| 0.00723710686141 | Unwinding of DNA | R-HSA-176974 |
| 0.00723948136558 | Deposition of new CENPA-containing nucleosomes at the centromere | R-HSA-606279 |
| 0.00723948136558 | Nucleosome assembly | R-HSA-774815 |
| 0.00753491368197 | Orc1 removal from chromatin | R-HSA-68949 |
| 0.00802353798341 | Human cytomegalovirus infection - Homo sapiens (human) | path:hsa05163 |
| 0.00855183839356 | Interleukin receptor SHC signaling | R-HSA-912526 |
| 0.00877107528153 | Chk1/Chk2(Cds1) mediated inactivation of Cyclin B:Cdk1 complex | R-HSA-75035 |
| 0.00879086588539 | Pathways in cancer - Homo sapiens (human) | path:hsa05200 |
| 0.00949296296244 | Golgi-to-ER retrograde transport | R-HSA-8856688 |
| 0.009813639905 | Influenza A - Homo sapiens (human) | path:hsa05164 |
| 0.009813639905 | HDR through Homologous Recombination (HRR) | R-HSA-5685942 |
| 0.0100106521468 | Small cell lung cancer - Homo sapiens (human) | path:hsa05222 |
| 0.0100912763672 | Phosphorylation of proteins involved in G1/S transition by active Cyclin E:Cdk2 complexes | R-HSA-69200 |
| 0.0101242700139 | TP53 regulates transcription of several additional cell death genes whose specific roles in p53-dependent apo | R-HSA-6803205 |
| 0.0101242700139 | Advanced glycosylation endproduct receptor signaling | R-HSA-879415 |
| 0.0101242700139 | TP53 Regulates Transcription of Genes Involved in G1 Cell Cycle Arrest | R-HSA-6804116 |
| 0.0101242700139 | CDK-mediated phosphorylation and removal of Cdc6 | R-HSA-69017 |
| 0.0101242700139 | Chaperonin-mediated protein folding | R-HSA-390466 |
| 0.0113123387219 | Asthma - Homo sapiens (human) | path:hsa05310 |
| 0.0113123387219 | ISG15 antiviral mechanism | R-HSA-1169408 |
| 0.0120361063213 | Interleukin-4 and Interleukin-13 signaling | R-HSA-6785807 |
| 0.0127897600375 | Transcriptional regulation by RUNX2 | R-HSA-8878166 |
| 0.0127897600375 | NOD-like receptor signaling pathway - Homo sapiens (human) | path:hsa04621 |
| 0.0128737273089 | Protein folding | R-HSA-391251 |
| 0.0128737273089 | Hematopoietic cell lineage - Homo sapiens (human) | path:hsa04640 |
| 0.0133188487085 | Innate Immune System | R-HSA-168249 |
| 0.0136257736375 | DNA strand elongation | R-HSA-69190 |
| 0.013722009948 | Downstream TCR signaling | R-HSA-202424 |
| 0.013722009948 | Transcription of E2F targets under negative control by DREAM complex | R-HSA-1362277 |
| 0.0184454127234 | Hepatitis B - Homo sapiens (human) | path:hsa05161 |
| 0.0184454127234 | Chromosome Maintenance | R-HSA-73886 |
| 0.0187199433087 | Kaposi sarcoma-associated herpesvirus infection - Homo sapiens (human) | path:hsa05167 |
| 0.0187199433087 | Deregulated CDK5 triggers multiple neurodegenerative pathways in Alzheimer,s disease models | R-HSA-8862803 |
| 0.0187199433087 | Neurodegenerative Diseases | R-HSA-8863678 |
| 0.0197799605771 | Transcriptional Regulation by TP53 | R-HSA-3700989 |
| 0.0203952709589 | PTK6 Regulates Cell Cycle | R-HSA-8849470 |
| 0.0203952709589 | Phosphorylation of Emi1 | R-HSA-176417 |
| 0.0209428226105 | p53-Dependent G1 DNA Damage Response | R-HSA-69563 |
| 0.0209428226105 | p53-Dependent G1/S DNA damage checkpoint | R-HSA-69580 |
| 0.0209428226105 | Other semaphorin interactions | R-HSA-416700 |
| 0.0209428226105 | Regulation of APC/C activators between G1/S and early anaphase | R-HSA-176408 |
| 0.0212785651183 | Human papillomavirus infection - Homo sapiens (human) | path:hsa05165 |
| 0.0219119731749 | Signaling by Rho GTPases, Miro GTPases and RHOBTB3 | R-HSA-9716542 |
| 0.0244680845015 | Viral carcinogenesis - Homo sapiens (human) | path:hsa05203 |
| 0.0246178682605 | Presynaptic phase of homologous DNA pairing and strand exchange | R-HSA-5693616 |
| 0.0264933846303 | Activation of NIMA Kinases NEK9, NEK6, NEK7 | R-HSA-2980767 |
| 0.0265256930537 | Fanconi Anemia Pathway | R-HSA-6783310 |
| 0.0265256930537 | Phosphorylation of the APC/C | R-HSA-176412 |
| 0.028630945136 | Homologous recombination - Homo sapiens (human) | path:hsa03440 |
| 0.0296880786801 | E2F mediated regulation of DNA replication | R-HSA-113510 |
| 0.0296880786801 | Diseases of programmed cell death | R-HSA-9645723 |
| 0.030496830075 | Signaling by Rho GTPases | R-HSA-194315 |
| 0.0326895351913 | CDC6 association with the ORC:origin complex | R-HSA-68689 |
| 0.0326895351913 | Interleukin-15 signaling | R-HSA-8983432 |
| 0.0326895351913 | Linoleic acid (LA) metabolism | R-HSA-2046105 |
| 0.0326895351913 | OAS antiviral response | R-HSA-8983711 |

#### clinTCMR downregulated

| q-value | Pathway name | Pathway ID | | q-value | pathway | external_id |
|---|---|---|---|---|---|---|
| 0.026722741 | positive regula | GO:0003099 | | 0.0828013387611 | Adrenoceptors | R-HSA-390696 |
| 0.027559630 | regulation of r | GO:0002090 | | 0.0828013387611 | Amine ligand-binding receptors | R-HSA-375280 |
| 0.027559630 | adenylate cycl | GO:0071880 | | 0.0828013387611 | Cocaine addiction - Homo sapiens (human) | path:hsa05030 |
| 0.027559630 | positive regula | GO:0001996 | | 0.126726264254 | Phase 0 - rapid depolarisation | R-HSA-5576892 |
| 0.027559630 | receptor inter | GO:0031623 | | 0.126726264254 | GPCR downstream signalling | R-HSA-388396 |
| 0.038329716 | striated musc | GO:0014706 | | | | |
| 0.112354967 | negative regul | GO:0003085 | | | | |
| 0.112354967 | heart field spe | GO:0003128 | | | | |
| 0.112354967 | embryonic lin | GO:0030326 | | | | |
| 0.112354967 | regulation of s | GO:2001014 | | | | |
| 0.112354967 | positive regula | GO:0045597 | | | | |
| 0.112354967 | regulation of t | GO:0035296 | | | | |
| 0.112354967 | regulation of l | GO:0050880 | | | | |
| 0.112354967 | regulation of s | GO:0016202 | | | | |
| 0.112354967 | terpenoid me | GO:0006721 | | | | |
| 0.112354967 | regulation of r | GO:0048634 | | | | |
| 0.112354967 | bone maturat | GO:0070977 | | | | |
| 0.119436518 | cellular polysa | GO:0033692 | | | | |
| 0.119436518 | physiological | GO:0003298 | | | | |
| 0.119436518 | myelination ir | GO:0022011 | | | | |
| 0.119436518 | negative regul | GO:0030279 | | | | |

#### Suppl. Table 6: Shared genes of cABMR and clinTCMR

Adjustment of p-value by false discovery rate.

##### Upregulated

| gene name | log2fold change cABMR | padj cABMR | log2fold change clinTCMR | padj clinTCMR |
|---|---|---|---|---|
| UBD | 3,478071266 | 6,29438E-14 | 3,781778143 | 3,60589E-16 |
| MMP7 | 4,403777448 | 2,80093E-12 | 1,78053712 | 0,014538139 |
| LOXL4 | 4,070652531 | 1,39048E-09 | 3,113830085 | 1,10021E-05 |
| SPATA21 | 3,332557182 | 1,45183E-09 | 2,130287476 | 0,000345697 |
| STMN2 | 6,086352207 | 1,60189E-09 | 3,289831984 | 0,003471866 |
| APOBEC3B | 3,017573296 | 1,03952E-08 | 3,457196343 | 2,61673E-11 |
| MUC13 | 4,364393334 | 1,34524E-08 | 4,156401677 | 7,32968E-08 |
| GPRC5B | 2,024271343 | 1,73955E-08 | 1,075596551 | 0,007530472 |
| CXCL10 | 2,69763015 | 1,81502E-08 | 3,984378996 | 1,7596E-17 |
| LCN2 | 2,800194736 | 6,15533E-08 | 2,095652015 | 0,000159748 |
| NFKB2 | 1,202682533 | 9,09875E-08 | 1,115708991 | 1,75446E-06 |
| RELB | 1,641532968 | 9,31485E-08 | 1,611925756 | 3,39958E-07 |
| RGCC | 2,125389619 | 1,17097E-07 | 1,494946986 | 0,000654531 |
| CCR7 | 2,651033603 | 1,34957E-07 | 2,314413461 | 8,87292E-06 |
| IL8 | 4,782511474 | 1,5033E-07 | 2,979095375 | 0,003257934 |
| HLA-G | 2,209051365 | 2,12474E-07 | 1,310898524 | 0,005725693 |
| SELM | 1,374586356 | 2,20082E-07 | 1,005000117 | 0,000470049 |
| MCM5 | 1,29174774 | 2,40494E-07 | 1,635851129 | 4,66755E-11 |
| BIRC3 | 1,576832626 | 2,4664E-07 | 1,724613323 | 2,91402E-08 |
| TTYH3 | 1,157001962 | 2,89735E-07 | 1,539182966 | 5,48416E-12 |
| NFE2L3 | 1,586860013 | 3,00355E-07 | 1,118999648 | 0,000902496 |
| MVP | 1,235585475 | 3,18148E-07 | 1,074699366 | 2,79158E-05 |
| CCL2 | 2,134975442 | 3,33019E-07 | 1,160576026 | 0,014600688 |
| TNFRSF9 | 3,061093096 | 3,61164E-07 | 2,938293077 | 1,65667E-06 |
| FSTL3 | 2,081769698 | 4,85788E-07 | 1,587311204 | 0,000398036 |

| | | | | |
|---|---|---|---|---|
| CORO1A | 1,258118187 | 5,00489E-07 | 1,169471078 | 8,9E-06 |
| KIAA1644 | 2,854327165 | 5,28243E-07 | 1,697445229 | 0,008172096 |
| DTL | 2,404634977 | 5,85467E-07 | 3,948238297 | 6,14311E-18 |
| PFKP | 1,848734859 | 6,79363E-07 | 1,016950306 | 0,016468199 |
| ITGA6 | 1,133626738 | 7,26827E-07 | 1,1123676 | 3,28673E-06 |
| MICAL1 | 1,477703316 | 1,01328E-06 | 1,195095613 | 0,000233741 |
| CDC20 | 2,558102848 | 1,08297E-06 | 3,744134277 | 1,18307E-14 |
| PCDP1 | 2,658179516 | 1,44903E-06 | 1,907570771 | 0,001592903 |
| LSP1 | 1,502772216 | 1,62127E-06 | 1,260936313 | 0,000177922 |
| CXCL11 | 2,241861021 | 2,04847E-06 | 2,846042806 | 1,72991E-09 |
| SPON2 | 1,540881942 | 2,16057E-06 | 1,641929877 | 1,08516E-06 |
| JUN | 1,37058197 | 2,2775E-06 | 1,018208646 | 0,001321517 |
| TAP1 | 1,048565738 | 2,2775E-06 | 1,580904233 | 5,24129E-13 |
| IGJ | 2,118633124 | 2,29946E-06 | 1,769280373 | 0,000250386 |
| SHISA3 | 2,648406022 | 2,31026E-06 | 1,422455832 | 0,029948973 |
| PRDM1 | 1,217031116 | 2,45498E-06 | 1,318782607 | 6,53133E-07 |
| LIF | 2,831463267 | 2,50046E-06 | 1,716267728 | 0,011553621 |
| CCL20 | 4,532267875 | 2,56854E-06 | 3,006962302 | 0,004692085 |
| CXCR3 | 2,193183529 | 3,02335E-06 | 1,893811968 | 0,000163954 |
| LOC102723407 | 2,58936578 | 3,56554E-06 | 2,070084222 | 0,000628092 |
| ANXA2 | 1,32794301 | 3,82169E-06 | 1,558219802 | 1,20976E-07 |
| NCAPH | 1,665623356 | 4,11962E-06 | 2,451206099 | 2,75527E-12 |
| DRAM1 | 1,043625783 | 4,26858E-06 | 1,003902398 | 2,85085E-05 |
| ECT2 | 1,17803861 | 5,15685E-06 | 1,965534688 | 2,17753E-15 |
| SLC51B | 4,472658997 | 5,45318E-06 | 5,034533437 | 6,81904E-07 |
| TYMP | 1,229263804 | 5,68123E-06 | 1,491897706 | 6,64418E-08 |
| STK17B | 1,060631236 | 6,20094E-06 | 1,082757678 | 1,06579E-05 |
| CXCR4 | 1,500920693 | 6,43294E-06 | 1,647860165 | 1,60926E-06 |
| GPR171 | 1,856414402 | 6,73125E-06 | 1,500858257 | 0,000680358 |
| ITGAX | 1,407229245 | 6,94467E-06 | 1,463070417 | 7,50639E-06 |
| IL32 | 1,863153157 | 7,70835E-06 | 2,471562351 | 3,9888E-09 |
| LEF1 | 2,000975308 | 8,11698E-06 | 1,821622098 | 0,000124069 |
| TYMS | 1,636917277 | 8,20754E-06 | 2,406058933 | 4,04841E-11 |
| KLHL29 | 2,024659887 | 8,27202E-06 | 1,372070582 | 0,006674259 |
| DBNDD1 | 2,592264873 | 9,25483E-06 | 2,203818522 | 0,00049564 |
| LOC102724895 | 4,256684595 | 9,27708E-06 | 4,060847198 | 5,42945E-05 |
| FAT1 | 1,63710553 | 9,88357E-06 | 1,552787708 | 8,84176E-05 |
| CCL21 | 1,887027511 | 1,07396E-05 | 1,479712014 | 0,001616615 |
| GDF15 | 2,719603704 | 1,12796E-05 | 2,245833676 | 0,000809857 |
| DPEP1 | 4,239925311 | 1,14569E-05 | 2,479369717 | 0,028079597 |
| SPP1 | 1,991011097 | 1,15612E-05 | 1,332950918 | 0,008791141 |
| CD2 | 1,350376628 | 1,20282E-05 | 1,423489957 | 1,10259E-05 |
| CD96 | 1,192826325 | 1,21904E-05 | 1,005020563 | 0,000642977 |
| CDC45 | 2,28991136 | 1,26949E-05 | 4,071329182 | 2,57077E-16 |
| FCAMR | 2,079291061 | 1,453E-05 | 2,20466832 | 1,22002E-05 |
| PDGFA | 1,894568301 | 1,52616E-05 | 1,525919146 | 0,001444197 |
| RHOF | 1,415764295 | 1,88416E-05 | 1,117439259 | 0,001948224 |

| | | | | |
|---|---|---|---|---|
| TNFRSF12A | 2,504023698 | 2,0274E-05 | 2,566885397 | 3,64356E-05 |
| CD3E | 1,334105103 | 2,20606E-05 | 1,164919715 | 0,000610802 |
| SEMA4A | 1,469450261 | 2,48258E-05 | 1,369525077 | 0,000226684 |
| ATP10A | 1,74020792 | 2,53016E-05 | 1,539217205 | 0,000516906 |
| CCL19 | 2,152887251 | 2,69109E-05 | 2,490912815 | 3,22181E-06 |
| HKDC1 | 3,220455981 | 2,85415E-05 | 2,873353748 | 0,000529451 |
| DCDC2 | 2,084961018 | 2,93074E-05 | 1,1980685 | 0,037666871 |
| CIITA | 1,167813493 | 3,00319E-05 | 1,195187839 | 5,62251E-05 |
| MCM6 | 1,179187812 | 3,00319E-05 | 1,919433512 | 5,58778E-12 |
| MTHFD2 | 1,550878988 | 3,11586E-05 | 1,7389942 | 7,60717E-06 |
| TNFRSF17 | 2,988464598 | 3,22074E-05 | 2,240838827 | 0,004448589 |
| GZMK | 1,432370331 | 3,28615E-05 | 1,270186912 | 0,000626297 |
| LIMD2 | 1,081493757 | 3,29241E-05 | 1,036511119 | 0,00019824 |
| FAM111B | 2,05345095 | 3,79274E-05 | 4,247248403 | 8,45992E-19 |
| PTPRCAP | 1,231351079 | 3,91424E-05 | 1,394304361 | 7,29189E-06 |
| PHLDA3 | 1,836436636 | 4,08205E-05 | 1,162333688 | 0,0227225 |
| CTSE | 2,918133004 | 4,11827E-05 | 1,841023921 | 0,024953621 |
| MYO1G | 1,394342038 | 4,40333E-05 | 1,228094115 | 0,000862249 |
| MCM2 | 1,312105451 | 4,43989E-05 | 2,051380644 | 8,83447E-11 |
| CD28 | 1,197459506 | 4,44245E-05 | 1,402599954 | 3,24736E-06 |
| GPRIN3 | 1,41674545 | 4,74667E-05 | 1,070789838 | 0,005432592 |
| BUB1B | 1,658126936 | 5,06235E-05 | 3,104666573 | 3,05676E-16 |
| SOX9-AS1 | 1,869457699 | 5,33796E-05 | 1,75019366 | 0,000394116 |
| EYS | 2,32984681 | 5,43817E-05 | 2,244564361 | 0,000226047 |
| CD37 | 1,179551561 | 5,52617E-05 | 1,312389202 | 1,83558E-05 |
| MMP19 | 1,305776772 | 5,53004E-05 | 1,389068636 | 4,6232E-05 |
| SPECC1 | 1,419385072 | 5,60198E-05 | 1,039627128 | 0,008090188 |
| F2RL1 | 1,28622218 | 5,65717E-05 | 1,251415647 | 0,000258144 |
| NPTX2 | 2,636342303 | 5,73593E-05 | 2,564322856 | 0,000213098 |
| KCNJ5 | 2,200116207 | 5,85234E-05 | 2,614479943 | 3,04949E-06 |
| SLC7A6 | 1,203407056 | 6,03832E-05 | 1,088258202 | 0,00073444 |
| SPOCK2 | 1,354669197 | 6,23383E-05 | 1,168536943 | 0,001519315 |
| CD5 | 1,463177695 | 6,50817E-05 | 1,470911787 | 0,000151844 |
| PTPN7 | 1,500536344 | 6,6139E-05 | 1,396016203 | 0,000550479 |
| RAB3B | 3,298992774 | 8,21927E-05 | 3,887374274 | 4,70663E-06 |
| ISG20 | 1,213764253 | 8,42664E-05 | 1,964543731 | 9,63143E-11 |
| EGR3 | 1,947113831 | 8,45896E-05 | 1,909328256 | 0,000256818 |
| LGALS2 | 1,935568426 | 8,93356E-05 | 1,606878466 | 0,003029369 |
| EPHB2 | 1,487852767 | 8,9717E-05 | 1,196030638 | 0,00415099 |
| SLAMF8 | 1,514924682 | 0,000104602 | 2,394996545 | 6,62754E-10 |
| IKBKE | 1,222157173 | 0,000110145 | 1,216879737 | 0,000274622 |
| VAMP5 | 1,066379769 | 0,000110208 | 1,446028218 | 3,02706E-07 |
| LAX1 | 1,325055657 | 0,000114482 | 1,062428224 | 0,005088885 |
| RGS1 | 1,27311525 | 0,000114482 | 1,690282516 | 5,04492E-07 |
| MELK | 2,029513645 | 0,000123675 | 3,946220899 | 2,0257E-15 |
| DEPDC1B | 1,898123958 | 0,0001238 | 1,98979446 | 8,68398E-05 |
| CD69 | 1,352597052 | 0,000127642 | 1,365679264 | 0,000272805 |

| | | | | |
|---|---|---|---|---|
| SLC7A1 | 1,368915898 | 0,000128576 | 1,038144216 | 0,009086082 |
| HLA-DOA | 1,033036494 | 0,000131013 | 1,02060825 | 0,000440334 |
| CXCL9 | 1,722679939 | 0,000137008 | 3,311142266 | 8,9897E-14 |
| TLR7 | 1,490454095 | 0,000146805 | 1,22945861 | 0,004325763 |
| MND1 | 2,283768218 | 0,000161964 | 3,685279833 | 7,34569E-10 |
| TCF19 | 1,114153119 | 0,000169646 | 2,066223433 | 5,56918E-13 |
| THEMIS | 1,248203534 | 0,000176254 | 1,23375515 | 0,000484839 |
| IL21R | 1,424599493 | 0,000188236 | 1,613385293 | 5,52268E-05 |
| HLA-H | 1,303215936 | 0,000188354 | 1,279003228 | 0,000676876 |
| IGLL1 | 2,170417598 | 0,00018886 | 1,874059676 | 0,003307931 |
| DERL3 | 2,081300634 | 0,00019039 | 2,380518148 | 4,33712E-05 |
| KRT23 | 4,674357245 | 0,000191614 | 4,955867246 | 0,000213098 |
| FCRL5 | 2,525030983 | 0,000199748 | 2,235926402 | 0,002268064 |
| MTHFD1L | 1,218296987 | 0,000203496 | 1,296757007 | 0,000208491 |
| CD8A | 1,454664504 | 0,000214552 | 1,78782839 | 1,29499E-05 |
| RASGRP1 | 1,259806195 | 0,000227301 | 1,134752636 | 0,002257464 |
| CD7 | 1,262110558 | 0,000239622 | 1,336873494 | 0,000227987 |
| NRIP2 | 1,941551812 | 0,000241117 | 1,227970489 | 0,045061612 |
| TTC9 | 1,950300057 | 0,000259696 | 1,295066199 | 0,034316216 |
| MIR155HG | 2,336043859 | 0,000265802 | 2,127997068 | 0,001799145 |
| PLXNC1 | 1,110201122 | 0,000266659 | 1,546739504 | 6,81904E-07 |
| LYZ | 1,121834786 | 0,000274317 | 1,905113576 | 5,3745E-10 |
| KLHL6 | 1,096153638 | 0,000285062 | 1,288609071 | 4,69239E-05 |
| ITK | 1,283333526 | 0,000286287 | 1,377271309 | 0,000253687 |
| CHEK1 | 1,521714173 | 0,000289523 | 2,661917634 | 5,9029E-11 |
| ADAMDEC1 | 2,785653137 | 0,000291111 | 4,424231152 | 3,00318E-09 |
| IDO1 | 1,643068531 | 0,000291217 | 2,22385724 | 1,52165E-06 |
| EGR2 | 1,740714248 | 0,000328086 | 2,284478655 | 3,79372E-06 |
| IL4I1 | 2,138915112 | 0,000330138 | 1,910295437 | 0,00337346 |
| KDELR3 | 1,242078617 | 0,000334771 | 1,508498745 | 3,34199E-05 |
| HELLS | 1,212123049 | 0,00033774 | 2,384116838 | 1,19303E-13 |
| TOP2A | 1,207401069 | 0,000339936 | 3,007207166 | 2,75925E-20 |
| LCK | 1,147301724 | 0,000353104 | 1,121748163 | 0,001174176 |
| IL1RL1 | 1,954399741 | 0,000353846 | 1,641590924 | 0,006859923 |
| RLTPR | 1,434886296 | 0,000353846 | 1,293219256 | 0,003237546 |
| NETO2 | 1,547822605 | 0,000356835 | 1,542769456 | 0,000965697 |
| RTKN2 | 2,438371066 | 0,000364845 | 1,893651509 | 0,013226812 |
| CD27 | 1,557548262 | 0,000368668 | 1,794878755 | 9,5204E-05 |
| KRT8P41 | 2,71913147 | 0,000369982 | 1,968800326 | 0,024285594 |
| CCR5 | 1,024551293 | 0,000374781 | 1,282502672 | 2,01551E-05 |
| ARHGAP11A | 1,063192705 | 0,00037939 | 1,846695409 | 2,53733E-10 |
| BTLA | 1,440639258 | 0,000394608 | 1,364359439 | 0,001726042 |
| PSTPIP1 | 1,177880353 | 0,00040347 | 1,010686358 | 0,005697293 |
| ZWINT | 1,143165779 | 0,000413532 | 2,66115725 | 4,72699E-18 |
| SLC2A6 | 1,093656538 | 0,000450513 | 1,040163984 | 0,002003501 |
| SLC35C1 | 1,20625989 | 0,000452105 | 1,103327127 | 0,003357749 |
| GDNF | 2,229459455 | 0,000459528 | 1,443332505 | 0,048658983 |

| | | | | |
|---|---|---|---|---|
| RARRES1 | 1,357432673 | 0,000471488 | 1,16887335 | 0,006448949 |
| FAM129A | 1,16691849 | 0,000478542 | 1,105846222 | 0,002329256 |
| AMICA1 | 1,059549273 | 0,00049102 | 1,055628357 | 0,001297752 |
| MYC | 1,09716973 | 0,000496754 | 1,148854403 | 0,000648841 |
| CRIP1 | 1,242225461 | 0,000499293 | 1,321839757 | 0,000516906 |
| KRT18 | 1,003216054 | 0,000500834 | 1,106667719 | 0,000325289 |
| GPNMB | 1,21575287 | 0,000515156 | 1,723682057 | 1,71051E-06 |
| CDCP1 | 1,284474867 | 0,000517413 | 1,316627088 | 0,000839442 |
| CLDN7 | 1,284361346 | 0,00052701 | 1,141790545 | 0,005068029 |
| MTMR11 | 1,687218343 | 0,000544936 | 1,240457454 | 0,024953621 |
| APOL3 | 1,059895908 | 0,000553231 | 2,145375492 | 9,60161E-13 |
| RRM2 | 1,319302001 | 0,000555183 | 3,279421268 | 2,78515E-19 |
| CD6 | 1,33732172 | 0,000569602 | 1,27248169 | 0,002480555 |
| PLAUR | 1,177205814 | 0,000596801 | 1,473538218 | 3,44614E-05 |
| E2F1 | 1,981224451 | 0,000616033 | 3,709952716 | 1,54814E-11 |
| UBASH3A | 1,410853825 | 0,000648295 | 1,127216861 | 0,013947635 |
| PAGE4 | 5,086138253 | 0,000662871 | 4,825823958 | 0,002868045 |
| FOXM1 | 1,27580658 | 0,000745851 | 3,166241378 | 2,78515E-19 |
| MARCH3 | 1,294154661 | 0,000752413 | 1,264316956 | 0,002053756 |
| TP53I3 | 1,058758426 | 0,000760174 | 1,571136253 | 9,4108E-07 |
| UBXN10 | 1,662786175 | 0,000769811 | 2,026574643 | 0,000105568 |
| HAPLN3 | 1,708975549 | 0,00078557 | 2,142299766 | 4,58603E-05 |
| CAPG | 1,344150371 | 0,000805826 | 2,043234724 | 5,03166E-07 |
| C9orf172 | 2,512552798 | 0,000841687 | 1,99978122 | 0,018238234 |
| ARNTL2 | 1,131035347 | 0,000878282 | 1,473808529 | 3,34199E-05 |
| LAMA3 | 1,2988547 | 0,000915566 | 1,400627782 | 0,000847213 |
| PLK1 | 1,265096619 | 0,000923642 | 2,273174059 | 2,63595E-10 |
| FAM19A5 | 2,048764824 | 0,000946001 | 1,947133257 | 0,004062819 |
| LOC642131 | 2,277048981 | 0,000946645 | 1,629595085 | 0,038018201 |
| SLAMF7 | 1,195948953 | 0,000947108 | 2,028554082 | 2,21146E-08 |
| LILRB3 | 1,012665659 | 0,000947313 | 1,250187667 | 0,000104505 |
| SNX20 | 1,252501709 | 0,00096315 | 1,562929744 | 6,52115E-05 |
| FAM169A | 1,327200959 | 0,000973148 | 1,319582865 | 0,002525685 |
| TRPM2 | 1,243498604 | 0,000981505 | 1,387827537 | 0,000535889 |
| CPNE5 | 1,828894912 | 0,000987783 | 1,315311991 | 0,038564921 |
| LINC00892 | 2,020748503 | 0,000992003 | 1,398172301 | 0,04512151 |
| NCAPG | 1,289505323 | 0,001015834 | 3,267074727 | 5,78565E-19 |
| DUSP8 | 2,55871891 | 0,001030208 | 2,489785492 | 0,003018924 |
| CD79A | 1,553730809 | 0,001034846 | 1,89605809 | 0,000130058 |
| SAMD11 | 1,342231518 | 0,001041319 | 1,031796595 | 0,025834481 |
| HLA-DOB | 1,590021553 | 0,001057979 | 1,991150547 | 7,4291E-05 |
| TESPA1 | 1,160907358 | 0,001153071 | 1,188232663 | 0,002072036 |
| RHOH | 1,039676376 | 0,00120249 | 1,336437836 | 5,55649E-05 |
| SEPT5-GP1BB | 2,672829145 | 0,001221666 | 2,123585427 | 0,02149629 |
| CCL18 | 2,000944891 | 0,001231679 | 2,404504727 | 0,000232122 |
| MYEOV | 3,181691996 | 0,001252014 | 2,833673188 | 0,009564744 |
| POU2AF1 | 1,647105075 | 0,001272953 | 1,952525336 | 0,000296413 |

| | | | | |
|---|---|---|---|---|
| HRH2 | 1,355003603 | 0,001303126 | 1,072576987 | 0,025592202 |
| PARPBP | 1,040295914 | 0,001328409 | 2,109878629 | 1,47077E-12 |
| KIFC1 | 1,205704468 | 0,001345041 | 2,670350465 | 1,58318E-14 |
| ZBP1 | 1,536663625 | 0,001345909 | 1,758428481 | 0,000560225 |
| MYO7B | 1,874431216 | 0,001350244 | 1,991474492 | 0,001454343 |
| GRIN2D | 1,414305446 | 0,001359939 | 1,188413685 | 0,016270665 |
| E2F8 | 1,78489187 | 0,001360947 | 3,38884905 | 6,36761E-11 |
| LOC101929475 | 3,429708596 | 0,001363518 | 3,440449203 | 0,002640149 |
| ERCC6L | 2,167807975 | 0,001449739 | 3,356636004 | 1,40624E-07 |
| BATF | 1,286508827 | 0,001470213 | 1,646162417 | 9,5204E-05 |
| CCRL2 | 1,086804541 | 0,001490149 | 1,824789945 | 4,75866E-08 |
| CCL5 | 1,08771587 | 0,001492225 | 1,220862474 | 0,000868397 |
| RASGRF1 | 2,671674541 | 0,001520586 | 2,264173666 | 0,016035252 |
| EMILIN2 | 1,329008444 | 0,001574009 | 1,490063394 | 0,000902496 |
| RAB7B | 1,418012076 | 0,001624101 | 1,411276458 | 0,003387759 |
| C15orf48 | 2,729358638 | 0,001638989 | 2,711204942 | 0,003808632 |
| TRAT1 | 1,247861857 | 0,001655932 | 1,510943432 | 0,000280259 |
| HAR1A | 2,061429282 | 0,001702762 | 2,298711991 | 0,000641834 |
| PTPN22 | 1,017216219 | 0,001715843 | 1,009219815 | 0,003967921 |
| KIF20A | 1,333724748 | 0,001772888 | 2,950604718 | 9,03806E-14 |
| AURKB | 1,395762684 | 0,001799597 | 2,856833543 | 2,0246E-12 |
| GAS7 | 1,006298497 | 0,001824627 | 1,029413229 | 0,003247673 |
| COL8A2 | 1,326496672 | 0,001841833 | 1,002217176 | 0,040559214 |
| TRIM59 | 1,175492003 | 0,001851231 | 1,066527089 | 0,009609828 |
| GINS4 | 1,169750927 | 0,001876145 | 1,843882274 | 9,37803E-07 |
| MZB1 | 1,812644366 | 0,001931503 | 2,110464815 | 0,000671446 |
| BCAS1 | 1,860342568 | 0,002191105 | 1,476100432 | 0,030411771 |
| FCN1 | 1,327448492 | 0,002192011 | 1,364525883 | 0,003782074 |
| AKR1B10 | 3,057945093 | 0,002221252 | 4,521062384 | 1,25131E-05 |
| ZNF541 | 2,246931061 | 0,002355296 | 2,13507815 | 0,008942473 |
| BCAT1 | 1,187642232 | 0,002437762 | 1,749136028 | 1,38485E-05 |
| LOC100126784 | 2,740016361 | 0,002475898 | 3,877825883 | 1,862E-05 |
| PLA2G2D | 1,682220803 | 0,002535249 | 2,225554672 | 0,000102329 |
| TRABD2A | 1,82755965 | 0,002545298 | 1,402434615 | 0,042429452 |
| BAIAP2L2 | 1,377639244 | 0,002550184 | 1,532328901 | 0,001648823 |
| CD3G | 1,091793905 | 0,002642897 | 1,096960182 | 0,005197224 |
| FABP5 | 1,070137425 | 0,002679641 | 2,727014185 | 1,66285E-15 |
| MYBL2 | 1,599361631 | 0,002717123 | 3,352882106 | 1,33305E-11 |
| CCNA2 | 1,082892784 | 0,002763254 | 2,724742889 | 5,08252E-16 |
| MCOLN2 | 1,562060769 | 0,002780048 | 1,5624968 | 0,005538621 |
| ABCD2 | 1,430754998 | 0,002801825 | 1,613361617 | 0,001394429 |
| NRIP3 | 2,434702248 | 0,003009775 | 2,560166322 | 0,003069827 |
| GINS2 | 1,14543027 | 0,003062455 | 2,11452043 | 2,27428E-08 |
| HLA-L | 1,189751156 | 0,00309299 | 1,007138112 | 0,026014351 |
| E2F7 | 2,017135323 | 0,003179216 | 3,742960176 | 4,28895E-09 |
| LPAR3 | 3,232069012 | 0,003193952 | 2,637436848 | 0,032240181 |
| ETV7 | 1,33932407 | 0,00319962 | 1,984149921 | 2,23014E-05 |

| | | | | |
|---|---|---|---|---|
| IL18 | 1,035689664 | 0,003239694 | 1,341954887 | 0,000283062 |
| C16orf54 | 1,229765875 | 0,003276712 | 1,118193444 | 0,01572365 |
| ICOS | 1,622613838 | 0,003300554 | 2,848607542 | 1,04767E-07 |
| LOC101060038 | 2,167527347 | 0,003305495 | 1,906738063 | 0,019835736 |
| TUBB2B | 1,483193474 | 0,003599797 | 1,179857389 | 0,041982916 |
| SIGLEC10 | 1,052531768 | 0,00379284 | 1,441014612 | 0,0001518 |
| CLSPN | 1,289927374 | 0,003824463 | 3,062245659 | 4,30326E-13 |
| METTL24 | 1,411681544 | 0,003913039 | 1,11464579 | 0,046426783 |
| LOC101059954 | 2,757175331 | 0,003922336 | 2,847247428 | 0,005116968 |
| PBK | 1,914959145 | 0,003965317 | 4,355382932 | 2,21926E-12 |
| CILP2 | 2,48045141 | 0,004028432 | 2,417271671 | 0,010505892 |
| PANDAR | 1,348371347 | 0,004113507 | 1,254531432 | 0,015311693 |
| PDCD1 | 1,943466965 | 0,00414248 | 2,84104123 | 2,75996E-05 |
| DUSP2 | 1,441091171 | 0,00415419 | 2,314958552 | 2,45244E-06 |
| MNS1 | 1,353544111 | 0,004193687 | 1,385903391 | 0,007340177 |
| TRIM31 | 2,393332412 | 0,004304372 | 3,346800916 | 0,000102717 |
| FZD2 | 1,606537507 | 0,004328729 | 1,485143218 | 0,016468199 |
| DEPDC1 | 1,694151048 | 0,004356465 | 3,316166043 | 5,14139E-09 |
| CD19 | 2,539505634 | 0,004531003 | 2,784282979 | 0,003247673 |
| BCL2A1 | 1,139616928 | 0,004613792 | 1,880386695 | 3,82712E-06 |
| SYTL1 | 1,219307011 | 0,004996385 | 1,138875413 | 0,017254075 |
| C5orf20 | 1,51298807 | 0,005170565 | 1,978757779 | 0,000406912 |
| LRRC43 | 1,902051987 | 0,005181892 | 2,587336409 | 0,000222275 |
| ORC1 | 1,416618539 | 0,005192968 | 2,455721395 | 4,94178E-07 |
| KIF18A | 1,45220738 | 0,005248192 | 2,848953206 | 2,63889E-09 |
| IQGAP3 | 1,829732184 | 0,005265089 | 3,023676544 | 3,50429E-06 |
| TTK | 1,263557124 | 0,005495975 | 2,553197489 | 1,44967E-09 |
| RPS6KL1 | 1,401316638 | 0,00554122 | 1,568329037 | 0,003495258 |
| SLAMF1 | 1,089717595 | 0,005998043 | 1,038691787 | 0,016846826 |
| CDT1 | 1,395474038 | 0,006028052 | 2,423722371 | 7,3311E-07 |
| FBN3 | 1,918303229 | 0,006196499 | 1,60622526 | 0,044830895 |
| KRT86 | 4,138966052 | 0,006304838 | 3,493811836 | 0,041330611 |
| ASF1B | 1,252242481 | 0,006372158 | 2,801041096 | 1,36181E-11 |
| UBE2C | 1,530546114 | 0,006502351 | 3,494683058 | 1,09853E-11 |
| SIT1 | 1,275491034 | 0,006527425 | 1,412704617 | 0,004337377 |
| GPR18 | 1,125357337 | 0,006904907 | 1,013199313 | 0,028405373 |
| TRNP1 | 1,297196102 | 0,007012811 | 1,494897869 | 0,003740119 |
| GNB1L | 1,348481732 | 0,007056478 | 1,297900308 | 0,018210931 |
| RMI2 | 1,468701055 | 0,007209603 | 2,970037435 | 2,4554E-08 |
| CXCR6 | 1,017893794 | 0,007246637 | 1,414082539 | 0,00032045 |
| YPEL4 | 2,167255691 | 0,007259352 | 1,98035181 | 0,025634646 |
| CCL4 | 1,065561232 | 0,007300456 | 1,165348491 | 0,006471396 |
| CENPK | 1,351171912 | 0,007450496 | 2,999463793 | 3,60391E-10 |
| UHRF1 | 1,219276462 | 0,007503594 | 3,063826777 | 5,2132E-13 |
| LINC00313 | 3,287736488 | 0,007517124 | 2,755114003 | 0,047479119 |
| LINC00152 | 1,153432963 | 0,007745972 | 1,658445184 | 0,000214392 |
| NRXN2 | 1,684802359 | 0,008130487 | 1,483180841 | 0,03841545 |

| | | | | |
|---|---|---|---|---|
| LINC00925 | 1,627385644 | 0,008175807 | 1,516197547 | 0,024800406 |
| P2RY10 | 1,101571121 | 0,008314827 | 1,135443555 | 0,012067956 |
| CDKN3 | 1,311248597 | 0,008629092 | 3,173533111 | 9,67143E-12 |
| RASD1 | 1,593601458 | 0,008660401 | 1,584844865 | 0,017898503 |
| LRCOL1 | 1,028930835 | 0,008792854 | 1,695947774 | 1,67476E-05 |
| EPHX3 | 2,178902456 | 0,008911988 | 1,940870501 | 0,035448912 |
| OIP5 | 1,584935585 | 0,008928381 | 3,07129429 | 5,72378E-08 |
| RIBC2 | 2,351124467 | 0,009336193 | 3,251718498 | 0,000300793 |
| HJURP | 1,167357999 | 0,009496755 | 2,942842112 | 4,60657E-13 |
| GPRIN1 | 1,836418149 | 0,009554678 | 2,729899667 | 0,000125908 |
| MS4A1 | 1,123437922 | 0,009630795 | 1,035934655 | 0,031915501 |
| ADD2 | 1,342189182 | 0,009698429 | 1,184452592 | 0,038218281 |
| SPC25 | 1,385417658 | 0,010654837 | 3,082214262 | 9,5561E-10 |
| C16orf59 | 1,516260228 | 0,010707686 | 2,223614922 | 0,000130643 |
| EXO1 | 1,474845684 | 0,010792522 | 3,568281383 | 3,38838E-11 |
| PHLDA2 | 2,253176576 | 0,010922279 | 2,221402237 | 0,02149629 |
| MCM10 | 1,46582539 | 0,011681674 | 3,421472716 | 2,20064E-10 |
| FAM154B | 1,808006911 | 0,01170482 | 2,512632545 | 0,000642977 |
| SHCBP1 | 1,07201637 | 0,011746632 | 2,476904631 | 3,66622E-10 |
| POLQ | 1,370923614 | 0,011864442 | 2,619870288 | 6,46773E-07 |
| SNAP25-AS1 | 1,21900781 | 0,01205585 | 1,486602642 | 0,003836648 |
| GZMM | 1,482694937 | 0,013590963 | 1,528103498 | 0,01643856 |
| AC020571.3 | 2,288976043 | 0,013925402 | 2,059298115 | 0,047394824 |
| MMP9 | 1,945627715 | 0,014588786 | 2,409793769 | 0,004689941 |
| ANLN | 1,093073287 | 0,015259733 | 2,856077879 | 1,25628E-11 |
| CENPI | 1,505295817 | 0,015308566 | 1,726996508 | 0,007082074 |
| ASPHD2 | 1,13841905 | 0,015334399 | 1,070217125 | 0,038475531 |
| CCNB2 | 1,330620051 | 0,015770183 | 3,285531296 | 7,0513E-11 |
| LTA | 1,483467601 | 0,015910348 | 1,566939035 | 0,01766004 |
| CCL3L3 | 2,195799522 | 0,01625686 | 2,091376417 | 0,036478813 |
| NEIL3 | 2,123451841 | 0,016363807 | 1,976487809 | 0,046988445 |
| DLGAP5 | 1,151436927 | 0,017354265 | 3,248667632 | 8,9897E-14 |
| SOCS1 | 1,070320452 | 0,017976276 | 1,596153106 | 0,000606556 |
| HS3ST2 | 1,882648509 | 0,018781187 | 3,099674162 | 0,000131602 |
| BUB1 | 1,104604373 | 0,019782465 | 2,846715311 | 5,73789E-11 |
| NEK2 | 1,108957119 | 0,019782465 | 2,71656408 | 1,85355E-10 |
| FCGR1A | 1,270840532 | 0,021211065 | 2,384736269 | 1,77848E-05 |
| F2RL3 | 1,758011282 | 0,021220733 | 1,674939148 | 0,048375726 |
| GSG2 | 1,981472737 | 0,021294071 | 3,118266898 | 0,000173554 |
| BHLHE41 | 1,160291779 | 0,024742508 | 1,358278251 | 0,014834116 |
| LOC100506098 | 1,590780044 | 0,025396676 | 1,906996785 | 0,011211624 |
| STMND1 | 1,915461201 | 0,026390963 | 2,45689292 | 0,005225193 |
| LOC102724385 | 2,171195806 | 0,026645403 | 2,380539781 | 0,024466563 |
| LINC00639 | 1,729027308 | 0,02843855 | 1,77855715 | 0,037923937 |
| ANKRD36BP2 | 1,662691507 | 0,028544422 | 1,786587517 | 0,030451437 |
| C2CD4A | 2,226560143 | 0,028956116 | 2,644106967 | 0,015867713 |
| SIGLEC8 | 1,160171978 | 0,029447608 | 2,179292976 | 3,89796E-05 |

| | | | | |
|---|---|---|---|---|
| IL2RA | 1,259967577 | 0,029455984 | 1,752563165 | 0,00377791 |
| CCAT1 | 1,634771397 | 0,029788612 | 2,702515619 | 0,000389087 |
| OPRL1 | 1,068073734 | 0,030568236 | 1,450677474 | 0,003953451 |
| FCRLA | 1,455360153 | 0,030754093 | 1,510429043 | 0,037096291 |
| CENPM | 1,978567191 | 0,030922781 | 3,203049404 | 0,000368037 |
| NUF2 | 1,00478598 | 0,032818265 | 3,050269978 | 2,30306E-13 |
| HS1BP3-IT1 | 1,0340911 | 0,033375663 | 1,445879233 | 0,004555987 |
| RPL39L | 1,027675427 | 0,035267086 | 1,369769317 | 0,006466561 |
| HSPA6 | 1,04520871 | 0,035786415 | 1,28465985 | 0,015125646 |
| DIAPH3 | 1,035585131 | 0,036412332 | 2,567143683 | 9,46099E-09 |
| POU5F1B | 2,061259504 | 0,036656801 | 2,192631288 | 0,041447751 |
| TROAP | 1,535750017 | 0,040017321 | 2,768976251 | 0,000145482 |
| SCIN | 1,429903072 | 0,04229904 | 2,211328948 | 0,001796833 |
| IFNG | 1,273824673 | 0,04507788 | 1,802530619 | 0,005101982 |

**Downregulated**

| gene name | log2fold change cABMR | padj cABMR | log2fold change clinTCMR | padj clinTCMR |
|---|---|---|---|---|
| ACSM3 | -1,252803454 | 1,44719E-09 | -1,001235654 | 4,1609E-06 |
| LOC102723845 | -1,153436192 | 6,86696E-06 | -1,042818785 | 0,000143443 |
| RP11-436I24.1 | -4,174521499 | 1,11227E-05 | -2,098111383 | 0,030532535 |
| C21orf37 | -2,295855993 | 1,32173E-05 | -1,445067725 | 0,010849794 |
| UXT-AS1 | -1,564271843 | 4,44245E-05 | -1,029623774 | 0,012372137 |
| CYP2C19 | -2,182207718 | 5,62142E-05 | -1,375076764 | 0,026328559 |
| RP11-12A2.3 | -1,401232076 | 9,94562E-05 | -1,473479775 | 0,000126047 |
| CUX2 | -1,307258666 | 0,000109582 | -1,359484188 | 0,000171218 |
| LINC01093 | -1,406685485 | 0,000118318 | -1,361141119 | 0,000553594 |
| RP11-458D21.1 | -1,269266589 | 0,000130396 | -1,63439327 | 3,80519E-06 |
| HSD17B14 | -1,581925439 | 0,00013739 | -1,346956925 | 0,003111377 |
| GPC5 | -1,421094224 | 0,000181756 | -1,432643253 | 0,000444185 |
| RP11-242D8.1 | -1,157359174 | 0,000310816 | -1,226971443 | 0,000386846 |
| LOC101929633 | -1,051407577 | 0,000332259 | -1,264790379 | 4,58603E-05 |
| GNMT | -1,224669344 | 0,000427014 | -1,269928799 | 0,000678416 |
| TRHDE-AS1 | -1,980038861 | 0,00058193 | -2,097608233 | 0,000689404 |
| HRASLS2 | -1,901151255 | 0,000650285 | -1,457895618 | 0,018961217 |
| IL1RAP | -1,262190265 | 0,000758884 | -1,353674423 | 0,000775782 |
| ENHO | -1,428187261 | 0,000779484 | -1,531353364 | 0,000828707 |
| MEIOB | -2,636448986 | 0,000842139 | -1,928870878 | 0,026565915 |
| DNM1 | -1,162358059 | 0,000849039 | -2,345474466 | 5,38405E-11 |
| AC097468.4 | -1,434993678 | 0,001104253 | -1,368103994 | 0,004211538 |
| DCDC5 | -1,095493239 | 0,001373166 | -1,305905365 | 0,000336894 |
| NCAM2 | -2,017602023 | 0,001976532 | -2,23989499 | 0,001427174 |
| LOC101927595 | -1,085633802 | 0,002836321 | -1,035229846 | 0,009790844 |
| LOC149684 | -2,096148677 | 0,003011335 | -2,487923715 | 0,002072036 |
| LOC101927170 | -2,42560413 | 0,004314503 | -2,953071426 | 0,001459957 |
| TRHDE | -1,602180085 | 0,004754805 | -1,357202715 | 0,033650991 |
| EFCAB1 | -1,137079031 | 0,005955513 | -1,489543037 | 0,000964208 |
| BCRP3 | -1,390481467 | 0,00595807 | -1,164212473 | 0,041523111 |

| | | | | |
|---|---|---|---|---|
| HSPB7 | -2,961954778 | 0,005965794 | -2,691874013 | 0,02544569 |
| SCG5 | -1,101753495 | 0,006859866 | -1,38062077 | 0,001501439 |
| FAM9B | -1,24901151 | 0,007771971 | -1,383179479 | 0,006756003 |
| RP11-75N4.2 | -1,074801146 | 0,01121742 | -1,659124478 | 0,000532198 |
| MYH3 | -1,219049654 | 0,012867888 | -1,737015279 | 0,000755323 |
| LOC102724208 | -1,808812813 | 0,013202697 | -2,341502637 | 0,007831425 |
| LGI1 | -1,415923821 | 0,013493644 | -1,498993141 | 0,016939338 |
| RFPL2 | -1,914648 | 0,015018695 | -1,963822655 | 0,023813124 |
| TNNT2 | -2,187106206 | 0,015259733 | -2,64466704 | 0,007616952 |
| DIO3OS | -1,508257939 | 0,015339764 | -1,52874725 | 0,026008198 |
| LOC101928816 | -1,025575792 | 0,015399723 | -1,030905165 | 0,03143244 |
| MGAM | -1,318601237 | 0,016062605 | -1,485361673 | 0,014729448 |
| CYP1A1 | -1,340455452 | 0,016372342 | -1,503868064 | 0,013238841 |
| DKFZp434J0226 | -2,18718257 | 0,016526147 | -2,154853943 | 0,029509963 |
| FAM9A | -2,089750337 | 0,018798889 | -2,017640745 | 0,046478164 |
| RP11-15A1.7 | -1,011113638 | 0,018861014 | -1,374241806 | 0,003228005 |
| LOC102724603 | -1,215345651 | 0,019487376 | -1,2743398 | 0,025564249 |
| DOK6 | -1,012454752 | 0,021688607 | -1,186928534 | 0,013238841 |
| LOC102724645 | -1,015622025 | 0,024863786 | -1,254805193 | 0,010786821 |
| AVPR1A | -1,232150571 | 0,025454844 | -1,627376034 | 0,005519469 |
| NPIPB6 | -1,117677519 | 0,027565639 | -1,54188828 | 0,004198778 |
| RP11-10L7.1 | -1,903817428 | 0,030712949 | -4,168624292 | 9,16044E-05 |
| CALR3 | -1,07619641 | 0,036342068 | -1,386306258 | 0,014729448 |
| THBS4 | -1,899286379 | 0,043443548 | -2,844640789 | 0,004122176 |

**Suppl. Table 7: Shared pathways of cABMR and clinTCMR in the discovery cohort**
**Upregulated**

| q-value | Pathway name | Pathway ID | | | q-value | Pathway name |
|---|---|---|---|---|---|---|
| 2.6101449866 | positive regula | GO:0022409 | | | 1.0861818489 | Chemokine rec |
| 2.6101449866 | regulation of ly | GO:0051249 | | | 6.1168076113 | Cytokine-cytok |
| 2.6101449866 | regulation of le | GO:1903037 | | | 6.1168076113 | Viral protein in |
| 9.5067672321 | cellular respon | GO:0071345 | | | 6.1168076113 | Cell Cycle |
| 9.5067672321 | chemokine-me | GO:0070098 | | | 1.3089473569 | Cell Cycle Chec |
| 9.5067672321 | hemopoiesis | GO:0030097 | | | 1.3929283131 | Cell Cycle, Mito |
| 3.6021126048 | response to ch | GO:1990868 | | | 3.0365592732 | Hematopoietic |
| 6.6698383869 | positive regula | GO:0002696 | | | 3.9998904968 | Human T-cell le |
| 1.2936869126 | positive regula | GO:0050867 | | | 6.7880884121 | Cell cycle - Hon |
| 1.2735819898 | neutrophil mig | GO:1990266 | | | 7.1291427503 | Peptide ligand- |
| 4.0172619007 | regulation of le | GO:1902105 | | | 7.8864232618 | Resolution of S |
| 4.0172619007 | DNA replicatio | GO:0006260 | | | 8.8373437060 | Primary immun |
| 4.8094635086 | nuclear divisio | GO:0000280 | | | 1.0364370682 | Amplification |
| 5.9345582041 | neutrophil che | GO:0030593 | | | 1.0364370682 | Amplification c |
| 1.1000216537 | G1/S transition | GO:0000082 | | | 1.0364370682 | Mitotic Spindle |
| 1.1874479419 | response to int | GO:0034341 | | | 1.1298288079 | Unwinding of D |
| 1.2339547296 | regulation of ir | GO:1902531 | | | 1.4399684381 | EML4 and NUD |
| 1.2339547296 | calcium ion tra | GO:0097553 | | | 1.9954484588 | Signal Transdu |
| 2.8792220668 | positive regula | GO:0050778 | | | 2.0612205030 | Mitotic G1 pha |
| 3.2074038059 | phosphorylatio | GO:0016310 | | | 2.0612205030 | RHO GTPases A |
| 3.2074038059 | positive regula | GO:0001819 | | | 3.2222340199 | Chemokine sig |
| 3.2074038059 | mitotic spindle | GO:0007094 | | | 3.2222340199 | Mitotic Prome |
| 8.1162545890 | positive regula | GO:0032103 | | | 3.6083702932 | Separation of S |
| 1.1853059837 | positive regula | GO:0009967 | | | 3.6188521776 | G1/S Transition |
| 1.9447357297 | positive regula | GO:0010647 | | | 3.6188521776 | Class A/1 (Rhod |
| 2.2643893944 | dendritic cell d | GO:0097028 | | | 4.3791096442 | Immune Systen |
| 2.3916490564 | ERK1 and ERK2 | GO:0070371 | | | 6.3003631245 | DNA Replicatio |
| 2.3916490564 | positive regula | GO:1903708 | | | 6.3114624206 | Activation of A |
| 2.3916490564 | negative regula | GO:0051985 | | | 1.6040828543 | Mitotic Anapha |
| 2.7804408817 | myeloid leukoc | GO:0002573 | | | 1.6601444845 | Mitotic Metapl |
| 4.0010978178 | regulation of p | GO:0043067 | | | 2.5453283409 | Synthesis of DN |
| 4.2750481977 | negative regula | GO:0045786 | | | 3.4941536063 | Activation of th |
| 5.6250050698 | regulation of c | GO:0010564 | | | 6.0170297498 | TNFR2 non-can |
| 5.8791113101 | DNA replicatio | GO:0006270 | | | 6.2662406484 | T cell receptor |
| 6.8074204717 | cellular cation | GO:0030003 | | | 7.2159067927 | DNA Replicatio |
| 9.3257788779 | regulation of o | GO:0033043 | | | 9.1622867752 | Epstein-Barr vi |
| 9.5743802718 | regulation of p | GO:0051174 | | | 9.7352575889 | Orc1 removal f |
| 0.0001052393 | leukocyte apop | GO:0071887 | | | 0.0001202267 | Rheumatoid ar |
| 0.0001446603 | regulation of ir | GO:0032649 | | | 0.0001522172 | G1/S-Specific T |
| 0.0001459865 | regulation of le | GO:0070663 | | | 0.0001593171 | GPCR ligand bi |
| 0.0001949551 | regulation of ir | GO:0050727 | | | 0.0001912205 | G2/M Checkpo |
| 0.0002086686 | regulation of d | GO:0031347 | | | 0.0001952028 | APC/C-mediate |
| 0.0002184430 | regulation of G | GO:0043087 | | | 0.0001952028 | Regulation of n |
| 0.0002297490 | response to tu | GO:0034612 | | | 0.0002782642 | Cell adhesion n |
| 0.0002297490 | positive regula | GO:0031349 | | | 0.0002823086 | DNA strand elo |
| 0.0003191471 | positive regula | GO:0051347 | | | 0.0002823086 | Switching of or |
| 0.0003756136 | regulation of T | GO:0002664 | | | 0.0002823086 | S Phase |
| 0.0004642300 | centromere co | GO:0034508 | | | 0.0002823086 | Toll-like recept |
| 0.0004677744 | positive regula | GO:0008284 | | | 0.0002897081 | Assembly of the |
| 0.0004691184 | antigen recept | GO:0050851 | | | 0.0003100278 | Activation of A |
| 0.0005367776 | DNA recombin | GO:0006310 | | | 0.0003151712 | Cytokine Signa |
| 0.0005598619 | T-helper 1 type | GO:0042088 | | | 0.0003908059 | M Phase |
| 0.0007310372 | T cell chemota | GO:0010818 | | | 0.0004329208 | PD-L1 expressi |
| 0.0007933783 | regulation of a | GO:0050854 | | | 0.0004765227 | Toxoplasmosis |
| 0.0008907310 | positive regula | GO:0010942 | | | 0.0009088142 | Adaptive Immu |
| 0.0010999223 | negative regula | GO:0051782 | | | 0.0009520947 | TP53 Regulates |
| 0.0011534124 | regulation of m | GO:0007346 | | | 0.0009588355 | Antigen proces |
| 0.0012430589 | chromatin rem | GO:0031055 | | | 0.0009588355 | G alpha (i) signa |
| 0.0013061338 | positive regula | GO:0051130 | | | 0.0012133405 | NF-kappa B sign |
| 0.0013061338 | positive regula | GO:0051272 | | | 0.0012133405 | Type I diabetes |
| 0.0014507871 | positive regula | GO:0045597 | | | 0.0015423086 | Polo-like kinas |
| 0.0015127198 | regulation of le | GO:0002688 | | | 0.0015526896 | Th17 cell differ |
| 0.0015699446 | signal transduc | GO:0072401 | | | 0.0017735291 | APC:Cdc20 me |
| 0.0015699446 | signal transduc | GO:0072422 | | | 0.0018515400 | Assembly of co |
| 0.0016864894 | ion homeostas | GO:0050801 | | | 0.0018515400 | Intestinal imm |
| 0.0018422138 | calcium-media | GO:0019722 | | | 0.0020305999 | TNF signaling p |
| 0.0021534616 | actin polymeri | GO:0008154 | | | 0.0020305999 | Signaling by Rh |
| 0.0021534616 | negative regula | GO:0001818 | | | 0.0022511561 | APC/C:Cdc20 m |
| 0.0022231448 | DNA replicatio | GO:0034724 | | | 0.0025139187 | Th1 and Th2 ce |
| 0.0022231448 | eosinophil che | GO:0048245 | | | 0.0027896689 | Signaling by Rh |
| 0.0023487970 | protein-DNA c | GO:0065004 | | | 0.0028282656 | IL-17 signaling |
| 0.0024702434 | positive regula | GO:0002699 | | | 0.0028345686 | Signaling by GF |
| 0.0025641030 | positive regula | GO:0002579 | | | 0.0041280788 | Allograft reject |
| 0.0025641030 | positive regula | GO:0033630 | | | 0.0041280788 | Regulation of A |
| 0.0026838256 | DNA conforma | GO:0071103 | | | 0.0041280788 | Costimulation |
| 0.0027524774 | regulation of vi | GO:0050792 | | | 0.0053787026 | Immunoregula |
| 0.0028405402 | negative regula | GO:0048525 | | | 0.0060627848 | RHO GTPase Eff |
| 0.0029144427 | regulation of ir | GO:0032655 | | | 0.0061703975 | Transcription c |
| 0.0034620696 | regulation of re | GO:1900120 | | | 0.0061703975 | Cyclin A/B1/B2 |
| 0.0035316325 | regulation of c | GO:0032268 | | | 0.0068417181 | Graft-versus-ho |
| 0.0037212959 | positive regula | GO:0051345 | | | 0.0068417181 | RHOF GTPase c |
| 0.0039506685 | eosinophil mig | GO:0072677 | | | 0.0069724070 | G0 and Early G |
| 0.0040154986 | regulation of n | GO:1902622 | | | 0.0071272365 | Cytosolic DNA- |
| 0.0041997061 | regulation of ly | GO:2000401 | | | 0.0072649233 | Deposition of n |
| 0.0041997061 | positive regula | GO:0002645 | | | 0.0072649233 | Nucleosome as |
| 0.0041997061 | regulation of d | GO:0002604 | | | 0.0074323702 | Condensation c |
| 0.0046824709 | regulation of a | GO:0032956 | | | 0.0074874327 | Measles - Homo |
| 0.0047642310 | interferon-gam | GO:0060333 | | | 0.0079187978 | Inflammatory b |
| 0.0047642310 | negative regula | GO:0022408 | | | 0.0086512059 | APC-Cdc20 me |
| 0.0048169160 | negative regula | GO:0071930 | | | 0.0089759149 | Influenza A - Ho |
| 0.0048169160 | positive regula | GO:0010604 | | | 0.0097256788 | Translocation c |
| 0.0051102459 | establishment | GO:0001768 | | | 0.0108764792 | Cdc20:Phosph |
| 0.0051102459 | natural killer c | GO:0035747 | | | 0.0108764792 | TNFs bind their |
| 0.0051102459 | negative regula | GO:0051129 | | | 0.0113502098 | GPCR downstre |
| 0.0062374552 | regulation of ir | GO:0002861 | | | 0.0119196255 | Interferon gam |
| 0.0070494618 | positive regula | GO:0031325 | | | 0.0119785447 | APC/C:Cdh1 m |
| 0.0073443369 | positive regula | GO:0051173 | | | 0.0130571170 | TCR signaling |
| 0.0094783613 | regulation of p | GO:0051246 | | | 0.0132741704 | Phosphorylatio |
| 0.0097941761 | regulation of ki | GO:0090183 | | | 0.0134615253 | TP53 Regulates |
| 0.0106482204 | regulation of ir | GO:0045088 | | | 0.0141714449 | Osteoclast diffe |
| 0.0114142046 | hepatocyte dif | GO:0070365 | | | 0.0145033536 | PD-1 signaling |
| 0.0116123253 | stress-activate | GO:0051403 | | | 0.0156314611 | Chagas disease |
| 0.0116502814 | positive regula | GO:0002687 | | | 0.0162159899 | G2 Phase |
| 0.0130328058 | pseudopodium | GO:0031268 | | | 0.0162159899 | Binding and en |
| 0.0130328058 | antigen proces | GO:0019885 | | | 0.0169203445 | C-type lectin re |
| 0.0130328058 | G2 DNA damag | GO:0031572 | | | 0.0173756945 | TNF receptor su |
| 0.0130328058 | regulation of D | GO:0051052 | | | 0.0175933532 | Constitutive Si |
| 0.0130328058 | positive regula | GO:0051962 | | | 0.0209410273 | African trypano |
| 0.0130382644 | regulation of a | GO:0002580 | | | 0.0229181236 | Phosphorylatio |
| 0.0141891897 | histone exchan | GO:0043486 | | | 0.0241449883 | Viral carcinoge |
| 0.0147677833 | morphogenesis | GO:0003159 | | | 0.0266754634 | One carbon po |
| 0.0151443063 | regulation of p | GO:0002700 | | | 0.0295336696 | Antigen proces |
| 0.0168677942 | chronic inflam | GO:0002544 | | | 0.0302952664 | Binding of TCF/ |
| 0.0168677942 | tetrahydrofola | GO:0046653 | | | 0.0302952664 | Activation of N |
| 0.0177182814 | positive regula | GO:0090184 | | | 0.0305599912 | Tuberculosis - I |
| 0.0177182814 | G2/M transitio | GO:0000086 | | | 0.0307969028 | Human immun |
| 0.0177182814 | regulation of a | GO:0002583 | | | 0.0308698274 | Bladder cancer |
| 0.0177182814 | antigen proces | GO:0002476 | | | 0.0317182566 | Collagen forma |
| 0.0177182814 | regulation of D | GO:2000371 | | | 0.0324160032 | Inactivation of |
| 0.0186233875 | myeloid dendr | GO:0043011 | | | 0.0324160032 | Inhibition of th |
| 0.0196408914 | regulation of gl | GO:1900449 | | | 0.0325092682 | Generation of s |
| 0.0209137373 | cation transpo | GO:0006812 | | | 0.0327042420 | Phagosome - H |
| 0.0224810385 | negative regula | GO:0050866 | | | 0.0346065026 | Signaling by SC |
| 0.0226706889 | viral genome re | GO:0019079 | | | 0.0346065026 | Acute myeloid |
| 0.0226706889 | viral life cycle | GO:0019058 | | | 0.0358071081 | RUNX3 regulat |
| 0.0228583715 | negative regula | GO:0060548 | | | 0.0358071081 | Dectin-1 media |
| 0.0229895474 | maintenance o | GO:0010216 | | | 0.0358071081 | NIK-->noncano |
| 0.0237684692 | regulation of cy | GO:0001959 | | | 0.0375672100 | Cellular senesc |
| 0.0267249681 | response to int | GO:0070555 | | | | |
| 0.0267249681 | regulation of ir | GO:0032675 | | | | |
| 0.0285638745 | regulation of gl | GO:1903018 | | | | |
| 0.0291704448 | regulation of ca | GO:0051940 | | | | |
| 0.0313603161 | tumor necrosis | GO:0033209 | | | | |
| 0.0331098454 | T cell mediated | GO:0002456 | | | | |
| 0.0331098454 | positive regula | GO:0120034 | | | | |
| 0.0331098454 | DNA damage re | GO:0030330 | | | | |
| 0.0331098454 | cellular protei | GO:0006464 | | | | |
| 0.0340849285 | negative regula | GO:0051348 | | | | |
| 0.0340849285 | regulation of sy | GO:0060142 | | | | |
| 0.0344844876 | regulation of ir | GO:0032677 | | | | |
| 0.0344844876 | negative regula | GO:0002578 | | | | |
| 0.0344844876 | positive regula | GO:0010912 | | | | |
| 0.0354747504 | negative regula | GO:0002695 | | | | |
| 0.0354747504 | regulation of fi | GO:0048145 | | | | |
| 0.0369261197 | negative regula | GO:0009968 | | | | |

**Downregulated**

| q-value | Pathway name | Pathway ID | | q-value | Pathway name | Pathway ID |
|---|---|---|---|---|---|---|
| 0.0147856455 | unsaturated fa | GO:0033559 | | 0.0005397953 | Synthesis of ep | R-HSA-214267 |
| 0.0147856455 | long-chain fatt | GO:0001676 | | 0.0005397953 | Synthesis of (16 | R-HSA-214281 |
| 0.0147856455 | terpenoid meta | GO:0006721 | | 0.0020805355 | Xenobiotics | R-HSA-211981 |
| 0.0152613293 | sarcomere orga | GO:0045214 | | 0.0043753232 | Striated Muscl | R-HSA-390522 |
| 0.0152613293 | actin-myosin fi | GO:0033275 | | 0.0052523975 | Fatty acid meta | R-HSA-897886 |
| 0.0152613293 | fatty acid biosy | GO:0006633 | | 0.0073005574 | Cytochrome P4 | R-HSA-211897 |
| 0.0241907678 | positive regula | GO:0051965 | | 0.0073005574 | Arachidonic ac | R-HSA-214275 |
| 0.0241907678 | arachidonic ac | GO:0019369 | | 0.0120734995 | Chemical carci | path:hsa05204 |
| 0.0299063637 | cellular glucan | GO:0006073 | | 0.0151909532 | Phase I - Functi | R-HSA-211945 |
| 0.0299063637 | glucan metabo | GO:0044042 | | 0.0151909532 | Metabolism of | R-HSA-556833 |
| 0.0299063637 | striated muscl | GO:0051146 | | | | |
| 0.0324288779 | heart contracti | GO:0060047 | | | | |
| 0.0347054094 | regulation of b | GO:1903522 | | | | |

**Suppl. Table 8: Differentially expressed genes of the validation cohort cABMR vs. NHR**

| log2FoldChange | padj | gene_names | significance | direction |
|---|---|---|---|---|
| 2,015723806 | 4,81871E-08 | GLS | TRUE | Upregulated |
| 4,220834716 | 4,81871E-08 | SLC7A14 | TRUE | Upregulated |
| 2,713994837 | 6,60574E-08 | C1orf204 | TRUE | Upregulated |
| 2,935567848 | 1,77857E-07 | FGF14 | TRUE | Upregulated |
| 2,928053701 | 1,77857E-07 | KCNA3 | TRUE | Upregulated |
| 3,746335609 | 1,77857E-07 | UBD | TRUE | Upregulated |
| 2,745080466 | 1,09512E-06 | PAPPA-AS1 | TRUE | Upregulated |
| 0,921870976 | 1,20156E-06 | CRYBG3 | FALSE | Upregulated |
| 1,921310999 | 1,34775E-06 | APOBEC4 | TRUE | Upregulated |
| 1,532955726 | 1,34775E-06 | COA1 | TRUE | Upregulated |
| 1,401015699 | 1,34775E-06 | DEXI | TRUE | Upregulated |
| 3,956344279 | 1,40505E-06 | LOC102723401 | TRUE | Upregulated |
| 2,212179843 | 1,41733E-06 | SLC1A7 | TRUE | Upregulated |
| 4,470939161 | 1,98722E-06 | NRSN1 | TRUE | Upregulated |
| 3,144271898 | 2,02652E-06 | RP11-77C3.3 | TRUE | Upregulated |
| 1,725384899 | 2,63182E-06 | ARSG | TRUE | Upregulated |
| 1,4789798 | 3,36553E-06 | FAM196B | TRUE | Upregulated |
| 1,725133873 | 3,36553E-06 | LOC102724536 | TRUE | Upregulated |
| 1,700135898 | 3,36553E-06 | PHOSPHO1 | TRUE | Upregulated |
| 2,146447248 | 3,848E-06 | ZBED2 | TRUE | Upregulated |
| 1,654153897 | 3,95917E-06 | CELF2-AS1 | TRUE | Upregulated |
| 2,018137379 | 4,44881E-06 | FAT1 | TRUE | Upregulated |
| 2,2089382 | 7,83497E-06 | HLA-F-AS1 | TRUE | Upregulated |
| 1,839441622 | 9,07596E-06 | AEBP1 | TRUE | Upregulated |
| 4,394940508 | 9,07596E-06 | IRF4 | TRUE | Upregulated |
| 1,943556929 | 9,13518E-06 | TMEM75 | TRUE | Upregulated |
| -0,998720423 | 1,12187E-05 | ZKSCAN1 | FALSE | Downregulated |
| -0,750157584 | 1,42079E-05 | FURIN | FALSE | Downregulated |
| 1,736729846 | 1,60326E-05 | TAPSAR1 | TRUE | Upregulated |
| 2,535535868 | 1,68827E-05 | MUC12 | TRUE | Upregulated |
| 1,342502232 | 2,44586E-05 | LOC100506990 | TRUE | Upregulated |

| | | | | |
|---|---|---|---|---|
| 1,403794159 | 2,47164E-05 | LIMK2 | TRUE | Upregulated |
| -0,790145021 | 2,47164E-05 | TFR2 | FALSE | Downregulated |
| 1,310437541 | 2,47164E-05 | TLDC2 | TRUE | Upregulated |
| 2,002708374 | 2,6018E-05 | IL32 | TRUE | Upregulated |
| 2,216441366 | 2,6018E-05 | LOC101928152 | TRUE | Upregulated |
| -1,026051544 | 3,15526E-05 | APOC3 | TRUE | Downregulated |
| -0,73976431 | 3,17772E-05 | CDHR5 | FALSE | Downregulated |
| 1,634551248 | 3,17772E-05 | LOC101927954 | TRUE | Upregulated |
| -1,297909525 | 3,39131E-05 | PTH1R | TRUE | Downregulated |
| 2,007767544 | 3,74818E-05 | AQP4-AS1 | TRUE | Upregulated |
| 2,786666092 | 3,7778E-05 | EFEMP1 | TRUE | Upregulated |
| 1,031306168 | 3,7778E-05 | KCNQ1OT1 | TRUE | Upregulated |
| 4,590354477 | 4,26497E-05 | CXCL10 | TRUE | Upregulated |
| -0,856115709 | 4,26497E-05 | HAGH | FALSE | Downregulated |
| 1,357242447 | 4,626E-05 | PIGF | TRUE | Upregulated |
| -0,747567805 | 5,0477E-05 | TJP2 | FALSE | Downregulated |
| 1,20049366 | 5,0477E-05 | ZNF433 | TRUE | Upregulated |
| 2,025052754 | 5,18423E-05 | COL1A1 | TRUE | Upregulated |
| 1,799899495 | 5,72062E-05 | FAM115C | TRUE | Upregulated |
| 1,266352643 | 5,72062E-05 | SAP30L-AS1 | TRUE | Upregulated |
| 1,427232915 | 6,09991E-05 | VIM-AS1 | TRUE | Upregulated |
| 1,364114359 | 6,64463E-05 | MBNL1-AS1 | TRUE | Upregulated |
| 3,662833608 | 7,17608E-05 | BCAT1 | TRUE | Upregulated |
| 1,241307539 | 7,17608E-05 | LOC441454 | TRUE | Upregulated |
| -0,710783861 | 7,49816E-05 | ASGR2 | FALSE | Downregulated |
| -0,877623299 | 8,74985E-05 | ACACB | FALSE | Downregulated |
| -1,819358544 | 9,99001E-05 | G6PC | TRUE | Downregulated |
| -2,990703425 | 0,00010418 | AVPR1A | TRUE | Downregulated |
| -0,626782419 | 0,00010418 | CLUH | FALSE | Downregulated |
| 2,346984977 | 0,00010418 | JAK3 | TRUE | Upregulated |
| 4,149665906 | 0,00010418 | LINC00900 | TRUE | Upregulated |
| 2,273532745 | 0,00010418 | LOC101928173 | TRUE | Upregulated |
| 2,216993085 | 0,00010418 | SPOCK2 | TRUE | Upregulated |
| -0,66896688 | 0,00010778 | ASGR1 | FALSE | Downregulated |
| 0,793263641 | 0,00010778 | NFE2L2 | FALSE | Upregulated |
| -1,038783164 | 0,000112488 | ALDOB | TRUE | Downregulated |
| -1,657580547 | 0,000112488 | PPP1R1A | TRUE | Downregulated |
| 2,655007156 | 0,000135651 | DTNA | TRUE | Upregulated |
| 1,550884374 | 0,000135651 | ITGA6 | TRUE | Upregulated |
| -0,748089384 | 0,000136388 | F2 | FALSE | Downregulated |
| 3,586416692 | 0,000136978 | CXCL9 | TRUE | Upregulated |
| 0,900592154 | 0,000137477 | PAPLN | FALSE | Upregulated |
| -0,967494394 | 0,000138998 | AGT | FALSE | Downregulated |
| -1,448614718 | 0,000139676 | CBS | TRUE | Downregulated |
| 1,449297875 | 0,000139676 | NLGN3 | TRUE | Upregulated |
| 1,120040467 | 0,000154854 | PVRIG | TRUE | Upregulated |
| -0,957450335 | 0,000169846 | ACADS | FALSE | Downregulated |
| 1,028644604 | 0,000169846 | C10orf10 | TRUE | Upregulated |
| 1,698719435 | 0,000169846 | COL1A2 | TRUE | Upregulated |
| 0,999633081 | 0,000169846 | DMTF1 | FALSE | Upregulated |
| -0,674279052 | 0,000169846 | IGFBP4 | FALSE | Downregulated |

| | | | | |
|---|---|---|---|---|
| 1,362344882 | 0,000169846 | LOC100506142 | TRUE | Upregulated |
| 3,668994054 | 0,000171879 | LOC101927734 | TRUE | Upregulated |
| 1,408416765 | 0,000171879 | SLC7A6OS | TRUE | Upregulated |
| 2,086313471 | 0,000183056 | IL21R-AS1 | TRUE | Upregulated |
| 0,885457535 | 0,000183056 | PYGB | FALSE | Upregulated |
| 1,245563464 | 0,000189564 | LOC100294362 | TRUE | Upregulated |
| 1,805948181 | 0,000189659 | APCDD1L | TRUE | Upregulated |
| -1,065399307 | 0,000196695 | PCDH1 | TRUE | Downregulated |
| 1,584458033 | 0,00021334 | STAT1 | TRUE | Upregulated |
| 1,670188885 | 0,000215093 | BYSL | TRUE | Upregulated |
| 0,95419381 | 0,000215093 | FAM168A | FALSE | Upregulated |
| 1,44749902 | 0,000215093 | RP11-436I24.1 | TRUE | Upregulated |
| 1,350227918 | 0,000221542 | FMNL1 | TRUE | Upregulated |
| -0,66836424 | 0,000221542 | HPN | FALSE | Downregulated |
| 1,02260617 | 0,000221542 | KLHL29 | TRUE | Upregulated |
| 2,6240642 | 0,000221542 | LOC101927085 | TRUE | Upregulated |
| 2,957288222 | 0,000221542 | RP11-44F21.3 | TRUE | Upregulated |
| 1,999701312 | 0,000223015 | FSTL3 | TRUE | Upregulated |
| 2,720833314 | 0,000223468 | IGJ | TRUE | Upregulated |
| 2,050480538 | 0,000236944 | PCNXL2 | TRUE | Upregulated |
| 1,876823074 | 0,000236944 | RUNX3 | TRUE | Upregulated |
| 3,108976889 | 0,00024271 | GCSAM | TRUE | Upregulated |
| 1,10933998 | 0,000253429 | GSN | TRUE | Upregulated |
| 1,670903583 | 0,000257895 | SLC4A5 | TRUE | Upregulated |
| 0,943545048 | 0,000274301 | FBXL2 | FALSE | Upregulated |
| 3,413040942 | 0,000274417 | PRDM8 | TRUE | Upregulated |
| 3,114597943 | 0,000283051 | GSG2 | TRUE | Upregulated |
| -0,773382443 | 0,000292848 | ARG1 | FALSE | Downregulated |
| 1,547169323 | 0,000292848 | CD84 | TRUE | Upregulated |
| 1,172432237 | 0,000292848 | KCTD12 | TRUE | Upregulated |
| 2,236018191 | 0,00029536 | DDI1 | TRUE | Upregulated |
| 1,283015593 | 0,000299252 | PLAC4 | TRUE | Upregulated |
| -0,843866658 | 0,000319332 | PROX1 | FALSE | Downregulated |
| 1,499059565 | 0,000321242 | LRRC18 | TRUE | Upregulated |
| -0,689493905 | 0,000323502 | C19orf66 | FALSE | Downregulated |
| 1,169895534 | 0,000333532 | PRPF40B | TRUE | Upregulated |
| -1,572020776 | 0,000345922 | PPP1R3C | TRUE | Downregulated |
| 1,602152425 | 0,000351153 | TMC8 | TRUE | Upregulated |
| -0,880547551 | 0,000352635 | ECHDC2 | FALSE | Downregulated |
| 1,027849486 | 0,000352635 | LOC100128653 | TRUE | Upregulated |
| 1,260920634 | 0,000360821 | TMEM120B | TRUE | Upregulated |
| 1,900618274 | 0,000386073 | ATP2A3 | TRUE | Upregulated |
| 1,08437144 | 0,000386073 | RP6-206I17.2 | TRUE | Upregulated |
| 1,969624851 | 0,000395353 | LOC101927848 | TRUE | Upregulated |
| 2,524919442 | 0,000397849 | BCL11B | TRUE | Upregulated |
| 0,927130335 | 0,000414805 | PDAP1 | FALSE | Upregulated |
| 2,516354379 | 0,000417815 | C11orf45 | TRUE | Upregulated |
| 1,38403983 | 0,000417815 | TSPAN3 | TRUE | Upregulated |
| -2,88955532 | 0,000436864 | GJB5 | TRUE | Downregulated |
| 0,96985593 | 0,000451184 | IRF1 | FALSE | Upregulated |
| -0,511982456 | 0,000456061 | MST1 | FALSE | Downregulated |

| | | | | |
|---|---|---|---|---|
| -0,970384483 | 0,000456061 | TMEM176A | FALSE | Downregulated |
| 1,012212548 | 0,000462896 | KRR1 | TRUE | Upregulated |
| 1,357012899 | 0,000466038 | LAMA2 | TRUE | Upregulated |
| 1,446117163 | 0,000466038 | LOC102724584 | TRUE | Upregulated |
| 3,72998583 | 0,00046815 | AC007750.5 | TRUE | Upregulated |
| -0,710499147 | 0,000469374 | AGMAT | FALSE | Downregulated |
| 3,073743399 | 0,000469374 | B3GNT5 | TRUE | Upregulated |
| 1,246738667 | 0,000469374 | TRIM17 | TRUE | Upregulated |
| 0,906502554 | 0,00047886 | C1orf56 | FALSE | Upregulated |
| 1,350368242 | 0,00047886 | CCND1 | TRUE | Upregulated |
| 2,151650019 | 0,00047886 | KAAG1 | TRUE | Upregulated |
| 0,896583473 | 0,000494155 | ASF1A | FALSE | Upregulated |
| 3,26131342 | 0,000494155 | LINC00691 | TRUE | Upregulated |
| -0,871016037 | 0,000507316 | MAT1A | FALSE | Downregulated |
| -1,288487671 | 0,000510315 | PACSIN3 | TRUE | Downregulated |
| -0,596713087 | 0,000522879 | RXRA | FALSE | Downregulated |
| 2,678289188 | 0,000529295 | AC007461.2 | TRUE | Upregulated |
| -0,776371716 | 0,000529295 | OAF | FALSE | Downregulated |
| 1,595284424 | 0,000532369 | RP11-401P9.5 | TRUE | Upregulated |
| -0,722300219 | 0,0005422 | ECE1 | FALSE | Downregulated |
| 2,790126363 | 0,000598201 | BHLHE41 | TRUE | Upregulated |
| 1,930991769 | 0,000598201 | XYLT1 | TRUE | Upregulated |
| 2,572718398 | 0,000611455 | CASC14 | TRUE | Upregulated |
| 1,707086545 | 0,000611455 | THBS2 | TRUE | Upregulated |
| -1,149100704 | 0,000615493 | HAO2 | TRUE | Downregulated |
| 3,999215077 | 0,00066364 | GRAP2 | TRUE | Upregulated |
| 1,617635699 | 0,000665355 | CLDN18 | TRUE | Upregulated |
| -1,377713632 | 0,000687318 | MOCOS | TRUE | Downregulated |
| 1,7249586 | 0,000731724 | LYZ | TRUE | Upregulated |
| -1,24970643 | 0,000731724 | MLPH | TRUE | Downregulated |
| 0,922354068 | 0,000731724 | OGFRL1 | FALSE | Upregulated |
| 1,564727669 | 0,000739254 | THY1 | TRUE | Upregulated |
| -1,878138301 | 0,000741847 | CFHR3 | TRUE | Downregulated |
| 1,493549735 | 0,000760984 | ATP8B2 | TRUE | Upregulated |
| -0,943179759 | 0,000761212 | GYS2 | FALSE | Downregulated |
| -0,811597359 | 0,000774029 | ABCC9 | FALSE | Downregulated |
| 0,767312834 | 0,000780055 | C12orf76 | FALSE | Upregulated |
| 1,983117123 | 0,000790574 | LOC100289455 | TRUE | Upregulated |
| 2,421010648 | 0,000790574 | LOC101928123 | TRUE | Upregulated |
| 1,378010263 | 0,000791871 | LYSMD1 | TRUE | Upregulated |
| 1,832276263 | 0,000807288 | AVPR2 | TRUE | Upregulated |
| -0,992586846 | 0,000807288 | TBX3 | FALSE | Downregulated |
| 1,78658808 | 0,000812961 | PLEKHG4 | TRUE | Upregulated |
| 3,333320168 | 0,000828615 | COL15A1 | TRUE | Upregulated |
| 1,99881676 | 0,000828779 | CD48 | TRUE | Upregulated |
| 0,97854748 | 0,000833633 | IGFBP7 | FALSE | Upregulated |
| -0,595862727 | 0,000835799 | LRP5 | FALSE | Downregulated |
| 1,791611384 | 0,000846284 | C1orf198 | TRUE | Upregulated |
| 1,664109743 | 0,000846284 | HMGN4 | TRUE | Upregulated |
| 1,462756359 | 0,000846284 | LOC100996723 | TRUE | Upregulated |
| 2,573057993 | 0,000846284 | POM121L9P | TRUE | Upregulated |

| | | | | |
|---|---|---|---|---|
| -0,882482819 | 0,000876878 | ACAA1 | FALSE | Downregulated |
| -0,838759545 | 0,000876878 | ALDH2 | FALSE | Downregulated |
| -0,808160972 | 0,000876878 | ITIH1 | FALSE | Downregulated |
| 1,313972853 | 0,000880887 | COL5A1 | TRUE | Upregulated |
| 1,882852354 | 0,000896516 | ALK | TRUE | Upregulated |
| -0,742985351 | 0,000901608 | ANPEP | FALSE | Downregulated |
| -0,854617407 | 0,000901608 | GC | FALSE | Downregulated |
| -0,880729013 | 0,000960633 | HAAO | FALSE | Downregulated |
| -0,682950827 | 0,000960633 | KANK1 | FALSE | Downregulated |
| 1,004314856 | 0,000960633 | LOC153684 | TRUE | Upregulated |
| 1,372318609 | 0,000960633 | MDFIC | TRUE | Upregulated |
| -0,963132995 | 0,000965709 | HADH | FALSE | Downregulated |
| 1,00936074 | 0,000965709 | STK10 | TRUE | Upregulated |
| -0,947558229 | 0,000971172 | CHCHD10 | FALSE | Downregulated |
| -0,794855991 | 0,000992333 | ADCY1 | FALSE | Downregulated |
| 1,352394336 | 0,000992557 | SERAC1 | TRUE | Upregulated |
| -0,807466008 | 0,000996196 | ALAD | FALSE | Downregulated |
| 2,544999108 | 0,001047712 | LOC101928352 | TRUE | Upregulated |
| -0,96209822 | 0,001057926 | ACADSB | FALSE | Downregulated |
| 1,726185217 | 0,001057926 | C1orf195 | TRUE | Upregulated |
| -1,338249163 | 0,001057926 | CD82 | TRUE | Downregulated |
| -1,385616863 | 0,001057926 | CNDP1 | TRUE | Downregulated |
| 1,325623459 | 0,001057926 | DOCK2 | TRUE | Upregulated |
| 0,832839844 | 0,001058148 | CLOCK | FALSE | Upregulated |
| 1,278070229 | 0,001062372 | CERKL | TRUE | Upregulated |
| -1,009789205 | 0,001067611 | HP | TRUE | Downregulated |
| 1,435558574 | 0,001067611 | RP11-379J5.5 | TRUE | Upregulated |
| 1,104409356 | 0,001071247 | SPRTN | TRUE | Upregulated |
| 1,131504329 | 0,001078174 | ZNF124 | TRUE | Upregulated |
| 1,054324749 | 0,00109033 | ZNF469 | TRUE | Upregulated |
| 1,846139779 | 0,001105176 | SLC38A1 | TRUE | Upregulated |
| 1,686783427 | 0,001119339 | SSC5D | TRUE | Upregulated |
| 1,370092463 | 0,001119339 | TMEM132A | TRUE | Upregulated |
| -0,84668004 | 0,001145484 | CGN | FALSE | Downregulated |
| 1,592628407 | 0,001163472 | ENTPD1-AS1 | TRUE | Upregulated |
| 3,194425673 | 0,001179745 | F13A1 | TRUE | Upregulated |
| 0,991437439 | 0,001179745 | LOC102724814 | FALSE | Upregulated |
| -0,581661824 | 0,001179745 | RPL3 | FALSE | Downregulated |
| -1,031185779 | 0,001212695 | HAL | TRUE | Downregulated |
| -0,748195389 | 0,001294252 | PIK3R1 | FALSE | Downregulated |
| -1,019889167 | 0,00136474 | ATP5D | TRUE | Downregulated |
| 1,121124335 | 0,00136474 | EVA1B | TRUE | Upregulated |
| 2,723661546 | 0,00136474 | LOC101928491 | TRUE | Upregulated |
| 2,186425562 | 0,00136474 | ZNF80 | TRUE | Upregulated |
| 0,823756711 | 0,001366662 | TMEM14E | FALSE | Upregulated |
| -1,046645047 | 0,001395254 | PKLR | TRUE | Downregulated |
| 2,064755802 | 0,001401728 | RYR2 | TRUE | Upregulated |
| -0,987423353 | 0,001401728 | TMUB1 | FALSE | Downregulated |
| 2,649194714 | 0,001426538 | SEMA4A | TRUE | Upregulated |
| -1,006015053 | 0,001505014 | APCS | TRUE | Downregulated |
| -0,783594581 | 0,001540587 | LMAN1 | FALSE | Downregulated |

| | | | | |
|---|---|---|---|---|
| -0,716095905 | 0,001617282 | RPL37A | FALSE | Downregulated |
| 1,752714532 | 0,001636903 | LAMC3 | TRUE | Upregulated |
| -1,170779027 | 0,00163824 | ECSIT | TRUE | Downregulated |
| 2,026609664 | 0,001660242 | CD6 | TRUE | Upregulated |
| 0,914714831 | 0,001669545 | RP5-1142A6.2 | FALSE | Upregulated |
| -1,182169953 | 0,001711866 | THSD4 | TRUE | Downregulated |
| -1,208613106 | 0,001753649 | ORM1 | TRUE | Downregulated |
| 1,340150496 | 0,001782887 | ANXA2 | TRUE | Upregulated |
| 3,27211598 | 0,001782887 | BCAN | TRUE | Upregulated |
| 0,529571386 | 0,001782887 | TBC1D8 | FALSE | Upregulated |
| 1,954022664 | 0,001799761 | ANXA1 | TRUE | Upregulated |
| 1,805353409 | 0,00184127 | STK17B | TRUE | Upregulated |
| -0,91087001 | 0,001844977 | SPTBN2 | FALSE | Downregulated |
| 1,233904389 | 0,001897558 | LOC642441 | TRUE | Upregulated |
| 1,110038468 | 0,001914208 | PREX1 | TRUE | Upregulated |
| -0,65690569 | 0,001914208 | RNF130 | FALSE | Downregulated |
| 0,79255906 | 0,001921912 | MARCKS | FALSE | Upregulated |
| 2,130861575 | 0,001937539 | ITGBL1 | TRUE | Upregulated |
| 3,70003665 | 0,001937539 | LOC102724585 | TRUE | Upregulated |
| 1,461671804 | 0,001963354 | LBH | TRUE | Upregulated |
| -0,772249961 | 0,00199878 | AR | FALSE | Downregulated |
| 1,271056928 | 0,002006647 | THBS1 | TRUE | Upregulated |
| 1,544516736 | 0,002006952 | SRGN | TRUE | Upregulated |
| 1,709617448 | 0,002043833 | ANTXR1 | TRUE | Upregulated |
| 1,027331799 | 0,002043833 | GPR82 | TRUE | Upregulated |
| 3,001417828 | 0,002053635 | LOC101927071 | TRUE | Upregulated |
| 1,520824291 | 0,002062824 | NEDD4L | TRUE | Upregulated |
| -1,221182604 | 0,002121802 | ATF5 | TRUE | Downregulated |
| 2,563651379 | 0,00215148 | LOC101928760 | TRUE | Upregulated |
| -0,743475849 | 0,002168879 | ACAT1 | FALSE | Downregulated |
| 2,776493306 | 0,002206418 | SCML4 | TRUE | Upregulated |
| 1,564310231 | 0,002206418 | SOX4 | TRUE | Upregulated |
| 0,675559834 | 0,002206418 | TMEM184A | FALSE | Upregulated |
| 1,384522357 | 0,002234858 | LOC101929732 | TRUE | Upregulated |
| -0,712755148 | 0,002259388 | ACAD11 | FALSE | Downregulated |
| 1,470898497 | 0,002259388 | DOCK11 | TRUE | Upregulated |
| 1,737989475 | 0,002266994 | LINGO3 | TRUE | Upregulated |
| 1,607294793 | 0,002268962 | LINC00612 | TRUE | Upregulated |
| 3,091600536 | 0,002290885 | TNFRSF12A | TRUE | Upregulated |
| 1,018297392 | 0,002296259 | VIM | TRUE | Upregulated |
| 1,316697055 | 0,002316703 | C7 | TRUE | Upregulated |
| 0,725933223 | 0,002336659 | MAB21L2 | FALSE | Upregulated |
| -1,587029263 | 0,002353067 | MT2A | TRUE | Downregulated |
| -0,590663045 | 0,002354025 | SHMT2 | FALSE | Downregulated |
| 1,672467465 | 0,002360353 | LOC101927955 | TRUE | Upregulated |
| 1,121663957 | 0,002370711 | ABCC1 | TRUE | Upregulated |
| -1,71259881 | 0,002372783 | CACNA1H | TRUE | Downregulated |
| -0,900788504 | 0,002393902 | CES1 | FALSE | Downregulated |
| 0,928825789 | 0,002420003 | FLNA | FALSE | Upregulated |
| -0,826399735 | 0,002435195 | CXXC5 | FALSE | Downregulated |
| 1,617982717 | 0,002435195 | TCTEX1D4 | TRUE | Upregulated |

| | | | | |
|---|---|---|---|---|
| -0,729211593 | 0,002446323 | F11 | FALSE | Downregulated |
| 1,573597245 | 0,002446323 | MIAT | TRUE | Upregulated |
| -0,795377516 | 0,002449048 | GLUD1 | FALSE | Downregulated |
| 1,830445526 | 0,002449048 | VWF | TRUE | Upregulated |
| 2,448588623 | 0,00247992 | LOC102723604 | TRUE | Upregulated |
| -1,098747169 | 0,002495111 | ADRA1A | TRUE | Downregulated |
| 2,445079927 | 0,002495111 | GEM | TRUE | Upregulated |
| 3,583048243 | 0,002499685 | LOC101927751 | TRUE | Upregulated |
| 0,766834716 | 0,002499685 | SLC2A12 | FALSE | Upregulated |
| 2,73452359 | 0,002546228 | LAX1 | TRUE | Upregulated |
| 1,450716783 | 0,002562875 | IQCK | TRUE | Upregulated |
| 0,837704548 | 0,002574413 | RAVER2 | FALSE | Upregulated |
| -0,633435883 | 0,00258218 | CLU | FALSE | Downregulated |
| -1,759747037 | 0,00258218 | MT1E | TRUE | Downregulated |
| 1,8771033 | 0,002599356 | C17orf97 | TRUE | Upregulated |
| 1,653000471 | 0,002808164 | PRRT2 | TRUE | Upregulated |
| -0,779634121 | 0,00290988 | ST13 | FALSE | Downregulated |
| -0,759700659 | 0,002941501 | KHK | FALSE | Downregulated |
| 1,41471215 | 0,00299811 | WDFY4 | TRUE | Upregulated |
| 1,451335355 | 0,0030296 | LOC101929786 | TRUE | Upregulated |
| -0,711694687 | 0,00314995 | C16orf58 | FALSE | Downregulated |
| -0,994011432 | 0,003230611 | PANK1 | FALSE | Downregulated |
| 1,15263336 | 0,003238219 | PTPRC | TRUE | Upregulated |
| -1,100614359 | 0,003257886 | LINC01018 | TRUE | Downregulated |
| 0,812975121 | 0,003291457 | MINA | FALSE | Upregulated |
| 1,338669353 | 0,003291457 | SEL1L3 | TRUE | Upregulated |
| 1,680715352 | 0,003319852 | CDKN2B | TRUE | Upregulated |
| 2,265347435 | 0,003319852 | DCDC2 | TRUE | Upregulated |
| -1,064667486 | 0,003319852 | SH3PXD2A | TRUE | Downregulated |
| -0,698019879 | 0,003332448 | DUS1L | FALSE | Downregulated |
| 1,25204513 | 0,003378269 | C1orf200 | TRUE | Upregulated |
| 1,508544 | 0,003401495 | CRMP1 | TRUE | Upregulated |
| 2,60066559 | 0,003401495 | ENAM | TRUE | Upregulated |
| 2,696595144 | 0,003401495 | GFI1 | TRUE | Upregulated |
| 1,159079184 | 0,003425513 | LOC102724466 | TRUE | Upregulated |
| -0,596405685 | 0,003528513 | MAN1A1 | FALSE | Downregulated |
| 1,575590969 | 0,003542415 | DOK1 | TRUE | Upregulated |
| 1,789868583 | 0,00357376 | BIRC3 | TRUE | Upregulated |
| -0,717514659 | 0,00357376 | PHLPP1 | FALSE | Downregulated |
| 1,111694381 | 0,003604179 | GPR34 | TRUE | Upregulated |
| 1,367362437 | 0,003610328 | FAM98B | TRUE | Upregulated |
| 1,548801889 | 0,003610328 | IKZF3 | TRUE | Upregulated |
| -0,888363791 | 0,003610328 | PXMP2 | FALSE | Downregulated |
| 2,525005232 | 0,003614358 | GABRE | TRUE | Upregulated |
| -1,096587892 | 0,00361461 | LIPC | TRUE | Downregulated |
| -0,637784102 | 0,003637859 | SERPINF2 | FALSE | Downregulated |
| -0,86688341 | 0,003649412 | NTN4 | FALSE | Downregulated |
| -0,469953343 | 0,003668468 | RPL41 | FALSE | Downregulated |
| -0,808703055 | 0,003697932 | DBH-AS1 | FALSE | Downregulated |
| 0,639833688 | 0,003697932 | VASN | FALSE | Upregulated |
| 3,172875852 | 0,003705605 | CACNA1C-AS4 | TRUE | Upregulated |

| | | | | |
|---|---|---|---|---|
| -0,66042828 | 0,003705605 | DPYS | FALSE | Downregulated |
| 1,471257023 | 0,003705605 | HLA-DOA | TRUE | Upregulated |
| 1,149705238 | 0,003705605 | PDGFRA | TRUE | Upregulated |
| -0,634974103 | 0,003705605 | RPS5 | FALSE | Downregulated |
| 1,070704725 | 0,003705605 | SH3RF3-AS1 | TRUE | Upregulated |
| 1,488239489 | 0,003705605 | SNX20 | TRUE | Upregulated |
| 1,848051503 | 0,003705605 | ZAP70 | TRUE | Upregulated |
| -0,739066019 | 0,003725266 | TMEM56 | FALSE | Downregulated |
| 3,848089693 | 0,003761997 | AC002480.3 | TRUE | Upregulated |
| -1,225948513 | 0,003761997 | MME | TRUE | Downregulated |
| 1,6986069 | 0,003761997 | NRSN2 | TRUE | Upregulated |
| 0,762572219 | 0,003761997 | SESN3 | FALSE | Upregulated |
| -1,308229722 | 0,003770057 | CFHR1 | TRUE | Downregulated |
| -0,966607438 | 0,003770057 | NME4 | FALSE | Downregulated |
| 1,481800353 | 0,003785234 | MICAL1 | TRUE | Upregulated |
| -0,927462639 | 0,003792492 | HOMER2 | FALSE | Downregulated |
| -0,705895472 | 0,003795222 | APOC1 | FALSE | Downregulated |
| 2,715784184 | 0,003802051 | SPATA12 | TRUE | Upregulated |
| 3,078383704 | 0,003813218 | VGLL3 | TRUE | Upregulated |
| 2,575670397 | 0,003843919 | CD96 | TRUE | Upregulated |
| 0,821273071 | 0,003843919 | SLC35G2 | FALSE | Upregulated |
| 1,054813923 | 0,003849574 | HLA-DRA | TRUE | Upregulated |
| 0,833560329 | 0,003901332 | LSM4 | FALSE | Upregulated |
| -0,778575461 | 0,003902103 | ECHS1 | FALSE | Downregulated |
| 1,136295903 | 0,00391932 | COL4A2 | TRUE | Upregulated |
| 1,126308108 | 0,00394956 | HCP5 | TRUE | Upregulated |
| 1,060678785 | 0,00402243 | MGC16275 | TRUE | Upregulated |
| -0,799624026 | 0,004033683 | ACSL1 | FALSE | Downregulated |
| -0,622003863 | 0,004057295 | C1R | FALSE | Downregulated |
| 1,697140937 | 0,004057295 | DPYSL3 | TRUE | Upregulated |
| 2,427910539 | 0,004057295 | FOXC1 | TRUE | Upregulated |
| -0,771397644 | 0,004057295 | HSD17B4 | FALSE | Downregulated |
| 0,42929118 | 0,004057295 | MALAT1 | FALSE | Upregulated |
| -0,643005 | 0,004057295 | RBP4 | FALSE | Downregulated |
| -0,798150061 | 0,004057295 | SERPINA5 | FALSE | Downregulated |
| -1,140495814 | 0,004057295 | SLC30A10 | TRUE | Downregulated |
| 0,888229804 | 0,004057295 | TGFB1 | FALSE | Upregulated |
| 1,780265556 | 0,004057295 | TSC22D1-AS1 | TRUE | Upregulated |
| 1,537134758 | 0,004081151 | RP11-277P12.20 | TRUE | Upregulated |
| -0,66928451 | 0,004122929 | ECH1 | FALSE | Downregulated |
| 0,748865312 | 0,004131765 | B3GALT2 | FALSE | Upregulated |
| -0,45753892 | 0,004131765 | ZC3H7B | FALSE | Downregulated |
| 3,642765147 | 0,004166834 | ACAN | TRUE | Upregulated |
| 1,047481043 | 0,004173021 | BTN3A2 | TRUE | Upregulated |
| -1,580212924 | 0,004173021 | KCNN2 | TRUE | Downregulated |
| -0,510262896 | 0,00420426 | HDLBP | FALSE | Downregulated |
| 1,237781454 | 0,004293617 | VTI1A | TRUE | Upregulated |
| 2,507219136 | 0,004337066 | TTC9 | TRUE | Upregulated |
| 0,961696628 | 0,004369238 | LPHN1 | FALSE | Upregulated |
| 3,279356785 | 0,004417906 | LINC00707 | TRUE | Upregulated |
| 2,427064944 | 0,004493482 | SSPN | TRUE | Upregulated |

| | | | | |
|---|---|---|---|---|
| 0,783800915 | 0,004494627 | CAPN2 | FALSE | Upregulated |
| 1,172271357 | 0,004494627 | RELL2 | TRUE | Upregulated |
| 1,363775521 | 0,004507867 | LOC80154 | TRUE | Upregulated |
| -0,544013524 | 0,004557409 | NFIA | FALSE | Downregulated |
| 1,192023724 | 0,004557409 | TRNAU1AP | TRUE | Upregulated |
| 0,984102197 | 0,004557409 | ZC3H12B | FALSE | Upregulated |
| -0,627529257 | 0,00460482 | CAMSAP3 | FALSE | Downregulated |
| 1,983944526 | 0,004637094 | SLAMF7 | TRUE | Upregulated |
| -0,708843853 | 0,00466031 | ADI1 | FALSE | Downregulated |
| -0,701763127 | 0,004712562 | C8A | FALSE | Downregulated |
| 3,0845787 | 0,004712562 | SMOC2 | TRUE | Upregulated |
| -0,775055388 | 0,004712562 | TMEM176B | FALSE | Downregulated |
| -0,667521911 | 0,004712562 | USF2 | FALSE | Downregulated |
| 0,81241262 | 0,004712562 | ZNF562 | FALSE | Upregulated |
| 0,757232682 | 0,004784629 | COL4A4 | FALSE | Upregulated |
| 1,395467101 | 0,004835772 | MMEL1 | TRUE | Upregulated |
| 0,956829828 | 0,004856958 | CENPO | FALSE | Upregulated |
| -0,879573171 | 0,004856958 | F9 | FALSE | Downregulated |
| -2,664171003 | 0,004874893 | NCAM2 | TRUE | Downregulated |
| -0,874829864 | 0,004905418 | ENPEP | FALSE | Downregulated |
| -0,811058227 | 0,004905418 | TCEA3 | FALSE | Downregulated |
| -0,983774091 | 0,004917835 | ALDH1L1 | FALSE | Downregulated |
| 1,126213484 | 0,004946203 | TAP1 | TRUE | Upregulated |
| 0,602887168 | 0,005028382 | ATG7 | FALSE | Upregulated |
| 1,5346228 | 0,005028382 | EVC | TRUE | Upregulated |
| 2,161840422 | 0,005067003 | CCDC80 | TRUE | Upregulated |
| 0,896411162 | 0,005165308 | CLSTN1 | FALSE | Upregulated |
| 1,213770184 | 0,005280242 | LOC100132781 | TRUE | Upregulated |
| -1,547196424 | 0,005280242 | LYRM1 | TRUE | Downregulated |
| 1,069736167 | 0,005280242 | NFKB2 | TRUE | Upregulated |
| 0,782837881 | 0,005280242 | WDFY2 | FALSE | Upregulated |
| -1,079582526 | 0,005460912 | C11orf31 | TRUE | Downregulated |
| -1,097611848 | 0,00547018 | ASPG | TRUE | Downregulated |
| -1,142825274 | 0,00547018 | ERRFI1 | TRUE | Downregulated |
| -0,893499495 | 0,00547018 | KBTBD11 | FALSE | Downregulated |
| 0,742067503 | 0,00547018 | LOC100128818 | FALSE | Upregulated |
| 1,809203568 | 0,00547018 | PDK1 | TRUE | Upregulated |
| 1,561672009 | 0,005482073 | BCL2 | TRUE | Upregulated |
| 0,706604745 | 0,005482073 | OGT | FALSE | Upregulated |
| -0,90233434 | 0,005513512 | CYP4A11 | FALSE | Downregulated |
| 1,82377519 | 0,005525354 | LOC101928373 | TRUE | Upregulated |
| 2,036286785 | 0,005597984 | VCAN | TRUE | Upregulated |
| 0,656237462 | 0,005643889 | SLC26A1 | FALSE | Upregulated |
| 0,70276871 | 0,00569509 | NAV1 | FALSE | Upregulated |
| 1,295093277 | 0,005775762 | ABCC5 | TRUE | Upregulated |
| 0,974492703 | 0,005795433 | MICALL2 | FALSE | Upregulated |
| -0,883585479 | 0,005796985 | MFAP3L | FALSE | Downregulated |
| -0,977504853 | 0,005844795 | IGFALS | FALSE | Downregulated |
| 2,736026184 | 0,005844795 | LOC101927069 | TRUE | Upregulated |
| 0,627446986 | 0,005858235 | SEPT11 | FALSE | Upregulated |
| -0,801144671 | 0,005892624 | HPX | FALSE | Downregulated |

| | | | | |
|---|---|---|---|---|
| -0,751379345 | 0,005931446 | SARDH | FALSE | Downregulated |
| -0,752395213 | 0,005942151 | CDO1 | FALSE | Downregulated |
| -1,22948924 | 0,005942151 | RHEBP2 | TRUE | Downregulated |
| 1,684815919 | 0,006156944 | DZIP1 | TRUE | Upregulated |
| 0,767095818 | 0,006193953 | ACTR3 | FALSE | Upregulated |
| 2,292607278 | 0,006197999 | LOC101927596 | TRUE | Upregulated |
| -2,122297848 | 0,006203321 | GJB4 | TRUE | Downregulated |
| -0,528748695 | 0,006248355 | CNBP | FALSE | Downregulated |
| 1,197896443 | 0,006265517 | NFASC | TRUE | Upregulated |
| 0,823386259 | 0,006274379 | PCDH12 | FALSE | Upregulated |
| -0,839818944 | 0,00627968 | HSD17B6 | FALSE | Downregulated |
| -0,898631938 | 0,00630607 | GHR | FALSE | Downregulated |
| 1,681303592 | 0,006317444 | MTRNR2L8 | TRUE | Upregulated |
| -0,59768132 | 0,006323162 | IQGAP2 | FALSE | Downregulated |
| 1,793025986 | 0,006326149 | FUT8-AS1 | TRUE | Upregulated |
| 1,599628047 | 0,006370427 | PHYHIPL | TRUE | Upregulated |
| -1,452082679 | 0,006373766 | CLEC4M | TRUE | Downregulated |
| 1,556888894 | 0,006392205 | PIN4P1 | TRUE | Upregulated |
| 1,656333498 | 0,006392205 | PODN | TRUE | Upregulated |
| 3,26915546 | 0,00640849 | LOC102723537 | TRUE | Upregulated |
| 1,025346926 | 0,006489009 | SLC48A1 | TRUE | Upregulated |
| -0,896589525 | 0,006659315 | SLC22A1 | FALSE | Downregulated |
| 2,838060657 | 0,006667503 | IFNG | TRUE | Upregulated |
| 3,073314866 | 0,006694947 | PIP5KL1 | TRUE | Upregulated |
| 0,674014367 | 0,006734439 | ZNF233 | FALSE | Upregulated |
| 2,260292941 | 0,006787332 | C6orf147 | TRUE | Upregulated |
| -0,824514537 | 0,006787332 | CTSF | FALSE | Downregulated |
| 1,122650854 | 0,006787332 | FBN1 | TRUE | Upregulated |
| -0,695155108 | 0,006793281 | EPHX2 | FALSE | Downregulated |
| -0,802206706 | 0,006826002 | SLC39A5 | FALSE | Downregulated |
| -0,715089547 | 0,006877399 | MAOA | FALSE | Downregulated |
| 1,845324855 | 0,006907773 | KCNS1 | TRUE | Upregulated |
| 2,354755242 | 0,006907773 | LOC101927255 | TRUE | Upregulated |
| -0,580989977 | 0,006951305 | ITIH4 | FALSE | Downregulated |
| -0,526902294 | 0,006951305 | SIGMAR1 | FALSE | Downregulated |
| 2,244864721 | 0,006958953 | LOC101927256 | TRUE | Upregulated |
| -0,772934654 | 0,006977082 | C8B | FALSE | Downregulated |
| -0,60212362 | 0,006981927 | PSMB7 | FALSE | Downregulated |
| -0,62706677 | 0,007033377 | SERPINF1 | FALSE | Downregulated |
| 1,112252554 | 0,007156157 | LOC728805 | TRUE | Upregulated |
| -0,599576518 | 0,007159047 | GLS2 | FALSE | Downregulated |
| 0,985388028 | 0,007161028 | LOC400553 | FALSE | Upregulated |
| 3,186416548 | 0,007161028 | MUSK | TRUE | Upregulated |
| 0,627873748 | 0,007161028 | RNF19A | FALSE | Upregulated |
| -0,619421689 | 0,007239616 | AZGP1 | FALSE | Downregulated |
| -2,754314908 | 0,007274592 | DNAJC12 | TRUE | Downregulated |
| 0,580885163 | 0,00728276 | JUND | FALSE | Upregulated |
| 2,326182576 | 0,007362006 | C16orf54 | TRUE | Upregulated |
| -1,820088461 | 0,007371192 | CCL14 | TRUE | Downregulated |
| 1,52325343 | 0,007404069 | ITGAM | TRUE | Upregulated |
| 1,179818677 | 0,00749908 | COL4A1 | TRUE | Upregulated |

| | | | | |
|---|---|---|---|---|
| 1,138169536 | 0,00749908 | CTB-113P19.1 | TRUE | Upregulated |
| 1,121382673 | 0,00749908 | MRC2 | TRUE | Upregulated |
| 1,085390211 | 0,00749908 | NLRC5 | TRUE | Upregulated |
| -0,794724739 | 0,00749908 | SLC27A2 | FALSE | Downregulated |
| -0,558399571 | 0,007499524 | SLC9A3R2 | FALSE | Downregulated |
| 1,423647223 | 0,007548149 | PAQR8 | TRUE | Upregulated |
| -0,875400789 | 0,007548149 | SORD | FALSE | Downregulated |
| 1,669881605 | 0,007565072 | ZMAT3 | TRUE | Upregulated |
| 1,392069325 | 0,007670825 | FRMD5 | TRUE | Upregulated |
| 1,748350278 | 0,007670825 | PRF1 | TRUE | Upregulated |
| -0,996116489 | 0,007784982 | MDN1 | FALSE | Downregulated |
| -1,616059464 | 0,007865033 | AOC4P | TRUE | Downregulated |
| -0,878629313 | 0,007865033 | ETFDH | FALSE | Downregulated |
| 0,736869448 | 0,007865033 | VAMP1 | FALSE | Upregulated |
| -0,801816335 | 0,007950858 | GCKR | FALSE | Downregulated |
| -0,650949484 | 0,008049622 | PLG | FALSE | Downregulated |
| 3,157244908 | 0,0080615 | RUNX1-IT1 | TRUE | Upregulated |
| 0,792253105 | 0,008114335 | LOC440300 | FALSE | Upregulated |
| -1,57393097 | 0,00817191 | MT1X | TRUE | Downregulated |
| 0,771849853 | 0,008216679 | KLHL5 | FALSE | Upregulated |
| 0,915799133 | 0,008243573 | PRC1 | FALSE | Upregulated |
| 1,872581225 | 0,008243573 | SLC9A5 | TRUE | Upregulated |
| 1,265797937 | 0,008257124 | HLA-F | TRUE | Upregulated |
| 0,67659561 | 0,008257124 | NUTM2G | FALSE | Upregulated |
| 0,895025321 | 0,008257124 | SPON2 | FALSE | Upregulated |
| -1,02592237 | 0,008404614 | APOA5 | TRUE | Downregulated |
| 1,856056431 | 0,008404614 | FBLN1 | TRUE | Upregulated |
| -1,475331381 | 0,008404614 | LINC00671 | TRUE | Downregulated |
| 1,074095805 | 0,008404614 | SLC7A5P2 | TRUE | Upregulated |
| -0,994319275 | 0,008493027 | FAHD2A | FALSE | Downregulated |
| 4,568789853 | 0,008588342 | AKR1B10 | TRUE | Upregulated |
| -0,653730473 | 0,008588342 | COMT | FALSE | Downregulated |
| -0,587704209 | 0,008659936 | AQP9 | FALSE | Downregulated |
| -0,887121575 | 0,008659936 | COX5B | FALSE | Downregulated |
| 2,71002498 | 0,008706962 | POU2AF1 | TRUE | Upregulated |
| -0,646933337 | 0,00875766 | BCKDHA | FALSE | Downregulated |
| 0,81015523 | 0,008762848 | LOC101927178 | FALSE | Upregulated |
| -0,816802384 | 0,008793124 | ADH1B | FALSE | Downregulated |
| 1,240997376 | 0,008793124 | FCHSD1 | TRUE | Upregulated |
| 0,717861663 | 0,008793124 | LOC644083 | FALSE | Upregulated |
| 1,088697588 | 0,008793124 | PPP1R16B | TRUE | Upregulated |
| -0,657488397 | 0,008812012 | IRS1 | FALSE | Downregulated |
| 0,738019516 | 0,008812012 | TMEM181 | FALSE | Upregulated |
| -1,393240789 | 0,008825408 | OCEL1 | TRUE | Downregulated |
| 0,537017991 | 0,008842449 | DTX4 | FALSE | Upregulated |
| 0,990708493 | 0,008864951 | FAT2 | FALSE | Upregulated |
| 2,270398403 | 0,008864951 | LOC100132891 | TRUE | Upregulated |
| -0,63297046 | 0,008893751 | REXO1 | FALSE | Downregulated |
| 1,181078225 | 0,008902043 | CBFB | TRUE | Upregulated |
| 0,813950818 | 0,008934361 | LOC100506603 | FALSE | Upregulated |
| -1,702234961 | 0,008934361 | NDUFB4 | TRUE | Downregulated |

| | | | | |
|---|---|---|---|---|
| -0,766996265 | 0,008934361 | NR1H3 | FALSE | Downregulated |
| 1,247157278 | 0,009105799 | TIMP1 | TRUE | Upregulated |
| 0,999803502 | 0,009197003 | PPAPDC2 | FALSE | Upregulated |
| -0,719189566 | 0,009214365 | EGFR | FALSE | Downregulated |
| 2,561958614 | 0,00922475 | CAP2 | TRUE | Upregulated |
| 0,634304275 | 0,00922475 | GUCD1 | FALSE | Upregulated |
| -0,995792713 | 0,00922475 | RORC | FALSE | Downregulated |
| 1,098381226 | 0,00922475 | TTC33 | TRUE | Upregulated |
| 0,945294771 | 0,009340022 | FAS-AS1 | FALSE | Upregulated |
| 1,890960848 | 0,009395271 | OSBPL3 | TRUE | Upregulated |
| -0,575634275 | 0,009420296 | PGM1 | FALSE | Downregulated |
| 0,851545236 | 0,009557578 | LOC101929240 | FALSE | Upregulated |
| 0,621987011 | 0,009559106 | NPC1 | FALSE | Upregulated |
| 2,596097503 | 0,009579573 | CACNA1C-AS1 | TRUE | Upregulated |
| 1,966937664 | 0,00961305 | BICC1 | TRUE | Upregulated |
| 1,1198918 | 0,00961305 | KCTD10 | TRUE | Upregulated |
| 1,432447286 | 0,00961305 | LMOD3 | TRUE | Upregulated |
| 1,316272224 | 0,00961305 | LOC100507346 | TRUE | Upregulated |
| 0,734465705 | 0,00961305 | PPAP2A | FALSE | Upregulated |
| -0,543086231 | 0,00980662 | NFIC | FALSE | Downregulated |
| 2,327912218 | 0,00980662 | PKMYT1 | TRUE | Upregulated |
| -0,854593008 | 0,00980662 | SLC37A4 | FALSE | Downregulated |
| -0,898461021 | 0,009870082 | XYLB | FALSE | Downregulated |
| 1,018412877 | 0,009877428 | LOC101928268 | TRUE | Upregulated |
| 2,389189607 | 0,009877428 | LOC101928409 | TRUE | Upregulated |
| 1,698491023 | 0,009882901 | LOC100129697 | TRUE | Upregulated |
| 1,165558043 | 0,009939696 | MAP3K19 | TRUE | Upregulated |
| -0,916010166 | 0,009940857 | C4B | FALSE | Downregulated |
| 0,748594689 | 0,009940857 | PYROXD1 | FALSE | Upregulated |
| 0,916123644 | 0,009968705 | ENO4 | FALSE | Upregulated |
| 1,429326709 | 0,010123211 | GPNMB | TRUE | Upregulated |
| -0,814450183 | 0,010123211 | NR1I2 | FALSE | Downregulated |
| 3,168929148 | 0,010140231 | AC145110.1 | TRUE | Upregulated |
| -0,528540846 | 0,010149533 | RPL8 | FALSE | Downregulated |
| 0,925957467 | 0,010179132 | ZNF497 | FALSE | Upregulated |
| 0,682547532 | 0,010235007 | FMNL2 | FALSE | Upregulated |
| -1,343438384 | 0,01028266 | TTLL4 | TRUE | Downregulated |
| 0,751290353 | 0,010361242 | IQCC | FALSE | Upregulated |
| 0,876166768 | 0,010394565 | FLJ32756 | FALSE | Upregulated |
| 1,820154403 | 0,010444506 | IL7R | TRUE | Upregulated |
| 1,875829013 | 0,010607128 | APBA2 | TRUE | Upregulated |
| 2,541351959 | 0,010607128 | FER1L4 | TRUE | Upregulated |
| 1,485044044 | 0,010607128 | LOC101928926 | TRUE | Upregulated |
| 2,148654132 | 0,010607128 | PDCD1 | TRUE | Upregulated |
| 2,925465325 | 0,010610884 | LOC100996318 | TRUE | Upregulated |
| 1,480148176 | 0,010657997 | PIGZ | TRUE | Upregulated |
| 0,875831052 | 0,010698131 | SAMHD1 | FALSE | Upregulated |
| 1,411677872 | 0,010714224 | GLI1 | TRUE | Upregulated |
| -1,243462785 | 0,010714224 | SLC19A2 | TRUE | Downregulated |
| -1,219670675 | 0,010720631 | RNF152 | TRUE | Downregulated |
| 0,762798203 | 0,010887866 | LOC101928120 | FALSE | Upregulated |

| | | | | |
|---|---|---|---|---|
| 0,66898877 | 0,010914821 | PLXNC1 | FALSE | Upregulated |
| 0,640586353 | 0,010941829 | RHCE | FALSE | Upregulated |
| -1,142228328 | 0,011045487 | PCK1 | TRUE | Downregulated |
| -0,738054995 | 0,011122959 | ACAA2 | FALSE | Downregulated |
| 1,19550743 | 0,011122959 | LOC101928240 | TRUE | Upregulated |
| 1,605395562 | 0,011208642 | BTG2 | TRUE | Upregulated |
| 1,326805575 | 0,011224367 | KIRREL | TRUE | Upregulated |
| 1,530671831 | 0,011377654 | FAM53B-AS1 | TRUE | Upregulated |
| -0,564559355 | 0,011554763 | VTN | FALSE | Downregulated |
| 0,991318408 | 0,01157188 | LOC100506497 | FALSE | Upregulated |
| 1,784348668 | 0,011574345 | PSTPIP1 | TRUE | Upregulated |
| 1,268057169 | 0,011692796 | SLA | TRUE | Upregulated |
| -0,858720178 | 0,011698992 | FAM213A | FALSE | Downregulated |
| 1,742847286 | 0,011698992 | IL24 | TRUE | Upregulated |
| 0,935922527 | 0,011723104 | IFNK | FALSE | Upregulated |
| 0,739167803 | 0,01175123 | ATP8B1 | FALSE | Upregulated |
| -0,487276548 | 0,011798149 | AKT2 | FALSE | Downregulated |
| 1,726612477 | 0,011798149 | KIAA1211 | TRUE | Upregulated |
| 1,166620231 | 0,01188502 | MMP25 | TRUE | Upregulated |
| 0,822524962 | 0,01188502 | SEMA4D | FALSE | Upregulated |
| 1,379290301 | 0,011949416 | FAM212A | TRUE | Upregulated |
| 3,321835976 | 0,011949416 | RASL12 | TRUE | Upregulated |
| 1,468585069 | 0,011949416 | SLC52A1 | TRUE | Upregulated |
| 0,746725085 | 0,011949416 | XRCC3 | FALSE | Upregulated |
| 0,689763544 | 0,012083914 | MAP3K3 | FALSE | Upregulated |
| 1,199932115 | 0,012098865 | LOC100507156 | TRUE | Upregulated |
| -0,842371286 | 0,012098865 | PCBD1 | FALSE | Downregulated |
| 1,399411056 | 0,012098865 | STX19 | TRUE | Upregulated |
| 2,293324447 | 0,01218172 | RP5-1027G4.3 | TRUE | Upregulated |
| 1,655184208 | 0,012212089 | COL16A1 | TRUE | Upregulated |
| 0,730190841 | 0,012336515 | MSN | FALSE | Upregulated |
| -0,673019872 | 0,0123834 | LCAT | FALSE | Downregulated |
| -0,728464451 | 0,012398156 | POFUT1 | FALSE | Downregulated |
| 1,038598378 | 0,012472251 | CCDC88B | TRUE | Upregulated |
| -0,574885454 | 0,012496694 | MPST | FALSE | Downregulated |
| -0,688028473 | 0,012557414 | AGXT | FALSE | Downregulated |
| -1,044709988 | 0,012594035 | DGAT2 | TRUE | Downregulated |
| -0,987613009 | 0,012638048 | TTR | FALSE | Downregulated |
| 0,65721469 | 0,012739559 | ENTPD7 | FALSE | Upregulated |
| -0,974338757 | 0,012739559 | HMGCS2 | FALSE | Downregulated |
| -1,083742746 | 0,012787438 | ANKRD24 | TRUE | Downregulated |
| 0,833775074 | 0,012920655 | LOC653513 | FALSE | Upregulated |
| 1,517053738 | 0,012967408 | CD248 | TRUE | Upregulated |
| -0,649819462 | 0,013265913 | PBLD | FALSE | Downregulated |
| -0,613883081 | 0,013265913 | PLIN5 | FALSE | Downregulated |
| 0,604960619 | 0,013265913 | RHOQ | FALSE | Upregulated |
| -0,508571961 | 0,013277177 | UBXN6 | FALSE | Downregulated |
| -1,383580556 | 0,013473128 | UQCRB | TRUE | Downregulated |
| 1,249266117 | 0,013490283 | CFD | TRUE | Upregulated |
| -0,750893712 | 0,013591787 | IRS2 | FALSE | Downregulated |
| -0,805732504 | 0,013591787 | STAB2 | FALSE | Downregulated |

| | | | | |
|---|---|---|---|---|
| -0,518337029 | 0,013653096 | RPL36 | FALSE | Downregulated |
| -0,477087954 | 0,013760337 | EIF4EBP2 | FALSE | Downregulated |
| -0,738322524 | 0,013760337 | NUPR1 | FALSE | Downregulated |
| 1,82878617 | 0,013787053 | KPNA5 | TRUE | Upregulated |
| 0,992618177 | 0,013787053 | SUV39H2 | FALSE | Upregulated |
| 0,925848954 | 0,01383244 | ITGA7 | FALSE | Upregulated |
| -0,621000639 | 0,01383244 | UQCRC1 | FALSE | Downregulated |
| 0,66914244 | 0,014034218 | ZNF544 | FALSE | Upregulated |
| -0,377248408 | 0,014084993 | TPT1 | FALSE | Downregulated |
| -0,840893918 | 0,014100247 | BHMT | FALSE | Downregulated |
| 1,37807503 | 0,014162096 | FAM111B | TRUE | Upregulated |
| 0,97389651 | 0,014162096 | LAPTM5 | FALSE | Upregulated |
| -0,906790314 | 0,014162096 | MARC1 | FALSE | Downregulated |
| 0,845529838 | 0,014162096 | SMURF2 | FALSE | Upregulated |
| 2,213729063 | 0,014220098 | CD24 | TRUE | Upregulated |
| -0,99642887 | 0,014220098 | DNASE1L3 | FALSE | Downregulated |
| 0,971623005 | 0,014220098 | JAK2 | FALSE | Upregulated |
| -0,846495139 | 0,014220098 | PRDX6 | FALSE | Downregulated |
| 1,53716864 | 0,014220098 | SYNGR1 | TRUE | Upregulated |
| 1,012059493 | 0,01426327 | ITGB2 | TRUE | Upregulated |
| -1,80494518 | 0,014318188 | HAPLN4 | TRUE | Downregulated |
| 1,237810323 | 0,014339862 | GRIP1 | TRUE | Upregulated |
| -0,576418845 | 0,014429302 | AHSG | FALSE | Downregulated |
| 1,087394153 | 0,014632395 | FLJ13224 | TRUE | Upregulated |
| 1,107398536 | 0,014647563 | LSP1 | TRUE | Upregulated |
| -1,044010801 | 0,014895525 | RHEB | TRUE | Downregulated |
| 0,607518079 | 0,014918679 | FANCC | FALSE | Upregulated |
| 3,068832432 | 0,014939296 | SPDL1 | TRUE | Upregulated |
| 0,599844939 | 0,014949664 | ZNF276 | FALSE | Upregulated |
| -0,630198287 | 0,015005278 | APOA1 | FALSE | Downregulated |
| 1,139572551 | 0,015005278 | LOC729683 | TRUE | Upregulated |
| 1,259365109 | 0,015005278 | UQCRBP1 | TRUE | Upregulated |
| 1,207843929 | 0,015005424 | CHRNG | TRUE | Upregulated |
| -2,850051036 | 0,015005424 | GOLGA6A | TRUE | Downregulated |
| 0,450948325 | 0,015008992 | EPB41 | FALSE | Upregulated |
| -0,633822424 | 0,015086435 | AMBP | FALSE | Downregulated |
| 1,829862446 | 0,015086435 | FCRL6 | TRUE | Upregulated |
| -0,872791331 | 0,015241528 | ACADM | FALSE | Downregulated |
| -0,685883175 | 0,015241528 | COX6A1 | FALSE | Downregulated |
| -0,548138187 | 0,015241528 | FKBP8 | FALSE | Downregulated |
| 1,274118217 | 0,015241528 | NFAM1 | TRUE | Upregulated |
| 1,179998017 | 0,015250489 | MAP1B | TRUE | Upregulated |
| -0,446598882 | 0,015263071 | CERS2 | FALSE | Downregulated |
| 1,508335064 | 0,01537771 | MYO1G | TRUE | Upregulated |
| 0,734819499 | 0,01544153 | RP11-231I16.1 | FALSE | Upregulated |
| 0,948413008 | 0,015451612 | SYTL4 | FALSE | Upregulated |
| -0,581732798 | 0,015621277 | KLC4 | FALSE | Downregulated |
| 0,868864817 | 0,015668409 | LTBP2 | FALSE | Upregulated |
| 0,574748018 | 0,015757667 | ADCY7 | FALSE | Upregulated |
| 1,682577973 | 0,015769891 | SLC16A5 | TRUE | Upregulated |
| 1,048197834 | 0,015803229 | AFAP1-AS1 | TRUE | Upregulated |

| | | | | |
|---|---|---|---|---|
| 1,058968923 | 0,015803229 | DBNDD1 | TRUE | Upregulated |
| -0,705750449 | 0,015908633 | ADAMTS13 | FALSE | Downregulated |
| 1,99489741 | 0,015958212 | EVC2 | TRUE | Upregulated |
| 1,745560248 | 0,015958212 | STARD3NL | TRUE | Upregulated |
| -0,401316169 | 0,015979337 | COL18A1 | FALSE | Downregulated |
| -0,541811318 | 0,015983036 | SERPINA1 | FALSE | Downregulated |
| 0,730402793 | 0,016000064 | PPT1 | FALSE | Upregulated |
| 1,119515467 | 0,016047232 | PKM | TRUE | Upregulated |
| -0,408424542 | 0,016053885 | PCBP2 | FALSE | Downregulated |
| -0,84738516 | 0,016070138 | ORM2 | FALSE | Downregulated |
| -0,983601727 | 0,01609112 | PEG3 | FALSE | Downregulated |
| 1,090069143 | 0,016149932 | LOC101928502 | TRUE | Upregulated |
| -1,065095557 | 0,016201874 | CYP2E1 | TRUE | Downregulated |
| 1,550881521 | 0,016201874 | FAM46C | TRUE | Upregulated |
| -0,882891064 | 0,016294396 | MRPL38 | FALSE | Downregulated |
| 1,940910497 | 0,016307997 | GPR176 | TRUE | Upregulated |
| 0,636689275 | 0,016307997 | LRRC37A4P | FALSE | Upregulated |
| -1,072286573 | 0,016414774 | CLEC4G | TRUE | Downregulated |
| 1,135671861 | 0,016414774 | CLUAP1 | TRUE | Upregulated |
| -0,648218975 | 0,016414774 | CYP4F3 | FALSE | Downregulated |
| 0,71227158 | 0,016414774 | ZBTB38 | FALSE | Upregulated |
| 1,235606866 | 0,016449579 | ZNF589 | TRUE | Upregulated |
| -0,80525087 | 0,016541854 | FOXA1 | FALSE | Downregulated |
| 1,546829248 | 0,016541854 | SOX9 | TRUE | Upregulated |
| 0,729441932 | 0,016568561 | LIMD2 | FALSE | Upregulated |
| -0,706416503 | 0,016720027 | COBLL1 | FALSE | Downregulated |
| -0,670014104 | 0,016720027 | KIAA1217 | FALSE | Downregulated |
| 1,990645729 | 0,016720027 | LOXL1 | TRUE | Upregulated |
| 0,892435241 | 0,016720027 | MCTP1 | FALSE | Upregulated |
| 1,00483034 | 0,016720027 | SCXB | TRUE | Upregulated |
| 3,152358137 | 0,016772178 | WISP2 | TRUE | Upregulated |
| -0,659315559 | 0,016837838 | KLKB1 | FALSE | Downregulated |
| 0,589797959 | 0,016908143 | SLC7A6 | FALSE | Upregulated |
| 1,033687714 | 0,017031668 | C3orf62 | TRUE | Upregulated |
| 0,726803189 | 0,017031668 | NRON | FALSE | Upregulated |
| -0,772264533 | 0,017031668 | SLC38A4 | FALSE | Downregulated |
| -1,015477108 | 0,01708689 | AKR1D1 | TRUE | Downregulated |
| 1,314328317 | 0,017196828 | C14orf79 | TRUE | Upregulated |
| 3,126377309 | 0,0172748 | DLGAP1-AS5 | TRUE | Upregulated |
| -0,762278806 | 0,01728801 | PNKD | FALSE | Downregulated |
| 1,099322575 | 0,017304846 | LOC101928212 | TRUE | Upregulated |
| 1,415756023 | 0,017381089 | LOC100506985 | TRUE | Upregulated |
| 1,146434705 | 0,017439558 | ELP5 | TRUE | Upregulated |
| -1,233065332 | 0,017483548 | NYNRIN | TRUE | Downregulated |
| -0,40369015 | 0,01750559 | EEF2 | FALSE | Downregulated |
| 1,265434561 | 0,017513057 | LOC100287934 | TRUE | Upregulated |
| 1,391679464 | 0,017522227 | GAS7 | TRUE | Upregulated |
| 1,083015297 | 0,017559697 | KLF2 | TRUE | Upregulated |
| 0,573610297 | 0,017592499 | CIITA | FALSE | Upregulated |
| -0,632869895 | 0,017617377 | DCAF11 | FALSE | Downregulated |
| -0,780062619 | 0,017641 | IYD | FALSE | Downregulated |

| | | | | |
|---|---|---|---|---|
| -0,544182377 | 0,01774418 | APOB | FALSE | Downregulated |
| 1,614486246 | 0,01774418 | HELB | TRUE | Upregulated |
| -1,366498204 | 0,01774418 | LINC01093 | TRUE | Downregulated |
| 1,968256022 | 0,017758574 | GPR132 | TRUE | Upregulated |
| 0,898290876 | 0,017946933 | BOLA3-AS1 | FALSE | Upregulated |
| 0,8186173 | 0,017946933 | C5orf66 | FALSE | Upregulated |
| 1,393011766 | 0,017946933 | CLEC2D | TRUE | Upregulated |
| -0,587632516 | 0,017946933 | EPHB4 | FALSE | Downregulated |
| -1,25621698 | 0,017946933 | GNMT | TRUE | Downregulated |
| -0,738166964 | 0,017946933 | PTGR1 | FALSE | Downregulated |
| 1,334783657 | 0,018006658 | AHNAK2 | TRUE | Upregulated |
| 0,573738436 | 0,0180329 | APPL2 | FALSE | Upregulated |
| 1,036569129 | 0,0180329 | IFI16 | TRUE | Upregulated |
| 1,496051241 | 0,018124196 | DPT | TRUE | Upregulated |
| 1,016400301 | 0,018124196 | FEZ2 | TRUE | Upregulated |
| 2,019479295 | 0,018124196 | LOC101929687 | TRUE | Upregulated |
| -0,582089764 | 0,018148955 | COBL | FALSE | Downregulated |
| 0,828375206 | 0,018175347 | ASAP1 | FALSE | Upregulated |
| 1,081412563 | 0,01824134 | OGN | TRUE | Upregulated |
| 1,039559754 | 0,018318786 | DOK6 | TRUE | Upregulated |
| 0,661901364 | 0,018318786 | M6PR | FALSE | Upregulated |
| -0,626807573 | 0,018351481 | MPC2 | FALSE | Downregulated |
| 1,263324049 | 0,018457394 | SFXN3 | TRUE | Upregulated |
| -0,656470636 | 0,018483856 | PGRMC1 | FALSE | Downregulated |
| 0,935829597 | 0,018504767 | INPP5D | FALSE | Upregulated |
| 0,719146525 | 0,018504767 | PCSK7 | FALSE | Upregulated |
| 1,605082049 | 0,018504767 | POU5F1 | TRUE | Upregulated |
| 0,949493338 | 0,018558314 | ANGPTL2 | FALSE | Upregulated |
| -0,614004687 | 0,018878471 | ACSL5 | FALSE | Downregulated |
| 1,065671227 | 0,018878471 | MYOF | TRUE | Upregulated |
| -1,02120576 | 0,018880797 | HSD17B8 | TRUE | Downregulated |
| 1,239002685 | 0,018880797 | LOC285762 | TRUE | Upregulated |
| 1,288304122 | 0,018928233 | LOC101928512 | TRUE | Upregulated |
| 1,014539403 | 0,018930619 | GJA9-MYCBP | TRUE | Upregulated |
| 1,881083754 | 0,019061214 | LMTK3 | TRUE | Upregulated |
| -0,51279171 | 0,019084781 | FAM20C | FALSE | Downregulated |
| 2,109199592 | 0,01908487 | TNFRSF11B | TRUE | Upregulated |
| -1,015260002 | 0,019156426 | SLCO1B3 | TRUE | Downregulated |
| 0,78500146 | 0,01923489 | CSTF2T | FALSE | Upregulated |
| 1,916987123 | 0,019590039 | LINC00957 | TRUE | Upregulated |
| 0,697123325 | 0,019682958 | GSN-AS1 | FALSE | Upregulated |
| 2,706192483 | 0,019682958 | NMNAT2 | TRUE | Upregulated |
| -0,756167777 | 0,019682958 | PRG4 | FALSE | Downregulated |
| -0,469205522 | 0,019682958 | RPL14 | FALSE | Downregulated |
| -0,698856099 | 0,019682958 | ZNF395 | FALSE | Downregulated |
| -1,06465702 | 0,019751033 | ARSB | TRUE | Downregulated |
| 1,552167692 | 0,019767468 | PIM2 | TRUE | Upregulated |
| 1,363054834 | 0,019798025 | FRMD6 | TRUE | Upregulated |
| 1,387642653 | 0,020025923 | PLEKHG5 | TRUE | Upregulated |
| 2,339682327 | 0,020257364 | CFTR | TRUE | Upregulated |
| -0,526128364 | 0,020257364 | CRAT | FALSE | Downregulated |

| | | | | |
|---|---|---|---|---|
| 0,689712611 | 0,020311271 | LPAR6 | FALSE | Upregulated |
| 1,389406949 | 0,020343672 | CHRNA2 | TRUE | Upregulated |
| -0,433994578 | 0,020348677 | CDC42BPB | FALSE | Downregulated |
| 1,222913142 | 0,020348677 | TAP2 | TRUE | Upregulated |
| 1,903304693 | 0,020456426 | SEPT1 | TRUE | Upregulated |
| 1,753129649 | 0,020484166 | ATP10A | TRUE | Upregulated |
| 2,860030579 | 0,02070049 | LOC100996535 | TRUE | Upregulated |
| 0,550489481 | 0,020703991 | LOC100379224 | FALSE | Upregulated |
| 1,172338168 | 0,020924578 | ZNF473 | TRUE | Upregulated |
| 1,386434228 | 0,020967167 | GBP5 | TRUE | Upregulated |
| -0,618453471 | 0,021025637 | MCFD2 | FALSE | Downregulated |
| -1,564785569 | 0,021179198 | IGSF9 | TRUE | Downregulated |
| 0,708295592 | 0,021232462 | MCMBP | FALSE | Upregulated |
| -0,607950307 | 0,021232462 | PHKA2 | FALSE | Downregulated |
| 1,132822674 | 0,021234616 | EVA1C | TRUE | Upregulated |
| 1,01492919 | 0,021234616 | PRKAG1 | TRUE | Upregulated |
| 2,998211173 | 0,021402522 | LOC100996758 | TRUE | Upregulated |
| -0,902404767 | 0,021447712 | FBXO17 | FALSE | Downregulated |
| 1,290430946 | 0,021527858 | GPRIN3 | TRUE | Upregulated |
| -0,523150898 | 0,02155624 | AKAP1 | FALSE | Downregulated |
| -1,061392962 | 0,02155624 | EXPH5 | TRUE | Downregulated |
| 1,752522509 | 0,02155624 | PTAFR | TRUE | Upregulated |
| 0,581165069 | 0,02155624 | RGL4 | FALSE | Upregulated |
| 0,65971071 | 0,021658918 | FAM45A | FALSE | Upregulated |
| -0,485751086 | 0,021688269 | MAN2C1 | FALSE | Downregulated |
| 0,789760332 | 0,021688269 | PIK3IP1 | FALSE | Upregulated |
| 1,188253649 | 0,021702743 | ABCA10 | TRUE | Upregulated |
| 1,317592714 | 0,021739183 | CRISPLD2 | TRUE | Upregulated |
| 0,637071813 | 0,021739183 | VGLL4 | FALSE | Upregulated |
| 0,544468869 | 0,021789595 | CSNK1D | FALSE | Upregulated |
| 0,578755363 | 0,021789595 | LRTOMT | FALSE | Upregulated |
| 0,732028777 | 0,021842492 | PKD2L2 | FALSE | Upregulated |
| -0,799131284 | 0,021918882 | CFHR2 | FALSE | Downregulated |
| 1,739543255 | 0,021966251 | LOC101927308 | TRUE | Upregulated |
| 0,581189235 | 0,022096713 | SPNS2 | FALSE | Upregulated |
| -0,914348114 | 0,022172073 | BCHE | FALSE | Downregulated |
| 0,953576771 | 0,022185003 | TRAF1 | FALSE | Upregulated |
| 0,737799311 | 0,02222488 | SPDYE6 | FALSE | Upregulated |
| -0,936780819 | 0,022288415 | PRLR | FALSE | Downregulated |
| 0,823170876 | 0,022356742 | GPR68 | FALSE | Upregulated |
| 1,162844311 | 0,022360386 | TRIM22 | TRUE | Upregulated |
| 0,697080642 | 0,022464604 | ADPGK | FALSE | Upregulated |
| 2,413283832 | 0,022592251 | NALCN-AS1 | TRUE | Upregulated |
| -0,890784387 | 0,022748303 | RNF5 | FALSE | Downregulated |
| 0,930009029 | 0,022782645 | CDC25B | FALSE | Upregulated |
| -0,845323745 | 0,022791721 | PDK4 | FALSE | Downregulated |
| 1,733136926 | 0,02283585 | RCOR2 | TRUE | Upregulated |
| 0,691922812 | 0,02283585 | RECQL | FALSE | Upregulated |
| 1,721321025 | 0,022871421 | HSPA7 | TRUE | Upregulated |
| -1,926974722 | 0,022932463 | TPPP2 | TRUE | Downregulated |
| 0,935425254 | 0,022972413 | LOC100996740 | FALSE | Upregulated |

| | | | | |
|---|---|---|---|---|
| 1,700908808 | 0,022972413 | S1PR4 | TRUE | Upregulated |
| 0,865156417 | 0,023098152 | ARHGAP4 | FALSE | Upregulated |
| 1,213324113 | 0,023098152 | GRIP2 | TRUE | Upregulated |
| -1,293118414 | 0,023165607 | HSD17B14 | TRUE | Downregulated |
| 2,156747157 | 0,023165607 | PART1 | TRUE | Upregulated |
| 0,817694102 | 0,023173832 | AP4B1-AS1 | FALSE | Upregulated |
| 0,659088185 | 0,023457652 | ABR | FALSE | Upregulated |
| -0,588576655 | 0,023457652 | SERPING1 | FALSE | Downregulated |
| 1,643971686 | 0,023572266 | LOC388906 | TRUE | Upregulated |
| 1,621676188 | 0,023651325 | B4GALNT1 | TRUE | Upregulated |
| 1,986712687 | 0,023651325 | PERM1 | TRUE | Upregulated |
| -0,486910873 | 0,023672397 | MYO1B | FALSE | Downregulated |
| -0,990762317 | 0,02372185 | GALK1 | FALSE | Downregulated |
| -0,455501163 | 0,023757679 | RPLP0 | FALSE | Downregulated |
| 1,41111398 | 0,023766399 | COL4A2-AS2 | TRUE | Upregulated |
| -0,610069587 | 0,023766399 | SDC2 | FALSE | Downregulated |
| -0,657246369 | 0,024038724 | ECI1 | FALSE | Downregulated |
| 1,348072135 | 0,024038724 | IER5L | TRUE | Upregulated |
| 0,978976295 | 0,024076176 | A2M-AS1 | FALSE | Upregulated |
| 0,824396886 | 0,024076176 | LOC102724965 | FALSE | Upregulated |
| 1,286268148 | 0,024076176 | ST14 | TRUE | Upregulated |
| -2,008101991 | 0,0241013 | BLOC1S5 | TRUE | Downregulated |
| 1,145190444 | 0,0241013 | FBXW8 | TRUE | Upregulated |
| 2,005681257 | 0,02410737 | STAMBPL1 | TRUE | Upregulated |
| 0,800163549 | 0,024195396 | SMC4 | FALSE | Upregulated |
| 1,779351183 | 0,024202658 | PDGFRL | TRUE | Upregulated |
| -0,652596172 | 0,024291529 | SLC27A5 | FALSE | Downregulated |
| -0,984151788 | 0,024354193 | MGMT | FALSE | Downregulated |
| 1,986843304 | 0,024533171 | GLIPR2 | TRUE | Upregulated |
| 0,506381358 | 0,024668829 | ADRBK1 | FALSE | Upregulated |
| 0,54439185 | 0,024668829 | SMAD3 | FALSE | Upregulated |
| -1,638144291 | 0,024673021 | BBOX1 | TRUE | Downregulated |
| 0,807344181 | 0,024673021 | LOC100293704 | FALSE | Upregulated |
| 0,838056838 | 0,024673021 | STRADA | FALSE | Upregulated |
| -0,59703077 | 0,024728833 | APMAP | FALSE | Downregulated |
| 1,830118738 | 0,024785005 | ARNTL2 | TRUE | Upregulated |
| 1,829302109 | 0,024966803 | DCK | TRUE | Upregulated |
| -0,642332221 | 0,024966803 | SNTB1 | FALSE | Downregulated |
| -0,624615183 | 0,024999602 | FIS1 | FALSE | Downregulated |
| 1,958900822 | 0,025026372 | LINC01152 | TRUE | Upregulated |
| 0,465657162 | 0,025026372 | VPS13C | FALSE | Upregulated |
| 0,70339735 | 0,025027383 | ARHGAP31 | FALSE | Upregulated |
| -0,454931229 | 0,02507225 | OS9 | FALSE | Downregulated |
| 2,595445144 | 0,025119114 | CPZ | TRUE | Upregulated |
| 2,905984945 | 0,025119442 | HKDC1 | TRUE | Upregulated |
| 0,750487082 | 0,025238663 | PIK3R3 | FALSE | Upregulated |
| -0,751310614 | 0,02529907 | AAMP | FALSE | Downregulated |
| -1,237131438 | 0,02529907 | ATP5I | TRUE | Downregulated |
| 0,688086522 | 0,025395657 | USP45 | FALSE | Upregulated |
| 1,444640558 | 0,025407084 | PIK3CG | TRUE | Upregulated |
| 0,996561116 | 0,025407084 | RFX5 | FALSE | Upregulated |

| | | | | |
|---|---|---|---|---|
| 0,967280798 | 0,025422979 | GBP4 | FALSE | Upregulated |
| 0,911847362 | 0,025598004 | FCGR3A | FALSE | Upregulated |
| 0,992664833 | 0,025598004 | GNRH1 | FALSE | Upregulated |
| -0,78956826 | 0,025626922 | ADH6 | FALSE | Downregulated |
| 1,186013708 | 0,02565352 | LOC390937 | TRUE | Upregulated |
| 1,807458623 | 0,02574147 | TDRKH | TRUE | Upregulated |
| 2,371779898 | 0,025768605 | SLC7A1 | TRUE | Upregulated |
| 1,094604224 | 0,02579982 | RHBDF2 | TRUE | Upregulated |
| -0,734613374 | 0,02579982 | SMOC1 | FALSE | Downregulated |
| -0,546268622 | 0,025819714 | ZFHX4 | FALSE | Downregulated |
| -0,744935806 | 0,025964132 | ITCH | FALSE | Downregulated |
| -1,721226256 | 0,025964132 | RP11-701P16.5 | TRUE | Downregulated |
| 1,203733765 | 0,025964132 | SGK223 | TRUE | Upregulated |
| 0,786691841 | 0,025964132 | TRIM52 | FALSE | Upregulated |
| 1,487289782 | 0,026044082 | C1orf210 | TRUE | Upregulated |
| 0,775657069 | 0,026044082 | CD74 | FALSE | Upregulated |
| 3,625754959 | 0,026051502 | PRR15 | TRUE | Upregulated |
| 0,88003569 | 0,026171888 | CYB5D1 | FALSE | Upregulated |
| 0,794330857 | 0,026214961 | AKAP5 | FALSE | Upregulated |
| -0,477945446 | 0,026214961 | HADHA | FALSE | Downregulated |
| 1,469541127 | 0,026214961 | LOC101927051 | TRUE | Upregulated |
| 1,927371943 | 0,026214961 | RP11-856M7.1 | TRUE | Upregulated |
| 0,813192558 | 0,026497596 | KLF6 | FALSE | Upregulated |
| 1,845672787 | 0,026497596 | PLXNB3 | TRUE | Upregulated |
| 1,475545763 | 0,026543826 | TC2N | TRUE | Upregulated |
| 0,975713605 | 0,026551983 | CENPU | FALSE | Upregulated |
| 1,692678396 | 0,026698388 | SSBP2 | TRUE | Upregulated |
| 0,761062047 | 0,026776615 | ATAD2B | FALSE | Upregulated |
| 2,279769703 | 0,026776615 | BHLHE22 | TRUE | Upregulated |
| -0,65635779 | 0,026776615 | RGN | FALSE | Downregulated |
| -0,721543855 | 0,026776615 | SLC25A10 | FALSE | Downregulated |
| -0,477535625 | 0,026811142 | PPP1R10 | FALSE | Downregulated |
| 1,122865222 | 0,026835765 | LOC388692 | TRUE | Upregulated |
| 0,940509518 | 0,026858664 | ZNF660 | FALSE | Upregulated |
| -1,016641481 | 0,026881145 | FCN3 | TRUE | Downregulated |
| -0,770866868 | 0,026919457 | HMGCL | FALSE | Downregulated |
| 0,75969395 | 0,026953652 | SOGA1 | FALSE | Upregulated |
| 2,845500849 | 0,027104448 | ACTBL2 | TRUE | Upregulated |
| 2,524586037 | 0,027192616 | CDCP1 | TRUE | Upregulated |
| -0,531825171 | 0,027233933 | F7 | FALSE | Downregulated |
| -0,804468125 | 0,027233933 | PLEKHA7 | FALSE | Downregulated |
| -0,597315525 | 0,02731441 | TRAP1 | FALSE | Downregulated |
| 1,931218694 | 0,027358673 | SGCD | TRUE | Upregulated |
| 0,928037179 | 0,027417296 | ARID5B | FALSE | Upregulated |
| 2,581616969 | 0,027764574 | LOC102724371 | TRUE | Upregulated |
| -0,472623744 | 0,027764574 | MAF | FALSE | Downregulated |
| 0,969397706 | 0,028050354 | CCDC64 | FALSE | Upregulated |
| 0,918194449 | 0,028050354 | KCNH1 | FALSE | Upregulated |
| -0,56318708 | 0,028050354 | MIEF1 | FALSE | Downregulated |
| 0,842503426 | 0,028119325 | FAS | FALSE | Upregulated |
| 1,33893085 | 0,028119325 | RP11-47I22.2 | TRUE | Upregulated |

| | | | | |
|---|---|---|---|---|
| -1,066830599 | 0,02813605 | APOC4 | TRUE | Downregulated |
| 0,644444614 | 0,028252841 | RAB15 | FALSE | Upregulated |
| -1,573379757 | 0,028380064 | CHRNA4 | TRUE | Downregulated |
| -0,707080619 | 0,028550619 | PGLYRP2 | FALSE | Downregulated |
| -0,443661531 | 0,028550619 | RPL29 | FALSE | Downregulated |
| 0,70679615 | 0,028550619 | VASP | FALSE | Upregulated |
| -0,504614322 | 0,028578815 | SERPIND1 | FALSE | Downregulated |
| 2,318847073 | 0,028655216 | CD2 | TRUE | Upregulated |
| 0,386482504 | 0,028663316 | LENG8 | FALSE | Upregulated |
| -0,718793155 | 0,028847577 | SELENBP1 | FALSE | Downregulated |
| -0,519497517 | 0,028847577 | STAB1 | FALSE | Downregulated |
| 0,835045141 | 0,028979623 | PLEKHO2 | FALSE | Upregulated |
| -0,394378424 | 0,028979623 | PTPRF | FALSE | Downregulated |
| 1,150483704 | 0,029263617 | TICRR | TRUE | Upregulated |
| -0,720796271 | 0,029295499 | ALDH7A1 | FALSE | Downregulated |
| -0,923134892 | 0,029295499 | C9orf78 | FALSE | Downregulated |
| 1,146184459 | 0,0292957 | C21orf67 | TRUE | Upregulated |
| 0,875257123 | 0,0293928 | LCP1 | FALSE | Upregulated |
| 0,660608463 | 0,029461121 | CHRNB2 | FALSE | Upregulated |
| 2,389664317 | 0,029534148 | CDH19 | TRUE | Upregulated |
| -0,517952989 | 0,029536142 | ADIPOR1 | FALSE | Downregulated |
| -0,616524168 | 0,029536142 | MARC2 | FALSE | Downregulated |
| 0,944695773 | 0,029536142 | ZNF333 | FALSE | Upregulated |
| -0,454468221 | 0,029538533 | ZFP36L1 | FALSE | Downregulated |
| 0,602906482 | 0,029554649 | APC2 | FALSE | Upregulated |
| 1,374920121 | 0,029554649 | DKK3 | TRUE | Upregulated |
| -0,506765839 | 0,029586651 | PQLC1 | FALSE | Downregulated |
| 0,524948294 | 0,029592416 | GGA1 | FALSE | Upregulated |
| -1,210000073 | 0,029592416 | PPM1H | TRUE | Downregulated |
| -0,455968587 | 0,029629188 | FCGRT | FALSE | Downregulated |
| 1,207410609 | 0,029629188 | GPSM1 | TRUE | Upregulated |
| 0,723506891 | 0,029629188 | NCAPH2 | FALSE | Upregulated |
| -0,758503209 | 0,029649496 | VIPR1 | FALSE | Downregulated |
| -0,540241603 | 0,02977379 | MTHFD1 | FALSE | Downregulated |
| 1,549069677 | 0,029777804 | FMN1 | TRUE | Upregulated |
| 0,632237223 | 0,029777804 | HMGXB3 | FALSE | Upregulated |
| 1,296603759 | 0,029856477 | FNDC9 | TRUE | Upregulated |
| 1,757706597 | 0,029882408 | STK39 | TRUE | Upregulated |
| 0,668992335 | 0,029930315 | MAP7D1 | FALSE | Upregulated |
| -1,863513435 | 0,029930315 | RFC2 | TRUE | Downregulated |
| 1,011874227 | 0,030143421 | IKZF1 | TRUE | Upregulated |
| 0,481973478 | 0,030194477 | VCL | FALSE | Upregulated |
| -0,766456952 | 0,030325432 | GCDH | FALSE | Downregulated |
| -1,342055646 | 0,030327575 | SSTR1 | TRUE | Downregulated |
| -0,52715035 | 0,030394251 | PPARA | FALSE | Downregulated |
| -0,677163853 | 0,030394251 | TOMM20 | FALSE | Downregulated |
| 0,753775284 | 0,030494622 | LINC01004 | FALSE | Upregulated |
| 1,22225082 | 0,030695469 | CCDC41 | TRUE | Upregulated |
| 0,717176372 | 0,030695469 | COX15 | FALSE | Upregulated |
| 0,776059598 | 0,030800887 | TNRC6C-AS1 | FALSE | Upregulated |
| 1,034201682 | 0,030938784 | RALGDS | TRUE | Upregulated |

| | | | | |
|---|---|---|---|---|
| 1,252782488 | 0,031195791 | KCNJ4 | TRUE | Upregulated |
| -0,73849261 | 0,031264312 | KLF15 | FALSE | Downregulated |
| 1,990627318 | 0,031264312 | MGAT3 | TRUE | Upregulated |
| 1,325616674 | 0,031268816 | LOC101927479 | TRUE | Upregulated |
| 0,89272245 | 0,031268816 | NAALAD2 | FALSE | Upregulated |
| 1,592433726 | 0,031268816 | PROB1 | TRUE | Upregulated |
| 0,483764786 | 0,031422792 | MYO1F | FALSE | Upregulated |
| 0,828537003 | 0,031470713 | VPS37B | FALSE | Upregulated |
| 1,787516983 | 0,031487997 | N4BP3 | TRUE | Upregulated |
| 0,577791694 | 0,031490702 | DNAJC14 | FALSE | Upregulated |
| 0,978081435 | 0,031490702 | DOCK8 | FALSE | Upregulated |
| 2,461198299 | 0,031503774 | CLSTN2-AS1 | TRUE | Upregulated |
| 1,971447128 | 0,031503774 | LOC102659288 | TRUE | Upregulated |
| -0,953983261 | 0,031524235 | FCN2 | FALSE | Downregulated |
| 0,293511229 | 0,03155533 | DDX17 | FALSE | Upregulated |
| 1,22130942 | 0,031569712 | PRR7-AS1 | TRUE | Upregulated |
| 0,447663655 | 0,031578078 | LGALS8 | FALSE | Upregulated |
| 0,816348292 | 0,031578078 | PARP12 | FALSE | Upregulated |
| -0,704055397 | 0,031578078 | PTPN3 | FALSE | Downregulated |
| 0,824766544 | 0,031578078 | RNF141 | FALSE | Upregulated |
| 2,24505279 | 0,031578078 | SPINT1 | TRUE | Upregulated |
| 0,747982828 | 0,031578078 | ZNF550 | FALSE | Upregulated |
| 1,353113904 | 0,031644319 | ITGA4 | TRUE | Upregulated |
| -0,649576745 | 0,031689221 | APOH | FALSE | Downregulated |
| 1,230100416 | 0,031812498 | CELSR3 | TRUE | Upregulated |
| 0,926642232 | 0,031843085 | ARL4C | FALSE | Upregulated |
| 2,840919445 | 0,031843085 | LOC101929444 | TRUE | Upregulated |
| 0,729321552 | 0,031873294 | DMXL2 | FALSE | Upregulated |
| 1,084218597 | 0,031873294 | RELT | TRUE | Upregulated |
| 0,783704571 | 0,03187801 | LOC101928061 | FALSE | Upregulated |
| -0,581448861 | 0,03193686 | PCK2 | FALSE | Downregulated |
| 1,831168235 | 0,031937566 | LINC00943 | TRUE | Upregulated |
| -0,645638859 | 0,032042586 | MTMR4 | FALSE | Downregulated |
| -1,110460052 | 0,032192872 | COQ5 | TRUE | Downregulated |
| -0,996104085 | 0,032287625 | MRPS5 | FALSE | Downregulated |
| 1,547744357 | 0,032648056 | CD27 | TRUE | Upregulated |
| 0,906480423 | 0,032730726 | FAM198B | FALSE | Upregulated |
| -0,591738564 | 0,032730726 | SEC62 | FALSE | Downregulated |
| 1,287676139 | 0,032730726 | UHRF1 | TRUE | Upregulated |
| -0,540261903 | 0,032735072 | PIPOX | FALSE | Downregulated |
| 0,619946103 | 0,032767356 | CRIM1 | FALSE | Upregulated |
| -0,771515002 | 0,03280762 | CES3 | FALSE | Downregulated |
| 1,002693943 | 0,032858968 | LOC101928604 | TRUE | Upregulated |
| -0,467133777 | 0,032898399 | A1CF | FALSE | Downregulated |
| 1,07523133 | 0,032898399 | LRIF1 | TRUE | Upregulated |
| 1,059673191 | 0,032898399 | MYL5 | TRUE | Upregulated |
| -0,613593626 | 0,032999041 | LRP6 | FALSE | Downregulated |
| 0,871433415 | 0,033071419 | HLA-B | FALSE | Upregulated |
| 1,370716064 | 0,033088594 | SLAMF8 | TRUE | Upregulated |
| 0,801156794 | 0,03314118 | LOC100506860 | FALSE | Upregulated |
| -0,395126692 | 0,033406173 | RPL13 | FALSE | Downregulated |

| | | | | |
|---|---|---|---|---|
| -0,568914685 | 0,033470675 | GOT2 | FALSE | Downregulated |
| 1,000134654 | 0,033470675 | NCKAP1L | TRUE | Upregulated |
| 0,961121398 | 0,033470675 | OR2A1 | FALSE | Upregulated |
| 3,147444108 | 0,033519347 | FCRL5 | TRUE | Upregulated |
| 1,635713659 | 0,033519347 | LUM | TRUE | Upregulated |
| 2,6412797 | 0,033519347 | PBOV1 | TRUE | Upregulated |
| -0,687059385 | 0,033519347 | SERPINA3 | FALSE | Downregulated |
| 0,967432016 | 0,033519347 | WIPF1 | FALSE | Upregulated |
| 3,248766505 | 0,033563838 | LOC101929133 | TRUE | Upregulated |
| 1,523651079 | 0,033847648 | HOXA2 | TRUE | Upregulated |
| -0,620505894 | 0,033891349 | CTSL | FALSE | Downregulated |
| 0,773457295 | 0,033891349 | RHBDD1 | FALSE | Upregulated |
| 0,499802514 | 0,033976367 | CREB1 | FALSE | Upregulated |
| 0,900020989 | 0,034048178 | CTC-338M12.4 | FALSE | Upregulated |
| 1,355499223 | 0,034048178 | LOC100499489 | TRUE | Upregulated |
| -0,517536782 | 0,034678351 | C2 | FALSE | Downregulated |
| 0,44394056 | 0,034842367 | EMC1 | FALSE | Upregulated |
| 0,936975613 | 0,034842367 | SPOPL | FALSE | Upregulated |
| 0,635451249 | 0,034903239 | SMA5 | FALSE | Upregulated |
| 0,526478347 | 0,034985007 | RNF213 | FALSE | Upregulated |
| 1,466766355 | 0,035086378 | CARD11 | TRUE | Upregulated |
| 0,983874207 | 0,035086378 | ZNF790 | FALSE | Upregulated |
| 1,041331214 | 0,035380145 | VPS9D1-AS1 | TRUE | Upregulated |
| 1,026588844 | 0,035381837 | RASSF1-AS1 | TRUE | Upregulated |
| 0,950740775 | 0,035494326 | PAGR1 | FALSE | Upregulated |
| -0,682414566 | 0,035494326 | RELN | FALSE | Downregulated |
| 0,527692196 | 0,035527422 | SYNC | FALSE | Upregulated |
| -0,927111637 | 0,035527817 | MACROD1 | FALSE | Downregulated |
| 0,855367541 | 0,035560767 | COL3A1 | FALSE | Upregulated |
| -0,577365402 | 0,035707531 | NT5E | FALSE | Downregulated |
| 0,465156031 | 0,035866593 | QKI | FALSE | Upregulated |
| -0,505550701 | 0,036025571 | DHTKD1 | FALSE | Downregulated |
| -0,690806384 | 0,036025571 | TXNRD2 | FALSE | Downregulated |
| -2,934323657 | 0,036155975 | LOC100506518 | TRUE | Downregulated |
| 1,165987556 | 0,036155975 | LOC101929668 | TRUE | Upregulated |
| 0,603432851 | 0,036163781 | NME7 | FALSE | Upregulated |
| -0,450892061 | 0,036220204 | MASP1 | FALSE | Downregulated |
| -0,546856772 | 0,03625652 | EHHADH | FALSE | Downregulated |
| 2,762825683 | 0,036272175 | IDO1 | TRUE | Upregulated |
| -0,701470247 | 0,036277632 | SH3BGRL2 | FALSE | Downregulated |
| 2,062352016 | 0,036304551 | NPAS1 | TRUE | Upregulated |
| -0,419446917 | 0,036350756 | HECTD1 | FALSE | Downregulated |
| -0,560105264 | 0,036408943 | CTAGE5 | FALSE | Downregulated |
| 0,596323892 | 0,036533316 | ELK3 | FALSE | Upregulated |
| 2,125068756 | 0,036672908 | CCR2 | TRUE | Upregulated |
| 0,57486328 | 0,036680029 | DGKQ | FALSE | Upregulated |
| 0,907446134 | 0,036881218 | ADAMTS2 | FALSE | Upregulated |
| 1,094866285 | 0,03692222 | AMPD3 | TRUE | Upregulated |
| -0,726325664 | 0,03692222 | SOWAHB | FALSE | Downregulated |
| 0,768456069 | 0,03692222 | TYW5 | FALSE | Upregulated |
| 0,997084901 | 0,037445085 | EMC3-AS1 | FALSE | Upregulated |

| | | | | |
|---|---|---|---|---|
| -0,409242699 | 0,037445085 | FOXN3 | FALSE | Downregulated |
| -0,70876754 | 0,037448529 | DCXR | FALSE | Downregulated |
| -0,636650191 | 0,037662123 | BDH1 | FALSE | Downregulated |
| -0,66212337 | 0,037855622 | FAH | FALSE | Downregulated |
| 1,054780725 | 0,037855622 | PDE4B | TRUE | Upregulated |
| 1,891124984 | 0,037888098 | LGALS3 | TRUE | Upregulated |
| 1,534421658 | 0,037888098 | PTGER2 | TRUE | Upregulated |
| 1,30314176 | 0,038001062 | MPL | TRUE | Upregulated |
| 1,581930412 | 0,038001062 | S100A1 | TRUE | Upregulated |
| -0,386029967 | 0,038001062 | UBR4 | FALSE | Downregulated |
| -1,150306948 | 0,038057955 | CFHR4 | TRUE | Downregulated |
| -0,567645259 | 0,038223022 | ERGIC1 | FALSE | Downregulated |
| 1,202176935 | 0,038223022 | SNPH | TRUE | Upregulated |
| -0,921985386 | 0,038250829 | MPND | FALSE | Downregulated |
| 1,185372047 | 0,038250829 | RUNX1 | TRUE | Upregulated |
| -0,725140374 | 0,038489773 | BTD | FALSE | Downregulated |
| 1,615883156 | 0,038489773 | DGKA | TRUE | Upregulated |
| 1,453968469 | 0,038585532 | LOC101927321 | TRUE | Upregulated |
| 0,93367233 | 0,038737919 | SERPINB1 | FALSE | Upregulated |
| 1,543633095 | 0,038737919 | SLC16A14 | TRUE | Upregulated |
| 2,145837839 | 0,038830724 | EMP1 | TRUE | Upregulated |
| 0,574396325 | 0,038830724 | LOC101927469 | FALSE | Upregulated |
| 2,24798301 | 0,038830724 | WIPF3 | TRUE | Upregulated |
| -0,527711068 | 0,038889006 | POLR2E | FALSE | Downregulated |
| 1,072116827 | 0,039262395 | LOC100996571 | TRUE | Upregulated |
| 1,727619121 | 0,039262395 | PLCXD3 | TRUE | Upregulated |
| 0,877683177 | 0,039353578 | CD160 | FALSE | Upregulated |
| 0,887335481 | 0,039353578 | SEPT8 | FALSE | Upregulated |
| 2,628537021 | 0,039427415 | MAMDC2-AS1 | TRUE | Upregulated |
| 0,977906792 | 0,039536548 | FGD2 | FALSE | Upregulated |
| 1,73313498 | 0,039536548 | LINC00319 | TRUE | Upregulated |
| 0,548380196 | 0,039536548 | LOC101927838 | FALSE | Upregulated |
| 0,778098616 | 0,039536548 | LOC101929396 | FALSE | Upregulated |
| -0,488207978 | 0,039536548 | RARRES2 | FALSE | Downregulated |
| 0,939101144 | 0,039536548 | SEZ6 | FALSE | Upregulated |
| 0,698477628 | 0,039687672 | GPN1 | FALSE | Upregulated |
| 1,115040297 | 0,039722355 | LYG2 | TRUE | Upregulated |
| 0,574002135 | 0,039727243 | ZNF7 | FALSE | Upregulated |
| 1,263763226 | 0,039756743 | SLC6A6 | TRUE | Upregulated |
| -0,714096445 | 0,039776709 | NR1H4 | FALSE | Downregulated |
| -1,711290757 | 0,039947942 | C5orf27 | TRUE | Downregulated |
| -0,948052116 | 0,040065116 | CCDC90B | FALSE | Downregulated |
| -0,848376128 | 0,040065116 | GNAO1 | FALSE | Downregulated |
| 2,183308952 | 0,0400787 | LOC102724233 | TRUE | Upregulated |
| 1,384103774 | 0,040091266 | FAM129A | TRUE | Upregulated |
| 1,889112279 | 0,040091266 | PGR | TRUE | Upregulated |
| 1,351361335 | 0,040093804 | FBLIM1 | TRUE | Upregulated |
| 1,365697208 | 0,040095319 | HLA-DMB | TRUE | Upregulated |
| 0,569977224 | 0,040095319 | PRH2 | FALSE | Upregulated |
| 1,306442347 | 0,040095319 | RELB | TRUE | Upregulated |
| 0,831989453 | 0,040147251 | LANCL2 | FALSE | Upregulated |

| | | | | |
|---|---|---|---|---|
| 1,311846733 | 0,040147251 | RASAL3 | TRUE | Upregulated |
| -0,372539322 | 0,040147251 | RPL19 | FALSE | Downregulated |
| 0,430899659 | 0,040235305 | ATM | FALSE | Upregulated |
| 1,211601752 | 0,040235305 | G6PD | TRUE | Upregulated |
| 1,744196831 | 0,040239798 | MIR3648 | TRUE | Upregulated |
| -0,72269285 | 0,040239798 | SLCO1B1 | FALSE | Downregulated |
| 2,513630042 | 0,040243083 | SPP1 | TRUE | Upregulated |
| -0,658918286 | 0,040377613 | ACOX2 | FALSE | Downregulated |
| -0,591158388 | 0,040377613 | FGA | FALSE | Downregulated |
| 0,56436447 | 0,040387635 | GPR18 | FALSE | Upregulated |
| 1,432435105 | 0,040430907 | LOC102723317 | TRUE | Upregulated |
| -0,579428377 | 0,040430907 | ZGPAT | FALSE | Downregulated |
| 2,777703176 | 0,040813695 | RGS4 | TRUE | Upregulated |
| -0,528383076 | 0,040813695 | USP19 | FALSE | Downregulated |
| 1,116805832 | 0,04081911 | DISC2 | TRUE | Upregulated |
| 1,169885971 | 0,041046115 | MAP1A | TRUE | Upregulated |
| -0,627454114 | 0,041053132 | HAO1 | FALSE | Downregulated |
| 1,079050995 | 0,041055081 | SLIT1 | TRUE | Upregulated |
| 2,612189128 | 0,041083666 | CACNG4 | TRUE | Upregulated |
| -0,662914448 | 0,041097068 | ANG | FALSE | Downregulated |
| -0,717463327 | 0,041097068 | CAT | FALSE | Downregulated |
| -0,486409909 | 0,041116282 | C1S | FALSE | Downregulated |
| 0,635027414 | 0,041116282 | MIR600HG | FALSE | Upregulated |
| 1,166451499 | 0,041116282 | SYTL1 | TRUE | Upregulated |
| -1,073158057 | 0,041116282 | TBCA | TRUE | Downregulated |
| -0,473961876 | 0,041116282 | VPS26B | FALSE | Downregulated |
| 1,036276326 | 0,041504695 | TAGAP | TRUE | Upregulated |
| -0,367521318 | 0,041547534 | ARHGAP35 | FALSE | Downregulated |
| 0,543680784 | 0,041547534 | FAM160B2 | FALSE | Upregulated |
| 2,732568474 | 0,041580021 | FRG2C | TRUE | Upregulated |
| -0,572231951 | 0,041583686 | DPYD | FALSE | Downregulated |
| 1,507350144 | 0,041583686 | SOD3 | TRUE | Upregulated |
| 0,59425089 | 0,041604802 | MIRLET7DHG | FALSE | Upregulated |
| 0,602127326 | 0,041637913 | FAM149B1 | FALSE | Upregulated |
| 0,793556096 | 0,041637913 | PPFIA2 | FALSE | Upregulated |
| 1,388878655 | 0,041675121 | LOC102723726 | TRUE | Upregulated |
| 0,966056417 | 0,041693817 | LOC102723735 | FALSE | Upregulated |
| 0,755305038 | 0,041707696 | LOC101927385 | FALSE | Upregulated |
| 0,521380743 | 0,041796689 | ITPR3 | FALSE | Upregulated |
| -0,726594992 | 0,041839957 | ELL2 | FALSE | Downregulated |
| 1,10994202 | 0,04197796 | KIF21B | TRUE | Upregulated |
| 2,166243341 | 0,04197796 | RP4-553F4.6 | TRUE | Upregulated |
| -0,484978312 | 0,04197796 | SELO | FALSE | Downregulated |
| -0,441042921 | 0,042059636 | SERBP1 | FALSE | Downregulated |
| 1,891408622 | 0,042122415 | RAPGEF4-AS1 | TRUE | Upregulated |
| 2,679507261 | 0,042246658 | C1orf106 | TRUE | Upregulated |
| -0,733053966 | 0,042246658 | CROCCP2 | FALSE | Downregulated |
| -0,548319132 | 0,042246658 | METTL7A | FALSE | Downregulated |
| 0,700979262 | 0,042355254 | PTOV1-AS1 | FALSE | Upregulated |
| 0,752198391 | 0,042415158 | TMSB10 | FALSE | Upregulated |
| 0,809466344 | 0,042449751 | NIN | FALSE | Upregulated |

| | | | | |
|---|---|---|---|---|
| 0,560848611 | 0,042530298 | PODXL | FALSE | Upregulated |
| 0,412021729 | 0,042530298 | UHRF1BP1 | FALSE | Upregulated |
| -0,478779091 | 0,042532534 | C3 | FALSE | Downregulated |
| 1,413273501 | 0,042532534 | CD27-AS1 | TRUE | Upregulated |
| 0,874435882 | 0,042594 | PELI3 | FALSE | Upregulated |
| -0,608560368 | 0,042653837 | NGEF | FALSE | Downregulated |
| -0,489000465 | 0,042653837 | ZBTB20 | FALSE | Downregulated |
| 1,493468572 | 0,042727812 | MMP2 | TRUE | Upregulated |
| -1,000616239 | 0,04274357 | DHODH | TRUE | Downregulated |
| 0,651973973 | 0,042850724 | SCUBE3 | FALSE | Upregulated |
| -0,474453212 | 0,042915223 | MAOB | FALSE | Downregulated |
| 0,840468444 | 0,042921908 | LENG9 | FALSE | Upregulated |
| 1,050325317 | 0,042921908 | SLC6A8 | TRUE | Upregulated |
| 1,239867798 | 0,042979694 | POLG2 | TRUE | Upregulated |
| 1,682082977 | 0,043093293 | KRT7 | TRUE | Upregulated |
| -0,517287888 | 0,043145659 | ABAT | FALSE | Downregulated |
| 0,970908663 | 0,043145878 | LOC101926887 | FALSE | Upregulated |
| 0,511789234 | 0,043733117 | FDPSP2 | FALSE | Upregulated |
| -0,784409799 | 0,043786045 | C4A | FALSE | Downregulated |
| 1,931728198 | 0,043828611 | CAPG | TRUE | Upregulated |
| 2,061555966 | 0,043882584 | CLDN11 | TRUE | Upregulated |
| 1,180889662 | 0,043882584 | RP11-356I2.4 | TRUE | Upregulated |
| 0,661878019 | 0,043917708 | LOC100130744 | FALSE | Upregulated |
| -0,532613694 | 0,044513604 | ALDH4A1 | FALSE | Downregulated |
| -0,598118492 | 0,044513604 | CAPN5 | FALSE | Downregulated |
| 2,6599246 | 0,044513604 | LOC102723511 | TRUE | Upregulated |
| 0,603141199 | 0,044513604 | RHOBTB2 | FALSE | Upregulated |
| 0,528448699 | 0,044513604 | USP24 | FALSE | Upregulated |
| -0,765378265 | 0,044648891 | ATP5G2 | FALSE | Downregulated |
| 0,610629403 | 0,044907657 | LOC101929290 | FALSE | Upregulated |
| -0,523500807 | 0,044973612 | IL13RA1 | FALSE | Downregulated |
| -0,77460677 | 0,044992903 | NAGS | FALSE | Downregulated |
| 1,469713798 | 0,045030841 | RAB31 | TRUE | Upregulated |
| 1,373697017 | 0,045054017 | PIPSL | TRUE | Upregulated |
| 1,555342461 | 0,045234967 | LOC101927529 | TRUE | Upregulated |
| 2,405476706 | 0,0452387 | CELF2-AS2 | TRUE | Upregulated |
| 1,542323827 | 0,045324996 | MARCH3 | TRUE | Upregulated |
| 1,377034344 | 0,045541931 | FBLN2 | TRUE | Upregulated |
| 0,721096105 | 0,045541931 | LDLRAD2 | FALSE | Upregulated |
| 2,442746132 | 0,045541931 | THEMIS | TRUE | Upregulated |
| 1,638033883 | 0,045609003 | PTGDS | TRUE | Upregulated |
| -0,533774525 | 0,045683802 | CYP27A1 | FALSE | Downregulated |
| 0,964628125 | 0,045713208 | TUBA1A | FALSE | Upregulated |
| 0,673316014 | 0,045714305 | UBE2Z | FALSE | Upregulated |
| 1,652682089 | 0,045735434 | C20orf144 | TRUE | Upregulated |
| 0,693283482 | 0,045854778 | LOC102724499 | FALSE | Upregulated |
| -1,940190675 | 0,045858962 | TMEM179 | TRUE | Downregulated |
| -0,547251708 | 0,04588027 | FUBP3 | FALSE | Downregulated |
| -1,288794411 | 0,04588027 | GLUD1P7 | TRUE | Downregulated |
| 0,845346241 | 0,04588027 | JPH3 | FALSE | Upregulated |
| -0,576896724 | 0,04588027 | SERPINC1 | FALSE | Downregulated |

| | | | | |
|---|---|---|---|---|
| -0,696891436 | 0,04588027 | SMO | FALSE | Downregulated |
| 1,569737691 | 0,045938697 | CD86 | TRUE | Upregulated |
| 0,640263386 | 0,045956575 | FAM86A | FALSE | Upregulated |
| 1,002918553 | 0,045956575 | TMEM243 | TRUE | Upregulated |
| 1,199631362 | 0,046006379 | DISP2 | TRUE | Upregulated |
| 1,007099539 | 0,046007252 | LOC100506621 | TRUE | Upregulated |
| -2,769491125 | 0,046177733 | LOC101926934 | TRUE | Downregulated |
| -0,523144095 | 0,046262123 | ZBTB7B | FALSE | Downregulated |
| -0,471716916 | 0,046278503 | GRINA | FALSE | Downregulated |
| -0,504044459 | 0,04629001 | CPT2 | FALSE | Downregulated |
| -0,646431482 | 0,04629001 | DGCR6L | FALSE | Downregulated |
| 0,789013144 | 0,04629001 | FUT2 | FALSE | Upregulated |
| 1,422815375 | 0,04629001 | MYO1H | TRUE | Upregulated |
| 1,758635094 | 0,04629001 | SH3TC2 | TRUE | Upregulated |
| 0,737369793 | 0,04629001 | TNFRSF6B | FALSE | Upregulated |
| 0,770651768 | 0,04629001 | TUBB1 | FALSE | Upregulated |
| -0,645785308 | 0,046319041 | HDGF | FALSE | Downregulated |
| -0,510079141 | 0,046605018 | CPN2 | FALSE | Downregulated |
| -2,31379237 | 0,046605018 | MT1F | TRUE | Downregulated |
| 1,165072292 | 0,046680707 | ELF4 | TRUE | Upregulated |
| -0,582234498 | 0,046871457 | SOWAHC | FALSE | Downregulated |
| 0,948665432 | 0,046871457 | ZNF830 | FALSE | Upregulated |
| 1,585089673 | 0,046889646 | FCN1 | TRUE | Upregulated |
| 1,186559947 | 0,047290889 | FAM126A | TRUE | Upregulated |
| 1,0228568 | 0,047418564 | PPP3CC | TRUE | Upregulated |
| 1,250958992 | 0,04758834 | CORO1A | TRUE | Upregulated |
| -0,576583806 | 0,04758834 | IL6R | FALSE | Downregulated |
| -0,717760311 | 0,04758834 | LRRC3 | FALSE | Downregulated |
| -0,798797104 | 0,047668194 | EBP | FALSE | Downregulated |
| 0,657616744 | 0,047824294 | RARB | FALSE | Upregulated |
| 1,185639708 | 0,047824294 | SPTBN5 | TRUE | Upregulated |
| 1,016319336 | 0,047824294 | SVEP1 | TRUE | Upregulated |
| 2,740075425 | 0,047824294 | TNFRSF17 | TRUE | Upregulated |
| 1,116995073 | 0,048012442 | RP11-118E18.4 | TRUE | Upregulated |
| 0,93373338 | 0,048313983 | ICAM1 | FALSE | Upregulated |
| 1,615154371 | 0,048313983 | LOC101927442 | TRUE | Upregulated |
| 1,551129681 | 0,048535797 | GDF1 | TRUE | Upregulated |
| -0,653298196 | 0,048739473 | CPB2 | FALSE | Downregulated |
| -0,69137864 | 0,048739473 | GATM | FALSE | Downregulated |
| 0,607616469 | 0,048942395 | RNF44 | FALSE | Upregulated |
| 3,206702407 | 0,049131405 | MGC50722 | TRUE | Upregulated |
| -0,67748499 | 0,049131405 | SAT2 | FALSE | Downregulated |
| 1,50405012 | 0,049223846 | RAB40A | TRUE | Upregulated |
| 0,466665166 | 0,049223846 | SLC12A4 | FALSE | Upregulated |
| -0,900129404 | 0,049231737 | GDF2 | FALSE | Downregulated |
| 0,985601348 | 0,049231737 | XPR1 | FALSE | Upregulated |
| -0,756219054 | 0,04925291 | ALB | FALSE | Downregulated |
| 0,797627487 | 0,04925291 | ASB14 | FALSE | Upregulated |
| -0,502495759 | 0,04925291 | GNB2L1 | FALSE | Downregulated |
| 2,513698659 | 0,04925291 | LHCGR | TRUE | Upregulated |
| 0,63163351 | 0,049502327 | C21orf2 | FALSE | Upregulated |

| | | | | |
|---|---|---|---|---|
| 1,154978213 | 0,049502327 | UCP2 | TRUE | Upregulated |
| 1,757443949 | 0,049552652 | ADRA2A | TRUE | Upregulated |
| 0,835250887 | 0,049552652 | LINC00854 | FALSE | Upregulated |
| -0,480607535 | 0,04970652 | PROC | FALSE | Downregulated |
| -0,831819353 | 0,049807397 | CMBL | FALSE | Downregulated |
| 1,146506762 | 0,049807397 | HIGD1B | TRUE | Upregulated |
| 0,654952636 | 0,049807397 | ITPR1 | FALSE | Upregulated |
| -1,433620306 | 0,050014221 | PLGLA | FALSE | Downregulated |
| -0,63299997 | 0,050056959 | KDR | FALSE | Downregulated |
| -0,537434893 | 0,05018365 | SYVN1 | FALSE | Downregulated |
| -0,624481581 | 0,050223194 | AGL | FALSE | Downregulated |
| 1,923766132 | 0,050223877 | CCR5 | FALSE | Upregulated |
| 0,774991432 | 0,050223877 | FGFR2 | FALSE | Upregulated |
| 1,015059163 | 0,050223877 | GBP1 | FALSE | Upregulated |
| 0,755871256 | 0,050274104 | C17orf72 | FALSE | Upregulated |
| -0,481275496 | 0,050274104 | PROS1 | FALSE | Downregulated |
| 1,007816312 | 0,050393978 | RP11-54O15.3 | FALSE | Upregulated |
| -0,427354806 | 0,050422793 | C8orf82 | FALSE | Downregulated |
| 0,961015749 | 0,050426575 | KIAA1614 | FALSE | Upregulated |
| -0,638783484 | 0,05051528 | AOX1 | FALSE | Downregulated |
| 0,849971934 | 0,05051528 | CD97 | FALSE | Upregulated |
| -0,80345128 | 0,050548977 | BPHL | FALSE | Downregulated |
| 0,889975398 | 0,050548977 | DPCD | FALSE | Upregulated |
| -1,175635492 | 0,050548977 | SHH | FALSE | Downregulated |
| 1,024534785 | 0,05059948 | IFI30 | FALSE | Upregulated |
| 0,800333471 | 0,050601113 | TTYH3 | FALSE | Upregulated |
| 0,665258674 | 0,050630826 | ALG1 | FALSE | Upregulated |
| 0,778665911 | 0,050630826 | FLJ32790 | FALSE | Upregulated |
| 0,936004786 | 0,050630826 | JUN | FALSE | Upregulated |
| 1,867948994 | 0,050630826 | LOC613038 | FALSE | Upregulated |
| -1,25299135 | 0,050670772 | CDA | FALSE | Downregulated |
| 0,698959759 | 0,050670772 | CDAN1 | FALSE | Upregulated |
| 1,038545676 | 0,050670772 | ZBTB33 | FALSE | Upregulated |
| 0,737003966 | 0,050930331 | CABLES2 | FALSE | Upregulated |
| 0,587937823 | 0,050942255 | LOC100996506 | FALSE | Upregulated |
| -0,621212859 | 0,050950921 | CPS1 | FALSE | Downregulated |
| -2,168349003 | 0,050950921 | LINC00884 | FALSE | Downregulated |
| 0,889337463 | 0,050950921 | SH3BGRL3 | FALSE | Upregulated |
| 1,537273396 | 0,050960217 | PLEKHG4B | FALSE | Upregulated |
| -0,580730559 | 0,050960217 | PTMS | FALSE | Downregulated |
| 4,600273966 | 0,05151926 | LOC102724370 | FALSE | Upregulated |
| 0,819058879 | 0,0518769 | C7orf43 | FALSE | Upregulated |
| 0,895207069 | 0,051945583 | INMT | FALSE | Upregulated |
| 1,271202565 | 0,052013552 | ARHGAP18 | FALSE | Upregulated |
| -0,55982142 | 0,052013552 | SMIM14 | FALSE | Downregulated |
| 1,784052994 | 0,05207518 | LOC101927756 | FALSE | Upregulated |
| 1,451025879 | 0,052175036 | SLC9A6 | FALSE | Upregulated |
| -0,584864822 | 0,052175036 | XDH | FALSE | Downregulated |
| 0,838642374 | 0,05222937 | BMP6 | FALSE | Upregulated |
| -0,637308849 | 0,05236083 | ENPP1 | FALSE | Downregulated |
| -0,717114964 | 0,05236083 | GOLIM4 | FALSE | Downregulated |

| | | | | |
|---|---|---|---|---|
| 0,63758832 | 0,05236083 | TIGD6 | FALSE | Upregulated |
| 1,105815333 | 0,052630863 | VAV1 | FALSE | Upregulated |
| 0,613268596 | 0,052679576 | ADNP | FALSE | Upregulated |
| -0,621546341 | 0,052817497 | HSD11B1 | FALSE | Downregulated |
| 0,9963772 | 0,05315098 | EPB41L3 | FALSE | Upregulated |
| 1,192291598 | 0,05321646 | PP14571 | FALSE | Upregulated |
| -0,852602758 | 0,053532057 | GBA3 | FALSE | Downregulated |
| 1,385601305 | 0,053631799 | RAX2 | FALSE | Upregulated |
| 0,747709864 | 0,053653895 | CROCC | FALSE | Upregulated |
| -0,453005415 | 0,053653895 | PRAP1 | FALSE | Downregulated |
| -0,906933137 | 0,053653895 | RP11-355N15.1 | FALSE | Downregulated |
| -0,521537203 | 0,053892113 | MPDZ | FALSE | Downregulated |
| 0,603156556 | 0,053908034 | PACSIN2 | FALSE | Upregulated |
| 0,579154875 | 0,053967971 | HEG1 | FALSE | Upregulated |
| -0,875905442 | 0,053967971 | HPR | FALSE | Downregulated |
| 0,668084647 | 0,054747906 | C19orf71 | FALSE | Upregulated |
| 0,661271799 | 0,054747906 | LOC100128374 | FALSE | Upregulated |
| 2,380581983 | 0,054747906 | SEMA7A | FALSE | Upregulated |
| -0,651655432 | 0,054747906 | TOB1 | FALSE | Downregulated |
| -0,574917324 | 0,054762702 | ASMTL | FALSE | Downregulated |
| -0,548829919 | 0,054779572 | GAMT | FALSE | Downregulated |
| 0,756095064 | 0,054779572 | NBPF15 | FALSE | Upregulated |
| 1,4739896 | 0,054779572 | PDZRN3 | FALSE | Upregulated |
| -0,773014391 | 0,054869508 | MTHFS | FALSE | Downregulated |
| 1,434060894 | 0,054869508 | TCEA2 | FALSE | Upregulated |
| 0,86455716 | 0,054902714 | NLRP1 | FALSE | Upregulated |
| 0,828517861 | 0,054920203 | STK16 | FALSE | Upregulated |
| 1,014633583 | 0,055094999 | LPPR3 | FALSE | Upregulated |
| -0,512136876 | 0,055094999 | PHB2 | FALSE | Downregulated |
| 1,502334875 | 0,055094999 | SMARCD3 | FALSE | Upregulated |
| -0,522424189 | 0,055235253 | ERBB3 | FALSE | Downregulated |
| 0,83176056 | 0,055235253 | PIK3CD | FALSE | Upregulated |
| 1,15140148 | 0,055235253 | SLC22A18AS | FALSE | Upregulated |
| 0,622044854 | 0,055248812 | GPR22 | FALSE | Upregulated |
| -0,356972369 | 0,055260288 | ACADVL | FALSE | Downregulated |
| 1,003044868 | 0,055325405 | FKBP14 | FALSE | Upregulated |
| -0,435173127 | 0,055325405 | LRP3 | FALSE | Downregulated |
| -0,594308002 | 0,055349506 | DGCR8 | FALSE | Downregulated |
| 2,33616437 | 0,055361258 | DNAJB13 | FALSE | Upregulated |
| 2,861933006 | 0,055361258 | TRIM46 | FALSE | Upregulated |
| 1,030167673 | 0,055361258 | TRIM59 | FALSE | Upregulated |
| -0,730781278 | 0,055383829 | SNX29P2 | FALSE | Downregulated |
| 0,62241179 | 0,055443588 | ASB16 | FALSE | Upregulated |
| 1,045509294 | 0,055443588 | C16orf3 | FALSE | Upregulated |
| 1,517275442 | 0,055443588 | LOC101241902 | FALSE | Upregulated |
| 2,625796426 | 0,055443588 | LSAMP | FALSE | Upregulated |
| 1,216641953 | 0,055528256 | EMR2 | FALSE | Upregulated |
| -0,419367602 | 0,05572338 | ST6GAL1 | FALSE | Downregulated |
| -0,980017985 | 0,05572338 | TMEM141 | FALSE | Downregulated |
| 0,567106945 | 0,056131739 | ADAMTSL4 | FALSE | Upregulated |
| -0,493456246 | 0,05617345 | NDUFV1 | FALSE | Downregulated |

| | | | | |
|---|---|---|---|---|
| 1,228440361 | 0,05620026 | LOC101929188 | FALSE | Upregulated |
| 1,611103141 | 0,05620026 | PLVAP | FALSE | Upregulated |
| 2,09049619 | 0,056218931 | RPS16P5 | FALSE | Upregulated |
| 0,689866714 | 0,056424665 | TNFAIP2 | FALSE | Upregulated |
| -0,618888111 | 0,056444943 | GTPBP6 | FALSE | Downregulated |
| -0,39389048 | 0,056444943 | HSPA9 | FALSE | Downregulated |
| -1,885443221 | 0,056526452 | MT1G | FALSE | Downregulated |
| -0,56888137 | 0,056526452 | NFKBIA | FALSE | Downregulated |
| -0,710925286 | 0,056526452 | NSUN6 | FALSE | Downregulated |
| -0,528023221 | 0,056550356 | C6 | FALSE | Downregulated |
| 3,217159812 | 0,056701393 | HS3ST2 | FALSE | Upregulated |
| 0,692335069 | 0,056701393 | LOC102724342 | FALSE | Upregulated |
| -0,465135497 | 0,056908492 | TOLLIP | FALSE | Downregulated |
| 0,595400188 | 0,056989776 | FANCI | FALSE | Upregulated |
| 1,696200001 | 0,057018107 | PAQR5 | FALSE | Upregulated |
| -0,483953805 | 0,05704781 | PEBP1 | FALSE | Downregulated |
| 0,649509931 | 0,057111105 | ZYX | FALSE | Upregulated |
| -0,530231066 | 0,057358266 | LDHD | FALSE | Downregulated |
| 1,00734626 | 0,057358266 | LOC100506731 | FALSE | Upregulated |
| -1,052582906 | 0,057500668 | PITPNM3 | FALSE | Downregulated |
| 3,381341696 | 0,057547337 | LOC101927055 | FALSE | Upregulated |
| 0,916069592 | 0,057659015 | CASP3 | FALSE | Upregulated |
| 0,877664637 | 0,057689955 | FBXO41 | FALSE | Upregulated |
| -0,633382274 | 0,057689955 | SLC25A23 | FALSE | Downregulated |
| 0,867871043 | 0,05788254 | GRK6 | FALSE | Upregulated |
| -1,932076383 | 0,057913949 | COL11A2 | FALSE | Downregulated |
| 0,647377357 | 0,058142147 | RP4-591N18.2 | FALSE | Upregulated |
| 1,418832146 | 0,058237522 | LAIR1 | FALSE | Upregulated |
| 1,04811638 | 0,058368141 | NIPAL2 | FALSE | Upregulated |
| -0,462107348 | 0,058394458 | REEP6 | FALSE | Downregulated |
| 0,411403571 | 0,058629518 | RBBP4 | FALSE | Upregulated |
| 0,859529858 | 0,058629518 | RECQL4 | FALSE | Upregulated |
| -0,606514343 | 0,058755056 | SHMT1 | FALSE | Downregulated |
| 2,695985084 | 0,0587956 | MOXD1 | FALSE | Upregulated |
| -0,361605384 | 0,058862026 | EEF1G | FALSE | Downregulated |
| 0,623511889 | 0,058957303 | LOC101929533 | FALSE | Upregulated |
| 1,598128048 | 0,059062525 | ACSL4 | FALSE | Upregulated |
| 0,896560664 | 0,059062525 | C9orf66 | FALSE | Upregulated |
| -0,742527237 | 0,059062525 | MICU1 | FALSE | Downregulated |
| 0,899241216 | 0,059070461 | FBXO34 | FALSE | Upregulated |
| 2,316055962 | 0,059070461 | PLCL1 | FALSE | Upregulated |
| 0,597729018 | 0,059161962 | LOC100505736 | FALSE | Upregulated |
| -0,367962321 | 0,059368563 | GNE | FALSE | Downregulated |
| -0,671328116 | 0,059368563 | LIPG | FALSE | Downregulated |
| 0,513594955 | 0,059368563 | MPHOSPH8 | FALSE | Upregulated |
| 1,544057078 | 0,059368563 | P2RY8 | FALSE | Upregulated |
| -0,873731317 | 0,059396051 | SLC17A2 | FALSE | Downregulated |
| 0,843784056 | 0,059422057 | C12orf60 | FALSE | Upregulated |
| 1,710387002 | 0,059700055 | FAM90A1 | FALSE | Upregulated |
| -0,55972977 | 0,059700055 | FGB | FALSE | Downregulated |
| -1,582545401 | 0,059700055 | LOC102724175 | FALSE | Downregulated |

| | | | | |
|---|---|---|---|---|
| 0,516411416 | 0,059764859 | BTBD9 | FALSE | Upregulated |
| 0,770084931 | 0,059831001 | ARHGEF39 | FALSE | Upregulated |
| -0,636886134 | 0,059831001 | EFNB1 | FALSE | Downregulated |
| 0,55937408 | 0,060086793 | NIPA2 | FALSE | Upregulated |
| 0,791046248 | 0,060402044 | LOXL4 | FALSE | Upregulated |
| 0,939960847 | 0,060411878 | AMMECR1 | FALSE | Upregulated |
| 0,871717397 | 0,060783187 | PTPLAD2 | FALSE | Upregulated |
| -0,720905714 | 0,060956181 | ARMC6 | FALSE | Downregulated |
| 1,678925364 | 0,060956181 | KCNC1 | FALSE | Upregulated |
| 0,845559855 | 0,060956181 | TPM1 | FALSE | Upregulated |
| 0,94952552 | 0,060984431 | IL3RA | FALSE | Upregulated |
| 0,719362916 | 0,060984431 | SSBP3-AS1 | FALSE | Upregulated |
| 0,546800225 | 0,061002535 | NEXN | FALSE | Upregulated |
| -0,391465239 | 0,061180621 | ALKBH5 | FALSE | Downregulated |
| -1,088689961 | 0,061180621 | APLNR | FALSE | Downregulated |
| -0,665037869 | 0,061180621 | BCAM | FALSE | Downregulated |
| 2,102510568 | 0,061180621 | EPHB2 | FALSE | Upregulated |
| -2,132356401 | 0,061180621 | LOC101930375 | FALSE | Downregulated |
| -1,275628598 | 0,061180621 | MANSC1 | FALSE | Downregulated |
| 1,512014513 | 0,061180621 | MMP23B | FALSE | Upregulated |
| 1,18483047 | 0,061180621 | NEXN-AS1 | FALSE | Upregulated |
| -0,963400674 | 0,061180621 | PALM3 | FALSE | Downregulated |
| -1,666193024 | 0,061180621 | PPIC | FALSE | Downregulated |
| 2,400398097 | 0,061180621 | SH2D2A | FALSE | Upregulated |
| 0,577600129 | 0,061180621 | USP35 | FALSE | Upregulated |
| 1,367184304 | 0,061218595 | AC006145.4 | FALSE | Upregulated |
| 1,441578789 | 0,061218595 | ERI1 | FALSE | Upregulated |
| -1,108003747 | 0,061218595 | FAM103A1 | FALSE | Downregulated |
| -0,446524204 | 0,061364456 | P4HB | FALSE | Downregulated |
| -0,51356457 | 0,061651885 | VPS51 | FALSE | Downregulated |
| 1,031207087 | 0,061837779 | FAM163B | FALSE | Upregulated |
| 0,49953946 | 0,061837779 | PIEZO1 | FALSE | Upregulated |
| 0,717858525 | 0,061876497 | CTNS | FALSE | Upregulated |
| -0,555265767 | 0,061900734 | LMAN2 | FALSE | Downregulated |
| -0,530485658 | 0,061941153 | KLHDC10 | FALSE | Downregulated |
| 0,497293453 | 0,062231082 | RNASEH2C | FALSE | Upregulated |
| 0,490176259 | 0,062258075 | ITPK1-AS1 | FALSE | Upregulated |
| -0,626104123 | 0,062258075 | SLC22A3 | FALSE | Downregulated |
| -0,919727138 | 0,062516765 | DHRS4 | FALSE | Downregulated |
| 0,683082012 | 0,062672917 | GK3P | FALSE | Upregulated |
| -0,784437668 | 0,062725231 | HTR2A | FALSE | Downregulated |
| 0,567397543 | 0,06273037 | GNB4 | FALSE | Upregulated |
| 1,174583234 | 0,062850531 | TP53TG5 | FALSE | Upregulated |
| -0,735740819 | 0,063638264 | DAK | FALSE | Downregulated |
| 1,303062923 | 0,063638264 | LOC101929352 | FALSE | Upregulated |
| 1,59803423 | 0,063638264 | LOC389332 | FALSE | Upregulated |
| 0,678567024 | 0,063638264 | LRP2BP | FALSE | Upregulated |
| 1,566655606 | 0,063638264 | PAQR4 | FALSE | Upregulated |
| -0,802392006 | 0,063638264 | SRCIN1 | FALSE | Downregulated |
| 1,104348557 | 0,063970102 | LOC101928482 | FALSE | Upregulated |
| -0,592625191 | 0,064044333 | AFMID | FALSE | Downregulated |

| | | | | |
|---|---|---|---|---|
| -0,559719977 | 0,064044333 | BRPF3 | FALSE | Downregulated |
| 0,663172736 | 0,064044333 | PISD | FALSE | Upregulated |
| -0,674258485 | 0,064069467 | ABCG8 | FALSE | Downregulated |
| -0,687519488 | 0,064129137 | MPC1 | FALSE | Downregulated |
| 1,433078005 | 0,064467733 | ZNF107 | FALSE | Upregulated |
| -2,477897363 | 0,064484263 | OPCML | FALSE | Downregulated |
| 0,600908801 | 0,064484263 | POLR2B | FALSE | Upregulated |
| 0,746162994 | 0,064503834 | IL10RA | FALSE | Upregulated |
| 2,915970058 | 0,064512151 | TNFRSF9 | FALSE | Upregulated |
| 0,581443799 | 0,064513439 | DOCK5 | FALSE | Upregulated |
| 1,017107012 | 0,064700662 | ZBTB22 | FALSE | Upregulated |
| 0,486941123 | 0,06471258 | FGL2 | FALSE | Upregulated |
| 1,819172616 | 0,06471258 | FOXO6 | FALSE | Upregulated |
| 0,905569245 | 0,06471258 | RTL1 | FALSE | Upregulated |
| -1,03505422 | 0,064878464 | CXCL2 | FALSE | Downregulated |
| 0,604499445 | 0,064878464 | LOC101928958 | FALSE | Upregulated |
| 0,635436061 | 0,064956118 | PDZD9 | FALSE | Upregulated |
| 0,681035954 | 0,065027509 | DNMT1 | FALSE | Upregulated |
| 0,56058652 | 0,065084463 | FCHSD2 | FALSE | Upregulated |
| -0,871200099 | 0,065093505 | OCIAD1 | FALSE | Downregulated |
| 0,368623726 | 0,065532675 | FAM13A | FALSE | Upregulated |
| 1,654620537 | 0,065532675 | LOC100996574 | FALSE | Upregulated |
| -0,797976688 | 0,065636449 | GALE | FALSE | Downregulated |
| -0,912594065 | 0,065853887 | COQ4 | FALSE | Downregulated |
| 0,839937899 | 0,06590313 | ITPKB | FALSE | Upregulated |
| 1,154598684 | 0,06590313 | MAP3K14-AS1 | FALSE | Upregulated |
| 2,72409144 | 0,065936809 | KIFC1 | FALSE | Upregulated |
| 0,496981898 | 0,065949539 | ANGPT2 | FALSE | Upregulated |
| 0,615820678 | 0,065949539 | LOC102724559 | FALSE | Upregulated |
| 0,610801552 | 0,065949539 | RILPL1 | FALSE | Upregulated |
| 0,435862528 | 0,065949539 | TMEM200B | FALSE | Upregulated |
| 1,346284039 | 0,065949539 | ZNF737 | FALSE | Upregulated |
| -0,672066746 | 0,066022862 | SHF | FALSE | Downregulated |
| 1,076000823 | 0,066157324 | NABP1 | FALSE | Upregulated |
| 1,180188952 | 0,066159012 | LOC101929837 | FALSE | Upregulated |
| 0,753837678 | 0,066192938 | ADAM22 | FALSE | Upregulated |
| 2,894225386 | 0,066213445 | RP11-506B6.6 | FALSE | Upregulated |
| 0,467864488 | 0,066482442 | FAM65C | FALSE | Upregulated |
| 1,121552838 | 0,066482442 | MLLT10P1 | FALSE | Upregulated |
| 0,77885948 | 0,066762801 | CACNA1C | FALSE | Upregulated |
| 1,096477018 | 0,066762801 | PHTF2 | FALSE | Upregulated |
| 0,594474522 | 0,066762801 | PITRM1-AS1 | FALSE | Upregulated |
| -1,878618437 | 0,066812324 | PPP1R3G | FALSE | Downregulated |
| 0,941481884 | 0,066871278 | FYB | FALSE | Upregulated |
| 1,686929678 | 0,066871278 | LOC101929074 | FALSE | Upregulated |
| 0,61976261 | 0,066922861 | SPDYE2 | FALSE | Upregulated |
| 0,345558505 | 0,066922861 | TRIM8 | FALSE | Upregulated |
| -0,697070993 | 0,066960563 | RPS15A | FALSE | Downregulated |
| 0,703438996 | 0,06709413 | ZNF571-AS1 | FALSE | Upregulated |
| -0,501982954 | 0,067253339 | RPL11 | FALSE | Downregulated |
| 1,80919668 | 0,067460072 | ITK | FALSE | Upregulated |

| | | | | |
|---|---|---|---|---|
| -2,117037549 | 0,0676598 | FAM64A | FALSE | Downregulated |
| 1,635624227 | 0,067804865 | FAIM3 | FALSE | Upregulated |
| 1,010419255 | 0,067804865 | LOC100130992 | FALSE | Upregulated |
| 0,980003551 | 0,067804865 | MAFF | FALSE | Upregulated |
| -0,461558556 | 0,067804865 | SORBS2 | FALSE | Downregulated |
| -0,684823866 | 0,067962202 | ASS1 | FALSE | Downregulated |
| 1,231325207 | 0,067983588 | TWIST1 | FALSE | Upregulated |
| -0,483078795 | 0,068049844 | APOA2 | FALSE | Downregulated |
| 0,915885127 | 0,0680772 | AGAP2 | FALSE | Upregulated |
| 1,816739326 | 0,0680772 | CD300LB | FALSE | Upregulated |
| 0,642731094 | 0,0680772 | KCNIP4 | FALSE | Upregulated |
| -1,266956111 | 0,0680772 | LRRTM3 | FALSE | Downregulated |
| 0,988100806 | 0,0680772 | PDGFB | FALSE | Upregulated |
| 1,436122672 | 0,0680772 | PFKP | FALSE | Upregulated |
| 0,637829621 | 0,0680772 | ZNF529 | FALSE | Upregulated |
| 1,0211437 | 0,068252342 | LOC100507373 | FALSE | Upregulated |
| -0,675528813 | 0,068252342 | RDH16 | FALSE | Downregulated |
| 0,682650754 | 0,068268921 | LOC727978 | FALSE | Upregulated |
| 0,96904417 | 0,068356974 | OSBPL10 | FALSE | Upregulated |
| 0,932932002 | 0,068991705 | ARHGEF33 | FALSE | Upregulated |
| 1,04676311 | 0,068991705 | FLJ32255 | FALSE | Upregulated |
| 1,786679344 | 0,068991705 | HVCN1 | FALSE | Upregulated |
| 1,186658229 | 0,068991705 | LOC102723555 | FALSE | Upregulated |
| 0,394004159 | 0,068991705 | NKTR | FALSE | Upregulated |
| -0,686303342 | 0,068991705 | TMEM192 | FALSE | Downregulated |
| 2,721468923 | 0,068997812 | ILDR1 | FALSE | Upregulated |
| 1,896199822 | 0,069423568 | FNDC1 | FALSE | Upregulated |
| 1,364889034 | 0,069423568 | SLFN13 | FALSE | Upregulated |
| 1,751205836 | 0,069476129 | NMUR1 | FALSE | Upregulated |
| 0,685958328 | 0,069515391 | JPX | FALSE | Upregulated |
| 1,148282278 | 0,069515391 | SECTM1 | FALSE | Upregulated |
| 2,15054539 | 0,069681267 | SEMA3D | FALSE | Upregulated |
| 1,667902762 | 0,069737764 | FAM105A | FALSE | Upregulated |
| 0,996853138 | 0,069737764 | LOC100128573 | FALSE | Upregulated |
| 0,678383908 | 0,069855058 | HIF1A-AS2 | FALSE | Upregulated |
| 1,907352938 | 0,069874751 | A2MP1 | FALSE | Upregulated |
| 1,347573637 | 0,069893533 | VSIG10L | FALSE | Upregulated |
| 1,667304933 | 0,069963648 | ABCC4 | FALSE | Upregulated |
| 1,362050974 | 0,069963648 | CEP89 | FALSE | Upregulated |
| -0,690018423 | 0,069963648 | IL17RC | FALSE | Downregulated |
| -0,515159006 | 0,069963648 | RPS16 | FALSE | Downregulated |
| -0,781545661 | 0,069963648 | SNAPC5 | FALSE | Downregulated |
| 0,922034595 | 0,070075289 | ACVRL1 | FALSE | Upregulated |
| 0,729924941 | 0,070075289 | PLCE1 | FALSE | Upregulated |
| 0,762883009 | 0,070075289 | RP11-864J10.4 | FALSE | Upregulated |
| -0,561012255 | 0,070171323 | DMGDH | FALSE | Downregulated |
| 2,090151052 | 0,070610209 | SP140 | FALSE | Upregulated |
| -0,404269458 | 0,070735002 | CALM1 | FALSE | Downregulated |
| 0,500947968 | 0,070735002 | GREB1L | FALSE | Upregulated |
| 0,54824028 | 0,070735002 | RUFY3 | FALSE | Upregulated |
| 0,635754715 | 0,070808107 | NCKAP5L | FALSE | Upregulated |

| | | | | |
|---|---|---|---|---|
| 0,641972761 | 0,070808107 | NICN1 | FALSE | Upregulated |
| 0,588198066 | 0,070808107 | SNN | FALSE | Upregulated |
| 0,804178114 | 0,070810416 | CPM | FALSE | Upregulated |
| -0,483190049 | 0,070835199 | AGFG2 | FALSE | Downregulated |
| 1,082286462 | 0,070947328 | RASSF2 | FALSE | Upregulated |
| -0,469126574 | 0,071024536 | FAM168B | FALSE | Downregulated |
| 2,089019593 | 0,071036854 | RBMXL3 | FALSE | Upregulated |
| 1,114404933 | 0,071138224 | FUT4 | FALSE | Upregulated |
| 0,511354279 | 0,071317714 | SPDYE1 | FALSE | Upregulated |
| 0,837032338 | 0,071371591 | ARHGAP9 | FALSE | Upregulated |
| -0,328986748 | 0,071371591 | EPN1 | FALSE | Downregulated |
| 0,711864048 | 0,071371591 | JAG1 | FALSE | Upregulated |
| 1,451855498 | 0,071371591 | LOC101927911 | FALSE | Upregulated |
| 1,294560866 | 0,071371591 | LOC102724705 | FALSE | Upregulated |
| -0,367491922 | 0,071371591 | RPS15 | FALSE | Downregulated |
| 0,378943565 | 0,07137249 | BAIAP2L1 | FALSE | Upregulated |
| 0,876363261 | 0,07137249 | CD44 | FALSE | Upregulated |
| 1,929906987 | 0,07137249 | CUBN | FALSE | Upregulated |
| 0,75073028 | 0,07137249 | LCP2 | FALSE | Upregulated |
| 0,540013405 | 0,071630776 | FANCA | FALSE | Upregulated |
| 0,681811731 | 0,071826511 | CTSS | FALSE | Upregulated |
| 0,799562052 | 0,071826511 | SGTB | FALSE | Upregulated |
| 1,745064297 | 0,071826511 | ZNF300 | FALSE | Upregulated |
| 0,734186082 | 0,072090451 | PSMB8 | FALSE | Upregulated |
| 1,143439692 | 0,072104686 | RP11-162J8.2 | FALSE | Upregulated |
| 1,343293884 | 0,072104686 | SLIT2 | FALSE | Upregulated |
| -0,675142156 | 0,072187848 | NDUFB7 | FALSE | Downregulated |
| -0,606909163 | 0,072484155 | ZHX1 | FALSE | Downregulated |
| -0,994490816 | 0,072829227 | RP11-478P10.1 | FALSE | Downregulated |
| -0,703209747 | 0,072875464 | RTKN | FALSE | Downregulated |
| 2,414566316 | 0,073062949 | RP11-594N15.2 | FALSE | Upregulated |
| 1,222291918 | 0,073062949 | SLC25A19 | FALSE | Upregulated |
| 1,714991965 | 0,073063446 | LILRB4 | FALSE | Upregulated |
| 1,982851759 | 0,073092869 | LOC101928738 | FALSE | Upregulated |
| -1,228474139 | 0,073103301 | DHRSX | FALSE | Downregulated |
| -0,664394605 | 0,073103301 | QPRT | FALSE | Downregulated |
| 2,011435506 | 0,073109091 | PTCHD1 | FALSE | Upregulated |
| 0,492384771 | 0,073280049 | PPAPDC1B | FALSE | Upregulated |
| 2,097786686 | 0,073622215 | FUT7 | FALSE | Upregulated |
| -0,676345534 | 0,07362611 | CFHR5 | FALSE | Downregulated |
| 0,780458331 | 0,07362611 | CYBB | FALSE | Upregulated |
| -0,581135693 | 0,07362611 | CYP4F2 | FALSE | Downregulated |
| 1,169567722 | 0,07362611 | GPRC5B | FALSE | Upregulated |
| -0,668317621 | 0,07362611 | HIBADH | FALSE | Downregulated |
| -0,485405929 | 0,07362611 | RN7SL1 | FALSE | Downregulated |
| 1,640005817 | 0,073721047 | IL2RG | FALSE | Upregulated |
| 2,65325173 | 0,073729519 | LOC101928985 | FALSE | Upregulated |
| 1,065190171 | 0,073770042 | LMBR1L | FALSE | Upregulated |
| -0,582229143 | 0,073800649 | DBI | FALSE | Downregulated |
| -0,610873821 | 0,073879605 | ACAD10 | FALSE | Downregulated |
| 0,819042523 | 0,073879605 | MFAP4 | FALSE | Upregulated |

| | | | | |
|---|---|---|---|---|
| 1,83015381 | 0,074219337 | LOC102724554 | FALSE | Upregulated |
| 0,743895015 | 0,074258461 | CSF1 | FALSE | Upregulated |
| -0,604138634 | 0,074258461 | HEBP2 | FALSE | Downregulated |
| -1,279193624 | 0,074258461 | MRPL55 | FALSE | Downregulated |
| -2,707158872 | 0,074346768 | LSM5 | FALSE | Downregulated |
| 0,492584988 | 0,074346768 | MMP24 | FALSE | Upregulated |
| 0,665203043 | 0,074346768 | ZNF227 | FALSE | Upregulated |
| -1,849613247 | 0,074671544 | C14orf180 | FALSE | Downregulated |
| 1,041075907 | 0,074810093 | CELF2 | FALSE | Upregulated |
| -0,833138815 | 0,074826031 | CCS | FALSE | Downregulated |
| 2,3823598 | 0,074826031 | IGSF21 | FALSE | Upregulated |
| -0,629015087 | 0,074826031 | RFNG | FALSE | Downregulated |
| -0,526628169 | 0,074826031 | RPLP2 | FALSE | Downregulated |
| 1,025357596 | 0,074855046 | ROBO1 | FALSE | Upregulated |
| 0,8019074 | 0,075111196 | C12orf65 | FALSE | Upregulated |
| -1,008911977 | 0,075111196 | FOLH1 | FALSE | Downregulated |
| 1,425378471 | 0,075111196 | LOC101927897 | FALSE | Upregulated |
| 2,053100013 | 0,075111196 | MGC39372 | FALSE | Upregulated |
| 0,749339422 | 0,075766157 | AKNA | FALSE | Upregulated |
| 1,904329928 | 0,075766157 | CD8A | FALSE | Upregulated |
| 2,202877155 | 0,075766157 | CECR6 | FALSE | Upregulated |
| 0,538568957 | 0,075905673 | SMG6 | FALSE | Upregulated |
| 1,483067555 | 0,075936001 | GPX7 | FALSE | Upregulated |
| 0,602656731 | 0,07595566 | AC093690.1 | FALSE | Upregulated |
| 1,330777745 | 0,07595566 | FLG2 | FALSE | Upregulated |
| 0,895142784 | 0,075992147 | LOC101928927 | FALSE | Upregulated |
| -0,464674815 | 0,075992147 | SMEK1 | FALSE | Downregulated |
| -0,622992073 | 0,076027189 | SMTN | FALSE | Downregulated |
| -0,392825799 | 0,076031421 | BHMT2 | FALSE | Downregulated |
| -2,400582455 | 0,076031421 | BRIP1 | FALSE | Downregulated |
| -1,395444495 | 0,076031421 | CES1P1 | FALSE | Downregulated |
| -0,594172955 | 0,076031421 | POLD2 | FALSE | Downregulated |
| 0,602929523 | 0,076031421 | TYMP | FALSE | Upregulated |
| 0,629286574 | 0,076153196 | LRRK1 | FALSE | Upregulated |
| 1,108281198 | 0,076188757 | FHOD3 | FALSE | Upregulated |
| -0,562969386 | 0,076214084 | ETNK2 | FALSE | Downregulated |
| 1,80346848 | 0,07638659 | FAM90A25P | FALSE | Upregulated |
| -0,854240835 | 0,076394114 | SLC33A1 | FALSE | Downregulated |
| 0,471737397 | 0,076451444 | DLGAP4 | FALSE | Upregulated |
| -1,165422931 | 0,076810934 | B3GNT1 | FALSE | Downregulated |
| -0,541356993 | 0,07682139 | APOF | FALSE | Downregulated |
| 2,108428788 | 0,076853372 | STK32B | FALSE | Upregulated |
| -0,539281782 | 0,076926313 | ZNF275 | FALSE | Downregulated |
| 1,690594546 | 0,07710639 | NAV3 | FALSE | Upregulated |
| 2,629360618 | 0,077270372 | JAKMIP1 | FALSE | Upregulated |
| 0,395400124 | 0,0773127 | MAP3K13 | FALSE | Upregulated |
| -0,397322461 | 0,0773127 | PCYOX1 | FALSE | Downregulated |
| 0,956830572 | 0,077538875 | TMEM74B | FALSE | Upregulated |
| 1,169366442 | 0,077562225 | FBLN5 | FALSE | Upregulated |
| 2,67922237 | 0,077584295 | MTMR11 | FALSE | Upregulated |
| 0,445069462 | 0,077597022 | CC2D1B | FALSE | Upregulated |

| | | | | |
|---|---|---|---|---|
| -0,382174242 | 0,077597022 | DIAPH1 | FALSE | Downregulated |
| 0,489366144 | 0,077685204 | LRRTM2 | FALSE | Upregulated |
| -0,836949751 | 0,077815326 | COX8A | FALSE | Downregulated |
| -0,640200186 | 0,077815326 | GFRA1 | FALSE | Downregulated |
| 1,433312191 | 0,077815326 | MDM1 | FALSE | Upregulated |
| -0,976231589 | 0,077815326 | MRPL14 | FALSE | Downregulated |
| -0,723256507 | 0,077815326 | SESN2 | FALSE | Downregulated |
| 0,549322799 | 0,077818697 | BBS1 | FALSE | Upregulated |
| 1,063764711 | 0,077818697 | STXBP1 | FALSE | Upregulated |
| 0,618870328 | 0,077818697 | TAGLN2 | FALSE | Upregulated |
| 2,303657916 | 0,077923938 | KIAA1211L | FALSE | Upregulated |
| -1,101776675 | 0,078182876 | PDE1B | FALSE | Downregulated |
| -0,77877624 | 0,078516252 | ADH4 | FALSE | Downregulated |
| -0,813979805 | 0,078516252 | HSD17B13 | FALSE | Downregulated |
| 1,206775188 | 0,079018611 | TMEM255A | FALSE | Upregulated |
| -1,246909461 | 0,079069049 | RP11-492A10.1 | FALSE | Downregulated |
| 0,539907546 | 0,079241858 | SPDYE3 | FALSE | Upregulated |
| 1,504113735 | 0,079267892 | EMB | FALSE | Upregulated |
| -0,556927798 | 0,079267892 | RNF187 | FALSE | Downregulated |
| 0,776321544 | 0,079279621 | LOC100506374 | FALSE | Upregulated |
| 2,032468947 | 0,079279621 | RLTPR | FALSE | Upregulated |
| -1,027895409 | 0,079279621 | TRABD2B | FALSE | Downregulated |
| 0,547274525 | 0,079304629 | MDM2 | FALSE | Upregulated |
| 0,447585687 | 0,079407961 | TMOD1 | FALSE | Upregulated |
| 1,302199459 | 0,079558437 | PAPSS1 | FALSE | Upregulated |
| -2,902864799 | 0,079870082 | GPC5 | FALSE | Downregulated |
| -0,538479384 | 0,079870082 | SMARCA1 | FALSE | Downregulated |
| 0,523940328 | 0,079926716 | NELFCD | FALSE | Upregulated |
| 0,601216792 | 0,0799867 | ASH1L-AS1 | FALSE | Upregulated |
| 1,968911838 | 0,080166211 | LOC102723311 | FALSE | Upregulated |
| 1,560870449 | 0,080166211 | NPNT | FALSE | Upregulated |
| 2,114047396 | 0,080166211 | PLD4 | FALSE | Upregulated |
| -0,556196319 | 0,080166211 | PPP1R15B | FALSE | Downregulated |
| 1,155718896 | 0,080166211 | TDRD7 | FALSE | Upregulated |
| 0,532774916 | 0,080540604 | LOC100996746 | FALSE | Upregulated |
| 2,713551518 | 0,080540604 | LOC101927840 | FALSE | Upregulated |
| -0,501206021 | 0,080540604 | OGDHL | FALSE | Downregulated |
| -0,643744504 | 0,080540604 | PRDX5 | FALSE | Downregulated |
| -0,654137228 | 0,080664672 | EIF1AX | FALSE | Downregulated |
| 1,508033051 | 0,080852146 | ZCCHC18 | FALSE | Upregulated |
| 0,754661393 | 0,080972345 | PTPRCAP | FALSE | Upregulated |
| 2,410681669 | 0,081190523 | HIF3A | FALSE | Upregulated |
| 1,592840995 | 0,081221445 | LOC101927963 | FALSE | Upregulated |
| -0,531215061 | 0,081246461 | C5 | FALSE | Downregulated |
| 0,933850286 | 0,081246461 | FABP5 | FALSE | Upregulated |
| 1,05730282 | 0,081246461 | PHF19 | FALSE | Upregulated |
| -0,353146694 | 0,081295156 | NFE2L1 | FALSE | Downregulated |
| -0,37170934 | 0,081295156 | NUCB1 | FALSE | Downregulated |
| 0,6405012 | 0,081335618 | ADAMTSL2 | FALSE | Upregulated |
| 2,208178541 | 0,081335618 | KCNA6 | FALSE | Upregulated |
| 0,996941407 | 0,081399632 | UNC5A | FALSE | Upregulated |

| | | | | |
|---|---|---|---|---|
| -0,800723323 | 0,081438015 | FUOM | FALSE | Downregulated |
| -0,551477455 | 0,081511581 | THRB | FALSE | Downregulated |
| 1,020547893 | 0,081632546 | LOC101927793 | FALSE | Upregulated |
| -0,706240463 | 0,08165066 | DHRS4-AS1 | FALSE | Downregulated |
| 0,885343787 | 0,081719898 | LOC101929633 | FALSE | Upregulated |
| -1,283358296 | 0,081719898 | NBPF13P | FALSE | Downregulated |
| 0,597516779 | 0,081985934 | LOC101928862 | FALSE | Upregulated |
| -0,470385269 | 0,082005529 | SAR1B | FALSE | Downregulated |
| -0,596499959 | 0,082005529 | TBL1X | FALSE | Downregulated |
| -0,53661585 | 0,082005529 | ZADH2 | FALSE | Downregulated |
| -0,600957166 | 0,082152494 | LY6E | FALSE | Downregulated |
| 0,532778526 | 0,082397822 | LOC101928323 | FALSE | Upregulated |
| 0,735645105 | 0,082397822 | PLXNA1 | FALSE | Upregulated |
| -0,303352567 | 0,082713399 | FAM8A1 | FALSE | Downregulated |
| -0,377154783 | 0,082841978 | RPL35 | FALSE | Downregulated |
| 0,554324057 | 0,082847542 | LOC441081 | FALSE | Upregulated |
| 0,618116257 | 0,082918082 | AC003665.1 | FALSE | Upregulated |
| 2,369206477 | 0,082918082 | CASC1 | FALSE | Upregulated |
| 0,747025858 | 0,082918082 | CBL | FALSE | Upregulated |
| 0,741335341 | 0,082918082 | COPG2IT1 | FALSE | Upregulated |
| -0,710581917 | 0,082918082 | FOXRED1 | FALSE | Downregulated |
| 2,753174952 | 0,082918082 | LOC101928554 | FALSE | Upregulated |
| 0,738862298 | 0,082918082 | PIGR | FALSE | Upregulated |
| -0,408079207 | 0,082918082 | RPL18 | FALSE | Downregulated |
| 0,922205711 | 0,082918082 | TDRD6 | FALSE | Upregulated |
| 0,47405014 | 0,082918082 | YPEL1 | FALSE | Upregulated |
| 1,308583298 | 0,082918082 | ZNF799 | FALSE | Upregulated |
| -1,45506731 | 0,083079458 | GPR98 | FALSE | Downregulated |
| -1,022540306 | 0,083085396 | CSNK2A2 | FALSE | Downregulated |
| 1,621042488 | 0,083085396 | GPRIN1 | FALSE | Upregulated |
| 1,191542005 | 0,083085396 | LOC101928699 | FALSE | Upregulated |
| -0,511242324 | 0,083085396 | MTERFD2 | FALSE | Downregulated |
| 0,56521156 | 0,083085396 | RASGRP1 | FALSE | Upregulated |
| 1,039471828 | 0,083085396 | SIGLEC10 | FALSE | Upregulated |
| 1,034173013 | 0,083091037 | CYR61 | FALSE | Upregulated |
| 1,186818577 | 0,083091037 | LOC101928363 | FALSE | Upregulated |
| 0,734754277 | 0,083268203 | COL6A3 | FALSE | Upregulated |
| -0,378212537 | 0,083268203 | FAU | FALSE | Downregulated |
| 1,589296428 | 0,083268203 | LOC101928465 | FALSE | Upregulated |
| 0,859628307 | 0,083268203 | ZNF385A | FALSE | Upregulated |
| 1,9825194 | 0,083548962 | LOC101929741 | FALSE | Upregulated |
| -0,466578764 | 0,083763614 | C1R-2 | FALSE | Downregulated |
| -0,69821366 | 0,083763614 | PEMT | FALSE | Downregulated |
| -0,613127099 | 0,083918537 | GPR125 | FALSE | Downregulated |
| 2,725441494 | 0,083918537 | RP11-93B14.5 | FALSE | Upregulated |
| 0,469237626 | 0,084270619 | CRLF3 | FALSE | Upregulated |
| 0,850028058 | 0,084270619 | PLEKHG2 | FALSE | Upregulated |
| 0,500452259 | 0,084328589 | CNTD1 | FALSE | Upregulated |
| 0,553932533 | 0,084644796 | GALNT10 | FALSE | Upregulated |
| 1,580470263 | 0,08466183 | TCEAL1 | FALSE | Upregulated |
| -0,403596076 | 0,084708639 | GPD1 | FALSE | Downregulated |

| | | | | |
|---|---|---|---|---|
| -1,178997494 | 0,084861295 | KCNK5 | FALSE | Downregulated |
| -0,605160456 | 0,084861295 | SEC16B | FALSE | Downregulated |
| 0,650911287 | 0,084861295 | SOX12 | FALSE | Upregulated |
| 0,720002237 | 0,084861295 | WDR20 | FALSE | Upregulated |
| -0,634203712 | 0,08491507 | EDC4 | FALSE | Downregulated |
| 1,732031923 | 0,084950437 | ARHGAP22 | FALSE | Upregulated |
| -2,550285896 | 0,084950437 | NLGN4X | FALSE | Downregulated |
| 2,304228109 | 0,085137314 | E2F1 | FALSE | Upregulated |
| 0,540602682 | 0,085137314 | EIF2S2 | FALSE | Upregulated |
| -1,127052722 | 0,085137314 | FXN | FALSE | Downregulated |
| 1,212065746 | 0,085137314 | KIAA1841 | FALSE | Upregulated |
| 1,666076696 | 0,085137314 | SLC1A3 | FALSE | Upregulated |
| -0,403599686 | 0,085218195 | NDST1 | FALSE | Downregulated |
| 0,615874172 | 0,08525774 | ABCB1 | FALSE | Upregulated |
| -0,973278522 | 0,08525774 | IBA57 | FALSE | Downregulated |
| -1,008602785 | 0,08525774 | LOC101927031 | FALSE | Downregulated |
| -0,482779335 | 0,085375695 | CS | FALSE | Downregulated |
| -0,719018691 | 0,085437215 | C1orf168 | FALSE | Downregulated |
| -0,530329646 | 0,085671078 | C4BPA | FALSE | Downregulated |
| 0,615097906 | 0,085671078 | FLJ38668 | FALSE | Upregulated |
| 0,946957679 | 0,085808824 | FIGNL2 | FALSE | Upregulated |
| -0,70113092 | 0,085992832 | RPS26 | FALSE | Downregulated |
| -0,674902731 | 0,086029246 | CCDC125 | FALSE | Downregulated |
| 0,91750555 | 0,086029246 | GVINP1 | FALSE | Upregulated |
| 0,901445638 | 0,086199146 | DFNB31 | FALSE | Upregulated |
| -0,502045398 | 0,086351145 | RPPH1 | FALSE | Downregulated |
| 0,997368057 | 0,086587346 | EPB41L4A-AS2 | FALSE | Upregulated |
| 0,914218535 | 0,086587346 | FZD6 | FALSE | Upregulated |
| 0,730878944 | 0,086587346 | LDHAL6B | FALSE | Upregulated |
| -1,112230314 | 0,08670999 | CNTFR | FALSE | Downregulated |
| 1,10752779 | 0,086906258 | PLEK | FALSE | Upregulated |
| 0,585339593 | 0,086949578 | LOC101928793 | FALSE | Upregulated |
| -0,557212034 | 0,086956841 | ANO1 | FALSE | Downregulated |
| 0,785667756 | 0,087026148 | OMD | FALSE | Upregulated |
| 0,712483889 | 0,087026148 | PARP8 | FALSE | Upregulated |
| -0,711868916 | 0,087034117 | EIF2B5 | FALSE | Downregulated |
| 0,822089331 | 0,087207716 | CYBA | FALSE | Upregulated |
| 1,539996407 | 0,087207716 | GLI2 | FALSE | Upregulated |
| -0,457009013 | 0,087207716 | PTPRK | FALSE | Downregulated |
| -0,4274872 | 0,087207716 | VEGFA | FALSE | Downregulated |
| -2,415206202 | 0,087413641 | CRHR1-IT1 | FALSE | Downregulated |
| 1,230025375 | 0,087413641 | GLT8D2 | FALSE | Upregulated |
| 0,522871237 | 0,087413641 | LOC101929333 | FALSE | Upregulated |
| -0,352281964 | 0,087413641 | RN7SK | FALSE | Downregulated |
| -0,416671073 | 0,087413641 | RPS7 | FALSE | Downregulated |
| -0,75119335 | 0,087413641 | SYNE3 | FALSE | Downregulated |
| -0,541647576 | 0,087413641 | TTC17 | FALSE | Downregulated |
| 0,539438294 | 0,087466171 | LOC100288162 | FALSE | Upregulated |
| 2,030294298 | 0,087703723 | PDGFD | FALSE | Upregulated |
| 0,813887571 | 0,087911639 | PCNA-AS1 | FALSE | Upregulated |
| -0,549437179 | 0,087962809 | SLC17A9 | FALSE | Downregulated |

| | | | | |
|---|---|---|---|---|
| 1,863195978 | 0,088099895 | SCAMP5 | FALSE | Upregulated |
| 0,410135316 | 0,088266629 | FMNL3 | FALSE | Upregulated |
| -1,079322652 | 0,088487597 | ADCY10 | FALSE | Downregulated |
| 1,333335506 | 0,088487597 | GALNT16 | FALSE | Upregulated |
| 0,730766437 | 0,08873 | ALOX12 | FALSE | Upregulated |
| -0,506776459 | 0,08873 | CNKSR3 | FALSE | Downregulated |
| 0,735166196 | 0,08873 | LOC101927732 | FALSE | Upregulated |
| 0,837994343 | 0,089393093 | AJUBA | FALSE | Upregulated |
| -0,762749509 | 0,089393093 | BDH2 | FALSE | Downregulated |
| -0,611867708 | 0,089393093 | C3P1 | FALSE | Downregulated |
| 1,55815638 | 0,089393093 | CYCSP52 | FALSE | Upregulated |
| 1,887038091 | 0,089393093 | DDX53 | FALSE | Upregulated |
| -0,826950003 | 0,089393093 | DHRS4L2 | FALSE | Downregulated |
| 0,781451203 | 0,089393093 | DNAJC24 | FALSE | Upregulated |
| -0,828340486 | 0,089393093 | DUSP10 | FALSE | Downregulated |
| 1,041342862 | 0,089393093 | FAM205B | FALSE | Upregulated |
| 0,997230294 | 0,089393093 | LOC102724243 | FALSE | Upregulated |
| 0,699993223 | 0,089393093 | LOC646127 | FALSE | Upregulated |
| 0,836118903 | 0,089393093 | LOXL2 | FALSE | Upregulated |
| 1,594735988 | 0,089393093 | MS4A4A | FALSE | Upregulated |
| -0,619927197 | 0,089393093 | RP11-713M15.1 | FALSE | Downregulated |
| 0,622669021 | 0,089393093 | SRGAP2-AS1 | FALSE | Upregulated |
| 0,645869614 | 0,089393093 | UNC13D | FALSE | Upregulated |
| 2,511088044 | 0,089503632 | PYY2 | FALSE | Upregulated |
| 1,306273747 | 0,08959775 | RPS6KA2-AS1 | FALSE | Upregulated |
| -0,415442274 | 0,08959775 | UBB | FALSE | Downregulated |
| 0,718559933 | 0,089597788 | CHD3 | FALSE | Upregulated |
| 0,665198066 | 0,089698526 | APOL3 | FALSE | Upregulated |
| 1,706969878 | 0,089698526 | ISG20 | FALSE | Upregulated |
| 0,688132859 | 0,089763541 | COTL1 | FALSE | Upregulated |
| 0,739758646 | 0,089763541 | KRT8 | FALSE | Upregulated |
| 1,029892936 | 0,089763541 | LINC00896 | FALSE | Upregulated |
| -0,473254075 | 0,089763541 | UBR3 | FALSE | Downregulated |
| -0,32814582 | 0,089904273 | REPIN1 | FALSE | Downregulated |
| -0,716893664 | 0,090056671 | COL4A3BP | FALSE | Downregulated |
| 0,541950117 | 0,09016306 | PDCD2 | FALSE | Upregulated |
| -0,704926482 | 0,09016306 | UQCR11 | FALSE | Downregulated |
| 0,747068325 | 0,090360457 | SLC5A9 | FALSE | Upregulated |
| -0,510716468 | 0,090473625 | DLC1 | FALSE | Downregulated |
| 0,537104338 | 0,090473625 | HDAC7 | FALSE | Upregulated |
| -0,316457967 | 0,090473625 | RPS14 | FALSE | Downregulated |
| 0,993673924 | 0,090473625 | ZRANB2-AS1 | FALSE | Upregulated |
| 2,044491894 | 0,090873466 | FCAMR | FALSE | Upregulated |
| -0,81183104 | 0,090873466 | PLA2G16 | FALSE | Downregulated |
| -0,360573586 | 0,090931299 | APP | FALSE | Downregulated |
| -0,275669049 | 0,091201354 | HIPK2 | FALSE | Downregulated |
| -0,505650346 | 0,091518899 | C19orf43 | FALSE | Downregulated |
| -0,754102532 | 0,091518899 | EIF3K | FALSE | Downregulated |
| -0,756835019 | 0,091518899 | KDM8 | FALSE | Downregulated |
| -0,587509671 | 0,091518899 | SLC2A2 | FALSE | Downregulated |
| 0,476695908 | 0,091553544 | PHC1 | FALSE | Upregulated |

| | | | | |
|---|---|---|---|---|
| -0,593862921 | 0,091608295 | HGD | FALSE | Downregulated |
| 0,586615409 | 0,091608295 | SMARCA5-AS1 | FALSE | Upregulated |
| -0,495925178 | 0,091709327 | SOD1 | FALSE | Downregulated |
| 0,526626905 | 0,091721674 | AZI2 | FALSE | Upregulated |
| 0,560760689 | 0,091721674 | DSCR3 | FALSE | Upregulated |
| -0,776918383 | 0,091721674 | RPS29 | FALSE | Downregulated |
| -0,53157878 | 0,091778739 | ACSM2B | FALSE | Downregulated |
| -0,61565157 | 0,091784284 | PIK3C2G | FALSE | Downregulated |
| 1,561814522 | 0,091818701 | TGM4 | FALSE | Upregulated |
| 1,5517596 | 0,091874162 | LOC100130539 | FALSE | Upregulated |
| 0,844717063 | 0,091917745 | WNT2B | FALSE | Upregulated |
| 0,75319059 | 0,091988929 | CMTM4 | FALSE | Upregulated |
| -0,300094492 | 0,091988929 | MTSS1 | FALSE | Downregulated |
| -1,430828284 | 0,092241454 | FBXL21 | FALSE | Downregulated |
| 0,824944694 | 0,092381294 | ATP1A1OS | FALSE | Upregulated |
| 2,056293979 | 0,092529221 | IL12RB1 | FALSE | Upregulated |
| 1,100293622 | 0,092556811 | FLG | FALSE | Upregulated |
| 0,395565217 | 0,092900676 | FAM208A | FALSE | Upregulated |
| -1,599070506 | 0,092900676 | PATE4 | FALSE | Downregulated |
| 0,686200576 | 0,092900676 | RAD52 | FALSE | Upregulated |
| -0,35116164 | 0,093149405 | ORMDL3 | FALSE | Downregulated |
| 0,79299529 | 0,093149405 | SPI1 | FALSE | Upregulated |
| 0,495280456 | 0,093225846 | FLJ31306 | FALSE | Upregulated |
| 0,600579543 | 0,093225846 | OSMR | FALSE | Upregulated |
| -0,487967912 | 0,093375794 | AHCY | FALSE | Downregulated |
| 0,717154678 | 0,093375794 | DENND3 | FALSE | Upregulated |
| -0,653140916 | 0,093375794 | VPS28 | FALSE | Downregulated |
| 1,308527927 | 0,093616172 | MBOAT1 | FALSE | Upregulated |
| -0,500017259 | 0,093640271 | ST3GAL1 | FALSE | Downregulated |
| 1,064420642 | 0,094267924 | RNASE1 | FALSE | Upregulated |
| 0,77255909 | 0,094499005 | DUSP14 | FALSE | Upregulated |
| 0,743713266 | 0,094499005 | LHFPL4 | FALSE | Upregulated |
| -0,418861959 | 0,094499005 | RALGAPA2 | FALSE | Downregulated |
| -1,528385832 | 0,094499005 | RPP25L | FALSE | Downregulated |
| 0,91094648 | 0,094525507 | FGD3 | FALSE | Upregulated |
| 2,213650969 | 0,094742612 | DCLK1 | FALSE | Upregulated |
| 2,092263646 | 0,094742612 | RIPK3 | FALSE | Upregulated |
| 1,4336433 | 0,094919279 | AL592528.1 | FALSE | Upregulated |
| -0,432191368 | 0,095101912 | BOK | FALSE | Downregulated |
| 1,009511695 | 0,095101912 | MEIS1-AS2 | FALSE | Upregulated |
| -0,413968679 | 0,095298634 | APOE | FALSE | Downregulated |
| -0,952728672 | 0,09534952 | ARL4D | FALSE | Downregulated |
| 0,49162255 | 0,095510907 | DNM1L | FALSE | Upregulated |
| 0,796250641 | 0,095525502 | LIMK1 | FALSE | Upregulated |
| 2,116420751 | 0,095667012 | SLC24A5 | FALSE | Upregulated |
| 2,137719191 | 0,095667012 | SMC5-AS1 | FALSE | Upregulated |
| -1,068287335 | 0,095777633 | HS2ST1 | FALSE | Downregulated |
| -0,518833047 | 0,095777633 | RPN1 | FALSE | Downregulated |
| 1,785845753 | 0,095944566 | RP1-102G20.5 | FALSE | Upregulated |
| -0,512470933 | 0,096117832 | ALDH6A1 | FALSE | Downregulated |
| -0,352535348 | 0,096117832 | GLTSCR2 | FALSE | Downregulated |

| | | | | |
|---|---|---|---|---|
| 0,602772909 | 0,096117832 | LRRC24 | FALSE | Upregulated |
| -0,443320383 | 0,096155557 | SLC25A13 | FALSE | Downregulated |
| 1,739978624 | 0,096502227 | PPM1N | FALSE | Upregulated |
| -0,717973644 | 0,096813669 | COX6B1 | FALSE | Downregulated |
| -0,507183749 | 0,097420935 | ABCA8 | FALSE | Downregulated |
| 0,751394367 | 0,09748945 | B4GALT3 | FALSE | Upregulated |
| 0,790411125 | 0,097579806 | LOC100272216 | FALSE | Upregulated |
| -1,178115676 | 0,097579806 | RRM1 | FALSE | Downregulated |
| 1,040870838 | 0,097899859 | LOC100996761 | FALSE | Upregulated |
| 0,876658273 | 0,097899859 | ZC3H12A | FALSE | Upregulated |
| -0,431814588 | 0,0981699 | FAM83H | FALSE | Downregulated |
| 0,973911903 | 0,098558112 | CCBE1 | FALSE | Upregulated |
| -0,738067077 | 0,098558112 | ZBTB39 | FALSE | Downregulated |
| -0,684312808 | 0,098615467 | GLYAT | FALSE | Downregulated |
| -0,612260236 | 0,098622706 | C4BPB | FALSE | Downregulated |
| -0,425362372 | 0,098622706 | GPX4 | FALSE | Downregulated |
| -0,407602638 | 0,098622706 | RPL30 | FALSE | Downregulated |
| 0,659384442 | 0,098809261 | ARHGAP23 | FALSE | Upregulated |
| -0,677582368 | 0,098809261 | PPIF | FALSE | Downregulated |
| -0,796532378 | 0,098809261 | RPL36AL | FALSE | Downregulated |
| -0,432127571 | 0,098906849 | EPB41L5 | FALSE | Downregulated |
| -0,522825911 | 0,098906849 | SEC23A | FALSE | Downregulated |
| 0,794576671 | 0,099070931 | LOC101927371 | FALSE | Upregulated |
| -0,673954907 | 0,099361594 | SFSWAP | FALSE | Downregulated |
| -0,521202293 | 0,099626488 | AURKAIP1 | FALSE | Downregulated |
| 0,919349841 | 0,099626488 | B3GNT4 | FALSE | Upregulated |
| -0,425407706 | 0,099626488 | MYH14 | FALSE | Downregulated |
| 0,5095469 | 0,099736404 | ERMP1 | FALSE | Upregulated |
| -0,56527988 | 0,099736404 | LLGL2 | FALSE | Downregulated |
| 0,625107975 | 0,099736404 | LOC101927066 | FALSE | Upregulated |
| 0,836715324 | 0,099736404 | LOC101928188 | FALSE | Upregulated |
| 1,399159832 | 0,099736404 | NXPE3 | FALSE | Upregulated |
| -2,359990172 | 0,099736404 | PSENEN | FALSE | Downregulated |
| 0,870736645 | 0,099736404 | PTGIS | FALSE | Upregulated |
| -0,469360244 | 0,099736404 | SEPP1 | FALSE | Downregulated |
| 1,822231687 | 0,099736404 | ZNF711 | FALSE | Upregulated |
| -1,329175476 | 0,099896113 | TSNAX | FALSE | Downregulated |
| -0,23644016 | 0,099937223 | ARHGEF12 | FALSE | Downregulated |
| -0,471773785 | 0,099972116 | MCCC2 | FALSE | Downregulated |
| 0,518769031 | 0,100075347 | AAK1 | FALSE | Upregulated |
| -1,134331062 | 0,100075347 | DUSP23 | FALSE | Downregulated |
| -0,43303276 | 0,100075347 | NMT1 | FALSE | Downregulated |
| -0,636318585 | 0,10020694 | EFNA1 | FALSE | Downregulated |
| 0,537314972 | 0,100291442 | CDC42SE2 | FALSE | Upregulated |
| -1,212754325 | 0,100291442 | MRPL15 | FALSE | Downregulated |
| 0,710308229 | 0,100436018 | CCDC18 | FALSE | Upregulated |
| -1,161491549 | 0,100926582 | ADRA2B | FALSE | Downregulated |
| -0,593801078 | 0,101329056 | ECI2 | FALSE | Downregulated |
| -0,549655274 | 0,101329056 | FGG | FALSE | Downregulated |
| 2,151913831 | 0,101548571 | ZBP1 | FALSE | Upregulated |
| 0,579089989 | 0,10184603 | LHFPL2 | FALSE | Upregulated |

| | | | | |
|---|---|---|---|---|
| -0,843666333 | 0,101956359 | YPEL2 | FALSE | Downregulated |
| 0,752270644 | 0,101989867 | ACAP1 | FALSE | Upregulated |
| 1,003462953 | 0,102142003 | COL4A3 | FALSE | Upregulated |
| 2,579696077 | 0,102172933 | LOC102724049 | FALSE | Upregulated |
| -1,455898132 | 0,102176581 | OAT | FALSE | Downregulated |
| 1,446931769 | 0,102195287 | MTHFD1L | FALSE | Upregulated |
| 0,946358101 | 0,102349862 | RANGRF | FALSE | Upregulated |
| -0,628702881 | 0,102813136 | FERMT2 | FALSE | Downregulated |
| -0,632874654 | 0,102813136 | FOXRED2 | FALSE | Downregulated |
| -0,294137114 | 0,102881327 | LRP1 | FALSE | Downregulated |
| 2,222960009 | 0,102898777 | C3orf30 | FALSE | Upregulated |
| 0,689369725 | 0,102931257 | ZNF410 | FALSE | Upregulated |
| 1,101407211 | 0,103031275 | HSPB6 | FALSE | Upregulated |
| 1,122148039 | 0,103031275 | MAP6D1 | FALSE | Upregulated |
| -1,514436006 | 0,103031275 | PCBD2 | FALSE | Downregulated |
| 0,765377159 | 0,103031275 | PCP4L1 | FALSE | Upregulated |
| 1,79941395 | 0,10311958 | LOC646214 | FALSE | Upregulated |
| -1,467911428 | 0,103151081 | IL1RN | FALSE | Downregulated |
| -0,479125123 | 0,103226831 | ATF4 | FALSE | Downregulated |
| 0,65524466 | 0,103724225 | LOC102723566 | FALSE | Upregulated |
| 1,323154334 | 0,103724225 | LOC102724791 | FALSE | Upregulated |
| 1,758361028 | 0,103857913 | LOC101928214 | FALSE | Upregulated |
| -0,526796454 | 0,104011287 | ETFA | FALSE | Downregulated |
| 0,483879012 | 0,104011287 | PDZD2 | FALSE | Upregulated |
| -0,690047167 | 0,104029231 | SNX14 | FALSE | Downregulated |
| 0,597120389 | 0,104029231 | WNT4 | FALSE | Upregulated |
| 0,503498581 | 0,104093663 | AGPAT4 | FALSE | Upregulated |
| -0,405102548 | 0,104093663 | RPL12 | FALSE | Downregulated |
| 2,303997168 | 0,10436471 | IGFBP7-AS1 | FALSE | Upregulated |
| 0,939275631 | 0,10436471 | STARD4 | FALSE | Upregulated |
| 0,663077792 | 0,104407672 | COA5 | FALSE | Upregulated |
| 2,257658735 | 0,104605242 | FAM46B | FALSE | Upregulated |
| -0,316681897 | 0,104605242 | SRRM2 | FALSE | Downregulated |
| 0,798392612 | 0,104681482 | LOC100131564 | FALSE | Upregulated |
| 0,824847846 | 0,104683938 | ABHD14A | FALSE | Upregulated |
| -0,423961063 | 0,104776525 | DHRS3 | FALSE | Downregulated |
| 1,150831549 | 0,104884099 | LOC101927340 | FALSE | Upregulated |
| 0,609436484 | 0,104964297 | KDELR3 | FALSE | Upregulated |
| 0,392477357 | 0,105044866 | DDX5 | FALSE | Upregulated |
| 0,406447938 | 0,105044866 | VAT1 | FALSE | Upregulated |
| -0,244930566 | 0,105549154 | HUWE1 | FALSE | Downregulated |
| -0,647783914 | 0,105549154 | NDUFA4 | FALSE | Downregulated |
| 0,717802425 | 0,10565046 | HMGA1 | FALSE | Upregulated |
| 0,748998015 | 0,10565046 | SLC46A2 | FALSE | Upregulated |
| 0,630379222 | 0,105657854 | FAM184B | FALSE | Upregulated |
| 0,55566723 | 0,105680543 | MAPT | FALSE | Upregulated |
| -0,433789417 | 0,105680543 | MAST3 | FALSE | Downregulated |
| 1,649974278 | 0,106067462 | HEXA-AS1 | FALSE | Upregulated |
| 0,734896342 | 0,106067462 | IKZF2 | FALSE | Upregulated |
| 0,615396908 | 0,106067462 | ITGAV | FALSE | Upregulated |
| -0,88163108 | 0,106067462 | LPA | FALSE | Downregulated |

| | | | | |
|---|---|---|---|---|
| 1,634185652 | 0,106067462 | PKIA | FALSE | Upregulated |
| 2,312909785 | 0,106067462 | TMEM55A | FALSE | Upregulated |
| -0,416430683 | 0,106556452 | PGRMC2 | FALSE | Downregulated |
| -0,632469862 | 0,106582173 | C9 | FALSE | Downregulated |
| 1,119821986 | 0,106582173 | TRPM2 | FALSE | Upregulated |
| 1,837921578 | 0,106631936 | LOC100653515 | FALSE | Upregulated |
| -1,309167107 | 0,106667707 | LAMTOR5 | FALSE | Downregulated |
| -0,562517446 | 0,10680482 | SLC13A5 | FALSE | Downregulated |
| 2,144213338 | 0,106854945 | LOC102724476 | FALSE | Upregulated |
| -0,427665896 | 0,106901562 | GRHPR | FALSE | Downregulated |
| 2,464882504 | 0,107084337 | LGR6 | FALSE | Upregulated |
| -0,475578119 | 0,10710597 | CRYL1 | FALSE | Downregulated |
| 0,426910517 | 0,107250093 | FLJ42627 | FALSE | Upregulated |
| 2,503971696 | 0,107375434 | LOC101929824 | FALSE | Upregulated |
| 0,344047969 | 0,107440894 | SPTAN1 | FALSE | Upregulated |
| -1,522823461 | 0,107440894 | TERF1 | FALSE | Downregulated |
| -0,804401468 | 0,107529402 | DNAJB9 | FALSE | Downregulated |
| -0,764849139 | 0,107543608 | APRT | FALSE | Downregulated |
| 0,769334067 | 0,10755616 | CCDC137 | FALSE | Upregulated |
| 1,82607246 | 0,10755616 | FAM157C | FALSE | Upregulated |
| 1,123357458 | 0,10755616 | PNMA1 | FALSE | Upregulated |
| 0,696734414 | 0,10755616 | ZNF83 | FALSE | Upregulated |
| -0,460440538 | 0,107735324 | SLC20A2 | FALSE | Downregulated |
| -0,523006101 | 0,108161715 | ADH1A | FALSE | Downregulated |
| 0,600096588 | 0,108161715 | FLJ40448 | FALSE | Upregulated |
| -0,382664974 | 0,108161715 | SORL1 | FALSE | Downregulated |
| 1,355532638 | 0,108325554 | FAM131B | FALSE | Upregulated |
| 1,368435254 | 0,108419695 | LEPREL2 | FALSE | Upregulated |
| 1,000557609 | 0,108464509 | SMC1B | FALSE | Upregulated |
| -1,227999945 | 0,108659077 | DAB1 | FALSE | Downregulated |
| 2,236178722 | 0,108716644 | GAS2L3 | FALSE | Upregulated |
| 0,777800595 | 0,10878459 | LOC101927161 | FALSE | Upregulated |
| -0,706136163 | 0,10878459 | REPS1 | FALSE | Downregulated |
| 2,160901061 | 0,10879563 | LOC100131825 | FALSE | Upregulated |
| 0,625073215 | 0,108839714 | GPSM2 | FALSE | Upregulated |
| -0,761762621 | 0,109060237 | TMEM109 | FALSE | Downregulated |
| 0,751928905 | 0,109339261 | LRRC37A6P | FALSE | Upregulated |
| 0,427187332 | 0,109362969 | TRMT112 | FALSE | Upregulated |
| 0,525358769 | 0,109393496 | SHANK2-AS1 | FALSE | Upregulated |
| -0,400327652 | 0,109560601 | AUTS2 | FALSE | Downregulated |
| -0,381993146 | 0,109563046 | FGFRL1 | FALSE | Downregulated |
| 0,743792472 | 0,109563046 | HLA-DPA1 | FALSE | Upregulated |
| -1,265781241 | 0,109563046 | SLCO4C1 | FALSE | Downregulated |
| 0,841294697 | 0,109695623 | HIVEP3 | FALSE | Upregulated |
| 0,69722647 | 0,109712133 | ASAP2 | FALSE | Upregulated |
| 0,928249491 | 0,109885626 | RP13-143G15.3 | FALSE | Upregulated |
| -0,640523901 | 0,109890443 | TCEA1 | FALSE | Downregulated |
| -0,692156657 | 0,110043726 | AQP3 | FALSE | Downregulated |
| 1,251100443 | 0,110043726 | LOC102724976 | FALSE | Upregulated |
| 0,508483176 | 0,110043726 | SCAND2P | FALSE | Upregulated |
| 1,598260994 | 0,110058319 | FCGR3B | FALSE | Upregulated |

| | | | | |
|---|---|---|---|---|
| 1,557804067 | 0,110058319 | GRK5-IT1 | FALSE | Upregulated |
| 0,811611538 | 0,110058319 | KIAA0513 | FALSE | Upregulated |
| -0,759577563 | 0,110058319 | NMRK1 | FALSE | Downregulated |
| -0,607601013 | 0,110058319 | PPID | FALSE | Downregulated |
| -0,370058565 | 0,110109976 | RNF40 | FALSE | Downregulated |
| -0,39750542 | 0,110159212 | OSBP | FALSE | Downregulated |
| -0,520462246 | 0,110250028 | GLYCTK | FALSE | Downregulated |
| 1,050815053 | 0,110344107 | HES4 | FALSE | Upregulated |
| 1,036745249 | 0,110498961 | CCL21 | FALSE | Upregulated |
| -0,427144894 | 0,110566707 | NUDT4 | FALSE | Downregulated |
| 0,545119558 | 0,11084227 | FSD2 | FALSE | Upregulated |
| -0,792418975 | 0,110910066 | AASS | FALSE | Downregulated |
| 0,498668134 | 0,110910066 | AGBL5 | FALSE | Upregulated |
| -0,564865205 | 0,110910066 | AKR1C4 | FALSE | Downregulated |
| 0,372132399 | 0,110910066 | EIF2S3 | FALSE | Upregulated |
| -0,536127227 | 0,110910066 | GPHN | FALSE | Downregulated |
| 0,495705448 | 0,110910066 | SRR | FALSE | Upregulated |
| -0,621129721 | 0,111448403 | ELFN1 | FALSE | Downregulated |
| -0,506131146 | 0,111627762 | TMEM63B | FALSE | Downregulated |
| 0,545741958 | 0,11186948 | ZNF805 | FALSE | Upregulated |
| -0,438356706 | 0,111966901 | C1orf115 | FALSE | Downregulated |
| -2,362165045 | 0,111966901 | LINC00263 | FALSE | Downregulated |
| 0,644954106 | 0,112147057 | ARRDC2 | FALSE | Upregulated |
| -0,363436544 | 0,112147057 | RPL31 | FALSE | Downregulated |
| 2,156407315 | 0,112147057 | ZWINT | FALSE | Upregulated |
| 0,502383206 | 0,112463778 | AIRN | FALSE | Upregulated |
| -1,061139472 | 0,112463778 | RP11-115J16.1 | FALSE | Downregulated |
| 1,842418348 | 0,112979019 | AC005264.2 | FALSE | Upregulated |
| -0,42115294 | 0,112981341 | ALDH5A1 | FALSE | Downregulated |
| -0,971812219 | 0,112981341 | BCO2 | FALSE | Downregulated |
| -0,616736846 | 0,112981341 | F8 | FALSE | Downregulated |
| 0,934190212 | 0,112981341 | GP6 | FALSE | Upregulated |
| -0,64723812 | 0,112981341 | IP6K1 | FALSE | Downregulated |
| 1,405210616 | 0,113098664 | CCND2 | FALSE | Upregulated |
| -2,303449068 | 0,113221604 | LOC101928961 | FALSE | Downregulated |
| 0,702693265 | 0,113338207 | ANKDD1A | FALSE | Upregulated |
| 0,312992343 | 0,113385188 | VPS13B | FALSE | Upregulated |
| 0,547856597 | 0,113426673 | KAZN | FALSE | Upregulated |
| 2,046046981 | 0,113450397 | HAPLN3 | FALSE | Upregulated |
| -2,048357918 | 0,113450397 | HYDIN | FALSE | Downregulated |
| -0,499017297 | 0,113450397 | IDH1 | FALSE | Downregulated |
| 0,71531614 | 0,113450397 | MYH11 | FALSE | Upregulated |
| 1,162189546 | 0,113450397 | SH3D21 | FALSE | Upregulated |
| 0,666947862 | 0,113627435 | CCNE2 | FALSE | Upregulated |
| 0,937034585 | 0,113854446 | ACSS1 | FALSE | Upregulated |
| 0,780513173 | 0,114420714 | RALB | FALSE | Upregulated |
| -0,925034946 | 0,114442076 | GPR116 | FALSE | Downregulated |
| 0,504532982 | 0,114499821 | SDCBP2-AS1 | FALSE | Upregulated |
| 0,523156807 | 0,11452141 | AQP11 | FALSE | Upregulated |
| 0,716096056 | 0,11474388 | CALB1 | FALSE | Upregulated |
| 0,913716351 | 0,114785987 | TRERF1 | FALSE | Upregulated |

| | | | | |
|---|---|---|---|---|
| 0,669423769 | 0,114851585 | COL18A1-AS1 | FALSE | Upregulated |
| -0,515655332 | 0,114851585 | PECR | FALSE | Downregulated |
| -0,426286057 | 0,114851585 | RN7SL2 | FALSE | Downregulated |
| -0,373715735 | 0,114851585 | SERPINA10 | FALSE | Downregulated |
| -0,494371552 | 0,114851585 | SLC25A22 | FALSE | Downregulated |
| 0,630384049 | 0,114851585 | TBC1D9 | FALSE | Upregulated |
| 1,464748025 | 0,114851585 | TM6SF1 | FALSE | Upregulated |
| 0,38866445 | 0,114851585 | XRRA1 | FALSE | Upregulated |
| 0,56266428 | 0,11515193 | AP3M1 | FALSE | Upregulated |
| -0,487214941 | 0,11515193 | MLXIPL | FALSE | Downregulated |
| -0,476299063 | 0,115316861 | IRF2BP1 | FALSE | Downregulated |
| -0,413466251 | 0,115363755 | NFX1 | FALSE | Downregulated |
| -0,528934917 | 0,115363755 | RPS28 | FALSE | Downregulated |
| -2,521597069 | 0,115506376 | LOC100289561 | FALSE | Downregulated |
| 0,753702982 | 0,115604379 | CD5 | FALSE | Upregulated |
| -0,885905339 | 0,115604379 | CTBS | FALSE | Downregulated |
| -2,473770193 | 0,115604379 | NEUROD6 | FALSE | Downregulated |
| 2,114684045 | 0,115604379 | ROR2 | FALSE | Upregulated |
| 1,151857131 | 0,115619346 | FOXP3 | FALSE | Upregulated |
| -0,379370963 | 0,115950034 | ESYT1 | FALSE | Downregulated |
| -0,299390039 | 0,115950034 | SLC38A10 | FALSE | Downregulated |
| -0,320227359 | 0,115953988 | BAG6 | FALSE | Downregulated |
| 0,378338636 | 0,116351072 | PTPMT1 | FALSE | Upregulated |
| 0,868375361 | 0,116500946 | SLC44A3 | FALSE | Upregulated |
| 0,477337277 | 0,116634267 | ATP11A-AS1 | FALSE | Upregulated |
| 0,821924555 | 0,116634267 | RP11-14D22.5 | FALSE | Upregulated |
| -0,54492071 | 0,116856094 | FBXL5 | FALSE | Downregulated |
| 1,285722338 | 0,117001295 | LOC154761 | FALSE | Upregulated |
| -0,735550197 | 0,117001295 | ZNRF3 | FALSE | Downregulated |
| 2,159568877 | 0,117385436 | LINC00941 | FALSE | Upregulated |
| -1,460767991 | 0,117831029 | OSBPL6 | FALSE | Downregulated |
| -0,326464073 | 0,117988151 | EEF1D | FALSE | Downregulated |
| -0,926776441 | 0,118011559 | ANAPC11 | FALSE | Downregulated |
| -0,478392708 | 0,118011559 | ARHGEF40 | FALSE | Downregulated |
| 1,654134267 | 0,118108836 | LINC00639 | FALSE | Upregulated |
| 1,080319117 | 0,118132388 | LAT | FALSE | Upregulated |
| 2,526213414 | 0,118213826 | LOC101929523 | FALSE | Upregulated |
| -0,658265986 | 0,118311669 | NDUFV3 | FALSE | Downregulated |
| -0,35889082 | 0,118544432 | INTS1 | FALSE | Downregulated |
| -0,406680978 | 0,118544432 | RPL5 | FALSE | Downregulated |
| 0,457226767 | 0,118767668 | RRN3P3 | FALSE | Upregulated |
| -0,612354985 | 0,118783161 | PARK7 | FALSE | Downregulated |
| 0,945894288 | 0,118875196 | CSF2RB | FALSE | Upregulated |
| 0,49241244 | 0,118875196 | JAZF1 | FALSE | Upregulated |
| -0,635171998 | 0,118880761 | GOLGA5 | FALSE | Downregulated |
| -0,645892059 | 0,118880761 | PHGDH | FALSE | Downregulated |
| 0,440801969 | 0,118880761 | RFTN1 | FALSE | Upregulated |
| 1,176819224 | 0,118880761 | WDR47 | FALSE | Upregulated |
| -0,571163525 | 0,118995728 | ALPL | FALSE | Downregulated |
| 0,702783732 | 0,118995728 | SPRYD3 | FALSE | Upregulated |
| 2,163800269 | 0,119084727 | IFNG-AS1 | FALSE | Upregulated |

| | | | | |
|---|---|---|---|---|
| 1,979934851 | 0,119356514 | ANXA2R | FALSE | Upregulated |
| -0,463036405 | 0,119356514 | ATP5B | FALSE | Downregulated |
| 0,744491779 | 0,119356514 | BTRC | FALSE | Upregulated |
| 0,368508133 | 0,119356514 | GATC | FALSE | Upregulated |
| -0,473505287 | 0,119356514 | ONECUT1 | FALSE | Downregulated |
| 0,584515381 | 0,119356514 | RPL13AP6 | FALSE | Upregulated |
| 1,43779083 | 0,119356514 | SLC25A33 | FALSE | Upregulated |
| 1,005372853 | 0,119356514 | SPACA6P-AS | FALSE | Upregulated |
| 0,957803753 | 0,119356514 | THRA | FALSE | Upregulated |
| -0,585246174 | 0,11969283 | IGFBP3 | FALSE | Downregulated |
| 1,947978928 | 0,11974648 | NALCN | FALSE | Upregulated |
| 1,164702837 | 0,11987695 | LOC102723741 | FALSE | Upregulated |
| -1,486761088 | 0,119913719 | MIR17HG | FALSE | Downregulated |
| -0,767658348 | 0,120198464 | PLIN2 | FALSE | Downregulated |
| -0,516066824 | 0,120211888 | NR1I3 | FALSE | Downregulated |
| 0,3793425 | 0,120333122 | GON4L | FALSE | Upregulated |
| 0,415829416 | 0,120578865 | AIDA | FALSE | Upregulated |
| 0,686876615 | 0,120650673 | GPCPD1 | FALSE | Upregulated |
| -0,419253735 | 0,121110688 | TSC2 | FALSE | Downregulated |
| 0,70407079 | 0,121122085 | IFNGR1 | FALSE | Upregulated |
| 0,919437633 | 0,121122085 | LOC100287852 | FALSE | Upregulated |
| 1,026352977 | 0,121195941 | LOC102724974 | FALSE | Upregulated |
| -0,549331203 | 0,12163005 | FTCD | FALSE | Downregulated |
| 0,936710101 | 0,121688333 | LINC00176 | FALSE | Upregulated |
| 2,661658796 | 0,121777682 | AQP6 | FALSE | Upregulated |
| 1,762587521 | 0,121810403 | CDCA4 | FALSE | Upregulated |
| 0,338014354 | 0,12181754 | MINK1 | FALSE | Upregulated |
| 0,367304095 | 0,12181754 | SMURF1 | FALSE | Upregulated |
| 0,546531011 | 0,122072015 | TEP1 | FALSE | Upregulated |
| 0,919807261 | 0,122899838 | GRIK1 | FALSE | Upregulated |
| 1,594473027 | 0,122899838 | LOC100505942 | FALSE | Upregulated |
| 0,562296366 | 0,122899838 | MFSD6 | FALSE | Upregulated |
| -1,702956226 | 0,122899838 | RP11-290F20.1 | FALSE | Downregulated |
| 0,52259076 | 0,12293683 | FAM175A | FALSE | Upregulated |
| 0,617766819 | 0,12293683 | ZNF789 | FALSE | Upregulated |
| 0,461353794 | 0,123030626 | TANC1 | FALSE | Upregulated |
| 2,104465288 | 0,123166609 | CDX2 | FALSE | Upregulated |
| 1,159962795 | 0,12347191 | C15orf54 | FALSE | Upregulated |
| 0,460321523 | 0,123481158 | DMPK | FALSE | Upregulated |
| -0,763618168 | 0,123647608 | SAA4 | FALSE | Downregulated |
| 1,877367882 | 0,123828328 | DAPP1 | FALSE | Upregulated |
| 1,310208062 | 0,123884972 | ZC3H12D | FALSE | Upregulated |
| 0,639357453 | 0,123939651 | DYNLL1 | FALSE | Upregulated |
| 0,754625712 | 0,123939651 | TRIM65 | FALSE | Upregulated |
| 0,725950622 | 0,124323979 | CALHM2 | FALSE | Upregulated |
| 0,782699399 | 0,124323979 | KCNAB2 | FALSE | Upregulated |
| 0,339792949 | 0,124323979 | KMT2E | FALSE | Upregulated |
| -0,936007727 | 0,124323979 | MOGAT2 | FALSE | Downregulated |
| -0,492656247 | 0,124323979 | NDRG2 | FALSE | Downregulated |
| -0,426820592 | 0,124418471 | PCNXL3 | FALSE | Downregulated |
| -0,575565994 | 0,124505469 | GPAA1 | FALSE | Downregulated |

| | | | | |
|---|---|---|---|---|
| 0,478005836 | 0,1245966 | LIMD1-AS1 | FALSE | Upregulated |
| 0,742261634 | 0,124764582 | CSNK1G2-AS1 | FALSE | Upregulated |
| 0,774240548 | 0,125103115 | UBE2L6 | FALSE | Upregulated |
| 2,201067845 | 0,125113919 | BMS1P18 | FALSE | Upregulated |
| 0,51342961 | 0,125134049 | DPYSL2 | FALSE | Upregulated |
| -2,19043555 | 0,125134049 | ESPNP | FALSE | Downregulated |
| -0,576123466 | 0,125302467 | ELMO1 | FALSE | Downregulated |
| 0,766992277 | 0,125439161 | KCTD4 | FALSE | Upregulated |
| 0,318738172 | 0,125604558 | SLC46A1 | FALSE | Upregulated |
| 0,406382453 | 0,125952237 | HEXIM2 | FALSE | Upregulated |
| -0,644609917 | 0,126081688 | PON1 | FALSE | Downregulated |
| 0,701387421 | 0,126219211 | BTN3A1 | FALSE | Upregulated |
| -0,607519692 | 0,126323258 | PID1 | FALSE | Downregulated |
| 0,40976624 | 0,126347579 | POLH | FALSE | Upregulated |
| -0,539933949 | 0,126375839 | LARP1B | FALSE | Downregulated |
| 1,899767776 | 0,126375839 | NRGN | FALSE | Upregulated |
| 0,570386937 | 0,126375839 | PLTP | FALSE | Upregulated |
| -0,697559678 | 0,126377006 | NEU4 | FALSE | Downregulated |
| -0,484884844 | 0,126487934 | F12 | FALSE | Downregulated |
| 0,831595902 | 0,126487934 | HCG4B | FALSE | Upregulated |
| 1,302212587 | 0,127017628 | CCL5 | FALSE | Upregulated |
| 0,482342393 | 0,127066682 | CALML4 | FALSE | Upregulated |
| 0,507280325 | 0,127247069 | FKBP1AP1 | FALSE | Upregulated |
| -0,620939534 | 0,127360705 | ACSM5 | FALSE | Downregulated |
| 0,512285138 | 0,127360705 | DAB2 | FALSE | Upregulated |
| 1,06671549 | 0,127360705 | FCGBP | FALSE | Upregulated |
| -0,446546185 | 0,127360705 | HSPD1 | FALSE | Downregulated |
| -2,189746317 | 0,127360705 | OVGP1 | FALSE | Downregulated |
| 0,392137894 | 0,127360705 | SERINC4 | FALSE | Upregulated |
| -0,619500541 | 0,127429778 | NDUFB9 | FALSE | Downregulated |
| -0,68227675 | 0,12747756 | PRDX4 | FALSE | Downregulated |
| 2,089105418 | 0,127669254 | CREB3L1 | FALSE | Upregulated |
| 0,683532533 | 0,127669254 | CYCS | FALSE | Upregulated |
| 0,635091417 | 0,127797871 | LOC101929599 | FALSE | Upregulated |
| 1,021755033 | 0,127898591 | NHS | FALSE | Upregulated |
| 0,403258461 | 0,127971383 | LASP1 | FALSE | Upregulated |
| 0,352713841 | 0,127971383 | TMEM140 | FALSE | Upregulated |
| -0,561538833 | 0,128068169 | QDPR | FALSE | Downregulated |
| 0,714123686 | 0,128146574 | B4GALT2 | FALSE | Upregulated |
| 0,834927862 | 0,128146574 | LOC102724172 | FALSE | Upregulated |
| -0,477639515 | 0,128146574 | PRKAG2 | FALSE | Downregulated |
| -0,452137732 | 0,128146574 | SEC63 | FALSE | Downregulated |
| 0,635468454 | 0,128345202 | SLC1A4 | FALSE | Upregulated |
| -0,432179619 | 0,128409539 | TM9SF3 | FALSE | Downregulated |
| 1,003898309 | 0,128409539 | TMOD2 | FALSE | Upregulated |
| -0,479790146 | 0,128577463 | CYC1 | FALSE | Downregulated |
| -0,561736248 | 0,12861479 | PHYH | FALSE | Downregulated |
| 1,268458761 | 0,128681413 | LOC101928424 | FALSE | Upregulated |
| 0,451294479 | 0,128681413 | SPRN | FALSE | Upregulated |
| -1,549665511 | 0,128681413 | TM6SF2 | FALSE | Downregulated |
| -0,474375732 | 0,128801341 | PDK2 | FALSE | Downregulated |

| | | | | |
|---|---|---|---|---|
| 1,20486951 | 0,129121381 | MIR100HG | FALSE | Upregulated |
| 1,888137046 | 0,12914519 | TRPV6 | FALSE | Upregulated |
| -0,931212351 | 0,129470272 | BTBD10 | FALSE | Downregulated |
| -1,019704462 | 0,129733734 | UNG | FALSE | Downregulated |
| 0,416286624 | 0,129873205 | KPNA1 | FALSE | Upregulated |
| 1,571735523 | 0,129873205 | LINC01108 | FALSE | Upregulated |
| -0,404157969 | 0,129873205 | RPS20 | FALSE | Downregulated |
| -0,431908943 | 0,129873205 | TMPRSS6 | FALSE | Downregulated |
| 1,231163724 | 0,130142059 | GINS4 | FALSE | Upregulated |
| -0,762656726 | 0,130142059 | LINC01061 | FALSE | Downregulated |
| 2,006068798 | 0,130142059 | LOC101926931 | FALSE | Upregulated |
| -0,710639756 | 0,130173319 | TSPAN33 | FALSE | Downregulated |
| 0,804945703 | 0,130173319 | ZNF35 | FALSE | Upregulated |
| 0,781069696 | 0,130204662 | LOC100131395 | FALSE | Upregulated |
| 0,617181701 | 0,130204662 | PACS1 | FALSE | Upregulated |
| -0,604475997 | 0,130204662 | UGT2B7 | FALSE | Downregulated |
| 0,40209081 | 0,13038664 | CYFIP1 | FALSE | Upregulated |
| 1,253065907 | 0,13038664 | MCM2 | FALSE | Upregulated |
| 0,607255387 | 0,13038948 | CFLAR-AS1 | FALSE | Upregulated |
| 0,526543479 | 0,13038948 | CNTRL | FALSE | Upregulated |
| -0,613839963 | 0,13038948 | THRB-AS1 | FALSE | Downregulated |
| 1,446758606 | 0,130397029 | LOC101928019 | FALSE | Upregulated |
| 0,401080287 | 0,130433263 | PNRC2 | FALSE | Upregulated |
| -0,702172356 | 0,130611767 | UROD | FALSE | Downregulated |
| 1,704627769 | 0,130662018 | C12orf71 | FALSE | Upregulated |
| -0,405278217 | 0,131413604 | HGS | FALSE | Downregulated |
| 0,359384184 | 0,131597868 | SYNPO | FALSE | Upregulated |
| 0,597469964 | 0,131811183 | CPSF4 | FALSE | Upregulated |
| 0,640200355 | 0,131839473 | DERL3 | FALSE | Upregulated |
| 0,51028896 | 0,131884972 | HTRA1 | FALSE | Upregulated |
| -0,400218122 | 0,131884972 | PTPLAD1 | FALSE | Downregulated |
| 0,671652477 | 0,131884972 | TXNRD1 | FALSE | Upregulated |
| 0,441728663 | 0,132149764 | SZRD1 | FALSE | Upregulated |
| -0,744475761 | 0,132242125 | ACAD8 | FALSE | Downregulated |
| 0,509152783 | 0,132435089 | ZNF850 | FALSE | Upregulated |
| 1,150995023 | 0,132640299 | HPDL | FALSE | Upregulated |
| 0,510077759 | 0,132640299 | POU2F1 | FALSE | Upregulated |
| 0,723617232 | 0,132640299 | ZNF720 | FALSE | Upregulated |
| 0,710701681 | 0,132659029 | CCNH | FALSE | Upregulated |
| -1,398938869 | 0,132659029 | SLC38A11 | FALSE | Downregulated |
| -0,650847788 | 0,132703558 | PLA2G15 | FALSE | Downregulated |
| -1,03521314 | 0,132703558 | RNF214 | FALSE | Downregulated |
| -0,874929202 | 0,132849606 | HOOK1 | FALSE | Downregulated |
| 0,596488452 | 0,132849606 | ICA1L | FALSE | Upregulated |
| 0,723795845 | 0,132849606 | PPAT | FALSE | Upregulated |
| 0,932262718 | 0,132849606 | SPARCL1 | FALSE | Upregulated |
| -0,426974154 | 0,132865621 | CFH | FALSE | Downregulated |
| 1,267738643 | 0,132865621 | LINC00999 | FALSE | Upregulated |
| 1,417056588 | 0,132910628 | SLC6A20 | FALSE | Upregulated |
| 1,0493521 | 0,132971924 | FDXR | FALSE | Upregulated |
| -0,567564277 | 0,132989475 | STRN3 | FALSE | Downregulated |

| | | | | |
|---|---|---|---|---|
| 1,015534198 | 0,133091314 | RP11-1H15.2 | FALSE | Upregulated |
| 1,260428034 | 0,133490794 | FDXACB1 | FALSE | Upregulated |
| 0,625339032 | 0,13352361 | ZNF606 | FALSE | Upregulated |
| 0,954647884 | 0,133962093 | FBXL18 | FALSE | Upregulated |
| 0,992593016 | 0,133962093 | PDE6B | FALSE | Upregulated |
| 0,608079469 | 0,133962093 | PGAM2 | FALSE | Upregulated |
| 0,644817915 | 0,133962093 | RP5-914P20.5 | FALSE | Upregulated |
| 0,988082205 | 0,134020212 | LOC101927028 | FALSE | Upregulated |
| -0,584350353 | 0,13413212 | LOC100507033 | FALSE | Downregulated |
| 0,647725045 | 0,134169332 | VAC14-AS1 | FALSE | Upregulated |
| 1,265865904 | 0,134227719 | CTC-558O2.1 | FALSE | Upregulated |
| 0,415745524 | 0,134227719 | SNRK-AS1 | FALSE | Upregulated |
| 0,514082259 | 0,134361791 | CCSAP | FALSE | Upregulated |
| 1,663805058 | 0,134361791 | RP11-379K22.2 | FALSE | Upregulated |
| 2,072795069 | 0,134460775 | FRMD7 | FALSE | Upregulated |
| -1,129204674 | 0,134460775 | NAIF1 | FALSE | Downregulated |
| -0,326339035 | 0,134460775 | SLC25A1 | FALSE | Downregulated |
| 0,863927642 | 0,134533543 | CCR9 | FALSE | Upregulated |
| -2,479427123 | 0,134555292 | FAM35DP | FALSE | Downregulated |
| 0,856295066 | 0,134578398 | B3GAT3 | FALSE | Upregulated |
| 0,505392724 | 0,134671602 | C12orf61 | FALSE | Upregulated |
| -0,407565724 | 0,134940981 | ACOX1 | FALSE | Downregulated |
| 0,947225307 | 0,134940981 | C6orf25 | FALSE | Upregulated |
| 0,690747804 | 0,134984097 | SNRPB | FALSE | Upregulated |
| 0,48659099 | 0,135314472 | ADD3 | FALSE | Upregulated |
| -0,662717286 | 0,135314472 | C8G | FALSE | Downregulated |
| -0,568623325 | 0,135314472 | ECHDC3 | FALSE | Downregulated |
| 0,49694169 | 0,135314472 | EPB41L4A | FALSE | Upregulated |
| 0,532133233 | 0,135314472 | ERVFRD-1 | FALSE | Upregulated |
| -0,328619509 | 0,135314472 | RETSAT | FALSE | Downregulated |
| 0,673694105 | 0,13555176 | RAPGEF6 | FALSE | Upregulated |
| 0,832040582 | 0,135579655 | ABCA7 | FALSE | Upregulated |
| -0,514045672 | 0,135588517 | HSD17B10 | FALSE | Downregulated |
| 1,300336911 | 0,13593781 | NFATC4 | FALSE | Upregulated |
| 0,413571788 | 0,136011442 | PHKG1 | FALSE | Upregulated |
| 1,439594265 | 0,136198534 | A4GNT | FALSE | Upregulated |
| -0,492954403 | 0,136198534 | GRB14 | FALSE | Downregulated |
| -0,380913248 | 0,136598625 | HABP2 | FALSE | Downregulated |
| -0,678255181 | 0,136598625 | RPRD1B | FALSE | Downregulated |
| 1,031844037 | 0,136598625 | TCF19 | FALSE | Upregulated |
| -0,8930504 | 0,136645663 | BANF1 | FALSE | Downregulated |
| 1,003528717 | 0,136645663 | FAM102B | FALSE | Upregulated |
| 0,807838037 | 0,136645663 | PBX1 | FALSE | Upregulated |
| -0,55781315 | 0,136871023 | BAIAP2 | FALSE | Downregulated |
| 0,788211932 | 0,137322674 | HSPA4L | FALSE | Upregulated |
| 0,974424694 | 0,137322674 | ITGAX | FALSE | Upregulated |
| -0,853397484 | 0,137374771 | PDSS2 | FALSE | Downregulated |
| 1,386480686 | 0,137374771 | TNFRSF10A | FALSE | Upregulated |
| 0,752374302 | 0,137535273 | KIAA1462 | FALSE | Upregulated |
| -1,321771576 | 0,137535273 | MIS12 | FALSE | Downregulated |
| 0,954626101 | 0,137594798 | PIANP | FALSE | Upregulated |

| | | | | |
|---|---|---|---|---|
| 1,012102996 | 0,13771744 | FCGR2A | FALSE | Upregulated |
| 0,535713882 | 0,13780431 | NPHP3 | FALSE | Upregulated |
| 0,309220074 | 0,138011997 | RBPMS | FALSE | Upregulated |
| 0,672032323 | 0,138023002 | A2M | FALSE | Upregulated |
| -0,599001602 | 0,13859023 | AUH | FALSE | Downregulated |
| -0,730803325 | 0,13934218 | CPOX | FALSE | Downregulated |
| 0,781945139 | 0,13934218 | CYB5R2 | FALSE | Upregulated |
| -0,720819064 | 0,13934218 | TTBK1 | FALSE | Downregulated |
| -1,092238808 | 0,139506567 | CLCN4 | FALSE | Downregulated |
| -0,354075927 | 0,139506567 | NHSL1 | FALSE | Downregulated |
| 0,491299574 | 0,13999528 | MARCH1 | FALSE | Upregulated |
| 0,564936256 | 0,140023797 | GPX2 | FALSE | Upregulated |
| 0,529439954 | 0,140023797 | MTRF1L | FALSE | Upregulated |
| 1,480086612 | 0,140023797 | NBL1 | FALSE | Upregulated |
| 0,502317087 | 0,140256904 | PTRF | FALSE | Upregulated |
| 0,658860959 | 0,140349161 | TP53I3 | FALSE | Upregulated |
| -0,978851702 | 0,140555067 | FUNDC2 | FALSE | Downregulated |
| -0,551431235 | 0,140555067 | KAT6B | FALSE | Downregulated |
| 1,476987169 | 0,140555067 | LOC101929937 | FALSE | Upregulated |
| 0,682870601 | 0,140574437 | LOC101929005 | FALSE | Upregulated |
| 1,473306421 | 0,140881169 | FOSL1 | FALSE | Upregulated |
| 0,42453366 | 0,141248175 | RABGAP1L | FALSE | Upregulated |
| 0,68998385 | 0,14130694 | HLA-DPB1 | FALSE | Upregulated |
| 0,599087027 | 0,141404975 | TRPC5 | FALSE | Upregulated |
| 1,295372407 | 0,141649787 | HOXA3 | FALSE | Upregulated |
| 1,676583871 | 0,141833244 | HOXB2 | FALSE | Upregulated |
| 0,951835093 | 0,141833244 | LOC100129148 | FALSE | Upregulated |
| 0,860248463 | 0,141833244 | PLBD1 | FALSE | Upregulated |
| -0,505722925 | 0,141833244 | PRDX2 | FALSE | Downregulated |
| -0,520747338 | 0,141833244 | RPS21 | FALSE | Downregulated |
| -0,432259417 | 0,141833244 | SCARB1 | FALSE | Downregulated |
| -0,401207965 | 0,141833244 | SORBS1 | FALSE | Downregulated |
| 0,704438539 | 0,141833244 | TAGLN | FALSE | Upregulated |
| 0,439106557 | 0,142206489 | IL17RB | FALSE | Upregulated |
| 1,575451323 | 0,142206489 | RP4-639F20.1 | FALSE | Upregulated |
| 1,860927713 | 0,142381075 | LINC00966 | FALSE | Upregulated |
| -0,404219482 | 0,142638024 | SLC39A14 | FALSE | Downregulated |
| 0,588817929 | 0,142967896 | FBXL16 | FALSE | Upregulated |
| -0,589625622 | 0,143026861 | TTC36 | FALSE | Downregulated |
| 1,588635083 | 0,143035113 | LOC101927183 | FALSE | Upregulated |
| -0,596540484 | 0,143035113 | ZNF281 | FALSE | Downregulated |
| 0,401072601 | 0,143092895 | SPATA21 | FALSE | Upregulated |
| -0,366179181 | 0,143242614 | CLPTM1 | FALSE | Downregulated |
| 1,143372339 | 0,143242614 | LGALS4 | FALSE | Upregulated |
| -0,29524203 | 0,143242614 | LONP2 | FALSE | Downregulated |
| 0,814260695 | 0,143242614 | MEGF6 | FALSE | Upregulated |
| -0,848430193 | 0,143242614 | NPR1 | FALSE | Downregulated |
| 0,903928387 | 0,143242614 | SACS | FALSE | Upregulated |
| -0,646637527 | 0,143242614 | STRAP | FALSE | Downregulated |
| -1,01815677 | 0,143646895 | TPSG1 | FALSE | Downregulated |
| -0,45562756 | 0,143739029 | STK40 | FALSE | Downregulated |

| | | | | |
|---|---|---|---|---|
| -0,480579788 | 0,143849583 | ERF | FALSE | Downregulated |
| -0,568567885 | 0,144042086 | KCND3 | FALSE | Downregulated |
| 1,677950647 | 0,144046117 | CDSN | FALSE | Upregulated |
| -1,528062477 | 0,144202534 | C17orf89 | FALSE | Downregulated |
| 2,12780151 | 0,144283881 | RP11-475N22.4 | FALSE | Upregulated |
| 2,350758906 | 0,144367171 | LOC729162 | FALSE | Upregulated |
| 1,960668014 | 0,144462074 | AC074138.3 | FALSE | Upregulated |
| -0,992567198 | 0,144462074 | AQP7P1 | FALSE | Downregulated |
| 0,697790967 | 0,144476803 | MGAT5 | FALSE | Upregulated |
| -0,791928078 | 0,144476803 | TBC1D2 | FALSE | Downregulated |
| -0,583660058 | 0,144584499 | TADA2B | FALSE | Downregulated |
| -0,495791047 | 0,144589768 | ABCA5 | FALSE | Downregulated |
| 2,168363007 | 0,14479471 | ITPKB-IT1 | FALSE | Upregulated |
| 0,524210317 | 0,144817429 | AGFG1 | FALSE | Upregulated |
| 1,132057984 | 0,144817429 | CHST4 | FALSE | Upregulated |
| -2,218020208 | 0,144817429 | MT1H | FALSE | Downregulated |
| -0,649168665 | 0,144817429 | SPRED2 | FALSE | Downregulated |
| -0,45857945 | 0,145105374 | GALNT2 | FALSE | Downregulated |
| 0,886191113 | 0,145105374 | TIMM22 | FALSE | Upregulated |
| 1,339180138 | 0,145124447 | HSH2D | FALSE | Upregulated |
| -1,009005854 | 0,145186099 | NOTUM | FALSE | Downregulated |
| 1,608302592 | 0,145376546 | EOMES | FALSE | Upregulated |
| 1,277458857 | 0,145988471 | ZSCAN12P1 | FALSE | Upregulated |
| -1,085825341 | 0,146134065 | ZNF248 | FALSE | Downregulated |
| 1,349908065 | 0,146367284 | RP11-260E18.1 | FALSE | Upregulated |
| 0,459273433 | 0,146773528 | GPR183 | FALSE | Upregulated |
| -0,834076737 | 0,146791113 | MRPS18A | FALSE | Downregulated |
| 1,057266579 | 0,147007773 | ITGB2-AS1 | FALSE | Upregulated |
| -0,606749272 | 0,147106891 | NAT8 | FALSE | Downregulated |
| 0,843438022 | 0,147106891 | NME6 | FALSE | Upregulated |
| 1,159701775 | 0,147106891 | SHISA6 | FALSE | Upregulated |
| -0,543087021 | 0,147237349 | F13B | FALSE | Downregulated |
| -0,759120701 | 0,147238387 | MRPL54 | FALSE | Downregulated |
| 0,798401329 | 0,147331991 | PTGER4 | FALSE | Upregulated |
| 0,495395798 | 0,147331991 | UBE2L3 | FALSE | Upregulated |
| 1,286722392 | 0,147668937 | CCDC181 | FALSE | Upregulated |
| 0,951208088 | 0,147668937 | NXPH3 | FALSE | Upregulated |
| 1,809585585 | 0,147798519 | KLHDC7B | FALSE | Upregulated |
| -0,490954327 | 0,147798519 | KTN1 | FALSE | Downregulated |
| 1,06779861 | 0,147919817 | PTPRE | FALSE | Upregulated |
| 0,438270217 | 0,147919817 | SART3 | FALSE | Upregulated |
| -0,481441668 | 0,148063211 | CECR2 | FALSE | Downregulated |
| -1,175661753 | 0,148063211 | GRIN2A | FALSE | Downregulated |
| 0,951701138 | 0,148063211 | RP3-395M20.9 | FALSE | Upregulated |
| 1,578190878 | 0,148063211 | RP4-544H6.2 | FALSE | Upregulated |
| 1,557881457 | 0,148234649 | LOC100506454 | FALSE | Upregulated |
| 0,575192281 | 0,148234649 | TYMS | FALSE | Upregulated |
| 0,997844952 | 0,148347896 | CLIP3 | FALSE | Upregulated |
| 0,36556089 | 0,148347896 | DUSP16 | FALSE | Upregulated |
| 1,068737725 | 0,148347896 | HMGCS1 | FALSE | Upregulated |
| 1,785495042 | 0,148424188 | LOC100131347 | FALSE | Upregulated |

| | | | | |
|---|---|---|---|---|
| -0,463899392 | 0,148424188 | SLC22A7 | FALSE | Downregulated |
| -0,471290836 | 0,148424188 | ZCCHC24 | FALSE | Downregulated |
| -0,429402408 | 0,148428143 | MTTP | FALSE | Downregulated |
| 1,19005729 | 0,148612333 | ARHGEF38 | FALSE | Upregulated |
| 0,4873674 | 0,149287803 | AVL9 | FALSE | Upregulated |
| 0,512754383 | 0,149664876 | FAR2 | FALSE | Upregulated |
| -0,400619214 | 0,149855892 | HNRNPM | FALSE | Downregulated |
| -0,641369225 | 0,149984195 | COX7C | FALSE | Downregulated |
| 0,543380118 | 0,150149312 | GPM6B | FALSE | Upregulated |
| -0,486362327 | 0,150149312 | RPLP1 | FALSE | Downregulated |
| 0,88448068 | 0,150269161 | CCDC146 | FALSE | Upregulated |
| 2,104636746 | 0,150269161 | RP11-141O11.2 | FALSE | Upregulated |
| 0,758510765 | 0,15035891 | LOC166994 | FALSE | Upregulated |
| 1,903212099 | 0,150657058 | LOC102724100 | FALSE | Upregulated |
| -0,548249225 | 0,150872722 | CGREF1 | FALSE | Downregulated |
| 0,774437581 | 0,150877294 | MCAM | FALSE | Upregulated |
| -0,43213909 | 0,150922524 | ATP11C | FALSE | Downregulated |
| -0,689478643 | 0,150922524 | CDK14 | FALSE | Downregulated |
| 0,466872269 | 0,15119978 | CELSR1 | FALSE | Upregulated |
| 1,44802944 | 0,151256181 | LOC648987 | FALSE | Upregulated |
| -0,344625591 | 0,151256181 | PAPSS2 | FALSE | Downregulated |
| 0,438892114 | 0,151256181 | RBM5-AS1 | FALSE | Upregulated |
| -0,444003883 | 0,151322416 | DDI2 | FALSE | Downregulated |
| 0,442304839 | 0,151335969 | SENP5 | FALSE | Upregulated |
| 0,639035455 | 0,151431155 | C5orf45 | FALSE | Upregulated |
| 0,345784547 | 0,151431155 | TMBIM1 | FALSE | Upregulated |
| 1,123616023 | 0,151491585 | AC159540.1-2 | FALSE | Upregulated |
| 0,530053961 | 0,151507661 | CORO1C | FALSE | Upregulated |
| 0,678923541 | 0,151507661 | RARRES1 | FALSE | Upregulated |
| -0,378113388 | 0,151522667 | RPL23A | FALSE | Downregulated |
| -0,723879018 | 0,151746899 | LYVE1 | FALSE | Downregulated |
| -0,46257269 | 0,151878346 | FGFR4 | FALSE | Downregulated |
| 0,808576845 | 0,151990076 | MUC5B | FALSE | Upregulated |
| 1,05024153 | 0,152108778 | LUZP6 | FALSE | Upregulated |
| 0,81997513 | 0,152235253 | ARL2BP | FALSE | Upregulated |
| -0,773626444 | 0,152269767 | DMRTA1 | FALSE | Downregulated |
| -1,11391518 | 0,152269767 | MTMR6 | FALSE | Downregulated |
| 0,636588627 | 0,152269767 | RAP2B | FALSE | Upregulated |
| 0,907713328 | 0,152280255 | CCNDBP1 | FALSE | Upregulated |
| 0,971416684 | 0,152324588 | CDKN1A | FALSE | Upregulated |
| 0,601620048 | 0,152517623 | SNORA73A | FALSE | Upregulated |
| 0,390617629 | 0,152844828 | SIDT2 | FALSE | Upregulated |
| 1,182437883 | 0,153012753 | SPINT2 | FALSE | Upregulated |
| 1,714834037 | 0,153012753 | TMEM92 | FALSE | Upregulated |
| 0,88729752 | 0,153388577 | PPP1R13L | FALSE | Upregulated |
| 0,697921186 | 0,153638039 | MLK7-AS1 | FALSE | Upregulated |
| -0,594653267 | 0,153775607 | PALM2 | FALSE | Downregulated |
| -0,629485464 | 0,153831249 | HS3ST3B1 | FALSE | Downregulated |
| -0,341397789 | 0,153832547 | NIPSNAP1 | FALSE | Downregulated |
| 0,813356221 | 0,153990016 | BLMH | FALSE | Upregulated |
| 0,794900843 | 0,153990016 | RIT1 | FALSE | Upregulated |

| | | | | |
|---|---|---|---|---|
| -0,514066775 | 0,154003545 | FLRT3 | FALSE | Downregulated |
| 1,861276161 | 0,154003545 | RNF157-AS1 | FALSE | Upregulated |
| 0,74230446 | 0,154003545 | SUPT20H | FALSE | Upregulated |
| 0,409200823 | 0,154027938 | LOC652276 | FALSE | Upregulated |
| -1,078740225 | 0,154090857 | SLC35B1 | FALSE | Downregulated |
| 0,434579701 | 0,154262083 | GNPTAB | FALSE | Upregulated |
| -0,555273929 | 0,154363539 | HLCS | FALSE | Downregulated |
| 0,458534409 | 0,154435278 | C15orf60 | FALSE | Upregulated |
| 0,585558658 | 0,154466534 | KCNAB1 | FALSE | Upregulated |
| -0,859901753 | 0,154476546 | UBAC1 | FALSE | Downregulated |
| 0,931498825 | 0,154608792 | NRG1 | FALSE | Upregulated |
| -0,612190682 | 0,154721082 | ZFP41 | FALSE | Downregulated |
| -0,703954465 | 0,154905678 | CSAD | FALSE | Downregulated |
| -0,516386955 | 0,154905678 | FH | FALSE | Downregulated |
| -0,78536119 | 0,154905678 | LOC101929752 | FALSE | Downregulated |
| -0,791993576 | 0,154905678 | RCL1 | FALSE | Downregulated |
| -1,115048979 | 0,154905678 | RNU4-1 | FALSE | Downregulated |
| -0,445669415 | 0,155036942 | ACSM2A | FALSE | Downregulated |
| -0,465460268 | 0,155104306 | GPS1 | FALSE | Downregulated |
| 0,590700807 | 0,155290097 | PRICKLE4 | FALSE | Upregulated |
| -0,250945001 | 0,155290097 | SUN2 | FALSE | Downregulated |
| 0,58788203 | 0,1553029 | RAB6B | FALSE | Upregulated |
| 1,425179371 | 0,1553029 | SIT1 | FALSE | Upregulated |
| 0,407938086 | 0,155407046 | CORO1B | FALSE | Upregulated |
| 0,687137433 | 0,155531032 | C9orf172 | FALSE | Upregulated |
| 0,59155411 | 0,155651402 | LOC102723328 | FALSE | Upregulated |
| -0,334876197 | 0,155651402 | MYO18A | FALSE | Downregulated |
| 1,187309659 | 0,155884407 | MAP4K1 | FALSE | Upregulated |
| 0,625914462 | 0,155884407 | REEP1 | FALSE | Upregulated |
| 0,897338154 | 0,156052933 | SH3PXD2B | FALSE | Upregulated |
| 0,351826695 | 0,156052933 | TNS3 | FALSE | Upregulated |
| 0,999631765 | 0,156052933 | ZNF565 | FALSE | Upregulated |
| 1,299983713 | 0,156052933 | ZNF831 | FALSE | Upregulated |
| 0,671806077 | 0,156069816 | LOC102723422 | FALSE | Upregulated |
| 0,549156673 | 0,156092079 | CCNC | FALSE | Upregulated |
| 0,955357969 | 0,156140297 | TNFSF14 | FALSE | Upregulated |
| 2,135205994 | 0,156230274 | CCDC33 | FALSE | Upregulated |
| -0,466002457 | 0,156488311 | PNPLA2 | FALSE | Downregulated |
| -0,859581794 | 0,156488311 | WDR34 | FALSE | Downregulated |
| -0,433844357 | 0,156512199 | ITIH2 | FALSE | Downregulated |
| 0,820138514 | 0,156512199 | SH2D3C | FALSE | Upregulated |
| 0,942689737 | 0,156512199 | TLCD1 | FALSE | Upregulated |
| -0,510412784 | 0,157106164 | EPHX1 | FALSE | Downregulated |
| -1,026767875 | 0,157125596 | WDR83OS | FALSE | Downregulated |
| 0,684163216 | 0,157144306 | PRRT3 | FALSE | Upregulated |
| -0,495992571 | 0,157340676 | FAM195A | FALSE | Downregulated |
| -0,671900251 | 0,157361353 | FBXO28 | FALSE | Downregulated |
| 0,456124027 | 0,157514278 | NANOS1 | FALSE | Upregulated |
| 1,002240479 | 0,15751877 | KCNG2 | FALSE | Upregulated |
| 0,64265474 | 0,157613519 | HLA-A | FALSE | Upregulated |
| -0,663837402 | 0,157613519 | RTF1 | FALSE | Downregulated |

| | | | | |
|---|---|---|---|---|
| 0,487607117 | 0,157673304 | H2AFY | FALSE | Upregulated |
| 0,421589712 | 0,157673304 | ICMT | FALSE | Upregulated |
| 0,558668324 | 0,157673304 | PITPNC1 | FALSE | Upregulated |
| 0,800073559 | 0,157673304 | STAG3 | FALSE | Upregulated |
| 0,44366531 | 0,15779673 | CACYBP | FALSE | Upregulated |
| -0,366619968 | 0,157861988 | RIN3 | FALSE | Downregulated |
| 2,187499198 | 0,158141117 | LOC100506737 | FALSE | Upregulated |
| 0,575323572 | 0,159031427 | RFC5 | FALSE | Upregulated |
| -2,156389064 | 0,159097801 | ZFHX4-AS1 | FALSE | Downregulated |
| -0,761313947 | 0,159191343 | SNX13 | FALSE | Downregulated |
| 0,622385647 | 0,159191343 | UCHL5 | FALSE | Upregulated |
| 0,810428221 | 0,159230808 | SYK | FALSE | Upregulated |
| -1,535012488 | 0,159347812 | C16orf87 | FALSE | Downregulated |
| 0,707905713 | 0,159471427 | CYP21A2 | FALSE | Upregulated |
| 0,912727373 | 0,159471427 | DKFZP434K028 | FALSE | Upregulated |
| 0,806382515 | 0,159471427 | MOB3A | FALSE | Upregulated |
| 0,406559823 | 0,159585637 | DALRD3 | FALSE | Upregulated |
| 0,510562404 | 0,159585637 | FAM196A | FALSE | Upregulated |
| -1,051001817 | 0,159585637 | POLR3GL | FALSE | Downregulated |
| 0,467399998 | 0,159585637 | PPP1R9B | FALSE | Upregulated |
| -0,354214899 | 0,15959738 | ABCC3 | FALSE | Downregulated |
| -0,641171338 | 0,159901683 | ALKBH7 | FALSE | Downregulated |
| 0,692225666 | 0,159901683 | LOC641367 | FALSE | Upregulated |
| 0,55587924 | 0,159901683 | VOPP1 | FALSE | Upregulated |
| 0,502803512 | 0,159926615 | CROCCP3 | FALSE | Upregulated |
| 0,752782755 | 0,159979843 | COL8A2 | FALSE | Upregulated |
| -0,663264182 | 0,160341922 | GCAT | FALSE | Downregulated |
| 0,550711624 | 0,160571262 | ARHGEF2 | FALSE | Upregulated |
| 1,000026051 | 0,160571262 | SFMBT2 | FALSE | Upregulated |
| 0,623454779 | 0,16065758 | IPPK | FALSE | Upregulated |
| 1,013962001 | 0,160720494 | FLJ46836 | FALSE | Upregulated |
| -0,425819886 | 0,160734771 | SLC7A2 | FALSE | Downregulated |
| 2,309894631 | 0,161334399 | DTHD1 | FALSE | Upregulated |
| 0,636189487 | 0,161337452 | ORMDL2 | FALSE | Upregulated |
| -0,555166822 | 0,161634874 | FBP1 | FALSE | Downregulated |
| -0,554149581 | 0,161921794 | AKR7A3 | FALSE | Downregulated |
| 0,625783767 | 0,161921794 | TMEM41A | FALSE | Upregulated |
| -1,46117873 | 0,161921794 | ZNF34 | FALSE | Downregulated |
| -0,519478996 | 0,162245362 | GET4 | FALSE | Downregulated |
| 0,616676619 | 0,162337551 | FOXN3-AS1 | FALSE | Upregulated |
| 0,960170387 | 0,162337551 | PCDH18 | FALSE | Upregulated |
| 1,094337586 | 0,162485007 | FIGNL1 | FALSE | Upregulated |
| 0,545624615 | 0,162485007 | KIAA0556 | FALSE | Upregulated |
| 0,88322856 | 0,162485007 | LOC101928281 | FALSE | Upregulated |
| -0,330246295 | 0,162485007 | RPL9 | FALSE | Downregulated |
| 0,738470045 | 0,162485007 | STX6 | FALSE | Upregulated |
| -0,49951587 | 0,162485007 | SYAP1 | FALSE | Downregulated |
| -0,606485218 | 0,162485007 | VKORC1L1 | FALSE | Downregulated |
| 0,691394692 | 0,162586323 | CABYR | FALSE | Upregulated |
| -1,254754474 | 0,162697546 | CYP2W1 | FALSE | Downregulated |
| 0,58601279 | 0,162697546 | FAM161B | FALSE | Upregulated |

| | | | | |
|---|---|---|---|---|
| 0,458350838 | 0,162697546 | GOLM1 | FALSE | Upregulated |
| 0,915118537 | 0,162697546 | RP11-234K24.3 | FALSE | Upregulated |
| 0,524859602 | 0,162923136 | MIER3 | FALSE | Upregulated |
| -0,51922449 | 0,162995735 | ERLIN1 | FALSE | Downregulated |
| 0,693682837 | 0,162995735 | FAM50A | FALSE | Upregulated |
| -0,701600504 | 0,162995735 | GANC | FALSE | Downregulated |
| -0,596178622 | 0,162995735 | GPR89A | FALSE | Downregulated |
| -0,566630456 | 0,162995735 | GPT2 | FALSE | Downregulated |
| 0,512413244 | 0,162995735 | IFT140 | FALSE | Upregulated |
| 0,601309265 | 0,162995735 | KCNJ10 | FALSE | Upregulated |
| 1,927246511 | 0,162995735 | MZB1 | FALSE | Upregulated |
| -0,389925853 | 0,162995735 | NID1 | FALSE | Downregulated |
| 0,824670436 | 0,162995735 | TSPO | FALSE | Upregulated |
| 0,581965726 | 0,163381475 | ECM2 | FALSE | Upregulated |
| -0,780246755 | 0,163391658 | CYP4F12 | FALSE | Downregulated |
| 0,43389838 | 0,163391658 | SRA1 | FALSE | Upregulated |
| 0,440877731 | 0,163412847 | RSBN1L-AS1 | FALSE | Upregulated |
| 0,462935797 | 0,163635172 | RICTOR | FALSE | Upregulated |
| 0,462396772 | 0,163672784 | EIF3J | FALSE | Upregulated |
| 1,744071729 | 0,163754405 | BHLHA15 | FALSE | Upregulated |
| -0,521481863 | 0,16380645 | FLCN | FALSE | Downregulated |
| -0,639475848 | 0,16380645 | RAB2A | FALSE | Downregulated |
| 1,171926365 | 0,164096147 | LOC100129083 | FALSE | Upregulated |
| 0,529307248 | 0,164786277 | CCDC97 | FALSE | Upregulated |
| -0,479282573 | 0,164786277 | CFI | FALSE | Downregulated |
| -0,783142886 | 0,164786277 | CYP2D7P | FALSE | Downregulated |
| 0,694138113 | 0,164786277 | TRIB2 | FALSE | Upregulated |
| 0,372096895 | 0,164786277 | ZNFX1 | FALSE | Upregulated |
| -0,497440815 | 0,165109678 | ABCD3 | FALSE | Downregulated |
| -0,712665509 | 0,165109678 | RWDD2B | FALSE | Downregulated |
| 1,467184476 | 0,16515013 | GJC1 | FALSE | Upregulated |
| -0,410700841 | 0,16515013 | SOX13 | FALSE | Downregulated |
| -1,110634544 | 0,165227948 | LOC101927870 | FALSE | Downregulated |
| -0,456934253 | 0,165378756 | GLYATL1 | FALSE | Downregulated |
| 0,270000085 | 0,165378756 | USP34 | FALSE | Upregulated |
| 0,557079553 | 0,165488721 | FAM186B | FALSE | Upregulated |
| 0,646203205 | 0,165488721 | TCF4 | FALSE | Upregulated |
| 1,774263953 | 0,165488721 | TMEM246 | FALSE | Upregulated |
| 0,849792938 | 0,165577598 | RPS6KA1 | FALSE | Upregulated |
| 2,782341371 | 0,1657506 | FAM133DP | FALSE | Upregulated |
| -1,506476343 | 0,165759829 | BCAS2 | FALSE | Downregulated |
| -0,535400438 | 0,165771522 | CPED1 | FALSE | Downregulated |
| -0,334633309 | 0,165771522 | SMARCA2 | FALSE | Downregulated |
| 0,493074973 | 0,16595283 | ZNF397 | FALSE | Upregulated |
| -0,485646387 | 0,166146892 | CNPY2 | FALSE | Downregulated |
| -0,524626654 | 0,166262828 | ADAMTSL3 | FALSE | Downregulated |
| 0,741386896 | 0,166262828 | PDGFRB | FALSE | Upregulated |
| 0,736928468 | 0,16679073 | EME1 | FALSE | Upregulated |
| 1,56376556 | 0,16679073 | ITGB8 | FALSE | Upregulated |
| 0,814770946 | 0,16679073 | RP1-1J6.2 | FALSE | Upregulated |
| 1,917674199 | 0,16679073 | SCN7A | FALSE | Upregulated |

| | | | | |
|---|---|---|---|---|
| -0,643611946 | 0,166882106 | CHID1 | FALSE | Downregulated |
| 1,199246415 | 0,166882106 | LPXN | FALSE | Upregulated |
| -0,545788896 | 0,167016224 | ASMTL-AS1 | FALSE | Downregulated |
| 0,54489705 | 0,16703747 | CASP8 | FALSE | Upregulated |
| 2,369771566 | 0,167069577 | FAM169B | FALSE | Upregulated |
| 0,675937925 | 0,167069577 | IFFO2 | FALSE | Upregulated |
| 0,765670454 | 0,167069577 | TGFB3 | FALSE | Upregulated |
| 1,228230984 | 0,167069577 | TRIM15 | FALSE | Upregulated |
| 0,626448596 | 0,167131912 | ABHD8 | FALSE | Upregulated |
| -0,302881257 | 0,167131912 | AES | FALSE | Downregulated |
| -0,390631116 | 0,167131912 | ALDH3A2 | FALSE | Downregulated |
| 0,712566869 | 0,167131912 | CREB3L2 | FALSE | Upregulated |
| 0,531575015 | 0,167131912 | FAM129B | FALSE | Upregulated |
| 2,143057438 | 0,167131912 | FBN3 | FALSE | Upregulated |
| 1,866638383 | 0,167131912 | LOC102724135 | FALSE | Upregulated |
| -0,78837261 | 0,167131912 | PIM1 | FALSE | Downregulated |
| 1,458746929 | 0,167131912 | PPIL3 | FALSE | Upregulated |
| -0,512066079 | 0,167131912 | RDH10 | FALSE | Downregulated |
| 0,854952521 | 0,16720303 | LACC1 | FALSE | Upregulated |
| -0,492130424 | 0,167291732 | CEBPA | FALSE | Downregulated |
| -0,271604606 | 0,167291732 | LSR | FALSE | Downregulated |
| -0,692422161 | 0,167291732 | MAPK14 | FALSE | Downregulated |
| 0,628854656 | 0,167291732 | MVP | FALSE | Upregulated |
| 1,101446028 | 0,167303437 | GAS6-AS1 | FALSE | Upregulated |
| -0,81858204 | 0,167344457 | LOC730102 | FALSE | Downregulated |
| 0,555676238 | 0,167398931 | TRAF3IP2 | FALSE | Upregulated |
| 0,687458834 | 0,167675224 | USP3-AS1 | FALSE | Upregulated |
| 1,51797361 | 0,167762647 | SNHG1 | FALSE | Upregulated |
| 0,443169016 | 0,167791021 | GPR107 | FALSE | Upregulated |
| 0,602668681 | 0,167791021 | RP11-450K4.1 | FALSE | Upregulated |
| -0,480056416 | 0,167877294 | ACBD4 | FALSE | Downregulated |
| -0,693455776 | 0,168499797 | LRG1 | FALSE | Downregulated |
| -0,467119816 | 0,168565094 | ZNF358 | FALSE | Downregulated |
| -0,575575787 | 0,168599079 | TCEB2 | FALSE | Downregulated |
| 0,558799767 | 0,169166037 | BNIP3L | FALSE | Upregulated |
| 0,769972171 | 0,169189223 | RNFT1 | FALSE | Upregulated |
| -0,912241723 | 0,169360494 | NNMT | FALSE | Downregulated |
| 2,11922919 | 0,169363634 | CD79A | FALSE | Upregulated |
| -0,591360063 | 0,169439382 | PLA2G12A | FALSE | Downregulated |
| 0,98262752 | 0,169736608 | PRNCR1 | FALSE | Upregulated |
| -0,642944297 | 0,169736608 | SLC9B2 | FALSE | Downregulated |
| -0,439056981 | 0,169830931 | ASPH | FALSE | Downregulated |
| -0,581579767 | 0,169830931 | ERI2 | FALSE | Downregulated |
| -1,270176641 | 0,169830931 | ESPNL | FALSE | Downregulated |
| 2,317147924 | 0,169830931 | LAMC2 | FALSE | Upregulated |
| 1,659638777 | 0,169830931 | LOC729737 | FALSE | Upregulated |
| 0,708239972 | 0,169830931 | OSBPL1A | FALSE | Upregulated |
| -0,437990398 | 0,169849228 | LRPPRC | FALSE | Downregulated |
| -0,50044653 | 0,169864966 | SEPSECS | FALSE | Downregulated |
| -0,54034311 | 0,16986534 | RPL37 | FALSE | Downregulated |
| -0,266786919 | 0,169920025 | RPL18A | FALSE | Downregulated |

| | | | | |
|---|---|---|---|---|
| 0,434420606 | 0,170007708 | ANKRD36BP1 | FALSE | Upregulated |
| -0,352536983 | 0,170016105 | MOV10 | FALSE | Downregulated |
| 0,597703205 | 0,170049894 | CRIP2 | FALSE | Upregulated |
| 0,437751381 | 0,170049894 | RBM4 | FALSE | Upregulated |
| 0,638482777 | 0,170091708 | NBEAL2 | FALSE | Upregulated |
| 1,53952991 | 0,170151837 | LOC101060195 | FALSE | Upregulated |
| -0,437230762 | 0,170151837 | MRPL12 | FALSE | Downregulated |
| 0,430381741 | 0,170554447 | DHDDS | FALSE | Upregulated |
| -0,867434093 | 0,170554447 | LOC102724852 | FALSE | Downregulated |
| -0,521424634 | 0,170554447 | RBM26 | FALSE | Downregulated |
| -0,431516449 | 0,170672524 | CDK5RAP3 | FALSE | Downregulated |
| 1,61159523 | 0,170681794 | GPR150 | FALSE | Upregulated |
| 2,170221994 | 0,170760144 | CLIC6 | FALSE | Upregulated |
| -1,403313474 | 0,170760144 | LNP1 | FALSE | Downregulated |
| -0,359527238 | 0,170831527 | CDK10 | FALSE | Downregulated |
| -0,631069495 | 0,170831527 | FRRS1 | FALSE | Downregulated |
| 0,746585095 | 0,171260881 | C19orf68 | FALSE | Upregulated |
| -2,331874244 | 0,171283006 | SLC45A2 | FALSE | Downregulated |
| 1,093423478 | 0,171408255 | GNG12-AS1 | FALSE | Upregulated |
| 0,44410528 | 0,171553653 | JADE2 | FALSE | Upregulated |
| 0,422343773 | 0,171553653 | LRRN3 | FALSE | Upregulated |
| 1,28908943 | 0,171587347 | OSCP1 | FALSE | Upregulated |
| 0,383264673 | 0,171822914 | MPEG1 | FALSE | Upregulated |
| 0,707315235 | 0,172048511 | ZNF518B | FALSE | Upregulated |
| 0,52897449 | 0,17205413 | FILIP1L | FALSE | Upregulated |
| 0,365158874 | 0,172280664 | LRP10 | FALSE | Upregulated |
| 1,11527689 | 0,17239666 | E2F3 | FALSE | Upregulated |
| 0,527392132 | 0,172470814 | ATP2B4 | FALSE | Upregulated |
| -0,552983398 | 0,172560555 | C11orf54 | FALSE | Downregulated |
| 0,500670853 | 0,172623701 | ZNF79 | FALSE | Upregulated |
| 0,439635294 | 0,172720373 | EYS | FALSE | Upregulated |
| 0,872061365 | 0,172720373 | HLA-H | FALSE | Upregulated |
| 1,3413778 | 0,172753528 | FSCN2 | FALSE | Upregulated |
| 0,459981091 | 0,172997569 | AGK | FALSE | Upregulated |
| 0,662759397 | 0,173033713 | TACR1 | FALSE | Upregulated |
| 0,980265566 | 0,173440481 | KCNH3 | FALSE | Upregulated |
| 1,781288511 | 0,173540263 | LPAR1 | FALSE | Upregulated |
| 0,679335303 | 0,17360704 | GATS | FALSE | Upregulated |
| -0,280441105 | 0,17360704 | KIF1C | FALSE | Downregulated |
| 0,807872446 | 0,173731876 | TMEM8B | FALSE | Upregulated |
| 0,871334097 | 0,173802736 | LCTL | FALSE | Upregulated |
| -2,111597331 | 0,173802736 | SMN1 | FALSE | Downregulated |
| -1,451728955 | 0,173802736 | TEC | FALSE | Downregulated |
| -0,681855773 | 0,173922153 | EARS2 | FALSE | Downregulated |
| 0,521738496 | 0,174018241 | BCL2L1 | FALSE | Upregulated |
| -0,613054251 | 0,174018241 | FAM205A | FALSE | Downregulated |
| 1,087357117 | 0,174018241 | PLEKHB2 | FALSE | Upregulated |
| 0,996349342 | 0,174018241 | SLFN11 | FALSE | Upregulated |
| -0,450740269 | 0,174028044 | BAAT | FALSE | Downregulated |
| 0,490456818 | 0,174028044 | SPTY2D1-AS1 | FALSE | Upregulated |
| 0,632625727 | 0,174519142 | DDX28 | FALSE | Upregulated |

| | | | | |
|---|---|---|---|---|
| -0,422849857 | 0,174556027 | C1RL | FALSE | Downregulated |
| -0,533810708 | 0,174556027 | CYP2C8 | FALSE | Downregulated |
| -2,517631153 | 0,174556027 | LOC389602 | FALSE | Downregulated |
| -0,528656609 | 0,174556027 | NET1 | FALSE | Downregulated |
| 1,857892085 | 0,174611722 | OXCT1 | FALSE | Upregulated |
| 0,585315456 | 0,174772439 | DBP | FALSE | Upregulated |
| -0,394422506 | 0,174982527 | RPS25 | FALSE | Downregulated |
| 0,463553459 | 0,175053802 | PTPN14 | FALSE | Upregulated |
| 0,568582941 | 0,175119703 | FAM227B | FALSE | Upregulated |
| 0,739905095 | 0,175119703 | HMGN3-AS1 | FALSE | Upregulated |
| -1,790259323 | 0,175119703 | LOC101926894 | FALSE | Downregulated |
| 1,080471193 | 0,175119703 | LOC101929698 | FALSE | Upregulated |
| 0,741015985 | 0,175502003 | CCDC3 | FALSE | Upregulated |
| -0,293495298 | 0,175545662 | CTDSP2 | FALSE | Downregulated |
| 0,660229536 | 0,17571591 | RFXANK | FALSE | Upregulated |
| -0,458862145 | 0,176000245 | RPS4X | FALSE | Downregulated |
| 0,717709801 | 0,176000245 | SCG3 | FALSE | Upregulated |
| -0,476233881 | 0,176000245 | SRSF11 | FALSE | Downregulated |
| 0,565102089 | 0,176000245 | UNC93B1 | FALSE | Upregulated |
| 0,65728525 | 0,176002567 | APOL4 | FALSE | Upregulated |
| 0,928762427 | 0,176093384 | CSRP2 | FALSE | Upregulated |
| 0,578548442 | 0,176093384 | PFDN1 | FALSE | Upregulated |
| -0,751845645 | 0,176093384 | ZFP1 | FALSE | Downregulated |
| -0,721087766 | 0,176219964 | ABLIM3 | FALSE | Downregulated |
| 0,458493326 | 0,176394771 | ARHGEF1 | FALSE | Upregulated |
| 1,251039092 | 0,17652689 | BICD1 | FALSE | Upregulated |
| 0,7858286 | 0,176785936 | PLEKHO1 | FALSE | Upregulated |
| 0,406299033 | 0,176790351 | ZACN | FALSE | Upregulated |
| 1,028968238 | 0,176812857 | CD81-AS1 | FALSE | Upregulated |
| 0,393622534 | 0,177173653 | LINC00094 | FALSE | Upregulated |
| -2,134840927 | 0,177380246 | LOC100507670 | FALSE | Downregulated |
| -0,424880902 | 0,177454633 | AKR7A2 | FALSE | Downregulated |
| 1,601908905 | 0,177454633 | GPR84 | FALSE | Upregulated |
| -1,007533882 | 0,177454633 | IER3IP1 | FALSE | Downregulated |
| -0,442604764 | 0,177454633 | LIME1 | FALSE | Downregulated |
| 1,650426344 | 0,177454633 | RRM2 | FALSE | Upregulated |
| -0,325829797 | 0,177454633 | SF3A1 | FALSE | Downregulated |
| -0,540895534 | 0,177454633 | SRD5A1 | FALSE | Downregulated |
| 0,833703268 | 0,177454633 | VAMP5 | FALSE | Upregulated |
| 0,822953963 | 0,177454633 | WRN | FALSE | Upregulated |
| 0,561571682 | 0,177632493 | DUSP28 | FALSE | Upregulated |
| 0,606505725 | 0,177632493 | GIMAP8 | FALSE | Upregulated |
| 0,65265141 | 0,177819062 | CBLB | FALSE | Upregulated |
| 0,616456537 | 0,177819062 | FAM63B | FALSE | Upregulated |
| 0,58228202 | 0,177819062 | SCRN3 | FALSE | Upregulated |
| -1,424563823 | 0,177819062 | TPRG1 | FALSE | Downregulated |
| 0,665962931 | 0,178011642 | ITGAL | FALSE | Upregulated |
| 0,867519219 | 0,178073962 | PRKG1 | FALSE | Upregulated |
| 0,604513221 | 0,178151023 | PEX12 | FALSE | Upregulated |
| 2,888244251 | 0,178210654 | ANKRD22 | FALSE | Upregulated |
| 0,498214887 | 0,178210654 | ATP5SL | FALSE | Upregulated |

| | | | | |
|---|---|---|---|---|
| 0,640738057 | 0,178210654 | TTL | FALSE | Upregulated |
| 1,611063965 | 0,178217188 | MEPE | FALSE | Upregulated |
| 1,109579024 | 0,178217188 | RAC2 | FALSE | Upregulated |
| 1,161154169 | 0,178299126 | LOC101928215 | FALSE | Upregulated |
| -2,132923541 | 0,178568271 | RTN4RL1 | FALSE | Downregulated |
| 0,390390648 | 0,178568271 | SH3BP5-AS1 | FALSE | Upregulated |
| -0,278547701 | 0,178709304 | XBP1 | FALSE | Downregulated |
| -1,050703666 | 0,178789535 | ZNF212 | FALSE | Downregulated |
| 0,861625255 | 0,179303695 | ZNF367 | FALSE | Upregulated |
| 0,557942883 | 0,179503362 | TBX2 | FALSE | Upregulated |
| 0,63976971 | 0,17956099 | KCNC3 | FALSE | Upregulated |
| 0,427042133 | 0,17956099 | MED26 | FALSE | Upregulated |
| 1,122952926 | 0,179721326 | AMPH | FALSE | Upregulated |
| -0,880041146 | 0,179755855 | CLEC3B | FALSE | Downregulated |
| 0,662991296 | 0,179755855 | LOC102724364 | FALSE | Upregulated |
| 1,957201576 | 0,179805494 | KCNJ5 | FALSE | Upregulated |
| -1,005679722 | 0,179812489 | RP11-475O6.1-2 | FALSE | Downregulated |
| -0,590564184 | 0,179847301 | PDIA6 | FALSE | Downregulated |
| 1,269917276 | 0,180024403 | PM20D1 | FALSE | Upregulated |
| 0,490179427 | 0,180024403 | SLC35B2 | FALSE | Upregulated |
| 0,713742349 | 0,18010926 | RNF24 | FALSE | Upregulated |
| -0,690925644 | 0,180233526 | SENP2 | FALSE | Downregulated |
| 0,531624169 | 0,180320012 | PTPN12 | FALSE | Upregulated |
| 2,185439692 | 0,180353603 | RUNX2 | FALSE | Upregulated |
| -0,483599032 | 0,180642188 | SERPINA4 | FALSE | Downregulated |
| -1,015942172 | 0,180914785 | LOC101928938 | FALSE | Downregulated |
| -0,519573751 | 0,181075987 | ATP13A1 | FALSE | Downregulated |
| -0,918198329 | 0,181162511 | ZNF408 | FALSE | Downregulated |
| 0,633773102 | 0,181221114 | LOC102723824 | FALSE | Upregulated |
| 0,994534468 | 0,181492417 | LOC102724518 | FALSE | Upregulated |
| 0,404400339 | 0,181838914 | DOPEY1 | FALSE | Upregulated |
| 1,121132605 | 0,181838914 | MB21D1 | FALSE | Upregulated |
| -0,393433566 | 0,181838914 | PPIB | FALSE | Downregulated |
| -0,921598256 | 0,181853013 | ZDBF2 | FALSE | Downregulated |
| -0,2998542 | 0,182198401 | KHSRP | FALSE | Downregulated |
| -0,484724636 | 0,182626543 | PNPLA7 | FALSE | Downregulated |
| 0,568101267 | 0,182741972 | LOC100128361 | FALSE | Upregulated |
| -0,442545382 | 0,182886467 | ACP2 | FALSE | Downregulated |
| 0,392949272 | 0,182952142 | GDF11 | FALSE | Upregulated |
| 0,67211172 | 0,182976672 | EMG1 | FALSE | Upregulated |
| 1,125158131 | 0,182979152 | IKBKE | FALSE | Upregulated |
| -0,987839734 | 0,182979152 | KCNMA1 | FALSE | Downregulated |
| -0,32422218 | 0,182979152 | STARD10 | FALSE | Downregulated |
| 1,126499617 | 0,183286178 | ADAM12 | FALSE | Upregulated |
| -0,911438794 | 0,183286178 | BRCA1 | FALSE | Downregulated |
| 0,651510729 | 0,183286178 | DLAT | FALSE | Upregulated |
| 0,939913339 | 0,183386707 | WNK2 | FALSE | Upregulated |
| -0,474575202 | 0,183539803 | LUC7L2 | FALSE | Downregulated |
| 0,446523202 | 0,183600585 | KDM1B | FALSE | Upregulated |
| 0,434418141 | 0,183643343 | CRBN | FALSE | Upregulated |
| -0,363311682 | 0,183649676 | KNG1 | FALSE | Downregulated |

| | | | | |
|---|---|---|---|---|
| 2,227469699 | 0,183649676 | LOC102724235 | FALSE | Upregulated |
| -0,465041409 | 0,183670656 | PDIA4 | FALSE | Downregulated |
| 2,36828808 | 0,183892762 | LGI2 | FALSE | Upregulated |
| 0,705674175 | 0,184087059 | C1QTNF6 | FALSE | Upregulated |
| 2,222498685 | 0,184087059 | TNF | FALSE | Upregulated |
| 0,595860745 | 0,184364809 | FAM212B | FALSE | Upregulated |
| 2,191261084 | 0,184364809 | GPRIN2 | FALSE | Upregulated |
| -0,334522673 | 0,184364809 | H6PD | FALSE | Downregulated |
| 0,554599764 | 0,184364809 | OPA3 | FALSE | Upregulated |
| -0,733073966 | 0,184364809 | PKIG | FALSE | Downregulated |
| -2,614636793 | 0,184381483 | H2AFB3 | FALSE | Downregulated |
| 1,829227791 | 0,184381483 | HUNK | FALSE | Upregulated |
| 0,512280457 | 0,184381483 | KDM4A-AS1 | FALSE | Upregulated |
| 0,380494605 | 0,184569451 | RBMS2 | FALSE | Upregulated |
| 0,52607476 | 0,184603688 | INSIG2 | FALSE | Upregulated |
| -0,354741911 | 0,184603688 | PARD3 | FALSE | Downregulated |
| 0,667811991 | 0,185146745 | LOC102724549 | FALSE | Upregulated |
| -0,745805367 | 0,185146745 | OST4 | FALSE | Downregulated |
| -1,025032275 | 0,185146745 | PZP | FALSE | Downregulated |
| -0,569021472 | 0,185146745 | SMPDL3A | FALSE | Downregulated |
| 1,103862451 | 0,18522338 | MAB21L1 | FALSE | Upregulated |
| 0,591361306 | 0,18522338 | RPP38 | FALSE | Upregulated |
| 1,123612828 | 0,185228875 | HMGB2 | FALSE | Upregulated |
| 0,585077987 | 0,185327353 | ARHGEF6 | FALSE | Upregulated |
| -0,514068438 | 0,185327353 | MSRB1 | FALSE | Downregulated |
| 0,418214858 | 0,185327353 | NUMB | FALSE | Upregulated |
| -0,334763997 | 0,185327353 | RBP5 | FALSE | Downregulated |
| 1,757747372 | 0,18542657 | EFCAB12 | FALSE | Upregulated |
| -0,668343585 | 0,185717158 | CYP4A22 | FALSE | Downregulated |
| -0,424741918 | 0,185717158 | IVD | FALSE | Downregulated |
| 2,011681279 | 0,185717158 | LOC101929442 | FALSE | Upregulated |
| 0,851907835 | 0,185717158 | MAP6 | FALSE | Upregulated |
| -0,558236032 | 0,185717158 | SEC22B | FALSE | Downregulated |
| -0,367340626 | 0,185717158 | SFXN5 | FALSE | Downregulated |
| 5,981850506 | 0,185717158 | TNNT3 | FALSE | Upregulated |
| -2,269476165 | 0,186111766 | LOC102723965 | FALSE | Downregulated |
| -0,536905597 | 0,186153684 | LOC100996492 | FALSE | Downregulated |
| -0,624807645 | 0,186153684 | RNU4-2 | FALSE | Downregulated |
| -0,513387943 | 0,186246212 | HFE2 | FALSE | Downregulated |
| 1,887951633 | 0,186246212 | S100A4 | FALSE | Upregulated |
| 1,171624159 | 0,186325253 | TFEC | FALSE | Upregulated |
| -0,596786898 | 0,186432286 | NDUFV2 | FALSE | Downregulated |
| 0,839226395 | 0,186685925 | CR1 | FALSE | Upregulated |
| 0,37211786 | 0,186685925 | KANSL1L | FALSE | Upregulated |
| 0,599127894 | 0,186766685 | CAND2 | FALSE | Upregulated |
| 0,640169381 | 0,186766685 | GPR113 | FALSE | Upregulated |
| -0,367525632 | 0,186766685 | GUSB | FALSE | Downregulated |
| -0,884646735 | 0,186766685 | IFITM10 | FALSE | Downregulated |
| -0,559383664 | 0,186850177 | MRPS34 | FALSE | Downregulated |
| -0,317434577 | 0,186850177 | MYLK | FALSE | Downregulated |
| 0,826817505 | 0,187220033 | RAB36 | FALSE | Upregulated |

| | | | | |
|---|---|---|---|---|
| 0,692020404 | 0,18733529 | VPS8 | FALSE | Upregulated |
| -0,50757695 | 0,18766858 | NHLRC2 | FALSE | Downregulated |
| 0,806838248 | 0,187858178 | LOC101929636 | FALSE | Upregulated |
| 0,590055132 | 0,188027124 | ARL17A | FALSE | Upregulated |
| 0,809700178 | 0,188027124 | LOC101928069 | FALSE | Upregulated |
| 1,952165686 | 0,188027124 | STOX1 | FALSE | Upregulated |
| 1,015289255 | 0,188045026 | SEMA3E | FALSE | Upregulated |
| -0,717519335 | 0,18824097 | IL1RAP | FALSE | Downregulated |
| 1,086867135 | 0,18824097 | LOC101927675 | FALSE | Upregulated |
| 0,354058796 | 0,18824097 | MPP5 | FALSE | Upregulated |
| -0,697989204 | 0,18824097 | NRK | FALSE | Downregulated |
| -0,366181003 | 0,188251816 | GCGR | FALSE | Downregulated |
| 0,411152942 | 0,188360015 | GZF1 | FALSE | Upregulated |
| 1,545518044 | 0,188360015 | LOC102724147 | FALSE | Upregulated |
| -0,539542367 | 0,188360015 | PRDX3 | FALSE | Downregulated |
| -1,01298884 | 0,188372705 | RP11-7F17.5 | FALSE | Downregulated |
| 0,750387118 | 0,188372705 | ZDHHC15 | FALSE | Upregulated |
| 0,975931932 | 0,188377262 | LONRF2 | FALSE | Upregulated |
| 0,68311541 | 0,188539055 | RUNDC3B | FALSE | Upregulated |
| 0,502015034 | 0,188643064 | LOC101929340 | FALSE | Upregulated |
| 0,589787607 | 0,188643064 | LTBP4 | FALSE | Upregulated |
| -0,704366975 | 0,188643064 | RPUSD4 | FALSE | Downregulated |
| 0,759943561 | 0,188920215 | ALDH1A3 | FALSE | Upregulated |
| -0,397102274 | 0,188920215 | CLDN1 | FALSE | Downregulated |
| 0,506712537 | 0,188920215 | PPP1R18 | FALSE | Upregulated |
| -1,621787015 | 0,188920215 | PRKAR2B | FALSE | Downregulated |
| -0,379043835 | 0,18892309 | IARS2 | FALSE | Downregulated |
| 0,409242213 | 0,189111586 | CD3EAP | FALSE | Upregulated |
| 0,735943601 | 0,189111586 | FAM206A | FALSE | Upregulated |
| -0,911787833 | 0,189111586 | GSPT2 | FALSE | Downregulated |
| -1,863828167 | 0,189111586 | HAMP | FALSE | Downregulated |
| 0,740484534 | 0,189111586 | LOC100499194 | FALSE | Upregulated |
| 1,027123233 | 0,189111586 | MED10 | FALSE | Upregulated |
| 1,960783778 | 0,189111586 | TCHH | FALSE | Upregulated |
| -0,396839716 | 0,189376753 | ESPN | FALSE | Downregulated |
| 0,456931305 | 0,18943601 | TUBGCP4 | FALSE | Upregulated |
| 0,511905753 | 0,189480737 | CCNK | FALSE | Upregulated |
| 1,239590922 | 0,189613413 | G6PC2 | FALSE | Upregulated |
| 0,432763318 | 0,189841183 | LOC100303749 | FALSE | Upregulated |
| -1,011887495 | 0,189841183 | TEDDM1 | FALSE | Downregulated |
| 1,113468055 | 0,189888676 | USHBP1 | FALSE | Upregulated |
| -0,383473453 | 0,190001935 | MAP2K7 | FALSE | Downregulated |
| 0,802473954 | 0,190001935 | PKDCC | FALSE | Upregulated |
| 0,508572381 | 0,190001935 | SPON1 | FALSE | Upregulated |
| 1,043167028 | 0,190001935 | SRPX | FALSE | Upregulated |
| 0,661239663 | 0,190001935 | ST7L | FALSE | Upregulated |
| 0,401264721 | 0,190001935 | WSB2 | FALSE | Upregulated |
| -0,943166219 | 0,190103432 | LOC400548 | FALSE | Downregulated |
| 1,791458971 | 0,190209474 | HPSE | FALSE | Upregulated |
| -0,397033447 | 0,190281271 | PPP1R16A | FALSE | Downregulated |
| 0,855987932 | 0,190417509 | PRCD | FALSE | Upregulated |

| | | | | |
|---|---|---|---|---|
| -2,505853024 | 0,190769419 | KCNK3 | FALSE | Downregulated |
| 0,907592654 | 0,191121394 | LOC101928038 | FALSE | Upregulated |
| -0,390395715 | 0,191294378 | EEF1B2 | FALSE | Downregulated |
| 0,603749792 | 0,191294378 | LOC101928658 | FALSE | Upregulated |
| 1,046529437 | 0,191306835 | METRNL | FALSE | Upregulated |
| -0,792346366 | 0,19132085 | DNAJC30 | FALSE | Downregulated |
| -0,380388986 | 0,19132085 | RPL24 | FALSE | Downregulated |
| -1,020258549 | 0,191847416 | RP11-475O6.1 | FALSE | Downregulated |
| 1,311716439 | 0,192019157 | RNF157 | FALSE | Upregulated |
| -0,366793969 | 0,192046203 | GRB10 | FALSE | Downregulated |
| 0,640701774 | 0,192046203 | SGPP2 | FALSE | Upregulated |
| 0,485155895 | 0,192415303 | LOC100128882 | FALSE | Upregulated |
| 1,086510708 | 0,192723859 | SULT1C2 | FALSE | Upregulated |
| -0,847866681 | 0,192750055 | ARNTL | FALSE | Downregulated |
| 1,543957006 | 0,193047859 | RASEF | FALSE | Upregulated |
| -2,972800987 | 0,193382004 | AC004053.1 | FALSE | Downregulated |
| -0,485112046 | 0,193528885 | ST3GAL2 | FALSE | Downregulated |
| 1,869590117 | 0,193726834 | HIST1H3I | FALSE | Upregulated |
| -0,376069015 | 0,193784366 | TOM1L1 | FALSE | Downregulated |
| 1,046971186 | 0,193879776 | LFNG | FALSE | Upregulated |
| -0,300503265 | 0,194033611 | PNPO | FALSE | Downregulated |
| -0,755105215 | 0,194149702 | SLC22A18 | FALSE | Downregulated |
| -0,940721042 | 0,194229845 | FAM110B | FALSE | Downregulated |
| 0,631009995 | 0,194289556 | NEDD9 | FALSE | Upregulated |
| -0,445111552 | 0,194480318 | CLCN5 | FALSE | Downregulated |
| -0,63530266 | 0,194524885 | EXOSC10 | FALSE | Downregulated |
| 1,132974388 | 0,194524885 | HCG26 | FALSE | Upregulated |
| 3,708198532 | 0,194524885 | KRT23 | FALSE | Upregulated |
| -1,584948589 | 0,194556826 | AC000124.1 | FALSE | Downregulated |
| 1,189667039 | 0,194675362 | MX2 | FALSE | Upregulated |
| -0,619178326 | 0,194675362 | SLC25A11 | FALSE | Downregulated |
| 0,645415058 | 0,194815443 | BCL2L11 | FALSE | Upregulated |
| -1,277270468 | 0,195201329 | CRYM | FALSE | Downregulated |
| 1,272223539 | 0,195201329 | LOC102723331 | FALSE | Upregulated |
| 0,512423425 | 0,195201329 | RANGAP1 | FALSE | Upregulated |
| -0,375762253 | 0,195353889 | AMOTL2 | FALSE | Downregulated |
| 0,698924804 | 0,195678195 | SLFN5 | FALSE | Upregulated |
| -0,449128697 | 0,195730307 | SECISBP2L | FALSE | Downregulated |
| -0,597389279 | 0,195805761 | C10orf105 | FALSE | Downregulated |
| 1,187833445 | 0,195805761 | INSM2 | FALSE | Upregulated |
| 0,943816771 | 0,195805761 | LOC101927296 | FALSE | Upregulated |
| 1,396813553 | 0,195847347 | RTN1 | FALSE | Upregulated |
| 0,948664891 | 0,195847347 | TMA16 | FALSE | Upregulated |
| 0,243828846 | 0,195968666 | SQSTM1 | FALSE | Upregulated |
| -0,835185879 | 0,196284607 | LOC102724562 | FALSE | Downregulated |
| -1,890338262 | 0,196313612 | EPHB1 | FALSE | Downregulated |
| -0,776259471 | 0,196604438 | LOC101928431 | FALSE | Downregulated |
| 0,898349161 | 0,196886514 | AGBL3 | FALSE | Upregulated |
| 0,433866197 | 0,196886514 | CEP95 | FALSE | Upregulated |
| 0,43164698 | 0,196886514 | GPR146 | FALSE | Upregulated |
| 0,806354588 | 0,196886514 | IFT80 | FALSE | Upregulated |

| | | | | |
|---|---|---|---|---|
| 1,127315585 | 0,196886514 | LMCD1 | FALSE | Upregulated |
| -0,295483984 | 0,196886514 | PLIN4 | FALSE | Downregulated |
| -0,721523404 | 0,196886514 | POLR2F | FALSE | Downregulated |
| 0,460691853 | 0,196895503 | SLC35A1 | FALSE | Upregulated |
| -0,464266278 | 0,197094908 | LMO7 | FALSE | Downregulated |
| -0,335815775 | 0,197215318 | TPI1 | FALSE | Downregulated |
| 1,948846182 | 0,19743159 | PXT1 | FALSE | Upregulated |
| 0,676706888 | 0,197465228 | HMGCR | FALSE | Upregulated |
| 0,477894216 | 0,197465228 | LOC101928719 | FALSE | Upregulated |
| 1,404881527 | 0,197471169 | MEFV | FALSE | Upregulated |
| 0,907634929 | 0,197499028 | AQP1 | FALSE | Upregulated |
| 0,83770189 | 0,197693186 | SGCE | FALSE | Upregulated |
| -1,109126468 | 0,197814977 | HEPACAM | FALSE | Downregulated |
| 0,712181788 | 0,198014436 | COL6A2 | FALSE | Upregulated |
| 0,826639688 | 0,198046207 | ENAH | FALSE | Upregulated |
| -0,988362049 | 0,198050357 | PLIN1 | FALSE | Downregulated |
| 0,720653335 | 0,198089029 | EMC9 | FALSE | Upregulated |
| -0,419421517 | 0,19816497 | CHP1 | FALSE | Downregulated |
| -0,52518791 | 0,19823863 | GNL1 | FALSE | Downregulated |
| 0,549166493 | 0,19823863 | PITPNM1 | FALSE | Upregulated |
| 0,487701838 | 0,198354491 | FAM111A | FALSE | Upregulated |
| -0,579096988 | 0,198471354 | TMEM230 | FALSE | Downregulated |
| -0,344879736 | 0,198471354 | ZC3H11A | FALSE | Downregulated |
| 0,69897096 | 0,198481999 | LYN | FALSE | Upregulated |
| -0,598603926 | 0,198636947 | TPCN2 | FALSE | Downregulated |
| 0,87650998 | 0,198804041 | RASSF3 | FALSE | Upregulated |
| 1,09324988 | 0,19880904 | FAM49B | FALSE | Upregulated |
| 2,181789208 | 0,19880904 | OR2D3 | FALSE | Upregulated |
| -0,599646298 | 0,19880904 | SLC44A1 | FALSE | Downregulated |
| 0,701798798 | 0,199068494 | COL12A1 | FALSE | Upregulated |
| -2,088959081 | 0,199303078 | C22orf31 | FALSE | Downregulated |
| 2,9000976 | 0,199303078 | LNX1-AS2 | FALSE | Upregulated |
| -0,77300553 | 0,199303244 | ATP6AP1 | FALSE | Downregulated |
| -0,585846266 | 0,199327714 | AMDHD1 | FALSE | Downregulated |
| -0,859739568 | 0,199327714 | C3orf58 | FALSE | Downregulated |
| 0,528558766 | 0,199327714 | CXCR6 | FALSE | Upregulated |
| -0,431715233 | 0,199327714 | CYP4F11 | FALSE | Downregulated |
| 1,288137367 | 0,19946618 | EHF | FALSE | Upregulated |
| 0,581212466 | 0,199577939 | LOC100287467 | FALSE | Upregulated |
| 1,088772436 | 0,199708063 | SQLE | FALSE | Upregulated |
| 0,581748642 | 0,200002268 | DDHD2 | FALSE | Upregulated |
| 0,590786949 | 0,200002268 | LOC730257 | FALSE | Upregulated |
| -0,488857737 | 0,200002268 | VAPB | FALSE | Downregulated |
| 0,483335901 | 0,200015703 | SYNE2 | FALSE | Upregulated |
| 0,862066099 | 0,20001953 | OSBPL5 | FALSE | Upregulated |
| -0,325749492 | 0,200181508 | A1BG | FALSE | Downregulated |
| -0,638287309 | 0,200181508 | ABHD6 | FALSE | Downregulated |
| 0,486020445 | 0,200181508 | CECR1 | FALSE | Upregulated |
| 0,76771262 | 0,200181508 | IDI2-AS1 | FALSE | Upregulated |
| 0,804472407 | 0,200181508 | MCM4 | FALSE | Upregulated |
| -0,295759559 | 0,200181508 | SNRNP200 | FALSE | Downregulated |

| | | | | |
|---|---|---|---|---|
| 0,710097902 | 0,200181508 | TMC6 | FALSE | Upregulated |
| 0,644326309 | 0,200181508 | ZNF57 | FALSE | Upregulated |
| -1,376815694 | 0,200181888 | DDO | FALSE | Downregulated |
| 0,396040962 | 0,200406706 | ACTB | FALSE | Upregulated |
| 1,508241722 | 0,200501231 | LOC101927098 | FALSE | Upregulated |
| -2,02790445 | 0,200783257 | MTERFD1 | FALSE | Downregulated |
| 0,975136349 | 0,200783257 | SLC30A4 | FALSE | Upregulated |
| -1,10763091 | 0,200866331 | NDUFB3 | FALSE | Downregulated |
| 0,846802501 | 0,201001465 | LOC729451 | FALSE | Upregulated |
| 0,710737242 | 0,2010358 | PLXDC2 | FALSE | Upregulated |
| -0,646584844 | 0,201130876 | PTPRD | FALSE | Downregulated |
| 0,432615965 | 0,201234923 | HLA-E | FALSE | Upregulated |
| 1,004005108 | 0,201671629 | NUSAP1 | FALSE | Upregulated |
| 0,67489131 | 0,20178252 | RBFOX3 | FALSE | Upregulated |
| 2,133119099 | 0,201896601 | ATP6V1G2 | FALSE | Upregulated |
| 0,567325506 | 0,201896601 | PARP6 | FALSE | Upregulated |
| 0,606684781 | 0,202208984 | LOC102724345 | FALSE | Upregulated |
| 0,944421055 | 0,202299587 | FAM120C | FALSE | Upregulated |
| -0,41483039 | 0,202299587 | MZT2B | FALSE | Downregulated |
| 0,87698434 | 0,202539853 | EMC2 | FALSE | Upregulated |
| 1,159267522 | 0,202807328 | RRN3P2 | FALSE | Upregulated |
| 0,251635451 | 0,203559874 | CD302 | FALSE | Upregulated |
| 0,503082007 | 0,203559874 | FAM219A | FALSE | Upregulated |
| 0,560268141 | 0,203624162 | LOC101927266 | FALSE | Upregulated |
| 0,567704152 | 0,203676364 | LOC101929451 | FALSE | Upregulated |
| 0,497505613 | 0,203786306 | CLTB | FALSE | Upregulated |
| 0,715654562 | 0,203946496 | NFATC1 | FALSE | Upregulated |
| 0,555559141 | 0,20428168 | FCHO2 | FALSE | Upregulated |
| -0,345808781 | 0,204532174 | JMJD1C | FALSE | Downregulated |
| 1,800276313 | 0,204532174 | LOC101928872 | FALSE | Upregulated |
| 0,857431828 | 0,20463968 | LOC101929657 | FALSE | Upregulated |
| -0,571731921 | 0,204786504 | NFKBIZ | FALSE | Downregulated |
| 0,652114098 | 0,204786504 | TRIM37 | FALSE | Upregulated |
| 1,707363958 | 0,204831683 | RHPN1 | FALSE | Upregulated |
| 0,709969903 | 0,204913784 | TLR9 | FALSE | Upregulated |
| 0,401860883 | 0,204973961 | SNX21 | FALSE | Upregulated |
| 0,620433646 | 0,204973961 | TWF1 | FALSE | Upregulated |
| 0,645351686 | 0,20501047 | NSMAF | FALSE | Upregulated |
| 1,290408727 | 0,205131025 | OR7E12P | FALSE | Upregulated |
| 0,924059871 | 0,205211201 | DUSP11 | FALSE | Upregulated |
| -1,047679583 | 0,205222019 | EMD | FALSE | Downregulated |
| 1,064985644 | 0,205470392 | LOC101927585 | FALSE | Upregulated |
| 0,336287907 | 0,205486393 | SUPT7L | FALSE | Upregulated |
| -0,501607307 | 0,205534075 | AP2S1 | FALSE | Downregulated |
| 1,27167146 | 0,205534075 | BOLA3 | FALSE | Upregulated |
| -0,508411766 | 0,205534075 | JMJD4 | FALSE | Downregulated |
| 0,534002021 | 0,20566325 | LOC101928291 | FALSE | Upregulated |
| 1,659926071 | 0,20582315 | LOC101930129 | FALSE | Upregulated |
| -1,303254882 | 0,20582315 | LRRC55 | FALSE | Downregulated |
| 1,698493806 | 0,20582315 | RP11-778J15.1 | FALSE | Upregulated |
| 0,446221101 | 0,205972288 | MPI | FALSE | Upregulated |

| | | | | |
|---|---|---|---|---|
| -0,368595883 | 0,205972288 | RAB43 | FALSE | Downregulated |
| -0,61515831 | 0,206084068 | CLDN14 | FALSE | Downregulated |
| 0,455836288 | 0,206084068 | NOC2L | FALSE | Upregulated |
| 0,810808653 | 0,206084068 | ZFAT-AS1 | FALSE | Upregulated |
| -0,424386276 | 0,20610223 | CDH5 | FALSE | Downregulated |
| 0,705058206 | 0,206265549 | LGALS9 | FALSE | Upregulated |
| -0,704803641 | 0,206265549 | NTHL1 | FALSE | Downregulated |
| 1,268406233 | 0,206265549 | NUTM1 | FALSE | Upregulated |
| 0,561540205 | 0,206265549 | TGFBI | FALSE | Upregulated |
| 0,669111478 | 0,206336959 | CNTNAP1 | FALSE | Upregulated |
| 0,74248704 | 0,206336959 | TAS1R3 | FALSE | Upregulated |
| -0,266366708 | 0,206382407 | MGAT4B | FALSE | Downregulated |
| -0,480905309 | 0,206668848 | CHPF | FALSE | Downregulated |
| 0,535549413 | 0,206719936 | B3GNTL1 | FALSE | Upregulated |
| 0,45977862 | 0,206719936 | ITGB3 | FALSE | Upregulated |
| 0,981163906 | 0,206719936 | LOC101928641 | FALSE | Upregulated |
| 0,548862977 | 0,207221253 | NRAP | FALSE | Upregulated |
| 0,419906977 | 0,207221253 | PLEKHG6 | FALSE | Upregulated |
| 0,505818181 | 0,207221253 | YEATS2 | FALSE | Upregulated |
| 1,79809392 | 0,207466228 | CADM3 | FALSE | Upregulated |
| 0,928177745 | 0,207618588 | LETM2 | FALSE | Upregulated |
| 0,495900937 | 0,207618588 | OGFOD1 | FALSE | Upregulated |
| 0,249699692 | 0,207618588 | PDXDC1 | FALSE | Upregulated |
| -0,474806716 | 0,207618588 | TOM1L2 | FALSE | Downregulated |
| 1,2262712 | 0,207757269 | PRC1-AS1 | FALSE | Upregulated |
| -0,519123527 | 0,208058305 | SIGIRR | FALSE | Downregulated |
| -0,428943621 | 0,208105369 | FMO5 | FALSE | Downregulated |
| -0,656028669 | 0,208105369 | INTS5 | FALSE | Downregulated |
| 0,718871236 | 0,208105369 | MCM5 | FALSE | Upregulated |
| -0,353767443 | 0,208105369 | NUCKS1 | FALSE | Downregulated |
| 0,383277927 | 0,208105369 | RASA1 | FALSE | Upregulated |
| 0,394963094 | 0,208105369 | RP11-374P20.4 | FALSE | Upregulated |
| 0,445808501 | 0,208105369 | THUMPD3 | FALSE | Upregulated |
| -2,211255857 | 0,208105369 | ZNF658B | FALSE | Downregulated |
| 0,876699375 | 0,208146294 | RAD1 | FALSE | Upregulated |
| 0,234030912 | 0,208170748 | ZFHX3 | FALSE | Upregulated |
| 0,49601618 | 0,208226124 | ZSCAN32 | FALSE | Upregulated |
| 0,429657518 | 0,208285219 | CNOT11 | FALSE | Upregulated |
| 0,413664259 | 0,208371061 | MTMR7 | FALSE | Upregulated |
| 1,390003261 | 0,208427583 | SLCO2A1 | FALSE | Upregulated |
| 0,522544735 | 0,208523289 | HIC1 | FALSE | Upregulated |
| 1,196498378 | 0,208523289 | TNFAIP8 | FALSE | Upregulated |
| 1,906793556 | 0,208674374 | ASNS | FALSE | Upregulated |
| 0,438630061 | 0,208674374 | EIF4G3 | FALSE | Upregulated |
| 1,150731505 | 0,208674374 | IER3 | FALSE | Upregulated |
| 0,677651422 | 0,208674374 | RP11-15A1.7 | FALSE | Upregulated |
| -0,344422458 | 0,208674374 | RPS9 | FALSE | Downregulated |
| 1,558276732 | 0,208674374 | RYR1 | FALSE | Upregulated |
| 0,539937255 | 0,208674374 | SERHL2 | FALSE | Upregulated |
| -0,542363754 | 0,208674374 | TLK1 | FALSE | Downregulated |
| -1,28016237 | 0,208873339 | GSTA2 | FALSE | Downregulated |

| | | | | |
|---|---|---|---|---|
| 0,930150561 | 0,208894373 | MOB3B | FALSE | Upregulated |
| 1,124125285 | 0,209321386 | JKAMP | FALSE | Upregulated |
| 0,570753337 | 0,209325132 | CSPG4 | FALSE | Upregulated |
| 0,399532531 | 0,209325132 | FAM20B | FALSE | Upregulated |
| -0,436506171 | 0,209325132 | PRODH2 | FALSE | Downregulated |
| 0,463684288 | 0,209325132 | TAF9B | FALSE | Upregulated |
| 0,838057547 | 0,209524216 | TMEM11 | FALSE | Upregulated |
| 1,009160578 | 0,209762829 | GSTM3 | FALSE | Upregulated |
| 1,170074373 | 0,209762829 | M1AP | FALSE | Upregulated |
| 0,4795848 | 0,209762829 | TSHZ2 | FALSE | Upregulated |
| 0,98390925 | 0,209762829 | TTF2 | FALSE | Upregulated |
| -0,570345482 | 0,210078363 | MARK4 | FALSE | Downregulated |
| 0,964615873 | 0,210128249 | LOC101927201 | FALSE | Upregulated |
| -0,756183322 | 0,210128249 | LSM12 | FALSE | Downregulated |
| 0,822643541 | 0,210128249 | ZBED5 | FALSE | Upregulated |
| -0,309347423 | 0,210174201 | EEF1A1 | FALSE | Downregulated |
| -0,515461798 | 0,210291836 | SLC25A20 | FALSE | Downregulated |
| 0,697675601 | 0,21037428 | HOXB3 | FALSE | Upregulated |
| 0,451182155 | 0,210404574 | DGKZ | FALSE | Upregulated |
| -0,29967923 | 0,210437889 | AKAP9 | FALSE | Downregulated |
| -0,335422128 | 0,210437889 | HNRNPC | FALSE | Downregulated |
| 0,367261132 | 0,210437889 | KCNJ13 | FALSE | Upregulated |
| -0,646618489 | 0,210742795 | INS-IGF2 | FALSE | Downregulated |
| 0,513181698 | 0,210871339 | MARS | FALSE | Upregulated |
| 0,783991352 | 0,210871339 | SFRP5 | FALSE | Upregulated |
| -0,782065765 | 0,211040731 | AGAP9 | FALSE | Downregulated |
| 0,84965297 | 0,211040731 | ARHGAP25 | FALSE | Upregulated |
| 0,522167875 | 0,211040731 | CCDC73 | FALSE | Upregulated |
| -0,422162431 | 0,211040731 | CXCL12 | FALSE | Downregulated |
| 1,020495346 | 0,211040731 | MRVI1 | FALSE | Upregulated |
| -0,629390555 | 0,211040731 | NDUFA11 | FALSE | Downregulated |
| -0,322365323 | 0,211040731 | NR2F6 | FALSE | Downregulated |
| -0,521883462 | 0,211040731 | RGS12 | FALSE | Downregulated |
| 0,401888964 | 0,211040731 | TACC1 | FALSE | Upregulated |
| 0,671934403 | 0,211040731 | TCEB3B | FALSE | Upregulated |
| -0,767753641 | 0,211040731 | TDO2 | FALSE | Downregulated |
| -3,023754857 | 0,211040731 | TSNAX-DISC1 | FALSE | Downregulated |
| -0,291222393 | 0,211221064 | F10 | FALSE | Downregulated |
| 0,590927522 | 0,211342973 | SASH3 | FALSE | Upregulated |
| 1,571551851 | 0,21160315 | ERCC8 | FALSE | Upregulated |
| 2,144117746 | 0,21169133 | CPEB1 | FALSE | Upregulated |
| 0,324992196 | 0,21169133 | TCF7 | FALSE | Upregulated |
| 0,469464433 | 0,211776011 | ERO1LB | FALSE | Upregulated |
| 1,702732977 | 0,211776011 | KIAA1324L | FALSE | Upregulated |
| 0,788226492 | 0,211776011 | LOC101927940 | FALSE | Upregulated |
| 1,119226657 | 0,211776011 | LOC102724626 | FALSE | Upregulated |
| 1,552230936 | 0,211776011 | ROBO2 | FALSE | Upregulated |
| -1,902666517 | 0,212002017 | AKR1C6P | FALSE | Downregulated |
| 0,333215209 | 0,212002017 | CTBP1-AS | FALSE | Upregulated |
| 0,517734814 | 0,212200981 | LYPD1 | FALSE | Upregulated |
| -0,791566417 | 0,212226731 | KCNC4 | FALSE | Downregulated |

| | | | | |
|---|---|---|---|---|
| 1,139509771 | 0,212226731 | LOC101059976 | FALSE | Upregulated |
| 1,690342113 | 0,212226731 | TMC4 | FALSE | Upregulated |
| -0,444753359 | 0,212499054 | HADHB | FALSE | Downregulated |
| -0,625608566 | 0,21261659 | AGTR1 | FALSE | Downregulated |
| -0,290053888 | 0,21261659 | APLP2 | FALSE | Downregulated |
| 0,440678347 | 0,21261659 | FUBP1 | FALSE | Upregulated |
| 0,844982713 | 0,21261659 | GALNT7 | FALSE | Upregulated |
| 0,639287394 | 0,21261659 | PTCHD3P1 | FALSE | Upregulated |
| 1,930526062 | 0,21264151 | LNX1-AS1 | FALSE | Upregulated |
| 0,417783186 | 0,212645489 | EMILIN1 | FALSE | Upregulated |
| -0,667914472 | 0,212987007 | PIAS4 | FALSE | Downregulated |
| -0,574220939 | 0,213153844 | C19orf60 | FALSE | Downregulated |
| 0,709567912 | 0,213153844 | LINC00984 | FALSE | Upregulated |
| 0,503350966 | 0,213159277 | LRRC19 | FALSE | Upregulated |
| 1,662163992 | 0,213184876 | LOC100132249 | FALSE | Upregulated |
| -1,779593722 | 0,213257545 | TMEM256 | FALSE | Downregulated |
| 0,627545445 | 0,213429498 | IQCG | FALSE | Upregulated |
| 1,622388389 | 0,213795041 | CA5BP1 | FALSE | Upregulated |
| -0,739323224 | 0,214068934 | BCL7C | FALSE | Downregulated |
| -0,950864909 | 0,214176803 | WDR72 | FALSE | Downregulated |
| -0,465020008 | 0,214230118 | GLUL | FALSE | Downregulated |
| -0,568538243 | 0,214306898 | POLR1D | FALSE | Downregulated |
| 0,400047223 | 0,21467309 | STARD7-AS1 | FALSE | Upregulated |
| 5,820581863 | 0,214727487 | ADIPOQ-AS1 | FALSE | Upregulated |
| -0,242001194 | 0,214727487 | COPA | FALSE | Downregulated |
| 0,866805453 | 0,214727487 | IL10RB-AS1 | FALSE | Upregulated |
| 0,75529852 | 0,214727487 | LOC100129518 | FALSE | Upregulated |
| -0,352241254 | 0,215047883 | GPBP1L1 | FALSE | Downregulated |
| 0,982087929 | 0,215047883 | LAMA3 | FALSE | Upregulated |
| 1,538867873 | 0,215047883 | PTGDR | FALSE | Upregulated |
| 1,01235115 | 0,215047883 | TMEM51 | FALSE | Upregulated |
| -0,434901828 | 0,215047883 | ZCCHC6 | FALSE | Downregulated |
| 1,39303853 | 0,215050606 | ARL2 | FALSE | Upregulated |
| -0,391946415 | 0,215138928 | BCKDK | FALSE | Downregulated |
| 0,293546309 | 0,215165713 | MAP4 | FALSE | Upregulated |
| -1,143379575 | 0,215306981 | C9orf72 | FALSE | Downregulated |
| -0,556940474 | 0,215315589 | CCDC117 | FALSE | Downregulated |
| -0,502298439 | 0,215316688 | ATG13 | FALSE | Downregulated |
| -2,136536344 | 0,215354123 | CCM2L | FALSE | Downregulated |
| 0,913972562 | 0,215354123 | DPM2 | FALSE | Upregulated |
| -0,629555158 | 0,215559856 | KLHL36 | FALSE | Downregulated |
| -1,815664067 | 0,215659593 | MFSD5 | FALSE | Downregulated |
| 1,863734134 | 0,216087431 | C5orf58 | FALSE | Upregulated |
| 0,509438219 | 0,216371328 | PXK | FALSE | Upregulated |
| 1,067057262 | 0,216654874 | CIT | FALSE | Upregulated |
| 1,099787123 | 0,216901298 | GOLGA2P7 | FALSE | Upregulated |
| 0,486946935 | 0,217140075 | SPNS1 | FALSE | Upregulated |
| 1,434485121 | 0,21718696 | LOC101930275 | FALSE | Upregulated |
| 1,130004831 | 0,217200977 | HDGFRP3 | FALSE | Upregulated |
| -0,5085306 | 0,217200977 | NAA60 | FALSE | Downregulated |
| 1,581820372 | 0,217307722 | LOC102724240 | FALSE | Upregulated |

| | | | | |
|---|---|---|---|---|
| 0,745664264 | 0,217632 | TIGIT | FALSE | Upregulated |
| 0,718393159 | 0,217662502 | LONRF1 | FALSE | Upregulated |
| -0,294606486 | 0,218018025 | NACA | FALSE | Downregulated |
| -0,618656068 | 0,218047394 | SCN9A | FALSE | Downregulated |
| 0,806355613 | 0,21813549 | HCFC1R1 | FALSE | Upregulated |
| 0,407123726 | 0,218219706 | ADPGK-AS1 | FALSE | Upregulated |
| -0,61573727 | 0,218470206 | NMRAL1 | FALSE | Downregulated |
| 1,346823909 | 0,219192051 | ATF3 | FALSE | Upregulated |
| 0,623473732 | 0,219192051 | C1QA | FALSE | Upregulated |
| 1,15189283 | 0,219192051 | CCL19 | FALSE | Upregulated |
| 1,064546929 | 0,219192051 | CDR1 | FALSE | Upregulated |
| 1,057720991 | 0,219192051 | GPR162 | FALSE | Upregulated |
| 2,336375311 | 0,219192051 | LOC101928163 | FALSE | Upregulated |
| -0,358063384 | 0,219192051 | NAPA | FALSE | Downregulated |
| 0,514711103 | 0,219192051 | SLC38A2 | FALSE | Upregulated |
| 1,847554489 | 0,219192051 | TPO | FALSE | Upregulated |
| 0,664095741 | 0,219192051 | TRAFD1 | FALSE | Upregulated |
| 0,385464447 | 0,219192051 | YWHAZ | FALSE | Upregulated |
| 0,444021676 | 0,219234318 | LOC100505502 | FALSE | Upregulated |
| -0,333225343 | 0,21929916 | C1orf43 | FALSE | Downregulated |
| -0,348527923 | 0,21929916 | UBN1 | FALSE | Downregulated |
| -0,386532379 | 0,219403753 | CD14 | FALSE | Downregulated |
| 1,469680499 | 0,219403753 | DHRS13 | FALSE | Upregulated |
| 0,940631479 | 0,219403753 | ERV3-1 | FALSE | Upregulated |
| 0,451440799 | 0,219403753 | NHP2 | FALSE | Upregulated |
| -0,52706363 | 0,219403753 | NRBP1 | FALSE | Downregulated |
| 1,426806563 | 0,219403753 | TMTC2 | FALSE | Upregulated |
| 0,563219854 | 0,219647458 | LOC102724811 | FALSE | Upregulated |
| 1,445463935 | 0,219825617 | TRNP1 | FALSE | Upregulated |
| -0,336841303 | 0,220002515 | SAT1 | FALSE | Downregulated |
| 1,235162173 | 0,220051414 | KIAA1549L | FALSE | Upregulated |
| 1,204346683 | 0,22023015 | RCAN2 | FALSE | Upregulated |
| -0,434761254 | 0,220527324 | ABCC6 | FALSE | Downregulated |
| -0,440838515 | 0,220527324 | RPL27 | FALSE | Downregulated |
| 0,314410572 | 0,220527324 | TMED5 | FALSE | Upregulated |
| 0,511544383 | 0,220789752 | GHRL | FALSE | Upregulated |
| -0,393192826 | 0,220933127 | SPRYD4 | FALSE | Downregulated |
| 0,423386299 | 0,220933127 | TMEM80 | FALSE | Upregulated |
| -0,434495533 | 0,220978784 | JTB | FALSE | Downregulated |
| 1,213790829 | 0,220978784 | SERPINB8 | FALSE | Upregulated |
| -1,83579144 | 0,220978784 | TMEM105 | FALSE | Downregulated |
| 0,476974715 | 0,221032432 | ERMAP | FALSE | Upregulated |
| 0,343664756 | 0,221032432 | NCBP1 | FALSE | Upregulated |
| 1,377446057 | 0,221072874 | LOC102724601 | FALSE | Upregulated |
| -0,38943281 | 0,221072874 | OPLAH | FALSE | Downregulated |
| 2,063139139 | 0,22148184 | HR | FALSE | Upregulated |
| 0,437690209 | 0,221485537 | LOC100132356 | FALSE | Upregulated |
| -0,689712335 | 0,221655151 | GJB3 | FALSE | Downregulated |
| -0,290333732 | 0,221783477 | KIAA1161 | FALSE | Downregulated |
| 0,45651054 | 0,221827561 | MAP3K14 | FALSE | Upregulated |
| 0,659128354 | 0,221827561 | NT5C | FALSE | Upregulated |

| | | | | |
|---|---|---|---|---|
| 1,589869946 | 0,221827561 | RSPH10B2 | FALSE | Upregulated |
| -0,4199703 | 0,221834281 | USO1 | FALSE | Downregulated |
| -0,558323561 | 0,2218883 | SLC39A11 | FALSE | Downregulated |
| 0,522676352 | 0,222096698 | RP11-266L9.2 | FALSE | Upregulated |
| 0,391450723 | 0,222096698 | SPARC | FALSE | Upregulated |
| -0,463048217 | 0,222096698 | VARS2 | FALSE | Downregulated |
| 0,6049884 | 0,2225348 | CKS1B | FALSE | Upregulated |
| 1,135811339 | 0,222615905 | ACER3 | FALSE | Upregulated |
| 1,803691604 | 0,222615905 | HIST2H3D | FALSE | Upregulated |
| 1,121251527 | 0,222615905 | RASAL2-AS1 | FALSE | Upregulated |
| 1,712459256 | 0,222652066 | LTB | FALSE | Upregulated |
| 0,749441889 | 0,22266538 | MYLK-AS1 | FALSE | Upregulated |
| 0,730898403 | 0,223085539 | DYNC2H1 | FALSE | Upregulated |
| -0,611264327 | 0,223168735 | TPRN | FALSE | Downregulated |
| 0,501293401 | 0,223545523 | CHST15 | FALSE | Upregulated |
| 0,222157159 | 0,223545523 | KIAA1147 | FALSE | Upregulated |
| 0,540193333 | 0,223545523 | MYH7B | FALSE | Upregulated |
| 1,117554842 | 0,223784777 | SLC8A3 | FALSE | Upregulated |
| -0,404100185 | 0,224012473 | F11-AS1 | FALSE | Downregulated |
| 0,52772966 | 0,224158093 | MRS2 | FALSE | Upregulated |
| -1,021616737 | 0,224444504 | ACP6 | FALSE | Downregulated |
| -0,754915457 | 0,224444504 | METTL14 | FALSE | Downregulated |
| 0,553746407 | 0,224482316 | CCT6P3 | FALSE | Upregulated |
| 0,861090003 | 0,224482316 | LOC101593348 | FALSE | Upregulated |
| 0,711552006 | 0,224723125 | KIF26B | FALSE | Upregulated |
| -0,632658111 | 0,224762579 | DHX34 | FALSE | Downregulated |
| -1,298423582 | 0,224762579 | ENHO | FALSE | Downregulated |
| 1,872340148 | 0,224762579 | LOC101928292 | FALSE | Upregulated |
| -0,459484765 | 0,224762579 | MET | FALSE | Downregulated |
| -0,768250472 | 0,224762579 | TAT | FALSE | Downregulated |
| -0,381364467 | 0,225032735 | AGPAT2 | FALSE | Downregulated |
| -0,765345258 | 0,225127868 | SKP2 | FALSE | Downregulated |
| 1,438658866 | 0,225129823 | ANKLE1 | FALSE | Upregulated |
| -0,422227453 | 0,225129823 | DNAJC11 | FALSE | Downregulated |
| -1,173032444 | 0,225129823 | LHX2 | FALSE | Downregulated |
| 1,023138852 | 0,225129823 | NAF1 | FALSE | Upregulated |
| -0,40749488 | 0,225129823 | NDUFS2 | FALSE | Downregulated |
| -0,849292263 | 0,225129823 | SNRNP25 | FALSE | Downregulated |
| 0,522059739 | 0,225762055 | LOC100506504 | FALSE | Upregulated |
| -1,577493171 | 0,225762055 | RPL23AP53 | FALSE | Downregulated |
| 0,435260463 | 0,225768189 | GDAP1L1 | FALSE | Upregulated |
| -0,712306098 | 0,225768189 | HLX | FALSE | Downregulated |
| -0,927348006 | 0,225768189 | SCART1 | FALSE | Downregulated |
| 0,357985143 | 0,225795408 | BRI3 | FALSE | Upregulated |
| 0,577716337 | 0,225795408 | TPM4 | FALSE | Upregulated |
| -0,526798813 | 0,226183737 | NDUFB11 | FALSE | Downregulated |
| 1,349012284 | 0,226272363 | CDH6 | FALSE | Upregulated |
| 0,731067996 | 0,226558129 | LOC100131270 | FALSE | Upregulated |
| -0,627184246 | 0,226558129 | TMEM25 | FALSE | Downregulated |
| -0,541514717 | 0,226802963 | GRPEL1 | FALSE | Downregulated |
| -0,428237186 | 0,226935357 | APOC2 | FALSE | Downregulated |

| | | | | |
|---|---|---|---|---|
| 1,479203044 | 0,226935357 | SPAG17 | FALSE | Upregulated |
| 1,763377481 | 0,227123487 | C5orf54 | FALSE | Upregulated |
| -0,233081163 | 0,227201235 | CREBBP | FALSE | Downregulated |
| -0,464327512 | 0,227201235 | NGFR | FALSE | Downregulated |
| 0,878294491 | 0,227201235 | SYT1 | FALSE | Upregulated |
| 0,295767493 | 0,227201235 | UTRN | FALSE | Upregulated |
| 1,664623133 | 0,227275425 | MCOLN3 | FALSE | Upregulated |
| 0,548067369 | 0,227275425 | RNF207 | FALSE | Upregulated |
| 0,412835171 | 0,227275425 | SENP6 | FALSE | Upregulated |
| -0,58754364 | 0,227636895 | MPDU1 | FALSE | Downregulated |
| 1,019437409 | 0,227946116 | ASF1B | FALSE | Upregulated |
| 1,529497401 | 0,227946116 | HIST2H4A | FALSE | Upregulated |
| -0,38312518 | 0,228001311 | CP | FALSE | Downregulated |
| 0,457191208 | 0,22803415 | PCGF3 | FALSE | Upregulated |
| 0,934902946 | 0,228216426 | AIF1 | FALSE | Upregulated |
| -0,40157289 | 0,228216426 | C2orf72 | FALSE | Downregulated |
| -0,436696804 | 0,228248683 | QARS | FALSE | Downregulated |
| 0,558333181 | 0,228268457 | C1QC | FALSE | Upregulated |
| -0,86012826 | 0,228273376 | GINM1 | FALSE | Downregulated |
| 0,917538841 | 0,22828261 | DSCR9 | FALSE | Upregulated |
| -0,40886419 | 0,22828261 | SCP2 | FALSE | Downregulated |
| 0,257569744 | 0,22828261 | SMARCC2 | FALSE | Upregulated |
| 0,48767203 | 0,228575178 | ZNHIT3 | FALSE | Upregulated |
| 0,89579806 | 0,228697249 | ANKS6 | FALSE | Upregulated |
| 0,457632404 | 0,228697249 | TVP23A | FALSE | Upregulated |
| 0,769927764 | 0,22888787 | BRICD5 | FALSE | Upregulated |
| 1,509833619 | 0,22925506 | LINC00967 | FALSE | Upregulated |
| -0,356621557 | 0,229356043 | ALDH8A1 | FALSE | Downregulated |
| 0,271807025 | 0,229356043 | SEPT9 | FALSE | Upregulated |
| 0,916814964 | 0,229524492 | CNRIP1 | FALSE | Upregulated |
| -0,416665475 | 0,230100047 | ANKS1A | FALSE | Downregulated |
| -0,741150465 | 0,230100047 | ATP5E | FALSE | Downregulated |
| -0,51846569 | 0,230800025 | PFKFB1 | FALSE | Downregulated |
| 1,417511053 | 0,2311611 | OR7E2P | FALSE | Upregulated |
| 0,486088047 | 0,231202033 | VASH2 | FALSE | Upregulated |
| 0,314287201 | 0,231235269 | FAM134A | FALSE | Upregulated |
| -0,277894269 | 0,231739987 | RPS2 | FALSE | Downregulated |
| 0,807278133 | 0,23178347 | ARHGEF3 | FALSE | Upregulated |
| -0,521133314 | 0,23178347 | ARID4A | FALSE | Downregulated |
| 1,724881077 | 0,23178347 | DBF4 | FALSE | Upregulated |
| -0,923763311 | 0,23178347 | DCDC5 | FALSE | Downregulated |
| -0,353933892 | 0,23178347 | GAA | FALSE | Downregulated |
| -1,249251122 | 0,23178347 | KIAA1652 | FALSE | Downregulated |
| 0,498504396 | 0,23178347 | SNTB2 | FALSE | Upregulated |
| 1,394583952 | 0,23178347 | ZNF396 | FALSE | Upregulated |
| 0,507335978 | 0,231791978 | GNPDA2 | FALSE | Upregulated |
| 0,927791226 | 0,231855107 | GRPEL2 | FALSE | Upregulated |
| 0,594696372 | 0,231866062 | ARHGAP27 | FALSE | Upregulated |
| 0,468912873 | 0,231866062 | C11orf49 | FALSE | Upregulated |
| 1,044966514 | 0,231866062 | CCDC36 | FALSE | Upregulated |
| 0,454087717 | 0,231866062 | GPBP1 | FALSE | Upregulated |

| | | | | |
|---|---|---|---|---|
| 0,960802715 | 0,231866062 | IPO9-AS1 | FALSE | Upregulated |
| 0,313402169 | 0,231866062 | SLC25A29 | FALSE | Upregulated |
| -0,729237067 | 0,232438521 | SYBU | FALSE | Downregulated |
| 1,352958973 | 0,232447504 | DPYSL4 | FALSE | Upregulated |
| 0,548336917 | 0,232627299 | EXOC8 | FALSE | Upregulated |
| -0,952502931 | 0,232637838 | IL17D | FALSE | Downregulated |
| 0,831030342 | 0,232751541 | ANKRD44 | FALSE | Upregulated |
| 1,141713552 | 0,232751541 | TMEM117 | FALSE | Upregulated |
| -0,572321175 | 0,232751541 | TTPA | FALSE | Downregulated |
| 1,7589914 | 0,232865106 | CD28 | FALSE | Upregulated |
| 0,422037644 | 0,232865106 | LYRM2 | FALSE | Upregulated |
| -0,898041495 | 0,232865106 | MRPL32 | FALSE | Downregulated |
| 0,362892483 | 0,233607806 | CEMP1 | FALSE | Upregulated |
| 1,230662802 | 0,233607806 | CHST9 | FALSE | Upregulated |
| -0,382559127 | 0,233607806 | DCAF6 | FALSE | Downregulated |
| 0,529851394 | 0,233795656 | PFKFB3 | FALSE | Upregulated |
| -0,414914438 | 0,233884303 | EFCAB14 | FALSE | Downregulated |
| -0,697651859 | 0,234072913 | REPS2 | FALSE | Downregulated |
| -0,63231684 | 0,234106303 | ACBD6 | FALSE | Downregulated |
| -0,482822331 | 0,234106303 | COG3 | FALSE | Downregulated |
| 0,694188402 | 0,234106303 | RAPGEF5 | FALSE | Upregulated |
| 0,438567537 | 0,234106303 | SAMD11 | FALSE | Upregulated |
| -0,53808803 | 0,234106303 | ZC3H13 | FALSE | Downregulated |
| -0,346629631 | 0,234170628 | RAF1 | FALSE | Downregulated |
| 0,537176533 | 0,234323801 | C18orf25 | FALSE | Upregulated |
| -0,440106236 | 0,234411243 | EXTL3 | FALSE | Downregulated |
| 1,367077983 | 0,23505928 | BRE-AS1 | FALSE | Upregulated |
| -0,897280979 | 0,235080784 | C2orf47 | FALSE | Downregulated |
| 0,485108028 | 0,235080784 | FAM69A | FALSE | Upregulated |
| 0,606977228 | 0,235080784 | HP09025 | FALSE | Upregulated |
| 0,558066921 | 0,235126611 | ASB9P1 | FALSE | Upregulated |
| 0,850792731 | 0,235126611 | C8orf59 | FALSE | Upregulated |
| 0,425384642 | 0,235819586 | GPX3 | FALSE | Upregulated |
| -1,275782093 | 0,235819586 | HIST1H4B | FALSE | Downregulated |
| -0,378820922 | 0,235819586 | NCLN | FALSE | Downregulated |
| 0,58594853 | 0,235819586 | UBE2Q1 | FALSE | Upregulated |
| -0,638789599 | 0,235851333 | PEX16 | FALSE | Downregulated |
| 1,266570594 | 0,236246926 | KCNK7 | FALSE | Upregulated |
| -0,499188453 | 0,236282863 | RRP1B | FALSE | Downregulated |
| -0,425491931 | 0,236329375 | LINC00261 | FALSE | Downregulated |
| 0,412063726 | 0,236329375 | RNF219-AS1 | FALSE | Upregulated |
| -0,366705668 | 0,236342347 | PACS2 | FALSE | Downregulated |
| -0,585556036 | 0,236381441 | IRF2 | FALSE | Downregulated |
| 0,440643101 | 0,236512591 | LOC101927647 | FALSE | Upregulated |
| -0,380693863 | 0,236735283 | CFB | FALSE | Downregulated |
| 1,481021634 | 0,236735283 | PAFAH1B3 | FALSE | Upregulated |
| 1,501851069 | 0,236735283 | PYHIN1 | FALSE | Upregulated |
| -0,280000639 | 0,236848181 | UPF1 | FALSE | Downregulated |
| 1,696266706 | 0,236876723 | C17orf82 | FALSE | Upregulated |
| -0,669316835 | 0,236876723 | DNAJC8 | FALSE | Downregulated |
| 1,3671509 | 0,236876723 | LOC101927131 | FALSE | Upregulated |

| | | | | |
|---|---|---|---|---|
| -1,240423536 | 0,236876723 | SAC3D1 | FALSE | Downregulated |
| -0,708805476 | 0,237109795 | TIGD2 | FALSE | Downregulated |
| 0,36580397 | 0,237151884 | TCOF1 | FALSE | Upregulated |
| -0,379624496 | 0,23716267 | GPI | FALSE | Downregulated |
| 1,961964285 | 0,237425577 | CDHR1 | FALSE | Upregulated |
| 0,461117703 | 0,237472041 | CASKIN1 | FALSE | Upregulated |
| -0,496335949 | 0,237472041 | GATA4 | FALSE | Downregulated |
| 1,428235838 | 0,237472041 | HSPA2 | FALSE | Upregulated |
| 0,670811959 | 0,237472041 | NUP107 | FALSE | Upregulated |
| -0,431645161 | 0,237472041 | PAH | FALSE | Downregulated |
| -0,445861417 | 0,237472041 | RHOBTB3 | FALSE | Downregulated |
| 0,856376902 | 0,237472041 | SELPLG | FALSE | Upregulated |
| 1,511632572 | 0,237472041 | SLAMF6 | FALSE | Upregulated |
| -1,461965398 | 0,237472041 | SNORD94 | FALSE | Downregulated |
| 0,353114293 | 0,237699357 | ANKRD13A | FALSE | Upregulated |
| 0,858020687 | 0,237699357 | DTWD1 | FALSE | Upregulated |
| 0,37215823 | 0,237699357 | MCTS1 | FALSE | Upregulated |
| -0,361620291 | 0,237699357 | SDHA | FALSE | Downregulated |
| 0,900839944 | 0,237792492 | FAM86B1 | FALSE | Upregulated |
| 0,553593643 | 0,237839871 | CYTH3 | FALSE | Upregulated |
| 1,413370695 | 0,237889982 | CORO2B | FALSE | Upregulated |
| 0,396164964 | 0,237889982 | SRRM5 | FALSE | Upregulated |
| -0,53111345 | 0,237937045 | TMEM33 | FALSE | Downregulated |
| 0,325345396 | 0,237995077 | DYRK1A | FALSE | Upregulated |
| -0,401474842 | 0,238356004 | BLVRB | FALSE | Downregulated |
| -1,380898223 | 0,238356004 | SALL4 | FALSE | Downregulated |
| -1,010189935 | 0,238389974 | BMPER | FALSE | Downregulated |
| 1,527546906 | 0,238389974 | PLAU | FALSE | Upregulated |
| 0,382863442 | 0,238389974 | PSIP1 | FALSE | Upregulated |
| 0,764032711 | 0,238397473 | EGOT | FALSE | Upregulated |
| 0,821447703 | 0,238397473 | LOC285095 | FALSE | Upregulated |
| 1,531324388 | 0,238397473 | PHOX2A | FALSE | Upregulated |
| -0,62908675 | 0,238397473 | POC1B | FALSE | Downregulated |
| 0,603632074 | 0,238474986 | MRPS18C | FALSE | Upregulated |
| 0,858595345 | 0,238532363 | GNG2 | FALSE | Upregulated |
| -1,336782832 | 0,238661633 | TRHDE | FALSE | Downregulated |
| 0,504501451 | 0,238707133 | PAPPA | FALSE | Upregulated |
| -0,689632079 | 0,238829113 | DEPDC7 | FALSE | Downregulated |
| -0,735491486 | 0,23906327 | SH3RF2 | FALSE | Downregulated |
| -0,418745473 | 0,239662673 | EHD3 | FALSE | Downregulated |
| -0,641432927 | 0,239662673 | MAMDC4 | FALSE | Downregulated |
| 0,79049083 | 0,239662673 | TMEM173 | FALSE | Upregulated |
| 0,446200351 | 0,239743436 | IAH1 | FALSE | Upregulated |
| 1,342638662 | 0,239743436 | LIN52 | FALSE | Upregulated |
| 0,67840422 | 0,239743436 | LLGL1 | FALSE | Upregulated |
| 0,547171699 | 0,240189791 | STX3 | FALSE | Upregulated |
| -1,680182308 | 0,240289183 | ATAD3C | FALSE | Downregulated |
| -0,418314912 | 0,240289183 | MDC1 | FALSE | Downregulated |
| 0,528765795 | 0,240289183 | OSGIN2 | FALSE | Upregulated |
| -0,393420171 | 0,240473211 | ERGIC3 | FALSE | Downregulated |
| 0,602310027 | 0,240477277 | ABCC10 | FALSE | Upregulated |

| | | | | |
|---|---|---|---|---|
| 0,375865977 | 0,240477277 | MMP19 | FALSE | Upregulated |
| 0,309784269 | 0,240860198 | MGEA5 | FALSE | Upregulated |
| -0,590264553 | 0,241360324 | SRP68 | FALSE | Downregulated |
| 1,709864279 | 0,241462033 | ADAM33 | FALSE | Upregulated |
| -0,412688122 | 0,241462033 | TMX4 | FALSE | Downregulated |
| -0,593830307 | 0,241481214 | MRFAP1L1 | FALSE | Downregulated |
| 1,172757988 | 0,241488845 | SPATA32 | FALSE | Upregulated |
| 0,532936597 | 0,241785913 | LOC101929040 | FALSE | Upregulated |
| 1,13453694 | 0,24185553 | C5orf63 | FALSE | Upregulated |
| 1,620022648 | 0,241862433 | CXCR4 | FALSE | Upregulated |
| 0,913732494 | 0,241862433 | DUS4L | FALSE | Upregulated |
| -0,30730329 | 0,241862433 | MAGI1 | FALSE | Downregulated |
| 0,680119855 | 0,241911329 | CCDC120 | FALSE | Upregulated |
| -1,213921778 | 0,242041546 | PHYHIP | FALSE | Downregulated |
| -1,527544392 | 0,242082095 | MEP1B | FALSE | Downregulated |
| -0,669516038 | 0,242152536 | CEP120 | FALSE | Downregulated |
| 0,639706466 | 0,242152536 | SMARCAL1 | FALSE | Upregulated |
| -0,307111642 | 0,242350589 | MDH2 | FALSE | Downregulated |
| 0,362183369 | 0,242452491 | GTF2IRD1P1 | FALSE | Upregulated |
| -1,390489823 | 0,243026589 | FOXN4 | FALSE | Downregulated |
| 0,960309505 | 0,243116817 | SASS6 | FALSE | Upregulated |
| -0,500166485 | 0,243116817 | VEGFB | FALSE | Downregulated |
| -1,567096056 | 0,243116817 | ZNF16 | FALSE | Downregulated |
| -0,815681645 | 0,243143801 | MCUR1 | FALSE | Downregulated |
| -0,422532509 | 0,243582459 | EXOC3 | FALSE | Downregulated |
| 1,388508025 | 0,243582459 | TBX6 | FALSE | Upregulated |
| 0,871947135 | 0,243671655 | GGN | FALSE | Upregulated |
| 1,232751101 | 0,243671655 | RP11-426L16.8 | FALSE | Upregulated |
| 0,530952956 | 0,243671655 | RP5-1033H22.2 | FALSE | Upregulated |
| -1,75868741 | 0,243671655 | TMEM220-AS1 | FALSE | Downregulated |
| 0,87912943 | 0,243788784 | SWSAP1 | FALSE | Upregulated |
| -0,388541116 | 0,24382321 | CDC42EP4 | FALSE | Downregulated |
| 0,348114679 | 0,24382321 | GNAL | FALSE | Upregulated |
| 0,460528882 | 0,24382321 | HPS4 | FALSE | Upregulated |
| 0,379697887 | 0,24382321 | JADE1 | FALSE | Upregulated |
| 1,260487442 | 0,24382321 | KIAA1755 | FALSE | Upregulated |
| -0,829609451 | 0,243824536 | APOA4 | FALSE | Downregulated |
| 0,499633506 | 0,243969733 | NAIP | FALSE | Upregulated |
| 0,427750934 | 0,243969733 | PIP5K1C | FALSE | Upregulated |
| 0,703277421 | 0,244013213 | MFSD10 | FALSE | Upregulated |
| 0,897375195 | 0,244193975 | FAR1 | FALSE | Upregulated |
| 0,405935919 | 0,244218314 | NUP188 | FALSE | Upregulated |
| -0,469007168 | 0,244218314 | PFDN5 | FALSE | Downregulated |
| -1,474565612 | 0,244642707 | ABCC11 | FALSE | Downregulated |
| -0,364503139 | 0,244642707 | GJB1 | FALSE | Downregulated |
| -0,60752695 | 0,244642707 | GRAMD3 | FALSE | Downregulated |
| -0,43296693 | 0,244642707 | HERC2P2 | FALSE | Downregulated |
| -1,194133578 | 0,244642707 | LOC101060212 | FALSE | Downregulated |
| 0,42372213 | 0,244642707 | SPDYE5 | FALSE | Upregulated |
| 1,642447412 | 0,244664718 | LOC101929578 | FALSE | Upregulated |
| 0,590987186 | 0,245006996 | CD47 | FALSE | Upregulated |

| | | | | |
|---|---|---|---|---|
| -0,506011051 | 0,24523586 | SURF1 | FALSE | Downregulated |
| 0,608285228 | 0,24567637 | DOLK | FALSE | Upregulated |
| 0,424038451 | 0,245771008 | OMG | FALSE | Upregulated |
| 0,991879579 | 0,246022306 | ASXL3 | FALSE | Upregulated |
| 1,601952749 | 0,246022306 | LOC102723625 | FALSE | Upregulated |
| -0,264288663 | 0,246022306 | RERE | FALSE | Downregulated |
| -0,913367906 | 0,246200959 | GID4 | FALSE | Downregulated |
| 0,603923152 | 0,246200959 | KLHL31 | FALSE | Upregulated |
| 0,891704024 | 0,246200959 | LOC101928439 | FALSE | Upregulated |
| 0,910967617 | 0,246326043 | GOLGA6L9 | FALSE | Upregulated |
| 0,805990807 | 0,246326043 | LOC102723708 | FALSE | Upregulated |
| -0,40670561 | 0,246326043 | MAP2K3 | FALSE | Downregulated |
| -0,581528578 | 0,246326043 | MRP63 | FALSE | Downregulated |
| -1,84398318 | 0,246326043 | ZNF229 | FALSE | Downregulated |
| -0,506344986 | 0,246734717 | DDX56 | FALSE | Downregulated |
| 0,287611775 | 0,246734717 | TRIO | FALSE | Upregulated |
| -2,013416115 | 0,246759235 | RPL21P28 | FALSE | Downregulated |
| 0,506312308 | 0,247254864 | CHPF2 | FALSE | Upregulated |
| 0,607898429 | 0,24764481 | NCOA7 | FALSE | Upregulated |
| 1,133861476 | 0,247706735 | ACHE | FALSE | Upregulated |
| 0,358076044 | 0,247706735 | CRAMP1L | FALSE | Upregulated |
| 1,290594112 | 0,247706735 | IL4I1 | FALSE | Upregulated |
| 0,532131099 | 0,247706735 | PSEN1 | FALSE | Upregulated |
| 1,212134233 | 0,247706735 | RDH12 | FALSE | Upregulated |
| 0,525507571 | 0,247706735 | RFX7 | FALSE | Upregulated |
| -0,739696461 | 0,248042859 | C19orf80 | FALSE | Downregulated |
| 0,546310595 | 0,248042859 | PECAM1 | FALSE | Upregulated |
| 2,763499799 | 0,248042859 | PYGM | FALSE | Upregulated |
| 0,591591736 | 0,248042859 | SPSB2 | FALSE | Upregulated |
| 0,51571184 | 0,248115103 | AC104134.2 | FALSE | Upregulated |
| 0,599290041 | 0,248228604 | ANKRD36 | FALSE | Upregulated |
| 0,361956493 | 0,248528571 | ATP11B | FALSE | Upregulated |
| -0,285636686 | 0,248528571 | CNNM3 | FALSE | Downregulated |
| -0,770050445 | 0,248640249 | FST | FALSE | Downregulated |
| -0,365790312 | 0,248953276 | GPR126 | FALSE | Downregulated |
| 0,409664779 | 0,248953276 | KCNRG | FALSE | Upregulated |
| -0,689024779 | 0,248953276 | TACO1 | FALSE | Downregulated |
| -1,068560412 | 0,248989015 | ACOT11 | FALSE | Downregulated |
| 0,651162806 | 0,248989015 | ADAMTS10 | FALSE | Upregulated |
| 1,308411859 | 0,248989015 | HCG11 | FALSE | Upregulated |
| 1,855313295 | 0,248989015 | LOC100506113 | FALSE | Upregulated |
| -0,643119732 | 0,248989015 | PEF1 | FALSE | Downregulated |
| 0,608393842 | 0,248994847 | TOR4A | FALSE | Upregulated |
| 1,016487904 | 0,24900721 | LOC101928745 | FALSE | Upregulated |
| 0,954246821 | 0,249022959 | F2RL3 | FALSE | Upregulated |
| 0,872474657 | 0,249022959 | LOC101928244 | FALSE | Upregulated |
| 0,487738581 | 0,24912993 | NUTM2A | FALSE | Upregulated |
| 1,090712998 | 0,24912993 | PLEKHB1 | FALSE | Upregulated |
| -0,739661591 | 0,24912993 | SAP130 | FALSE | Downregulated |
| -0,474710122 | 0,24912993 | TRPV1 | FALSE | Downregulated |
| 0,372576392 | 0,24923996 | TBC1D10B | FALSE | Upregulated |

| | | | | |
|---|---|---|---|---|
| -0,72390506 | 0,249506389 | HAUS4 | FALSE | Downregulated |
| -0,323159808 | 0,249644989 | RPS17 | FALSE | Downregulated |
| -0,920232798 | 0,249890952 | SRGAP3 | FALSE | Downregulated |
| 0,616080687 | 0,249980137 | CHSY1 | FALSE | Upregulated |
| 0,691209882 | 0,250027492 | LOC101928059 | FALSE | Upregulated |
| 2,225709792 | 0,25022688 | PLA2G2D | FALSE | Upregulated |
| 0,590515111 | 0,250331578 | ARIH2OS | FALSE | Upregulated |
| 0,295445861 | 0,250331578 | ATN1 | FALSE | Upregulated |
| 1,750431806 | 0,250331578 | CGB7 | FALSE | Upregulated |
| 1,913177033 | 0,250331578 | DUSP8 | FALSE | Upregulated |
| 1,167542242 | 0,250331578 | EPS8L1 | FALSE | Upregulated |
| 0,507567918 | 0,250331578 | MFSD4 | FALSE | Upregulated |
| 0,575576302 | 0,250331578 | NUDT15 | FALSE | Upregulated |
| -2,157271114 | 0,250331578 | PCNAP1 | FALSE | Downregulated |
| 2,112751595 | 0,250331578 | SCNN1B | FALSE | Upregulated |
| 0,70082376 | 0,250331578 | SNX12 | FALSE | Upregulated |
| -0,699883817 | 0,250331578 | USP37 | FALSE | Downregulated |
| -0,652842097 | 0,250331578 | WDR19 | FALSE | Downregulated |
| 0,524161208 | 0,250640403 | BMPR1A | FALSE | Upregulated |
| 1,323480826 | 0,250640403 | RP11-158M2.5 | FALSE | Upregulated |
| -0,594340801 | 0,250691993 | KLHL2 | FALSE | Downregulated |
| 0,831513692 | 0,250691993 | STMN3 | FALSE | Upregulated |
| -1,627771472 | 0,250731401 | XPNPEP2 | FALSE | Downregulated |
| 1,029817477 | 0,251052955 | MSRB3 | FALSE | Upregulated |
| -1,078632386 | 0,251265715 | RNF215 | FALSE | Downregulated |
| 0,516573821 | 0,251374903 | RNF139-AS1 | FALSE | Upregulated |
| 1,071471003 | 0,251474927 | OR2A1-AS1 | FALSE | Upregulated |
| -0,6318727 | 0,251474927 | RCOR3 | FALSE | Downregulated |
| -0,637123201 | 0,251474927 | VPS37A | FALSE | Downregulated |
| 0,648433142 | 0,251691547 | PSMD5-AS1 | FALSE | Upregulated |
| -0,425136781 | 0,251841239 | AKR1A1 | FALSE | Downregulated |
| 0,905245786 | 0,251841239 | HLA-DMA | FALSE | Upregulated |
| 1,596349679 | 0,251841239 | IL15 | FALSE | Upregulated |
| 1,497975162 | 0,251841239 | SERPINA12 | FALSE | Upregulated |
| 0,662353496 | 0,251841239 | SESN1 | FALSE | Upregulated |
| 0,385536446 | 0,252463909 | TTC28-AS1 | FALSE | Upregulated |
| 0,626476348 | 0,25252083 | EPX | FALSE | Upregulated |
| 0,381859965 | 0,252521569 | NFE2L3 | FALSE | Upregulated |
| -1,769933958 | 0,252713843 | SPRNP1 | FALSE | Downregulated |
| 0,899978218 | 0,252732577 | CSNK1A1P1 | FALSE | Upregulated |
| -0,26642518 | 0,252732577 | RPS19 | FALSE | Downregulated |
| 0,461696628 | 0,252971219 | MRPS14 | FALSE | Upregulated |
| -0,461500114 | 0,253189247 | TELO2 | FALSE | Downregulated |
| 1,323701806 | 0,253189247 | TSPAN5 | FALSE | Upregulated |
| 0,626763846 | 0,253402884 | GMIP | FALSE | Upregulated |
| 1,65009226 | 0,25352329 | LOC101927767 | FALSE | Upregulated |
| 0,640234331 | 0,253533541 | ADRBK2 | FALSE | Upregulated |
| 0,520905485 | 0,253639118 | ENDOU | FALSE | Upregulated |
| -0,358581628 | 0,253639118 | MEGF9 | FALSE | Downregulated |
| 1,672211013 | 0,253639118 | TLR5 | FALSE | Upregulated |
| -0,883873647 | 0,253675092 | RPUSD1 | FALSE | Downregulated |

| | | | | |
|---|---|---|---|---|
| 0,562431836 | 0,253683044 | CMTM3 | FALSE | Upregulated |
| -0,497162366 | 0,253764497 | SLMAP | FALSE | Downregulated |
| -0,364872436 | 0,254067617 | FN1 | FALSE | Downregulated |
| -0,892298517 | 0,254145466 | TSPAN7 | FALSE | Downregulated |
| 0,706990774 | 0,254327797 | TNC | FALSE | Upregulated |
| 0,582436862 | 0,254418881 | PRKCH | FALSE | Upregulated |
| 0,544814956 | 0,254449722 | NPB | FALSE | Upregulated |
| -0,82895383 | 0,254453267 | RBM41 | FALSE | Downregulated |
| -0,47690536 | 0,254605139 | FAM149A | FALSE | Downregulated |
| -0,390758935 | 0,254724754 | HINT1 | FALSE | Downregulated |
| 0,848761446 | 0,254724754 | TNFSF13 | FALSE | Upregulated |
| 0,630812678 | 0,255048656 | LOC100996813 | FALSE | Upregulated |
| 0,795974889 | 0,255048656 | SLC9A9 | FALSE | Upregulated |
| -0,9878647 | 0,255343666 | MLIP | FALSE | Downregulated |
| -2,320998332 | 0,255343666 | TAAR3 | FALSE | Downregulated |
| -0,311178536 | 0,255381399 | PRPF6 | FALSE | Downregulated |
| 0,451353711 | 0,255381399 | RPF1 | FALSE | Upregulated |
| 0,525258461 | 0,255381399 | SLC51A | FALSE | Upregulated |
| 0,394403377 | 0,255506327 | MEGF11 | FALSE | Upregulated |
| 0,476919252 | 0,255557264 | TNFAIP3 | FALSE | Upregulated |
| 0,635985082 | 0,255610945 | IDI2 | FALSE | Upregulated |
| 1,083562905 | 0,255610945 | VLDLR | FALSE | Upregulated |
| 0,81796718 | 0,255711595 | TRPA1 | FALSE | Upregulated |
| -0,723969284 | 0,255934605 | ADSSL1 | FALSE | Downregulated |
| -1,862267317 | 0,255934605 | KCNH8 | FALSE | Downregulated |
| -0,358979844 | 0,255934605 | TMEM57 | FALSE | Downregulated |
| -0,629671222 | 0,255934605 | ZC3H3 | FALSE | Downregulated |
| 0,356337278 | 0,256681549 | CHKA | FALSE | Upregulated |
| 0,363145268 | 0,25699545 | CARHSP1 | FALSE | Upregulated |
| -0,616333668 | 0,257180115 | NUDT5 | FALSE | Downregulated |
| -0,424483063 | 0,257182144 | LRRC8A | FALSE | Downregulated |
| -0,507927815 | 0,257270177 | DUSP6 | FALSE | Downregulated |
| 1,072556674 | 0,257625567 | CD3E | FALSE | Upregulated |
| 0,499089867 | 0,257737585 | SSH3 | FALSE | Upregulated |
| -0,357064556 | 0,257833825 | HSDL2 | FALSE | Downregulated |
| -0,305592192 | 0,257865047 | SEPHS2 | FALSE | Downregulated |
| 0,535874796 | 0,257923261 | ARL5B | FALSE | Upregulated |
| 0,459145695 | 0,257923261 | STARD9 | FALSE | Upregulated |
| 0,571717097 | 0,258133674 | BZRAP1-AS1 | FALSE | Upregulated |
| 0,332900754 | 0,25829303 | PPP1CB | FALSE | Upregulated |
| 1,036033102 | 0,258312339 | SCGN | FALSE | Upregulated |
| 0,919737266 | 0,258345861 | HID1 | FALSE | Upregulated |
| -0,451345502 | 0,25837799 | HIATL1 | FALSE | Downregulated |
| 0,771459207 | 0,25837799 | PANO | FALSE | Upregulated |
| 0,460467023 | 0,258472783 | MTRNR2L3 | FALSE | Upregulated |
| 1,199623244 | 0,258474136 | S100A6 | FALSE | Upregulated |
| -0,397013244 | 0,25873238 | CTSO | FALSE | Downregulated |
| 0,586010291 | 0,25873238 | LOC100126784 | FALSE | Upregulated |
| 0,327269787 | 0,258954194 | ZBED4 | FALSE | Upregulated |
| 2,839872948 | 0,259100287 | DUOX2 | FALSE | Upregulated |
| 1,357787776 | 0,259100287 | FAM227A | FALSE | Upregulated |

| | | | | |
|---|---|---|---|---|
| 1,756844172 | 0,259350264 | ASRGL1 | FALSE | Upregulated |
| -0,422682039 | 0,259421629 | GSDMB | FALSE | Downregulated |
| -0,398571196 | 0,259551031 | EGFR-AS1 | FALSE | Downregulated |
| -0,287192283 | 0,259551031 | RPL32 | FALSE | Downregulated |
| 0,360624103 | 0,259612657 | ANKRD6 | FALSE | Upregulated |
| -0,495723444 | 0,259655999 | ICAM3 | FALSE | Downregulated |
| -0,357079381 | 0,259655999 | SLC30A1 | FALSE | Downregulated |
| 0,623345662 | 0,259655999 | UVRAG | FALSE | Upregulated |
| -0,422093783 | 0,259904457 | MCCC1 | FALSE | Downregulated |
| 0,68447958 | 0,259953253 | AKAP3 | FALSE | Upregulated |
| -1,87494191 | 0,259953253 | LOC101929007 | FALSE | Downregulated |
| 0,463396344 | 0,259953253 | NOL8 | FALSE | Upregulated |
| -0,294049623 | 0,259953253 | SLCO2B1 | FALSE | Downregulated |
| -0,829316625 | 0,260081117 | CNTFR-AS1 | FALSE | Downregulated |
| 0,393969479 | 0,260098136 | CSPP1 | FALSE | Upregulated |
| 0,400182006 | 0,260098136 | EZR | FALSE | Upregulated |
| 1,2786367 | 0,260212065 | LINC00630 | FALSE | Upregulated |
| 1,413240509 | 0,260212065 | RP3-437I16.1 | FALSE | Upregulated |
| -0,452624505 | 0,260212065 | USP31 | FALSE | Downregulated |
| 0,374507541 | 0,260766987 | NR2F1 | FALSE | Upregulated |
| -0,366344842 | 0,261076086 | CYB5R3 | FALSE | Downregulated |
| -0,760850607 | 0,261117917 | ART4 | FALSE | Downregulated |
| -0,696246141 | 0,261117917 | SLC6A13 | FALSE | Downregulated |
| 0,448007833 | 0,261515795 | C7orf49 | FALSE | Upregulated |
| -0,456438383 | 0,261686058 | ACVR1B | FALSE | Downregulated |
| -0,500577109 | 0,261686058 | COPS6 | FALSE | Downregulated |
| -0,536448789 | 0,261686058 | GCC1 | FALSE | Downregulated |
| 0,944117564 | 0,261686058 | LOC101927730 | FALSE | Upregulated |
| 0,760520412 | 0,261772565 | MYO5A | FALSE | Upregulated |
| 0,563886085 | 0,261772565 | TRPM4 | FALSE | Upregulated |
| -0,349441287 | 0,261873722 | MTDH | FALSE | Downregulated |
| -0,60381954 | 0,262023728 | HCN3 | FALSE | Downregulated |
| -0,408056518 | 0,262135788 | MAGT1 | FALSE | Downregulated |
| 0,322172935 | 0,262135788 | MYO9B | FALSE | Upregulated |
| -0,325316293 | 0,262269668 | JUP | FALSE | Downregulated |
| 0,487953068 | 0,262279125 | ARL8B | FALSE | Upregulated |
| -0,485701156 | 0,262279125 | ATXN10 | FALSE | Downregulated |
| 0,470701555 | 0,262451955 | IFNAR2 | FALSE | Upregulated |
| -1,888323753 | 0,262909992 | N6AMT1 | FALSE | Downregulated |
| 1,012301393 | 0,262949491 | ARMC9 | FALSE | Upregulated |
| 0,483887326 | 0,263237026 | IFFO1 | FALSE | Upregulated |
| 0,297884746 | 0,263237026 | MBD6 | FALSE | Upregulated |
| 0,38824141 | 0,263510371 | ATG14 | FALSE | Upregulated |
| 0,562316271 | 0,263575433 | NCF2 | FALSE | Upregulated |
| -0,93437047 | 0,263616172 | FAM151A | FALSE | Downregulated |
| 1,758491336 | 0,263616172 | LOC102724330 | FALSE | Upregulated |
| 0,543318367 | 0,264117436 | LOC101928917 | FALSE | Upregulated |
| 0,477989396 | 0,264144457 | SUMO1P3 | FALSE | Upregulated |
| -0,575284359 | 0,264167168 | AC011343.1 | FALSE | Downregulated |
| 2,983503175 | 0,264188979 | RAB3B | FALSE | Upregulated |
| -1,593234481 | 0,264222833 | RAG1 | FALSE | Downregulated |

| | | | | |
|---|---|---|---|---|
| -0,417083651 | 0,264398933 | LETM1 | FALSE | Downregulated |
| 0,433493647 | 0,264691752 | LRRC4 | FALSE | Upregulated |
| 1,338965803 | 0,264691752 | MEIS3 | FALSE | Upregulated |
| -1,628780512 | 0,264691752 | PRB2 | FALSE | Downregulated |
| -0,280664445 | 0,264691752 | RPS8 | FALSE | Downregulated |
| 1,405256259 | 0,264691752 | ZNF215 | FALSE | Upregulated |
| -0,880774827 | 0,264974716 | ZNF234 | FALSE | Downregulated |
| 0,416973621 | 0,265027953 | EIF4ENIF1 | FALSE | Upregulated |
| 0,420549879 | 0,265027953 | LOC100133445 | FALSE | Upregulated |
| -0,753407383 | 0,265027953 | PHYHD1 | FALSE | Downregulated |
| -0,391082215 | 0,265027953 | SLC6A1 | FALSE | Downregulated |
| -0,494236421 | 0,26543504 | GOT1 | FALSE | Downregulated |
| 0,835929927 | 0,265750514 | C17orf100 | FALSE | Upregulated |
| -0,365219918 | 0,265791802 | SERP1 | FALSE | Downregulated |
| -0,793591136 | 0,265793798 | SHFM1 | FALSE | Downregulated |
| -0,53769737 | 0,26621321 | ROBO4 | FALSE | Downregulated |
| 0,645650152 | 0,267392312 | ZNF620 | FALSE | Upregulated |
| 0,921173166 | 0,267541704 | FAM150B | FALSE | Upregulated |
| 0,859709069 | 0,267541704 | HOXA-AS2 | FALSE | Upregulated |
| 0,455509288 | 0,267688564 | GOLGA8H | FALSE | Upregulated |
| -0,86942583 | 0,267688564 | GTF2H3 | FALSE | Downregulated |
| 0,480762972 | 0,267688564 | LOC102723863 | FALSE | Upregulated |
| -0,646996481 | 0,267808634 | HPS6 | FALSE | Downregulated |
| 0,876702413 | 0,267808634 | MROH6 | FALSE | Upregulated |
| 1,556766363 | 0,267976057 | LOC102723500 | FALSE | Upregulated |
| -0,421715102 | 0,268263397 | CHTF8 | FALSE | Downregulated |
| 1,703858941 | 0,268315699 | HENMT1 | FALSE | Upregulated |
| 0,734215932 | 0,268315699 | NBEA | FALSE | Upregulated |
| 1,204925766 | 0,268315699 | PIK3R6 | FALSE | Upregulated |
| 1,196042347 | 0,268387189 | C11orf16 | FALSE | Upregulated |
| 0,852628198 | 0,268479879 | KLF4 | FALSE | Upregulated |
| 0,53735147 | 0,268479879 | ZFYVE21 | FALSE | Upregulated |
| -0,713593236 | 0,26857987 | STRA13 | FALSE | Downregulated |
| 0,560991087 | 0,268631783 | LRRC37A2 | FALSE | Upregulated |
| -0,597838999 | 0,268662879 | CRTC2 | FALSE | Downregulated |
| 0,844434525 | 0,268667044 | AKAP7 | FALSE | Upregulated |
| -0,317905673 | 0,268676897 | BRD1 | FALSE | Downregulated |
| -1,141147242 | 0,268676897 | NDUFAB1 | FALSE | Downregulated |
| 0,58085236 | 0,268676897 | SLC35D2 | FALSE | Upregulated |
| 0,415091107 | 0,268968914 | ABI3BP | FALSE | Upregulated |
| 0,344798482 | 0,269081937 | SASH1 | FALSE | Upregulated |
| 0,224081205 | 0,269212723 | SON | FALSE | Upregulated |
| -1,053835005 | 0,269315414 | EMC6 | FALSE | Downregulated |
| 0,571189013 | 0,269315414 | WIPI1 | FALSE | Upregulated |
| -0,693330097 | 0,269341586 | IL10RB | FALSE | Downregulated |
| 0,725194601 | 0,269341586 | NDE1 | FALSE | Upregulated |
| 2,808304378 | 0,269341586 | RP11-2O17.2 | FALSE | Upregulated |
| -0,392567924 | 0,269341586 | ZNHIT1 | FALSE | Downregulated |
| 0,351532434 | 0,269521393 | GLTSCR1 | FALSE | Upregulated |
| -0,630280647 | 0,269615788 | CPEB3 | FALSE | Downregulated |
| 1,277453701 | 0,269615788 | GDAP1 | FALSE | Upregulated |

| | | | | |
|---|---|---|---|---|
| 0,616464178 | 0,269615788 | SUPT4H1 | FALSE | Upregulated |
| -1,240944688 | 0,269681103 | MIPOL1 | FALSE | Downregulated |
| 0,919974721 | 0,270685393 | C5AR1 | FALSE | Upregulated |
| 0,387429259 | 0,270685393 | STAT5A | FALSE | Upregulated |
| -0,671899107 | 0,270687491 | GJB2 | FALSE | Downregulated |
| 0,975636369 | 0,270687491 | WDSUB1 | FALSE | Upregulated |
| 0,27946022 | 0,270814011 | MSH6 | FALSE | Upregulated |
| 0,278676931 | 0,270838554 | AHNAK | FALSE | Upregulated |
| -0,281146817 | 0,271160255 | DST | FALSE | Downregulated |
| -0,81403883 | 0,271161662 | GRB7 | FALSE | Downregulated |
| -0,842812129 | 0,271161662 | TMEM177 | FALSE | Downregulated |
| 0,88516049 | 0,271200807 | OR2A9P | FALSE | Upregulated |
| -0,515815184 | 0,271200807 | PMVK | FALSE | Downregulated |
| -1,818362272 | 0,27128005 | CEACAM8 | FALSE | Downregulated |
| 0,364844774 | 0,271514382 | DNHD1 | FALSE | Upregulated |
| 0,601713349 | 0,271621402 | GAS5-AS1 | FALSE | Upregulated |
| -1,691661726 | 0,271621402 | LOC100499484 | FALSE | Downregulated |
| 0,94232633 | 0,271731562 | AC010226.4 | FALSE | Upregulated |
| 0,555536161 | 0,271731562 | SMAD1-AS2 | FALSE | Upregulated |
| 1,109136865 | 0,271743888 | CD34 | FALSE | Upregulated |
| 0,57241801 | 0,271743888 | YRDC | FALSE | Upregulated |
| 1,619907567 | 0,271982283 | ATP2A1 | FALSE | Upregulated |
| 0,830791113 | 0,271982283 | ITGB4 | FALSE | Upregulated |
| -1,298650539 | 0,272051579 | LINC00652 | FALSE | Downregulated |
| 0,925859102 | 0,272384296 | ARL6IP6 | FALSE | Upregulated |
| -0,637039706 | 0,272384296 | C22orf15 | FALSE | Downregulated |
| 0,842471183 | 0,272384296 | GCNT1 | FALSE | Upregulated |
| -0,26837105 | 0,272384296 | RBM33 | FALSE | Downregulated |
| 1,498829948 | 0,272614472 | ADAM28 | FALSE | Upregulated |
| -0,366415288 | 0,272614472 | YME1L1 | FALSE | Downregulated |
| -0,306166835 | 0,272916405 | CD81 | FALSE | Downregulated |
| 0,414935752 | 0,273083068 | SLC24A1 | FALSE | Upregulated |
| 1,433364404 | 0,273157012 | KCNMB1 | FALSE | Upregulated |
| 0,767018441 | 0,273157012 | PAX8-AS1 | FALSE | Upregulated |
| -0,382722855 | 0,273272008 | TF | FALSE | Downregulated |
| 0,44232842 | 0,273543231 | IL7 | FALSE | Upregulated |
| -0,432984652 | 0,273605437 | KDM4C | FALSE | Downregulated |
| 0,716409217 | 0,273605437 | PIP4K2C | FALSE | Upregulated |
| -0,419099169 | 0,27362571 | NBPF9 | FALSE | Downregulated |
| -0,688123011 | 0,273668603 | HUS1 | FALSE | Downregulated |
| 0,382862302 | 0,273668603 | PPP1R13B | FALSE | Upregulated |
| 0,473808218 | 0,273920001 | CD93 | FALSE | Upregulated |
| 0,389934965 | 0,274771442 | DHRS12 | FALSE | Upregulated |
| 0,889941879 | 0,2750552 | PARVG | FALSE | Upregulated |
| 0,981820902 | 0,275172142 | ALG1L9P | FALSE | Upregulated |
| 0,567103945 | 0,275697275 | DNAJA4 | FALSE | Upregulated |
| 1,185073301 | 0,275910804 | APOBEC3D | FALSE | Upregulated |
| -0,367612311 | 0,275935486 | KIAA2026 | FALSE | Downregulated |
| -0,397827767 | 0,276091251 | HDHD3 | FALSE | Downregulated |
| 1,432572684 | 0,276091251 | KCND1 | FALSE | Upregulated |
| 0,672125312 | 0,276226727 | SOGA2 | FALSE | Upregulated |

| | | | | |
|---|---|---|---|---|
| -0,397863531 | 0,276820049 | AIG1 | FALSE | Downregulated |
| -0,306451537 | 0,276820049 | SLC2A4RG | FALSE | Downregulated |
| 0,403707085 | 0,276939967 | TSSK3 | FALSE | Upregulated |
| -0,631236333 | 0,277324423 | ALDOC | FALSE | Downregulated |
| -0,259723303 | 0,277439155 | EIF4H | FALSE | Downregulated |
| -0,942611654 | 0,277463134 | LOC102724238 | FALSE | Downregulated |
| 0,741971854 | 0,277671419 | NOTCH3 | FALSE | Upregulated |
| -0,573217326 | 0,277671419 | SKIDA1 | FALSE | Downregulated |
| 0,35605158 | 0,277671419 | TXNDC5 | FALSE | Upregulated |
| 0,961493316 | 0,278466665 | PRAF2 | FALSE | Upregulated |
| 1,355685792 | 0,278544266 | ZNF267 | FALSE | Upregulated |
| -0,255954177 | 0,27859925 | GOLGA4 | FALSE | Downregulated |
| -0,347551838 | 0,278686412 | MAP2K1 | FALSE | Downregulated |
| -0,632838387 | 0,278686412 | TMEM219 | FALSE | Downregulated |
| 0,529052003 | 0,279264257 | ARMC7 | FALSE | Upregulated |
| -0,844048969 | 0,279264257 | JAGN1 | FALSE | Downregulated |
| 1,065630867 | 0,279264257 | LOC101927521 | FALSE | Upregulated |
| -0,347020434 | 0,279264257 | PCYT2 | FALSE | Downregulated |
| -0,341588493 | 0,279264257 | RPS27A | FALSE | Downregulated |
| 0,301490351 | 0,279264257 | TOP2B | FALSE | Upregulated |
| 1,255881872 | 0,279329918 | MIR143HG | FALSE | Upregulated |
| 0,956913424 | 0,279697336 | SDR42E1 | FALSE | Upregulated |
| 1,819555355 | 0,27974229 | AIM2 | FALSE | Upregulated |
| 2,112260274 | 0,27974229 | C10orf55 | FALSE | Upregulated |
| 0,316818298 | 0,27974229 | C15orf62 | FALSE | Upregulated |
| 0,802356518 | 0,27974229 | CHORDC1 | FALSE | Upregulated |
| 0,430814562 | 0,27974229 | INPP4A | FALSE | Upregulated |
| -0,399257406 | 0,27974229 | LDHA | FALSE | Downregulated |
| 0,541273413 | 0,27974229 | LRRC37A | FALSE | Upregulated |
| -0,598021825 | 0,27974229 | MTIF2 | FALSE | Downregulated |
| 0,636932537 | 0,27974229 | MYO5C | FALSE | Upregulated |
| -0,46779347 | 0,27974229 | NAPRT1 | FALSE | Downregulated |
| 0,986755203 | 0,27974229 | SRGAP2C | FALSE | Upregulated |
| -0,378192743 | 0,27974229 | TMEM110 | FALSE | Downregulated |
| 0,327447493 | 0,27974229 | TNFRSF10B | FALSE | Upregulated |
| 0,339376608 | 0,27974229 | UCKL1-AS1 | FALSE | Upregulated |
| -0,430826782 | 0,27974229 | UGT2B4 | FALSE | Downregulated |
| -0,359063328 | 0,27974229 | ZMYM4 | FALSE | Downregulated |
| 1,551126964 | 0,27974229 | ZNF165 | FALSE | Upregulated |
| -0,362619983 | 0,27974229 | ZNF91 | FALSE | Downregulated |
| -1,971501679 | 0,279757383 | C21orf90 | FALSE | Downregulated |
| -0,495638846 | 0,279757383 | PAPD5 | FALSE | Downregulated |
| 0,868405788 | 0,279757383 | PRKCQ | FALSE | Upregulated |
| -0,515552142 | 0,279960439 | INPP5B | FALSE | Downregulated |
| -0,320303126 | 0,280010336 | F5 | FALSE | Downregulated |
| 0,920318749 | 0,280504282 | LXN | FALSE | Upregulated |
| 0,404875005 | 0,280504282 | NUDT13 | FALSE | Upregulated |
| -0,449813108 | 0,280504282 | UCK1 | FALSE | Downregulated |
| -0,51161755 | 0,280916139 | NMD3 | FALSE | Downregulated |
| -0,428014478 | 0,281221027 | TALDO1 | FALSE | Downregulated |
| 0,811445352 | 0,281364388 | OAS2 | FALSE | Upregulated |

| | | | | |
|---|---|---|---|---|
| 1,123192948 | 0,281523502 | ZNF596 | FALSE | Upregulated |
| 0,873347859 | 0,281970843 | HN1 | FALSE | Upregulated |
| 0,548611126 | 0,281979891 | C19orf54 | FALSE | Upregulated |
| 0,867636448 | 0,281979891 | DBN1 | FALSE | Upregulated |
| 0,415725144 | 0,282525009 | B2M | FALSE | Upregulated |
| -0,375264508 | 0,282525009 | SIRT5 | FALSE | Downregulated |
| 0,584403622 | 0,282534486 | NMNAT3 | FALSE | Upregulated |
| 0,640333414 | 0,282534486 | PGAM4 | FALSE | Upregulated |
| 1,232518535 | 0,282582416 | IPCEF1 | FALSE | Upregulated |
| 0,58055668 | 0,28286874 | AMZ1 | FALSE | Upregulated |
| -0,340580872 | 0,282926469 | FBXW2 | FALSE | Downregulated |
| 0,42194575 | 0,283043989 | KIAA1551 | FALSE | Upregulated |
| 1,059021403 | 0,283046202 | AGRP | FALSE | Upregulated |
| 1,081905325 | 0,283046202 | PFN4 | FALSE | Upregulated |
| -0,607859714 | 0,28307398 | LOC101927345 | FALSE | Downregulated |
| 0,721322694 | 0,283092229 | CMKLR1 | FALSE | Upregulated |
| -0,657716702 | 0,283092229 | GLT1D1 | FALSE | Downregulated |
| 0,471622396 | 0,283092229 | HIF1A | FALSE | Upregulated |
| 0,461437796 | 0,283092229 | ZBTB46 | FALSE | Upregulated |
| 0,47857742 | 0,283291802 | LOC102724552 | FALSE | Upregulated |
| 0,321479956 | 0,283413096 | ACTN1 | FALSE | Upregulated |
| 1,402926391 | 0,283413096 | FZD7 | FALSE | Upregulated |
| 0,294837463 | 0,283451158 | CDK5RAP2 | FALSE | Upregulated |
| -0,526081006 | 0,283508486 | SEMA6C | FALSE | Downregulated |
| -0,589986096 | 0,283580245 | LONRF3 | FALSE | Downregulated |
| -1,762255556 | 0,283619433 | FRG1B | FALSE | Downregulated |
| -0,305619357 | 0,283682641 | FETUB | FALSE | Downregulated |
| 0,935823861 | 0,283947896 | SLC26A8 | FALSE | Upregulated |
| -1,381449439 | 0,284031578 | FAM53A | FALSE | Downregulated |
| -0,511104479 | 0,284031578 | GLRX5 | FALSE | Downregulated |
| -1,42785722 | 0,284031578 | PKD1L2 | FALSE | Downregulated |
| 0,661966743 | 0,2842419 | RGS20 | FALSE | Upregulated |
| 0,861319686 | 0,284402785 | NCS1 | FALSE | Upregulated |
| 1,151340024 | 0,284629496 | FAM83C | FALSE | Upregulated |
| -0,823041446 | 0,284629496 | GTPBP4 | FALSE | Downregulated |
| 0,399205735 | 0,284629496 | TMSB4X | FALSE | Upregulated |
| 1,102807182 | 0,284629496 | UPK1B | FALSE | Upregulated |
| 0,607294484 | 0,284756435 | DXO | FALSE | Upregulated |
| 1,601952058 | 0,284756435 | LOC101927608 | FALSE | Upregulated |
| -1,564644573 | 0,284756435 | LOC102724720 | FALSE | Downregulated |
| 0,336073097 | 0,284756435 | LRRFIP2 | FALSE | Upregulated |
| 1,278124146 | 0,284888444 | KPNA2 | FALSE | Upregulated |
| 0,772813547 | 0,284888444 | PTPN13 | FALSE | Upregulated |
| 0,606086985 | 0,284943686 | CXCR5 | FALSE | Upregulated |
| -0,306066231 | 0,284943686 | FTH1 | FALSE | Downregulated |
| -0,266937396 | 0,284943686 | VPS4A | FALSE | Downregulated |
| 0,555222896 | 0,285004336 | SH3BP4 | FALSE | Upregulated |
| 0,564999957 | 0,285049978 | OCLN | FALSE | Upregulated |
| -0,435574909 | 0,285106454 | TMEM8A | FALSE | Downregulated |
| -0,60387011 | 0,28569087 | WFS1 | FALSE | Downregulated |
| 0,741074577 | 0,285817179 | FAM127A | FALSE | Upregulated |

| | | | | |
|---|---|---|---|---|
| -0,724226733 | 0,285817179 | TMEM254 | FALSE | Downregulated |
| 0,37447875 | 0,285896446 | MFSD11 | FALSE | Upregulated |
| 0,379068302 | 0,285896446 | ZNF814 | FALSE | Upregulated |
| -0,310698162 | 0,286078534 | TUFM | FALSE | Downregulated |
| 0,754725361 | 0,286326383 | AFAP1 | FALSE | Upregulated |
| 0,472763518 | 0,286352608 | TLE3 | FALSE | Upregulated |
| 0,304914406 | 0,286370535 | TRABD | FALSE | Upregulated |
| -1,582488015 | 0,286538765 | TECTA | FALSE | Downregulated |
| 0,881426618 | 0,286757723 | PDE4D | FALSE | Upregulated |
| 0,366467577 | 0,286787545 | LOC100506634 | FALSE | Upregulated |
| 0,424519875 | 0,28691807 | C14orf164 | FALSE | Upregulated |
| 0,889941652 | 0,28691807 | CD163L1 | FALSE | Upregulated |
| 1,187952436 | 0,28691807 | HCG27 | FALSE | Upregulated |
| -0,296813004 | 0,28691807 | HIPK3 | FALSE | Downregulated |
| -0,325917522 | 0,28691807 | MED13 | FALSE | Downregulated |
| 0,394769654 | 0,287099324 | CD37 | FALSE | Upregulated |
| 0,634180472 | 0,287099324 | SLC1A5 | FALSE | Upregulated |
| 0,286337016 | 0,287270039 | KIAA1432 | FALSE | Upregulated |
| 1,351827753 | 0,287270039 | KIF2C | FALSE | Upregulated |
| 1,053238219 | 0,28729558 | APOBEC3F | FALSE | Upregulated |
| 0,444334133 | 0,28729558 | RBM25 | FALSE | Upregulated |
| 1,529243001 | 0,28729558 | TMEM108 | FALSE | Upregulated |
| -0,482167718 | 0,287358296 | HINT2 | FALSE | Downregulated |
| 0,446696124 | 0,287444938 | MYSM1 | FALSE | Upregulated |
| -1,021580287 | 0,287918502 | SPATA31E1 | FALSE | Downregulated |
| 1,573609778 | 0,288483056 | LOC100129215 | FALSE | Upregulated |
| 1,898850267 | 0,288483056 | LOC550643 | FALSE | Upregulated |
| 0,925820619 | 0,288483056 | SFTPB | FALSE | Upregulated |
| 0,313776918 | 0,288483056 | SRPK2 | FALSE | Upregulated |
| 1,03430471 | 0,288733252 | LOC102724699 | FALSE | Upregulated |
| -0,714416146 | 0,288838354 | SLC1A2 | FALSE | Downregulated |
| 0,471305657 | 0,288902922 | TSSK4 | FALSE | Upregulated |
| -0,491020045 | 0,288902922 | UGDH | FALSE | Downregulated |
| 0,688859227 | 0,288902922 | ZNF23 | FALSE | Upregulated |
| -0,555098471 | 0,289086991 | SGK2 | FALSE | Downregulated |
| 0,930291105 | 0,2891203 | DEF6 | FALSE | Upregulated |
| 0,417070973 | 0,28916905 | NTMT1 | FALSE | Upregulated |
| -0,983442225 | 0,289435029 | ARID3C | FALSE | Downregulated |
| -2,153043085 | 0,289499651 | BSCL2 | FALSE | Downregulated |
| -0,374695824 | 0,289499651 | FXYD1 | FALSE | Downregulated |
| 0,381345004 | 0,289499651 | GNPDA1 | FALSE | Upregulated |
| 1,462514142 | 0,289499651 | SLC24A4 | FALSE | Upregulated |
| -0,344416805 | 0,289574241 | CYB5A | FALSE | Downregulated |
| 0,549336184 | 0,289683344 | LOC101929204 | FALSE | Upregulated |
| 1,965610567 | 0,290055471 | BATF | FALSE | Upregulated |
| -0,446230529 | 0,290110978 | CHMP1A | FALSE | Downregulated |
| 0,849807244 | 0,290390146 | RP11-692N5.1 | FALSE | Upregulated |
| 0,343282712 | 0,290643574 | HEATR1 | FALSE | Upregulated |
| -0,975906558 | 0,290643574 | PAAF1 | FALSE | Downregulated |
| -0,299846192 | 0,291040442 | CYP4V2 | FALSE | Downregulated |
| -0,365206822 | 0,291040442 | TOMM40L | FALSE | Downregulated |

| | | | | |
|---|---|---|---|---|
| -1,68664447 | 0,291181846 | GUCA1B | FALSE | Downregulated |
| -0,512475567 | 0,291181846 | TFDP2 | FALSE | Downregulated |
| -1,409676428 | 0,291344438 | TREML1 | FALSE | Downregulated |
| 0,447891309 | 0,291389113 | ATAD5 | FALSE | Upregulated |
| 0,822330829 | 0,291624942 | LOC101927227 | FALSE | Upregulated |
| -1,902852414 | 0,291624942 | LOC101929296 | FALSE | Downregulated |
| 0,890723533 | 0,291624942 | SPATA18 | FALSE | Upregulated |
| 0,470803319 | 0,291624942 | TESK1 | FALSE | Upregulated |
| -1,840918701 | 0,291826576 | OR10J1 | FALSE | Downregulated |
| -0,448212191 | 0,291826576 | TMEM51-AS1 | FALSE | Downregulated |
| 2,263964543 | 0,292181266 | CTAGE15 | FALSE | Upregulated |
| 1,624849981 | 0,292181266 | KIAA0087 | FALSE | Upregulated |
| -1,148991285 | 0,292181266 | LOC101927105 | FALSE | Downregulated |
| -0,31775375 | 0,292181266 | MBTPS1 | FALSE | Downregulated |
| -0,82773544 | 0,292181266 | TMEM208 | FALSE | Downregulated |
| 0,754516976 | 0,292246381 | ALPK3 | FALSE | Upregulated |
| -0,589692176 | 0,292322667 | FAM200B | FALSE | Downregulated |
| -0,659450664 | 0,292591917 | LDOC1L | FALSE | Downregulated |
| 0,989390383 | 0,292631612 | DTX3 | FALSE | Upregulated |
| 1,415763358 | 0,292983485 | LOC100131107 | FALSE | Upregulated |
| 0,439709358 | 0,293164005 | RP11-197N18.7 | FALSE | Upregulated |
| 1,621016771 | 0,293230413 | LOC440173 | FALSE | Upregulated |
| 1,309889337 | 0,293363465 | MALL | FALSE | Upregulated |
| -0,519723467 | 0,293570986 | OXR1 | FALSE | Downregulated |
| 0,551469323 | 0,293786999 | C1QB | FALSE | Upregulated |
| -0,335383058 | 0,293786999 | PGPEP1 | FALSE | Downregulated |
| 0,437064473 | 0,293786999 | USP25 | FALSE | Upregulated |
| 0,817951399 | 0,293877405 | C9orf116 | FALSE | Upregulated |
| -0,536587994 | 0,293877405 | TRMT44 | FALSE | Downregulated |
| 0,76199973 | 0,294325442 | NECAB1 | FALSE | Upregulated |
| -0,534585955 | 0,294330479 | CREB3L3 | FALSE | Downregulated |
| 1,048119546 | 0,294662576 | IQCB1 | FALSE | Upregulated |
| 1,10228274 | 0,295067656 | LINC01123 | FALSE | Upregulated |
| -0,810273718 | 0,295102782 | LOC100133331 | FALSE | Downregulated |
| -1,116274452 | 0,295133087 | RFPL1S | FALSE | Downregulated |
| 0,524800905 | 0,295287405 | PTPN7 | FALSE | Upregulated |
| 1,227056706 | 0,295381097 | AC022007.5 | FALSE | Upregulated |
| 1,133577952 | 0,295418864 | PM20D2 | FALSE | Upregulated |
| 0,747404191 | 0,29542647 | HCK | FALSE | Upregulated |
| -0,314408523 | 0,29542647 | NISCH | FALSE | Downregulated |
| -2,106480233 | 0,29542647 | NPTXR | FALSE | Downregulated |
| 0,37724553 | 0,295453178 | CYLD | FALSE | Upregulated |
| -0,440393647 | 0,295533418 | SALL1 | FALSE | Downregulated |
| -0,37004442 | 0,295810221 | APOL1 | FALSE | Downregulated |
| 0,436187705 | 0,295810221 | PDE5A | FALSE | Upregulated |
| 0,752445763 | 0,295854673 | ANKEF1 | FALSE | Upregulated |
| -0,348468342 | 0,295854673 | CUX1 | FALSE | Downregulated |
| 0,464186711 | 0,296439953 | DNAJC10 | FALSE | Upregulated |
| 0,407786317 | 0,296439953 | MARCKSL1 | FALSE | Upregulated |
| -0,278142964 | 0,296439953 | STAU1 | FALSE | Downregulated |
| -0,450060459 | 0,296439953 | TIAM1 | FALSE | Downregulated |

| | | | | |
|---|---|---|---|---|
| -0,312309262 | 0,296439953 | TOR1AIP1 | FALSE | Downregulated |
| -0,465200332 | 0,296507106 | CD2BP2 | FALSE | Downregulated |
| 1,133099515 | 0,296507106 | COX6B2 | FALSE | Upregulated |
| -0,526410428 | 0,296507106 | PREB | FALSE | Downregulated |
| 1,115219337 | 0,296507106 | ST8SIA4 | FALSE | Upregulated |
| 0,344597485 | 0,296507106 | TRAF4 | FALSE | Upregulated |
| -0,511746495 | 0,29687549 | PHKA2-AS1 | FALSE | Downregulated |
| -0,281205431 | 0,29687549 | ZBTB7A | FALSE | Downregulated |
| 1,270184425 | 0,297182905 | CDH11 | FALSE | Upregulated |
| -1,631865285 | 0,297230321 | C9orf85 | FALSE | Downregulated |
| -3,913553714 | 0,297247577 | HIST1H4J | FALSE | Downregulated |
| 0,45327505 | 0,297247577 | SIGLEC1 | FALSE | Upregulated |
| -0,497516414 | 0,297321221 | CYB5D2 | FALSE | Downregulated |
| 0,525734491 | 0,297321221 | DNM3 | FALSE | Upregulated |
| 0,673115814 | 0,297321221 | ISPD-AS1 | FALSE | Upregulated |
| 1,553142184 | 0,29735153 | LOC101929042 | FALSE | Upregulated |
| 0,736599079 | 0,297539635 | CYBRD1 | FALSE | Upregulated |
| 1,294297363 | 0,297539635 | HERC2P3 | FALSE | Upregulated |
| 0,330953968 | 0,297960153 | LOC100505658 | FALSE | Upregulated |
| 1,447058999 | 0,298040111 | ESRRB | FALSE | Upregulated |
| 0,457632165 | 0,298176223 | DCUN1D1 | FALSE | Upregulated |
| 0,263102813 | 0,298394923 | ATXN7 | FALSE | Upregulated |
| 0,390395591 | 0,298394923 | BTBD18 | FALSE | Upregulated |
| -0,307603138 | 0,298394923 | GLTPD2 | FALSE | Downregulated |
| 0,851609263 | 0,298394923 | GPR39 | FALSE | Upregulated |
| -0,278199805 | 0,298394923 | HECTD4 | FALSE | Downregulated |
| -0,506778905 | 0,298394923 | HPGD | FALSE | Downregulated |
| 1,025014043 | 0,298394923 | INPP4B | FALSE | Upregulated |
| 0,346844001 | 0,298394923 | ITGB5 | FALSE | Upregulated |
| 0,809285003 | 0,298394923 | LOC100507530 | FALSE | Upregulated |
| 0,466479713 | 0,298394923 | LOC102723990 | FALSE | Upregulated |
| -0,528362989 | 0,298394923 | MERTK | FALSE | Downregulated |
| -0,977262396 | 0,298394923 | MRO | FALSE | Downregulated |
| 0,268792776 | 0,298394923 | MYO1C | FALSE | Upregulated |
| -0,732923897 | 0,298394923 | NLRP6 | FALSE | Downregulated |
| 0,680232395 | 0,298394923 | PDCD1LG2 | FALSE | Upregulated |
| -0,247684144 | 0,298394923 | PFKL | FALSE | Downregulated |
| -1,828976293 | 0,298394923 | PRB3 | FALSE | Downregulated |
| -0,628299624 | 0,298394923 | RARS | FALSE | Downregulated |
| 0,405026296 | 0,298394923 | RASAL2 | FALSE | Upregulated |
| 0,442215973 | 0,298394923 | RASSF4 | FALSE | Upregulated |
| 0,925723639 | 0,298394923 | RP1-257A7.4 | FALSE | Upregulated |
| -0,416223814 | 0,298394923 | TBC1D24 | FALSE | Downregulated |
| -1,055336619 | 0,298394923 | TXNDC16 | FALSE | Downregulated |
| 1,151286952 | 0,298394923 | VILL | FALSE | Upregulated |
| 0,357133719 | 0,298394923 | ZFYVE26 | FALSE | Upregulated |
| 0,77116741 | 0,298394923 | ZNF441 | FALSE | Upregulated |
| 0,610095766 | 0,298394923 | ZNF532 | FALSE | Upregulated |
| -0,782847075 | 0,298617869 | KIF13A | FALSE | Downregulated |
| -0,41366423 | 0,298617869 | NDUFB10 | FALSE | Downregulated |
| 0,440638446 | 0,298617869 | TNFRSF1B | FALSE | Upregulated |

| | | | | |
|---|---|---|---|---|
| 0,64817481 | 0,298637012 | C1QL3 | FALSE | Upregulated |
| 1,538594041 | 0,298639236 | LINC00959 | FALSE | Upregulated |
| 0,987410109 | 0,298639236 | LOC101926994 | FALSE | Upregulated |
| 1,827343489 | 0,298639236 | RP11-572M11.3 | FALSE | Upregulated |
| 1,368118163 | 0,298664529 | CYP2F1 | FALSE | Upregulated |
| -1,116172521 | 0,298664529 | IFT27 | FALSE | Downregulated |
| 0,288360825 | 0,298664529 | INF2 | FALSE | Upregulated |
| 0,683715859 | 0,298664529 | MAPK7 | FALSE | Upregulated |
| 0,985607644 | 0,298664529 | TAF1B | FALSE | Upregulated |
| 0,714835735 | 0,298690843 | BASP1 | FALSE | Upregulated |
| -0,486396448 | 0,299022299 | ZDHHC3 | FALSE | Downregulated |
| 0,65084987 | 0,299077734 | GHRLOS | FALSE | Upregulated |
| 1,66375082 | 0,299182923 | RP11-329B9.1 | FALSE | Upregulated |
| 0,30892657 | 0,299182923 | TULP4 | FALSE | Upregulated |
| -0,769568046 | 0,29938395 | LOC101927645 | FALSE | Downregulated |
| 0,315748077 | 0,299492899 | COX20 | FALSE | Upregulated |
| 0,732276785 | 0,299492899 | DIEXF | FALSE | Upregulated |
| -0,425873902 | 0,299492899 | SLC25A47 | FALSE | Downregulated |
| -0,974743039 | 0,299695058 | BMS1P4 | FALSE | Downregulated |
| 0,334471669 | 0,299695058 | POU5F1P4 | FALSE | Upregulated |
| -1,363337572 | 0,299695058 | ZNF311 | FALSE | Downregulated |
| -0,451023246 | 0,300129719 | TRIB1 | FALSE | Downregulated |
| 0,680983786 | 0,300345517 | ACTA2 | FALSE | Upregulated |
| 0,634697945 | 0,300464101 | ADAMTS16 | FALSE | Upregulated |
| 1,227887528 | 0,300464101 | CXorf36 | FALSE | Upregulated |
| 0,311737848 | 0,300464101 | KLC1 | FALSE | Upregulated |
| 1,35195986 | 0,300464101 | LOC388242 | FALSE | Upregulated |
| 0,870468218 | 0,300464101 | PRSS54 | FALSE | Upregulated |
| 0,739641867 | 0,300464101 | SLC5A10 | FALSE | Upregulated |
| 0,34186589 | 0,300464101 | SUDS3 | FALSE | Upregulated |
| -2,229724957 | 0,300663597 | ANKRD20A12P | FALSE | Downregulated |
| 0,416366921 | 0,300663597 | DESI1 | FALSE | Upregulated |
| -0,706958679 | 0,300663597 | SUZ12 | FALSE | Downregulated |
| 0,974152077 | 0,300909272 | WDR86 | FALSE | Upregulated |
| 1,036411808 | 0,300953778 | ZNF71 | FALSE | Upregulated |
| -0,219442516 | 0,301009851 | CSDE1 | FALSE | Downregulated |
| 0,517017591 | 0,301062876 | JUNB | FALSE | Upregulated |
| 0,469218421 | 0,301150681 | TIMM8B | FALSE | Upregulated |
| 0,640594253 | 0,301394284 | IGLON5 | FALSE | Upregulated |
| 0,633103815 | 0,301394284 | ZBTB32 | FALSE | Upregulated |
| 0,315679787 | 0,301401929 | BTBD6 | FALSE | Upregulated |
| 0,337902544 | 0,301401929 | HSD17B1 | FALSE | Upregulated |
| -0,40858907 | 0,301412302 | DNAJA3 | FALSE | Downregulated |
| 0,75369661 | 0,301412302 | FAM78A | FALSE | Upregulated |
| 0,408013606 | 0,301412302 | SCO2 | FALSE | Upregulated |
| 1,036700753 | 0,301580972 | C1orf145 | FALSE | Upregulated |
| 2,144396747 | 0,301580972 | FAM72D | FALSE | Upregulated |
| -0,391199218 | 0,301580972 | HGFAC | FALSE | Downregulated |
| 0,320617901 | 0,301580972 | IMPAD1 | FALSE | Upregulated |
| 1,823669144 | 0,301580972 | LOC102031319 | FALSE | Upregulated |
| -0,436452973 | 0,301580972 | RPL35A | FALSE | Downregulated |

| | | | | |
|---|---|---|---|---|
| 0,491013423 | 0,301686324 | ANGPTL7 | FALSE | Upregulated |
| 0,496532967 | 0,301830416 | MCMDC2 | FALSE | Upregulated |
| -0,756464931 | 0,302043583 | DIABLO | FALSE | Downregulated |
| -0,509355827 | 0,302043583 | HEATR4 | FALSE | Downregulated |
| -0,695296957 | 0,302098485 | HIST1H2BC | FALSE | Downregulated |
| -1,04782647 | 0,302112511 | SF3B14 | FALSE | Downregulated |
| -0,405844239 | 0,302419137 | ENTPD5 | FALSE | Downregulated |
| 0,753419481 | 0,302419137 | PPM1D | FALSE | Upregulated |
| -0,541546961 | 0,302419137 | TOP1MT | FALSE | Downregulated |
| 0,998343807 | 0,302419137 | TSHZ3 | FALSE | Upregulated |
| 0,420574163 | 0,302419137 | XPA | FALSE | Upregulated |
| -1,105335804 | 0,302530096 | MBL1P | FALSE | Downregulated |
| 0,320600366 | 0,3026885 | ACOT13 | FALSE | Upregulated |
| 1,382622393 | 0,3026885 | HIST1H1B | FALSE | Upregulated |
| 0,960451788 | 0,3026885 | LOC102723506 | FALSE | Upregulated |
| 0,374609242 | 0,303085946 | MAFK | FALSE | Upregulated |
| -0,620320695 | 0,303303418 | PROX1-AS1 | FALSE | Downregulated |
| 0,561938145 | 0,303397917 | LOC101927420 | FALSE | Upregulated |
| 0,854659531 | 0,303406742 | CHST2 | FALSE | Upregulated |
| 1,057879854 | 0,303582258 | ACE | FALSE | Upregulated |
| -0,450155474 | 0,303582258 | ADHFE1 | FALSE | Downregulated |
| 0,510819247 | 0,303582258 | PRR15L | FALSE | Upregulated |
| 0,614258552 | 0,303582258 | UGCG | FALSE | Upregulated |
| 0,443223991 | 0,303582258 | USP42 | FALSE | Upregulated |
| 0,267763619 | 0,303615125 | TAOK2 | FALSE | Upregulated |
| -0,379635403 | 0,30382499 | FAM53B | FALSE | Downregulated |
| 1,645729072 | 0,30382499 | FHIT | FALSE | Upregulated |
| 0,811832386 | 0,30382499 | GRIN2B | FALSE | Upregulated |
| 1,684415767 | 0,30382499 | MTRNR2L6 | FALSE | Upregulated |
| 0,678600972 | 0,30382499 | TMEM86A | FALSE | Upregulated |
| 0,684119223 | 0,303849047 | LOC102724335 | FALSE | Upregulated |
| 0,68420521 | 0,303867701 | ADAMTS7 | FALSE | Upregulated |
| 0,415163533 | 0,303867701 | OTUD4 | FALSE | Upregulated |
| -0,299318251 | 0,303867701 | RPS23 | FALSE | Downregulated |
| 0,424772645 | 0,303921861 | MTMR2 | FALSE | Upregulated |
| 0,338837199 | 0,303921861 | RP11-133O22.6 | FALSE | Upregulated |
| -0,391144072 | 0,303944277 | ZNF787 | FALSE | Downregulated |
| -0,510149947 | 0,304343118 | SULT1A1 | FALSE | Downregulated |
| 1,363113109 | 0,304554717 | GABARAPL3 | FALSE | Upregulated |
| -1,666173073 | 0,304586125 | BNIP1 | FALSE | Downregulated |
| -0,86815989 | 0,304586125 | TOMM6 | FALSE | Downregulated |
| 0,667599511 | 0,304775143 | CCDC173 | FALSE | Upregulated |
| -0,683178074 | 0,3047948 | ADH1C | FALSE | Downregulated |
| -0,428707858 | 0,3047948 | DNAJC16 | FALSE | Downregulated |
| -1,492156601 | 0,3047948 | LOC102723788 | FALSE | Downregulated |
| -0,749935184 | 0,305049474 | TMEM143 | FALSE | Downregulated |
| 0,56071558 | 0,305181262 | PARD6G-AS1 | FALSE | Upregulated |
| -0,296530121 | 0,305262448 | DCAF5 | FALSE | Downregulated |
| -0,393646529 | 0,305262448 | PON3 | FALSE | Downregulated |
| -0,364673992 | 0,305262448 | RANBP1 | FALSE | Downregulated |
| -0,637459244 | 0,305262448 | TOMM7 | FALSE | Downregulated |

| | | | | |
|---|---|---|---|---|
| 0,366610144 | 0,305264995 | LOC100131496 | FALSE | Upregulated |
| 0,8110916 | 0,305446334 | DTNBP1 | FALSE | Upregulated |
| -0,565731882 | 0,305446334 | LBP | FALSE | Downregulated |
| 1,108643222 | 0,305446334 | WNT16 | FALSE | Upregulated |
| 1,139663976 | 0,306024651 | SEMA3G | FALSE | Upregulated |
| 1,234071668 | 0,306024651 | THSD1 | FALSE | Upregulated |
| 0,678150528 | 0,30611214 | LOC102723369 | FALSE | Upregulated |
| 1,107741752 | 0,30611214 | RP1-234P15.4 | FALSE | Upregulated |
| 0,522460065 | 0,306175816 | ZNF503-AS2 | FALSE | Upregulated |
| -0,35504977 | 0,306233948 | ACY1 | FALSE | Downregulated |
| 1,725404756 | 0,306233948 | C11orf48 | FALSE | Upregulated |
| 1,567015856 | 0,306517043 | CREB5 | FALSE | Upregulated |
| 1,215455716 | 0,306517043 | LOC728392 | FALSE | Upregulated |
| 0,954405094 | 0,306627368 | TMEM251 | FALSE | Upregulated |
| 0,593846355 | 0,306664428 | OAS3 | FALSE | Upregulated |
| -0,318217601 | 0,306940903 | ZCCHC14 | FALSE | Downregulated |
| -0,624411822 | 0,306997077 | LOC102723769 | FALSE | Downregulated |
| 1,046294718 | 0,306997077 | MITF | FALSE | Upregulated |
| 1,100277293 | 0,307048501 | SAMSN1 | FALSE | Upregulated |
| 0,56055134 | 0,307168877 | DYRK2 | FALSE | Upregulated |
| 0,442311553 | 0,307168877 | ST7-AS2 | FALSE | Upregulated |
| 0,268674828 | 0,307301458 | MCC | FALSE | Upregulated |
| 0,642979176 | 0,307371923 | LOC102723309 | FALSE | Upregulated |
| 0,430568786 | 0,307511912 | LOC101927018 | FALSE | Upregulated |
| -0,291560629 | 0,307583206 | AMFR | FALSE | Downregulated |
| -0,34287928 | 0,307604206 | DECR2 | FALSE | Downregulated |
| 0,39645483 | 0,307604206 | MRS2P2 | FALSE | Upregulated |
| -0,403520286 | 0,307647287 | NOP9 | FALSE | Downregulated |
| 0,611146842 | 0,307651855 | ALKBH4 | FALSE | Upregulated |
| -0,390792723 | 0,308037724 | CPT1A | FALSE | Downregulated |
| 0,669915431 | 0,308037724 | SYTL2 | FALSE | Upregulated |
| 0,337449994 | 0,308037724 | WDR48 | FALSE | Upregulated |
| -0,613743936 | 0,308113094 | DRG2 | FALSE | Downregulated |
| 0,907443803 | 0,308113094 | HLA-DQA1 | FALSE | Upregulated |
| -0,789091662 | 0,308113094 | IQSEC2 | FALSE | Downregulated |
| 0,617718225 | 0,308113094 | TCP11L1 | FALSE | Upregulated |
| 0,9064427 | 0,308160474 | INE1 | FALSE | Upregulated |
| 0,604495624 | 0,308160474 | LOC115110 | FALSE | Upregulated |
| -1,850415253 | 0,308160474 | TENM3 | FALSE | Downregulated |
| 0,3365765 | 0,308757579 | ARMC5 | FALSE | Upregulated |
| 0,573315852 | 0,308757579 | DUSP22 | FALSE | Upregulated |
| -0,380092537 | 0,308757579 | GAPVD1 | FALSE | Downregulated |
| 1,642388932 | 0,308757579 | HIST1H3C | FALSE | Upregulated |
| 0,365841822 | 0,308757579 | LOC100507520 | FALSE | Upregulated |
| -0,354861893 | 0,308757579 | NUDCD3 | FALSE | Downregulated |
| -0,872442576 | 0,309046278 | SULT1E1 | FALSE | Downregulated |
| -0,435823397 | 0,309098451 | ACADL | FALSE | Downregulated |
| 0,543023631 | 0,309485767 | TAC4 | FALSE | Upregulated |
| 0,632766563 | 0,309807496 | KSR1 | FALSE | Upregulated |
| -0,58474738 | 0,310004442 | EVI5 | FALSE | Downregulated |
| 0,586503645 | 0,310037682 | ATP13A2 | FALSE | Upregulated |

| | | | | |
|---|---|---|---|---|
| 0,341122356 | 0,310045376 | EVI2B | FALSE | Upregulated |
| 0,728437197 | 0,310105954 | ANK2 | FALSE | Upregulated |
| 0,986163915 | 0,310105954 | CEP128 | FALSE | Upregulated |
| 0,420284903 | 0,310105954 | VSIG10 | FALSE | Upregulated |
| 0,46128765 | 0,310218039 | B9D2 | FALSE | Upregulated |
| -1,157861768 | 0,310218039 | HCAR3 | FALSE | Downregulated |
| 1,074251688 | 0,310218039 | MKI67 | FALSE | Upregulated |
| -0,274160361 | 0,310218039 | RNF123 | FALSE | Downregulated |
| -0,609643468 | 0,310218039 | SLC19A1 | FALSE | Downregulated |
| 0,313612325 | 0,310218039 | TNKS2 | FALSE | Upregulated |
| 0,52451688 | 0,310295932 | AP5Z1 | FALSE | Upregulated |
| -0,53952453 | 0,310295932 | FGD4 | FALSE | Downregulated |
| -0,586717726 | 0,310295932 | RTFDC1 | FALSE | Downregulated |
| -0,34545694 | 0,310295932 | UROC1 | FALSE | Downregulated |
| -0,487394591 | 0,310405069 | FNIP2 | FALSE | Downregulated |
| 0,94716765 | 0,310645979 | OR1L4 | FALSE | Upregulated |
| 0,502164056 | 0,310659537 | PTCH1 | FALSE | Upregulated |
| -0,328027988 | 0,31104738 | HPD | FALSE | Downregulated |
| -0,793142115 | 0,311085768 | CYP3A43 | FALSE | Downregulated |
| 0,957948028 | 0,311085768 | PTGIR | FALSE | Upregulated |
| 0,583707965 | 0,311169706 | UBALD2 | FALSE | Upregulated |
| -0,311047798 | 0,311693765 | ARHGAP21 | FALSE | Downregulated |
| -0,329940214 | 0,311693765 | SCN8A | FALSE | Downregulated |
| -0,329867445 | 0,311912368 | AP1M1 | FALSE | Downregulated |
| 0,620347741 | 0,311912368 | LOC101928098 | FALSE | Upregulated |
| -0,285585965 | 0,311912368 | SLC16A2 | FALSE | Downregulated |
| 0,34208868 | 0,311958859 | PRDM2 | FALSE | Upregulated |
| 0,923282618 | 0,312341491 | ZNF600 | FALSE | Upregulated |
| -1,231517132 | 0,312392268 | ACTR3B | FALSE | Downregulated |
| 0,968774393 | 0,312392268 | FMOD | FALSE | Upregulated |
| -0,492834536 | 0,312749998 | GTF3C2 | FALSE | Downregulated |
| -1,789707508 | 0,312874907 | A2ML1 | FALSE | Downregulated |
| -0,659927922 | 0,312884563 | SRPK1 | FALSE | Downregulated |
| -2,078307847 | 0,313594819 | GUSBP3 | FALSE | Downregulated |
| 0,77507539 | 0,313594819 | VWA2 | FALSE | Upregulated |
| 0,859718953 | 0,314041008 | PPARG | FALSE | Upregulated |
| 0,583665708 | 0,314568015 | ARPC5L | FALSE | Upregulated |
| 0,551343959 | 0,314706834 | AHI1 | FALSE | Upregulated |
| 0,999625669 | 0,31471619 | RP1-27K12.2 | FALSE | Upregulated |
| -0,352460242 | 0,314906411 | TSHZ1 | FALSE | Downregulated |
| -0,288708387 | 0,31493781 | RAB11B | FALSE | Downregulated |
| -1,663908331 | 0,31499494 | LOC102724603 | FALSE | Downregulated |
| 0,656931072 | 0,315340394 | DDX11 | FALSE | Upregulated |
| 0,461653867 | 0,315340394 | DNASE1L1 | FALSE | Upregulated |
| -0,53984348 | 0,315340394 | RAB11B-AS1 | FALSE | Downregulated |
| -0,586997979 | 0,315340394 | TRIM55 | FALSE | Downregulated |
| -0,447294993 | 0,315356337 | ZBTB45 | FALSE | Downregulated |
| 1,718236212 | 0,315633119 | P2RY14 | FALSE | Upregulated |
| -0,523275623 | 0,316204485 | TPMT | FALSE | Downregulated |
| -0,311245216 | 0,316225469 | RPS10 | FALSE | Downregulated |
| -0,418692712 | 0,316324751 | FAM3A | FALSE | Downregulated |

| | | | | |
|---|---|---|---|---|
| -0,284993563 | 0,316547629 | SCYL1 | FALSE | Downregulated |
| 0,459629277 | 0,316675778 | DNALI1 | FALSE | Upregulated |
| -0,556710809 | 0,316697074 | RBM23 | FALSE | Downregulated |
| -0,338610671 | 0,316850048 | ADH5 | FALSE | Downregulated |
| -0,50992447 | 0,316850048 | GSTA1 | FALSE | Downregulated |
| 0,627136761 | 0,316850048 | IER5 | FALSE | Upregulated |
| 0,593116386 | 0,316915589 | TNFRSF25 | FALSE | Upregulated |
| 0,640688308 | 0,316991633 | ETV3 | FALSE | Upregulated |
| 0,60338946 | 0,3170631 | ZNF841 | FALSE | Upregulated |
| 1,005473603 | 0,317181611 | GRM6 | FALSE | Upregulated |
| 0,45180317 | 0,317181611 | SDCCAG3 | FALSE | Upregulated |
| 1,656535563 | 0,317206895 | PLEKHN1 | FALSE | Upregulated |
| -0,298734347 | 0,317258263 | EIF5A | FALSE | Downregulated |
| 1,440428044 | 0,317258263 | GRM7 | FALSE | Upregulated |
| -0,235523897 | 0,317258263 | RPS6 | FALSE | Downregulated |
| 0,484082112 | 0,317286012 | FOXH1 | FALSE | Upregulated |
| 2,077280451 | 0,317454366 | FCGR1B | FALSE | Upregulated |
| -0,313637644 | 0,317519219 | TRIM26 | FALSE | Downregulated |
| -0,340603463 | 0,317554628 | GALM | FALSE | Downregulated |
| 0,747814507 | 0,317561067 | LOC101929512 | FALSE | Upregulated |
| 1,118366031 | 0,317561067 | SPATA24 | FALSE | Upregulated |
| 1,421419812 | 0,317561067 | ZNF100 | FALSE | Upregulated |
| 0,685097018 | 0,317610705 | CLEC2B | FALSE | Upregulated |
| 0,982370993 | 0,317639026 | SULF1 | FALSE | Upregulated |
| -0,396555289 | 0,318097719 | DAAM2 | FALSE | Downregulated |
| -0,348975646 | 0,318101492 | GPC6 | FALSE | Downregulated |
| 0,729270506 | 0,318205409 | SMTNL1 | FALSE | Upregulated |
| -0,830456735 | 0,318277355 | CCDC159 | FALSE | Downregulated |
| 0,443233893 | 0,318319263 | BRD7P3 | FALSE | Upregulated |
| 2,311596387 | 0,318319263 | GTF2IP1 | FALSE | Upregulated |
| -0,340976238 | 0,318319263 | MOCS1 | FALSE | Downregulated |
| 0,653860628 | 0,318319263 | RABGGTB | FALSE | Upregulated |
| 0,9788764 | 0,318319263 | RMI1 | FALSE | Upregulated |
| 0,476782382 | 0,318336508 | ZNF394 | FALSE | Upregulated |
| 1,23191709 | 0,318366119 | EIF3J-AS1 | FALSE | Upregulated |
| 1,54859032 | 0,318607245 | CD8B | FALSE | Upregulated |
| 1,69016546 | 0,318657742 | ENPP5 | FALSE | Upregulated |
| -1,035748165 | 0,318666262 | CISD1 | FALSE | Downregulated |
| -0,277278156 | 0,318666262 | ETFB | FALSE | Downregulated |
| 1,576695741 | 0,319114988 | HSD3B1 | FALSE | Upregulated |
| -0,403297907 | 0,319114988 | PDE2A | FALSE | Downregulated |
| 0,341542658 | 0,319349799 | GNRHR2 | FALSE | Upregulated |
| 0,802874722 | 0,319956913 | SCPEP1 | FALSE | Upregulated |
| 0,626924864 | 0,319956913 | SKA2 | FALSE | Upregulated |
| 0,523735591 | 0,320030659 | PTPRH | FALSE | Upregulated |
| 0,409580307 | 0,320070714 | ITGA11 | FALSE | Upregulated |
| -1,140105829 | 0,320131569 | PTPRT | FALSE | Downregulated |
| -0,879364728 | 0,320131569 | ZNF334 | FALSE | Downregulated |
| 1,913482943 | 0,320352404 | RIMBP3B | FALSE | Upregulated |
| -0,602021866 | 0,320454985 | FASTKD2 | FALSE | Downregulated |
| 1,24938241 | 0,320511662 | ADAMTS6 | FALSE | Upregulated |

| | | | | |
|---|---|---|---|---|
| -1,190461395 | 0,32074346 | WDR92 | FALSE | Downregulated |
| 1,644845406 | 0,320762075 | TBC1D30 | FALSE | Upregulated |
| -0,442237073 | 0,320860985 | EVA1A | FALSE | Downregulated |
| 0,631795614 | 0,320919186 | SAMD9L | FALSE | Upregulated |
| 1,397264558 | 0,321049599 | CASP16 | FALSE | Upregulated |
| 0,572627345 | 0,321084228 | CAMSAP2 | FALSE | Upregulated |
| -0,621061958 | 0,321667262 | CENPT | FALSE | Downregulated |
| -0,221569593 | 0,321667262 | CIDEB | FALSE | Downregulated |
| -1,064706304 | 0,321667262 | ING3 | FALSE | Downregulated |
| 0,739125447 | 0,321667262 | LOC101929012 | FALSE | Upregulated |
| 0,420364353 | 0,321667262 | PARD3B | FALSE | Upregulated |
| 1,01861253 | 0,32173556 | CCDC141 | FALSE | Upregulated |
| -0,494320136 | 0,321844473 | AMACR | FALSE | Downregulated |
| -1,021923187 | 0,321844473 | ETV1 | FALSE | Downregulated |
| 0,800578391 | 0,321844473 | EXOSC4 | FALSE | Upregulated |
| -1,10516024 | 0,321844473 | JAKMIP2 | FALSE | Downregulated |
| 1,525386008 | 0,321844473 | LOC102724196 | FALSE | Upregulated |
| -0,484620911 | 0,321844473 | SLC23A2 | FALSE | Downregulated |
| 0,99504382 | 0,321844473 | SRFBP1 | FALSE | Upregulated |
| 0,786922747 | 0,322437062 | CMAHP | FALSE | Upregulated |
| 1,162949422 | 0,322437062 | EYA4 | FALSE | Upregulated |
| 0,791344521 | 0,322437062 | WAS | FALSE | Upregulated |
| 0,409968679 | 0,322465804 | ZSCAN21 | FALSE | Upregulated |
| 0,766607765 | 0,322511972 | CDKN1C | FALSE | Upregulated |
| 1,626033009 | 0,322511972 | KBTBD8 | FALSE | Upregulated |
| 1,270019281 | 0,322617071 | FLJ46552 | FALSE | Upregulated |
| 1,358275809 | 0,322617071 | SNORA7B | FALSE | Upregulated |
| -0,398576167 | 0,322637939 | ATP2B2 | FALSE | Downregulated |
| 0,544889755 | 0,322637939 | LOX | FALSE | Upregulated |
| 1,299714699 | 0,322654625 | FAM27E2 | FALSE | Upregulated |
| -1,437892463 | 0,322654625 | RP11-101E14.2 | FALSE | Downregulated |
| -1,594805926 | 0,322854362 | LOC102724643 | FALSE | Downregulated |
| 1,061586138 | 0,322989632 | CLEC11A | FALSE | Upregulated |
| 0,817703334 | 0,322989632 | FRMD3 | FALSE | Upregulated |
| 0,857588484 | 0,322989632 | HACE1 | FALSE | Upregulated |
| 1,780615166 | 0,322989632 | HLA-DQB2 | FALSE | Upregulated |
| 0,466712981 | 0,322989632 | LOC102724611 | FALSE | Upregulated |
| -0,469994896 | 0,322989632 | PEX13 | FALSE | Downregulated |
| 0,461291902 | 0,322989632 | PRMT1 | FALSE | Upregulated |
| -1,376422476 | 0,322989632 | RAB27B | FALSE | Downregulated |
| -0,282768238 | 0,322989632 | SRSF4 | FALSE | Downregulated |
| 0,508262473 | 0,322989632 | USP6 | FALSE | Upregulated |
| 1,512034901 | 0,323254944 | CD300LF | FALSE | Upregulated |
| -0,605651999 | 0,323551306 | REXO4 | FALSE | Downregulated |
| -0,491406997 | 0,323631444 | HNF1A-AS1 | FALSE | Downregulated |
| 0,398395358 | 0,324003053 | GAS6 | FALSE | Upregulated |
| -0,363633482 | 0,324104845 | BZRAP1 | FALSE | Downregulated |
| -0,576025347 | 0,324104845 | CPS1-IT1 | FALSE | Downregulated |
| -0,986591615 | 0,324109223 | FAM172A | FALSE | Downregulated |
| 0,503858395 | 0,324363413 | REL | FALSE | Upregulated |
| 0,558907607 | 0,324506295 | INIP | FALSE | Upregulated |

| | | | | |
|---|---|---|---|---|
| 1,324726336 | 0,324574761 | NPIPA2 | FALSE | Upregulated |
| 0,699802895 | 0,324574761 | ZNF225 | FALSE | Upregulated |
| -0,322937731 | 0,324597644 | ACBD5 | FALSE | Downregulated |
| -0,922565617 | 0,324619575 | HERC2P9 | FALSE | Downregulated |
| 0,259435232 | 0,324653816 | CIAO1 | FALSE | Upregulated |
| 0,628628522 | 0,324674542 | AKAP10 | FALSE | Upregulated |
| 0,652025027 | 0,324674542 | BRWD1-IT2 | FALSE | Upregulated |
| 0,717430588 | 0,324674542 | EDRF1-AS1 | FALSE | Upregulated |
| 0,35818544 | 0,324674542 | KIAA0754 | FALSE | Upregulated |
| 1,105484252 | 0,324674542 | LOC102723589 | FALSE | Upregulated |
| -0,384732367 | 0,324674542 | MSH3 | FALSE | Downregulated |
| -0,675167141 | 0,324674542 | NFIL3 | FALSE | Downregulated |
| -1,480996852 | 0,324674542 | PINX1 | FALSE | Downregulated |
| -0,755254889 | 0,324674542 | PLEKHA3 | FALSE | Downregulated |
| 0,524343538 | 0,324674542 | PLK2 | FALSE | Upregulated |
| -1,246761541 | 0,324674542 | POSTN | FALSE | Downregulated |
| -0,271706947 | 0,324674542 | ROCK2 | FALSE | Downregulated |
| 0,765960704 | 0,324674542 | SLC16A3 | FALSE | Upregulated |
| 0,537615857 | 0,324674542 | SLC27A4 | FALSE | Upregulated |
| 0,305739789 | 0,324674542 | TRIM13 | FALSE | Upregulated |
| 1,277050706 | 0,324674542 | WDR38 | FALSE | Upregulated |
| -1,064732128 | 0,324827031 | ANKMY2 | FALSE | Downregulated |
| -0,542126025 | 0,324827031 | ETNPPL | FALSE | Downregulated |
| 0,48627153 | 0,324827031 | GLIPR1L1 | FALSE | Upregulated |
| -0,566313576 | 0,324827031 | MYCL | FALSE | Downregulated |
| -0,506843044 | 0,324827031 | RBBP7 | FALSE | Downregulated |
| -0,424852881 | 0,324827031 | TAF6L | FALSE | Downregulated |
| -0,400599802 | 0,324886437 | LMF2 | FALSE | Downregulated |
| 0,795393017 | 0,324886437 | NOC3L | FALSE | Upregulated |
| 0,755056635 | 0,324886437 | RP11-890B15.2 | FALSE | Upregulated |
| -1,632646892 | 0,324886437 | RP5-1180C10.2 | FALSE | Downregulated |
| -0,610527215 | 0,324916985 | UQCRFS1 | FALSE | Downregulated |
| -0,424736549 | 0,325150702 | SEC24B | FALSE | Downregulated |
| -0,515111506 | 0,325156767 | TTC7B | FALSE | Downregulated |
| 0,664068643 | 0,325475112 | DCTN5 | FALSE | Upregulated |
| -0,338224541 | 0,325475112 | SLC4A4 | FALSE | Downregulated |
| 0,760670501 | 0,325697499 | RNF8 | FALSE | Upregulated |
| 1,07514608 | 0,325716204 | DBR1 | FALSE | Upregulated |
| 0,703325251 | 0,325716204 | RAB8B | FALSE | Upregulated |
| 1,659183628 | 0,325738129 | AC018647.3 | FALSE | Upregulated |
| -0,623675179 | 0,325767712 | PIGO | FALSE | Downregulated |
| 0,971933606 | 0,325767712 | SLC2A1 | FALSE | Upregulated |
| 0,971042409 | 0,325802967 | RP11-16E12.2 | FALSE | Upregulated |
| -2,331051753 | 0,326090168 | AP3S2 | FALSE | Downregulated |
| -0,556471413 | 0,326717258 | AGAP1 | FALSE | Downregulated |
| -0,60880923 | 0,326717258 | IL11RA | FALSE | Downregulated |
| 1,341475555 | 0,326945802 | DDX11L5 | FALSE | Upregulated |
| 0,816670911 | 0,326945802 | NR1D1 | FALSE | Upregulated |
| -0,296167551 | 0,326945802 | R3HDM2 | FALSE | Downregulated |
| 1,510685715 | 0,32703244 | FLJ46026 | FALSE | Upregulated |
| 0,432617104 | 0,327064655 | DLG5 | FALSE | Upregulated |

| | | | | |
|---|---|---|---|---|
| 0,98051218 | 0,327064655 | LOC101928284 | FALSE | Upregulated |
| -0,909666343 | 0,327064655 | PIGY | FALSE | Downregulated |
| 1,446789384 | 0,327232469 | ATP6V1E2 | FALSE | Upregulated |
| 0,688708498 | 0,327232469 | UBTD2 | FALSE | Upregulated |
| 0,577814639 | 0,327260176 | PLXNA3 | FALSE | Upregulated |
| -0,468383564 | 0,327665664 | EPS15L1 | FALSE | Downregulated |
| -0,33904882 | 0,327688576 | YWHAE | FALSE | Downregulated |
| 0,601809913 | 0,328020565 | CDK18 | FALSE | Upregulated |
| 1,54760108 | 0,328116266 | DKFZp667F0711 | FALSE | Upregulated |
| 0,52379939 | 0,328116266 | TNK1 | FALSE | Upregulated |
| 0,452030363 | 0,328452235 | MYL12A | FALSE | Upregulated |
| -0,526794936 | 0,328555777 | MAP3K5 | FALSE | Downregulated |
| -0,865109551 | 0,328670001 | CXorf40A | FALSE | Downregulated |
| -1,648057281 | 0,328699734 | LOC100132813 | FALSE | Downregulated |
| -0,501023702 | 0,329184873 | NPAT | FALSE | Downregulated |
| 0,506984257 | 0,329184873 | SERPINH1 | FALSE | Upregulated |
| 0,718067493 | 0,329312615 | ADAMTS1 | FALSE | Upregulated |
| -0,348885338 | 0,329312615 | NUGGC | FALSE | Downregulated |
| -0,429977016 | 0,329312615 | PROSC | FALSE | Downregulated |
| -0,289799176 | 0,329312615 | WWP1 | FALSE | Downregulated |
| -0,275715857 | 0,329312615 | ZBED1 | FALSE | Downregulated |
| 0,521600552 | 0,329619679 | TOMM34 | FALSE | Upregulated |
| -0,651951332 | 0,329799897 | AMZ2 | FALSE | Downregulated |
| 0,751850811 | 0,329799897 | ZNF10 | FALSE | Upregulated |
| 0,299275689 | 0,330276396 | YAP1 | FALSE | Upregulated |
| 0,563217255 | 0,330276396 | ZSWIM4 | FALSE | Upregulated |
| 0,367925943 | 0,330347902 | PTPN21 | FALSE | Upregulated |
| -1,262024441 | 0,330806273 | DSCAML1 | FALSE | Downregulated |
| -0,427002916 | 0,330806273 | ECM1 | FALSE | Downregulated |
| 1,450983109 | 0,330806273 | SEMA3C | FALSE | Upregulated |
| -1,536232981 | 0,330919468 | EGFLAM-AS4 | FALSE | Downregulated |
| 0,37412699 | 0,330935599 | ABHD16B | FALSE | Upregulated |
| -0,549009795 | 0,330935599 | LEAP2 | FALSE | Downregulated |
| 0,915175225 | 0,330935599 | NCF4 | FALSE | Upregulated |
| -0,447730914 | 0,330935599 | PHC3 | FALSE | Downregulated |
| -0,367612247 | 0,330935599 | PIK3R4 | FALSE | Downregulated |
| 0,849192467 | 0,330992494 | CDH13 | FALSE | Upregulated |
| 0,499989708 | 0,330992494 | TAF5 | FALSE | Upregulated |
| -1,395634221 | 0,331117026 | LRP2 | FALSE | Downregulated |
| -0,497897076 | 0,331138555 | ATPAF1 | FALSE | Downregulated |
| -0,299085406 | 0,331153045 | HRG | FALSE | Downregulated |
| -0,67918451 | 0,331202854 | LAMTOR2 | FALSE | Downregulated |
| -0,251470812 | 0,331406742 | ARFGEF2 | FALSE | Downregulated |
| 0,574660842 | 0,33155282 | LIPE | FALSE | Upregulated |
| 0,452330199 | 0,331584902 | LOC101928070 | FALSE | Upregulated |
| 1,155590062 | 0,331690402 | HS3ST1 | FALSE | Upregulated |
| 0,647273872 | 0,331690402 | IFIT1B | FALSE | Upregulated |
| 2,166068255 | 0,331690402 | ITIH6 | FALSE | Upregulated |
| 0,774441888 | 0,331690402 | LINC00102 | FALSE | Upregulated |
| -0,707172212 | 0,331690402 | MTPAP | FALSE | Downregulated |
| 0,290223397 | 0,331690402 | PARP14 | FALSE | Upregulated |

| | | | | |
|---|---|---|---|---|
| 0,671302202 | 0,331690402 | TM2D2 | FALSE | Upregulated |
| -0,454978618 | 0,331702081 | LYNX1 | FALSE | Downregulated |
| 0,834372748 | 0,331886707 | FLJ42258 | FALSE | Upregulated |
| -1,47794583 | 0,331886707 | IL1RAPL2 | FALSE | Downregulated |
| -1,600096481 | 0,331886707 | LOC100129473 | FALSE | Downregulated |
| -1,343554025 | 0,331886707 | RSG1 | FALSE | Downregulated |
| -1,559241059 | 0,331886707 | SPTSSB | FALSE | Downregulated |
| -0,404553634 | 0,332333546 | D2HGDH | FALSE | Downregulated |
| 0,430698832 | 0,332370638 | HIVEP2 | FALSE | Upregulated |
| 0,488441779 | 0,332814316 | ANK3 | FALSE | Upregulated |
| 0,902428278 | 0,332814316 | CCDC65 | FALSE | Upregulated |
| 0,768498402 | 0,332863768 | LOC101929479 | FALSE | Upregulated |
| -0,873190755 | 0,333138137 | P2RY2 | FALSE | Downregulated |
| 0,685977185 | 0,333225367 | LOC102724490 | FALSE | Upregulated |
| 0,46290946 | 0,333854529 | IP6K2 | FALSE | Upregulated |
| 0,423230021 | 0,334461657 | GAB2 | FALSE | Upregulated |
| 0,350554148 | 0,334713847 | OTUB2 | FALSE | Upregulated |
| 0,523627795 | 0,334741884 | MED22 | FALSE | Upregulated |
| 0,9836845 | 0,334853646 | AC022173.2 | FALSE | Upregulated |
| 0,725864287 | 0,334870291 | TMEM60 | FALSE | Upregulated |
| -0,844660492 | 0,335432232 | ZNF18 | FALSE | Downregulated |
| -0,727240309 | 0,335619932 | PRSS53 | FALSE | Downregulated |
| -0,720042252 | 0,335684453 | NKAIN2 | FALSE | Downregulated |
| -0,20228368 | 0,335684453 | SRCAP | FALSE | Downregulated |
| 0,353239998 | 0,335738172 | CAB39 | FALSE | Upregulated |
| 0,329907604 | 0,335738172 | HEXDC | FALSE | Upregulated |
| -0,668425551 | 0,335738172 | SLC25A28 | FALSE | Downregulated |
| -0,376599822 | 0,335738172 | SNX18 | FALSE | Downregulated |
| 1,83124768 | 0,33619871 | SLFN12 | FALSE | Upregulated |
| -1,696321175 | 0,336400447 | CNGB3 | FALSE | Downregulated |
| 0,632443485 | 0,336400447 | LOC100507071 | FALSE | Upregulated |
| -0,404187188 | 0,336400447 | TM9SF2 | FALSE | Downregulated |
| 0,42847646 | 0,336400447 | WARS | FALSE | Upregulated |
| 0,41214639 | 0,336423531 | AC009501.4 | FALSE | Upregulated |
| -0,503162366 | 0,336885775 | FAM99A | FALSE | Downregulated |
| -0,551932083 | 0,337682711 | BLOC1S1 | FALSE | Downregulated |
| -0,454102391 | 0,337682711 | GCSH | FALSE | Downregulated |
| 0,96430648 | 0,337682711 | GLI3 | FALSE | Upregulated |
| -0,461318238 | 0,337682711 | IRF6 | FALSE | Downregulated |
| -0,380836863 | 0,337682711 | TMPO | FALSE | Downregulated |
| -0,585474254 | 0,337736612 | CSNK2B | FALSE | Downregulated |
| 0,472445231 | 0,337736612 | PLCB2 | FALSE | Upregulated |
| -0,358594838 | 0,337736612 | SNRPD3 | FALSE | Downregulated |
| 0,560785152 | 0,337927542 | C20orf27 | FALSE | Upregulated |
| 0,578994856 | 0,337927542 | VPS35 | FALSE | Upregulated |
| 1,909746955 | 0,337927625 | AP001258.4 | FALSE | Upregulated |
| -0,92025846 | 0,337927625 | FAM96B | FALSE | Downregulated |
| 0,492605136 | 0,337927625 | LOC101927831 | FALSE | Upregulated |
| -0,290602565 | 0,337927625 | TMEM66 | FALSE | Downregulated |
| 0,46562123 | 0,337993691 | EML6 | FALSE | Upregulated |
| 1,60749936 | 0,338576773 | LOC101927082 | FALSE | Upregulated |

| | | | | |
|---|---|---|---|---|
| 1,384219449 | 0,338774457 | AC097662.2 | FALSE | Upregulated |
| 0,370392918 | 0,338774457 | NSUN5 | FALSE | Upregulated |
| 0,259916557 | 0,338774457 | VPS13A | FALSE | Upregulated |
| 0,32629738 | 0,338860095 | HIP1R | FALSE | Upregulated |
| -0,269066692 | 0,338887454 | NADK2 | FALSE | Downregulated |
| 1,742616656 | 0,339343267 | AC011738.4 | FALSE | Upregulated |
| 1,817766844 | 0,33966683 | C4orf6 | FALSE | Upregulated |
| -0,647960438 | 0,33966683 | ZNF574 | FALSE | Downregulated |
| -0,612430572 | 0,339887936 | NUPL1 | FALSE | Downregulated |
| 0,730497514 | 0,339887936 | TMEM126A | FALSE | Upregulated |
| 0,558938306 | 0,340501124 | LINC00963 | FALSE | Upregulated |
| 0,435158871 | 0,340569662 | PHLDB1 | FALSE | Upregulated |
| -0,68049814 | 0,340815604 | TAF3 | FALSE | Downregulated |
| 0,456095939 | 0,3408441 | UBA6 | FALSE | Upregulated |
| -0,461861123 | 0,340880847 | BRD8 | FALSE | Downregulated |
| 0,579135007 | 0,340880847 | TUBA8 | FALSE | Upregulated |
| 1,318460023 | 0,340889635 | LOC101927583 | FALSE | Upregulated |
| 0,494023814 | 0,340889635 | LOC101929046 | FALSE | Upregulated |
| 0,465627897 | 0,340975523 | ACLY | FALSE | Upregulated |
| 0,26258612 | 0,340975523 | ANKH | FALSE | Upregulated |
| -1,489561162 | 0,340975523 | CAMK2N2 | FALSE | Downregulated |
| 0,481235277 | 0,340975523 | FERMT3 | FALSE | Upregulated |
| -1,03948405 | 0,340975523 | KLHDC8B | FALSE | Downregulated |
| -1,868548515 | 0,340975523 | LINC01133 | FALSE | Downregulated |
| -0,363653786 | 0,340975523 | POGK | FALSE | Downregulated |
| -1,714117183 | 0,341089413 | C4orf33 | FALSE | Downregulated |
| -0,695568135 | 0,341095029 | NAA16 | FALSE | Downregulated |
| 0,827600874 | 0,34152986 | NEK8 | FALSE | Upregulated |
| -0,36127554 | 0,341584486 | RNF185 | FALSE | Downregulated |
| -0,910806229 | 0,341607623 | DCTN3 | FALSE | Downregulated |
| -0,600576023 | 0,341690383 | SRD5A2 | FALSE | Downregulated |
| 0,578261147 | 0,341877031 | ZNF540 | FALSE | Upregulated |
| -0,909471338 | 0,341961695 | ARHGAP20 | FALSE | Downregulated |
| 1,360717427 | 0,341997734 | SRD5A3-AS1 | FALSE | Upregulated |
| 0,587803266 | 0,342493315 | CEACAM16 | FALSE | Upregulated |
| 0,477566609 | 0,342664571 | ASAP3 | FALSE | Upregulated |
| 0,301873389 | 0,343011657 | C19orf44 | FALSE | Upregulated |
| -0,541244687 | 0,343068845 | PHLDB3 | FALSE | Downregulated |
| 0,538492395 | 0,34320266 | CCDC28B | FALSE | Upregulated |
| -0,271700643 | 0,343292777 | IRF2BPL | FALSE | Downregulated |
| 0,312250271 | 0,343292777 | PRR26 | FALSE | Upregulated |
| 0,757971985 | 0,343371044 | SLC35A5 | FALSE | Upregulated |
| 0,660474671 | 0,343451425 | LOC100287808 | FALSE | Upregulated |
| 1,013356767 | 0,343466808 | GPR56 | FALSE | Upregulated |
| -0,658240226 | 0,34348116 | PNP | FALSE | Downregulated |
| 0,562251596 | 0,343592359 | DDR2 | FALSE | Upregulated |
| 1,676295733 | 0,343592359 | TLR7 | FALSE | Upregulated |
| 0,470302737 | 0,343708256 | COL5A2 | FALSE | Upregulated |
| 0,34502553 | 0,343708256 | MYO9A | FALSE | Upregulated |
| 1,124563632 | 0,343751478 | CLSTN2 | FALSE | Upregulated |
| 0,998254846 | 0,343935679 | PIGC | FALSE | Upregulated |

| | | | | |
|---|---|---|---|---|
| 1,328164395 | 0,343975476 | KCNMB2 | FALSE | Upregulated |
| 0,818385824 | 0,343975476 | MED12L | FALSE | Upregulated |
| 1,451272685 | 0,344013811 | RP11-883G14.2 | FALSE | Upregulated |
| 1,682915219 | 0,344102539 | TACSTD2 | FALSE | Upregulated |
| 1,00925423 | 0,34422262 | ZEB2-AS1 | FALSE | Upregulated |
| -0,984150963 | 0,344354765 | PTRHD1 | FALSE | Downregulated |
| -0,843747715 | 0,344600397 | LGSN | FALSE | Downregulated |
| -0,320678302 | 0,344600397 | MTUS1 | FALSE | Downregulated |
| 0,567044721 | 0,344699704 | COL14A1 | FALSE | Upregulated |
| -0,422251176 | 0,344883043 | CAMK1 | FALSE | Downregulated |
| 0,30689132 | 0,344883043 | KIF5B | FALSE | Upregulated |
| -0,453245284 | 0,345265381 | ANXA7 | FALSE | Downregulated |
| -0,799606992 | 0,345265381 | CYP1A2 | FALSE | Downregulated |
| 1,897200132 | 0,345265381 | HIST1H2AI | FALSE | Upregulated |
| -0,295613827 | 0,345265381 | RMND5A | FALSE | Downregulated |
| 1,968852918 | 0,345301201 | LOC102724754 | FALSE | Upregulated |
| 0,254548174 | 0,345301201 | POM121 | FALSE | Upregulated |
| -0,680327997 | 0,345841482 | KIAA1804 | FALSE | Downregulated |
| 0,704123597 | 0,345841482 | KIRREL3 | FALSE | Upregulated |
| 0,391408978 | 0,345930568 | AGPAT6 | FALSE | Upregulated |
| 0,281967962 | 0,346407316 | DAZAP2 | FALSE | Upregulated |
| -0,324997861 | 0,346581661 | YWHAQ | FALSE | Downregulated |
| 0,500560329 | 0,346605822 | LMO4 | FALSE | Upregulated |
| -1,205151907 | 0,346605822 | TSPAN1 | FALSE | Downregulated |
| 0,297304496 | 0,346690286 | MTOR | FALSE | Upregulated |
| 0,73896001 | 0,346915969 | NR2C2AP | FALSE | Upregulated |
| -0,308449709 | 0,34700622 | CMPK1 | FALSE | Downregulated |
| 1,332457771 | 0,347103558 | GJC2 | FALSE | Upregulated |
| 0,552543935 | 0,347362284 | PCED1B-AS1 | FALSE | Upregulated |
| -0,380689586 | 0,347474807 | PLEKHG3 | FALSE | Downregulated |
| -1,693585764 | 0,347526562 | LOC100652824 | FALSE | Downregulated |
| -0,329775287 | 0,347526562 | PPFIBP2 | FALSE | Downregulated |
| 0,949647522 | 0,347608149 | ATP10B | FALSE | Upregulated |
| -0,512069733 | 0,347608149 | CEP70 | FALSE | Downregulated |
| -1,535040811 | 0,347608149 | PDZRN4 | FALSE | Downregulated |
| -1,250567028 | 0,347608149 | RP11-261N11.8 | FALSE | Downregulated |
| 1,571200082 | 0,347608149 | RP11-789C17.3 | FALSE | Upregulated |
| 0,607558684 | 0,347608149 | THOC1 | FALSE | Upregulated |
| -0,994758802 | 0,348046481 | SP5 | FALSE | Downregulated |
| -1,422228498 | 0,348046481 | TSNAXIP1 | FALSE | Downregulated |
| 0,465048761 | 0,348132974 | PRPH2 | FALSE | Upregulated |
| 0,35789925 | 0,348168672 | LOC101929291 | FALSE | Upregulated |
| 0,608153082 | 0,348168672 | PWWP2A | FALSE | Upregulated |
| 1,82698858 | 0,34819485 | CD3G | FALSE | Upregulated |
| 0,653771712 | 0,348584661 | LHFP | FALSE | Upregulated |
| 0,581527623 | 0,348584661 | SRC | FALSE | Upregulated |
| 0,422074293 | 0,348731311 | RNF32 | FALSE | Upregulated |
| -0,505046953 | 0,348779452 | EID1 | FALSE | Downregulated |
| -0,687819393 | 0,348830835 | SEC31B | FALSE | Downregulated |
| -0,626379284 | 0,349609015 | PLAGL2 | FALSE | Downregulated |
| 0,473540046 | 0,349609015 | RRS1 | FALSE | Upregulated |

| | | | | |
|---|---|---|---|---|
| 0,847745708 | 0,349609015 | RTN4R | FALSE | Upregulated |
| -0,790475713 | 0,349609015 | TRPT1 | FALSE | Downregulated |
| 0,511809331 | 0,349710184 | LOC101927681 | FALSE | Upregulated |
| 0,625106879 | 0,350299591 | CHST11 | FALSE | Upregulated |
| 0,370445631 | 0,350371172 | BAIAP3 | FALSE | Upregulated |
| 0,642042942 | 0,350371172 | CTHRC1 | FALSE | Upregulated |
| -0,42188715 | 0,350371172 | PCTP | FALSE | Downregulated |
| 0,356752247 | 0,350371172 | WDR7 | FALSE | Upregulated |
| -0,543101268 | 0,350457916 | LNX2 | FALSE | Downregulated |
| 1,069577541 | 0,350636809 | WNT11 | FALSE | Upregulated |
| 0,739375222 | 0,350831333 | LOC101928595 | FALSE | Upregulated |
| -0,313908465 | 0,350831333 | PSMD3 | FALSE | Downregulated |
| 0,612819967 | 0,350831333 | SGK1 | FALSE | Upregulated |
| -0,419964818 | 0,350831333 | TKT | FALSE | Downregulated |
| 0,263321142 | 0,351050658 | ZNF687 | FALSE | Upregulated |
| 0,992732694 | 0,351260195 | CDCA5 | FALSE | Upregulated |
| -1,367804068 | 0,351260195 | FRG1 | FALSE | Downregulated |
| 0,524673123 | 0,351260195 | LOC101928657 | FALSE | Upregulated |
| 1,071481712 | 0,351260195 | MC1R | FALSE | Upregulated |
| 0,435724112 | 0,351260195 | SERPINB6 | FALSE | Upregulated |
| 0,477198244 | 0,351282429 | LNPEP | FALSE | Upregulated |
| -0,606967743 | 0,351562631 | RANBP3L | FALSE | Downregulated |
| 0,873918362 | 0,351722108 | LOC101928510 | FALSE | Upregulated |
| -0,448006884 | 0,351741297 | PCOLCE | FALSE | Downregulated |
| 0,874056406 | 0,351821618 | FEN1 | FALSE | Upregulated |
| -1,266026328 | 0,351821618 | NDUFAF4 | FALSE | Downregulated |
| -0,200466073 | 0,351821618 | RPL4 | FALSE | Downregulated |
| -0,424674805 | 0,352016625 | DNAJC4 | FALSE | Downregulated |
| 0,824581589 | 0,352016625 | PAPOLG | FALSE | Upregulated |
| -0,542278217 | 0,352016625 | SIVA1 | FALSE | Downregulated |
| 0,990668654 | 0,35238522 | CILP | FALSE | Upregulated |
| -0,347793534 | 0,35238522 | EI24 | FALSE | Downregulated |
| -0,890335171 | 0,35238522 | HNF4A-AS1 | FALSE | Downregulated |
| 0,528144382 | 0,35265511 | NOL3 | FALSE | Upregulated |
| -0,738995956 | 0,35265511 | TEX10 | FALSE | Downregulated |
| -0,301494106 | 0,352698382 | PLOD1 | FALSE | Downregulated |
| -0,276098265 | 0,352729405 | CPSF1 | FALSE | Downregulated |
| -0,562020453 | 0,35275168 | B3GNT2 | FALSE | Downregulated |
| -1,56656756 | 0,35275168 | NHLRC1 | FALSE | Downregulated |
| -0,339784051 | 0,35275168 | STT3B | FALSE | Downregulated |
| -0,448851401 | 0,352962761 | C10orf54 | FALSE | Downregulated |
| 0,43169009 | 0,352962761 | IQGAP1 | FALSE | Upregulated |
| -0,439057124 | 0,352962761 | N4BP2L1 | FALSE | Downregulated |
| -0,291516724 | 0,352962761 | PTPRB | FALSE | Downregulated |
| 1,641501123 | 0,352962761 | TESPA1 | FALSE | Upregulated |
| 0,716448238 | 0,353073017 | FLYWCH2 | FALSE | Upregulated |
| 1,510038499 | 0,353181947 | RNU12 | FALSE | Upregulated |
| 0,426264194 | 0,35381165 | KRT18 | FALSE | Upregulated |
| 0,813790813 | 0,353970199 | EXOC3L1 | FALSE | Upregulated |
| -0,898667957 | 0,354048917 | LOC102723694 | FALSE | Downregulated |
| -0,836048473 | 0,354048917 | TFAP4 | FALSE | Downregulated |

| | | | | |
|---|---|---|---|---|
| 0,703848488 | 0,354141647 | POLR3C | FALSE | Upregulated |
| -0,492993799 | 0,354215732 | GFOD2 | FALSE | Downregulated |
| 1,207853541 | 0,354215732 | RP11-219G17.4 | FALSE | Upregulated |
| 0,509526474 | 0,354215732 | YWHAH | FALSE | Upregulated |
| 0,613601516 | 0,354451475 | LOC101927727 | FALSE | Upregulated |
| 0,789525224 | 0,354451475 | TM4SF1 | FALSE | Upregulated |
| 1,114034697 | 0,354480552 | LOC101927355 | FALSE | Upregulated |
| -0,574619279 | 0,354549312 | NLK | FALSE | Downregulated |
| 1,134080525 | 0,354770192 | C9orf169 | FALSE | Upregulated |
| -0,510089233 | 0,354770192 | KYNU | FALSE | Downregulated |
| -0,435419657 | 0,354770192 | MED24 | FALSE | Downregulated |
| 0,431034873 | 0,354839042 | VPS41 | FALSE | Upregulated |
| 0,230844033 | 0,355919904 | AHSA2 | FALSE | Upregulated |
| -0,842248444 | 0,355919904 | COMMD7 | FALSE | Downregulated |
| 0,467638428 | 0,355919904 | PITHD1 | FALSE | Upregulated |
| -0,374599381 | 0,355984269 | MAPRE2 | FALSE | Downregulated |
| 0,357947597 | 0,35628041 | FAM211A | FALSE | Upregulated |
| 1,764308473 | 0,356289839 | IQGAP3 | FALSE | Upregulated |
| 0,465392169 | 0,356300612 | SERHL | FALSE | Upregulated |
| 0,28847067 | 0,356318555 | KCTD20 | FALSE | Upregulated |
| 0,473234479 | 0,356642006 | DCUN1D3 | FALSE | Upregulated |
| 1,309432081 | 0,35669941 | ABCA17P | FALSE | Upregulated |
| -0,425606994 | 0,35669941 | ETS2 | FALSE | Downregulated |
| 0,714329724 | 0,356824661 | GOLGA6L5P | FALSE | Upregulated |
| 0,405877789 | 0,356824661 | MTERFD3 | FALSE | Upregulated |
| -0,47639484 | 0,357045116 | CENPV | FALSE | Downregulated |
| -1,397239097 | 0,357045116 | LOC101928706 | FALSE | Downregulated |
| -1,751230732 | 0,357045116 | MICALCL | FALSE | Downregulated |
| 2,873255101 | 0,357045116 | TRIM73 | FALSE | Upregulated |
| 1,72590505 | 0,357221147 | CA3 | FALSE | Upregulated |
| -0,814969342 | 0,357388259 | LCLAT1 | FALSE | Downregulated |
| 0,687839798 | 0,357644109 | CENPBD1 | FALSE | Upregulated |
| 0,637398475 | 0,357644109 | GPRC5D | FALSE | Upregulated |
| -1,242327891 | 0,357644109 | OR13C8 | FALSE | Downregulated |
| 1,596708574 | 0,357644109 | SLC5A1 | FALSE | Upregulated |
| 0,934131057 | 0,357652415 | RASD1 | FALSE | Upregulated |
| -0,259731614 | 0,357739143 | MARCH6 | FALSE | Downregulated |
| 1,366265499 | 0,357739143 | OSCAR | FALSE | Upregulated |
| 0,398955878 | 0,357739143 | SETMAR | FALSE | Upregulated |
| -0,245450422 | 0,357739143 | ZNF592 | FALSE | Downregulated |
| 0,482170995 | 0,357756648 | RHOG | FALSE | Upregulated |
| -0,293751893 | 0,357844677 | ARAF | FALSE | Downregulated |
| 1,305125528 | 0,357844677 | GPR173 | FALSE | Upregulated |
| -0,350376679 | 0,357844677 | LRRC3DN | FALSE | Downregulated |
| 0,563325424 | 0,357844677 | SUCO | FALSE | Upregulated |
| 0,437269191 | 0,358050943 | LOC101929387 | FALSE | Upregulated |
| 0,74531048 | 0,358159984 | ZNF575 | FALSE | Upregulated |
| 0,98765049 | 0,358191069 | FAM26F | FALSE | Upregulated |
| -0,292809057 | 0,358476077 | LDLRAD4 | FALSE | Downregulated |
| 0,706740364 | 0,358583152 | ABCA3 | FALSE | Upregulated |
| 0,466376107 | 0,358583152 | IFNGR2 | FALSE | Upregulated |

| | | | | |
|---|---|---|---|---|
| 0,543457287 | 0,358583152 | LOC101928830 | FALSE | Upregulated |
| 0,613333382 | 0,358879542 | NIF3L1 | FALSE | Upregulated |
| -0,777203092 | 0,358879542 | SCN3A | FALSE | Downregulated |
| 0,321378038 | 0,359048957 | WDR24 | FALSE | Upregulated |
| -0,793364399 | 0,35912671 | LOC101929195 | FALSE | Downregulated |
| -0,968954096 | 0,359226421 | ZNF134 | FALSE | Downregulated |
| -0,364438506 | 0,359329939 | LRRC45 | FALSE | Downregulated |
| -0,366833163 | 0,360074766 | ARID2 | FALSE | Downregulated |
| 1,070144861 | 0,360074766 | LOC100130705 | FALSE | Upregulated |
| -0,317614208 | 0,360162458 | PEPD | FALSE | Downregulated |
| -1,405436222 | 0,360162458 | SLC5A4 | FALSE | Downregulated |
| 0,361402091 | 0,360171364 | LOC102723666 | FALSE | Upregulated |
| 0,442201116 | 0,360319948 | LOC100996741 | FALSE | Upregulated |
| -0,654020072 | 0,360644336 | LOC730101 | FALSE | Downregulated |
| -0,607047261 | 0,360940494 | AC007099.1 | FALSE | Downregulated |
| 1,103427086 | 0,361032769 | CACNA2D1 | FALSE | Upregulated |
| 0,537066954 | 0,361032769 | CITED4 | FALSE | Upregulated |
| -0,565104372 | 0,361032769 | CUX2 | FALSE | Downregulated |
| 0,798064179 | 0,361032769 | LY9 | FALSE | Upregulated |
| 0,54903342 | 0,361032769 | PODNL1 | FALSE | Upregulated |
| 0,957583086 | 0,361032769 | SYNPO2L | FALSE | Upregulated |
| 1,298853046 | 0,361247336 | RASGEF1A | FALSE | Upregulated |
| -0,445285697 | 0,361643773 | FAM73B | FALSE | Downregulated |
| 0,361911699 | 0,361866883 | TMED1 | FALSE | Upregulated |
| -0,413072434 | 0,361917229 | ATF7 | FALSE | Downregulated |
| 0,705212857 | 0,361917229 | BRMS1 | FALSE | Upregulated |
| 0,740239891 | 0,361917229 | FICD | FALSE | Upregulated |
| -0,338832528 | 0,361917229 | RHOU | FALSE | Downregulated |
| -0,298171338 | 0,361917229 | TARDBP | FALSE | Downregulated |
| -1,212789113 | 0,361940754 | GP5 | FALSE | Downregulated |
| -0,685601641 | 0,362075951 | C8orf4 | FALSE | Downregulated |
| -0,372716135 | 0,362202898 | C11orf58 | FALSE | Downregulated |
| 0,319247497 | 0,362657077 | FSIP2 | FALSE | Upregulated |
| 0,66259185 | 0,363142011 | MTMR9LP | FALSE | Upregulated |
| 0,711604982 | 0,363197133 | RSPH9 | FALSE | Upregulated |
| 0,541389004 | 0,363464177 | C6orf136 | FALSE | Upregulated |
| 0,650830299 | 0,363464177 | CASP4 | FALSE | Upregulated |
| 1,28235566 | 0,363464177 | FRMD6-AS1 | FALSE | Upregulated |
| -0,256052924 | 0,363464177 | ITPR2 | FALSE | Downregulated |
| -0,364502565 | 0,363464177 | NUDT16 | FALSE | Downregulated |
| -0,346269866 | 0,363464177 | ZNF444 | FALSE | Downregulated |
| 1,116657512 | 0,363498579 | SULT1C4 | FALSE | Upregulated |
| 1,301429247 | 0,363507925 | BDNF-AS | FALSE | Upregulated |
| 0,575467172 | 0,363507925 | PRICKLE2-AS3 | FALSE | Upregulated |
| 0,316611242 | 0,36381469 | CERS6-AS1 | FALSE | Upregulated |
| 0,308703452 | 0,363818593 | RHOC | FALSE | Upregulated |
| 1,327522397 | 0,363818593 | TCTN2 | FALSE | Upregulated |
| 0,316214733 | 0,363818593 | TIMP3 | FALSE | Upregulated |
| -0,289995006 | 0,363883126 | OGDH | FALSE | Downregulated |
| 0,350393712 | 0,363994105 | FAM104A | FALSE | Upregulated |
| -0,731009436 | 0,364046428 | RABL5 | FALSE | Downregulated |

| | | | | |
|---|---|---|---|---|
| 0,442932461 | 0,364047819 | APPL1 | FALSE | Upregulated |
| -0,525703664 | 0,364047819 | PMF1 | FALSE | Downregulated |
| 0,866529377 | 0,364149136 | KLHL6 | FALSE | Upregulated |
| 0,538876484 | 0,364149136 | ZNF37BP | FALSE | Upregulated |
| -0,273149872 | 0,364228781 | ABHD2 | FALSE | Downregulated |
| 0,384752481 | 0,364228781 | NUTM2D | FALSE | Upregulated |
| -0,323792889 | 0,364228781 | SIK3 | FALSE | Downregulated |
| -0,393251498 | 0,364303448 | ATP7B | FALSE | Downregulated |
| 0,363543182 | 0,364333483 | ESYT3 | FALSE | Upregulated |
| 0,604337718 | 0,364375068 | ME3 | FALSE | Upregulated |
| -0,530480966 | 0,364480901 | GEMIN5 | FALSE | Downregulated |
| 0,227885636 | 0,36459903 | LOC100190986 | FALSE | Upregulated |
| -0,636176805 | 0,364622489 | AZGP1P1 | FALSE | Downregulated |
| -0,499280476 | 0,364622489 | DNAJC19 | FALSE | Downregulated |
| 0,430890294 | 0,364622489 | LOC100271836 | FALSE | Upregulated |
| -0,183897598 | 0,364622489 | SCAF1 | FALSE | Downregulated |
| -0,277969488 | 0,365095652 | ARHGEF10L | FALSE | Downregulated |
| -0,667821852 | 0,365095652 | ESAM | FALSE | Downregulated |
| -0,284960663 | 0,365095652 | RPL26 | FALSE | Downregulated |
| -1,813142666 | 0,365095652 | TBX19 | FALSE | Downregulated |
| -1,984638952 | 0,365138475 | STRC | FALSE | Downregulated |
| -1,038674536 | 0,365138475 | SWI5 | FALSE | Downregulated |
| 0,313388281 | 0,365164723 | NRIP2 | FALSE | Upregulated |
| -0,267113192 | 0,365164723 | SHANK2 | FALSE | Downregulated |
| 1,522019131 | 0,365325164 | SCN1A | FALSE | Upregulated |
| -0,23781279 | 0,365329957 | RPL13A | FALSE | Downregulated |
| -0,534188347 | 0,365493443 | GAREM | FALSE | Downregulated |
| 0,438085925 | 0,365542138 | LOC102723806 | FALSE | Upregulated |
| -0,217459492 | 0,365542138 | PRKCSH | FALSE | Downregulated |
| -0,371212595 | 0,365579715 | ATP5G3 | FALSE | Downregulated |
| -1,799548105 | 0,365579715 | HYKK | FALSE | Downregulated |
| 1,097209013 | 0,365649192 | HRH2 | FALSE | Upregulated |
| -0,362865107 | 0,365716028 | APEX1 | FALSE | Downregulated |
| -0,455060292 | 0,365716028 | MST1L | FALSE | Downregulated |
| -1,101644224 | 0,365729927 | IL27 | FALSE | Downregulated |
| -0,539556727 | 0,365760664 | GPAM | FALSE | Downregulated |
| 1,345304891 | 0,365813856 | CENPF | FALSE | Upregulated |
| -0,619825914 | 0,365826172 | PRKAR1B | FALSE | Downregulated |
| -1,058044442 | 0,365910711 | COMMD1 | FALSE | Downregulated |
| 0,601092665 | 0,365910711 | LIPT1 | FALSE | Upregulated |
| -0,938453847 | 0,365966252 | USP43 | FALSE | Downregulated |
| 0,700333072 | 0,366099498 | PRKG1-AS1 | FALSE | Upregulated |
| 0,766384513 | 0,366099498 | RAB9A | FALSE | Upregulated |
| -0,436120864 | 0,36621596 | GATAD1 | FALSE | Downregulated |
| 0,547388886 | 0,36621596 | PPARD | FALSE | Upregulated |
| -0,303173403 | 0,36621596 | RPS27 | FALSE | Downregulated |
| 1,532776123 | 0,366588761 | NEURL3 | FALSE | Upregulated |
| 0,855637982 | 0,366702256 | IL23A | FALSE | Upregulated |
| 0,404659394 | 0,366702256 | LRRC39 | FALSE | Upregulated |
| -1,017501629 | 0,366813653 | DSC1 | FALSE | Downregulated |
| 1,45816409 | 0,366813653 | KRT19 | FALSE | Upregulated |

| | | | | |
|---|---|---|---|---|
| -0,483517876 | 0,366813653 | LIG4 | FALSE | Downregulated |
| -0,600445038 | 0,366813653 | SRM | FALSE | Downregulated |
| 0,493261587 | 0,366821965 | ARHGAP26 | FALSE | Upregulated |
| 0,247664103 | 0,366821965 | KIAA0430 | FALSE | Upregulated |
| 0,533118617 | 0,366821965 | LSM8 | FALSE | Upregulated |
| -0,709925272 | 0,366997654 | YOD1 | FALSE | Downregulated |
| 0,650295136 | 0,367027085 | LMO3 | FALSE | Upregulated |
| 0,281492736 | 0,367027085 | LOC102724445 | FALSE | Upregulated |
| 0,489936198 | 0,367027085 | LOH12CR1 | FALSE | Upregulated |
| -0,265531056 | 0,367027085 | MGAT2 | FALSE | Downregulated |
| -0,341507144 | 0,367091658 | LGR4 | FALSE | Downregulated |
| 0,401313152 | 0,367097203 | DLG2 | FALSE | Upregulated |
| -0,597188397 | 0,367097203 | KLHL15 | FALSE | Downregulated |
| 1,818351122 | 0,367097203 | LOC102723678 | FALSE | Upregulated |
| 0,2896012 | 0,367097203 | THAP3 | FALSE | Upregulated |
| -0,918530137 | 0,367097203 | ZNF180 | FALSE | Downregulated |
| 0,514985474 | 0,367097203 | ZNF510 | FALSE | Upregulated |
| 1,668120106 | 0,367314708 | DUSP2 | FALSE | Upregulated |
| 1,285930035 | 0,367344676 | STON1 | FALSE | Upregulated |
| 0,310532131 | 0,36735496 | SLC31A2 | FALSE | Upregulated |
| 0,38414867 | 0,367370486 | PSMD6-AS2 | FALSE | Upregulated |
| 1,26431638 | 0,36743116 | CSF2RA | FALSE | Upregulated |
| -0,213576085 | 0,36743116 | IQSEC1 | FALSE | Downregulated |
| -0,834341181 | 0,367596787 | MINOS1P1 | FALSE | Downregulated |
| -0,262491305 | 0,367688337 | ADCK3 | FALSE | Downregulated |
| 1,012058724 | 0,36772433 | LOC101926895 | FALSE | Upregulated |
| -0,444506795 | 0,367782839 | TAPT1 | FALSE | Downregulated |
| 0,460625324 | 0,367782839 | TREX1 | FALSE | Upregulated |
| 0,479128929 | 0,367887285 | EMILIN2 | FALSE | Upregulated |
| 0,421440115 | 0,367887285 | KDM6A | FALSE | Upregulated |
| -0,739016851 | 0,367887285 | RBP2 | FALSE | Downregulated |
| -0,46868617 | 0,367973598 | HDHD2 | FALSE | Downregulated |
| 1,577672752 | 0,368147579 | C5orf28 | FALSE | Upregulated |
| -0,191064535 | 0,368147579 | KIAA1671 | FALSE | Downregulated |
| -1,025567013 | 0,368148733 | DNM1 | FALSE | Downregulated |
| 0,484578191 | 0,368148733 | RGMB | FALSE | Upregulated |
| 0,614139463 | 0,368605884 | SLC26A2 | FALSE | Upregulated |
| 0,772480991 | 0,368750354 | AKR1B1 | FALSE | Upregulated |
| -0,896347243 | 0,369367198 | AMN | FALSE | Downregulated |
| -0,730787601 | 0,36957445 | USE1 | FALSE | Downregulated |
| -0,460049472 | 0,369606265 | FGD5 | FALSE | Downregulated |
| -0,357329545 | 0,369606265 | FMO3 | FALSE | Downregulated |
| -0,340443642 | 0,369606265 | SEC24D | FALSE | Downregulated |
| 0,667812398 | 0,369615796 | LOC102724077 | FALSE | Upregulated |
| -0,298562427 | 0,369702508 | SHC2 | FALSE | Downregulated |
| -0,307912502 | 0,369712895 | SHPK | FALSE | Downregulated |
| 0,506841235 | 0,369874448 | SNX11 | FALSE | Upregulated |
| 1,741070241 | 0,370520924 | NCAM1-AS1 | FALSE | Upregulated |
| 1,200406534 | 0,3708492 | KLRD1 | FALSE | Upregulated |
| 0,463774026 | 0,370872165 | DOCK3 | FALSE | Upregulated |
| 0,522289711 | 0,370872165 | EXOC1 | FALSE | Upregulated |

| | | | | |
|---|---|---|---|---|
| 1,270347314 | 0,370872165 | HCG25 | FALSE | Upregulated |
| 0,710207252 | 0,370872165 | NFKBIE | FALSE | Upregulated |
| 1,002292347 | 0,370872165 | OLFM1 | FALSE | Upregulated |
| -0,470866162 | 0,370901798 | PAFAH2 | FALSE | Downregulated |
| 0,251637908 | 0,371042586 | TMEM127 | FALSE | Upregulated |
| 1,23618757 | 0,371208247 | LOC100130987 | FALSE | Upregulated |
| 0,601213225 | 0,371267786 | CYTH2 | FALSE | Upregulated |
| -0,414711844 | 0,371524578 | GLDC | FALSE | Downregulated |
| 0,321462006 | 0,371591094 | MCM3AP-AS1 | FALSE | Upregulated |
| 1,205375128 | 0,371671699 | C1QTNF3 | FALSE | Upregulated |
| 0,222843664 | 0,372064767 | SET | FALSE | Upregulated |
| -0,73345227 | 0,372064767 | THYN1 | FALSE | Downregulated |
| 1,109563932 | 0,372139713 | HCAR2 | FALSE | Upregulated |
| 1,265670723 | 0,372271187 | UST | FALSE | Upregulated |
| -0,285154996 | 0,372436271 | ARHGAP29 | FALSE | Downregulated |
| -0,379532229 | 0,372436271 | DNAJB12 | FALSE | Downregulated |
| 1,185602822 | 0,372436271 | FAM86HP | FALSE | Upregulated |
| 1,122403677 | 0,372436271 | MPP7 | FALSE | Upregulated |
| -0,39182574 | 0,372436271 | POMT1 | FALSE | Downregulated |
| -0,564195252 | 0,372564392 | SLC5A6 | FALSE | Downregulated |
| 0,391342222 | 0,372703969 | C16orf62 | FALSE | Upregulated |
| 0,597021427 | 0,372880198 | CHMP4A | FALSE | Upregulated |
| 0,962793433 | 0,373159708 | CNIH2 | FALSE | Upregulated |
| 0,352326607 | 0,373159708 | EVI2A | FALSE | Upregulated |
| 0,507741779 | 0,373208191 | DAPK3 | FALSE | Upregulated |
| 0,744508061 | 0,373208191 | ZNF571 | FALSE | Upregulated |
| -0,577120107 | 0,373254895 | CCDC25 | FALSE | Downregulated |
| 0,742031031 | 0,3734911 | PDSS1 | FALSE | Upregulated |
| 0,703397638 | 0,373591851 | ADAP1 | FALSE | Upregulated |
| 1,242110626 | 0,373591851 | LOC283887 | FALSE | Upregulated |
| 0,557171859 | 0,373921416 | RNF166 | FALSE | Upregulated |
| -0,220094258 | 0,374040237 | GLG1 | FALSE | Downregulated |
| -0,868226986 | 0,374263431 | AKR7A2P1 | FALSE | Downregulated |
| 1,642151468 | 0,374263431 | GDNF | FALSE | Upregulated |
| -1,268626664 | 0,374263431 | LOC101060632 | FALSE | Downregulated |
| -0,324288389 | 0,374263431 | RAB17 | FALSE | Downregulated |
| -0,449057742 | 0,374263431 | SCAPER | FALSE | Downregulated |
| 1,610029897 | 0,374263431 | UBXN10-AS1 | FALSE | Upregulated |
| 0,730019573 | 0,374286723 | GTF2IRD1 | FALSE | Upregulated |
| 0,932837624 | 0,374286723 | LZTS1 | FALSE | Upregulated |
| -1,240466333 | 0,374309568 | TIMD4 | FALSE | Downregulated |
| 1,007327557 | 0,374313894 | LOC392196 | FALSE | Upregulated |
| -0,322646814 | 0,374313894 | SRRM1 | FALSE | Downregulated |
| 0,444420238 | 0,374367704 | LOC101927057 | FALSE | Upregulated |
| 1,026978231 | 0,374397064 | PMP22 | FALSE | Upregulated |
| -1,788150345 | 0,374415578 | APOL5 | FALSE | Downregulated |
| -0,410646076 | 0,374550334 | GOSR1 | FALSE | Downregulated |
| 0,759800796 | 0,374550334 | LOC101927382 | FALSE | Upregulated |
| -0,83524634 | 0,374664685 | BRI3BP | FALSE | Downregulated |
| -0,668821701 | 0,374664685 | F8A1 | FALSE | Downregulated |
| -1,299182965 | 0,374664685 | LLPH | FALSE | Downregulated |

| | | | | |
|---|---|---|---|---|
| -0,261342146 | 0,37473412 | PC | FALSE | Downregulated |
| 0,43791983 | 0,374962676 | LOC101929747 | FALSE | Upregulated |
| 0,685964366 | 0,375000615 | PDP2 | FALSE | Upregulated |
| 0,493108021 | 0,375023753 | NPY6R | FALSE | Upregulated |
| 0,60679273 | 0,375131478 | CHAC2 | FALSE | Upregulated |
| 0,5305592 | 0,375193836 | TNFRSF21 | FALSE | Upregulated |
| -0,317609187 | 0,375250016 | DDAH1 | FALSE | Downregulated |
| 0,235894432 | 0,375250016 | MCL1 | FALSE | Upregulated |
| 0,535151109 | 0,375250016 | RP3-325F22.3 | FALSE | Upregulated |
| 0,392471102 | 0,375402913 | AP000688.14 | FALSE | Upregulated |
| -0,39784326 | 0,375402913 | TEX261 | FALSE | Downregulated |
| 1,272033359 | 0,375499201 | FAM86JP | FALSE | Upregulated |
| -0,863013723 | 0,375653816 | LOC100288102 | FALSE | Downregulated |
| -0,421251515 | 0,375662722 | RITA1 | FALSE | Downregulated |
| -1,659085239 | 0,375785728 | CDC20 | FALSE | Downregulated |
| 0,551631858 | 0,375785728 | DPH7 | FALSE | Upregulated |
| -0,316805691 | 0,375785728 | MBNL3 | FALSE | Downregulated |
| 0,636798973 | 0,375888215 | YBX3 | FALSE | Upregulated |
| -0,411992332 | 0,375898072 | PRPSAP1 | FALSE | Downregulated |
| -0,358358796 | 0,376081942 | GPT | FALSE | Downregulated |
| -0,369748756 | 0,376081942 | HRSP12 | FALSE | Downregulated |
| -0,320569416 | 0,376081942 | ITIH3 | FALSE | Downregulated |
| -1,027421365 | 0,376081942 | LOC101928158 | FALSE | Downregulated |
| 0,543639771 | 0,376081942 | LOC642648 | FALSE | Upregulated |
| 0,470606184 | 0,376081942 | WDR52 | FALSE | Upregulated |
| -0,780493524 | 0,376142072 | CBWD5 | FALSE | Downregulated |
| 1,549443742 | 0,376306067 | LINC00865 | FALSE | Upregulated |
| -1,646815906 | 0,376306067 | LYAR | FALSE | Downregulated |
| 0,892470872 | 0,376366436 | SRI | FALSE | Upregulated |
| -0,536856897 | 0,376480497 | MGC12916 | FALSE | Downregulated |
| -0,370020982 | 0,376535025 | TFPI | FALSE | Downregulated |
| -0,692330835 | 0,376757903 | DGKE | FALSE | Downregulated |
| 0,434485162 | 0,376757903 | FBXL22 | FALSE | Upregulated |
| -0,306723214 | 0,376757903 | RAPH1 | FALSE | Downregulated |
| 0,714355816 | 0,376757903 | TES | FALSE | Upregulated |
| -0,766898602 | 0,376877199 | GPR133 | FALSE | Downregulated |
| 1,191918402 | 0,376959352 | HIST1H2AG | FALSE | Upregulated |
| -1,817709847 | 0,376959352 | LOC101928886 | FALSE | Downregulated |
| -0,377465442 | 0,376959352 | PDDC1 | FALSE | Downregulated |
| 1,677177192 | 0,376959352 | RFX4 | FALSE | Upregulated |
| 0,48886121 | 0,377030543 | C10orf12 | FALSE | Upregulated |
| 0,319636209 | 0,377219498 | FOXJ3 | FALSE | Upregulated |
| 0,287280295 | 0,377329587 | GPR124 | FALSE | Upregulated |
| -0,948317256 | 0,377329587 | RINT1 | FALSE | Downregulated |
| -1,981874035 | 0,377541633 | DCHS2 | FALSE | Downregulated |
| 0,563422975 | 0,377558221 | SPIN3 | FALSE | Upregulated |
| 0,441766688 | 0,37757101 | CCAT1 | FALSE | Upregulated |
| 0,450937612 | 0,37757101 | PCYT1A | FALSE | Upregulated |
| 0,701309934 | 0,377620023 | LOC101928794 | FALSE | Upregulated |
| -0,591754349 | 0,378257392 | ADAT1 | FALSE | Downregulated |
| 1,169108817 | 0,378257392 | C10orf88 | FALSE | Upregulated |

| | | | | |
|---|---|---|---|---|
| 1,634496598 | 0,378257392 | LOC101928553 | FALSE | Upregulated |
| -0,330785727 | 0,378257392 | NAGLU | FALSE | Downregulated |
| -1,363054064 | 0,378387507 | GPR143 | FALSE | Downregulated |
| 1,144072768 | 0,378528623 | TRPM5 | FALSE | Upregulated |
| 0,477740812 | 0,378547047 | SLC43A2 | FALSE | Upregulated |
| -0,523682566 | 0,378720195 | RAPGEF4 | FALSE | Downregulated |
| -0,416528413 | 0,378820527 | NUP88 | FALSE | Downregulated |
| 1,604112282 | 0,378891797 | GALNT9 | FALSE | Upregulated |
| 0,323151865 | 0,378891797 | PAN3-AS1 | FALSE | Upregulated |
| 0,520077253 | 0,378891797 | SHKBP1 | FALSE | Upregulated |
| -1,137046685 | 0,378891797 | TGDS | FALSE | Downregulated |
| -0,69756791 | 0,378891797 | ZFAND2A | FALSE | Downregulated |
| 0,671821425 | 0,378899686 | VSTM4 | FALSE | Upregulated |
| -0,88021917 | 0,378934341 | MTUS2 | FALSE | Downregulated |
| 0,360150847 | 0,379002473 | EHBP1L1 | FALSE | Upregulated |
| -1,711884134 | 0,379014392 | LOC101929057 | FALSE | Downregulated |
| 0,724538513 | 0,379712918 | PPIL1 | FALSE | Upregulated |
| 0,510745468 | 0,38019264 | ANKRD32 | FALSE | Upregulated |
| 0,41164064 | 0,380468688 | MAK | FALSE | Upregulated |
| 1,354684614 | 0,380543393 | LOC101928993 | FALSE | Upregulated |
| 0,412815775 | 0,380560886 | FAM21A | FALSE | Upregulated |
| -0,391887578 | 0,380560886 | TK2 | FALSE | Downregulated |
| -0,821784655 | 0,380745 | CORO2A | FALSE | Downregulated |
| -0,20445877 | 0,381044723 | MASP2 | FALSE | Downregulated |
| -0,85452865 | 0,381302495 | THOC6 | FALSE | Downregulated |
| -0,756249054 | 0,381546918 | COL26A1 | FALSE | Downregulated |
| 0,282318509 | 0,381546918 | TRIM41 | FALSE | Upregulated |
| -0,309895703 | 0,381698063 | FTL | FALSE | Downregulated |
| 1,394811217 | 0,381758266 | GLRX2 | FALSE | Upregulated |
| 1,387271849 | 0,381768798 | CCDC30 | FALSE | Upregulated |
| -0,701796302 | 0,381768798 | FGF11 | FALSE | Downregulated |
| 0,323894527 | 0,381768798 | MED15 | FALSE | Upregulated |
| -0,266697083 | 0,381768798 | WDR81 | FALSE | Downregulated |
| 0,49890413 | 0,381842265 | CEACAM19 | FALSE | Upregulated |
| -0,520361095 | 0,381842265 | MARVELD3 | FALSE | Downregulated |
| 0,60336134 | 0,381886485 | TSPO2 | FALSE | Upregulated |
| 0,458634958 | 0,381894219 | ANKRD36C | FALSE | Upregulated |
| -0,51236578 | 0,381894219 | ARRDC3 | FALSE | Downregulated |
| 0,735985865 | 0,382021435 | LECT1 | FALSE | Upregulated |
| 0,355071494 | 0,382021435 | MXRA7 | FALSE | Upregulated |
| -0,30216142 | 0,382344676 | MTCH1 | FALSE | Downregulated |
| 0,993099673 | 0,382480969 | C1orf74 | FALSE | Upregulated |
| -0,389130398 | 0,382660929 | INTS10 | FALSE | Downregulated |
| 0,544175177 | 0,382709499 | IGFBP2 | FALSE | Upregulated |
| 1,374858215 | 0,382737998 | NGF | FALSE | Upregulated |
| -0,643486829 | 0,382874876 | C1orf123 | FALSE | Downregulated |
| -1,536165327 | 0,382874876 | DRD1 | FALSE | Downregulated |
| 0,985433844 | 0,382874876 | ZDHHC1 | FALSE | Upregulated |
| -0,583457886 | 0,3829453 | DRG1 | FALSE | Downregulated |
| -0,266766989 | 0,3829453 | SERF2 | FALSE | Downregulated |
| -0,430479885 | 0,3829453 | TANGO6 | FALSE | Downregulated |

| | | | | |
|---|---|---|---|---|
| 1,712692235 | 0,383054225 | PLA2G4E | FALSE | Upregulated |
| 0,834291899 | 0,383101947 | C11orf84 | FALSE | Upregulated |
| 0,356560656 | 0,383101947 | LOC101929545 | FALSE | Upregulated |
| 0,350571279 | 0,383101947 | SNX22 | FALSE | Upregulated |
| 1,333736931 | 0,383155859 | CD300E | FALSE | Upregulated |
| -0,307720378 | 0,383387354 | SAP30L | FALSE | Downregulated |
| -0,303820776 | 0,383636795 | ASL | FALSE | Downregulated |
| -0,502794782 | 0,384005953 | NTSR1 | FALSE | Downregulated |
| 0,605574088 | 0,384005953 | SLCO3A1 | FALSE | Upregulated |
| -0,301056086 | 0,384005953 | UBA52 | FALSE | Downregulated |
| 1,295160983 | 0,384208283 | LPAR2 | FALSE | Upregulated |
| -1,097244485 | 0,384208283 | SULT1B1 | FALSE | Downregulated |
| 0,575962592 | 0,384544616 | CSGALNACT2 | FALSE | Upregulated |
| -0,578602785 | 0,384587427 | THRSP | FALSE | Downregulated |
| 0,829769416 | 0,384839191 | PI4KAP1 | FALSE | Upregulated |
| -0,291178965 | 0,384860507 | ABCA1 | FALSE | Downregulated |
| -0,540469908 | 0,384860507 | SUGP1 | FALSE | Downregulated |
| -2,073723081 | 0,385065245 | FLJ33581 | FALSE | Downregulated |
| 0,27003987 | 0,385153835 | ABHD17A | FALSE | Upregulated |
| 0,965721284 | 0,385153835 | HILS1 | FALSE | Upregulated |
| -1,251695553 | 0,385153835 | METTL21C | FALSE | Downregulated |
| 0,366813015 | 0,385153835 | RALGAPB | FALSE | Upregulated |
| 0,504173626 | 0,385153835 | RP5-977B1.7 | FALSE | Upregulated |
| 1,842083832 | 0,385153835 | WDR11-AS1 | FALSE | Upregulated |
| -0,70422851 | 0,385155565 | POP7 | FALSE | Downregulated |
| -0,361025692 | 0,385163521 | ZNF407 | FALSE | Downregulated |
| 0,53682188 | 0,385282025 | CALCRL | FALSE | Upregulated |
| 0,519790095 | 0,385403301 | FBXL14 | FALSE | Upregulated |
| 1,067537992 | 0,385403301 | GNAZ | FALSE | Upregulated |
| 1,100645441 | 0,385403301 | LARP6 | FALSE | Upregulated |
| 1,62354093 | 0,385403301 | LOC100288175 | FALSE | Upregulated |
| -0,904005769 | 0,385403301 | LRRC16B | FALSE | Downregulated |
| -0,23079714 | 0,385403301 | PRPF8 | FALSE | Downregulated |
| -0,428475807 | 0,385403301 | SEC24A | FALSE | Downregulated |
| -0,250883757 | 0,385403301 | TENC1 | FALSE | Downregulated |
| 0,690646725 | 0,385473147 | ALOX5 | FALSE | Upregulated |
| 0,749754154 | 0,385473147 | CCL22 | FALSE | Upregulated |
| -0,52525365 | 0,385688298 | TMEM69 | FALSE | Downregulated |
| 1,224553551 | 0,385728039 | ATP2C2 | FALSE | Upregulated |
| 0,237463216 | 0,385776814 | DDX6 | FALSE | Upregulated |
| -0,244225985 | 0,38607427 | TMEM248 | FALSE | Downregulated |
| -0,914474913 | 0,38608075 | C1orf174 | FALSE | Downregulated |
| 1,298955801 | 0,38608075 | C20orf173 | FALSE | Upregulated |
| 0,403529718 | 0,38608075 | C2CD3 | FALSE | Upregulated |
| 0,631274727 | 0,38608075 | C9orf139 | FALSE | Upregulated |
| -0,492149536 | 0,38608075 | ERCC6 | FALSE | Downregulated |
| -0,362032197 | 0,38608075 | GBE1 | FALSE | Downregulated |
| 0,782808156 | 0,38608075 | GINS1 | FALSE | Upregulated |
| 1,435707012 | 0,38608075 | HS3ST5 | FALSE | Upregulated |
| 1,389861867 | 0,38608075 | LOC102724515 | FALSE | Upregulated |
| -0,866350773 | 0,38608075 | MEX3A | FALSE | Downregulated |

| | | | | |
|---|---|---|---|---|
| 1,437901352 | 0,38608075 | ODF3L1 | FALSE | Upregulated |
| -0,409525575 | 0,38608075 | SEPT10 | FALSE | Downregulated |
| -0,50099666 | 0,38608075 | SPAG7 | FALSE | Downregulated |
| 0,714152976 | 0,386329693 | ARHGEF16 | FALSE | Upregulated |
| 0,720496379 | 0,386329693 | C17orf78 | FALSE | Upregulated |
| 0,275917016 | 0,386329693 | CACFD1 | FALSE | Upregulated |
| 0,446710388 | 0,386329693 | PAXBP1-AS1 | FALSE | Upregulated |
| -0,531899495 | 0,386539862 | BMS1P6 | FALSE | Downregulated |
| 0,940816473 | 0,386700401 | KCNJ15 | FALSE | Upregulated |
| 0,546879888 | 0,386700401 | USP39 | FALSE | Upregulated |
| 0,598748703 | 0,386700401 | ZNF69 | FALSE | Upregulated |
| 0,722534568 | 0,386842597 | LMNB1 | FALSE | Upregulated |
| 1,015536067 | 0,386868941 | HSPA6 | FALSE | Upregulated |
| 0,633189067 | 0,386930855 | LOC100129203 | FALSE | Upregulated |
| -1,291977727 | 0,387260358 | LOC257396 | FALSE | Downregulated |
| 0,422818596 | 0,387260358 | PGLS | FALSE | Upregulated |
| -0,304442551 | 0,387260358 | TST | FALSE | Downregulated |
| -0,544905109 | 0,387265401 | C9orf64 | FALSE | Downregulated |
| 0,788273819 | 0,387619023 | SLC45A4 | FALSE | Upregulated |
| 0,510019477 | 0,387619023 | TRIM35 | FALSE | Upregulated |
| -0,763253393 | 0,387672908 | B3GAT1 | FALSE | Downregulated |
| 0,43982277 | 0,388015657 | ADAM1A | FALSE | Upregulated |
| 0,78433988 | 0,388092566 | LOC101926980 | FALSE | Upregulated |
| 0,456239799 | 0,388191974 | ANXA2P1 | FALSE | Upregulated |
| 0,3325338 | 0,388191974 | ATP6V0A1 | FALSE | Upregulated |
| -0,365402646 | 0,388191974 | EHHADH-AS1 | FALSE | Downregulated |
| -0,370333183 | 0,388191974 | LOC101929648 | FALSE | Downregulated |
| -1,343206231 | 0,388191974 | PET117 | FALSE | Downregulated |
| 1,028633482 | 0,388191974 | PTCHD4 | FALSE | Upregulated |
| 0,556930235 | 0,388200846 | IRF8 | FALSE | Upregulated |
| 1,028519681 | 0,388481233 | SPATA6 | FALSE | Upregulated |
| -0,344717808 | 0,388517832 | FLOT1 | FALSE | Downregulated |
| -0,305151987 | 0,388517832 | SYNJ2BP | FALSE | Downregulated |
| -0,56597688 | 0,388622604 | C19orf53 | FALSE | Downregulated |
| 1,256516796 | 0,388622604 | C9orf171 | FALSE | Upregulated |
| 0,366716145 | 0,388695044 | METTL12 | FALSE | Upregulated |
| -1,489322923 | 0,388965104 | CDK5 | FALSE | Downregulated |
| 0,65197599 | 0,388965104 | ZNF182 | FALSE | Upregulated |
| -0,420076441 | 0,389057533 | FAM120B | FALSE | Downregulated |
| 0,582554973 | 0,389078584 | CLYBL-AS2 | FALSE | Upregulated |
| -0,565197658 | 0,38920151 | NR0B2 | FALSE | Downregulated |
| -2,22107408 | 0,389263562 | UGT2A2 | FALSE | Downregulated |
| 1,551939105 | 0,389566485 | CD300C | FALSE | Upregulated |
| 1,400699044 | 0,389566485 | FLJ22184 | FALSE | Upregulated |
| 1,070722799 | 0,389566485 | KCNK12 | FALSE | Upregulated |
| 1,017067634 | 0,389631913 | EIF5A2 | FALSE | Upregulated |
| 0,318724781 | 0,389631913 | ZBTB11-AS1 | FALSE | Upregulated |
| -0,286837839 | 0,389638426 | OIT3 | FALSE | Downregulated |
| -0,285409815 | 0,38981725 | ANGPTL3 | FALSE | Downregulated |
| 0,344948687 | 0,389909369 | RUFY2 | FALSE | Upregulated |
| 0,60661562 | 0,39010114 | EDNRB-AS1 | FALSE | Upregulated |

| | | | | |
|---|---|---|---|---|
| -0,525544005 | 0,390161442 | TSKU | FALSE | Downregulated |
| 0,731189221 | 0,390168044 | CCDC78 | FALSE | Upregulated |
| 0,328698951 | 0,390168044 | CEP68 | FALSE | Upregulated |
| 0,898987448 | 0,390168044 | FAM86FP | FALSE | Upregulated |
| 0,321809742 | 0,390168044 | IRF3 | FALSE | Upregulated |
| 0,495102263 | 0,390168044 | XRCC1 | FALSE | Upregulated |
| 0,175834974 | 0,390168044 | ZNF609 | FALSE | Upregulated |
| 0,399732589 | 0,390690874 | TADA1 | FALSE | Upregulated |
| 1,075134762 | 0,390932873 | BARD1 | FALSE | Upregulated |
| 0,724357171 | 0,391019301 | LEPROTL1 | FALSE | Upregulated |
| 0,354713968 | 0,391172922 | LOC102723929 | FALSE | Upregulated |
| -1,05149848 | 0,391307276 | CA13 | FALSE | Downregulated |
| -0,530406215 | 0,391307276 | FAM162A | FALSE | Downregulated |
| 0,597521704 | 0,391307276 | LOC728613 | FALSE | Upregulated |
| 0,26149851 | 0,391364751 | SEPT2 | FALSE | Upregulated |
| -0,991626161 | 0,391718367 | USP51 | FALSE | Downregulated |
| 0,466411185 | 0,39176768 | INTS8 | FALSE | Upregulated |
| 0,647110166 | 0,39176768 | NRADDP | FALSE | Upregulated |
| -0,54588192 | 0,39176768 | STAM2 | FALSE | Downregulated |
| -1,408770337 | 0,391983542 | EMX2OS | FALSE | Downregulated |
| 0,683091096 | 0,392006637 | DOK7 | FALSE | Upregulated |
| -1,155990402 | 0,392053164 | C4orf32 | FALSE | Downregulated |
| -1,684075638 | 0,392069503 | AC074286.1 | FALSE | Downregulated |
| -0,6652167 | 0,392069503 | CENPBD1P1 | FALSE | Downregulated |
| -1,007523554 | 0,392278716 | LHX9 | FALSE | Downregulated |
| 0,509565846 | 0,392377012 | DYNLL1-AS1 | FALSE | Upregulated |
| -0,957187008 | 0,392406899 | MAGEH1 | FALSE | Downregulated |
| -0,344937825 | 0,392586794 | IBTK | FALSE | Downregulated |
| 0,749612386 | 0,392586794 | LOC102724133 | FALSE | Upregulated |
| -0,371569598 | 0,392670194 | EIF3E | FALSE | Downregulated |
| 0,704244566 | 0,39290956 | C1QTNF5 | FALSE | Upregulated |
| -0,501128861 | 0,39290956 | MRPS27 | FALSE | Downregulated |
| 0,561273265 | 0,39307141 | IFI35 | FALSE | Upregulated |
| 0,364352217 | 0,39307141 | MSTO2P | FALSE | Upregulated |
| 0,340471326 | 0,39307141 | PI4K2A | FALSE | Upregulated |
| 0,496950066 | 0,39307141 | RSPH3 | FALSE | Upregulated |
| 0,626764813 | 0,393076151 | C21orf91 | FALSE | Upregulated |
| 1,329274904 | 0,393076151 | LOC101929805 | FALSE | Upregulated |
| -0,286869208 | 0,393076151 | SLC25A42 | FALSE | Downregulated |
| -1,375172133 | 0,393279516 | GRHL3 | FALSE | Downregulated |
| 0,762852935 | 0,393351843 | EPB41L4A-AS1 | FALSE | Upregulated |
| -0,707981894 | 0,39347677 | ABHD10 | FALSE | Downregulated |
| 0,32500835 | 0,393592175 | TMEM2 | FALSE | Upregulated |
| 1,545790524 | 0,393621207 | CLIC5 | FALSE | Upregulated |
| -1,192219213 | 0,393621207 | NBPF3 | FALSE | Downregulated |
| -0,413173729 | 0,393636989 | FES | FALSE | Downregulated |
| 0,332101246 | 0,393636989 | IRAK1 | FALSE | Upregulated |
| 0,29050298 | 0,393636989 | KDM1A | FALSE | Upregulated |
| 0,307525765 | 0,393733004 | ATE1 | FALSE | Upregulated |
| -0,415388356 | 0,393801203 | CYP20A1 | FALSE | Downregulated |
| -0,312917446 | 0,393801203 | RPL22 | FALSE | Downregulated |

| | | | | |
|---|---|---|---|---|
| 0,242230132 | 0,393991916 | KPNB1 | FALSE | Upregulated |
| -0,377308551 | 0,393991916 | SETBP1 | FALSE | Downregulated |
| 1,263924141 | 0,394054608 | PLAUR | FALSE | Upregulated |
| 0,349744485 | 0,394056196 | ZNF671 | FALSE | Upregulated |
| 0,760924423 | 0,394326167 | FLRT2 | FALSE | Upregulated |
| -0,330033772 | 0,394332654 | MTHFR | FALSE | Downregulated |
| 0,80227353 | 0,394332654 | PDP1 | FALSE | Upregulated |
| 0,203997795 | 0,394332654 | PDPK1 | FALSE | Upregulated |
| 0,726379512 | 0,39438098 | GSG1 | FALSE | Upregulated |
| 0,363985607 | 0,394603741 | BRAT1 | FALSE | Upregulated |
| 0,460791213 | 0,39485765 | DOCK9 | FALSE | Upregulated |
| 1,242016872 | 0,39485765 | LOC101930452 | FALSE | Upregulated |
| 0,376437748 | 0,39485765 | LOC146880 | FALSE | Upregulated |
| -0,306190388 | 0,39485765 | PANK3 | FALSE | Downregulated |
| 0,79460307 | 0,394993536 | METTL10 | FALSE | Upregulated |
| 0,26431589 | 0,395069899 | TPPP | FALSE | Upregulated |
| -1,680397071 | 0,395189714 | RP9 | FALSE | Downregulated |
| 1,324744824 | 0,39533289 | FBN2 | FALSE | Upregulated |
| 1,003085532 | 0,395439732 | PPP1R14A | FALSE | Upregulated |
| -0,417777715 | 0,395439732 | TAF10 | FALSE | Downregulated |
| 0,358500422 | 0,395479335 | HMGN1 | FALSE | Upregulated |
| 0,774978719 | 0,395479335 | TMEM218 | FALSE | Upregulated |
| -0,692196576 | 0,395479335 | ZFPL1 | FALSE | Downregulated |
| 0,422666235 | 0,395509477 | SSBP4 | FALSE | Upregulated |
| 0,41673656 | 0,395510406 | LOC102724383 | FALSE | Upregulated |
| 0,962585679 | 0,395510406 | RP11-64D24.4 | FALSE | Upregulated |
| -1,394966225 | 0,395510406 | TMPRSS11E | FALSE | Downregulated |
| -0,587405179 | 0,395510406 | ZXDA | FALSE | Downregulated |
| -0,30614511 | 0,395644653 | XRCC5 | FALSE | Downregulated |
| 0,307340114 | 0,396186632 | TMPRSS9 | FALSE | Upregulated |
| -0,773324064 | 0,396291567 | METTL6 | FALSE | Downregulated |
| 0,297348448 | 0,396371346 | AKAP8 | FALSE | Upregulated |
| 0,331944744 | 0,396663026 | SPG21 | FALSE | Upregulated |
| 0,629100625 | 0,396838677 | PRMT6 | FALSE | Upregulated |
| -0,791983583 | 0,396838677 | SAP25 | FALSE | Downregulated |
| -0,217554416 | 0,396838677 | SETD7 | FALSE | Downregulated |
| -2,144462472 | 0,397375124 | CDH15 | FALSE | Downregulated |
| -1,570197679 | 0,397375124 | LOC102723383 | FALSE | Downregulated |
| -0,498707305 | 0,397375124 | MRPL23-AS1 | FALSE | Downregulated |
| -1,456544087 | 0,397375124 | TSPAN8 | FALSE | Downregulated |
| -1,537831842 | 0,397588324 | LSMEM1 | FALSE | Downregulated |
| 0,576463518 | 0,397588324 | RP11-508N12.2 | FALSE | Upregulated |
| 1,352374373 | 0,397607586 | H1FNT | FALSE | Upregulated |
| 0,66780735 | 0,397607586 | NREP | FALSE | Upregulated |
| 0,851267051 | 0,397633576 | TSPAN32 | FALSE | Upregulated |
| 0,35828184 | 0,397716105 | CDKL1 | FALSE | Upregulated |
| 0,638591168 | 0,397777545 | LRRC4B | FALSE | Upregulated |
| 0,203528502 | 0,397926193 | ATXN1 | FALSE | Upregulated |
| 0,342105022 | 0,397981134 | NCAPG | FALSE | Upregulated |
| -0,977620816 | 0,397981134 | NOC4L | FALSE | Downregulated |
| -0,312648387 | 0,398041164 | AKAP12 | FALSE | Downregulated |

| | | | | |
|---|---|---|---|---|
| 0,522212831 | 0,398041164 | BCDIN3D | FALSE | Upregulated |
| 1,02145145 | 0,398041164 | C12orf5 | FALSE | Upregulated |
| 0,329630999 | 0,398041164 | CCPG1 | FALSE | Upregulated |
| -0,568623255 | 0,398041164 | SDHAF1 | FALSE | Downregulated |
| 1,55870478 | 0,39806485 | LOC101928221 | FALSE | Upregulated |
| 0,571251714 | 0,398121753 | ELN | FALSE | Upregulated |
| -0,322764334 | 0,398571619 | RPL21 | FALSE | Downregulated |
| -0,747847293 | 0,39914546 | TMTC1 | FALSE | Downregulated |
| 0,645592259 | 0,399200478 | C16orf86 | FALSE | Upregulated |
| 0,551014542 | 0,399200478 | DENR | FALSE | Upregulated |
| -0,472885893 | 0,399200478 | DERA | FALSE | Downregulated |
| -0,317007578 | 0,399200478 | FDX1 | FALSE | Downregulated |
| -0,60304497 | 0,399200478 | HIST1H4E | FALSE | Downregulated |
| 0,684268282 | 0,399200478 | KNTC1 | FALSE | Upregulated |
| 1,410812553 | 0,399200478 | LDB3 | FALSE | Upregulated |
| 0,298502294 | 0,399200478 | PANK2 | FALSE | Upregulated |
| 0,413700083 | 0,399200478 | PKHD1 | FALSE | Upregulated |
| -0,358692153 | 0,399200478 | PSMF1 | FALSE | Downregulated |
| 0,670354034 | 0,399200478 | SH3GL1P2 | FALSE | Upregulated |
| -0,537480496 | 0,399200478 | SMDT1 | FALSE | Downregulated |
| -0,433936332 | 0,399200478 | SUMF1 | FALSE | Downregulated |
| 0,253845826 | 0,399341138 | AGRN | FALSE | Upregulated |
| -0,283808415 | 0,399341138 | CASC4 | FALSE | Downregulated |
| 0,586972984 | 0,399341138 | GRIN2C | FALSE | Upregulated |
| 0,368764494 | 0,399341138 | GUF1 | FALSE | Upregulated |
| -0,337318085 | 0,399341138 | RAB5C | FALSE | Downregulated |
| 0,420522353 | 0,399376986 | ANKRD50 | FALSE | Upregulated |
| -2,195191718 | 0,399376986 | GUSBP2 | FALSE | Downregulated |
| 0,823201315 | 0,399376986 | PMEPA1 | FALSE | Upregulated |
| -0,739472262 | 0,399376986 | PPIE | FALSE | Downregulated |
| 0,980731909 | 0,399376986 | SAMD15 | FALSE | Upregulated |
| -0,657577176 | 0,399578536 | WNT5A | FALSE | Downregulated |
| 0,388051372 | 0,399578536 | ZSCAN22 | FALSE | Upregulated |
| 0,428494181 | 0,399779372 | ASTE1 | FALSE | Upregulated |
| -0,353977485 | 0,399896516 | FECH | FALSE | Downregulated |
| -0,368382398 | 0,399966482 | SLC22A25 | FALSE | Downregulated |
| 0,305864126 | 0,400000922 | ASCC1 | FALSE | Upregulated |
| -0,442786905 | 0,400000922 | CA2 | FALSE | Downregulated |
| 0,464180045 | 0,400000922 | FABP3 | FALSE | Upregulated |
| -1,771071634 | 0,400000922 | LYRM9 | FALSE | Downregulated |
| -0,814471089 | 0,400000922 | SEPT7P2 | FALSE | Downregulated |
| -0,693128653 | 0,40008887 | PHKG2 | FALSE | Downregulated |
| -1,556326514 | 0,400154156 | MYEOV2 | FALSE | Downregulated |
| -0,495311372 | 0,400472236 | RNR2 | FALSE | Downregulated |
| 0,969084174 | 0,400472236 | SLC9B1 | FALSE | Upregulated |
| -0,34362488 | 0,400472236 | WNK3 | FALSE | Downregulated |
| 0,39181614 | 0,400501689 | RCC2 | FALSE | Upregulated |
| 1,800584344 | 0,400660495 | BAI1 | FALSE | Upregulated |
| 1,003734562 | 0,400815358 | LIX1 | FALSE | Upregulated |
| -0,559129993 | 0,400846527 | S1PR1 | FALSE | Downregulated |
| -0,241141863 | 0,40144274 | AFF1 | FALSE | Downregulated |

| | | | | |
|---|---|---|---|---|
| 1,912326373 | 0,401638049 | TMEM183B | FALSE | Upregulated |
| 0,559896498 | 0,40170658 | LYRM5 | FALSE | Upregulated |
| 0,361592173 | 0,401745166 | KIAA0930 | FALSE | Upregulated |
| 1,243170747 | 0,401745166 | PARM1 | FALSE | Upregulated |
| -0,751069301 | 0,402018935 | C16orf59 | FALSE | Downregulated |
| 0,354147328 | 0,402172725 | KBTBD2 | FALSE | Upregulated |
| 0,669942647 | 0,402300002 | TK1 | FALSE | Upregulated |
| 1,141685228 | 0,402613486 | ANK1 | FALSE | Upregulated |
| 0,582299886 | 0,402784027 | TSPAN12 | FALSE | Upregulated |
| 0,435479758 | 0,402815095 | KIAA0895L | FALSE | Upregulated |
| -0,267843325 | 0,402815095 | PSD4 | FALSE | Downregulated |
| 0,321060882 | 0,402920349 | LOC101928042 | FALSE | Upregulated |
| -0,637167428 | 0,402920349 | NECAB3 | FALSE | Downregulated |
| 0,400884381 | 0,402920349 | RP1-232P20.1 | FALSE | Upregulated |
| 0,626127239 | 0,402920349 | SLC12A2 | FALSE | Upregulated |
| 0,708688851 | 0,402920349 | SNX32 | FALSE | Upregulated |
| 0,845684264 | 0,402974802 | PRKCDBP | FALSE | Upregulated |
| 0,571273898 | 0,402974802 | RPS2P32 | FALSE | Upregulated |
| -0,587557732 | 0,403133233 | MRPL45 | FALSE | Downregulated |
| 0,463342796 | 0,403304706 | FTX | FALSE | Upregulated |
| 1,552164732 | 0,403304706 | TMEM231 | FALSE | Upregulated |
| 0,288428262 | 0,403517295 | SKIV2L2 | FALSE | Upregulated |
| -0,710501309 | 0,403530555 | GPR45 | FALSE | Downregulated |
| -1,019899912 | 0,403530555 | SCARNA9 | FALSE | Downregulated |
| -0,814687128 | 0,403942524 | KCNE2 | FALSE | Downregulated |
| -0,269324735 | 0,404194264 | KCTD3 | FALSE | Downregulated |
| 1,166412657 | 0,404194264 | TEAD4 | FALSE | Upregulated |
| 0,704291882 | 0,404359086 | PDZD4 | FALSE | Upregulated |
| -0,282948482 | 0,404518531 | ZNF3 | FALSE | Downregulated |
| 0,324291354 | 0,404545441 | PHACTR1 | FALSE | Upregulated |
| 0,394710121 | 0,404926999 | AC137932.6 | FALSE | Upregulated |
| -0,270921851 | 0,404926999 | C6orf89 | FALSE | Downregulated |
| 0,843106644 | 0,404926999 | LOC102723335 | FALSE | Upregulated |
| -0,772555679 | 0,404926999 | MVB12A | FALSE | Downregulated |
| -0,300630266 | 0,404926999 | NDFIP1 | FALSE | Downregulated |
| 1,937937634 | 0,404926999 | OR10V2P | FALSE | Upregulated |
| -0,510112809 | 0,404926999 | PHAX | FALSE | Downregulated |
| -0,299246435 | 0,405099896 | WDR59 | FALSE | Downregulated |
| 0,380836239 | 0,405163129 | DLK2 | FALSE | Upregulated |
| 1,501325961 | 0,40528855 | OR10A2 | FALSE | Upregulated |
| 0,312119487 | 0,405305244 | LOC286190 | FALSE | Upregulated |
| 0,88076797 | 0,405532707 | UBASH3B | FALSE | Upregulated |
| 0,211076405 | 0,405697783 | AIM1 | FALSE | Upregulated |
| -0,4516839 | 0,406002779 | PPP1R11 | FALSE | Downregulated |
| 1,005647446 | 0,406002779 | RP11-473M20.5 | FALSE | Upregulated |
| -0,417452856 | 0,406002779 | VKORC1 | FALSE | Downregulated |
| 1,718743917 | 0,406069683 | SEPT5 | FALSE | Upregulated |
| 0,778314797 | 0,406190586 | LOC101928263 | FALSE | Upregulated |
| -0,969159594 | 0,406190586 | LOC440570 | FALSE | Downregulated |
| 1,034008907 | 0,406190586 | PTRH1 | FALSE | Upregulated |
| 0,438272379 | 0,406190586 | SOAT1 | FALSE | Upregulated |

| | | | | |
|---|---|---|---|---|
| 0,808597747 | 0,406190586 | TMED6 | FALSE | Upregulated |
| 0,525390457 | 0,406190586 | ZNF821 | FALSE | Upregulated |
| 0,499091649 | 0,406212106 | MRPS12 | FALSE | Upregulated |
| -0,336168203 | 0,406252272 | RIPK1 | FALSE | Downregulated |
| -1,496606432 | 0,406288039 | OTUD7A | FALSE | Downregulated |
| 0,588579394 | 0,406577971 | LCA5L | FALSE | Upregulated |
| -0,465528356 | 0,406577971 | OTUD1 | FALSE | Downregulated |
| -1,002174877 | 0,406670054 | ZNF684 | FALSE | Downregulated |
| 0,734583093 | 0,406742954 | ARHGAP15 | FALSE | Upregulated |
| 0,727388065 | 0,406941773 | SLC52A2 | FALSE | Upregulated |
| 1,593793469 | 0,407112666 | CD52 | FALSE | Upregulated |
| 0,548956403 | 0,407112666 | IGSF6 | FALSE | Upregulated |
| 0,690123585 | 0,407200848 | COL8A1 | FALSE | Upregulated |
| 0,488847133 | 0,407447945 | UBE2W | FALSE | Upregulated |
| 0,350129125 | 0,407649806 | GPRASP1 | FALSE | Upregulated |
| 0,512084163 | 0,40785076 | IMPG1 | FALSE | Upregulated |
| -0,583949128 | 0,40785076 | MRPL24 | FALSE | Downregulated |
| -0,215453183 | 0,40790941 | KIAA2013 | FALSE | Downregulated |
| -1,436948966 | 0,407932517 | AKR1E2 | FALSE | Downregulated |
| 1,384555397 | 0,408159143 | PROM1 | FALSE | Upregulated |
| 0,734857491 | 0,408284701 | TRIM21 | FALSE | Upregulated |
| -0,891599422 | 0,408363808 | HAUS2 | FALSE | Downregulated |
| -0,235987398 | 0,408633265 | TMBIM6 | FALSE | Downregulated |
| 0,40475212 | 0,408638179 | TGFBR1 | FALSE | Upregulated |
| 0,764765016 | 0,40869913 | TRIM34 | FALSE | Upregulated |
| 0,406830228 | 0,408812603 | PSMC2 | FALSE | Upregulated |
| -1,549702418 | 0,408885628 | RP11-695J4.2 | FALSE | Downregulated |
| -0,500617842 | 0,408970259 | SPP2 | FALSE | Downregulated |
| 1,581163795 | 0,408977612 | GOLGA6L10 | FALSE | Upregulated |
| 0,539868965 | 0,408977612 | MSMO1 | FALSE | Upregulated |
| 0,349546316 | 0,409254451 | OXNAD1 | FALSE | Upregulated |
| 0,642230162 | 0,409254451 | ZFAS1 | FALSE | Upregulated |
| -1,252618272 | 0,409422808 | ADCY10P1 | FALSE | Downregulated |
| -0,335149492 | 0,409422808 | DDX23 | FALSE | Downregulated |
| 0,405409585 | 0,409422808 | ZNF551 | FALSE | Upregulated |
| 0,402954197 | 0,409447796 | RAP2C-AS1 | FALSE | Upregulated |
| -1,47620153 | 0,409584079 | LOC102723766 | FALSE | Downregulated |
| 0,319442718 | 0,409584079 | RLIM | FALSE | Upregulated |
| -0,810447633 | 0,409646031 | PMS2 | FALSE | Downregulated |
| 0,408926064 | 0,410190298 | GLT8D1 | FALSE | Upregulated |
| 0,410974663 | 0,410190298 | SPATA1 | FALSE | Upregulated |
| 0,324323101 | 0,410264431 | PROX2 | FALSE | Upregulated |
| 0,813318319 | 0,410483472 | EPHA3 | FALSE | Upregulated |
| -0,589479137 | 0,410488561 | PARP16 | FALSE | Downregulated |
| -0,348826548 | 0,4106498 | SHROOM1 | FALSE | Downregulated |
| 0,77302878 | 0,410896997 | CDYL2 | FALSE | Upregulated |
| -0,757768687 | 0,411102585 | SMA4 | FALSE | Downregulated |
| 0,300326443 | 0,411164073 | PLCG1 | FALSE | Upregulated |
| 0,387393804 | 0,411288075 | PFN1P2 | FALSE | Upregulated |
| -1,544261836 | 0,411288075 | PWP2 | FALSE | Downregulated |
| -0,810457463 | 0,411390913 | LOC101928139 | FALSE | Downregulated |

| | | | | |
|---|---|---|---|---|
| -1,537930285 | 0,411542675 | RP11-328K4.1 | FALSE | Downregulated |
| 0,287092316 | 0,411745742 | LARP4B | FALSE | Upregulated |
| -0,279847414 | 0,411833352 | GNA11 | FALSE | Downregulated |
| -0,633725483 | 0,411849351 | FLJ44342 | FALSE | Downregulated |
| -0,890862792 | 0,411936476 | ANAPC15 | FALSE | Downregulated |
| 0,423744753 | 0,411936476 | HRNR | FALSE | Upregulated |
| 0,893457811 | 0,411936476 | LOC101928473 | FALSE | Upregulated |
| -0,32251964 | 0,411936476 | MLYCD | FALSE | Downregulated |
| -0,274061235 | 0,411936476 | POR | FALSE | Downregulated |
| -1,267277257 | 0,411936476 | PP13 | FALSE | Downregulated |
| 1,075675551 | 0,411936476 | RACGAP1 | FALSE | Upregulated |
| 0,597681281 | 0,411936476 | SLC15A4 | FALSE | Upregulated |
| -0,650235194 | 0,411936476 | ZKSCAN2 | FALSE | Downregulated |
| 1,097230716 | 0,411936476 | ZNF763 | FALSE | Upregulated |
| -1,188784687 | 0,411953425 | LOC101928558 | FALSE | Downregulated |
| 1,380032936 | 0,41208637 | RP11-318M2.2 | FALSE | Upregulated |
| -0,345973899 | 0,412321199 | CYP2J2 | FALSE | Downregulated |
| 0,277433303 | 0,412321199 | RHOA | FALSE | Upregulated |
| 0,319118046 | 0,41275997 | LOC102723600 | FALSE | Upregulated |
| 0,430751057 | 0,412820279 | ZNF582 | FALSE | Upregulated |
| -0,280651625 | 0,412975684 | ABCG5 | FALSE | Downregulated |
| -1,29082717 | 0,412975684 | PEG10 | FALSE | Downregulated |
| -0,419776214 | 0,412975684 | SRSF3 | FALSE | Downregulated |
| -0,701282779 | 0,412975684 | UBE2F | FALSE | Downregulated |
| 1,226223929 | 0,412975684 | ZNF570 | FALSE | Upregulated |
| 2,205092638 | 0,413026702 | ACTN3 | FALSE | Upregulated |
| 0,823383559 | 0,413137754 | CD7 | FALSE | Upregulated |
| -0,554197663 | 0,413137754 | CHMP5 | FALSE | Downregulated |
| 0,817316224 | 0,413137754 | CTGF | FALSE | Upregulated |
| 0,367988662 | 0,413137754 | ETV6 | FALSE | Upregulated |
| -0,285689544 | 0,413137754 | FAM105B | FALSE | Downregulated |
| -0,525552789 | 0,413137754 | HDAC3 | FALSE | Downregulated |
| 0,500439521 | 0,413137754 | INPP5F | FALSE | Upregulated |
| 1,469007843 | 0,413137754 | LOC100131303 | FALSE | Upregulated |
| -0,565150196 | 0,413137754 | UROS | FALSE | Downregulated |
| 0,430094754 | 0,413191128 | C2orf15 | FALSE | Upregulated |
| 0,305945897 | 0,413294227 | B3GALNT2 | FALSE | Upregulated |
| 0,461721341 | 0,413294227 | LOC102724748 | FALSE | Upregulated |
| -0,713129057 | 0,413294227 | LOC729652 | FALSE | Downregulated |
| 0,447580933 | 0,41368538 | ARPC1B | FALSE | Upregulated |
| -0,950618066 | 0,41368538 | CYP3A7 | FALSE | Downregulated |
| 0,58699209 | 0,41368538 | KCNQ1 | FALSE | Upregulated |
| 0,599270552 | 0,41368538 | LINC00472 | FALSE | Upregulated |
| 0,870517458 | 0,41368538 | LOC101927030 | FALSE | Upregulated |
| 0,290223217 | 0,41368538 | ZNF584 | FALSE | Upregulated |
| 0,451114376 | 0,413858469 | PIP4K2A | FALSE | Upregulated |
| -0,317511225 | 0,414430459 | KLHL24 | FALSE | Downregulated |
| 0,740390089 | 0,414602436 | DGKH | FALSE | Upregulated |
| 0,435411874 | 0,41460619 | ALKBH3-AS1 | FALSE | Upregulated |
| 0,607901846 | 0,414789032 | TMEM164 | FALSE | Upregulated |
| -0,321183457 | 0,415146306 | SART1 | FALSE | Downregulated |

| | | | | |
|---|---|---|---|---|
| 0,468871047 | 0,415222776 | ADARB1 | FALSE | Upregulated |
| -0,284118658 | 0,415295604 | CHRD | FALSE | Downregulated |
| -0,595816184 | 0,415295604 | PEX11A | FALSE | Downregulated |
| -0,267177667 | 0,41573345 | CD99 | FALSE | Downregulated |
| 0,449283344 | 0,41573345 | GPATCH4 | FALSE | Upregulated |
| 0,294822397 | 0,41573345 | SYT2 | FALSE | Upregulated |
| 0,328834152 | 0,41573345 | ZC2HC1C | FALSE | Upregulated |
| -0,5787268 | 0,4160037 | CYB5R1 | FALSE | Downregulated |
| 1,367159888 | 0,416017221 | ITGA8 | FALSE | Upregulated |
| -0,607407839 | 0,416017221 | PIGN | FALSE | Downregulated |
| 0,591048976 | 0,416290435 | METTL15 | FALSE | Upregulated |
| 0,574666086 | 0,416290435 | TYW1B | FALSE | Upregulated |
| 0,453041514 | 0,416918103 | CA5A | FALSE | Upregulated |
| -0,67123671 | 0,417081196 | C2orf42 | FALSE | Downregulated |
| 0,489595685 | 0,417096904 | CCDC39 | FALSE | Upregulated |
| -0,204884103 | 0,417096904 | PLXNB1 | FALSE | Downregulated |
| -0,594449405 | 0,41714019 | RP11-622A1.2 | FALSE | Downregulated |
| 0,615435263 | 0,417441217 | MX1 | FALSE | Upregulated |
| 1,521345193 | 0,417441217 | NELL2 | FALSE | Upregulated |
| 0,36383337 | 0,417441217 | UEVLD | FALSE | Upregulated |
| 0,70518273 | 0,417498173 | TRAPPC13 | FALSE | Upregulated |
| -0,486292008 | 0,417525412 | ZNF44 | FALSE | Downregulated |
| 0,535586649 | 0,418045171 | EFCAB2 | FALSE | Upregulated |
| 0,598138046 | 0,418050745 | DSEL | FALSE | Upregulated |
| 0,228053624 | 0,418294036 | RRBP1 | FALSE | Upregulated |
| -0,790929939 | 0,418538638 | LOC100507131 | FALSE | Downregulated |
| -0,334893801 | 0,418784283 | CLPX | FALSE | Downregulated |
| 0,52546073 | 0,418784283 | LOC101927124 | FALSE | Upregulated |
| -0,435987831 | 0,418784283 | NDUFS7 | FALSE | Downregulated |
| -0,459105775 | 0,418974312 | SDHB | FALSE | Downregulated |
| -0,509034784 | 0,419085684 | CWF19L2 | FALSE | Downregulated |
| 0,597608631 | 0,419085684 | MAP3K6 | FALSE | Upregulated |
| -0,408374205 | 0,419085684 | RPS19BP1 | FALSE | Downregulated |
| 0,903383747 | 0,419275102 | C14orf182 | FALSE | Upregulated |
| 0,293403981 | 0,419337382 | FBXL17 | FALSE | Upregulated |
| 1,033004008 | 0,419337382 | SIGLEC14 | FALSE | Upregulated |
| 0,273631846 | 0,419376824 | LRRFIP1 | FALSE | Upregulated |
| -0,542122464 | 0,419376824 | TIMM50 | FALSE | Downregulated |
| 0,341174464 | 0,419389004 | ARGFXP2 | FALSE | Upregulated |
| 0,553085934 | 0,41956082 | CCDC104 | FALSE | Upregulated |
| -0,623647623 | 0,41956082 | GLCE | FALSE | Downregulated |
| 0,349394246 | 0,419572737 | LOC100506922 | FALSE | Upregulated |
| 0,571870837 | 0,419806597 | QTRTD1 | FALSE | Upregulated |
| -0,409878273 | 0,419908302 | COA4 | FALSE | Downregulated |
| 0,432641551 | 0,419908302 | TRPS1 | FALSE | Upregulated |
| 0,660197661 | 0,420670129 | ASUN | FALSE | Upregulated |
| 0,332965399 | 0,420670129 | LOC101928445 | FALSE | Upregulated |
| 0,986104493 | 0,420670129 | TBX21 | FALSE | Upregulated |
| 0,304431626 | 0,420880839 | COL6A1 | FALSE | Upregulated |
| -0,410371905 | 0,420880839 | ZBED3 | FALSE | Downregulated |
| 0,880090707 | 0,420934816 | C9orf9 | FALSE | Upregulated |

| | | | | |
|---|---|---|---|---|
| 0,926454048 | 0,420934816 | DLG3 | FALSE | Upregulated |
| -0,818984183 | 0,420934816 | MTPN | FALSE | Downregulated |
| -1,247253756 | 0,420934816 | RP11-418J17.1 | FALSE | Downregulated |
| -0,175114593 | 0,420934816 | SEC14L2 | FALSE | Downregulated |
| -1,254335032 | 0,421056507 | SEC14L3 | FALSE | Downregulated |
| 0,554057253 | 0,421264058 | MTURN | FALSE | Upregulated |
| 0,672030371 | 0,421286191 | H1FX-AS1 | FALSE | Upregulated |
| -0,460428062 | 0,421479995 | TMEM222 | FALSE | Downregulated |
| 0,361248372 | 0,421587491 | HDAC1 | FALSE | Upregulated |
| 1,088777438 | 0,421587491 | IGSF10 | FALSE | Upregulated |
| -0,857426058 | 0,421587491 | MROH8 | FALSE | Downregulated |
| -0,21051186 | 0,421587491 | PABPC1 | FALSE | Downregulated |
| 0,969189885 | 0,421587491 | PKNOX2 | FALSE | Upregulated |
| -0,217917077 | 0,421587491 | TBC1D9B | FALSE | Downregulated |
| 0,494312022 | 0,421587491 | ZNF322 | FALSE | Upregulated |
| -0,336820285 | 0,421635539 | IGF2-AS | FALSE | Downregulated |
| 0,924001721 | 0,421788541 | C8orf37 | FALSE | Upregulated |
| 0,294436161 | 0,421938547 | MSMP | FALSE | Upregulated |
| -0,75456574 | 0,421938547 | PTER | FALSE | Downregulated |
| 1,021180671 | 0,421938547 | ZNF135 | FALSE | Upregulated |
| 0,478403086 | 0,422096997 | PTTG3P | FALSE | Upregulated |
| -0,322215855 | 0,422157975 | CUL3 | FALSE | Downregulated |
| -0,59798586 | 0,422157975 | DUSP1 | FALSE | Downregulated |
| 0,240780716 | 0,422157975 | SETX | FALSE | Upregulated |
| -1,32605236 | 0,422400621 | METTL21B | FALSE | Downregulated |
| 1,029414515 | 0,422548246 | INTU | FALSE | Upregulated |
| 0,335370399 | 0,422548246 | NRP1 | FALSE | Upregulated |
| -0,219144914 | 0,422548246 | PDE8A | FALSE | Downregulated |
| -0,311538794 | 0,422548246 | PIK3R2 | FALSE | Downregulated |
| 1,210588406 | 0,422772573 | SLC4A8 | FALSE | Upregulated |
| 0,366706015 | 0,42278686 | LOC101928892 | FALSE | Upregulated |
| -0,423618485 | 0,422839483 | DOCK7 | FALSE | Downregulated |
| 1,258514032 | 0,422839483 | EFCAB10 | FALSE | Upregulated |
| 0,378242897 | 0,42293396 | KIFC2 | FALSE | Upregulated |
| -0,597465878 | 0,422987155 | DFFA | FALSE | Downregulated |
| -0,384242483 | 0,422993758 | TMED9 | FALSE | Downregulated |
| 5,055596975 | 0,423186606 | TNNC2 | FALSE | Upregulated |
| 0,291146997 | 0,423443308 | PEAK1 | FALSE | Upregulated |
| 1,4553591 | 0,423443308 | TTLL13 | FALSE | Upregulated |
| -2,190560988 | 0,423443308 | ZFP91-CNTF | FALSE | Downregulated |
| -1,448593133 | 0,423443986 | POC1A | FALSE | Downregulated |
| 0,555942612 | 0,423443986 | SPATC1 | FALSE | Upregulated |
| 0,276538891 | 0,423443986 | ZNF337 | FALSE | Upregulated |
| 1,216449582 | 0,423670539 | VDR | FALSE | Upregulated |
| -0,712023226 | 0,42368794 | KIF27 | FALSE | Downregulated |
| -0,268678006 | 0,423850384 | CES2 | FALSE | Downregulated |
| -1,033277581 | 0,423860779 | ANKRD39 | FALSE | Downregulated |
| 3,358630119 | 0,424052708 | LOC440354 | FALSE | Upregulated |
| 1,474184879 | 0,424052708 | OR2A12 | FALSE | Upregulated |
| -0,57020875 | 0,42407185 | LOC150776 | FALSE | Downregulated |
| 0,273019999 | 0,425018174 | HEXIM1 | FALSE | Upregulated |

| | | | | |
|---|---|---|---|---|
| 0,582458784 | 0,425117199 | CELF4 | FALSE | Upregulated |
| 0,32466168 | 0,425362477 | C22orf23 | FALSE | Upregulated |
| -0,350061846 | 0,425362477 | MRPL49 | FALSE | Downregulated |
| -0,309055562 | 0,425618741 | RANBP10 | FALSE | Downregulated |
| 0,704091479 | 0,425639094 | IMPDH1 | FALSE | Upregulated |
| 0,588266992 | 0,425639094 | SLFN14 | FALSE | Upregulated |
| 0,37706879 | 0,425725779 | TP53BP2 | FALSE | Upregulated |
| -0,481336087 | 0,425725779 | ZNF160 | FALSE | Downregulated |
| 0,714246539 | 0,425725779 | ZNF75A | FALSE | Upregulated |
| 0,476525018 | 0,426009605 | EIF2B2 | FALSE | Upregulated |
| -0,398583318 | 0,426109425 | FOXA2 | FALSE | Downregulated |
| 0,44426095 | 0,426109425 | LOC101927437 | FALSE | Upregulated |
| 1,398291634 | 0,426135268 | HIST1H4I | FALSE | Upregulated |
| -0,487132912 | 0,42641739 | MRPL16 | FALSE | Downregulated |
| -1,200159506 | 0,426546267 | CALML3 | FALSE | Downregulated |
| -0,280439732 | 0,426565123 | DAP | FALSE | Downregulated |
| 0,730293256 | 0,426614923 | LOC102723665 | FALSE | Upregulated |
| -0,583144929 | 0,426686351 | MAP10 | FALSE | Downregulated |
| -0,254078347 | 0,426821466 | GLYR1 | FALSE | Downregulated |
| -0,314150034 | 0,426821466 | SPPL2B | FALSE | Downregulated |
| -0,481144403 | 0,426950823 | BMS1P5 | FALSE | Downregulated |
| 0,370590878 | 0,42715707 | ASB8 | FALSE | Upregulated |
| -1,354971249 | 0,427240465 | LOC101928303 | FALSE | Downregulated |
| 0,94017403 | 0,427526819 | AIFM3 | FALSE | Upregulated |
| 0,41706783 | 0,427526819 | ATP6V1C1 | FALSE | Upregulated |
| 0,562993844 | 0,427526819 | EPC1 | FALSE | Upregulated |
| 1,856190508 | 0,427526819 | RP11-569G13.2 | FALSE | Upregulated |
| -0,709624622 | 0,427545388 | C12orf66 | FALSE | Downregulated |
| -0,633436806 | 0,427545388 | LOC102723772 | FALSE | Downregulated |
| 0,892248134 | 0,427545388 | RP11-27K13.3 | FALSE | Upregulated |
| -0,379770673 | 0,427549259 | IL12RB2 | FALSE | Downregulated |
| -0,290689443 | 0,427549259 | PPP6R3 | FALSE | Downregulated |
| 0,8875678 | 0,427549259 | TTC5 | FALSE | Upregulated |
| -0,57630445 | 0,427947375 | TMEM55B | FALSE | Downregulated |
| -0,970392057 | 0,428004315 | MOGAT1 | FALSE | Downregulated |
| 0,456747655 | 0,428004315 | TTC3-AS1 | FALSE | Upregulated |
| -1,220307983 | 0,428004315 | ZCCHC4 | FALSE | Downregulated |
| 0,781402319 | 0,428661694 | DEGS2 | FALSE | Upregulated |
| -0,690203321 | 0,428661694 | KHDRBS3 | FALSE | Downregulated |
| 0,67204193 | 0,428789507 | AEBP2 | FALSE | Upregulated |
| -0,385923556 | 0,428812002 | CLN8 | FALSE | Downregulated |
| -0,458687318 | 0,428812002 | LIMCH1 | FALSE | Downregulated |
| -0,455324469 | 0,428812002 | TMEM37 | FALSE | Downregulated |
| 0,829297825 | 0,428850883 | COL9A2 | FALSE | Upregulated |
| 0,288982325 | 0,428850883 | CUL1 | FALSE | Upregulated |
| -0,927529861 | 0,428850883 | DNA2 | FALSE | Downregulated |
| -0,837621213 | 0,428850883 | ING2 | FALSE | Downregulated |
| 1,049057698 | 0,428850883 | ITPRIPL1 | FALSE | Upregulated |
| 0,694995278 | 0,428850883 | P2RX2 | FALSE | Upregulated |
| 1,215508609 | 0,428850883 | PRRX1 | FALSE | Upregulated |
| 0,699360888 | 0,428850883 | RP11-333E1.1 | FALSE | Upregulated |

| | | | | |
|---|---|---|---|---|
| -0,234628149 | 0,428857038 | PHLDB2 | FALSE | Downregulated |
| 0,966288966 | 0,428857038 | SCN3B | FALSE | Upregulated |
| -0,191597701 | 0,428900719 | DCAF8 | FALSE | Downregulated |
| -0,762785856 | 0,428965846 | NDUFAF2 | FALSE | Downregulated |
| 0,304251542 | 0,429026867 | USP3 | FALSE | Upregulated |
| -0,330408671 | 0,429100275 | SUCLG2 | FALSE | Downregulated |
| 1,117811407 | 0,429265695 | LOC100506379 | FALSE | Upregulated |
| 0,965646869 | 0,429265695 | TMC2 | FALSE | Upregulated |
| -0,390908194 | 0,429325062 | CRLS1 | FALSE | Downregulated |
| 0,521103666 | 0,429325062 | LDLR | FALSE | Upregulated |
| -0,2553754 | 0,429325062 | PCSK6 | FALSE | Downregulated |
| 0,417945203 | 0,429325062 | TSPAN18 | FALSE | Upregulated |
| -0,290651942 | 0,429365693 | ATP13A3 | FALSE | Downregulated |
| 0,247479027 | 0,429365693 | TCF20 | FALSE | Upregulated |
| -0,356221754 | 0,42979985 | C11orf24 | FALSE | Downregulated |
| 0,724565256 | 0,429980494 | SLC39A10 | FALSE | Upregulated |
| 1,497927545 | 0,430089755 | C1orf51 | FALSE | Upregulated |
| -0,932920809 | 0,430195989 | GRHL1 | FALSE | Downregulated |
| -0,503407203 | 0,430195989 | PBX3 | FALSE | Downregulated |
| 0,99617907 | 0,430195989 | RNF224 | FALSE | Upregulated |
| 0,498664279 | 0,430295419 | GABPB2 | FALSE | Upregulated |
| -0,623839217 | 0,430295419 | LRRC28 | FALSE | Downregulated |
| 0,899099876 | 0,430379243 | ODF3 | FALSE | Upregulated |
| 0,476904015 | 0,430379243 | SPTLC3 | FALSE | Upregulated |
| -0,5586544 | 0,430439843 | CLK4 | FALSE | Downregulated |
| 0,410988836 | 0,430439843 | LIMA1 | FALSE | Upregulated |
| 0,715590472 | 0,430439843 | RPL32P3 | FALSE | Upregulated |
| -0,266917318 | 0,430439843 | TEAD1 | FALSE | Downregulated |
| 0,420699535 | 0,430621432 | TMEM206 | FALSE | Upregulated |
| 1,925559295 | 0,430648922 | E2F2 | FALSE | Upregulated |
| -1,125988191 | 0,430964004 | OXLD1 | FALSE | Downregulated |
| 0,364844294 | 0,431415542 | PANX1 | FALSE | Upregulated |
| 0,506761986 | 0,431434777 | DIS3L2 | FALSE | Upregulated |
| -0,484546984 | 0,431434777 | GCNT2 | FALSE | Downregulated |
| 0,555767773 | 0,431434777 | LILRB1 | FALSE | Upregulated |
| 0,516624262 | 0,431434777 | RDH13 | FALSE | Upregulated |
| 0,924774332 | 0,431733862 | FJX1 | FALSE | Upregulated |
| 0,407166489 | 0,431733862 | PLAT | FALSE | Upregulated |
| 0,99902017 | 0,431733862 | PTK7 | FALSE | Upregulated |
| -0,496271276 | 0,431733862 | QRICH2 | FALSE | Downregulated |
| 0,632301818 | 0,431924135 | C16orf98 | FALSE | Upregulated |
| -1,437071888 | 0,431924135 | LOC101928669 | FALSE | Downregulated |
| -0,441504662 | 0,431927331 | AADAC | FALSE | Downregulated |
| 0,267120047 | 0,431927331 | MAN2B1 | FALSE | Upregulated |
| 1,412108605 | 0,431927331 | MATN1-AS1 | FALSE | Upregulated |
| 0,227794545 | 0,432116306 | SMLR1 | FALSE | Upregulated |
| -0,488991726 | 0,43291731 | KIAA1919 | FALSE | Downregulated |
| -1,084878436 | 0,433362166 | TMEM68 | FALSE | Downregulated |
| -0,36416953 | 0,433811819 | DECR1 | FALSE | Downregulated |
| 1,232019927 | 0,433839509 | CPNE5 | FALSE | Upregulated |
| -0,273944474 | 0,433843823 | KANSL3 | FALSE | Downregulated |

| | | | | |
|---|---|---|---|---|
| -1,379932567 | 0,433843823 | VSTM2L | FALSE | Downregulated |
| -1,394152092 | 0,433843823 | YBX2 | FALSE | Downregulated |
| 0,986821728 | 0,434623664 | GJA1 | FALSE | Upregulated |
| -0,498639848 | 0,434623664 | PAPD4 | FALSE | Downregulated |
| 0,682997575 | 0,434771829 | TMEM41B | FALSE | Upregulated |
| 0,314008848 | 0,434785622 | ZNF506 | FALSE | Upregulated |
| -0,43738227 | 0,435059788 | ZBTB48 | FALSE | Downregulated |
| 0,290116215 | 0,435193797 | TIMP2 | FALSE | Upregulated |
| 0,849291855 | 0,435260341 | GPIHBP1 | FALSE | Upregulated |
| 0,442643991 | 0,435260341 | NEDD8 | FALSE | Upregulated |
| -0,575639886 | 0,435326244 | ZDHHC4 | FALSE | Downregulated |
| 0,340575034 | 0,435359009 | CRYZ | FALSE | Upregulated |
| -0,216766578 | 0,435359009 | IRF2BP2 | FALSE | Downregulated |
| 0,346112937 | 0,435359009 | OSGEPL1 | FALSE | Upregulated |
| -0,32713266 | 0,435626621 | PPP1R26 | FALSE | Downregulated |
| -1,461269561 | 0,435828972 | APOBEC3G | FALSE | Downregulated |
| -0,318414188 | 0,43587482 | DCBLD2 | FALSE | Downregulated |
| 1,000699101 | 0,43587482 | SPEG | FALSE | Upregulated |
| 0,416160032 | 0,435993414 | NME9 | FALSE | Upregulated |
| -0,212865815 | 0,435993414 | RPL7A | FALSE | Downregulated |
| -0,359912574 | 0,43601096 | AKAP11 | FALSE | Downregulated |
| -0,473392332 | 0,43601096 | ARSA | FALSE | Downregulated |
| 1,131459574 | 0,43601096 | ASAP1-IT2 | FALSE | Upregulated |
| 0,672456774 | 0,43601096 | CHTF18 | FALSE | Upregulated |
| 0,527989571 | 0,43601096 | ELOVL2 | FALSE | Upregulated |
| 0,383326261 | 0,43601096 | ERCC6L2 | FALSE | Upregulated |
| 0,45446196 | 0,43601096 | FHOD1 | FALSE | Upregulated |
| 0,291749993 | 0,43601096 | KDM2B | FALSE | Upregulated |
| 0,506777979 | 0,43601096 | LOC102724262 | FALSE | Upregulated |
| 0,990536522 | 0,43601096 | LOC81691 | FALSE | Upregulated |
| -0,290780937 | 0,43601096 | MRPL20 | FALSE | Downregulated |
| -0,470765046 | 0,43601096 | MRPL28 | FALSE | Downregulated |
| 0,933237241 | 0,43601096 | NUFIP1 | FALSE | Upregulated |
| 0,287696411 | 0,43601096 | ORC4 | FALSE | Upregulated |
| -1,315770171 | 0,43601096 | PHOSPHO2 | FALSE | Downregulated |
| 0,430929642 | 0,43601096 | RQCD1 | FALSE | Upregulated |
| 0,355914659 | 0,43601096 | SDCBP | FALSE | Upregulated |
| 0,173732927 | 0,43601096 | SPTBN1 | FALSE | Upregulated |
| -0,69097294 | 0,43601096 | TBXA2R | FALSE | Downregulated |
| -0,802421097 | 0,436129062 | RP11-1096D5.1 | FALSE | Downregulated |
| -0,580458192 | 0,436145609 | UBALD1 | FALSE | Downregulated |
| 0,386395221 | 0,436145609 | ZNF224 | FALSE | Upregulated |
| 0,569545842 | 0,436172108 | CCHCR1 | FALSE | Upregulated |
| 0,592603851 | 0,436172108 | COL17A1 | FALSE | Upregulated |
| 0,544066978 | 0,436172108 | FUT8 | FALSE | Upregulated |
| -0,485113756 | 0,436172108 | KIF12 | FALSE | Downregulated |
| 0,420067797 | 0,436172108 | LOC102724930 | FALSE | Upregulated |
| 1,186916888 | 0,436172108 | MOSPD1 | FALSE | Upregulated |
| -1,261142625 | 0,436172108 | MROH2A | FALSE | Downregulated |
| 0,335115454 | 0,436172108 | MYL12B | FALSE | Upregulated |
| 0,860643938 | 0,436172108 | NCR3LG1 | FALSE | Upregulated |

| | | | | |
|---|---|---|---|---|
| -1,368853497 | 0,436172108 | PACRG-AS1 | FALSE | Downregulated |
| -0,774637593 | 0,436172108 | SLC23A3 | FALSE | Downregulated |
| -0,817575536 | 0,436172108 | SPTSSA | FALSE | Downregulated |
| -1,02999008 | 0,436172108 | ZFP30 | FALSE | Downregulated |
| 0,266664942 | 0,436356842 | AKAP17A | FALSE | Upregulated |
| 0,868319719 | 0,436570869 | COLGALT2 | FALSE | Upregulated |
| 0,251941512 | 0,436570869 | CSNK1G2 | FALSE | Upregulated |
| 0,573509864 | 0,436570869 | PRR5L | FALSE | Upregulated |
| 0,318691524 | 0,436570869 | PRSS12 | FALSE | Upregulated |
| 1,420241638 | 0,436570869 | RNASE8 | FALSE | Upregulated |
| 0,284649917 | 0,436570869 | SIK2 | FALSE | Upregulated |
| -0,158939983 | 0,436665171 | EIF4B | FALSE | Downregulated |
| -1,41041745 | 0,436665171 | IL1R2 | FALSE | Downregulated |
| -0,646830553 | 0,436885149 | FOLH1B | FALSE | Downregulated |
| -0,411442252 | 0,436902513 | PEG3-AS1 | FALSE | Downregulated |
| 0,447708992 | 0,436950138 | RGS3 | FALSE | Upregulated |
| 0,375308607 | 0,436962678 | ATP6AP2 | FALSE | Upregulated |
| 1,262954641 | 0,437268155 | SGOL2 | FALSE | Upregulated |
| -1,353250822 | 0,437532945 | SCUBE1 | FALSE | Downregulated |
| -0,310046573 | 0,437852145 | PYGL | FALSE | Downregulated |
| 0,297889161 | 0,437852145 | RPL23AP64 | FALSE | Upregulated |
| 0,396805562 | 0,437858193 | TMCC1 | FALSE | Upregulated |
| -0,479450108 | 0,437873207 | CAMK1D | FALSE | Downregulated |
| 1,211604913 | 0,437873207 | PFKFB4 | FALSE | Upregulated |
| 1,713468491 | 0,438087239 | OR2A20P | FALSE | Upregulated |
| 0,778412866 | 0,438087239 | SEBOX | FALSE | Upregulated |
| -0,322224869 | 0,43822639 | ASPDH | FALSE | Downregulated |
| 0,900320753 | 0,438680491 | LOC729930 | FALSE | Upregulated |
| -1,232342422 | 0,438692961 | NUPL2 | FALSE | Downregulated |
| -0,375204146 | 0,438733288 | RUVBL2 | FALSE | Downregulated |
| -0,348948923 | 0,438868673 | MAP7 | FALSE | Downregulated |
| -0,594413555 | 0,439089023 | LINC00478 | FALSE | Downregulated |
| -0,356994768 | 0,439089023 | RAI14 | FALSE | Downregulated |
| -0,493763254 | 0,439089023 | SAMD5 | FALSE | Downregulated |
| -0,773784034 | 0,439224857 | GMFB | FALSE | Downregulated |
| 0,440570828 | 0,439224857 | JDP2 | FALSE | Upregulated |
| -0,148342325 | 0,439224857 | TNRC6B | FALSE | Downregulated |
| -0,360486519 | 0,439487933 | ZFYVE9 | FALSE | Downregulated |
| 0,567649991 | 0,439488327 | LOC101928318 | FALSE | Upregulated |
| 0,641217831 | 0,439547738 | ZNRF2P2 | FALSE | Upregulated |
| 0,972370089 | 0,439796103 | AC011899.9 | FALSE | Upregulated |
| 0,636929893 | 0,4398535 | IGSF22 | FALSE | Upregulated |
| -0,966944043 | 0,440350195 | ELTD1 | FALSE | Downregulated |
| 0,297529996 | 0,440416237 | PPP2R3A | FALSE | Upregulated |
| 0,37773802 | 0,440451816 | HIGD1A | FALSE | Upregulated |
| -1,553976836 | 0,440451816 | LOC102724064 | FALSE | Downregulated |
| -0,497514919 | 0,440452074 | MTRR | FALSE | Downregulated |
| -0,697975746 | 0,440529747 | PDCD5 | FALSE | Downregulated |
| -0,483383125 | 0,440621284 | LINC01128 | FALSE | Downregulated |
| -0,8573286 | 0,440868072 | UBL7 | FALSE | Downregulated |
| -0,460394505 | 0,440905501 | SLC45A3 | FALSE | Downregulated |

| | | | | |
|---|---|---|---|---|
| -0,266626545 | 0,441006194 | DOCK4 | FALSE | Downregulated |
| 0,298724367 | 0,441076133 | XPO5 | FALSE | Upregulated |
| -0,430433583 | 0,441153243 | IFNLR1 | FALSE | Downregulated |
| 0,246184616 | 0,441202969 | C6orf120 | FALSE | Upregulated |
| 0,822731475 | 0,441202969 | PDGFA | FALSE | Upregulated |
| 0,228320511 | 0,441419745 | CNP | FALSE | Upregulated |
| -0,378619869 | 0,441419745 | FAM222B | FALSE | Downregulated |
| 0,433106209 | 0,441419745 | PIK3CB | FALSE | Upregulated |
| -0,322114592 | 0,441598121 | CCT4 | FALSE | Downregulated |
| 0,288904543 | 0,441598121 | QRICH1 | FALSE | Upregulated |
| -2,043600664 | 0,441598121 | SYCE1 | FALSE | Downregulated |
| 0,39496167 | 0,441598121 | THEMIS2 | FALSE | Upregulated |
| 0,441418089 | 0,4421896 | LRRC32 | FALSE | Upregulated |
| -0,52787486 | 0,4421896 | TEK | FALSE | Downregulated |
| 1,080271788 | 0,442381698 | NRG3 | FALSE | Upregulated |
| -0,334790778 | 0,442471596 | ACMSD | FALSE | Downregulated |
| -1,444265279 | 0,442527116 | CCDC15 | FALSE | Downregulated |
| -1,153184563 | 0,442527116 | PRR22 | FALSE | Downregulated |
| -0,294040279 | 0,442527116 | RFC1 | FALSE | Downregulated |
| 0,383279315 | 0,442527116 | TUB | FALSE | Upregulated |
| 0,469465915 | 0,442596102 | ANKRD27 | FALSE | Upregulated |
| 0,911141019 | 0,442596102 | DAPK2 | FALSE | Upregulated |
| 0,54660928 | 0,442596102 | DIMT1 | FALSE | Upregulated |
| -0,342036547 | 0,442596102 | DNPEP | FALSE | Downregulated |
| 0,390228234 | 0,442596102 | EPN2 | FALSE | Upregulated |
| -0,974818769 | 0,442596102 | FRA10AC1 | FALSE | Downregulated |
| -0,785906031 | 0,442596102 | LOC220729 | FALSE | Downregulated |
| -0,508702189 | 0,442596102 | MAPK9 | FALSE | Downregulated |
| -0,229586851 | 0,442596102 | MAT2A | FALSE | Downregulated |
| 0,459720267 | 0,442596102 | P4HTM | FALSE | Upregulated |
| -0,708808107 | 0,442596102 | PDIA3P1 | FALSE | Downregulated |
| 0,333770482 | 0,442596102 | PLCL2 | FALSE | Upregulated |
| 0,792218675 | 0,442596102 | PPT2 | FALSE | Upregulated |
| 0,427412767 | 0,442596102 | RP11-61I13.3 | FALSE | Upregulated |
| 0,26168644 | 0,442596102 | TNPO2 | FALSE | Upregulated |
| 0,721831152 | 0,443070344 | AC058791.1 | FALSE | Upregulated |
| 0,51481525 | 0,443241867 | LOC101928998 | FALSE | Upregulated |
| 0,210616078 | 0,443241867 | NEAT1 | FALSE | Upregulated |
| -0,400161907 | 0,443428668 | VRK3 | FALSE | Downregulated |
| -0,432913601 | 0,443428819 | PPP1R12B | FALSE | Downregulated |
| 0,394436209 | 0,44344137 | DCN | FALSE | Upregulated |
| 1,238117078 | 0,44344137 | GTSE1 | FALSE | Upregulated |
| -0,385417576 | 0,44344137 | LOC102724607 | FALSE | Downregulated |
| -0,345176025 | 0,44344137 | MEN1 | FALSE | Downregulated |
| 0,493298681 | 0,44344137 | SLC12A5 | FALSE | Upregulated |
| -0,696252591 | 0,443680031 | ZBTB16 | FALSE | Downregulated |
| 0,437052148 | 0,443705862 | TIMMDC1 | FALSE | Upregulated |
| 0,55781779 | 0,443705862 | YIPF5 | FALSE | Upregulated |
| 0,651579036 | 0,44375534 | CXCR3 | FALSE | Upregulated |
| 0,351260864 | 0,44375534 | FNBP1 | FALSE | Upregulated |
| 1,07675554 | 0,44375534 | MYLK4 | FALSE | Upregulated |

| | | | | |
|---|---|---|---|---|
| -0,55171896 | 0,444294226 | LOC401286 | FALSE | Downregulated |
| 0,335507282 | 0,444387641 | ATP6V1B2 | FALSE | Upregulated |
| -0,301218072 | 0,444387641 | TCF12 | FALSE | Downregulated |
| 0,254264997 | 0,444387641 | TRAPPC12 | FALSE | Upregulated |
| 0,325897402 | 0,444426602 | ACTG1 | FALSE | Upregulated |
| -0,685039956 | 0,444426602 | SENP7 | FALSE | Downregulated |
| 0,289366319 | 0,444802816 | CERS5 | FALSE | Upregulated |
| 0,268812494 | 0,444869114 | DTX3L | FALSE | Upregulated |
| -1,383807316 | 0,445134428 | CADM2-AS1 | FALSE | Downregulated |
| -0,333940121 | 0,445274467 | CPNE3 | FALSE | Downregulated |
| 1,444960615 | 0,445274467 | LOC102724756 | FALSE | Upregulated |
| -0,404469907 | 0,445274467 | SLC30A9 | FALSE | Downregulated |
| -1,211354174 | 0,445343523 | PRRG2 | FALSE | Downregulated |
| -0,483053268 | 0,445343523 | WDR45 | FALSE | Downregulated |
| 1,254045387 | 0,445644973 | LINC00035 | FALSE | Upregulated |
| 0,387776733 | 0,445791227 | LOC729732 | FALSE | Upregulated |
| 0,226754688 | 0,445791227 | ZNRF2 | FALSE | Upregulated |
| 0,317471859 | 0,445916717 | BCDIN3D-AS1 | FALSE | Upregulated |
| -0,381293391 | 0,445921368 | NDEL1 | FALSE | Downregulated |
| 0,339423254 | 0,44595663 | SMCR5 | FALSE | Upregulated |
| -1,350425027 | 0,44595663 | TMEM255B | FALSE | Downregulated |
| 0,992130928 | 0,44595663 | ZNF354A | FALSE | Upregulated |
| 0,314941044 | 0,446270378 | NINJ1 | FALSE | Upregulated |
| -1,017408189 | 0,446270378 | RPS6KA5 | FALSE | Downregulated |
| 0,264740778 | 0,446365257 | COMMD5 | FALSE | Upregulated |
| -0,212275162 | 0,446365257 | EIF5 | FALSE | Downregulated |
| -0,468443938 | 0,446365257 | FCF1 | FALSE | Downregulated |
| 0,388823152 | 0,446365257 | PLEKHH1 | FALSE | Upregulated |
| 0,387807574 | 0,446420783 | TMEM167A | FALSE | Upregulated |
| -0,328260322 | 0,446592372 | DNAJC22 | FALSE | Downregulated |
| 0,292675383 | 0,446643218 | DNAAF2 | FALSE | Upregulated |
| 0,860319738 | 0,44680736 | ARSK | FALSE | Upregulated |
| 0,733120512 | 0,44683956 | PLK3 | FALSE | Upregulated |
| -0,623085142 | 0,44683956 | TSPAN17 | FALSE | Downregulated |
| 0,19228797 | 0,44683956 | ZNF106 | FALSE | Upregulated |
| -0,273447073 | 0,446848633 | TMEM245 | FALSE | Downregulated |
| -0,604202607 | 0,447508326 | FADS3 | FALSE | Downregulated |
| -0,476737933 | 0,447508326 | GADD45A | FALSE | Downregulated |
| -0,732597034 | 0,447508326 | IQCJ-SCHIP1 | FALSE | Downregulated |
| 0,187770664 | 0,447508326 | NCOR2 | FALSE | Upregulated |
| 0,258292737 | 0,447508326 | ORAI2 | FALSE | Upregulated |
| -0,260884961 | 0,448030473 | IFITM3 | FALSE | Downregulated |
| 0,54469305 | 0,448030473 | PRDM15 | FALSE | Upregulated |
| -0,312319292 | 0,448476614 | SATB1 | FALSE | Downregulated |
| -0,369315942 | 0,448717331 | ANKRD28 | FALSE | Downregulated |
| -0,343799301 | 0,448827188 | ELK1 | FALSE | Downregulated |
| 0,69189555 | 0,448869736 | FAM13C | FALSE | Upregulated |
| 1,261989217 | 0,448869736 | LOC101929039 | FALSE | Upregulated |
| -0,365370243 | 0,448869736 | RMDN3 | FALSE | Downregulated |
| 1,163030995 | 0,448906435 | LOC101927086 | FALSE | Upregulated |
| -0,262280458 | 0,448935347 | ACSM3 | FALSE | Downregulated |

| | | | | |
|---|---|---|---|---|
| 0,773083069 | 0,448983636 | SORT1 | FALSE | Upregulated |
| -0,834897538 | 0,449064976 | EXTL2 | FALSE | Downregulated |
| -1,175991145 | 0,449366421 | AOC2 | FALSE | Downregulated |
| 0,737390839 | 0,449366421 | CCNI2 | FALSE | Upregulated |
| -0,342098866 | 0,449366421 | EPB41L4B | FALSE | Downregulated |
| -1,257848573 | 0,449366421 | LOC101927407 | FALSE | Downregulated |
| -1,887362768 | 0,449366421 | NDUFC2-KCTD14 | FALSE | Downregulated |
| 1,375109708 | 0,449366421 | NLGN4Y | FALSE | Upregulated |
| 0,612500277 | 0,449366421 | ZFAT | FALSE | Upregulated |
| 0,515623138 | 0,449415226 | XKR8 | FALSE | Upregulated |
| -0,494386022 | 0,44963341 | GREM2 | FALSE | Downregulated |
| -1,193796829 | 0,44963341 | MST4 | FALSE | Downregulated |
| -0,478167978 | 0,44963341 | R3HDM1 | FALSE | Downregulated |
| 0,475016584 | 0,449999373 | SMARCAD1 | FALSE | Upregulated |
| 0,305940457 | 0,45001751 | RAD9A | FALSE | Upregulated |
| 0,371916297 | 0,450460269 | WDR13 | FALSE | Upregulated |
| 0,725260587 | 0,450623204 | CCDC102A | FALSE | Upregulated |
| -0,65873979 | 0,451267999 | VANGL1 | FALSE | Downregulated |
| -0,218425481 | 0,451438376 | RANBP2 | FALSE | Downregulated |
| 1,012956556 | 0,452092024 | HIST1H1D | FALSE | Upregulated |
| 0,725728288 | 0,452099896 | DUSP12 | FALSE | Upregulated |
| -0,279821358 | 0,452099896 | GCLC | FALSE | Downregulated |
| 0,37790543 | 0,452099896 | IFIT3 | FALSE | Upregulated |
| 0,63303553 | 0,452099896 | LINC01043 | FALSE | Upregulated |
| -0,334443832 | 0,452099896 | MTMR12 | FALSE | Downregulated |
| 0,433261235 | 0,452099896 | NDUFS5 | FALSE | Upregulated |
| 0,715180687 | 0,452099896 | PHLDA3 | FALSE | Upregulated |
| 0,859007155 | 0,452099896 | SAP30 | FALSE | Upregulated |
| 0,349479015 | 0,452099896 | UACA | FALSE | Upregulated |
| 0,504361259 | 0,452099896 | VSIG4 | FALSE | Upregulated |
| -0,350770302 | 0,452266199 | ACVR2B | FALSE | Downregulated |
| -0,271829522 | 0,452266199 | NNT | FALSE | Downregulated |
| 0,312545081 | 0,452266199 | PTPRM | FALSE | Upregulated |
| 0,307639482 | 0,452266199 | SREK1 | FALSE | Upregulated |
| 0,423496704 | 0,452329517 | FLVCR2 | FALSE | Upregulated |
| 0,692537657 | 0,452579 | MAGI1-IT1 | FALSE | Upregulated |
| -1,072897524 | 0,452639843 | NDUFAF5 | FALSE | Downregulated |
| 0,399253886 | 0,452852251 | IKZF4 | FALSE | Upregulated |
| 1,011997544 | 0,452852251 | PRDM16 | FALSE | Upregulated |
| 1,231216035 | 0,452852251 | QRFP | FALSE | Upregulated |
| 0,82594379 | 0,452852251 | RNF2 | FALSE | Upregulated |
| -0,907694838 | 0,452852251 | WNT8B | FALSE | Downregulated |
| 0,838214177 | 0,452958153 | RP11-118E18.2 | FALSE | Upregulated |
| 1,131113414 | 0,453351337 | PAPPA2 | FALSE | Upregulated |
| 0,372277132 | 0,453436363 | FGD1 | FALSE | Upregulated |
| 0,33549429 | 0,453789033 | PPM1M | FALSE | Upregulated |
| 0,216017008 | 0,453789033 | TCIRG1 | FALSE | Upregulated |
| -0,332466183 | 0,453825384 | ANAPC16 | FALSE | Downregulated |
| 0,342681654 | 0,454153219 | ARHGDIB | FALSE | Upregulated |
| -0,287985108 | 0,454163126 | CYP2C9 | FALSE | Downregulated |
| -0,406086698 | 0,454215201 | HPCAL1 | FALSE | Downregulated |

| | | | | |
|---|---|---|---|---|
| 0,296764477 | 0,454215201 | URAHP | FALSE | Upregulated |
| 0,502579291 | 0,454254086 | NKPD1 | FALSE | Upregulated |
| -0,469603978 | 0,454356001 | EIF1B-AS1 | FALSE | Downregulated |
| -0,475827319 | 0,454362008 | PRR14 | FALSE | Downregulated |
| -1,139005822 | 0,454377346 | ACSL6 | FALSE | Downregulated |
| -0,218542245 | 0,454377346 | GCN1L1 | FALSE | Downregulated |
| -0,433912803 | 0,454377346 | HSD17B12 | FALSE | Downregulated |
| 0,43826414 | 0,454377346 | LOC101927898 | FALSE | Upregulated |
| 0,309470931 | 0,454377346 | LRRC14 | FALSE | Upregulated |
| -0,487279827 | 0,454377346 | PIGG | FALSE | Downregulated |
| 0,670309788 | 0,454377346 | PRKX | FALSE | Upregulated |
| 0,348521073 | 0,454377346 | RBM12B | FALSE | Upregulated |
| -0,446916011 | 0,454467593 | KPNA3 | FALSE | Downregulated |
| -0,278495881 | 0,454647599 | CLIC4 | FALSE | Downregulated |
| -0,442612654 | 0,454647599 | IMP4 | FALSE | Downregulated |
| -1,103117249 | 0,454647599 | SLC12A8 | FALSE | Downregulated |
| 0,374817093 | 0,454647599 | ZNF692 | FALSE | Upregulated |
| -0,81626329 | 0,454772072 | PYURF | FALSE | Downregulated |
| 0,426732947 | 0,454831798 | CLIP2 | FALSE | Upregulated |
| 0,391590207 | 0,454845435 | PDE8B | FALSE | Upregulated |
| 0,678848696 | 0,454849766 | RIPK2 | FALSE | Upregulated |
| 0,872086093 | 0,455000922 | NCF1C | FALSE | Upregulated |
| -0,290184241 | 0,455070271 | POLE | FALSE | Downregulated |
| 0,542911559 | 0,455070271 | SPSB1 | FALSE | Upregulated |
| 0,347371929 | 0,45510461 | DAGLB | FALSE | Upregulated |
| 0,443915724 | 0,45510461 | TIGD7 | FALSE | Upregulated |
| 1,043195157 | 0,455289028 | BMP5 | FALSE | Upregulated |
| -1,499762206 | 0,455293989 | APOO | FALSE | Downregulated |
| 0,285881292 | 0,455520551 | RBM15 | FALSE | Upregulated |
| -1,160741807 | 0,455778315 | C17orf49 | FALSE | Downregulated |
| 0,996504191 | 0,455778315 | CHEK2 | FALSE | Upregulated |
| 0,498535755 | 0,455778315 | DNAJC9 | FALSE | Upregulated |
| 0,389970637 | 0,455778315 | NAP1L1 | FALSE | Upregulated |
| 0,275142358 | 0,455778315 | NMT2 | FALSE | Upregulated |
| 0,514333516 | 0,455778315 | POMT2 | FALSE | Upregulated |
| 0,335980798 | 0,455778315 | RBM4B | FALSE | Upregulated |
| 0,500840478 | 0,455778315 | ZNF319 | FALSE | Upregulated |
| 0,583460895 | 0,455799986 | TMEM209 | FALSE | Upregulated |
| 0,409651494 | 0,456100471 | ARAP2 | FALSE | Upregulated |
| 0,266316698 | 0,456259323 | RANBP9 | FALSE | Upregulated |
| 0,782268224 | 0,456386984 | FAM71E1 | FALSE | Upregulated |
| 0,212536861 | 0,456386984 | GGA2 | FALSE | Upregulated |
| -0,318581092 | 0,456386984 | PPFIBP1 | FALSE | Downregulated |
| -0,370941955 | 0,456474179 | DDX41 | FALSE | Downregulated |
| -0,550368601 | 0,456542221 | COPS3 | FALSE | Downregulated |
| 0,382871428 | 0,456542221 | EFHD2 | FALSE | Upregulated |
| -0,719780131 | 0,456542221 | NEGR1 | FALSE | Downregulated |
| 0,594129533 | 0,456542221 | TRIM32 | FALSE | Upregulated |
| -0,256658234 | 0,456684886 | PRPF31 | FALSE | Downregulated |
| 0,290739607 | 0,456835562 | ARHGAP30 | FALSE | Upregulated |
| -0,851619958 | 0,45699185 | LINC00116 | FALSE | Downregulated |

| | | | | |
|---|---|---|---|---|
| -1,023436328 | 0,45699185 | LINC00998 | FALSE | Downregulated |
| 0,685949497 | 0,45699185 | PCSK5 | FALSE | Upregulated |
| -0,521288139 | 0,45699185 | PPP6C | FALSE | Downregulated |
| 0,639282308 | 0,45699185 | RGS19 | FALSE | Upregulated |
| -0,203944602 | 0,45699185 | SLC10A1 | FALSE | Downregulated |
| 0,30114682 | 0,45699185 | SP2 | FALSE | Upregulated |
| -0,429173293 | 0,45699185 | SYNGR2 | FALSE | Downregulated |
| 0,298756828 | 0,45699185 | WDR26 | FALSE | Upregulated |
| 0,312271845 | 0,456999726 | BBIP1 | FALSE | Upregulated |
| 0,381192695 | 0,457114499 | ELOVL5 | FALSE | Upregulated |
| -0,266597078 | 0,45725586 | ABHD15 | FALSE | Downregulated |
| -0,228835577 | 0,45725586 | DHFR | FALSE | Downregulated |
| -0,364001004 | 0,458134274 | SCARNA17 | FALSE | Downregulated |
| 0,303613861 | 0,45825151 | TECPR1 | FALSE | Upregulated |
| 0,294298778 | 0,458474985 | ATF7IP | FALSE | Upregulated |
| -1,003787623 | 0,458988211 | C11orf52 | FALSE | Downregulated |
| 0,345673027 | 0,459210835 | AC009948.5 | FALSE | Upregulated |
| 0,257923838 | 0,459210835 | ACTR2 | FALSE | Upregulated |
| 1,020815612 | 0,459258924 | ANO3 | FALSE | Upregulated |
| 0,287628433 | 0,459258924 | ATP9A | FALSE | Upregulated |
| 0,685523935 | 0,459258924 | C1orf216 | FALSE | Upregulated |
| -0,628158219 | 0,459258924 | GMPPA | FALSE | Downregulated |
| -0,632562127 | 0,459258924 | LYPLAL1 | FALSE | Downregulated |
| 0,252926632 | 0,459258924 | RALBP1 | FALSE | Upregulated |
| -1,398577848 | 0,459258924 | SUCNR1 | FALSE | Downregulated |
| 1,283793024 | 0,459258924 | TLR8 | FALSE | Upregulated |
| 0,907271545 | 0,459258924 | ULK4P1 | FALSE | Upregulated |
| 0,724794291 | 0,459623913 | CLCN2 | FALSE | Upregulated |
| -0,373926035 | 0,459623913 | PGAM1P5 | FALSE | Downregulated |
| -1,597857699 | 0,459769865 | LOC100507316 | FALSE | Downregulated |
| -0,921671857 | 0,459817693 | PARD6A | FALSE | Downregulated |
| -0,364400363 | 0,459817693 | SEC14L4 | FALSE | Downregulated |
| 0,340791406 | 0,459843574 | ABI1 | FALSE | Upregulated |
| 0,764318612 | 0,459843574 | AMICA1 | FALSE | Upregulated |
| 0,610281813 | 0,459843574 | C17orf59 | FALSE | Upregulated |
| 0,399226072 | 0,459843574 | LOC101928552 | FALSE | Upregulated |
| -0,414036234 | 0,459843574 | ZNF292 | FALSE | Downregulated |
| 0,301104153 | 0,459868135 | MFHAS1 | FALSE | Upregulated |
| 0,620297009 | 0,460274323 | LMOD1 | FALSE | Upregulated |
| 1,28220805 | 0,460274323 | LOC102724867 | FALSE | Upregulated |
| -0,624130118 | 0,460345147 | MED4 | FALSE | Downregulated |
| 1,152774242 | 0,460496787 | GPR171 | FALSE | Upregulated |
| 0,286096413 | 0,460496787 | LOC102724391 | FALSE | Upregulated |
| 0,453685695 | 0,460496787 | PTGFRN | FALSE | Upregulated |
| 0,312686131 | 0,460555039 | LBX2-AS1 | FALSE | Upregulated |
| -0,28584508 | 0,460645161 | ZFPM1 | FALSE | Downregulated |
| 0,920234504 | 0,460810001 | CACNA1G-AS1 | FALSE | Upregulated |
| 1,179849794 | 0,460810001 | HIST1H2BG | FALSE | Upregulated |
| 0,317166898 | 0,46088213 | AKT3 | FALSE | Upregulated |
| -1,182620786 | 0,46088213 | EHMT1-IT1 | FALSE | Downregulated |
| -0,411377676 | 0,46112209 | DHX33 | FALSE | Downregulated |

| | | | | |
|---|---|---|---|---|
| 0,342359084 | 0,46112209 | LOC100505767 | FALSE | Upregulated |
| 0,670852543 | 0,46112209 | LOC102724782 | FALSE | Upregulated |
| 0,663288487 | 0,46112209 | PKN3 | FALSE | Upregulated |
| -1,55193377 | 0,46112209 | SUSD4 | FALSE | Downregulated |
| 0,273377429 | 0,461616042 | RB1CC1 | FALSE | Upregulated |
| 1,040226236 | 0,461707273 | NRXN2 | FALSE | Upregulated |
| 0,292441965 | 0,461809501 | JOSD1 | FALSE | Upregulated |
| 0,781851426 | 0,461809501 | PPP1R2 | FALSE | Upregulated |
| 0,404546684 | 0,461809501 | RP11-31F15.2 | FALSE | Upregulated |
| 0,670545708 | 0,462458872 | C16orf52 | FALSE | Upregulated |
| -1,110298107 | 0,462458872 | KIF1A | FALSE | Downregulated |
| -0,48900526 | 0,462458872 | LOC101929792 | FALSE | Downregulated |
| 0,236252749 | 0,462458872 | TCF25 | FALSE | Upregulated |
| 0,351876658 | 0,4626169 | INPP5K | FALSE | Upregulated |
| 0,637279823 | 0,4626169 | RP11-346I3.4 | FALSE | Upregulated |
| 0,217163353 | 0,4626169 | TXLNA | FALSE | Upregulated |
| 0,602304037 | 0,462713008 | PTK6 | FALSE | Upregulated |
| 0,391238135 | 0,462713008 | ZNF829 | FALSE | Upregulated |
| 1,373140015 | 0,463354304 | HIST1H2AL | FALSE | Upregulated |
| 0,296921988 | 0,463354304 | MLANA | FALSE | Upregulated |
| -0,427135473 | 0,463354304 | RABEPK | FALSE | Downregulated |
| 1,090965165 | 0,463564349 | LOC728819 | FALSE | Upregulated |
| -1,180256464 | 0,463564349 | SHD | FALSE | Downregulated |
| 0,600029663 | 0,463667803 | DTYMK | FALSE | Upregulated |
| 1,068444882 | 0,463667803 | LOC100129534 | FALSE | Upregulated |
| 0,29280415 | 0,463667803 | ZBTB5 | FALSE | Upregulated |
| -0,629824827 | 0,463814939 | TIMM10 | FALSE | Downregulated |
| -1,154249698 | 0,463915202 | C12orf45 | FALSE | Downregulated |
| -0,656349319 | 0,463915202 | CARF | FALSE | Downregulated |
| 0,622633194 | 0,463949401 | LOC101928466 | FALSE | Upregulated |
| -0,325599446 | 0,463949401 | TAF15 | FALSE | Downregulated |
| 0,512466675 | 0,46421361 | LRRC8B | FALSE | Upregulated |
| 0,506819369 | 0,464260343 | CTSC | FALSE | Upregulated |
| -0,275573497 | 0,464260343 | DLG1 | FALSE | Downregulated |
| 0,45176236 | 0,464434691 | RASA3 | FALSE | Upregulated |
| -0,354086989 | 0,464543009 | LTBP1 | FALSE | Downregulated |
| 0,269133222 | 0,464786186 | ALS2 | FALSE | Upregulated |
| -0,674441542 | 0,464940687 | RPUSD3 | FALSE | Downregulated |
| 0,643070888 | 0,465497895 | PLEKHA8 | FALSE | Upregulated |
| 0,680763417 | 0,465580916 | C7orf31 | FALSE | Upregulated |
| -1,453822014 | 0,465580916 | CBLN4 | FALSE | Downregulated |
| 0,242422853 | 0,465580916 | LOC101927509 | FALSE | Upregulated |
| -0,272606576 | 0,465580916 | SSU72 | FALSE | Downregulated |
| 0,362227331 | 0,465580916 | SULF2 | FALSE | Upregulated |
| 0,348415393 | 0,465580916 | ZSCAN16 | FALSE | Upregulated |
| 0,501233388 | 0,46563402 | SPRY1 | FALSE | Upregulated |
| -0,430025323 | 0,466137219 | B9D1 | FALSE | Downregulated |
| -1,158481567 | 0,46629166 | STAG3L4 | FALSE | Downregulated |
| 0,81714241 | 0,466383809 | LOC102724938 | FALSE | Upregulated |
| -0,457296407 | 0,467107851 | CUTA | FALSE | Downregulated |
| 1,257300056 | 0,467107851 | NR2F2-AS1 | FALSE | Upregulated |

| | | | | |
|---|---|---|---|---|
| 0,429524291 | 0,467197874 | ANKS1B | FALSE | Upregulated |
| -0,47719066 | 0,467197874 | DNPH1 | FALSE | Downregulated |
| 0,381644846 | 0,467197874 | IGFBP5 | FALSE | Upregulated |
| 0,476161888 | 0,467197874 | MFAP3 | FALSE | Upregulated |
| 0,421280707 | 0,467197874 | SLA2 | FALSE | Upregulated |
| 0,827449153 | 0,467207486 | LOC101928289 | FALSE | Upregulated |
| -0,266037496 | 0,467207486 | PLEKHA6 | FALSE | Downregulated |
| -0,215582916 | 0,467240963 | PRKACA | FALSE | Downregulated |
| -0,521895733 | 0,467671647 | SYNJ1 | FALSE | Downregulated |
| -0,776756034 | 0,46767677 | SSTR2 | FALSE | Downregulated |
| -0,619699419 | 0,467824867 | GPALPP1 | FALSE | Downregulated |
| -0,226578564 | 0,467888358 | BRD2 | FALSE | Downregulated |
| 0,536849279 | 0,467888358 | LOC101928206 | FALSE | Upregulated |
| 0,578156135 | 0,467888358 | OR10AD1 | FALSE | Upregulated |
| 0,345225605 | 0,467888358 | RPS6KL1 | FALSE | Upregulated |
| 0,481746049 | 0,467888358 | SETDB2 | FALSE | Upregulated |
| 0,222820618 | 0,467888358 | TRIM14 | FALSE | Upregulated |
| -0,451534153 | 0,468375074 | FAM198A | FALSE | Downregulated |
| 0,316108934 | 0,468375074 | KIAA0247 | FALSE | Upregulated |
| -1,388759577 | 0,468375074 | LOC102723799 | FALSE | Downregulated |
| -0,375160631 | 0,468375074 | MS4A6A | FALSE | Downregulated |
| 0,406791839 | 0,468375074 | MTMR14 | FALSE | Upregulated |
| 0,236298092 | 0,468691795 | CCAR2 | FALSE | Upregulated |
| -0,300791727 | 0,468691795 | EEA1 | FALSE | Downregulated |
| -0,38514894 | 0,468957006 | HNMT | FALSE | Downregulated |
| -0,426017244 | 0,469411188 | AADAT | FALSE | Downregulated |
| -0,517464936 | 0,469411188 | ATHL1 | FALSE | Downregulated |
| -0,392211179 | 0,469411188 | ATP5H | FALSE | Downregulated |
| 0,390564599 | 0,469411188 | ATP5J | FALSE | Upregulated |
| 0,56361134 | 0,469411188 | ATP6V1F | FALSE | Upregulated |
| 0,568950641 | 0,469411188 | CCDC68 | FALSE | Upregulated |
| 0,324557956 | 0,469411188 | CEP57 | FALSE | Upregulated |
| 0,346283295 | 0,469411188 | CIRBP-AS1 | FALSE | Upregulated |
| 0,681055368 | 0,469411188 | GEMIN2 | FALSE | Upregulated |
| 0,44066523 | 0,469411188 | GPR137B | FALSE | Upregulated |
| 0,868151648 | 0,469411188 | HAVCR2 | FALSE | Upregulated |
| 0,619248547 | 0,469411188 | HLA-DQB1 | FALSE | Upregulated |
| 0,361511279 | 0,469411188 | KMT2E-AS1 | FALSE | Upregulated |
| 1,988929006 | 0,469411188 | LOC101928344 | FALSE | Upregulated |
| 0,474617903 | 0,469411188 | LPP-AS1 | FALSE | Upregulated |
| -0,990280227 | 0,469411188 | NXPH4 | FALSE | Downregulated |
| 0,646493308 | 0,469411188 | POLR1C | FALSE | Upregulated |
| 0,456757983 | 0,469411188 | PRORSD1P | FALSE | Upregulated |
| 0,654749048 | 0,469411188 | RIMKLB | FALSE | Upregulated |
| 0,929144009 | 0,469411188 | RP11-264C15.2 | FALSE | Upregulated |
| 0,480063066 | 0,469411188 | SIPA1L2 | FALSE | Upregulated |
| -0,243842768 | 0,469411188 | SND1 | FALSE | Downregulated |
| -0,330017283 | 0,469411188 | TGFBR3L | FALSE | Downregulated |
| -0,412017576 | 0,469411188 | TMEM183A | FALSE | Downregulated |
| -0,764447802 | 0,469411188 | TSTD1 | FALSE | Downregulated |
| 0,339669914 | 0,469411188 | TTI2 | FALSE | Upregulated |

| | | | | |
|---|---|---|---|---|
| -0,335832625 | 0,469411188 | UBE2E1 | FALSE | Downregulated |
| 0,950676981 | 0,469411188 | ZNF354B | FALSE | Upregulated |
| 0,564071101 | 0,469497432 | LINC01126 | FALSE | Upregulated |
| -1,278689375 | 0,469600178 | ZNF415 | FALSE | Downregulated |
| -1,428264018 | 0,469619447 | PRIM2 | FALSE | Downregulated |
| -0,352773353 | 0,469784886 | DPP7 | FALSE | Downregulated |
| 0,618583797 | 0,469948824 | EML2 | FALSE | Upregulated |
| -0,270232868 | 0,469948824 | SKIV2L | FALSE | Downregulated |
| 0,88813252 | 0,470428446 | ASTN2 | FALSE | Upregulated |
| -0,442628638 | 0,470428446 | SSH2 | FALSE | Downregulated |
| 0,333719538 | 0,470445586 | SNX30 | FALSE | Upregulated |
| 0,379116918 | 0,470495064 | CREB3 | FALSE | Upregulated |
| 0,455871241 | 0,470522684 | IGSF23 | FALSE | Upregulated |
| -0,234097521 | 0,470654461 | PEX19 | FALSE | Downregulated |
| -0,493053012 | 0,470654461 | ZNF259 | FALSE | Downregulated |
| 0,50931632 | 0,470898526 | TTC30B | FALSE | Upregulated |
| 0,311272123 | 0,471271029 | AIMP1 | FALSE | Upregulated |
| -1,180576338 | 0,471271029 | AK6 | FALSE | Downregulated |
| 0,581779274 | 0,471271029 | BTBD3 | FALSE | Upregulated |
| -0,184641317 | 0,471271029 | CTDSP1 | FALSE | Downregulated |
| 0,472690767 | 0,471271029 | FIP1L1 | FALSE | Upregulated |
| 0,813448305 | 0,471271029 | LOC102724548 | FALSE | Upregulated |
| -0,445188685 | 0,471271029 | LOC389906 | FALSE | Downregulated |
| -0,810754289 | 0,471271029 | RNF113A | FALSE | Downregulated |
| -0,899975615 | 0,471271029 | RNF208 | FALSE | Downregulated |
| -0,429928602 | 0,471271029 | SEPT4 | FALSE | Downregulated |
| 0,32757135 | 0,471271029 | SPCS3 | FALSE | Upregulated |
| 0,792908396 | 0,471271029 | SPECC1 | FALSE | Upregulated |
| 0,616840804 | 0,471271029 | TRPV2 | FALSE | Upregulated |
| -0,594557537 | 0,471271029 | UCK2 | FALSE | Downregulated |
| 0,671089814 | 0,471313465 | ACOT9 | FALSE | Upregulated |
| 0,393841194 | 0,471313465 | KLHL12 | FALSE | Upregulated |
| -0,24282371 | 0,471502358 | TPR | FALSE | Downregulated |
| 1,351217197 | 0,471712898 | GOLGA8S | FALSE | Upregulated |
| -0,448853611 | 0,471712898 | UBE2N | FALSE | Downregulated |
| 0,271221161 | 0,472052158 | ZNF552 | FALSE | Upregulated |
| 0,392787181 | 0,472072063 | LOC102724672 | FALSE | Upregulated |
| -0,208664396 | 0,472228646 | MAN2A2 | FALSE | Downregulated |
| 1,095107817 | 0,472384017 | DKFZP434I0714 | FALSE | Upregulated |
| -0,903179732 | 0,472384017 | LOC102723864 | FALSE | Downregulated |
| 0,487796159 | 0,472384017 | PPP2R3C | FALSE | Upregulated |
| -0,340898736 | 0,472384017 | RBBP6 | FALSE | Downregulated |
| -0,346065846 | 0,472384017 | YIPF3 | FALSE | Downregulated |
| -0,779014893 | 0,472452173 | MIPEP | FALSE | Downregulated |
| -1,130686243 | 0,47257504 | LINC00167 | FALSE | Downregulated |
| 0,217103904 | 0,472613346 | LOC100507577 | FALSE | Upregulated |
| 0,462240479 | 0,472613346 | U2AF1 | FALSE | Upregulated |
| -0,275599732 | 0,47265753 | DDX54 | FALSE | Downregulated |
| 0,345259134 | 0,47265753 | KIAA0226 | FALSE | Upregulated |
| 3,843909575 | 0,47265753 | MYBPC2 | FALSE | Upregulated |
| -0,413507376 | 0,47265753 | RRNAD1 | FALSE | Downregulated |

| | | | | |
|---|---|---|---|---|
| 0,251285747 | 0,473104725 | ITPRIPL2 | FALSE | Upregulated |
| -0,628557176 | 0,473163457 | RRP8 | FALSE | Downregulated |
| 0,473825452 | 0,473163457 | TMEM139 | FALSE | Upregulated |
| 0,370891942 | 0,473175582 | RNF4 | FALSE | Upregulated |
| -0,234098723 | 0,473238842 | ANKRD52 | FALSE | Downregulated |
| -0,743939794 | 0,473238842 | BLZF1 | FALSE | Downregulated |
| 0,6611072 | 0,473238842 | CXorf23 | FALSE | Upregulated |
| -0,359584033 | 0,473238842 | DERL1 | FALSE | Downregulated |
| 0,295956879 | 0,473238842 | FLJ31945 | FALSE | Upregulated |
| -0,88792964 | 0,473238842 | FOXF1 | FALSE | Downregulated |
| -0,209007787 | 0,473238842 | KIAA0319L | FALSE | Downregulated |
| 0,472028719 | 0,473238842 | LOC101930171 | FALSE | Upregulated |
| 0,37240618 | 0,473238842 | MUC6 | FALSE | Upregulated |
| -0,273218365 | 0,473238842 | NBPF14 | FALSE | Downregulated |
| -0,324001176 | 0,473238842 | PSMD8 | FALSE | Downregulated |
| 1,339833996 | 0,473238842 | RP11-17A4.2 | FALSE | Upregulated |
| 0,60563906 | 0,473238842 | RP11-506O24.1 | FALSE | Upregulated |
| -0,684018013 | 0,473238842 | THUMPD3-AS1 | FALSE | Downregulated |
| 0,166460147 | 0,473238842 | TMEM123 | FALSE | Upregulated |
| 0,390126622 | 0,473238842 | UBAP2 | FALSE | Upregulated |
| -0,490400552 | 0,473350553 | MLST8 | FALSE | Downregulated |
| 0,627015161 | 0,473404208 | TVP23C | FALSE | Upregulated |
| -1,286277998 | 0,473532582 | CD99P1 | FALSE | Downregulated |
| 0,302718939 | 0,473577327 | C9orf84 | FALSE | Upregulated |
| 0,424761433 | 0,473577327 | ZNHIT2 | FALSE | Upregulated |
| -0,877597381 | 0,473716713 | C17orf96 | FALSE | Downregulated |
| -0,1791585 | 0,473824535 | CTBP1 | FALSE | Downregulated |
| -0,427995085 | 0,473851069 | FRAT2 | FALSE | Downregulated |
| 0,564408441 | 0,473851069 | ULK4 | FALSE | Upregulated |
| 0,584372339 | 0,473881965 | BAALC | FALSE | Upregulated |
| 0,333005552 | 0,473881965 | FAM118B | FALSE | Upregulated |
| -0,987808302 | 0,473881965 | KRTCAP2 | FALSE | Downregulated |
| -0,130400744 | 0,474038072 | LINC00657 | FALSE | Downregulated |
| 0,662319638 | 0,474038072 | LOC102723798 | FALSE | Upregulated |
| 0,311996474 | 0,474038072 | PPP1R37 | FALSE | Upregulated |
| 0,260908625 | 0,474533294 | C9orf131 | FALSE | Upregulated |
| 1,426969472 | 0,475016234 | C5orf20 | FALSE | Upregulated |
| 0,231307638 | 0,475016234 | DENND4B | FALSE | Upregulated |
| 0,430789699 | 0,475637262 | ARAP3 | FALSE | Upregulated |
| 0,703971611 | 0,475637262 | HIPK4 | FALSE | Upregulated |
| 0,27963803 | 0,475637262 | ILK | FALSE | Upregulated |
| -0,52385236 | 0,475713438 | PAXIP1 | FALSE | Downregulated |
| 1,088254538 | 0,476178107 | CD58 | FALSE | Upregulated |
| 0,286733791 | 0,476178107 | LAMP2 | FALSE | Upregulated |
| 0,49246193 | 0,476178107 | TUSC2 | FALSE | Upregulated |
| -1,200998608 | 0,476269287 | HAUS8 | FALSE | Downregulated |
| 0,155837145 | 0,476359241 | CLTC | FALSE | Upregulated |
| 0,402278633 | 0,476359241 | FILIP1 | FALSE | Upregulated |
| -0,280787996 | 0,476359241 | INVS | FALSE | Downregulated |
| 0,299977917 | 0,476359241 | LEPROT | FALSE | Upregulated |
| 0,497171028 | 0,476359241 | PGM5P2 | FALSE | Upregulated |

| | | | | |
|---|---|---|---|---|
| -0,374658273 | 0,476359241 | PTEN | FALSE | Downregulated |
| 0,382240177 | 0,476400214 | XAF1 | FALSE | Upregulated |
| -0,286384082 | 0,476458331 | ALDH9A1 | FALSE | Downregulated |
| 0,467450438 | 0,476529567 | CCDC12 | FALSE | Upregulated |
| -0,296302324 | 0,476994394 | FAM210B | FALSE | Downregulated |
| -0,212850415 | 0,476994394 | FZD5 | FALSE | Downregulated |
| 0,607863462 | 0,477059792 | TRIL | FALSE | Upregulated |
| 0,258744002 | 0,477069454 | ZNF140 | FALSE | Upregulated |
| -0,468073322 | 0,477446595 | ESD | FALSE | Downregulated |
| 1,244295183 | 0,477624689 | GAL3ST4 | FALSE | Upregulated |
| 0,408232585 | 0,477624689 | KIAA1407 | FALSE | Upregulated |
| -0,464811417 | 0,477624689 | SSNA1 | FALSE | Downregulated |
| 0,319491364 | 0,477790758 | C19orf10 | FALSE | Upregulated |
| -0,848205792 | 0,477819983 | XKR9 | FALSE | Downregulated |
| 0,213384061 | 0,477834565 | FRMD4A | FALSE | Upregulated |
| -0,282519883 | 0,478130987 | LUC7L3 | FALSE | Downregulated |
| -0,26844548 | 0,478229305 | LRRC47 | FALSE | Downregulated |
| -0,465888812 | 0,478487727 | BLOC1S6 | FALSE | Downregulated |
| 0,922514495 | 0,478545717 | C11orf21 | FALSE | Upregulated |
| 0,551293678 | 0,478545717 | EIF1AD | FALSE | Upregulated |
| 0,305866054 | 0,478545717 | LBR | FALSE | Upregulated |
| 1,437418372 | 0,478545717 | MSH5-SAPCD1 | FALSE | Upregulated |
| 0,50040375 | 0,478545717 | RNF25 | FALSE | Upregulated |
| -0,258852789 | 0,478545717 | SETD1A | FALSE | Downregulated |
| 1,226282028 | 0,478601526 | ASB2 | FALSE | Upregulated |
| -0,291306975 | 0,47873795 | TAB3 | FALSE | Downregulated |
| -0,286190801 | 0,47884111 | AK4 | FALSE | Downregulated |
| 0,947162673 | 0,47884111 | F3 | FALSE | Upregulated |
| -0,95791498 | 0,47884111 | KCNJ3 | FALSE | Downregulated |
| -0,252381654 | 0,47884111 | RBM10 | FALSE | Downregulated |
| -0,411581 | 0,479018079 | IMPA1 | FALSE | Downregulated |
| 0,484271806 | 0,479018079 | LOC101928517 | FALSE | Upregulated |
| 0,38741667 | 0,479108907 | IL18BP | FALSE | Upregulated |
| -1,260408116 | 0,479146354 | ABCC6P2 | FALSE | Downregulated |
| 0,189320087 | 0,479423256 | HNRNPK | FALSE | Upregulated |
| -0,384381586 | 0,479632523 | EMC10 | FALSE | Downregulated |
| 0,549707609 | 0,479651512 | RRAS | FALSE | Upregulated |
| 0,456309488 | 0,479978752 | CNGA1 | FALSE | Upregulated |
| 0,439923664 | 0,47999308 | FAM185A | FALSE | Upregulated |
| -0,222943673 | 0,479995671 | ESRP2 | FALSE | Downregulated |
| 0,238683318 | 0,479995671 | LOC102723521 | FALSE | Upregulated |
| -0,2019688 | 0,479995671 | TCEB3 | FALSE | Downregulated |
| 0,427167102 | 0,480066562 | DKFZP434H168 | FALSE | Upregulated |
| 0,358310719 | 0,480178333 | CDK11B | FALSE | Upregulated |
| 3,628222282 | 0,480202449 | ACTN2 | FALSE | Upregulated |
| 0,731349123 | 0,480202449 | CYP2B7P | FALSE | Upregulated |
| 0,349319669 | 0,480202449 | KIF2A | FALSE | Upregulated |
| -0,491527808 | 0,480202449 | TXNRD3 | FALSE | Downregulated |
| -1,716793834 | 0,480507506 | RP11-534N16.1 | FALSE | Downregulated |
| 1,016677392 | 0,480587813 | ADPRH | FALSE | Upregulated |
| -0,322166819 | 0,480587813 | CHCHD2 | FALSE | Downregulated |

| | | | | |
|---|---|---|---|---|
| 0,372873475 | 0,480587813 | HLTF | FALSE | Upregulated |
| -0,215573262 | 0,480587813 | MLEC | FALSE | Downregulated |
| 0,238488854 | 0,480587813 | NAPEPLD | FALSE | Upregulated |
| 0,829031494 | 0,480818394 | USP17L7 | FALSE | Upregulated |
| -0,693754709 | 0,480840395 | QRSL1 | FALSE | Downregulated |
| 0,51149178 | 0,480840395 | SLC35C1 | FALSE | Upregulated |
| 1,219479081 | 0,480840395 | STAP1 | FALSE | Upregulated |
| -0,992936466 | 0,4810031 | KCNJ2 | FALSE | Downregulated |
| -0,566837492 | 0,48122966 | ZG16 | FALSE | Downregulated |
| 0,263212453 | 0,481476545 | EIF3I | FALSE | Upregulated |
| 0,417640924 | 0,481476545 | LOC101927685 | FALSE | Upregulated |
| -1,00281117 | 0,481718732 | FADS6 | FALSE | Downregulated |
| -0,190634857 | 0,48183181 | EPAS1 | FALSE | Downregulated |
| 0,179658818 | 0,481838775 | RBM15B | FALSE | Upregulated |
| 0,977806838 | 0,481916199 | KLRK1 | FALSE | Upregulated |
| -0,517552306 | 0,481916199 | LOC101927790 | FALSE | Downregulated |
| -0,769709546 | 0,481916199 | MED28 | FALSE | Downregulated |
| 0,306332313 | 0,481916199 | ZEB2 | FALSE | Upregulated |
| -0,357578544 | 0,481977146 | MAGED2 | FALSE | Downregulated |
| -0,223783146 | 0,481990871 | RPL7 | FALSE | Downregulated |
| -0,389313299 | 0,482142978 | RNF38 | FALSE | Downregulated |
| 1,04002807 | 0,482177774 | YJEFN3 | FALSE | Upregulated |
| 0,647957237 | 0,482177774 | ZNF701 | FALSE | Upregulated |
| -0,411230535 | 0,482223813 | ABCB10 | FALSE | Downregulated |
| -0,420958519 | 0,482250924 | RASGEF1B | FALSE | Downregulated |
| 0,465135684 | 0,482386068 | CYTH4 | FALSE | Upregulated |
| 0,248806374 | 0,482386068 | UBA5 | FALSE | Upregulated |
| -1,054783676 | 0,482410092 | TEN1-CDK3 | FALSE | Downregulated |
| 0,517499188 | 0,482445566 | ANO1-AS2 | FALSE | Upregulated |
| 0,283457084 | 0,482445566 | NOTCH2NL | FALSE | Upregulated |
| 0,44281009 | 0,482459431 | ZNF37A | FALSE | Upregulated |
| -0,449347019 | 0,482502773 | DPP4 | FALSE | Downregulated |
| -0,384281725 | 0,482558801 | FAM214A | FALSE | Downregulated |
| -0,272270697 | 0,482558801 | PSMC3 | FALSE | Downregulated |
| 0,873238372 | 0,482589526 | FBXO5 | FALSE | Upregulated |
| -0,219017991 | 0,482589526 | KIAA2018 | FALSE | Downregulated |
| -0,39568751 | 0,482695756 | AKR1CL1 | FALSE | Downregulated |
| 0,206774124 | 0,482695756 | GTPBP1 | FALSE | Upregulated |
| 0,824104173 | 0,482695756 | LOC102724528 | FALSE | Upregulated |
| -0,55579172 | 0,482695756 | PRRG4 | FALSE | Downregulated |
| -0,790109876 | 0,482695756 | RND2 | FALSE | Downregulated |
| 0,393724305 | 0,482695756 | SNAI3 | FALSE | Upregulated |
| 0,271857674 | 0,482695756 | ZNF672 | FALSE | Upregulated |
| -0,820656613 | 0,482712751 | ASB16-AS1 | FALSE | Downregulated |
| -0,513776227 | 0,482716187 | FBXL6 | FALSE | Downregulated |
| -0,247867814 | 0,482760972 | SNX9 | FALSE | Downregulated |
| -0,481992538 | 0,48277393 | MAP3K9 | FALSE | Downregulated |
| -0,247044501 | 0,482784758 | KAT2B | FALSE | Downregulated |
| -0,736257089 | 0,482908535 | CBWD2 | FALSE | Downregulated |
| 0,818267296 | 0,483041444 | LRRC37B | FALSE | Upregulated |
| 0,712952836 | 0,483458667 | SPATA9 | FALSE | Upregulated |

| | | | | |
|---|---|---|---|---|
| 1,507691876 | 0,483477047 | FUNDC2P2 | FALSE | Upregulated |
| -0,51152328 | 0,483477047 | NDUFA3 | FALSE | Downregulated |
| 0,510871423 | 0,483477047 | NRF1 | FALSE | Upregulated |
| -0,428041894 | 0,483591276 | TSTA3 | FALSE | Downregulated |
| -0,617079848 | 0,483621646 | TMEM161A | FALSE | Downregulated |
| 0,446020232 | 0,483762254 | TNIK | FALSE | Upregulated |
| -0,858110998 | 0,483818333 | LOC101928997 | FALSE | Downregulated |
| 0,779082935 | 0,483837515 | GPR114 | FALSE | Upregulated |
| 0,479103815 | 0,484311693 | ANAPC7 | FALSE | Upregulated |
| -0,569353962 | 0,484311693 | CDK2 | FALSE | Downregulated |
| -0,312710534 | 0,484340742 | DSG2 | FALSE | Downregulated |
| 1,067119129 | 0,484340742 | DUSP4 | FALSE | Upregulated |
| 0,60504198 | 0,484340742 | EFR3B | FALSE | Upregulated |
| 0,649744236 | 0,484340742 | FAM213B | FALSE | Upregulated |
| 0,635891234 | 0,484340742 | FAM78B | FALSE | Upregulated |
| -0,497452439 | 0,484340742 | L2HGDH | FALSE | Downregulated |
| 0,348538814 | 0,484340742 | PDE4DIP | FALSE | Upregulated |
| -0,354123762 | 0,484340742 | PUS3 | FALSE | Downregulated |
| -0,224292287 | 0,484340742 | STARD7 | FALSE | Downregulated |
| -0,218068668 | 0,484351179 | FBXW5 | FALSE | Downregulated |
| 0,327164294 | 0,484427218 | HNRNPR | FALSE | Upregulated |
| 0,328056321 | 0,484531117 | TP73 | FALSE | Upregulated |
| -0,278899764 | 0,484583844 | CKAP5 | FALSE | Downregulated |
| -0,155228684 | 0,484583844 | EIF4G1 | FALSE | Downregulated |
| -0,594550103 | 0,484583844 | FAM118A | FALSE | Downregulated |
| 1,119203565 | 0,484583844 | LINC00353 | FALSE | Upregulated |
| -0,719111987 | 0,484583844 | NRARP | FALSE | Downregulated |
| 0,640083716 | 0,484583844 | RHD | FALSE | Upregulated |
| 0,795063712 | 0,484583844 | SSSCA1-AS1 | FALSE | Upregulated |
| -0,819519855 | 0,484719209 | DHRS4L1 | FALSE | Downregulated |
| -0,615185609 | 0,484793249 | CRB3 | FALSE | Downregulated |
| 0,631008096 | 0,484793249 | DZIP1L | FALSE | Upregulated |
| 0,562717704 | 0,484793249 | HCRTR1 | FALSE | Upregulated |
| -0,877944414 | 0,484793249 | HDC | FALSE | Downregulated |
| -0,70371929 | 0,484793249 | HTR7P1 | FALSE | Downregulated |
| 0,235687718 | 0,484793249 | KDSR | FALSE | Upregulated |
| 0,243006669 | 0,484793249 | LOC100130093 | FALSE | Upregulated |
| -0,393065567 | 0,484793249 | LOC100130872 | FALSE | Downregulated |
| 0,300871882 | 0,484793249 | LSMEM2 | FALSE | Upregulated |
| 0,639872573 | 0,484793249 | MFI2 | FALSE | Upregulated |
| -1,231615403 | 0,484793249 | PSTK | FALSE | Downregulated |
| 1,138215451 | 0,484793249 | ZNF804A | FALSE | Upregulated |
| 0,357576909 | 0,48497821 | B4GALT5 | FALSE | Upregulated |
| -0,6694553 | 0,48497821 | ELP6 | FALSE | Downregulated |
| -0,301389842 | 0,485064042 | NOTCH4 | FALSE | Downregulated |
| 0,783787627 | 0,485176416 | APH1B | FALSE | Upregulated |
| 0,285217785 | 0,485176416 | BBX | FALSE | Upregulated |
| 0,288599684 | 0,485176416 | SRP19 | FALSE | Upregulated |
| -0,354386337 | 0,48538374 | COLEC10 | FALSE | Downregulated |
| 1,808294469 | 0,485519278 | LOC101929901 | FALSE | Upregulated |
| -0,377009352 | 0,485709712 | LINC00538 | FALSE | Downregulated |

| | | | | |
|---|---|---|---|---|
| -0,18802961 | 0,485724214 | GNAI2 | FALSE | Downregulated |
| 0,547769596 | 0,485842213 | KDELC2 | FALSE | Upregulated |
| -0,77120451 | 0,486088519 | COL4A5 | FALSE | Downregulated |
| -0,820951848 | 0,486088519 | SLC26A6 | FALSE | Downregulated |
| 0,4743991 | 0,486487667 | KLF16 | FALSE | Upregulated |
| -0,38086187 | 0,486648638 | FASTK | FALSE | Downregulated |
| -0,356406838 | 0,486648638 | HEATR5B | FALSE | Downregulated |
| -0,386821115 | 0,48671411 | NDRG4 | FALSE | Downregulated |
| -0,24195338 | 0,48671411 | ZFAND3 | FALSE | Downregulated |
| -0,383261161 | 0,486774915 | UBQLN4 | FALSE | Downregulated |
| 0,304074724 | 0,486890912 | ATG16L2 | FALSE | Upregulated |
| 0,68864753 | 0,486922319 | C16orf93 | FALSE | Upregulated |
| 0,603930227 | 0,486967473 | ZFP90 | FALSE | Upregulated |
| -0,839580186 | 0,487262392 | CBLN3 | FALSE | Downregulated |
| -0,520554965 | 0,487346685 | CDK6 | FALSE | Downregulated |
| -0,431455908 | 0,487462982 | CEP63 | FALSE | Downregulated |
| 0,600251119 | 0,487570097 | ACSBG2 | FALSE | Upregulated |
| -0,219782797 | 0,487571342 | RPL6 | FALSE | Downregulated |
| 0,347860772 | 0,487578759 | ACTL10 | FALSE | Upregulated |
| 0,186365593 | 0,487578759 | GSPT1 | FALSE | Upregulated |
| 0,984977983 | 0,487578759 | IL34 | FALSE | Upregulated |
| 0,775859496 | 0,487720749 | IL21R | FALSE | Upregulated |
| -0,305469516 | 0,487720749 | N4BP2 | FALSE | Downregulated |
| 0,608122315 | 0,487839424 | GTF2A2 | FALSE | Upregulated |
| 0,259298838 | 0,48791937 | CCRN4L | FALSE | Upregulated |
| 0,42987995 | 0,48791937 | EFHC1 | FALSE | Upregulated |
| -0,398302143 | 0,48791937 | FGL1 | FALSE | Downregulated |
| 1,203539125 | 0,48791937 | LOC101927249 | FALSE | Upregulated |
| 0,326450697 | 0,48791937 | LOC102723520 | FALSE | Upregulated |
| 0,646115673 | 0,48791937 | MRAS | FALSE | Upregulated |
| 0,538964434 | 0,48791937 | POMK | FALSE | Upregulated |
| 0,238229473 | 0,48791937 | SEPN1 | FALSE | Upregulated |
| -0,256483112 | 0,48791937 | ZER1 | FALSE | Downregulated |
| 0,788623375 | 0,488321135 | PRIM1 | FALSE | Upregulated |
| -0,32365281 | 0,488402487 | RNF216 | FALSE | Downregulated |
| -0,392407185 | 0,488481965 | TMEM106B | FALSE | Downregulated |
| 1,262234846 | 0,488614712 | AC092295.7 | FALSE | Upregulated |
| -0,857832623 | 0,488614712 | ALKBH2 | FALSE | Downregulated |
| -0,366483811 | 0,488614712 | CRCP | FALSE | Downregulated |
| -1,051459201 | 0,488614922 | PDE6D | FALSE | Downregulated |
| 0,849144099 | 0,488668989 | LOC101927952 | FALSE | Upregulated |
| 0,425827723 | 0,488680481 | DPY19L2P3 | FALSE | Upregulated |
| -0,745803958 | 0,488680481 | LOC101928058 | FALSE | Downregulated |
| 0,565076534 | 0,488745019 | LRRC15 | FALSE | Upregulated |
| 0,498028094 | 0,488798674 | TMA7 | FALSE | Upregulated |
| -0,994280171 | 0,488798674 | UTP23 | FALSE | Downregulated |
| 0,368743797 | 0,488897929 | NAA20 | FALSE | Upregulated |
| -0,176600433 | 0,488915219 | ACO2 | FALSE | Downregulated |
| 0,848607149 | 0,488915219 | CCDC23 | FALSE | Upregulated |
| 0,765827747 | 0,488915219 | CCDC40 | FALSE | Upregulated |
| -0,319836953 | 0,488915219 | CHPT1 | FALSE | Downregulated |

| | | | | |
|---|---|---|---|---|
| 1,397163771 | 0,488915219 | GZMA | FALSE | Upregulated |
| 0,789534559 | 0,488915219 | KCNH7 | FALSE | Upregulated |
| -0,379181934 | 0,488915219 | LOC100996724 | FALSE | Downregulated |
| -1,378513633 | 0,488915219 | LOC101927279 | FALSE | Downregulated |
| 0,775835458 | 0,488915219 | LOC102724163 | FALSE | Upregulated |
| -1,291044378 | 0,488915219 | LOC102724483 | FALSE | Downregulated |
| 0,494972288 | 0,488915219 | LRRC57 | FALSE | Upregulated |
| 0,304107342 | 0,488915219 | MRPL37 | FALSE | Upregulated |
| 0,793152585 | 0,488915219 | NTRK2 | FALSE | Upregulated |
| -0,30140327 | 0,488915219 | ORAI3 | FALSE | Downregulated |
| -0,466869597 | 0,488915219 | POLR2J | FALSE | Downregulated |
| 0,597819639 | 0,488915219 | RPP25 | FALSE | Upregulated |
| 0,514881344 | 0,488915219 | SPDYE7P | FALSE | Upregulated |
| -0,209935272 | 0,488915219 | SRPR | FALSE | Downregulated |
| 0,437369801 | 0,488915219 | STK4 | FALSE | Upregulated |
| 0,561379717 | 0,488915219 | ZNF703 | FALSE | Upregulated |
| -0,419922018 | 0,489351366 | TSEN54 | FALSE | Downregulated |
| -0,247466269 | 0,489382671 | EIF3L | FALSE | Downregulated |
| -1,565595632 | 0,489505347 | EIF1AY | FALSE | Downregulated |
| -0,28504595 | 0,489505347 | PXDN | FALSE | Downregulated |
| 0,259522336 | 0,489505347 | STK17A | FALSE | Upregulated |
| -0,366971867 | 0,489505347 | TTLL12 | FALSE | Downregulated |
| 0,243519042 | 0,489519201 | SLK | FALSE | Upregulated |
| -1,338033757 | 0,489519201 | ZNF391 | FALSE | Downregulated |
| 0,327805408 | 0,489769803 | PPM1E | FALSE | Upregulated |
| 0,510500565 | 0,489769803 | RNF7 | FALSE | Upregulated |
| 0,496823175 | 0,489771903 | ATG9B | FALSE | Upregulated |
| -0,70365174 | 0,489771903 | CLRN3 | FALSE | Downregulated |
| -0,32888457 | 0,489771903 | PDIA5 | FALSE | Downregulated |
| 0,360436204 | 0,489771903 | PEX5L | FALSE | Upregulated |
| -0,350671539 | 0,489771903 | PPP1CC | FALSE | Downregulated |
| -1,370390109 | 0,489771903 | RP11-118B18.1 | FALSE | Downregulated |
| -0,44490747 | 0,489771903 | RYK | FALSE | Downregulated |
| 0,591314173 | 0,489771903 | ZNF136 | FALSE | Upregulated |
| 0,647843716 | 0,489774997 | MRPS6 | FALSE | Upregulated |
| -0,276690611 | 0,489851598 | DNAJC13 | FALSE | Downregulated |
| 0,296979559 | 0,489851598 | SEC14L1 | FALSE | Upregulated |
| 1,040307848 | 0,490067699 | COL7A1 | FALSE | Upregulated |
| 0,288382885 | 0,490067699 | FOXN3-AS2 | FALSE | Upregulated |
| 3,043639971 | 0,490136937 | LOC101060216 | FALSE | Upregulated |
| -0,492055858 | 0,490938086 | FGFR1OP2 | FALSE | Downregulated |
| -1,061925023 | 0,490965965 | ZBTB6 | FALSE | Downregulated |
| -0,309671378 | 0,491037351 | CEBPB | FALSE | Downregulated |
| -0,618781886 | 0,491087584 | ARMCX5 | FALSE | Downregulated |
| 0,501070733 | 0,491087584 | ATP8A1 | FALSE | Upregulated |
| 0,436448942 | 0,491087584 | LOC101929347 | FALSE | Upregulated |
| -0,531643183 | 0,491087584 | NUDT22 | FALSE | Downregulated |
| 1,057001904 | 0,491264742 | CLCA3P | FALSE | Upregulated |
| -0,167691351 | 0,491264742 | GOLGB1 | FALSE | Downregulated |
| 0,675859038 | 0,491328081 | AGAP2-AS1 | FALSE | Upregulated |
| 1,11208311 | 0,49135303 | NAALADL1 | FALSE | Upregulated |

| | | | | |
|---|---|---|---|---|
| 0,618132388 | 0,49135303 | RP11-521B24.3 | FALSE | Upregulated |
| -0,518908284 | 0,491378268 | ATP6V1G1 | FALSE | Downregulated |
| 1,500612125 | 0,491505776 | CDC7 | FALSE | Upregulated |
| 0,595197628 | 0,491505776 | DDX39A | FALSE | Upregulated |
| 0,614614135 | 0,491505776 | DHX35 | FALSE | Upregulated |
| 0,804857508 | 0,491505776 | GSTCD | FALSE | Upregulated |
| 0,942577848 | 0,491505776 | MYBL1 | FALSE | Upregulated |
| 0,571473815 | 0,491505776 | NCBP2-AS2 | FALSE | Upregulated |
| -0,352059361 | 0,491505776 | RAB40C | FALSE | Downregulated |
| 0,6039857 | 0,491505776 | RP11-35G9.3 | FALSE | Upregulated |
| 0,672718915 | 0,491505776 | SIGLEC9 | FALSE | Upregulated |
| -0,41319915 | 0,491505776 | TBC1D16 | FALSE | Downregulated |
| -0,465930963 | 0,491505776 | TXNDC12 | FALSE | Downregulated |
| 0,411563531 | 0,491505776 | YPEL4 | FALSE | Upregulated |
| -0,489463412 | 0,491505776 | ZNF697 | FALSE | Downregulated |
| 1,949159431 | 0,491680384 | HOXA-AS3 | FALSE | Upregulated |
| 0,425018568 | 0,491682907 | C9orf96 | FALSE | Upregulated |
| -0,567981994 | 0,491696615 | FBXL12 | FALSE | Downregulated |
| -0,531649762 | 0,49173402 | NR6A1 | FALSE | Downregulated |
| -0,383352703 | 0,491762658 | BCL7B | FALSE | Downregulated |
| 0,296895377 | 0,491762658 | ERCC6L | FALSE | Upregulated |
| -0,2287201 | 0,491779958 | GSTK1 | FALSE | Downregulated |
| 0,59057182 | 0,491779958 | KCTD13 | FALSE | Upregulated |
| 1,014913011 | 0,491863567 | C12orf75 | FALSE | Upregulated |
| 1,241528248 | 0,491863567 | PRKAG2-AS1 | FALSE | Upregulated |
| -0,407677046 | 0,491863567 | RAB33B | FALSE | Downregulated |
| -0,210976709 | 0,492231104 | ILF3 | FALSE | Downregulated |
| 0,262596632 | 0,492231104 | PIK3C2A | FALSE | Upregulated |
| -0,427192167 | 0,492239631 | ZMYND19 | FALSE | Downregulated |
| 0,48164852 | 0,492592668 | ABHD13 | FALSE | Upregulated |
| 1,086980915 | 0,492592668 | LIMS3 | FALSE | Upregulated |
| 0,54607524 | 0,492592668 | SCN1B | FALSE | Upregulated |
| -0,48429767 | 0,492681195 | ARHGEF28 | FALSE | Downregulated |
| 0,778599215 | 0,492886989 | MED20 | FALSE | Upregulated |
| -0,309916114 | 0,492886989 | SGK494 | FALSE | Downregulated |
| 0,461869779 | 0,492958143 | CLDN7 | FALSE | Upregulated |
| -0,599085313 | 0,493059173 | NAT1 | FALSE | Downregulated |
| 0,321123546 | 0,493086611 | NUP62 | FALSE | Upregulated |
| -0,441255133 | 0,493089538 | CPNE2 | FALSE | Downregulated |
| -0,338073945 | 0,493182917 | DMD | FALSE | Downregulated |
| -0,249678408 | 0,493256475 | RNF103 | FALSE | Downregulated |
| -0,600906988 | 0,493256475 | THAP2 | FALSE | Downregulated |
| 0,320494045 | 0,493492918 | MLX | FALSE | Upregulated |
| 0,784227385 | 0,493502056 | ZMAT5 | FALSE | Upregulated |
| -1,9411885 | 0,493580309 | LOC102723360 | FALSE | Downregulated |
| 0,352097674 | 0,493580309 | ZDHHC23 | FALSE | Upregulated |
| 0,271770877 | 0,493632375 | SMARCD2 | FALSE | Upregulated |
| 0,685222934 | 0,493666534 | FEZ1 | FALSE | Upregulated |
| -1,273048952 | 0,493666534 | GALNT3 | FALSE | Downregulated |
| 0,233760638 | 0,493666534 | LOC101929264 | FALSE | Upregulated |
| 0,897335873 | 0,493666534 | ZNF431 | FALSE | Upregulated |

| | | | | |
|---|---|---|---|---|
| 0,550067548 | 0,493725809 | EIF4E | FALSE | Upregulated |
| 0,558359184 | 0,493960346 | LOC102724141 | FALSE | Upregulated |
| 0,267698658 | 0,494257668 | PGAM5 | FALSE | Upregulated |
| 0,298541331 | 0,494375355 | SLC30A7 | FALSE | Upregulated |
| -0,496052722 | 0,494410779 | CDK8 | FALSE | Downregulated |
| 0,972130793 | 0,494410779 | CHIC1 | FALSE | Upregulated |
| 1,081583852 | 0,494410779 | IGFBP6 | FALSE | Upregulated |
| -0,943940361 | 0,494410779 | TP53TG1 | FALSE | Downregulated |
| -0,691837834 | 0,494512384 | GCSHP3 | FALSE | Downregulated |
| 0,278957808 | 0,494520413 | NBPF8 | FALSE | Upregulated |
| -0,264330224 | 0,494520413 | PBX2 | FALSE | Downregulated |
| 0,718807022 | 0,494520413 | PRDM1 | FALSE | Upregulated |
| -0,299806437 | 0,494520413 | SAMD14 | FALSE | Downregulated |
| 0,686973881 | 0,494536273 | LOC101059906 | FALSE | Upregulated |
| 0,331865367 | 0,494536273 | ZMIZ2 | FALSE | Upregulated |
| -0,194304301 | 0,494593763 | SEMA6A | FALSE | Downregulated |
| 0,438292932 | 0,495231852 | ALPK1 | FALSE | Upregulated |
| -0,432618956 | 0,495241717 | KIAA0907 | FALSE | Downregulated |
| 0,425493145 | 0,495307525 | NBPF1 | FALSE | Upregulated |
| -0,447635876 | 0,495307525 | TGIF1 | FALSE | Downregulated |
| 0,34156465 | 0,495319778 | TAF2 | FALSE | Upregulated |
| -0,1244205 | 0,495427071 | PLEC | FALSE | Downregulated |
| -0,260001921 | 0,496030652 | ARL6IP4 | FALSE | Downregulated |
| 0,307318948 | 0,496193719 | PPP3R1 | FALSE | Upregulated |
| 0,890339022 | 0,496407018 | HIST2H3A | FALSE | Upregulated |
| -0,883241936 | 0,49643341 | EDN1 | FALSE | Downregulated |
| 0,324071205 | 0,496539553 | ADORA2A | FALSE | Upregulated |
| -0,393595867 | 0,496612362 | LAMTOR1 | FALSE | Downregulated |
| 0,302070077 | 0,496612362 | ZBTB12 | FALSE | Upregulated |
| -0,54243898 | 0,496906366 | FAM184A | FALSE | Downregulated |
| 0,43731747 | 0,497136915 | C12orf29 | FALSE | Upregulated |
| -0,484895961 | 0,497210642 | ATRAID | FALSE | Downregulated |
| -0,602369799 | 0,497210642 | MIR181A2HG | FALSE | Downregulated |
| 0,293276133 | 0,497210642 | MXD3 | FALSE | Upregulated |
| 0,340279635 | 0,497277106 | ZNF546 | FALSE | Upregulated |
| -0,277265425 | 0,497486368 | SERPINA6 | FALSE | Downregulated |
| 0,598612785 | 0,497776137 | ZNF559 | FALSE | Upregulated |
| 0,225339716 | 0,498033658 | USP47 | FALSE | Upregulated |
| 0,370898556 | 0,498105516 | RIPK4 | FALSE | Upregulated |
| -0,417262492 | 0,498105516 | SPRY2 | FALSE | Downregulated |
| 0,465003582 | 0,498105516 | TNXB | FALSE | Upregulated |
| 0,50905619 | 0,498248069 | CCNG2 | FALSE | Upregulated |
| 0,512159374 | 0,49842221 | ABHD4 | FALSE | Upregulated |
| 1,518766181 | 0,49842221 | CCDC168 | FALSE | Upregulated |
| -0,941871924 | 0,49842221 | IGF2 | FALSE | Downregulated |
| 0,343886948 | 0,498536844 | BOP1 | FALSE | Upregulated |
| 0,244509772 | 0,498536844 | C20orf112 | FALSE | Upregulated |
| 1,286302923 | 0,498536844 | FAM222A-AS1 | FALSE | Upregulated |
| 0,51969309 | 0,498536844 | LOC101928408 | FALSE | Upregulated |
| 0,865804077 | 0,498536844 | LOC102723614 | FALSE | Upregulated |
| 1,04075641 | 0,498536844 | LOC728769 | FALSE | Upregulated |

| | | | | |
|---|---|---|---|---|
| 0,743787028 | 0,498536844 | NDN | FALSE | Upregulated |
| 0,311580761 | 0,498536844 | NUDT21 | FALSE | Upregulated |
| 0,406857581 | 0,498536844 | SEPT7 | FALSE | Upregulated |
| 0,713783084 | 0,498536844 | SLC4A7 | FALSE | Upregulated |
| 0,506877364 | 0,498604526 | LPPR1 | FALSE | Upregulated |
| 0,42434806 | 0,498645768 | CLCF1 | FALSE | Upregulated |
| -0,293522988 | 0,498645768 | FGF2 | FALSE | Downregulated |
| 0,895757671 | 0,498645768 | HAR1A | FALSE | Upregulated |
| 0,721710054 | 0,498645768 | LOC102724808 | FALSE | Upregulated |
| -0,267717284 | 0,498645768 | NCOA6 | FALSE | Downregulated |
| -0,303108345 | 0,498762376 | PAXBP1 | FALSE | Downregulated |
| 0,386910765 | 0,498864542 | MED19 | FALSE | Upregulated |
| 0,836238837 | 0,498872205 | TTYH2 | FALSE | Upregulated |
| -0,360484616 | 0,498887008 | NOP56 | FALSE | Downregulated |
| 0,398797645 | 0,498887008 | TTN | FALSE | Upregulated |
| 0,552416928 | 0,498931422 | AC011747.4 | FALSE | Upregulated |
| -0,521690443 | 0,498931422 | NSUN7 | FALSE | Downregulated |
| -1,379554106 | 0,498931422 | PIF1 | FALSE | Downregulated |
| -1,235875123 | 0,498931422 | ZNF491 | FALSE | Downregulated |
| 0,474475516 | 0,498938831 | CTAGE8 | FALSE | Upregulated |
| 0,249694977 | 0,49900702 | APOL6 | FALSE | Upregulated |
| -0,163060751 | 0,499035351 | RBL2 | FALSE | Downregulated |
| 0,862100095 | 0,499106757 | AC009264.1 | FALSE | Upregulated |
| -0,481167243 | 0,499106757 | ATF2 | FALSE | Downregulated |
| 0,971814579 | 0,499106757 | HERC2P7 | FALSE | Upregulated |
| 0,23116016 | 0,499132488 | CARM1 | FALSE | Upregulated |
| -0,30539445 | 0,499350837 | ZDHHC8 | FALSE | Downregulated |
| 0,439580569 | 0,499372062 | ACOT6 | FALSE | Upregulated |
| 0,262865079 | 0,499759076 | KRI1 | FALSE | Upregulated |
| 0,429360752 | 0,499781212 | FXYD6 | FALSE | Upregulated |
| -0,278446712 | 0,499781212 | ZBTB40 | FALSE | Downregulated |
| 0,604694807 | 0,499781212 | ZNF331 | FALSE | Upregulated |
| 0,255560088 | 0,499781212 | ZNF750 | FALSE | Upregulated |
| 0,491555542 | 0,499795104 | C9orf163 | FALSE | Upregulated |
| -0,560180488 | 0,499795104 | CLDN5 | FALSE | Downregulated |
| 0,247259522 | 0,499795104 | CMTR1 | FALSE | Upregulated |
| 0,537540602 | 0,499795104 | CXCR2P1 | FALSE | Upregulated |
| 0,405947962 | 0,499795104 | KIAA0391 | FALSE | Upregulated |
| 1,136438814 | 0,499795104 | KLHL33 | FALSE | Upregulated |
| 1,382156928 | 0,499795104 | LOC101929949 | FALSE | Upregulated |
| 0,625726171 | 0,499795104 | LOC102723381 | FALSE | Upregulated |
| 0,320425866 | 0,499795104 | LOC102723451 | FALSE | Upregulated |
| -0,187387882 | 0,499795104 | OBSL1 | FALSE | Downregulated |
| -0,362749381 | 0,499795104 | PSMB3 | FALSE | Downregulated |
| -0,274733761 | 0,499795104 | RELA | FALSE | Downregulated |
| 1,131331916 | 0,499795104 | RNASE6 | FALSE | Upregulated |
| 0,288564119 | 0,499795104 | SLC16A4 | FALSE | Upregulated |
| 0,788973737 | 0,499795104 | SPHK1 | FALSE | Upregulated |
| 0,653251725 | 0,499795104 | ST6GALNAC4 | FALSE | Upregulated |
| 0,59744621 | 0,499837351 | LOC100507191 | FALSE | Upregulated |
| 0,761740481 | 0,49989381 | SRCRB4D | FALSE | Upregulated |

| | | | | |
|---|---|---|---|---|
| 0,867157374 | 0,500150042 | CCDC64B | FALSE | Upregulated |
| -0,725186908 | 0,500150042 | PCSK4 | FALSE | Downregulated |
| -0,269291487 | 0,500150042 | PITRM1 | FALSE | Downregulated |
| -0,341296658 | 0,500202277 | NDUFC2 | FALSE | Downregulated |
| 0,492736919 | 0,500497068 | CHMP2B | FALSE | Upregulated |
| 0,373188742 | 0,500497068 | MAD1L1 | FALSE | Upregulated |
| 0,236588472 | 0,500536132 | GRB2 | FALSE | Upregulated |
| -0,760644263 | 0,501058521 | ERICH1 | FALSE | Downregulated |
| 2,435082776 | 0,501058521 | GTF2H2 | FALSE | Upregulated |
| -0,593811913 | 0,501058521 | PLEKHF2 | FALSE | Downregulated |
| 0,379319124 | 0,501109046 | DFNA5 | FALSE | Upregulated |
| 1,346885632 | 0,501109046 | RAB7B | FALSE | Upregulated |
| -0,688029594 | 0,501172822 | CDRT15 | FALSE | Downregulated |
| 0,274868965 | 0,501182602 | PUS10 | FALSE | Upregulated |
| 0,356237099 | 0,501211258 | ALDOA | FALSE | Upregulated |
| 0,684540544 | 0,501211258 | CPLX2 | FALSE | Upregulated |
| 0,363684598 | 0,501211258 | KCTD2 | FALSE | Upregulated |
| 0,227899298 | 0,501211258 | PPP2R5A | FALSE | Upregulated |
| -0,623276771 | 0,501211258 | ROGDI | FALSE | Downregulated |
| 0,488795886 | 0,501211258 | SMCO4 | FALSE | Upregulated |
| -1,363372395 | 0,501211258 | UPK3BL | FALSE | Downregulated |
| 1,126757521 | 0,501211258 | WDR16 | FALSE | Upregulated |
| -1,18543617 | 0,501455243 | RMDN2-AS1 | FALSE | Downregulated |
| -0,387671649 | 0,501672864 | LOC101929330 | FALSE | Downregulated |
| -1,032540503 | 0,501682026 | UFSP1 | FALSE | Downregulated |
| 0,407644405 | 0,501764879 | ARL13A | FALSE | Upregulated |
| 0,406220754 | 0,501764879 | LOC730183 | FALSE | Upregulated |
| 0,663005031 | 0,501764879 | TRAF5 | FALSE | Upregulated |
| 0,314431589 | 0,501867844 | SNAPC3 | FALSE | Upregulated |
| -0,898985313 | 0,501873269 | TRIM16L | FALSE | Downregulated |
| -0,143479741 | 0,501896424 | ZSWIM8 | FALSE | Downregulated |
| 0,988629854 | 0,501965285 | LOC101929440 | FALSE | Upregulated |
| 0,34845507 | 0,501965285 | NCAPD2 | FALSE | Upregulated |
| 0,908669435 | 0,501965285 | POU5F2 | FALSE | Upregulated |
| 0,849224068 | 0,501965285 | VEGFC | FALSE | Upregulated |
| 0,707695516 | 0,502032461 | TMEM125 | FALSE | Upregulated |
| 0,302189222 | 0,502098665 | ADAM17 | FALSE | Upregulated |
| -0,658941597 | 0,502098665 | CTTNBP2 | FALSE | Downregulated |
| -0,201621685 | 0,502098665 | DOT1L | FALSE | Downregulated |
| -0,383245549 | 0,502098665 | EXOC6B | FALSE | Downregulated |
| -0,342360267 | 0,502098665 | SLC29A1 | FALSE | Downregulated |
| -0,285404432 | 0,502098665 | TRIM62 | FALSE | Downregulated |
| 0,443534493 | 0,502210544 | ZBTB47 | FALSE | Upregulated |
| -0,564373315 | 0,502288389 | IQCH-AS1 | FALSE | Downregulated |
| -0,192082073 | 0,502398224 | HERC2 | FALSE | Downregulated |
| -0,300871992 | 0,502398224 | LOC101928716 | FALSE | Downregulated |
| 0,200355559 | 0,502398224 | PPP1R12C | FALSE | Upregulated |
| 0,451992729 | 0,502398224 | YIPF1 | FALSE | Upregulated |
| 0,570479417 | 0,502875887 | LGALSL | FALSE | Upregulated |
| -1,200039373 | 0,502875887 | RPL13AP17 | FALSE | Downregulated |
| -1,015357502 | 0,502965373 | AC079117.1 | FALSE | Downregulated |

| | | | | |
|---|---|---|---|---|
| -0,248670948 | 0,503040772 | SLC38A3 | FALSE | Downregulated |
| 0,627739433 | 0,50317856 | RP11-127L20.3 | FALSE | Upregulated |
| 1,651139328 | 0,503826145 | ESP33 | FALSE | Upregulated |
| 0,97176559 | 0,504041761 | NAT8L | FALSE | Upregulated |
| -0,296249107 | 0,504113098 | CXXC1 | FALSE | Downregulated |
| -0,342941113 | 0,504122217 | TBC1D13 | FALSE | Downregulated |
| -0,225921565 | 0,504547566 | CCDC50 | FALSE | Downregulated |
| 0,22053711 | 0,504672892 | CAPRIN1 | FALSE | Upregulated |
| 1,304953766 | 0,504820195 | RP11-630D6.5 | FALSE | Upregulated |
| 0,364641983 | 0,504991563 | PKD2 | FALSE | Upregulated |
| 1,266283609 | 0,505041615 | CDH24 | FALSE | Upregulated |
| -0,248736806 | 0,505041615 | DCHS1 | FALSE | Downregulated |
| 0,324061606 | 0,505041615 | FAM134B | FALSE | Upregulated |
| 0,476373662 | 0,505041615 | SLC8A1 | FALSE | Upregulated |
| 1,129942184 | 0,505041615 | TGFB2 | FALSE | Upregulated |
| -0,272951294 | 0,505071306 | PALMD | FALSE | Downregulated |
| 1,056176159 | 0,505236643 | NSUN3 | FALSE | Upregulated |
| -0,384712861 | 0,505376844 | AHSA1 | FALSE | Downregulated |
| -0,254050513 | 0,505376844 | LOC102724129 | FALSE | Downregulated |
| 0,333970379 | 0,505906344 | MEX3C | FALSE | Upregulated |
| 0,583003233 | 0,505906344 | NR1D2 | FALSE | Upregulated |
| -0,323368214 | 0,506039852 | ADAM10 | FALSE | Downregulated |
| 0,406705114 | 0,506039852 | KCTD9 | FALSE | Upregulated |
| 0,448680751 | 0,506555719 | ME2 | FALSE | Upregulated |
| -0,253044152 | 0,506904991 | INHBC | FALSE | Downregulated |
| -0,229825103 | 0,507173575 | RPL10A | FALSE | Downregulated |
| 0,569813937 | 0,507215217 | GPAT2 | FALSE | Upregulated |
| 0,284366725 | 0,507364691 | ATP6V1C2 | FALSE | Upregulated |
| 0,700905853 | 0,507364691 | RP11-540A21.2 | FALSE | Upregulated |
| -0,611046081 | 0,507497587 | GTPBP3 | FALSE | Downregulated |
| -0,452635175 | 0,507515251 | KIF26A | FALSE | Downregulated |
| -0,323579869 | 0,507572336 | DDRGK1 | FALSE | Downregulated |
| 0,925511295 | 0,507572336 | OR5C1 | FALSE | Upregulated |
| -0,608413628 | 0,507572336 | SLC13A3 | FALSE | Downregulated |
| -0,477264318 | 0,507630968 | MRPS22 | FALSE | Downregulated |
| -0,280264348 | 0,507630968 | TXN2 | FALSE | Downregulated |
| -0,297961427 | 0,507724072 | MTFR1L | FALSE | Downregulated |
| 0,626758616 | 0,508115127 | CASP1 | FALSE | Upregulated |
| -0,592890438 | 0,508115127 | EBPL | FALSE | Downregulated |
| -1,157874437 | 0,508115127 | SOSTDC1 | FALSE | Downregulated |
| 0,238163468 | 0,508175724 | EHBP1 | FALSE | Upregulated |
| 0,631603962 | 0,508175724 | TAF9 | FALSE | Upregulated |
| 0,293140972 | 0,508249667 | GDI1 | FALSE | Upregulated |
| 0,454095562 | 0,508262962 | ZNF843 | FALSE | Upregulated |
| -0,465341361 | 0,50840893 | GPKOW | FALSE | Downregulated |
| 0,258413799 | 0,508801779 | BDP1 | FALSE | Upregulated |
| -0,332851308 | 0,508801779 | BZW1 | FALSE | Downregulated |
| 1,441576958 | 0,508965508 | LOC101927188 | FALSE | Upregulated |
| -0,143921761 | 0,509001861 | SPEN | FALSE | Downregulated |
| -0,340621306 | 0,509162658 | ARF5 | FALSE | Downregulated |
| 0,601388393 | 0,509162658 | CEP152 | FALSE | Upregulated |

| | | | | |
|---|---|---|---|---|
| -0,822400306 | 0,509162658 | CTNNA3 | FALSE | Downregulated |
| -1,329043358 | 0,509162658 | LINC00659 | FALSE | Downregulated |
| 1,171268805 | 0,509162658 | LOC101927474 | FALSE | Upregulated |
| 0,993686722 | 0,509162658 | LOC729739 | FALSE | Upregulated |
| -0,212021317 | 0,509162658 | MED13L | FALSE | Downregulated |
| 0,265880041 | 0,509162658 | MORC2-AS1 | FALSE | Upregulated |
| 0,306702922 | 0,509162658 | RNF139 | FALSE | Upregulated |
| 0,677650497 | 0,509162658 | RP4-777D9.2 | FALSE | Upregulated |
| 0,36661152 | 0,509589403 | TMEM87A | FALSE | Upregulated |
| -0,223145156 | 0,5096644 | ARHGAP5 | FALSE | Downregulated |
| 0,622686222 | 0,5096644 | RHBDL2 | FALSE | Upregulated |
| 0,205741286 | 0,5096644 | TNRC6C | FALSE | Upregulated |
| -0,347086717 | 0,50969956 | HOGA1 | FALSE | Downregulated |
| -0,261564563 | 0,50969956 | IER2 | FALSE | Downregulated |
| -0,311131491 | 0,509743117 | FABP1 | FALSE | Downregulated |
| 1,554432724 | 0,509743117 | GABRP | FALSE | Upregulated |
| 1,26457677 | 0,509743117 | LOC100335030 | FALSE | Upregulated |
| 0,616376712 | 0,509743117 | NAA40 | FALSE | Upregulated |
| -0,387288357 | 0,509743117 | SMIM15 | FALSE | Downregulated |
| 0,567355084 | 0,509835415 | MSH2 | FALSE | Upregulated |
| 0,475780388 | 0,50986709 | MSR1 | FALSE | Upregulated |
| -0,394345803 | 0,509908535 | SPR | FALSE | Downregulated |
| 0,223741391 | 0,509913626 | TINF2 | FALSE | Upregulated |
| 0,384190905 | 0,510146159 | MAP3K10 | FALSE | Upregulated |
| -0,277640549 | 0,510211611 | TBC1D5 | FALSE | Downregulated |
| 0,384535743 | 0,510372987 | ZNF777 | FALSE | Upregulated |
| -0,193582718 | 0,510502798 | HTT | FALSE | Downregulated |
| 1,014449152 | 0,510837442 | ACTL7A | FALSE | Upregulated |
| -0,503646495 | 0,511018445 | MBLAC2 | FALSE | Downregulated |
| 0,899119753 | 0,511126606 | HIST2H3C | FALSE | Upregulated |
| -1,041077516 | 0,511344155 | HCN2 | FALSE | Downregulated |
| 0,418796563 | 0,511344155 | TMEM184B | FALSE | Upregulated |
| -0,278862138 | 0,511419761 | PPP5C | FALSE | Downregulated |
| 0,985389339 | 0,511425969 | ADAMTS9-AS2 | FALSE | Upregulated |
| 0,239694393 | 0,511425969 | HSPG2 | FALSE | Upregulated |
| -0,929497974 | 0,511425969 | KIAA1586 | FALSE | Downregulated |
| -0,257286134 | 0,511425969 | MORC2 | FALSE | Downregulated |
| -1,255035223 | 0,511425969 | SLC4A9 | FALSE | Downregulated |
| -0,489801819 | 0,511495757 | MFSD9 | FALSE | Downregulated |
| 0,483071197 | 0,511495757 | PWWP2B | FALSE | Upregulated |
| 0,772230948 | 0,511499311 | REC8 | FALSE | Upregulated |
| 0,711759916 | 0,511521979 | C17orf53 | FALSE | Upregulated |
| 0,332718451 | 0,511521979 | TTC7A | FALSE | Upregulated |
| 1,257231094 | 0,511561814 | FAM225A | FALSE | Upregulated |
| 0,5730099 | 0,511601077 | LILRB2 | FALSE | Upregulated |
| 0,347893387 | 0,511629849 | CNN2 | FALSE | Upregulated |
| -1,755583552 | 0,511661817 | KIAA1751 | FALSE | Downregulated |
| 0,654863415 | 0,511870444 | RASA4 | FALSE | Upregulated |
| 0,393062447 | 0,51205823 | LINC00514 | FALSE | Upregulated |
| 0,889072052 | 0,51205823 | LOC100128006 | FALSE | Upregulated |
| 1,019017767 | 0,51205823 | LOC101928854 | FALSE | Upregulated |

| | | | | |
|---|---|---|---|---|
| 0,245770622 | 0,51205823 | MYRF | FALSE | Upregulated |
| 1,120955459 | 0,51205823 | RALGAPA1P | FALSE | Upregulated |
| -0,267403465 | 0,51205823 | SARM1 | FALSE | Downregulated |
| -0,1503295 | 0,512155875 | EP300 | FALSE | Downregulated |
| 0,244471274 | 0,512155875 | GABPB1-AS1 | FALSE | Upregulated |
| -0,313590124 | 0,512155875 | KCNJ8 | FALSE | Downregulated |
| 0,90041553 | 0,512155875 | WDYHV1 | FALSE | Upregulated |
| -1,296817747 | 0,512319683 | CSTA | FALSE | Downregulated |
| -0,211120768 | 0,512319683 | HNRNPA2B1 | FALSE | Downregulated |
| -1,407805862 | 0,512319683 | LOC101929715 | FALSE | Downregulated |
| -1,221883839 | 0,512319683 | NRN1 | FALSE | Downregulated |
| -2,518806603 | 0,512678211 | LOC102724258 | FALSE | Downregulated |
| 0,422211264 | 0,512701916 | IL15RA | FALSE | Upregulated |
| 0,712152937 | 0,512701916 | STXBP5-AS1 | FALSE | Upregulated |
| -0,536886031 | 0,512971404 | CBLC | FALSE | Downregulated |
| -0,37930408 | 0,512971404 | DDX21 | FALSE | Downregulated |
| 0,311792706 | 0,512971404 | LOC101928621 | FALSE | Upregulated |
| 0,557831376 | 0,512971404 | MARVELD1 | FALSE | Upregulated |
| -0,991898118 | 0,512971404 | MEIS3P1 | FALSE | Downregulated |
| 0,983980262 | 0,512971404 | POC5 | FALSE | Upregulated |
| 0,805434509 | 0,512971404 | ZNF137P | FALSE | Upregulated |
| 0,911098285 | 0,512985774 | C1orf116 | FALSE | Upregulated |
| 0,321115773 | 0,512985774 | MLLT10 | FALSE | Upregulated |
| 0,667668172 | 0,513031314 | GTPBP8 | FALSE | Upregulated |
| 0,35329654 | 0,513062109 | HIRIP3 | FALSE | Upregulated |
| 0,350305299 | 0,513062109 | SPIRE2 | FALSE | Upregulated |
| -0,897719137 | 0,513110335 | FAM86B3P | FALSE | Downregulated |
| 0,603728319 | 0,513203425 | HAGHL | FALSE | Upregulated |
| -1,432649955 | 0,513203425 | POU3F2 | FALSE | Downregulated |
| -0,728855601 | 0,513203425 | ST6GAL2 | FALSE | Downregulated |
| -0,375791909 | 0,513242217 | DAO | FALSE | Downregulated |
| -0,848155856 | 0,513242217 | EIF3CL | FALSE | Downregulated |
| 0,988055678 | 0,513242217 | LOC101929571 | FALSE | Upregulated |
| 0,331198922 | 0,513242217 | PP2D1 | FALSE | Upregulated |
| 0,228759544 | 0,513242217 | SIPA1L3 | FALSE | Upregulated |
| -0,503502152 | 0,513242217 | SLC25A34 | FALSE | Downregulated |
| -0,330858125 | 0,513242217 | UTP18 | FALSE | Downregulated |
| 0,277308668 | 0,513258378 | LOC101928524 | FALSE | Upregulated |
| -0,460428636 | 0,513262506 | DNAJC25 | FALSE | Downregulated |
| 0,450067617 | 0,513262506 | PRELP | FALSE | Upregulated |
| -0,419604516 | 0,513453869 | C14orf105 | FALSE | Downregulated |
| 0,70114113 | 0,513536594 | ACTR6 | FALSE | Upregulated |
| -0,613297395 | 0,513536594 | FUT10 | FALSE | Downregulated |
| -1,402307892 | 0,513617026 | SNORA54 | FALSE | Downregulated |
| -0,388099937 | 0,513797825 | LOC101927514 | FALSE | Downregulated |
| 1,142547378 | 0,514014356 | GATA2 | FALSE | Upregulated |
| 0,242676059 | 0,514014356 | SH3BP5 | FALSE | Upregulated |
| 0,301183985 | 0,514014356 | SMAD5 | FALSE | Upregulated |
| -0,268240299 | 0,514267007 | HERPUD1 | FALSE | Downregulated |
| 0,173681824 | 0,514346172 | SP1 | FALSE | Upregulated |
| -0,263741366 | 0,514566108 | AMOTL1 | FALSE | Downregulated |

| | | | | |
|---|---|---|---|---|
| -0,866250456 | 0,514566108 | LOC100272217 | FALSE | Downregulated |
| -1,524548771 | 0,514835379 | TMEM233 | FALSE | Downregulated |
| 0,261123376 | 0,514938381 | TMED10P1 | FALSE | Upregulated |
| 0,687716427 | 0,514948069 | PIGU | FALSE | Upregulated |
| -0,91152741 | 0,515052679 | GKAP1 | FALSE | Downregulated |
| 2,392061042 | 0,515103628 | FLG-AS1 | FALSE | Upregulated |
| 0,539331593 | 0,515103628 | MRPS31P5 | FALSE | Upregulated |
| 0,327775609 | 0,515103628 | RPAP1 | FALSE | Upregulated |
| -0,427543446 | 0,515103628 | ZEB1 | FALSE | Downregulated |
| 0,282576048 | 0,515204818 | HSPB1 | FALSE | Upregulated |
| -1,276697109 | 0,515204818 | TEX22 | FALSE | Downregulated |
| 0,466802455 | 0,515275639 | RABEP2 | FALSE | Upregulated |
| 0,2068831 | 0,51550647 | SHC1 | FALSE | Upregulated |
| 0,55810653 | 0,515682015 | ENO2 | FALSE | Upregulated |
| 0,328344902 | 0,515682015 | NUTM2B | FALSE | Upregulated |
| -0,34934194 | 0,515704123 | CDK2AP2 | FALSE | Downregulated |
| 0,563055736 | 0,515704123 | MBOAT4 | FALSE | Upregulated |
| 0,89000268 | 0,515803586 | LINC00869 | FALSE | Upregulated |
| -0,242879359 | 0,515843614 | FAM193A | FALSE | Downregulated |
| 0,306032128 | 0,516023932 | C1orf122 | FALSE | Upregulated |
| -0,254797745 | 0,516086402 | PDIA3 | FALSE | Downregulated |
| -0,273402522 | 0,516145967 | ATF6 | FALSE | Downregulated |
| 2,147624555 | 0,516145967 | PRH1 | FALSE | Upregulated |
| -0,279789619 | 0,516273463 | AIFM1 | FALSE | Downregulated |
| 0,383929582 | 0,516273463 | CLK2 | FALSE | Upregulated |
| 0,722748084 | 0,516273463 | DOHH | FALSE | Upregulated |
| 1,389366306 | 0,516273463 | KGFLP1 | FALSE | Upregulated |
| 1,08890878 | 0,516273463 | LRRC14B | FALSE | Upregulated |
| 0,542458773 | 0,516273463 | POU2F2 | FALSE | Upregulated |
| 0,342145214 | 0,516273463 | RAP1B | FALSE | Upregulated |
| 0,953505692 | 0,516273463 | RP11-575L7.4 | FALSE | Upregulated |
| 0,49724352 | 0,516315397 | SLC6A4 | FALSE | Upregulated |
| -0,562569223 | 0,516322126 | DTNB | FALSE | Downregulated |
| 0,816306957 | 0,516620077 | ACTRT3 | FALSE | Upregulated |
| 0,454026506 | 0,516620077 | FAM179B | FALSE | Upregulated |
| 1,017619404 | 0,516620077 | PLA2G4A | FALSE | Upregulated |
| 0,660257908 | 0,517128643 | ARL10 | FALSE | Upregulated |
| -0,405844597 | 0,517128643 | BMS1 | FALSE | Downregulated |
| -0,18260282 | 0,517301409 | AP3D1 | FALSE | Downregulated |
| 0,368759445 | 0,517301409 | DONSON | FALSE | Upregulated |
| -0,226980012 | 0,517316853 | DAPK1 | FALSE | Downregulated |
| -0,261879801 | 0,517316853 | LBX2 | FALSE | Downregulated |
| -0,544790519 | 0,517316853 | LOC100129722 | FALSE | Downregulated |
| -0,215179096 | 0,517316853 | RAB7A | FALSE | Downregulated |
| -0,869790382 | 0,517316853 | TMEM42 | FALSE | Downregulated |
| -0,598100488 | 0,517316853 | TMPRSS11D | FALSE | Downregulated |
| 0,306597632 | 0,517549898 | ASAH1 | FALSE | Upregulated |
| -0,80234775 | 0,517556189 | FAM27E3 | FALSE | Downregulated |
| -0,41173898 | 0,517949138 | EMX1 | FALSE | Downregulated |
| 0,339771375 | 0,517949138 | FAM87B | FALSE | Upregulated |
| -0,359861567 | 0,517949138 | LIN7C | FALSE | Downregulated |

| | | | | |
|---|---|---|---|---|
| 1,517562534 | 0,517949138 | NTF4 | FALSE | Upregulated |
| 0,500243732 | 0,517949138 | POP4 | FALSE | Upregulated |
| -0,307202912 | 0,518349502 | NPIPB11 | FALSE | Downregulated |
| 0,225143769 | 0,518374949 | KAT2A | FALSE | Upregulated |
| 0,430561917 | 0,518374949 | ZNF436 | FALSE | Upregulated |
| 0,412052536 | 0,518396858 | RASGRP3 | FALSE | Upregulated |
| 0,395435428 | 0,519118665 | BTN2A1 | FALSE | Upregulated |
| 0,675881305 | 0,519236667 | NCF1 | FALSE | Upregulated |
| -0,39150661 | 0,519236667 | SLC22A9 | FALSE | Downregulated |
| 0,308415548 | 0,519236667 | UBA7 | FALSE | Upregulated |
| 0,54290362 | 0,520053937 | SLC7A5 | FALSE | Upregulated |
| -0,282244389 | 0,520118334 | DHX16 | FALSE | Downregulated |
| 0,321434955 | 0,520134424 | VAMP2 | FALSE | Upregulated |
| 0,665636184 | 0,520290153 | RSPH4A | FALSE | Upregulated |
| 0,745266247 | 0,520459107 | CLIP4 | FALSE | Upregulated |
| 0,359253311 | 0,520480316 | TIRAP | FALSE | Upregulated |
| 0,25504494 | 0,520594327 | CTDSPL2 | FALSE | Upregulated |
| -0,316526138 | 0,520594327 | PHACTR4 | FALSE | Downregulated |
| 1,3781299 | 0,520766882 | LOC102723917 | FALSE | Upregulated |
| 0,825837822 | 0,521120535 | GMPR | FALSE | Upregulated |
| 0,458088522 | 0,521315257 | RSU1 | FALSE | Upregulated |
| 0,601937799 | 0,521488928 | ANKRD53 | FALSE | Upregulated |
| -0,63879964 | 0,521488928 | BBS10 | FALSE | Downregulated |
| -0,273517277 | 0,521488928 | BCLAF1 | FALSE | Downregulated |
| -0,51696245 | 0,521488928 | CEBPA-AS1 | FALSE | Downregulated |
| 0,666506 | 0,521488928 | CEP97 | FALSE | Upregulated |
| 0,628518705 | 0,521488928 | CLDN4 | FALSE | Upregulated |
| 0,316288357 | 0,521488928 | COG7 | FALSE | Upregulated |
| -0,571498214 | 0,521488928 | CYP2C19 | FALSE | Downregulated |
| -0,469935504 | 0,521488928 | CYP7B1 | FALSE | Downregulated |
| -0,289557025 | 0,521488928 | EXOC3L4 | FALSE | Downregulated |
| 0,232067149 | 0,521488928 | GATAD2B | FALSE | Upregulated |
| -0,396620903 | 0,521488928 | GFER | FALSE | Downregulated |
| -0,8457397 | 0,521488928 | GSTA4 | FALSE | Downregulated |
| 0,679795501 | 0,521488928 | HCCS | FALSE | Upregulated |
| -0,369779748 | 0,521488928 | KCTD21 | FALSE | Downregulated |
| 0,461192101 | 0,521488928 | KLC3 | FALSE | Upregulated |
| 0,516464152 | 0,521488928 | LCORL | FALSE | Upregulated |
| -0,503340944 | 0,521488928 | LOC100507616 | FALSE | Downregulated |
| -1,288974117 | 0,521488928 | LOC101927372 | FALSE | Downregulated |
| -0,450168698 | 0,521488928 | LOC102724455 | FALSE | Downregulated |
| -0,894112547 | 0,521488928 | LOC102724873 | FALSE | Downregulated |
| 0,417238243 | 0,521488928 | LOC643072 | FALSE | Upregulated |
| 1,148657857 | 0,521488928 | MEF2BNB-MEF2B | FALSE | Upregulated |
| 0,275088452 | 0,521488928 | MKNK1-AS1 | FALSE | Upregulated |
| 0,400587556 | 0,521488928 | MRPL40 | FALSE | Upregulated |
| 1,280305735 | 0,521488928 | NXPE4 | FALSE | Upregulated |
| -0,364350966 | 0,521488928 | PABPC1L | FALSE | Downregulated |
| 0,363191569 | 0,521488928 | PIKFYVE | FALSE | Upregulated |
| 0,217380934 | 0,521488928 | PIP4K2B | FALSE | Upregulated |
| -0,570654357 | 0,521488928 | PMS2P5 | FALSE | Downregulated |

| | | | | |
|---|---|---|---|---|
| 0,37641628 | 0,521488928 | POM121L10P | FALSE | Upregulated |
| 0,125948269 | 0,521488928 | PRRC2A | FALSE | Upregulated |
| 0,451185199 | 0,521488928 | PTPN2 | FALSE | Upregulated |
| 0,726292207 | 0,521488928 | RBMS3-AS2 | FALSE | Upregulated |
| -0,373704289 | 0,521488928 | SC5D | FALSE | Downregulated |
| -0,400130136 | 0,521488928 | SCRN2 | FALSE | Downregulated |
| -0,240933511 | 0,521488928 | SF3B3 | FALSE | Downregulated |
| 0,860661833 | 0,521488928 | SLC15A2 | FALSE | Upregulated |
| -0,541065059 | 0,521488928 | SLC35E3 | FALSE | Downregulated |
| 0,18680237 | 0,521488928 | STAT6 | FALSE | Upregulated |
| -0,273722765 | 0,521488928 | STT3A | FALSE | Downregulated |
| 0,277681997 | 0,521488928 | TFE3 | FALSE | Upregulated |
| -0,238710436 | 0,521488928 | TOR1AIP2 | FALSE | Downregulated |
| -0,699803931 | 0,521488928 | USP21 | FALSE | Downregulated |
| 0,195371705 | 0,521488928 | WDR1 | FALSE | Upregulated |
| 0,215977573 | 0,521488928 | ZFYVE16 | FALSE | Upregulated |
| 0,317855116 | 0,521524944 | KATNAL2 | FALSE | Upregulated |
| 0,22970917 | 0,521547457 | PMPCB | FALSE | Upregulated |
| 0,422660702 | 0,521591743 | MCM3 | FALSE | Upregulated |
| -0,525118005 | 0,521656908 | CCDC86 | FALSE | Downregulated |
| 0,21475812 | 0,521656908 | PGM3 | FALSE | Upregulated |
| 0,376999569 | 0,521656908 | ZNF462 | FALSE | Upregulated |
| -0,427692789 | 0,521660987 | INHBE | FALSE | Downregulated |
| -0,706723751 | 0,52172148 | DBIL5P | FALSE | Downregulated |
| -0,198759607 | 0,52172148 | SMEK2 | FALSE | Downregulated |
| -0,521060471 | 0,521768164 | POMP | FALSE | Downregulated |
| 0,395311698 | 0,521768164 | RCSD1 | FALSE | Upregulated |
| 0,781393717 | 0,522015029 | LOC101928498 | FALSE | Upregulated |
| -0,596683194 | 0,522015029 | LOC646762 | FALSE | Downregulated |
| -1,159060322 | 0,522256847 | CA11 | FALSE | Downregulated |
| -0,946651978 | 0,522256847 | DPM3 | FALSE | Downregulated |
| -0,230348526 | 0,522539901 | UPB1 | FALSE | Downregulated |
| 1,722962253 | 0,522579345 | TRIM39-RPP21 | FALSE | Upregulated |
| -0,277468798 | 0,522579345 | UBE2G2 | FALSE | Downregulated |
| 0,819818114 | 0,522807119 | PTGES3L | FALSE | Upregulated |
| 0,176903267 | 0,522941099 | ASXL1 | FALSE | Upregulated |
| -0,895406448 | 0,523097994 | TADA2A | FALSE | Downregulated |
| 0,313711515 | 0,523235757 | JAM3 | FALSE | Upregulated |
| 0,66478083 | 0,523235757 | TRIM74 | FALSE | Upregulated |
| -0,442554016 | 0,523286767 | MRPS10 | FALSE | Downregulated |
| 0,238696273 | 0,523346904 | C1orf86 | FALSE | Upregulated |
| 0,334268053 | 0,523346904 | RP11-103C16.2 | FALSE | Upregulated |
| 0,22559323 | 0,523365757 | FKBP15 | FALSE | Upregulated |
| -0,435109909 | 0,523508136 | KDM5B | FALSE | Downregulated |
| -0,308332999 | 0,523508136 | MARVELD2 | FALSE | Downregulated |
| 0,173465609 | 0,523908071 | HNRNPA1 | FALSE | Upregulated |
| 0,33350511 | 0,523908071 | LGALS3BP | FALSE | Upregulated |
| -0,17131401 | 0,523908071 | SUPT6H | FALSE | Downregulated |
| -0,253179261 | 0,524168733 | CALCOCO2 | FALSE | Downregulated |
| 0,498140077 | 0,524168733 | ELMOD2 | FALSE | Upregulated |
| -1,068283485 | 0,524168733 | LOC643355 | FALSE | Downregulated |

| | | | | |
|---|---|---|---|---|
| -0,377897153 | 0,524168733 | YIPF4 | FALSE | Downregulated |
| -0,803073064 | 0,524231241 | ASB9 | FALSE | Downregulated |
| -0,713093576 | 0,524231241 | LOC101928615 | FALSE | Downregulated |
| 0,615681493 | 0,524695568 | RHBDF1 | FALSE | Upregulated |
| 0,258056765 | 0,524950996 | IRGQ | FALSE | Upregulated |
| 0,791585502 | 0,524950996 | ZNF827 | FALSE | Upregulated |
| -2,054661948 | 0,525209415 | APITD1-CORT | FALSE | Downregulated |
| -0,199246622 | 0,525333053 | GPLD1 | FALSE | Downregulated |
| -0,52768537 | 0,52539342 | CUL2 | FALSE | Downregulated |
| -0,928058213 | 0,525616915 | LOC102724255 | FALSE | Downregulated |
| 1,106809894 | 0,525651214 | LOC102724916 | FALSE | Upregulated |
| 0,230545993 | 0,525651214 | ZMYM2 | FALSE | Upregulated |
| 0,336667155 | 0,525779722 | RAB35 | FALSE | Upregulated |
| 0,562978975 | 0,525779722 | SNORD10 | FALSE | Upregulated |
| 0,861903579 | 0,525928082 | CACNB1 | FALSE | Upregulated |
| 0,233608119 | 0,526167642 | CPEB4 | FALSE | Upregulated |
| 0,293807576 | 0,52626618 | PRICKLE2-AS1 | FALSE | Upregulated |
| -0,592040562 | 0,526301297 | NUDT8 | FALSE | Downregulated |
| -0,756431599 | 0,526343032 | CCDC59 | FALSE | Downregulated |
| -0,563447258 | 0,526343032 | CDHR2 | FALSE | Downregulated |
| 0,433323879 | 0,526343032 | DOCK10 | FALSE | Upregulated |
| -0,48145235 | 0,526453687 | SDSL | FALSE | Downregulated |
| -0,282731111 | 0,526457637 | PEX5 | FALSE | Downregulated |
| 0,936838646 | 0,526532279 | C16orf90 | FALSE | Upregulated |
| -0,200012012 | 0,526532279 | CAPN15 | FALSE | Downregulated |
| -1,411654637 | 0,526532279 | FAM71A | FALSE | Downregulated |
| 0,340022621 | 0,526532279 | IDUA | FALSE | Upregulated |
| 0,646649412 | 0,526532279 | ITGA3 | FALSE | Upregulated |
| 0,756478669 | 0,526532279 | KIAA1549 | FALSE | Upregulated |
| 1,002714802 | 0,526532279 | LINC00961 | FALSE | Upregulated |
| -0,574789503 | 0,526532279 | LMAN2L | FALSE | Downregulated |
| 0,377682827 | 0,526532279 | LNX1 | FALSE | Upregulated |
| -0,764415686 | 0,526532279 | POLB | FALSE | Downregulated |
| -0,587098671 | 0,526532279 | VWCE | FALSE | Downregulated |
| -0,695461923 | 0,526532279 | ZNF555 | FALSE | Downregulated |
| 0,850291628 | 0,52673399 | CHRND | FALSE | Upregulated |
| -0,237004402 | 0,526902687 | MRPL43 | FALSE | Downregulated |
| 0,588444587 | 0,527137301 | OARD1 | FALSE | Upregulated |
| 0,29696127 | 0,527137301 | RNF149 | FALSE | Upregulated |
| -0,553245681 | 0,527140426 | PDE11A | FALSE | Downregulated |
| 0,361216579 | 0,527388342 | DCUN1D4 | FALSE | Upregulated |
| 0,870380717 | 0,527388342 | DNAH14 | FALSE | Upregulated |
| 0,224543433 | 0,527388342 | FAM120AOS | FALSE | Upregulated |
| -1,280789093 | 0,527388342 | LOC100506124 | FALSE | Downregulated |
| 0,405827275 | 0,527388342 | MANEAL | FALSE | Upregulated |
| 0,503077077 | 0,527388342 | RP11-307E17.8 | FALSE | Upregulated |
| 0,722655154 | 0,527388342 | S100A11 | FALSE | Upregulated |
| 0,53275213 | 0,527422869 | RP11-536C5.7 | FALSE | Upregulated |
| 0,67395339 | 0,527491003 | CERS1 | FALSE | Upregulated |
| -0,505733075 | 0,527491003 | GREB1 | FALSE | Downregulated |
| -0,448410503 | 0,527491003 | MRPS30 | FALSE | Downregulated |

| | | | | |
|---|---|---|---|---|
| 0,240929715 | 0,527491003 | SLC9A7 | FALSE | Upregulated |
| 0,543858967 | 0,527491003 | TRIM45 | FALSE | Upregulated |
| 0,266933515 | 0,527589661 | LOC102723489 | FALSE | Upregulated |
| -0,344594984 | 0,527589661 | SUMO1 | FALSE | Downregulated |
| -0,279318006 | 0,527675972 | NDUFB8 | FALSE | Downregulated |
| -0,303143351 | 0,527784449 | PRPF19 | FALSE | Downregulated |
| 0,421294945 | 0,527784449 | USP50 | FALSE | Upregulated |
| 0,800614138 | 0,527833218 | HEPH | FALSE | Upregulated |
| -0,272661993 | 0,527833218 | RAB14 | FALSE | Downregulated |
| -0,426896318 | 0,527968868 | YPEL5 | FALSE | Downregulated |
| 0,316544607 | 0,528506561 | ZNF518A | FALSE | Upregulated |
| -0,278690937 | 0,528586442 | GSDMD | FALSE | Downregulated |
| -0,389243503 | 0,528619933 | PPP2R5C | FALSE | Downregulated |
| -0,809426973 | 0,528655453 | CNIH4 | FALSE | Downregulated |
| -0,848861437 | 0,528670079 | CRH | FALSE | Downregulated |
| 0,74346975 | 0,528670079 | PYCARD | FALSE | Upregulated |
| 0,637700163 | 0,528860387 | CMPK2 | FALSE | Upregulated |
| -0,436709312 | 0,528860387 | IFRD1 | FALSE | Downregulated |
| 0,741131835 | 0,529248241 | C1orf95 | FALSE | Upregulated |
| -1,29866629 | 0,529248241 | LOC100128276 | FALSE | Downregulated |
| -0,20819267 | 0,529248241 | SLC5A2 | FALSE | Downregulated |
| 0,313642147 | 0,529248241 | WDR37 | FALSE | Upregulated |
| -1,320679541 | 0,52940895 | OR51Q1 | FALSE | Downregulated |
| 0,725223236 | 0,529428241 | CCDC102B | FALSE | Upregulated |
| -0,371479885 | 0,529472016 | BAIAP2-AS1 | FALSE | Downregulated |
| 0,901123787 | 0,529801057 | SYT11 | FALSE | Upregulated |
| 0,652298533 | 0,529998361 | LOC102724757 | FALSE | Upregulated |
| -0,295763464 | 0,530020199 | EIF3F | FALSE | Downregulated |
| 0,788066216 | 0,530115925 | MLLT3 | FALSE | Upregulated |
| -0,26247011 | 0,530274251 | UTP14C | FALSE | Downregulated |
| 0,289217017 | 0,530291395 | DARS2 | FALSE | Upregulated |
| 1,309992824 | 0,530291395 | TUBB2B | FALSE | Upregulated |
| 0,469203408 | 0,53043793 | PXN-AS1 | FALSE | Upregulated |
| 0,215443179 | 0,530478362 | ANXA9 | FALSE | Upregulated |
| 0,493305797 | 0,530478362 | IDH3A | FALSE | Upregulated |
| -0,266576121 | 0,530592299 | BCOR | FALSE | Downregulated |
| 0,405064149 | 0,530614053 | ENPP7 | FALSE | Upregulated |
| 0,99463117 | 0,530614053 | KCNJ1 | FALSE | Upregulated |
| -0,271514225 | 0,530630504 | PHF8 | FALSE | Downregulated |
| 0,322669546 | 0,530645691 | ARMC10 | FALSE | Upregulated |
| -0,393430316 | 0,530645691 | DHX29 | FALSE | Downregulated |
| -0,478455845 | 0,530645691 | HLX-AS1 | FALSE | Downregulated |
| 1,087786193 | 0,530645691 | KAL1 | FALSE | Upregulated |
| -0,387261147 | 0,530645691 | NFKBIB | FALSE | Downregulated |
| 0,383676812 | 0,530645691 | ORMDL1 | FALSE | Upregulated |
| -0,302994325 | 0,530645691 | SKP1 | FALSE | Downregulated |
| -0,445194032 | 0,530645691 | VPS11 | FALSE | Downregulated |
| -0,970185977 | 0,530645691 | ZNRD1 | FALSE | Downregulated |
| 0,426779256 | 0,530652992 | ABT1 | FALSE | Upregulated |
| 0,827105279 | 0,530652992 | AK9 | FALSE | Upregulated |
| 0,416685049 | 0,530652992 | LOC102724153 | FALSE | Upregulated |

| | | | | |
|---|---|---|---|---|
| 0,550040774 | 0,530860309 | LOC101929425 | FALSE | Upregulated |
| -0,219926131 | 0,53094732 | ZBTB44 | FALSE | Downregulated |
| 0,234142062 | 0,530949785 | POLR3H | FALSE | Upregulated |
| -0,805632629 | 0,531059921 | LOC101927559 | FALSE | Downregulated |
| -0,655747394 | 0,531059921 | SIRT6 | FALSE | Downregulated |
| -1,289258809 | 0,531349442 | NXT2 | FALSE | Downregulated |
| 0,193312798 | 0,531349442 | TENM1 | FALSE | Upregulated |
| 0,957772257 | 0,531349442 | ZNF714 | FALSE | Upregulated |
| -1,232602612 | 0,532037667 | AADACL2 | FALSE | Downregulated |
| 0,924309621 | 0,532037667 | ACE2 | FALSE | Upregulated |
| 0,842245878 | 0,532427307 | GCM2 | FALSE | Upregulated |
| 0,415007676 | 0,532527354 | FAM73A | FALSE | Upregulated |
| -1,335329654 | 0,532842682 | SRGAP2B | FALSE | Downregulated |
| -0,880125677 | 0,533105618 | KDM5D | FALSE | Downregulated |
| 0,719775157 | 0,533253786 | B3GALT4 | FALSE | Upregulated |
| -1,169029192 | 0,533578799 | ATG4C | FALSE | Downregulated |
| 0,438235843 | 0,533578799 | C17orf104 | FALSE | Upregulated |
| 1,423204119 | 0,533617386 | NDUFA6-AS1 | FALSE | Upregulated |
| -0,310556514 | 0,533658281 | DDOST | FALSE | Downregulated |
| 0,381791576 | 0,533658281 | GRAMD1A | FALSE | Upregulated |
| -0,923415717 | 0,533658281 | HRC | FALSE | Downregulated |
| -0,345224477 | 0,533658281 | SFMBT1 | FALSE | Downregulated |
| 0,245466634 | 0,533668731 | HIF1AN | FALSE | Upregulated |
| -0,350686063 | 0,533719497 | PFAS | FALSE | Downregulated |
| 0,618873241 | 0,533752396 | CEP78 | FALSE | Upregulated |
| 0,260533746 | 0,533913891 | DCAF10 | FALSE | Upregulated |
| 0,207577285 | 0,533958544 | MIB2 | FALSE | Upregulated |
| -0,641102087 | 0,534704063 | GPR37 | FALSE | Downregulated |
| 0,246783534 | 0,535173474 | RAD50 | FALSE | Upregulated |
| 0,268315287 | 0,53519924 | PRKCZ | FALSE | Upregulated |
| 0,914614933 | 0,535633222 | RFC3 | FALSE | Upregulated |
| 1,065294366 | 0,535692397 | WBSCR27 | FALSE | Upregulated |
| 0,324696577 | 0,535708193 | WWC3 | FALSE | Upregulated |
| 0,811440683 | 0,535774689 | PDZD7 | FALSE | Upregulated |
| -0,290665081 | 0,536034456 | CRTAP | FALSE | Downregulated |
| 0,278815012 | 0,5361384 | PVR | FALSE | Upregulated |
| -0,384581042 | 0,536286251 | CC2D2B | FALSE | Downregulated |
| -1,309724326 | 0,536286251 | LOC102723466 | FALSE | Downregulated |
| -0,226301699 | 0,536320871 | MKNK2 | FALSE | Downregulated |
| -0,381887589 | 0,536320871 | MTO1 | FALSE | Downregulated |
| 0,691925598 | 0,536320871 | RP11-430B1.2 | FALSE | Upregulated |
| -0,305618194 | 0,536543694 | HARS | FALSE | Downregulated |
| 1,313830248 | 0,536563522 | CCL2 | FALSE | Upregulated |
| 0,355389709 | 0,536563522 | MAPKAPK3 | FALSE | Upregulated |
| 0,546640564 | 0,536563522 | NIFK-AS1 | FALSE | Upregulated |
| 0,28317749 | 0,536633312 | DOC2A | FALSE | Upregulated |
| 1,111637093 | 0,536633312 | SENCR | FALSE | Upregulated |
| 0,371242869 | 0,536633312 | SLC22A5 | FALSE | Upregulated |
| 0,415370211 | 0,536633312 | TAPT1-AS1 | FALSE | Upregulated |
| 1,258345533 | 0,536866378 | DDX43 | FALSE | Upregulated |
| -0,213845832 | 0,536866378 | DPP9 | FALSE | Downregulated |

| | | | | |
|---|---|---|---|---|
| -0,388256663 | 0,536866378 | FAM101B | FALSE | Downregulated |
| 0,556882578 | 0,536866378 | GAS5 | FALSE | Upregulated |
| -0,238781136 | 0,536866378 | HSD3B7 | FALSE | Downregulated |
| -0,208048999 | 0,536866378 | MLF2 | FALSE | Downregulated |
| -0,195757494 | 0,536866378 | SCAP | FALSE | Downregulated |
| 0,476228286 | 0,536866378 | SMPD5 | FALSE | Upregulated |
| -0,290609563 | 0,536866378 | SSRP1 | FALSE | Downregulated |
| -0,397892456 | 0,536866378 | ZNF446 | FALSE | Downregulated |
| 0,480349253 | 0,536992037 | ATL1 | FALSE | Upregulated |
| 0,749615294 | 0,536992037 | FREM2 | FALSE | Upregulated |
| 0,525934063 | 0,536992037 | TSSK2 | FALSE | Upregulated |
| -0,244429263 | 0,537046357 | ULK1 | FALSE | Downregulated |
| -0,390153574 | 0,53711623 | STX1B | FALSE | Downregulated |
| -0,761754553 | 0,537257434 | PFDN6 | FALSE | Downregulated |
| 0,250814418 | 0,537257434 | PHF12 | FALSE | Upregulated |
| -0,737682029 | 0,5376303 | LOC100287896 | FALSE | Downregulated |
| 0,718063872 | 0,537646009 | C19orf57 | FALSE | Upregulated |
| 0,78259385 | 0,537646009 | CERCAM | FALSE | Upregulated |
| 0,380957034 | 0,537646009 | DSERG1 | FALSE | Upregulated |
| 0,333164819 | 0,537646009 | SOX18 | FALSE | Upregulated |
| -0,574832274 | 0,537646009 | TCN2 | FALSE | Downregulated |
| 0,427958185 | 0,537646009 | UNC5B | FALSE | Upregulated |
| 0,821740448 | 0,537718107 | MDK | FALSE | Upregulated |
| -0,855707617 | 0,537724113 | CCDC53 | FALSE | Downregulated |
| -0,247429937 | 0,537724113 | IDH2 | FALSE | Downregulated |
| 0,746947917 | 0,537724113 | KLF8 | FALSE | Upregulated |
| -0,925185787 | 0,537724113 | TCP10L | FALSE | Downregulated |
| 0,599857624 | 0,537734315 | BLOC1S4 | FALSE | Upregulated |
| -0,635948558 | 0,537734315 | C1orf226 | FALSE | Downregulated |
| -0,411474318 | 0,537891224 | TMEM50B | FALSE | Downregulated |
| -0,255368049 | 0,538216095 | CAPN1 | FALSE | Downregulated |
| -1,270608411 | 0,538216095 | LOC284581 | FALSE | Downregulated |
| -0,452549908 | 0,538238088 | MKKS | FALSE | Downregulated |
| 0,326923757 | 0,538325235 | GLTSCR1L | FALSE | Upregulated |
| 0,592658414 | 0,538325235 | LOC100507547 | FALSE | Upregulated |
| -0,33269644 | 0,538325235 | PKP2 | FALSE | Downregulated |
| 0,24782511 | 0,538325235 | SIRT1 | FALSE | Upregulated |
| -0,460606366 | 0,538376026 | CYP3A4 | FALSE | Downregulated |
| -1,792779003 | 0,538376026 | LOC101927374 | FALSE | Downregulated |
| 0,322794441 | 0,538396597 | DCAF12 | FALSE | Upregulated |
| -1,001070839 | 0,538413908 | ANKRD35 | FALSE | Downregulated |
| 1,114810771 | 0,538458474 | XKR6 | FALSE | Upregulated |
| 0,689744629 | 0,538861018 | DEPDC4 | FALSE | Upregulated |
| 0,88994448 | 0,538861018 | IL5 | FALSE | Upregulated |
| 1,000029484 | 0,538861018 | LOC90834 | FALSE | Upregulated |
| -0,299134221 | 0,538861018 | PIGQ | FALSE | Downregulated |
| -0,272224986 | 0,538861018 | PRPF3 | FALSE | Downregulated |
| -0,642389074 | 0,538861018 | ZNF595 | FALSE | Downregulated |
| 0,918339793 | 0,538872825 | IGSF3 | FALSE | Upregulated |
| 0,229676867 | 0,538968794 | NCOA3 | FALSE | Upregulated |
| -0,472436675 | 0,539078764 | STOX2 | FALSE | Downregulated |

| | | | | |
|---|---|---|---|---|
| 0,638685015 | 0,539493357 | HYAL3 | FALSE | Upregulated |
| -0,657632299 | 0,539493357 | LOC101060494 | FALSE | Downregulated |
| -0,348327469 | 0,539493357 | SERTAD2 | FALSE | Downregulated |
| 0,658088561 | 0,539610183 | CKAP2 | FALSE | Upregulated |
| -0,168474193 | 0,539696814 | SETD2 | FALSE | Downregulated |
| -0,36219723 | 0,540031586 | PET112 | FALSE | Downregulated |
| -0,577122466 | 0,540044874 | AGPAT5 | FALSE | Downregulated |
| 0,214953631 | 0,540044874 | OAZ2 | FALSE | Upregulated |
| -0,312151411 | 0,540238633 | THBS3 | FALSE | Downregulated |
| -0,314972625 | 0,540414709 | SLC25A44 | FALSE | Downregulated |
| -0,268075255 | 0,540431715 | AFM | FALSE | Downregulated |
| -0,680550523 | 0,540431715 | CEP112 | FALSE | Downregulated |
| -0,882465776 | 0,540431715 | LOC101927752 | FALSE | Downregulated |
| 0,414799009 | 0,540431715 | ZNF622 | FALSE | Upregulated |
| 0,90806819 | 0,540534669 | C19orf84 | FALSE | Upregulated |
| -0,333716357 | 0,540641243 | BST2 | FALSE | Downregulated |
| 0,25865344 | 0,540641243 | CNOT7 | FALSE | Upregulated |
| 0,737902185 | 0,540641243 | LEF1 | FALSE | Upregulated |
| -0,628554772 | 0,540641243 | LINC00886 | FALSE | Downregulated |
| 0,927355756 | 0,540641243 | NLRP14 | FALSE | Upregulated |
| 0,659710677 | 0,540641243 | VAV3 | FALSE | Upregulated |
| -0,379668614 | 0,540658361 | GNAI1 | FALSE | Downregulated |
| 0,400982234 | 0,540897757 | ELMOD3 | FALSE | Upregulated |
| -0,699669862 | 0,540933979 | TMEM5 | FALSE | Downregulated |
| 0,338678106 | 0,541033449 | HTRA2 | FALSE | Upregulated |
| 0,59573407 | 0,541033449 | SLC35B3 | FALSE | Upregulated |
| -0,600928178 | 0,541047215 | FCGR2B | FALSE | Downregulated |
| 0,329958002 | 0,541047215 | LOC101929004 | FALSE | Upregulated |
| 0,290413036 | 0,541047215 | PHPT1 | FALSE | Upregulated |
| 0,318799412 | 0,541056152 | HMGN3 | FALSE | Upregulated |
| -0,244086031 | 0,541056152 | HSF1 | FALSE | Downregulated |
| -0,269096829 | 0,541100953 | IMPDH2 | FALSE | Downregulated |
| 0,861734502 | 0,541168828 | GPR26 | FALSE | Upregulated |
| 0,197004726 | 0,541168828 | XPO1 | FALSE | Upregulated |
| 0,651807395 | 0,54122121 | GMFG | FALSE | Upregulated |
| 0,797687091 | 0,541312249 | RPSAP58 | FALSE | Upregulated |
| 0,895915814 | 0,541312299 | DGAT2L6 | FALSE | Upregulated |
| 0,295663218 | 0,541362334 | CERS6 | FALSE | Upregulated |
| -0,760160258 | 0,541362334 | LOC113230 | FALSE | Downregulated |
| -0,326226608 | 0,541362334 | YY1AP1 | FALSE | Downregulated |
| -0,141372993 | 0,54146351 | PINK1 | FALSE | Downregulated |
| 0,381995121 | 0,541783224 | TPTE2P5 | FALSE | Upregulated |
| -0,484895592 | 0,541783224 | TTC1 | FALSE | Downregulated |
| 0,311683101 | 0,541974469 | BRPF1 | FALSE | Upregulated |
| 0,928848152 | 0,541974469 | CHCHD6 | FALSE | Upregulated |
| 0,444706758 | 0,541974469 | DGCR9 | FALSE | Upregulated |
| 0,346102286 | 0,541974469 | GIPC1 | FALSE | Upregulated |
| 0,540857975 | 0,541974469 | SMC2 | FALSE | Upregulated |
| -0,534438015 | 0,542017276 | C15orf40 | FALSE | Downregulated |
| -0,413122167 | 0,542017276 | FBXO44 | FALSE | Downregulated |
| 1,07081788 | 0,542017276 | LOC101929490 | FALSE | Upregulated |

| | | | | |
|---|---|---|---|---|
| 0,548847428 | 0,542017276 | LOC102723457 | FALSE | Upregulated |
| -0,477398796 | 0,542017276 | MRPL46 | FALSE | Downregulated |
| 0,219153084 | 0,542017276 | RBM19 | FALSE | Upregulated |
| 0,49157277 | 0,542017276 | TFRC | FALSE | Upregulated |
| -0,529539279 | 0,542150792 | EP400NL | FALSE | Downregulated |
| 0,959927603 | 0,542312798 | JAKMIP3 | FALSE | Upregulated |
| 0,531677299 | 0,542321734 | C6orf211 | FALSE | Upregulated |
| -0,186446979 | 0,542321734 | EIF3A | FALSE | Downregulated |
| -0,346490356 | 0,542321734 | RPS12 | FALSE | Downregulated |
| 0,34021389 | 0,542325815 | TRIM23 | FALSE | Upregulated |
| -0,493212678 | 0,542555071 | NDUFA9 | FALSE | Downregulated |
| 0,312485946 | 0,542820489 | LOC101927903 | FALSE | Upregulated |
| 0,404682663 | 0,542820489 | SLFNL1 | FALSE | Upregulated |
| -0,221417907 | 0,542820489 | TADA3 | FALSE | Downregulated |
| -0,512432246 | 0,542820489 | TTLL11 | FALSE | Downregulated |
| 0,403253423 | 0,542820489 | WWC1 | FALSE | Upregulated |
| 0,255367318 | 0,543091802 | ATP2C1 | FALSE | Upregulated |
| -0,253675825 | 0,543091802 | KLHDC3 | FALSE | Downregulated |
| 0,282585264 | 0,543091802 | RBMS1 | FALSE | Upregulated |
| 0,293817466 | 0,543091802 | SPATA20 | FALSE | Upregulated |
| -0,440792059 | 0,543091802 | TSSC4 | FALSE | Downregulated |
| 0,237614618 | 0,543091802 | WIPI2 | FALSE | Upregulated |
| -0,241288021 | 0,543202109 | ATF6B | FALSE | Downregulated |
| 0,263720846 | 0,543202109 | GMPR2 | FALSE | Upregulated |
| -0,358593335 | 0,543202109 | KIAA1429 | FALSE | Downregulated |
| -0,635797172 | 0,543202109 | MRPL21 | FALSE | Downregulated |
| 0,30318132 | 0,543202109 | PTRH2 | FALSE | Upregulated |
| 0,251259005 | 0,543202109 | USP36 | FALSE | Upregulated |
| 0,528515682 | 0,543425954 | LOC102724164 | FALSE | Upregulated |
| 0,342373408 | 0,543425954 | RSBN1L | FALSE | Upregulated |
| 0,344016618 | 0,543687268 | CCT8 | FALSE | Upregulated |
| 0,447759243 | 0,543687268 | CDT1 | FALSE | Upregulated |
| 0,693358467 | 0,543918481 | CA5B | FALSE | Upregulated |
| -0,308586128 | 0,543918481 | SLC39A13 | FALSE | Downregulated |
| -0,623959053 | 0,543918481 | SOWAHA | FALSE | Downregulated |
| -0,441729951 | 0,543918481 | TRMT2B | FALSE | Downregulated |
| 0,971720517 | 0,543918481 | ZNF284 | FALSE | Upregulated |
| 0,660392977 | 0,543984061 | MKLN1-AS | FALSE | Upregulated |
| 0,914640958 | 0,544163413 | ANXA13 | FALSE | Upregulated |
| 0,936522994 | 0,544163413 | C22orf34 | FALSE | Upregulated |
| 0,478586911 | 0,544163413 | CACNB3 | FALSE | Upregulated |
| 0,831773669 | 0,544163413 | TMEM150B | FALSE | Upregulated |
| -0,219800874 | 0,544163413 | TSPAN9 | FALSE | Downregulated |
| 0,209473879 | 0,544242655 | PRKD3 | FALSE | Upregulated |
| 0,696874316 | 0,544242655 | RIMS3 | FALSE | Upregulated |
| -0,42513409 | 0,544573496 | ATAD3B | FALSE | Downregulated |
| 0,456557741 | 0,544573496 | ATP8B4 | FALSE | Upregulated |
| 0,452827877 | 0,544573496 | CCDC121 | FALSE | Upregulated |
| 0,883193837 | 0,544573496 | DENND6B | FALSE | Upregulated |
| -1,226072879 | 0,544573496 | LOC344967 | FALSE | Downregulated |
| -0,405076486 | 0,544573496 | SAPCD2 | FALSE | Downregulated |

| | | | | |
|---|---|---|---|---|
| 0,29304361 | 0,544596676 | MAK16 | FALSE | Upregulated |
| 0,302529751 | 0,544634604 | C6orf201 | FALSE | Upregulated |
| -0,21486276 | 0,544634604 | GTF2I | FALSE | Downregulated |
| 1,004610121 | 0,544634604 | LOC101929165 | FALSE | Upregulated |
| 0,964686649 | 0,544724537 | P2RY6 | FALSE | Upregulated |
| 0,413054076 | 0,544812359 | FAM86C1 | FALSE | Upregulated |
| 0,520104293 | 0,544812359 | HOMER3 | FALSE | Upregulated |
| -0,374425699 | 0,544812359 | POLL | FALSE | Downregulated |
| 0,22594618 | 0,544812359 | RHOF | FALSE | Upregulated |
| -0,233765339 | 0,544936135 | CDH2 | FALSE | Downregulated |
| -0,456154071 | 0,544936135 | FBXO42 | FALSE | Downregulated |
| -0,265887243 | 0,544936135 | RXRB | FALSE | Downregulated |
| -0,489059995 | 0,544956832 | EFCAB4A | FALSE | Downregulated |
| 0,300426296 | 0,545169028 | RP4-724E16.2 | FALSE | Upregulated |
| -1,452278644 | 0,545185185 | LOC101928841 | FALSE | Downregulated |
| -0,267027561 | 0,545471157 | UBE2O | FALSE | Downregulated |
| 0,163626865 | 0,545600543 | HBP1 | FALSE | Upregulated |
| -1,155880642 | 0,54564465 | SNHG19 | FALSE | Downregulated |
| -0,44273604 | 0,545807436 | LTN1 | FALSE | Downregulated |
| 0,429604288 | 0,545807436 | SERPINB9 | FALSE | Upregulated |
| 0,542074877 | 0,545898229 | ANKRD36B | FALSE | Upregulated |
| -0,850290832 | 0,545898229 | RP11-513G11.3 | FALSE | Downregulated |
| -0,337886502 | 0,545898229 | VTI1B | FALSE | Downregulated |
| 0,365298478 | 0,545898229 | ZC3H15 | FALSE | Upregulated |
| 1,263759133 | 0,545920337 | GRM3 | FALSE | Upregulated |
| 0,727882096 | 0,546061422 | B3GALTL | FALSE | Upregulated |
| 0,763816389 | 0,546061422 | CEP19 | FALSE | Upregulated |
| 0,258452944 | 0,546061422 | CRB2 | FALSE | Upregulated |
| 0,543924364 | 0,546061422 | LPP-AS2 | FALSE | Upregulated |
| 0,41126115 | 0,546061422 | MS4A7 | FALSE | Upregulated |
| 0,41541621 | 0,546061422 | ZNF354C | FALSE | Upregulated |
| 0,487789398 | 0,546218203 | CENPN | FALSE | Upregulated |
| -0,208030245 | 0,546218203 | CYP8B1 | FALSE | Downregulated |
| 0,834912595 | 0,546218203 | GJC3 | FALSE | Upregulated |
| 0,813145945 | 0,546218203 | LOC102724368 | FALSE | Upregulated |
| -0,575817659 | 0,546218203 | MTA3 | FALSE | Downregulated |
| -0,692793384 | 0,54643515 | ARHGAP19 | FALSE | Downregulated |
| 0,893782095 | 0,54643515 | LSM6 | FALSE | Upregulated |
| -0,307602185 | 0,54643515 | MESTIT1 | FALSE | Downregulated |
| 0,32996368 | 0,54643515 | TLE4 | FALSE | Upregulated |
| 0,451349394 | 0,54643515 | WNK4 | FALSE | Upregulated |
| -0,224432956 | 0,546469175 | FPGS | FALSE | Downregulated |
| -0,955655246 | 0,546481013 | TBC1D7 | FALSE | Downregulated |
| -0,298556586 | 0,546649343 | GOLGA2P5 | FALSE | Downregulated |
| -0,880653776 | 0,546689729 | CYP1A1 | FALSE | Downregulated |
| -0,388288099 | 0,546689729 | PANX2 | FALSE | Downregulated |
| 0,489531196 | 0,546689729 | ZNF324B | FALSE | Upregulated |
| 0,594175847 | 0,546753547 | PNMA2 | FALSE | Upregulated |
| 1,098717815 | 0,546753547 | SLC35F1 | FALSE | Upregulated |
| 0,874765945 | 0,546753547 | ZNF185 | FALSE | Upregulated |
| 0,301982039 | 0,546905483 | MAP3K1 | FALSE | Upregulated |

| | | | | |
|---|---|---|---|---|
| 0,330410662 | 0,547009275 | DNMT3A | FALSE | Upregulated |
| 0,457211249 | 0,547205646 | EMC8 | FALSE | Upregulated |
| -0,192806945 | 0,547382626 | TRAK1 | FALSE | Downregulated |
| -0,353979487 | 0,547455061 | SAMD10 | FALSE | Downregulated |
| 0,254258721 | 0,547500378 | C14orf93 | FALSE | Upregulated |
| -0,323638806 | 0,547500378 | DEK | FALSE | Downregulated |
| 1,335527353 | 0,547500378 | EGLN3 | FALSE | Upregulated |
| -0,22670506 | 0,547500378 | SLC35D1 | FALSE | Downregulated |
| 0,579285057 | 0,547545813 | PIGS | FALSE | Upregulated |
| -0,224173543 | 0,547621554 | HNRNPDL | FALSE | Downregulated |
| -0,355969234 | 0,547797209 | FDPS | FALSE | Downregulated |
| 0,435119382 | 0,547797209 | POLD1 | FALSE | Upregulated |
| 0,499904418 | 0,547871124 | LRP12 | FALSE | Upregulated |
| -0,197395414 | 0,547871124 | SUPT5H | FALSE | Downregulated |
| 0,340843853 | 0,547891202 | COCH | FALSE | Upregulated |
| 0,434697251 | 0,547891202 | SWAP70 | FALSE | Upregulated |
| -1,004062814 | 0,548242088 | ACPL2 | FALSE | Downregulated |
| -0,216554667 | 0,548242088 | ITSN2 | FALSE | Downregulated |
| 0,68763432 | 0,548242088 | PIGA | FALSE | Upregulated |
| 0,415598762 | 0,54843218 | AK5 | FALSE | Upregulated |
| -0,426093209 | 0,54843218 | CYGB | FALSE | Downregulated |
| 0,783285176 | 0,54843218 | RALGPS1 | FALSE | Upregulated |
| -0,304746187 | 0,54843218 | TEN1 | FALSE | Downregulated |
| 0,586879805 | 0,548455581 | TSPYL5 | FALSE | Upregulated |
| 0,295169675 | 0,54853502 | CTDSPL | FALSE | Upregulated |
| 0,51986557 | 0,548671286 | RARG | FALSE | Upregulated |
| 0,540297374 | 0,548698951 | APBB3 | FALSE | Upregulated |
| -0,728649816 | 0,548698951 | HSDL1 | FALSE | Downregulated |
| 0,614961542 | 0,548698951 | UTP15 | FALSE | Upregulated |
| -1,17669183 | 0,548776382 | MIOX | FALSE | Downregulated |
| 0,18402898 | 0,548796065 | PTCD3 | FALSE | Upregulated |
| -1,111831025 | 0,548848191 | ESPL1 | FALSE | Downregulated |
| -0,332902782 | 0,548848191 | HIC2 | FALSE | Downregulated |
| -0,360327124 | 0,548848191 | KCNS2 | FALSE | Downregulated |
| -0,745322743 | 0,548848191 | NIPSNAP3B | FALSE | Downregulated |
| 1,095859604 | 0,548848191 | PDLIM3 | FALSE | Upregulated |
| 0,547450234 | 0,548848191 | TMEM147 | FALSE | Upregulated |
| -0,343387073 | 0,548848191 | TTC39C | FALSE | Downregulated |
| 0,527186133 | 0,548848191 | UBE2Q2 | FALSE | Upregulated |
| -0,178946353 | 0,548848191 | ZNF516 | FALSE | Downregulated |
| 1,018516452 | 0,549233831 | LOC102724428 | FALSE | Upregulated |
| -0,716829779 | 0,549330687 | NGDN | FALSE | Downregulated |
| -1,470577395 | 0,549407952 | GPR75-ASB3 | FALSE | Downregulated |
| 0,462882305 | 0,549407952 | ZNF141 | FALSE | Upregulated |
| 0,680389833 | 0,549616241 | CDK5R1 | FALSE | Upregulated |
| -0,849400201 | 0,549620079 | LRP4-AS1 | FALSE | Downregulated |
| 0,551302359 | 0,549662198 | LOC102723405 | FALSE | Upregulated |
| -0,293036083 | 0,549946535 | LOC101929109 | FALSE | Downregulated |
| -1,315772459 | 0,550122455 | LOC100287477 | FALSE | Downregulated |
| 0,372081823 | 0,550202776 | RP11-300J18.1 | FALSE | Upregulated |
| 0,268516067 | 0,550435279 | RBM3 | FALSE | Upregulated |

| | | | | |
|---|---|---|---|---|
| 0,38380976 | 0,550561927 | C15orf52 | FALSE | Upregulated |
| 1,420502462 | 0,550561927 | OR10A5 | FALSE | Upregulated |
| 0,213621345 | 0,550561927 | TGFBR2 | FALSE | Upregulated |
| -0,754892792 | 0,55062485 | PRB1 | FALSE | Downregulated |
| 0,71202421 | 0,55081126 | ZNF845 | FALSE | Upregulated |
| -0,524546164 | 0,551110178 | DNAJB14 | FALSE | Downregulated |
| 0,75893916 | 0,551110178 | SLC9A3 | FALSE | Upregulated |
| 0,655502466 | 0,551316258 | AC147651.4 | FALSE | Upregulated |
| 1,375443555 | 0,551542162 | FLNC | FALSE | Upregulated |
| 0,313916461 | 0,551542162 | WSB1 | FALSE | Upregulated |
| -0,438845157 | 0,551545454 | DDX31 | FALSE | Downregulated |
| -0,26434427 | 0,552075319 | FBXO7 | FALSE | Downregulated |
| 0,464372869 | 0,552091003 | LOC100506114 | FALSE | Upregulated |
| -0,924690436 | 0,552091003 | SNORD118 | FALSE | Downregulated |
| -0,25079286 | 0,552091003 | TGFBR3 | FALSE | Downregulated |
| 0,274306012 | 0,552539966 | DAB2IP | FALSE | Upregulated |
| 0,189264652 | 0,552539966 | FAM208B | FALSE | Upregulated |
| -0,557415566 | 0,552660064 | NLE1 | FALSE | Downregulated |
| -0,7825998 | 0,552665515 | RPP30 | FALSE | Downregulated |
| 0,527437009 | 0,552889668 | PCDH17 | FALSE | Upregulated |
| -0,205049215 | 0,552923762 | ATP5A1 | FALSE | Downregulated |
| -1,200017243 | 0,552923762 | KCNJ14 | FALSE | Downregulated |
| -0,265955802 | 0,552923762 | NOP14 | FALSE | Downregulated |
| -0,259013578 | 0,552923762 | SCAND1 | FALSE | Downregulated |
| -0,464343594 | 0,552923762 | SIRT4 | FALSE | Downregulated |
| 0,293275597 | 0,552973709 | NEO1 | FALSE | Upregulated |
| 0,591626334 | 0,552995775 | ALG6 | FALSE | Upregulated |
| -0,618514769 | 0,552995775 | AOC1 | FALSE | Downregulated |
| 0,19023048 | 0,552995775 | COG5 | FALSE | Upregulated |
| 0,502669254 | 0,552995775 | COQ10B | FALSE | Upregulated |
| -1,281364018 | 0,552995775 | LINC00202-1 | FALSE | Downregulated |
| 0,590907331 | 0,552995775 | LOC102724061 | FALSE | Upregulated |
| -0,561455778 | 0,552995775 | RNGTT | FALSE | Downregulated |
| -0,342534586 | 0,552995775 | SCARNA12 | FALSE | Downregulated |
| 0,532872509 | 0,552995775 | UTP6 | FALSE | Upregulated |
| 0,680415953 | 0,552995775 | WDPCP | FALSE | Upregulated |
| -0,638105497 | 0,553021159 | NUCB2 | FALSE | Downregulated |
| -0,291485714 | 0,553099158 | SMAD6 | FALSE | Downregulated |
| 0,448029523 | 0,553099158 | STS | FALSE | Upregulated |
| -0,501132368 | 0,553324108 | CCDC106 | FALSE | Downregulated |
| 0,248066677 | 0,553324108 | CKAP4 | FALSE | Upregulated |
| 0,539217238 | 0,553324108 | EXO5 | FALSE | Upregulated |
| 0,583251927 | 0,553324108 | LYSMD4 | FALSE | Upregulated |
| -0,543891337 | 0,553324108 | NVL | FALSE | Downregulated |
| -1,275661798 | 0,553324108 | PFDN4 | FALSE | Downregulated |
| 0,521674659 | 0,553437841 | LOC101929018 | FALSE | Upregulated |
| -0,411970939 | 0,553582772 | FUT6 | FALSE | Downregulated |
| -1,24211983 | 0,553582772 | LOC100129940 | FALSE | Downregulated |
| 0,805511542 | 0,553582772 | LOC102724387 | FALSE | Upregulated |
| -0,496745976 | 0,553735686 | COX6C | FALSE | Downregulated |
| 0,951166062 | 0,553735686 | LOC101928571 | FALSE | Upregulated |

| | | | | |
|---|---|---|---|---|
| 0,204760655 | 0,553735686 | NUP50 | FALSE | Upregulated |
| 0,633265551 | 0,553735686 | RBPMS2 | FALSE | Upregulated |
| -0,859593757 | 0,553735686 | SNHG11 | FALSE | Downregulated |
| 0,449534456 | 0,553735686 | SRPX2 | FALSE | Upregulated |
| 0,665077552 | 0,553735686 | TSPAN13 | FALSE | Upregulated |
| -0,433328336 | 0,553735686 | UBL5 | FALSE | Downregulated |
| 0,490608934 | 0,554224602 | EML1 | FALSE | Upregulated |
| 0,433873936 | 0,554233365 | LOC101929072 | FALSE | Upregulated |
| -1,169299972 | 0,554317594 | DSCAM | FALSE | Downregulated |
| 0,273724801 | 0,554317594 | GAB1 | FALSE | Upregulated |
| -0,378478594 | 0,554317594 | LOC102724790 | FALSE | Downregulated |
| -0,381304523 | 0,554317594 | NBPF12 | FALSE | Downregulated |
| -1,063768662 | 0,554317594 | NOL4 | FALSE | Downregulated |
| 0,5005175 | 0,554317594 | NPR3 | FALSE | Upregulated |
| 1,286751777 | 0,554317594 | NRG2 | FALSE | Upregulated |
| -0,311930243 | 0,554317594 | PDHA1 | FALSE | Downregulated |
| 0,501385489 | 0,554317594 | PSMD14 | FALSE | Upregulated |
| -0,707565543 | 0,554317594 | SNRPF | FALSE | Downregulated |
| -0,267194002 | 0,554317594 | STRADB | FALSE | Downregulated |
| 0,28313983 | 0,554317594 | TPI1P3 | FALSE | Upregulated |
| 0,337759042 | 0,554317594 | VAMP3 | FALSE | Upregulated |
| -0,209854281 | 0,554619058 | MAML3 | FALSE | Downregulated |
| -0,656494128 | 0,554619058 | TRHDE-AS1 | FALSE | Downregulated |
| -0,46757844 | 0,555679661 | C12orf57 | FALSE | Downregulated |
| -0,471032102 | 0,555679661 | DPH2 | FALSE | Downregulated |
| -0,659026136 | 0,555679661 | GMCL1 | FALSE | Downregulated |
| -0,988701975 | 0,555679661 | TNRC18P1 | FALSE | Downregulated |
| -0,442114649 | 0,555768285 | LOC389834 | FALSE | Downregulated |
| 0,392402073 | 0,555768285 | RNF34 | FALSE | Upregulated |
| 0,279093869 | 0,555799173 | FLJ37453 | FALSE | Upregulated |
| 0,520242316 | 0,555799173 | LINC00843 | FALSE | Upregulated |
| 0,424498499 | 0,555799173 | MFSD1 | FALSE | Upregulated |
| 0,305219255 | 0,555799173 | PRR13 | FALSE | Upregulated |
| 0,3339402 | 0,555799173 | QSOX1 | FALSE | Upregulated |
| -1,137894457 | 0,555923011 | RP11-563J2.2 | FALSE | Downregulated |
| 0,390188386 | 0,55614926 | ZNF808 | FALSE | Upregulated |
| 0,376618296 | 0,556164834 | IPO4 | FALSE | Upregulated |
| -0,928368893 | 0,556251095 | LOC101927032 | FALSE | Downregulated |
| 0,398417798 | 0,556251095 | LPCAT1 | FALSE | Upregulated |
| 0,727549851 | 0,556251095 | SELM | FALSE | Upregulated |
| 0,482872749 | 0,556265344 | DDHD1 | FALSE | Upregulated |
| -0,790730294 | 0,556265344 | LINC00852 | FALSE | Downregulated |
| -0,25509826 | 0,556265344 | SKIL | FALSE | Downregulated |
| 0,228967282 | 0,556487849 | NR2C2 | FALSE | Upregulated |
| -0,28443945 | 0,556487849 | PATZ1 | FALSE | Downregulated |
| -0,204810697 | 0,556660932 | SMPD4 | FALSE | Downregulated |
| 0,352354886 | 0,556741986 | TPGS2 | FALSE | Upregulated |
| -0,22330126 | 0,556989092 | SLC25A5 | FALSE | Downregulated |
| 0,755019907 | 0,557319782 | RUNDC3A | FALSE | Upregulated |
| -0,695068954 | 0,557492473 | TMEM14A | FALSE | Downregulated |
| 0,727942839 | 0,557493447 | CTSK | FALSE | Upregulated |

| | | | | |
|---|---|---|---|---|
| 0,301550628 | 0,557533095 | C21orf62 | FALSE | Upregulated |
| -0,471355355 | 0,557668034 | MAD2L2 | FALSE | Downregulated |
| -0,199508462 | 0,557890533 | SYNE1 | FALSE | Downregulated |
| -1,332814895 | 0,558010411 | BEST2 | FALSE | Downregulated |
| -0,736641024 | 0,558010411 | LOC283177 | FALSE | Downregulated |
| 0,328594768 | 0,558070411 | RAVER1 | FALSE | Upregulated |
| 0,695582687 | 0,558769425 | FLJ46363 | FALSE | Upregulated |
| 0,448674473 | 0,558786364 | APBB1 | FALSE | Upregulated |
| -0,325920168 | 0,558786364 | FN3K | FALSE | Downregulated |
| 0,223793658 | 0,558978764 | ARHGEF26 | FALSE | Upregulated |
| -0,315547257 | 0,558978764 | KXD1 | FALSE | Downregulated |
| -0,527190823 | 0,559078959 | CCP110 | FALSE | Downregulated |
| -0,650586015 | 0,559159199 | TTC18 | FALSE | Downregulated |
| 0,865766789 | 0,559248308 | ASCL2 | FALSE | Upregulated |
| -0,379569974 | 0,559248308 | EIF4EBP1 | FALSE | Downregulated |
| 0,834601373 | 0,559248308 | KCNIP2-AS1 | FALSE | Upregulated |
| -1,149972439 | 0,559248308 | RP11-637A17.2 | FALSE | Downregulated |
| -0,17234102 | 0,559248308 | TNKS1BP1 | FALSE | Downregulated |
| -0,620403139 | 0,559248308 | TVP23B | FALSE | Downregulated |
| -0,229132361 | 0,559248308 | UBE3C | FALSE | Downregulated |
| 0,536949161 | 0,559536242 | FLJ38576 | FALSE | Upregulated |
| 0,947412873 | 0,559561742 | FBXW9 | FALSE | Upregulated |
| 0,554839379 | 0,559813841 | P4HA2 | FALSE | Upregulated |
| -0,3691815 | 0,559831835 | SH2D3A | FALSE | Downregulated |
| 0,274932591 | 0,559931357 | RAPGEF3 | FALSE | Upregulated |
| -0,331025252 | 0,559949092 | SMC6 | FALSE | Downregulated |
| -0,365985699 | 0,560298479 | PLA2G6 | FALSE | Downregulated |
| -0,56801525 | 0,560368539 | CPAMD8 | FALSE | Downregulated |
| 0,489125998 | 0,560368539 | DCUN1D2 | FALSE | Upregulated |
| 0,236311833 | 0,560368539 | KANK3 | FALSE | Upregulated |
| -0,31572779 | 0,560584823 | NOL9 | FALSE | Downregulated |
| 0,300747607 | 0,560894738 | ZDHHC20 | FALSE | Upregulated |
| 0,338307896 | 0,560909824 | ZNF41 | FALSE | Upregulated |
| 0,693673479 | 0,561023132 | RFPL1 | FALSE | Upregulated |
| 0,437701843 | 0,561137372 | ALX4 | FALSE | Upregulated |
| -0,26155597 | 0,561156054 | SSH1 | FALSE | Downregulated |
| -0,256346219 | 0,561209368 | CLK3 | FALSE | Downregulated |
| 1,800310454 | 0,561234581 | LOC102723584 | FALSE | Upregulated |
| 0,333254156 | 0,561341823 | ADCY3 | FALSE | Upregulated |
| -0,254640841 | 0,561341823 | SEMA5A | FALSE | Downregulated |
| 0,268915678 | 0,561341823 | SPIDR | FALSE | Upregulated |
| -0,631165439 | 0,561350301 | LOC102723954 | FALSE | Downregulated |
| -0,240678532 | 0,561350301 | NDUFS1 | FALSE | Downregulated |
| -0,535096661 | 0,561350301 | PGBD5 | FALSE | Downregulated |
| 0,46245908 | 0,561350301 | TRAF3IP1 | FALSE | Upregulated |
| 0,373186559 | 0,561427238 | NR2C1 | FALSE | Upregulated |
| -0,553623322 | 0,561698856 | ATP5J2 | FALSE | Downregulated |
| -0,244007815 | 0,561698856 | BTF3 | FALSE | Downregulated |
| 0,370806467 | 0,561698856 | GPANK1 | FALSE | Upregulated |
| -0,186559942 | 0,561698856 | GPRC5C | FALSE | Downregulated |
| 0,384322791 | 0,561698856 | LOC100506388 | FALSE | Upregulated |

| | | | | |
|---|---|---|---|---|
| -0,299743554 | 0,561698856 | LOC101927339 | FALSE | Downregulated |
| 0,491689113 | 0,561698856 | NHSL2 | FALSE | Upregulated |
| 0,374500894 | 0,561698856 | SEC22A | FALSE | Upregulated |
| -0,624014006 | 0,561698856 | SNX24 | FALSE | Downregulated |
| -0,723288197 | 0,561698856 | ZNF20 | FALSE | Downregulated |
| 0,298083037 | 0,56171135 | SMAD9 | FALSE | Upregulated |
| 0,741257324 | 0,561778632 | RTP4 | FALSE | Upregulated |
| 0,243185864 | 0,562006994 | ANXA4 | FALSE | Upregulated |
| 1,278079109 | 0,562217549 | FASLG | FALSE | Upregulated |
| -1,186040661 | 0,562217549 | LINC00314 | FALSE | Downregulated |
| 0,365594812 | 0,562217549 | PSMD9 | FALSE | Upregulated |
| 0,581055315 | 0,562217549 | TRMT61B | FALSE | Upregulated |
| -0,343635473 | 0,562329994 | ALG13 | FALSE | Downregulated |
| -0,194509046 | 0,562807135 | GAPDH | FALSE | Downregulated |
| 0,530197205 | 0,562979038 | AC037193.1 | FALSE | Upregulated |
| 0,268234474 | 0,562979038 | CENPP | FALSE | Upregulated |
| -0,283069605 | 0,562979038 | ESR1 | FALSE | Downregulated |
| -0,26819619 | 0,562979038 | UQCRC2 | FALSE | Downregulated |
| 0,236677436 | 0,56321876 | WDR6 | FALSE | Upregulated |
| -0,354050007 | 0,563574403 | THUMPD1 | FALSE | Downregulated |
| -0,2384874 | 0,563627363 | HSP90AA1 | FALSE | Downregulated |
| 0,189250752 | 0,563627363 | RNPEPL1 | FALSE | Upregulated |
| 0,365736416 | 0,563631194 | ADNP2 | FALSE | Upregulated |
| 0,694980273 | 0,563631194 | DACT3 | FALSE | Upregulated |
| 0,175085857 | 0,563631194 | FOXK1 | FALSE | Upregulated |
| -0,370574444 | 0,563631194 | PPP1R7 | FALSE | Downregulated |
| -0,367568381 | 0,563631194 | SLU7 | FALSE | Downregulated |
| 0,209688754 | 0,563643127 | CCNL1 | FALSE | Upregulated |
| -0,33940826 | 0,563643171 | SGSH | FALSE | Downregulated |
| 0,41440801 | 0,563733769 | TRIM5 | FALSE | Upregulated |
| -0,340282543 | 0,563942935 | ALX3 | FALSE | Downregulated |
| 0,431111929 | 0,563942935 | LOC100507412 | FALSE | Upregulated |
| 0,795131929 | 0,563942935 | LOC729040 | FALSE | Upregulated |
| 0,203792365 | 0,564000528 | DAG1 | FALSE | Upregulated |
| -0,360045951 | 0,564000528 | ZNF197 | FALSE | Downregulated |
| -0,215185807 | 0,564117815 | STOM | FALSE | Downregulated |
| -0,292420821 | 0,564200422 | APOC4-APOC2 | FALSE | Downregulated |
| 0,439584799 | 0,564200422 | C1orf111 | FALSE | Upregulated |
| -0,360729699 | 0,564200422 | DBT | FALSE | Downregulated |
| 0,376885507 | 0,564200422 | DNM1P41 | FALSE | Upregulated |
| -1,139619552 | 0,564200422 | GPR151 | FALSE | Downregulated |
| -0,227912586 | 0,564200422 | HIP1 | FALSE | Downregulated |
| 0,537356983 | 0,564200422 | KCNK6 | FALSE | Upregulated |
| -0,506442577 | 0,564200422 | MPP6 | FALSE | Downregulated |
| -0,398313011 | 0,564200422 | MRPS35 | FALSE | Downregulated |
| 0,345309008 | 0,564200422 | PPIP5K1 | FALSE | Upregulated |
| -0,269349148 | 0,564200422 | ST3GAL6 | FALSE | Downregulated |
| 0,638217956 | 0,564200422 | SUSD5 | FALSE | Upregulated |
| -0,362712333 | 0,564200422 | UBE2D2 | FALSE | Downregulated |
| -0,189759648 | 0,564200422 | ZRANB1 | FALSE | Downregulated |
| -0,408814689 | 0,564206392 | COMMD9 | FALSE | Downregulated |

| | | | | |
|---|---|---|---|---|
| 0,481317971 | 0,564206392 | EED | FALSE | Upregulated |
| 0,72383939 | 0,564206392 | LOC101928025 | FALSE | Upregulated |
| -0,177508117 | 0,564206392 | MAP3K11 | FALSE | Downregulated |
| -0,248529909 | 0,564206392 | PRKAA1 | FALSE | Downregulated |
| -0,26396211 | 0,564206392 | PSMB2 | FALSE | Downregulated |
| 1,059241998 | 0,564206392 | SLC51B | FALSE | Upregulated |
| -0,311790514 | 0,564406832 | TRAF6 | FALSE | Downregulated |
| -0,628862444 | 0,564846853 | GPR153 | FALSE | Downregulated |
| 0,336135481 | 0,564986632 | VEZT | FALSE | Upregulated |
| 0,331655944 | 0,565160664 | SMYD4 | FALSE | Upregulated |
| 0,274808555 | 0,565160664 | UBE2J1 | FALSE | Upregulated |
| 0,269001403 | 0,56535462 | MKL1 | FALSE | Upregulated |
| -1,4541636 | 0,56577945 | LTF | FALSE | Downregulated |
| 0,16909353 | 0,56577945 | SPG11 | FALSE | Upregulated |
| 0,183458432 | 0,565796954 | GSE1 | FALSE | Upregulated |
| -0,291594698 | 0,565796954 | PAIP2 | FALSE | Downregulated |
| -0,927336495 | 0,565843149 | LINC01168 | FALSE | Downregulated |
| -0,698281352 | 0,565909903 | RP11-761I4.3 | FALSE | Downregulated |
| 0,362787578 | 0,565937318 | B4GALT7 | FALSE | Upregulated |
| 0,358903898 | 0,566197309 | ABI3 | FALSE | Upregulated |
| 0,20013759 | 0,566197309 | CDC42SE1 | FALSE | Upregulated |
| 0,473784893 | 0,566197309 | DEGS1 | FALSE | Upregulated |
| -0,414618297 | 0,566197309 | NEK1 | FALSE | Downregulated |
| -1,21035106 | 0,566223919 | TMPRSS4 | FALSE | Downregulated |
| 0,549588134 | 0,56649963 | SNRNP27 | FALSE | Upregulated |
| -0,842887438 | 0,566523366 | LOC340074 | FALSE | Downregulated |
| -0,252793116 | 0,566560615 | CBR1 | FALSE | Downregulated |
| -0,510956011 | 0,566875972 | BCL6B | FALSE | Downregulated |
| -0,377900808 | 0,566886674 | C19orf26 | FALSE | Downregulated |
| -0,484777895 | 0,566886674 | MAP4K2 | FALSE | Downregulated |
| -0,199147323 | 0,566886674 | NFIB | FALSE | Downregulated |
| 0,307347626 | 0,566886674 | SCYL2 | FALSE | Upregulated |
| 1,272274108 | 0,566886674 | STXBP5L | FALSE | Upregulated |
| -0,855695841 | 0,567108311 | CCDC150 | FALSE | Downregulated |
| -0,408155231 | 0,567108311 | EIF4A3 | FALSE | Downregulated |
| 0,386416338 | 0,567108311 | EVI5L | FALSE | Upregulated |
| 0,249835504 | 0,567108311 | GFPT1 | FALSE | Upregulated |
| 1,252652421 | 0,567108311 | GUSBP4 | FALSE | Upregulated |
| -0,339073989 | 0,567108311 | SYDE2 | FALSE | Downregulated |
| 0,30727493 | 0,567108311 | TRAF3 | FALSE | Upregulated |
| 0,710483317 | 0,567193271 | NKD2 | FALSE | Upregulated |
| 0,494699775 | 0,567193271 | NT5DC2 | FALSE | Upregulated |
| -0,354758814 | 0,567193271 | RNASE13 | FALSE | Downregulated |
| 0,877736269 | 0,567193271 | TBX10 | FALSE | Upregulated |
| 1,184302707 | 0,567213952 | VNN2 | FALSE | Upregulated |
| 0,404567612 | 0,567398786 | MOB1A | FALSE | Upregulated |
| 0,393036997 | 0,567398786 | RNF138P1 | FALSE | Upregulated |
| 0,20772937 | 0,567398786 | RP11-545I5.3 | FALSE | Upregulated |
| 0,700531606 | 0,567435667 | USP17L2 | FALSE | Upregulated |
| -0,465825957 | 0,567467625 | HDDC3 | FALSE | Downregulated |
| 0,781069734 | 0,567566255 | PLCZ1 | FALSE | Upregulated |

| | | | | |
|---|---|---|---|---|
| 0,656139475 | 0,567634951 | LOC101927895 | FALSE | Upregulated |
| 0,357625554 | 0,567634951 | RAP1A | FALSE | Upregulated |
| 0,249903439 | 0,567634951 | TRMT10B | FALSE | Upregulated |
| 0,629054406 | 0,567709931 | TPM3P9 | FALSE | Upregulated |
| 0,513586992 | 0,567960505 | RNF39 | FALSE | Upregulated |
| -0,56129067 | 0,567960505 | ZNF257 | FALSE | Downregulated |
| 0,229448568 | 0,568030408 | INO80D | FALSE | Upregulated |
| -0,963638625 | 0,568203127 | ENPP3 | FALSE | Downregulated |
| -0,603070404 | 0,568439933 | PPIH | FALSE | Downregulated |
| 1,020121179 | 0,568439933 | SPIC | FALSE | Upregulated |
| 0,327220186 | 0,568836006 | ZNF132 | FALSE | Upregulated |
| 0,875733356 | 0,569235871 | MPZ | FALSE | Upregulated |
| 0,277107567 | 0,569235871 | NOD1 | FALSE | Upregulated |
| 0,246609063 | 0,569235871 | RFFL | FALSE | Upregulated |
| -0,843660121 | 0,569235871 | RGPD5 | FALSE | Downregulated |
| -0,206568079 | 0,56938514 | RAD23A | FALSE | Downregulated |
| 0,544998495 | 0,569424236 | PLD6 | FALSE | Upregulated |
| -1,06860862 | 0,569424236 | SLC5A12 | FALSE | Downregulated |
| -0,282235904 | 0,569500769 | FRK | FALSE | Downregulated |
| -1,545838537 | 0,569500769 | LINC00332 | FALSE | Downregulated |
| 0,344481145 | 0,569500769 | PSME3 | FALSE | Upregulated |
| 0,451350536 | 0,569500769 | SFRP1 | FALSE | Upregulated |
| 0,273408179 | 0,569517152 | RP11-178C3.2 | FALSE | Upregulated |
| -0,814240801 | 0,569834592 | CAB39L | FALSE | Downregulated |
| 0,430859898 | 0,569834592 | CLUHP3 | FALSE | Upregulated |
| -0,215169469 | 0,569834592 | IPO5 | FALSE | Downregulated |
| -0,52091658 | 0,569834592 | LOC100130285 | FALSE | Downregulated |
| -0,358864847 | 0,56991736 | LOC101928185 | FALSE | Downregulated |
| -0,268177331 | 0,570071083 | SNX2 | FALSE | Downregulated |
| -0,3527554 | 0,570071083 | SPPL3 | FALSE | Downregulated |
| -0,310291104 | 0,570459975 | HERC4 | FALSE | Downregulated |
| -0,198568906 | 0,570459975 | RPS24 | FALSE | Downregulated |
| -0,433396243 | 0,570533979 | ZNF417 | FALSE | Downregulated |
| 0,209517999 | 0,570881571 | LOC100128494 | FALSE | Upregulated |
| -0,596135375 | 0,570881571 | METTL8 | FALSE | Downregulated |
| -0,351151825 | 0,570881571 | SPATA6L | FALSE | Downregulated |
| -0,28633997 | 0,570888423 | AZIN1 | FALSE | Downregulated |
| -0,369895426 | 0,570888423 | BSN | FALSE | Downregulated |
| -0,454085018 | 0,570888423 | C11orf71 | FALSE | Downregulated |
| -0,416819679 | 0,570888423 | EMC7 | FALSE | Downregulated |
| 0,248899884 | 0,570888423 | KIAA1598 | FALSE | Upregulated |
| -0,394210088 | 0,570888423 | LOC388849 | FALSE | Downregulated |
| 0,379233408 | 0,570888423 | MYO7B | FALSE | Upregulated |
| -0,751421766 | 0,570888423 | NDUFA7 | FALSE | Downregulated |
| 0,763032375 | 0,570888423 | NOD2 | FALSE | Upregulated |
| -0,540179891 | 0,570888423 | OLA1 | FALSE | Downregulated |
| 0,182699898 | 0,570888423 | PDCD4 | FALSE | Upregulated |
| -0,580399003 | 0,570888423 | POLA2 | FALSE | Downregulated |
| -0,479401814 | 0,570888423 | RNF165 | FALSE | Downregulated |
| -0,479556434 | 0,570888423 | SDF2L1 | FALSE | Downregulated |
| 0,312956997 | 0,570888423 | SIM2 | FALSE | Upregulated |

| | | | | |
|---|---|---|---|---|
| 0,144050514 | 0,570888423 | TAPBP | FALSE | Upregulated |
| -0,833270612 | 0,570888423 | TRPC1 | FALSE | Downregulated |
| -0,986433176 | 0,570888423 | TYRO3P | FALSE | Downregulated |
| -0,685413941 | 0,570888423 | WDR89 | FALSE | Downregulated |
| 0,352144914 | 0,570984316 | ANGEL2 | FALSE | Upregulated |
| -0,216707719 | 0,570984316 | KEAP1 | FALSE | Downregulated |
| 0,198788058 | 0,571251308 | CD59 | FALSE | Upregulated |
| -0,194982723 | 0,571251308 | DDB1 | FALSE | Downregulated |
| -0,539970536 | 0,571251308 | KCNIP2 | FALSE | Downregulated |
| -0,924286548 | 0,571251308 | THBS4 | FALSE | Downregulated |
| -0,234365931 | 0,571339166 | RTN3 | FALSE | Downregulated |
| 0,255184108 | 0,572076553 | TRAPPC11 | FALSE | Upregulated |
| 3,368597122 | 0,572159685 | MYO18B | FALSE | Upregulated |
| -0,184851222 | 0,572159685 | RBCK1 | FALSE | Downregulated |
| -0,166567359 | 0,572159685 | SDF4 | FALSE | Downregulated |
| -0,335761962 | 0,572276188 | CD2AP | FALSE | Downregulated |
| 0,373304653 | 0,572320125 | AC004019.13 | FALSE | Upregulated |
| 0,354612789 | 0,572320125 | NKIRAS2 | FALSE | Upregulated |
| 0,440281802 | 0,572320125 | SLC2A11 | FALSE | Upregulated |
| -0,821936987 | 0,572320125 | TCEANC | FALSE | Downregulated |
| -0,222437144 | 0,572320125 | XRCC6 | FALSE | Downregulated |
| 0,300908506 | 0,572352297 | LOC102723554 | FALSE | Upregulated |
| -2,203278701 | 0,572381202 | RP4-669L17.10 | FALSE | Downregulated |
| -0,247303424 | 0,572511079 | CCT7 | FALSE | Downregulated |
| -0,261924939 | 0,573594779 | ATP5C1 | FALSE | Downregulated |
| 0,353333747 | 0,573594779 | DPH3P1 | FALSE | Upregulated |
| 0,36416831 | 0,573594779 | FAM117B | FALSE | Upregulated |
| 0,321881211 | 0,573594779 | LOC101927839 | FALSE | Upregulated |
| -0,323709 | 0,573594779 | MTFR1 | FALSE | Downregulated |
| 0,185446675 | 0,573594779 | PLXND1 | FALSE | Upregulated |
| 0,843665552 | 0,573596589 | CASQ1 | FALSE | Upregulated |
| -0,188911849 | 0,573596589 | DGCR2 | FALSE | Downregulated |
| 0,840419955 | 0,573596589 | KCNN3 | FALSE | Upregulated |
| -0,979426011 | 0,573596589 | LRFN1 | FALSE | Downregulated |
| 0,342047544 | 0,573596589 | MBD4 | FALSE | Upregulated |
| -0,393004307 | 0,573596589 | SPRY4 | FALSE | Downregulated |
| 0,490710303 | 0,573596589 | TIPRL | FALSE | Upregulated |
| 0,206570113 | 0,573762058 | CCDC85C | FALSE | Upregulated |
| 0,32972535 | 0,573881714 | LOC642852 | FALSE | Upregulated |
| -0,269297823 | 0,573925599 | MAST4 | FALSE | Downregulated |
| -1,083397751 | 0,573946514 | GATA1 | FALSE | Downregulated |
| -0,29593773 | 0,573976158 | SLC25A46 | FALSE | Downregulated |
| -0,558503007 | 0,573992413 | ADORA2B | FALSE | Downregulated |
| -1,188505024 | 0,574095079 | HSFX1 | FALSE | Downregulated |
| -0,575849938 | 0,574200956 | LINC00641 | FALSE | Downregulated |
| 0,359577509 | 0,574217463 | CHST12 | FALSE | Upregulated |
| -0,266229796 | 0,574217463 | ISOC2 | FALSE | Downregulated |
| -1,110206348 | 0,574227667 | CYP2S1 | FALSE | Downregulated |
| 0,223817026 | 0,574516801 | SNW1 | FALSE | Upregulated |
| -0,593223211 | 0,574565909 | ZNF577 | FALSE | Downregulated |
| -1,31417848 | 0,575088759 | BNIPL | FALSE | Downregulated |

| | | | | |
|---|---|---|---|---|
| 0,362779595 | 0,575088759 | DHFRL1 | FALSE | Upregulated |
| -0,221585895 | 0,575418481 | APH1A | FALSE | Downregulated |
| -1,135765807 | 0,575891776 | GPR123 | FALSE | Downregulated |
| 0,490480794 | 0,575894857 | TMC5 | FALSE | Upregulated |
| 0,194155874 | 0,576184832 | COL10A1 | FALSE | Upregulated |
| 0,540148481 | 0,576184832 | RASSF6 | FALSE | Upregulated |
| 1,231292114 | 0,576184832 | RERG | FALSE | Upregulated |
| 0,265978615 | 0,576184832 | TMED8 | FALSE | Upregulated |
| 0,57693393 | 0,57629076 | BACE2 | FALSE | Upregulated |
| -0,292466646 | 0,57629076 | CASZ1 | FALSE | Downregulated |
| 0,895373277 | 0,57629076 | LZTS1-AS1 | FALSE | Upregulated |
| 0,506088279 | 0,57629076 | SRD5A3 | FALSE | Upregulated |
| 0,327217174 | 0,576380339 | GXYLT1 | FALSE | Upregulated |
| 0,319666218 | 0,576380339 | HAUS6 | FALSE | Upregulated |
| 0,420898626 | 0,576380339 | SNX29 | FALSE | Upregulated |
| 0,207128434 | 0,576791149 | EPS8L2 | FALSE | Upregulated |
| -0,308859052 | 0,576791149 | PLSCR4 | FALSE | Downregulated |
| -0,18431983 | 0,577022644 | GDI2 | FALSE | Downregulated |
| -0,965674789 | 0,57706755 | EXOSC9 | FALSE | Downregulated |
| 0,390557361 | 0,57706755 | SOBP | FALSE | Upregulated |
| 0,533745 | 0,577285596 | SCRN1 | FALSE | Upregulated |
| -0,751899414 | 0,577303165 | RIOK1 | FALSE | Downregulated |
| 0,343815367 | 0,577514591 | FIGN | FALSE | Upregulated |
| 0,330036793 | 0,577742805 | TANGO2 | FALSE | Upregulated |
| 0,463910192 | 0,577810203 | WWOX | FALSE | Upregulated |
| -0,555209129 | 0,577894513 | RAPGEFL1 | FALSE | Downregulated |
| 0,527323762 | 0,577894513 | TACC3 | FALSE | Upregulated |
| 0,298435491 | 0,578309874 | ARPC2 | FALSE | Upregulated |
| 0,48816864 | 0,578309874 | PSD | FALSE | Upregulated |
| -0,58626845 | 0,578318875 | CDK11A | FALSE | Downregulated |
| 0,893266579 | 0,578318875 | DEAR | FALSE | Upregulated |
| -0,376551549 | 0,578524886 | MUC20 | FALSE | Downregulated |
| -0,302069079 | 0,578832374 | NBAS | FALSE | Downregulated |
| -0,39525122 | 0,578832374 | ZFP64 | FALSE | Downregulated |
| -0,690160932 | 0,579250556 | KIF19 | FALSE | Downregulated |
| -0,884461344 | 0,579250556 | TMEM116 | FALSE | Downregulated |
| -0,514134511 | 0,579250556 | ZNF226 | FALSE | Downregulated |
| 0,764824976 | 0,57972116 | CD72 | FALSE | Upregulated |
| 0,612843498 | 0,57972116 | DTD1 | FALSE | Upregulated |
| -0,806393516 | 0,579816515 | GYLTL1B | FALSE | Downregulated |
| 0,429970631 | 0,579816515 | RABIF | FALSE | Upregulated |
| 0,928960788 | 0,579831771 | LOC440934 | FALSE | Upregulated |
| 0,389101397 | 0,579972658 | PCBP4 | FALSE | Upregulated |
| -0,39847808 | 0,580043249 | SNORD3A | FALSE | Downregulated |
| 0,223959231 | 0,580086969 | TDRP | FALSE | Upregulated |
| 1,002750287 | 0,580093414 | GREM1 | FALSE | Upregulated |
| 0,343335894 | 0,580102478 | LOC642361 | FALSE | Upregulated |
| 0,211573602 | 0,580129592 | AP1G2 | FALSE | Upregulated |
| 1,176003506 | 0,580135345 | JAKMIP2-AS1 | FALSE | Upregulated |
| -1,055207961 | 0,580135345 | ZBTB26 | FALSE | Downregulated |
| -0,678574656 | 0,580175837 | PIGK | FALSE | Downregulated |

| | | | | |
|---|---|---|---|---|
| 0,593299234 | 0,580175837 | VMO1 | FALSE | Upregulated |
| -0,326900881 | 0,580264585 | PRMT7 | FALSE | Downregulated |
| -0,286234082 | 0,58031354 | ALDH16A1 | FALSE | Downregulated |
| -0,712170549 | 0,580346868 | C8orf58 | FALSE | Downregulated |
| 0,338255705 | 0,580408445 | NPIPB6 | FALSE | Upregulated |
| -0,232775277 | 0,580408445 | RPL17 | FALSE | Downregulated |
| -0,407569561 | 0,580546002 | LOC101927809 | FALSE | Downregulated |
| -0,503505213 | 0,580736266 | RAMP3 | FALSE | Downregulated |
| -0,44827547 | 0,580736266 | SUGCT | FALSE | Downregulated |
| 0,525921043 | 0,580782986 | BACH2 | FALSE | Upregulated |
| 0,589563825 | 0,580782986 | NIPAL1 | FALSE | Upregulated |
| 0,216018537 | 0,580782986 | ZMIZ1 | FALSE | Upregulated |
| 0,203167998 | 0,580782986 | ZNF587 | FALSE | Upregulated |
| 0,434158396 | 0,580899509 | ARHGEF10 | FALSE | Upregulated |
| -0,341183541 | 0,581024437 | TIE1 | FALSE | Downregulated |
| 0,256705075 | 0,58118702 | NLRC3 | FALSE | Upregulated |
| -0,180460867 | 0,58118702 | RPL23 | FALSE | Downregulated |
| 1,199539819 | 0,581324882 | SPAG4 | FALSE | Upregulated |
| -0,197658708 | 0,581579817 | COL27A1 | FALSE | Downregulated |
| -0,162397871 | 0,581579817 | H1FX | FALSE | Downregulated |
| -1,290719377 | 0,581834657 | MROH7-TTC4 | FALSE | Downregulated |
| 0,587528998 | 0,58196162 | TATDN3 | FALSE | Upregulated |
| 0,470448907 | 0,581995643 | PSAPL1 | FALSE | Upregulated |
| 0,176988096 | 0,58225378 | STX16 | FALSE | Upregulated |
| 0,218877679 | 0,582347924 | ANGPTL1 | FALSE | Upregulated |
| -0,817971889 | 0,582347924 | DBF4B | FALSE | Downregulated |
| -0,235695013 | 0,582347924 | GRIN3A | FALSE | Downregulated |
| 0,363543769 | 0,582347924 | LOC101928355 | FALSE | Upregulated |
| -0,808737145 | 0,582347924 | MAP2K6 | FALSE | Downregulated |
| 0,26215674 | 0,582347924 | MROH1 | FALSE | Upregulated |
| 0,255841034 | 0,582347924 | MYL9 | FALSE | Upregulated |
| 0,322366804 | 0,582347924 | NUP133 | FALSE | Upregulated |
| -0,255244726 | 0,582347924 | PPAP2B | FALSE | Downregulated |
| -0,441350789 | 0,582347924 | SLC17A1 | FALSE | Downregulated |
| 0,697555656 | 0,582347924 | ZFP2 | FALSE | Upregulated |
| -0,459179419 | 0,58246999 | GPS2 | FALSE | Downregulated |
| 0,211460755 | 0,58246999 | PHIP | FALSE | Upregulated |
| -0,249861765 | 0,58246999 | UQCRQ | FALSE | Downregulated |
| -0,298705823 | 0,582711117 | FGGY | FALSE | Downregulated |
| -0,488260666 | 0,582887607 | ATPAF2 | FALSE | Downregulated |
| -0,32221735 | 0,582887607 | FAM211A-AS1 | FALSE | Downregulated |
| -0,379046667 | 0,582887607 | GNB5 | FALSE | Downregulated |
| 0,307376629 | 0,582887607 | IGF2BP3 | FALSE | Upregulated |
| -0,241456652 | 0,582887607 | ITGA5 | FALSE | Downregulated |
| 0,457544278 | 0,582887607 | MRPL27 | FALSE | Upregulated |
| -0,178955582 | 0,582887607 | SLC31A1 | FALSE | Downregulated |
| -0,616856466 | 0,582971026 | GCA | FALSE | Downregulated |
| -0,286280864 | 0,582971026 | KIAA1468 | FALSE | Downregulated |
| 0,175881234 | 0,58304212 | SLC12A7 | FALSE | Upregulated |
| -0,48534405 | 0,583142502 | AGBL2 | FALSE | Downregulated |
| 0,643757922 | 0,583142502 | HIST1H4D | FALSE | Upregulated |

| | | | | |
|---|---|---|---|---|
| -0,683013909 | 0,583142502 | IFT74 | FALSE | Downregulated |
| 0,501999931 | 0,583142502 | LOC102724217 | FALSE | Upregulated |
| -0,327705964 | 0,583142502 | LUC7L | FALSE | Downregulated |
| 0,445940842 | 0,583142502 | MTRF1 | FALSE | Upregulated |
| -0,300820938 | 0,583142502 | NPTN | FALSE | Downregulated |
| -0,282801518 | 0,583142502 | PIM3 | FALSE | Downregulated |
| 0,373116077 | 0,583142502 | SCMH1 | FALSE | Upregulated |
| -0,290006177 | 0,583142502 | SH3BP5L | FALSE | Downregulated |
| 0,232915112 | 0,583149935 | PNPLA6 | FALSE | Upregulated |
| -0,223632834 | 0,583409196 | PLS3 | FALSE | Downregulated |
| -0,700467279 | 0,583488665 | C7orf73 | FALSE | Downregulated |
| -0,619683885 | 0,583488665 | EXOC6 | FALSE | Downregulated |
| 0,440284178 | 0,583488665 | LOC102723543 | FALSE | Upregulated |
| 0,800741568 | 0,583488665 | NAT14 | FALSE | Upregulated |
| 0,686488436 | 0,583488665 | PPP1R3D | FALSE | Upregulated |
| -0,236190974 | 0,583488665 | RPL38 | FALSE | Downregulated |
| -0,432046065 | 0,583488665 | VIPAS39 | FALSE | Downregulated |
| -0,916252588 | 0,583496249 | IHH | FALSE | Downregulated |
| -0,454210405 | 0,583600683 | BUD31 | FALSE | Downregulated |
| 0,247803869 | 0,583600683 | C22orf26 | FALSE | Upregulated |
| 0,997438148 | 0,583600683 | CD101 | FALSE | Upregulated |
| -1,127511003 | 0,583600683 | CPXM2 | FALSE | Downregulated |
| -0,396578914 | 0,583600683 | CTH | FALSE | Downregulated |
| 0,259070894 | 0,583600683 | DCTN2 | FALSE | Upregulated |
| 0,340222624 | 0,583600683 | FAM132B | FALSE | Upregulated |
| -0,494132389 | 0,583600683 | GUSBP1 | FALSE | Downregulated |
| 0,938015236 | 0,583600683 | INGX | FALSE | Upregulated |
| -0,266752397 | 0,583600683 | IPO8 | FALSE | Downregulated |
| -1,263695837 | 0,583600683 | LOC100653233 | FALSE | Downregulated |
| 0,813269577 | 0,583600683 | MSS51 | FALSE | Upregulated |
| 0,303089821 | 0,583600683 | PTPN6 | FALSE | Upregulated |
| -0,51263051 | 0,583600683 | RP11-279F6.1 | FALSE | Downregulated |
| -0,318337784 | 0,583600683 | ZMYM3 | FALSE | Downregulated |
| -0,26493591 | 0,583725243 | FBXL15 | FALSE | Downregulated |
| -0,768828651 | 0,583725243 | LOC102724770 | FALSE | Downregulated |
| 0,306317386 | 0,583725243 | OSGIN1 | FALSE | Upregulated |
| -1,248699225 | 0,583725243 | SNHG6 | FALSE | Downregulated |
| -0,392646714 | 0,583733784 | NBPF10 | FALSE | Downregulated |
| 0,353995201 | 0,583789758 | ATAD2 | FALSE | Upregulated |
| 0,310362482 | 0,583789758 | IKBKG | FALSE | Upregulated |
| 0,696933549 | 0,583789758 | MAGOHB | FALSE | Upregulated |
| 0,238708284 | 0,583789758 | TMEM43 | FALSE | Upregulated |
| 0,958329411 | 0,583872461 | C8orf49 | FALSE | Upregulated |
| -0,293261023 | 0,583897739 | NBPF11 | FALSE | Downregulated |
| -1,345261099 | 0,583897739 | PLA2G4B | FALSE | Downregulated |
| -0,280363987 | 0,583897739 | STIP1 | FALSE | Downregulated |
| 0,286426413 | 0,583997646 | ARHGAP17 | FALSE | Upregulated |
| -0,458440194 | 0,583997646 | PLS1 | FALSE | Downregulated |
| -0,275003879 | 0,583997646 | ZNF33B | FALSE | Downregulated |
| 0,390295451 | 0,584103589 | PGS1 | FALSE | Upregulated |
| -1,142132946 | 0,584267447 | MFSD2B | FALSE | Downregulated |

| | | | | |
|---|---|---|---|---|
| 0,274080455 | 0,584267447 | USP20 | FALSE | Upregulated |
| 0,488253701 | 0,584766671 | LRRC25 | FALSE | Upregulated |
| 0,504165581 | 0,584802622 | CCR1 | FALSE | Upregulated |
| 0,404204918 | 0,584802622 | ETNK1 | FALSE | Upregulated |
| 0,507047766 | 0,584802622 | TAF8 | FALSE | Upregulated |
| 0,485580612 | 0,584969582 | DOK2 | FALSE | Upregulated |
| 0,37239782 | 0,584969582 | TMEM50A | FALSE | Upregulated |
| -0,301370048 | 0,584969582 | UFL1 | FALSE | Downregulated |
| -0,336054147 | 0,585050601 | GBP7 | FALSE | Downregulated |
| 0,997681518 | 0,585149926 | DDX11L1 | FALSE | Upregulated |
| -0,197768593 | 0,585234353 | TNFRSF1A | FALSE | Downregulated |
| 0,377162056 | 0,585266846 | OGFOD2 | FALSE | Upregulated |
| -0,346689866 | 0,585290289 | C17orf85 | FALSE | Downregulated |
| 0,344485814 | 0,585290289 | DKC1 | FALSE | Upregulated |
| -0,353385507 | 0,585290289 | JRK | FALSE | Downregulated |
| -0,259451379 | 0,585290289 | LOC101928946 | FALSE | Downregulated |
| 1,023292813 | 0,585290289 | MATN1 | FALSE | Upregulated |
| -0,185924486 | 0,585290289 | SEC61A1 | FALSE | Downregulated |
| 0,46385091 | 0,585290289 | TEAD2 | FALSE | Upregulated |
| 0,944964122 | 0,585569867 | RAG2 | FALSE | Upregulated |
| 0,716208048 | 0,585569867 | TMEM169 | FALSE | Upregulated |
| 0,749571802 | 0,585601282 | LOC101929519 | FALSE | Upregulated |
| -0,324817198 | 0,585601282 | SH3TC1 | FALSE | Downregulated |
| 0,291548219 | 0,585756322 | RC3H2 | FALSE | Upregulated |
| 0,417792011 | 0,586074946 | ARID3B | FALSE | Upregulated |
| 1,028836532 | 0,586133193 | PRTG | FALSE | Upregulated |
| -0,911935633 | 0,586256432 | FLJ37035 | FALSE | Downregulated |
| 0,256766158 | 0,586463445 | DUOX1 | FALSE | Upregulated |
| -0,278439415 | 0,586542117 | LHPP | FALSE | Downregulated |
| 0,328482783 | 0,586675716 | ELOF1 | FALSE | Upregulated |
| 0,277470174 | 0,586675716 | KIAA1731 | FALSE | Upregulated |
| -0,976918051 | 0,586931835 | ADH7 | FALSE | Downregulated |
| -0,575249807 | 0,587046826 | LOC102723701 | FALSE | Downregulated |
| -0,183575735 | 0,587046826 | MGAT1 | FALSE | Downregulated |
| 1,062829014 | 0,587092872 | BCL11A | FALSE | Upregulated |
| -0,255202029 | 0,587092872 | IPO7 | FALSE | Downregulated |
| -0,36136958 | 0,587093267 | USP38 | FALSE | Downregulated |
| 0,616597673 | 0,587181742 | CLEC7A | FALSE | Upregulated |
| 0,342670529 | 0,587391184 | FZD1 | FALSE | Upregulated |
| -0,282768742 | 0,58765215 | FAM160A2 | FALSE | Downregulated |
| 0,982458394 | 0,587727918 | RP11-486O12.2 | FALSE | Upregulated |
| 0,576513355 | 0,587757605 | BCL10 | FALSE | Upregulated |
| -0,42844404 | 0,587757605 | LOC729173 | FALSE | Downregulated |
| -0,351816999 | 0,587757605 | PQBP1 | FALSE | Downregulated |
| -1,139835664 | 0,587757605 | TBC1D19 | FALSE | Downregulated |
| 0,760175867 | 0,588038074 | CBX2 | FALSE | Upregulated |
| -0,577379243 | 0,588062974 | OBFC1 | FALSE | Downregulated |
| -0,489041779 | 0,588114586 | CETP | FALSE | Downregulated |
| 0,184020455 | 0,588605136 | HAS3 | FALSE | Upregulated |
| -0,379677965 | 0,588605136 | RAB5A | FALSE | Downregulated |
| 0,148002768 | 0,588605136 | RNF10 | FALSE | Upregulated |

| | | | | |
|---|---|---|---|---|
| -0,306607421 | 0,588695874 | FARSA | FALSE | Downregulated |
| -0,591622652 | 0,588695874 | LOC101060376 | FALSE | Downregulated |
| 0,419879968 | 0,588695874 | WAC-AS1 | FALSE | Upregulated |
| -0,264422002 | 0,588711302 | PARP3 | FALSE | Downregulated |
| -1,168525011 | 0,588718857 | ITPKA | FALSE | Downregulated |
| 0,261529164 | 0,588718857 | PTBP3 | FALSE | Upregulated |
| 0,292580049 | 0,588718857 | TRIM38 | FALSE | Upregulated |
| -1,047109344 | 0,588806078 | PACRG | FALSE | Downregulated |
| -0,386262845 | 0,588937544 | CDK19 | FALSE | Downregulated |
| 1,009150976 | 0,589533515 | PATL2 | FALSE | Upregulated |
| 1,366094267 | 0,589677343 | ADARB2 | FALSE | Upregulated |
| -0,358539996 | 0,589677343 | EMP2 | FALSE | Downregulated |
| 0,262352909 | 0,589677343 | GALNS | FALSE | Upregulated |
| -1,349485652 | 0,589677343 | MROH7 | FALSE | Downregulated |
| -0,360078461 | 0,58971337 | JMY | FALSE | Downregulated |
| 0,364730882 | 0,58971337 | MGAT4A | FALSE | Upregulated |
| 0,282190287 | 0,58971337 | PDE12 | FALSE | Upregulated |
| -0,158092423 | 0,589901053 | RPL27A | FALSE | Downregulated |
| -0,178385721 | 0,590204621 | MAZ | FALSE | Downregulated |
| -0,381772718 | 0,590204621 | PPIP5K2 | FALSE | Downregulated |
| 0,397345803 | 0,590204621 | SLC37A1 | FALSE | Upregulated |
| -0,266533855 | 0,590378156 | CPQ | FALSE | Downregulated |
| 0,334064577 | 0,590487481 | GNAT1 | FALSE | Upregulated |
| 0,747871041 | 0,590497854 | HIST1H2AE | FALSE | Upregulated |
| -0,387711596 | 0,590497854 | PXMP4 | FALSE | Downregulated |
| -0,474797162 | 0,590497854 | SLC25A45 | FALSE | Downregulated |
| 0,310949962 | 0,590497854 | SPTLC2 | FALSE | Upregulated |
| -0,381461228 | 0,590497854 | TMEM47 | FALSE | Downregulated |
| -0,874966945 | 0,590497854 | UBL7-AS1 | FALSE | Downregulated |
| 0,90386194 | 0,590601179 | FUT1 | FALSE | Upregulated |
| -0,200235929 | 0,590601179 | UFD1L | FALSE | Downregulated |
| -0,534109763 | 0,590601179 | ZSCAN26 | FALSE | Downregulated |
| 0,325104182 | 0,590905733 | UPF3A | FALSE | Upregulated |
| 0,89410953 | 0,591119783 | RXFP4 | FALSE | Upregulated |
| -1,072022894 | 0,591347986 | ABCA13 | FALSE | Downregulated |
| -0,544234386 | 0,591347986 | C14orf80 | FALSE | Downregulated |
| 0,255607395 | 0,591347986 | CTAGE7P | FALSE | Upregulated |
| -0,444562762 | 0,591347986 | ZNF324 | FALSE | Downregulated |
| -0,271113363 | 0,591948078 | CTR9 | FALSE | Downregulated |
| 0,651269158 | 0,591948078 | EDNRA | FALSE | Upregulated |
| 0,298763631 | 0,591948078 | FOXD2 | FALSE | Upregulated |
| -0,250978828 | 0,591948078 | ITPKC | FALSE | Downregulated |
| 0,685923095 | 0,591948078 | TBC1D32 | FALSE | Upregulated |
| -1,172835034 | 0,591948078 | TXK | FALSE | Downregulated |
| -0,237440057 | 0,591948078 | ZC3H18 | FALSE | Downregulated |
| 0,251005992 | 0,591997768 | DGAT1 | FALSE | Upregulated |
| -0,230207254 | 0,592026359 | LINC00674 | FALSE | Downregulated |
| 0,363344141 | 0,592054379 | HK1 | FALSE | Upregulated |
| 0,563290589 | 0,592111679 | LRRC42 | FALSE | Upregulated |
| -1,100762306 | 0,592111679 | PROM2 | FALSE | Downregulated |
| -0,21100293 | 0,592180819 | SMAP2 | FALSE | Downregulated |

| | | | | |
|---|---|---|---|---|
| -0,274684352 | 0,592180819 | WDR5 | FALSE | Downregulated |
| -0,949666037 | 0,592222778 | ADAM20P1 | FALSE | Downregulated |
| 0,282103056 | 0,592292861 | CEPT1 | FALSE | Upregulated |
| 0,782728752 | 0,592303489 | LOC102723611 | FALSE | Upregulated |
| 0,314898559 | 0,592466201 | CBX1 | FALSE | Upregulated |
| 0,355080571 | 0,59266588 | PLIN3 | FALSE | Upregulated |
| 1,229283793 | 0,593197419 | GDNF-AS1 | FALSE | Upregulated |
| 0,531947838 | 0,593269292 | DACT1 | FALSE | Upregulated |
| 0,602377615 | 0,593269292 | LRMP | FALSE | Upregulated |
| -0,168398899 | 0,593390952 | SDC1 | FALSE | Downregulated |
| -0,251849555 | 0,593634037 | INTS6 | FALSE | Downregulated |
| 0,908169483 | 0,593923082 | ANKRD20A2 | FALSE | Upregulated |
| -1,098504103 | 0,594001827 | CES5A | FALSE | Downregulated |
| 0,332845166 | 0,594253373 | LOC101928789 | FALSE | Upregulated |
| 0,664417776 | 0,594294664 | TRPC5OS | FALSE | Upregulated |
| 0,401364765 | 0,594558511 | IFT46 | FALSE | Upregulated |
| 0,407546575 | 0,594626163 | SRGAP3-AS3 | FALSE | Upregulated |
| 0,325329889 | 0,594655561 | PFN2 | FALSE | Upregulated |
| -0,340477383 | 0,594655561 | SYT17 | FALSE | Downregulated |
| -0,25902215 | 0,594758013 | SLC8B1 | FALSE | Downregulated |
| 0,207140382 | 0,594784427 | HNRNPAB | FALSE | Upregulated |
| 0,259403352 | 0,594839248 | ABCE1 | FALSE | Upregulated |
| -0,241114784 | 0,594839248 | ADRM1 | FALSE | Downregulated |
| -0,204199077 | 0,594839248 | GHITM | FALSE | Downregulated |
| -0,259360302 | 0,594839248 | LYPLA2 | FALSE | Downregulated |
| 0,492721175 | 0,594839248 | P4HA2-AS1 | FALSE | Upregulated |
| 0,251825148 | 0,594886796 | HERC3 | FALSE | Upregulated |
| -0,278151028 | 0,595092884 | ASB13 | FALSE | Downregulated |
| 0,410890253 | 0,595117337 | LOC102724730 | FALSE | Upregulated |
| -0,985752018 | 0,595370952 | NPHP3-ACAD11 | FALSE | Downregulated |
| 0,317197779 | 0,595538376 | DNM3OS | FALSE | Upregulated |
| 0,334890579 | 0,595538376 | LRWD1 | FALSE | Upregulated |
| 0,596438591 | 0,595538376 | MCM6 | FALSE | Upregulated |
| 0,180935485 | 0,595538376 | RP11-793H13.3 | FALSE | Upregulated |
| -0,679954526 | 0,595577156 | GPR157 | FALSE | Downregulated |
| -1,008586861 | 0,595657582 | C5orf55 | FALSE | Downregulated |
| 1,080508089 | 0,595782219 | B3GALNT1 | FALSE | Upregulated |
| 0,362838993 | 0,59591245 | CACNA2D4 | FALSE | Upregulated |
| 0,494317287 | 0,59591245 | IRAK3 | FALSE | Upregulated |
| -0,603912057 | 0,59591245 | RP11-372E1.4 | FALSE | Downregulated |
| 0,88579874 | 0,596142093 | ZNF92 | FALSE | Upregulated |
| 0,650380051 | 0,596150758 | LOC439994 | FALSE | Upregulated |
| -1,01624019 | 0,596150758 | LRRCC1 | FALSE | Downregulated |
| 0,62296823 | 0,596201982 | AGA | FALSE | Upregulated |
| -0,492661135 | 0,596201982 | NAPB | FALSE | Downregulated |
| 0,255129501 | 0,596201982 | RP11-392P7.6 | FALSE | Upregulated |
| -0,386195204 | 0,596201982 | SEPW1 | FALSE | Downregulated |
| -0,254692773 | 0,596201982 | SNX5 | FALSE | Downregulated |
| -0,454784761 | 0,596393373 | ANO7 | FALSE | Downregulated |
| 0,340931787 | 0,596393373 | DCPS | FALSE | Upregulated |
| -0,471117006 | 0,596393373 | DDT | FALSE | Downregulated |

| | | | | |
|---|---|---|---|---|
| 0,296267013 | 0,596393373 | DHRS7 | FALSE | Upregulated |
| -0,33429692 | 0,596393373 | FRY-AS1 | FALSE | Downregulated |
| -0,32164867 | 0,596393373 | SLC16A1 | FALSE | Downregulated |
| 0,694892905 | 0,596625331 | LOC389765 | FALSE | Upregulated |
| 0,546768969 | 0,596657528 | MTHFD2 | FALSE | Upregulated |
| 0,527679946 | 0,596657528 | SPAG8 | FALSE | Upregulated |
| 0,624501774 | 0,596845847 | KIF3C | FALSE | Upregulated |
| -0,383230157 | 0,596845847 | TPD52 | FALSE | Downregulated |
| -0,331990578 | 0,596855693 | IFT122 | FALSE | Downregulated |
| -0,156132569 | 0,596971678 | ERP29 | FALSE | Downregulated |
| -0,338069387 | 0,596980125 | BFAR | FALSE | Downregulated |
| 0,418677225 | 0,596980125 | C1orf159 | FALSE | Upregulated |
| 0,447616017 | 0,596980125 | LRRC16A | FALSE | Upregulated |
| -0,384116975 | 0,596980125 | MAP2K5 | FALSE | Downregulated |
| 0,237617958 | 0,596980125 | MAPKAPK5-AS1 | FALSE | Upregulated |
| 0,458477225 | 0,596980125 | UBXN2B | FALSE | Upregulated |
| 0,907392866 | 0,597249173 | RAMP2-AS1 | FALSE | Upregulated |
| -0,399510842 | 0,59750748 | CBFA2T3 | FALSE | Downregulated |
| -0,412594791 | 0,597541869 | CLN3 | FALSE | Downregulated |
| 0,222459138 | 0,597541869 | LOC100506710 | FALSE | Upregulated |
| 0,728653376 | 0,59764938 | ENKUR | FALSE | Upregulated |
| -0,3825015 | 0,59764938 | NCALD | FALSE | Downregulated |
| -0,276158041 | 0,59764938 | OGG1 | FALSE | Downregulated |
| 0,294150557 | 0,59764938 | PLCH2 | FALSE | Upregulated |
| 0,371561829 | 0,597925752 | GRK5 | FALSE | Upregulated |
| 0,256636056 | 0,598267798 | C9orf114 | FALSE | Upregulated |
| -0,285508754 | 0,598267798 | CLYBL | FALSE | Downregulated |
| 0,268088887 | 0,598267798 | RNA45S5 | FALSE | Upregulated |
| 0,2104155 | 0,598284123 | BAZ1A | FALSE | Upregulated |
| -0,287943318 | 0,598284123 | MBTD1 | FALSE | Downregulated |
| -0,298305408 | 0,598284123 | THAP4 | FALSE | Downregulated |
| -0,270208706 | 0,598435288 | BET1L | FALSE | Downregulated |
| -0,421633185 | 0,598435288 | CCDC115 | FALSE | Downregulated |
| -0,515290492 | 0,598435288 | CDKN2AIP | FALSE | Downregulated |
| 0,279336796 | 0,598435288 | CTRL | FALSE | Upregulated |
| 0,32686097 | 0,598435288 | LOC101927989 | FALSE | Upregulated |
| -0,21701141 | 0,598435288 | ZNF598 | FALSE | Downregulated |
| 0,308799673 | 0,598458051 | SYNPO2 | FALSE | Upregulated |
| -0,504147533 | 0,598458051 | WBSCR22 | FALSE | Downregulated |
| -0,622377833 | 0,598629933 | NOP10 | FALSE | Downregulated |
| -0,335616143 | 0,598688933 | APBA3 | FALSE | Downregulated |
| -0,429434685 | 0,598688933 | PREPL | FALSE | Downregulated |
| -0,318188761 | 0,598688933 | SNX4 | FALSE | Downregulated |
| 1,337847157 | 0,598784263 | TRIM29 | FALSE | Upregulated |
| -0,189690821 | 0,59920392 | MAN1A2 | FALSE | Downregulated |
| -0,221862844 | 0,59920392 | RNASE4 | FALSE | Downregulated |
| 0,32261897 | 0,599522201 | ASPN | FALSE | Upregulated |
| 0,283655481 | 0,599522201 | INE2 | FALSE | Upregulated |
| 0,344268372 | 0,599876623 | POU5F1P3 | FALSE | Upregulated |
| -0,218618246 | 0,599889393 | RHOT2 | FALSE | Downregulated |
| -0,212364903 | 0,599905855 | HSP90B1 | FALSE | Downregulated |

| | | | | |
|---|---|---|---|---|
| -0,182632211 | 0,599905855 | NFIX | FALSE | Downregulated |
| 0,171079231 | 0,600013358 | SBF2 | FALSE | Upregulated |
| -0,328127157 | 0,600226297 | PHF23 | FALSE | Downregulated |
| -0,920764844 | 0,600226297 | WDR53 | FALSE | Downregulated |
| 0,130965118 | 0,600548026 | KMT2A | FALSE | Upregulated |
| -0,502630566 | 0,600548026 | PIN4 | FALSE | Downregulated |
| -0,514255669 | 0,600548026 | STL | FALSE | Downregulated |
| -0,34101238 | 0,600949293 | PCGF5 | FALSE | Downregulated |
| -0,461894699 | 0,6010592 | PWP1 | FALSE | Downregulated |
| 0,246199019 | 0,601109469 | ALG12 | FALSE | Upregulated |
| 0,232673347 | 0,601109469 | NBEAL1 | FALSE | Upregulated |
| 0,283259346 | 0,601109469 | RPA1 | FALSE | Upregulated |
| -0,190200506 | 0,601109469 | SMC1A | FALSE | Downregulated |
| 1,059909426 | 0,601128183 | KREMEN2 | FALSE | Upregulated |
| -1,791652277 | 0,601299682 | ANKRD20A1 | FALSE | Downregulated |
| 0,296603942 | 0,601299682 | MRPL4 | FALSE | Upregulated |
| -0,230310161 | 0,60134907 | ABHD14B | FALSE | Downregulated |
| 0,513366552 | 0,60134907 | C1orf213 | FALSE | Upregulated |
| -0,329247343 | 0,601373794 | GORASP2 | FALSE | Downregulated |
| 0,396466206 | 0,601425284 | LOC100130075 | FALSE | Upregulated |
| 0,221454825 | 0,601425284 | LYL1 | FALSE | Upregulated |
| 0,340610155 | 0,601509989 | C22orf46 | FALSE | Upregulated |
| -0,292954131 | 0,601509989 | CTB-118N6.3 | FALSE | Downregulated |
| -0,350677035 | 0,601509989 | CYHR1 | FALSE | Downregulated |
| -0,962652921 | 0,601509989 | DLEU2L | FALSE | Downregulated |
| 0,261964194 | 0,601509989 | GRIPAP1 | FALSE | Upregulated |
| -0,860478114 | 0,601509989 | LINC00637 | FALSE | Downregulated |
| -0,372270248 | 0,601509989 | MCOLN1 | FALSE | Downregulated |
| 0,327136421 | 0,601509989 | ZFYVE27 | FALSE | Upregulated |
| -0,867304568 | 0,601509989 | ZGLP1 | FALSE | Downregulated |
| -0,699230733 | 0,601509989 | ZNF253 | FALSE | Downregulated |
| 0,467494317 | 0,601793209 | AOAH | FALSE | Upregulated |
| -0,558020541 | 0,601920671 | GLUD2 | FALSE | Downregulated |
| 0,237096137 | 0,601920671 | HTR2B | FALSE | Upregulated |
| -0,534315262 | 0,602033778 | ACSM1 | FALSE | Downregulated |
| -0,210623795 | 0,60214271 | BAHCC1 | FALSE | Downregulated |
| -0,399979601 | 0,60214271 | HIST1H2BK | FALSE | Downregulated |
| 0,161401633 | 0,60214271 | LRBA | FALSE | Upregulated |
| -0,804723729 | 0,60214271 | SLC2A4 | FALSE | Downregulated |
| 0,253849881 | 0,60214271 | TCTA | FALSE | Upregulated |
| -0,273573397 | 0,60214271 | ZNF146 | FALSE | Downregulated |
| 0,340375338 | 0,602205695 | SBSPON | FALSE | Upregulated |
| 0,573503817 | 0,602209949 | TMEM185A | FALSE | Upregulated |
| -0,313925476 | 0,602340663 | CDC42EP3 | FALSE | Downregulated |
| 0,727108088 | 0,602563853 | AMZ2P1 | FALSE | Upregulated |
| 0,346241554 | 0,602563853 | GOLGA8O | FALSE | Upregulated |
| -0,39419997 | 0,602641138 | APOOL | FALSE | Downregulated |
| 0,251788014 | 0,602713701 | EFCAB13 | FALSE | Upregulated |
| 0,220479077 | 0,602829246 | CCDC93 | FALSE | Upregulated |
| -0,624112329 | 0,602950665 | IPO11 | FALSE | Downregulated |
| -0,187846925 | 0,603476283 | CD99L2 | FALSE | Downregulated |

| | | | | |
|---|---|---|---|---|
| 0,934181627 | 0,603476283 | RP1-69M21.2 | FALSE | Upregulated |
| 1,297623906 | 0,603570862 | FAM19A5 | FALSE | Upregulated |
| 0,388744173 | 0,603570862 | LOC100289230 | FALSE | Upregulated |
| 0,720547425 | 0,603570862 | LOC285819 | FALSE | Upregulated |
| -0,484666431 | 0,604150421 | MYH3 | FALSE | Downregulated |
| -0,841073328 | 0,604298097 | MT1M | FALSE | Downregulated |
| 0,199379187 | 0,604362531 | RAB3GAP2 | FALSE | Upregulated |
| 0,526022346 | 0,604399775 | PLA2R1 | FALSE | Upregulated |
| 0,530913688 | 0,604485193 | PLEKHH2 | FALSE | Upregulated |
| -0,261974689 | 0,604590705 | KLF9 | FALSE | Downregulated |
| 0,922083791 | 0,604590705 | KLHL41 | FALSE | Upregulated |
| 0,171394652 | 0,60480529 | DEDD | FALSE | Upregulated |
| -0,492790632 | 0,60489526 | COPS8 | FALSE | Downregulated |
| 0,549870035 | 0,60489526 | EPHB3 | FALSE | Upregulated |
| 0,248923759 | 0,605013598 | CHTOP | FALSE | Upregulated |
| 0,201740595 | 0,605013598 | DYNC1LI2 | FALSE | Upregulated |
| 0,403817784 | 0,605013598 | MBD5 | FALSE | Upregulated |
| -0,22976512 | 0,605013598 | MRPS26 | FALSE | Downregulated |
| 0,455948807 | 0,605013598 | NAA38 | FALSE | Upregulated |
| 0,97835501 | 0,605013598 | RP11-180I22.2 | FALSE | Upregulated |
| 1,141413999 | 0,605013598 | SMIM4 | FALSE | Upregulated |
| 0,534328706 | 0,605024028 | NUP93 | FALSE | Upregulated |
| 0,516018169 | 0,605097861 | ALG14 | FALSE | Upregulated |
| -0,331874328 | 0,605097861 | LRRK2 | FALSE | Downregulated |
| 0,234562403 | 0,605097861 | MVB12B | FALSE | Upregulated |
| 1,126746814 | 0,605393574 | IGSF9B | FALSE | Upregulated |
| 0,602645462 | 0,605616894 | AACS | FALSE | Upregulated |
| -0,260454012 | 0,605616894 | FAM84B | FALSE | Downregulated |
| 0,419418881 | 0,605616894 | GRIN3B | FALSE | Upregulated |
| 1,521220593 | 0,605616894 | HNRNPH2 | FALSE | Upregulated |
| -0,888430009 | 0,605616894 | HSPB11 | FALSE | Downregulated |
| 0,483256269 | 0,605616894 | MYCT1 | FALSE | Upregulated |
| -0,20619009 | 0,605616894 | PCCA | FALSE | Downregulated |
| -1,114771028 | 0,605616894 | RP11-421P23.1 | FALSE | Downregulated |
| 0,152594738 | 0,605616894 | SHISA5 | FALSE | Upregulated |
| -0,250296994 | 0,605616894 | SLC41A2 | FALSE | Downregulated |
| 0,389039827 | 0,605616894 | SPN | FALSE | Upregulated |
| 0,61534292 | 0,605911391 | B3GNT7 | FALSE | Upregulated |
| -0,247098254 | 0,606452112 | CLASRP | FALSE | Downregulated |
| 0,280332612 | 0,606452112 | E2F4 | FALSE | Upregulated |
| 0,827850356 | 0,606452112 | RHCG | FALSE | Upregulated |
| 0,3548247 | 0,606559353 | LOC102467073 | FALSE | Upregulated |
| -0,467311142 | 0,606610828 | PCOLCE2 | FALSE | Downregulated |
| 0,610336813 | 0,60715728 | BRMS1L | FALSE | Upregulated |
| -0,323076992 | 0,60715728 | GM2A | FALSE | Downregulated |
| 0,25250938 | 0,60715728 | XRN1 | FALSE | Upregulated |
| 0,390559647 | 0,607241808 | ID3 | FALSE | Upregulated |
| 0,330381034 | 0,607241808 | LARS2-AS1 | FALSE | Upregulated |
| 0,598714867 | 0,607273485 | LRCOL1 | FALSE | Upregulated |
| 0,264974384 | 0,607273485 | PIP5K1A | FALSE | Upregulated |
| 0,324618059 | 0,607278328 | TBCE | FALSE | Upregulated |

| | | | | |
|---|---|---|---|---|
| 0,144865535 | 0,60737734 | PBRM1 | FALSE | Upregulated |
| -1,059026703 | 0,60750285 | LINC00950 | FALSE | Downregulated |
| 0,247896633 | 0,607822261 | LOC101927393 | FALSE | Upregulated |
| 0,168002612 | 0,607822261 | LOC101927556 | FALSE | Upregulated |
| -0,220426716 | 0,607930059 | MED23 | FALSE | Downregulated |
| -0,555080437 | 0,6079556 | HSD17B3 | FALSE | Downregulated |
| 0,160777054 | 0,608206048 | MCM3AP | FALSE | Upregulated |
| 0,223912157 | 0,608240563 | CANT1 | FALSE | Upregulated |
| -0,473539946 | 0,608240563 | COX7B | FALSE | Downregulated |
| 0,832158462 | 0,608240563 | RP11-638L3.1 | FALSE | Upregulated |
| -0,380867233 | 0,608240563 | RRAGA | FALSE | Downregulated |
| -0,255961818 | 0,608424378 | SUMO3 | FALSE | Downregulated |
| -1,000192447 | 0,608429555 | RP11-342C23.4 | FALSE | Downregulated |
| 0,323658079 | 0,608626544 | LOC101929986 | FALSE | Upregulated |
| -0,411395793 | 0,608694852 | COPS7B | FALSE | Downregulated |
| -0,308686644 | 0,608694852 | HSD17B11 | FALSE | Downregulated |
| -0,236463269 | 0,608694852 | TSR3 | FALSE | Downregulated |
| -0,410322079 | 0,608812774 | RSC1A1 | FALSE | Downregulated |
| 0,334257485 | 0,608887912 | EFEMP2 | FALSE | Upregulated |
| 0,649356419 | 0,609172597 | TPM2 | FALSE | Upregulated |
| 0,285557309 | 0,609293444 | TBC1D22A | FALSE | Upregulated |
| -0,498186083 | 0,609346061 | DDX52 | FALSE | Downregulated |
| 0,215484729 | 0,609346061 | LOC100288123 | FALSE | Upregulated |
| 0,191878709 | 0,609346061 | PIK3AP1 | FALSE | Upregulated |
| 0,72589224 | 0,609416602 | SEPT3 | FALSE | Upregulated |
| -0,719898811 | 0,609505944 | LINC01089 | FALSE | Downregulated |
| 0,522177746 | 0,60952257 | TLR1 | FALSE | Upregulated |
| -1,197962048 | 0,609602686 | TTTY14 | FALSE | Downregulated |
| -0,258060368 | 0,610148961 | RCN3 | FALSE | Downregulated |
| 0,227792173 | 0,610204159 | RGL2 | FALSE | Upregulated |
| 0,2114549 | 0,610205019 | GALNT11 | FALSE | Upregulated |
| -0,352137091 | 0,610205019 | RHPN2 | FALSE | Downregulated |
| 0,229757595 | 0,610205019 | TNFRSF14 | FALSE | Upregulated |
| 0,153023225 | 0,610239063 | AKAP13 | FALSE | Upregulated |
| -0,19882019 | 0,610239063 | BGN | FALSE | Downregulated |
| -1,289944959 | 0,610239063 | LOC101929950 | FALSE | Downregulated |
| 0,440962318 | 0,610603495 | EPDR1 | FALSE | Upregulated |
| 0,791601633 | 0,610603495 | LOC100507424 | FALSE | Upregulated |
| -0,445147751 | 0,610603495 | ZNF329 | FALSE | Downregulated |
| 0,249506581 | 0,610667646 | KGFLP2 | FALSE | Upregulated |
| 0,236118867 | 0,610833217 | FAM21C | FALSE | Upregulated |
| -0,299153716 | 0,610833217 | LRRN4 | FALSE | Downregulated |
| 0,393891601 | 0,610833217 | MEMO1 | FALSE | Upregulated |
| -0,180896995 | 0,610833217 | SAFB | FALSE | Downregulated |
| -0,1620072 | 0,610938955 | NBR1 | FALSE | Downregulated |
| -0,217314219 | 0,610938955 | RELL1 | FALSE | Downregulated |
| 0,295658689 | 0,610938955 | TSPAN11 | FALSE | Upregulated |
| 0,828531807 | 0,611175904 | MILR1 | FALSE | Upregulated |
| -0,813667816 | 0,611250663 | LINC01184 | FALSE | Downregulated |
| 0,193487425 | 0,611250663 | MZF1 | FALSE | Upregulated |
| 0,185137971 | 0,611250663 | PAN3 | FALSE | Upregulated |

| | | | | |
|---|---|---|---|---|
| -0,21032371 | 0,611250663 | SAMD4B | FALSE | Downregulated |
| 0,263109548 | 0,611250663 | ZFP69 | FALSE | Upregulated |
| -0,397456761 | 0,612120803 | PSMG3-AS1 | FALSE | Downregulated |
| 0,189424831 | 0,612271889 | CISD3 | FALSE | Upregulated |
| -0,20961879 | 0,612271889 | TRIM66 | FALSE | Downregulated |
| -0,435574318 | 0,612282159 | TIMM17A | FALSE | Downregulated |
| 0,914786469 | 0,612374622 | FBXW4P1 | FALSE | Upregulated |
| -0,636716522 | 0,612374622 | LOC100506302 | FALSE | Downregulated |
| -0,304384643 | 0,612374622 | NUP160 | FALSE | Downregulated |
| 0,526031577 | 0,612441818 | CD244 | FALSE | Upregulated |
| -0,408459592 | 0,612450974 | ADAM9 | FALSE | Downregulated |
| 0,432396207 | 0,612450974 | RCBTB2 | FALSE | Upregulated |
| 0,161407108 | 0,612450974 | TET3 | FALSE | Upregulated |
| 0,302439225 | 0,612671938 | ACOT12 | FALSE | Upregulated |
| 0,876202642 | 0,612671938 | SAA2 | FALSE | Upregulated |
| 0,443962118 | 0,612691755 | CHAF1A | FALSE | Upregulated |
| -0,294052575 | 0,612691755 | CNNM2 | FALSE | Downregulated |
| -1,461464242 | 0,612691755 | GNG10 | FALSE | Downregulated |
| 0,447826479 | 0,612691755 | LOC100126582 | FALSE | Upregulated |
| 1,082706666 | 0,612691755 | NPM3 | FALSE | Upregulated |
| -0,917648925 | 0,612691755 | PGBD1 | FALSE | Downregulated |
| 0,516683521 | 0,612691755 | SYTL5 | FALSE | Upregulated |
| -0,422323085 | 0,612936376 | COA3 | FALSE | Downregulated |
| 0,458412628 | 0,612976563 | ETV4 | FALSE | Upregulated |
| -0,140602389 | 0,613028509 | PRRC2C | FALSE | Downregulated |
| 0,337923152 | 0,613032115 | SLX4 | FALSE | Upregulated |
| -0,210741448 | 0,613032115 | TPST2 | FALSE | Downregulated |
| -0,867001672 | 0,613034123 | FAM47E | FALSE | Downregulated |
| -0,562023583 | 0,613056185 | NUDT14 | FALSE | Downregulated |
| 0,67186953 | 0,613875005 | ALKBH3 | FALSE | Upregulated |
| -0,553142876 | 0,613877186 | BMS1P2 | FALSE | Downregulated |
| -0,498686931 | 0,613902647 | VPS72 | FALSE | Downregulated |
| 0,499757717 | 0,614065438 | QPCTL | FALSE | Upregulated |
| -0,236037783 | 0,614065438 | UGT2B15 | FALSE | Downregulated |
| 0,520931799 | 0,614118387 | ISY1 | FALSE | Upregulated |
| 0,588515958 | 0,614289136 | P4HA3 | FALSE | Upregulated |
| 0,990833868 | 0,614289136 | SLC35G1 | FALSE | Upregulated |
| 0,245259217 | 0,614289136 | TYW1 | FALSE | Upregulated |
| -0,480081451 | 0,614300329 | SEH1L | FALSE | Downregulated |
| 0,548819112 | 0,614300329 | SNAP25 | FALSE | Upregulated |
| 0,322200684 | 0,614300329 | TANC2 | FALSE | Upregulated |
| 0,173224106 | 0,614300329 | WBSCR16 | FALSE | Upregulated |
| 0,561325654 | 0,614364918 | LOC101928689 | FALSE | Upregulated |
| -1,080048521 | 0,614364918 | ZNF239 | FALSE | Downregulated |
| -0,433640334 | 0,614379925 | ZNF254 | FALSE | Downregulated |
| -0,153128501 | 0,614681907 | EIF3B | FALSE | Downregulated |
| 0,449365444 | 0,614681907 | EPPK1 | FALSE | Upregulated |
| -0,133702644 | 0,614681907 | TRIP12 | FALSE | Downregulated |
| 0,744707628 | 0,614681907 | ZSCAN1 | FALSE | Upregulated |
| 0,970325006 | 0,61480009 | SLC6A17 | FALSE | Upregulated |
| 0,902859662 | 0,614852558 | ACRBP | FALSE | Upregulated |

| | | | | |
|---|---|---|---|---|
| 0,28090817 | 0,61493976 | CACNA1D | FALSE | Upregulated |
| -0,856942609 | 0,61493976 | KLHL13 | FALSE | Downregulated |
| 0,305504241 | 0,61493976 | TCHP | FALSE | Upregulated |
| -0,300957118 | 0,61493976 | YIPF6 | FALSE | Downregulated |
| -0,766785895 | 0,614962935 | LOC102724314 | FALSE | Downregulated |
| -0,51507178 | 0,614962935 | RP5-1013A22.2 | FALSE | Downregulated |
| 0,201026584 | 0,615762264 | POLD4 | FALSE | Upregulated |
| -0,159485392 | 0,615762264 | UBE3A | FALSE | Downregulated |
| 0,445291028 | 0,615883137 | RAD51B | FALSE | Upregulated |
| -0,519419541 | 0,6159796 | ALG3 | FALSE | Downregulated |
| 0,18676461 | 0,616014346 | FBXL3 | FALSE | Upregulated |
| 0,516460485 | 0,61616484 | OSGEPL1-AS1 | FALSE | Upregulated |
| 0,332282678 | 0,61616484 | RPRD1A | FALSE | Upregulated |
| 0,613186326 | 0,61616484 | TNFSF13B | FALSE | Upregulated |
| -0,272916889 | 0,616171215 | RGS14 | FALSE | Downregulated |
| 0,159639171 | 0,616171215 | SH2B3 | FALSE | Upregulated |
| -0,539934708 | 0,616171215 | ZNF346 | FALSE | Downregulated |
| -0,234173447 | 0,616201921 | LINC00667 | FALSE | Downregulated |
| -0,304331746 | 0,616265564 | PHF20L1 | FALSE | Downregulated |
| -0,286243484 | 0,616265564 | RNF125 | FALSE | Downregulated |
| 0,342353371 | 0,616271178 | AFF4 | FALSE | Upregulated |
| 0,872344204 | 0,616342731 | ZBTB7C | FALSE | Upregulated |
| 0,792976389 | 0,616430533 | FLVCR1 | FALSE | Upregulated |
| 0,799935207 | 0,616641072 | BBS7 | FALSE | Upregulated |
| 0,43717301 | 0,616792664 | SUSD1 | FALSE | Upregulated |
| 0,632016973 | 0,617086367 | KLHDC9 | FALSE | Upregulated |
| -0,322828747 | 0,617086367 | PES1 | FALSE | Downregulated |
| 0,237248279 | 0,617086367 | RAB23 | FALSE | Upregulated |
| -0,285642896 | 0,617107269 | GSTZ1 | FALSE | Downregulated |
| 0,350298003 | 0,617155377 | LOC102724026 | FALSE | Upregulated |
| 0,278745639 | 0,61724849 | DNAH17 | FALSE | Upregulated |
| -0,204603415 | 0,61724849 | EXT1 | FALSE | Downregulated |
| 0,689142886 | 0,61724849 | NEUROD2 | FALSE | Upregulated |
| 0,469452818 | 0,617454772 | DCAF17 | FALSE | Upregulated |
| 0,301991717 | 0,617454772 | FAM53C | FALSE | Upregulated |
| -0,54057057 | 0,617460772 | PALB2 | FALSE | Downregulated |
| 0,145455716 | 0,617460772 | ZFP36L2 | FALSE | Upregulated |
| -0,221804989 | 0,617503896 | NAV2 | FALSE | Downregulated |
| 0,412972993 | 0,617503896 | PASK | FALSE | Upregulated |
| 0,44657901 | 0,617503896 | RAP1GAP2 | FALSE | Upregulated |
| 0,472400503 | 0,617667201 | PRKCB | FALSE | Upregulated |
| 0,820785601 | 0,617808913 | LOC339874 | FALSE | Upregulated |
| -0,590527321 | 0,617959922 | HS3ST3A1 | FALSE | Downregulated |
| 0,21017568 | 0,617978306 | FBXO18 | FALSE | Upregulated |
| -0,30119336 | 0,618079235 | BRWD3 | FALSE | Downregulated |
| -0,198556394 | 0,618079235 | EIF3H | FALSE | Downregulated |
| 0,548958991 | 0,618079235 | GRIN2D | FALSE | Upregulated |
| -0,30684801 | 0,618079235 | MTG1 | FALSE | Downregulated |
| -0,364253626 | 0,618079235 | TRPM8 | FALSE | Downregulated |
| -0,920595081 | 0,618117994 | RAB30-AS1 | FALSE | Downregulated |
| -0,276282765 | 0,618117994 | RP11-696N14.1 | FALSE | Downregulated |

| | | | | |
|---|---|---|---|---|
| -0,885305092 | 0,618117994 | ZFP92 | FALSE | Downregulated |
| 0,380349919 | 0,618304778 | CASP10 | FALSE | Upregulated |
| -0,508992446 | 0,618304778 | EXOSC7 | FALSE | Downregulated |
| 0,23937869 | 0,618503846 | C9orf37 | FALSE | Upregulated |
| -0,62406077 | 0,618503846 | MINOS1 | FALSE | Downregulated |
| -0,235585425 | 0,618555012 | ATP6V0E2 | FALSE | Downregulated |
| 0,193931508 | 0,618555012 | MMRN2 | FALSE | Upregulated |
| -0,268403491 | 0,618555012 | MRPL34 | FALSE | Downregulated |
| -0,848939357 | 0,618555012 | RHBDL1 | FALSE | Downregulated |
| 0,305362534 | 0,618555012 | SLC35F6 | FALSE | Upregulated |
| -0,264051393 | 0,618555012 | TMX3 | FALSE | Downregulated |
| 0,789692862 | 0,618687034 | SPIB | FALSE | Upregulated |
| 1,237642104 | 0,61875952 | ATP1A2 | FALSE | Upregulated |
| 0,323558122 | 0,618835841 | C14orf166 | FALSE | Upregulated |
| 0,953388823 | 0,618946795 | SUPT3H | FALSE | Upregulated |
| 0,391062014 | 0,619035355 | GLTP | FALSE | Upregulated |
| 0,530602501 | 0,619125813 | LACE1 | FALSE | Upregulated |
| 0,188002253 | 0,619125813 | MGA | FALSE | Upregulated |
| -0,469260948 | 0,619414649 | FBXO27 | FALSE | Downregulated |
| 0,703011839 | 0,619414649 | LOC101927125 | FALSE | Upregulated |
| -0,280248838 | 0,619478802 | ABHD16A | FALSE | Downregulated |
| -0,39855346 | 0,619478802 | CACNA2D2 | FALSE | Downregulated |
| -0,446446631 | 0,619478802 | CFDP1 | FALSE | Downregulated |
| 0,248873313 | 0,619478802 | LCOR | FALSE | Upregulated |
| -0,590677516 | 0,619478802 | MAST1 | FALSE | Downregulated |
| 0,290378291 | 0,619478802 | PLEKHA2 | FALSE | Upregulated |
| -0,844368402 | 0,619573923 | LOC101928561 | FALSE | Downregulated |
| 0,868702109 | 0,619618642 | MCHR1 | FALSE | Upregulated |
| 1,123385077 | 0,619618642 | SOCS1 | FALSE | Upregulated |
| -1,083569212 | 0,619618642 | USP6NL-IT1 | FALSE | Downregulated |
| -0,829435931 | 0,619638364 | LOC389831 | FALSE | Downregulated |
| 0,37598335 | 0,619725569 | ZSCAN25 | FALSE | Upregulated |
| 0,590128837 | 0,619790325 | LOC102724587 | FALSE | Upregulated |
| 0,503167442 | 0,619790325 | XXYLT1-AS2 | FALSE | Upregulated |
| -0,711393555 | 0,620052062 | FREM1 | FALSE | Downregulated |
| 0,232927886 | 0,620167112 | MXRA8 | FALSE | Upregulated |
| 0,35061727 | 0,620167112 | PLEKHF1 | FALSE | Upregulated |
| 0,690766505 | 0,620167112 | PRICKLE2-AS2 | FALSE | Upregulated |
| -0,575586387 | 0,620167112 | RAI2 | FALSE | Downregulated |
| -0,21038247 | 0,620167112 | SHROOM2 | FALSE | Downregulated |
| 0,160472115 | 0,620167112 | SNED1 | FALSE | Upregulated |
| 0,404705933 | 0,620167112 | UBOX5 | FALSE | Upregulated |
| -0,19904589 | 0,620167112 | WBP2 | FALSE | Downregulated |
| 0,208810193 | 0,620636469 | GNS | FALSE | Upregulated |
| 0,290054265 | 0,620778598 | RNF41 | FALSE | Upregulated |
| 0,372550835 | 0,620998433 | SCARNA13 | FALSE | Upregulated |
| 0,391440516 | 0,621081837 | BBS9 | FALSE | Upregulated |
| -0,263670754 | 0,621085278 | ACSL3 | FALSE | Downregulated |
| 0,26460926 | 0,621085278 | AMDHD2 | FALSE | Upregulated |
| -0,351630156 | 0,621085278 | ATOX1 | FALSE | Downregulated |
| -0,27597844 | 0,621085278 | BCAR3 | FALSE | Downregulated |

| | | | | |
|---|---|---|---|---|
| 0,247986129 | 0,621085278 | C22orf29 | FALSE | Upregulated |
| -0,214415428 | 0,621085278 | CARNS1 | FALSE | Downregulated |
| 0,428983445 | 0,621085278 | FAM135A | FALSE | Upregulated |
| 0,910758118 | 0,621085278 | FANCE | FALSE | Upregulated |
| 0,533901532 | 0,621085278 | GLIDR | FALSE | Upregulated |
| -0,30511523 | 0,621085278 | GMEB2 | FALSE | Downregulated |
| 0,298456196 | 0,621085278 | GNG12 | FALSE | Upregulated |
| -0,534222695 | 0,621085278 | GTF2H5 | FALSE | Downregulated |
| -0,145005612 | 0,621085278 | HELZ | FALSE | Downregulated |
| -0,271880486 | 0,621085278 | IKZF5 | FALSE | Downregulated |
| -0,883080991 | 0,621085278 | LOC100505771 | FALSE | Downregulated |
| 0,276241349 | 0,621085278 | LRRN2 | FALSE | Upregulated |
| -0,222438953 | 0,621085278 | MTR | FALSE | Downregulated |
| 0,561264818 | 0,621085278 | OTUD6B | FALSE | Upregulated |
| -0,239959532 | 0,621085278 | RNF170 | FALSE | Downregulated |
| -0,523319663 | 0,621085278 | SLC41A3 | FALSE | Downregulated |
| -0,197337089 | 0,621085278 | TECR | FALSE | Downregulated |
| 0,714995882 | 0,621085278 | TMEM229B | FALSE | Upregulated |
| 0,748792914 | 0,621085278 | ULK4P2 | FALSE | Upregulated |
| -0,251534478 | 0,621085278 | URI1 | FALSE | Downregulated |
| -0,590766524 | 0,621085278 | VPS33B | FALSE | Downregulated |
| 0,170447437 | 0,621362062 | SNRK | FALSE | Upregulated |
| 0,602732459 | 0,621502723 | FAM200A | FALSE | Upregulated |
| -1,258974732 | 0,621502723 | LOC100509620 | FALSE | Downregulated |
| -1,073940759 | 0,621502723 | RP11-115H18.1 | FALSE | Downregulated |
| -0,367421701 | 0,621502723 | TEX264 | FALSE | Downregulated |
| -0,299523914 | 0,621533405 | ANKS4B | FALSE | Downregulated |
| -0,384568729 | 0,621533405 | ELOVL6 | FALSE | Downregulated |
| 0,893583353 | 0,621570507 | LOC101928936 | FALSE | Upregulated |
| -0,52097917 | 0,621570507 | MTFMT | FALSE | Downregulated |
| 0,194975967 | 0,62186367 | ELMSAN1 | FALSE | Upregulated |
| -0,168857592 | 0,62186367 | LMTK2 | FALSE | Downregulated |
| -0,603877884 | 0,62186367 | NOVA2 | FALSE | Downregulated |
| -0,25102082 | 0,62186367 | PSMD5 | FALSE | Downregulated |
| -0,41626774 | 0,62186367 | ST3GAL5 | FALSE | Downregulated |
| -0,383697034 | 0,621894502 | SEC11C | FALSE | Downregulated |
| 0,780276144 | 0,622100158 | LOC101927693 | FALSE | Upregulated |
| 0,810431553 | 0,622163698 | EBF1 | FALSE | Upregulated |
| -0,234299928 | 0,622163698 | TOR3A | FALSE | Downregulated |
| 0,743383106 | 0,622201436 | GDF15 | FALSE | Upregulated |
| 0,442284566 | 0,622201436 | GPD2 | FALSE | Upregulated |
| 0,60685147 | 0,622201436 | IGF2BP2 | FALSE | Upregulated |
| -0,395196237 | 0,622201436 | LINC00909 | FALSE | Downregulated |
| 0,603410797 | 0,622201436 | LST1 | FALSE | Upregulated |
| -0,533611732 | 0,622201436 | MPPED1 | FALSE | Downregulated |
| 0,245478274 | 0,622201436 | NFYB | FALSE | Upregulated |
| 0,438120258 | 0,622201436 | SKAP2 | FALSE | Upregulated |
| -0,50188488 | 0,622201436 | TYW3 | FALSE | Downregulated |
| 0,534975539 | 0,622253127 | PRKD1 | FALSE | Upregulated |
| 0,641979811 | 0,622377271 | CD40LG | FALSE | Upregulated |
| 0,224079596 | 0,622491482 | DHX8 | FALSE | Upregulated |

| | | | | |
|---|---|---|---|---|
| -0,150506927 | 0,622491482 | LAMP1 | FALSE | Downregulated |
| -0,744561209 | 0,622491482 | LOC439951 | FALSE | Downregulated |
| 0,269296088 | 0,622677995 | CBX6 | FALSE | Upregulated |
| 0,18674114 | 0,622677995 | CDS2 | FALSE | Upregulated |
| 0,372573592 | 0,622750863 | FAM214B | FALSE | Upregulated |
| -0,323979972 | 0,622750863 | KCNT2 | FALSE | Downregulated |
| -0,986158583 | 0,622750863 | LGR5 | FALSE | Downregulated |
| -0,320761827 | 0,622755063 | ATG16L1 | FALSE | Downregulated |
| 1,549407999 | 0,622755063 | RIMBP3C | FALSE | Upregulated |
| 0,52148847 | 0,622755063 | ZNF341 | FALSE | Upregulated |
| -0,217228091 | 0,622807304 | SNAP47 | FALSE | Downregulated |
| -1,065397138 | 0,622831162 | LOC101929656 | FALSE | Downregulated |
| 0,276890749 | 0,622831162 | ZCCHC3 | FALSE | Upregulated |
| 0,244109177 | 0,622874981 | HTATSF1P2 | FALSE | Upregulated |
| 1,000674187 | 0,622874981 | PCYOX1L | FALSE | Upregulated |
| 0,563824777 | 0,623042109 | LOC101929449 | FALSE | Upregulated |
| 0,521342293 | 0,623091847 | C12orf4 | FALSE | Upregulated |
| 0,29978709 | 0,623183233 | SAMD8 | FALSE | Upregulated |
| -0,266379989 | 0,623239105 | ABCB8 | FALSE | Downregulated |
| -0,37454587 | 0,623290131 | FAM203A | FALSE | Downregulated |
| -0,705263686 | 0,623290131 | LOC100289361 | FALSE | Downregulated |
| -0,406189739 | 0,623386853 | ZNF688 | FALSE | Downregulated |
| -0,22357809 | 0,623478881 | CYP2C18 | FALSE | Downregulated |
| 0,188895001 | 0,623478881 | LRCH4 | FALSE | Upregulated |
| 0,582194841 | 0,623478881 | PKHD1L1 | FALSE | Upregulated |
| -0,292837915 | 0,623478881 | UMPS | FALSE | Downregulated |
| -0,254117113 | 0,623478881 | ZNF800 | FALSE | Downregulated |
| 0,400115725 | 0,623573389 | ADCK5 | FALSE | Upregulated |
| -0,95264079 | 0,623579291 | ALMS1P | FALSE | Downregulated |
| 0,548289039 | 0,62373559 | AL022476.2 | FALSE | Upregulated |
| -0,25325658 | 0,62373559 | SOS2 | FALSE | Downregulated |
| 0,675476817 | 0,62373559 | TSPAN15 | FALSE | Upregulated |
| 0,521581928 | 0,62376038 | MRPL36 | FALSE | Upregulated |
| 0,374813507 | 0,62376038 | SSB | FALSE | Upregulated |
| -0,218120271 | 0,62378305 | POLR1A | FALSE | Downregulated |
| -0,19261612 | 0,624606313 | ZNF664 | FALSE | Downregulated |
| -0,490394862 | 0,624691077 | CRY1 | FALSE | Downregulated |
| 0,13640295 | 0,624691077 | DYNC1H1 | FALSE | Upregulated |
| 0,332504458 | 0,624691077 | LOC100287042 | FALSE | Upregulated |
| 0,776573854 | 0,624691077 | U2AF1L4 | FALSE | Upregulated |
| -0,271560268 | 0,624694413 | PCIF1 | FALSE | Downregulated |
| 0,240754868 | 0,62472581 | RGAG4 | FALSE | Upregulated |
| -0,481725939 | 0,62472581 | TRAPPC6A | FALSE | Downregulated |
| 0,499897304 | 0,624960644 | PDLIM7 | FALSE | Upregulated |
| -0,410076785 | 0,625109813 | CHEK1 | FALSE | Downregulated |
| -1,134874049 | 0,625109813 | LOC101929580 | FALSE | Downregulated |
| -0,27464728 | 0,625109813 | TSPAN31 | FALSE | Downregulated |
| 0,153700309 | 0,625358663 | TUBB | FALSE | Upregulated |
| 0,529975706 | 0,62544324 | RRAGC | FALSE | Upregulated |
| -0,20276193 | 0,625464431 | CEP164 | FALSE | Downregulated |
| -0,263534887 | 0,625464431 | RAD54L2 | FALSE | Downregulated |

| | | | | |
|---|---|---|---|---|
| 0,568903848 | 0,625555287 | CDC14A | FALSE | Upregulated |
| 0,271383504 | 0,625683367 | GNAI3 | FALSE | Upregulated |
| 0,291466442 | 0,625683367 | NEIL2 | FALSE | Upregulated |
| -0,350770919 | 0,625683367 | TMEM150C | FALSE | Downregulated |
| -0,301205072 | 0,625683367 | UFC1 | FALSE | Downregulated |
| 0,253012607 | 0,625712756 | GJD3 | FALSE | Upregulated |
| -0,400184731 | 0,625732476 | ABCF2 | FALSE | Downregulated |
| -0,135880177 | 0,625732476 | ADAR | FALSE | Downregulated |
| -0,316569485 | 0,625732476 | BCS1L | FALSE | Downregulated |
| 0,195272444 | 0,625732476 | CAPN12 | FALSE | Upregulated |
| -0,259556256 | 0,625732476 | CNPY3 | FALSE | Downregulated |
| 0,25226389 | 0,625732476 | DCLRE1C | FALSE | Upregulated |
| 0,372297134 | 0,625732476 | NIPAL4 | FALSE | Upregulated |
| 0,309025198 | 0,625732476 | RP11-475I24.8 | FALSE | Upregulated |
| 0,606569164 | 0,626104385 | IFI44 | FALSE | Upregulated |
| -0,502470222 | 0,626223871 | ZNF778 | FALSE | Downregulated |
| 0,187592998 | 0,626333714 | MAPKAPK2 | FALSE | Upregulated |
| -0,917500311 | 0,626527719 | KPTN | FALSE | Downregulated |
| -0,417302211 | 0,626922811 | ZNF155 | FALSE | Downregulated |
| -0,247828632 | 0,626934316 | MRPS18B | FALSE | Downregulated |
| -0,387370897 | 0,62714588 | TMEM135 | FALSE | Downregulated |
| -0,317537435 | 0,62714588 | ZCCHC8 | FALSE | Downregulated |
| -0,177864101 | 0,627196667 | SHROOM3 | FALSE | Downregulated |
| 0,285776644 | 0,627230278 | GBP2 | FALSE | Upregulated |
| 0,658435764 | 0,627266727 | KCNK10 | FALSE | Upregulated |
| -0,282988834 | 0,627295574 | USP14 | FALSE | Downregulated |
| -0,27964618 | 0,62749159 | EXOC5 | FALSE | Downregulated |
| 1,454830395 | 0,627632844 | DMKN | FALSE | Upregulated |
| 2,675634663 | 0,627632844 | MYLK2 | FALSE | Upregulated |
| -0,242082176 | 0,627632844 | SLC39A9 | FALSE | Downregulated |
| 0,325698421 | 0,627783269 | ACSS2 | FALSE | Upregulated |
| 0,316446801 | 0,627841203 | HCLS1 | FALSE | Upregulated |
| -0,319225229 | 0,628074603 | FUZ | FALSE | Downregulated |
| 0,551758707 | 0,628074603 | LRRC9 | FALSE | Upregulated |
| -0,302516011 | 0,628074603 | NME2 | FALSE | Downregulated |
| 0,233525324 | 0,628194071 | PSMC3IP | FALSE | Upregulated |
| -0,517071944 | 0,628329175 | PRH1-PRR4 | FALSE | Downregulated |
| -0,245984257 | 0,628329175 | RNF13 | FALSE | Downregulated |
| -0,330454197 | 0,628532827 | ACKR2 | FALSE | Downregulated |
| 0,484963843 | 0,628532827 | RP11-1072A3.2 | FALSE | Upregulated |
| -0,561398812 | 0,628532827 | TRPV3 | FALSE | Downregulated |
| -1,017468422 | 0,628532827 | WDHD1 | FALSE | Downregulated |
| -0,310892172 | 0,62859432 | RRN3 | FALSE | Downregulated |
| 0,305278412 | 0,62859432 | ZMYM6 | FALSE | Upregulated |
| -0,560261114 | 0,628883456 | NPRL2 | FALSE | Downregulated |
| 0,250650133 | 0,628883456 | WDR5B | FALSE | Upregulated |
| 0,758305848 | 0,629107956 | CAMKK1 | FALSE | Upregulated |
| 0,694090445 | 0,629418113 | AWAT1 | FALSE | Upregulated |
| -0,28624447 | 0,629418113 | RSAD1 | FALSE | Downregulated |
| 0,441368882 | 0,629418113 | VEPH1 | FALSE | Upregulated |
| -0,333682828 | 0,629491468 | SNHG7 | FALSE | Downregulated |

| | | | | |
|---|---|---|---|---|
| -0,2382496 | 0,629543508 | FZR1 | FALSE | Downregulated |
| 3,244611873 | 0,629543508 | LOC102724956 | FALSE | Upregulated |
| 0,630710736 | 0,629543508 | LPCAT2 | FALSE | Upregulated |
| -0,467717526 | 0,629543508 | MRPL45P2 | FALSE | Downregulated |
| -1,610097593 | 0,629557727 | LY75 | FALSE | Downregulated |
| 0,27043319 | 0,629720015 | ZFP28 | FALSE | Upregulated |
| -0,161996884 | 0,629739862 | MEF2A | FALSE | Downregulated |
| -0,237085373 | 0,630073204 | ABCA9 | FALSE | Downregulated |
| -0,912210926 | 0,630412605 | C4orf27 | FALSE | Downregulated |
| -0,614986072 | 0,630435439 | C2orf74 | FALSE | Downregulated |
| 0,989510863 | 0,630435439 | RGS16 | FALSE | Upregulated |
| 0,53962725 | 0,630670726 | MRM1 | FALSE | Upregulated |
| -0,416722839 | 0,630670726 | PPP2R5B | FALSE | Downregulated |
| 0,129383428 | 0,630841974 | MYH10 | FALSE | Upregulated |
| 0,163567472 | 0,631030866 | HNRNPU-AS1 | FALSE | Upregulated |
| -0,299945351 | 0,631046069 | AFTPH | FALSE | Downregulated |
| -0,162343571 | 0,631046069 | KAT6A | FALSE | Downregulated |
| -0,507156545 | 0,631046069 | UAP1L1 | FALSE | Downregulated |
| 0,371696401 | 0,631046069 | ZDHHC21 | FALSE | Upregulated |
| 0,388353528 | 0,631190434 | CASP2 | FALSE | Upregulated |
| 0,164779865 | 0,631190434 | PCF11 | FALSE | Upregulated |
| -0,218553494 | 0,631200275 | ANGPTL4 | FALSE | Downregulated |
| 0,490589179 | 0,631200275 | PARVB | FALSE | Upregulated |
| 0,460815713 | 0,631412146 | CHGA | FALSE | Upregulated |
| -0,169532091 | 0,631479366 | ARFGAP2 | FALSE | Downregulated |
| -0,356418241 | 0,631479366 | C17orf47 | FALSE | Downregulated |
| -0,874669467 | 0,631479366 | C8orf31 | FALSE | Downregulated |
| 0,298069584 | 0,631479366 | PRKAA2 | FALSE | Upregulated |
| -0,207012534 | 0,631479366 | STAG2 | FALSE | Downregulated |
| 0,323805859 | 0,631479366 | TIMELESS | FALSE | Upregulated |
| 0,43956517 | 0,631572958 | DIRC2 | FALSE | Upregulated |
| 0,177056333 | 0,631572958 | RTEL1 | FALSE | Upregulated |
| 0,561074625 | 0,631752754 | LOC389199 | FALSE | Upregulated |
| -0,21232318 | 0,631847228 | SERINC5 | FALSE | Downregulated |
| 0,510121013 | 0,631973809 | FAM98C | FALSE | Upregulated |
| -0,259404234 | 0,632149774 | CPB2-AS1 | FALSE | Downregulated |
| 0,614951769 | 0,632149774 | LRRC29 | FALSE | Upregulated |
| -0,591621033 | 0,632405415 | IMMP2L | FALSE | Downregulated |
| 0,378278473 | 0,63249469 | ERMARD | FALSE | Upregulated |
| -0,556585162 | 0,632530561 | CHST14 | FALSE | Downregulated |
| 0,254126478 | 0,632579794 | ATG4D | FALSE | Upregulated |
| -0,322026132 | 0,632579794 | CMAS | FALSE | Downregulated |
| -0,600661489 | 0,632782379 | EGR1 | FALSE | Downregulated |
| 0,182177905 | 0,632782379 | NNAT | FALSE | Upregulated |
| -0,719106801 | 0,632782379 | PBDC1 | FALSE | Downregulated |
| -0,560570473 | 0,632783975 | ZNF658 | FALSE | Downregulated |
| -0,325798026 | 0,632891817 | POLR3B | FALSE | Downregulated |
| 0,430314488 | 0,632933923 | WASF3 | FALSE | Upregulated |
| 0,219778533 | 0,632947919 | SEC61A2 | FALSE | Upregulated |
| -0,440187328 | 0,632966947 | CHM | FALSE | Downregulated |
| -0,270724609 | 0,633007997 | METAP2 | FALSE | Downregulated |

| | | | | |
|---|---|---|---|---|
| -0,219787413 | 0,633079721 | PSMC1 | FALSE | Downregulated |
| 0,26487501 | 0,633079721 | TRAPPC2 | FALSE | Upregulated |
| -0,266636033 | 0,633232852 | LOC101928319 | FALSE | Downregulated |
| 0,226177616 | 0,633548565 | POLR2L | FALSE | Upregulated |
| -0,48038866 | 0,633548565 | RNASEK | FALSE | Downregulated |
| -0,266019541 | 0,633581212 | CEP104 | FALSE | Downregulated |
| -0,521993723 | 0,633581212 | DPY19L2P4 | FALSE | Downregulated |
| -0,356631311 | 0,633663736 | MGC27345 | FALSE | Downregulated |
| 0,692714941 | 0,633663736 | PRELID2 | FALSE | Upregulated |
| -0,32395637 | 0,633691662 | BTBD7 | FALSE | Downregulated |
| -0,159859235 | 0,633691662 | DDX42 | FALSE | Downregulated |
| -0,434463276 | 0,633691662 | HIST1H2BD | FALSE | Downregulated |
| 0,375970137 | 0,633691662 | HRAS | FALSE | Upregulated |
| 0,52065922 | 0,633691662 | MAPK11 | FALSE | Upregulated |
| -0,58895395 | 0,633691662 | MEIOB | FALSE | Downregulated |
| -0,514181962 | 0,633691662 | PAXIP1-AS1 | FALSE | Downregulated |
| -0,267593569 | 0,633713824 | NQO2 | FALSE | Downregulated |
| -0,833229873 | 0,633715747 | LOC728989 | FALSE | Downregulated |
| -0,193987831 | 0,633715747 | SMG5 | FALSE | Downregulated |
| 0,310219793 | 0,63399304 | XPNPEP1 | FALSE | Upregulated |
| 0,684379738 | 0,634153049 | LINC00493 | FALSE | Upregulated |
| 0,340187478 | 0,634212679 | CARD8 | FALSE | Upregulated |
| -0,631549697 | 0,634216917 | C3orf35 | FALSE | Downregulated |
| 0,27368294 | 0,634216917 | CC2D2A | FALSE | Upregulated |
| -0,400927091 | 0,634216917 | SLC25A26 | FALSE | Downregulated |
| -0,174608508 | 0,634499182 | TET2 | FALSE | Downregulated |
| -0,26256851 | 0,634559405 | BANP | FALSE | Downregulated |
| 1,016089967 | 0,634559405 | IL1B | FALSE | Upregulated |
| -0,503355447 | 0,634559405 | NNT-AS1 | FALSE | Downregulated |
| -0,271516482 | 0,634559405 | TUSC1 | FALSE | Downregulated |
| 0,28484122 | 0,634559405 | ULK3 | FALSE | Upregulated |
| 0,132066632 | 0,634651593 | NIPBL | FALSE | Upregulated |
| 0,29648797 | 0,63468417 | LOC101929714 | FALSE | Upregulated |
| -0,199750543 | 0,63468417 | MFSD12 | FALSE | Downregulated |
| 0,288906136 | 0,634843104 | PDHB | FALSE | Upregulated |
| 0,600214762 | 0,634855611 | COMMD6 | FALSE | Upregulated |
| -0,274776305 | 0,635077619 | AKAP6 | FALSE | Downregulated |
| 0,548383421 | 0,635077619 | ALG10B | FALSE | Upregulated |
| -0,206733111 | 0,635077619 | ARHGAP32 | FALSE | Downregulated |
| -0,944156306 | 0,635077619 | C8orf44 | FALSE | Downregulated |
| 0,507490269 | 0,635077619 | CLRN1 | FALSE | Upregulated |
| 0,680386535 | 0,635077619 | DISC1 | FALSE | Upregulated |
| -0,439782147 | 0,635077619 | ENO3 | FALSE | Downregulated |
| -1,362665372 | 0,635077619 | GRID1 | FALSE | Downregulated |
| 0,294432741 | 0,635077619 | HCFC2 | FALSE | Upregulated |
| 0,890078445 | 0,635077619 | IRAK1BP1 | FALSE | Upregulated |
| -0,228794543 | 0,635077619 | IWS1 | FALSE | Downregulated |
| 0,308271498 | 0,635077619 | KLF7 | FALSE | Upregulated |
| 0,589321821 | 0,635077619 | KRT10 | FALSE | Upregulated |
| 0,812301993 | 0,635077619 | LINC00662 | FALSE | Upregulated |
| -1,016899372 | 0,635077619 | LOC100506714 | FALSE | Downregulated |

| | | | | |
|---|---|---|---|---|
| 0,625325838 | 0,635077619 | LOC100507507 | FALSE | Upregulated |
| 0,209139007 | 0,635077619 | LOC101930421 | FALSE | Upregulated |
| 1,027029625 | 0,635077619 | LOC102724621 | FALSE | Upregulated |
| 0,472043143 | 0,635077619 | LRRC37A3 | FALSE | Upregulated |
| 0,438907938 | 0,635077619 | NKRF | FALSE | Upregulated |
| 0,847695219 | 0,635077619 | PCLO | FALSE | Upregulated |
| -0,333738577 | 0,635077619 | PPP2R3B | FALSE | Downregulated |
| 0,448831673 | 0,635077619 | RAB42 | FALSE | Upregulated |
| 0,175021544 | 0,635077619 | SH3D19 | FALSE | Upregulated |
| -0,180077417 | 0,635077619 | SS18L1 | FALSE | Downregulated |
| -0,369700268 | 0,635077619 | TBL2 | FALSE | Downregulated |
| 0,292974878 | 0,635077619 | VAMP8 | FALSE | Upregulated |
| -0,175574491 | 0,635077619 | ZNF33A | FALSE | Downregulated |
| 0,325982009 | 0,635116975 | ECD | FALSE | Upregulated |
| 0,299113371 | 0,635328504 | ATAT1 | FALSE | Upregulated |
| 0,506274371 | 0,635328504 | STXBP5 | FALSE | Upregulated |
| 0,754390538 | 0,635329227 | LOC101929112 | FALSE | Upregulated |
| -0,943863471 | 0,635673488 | CCL3 | FALSE | Downregulated |
| -0,324410455 | 0,635673488 | CXADR | FALSE | Downregulated |
| 0,162450932 | 0,635673488 | H3F3A | FALSE | Upregulated |
| -0,979571442 | 0,63576788 | C2CD4B | FALSE | Downregulated |
| -0,261622482 | 0,635800827 | LECT2 | FALSE | Downregulated |
| -0,964842544 | 0,635800827 | PRG2 | FALSE | Downregulated |
| 0,234886248 | 0,635800827 | TOR1A | FALSE | Upregulated |
| -0,351326734 | 0,635974591 | AMOT | FALSE | Downregulated |
| 0,465189818 | 0,635974591 | CAAP1 | FALSE | Upregulated |
| -0,240489626 | 0,635974591 | HN1L | FALSE | Downregulated |
| 0,332965885 | 0,635974591 | KIF16B | FALSE | Upregulated |
| 0,665548166 | 0,635974591 | LOC101929015 | FALSE | Upregulated |
| 0,319259159 | 0,635974591 | LOC101929270 | FALSE | Upregulated |
| 0,803527396 | 0,635974591 | POM121L8P | FALSE | Upregulated |
| 0,727412157 | 0,635974591 | RP11-111E14.1 | FALSE | Upregulated |
| -0,187763312 | 0,635974591 | TMEM30B | FALSE | Downregulated |
| 0,417743779 | 0,635974591 | ZNF440 | FALSE | Upregulated |
| 0,956117145 | 0,636006027 | FAIM | FALSE | Upregulated |
| -0,239850194 | 0,636022617 | ARPC1A | FALSE | Downregulated |
| 0,45483836 | 0,636063792 | DTD2 | FALSE | Upregulated |
| 0,317124854 | 0,636063792 | GOLT1A | FALSE | Upregulated |
| -0,394447633 | 0,636063792 | GPR180 | FALSE | Downregulated |
| 0,735577355 | 0,636063792 | HSPB7 | FALSE | Upregulated |
| -0,883380234 | 0,636063792 | MAN1B1-AS1 | FALSE | Downregulated |
| 0,416820728 | 0,636063792 | RAMP1 | FALSE | Upregulated |
| -0,472914689 | 0,636063792 | SELK | FALSE | Downregulated |
| 0,656849073 | 0,63606421 | NLRP3 | FALSE | Upregulated |
| 1,015235932 | 0,63606421 | PLA2G2A | FALSE | Upregulated |
| -0,979156141 | 0,636555705 | HDX | FALSE | Downregulated |
| 0,327107787 | 0,636617927 | ARHGAP44 | FALSE | Upregulated |
| -0,177451649 | 0,636617927 | CCNI | FALSE | Downregulated |
| -0,181589084 | 0,636617927 | CELF1 | FALSE | Downregulated |
| 0,396990762 | 0,636617927 | ELAC1 | FALSE | Upregulated |
| -0,477744616 | 0,636617927 | LIAS | FALSE | Downregulated |

| | | | | |
|---|---|---|---|---|
| -0,693127842 | 0,636617927 | LOC102724423 | FALSE | Downregulated |
| -0,510443304 | 0,636617927 | LOC102724788 | FALSE | Downregulated |
| -0,318425195 | 0,636617927 | NOL7 | FALSE | Downregulated |
| 0,348389724 | 0,636617927 | NT5C1B | FALSE | Upregulated |
| 0,427750343 | 0,636617927 | PMEL | FALSE | Upregulated |
| 0,449062957 | 0,636617927 | RP3-400N23.6 | FALSE | Upregulated |
| -0,227347263 | 0,636617927 | RPL39 | FALSE | Downregulated |
| 0,273901607 | 0,636617927 | SLC35E2 | FALSE | Upregulated |
| -1,16809849 | 0,636617927 | SPIN2B | FALSE | Downregulated |
| 0,253045136 | 0,636617927 | SYNGAP1 | FALSE | Upregulated |
| -0,71964252 | 0,636617927 | TMEM200C | FALSE | Downregulated |
| -0,925983842 | 0,636617927 | ZNF709 | FALSE | Downregulated |
| -1,073292891 | 0,636628073 | NPIPB8 | FALSE | Downregulated |
| -0,880596811 | 0,636712612 | LOC102724598 | FALSE | Downregulated |
| 0,875520483 | 0,636712612 | RNASEK-C17orf49 | FALSE | Upregulated |
| -0,417578353 | 0,636804769 | TMEM82 | FALSE | Downregulated |
| 0,243215111 | 0,636893943 | ACACA | FALSE | Upregulated |
| 0,899602522 | 0,636893943 | SCG5 | FALSE | Upregulated |
| 0,569759342 | 0,637052486 | COMMD2 | FALSE | Upregulated |
| -0,422557977 | 0,637110142 | CSTF1 | FALSE | Downregulated |
| 0,220956026 | 0,637110142 | SBF2-AS1 | FALSE | Upregulated |
| 0,432227233 | 0,637204102 | GAS1 | FALSE | Upregulated |
| 0,268363735 | 0,637570276 | GCNT7 | FALSE | Upregulated |
| 2,37218163 | 0,637570276 | LOC102724516 | FALSE | Upregulated |
| -1,026064152 | 0,637680054 | C1orf229 | FALSE | Downregulated |
| -0,314228463 | 0,63789496 | AFG3L1P | FALSE | Downregulated |
| 0,464642075 | 0,63789496 | HEPHL1 | FALSE | Upregulated |
| -0,277569489 | 0,63789496 | IDH3G | FALSE | Downregulated |
| -0,281407193 | 0,63789496 | NELFA | FALSE | Downregulated |
| -0,305511962 | 0,637898734 | ZFAND6 | FALSE | Downregulated |
| -1,026326412 | 0,637991993 | RFESD | FALSE | Downregulated |
| -0,632863716 | 0,638072843 | KLHL23 | FALSE | Downregulated |
| -0,270568218 | 0,638157228 | TMX2 | FALSE | Downregulated |
| 0,399938961 | 0,638157228 | TTLL3 | FALSE | Upregulated |
| 0,51346527 | 0,63824203 | ARNT2 | FALSE | Upregulated |
| 0,191030339 | 0,63824203 | ENTPD1 | FALSE | Upregulated |
| -0,433522176 | 0,63824203 | LOC101929384 | FALSE | Downregulated |
| 1,03327364 | 0,63824203 | LOC653653 | FALSE | Upregulated |
| -0,256844803 | 0,63824203 | METRN | FALSE | Downregulated |
| -0,269678372 | 0,63824203 | TIA1 | FALSE | Downregulated |
| -0,271793742 | 0,638257032 | SDHD | FALSE | Downregulated |
| -0,330650151 | 0,638325725 | MRPS15 | FALSE | Downregulated |
| -0,376434732 | 0,638803222 | LOC731656 | FALSE | Downregulated |
| 0,273439426 | 0,638893228 | NTN3 | FALSE | Upregulated |
| -0,820548744 | 0,638943005 | PTPRO | FALSE | Downregulated |
| 0,213534 | 0,639055354 | HMOX2 | FALSE | Upregulated |
| 0,482607047 | 0,639155145 | LENEP | FALSE | Upregulated |
| -0,269980715 | 0,639155145 | PRCC | FALSE | Downregulated |
| -0,267659645 | 0,639205865 | C9orf173 | FALSE | Downregulated |
| 0,207118056 | 0,639231615 | ARID5A | FALSE | Upregulated |
| 0,211862845 | 0,639239096 | AC012314.8 | FALSE | Upregulated |

| | | | | |
|---|---|---|---|---|
| 0,181324713 | 0,639239096 | FGD6 | FALSE | Upregulated |
| -0,243063551 | 0,639263898 | EIF6 | FALSE | Downregulated |
| 0,283670485 | 0,639263898 | FAM126B | FALSE | Upregulated |
| -0,237646548 | 0,639263898 | LIMS2 | FALSE | Downregulated |
| 0,247641975 | 0,639300325 | HLA-C | FALSE | Upregulated |
| -0,842668645 | 0,639300325 | MAP3K7CL | FALSE | Downregulated |
| 0,255616269 | 0,639300325 | OFD1 | FALSE | Upregulated |
| -0,434817003 | 0,639300325 | SYPL2 | FALSE | Downregulated |
| 0,243700659 | 0,639332675 | C12orf43 | FALSE | Upregulated |
| -0,70355679 | 0,639381378 | IFT43 | FALSE | Downregulated |
| -0,245784378 | 0,639381378 | RP11-57H12.3 | FALSE | Downregulated |
| -0,416262658 | 0,639454649 | FAM65B | FALSE | Downregulated |
| 0,564893557 | 0,639454649 | ZNF2 | FALSE | Upregulated |
| 0,344040718 | 0,639610203 | C19orf25 | FALSE | Upregulated |
| 0,187790619 | 0,639610203 | HMHA1 | FALSE | Upregulated |
| -0,504577805 | 0,639610203 | LOC101928668 | FALSE | Downregulated |
| -1,130755471 | 0,639610203 | RP11-362F19.1 | FALSE | Downregulated |
| 0,383584835 | 0,639784941 | PROSER1 | FALSE | Upregulated |
| -0,306517151 | 0,63978802 | CLNS1A | FALSE | Downregulated |
| -0,191110844 | 0,63997259 | KLF12 | FALSE | Downregulated |
| -0,565676169 | 0,640088274 | DCP1B | FALSE | Downregulated |
| -0,3384275 | 0,640088274 | SEC22C | FALSE | Downregulated |
| -0,110255121 | 0,64022009 | ARID1A | FALSE | Downregulated |
| -0,265307558 | 0,640253031 | PSMD1 | FALSE | Downregulated |
| 0,509965168 | 0,640697317 | TASP1 | FALSE | Upregulated |
| 0,187369572 | 0,640739952 | C3orf17 | FALSE | Upregulated |
| 0,238606425 | 0,640739952 | DND1 | FALSE | Upregulated |
| -0,279235653 | 0,640739952 | NSMF | FALSE | Downregulated |
| 0,372452672 | 0,640739952 | PCDH9 | FALSE | Upregulated |
| -0,17054611 | 0,640852206 | ATXN1L | FALSE | Downregulated |
| -0,165370482 | 0,640852206 | EIF3G | FALSE | Downregulated |
| 0,213517589 | 0,640920809 | FAM178A | FALSE | Upregulated |
| 0,230303024 | 0,640920809 | PHLPP2 | FALSE | Upregulated |
| 0,486937444 | 0,641014991 | BIN2 | FALSE | Upregulated |
| -1,20312565 | 0,641014991 | HTT-AS | FALSE | Downregulated |
| 0,53046021 | 0,641098718 | GIMAP7 | FALSE | Upregulated |
| -0,789410795 | 0,641139985 | CDNF | FALSE | Downregulated |
| -0,311757944 | 0,641192087 | CCL16 | FALSE | Downregulated |
| -0,203194648 | 0,641192087 | GOLGA1 | FALSE | Downregulated |
| -0,332238984 | 0,641192087 | RNR1 | FALSE | Downregulated |
| -0,640064588 | 0,641192087 | ZBTB9 | FALSE | Downregulated |
| 0,612469748 | 0,641459587 | GPR89B | FALSE | Upregulated |
| 0,20114365 | 0,64196632 | BEST1 | FALSE | Upregulated |
| 0,250069022 | 0,641978962 | GPR17 | FALSE | Upregulated |
| -0,261029119 | 0,641978962 | LPIN3 | FALSE | Downregulated |
| -0,321674513 | 0,641978962 | MED25 | FALSE | Downregulated |
| 0,34631348 | 0,641978962 | PRPF39 | FALSE | Upregulated |
| -0,243712535 | 0,642088417 | ACOT2 | FALSE | Downregulated |
| -2,551325937 | 0,642088417 | COMMD3-BMI1 | FALSE | Downregulated |
| 0,497885661 | 0,642088417 | KIF20A | FALSE | Upregulated |
| 0,245210513 | 0,642088417 | LOC101929017 | FALSE | Upregulated |

| | | | | |
|---|---|---|---|---|
| -0,190396034 | 0,642088417 | PARP10 | FALSE | Downregulated |
| -0,192788052 | 0,642088417 | RPSA | FALSE | Downregulated |
| 0,473214845 | 0,642088417 | STAG3L5P | FALSE | Upregulated |
| -0,221553185 | 0,642097706 | ATMIN | FALSE | Downregulated |
| -0,925742605 | 0,642097706 | GDPGP1 | FALSE | Downregulated |
| 0,150780673 | 0,642175363 | LAMB2 | FALSE | Upregulated |
| -0,340584399 | 0,642175363 | PEX2 | FALSE | Downregulated |
| -0,430595031 | 0,642175363 | PSMD12 | FALSE | Downregulated |
| 0,45456664 | 0,642175363 | STARD4-AS1 | FALSE | Upregulated |
| -0,36356406 | 0,642175363 | TUBGCP5 | FALSE | Downregulated |
| -0,194849715 | 0,642175363 | WBP1L | FALSE | Downregulated |
| 0,128165083 | 0,642348338 | CTSB | FALSE | Upregulated |
| 0,801658296 | 0,642348338 | EBI3 | FALSE | Upregulated |
| -0,659038852 | 0,642368983 | RP11-81N13.1 | FALSE | Downregulated |
| -2,775353755 | 0,642368983 | SLX1B-SULT1A4 | FALSE | Downregulated |
| 0,254146601 | 0,642368983 | ZC3HAV1 | FALSE | Upregulated |
| 1,386189255 | 0,64239813 | TBC1D3H | FALSE | Upregulated |
| -0,392502508 | 0,64239813 | TMEM203 | FALSE | Downregulated |
| 0,376686301 | 0,642422333 | LOC101929401 | FALSE | Upregulated |
| -0,210546049 | 0,642444923 | CTIF | FALSE | Downregulated |
| -0,665094561 | 0,642532369 | C10orf62 | FALSE | Downregulated |
| -0,308354571 | 0,642532369 | CLDN15 | FALSE | Downregulated |
| -0,348769719 | 0,642532369 | INS | FALSE | Downregulated |
| -0,965898437 | 0,642532369 | RGS6 | FALSE | Downregulated |
| 0,248654341 | 0,642805715 | PIK3CA | FALSE | Upregulated |
| -0,808186411 | 0,64283731 | SLMO2-ATP5E | FALSE | Downregulated |
| -0,357295802 | 0,643082135 | TMCO4 | FALSE | Downregulated |
| -0,30406802 | 0,64319402 | DDC | FALSE | Downregulated |
| -0,406651404 | 0,64319402 | TM2D3 | FALSE | Downregulated |
| -0,262907481 | 0,643457152 | ACVR2B-AS1 | FALSE | Downregulated |
| -0,358061302 | 0,643457152 | ERCC2 | FALSE | Downregulated |
| 0,392353483 | 0,643457152 | FKTN | FALSE | Upregulated |
| -0,208714957 | 0,643682092 | SNX33 | FALSE | Downregulated |
| -0,309844849 | 0,643682092 | ZFAND2B | FALSE | Downregulated |
| -0,201220301 | 0,643731816 | RPS3 | FALSE | Downregulated |
| 0,67318729 | 0,644084886 | RGS7BP | FALSE | Upregulated |
| 0,284990285 | 0,644333354 | MICAL2 | FALSE | Upregulated |
| 0,745615608 | 0,644333354 | SPATA13-AS1 | FALSE | Upregulated |
| 0,155524903 | 0,644406158 | PAFAH1B1 | FALSE | Upregulated |
| 0,138681136 | 0,644500636 | PRKDC | FALSE | Upregulated |
| -0,177786373 | 0,644702486 | TNKS | FALSE | Downregulated |
| -0,14507359 | 0,645322319 | ARF1 | FALSE | Downregulated |
| 0,238711505 | 0,645443666 | HEIH | FALSE | Upregulated |
| 0,892664675 | 0,645443666 | LOC100287015 | FALSE | Upregulated |
| 0,23984192 | 0,645498408 | PGD | FALSE | Upregulated |
| -0,722566618 | 0,645498408 | POLR2M | FALSE | Downregulated |
| -0,190563376 | 0,645498408 | PRELID1 | FALSE | Downregulated |
| 0,668358406 | 0,645498408 | SLC22A17 | FALSE | Upregulated |
| 0,681674306 | 0,645670848 | AGAP11 | FALSE | Upregulated |
| -0,500840512 | 0,645670848 | CCBL2 | FALSE | Downregulated |
| -0,212754397 | 0,645670848 | IL6ST | FALSE | Downregulated |

| | | | | |
|---|---|---|---|---|
| -0,509621804 | 0,645670848 | KATNA1 | FALSE | Downregulated |
| 0,265333096 | 0,645670848 | LINC00476 | FALSE | Upregulated |
| 0,488020177 | 0,645670848 | LOC102724597 | FALSE | Upregulated |
| -1,008501289 | 0,645670848 | LOC648044 | FALSE | Downregulated |
| -0,303320881 | 0,645670848 | OGFOD3 | FALSE | Downregulated |
| -0,428417967 | 0,645670848 | PIWIL2 | FALSE | Downregulated |
| -0,476597748 | 0,645670848 | PUS7L | FALSE | Downregulated |
| -0,665468364 | 0,645670848 | RILPL2 | FALSE | Downregulated |
| -0,182179668 | 0,645670848 | TTC28 | FALSE | Downregulated |
| 0,542369961 | 0,645670848 | WASH3P | FALSE | Upregulated |
| -0,296298705 | 0,645767705 | HHEX | FALSE | Downregulated |
| 0,179582368 | 0,64585531 | DICER1 | FALSE | Upregulated |
| -0,121805283 | 0,64585531 | KMT2D | FALSE | Downregulated |
| -0,266208578 | 0,64585531 | RPP14 | FALSE | Downregulated |
| -0,763032458 | 0,645861462 | SLC29A4 | FALSE | Downregulated |
| -0,284895483 | 0,646163762 | EDEM2 | FALSE | Downregulated |
| 0,725200957 | 0,646163762 | TUSC3 | FALSE | Upregulated |
| 0,245810906 | 0,646163762 | ZAK | FALSE | Upregulated |
| 0,216021819 | 0,646180472 | PTP4A2 | FALSE | Upregulated |
| -0,932048622 | 0,646316703 | OPN1SW | FALSE | Downregulated |
| 0,240880266 | 0,646501601 | AC005594.3 | FALSE | Upregulated |
| -0,197513267 | 0,646501601 | ADCY5 | FALSE | Downregulated |
| 0,995669313 | 0,646501601 | DHRS9 | FALSE | Upregulated |
| -0,287098706 | 0,646501601 | HIST2H2AC | FALSE | Downregulated |
| -0,519110517 | 0,646501601 | MRPL17 | FALSE | Downregulated |
| -0,524654638 | 0,646501601 | RPL13AP5 | FALSE | Downregulated |
| -0,207707518 | 0,646501601 | TRAPPC8 | FALSE | Downregulated |
| -0,217904345 | 0,646711107 | CTDP1 | FALSE | Downregulated |
| -0,243755186 | 0,646711107 | KIAA0922 | FALSE | Downregulated |
| 0,355230141 | 0,646711107 | LOC101928443 | FALSE | Upregulated |
| -0,418277695 | 0,647013302 | EPB41L1 | FALSE | Downregulated |
| -0,495323008 | 0,647258516 | LEPREL1 | FALSE | Downregulated |
| -0,376241431 | 0,647394235 | ADIRF | FALSE | Downregulated |
| 0,126152697 | 0,647394235 | BACE1 | FALSE | Upregulated |
| 1,008421797 | 0,647394235 | LOC102724124 | FALSE | Upregulated |
| 0,359224037 | 0,647394235 | MID2 | FALSE | Upregulated |
| -0,281909966 | 0,647394235 | PTPN4 | FALSE | Downregulated |
| -0,967516976 | 0,647394235 | RP11-261C10.3 | FALSE | Downregulated |
| 0,304223206 | 0,647394235 | SENP8 | FALSE | Upregulated |
| 0,881394454 | 0,647394235 | TRHR | FALSE | Upregulated |
| 0,468447501 | 0,647394235 | ZNF639 | FALSE | Upregulated |
| 0,268683751 | 0,647726838 | ELL3 | FALSE | Upregulated |
| 0,29865458 | 0,647744462 | ABHD12 | FALSE | Upregulated |
| 0,798493127 | 0,647904296 | CCDC77 | FALSE | Upregulated |
| 0,165635865 | 0,64806262 | USP54 | FALSE | Upregulated |
| -0,559604243 | 0,648169351 | LOC101060398 | FALSE | Downregulated |
| 0,458129149 | 0,648271535 | BBC3 | FALSE | Upregulated |
| -0,540398128 | 0,648271535 | C6orf123 | FALSE | Downregulated |
| -0,203325859 | 0,648488946 | CLCN3 | FALSE | Downregulated |
| 0,490246991 | 0,648488946 | DOPEY2 | FALSE | Upregulated |
| 0,221017463 | 0,648631079 | CYB561A3 | FALSE | Upregulated |

| | | | | |
|---|---|---|---|---|
| -0,773151941 | 0,648631079 | LILRA4 | FALSE | Downregulated |
| 0,455015903 | 0,648631079 | NUDT7 | FALSE | Upregulated |
| 0,170987792 | 0,648631079 | SMIM5 | FALSE | Upregulated |
| -0,244451369 | 0,648631079 | SNHG16 | FALSE | Downregulated |
| 0,516691934 | 0,648727994 | SH3RF3 | FALSE | Upregulated |
| 0,391952959 | 0,648727994 | SLC16A6 | FALSE | Upregulated |
| 0,173856439 | 0,648789159 | F11R | FALSE | Upregulated |
| 0,200968402 | 0,64902144 | ACVR2A | FALSE | Upregulated |
| -0,140674313 | 0,649166274 | AP2M1 | FALSE | Downregulated |
| -0,490452996 | 0,649166274 | NUAK2 | FALSE | Downregulated |
| 0,231555768 | 0,649329582 | KDELR1 | FALSE | Upregulated |
| 0,581610334 | 0,649359187 | SV2A | FALSE | Upregulated |
| 0,372958277 | 0,64951927 | SH3BP1 | FALSE | Upregulated |
| 0,315534503 | 0,64951927 | ZDHHC6 | FALSE | Upregulated |
| 0,315503686 | 0,64954123 | MTX3 | FALSE | Upregulated |
| 0,551640693 | 0,649728524 | ABLIM2 | FALSE | Upregulated |
| -0,558426816 | 0,649728524 | ATP1B3 | FALSE | Downregulated |
| 0,186198122 | 0,649728524 | AXL | FALSE | Upregulated |
| 0,248407495 | 0,649728524 | LDLRAD4-AS1 | FALSE | Upregulated |
| -0,273815272 | 0,649728524 | LZTS3 | FALSE | Downregulated |
| 0,447733563 | 0,649728524 | MAP2 | FALSE | Upregulated |
| 0,402696802 | 0,649728524 | USP44 | FALSE | Upregulated |
| 0,855027462 | 0,65000556 | FCRL3 | FALSE | Upregulated |
| 0,565745059 | 0,650043328 | H3F3AP4 | FALSE | Upregulated |
| -0,301645407 | 0,650058592 | FAM107A | FALSE | Downregulated |
| 0,392750901 | 0,650058592 | SKAP1 | FALSE | Upregulated |
| -0,536892043 | 0,650196422 | VPS37D | FALSE | Downregulated |
| 0,311766631 | 0,650449222 | FAM3C | FALSE | Upregulated |
| -0,759115428 | 0,650449222 | UBXN7-AS1 | FALSE | Downregulated |
| -0,761587218 | 0,650755584 | LEFTY1 | FALSE | Downregulated |
| 0,285251614 | 0,650755584 | MRPS11 | FALSE | Upregulated |
| -0,214860089 | 0,650755584 | POLG | FALSE | Downregulated |
| -0,238318643 | 0,650755584 | RGL1 | FALSE | Downregulated |
| 0,221201371 | 0,650755584 | TRUB2 | FALSE | Upregulated |
| 0,444028505 | 0,650775003 | NOXO1 | FALSE | Upregulated |
| 0,756734822 | 0,650775003 | TBX1 | FALSE | Upregulated |
| 0,430078087 | 0,650815288 | SNRNP40 | FALSE | Upregulated |
| -0,650859348 | 0,650815288 | TMEM258 | FALSE | Downregulated |
| -0,188894756 | 0,650860651 | USP32 | FALSE | Downregulated |
| -0,870073176 | 0,650867618 | SDHAP3 | FALSE | Downregulated |
| -0,437817203 | 0,65094347 | SHBG | FALSE | Downregulated |
| 0,225880359 | 0,650977865 | TOR1B | FALSE | Upregulated |
| -0,290954438 | 0,651008586 | NEU1 | FALSE | Downregulated |
| 0,191680922 | 0,651180561 | UBP1 | FALSE | Upregulated |
| -0,273518498 | 0,651280343 | GLB1 | FALSE | Downregulated |
| 0,455597972 | 0,651782514 | LOC101927180 | FALSE | Upregulated |
| 0,839043201 | 0,652017166 | EFCAB11 | FALSE | Upregulated |
| 0,628237823 | 0,652017166 | LOC374443 | FALSE | Upregulated |
| -0,187899827 | 0,652017166 | NSUN2 | FALSE | Downregulated |
| -0,350117022 | 0,652017166 | USP12 | FALSE | Downregulated |
| 0,625949827 | 0,65207616 | TMLHE-AS1 | FALSE | Upregulated |

| | | | | |
|---|---|---|---|---|
| 0,3430507 | 0,652076693 | LRIG2 | FALSE | Upregulated |
| 0,300603028 | 0,652094602 | B3GAT2 | FALSE | Upregulated |
| 0,19675687 | 0,652317889 | ANKRD23 | FALSE | Upregulated |
| 0,730852321 | 0,652317889 | GNA15 | FALSE | Upregulated |
| 0,231443711 | 0,652317889 | LAP3 | FALSE | Upregulated |
| 0,900180511 | 0,652317889 | LOC101928049 | FALSE | Upregulated |
| -0,314426783 | 0,652317889 | RSPRY1 | FALSE | Downregulated |
| 0,474573888 | 0,652317889 | TAS1R1 | FALSE | Upregulated |
| 0,247065586 | 0,652317889 | TCF7L1 | FALSE | Upregulated |
| 0,234166886 | 0,652317889 | TUBA4A | FALSE | Upregulated |
| 0,297305362 | 0,652317889 | ZNF317 | FALSE | Upregulated |
| -0,555216811 | 0,652377726 | WRAP73 | FALSE | Downregulated |
| 0,322757292 | 0,652385361 | C11orf96 | FALSE | Upregulated |
| -0,145328333 | 0,652573156 | ACO1 | FALSE | Downregulated |
| 0,127782656 | 0,652712864 | CDK12 | FALSE | Upregulated |
| 0,26047192 | 0,652712864 | SQRDL | FALSE | Upregulated |
| 0,268807695 | 0,652712864 | ZBTB43 | FALSE | Upregulated |
| -0,768790989 | 0,652813749 | CCDC144A | FALSE | Downregulated |
| 0,797874296 | 0,652813749 | ECE2 | FALSE | Upregulated |
| -1,449660404 | 0,652813749 | GOLGA8T | FALSE | Downregulated |
| 0,342951579 | 0,652813749 | HSBP1L1 | FALSE | Upregulated |
| 0,112836689 | 0,652813749 | PCBP1 | FALSE | Upregulated |
| 0,179066083 | 0,652813749 | SHB | FALSE | Upregulated |
| -0,524837402 | 0,652813749 | TMEM161B | FALSE | Downregulated |
| -0,412504605 | 0,652976311 | FAM222A | FALSE | Downregulated |
| -0,332876907 | 0,653111432 | PIAS2 | FALSE | Downregulated |
| 0,272712912 | 0,653111432 | THOC5 | FALSE | Upregulated |
| -0,276725237 | 0,65316362 | DDX1 | FALSE | Downregulated |
| 0,267262547 | 0,65351469 | STX10 | FALSE | Upregulated |
| 0,41578514 | 0,653636716 | DHRS7B | FALSE | Upregulated |
| 0,503458347 | 0,653636716 | FAM109B | FALSE | Upregulated |
| -0,355371792 | 0,653636716 | GADD45B | FALSE | Downregulated |
| -0,51197363 | 0,653636716 | HEY2 | FALSE | Downregulated |
| -0,656498061 | 0,653636716 | MTHFD2L | FALSE | Downregulated |
| -0,857163365 | 0,653636716 | PPP1R27 | FALSE | Downregulated |
| 0,180876659 | 0,653636716 | SETD6 | FALSE | Upregulated |
| 0,186740555 | 0,653636716 | TSPAN4 | FALSE | Upregulated |
| -0,634357188 | 0,65366373 | TGFA | FALSE | Downregulated |
| 0,388469005 | 0,653782255 | THAP8 | FALSE | Upregulated |
| 0,157522962 | 0,653794821 | DIO1 | FALSE | Upregulated |
| -0,397491074 | 0,653862308 | REN | FALSE | Downregulated |
| -0,228463401 | 0,653920329 | FAM35A | FALSE | Downregulated |
| -0,201107504 | 0,65395778 | ERBB2 | FALSE | Downregulated |
| 0,303464214 | 0,654025201 | ADAMTS9 | FALSE | Upregulated |
| -0,176521064 | 0,654048137 | RMRP | FALSE | Downregulated |
| 0,508584447 | 0,65430443 | JAG2 | FALSE | Upregulated |
| -0,157552824 | 0,654565228 | GLTPD1 | FALSE | Downregulated |
| 0,203833461 | 0,655133483 | CARD14 | FALSE | Upregulated |
| 0,371707358 | 0,655666452 | PDE6A | FALSE | Upregulated |
| -0,81819348 | 0,655872582 | CHST1 | FALSE | Downregulated |
| 0,191545951 | 0,655872582 | CPD | FALSE | Upregulated |

| | | | | |
|---|---|---|---|---|
| 0,201952403 | 0,655872582 | DDX58 | FALSE | Upregulated |
| -0,929990045 | 0,655872582 | LOC102724192 | FALSE | Downregulated |
| 1,04856261 | 0,655872582 | MIR4435-1HG | FALSE | Upregulated |
| 0,277792112 | 0,655872582 | RPS6KA4 | FALSE | Upregulated |
| -0,301104881 | 0,656008165 | DPY19L4 | FALSE | Downregulated |
| -0,816147171 | 0,656008165 | LOC100652901 | FALSE | Downregulated |
| 1,10256331 | 0,656008165 | RP11-290F20.3 | FALSE | Upregulated |
| -0,179520887 | 0,656008165 | TRIM24 | FALSE | Downregulated |
| 0,325823109 | 0,656069175 | SLC39A3 | FALSE | Upregulated |
| 0,264528784 | 0,656115801 | FAM57A | FALSE | Upregulated |
| -0,165200647 | 0,656182657 | ABTB2 | FALSE | Downregulated |
| 0,666991185 | 0,656182657 | LOC729083 | FALSE | Upregulated |
| 0,565472899 | 0,656182657 | TOR2A | FALSE | Upregulated |
| -0,5427159 | 0,656185745 | FAM229B | FALSE | Downregulated |
| -0,317621428 | 0,656185745 | NLN | FALSE | Downregulated |
| 0,433857987 | 0,656186813 | LSM10 | FALSE | Upregulated |
| 0,347711786 | 0,656537041 | TDG | FALSE | Upregulated |
| 0,734599317 | 0,656548409 | WRAP53 | FALSE | Upregulated |
| 0,679500429 | 0,656651265 | MGP | FALSE | Upregulated |
| 0,297584615 | 0,656651265 | NSMCE4A | FALSE | Upregulated |
| -0,484648129 | 0,656651265 | SHQ1 | FALSE | Downregulated |
| -0,152311723 | 0,656779948 | AK3 | FALSE | Downregulated |
| 0,561517597 | 0,65679498 | RP6-191P20.4 | FALSE | Upregulated |
| 0,220864027 | 0,657035843 | CEBPZ-AS1 | FALSE | Upregulated |
| 0,205840067 | 0,657144908 | KIF24 | FALSE | Upregulated |
| -0,184176677 | 0,657144908 | OGFR | FALSE | Downregulated |
| -1,034942795 | 0,657144908 | PCCA-AS1 | FALSE | Downregulated |
| 0,492242387 | 0,657377662 | KLHL10 | FALSE | Upregulated |
| -0,195512269 | 0,657767799 | XAB2 | FALSE | Downregulated |
| -0,230600347 | 0,657767799 | ZFP91 | FALSE | Downregulated |
| 0,740298002 | 0,657913239 | RAB34 | FALSE | Upregulated |
| -0,579624175 | 0,657950075 | CHCHD7 | FALSE | Downregulated |
| 0,195988156 | 0,658038865 | CDIP1 | FALSE | Upregulated |
| 0,357190613 | 0,658038865 | EHD2 | FALSE | Upregulated |
| -0,139400205 | 0,658038865 | EWSR1 | FALSE | Downregulated |
| 0,319409121 | 0,658038865 | FLJ46906 | FALSE | Upregulated |
| 0,563130093 | 0,658038865 | GPR152 | FALSE | Upregulated |
| -0,489588684 | 0,658038865 | HPRT1 | FALSE | Downregulated |
| -0,865368659 | 0,658038865 | LINC00885 | FALSE | Downregulated |
| 0,423121111 | 0,658038865 | LOC101928762 | FALSE | Upregulated |
| 0,250880989 | 0,658038865 | LRFN4 | FALSE | Upregulated |
| -0,303787919 | 0,658038865 | PAQR7 | FALSE | Downregulated |
| 0,178579728 | 0,658038865 | RBM12 | FALSE | Upregulated |
| 0,196598981 | 0,658038865 | SCARNA10 | FALSE | Upregulated |
| 0,221740694 | 0,658038865 | SRSF10 | FALSE | Upregulated |
| 0,303531066 | 0,658038865 | TPT1-AS1 | FALSE | Upregulated |
| -0,185772797 | 0,658038865 | UGDH-AS1 | FALSE | Downregulated |
| -0,403306376 | 0,658168513 | PGP | FALSE | Downregulated |
| 0,286309304 | 0,658218845 | MUS81 | FALSE | Upregulated |
| -0,19872192 | 0,658218845 | SAFB2 | FALSE | Downregulated |
| 0,341646771 | 0,658444631 | MAP4K5 | FALSE | Upregulated |

| | | | | |
|---|---|---|---|---|
| -0,933224282 | 0,658499991 | RHOJ | FALSE | Downregulated |
| -0,455002658 | 0,658511122 | CENPJ | FALSE | Downregulated |
| -0,148607801 | 0,658529819 | CREBZF | FALSE | Downregulated |
| -0,918482984 | 0,658849986 | LOC100630918 | FALSE | Downregulated |
| 0,749369894 | 0,6589373 | FIBIN | FALSE | Upregulated |
| 0,845146969 | 0,658951613 | LINC00346 | FALSE | Upregulated |
| -0,239892308 | 0,65908252 | DNMBP | FALSE | Downregulated |
| 0,219497084 | 0,65908252 | GGACT | FALSE | Upregulated |
| 0,341623303 | 0,65908252 | GPN2 | FALSE | Upregulated |
| 0,540694295 | 0,65908252 | PTPRR | FALSE | Upregulated |
| 0,2000149 | 0,65908252 | SUPT16H | FALSE | Upregulated |
| 0,668379459 | 0,659454707 | RP11-381O7.3 | FALSE | Upregulated |
| -0,239622509 | 0,659519565 | CCM2 | FALSE | Downregulated |
| 0,663848421 | 0,659524985 | CD22 | FALSE | Upregulated |
| 0,254153147 | 0,659524985 | MFSD7 | FALSE | Upregulated |
| -0,421428499 | 0,659666108 | C8orf46 | FALSE | Downregulated |
| 0,287570304 | 0,659666108 | SMYD5 | FALSE | Upregulated |
| -0,178745693 | 0,659666108 | TBC1D14 | FALSE | Downregulated |
| 0,232081235 | 0,659666108 | TRMT5 | FALSE | Upregulated |
| -0,455172699 | 0,659856876 | C17orf51 | FALSE | Downregulated |
| 0,429364689 | 0,659856876 | NCAM1 | FALSE | Upregulated |
| -0,235660375 | 0,659961026 | C5orf42 | FALSE | Downregulated |
| -0,363731223 | 0,659961026 | FBXL19 | FALSE | Downregulated |
| 0,854362004 | 0,659961026 | MICB | FALSE | Upregulated |
| 0,269178103 | 0,659961026 | TARBP1 | FALSE | Upregulated |
| 0,297369345 | 0,660053327 | E2F5 | FALSE | Upregulated |
| 0,377278088 | 0,660053327 | LOC102723809 | FALSE | Upregulated |
| -0,181460452 | 0,660147032 | ANGPTL6 | FALSE | Downregulated |
| -0,531370733 | 0,660176686 | OR51M1 | FALSE | Downregulated |
| -0,258909797 | 0,660356024 | MSRA | FALSE | Downregulated |
| 0,256846276 | 0,660356024 | ZDHHC16 | FALSE | Upregulated |
| 1,211277252 | 0,660368093 | JPH2 | FALSE | Upregulated |
| 0,567289434 | 0,660368093 | ZNF878 | FALSE | Upregulated |
| 0,336778382 | 0,660990324 | LOC100652768 | FALSE | Upregulated |
| 0,495689996 | 0,66100501 | FHL2 | FALSE | Upregulated |
| 0,286187344 | 0,661341049 | PTDSS2 | FALSE | Upregulated |
| 0,449255203 | 0,661659493 | GJB7 | FALSE | Upregulated |
| -0,54220491 | 0,661659493 | KLHL17 | FALSE | Downregulated |
| -0,811644312 | 0,661659493 | LOC102724881 | FALSE | Downregulated |
| -0,434500838 | 0,661659493 | SMIM2 | FALSE | Downregulated |
| 2,682574653 | 0,661659493 | TNNI2 | FALSE | Upregulated |
| 0,171820234 | 0,661690634 | CCDC88A | FALSE | Upregulated |
| -0,313216123 | 0,661690634 | MOAP1 | FALSE | Downregulated |
| -0,418172195 | 0,661690634 | SMIM13 | FALSE | Downregulated |
| -0,376201046 | 0,661690634 | ZNF558 | FALSE | Downregulated |
| 0,670677886 | 0,661718991 | HIST1H3D | FALSE | Upregulated |
| 0,336096934 | 0,661919892 | BAX | FALSE | Upregulated |
| 0,152866306 | 0,661919892 | CABIN1 | FALSE | Upregulated |
| 0,724069893 | 0,661919892 | LIPH | FALSE | Upregulated |
| 0,23206992 | 0,661919892 | LOC646652 | FALSE | Upregulated |
| 0,228481319 | 0,661919892 | PMS2P1 | FALSE | Upregulated |

| | | | | |
|---|---|---|---|---|
| 0,804771826 | 0,661919892 | RIN1 | FALSE | Upregulated |
| -0,348352793 | 0,661919892 | RYBP | FALSE | Downregulated |
| 0,254938493 | 0,661919892 | WLS | FALSE | Upregulated |
| 0,181568629 | 0,661919892 | XPO6 | FALSE | Upregulated |
| -0,195807636 | 0,661985999 | MUT | FALSE | Downregulated |
| -0,296104237 | 0,662133677 | ZNF175 | FALSE | Downregulated |
| -0,753482841 | 0,662203619 | CNTNAP3 | FALSE | Downregulated |
| -0,115180331 | 0,662282163 | BIRC6 | FALSE | Downregulated |
| 0,723346389 | 0,662282163 | FAM83F | FALSE | Upregulated |
| -0,170493566 | 0,662282163 | LTBR | FALSE | Downregulated |
| -0,825932996 | 0,662282163 | TNFSF4 | FALSE | Downregulated |
| -0,642470499 | 0,66270595 | FAM229A | FALSE | Downregulated |
| -0,162203245 | 0,66272079 | PNRC1 | FALSE | Downregulated |
| 0,259333456 | 0,663126739 | ZNF84 | FALSE | Upregulated |
| 0,244795853 | 0,663279992 | ATXN3 | FALSE | Upregulated |
| -0,556177456 | 0,663325919 | TGFB1I1 | FALSE | Downregulated |
| -0,509081478 | 0,663342101 | CTD-3080P12.3 | FALSE | Downregulated |
| 0,360733164 | 0,663425243 | HELQ | FALSE | Upregulated |
| -0,562790521 | 0,663425243 | LOC101929548 | FALSE | Downregulated |
| 0,316687616 | 0,663454626 | BCL7A | FALSE | Upregulated |
| 0,853476798 | 0,663650171 | ADAMTS15 | FALSE | Upregulated |
| -1,282217558 | 0,663650171 | AGAP7 | FALSE | Downregulated |
| 0,227447403 | 0,663650171 | DDB2 | FALSE | Upregulated |
| -0,123396048 | 0,663650171 | HMBOX1 | FALSE | Downregulated |
| 0,162451161 | 0,663650171 | LOC148696 | FALSE | Upregulated |
| -0,705968473 | 0,663650171 | MOGAT3 | FALSE | Downregulated |
| 0,883689029 | 0,663650171 | NACAD | FALSE | Upregulated |
| -0,146659842 | 0,663650171 | UBC | FALSE | Downregulated |
| 0,237967302 | 0,663769807 | CALU | FALSE | Upregulated |
| 0,871175104 | 0,663769807 | F2RL1 | FALSE | Upregulated |
| -0,480336565 | 0,663769807 | MICU3 | FALSE | Downregulated |
| 0,310048009 | 0,663796639 | CCDC61 | FALSE | Upregulated |
| 0,739296292 | 0,663796639 | LINC00638 | FALSE | Upregulated |
| -0,138527578 | 0,663796639 | PTBP1 | FALSE | Downregulated |
| 0,485339885 | 0,663796639 | SLC25A36 | FALSE | Upregulated |
| -1,086879376 | 0,663826636 | PRSS55 | FALSE | Downregulated |
| -0,174501107 | 0,663898173 | AHCTF1 | FALSE | Downregulated |
| 0,236625572 | 0,663898173 | CLASP2 | FALSE | Upregulated |
| 0,530612339 | 0,663898173 | LOC101928982 | FALSE | Upregulated |
| 0,288932892 | 0,663898173 | LOC388282 | FALSE | Upregulated |
| -0,628145909 | 0,663898173 | MIR497HG | FALSE | Downregulated |
| 0,201829859 | 0,663898173 | VWA1 | FALSE | Upregulated |
| 0,300447704 | 0,663898173 | ZFHX2 | FALSE | Upregulated |
| -0,326721601 | 0,663898173 | ZNF252P | FALSE | Downregulated |
| 0,672578772 | 0,664123043 | TNFRSF11A | FALSE | Upregulated |
| -0,188730012 | 0,664147873 | DBH | FALSE | Downregulated |
| 0,681923231 | 0,664164561 | LKAAEAR1 | FALSE | Upregulated |
| 0,320124505 | 0,664164561 | SLC2A9 | FALSE | Upregulated |
| -1,380933485 | 0,664195336 | GOLGA6L4 | FALSE | Downregulated |
| -0,680685037 | 0,664195336 | NDUFAF6 | FALSE | Downregulated |
| 0,710924248 | 0,664195336 | TRMT6 | FALSE | Upregulated |

| | | | | |
|---|---|---|---|---|
| -0,414320789 | 0,664300191 | ALG8 | FALSE | Downregulated |
| -0,550782622 | 0,664300191 | FAM220A | FALSE | Downregulated |
| 0,734277928 | 0,664300191 | FCGR1A | FALSE | Upregulated |
| -0,672020235 | 0,664300191 | IL1RL2 | FALSE | Downregulated |
| 0,83734528 | 0,664300191 | LOC100506161 | FALSE | Upregulated |
| 0,521083289 | 0,664300191 | SNORA48 | FALSE | Upregulated |
| -0,222046765 | 0,664402028 | LOC101928897 | FALSE | Downregulated |
| 0,204909804 | 0,664474932 | GGT5 | FALSE | Upregulated |
| 0,214644006 | 0,664484644 | ALAS1 | FALSE | Upregulated |
| 0,518606422 | 0,664484644 | BHLHB9 | FALSE | Upregulated |
| 0,450795209 | 0,664484644 | HULC | FALSE | Upregulated |
| 0,25185194 | 0,664484644 | NSUN5P1 | FALSE | Upregulated |
| 0,346086221 | 0,664484644 | SNHG3 | FALSE | Upregulated |
| 0,457788219 | 0,664484644 | ZNF493 | FALSE | Upregulated |
| 0,350717725 | 0,664543049 | CSRNP1 | FALSE | Upregulated |
| 0,64656635 | 0,664543049 | DSCC1 | FALSE | Upregulated |
| 0,306279878 | 0,664543049 | EIF2S1 | FALSE | Upregulated |
| -0,288906357 | 0,664543049 | GRAMD1C | FALSE | Downregulated |
| 0,545192798 | 0,664543049 | H2AFX | FALSE | Upregulated |
| 0,375450348 | 0,664543049 | HK2 | FALSE | Upregulated |
| 1,067098854 | 0,664543049 | MST1R | FALSE | Upregulated |
| 0,293721035 | 0,664543049 | PALM | FALSE | Upregulated |
| 0,430189668 | 0,664543049 | PIGH | FALSE | Upregulated |
| 0,429742838 | 0,664543049 | RBM7 | FALSE | Upregulated |
| -0,244704187 | 0,664543049 | SPATA2L | FALSE | Downregulated |
| 0,24208042 | 0,664564591 | CCDC14 | FALSE | Upregulated |
| -0,403264813 | 0,664564591 | LOC101928384 | FALSE | Downregulated |
| 0,176845009 | 0,665084776 | SUOX | FALSE | Upregulated |
| 0,367419599 | 0,665248374 | SUGT1 | FALSE | Upregulated |
| 0,373323719 | 0,665339541 | C2orf44 | FALSE | Upregulated |
| 0,868213115 | 0,665339541 | GPR156 | FALSE | Upregulated |
| 0,273266881 | 0,665339541 | L3MBTL4 | FALSE | Upregulated |
| 0,291965093 | 0,665339541 | PAQR9 | FALSE | Upregulated |
| 0,403285485 | 0,665339541 | POMZP3 | FALSE | Upregulated |
| -0,294170837 | 0,665339541 | RAB7L1 | FALSE | Downregulated |
| 0,139798785 | 0,665339541 | REV3L | FALSE | Upregulated |
| -0,302028654 | 0,665339541 | RHBDD2 | FALSE | Downregulated |
| -0,550145054 | 0,665433714 | NOSTRIN | FALSE | Downregulated |
| 0,229436279 | 0,665433714 | PML | FALSE | Upregulated |
| -0,229836668 | 0,665433714 | SLC13A4 | FALSE | Downregulated |
| 0,262134709 | 0,665433714 | USP16 | FALSE | Upregulated |
| 0,24094666 | 0,665550679 | TP53I11 | FALSE | Upregulated |
| 0,330820001 | 0,665589849 | THAP7-AS1 | FALSE | Upregulated |
| -0,325766208 | 0,665900201 | LOC101928798 | FALSE | Downregulated |
| 1,1268944 | 0,665910537 | GCOM1 | FALSE | Upregulated |
| 0,518015391 | 0,665910537 | MRPL33 | FALSE | Upregulated |
| -0,298824675 | 0,665910537 | ZCCHC7 | FALSE | Downregulated |
| 0,450976826 | 0,666068101 | ODF2L | FALSE | Upregulated |
| -0,178720643 | 0,666068101 | STAG3L5P-PVRIG2P-PILRB | FALSE | Downregulated |
| -0,349412924 | 0,666431143 | CLEC1B | FALSE | Downregulated |

| | | | | |
|---|---|---|---|---|
| -0,217422145 | 0,666709452 | NAT10 | FALSE | Downregulated |
| 0,355999236 | 0,666709452 | PRKRA | FALSE | Upregulated |
| 0,199131873 | 0,666748414 | CSF1R | FALSE | Upregulated |
| 0,737401895 | 0,666748414 | LINC00294 | FALSE | Upregulated |
| -0,677984306 | 0,666748414 | LINC01010 | FALSE | Downregulated |
| -0,204201972 | 0,666748414 | MED29 | FALSE | Downregulated |
| -0,261787979 | 0,666748414 | MRPS24 | FALSE | Downregulated |
| -0,609700156 | 0,666748414 | NDUFB2-AS1 | FALSE | Downregulated |
| -0,329739622 | 0,666748414 | NOP58 | FALSE | Downregulated |
| 0,218866511 | 0,666748414 | SLC37A2 | FALSE | Upregulated |
| 0,222221483 | 0,666748414 | TUBA1B | FALSE | Upregulated |
| -0,397078532 | 0,666748414 | USP28 | FALSE | Downregulated |
| 0,641383333 | 0,666748414 | ZNF439 | FALSE | Upregulated |
| -0,266806536 | 0,667148922 | ZNF696 | FALSE | Downregulated |
| 0,530442827 | 0,667325197 | MNAT1 | FALSE | Upregulated |
| 0,585303151 | 0,667350254 | EXOG | FALSE | Upregulated |
| -0,216483267 | 0,667350254 | UBE2K | FALSE | Downregulated |
| 0,183136887 | 0,667493403 | LOC101928853 | FALSE | Upregulated |
| 0,301491458 | 0,667493403 | ZNF512 | FALSE | Upregulated |
| -0,195255928 | 0,66767628 | EIF1 | FALSE | Downregulated |
| -0,514288988 | 0,668184817 | DFFB | FALSE | Downregulated |
| 0,795346651 | 0,668184817 | KPNA7 | FALSE | Upregulated |
| -0,300647471 | 0,668549759 | ATP5S | FALSE | Downregulated |
| -0,177170087 | 0,668571164 | RDX | FALSE | Downregulated |
| -0,181804343 | 0,668741466 | CALR | FALSE | Downregulated |
| 0,482453404 | 0,668741466 | FLJ23867 | FALSE | Upregulated |
| -0,205445548 | 0,668741466 | HNRNPH3 | FALSE | Downregulated |
| -0,327199669 | 0,668741466 | LAD1 | FALSE | Downregulated |
| 0,585583054 | 0,668741466 | LOC101929506 | FALSE | Upregulated |
| -0,226916897 | 0,668851243 | PPP3CA | FALSE | Downregulated |
| -0,246088906 | 0,668851243 | WBP11 | FALSE | Downregulated |
| 0,238775527 | 0,66892681 | TTC23 | FALSE | Upregulated |
| 0,390267494 | 0,66934305 | DPEP2 | FALSE | Upregulated |
| 0,168804579 | 0,66934305 | RAB13 | FALSE | Upregulated |
| 0,257872649 | 0,669371352 | FAM173A | FALSE | Upregulated |
| -0,214983349 | 0,669663553 | ATP2B1 | FALSE | Downregulated |
| 0,151064726 | 0,669663553 | MBNL1 | FALSE | Upregulated |
| 0,182113941 | 0,669663553 | RP5-886K2.3 | FALSE | Upregulated |
| -0,519724088 | 0,66967378 | ADAMTSL1 | FALSE | Downregulated |
| -0,205925048 | 0,66967378 | CCDC176 | FALSE | Downregulated |
| -0,24520022 | 0,66967378 | CTNNBL1 | FALSE | Downregulated |
| -0,22537876 | 0,66967378 | DLG4 | FALSE | Downregulated |
| -0,335934788 | 0,66967378 | GEN1 | FALSE | Downregulated |
| -0,206222588 | 0,66967378 | MAP3K4 | FALSE | Downregulated |
| 0,169645678 | 0,66967378 | PCM1 | FALSE | Upregulated |
| -0,2950076 | 0,66967378 | SLC25A37 | FALSE | Downregulated |
| 0,220265779 | 0,66967378 | TAF11 | FALSE | Upregulated |
| 0,271006648 | 0,66967378 | TYROBP | FALSE | Upregulated |
| 0,541634303 | 0,669766024 | LOC101929185 | FALSE | Upregulated |
| 0,408895259 | 0,669766024 | OBSCN | FALSE | Upregulated |
| 0,285884577 | 0,669766024 | TGIF2 | FALSE | Upregulated |

| | | | | |
|---|---|---|---|---|
| -0,469310496 | 0,669842025 | ZNF25 | FALSE | Downregulated |
| 0,569737022 | 0,669953905 | CASC5 | FALSE | Upregulated |
| 0,58677144 | 0,669953905 | CRLF1 | FALSE | Upregulated |
| 0,195426961 | 0,669953905 | CTBP2 | FALSE | Upregulated |
| -0,652973509 | 0,669953905 | CYP4F22 | FALSE | Downregulated |
| 0,33565612 | 0,669953905 | DENND6A | FALSE | Upregulated |
| -0,241987355 | 0,669953905 | FOXA3 | FALSE | Downregulated |
| -0,55900103 | 0,669953905 | MANBAL | FALSE | Downregulated |
| 0,777038877 | 0,669953905 | NAPSA | FALSE | Upregulated |
| -0,175031327 | 0,669953905 | PUF60 | FALSE | Downregulated |
| -0,3135856 | 0,669953905 | PVRL3 | FALSE | Downregulated |
| -0,17031077 | 0,669953905 | ST5 | FALSE | Downregulated |
| -0,109020903 | 0,669953905 | TMED10 | FALSE | Downregulated |
| 0,226414403 | 0,669953905 | TMEM175 | FALSE | Upregulated |
| -0,3395012 | 0,669991338 | C1orf27 | FALSE | Downregulated |
| -0,257430687 | 0,67041703 | DNASE2 | FALSE | Downregulated |
| 0,167098343 | 0,670560014 | EIF4A1 | FALSE | Upregulated |
| 0,256866552 | 0,670604216 | ITM2C | FALSE | Upregulated |
| -0,56601098 | 0,670604216 | LOC101928230 | FALSE | Downregulated |
| 0,968902023 | 0,670604216 | SLC34A2 | FALSE | Upregulated |
| -0,543030973 | 0,670934186 | ARV1 | FALSE | Downregulated |
| -0,407012135 | 0,670934186 | FAM99B | FALSE | Downregulated |
| 0,206658695 | 0,670934186 | NDST2 | FALSE | Upregulated |
| 0,266134188 | 0,670934186 | SURF6 | FALSE | Upregulated |
| -0,230194479 | 0,67094851 | AGO3 | FALSE | Downregulated |
| 0,273135527 | 0,67094851 | C16orf71 | FALSE | Upregulated |
| 0,146466773 | 0,67094851 | LIMD1 | FALSE | Upregulated |
| -0,485979901 | 0,67094851 | WASH2P | FALSE | Downregulated |
| 0,735326104 | 0,67094851 | ZNF169 | FALSE | Upregulated |
| -0,51897309 | 0,670961347 | MPZL2 | FALSE | Downregulated |
| 0,22545185 | 0,670961347 | RNF113B | FALSE | Upregulated |
| -0,288938458 | 0,671168011 | COX5A | FALSE | Downregulated |
| -0,322824762 | 0,671168011 | DNTTIP2 | FALSE | Downregulated |
| 0,184413045 | 0,671168011 | MTCH2 | FALSE | Upregulated |
| 0,255474539 | 0,671168011 | PARN | FALSE | Upregulated |
| 0,179002962 | 0,671168011 | PDCD11 | FALSE | Upregulated |
| -0,319893986 | 0,671168011 | SDE2 | FALSE | Downregulated |
| 0,350937971 | 0,671168011 | UBR7 | FALSE | Upregulated |
| -0,559158134 | 0,671180768 | CYP21A1P | FALSE | Downregulated |
| 0,161104204 | 0,671180768 | PAQR3 | FALSE | Upregulated |
| -0,219688834 | 0,67137036 | GOLGA2 | FALSE | Downregulated |
| -0,958390129 | 0,67141058 | LOC101929227 | FALSE | Downregulated |
| -0,142092642 | 0,67141058 | SP3 | FALSE | Downregulated |
| -0,275702866 | 0,67141058 | TCEAL4 | FALSE | Downregulated |
| 0,429238303 | 0,671421244 | TRIT1 | FALSE | Upregulated |
| -0,364882637 | 0,671442359 | AGAP5 | FALSE | Downregulated |
| 0,553383873 | 0,671626578 | KCNK16 | FALSE | Upregulated |
| 0,362741445 | 0,671663422 | PARP2 | FALSE | Upregulated |
| -0,79104079 | 0,671789005 | SMIM11 | FALSE | Downregulated |
| 0,546830705 | 0,671939816 | RPGR | FALSE | Upregulated |
| 0,626824608 | 0,671954838 | ZNF468 | FALSE | Upregulated |

| | | | | |
|---|---|---|---|---|
| -0,196362995 | 0,672183611 | ONECUT2 | FALSE | Downregulated |
| 0,462943197 | 0,672217861 | MOCS3 | FALSE | Upregulated |
| 0,595970906 | 0,672357179 | WISP1 | FALSE | Upregulated |
| 0,241374924 | 0,672701287 | MCM9 | FALSE | Upregulated |
| -0,228971373 | 0,67282805 | MTA2 | FALSE | Downregulated |
| -1,016149144 | 0,672971406 | PPP1R32 | FALSE | Downregulated |
| 0,241103368 | 0,673123351 | ZNF770 | FALSE | Upregulated |
| 0,479343483 | 0,673217427 | C19orf67 | FALSE | Upregulated |
| 0,591422345 | 0,673217427 | CCDC8 | FALSE | Upregulated |
| -0,317532637 | 0,673217427 | CCDC85B | FALSE | Downregulated |
| 0,227003558 | 0,673217427 | CLVS1 | FALSE | Upregulated |
| 0,379161443 | 0,673217427 | LMLN | FALSE | Upregulated |
| 0,278421907 | 0,673217427 | MOK | FALSE | Upregulated |
| 2,314271221 | 0,673217427 | MYLPF | FALSE | Upregulated |
| 0,178414204 | 0,673217427 | RBM8A | FALSE | Upregulated |
| 0,307173246 | 0,673217427 | SPDYE2B | FALSE | Upregulated |
| 0,17234695 | 0,673217427 | TMEM131 | FALSE | Upregulated |
| -0,533884631 | 0,673225197 | SERTAD1 | FALSE | Downregulated |
| -0,239416066 | 0,673313307 | DNASE1 | FALSE | Downregulated |
| -0,411264774 | 0,6733631 | ATP5L | FALSE | Downregulated |
| -0,210881208 | 0,673578993 | SULT2A1 | FALSE | Downregulated |
| -0,803363912 | 0,673578993 | ZNF442 | FALSE | Downregulated |
| 0,387450801 | 0,673748872 | DNASE1L2 | FALSE | Upregulated |
| -0,201199751 | 0,673748872 | MAN1C1 | FALSE | Downregulated |
| -0,320237257 | 0,673748872 | NDUFB2 | FALSE | Downregulated |
| 0,436706066 | 0,673748872 | SLC7A9 | FALSE | Upregulated |
| 0,166611599 | 0,67374956 | FEM1B | FALSE | Upregulated |
| 0,235665818 | 0,673772194 | AKR7L | FALSE | Upregulated |
| 0,376185018 | 0,673772194 | C7orf71 | FALSE | Upregulated |
| 0,194865005 | 0,673772194 | CGGBP1 | FALSE | Upregulated |
| 0,258173159 | 0,673772194 | MED31 | FALSE | Upregulated |
| -0,414758075 | 0,673772194 | ZNF414 | FALSE | Downregulated |
| -0,214636211 | 0,673846177 | COPE | FALSE | Downregulated |
| 0,104850047 | 0,673920267 | ACTN4 | FALSE | Upregulated |
| -0,588854055 | 0,673920267 | NAE1 | FALSE | Downregulated |
| -0,857672095 | 0,673920267 | XRCC6BP1 | FALSE | Downregulated |
| 0,776876559 | 0,673931 | PRB4 | FALSE | Upregulated |
| -0,418370837 | 0,674055567 | COX18 | FALSE | Downregulated |
| 0,541690625 | 0,674232774 | AP4M1 | FALSE | Upregulated |
| -0,340261247 | 0,674232774 | CEP44 | FALSE | Downregulated |
| -0,932924648 | 0,674232774 | RPL36A-HNRNPH2 | FALSE | Downregulated |
| 0,219693995 | 0,674232774 | SHANK2-AS3 | FALSE | Upregulated |
| -0,324660258 | 0,674232774 | SLC4A1AP | FALSE | Downregulated |
| 0,294633212 | 0,674232774 | SPRED1 | FALSE | Upregulated |
| -0,168958185 | 0,674369188 | BMP1 | FALSE | Downregulated |
| 0,290609688 | 0,674369188 | DUT | FALSE | Upregulated |
| -0,26525309 | 0,674369188 | EIF3M | FALSE | Downregulated |
| 0,495381867 | 0,674549756 | LINC01107 | FALSE | Upregulated |
| -0,281832852 | 0,674830421 | CENPC | FALSE | Downregulated |
| -0,94651756 | 0,674842246 | C14orf37 | FALSE | Downregulated |
| 0,399229622 | 0,674842246 | PLA2G4C | FALSE | Upregulated |

| | | | | |
|---|---|---|---|---|
| -0,272783625 | 0,674995201 | PPARGC1A | FALSE | Downregulated |
| -0,518420568 | 0,67508711 | DERL2 | FALSE | Downregulated |
| 0,169168527 | 0,67508711 | ERAP1 | FALSE | Upregulated |
| 0,931734187 | 0,67508711 | GATSL2 | FALSE | Upregulated |
| -0,967418631 | 0,67508711 | IP6K3 | FALSE | Downregulated |
| -0,795526694 | 0,67508711 | LOC101929482 | FALSE | Downregulated |
| 0,704602286 | 0,67508711 | LTC4S | FALSE | Upregulated |
| 0,289197722 | 0,67508711 | NHP2L1 | FALSE | Upregulated |
| -0,204732995 | 0,67508711 | RAB10 | FALSE | Downregulated |
| -0,231231887 | 0,67508711 | RCAN1 | FALSE | Downregulated |
| 0,278819041 | 0,67508711 | TLE2 | FALSE | Upregulated |
| 0,309379929 | 0,67508711 | TOPORS-AS1 | FALSE | Upregulated |
| -0,430023711 | 0,67508711 | TSSC1 | FALSE | Downregulated |
| 0,302676518 | 0,675156273 | ACSS3 | FALSE | Upregulated |
| -0,223297211 | 0,675156273 | AKR1C1 | FALSE | Downregulated |
| -0,622296522 | 0,675156273 | ASIC1 | FALSE | Downregulated |
| 0,436536622 | 0,675156273 | CD274 | FALSE | Upregulated |
| 0,132420612 | 0,675156273 | CSNK1A1 | FALSE | Upregulated |
| 0,279793642 | 0,675156273 | ERAL1 | FALSE | Upregulated |
| -0,232081471 | 0,675156273 | HIBCH | FALSE | Downregulated |
| -0,940473813 | 0,675156273 | LINC00705 | FALSE | Downregulated |
| 0,565540949 | 0,675156273 | LOC100507599 | FALSE | Upregulated |
| -0,740107505 | 0,675156273 | LOC101927009 | FALSE | Downregulated |
| -0,337309359 | 0,675156273 | LOC101927894 | FALSE | Downregulated |
| -0,24626024 | 0,675156273 | LOC102724281 | FALSE | Downregulated |
| 0,655910566 | 0,675156273 | OLFML2A | FALSE | Upregulated |
| 0,467837731 | 0,675156273 | SCRG1 | FALSE | Upregulated |
| 0,308781741 | 0,675156273 | TSFM | FALSE | Upregulated |
| -0,563416298 | 0,675156273 | UBLCP1 | FALSE | Downregulated |
| -0,86920615 | 0,675156273 | ZNF879 | FALSE | Downregulated |
| 0,310308878 | 0,675174732 | CWC25 | FALSE | Upregulated |
| 0,900886187 | 0,675175718 | NPAS3 | FALSE | Upregulated |
| 0,275650153 | 0,675291583 | AIMP2 | FALSE | Upregulated |
| -0,193828472 | 0,675291583 | DAAM1 | FALSE | Downregulated |
| -0,097942339 | 0,675291583 | TNRC18 | FALSE | Downregulated |
| -0,112445239 | 0,675291583 | USP9X | FALSE | Downregulated |
| 0,185811901 | 0,675397039 | C16orf70 | FALSE | Upregulated |
| 0,488658301 | 0,675397039 | RBPMS-AS1 | FALSE | Upregulated |
| 0,753240097 | 0,675397039 | UNC5B-AS1 | FALSE | Upregulated |
| -0,231296768 | 0,675397039 | USP30 | FALSE | Downregulated |
| 0,298953647 | 0,675397039 | YARS2 | FALSE | Upregulated |
| 0,441167636 | 0,67570831 | KIAA1244 | FALSE | Upregulated |
| -0,166670783 | 0,67570831 | PRKAR1A | FALSE | Downregulated |
| -0,426152815 | 0,675728639 | GALC | FALSE | Downregulated |
| 0,299900452 | 0,675769534 | ARMC8 | FALSE | Upregulated |
| -0,144710262 | 0,675769534 | SORBS3 | FALSE | Downregulated |
| 0,498906765 | 0,675814072 | C5orf56 | FALSE | Upregulated |
| 0,661290185 | 0,675814072 | CD79B | FALSE | Upregulated |
| 0,270625009 | 0,675814072 | HPS1 | FALSE | Upregulated |
| 0,297513736 | 0,675814072 | SAV1 | FALSE | Upregulated |
| 0,386820822 | 0,675814072 | SLC37A3 | FALSE | Upregulated |

| | | | | |
|---|---|---|---|---|
| -0,149748038 | 0,675814072 | SRSF5 | FALSE | Downregulated |
| -0,241794719 | 0,675949196 | COPZ1 | FALSE | Downregulated |
| -0,190102411 | 0,676089969 | ARHGEF11 | FALSE | Downregulated |
| 0,259181472 | 0,676089969 | GNA12 | FALSE | Upregulated |
| -0,474659178 | 0,676089969 | LOC101928714 | FALSE | Downregulated |
| 0,356539494 | 0,676089969 | RARRES3 | FALSE | Upregulated |
| 0,227458774 | 0,676136241 | ARPC4 | FALSE | Upregulated |
| 0,302407851 | 0,676136241 | SLC20A1 | FALSE | Upregulated |
| 0,293056832 | 0,676205581 | ATL3 | FALSE | Upregulated |
| -0,335392603 | 0,676205581 | DOLPP1 | FALSE | Downregulated |
| -0,192669474 | 0,676205581 | SLC35A3 | FALSE | Downregulated |
| 0,569653734 | 0,676205581 | ZNF525 | FALSE | Upregulated |
| -0,965672211 | 0,676226271 | AGR2 | FALSE | Downregulated |
| 0,259832354 | 0,676405197 | ADAM15 | FALSE | Upregulated |
| 0,332340761 | 0,676405197 | ARHGAP10 | FALSE | Upregulated |
| -0,161681655 | 0,676405197 | FBXO31 | FALSE | Downregulated |
| -0,433792265 | 0,676405197 | LOC102723539 | FALSE | Downregulated |
| 0,157770965 | 0,676405197 | ROCK1 | FALSE | Upregulated |
| 0,308013326 | 0,676505 | MAL2 | FALSE | Upregulated |
| 0,149556593 | 0,676570746 | FAM134C | FALSE | Upregulated |
| 0,204807484 | 0,676570746 | PARP9 | FALSE | Upregulated |
| 0,811739 | 0,67657506 | LOC102724889 | FALSE | Upregulated |
| -0,888596733 | 0,676712895 | NANP | FALSE | Downregulated |
| 0,213198954 | 0,676747199 | LOC284837 | FALSE | Upregulated |
| -0,336242695 | 0,676780827 | LOC728763 | FALSE | Downregulated |
| -0,175394203 | 0,676780827 | PTK2 | FALSE | Downregulated |
| 0,136971748 | 0,676780827 | RAPGEF1 | FALSE | Upregulated |
| 0,81393409 | 0,676852713 | LOC102724850 | FALSE | Upregulated |
| -0,462673351 | 0,676903092 | CTU2 | FALSE | Downregulated |
| -0,161416837 | 0,676903092 | PKD1 | FALSE | Downregulated |
| 0,150750806 | 0,676994484 | FAM65A | FALSE | Upregulated |
| 0,905823293 | 0,676994484 | STAC3 | FALSE | Upregulated |
| -1,400325257 | 0,676994484 | TTC4 | FALSE | Downregulated |
| -0,375413479 | 0,677043144 | FLJ30403 | FALSE | Downregulated |
| -0,364778345 | 0,677045805 | TRAF3IP3 | FALSE | Downregulated |
| 0,254688378 | 0,677134518 | SOD2 | FALSE | Upregulated |
| 0,206604023 | 0,677137202 | LOC102723342 | FALSE | Upregulated |
| -0,142441457 | 0,67725063 | GANAB | FALSE | Downregulated |
| -0,613966601 | 0,67725063 | PAK1IP1 | FALSE | Downregulated |
| -0,237680323 | 0,677376288 | CHN2 | FALSE | Downregulated |
| -0,507372284 | 0,677376288 | COX10-AS1 | FALSE | Downregulated |
| -0,153582775 | 0,677503375 | ADIPOR2 | FALSE | Downregulated |
| -0,867126245 | 0,677668158 | BARHL1 | FALSE | Downregulated |
| 0,525362844 | 0,677668158 | ISG15 | FALSE | Upregulated |
| 0,231124974 | 0,677668158 | NEDD4 | FALSE | Upregulated |
| -0,644256799 | 0,677676845 | AGTRAP | FALSE | Downregulated |
| -0,362687438 | 0,677676845 | EFNB2 | FALSE | Downregulated |
| 0,211135856 | 0,677676845 | LIX1L | FALSE | Upregulated |
| 0,256516849 | 0,677708637 | MRAP | FALSE | Upregulated |
| -0,265384839 | 0,677708637 | SMIM7 | FALSE | Downregulated |
| -0,159852713 | 0,677803094 | ADCY6 | FALSE | Downregulated |

| | | | | |
|---|---|---|---|---|
| -0,279150036 | 0,677803094 | GNRHR | FALSE | Downregulated |
| -0,535867609 | 0,677918388 | LOC102724833 | FALSE | Downregulated |
| 0,338646639 | 0,677927519 | MAPK3 | FALSE | Upregulated |
| -0,198632262 | 0,677927519 | MORF4L2 | FALSE | Downregulated |
| 0,636431802 | 0,678035683 | SNX15 | FALSE | Upregulated |
| 0,160541152 | 0,678093772 | TBCD | FALSE | Upregulated |
| 0,502472078 | 0,678251224 | RNF150 | FALSE | Upregulated |
| -1,020157342 | 0,678304934 | FXYD2 | FALSE | Downregulated |
| -0,338242102 | 0,678304934 | L3MBTL3 | FALSE | Downregulated |
| 0,272220892 | 0,678304934 | MAPK8IP1 | FALSE | Upregulated |
| -0,179880463 | 0,678304934 | RPL28 | FALSE | Downregulated |
| -0,155769153 | 0,678304934 | TRAF7 | FALSE | Downregulated |
| -0,951691033 | 0,678363656 | SCGB2B3P | FALSE | Downregulated |
| 0,569892502 | 0,678559232 | GTF2E1 | FALSE | Upregulated |
| 0,487585193 | 0,678566306 | HHAT | FALSE | Upregulated |
| 0,210511397 | 0,67864852 | PHF20 | FALSE | Upregulated |
| 0,344172009 | 0,67864852 | TMPPE | FALSE | Upregulated |
| -0,313680153 | 0,678653188 | MBTPS2 | FALSE | Downregulated |
| -0,505623797 | 0,678994117 | MED9 | FALSE | Downregulated |
| 0,381167406 | 0,67907239 | ZNF823 | FALSE | Upregulated |
| 0,296633289 | 0,679156901 | TCFL5 | FALSE | Upregulated |
| -0,951512702 | 0,67941994 | LOC728752 | FALSE | Downregulated |
| -0,694156509 | 0,679917427 | ARHGAP5-AS1 | FALSE | Downregulated |
| 0,577818887 | 0,679917427 | KIF3A | FALSE | Upregulated |
| 0,703003908 | 0,679917427 | LINC00528 | FALSE | Upregulated |
| -0,258072643 | 0,679917427 | LOC101928728 | FALSE | Downregulated |
| 0,217710264 | 0,679917427 | MAPKAP1 | FALSE | Upregulated |
| -0,236557848 | 0,680130537 | ERLIN2 | FALSE | Downregulated |
| -0,3163124 | 0,680130537 | GTPBP10 | FALSE | Downregulated |
| -0,433897137 | 0,680151532 | BPNT1 | FALSE | Downregulated |
| -0,205898872 | 0,680181307 | FRMD8 | FALSE | Downregulated |
| 0,579776417 | 0,680181307 | PARK2 | FALSE | Upregulated |
| -0,557549601 | 0,680353334 | PPP1R1C | FALSE | Downregulated |
| 0,232493985 | 0,680540777 | C11orf42 | FALSE | Upregulated |
| 0,477271059 | 0,680540777 | C1orf109 | FALSE | Upregulated |
| 0,239160949 | 0,680540777 | FRY | FALSE | Upregulated |
| -0,27044908 | 0,680540777 | GLOD4 | FALSE | Downregulated |
| -0,360245261 | 0,680540777 | HSPA1B | FALSE | Downregulated |
| 0,293133408 | 0,680540777 | LDHAL6A | FALSE | Upregulated |
| -0,440086585 | 0,680540777 | REEP3 | FALSE | Downregulated |
| 0,497568202 | 0,680639515 | ATP1B2 | FALSE | Upregulated |
| 0,371609047 | 0,680874831 | ISLR | FALSE | Upregulated |
| -0,647764682 | 0,680874831 | LINGO4 | FALSE | Downregulated |
| 0,194739716 | 0,680942047 | RP11-143J12.2 | FALSE | Upregulated |
| 0,203698722 | 0,681092661 | CNOT2 | FALSE | Upregulated |
| 0,678300646 | 0,681092661 | CORO7 | FALSE | Upregulated |
| 0,44062177 | 0,681092661 | ZNF521 | FALSE | Upregulated |
| 0,524741172 | 0,681106571 | RP11-863K10.2 | FALSE | Upregulated |
| 0,163124492 | 0,681106571 | YWHAB | FALSE | Upregulated |
| 0,302000982 | 0,681184877 | ANXA5 | FALSE | Upregulated |
| -0,120913824 | 0,681198304 | RPS11 | FALSE | Downregulated |

| | | | | |
|---|---|---|---|---|
| 0,227962365 | 0,6813134 | RPS6KA2 | FALSE | Upregulated |
| -0,352271312 | 0,681464013 | MIIP | FALSE | Downregulated |
| 0,607908144 | 0,681842229 | SMOX | FALSE | Upregulated |
| 0,254222626 | 0,681842229 | TRMT1L | FALSE | Upregulated |
| 0,493131778 | 0,681996607 | GALNT6 | FALSE | Upregulated |
| -0,596116624 | 0,681996607 | JMJD7 | FALSE | Downregulated |
| -0,644711669 | 0,681996607 | LOC100506571 | FALSE | Downregulated |
| 0,438347381 | 0,681996607 | PGAP1 | FALSE | Upregulated |
| -0,140712796 | 0,681996607 | PPP2R1B | FALSE | Downregulated |
| 0,278726767 | 0,681996607 | TRA2B | FALSE | Upregulated |
| -0,34242992 | 0,682344251 | ATP10D | FALSE | Downregulated |
| 0,249623322 | 0,682344251 | PSMB10 | FALSE | Upregulated |
| -0,174997562 | 0,682344251 | RABEP1 | FALSE | Downregulated |
| -0,126461104 | 0,682344251 | RPL10 | FALSE | Downregulated |
| 0,741534404 | 0,682344251 | ZNF14 | FALSE | Upregulated |
| -0,84656817 | 0,682436501 | CETN2 | FALSE | Downregulated |
| 0,163126128 | 0,682440421 | P2RX5-TAX1BP3 | FALSE | Upregulated |
| 1,083033797 | 0,682768529 | HSD3B2 | FALSE | Upregulated |
| 0,517204944 | 0,682768529 | LOC389607 | FALSE | Upregulated |
| 0,185155054 | 0,682768529 | TAF4 | FALSE | Upregulated |
| 0,710104655 | 0,683094528 | KCNT1 | FALSE | Upregulated |
| -0,533845076 | 0,683227172 | LOC155060 | FALSE | Downregulated |
| -0,218245453 | 0,683312991 | ATP6V0C | FALSE | Downregulated |
| -0,130881918 | 0,683312991 | CENPB | FALSE | Downregulated |
| 0,483356698 | 0,683312991 | HIST1H2APS1 | FALSE | Upregulated |
| 0,311693435 | 0,683312991 | MXRA5 | FALSE | Upregulated |
| -0,773484891 | 0,683312991 | NIM1K | FALSE | Downregulated |
| -0,308203193 | 0,683312991 | TSR2 | FALSE | Downregulated |
| -0,37263184 | 0,683312991 | WIBG | FALSE | Downregulated |
| -0,404578889 | 0,683366644 | HSF4 | FALSE | Downregulated |
| 0,213171489 | 0,683374576 | ABCB4 | FALSE | Upregulated |
| 0,181383849 | 0,683374576 | LDB1 | FALSE | Upregulated |
| -0,153906833 | 0,683374576 | MATR3 | FALSE | Downregulated |
| -0,163357784 | 0,683374576 | SH3GL1 | FALSE | Downregulated |
| 0,418787888 | 0,68362478 | ATXN7L2 | FALSE | Upregulated |
| 0,475386808 | 0,68362478 | C14orf119 | FALSE | Upregulated |
| -0,184306925 | 0,68362478 | CALCOCO1 | FALSE | Downregulated |
| 0,498135799 | 0,68362478 | CCDC43 | FALSE | Upregulated |
| 0,337154224 | 0,68362478 | CHST3 | FALSE | Upregulated |
| 0,52425998 | 0,68362478 | FOS | FALSE | Upregulated |
| 0,185251856 | 0,68362478 | LINC00242 | FALSE | Upregulated |
| -0,150230215 | 0,68362478 | SIPA1L1 | FALSE | Downregulated |
| 0,455394704 | 0,68362478 | SURF2 | FALSE | Upregulated |
| -0,355255361 | 0,68362478 | TEFM | FALSE | Downregulated |
| 0,350773005 | 0,68362478 | TMEM194B | FALSE | Upregulated |
| 0,195116401 | 0,68362478 | TMEM79 | FALSE | Upregulated |
| 0,600548493 | 0,68362478 | USP9Y | FALSE | Upregulated |
| 0,678965091 | 0,68362478 | ZNF667 | FALSE | Upregulated |
| 0,519863886 | 0,683693861 | VN1R1 | FALSE | Upregulated |
| 0,152862836 | 0,683705902 | ATG2A | FALSE | Upregulated |
| 1,163252848 | 0,683814916 | TPSAB1 | FALSE | Upregulated |

| | | | | |
|---|---|---|---|---|
| 0,325292114 | 0,683859208 | FTSJ1 | FALSE | Upregulated |
| -0,238623067 | 0,68391953 | PER2 | FALSE | Downregulated |
| 0,237007794 | 0,68391953 | PSMB5 | FALSE | Upregulated |
| 0,187265678 | 0,68391953 | ZFX | FALSE | Upregulated |
| 0,204521596 | 0,684098611 | GCLM | FALSE | Upregulated |
| -0,250849476 | 0,684098611 | TP53 | FALSE | Downregulated |
| 1,023502197 | 0,684163673 | FAM180B | FALSE | Upregulated |
| 0,330017902 | 0,684311242 | PTCD1 | FALSE | Upregulated |
| 0,795237215 | 0,684311242 | SBK1 | FALSE | Upregulated |
| -0,708431549 | 0,685088644 | CNEP1R1 | FALSE | Downregulated |
| 0,179467538 | 0,685088644 | INO80 | FALSE | Upregulated |
| 0,216058122 | 0,685088644 | MTF1 | FALSE | Upregulated |
| 0,869580549 | 0,685088644 | TMCC2 | FALSE | Upregulated |
| -0,243985948 | 0,685320953 | FAM211B | FALSE | Downregulated |
| -0,541865291 | 0,685355837 | LOC101928875 | FALSE | Downregulated |
| 0,158597727 | 0,685355837 | YWHAG | FALSE | Upregulated |
| -0,265173853 | 0,685456437 | LYRM7 | FALSE | Downregulated |
| -0,355087847 | 0,685664659 | C5orf15 | FALSE | Downregulated |
| 0,872465519 | 0,685664659 | KIAA1239 | FALSE | Upregulated |
| -0,363772062 | 0,685937985 | AP5S1 | FALSE | Downregulated |
| 0,649244362 | 0,686048242 | RSPH10B | FALSE | Upregulated |
| 0,345207876 | 0,686048242 | SLC36A1 | FALSE | Upregulated |
| 0,344056027 | 0,68620123 | LOC101927181 | FALSE | Upregulated |
| 0,460365814 | 0,68620123 | PLAC8 | FALSE | Upregulated |
| 0,547838227 | 0,686346497 | CCDC170 | FALSE | Upregulated |
| 0,444874593 | 0,686346497 | CEP85L | FALSE | Upregulated |
| 0,25998461 | 0,686346497 | LOC100506730 | FALSE | Upregulated |
| -0,219044855 | 0,686346497 | PITPNB | FALSE | Downregulated |
| -0,474039947 | 0,686346497 | PLGLB1 | FALSE | Downregulated |
| 1,102061406 | 0,686468652 | EGFL8 | FALSE | Upregulated |
| -0,359271485 | 0,686553271 | ADAMTS8 | FALSE | Downregulated |
| 0,402091336 | 0,686553271 | MEG3 | FALSE | Upregulated |
| -0,324693609 | 0,687157392 | ANKRD42 | FALSE | Downregulated |
| 0,31968678 | 0,687157392 | CISD2 | FALSE | Upregulated |
| 0,304417329 | 0,687157392 | EDRF1 | FALSE | Upregulated |
| 0,120150289 | 0,687157392 | HCFC1 | FALSE | Upregulated |
| -0,419287499 | 0,687157392 | LOC100506083 | FALSE | Downregulated |
| -1,960617686 | 0,687157392 | LOC102724828 | FALSE | Downregulated |
| -0,15022984 | 0,687157392 | PCNT | FALSE | Downregulated |
| -0,169615273 | 0,687157392 | SLTM | FALSE | Downregulated |
| -0,682744218 | 0,687157392 | SMPD2 | FALSE | Downregulated |
| 0,294321737 | 0,687269539 | AIFM2 | FALSE | Upregulated |
| -0,297527593 | 0,687269539 | COPS5 | FALSE | Downregulated |
| 0,2391457 | 0,687269539 | DDX19A | FALSE | Upregulated |
| -0,973434072 | 0,687269539 | FAM87A | FALSE | Downregulated |
| 0,159280861 | 0,687269539 | FGFR3 | FALSE | Upregulated |
| 0,985225767 | 0,687269539 | LOC102723675 | FALSE | Upregulated |
| -0,371353396 | 0,687269539 | LOC286083 | FALSE | Downregulated |
| -0,299694793 | 0,687269539 | NAAA | FALSE | Downregulated |
| -0,135064304 | 0,687269539 | PKD1P1 | FALSE | Downregulated |
| 0,285526839 | 0,687269539 | RBM28 | FALSE | Upregulated |

| | | | | |
|---|---|---|---|---|
| -0,264888251 | 0,687269539 | SCNN1D | FALSE | Downregulated |
| -0,200650161 | 0,687269539 | SLC25A25 | FALSE | Downregulated |
| -0,228221529 | 0,687269539 | SLC35A4 | FALSE | Downregulated |
| -0,286492492 | 0,687269539 | ZNF22 | FALSE | Downregulated |
| -0,183090234 | 0,687469772 | CLSTN3 | FALSE | Downregulated |
| -0,167250963 | 0,687469772 | DIP2B | FALSE | Downregulated |
| 0,745656076 | 0,687606422 | ASIP | FALSE | Upregulated |
| 0,5117466 | 0,687606422 | NCEH1 | FALSE | Upregulated |
| 0,123759239 | 0,687606422 | TNIP1 | FALSE | Upregulated |
| 0,806484789 | 0,687825749 | ID4 | FALSE | Upregulated |
| -0,366110642 | 0,687990048 | B4GALT6 | FALSE | Downregulated |
| -1,329323374 | 0,687992975 | CTGLF12P | FALSE | Downregulated |
| 0,193534797 | 0,687992975 | LRRC61 | FALSE | Upregulated |
| 0,31468771 | 0,687992975 | PRKCI | FALSE | Upregulated |
| 0,673387123 | 0,687992975 | SLC6A9 | FALSE | Upregulated |
| 0,215541425 | 0,687992975 | SNF8 | FALSE | Upregulated |
| 0,339340488 | 0,688053509 | GPBAR1 | FALSE | Upregulated |
| -0,11187987 | 0,688129413 | NEK6 | FALSE | Downregulated |
| 0,139931742 | 0,688129413 | TJP1 | FALSE | Upregulated |
| -0,160610283 | 0,688267313 | TPRG1L | FALSE | Downregulated |
| 0,686247039 | 0,688267313 | TUBA3E | FALSE | Upregulated |
| 0,175441957 | 0,688413005 | CAP1 | FALSE | Upregulated |
| 0,194899154 | 0,688413005 | JMJD1C-AS1 | FALSE | Upregulated |
| -0,223817442 | 0,688413005 | PNPLA3 | FALSE | Downregulated |
| 0,346075311 | 0,689168634 | SOCS5 | FALSE | Upregulated |
| -0,283018375 | 0,689214842 | C16orf13 | FALSE | Downregulated |
| -0,253254515 | 0,689215778 | NDRG1 | FALSE | Downregulated |
| -0,215141529 | 0,689218276 | NF2 | FALSE | Downregulated |
| -0,340615378 | 0,689218276 | RFK | FALSE | Downregulated |
| -0,987871731 | 0,689218276 | TSACC | FALSE | Downregulated |
| -0,544307545 | 0,689289474 | ZNF527 | FALSE | Downregulated |
| -0,818078909 | 0,689641949 | ARL11 | FALSE | Downregulated |
| -0,229235111 | 0,689641949 | CHMP7 | FALSE | Downregulated |
| -0,631051008 | 0,689641949 | LOC100129617 | FALSE | Downregulated |
| -0,432881938 | 0,689641949 | TUBA1C | FALSE | Downregulated |
| -0,478204458 | 0,689644943 | IL22RA1 | FALSE | Downregulated |
| 0,824757981 | 0,689805312 | UCP3 | FALSE | Upregulated |
| 0,190693754 | 0,689882893 | LOC101928874 | FALSE | Upregulated |
| -0,184088684 | 0,690042315 | BCAT2 | FALSE | Downregulated |
| -0,194232372 | 0,690042315 | BTG1 | FALSE | Downregulated |
| -0,336228144 | 0,690042315 | PNPLA8 | FALSE | Downregulated |
| -0,27818051 | 0,690042315 | RPH3AL | FALSE | Downregulated |
| -0,840995458 | 0,690369063 | COL28A1 | FALSE | Downregulated |
| 0,311582798 | 0,690429639 | NECAP1 | FALSE | Upregulated |
| 0,522797757 | 0,690727223 | AC024592.9 | FALSE | Upregulated |
| 0,478948108 | 0,690727223 | ADA | FALSE | Upregulated |
| 0,275747845 | 0,690727223 | BBS4 | FALSE | Upregulated |
| 0,300568218 | 0,690727223 | CCND3 | FALSE | Upregulated |
| -0,510584344 | 0,690727223 | CSTF2 | FALSE | Downregulated |
| 0,541923849 | 0,690727223 | ERBB4 | FALSE | Upregulated |
| 0,538267342 | 0,690727223 | LILRA1 | FALSE | Upregulated |

| | | | | |
|---|---|---|---|---|
| -0,153610322 | 0,690727223 | PUM1 | FALSE | Downregulated |
| -0,310830986 | 0,690727223 | TPD52L1 | FALSE | Downregulated |
| 0,120281845 | 0,690727223 | WHSC1L1 | FALSE | Upregulated |
| 0,404773367 | 0,690727223 | ZNF211 | FALSE | Upregulated |
| 0,254453862 | 0,690868808 | MLH1 | FALSE | Upregulated |
| -0,526028111 | 0,690912521 | AKIP1 | FALSE | Downregulated |
| 0,349380019 | 0,690912521 | ARHGEF15 | FALSE | Upregulated |
| -0,400441338 | 0,690912521 | CES4A | FALSE | Downregulated |
| 0,400568739 | 0,690912521 | CNOT6 | FALSE | Upregulated |
| 0,416230103 | 0,690912521 | CTTNBP2NL | FALSE | Upregulated |
| 0,315652265 | 0,690912521 | DDX47 | FALSE | Upregulated |
| 0,190942913 | 0,690912521 | GRN | FALSE | Upregulated |
| -0,512008158 | 0,690912521 | GYPE | FALSE | Downregulated |
| -0,19085189 | 0,690912521 | MEPCE | FALSE | Downregulated |
| 0,685189848 | 0,690912521 | NFKBID | FALSE | Upregulated |
| 0,338399322 | 0,690912521 | PFDN2 | FALSE | Upregulated |
| 0,802365832 | 0,690912521 | PTGES2-AS1 | FALSE | Upregulated |
| -0,222619554 | 0,690912521 | SERPINA7 | FALSE | Downregulated |
| -0,366432353 | 0,690912521 | UQCRH | FALSE | Downregulated |
| -0,186520798 | 0,690970729 | LPHN2 | FALSE | Downregulated |
| 0,366265218 | 0,691003515 | LOC727710 | FALSE | Upregulated |
| 0,158075326 | 0,691003515 | NTN1 | FALSE | Upregulated |
| 0,227953683 | 0,691043334 | AC004893.11 | FALSE | Upregulated |
| -0,361597005 | 0,691295634 | NPIPA5 | FALSE | Downregulated |
| 0,125122622 | 0,691297514 | MAPK8IP3 | FALSE | Upregulated |
| 0,730393763 | 0,69130841 | KCNS3 | FALSE | Upregulated |
| 0,157021457 | 0,691453602 | CC2D1A | FALSE | Upregulated |
| -0,217019847 | 0,691453602 | NUDC | FALSE | Downregulated |
| 0,306525285 | 0,691453602 | ZNF767 | FALSE | Upregulated |
| 0,308310941 | 0,691474089 | RBM22 | FALSE | Upregulated |
| -0,377219746 | 0,691476706 | DISP1 | FALSE | Downregulated |
| -0,808898533 | 0,691476706 | RPL23AP82 | FALSE | Downregulated |
| -0,294200374 | 0,691588153 | NDUFAF3 | FALSE | Downregulated |
| 0,437083144 | 0,691604669 | ZMYM1 | FALSE | Upregulated |
| 0,17280729 | 0,691860224 | CUL4A | FALSE | Upregulated |
| 0,191102953 | 0,691860224 | SOCS6 | FALSE | Upregulated |
| 0,783353445 | 0,691867937 | LOC101929336 | FALSE | Upregulated |
| 0,184112736 | 0,691867937 | NECAP2 | FALSE | Upregulated |
| -0,243838998 | 0,691867937 | SDPR | FALSE | Downregulated |
| -0,554615857 | 0,692208404 | TXNDC9 | FALSE | Downregulated |
| 0,751593716 | 0,692224564 | EPN3 | FALSE | Upregulated |
| 0,254568407 | 0,69226439 | ATP6V1D | FALSE | Upregulated |
| 0,231137042 | 0,69226439 | CDIPT | FALSE | Upregulated |
| 0,246964459 | 0,69226439 | FUS | FALSE | Upregulated |
| 0,490916041 | 0,69226439 | LIN28A | FALSE | Upregulated |
| 0,320532154 | 0,69226439 | MLLT11 | FALSE | Upregulated |
| -0,558565328 | 0,69226439 | THG1L | FALSE | Downregulated |
| -0,496408782 | 0,692426113 | LILRA6 | FALSE | Downregulated |
| -0,648277202 | 0,692520563 | RSAD2 | FALSE | Downregulated |
| 0,381539036 | 0,692553012 | KCTD11 | FALSE | Upregulated |
| -0,266279665 | 0,693262046 | ACP1 | FALSE | Downregulated |

| | | | | |
|---|---|---|---|---|
| -0,249365551 | 0,693262046 | MMAB | FALSE | Downregulated |
| 0,347400126 | 0,693479726 | BRF2 | FALSE | Upregulated |
| 0,459138377 | 0,693479726 | PDE1A | FALSE | Upregulated |
| 0,407207052 | 0,693479726 | RP11-466A17.1 | FALSE | Upregulated |
| -0,145857553 | 0,693699897 | WDTC1 | FALSE | Downregulated |
| -0,235352738 | 0,693745035 | EEPD1 | FALSE | Downregulated |
| 0,364353191 | 0,693745035 | MLKL | FALSE | Upregulated |
| 0,142881217 | 0,693745035 | SLC25A6 | FALSE | Upregulated |
| -1,063547816 | 0,693833243 | GALNT4 | FALSE | Downregulated |
| -0,845222509 | 0,693833243 | LOC102723512 | FALSE | Downregulated |
| -0,352696712 | 0,693833243 | RUNDC1 | FALSE | Downregulated |
| 0,500589277 | 0,693875207 | WTIP | FALSE | Upregulated |
| 0,590355625 | 0,693973751 | C14orf178 | FALSE | Upregulated |
| -0,177610721 | 0,693973751 | ITSN1 | FALSE | Downregulated |
| -0,395860988 | 0,693989903 | NPY1R | FALSE | Downregulated |
| -0,172233075 | 0,693996599 | CSRP1 | FALSE | Downregulated |
| -0,667651591 | 0,694053613 | C16orf46 | FALSE | Downregulated |
| -0,148001876 | 0,694069308 | PBXIP1 | FALSE | Downregulated |
| -0,286334672 | 0,694265109 | FKBP3 | FALSE | Downregulated |
| 0,435916544 | 0,694280625 | CYS1 | FALSE | Upregulated |
| 0,174525656 | 0,694378215 | C17orf103 | FALSE | Upregulated |
| -0,781314069 | 0,694378215 | CCDC67 | FALSE | Downregulated |
| 0,239440229 | 0,694378215 | GPR160 | FALSE | Upregulated |
| 0,217162851 | 0,694378215 | NOP14-AS1 | FALSE | Upregulated |
| -0,174541977 | 0,694378215 | RAB26 | FALSE | Downregulated |
| 0,127829245 | 0,694378215 | SLC22A23 | FALSE | Upregulated |
| -0,916137984 | 0,694487382 | LINC00890 | FALSE | Downregulated |
| -0,365125335 | 0,694487382 | NUP54 | FALSE | Downregulated |
| 0,317133639 | 0,694720086 | LAMTOR3 | FALSE | Upregulated |
| -0,280847294 | 0,69484806 | ERO1L | FALSE | Downregulated |
| -0,632331532 | 0,695066484 | CSRNP3 | FALSE | Downregulated |
| -0,30739522 | 0,695346843 | GALK2 | FALSE | Downregulated |
| -0,405074489 | 0,695346843 | TMEM194A | FALSE | Downregulated |
| -0,465060637 | 0,695346843 | USP18 | FALSE | Downregulated |
| -0,309808803 | 0,695671449 | TIFA | FALSE | Downregulated |
| -0,3538656 | 0,695858516 | PMS1 | FALSE | Downregulated |
| 0,507942084 | 0,695926115 | LSM11 | FALSE | Upregulated |
| -0,260740225 | 0,696012906 | ATP5O | FALSE | Downregulated |
| 0,52816899 | 0,696012906 | HDAC11 | FALSE | Upregulated |
| 0,398155161 | 0,696012906 | OSTM1 | FALSE | Upregulated |
| -0,1277056 | 0,696012906 | PHF3 | FALSE | Downregulated |
| -0,300480604 | 0,696012906 | PLCB1 | FALSE | Downregulated |
| -0,327305789 | 0,696012906 | RNF121 | FALSE | Downregulated |
| 0,339568691 | 0,696012906 | RPAP3 | FALSE | Upregulated |
| -0,423286286 | 0,696012906 | RPUSD2 | FALSE | Downregulated |
| -0,11004784 | 0,696012906 | SF1 | FALSE | Downregulated |
| -0,212446142 | 0,696012906 | SRP72 | FALSE | Downregulated |
| 0,209941369 | 0,696012906 | TAPBPL | FALSE | Upregulated |
| 0,648611827 | 0,696012906 | TPBG | FALSE | Upregulated |
| 0,266769546 | 0,696012906 | TRAPPC6B | FALSE | Upregulated |
| 0,343827264 | 0,696012906 | TRIM67 | FALSE | Upregulated |

| | | | | |
|---|---|---|---|---|
| -0,14879803 | 0,696012906 | TRMT2A | FALSE | Downregulated |
| 0,453546383 | 0,696012906 | ZNF438 | FALSE | Upregulated |
| -0,344732758 | 0,69614939 | CDC23 | FALSE | Downregulated |
| 0,541358139 | 0,69614939 | FBP2 | FALSE | Upregulated |
| 0,487112382 | 0,69614939 | RP11-379B18.5 | FALSE | Upregulated |
| -0,355704112 | 0,696287431 | ESF1 | FALSE | Downregulated |
| -0,983334832 | 0,696287431 | LOC101927609 | FALSE | Downregulated |
| 0,587396377 | 0,696287431 | SLC9A2 | FALSE | Upregulated |
| -0,465514934 | 0,696342207 | ISPD | FALSE | Downregulated |
| -0,437019558 | 0,696358879 | ZDHHC11 | FALSE | Downregulated |
| 0,276564918 | 0,696662792 | CLDN24 | FALSE | Upregulated |
| 0,403003785 | 0,69673062 | RP1-149A16.3 | FALSE | Upregulated |
| -0,134777983 | 0,69673062 | SF3B2 | FALSE | Downregulated |
| -0,213175485 | 0,696968196 | LRRC58 | FALSE | Downregulated |
| 0,152517587 | 0,697042819 | DNAJC5 | FALSE | Upregulated |
| 0,312915218 | 0,697053827 | LOC101927719 | FALSE | Upregulated |
| 0,378838257 | 0,697231199 | LOC102723908 | FALSE | Upregulated |
| 0,277871299 | 0,6973241 | STEAP4 | FALSE | Upregulated |
| -0,493013453 | 0,6973241 | TIMM8A | FALSE | Downregulated |
| 0,314290376 | 0,697491452 | LMNB2 | FALSE | Upregulated |
| 0,502380132 | 0,697491452 | NAT6 | FALSE | Upregulated |
| -0,258808391 | 0,697491452 | PSAT1 | FALSE | Downregulated |
| -0,370417006 | 0,697628882 | KIAA1009 | FALSE | Downregulated |
| 0,208262527 | 0,69786906 | SNCAIP | FALSE | Upregulated |
| -0,518362374 | 0,698070988 | LOC100505727 | FALSE | Downregulated |
| -0,279920057 | 0,698109826 | PRADC1 | FALSE | Downregulated |
| 0,216907512 | 0,69815033 | DNAH11 | FALSE | Upregulated |
| 0,499295331 | 0,69815033 | DUSP18 | FALSE | Upregulated |
| -0,216420525 | 0,698272876 | FNDC4 | FALSE | Downregulated |
| 0,567394098 | 0,698293863 | SLC2A3 | FALSE | Upregulated |
| -0,240911988 | 0,698384308 | CUEDC2 | FALSE | Downregulated |
| 0,548450795 | 0,698384308 | EXTL3-AS1 | FALSE | Upregulated |
| 0,959040452 | 0,698524261 | LOC101927745 | FALSE | Upregulated |
| 0,470687084 | 0,698680389 | C15orf61 | FALSE | Upregulated |
| 0,489577798 | 0,698680389 | CDC42BPG | FALSE | Upregulated |
| 0,10310258 | 0,698680389 | EGLN1 | FALSE | Upregulated |
| -0,333821194 | 0,698680389 | INPP1 | FALSE | Downregulated |
| 0,675400893 | 0,698680389 | LOC101929500 | FALSE | Upregulated |
| 0,841818795 | 0,698680389 | LOC339192 | FALSE | Upregulated |
| -0,653409632 | 0,698680389 | NSMCE2 | FALSE | Downregulated |
| -0,483525107 | 0,698680389 | TIMM21 | FALSE | Downregulated |
| -0,310859123 | 0,698680389 | TMEM53 | FALSE | Downregulated |
| 0,640920895 | 0,698680389 | YAE1D1 | FALSE | Upregulated |
| 0,569337422 | 0,698680389 | ZNF214 | FALSE | Upregulated |
| -0,243202036 | 0,698680389 | ZNF746 | FALSE | Downregulated |
| -0,132247972 | 0,698722425 | SPATA13 | FALSE | Downregulated |
| 0,408525114 | 0,698749839 | CYP2A7 | FALSE | Upregulated |
| -1,248690283 | 0,69907874 | C19orf77 | FALSE | Downregulated |
| -0,5249915 | 0,69907874 | PSPH | FALSE | Downregulated |
| -0,43798874 | 0,699093847 | NPW | FALSE | Downregulated |
| 0,187447954 | 0,699093847 | RUSC1 | FALSE | Upregulated |

| | | | | |
|---|---|---|---|---|
| -0,260692056 | 0,699169993 | NLGN2 | FALSE | Downregulated |
| 0,289367577 | 0,699743056 | PPP1R35 | FALSE | Upregulated |
| 0,508136489 | 0,699828504 | GSKIP | FALSE | Upregulated |
| 0,686049411 | 0,699828504 | HELLS | FALSE | Upregulated |
| 0,618311918 | 0,699828504 | ZNF607 | FALSE | Upregulated |
| 0,355820482 | 0,700028294 | LOC102724254 | FALSE | Upregulated |
| 0,721529129 | 0,700028294 | MACC1 | FALSE | Upregulated |
| 0,401625947 | 0,700028294 | RPL31P11 | FALSE | Upregulated |
| -0,467809547 | 0,700028294 | TRIM68 | FALSE | Downregulated |
| -0,248520401 | 0,700095691 | NCKIPSD | FALSE | Downregulated |
| 0,261955834 | 0,700199766 | FSTL1 | FALSE | Upregulated |
| 0,470521342 | 0,700199766 | IFI6 | FALSE | Upregulated |
| -0,373987223 | 0,700439063 | MRPS31 | FALSE | Downregulated |
| -0,955587786 | 0,700439063 | PDCL3P4 | FALSE | Downregulated |
| -0,21480401 | 0,700565987 | AKR1C3 | FALSE | Downregulated |
| -0,480539898 | 0,70081155 | LINC00211 | FALSE | Downregulated |
| -0,151211982 | 0,700903886 | B4GALT1 | FALSE | Downregulated |
| -0,259019011 | 0,700903886 | GPR182 | FALSE | Downregulated |
| 0,254435936 | 0,700903886 | PER3 | FALSE | Upregulated |
| 0,39013813 | 0,700903886 | RBBP8 | FALSE | Upregulated |
| -0,513308957 | 0,700903886 | SDK2 | FALSE | Downregulated |
| 0,248390921 | 0,700903886 | SH2D4A | FALSE | Upregulated |
| 0,164436481 | 0,700903886 | SNX17 | FALSE | Upregulated |
| 0,198949273 | 0,700903886 | TESK2 | FALSE | Upregulated |
| -0,658283167 | 0,700911211 | SEMA3B-AS1 | FALSE | Downregulated |
| 0,282759784 | 0,701133699 | GNL2 | FALSE | Upregulated |
| 0,347882093 | 0,701133699 | RTCA | FALSE | Upregulated |
| -0,232456651 | 0,701188503 | ACOX3 | FALSE | Downregulated |
| 0,19915152 | 0,701244627 | CLCC1 | FALSE | Upregulated |
| 0,1818731 | 0,701244627 | CLDN20 | FALSE | Upregulated |
| 0,442107693 | 0,701244627 | DGCR11 | FALSE | Upregulated |
| -0,259313003 | 0,701244627 | DLEC1 | FALSE | Downregulated |
| -0,240379252 | 0,701244627 | DVL2 | FALSE | Downregulated |
| 0,456755116 | 0,701244627 | EFCAB4B | FALSE | Upregulated |
| -0,256264702 | 0,701244627 | ENC1 | FALSE | Downregulated |
| 0,284264499 | 0,701244627 | GDPD4 | FALSE | Upregulated |
| -0,149223807 | 0,701244627 | GUK1 | FALSE | Downregulated |
| -0,405713874 | 0,701244627 | LOC102724023 | FALSE | Downregulated |
| 0,396850803 | 0,701244627 | P2RY13 | FALSE | Upregulated |
| -0,478040812 | 0,701244627 | PALM2-AKAP2 | FALSE | Downregulated |
| 0,544206996 | 0,701244627 | RP1-249H1.4 | FALSE | Upregulated |
| -0,831722988 | 0,701244627 | RP5-855F14.1 | FALSE | Downregulated |
| 0,660514617 | 0,701244627 | S100P | FALSE | Upregulated |
| 0,405009713 | 0,701244627 | TMEM189 | FALSE | Upregulated |
| -0,280194768 | 0,701244627 | YPEL3 | FALSE | Downregulated |
| 0,520806341 | 0,701244627 | ZNF648 | FALSE | Upregulated |
| 0,93617765 | 0,70126037 | LOC102723927 | FALSE | Upregulated |
| 0,288691297 | 0,701616768 | SMAP1 | FALSE | Upregulated |
| -0,814149204 | 0,701649093 | TAS2R14 | FALSE | Downregulated |
| 0,498489357 | 0,701649093 | WASF1 | FALSE | Upregulated |
| 0,258089257 | 0,702048067 | C15orf39 | FALSE | Upregulated |

| | | | | |
|---|---|---|---|---|
| 0,787726741 | 0,702048067 | COL18A1-AS2 | FALSE | Upregulated |
| -0,231907057 | 0,702048067 | EBNA1BP2 | FALSE | Downregulated |
| -0,175381889 | 0,702048067 | GABARAP | FALSE | Downregulated |
| -0,254644783 | 0,702048067 | IGF1 | FALSE | Downregulated |
| -0,332192172 | 0,702048067 | SCLY | FALSE | Downregulated |
| -0,148083373 | 0,702048067 | TNS1 | FALSE | Downregulated |
| -0,233046801 | 0,702129783 | DRAP1 | FALSE | Downregulated |
| 0,295529082 | 0,702129783 | DYNC1LI1 | FALSE | Upregulated |
| -0,73469617 | 0,702129783 | GPR21 | FALSE | Downregulated |
| -0,743231761 | 0,702129783 | GTF2H2C | FALSE | Downregulated |
| -0,352076722 | 0,702129783 | UQCC2 | FALSE | Downregulated |
| -0,197564658 | 0,702250046 | GAS8 | FALSE | Downregulated |
| -1,191989135 | 0,702250046 | RBM14-RBM4 | FALSE | Downregulated |
| -0,32490203 | 0,702250046 | TRMT11 | FALSE | Downregulated |
| 0,737218513 | 0,70254534 | CDS1 | FALSE | Upregulated |
| -0,511351014 | 0,703017987 | LOC100270746 | FALSE | Downregulated |
| -0,327409587 | 0,703131857 | SLC35A2 | FALSE | Downregulated |
| 0,341793618 | 0,703279184 | CAV2 | FALSE | Upregulated |
| 0,314441831 | 0,703631952 | CRIPT | FALSE | Upregulated |
| -0,248958886 | 0,703631952 | DNAJB6 | FALSE | Downregulated |
| -0,166894454 | 0,703631952 | UBR2 | FALSE | Downregulated |
| -0,772345045 | 0,703932437 | LOC101927138 | FALSE | Downregulated |
| 0,685982152 | 0,7039739 | CCDC24 | FALSE | Upregulated |
| 0,203232047 | 0,704049789 | C19orf12 | FALSE | Upregulated |
| 0,352551696 | 0,704205486 | GGCT | FALSE | Upregulated |
| 0,228285258 | 0,704297064 | ZDHHC9 | FALSE | Upregulated |
| 0,380904787 | 0,704339885 | CBR3 | FALSE | Upregulated |
| -0,210857904 | 0,704339885 | CNPPD1 | FALSE | Downregulated |
| 0,472871795 | 0,704339885 | METTL25 | FALSE | Upregulated |
| 0,122902417 | 0,704339885 | MKLN1 | FALSE | Upregulated |
| 0,767351869 | 0,704346503 | TAS2R39 | FALSE | Upregulated |
| -0,230383853 | 0,704346503 | ZRANB3 | FALSE | Downregulated |
| 0,88152045 | 0,704739796 | CSF3 | FALSE | Upregulated |
| 0,203699267 | 0,704739796 | SRF | FALSE | Upregulated |
| -0,302788615 | 0,704894034 | LOC101928068 | FALSE | Downregulated |
| 0,234097378 | 0,704895964 | BRD7 | FALSE | Upregulated |
| -0,226658808 | 0,705091994 | ACAT2 | FALSE | Downregulated |
| -0,266962929 | 0,705126814 | MIR210HG | FALSE | Downregulated |
| 0,391962534 | 0,705126814 | SLC38A9 | FALSE | Upregulated |
| 2,326085791 | 0,705264207 | RNF103-CHMP3 | FALSE | Upregulated |
| 0,177163614 | 0,705275222 | ATXN2 | FALSE | Upregulated |
| 0,628191826 | 0,705275222 | DFNB59 | FALSE | Upregulated |
| -0,86974479 | 0,705275222 | GABRB3 | FALSE | Downregulated |
| 0,306223398 | 0,705336182 | NPR2 | FALSE | Upregulated |
| 0,557913912 | 0,705445706 | HSPA12A | FALSE | Upregulated |
| 0,69755118 | 0,705715686 | RPL22L1 | FALSE | Upregulated |
| -0,614383829 | 0,705715686 | ZNF200 | FALSE | Downregulated |
| 2,425816657 | 0,705732809 | MB | FALSE | Upregulated |
| -0,259135443 | 0,705732809 | ZYG11A | FALSE | Downregulated |
| -0,611258886 | 0,705755109 | LOC100506472 | FALSE | Downregulated |
| -0,27400585 | 0,705769509 | ESCO1 | FALSE | Downregulated |

| | | | | |
|---|---|---|---|---|
| -0,18782614 | 0,705769509 | ZBTB10 | FALSE | Downregulated |
| 0,198202153 | 0,706310106 | DOCK4-AS1 | FALSE | Upregulated |
| -0,679779106 | 0,706348252 | RPS4Y1 | FALSE | Downregulated |
| -0,25501026 | 0,706406621 | RBAK | FALSE | Downregulated |
| 0,207732502 | 0,706786309 | BEND3P3 | FALSE | Upregulated |
| 0,576651057 | 0,706786309 | FXYD3 | FALSE | Upregulated |
| 0,141621237 | 0,706791211 | RC3H1 | FALSE | Upregulated |
| 0,224413007 | 0,706791211 | SLC1A1 | FALSE | Upregulated |
| -0,443008104 | 0,706807656 | LINC00844 | FALSE | Downregulated |
| 0,24093304 | 0,706842634 | TOPBP1 | FALSE | Upregulated |
| -0,19614367 | 0,707014094 | ZNF362 | FALSE | Downregulated |
| 0,752777628 | 0,707022799 | FRZB | FALSE | Upregulated |
| -0,174327238 | 0,707595838 | STAT3 | FALSE | Downregulated |
| 0,891157892 | 0,707644558 | ANXA8 | FALSE | Upregulated |
| -0,123900092 | 0,707644558 | FNDC3A | FALSE | Downregulated |
| 0,260303258 | 0,707644558 | SLC35C2 | FALSE | Upregulated |
| 0,475968249 | 0,707644558 | ZSCAN9 | FALSE | Upregulated |
| 0,244875985 | 0,707702448 | GDE1 | FALSE | Upregulated |
| 0,513000819 | 0,70806235 | ANKRD49 | FALSE | Upregulated |
| 0,446008285 | 0,70806235 | ARHGEF25 | FALSE | Upregulated |
| -0,255444352 | 0,70806235 | DARS | FALSE | Downregulated |
| -0,813714511 | 0,70806235 | FAM95C | FALSE | Downregulated |
| -0,26222132 | 0,70806235 | HOOK2 | FALSE | Downregulated |
| 0,691679732 | 0,70806235 | LINC00338 | FALSE | Upregulated |
| -0,404848855 | 0,70806235 | LOC102723382 | FALSE | Downregulated |
| -0,65324583 | 0,70806235 | MNDA | FALSE | Downregulated |
| -0,184580301 | 0,70806235 | PAN2 | FALSE | Downregulated |
| -0,189937053 | 0,70806235 | PDLIM2 | FALSE | Downregulated |
| -0,58296172 | 0,70806235 | RP5-1074L1.1 | FALSE | Downregulated |
| -0,194058911 | 0,70806235 | SLC43A3 | FALSE | Downregulated |
| -0,440264889 | 0,70806235 | VIMP | FALSE | Downregulated |
| 0,434193251 | 0,70806235 | ZNF678 | FALSE | Upregulated |
| -0,429673794 | 0,708201993 | THOC7 | FALSE | Downregulated |
| -0,135424827 | 0,708252902 | TUBGCP2 | FALSE | Downregulated |
| -0,281705146 | 0,708661749 | HIGD2A | FALSE | Downregulated |
| 0,248811007 | 0,70868121 | P4HA1 | FALSE | Upregulated |
| 0,195805651 | 0,708785533 | FAM92A1 | FALSE | Upregulated |
| -4,013133575 | 0,708785533 | GJA9 | FALSE | Downregulated |
| 0,221718558 | 0,708954309 | SNTA1 | FALSE | Upregulated |
| -0,269437344 | 0,70906268 | SREBF1 | FALSE | Downregulated |
| -0,200295993 | 0,70922478 | CHRM5 | FALSE | Downregulated |
| 0,238560266 | 0,70922478 | GOLGA8N | FALSE | Upregulated |
| -2,243962048 | 0,70932819 | LOC401589 | FALSE | Downregulated |
| 0,276509696 | 0,70932819 | MAX | FALSE | Upregulated |
| -0,249757658 | 0,70932819 | NPC1L1 | FALSE | Downregulated |
| -0,307903727 | 0,70932819 | TOPORS | FALSE | Downregulated |
| 0,227065495 | 0,709499395 | HYAL2 | FALSE | Upregulated |
| -0,175020887 | 0,709499395 | IGSF8 | FALSE | Downregulated |
| -0,158770388 | 0,709499395 | MYO10 | FALSE | Downregulated |
| -0,456608692 | 0,709499395 | STX1A | FALSE | Downregulated |
| 0,509284838 | 0,709499395 | USP32P2 | FALSE | Upregulated |

| | | | | |
|---|---|---|---|---|
| 0,255178288 | 0,709556524 | CCDC66 | FALSE | Upregulated |
| 0,186967617 | 0,709556524 | TLK2 | FALSE | Upregulated |
| 0,220927854 | 0,709556524 | TUBB6 | FALSE | Upregulated |
| -0,345719296 | 0,709572086 | DYNLT3 | FALSE | Downregulated |
| 0,21702428 | 0,709572086 | PEX1 | FALSE | Upregulated |
| -0,308034964 | 0,709624838 | ZNF511 | FALSE | Downregulated |
| 0,251418705 | 0,709684793 | FOXO4 | FALSE | Upregulated |
| 0,409878975 | 0,709707378 | ZNF528 | FALSE | Upregulated |
| -0,254267458 | 0,709708208 | CHD1L | FALSE | Downregulated |
| -0,246412357 | 0,709708208 | EXOC4 | FALSE | Downregulated |
| -0,689299433 | 0,709708208 | FAM104B | FALSE | Downregulated |
| -0,179321108 | 0,709708208 | H2AFV | FALSE | Downregulated |
| -0,246375766 | 0,709708208 | SBDS | FALSE | Downregulated |
| 0,504834899 | 0,709890646 | LOC102724550 | FALSE | Upregulated |
| -0,235850974 | 0,709890646 | SNX3 | FALSE | Downregulated |
| 0,254169319 | 0,709950576 | FAM178B | FALSE | Upregulated |
| -0,257338926 | 0,709950576 | SOX5 | FALSE | Downregulated |
| -0,262697515 | 0,710068406 | CUEDC1 | FALSE | Downregulated |
| -0,240824282 | 0,71015706 | AGXT2 | FALSE | Downregulated |
| -0,32769778 | 0,71015706 | GPR75 | FALSE | Downregulated |
| -0,167126246 | 0,71015706 | NAGA | FALSE | Downregulated |
| -0,166595767 | 0,71015706 | PCCB | FALSE | Downregulated |
| -0,269710516 | 0,71015706 | TH | FALSE | Downregulated |
| 0,533423907 | 0,710525843 | FAM86DP | FALSE | Upregulated |
| 0,397987485 | 0,710793541 | CST7 | FALSE | Upregulated |
| -0,217411441 | 0,710831057 | FAF2 | FALSE | Downregulated |
| 0,483985608 | 0,710884181 | COL5A3 | FALSE | Upregulated |
| -0,255061817 | 0,710909635 | C2orf43 | FALSE | Downregulated |
| -0,639424704 | 0,710938651 | LINC01181 | FALSE | Downregulated |
| 0,355469956 | 0,710938651 | LOC102723877 | FALSE | Upregulated |
| -0,574353206 | 0,710938651 | UBAP1L | FALSE | Downregulated |
| 0,431106362 | 0,710946645 | CCDC84 | FALSE | Upregulated |
| 0,23042215 | 0,710946645 | TSTD2 | FALSE | Upregulated |
| 0,943403045 | 0,711161684 | KIAA1644 | FALSE | Upregulated |
| -0,372985024 | 0,711161684 | NDUFA12 | FALSE | Downregulated |
| 0,311730885 | 0,711225434 | PEAR1 | FALSE | Upregulated |
| 0,347070171 | 0,711421718 | TRAM2-AS1 | FALSE | Upregulated |
| 0,535686884 | 0,711651627 | SLC25A24 | FALSE | Upregulated |
| -0,249347854 | 0,711708743 | ISG20L2 | FALSE | Downregulated |
| -0,821562901 | 0,711709647 | SOAT2 | FALSE | Downregulated |
| 0,172265474 | 0,712028844 | CALM2 | FALSE | Upregulated |
| -0,185070548 | 0,712340902 | CLTA | FALSE | Downregulated |
| 0,480590313 | 0,712340902 | DNAJC3-AS1 | FALSE | Upregulated |
| -0,406919732 | 0,712340902 | LOC102288414 | FALSE | Downregulated |
| -0,493704459 | 0,712340902 | SLC10A3 | FALSE | Downregulated |
| 0,208870575 | 0,712340902 | TMEM39A | FALSE | Upregulated |
| 0,397047996 | 0,712340902 | ZC2HC1A | FALSE | Upregulated |
| -0,233615926 | 0,712360011 | BABAM1 | FALSE | Downregulated |
| 0,541537295 | 0,712360011 | LOC100506538 | FALSE | Upregulated |
| 0,134959677 | 0,712360011 | PI4KB | FALSE | Upregulated |
| 0,344166495 | 0,712360011 | SLC11A1 | FALSE | Upregulated |

| | | | | |
|---|---|---|---|---|
| 0,72590768 | 0,712360011 | ZNF430 | FALSE | Upregulated |
| 0,25281594 | 0,712360011 | ZNF668 | FALSE | Upregulated |
| -0,728330788 | 0,712486535 | GPHA2 | FALSE | Downregulated |
| -0,392548097 | 0,712516474 | TRIM39 | FALSE | Downregulated |
| -0,447245376 | 0,712523085 | TRAPPC2L | FALSE | Downregulated |
| 0,244625161 | 0,712527346 | FEM1C | FALSE | Upregulated |
| -0,624006908 | 0,712527346 | ITGA10 | FALSE | Downregulated |
| 0,632085483 | 0,712527346 | LOC102724442 | FALSE | Upregulated |
| 0,154138913 | 0,712527346 | NOTCH2 | FALSE | Upregulated |
| -0,311874025 | 0,712527346 | NRBF2 | FALSE | Downregulated |
| -0,550608686 | 0,712527346 | SAPCD1 | FALSE | Downregulated |
| 0,146194669 | 0,712571014 | CST3 | FALSE | Upregulated |
| 0,71097793 | 0,712571014 | ZIK1 | FALSE | Upregulated |
| 0,227946773 | 0,712607357 | FAM177A1 | FALSE | Upregulated |
| 0,194838746 | 0,712607357 | FBXL13 | FALSE | Upregulated |
| 0,257416426 | 0,712607357 | LOC606724 | FALSE | Upregulated |
| 0,328130827 | 0,712607357 | TM4SF20 | FALSE | Upregulated |
| -0,550133432 | 0,712607357 | ZSCAN16-AS1 | FALSE | Downregulated |
| 0,40886497 | 0,71262591 | IFI27 | FALSE | Upregulated |
| 0,504254556 | 0,71262591 | LOC101928055 | FALSE | Upregulated |
| 0,236322859 | 0,71262591 | MIRLET7BHG | FALSE | Upregulated |
| 0,297000045 | 0,71262591 | S100A14 | FALSE | Upregulated |
| 0,763484238 | 0,71262591 | SAAL1 | FALSE | Upregulated |
| 0,128482678 | 0,71262591 | SCAF11 | FALSE | Upregulated |
| -0,237736511 | 0,71262591 | TM7SF2 | FALSE | Downregulated |
| 0,133422661 | 0,712673628 | FBXO11 | FALSE | Upregulated |
| -0,5675575 | 0,712706725 | ITGAD | FALSE | Downregulated |
| 0,80073987 | 0,712837355 | ARL6 | FALSE | Upregulated |
| 0,382402842 | 0,712911623 | LOC102724316 | FALSE | Upregulated |
| 0,669478587 | 0,713011908 | SNORD89 | FALSE | Upregulated |
| -0,204843452 | 0,713604547 | OPN3 | FALSE | Downregulated |
| -0,194337052 | 0,713711426 | SUV39H1 | FALSE | Downregulated |
| 0,281961339 | 0,713801 | RAB27A | FALSE | Upregulated |
| 0,190500325 | 0,713980204 | FBXO22-AS1 | FALSE | Upregulated |
| 0,197611541 | 0,713986622 | C5orf24 | FALSE | Upregulated |
| 0,164337004 | 0,713986622 | CADM1 | FALSE | Upregulated |
| 0,164301815 | 0,713986622 | CSRNP2 | FALSE | Upregulated |
| 0,228666289 | 0,713986622 | RP11-84C10.4 | FALSE | Upregulated |
| 1,203033167 | 0,713986622 | SKA3 | FALSE | Upregulated |
| -0,18503946 | 0,713986622 | TINAGL1 | FALSE | Downregulated |
| -0,259186193 | 0,713986622 | ZKSCAN5 | FALSE | Downregulated |
| -0,377877684 | 0,714046414 | ZNF706 | FALSE | Downregulated |
| -0,5320661 | 0,71405497 | CCDC51 | FALSE | Downregulated |
| -0,211739836 | 0,71405497 | KCTD6 | FALSE | Downregulated |
| 0,892812507 | 0,71405497 | PPFIA4 | FALSE | Upregulated |
| 0,267893766 | 0,71413993 | SMS | FALSE | Upregulated |
| 0,22333538 | 0,71413993 | ZBTB37 | FALSE | Upregulated |
| -0,201981413 | 0,714259296 | ATL2 | FALSE | Downregulated |
| -0,518309692 | 0,714259296 | C1orf50 | FALSE | Downregulated |
| 0,16351752 | 0,714259296 | LYST | FALSE | Upregulated |
| 0,39542933 | 0,714259296 | NCMAP | FALSE | Upregulated |

| | | | | |
|---|---|---|---|---|
| -0,126677403 | 0,714259296 | PAICS | FALSE | Downregulated |
| 0,272301338 | 0,714259296 | TMEM223 | FALSE | Upregulated |
| 0,178986162 | 0,714264752 | ZNF217 | FALSE | Upregulated |
| 0,39256138 | 0,71429005 | DGKG | FALSE | Upregulated |
| -0,177434899 | 0,714415155 | CDH1 | FALSE | Downregulated |
| -0,310577955 | 0,714415155 | SMIM12 | FALSE | Downregulated |
| 0,167114849 | 0,714497449 | SLC25A4 | FALSE | Upregulated |
| 0,319099725 | 0,714534465 | SLC25A35 | FALSE | Upregulated |
| 0,377905918 | 0,714629472 | PALD1 | FALSE | Upregulated |
| -0,216655104 | 0,714688077 | ILVBL | FALSE | Downregulated |
| 0,269653556 | 0,714839539 | SLC25A30 | FALSE | Upregulated |
| 0,157738852 | 0,714871275 | COQ6 | FALSE | Upregulated |
| 0,176700376 | 0,714871275 | FUCA2 | FALSE | Upregulated |
| 0,452231163 | 0,714871275 | HCG9 | FALSE | Upregulated |
| 0,305901754 | 0,714871275 | HKR1 | FALSE | Upregulated |
| 0,662671382 | 0,714871275 | LOC101928879 | FALSE | Upregulated |
| 0,194535867 | 0,714871275 | MAGI1-AS1 | FALSE | Upregulated |
| 0,372104586 | 0,714871275 | METTL20 | FALSE | Upregulated |
| 0,288603637 | 0,715280892 | CEP85 | FALSE | Upregulated |
| -0,200443183 | 0,715280892 | NDUFA13 | FALSE | Downregulated |
| 0,725692194 | 0,715356565 | AC016582.2 | FALSE | Upregulated |
| 0,582361667 | 0,715356565 | DNMT3B | FALSE | Upregulated |
| -0,198833094 | 0,715463006 | ARSD | FALSE | Downregulated |
| -0,691145342 | 0,715463006 | LINC00933 | FALSE | Downregulated |
| 0,415986678 | 0,715463006 | PET100 | FALSE | Upregulated |
| 0,222259028 | 0,715463006 | RSF1 | FALSE | Upregulated |
| 0,589547568 | 0,715522788 | ANKRD18A | FALSE | Upregulated |
| 0,119825522 | 0,715522788 | CLASP1 | FALSE | Upregulated |
| -0,159213542 | 0,715522788 | KLF13 | FALSE | Downregulated |
| 0,163253272 | 0,715522788 | SLC18A2 | FALSE | Upregulated |
| -0,802652187 | 0,715569159 | TIGD1 | FALSE | Downregulated |
| 0,338496133 | 0,715579737 | SNAPIN | FALSE | Upregulated |
| 0,640009874 | 0,715810679 | HIST1H2AK | FALSE | Upregulated |
| -0,303149259 | 0,715810679 | RCAN3 | FALSE | Downregulated |
| 0,188444312 | 0,715810679 | ZNF557 | FALSE | Upregulated |
| -0,316234842 | 0,716157608 | CRADD | FALSE | Downregulated |
| 0,264574183 | 0,716157608 | FMO4 | FALSE | Upregulated |
| -0,452499347 | 0,716157608 | GAN | FALSE | Downregulated |
| 0,548288852 | 0,716157608 | HNRNPA1L2 | FALSE | Upregulated |
| 0,184553283 | 0,716157608 | KCTD21-AS1 | FALSE | Upregulated |
| 0,121916309 | 0,716157608 | NUFIP2 | FALSE | Upregulated |
| 0,150642743 | 0,716157608 | STAT2 | FALSE | Upregulated |
| 0,154975422 | 0,716157608 | ZNF507 | FALSE | Upregulated |
| 0,225019083 | 0,71617696 | ACTR8 | FALSE | Upregulated |
| -0,441482998 | 0,71617696 | MOSPD2 | FALSE | Downregulated |
| -0,341639354 | 0,716264765 | CDC73 | FALSE | Downregulated |
| -0,195818895 | 0,716264765 | FAM160A1 | FALSE | Downregulated |
| 0,320681847 | 0,716264765 | LOC100996405 | FALSE | Upregulated |
| -0,159823978 | 0,716264765 | SYNCRIP | FALSE | Downregulated |
| -0,384770858 | 0,716585001 | RAB3IP | FALSE | Downregulated |
| 1,06534857 | 0,716585001 | RNVU1-7 | FALSE | Upregulated |

| | | | | |
|---|---|---|---|---|
| 0,248666879 | 0,716585001 | RP11-809M12.1 | FALSE | Upregulated |
| 0,202738486 | 0,716585001 | SERINC2 | FALSE | Upregulated |
| -0,52555605 | 0,716585001 | ZNF461 | FALSE | Downregulated |
| 0,158051016 | 0,716605038 | DHX38 | FALSE | Upregulated |
| 0,264048776 | 0,716667125 | BRK1 | FALSE | Upregulated |
| 0,174930902 | 0,716667125 | PIAS1 | FALSE | Upregulated |
| -0,371222091 | 0,716667125 | ZNHIT6 | FALSE | Downregulated |
| -0,177091567 | 0,716750141 | AHR | FALSE | Downregulated |
| 0,596289601 | 0,716750141 | ANKRD65 | FALSE | Upregulated |
| 0,31524006 | 0,716750141 | CCDC91 | FALSE | Upregulated |
| -0,234886006 | 0,716750141 | DYNC1I2 | FALSE | Downregulated |
| -0,451482307 | 0,716750141 | PILRA | FALSE | Downregulated |
| 0,111868092 | 0,716750141 | SZT2 | FALSE | Upregulated |
| 0,353562708 | 0,717020725 | C9orf142 | FALSE | Upregulated |
| 0,648711253 | 0,717020725 | LOC101929410 | FALSE | Upregulated |
| 0,423493704 | 0,717020725 | UNC50 | FALSE | Upregulated |
| 0,518982651 | 0,717115768 | MBIP | FALSE | Upregulated |
| -0,253704549 | 0,717115768 | PKDREJ | FALSE | Downregulated |
| 0,361599472 | 0,717128699 | LINC00936 | FALSE | Upregulated |
| 0,13060215 | 0,717140978 | KMO | FALSE | Upregulated |
| -0,74560654 | 0,717185137 | GCK | FALSE | Downregulated |
| -0,127386715 | 0,717185137 | TYK2 | FALSE | Downregulated |
| -0,611580567 | 0,717348658 | TUFT1 | FALSE | Downregulated |
| -0,129507084 | 0,717353173 | MED1 | FALSE | Downregulated |
| -0,46482574 | 0,717423052 | PSMG4 | FALSE | Downregulated |
| -0,88037996 | 0,717585054 | FAM171A2 | FALSE | Downregulated |
| 0,157837944 | 0,717585054 | LOXL3 | FALSE | Upregulated |
| 0,297676668 | 0,717585054 | PPIL6 | FALSE | Upregulated |
| 0,559287991 | 0,717644775 | MNS1 | FALSE | Upregulated |
| -0,197938957 | 0,717644775 | NR3C2 | FALSE | Downregulated |
| -0,319090118 | 0,717644775 | VPS25 | FALSE | Downregulated |
| -0,796166804 | 0,717698818 | CPNE6 | FALSE | Downregulated |
| 0,206781294 | 0,717698818 | FAM195B | FALSE | Upregulated |
| 1,003408042 | 0,717698818 | HTR2C | FALSE | Upregulated |
| 0,231544853 | 0,717698818 | LOC102724006 | FALSE | Upregulated |
| 0,282929063 | 0,717698818 | RIBC1 | FALSE | Upregulated |
| 0,31595496 | 0,717698818 | SMCO3 | FALSE | Upregulated |
| -0,129632361 | 0,717698818 | ZNF316 | FALSE | Downregulated |
| -0,303368819 | 0,717700965 | GMPS | FALSE | Downregulated |
| 0,697591195 | 0,717700965 | RP11-677M14.7 | FALSE | Upregulated |
| 0,254461554 | 0,718099496 | NIPA1 | FALSE | Upregulated |
| -0,301623058 | 0,718167365 | ING4 | FALSE | Downregulated |
| -0,329764659 | 0,718207152 | CAPN10 | FALSE | Downregulated |
| 0,24290521 | 0,718207152 | RCE1 | FALSE | Upregulated |
| 0,538346172 | 0,718207152 | SPATA33 | FALSE | Upregulated |
| -0,139739619 | 0,718258573 | ANO6 | FALSE | Downregulated |
| 0,361281024 | 0,718258573 | NRIP3 | FALSE | Upregulated |
| -0,460056208 | 0,718274134 | APIP | FALSE | Downregulated |
| 0,276894604 | 0,718274134 | GBP3 | FALSE | Upregulated |
| -0,620770252 | 0,718291846 | LOC101930307 | FALSE | Downregulated |
| -0,273452405 | 0,718349597 | LOC285593 | FALSE | Downregulated |

| | | | | |
|---|---|---|---|---|
| -0,607140072 | 0,718456581 | NUDT1 | FALSE | Downregulated |
| -0,266324438 | 0,718456581 | ZNF467 | FALSE | Downregulated |
| -0,206873235 | 0,718687856 | LOC102724696 | FALSE | Downregulated |
| -0,361603558 | 0,71877973 | MUSTN1 | FALSE | Downregulated |
| 0,17358169 | 0,71877973 | PPM1F | FALSE | Upregulated |
| -0,399453855 | 0,718929228 | RBM43 | FALSE | Downregulated |
| -0,125418105 | 0,718929228 | SNX1 | FALSE | Downregulated |
| -0,109302077 | 0,718929228 | YBX1 | FALSE | Downregulated |
| -0,253752872 | 0,718948108 | CCT2 | FALSE | Downregulated |
| -0,283976428 | 0,719057888 | APAF1 | FALSE | Downregulated |
| 0,141019813 | 0,719057888 | ARIH2 | FALSE | Upregulated |
| -0,771126781 | 0,719057888 | KBTBD3 | FALSE | Downregulated |
| 0,156686643 | 0,719057888 | PALLD | FALSE | Upregulated |
| -0,495633745 | 0,719057888 | TRMT10A | FALSE | Downregulated |
| 0,351399157 | 0,719306683 | LGI4 | FALSE | Upregulated |
| -0,481823429 | 0,719306683 | LOC101927160 | FALSE | Downregulated |
| -0,296464685 | 0,719306683 | THAP7 | FALSE | Downregulated |
| -1,003723603 | 0,719306683 | UBE2QL1 | FALSE | Downregulated |
| 0,240074195 | 0,719374601 | CDK2AP1 | FALSE | Upregulated |
| 0,308984087 | 0,719721262 | DDR1 | FALSE | Upregulated |
| -0,164226319 | 0,719721262 | PPP2R2D | FALSE | Downregulated |
| -0,219980993 | 0,719723473 | KDM7A | FALSE | Downregulated |
| 0,469886136 | 0,719951565 | CLP1 | FALSE | Upregulated |
| 0,582307685 | 0,719964979 | LOC554223 | FALSE | Upregulated |
| -0,274664106 | 0,720029372 | GHDC | FALSE | Downregulated |
| 0,272705434 | 0,720095039 | C2CD4D | FALSE | Upregulated |
| -0,460869274 | 0,720095039 | CCDC70 | FALSE | Downregulated |
| -0,154252258 | 0,720095039 | HIST1H1E | FALSE | Downregulated |
| -0,247518979 | 0,720095039 | METAP1 | FALSE | Downregulated |
| 0,129332572 | 0,720095039 | SDHC | FALSE | Upregulated |
| 0,253820683 | 0,720095039 | SEPT6 | FALSE | Upregulated |
| -0,189271426 | 0,720095039 | SSR4 | FALSE | Downregulated |
| 0,450902983 | 0,720095039 | ZNF485 | FALSE | Upregulated |
| -0,131395174 | 0,720156623 | DUSP3 | FALSE | Downregulated |
| 0,526160141 | 0,720168057 | AMIGO2 | FALSE | Upregulated |
| -0,286993401 | 0,720168057 | CNTROB | FALSE | Downregulated |
| -0,255739405 | 0,720168057 | LOC101927513 | FALSE | Downregulated |
| 0,593424588 | 0,720168057 | WDR62 | FALSE | Upregulated |
| 0,334802903 | 0,720304188 | CCDC132 | FALSE | Upregulated |
| -0,458223667 | 0,720418631 | MYL6B | FALSE | Downregulated |
| -0,530185246 | 0,720418631 | UCHL3 | FALSE | Downregulated |
| 0,20713507 | 0,72061426 | NAT9 | FALSE | Upregulated |
| 0,278224832 | 0,721320314 | TRIAP1 | FALSE | Upregulated |
| -0,404853923 | 0,721726925 | MTIF3 | FALSE | Downregulated |
| -0,417603897 | 0,721776174 | ATF1 | FALSE | Downregulated |
| 0,111425487 | 0,721776174 | CDC14B | FALSE | Upregulated |
| 0,534258924 | 0,721929889 | LINC00960 | FALSE | Upregulated |
| 0,279153514 | 0,721929889 | RCHY1 | FALSE | Upregulated |
| 0,350042703 | 0,721934142 | SLC16A7 | FALSE | Upregulated |
| -0,18582993 | 0,722019924 | CBX4 | FALSE | Downregulated |
| 0,326244089 | 0,722019924 | CCT6P1 | FALSE | Upregulated |

| | | | | |
|---|---|---|---|---|
| 0,398664739 | 0,722019924 | DCDC2B | FALSE | Upregulated |
| 0,8466851 | 0,722019924 | DGKB | FALSE | Upregulated |
| 0,413193072 | 0,722019924 | DRAM1 | FALSE | Upregulated |
| -0,533420525 | 0,722019924 | LOC100289097 | FALSE | Downregulated |
| -0,296823293 | 0,722019924 | POMGNT2 | FALSE | Downregulated |
| 0,769497191 | 0,722019924 | RAB41 | FALSE | Upregulated |
| 0,382602289 | 0,722019924 | SFXN4 | FALSE | Upregulated |
| 0,402734203 | 0,722019924 | WASH1 | FALSE | Upregulated |
| -1,007315524 | 0,722019924 | ZNF815P | FALSE | Downregulated |
| -0,189485658 | 0,722019924 | ZZZ3 | FALSE | Downregulated |
| -0,650039758 | 0,722120969 | XIRP1 | FALSE | Downregulated |
| 0,549651381 | 0,722249996 | CLDN9 | FALSE | Upregulated |
| -0,238272909 | 0,722249996 | RNF114 | FALSE | Downregulated |
| 0,333073992 | 0,722347311 | SH3YL1 | FALSE | Upregulated |
| -0,24166337 | 0,722347311 | SMIM19 | FALSE | Downregulated |
| -0,197735787 | 0,7225627 | GLUD1P3 | FALSE | Downregulated |
| -0,70651704 | 0,722633912 | AC007163.3 | FALSE | Downregulated |
| 0,689298956 | 0,722633912 | AC018742.1 | FALSE | Upregulated |
| -0,583043768 | 0,722633912 | AGPAT9 | FALSE | Downregulated |
| -0,255596824 | 0,722633912 | AOC3 | FALSE | Downregulated |
| 0,130228439 | 0,722633912 | CEP170B | FALSE | Upregulated |
| -0,157831492 | 0,722633912 | CLINT1 | FALSE | Downregulated |
| -0,494486343 | 0,722633912 | GSTA7P | FALSE | Downregulated |
| -0,206584992 | 0,722633912 | GTF2F1 | FALSE | Downregulated |
| -0,400883024 | 0,722633912 | MAGI2-AS3 | FALSE | Downregulated |
| -0,097013456 | 0,722633912 | SEMA4G | FALSE | Downregulated |
| 0,261076167 | 0,722633912 | SPDYA | FALSE | Upregulated |
| 2,986306113 | 0,722633912 | TNNT1 | FALSE | Upregulated |
| -0,142218276 | 0,722681901 | CTSH | FALSE | Downregulated |
| -0,273088892 | 0,722681901 | FBXO22 | FALSE | Downregulated |
| -0,601962129 | 0,722829709 | LOC401127 | FALSE | Downregulated |
| 0,453581643 | 0,722829709 | LRR1 | FALSE | Upregulated |
| -0,144969133 | 0,722829709 | PMPCA | FALSE | Downregulated |
| 0,233957731 | 0,723149575 | C11orf83 | FALSE | Upregulated |
| 0,51495073 | 0,723174272 | LINC00997 | FALSE | Upregulated |
| 0,300257118 | 0,723216746 | TRAF2 | FALSE | Upregulated |
| -0,165152484 | 0,723276325 | CITED2 | FALSE | Downregulated |
| -0,136501706 | 0,723276325 | HYAL1 | FALSE | Downregulated |
| -0,798996328 | 0,723276325 | RRAD | FALSE | Downregulated |
| 0,122436045 | 0,723276325 | UBR5 | FALSE | Upregulated |
| 0,29544063 | 0,723276325 | ZNF343 | FALSE | Upregulated |
| 0,271715719 | 0,723306863 | FARS2 | FALSE | Upregulated |
| 0,413415467 | 0,723306863 | PSMB9 | FALSE | Upregulated |
| -0,200300794 | 0,723337303 | USP1 | FALSE | Downregulated |
| -0,39005733 | 0,72339354 | LOC90784 | FALSE | Downregulated |
| 0,83754782 | 0,723659557 | CYP3A7-CYP3AP1 | FALSE | Upregulated |
| 0,631987743 | 0,723880218 | CAMKMT | FALSE | Upregulated |
| -0,28475716 | 0,723916192 | COX7A2L | FALSE | Downregulated |
| -0,171867356 | 0,723916192 | DROSHA | FALSE | Downregulated |
| 0,163483965 | 0,723916192 | FAM186A | FALSE | Upregulated |
| 0,184978696 | 0,723916192 | FAM69B | FALSE | Upregulated |

| | | | | |
|---|---|---|---|---|
| -0,391017748 | 0,723916192 | GRASP | FALSE | Downregulated |
| 0,201967712 | 0,723916192 | KIF28P | FALSE | Upregulated |
| 0,761320595 | 0,723916192 | LOC101928040 | FALSE | Upregulated |
| 0,300707468 | 0,723916192 | MTHFSD | FALSE | Upregulated |
| 0,147734506 | 0,723916192 | SLC47A1 | FALSE | Upregulated |
| 0,297395344 | 0,723916192 | TBKBP1 | FALSE | Upregulated |
| 0,404494507 | 0,723916192 | TEX9 | FALSE | Upregulated |
| 0,302487793 | 0,723916192 | TRIM11 | FALSE | Upregulated |
| 0,177324937 | 0,723916192 | WWP2 | FALSE | Upregulated |
| -0,298181037 | 0,724003923 | SLC2A8 | FALSE | Downregulated |
| 0,146310218 | 0,724003923 | TMED7 | FALSE | Upregulated |
| 0,423051611 | 0,724171889 | FGF7 | FALSE | Upregulated |
| -0,261725938 | 0,724181958 | ABCC6P1 | FALSE | Downregulated |
| -0,107569911 | 0,724181958 | AKT1 | FALSE | Downregulated |
| -0,184965295 | 0,724181958 | C1QTNF1 | FALSE | Downregulated |
| 0,459310397 | 0,724181958 | SYTL3 | FALSE | Upregulated |
| -0,213248262 | 0,7242582 | ANKZF1 | FALSE | Downregulated |
| 0,308865911 | 0,7242582 | BUD13 | FALSE | Upregulated |
| 0,169563417 | 0,7242582 | CHAD | FALSE | Upregulated |
| -0,291318726 | 0,7242582 | LINC00910 | FALSE | Downregulated |
| 0,517428504 | 0,7242582 | LOC100101148 | FALSE | Upregulated |
| 0,77865844 | 0,7242582 | LOC101928236 | FALSE | Upregulated |
| 0,352291863 | 0,7242582 | OIP5 | FALSE | Upregulated |
| 0,229447254 | 0,7242582 | RP11-538P18.2 | FALSE | Upregulated |
| 0,121435182 | 0,7242582 | UBE2D3 | FALSE | Upregulated |
| -0,329058366 | 0,724358546 | NUDT9 | FALSE | Downregulated |
| 0,322831322 | 0,724648943 | DGCR14 | FALSE | Upregulated |
| -0,740425545 | 0,724648943 | MED27 | FALSE | Downregulated |
| 0,214815499 | 0,724648943 | NUP205 | FALSE | Upregulated |
| -0,20510044 | 0,724648943 | PDZD8 | FALSE | Downregulated |
| -0,213263975 | 0,724648943 | UHRF1BP1L | FALSE | Downregulated |
| 0,115093584 | 0,724772981 | MAGI3 | FALSE | Upregulated |
| -0,266000176 | 0,724772981 | NTAN1 | FALSE | Downregulated |
| -0,501792863 | 0,724785182 | CPE | FALSE | Downregulated |
| 0,536045826 | 0,724785182 | LOC642423 | FALSE | Upregulated |
| 0,326073248 | 0,725101562 | ACOT1 | FALSE | Upregulated |
| 0,649681372 | 0,725101562 | CD38 | FALSE | Upregulated |
| 0,802875192 | 0,725101562 | MARK1 | FALSE | Upregulated |
| 0,209115412 | 0,72519697 | THNSL1 | FALSE | Upregulated |
| -0,109383244 | 0,72520908 | MIA3 | FALSE | Downregulated |
| 0,1439854 | 0,725223998 | AFG3L2 | FALSE | Upregulated |
| 0,81072012 | 0,725292959 | HIST1H2AM | FALSE | Upregulated |
| -0,121733217 | 0,725292959 | PNISR | FALSE | Downregulated |
| 0,687264264 | 0,725537347 | ICOSLG | FALSE | Upregulated |
| -0,403714914 | 0,725960523 | BOLA1 | FALSE | Downregulated |
| -0,242428472 | 0,725960523 | EDNRB | FALSE | Downregulated |
| -0,407787792 | 0,725960523 | ZNF649 | FALSE | Downregulated |
| 0,161556395 | 0,725981498 | CASC3 | FALSE | Upregulated |
| 0,170621859 | 0,726052848 | GTF3C5 | FALSE | Upregulated |
| -1,387251525 | 0,726052848 | LOC101060389 | FALSE | Downregulated |
| -0,702431313 | 0,726052848 | LOC102724475 | FALSE | Downregulated |

| | | | | |
|---|---|---|---|---|
| 0,592175697 | 0,726052848 | RP11-372E1.6 | FALSE | Upregulated |
| -0,234355258 | 0,726052848 | SLC19A3 | FALSE | Downregulated |
| -0,349204044 | 0,726052848 | TARSL2 | FALSE | Downregulated |
| -0,500780957 | 0,726052848 | TCEAL8 | FALSE | Downregulated |
| -0,487856959 | 0,726052848 | TMEM170A | FALSE | Downregulated |
| -0,197751277 | 0,726065768 | CD276 | FALSE | Downregulated |
| -0,303025865 | 0,726065768 | MPHOSPH10 | FALSE | Downregulated |
| 0,349334913 | 0,726088132 | EMID1 | FALSE | Upregulated |
| -0,175203891 | 0,726707505 | HEATR2 | FALSE | Downregulated |
| 0,603361727 | 0,726707505 | LOC101927861 | FALSE | Upregulated |
| 0,261313718 | 0,726707505 | MAML2 | FALSE | Upregulated |
| -0,314631033 | 0,726799997 | SF3B5 | FALSE | Downregulated |
| 0,665345959 | 0,726799997 | UBASH3A | FALSE | Upregulated |
| -0,15667151 | 0,727351253 | ARHGEF7 | FALSE | Downregulated |
| -0,933410832 | 0,727440092 | CCL15 | FALSE | Downregulated |
| 0,424678987 | 0,727440092 | GPATCH2 | FALSE | Upregulated |
| 0,16901266 | 0,727440092 | MTX1 | FALSE | Upregulated |
| -0,300477909 | 0,727509928 | HARS2 | FALSE | Downregulated |
| -0,252670408 | 0,727509928 | SLC39A8 | FALSE | Downregulated |
| 0,385203661 | 0,727723321 | KLHL18 | FALSE | Upregulated |
| 0,196494891 | 0,727765792 | TSC22D4 | FALSE | Upregulated |
| 0,139715949 | 0,727824006 | FGD5-AS1 | FALSE | Upregulated |
| -0,127429153 | 0,727824006 | GAK | FALSE | Downregulated |
| -0,229112866 | 0,727824006 | PLOD2 | FALSE | Downregulated |
| 0,169164013 | 0,727824006 | UFM1 | FALSE | Upregulated |
| 0,222585087 | 0,727938238 | CROT | FALSE | Upregulated |
| 0,741344465 | 0,728146273 | HIST2H2AA4 | FALSE | Upregulated |
| -0,454403332 | 0,728146273 | LOC440390 | FALSE | Downregulated |
| -0,950812141 | 0,728146273 | PCDH20 | FALSE | Downregulated |
| -0,131711635 | 0,728164767 | FAM193B | FALSE | Downregulated |
| -0,757204457 | 0,728294894 | C3orf67 | FALSE | Downregulated |
| 0,412523813 | 0,728294894 | MSC | FALSE | Upregulated |
| 0,174169278 | 0,728408663 | BMP8B | FALSE | Upregulated |
| -0,982113261 | 0,728551083 | LOC102723580 | FALSE | Downregulated |
| 0,462942928 | 0,728551083 | TM4SF18 | FALSE | Upregulated |
| -0,512910454 | 0,728551083 | ZNF283 | FALSE | Downregulated |
| 0,186309931 | 0,728612072 | NUP210 | FALSE | Upregulated |
| -0,467517503 | 0,728612072 | SPDYE4 | FALSE | Downregulated |
| -0,470091797 | 0,728672646 | ZDHHC11B | FALSE | Downregulated |
| -0,135393497 | 0,7287772 | LINC01000 | FALSE | Downregulated |
| -0,1519863 | 0,728790676 | ERC1 | FALSE | Downregulated |
| 0,11279542 | 0,728790676 | SEC31A | FALSE | Upregulated |
| 0,330195082 | 0,729062616 | DDX11L2 | FALSE | Upregulated |
| -0,471153207 | 0,729087928 | SLC36A4 | FALSE | Downregulated |
| -0,206915688 | 0,729154997 | SCARNA7 | FALSE | Downregulated |
| 0,167673295 | 0,72917645 | ZNF142 | FALSE | Upregulated |
| -0,190600513 | 0,729195523 | NAA15 | FALSE | Downregulated |
| 2,589495673 | 0,729341425 | CACNA1S | FALSE | Upregulated |
| -0,598000024 | 0,729405115 | HIST4H4 | FALSE | Downregulated |
| 0,147676198 | 0,729405115 | WDR82 | FALSE | Upregulated |
| -0,49776756 | 0,729479221 | TAL1 | FALSE | Downregulated |

| | | | | |
|---|---|---|---|---|
| -0,141082244 | 0,729702358 | ARVCF | FALSE | Downregulated |
| 0,141733966 | 0,729702358 | CLCN7 | FALSE | Upregulated |
| 0,238020062 | 0,729702358 | DTX2 | FALSE | Upregulated |
| 0,538241327 | 0,729702358 | LOC101929433 | FALSE | Upregulated |
| 0,236459972 | 0,729702358 | PER1 | FALSE | Upregulated |
| -0,420810435 | 0,729702358 | SRL | FALSE | Downregulated |
| -0,243985681 | 0,72992428 | CDC5L | FALSE | Downregulated |
| -0,179250387 | 0,72992428 | GALT | FALSE | Downregulated |
| -0,424090729 | 0,72992428 | NUDT19 | FALSE | Downregulated |
| -0,280572391 | 0,73006089 | CBR4 | FALSE | Downregulated |
| -0,7482154 | 0,73006089 | LOC101929621 | FALSE | Downregulated |
| -0,281896121 | 0,73006089 | NDUFS4 | FALSE | Downregulated |
| -0,211601101 | 0,73006089 | SLC25A39 | FALSE | Downregulated |
| 0,688948759 | 0,73006089 | TOP2A | FALSE | Upregulated |
| 0,568483451 | 0,730095126 | LOC728290 | FALSE | Upregulated |
| 0,218974331 | 0,730112363 | ZNF483 | FALSE | Upregulated |
| 0,49757018 | 0,730275158 | SLC34A3 | FALSE | Upregulated |
| 0,176930533 | 0,73029537 | LOC101928660 | FALSE | Upregulated |
| -0,221131478 | 0,730316986 | AARS2 | FALSE | Downregulated |
| 0,470316079 | 0,730316986 | LINC01138 | FALSE | Upregulated |
| 0,379155514 | 0,730392872 | BMP2 | FALSE | Upregulated |
| 0,155872573 | 0,730449804 | ANXA11 | FALSE | Upregulated |
| 0,245099139 | 0,730478817 | MPZL1 | FALSE | Upregulated |
| -0,168405516 | 0,730478817 | SMPD1 | FALSE | Downregulated |
| -0,462112924 | 0,730535162 | MRPL13 | FALSE | Downregulated |
| 1,007322818 | 0,730535162 | RIMBP3 | FALSE | Upregulated |
| -0,174274834 | 0,730563721 | GSK3B | FALSE | Downregulated |
| -0,302366741 | 0,730735107 | EBF3 | FALSE | Downregulated |
| -0,129952996 | 0,730805585 | EXOC7 | FALSE | Downregulated |
| 0,201482239 | 0,73102888 | AP3B1 | FALSE | Upregulated |
| 0,670160691 | 0,73102888 | C9orf129 | FALSE | Upregulated |
| -0,181815164 | 0,73102888 | CPEB2 | FALSE | Downregulated |
| 0,211957385 | 0,73102888 | LOC101930137 | FALSE | Upregulated |
| 0,593594196 | 0,73102888 | NEB | FALSE | Upregulated |
| -0,210069862 | 0,73102888 | NODAL | FALSE | Downregulated |
| -0,325075484 | 0,73102888 | RASIP1 | FALSE | Downregulated |
| -0,117687904 | 0,73102888 | RBM47 | FALSE | Downregulated |
| -0,164605233 | 0,73102888 | TAB2 | FALSE | Downregulated |
| -0,511526292 | 0,731103121 | ZBTB8OS | FALSE | Downregulated |
| 0,226469422 | 0,731175551 | CCDC157 | FALSE | Upregulated |
| 0,178795588 | 0,73142397 | DMWD | FALSE | Upregulated |
| 0,246784245 | 0,73146857 | ABCB7 | FALSE | Upregulated |
| -0,848114458 | 0,73146857 | FAM231A | FALSE | Downregulated |
| 0,57800504 | 0,73146857 | LOC101929309 | FALSE | Upregulated |
| 0,511878461 | 0,73146857 | OAZ3 | FALSE | Upregulated |
| 0,408044193 | 0,73146857 | UXS1 | FALSE | Upregulated |
| 0,386072782 | 0,73146857 | ZNF423 | FALSE | Upregulated |
| -0,154592338 | 0,731816656 | TSPAN14 | FALSE | Downregulated |
| -0,312179342 | 0,73184448 | C19orf47 | FALSE | Downregulated |
| -0,576379754 | 0,73184448 | DHRS2 | FALSE | Downregulated |
| -0,418614981 | 0,73184448 | LOC100996763 | FALSE | Downregulated |

| | | | | |
|---|---|---|---|---|
| -0,107275409 | 0,73184448 | MAN2B2 | FALSE | Downregulated |
| -0,142838846 | 0,73184448 | RAB5B | FALSE | Downregulated |
| 0,715862509 | 0,73184448 | SGCA | FALSE | Upregulated |
| 0,444468093 | 0,73184448 | TTC30A | FALSE | Upregulated |
| -0,195256363 | 0,73184448 | UBFD1 | FALSE | Downregulated |
| 0,192892283 | 0,731890264 | C19orf24 | FALSE | Upregulated |
| 0,289300579 | 0,731890264 | LOC101926943 | FALSE | Upregulated |
| -0,484954715 | 0,731891592 | ASAH2B | FALSE | Downregulated |
| 0,260933742 | 0,731891592 | INTS9 | FALSE | Upregulated |
| 0,667136292 | 0,73189185 | FPR1 | FALSE | Upregulated |
| -0,256801012 | 0,731969352 | DEPTOR | FALSE | Downregulated |
| -0,180785894 | 0,731969352 | KARS | FALSE | Downregulated |
| -0,31756619 | 0,732135251 | B4GALT4 | FALSE | Downregulated |
| 0,173258253 | 0,732176433 | DOK4 | FALSE | Upregulated |
| -0,161879747 | 0,732177897 | KIF21A | FALSE | Downregulated |
| -0,773612827 | 0,732177897 | LOC101927595 | FALSE | Downregulated |
| 0,348959717 | 0,732177897 | TTF1 | FALSE | Upregulated |
| 0,204285497 | 0,732291891 | ABHD1 | FALSE | Upregulated |
| 0,193956428 | 0,732291891 | CCDC71L | FALSE | Upregulated |
| -0,197514468 | 0,732291891 | CEBPD | FALSE | Downregulated |
| 0,554525003 | 0,732291891 | CMC1 | FALSE | Upregulated |
| -0,278732389 | 0,732291891 | GLIS3 | FALSE | Downregulated |
| 0,162069408 | 0,732325879 | FLII | FALSE | Upregulated |
| -0,392046265 | 0,732325879 | GINS2 | FALSE | Downregulated |
| 0,115004821 | 0,732325879 | MYO5B | FALSE | Upregulated |
| -0,517124336 | 0,732325879 | POLE2 | FALSE | Downregulated |
| 0,350279436 | 0,732325879 | PTCD2 | FALSE | Upregulated |
| 0,228862178 | 0,73250392 | MIR22HG | FALSE | Upregulated |
| 0,19846642 | 0,73250392 | MTMR1 | FALSE | Upregulated |
| 0,412948673 | 0,73250392 | OPHN1 | FALSE | Upregulated |
| 0,363988999 | 0,73250392 | PUSL1 | FALSE | Upregulated |
| 0,841200818 | 0,73250392 | SMN2 | FALSE | Upregulated |
| -0,154359546 | 0,73250392 | TTC31 | FALSE | Downregulated |
| -0,13945715 | 0,73268787 | MGLL | FALSE | Downregulated |
| -0,393176322 | 0,732730134 | CUTC | FALSE | Downregulated |
| 0,392649757 | 0,732849483 | C6orf132 | FALSE | Upregulated |
| -0,589714528 | 0,732849483 | C9orf152 | FALSE | Downregulated |
| -0,343579874 | 0,732849483 | CCDC28A | FALSE | Downregulated |
| 0,266953789 | 0,732849483 | CLEC12A | FALSE | Upregulated |
| -0,189425421 | 0,732849483 | IARS | FALSE | Downregulated |
| 0,268956011 | 0,732849483 | LINC00342 | FALSE | Upregulated |
| -0,421658116 | 0,732849483 | TAF1D | FALSE | Downregulated |
| 0,234503816 | 0,732849483 | UBE2M | FALSE | Upregulated |
| 0,149202035 | 0,733115574 | ZBTB4 | FALSE | Upregulated |
| -0,114501931 | 0,733205942 | CAST | FALSE | Downregulated |
| -0,373369072 | 0,733303373 | WEE2 | FALSE | Downregulated |
| -0,221614178 | 0,733414103 | C22orf39 | FALSE | Downregulated |
| -0,190763741 | 0,733414103 | ELF2 | FALSE | Downregulated |
| 0,347505134 | 0,733414103 | GMDS | FALSE | Upregulated |
| -0,856081388 | 0,733414103 | MT1L | FALSE | Downregulated |
| 0,255507438 | 0,733414103 | PRPF38B | FALSE | Upregulated |

| | | | | |
|---|---|---|---|---|
| 0,401120877 | 0,733414103 | TTC39A | FALSE | Upregulated |
| -0,16948108 | 0,733414103 | USP4 | FALSE | Downregulated |
| 0,121604607 | 0,733485308 | BRWD1 | FALSE | Upregulated |
| -0,49005642 | 0,733485308 | GYG1 | FALSE | Downregulated |
| 0,239757195 | 0,733485308 | PUS1 | FALSE | Upregulated |
| 0,223573804 | 0,733489831 | AP1AR | FALSE | Upregulated |
| -0,267828002 | 0,733636971 | HDAC8 | FALSE | Downregulated |
| 0,76388847 | 0,733641796 | CMTM1 | FALSE | Upregulated |
| -0,223958895 | 0,7340789 | LOC102724975 | FALSE | Downregulated |
| 0,149323918 | 0,734198924 | DENND2D | FALSE | Upregulated |
| -0,406204798 | 0,734198924 | MAGIX | FALSE | Downregulated |
| -0,352805191 | 0,734198924 | METTL22 | FALSE | Downregulated |
| 0,644888821 | 0,734198924 | OR1K1 | FALSE | Upregulated |
| 0,451987385 | 0,734198924 | PLCD3 | FALSE | Upregulated |
| 0,190642515 | 0,734264402 | CBFA2T2 | FALSE | Upregulated |
| 0,410778484 | 0,734470171 | CDK7 | FALSE | Upregulated |
| 0,543420098 | 0,734470171 | GBGT1 | FALSE | Upregulated |
| 1,106831061 | 0,734470171 | KIAA0408 | FALSE | Upregulated |
| 0,502569533 | 0,734470171 | VWA5B2 | FALSE | Upregulated |
| 0,27115463 | 0,734503301 | METTL23 | FALSE | Upregulated |
| 0,763719067 | 0,734568364 | GDF6 | FALSE | Upregulated |
| -0,493937059 | 0,734662133 | C7orf63 | FALSE | Downregulated |
| 0,4746427 | 0,734844002 | TSEN15 | FALSE | Upregulated |
| 0,223596042 | 0,734878186 | DMTN | FALSE | Upregulated |
| -0,154932826 | 0,734987705 | ARL6IP1 | FALSE | Downregulated |
| 0,841343636 | 0,735306045 | AJAP1 | FALSE | Upregulated |
| 0,160030473 | 0,735306045 | COG1 | FALSE | Upregulated |
| 0,349354778 | 0,735306045 | GTDC1 | FALSE | Upregulated |
| -0,255902614 | 0,735306045 | HIST2H2BE | FALSE | Downregulated |
| 0,165929307 | 0,735306045 | HSPA8 | FALSE | Upregulated |
| -0,770907097 | 0,735306045 | LOC101927536 | FALSE | Downregulated |
| 0,588717505 | 0,735306045 | LOC102724293 | FALSE | Upregulated |
| 0,317101562 | 0,735306045 | LOC102724784 | FALSE | Upregulated |
| 0,517673028 | 0,735306045 | MUC15 | FALSE | Upregulated |
| 0,187260828 | 0,735306045 | NSFL1C | FALSE | Upregulated |
| 0,716655588 | 0,735306045 | PRR7 | FALSE | Upregulated |
| -0,579442398 | 0,735306045 | SLC7A10 | FALSE | Downregulated |
| 0,205508432 | 0,735306045 | TUBB2A | FALSE | Upregulated |
| -0,226655661 | 0,735306045 | VTA1 | FALSE | Downregulated |
| -0,316008762 | 0,735306045 | ZNF766 | FALSE | Downregulated |
| -2,778400756 | 0,735398387 | LOC102724117 | FALSE | Downregulated |
| 0,153388843 | 0,735657929 | DHX9 | FALSE | Upregulated |
| -0,511805462 | 0,736046958 | CD247 | FALSE | Downregulated |
| 0,339539372 | 0,736046958 | ZNF117 | FALSE | Upregulated |
| -0,461329713 | 0,736124518 | LINC00987 | FALSE | Downregulated |
| 0,324498711 | 0,736124518 | LOC100128364 | FALSE | Upregulated |
| 0,379055298 | 0,736124518 | RP11-43N5.1 | FALSE | Upregulated |
| 0,370649887 | 0,736124518 | ZNF569 | FALSE | Upregulated |
| 0,1520454 | 0,736146044 | LSM14B | FALSE | Upregulated |
| 0,176117978 | 0,736259718 | TARS | FALSE | Upregulated |
| -0,134879961 | 0,736410051 | ZNF207 | FALSE | Downregulated |

| | | | | |
|---|---|---|---|---|
| 0,336837142 | 0,736696049 | NR2F1-AS1 | FALSE | Upregulated |
| -0,298167302 | 0,737499813 | SNRNP48 | FALSE | Downregulated |
| -0,193105856 | 0,73753466 | TNFSF10 | FALSE | Downregulated |
| 0,301255867 | 0,737742313 | AAR2 | FALSE | Upregulated |
| -0,250994303 | 0,737742313 | CYP39A1 | FALSE | Downregulated |
| 0,215969093 | 0,737742313 | HLA-L | FALSE | Upregulated |
| 0,340171843 | 0,737742313 | NMI | FALSE | Upregulated |
| -0,322170332 | 0,737742313 | PLLP | FALSE | Downregulated |
| 0,317733892 | 0,737753586 | LOC101060553 | FALSE | Upregulated |
| 0,154867721 | 0,737753586 | SYNJ2 | FALSE | Upregulated |
| 0,223179181 | 0,737759273 | NDST3 | FALSE | Upregulated |
| 0,280681167 | 0,737839658 | BROX | FALSE | Upregulated |
| -0,165689195 | 0,737839658 | PRPF4B | FALSE | Downregulated |
| -0,219032603 | 0,737839658 | RPS13 | FALSE | Downregulated |
| 0,714937308 | 0,737839658 | SLAMF1 | FALSE | Upregulated |
| 0,20130862 | 0,737850129 | ARL5A | FALSE | Upregulated |
| -0,317472531 | 0,737892809 | PRR18 | FALSE | Downregulated |
| -0,185847429 | 0,737892809 | STAR | FALSE | Downregulated |
| 0,180261074 | 0,738091658 | DNAJB7 | FALSE | Upregulated |
| -0,284830362 | 0,738091658 | DYNLRB1 | FALSE | Downregulated |
| 0,711584781 | 0,738091658 | FLJ45872 | FALSE | Upregulated |
| -0,310076254 | 0,738093653 | C8orf33 | FALSE | Downregulated |
| -0,267446444 | 0,738093653 | ICK | FALSE | Downregulated |
| 0,142648393 | 0,738153827 | CAMKK2 | FALSE | Upregulated |
| 0,21944146 | 0,73833619 | ATP6V0D1 | FALSE | Upregulated |
| 0,172563582 | 0,738451985 | PLOD3 | FALSE | Upregulated |
| 0,369201099 | 0,73850245 | CORT | FALSE | Upregulated |
| -0,489039487 | 0,73850245 | GFRA2 | FALSE | Downregulated |
| 0,160128695 | 0,73850245 | GLIS2 | FALSE | Upregulated |
| -1,075773803 | 0,73850245 | LOC647859 | FALSE | Downregulated |
| 0,281811561 | 0,738775797 | ATXN7L1 | FALSE | Upregulated |
| -0,178802481 | 0,738775797 | DHX15 | FALSE | Downregulated |
| -0,20035051 | 0,738775797 | DIAPH2 | FALSE | Downregulated |
| 0,267937599 | 0,738775797 | ERH | FALSE | Upregulated |
| 0,61110716 | 0,738775797 | FAM83D | FALSE | Upregulated |
| -0,22804973 | 0,738775797 | LDB2 | FALSE | Downregulated |
| -0,230400874 | 0,738775797 | TMUB2 | FALSE | Downregulated |
| -0,099571 | 0,738890559 | STK11 | FALSE | Downregulated |
| 0,301157159 | 0,738996661 | FAM180A | FALSE | Upregulated |
| 0,264858304 | 0,73902683 | ANKRD46 | FALSE | Upregulated |
| 0,445079197 | 0,739535145 | ARL4A | FALSE | Upregulated |
| -0,220113773 | 0,739679748 | FLAD1 | FALSE | Downregulated |
| 0,190994886 | 0,739783695 | ACVR1 | FALSE | Upregulated |
| -0,251551622 | 0,740012707 | DCBLD1 | FALSE | Downregulated |
| 0,174869766 | 0,740098427 | MAP1S | FALSE | Upregulated |
| -0,165852417 | 0,740221725 | DHRS1 | FALSE | Downregulated |
| -0,394159467 | 0,740221725 | LOC399900 | FALSE | Downregulated |
| -0,108689926 | 0,740221725 | TNFAIP1 | FALSE | Downregulated |
| -0,197213605 | 0,740221725 | TOMM40 | FALSE | Downregulated |
| 0,262959347 | 0,740286099 | CXorf38 | FALSE | Upregulated |
| -0,159050245 | 0,740312438 | CCDC152 | FALSE | Downregulated |

| | | | | |
|---|---|---|---|---|
| 0,30081002 | 0,740582852 | HMCN2 | FALSE | Upregulated |
| 0,244255094 | 0,740685466 | NUMBL | FALSE | Upregulated |
| 0,217053942 | 0,740685466 | RP11-4F22.2 | FALSE | Upregulated |
| -0,165612898 | 0,741191804 | SECISBP2 | FALSE | Downregulated |
| -0,26655371 | 0,741211917 | LOC101929722 | FALSE | Downregulated |
| -0,442670912 | 0,741211917 | STYXL1 | FALSE | Downregulated |
| -0,171241617 | 0,742296521 | HPN-AS1 | FALSE | Downregulated |
| -0,170983334 | 0,742296521 | TSHR | FALSE | Downregulated |
| -0,381043328 | 0,742449006 | BRE | FALSE | Downregulated |
| 0,291040851 | 0,742483681 | CCDC158 | FALSE | Upregulated |
| -0,151775264 | 0,742620111 | KPNA6 | FALSE | Downregulated |
| 0,15596529 | 0,742620111 | LIG3 | FALSE | Upregulated |
| -0,23583817 | 0,742647689 | GPATCH2L | FALSE | Downregulated |
| -0,22193157 | 0,742747283 | STAG3L2 | FALSE | Downregulated |
| 0,616222715 | 0,742821268 | C14orf166B | FALSE | Upregulated |
| 0,724303125 | 0,742821268 | DNAJC18 | FALSE | Upregulated |
| 0,867827081 | 0,742848156 | KCNK13 | FALSE | Upregulated |
| 0,295901344 | 0,742848156 | LOC101927151 | FALSE | Upregulated |
| 0,930963888 | 0,742848156 | SOX9-AS1 | FALSE | Upregulated |
| -0,749279621 | 0,742901905 | CCDC34 | FALSE | Downregulated |
| -0,221799765 | 0,742995889 | AHCYL2 | FALSE | Downregulated |
| 0,128172896 | 0,743244266 | CMTM6 | FALSE | Upregulated |
| -0,249103867 | 0,743244266 | DDX60L | FALSE | Downregulated |
| -0,187369117 | 0,743244266 | L3MBTL2 | FALSE | Downregulated |
| 0,221740402 | 0,743244266 | MAPK8 | FALSE | Upregulated |
| -0,273999435 | 0,743244266 | NAV2-AS4 | FALSE | Downregulated |
| 0,316026128 | 0,743346735 | SPATA2 | FALSE | Upregulated |
| -0,482490986 | 0,743489368 | C14orf142 | FALSE | Downregulated |
| 0,116613123 | 0,743489368 | MAML1 | FALSE | Upregulated |
| 0,159398045 | 0,743889129 | FBXO38 | FALSE | Upregulated |
| -0,261645703 | 0,743889129 | WDR74 | FALSE | Downregulated |
| 0,204645301 | 0,743993394 | IL16 | FALSE | Upregulated |
| -0,177144402 | 0,744005698 | YTHDC1 | FALSE | Downregulated |
| 0,482533265 | 0,744089826 | CLEC5A | FALSE | Upregulated |
| -0,213214188 | 0,744089826 | MAP4K3 | FALSE | Downregulated |
| -0,18828242 | 0,744206613 | DOCK6 | FALSE | Downregulated |
| -0,564702566 | 0,744275554 | LINC00939 | FALSE | Downregulated |
| 0,421825856 | 0,744359635 | CNTNAP2 | FALSE | Upregulated |
| 0,203236968 | 0,744359635 | EPHA2 | FALSE | Upregulated |
| -0,381514244 | 0,744359635 | LOC644794 | FALSE | Downregulated |
| 0,772702357 | 0,744772542 | IL17REL | FALSE | Upregulated |
| -0,482856155 | 0,744805495 | LRRC34 | FALSE | Downregulated |
| -0,300161653 | 0,745023531 | NAT8B | FALSE | Downregulated |
| 0,109652602 | 0,745023531 | NPLOC4 | FALSE | Upregulated |
| 0,440751546 | 0,745230798 | CCDC127 | FALSE | Upregulated |
| 0,232606493 | 0,745238836 | ANKRD37 | FALSE | Upregulated |
| -0,44610686 | 0,745280502 | ADM | FALSE | Downregulated |
| 0,521031986 | 0,745381993 | LOC102723583 | FALSE | Upregulated |
| -0,215019981 | 0,745381993 | PEX14 | FALSE | Downregulated |
| 0,370331367 | 0,745381993 | SOCS2 | FALSE | Upregulated |
| -0,389292048 | 0,745440213 | PGBD2 | FALSE | Downregulated |

| | | | | |
|---|---|---|---|---|
| 0,801248128 | 0,745469346 | LOC101929141 | FALSE | Upregulated |
| 0,181440462 | 0,745469346 | LOC102723743 | FALSE | Upregulated |
| -0,15013079 | 0,746110686 | DCST2 | FALSE | Downregulated |
| 0,43350374 | 0,746110686 | LOC100506023 | FALSE | Upregulated |
| -0,346394876 | 0,746110686 | PNMA6A | FALSE | Downregulated |
| 0,182320436 | 0,746110686 | RNF111 | FALSE | Upregulated |
| 0,35527156 | 0,746110686 | ZNF205-AS1 | FALSE | Upregulated |
| 0,357812141 | 0,746281848 | DRD5 | FALSE | Upregulated |
| -0,210189173 | 0,746285252 | PIGT | FALSE | Downregulated |
| 0,775489431 | 0,746451173 | GRAPL | FALSE | Upregulated |
| 0,304943059 | 0,746451173 | RNF115 | FALSE | Upregulated |
| 0,28696812 | 0,746451173 | ZNF717 | FALSE | Upregulated |
| 0,380052243 | 0,746474524 | C15orf57 | FALSE | Upregulated |
| -0,30251674 | 0,746474524 | CNOT10 | FALSE | Downregulated |
| -0,142754754 | 0,746474524 | GRTP1 | FALSE | Downregulated |
| -0,231750546 | 0,746474524 | HEATR5A | FALSE | Downregulated |
| 0,232845918 | 0,746474524 | HTR6 | FALSE | Upregulated |
| 0,408994742 | 0,746474524 | LOC101927163 | FALSE | Upregulated |
| -0,398204736 | 0,746474524 | LOC101929065 | FALSE | Downregulated |
| 0,304823547 | 0,746474524 | MAP7D3 | FALSE | Upregulated |
| 0,17736219 | 0,746474524 | NFYA | FALSE | Upregulated |
| 0,193845767 | 0,746474524 | NFYC-AS1 | FALSE | Upregulated |
| 0,136498106 | 0,746474524 | PRR14L | FALSE | Upregulated |
| 0,132410766 | 0,746474524 | PTPN11 | FALSE | Upregulated |
| 0,270125024 | 0,746474524 | RAP1GAP | FALSE | Upregulated |
| 0,290292517 | 0,746474524 | TBPL1 | FALSE | Upregulated |
| -0,201915943 | 0,746474524 | WDR90 | FALSE | Downregulated |
| 0,180719235 | 0,746688956 | ENTPD4 | FALSE | Upregulated |
| 0,173744436 | 0,746688956 | HCG18 | FALSE | Upregulated |
| 0,332006322 | 0,746841962 | DICER1-AS1 | FALSE | Upregulated |
| 0,21714459 | 0,746841962 | LOC102724724 | FALSE | Upregulated |
| -0,294397204 | 0,746841962 | TCTN3 | FALSE | Downregulated |
| -0,167450702 | 0,746848829 | ARHGAP1 | FALSE | Downregulated |
| 0,4065597 | 0,746848829 | ZNF17 | FALSE | Upregulated |
| 0,37413203 | 0,74708725 | CD55 | FALSE | Upregulated |
| 0,13421574 | 0,747257168 | AGO1 | FALSE | Upregulated |
| -0,223891991 | 0,747257168 | MTAP | FALSE | Downregulated |
| 0,207016885 | 0,747257168 | SF3A3 | FALSE | Upregulated |
| -0,347954079 | 0,74738131 | MRPL42 | FALSE | Downregulated |
| 0,195771769 | 0,747384645 | RRAS2 | FALSE | Upregulated |
| 0,16655662 | 0,747384645 | STARD8 | FALSE | Upregulated |
| -0,700889663 | 0,747415104 | FAM3D | FALSE | Downregulated |
| 0,253112381 | 0,747710732 | QSOX2 | FALSE | Upregulated |
| 0,22159054 | 0,747710732 | TNIP2 | FALSE | Upregulated |
| -0,216779737 | 0,747931191 | LPAL2 | FALSE | Downregulated |
| -0,221325873 | 0,747935334 | PHF14 | FALSE | Downregulated |
| 0,187690611 | 0,748019868 | NOL6 | FALSE | Upregulated |
| 0,234652236 | 0,748207261 | MID1IP1 | FALSE | Upregulated |
| 0,153317281 | 0,748207261 | SLC29A2 | FALSE | Upregulated |
| -0,70756146 | 0,748516304 | PDE9A | FALSE | Downregulated |
| 0,54981309 | 0,748534813 | PPP1R1B | FALSE | Upregulated |

| | | | | |
|---|---|---|---|---|
| 0,323661492 | 0,748654271 | CAV1 | FALSE | Upregulated |
| 0,341442448 | 0,748654271 | COMMD4 | FALSE | Upregulated |
| 0,150092097 | 0,748654271 | PSPC1 | FALSE | Upregulated |
| 0,137897154 | 0,748774888 | LOC101929469 | FALSE | Upregulated |
| -0,550914224 | 0,748774888 | RP11-680F20.6 | FALSE | Downregulated |
| -0,309580004 | 0,748774888 | ZNF350 | FALSE | Downregulated |
| -0,17687297 | 0,748891035 | EHMT2 | FALSE | Downregulated |
| 0,37793572 | 0,748891035 | RDH5 | FALSE | Upregulated |
| 0,416851178 | 0,748939596 | C1orf21 | FALSE | Upregulated |
| 0,277233317 | 0,748945643 | INCENP | FALSE | Upregulated |
| 0,447853911 | 0,748945643 | RNF219 | FALSE | Upregulated |
| 0,121404787 | 0,748945643 | SRSF2 | FALSE | Upregulated |
| -0,292604929 | 0,749077893 | ANP32E | FALSE | Downregulated |
| -0,641900595 | 0,749077893 | RNASEH1-AS1 | FALSE | Downregulated |
| -0,797807073 | 0,749215943 | MYH16 | FALSE | Downregulated |
| 0,254031167 | 0,749235981 | IPP | FALSE | Upregulated |
| 0,215628532 | 0,749260785 | NUDT16L1 | FALSE | Upregulated |
| -0,120571114 | 0,749389503 | APEH | FALSE | Downregulated |
| -1,538723858 | 0,749399375 | C21orf140 | FALSE | Downregulated |
| -0,407428144 | 0,749399375 | NKAP | FALSE | Downregulated |
| 0,186262313 | 0,749399375 | PELP1 | FALSE | Upregulated |
| 0,305064929 | 0,749399375 | PRICKLE2 | FALSE | Upregulated |
| -0,142370736 | 0,749399375 | SARS | FALSE | Downregulated |
| -0,200070587 | 0,749644683 | ANP32B | FALSE | Downregulated |
| 0,185925767 | 0,749644683 | CDRT15P1 | FALSE | Upregulated |
| -0,350809348 | 0,749644683 | DNAJC15 | FALSE | Downregulated |
| -0,475906345 | 0,749644683 | FRS3 | FALSE | Downregulated |
| -0,209464346 | 0,749644683 | LTA4H | FALSE | Downregulated |
| 2,505658124 | 0,749644683 | MYL2 | FALSE | Upregulated |
| -0,229420703 | 0,749644683 | NDUFA1 | FALSE | Downregulated |
| -0,417080559 | 0,749644683 | SNORA7A | FALSE | Downregulated |
| -0,150061137 | 0,749644683 | ZNF318 | FALSE | Downregulated |
| -0,202797865 | 0,749816191 | CYP51A1 | FALSE | Downregulated |
| -0,392513936 | 0,749816191 | LYSMD2 | FALSE | Downregulated |
| 3,111425518 | 0,749859432 | KRT5 | FALSE | Upregulated |
| -0,246210499 | 0,749935372 | TMEM167B | FALSE | Downregulated |
| 0,375039718 | 0,749935372 | UPRT | FALSE | Upregulated |
| -0,197111199 | 0,750019943 | RNF128 | FALSE | Downregulated |
| -0,792652176 | 0,750019943 | UGT3A2 | FALSE | Downregulated |
| 0,594514705 | 0,750063842 | ADAMTSL4-AS1 | FALSE | Upregulated |
| -0,192990211 | 0,750063842 | AGO4 | FALSE | Downregulated |
| 0,267201939 | 0,750063842 | LOC102724634 | FALSE | Upregulated |
| 0,333478717 | 0,750063842 | PRIMPOL | FALSE | Upregulated |
| -0,164361339 | 0,750107408 | COG4 | FALSE | Downregulated |
| -0,202613944 | 0,750107408 | GOSR2 | FALSE | Downregulated |
| -0,420117144 | 0,750107408 | MEAF6 | FALSE | Downregulated |
| 0,353321692 | 0,750182501 | ZNF32-AS2 | FALSE | Upregulated |
| 0,201155115 | 0,750333485 | C20orf24 | FALSE | Upregulated |
| 0,269773947 | 0,750333485 | CD36 | FALSE | Upregulated |
| -0,713889254 | 0,750333485 | LINC00311 | FALSE | Downregulated |
| -0,231714337 | 0,750333485 | RGP1 | FALSE | Downregulated |

| | | | | |
|---|---|---|---|---|
| -0,344749843 | 0,750333485 | RMDN2 | FALSE | Downregulated |
| 0,488635714 | 0,750333485 | SIRPB2 | FALSE | Upregulated |
| 0,143048029 | 0,750333485 | VHL | FALSE | Upregulated |
| -0,300735014 | 0,750433537 | CCL23 | FALSE | Downregulated |
| 0,432158199 | 0,750445548 | ETV2 | FALSE | Upregulated |
| -0,582463285 | 0,750445548 | RASL10B | FALSE | Downregulated |
| 0,248787062 | 0,750445548 | TMX1 | FALSE | Upregulated |
| 0,260883947 | 0,750451874 | ANKRD34A | FALSE | Upregulated |
| -0,198388468 | 0,750451874 | FSCN1 | FALSE | Downregulated |
| 0,978152285 | 0,750451874 | MYBL2 | FALSE | Upregulated |
| -0,202349285 | 0,750451874 | POLR1B | FALSE | Downregulated |
| 0,418383667 | 0,750559594 | HSD17B7 | FALSE | Upregulated |
| -0,127368742 | 0,750559594 | KANK2 | FALSE | Downregulated |
| -0,342799239 | 0,750559594 | SP7 | FALSE | Downregulated |
| 0,875543583 | 0,75108202 | RGS9 | FALSE | Upregulated |
| 0,510731519 | 0,751125684 | ZNF682 | FALSE | Upregulated |
| -0,211901233 | 0,751175027 | MAT2B | FALSE | Downregulated |
| 0,277677836 | 0,751187887 | ITPRIP | FALSE | Upregulated |
| -0,226909976 | 0,751187887 | ZMAT1 | FALSE | Downregulated |
| 0,248180373 | 0,751221698 | HLA-J | FALSE | Upregulated |
| -0,124329281 | 0,751221698 | MAP4K4 | FALSE | Downregulated |
| 0,336468661 | 0,751372292 | CCDC167 | FALSE | Upregulated |
| 0,42977497 | 0,751372292 | LOC101929673 | FALSE | Upregulated |
| -0,591005451 | 0,751372292 | LOC102723779 | FALSE | Downregulated |
| 1,717411983 | 0,751372292 | LOC102724398 | FALSE | Upregulated |
| 0,329310711 | 0,751372292 | ZFAND1 | FALSE | Upregulated |
| -0,618753067 | 0,751372292 | ZNF630 | FALSE | Downregulated |
| 0,717964373 | 0,751787008 | LOC440434 | FALSE | Upregulated |
| 0,13960124 | 0,751787008 | N4BP2L2 | FALSE | Upregulated |
| 0,733384009 | 0,751789637 | AC007551.3 | FALSE | Upregulated |
| -0,104527678 | 0,751789637 | PUM2 | FALSE | Downregulated |
| 0,731872436 | 0,751823513 | RFC4 | FALSE | Upregulated |
| -0,161399556 | 0,75186205 | CDK9 | FALSE | Downregulated |
| 0,173987084 | 0,75186205 | SMC3 | FALSE | Upregulated |
| -0,109465471 | 0,751883415 | ZC3H4 | FALSE | Downregulated |
| 0,18126602 | 0,752325951 | HEXA | FALSE | Upregulated |
| 0,134690443 | 0,752431011 | POM121C | FALSE | Upregulated |
| 0,122766124 | 0,752475436 | NPEPPS | FALSE | Upregulated |
| -0,241977436 | 0,752475436 | WDR60 | FALSE | Downregulated |
| -1,310974336 | 0,752491258 | GBAP1 | FALSE | Downregulated |
| -0,540969155 | 0,752720897 | USP32P1 | FALSE | Downregulated |
| 0,1459108 | 0,752726818 | LOC100505564 | FALSE | Upregulated |
| 0,197469063 | 0,752726818 | RAB3GAP1 | FALSE | Upregulated |
| -0,244223198 | 0,752726818 | SYT7 | FALSE | Downregulated |
| -0,3224475 | 0,752726818 | WHAMMP2 | FALSE | Downregulated |
| 0,809894866 | 0,752847528 | KRT19P2 | FALSE | Upregulated |
| -0,727244952 | 0,752856372 | LOC102723671 | FALSE | Downregulated |
| 0,17693073 | 0,752856372 | SMAD5-AS1 | FALSE | Upregulated |
| -0,120978684 | 0,752856524 | MAP2K2 | FALSE | Downregulated |
| -0,295176092 | 0,752949189 | CSRP2BP | FALSE | Downregulated |
| -0,255614862 | 0,753127179 | VPS16 | FALSE | Downregulated |

| | | | | |
|---|---|---|---|---|
| -1,682431704 | 0,753208426 | LOC102724843 | FALSE | Downregulated |
| 0,348866913 | 0,753339222 | URB2 | FALSE | Upregulated |
| 1,018457138 | 0,753623094 | HIST1H2AD | FALSE | Upregulated |
| -0,193395452 | 0,753646551 | CFP | FALSE | Downregulated |
| 0,186078465 | 0,753646551 | CSGALNACT1 | FALSE | Upregulated |
| 0,179042268 | 0,753646551 | GAS2 | FALSE | Upregulated |
| -0,543101318 | 0,753646551 | LOC101928628 | FALSE | Downregulated |
| -0,279848157 | 0,753646551 | OR52W1 | FALSE | Downregulated |
| -0,916181824 | 0,753646551 | PAM16 | FALSE | Downregulated |
| 0,224401486 | 0,753646551 | PDGFC | FALSE | Upregulated |
| -0,116250816 | 0,753646551 | RAPGEF2 | FALSE | Downregulated |
| 0,136068775 | 0,753646551 | RMDN1 | FALSE | Upregulated |
| 0,167787242 | 0,753646551 | SMAD2 | FALSE | Upregulated |
| -0,698562193 | 0,753646551 | SRRM2-AS1 | FALSE | Downregulated |
| -0,156230646 | 0,753646551 | TIMM13 | FALSE | Downregulated |
| 0,146952936 | 0,753646551 | TSPYL6 | FALSE | Upregulated |
| -0,61232743 | 0,753646551 | ZNF773 | FALSE | Downregulated |
| 0,477869716 | 0,753646551 | ZNF853 | FALSE | Upregulated |
| -0,364179434 | 0,753646551 | ZSWIM5 | FALSE | Downregulated |
| 0,241912201 | 0,753653006 | AP1S2 | FALSE | Upregulated |
| 0,120267614 | 0,753720478 | GLIPR1 | FALSE | Upregulated |
| 0,231002104 | 0,753754304 | CD53 | FALSE | Upregulated |
| 0,342665189 | 0,753754304 | ZNF8 | FALSE | Upregulated |
| 0,226337886 | 0,753960153 | PAXIP1-AS2 | FALSE | Upregulated |
| 0,405508493 | 0,754069481 | BEGAIN | FALSE | Upregulated |
| 0,136144358 | 0,754272649 | NFYC | FALSE | Upregulated |
| 0,130557344 | 0,75428589 | G3BP1 | FALSE | Upregulated |
| -0,406297726 | 0,754465637 | HSP90B2P | FALSE | Downregulated |
| -0,30325316 | 0,754486158 | AC018705.5 | FALSE | Downregulated |
| -0,211402684 | 0,754486158 | ZNF608 | FALSE | Downregulated |
| -0,73821898 | 0,754731862 | RP11-12A2.3 | FALSE | Downregulated |
| -0,160052946 | 0,754870856 | ZNF740 | FALSE | Downregulated |
| 0,328972451 | 0,755231283 | CASP9 | FALSE | Upregulated |
| 0,200067042 | 0,755231283 | SLC30A5 | FALSE | Upregulated |
| -0,21071635 | 0,755231283 | SLC3A2 | FALSE | Downregulated |
| 1,357955398 | 0,755265216 | RHOXF2B | FALSE | Upregulated |
| 0,187587532 | 0,755443405 | BRF1 | FALSE | Upregulated |
| 0,151819607 | 0,755486259 | KRAS | FALSE | Upregulated |
| 0,260542942 | 0,755505403 | C2orf69 | FALSE | Upregulated |
| -0,49214916 | 0,755505403 | LOC101927941 | FALSE | Downregulated |
| 0,353853775 | 0,755505403 | MAPK10 | FALSE | Upregulated |
| 0,584467993 | 0,755505403 | SMIM3 | FALSE | Upregulated |
| -0,371676427 | 0,755601154 | HAUS7 | FALSE | Downregulated |
| 0,175471989 | 0,755601154 | N4BP1 | FALSE | Upregulated |
| -0,165730403 | 0,755601154 | PA2G4 | FALSE | Downregulated |
| 0,746626907 | 0,755691341 | SCN4A | FALSE | Upregulated |
| -0,210632269 | 0,755833161 | ENTHD2 | FALSE | Downregulated |
| -0,13373459 | 0,755833161 | RTN4 | FALSE | Downregulated |
| -0,280522888 | 0,756118769 | VPS29 | FALSE | Downregulated |
| 0,275203825 | 0,756649212 | LOC100132731 | FALSE | Upregulated |
| 0,619099099 | 0,756667355 | HOXB6 | FALSE | Upregulated |

| | | | | |
|---|---|---|---|---|
| 0,257881344 | 0,756667355 | KIAA0586 | FALSE | Upregulated |
| 0,355671682 | 0,756667355 | LOC102724286 | FALSE | Upregulated |
| 0,149631248 | 0,756667355 | RAC1 | FALSE | Upregulated |
| -0,276877954 | 0,756667355 | TXN | FALSE | Downregulated |
| -0,27133279 | 0,756667355 | ZNF619 | FALSE | Downregulated |
| -0,430438867 | 0,756667355 | ZNF653 | FALSE | Downregulated |
| -0,374606349 | 0,757005857 | LSM1 | FALSE | Downregulated |
| 0,406934662 | 0,757111331 | RP11-797H7.5 | FALSE | Upregulated |
| -0,244063628 | 0,757111331 | SYCE2 | FALSE | Downregulated |
| 0,323508447 | 0,757163629 | THOC7-AS1 | FALSE | Upregulated |
| -0,234599042 | 0,757181342 | LOC100506548 | FALSE | Downregulated |
| -0,26147116 | 0,757181342 | LOC150051 | FALSE | Downregulated |
| 0,773185683 | 0,757226883 | LOC101929689 | FALSE | Upregulated |
| 0,304765391 | 0,757255112 | C19orf82 | FALSE | Upregulated |
| -0,229242656 | 0,757255112 | LARP7 | FALSE | Downregulated |
| 0,496487435 | 0,757255112 | MUC3A | FALSE | Upregulated |
| 0,277180085 | 0,757255112 | NAT2 | FALSE | Upregulated |
| 0,331746559 | 0,757255112 | RPF2 | FALSE | Upregulated |
| -0,2471527 | 0,757340346 | WDR18 | FALSE | Downregulated |
| -0,41097646 | 0,757452684 | MRPS36 | FALSE | Downregulated |
| -0,222307819 | 0,757467613 | HSPH1 | FALSE | Downregulated |
| 0,417870803 | 0,757467613 | ZC3H8 | FALSE | Upregulated |
| -0,182901035 | 0,757744617 | AQR | FALSE | Downregulated |
| 0,256305167 | 0,757744617 | CBX3 | FALSE | Upregulated |
| -0,306853144 | 0,757744617 | CMTR2 | FALSE | Downregulated |
| 0,199857667 | 0,757744617 | DIP2A | FALSE | Upregulated |
| -0,279996214 | 0,757744617 | LGALS1 | FALSE | Downregulated |
| -0,388732738 | 0,757744617 | LIFR-AS1 | FALSE | Downregulated |
| 0,436068907 | 0,757744617 | LOC642846 | FALSE | Upregulated |
| -0,130365437 | 0,757744617 | MAN2A1 | FALSE | Downregulated |
| 0,468850991 | 0,757842226 | OR52I1 | FALSE | Upregulated |
| 0,64895939 | 0,757842226 | PTPN22 | FALSE | Upregulated |
| 0,184129842 | 0,757842226 | TBRG1 | FALSE | Upregulated |
| -0,618054511 | 0,757881945 | SRP14-AS1 | FALSE | Downregulated |
| -1,105929207 | 0,757881945 | TNXA | FALSE | Downregulated |
| -3,277434576 | 0,75798201 | RAD51L3-RFFL | FALSE | Downregulated |
| 0,305543728 | 0,758066062 | LINC00861 | FALSE | Upregulated |
| 0,233453037 | 0,758111021 | TRIM7 | FALSE | Upregulated |
| 0,416353276 | 0,758209858 | WDR4 | FALSE | Upregulated |
| -0,23522532 | 0,758328023 | DPP3 | FALSE | Downregulated |
| 0,225415428 | 0,758336439 | SNIP1 | FALSE | Upregulated |
| -0,617450833 | 0,758489826 | ARMC2-AS1 | FALSE | Downregulated |
| -0,284162657 | 0,758555371 | RNF181 | FALSE | Downregulated |
| -0,320621134 | 0,758656672 | HERC5 | FALSE | Downregulated |
| -0,347958474 | 0,758713499 | JRKL | FALSE | Downregulated |
| 0,560474154 | 0,758865209 | SALL2 | FALSE | Upregulated |
| -0,346442701 | 0,758999955 | SNHG8 | FALSE | Downregulated |
| -0,146375639 | 0,759167338 | HMG20B | FALSE | Downregulated |
| -0,186548572 | 0,759167338 | MXD1 | FALSE | Downregulated |
| -0,177493209 | 0,759167338 | PAIP1 | FALSE | Downregulated |
| 0,191837358 | 0,759167338 | SATL1 | FALSE | Upregulated |

| | | | | |
|---|---|---|---|---|
| -0,319923534 | 0,759167338 | TMEM17 | FALSE | Downregulated |
| -0,94192576 | 0,759167338 | TMEM56-RWDD3 | FALSE | Downregulated |
| -0,550106812 | 0,759361556 | ACD | FALSE | Downregulated |
| -0,293071866 | 0,759361556 | ADAM19 | FALSE | Downregulated |
| -0,617295099 | 0,759361556 | CATSPER2P1 | FALSE | Downregulated |
| 0,288537535 | 0,759361556 | COIL | FALSE | Upregulated |
| -0,133887933 | 0,759361556 | PHC2 | FALSE | Downregulated |
| -0,387595468 | 0,759361556 | PPM1L | FALSE | Downregulated |
| 0,304377697 | 0,759361556 | SOCS4 | FALSE | Upregulated |
| -0,155689487 | 0,75948302 | GID8 | FALSE | Downregulated |
| 0,229239521 | 0,75948302 | IRF9 | FALSE | Upregulated |
| 0,226967559 | 0,75948302 | MRPS21 | FALSE | Upregulated |
| 0,203419845 | 0,75948302 | SPAG1 | FALSE | Upregulated |
| -0,136971446 | 0,75948302 | TRIM27 | FALSE | Downregulated |
| -0,251072519 | 0,75948302 | UG0898H09 | FALSE | Downregulated |
| 0,158076617 | 0,75948302 | ZBTB18 | FALSE | Upregulated |
| 0,414647821 | 0,759614879 | SH2B2 | FALSE | Upregulated |
| -0,155503925 | 0,759614879 | ZSWIM1 | FALSE | Downregulated |
| 0,309295647 | 0,759670782 | HSPA13 | FALSE | Upregulated |
| 0,194331679 | 0,759799211 | SCAMP2 | FALSE | Upregulated |
| 0,121456644 | 0,759875749 | FOXK2 | FALSE | Upregulated |
| 0,126343112 | 0,759909721 | SMARCC1 | FALSE | Upregulated |
| 0,094832413 | 0,761029933 | CHERP | FALSE | Upregulated |
| 0,18740094 | 0,761029933 | DHX32 | FALSE | Upregulated |
| -0,143329325 | 0,761122661 | AUP1 | FALSE | Downregulated |
| -0,204408155 | 0,761122661 | CYP2A6 | FALSE | Downregulated |
| -0,121141135 | 0,761122661 | HNF4A | FALSE | Downregulated |
| -0,122392274 | 0,761122661 | IL1R1 | FALSE | Downregulated |
| -0,712293592 | 0,761122661 | LOC101930567 | FALSE | Downregulated |
| 0,469399933 | 0,761122661 | NKG7 | FALSE | Upregulated |
| 0,470157566 | 0,761122661 | PARS2 | FALSE | Upregulated |
| -0,144942928 | 0,761122661 | PDS5A | FALSE | Downregulated |
| -0,141761389 | 0,761122661 | RNF217 | FALSE | Downregulated |
| 0,334926138 | 0,761122661 | UGGT2 | FALSE | Upregulated |
| -0,47519109 | 0,761122661 | WBP1 | FALSE | Downregulated |
| 0,227068655 | 0,761205868 | ABTB1 | FALSE | Upregulated |
| -0,165229524 | 0,761205868 | NEURL4 | FALSE | Downregulated |
| 0,259491672 | 0,761205868 | PLEK2 | FALSE | Upregulated |
| -0,850434509 | 0,761205868 | TNFRSF10C | FALSE | Downregulated |
| -0,307551746 | 0,761396619 | ETV5 | FALSE | Downregulated |
| 0,179192751 | 0,761396619 | MOB3C | FALSE | Upregulated |
| -0,238099502 | 0,761396619 | PPTC7 | FALSE | Downregulated |
| -0,229246411 | 0,761586221 | BNIP2 | FALSE | Downregulated |
| -0,606257412 | 0,761769228 | SULT1A4 | FALSE | Downregulated |
| 0,125567293 | 0,761769228 | TRAM2 | FALSE | Upregulated |
| -0,341771982 | 0,761839325 | CWF19L1 | FALSE | Downregulated |
| 0,185633499 | 0,761839325 | FOXJ2 | FALSE | Upregulated |
| 0,369246069 | 0,761839325 | UPF3B | FALSE | Upregulated |
| 0,601715999 | 0,761903088 | NPFF | FALSE | Upregulated |
| -0,434817794 | 0,761903088 | SCLT1 | FALSE | Downregulated |
| -0,135211068 | 0,761964071 | CHD9 | FALSE | Downregulated |

| | | | | |
|---|---|---|---|---|
| 0,619356802 | 0,761964071 | LOC102724292 | FALSE | Upregulated |
| 0,211541414 | 0,761964071 | SETD8 | FALSE | Upregulated |
| 0,280143129 | 0,761964071 | ZNF429 | FALSE | Upregulated |
| 0,281481577 | 0,761964071 | ZNF704 | FALSE | Upregulated |
| -0,59013913 | 0,762045588 | LINC00539 | FALSE | Downregulated |
| 0,220118206 | 0,762045588 | LRSAM1 | FALSE | Upregulated |
| -0,167865022 | 0,762045588 | SERPINA11 | FALSE | Downregulated |
| 0,156835489 | 0,762070789 | DNAJB5 | FALSE | Upregulated |
| 0,099895531 | 0,762099899 | EIF2AK1 | FALSE | Upregulated |
| -0,315353409 | 0,762136299 | MRPL11 | FALSE | Downregulated |
| 0,139400704 | 0,762136299 | SCAF4 | FALSE | Upregulated |
| 0,235670246 | 0,762136299 | ZNF213 | FALSE | Upregulated |
| 0,690661875 | 0,762446094 | LOC101929047 | FALSE | Upregulated |
| 0,279288533 | 0,762446094 | RIC3 | FALSE | Upregulated |
| -0,472313612 | 0,762446094 | ZNF583 | FALSE | Downregulated |
| -0,815288338 | 0,762497279 | SLC34A1 | FALSE | Downregulated |
| -0,590979037 | 0,762520877 | P2RX6 | FALSE | Downregulated |
| 0,169062828 | 0,762520877 | RNF144B | FALSE | Upregulated |
| -0,286430284 | 0,762520877 | ZFAND4 | FALSE | Downregulated |
| -0,192781515 | 0,762520877 | ZRANB2 | FALSE | Downregulated |
| -0,69597433 | 0,762602639 | OR1L6 | FALSE | Downregulated |
| 0,407576765 | 0,762618131 | EXOSC1 | FALSE | Upregulated |
| -0,764898141 | 0,762618131 | LOC101928030 | FALSE | Downregulated |
| 0,63062429 | 0,762618131 | LOC102724920 | FALSE | Upregulated |
| 0,301758286 | 0,762618131 | MYOM1 | FALSE | Upregulated |
| 0,246105887 | 0,762618131 | NEU3 | FALSE | Upregulated |
| -0,235910377 | 0,762642221 | BCKDHB | FALSE | Downregulated |
| -0,209286252 | 0,762642221 | DR1 | FALSE | Downregulated |
| -0,146697068 | 0,762642221 | ETF1 | FALSE | Downregulated |
| -1,166206502 | 0,762642221 | GOLGA6D | FALSE | Downregulated |
| -0,121808828 | 0,762642221 | KIDINS220 | FALSE | Downregulated |
| 0,24970565 | 0,762642221 | LOC101929173 | FALSE | Upregulated |
| -0,26267665 | 0,762642221 | LOC729658 | FALSE | Downregulated |
| 0,233523808 | 0,762642221 | MLLT4-AS1 | FALSE | Upregulated |
| -0,157167042 | 0,762642221 | MRPL41 | FALSE | Downregulated |
| 0,444146975 | 0,762642221 | SNAI2 | FALSE | Upregulated |
| -0,085616396 | 0,762736271 | CALD1 | FALSE | Downregulated |
| -0,883794351 | 0,762736271 | FRG2 | FALSE | Downregulated |
| -0,196553154 | 0,762736271 | IST1 | FALSE | Downregulated |
| -0,291977966 | 0,762736271 | KCNIP3 | FALSE | Downregulated |
| -0,212312045 | 0,762736271 | LOC101927854 | FALSE | Downregulated |
| -0,375851321 | 0,762736271 | LOC102724895 | FALSE | Downregulated |
| 0,283083458 | 0,762736271 | RASGRP2 | FALSE | Upregulated |
| 0,318954465 | 0,762736271 | THAP6 | FALSE | Upregulated |
| -0,15481242 | 0,762736271 | ZNRF1 | FALSE | Downregulated |
| -0,272929211 | 0,762867624 | JMJD6 | FALSE | Downregulated |
| 0,075228901 | 0,762867624 | MYH9 | FALSE | Upregulated |
| -0,201926974 | 0,762867624 | SLC25A43 | FALSE | Downregulated |
| -0,47411216 | 0,76312943 | SAMD3 | FALSE | Downregulated |
| 0,330142608 | 0,76312943 | ZNF432 | FALSE | Upregulated |
| -0,276879544 | 0,763613003 | ATP7A | FALSE | Downregulated |

| | | | | |
|---|---|---|---|---|
| 0,537481319 | 0,763613003 | LOC645485 | FALSE | Upregulated |
| 0,517939872 | 0,76363996 | APCDD1 | FALSE | Upregulated |
| 0,27607757 | 0,76363996 | CCDC109B | FALSE | Upregulated |
| 0,337139218 | 0,76363996 | LOC102724872 | FALSE | Upregulated |
| -0,188234202 | 0,76363996 | SMC5 | FALSE | Downregulated |
| 0,349947355 | 0,76363996 | TLR6 | FALSE | Upregulated |
| -0,204639666 | 0,76363996 | ZNF641 | FALSE | Downregulated |
| 0,780636728 | 0,763666358 | DDX11L9 | FALSE | Upregulated |
| 0,253007742 | 0,763666358 | LOC101059949 | FALSE | Upregulated |
| 0,12790897 | 0,763666358 | NSD1 | FALSE | Upregulated |
| -0,221786253 | 0,763776298 | MTF2 | FALSE | Downregulated |
| -0,210430201 | 0,764284645 | OSBPL11 | FALSE | Downregulated |
| -0,213318391 | 0,764624356 | ARHGAP33 | FALSE | Downregulated |
| 0,72625477 | 0,764624356 | LINC00880 | FALSE | Upregulated |
| -0,441159734 | 0,764624356 | MN1 | FALSE | Downregulated |
| -0,329968236 | 0,764624356 | STAM | FALSE | Downregulated |
| 0,163258673 | 0,764793808 | RABGAP1 | FALSE | Upregulated |
| 0,170907217 | 0,764897803 | ZBTB8A | FALSE | Upregulated |
| -0,377430428 | 0,76495347 | APEX2 | FALSE | Downregulated |
| -0,228066518 | 0,76498681 | ERI3 | FALSE | Downregulated |
| -0,17567705 | 0,76498681 | OCRL | FALSE | Downregulated |
| -0,126125272 | 0,76498681 | PDE3B | FALSE | Downregulated |
| -0,14240507 | 0,76498681 | RPN2 | FALSE | Downregulated |
| 0,605951414 | 0,76498681 | SIGLEC5 | FALSE | Upregulated |
| 0,103280954 | 0,76507805 | SVIL | FALSE | Upregulated |
| -0,179760551 | 0,765089042 | FLJ35282 | FALSE | Downregulated |
| -0,21481242 | 0,765122605 | DYNC2LI1 | FALSE | Downregulated |
| 0,347536349 | 0,765122605 | HDAC9 | FALSE | Upregulated |
| 0,302952698 | 0,765122605 | HINFP | FALSE | Upregulated |
| -0,618125455 | 0,765122605 | LOC101927119 | FALSE | Downregulated |
| -0,256818165 | 0,765122605 | MFF | FALSE | Downregulated |
| 0,228686527 | 0,765282075 | PPA1 | FALSE | Upregulated |
| -0,129206163 | 0,765283319 | PAPD7 | FALSE | Downregulated |
| 0,365385084 | 0,765345415 | OR10J5 | FALSE | Upregulated |
| -0,589982155 | 0,765473434 | OR2A7 | FALSE | Downregulated |
| 0,304805182 | 0,765473434 | RABL2B | FALSE | Upregulated |
| 0,293579177 | 0,765473434 | STAU2-AS1 | FALSE | Upregulated |
| 0,762591562 | 0,765473434 | ZIM2 | FALSE | Upregulated |
| -0,441294865 | 0,765733281 | C11orf73 | FALSE | Downregulated |
| 0,244078642 | 0,765733281 | CELF3 | FALSE | Upregulated |
| 0,388152278 | 0,765733281 | CLDN10 | FALSE | Upregulated |
| 0,724311884 | 0,765733281 | FAM101A | FALSE | Upregulated |
| -0,674437169 | 0,765733281 | FAM124B | FALSE | Downregulated |
| 0,49637504 | 0,765733281 | GPR135 | FALSE | Upregulated |
| -0,20197603 | 0,765733281 | HINT3 | FALSE | Downregulated |
| -0,335299642 | 0,765733281 | LOC101928433 | FALSE | Downregulated |
| 0,200727146 | 0,765733281 | NOB1 | FALSE | Upregulated |
| 0,217070776 | 0,765733281 | RAB21 | FALSE | Upregulated |
| -0,148611398 | 0,765733281 | SFT2D3 | FALSE | Downregulated |
| 0,213914438 | 0,765733281 | STARD5 | FALSE | Upregulated |
| -0,196651549 | 0,765733281 | ZNF45 | FALSE | Downregulated |

| | | | | |
|---|---|---|---|---|
| 0,176016855 | 0,766026482 | MPLKIP | FALSE | Upregulated |
| -0,324547256 | 0,766026482 | RAC3 | FALSE | Downregulated |
| 0,323187202 | 0,766026482 | TRMT13 | FALSE | Upregulated |
| -0,325647314 | 0,766026482 | TTC27 | FALSE | Downregulated |
| -0,085695072 | 0,766132331 | CTNNA1 | FALSE | Downregulated |
| 0,107227173 | 0,766207408 | IMMT | FALSE | Upregulated |
| -0,428187933 | 0,766207408 | LINC01024 | FALSE | Downregulated |
| -0,590367179 | 0,766220603 | HOXA4 | FALSE | Downregulated |
| 0,111676452 | 0,766220603 | VPRBP | FALSE | Upregulated |
| 0,223510693 | 0,766282933 | ATP6V0A2 | FALSE | Upregulated |
| -0,344770443 | 0,766282933 | PRPF18 | FALSE | Downregulated |
| -0,166660647 | 0,766282933 | SEPHS1 | FALSE | Downregulated |
| 0,164070733 | 0,766354295 | ZNF542 | FALSE | Upregulated |
| -0,379543674 | 0,766587187 | DYNLT1 | FALSE | Downregulated |
| 0,36178807 | 0,766708182 | LURAP1L | FALSE | Upregulated |
| -0,289506814 | 0,766725448 | TATDN1 | FALSE | Downregulated |
| 0,147342775 | 0,767007252 | TOP1 | FALSE | Upregulated |
| 0,554353362 | 0,767010095 | LCK | FALSE | Upregulated |
| -0,16430201 | 0,767010095 | RP11-778H2.1 | FALSE | Downregulated |
| -0,078165205 | 0,767052832 | HNRNPUL1 | FALSE | Downregulated |
| 0,134877403 | 0,767156282 | LRP11 | FALSE | Upregulated |
| -0,313369507 | 0,767268887 | HMGB3 | FALSE | Downregulated |
| -0,760391985 | 0,767268887 | LOC100505909 | FALSE | Downregulated |
| -0,150279909 | 0,767334102 | NUDT3 | FALSE | Downregulated |
| 0,651177116 | 0,767344264 | PLA2G5 | FALSE | Upregulated |
| -0,57234619 | 0,767344264 | PTP4A3 | FALSE | Downregulated |
| 0,381924319 | 0,767418459 | SBDSP1 | FALSE | Upregulated |
| 0,151800716 | 0,767435567 | CCNJ | FALSE | Upregulated |
| -0,174947306 | 0,767450478 | GALNT1 | FALSE | Downregulated |
| 0,204161243 | 0,767450478 | GOLPH3L | FALSE | Upregulated |
| -0,282680514 | 0,767450478 | LRRC40 | FALSE | Downregulated |
| -0,246397906 | 0,767450478 | NARFL | FALSE | Downregulated |
| 0,156332047 | 0,767728883 | IDNK | FALSE | Upregulated |
| 0,192176231 | 0,767728883 | SPECC1L | FALSE | Upregulated |
| 0,506953237 | 0,767808939 | BAIAP2L2 | FALSE | Upregulated |
| -0,382474265 | 0,767808939 | IRAK2 | FALSE | Downregulated |
| -0,598535819 | 0,767808939 | OR51B6 | FALSE | Downregulated |
| 0,634939831 | 0,767808939 | RANBP17 | FALSE | Upregulated |
| 0,445292455 | 0,768227129 | LOC102723306 | FALSE | Upregulated |
| -0,431420547 | 0,768350138 | C3orf18 | FALSE | Downregulated |
| -0,608906133 | 0,768350138 | ZNRD1-AS1 | FALSE | Downregulated |
| 0,539800772 | 0,768513403 | HIST1H4H | FALSE | Upregulated |
| 0,188797155 | 0,768557615 | CDK17 | FALSE | Upregulated |
| -0,786336358 | 0,768557615 | CTSG | FALSE | Downregulated |
| -0,120277781 | 0,768557615 | GBF1 | FALSE | Downregulated |
| -0,194972107 | 0,768557615 | IFITM2 | FALSE | Downregulated |
| -0,127669934 | 0,768557615 | KLB | FALSE | Downregulated |
| 0,23938876 | 0,768557615 | NAA10 | FALSE | Upregulated |
| 0,831380566 | 0,768557615 | OR2A4 | FALSE | Upregulated |
| -0,245557815 | 0,768557615 | PDK3 | FALSE | Downregulated |
| 0,221311964 | 0,768557615 | THAP9 | FALSE | Upregulated |

| | | | | |
|---|---|---|---|---|
| -0,183354442 | 0,768557615 | THSD7A | FALSE | Downregulated |
| -0,303158246 | 0,768557615 | TMEM260 | FALSE | Downregulated |
| 0,127814351 | 0,768557615 | VAC14 | FALSE | Upregulated |
| 0,268233742 | 0,768779486 | FXYD5 | FALSE | Upregulated |
| 0,246493376 | 0,768779486 | KCNV2 | FALSE | Upregulated |
| 0,151221534 | 0,768779486 | UAP1 | FALSE | Upregulated |
| -0,094664651 | 0,768858824 | ZZEF1 | FALSE | Downregulated |
| 0,473688531 | 0,76902234 | KIAA0226L | FALSE | Upregulated |
| -0,147927624 | 0,769037601 | MAU2 | FALSE | Downregulated |
| 0,203491753 | 0,769037601 | UHRF2 | FALSE | Upregulated |
| -0,235970911 | 0,76905947 | KIAA0319 | FALSE | Downregulated |
| 0,431135345 | 0,769356548 | LOC729870 | FALSE | Upregulated |
| 0,791482282 | 0,769420807 | IGFN1 | FALSE | Upregulated |
| -0,185418934 | 0,769482228 | GLE1 | FALSE | Downregulated |
| -0,393275168 | 0,769482228 | LOC645513 | FALSE | Downregulated |
| 0,529946379 | 0,769532214 | KCNK17 | FALSE | Upregulated |
| 0,402087311 | 0,769532214 | LOC101927054 | FALSE | Upregulated |
| 0,498968035 | 0,769532214 | LOC102546226 | FALSE | Upregulated |
| 0,523102054 | 0,769532214 | LOC102724193 | FALSE | Upregulated |
| -0,139113877 | 0,769532214 | PATL1 | FALSE | Downregulated |
| -0,255489691 | 0,769532214 | SSPO | FALSE | Downregulated |
| 0,262608974 | 0,769851711 | AP5M1 | FALSE | Upregulated |
| 0,241733716 | 0,769851711 | CHFR | FALSE | Upregulated |
| 0,437114315 | 0,769851711 | CMSS1 | FALSE | Upregulated |
| 0,15211567 | 0,769851711 | ST7-AS1 | FALSE | Upregulated |
| -0,291359048 | 0,769861102 | PDE3A | FALSE | Downregulated |
| -0,141693359 | 0,770347126 | DBNL | FALSE | Downregulated |
| 0,261027052 | 0,770359269 | BBS5 | FALSE | Upregulated |
| 0,131561696 | 0,770359269 | CCT5 | FALSE | Upregulated |
| -0,179581198 | 0,770359269 | EIF2D | FALSE | Downregulated |
| -0,224298396 | 0,770359269 | HIST1H4C | FALSE | Downregulated |
| 0,420023694 | 0,770359269 | LOC100996419 | FALSE | Upregulated |
| -0,680391125 | 0,770359269 | LOC102724851 | FALSE | Downregulated |
| -0,536771869 | 0,770359269 | SELP | FALSE | Downregulated |
| 0,614917725 | 0,770359269 | SGSM1 | FALSE | Upregulated |
| 0,272923115 | 0,770359269 | STIM2 | FALSE | Upregulated |
| -0,458438263 | 0,770359269 | TTC3P1 | FALSE | Downregulated |
| 0,106441888 | 0,770469944 | UBE2R2 | FALSE | Upregulated |
| 0,406370782 | 0,77060634 | FAM217A | FALSE | Upregulated |
| -0,331712997 | 0,770634018 | PRX | FALSE | Downregulated |
| -0,158753328 | 0,770634018 | SCAI | FALSE | Downregulated |
| -0,246313065 | 0,770634018 | SLC16A13 | FALSE | Downregulated |
| 0,206662795 | 0,770664842 | LATS1 | FALSE | Upregulated |
| -0,127264296 | 0,770667803 | ATP2A2 | FALSE | Downregulated |
| -0,222249271 | 0,770667803 | C1QBP | FALSE | Downregulated |
| 0,179157494 | 0,770667803 | CERS4 | FALSE | Upregulated |
| -0,126607932 | 0,770667803 | DHCR24 | FALSE | Downregulated |
| 0,584054991 | 0,770667803 | LOC102724556 | FALSE | Upregulated |
| 0,330816917 | 0,770667803 | MICU2 | FALSE | Upregulated |
| 0,438478076 | 0,770667803 | RAD54B | FALSE | Upregulated |
| 0,398865781 | 0,770667803 | RP11-147N17.1 | FALSE | Upregulated |

| | | | | |
|---|---|---|---|---|
| 0,281980741 | 0,770667803 | TRADD | FALSE | Upregulated |
| 0,599724446 | 0,770793993 | RP11-478K7.2 | FALSE | Upregulated |
| 0,474295542 | 0,770950011 | CD226 | FALSE | Upregulated |
| 0,368042534 | 0,770950011 | CYP11A1 | FALSE | Upregulated |
| 0,319075114 | 0,770950011 | RP11-996F15.2 | FALSE | Upregulated |
| 0,188208915 | 0,771024494 | PYCRL | FALSE | Upregulated |
| 0,324041608 | 0,771335066 | ARHGEF34P | FALSE | Upregulated |
| -0,220293015 | 0,771335066 | RBBP5 | FALSE | Downregulated |
| 0,827364503 | 0,771335066 | SFN | FALSE | Upregulated |
| -0,13677986 | 0,771421882 | PDPR | FALSE | Downregulated |
| -0,430323677 | 0,771516798 | NPPA | FALSE | Downregulated |
| -0,648533498 | 0,771728614 | CTD-2201I18.1 | FALSE | Downregulated |
| 0,13542538 | 0,771728614 | PPP1R14B | FALSE | Upregulated |
| 0,186036447 | 0,771786599 | ANAPC1 | FALSE | Upregulated |
| 0,247365209 | 0,771786599 | ARHGEF37 | FALSE | Upregulated |
| 0,26929811 | 0,771786599 | TMEM242 | FALSE | Upregulated |
| 0,478157865 | 0,771786599 | TNFAIP8L2-SCNM1 | FALSE | Upregulated |
| 0,507284624 | 0,771786646 | TPK1 | FALSE | Upregulated |
| -0,306320416 | 0,77184764 | WASH7P | FALSE | Downregulated |
| -0,140208463 | 0,772011414 | APOM | FALSE | Downregulated |
| -0,313608659 | 0,772011414 | DANCR | FALSE | Downregulated |
| 0,63933432 | 0,772011414 | GATA5 | FALSE | Upregulated |
| 0,554672732 | 0,772011414 | ITM2A | FALSE | Upregulated |
| -0,226170765 | 0,77207475 | NAMPT | FALSE | Downregulated |
| 0,442452325 | 0,772076103 | CTSW | FALSE | Upregulated |
| -0,165471214 | 0,772091396 | RNF43 | FALSE | Downregulated |
| 0,28496649 | 0,772131773 | GNAT2 | FALSE | Upregulated |
| -0,234286855 | 0,772131773 | POLE3 | FALSE | Downregulated |
| 0,271823156 | 0,772131773 | TTLL9 | FALSE | Upregulated |
| -0,215793598 | 0,772131773 | ZNF75D | FALSE | Downregulated |
| -0,19576329 | 0,772192784 | LOC595101 | FALSE | Downregulated |
| -0,270479413 | 0,772192784 | SUPV3L1 | FALSE | Downregulated |
| 0,231860522 | 0,772236836 | DDX60 | FALSE | Upregulated |
| -0,117406309 | 0,772264944 | MAFB | FALSE | Downregulated |
| -0,144350277 | 0,772264944 | SLC9A8 | FALSE | Downregulated |
| 0,144968293 | 0,772269481 | SDC3 | FALSE | Upregulated |
| 0,171185966 | 0,772419134 | ABHD12B | FALSE | Upregulated |
| 0,599771363 | 0,772419134 | ARMC2 | FALSE | Upregulated |
| -0,236798022 | 0,772419134 | PDCD6 | FALSE | Downregulated |
| 0,27090153 | 0,772602351 | MSL3 | FALSE | Upregulated |
| -0,717757558 | 0,772602351 | NCR3 | FALSE | Downregulated |
| 0,36943035 | 0,772672888 | C17orf58 | FALSE | Upregulated |
| 0,227528441 | 0,772672888 | C7orf50 | FALSE | Upregulated |
| 0,093141922 | 0,772672888 | CHD4 | FALSE | Upregulated |
| 0,279945327 | 0,772672888 | CNTN3 | FALSE | Upregulated |
| 0,498867985 | 0,772672888 | DDX26B | FALSE | Upregulated |
| 0,593078139 | 0,772672888 | EVPL | FALSE | Upregulated |
| 0,175474599 | 0,772672888 | LOC100996873 | FALSE | Upregulated |
| 0,572056105 | 0,772672888 | LOC101927258 | FALSE | Upregulated |
| 0,49835066 | 0,772672888 | LOC101928054 | FALSE | Upregulated |
| -0,516506319 | 0,772672888 | LOC102724933 | FALSE | Downregulated |

| | | | | |
|---|---|---|---|---|
| -0,167331291 | 0,772672888 | LSG1 | FALSE | Downregulated |
| -0,209498676 | 0,772672888 | MRPS2 | FALSE | Downregulated |
| -0,22927158 | 0,772672888 | NARG2 | FALSE | Downregulated |
| -0,151958604 | 0,772672888 | NR5A2 | FALSE | Downregulated |
| 0,342069102 | 0,772672888 | PEX6 | FALSE | Upregulated |
| -0,099016187 | 0,772672888 | PJA2 | FALSE | Downregulated |
| -0,083368701 | 0,772672888 | POLR2A | FALSE | Downregulated |
| -0,47334742 | 0,772672888 | RLBP1 | FALSE | Downregulated |
| -0,605499235 | 0,772672888 | RP11-554A11.8 | FALSE | Downregulated |
| 0,295190697 | 0,772672888 | SRXN1 | FALSE | Upregulated |
| 0,212511882 | 0,772672888 | TMEM45B | FALSE | Upregulated |
| -0,309838592 | 0,772672888 | TUBG1 | FALSE | Downregulated |
| 0,325308999 | 0,772672888 | ZNF732 | FALSE | Upregulated |
| -0,223125393 | 0,772743471 | ANKRD26 | FALSE | Downregulated |
| -0,375840208 | 0,772743471 | MKS1 | FALSE | Downregulated |
| 0,330560617 | 0,772798868 | ATE1-AS1 | FALSE | Upregulated |
| -0,485101616 | 0,772798868 | C17orf67 | FALSE | Downregulated |
| -0,725122188 | 0,772798868 | C3orf84 | FALSE | Downregulated |
| -0,432226938 | 0,772798868 | CBX8 | FALSE | Downregulated |
| 0,469997467 | 0,772798868 | CYTIP | FALSE | Upregulated |
| -0,262695658 | 0,772798868 | DSG1 | FALSE | Downregulated |
| 0,285351313 | 0,772798868 | ELF3 | FALSE | Upregulated |
| 0,364894667 | 0,772798868 | EZH2 | FALSE | Upregulated |
| -0,131899903 | 0,772798868 | FAM63A | FALSE | Downregulated |
| -0,282152599 | 0,772798868 | GTF2E2 | FALSE | Downregulated |
| 0,99874883 | 0,772798868 | JMJD7-PLA2G4B | FALSE | Upregulated |
| 0,663460044 | 0,772798868 | LOC101929160 | FALSE | Upregulated |
| 0,199637614 | 0,772798868 | PFKM | FALSE | Upregulated |
| -0,106479737 | 0,772798868 | RPS3A | FALSE | Downregulated |
| 0,279051876 | 0,772798868 | SEC1P | FALSE | Upregulated |
| 0,292099342 | 0,772798868 | SERGEF | FALSE | Upregulated |
| 0,217660006 | 0,772798868 | SLC35G6 | FALSE | Upregulated |
| 0,193068738 | 0,772798868 | USP11 | FALSE | Upregulated |
| 0,201534923 | 0,772798868 | VMAC | FALSE | Upregulated |
| -0,466339307 | 0,772909542 | ARMCX2 | FALSE | Downregulated |
| 1,257366338 | 0,772909542 | RASA4B | FALSE | Upregulated |
| -0,311724408 | 0,773016534 | CYB5RL | FALSE | Downregulated |
| -0,617906192 | 0,773157836 | LOC375196 | FALSE | Downregulated |
| -0,532051301 | 0,773336569 | GSTO2 | FALSE | Downregulated |
| -0,140533336 | 0,773336569 | NF1 | FALSE | Downregulated |
| -0,180524445 | 0,773336569 | NFKB1 | FALSE | Downregulated |
| 0,485912353 | 0,773336569 | ZUFSP | FALSE | Upregulated |
| -0,397867179 | 0,773561533 | KCNE3 | FALSE | Downregulated |
| 0,497797833 | 0,773923 | LOC101928969 | FALSE | Upregulated |
| 0,134868927 | 0,774053672 | MXD4 | FALSE | Upregulated |
| 0,139805318 | 0,774053672 | PLD1 | FALSE | Upregulated |
| -0,16501742 | 0,774060984 | ELL | FALSE | Downregulated |
| 0,196343847 | 0,774060984 | FER | FALSE | Upregulated |
| -0,379744721 | 0,774060984 | KCTD1 | FALSE | Downregulated |
| 0,229759714 | 0,774060984 | KLHDC4 | FALSE | Upregulated |
| -0,174397072 | 0,774060984 | RGPD4 | FALSE | Downregulated |

| | | | | |
|---|---|---|---|---|
| -0,542442229 | 0,774060984 | RRP9 | FALSE | Downregulated |
| 0,189720742 | 0,774060984 | SIRPA | FALSE | Upregulated |
| -0,220141145 | 0,774213403 | ANKS3 | FALSE | Downregulated |
| -0,191417648 | 0,774213403 | PIRT | FALSE | Downregulated |
| 0,246380525 | 0,774233403 | RNF135 | FALSE | Upregulated |
| 0,317321985 | 0,774239267 | FAM122C | FALSE | Upregulated |
| 0,231180824 | 0,774357026 | ADCK4 | FALSE | Upregulated |
| 0,183576359 | 0,774504202 | ABL2 | FALSE | Upregulated |
| 0,968123145 | 0,774504202 | CCDC180 | FALSE | Upregulated |
| -0,268089141 | 0,774504202 | CDKAL1 | FALSE | Downregulated |
| -0,531160105 | 0,774504202 | GEMIN6 | FALSE | Downregulated |
| -0,187626284 | 0,774504202 | MRPL51 | FALSE | Downregulated |
| 0,400991341 | 0,774504202 | PGM2L1 | FALSE | Upregulated |
| -0,235094744 | 0,774504202 | PHF13 | FALSE | Downregulated |
| -0,382947 | 0,774512758 | BZW2 | FALSE | Downregulated |
| -0,639568311 | 0,774512758 | LOC101929225 | FALSE | Downregulated |
| 0,219611214 | 0,774512758 | LOC102723406 | FALSE | Upregulated |
| 0,728174276 | 0,774512758 | LPHN3 | FALSE | Upregulated |
| -1,005653701 | 0,774512758 | MYL1 | FALSE | Downregulated |
| 0,133553128 | 0,774524297 | CDC37 | FALSE | Upregulated |
| 0,127243151 | 0,774660069 | GOLPH3 | FALSE | Upregulated |
| 0,445956022 | 0,774753141 | TRDMT1 | FALSE | Upregulated |
| 0,449578759 | 0,774845254 | LOC101929360 | FALSE | Upregulated |
| 0,35096824 | 0,774845254 | LOC92249 | FALSE | Upregulated |
| -0,251970275 | 0,774845254 | TMEM238 | FALSE | Downregulated |
| -0,271464507 | 0,775005332 | SP140L | FALSE | Downregulated |
| 0,214977691 | 0,775154066 | TRIP10 | FALSE | Upregulated |
| 0,223773088 | 0,775717949 | NCOA5 | FALSE | Upregulated |
| 0,294908718 | 0,77572545 | RAB32 | FALSE | Upregulated |
| -0,091735897 | 0,77572545 | SSFA2 | FALSE | Downregulated |
| -0,458654327 | 0,775822753 | TMEM254-AS1 | FALSE | Downregulated |
| -0,471989052 | 0,775844883 | MMD | FALSE | Downregulated |
| 0,447125312 | 0,77590742 | A4GALT | FALSE | Upregulated |
| 0,220291405 | 0,77590742 | ADSS | FALSE | Upregulated |
| 0,284186638 | 0,77590742 | HMBS | FALSE | Upregulated |
| 0,137883911 | 0,775915856 | SYNM | FALSE | Upregulated |
| -0,288899603 | 0,775923507 | PRPSAP2 | FALSE | Downregulated |
| 0,330764044 | 0,775923507 | STAT4 | FALSE | Upregulated |
| -0,76785676 | 0,775957085 | LOC102724010 | FALSE | Downregulated |
| 0,26261244 | 0,776191939 | LDHB | FALSE | Upregulated |
| -0,23417005 | 0,776191939 | LIMS1 | FALSE | Downregulated |
| -0,434256277 | 0,776191939 | LOC553103 | FALSE | Downregulated |
| 0,313509079 | 0,776191939 | MAPT-IT1 | FALSE | Upregulated |
| 0,303294514 | 0,776191939 | MYOZ3 | FALSE | Upregulated |
| 0,172756039 | 0,776191939 | SNX6 | FALSE | Upregulated |
| 0,19974218 | 0,776191939 | TOMM22 | FALSE | Upregulated |
| 3,865246165 | 0,776215728 | KRT1 | FALSE | Upregulated |
| 0,65434152 | 0,776511866 | CNKSR1 | FALSE | Upregulated |
| 0,149131674 | 0,776604381 | GGNBP2 | FALSE | Upregulated |
| 0,141686854 | 0,776617813 | AKR1C2 | FALSE | Upregulated |
| 0,240020045 | 0,776617813 | FN3KRP | FALSE | Upregulated |

| | | | | |
|---|---|---|---|---|
| -0,252163756 | 0,776617813 | RP11-383C5.3 | FALSE | Downregulated |
| -0,293916176 | 0,776660493 | TICAM2 | FALSE | Downregulated |
| 0,144588005 | 0,77671121 | TRAK2 | FALSE | Upregulated |
| -0,116440583 | 0,776827327 | SEC13 | FALSE | Downregulated |
| 0,30720224 | 0,776847132 | ADCY4 | FALSE | Upregulated |
| -0,187628742 | 0,776847132 | WWTR1 | FALSE | Downregulated |
| -0,243275295 | 0,776869495 | EMBP1 | FALSE | Downregulated |
| 0,493677391 | 0,77688025 | CCDC83 | FALSE | Upregulated |
| -0,189157867 | 0,777095904 | RGL3 | FALSE | Downregulated |
| -0,539484017 | 0,77711442 | MIR4500HG | FALSE | Downregulated |
| 0,63591339 | 0,777120215 | FENDRR | FALSE | Upregulated |
| 0,158794323 | 0,777120215 | MAEA | FALSE | Upregulated |
| 0,184350798 | 0,777120215 | PLEKHA4 | FALSE | Upregulated |
| -0,526067825 | 0,777120215 | RAB4B | FALSE | Downregulated |
| 0,224505911 | 0,777482858 | RARS2 | FALSE | Upregulated |
| 0,189552799 | 0,777643372 | TMPO-AS1 | FALSE | Upregulated |
| -0,754124743 | 0,777651982 | LINC00598 | FALSE | Downregulated |
| 0,348658156 | 0,777680986 | APOPT1 | FALSE | Upregulated |
| -0,455152193 | 0,777680986 | LEPREL4 | FALSE | Downregulated |
| -0,191252515 | 0,777680986 | LILRB3 | FALSE | Downregulated |
| 0,431563884 | 0,777680986 | LOC101927300 | FALSE | Upregulated |
| -0,315793888 | 0,777680986 | LZTFL1 | FALSE | Downregulated |
| -0,307051758 | 0,777680986 | SARS2 | FALSE | Downregulated |
| 0,205035871 | 0,777680986 | SYS1 | FALSE | Upregulated |
| 0,420498676 | 0,777680986 | TAMM41 | FALSE | Upregulated |
| -0,194676415 | 0,777680986 | THAP11 | FALSE | Downregulated |
| 0,11535996 | 0,777680986 | TUG1 | FALSE | Upregulated |
| 0,139530753 | 0,777680986 | UNK | FALSE | Upregulated |
| 1,214312563 | 0,777802229 | LOC102724200 | FALSE | Upregulated |
| -0,117229591 | 0,777827779 | PRDX1 | FALSE | Downregulated |
| -0,097550551 | 0,777842078 | ADCY9 | FALSE | Downregulated |
| 0,318583354 | 0,777842078 | RRP15 | FALSE | Upregulated |
| 0,108876294 | 0,77789843 | ARGLU1 | FALSE | Upregulated |
| -0,7123765 | 0,77789843 | NR4A2 | FALSE | Downregulated |
| -0,933789384 | 0,778081283 | AOAH-IT1 | FALSE | Downregulated |
| -0,258239707 | 0,778081283 | PSMA3 | FALSE | Downregulated |
| -0,336979578 | 0,778081283 | ZNF296 | FALSE | Downregulated |
| -0,580892168 | 0,778231736 | LOC283731 | FALSE | Downregulated |
| -0,379067321 | 0,778231736 | ZNF818P | FALSE | Downregulated |
| 0,125807643 | 0,778327221 | BMP8A | FALSE | Upregulated |
| -0,587035093 | 0,778327221 | CCDC85A | FALSE | Downregulated |
| 0,202747415 | 0,778327221 | CCDC88C | FALSE | Upregulated |
| -0,180016029 | 0,778327221 | CRHBP | FALSE | Downregulated |
| -0,178061509 | 0,778327221 | GFM2 | FALSE | Downregulated |
| 0,453846672 | 0,778327221 | LOC100506859 | FALSE | Upregulated |
| -0,223597733 | 0,778327221 | SPG20 | FALSE | Downregulated |
| -0,273553804 | 0,778347245 | C4orf29 | FALSE | Downregulated |
| 0,815180245 | 0,778390172 | CRYAB | FALSE | Upregulated |
| 0,178143434 | 0,778390172 | FLYWCH1 | FALSE | Upregulated |
| -0,109753039 | 0,778459108 | SPG7 | FALSE | Downregulated |
| 0,1848445 | 0,778668479 | ALMS1 | FALSE | Upregulated |

| | | | | |
|---|---|---|---|---|
| 0,186448178 | 0,778668479 | FBXO25 | FALSE | Upregulated |
| 0,215083513 | 0,778668479 | LOC101928576 | FALSE | Upregulated |
| -0,756173769 | 0,778668479 | POF1B | FALSE | Downregulated |
| 0,155258106 | 0,778668479 | TJP3 | FALSE | Upregulated |
| 0,521226632 | 0,778761706 | DOC2B | FALSE | Upregulated |
| 0,454851738 | 0,778761706 | FAM71C | FALSE | Upregulated |
| 0,858887666 | 0,778761706 | KCNE1 | FALSE | Upregulated |
| -0,12566627 | 0,778761706 | LOC101927377 | FALSE | Downregulated |
| 0,182590549 | 0,778761706 | LRRC8E | FALSE | Upregulated |
| -0,217869762 | 0,778761706 | PLCD1 | FALSE | Downregulated |
| -0,331743535 | 0,778761706 | RALA | FALSE | Downregulated |
| 0,21274546 | 0,778761706 | SOCS7 | FALSE | Upregulated |
| 0,432613002 | 0,778980969 | IL3 | FALSE | Upregulated |
| -0,506205702 | 0,779005211 | PITX1 | FALSE | Downregulated |
| -0,109567709 | 0,77916121 | TAOK1 | FALSE | Downregulated |
| 0,342653602 | 0,779351146 | AC007246.3 | FALSE | Upregulated |
| 0,132755068 | 0,779351146 | ACKR4 | FALSE | Upregulated |
| -0,646748549 | 0,779351146 | ASCL3 | FALSE | Downregulated |
| 0,223791248 | 0,779351146 | C10orf118 | FALSE | Upregulated |
| -0,539043021 | 0,779351146 | C10orf128 | FALSE | Downregulated |
| 0,247509063 | 0,779351146 | CDON | FALSE | Upregulated |
| 0,176177868 | 0,779351146 | CPT1B | FALSE | Upregulated |
| 0,651639233 | 0,779351146 | DNAJC25-GNG10 | FALSE | Upregulated |
| 0,254812849 | 0,779351146 | FAM122B | FALSE | Upregulated |
| 0,386727483 | 0,779351146 | FBXL19-AS1 | FALSE | Upregulated |
| -0,125195681 | 0,779351146 | FNDC3B | FALSE | Downregulated |
| -0,160572313 | 0,779351146 | GABPA | FALSE | Downregulated |
| -0,39529828 | 0,779351146 | IFT81 | FALSE | Downregulated |
| -0,184276769 | 0,779351146 | LOC100996573 | FALSE | Downregulated |
| 0,214542482 | 0,779351146 | LOC101929140 | FALSE | Upregulated |
| -0,212693143 | 0,779351146 | MEA1 | FALSE | Downregulated |
| 0,37900365 | 0,779351146 | MRPS28 | FALSE | Upregulated |
| -0,442614715 | 0,779351146 | MTOR-AS1 | FALSE | Downregulated |
| -0,107681269 | 0,779351146 | NADK | FALSE | Downregulated |
| -0,201592771 | 0,779351146 | NLRX1 | FALSE | Downregulated |
| 0,101553878 | 0,779351146 | NPIPB4 | FALSE | Upregulated |
| -0,529426165 | 0,779351146 | ODF3L2 | FALSE | Downregulated |
| -0,315195148 | 0,779351146 | PRAM1 | FALSE | Downregulated |
| 0,248541929 | 0,779351146 | RGS17 | FALSE | Upregulated |
| 0,185256903 | 0,779351146 | SH3KBP1 | FALSE | Upregulated |
| 0,214810517 | 0,779351146 | TANK | FALSE | Upregulated |
| 0,256714654 | 0,779351146 | TMEM201 | FALSE | Upregulated |
| -0,118951623 | 0,779351146 | TRIM56 | FALSE | Downregulated |
| 0,161019547 | 0,779351146 | TSPYL4 | FALSE | Upregulated |
| -0,174439169 | 0,779351146 | ZBED6 | FALSE | Downregulated |
| -0,533592538 | 0,779351146 | ZWILCH | FALSE | Downregulated |
| -0,301794351 | 0,779434842 | COMMD3 | FALSE | Downregulated |
| 0,440016151 | 0,779470936 | ZNF599 | FALSE | Upregulated |
| 0,150086212 | 0,779492532 | NFRKB | FALSE | Upregulated |
| -0,229601634 | 0,779544341 | LOC644662 | FALSE | Downregulated |
| 2,399130619 | 0,779579452 | EEF1E1-BLOC1S5 | FALSE | Upregulated |

| | | | | |
|---|---|---|---|---|
| 0,496309731 | 0,779579452 | EFCAB6 | FALSE | Upregulated |
| 0,170198159 | 0,779579452 | FBXW7 | FALSE | Upregulated |
| 0,479173304 | 0,779718297 | LOC102723683 | FALSE | Upregulated |
| -0,706092043 | 0,779751654 | PRR19 | FALSE | Downregulated |
| 0,643060977 | 0,779762078 | LOC100506753 | FALSE | Upregulated |
| -0,098010067 | 0,779762078 | SPAG9 | FALSE | Downregulated |
| -0,38756651 | 0,780282052 | MGC70870 | FALSE | Downregulated |
| -0,416757048 | 0,780515396 | LRRC2-AS1 | FALSE | Downregulated |
| 0,182658596 | 0,780622539 | NSG1 | FALSE | Upregulated |
| 0,30210047 | 0,780622539 | STX2 | FALSE | Upregulated |
| 0,143734258 | 0,780718112 | ARCN1 | FALSE | Upregulated |
| 0,394655714 | 0,780800178 | LOC102724005 | FALSE | Upregulated |
| 0,222590025 | 0,780840306 | DDX55 | FALSE | Upregulated |
| 0,32184047 | 0,780840306 | NBR2 | FALSE | Upregulated |
| 0,251383519 | 0,780840306 | ZNF638-IT1 | FALSE | Upregulated |
| -0,176949015 | 0,78143973 | CNST | FALSE | Downregulated |
| -0,201582088 | 0,78143973 | NOSIP | FALSE | Downregulated |
| 0,26024113 | 0,781539984 | SPCS1 | FALSE | Upregulated |
| 0,495569854 | 0,781723519 | BRSK1 | FALSE | Upregulated |
| -0,262895338 | 0,781723519 | PMS2CL | FALSE | Downregulated |
| -0,316373457 | 0,781723519 | TSEN2 | FALSE | Downregulated |
| -0,358323431 | 0,781723519 | ZNF844 | FALSE | Downregulated |
| -0,167402005 | 0,781829874 | RP11-219B17.1 | FALSE | Downregulated |
| -0,425205669 | 0,781829874 | RP11-97O12.2 | FALSE | Downregulated |
| 0,416543723 | 0,781964532 | LRRC31 | FALSE | Upregulated |
| -0,352503318 | 0,782172916 | NUDT18 | FALSE | Downregulated |
| 0,252954311 | 0,782180274 | KIAA1045 | FALSE | Upregulated |
| 0,158606366 | 0,782245997 | DIS3 | FALSE | Upregulated |
| 1,066418133 | 0,782245997 | FNTB | FALSE | Upregulated |
| -0,516518534 | 0,782245997 | IFITM5 | FALSE | Downregulated |
| 0,25565493 | 0,782245997 | KCNMB3 | FALSE | Upregulated |
| -0,157936538 | 0,782245997 | MED14 | FALSE | Downregulated |
| -0,135563488 | 0,782245997 | PDLIM5 | FALSE | Downregulated |
| -0,141648184 | 0,782245997 | PURA | FALSE | Downregulated |
| -0,197010142 | 0,782245997 | RPL34 | FALSE | Downregulated |
| -0,161394235 | 0,782245997 | TBC1D4 | FALSE | Downregulated |
| -0,129824265 | 0,782263864 | NFAT5 | FALSE | Downregulated |
| 0,487859496 | 0,782263864 | TMEM163 | FALSE | Upregulated |
| -0,661803718 | 0,782525834 | HILPDA | FALSE | Downregulated |
| 0,799462024 | 0,782525834 | LIF | FALSE | Upregulated |
| 0,12787615 | 0,782525834 | PURB | FALSE | Upregulated |
| -0,106856846 | 0,782525834 | RBM39 | FALSE | Downregulated |
| -0,243170526 | 0,782525834 | RP11-983P16.4 | FALSE | Downregulated |
| 0,228296925 | 0,782525834 | TNFSF12 | FALSE | Upregulated |
| -0,225433479 | 0,782813023 | METTL16 | FALSE | Downregulated |
| 0,203713573 | 0,782850541 | FBXO32 | FALSE | Upregulated |
| 0,305577634 | 0,782850541 | WDR75 | FALSE | Upregulated |
| -0,273930878 | 0,783004504 | ZNF616 | FALSE | Downregulated |
| 0,275390936 | 0,783009267 | FASN | FALSE | Upregulated |
| 0,219581108 | 0,783009267 | PHTF1 | FALSE | Upregulated |
| 0,250676243 | 0,783009267 | SLC25A51 | FALSE | Upregulated |

| | | | | |
|---|---|---|---|---|
| 0,204330013 | 0,783009267 | WASH5P | FALSE | Upregulated |
| 0,269606108 | 0,783009267 | ZNF761 | FALSE | Upregulated |
| 0,359084016 | 0,783047031 | CARD9 | FALSE | Upregulated |
| -0,246480495 | 0,783123537 | CDKN2AIPNL | FALSE | Downregulated |
| -0,177953892 | 0,783123537 | COPS7A | FALSE | Downregulated |
| 0,231298529 | 0,783123537 | DYRK1B | FALSE | Upregulated |
| -0,403411992 | 0,783123537 | LOC102723980 | FALSE | Downregulated |
| 0,137802165 | 0,783123537 | LPPR2 | FALSE | Upregulated |
| -0,36534057 | 0,783123537 | OR13F1 | FALSE | Downregulated |
| 0,179064055 | 0,783123537 | RNASET2 | FALSE | Upregulated |
| -0,60520895 | 0,783123537 | SELE | FALSE | Downregulated |
| 0,131649982 | 0,783123537 | URGCP | FALSE | Upregulated |
| 0,461031695 | 0,783231956 | SH2D1A | FALSE | Upregulated |
| -0,214620153 | 0,783320242 | MIOS | FALSE | Downregulated |
| -0,300553783 | 0,783391545 | MPV17L | FALSE | Downregulated |
| 0,127100642 | 0,78345191 | PLD3 | FALSE | Upregulated |
| -0,336900049 | 0,78368213 | LOC101927206 | FALSE | Downregulated |
| -0,134743645 | 0,783810669 | ZNF219 | FALSE | Downregulated |
| 0,162783324 | 0,783940547 | KDM3A | FALSE | Upregulated |
| -0,3315477 | 0,783940547 | TLR3 | FALSE | Downregulated |
| 0,240809931 | 0,784016166 | TMCO3 | FALSE | Upregulated |
| -0,612104558 | 0,784075044 | OR56B4 | FALSE | Downregulated |
| 0,19218746 | 0,784075044 | STAG3L3 | FALSE | Upregulated |
| -0,643352562 | 0,784097042 | LOC101929480 | FALSE | Downregulated |
| 0,377090685 | 0,784101299 | PIGX | FALSE | Upregulated |
| -0,315392224 | 0,784242111 | LAMB3 | FALSE | Downregulated |
| -0,225967493 | 0,784849935 | CCDC71 | FALSE | Downregulated |
| 0,241144545 | 0,784849935 | NKD1 | FALSE | Upregulated |
| -0,241417835 | 0,784849935 | RPS27L | FALSE | Downregulated |
| 0,652150874 | 0,784849935 | SLC22A15 | FALSE | Upregulated |
| 0,285271688 | 0,784849935 | TMEM186 | FALSE | Upregulated |
| 0,317259346 | 0,784849935 | VBP1 | FALSE | Upregulated |
| -0,163115606 | 0,784908656 | BCORL1 | FALSE | Downregulated |
| -0,234465698 | 0,784931523 | ADO | FALSE | Downregulated |
| -0,108057871 | 0,784931523 | KLHDC2 | FALSE | Downregulated |
| -0,179944314 | 0,784931523 | POLM | FALSE | Downregulated |
| -0,138818159 | 0,784946345 | DNMBP-AS1 | FALSE | Downregulated |
| 0,124292947 | 0,7850076 | PSMD7 | FALSE | Upregulated |
| 0,253042602 | 0,785048569 | NOS1AP | FALSE | Upregulated |
| -0,284996712 | 0,78516828 | ANO10 | FALSE | Downregulated |
| 0,23119799 | 0,78516828 | PRDM4 | FALSE | Upregulated |
| 0,298214563 | 0,78516828 | VMA21 | FALSE | Upregulated |
| 0,689607531 | 0,785172558 | RP11-402L6.1 | FALSE | Upregulated |
| 0,106650019 | 0,785172558 | STX17 | FALSE | Upregulated |
| 0,291856494 | 0,785172558 | TLR2 | FALSE | Upregulated |
| 0,305256212 | 0,785188889 | DLL1 | FALSE | Upregulated |
| -0,138950014 | 0,785248703 | EGFL7 | FALSE | Downregulated |
| -0,121926751 | 0,785274253 | RNF11 | FALSE | Downregulated |
| 0,244140445 | 0,785284811 | ARFIP1 | FALSE | Upregulated |
| 0,308591018 | 0,785284811 | CAPN10-AS1 | FALSE | Upregulated |
| 0,305914304 | 0,785284811 | LOC101060181 | FALSE | Upregulated |

| | | | | |
|---|---|---|---|---|
| 0,270852368 | 0,785284811 | SLBP | FALSE | Upregulated |
| 0,561248264 | 0,785670121 | LOC284023 | FALSE | Upregulated |
| 0,227375121 | 0,785883448 | C1orf177 | FALSE | Upregulated |
| 0,298626939 | 0,786047427 | ENPP4 | FALSE | Upregulated |
| -0,256405593 | 0,786047427 | GLRX3 | FALSE | Downregulated |
| 0,130394724 | 0,786047427 | LUZP1 | FALSE | Upregulated |
| -0,155845557 | 0,786074675 | MAPK6 | FALSE | Downregulated |
| 0,401286529 | 0,78649301 | WDR66 | FALSE | Upregulated |
| 0,116346031 | 0,786569862 | AARS | FALSE | Upregulated |
| 0,178228889 | 0,786585119 | LARGE-AS1 | FALSE | Upregulated |
| -0,140164668 | 0,786703178 | PAK2 | FALSE | Downregulated |
| 0,188400515 | 0,786714221 | PLCD4 | FALSE | Upregulated |
| 0,253967259 | 0,786926781 | KIF11 | FALSE | Upregulated |
| -0,573027607 | 0,786962332 | PPIEL | FALSE | Downregulated |
| -0,577676167 | 0,787021954 | LOC101929608 | FALSE | Downregulated |
| 0,284011347 | 0,787283139 | ANKAR | FALSE | Upregulated |
| -0,122483511 | 0,78738991 | EPB41L2 | FALSE | Downregulated |
| 0,615519095 | 0,78738991 | KCNH4 | FALSE | Upregulated |
| 0,215920599 | 0,78738991 | PNMAL2 | FALSE | Upregulated |
| -0,697201533 | 0,787397557 | TEX13A | FALSE | Downregulated |
| -0,123648888 | 0,787397557 | UBL3 | FALSE | Downregulated |
| -0,186361587 | 0,787429683 | FAM192A | FALSE | Downregulated |
| 0,387124199 | 0,787470844 | FLJ12825 | FALSE | Upregulated |
| -0,107799046 | 0,787470844 | MAP3K2 | FALSE | Downregulated |
| 0,530447871 | 0,787551126 | LOC101928189 | FALSE | Upregulated |
| 0,286379021 | 0,787585935 | STXBP3 | FALSE | Upregulated |
| 0,3100583 | 0,78794448 | DENND2C | FALSE | Upregulated |
| -0,195137236 | 0,78794448 | ZNF652 | FALSE | Downregulated |
| -0,139807441 | 0,788003384 | PI4K2B | FALSE | Downregulated |
| -0,229179321 | 0,788003384 | SOX6 | FALSE | Downregulated |
| 0,400296599 | 0,788015492 | HSF2 | FALSE | Upregulated |
| 0,66803441 | 0,788132161 | BANK1 | FALSE | Upregulated |
| 0,073425011 | 0,788132161 | FAXDC2 | FALSE | Upregulated |
| 0,206807551 | 0,788132161 | FKBP10 | FALSE | Upregulated |
| 0,199830449 | 0,788132161 | HSPA14 | FALSE | Upregulated |
| 0,466167269 | 0,788132161 | HTRA3 | FALSE | Upregulated |
| -0,257267463 | 0,788132161 | IL17RE | FALSE | Downregulated |
| -0,116766425 | 0,788132161 | LIFR | FALSE | Downregulated |
| 0,32732846 | 0,788132161 | LOC100132352 | FALSE | Upregulated |
| 0,602314707 | 0,788132161 | LOC100505938 | FALSE | Upregulated |
| 0,655352195 | 0,788132161 | LOC101928557 | FALSE | Upregulated |
| -0,969053812 | 0,788132161 | LOC728554 | FALSE | Downregulated |
| 0,111196237 | 0,788132161 | MAP3K12 | FALSE | Upregulated |
| 0,151295931 | 0,788132161 | MICA | FALSE | Upregulated |
| -0,112175498 | 0,788132161 | NACC2 | FALSE | Downregulated |
| 0,139513491 | 0,788132161 | NCBP2 | FALSE | Upregulated |
| -0,259953349 | 0,788132161 | OAS1 | FALSE | Downregulated |
| -0,627262337 | 0,788132161 | SLC14A2 | FALSE | Downregulated |
| -0,201041417 | 0,788132161 | SLC38A7 | FALSE | Downregulated |
| -0,271718229 | 0,788132161 | SRBD1 | FALSE | Downregulated |
| -0,155007573 | 0,788132161 | SRGAP1 | FALSE | Downregulated |

| | | | | |
|---|---|---|---|---|
| 0,16570624 | 0,788132161 | SYCP3 | FALSE | Upregulated |
| -0,138786852 | 0,788280581 | EYA3 | FALSE | Downregulated |
| -0,26511121 | 0,788292683 | NAPSB | FALSE | Downregulated |
| -0,17524151 | 0,788393708 | POFUT2 | FALSE | Downregulated |
| -0,995816119 | 0,788393708 | SIK1 | FALSE | Downregulated |
| 0,156211348 | 0,788426091 | CTDNEP1 | FALSE | Upregulated |
| -0,216809222 | 0,788612611 | ERCC3 | FALSE | Downregulated |
| 0,161410773 | 0,788683271 | TRIP6 | FALSE | Upregulated |
| 0,374693725 | 0,78937297 | STX11 | FALSE | Upregulated |
| -0,661155149 | 0,789391454 | OXT | FALSE | Downregulated |
| 0,199308642 | 0,789647798 | NCK2 | FALSE | Upregulated |
| 0,293145797 | 0,789767352 | EBF4 | FALSE | Upregulated |
| -0,196429072 | 0,79001322 | CADPS2 | FALSE | Downregulated |
| 0,391955204 | 0,79001322 | RP11-85F14.5 | FALSE | Upregulated |
| -0,133508031 | 0,790248437 | APBB2 | FALSE | Downregulated |
| 0,659574491 | 0,790248437 | BIK | FALSE | Upregulated |
| 0,195706613 | 0,790248437 | CCSER2 | FALSE | Upregulated |
| 0,441054614 | 0,790248437 | CNKSR2 | FALSE | Upregulated |
| -0,124443199 | 0,790248437 | COASY | FALSE | Downregulated |
| 0,327920181 | 0,790248437 | FAM216A | FALSE | Upregulated |
| -0,162574329 | 0,790248437 | HABP4 | FALSE | Downregulated |
| -0,133298088 | 0,790248437 | INPP5A | FALSE | Downregulated |
| 0,564180658 | 0,790248437 | LOC100996517 | FALSE | Upregulated |
| 0,137919843 | 0,790248437 | PAWR | FALSE | Upregulated |
| 0,223158212 | 0,790248437 | RWDD2A | FALSE | Upregulated |
| -0,083465628 | 0,790248437 | SCARB2 | FALSE | Downregulated |
| -0,172881876 | 0,790248437 | SSBP3 | FALSE | Downregulated |
| 0,122615246 | 0,790248437 | TP53INP2 | FALSE | Upregulated |
| 0,253242647 | 0,790248437 | ZSCAN12 | FALSE | Upregulated |
| -0,161075626 | 0,790425962 | FAHD1 | FALSE | Downregulated |
| 0,240820262 | 0,790686085 | SMAGP | FALSE | Upregulated |
| 0,324269226 | 0,790719572 | C14orf2 | FALSE | Upregulated |
| 0,204802653 | 0,790780893 | NES | FALSE | Upregulated |
| -0,107687659 | 0,790948614 | SCAF8 | FALSE | Downregulated |
| -0,448772042 | 0,791406395 | GPC6-AS1 | FALSE | Downregulated |
| 0,330281787 | 0,791406395 | NSUN5P2 | FALSE | Upregulated |
| 0,661637548 | 0,791531302 | PLB1 | FALSE | Upregulated |
| 0,213116741 | 0,791531302 | RRAGD | FALSE | Upregulated |
| 0,566831305 | 0,791838362 | METTL4 | FALSE | Upregulated |
| -0,376279012 | 0,792020653 | UTP11L | FALSE | Downregulated |
| -0,247623234 | 0,792494373 | LOC100132832 | FALSE | Downregulated |
| 0,494244512 | 0,792556702 | EFCAB6-AS1 | FALSE | Upregulated |
| -0,246473123 | 0,792556702 | IFITM1 | FALSE | Downregulated |
| 0,100666657 | 0,792556702 | JMJD8 | FALSE | Upregulated |
| -0,130285596 | 0,792556702 | MKL2 | FALSE | Downregulated |
| -0,085565233 | 0,792556702 | SURF4 | FALSE | Downregulated |
| -0,660292216 | 0,792747292 | LOC729164 | FALSE | Downregulated |
| -0,091822406 | 0,792757144 | PTP4A1 | FALSE | Downregulated |
| 0,577696634 | 0,792757144 | TMEM72 | FALSE | Upregulated |
| 0,468777838 | 0,792887211 | FANCD2 | FALSE | Upregulated |
| 0,345270962 | 0,792924371 | RAD18 | FALSE | Upregulated |

| | | | | |
|---|---|---|---|---|
| 0,410961623 | 0,793029427 | BOLL | FALSE | Upregulated |
| 0,163612296 | 0,793029427 | CLDN3 | FALSE | Upregulated |
| -0,339950882 | 0,793029427 | LOC101928734 | FALSE | Downregulated |
| -0,459509899 | 0,79308696 | BMP10 | FALSE | Downregulated |
| -0,11924395 | 0,79308696 | BSDC1 | FALSE | Downregulated |
| -0,15220769 | 0,79308696 | CAPZB | FALSE | Downregulated |
| -0,143284301 | 0,79308696 | CD151 | FALSE | Downregulated |
| -0,406952422 | 0,79308696 | COX17 | FALSE | Downregulated |
| 0,619847962 | 0,79308696 | EPB42 | FALSE | Upregulated |
| 0,151732339 | 0,79308696 | FRMD4B | FALSE | Upregulated |
| -0,180008807 | 0,79308696 | GSTM4 | FALSE | Downregulated |
| -0,694617723 | 0,79308696 | NBPF25P | FALSE | Downregulated |
| 0,900116484 | 0,79308696 | NPIPB3 | FALSE | Upregulated |
| 0,827853839 | 0,79308696 | PPAN-P2RY11 | FALSE | Upregulated |
| -0,190296613 | 0,79308696 | RBBP9 | FALSE | Downregulated |
| 0,5038612 | 0,79308696 | STK32C | FALSE | Upregulated |
| -0,191673497 | 0,79308696 | ZNF404 | FALSE | Downregulated |
| -0,225027646 | 0,793128973 | NSMCE1 | FALSE | Downregulated |
| -0,262419331 | 0,793254081 | ZNF484 | FALSE | Downregulated |
| -0,169773534 | 0,793413037 | PPP1R3B | FALSE | Downregulated |
| -0,177266075 | 0,793482964 | GDA | FALSE | Downregulated |
| -0,22382652 | 0,793482964 | VPS53 | FALSE | Downregulated |
| 0,193055595 | 0,793513163 | ACAD9 | FALSE | Upregulated |
| -0,536925027 | 0,793723696 | SELL | FALSE | Downregulated |
| -0,743848438 | 0,793723696 | SLCO1C1 | FALSE | Downregulated |
| -0,287472192 | 0,793775664 | RRP12 | FALSE | Downregulated |
| -0,198195295 | 0,793943224 | GJA4 | FALSE | Downregulated |
| -0,419916091 | 0,79406223 | MAGOH | FALSE | Downregulated |
| -0,246326204 | 0,794064952 | C12orf44 | FALSE | Downregulated |
| -0,514562238 | 0,794098244 | BTNL3 | FALSE | Downregulated |
| 0,150022091 | 0,794098244 | PERP | FALSE | Upregulated |
| 0,214243771 | 0,794098244 | PLK1S1 | FALSE | Upregulated |
| 0,378800274 | 0,794173969 | EFCC1 | FALSE | Upregulated |
| 0,390705296 | 0,794173969 | GLB1L2 | FALSE | Upregulated |
| -0,204679472 | 0,794177833 | ARHGEF9 | FALSE | Downregulated |
| -0,135797255 | 0,794212323 | HDGFRP2 | FALSE | Downregulated |
| -0,111295289 | 0,794483482 | DYSF | FALSE | Downregulated |
| -0,627596942 | 0,794712684 | LOC102606465 | FALSE | Downregulated |
| -0,170757617 | 0,794873576 | MST1P2 | FALSE | Downregulated |
| 0,156836194 | 0,794897939 | LAMA5 | FALSE | Upregulated |
| 0,210546125 | 0,794906441 | HERC6 | FALSE | Upregulated |
| 0,241390388 | 0,795005673 | ACTR3C | FALSE | Upregulated |
| -0,130171526 | 0,795005673 | EAF1 | FALSE | Downregulated |
| 0,188677258 | 0,795005673 | LOC101928045 | FALSE | Upregulated |
| 0,609375636 | 0,795005673 | LOC102724413 | FALSE | Upregulated |
| -0,6019623 | 0,795005673 | POU6F2 | FALSE | Downregulated |
| -0,176547029 | 0,795113982 | ACOT8 | FALSE | Downregulated |
| 0,137323326 | 0,795113982 | ANKRD12 | FALSE | Upregulated |
| 0,438315178 | 0,79518996 | SLC39A4 | FALSE | Upregulated |
| -0,274418627 | 0,79518996 | ZCCHC17 | FALSE | Downregulated |
| -0,435347029 | 0,795437947 | CA1 | FALSE | Downregulated |

| | | | | |
|---|---|---|---|---|
| 0,08673237 | 0,795592671 | LPP | FALSE | Upregulated |
| 0,250054673 | 0,795701791 | RP11-395P17.3 | FALSE | Upregulated |
| 0,091070456 | 0,795701791 | ZDHHC5 | FALSE | Upregulated |
| 0,473481995 | 0,795720242 | C8orf76 | FALSE | Upregulated |
| 0,248398053 | 0,795720242 | FRAS1 | FALSE | Upregulated |
| -0,275996239 | 0,795720242 | ID1 | FALSE | Downregulated |
| -0,239858852 | 0,795720242 | LOC101928170 | FALSE | Downregulated |
| -0,188738929 | 0,795720242 | SIDT1 | FALSE | Downregulated |
| 0,57015281 | 0,795720242 | TEKT4P2 | FALSE | Upregulated |
| -0,212175167 | 0,795720242 | UNC5CL | FALSE | Downregulated |
| -0,489116496 | 0,795867626 | SAA1 | FALSE | Downregulated |
| 0,157738641 | 0,795895194 | BTBD1 | FALSE | Upregulated |
| 0,166249038 | 0,795895194 | ISCU | FALSE | Upregulated |
| -0,237182239 | 0,796246922 | NENF | FALSE | Downregulated |
| -0,131384136 | 0,796313433 | STOML2 | FALSE | Downregulated |
| 0,113307243 | 0,796343711 | KIAA0355 | FALSE | Upregulated |
| 0,193800153 | 0,79636941 | PTPRG-AS1 | FALSE | Upregulated |
| 0,224799499 | 0,796441346 | DAXX | FALSE | Upregulated |
| 0,173303906 | 0,796523227 | RP11-98I9.4 | FALSE | Upregulated |
| 0,130262603 | 0,796652182 | ARHGEF17 | FALSE | Upregulated |
| 0,327329931 | 0,796680883 | MUTYH | FALSE | Upregulated |
| 0,277528409 | 0,796692265 | DNAH6 | FALSE | Upregulated |
| -0,236379325 | 0,796705936 | THAP9-AS1 | FALSE | Downregulated |
| 0,596674458 | 0,796774294 | ADCYAP1R1 | FALSE | Upregulated |
| -0,268430665 | 0,796774294 | GDPD5 | FALSE | Downregulated |
| 0,569285809 | 0,796774294 | LOC102724691 | FALSE | Upregulated |
| 0,389787061 | 0,796774294 | SPEF2 | FALSE | Upregulated |
| -0,092822892 | 0,796871219 | NCL | FALSE | Downregulated |
| 0,202370351 | 0,796871219 | PNKP | FALSE | Upregulated |
| 0,469679234 | 0,796979004 | LOC102723425 | FALSE | Upregulated |
| -0,101973251 | 0,797730249 | ABCA2 | FALSE | Downregulated |
| -0,146303046 | 0,797730249 | USP6NL | FALSE | Downregulated |
| 0,147408103 | 0,797895594 | COQ9 | FALSE | Upregulated |
| 0,17507736 | 0,797895594 | FUT11 | FALSE | Upregulated |
| 0,440090711 | 0,797895594 | ROPN1L-AS1 | FALSE | Upregulated |
| 0,237113488 | 0,797895594 | ZNF605 | FALSE | Upregulated |
| 0,161147248 | 0,79790084 | C3orf22 | FALSE | Upregulated |
| -0,186413529 | 0,79790084 | FGFR1OP | FALSE | Downregulated |
| -0,170774734 | 0,798303912 | AGMO | FALSE | Downregulated |
| 0,347926249 | 0,798303912 | CCDC7 | FALSE | Upregulated |
| -0,166785513 | 0,798303912 | GCH1 | FALSE | Downregulated |
| -0,105995706 | 0,798303912 | MICAL3 | FALSE | Downregulated |
| -0,072628589 | 0,798303912 | PPP6R2 | FALSE | Downregulated |
| 1,35425703 | 0,798303912 | RAB4B-EGLN2 | FALSE | Upregulated |
| -0,198510938 | 0,798394415 | AMY2B | FALSE | Downregulated |
| -0,17148723 | 0,798663995 | CASK | FALSE | Downregulated |
| -0,839242851 | 0,798663995 | CDK3 | FALSE | Downregulated |
| -0,079287935 | 0,798663995 | EP400 | FALSE | Downregulated |
| -0,15477675 | 0,798663995 | LOC285074 | FALSE | Downregulated |
| -0,280493666 | 0,798663995 | SOX15 | FALSE | Downregulated |
| 0,189708216 | 0,798954346 | GRWD1 | FALSE | Upregulated |

| | | | | |
|---|---|---|---|---|
| -0,284774411 | 0,798954346 | MIR155HG | FALSE | Downregulated |
| -0,101607604 | 0,798954346 | UHMK1 | FALSE | Downregulated |
| 0,149918978 | 0,798954346 | UNC119B | FALSE | Upregulated |
| 0,17083561 | 0,798985764 | GLO1 | FALSE | Upregulated |
| -0,178132331 | 0,798985764 | SGK3 | FALSE | Downregulated |
| 0,223731201 | 0,799029733 | PTGR2 | FALSE | Upregulated |
| 0,144332249 | 0,799029733 | TSC22D1 | FALSE | Upregulated |
| 0,227994351 | 0,799029733 | WDR70 | FALSE | Upregulated |
| -0,156881625 | 0,79906852 | TCF7L2 | FALSE | Downregulated |
| 0,484420708 | 0,79906852 | ZNF19 | FALSE | Upregulated |
| -0,557992649 | 0,799158724 | C9orf40 | FALSE | Downregulated |
| 0,235515959 | 0,799158724 | JARID2-AS1 | FALSE | Upregulated |
| -0,141345355 | 0,799182552 | CACNB2 | FALSE | Downregulated |
| -0,11895667 | 0,799496732 | TAOK3 | FALSE | Downregulated |
| 0,146993761 | 0,799868044 | RUFY1 | FALSE | Upregulated |
| 0,195375394 | 0,799945641 | STARD13-AS | FALSE | Upregulated |
| 0,205300651 | 0,79998185 | RASGRF2 | FALSE | Upregulated |
| 0,167548275 | 0,800036449 | PROCR | FALSE | Upregulated |
| -0,112143115 | 0,800036449 | TFIP11 | FALSE | Downregulated |
| 0,160082151 | 0,800036449 | ZCWPW1 | FALSE | Upregulated |
| -0,18630203 | 0,800068448 | AMER1 | FALSE | Downregulated |
| 0,258175414 | 0,800251395 | ATOH8 | FALSE | Upregulated |
| -1,105291047 | 0,800251395 | ISY1-RAB43 | FALSE | Downregulated |
| -0,338374076 | 0,800251395 | KCNC2 | FALSE | Downregulated |
| -0,203248893 | 0,800251395 | OR13D1 | FALSE | Downregulated |
| 0,151245962 | 0,800251395 | SF3A2 | FALSE | Upregulated |
| -0,274821938 | 0,800424947 | DTWD2 | FALSE | Downregulated |
| -0,171100778 | 0,800765977 | PRCP | FALSE | Downregulated |
| 0,224007387 | 0,800782003 | CCDC22 | FALSE | Upregulated |
| -0,151511196 | 0,800782003 | ID2 | FALSE | Downregulated |
| -0,098915429 | 0,800782003 | SHANK3 | FALSE | Downregulated |
| -0,224790293 | 0,800789816 | C1orf85 | FALSE | Downregulated |
| -0,317190024 | 0,800789816 | MED6 | FALSE | Downregulated |
| 0,252782336 | 0,800789816 | SPATA5 | FALSE | Upregulated |
| -0,349041155 | 0,801094025 | NDC1 | FALSE | Downregulated |
| -0,346477547 | 0,801291573 | GATM-AS1 | FALSE | Downregulated |
| 0,544111402 | 0,801291573 | GRIA4 | FALSE | Upregulated |
| 1,92043178 | 0,801291573 | NPIPA8 | FALSE | Upregulated |
| -0,173647919 | 0,801291573 | OXA1L | FALSE | Downregulated |
| -0,292185787 | 0,801317698 | AURKA | FALSE | Downregulated |
| 0,549808407 | 0,80140081 | LOC101927699 | FALSE | Upregulated |
| 0,282074779 | 0,80140081 | PLP2 | FALSE | Upregulated |
| -0,149236267 | 0,80140081 | UGT2B10 | FALSE | Downregulated |
| 0,146372336 | 0,801402899 | FLJ13197 | FALSE | Upregulated |
| 0,583824878 | 0,801402899 | POLR2J3 | FALSE | Upregulated |
| -0,122802821 | 0,801575897 | GNPTG | FALSE | Downregulated |
| 0,519918487 | 0,801575897 | LOC100507258 | FALSE | Upregulated |
| 0,536048283 | 0,801648326 | RHOH | FALSE | Upregulated |
| 0,092114104 | 0,801666446 | BTBD2 | FALSE | Upregulated |
| 0,140488349 | 0,801666446 | GP1BA | FALSE | Upregulated |
| -0,14997507 | 0,801666446 | HGSNAT | FALSE | Downregulated |

| | | | | |
|---|---|---|---|---|
| 0,11804496 | 0,801824502 | B3GNT9 | FALSE | Upregulated |
| 1,778733587 | 0,801945947 | TNNC1 | FALSE | Upregulated |
| -0,189450058 | 0,802320873 | DDX19B | FALSE | Downregulated |
| -0,151957325 | 0,802320873 | NCAPD3 | FALSE | Downregulated |
| 0,298529816 | 0,802687305 | KLC2 | FALSE | Upregulated |
| -0,146156279 | 0,802687305 | TBC1D1 | FALSE | Downregulated |
| 0,330832807 | 0,80270437 | EOGT | FALSE | Upregulated |
| -0,250994281 | 0,80270437 | SLC10A5 | FALSE | Downregulated |
| -0,267267869 | 0,802715685 | C7orf25 | FALSE | Downregulated |
| -0,131505357 | 0,802715685 | FAM32A | FALSE | Downregulated |
| 0,407434902 | 0,802802237 | CDKN3 | FALSE | Upregulated |
| 0,188960362 | 0,802802237 | DHX37 | FALSE | Upregulated |
| 0,165972788 | 0,802802237 | LILRB5 | FALSE | Upregulated |
| 0,141259348 | 0,802802237 | WDFY1 | FALSE | Upregulated |
| 0,367276554 | 0,803059745 | PLAGL1 | FALSE | Upregulated |
| -0,176515374 | 0,803191118 | HSPE1 | FALSE | Downregulated |
| -0,524587993 | 0,803413614 | CYP26A1 | FALSE | Downregulated |
| -0,12664519 | 0,803747397 | CLMN | FALSE | Downregulated |
| -0,306503323 | 0,803855878 | SREK1IP1 | FALSE | Downregulated |
| 0,186334681 | 0,803855878 | ZNF330 | FALSE | Upregulated |
| 0,457955814 | 0,803888627 | OR7E14P | FALSE | Upregulated |
| -0,199266966 | 0,80410812 | HFE | FALSE | Downregulated |
| 0,457688073 | 0,804175625 | LOC102723581 | FALSE | Upregulated |
| 0,640179131 | 0,804200638 | LOC102723329 | FALSE | Upregulated |
| 0,297945875 | 0,804406041 | ATRIP | FALSE | Upregulated |
| 0,250343709 | 0,804614012 | CDCA7L | FALSE | Upregulated |
| -0,073523593 | 0,804614012 | CDK13 | FALSE | Downregulated |
| -0,106793973 | 0,804614012 | KIF3B | FALSE | Downregulated |
| 0,18755768 | 0,804614012 | PIK3C3 | FALSE | Upregulated |
| -0,227606195 | 0,804614012 | ZNF385B | FALSE | Downregulated |
| -0,501951736 | 0,804631104 | C1orf162 | FALSE | Downregulated |
| -0,471021983 | 0,804681483 | LIMS3L | FALSE | Downregulated |
| 0,177171823 | 0,804681483 | PLN | FALSE | Upregulated |
| 0,332754333 | 0,80494505 | CD180 | FALSE | Upregulated |
| -0,154493964 | 0,80494505 | ILF2 | FALSE | Downregulated |
| -0,191321705 | 0,805025883 | AIP | FALSE | Downregulated |
| 0,236971388 | 0,805025883 | HHIPL1 | FALSE | Upregulated |
| -0,277017723 | 0,805025883 | MGST3 | FALSE | Downregulated |
| -0,131753431 | 0,805030894 | DNAJC21 | FALSE | Downregulated |
| 0,16320223 | 0,80507528 | GBA | FALSE | Upregulated |
| 0,19190412 | 0,80548389 | CAPZA2 | FALSE | Upregulated |
| -0,196631802 | 0,80548389 | FRAT1 | FALSE | Downregulated |
| 0,473973465 | 0,80548389 | LRRC7 | FALSE | Upregulated |
| -0,137402521 | 0,80548389 | MOCS2 | FALSE | Downregulated |
| -0,086867827 | 0,80548389 | STK25 | FALSE | Downregulated |
| 0,143769786 | 0,80548389 | SUMO2 | FALSE | Upregulated |
| 0,189679352 | 0,80548389 | UBE2B | FALSE | Upregulated |
| -0,548338492 | 0,805613625 | HIST1H2BN | FALSE | Downregulated |
| -0,505167525 | 0,805613625 | LOC100289026 | FALSE | Downregulated |
| 0,127557012 | 0,805613625 | TARBP2 | FALSE | Upregulated |
| -0,129570192 | 0,805614091 | C9orf69 | FALSE | Downregulated |

| | | | | |
|---|---|---|---|---|
| -0,095815955 | 0,805614091 | CPSF7 | FALSE | Downregulated |
| 0,255426946 | 0,805614091 | DUSP5 | FALSE | Upregulated |
| -0,259521504 | 0,805614091 | GPATCH3 | FALSE | Downregulated |
| 0,138489855 | 0,805614091 | SLC23A1 | FALSE | Upregulated |
| -0,176538508 | 0,805614091 | TFDP1 | FALSE | Downregulated |
| -0,120022683 | 0,805796789 | CCNY | FALSE | Downregulated |
| -0,204591611 | 0,805802866 | GMPPB | FALSE | Downregulated |
| -0,156128454 | 0,805882646 | BACH1 | FALSE | Downregulated |
| 0,205488723 | 0,805882646 | BAG3 | FALSE | Upregulated |
| -0,118198881 | 0,805882646 | C17orf70 | FALSE | Downregulated |
| 0,247324046 | 0,805882646 | CCNB1IP1 | FALSE | Upregulated |
| 0,40924434 | 0,805882646 | LOC101927623 | FALSE | Upregulated |
| -0,377936594 | 0,805882646 | PSMG1 | FALSE | Downregulated |
| 0,289874499 | 0,805882646 | TTC9C | FALSE | Upregulated |
| 0,708502093 | 0,805895499 | CARD16 | FALSE | Upregulated |
| -0,084111551 | 0,805895499 | LRPAP1 | FALSE | Downregulated |
| -0,218069862 | 0,806015424 | CTBP1-AS2 | FALSE | Downregulated |
| 0,222953006 | 0,806079926 | SNORD17 | FALSE | Upregulated |
| -0,193255409 | 0,806285629 | RHOB | FALSE | Downregulated |
| 0,183555354 | 0,806285629 | TWF2 | FALSE | Upregulated |
| 0,435966344 | 0,806292057 | POLR3K | FALSE | Upregulated |
| 0,297862264 | 0,806405208 | EIF1B | FALSE | Upregulated |
| -0,37795363 | 0,806405208 | HM13-AS1 | FALSE | Downregulated |
| 0,49887747 | 0,806405208 | LOC102723501 | FALSE | Upregulated |
| -0,183570669 | 0,806405208 | LYPLA1 | FALSE | Downregulated |
| 0,294194695 | 0,806405208 | RIOK2 | FALSE | Upregulated |
| -0,299496127 | 0,806505097 | ACTR5 | FALSE | Downregulated |
| 0,856768788 | 0,806738026 | CORO7-PAM16 | FALSE | Upregulated |
| -0,125945678 | 0,806777771 | AP2A2 | FALSE | Downregulated |
| -0,422477915 | 0,806777771 | GTF2IRD2 | FALSE | Downregulated |
| 0,67741777 | 0,806777771 | MYPN | FALSE | Upregulated |
| 0,328120455 | 0,806777771 | ZNF232 | FALSE | Upregulated |
| 0,156831486 | 0,806819211 | CAMK2G | FALSE | Upregulated |
| -0,687114689 | 0,806819211 | LINC00582 | FALSE | Downregulated |
| 0,244007066 | 0,806819211 | ZBTB34 | FALSE | Upregulated |
| -0,301083215 | 0,806937596 | FOPNL | FALSE | Downregulated |
| 0,261621203 | 0,806937596 | RBM38 | FALSE | Upregulated |
| 0,13621484 | 0,807045285 | APC | FALSE | Upregulated |
| -0,070797229 | 0,807045285 | BRD4 | FALSE | Downregulated |
| 0,1890857 | 0,807045285 | BTN3A3 | FALSE | Upregulated |
| 0,134135158 | 0,807045285 | CAMK2D | FALSE | Upregulated |
| -0,180142799 | 0,807045285 | CASC10 | FALSE | Downregulated |
| 0,365860249 | 0,807045285 | DLEU7 | FALSE | Upregulated |
| 0,26060766 | 0,807045285 | GK5 | FALSE | Upregulated |
| -0,301701215 | 0,807045285 | HSD11B2 | FALSE | Downregulated |
| 0,127837251 | 0,807045285 | MBP | FALSE | Upregulated |
| 0,127629545 | 0,807045285 | NEK4 | FALSE | Upregulated |
| -0,279589144 | 0,807045285 | PFKFB2 | FALSE | Downregulated |
| -0,230172216 | 0,807045285 | RP5-1029K10.2 | FALSE | Downregulated |
| -0,1854165 | 0,807045285 | SEP15 | FALSE | Downregulated |
| -0,185433594 | 0,807045285 | STARD3 | FALSE | Downregulated |

| | | | | |
|---|---|---|---|---|
| -0,167139029 | 0,807045285 | ZFYVE20 | FALSE | Downregulated |
| -0,179815934 | 0,807050768 | SNX25 | FALSE | Downregulated |
| -0,313046778 | 0,80712844 | CNTNAP3B | FALSE | Downregulated |
| 0,132187662 | 0,80712844 | NAB2 | FALSE | Upregulated |
| -0,222511792 | 0,80712844 | ROMO1 | FALSE | Downregulated |
| 0,199906314 | 0,807145019 | ALYREF | FALSE | Upregulated |
| 0,116032414 | 0,807145019 | OLFML1 | FALSE | Upregulated |
| 0,090815299 | 0,807225712 | PKP4 | FALSE | Upregulated |
| 0,575331761 | 0,807454801 | CECR5-AS1 | FALSE | Upregulated |
| -0,14026776 | 0,807523653 | ZFYVE1 | FALSE | Downregulated |
| -0,150540531 | 0,807534648 | DCAF13 | FALSE | Downregulated |
| -0,172353445 | 0,807534648 | DUS3L | FALSE | Downregulated |
| -0,376281806 | 0,807557723 | FCER1G | FALSE | Downregulated |
| 0,194017556 | 0,807819337 | ZNF700 | FALSE | Upregulated |
| -0,388279415 | 0,80797195 | NUP35 | FALSE | Downregulated |
| -0,179846416 | 0,808029795 | LZTS2 | FALSE | Downregulated |
| -0,21495699 | 0,808029795 | SNRPG | FALSE | Downregulated |
| -0,355373454 | 0,808088401 | DAD1 | FALSE | Downregulated |
| -0,572622155 | 0,808088401 | RP11-736K20.6 | FALSE | Downregulated |
| -0,227884468 | 0,808088401 | RPS18P9 | FALSE | Downregulated |
| -0,192666704 | 0,808088401 | UQCC1 | FALSE | Downregulated |
| -0,197503197 | 0,808088401 | ZNF721 | FALSE | Downregulated |
| 0,24603185 | 0,808088401 | ZNF833P | FALSE | Upregulated |
| -0,295426395 | 0,808110396 | CRX | FALSE | Downregulated |
| 0,115117302 | 0,808110396 | EML5 | FALSE | Upregulated |
| 0,373069434 | 0,808389047 | BPGM | FALSE | Upregulated |
| 0,40350575 | 0,808389047 | HS1BP3-IT1 | FALSE | Upregulated |
| 0,300684277 | 0,808641692 | MYC | FALSE | Upregulated |
| -0,158639341 | 0,808641692 | UBXN1 | FALSE | Downregulated |
| -0,166580512 | 0,808729819 | CCDC57 | FALSE | Downregulated |
| -0,192746253 | 0,80877283 | ZNF623 | FALSE | Downregulated |
| 0,379269324 | 0,808813396 | ICAM2 | FALSE | Upregulated |
| 0,112511111 | 0,808813396 | PABPN1 | FALSE | Upregulated |
| -0,154185246 | 0,808813396 | RNMT | FALSE | Downregulated |
| -0,109303624 | 0,808813396 | STAT5B | FALSE | Downregulated |
| -0,321456085 | 0,808813396 | ZNF235 | FALSE | Downregulated |
| 0,422806536 | 0,808821361 | UPK3B | FALSE | Upregulated |
| 0,102376983 | 0,808878151 | BAZ1B | FALSE | Upregulated |
| 0,323063318 | 0,808878151 | ITIH5 | FALSE | Upregulated |
| 0,523416064 | 0,808878151 | LOC101929758 | FALSE | Upregulated |
| 0,090794387 | 0,808944194 | HMGN2 | FALSE | Upregulated |
| -0,244832124 | 0,809114169 | HAUS5 | FALSE | Downregulated |
| 0,346167541 | 0,809114169 | MTFP1 | FALSE | Upregulated |
| 0,20408373 | 0,809114169 | PAK1 | FALSE | Upregulated |
| -0,151459793 | 0,809114169 | SLC25A38 | FALSE | Downregulated |
| -0,385914268 | 0,809114169 | SLED1 | FALSE | Downregulated |
| -0,188862121 | 0,809114169 | UBE2A | FALSE | Downregulated |
| 0,109596123 | 0,809275327 | IVNS1ABP | FALSE | Upregulated |
| 0,162884541 | 0,809275327 | MMP14 | FALSE | Upregulated |
| -0,16916619 | 0,809332458 | ITGB7 | FALSE | Downregulated |
| 0,177342313 | 0,809332458 | UBOX5-AS1 | FALSE | Upregulated |

| | | | | |
|---|---|---|---|---|
| -0,300860317 | 0,80934858 | RP11-106M7.1 | FALSE | Downregulated |
| 0,183465618 | 0,809498267 | CCNF | FALSE | Upregulated |
| 0,242185772 | 0,809498267 | KLHL26 | FALSE | Upregulated |
| -0,116535818 | 0,809498267 | PSME4 | FALSE | Downregulated |
| 0,570147801 | 0,809498267 | RP11-295M18.2 | FALSE | Upregulated |
| -0,215417359 | 0,809498267 | SEMA6D | FALSE | Downregulated |
| -0,402928742 | 0,809498267 | TMEM121 | FALSE | Downregulated |
| -0,104044882 | 0,809512892 | ESRRA | FALSE | Downregulated |
| -0,150721086 | 0,809529201 | MKRN1 | FALSE | Downregulated |
| 0,951195782 | 0,809815851 | LOC102724560 | FALSE | Upregulated |
| 0,178419801 | 0,809879478 | ZNF791 | FALSE | Upregulated |
| -0,136680813 | 0,810186001 | CLK1 | FALSE | Downregulated |
| -0,36534088 | 0,810242219 | KANK4 | FALSE | Downregulated |
| 0,630825997 | 0,810242219 | PSRC1 | FALSE | Upregulated |
| -0,168415261 | 0,810621534 | PSMD13 | FALSE | Downregulated |
| 0,221070152 | 0,810712278 | AC011747.7 | FALSE | Upregulated |
| 0,138865055 | 0,810866263 | ZFP36 | FALSE | Upregulated |
| 0,099326222 | 0,810990959 | DLEU1 | FALSE | Upregulated |
| 0,361503085 | 0,810990959 | FANCG | FALSE | Upregulated |
| 0,48886737 | 0,810990959 | SLC26A5 | FALSE | Upregulated |
| 0,270370511 | 0,811015413 | MORN1 | FALSE | Upregulated |
| 0,133845482 | 0,811111948 | TMEM184C | FALSE | Upregulated |
| 0,138663267 | 0,811290058 | RP11-28G8.1 | FALSE | Upregulated |
| 0,309776719 | 0,81134577 | FBF1 | FALSE | Upregulated |
| 0,116640703 | 0,811606409 | ACTR1A | FALSE | Upregulated |
| -0,12191449 | 0,811785532 | HIRA | FALSE | Downregulated |
| 0,547523006 | 0,811785532 | LOC101927934 | FALSE | Upregulated |
| -0,242724111 | 0,811785532 | LOC102723773 | FALSE | Downregulated |
| -0,203502665 | 0,811785532 | TRAPPC3 | FALSE | Downregulated |
| 0,255322544 | 0,811849497 | AC020571.3 | FALSE | Upregulated |
| 0,12435009 | 0,812035304 | ACAP3 | FALSE | Upregulated |
| 0,296330997 | 0,812035304 | BTN2A2 | FALSE | Upregulated |
| -0,12101731 | 0,812035304 | CBX7 | FALSE | Downregulated |
| 0,343597549 | 0,812035304 | MADCAM1 | FALSE | Upregulated |
| -0,409968579 | 0,812035304 | POP1 | FALSE | Downregulated |
| -0,722609061 | 0,81221639 | COL19A1 | FALSE | Downregulated |
| 0,360748612 | 0,81227124 | MRPL47 | FALSE | Upregulated |
| 0,144116853 | 0,812338846 | EPHA1 | FALSE | Upregulated |
| -0,160938649 | 0,812338846 | LSM14A | FALSE | Downregulated |
| -0,467368989 | 0,812338846 | LY86 | FALSE | Downregulated |
| -0,134598882 | 0,812338846 | PDXP | FALSE | Downregulated |
| -0,618137847 | 0,812338846 | RGS10 | FALSE | Downregulated |
| -0,256949529 | 0,812338846 | TRIP4 | FALSE | Downregulated |
| 0,228363657 | 0,812989143 | ABCA4 | FALSE | Upregulated |
| -0,29596307 | 0,812989143 | DLGAP2 | FALSE | Downregulated |
| -0,285623505 | 0,812989143 | VIL1 | FALSE | Downregulated |
| 0,068762623 | 0,813010184 | C6orf62 | FALSE | Upregulated |
| -0,115319538 | 0,813010184 | TMEM97 | FALSE | Downregulated |
| -0,292224503 | 0,813019739 | MCCD1 | FALSE | Downregulated |
| 0,139613192 | 0,813092761 | C15orf27 | FALSE | Upregulated |
| 0,114471709 | 0,813092761 | MARK3 | FALSE | Upregulated |

| | | | | |
|---|---|---|---|---|
| -0,599835001 | 0,813163998 | CGRRF1 | FALSE | Downregulated |
| 0,106473156 | 0,813163998 | EPS15 | FALSE | Upregulated |
| 0,216285261 | 0,813163998 | NPIPB9 | FALSE | Upregulated |
| -0,175848216 | 0,813163998 | RSRC2 | FALSE | Downregulated |
| 0,105832855 | 0,813163998 | SMG7 | FALSE | Upregulated |
| 0,351774946 | 0,813163998 | TMEM102 | FALSE | Upregulated |
| 0,315756688 | 0,813163998 | VCPKMT | FALSE | Upregulated |
| 0,332099443 | 0,813317686 | AP4B1 | FALSE | Upregulated |
| -0,472386622 | 0,813317686 | LINC01127 | FALSE | Downregulated |
| 0,145696448 | 0,813317686 | NELFE | FALSE | Upregulated |
| 0,59682211 | 0,813473952 | PLK1 | FALSE | Upregulated |
| 0,41170458 | 0,813677586 | ABRACL | FALSE | Upregulated |
| 0,179220173 | 0,813677586 | LITAF | FALSE | Upregulated |
| 0,282760101 | 0,813677586 | RP11-24J23.2 | FALSE | Upregulated |
| -0,191511797 | 0,813677586 | STK36 | FALSE | Downregulated |
| -0,182103927 | 0,813677586 | WHAMM | FALSE | Downregulated |
| 0,164567364 | 0,813717577 | TTBK2 | FALSE | Upregulated |
| -0,167400942 | 0,813717577 | ZNF76 | FALSE | Downregulated |
| -0,121769191 | 0,813927688 | ZDHHC18 | FALSE | Downregulated |
| -0,145905372 | 0,813977348 | PHF10 | FALSE | Downregulated |
| -0,156481385 | 0,814117673 | CLCN6 | FALSE | Downregulated |
| -0,555266222 | 0,814117673 | LINC00907 | FALSE | Downregulated |
| 0,55847049 | 0,814117673 | LOC100131257 | FALSE | Upregulated |
| -0,095503827 | 0,814117673 | PAPOLA | FALSE | Downregulated |
| 0,086047661 | 0,814117673 | UBQLN1 | FALSE | Upregulated |
| -0,318722892 | 0,814170896 | CCDC87 | FALSE | Downregulated |
| 0,494745636 | 0,814170896 | LPCAT4 | FALSE | Upregulated |
| 0,14436302 | 0,814170896 | PSMA6 | FALSE | Upregulated |
| -0,154356283 | 0,814238664 | LATS2 | FALSE | Downregulated |
| 0,249598236 | 0,814238664 | LOC93622 | FALSE | Upregulated |
| -0,204009439 | 0,814241815 | SYNE4 | FALSE | Downregulated |
| -0,283376484 | 0,814390249 | MRPL44 | FALSE | Downregulated |
| -0,414905415 | 0,814390249 | TMPRSS11A | FALSE | Downregulated |
| -0,188882966 | 0,814632004 | HACL1 | FALSE | Downregulated |
| 0,623657471 | 0,814632004 | LOC101930033 | FALSE | Upregulated |
| 0,227148472 | 0,814947981 | ZXDB | FALSE | Upregulated |
| -0,186679107 | 0,814955942 | IFT88 | FALSE | Downregulated |
| 0,104440901 | 0,814955942 | KDM5C | FALSE | Upregulated |
| -0,242500221 | 0,815148503 | DDX50 | FALSE | Downregulated |
| 0,199740217 | 0,815148503 | DTX2P1-UPK3BP1-PMS2P11 | FALSE | Upregulated |
| -0,146607752 | 0,81543037 | NIT2 | FALSE | Downregulated |
| -0,488384358 | 0,81543037 | SLC16A1-AS1 | FALSE | Downregulated |
| -0,176521513 | 0,815851519 | SP110 | FALSE | Downregulated |
| -0,529219536 | 0,815858929 | FAM26E | FALSE | Downregulated |
| 0,125184587 | 0,815858929 | NEURL1B | FALSE | Upregulated |
| 0,126477478 | 0,816006109 | NFATC2 | FALSE | Upregulated |
| 0,194363584 | 0,816427301 | MARCH5 | FALSE | Upregulated |
| 0,264022585 | 0,816519468 | LOC100268168 | FALSE | Upregulated |
| 0,517280396 | 0,816697608 | LINC00265 | FALSE | Upregulated |
| 0,187364942 | 0,816697608 | NOM1 | FALSE | Upregulated |

| | | | | |
|---|---|---|---|---|
| 0,285116567 | 0,816781571 | FAM173B | FALSE | Upregulated |
| 0,396121706 | 0,816781571 | STXBP4 | FALSE | Upregulated |
| -0,128780845 | 0,817265913 | PRKAB2 | FALSE | Downregulated |
| -0,145549704 | 0,817265913 | SEC61B | FALSE | Downregulated |
| 0,159027507 | 0,817359453 | CAPZA1 | FALSE | Upregulated |
| -0,152380813 | 0,817359453 | RP11-290F20.2 | FALSE | Downregulated |
| 0,056852345 | 0,817359453 | SBF1 | FALSE | Upregulated |
| -0,312316731 | 0,817359453 | YBEY | FALSE | Downregulated |
| -0,126285141 | 0,817392742 | AMPD2 | FALSE | Downregulated |
| 0,497740249 | 0,817392742 | CHI3L1 | FALSE | Upregulated |
| 0,138190417 | 0,817392742 | EPN2-AS1 | FALSE | Upregulated |
| 0,144567899 | 0,817481421 | PROSER2-AS1 | FALSE | Upregulated |
| 0,449711187 | 0,817707345 | ASIC4 | FALSE | Upregulated |
| -0,752934541 | 0,817707345 | LOC100996442 | FALSE | Downregulated |
| 0,405352752 | 0,817737064 | ISM1-AS1 | FALSE | Upregulated |
| -0,408590125 | 0,817882931 | SULT1C2P1 | FALSE | Downregulated |
| 0,157632861 | 0,817937408 | ADRA1B | FALSE | Upregulated |
| 0,329013748 | 0,817937408 | ATRNL1 | FALSE | Upregulated |
| -0,124854402 | 0,817937408 | PRKCA | FALSE | Downregulated |
| -0,210522918 | 0,817937408 | ZNF680 | FALSE | Downregulated |
| 0,19480403 | 0,817948943 | IMPACT | FALSE | Upregulated |
| 0,231431583 | 0,817948943 | LOC102724733 | FALSE | Upregulated |
| -0,211291952 | 0,817948943 | PSMC6 | FALSE | Downregulated |
| 0,287821042 | 0,817948943 | RAE1 | FALSE | Upregulated |
| 0,108630382 | 0,818013703 | ZCCHC11 | FALSE | Upregulated |
| -0,229512413 | 0,818056001 | FOLR2 | FALSE | Downregulated |
| -0,259447114 | 0,818454762 | ARID3A | FALSE | Downregulated |
| -0,134308392 | 0,818454762 | CYP1B1 | FALSE | Downregulated |
| 0,597918565 | 0,818454762 | ERI3-IT1 | FALSE | Upregulated |
| -1,23576088 | 0,818454762 | GOLGA8K | FALSE | Downregulated |
| 0,191409362 | 0,818454762 | GTF3C3 | FALSE | Upregulated |
| 0,139623484 | 0,818454762 | JARID2 | FALSE | Upregulated |
| 0,131137005 | 0,818454762 | LOC102724177 | FALSE | Upregulated |
| -0,266729726 | 0,818454762 | PNPLA4 | FALSE | Downregulated |
| 0,104630434 | 0,818454762 | TSR1 | FALSE | Upregulated |
| -0,080819834 | 0,818504321 | KIF1B | FALSE | Downregulated |
| 0,41042874 | 0,818533451 | LYG1 | FALSE | Upregulated |
| -0,12292651 | 0,818558169 | POLDIP3 | FALSE | Downregulated |
| 0,499533496 | 0,818582494 | C2CD4A | FALSE | Upregulated |
| -0,521288024 | 0,818582494 | C7orf55 | FALSE | Downregulated |
| 0,179040572 | 0,818582494 | DDIT3 | FALSE | Upregulated |
| -0,125645434 | 0,818582494 | DLST | FALSE | Downregulated |
| -0,130429589 | 0,818582494 | ELF1 | FALSE | Downregulated |
| 0,445750434 | 0,818582494 | ELMO1-AS1 | FALSE | Upregulated |
| -0,10936053 | 0,818582494 | F2RL2 | FALSE | Downregulated |
| -0,351668006 | 0,818582494 | GUCY2C | FALSE | Downregulated |
| -0,127946929 | 0,818787595 | PROSER2 | FALSE | Downregulated |
| 0,190108886 | 0,819053192 | LOC100996273 | FALSE | Upregulated |
| 0,586185918 | 0,81909025 | MS4A1 | FALSE | Upregulated |
| -0,155814252 | 0,819173972 | VPS4B | FALSE | Downregulated |
| 0,610746533 | 0,819228944 | TMEM54 | FALSE | Upregulated |

| | | | | |
|---|---|---|---|---|
| 0,368364025 | 0,819263397 | FOXQ1 | FALSE | Upregulated |
| 0,086835028 | 0,819305905 | MSL1 | FALSE | Upregulated |
| -0,408010085 | 0,819395599 | MPZL3 | FALSE | Downregulated |
| -0,11749787 | 0,819553077 | CMIP | FALSE | Downregulated |
| 0,188738765 | 0,819553077 | DSTYK | FALSE | Upregulated |
| 0,372933006 | 0,819553077 | RPA3-AS1 | FALSE | Upregulated |
| -0,221105386 | 0,819553077 | ZNF250 | FALSE | Downregulated |
| 0,407758021 | 0,819671844 | MGC34796 | FALSE | Upregulated |
| 0,355913572 | 0,819700529 | SOGA3 | FALSE | Upregulated |
| -0,115328911 | 0,819790163 | DNAJB1 | FALSE | Downregulated |
| 0,193944872 | 0,820208439 | GUCY1A2 | FALSE | Upregulated |
| 0,101286316 | 0,820649935 | ZNF638 | FALSE | Upregulated |
| 0,31036719 | 0,820707588 | LOC102723782 | FALSE | Upregulated |
| 0,562581793 | 0,82075945 | LINC00152 | FALSE | Upregulated |
| 0,56161353 | 0,82075945 | RP11-234B24.2 | FALSE | Upregulated |
| -0,195994459 | 0,82075945 | SLC25A32 | FALSE | Downregulated |
| 0,287166329 | 0,82075945 | TNFSF8 | FALSE | Upregulated |
| 0,13129218 | 0,820813616 | BAHD1 | FALSE | Upregulated |
| 0,551912423 | 0,820813616 | BST1 | FALSE | Upregulated |
| -0,178851891 | 0,820813616 | EIF2B4 | FALSE | Downregulated |
| 0,439161641 | 0,820813616 | EIF3C | FALSE | Upregulated |
| -0,215971435 | 0,820813616 | FAM117A | FALSE | Downregulated |
| 3,268624631 | 0,820813616 | KRT14 | FALSE | Upregulated |
| -0,378121924 | 0,820813616 | MAGEA8 | FALSE | Downregulated |
| -0,16104028 | 0,820813616 | MESDC2 | FALSE | Downregulated |
| 0,200807282 | 0,820813616 | MRPL10 | FALSE | Upregulated |
| 0,236456285 | 0,820813616 | NME3 | FALSE | Upregulated |
| -0,12404471 | 0,820813616 | SIL1 | FALSE | Downregulated |
| 0,092409581 | 0,820813616 | SMAD4 | FALSE | Upregulated |
| -0,533653119 | 0,820813616 | TBX20 | FALSE | Downregulated |
| -0,411664928 | 0,820820428 | KCNK1 | FALSE | Downregulated |
| -0,231961025 | 0,820820428 | LINC00574 | FALSE | Downregulated |
| 0,153872533 | 0,820820428 | PLEKHA5 | FALSE | Upregulated |
| -0,093562391 | 0,820846892 | DVL3 | FALSE | Downregulated |
| -0,429074922 | 0,820846892 | LINC01003 | FALSE | Downregulated |
| -0,13201487 | 0,820846892 | MAN1B1 | FALSE | Downregulated |
| 0,303007314 | 0,820871034 | LOC100862671 | FALSE | Upregulated |
| 0,700365245 | 0,820871034 | LY75-CD302 | FALSE | Upregulated |
| 0,501810614 | 0,820871034 | MGC72080 | FALSE | Upregulated |
| -0,110603682 | 0,820871034 | MGST1 | FALSE | Downregulated |
| -0,178136817 | 0,820871034 | MRPL3 | FALSE | Downregulated |
| -0,108980377 | 0,820871034 | PON2 | FALSE | Downregulated |
| 0,169158469 | 0,820871034 | RAB11FIP4 | FALSE | Upregulated |
| 0,18010405 | 0,820871034 | SPAG5-AS1 | FALSE | Upregulated |
| 0,258770851 | 0,820871034 | ZNF561 | FALSE | Upregulated |
| 0,147440811 | 0,820964242 | FBXO8 | FALSE | Upregulated |
| -0,274956977 | 0,820964242 | HIST1H1A | FALSE | Downregulated |
| 0,105169449 | 0,820964242 | KDM4A | FALSE | Upregulated |
| -0,931024968 | 0,820964242 | LINC00520 | FALSE | Downregulated |
| -0,207854595 | 0,820964242 | LOC101928143 | FALSE | Downregulated |
| 0,299580962 | 0,820964242 | MPV17L2 | FALSE | Upregulated |

| | | | | |
|---|---|---|---|---|
| 0,117713792 | 0,820964242 | PHKB | FALSE | Upregulated |
| -0,158651141 | 0,821204991 | CCNT1 | FALSE | Downregulated |
| -0,181997435 | 0,821204991 | LOC102723722 | FALSE | Downregulated |
| -0,079332828 | 0,821204991 | SSR1 | FALSE | Downregulated |
| 0,513780604 | 0,821204991 | TMEM99 | FALSE | Upregulated |
| 0,220290083 | 0,821411151 | GABBR1 | FALSE | Upregulated |
| 0,109829598 | 0,82151348 | UBE2D4 | FALSE | Upregulated |
| 0,420755515 | 0,821673979 | ANKDD1B | FALSE | Upregulated |
| -0,119251363 | 0,821673979 | INTS3 | FALSE | Downregulated |
| -0,485794448 | 0,821673979 | RBM45 | FALSE | Downregulated |
| -0,14307969 | 0,821803801 | ANGEL1 | FALSE | Downregulated |
| 0,141821629 | 0,821803801 | C15orf37 | FALSE | Upregulated |
| 0,388031546 | 0,821803801 | C6orf10 | FALSE | Upregulated |
| 0,191541918 | 0,821803801 | CDR2 | FALSE | Upregulated |
| -0,132127557 | 0,821803801 | CPPED1 | FALSE | Downregulated |
| -0,273998888 | 0,821803801 | GPN3 | FALSE | Downregulated |
| 0,432066629 | 0,821803801 | LOC388820 | FALSE | Upregulated |
| -0,33512499 | 0,821803801 | METTL2A | FALSE | Downregulated |
| 0,170313799 | 0,821803801 | RAB4A | FALSE | Upregulated |
| 0,296056362 | 0,821960723 | SLC29A3 | FALSE | Upregulated |
| -0,167956654 | 0,822036253 | ARMCX4 | FALSE | Downregulated |
| 0,14218357 | 0,822055098 | ACP5 | FALSE | Upregulated |
| 0,10574456 | 0,822055098 | ATP1B1 | FALSE | Upregulated |
| -0,1392893 | 0,822055098 | CAPNS1 | FALSE | Downregulated |
| -0,549786749 | 0,822055098 | PSPN | FALSE | Downregulated |
| 0,279278121 | 0,822055098 | RWDD4 | FALSE | Upregulated |
| -0,199755734 | 0,822055098 | SDAD1 | FALSE | Downregulated |
| -0,175697779 | 0,822055098 | ZNF579 | FALSE | Downregulated |
| 0,132771231 | 0,822127099 | THEM5 | FALSE | Upregulated |
| 0,100578257 | 0,822258157 | BSG | FALSE | Upregulated |
| 0,261435374 | 0,822258157 | C3orf38 | FALSE | Upregulated |
| -0,133006942 | 0,822268774 | IKBKAP | FALSE | Downregulated |
| -0,259639348 | 0,822268774 | NDUFAF1 | FALSE | Downregulated |
| 1,548988208 | 0,822268774 | PRODH | FALSE | Upregulated |
| -0,223673396 | 0,822268774 | UQCR10 | FALSE | Downregulated |
| 0,130877246 | 0,822268774 | WDR88 | FALSE | Upregulated |
| -0,118900426 | 0,822351805 | DCTD | FALSE | Downregulated |
| -0,293615756 | 0,822351805 | LOC339803 | FALSE | Downregulated |
| -0,240699824 | 0,822351805 | MRPL23 | FALSE | Downregulated |
| 0,08853532 | 0,822682689 | FKBP4 | FALSE | Upregulated |
| -0,089437259 | 0,822682689 | GTF3C1 | FALSE | Downregulated |
| 0,137485272 | 0,822682689 | HIST1H2AC | FALSE | Upregulated |
| -0,393464748 | 0,822682689 | LOC101929513 | FALSE | Downregulated |
| 0,38519251 | 0,822682689 | LOC102724578 | FALSE | Upregulated |
| 0,12146984 | 0,822753799 | LAMB1 | FALSE | Upregulated |
| 0,308629094 | 0,823142049 | ARR3 | FALSE | Upregulated |
| 0,201376443 | 0,823142049 | DDX27 | FALSE | Upregulated |
| 0,155535359 | 0,823142049 | LHX4 | FALSE | Upregulated |
| -0,271151345 | 0,823142049 | MRPL39 | FALSE | Downregulated |
| -0,363949008 | 0,823142049 | OSBPL7 | FALSE | Downregulated |
| -1,042926567 | 0,823142049 | RNU1-2 | FALSE | Downregulated |

| | | | | |
|---|---|---|---|---|
| 0,194837682 | 0,823142049 | SMG7-AS1 | FALSE | Upregulated |
| 0,101636665 | 0,823142049 | TRANK1 | FALSE | Upregulated |
| 0,155250789 | 0,823254283 | RRP7A | FALSE | Upregulated |
| 0,478243544 | 0,823270678 | TAS2R19 | FALSE | Upregulated |
| 0,098596012 | 0,823292809 | ZBTB11 | FALSE | Upregulated |
| -0,100603467 | 0,823384474 | ATXN7L3B | FALSE | Downregulated |
| 0,302531532 | 0,823384474 | RCN2 | FALSE | Upregulated |
| 0,12554443 | 0,823402121 | PSME2 | FALSE | Upregulated |
| 0,081698833 | 0,823590046 | ADD1 | FALSE | Upregulated |
| 0,147483939 | 0,823809268 | FRS2 | FALSE | Upregulated |
| 0,328585371 | 0,823809268 | GADD45G | FALSE | Upregulated |
| -0,61801638 | 0,823809268 | RGS1 | FALSE | Downregulated |
| 0,288714016 | 0,823917383 | SNORD22 | FALSE | Upregulated |
| -0,201216715 | 0,824061568 | HTATIP2 | FALSE | Downregulated |
| 0,19984318 | 0,82429946 | AP3S1 | FALSE | Upregulated |
| 0,289355955 | 0,82429946 | APLF | FALSE | Upregulated |
| 0,174875156 | 0,82429946 | CSTF3 | FALSE | Upregulated |
| -0,161376026 | 0,82429946 | FANCF | FALSE | Downregulated |
| 0,707048895 | 0,82429946 | LOC102724374 | FALSE | Upregulated |
| 0,246896918 | 0,82429946 | LOC284412 | FALSE | Upregulated |
| 0,205396616 | 0,82429946 | NBPF20 | FALSE | Upregulated |
| -0,117987406 | 0,82429946 | PGK1 | FALSE | Downregulated |
| -0,25975864 | 0,82429946 | PGM2 | FALSE | Downregulated |
| -0,114931895 | 0,82429946 | SNRNP70 | FALSE | Downregulated |
| 0,257179902 | 0,82429946 | TMCC3 | FALSE | Upregulated |
| 0,36358573 | 0,824423838 | MCTP2 | FALSE | Upregulated |
| -0,521246885 | 0,824423838 | METAP1D | FALSE | Downregulated |
| -0,124462029 | 0,824665438 | CD4 | FALSE | Downregulated |
| 0,291554065 | 0,824732718 | LRP4 | FALSE | Upregulated |
| -1,772482769 | 0,824732718 | TRIM6-TRIM34 | FALSE | Downregulated |
| -0,130552226 | 0,82475937 | ATP9B | FALSE | Downregulated |
| 0,591006654 | 0,82475937 | ERCC6-PGBD3 | FALSE | Upregulated |
| 0,783464601 | 0,82475937 | FAM156A | FALSE | Upregulated |
| 0,148151827 | 0,82475937 | FAM169A | FALSE | Upregulated |
| 0,177567881 | 0,82475937 | FNDC5 | FALSE | Upregulated |
| 0,30926702 | 0,82475937 | GOLT1B | FALSE | Upregulated |
| 0,246966984 | 0,82475937 | GSTP1 | FALSE | Upregulated |
| -0,488696044 | 0,82475937 | LOC100233156 | FALSE | Downregulated |
| 0,140993382 | 0,82475937 | LPIN1 | FALSE | Upregulated |
| 0,268636448 | 0,82475937 | MED17 | FALSE | Upregulated |
| 0,50863653 | 0,82475937 | MMP17 | FALSE | Upregulated |
| -0,088296812 | 0,82475937 | NONO | FALSE | Downregulated |
| 0,225754386 | 0,82475937 | RP11-840I19.3 | FALSE | Upregulated |
| -0,095306967 | 0,82475937 | STK24 | FALSE | Downregulated |
| -0,159154877 | 0,82475937 | TARS2 | FALSE | Downregulated |
| 0,524694821 | 0,82475937 | TCEB3CL | FALSE | Upregulated |
| 0,139643047 | 0,82475937 | ZNF618 | FALSE | Upregulated |
| -0,252412107 | 0,82475937 | ZNF74 | FALSE | Downregulated |
| 0,076240347 | 0,824776112 | MYL6 | FALSE | Upregulated |
| -0,104992018 | 0,824837644 | TRIM33 | FALSE | Downregulated |
| -0,537959678 | 0,824880718 | CCDC60 | FALSE | Downregulated |

| | | | | |
|---|---|---|---|---|
| 0,440934171 | 0,824880718 | CRYAA | FALSE | Upregulated |
| -0,121197244 | 0,824880718 | CSK | FALSE | Downregulated |
| -0,264684508 | 0,824880718 | DCLRE1A | FALSE | Downregulated |
| 0,123446281 | 0,824880718 | FDFT1 | FALSE | Upregulated |
| -0,109414036 | 0,824880718 | KDELR2 | FALSE | Downregulated |
| 0,149284364 | 0,824880718 | PEX11G | FALSE | Upregulated |
| -0,082192265 | 0,824880718 | SCARNA2 | FALSE | Downregulated |
| 0,405307336 | 0,824880718 | SERP2 | FALSE | Upregulated |
| -0,130527251 | 0,824880718 | SMCR8 | FALSE | Downregulated |
| -0,14650914 | 0,824967295 | ARF3 | FALSE | Downregulated |
| 0,362929954 | 0,824967295 | FOXD2-AS1 | FALSE | Upregulated |
| -0,201028809 | 0,824967295 | LGALS8-AS1 | FALSE | Downregulated |
| 0,420645234 | 0,825000066 | WFDC1 | FALSE | Upregulated |
| 0,147493775 | 0,825121563 | RAB24 | FALSE | Upregulated |
| 0,242949417 | 0,825205917 | LINC00092 | FALSE | Upregulated |
| -0,257355584 | 0,825374576 | GALNT18 | FALSE | Downregulated |
| 0,157643288 | 0,825549796 | ANAPC13 | FALSE | Upregulated |
| 0,303870273 | 0,825549796 | DPY19L1P1 | FALSE | Upregulated |
| -0,058471837 | 0,825549796 | HNRNPU | FALSE | Downregulated |
| 0,387943881 | 0,825549796 | LOC101928895 | FALSE | Upregulated |
| 0,23643937 | 0,825549796 | LOC102723820 | FALSE | Upregulated |
| -0,174234859 | 0,825549796 | NFS1 | FALSE | Downregulated |
| 0,359990473 | 0,825549796 | RP11-75N4.2 | FALSE | Upregulated |
| -0,283811384 | 0,825549796 | TRIQK | FALSE | Downregulated |
| 0,214163929 | 0,825811047 | MCU | FALSE | Upregulated |
| 0,151787142 | 0,825811047 | PQLC2 | FALSE | Upregulated |
| 0,167347563 | 0,825962846 | SATB2 | FALSE | Upregulated |
| -0,260138017 | 0,826040652 | ARMC1 | FALSE | Downregulated |
| -0,085429112 | 0,826040652 | KDM3B | FALSE | Downregulated |
| -0,245922353 | 0,826044466 | FABP2 | FALSE | Downregulated |
| 0,181051072 | 0,826163706 | PHF21A | FALSE | Upregulated |
| 0,156841845 | 0,826267425 | METTL7B | FALSE | Upregulated |
| -0,11762127 | 0,826339565 | AKT1S1 | FALSE | Downregulated |
| -0,541758499 | 0,826339565 | EGFLAM-AS2 | FALSE | Downregulated |
| 0,264830899 | 0,826339565 | FMO2 | FALSE | Upregulated |
| -0,248095012 | 0,826339565 | ORAOV1 | FALSE | Downregulated |
| -0,11927831 | 0,826339565 | PXDC1 | FALSE | Downregulated |
| 0,510103299 | 0,826339565 | ROR1 | FALSE | Upregulated |
| 0,160309393 | 0,826489299 | ZNF205 | FALSE | Upregulated |
| 0,432733193 | 0,826549763 | FERMT1 | FALSE | Upregulated |
| 0,289632519 | 0,826652029 | PSTPIP2 | FALSE | Upregulated |
| 1,255139654 | 0,826713476 | F8A2 | FALSE | Upregulated |
| -0,10641787 | 0,826713476 | INPPL1 | FALSE | Downregulated |
| -0,561648882 | 0,826713476 | LOC101928947 | FALSE | Downregulated |
| -0,635193709 | 0,826713476 | RPL19P12 | FALSE | Downregulated |
| -0,127352952 | 0,826715046 | OTUB1 | FALSE | Downregulated |
| 0,228557178 | 0,826908903 | ABRA | FALSE | Upregulated |
| -0,297526759 | 0,826908903 | ADM5 | FALSE | Downregulated |
| -0,493572798 | 0,827004998 | GRM8 | FALSE | Downregulated |
| -0,106535251 | 0,82703233 | PPP2R1A | FALSE | Downregulated |
| 0,422755656 | 0,827070132 | ASPRV1 | FALSE | Upregulated |

| | | | | |
|---|---|---|---|---|
| 0,091268089 | 0,827070132 | CALM3 | FALSE | Upregulated |
| 0,18638216 | 0,827070132 | CCT6A | FALSE | Upregulated |
| 0,363135649 | 0,827070132 | CHSY3 | FALSE | Upregulated |
| 0,402920981 | 0,827070132 | FCHO1 | FALSE | Upregulated |
| -0,549838463 | 0,827070132 | FLJ20712 | FALSE | Downregulated |
| 0,152046523 | 0,827070132 | LOC100128531 | FALSE | Upregulated |
| 0,560596387 | 0,827070132 | POLR2J2 | FALSE | Upregulated |
| 0,305601581 | 0,827070132 | RNF216P1 | FALSE | Upregulated |
| 0,09269895 | 0,827070132 | RORA | FALSE | Upregulated |
| 0,238446135 | 0,827070132 | RP5-1057J7.6 | FALSE | Upregulated |
| 0,123515302 | 0,827070132 | SIKE1 | FALSE | Upregulated |
| -0,259122327 | 0,827070132 | TBC1D22B | FALSE | Downregulated |
| -0,175854338 | 0,827070132 | TTLL2 | FALSE | Downregulated |
| 0,469986366 | 0,827070132 | ZNF280C | FALSE | Upregulated |
| -0,180740038 | 0,827070132 | ZNF628 | FALSE | Downregulated |
| -0,324015573 | 0,827131939 | HAO2-IT1 | FALSE | Downregulated |
| 0,2602524 | 0,82737944 | FBLN7 | FALSE | Upregulated |
| 0,277513212 | 0,827479835 | CCZ1 | FALSE | Upregulated |
| -0,156118746 | 0,827495287 | NCSTN | FALSE | Downregulated |
| -0,189065275 | 0,827495287 | NDUFA5 | FALSE | Downregulated |
| 0,166817528 | 0,827495287 | RAB3IL1 | FALSE | Upregulated |
| -0,092619981 | 0,827590452 | RARA | FALSE | Downregulated |
| -0,419916593 | 0,827658392 | DNAH5 | FALSE | Downregulated |
| -0,304136461 | 0,827955678 | C11orf1 | FALSE | Downregulated |
| -0,155690708 | 0,828040493 | DNAJA2 | FALSE | Downregulated |
| 0,07359602 | 0,828040493 | MYCBP2 | FALSE | Upregulated |
| -0,250063053 | 0,828040493 | NIPSNAP3A | FALSE | Downregulated |
| 0,111061388 | 0,828040493 | PLCB3 | FALSE | Upregulated |
| 0,196384504 | 0,828040493 | SESTD1 | FALSE | Upregulated |
| -1,529228005 | 0,828040493 | TBC1D3C | FALSE | Downregulated |
| -0,207023021 | 0,828134993 | EBAG9 | FALSE | Downregulated |
| -0,12143591 | 0,828134993 | EFR3A | FALSE | Downregulated |
| -0,232397174 | 0,828203962 | PRRG1 | FALSE | Downregulated |
| 0,319586647 | 0,82829932 | CRYBA1 | FALSE | Upregulated |
| 0,38962702 | 0,82829932 | LOC101928527 | FALSE | Upregulated |
| -0,232366916 | 0,82829932 | SLC17A3 | FALSE | Downregulated |
| -0,365475616 | 0,828429202 | LOC101927663 | FALSE | Downregulated |
| 0,111862565 | 0,828491477 | SLC35E1 | FALSE | Upregulated |
| -0,285754398 | 0,828607785 | KIAA1875 | FALSE | Downregulated |
| -0,320293274 | 0,828607785 | LOC100506319 | FALSE | Downregulated |
| 0,199136334 | 0,828627634 | EXOC3L2 | FALSE | Upregulated |
| -0,174696112 | 0,828637351 | LOC101927157 | FALSE | Downregulated |
| -0,063062979 | 0,828637351 | POLDIP2 | FALSE | Downregulated |
| -0,160687566 | 0,828699685 | MFSD3 | FALSE | Downregulated |
| 0,170110806 | 0,828716808 | NRTN | FALSE | Upregulated |
| -0,154958397 | 0,828716808 | SND1-IT1 | FALSE | Downregulated |
| -0,227924838 | 0,828716808 | TMEM45A | FALSE | Downregulated |
| -0,80455117 | 0,828935655 | GSTM1 | FALSE | Downregulated |
| 0,145703191 | 0,828935655 | ZNF496 | FALSE | Upregulated |
| -0,175381697 | 0,82909188 | ST7 | FALSE | Downregulated |
| 0,142199774 | 0,829218733 | AGO2 | FALSE | Upregulated |

| | | | | |
|---|---|---|---|---|
| 0,126885835 | 0,829218733 | ANKIB1 | FALSE | Upregulated |
| -0,153142774 | 0,829218733 | GNA14 | FALSE | Downregulated |
| -0,289510339 | 0,829218733 | KCTD5 | FALSE | Downregulated |
| 0,313609996 | 0,829218733 | LOC102724047 | FALSE | Upregulated |
| 0,115072724 | 0,829218733 | LPGAT1 | FALSE | Upregulated |
| -0,134965624 | 0,829218733 | SERINC3 | FALSE | Downregulated |
| -0,193969407 | 0,829218733 | TRAPPC4 | FALSE | Downregulated |
| -0,251607214 | 0,829218733 | TRIM6 | FALSE | Downregulated |
| -0,201068019 | 0,829218733 | ZNF470 | FALSE | Downregulated |
| 0,256604405 | 0,829323755 | HSPA12B | FALSE | Upregulated |
| 0,11534479 | 0,829323755 | SMU1 | FALSE | Upregulated |
| -0,128510988 | 0,829408601 | FBRS | FALSE | Downregulated |
| -0,388800684 | 0,829470979 | NUDT4P1 | FALSE | Downregulated |
| 0,111863049 | 0,829470979 | PSMD2 | FALSE | Upregulated |
| 0,165531425 | 0,829470979 | RP11-85B7.2 | FALSE | Upregulated |
| -0,195844074 | 0,829470979 | TBC1D10A | FALSE | Downregulated |
| -0,13027442 | 0,829571151 | CHST13 | FALSE | Downregulated |
| 0,157472575 | 0,829571151 | STX7 | FALSE | Upregulated |
| -0,231518502 | 0,829696022 | C21orf59 | FALSE | Downregulated |
| 0,147437868 | 0,829696022 | GPATCH11 | FALSE | Upregulated |
| 0,299076148 | 0,829696022 | GRHL2 | FALSE | Upregulated |
| -0,170863954 | 0,829696022 | S100A16 | FALSE | Downregulated |
| -0,115821312 | 0,829815177 | RNPS1 | FALSE | Downregulated |
| -0,15659487 | 0,829815177 | TMEM198B | FALSE | Downregulated |
| 0,484341995 | 0,8300554 | C3orf70 | FALSE | Upregulated |
| -0,127518238 | 0,830067922 | PSMB4 | FALSE | Downregulated |
| -0,090561034 | 0,830164109 | CDC42EP1 | FALSE | Downregulated |
| -0,263162715 | 0,830164109 | NDNL2 | FALSE | Downregulated |
| 0,103917514 | 0,830164109 | SYT9 | FALSE | Upregulated |
| -0,689592084 | 0,83026378 | FAM47E-STBD1 | FALSE | Downregulated |
| 0,15586641 | 0,830585808 | LOC102723851 | FALSE | Upregulated |
| -0,135259499 | 0,830640556 | TAF1C | FALSE | Downregulated |
| 0,196390188 | 0,830720335 | LOC102724350 | FALSE | Upregulated |
| 0,123220732 | 0,830732822 | PTPN1 | FALSE | Upregulated |
| -0,281826825 | 0,830732822 | ZNF765 | FALSE | Downregulated |
| -0,289723275 | 0,830747288 | FAM124A | FALSE | Downregulated |
| 0,114247653 | 0,830902191 | POMGNT1 | FALSE | Upregulated |
| 0,263819317 | 0,831113336 | TIMM9 | FALSE | Upregulated |
| -0,305386528 | 0,831360288 | ANAPC4 | FALSE | Downregulated |
| 0,260157448 | 0,831625112 | FASTKD5 | FALSE | Upregulated |
| 0,143267737 | 0,831657771 | FLJ38717 | FALSE | Upregulated |
| 0,139868242 | 0,831710292 | CYP2R1 | FALSE | Upregulated |
| -0,082326118 | 0,831784046 | WDR33 | FALSE | Downregulated |
| -0,122177703 | 0,831786563 | CAD | FALSE | Downregulated |
| -0,30382831 | 0,831839233 | PCDH7 | FALSE | Downregulated |
| -0,129732462 | 0,831839233 | ZNF710 | FALSE | Downregulated |
| 0,11536462 | 0,831993754 | GNL3L | FALSE | Upregulated |
| 0,269387687 | 0,832010873 | CDKN2C | FALSE | Upregulated |
| -0,124531513 | 0,832010873 | CLEC16A | FALSE | Downregulated |
| 0,364584647 | 0,832010873 | DNAL4 | FALSE | Upregulated |
| -0,111972224 | 0,832010873 | EPOR | FALSE | Downregulated |

| | | | | |
|---|---|---|---|---|
| -0,154817525 | 0,832010873 | ING5 | FALSE | Downregulated |
| 0,215611637 | 0,832010873 | KLHL25 | FALSE | Upregulated |
| 0,408109124 | 0,832010873 | LINC00563 | FALSE | Upregulated |
| -0,265507338 | 0,832010873 | LOC101929130 | FALSE | Downregulated |
| 0,16672994 | 0,832010873 | MCF2L | FALSE | Upregulated |
| -0,213878056 | 0,832010873 | OLFML3 | FALSE | Downregulated |
| 0,079153981 | 0,832010873 | PICALM | FALSE | Upregulated |
| 0,277642342 | 0,832010873 | POLR2G | FALSE | Upregulated |
| -0,336538513 | 0,832010873 | RPL36A | FALSE | Downregulated |
| 0,163289137 | 0,832010873 | STX5 | FALSE | Upregulated |
| 1,091469423 | 0,832010873 | TTN-AS1 | FALSE | Upregulated |
| -0,18503411 | 0,832010873 | ZNF271 | FALSE | Downregulated |
| -0,091711405 | 0,832039809 | ATXN7L3 | FALSE | Downregulated |
| 0,127149439 | 0,832039809 | GSR | FALSE | Upregulated |
| 0,379187331 | 0,832039809 | RS1 | FALSE | Upregulated |
| -0,247051185 | 0,832039809 | RWDD1 | FALSE | Downregulated |
| -0,217222122 | 0,832039809 | ZNF593 | FALSE | Downregulated |
| 0,550652055 | 0,832040725 | ARHGEF19 | FALSE | Upregulated |
| -0,166651939 | 0,832040725 | FAM89B | FALSE | Downregulated |
| -0,074761554 | 0,832040725 | KMT2B | FALSE | Downregulated |
| 0,801697479 | 0,832040725 | LOC102724159 | FALSE | Upregulated |
| 0,210380533 | 0,832040725 | SFTPD | FALSE | Upregulated |
| -0,308903567 | 0,832040725 | SNORA63 | FALSE | Downregulated |
| 0,289651318 | 0,832040725 | TEX30 | FALSE | Upregulated |
| 0,080314302 | 0,832040725 | TMEM63A | FALSE | Upregulated |
| -0,076506474 | 0,832040725 | TRIM25 | FALSE | Downregulated |
| -0,256926274 | 0,832040725 | ZNF143 | FALSE | Downregulated |
| 0,351976947 | 0,832048723 | DPYD-AS1 | FALSE | Upregulated |
| 0,249969827 | 0,832048723 | ELOVL1 | FALSE | Upregulated |
| -0,551263051 | 0,832048723 | OR2B2 | FALSE | Downregulated |
| -0,207188405 | 0,832048723 | ZNF26 | FALSE | Downregulated |
| 0,080198665 | 0,832051393 | SYMPK | FALSE | Upregulated |
| 0,12295079 | 0,832234356 | ZNF865 | FALSE | Upregulated |
| -0,291825409 | 0,832268161 | CCDC149 | FALSE | Downregulated |
| -0,329808782 | 0,832296591 | DDX12P | FALSE | Downregulated |
| -0,147424902 | 0,832296591 | IPO13 | FALSE | Downregulated |
| 0,099974121 | 0,832296591 | MAF1 | FALSE | Upregulated |
| 0,11692986 | 0,832296591 | PSMC4 | FALSE | Upregulated |
| 0,105757944 | 0,832296591 | RASSF5 | FALSE | Upregulated |
| 0,175291955 | 0,83233812 | PHF11 | FALSE | Upregulated |
| 0,564782516 | 0,832502558 | AANAT | FALSE | Upregulated |
| 0,363545653 | 0,832502558 | ARMCX1 | FALSE | Upregulated |
| 0,13944953 | 0,832502558 | ASCC3 | FALSE | Upregulated |
| 0,154812833 | 0,832502558 | ATG12 | FALSE | Upregulated |
| -0,123078552 | 0,832502558 | BCAP31 | FALSE | Downregulated |
| -0,53860899 | 0,832502558 | C13orf35 | FALSE | Downregulated |
| 0,293958313 | 0,832502558 | CACHD1 | FALSE | Upregulated |
| -0,142963986 | 0,832502558 | CEP290 | FALSE | Downregulated |
| -0,317469747 | 0,832502558 | CHCHD5 | FALSE | Downregulated |
| 0,129534691 | 0,832502558 | DENND5A | FALSE | Upregulated |
| -0,384789384 | 0,832502558 | DYRK3 | FALSE | Downregulated |

| | | | | |
|---|---|---|---|---|
| 0,633634494 | 0,832502558 | GIMAP1-GIMAP5 | FALSE | Upregulated |
| 0,375686237 | 0,832502558 | LDLRAD3 | FALSE | Upregulated |
| 0,374806129 | 0,832502558 | LOC100287497 | FALSE | Upregulated |
| 0,507748942 | 0,832502558 | LOC102724312 | FALSE | Upregulated |
| 0,218926226 | 0,832502558 | RBKS | FALSE | Upregulated |
| -0,131004879 | 0,832502558 | RBM27 | FALSE | Downregulated |
| -0,119862187 | 0,832502558 | RILP | FALSE | Downregulated |
| 0,470093917 | 0,832502558 | RP11-661P17.1 | FALSE | Upregulated |
| 0,182377341 | 0,832502558 | SGOL1 | FALSE | Upregulated |
| -0,276595279 | 0,832502558 | SUV420H2 | FALSE | Downregulated |
| 0,257488699 | 0,832502558 | TLCD2 | FALSE | Upregulated |
| -0,231421222 | 0,832502558 | WDR27 | FALSE | Downregulated |
| 0,077658461 | 0,832502558 | YY1 | FALSE | Upregulated |
| -0,267854086 | 0,832502558 | ZDHHC24 | FALSE | Downregulated |
| 0,178110458 | 0,832508411 | CYB561 | FALSE | Upregulated |
| 0,102289865 | 0,832508411 | ENDOG | FALSE | Upregulated |
| -0,546581253 | 0,832508411 | GPX8 | FALSE | Downregulated |
| 0,144456778 | 0,832508411 | LEPRE1 | FALSE | Upregulated |
| 0,360859397 | 0,832508411 | LOC101929409 | FALSE | Upregulated |
| 0,205332138 | 0,832508411 | LOC283788 | FALSE | Upregulated |
| -0,212558799 | 0,832508411 | MUC4 | FALSE | Downregulated |
| 0,118762568 | 0,832508411 | NBN | FALSE | Upregulated |
| 0,524552685 | 0,832508411 | PAK6 | FALSE | Upregulated |
| -0,111445738 | 0,832508411 | PCGF2 | FALSE | Downregulated |
| -0,379501665 | 0,832508411 | SNX7 | FALSE | Downregulated |
| 0,240188946 | 0,832508411 | UGT2A3 | FALSE | Upregulated |
| -0,435027089 | 0,832508411 | ZFY | FALSE | Downregulated |
| -0,137313815 | 0,832560375 | RALGPS2 | FALSE | Downregulated |
| -0,275873994 | 0,832671208 | ZNF783 | FALSE | Downregulated |
| -0,088125358 | 0,832852533 | BAZ2B | FALSE | Downregulated |
| -0,1475087 | 0,832852533 | ZNF764 | FALSE | Downregulated |
| -0,194231283 | 0,832965991 | LOC102724921 | FALSE | Downregulated |
| -0,285997922 | 0,833008998 | MRPL22 | FALSE | Downregulated |
| 0,178570801 | 0,833038176 | GAPDHS | FALSE | Upregulated |
| 0,14832367 | 0,833038176 | PCSK9 | FALSE | Upregulated |
| 0,490858451 | 0,833136469 | C2orf81 | FALSE | Upregulated |
| 0,096699437 | 0,833136469 | LTBP3 | FALSE | Upregulated |
| -0,203666246 | 0,833136469 | TRIM3 | FALSE | Downregulated |
| -0,297977177 | 0,833364763 | SDHAF2 | FALSE | Downregulated |
| 0,329353519 | 0,83351543 | CD209 | FALSE | Upregulated |
| 0,113553534 | 0,83351543 | FBXW4 | FALSE | Upregulated |
| -0,555485837 | 0,83351543 | KLHDC8A | FALSE | Downregulated |
| 0,190949208 | 0,83351543 | ZBTB3 | FALSE | Upregulated |
| -0,099416856 | 0,83351543 | ZNF24 | FALSE | Downregulated |
| -0,488153017 | 0,83351543 | ZNF876P | FALSE | Downregulated |
| 0,329889395 | 0,833563415 | BMI1 | FALSE | Upregulated |
| 0,118656725 | 0,833563415 | CHST7 | FALSE | Upregulated |
| -0,273598551 | 0,833563415 | NPAS2 | FALSE | Downregulated |
| -0,230098213 | 0,833563415 | SDS | FALSE | Downregulated |
| 0,134887 | 0,83360184 | LOC102724273 | FALSE | Upregulated |
| 0,119341036 | 0,83360184 | PHF1 | FALSE | Upregulated |

| | | | | |
|---|---|---|---|---|
| 0,495857987 | 0,83360184 | PINLYP | FALSE | Upregulated |
| -0,18850286 | 0,83360184 | RAP1GDS1 | FALSE | Downregulated |
| 0,207777114 | 0,83360184 | TFAP2E | FALSE | Upregulated |
| 0,203271239 | 0,83360184 | TMEM87B | FALSE | Upregulated |
| -0,951648655 | 0,83360184 | TNFSF12-TNFSF13 | FALSE | Downregulated |
| -0,145693057 | 0,83360184 | ZC3H6 | FALSE | Downregulated |
| 0,532166665 | 0,833640753 | RP11-87E22.2 | FALSE | Upregulated |
| -0,397111541 | 0,833883302 | LOC101928182 | FALSE | Downregulated |
| 0,300147301 | 0,834037282 | ELP4 | FALSE | Upregulated |
| -0,085499438 | 0,834361548 | DIS3L | FALSE | Downregulated |
| 0,096250051 | 0,834374005 | GPR161 | FALSE | Upregulated |
| -0,201166668 | 0,834374005 | SAMM50 | FALSE | Downregulated |
| 0,442189514 | 0,834483446 | LOC102723960 | FALSE | Upregulated |
| -0,217677555 | 0,834498253 | RP2 | FALSE | Downregulated |
| -0,175153148 | 0,834640179 | ZNF576 | FALSE | Downregulated |
| 0,239958097 | 0,834799191 | AIF1L | FALSE | Upregulated |
| 0,127478448 | 0,8351852 | TP53BP1 | FALSE | Upregulated |
| -0,144256031 | 0,835464913 | BECN1 | FALSE | Downregulated |
| 0,117884007 | 0,835740829 | ANXA6 | FALSE | Upregulated |
| -0,495167416 | 0,835740829 | C20orf96 | FALSE | Downregulated |
| 0,154085697 | 0,835740829 | DSC2 | FALSE | Upregulated |
| -0,133613549 | 0,835740829 | NAP1L4 | FALSE | Downregulated |
| 0,094141228 | 0,835740829 | RPE | FALSE | Upregulated |
| 0,428546365 | 0,835832457 | CASS4 | FALSE | Upregulated |
| 0,337432991 | 0,836221816 | ARL15 | FALSE | Upregulated |
| 0,273646222 | 0,836234932 | NABP2 | FALSE | Upregulated |
| -0,068897572 | 0,836280655 | GIGYF1 | FALSE | Downregulated |
| -0,085812034 | 0,83628099 | ABL1 | FALSE | Downregulated |
| -0,169140905 | 0,83628099 | AC144449.1 | FALSE | Downregulated |
| -0,132946202 | 0,83628099 | TNNI1 | FALSE | Downregulated |
| 0,26317765 | 0,836301131 | LOC286367 | FALSE | Upregulated |
| 0,123867056 | 0,836301131 | NOLC1 | FALSE | Upregulated |
| 0,248817225 | 0,836301131 | NPHP4 | FALSE | Upregulated |
| -0,174982731 | 0,836331523 | TIPARP-AS1 | FALSE | Downregulated |
| -0,144430644 | 0,836399006 | DHPS | FALSE | Downregulated |
| -0,217067973 | 0,836399006 | DLL4 | FALSE | Downregulated |
| 0,507278555 | 0,836504468 | LOC102724682 | FALSE | Upregulated |
| -0,173296415 | 0,836612866 | ARRB1 | FALSE | Downregulated |
| -0,279284493 | 0,836663708 | RBM34 | FALSE | Downregulated |
| -0,2513205 | 0,83671699 | NDUFC1 | FALSE | Downregulated |
| 0,142772636 | 0,836721468 | DDX20 | FALSE | Upregulated |
| 0,098563411 | 0,836721468 | MOGS | FALSE | Upregulated |
| -0,144143202 | 0,836768867 | ABCD4 | FALSE | Downregulated |
| -0,507607118 | 0,836768867 | AHRR | FALSE | Downregulated |
| -0,29941268 | 0,836768867 | C1D | FALSE | Downregulated |
| -0,290525927 | 0,836768867 | OXSM | FALSE | Downregulated |
| 0,200810646 | 0,836768867 | RASGRP4 | FALSE | Upregulated |
| 0,224449155 | 0,836768867 | SCARA3 | FALSE | Upregulated |
| 0,220580381 | 0,836796003 | CDHR3 | FALSE | Upregulated |
| -0,189330255 | 0,836796003 | HNF4G | FALSE | Downregulated |

| | | | | |
|---|---|---|---|---|
| 0,171671417 | 0,836796003 | NRAS | FALSE | Upregulated |
| 0,18856656 | 0,836796003 | SAMD1 | FALSE | Upregulated |
| -0,175842473 | 0,836796003 | USP30-AS1 | FALSE | Downregulated |
| 0,203194659 | 0,836846792 | RASSF1 | FALSE | Upregulated |
| -0,149956735 | 0,837083149 | HES1 | FALSE | Downregulated |
| -0,146611439 | 0,837086958 | PRRC1 | FALSE | Downregulated |
| 0,171710432 | 0,837164192 | POLR3F | FALSE | Upregulated |
| -0,25681899 | 0,837274244 | DPY19L2P2 | FALSE | Downregulated |
| -0,556529446 | 0,837274244 | LOC102723794 | FALSE | Downregulated |
| -0,17201123 | 0,837274244 | NAV2-AS5 | FALSE | Downregulated |
| 0,245205369 | 0,837274244 | RPA3 | FALSE | Upregulated |
| 0,115638967 | 0,837274244 | TAF1 | FALSE | Upregulated |
| -0,181723827 | 0,837696545 | OSTC | FALSE | Downregulated |
| -0,145004608 | 0,837825769 | PTPN9 | FALSE | Downregulated |
| 0,241981945 | 0,837975401 | FPR3 | FALSE | Upregulated |
| 0,235430958 | 0,837980363 | ADAMTS12 | FALSE | Upregulated |
| -0,208505869 | 0,837980363 | GIMAP2 | FALSE | Downregulated |
| 0,368065872 | 0,837980363 | LOC102724356 | FALSE | Upregulated |
| -0,385060503 | 0,837980363 | LOC102724448 | FALSE | Downregulated |
| 0,540654951 | 0,837980363 | TESC | FALSE | Upregulated |
| 0,368326595 | 0,837980363 | TXLNG2P | FALSE | Upregulated |
| 0,256946542 | 0,837980509 | RASA2 | FALSE | Upregulated |
| 0,178730238 | 0,838224506 | LOC101928429 | FALSE | Upregulated |
| 0,13506725 | 0,838224506 | LOC102723750 | FALSE | Upregulated |
| -0,424337643 | 0,838224506 | MRPL1 | FALSE | Downregulated |
| 0,411281171 | 0,838224506 | RAMP2 | FALSE | Upregulated |
| -0,527837308 | 0,838224506 | SIRPD | FALSE | Downregulated |
| -0,133704535 | 0,838224506 | TAZ | FALSE | Downregulated |
| 0,292472898 | 0,838522141 | VNN3 | FALSE | Upregulated |
| 0,43965797 | 0,838650484 | RP11-806K15.1 | FALSE | Upregulated |
| -0,292696454 | 0,838742635 | RAET1E | FALSE | Downregulated |
| -0,135101567 | 0,838930723 | LOC101929751 | FALSE | Downregulated |
| -0,207581016 | 0,839159895 | HECTD2 | FALSE | Downregulated |
| -0,115311233 | 0,83922767 | C1RL-AS1 | FALSE | Downregulated |
| 0,347793307 | 0,83922767 | KDELC1 | FALSE | Upregulated |
| 0,094983213 | 0,83922767 | ZNF460 | FALSE | Upregulated |
| -0,366693521 | 0,839246834 | ALDH1L1-AS1 | FALSE | Downregulated |
| -0,54296265 | 0,839246834 | BMS1P17 | FALSE | Downregulated |
| -0,549713886 | 0,839246834 | HIST1H2BE | FALSE | Downregulated |
| 0,209353515 | 0,839246834 | MGME1 | FALSE | Upregulated |
| -0,188553879 | 0,839246834 | MRPL9 | FALSE | Downregulated |
| 0,20661504 | 0,839246834 | PRKCD | FALSE | Upregulated |
| -0,23782708 | 0,839246834 | ZNF480 | FALSE | Downregulated |
| -0,099673208 | 0,839718903 | PHB | FALSE | Downregulated |
| 0,186837918 | 0,839839528 | DCAF15 | FALSE | Upregulated |
| -0,147063353 | 0,839983917 | UBXN7 | FALSE | Downregulated |
| 0,112017309 | 0,840207816 | PPP2CB | FALSE | Upregulated |
| 0,160661988 | 0,840226319 | CSNK1G1 | FALSE | Upregulated |
| -0,190790222 | 0,840404792 | NOL12 | FALSE | Downregulated |
| 0,172225552 | 0,840461769 | COPG2 | FALSE | Upregulated |
| 0,238370527 | 0,840461769 | MCAT | FALSE | Upregulated |

| | | | | |
|---|---|---|---|---|
| 0,148183777 | 0,840467311 | CHMP6 | FALSE | Upregulated |
| -0,611922226 | 0,840527798 | BTBD17 | FALSE | Downregulated |
| 0,193766623 | 0,840561326 | MYPOP | FALSE | Upregulated |
| 0,459210098 | 0,84058619 | ASB3 | FALSE | Upregulated |
| -0,12122755 | 0,84058619 | LEMD2 | FALSE | Downregulated |
| -0,184974883 | 0,840659363 | DPY19L3 | FALSE | Downregulated |
| -0,087109514 | 0,840659363 | ERP44 | FALSE | Downregulated |
| -0,456221224 | 0,840659363 | LINC01124 | FALSE | Downregulated |
| 0,100665244 | 0,840659363 | LPIN2 | FALSE | Upregulated |
| -0,131910319 | 0,840659363 | NDUFA6 | FALSE | Downregulated |
| 0,160897691 | 0,840659363 | TONSL | FALSE | Upregulated |
| 0,222170646 | 0,840957623 | ENDOD1 | FALSE | Upregulated |
| 0,094268438 | 0,840957623 | ERBB2IP | FALSE | Upregulated |
| 0,105539305 | 0,840957623 | FAM219B | FALSE | Upregulated |
| 0,314094007 | 0,840957623 | KIF7 | FALSE | Upregulated |
| -0,349300751 | 0,840957623 | RHEBL1 | FALSE | Downregulated |
| -0,318746418 | 0,840957623 | ST3GAL6-AS1 | FALSE | Downregulated |
| -0,195077227 | 0,840960727 | LOC101060581 | FALSE | Downregulated |
| 0,110844882 | 0,841004695 | LOC100289511 | FALSE | Upregulated |
| 0,143827094 | 0,841023704 | ADPRM | FALSE | Upregulated |
| 0,154668783 | 0,841023704 | CAPN3 | FALSE | Upregulated |
| 0,128424734 | 0,841023704 | LOC728024 | FALSE | Upregulated |
| 0,294045507 | 0,841023704 | MFGE8 | FALSE | Upregulated |
| -0,539897046 | 0,841023704 | RP11-503N18.1 | FALSE | Downregulated |
| 0,072878029 | 0,841145183 | SEC24C | FALSE | Upregulated |
| 0,189300887 | 0,841268734 | CRELD2 | FALSE | Upregulated |
| -0,397942999 | 0,841268734 | KLHL32 | FALSE | Downregulated |
| 0,267066761 | 0,841273473 | LINC00685 | FALSE | Upregulated |
| 0,167851275 | 0,841332738 | ST3GAL4 | FALSE | Upregulated |
| 0,13667839 | 0,841582375 | EPM2AIP1 | FALSE | Upregulated |
| -0,266158125 | 0,841582375 | TBCC | FALSE | Downregulated |
| 0,372936148 | 0,841645967 | BTN1A1 | FALSE | Upregulated |
| 0,239222491 | 0,841645967 | FBXO2 | FALSE | Upregulated |
| -0,161785575 | 0,841645967 | PRKAG3 | FALSE | Downregulated |
| -0,173205118 | 0,841645967 | TMEM104 | FALSE | Downregulated |
| -0,230896058 | 0,841799972 | RP11-432I5.1 | FALSE | Downregulated |
| 0,378933458 | 0,841896317 | CACNG1 | FALSE | Upregulated |
| -0,223187849 | 0,841896317 | GPR62 | FALSE | Downregulated |
| -0,059899813 | 0,842054632 | TMEM259 | FALSE | Downregulated |
| 0,180998958 | 0,842167586 | PELI1 | FALSE | Upregulated |
| 0,411241576 | 0,842273448 | AC078941.1 | FALSE | Upregulated |
| -0,219878002 | 0,842273448 | C21orf58 | FALSE | Downregulated |
| 0,297856818 | 0,842273448 | C3orf65 | FALSE | Upregulated |
| 0,180115802 | 0,842273448 | CCDC94 | FALSE | Upregulated |
| -0,209159153 | 0,842273448 | CXorf40B | FALSE | Downregulated |
| 0,214868549 | 0,842273448 | FAM204A | FALSE | Upregulated |
| 0,359986637 | 0,842273448 | HIST1H4K | FALSE | Upregulated |
| -0,235559179 | 0,842273448 | IGFLR1 | FALSE | Downregulated |
| -0,223642368 | 0,842273448 | LAPTM4B | FALSE | Downregulated |
| 0,245254734 | 0,842273448 | LOC101929051 | FALSE | Upregulated |
| -0,44899839 | 0,842273448 | METTL1 | FALSE | Downregulated |

| | | | | |
|---|---|---|---|---|
| 0,105925702 | 0,842273448 | MEX3D | FALSE | Upregulated |
| -0,358773059 | 0,842273448 | NUTM2F | FALSE | Downregulated |
| -0,296475678 | 0,842273448 | PDCD10 | FALSE | Downregulated |
| -0,22525143 | 0,842273448 | PTPLB | FALSE | Downregulated |
| -0,453905159 | 0,842273448 | RPS6KA6 | FALSE | Downregulated |
| 0,148065997 | 0,842273448 | SOX10 | FALSE | Upregulated |
| -0,476028504 | 0,842273448 | TRIM52-AS1 | FALSE | Downregulated |
| 0,175746141 | 0,842273448 | ZNRF2P1 | FALSE | Upregulated |
| 0,234166872 | 0,842390819 | THBD | FALSE | Upregulated |
| 0,079924099 | 0,842463794 | PTOV1 | FALSE | Upregulated |
| 0,152992963 | 0,842490068 | HNF1B | FALSE | Upregulated |
| -0,349625862 | 0,842547539 | TRIM16 | FALSE | Downregulated |
| 0,255513084 | 0,842647473 | KLHL7 | FALSE | Upregulated |
| 0,391682311 | 0,842647473 | MROH9 | FALSE | Upregulated |
| -0,080655474 | 0,842647473 | POLRMT | FALSE | Downregulated |
| 0,455964122 | 0,842647473 | RIMS4 | FALSE | Upregulated |
| 0,20496999 | 0,842647473 | ST6GALNAC2 | FALSE | Upregulated |
| 0,171576072 | 0,842647473 | TBC1D12 | FALSE | Upregulated |
| 0,536679473 | 0,842647473 | TFPI2 | FALSE | Upregulated |
| 0,32527184 | 0,842647473 | ZNF449 | FALSE | Upregulated |
| -0,157378576 | 0,842874498 | TMTC3 | FALSE | Downregulated |
| 0,132858545 | 0,84299911 | FBL | FALSE | Upregulated |
| 1,256717405 | 0,843290738 | SERF2-C15ORF63 | FALSE | Upregulated |
| -0,195849515 | 0,843581552 | GPR88 | FALSE | Downregulated |
| 0,178279333 | 0,8438324 | NDUFB5 | FALSE | Upregulated |
| -1,062066762 | 0,843974622 | GUSBP9 | FALSE | Downregulated |
| -0,220825295 | 0,84406573 | EDAR | FALSE | Downregulated |
| -0,11079366 | 0,84406573 | KPNA4 | FALSE | Downregulated |
| 0,26316978 | 0,844308999 | PPFIA3 | FALSE | Upregulated |
| 0,089385505 | 0,844343721 | GPER1 | FALSE | Upregulated |
| 0,125511858 | 0,844343721 | MAPK1 | FALSE | Upregulated |
| -0,165678527 | 0,844343721 | RHBDD3 | FALSE | Downregulated |
| 0,096674292 | 0,844371343 | STUB1 | FALSE | Upregulated |
| -0,081032533 | 0,844475479 | NOTCH1 | FALSE | Downregulated |
| -0,154324305 | 0,844525899 | TBL3 | FALSE | Downregulated |
| 0,258395655 | 0,844525899 | TTC8 | FALSE | Upregulated |
| -0,111703658 | 0,844525899 | VPS54 | FALSE | Downregulated |
| -0,100091815 | 0,844566493 | NXF1 | FALSE | Downregulated |
| -0,510378391 | 0,844566493 | TAS2R30 | FALSE | Downregulated |
| 0,282674747 | 0,844718942 | TYRO3 | FALSE | Upregulated |
| 0,507961292 | 0,84487364 | MAB21L3 | FALSE | Upregulated |
| 0,427119461 | 0,844876996 | LOC102724037 | FALSE | Upregulated |
| 0,20513576 | 0,844918633 | ACY3 | FALSE | Upregulated |
| 0,440118216 | 0,845157332 | CD109 | FALSE | Upregulated |
| 0,305787682 | 0,845157332 | COL4A6 | FALSE | Upregulated |
| 0,227188263 | 0,845157332 | ERCC4 | FALSE | Upregulated |
| 0,155514035 | 0,845157332 | FAM221B | FALSE | Upregulated |
| 0,23676989 | 0,845157332 | MCEE | FALSE | Upregulated |
| -0,192616642 | 0,845157332 | NUP43 | FALSE | Downregulated |
| 0,116356629 | 0,845157332 | TECPR2 | FALSE | Upregulated |
| -0,318225508 | 0,845157332 | TEX38 | FALSE | Downregulated |

| | | | | |
|---|---|---|---|---|
| -0,178008676 | 0,845157332 | TMEM170B | FALSE | Downregulated |
| 0,199694098 | 0,845157332 | ZNF195 | FALSE | Upregulated |
| 0,421934734 | 0,845166061 | FHL3 | FALSE | Upregulated |
| 0,124748619 | 0,845378309 | CNPY4 | FALSE | Upregulated |
| 0,134868147 | 0,84550409 | PRMT2 | FALSE | Upregulated |
| -0,204203956 | 0,84550409 | YAF2 | FALSE | Downregulated |
| 0,348302518 | 0,845557461 | LINC00485 | FALSE | Upregulated |
| 0,166895883 | 0,845557461 | LRRC46 | FALSE | Upregulated |
| 0,221342126 | 0,845557461 | RNU2-2P | FALSE | Upregulated |
| -0,333209052 | 0,845618478 | ME1 | FALSE | Downregulated |
| 0,216518975 | 0,845618478 | OR52B6 | FALSE | Upregulated |
| 0,245809339 | 0,845618478 | RP11-1078H9.5 | FALSE | Upregulated |
| 0,19567852 | 0,845890057 | PAQR6 | FALSE | Upregulated |
| -0,162814651 | 0,846025294 | ABCB6 | FALSE | Downregulated |
| 0,101109633 | 0,846025294 | BLCAP | FALSE | Upregulated |
| 0,389937 | 0,846025294 | CD33 | FALSE | Upregulated |
| 0,338927333 | 0,846025294 | GLIS3-AS1 | FALSE | Upregulated |
| -0,437117009 | 0,846025294 | LOC101928768 | FALSE | Downregulated |
| 0,345896052 | 0,846025294 | LOC102723651 | FALSE | Upregulated |
| 0,182699434 | 0,846025294 | PPP1R21 | FALSE | Upregulated |
| 0,146628887 | 0,846025294 | SPIRE1 | FALSE | Upregulated |
| -0,218130267 | 0,846466608 | CEP76 | FALSE | Downregulated |
| 0,1879651 | 0,846466608 | SPATA5L1 | FALSE | Upregulated |
| 0,152293163 | 0,846466608 | VPS18 | FALSE | Upregulated |
| 0,111069608 | 0,846475915 | A1BG-AS1 | FALSE | Upregulated |
| 0,107708027 | 0,846475915 | APOL2 | FALSE | Upregulated |
| 0,231892494 | 0,846475915 | GMEB1 | FALSE | Upregulated |
| -0,307615333 | 0,846475915 | MYOM2 | FALSE | Downregulated |
| -0,101617534 | 0,846475915 | TUT1 | FALSE | Downregulated |
| -0,084436087 | 0,846488236 | DVL1 | FALSE | Downregulated |
| 0,17133449 | 0,84652372 | MTMR9 | FALSE | Upregulated |
| -0,216351496 | 0,84652372 | ZNF563 | FALSE | Downregulated |
| -0,129467936 | 0,846608984 | ATP5F1 | FALSE | Downregulated |
| -0,185797929 | 0,84672512 | TNFRSF10D | FALSE | Downregulated |
| -0,430578759 | 0,846811493 | C16orf91 | FALSE | Downregulated |
| -0,124319242 | 0,846811493 | NUBP2 | FALSE | Downregulated |
| -0,270743252 | 0,847001272 | LOC101927402 | FALSE | Downregulated |
| -0,148586227 | 0,847001272 | MPP1 | FALSE | Downregulated |
| -0,10802851 | 0,847407551 | GCHFR | FALSE | Downregulated |
| 0,343046016 | 0,847407551 | HCCAT3 | FALSE | Upregulated |
| 0,412375183 | 0,847407551 | MKX | FALSE | Upregulated |
| 0,096483383 | 0,847407551 | SLC27A1 | FALSE | Upregulated |
| 0,112744803 | 0,847407551 | TCP1 | FALSE | Upregulated |
| -0,095935513 | 0,847428592 | ZSCAN29 | FALSE | Downregulated |
| 0,089511249 | 0,847518193 | VCP | FALSE | Upregulated |
| -0,071314857 | 0,847739539 | CHD2 | FALSE | Downregulated |
| 0,400605057 | 0,847739539 | TGM1 | FALSE | Upregulated |
| 0,171578203 | 0,847911328 | NFKBIL1 | FALSE | Upregulated |
| 0,082129398 | 0,847917305 | GPATCH8 | FALSE | Upregulated |
| 0,094471905 | 0,848060228 | SHPRH | FALSE | Upregulated |
| -0,080691823 | 0,84820888 | GBA2 | FALSE | Downregulated |

| | | | | |
|---|---|---|---|---|
| 0,237422277 | 0,848292002 | RP11-365O16.6 | FALSE | Upregulated |
| 0,158877769 | 0,848403044 | MEF2C | FALSE | Upregulated |
| 0,115590156 | 0,848533854 | HS6ST1 | FALSE | Upregulated |
| -0,263639074 | 0,848533854 | LOC100288778 | FALSE | Downregulated |
| 0,556035472 | 0,848533854 | LOC102724369 | FALSE | Upregulated |
| 0,102309136 | 0,848568109 | HELZ2 | FALSE | Upregulated |
| -0,299994747 | 0,848575185 | ZNF581 | FALSE | Downregulated |
| -0,171805564 | 0,848694526 | ABCG1 | FALSE | Downregulated |
| 0,331086677 | 0,848694526 | ASPM | FALSE | Upregulated |
| 0,237664944 | 0,848694526 | LOC100996643 | FALSE | Upregulated |
| 0,184840222 | 0,848694526 | PSEN2 | FALSE | Upregulated |
| 0,099964381 | 0,848694526 | SBNO1 | FALSE | Upregulated |
| 0,193948106 | 0,848796013 | C12orf73 | FALSE | Upregulated |
| 0,121546161 | 0,848796013 | PPP2CA | FALSE | Upregulated |
| 0,128585641 | 0,84898164 | C14orf159 | FALSE | Upregulated |
| -0,316057327 | 0,84898164 | FAM167B | FALSE | Downregulated |
| 0,194032941 | 0,84898164 | KIAA0040 | FALSE | Upregulated |
| -0,359395559 | 0,84898164 | LOC102724336 | FALSE | Downregulated |
| 0,161260486 | 0,849026 | ZC3H10 | FALSE | Upregulated |
| 0,062120301 | 0,849074809 | GGCX | FALSE | Upregulated |
| 0,41902083 | 0,849074809 | NUTM2A-AS1 | FALSE | Upregulated |
| -0,095728293 | 0,849107536 | LOC101927803 | FALSE | Downregulated |
| 0,111084031 | 0,849238326 | KAT5 | FALSE | Upregulated |
| -0,146532994 | 0,849238326 | TMEM214 | FALSE | Downregulated |
| -0,114510168 | 0,849287913 | U2SURP | FALSE | Downregulated |
| -0,126274385 | 0,849397894 | HERPUD2 | FALSE | Downregulated |
| 0,252950239 | 0,849397894 | IFI44L | FALSE | Upregulated |
| -0,316404054 | 0,849397894 | TUBD1 | FALSE | Downregulated |
| -0,507010247 | 0,84965191 | EIF5AL1 | FALSE | Downregulated |
| -0,178964351 | 0,84965191 | FREM3 | FALSE | Downregulated |
| -0,119135233 | 0,84965191 | RIN2 | FALSE | Downregulated |
| -0,366445547 | 0,84965191 | ZNF223 | FALSE | Downregulated |
| -0,675346362 | 0,84966022 | INO80B | FALSE | Downregulated |
| 0,181796232 | 0,849793099 | ANKRD13D | FALSE | Upregulated |
| 0,11515267 | 0,849793099 | CACUL1 | FALSE | Upregulated |
| -0,11956988 | 0,849793099 | CTCF | FALSE | Downregulated |
| -0,196877308 | 0,849793099 | CWC15 | FALSE | Downregulated |
| 0,167451189 | 0,849793099 | DNAJC9-AS1 | FALSE | Upregulated |
| -0,108791213 | 0,849793099 | GSTO1 | FALSE | Downregulated |
| -0,130583194 | 0,849793099 | LIPA | FALSE | Downregulated |
| 0,123274691 | 0,849793099 | PRNP | FALSE | Upregulated |
| -0,167730082 | 0,849793099 | R3HCC1L | FALSE | Downregulated |
| 0,101282439 | 0,849793099 | SMPD3 | FALSE | Upregulated |
| -0,114093903 | 0,849813405 | LOC101929079 | FALSE | Downregulated |
| 0,127330375 | 0,849813405 | LYSMD3 | FALSE | Upregulated |
| 0,132728003 | 0,849966231 | GIT2 | FALSE | Upregulated |
| 0,128422774 | 0,849994281 | LARS2 | FALSE | Upregulated |
| 0,326094646 | 0,849994281 | MEOX2 | FALSE | Upregulated |
| -0,565810105 | 0,849994281 | PRKRIP1 | FALSE | Downregulated |
| -0,149461395 | 0,849994281 | RBMXL1 | FALSE | Downregulated |
| 0,426960634 | 0,849994281 | RP11-626I20.3 | FALSE | Upregulated |

| | | | | |
|---|---|---|---|---|
| -0,119698601 | 0,849994281 | STX4 | FALSE | Downregulated |
| -0,133902107 | 0,850158599 | ACSF2 | FALSE | Downregulated |
| -0,176435311 | 0,850158599 | CNIH1 | FALSE | Downregulated |
| 0,259760847 | 0,850158599 | CRYZL1 | FALSE | Upregulated |
| 0,152939417 | 0,850158599 | GDAP2 | FALSE | Upregulated |
| 0,099430962 | 0,850158599 | MRPL19 | FALSE | Upregulated |
| 0,09951958 | 0,850158599 | RAB8A | FALSE | Upregulated |
| -0,089859355 | 0,850158599 | TP53INP1 | FALSE | Downregulated |
| 0,129032375 | 0,850438226 | QSER1 | FALSE | Upregulated |
| -0,158496451 | 0,85046181 | RP1-60O19.1 | FALSE | Downregulated |
| 0,092735991 | 0,85046181 | UBAP2L | FALSE | Upregulated |
| -0,124384173 | 0,850635636 | CARS2 | FALSE | Downregulated |
| -0,134039003 | 0,850750915 | CSNK2A1 | FALSE | Downregulated |
| -0,09486334 | 0,850858783 | HSP90AB1 | FALSE | Downregulated |
| -0,138814691 | 0,850858783 | LAS1L | FALSE | Downregulated |
| -0,092143211 | 0,850935751 | PLEKHM2 | FALSE | Downregulated |
| -0,101869418 | 0,851073814 | SCCPDH | FALSE | Downregulated |
| 0,308993616 | 0,851103575 | FBXW10 | FALSE | Upregulated |
| -0,125995782 | 0,851214423 | KIAA0232 | FALSE | Downregulated |
| 0,178704095 | 0,851275638 | MVD | FALSE | Upregulated |
| -0,352067643 | 0,851275638 | UBE2D1 | FALSE | Downregulated |
| 0,20972121 | 0,851646815 | BTNL9 | FALSE | Upregulated |
| -0,130124201 | 0,851646815 | ZNF384 | FALSE | Downregulated |
| -0,197651239 | 0,851753235 | AAGAB | FALSE | Downregulated |
| 0,14671208 | 0,851753235 | NEK11 | FALSE | Upregulated |
| -0,168112768 | 0,851753235 | PCMT1 | FALSE | Downregulated |
| -0,198192378 | 0,85180433 | CIAPIN1 | FALSE | Downregulated |
| -0,160679554 | 0,852020944 | KREMEN1 | FALSE | Downregulated |
| 0,161452958 | 0,852133803 | KLHDC7A | FALSE | Upregulated |
| -0,202205665 | 0,852195011 | SLC16A11 | FALSE | Downregulated |
| -0,407558086 | 0,852195011 | SNHG10 | FALSE | Downregulated |
| 0,099362608 | 0,852366407 | C1orf192 | FALSE | Upregulated |
| -0,479223086 | 0,852366407 | LOC100505650 | FALSE | Downregulated |
| 0,108676885 | 0,852366407 | RAB6A | FALSE | Upregulated |
| -0,247998941 | 0,852366407 | SLAIN1 | FALSE | Downregulated |
| -0,107898405 | 0,852366407 | SLC17A4 | FALSE | Downregulated |
| 0,21804666 | 0,852410876 | ABHD17B | FALSE | Upregulated |
| 0,108400137 | 0,852604895 | LOC101927190 | FALSE | Upregulated |
| 0,114727243 | 0,852604895 | NEIL1 | FALSE | Upregulated |
| 0,10470995 | 0,852616543 | MDH1 | FALSE | Upregulated |
| 0,116528592 | 0,852719721 | FBXO46 | FALSE | Upregulated |
| 0,276390436 | 0,852719721 | HDDC2 | FALSE | Upregulated |
| -0,108601152 | 0,852719721 | HSPA5 | FALSE | Downregulated |
| -0,12133169 | 0,852719721 | METTL13 | FALSE | Downregulated |
| -0,119545156 | 0,852719721 | SLC46A3 | FALSE | Downregulated |
| 0,156879773 | 0,852751779 | SCTR | FALSE | Upregulated |
| 0,186647832 | 0,853153263 | NSL1 | FALSE | Upregulated |
| -0,183286381 | 0,853153263 | TMEM86B | FALSE | Downregulated |
| -0,097630305 | 0,853153263 | VRK2 | FALSE | Downregulated |
| 0,064556484 | 0,853153263 | WAC | FALSE | Upregulated |
| 0,320154624 | 0,853153263 | ZNF816 | FALSE | Upregulated |

| | | | | |
|---|---|---|---|---|
| -0,143630676 | 0,853245403 | ADK | FALSE | Downregulated |
| 0,138698454 | 0,853245403 | ANO9 | FALSE | Upregulated |
| 0,18611184 | 0,853245403 | C2orf54 | FALSE | Upregulated |
| 0,217087963 | 0,853245403 | CCDC144B | FALSE | Upregulated |
| 0,087932735 | 0,853245403 | CTSA | FALSE | Upregulated |
| 0,286539149 | 0,853245403 | DACH1 | FALSE | Upregulated |
| 0,260591421 | 0,853245403 | FAM166A | FALSE | Upregulated |
| 0,099267186 | 0,853245403 | FLRT1 | FALSE | Upregulated |
| -0,201089741 | 0,853245403 | GPR128 | FALSE | Downregulated |
| -0,178868625 | 0,853245403 | HK3 | FALSE | Downregulated |
| -0,440886454 | 0,853245403 | KCNH6 | FALSE | Downregulated |
| -0,060418717 | 0,853245403 | KMT2C | FALSE | Downregulated |
| 0,461032668 | 0,853245403 | LINC00525 | FALSE | Upregulated |
| 0,393628199 | 0,853245403 | LOC100506746 | FALSE | Upregulated |
| -0,190715219 | 0,853245403 | LOC101060351 | FALSE | Downregulated |
| 0,209349692 | 0,853245403 | LOC102724709 | FALSE | Upregulated |
| -0,139921605 | 0,853245403 | LRRC2 | FALSE | Downregulated |
| -0,137049928 | 0,853245403 | MAPKAPK5 | FALSE | Downregulated |
| 0,23209173 | 0,853245403 | MCM8 | FALSE | Upregulated |
| 0,108830944 | 0,853245403 | MYO19 | FALSE | Upregulated |
| -0,149072634 | 0,853245403 | NARF | FALSE | Downregulated |
| -0,26124771 | 0,853245403 | NHEJ1 | FALSE | Downregulated |
| 0,458981768 | 0,853245403 | PLA2G7 | FALSE | Upregulated |
| 0,17771595 | 0,853245403 | POLR2I | FALSE | Upregulated |
| -0,405221798 | 0,853245403 | PRR11 | FALSE | Downregulated |
| 0,139762748 | 0,853245403 | RMND5B | FALSE | Upregulated |
| 0,24351764 | 0,853245403 | RNF144A | FALSE | Upregulated |
| -0,506658843 | 0,853245403 | RP11-222A5.1 | FALSE | Downregulated |
| -0,368840969 | 0,853245403 | RP11-673C5.2 | FALSE | Downregulated |
| 0,055997137 | 0,853245403 | SFPQ | FALSE | Upregulated |
| -0,256041937 | 0,853245403 | SNAPC2 | FALSE | Downregulated |
| 0,141976045 | 0,853245403 | STAU2 | FALSE | Upregulated |
| -0,22513793 | 0,853245403 | TFB2M | FALSE | Downregulated |
| -0,124522769 | 0,853245403 | TMEM220 | FALSE | Downregulated |
| -0,077268562 | 0,853245403 | TNFAIP8L1 | FALSE | Downregulated |
| 0,18179215 | 0,853245403 | UNC5C | FALSE | Upregulated |
| 0,315873749 | 0,853245403 | WDR93 | FALSE | Upregulated |
| 0,279460367 | 0,853378441 | GORAB | FALSE | Upregulated |
| -0,118427664 | 0,853378441 | PSME1 | FALSE | Downregulated |
| -0,192724625 | 0,853422023 | ABHD3 | FALSE | Downregulated |
| 0,212578076 | 0,853422023 | GPRASP2 | FALSE | Upregulated |
| 0,100096729 | 0,853422023 | LOC100129361 | FALSE | Upregulated |
| 0,424996089 | 0,853422023 | LOC101928047 | FALSE | Upregulated |
| -0,083676676 | 0,853422023 | PDXK | FALSE | Downregulated |
| 0,237100138 | 0,853570232 | C12orf49 | FALSE | Upregulated |
| 0,103468936 | 0,853570232 | CCDC92 | FALSE | Upregulated |
| 0,09514343 | 0,853570232 | MED12 | FALSE | Upregulated |
| -0,082692886 | 0,853570232 | PTPRJ | FALSE | Downregulated |
| -0,203458535 | 0,853570232 | TDRD10 | FALSE | Downregulated |
| 0,164483737 | 0,853953752 | CBLL1 | FALSE | Upregulated |
| 0,258244493 | 0,853953752 | COPS4 | FALSE | Upregulated |

| | | | | |
|---|---|---|---|---|
| 0,078651659 | 0,853953752 | CREBL2 | FALSE | Upregulated |
| 0,33086944 | 0,853953752 | KIAA1024 | FALSE | Upregulated |
| -0,479850067 | 0,853953752 | LOC100505985 | FALSE | Downregulated |
| 0,37041489 | 0,853953752 | POC1B-GALNT4 | FALSE | Upregulated |
| -0,192762009 | 0,853953752 | PRDM10 | FALSE | Downregulated |
| -0,09752573 | 0,853953752 | PTK2B | FALSE | Downregulated |
| -0,113536633 | 0,853953752 | SLC35B4 | FALSE | Downregulated |
| -0,125131224 | 0,853953752 | SLC35F5 | FALSE | Downregulated |
| -0,186338602 | 0,853953752 | TBP | FALSE | Downregulated |
| 0,188485375 | 0,853953752 | ULK2 | FALSE | Upregulated |
| -0,251399738 | 0,853953752 | ZNF273 | FALSE | Downregulated |
| -0,189785463 | 0,854099697 | AXIN2 | FALSE | Downregulated |
| -0,186710836 | 0,854099697 | PPA2 | FALSE | Downregulated |
| -0,307368536 | 0,854099697 | TMEM18 | FALSE | Downregulated |
| 0,234327898 | 0,854210433 | STOML1 | FALSE | Upregulated |
| 0,449554254 | 0,854210433 | TAF4B | FALSE | Upregulated |
| 0,279683075 | 0,854210433 | TREH | FALSE | Upregulated |
| -0,061935334 | 0,854210433 | TTPAL | FALSE | Downregulated |
| 0,094399836 | 0,854210433 | ZNF513 | FALSE | Upregulated |
| 0,236087609 | 0,854225303 | ZNF846 | FALSE | Upregulated |
| 0,198160147 | 0,854400083 | HEATR3 | FALSE | Upregulated |
| -0,118254989 | 0,85485966 | ARPC5 | FALSE | Downregulated |
| 0,14449307 | 0,85485966 | SIGLEC16 | FALSE | Upregulated |
| 0,207545503 | 0,854955319 | C5orf34 | FALSE | Upregulated |
| -0,102957971 | 0,855040943 | RPRD2 | FALSE | Downregulated |
| 0,191482553 | 0,855190967 | TERC | FALSE | Upregulated |
| 0,114761244 | 0,855315034 | CABP7 | FALSE | Upregulated |
| 0,344332039 | 0,855315034 | CDH20 | FALSE | Upregulated |
| -0,377796814 | 0,855315034 | CYP4X1 | FALSE | Downregulated |
| 0,474976709 | 0,85535808 | FAM129C | FALSE | Upregulated |
| -0,369905167 | 0,85535808 | FBXO48 | FALSE | Downregulated |
| 0,2743025 | 0,85535808 | LOC100289533 | FALSE | Upregulated |
| -0,099037999 | 0,85535808 | SH3GLB1 | FALSE | Downregulated |
| 0,173984848 | 0,855411665 | TTC13 | FALSE | Upregulated |
| -0,258799844 | 0,855587515 | FAM133B | FALSE | Downregulated |
| -0,147194016 | 0,855615588 | ARHGAP42 | FALSE | Downregulated |
| 0,048821345 | 0,855615588 | MLLT6 | FALSE | Upregulated |
| 0,218498672 | 0,855779172 | SNRPB2 | FALSE | Upregulated |
| 0,471408248 | 0,855897231 | ARHGAP11A | FALSE | Upregulated |
| -0,113920678 | 0,855897231 | NDUFA10 | FALSE | Downregulated |
| -0,218125599 | 0,855897231 | PIGM | FALSE | Downregulated |
| -0,157663912 | 0,856312435 | CCDC174 | FALSE | Downregulated |
| -0,214166491 | 0,856312435 | ERGIC2 | FALSE | Downregulated |
| -0,153756511 | 0,856312435 | TTC21B | FALSE | Downregulated |
| -0,138280932 | 0,856356823 | FGFR1 | FALSE | Downregulated |
| -0,32419677 | 0,856356823 | INO80C | FALSE | Downregulated |
| 2,580020169 | 0,856356823 | LOC102724441 | FALSE | Upregulated |
| 0,186171927 | 0,856356823 | LOC729291 | FALSE | Upregulated |
| 0,06712735 | 0,856356823 | NEK9 | FALSE | Upregulated |
| -0,145086969 | 0,856356823 | WDR43 | FALSE | Downregulated |
| -0,166957597 | 0,856356823 | ZNF48 | FALSE | Downregulated |

| | | | | |
|---|---|---|---|---|
| -0,087779675 | 0,856449555 | TRIP11 | FALSE | Downregulated |
| 0,232565185 | 0,85654642 | PTCH2 | FALSE | Upregulated |
| -0,485848075 | 0,856638245 | ZNF252P-AS1 | FALSE | Downregulated |
| 0,117513727 | 0,856806783 | DDX46 | FALSE | Upregulated |
| -0,115463674 | 0,856806783 | GTPBP2 | FALSE | Downregulated |
| -0,254380745 | 0,856806783 | RFT1 | FALSE | Downregulated |
| 0,151412626 | 0,856944169 | LOC284889 | FALSE | Upregulated |
| 0,089102845 | 0,857110547 | USP49 | FALSE | Upregulated |
| -0,164029508 | 0,857152929 | CPLX1 | FALSE | Downregulated |
| -0,074819262 | 0,857152929 | HNRNPF | FALSE | Downregulated |
| -0,211010151 | 0,857152929 | NDFIP2 | FALSE | Downregulated |
| 0,078463965 | 0,857248075 | DNAJC3 | FALSE | Upregulated |
| -0,113934002 | 0,857248075 | SMARCE1 | FALSE | Downregulated |
| -0,248761147 | 0,85739174 | C1orf220 | FALSE | Downregulated |
| 0,262190172 | 0,85739174 | EPSTI1 | FALSE | Upregulated |
| 0,133017778 | 0,85739174 | ESR2 | FALSE | Upregulated |
| -0,513589573 | 0,85740066 | AC004862.6 | FALSE | Downregulated |
| -0,103959402 | 0,85740066 | AHDC1 | FALSE | Downregulated |
| 0,349169485 | 0,85740066 | FGFBP3 | FALSE | Upregulated |
| -0,157610131 | 0,85740066 | RTP3 | FALSE | Downregulated |
| -0,153615669 | 0,85740066 | TBCEL | FALSE | Downregulated |
| -0,193604007 | 0,85740066 | ZIC4 | FALSE | Downregulated |
| 0,143767756 | 0,857744548 | GGT1 | FALSE | Upregulated |
| 0,440192064 | 0,857799507 | CCNB2 | FALSE | Upregulated |
| 0,133257605 | 0,858078489 | NEK7 | FALSE | Upregulated |
| -0,107530396 | 0,858078489 | SUV420H1 | FALSE | Downregulated |
| 0,066482888 | 0,858315127 | LONP1 | FALSE | Upregulated |
| 0,131174982 | 0,858370073 | SNRPD2 | FALSE | Upregulated |
| -0,061977374 | 0,858370073 | TRRAP | FALSE | Downregulated |
| 0,264622892 | 0,858485922 | CAMTA1 | FALSE | Upregulated |
| 0,61019055 | 0,858528027 | ACKR3 | FALSE | Upregulated |
| 0,4769592 | 0,858543798 | SLFN12L | FALSE | Upregulated |
| 0,370948737 | 0,858556412 | FGF17 | FALSE | Upregulated |
| 0,508514826 | 0,858906885 | CBWD6 | FALSE | Upregulated |
| 0,305556203 | 0,858906885 | FAM110A | FALSE | Upregulated |
| 0,265408172 | 0,858906885 | LPAR5 | FALSE | Upregulated |
| 0,166523945 | 0,858906885 | SPNS3 | FALSE | Upregulated |
| 0,160530816 | 0,858906885 | TMPRSS2 | FALSE | Upregulated |
| 0,170190661 | 0,859116665 | HDAC2 | FALSE | Upregulated |
| -0,117696266 | 0,859116665 | MIA2 | FALSE | Downregulated |
| 0,076543569 | 0,859116665 | RIC8A | FALSE | Upregulated |
| 0,159722984 | 0,859116665 | RP11-426A6.5 | FALSE | Upregulated |
| -0,12392139 | 0,859116665 | ZFYVE19 | FALSE | Downregulated |
| 0,478200312 | 0,85912045 | CD200 | FALSE | Upregulated |
| 0,215743185 | 0,85912045 | SLX1A | FALSE | Upregulated |
| 0,104441184 | 0,859251296 | ZNF148 | FALSE | Upregulated |
| -0,097295808 | 0,859251296 | ZYG11B | FALSE | Downregulated |
| -0,314876308 | 0,859267369 | GMNC | FALSE | Downregulated |
| 0,115814438 | 0,859350559 | STK19 | FALSE | Upregulated |
| -0,162634438 | 0,859536532 | C10orf32 | FALSE | Downregulated |
| 0,160625228 | 0,859536532 | C19orf35 | FALSE | Upregulated |

| | | | | |
|---|---|---|---|---|
| 0,112981362 | 0,859536532 | C2CD5 | FALSE | Upregulated |
| -0,103366145 | 0,859536532 | GIT1 | FALSE | Downregulated |
| 0,292215549 | 0,859536532 | ROBO3 | FALSE | Upregulated |
| 0,25664911 | 0,859536532 | TACR2 | FALSE | Upregulated |
| 0,170020383 | 0,859536532 | TET1 | FALSE | Upregulated |
| -0,120182366 | 0,859536532 | TIGD5 | FALSE | Downregulated |
| 0,35730955 | 0,859536532 | ZNF625 | FALSE | Upregulated |
| -0,144430751 | 0,859804035 | AATK | FALSE | Downregulated |
| -0,143412461 | 0,859804035 | LOC101928588 | FALSE | Downregulated |
| 0,087114165 | 0,859804035 | LPCAT3 | FALSE | Upregulated |
| -0,198284758 | 0,859804035 | RCBTB1 | FALSE | Downregulated |
| 0,279061989 | 0,859804035 | ZNF852 | FALSE | Upregulated |
| 0,165936446 | 0,859917008 | KLLN | FALSE | Upregulated |
| -0,110690291 | 0,859965894 | ATG4B | FALSE | Downregulated |
| -0,257888751 | 0,859989008 | LOC101928381 | FALSE | Downregulated |
| 0,362970619 | 0,860031545 | C1orf53 | FALSE | Upregulated |
| -0,100457517 | 0,860031545 | CNNM4 | FALSE | Downregulated |
| 0,131603284 | 0,860031545 | GNA13 | FALSE | Upregulated |
| -0,059127154 | 0,860031545 | INSR | FALSE | Downregulated |
| -0,337525058 | 0,860031545 | LOC100996497 | FALSE | Downregulated |
| -0,199750906 | 0,860031545 | MFSD8 | FALSE | Downregulated |
| 0,358251111 | 0,860031545 | OR2A2 | FALSE | Upregulated |
| -0,256582635 | 0,860031545 | RP11-473M20.7 | FALSE | Downregulated |
| 0,230512841 | 0,860031545 | SAMD12 | FALSE | Upregulated |
| -0,276025892 | 0,860031545 | SLX1B | FALSE | Downregulated |
| 0,085495838 | 0,860031545 | TSC22D2 | FALSE | Upregulated |
| -0,332524327 | 0,860100639 | LOC101928635 | FALSE | Downregulated |
| -0,17816234 | 0,860100639 | UXT | FALSE | Downregulated |
| 0,143932627 | 0,860185662 | DPY19L1 | FALSE | Upregulated |
| 0,119554346 | 0,860303939 | OSBP2 | FALSE | Upregulated |
| 0,184083885 | 0,860336831 | TUBA3FP | FALSE | Upregulated |
| 0,353791571 | 0,860445842 | BNC2 | FALSE | Upregulated |
| -0,113024639 | 0,860445842 | CEBPG | FALSE | Downregulated |
| 0,203632878 | 0,860445842 | PDE4C | FALSE | Upregulated |
| -0,123123115 | 0,860445842 | TACC2 | FALSE | Downregulated |
| -0,18467252 | 0,860466424 | HNRNPA1P10 | FALSE | Downregulated |
| 0,134182536 | 0,860523379 | CASP8AP2 | FALSE | Upregulated |
| 0,394170505 | 0,860599174 | CCDC171 | FALSE | Upregulated |
| -0,200127443 | 0,860599174 | LRFN3 | FALSE | Downregulated |
| -0,408072348 | 0,860599174 | RHNO1 | FALSE | Downregulated |
| 0,112728772 | 0,860617579 | G3BP2 | FALSE | Upregulated |
| 0,075096446 | 0,860858394 | ADORA2A-AS1 | FALSE | Upregulated |
| -0,081201443 | 0,860858394 | BNIP3 | FALSE | Downregulated |
| 0,168004608 | 0,860858394 | HGF | FALSE | Upregulated |
| 0,666880779 | 0,860858394 | LOC102723615 | FALSE | Upregulated |
| 0,172546789 | 0,860858394 | PRSS23 | FALSE | Upregulated |
| 0,196965525 | 0,860858394 | STEAP1 | FALSE | Upregulated |
| -0,295962898 | 0,860858394 | TTC22 | FALSE | Downregulated |
| 0,204269086 | 0,860858394 | ZNF347 | FALSE | Upregulated |
| 0,344332956 | 0,86086011 | APOBEC3C | FALSE | Upregulated |
| -0,47101617 | 0,86086011 | LOC102723603 | FALSE | Downregulated |

| | | | | |
|---|---|---|---|---|
| 0,281615797 | 0,860877424 | ZNF286A | FALSE | Upregulated |
| 0,32274579 | 0,861003788 | HCG8 | FALSE | Upregulated |
| -0,150829491 | 0,861003788 | LRP5L | FALSE | Downregulated |
| 0,148942259 | 0,861003788 | PREP | FALSE | Upregulated |
| -0,076641462 | 0,861009784 | CANX | FALSE | Downregulated |
| 0,351806492 | 0,861009784 | EFNB3 | FALSE | Upregulated |
| -0,055685129 | 0,861009784 | MACF1 | FALSE | Downregulated |
| 0,166519839 | 0,861009784 | S100A10 | FALSE | Upregulated |
| 0,125752769 | 0,861009784 | UBE2V1 | FALSE | Upregulated |
| -0,150943521 | 0,861009784 | UBIAD1 | FALSE | Downregulated |
| 0,151769733 | 0,861086664 | ATP6V0B | FALSE | Upregulated |
| -0,189715571 | 0,861162267 | C1orf35 | FALSE | Downregulated |
| 0,438101065 | 0,861162267 | FKBPL | FALSE | Upregulated |
| 0,183457969 | 0,861162267 | GLRA1 | FALSE | Upregulated |
| -0,438399814 | 0,861162267 | HCG4 | FALSE | Downregulated |
| -0,118658894 | 0,861162267 | KIFC3 | FALSE | Downregulated |
| 0,210557886 | 0,861162267 | LOC100126584 | FALSE | Upregulated |
| -0,29163378 | 0,861162267 | OR51I2 | FALSE | Downregulated |
| 0,156750593 | 0,861162267 | PLAA | FALSE | Upregulated |
| 0,103619395 | 0,861162267 | RIOK3 | FALSE | Upregulated |
| 0,251664584 | 0,861162267 | SFT2D2 | FALSE | Upregulated |
| -0,118753201 | 0,861162267 | STRN | FALSE | Downregulated |
| 0,273774503 | 0,861190784 | CCDC37 | FALSE | Upregulated |
| -0,087588644 | 0,861220094 | ZNF512B | FALSE | Downregulated |
| 0,457659429 | 0,861534846 | SOWAHD | FALSE | Upregulated |
| 0,230679378 | 0,861626987 | IL18 | FALSE | Upregulated |
| -0,278981797 | 0,861626987 | ZBTB24 | FALSE | Downregulated |
| -0,274571216 | 0,861698654 | SETD9 | FALSE | Downregulated |
| 0,214250745 | 0,861710216 | SPATA25 | FALSE | Upregulated |
| -0,702533716 | 0,861759921 | C1orf140 | FALSE | Downregulated |
| 0,240179939 | 0,862010023 | DNLZ | FALSE | Upregulated |
| -0,190305411 | 0,862010023 | ERG | FALSE | Downregulated |
| 0,475411699 | 0,862010023 | HRH1 | FALSE | Upregulated |
| 0,151901538 | 0,862010023 | WDR41 | FALSE | Upregulated |
| -0,187464723 | 0,862099274 | PARP15 | FALSE | Downregulated |
| 0,078834429 | 0,862099274 | YLPM1 | FALSE | Upregulated |
| 0,485170176 | 0,862099794 | SOX21 | FALSE | Upregulated |
| 0,153270614 | 0,862114932 | LAMA4 | FALSE | Upregulated |
| -0,074433771 | 0,862114932 | NRD1 | FALSE | Downregulated |
| -0,169496615 | 0,862114932 | OLFM2 | FALSE | Downregulated |
| -0,068673087 | 0,862114932 | PPL | FALSE | Downregulated |
| 0,232185058 | 0,862114932 | RFX3 | FALSE | Upregulated |
| 0,295360219 | 0,862114932 | RP11-307C12.11 | FALSE | Upregulated |
| 0,082171901 | 0,862114932 | SMG1P1 | FALSE | Upregulated |
| -0,137673162 | 0,862114932 | TMEM101 | FALSE | Downregulated |
| 0,199581747 | 0,862155352 | BEND7 | FALSE | Upregulated |
| 0,160560605 | 0,862155352 | MRRF | FALSE | Upregulated |
| -0,048587773 | 0,862155352 | NCOR1 | FALSE | Downregulated |
| 0,156187431 | 0,862155352 | PAFAH1B2 | FALSE | Upregulated |
| -0,102754726 | 0,862155352 | PCMTD2 | FALSE | Downregulated |
| -0,117700238 | 0,862155352 | SEC11A | FALSE | Downregulated |

| | | | | |
|---|---|---|---|---|
| -0,329982073 | 0,862155352 | SLIRP | FALSE | Downregulated |
| 0,18488288 | 0,862155352 | TCP11L2 | FALSE | Upregulated |
| -0,118903848 | 0,862155352 | TPM3 | FALSE | Downregulated |
| -0,254696667 | 0,862155352 | ZNF691 | FALSE | Downregulated |
| -0,463881771 | 0,862427346 | AC009299.3 | FALSE | Downregulated |
| -0,094580914 | 0,862427346 | ATG9A | FALSE | Downregulated |
| 0,241295173 | 0,862427346 | HYPK | FALSE | Upregulated |
| 0,305101838 | 0,862427346 | KITLG | FALSE | Upregulated |
| -0,104506363 | 0,862427346 | PROZ | FALSE | Downregulated |
| -0,106652493 | 0,862427346 | SREBF2 | FALSE | Downregulated |
| -0,194193235 | 0,862427346 | ZNF32-AS1 | FALSE | Downregulated |
| -0,329529601 | 0,862442328 | ABCA11P | FALSE | Downregulated |
| 0,139005057 | 0,86251439 | AMD1 | FALSE | Upregulated |
| 0,303370822 | 0,86251439 | LOC102724583 | FALSE | Upregulated |
| 0,100702474 | 0,862571813 | COLGALT1 | FALSE | Upregulated |
| -0,240314118 | 0,862571813 | LRAT | FALSE | Downregulated |
| -0,085691424 | 0,862571813 | MUM1 | FALSE | Downregulated |
| -0,130891383 | 0,862571813 | PDS5B | FALSE | Downregulated |
| 0,370970975 | 0,862905999 | RP11-829H16.3 | FALSE | Upregulated |
| 0,165125836 | 0,862965908 | CYP2B6 | FALSE | Upregulated |
| 0,067877462 | 0,862965908 | DENND4C | FALSE | Upregulated |
| -0,172079523 | 0,863047417 | PLXDC1 | FALSE | Downregulated |
| 0,125089537 | 0,863107153 | GSAP | FALSE | Upregulated |
| 0,150411884 | 0,863107153 | SCARNA6 | FALSE | Upregulated |
| -0,173593195 | 0,86316134 | TGS1 | FALSE | Downregulated |
| 0,176832448 | 0,863202277 | C9orf41 | FALSE | Upregulated |
| -0,122613155 | 0,863202277 | FOXO1 | FALSE | Downregulated |
| 0,192264361 | 0,863202277 | LOC100506596 | FALSE | Upregulated |
| 0,51170103 | 0,863202277 | LOC102723923 | FALSE | Upregulated |
| -0,11055402 | 0,863202277 | PLGLB2 | FALSE | Downregulated |
| 0,26476216 | 0,863202277 | STX8 | FALSE | Upregulated |
| -0,263898952 | 0,863202277 | TMEM217 | FALSE | Downregulated |
| 0,076817677 | 0,863202277 | TP53I13 | FALSE | Upregulated |
| 0,243867183 | 0,863202277 | ZNF573 | FALSE | Upregulated |
| 0,390684304 | 0,86340513 | AC007163.6 | FALSE | Upregulated |
| 0,144862768 | 0,86340513 | ACAP2 | FALSE | Upregulated |
| -2,593359595 | 0,86340513 | ATP5J2-PTCD1 | FALSE | Downregulated |
| 0,105460795 | 0,86340513 | CHURC1 | FALSE | Upregulated |
| 0,169156907 | 0,86340513 | CYB561D2 | FALSE | Upregulated |
| -0,089149927 | 0,86340513 | FAM115A | FALSE | Downregulated |
| 0,499605551 | 0,86340513 | GCNT4 | FALSE | Upregulated |
| -0,256344197 | 0,86340513 | HIATL2 | FALSE | Downregulated |
| 0,100159188 | 0,86340513 | HSPA4 | FALSE | Upregulated |
| -0,193606992 | 0,86340513 | KIAA0020 | FALSE | Downregulated |
| -0,122832059 | 0,86340513 | LOC102060414 | FALSE | Downregulated |
| -0,20555176 | 0,86340513 | POU6F1 | FALSE | Downregulated |
| -0,216713607 | 0,86340513 | PPHLN1 | FALSE | Downregulated |
| 0,277240246 | 0,86340513 | RGS11 | FALSE | Upregulated |
| -0,225935221 | 0,86340513 | UBTD1 | FALSE | Downregulated |
| -0,241416437 | 0,86340513 | ZNF585A | FALSE | Downregulated |
| 0,188512922 | 0,863452534 | C2orf49 | FALSE | Upregulated |

| | | | | |
|---|---|---|---|---|
| 0,176851094 | 0,863493041 | ARHGAP24 | FALSE | Upregulated |
| -0,230985278 | 0,863493041 | LOC101929078 | FALSE | Downregulated |
| -0,113339669 | 0,863509541 | USP33 | FALSE | Downregulated |
| -0,164369997 | 0,863535747 | PRUNE | FALSE | Downregulated |
| -0,093763756 | 0,863572824 | BMP2K | FALSE | Downregulated |
| 0,147202457 | 0,863572824 | C14orf1 | FALSE | Upregulated |
| -0,135465735 | 0,863761317 | SCO1 | FALSE | Downregulated |
| -0,256999233 | 0,863769784 | C10orf2 | FALSE | Downregulated |
| 0,086341473 | 0,863769784 | CDC42 | FALSE | Upregulated |
| -0,11445549 | 0,863769784 | ELAVL1 | FALSE | Downregulated |
| -0,149691666 | 0,863769784 | EXOC2 | FALSE | Downregulated |
| -0,088205258 | 0,863769784 | MED16 | FALSE | Downregulated |
| -0,11901583 | 0,863769784 | NDOR1 | FALSE | Downregulated |
| 0,240022978 | 0,863769784 | NRROS | FALSE | Upregulated |
| 0,084925136 | 0,863769784 | RUVBL1 | FALSE | Upregulated |
| -0,433258547 | 0,863809018 | LOC100996350 | FALSE | Downregulated |
| 0,150514769 | 0,863930933 | DEDD2 | FALSE | Upregulated |
| -0,209365675 | 0,863930933 | FAM210A | FALSE | Downregulated |
| 0,097975915 | 0,863930933 | TBC1D2B | FALSE | Upregulated |
| 0,250726149 | 0,863964118 | LOC101927023 | FALSE | Upregulated |
| -0,130119628 | 0,864000793 | MBOAT7 | FALSE | Downregulated |
| 0,30934913 | 0,864104535 | AAED1 | FALSE | Upregulated |
| -0,147219109 | 0,864104535 | AFF3 | FALSE | Downregulated |
| -0,086084681 | 0,864104535 | PHF2 | FALSE | Downregulated |
| 0,092272381 | 0,864104535 | XRN2 | FALSE | Upregulated |
| -0,225406863 | 0,864104535 | ZSWIM7 | FALSE | Downregulated |
| 0,185290372 | 0,864124544 | CDKL5 | FALSE | Upregulated |
| 0,078858977 | 0,864124544 | COPB2 | FALSE | Upregulated |
| 0,283081371 | 0,864124544 | GALNT15 | FALSE | Upregulated |
| 0,121552323 | 0,864124544 | LIG1 | FALSE | Upregulated |
| -0,11550854 | 0,864124544 | SGTA | FALSE | Downregulated |
| -0,194681547 | 0,864124544 | ST8SIA3 | FALSE | Downregulated |
| 0,246992578 | 0,864124544 | TMEM234 | FALSE | Upregulated |
| -0,094082952 | 0,864125986 | FMR1 | FALSE | Downregulated |
| 0,457703343 | 0,864136545 | HSP90AB4P | FALSE | Upregulated |
| 0,108106152 | 0,864161852 | ARRB2 | FALSE | Upregulated |
| 0,160258649 | 0,864161852 | NDUFS6 | FALSE | Upregulated |
| -0,177785855 | 0,864161852 | STBD1 | FALSE | Downregulated |
| 0,184528819 | 0,864163368 | HOMEZ | FALSE | Upregulated |
| -0,118486269 | 0,864165824 | RNF14 | FALSE | Downregulated |
| 0,195570713 | 0,864169161 | CWC22 | FALSE | Upregulated |
| -0,187467776 | 0,864169161 | NS3BP | FALSE | Downregulated |
| 0,323458966 | 0,864169161 | TIAF1 | FALSE | Upregulated |
| 0,196218136 | 0,864220153 | PPP1R3F | FALSE | Upregulated |
| -0,176730298 | 0,864618766 | LOC613037 | FALSE | Downregulated |
| 0,18517786 | 0,864704627 | FAM98A | FALSE | Upregulated |
| -0,140267558 | 0,864704627 | LMBRD1 | FALSE | Downregulated |
| 0,211438218 | 0,864704627 | PKI55 | FALSE | Upregulated |
| 0,43213749 | 0,864704627 | RASSF9 | FALSE | Upregulated |
| 0,145216356 | 0,864730701 | RAN | FALSE | Upregulated |
| 0,107350807 | 0,864786721 | TRNT1 | FALSE | Upregulated |

| | | | | |
|---|---|---|---|---|
| 0,062395759 | 0,864813434 | BAZ2A | FALSE | Upregulated |
| -0,102121382 | 0,86491283 | NUAK1 | FALSE | Downregulated |
| 0,151602325 | 0,86491283 | PNO1 | FALSE | Upregulated |
| 0,195688598 | 0,86491283 | TP53RK | FALSE | Upregulated |
| 0,185157315 | 0,864985462 | GPSM3 | FALSE | Upregulated |
| 0,413755867 | 0,864991975 | LOC101927733 | FALSE | Upregulated |
| -0,057189247 | 0,865148169 | RPS18 | FALSE | Downregulated |
| 0,115742034 | 0,865148169 | VEZF1 | FALSE | Upregulated |
| 0,356650754 | 0,865241922 | FITM2 | FALSE | Upregulated |
| -0,231768728 | 0,865241922 | POGLUT1 | FALSE | Downregulated |
| -0,048138166 | 0,865368375 | NCOA4 | FALSE | Downregulated |
| 0,168784071 | 0,865368375 | ZNF277 | FALSE | Upregulated |
| -0,164031278 | 0,865406392 | FIZ1 | FALSE | Downregulated |
| 0,25161642 | 0,865549198 | BAK1 | FALSE | Upregulated |
| -0,254414531 | 0,865549198 | EGFLAM | FALSE | Downregulated |
| 0,201359559 | 0,865549198 | GNG3 | FALSE | Upregulated |
| -0,157892402 | 0,865757213 | GGH | FALSE | Downregulated |
| -0,223752636 | 0,865757213 | NCAPG2 | FALSE | Downregulated |
| 0,127163827 | 0,865757213 | PPM1G | FALSE | Upregulated |
| 0,418162892 | 0,865757213 | ZNF888 | FALSE | Upregulated |
| 0,096129848 | 0,865796344 | ABI2 | FALSE | Upregulated |
| 0,084819601 | 0,865796344 | PHACTR2 | FALSE | Upregulated |
| 0,28258796 | 0,865796344 | SPRY3 | FALSE | Upregulated |
| 0,493952419 | 0,865962436 | HIST1H3B | FALSE | Upregulated |
| -0,12686767 | 0,866143384 | PRR5 | FALSE | Downregulated |
| 0,424103764 | 0,866172644 | LOC101929796 | FALSE | Upregulated |
| -0,138027344 | 0,866275848 | DDA1 | FALSE | Downregulated |
| 0,101654101 | 0,866275848 | IKBKB | FALSE | Upregulated |
| -0,463053895 | 0,866417613 | CAHM | FALSE | Downregulated |
| -0,268785166 | 0,866417613 | LOC102723845 | FALSE | Downregulated |
| -0,223976508 | 0,866417613 | METTL21A | FALSE | Downregulated |
| -0,167590951 | 0,866417613 | METTL9 | FALSE | Downregulated |
| 0,145816948 | 0,866417613 | PLET1 | FALSE | Upregulated |
| 0,244095327 | 0,866495526 | C9orf89 | FALSE | Upregulated |
| 1,733767949 | 0,866495526 | LOC102723728 | FALSE | Upregulated |
| 0,152605184 | 0,866495526 | PLEKHG7 | FALSE | Upregulated |
| -0,392583622 | 0,866975765 | AURKAPS1 | FALSE | Downregulated |
| -0,081486997 | 0,866975765 | DCTN1 | FALSE | Downregulated |
| -0,399872151 | 0,866980204 | UTY | FALSE | Downregulated |
| -0,098581885 | 0,866991081 | ABCF1 | FALSE | Downregulated |
| -0,183437316 | 0,867046183 | LRP8 | FALSE | Downregulated |
| 0,194918537 | 0,867046183 | SHC3 | FALSE | Upregulated |
| 0,092539113 | 0,867355628 | BCAR1 | FALSE | Upregulated |
| -0,151146231 | 0,867355628 | CKMT2-AS1 | FALSE | Downregulated |
| 0,116269803 | 0,867355628 | CLIC1 | FALSE | Upregulated |
| -0,108698208 | 0,867355628 | CUL4B | FALSE | Downregulated |
| 0,146850295 | 0,867355628 | DPYD-AS2 | FALSE | Upregulated |
| -0,110706518 | 0,867355628 | EIF2B1 | FALSE | Downregulated |
| 0,121855043 | 0,867355628 | FAAH | FALSE | Upregulated |
| -0,142973978 | 0,867355628 | FAM110C | FALSE | Downregulated |
| 0,086901438 | 0,867355628 | FOXO3 | FALSE | Upregulated |

| | | | | |
|---|---|---|---|---|
| 0,095548196 | 0,867355628 | GIGYF2 | FALSE | Upregulated |
| -0,07647815 | 0,867355628 | HNF1A | FALSE | Downregulated |
| 0,107373716 | 0,867355628 | KLHL21 | FALSE | Upregulated |
| 0,365357213 | 0,867355628 | LILRA2 | FALSE | Upregulated |
| -0,124766319 | 0,867355628 | LINC00847 | FALSE | Downregulated |
| 0,209760019 | 0,867355628 | LOC101927864 | FALSE | Upregulated |
| -0,406683733 | 0,867355628 | LOC102724841 | FALSE | Downregulated |
| -0,088187717 | 0,867355628 | MARCH7 | FALSE | Downregulated |
| -0,070611037 | 0,867355628 | MXI1 | FALSE | Downregulated |
| 0,149613559 | 0,867355628 | NAGK | FALSE | Upregulated |
| -0,171474905 | 0,867355628 | PPCS | FALSE | Downregulated |
| 0,067694152 | 0,867355628 | PPFIA1 | FALSE | Upregulated |
| 0,527112286 | 0,867355628 | RPP21 | FALSE | Upregulated |
| 0,096867102 | 0,867355628 | SAR1A | FALSE | Upregulated |
| -0,32959859 | 0,867355628 | TMEM241 | FALSE | Downregulated |
| -0,107985634 | 0,867355628 | TMF1 | FALSE | Downregulated |
| 0,060624892 | 0,867355628 | TSPYL1 | FALSE | Upregulated |
| 0,299464926 | 0,867355628 | TTC16 | FALSE | Upregulated |
| 0,139424164 | 0,867355628 | ZNF749 | FALSE | Upregulated |
| 0,173109185 | 0,867421249 | LAMTOR4 | FALSE | Upregulated |
| 0,316770328 | 0,867421249 | RP11-298I3.1 | FALSE | Upregulated |
| 0,063991927 | 0,867457702 | CCNL2 | FALSE | Upregulated |
| 0,192323716 | 0,867550834 | FPGT | FALSE | Upregulated |
| 0,073787637 | 0,86763669 | SRP9 | FALSE | Upregulated |
| 0,116905616 | 0,86773059 | ENOSF1 | FALSE | Upregulated |
| 0,101965181 | 0,86773059 | TRO | FALSE | Upregulated |
| -0,122108349 | 0,867770001 | JADE3 | FALSE | Downregulated |
| 0,135945196 | 0,867770001 | LOC102724043 | FALSE | Upregulated |
| 0,822688399 | 0,867770001 | MYOZ1 | FALSE | Upregulated |
| -0,243703451 | 0,867770001 | SLC25A52 | FALSE | Downregulated |
| -0,074031234 | 0,868032185 | CTSD | FALSE | Downregulated |
| -0,166851022 | 0,868032185 | LOC101926985 | FALSE | Downregulated |
| 0,376958717 | 0,868032185 | OPRM1 | FALSE | Upregulated |
| 0,12176988 | 0,868032185 | PDCD7 | FALSE | Upregulated |
| -0,160761558 | 0,868032185 | TMEM44-AS1 | FALSE | Downregulated |
| -0,185616524 | 0,868032185 | UBE2V2 | FALSE | Downregulated |
| -0,122926411 | 0,868032185 | UBQLN2 | FALSE | Downregulated |
| 0,145127886 | 0,868194278 | KIAA1143 | FALSE | Upregulated |
| 0,107488045 | 0,868194278 | PCED1A | FALSE | Upregulated |
| 0,08473856 | 0,868194278 | RP5-1071N3.1 | FALSE | Upregulated |
| -0,116132509 | 0,868194278 | SH3GLB2 | FALSE | Downregulated |
| -0,108727171 | 0,868194278 | YIF1A | FALSE | Downregulated |
| -0,146154192 | 0,868270613 | MED8 | FALSE | Downregulated |
| -0,350031316 | 0,868397336 | FTCDNL1 | FALSE | Downregulated |
| -0,090259184 | 0,868397336 | VWA8 | FALSE | Downregulated |
| 0,115526757 | 0,868468087 | ATR | FALSE | Upregulated |
| 0,069150148 | 0,868468087 | C6orf106 | FALSE | Upregulated |
| -0,17132043 | 0,868468087 | FAM189B | FALSE | Downregulated |
| 0,090643474 | 0,868468087 | KBTBD4 | FALSE | Upregulated |
| -0,150615544 | 0,868468087 | PCGF1 | FALSE | Downregulated |
| 0,11547697 | 0,868468087 | RAET1E-AS1 | FALSE | Upregulated |

| | | | | |
|---|---|---|---|---|
| 0,228064704 | 0,868468087 | TTC21B-AS1 | FALSE | Upregulated |
| -0,147176311 | 0,86859729 | HBS1L | FALSE | Downregulated |
| 0,106085978 | 0,86859729 | NFATC2IP | FALSE | Upregulated |
| -0,396257349 | 0,868601662 | HMGN5 | FALSE | Downregulated |
| 0,163374417 | 0,868601662 | PGM5 | FALSE | Upregulated |
| 0,214439172 | 0,868698515 | EXOSC5 | FALSE | Upregulated |
| -0,386352561 | 0,868698515 | OLFML2B | FALSE | Downregulated |
| 0,109528802 | 0,868698515 | ZBTB20-AS1 | FALSE | Upregulated |
| 0,354125331 | 0,868945797 | THAP1 | FALSE | Upregulated |
| 0,238868777 | 0,869097573 | CKB | FALSE | Upregulated |
| -0,13271018 | 0,869097573 | GIPC2 | FALSE | Downregulated |
| 0,244990526 | 0,869097573 | SSX2IP | FALSE | Upregulated |
| 0,379157433 | 0,869143909 | HSPB8 | FALSE | Upregulated |
| 0,450856859 | 0,869189423 | ZBED5-AS1 | FALSE | Upregulated |
| 0,06281375 | 0,869315985 | KDM5A | FALSE | Upregulated |
| -0,156726357 | 0,869315985 | SLC50A1 | FALSE | Downregulated |
| 0,097893114 | 0,869546115 | ANKLE2 | FALSE | Upregulated |
| -0,092725948 | 0,869546115 | EML3 | FALSE | Downregulated |
| 0,074244212 | 0,86960337 | CTNND1 | FALSE | Upregulated |
| 0,19187224 | 0,86960337 | GTF2B | FALSE | Upregulated |
| 0,150349017 | 0,86960337 | MARCH9 | FALSE | Upregulated |
| 0,123336623 | 0,86960337 | RP11-242D8.1 | FALSE | Upregulated |
| 0,256529642 | 0,86960337 | RP11-725G5.2 | FALSE | Upregulated |
| -0,17065666 | 0,869629304 | PSMA4 | FALSE | Downregulated |
| 0,12341433 | 0,869629304 | ZC3H7A | FALSE | Upregulated |
| -0,159261093 | 0,869664769 | NCDN | FALSE | Downregulated |
| -0,146576122 | 0,869664769 | PIGV | FALSE | Downregulated |
| -0,064300893 | 0,86976266 | RREB1 | FALSE | Downregulated |
| -0,084957791 | 0,870102024 | COX11 | FALSE | Downregulated |
| 0,257338454 | 0,870102024 | CRYBB2P1 | FALSE | Upregulated |
| 0,362005273 | 0,870102024 | LOC100505625 | FALSE | Upregulated |
| 0,072688 | 0,870102024 | RCN1 | FALSE | Upregulated |
| 0,238615515 | 0,870102024 | SPATA7 | FALSE | Upregulated |
| 0,144913001 | 0,870102024 | ZBTB42 | FALSE | Upregulated |
| 0,100653906 | 0,870162782 | ARF4 | FALSE | Upregulated |
| 0,175604697 | 0,870162782 | ARL16 | FALSE | Upregulated |
| -0,228793556 | 0,870162782 | C16orf80 | FALSE | Downregulated |
| -0,162914093 | 0,870162782 | CEP72 | FALSE | Downregulated |
| -0,100840025 | 0,870162782 | EXOSC6 | FALSE | Downregulated |
| -0,290453038 | 0,870162782 | FAM212B-AS1 | FALSE | Downregulated |
| 0,523905647 | 0,870162782 | FCGR2C | FALSE | Upregulated |
| 0,074274956 | 0,870162782 | FKBP9 | FALSE | Upregulated |
| -0,107688719 | 0,870162782 | ISOC1 | FALSE | Downregulated |
| 0,091222549 | 0,870162782 | LEMD3 | FALSE | Upregulated |
| -0,105651992 | 0,870162782 | RASSF7 | FALSE | Downregulated |
| -0,469613897 | 0,870162782 | RP11-314D7.2 | FALSE | Downregulated |
| 0,067613245 | 0,870162782 | SGSM2 | FALSE | Upregulated |
| 0,594082374 | 0,870342933 | CYP2A13 | FALSE | Upregulated |
| -0,078471227 | 0,870342933 | FAM107B | FALSE | Downregulated |
| 0,188532202 | 0,870342933 | HIGD1C | FALSE | Upregulated |
| 0,416667164 | 0,870342933 | LINC00506 | FALSE | Upregulated |

| | | | | |
|---|---|---|---|---|
| 0,162668286 | 0,870342933 | LMO2 | FALSE | Upregulated |
| -0,134348586 | 0,870342933 | POLR1E | FALSE | Downregulated |
| 0,158693983 | 0,870342933 | RRP1 | FALSE | Upregulated |
| -0,225507214 | 0,870374583 | IGFBP1 | FALSE | Downregulated |
| -0,099551659 | 0,870374583 | PTPRG | FALSE | Downregulated |
| 0,100773007 | 0,870409861 | BCCIP | FALSE | Upregulated |
| -0,169114875 | 0,87043367 | ACTL6A | FALSE | Downregulated |
| 0,094645019 | 0,87043367 | HNRNPA0 | FALSE | Upregulated |
| 0,179548337 | 0,870596374 | TOMM5 | FALSE | Upregulated |
| 0,304825801 | 0,870603263 | EFS | FALSE | Upregulated |
| 0,246123663 | 0,870670223 | GAB3 | FALSE | Upregulated |
| 0,310088876 | 0,870691565 | FAM188B | FALSE | Upregulated |
| 0,176957241 | 0,870691565 | FBXO40 | FALSE | Upregulated |
| -0,08782301 | 0,870790886 | COL6A6 | FALSE | Downregulated |
| 0,31564733 | 0,870790886 | LINGO1 | FALSE | Upregulated |
| -0,224273273 | 0,87084677 | TMEM252 | FALSE | Downregulated |
| -0,107647194 | 0,870909465 | CA14 | FALSE | Downregulated |
| -0,113185483 | 0,870909465 | DSTN | FALSE | Downregulated |
| -0,400273727 | 0,870909465 | LOC100130238 | FALSE | Downregulated |
| 0,060971401 | 0,870909465 | NR2F2 | FALSE | Upregulated |
| 0,166385911 | 0,870909465 | PAX8 | FALSE | Upregulated |
| 0,122313804 | 0,870909465 | PPP1R15A | FALSE | Upregulated |
| 0,077267029 | 0,870909465 | SLC12A6 | FALSE | Upregulated |
| -0,29324408 | 0,870909465 | SPAG16 | FALSE | Downregulated |
| -0,101269316 | 0,870909465 | SPPL2A | FALSE | Downregulated |
| 0,113114094 | 0,870909465 | UVSSA | FALSE | Upregulated |
| 0,089703795 | 0,870909465 | ZSCAN2 | FALSE | Upregulated |
| -0,179943296 | 0,870971717 | RP11-279D17.2 | FALSE | Downregulated |
| 0,060516309 | 0,870971717 | USP22 | FALSE | Upregulated |
| 0,132504249 | 0,871089675 | ANTXR2 | FALSE | Upregulated |
| 0,33524248 | 0,871089675 | ZNF699 | FALSE | Upregulated |
| 0,23184032 | 0,87117376 | MALSU1 | FALSE | Upregulated |
| 0,100148316 | 0,871178702 | LAT2 | FALSE | Upregulated |
| -0,186623572 | 0,871178702 | SLC2A13 | FALSE | Downregulated |
| -0,088585134 | 0,871178702 | ZKSCAN8 | FALSE | Downregulated |
| -0,446717321 | 0,871183051 | CCER2 | FALSE | Downregulated |
| 0,147661173 | 0,871183051 | CDH4 | FALSE | Upregulated |
| 0,249102921 | 0,87131644 | G2E3 | FALSE | Upregulated |
| 0,123550988 | 0,871461353 | SULT1A2 | FALSE | Upregulated |
| -0,125104975 | 0,871812387 | ARRDC3-AS1 | FALSE | Downregulated |
| 0,187357553 | 0,871895217 | SARNP | FALSE | Upregulated |
| 0,362904309 | 0,871969878 | LOC102724319 | FALSE | Upregulated |
| -0,287302812 | 0,871969878 | ZNF112 | FALSE | Downregulated |
| 0,075852222 | 0,872276196 | YTHDF3 | FALSE | Upregulated |
| -0,43096991 | 0,872355878 | KLHL35 | FALSE | Downregulated |
| 1,062708812 | 0,872361884 | LOC102724624 | FALSE | Upregulated |
| -0,427028489 | 0,872361884 | MEGF10 | FALSE | Downregulated |
| -0,063578452 | 0,872361884 | SMG1 | FALSE | Downregulated |
| 0,110738545 | 0,872361884 | SYT15 | FALSE | Upregulated |
| 0,07533704 | 0,872508966 | DDX3X | FALSE | Upregulated |
| 0,165946776 | 0,872508966 | ENOX2 | FALSE | Upregulated |

| | | | | |
|---|---|---|---|---|
| 0,104199864 | 0,872508966 | PNPT1 | FALSE | Upregulated |
| 0,074811609 | 0,872508966 | RIF1 | FALSE | Upregulated |
| -0,499717575 | 0,872700263 | LIPE-AS1 | FALSE | Downregulated |
| 0,292673297 | 0,872763502 | CXADRP2 | FALSE | Upregulated |
| -0,10230101 | 0,872968044 | NCKAP5 | FALSE | Downregulated |
| -0,456561746 | 0,873000689 | C11orf74 | FALSE | Downregulated |
| 0,170868578 | 0,873137678 | AK1 | FALSE | Upregulated |
| 0,1749207 | 0,873137678 | PREX2 | FALSE | Upregulated |
| -0,082616893 | 0,873162394 | ARHGAP12 | FALSE | Downregulated |
| 0,176209364 | 0,873162394 | GEMIN7 | FALSE | Upregulated |
| 0,091419273 | 0,873162394 | MATN2 | FALSE | Upregulated |
| -0,148856629 | 0,873326201 | RNU105A | FALSE | Downregulated |
| -0,146872347 | 0,873506596 | ADAM21 | FALSE | Downregulated |
| -0,246539386 | 0,873506596 | FAM221A | FALSE | Downregulated |
| -0,46392453 | 0,873506596 | FHDC1 | FALSE | Downregulated |
| -0,089689091 | 0,873506596 | NASP | FALSE | Downregulated |
| 0,109756041 | 0,873506596 | TCF3 | FALSE | Upregulated |
| -0,138834225 | 0,873506596 | TFCP2 | FALSE | Downregulated |
| -0,108572974 | 0,873530366 | HHIP | FALSE | Downregulated |
| 0,161659053 | 0,873530366 | RFWD3 | FALSE | Upregulated |
| 0,228893013 | 0,873623482 | LOC102723356 | FALSE | Upregulated |
| -0,250093235 | 0,873690909 | C1GALT1C1 | FALSE | Downregulated |
| 0,277752301 | 0,873690909 | LOC101928136 | FALSE | Upregulated |
| 0,205813037 | 0,873690909 | MANF | FALSE | Upregulated |
| 0,206467523 | 0,873690909 | RBL1 | FALSE | Upregulated |
| 0,202180607 | 0,873690909 | ZKSCAN7 | FALSE | Upregulated |
| 0,356603932 | 0,873690909 | ZMYND10 | FALSE | Upregulated |
| -0,155880788 | 0,873690909 | ZNF524 | FALSE | Downregulated |
| 0,207736784 | 0,873723326 | DET1 | FALSE | Upregulated |
| -0,06636378 | 0,873723326 | MBD3 | FALSE | Downregulated |
| -0,325036129 | 0,873827015 | RP3-510O8.4 | FALSE | Downregulated |
| -0,080085384 | 0,873895466 | ELK4 | FALSE | Downregulated |
| 0,175693325 | 0,873895466 | LOC729603 | FALSE | Upregulated |
| 0,399015139 | 0,873895466 | TAF1A | FALSE | Upregulated |
| 0,158069147 | 0,873899383 | LOC101929265 | FALSE | Upregulated |
| 0,211860359 | 0,874002352 | GIN1 | FALSE | Upregulated |
| 0,258165769 | 0,874002352 | ZNF28 | FALSE | Upregulated |
| 0,272815363 | 0,874277028 | FAM58A | FALSE | Upregulated |
| -0,223818581 | 0,874282824 | VPS33A | FALSE | Downregulated |
| 0,184195513 | 0,87438899 | LOC101928816 | FALSE | Upregulated |
| 0,156769323 | 0,87438899 | LOC145783 | FALSE | Upregulated |
| 0,090761043 | 0,87438899 | PDE7A | FALSE | Upregulated |
| 0,081364023 | 0,87438899 | PSMA7 | FALSE | Upregulated |
| 0,216094517 | 0,87438899 | RASSF8 | FALSE | Upregulated |
| -0,164727363 | 0,874542058 | IFT172 | FALSE | Downregulated |
| -0,107204508 | 0,874570027 | MAP1LC3A | FALSE | Downregulated |
| -0,236275345 | 0,874994396 | MRPS33 | FALSE | Downregulated |
| 0,18485337 | 0,875309344 | LOC102723597 | FALSE | Upregulated |
| -0,221553418 | 0,875428668 | LOC100506060 | FALSE | Downregulated |
| -0,115243377 | 0,875449822 | CDC37L1 | FALSE | Downregulated |
| -0,163169664 | 0,875449822 | SCRT1 | FALSE | Downregulated |

| | | | | |
|---|---|---|---|---|
| -0,085363155 | 0,875449822 | UBE4B | FALSE | Downregulated |
| -0,292190716 | 0,875449822 | UBXN10 | FALSE | Downregulated |
| -0,074548313 | 0,875683414 | RUSC2 | FALSE | Downregulated |
| 0,115057688 | 0,875722755 | DDX39B | FALSE | Upregulated |
| -0,128229961 | 0,875786253 | KLHL22 | FALSE | Downregulated |
| 0,159966653 | 0,875786253 | WDR77 | FALSE | Upregulated |
| 0,380710077 | 0,875802256 | FAM86C2P | FALSE | Upregulated |
| -0,0364127 | 0,875802256 | MAVS | FALSE | Downregulated |
| -0,283560913 | 0,875802256 | ZNF43 | FALSE | Downregulated |
| 0,142150997 | 0,876146539 | FAM96A | FALSE | Upregulated |
| -0,080501486 | 0,876174983 | GCC2 | FALSE | Downregulated |
| 0,210127153 | 0,876174983 | MRPL48 | FALSE | Upregulated |
| -0,128831034 | 0,876174983 | NDUFS8 | FALSE | Downregulated |
| 0,099349832 | 0,876174983 | ZNF587B | FALSE | Upregulated |
| -0,183296201 | 0,876200609 | ACYP2 | FALSE | Downregulated |
| -0,091662627 | 0,876200609 | CDK16 | FALSE | Downregulated |
| 0,315564344 | 0,876200609 | DNAH7 | FALSE | Upregulated |
| 0,17201169 | 0,876200609 | NUDT12 | FALSE | Upregulated |
| -0,402442982 | 0,876200609 | SNAPC1 | FALSE | Downregulated |
| 0,310322597 | 0,876200609 | ZNF230 | FALSE | Upregulated |
| -0,069198319 | 0,876274828 | POGZ | FALSE | Downregulated |
| 0,064694242 | 0,876307347 | CRKL | FALSE | Upregulated |
| 0,253305039 | 0,87633907 | CHCHD1 | FALSE | Upregulated |
| 0,162761122 | 0,876519054 | TBC1D8B | FALSE | Upregulated |
| -0,245068594 | 0,876639197 | LOC102724514 | FALSE | Downregulated |
| 0,099562856 | 0,876897565 | BAD | FALSE | Upregulated |
| -0,145299725 | 0,876897565 | LOC100506175 | FALSE | Downregulated |
| -0,262437018 | 0,877107344 | ORC5 | FALSE | Downregulated |
| -0,213768582 | 0,877107344 | SMG8 | FALSE | Downregulated |
| 0,200873756 | 0,877111486 | IFIT5 | FALSE | Upregulated |
| 0,23484699 | 0,877111486 | KIAA1107 | FALSE | Upregulated |
| 0,095919602 | 0,877111486 | LOC101928817 | FALSE | Upregulated |
| 0,191059487 | 0,877111486 | MOSPD3 | FALSE | Upregulated |
| -0,11325897 | 0,877111486 | PLCXD1 | FALSE | Downregulated |
| 0,246246208 | 0,877111486 | TOP1P1 | FALSE | Upregulated |
| 0,362886838 | 0,877215286 | FAM157A | FALSE | Upregulated |
| 0,102170272 | 0,877352553 | ATF7IP2 | FALSE | Upregulated |
| 0,373701289 | 0,877414811 | LOC100507656 | FALSE | Upregulated |
| -0,275228277 | 0,877414811 | RNF175 | FALSE | Downregulated |
| -0,099968003 | 0,877414811 | RNF31 | FALSE | Downregulated |
| 0,167748267 | 0,877414811 | SCD | FALSE | Upregulated |
| -0,135364099 | 0,877414811 | SSBP1 | FALSE | Downregulated |
| -0,07480509 | 0,877414811 | STAG1 | FALSE | Downregulated |
| 0,14619549 | 0,877463789 | S100PBP | FALSE | Upregulated |
| 0,118346254 | 0,877463789 | TMEM64 | FALSE | Upregulated |
| -0,190787465 | 0,877632764 | SPATA41 | FALSE | Downregulated |
| -0,051429266 | 0,877638939 | KIF13B | FALSE | Downregulated |
| -0,102378326 | 0,87782572 | FLT1 | FALSE | Downregulated |
| 0,095211103 | 0,87782572 | RPS6KB2 | FALSE | Upregulated |
| 0,129007687 | 0,877985212 | LOC102723992 | FALSE | Upregulated |
| 0,244310971 | 0,878028525 | TMED3 | FALSE | Upregulated |

| | | | | |
|---|---|---|---|---|
| -0,056037589 | 0,878028525 | ZHX3 | FALSE | Downregulated |
| -0,167066407 | 0,878064527 | ADSL | FALSE | Downregulated |
| 0,084917293 | 0,878064527 | ANKRD40 | FALSE | Upregulated |
| -0,126399562 | 0,878064527 | B3GALT6 | FALSE | Downregulated |
| 0,109610533 | 0,878064527 | LOC102724103 | FALSE | Upregulated |
| -0,106549589 | 0,878064527 | MBD2 | FALSE | Downregulated |
| 0,169918514 | 0,878064527 | MRPL18 | FALSE | Upregulated |
| 0,089110019 | 0,878064527 | MYBBP1A | FALSE | Upregulated |
| 0,159544145 | 0,878064527 | PEX3 | FALSE | Upregulated |
| -0,193700705 | 0,878064527 | RTN4IP1 | FALSE | Downregulated |
| -0,080344131 | 0,878064527 | SMARCA5 | FALSE | Downregulated |
| -0,070798394 | 0,878077752 | EIF4A2 | FALSE | Downregulated |
| 0,241863681 | 0,878077752 | LOC102723544 | FALSE | Upregulated |
| 0,17609331 | 0,878108953 | LOC101928707 | FALSE | Upregulated |
| 0,3672555 | 0,878379435 | ARPP21 | FALSE | Upregulated |
| 0,103497221 | 0,878379435 | EGLN2 | FALSE | Upregulated |
| -0,327205387 | 0,878379435 | EYA1 | FALSE | Downregulated |
| -0,417782539 | 0,878379435 | LOC101927143 | FALSE | Downregulated |
| -0,423613633 | 0,878379435 | LOC102723564 | FALSE | Downregulated |
| -0,109494888 | 0,878487277 | CNTN2 | FALSE | Downregulated |
| -0,44570884 | 0,878661529 | C16orf96 | FALSE | Downregulated |
| 0,117844799 | 0,878661529 | DMAP1 | FALSE | Upregulated |
| -0,060552126 | 0,878661529 | FAM120A | FALSE | Downregulated |
| 0,324963009 | 0,878661529 | KLF5 | FALSE | Upregulated |
| 0,32841207 | 0,878661529 | LOC101927999 | FALSE | Upregulated |
| -0,283494318 | 0,878661529 | LOC101929070 | FALSE | Downregulated |
| 0,129893935 | 0,878661529 | LOC730668 | FALSE | Upregulated |
| 0,198386165 | 0,878661529 | SDK1 | FALSE | Upregulated |
| -0,210887881 | 0,878720966 | KRBA1 | FALSE | Downregulated |
| -0,133313786 | 0,878904921 | ARL3 | FALSE | Downregulated |
| 0,405597076 | 0,878904921 | CYP27B1 | FALSE | Upregulated |
| -0,19054526 | 0,878904921 | DCSTAMP | FALSE | Downregulated |
| -0,219849854 | 0,878968575 | LOC100130876 | FALSE | Downregulated |
| 0,145798723 | 0,87898943 | TSPEAR | FALSE | Upregulated |
| 0,167974889 | 0,879379546 | COG2 | FALSE | Upregulated |
| 0,217990084 | 0,879499298 | ALG5 | FALSE | Upregulated |
| 0,337549946 | 0,879499298 | C18orf21 | FALSE | Upregulated |
| -0,146129091 | 0,879499298 | C20orf194 | FALSE | Downregulated |
| 0,321823192 | 0,879499298 | COLCA1 | FALSE | Upregulated |
| -0,112978298 | 0,879499298 | CTPS1 | FALSE | Downregulated |
| -0,291704371 | 0,879499298 | DOCK9-AS2 | FALSE | Downregulated |
| -0,177084662 | 0,879499298 | GPATCH1 | FALSE | Downregulated |
| -0,143627307 | 0,879499298 | GPR137 | FALSE | Downregulated |
| 0,182493893 | 0,879499298 | HEATR6 | FALSE | Upregulated |
| 0,235249282 | 0,879499298 | KCNB1 | FALSE | Upregulated |
| 0,265576224 | 0,879499298 | KIAA1467 | FALSE | Upregulated |
| -0,288226771 | 0,879499298 | LOC101927520 | FALSE | Downregulated |
| 0,123734222 | 0,879499298 | NKIRAS1 | FALSE | Upregulated |
| 0,124758227 | 0,879499298 | P2RX7 | FALSE | Upregulated |
| 0,097815722 | 0,879499298 | PGAP2 | FALSE | Upregulated |
| 0,173845307 | 0,879499298 | POP5 | FALSE | Upregulated |

| | | | | |
|---|---|---|---|---|
| -0,108760166 | 0,879499298 | RMND1 | FALSE | Downregulated |
| 0,183902236 | 0,879499298 | RNASEL | FALSE | Upregulated |
| -0,113520514 | 0,879499298 | RNF20 | FALSE | Downregulated |
| -2,312778862 | 0,879499298 | RNU1-1 | FALSE | Downregulated |
| 0,187154511 | 0,879499298 | RP11-266K4.9 | FALSE | Upregulated |
| 0,08793598 | 0,879499298 | RPAIN | FALSE | Upregulated |
| 0,178444367 | 0,879499298 | SENP1 | FALSE | Upregulated |
| 0,129663001 | 0,879499298 | THTPA | FALSE | Upregulated |
| 0,116298051 | 0,879499298 | TLN2 | FALSE | Upregulated |
| -0,129208616 | 0,879499298 | URM1 | FALSE | Downregulated |
| 0,135726822 | 0,87951864 | LINC00324 | FALSE | Upregulated |
| -0,095890387 | 0,879623161 | CYP2U1 | FALSE | Downregulated |
| 0,181685359 | 0,879623161 | GLI4 | FALSE | Upregulated |
| -0,099602765 | 0,879623161 | KLF11 | FALSE | Downregulated |
| -0,13259242 | 0,879623161 | LTB4R | FALSE | Downregulated |
| -0,068940231 | 0,879623161 | OAZ1 | FALSE | Downregulated |
| 0,151338344 | 0,879623161 | PIP5K1P1 | FALSE | Upregulated |
| -0,097276631 | 0,879623161 | ZNF274 | FALSE | Downregulated |
| 0,123420742 | 0,879699716 | KIAA0196 | FALSE | Upregulated |
| -0,156209178 | 0,879798975 | GIMAP4 | FALSE | Downregulated |
| 0,290057936 | 0,879844311 | LOC101928195 | FALSE | Upregulated |
| 0,247084602 | 0,879844311 | LOC101929719 | FALSE | Upregulated |
| -0,084000158 | 0,8798704 | FXR2 | FALSE | Downregulated |
| 0,222446765 | 0,879993013 | ALKBH6 | FALSE | Upregulated |
| 0,163468968 | 0,879993013 | PHKA1 | FALSE | Upregulated |
| -0,453330323 | 0,879993013 | RP11-434C1.1 | FALSE | Downregulated |
| 0,062951015 | 0,880032449 | GNPNAT1 | FALSE | Upregulated |
| 0,18599352 | 0,880109842 | ANKRA2 | FALSE | Upregulated |
| 0,1022947 | 0,880109842 | TM4SF4 | FALSE | Upregulated |
| 0,157135151 | 0,880151677 | AGER | FALSE | Upregulated |
| -0,066567459 | 0,880151677 | PGAM1 | FALSE | Downregulated |
| 0,233042062 | 0,880169805 | MMS22L | FALSE | Upregulated |
| 0,219589194 | 0,880169805 | SCT | FALSE | Upregulated |
| 0,113450022 | 0,88023093 | SLC25A27 | FALSE | Upregulated |
| -0,194789837 | 0,880376801 | UNC119 | FALSE | Downregulated |
| 0,080127973 | 0,880563125 | MESDC1 | FALSE | Upregulated |
| -0,113835535 | 0,880641397 | ZFAND5 | FALSE | Downregulated |
| -0,102250571 | 0,880752353 | PSMA5 | FALSE | Downregulated |
| 0,327398538 | 0,880752353 | SEC14L1P1 | FALSE | Upregulated |
| -0,07588954 | 0,880787164 | ARRDC1 | FALSE | Downregulated |
| 0,133329363 | 0,880787164 | DPF2 | FALSE | Upregulated |
| 0,313068211 | 0,880787164 | FSD1 | FALSE | Upregulated |
| -0,400924683 | 0,880787164 | LOC101929117 | FALSE | Downregulated |
| 0,128700784 | 0,880787164 | NLRP9 | FALSE | Upregulated |
| -0,083275157 | 0,880787164 | PAIP2B | FALSE | Downregulated |
| -0,120693832 | 0,880787164 | PICK1 | FALSE | Downregulated |
| 0,15832416 | 0,881064676 | CAMLG | FALSE | Upregulated |
| 0,147960721 | 0,881089624 | ARRDC4 | FALSE | Upregulated |
| -0,095917511 | 0,881089624 | DIXDC1 | FALSE | Downregulated |
| -0,120460325 | 0,881089624 | LMBR1 | FALSE | Downregulated |
| 0,412538048 | 0,881089624 | MSH5 | FALSE | Upregulated |

| | | | | |
|---|---|---|---|---|
| -0,477033448 | 0,881089624 | NTRK3-AS1 | FALSE | Downregulated |
| -0,069445468 | 0,881089624 | RAB1B | FALSE | Downregulated |
| 0,09617222 | 0,881089624 | SIRT3 | FALSE | Upregulated |
| 0,285314219 | 0,881089624 | ZNF77 | FALSE | Upregulated |
| 0,267738406 | 0,881089624 | ZNF793 | FALSE | Upregulated |
| 0,239424084 | 0,881094423 | YEATS4 | FALSE | Upregulated |
| 0,28771476 | 0,881185073 | ZNF736 | FALSE | Upregulated |
| -0,169774111 | 0,881239002 | ZNF554 | FALSE | Downregulated |
| 0,315219506 | 0,881262448 | DNAH3 | FALSE | Upregulated |
| -0,432369544 | 0,881262448 | FUNDC1 | FALSE | Downregulated |
| 0,141204603 | 0,881262448 | NKAIN3 | FALSE | Upregulated |
| 0,345790765 | 0,881262448 | SLC24A3 | FALSE | Upregulated |
| 0,237350763 | 0,881262448 | STMN1 | FALSE | Upregulated |
| 0,130193675 | 0,881309669 | TBK1 | FALSE | Upregulated |
| -0,106763802 | 0,881379191 | IK | FALSE | Downregulated |
| 0,219730227 | 0,881380639 | SIGLEC7 | FALSE | Upregulated |
| 0,076754937 | 0,881416139 | DENND1A | FALSE | Upregulated |
| 0,232556915 | 0,881416139 | LOC100996433 | FALSE | Upregulated |
| -0,337615206 | 0,881416139 | PPT2-EGFL8 | FALSE | Downregulated |
| 0,099054204 | 0,881525616 | LOC102724144 | FALSE | Upregulated |
| -0,259797063 | 0,881525616 | RP11-899L11.1 | FALSE | Downregulated |
| -0,141640029 | 0,881546485 | UFSP2 | FALSE | Downregulated |
| -0,131467472 | 0,881582328 | RHOBTB1 | FALSE | Downregulated |
| 0,407502467 | 0,881628682 | CRIP1 | FALSE | Upregulated |
| -0,179713755 | 0,881648807 | NOMO2 | FALSE | Downregulated |
| -0,213451076 | 0,881862663 | SYF2 | FALSE | Downregulated |
| -0,086450718 | 0,881882244 | HIPK1 | FALSE | Downregulated |
| 0,115828763 | 0,881887752 | CPNE8 | FALSE | Upregulated |
| 0,052215334 | 0,881887752 | MRFAP1 | FALSE | Upregulated |
| 0,174143388 | 0,882033081 | PDC | FALSE | Upregulated |
| 0,074091084 | 0,882088668 | TM7SF3 | FALSE | Upregulated |
| -0,428365352 | 0,882095392 | HIST1H2BF | FALSE | Downregulated |
| 1,56668935 | 0,882095392 | LOC102723399 | FALSE | Upregulated |
| -0,155613118 | 0,882102298 | ADCK2 | FALSE | Downregulated |
| -0,181598272 | 0,882102298 | CCDC107 | FALSE | Downregulated |
| -0,060221166 | 0,882102298 | CDC42BPA | FALSE | Downregulated |
| 0,146510203 | 0,882102298 | CLDN23 | FALSE | Upregulated |
| -0,150887878 | 0,882102298 | FHL1 | FALSE | Downregulated |
| -0,105940686 | 0,882102298 | GART | FALSE | Downregulated |
| 0,298000121 | 0,882102298 | GMNN | FALSE | Upregulated |
| -0,248167785 | 0,882102298 | MATK | FALSE | Downregulated |
| -0,054306892 | 0,882102298 | MORF4L1 | FALSE | Downregulated |
| 0,077578318 | 0,882102298 | MRC1 | FALSE | Upregulated |
| 0,072644193 | 0,882102298 | NACC1 | FALSE | Upregulated |
| -0,106912673 | 0,882102298 | RAB1A | FALSE | Downregulated |
| 0,22158978 | 0,882102298 | RP11-480I12.7 | FALSE | Upregulated |
| 0,127260381 | 0,882102298 | SCML1 | FALSE | Upregulated |
| -0,255543959 | 0,882105899 | BTBD19 | FALSE | Downregulated |
| -0,234614911 | 0,882105899 | COPZ2 | FALSE | Downregulated |
| -0,160149131 | 0,882105899 | TMEM14C | FALSE | Downregulated |
| -0,094088571 | 0,882113043 | SETD3 | FALSE | Downregulated |

| | | | | |
|---|---|---|---|---|
| 0,196335236 | 0,882319476 | POLD3 | FALSE | Upregulated |
| 0,194364147 | 0,882352105 | DZIP3 | FALSE | Upregulated |
| -0,134250649 | 0,882352105 | GABARAPL2 | FALSE | Downregulated |
| 0,097130941 | 0,882352105 | INO80E | FALSE | Upregulated |
| 0,238107868 | 0,882352105 | LOC101929115 | FALSE | Upregulated |
| 0,289530895 | 0,882352105 | LOC644961 | FALSE | Upregulated |
| -0,078333795 | 0,882352105 | MMP15 | FALSE | Downregulated |
| -0,068891542 | 0,882352105 | RABGEF1 | FALSE | Downregulated |
| -0,07052288 | 0,882352105 | SUMF2 | FALSE | Downregulated |
| -0,247622976 | 0,882352105 | TMEM128 | FALSE | Downregulated |
| 0,062549735 | 0,882352105 | TMEM129 | FALSE | Upregulated |
| -0,124399752 | 0,882382858 | EXD3 | FALSE | Downregulated |
| -0,166321777 | 0,882382858 | L3HYPDH | FALSE | Downregulated |
| 0,188060142 | 0,88257687 | GGTA1P | FALSE | Upregulated |
| 0,222581681 | 0,882614108 | DSN1 | FALSE | Upregulated |
| 0,21188907 | 0,882622154 | SNAI3-AS1 | FALSE | Upregulated |
| -0,287594499 | 0,882627968 | LOC101928351 | FALSE | Downregulated |
| -0,121675858 | 0,882627968 | STK3 | FALSE | Downregulated |
| 0,060246596 | 0,882635954 | FNBP4 | FALSE | Upregulated |
| 0,216476938 | 0,882644033 | TNN | FALSE | Upregulated |
| 0,07670628 | 0,882828824 | ZFC3H1 | FALSE | Upregulated |
| 0,201291263 | 0,882915034 | FASTKD1 | FALSE | Upregulated |
| -0,096320669 | 0,883013761 | CRTC1 | FALSE | Downregulated |
| 0,260746862 | 0,883018179 | RAD51D | FALSE | Upregulated |
| -0,134771617 | 0,883021146 | ZNF30 | FALSE | Downregulated |
| -0,121759369 | 0,883112911 | CCDC124 | FALSE | Downregulated |
| 0,177280994 | 0,883112911 | GRAMD1B | FALSE | Upregulated |
| 0,2068055 | 0,883112911 | RHBG | FALSE | Upregulated |
| -0,279755796 | 0,883112911 | RP5-968D22.1 | FALSE | Downregulated |
| 0,078323822 | 0,883112911 | ZNF644 | FALSE | Upregulated |
| 0,070804292 | 0,883112911 | ZNF839 | FALSE | Upregulated |
| -0,148550691 | 0,883151662 | NT5DC3 | FALSE | Downregulated |
| 0,161779133 | 0,883237415 | LOC101928741 | FALSE | Upregulated |
| 0,274974147 | 0,883237415 | TMEFF2 | FALSE | Upregulated |
| -0,122769918 | 0,88352605 | AVPI1 | FALSE | Downregulated |
| 0,295612233 | 0,88352605 | EYA2 | FALSE | Upregulated |
| -0,151424846 | 0,88352605 | LOC101927741 | FALSE | Downregulated |
| -0,117094437 | 0,88352605 | NUP85 | FALSE | Downregulated |
| -0,124964258 | 0,883615679 | CCDC82 | FALSE | Downregulated |
| -0,059592322 | 0,883615679 | MYO15B | FALSE | Downregulated |
| -0,094642845 | 0,883715943 | LAPTM4A | FALSE | Downregulated |
| -0,189510783 | 0,883848947 | NUDT2 | FALSE | Downregulated |
| 0,170026554 | 0,883958974 | AASDH | FALSE | Upregulated |
| 0,367590198 | 0,883958974 | LOC101927772 | FALSE | Upregulated |
| -0,127506167 | 0,883958974 | LRRN1 | FALSE | Downregulated |
| -0,193017857 | 0,883991248 | BLOC1S3 | FALSE | Downregulated |
| -0,070352644 | 0,883991248 | ESYT2 | FALSE | Downregulated |
| 0,174633575 | 0,883991248 | RP11-434D9.1 | FALSE | Upregulated |
| -0,114850394 | 0,883991248 | SUFU | FALSE | Downregulated |
| 0,234760168 | 0,884056753 | RAD51-AS1 | FALSE | Upregulated |
| -0,21879995 | 0,884090963 | HSPBAP1 | FALSE | Downregulated |

| | | | | |
|---|---|---|---|---|
| -0,206397578 | 0,884095523 | ETHE1 | FALSE | Downregulated |
| -0,42988434 | 0,884095523 | STXBP6 | FALSE | Downregulated |
| 0,087041564 | 0,884527805 | NUB1 | FALSE | Upregulated |
| -0,338159919 | 0,884886894 | LOC101927103 | FALSE | Downregulated |
| -0,138172623 | 0,884937003 | SRRD | FALSE | Downregulated |
| 0,106037735 | 0,885195626 | MFN1 | FALSE | Upregulated |
| -0,220029982 | 0,885195626 | PTS | FALSE | Downregulated |
| 0,140540649 | 0,885270341 | LOC100507472 | FALSE | Upregulated |
| 0,191916116 | 0,885270341 | ZSCAN31 | FALSE | Upregulated |
| -0,107290466 | 0,885292057 | APBA1 | FALSE | Downregulated |
| -0,120896585 | 0,885292057 | BIVM | FALSE | Downregulated |
| 0,080580673 | 0,885292057 | PAG1 | FALSE | Upregulated |
| 0,076261794 | 0,885447195 | MIB1 | FALSE | Upregulated |
| 0,12637581 | 0,885672282 | ALOX12P2 | FALSE | Upregulated |
| 0,531596805 | 0,885752332 | LOC644172 | FALSE | Upregulated |
| -0,088894181 | 0,88582985 | ARFRP1 | FALSE | Downregulated |
| 0,189170704 | 0,88582985 | ARHGAP6 | FALSE | Upregulated |
| -0,123724145 | 0,88582985 | BCL2L14 | FALSE | Downregulated |
| -0,206168776 | 0,88582985 | DCTN6 | FALSE | Downregulated |
| 0,136502887 | 0,88582985 | DNAJB11 | FALSE | Upregulated |
| -0,18832626 | 0,88582985 | DPM1 | FALSE | Downregulated |
| 0,07093747 | 0,88582985 | EDC3 | FALSE | Upregulated |
| 0,088977086 | 0,88582985 | FAF1 | FALSE | Upregulated |
| -0,128668923 | 0,88582985 | FKBP7 | FALSE | Downregulated |
| 0,318707162 | 0,88582985 | GPC4 | FALSE | Upregulated |
| -0,346528488 | 0,88582985 | GRK4 | FALSE | Downregulated |
| 0,178053074 | 0,88582985 | LOC101929787 | FALSE | Upregulated |
| 0,199832802 | 0,88582985 | LOC102723313 | FALSE | Upregulated |
| -0,058853392 | 0,88582985 | MMACHC | FALSE | Downregulated |
| 0,280414089 | 0,88582985 | MUM1L1 | FALSE | Upregulated |
| -0,068171956 | 0,88582985 | NFATC3 | FALSE | Downregulated |
| 0,07387485 | 0,88582985 | PAK4 | FALSE | Upregulated |
| 0,075216995 | 0,88582985 | PHYKPL | FALSE | Upregulated |
| -0,117499367 | 0,88582985 | PIH1D1 | FALSE | Downregulated |
| 0,116947629 | 0,88582985 | PQLC3 | FALSE | Upregulated |
| -0,052189137 | 0,88582985 | PRR12 | FALSE | Downregulated |
| 0,058515674 | 0,88582985 | RBMX | FALSE | Upregulated |
| 0,368693235 | 0,88582985 | RNF112 | FALSE | Upregulated |
| 0,131988785 | 0,88582985 | SCARF2 | FALSE | Upregulated |
| 0,08828392 | 0,88582985 | SEMA4B | FALSE | Upregulated |
| -0,150546544 | 0,88582985 | SH3BGRL | FALSE | Downregulated |
| 0,416900918 | 0,88582985 | SNCA | FALSE | Upregulated |
| 0,081497712 | 0,88582985 | STYX | FALSE | Upregulated |
| -0,068705977 | 0,88582985 | SYNRG | FALSE | Downregulated |
| 0,346733443 | 0,88582985 | TG | FALSE | Upregulated |
| -0,148724669 | 0,88582985 | ZMPSTE24 | FALSE | Downregulated |
| -0,092424166 | 0,88582985 | ZNF12 | FALSE | Downregulated |
| 0,192502946 | 0,885902438 | FAM76A | FALSE | Upregulated |
| -0,099680592 | 0,885902438 | SAP30BP | FALSE | Downregulated |
| -0,132463161 | 0,885902438 | SUB1 | FALSE | Downregulated |
| 0,098813713 | 0,885902438 | UBE3B | FALSE | Upregulated |

| | | | | |
|---|---|---|---|---|
| -0,191749487 | 0,885902438 | ZNF891 | FALSE | Downregulated |
| 0,165652779 | 0,885914332 | AEN | FALSE | Upregulated |
| -0,258615272 | 0,885914332 | LOC728407 | FALSE | Downregulated |
| 0,093238161 | 0,885929387 | GPR64 | FALSE | Upregulated |
| -0,229780477 | 0,885929387 | PTBP2 | FALSE | Downregulated |
| 0,273153341 | 0,886054579 | AC000099.1 | FALSE | Upregulated |
| 0,133790482 | 0,886054579 | ODC1 | FALSE | Upregulated |
| 0,080444309 | 0,886054579 | SGSM3 | FALSE | Upregulated |
| 0,153996665 | 0,886054579 | SLC8A1-AS1 | FALSE | Upregulated |
| 0,124608981 | 0,886092763 | NPC2 | FALSE | Upregulated |
| 0,15307686 | 0,886092763 | RFTN2 | FALSE | Upregulated |
| 0,117945215 | 0,886092763 | TBC1D15 | FALSE | Upregulated |
| -0,110095891 | 0,886092763 | UGT3A1 | FALSE | Downregulated |
| -0,101247896 | 0,886092763 | ZNF780A | FALSE | Downregulated |
| -0,187352948 | 0,886093924 | ACSBG1 | FALSE | Downregulated |
| 0,230194615 | 0,886093924 | LOC100287175 | FALSE | Upregulated |
| -0,095411392 | 0,886093924 | MRPS16 | FALSE | Downregulated |
| 2,169383482 | 0,886093924 | RNU1-4 | FALSE | Upregulated |
| -0,069031954 | 0,886093924 | SDC4 | FALSE | Downregulated |
| -0,091002238 | 0,886093924 | SIAH2 | FALSE | Downregulated |
| -0,107103632 | 0,886157823 | RBPJ | FALSE | Downregulated |
| -0,359948639 | 0,886220862 | PLAC8L1 | FALSE | Downregulated |
| -0,342228346 | 0,886384612 | LRRC37A5P | FALSE | Downregulated |
| 0,258491883 | 0,886448288 | LOC440157 | FALSE | Upregulated |
| -0,133485043 | 0,886622966 | MSANTD2 | FALSE | Downregulated |
| -0,32681186 | 0,886685418 | AL121656.5 | FALSE | Downregulated |
| 0,515137649 | 0,886685418 | C18orf32 | FALSE | Upregulated |
| 0,170519241 | 0,886685418 | INCA1 | FALSE | Upregulated |
| -0,160522514 | 0,886697565 | CIR1 | FALSE | Downregulated |
| 0,03918803 | 0,886940516 | WNK1 | FALSE | Upregulated |
| 0,068560344 | 0,886955312 | DDX24 | FALSE | Upregulated |
| 0,158503334 | 0,887025088 | ENY2 | FALSE | Upregulated |
| 0,308672468 | 0,887025088 | LOC100506281 | FALSE | Upregulated |
| -0,241599268 | 0,887025088 | LOC101928820 | FALSE | Downregulated |
| -0,103121145 | 0,887025088 | PELO | FALSE | Downregulated |
| 1,786513879 | 0,887025088 | PKP1 | FALSE | Upregulated |
| 0,099627132 | 0,887025088 | RNF169 | FALSE | Upregulated |
| 0,062113441 | 0,887025088 | SLC3A1 | FALSE | Upregulated |
| -0,113513063 | 0,887055029 | ALG2 | FALSE | Downregulated |
| 0,072331203 | 0,887055029 | API5 | FALSE | Upregulated |
| 0,279259571 | 0,887055029 | SLC9A9-AS1 | FALSE | Upregulated |
| 0,13427917 | 0,887068511 | GIMAP6 | FALSE | Upregulated |
| -0,070473333 | 0,887068511 | HNRNPH1 | FALSE | Downregulated |
| -0,233565495 | 0,887068511 | PUS7 | FALSE | Downregulated |
| 0,075678557 | 0,887068511 | WDR11 | FALSE | Upregulated |
| 0,140231892 | 0,887449819 | PIAS3 | FALSE | Upregulated |
| -0,082452462 | 0,887449819 | RALGAPA1 | FALSE | Downregulated |
| -0,118314492 | 0,887841592 | OCIAD2 | FALSE | Downregulated |
| -0,285271178 | 0,888064251 | DDX3Y | FALSE | Downregulated |
| -0,085307785 | 0,888165338 | CPSF6 | FALSE | Downregulated |
| -0,15373624 | 0,888165338 | DIRAS3 | FALSE | Downregulated |

| | | | | |
|---|---|---|---|---|
| -0,075584684 | 0,888165338 | FAM13A-AS1 | FALSE | Downregulated |
| 0,121701824 | 0,888165338 | SRGAP2 | FALSE | Upregulated |
| -0,081569106 | 0,888165338 | ZNF445 | FALSE | Downregulated |
| -0,169159689 | 0,888363824 | KLHL28 | FALSE | Downregulated |
| 0,122509443 | 0,888416176 | FAM83G | FALSE | Upregulated |
| 0,393137318 | 0,888457735 | LOC101927723 | FALSE | Upregulated |
| 0,264944845 | 0,888508989 | EDA | FALSE | Upregulated |
| -0,208747696 | 0,88879096 | BAMBI | FALSE | Downregulated |
| 0,250805198 | 0,888864086 | BTK | FALSE | Upregulated |
| 0,100221663 | 0,888864086 | CARS | FALSE | Upregulated |
| -0,104480115 | 0,889045021 | MREG | FALSE | Downregulated |
| 0,173166058 | 0,889152125 | NECAB2 | FALSE | Upregulated |
| 0,168252968 | 0,88921756 | FLJ39095 | FALSE | Upregulated |
| 0,05819127 | 0,88921756 | MON2 | FALSE | Upregulated |
| 0,340600211 | 0,889229943 | TBC1D31 | FALSE | Upregulated |
| 0,063237388 | 0,889239264 | PXN | FALSE | Upregulated |
| -0,27118408 | 0,889239264 | TCEB1 | FALSE | Downregulated |
| -0,071790165 | 0,889711405 | EXT2 | FALSE | Downregulated |
| 0,125712597 | 0,889711405 | LRRC48 | FALSE | Upregulated |
| -0,132639052 | 0,889711405 | SMYD2 | FALSE | Downregulated |
| -0,117586349 | 0,889711405 | SYPL1 | FALSE | Downregulated |
| 0,221559056 | 0,889711405 | TMEM199 | FALSE | Upregulated |
| 0,076699058 | 0,889714978 | UPF2 | FALSE | Upregulated |
| -0,182607579 | 0,889755517 | S100A13 | FALSE | Downregulated |
| -0,127154495 | 0,889794529 | LSS | FALSE | Downregulated |
| -0,203456096 | 0,889885315 | COX16 | FALSE | Downregulated |
| -0,21669522 | 0,889921513 | BATF2 | FALSE | Downregulated |
| 0,204635313 | 0,889921513 | CTU1 | FALSE | Upregulated |
| -0,285318353 | 0,889921513 | GATSL3 | FALSE | Downregulated |
| -0,294204935 | 0,889921513 | LOC101927198 | FALSE | Downregulated |
| 0,139782812 | 0,889921513 | LOC101928051 | FALSE | Upregulated |
| -1,411800069 | 0,889921513 | LOC102723719 | FALSE | Downregulated |
| 0,142066841 | 0,889921513 | OSGEP | FALSE | Upregulated |
| -0,16210783 | 0,889921513 | PIBF1 | FALSE | Downregulated |
| -0,385705719 | 0,889921513 | TMEM232 | FALSE | Downregulated |
| 0,231862197 | 0,889977917 | FBXO4 | FALSE | Upregulated |
| 0,238922137 | 0,890018096 | MMGT1 | FALSE | Upregulated |
| -0,114020994 | 0,890133582 | FOXP1 | FALSE | Downregulated |
| -0,095798042 | 0,890133582 | ZNF654 | FALSE | Downregulated |
| 0,295613869 | 0,890135982 | ACER2 | FALSE | Upregulated |
| 0,264581057 | 0,890164061 | CAPS2 | FALSE | Upregulated |
| 0,282419692 | 0,890164061 | LINC00087 | FALSE | Upregulated |
| 0,158967057 | 0,890164061 | PCOLCE-AS1 | FALSE | Upregulated |
| -0,220947695 | 0,890164061 | PELI2 | FALSE | Downregulated |
| -0,136869809 | 0,890774944 | NUDT6 | FALSE | Downregulated |
| -0,112682426 | 0,89117089 | PLEKHA1 | FALSE | Downregulated |
| 0,151346859 | 0,891176504 | PVRL1 | FALSE | Upregulated |
| -0,156199544 | 0,891652987 | MECR | FALSE | Downregulated |
| 0,03844981 | 0,891729779 | NUMA1 | FALSE | Upregulated |
| -0,400828391 | 0,891903449 | OVOL3 | FALSE | Downregulated |
| 0,091386888 | 0,892185961 | DNAJC1 | FALSE | Upregulated |

| | | | | |
|---|---|---|---|---|
| -0,137084234 | 0,892322693 | BRCC3 | FALSE | Downregulated |
| 0,095204135 | 0,892322693 | GOLGA8B | FALSE | Upregulated |
| 0,147916535 | 0,892322693 | GTF2H4 | FALSE | Upregulated |
| 0,163288164 | 0,892322693 | LOC100506734 | FALSE | Upregulated |
| -0,378846907 | 0,892322693 | MTUS2-AS1 | FALSE | Downregulated |
| -0,1408501 | 0,892322693 | NDUFB6 | FALSE | Downregulated |
| 0,129394661 | 0,892322693 | PTAR1 | FALSE | Upregulated |
| -0,106565122 | 0,892322693 | TM9SF1 | FALSE | Downregulated |
| 0,273198997 | 0,892435273 | SSSCA1 | FALSE | Upregulated |
| 0,364970387 | 0,892445708 | CALHM1 | FALSE | Upregulated |
| -0,103901517 | 0,892536628 | GIPC3 | FALSE | Downregulated |
| -0,098932235 | 0,892540913 | BIRC2 | FALSE | Downregulated |
| -0,056342878 | 0,892540913 | TTC3 | FALSE | Downregulated |
| -0,271085118 | 0,892540913 | ZNF624 | FALSE | Downregulated |
| -0,146424824 | 0,892550385 | ASB7 | FALSE | Downregulated |
| 0,048314325 | 0,892554563 | UBTF | FALSE | Upregulated |
| -0,452683204 | 0,892700941 | P2RY11 | FALSE | Downregulated |
| 0,156265924 | 0,892765991 | C3orf49 | FALSE | Upregulated |
| 0,13557432 | 0,892765991 | GOLGA7 | FALSE | Upregulated |
| -0,07370723 | 0,892765991 | ITGA9 | FALSE | Downregulated |
| 0,12107787 | 0,892765991 | TPGS1 | FALSE | Upregulated |
| -0,215855868 | 0,892766633 | PILRB | FALSE | Downregulated |
| 0,282939744 | 0,892876173 | FAM83A | FALSE | Upregulated |
| -0,284034789 | 0,893083549 | EMR1 | FALSE | Downregulated |
| 0,087194153 | 0,893172433 | RAB11A | FALSE | Upregulated |
| 0,301421486 | 0,893623638 | EPHA4 | FALSE | Upregulated |
| -0,085001589 | 0,893623638 | GTF3A | FALSE | Downregulated |
| -0,262492608 | 0,893623638 | LOC149351 | FALSE | Downregulated |
| 0,090018644 | 0,893714231 | RBM17 | FALSE | Upregulated |
| -0,072499255 | 0,89380779 | CNOT3 | FALSE | Downregulated |
| 0,141294273 | 0,89380779 | LOC102723998 | FALSE | Upregulated |
| 0,107982081 | 0,89380779 | SPAST | FALSE | Upregulated |
| -0,112431839 | 0,893815099 | USPL1 | FALSE | Downregulated |
| -0,208122317 | 0,893941751 | ALS2CR12 | FALSE | Downregulated |
| -0,064997687 | 0,893941751 | AP1B1 | FALSE | Downregulated |
| -0,191530624 | 0,893941751 | MFNG | FALSE | Downregulated |
| -0,199669553 | 0,893941751 | PDCL | FALSE | Downregulated |
| 0,069629892 | 0,893941751 | RABL6 | FALSE | Upregulated |
| -0,099409557 | 0,894166605 | ARL8A | FALSE | Downregulated |
| 0,065508895 | 0,894166605 | NT5C2 | FALSE | Upregulated |
| -0,709227037 | 0,89418231 | C1QTNF3-AMACR | FALSE | Downregulated |
| -0,197372772 | 0,89418231 | COL23A1 | FALSE | Downregulated |
| 0,076214032 | 0,89418231 | RGPD1 | FALSE | Upregulated |
| -0,06541421 | 0,89418231 | WAPAL | FALSE | Downregulated |
| 0,363476597 | 0,894291397 | LOC100505851 | FALSE | Upregulated |
| -0,173722366 | 0,894325559 | MIEF2 | FALSE | Downregulated |
| -0,093434208 | 0,894350796 | TAF5L | FALSE | Downregulated |
| 0,187147374 | 0,894362901 | APOC1P1 | FALSE | Upregulated |
| -0,131203779 | 0,894362901 | RAB20 | FALSE | Downregulated |
| 0,102294234 | 0,894362901 | RYR3 | FALSE | Upregulated |
| 0,206029886 | 0,894362901 | SLC25A12 | FALSE | Upregulated |

| | | | | |
|---|---|---|---|---|
| -0,20626716 | 0,894362901 | ZIC1 | FALSE | Downregulated |
| 0,126758482 | 0,894362901 | ZNF771 | FALSE | Upregulated |
| -0,062519918 | 0,894408016 | ZCCHC2 | FALSE | Downregulated |
| -0,09546291 | 0,894415437 | CSDC2 | FALSE | Downregulated |
| 0,156585131 | 0,89450894 | TBCCD1 | FALSE | Upregulated |
| 0,173805854 | 0,89450894 | TDP1 | FALSE | Upregulated |
| 0,167560753 | 0,894657535 | HIST2H2BF | FALSE | Upregulated |
| -0,095398292 | 0,894687111 | SRSF12 | FALSE | Downregulated |
| -0,146404689 | 0,894808723 | FAM188A | FALSE | Downregulated |
| -0,093428556 | 0,894808723 | MIF | FALSE | Downregulated |
| -0,162982201 | 0,894905336 | PDIK1L | FALSE | Downregulated |
| -0,373243979 | 0,895166709 | BMX | FALSE | Downregulated |
| 0,144871213 | 0,895166709 | BUB3 | FALSE | Upregulated |
| 0,210348118 | 0,895166709 | CHCHD4 | FALSE | Upregulated |
| -0,162191083 | 0,895166709 | DGCR5 | FALSE | Downregulated |
| 0,075072101 | 0,895166709 | EDEM1 | FALSE | Upregulated |
| 0,465873025 | 0,895166709 | EIF4EBP3 | FALSE | Upregulated |
| 0,314218638 | 0,895166709 | LOC100499484-C9ORF174 | FALSE | Upregulated |
| 0,373696973 | 0,895166709 | LOC101929664 | FALSE | Upregulated |
| 0,26934625 | 0,895166709 | MIXL1 | FALSE | Upregulated |
| -0,107571967 | 0,895166709 | RHOT1 | FALSE | Downregulated |
| -0,20558653 | 0,895166709 | S100A9 | FALSE | Downregulated |
| -0,110014469 | 0,895166709 | SLC28A1 | FALSE | Downregulated |
| 0,108636901 | 0,895166709 | SNRNP35 | FALSE | Upregulated |
| -0,225539953 | 0,895166709 | WBP4 | FALSE | Downregulated |
| -0,098828295 | 0,895166709 | XPNPEP3 | FALSE | Downregulated |
| -0,085463803 | 0,895166709 | YIPF2 | FALSE | Downregulated |
| -0,230701405 | 0,895306031 | ARMCX6 | FALSE | Downregulated |
| 0,121113352 | 0,895306031 | BOC | FALSE | Upregulated |
| 0,182039226 | 0,895306031 | C6orf1 | FALSE | Upregulated |
| -0,076036277 | 0,895306031 | CACTIN | FALSE | Downregulated |
| -0,085277741 | 0,895306031 | ERN1 | FALSE | Downregulated |
| -0,118673836 | 0,895306031 | FBXO6 | FALSE | Downregulated |
| 0,169457156 | 0,895306031 | KIAA1279 | FALSE | Upregulated |
| 0,295899098 | 0,895306031 | LOC101927818 | FALSE | Upregulated |
| 0,165064045 | 0,895306031 | LOC143666 | FALSE | Upregulated |
| -0,176611923 | 0,895306031 | PHF6 | FALSE | Downregulated |
| 0,076383652 | 0,895306031 | RBM12B-AS1 | FALSE | Upregulated |
| 0,131853417 | 0,895306031 | RNMTL1 | FALSE | Upregulated |
| 0,226602209 | 0,895306031 | RP6-109B7.3 | FALSE | Upregulated |
| 0,053455458 | 0,895306031 | TEX2 | FALSE | Upregulated |
| -0,087962822 | 0,895306031 | XPO7 | FALSE | Downregulated |
| 0,117048776 | 0,895338615 | ARFIP2 | FALSE | Upregulated |
| 0,142529073 | 0,895367801 | ANKRD16 | FALSE | Upregulated |
| -0,071288022 | 0,895367801 | KIAA0947 | FALSE | Downregulated |
| 0,399583854 | 0,895367801 | LOC102723692 | FALSE | Upregulated |
| -0,179164531 | 0,895367801 | RRP7B | FALSE | Downregulated |
| 0,224411887 | 0,895367801 | SUSD2 | FALSE | Upregulated |
| 0,512420675 | 0,895520804 | CHURC1-FNTB | FALSE | Upregulated |
| 0,089358354 | 0,895524146 | NAA50 | FALSE | Upregulated |

| | | | | |
|---|---|---|---|---|
| 0,145439898 | 0,895683271 | CABP4 | FALSE | Upregulated |
| -0,109462968 | 0,895683271 | MTM1 | FALSE | Downregulated |
| 0,121091692 | 0,895768599 | NRDE2 | FALSE | Upregulated |
| 0,145474631 | 0,895768599 | TMEM65 | FALSE | Upregulated |
| 0,074239377 | 0,89582974 | LOC101929844 | FALSE | Upregulated |
| -0,131688505 | 0,895943468 | ASNA1 | FALSE | Downregulated |
| -0,329298799 | 0,895943468 | C6orf141 | FALSE | Downregulated |
| -0,309578522 | 0,895943468 | KRTAP5-AS1 | FALSE | Downregulated |
| 0,0723218 | 0,895943468 | PKN1 | FALSE | Upregulated |
| 0,073713029 | 0,895943468 | SNX27 | FALSE | Upregulated |
| 0,099333045 | 0,89595282 | SDCCAG8 | FALSE | Upregulated |
| -0,103330409 | 0,896014527 | DPP8 | FALSE | Downregulated |
| -0,127329645 | 0,896014527 | TMC3 | FALSE | Downregulated |
| -0,083979776 | 0,89615076 | MTG2 | FALSE | Downregulated |
| -0,090672246 | 0,896165054 | CXCL16 | FALSE | Downregulated |
| -0,099911257 | 0,896165054 | LOC101927673 | FALSE | Downregulated |
| 0,241642137 | 0,896331751 | ANKRD55 | FALSE | Upregulated |
| -0,079909051 | 0,896331751 | ARID4B | FALSE | Downregulated |
| 0,091403454 | 0,896331751 | BICD2 | FALSE | Upregulated |
| 0,119889685 | 0,896331751 | C17orf107 | FALSE | Upregulated |
| 0,135149768 | 0,896331751 | C7orf26 | FALSE | Upregulated |
| 0,143895986 | 0,896331751 | CCR7 | FALSE | Upregulated |
| 0,115696552 | 0,896331751 | CDC42P3 | FALSE | Upregulated |
| 0,062721781 | 0,896331751 | CFLAR | FALSE | Upregulated |
| 0,297442279 | 0,896331751 | CLHC1 | FALSE | Upregulated |
| 0,340907956 | 0,896331751 | GLA | FALSE | Upregulated |
| -0,256293545 | 0,896331751 | GSTT2B | FALSE | Downregulated |
| 0,247533104 | 0,896331751 | LOC101927579 | FALSE | Upregulated |
| 0,169926796 | 0,896331751 | LOC101928678 | FALSE | Upregulated |
| 2,031525523 | 0,896331751 | MUC13 | FALSE | Upregulated |
| 0,115873005 | 0,896331751 | NIPAL3 | FALSE | Upregulated |
| -0,279433125 | 0,896331751 | NPIPB15 | FALSE | Downregulated |
| 0,119233241 | 0,896331751 | NSUN4 | FALSE | Upregulated |
| -0,29903997 | 0,896331751 | RP11-458D21.1 | FALSE | Downregulated |
| -0,254788579 | 0,896331751 | SLC6A19 | FALSE | Downregulated |
| -0,157468695 | 0,896331751 | TM4SF5 | FALSE | Downregulated |
| -0,090477195 | 0,896331751 | UBL4A | FALSE | Downregulated |
| 0,180985945 | 0,896331751 | UTP14A | FALSE | Upregulated |
| -0,219326059 | 0,896331751 | ZFP3 | FALSE | Downregulated |
| -0,385557385 | 0,896349624 | LOC100288181 | FALSE | Downregulated |
| -0,081642438 | 0,896349624 | SIX5 | FALSE | Downregulated |
| 0,332512506 | 0,896349624 | SPSB4 | FALSE | Upregulated |
| 0,158466799 | 0,896349624 | ZW10 | FALSE | Upregulated |
| 0,160331019 | 0,896357684 | ZNF568 | FALSE | Upregulated |
| -0,157310514 | 0,896433113 | JOSD2 | FALSE | Downregulated |
| 0,192445883 | 0,896433113 | SNRPE | FALSE | Upregulated |
| -0,39846657 | 0,896520818 | C1orf185 | FALSE | Downregulated |
| -0,135921366 | 0,896625265 | ASPHD2 | FALSE | Downregulated |
| 0,270039338 | 0,896627099 | RP11-598F7.4 | FALSE | Upregulated |
| -0,138408773 | 0,896627099 | TTC6 | FALSE | Downregulated |
| 0,096836129 | 0,896771712 | FAM199X | FALSE | Upregulated |

| | | | | |
|---|---|---|---|---|
| -0,078056606 | 0,896771712 | PSMD4 | FALSE | Downregulated |
| -0,119554836 | 0,89683058 | UBXN8 | FALSE | Downregulated |
| 0,318870421 | 0,896954675 | HYDIN2 | FALSE | Upregulated |
| 0,169381196 | 0,897008741 | ANXA10 | FALSE | Upregulated |
| 0,18010512 | 0,897008741 | C19orf52 | FALSE | Upregulated |
| -0,282534029 | 0,897008741 | CMTM2 | FALSE | Downregulated |
| -0,18917662 | 0,897008741 | COL25A1 | FALSE | Downregulated |
| 0,111107387 | 0,897008741 | PROCA1 | FALSE | Upregulated |
| -0,108945929 | 0,897008741 | SLC2A10 | FALSE | Downregulated |
| 0,103319496 | 0,897008741 | ZNF780B | FALSE | Upregulated |
| -0,070966077 | 0,897062979 | ACIN1 | FALSE | Downregulated |
| -0,1072677 | 0,897062979 | LEPR | FALSE | Downregulated |
| 0,109056689 | 0,897062979 | NEURL1 | FALSE | Upregulated |
| -0,122290026 | 0,897062979 | PSMB1 | FALSE | Downregulated |
| -0,091955123 | 0,897062979 | SIRT2 | FALSE | Downregulated |
| -0,20011881 | 0,897062979 | TCTN1 | FALSE | Downregulated |
| -0,088943209 | 0,897062979 | TLR4 | FALSE | Downregulated |
| 0,288629832 | 0,897080505 | AQP4 | FALSE | Upregulated |
| 0,156112127 | 0,897206257 | NDUFB1 | FALSE | Upregulated |
| -0,322222132 | 0,897248091 | BTN2A3P | FALSE | Downregulated |
| -0,119296322 | 0,897248091 | CHADL | FALSE | Downregulated |
| -0,126686167 | 0,897248091 | FLI1 | FALSE | Downregulated |
| -0,188775977 | 0,897248091 | TPRG1-AS2 | FALSE | Downregulated |
| -0,062836486 | 0,897297137 | ENSA | FALSE | Downregulated |
| 0,176784217 | 0,897297137 | NUDT17 | FALSE | Upregulated |
| -0,080585464 | 0,897304229 | EEF2K | FALSE | Downregulated |
| 0,12283305 | 0,897304229 | GNL3 | FALSE | Upregulated |
| 0,215709249 | 0,897304229 | LOC101060060 | FALSE | Upregulated |
| 0,102349493 | 0,897304229 | LRCH3 | FALSE | Upregulated |
| -0,069774028 | 0,897304229 | RBM14 | FALSE | Downregulated |
| -0,294087603 | 0,897304229 | SLC16A12 | FALSE | Downregulated |
| 0,053444825 | 0,897304229 | TLN1 | FALSE | Upregulated |
| -0,131688491 | 0,897304229 | ZFP14 | FALSE | Downregulated |
| 0,080697668 | 0,897304229 | ZNF646 | FALSE | Upregulated |
| -0,059054768 | 0,897391798 | CDC34 | FALSE | Downregulated |
| -0,178361026 | 0,897391798 | KY | FALSE | Downregulated |
| 0,141324226 | 0,897670339 | BIN3 | FALSE | Upregulated |
| 0,151394941 | 0,897670339 | CTNNAL1 | FALSE | Upregulated |
| -0,070631553 | 0,897670339 | GPX1 | FALSE | Downregulated |
| 0,045815619 | 0,897670339 | KIAA1109 | FALSE | Upregulated |
| -0,226543818 | 0,897670339 | LOC100132111 | FALSE | Downregulated |
| -0,07961216 | 0,897679635 | FUCA1 | FALSE | Downregulated |
| -0,092723477 | 0,897718413 | ASB1 | FALSE | Downregulated |
| -0,071964045 | 0,897881938 | TUBB4B | FALSE | Downregulated |
| -0,079103376 | 0,898309388 | APOBR | FALSE | Downregulated |
| 0,06482685 | 0,898309388 | MGRN1 | FALSE | Upregulated |
| 0,171393345 | 0,898348027 | LOC102723886 | FALSE | Upregulated |
| -0,145641655 | 0,898370791 | FLJ20464 | FALSE | Downregulated |
| 0,204960245 | 0,898370791 | LOC101928729 | FALSE | Upregulated |
| -0,410619336 | 0,898370791 | MANEA-AS1 | FALSE | Downregulated |
| -0,065325936 | 0,898370791 | PITPNM2 | FALSE | Downregulated |

| | | | | |
|---|---|---|---|---|
| -0,090960161 | 0,898370791 | RING1 | FALSE | Downregulated |
| -0,163472318 | 0,898370791 | SCOC | FALSE | Downregulated |
| 0,089888501 | 0,898370791 | SLC25A16 | FALSE | Upregulated |
| 0,098174784 | 0,898370791 | TXNDC15 | FALSE | Upregulated |
| 0,092574466 | 0,898370791 | YTHDC2 | FALSE | Upregulated |
| -0,077423538 | 0,898403458 | RNF167 | FALSE | Downregulated |
| -0,094562934 | 0,898451384 | ARMCX3 | FALSE | Downregulated |
| 0,204012354 | 0,898451384 | BAI3 | FALSE | Upregulated |
| 0,256827487 | 0,898451384 | BLK | FALSE | Upregulated |
| 0,116218446 | 0,898451384 | BTF3L4 | FALSE | Upregulated |
| 0,33688407 | 0,898451384 | KBTBD13 | FALSE | Upregulated |
| -0,337157178 | 0,898451384 | KRT222 | FALSE | Downregulated |
| 0,162349591 | 0,898451384 | MRPS9 | FALSE | Upregulated |
| -0,04915268 | 0,898451384 | MTMR10 | FALSE | Downregulated |
| 0,136057633 | 0,898451384 | SLC22A10 | FALSE | Upregulated |
| -0,066956882 | 0,898451384 | UNC13B | FALSE | Downregulated |
| 0,070007581 | 0,898451384 | WDR45B | FALSE | Upregulated |
| -0,112578593 | 0,898707689 | EIF2AK3 | FALSE | Downregulated |
| -0,093344995 | 0,898707689 | LOC101928081 | FALSE | Downregulated |
| 0,195273939 | 0,898707689 | PCGF6 | FALSE | Upregulated |
| 0,135487288 | 0,898888234 | AASDHPPT | FALSE | Upregulated |
| -0,18387928 | 0,898888234 | ANO5 | FALSE | Downregulated |
| -0,059640716 | 0,898888234 | ATG2B | FALSE | Downregulated |
| 0,080466942 | 0,898888234 | C12orf23 | FALSE | Upregulated |
| 0,088764583 | 0,898888234 | C17orf62 | FALSE | Upregulated |
| 0,175323542 | 0,898888234 | CARD6 | FALSE | Upregulated |
| 0,272834247 | 0,898888234 | EMP3 | FALSE | Upregulated |
| -0,164619365 | 0,898888234 | FGF14-AS2 | FALSE | Downregulated |
| 0,061786949 | 0,898888234 | GATAD2A | FALSE | Upregulated |
| -0,095366679 | 0,898888234 | GTSE1-AS1 | FALSE | Downregulated |
| -0,050771768 | 0,898888234 | KIAA0195 | FALSE | Downregulated |
| -0,284694825 | 0,898888234 | LOC201651 | FALSE | Downregulated |
| -0,063614817 | 0,898888234 | LRIG1 | FALSE | Downregulated |
| 0,122409011 | 0,898888234 | MANEA | FALSE | Upregulated |
| 0,281700233 | 0,898888234 | N6AMT2 | FALSE | Upregulated |
| -0,349785588 | 0,898888234 | PCAT19 | FALSE | Downregulated |
| 0,129635132 | 0,898888234 | RNASE7 | FALSE | Upregulated |
| -0,141179171 | 0,898888234 | SMUG1 | FALSE | Downregulated |
| 0,186410062 | 0,898888234 | SNORA74B | FALSE | Upregulated |
| -0,120547587 | 0,898888234 | SYT12 | FALSE | Downregulated |
| -0,140811892 | 0,898888234 | UBA3 | FALSE | Downregulated |
| -0,09898118 | 0,898888234 | UBE2I | FALSE | Downregulated |
| -0,095504544 | 0,898888234 | ZBTB1 | FALSE | Downregulated |
| 0,112873503 | 0,898888234 | ZDHHC2 | FALSE | Upregulated |
| 0,188847522 | 0,898888234 | ZNF582-AS1 | FALSE | Upregulated |
| 0,212356118 | 0,899428793 | AP000479.1 | FALSE | Upregulated |
| -0,056598056 | 0,899428793 | SRSF9 | FALSE | Downregulated |
| -0,236651902 | 0,899567898 | NFU1 | FALSE | Downregulated |
| 0,218710617 | 0,899567898 | PRKY | FALSE | Upregulated |
| -0,311539945 | 0,899608799 | TAS2R13 | FALSE | Downregulated |
| -0,186230736 | 0,899620005 | ATG4A | FALSE | Downregulated |

| | | | | |
|---|---|---|---|---|
| -0,314281495 | 0,899620005 | LOC102724024 | FALSE | Downregulated |
| 0,063666939 | 0,8996276 | CRK | FALSE | Upregulated |
| -0,110435972 | 0,899848632 | PDE7B | FALSE | Downregulated |
| 0,20387315 | 0,899848632 | PKD1L3 | FALSE | Upregulated |
| -0,090001404 | 0,899958844 | WDR46 | FALSE | Downregulated |
| 0,090535126 | 0,900082957 | GOLGA8M | FALSE | Upregulated |
| -0,223831635 | 0,900082957 | PDZD11 | FALSE | Downregulated |
| 0,149872056 | 0,900082957 | RUNX1T1 | FALSE | Upregulated |
| -0,133038603 | 0,900281134 | APTX | FALSE | Downregulated |
| -0,049398606 | 0,900281134 | ASH1L | FALSE | Downregulated |
| -0,300507557 | 0,900281134 | BCAS4 | FALSE | Downregulated |
| 0,063093924 | 0,900281134 | C1orf167 | FALSE | Upregulated |
| -0,121459247 | 0,900281134 | CELSR2 | FALSE | Downregulated |
| 0,031566479 | 0,900281134 | CNOT1 | FALSE | Upregulated |
| -0,140490373 | 0,900281134 | FADS2 | FALSE | Downregulated |
| 0,157406435 | 0,900281134 | FAM207A | FALSE | Upregulated |
| 0,094798393 | 0,900281134 | FBXL20 | FALSE | Upregulated |
| -0,083325066 | 0,900281134 | GEMIN4 | FALSE | Downregulated |
| 0,122859347 | 0,900281134 | HSPB9 | FALSE | Upregulated |
| -0,616484553 | 0,900281134 | NPY4R | FALSE | Downregulated |
| 0,156280544 | 0,900281134 | NTPCR | FALSE | Upregulated |
| -0,096761166 | 0,900281134 | ZSWIM6 | FALSE | Downregulated |
| -0,089591091 | 0,900338453 | ASPSCR1 | FALSE | Downregulated |
| 0,249297631 | 0,900338453 | LOC284454 | FALSE | Upregulated |
| 0,214314661 | 0,900652599 | OR5AK4P | FALSE | Upregulated |
| 0,19699048 | 0,900819024 | MTERF | FALSE | Upregulated |
| -0,065475915 | 0,900819024 | PPP1CA | FALSE | Downregulated |
| -0,227875312 | 0,900929279 | ADAM11 | FALSE | Downregulated |
| 0,271181145 | 0,900929279 | LINC00893 | FALSE | Upregulated |
| 0,082696937 | 0,900929279 | MAP1LC3B | FALSE | Upregulated |
| -0,295699974 | 0,900929279 | SCAND3 | FALSE | Downregulated |
| -0,33917227 | 0,901204277 | TXLNB | FALSE | Downregulated |
| -0,16388913 | 0,901291068 | LEKR1 | FALSE | Downregulated |
| -0,070420275 | 0,901291068 | SH2B1 | FALSE | Downregulated |
| 0,139570457 | 0,901312298 | CNTNAP4 | FALSE | Upregulated |
| 0,274059981 | 0,901382956 | NOMO3 | FALSE | Upregulated |
| 0,049578623 | 0,901415624 | ANKRD11 | FALSE | Upregulated |
| -0,251487882 | 0,901415624 | CDRT4 | FALSE | Downregulated |
| -0,25892687 | 0,901415624 | FASTKD3 | FALSE | Downregulated |
| -0,151532614 | 0,901415624 | GIMAP1 | FALSE | Downregulated |
| -0,153645124 | 0,901415624 | LOC102724157 | FALSE | Downregulated |
| -0,093773434 | 0,901415624 | MPPE1 | FALSE | Downregulated |
| -0,156716908 | 0,901415624 | NUTF2 | FALSE | Downregulated |
| 0,077675957 | 0,901415624 | PPP2R5E | FALSE | Upregulated |
| 0,093637577 | 0,901415624 | RAB11FIP5 | FALSE | Upregulated |
| 0,311175698 | 0,901415624 | RASL11A | FALSE | Upregulated |
| 0,163705313 | 0,901415624 | RSRC1 | FALSE | Upregulated |
| 0,099086678 | 0,901415624 | SLC17A5 | FALSE | Upregulated |
| -0,118718619 | 0,901415624 | TDP2 | FALSE | Downregulated |
| -0,150481743 | 0,901415624 | TTC12 | FALSE | Downregulated |
| 0,108096392 | 0,901415624 | ZNF514 | FALSE | Upregulated |

| | | | | |
|---|---|---|---|---|
| -0,073231755 | 0,9015085 | DENND5B | FALSE | Downregulated |
| -0,354335758 | 0,901863129 | CORO6 | FALSE | Downregulated |
| 0,228602333 | 0,901863129 | TRIM10 | FALSE | Upregulated |
| 0,114954995 | 0,901916905 | DUS2 | FALSE | Upregulated |
| 0,092956069 | 0,901916905 | EXOSC3 | FALSE | Upregulated |
| 0,286840894 | 0,901916905 | FLJ44511 | FALSE | Upregulated |
| 0,116960467 | 0,901916905 | MRGBP | FALSE | Upregulated |
| 0,202990761 | 0,901916905 | PTPDC1 | FALSE | Upregulated |
| -0,335946701 | 0,901916905 | TMEM100 | FALSE | Downregulated |
| 0,107257455 | 0,90193199 | CBY3 | FALSE | Upregulated |
| 0,070640803 | 0,901938655 | HECA | FALSE | Upregulated |
| 0,062883394 | 0,902135446 | CD46 | FALSE | Upregulated |
| -0,270227451 | 0,902135446 | VWA7 | FALSE | Downregulated |
| 0,091046571 | 0,902179265 | ZNF526 | FALSE | Upregulated |
| 0,206200009 | 0,90243316 | FUT3 | FALSE | Upregulated |
| -0,077725661 | 0,90243316 | RNPEP | FALSE | Downregulated |
| -0,234285803 | 0,902470086 | LOC643797 | FALSE | Downregulated |
| 0,120803223 | 0,902493892 | GGT7 | FALSE | Upregulated |
| 0,340964492 | 0,902493892 | MARK2P9 | FALSE | Upregulated |
| 0,063078855 | 0,902579834 | DENND4A | FALSE | Upregulated |
| -0,305612434 | 0,902579834 | NTRK3 | FALSE | Downregulated |
| -0,077510892 | 0,902579834 | PKN2 | FALSE | Downregulated |
| 1,90872035 | 0,902579834 | SNORD3C | FALSE | Upregulated |
| 0,154130208 | 0,902579834 | TMEM132D | FALSE | Upregulated |
| -0,079532832 | 0,902638412 | CHMP2A | FALSE | Downregulated |
| 0,17915599 | 0,902638412 | FOLR1 | FALSE | Upregulated |
| 0,126349444 | 0,902638412 | MYO15A | FALSE | Upregulated |
| -0,227042942 | 0,902638412 | ZNF383 | FALSE | Downregulated |
| 0,196825645 | 0,902672696 | LOC101927986 | FALSE | Upregulated |
| 0,124220467 | 0,902767458 | PRKCE | FALSE | Upregulated |
| 0,102092944 | 0,902932067 | TMEM185B | FALSE | Upregulated |
| -0,093922425 | 0,902998353 | KLF10 | FALSE | Downregulated |
| -0,094260668 | 0,902998353 | NRG4 | FALSE | Downregulated |
| 0,136897036 | 0,902998353 | SERTAD3 | FALSE | Upregulated |
| -0,158388906 | 0,903012918 | HEYL | FALSE | Downregulated |
| -0,123071313 | 0,903012918 | ZNF585B | FALSE | Downregulated |
| 0,06697096 | 0,903021707 | CABLES1 | FALSE | Upregulated |
| -0,092893924 | 0,903103717 | RAB11FIP1 | FALSE | Downregulated |
| 0,0671208 | 0,903228582 | LOC646471 | FALSE | Upregulated |
| 0,114021915 | 0,903228582 | LYRM4 | FALSE | Upregulated |
| -0,06994845 | 0,903228582 | SEMA3F | FALSE | Downregulated |
| -1,881766549 | 0,903724308 | RNU1-3 | FALSE | Downregulated |
| 0,126664631 | 0,903741268 | COX7A1 | FALSE | Upregulated |
| -0,255469759 | 0,903741268 | ZNF256 | FALSE | Downregulated |
| 0,068861393 | 0,903893405 | UNKL | FALSE | Upregulated |
| 0,141959 | 0,903960445 | IQSEC3 | FALSE | Upregulated |
| 0,048911572 | 0,903960445 | JAK1 | FALSE | Upregulated |
| 0,250424333 | 0,903960445 | LOC100131626 | FALSE | Upregulated |
| 0,274832642 | 0,903960445 | MFSD6L | FALSE | Upregulated |
| 0,036578647 | 0,903960445 | MLLT4 | FALSE | Upregulated |
| 0,147915584 | 0,903960445 | YIF1B | FALSE | Upregulated |

| | | | | |
|---|---|---|---|---|
| 0,400898622 | 0,903971221 | INO80B-WBP1 | FALSE | Upregulated |
| 0,188002033 | 0,904082388 | C19orf40 | FALSE | Upregulated |
| 0,236424754 | 0,904089907 | DCLRE1B | FALSE | Upregulated |
| 0,0761266 | 0,904089907 | FBXO21 | FALSE | Upregulated |
| 0,146391343 | 0,904089907 | ZRSR2 | FALSE | Upregulated |
| 0,258108978 | 0,90412072 | FLJ38122 | FALSE | Upregulated |
| -0,117643623 | 0,904173113 | MRPL30 | FALSE | Downregulated |
| -0,064540121 | 0,904294403 | IFNAR1 | FALSE | Downregulated |
| 0,06317441 | 0,904294403 | PTPN18 | FALSE | Upregulated |
| -0,091364845 | 0,904393753 | NDUFA2 | FALSE | Downregulated |
| -0,318822605 | 0,904393753 | SCD5 | FALSE | Downregulated |
| -0,269709039 | 0,904400615 | C19orf55 | FALSE | Downregulated |
| 0,121287443 | 0,904400615 | C9orf16 | FALSE | Upregulated |
| 0,069555863 | 0,904400615 | RARA-AS1 | FALSE | Upregulated |
| -0,152106392 | 0,90449343 | LTK | FALSE | Downregulated |
| -0,174211017 | 0,904543899 | ADPRHL2 | FALSE | Downregulated |
| -0,093870088 | 0,904712312 | COPB1 | FALSE | Downregulated |
| -0,141385151 | 0,904757802 | H2AFY2 | FALSE | Downregulated |
| 0,267060049 | 0,90476621 | ACN9 | FALSE | Upregulated |
| 0,072940685 | 0,90476621 | C6orf47 | FALSE | Upregulated |
| 0,126416219 | 0,90476621 | CD9 | FALSE | Upregulated |
| 0,064187385 | 0,90476621 | CLN5 | FALSE | Upregulated |
| -0,133647518 | 0,90476621 | CPSF3 | FALSE | Downregulated |
| -0,24508393 | 0,90476621 | CTAGE4 | FALSE | Downregulated |
| 0,088989612 | 0,90476621 | DDX49 | FALSE | Upregulated |
| 0,263947008 | 0,90476621 | FLJ45513 | FALSE | Upregulated |
| -0,10077671 | 0,90476621 | FYN | FALSE | Downregulated |
| 0,315922528 | 0,90476621 | GFAP | FALSE | Upregulated |
| -0,087236671 | 0,90476621 | HDAC10 | FALSE | Downregulated |
| 0,121913662 | 0,90476621 | IRAK4 | FALSE | Upregulated |
| -0,095836446 | 0,90476621 | KIAA1430 | FALSE | Downregulated |
| 0,205960008 | 0,90476621 | KIF20B | FALSE | Upregulated |
| -0,247749251 | 0,90476621 | LAG3 | FALSE | Downregulated |
| 0,351948921 | 0,90476621 | LOC102723444 | FALSE | Upregulated |
| 0,146021311 | 0,90476621 | MID1 | FALSE | Upregulated |
| 0,100939635 | 0,90476621 | NXN | FALSE | Upregulated |
| -0,215170208 | 0,90476621 | PITPNA-AS1 | FALSE | Downregulated |
| 0,090693629 | 0,90476621 | PRKAR2A | FALSE | Upregulated |
| -0,111835943 | 0,90476621 | PSMA1 | FALSE | Downregulated |
| -0,132748187 | 0,90476621 | TMEM221 | FALSE | Downregulated |
| 0,062875871 | 0,90476621 | VPS39 | FALSE | Upregulated |
| -0,056264835 | 0,90476621 | WWC2 | FALSE | Downregulated |
| -0,229903592 | 0,90476621 | ZNF138 | FALSE | Downregulated |
| 0,167523635 | 0,90476621 | ZNF174 | FALSE | Upregulated |
| -0,235236711 | 0,904912415 | APITD1 | FALSE | Downregulated |
| 0,184492347 | 0,904912415 | COMTD1 | FALSE | Upregulated |
| 0,05075058 | 0,904912415 | EDF1 | FALSE | Upregulated |
| 0,134084485 | 0,904912415 | PRSS37 | FALSE | Upregulated |
| 0,233979115 | 0,904912415 | SERF1B | FALSE | Upregulated |
| 0,22311089 | 0,904912415 | SERPINE1 | FALSE | Upregulated |
| 0,183596056 | 0,904912415 | ZNF366 | FALSE | Upregulated |

| | | | | |
|---|---|---|---|---|
| 0,112811041 | 0,90538652 | ZNF782 | FALSE | Upregulated |
| -0,085552946 | 0,905403634 | DDX51 | FALSE | Downregulated |
| -0,071556109 | 0,905403634 | ENGASE | FALSE | Downregulated |
| -0,095282742 | 0,905403634 | MRPS7 | FALSE | Downregulated |
| -0,115424643 | 0,905403634 | SIRT7 | FALSE | Downregulated |
| -0,108163427 | 0,905421969 | ATPIF1 | FALSE | Downregulated |
| -0,055227007 | 0,905509995 | HECTD3 | FALSE | Downregulated |
| -0,114649298 | 0,905792963 | SCARNA5 | FALSE | Downregulated |
| -0,132145862 | 0,905816099 | CLDND1 | FALSE | Downregulated |
| -0,129229194 | 0,906020581 | NOP2 | FALSE | Downregulated |
| -0,065257765 | 0,906072461 | CD164 | FALSE | Downregulated |
| -0,128747739 | 0,906130264 | RND3 | FALSE | Downregulated |
| 0,083061649 | 0,906130264 | TEAD3 | FALSE | Upregulated |
| -0,116449206 | 0,906414452 | ALG9 | FALSE | Downregulated |
| 0,282582724 | 0,906414452 | C5orf49 | FALSE | Upregulated |
| 0,157385777 | 0,906414452 | FGR | FALSE | Upregulated |
| 0,067809184 | 0,906473588 | LOC101929211 | FALSE | Upregulated |
| 0,102378281 | 0,906478442 | MSTO1 | FALSE | Upregulated |
| 0,06179548 | 0,906869954 | ATP11A | FALSE | Upregulated |
| -0,109274385 | 0,906869954 | CDC123 | FALSE | Downregulated |
| 0,100044082 | 0,906869954 | CDC16 | FALSE | Upregulated |
| -0,634539933 | 0,906869954 | EEF1A2 | FALSE | Downregulated |
| 0,176811116 | 0,906869954 | LOC102724488 | FALSE | Upregulated |
| -0,161638269 | 0,906869954 | NDRG3 | FALSE | Downregulated |
| 0,11009493 | 0,906869954 | ZNF549 | FALSE | Upregulated |
| -0,43151401 | 0,906975703 | H2AFB2 | FALSE | Downregulated |
| -0,160582921 | 0,906989131 | OR52D1 | FALSE | Downregulated |
| 0,108437366 | 0,907152869 | USB1 | FALSE | Upregulated |
| -0,284400304 | 0,907185276 | FRMPD1 | FALSE | Downregulated |
| -0,083978437 | 0,907185276 | PYGO2 | FALSE | Downregulated |
| 0,174232498 | 0,907185276 | WDR54 | FALSE | Upregulated |
| 0,126226901 | 0,907185276 | ZC3H12C | FALSE | Upregulated |
| -0,12794432 | 0,907185276 | ZDHHC12 | FALSE | Downregulated |
| 0,132682069 | 0,907185276 | ZNF385C | FALSE | Upregulated |
| -0,09151225 | 0,907349553 | BAG5 | FALSE | Downregulated |
| 0,247903727 | 0,907349553 | FMR1-AS1 | FALSE | Upregulated |
| 0,087679039 | 0,907721479 | WDR91 | FALSE | Upregulated |
| -0,150767482 | 0,907947913 | NSRP1 | FALSE | Downregulated |
| -0,069535653 | 0,907947913 | SLC5A3 | FALSE | Downregulated |
| -0,191677568 | 0,908009335 | TSPEAR-AS1 | FALSE | Downregulated |
| -0,230677692 | 0,908009335 | ZNF667-AS1 | FALSE | Downregulated |
| 0,056415913 | 0,90804942 | CBX5 | FALSE | Upregulated |
| 0,284119322 | 0,90804942 | LOC102723767 | FALSE | Upregulated |
| -0,089837359 | 0,90804942 | NOA1 | FALSE | Downregulated |
| -0,050064032 | 0,908115023 | BCR | FALSE | Downregulated |
| -0,189692169 | 0,908115023 | PLSCR3 | FALSE | Downregulated |
| 0,157650427 | 0,908115023 | RCCD1 | FALSE | Upregulated |
| 0,252431472 | 0,908115023 | VIPR2 | FALSE | Upregulated |
| 0,20465519 | 0,908139713 | SS18L2 | FALSE | Upregulated |
| 0,077637841 | 0,908166895 | TMED4 | FALSE | Upregulated |
| 0,042716243 | 0,908166895 | TPCN1 | FALSE | Upregulated |

| | | | | |
|---|---|---|---|---|
| 0,050601581 | 0,908204582 | CIRBP | FALSE | Upregulated |
| 0,051780043 | 0,908222045 | PABPC4 | FALSE | Upregulated |
| -0,063689952 | 0,908308297 | CHD7 | FALSE | Downregulated |
| -0,037615538 | 0,908378605 | FARP1 | FALSE | Downregulated |
| -0,124049921 | 0,908378605 | MON1A | FALSE | Downregulated |
| 0,131546982 | 0,908378605 | MYADM | FALSE | Upregulated |
| -0,077084669 | 0,908378605 | SGMS2 | FALSE | Downregulated |
| 0,205159577 | 0,908378605 | SLC14A1 | FALSE | Upregulated |
| 0,16155856 | 0,908378605 | WDR35 | FALSE | Upregulated |
| 0,096251256 | 0,908378605 | ZBTB14 | FALSE | Upregulated |
| -0,048315246 | 0,908408888 | ARIH1 | FALSE | Downregulated |
| 0,095396199 | 0,908951351 | ORC2 | FALSE | Upregulated |
| -0,036909304 | 0,908992939 | ARID1B | FALSE | Downregulated |
| 0,22134868 | 0,909006826 | TMEM198 | FALSE | Upregulated |
| 0,187053954 | 0,909009901 | C1QTNF1-AS1 | FALSE | Upregulated |
| 0,078078365 | 0,909009901 | GSK3A | FALSE | Upregulated |
| -0,126641286 | 0,909009901 | LOC101927850 | FALSE | Downregulated |
| 0,903065075 | 0,909009901 | LOC102724219 | FALSE | Upregulated |
| 0,137855768 | 0,909009901 | MTMR3 | FALSE | Upregulated |
| -0,059149485 | 0,909009901 | WASL | FALSE | Downregulated |
| -0,117019995 | 0,909060526 | DHX40 | FALSE | Downregulated |
| -0,119934971 | 0,909060526 | TSPYL2 | FALSE | Downregulated |
| 0,165561834 | 0,909336383 | LOC101928505 | FALSE | Upregulated |
| -0,199482911 | 0,909336383 | NR2E3 | FALSE | Downregulated |
| -0,088927035 | 0,909336383 | TGFBRAP1 | FALSE | Downregulated |
| -0,209783366 | 0,909347849 | LOC102724002 | FALSE | Downregulated |
| 0,161645191 | 0,909487094 | CYYR1 | FALSE | Upregulated |
| -0,218283727 | 0,909487094 | LINC00623 | FALSE | Downregulated |
| -0,059573044 | 0,909586663 | UGGT1 | FALSE | Downregulated |
| -0,049686341 | 0,909646782 | AK2 | FALSE | Downregulated |
| -0,118414708 | 0,909646782 | HNRNPLL | FALSE | Downregulated |
| -0,359167037 | 0,909646782 | STAC | FALSE | Downregulated |
| 0,131583274 | 0,909780785 | RAB30 | FALSE | Upregulated |
| 0,134921213 | 0,909802657 | AC091729.9 | FALSE | Upregulated |
| 0,113578056 | 0,909802657 | ARL13B | FALSE | Upregulated |
| -0,224057522 | 0,909802657 | C4orf21 | FALSE | Downregulated |
| 0,086015177 | 0,909815116 | UNC45A | FALSE | Upregulated |
| -0,134538178 | 0,909839999 | ETAA1 | FALSE | Downregulated |
| 0,057538675 | 0,909839999 | STK35 | FALSE | Upregulated |
| -0,054570373 | 0,909851724 | HMGB1 | FALSE | Downregulated |
| 0,164520163 | 0,909875719 | DCAF4 | FALSE | Upregulated |
| 0,051167884 | 0,909875719 | FARP2 | FALSE | Upregulated |
| 0,107094565 | 0,909875719 | KIAA1683 | FALSE | Upregulated |
| -0,057971261 | 0,909875719 | NCOA1 | FALSE | Downregulated |
| -0,156317141 | 0,909875719 | PMS2P3 | FALSE | Downregulated |
| 0,152931942 | 0,909875719 | SYN1 | FALSE | Upregulated |
| -0,088179571 | 0,909875719 | TSPAN6 | FALSE | Downregulated |
| 0,081024616 | 0,909875719 | ZBTB17 | FALSE | Upregulated |
| -0,149296875 | 0,909875719 | ZNF131 | FALSE | Downregulated |
| 0,093755482 | 0,910059364 | CLMP | FALSE | Upregulated |
| -0,068395852 | 0,910059364 | PSD3 | FALSE | Downregulated |

| | | | | |
|---|---|---|---|---|
| -0,054022872 | 0,910084951 | UBA1 | FALSE | Downregulated |
| 0,076615461 | 0,910171941 | GFOD1 | FALSE | Upregulated |
| -0,135008307 | 0,910171941 | IL33 | FALSE | Downregulated |
| -0,179014826 | 0,910171941 | LINC00649 | FALSE | Downregulated |
| -0,219048875 | 0,910171941 | LOC102723996 | FALSE | Downregulated |
| 0,080213287 | 0,910171941 | NID2 | FALSE | Upregulated |
| -0,066401477 | 0,910171941 | SLC43A1 | FALSE | Downregulated |
| 0,093961564 | 0,910171941 | TRMU | FALSE | Upregulated |
| 0,135010672 | 0,910171941 | ZNF548 | FALSE | Upregulated |
| -0,141602476 | 0,910331884 | EAPP | FALSE | Downregulated |
| -0,160984986 | 0,910351945 | LOC286437 | FALSE | Downregulated |
| 0,361267 | 0,910666921 | LOC102724545 | FALSE | Upregulated |
| -0,163073005 | 0,911015152 | OR5AK2 | FALSE | Downregulated |
| -0,119784955 | 0,911061543 | SNUPN | FALSE | Downregulated |
| 0,1560573 | 0,911107256 | ZMYM5 | FALSE | Upregulated |
| -0,194527589 | 0,911114013 | TMEM26 | FALSE | Downregulated |
| -0,391564211 | 0,911164608 | AKAP2 | FALSE | Downregulated |
| -0,197360685 | 0,911164608 | LOC101927759 | FALSE | Downregulated |
| -0,15013572 | 0,911164608 | MCF2L-AS1 | FALSE | Downregulated |
| -0,113973723 | 0,911164608 | NAA35 | FALSE | Downregulated |
| 0,044628728 | 0,911164608 | SCRIB | FALSE | Upregulated |
| -0,109609374 | 0,9112249 | IFT20 | FALSE | Downregulated |
| 0,122436818 | 0,911295431 | HIAT1 | FALSE | Upregulated |
| -0,098610225 | 0,911295431 | KCTD17 | FALSE | Downregulated |
| -0,096051349 | 0,911295431 | KLHL8 | FALSE | Downregulated |
| 0,197105906 | 0,911295431 | LSM2 | FALSE | Upregulated |
| 0,089359905 | 0,911295431 | NANS | FALSE | Upregulated |
| 0,062296806 | 0,911295431 | SLAIN2 | FALSE | Upregulated |
| -0,075750484 | 0,911295431 | STXBP2 | FALSE | Downregulated |
| -0,135490537 | 0,91132556 | AGTPBP1 | FALSE | Downregulated |
| -0,163878908 | 0,91132556 | EGR2 | FALSE | Downregulated |
| -0,162055889 | 0,91132556 | FKBP11 | FALSE | Downregulated |
| 0,063624112 | 0,91132556 | FLT4 | FALSE | Upregulated |
| 0,254119574 | 0,91132556 | PRSS50 | FALSE | Upregulated |
| -0,155884775 | 0,91132556 | RBM18 | FALSE | Downregulated |
| -0,125424373 | 0,91132556 | SACM1L | FALSE | Downregulated |
| -0,118342534 | 0,91132556 | TSEN34 | FALSE | Downregulated |
| 0,079491829 | 0,91132556 | ZNF768 | FALSE | Upregulated |
| -0,049546109 | 0,911453674 | KIAA0368 | FALSE | Downregulated |
| 0,223221028 | 0,911453674 | LOC101926935 | FALSE | Upregulated |
| -0,074540693 | 0,911453674 | MEIS2 | FALSE | Downregulated |
| -0,091221716 | 0,911453674 | PARVA | FALSE | Downregulated |
| 0,03566275 | 0,911613076 | ANKFY1 | FALSE | Upregulated |
| -0,066604266 | 0,911613076 | GNAQ | FALSE | Downregulated |
| 0,077050771 | 0,911613076 | ZNF263 | FALSE | Upregulated |
| 0,242677218 | 0,911639224 | EMCN | FALSE | Upregulated |
| -0,080826431 | 0,91168588 | XPOT | FALSE | Downregulated |
| 0,113168865 | 0,911710001 | CHCHD3 | FALSE | Upregulated |
| -0,305348969 | 0,911710001 | LOC102724348 | FALSE | Downregulated |
| -0,064129063 | 0,911710001 | REST | FALSE | Downregulated |
| 0,076227869 | 0,911765312 | ARL1 | FALSE | Upregulated |

| | | | | |
|---|---|---|---|---|
| 0,074058761 | 0,911765312 | C2orf68 | FALSE | Upregulated |
| 0,16934395 | 0,911765312 | CYB5R4 | FALSE | Upregulated |
| -0,150426768 | 0,911765312 | SNRPA1 | FALSE | Downregulated |
| -0,209579112 | 0,91193619 | CRP | FALSE | Downregulated |
| 0,130297018 | 0,912000467 | UBE2S | FALSE | Upregulated |
| 0,108872954 | 0,912159171 | FADD | FALSE | Upregulated |
| 0,073130555 | 0,912184954 | ITGA2 | FALSE | Upregulated |
| -0,129554046 | 0,912184954 | LOC100134822 | FALSE | Downregulated |
| 0,095413297 | 0,912231469 | ALDH3B1 | FALSE | Upregulated |
| 0,159593468 | 0,912231469 | C17orf75 | FALSE | Upregulated |
| -0,195382317 | 0,912377568 | CMC4 | FALSE | Downregulated |
| -0,175464113 | 0,912377568 | MSANTD3 | FALSE | Downregulated |
| 0,191300473 | 0,912377568 | RAB28 | FALSE | Upregulated |
| 0,094653155 | 0,912377568 | RHOD | FALSE | Upregulated |
| 0,074914482 | 0,912377568 | TBC1D10C | FALSE | Upregulated |
| -0,142898034 | 0,912377568 | TENM2 | FALSE | Downregulated |
| 0,155584668 | 0,912442903 | AP3M2 | FALSE | Upregulated |
| -0,125026912 | 0,912442903 | CD320 | FALSE | Downregulated |
| -0,120592172 | 0,912651643 | EID2 | FALSE | Downregulated |
| -0,201866947 | 0,912756862 | ARTN | FALSE | Downregulated |
| -0,065114675 | 0,912756862 | DHX30 | FALSE | Downregulated |
| 0,095025062 | 0,912756862 | ECT2L | FALSE | Upregulated |
| -0,074725109 | 0,912756862 | FNIP1 | FALSE | Downregulated |
| -0,034221328 | 0,912756862 | LOC100129034 | FALSE | Downregulated |
| -0,084339217 | 0,912756862 | PCNXL4 | FALSE | Downregulated |
| 0,077595173 | 0,912756862 | PGAP3 | FALSE | Upregulated |
| -0,100247307 | 0,912756862 | PIDD | FALSE | Downregulated |
| -0,129495759 | 0,912756862 | PRPF4 | FALSE | Downregulated |
| -0,124440723 | 0,912756862 | ZBTB25 | FALSE | Downregulated |
| 0,09771306 | 0,912756862 | ZNF500 | FALSE | Upregulated |
| 0,235314388 | 0,912863067 | LINC00339 | FALSE | Upregulated |
| 0,294748395 | 0,912863067 | OSTCP1 | FALSE | Upregulated |
| -0,160473147 | 0,912885683 | CWC27 | FALSE | Downregulated |
| -0,27012865 | 0,913070802 | LOC101929540 | FALSE | Downregulated |
| 0,076517881 | 0,913278361 | NRP2 | FALSE | Upregulated |
| -0,103495671 | 0,913420477 | NR4A3 | FALSE | Downregulated |
| -0,222871366 | 0,913420477 | PPAN | FALSE | Downregulated |
| -0,091834657 | 0,913458998 | DYM | FALSE | Downregulated |
| 0,170896617 | 0,913458998 | LOC400043 | FALSE | Upregulated |
| -0,390923429 | 0,913458998 | MYOZ2 | FALSE | Downregulated |
| 0,060124517 | 0,913578935 | CHD1 | FALSE | Upregulated |
| -0,058975672 | 0,913815666 | CRNKL1 | FALSE | Downregulated |
| 0,219653168 | 0,913815666 | LOC102723989 | FALSE | Upregulated |
| 0,106915581 | 0,913815666 | PRKACB | FALSE | Upregulated |
| 0,120376926 | 0,913815666 | TBCB | FALSE | Upregulated |
| 0,19350808 | 0,91388793 | CRHR2 | FALSE | Upregulated |
| -0,046806373 | 0,91388793 | ENG | FALSE | Downregulated |
| 0,175874191 | 0,91388793 | NPIPA1 | FALSE | Upregulated |
| 0,052097108 | 0,91388793 | RP11-367J11.2 | FALSE | Upregulated |
| -0,742571102 | 0,914198802 | BLOC1S1-RDH5 | FALSE | Downregulated |
| -0,081710579 | 0,914198802 | C15orf38 | FALSE | Downregulated |

| | | | | |
|---|---|---|---|---|
| -0,122193526 | 0,914198802 | CDK5RAP1 | FALSE | Downregulated |
| -0,082905894 | 0,914198802 | GLCCI1 | FALSE | Downregulated |
| -0,099594316 | 0,914198802 | OSTF1 | FALSE | Downregulated |
| -0,178405638 | 0,914198802 | PDRG1 | FALSE | Downregulated |
| -0,077052658 | 0,914198802 | SEMA4C | FALSE | Downregulated |
| 0,076500831 | 0,914198802 | SMCHD1 | FALSE | Upregulated |
| 0,039733636 | 0,914198802 | TGOLN2 | FALSE | Upregulated |
| 0,060118634 | 0,914198802 | TRAPPC9 | FALSE | Upregulated |
| 0,110389679 | 0,91434667 | CPVL | FALSE | Upregulated |
| -0,273560498 | 0,91434667 | IRX3 | FALSE | Downregulated |
| -0,105747258 | 0,91434667 | LOC101927185 | FALSE | Downregulated |
| -0,220254787 | 0,91434667 | SLC18B1 | FALSE | Downregulated |
| 0,193422979 | 0,91434667 | SLC25A21 | FALSE | Upregulated |
| -0,06413721 | 0,91434667 | ZNF655 | FALSE | Downregulated |
| 0,078367908 | 0,914347915 | NAP1L5 | FALSE | Upregulated |
| 0,035350344 | 0,914347915 | RAD23B | FALSE | Upregulated |
| 0,108041796 | 0,914347915 | SLC7A8 | FALSE | Upregulated |
| 0,140176872 | 0,914347915 | WDR61 | FALSE | Upregulated |
| 0,264653418 | 0,91435111 | RP11-875O11.2 | FALSE | Upregulated |
| 0,117031374 | 0,914428399 | DHX57 | FALSE | Upregulated |
| 0,367588735 | 0,914462882 | BOLA2 | FALSE | Upregulated |
| -0,076138235 | 0,914462882 | CNTLN | FALSE | Downregulated |
| 0,078038038 | 0,914462882 | MZT2A | FALSE | Upregulated |
| 0,141748138 | 0,914462882 | NDUFA8 | FALSE | Upregulated |
| 0,117898085 | 0,914462882 | NIP7 | FALSE | Upregulated |
| -0,105717484 | 0,914586385 | KATNB1 | FALSE | Downregulated |
| -0,082831494 | 0,914586385 | SPIN2A | FALSE | Downregulated |
| 0,202599599 | 0,914803822 | LOC100505761 | FALSE | Upregulated |
| -0,070278161 | 0,91482049 | FUK | FALSE | Downregulated |
| 0,124441972 | 0,91482049 | PKNOX1 | FALSE | Upregulated |
| -0,141024741 | 0,914861108 | CSTB | FALSE | Downregulated |
| 0,260014225 | 0,914861108 | FAHD2B | FALSE | Upregulated |
| -0,171566179 | 0,915005725 | STK11IP | FALSE | Downregulated |
| 0,06639846 | 0,915079258 | FKBP2 | FALSE | Upregulated |
| 0,170384877 | 0,915079258 | PPCDC | FALSE | Upregulated |
| -0,049965455 | 0,915107621 | ARNT | FALSE | Downregulated |
| -0,100183031 | 0,915107621 | METTL17 | FALSE | Downregulated |
| 0,04838003 | 0,915107621 | POLR2J4 | FALSE | Upregulated |
| -0,132694124 | 0,915107621 | PRKXP1 | FALSE | Downregulated |
| 0,105413831 | 0,91511445 | C1QTNF7 | FALSE | Upregulated |
| 0,095865781 | 0,915266135 | LOC101927644 | FALSE | Upregulated |
| -0,137583943 | 0,915340318 | CCDC183-AS1 | FALSE | Downregulated |
| 0,098994417 | 0,91542941 | C10orf76 | FALSE | Upregulated |
| 0,199000963 | 0,91542941 | C14orf28 | FALSE | Upregulated |
| 0,216028807 | 0,91542941 | CCDC113 | FALSE | Upregulated |
| -0,054157588 | 0,91542941 | DIP2C | FALSE | Downregulated |
| -0,118596957 | 0,91542941 | EFHD1 | FALSE | Downregulated |
| -0,144919104 | 0,91542941 | FANCM | FALSE | Downregulated |
| -0,059428186 | 0,91542941 | PLA2G12B | FALSE | Downregulated |
| -0,064333645 | 0,91542941 | RALY | FALSE | Downregulated |
| 0,055301078 | 0,91542941 | SOS1 | FALSE | Upregulated |

| | | | | |
|---|---|---|---|---|
| 0,316930586 | 0,91542941 | STK32A | FALSE | Upregulated |
| -0,090287583 | 0,91542941 | TAF6 | FALSE | Downregulated |
| 0,090516071 | 0,91542941 | TTI1 | FALSE | Upregulated |
| -0,065321802 | 0,915497864 | COG8 | FALSE | Downregulated |
| -0,047526716 | 0,915497864 | RPTOR | FALSE | Downregulated |
| -0,302946716 | 0,915644508 | PPP1R26-AS1 | FALSE | Downregulated |
| -0,07830336 | 0,915644508 | ZNF428 | FALSE | Downregulated |
| -0,034530865 | 0,916062718 | SETD5 | FALSE | Downregulated |
| 0,060912667 | 0,916200607 | MADD | FALSE | Upregulated |
| 0,121327735 | 0,916211116 | ZNF471 | FALSE | Upregulated |
| -0,071510332 | 0,916387458 | FAM91A1 | FALSE | Downregulated |
| -0,243402195 | 0,916387458 | SMPDL3B | FALSE | Downregulated |
| -0,289248271 | 0,916395458 | LINC01144 | FALSE | Downregulated |
| 0,103677194 | 0,916406294 | PPARGC1B | FALSE | Upregulated |
| -0,081269721 | 0,916484774 | MORN4 | FALSE | Downregulated |
| -0,094180442 | 0,916802497 | AREL1 | FALSE | Downregulated |
| 0,450088435 | 0,916802497 | CXCL13 | FALSE | Upregulated |
| -0,054119285 | 0,916802497 | DMXL1 | FALSE | Downregulated |
| -0,073983804 | 0,916802497 | ENTPD6 | FALSE | Downregulated |
| -0,060734947 | 0,916802497 | HEBP1 | FALSE | Downregulated |
| -0,270681098 | 0,916802497 | ILDR2 | FALSE | Downregulated |
| -0,240872236 | 0,916802497 | LOC100652758 | FALSE | Downregulated |
| -0,071019421 | 0,916802497 | MASTL | FALSE | Downregulated |
| 0,05699045 | 0,916802497 | PPP2R5D | FALSE | Upregulated |
| -0,093815844 | 0,916802497 | USP13 | FALSE | Downregulated |
| 0,112685042 | 0,916802497 | ZKSCAN4 | FALSE | Upregulated |
| -0,054595226 | 0,916802497 | ZXDC | FALSE | Downregulated |
| -0,086815383 | 0,916808154 | TOP3B | FALSE | Downregulated |
| 0,086995346 | 0,916848462 | LOC102723804 | FALSE | Upregulated |
| -0,081170873 | 0,916869812 | VMP1 | FALSE | Downregulated |
| 0,119813003 | 0,91688006 | ZNF287 | FALSE | Upregulated |
| -0,098775204 | 0,916904749 | RNASE10 | FALSE | Downregulated |
| -0,061464952 | 0,916909048 | MLLT1 | FALSE | Downregulated |
| 0,067166514 | 0,9170739 | ACSF3 | FALSE | Upregulated |
| -0,071284455 | 0,9170739 | BAG4 | FALSE | Downregulated |
| 0,040496164 | 0,9170739 | H3F3B | FALSE | Upregulated |
| 0,072148225 | 0,91726739 | PPP1R3E | FALSE | Upregulated |
| 0,18358074 | 0,91726739 | TLDC1 | FALSE | Upregulated |
| -0,061055047 | 0,91726739 | TM9SF4 | FALSE | Downregulated |
| -0,165090699 | 0,9173228 | FAAH2 | FALSE | Downregulated |
| 0,231242031 | 0,917370383 | NINJ2 | FALSE | Upregulated |
| -0,14468726 | 0,917370383 | RASL11B | FALSE | Downregulated |
| 0,075976544 | 0,91737359 | ACBD3 | FALSE | Upregulated |
| 0,09419489 | 0,917417057 | ARPC3 | FALSE | Upregulated |
| 0,074448554 | 0,917417057 | DHX36 | FALSE | Upregulated |
| -0,057638853 | 0,917417057 | FAT4 | FALSE | Downregulated |
| 0,096571647 | 0,917417057 | ZNF614 | FALSE | Upregulated |
| 0,143148059 | 0,917429324 | POLR3D | FALSE | Upregulated |
| -0,187180581 | 0,917436103 | ADAMTSL5 | FALSE | Downregulated |
| -0,064293754 | 0,917436103 | CEND1 | FALSE | Downregulated |
| 0,290967482 | 0,917436103 | CHRM3 | FALSE | Upregulated |

| | | | | |
|---|---|---|---|---|
| 0,083321539 | 0,917436103 | FBXO10 | FALSE | Upregulated |
| 0,118368955 | 0,917436103 | HAND2 | FALSE | Upregulated |
| -0,34884132 | 0,917436103 | ITLN1 | FALSE | Downregulated |
| 0,067234889 | 0,917436103 | LGMN | FALSE | Upregulated |
| 0,072943064 | 0,917436103 | LINC00174 | FALSE | Upregulated |
| -0,331937816 | 0,917436103 | LOC100506384 | FALSE | Downregulated |
| 0,155550677 | 0,917436103 | LOC102723542 | FALSE | Upregulated |
| -0,086774603 | 0,917436103 | NBPF19 | FALSE | Downregulated |
| 0,083756484 | 0,917436103 | NLRC4 | FALSE | Upregulated |
| -0,093208977 | 0,917436103 | ORAI1 | FALSE | Downregulated |
| -0,048677808 | 0,917436103 | PPIA | FALSE | Downregulated |
| -0,104820845 | 0,917436103 | PRMT5 | FALSE | Downregulated |
| -0,276075238 | 0,917436103 | SMIM2-AS1 | FALSE | Downregulated |
| -0,082716151 | 0,917436103 | TMEM120A | FALSE | Downregulated |
| -0,059679363 | 0,917436103 | TRPC4AP | FALSE | Downregulated |
| 0,082600848 | 0,917535915 | AKIRIN1 | FALSE | Upregulated |
| -0,080916813 | 0,917535915 | APBB1IP | FALSE | Downregulated |
| 0,381249138 | 0,917535915 | BIVM-ERCC5 | FALSE | Upregulated |
| -0,074038425 | 0,917535915 | RTCB | FALSE | Downregulated |
| -0,079894815 | 0,917535915 | USF1 | FALSE | Downregulated |
| 0,15711874 | 0,917592106 | DRD4 | FALSE | Upregulated |
| -0,06717836 | 0,917690943 | CD63 | FALSE | Downregulated |
| 0,141993509 | 0,917690943 | FAM131A | FALSE | Upregulated |
| -0,135223591 | 0,917690943 | FBXO45 | FALSE | Downregulated |
| 0,289227343 | 0,917690943 | IQCD | FALSE | Upregulated |
| 0,069572164 | 0,917690943 | SNAP23 | FALSE | Upregulated |
| 0,074694833 | 0,91774332 | HMG20A | FALSE | Upregulated |
| 0,065898428 | 0,91774332 | PLEKHM1 | FALSE | Upregulated |
| 0,085431608 | 0,91774332 | YKT6 | FALSE | Upregulated |
| 0,11894398 | 0,917756595 | GEMIN8P4 | FALSE | Upregulated |
| 0,087495715 | 0,917756595 | MAPRE1 | FALSE | Upregulated |
| -0,048177858 | 0,917756595 | SERINC1 | FALSE | Downregulated |
| -0,272740174 | 0,917787084 | LINC01146 | FALSE | Downregulated |
| -0,060500881 | 0,917787084 | UBN2 | FALSE | Downregulated |
| -0,196660495 | 0,917787084 | ZNF669 | FALSE | Downregulated |
| 0,066621231 | 0,917794749 | CAPS | FALSE | Upregulated |
| -0,115924746 | 0,917794749 | KRCC1 | FALSE | Downregulated |
| -0,082419325 | 0,917794749 | LOC100996255 | FALSE | Downregulated |
| -0,264496331 | 0,917794749 | NPAP1 | FALSE | Downregulated |
| 0,2147899 | 0,917794749 | SLC38A6 | FALSE | Upregulated |
| -0,069970382 | 0,917794749 | STK38 | FALSE | Downregulated |
| 0,186510461 | 0,918085611 | FKBP5 | FALSE | Upregulated |
| -0,139115626 | 0,918085611 | LRRC37BP1 | FALSE | Downregulated |
| 0,094554103 | 0,918085611 | RRM2B | FALSE | Upregulated |
| 0,066902981 | 0,918085611 | UBXN4 | FALSE | Upregulated |
| 0,062423297 | 0,918094398 | C1orf63 | FALSE | Upregulated |
| -0,097183708 | 0,918102579 | ALDH1L1-AS2 | FALSE | Downregulated |
| 0,088132067 | 0,918102579 | ANKRD54 | FALSE | Upregulated |
| -0,124513703 | 0,918102579 | FADS1 | FALSE | Downregulated |
| 0,080914061 | 0,918102579 | KCNJ9 | FALSE | Upregulated |
| 0,251113123 | 0,918102579 | LOC101928168 | FALSE | Upregulated |

| | | | | |
|---|---|---|---|---|
| -0,071936921 | 0,918102579 | LOC101928717 | FALSE | Downregulated |
| 0,118919914 | 0,918102579 | LOC101929354 | FALSE | Upregulated |
| -0,221053162 | 0,918102579 | LOC102723827 | FALSE | Downregulated |
| -0,147075479 | 0,918102579 | MMRN1 | FALSE | Downregulated |
| 0,095813888 | 0,918102579 | MRE11A | FALSE | Upregulated |
| -0,100476248 | 0,918102579 | MRPL35 | FALSE | Downregulated |
| 0,191593272 | 0,918102579 | PCED1B | FALSE | Upregulated |
| 0,103101764 | 0,918102579 | PPM1K | FALSE | Upregulated |
| -0,063381823 | 0,918102579 | SRSF8 | FALSE | Downregulated |
| 0,155199506 | 0,918102579 | TRIM69 | FALSE | Upregulated |
| 0,103727135 | 0,918102579 | ZDHHC17 | FALSE | Upregulated |
| 0,111343679 | 0,918210772 | CAMK2B | FALSE | Upregulated |
| 1,625292503 | 0,918210772 | KRT2 | FALSE | Upregulated |
| 0,075077445 | 0,918521736 | E4F1 | FALSE | Upregulated |
| -0,518127127 | 0,918534089 | MEF2B | FALSE | Downregulated |
| 0,076988795 | 0,918546871 | MAPRE3 | FALSE | Upregulated |
| -0,080753571 | 0,91859201 | LOC100506258 | FALSE | Downregulated |
| 0,104054217 | 0,91859201 | ORC3 | FALSE | Upregulated |
| 0,099987721 | 0,91859201 | YDJC | FALSE | Upregulated |
| 0,071174484 | 0,918613596 | MDM4 | FALSE | Upregulated |
| 0,14230145 | 0,918628217 | LOC101928609 | FALSE | Upregulated |
| 0,058548755 | 0,918628217 | MLH3 | FALSE | Upregulated |
| 0,166317824 | 0,918633041 | PTPRN2 | FALSE | Upregulated |
| -0,084986304 | 0,918717898 | FBXO9 | FALSE | Downregulated |
| 0,256375762 | 0,918717898 | THUMPD2 | FALSE | Upregulated |
| -0,155340568 | 0,918717898 | ZNF597 | FALSE | Downregulated |
| -0,067608446 | 0,918750289 | HMCN1 | FALSE | Downregulated |
| -0,0791647 | 0,918750289 | LEO1 | FALSE | Downregulated |
| -0,12078706 | 0,918750289 | ST3GAL4-AS1 | FALSE | Downregulated |
| -0,049065271 | 0,918750289 | UGP2 | FALSE | Downregulated |
| 0,550248707 | 0,918786231 | ARL2-SNX15 | FALSE | Upregulated |
| -0,072647128 | 0,918786231 | BCL2L2 | FALSE | Downregulated |
| 0,061795024 | 0,918786231 | CYP3A5 | FALSE | Upregulated |
| -0,115098714 | 0,918786231 | MRPL53 | FALSE | Downregulated |
| 0,076065093 | 0,918786231 | SLC27A3 | FALSE | Upregulated |
| 0,347401604 | 0,918893881 | LOC101927871 | FALSE | Upregulated |
| -0,126449882 | 0,918925437 | KATNAL1 | FALSE | Downregulated |
| -0,184739325 | 0,918991091 | ALG1L | FALSE | Downregulated |
| -0,056986382 | 0,918991091 | ETS1 | FALSE | Downregulated |
| -0,161381388 | 0,918991091 | WISP3 | FALSE | Downregulated |
| -0,048725547 | 0,919028014 | SAMD4A | FALSE | Downregulated |
| 0,044639329 | 0,919035041 | REV1 | FALSE | Upregulated |
| 0,139660655 | 0,919035041 | RP11-173B14.5 | FALSE | Upregulated |
| 0,3279638 | 0,919035041 | RP13-228J13.1 | FALSE | Upregulated |
| -0,286725367 | 0,919095367 | LOC102724615 | FALSE | Downregulated |
| -0,088410398 | 0,919312212 | FAM171A1 | FALSE | Downregulated |
| -0,077420222 | 0,919312212 | FOXN2 | FALSE | Downregulated |
| -0,059313515 | 0,919466193 | RNH1 | FALSE | Downregulated |
| 0,054351418 | 0,919466193 | TXNDC11 | FALSE | Upregulated |
| -0,050742311 | 0,919554086 | C16orf72 | FALSE | Downregulated |
| 0,053015174 | 0,919554086 | ITPK1 | FALSE | Upregulated |

| | | | | |
|---|---|---|---|---|
| -0,158277288 | 0,919554086 | KANSL1-AS1 | FALSE | Downregulated |
| -0,270945606 | 0,919554086 | LRRC49 | FALSE | Downregulated |
| 0,094857208 | 0,919697364 | LOC101928499 | FALSE | Upregulated |
| 0,055456965 | 0,919988381 | AKTIP | FALSE | Upregulated |
| -0,307786324 | 0,919988381 | CALN1 | FALSE | Downregulated |
| 0,109021402 | 0,919988381 | CMYA5 | FALSE | Upregulated |
| -0,112994971 | 0,919988381 | MBLAC1 | FALSE | Downregulated |
| 0,058579578 | 0,919988381 | NAA30 | FALSE | Upregulated |
| -0,324655646 | 0,920030795 | LOC100630923 | FALSE | Downregulated |
| 0,150274848 | 0,920030795 | POLR2K | FALSE | Upregulated |
| 0,072859626 | 0,920030795 | PPDPF | FALSE | Upregulated |
| 0,105410387 | 0,920030795 | SCAMP3 | FALSE | Upregulated |
| -0,086961576 | 0,920030795 | TFAM | FALSE | Downregulated |
| -0,039438488 | 0,920200753 | CGNL1 | FALSE | Downregulated |
| -0,141593558 | 0,920200753 | FAM174B | FALSE | Downregulated |
| 0,087161484 | 0,920200753 | KCTD18 | FALSE | Upregulated |
| 0,104447611 | 0,920200753 | LSM7 | FALSE | Upregulated |
| -0,051769534 | 0,920483551 | TMEM30A | FALSE | Downregulated |
| 0,059585837 | 0,920496798 | MLXIP | FALSE | Upregulated |
| -0,164319025 | 0,920496798 | TAF12 | FALSE | Downregulated |
| -0,054919024 | 0,920549218 | AGPAT1 | FALSE | Downregulated |
| -0,093318446 | 0,920549218 | IMPA2 | FALSE | Downregulated |
| -0,044809767 | 0,920551548 | SRRT | FALSE | Downregulated |
| 0,07723406 | 0,920711033 | AP5B1 | FALSE | Upregulated |
| -0,169015851 | 0,920711033 | CISH | FALSE | Downregulated |
| 0,078667017 | 0,920711033 | CSE1L | FALSE | Upregulated |
| 0,170015169 | 0,920711033 | HOXB4 | FALSE | Upregulated |
| -0,139813787 | 0,920711033 | SCNN1A | FALSE | Downregulated |
| -0,152536159 | 0,920711033 | WNT5B | FALSE | Downregulated |
| 0,160936991 | 0,920737002 | PSMG3 | FALSE | Upregulated |
| 0,166172761 | 0,920781373 | DNAJC28 | FALSE | Upregulated |
| -0,212555201 | 0,920989342 | CYP26B1 | FALSE | Downregulated |
| -0,114679853 | 0,920989342 | GYPC | FALSE | Downregulated |
| -0,160490469 | 0,920989342 | ST20 | FALSE | Downregulated |
| -0,15198424 | 0,92102177 | ALKBH8 | FALSE | Downregulated |
| 0,0694938 | 0,92102177 | CCDC6 | FALSE | Upregulated |
| 0,075470054 | 0,92102177 | CPSF2 | FALSE | Upregulated |
| 0,061135352 | 0,92102177 | TNFAIP8L2 | FALSE | Upregulated |
| 0,07666464 | 0,92102177 | UTP20 | FALSE | Upregulated |
| -0,095969688 | 0,92102177 | ZNF268 | FALSE | Downregulated |
| -0,096550054 | 0,921022463 | CCDC101 | FALSE | Downregulated |
| 0,17694131 | 0,921022463 | PLCXD2 | FALSE | Upregulated |
| -0,051953983 | 0,921023122 | CYTH1 | FALSE | Downregulated |
| -0,090528425 | 0,921047295 | RAB37 | FALSE | Downregulated |
| 0,197048023 | 0,921055637 | CBWD1 | FALSE | Upregulated |
| 0,207093705 | 0,921097086 | RFPL3 | FALSE | Upregulated |
| 0,122378639 | 0,921301303 | TMEM39B | FALSE | Upregulated |
| -0,113487351 | 0,921384365 | PLSCR1 | FALSE | Downregulated |
| -0,087134482 | 0,921430844 | CMC2 | FALSE | Downregulated |
| -0,059113625 | 0,921510588 | YES1 | FALSE | Downregulated |
| 0,054737433 | 0,921510588 | ZSCAN30 | FALSE | Upregulated |

| | | | | |
|---|---|---|---|---|
| 0,094265323 | 0,921527947 | ADAM8 | FALSE | Upregulated |
| 0,248020027 | 0,921527947 | LOC101059936 | FALSE | Upregulated |
| -0,276136091 | 0,921610159 | BEND4 | FALSE | Downregulated |
| 0,265568999 | 0,921610159 | BOLA2B | FALSE | Upregulated |
| -0,146945748 | 0,921610159 | UBE2E2 | FALSE | Downregulated |
| -0,128441969 | 0,921611426 | DRAM2 | FALSE | Downregulated |
| 0,099847604 | 0,92170916 | HPS5 | FALSE | Upregulated |
| -0,065052393 | 0,921758051 | AGGF1 | FALSE | Downregulated |
| -0,182101157 | 0,921758051 | CCL3L3 | FALSE | Downregulated |
| 0,136746059 | 0,921758051 | DDIT4 | FALSE | Upregulated |
| 0,128166594 | 0,921758051 | INTS12 | FALSE | Upregulated |
| -0,09207631 | 0,921758051 | KIF22 | FALSE | Downregulated |
| -0,41872654 | 0,921758051 | MYOM3 | FALSE | Downregulated |
| -0,146206047 | 0,921758051 | NUBPL | FALSE | Downregulated |
| 0,066816401 | 0,921758051 | PDXDC2P | FALSE | Upregulated |
| 0,037560162 | 0,921758051 | PFN1 | FALSE | Upregulated |
| 0,114880994 | 0,921758051 | RNF145 | FALSE | Upregulated |
| -0,163654441 | 0,921758051 | TMEM38A | FALSE | Downregulated |
| 0,103860672 | 0,921758051 | UBXN11 | FALSE | Upregulated |
| -0,107361324 | 0,921760234 | DPH5 | FALSE | Downregulated |
| -0,193565224 | 0,921795308 | NTF3 | FALSE | Downregulated |
| 0,064756499 | 0,921832849 | HAX1 | FALSE | Upregulated |
| 0,062625293 | 0,921832849 | MYD88 | FALSE | Upregulated |
| -0,09885952 | 0,921955252 | TSN | FALSE | Downregulated |
| 0,090627557 | 0,921955252 | TXNL4A | FALSE | Upregulated |
| -0,08188411 | 0,922102065 | BIN1 | FALSE | Downregulated |
| 0,161080371 | 0,922102065 | GNG11 | FALSE | Upregulated |
| 0,277428365 | 0,922108732 | MBOAT2 | FALSE | Upregulated |
| 0,098525303 | 0,922274462 | DDAH2 | FALSE | Upregulated |
| 0,06048801 | 0,922274462 | DYNLL2 | FALSE | Upregulated |
| 0,180791229 | 0,922274462 | LOC100128079 | FALSE | Upregulated |
| 0,253240417 | 0,922274462 | LOC101927011 | FALSE | Upregulated |
| -0,117433096 | 0,922274462 | MRGPRF | FALSE | Downregulated |
| 0,057068448 | 0,922274462 | ZFR | FALSE | Upregulated |
| 0,133196741 | 0,922341087 | FAM76B | FALSE | Upregulated |
| 0,073870682 | 0,922402257 | FTO | FALSE | Upregulated |
| -0,229140345 | 0,922767563 | C15orf38-AP3S2 | FALSE | Downregulated |
| 0,088911681 | 0,922767563 | WEE1 | FALSE | Upregulated |
| 0,060979232 | 0,922830887 | UBE2J2 | FALSE | Upregulated |
| 0,199522477 | 0,922889365 | CDK20 | FALSE | Upregulated |
| 0,218079878 | 0,922889365 | LOC100130899 | FALSE | Upregulated |
| 0,075394146 | 0,92292423 | ANP32A | FALSE | Upregulated |
| -0,052659946 | 0,92292423 | GTF2IRD2B | FALSE | Downregulated |
| 0,046396656 | 0,92292423 | NPM1 | FALSE | Upregulated |
| 0,040929482 | 0,922965432 | WASF2 | FALSE | Upregulated |
| 0,103966794 | 0,92308456 | NOXA1 | FALSE | Upregulated |
| -0,326594665 | 0,923217515 | LOC101929583 | FALSE | Downregulated |
| -0,094482193 | 0,923441567 | RAB11FIP2 | FALSE | Downregulated |
| -0,063924782 | 0,92355706 | IDH3B | FALSE | Downregulated |
| 0,052646888 | 0,92355706 | OPA1 | FALSE | Upregulated |
| -0,223626991 | 0,92355706 | TTTY15 | FALSE | Downregulated |

| | | | | |
|---|---|---|---|---|
| 0,053567166 | 0,923569615 | CHD6 | FALSE | Upregulated |
| 0,078305565 | 0,923615783 | PCNP | FALSE | Upregulated |
| -0,085817277 | 0,923675774 | ALS2CR11 | FALSE | Downregulated |
| -0,119828854 | 0,923675774 | FIG4 | FALSE | Downregulated |
| -0,071267207 | 0,923675774 | TIAL1 | FALSE | Downregulated |
| -0,133181299 | 0,923735479 | FOXE1 | FALSE | Downregulated |
| 0,253324789 | 0,92397456 | LOC100506127 | FALSE | Upregulated |
| -0,074088848 | 0,92397456 | SSR2 | FALSE | Downregulated |
| -0,049647585 | 0,92397456 | THRAP3 | FALSE | Downregulated |
| -0,058187366 | 0,924311198 | HPS3 | FALSE | Downregulated |
| -0,078957945 | 0,924434629 | LINC00675 | FALSE | Downregulated |
| 0,059531194 | 0,924524936 | TNPO3 | FALSE | Upregulated |
| -0,136138437 | 0,924559821 | POLI | FALSE | Downregulated |
| -0,192245321 | 0,92457315 | IFT57 | FALSE | Downregulated |
| 0,070602147 | 0,92457315 | KIAA1199 | FALSE | Upregulated |
| 0,084915404 | 0,92457315 | PPP4R1 | FALSE | Upregulated |
| -0,14669304 | 0,92457315 | RNF180 | FALSE | Downregulated |
| -0,128202232 | 0,924598462 | FBXO24 | FALSE | Downregulated |
| -0,056926373 | 0,924598462 | FYCO1 | FALSE | Downregulated |
| -0,083305592 | 0,924719743 | CXCL14 | FALSE | Downregulated |
| -0,222096689 | 0,924778374 | LOC441124 | FALSE | Downregulated |
| 0,065798534 | 0,924831694 | KIAA1522 | FALSE | Upregulated |
| 0,116810842 | 0,924831694 | LOC100294145 | FALSE | Upregulated |
| 0,053407726 | 0,924881618 | ARFGAP1 | FALSE | Upregulated |
| 0,229771394 | 0,925039323 | TPSB2 | FALSE | Upregulated |
| -0,140975241 | 0,92505059 | ALG11 | FALSE | Downregulated |
| 0,339162236 | 0,92505059 | CCL15-CCL14 | FALSE | Upregulated |
| 0,180693973 | 0,92505059 | FAM127B | FALSE | Upregulated |
| 0,045461019 | 0,92505059 | HDAC5 | FALSE | Upregulated |
| -0,144990241 | 0,92505059 | RINL | FALSE | Downregulated |
| 0,0854583 | 0,92505059 | SAE1 | FALSE | Upregulated |
| 0,054474885 | 0,92505059 | TAX1BP1 | FALSE | Upregulated |
| -0,077775523 | 0,92505059 | TERF2 | FALSE | Downregulated |
| 0,091531617 | 0,925073934 | SVIP | FALSE | Upregulated |
| -0,066843266 | 0,925081315 | PIK3C2B | FALSE | Downregulated |
| 0,127325282 | 0,925084458 | DNAH10 | FALSE | Upregulated |
| -0,064482756 | 0,925178435 | CLPP | FALSE | Downregulated |
| 0,079054593 | 0,925236657 | R3HCC1 | FALSE | Upregulated |
| -0,127384552 | 0,925551701 | MED18 | FALSE | Downregulated |
| 0,04441578 | 0,925687159 | CAMTA2 | FALSE | Upregulated |
| 0,082707743 | 0,925929584 | ADAMTS5 | FALSE | Upregulated |
| 0,061493851 | 0,925929584 | C16orf45 | FALSE | Upregulated |
| -0,185262687 | 0,925929584 | CALCA | FALSE | Downregulated |
| -0,157790472 | 0,925929584 | GPM6A | FALSE | Downregulated |
| -0,08165024 | 0,925929584 | HMOX1 | FALSE | Downregulated |
| 0,080476337 | 0,925929584 | HNRNPUL2-BSCL2 | FALSE | Upregulated |
| -0,060821299 | 0,925929584 | ITFG3 | FALSE | Downregulated |
| -0,067118704 | 0,925929584 | KAT7 | FALSE | Downregulated |
| -0,106655259 | 0,925929584 | KIF9 | FALSE | Downregulated |
| 0,089900089 | 0,925929584 | ZBTB41 | FALSE | Upregulated |
| -0,033174417 | 0,926318334 | FAM20A | FALSE | Downregulated |

| | | | | |
|---|---|---|---|---|
| 0,061937528 | 0,926407484 | METTL3 | FALSE | Upregulated |
| -0,046873013 | 0,926407484 | NUP214 | FALSE | Downregulated |
| 0,037723327 | 0,926596464 | AP1G1 | FALSE | Upregulated |
| -0,062267788 | 0,926596464 | BCL6 | FALSE | Downregulated |
| 0,102764879 | 0,926596464 | FBXO30 | FALSE | Upregulated |
| -0,047805619 | 0,926596464 | VAV2 | FALSE | Downregulated |
| -0,0697179 | 0,926649794 | DCP1A | FALSE | Downregulated |
| -0,149046797 | 0,926817046 | LCN12 | FALSE | Downregulated |
| 0,305313868 | 0,926817046 | SPECC1L-ADORA2A | FALSE | Upregulated |
| -0,048046495 | 0,926931864 | HNRNPL | FALSE | Downregulated |
| 0,217225575 | 0,92693929 | LOC101929092 | FALSE | Upregulated |
| 0,100390139 | 0,92693929 | PGGT1B | FALSE | Upregulated |
| -0,047241148 | 0,92693929 | ZNF862 | FALSE | Downregulated |
| -0,131585629 | 0,926986114 | CDADC1 | FALSE | Downregulated |
| 0,223209549 | 0,927027291 | B4GALNT3 | FALSE | Upregulated |
| -0,053730071 | 0,927027291 | MKRN2 | FALSE | Downregulated |
| 0,132914796 | 0,927076787 | MTCP1 | FALSE | Upregulated |
| -0,043306211 | 0,927076787 | PTMA | FALSE | Downregulated |
| -0,098983334 | 0,927076787 | ZCRB1 | FALSE | Downregulated |
| 0,173946205 | 0,927165863 | CEP135 | FALSE | Upregulated |
| -0,066852096 | 0,927165863 | NT5DC1 | FALSE | Downregulated |
| 0,088504426 | 0,927165863 | PRMT9 | FALSE | Upregulated |
| 0,117809055 | 0,927165863 | WFIKKN1 | FALSE | Upregulated |
| -0,072838837 | 0,92716802 | MORC3 | FALSE | Downregulated |
| -0,076493821 | 0,927174727 | BCL3 | FALSE | Downregulated |
| -0,259022531 | 0,92721484 | CEACAM21 | FALSE | Downregulated |
| -0,076849272 | 0,92721484 | CHKB | FALSE | Downregulated |
| -0,055593747 | 0,92721484 | CLDN22 | FALSE | Downregulated |
| -0,074408124 | 0,92721484 | DPH6 | FALSE | Downregulated |
| -0,052071072 | 0,92721484 | NUP98 | FALSE | Downregulated |
| 0,044969551 | 0,92721484 | SBNO2 | FALSE | Upregulated |
| 0,215528287 | 0,927305565 | BLVRA | FALSE | Upregulated |
| -0,043347137 | 0,927305565 | C11orf95 | FALSE | Downregulated |
| 0,169157164 | 0,927305565 | CCDC183 | FALSE | Upregulated |
| 0,051605807 | 0,927305565 | FRYL | FALSE | Upregulated |
| 0,214081286 | 0,927305565 | GAL3ST2 | FALSE | Upregulated |
| 0,053959463 | 0,927305565 | GPR155 | FALSE | Upregulated |
| -0,315416135 | 0,927305565 | IL18RAP | FALSE | Downregulated |
| -0,185765474 | 0,927305565 | KNSTRN | FALSE | Downregulated |
| 0,313693308 | 0,927305565 | MMP9 | FALSE | Upregulated |
| 0,07774018 | 0,927305565 | PARG | FALSE | Upregulated |
| -0,17958185 | 0,927305565 | PCBP1-AS1 | FALSE | Downregulated |
| 0,081596765 | 0,927305565 | PRPS1 | FALSE | Upregulated |
| 0,067686006 | 0,927305565 | PSMD11 | FALSE | Upregulated |
| 0,171965926 | 0,927305565 | RP11-611D20.2 | FALSE | Upregulated |
| 0,071099561 | 0,927305565 | RP11-685M7.3 | FALSE | Upregulated |
| 0,089219185 | 0,927305565 | SNRPC | FALSE | Upregulated |
| -0,077282814 | 0,927305565 | TYSND1 | FALSE | Downregulated |
| -0,063733912 | 0,927305565 | ZNF586 | FALSE | Downregulated |
| 0,086539673 | 0,927400565 | SPAG5 | FALSE | Upregulated |
| -0,036199267 | 0,927408422 | MTSS1L | FALSE | Downregulated |

| | | | | |
|---|---|---|---|---|
| 0,104367337 | 0,92742474 | AP4E1 | FALSE | Upregulated |
| 0,05575193 | 0,927505792 | SUCLG1 | FALSE | Upregulated |
| 0,074506234 | 0,927524873 | GLRX | FALSE | Upregulated |
| -0,097096005 | 0,927583261 | COX14 | FALSE | Downregulated |
| 0,07702581 | 0,927583261 | FNBP1L | FALSE | Upregulated |
| 0,254380489 | 0,927583261 | H2AFB1 | FALSE | Upregulated |
| 0,066200658 | 0,927583261 | HYLS1 | FALSE | Upregulated |
| -0,105135151 | 0,927583261 | LOC102724372 | FALSE | Downregulated |
| 0,069081903 | 0,927583261 | NGRN | FALSE | Upregulated |
| 0,051091378 | 0,927583261 | PTPRA | FALSE | Upregulated |
| 0,077379278 | 0,927583261 | SLC44A2 | FALSE | Upregulated |
| 0,163055939 | 0,927583261 | ZNF567 | FALSE | Upregulated |
| 0,26059029 | 0,927657244 | LINC00238 | FALSE | Upregulated |
| 0,045585428 | 0,927714674 | RAI1 | FALSE | Upregulated |
| -0,499538649 | 0,927714674 | SCNM1 | FALSE | Downregulated |
| -0,042538775 | 0,927714674 | TNRC6A | FALSE | Downregulated |
| -0,054005232 | 0,927747472 | AUNIP | FALSE | Downregulated |
| 0,129056913 | 0,927747472 | DCTPP1 | FALSE | Upregulated |
| -0,062199461 | 0,927747472 | ELP2 | FALSE | Downregulated |
| 0,1553405 | 0,927747472 | GNN | FALSE | Upregulated |
| 0,174707615 | 0,927747472 | KRTCAP3 | FALSE | Upregulated |
| -0,216182371 | 0,927747472 | LOC100093631 | FALSE | Downregulated |
| -0,052131035 | 0,927747472 | NAB1 | FALSE | Downregulated |
| -0,036416917 | 0,927747472 | PDZK1 | FALSE | Downregulated |
| -0,206081227 | 0,927747472 | RNASEH2A | FALSE | Downregulated |
| 0,095204552 | 0,927747472 | SLC15A1 | FALSE | Upregulated |
| -0,04948014 | 0,927747472 | SLC39A7 | FALSE | Downregulated |
| -0,190138318 | 0,927747472 | TRPM1 | FALSE | Downregulated |
| 0,120209052 | 0,927747472 | ZNF543 | FALSE | Upregulated |
| 0,065944037 | 0,927760746 | CCDC9 | FALSE | Upregulated |
| -0,129586828 | 0,927855175 | LIN37 | FALSE | Downregulated |
| 0,216524933 | 0,927855175 | LOC101929574 | FALSE | Upregulated |
| -0,078694263 | 0,927933279 | DENND5B-AS1 | FALSE | Downregulated |
| -0,068348617 | 0,927933279 | GRAMD4 | FALSE | Downregulated |
| -0,09155049 | 0,927933279 | PAM | FALSE | Downregulated |
| -0,045904728 | 0,927937618 | ASXL2 | FALSE | Downregulated |
| 0,310041172 | 0,927937618 | LINC01106 | FALSE | Upregulated |
| -0,09514479 | 0,927963901 | ACCS | FALSE | Downregulated |
| -0,106393003 | 0,927963901 | ARHGEF35 | FALSE | Downregulated |
| 0,046278213 | 0,927963901 | ASB6 | FALSE | Upregulated |
| 0,052540052 | 0,927963901 | C2orf88 | FALSE | Upregulated |
| 0,082135958 | 0,927963901 | CTD-2201E9.1 | FALSE | Upregulated |
| 0,062881532 | 0,927963901 | CYP2D6 | FALSE | Upregulated |
| 0,202562755 | 0,927963901 | EXTL1 | FALSE | Upregulated |
| 0,07164099 | 0,927963901 | PRKAR2A-AS1 | FALSE | Upregulated |
| -0,078200075 | 0,927963901 | RUSC1-AS1 | FALSE | Downregulated |
| 0,053374883 | 0,927963901 | WDR55 | FALSE | Upregulated |
| -0,115656018 | 0,927963901 | ZSWIM3 | FALSE | Downregulated |
| -0,148632066 | 0,928274713 | DCUN1D5 | FALSE | Downregulated |
| 0,044068478 | 0,928274713 | GORASP1 | FALSE | Upregulated |
| 0,057783815 | 0,928274713 | LOC100505915 | FALSE | Upregulated |

| | | | | |
|---|---|---|---|---|
| 0,041767873 | 0,928274713 | OTUD5 | FALSE | Upregulated |
| 0,065212904 | 0,928274713 | SH3RF1 | FALSE | Upregulated |
| -0,122250761 | 0,928274713 | SRMS | FALSE | Downregulated |
| -0,032446902 | 0,928301517 | BPTF | FALSE | Downregulated |
| 0,032380855 | 0,928309899 | AP2A1 | FALSE | Upregulated |
| 0,131040633 | 0,928398543 | TMEM70 | FALSE | Upregulated |
| -0,112583025 | 0,928657152 | FAM175B | FALSE | Downregulated |
| 0,2265256 | 0,928859011 | CD200R1 | FALSE | Upregulated |
| 0,06305446 | 0,929055042 | LOC100288846 | FALSE | Upregulated |
| -0,151493038 | 0,9291617 | BMS1P20 | FALSE | Downregulated |
| 0,114301568 | 0,9291617 | ZNF566 | FALSE | Upregulated |
| -0,066500664 | 0,929161719 | PPP4R4 | FALSE | Downregulated |
| 0,05333553 | 0,929330519 | LOC101929426 | FALSE | Upregulated |
| -0,050322126 | 0,929398568 | AMBRA1 | FALSE | Downregulated |
| 0,07655834 | 0,929398568 | ANKRD19P | FALSE | Upregulated |
| -0,048422756 | 0,929398568 | BRD3 | FALSE | Downregulated |
| 0,080429692 | 0,929398568 | CYB561D1 | FALSE | Upregulated |
| -0,185937099 | 0,929398568 | FAM174A | FALSE | Downregulated |
| 0,047046691 | 0,929398568 | GGA3 | FALSE | Upregulated |
| 0,074348806 | 0,929398568 | IDS | FALSE | Upregulated |
| -0,065114637 | 0,929398568 | IFIT2 | FALSE | Downregulated |
| 0,054041984 | 0,929398568 | LAMC1 | FALSE | Upregulated |
| -0,061154019 | 0,929398568 | LOC101928690 | FALSE | Downregulated |
| 0,032496007 | 0,929398568 | MAST2 | FALSE | Upregulated |
| -0,046172804 | 0,929398568 | SLC39A1 | FALSE | Downregulated |
| -0,102074003 | 0,929398568 | ZNF792 | FALSE | Downregulated |
| -0,036730923 | 0,929423127 | DIDO1 | FALSE | Downregulated |
| -0,315891083 | 0,929423127 | IL20RB | FALSE | Downregulated |
| -0,150097908 | 0,929423127 | LOC646626 | FALSE | Downregulated |
| 0,247823772 | 0,929423127 | RTN2 | FALSE | Upregulated |
| -0,297585927 | 0,929423127 | SMIM2-IT1 | FALSE | Downregulated |
| 0,070051596 | 0,929443366 | LTB4R2 | FALSE | Upregulated |
| 0,090014819 | 0,929443366 | TSTD3 | FALSE | Upregulated |
| -0,053598008 | 0,929558473 | ACTR1B | FALSE | Downregulated |
| -0,134513427 | 0,929722391 | AAMDC | FALSE | Downregulated |
| 0,057206861 | 0,929722391 | ANAPC2 | FALSE | Upregulated |
| -0,138328804 | 0,929722391 | C11orf80 | FALSE | Downregulated |
| 0,039445784 | 0,929722391 | CNDP2 | FALSE | Upregulated |
| -0,134744039 | 0,929722391 | E2F6 | FALSE | Downregulated |
| 0,164469966 | 0,929722391 | GPD1L | FALSE | Upregulated |
| -0,205212284 | 0,929722391 | HSD11B1L | FALSE | Downregulated |
| 0,090430911 | 0,929722391 | IDI1 | FALSE | Upregulated |
| 0,085419376 | 0,929722391 | MAGI2 | FALSE | Upregulated |
| 0,074910757 | 0,929722391 | MAP2K4 | FALSE | Upregulated |
| 0,19764908 | 0,929722391 | MITD1 | FALSE | Upregulated |
| -0,062343668 | 0,929722391 | NELFB | FALSE | Downregulated |
| -0,084623165 | 0,929722391 | NOL10 | FALSE | Downregulated |
| -0,082250835 | 0,929722391 | POLR2D | FALSE | Downregulated |
| -0,06613549 | 0,929722391 | RGPD3 | FALSE | Downregulated |
| 0,107865289 | 0,929722391 | RRAGB | FALSE | Upregulated |
| -0,169138744 | 0,929722391 | SDCBP2 | FALSE | Downregulated |

| | | | | |
|---|---|---|---|---|
| -0,179341842 | 0,929722391 | SEMA4F | FALSE | Downregulated |
| -0,069707496 | 0,929722391 | SFI1 | FALSE | Downregulated |
| -0,04183039 | 0,929722391 | STEAP3 | FALSE | Downregulated |
| 0,054824504 | 0,929722391 | UCKL1 | FALSE | Upregulated |
| 0,147512151 | 0,929722391 | ZNF611 | FALSE | Upregulated |
| -0,050189943 | 0,929879412 | FKBP1A | FALSE | Downregulated |
| 0,209897287 | 0,929983318 | LINC00654 | FALSE | Upregulated |
| 0,090937303 | 0,930040512 | ANKRD13C | FALSE | Upregulated |
| 0,078287759 | 0,930040512 | EPC2 | FALSE | Upregulated |
| -0,260351136 | 0,930211069 | C2orf73 | FALSE | Downregulated |
| -0,115397403 | 0,930211069 | PDE4A | FALSE | Downregulated |
| 0,042766213 | 0,930340407 | ARG2 | FALSE | Upregulated |
| 0,19341079 | 0,930394356 | LOC102723465 | FALSE | Upregulated |
| 0,278810716 | 0,930394356 | PAX5 | FALSE | Upregulated |
| -0,221761495 | 0,930446979 | ADC | FALSE | Downregulated |
| 0,222458942 | 0,930446979 | LIMS3-LOC440895 | FALSE | Upregulated |
| -0,25068699 | 0,930478782 | AC006272.1 | FALSE | Downregulated |
| 0,184480491 | 0,930478782 | CDC42EP5 | FALSE | Upregulated |
| 0,295236177 | 0,930478782 | PRDM6 | FALSE | Upregulated |
| 0,038322524 | 0,930478782 | PSAP | FALSE | Upregulated |
| 0,133222026 | 0,930478782 | TWISTNB | FALSE | Upregulated |
| -0,055575783 | 0,930548824 | ABCB11 | FALSE | Downregulated |
| -0,060289996 | 0,930548824 | AMN1 | FALSE | Downregulated |
| 0,109294418 | 0,930548824 | CCDC17 | FALSE | Upregulated |
| 0,037115055 | 0,930548824 | CTSZ | FALSE | Upregulated |
| -0,082066808 | 0,930548824 | GNG5 | FALSE | Downregulated |
| 0,069089905 | 0,930548824 | ITFG1 | FALSE | Upregulated |
| 0,066756265 | 0,930548824 | LOC102724016 | FALSE | Upregulated |
| -0,122706498 | 0,930548824 | PARP11 | FALSE | Downregulated |
| -0,040189953 | 0,930548824 | PITPNA | FALSE | Downregulated |
| 0,064271586 | 0,930548824 | ZNRF3-AS1 | FALSE | Upregulated |
| -0,069737765 | 0,930695265 | CTPS2 | FALSE | Downregulated |
| -0,094471034 | 0,930695265 | RPA2 | FALSE | Downregulated |
| -0,17046815 | 0,930713841 | TMEM187 | FALSE | Downregulated |
| -0,065999591 | 0,930791257 | TMEM44 | FALSE | Downregulated |
| -0,18455587 | 0,930807162 | DUSP19 | FALSE | Downregulated |
| -0,053357508 | 0,930812174 | CEP170 | FALSE | Downregulated |
| -0,093959141 | 0,930857102 | AZI1 | FALSE | Downregulated |
| -0,04645587 | 0,930857102 | C11orf57 | FALSE | Downregulated |
| -0,209492079 | 0,930857102 | C7orf60 | FALSE | Downregulated |
| -0,038903833 | 0,930857102 | CDV3 | FALSE | Downregulated |
| 0,217120219 | 0,930857102 | LINC00881 | FALSE | Upregulated |
| 0,050454061 | 0,930857102 | ZNF517 | FALSE | Upregulated |
| 0,226603714 | 0,930857818 | ASB4 | FALSE | Upregulated |
| 0,162446122 | 0,930857818 | EBLN2 | FALSE | Upregulated |
| -0,062829819 | 0,930857818 | EIF2AK2 | FALSE | Downregulated |
| 0,297814277 | 0,930857818 | GIMAP5 | FALSE | Upregulated |
| -0,067426312 | 0,930857818 | KIAA0141 | FALSE | Downregulated |
| 0,255772265 | 0,930857818 | LOC100996716 | FALSE | Upregulated |
| 0,058883345 | 0,930857818 | LOC102724535 | FALSE | Upregulated |
| 0,22718562 | 0,930857818 | LOC285556 | FALSE | Upregulated |

| | | | | |
|---|---|---|---|---|
| 0,079264481 | 0,930857818 | MAPK13 | FALSE | Upregulated |
| 0,049198393 | 0,930857818 | MIER1 | FALSE | Upregulated |
| -0,049814741 | 0,930857818 | PDCD6IP | FALSE | Downregulated |
| 0,044809857 | 0,930857818 | PTTG1IP | FALSE | Upregulated |
| -0,075322898 | 0,930857818 | RLF | FALSE | Downregulated |
| -0,103131671 | 0,930857818 | TMEM253 | FALSE | Downregulated |
| -0,13453182 | 0,931098174 | PRICKLE3 | FALSE | Downregulated |
| 0,219574271 | 0,931098174 | TMEM191A | FALSE | Upregulated |
| 0,133366401 | 0,931313415 | C1orf52 | FALSE | Upregulated |
| 0,095702678 | 0,931313415 | C9orf91 | FALSE | Upregulated |
| -0,093995493 | 0,931313415 | EFTUD1 | FALSE | Downregulated |
| -0,105499688 | 0,931313415 | HAUS3 | FALSE | Downregulated |
| 0,029315267 | 0,931313415 | IGF2R | FALSE | Upregulated |
| 0,225113451 | 0,931313415 | LOC102724378 | FALSE | Upregulated |
| -0,07396849 | 0,931322225 | ACOT7 | FALSE | Downregulated |
| -0,049062897 | 0,931322225 | C4orf19 | FALSE | Downregulated |
| 0,170487337 | 0,931322225 | IKBIP | FALSE | Upregulated |
| 0,140219272 | 0,931322225 | MAPK12 | FALSE | Upregulated |
| 0,091736269 | 0,931322225 | MEIS1 | FALSE | Upregulated |
| -0,062114936 | 0,931322225 | NOMO1 | FALSE | Downregulated |
| -0,067738735 | 0,931322225 | P2RX4 | FALSE | Downregulated |
| 0,08466502 | 0,931322225 | SIMC1 | FALSE | Upregulated |
| -0,119815311 | 0,931322225 | SLC41A1 | FALSE | Downregulated |
| -0,110101335 | 0,931322225 | SUCLA2 | FALSE | Downregulated |
| 0,080126847 | 0,931322225 | TIMM10B | FALSE | Upregulated |
| -0,161723651 | 0,931322225 | TMEM133 | FALSE | Downregulated |
| 0,100287883 | 0,931322225 | WHAMMP3 | FALSE | Upregulated |
| -0,231905954 | 0,931599525 | LOC441528 | FALSE | Downregulated |
| -0,160017816 | 0,931627562 | GTF2F2 | FALSE | Downregulated |
| -0,15731447 | 0,931627562 | NKX3-1 | FALSE | Downregulated |
| -0,147758537 | 0,931675993 | ADAMTS4 | FALSE | Downregulated |
| -0,039086067 | 0,931675993 | CDK4 | FALSE | Downregulated |
| -0,190766606 | 0,931675993 | MGC15885 | FALSE | Downregulated |
| 0,069396154 | 0,931675993 | TRAF3IP2-AS1 | FALSE | Upregulated |
| -0,133064303 | 0,931739457 | SOCS3 | FALSE | Downregulated |
| 0,05511807 | 0,931862927 | YTHDF2 | FALSE | Upregulated |
| 0,094443507 | 0,931945565 | DPAGT1 | FALSE | Upregulated |
| 0,13948723 | 0,931945565 | LTV1 | FALSE | Upregulated |
| -0,076643712 | 0,93199381 | R3HDM4 | FALSE | Downregulated |
| 0,068132952 | 0,93199507 | HMCES | FALSE | Upregulated |
| -0,09451748 | 0,93199507 | IDE | FALSE | Downregulated |
| -0,166806195 | 0,93199507 | LOC285389 | FALSE | Downregulated |
| -0,196372295 | 0,93199507 | RNU6ATAC | FALSE | Downregulated |
| -0,096466526 | 0,932629596 | RABGGTA | FALSE | Downregulated |
| 0,050889658 | 0,932777278 | DLD | FALSE | Upregulated |
| -0,07131883 | 0,932777278 | H2AFJ | FALSE | Downregulated |
| 0,072892774 | 0,932777278 | MINPP1 | FALSE | Upregulated |
| -0,079962564 | 0,932777278 | PLEKHM3 | FALSE | Downregulated |
| 0,094002573 | 0,932777278 | RHOXF2 | FALSE | Upregulated |
| 0,091528152 | 0,932777278 | RNF138 | FALSE | Upregulated |
| -0,186731649 | 0,932777278 | ZCCHC10 | FALSE | Downregulated |

| | | | | |
|---|---|---|---|---|
| -0,145964853 | 0,932813481 | EPHB6 | FALSE | Downregulated |
| 0,155259 | 0,932820822 | AC109828.1 | FALSE | Upregulated |
| -0,144131991 | 0,932820822 | C6orf203 | FALSE | Downregulated |
| 0,070725583 | 0,932820822 | F2R | FALSE | Upregulated |
| -0,077338836 | 0,932820822 | FOSL2 | FALSE | Downregulated |
| 0,056831121 | 0,932820822 | FTSJ3 | FALSE | Upregulated |
| -0,078689343 | 0,932820822 | GNPAT | FALSE | Downregulated |
| -0,213885256 | 0,932820822 | LOC101927060 | FALSE | Downregulated |
| -0,115939506 | 0,932820822 | REXO2 | FALSE | Downregulated |
| 0,125943326 | 0,932820822 | RFX2 | FALSE | Upregulated |
| -0,12723155 | 0,932820822 | TAS2R42 | FALSE | Downregulated |
| 0,078051102 | 0,932820822 | TMEM106A | FALSE | Upregulated |
| -0,108920746 | 0,932820822 | TRUB1 | FALSE | Downregulated |
| 0,069110207 | 0,932820822 | TSNARE1 | FALSE | Upregulated |
| 0,110786308 | 0,932820822 | UPP1 | FALSE | Upregulated |
| -0,095546364 | 0,932820822 | ZNF627 | FALSE | Downregulated |
| 0,223742317 | 0,932820822 | ZNF813 | FALSE | Upregulated |
| -0,256900047 | 0,932833832 | C1QTNF9B | FALSE | Downregulated |
| -0,058652244 | 0,932833832 | CDC26 | FALSE | Downregulated |
| -0,122523392 | 0,932833832 | CDR2L | FALSE | Downregulated |
| 0,069803531 | 0,932833832 | ISYNA1 | FALSE | Upregulated |
| 0,184883706 | 0,932906183 | GPR52 | FALSE | Upregulated |
| -0,151376489 | 0,932906183 | SEPSECS-AS1 | FALSE | Downregulated |
| 0,103759385 | 0,932911656 | ZNF101 | FALSE | Upregulated |
| -0,127042885 | 0,933043289 | C5orf22 | FALSE | Downregulated |
| 0,166898819 | 0,933043289 | C6orf226 | FALSE | Upregulated |
| 0,210217948 | 0,933043289 | RPGRIP1L | FALSE | Upregulated |
| -0,06807952 | 0,933043289 | SPCS2 | FALSE | Downregulated |
| 0,07924673 | 0,933348984 | MIF4GD | FALSE | Upregulated |
| 0,073151945 | 0,933394354 | SRPRB | FALSE | Upregulated |
| -0,037811393 | 0,933430178 | MEGF8 | FALSE | Downregulated |
| -0,11023821 | 0,933430178 | SOX7 | FALSE | Downregulated |
| -0,180579599 | 0,933511563 | FLT3LG | FALSE | Downregulated |
| 0,167994969 | 0,933511563 | TMEM237 | FALSE | Upregulated |
| -0,182032809 | 0,933664429 | JHDM1D-AS1 | FALSE | Downregulated |
| 0,101792351 | 0,933666752 | AFAP1L2 | FALSE | Upregulated |
| -0,061121481 | 0,933666752 | CX3CL1 | FALSE | Downregulated |
| 0,063215549 | 0,933666752 | KIAA1656 | FALSE | Upregulated |
| 0,107334419 | 0,933666752 | LOC101926918 | FALSE | Upregulated |
| 0,073611215 | 0,933666752 | SDHAP1 | FALSE | Upregulated |
| -0,189474062 | 0,933666752 | ST7-OT4 | FALSE | Downregulated |
| -0,038948549 | 0,933666752 | XYLT2 | FALSE | Downregulated |
| -0,137493931 | 0,933697458 | ARL14EP | FALSE | Downregulated |
| -0,125696907 | 0,933697458 | BRIX1 | FALSE | Downregulated |
| -0,047690457 | 0,933697458 | RAD21 | FALSE | Downregulated |
| -0,056084916 | 0,933765609 | SRP54 | FALSE | Downregulated |
| 0,098114849 | 0,933816605 | OSER1 | FALSE | Upregulated |
| -0,031479927 | 0,934064934 | FLNB | FALSE | Downregulated |
| -0,069763 | 0,934064934 | TROVE2 | FALSE | Downregulated |
| -0,148557604 | 0,934074878 | LCMT1 | FALSE | Downregulated |
| 0,105748753 | 0,934110015 | LINC00883 | FALSE | Upregulated |

| | | | | |
|---|---|---|---|---|
| 0,125623299 | 0,934116871 | LOC102724206 | FALSE | Upregulated |
| -0,195519784 | 0,934120167 | TMIE | FALSE | Downregulated |
| 0,203220038 | 0,934144702 | DLG5-AS1 | FALSE | Upregulated |
| 0,047610133 | 0,934144702 | OPTN | FALSE | Upregulated |
| -0,104553917 | 0,934310267 | ASPHD1 | FALSE | Downregulated |
| -0,038490385 | 0,934310267 | LZTR1 | FALSE | Downregulated |
| 0,109597345 | 0,934384518 | MTRNR2L4 | FALSE | Upregulated |
| -0,069224967 | 0,934384518 | POLR3G | FALSE | Downregulated |
| -0,095961649 | 0,934411972 | L3MBTL1 | FALSE | Downregulated |
| 0,199657145 | 0,93458828 | ANKRD13B | FALSE | Upregulated |
| 0,058508229 | 0,934636841 | BORA | FALSE | Upregulated |
| 0,081975687 | 0,934900466 | LOC101927044 | FALSE | Upregulated |
| 0,15951555 | 0,934900466 | LOC101928083 | FALSE | Upregulated |
| 0,068751237 | 0,934900466 | LRIG3 | FALSE | Upregulated |
| 0,041347306 | 0,934967856 | SRSF6 | FALSE | Upregulated |
| 0,068901823 | 0,93500342 | ATIC | FALSE | Upregulated |
| -0,170020537 | 0,93500342 | C1orf228 | FALSE | Downregulated |
| 0,116447301 | 0,93500342 | SPATS2 | FALSE | Upregulated |
| 0,050687059 | 0,93500342 | ZNF154 | FALSE | Upregulated |
| 0,0767971 | 0,935087082 | LMF1 | FALSE | Upregulated |
| -0,058272539 | 0,935271717 | NUS1 | FALSE | Downregulated |
| 0,039743904 | 0,935296178 | ECHDC1 | FALSE | Upregulated |
| -0,120378351 | 0,935296178 | ENKD1 | FALSE | Downregulated |
| 0,179708291 | 0,935296178 | LOC79999 | FALSE | Upregulated |
| -0,09125107 | 0,935296178 | SERPINE3 | FALSE | Downregulated |
| 0,095291322 | 0,935296178 | TSC22D3 | FALSE | Upregulated |
| -0,133759152 | 0,935312081 | PRDM7 | FALSE | Downregulated |
| -0,076480579 | 0,935378806 | TMEM134 | FALSE | Downregulated |
| 0,080971088 | 0,935385649 | DUSP7 | FALSE | Upregulated |
| 0,064819935 | 0,935606791 | TBC1D25 | FALSE | Upregulated |
| -0,23570112 | 0,935619703 | MYLK3 | FALSE | Downregulated |
| -0,191486288 | 0,935685186 | PABPN1L | FALSE | Downregulated |
| 0,072291028 | 0,935716781 | TGM2 | FALSE | Upregulated |
| 0,155822479 | 0,935751943 | LOC284825 | FALSE | Upregulated |
| -0,144869864 | 0,935760741 | PCP2 | FALSE | Downregulated |
| 0,067707352 | 0,935890841 | DCAF16 | FALSE | Upregulated |
| -0,057223716 | 0,936006465 | EHD1 | FALSE | Downregulated |
| -0,163223387 | 0,936006465 | PGF | FALSE | Downregulated |
| 0,059895677 | 0,936071444 | CKMT2 | FALSE | Upregulated |
| -0,062048887 | 0,93617008 | LOC101928192 | FALSE | Downregulated |
| 0,043396009 | 0,93617008 | PVRL2 | FALSE | Upregulated |
| -0,207537657 | 0,936285539 | ABCB9 | FALSE | Downregulated |
| -0,118469612 | 0,936285539 | DAGLA | FALSE | Downregulated |
| 0,322398698 | 0,936285539 | GGTLC2 | FALSE | Upregulated |
| 0,071806061 | 0,936285539 | PPM1B | FALSE | Upregulated |
| 0,035113439 | 0,936285539 | SLC9A3R1 | FALSE | Upregulated |
| 0,060549828 | 0,936294034 | UBR1 | FALSE | Upregulated |
| -0,120133088 | 0,936294034 | USMG5 | FALSE | Downregulated |
| -0,209937521 | 0,936357855 | TIPIN | FALSE | Downregulated |
| 0,071351883 | 0,936357855 | ZNF264 | FALSE | Upregulated |
| 0,081964179 | 0,936357855 | ZNF32 | FALSE | Upregulated |

| | | | | |
|---|---|---|---|---|
| -0,056372811 | 0,936488691 | PIGW | FALSE | Downregulated |
| 0,14393083 | 0,936488691 | SEMA3B | FALSE | Upregulated |
| -0,032545841 | 0,936535925 | BAP1 | FALSE | Downregulated |
| -0,042464196 | 0,936535925 | CIZ1 | FALSE | Downregulated |
| -0,087832968 | 0,936535925 | COQ7 | FALSE | Downregulated |
| -0,076794921 | 0,936535925 | COX7A2 | FALSE | Downregulated |
| 0,044330456 | 0,936535925 | FLJ10038 | FALSE | Upregulated |
| -0,178589717 | 0,936535925 | FZD3 | FALSE | Downregulated |
| -0,206734156 | 0,936535925 | LINC01137 | FALSE | Downregulated |
| 0,163291802 | 0,936535925 | LOC100287632 | FALSE | Upregulated |
| 0,116385022 | 0,936535925 | MAMLD1 | FALSE | Upregulated |
| -0,094219572 | 0,936535925 | PEX11B | FALSE | Downregulated |
| -0,084721627 | 0,936535925 | PPIL4 | FALSE | Downregulated |
| -0,054819193 | 0,936535925 | PRKAB1 | FALSE | Downregulated |
| 0,085388479 | 0,936535925 | RAB40B | FALSE | Upregulated |
| 0,111217254 | 0,936535925 | RGPD2 | FALSE | Upregulated |
| 0,102464139 | 0,936535925 | RP11-665G4.1 | FALSE | Upregulated |
| -0,056748965 | 0,936535925 | SMARCB1 | FALSE | Downregulated |
| 0,041585313 | 0,936535925 | SPTB | FALSE | Upregulated |
| 0,069150538 | 0,936535925 | ST3GAL3 | FALSE | Upregulated |
| 0,079089105 | 0,936535925 | STON2 | FALSE | Upregulated |
| -0,052054841 | 0,936535925 | TRIM2 | FALSE | Downregulated |
| -0,15795289 | 0,936535925 | TRMT10C | FALSE | Downregulated |
| -0,081010821 | 0,936535925 | WDR44 | FALSE | Downregulated |
| -0,08444032 | 0,936595916 | ABHD5 | FALSE | Downregulated |
| -0,05391577 | 0,936595916 | PPIG | FALSE | Downregulated |
| -0,104780323 | 0,936735775 | KIAA0895 | FALSE | Downregulated |
| 0,049621963 | 0,93678195 | MICALL1 | FALSE | Upregulated |
| 0,076532216 | 0,9368559 | BMF | FALSE | Upregulated |
| 0,084598131 | 0,936936618 | COX19 | FALSE | Upregulated |
| -0,039196415 | 0,936936618 | SLC25A3 | FALSE | Downregulated |
| 0,075196314 | 0,936994956 | ADAMTS17 | FALSE | Upregulated |
| 0,361356944 | 0,937376636 | TCAP | FALSE | Upregulated |
| 0,153347005 | 0,937432525 | LOC101926889 | FALSE | Upregulated |
| -0,055023124 | 0,937432525 | MEST | FALSE | Downregulated |
| 0,166346669 | 0,937432525 | RP11-278H7.1 | FALSE | Upregulated |
| 0,158109455 | 0,93750661 | GOLGA6B | FALSE | Upregulated |
| 0,068793631 | 0,937513688 | CD80 | FALSE | Upregulated |
| -0,032075992 | 0,937513688 | FBRSL1 | FALSE | Downregulated |
| -0,037263066 | 0,937513688 | FZD4 | FALSE | Downregulated |
| -0,051500141 | 0,937513688 | HSD17B2 | FALSE | Downregulated |
| -0,076608047 | 0,937513688 | LOC646719 | FALSE | Downregulated |
| 0,104927017 | 0,937513688 | MYLIP | FALSE | Upregulated |
| -0,087217433 | 0,937513688 | PHLDA1 | FALSE | Downregulated |
| -0,215356129 | 0,937513688 | RP11-466F5.8 | FALSE | Downregulated |
| 0,041595154 | 0,937513688 | SDR39U1 | FALSE | Upregulated |
| -0,075364596 | 0,937513688 | SMAD1 | FALSE | Downregulated |
| -0,261099156 | 0,937513688 | TRIM9 | FALSE | Downregulated |
| 0,033387241 | 0,937513688 | USP7 | FALSE | Upregulated |
| -0,052267597 | 0,937513688 | VPS52 | FALSE | Downregulated |
| 0,093220118 | 0,937513688 | ZNF790-AS1 | FALSE | Upregulated |

| | | | | |
|---|---|---|---|---|
| 0,155901574 | 0,937709288 | GMDS-AS1 | FALSE | Upregulated |
| 0,174111701 | 0,937709288 | HES6 | FALSE | Upregulated |
| -0,046456046 | 0,937709288 | KIAA1191 | FALSE | Downregulated |
| 0,069409899 | 0,937709288 | LINC00954 | FALSE | Upregulated |
| 0,100283249 | 0,937709288 | LOC101929441 | FALSE | Upregulated |
| -0,197724159 | 0,937709288 | RP11-527H14.2 | FALSE | Downregulated |
| 0,134588897 | 0,937709288 | ZNF775 | FALSE | Upregulated |
| -0,038581057 | 0,937840032 | ABCC2 | FALSE | Downregulated |
| -0,169945479 | 0,937840032 | ASB15 | FALSE | Downregulated |
| 0,031494282 | 0,937840032 | BCL9L | FALSE | Upregulated |
| -0,067340311 | 0,937840032 | C19orf70 | FALSE | Downregulated |
| -0,048962501 | 0,937840032 | C4orf3 | FALSE | Downregulated |
| 0,036687241 | 0,937840032 | KANSL1 | FALSE | Upregulated |
| -0,196112725 | 0,937840032 | LOC101928676 | FALSE | Downregulated |
| -0,057604339 | 0,937840032 | NEURL2 | FALSE | Downregulated |
| -0,075209004 | 0,937840032 | PPP1R8 | FALSE | Downregulated |
| 0,11018771 | 0,937840032 | PTGS1 | FALSE | Upregulated |
| 0,266689421 | 0,937840032 | SEPT5-GP1BB | FALSE | Upregulated |
| -0,158686462 | 0,937840032 | SPTBN4 | FALSE | Downregulated |
| 0,050729441 | 0,937840032 | TMLHE | FALSE | Upregulated |
| -0,081017134 | 0,937840032 | ZMAT2 | FALSE | Downregulated |
| -0,048168723 | 0,937944163 | TRIOBP | FALSE | Downregulated |
| 0,081470209 | 0,938111189 | FAM122A | FALSE | Upregulated |
| -0,068420603 | 0,938165212 | PTGDR2 | FALSE | Downregulated |
| -0,123120579 | 0,938236453 | RWDD3 | FALSE | Downregulated |
| -1,231509323 | 0,938456683 | RNVU1-18 | FALSE | Downregulated |
| 0,078011072 | 0,93853933 | CORIN | FALSE | Upregulated |
| 0,144789474 | 0,93853933 | FAM84A | FALSE | Upregulated |
| 0,112778529 | 0,93853933 | LOC102723997 | FALSE | Upregulated |
| -0,092673883 | 0,93866322 | C1orf172 | FALSE | Downregulated |
| 0,168113323 | 0,93866322 | CCDC54 | FALSE | Upregulated |
| 0,046672562 | 0,93866322 | CNOT6L | FALSE | Upregulated |
| 0,046657201 | 0,93866322 | DNAJC7 | FALSE | Upregulated |
| -0,056245947 | 0,93866322 | JAM2 | FALSE | Downregulated |
| 0,108319898 | 0,93866322 | LOC101928884 | FALSE | Upregulated |
| 0,111963061 | 0,93866322 | LOC102724322 | FALSE | Upregulated |
| 0,064583944 | 0,93866322 | NHLRC3 | FALSE | Upregulated |
| -0,057850458 | 0,93866322 | PLEKHH3 | FALSE | Downregulated |
| 0,088790434 | 0,938916173 | STEAP2 | FALSE | Upregulated |
| 0,185857496 | 0,938943246 | C16orf95 | FALSE | Upregulated |
| -0,219890128 | 0,938943246 | CLEC18A | FALSE | Downregulated |
| -0,051020326 | 0,938943246 | KIN | FALSE | Downregulated |
| -0,06027337 | 0,938943246 | KLHL20 | FALSE | Downregulated |
| -0,060379529 | 0,938943246 | LETMD1 | FALSE | Downregulated |
| -0,070303067 | 0,938943246 | LOC101927761 | FALSE | Downregulated |
| -0,140352434 | 0,938943246 | LOC102724652 | FALSE | Downregulated |
| 0,058059626 | 0,938943246 | RNF19B | FALSE | Upregulated |
| -0,030464102 | 0,938943246 | SF3B1 | FALSE | Downregulated |
| 0,196202673 | 0,939255887 | EFNA4 | FALSE | Upregulated |
| -0,080007748 | 0,939255887 | LOC101928089 | FALSE | Downregulated |
| 0,172463412 | 0,939255887 | NLRP7 | FALSE | Upregulated |

| | | | | |
|---|---|---|---|---|
| 0,058910237 | 0,939255887 | SHOC2 | FALSE | Upregulated |
| 0,170089218 | 0,939712352 | PDE10A | FALSE | Upregulated |
| 0,121527845 | 0,939733576 | SH2D1B | FALSE | Upregulated |
| -0,053021996 | 0,939997392 | LOC101929781 | FALSE | Downregulated |
| -0,062777606 | 0,939997392 | TMEM150A | FALSE | Downregulated |
| -0,071759095 | 0,940077472 | BCAP29 | FALSE | Downregulated |
| 0,106814397 | 0,940077472 | LENG1 | FALSE | Upregulated |
| -0,080520071 | 0,940121922 | OSER1-AS1 | FALSE | Downregulated |
| 0,060639537 | 0,940121922 | RTTN | FALSE | Upregulated |
| -0,057329137 | 0,9403665 | TCERG1 | FALSE | Downregulated |
| 0,031112293 | 0,940509164 | CD163 | FALSE | Upregulated |
| 0,150883478 | 0,940509164 | FLJ42393 | FALSE | Upregulated |
| 0,085194955 | 0,940509164 | ILF3-AS1 | FALSE | Upregulated |
| 0,057508584 | 0,940509164 | MAPKBP1 | FALSE | Upregulated |
| 0,069494534 | 0,940509164 | NRL | FALSE | Upregulated |
| -0,078240105 | 0,940509164 | TMEM14B | FALSE | Downregulated |
| -0,088941389 | 0,940509164 | UTS2B | FALSE | Downregulated |
| -0,08669923 | 0,940509164 | ZNF251 | FALSE | Downregulated |
| -0,094951864 | 0,940542088 | CELF6 | FALSE | Downregulated |
| 0,056577668 | 0,940542088 | CRELD1 | FALSE | Upregulated |
| 0,062628672 | 0,940542088 | CTNNBIP1 | FALSE | Upregulated |
| -0,037052377 | 0,940542088 | RAB11FIP3 | FALSE | Downregulated |
| -0,06463494 | 0,940542088 | SLC12A9 | FALSE | Downregulated |
| -0,070395432 | 0,940542088 | VPS45 | FALSE | Downregulated |
| -0,060378158 | 0,940617931 | GATA6 | FALSE | Downregulated |
| -0,045542142 | 0,94102897 | SPTY2D1 | FALSE | Downregulated |
| 0,130477743 | 0,941154961 | CX3CR1 | FALSE | Upregulated |
| -0,126605331 | 0,941170923 | OASL | FALSE | Downregulated |
| 0,18908867 | 0,941170923 | PLGRKT | FALSE | Upregulated |
| 0,068779811 | 0,941170923 | ZCCHC9 | FALSE | Upregulated |
| 0,052152421 | 0,941270608 | GFM1 | FALSE | Upregulated |
| -0,042967456 | 0,941279688 | CRY2 | FALSE | Downregulated |
| -0,162194984 | 0,941279688 | MED30 | FALSE | Downregulated |
| -0,141570653 | 0,941279688 | S1PR2 | FALSE | Downregulated |
| 0,142662964 | 0,941372516 | ADAT3 | FALSE | Upregulated |
| 0,135841611 | 0,941372516 | TNS4 | FALSE | Upregulated |
| -0,10740284 | 0,941372516 | UBXN2A | FALSE | Downregulated |
| -0,121733866 | 0,941372516 | ZNF320 | FALSE | Downregulated |
| -0,123315938 | 0,941571281 | HEPN1 | FALSE | Downregulated |
| -0,043000866 | 0,941866755 | HNRNPUL2 | FALSE | Downregulated |
| 0,100773725 | 0,941866755 | WRB | FALSE | Upregulated |
| -0,097315828 | 0,94212967 | ADAT2 | FALSE | Downregulated |
| 0,036584289 | 0,94212967 | COPG1 | FALSE | Upregulated |
| 0,039979482 | 0,94212967 | PSKH1 | FALSE | Upregulated |
| 0,066400162 | 0,94212967 | RNF26 | FALSE | Upregulated |
| 0,130781237 | 0,942158972 | CTAGE9 | FALSE | Upregulated |
| 0,063186292 | 0,942158972 | PAOX | FALSE | Upregulated |
| 0,032306317 | 0,942158972 | SMARCA4 | FALSE | Upregulated |
| -0,054892197 | 0,942158972 | TBC1D23 | FALSE | Downregulated |
| 0,054342893 | 0,942158972 | TIMM44 | FALSE | Upregulated |
| 0,067721007 | 0,942216546 | KIAA1958 | FALSE | Upregulated |

| | | | | |
|---|---|---|---|---|
| 0,072586861 | 0,942623555 | ARHGEF4 | FALSE | Upregulated |
| -0,128427235 | 0,942623555 | CCNA2 | FALSE | Downregulated |
| -0,210327495 | 0,942623555 | CMTM7 | FALSE | Downregulated |
| 0,142053765 | 0,942623555 | COL9A3 | FALSE | Upregulated |
| 0,116019794 | 0,942623555 | CXXC4 | FALSE | Upregulated |
| 0,093965369 | 0,942623555 | DPH3 | FALSE | Upregulated |
| 0,181857884 | 0,942623555 | FOXD4L4 | FALSE | Upregulated |
| -0,177838434 | 0,942623555 | LOC100128398 | FALSE | Downregulated |
| 0,053375491 | 0,942623555 | LOC102723535 | FALSE | Upregulated |
| 0,234202015 | 0,942623555 | LOC541473 | FALSE | Upregulated |
| -0,101985048 | 0,942623555 | LZIC | FALSE | Downregulated |
| -0,107122904 | 0,942623555 | MED4-AS1 | FALSE | Downregulated |
| 0,079963433 | 0,942623555 | MGST2 | FALSE | Upregulated |
| 0,0748817 | 0,942623555 | MVK | FALSE | Upregulated |
| -0,157026867 | 0,942623555 | TMEM136 | FALSE | Downregulated |
| -0,054721306 | 0,942623555 | TMOD3 | FALSE | Downregulated |
| -0,06986717 | 0,942623555 | ZNF564 | FALSE | Downregulated |
| 0,035588014 | 0,942916245 | DAZAP1 | FALSE | Upregulated |
| -0,208399455 | 0,942959933 | LOC100133920 | FALSE | Downregulated |
| -0,199979404 | 0,943054266 | C20orf62 | FALSE | Downregulated |
| -0,10457587 | 0,943054266 | CREM | FALSE | Downregulated |
| -0,168613576 | 0,943054266 | CSF2 | FALSE | Downregulated |
| -0,047051808 | 0,943054266 | TBC1D17 | FALSE | Downregulated |
| -0,190547506 | 0,94310836 | RRN3P1 | FALSE | Downregulated |
| 0,066160843 | 0,943191521 | POLR2H | FALSE | Upregulated |
| -0,062532742 | 0,943191521 | PORCN | FALSE | Downregulated |
| -0,074741554 | 0,943203375 | MYO16 | FALSE | Downregulated |
| -0,053713024 | 0,943210383 | FAM13B | FALSE | Downregulated |
| -0,113416656 | 0,943210383 | METTL18 | FALSE | Downregulated |
| 0,072271752 | 0,943210383 | NPEPL1 | FALSE | Upregulated |
| 0,02921899 | 0,943210383 | SRSF1 | FALSE | Upregulated |
| 0,048421671 | 0,943210383 | USP48 | FALSE | Upregulated |
| 0,096289538 | 0,943210383 | ZKSCAN3 | FALSE | Upregulated |
| -0,068366942 | 0,94321515 | VCAM1 | FALSE | Downregulated |
| -0,046676721 | 0,943285748 | ELMO2 | FALSE | Downregulated |
| -0,225006 | 0,943285748 | LINC00467 | FALSE | Downregulated |
| 0,144001241 | 0,943285748 | RNF122 | FALSE | Upregulated |
| 0,059350795 | 0,943285748 | ZNF398 | FALSE | Upregulated |
| 0,105165327 | 0,943347701 | RBM48 | FALSE | Upregulated |
| 0,184493536 | 0,943382028 | WDR78 | FALSE | Upregulated |
| -0,038190932 | 0,943489098 | ATRN | FALSE | Downregulated |
| 0,079617359 | 0,943489098 | LOC101929343 | FALSE | Upregulated |
| -0,052718268 | 0,943489098 | TMEM204 | FALSE | Downregulated |
| -0,037177445 | 0,943566535 | BCL2L13 | FALSE | Downregulated |
| 0,087801464 | 0,943566535 | CHIC2 | FALSE | Upregulated |
| -0,183343146 | 0,943566535 | PNOC | FALSE | Downregulated |
| 0,07519025 | 0,943566535 | UTP3 | FALSE | Upregulated |
| 0,187548886 | 0,943586351 | PPP1R14D | FALSE | Upregulated |
| -0,054283785 | 0,943802059 | BRAF | FALSE | Downregulated |
| 0,04045725 | 0,943802059 | CPSF3L | FALSE | Upregulated |
| 0,102724404 | 0,943802059 | SIGLEC15 | FALSE | Upregulated |

| | | | | |
|---|---|---|---|---|
| -0,217946661 | 0,943951957 | SRGAP2D | FALSE | Downregulated |
| 0,071647776 | 0,944025481 | GTF3C4 | FALSE | Upregulated |
| 0,055759041 | 0,944025481 | SHARPIN | FALSE | Upregulated |
| -0,073561652 | 0,944025481 | ZNF621 | FALSE | Downregulated |
| 0,137503198 | 0,944085726 | C21orf119 | FALSE | Upregulated |
| 0,202013077 | 0,944085726 | KRT8P41 | FALSE | Upregulated |
| 0,103360997 | 0,944085726 | METTL5 | FALSE | Upregulated |
| 0,060708249 | 0,944085726 | SEC23B | FALSE | Upregulated |
| 0,058386776 | 0,944085726 | TBC1D20 | FALSE | Upregulated |
| 0,189081201 | 0,94429968 | TMEM119 | FALSE | Upregulated |
| 0,044563974 | 0,944328167 | UBAP1 | FALSE | Upregulated |
| -0,034519228 | 0,944328167 | USP8 | FALSE | Downregulated |
| -0,043957617 | 0,944573501 | MAGED1 | FALSE | Downregulated |
| 0,068474668 | 0,944573501 | NGFRAP1 | FALSE | Upregulated |
| 0,110546619 | 0,944590598 | ACVR1C | FALSE | Upregulated |
| 0,0517136 | 0,944590598 | BRAP | FALSE | Upregulated |
| 0,051017618 | 0,944590598 | C9orf3 | FALSE | Upregulated |
| 0,050612364 | 0,944590598 | DDX18 | FALSE | Upregulated |
| 0,047560781 | 0,944590598 | EFTUD2 | FALSE | Upregulated |
| -0,284060141 | 0,944590598 | FOXD4L5 | FALSE | Downregulated |
| 0,076422461 | 0,944590598 | INHBB | FALSE | Upregulated |
| -0,173190268 | 0,944590598 | LOC102724126 | FALSE | Downregulated |
| -0,104361839 | 0,944590598 | RPS10P7 | FALSE | Downregulated |
| 0,05544868 | 0,944590598 | SCAMP4 | FALSE | Upregulated |
| -0,078450819 | 0,944590598 | TBXAS1 | FALSE | Downregulated |
| 0,094665287 | 0,944590598 | ZNF708 | FALSE | Upregulated |
| 0,067590499 | 0,944697875 | RNF222 | FALSE | Upregulated |
| -0,153638862 | 0,944812706 | TLX1NB | FALSE | Downregulated |
| -0,1146361 | 0,94498089 | ADAM20 | FALSE | Downregulated |
| 0,087269559 | 0,94498089 | NARS2 | FALSE | Upregulated |
| -0,033343306 | 0,94498089 | PPM1A | FALSE | Downregulated |
| -0,032823351 | 0,944999439 | OSBPL9 | FALSE | Downregulated |
| -0,04995881 | 0,945024998 | TPD52L2 | FALSE | Downregulated |
| -0,145088863 | 0,945048294 | GAL3ST1 | FALSE | Downregulated |
| -0,069276112 | 0,94505489 | ABCG2 | FALSE | Downregulated |
| 0,051214025 | 0,94505489 | BRD9 | FALSE | Upregulated |
| 0,112972431 | 0,94505489 | CYP7A1 | FALSE | Upregulated |
| 0,073297762 | 0,94507039 | C14orf169 | FALSE | Upregulated |
| -0,077515022 | 0,94507039 | COPRS | FALSE | Downregulated |
| 0,19410198 | 0,94507039 | ZNF675 | FALSE | Upregulated |
| -0,165049809 | 0,945175275 | ZNF572 | FALSE | Downregulated |
| 0,151502109 | 0,945241744 | YY2 | FALSE | Upregulated |
| 0,037505866 | 0,945455876 | NRBP2 | FALSE | Upregulated |
| 0,215999063 | 0,945455876 | SLC26A4-AS1 | FALSE | Upregulated |
| -0,036771128 | 0,945583925 | SLC25A15 | FALSE | Downregulated |
| -0,063298709 | 0,945607829 | DHX58 | FALSE | Downregulated |
| -0,128236932 | 0,945607829 | ZNF502 | FALSE | Downregulated |
| 0,019501882 | 0,945622735 | LARP1 | FALSE | Upregulated |
| 0,125068469 | 0,945622735 | LRRC27 | FALSE | Upregulated |
| -0,072731118 | 0,945786179 | C3orf83 | FALSE | Downregulated |
| -0,080635492 | 0,945786179 | CD40 | FALSE | Downregulated |

| | | | | |
|---|---|---|---|---|
| 0,055796508 | 0,945786179 | INTS4 | FALSE | Upregulated |
| -0,092160195 | 0,945786179 | KBTBD12 | FALSE | Downregulated |
| 0,055946281 | 0,945786179 | LANCL1 | FALSE | Upregulated |
| -0,061592192 | 0,945786179 | MIEN1 | FALSE | Downregulated |
| 0,043901039 | 0,945786179 | MYO6 | FALSE | Upregulated |
| -0,053791991 | 0,945786179 | SPOP | FALSE | Downregulated |
| -0,04415827 | 0,946015953 | SCARF1 | FALSE | Downregulated |
| 0,091973714 | 0,946214498 | ILKAP | FALSE | Upregulated |
| -0,121918096 | 0,946214498 | MMP21 | FALSE | Downregulated |
| 0,147613664 | 0,946214498 | MRPS17 | FALSE | Upregulated |
| 0,105217159 | 0,946214498 | SNHG5 | FALSE | Upregulated |
| -0,180601473 | 0,946321191 | CEACAM22P | FALSE | Downregulated |
| -0,06053913 | 0,946321191 | GARS | FALSE | Downregulated |
| 0,209142807 | 0,946321191 | LOC101929667 | FALSE | Upregulated |
| -0,098624455 | 0,946321191 | RPL26L1 | FALSE | Downregulated |
| 0,071685877 | 0,946321191 | TRPM3 | FALSE | Upregulated |
| -0,07726781 | 0,946321191 | WDR83 | FALSE | Downregulated |
| -0,1595311 | 0,946342027 | ST6GALNAC3 | FALSE | Downregulated |
| -0,155260971 | 0,946366427 | DYRK4 | FALSE | Downregulated |
| 0,065532765 | 0,946366427 | WARS2 | FALSE | Upregulated |
| -0,083757597 | 0,946374509 | FIBP | FALSE | Downregulated |
| -0,9799767 | 0,946374509 | NPIPA7 | FALSE | Downregulated |
| 0,080950576 | 0,946410198 | FFAR4 | FALSE | Upregulated |
| 0,05257037 | 0,946410198 | PTDSS1 | FALSE | Upregulated |
| -0,033017303 | 0,946410198 | SIPA1 | FALSE | Downregulated |
| 0,057758509 | 0,946410198 | TRPV4 | FALSE | Upregulated |
| 0,034295135 | 0,946581446 | CEP350 | FALSE | Upregulated |
| -0,046216783 | 0,946601726 | CCDC142 | FALSE | Downregulated |
| 0,05522062 | 0,946601726 | CDPF1 | FALSE | Upregulated |
| -0,079874349 | 0,946615114 | C12orf10 | FALSE | Downregulated |
| 0,056058141 | 0,946675406 | C1GALT1 | FALSE | Upregulated |
| 0,047063161 | 0,946675406 | PPP4R2 | FALSE | Upregulated |
| 0,0341 | 0,946748471 | LOC148413 | FALSE | Upregulated |
| 0,096780413 | 0,946783726 | LOC728743 | FALSE | Upregulated |
| -0,115160891 | 0,946902752 | C17orf66 | FALSE | Downregulated |
| 0,056456183 | 0,946921531 | BEAN1 | FALSE | Upregulated |
| -0,161720246 | 0,947071102 | ANKHD1 | FALSE | Downregulated |
| 0,040257553 | 0,947071102 | EPS8 | FALSE | Upregulated |
| -0,033909256 | 0,947071102 | MFN2 | FALSE | Downregulated |
| -0,068387685 | 0,947071102 | SLC30A6 | FALSE | Downregulated |
| 0,129589569 | 0,947156219 | AP000472.2 | FALSE | Upregulated |
| 0,044419049 | 0,947156219 | MPG | FALSE | Upregulated |
| 0,049478181 | 0,947156219 | SMAD7 | FALSE | Upregulated |
| 0,073811125 | 0,947156219 | TMEM19 | FALSE | Upregulated |
| -0,062802071 | 0,94736233 | MALT1 | FALSE | Downregulated |
| 0,041669362 | 0,947629855 | CFL2 | FALSE | Upregulated |
| 0,048219749 | 0,947762703 | ANO8 | FALSE | Upregulated |
| 0,032548527 | 0,947762703 | MYRIP | FALSE | Upregulated |
| -0,046612685 | 0,947762703 | USP53 | FALSE | Downregulated |
| 0,029037156 | 0,947862082 | AMT | FALSE | Upregulated |
| 0,197858205 | 0,947862082 | ATP4B | FALSE | Upregulated |

| | | | | |
|---|---|---|---|---|
| -0,180848361 | 0,947862082 | LIN9 | FALSE | Downregulated |
| -0,109900606 | 0,947862082 | NEBL | FALSE | Downregulated |
| 0,031738123 | 0,947862082 | POLR3E | FALSE | Upregulated |
| 0,178706978 | 0,947862082 | PSG1 | FALSE | Upregulated |
| -0,0322432 | 0,947862082 | TATDN2 | FALSE | Downregulated |
| 0,047753756 | 0,947862082 | TIAM2 | FALSE | Upregulated |
| 0,052598622 | 0,948229727 | ACOT4 | FALSE | Upregulated |
| -0,049361936 | 0,948229727 | CEP192 | FALSE | Downregulated |
| 0,063433033 | 0,948229727 | EHD4 | FALSE | Upregulated |
| 0,050122789 | 0,948229727 | GUSBP11 | FALSE | Upregulated |
| -0,024507461 | 0,948229727 | KHNYN | FALSE | Downregulated |
| -0,034801962 | 0,948229727 | MARK2 | FALSE | Downregulated |
| 0,107630606 | 0,948229727 | P2RY1 | FALSE | Upregulated |
| -0,042442187 | 0,948229727 | PRSS48 | FALSE | Downregulated |
| -0,058180616 | 0,948229727 | PSMD6 | FALSE | Downregulated |
| 0,039086114 | 0,948229727 | RECQL5 | FALSE | Upregulated |
| 0,050455173 | 0,948229727 | RNA28S5 | FALSE | Upregulated |
| -0,122117224 | 0,948229727 | RP4-669L17.10-2 | FALSE | Downregulated |
| 0,033140143 | 0,948229727 | SP100 | FALSE | Upregulated |
| 0,056040323 | 0,948229727 | SPTLC1 | FALSE | Upregulated |
| -0,125281674 | 0,948229727 | ZSCAN20 | FALSE | Downregulated |
| -0,081360459 | 0,948252116 | RIC8B | FALSE | Downregulated |
| -0,087311878 | 0,948252116 | ZNF202 | FALSE | Downregulated |
| 0,064331428 | 0,948272267 | LOC102724009 | FALSE | Upregulated |
| -1,053761532 | 0,94837793 | ARMCX5-GPRASP2 | FALSE | Downregulated |
| -0,155530478 | 0,948585488 | LOC100133182 | FALSE | Downregulated |
| -0,025800362 | 0,948901389 | TOMM70A | FALSE | Downregulated |
| -0,059656494 | 0,948909649 | NSF | FALSE | Downregulated |
| 0,18669946 | 0,948909649 | ST8SIA1 | FALSE | Upregulated |
| -0,125316779 | 0,948909649 | ZBTB49 | FALSE | Downregulated |
| 0,162190518 | 0,949065861 | NR4A1 | FALSE | Upregulated |
| -0,137140673 | 0,949108101 | FAM89A | FALSE | Downregulated |
| -0,092771183 | 0,949110113 | ENDOV | FALSE | Downregulated |
| 0,047840812 | 0,949174982 | FNTA | FALSE | Upregulated |
| 0,108213961 | 0,949174982 | LINC00526 | FALSE | Upregulated |
| -0,179335381 | 0,949174982 | LOC102723461 | FALSE | Downregulated |
| -0,048950956 | 0,949174982 | RDH11 | FALSE | Downregulated |
| -0,129857115 | 0,949255083 | CCDC126 | FALSE | Downregulated |
| -0,053991085 | 0,949255083 | CRIPAK | FALSE | Downregulated |
| 0,203804456 | 0,949255083 | FOXE3 | FALSE | Upregulated |
| -0,034365575 | 0,949255083 | H1F0 | FALSE | Downregulated |
| 0,089141705 | 0,949255083 | IPMK | FALSE | Upregulated |
| 0,048263458 | 0,949255083 | LINC-PINT | FALSE | Upregulated |
| -0,05357395 | 0,949255083 | LOC101927451 | FALSE | Downregulated |
| 0,060422787 | 0,949255083 | MECP2 | FALSE | Upregulated |
| 0,039981438 | 0,949255083 | NEMF | FALSE | Upregulated |
| -0,032764891 | 0,949255083 | NRIP1 | FALSE | Downregulated |
| -0,069387556 | 0,949255083 | RBMS3 | FALSE | Downregulated |
| 0,058665237 | 0,949255083 | RSL24D1 | FALSE | Upregulated |
| -0,142570007 | 0,949255083 | SHISA4 | FALSE | Downregulated |

| | | | | |
|---|---|---|---|---|
| -0,054959786 | 0,949255083 | TTYH1 | FALSE | Downregulated |
| -0,042244345 | 0,949255083 | USP10 | FALSE | Downregulated |
| 0,120631864 | 0,949255577 | BACE1-AS | FALSE | Upregulated |
| 0,135653875 | 0,949255577 | MTX2 | FALSE | Upregulated |
| 1,040558543 | 0,949255577 | MYH2 | FALSE | Upregulated |
| 0,138624388 | 0,949399772 | NCF1B | FALSE | Upregulated |
| 0,044482662 | 0,949399772 | OSBPL2 | FALSE | Upregulated |
| -0,050554604 | 0,949399772 | POLR3A | FALSE | Downregulated |
| 0,057833253 | 0,949399772 | PRSS8 | FALSE | Upregulated |
| -0,140237411 | 0,949410911 | LINC00665 | FALSE | Downregulated |
| -0,039583005 | 0,949410911 | POLR2C | FALSE | Downregulated |
| -0,038176322 | 0,949410911 | ZC3H14 | FALSE | Downregulated |
| -0,056331548 | 0,94951295 | ARFGAP3 | FALSE | Downregulated |
| -0,068250234 | 0,949552365 | THEM4 | FALSE | Downregulated |
| 0,038858691 | 0,949552365 | VAPA | FALSE | Upregulated |
| -0,119957628 | 0,94955572 | CHMP4C | FALSE | Downregulated |
| 0,072563144 | 0,94955572 | NOL11 | FALSE | Upregulated |
| 0,051289994 | 0,949704438 | NPDC1 | FALSE | Upregulated |
| -0,105757507 | 0,949704438 | ZC4H2 | FALSE | Downregulated |
| 0,077024754 | 0,94970504 | LOC493754 | FALSE | Upregulated |
| 0,03365357 | 0,94970504 | MEF2D | FALSE | Upregulated |
| -0,092565106 | 0,94975944 | BLOC1S2 | FALSE | Downregulated |
| 0,063453651 | 0,94975944 | CREB3L4 | FALSE | Upregulated |
| -0,045404954 | 0,94975944 | KRIT1 | FALSE | Downregulated |
| -0,066653114 | 0,94975944 | THNSL2 | FALSE | Downregulated |
| -0,030910173 | 0,949851968 | KDM6B | FALSE | Downregulated |
| 0,036227318 | 0,949851968 | MYO1D | FALSE | Upregulated |
| 0,06148318 | 0,949851968 | STX12 | FALSE | Upregulated |
| 0,093359984 | 0,949920306 | RAD54L | FALSE | Upregulated |
| -0,04743056 | 0,950319017 | GAS2L1 | FALSE | Downregulated |
| -0,050628329 | 0,950319017 | LOC100132815 | FALSE | Downregulated |
| -0,075304289 | 0,950451705 | LACTB2 | FALSE | Downregulated |
| -0,136806631 | 0,950451705 | LOC101927899 | FALSE | Downregulated |
| 0,073742559 | 0,950451705 | ZNF121 | FALSE | Upregulated |
| 0,052051601 | 0,950451705 | ZNF189 | FALSE | Upregulated |
| -0,416648845 | 0,950804007 | NPTN-IT1 | FALSE | Downregulated |
| 0,169901082 | 0,950842869 | PSIMCT-1 | FALSE | Upregulated |
| 0,094831792 | 0,951023035 | COA6 | FALSE | Upregulated |
| 0,072993045 | 0,951149624 | BCL2L10 | FALSE | Upregulated |
| 0,07649304 | 0,951149624 | HIST1H2BA | FALSE | Upregulated |
| 0,044778006 | 0,951197946 | TFG | FALSE | Upregulated |
| -0,035973304 | 0,951201046 | SSR3 | FALSE | Downregulated |
| 0,050939794 | 0,951201046 | ZDHHC7 | FALSE | Upregulated |
| -0,035777172 | 0,951341113 | LARGE | FALSE | Downregulated |
| -0,039005024 | 0,951341113 | THOP1 | FALSE | Downregulated |
| 0,07216504 | 0,951381743 | DENND2A | FALSE | Upregulated |
| 0,176891489 | 0,951490414 | RP9P | FALSE | Upregulated |
| 0,054961156 | 0,951490414 | SNRPA | FALSE | Upregulated |
| 0,094432985 | 0,951659217 | CIRH1A | FALSE | Upregulated |
| -0,087599557 | 0,951659217 | LOC101928574 | FALSE | Downregulated |
| 0,029654749 | 0,951659217 | SEL1L | FALSE | Upregulated |

| | | | | |
|---|---|---|---|---|
| -0,068214296 | 0,951728408 | WDR12 | FALSE | Downregulated |
| 0,057958993 | 0,951820572 | ARL6IP5 | FALSE | Upregulated |
| 0,066601926 | 0,951840613 | GRAMD2 | FALSE | Upregulated |
| -0,0628183 | 0,951890341 | MANBA | FALSE | Downregulated |
| -0,079837166 | 0,951892148 | C10orf11 | FALSE | Downregulated |
| 0,229268874 | 0,951892148 | C21orf33 | FALSE | Upregulated |
| 0,032376259 | 0,951892148 | FXR1 | FALSE | Upregulated |
| -0,028641152 | 0,951892148 | HOOK3 | FALSE | Downregulated |
| -0,094803401 | 0,951892148 | LRRC56 | FALSE | Downregulated |
| -0,021806652 | 0,951892148 | VPS13D | FALSE | Downregulated |
| 0,11339313 | 0,951892148 | ZMYM6NB | FALSE | Upregulated |
| -0,08505201 | 0,951896989 | LOC101927542 | FALSE | Downregulated |
| 0,062354241 | 0,951927876 | ASNSD1 | FALSE | Upregulated |
| 0,052482581 | 0,951991109 | PSMA2 | FALSE | Upregulated |
| -0,075117148 | 0,952133962 | FARSB | FALSE | Downregulated |
| 0,071943605 | 0,952242456 | RAD17 | FALSE | Upregulated |
| -0,053956788 | 0,952397771 | ARSE | FALSE | Downregulated |
| -0,126663665 | 0,952397771 | COL21A1 | FALSE | Downregulated |
| 0,028401174 | 0,952653296 | AGAP3 | FALSE | Upregulated |
| 0,045455237 | 0,952653296 | GRSF1 | FALSE | Upregulated |
| 0,050992686 | 0,952653296 | KBTBD7 | FALSE | Upregulated |
| 0,090702219 | 0,952653296 | UNC93A | FALSE | Upregulated |
| -0,055787896 | 0,952833149 | AAAS | FALSE | Downregulated |
| 0,080812886 | 0,952833149 | LIN7A | FALSE | Upregulated |
| 0,067787554 | 0,952833149 | LINC00341 | FALSE | Upregulated |
| 0,076669102 | 0,952833149 | LRRC8C | FALSE | Upregulated |
| -0,045298335 | 0,952833149 | WIPF2 | FALSE | Downregulated |
| -0,146081759 | 0,952885397 | CLDND2 | FALSE | Downregulated |
| -0,114707457 | 0,952885397 | WSCD1 | FALSE | Downregulated |
| -0,049016409 | 0,9529845 | TMEM59 | FALSE | Downregulated |
| -0,104872383 | 0,953074848 | MRPS23 | FALSE | Downregulated |
| -0,046798696 | 0,953196715 | CCAR1 | FALSE | Downregulated |
| 0,082057205 | 0,953196715 | LOC102723622 | FALSE | Upregulated |
| 0,125202655 | 0,953196715 | LOC102723679 | FALSE | Upregulated |
| 0,032053514 | 0,953274121 | ALCAM | FALSE | Upregulated |
| -0,040258542 | 0,953274121 | APOA1BP | FALSE | Downregulated |
| 0,188868211 | 0,953274121 | KDM4D | FALSE | Upregulated |
| 0,0688728 | 0,953274121 | LOC202181 | FALSE | Upregulated |
| -0,038163661 | 0,953274121 | SNAPC4 | FALSE | Downregulated |
| 0,155293393 | 0,953319165 | C1orf54 | FALSE | Upregulated |
| -0,035125647 | 0,953319165 | GOPC | FALSE | Downregulated |
| -0,056581433 | 0,953319165 | PYCR2 | FALSE | Downregulated |
| 0,0419072 | 0,953319165 | RPS6KB1 | FALSE | Upregulated |
| -0,039158346 | 0,953319165 | TTC19 | FALSE | Downregulated |
| 0,054081886 | 0,953331379 | IGBP1 | FALSE | Upregulated |
| -0,162352033 | 0,953331379 | TCTEX1D1 | FALSE | Downregulated |
| 0,100900534 | 0,953331379 | ZNF426 | FALSE | Upregulated |
| -0,058241378 | 0,953369754 | C2CD2L | FALSE | Downregulated |
| 0,031399243 | 0,953593553 | WDR73 | FALSE | Upregulated |
| 0,049330148 | 0,953598348 | IFIH1 | FALSE | Upregulated |
| -0,089959492 | 0,953669648 | TMEM126B | FALSE | Downregulated |

| | | | | |
|---|---|---|---|---|
| -0,052171244 | 0,953803346 | MYNN | FALSE | Downregulated |
| 0,030825969 | 0,953921921 | GOLGA3 | FALSE | Upregulated |
| 0,038989489 | 0,95407561 | ST6GALNAC6 | FALSE | Upregulated |
| -0,166484125 | 0,954078861 | RP11-620J15.2 | FALSE | Downregulated |
| 0,078324075 | 0,954102983 | GBAS | FALSE | Upregulated |
| -0,056865922 | 0,954185782 | EXOSC2 | FALSE | Downregulated |
| -0,042590241 | 0,954185782 | PRKD2 | FALSE | Downregulated |
| -0,060972686 | 0,954301405 | LOC102724187 | FALSE | Downregulated |
| -0,11651682 | 0,954324555 | MRPL50 | FALSE | Downregulated |
| -0,039988906 | 0,954461206 | CCDC69 | FALSE | Downregulated |
| 0,085395038 | 0,954461206 | COLQ | FALSE | Upregulated |
| 0,072354436 | 0,954461206 | DDX59 | FALSE | Upregulated |
| -0,03632073 | 0,954461206 | EPG5 | FALSE | Downregulated |
| -0,040949639 | 0,954461206 | FBXW11 | FALSE | Downregulated |
| 0,0386061 | 0,954461206 | FOXP4 | FALSE | Upregulated |
| 0,047571358 | 0,954461206 | GK | FALSE | Upregulated |
| -0,164559024 | 0,954461206 | KIT | FALSE | Downregulated |
| 0,096431559 | 0,954461206 | NME1 | FALSE | Upregulated |
| -0,067809344 | 0,954461206 | RRP36 | FALSE | Downregulated |
| 0,047276079 | 0,954461206 | SLC2A6 | FALSE | Upregulated |
| 0,025098936 | 0,954461206 | SUGP2 | FALSE | Upregulated |
| -0,031037656 | 0,954493196 | ABCA6 | FALSE | Downregulated |
| 0,088960258 | 0,954493196 | DPY30 | FALSE | Upregulated |
| 0,115120258 | 0,954493196 | EMR4P | FALSE | Upregulated |
| 0,108670485 | 0,954493196 | ERCC5 | FALSE | Upregulated |
| 0,140413 | 0,954493196 | MORF4L2-AS1 | FALSE | Upregulated |
| -0,054929139 | 0,954493196 | TOE1 | FALSE | Downregulated |
| -0,044011972 | 0,954493196 | ZNF280D | FALSE | Downregulated |
| -0,038039099 | 0,954580326 | ABCF3 | FALSE | Downregulated |
| -0,066200489 | 0,954580326 | ANP32A-IT1 | FALSE | Downregulated |
| 0,102578195 | 0,954580326 | BSPRY | FALSE | Upregulated |
| -0,042370954 | 0,954580326 | CECR5 | FALSE | Downregulated |
| -0,078411936 | 0,954580326 | FAM156B | FALSE | Downregulated |
| -0,148561636 | 0,954580326 | HIST2H4B | FALSE | Downregulated |
| 0,047865649 | 0,954580326 | LOC100134040 | FALSE | Upregulated |
| -0,094859197 | 0,954580326 | LOC101929280 | FALSE | Downregulated |
| -0,062131678 | 0,954580326 | LOC101929522 | FALSE | Downregulated |
| -0,1182234 | 0,954580326 | LOC102723703 | FALSE | Downregulated |
| 0,056126664 | 0,954580326 | MACROD2-AS1 | FALSE | Upregulated |
| 0,100019468 | 0,954580326 | MAD2L1BP | FALSE | Upregulated |
| -0,041085881 | 0,954580326 | NINL | FALSE | Downregulated |
| 0,037258882 | 0,954580326 | RP11-142C4.6 | FALSE | Upregulated |
| 0,052733085 | 0,954580326 | RP3-476K8.3 | FALSE | Upregulated |
| -0,020201336 | 0,954580326 | RPL15 | FALSE | Downregulated |
| 0,143594801 | 0,954580326 | SLC7A4 | FALSE | Upregulated |
| -0,147885579 | 0,954580326 | SNORA57 | FALSE | Downregulated |
| 0,066420484 | 0,954580326 | STX18 | FALSE | Upregulated |
| -0,054218663 | 0,954617636 | UBA2 | FALSE | Downregulated |
| 0,028714575 | 0,954688614 | AHCYL1 | FALSE | Upregulated |
| 0,037397205 | 0,954688614 | SNX19 | FALSE | Upregulated |
| -0,07640489 | 0,95471191 | LOC101928154 | FALSE | Downregulated |

| | | | | |
|---|---|---|---|---|
| 0,12584027 | 0,95471559 | SYP | FALSE | Upregulated |
| 0,083445073 | 0,954792336 | PRR24 | FALSE | Upregulated |
| 0,125704569 | 0,954792336 | SIRPB1 | FALSE | Upregulated |
| -0,032342015 | 0,955035974 | CFL1 | FALSE | Downregulated |
| 0,048734801 | 0,955122197 | SLIT3 | FALSE | Upregulated |
| -0,08798619 | 0,955122197 | WDR25 | FALSE | Downregulated |
| -0,064953642 | 0,955127624 | ATP6V0E1 | FALSE | Downregulated |
| 0,095135961 | 0,955127624 | CNTN4 | FALSE | Upregulated |
| 0,033995167 | 0,955127624 | EIF2A | FALSE | Upregulated |
| -0,18702746 | 0,955127624 | OPA1-AS1 | FALSE | Downregulated |
| -0,194437025 | 0,955127624 | RGS18 | FALSE | Downregulated |
| -0,062877624 | 0,955127624 | ZNF260 | FALSE | Downregulated |
| -0,142148102 | 0,955230035 | DBNDD2 | FALSE | Downregulated |
| -0,096319395 | 0,955230035 | LINC00938 | FALSE | Downregulated |
| 0,119965056 | 0,955240383 | LOC102724376 | FALSE | Upregulated |
| -0,112504705 | 0,955373278 | C1orf131 | FALSE | Downregulated |
| 0,024255802 | 0,955373278 | HNRNPA3 | FALSE | Upregulated |
| 0,053774023 | 0,955373278 | SCFD2 | FALSE | Upregulated |
| -0,252929093 | 0,955373278 | STX16-NPEPL1 | FALSE | Downregulated |
| 0,03708233 | 0,955441809 | CACTIN-AS1 | FALSE | Upregulated |
| 0,025996369 | 0,9557139 | CLIP1 | FALSE | Upregulated |
| 0,196960808 | 0,9557139 | FRMPD3 | FALSE | Upregulated |
| 0,023890614 | 0,955718233 | ANKRD17 | FALSE | Upregulated |
| -0,052912191 | 0,955718233 | COQ10A | FALSE | Downregulated |
| -0,094132675 | 0,955718233 | GAREML | FALSE | Downregulated |
| 0,133795409 | 0,955718233 | KIF17 | FALSE | Upregulated |
| 0,099526838 | 0,955718233 | LOC100506639 | FALSE | Upregulated |
| -0,108329104 | 0,955718233 | NEDD1 | FALSE | Downregulated |
| 0,064360895 | 0,955718233 | RCC1 | FALSE | Upregulated |
| -0,06434276 | 0,955718233 | RP11-41O4.2 | FALSE | Downregulated |
| -0,068815639 | 0,955718233 | SETD4 | FALSE | Downregulated |
| -0,087069426 | 0,955718233 | SNORA81 | FALSE | Downregulated |
| -0,029536297 | 0,955718233 | USP40 | FALSE | Downregulated |
| -0,053831089 | 0,955718233 | ZNF490 | FALSE | Downregulated |
| -0,114028019 | 0,955746351 | KLHDC1 | FALSE | Downregulated |
| 0,034451423 | 0,955746351 | OTC | FALSE | Upregulated |
| -0,078815566 | 0,955746351 | PLA1A | FALSE | Downregulated |
| -0,038975663 | 0,955746351 | RBM42 | FALSE | Downregulated |
| -0,045785309 | 0,955805766 | CNOT8 | FALSE | Downregulated |
| -0,062170166 | 0,955873183 | LOC101929691 | FALSE | Downregulated |
| 0,05709862 | 0,955898824 | LOC102724474 | FALSE | Upregulated |
| 0,077858048 | 0,955987759 | ISM1 | FALSE | Upregulated |
| -0,110185614 | 0,956013836 | MECOM | FALSE | Downregulated |
| 0,037685825 | 0,956194727 | PP7080 | FALSE | Upregulated |
| -0,033104112 | 0,956194727 | RBM5 | FALSE | Downregulated |
| 0,066506053 | 0,956344142 | RAB2B | FALSE | Upregulated |
| -0,054374691 | 0,956375536 | CYSTM1 | FALSE | Downregulated |
| 0,181433921 | 0,956431074 | GPR174 | FALSE | Upregulated |
| -0,14133206 | 0,956901282 | KCNAB3 | FALSE | Downregulated |
| 0,169034118 | 0,956901282 | LOC102724230 | FALSE | Upregulated |
| -0,047612191 | 0,956901282 | MAMDC2 | FALSE | Downregulated |

| | | | | |
|---|---|---|---|---|
| -0,058089503 | 0,956901282 | SCFD1 | FALSE | Downregulated |
| 0,077223311 | 0,956918143 | BAG2 | FALSE | Upregulated |
| -0,045173664 | 0,956918143 | CARD10 | FALSE | Downregulated |
| -0,038675419 | 0,956918143 | WRNIP1 | FALSE | Downregulated |
| -0,068990995 | 0,956937489 | DLEU2 | FALSE | Downregulated |
| -0,062774484 | 0,95694578 | MAFIP | FALSE | Downregulated |
| 0,030746354 | 0,956966456 | CASKIN2 | FALSE | Upregulated |
| -0,11089095 | 0,957038171 | CLIC2 | FALSE | Downregulated |
| 0,050812883 | 0,957132809 | CREBRF | FALSE | Upregulated |
| 0,029832286 | 0,957132809 | PCNX | FALSE | Upregulated |
| 0,086203436 | 0,957267682 | RP11-112H10.4 | FALSE | Upregulated |
| -0,090261243 | 0,957267682 | SCARA5 | FALSE | Downregulated |
| -0,039565088 | 0,957312398 | BHLHE40 | FALSE | Downregulated |
| 0,086455489 | 0,957509917 | MDGA1 | FALSE | Upregulated |
| -0,072122989 | 0,957509917 | NOP16 | FALSE | Downregulated |
| 0,118902019 | 0,957509917 | SYN3 | FALSE | Upregulated |
| -0,057064712 | 0,957545485 | DDX10 | FALSE | Downregulated |
| 0,053443861 | 0,957545485 | ERAP2 | FALSE | Upregulated |
| -0,046800694 | 0,957650807 | EFCAB7 | FALSE | Downregulated |
| -0,064046578 | 0,957650807 | LOC101929272 | FALSE | Downregulated |
| -0,02601561 | 0,957685941 | RBM6 | FALSE | Downregulated |
| -0,09039191 | 0,957933335 | MARS2 | FALSE | Downregulated |
| -0,036275673 | 0,957933335 | RANBP3 | FALSE | Downregulated |
| 0,132789785 | 0,957950411 | SPIN4 | FALSE | Upregulated |
| -0,038585198 | 0,958308709 | DNAJB2 | FALSE | Downregulated |
| -0,141292471 | 0,958378437 | ASIC3 | FALSE | Downregulated |
| -0,071525412 | 0,958394831 | INHBA | FALSE | Downregulated |
| -0,185412169 | 0,958534883 | EVPLL | FALSE | Downregulated |
| 0,098409497 | 0,958557389 | ICA1 | FALSE | Upregulated |
| -0,060821373 | 0,958557389 | ZNF594 | FALSE | Downregulated |
| -0,042854387 | 0,958589227 | CSNK1G3 | FALSE | Downregulated |
| 0,038937986 | 0,958678481 | PLEKHG1 | FALSE | Upregulated |
| -0,032709437 | 0,958773884 | DENND1C | FALSE | Downregulated |
| 0,047945898 | 0,958857434 | KLHL3 | FALSE | Upregulated |
| -0,080442872 | 0,958893101 | ART1 | FALSE | Downregulated |
| 0,030072002 | 0,958893101 | DNAH1 | FALSE | Upregulated |
| -0,07624765 | 0,958893101 | LINC00282 | FALSE | Downregulated |
| -0,111083721 | 0,958893101 | LOC100996693 | FALSE | Downregulated |
| 0,041620127 | 0,958893101 | LOC102723762 | FALSE | Upregulated |
| 0,120761338 | 0,958893101 | MACROD2 | FALSE | Upregulated |
| -0,034619681 | 0,958893101 | MNT | FALSE | Downregulated |
| 0,088015508 | 0,958893101 | NIFK | FALSE | Upregulated |
| -0,020735539 | 0,958893101 | RER1 | FALSE | Downregulated |
| 0,094393092 | 0,958893101 | SLMO2 | FALSE | Upregulated |
| -0,037007773 | 0,958893101 | STARD13 | FALSE | Downregulated |
| -0,110379593 | 0,958893101 | TENM4 | FALSE | Downregulated |
| -0,093604147 | 0,958893101 | TMEM27 | FALSE | Downregulated |
| 0,096037297 | 0,958893101 | TSSK6 | FALSE | Upregulated |
| 0,087734299 | 0,958893101 | VWA5A | FALSE | Upregulated |
| -0,031647969 | 0,958952466 | AXIN1 | FALSE | Downregulated |
| -0,071319312 | 0,959115311 | IL18R1 | FALSE | Downregulated |

| | | | | |
|---|---|---|---|---|
| -0,082403061 | 0,959177534 | SLC25A17 | FALSE | Downregulated |
| 0,03459902 | 0,95922511 | ERLEC1 | FALSE | Upregulated |
| 0,060753962 | 0,95922511 | FER1L5 | FALSE | Upregulated |
| 0,141566404 | 0,95922511 | LOC101929683 | FALSE | Upregulated |
| 0,034335934 | 0,95922511 | MCM7 | FALSE | Upregulated |
| -0,073277327 | 0,95922511 | PDF | FALSE | Downregulated |
| -0,049143219 | 0,95922511 | ZFYVE28 | FALSE | Downregulated |
| -0,05648354 | 0,959230028 | C19orf48 | FALSE | Downregulated |
| 0,10990898 | 0,959278389 | DGCR10 | FALSE | Upregulated |
| 0,0428562 | 0,959314376 | CLDN12 | FALSE | Upregulated |
| 0,135283963 | 0,959420537 | CKLF | FALSE | Upregulated |
| 0,119560261 | 0,959486739 | GATA3 | FALSE | Upregulated |
| -0,053080356 | 0,959687587 | PANK4 | FALSE | Downregulated |
| -0,037239276 | 0,959687587 | RBP1 | FALSE | Downregulated |
| 0,03522273 | 0,959898536 | TOP3A | FALSE | Upregulated |
| 0,066422378 | 0,960007651 | USP46 | FALSE | Upregulated |
| -0,020764672 | 0,960318149 | GNB1 | FALSE | Downregulated |
| 0,034644953 | 0,960330256 | ZNF236 | FALSE | Upregulated |
| -0,242516525 | 0,960835837 | CCZ1B | FALSE | Downregulated |
| 0,082234543 | 0,960944999 | LOC101928984 | FALSE | Upregulated |
| 0,210233658 | 0,960944999 | SNORD3B-2 | FALSE | Upregulated |
| -0,030537118 | 0,960944999 | VARS | FALSE | Downregulated |
| 0,168056478 | 0,961085793 | LOC100996634 | FALSE | Upregulated |
| 0,021728815 | 0,961102427 | NR3C1 | FALSE | Upregulated |
| -0,046538817 | 0,96111619 | C10orf25 | FALSE | Downregulated |
| -0,104935707 | 0,961267676 | GNB1L | FALSE | Downregulated |
| 0,028393669 | 0,961308379 | EZH1 | FALSE | Upregulated |
| -0,060358512 | 0,961308379 | STK38L | FALSE | Downregulated |
| 0,043096034 | 0,96134168 | KIAA0753 | FALSE | Upregulated |
| -0,034680859 | 0,961453076 | AGPAT3 | FALSE | Downregulated |
| 0,097706406 | 0,961460768 | CYP17A1 | FALSE | Upregulated |
| 0,128994286 | 0,961465962 | C6orf164 | FALSE | Upregulated |
| -0,267814018 | 0,961465962 | C7orf55-LUC7L2 | FALSE | Downregulated |
| -0,021513173 | 0,961465962 | CERK | FALSE | Downregulated |
| -0,289545743 | 0,961465962 | F8A3 | FALSE | Downregulated |
| 0,111464857 | 0,961465962 | HPGDS | FALSE | Upregulated |
| 0,126637838 | 0,961465962 | LINC01071 | FALSE | Upregulated |
| 0,090654138 | 0,961465962 | SERPINE2 | FALSE | Upregulated |
| 0,033241312 | 0,961465962 | VPS9D1 | FALSE | Upregulated |
| -0,033363811 | 0,961468391 | OXER1 | FALSE | Downregulated |
| -0,044125133 | 0,961577761 | TUBGCP3 | FALSE | Downregulated |
| 0,029319167 | 0,961595792 | AMIGO3 | FALSE | Upregulated |
| 0,072331061 | 0,961595792 | TMEM91 | FALSE | Upregulated |
| -0,038578469 | 0,961901753 | CHMP1B | FALSE | Downregulated |
| -0,087900818 | 0,961901753 | COQ2 | FALSE | Downregulated |
| 0,078197384 | 0,961901753 | GRIA3 | FALSE | Upregulated |
| -0,062697973 | 0,961901753 | MSI2 | FALSE | Downregulated |
| 0,082319793 | 0,961901753 | PRICKLE1 | FALSE | Upregulated |
| -0,031908619 | 0,961901753 | RNF126 | FALSE | Downregulated |
| 0,041803123 | 0,961901753 | SWT1 | FALSE | Upregulated |
| 0,038103176 | 0,961901753 | TIPARP | FALSE | Upregulated |

| | | | | |
|---|---|---|---|---|
| -0,031023234 | 0,961935134 | RNF168 | FALSE | Downregulated |
| 0,151598227 | 0,961935134 | RPL17-C18orf32 | FALSE | Upregulated |
| -0,029137386 | 0,961945928 | HIST1H1C | FALSE | Downregulated |
| -0,086514496 | 0,961945928 | LOC102724040 | FALSE | Downregulated |
| -0,017609612 | 0,962035536 | EML4 | FALSE | Downregulated |
| 0,029186708 | 0,962042699 | HM13 | FALSE | Upregulated |
| -0,109101826 | 0,962058086 | SMYD3 | FALSE | Downregulated |
| -0,04602703 | 0,962058086 | TUBE1 | FALSE | Downregulated |
| -0,050945573 | 0,962060096 | C14orf132 | FALSE | Downregulated |
| -0,030546878 | 0,962072076 | NR1H2 | FALSE | Downregulated |
| 0,089149182 | 0,962127754 | ASPA | FALSE | Upregulated |
| -0,031898486 | 0,962127754 | BCAS3 | FALSE | Downregulated |
| -0,032989963 | 0,962127754 | CPN1 | FALSE | Downregulated |
| -0,04554604 | 0,962127754 | CSF3R | FALSE | Downregulated |
| 0,070620447 | 0,962127754 | FAM49A | FALSE | Upregulated |
| 0,196725625 | 0,962127754 | HJURP | FALSE | Upregulated |
| 0,083302401 | 0,962127754 | LOC101928485 | FALSE | Upregulated |
| 0,033637554 | 0,962127754 | NARS | FALSE | Upregulated |
| 0,028309682 | 0,962127754 | SIAE | FALSE | Upregulated |
| 0,149737618 | 0,962127754 | TFCP2L1 | FALSE | Upregulated |
| -0,058665032 | 0,962127754 | TMEM165 | FALSE | Downregulated |
| -0,042577145 | 0,962127754 | TMEM9 | FALSE | Downregulated |
| 0,033192639 | 0,962127754 | TXLNG | FALSE | Upregulated |
| 0,055313479 | 0,962127754 | ZNF615 | FALSE | Upregulated |
| 0,037034703 | 0,962386423 | THOC2 | FALSE | Upregulated |
| 0,040816778 | 0,962547334 | GYG2 | FALSE | Upregulated |
| 0,045808629 | 0,962599261 | AC010127.3 | FALSE | Upregulated |
| 0,072640621 | 0,962599261 | DNAH9 | FALSE | Upregulated |
| 0,036277421 | 0,962675021 | CAPRIN2 | FALSE | Upregulated |
| -0,052013088 | 0,962675021 | DESI2 | FALSE | Downregulated |
| -0,019574449 | 0,962675021 | GNB2 | FALSE | Downregulated |
| 0,06562309 | 0,962675021 | GTF3C6 | FALSE | Upregulated |
| 0,056329125 | 0,962675021 | NAGPA | FALSE | Upregulated |
| -0,039077636 | 0,962675021 | NBPF26 | FALSE | Downregulated |
| 0,03647678 | 0,962675021 | PRR3 | FALSE | Upregulated |
| 0,086964156 | 0,962675021 | PYCR1 | FALSE | Upregulated |
| -0,05747498 | 0,962675021 | TIMM17B | FALSE | Downregulated |
| 0,084586557 | 0,962675021 | TMIGD2 | FALSE | Upregulated |
| -0,02524462 | 0,962675021 | VASH1 | FALSE | Downregulated |
| 0,68422348 | 0,962675021 | XIST | FALSE | Upregulated |
| -0,051425862 | 0,962837828 | BOD1 | FALSE | Downregulated |
| 0,019586362 | 0,962837828 | SKI | FALSE | Upregulated |
| 0,023593753 | 0,962887086 | ITGA1 | FALSE | Upregulated |
| -0,038011649 | 0,962992887 | HYOU1 | FALSE | Downregulated |
| 0,068870878 | 0,962992887 | ZNF443 | FALSE | Upregulated |
| -0,040623601 | 0,962992887 | ZNF503 | FALSE | Downregulated |
| -0,070876742 | 0,963148838 | DPF3 | FALSE | Downregulated |
| -0,111371321 | 0,963148838 | KTI12 | FALSE | Downregulated |
| 0,128843504 | 0,963330748 | HSPE1-MOB4 | FALSE | Upregulated |
| -0,061688458 | 0,963330748 | TEX14 | FALSE | Downregulated |
| 0,083632054 | 0,963341967 | CCDC122 | FALSE | Upregulated |

| | | | | |
|---|---|---|---|---|
| -0,026598378 | 0,963341967 | HP1BP3 | FALSE | Downregulated |
| -0,037719023 | 0,963341967 | PRKRIR | FALSE | Downregulated |
| 0,05268728 | 0,963341967 | PRUNE2 | FALSE | Upregulated |
| 0,032231126 | 0,963341967 | SMARCD1 | FALSE | Upregulated |
| 0,032512384 | 0,963341967 | USP15 | FALSE | Upregulated |
| 0,052363613 | 0,963382903 | NMNAT1 | FALSE | Upregulated |
| -0,055538194 | 0,963382903 | OMA1 | FALSE | Downregulated |
| 0,066147127 | 0,963382903 | ZNF418 | FALSE | Upregulated |
| 0,087142021 | 0,963606434 | CERS3 | FALSE | Upregulated |
| 0,07866546 | 0,963606434 | DGUOK | FALSE | Upregulated |
| 0,096204058 | 0,963606434 | IFT52 | FALSE | Upregulated |
| -0,103036724 | 0,963606434 | KIAA1324 | FALSE | Downregulated |
| -0,084913648 | 0,963606434 | KIAA1328 | FALSE | Downregulated |
| -0,05545143 | 0,963606434 | MRPL52 | FALSE | Downregulated |
| 0,078673422 | 0,963606434 | MYOT | FALSE | Upregulated |
| 0,04827099 | 0,963606434 | NSA2 | FALSE | Upregulated |
| -0,024958158 | 0,963606434 | PMM2 | FALSE | Downregulated |
| 0,068377657 | 0,963606434 | PPOX | FALSE | Upregulated |
| 0,084649782 | 0,963606434 | TMEM9B-AS1 | FALSE | Upregulated |
| 0,040550898 | 0,963606434 | TRIB3 | FALSE | Upregulated |
| -0,041311371 | 0,963606434 | TTLL5 | FALSE | Downregulated |
| -0,026979092 | 0,963606434 | ZMYND8 | FALSE | Downregulated |
| -0,022256342 | 0,963783843 | KDM4B | FALSE | Downregulated |
| -0,052883319 | 0,963909658 | ATP6V1H | FALSE | Downregulated |
| 0,065743336 | 0,963909658 | FBXL7 | FALSE | Upregulated |
| 0,030473507 | 0,963909658 | PPP2R4 | FALSE | Upregulated |
| 0,034385382 | 0,963909658 | TEF | FALSE | Upregulated |
| -0,041504548 | 0,963975698 | CCNT2-AS1 | FALSE | Downregulated |
| 0,134001047 | 0,964005823 | FAM35BP | FALSE | Upregulated |
| -0,021607191 | 0,964005823 | SIN3A | FALSE | Downregulated |
| 0,03172495 | 0,964034436 | LOC102723582 | FALSE | Upregulated |
| 0,034526132 | 0,9640907 | CEBPZ | FALSE | Upregulated |
| -0,052940374 | 0,964099381 | SH3PXD2A-AS1 | FALSE | Downregulated |
| 0,087656842 | 0,96413943 | ANAPC10 | FALSE | Upregulated |
| -0,037145416 | 0,964295325 | AGAP6 | FALSE | Downregulated |
| 0,087271062 | 0,964295325 | C7orf13 | FALSE | Upregulated |
| 0,030146328 | 0,964295325 | DEPDC5 | FALSE | Upregulated |
| 0,075838074 | 0,964295325 | DNAH2 | FALSE | Upregulated |
| -0,03045767 | 0,964295325 | FBXO33 | FALSE | Downregulated |
| 0,050302273 | 0,964295325 | HMGXB4 | FALSE | Upregulated |
| 0,068174166 | 0,964295325 | HSCB | FALSE | Upregulated |
| 0,08079885 | 0,964295325 | IL5RA | FALSE | Upregulated |
| 0,043425056 | 0,964295325 | IQCH | FALSE | Upregulated |
| 0,06055649 | 0,964295325 | ITGB1BP1 | FALSE | Upregulated |
| -0,157557141 | 0,964295325 | KCNA7 | FALSE | Downregulated |
| -0,111451253 | 0,964295325 | LOC101928398 | FALSE | Downregulated |
| 0,025018036 | 0,964295325 | MBNL2 | FALSE | Upregulated |
| -0,044731244 | 0,964295325 | MSRB2 | FALSE | Downregulated |
| 0,024853546 | 0,964295325 | MTA1 | FALSE | Upregulated |
| -0,072630555 | 0,964295325 | SLC16A10 | FALSE | Downregulated |
| -0,043739058 | 0,964295325 | UIMC1 | FALSE | Downregulated |

| | | | | |
|---|---|---|---|---|
| -0,054278519 | 0,964295325 | ZBTB2 | FALSE | Downregulated |
| -0,030957587 | 0,964300898 | SLC25A18 | FALSE | Downregulated |
| -0,044934622 | 0,964300898 | SMNDC1 | FALSE | Downregulated |
| 0,066215722 | 0,964387516 | ADM2 | FALSE | Upregulated |
| -0,029091913 | 0,964398943 | HIVEP1 | FALSE | Downregulated |
| 0,043156664 | 0,964398943 | RNPC3 | FALSE | Upregulated |
| -0,024503568 | 0,964620262 | PI4KA | FALSE | Downregulated |
| -0,050723784 | 0,964620262 | SEMA6B | FALSE | Downregulated |
| -0,048980138 | 0,964685507 | BCL2L12 | FALSE | Downregulated |
| -0,02677106 | 0,964793815 | ELAC2 | FALSE | Downregulated |
| -0,058065583 | 0,964793815 | ICT1 | FALSE | Downregulated |
| 0,04496616 | 0,964793815 | NGLY1 | FALSE | Upregulated |
| -0,043779806 | 0,964881576 | VAMP7 | FALSE | Downregulated |
| 0,06188677 | 0,9649665 | COX10 | FALSE | Upregulated |
| -0,026248842 | 0,9649665 | ZNF629 | FALSE | Downregulated |
| 0,078028796 | 0,964967262 | BMP4 | FALSE | Upregulated |
| 0,027374085 | 0,965111322 | URB1 | FALSE | Upregulated |
| 0,055356793 | 0,965173444 | GUCY1A3 | FALSE | Upregulated |
| 0,028264058 | 0,965173444 | IL2RB | FALSE | Upregulated |
| 0,088219914 | 0,965173444 | ISCA2 | FALSE | Upregulated |
| -0,184145299 | 0,965173444 | LOC102723433 | FALSE | Downregulated |
| -0,07226728 | 0,965173444 | TMEM98 | FALSE | Downregulated |
| -0,048456978 | 0,965173444 | ZNF81 | FALSE | Downregulated |
| -0,028342295 | 0,96526551 | ASH2L | FALSE | Downregulated |
| -0,099176247 | 0,96526551 | CEP41 | FALSE | Downregulated |
| 0,034894783 | 0,965282273 | ELP3 | FALSE | Upregulated |
| -0,018576491 | 0,965288622 | KDM2A | FALSE | Downregulated |
| 0,053280421 | 0,965479636 | EME2 | FALSE | Upregulated |
| -0,025847739 | 0,965785684 | ALDH1A1 | FALSE | Downregulated |
| 0,038360472 | 0,966020205 | MAP3K7 | FALSE | Upregulated |
| 0,018953326 | 0,966036669 | ATXN2L | FALSE | Upregulated |
| 0,039384948 | 0,966430796 | IGHMBP2 | FALSE | Upregulated |
| 0,03182792 | 0,966430796 | LOC102723853 | FALSE | Upregulated |
| 0,118372847 | 0,966430796 | RP11-182I10.3 | FALSE | Upregulated |
| -0,050731566 | 0,966520207 | C8orf47 | FALSE | Downregulated |
| 0,099945439 | 0,966520207 | HTR3E | FALSE | Upregulated |
| -0,027174686 | 0,966520207 | PEX26 | FALSE | Downregulated |
| 0,073162561 | 0,966520207 | SMIM20 | FALSE | Upregulated |
| 0,082305902 | 0,966559189 | CAMK4 | FALSE | Upregulated |
| 0,028083895 | 0,966559189 | CUL9 | FALSE | Upregulated |
| 0,038352848 | 0,966559189 | EMILIN3 | FALSE | Upregulated |
| 0,03891643 | 0,966559189 | KALRN | FALSE | Upregulated |
| 0,106029361 | 0,966559189 | KNDC1 | FALSE | Upregulated |
| -0,033006589 | 0,966559189 | LOC101928415 | FALSE | Downregulated |
| 0,198653729 | 0,966559189 | LOC102724504 | FALSE | Upregulated |
| -0,02817379 | 0,966559189 | PTPRS | FALSE | Downregulated |
| -0,05216337 | 0,966559189 | RP11-387D10.3 | FALSE | Downregulated |
| -0,023639098 | 0,966559189 | SPSB3 | FALSE | Downregulated |
| -0,039394953 | 0,966559189 | TMEM9B | FALSE | Downregulated |
| 0,021515349 | 0,966559189 | TSC1 | FALSE | Upregulated |
| -0,124851523 | 0,966559189 | VCX2 | FALSE | Downregulated |

| | | | | |
|---|---|---|---|---|
| -0,057444606 | 0,966740982 | DNAJC17 | FALSE | Downregulated |
| 0,047985101 | 0,966740982 | ENOPH1 | FALSE | Upregulated |
| 0,056942538 | 0,966740982 | MR1 | FALSE | Upregulated |
| -0,045205315 | 0,96686072 | MED11 | FALSE | Downregulated |
| 0,041467259 | 0,966884593 | C10orf95 | FALSE | Upregulated |
| 0,105255152 | 0,966884593 | FSD1L | FALSE | Upregulated |
| -0,134185698 | 0,966884593 | NUDT10 | FALSE | Downregulated |
| 0,120725378 | 0,966884593 | PTN | FALSE | Upregulated |
| -0,057674527 | 0,966884593 | SLPI | FALSE | Downregulated |
| 0,029731987 | 0,966884593 | ZNF674 | FALSE | Upregulated |
| 0,024116876 | 0,966915227 | CTC1 | FALSE | Upregulated |
| -0,036910068 | 0,966915227 | HNRNPD | FALSE | Downregulated |
| -0,026432161 | 0,966915227 | NUP153 | FALSE | Downregulated |
| -0,017145417 | 0,966968275 | ARFGEF1 | FALSE | Downregulated |
| 0,030848074 | 0,96700987 | SLC6A12 | FALSE | Upregulated |
| -0,028409738 | 0,967087322 | LOC646976 | FALSE | Downregulated |
| 0,02348035 | 0,967122714 | SETDB1 | FALSE | Upregulated |
| -0,017517081 | 0,967174446 | FAN1 | FALSE | Downregulated |
| 0,060782139 | 0,967174446 | PJA1 | FALSE | Upregulated |
| -0,023700558 | 0,967174446 | PPIL2 | FALSE | Downregulated |
| 0,024199297 | 0,967174446 | SUN1 | FALSE | Upregulated |
| -0,024527196 | 0,967205118 | RPS6KA3 | FALSE | Downregulated |
| 0,030417757 | 0,967276969 | FANCL | FALSE | Upregulated |
| 0,092303912 | 0,967276969 | ITGB3BP | FALSE | Upregulated |
| -0,037037341 | 0,967276969 | LOC101928555 | FALSE | Downregulated |
| 0,484345139 | 0,967276969 | UBE2F-SCLY | FALSE | Upregulated |
| -0,028859226 | 0,967446821 | RSL1D1 | FALSE | Downregulated |
| -0,053493133 | 0,967446821 | SLC25A40 | FALSE | Downregulated |
| 0,534832357 | 0,967475687 | CKM | FALSE | Upregulated |
| -0,113908535 | 0,967481848 | ABHD17C | FALSE | Downregulated |
| -0,026186805 | 0,967481848 | CRTC3 | FALSE | Downregulated |
| 0,059766273 | 0,967481848 | DHRS11 | FALSE | Upregulated |
| 0,024609461 | 0,967481848 | DOCK1 | FALSE | Upregulated |
| -0,095969781 | 0,967481848 | LOC91450 | FALSE | Downregulated |
| 0,035447673 | 0,967481848 | NQO1 | FALSE | Upregulated |
| 0,064302229 | 0,967481848 | POT1 | FALSE | Upregulated |
| 0,026085199 | 0,967481848 | SNAP29 | FALSE | Upregulated |
| 0,061498719 | 0,967481848 | TOX3 | FALSE | Upregulated |
| -0,032111888 | 0,967495653 | MMS19 | FALSE | Downregulated |
| -0,036473458 | 0,967495653 | STRIP1 | FALSE | Downregulated |
| -0,029653279 | 0,967584755 | IFRD2 | FALSE | Downregulated |
| -0,10057316 | 0,967632025 | DNAJC27 | FALSE | Downregulated |
| -0,077436251 | 0,967634333 | LOC441242 | FALSE | Downregulated |
| -0,025045905 | 0,967645161 | ZDHHC14 | FALSE | Downregulated |
| 0,027489925 | 0,967733924 | C2orf16 | FALSE | Upregulated |
| 0,079141401 | 0,967733924 | SNRPD1 | FALSE | Upregulated |
| -0,046602552 | 0,967771666 | EIF4E3 | FALSE | Downregulated |
| -0,041850727 | 0,967878933 | C11orf35 | FALSE | Downregulated |
| 0,028528496 | 0,967878933 | THADA | FALSE | Upregulated |
| -0,029384333 | 0,967986475 | LRCH1 | FALSE | Downregulated |
| 0,100898052 | 0,967986475 | UPP2 | FALSE | Upregulated |

| | | | | |
|---|---|---|---|---|
| 0,041988126 | 0,967994146 | ARHGEF5 | FALSE | Upregulated |
| -0,053696744 | 0,968020404 | ZNF70 | FALSE | Downregulated |
| -0,036021584 | 0,968022589 | TPRA1 | FALSE | Downregulated |
| 0,04589203 | 0,968048984 | DNAL1 | FALSE | Upregulated |
| 0,080289462 | 0,968048984 | FAM92A1P2 | FALSE | Upregulated |
| -0,035488762 | 0,968048984 | LOC102724250 | FALSE | Downregulated |
| -0,057379652 | 0,968048984 | ZNF486 | FALSE | Downregulated |
| -0,033097798 | 0,96819659 | CIB1 | FALSE | Downregulated |
| 0,053345612 | 0,96819659 | LMOD2 | FALSE | Upregulated |
| 0,072412535 | 0,968415039 | PRMT3 | FALSE | Upregulated |
| 0,03720721 | 0,968629738 | FAM109A | FALSE | Upregulated |
| 0,091349623 | 0,968629738 | LOC101929790 | FALSE | Upregulated |
| 0,05148883 | 0,968689469 | FOXP2 | FALSE | Upregulated |
| 0,035952776 | 0,968881305 | KLHL9 | FALSE | Upregulated |
| 0,050086153 | 0,968881305 | LOC100996876 | FALSE | Upregulated |
| -0,091813328 | 0,968881305 | MIR4458HG | FALSE | Downregulated |
| -0,031300169 | 0,969030837 | FEM1A | FALSE | Downregulated |
| -0,03270868 | 0,969062756 | CLDN2 | FALSE | Downregulated |
| -0,126248856 | 0,969062756 | LOC101927506 | FALSE | Downregulated |
| 0,105819538 | 0,969062756 | MGC57346 | FALSE | Upregulated |
| 0,04117162 | 0,969062756 | WNT3 | FALSE | Upregulated |
| 0,016658123 | 0,969307091 | ATP1A1 | FALSE | Upregulated |
| -0,053637245 | 0,969307091 | CD1D | FALSE | Downregulated |
| -0,021521114 | 0,969307091 | ERVK13-1 | FALSE | Downregulated |
| 0,075337172 | 0,969307091 | LOC101927073 | FALSE | Upregulated |
| -0,034299256 | 0,969307091 | LOC440386 | FALSE | Downregulated |
| 0,055478042 | 0,969307091 | MAP3K8 | FALSE | Upregulated |
| -0,031455838 | 0,969307091 | PSMB6 | FALSE | Downregulated |
| 0,038789509 | 0,969307091 | RPS6KC1 | FALSE | Upregulated |
| 0,017567367 | 0,969307091 | SEC16A | FALSE | Upregulated |
| -0,119577984 | 0,969307091 | SNX16 | FALSE | Downregulated |
| -0,040715374 | 0,969307091 | ZNF772 | FALSE | Downregulated |
| 0,107985211 | 0,969326506 | LOC100652736 | FALSE | Upregulated |
| 0,04465248 | 0,969567335 | AGAP4 | FALSE | Upregulated |
| -0,053622965 | 0,969567335 | CASP6 | FALSE | Downregulated |
| 0,112038572 | 0,969567335 | CCSER1 | FALSE | Upregulated |
| 0,016991619 | 0,969567335 | NCOA2 | FALSE | Upregulated |
| 0,051391611 | 0,969567335 | NSDHL | FALSE | Upregulated |
| 0,037945118 | 0,969567335 | NUDT16P1 | FALSE | Upregulated |
| 0,025076847 | 0,969567335 | SHE | FALSE | Upregulated |
| -0,0282376 | 0,96970136 | LOC102724761 | FALSE | Downregulated |
| -0,021879041 | 0,969713932 | TRAM1 | FALSE | Downregulated |
| 0,035096797 | 0,969825166 | QTRT1 | FALSE | Upregulated |
| 0,02248504 | 0,969934426 | EVL | FALSE | Upregulated |
| -0,028786668 | 0,969973457 | ATP6V1A | FALSE | Downregulated |
| -0,021883436 | 0,969973457 | CUL7 | FALSE | Downregulated |
| 0,044448022 | 0,969973457 | LINC00894 | FALSE | Upregulated |
| 0,057806345 | 0,969973457 | NEK3 | FALSE | Upregulated |
| -0,036453348 | 0,969973457 | NRM | FALSE | Downregulated |
| 0,024179249 | 0,969973457 | TNK2 | FALSE | Upregulated |
| 0,059640992 | 0,969973457 | WDR31 | FALSE | Upregulated |

| | | | | |
|---|---|---|---|---|
| 0,031363138 | 0,970058469 | FAM136A | FALSE | Upregulated |
| 0,026796128 | 0,970058469 | TAB1 | FALSE | Upregulated |
| -0,028353379 | 0,970160547 | AGPS | FALSE | Downregulated |
| -0,041142277 | 0,970162688 | IQCE | FALSE | Downregulated |
| 0,052332638 | 0,970162923 | DNAJB4 | FALSE | Upregulated |
| 0,021491575 | 0,970186951 | ARF6 | FALSE | Upregulated |
| -0,168565318 | 0,970250019 | ASCL1 | FALSE | Downregulated |
| -0,028764149 | 0,970250019 | CSNK1E | FALSE | Downregulated |
| -0,036082408 | 0,970250019 | TMBIM4 | FALSE | Downregulated |
| 0,023728731 | 0,970350007 | STRN4 | FALSE | Upregulated |
| -0,047059632 | 0,970422436 | NDUFA4L2 | FALSE | Downregulated |
| -0,030802979 | 0,970439153 | PCID2 | FALSE | Downregulated |
| -0,023724474 | 0,970499365 | CAMK2N1 | FALSE | Downregulated |
| 0,022383385 | 0,970499365 | CEP250 | FALSE | Upregulated |
| 0,041992741 | 0,970499365 | CYB5B | FALSE | Upregulated |
| -0,030238693 | 0,970499365 | DDTL | FALSE | Downregulated |
| -0,022403939 | 0,970499365 | GABARAPL1 | FALSE | Downregulated |
| 0,028306185 | 0,970499365 | LOC100507217 | FALSE | Upregulated |
| -0,063643736 | 0,970499365 | ZNF221 | FALSE | Downregulated |
| 0,094296516 | 0,970499365 | ZNF547 | FALSE | Upregulated |
| -0,064184238 | 0,970503055 | RPL21P44 | FALSE | Downregulated |
| 0,019523552 | 0,970503055 | WDFY3 | FALSE | Upregulated |
| 0,01489048 | 0,970517836 | DNM2 | FALSE | Upregulated |
| -0,044207388 | 0,970517836 | FBXL4 | FALSE | Downregulated |
| 0,094243387 | 0,970517836 | LILRA5 | FALSE | Upregulated |
| 0,026047158 | 0,970517836 | SLC11A2 | FALSE | Upregulated |
| 0,032652718 | 0,970594931 | DAP3 | FALSE | Upregulated |
| -0,055178204 | 0,970594931 | GPR65 | FALSE | Downregulated |
| -0,116822052 | 0,970594931 | LOC101929145 | FALSE | Downregulated |
| -0,029004775 | 0,970594931 | RAB12 | FALSE | Downregulated |
| 0,037445989 | 0,970594931 | RNLS | FALSE | Upregulated |
| 0,049060565 | 0,970651943 | CD300A | FALSE | Upregulated |
| -0,062131357 | 0,970651943 | LOC101928334 | FALSE | Downregulated |
| -0,035533367 | 0,970651943 | MPHOSPH9 | FALSE | Downregulated |
| -0,01306347 | 0,970651943 | PHRF1 | FALSE | Downregulated |
| -0,03336483 | 0,970860378 | CMTM8 | FALSE | Downregulated |
| -0,032760012 | 0,970860378 | MKNK1 | FALSE | Downregulated |
| 0,024890605 | 0,970860378 | PDLIM1 | FALSE | Upregulated |
| -0,044795498 | 0,970860378 | VAMP4 | FALSE | Downregulated |
| 0,058205487 | 0,970860378 | ZNF177 | FALSE | Upregulated |
| -0,020480727 | 0,970902672 | XPC | FALSE | Downregulated |
| -0,024748694 | 0,970936518 | LOC644090 | FALSE | Downregulated |
| 0,039501641 | 0,971020295 | LRRC20 | FALSE | Upregulated |
| -0,016794411 | 0,971020295 | PTPN23 | FALSE | Downregulated |
| 0,067165454 | 0,971020295 | ZNF416 | FALSE | Upregulated |
| -0,028506465 | 0,971107412 | SLC16A8 | FALSE | Downregulated |
| -0,068577835 | 0,971107412 | TMEM132E | FALSE | Downregulated |
| -0,05795667 | 0,971134389 | LCMT2 | FALSE | Downregulated |
| 0,088959718 | 0,971134389 | RPS15AP10 | FALSE | Upregulated |
| 0,060311851 | 0,971203349 | GUCY1B3 | FALSE | Upregulated |
| -0,022708022 | 0,971203349 | MIDN | FALSE | Downregulated |

| | | | | |
|---|---|---|---|---|
| 0,055990218 | 0,971259725 | FAM217B | FALSE | Upregulated |
| 0,017634799 | 0,971259725 | WHSC1 | FALSE | Upregulated |
| -0,078711659 | 0,971439611 | LY6K | FALSE | Downregulated |
| 0,026012087 | 0,971455556 | ZNF181 | FALSE | Upregulated |
| 0,028724813 | 0,971456958 | OIP5-AS1 | FALSE | Upregulated |
| 0,095513613 | 0,971456958 | TSLP | FALSE | Upregulated |
| 0,081797093 | 0,971461399 | C5AR2 | FALSE | Upregulated |
| 0,064348082 | 0,971664816 | DCAF4L1 | FALSE | Upregulated |
| 0,03794872 | 0,971675672 | ATG5 | FALSE | Upregulated |
| 0,027820775 | 0,971675672 | CAMSAP1 | FALSE | Upregulated |
| -0,09407388 | 0,971675672 | FAM154B | FALSE | Downregulated |
| -0,040957717 | 0,971675672 | HSPBP1 | FALSE | Downregulated |
| -0,081048067 | 0,971675672 | LOC102723893 | FALSE | Downregulated |
| -0,133020081 | 0,971675672 | PMF1-BGLAP | FALSE | Downregulated |
| 0,026771927 | 0,971675672 | ZNF837 | FALSE | Upregulated |
| 0,060597861 | 0,971893731 | ZNF626 | FALSE | Upregulated |
| 0,32117156 | 0,971930096 | TMEM189-UBE2V1 | FALSE | Upregulated |
| -0,101151484 | 0,971955413 | RP11-205M3.3 | FALSE | Downregulated |
| 0,260052415 | 0,972025619 | SLX1A-SULT1A3 | FALSE | Upregulated |
| -0,027816069 | 0,972250854 | MAFG | FALSE | Downregulated |
| 0,069008169 | 0,972250854 | RAD21-AS1 | FALSE | Upregulated |
| -0,025481889 | 0,972250854 | SLC25A5-AS1 | FALSE | Downregulated |
| 0,013788422 | 0,972250854 | TUBGCP6 | FALSE | Upregulated |
| -0,055738539 | 0,972250854 | ZNF613 | FALSE | Downregulated |
| 0,028520479 | 0,972348254 | B3GNT8 | FALSE | Upregulated |
| -0,071807811 | 0,972403904 | ANKRD33B | FALSE | Downregulated |
| 0,297005996 | 0,972403904 | C10orf32-ASMT | FALSE | Upregulated |
| -0,046680194 | 0,972403904 | CASD1 | FALSE | Downregulated |
| -0,079591593 | 0,972403904 | GATA6-AS1 | FALSE | Downregulated |
| 0,033896597 | 0,972403904 | HBCBP | FALSE | Upregulated |
| 0,021824022 | 0,972403904 | INADL | FALSE | Upregulated |
| 0,030034871 | 0,972403904 | IRF5 | FALSE | Upregulated |
| 0,03911327 | 0,972403904 | ISCA1 | FALSE | Upregulated |
| 0,036897422 | 0,972403904 | ITPA | FALSE | Upregulated |
| -0,019902261 | 0,972403904 | PARP1 | FALSE | Downregulated |
| 0,075394555 | 0,972403904 | PKD1L1 | FALSE | Upregulated |
| 0,078635663 | 0,972403904 | UXT-AS1 | FALSE | Upregulated |
| 0,029838193 | 0,972403904 | ZNF266 | FALSE | Upregulated |
| -0,019725113 | 0,97241181 | CUL5 | FALSE | Downregulated |
| -0,031942895 | 0,972425618 | LDLRAP1 | FALSE | Downregulated |
| -0,020452822 | 0,972593462 | SLC4A2 | FALSE | Downregulated |
| 0,017982926 | 0,972793152 | GPC1 | FALSE | Upregulated |
| -0,023836445 | 0,972830901 | BAG1 | FALSE | Downregulated |
| -0,022724002 | 0,972830901 | MARCH8 | FALSE | Downregulated |
| -0,011810329 | 0,972840252 | MPRIP | FALSE | Downregulated |
| -0,03307882 | 0,972840252 | PTGES2 | FALSE | Downregulated |
| -0,078452979 | 0,972840252 | STAG3L1 | FALSE | Downregulated |
| -0,061506381 | 0,973020134 | C2orf82 | FALSE | Downregulated |
| -0,014467766 | 0,973037782 | ARHGDIA | FALSE | Downregulated |
| 0,051727722 | 0,973037782 | GGPS1 | FALSE | Upregulated |
| 0,053221463 | 0,973070999 | DENND1B | FALSE | Upregulated |

| | | | | |
|---|---|---|---|---|
| 0,05320439 | 0,973361208 | ZNF345 | FALSE | Upregulated |
| -0,044902744 | 0,973438673 | SAMD9 | FALSE | Downregulated |
| -0,06512024 | 0,973563668 | CDC42EP2 | FALSE | Downregulated |
| -0,032114257 | 0,973563668 | CHRNE | FALSE | Downregulated |
| -0,018376442 | 0,973563668 | MBD1 | FALSE | Downregulated |
| 0,032149043 | 0,973563668 | TDRD3 | FALSE | Upregulated |
| 0,03037384 | 0,973626407 | ANKMY1 | FALSE | Upregulated |
| 0,035948927 | 0,973626407 | C11orf68 | FALSE | Upregulated |
| -0,022193461 | 0,973626407 | DEF8 | FALSE | Downregulated |
| -0,061185423 | 0,973626407 | FBXL8 | FALSE | Downregulated |
| -0,02249457 | 0,973626407 | FOXM1 | FALSE | Downregulated |
| 0,056088118 | 0,973626407 | LINC00899 | FALSE | Upregulated |
| -0,048875191 | 0,973626407 | LINC00921 | FALSE | Downregulated |
| 0,080404623 | 0,973626407 | LOC101929295 | FALSE | Upregulated |
| -0,052999302 | 0,973626407 | LOC727896 | FALSE | Downregulated |
| 0,033809587 | 0,973626407 | NOS3 | FALSE | Upregulated |
| 0,047369584 | 0,973626407 | RAB3D | FALSE | Upregulated |
| 0,091379502 | 0,973626407 | RET | FALSE | Upregulated |
| 0,159481457 | 0,973626407 | RGPD6 | FALSE | Upregulated |
| 0,022686116 | 0,973626407 | SCIMP | FALSE | Upregulated |
| 0,013880152 | 0,973626407 | TRIM44 | FALSE | Upregulated |
| -0,020924819 | 0,973626407 | YARS | FALSE | Downregulated |
| 0,079108589 | 0,973626407 | ZNF519 | FALSE | Upregulated |
| 0,027348645 | 0,973865318 | SYDE1 | FALSE | Upregulated |
| 0,030305379 | 0,973979305 | MMAA | FALSE | Upregulated |
| 0,027034845 | 0,974076672 | PPP1R9A | FALSE | Upregulated |
| 0,042068152 | 0,97421905 | FAM43A | FALSE | Upregulated |
| 0,091442491 | 0,97421905 | LINC00888 | FALSE | Upregulated |
| -0,01758783 | 0,97421905 | WIZ | FALSE | Downregulated |
| 0,017290994 | 0,974221347 | ENO1 | FALSE | Upregulated |
| 0,042909005 | 0,974259891 | INTS2 | FALSE | Upregulated |
| 0,034110396 | 0,974259891 | PMCHL2 | FALSE | Upregulated |
| 0,029305048 | 0,974259891 | SDHAP2 | FALSE | Upregulated |
| 0,101886015 | 0,974417146 | CLEC18C | FALSE | Upregulated |
| -0,044570909 | 0,974417146 | DKFZP586I1420 | FALSE | Downregulated |
| -0,02694016 | 0,974417146 | MMADHC | FALSE | Downregulated |
| 0,062672118 | 0,974417146 | MTMR8 | FALSE | Upregulated |
| 0,038790429 | 0,97449256 | HDHD1 | FALSE | Upregulated |
| -0,092054216 | 0,974554018 | RASA4CP | FALSE | Downregulated |
| -0,030930603 | 0,974554018 | SCYL3 | FALSE | Downregulated |
| 0,078984972 | 0,974554018 | TMEM154 | FALSE | Upregulated |
| 0,117117642 | 0,974554018 | UGT2B17 | FALSE | Upregulated |
| 0,018087707 | 0,974554018 | ZHX2 | FALSE | Upregulated |
| 0,073014621 | 0,974562839 | RASAL1 | FALSE | Upregulated |
| 0,04717159 | 0,974562839 | THOC3 | FALSE | Upregulated |
| 0,031765605 | 0,974607899 | C17orf80 | FALSE | Upregulated |
| 0,020846043 | 0,974607899 | ERCC1 | FALSE | Upregulated |
| 0,0171721 | 0,974607899 | HDAC6 | FALSE | Upregulated |
| 0,020825766 | 0,974802714 | BCL9 | FALSE | Upregulated |
| 0,082256687 | 0,974802714 | SLC25A21-AS1 | FALSE | Upregulated |
| 0,053223873 | 0,974899205 | ALPK2 | FALSE | Upregulated |

| | | | | |
|---|---|---|---|---|
| -0,018654957 | 0,974899205 | AP4S1 | FALSE | Downregulated |
| -0,025707181 | 0,974899205 | CLPTM1L | FALSE | Downregulated |
| 0,090766066 | 0,974899205 | GPR4 | FALSE | Upregulated |
| -0,055597462 | 0,974899205 | LOC100505570 | FALSE | Downregulated |
| 0,066556541 | 0,974899205 | TOB1-AS1 | FALSE | Upregulated |
| -0,039439957 | 0,974909103 | TMEM261 | FALSE | Downregulated |
| -0,064498376 | 0,975158597 | RPIA | FALSE | Downregulated |
| -0,026293232 | 0,975315555 | GYS1 | FALSE | Downregulated |
| -0,091870555 | 0,975379729 | ALDH1A2 | FALSE | Downregulated |
| -0,05596299 | 0,975379729 | CCR10 | FALSE | Downregulated |
| -0,034366172 | 0,975379729 | EPM2A | FALSE | Downregulated |
| -0,030982231 | 0,975379729 | LOC102723972 | FALSE | Downregulated |
| -0,030831093 | 0,975379729 | MBL2 | FALSE | Downregulated |
| 0,054535466 | 0,975379729 | ROM1 | FALSE | Upregulated |
| 0,018676948 | 0,975379729 | SS18 | FALSE | Upregulated |
| 0,034501043 | 0,975379729 | WWC2-AS2 | FALSE | Upregulated |
| 0,105536806 | 0,975592907 | HTR7 | FALSE | Upregulated |
| -0,020848187 | 0,97559507 | LARS | FALSE | Downregulated |
| -0,043401338 | 0,975606315 | EFNA2 | FALSE | Downregulated |
| -0,014103314 | 0,975606315 | NIT1 | FALSE | Downregulated |
| -0,042077222 | 0,975606315 | ZNF662 | FALSE | Downregulated |
| -0,024311484 | 0,975724495 | AC007038.7 | FALSE | Downregulated |
| 0,0288457 | 0,975724495 | CNNM1 | FALSE | Upregulated |
| -0,044827594 | 0,975724495 | DNAJC2 | FALSE | Downregulated |
| 0,021187491 | 0,975724495 | EPT1 | FALSE | Upregulated |
| -0,084039017 | 0,975724495 | HIST1H3H | FALSE | Downregulated |
| 0,133601695 | 0,975724495 | HIST2H2AA3 | FALSE | Upregulated |
| 0,023500948 | 0,975724495 | IMP3 | FALSE | Upregulated |
| 0,05866499 | 0,975724495 | LOC101928172 | FALSE | Upregulated |
| 0,051865807 | 0,975724495 | NUP210L | FALSE | Upregulated |
| -0,024109108 | 0,975724495 | PCMTD1 | FALSE | Downregulated |
| 0,017802875 | 0,975724495 | PLEKHJ1 | FALSE | Upregulated |
| 0,064126751 | 0,975724495 | PRO2852 | FALSE | Upregulated |
| -0,019834478 | 0,975724495 | PSMC5 | FALSE | Downregulated |
| -0,103519053 | 0,975724495 | SENP3 | FALSE | Downregulated |
| -0,020441847 | 0,975724495 | SFXN1 | FALSE | Downregulated |
| -0,031600567 | 0,975724495 | SGPP1 | FALSE | Downregulated |
| -0,035019117 | 0,975724495 | TMTC4 | FALSE | Downregulated |
| 0,02710559 | 0,975724495 | TRAPPC5 | FALSE | Upregulated |
| -0,017398627 | 0,975724495 | TRIM28 | FALSE | Downregulated |
| 0,041988546 | 0,975724495 | TULP3 | FALSE | Upregulated |
| -0,023798676 | 0,975724495 | VPS36 | FALSE | Downregulated |
| 0,031834693 | 0,975724495 | VSNL1 | FALSE | Upregulated |
| 0,016178415 | 0,975724582 | AP2B1 | FALSE | Upregulated |
| -0,103475681 | 0,975724582 | DAB1-AS1 | FALSE | Downregulated |
| -0,03953708 | 0,975724582 | FAM114A2 | FALSE | Downregulated |
| -0,040646656 | 0,975724582 | GLB1L | FALSE | Downregulated |
| -0,023703506 | 0,975724582 | GTF2A1 | FALSE | Downregulated |
| -0,018369321 | 0,975724582 | IRF7 | FALSE | Downregulated |
| 0,036889786 | 0,975724582 | KLHL11 | FALSE | Upregulated |
| 0,039871968 | 0,975724582 | OTUD3 | FALSE | Upregulated |

| | | | | |
|---|---|---|---|---|
| 0,042525738 | 0,975724582 | RP11-203B9.4 | FALSE | Upregulated |
| -0,019633908 | 0,975724582 | SIN3B | FALSE | Downregulated |
| 0,030526347 | 0,975724582 | ZNF420 | FALSE | Upregulated |
| -0,013493477 | 0,975737495 | SGPL1 | FALSE | Downregulated |
| 0,032846081 | 0,975737495 | ZNF785 | FALSE | Upregulated |
| -0,067024087 | 0,975753019 | BEND3 | FALSE | Downregulated |
| -0,01902966 | 0,975753019 | MYO7A | FALSE | Downregulated |
| 0,025357781 | 0,975770338 | ANKRD10 | FALSE | Upregulated |
| -0,050078413 | 0,975770338 | BTBD11 | FALSE | Downregulated |
| 0,024459768 | 0,975770338 | GADD45GIP1 | FALSE | Upregulated |
| 0,318919921 | 0,975770338 | LOC102723595 | FALSE | Upregulated |
| -0,064634883 | 0,975770338 | LOC728323 | FALSE | Downregulated |
| 0,049028794 | 0,975770338 | PCNA | FALSE | Upregulated |
| -0,042473093 | 0,975770338 | RSPO3 | FALSE | Downregulated |
| 0,039016938 | 0,975770338 | SLC9A1 | FALSE | Upregulated |
| -0,029948582 | 0,975770338 | SPICE1 | FALSE | Downregulated |
| -0,035739811 | 0,975770338 | TBX15 | FALSE | Downregulated |
| -0,025995787 | 0,976092076 | ATAD3A | FALSE | Downregulated |
| -0,030219817 | 0,976092076 | MCPH1 | FALSE | Downregulated |
| 0,023830754 | 0,976092076 | SLFNL1-AS1 | FALSE | Upregulated |
| 0,035659191 | 0,97625345 | FBXO3 | FALSE | Upregulated |
| 0,042220493 | 0,976309536 | CXorf56 | FALSE | Upregulated |
| 0,02309408 | 0,976309536 | LOC101927208 | FALSE | Upregulated |
| 0,077206434 | 0,976309536 | NTN5 | FALSE | Upregulated |
| -0,022172611 | 0,976348451 | APOBEC2 | FALSE | Downregulated |
| 0,091498119 | 0,976420224 | ECT2 | FALSE | Upregulated |
| -0,028551695 | 0,976420224 | LOC101926967 | FALSE | Downregulated |
| 0,035878432 | 0,976478771 | PIK3R5 | FALSE | Upregulated |
| 0,025429935 | 0,976787402 | ITFG2 | FALSE | Upregulated |
| 0,060428964 | 0,976838032 | LOC101927045 | FALSE | Upregulated |
| 0,014591303 | 0,976952869 | UBE2H | FALSE | Upregulated |
| 0,071609864 | 0,976980886 | ASIC5 | FALSE | Upregulated |
| -0,071912399 | 0,976980886 | GOLGA8R | FALSE | Downregulated |
| -0,018399989 | 0,976980886 | SPHK2 | FALSE | Downregulated |
| -0,101215411 | 0,977138309 | PROP1 | FALSE | Downregulated |
| -0,033525534 | 0,977138309 | ZBTB21 | FALSE | Downregulated |
| 0,049589822 | 0,977282764 | DCST1 | FALSE | Upregulated |
| 0,054642651 | 0,977282764 | NKAPP1 | FALSE | Upregulated |
| -0,02451152 | 0,977282764 | USP2 | FALSE | Downregulated |
| 0,033325077 | 0,977319321 | COA7 | FALSE | Upregulated |
| -0,077488854 | 0,977448214 | TTC21A | FALSE | Downregulated |
| 0,016307003 | 0,977532966 | CREG1 | FALSE | Upregulated |
| -0,030656213 | 0,977677957 | TMEM168 | FALSE | Downregulated |
| -0,027056858 | 0,977766038 | AC005029.1 | FALSE | Downregulated |
| -0,089325835 | 0,977766038 | CBWD3 | FALSE | Downregulated |
| 0,028097671 | 0,977766038 | CLN6 | FALSE | Upregulated |
| 0,048919073 | 0,977766038 | DACT2 | FALSE | Upregulated |
| -0,018456872 | 0,977766038 | FAM114A1 | FALSE | Downregulated |
| -0,023536609 | 0,977766038 | FAM160B1 | FALSE | Downregulated |
| 0,051289276 | 0,977766038 | GSTM2 | FALSE | Upregulated |
| 0,024540312 | 0,977766038 | HECW2 | FALSE | Upregulated |

| | | | | |
|---|---|---|---|---|
| -0,047071187 | 0,977766038 | KIF6 | FALSE | Downregulated |
| -0,013468403 | 0,977766038 | KLF3 | FALSE | Downregulated |
| 0,021468577 | 0,977766038 | OXSR1 | FALSE | Upregulated |
| -0,052554643 | 0,977766038 | PHF7 | FALSE | Downregulated |
| 0,022037465 | 0,977766038 | RCOR1 | FALSE | Upregulated |
| 0,03305946 | 0,977832741 | RABAC1 | FALSE | Upregulated |
| 0,043455607 | 0,977832741 | SRPK3 | FALSE | Upregulated |
| 0,016334386 | 0,977944581 | CCT3 | FALSE | Upregulated |
| -0,033798736 | 0,978202934 | KCTD7 | FALSE | Downregulated |
| 0,037343336 | 0,978202934 | NPL | FALSE | Upregulated |
| 0,032015472 | 0,978202934 | NUDCD2 | FALSE | Upregulated |
| -0,031897242 | 0,978202934 | PARD6B | FALSE | Downregulated |
| -0,023512587 | 0,978202934 | SAP18 | FALSE | Downregulated |
| 0,021186195 | 0,978202934 | STAP2 | FALSE | Upregulated |
| -0,051350404 | 0,978228855 | CCDC108 | FALSE | Downregulated |
| 0,031532504 | 0,978228855 | SHROOM4 | FALSE | Upregulated |
| 0,050355233 | 0,978228855 | ZNF419 | FALSE | Upregulated |
| 0,061485751 | 0,978251344 | RENBP | FALSE | Upregulated |
| -0,023642729 | 0,978514532 | PARD6G | FALSE | Downregulated |
| 0,05193409 | 0,978514532 | POU6F2-AS2 | FALSE | Upregulated |
| 0,021018575 | 0,978572811 | PLCG2 | FALSE | Upregulated |
| 0,479823786 | 0,978572811 | TSIX | FALSE | Upregulated |
| 0,024082023 | 0,978611458 | CADM4 | FALSE | Upregulated |
| 0,036423857 | 0,978611458 | GEMIN8 | FALSE | Upregulated |
| 0,020145455 | 0,978619179 | CHUK | FALSE | Upregulated |
| -0,020786988 | 0,978619179 | DCAKD | FALSE | Downregulated |
| 0,022706675 | 0,978619179 | HAVCR1P1 | FALSE | Upregulated |
| -0,018833775 | 0,978619179 | PAF1 | FALSE | Downregulated |
| 0,025300483 | 0,978820024 | ATAD1 | FALSE | Upregulated |
| -0,025871767 | 0,978820024 | GTF2H1 | FALSE | Downregulated |
| -0,012305411 | 0,979012538 | ARAP1 | FALSE | Downregulated |
| -0,030837102 | 0,979012538 | NDUFS3 | FALSE | Downregulated |
| -0,043798722 | 0,979144288 | MED21 | FALSE | Downregulated |
| -0,018495871 | 0,979144288 | SLCO1A2 | FALSE | Downregulated |
| -0,033532787 | 0,979171793 | C18orf8 | FALSE | Downregulated |
| -0,049233067 | 0,979171793 | KIAA1456 | FALSE | Downregulated |
| 0,022880036 | 0,979171793 | MARCH2 | FALSE | Upregulated |
| 0,025227677 | 0,979171793 | RANBP6 | FALSE | Upregulated |
| -0,06572621 | 0,979171793 | SEC61G | FALSE | Downregulated |
| -0,064905187 | 0,979171793 | SNORA23 | FALSE | Downregulated |
| 0,038288252 | 0,979171793 | SPRYD7 | FALSE | Upregulated |
| 0,017388354 | 0,979171793 | TMEM205 | FALSE | Upregulated |
| -0,034217113 | 0,979171793 | TXNL1 | FALSE | Downregulated |
| 0,018998428 | 0,979385276 | UBAC2 | FALSE | Upregulated |
| -0,029879859 | 0,979559817 | DPH1 | FALSE | Downregulated |
| -0,038920603 | 0,979590989 | ARHGAP39 | FALSE | Downregulated |
| -0,067703188 | 0,979590989 | ZNF425 | FALSE | Downregulated |
| 0,020654911 | 0,979675985 | C2CD2 | FALSE | Upregulated |
| -0,032639405 | 0,979675985 | LOC102723758 | FALSE | Downregulated |
| -0,056078739 | 0,979675985 | PLXNA4 | FALSE | Downregulated |
| 0,033035082 | 0,979675985 | RP1-293L8.2 | FALSE | Upregulated |

| | | | | |
|---|---|---|---|---|
| -0,024464685 | 0,979675985 | SNX10 | FALSE | Downregulated |
| 0,026298694 | 0,979675985 | ZBED6CL | FALSE | Upregulated |
| -0,036476628 | 0,979782041 | LOC102723469 | FALSE | Downregulated |
| -0,041400094 | 0,979782041 | SLC10A7 | FALSE | Downregulated |
| 0,013265479 | 0,979851953 | ASCC2 | FALSE | Upregulated |
| -0,024957921 | 0,979851953 | SFXN2 | FALSE | Downregulated |
| -0,080073182 | 0,979940746 | AC016910.1 | FALSE | Downregulated |
| 0,032572607 | 0,979940746 | AMIGO1 | FALSE | Upregulated |
| 0,030758533 | 0,979940746 | NUBP1 | FALSE | Upregulated |
| 0,03860925 | 0,979945783 | LOC101927610 | FALSE | Upregulated |
| 0,05756806 | 0,979956125 | DLGAP1-AS1 | FALSE | Upregulated |
| -0,00823639 | 0,979956125 | GNAS | FALSE | Downregulated |
| -0,081055212 | 0,979956125 | LOC102723874 | FALSE | Downregulated |
| -0,030850248 | 0,979956125 | MOB1B | FALSE | Downregulated |
| 0,053636161 | 0,979956125 | OGFR-AS1 | FALSE | Upregulated |
| 0,035134055 | 0,979956125 | PPME1 | FALSE | Upregulated |
| 0,020313026 | 0,979956125 | RAD9B | FALSE | Upregulated |
| 0,029417205 | 0,979956125 | RND1 | FALSE | Upregulated |
| -0,014226809 | 0,979956125 | THEM6 | FALSE | Downregulated |
| 0,012761328 | 0,979956125 | VDAC1 | FALSE | Upregulated |
| -0,021291206 | 0,979956125 | ZNF302 | FALSE | Downregulated |
| -0,031124886 | 0,9799766 | LOC101929328 | FALSE | Downregulated |
| 0,024007298 | 0,980088816 | STRBP | FALSE | Upregulated |
| -0,024693134 | 0,980196638 | AMMECR1L | FALSE | Downregulated |
| -0,015225075 | 0,980196638 | CNN3 | FALSE | Downregulated |
| 0,190392001 | 0,98024456 | C10orf71 | FALSE | Upregulated |
| -0,026389705 | 0,98024456 | MIS18BP1 | FALSE | Downregulated |
| 0,039597293 | 0,980252118 | SHISA9 | FALSE | Upregulated |
| 0,018561999 | 0,980322768 | ALDH1L2 | FALSE | Upregulated |
| -0,016882692 | 0,980322768 | EIF3D | FALSE | Downregulated |
| -0,019792016 | 0,980322768 | ENTPD3 | FALSE | Downregulated |
| 0,037347134 | 0,980322768 | LOC101928548 | FALSE | Upregulated |
| -0,017328321 | 0,980322768 | MRPL2 | FALSE | Downregulated |
| 0,033479981 | 0,980322768 | NEFH | FALSE | Upregulated |
| -0,034178355 | 0,980322768 | PA2G4P4 | FALSE | Downregulated |
| 0,025583218 | 0,980322768 | SELT | FALSE | Upregulated |
| -0,056431627 | 0,980578869 | MED7 | FALSE | Downregulated |
| -0,028370306 | 0,980672149 | EXD2 | FALSE | Downregulated |
| -0,032640082 | 0,980672149 | VPS37C | FALSE | Downregulated |
| -0,046362646 | 0,98086055 | EXOSC8 | FALSE | Downregulated |
| -0,028354671 | 0,980883941 | ANKRD9 | FALSE | Downregulated |
| -0,020255334 | 0,980883941 | LRRC59 | FALSE | Downregulated |
| 0,015897224 | 0,980923905 | CAPN7 | FALSE | Upregulated |
| -0,018847573 | 0,980923905 | CDKN1B | FALSE | Downregulated |
| -0,011563033 | 0,980923905 | CIC | FALSE | Downregulated |
| 0,030656919 | 0,980923905 | H2AFZ | FALSE | Upregulated |
| -0,030656093 | 0,980923905 | UCA1 | FALSE | Downregulated |
| 0,26364106 | 0,980923905 | ZNF559-ZNF177 | FALSE | Upregulated |
| 0,063504003 | 0,980959121 | AC012309.5 | FALSE | Upregulated |
| 0,026065081 | 0,980959121 | IPO9 | FALSE | Upregulated |
| 0,029306608 | 0,980959121 | RGPD8 | FALSE | Upregulated |

| | | | | |
|---|---|---|---|---|
| -0,028734721 | 0,980959121 | SMIM18 | FALSE | Downregulated |
| 0,016030067 | 0,980963946 | DCAF7 | FALSE | Upregulated |
| 0,021020881 | 0,980963946 | KCMF1 | FALSE | Upregulated |
| 0,01382796 | 0,980963946 | RNF220 | FALSE | Upregulated |
| 0,020962692 | 0,980980645 | CRIP3 | FALSE | Upregulated |
| -0,044371622 | 0,980980645 | RP11-15A1.3 | FALSE | Downregulated |
| -0,03133164 | 0,980987347 | LOC101929670 | FALSE | Downregulated |
| -0,029558885 | 0,981122201 | FAM50B | FALSE | Downregulated |
| 0,025107241 | 0,981163224 | HSBP1 | FALSE | Upregulated |
| -0,072808313 | 0,981186607 | LOC100510707 | FALSE | Downregulated |
| 0,014144152 | 0,981293892 | CAND1 | FALSE | Upregulated |
| 0,078448283 | 0,981367847 | LOC102724105 | FALSE | Upregulated |
| -0,025259169 | 0,981367847 | SORCS2 | FALSE | Downregulated |
| -0,019375849 | 0,981367847 | TMEM138 | FALSE | Downregulated |
| -0,017500104 | 0,981454877 | HEXB | FALSE | Downregulated |
| 0,037590807 | 0,981454877 | ZNF718 | FALSE | Upregulated |
| -0,028294845 | 0,981479183 | ADAL | FALSE | Downregulated |
| -0,025205438 | 0,981479183 | ZNF133 | FALSE | Downregulated |
| -0,014519314 | 0,981621397 | HDAC4 | FALSE | Downregulated |
| 0,086230714 | 0,981621397 | SLCO4A1 | FALSE | Upregulated |
| -0,0515517 | 0,981768421 | DLG1-AS1 | FALSE | Downregulated |
| 0,038111503 | 0,981768421 | LCT | FALSE | Upregulated |
| 0,046785667 | 0,981789946 | UBE2Q2P1 | FALSE | Upregulated |
| 0,01163858 | 0,98179956 | EIF4G2 | FALSE | Upregulated |
| 0,01435378 | 0,981813327 | LOC101928451 | FALSE | Upregulated |
| 0,028577459 | 0,981899335 | AVEN | FALSE | Upregulated |
| -0,021236146 | 0,981899335 | PDHX | FALSE | Downregulated |
| 0,173622428 | 0,981899335 | RNA18S5 | FALSE | Upregulated |
| -0,018415826 | 0,981899335 | SIAH1 | FALSE | Downregulated |
| -0,015835648 | 0,982000191 | INPP5E | FALSE | Downregulated |
| 0,011344058 | 0,982080937 | ITGB1 | FALSE | Upregulated |
| 0,010180988 | 0,982080937 | SETD1B | FALSE | Upregulated |
| 0,031054905 | 0,982174653 | ANP32AP1 | FALSE | Upregulated |
| 0,064069054 | 0,982174653 | FGF1 | FALSE | Upregulated |
| -0,016888416 | 0,982174653 | LOC102724484 | FALSE | Downregulated |
| 0,021004849 | 0,982174653 | PLEKHM1P | FALSE | Upregulated |
| 0,015550223 | 0,982255677 | SRP14 | FALSE | Upregulated |
| 0,021824995 | 0,982510401 | CNOT4 | FALSE | Upregulated |
| 0,063938026 | 0,982531624 | C20orf195 | FALSE | Upregulated |
| -0,103813212 | 0,982531624 | FAM72C | FALSE | Downregulated |
| -0,048991274 | 0,982531624 | LOC102724507 | FALSE | Downregulated |
| -0,021217002 | 0,982531624 | TRA2A | FALSE | Downregulated |
| 0,044953516 | 0,982587217 | ATG10 | FALSE | Upregulated |
| -0,013948541 | 0,982587217 | CCNT2 | FALSE | Downregulated |
| 0,020842483 | 0,982587217 | DTX1 | FALSE | Upregulated |
| -0,084315507 | 0,982587217 | MYCN | FALSE | Downregulated |
| -0,026698151 | 0,982587217 | PRPS2 | FALSE | Downregulated |
| -0,060634831 | 0,982587217 | RP11-473M20.14 | FALSE | Downregulated |
| 0,028962089 | 0,982587217 | ZNF304 | FALSE | Upregulated |
| -0,048220117 | 0,982712506 | NUDCD1 | FALSE | Downregulated |
| 0,018130192 | 0,982712506 | VDAC2 | FALSE | Upregulated |

| | | | | |
|---|---|---|---|---|
| -0,044587474 | 0,982808999 | LINC00663 | FALSE | Downregulated |
| 0,014673193 | 0,982808999 | ODF3B | FALSE | Upregulated |
| 0,01427415 | 0,98283781 | CEACAM1 | FALSE | Upregulated |
| -0,021468693 | 0,98283781 | LOC100506100 | FALSE | Downregulated |
| -0,014671408 | 0,98283781 | PLXNA2 | FALSE | Downregulated |
| 0,017659924 | 0,982895036 | CCNG1 | FALSE | Upregulated |
| -0,052132288 | 0,982895036 | LOC101448202 | FALSE | Downregulated |
| 0,099984256 | 0,982895036 | TCEB3C | FALSE | Upregulated |
| -0,013505797 | 0,982895036 | TLE1 | FALSE | Downregulated |
| 0,025125204 | 0,982895036 | TMEM106C | FALSE | Upregulated |
| -0,032883169 | 0,982895036 | TRMT12 | FALSE | Downregulated |
| -0,01897522 | 0,982926434 | GSS | FALSE | Downregulated |
| 0,040351993 | 0,982926434 | SLCO1B7 | FALSE | Upregulated |
| 0,017041051 | 0,983218897 | CDC40 | FALSE | Upregulated |
| -0,016659815 | 0,983218897 | FLOT2 | FALSE | Downregulated |
| 0,012925061 | 0,983218897 | PRPF40A | FALSE | Upregulated |
| -0,010067729 | 0,983218897 | TOB2 | FALSE | Downregulated |
| 0,051125761 | 0,983269985 | NEK10 | FALSE | Upregulated |
| 0,057636865 | 0,983413209 | RP11-364P22.1 | FALSE | Upregulated |
| 0,011534264 | 0,983448421 | DGKD | FALSE | Upregulated |
| 0,386191512 | 0,983448421 | MYH1 | FALSE | Upregulated |
| -0,047037765 | 0,983508353 | IMMP1L | FALSE | Downregulated |
| 0,022541108 | 0,983522664 | TSG101 | FALSE | Upregulated |
| -0,009234436 | 0,983545313 | EHMT1 | FALSE | Downregulated |
| -0,028232349 | 0,983545313 | LOC101929263 | FALSE | Downregulated |
| 0,012774507 | 0,983545313 | PINK1-AS | FALSE | Upregulated |
| 0,018472967 | 0,983545313 | RAB18 | FALSE | Upregulated |
| 0,057164214 | 0,983545313 | TRABD2A | FALSE | Upregulated |
| 0,043997885 | 0,983545313 | ZNF184 | FALSE | Upregulated |
| 0,111726257 | 0,983553616 | LOC284933 | FALSE | Upregulated |
| 0,020641298 | 0,9836143 | CASP7 | FALSE | Upregulated |
| -0,047495312 | 0,9836425 | ADCK1 | FALSE | Downregulated |
| -0,061354804 | 0,983809726 | ATOH7 | FALSE | Downregulated |
| -0,016867301 | 0,983925072 | DCTN4 | FALSE | Downregulated |
| -0,017256724 | 0,983925072 | EEFSEC | FALSE | Downregulated |
| -0,065894982 | 0,983925072 | SNORD15B | FALSE | Downregulated |
| -0,027917701 | 0,983925072 | TFB1M | FALSE | Downregulated |
| 0,018585244 | 0,983996996 | RBFOX2 | FALSE | Upregulated |
| 0,016241098 | 0,984145592 | CCDC130 | FALSE | Upregulated |
| 0,058924312 | 0,984164203 | SLC16A9 | FALSE | Upregulated |
| 0,046457103 | 0,984191038 | NOXRED1 | FALSE | Upregulated |
| 0,021876884 | 0,984191038 | ODF2 | FALSE | Upregulated |
| -0,04345204 | 0,984211217 | FLJ36777 | FALSE | Downregulated |
| 0,052519083 | 0,984221385 | CTSLP2 | FALSE | Upregulated |
| -0,026313553 | 0,984221385 | POLA1 | FALSE | Downregulated |
| -0,013701812 | 0,984543559 | TPP2 | FALSE | Downregulated |
| 0,024870507 | 0,984654355 | CCBL1 | FALSE | Upregulated |
| -0,014275232 | 0,984654355 | MON1B | FALSE | Downregulated |
| -0,023646507 | 0,984654355 | SIGLEC11 | FALSE | Downregulated |
| 0,025071791 | 0,984654355 | SLC15A3 | FALSE | Upregulated |
| 0,008694315 | 0,984654355 | SLC35E2B | FALSE | Upregulated |

| | | | | |
|---|---|---|---|---|
| 0,050289777 | 0,984654355 | TMEM178A | FALSE | Upregulated |
| -0,024511806 | 0,984702763 | TMCO6 | FALSE | Downregulated |
| -0,011273677 | 0,984950691 | RPL7L1 | FALSE | Downregulated |
| 0,029609488 | 0,985096801 | LOC100129550 | FALSE | Upregulated |
| -0,043107611 | 0,985096801 | LOC102724493 | FALSE | Downregulated |
| -0,058946565 | 0,985128427 | CHL1 | FALSE | Downregulated |
| 0,014465213 | 0,985128427 | GABPB1 | FALSE | Upregulated |
| 0,027016461 | 0,985128427 | IDO2 | FALSE | Upregulated |
| 0,031060617 | 0,985128427 | KIF9-AS1 | FALSE | Upregulated |
| -0,02525426 | 0,985128427 | LOC100129637 | FALSE | Downregulated |
| 0,051955562 | 0,985128427 | LOC100996717 | FALSE | Upregulated |
| -0,024857179 | 0,985128427 | LOC101927578 | FALSE | Downregulated |
| 0,035017519 | 0,985128427 | LOC101929573 | FALSE | Upregulated |
| 0,044795971 | 0,985128427 | LOC102723709 | FALSE | Upregulated |
| 0,018418869 | 0,985128427 | LOC401320 | FALSE | Upregulated |
| 0,034573897 | 0,985128427 | PSMD10 | FALSE | Upregulated |
| -0,017635598 | 0,985128427 | RBFA | FALSE | Downregulated |
| 0,073236811 | 0,985128427 | REM2 | FALSE | Upregulated |
| -0,013608264 | 0,985128427 | RFX1 | FALSE | Downregulated |
| 0,013620626 | 0,985128427 | RNF146 | FALSE | Upregulated |
| -0,01941562 | 0,985128427 | TBRG4 | FALSE | Downregulated |
| 0,011185271 | 0,985128427 | TMEM115 | FALSE | Upregulated |
| 0,012174098 | 0,985128427 | TNPO1 | FALSE | Upregulated |
| 0,04315412 | 0,985128427 | VPS13A-AS1 | FALSE | Upregulated |
| -0,011104713 | 0,985135052 | EIF2AK4 | FALSE | Downregulated |
| 0,023060451 | 0,985135052 | KRBOX4 | FALSE | Upregulated |
| 0,010390724 | 0,98518674 | VCPIP1 | FALSE | Upregulated |
| -0,041151507 | 0,985227896 | ACRC | FALSE | Downregulated |
| -0,024243927 | 0,985227896 | CACNA1A | FALSE | Downregulated |
| 0,01934495 | 0,985227896 | MCRS1 | FALSE | Upregulated |
| -0,008719267 | 0,985258549 | CTTN | FALSE | Downregulated |
| 0,059674336 | 0,985277999 | ARHGAP26-AS1 | FALSE | Upregulated |
| -0,024825071 | 0,985277999 | C1orf112 | FALSE | Downregulated |
| 0,013803189 | 0,985277999 | COLEC11 | FALSE | Upregulated |
| -0,025876496 | 0,985277999 | FTSJ2 | FALSE | Downregulated |
| 0,025721509 | 0,985277999 | LINC00161 | FALSE | Upregulated |
| -0,020471275 | 0,985277999 | PWRN2 | FALSE | Downregulated |
| -0,028568055 | 0,985277999 | PYROXD2 | FALSE | Downregulated |
| 0,017848617 | 0,985277999 | TIMM23 | FALSE | Upregulated |
| 0,013363429 | 0,985277999 | WDR65 | FALSE | Upregulated |
| 0,016428467 | 0,985386145 | RSBN1 | FALSE | Upregulated |
| 0,036132565 | 0,985390015 | ASB12 | FALSE | Upregulated |
| 0,050216909 | 0,985390015 | CCDC134 | FALSE | Upregulated |
| 0,025738238 | 0,985390015 | ENPP2 | FALSE | Upregulated |
| 0,018264491 | 0,985390015 | FYTTD1 | FALSE | Upregulated |
| 0,029904161 | 0,985390015 | KBTBD6 | FALSE | Upregulated |
| 0,013862929 | 0,985390015 | LOC101926920 | FALSE | Upregulated |
| 0,089865109 | 0,985390015 | LOC102723475 | FALSE | Upregulated |
| -0,032498467 | 0,985390015 | LOC200772 | FALSE | Downregulated |
| -0,056916076 | 0,985390015 | LOC494141 | FALSE | Downregulated |
| -0,040132382 | 0,985390015 | LOC644656 | FALSE | Downregulated |

| | | | | |
|---|---|---|---|---|
| -0,041543762 | 0,985390015 | PGLYRP1 | FALSE | Downregulated |
| -0,087388875 | 0,985390015 | RNU1-27P | FALSE | Downregulated |
| 0,053438009 | 0,985390015 | RPL23AP7 | FALSE | Upregulated |
| -0,019698411 | 0,985390015 | SMG9 | FALSE | Downregulated |
| 0,052666207 | 0,985390015 | SNHG17 | FALSE | Upregulated |
| 0,020562792 | 0,985390015 | ZNF786 | FALSE | Upregulated |
| -0,012701261 | 0,985438487 | SLC40A1 | FALSE | Downregulated |
| 0,014283696 | 0,985564228 | GPR179 | FALSE | Upregulated |
| 0,043039883 | 0,985564228 | TSSK1B | FALSE | Upregulated |
| -0,012603934 | 0,985564228 | TXNIP | FALSE | Downregulated |
| -0,062896461 | 0,98556678 | AC016738.3 | FALSE | Downregulated |
| -0,022752634 | 0,98556678 | FOCAD | FALSE | Downregulated |
| 0,018885353 | 0,98556678 | LOC101928783 | FALSE | Upregulated |
| 0,018036659 | 0,98556678 | MUL1 | FALSE | Upregulated |
| -0,014363599 | 0,98556678 | PIN1 | FALSE | Downregulated |
| 0,019403988 | 0,98556678 | RB1 | FALSE | Upregulated |
| -0,027448295 | 0,98556678 | STAMBP | FALSE | Downregulated |
| -0,012613364 | 0,98556678 | TBL1XR1 | FALSE | Downregulated |
| -0,013790775 | 0,98556678 | TCAIM | FALSE | Downregulated |
| 0,016297061 | 0,98556678 | ZNF580 | FALSE | Upregulated |
| -0,014836555 | 0,985695183 | NAA25 | FALSE | Downregulated |
| -0,01366188 | 0,985772021 | RDH14 | FALSE | Downregulated |
| -0,014893843 | 0,985772021 | TTC14 | FALSE | Downregulated |
| -0,060710578 | 0,985958695 | FMN2 | FALSE | Downregulated |
| 0,029315386 | 0,985958695 | TCEANC2 | FALSE | Upregulated |
| 0,018379244 | 0,985993647 | CD5L | FALSE | Upregulated |
| -0,025442788 | 0,985993647 | CHAMP1 | FALSE | Downregulated |
| 0,013164352 | 0,985993647 | IL17RA | FALSE | Upregulated |
| -0,015829577 | 0,985993647 | LOC100129917 | FALSE | Downregulated |
| -0,168858291 | 0,985993647 | LOC102724951 | FALSE | Downregulated |
| -0,013632192 | 0,985993647 | PPP2R2A | FALSE | Downregulated |
| -0,01483797 | 0,985993647 | SEC23IP | FALSE | Downregulated |
| 0,017409356 | 0,985993647 | SMIM6 | FALSE | Upregulated |
| -0,011219402 | 0,986007651 | BTAF1 | FALSE | Downregulated |
| 0,023966224 | 0,986106611 | MIER2 | FALSE | Upregulated |
| 0,029156304 | 0,986117306 | C3AR1 | FALSE | Upregulated |
| -0,014266096 | 0,986117306 | DNAJA1 | FALSE | Downregulated |
| 0,02771497 | 0,986117306 | KRBA2 | FALSE | Upregulated |
| 0,023314556 | 0,986117306 | PI4KAP2 | FALSE | Upregulated |
| 0,024137824 | 0,986117306 | SLC26A11 | FALSE | Upregulated |
| 0,022725094 | 0,986161688 | RPAP2 | FALSE | Upregulated |
| 0,02923948 | 0,986192915 | KDM4E | FALSE | Upregulated |
| -0,017261015 | 0,986192915 | VWA9 | FALSE | Downregulated |
| 0,043927679 | 0,986448327 | SMIM1 | FALSE | Upregulated |
| -0,014188539 | 0,986495216 | PIEZO2 | FALSE | Downregulated |
| -0,020525667 | 0,986512127 | GPR61 | FALSE | Downregulated |
| 0,018925725 | 0,986512127 | MFAP1 | FALSE | Upregulated |
| -0,030066689 | 0,986512127 | REEP4 | FALSE | Downregulated |
| -0,049933209 | 0,986512127 | REP15 | FALSE | Downregulated |
| -0,021167322 | 0,986512127 | SLC39A6 | FALSE | Downregulated |
| -0,014826661 | 0,986559422 | CDYL | FALSE | Downregulated |

| | | | | |
|---|---|---|---|---|
| 0,022232187 | 0,986559422 | CLEC14A | FALSE | Upregulated |
| 0,023314306 | 0,986559422 | COG6 | FALSE | Upregulated |
| 0,011418752 | 0,986559422 | MSL2 | FALSE | Upregulated |
| -0,020233776 | 0,986559422 | NPIPB5 | FALSE | Downregulated |
| -0,024982934 | 0,986570477 | RABL2A | FALSE | Downregulated |
| 0,045573791 | 0,986570477 | SPACA6P | FALSE | Upregulated |
| -0,019980038 | 0,986709735 | BBS2 | FALSE | Downregulated |
| 0,011055127 | 0,986760298 | CCNYL1 | FALSE | Upregulated |
| -0,0248818 | 0,986828214 | CCR8 | FALSE | Downregulated |
| -0,007943862 | 0,986828214 | CHD8 | FALSE | Downregulated |
| -0,009391577 | 0,986828214 | COX4I1 | FALSE | Downregulated |
| -0,031976545 | 0,986828214 | LINS | FALSE | Downregulated |
| 0,017016629 | 0,986828214 | LMBRD2 | FALSE | Upregulated |
| 0,030210378 | 0,986828214 | RBX1 | FALSE | Upregulated |
| -0,03896444 | 0,986828214 | RFXAP | FALSE | Downregulated |
| -0,007846164 | 0,986828214 | TMED2 | FALSE | Downregulated |
| 0,025838672 | 0,986828214 | UBA6-AS1 | FALSE | Upregulated |
| 0,015152934 | 0,986828214 | WDR36 | FALSE | Upregulated |
| -0,011941313 | 0,986828214 | XPO4 | FALSE | Downregulated |
| 0,012412612 | 0,986896534 | NPRL3 | FALSE | Upregulated |
| -0,020942078 | 0,986925645 | NDUFAF7 | FALSE | Downregulated |
| 0,052829303 | 0,986925645 | WDR76 | FALSE | Upregulated |
| -0,010870573 | 0,986941915 | ERVW-1 | FALSE | Downregulated |
| -0,014518702 | 0,986941915 | HEMK1 | FALSE | Downregulated |
| 0,025513073 | 0,986941915 | NAALADL2 | FALSE | Upregulated |
| -0,035937861 | 0,986941915 | RBMX2 | FALSE | Downregulated |
| -0,029348231 | 0,986941915 | RP11-96H19.1 | FALSE | Downregulated |
| -0,016631594 | 0,986941915 | THAP5 | FALSE | Downregulated |
| -0,021293542 | 0,986991734 | AP1S1 | FALSE | Downregulated |
| 0,020669994 | 0,986991734 | ATP5G1 | FALSE | Upregulated |
| -0,033629038 | 0,986991734 | CEP57L1 | FALSE | Downregulated |
| -0,015139143 | 0,986991734 | PAX6 | FALSE | Downregulated |
| -0,036110798 | 0,987036024 | FOXO3B | FALSE | Downregulated |
| 0,017594518 | 0,987036024 | VPS26A | FALSE | Upregulated |
| 0,014486528 | 0,987127908 | TJAP1 | FALSE | Upregulated |
| 0,013033723 | 0,987328252 | ALS2CL | FALSE | Upregulated |
| 0,01540224 | 0,987328252 | MAGEF1 | FALSE | Upregulated |
| 0,023996012 | 0,987328252 | TTLL7 | FALSE | Upregulated |
| -0,004409192 | 0,987395866 | PRRC2B | FALSE | Downregulated |
| 0,008671017 | 0,987465734 | KHDRBS1 | FALSE | Upregulated |
| -0,280361616 | 0,987465734 | RNU1-28P | FALSE | Downregulated |
| -0,009927984 | 0,987533626 | AKAP8L | FALSE | Downregulated |
| 0,022591659 | 0,987533626 | CHMP3 | FALSE | Upregulated |
| -0,011263458 | 0,987533626 | FAM102A | FALSE | Downregulated |
| -0,006590339 | 0,987533626 | MRPS25 | FALSE | Downregulated |
| 0,03678795 | 0,98754181 | DEFB1 | FALSE | Upregulated |
| -0,036443596 | 0,98754181 | PEX7 | FALSE | Downregulated |
| 0,04436675 | 0,987563262 | LRP1B | FALSE | Upregulated |
| 0,011968745 | 0,987563262 | MRTO4 | FALSE | Upregulated |
| -0,01487394 | 0,987563262 | SLC22A4 | FALSE | Downregulated |
| 0,012726217 | 0,98765529 | ING1 | FALSE | Upregulated |

| | | | | |
|---|---|---|---|---|
| 0,277198781 | 0,98765529 | MYBPC1 | FALSE | Upregulated |
| -0,018366653 | 0,987814081 | PAMR1 | FALSE | Downregulated |
| -0,013228687 | 0,987814193 | CDH23 | FALSE | Downregulated |
| -0,022532926 | 0,988172679 | S1PR3 | FALSE | Downregulated |
| -0,028707762 | 0,988192299 | FAT3 | FALSE | Downregulated |
| 0,021305002 | 0,988192299 | SGCB | FALSE | Upregulated |
| -0,019960624 | 0,988225602 | AARSD1 | FALSE | Downregulated |
| 0,009972886 | 0,988225602 | ANKRD61 | FALSE | Upregulated |
| -0,010860247 | 0,988225602 | HLF | FALSE | Downregulated |
| 0,010524691 | 0,988225602 | IFIT1 | FALSE | Upregulated |
| 0,035781009 | 0,988225602 | LOC101926901 | FALSE | Upregulated |
| -0,014264104 | 0,988225602 | LOC101928101 | FALSE | Downregulated |
| -0,018794094 | 0,988225602 | LOC102723897 | FALSE | Downregulated |
| -0,017664378 | 0,988225602 | LOC441179 | FALSE | Downregulated |
| 0,053579689 | 0,988225602 | NCAPH | FALSE | Upregulated |
| -0,004832666 | 0,988225602 | PLXNB2 | FALSE | Downregulated |
| 0,014559403 | 0,988225602 | RP11-760N9.1 | FALSE | Upregulated |
| -0,029220257 | 0,988225602 | RP3-365O12.2 | FALSE | Downregulated |
| 0,013622981 | 0,988225602 | TRMT61A | FALSE | Upregulated |
| 0,010323385 | 0,988225602 | TTC37 | FALSE | Upregulated |
| -0,011294105 | 0,988259901 | ALDH1B1 | FALSE | Downregulated |
| -0,044896604 | 0,988259901 | LOC101929819 | FALSE | Downregulated |
| -0,02682941 | 0,988301558 | MEF2BNB | FALSE | Downregulated |
| -0,023596691 | 0,988326979 | FDX1L | FALSE | Downregulated |
| -0,027266726 | 0,988326979 | HAT1 | FALSE | Downregulated |
| 0,013741027 | 0,988408794 | PLBD2 | FALSE | Upregulated |
| -0,010409429 | 0,988587758 | RFWD2 | FALSE | Downregulated |
| 0,01599446 | 0,988697858 | PHF5A | FALSE | Upregulated |
| 0,014517411 | 0,988731206 | TPST1 | FALSE | Upregulated |
| -0,013036652 | 0,988740169 | NT5C3B | FALSE | Downregulated |
| 0,012726793 | 0,988740169 | RP11-384K6.6 | FALSE | Upregulated |
| 0,013925935 | 0,988761566 | ENTPD8 | FALSE | Upregulated |
| 0,028739973 | 0,988775849 | C15orf41 | FALSE | Upregulated |
| -0,018707426 | 0,988798818 | CHML | FALSE | Downregulated |
| 0,021561079 | 0,988995663 | MANSC4 | FALSE | Upregulated |
| -0,030057356 | 0,988995663 | NXT1 | FALSE | Downregulated |
| -0,007704004 | 0,989195598 | PEX10 | FALSE | Downregulated |
| -0,018953238 | 0,989216371 | NCK1 | FALSE | Downregulated |
| 0,027978986 | 0,98945584 | RASL10A | FALSE | Upregulated |
| 0,021036505 | 0,989510003 | RNASEH2B | FALSE | Upregulated |
| -0,012444693 | 0,989536131 | BTG3 | FALSE | Downregulated |
| 0,039784473 | 0,989552162 | FAM166B | FALSE | Upregulated |
| -0,036784695 | 0,989622195 | AC104655.2 | FALSE | Downregulated |
| 0,019611807 | 0,989622195 | TFPT | FALSE | Upregulated |
| -0,008969871 | 0,989622195 | USP5 | FALSE | Downregulated |
| -0,014487993 | 0,989695789 | LOC102723811 | FALSE | Downregulated |
| 0,010325553 | 0,989695789 | PIGB | FALSE | Upregulated |
| 0,012223689 | 0,989790311 | CYFIP2 | FALSE | Upregulated |
| 0,010187961 | 0,989790311 | EMC3 | FALSE | Upregulated |
| -0,01410664 | 0,989790311 | LOC102724683 | FALSE | Downregulated |
| -0,014349361 | 0,989790311 | NUP155 | FALSE | Downregulated |

| | | | | |
|---|---|---|---|---|
| 0,012641001 | 0,989790311 | SLC6A1-AS1 | FALSE | Upregulated |
| 0,008484356 | 0,989790311 | THPO | FALSE | Upregulated |
| -0,041188439 | 0,989790311 | TNMD | FALSE | Downregulated |
| -0,014821665 | 0,989790311 | TRAPPC1 | FALSE | Downregulated |
| -0,018282413 | 0,989790311 | ZFPM2 | FALSE | Downregulated |
| -0,029760944 | 0,989790311 | ZNF836 | FALSE | Downregulated |
| -0,022836689 | 0,989800874 | LOC101927313 | FALSE | Downregulated |
| 0,010745993 | 0,989985149 | MORC4 | FALSE | Upregulated |
| -0,013632546 | 0,989985149 | ZFP62 | FALSE | Downregulated |
| -0,039964609 | 0,990082159 | ALAS2 | FALSE | Downregulated |
| 0,023691147 | 0,990082159 | ARSJ | FALSE | Upregulated |
| -0,030896928 | 0,990082159 | GS1-251I9.4 | FALSE | Downregulated |
| -0,04309704 | 0,990082159 | IL1A | FALSE | Downregulated |
| 0,008002579 | 0,990082159 | IL4R | FALSE | Upregulated |
| 0,011060993 | 0,990082159 | MRI1 | FALSE | Upregulated |
| 0,007036188 | 0,990082159 | PARP4 | FALSE | Upregulated |
| 0,006703308 | 0,990082159 | PTGES3 | FALSE | Upregulated |
| -0,028534833 | 0,990082159 | SLC28A2 | FALSE | Downregulated |
| 0,010413863 | 0,990082159 | TRAPPC10 | FALSE | Upregulated |
| -0,016196304 | 0,990082159 | TRIM47 | FALSE | Downregulated |
| 0,010774586 | 0,990082159 | ZNF747 | FALSE | Upregulated |
| -0,006928284 | 0,990155417 | ZNF451 | FALSE | Downregulated |
| 0,014241031 | 0,990275269 | GNG7 | FALSE | Upregulated |
| 0,039268091 | 0,990338683 | LOC388210 | FALSE | Upregulated |
| 0,012865055 | 0,990452636 | SLC7A7 | FALSE | Upregulated |
| -0,008247739 | 0,990597412 | SH3BP2 | FALSE | Downregulated |
| -0,019628809 | 0,990833913 | C9orf156 | FALSE | Downregulated |
| 0,009335162 | 0,990848224 | NADSYN1 | FALSE | Upregulated |
| -0,016113694 | 0,990906448 | PARL | FALSE | Downregulated |
| -0,006654765 | 0,990942679 | ZMYND11 | FALSE | Downregulated |
| 0,011697598 | 0,99096576 | BLNK | FALSE | Upregulated |
| -0,013797726 | 0,99096576 | GTF2H2B | FALSE | Downregulated |
| 0,011372871 | 0,99096576 | KLHL42 | FALSE | Upregulated |
| 0,009489596 | 0,99096576 | SNX8 | FALSE | Upregulated |
| 0,031870635 | 0,991022806 | LOC101928673 | FALSE | Upregulated |
| -0,014598808 | 0,991131589 | KCTD15 | FALSE | Downregulated |
| -0,009227605 | 0,991215232 | ANAPC5 | FALSE | Downregulated |
| -0,009500887 | 0,991215232 | BRCA2 | FALSE | Downregulated |
| -0,009003882 | 0,991215232 | CHMP4B | FALSE | Downregulated |
| 0,011147864 | 0,991215232 | CHRNB1 | FALSE | Upregulated |
| 0,011761622 | 0,991215232 | G0S2 | FALSE | Upregulated |
| -0,019270933 | 0,991215232 | KIAA0825 | FALSE | Downregulated |
| 0,017770892 | 0,991215232 | RNASEH1 | FALSE | Upregulated |
| 0,042859336 | 0,991215232 | TCEB3CL2 | FALSE | Upregulated |
| -0,01496517 | 0,991215232 | TUBA3D | FALSE | Downregulated |
| 0,012065977 | 0,991294852 | ATP6V1E1 | FALSE | Upregulated |
| 0,020834182 | 0,991294852 | COMMD10 | FALSE | Upregulated |
| -0,010532232 | 0,991294852 | LOC401296 | FALSE | Downregulated |
| 0,01732688 | 0,991441944 | AQP7 | FALSE | Upregulated |
| 0,004881126 | 0,991441944 | BOD1L1 | FALSE | Upregulated |
| -0,006066501 | 0,991441944 | DSP | FALSE | Downregulated |

| | | | | |
|---|---|---|---|---|
| 0,011987883 | 0,991441944 | INSIG1 | FALSE | Upregulated |
| 0,033612561 | 0,991441944 | NYAP1 | FALSE | Upregulated |
| -0,010448172 | 0,991441944 | ZNF689 | FALSE | Downregulated |
| -0,032266575 | 0,991443926 | AC006262.5 | FALSE | Downregulated |
| -0,009608044 | 0,991443926 | ADAP2 | FALSE | Downregulated |
| -0,029505592 | 0,991443926 | ADORA3 | FALSE | Downregulated |
| -0,011263401 | 0,991443926 | ALDH18A1 | FALSE | Downregulated |
| 0,021602819 | 0,991443926 | CTNND2 | FALSE | Upregulated |
| -0,010177552 | 0,991443926 | DSE | FALSE | Downregulated |
| 0,012542565 | 0,991443926 | EIF2B3 | FALSE | Upregulated |
| 0,008367978 | 0,991443926 | IGF1R | FALSE | Upregulated |
| -0,016753232 | 0,991443926 | IL27RA | FALSE | Downregulated |
| 0,006810072 | 0,991443926 | IREB2 | FALSE | Upregulated |
| -0,008565418 | 0,991443926 | KAT8 | FALSE | Downregulated |
| -0,008906072 | 0,991443926 | LIN54 | FALSE | Downregulated |
| 0,016087221 | 0,991443926 | LOC100288069 | FALSE | Upregulated |
| 0,010263487 | 0,991443926 | LOC645967 | FALSE | Upregulated |
| 0,014680223 | 0,991443926 | MAGOH2 | FALSE | Upregulated |
| 0,013475678 | 0,991443926 | MFSD2A | FALSE | Upregulated |
| 0,07278575 | 0,991443926 | MYCBP | FALSE | Upregulated |
| -0,005903605 | 0,991443926 | MYO1E | FALSE | Downregulated |
| 0,007412571 | 0,991443926 | OSBPL8 | FALSE | Upregulated |
| -0,006908687 | 0,991443926 | OTUD7B | FALSE | Downregulated |
| 0,011664013 | 0,991443926 | PLRG1 | FALSE | Upregulated |
| -0,020939704 | 0,991443926 | POLE4 | FALSE | Downregulated |
| 0,01198818 | 0,991443926 | PPP3CB | FALSE | Upregulated |
| -0,010661464 | 0,991443926 | RNF6 | FALSE | Downregulated |
| -0,018990801 | 0,991443926 | RP11-267N12.1 | FALSE | Downregulated |
| -0,010178863 | 0,991443926 | SDF2 | FALSE | Downregulated |
| 0,03458701 | 0,991443926 | SLC4A10 | FALSE | Upregulated |
| -0,007382318 | 0,991443926 | TERF2IP | FALSE | Downregulated |
| 0,014952999 | 0,991443926 | TMEM160 | FALSE | Upregulated |
| 0,02874002 | 0,991443926 | TMEM38B | FALSE | Upregulated |
| -0,005914959 | 0,991443926 | TOM1 | FALSE | Downregulated |
| -0,036180749 | 0,991443926 | TPTE2P1 | FALSE | Downregulated |
| 0,010648923 | 0,991443926 | WDR3 | FALSE | Upregulated |
| 0,014325046 | 0,991443926 | ZC3HC1 | FALSE | Upregulated |
| -0,00921448 | 0,991443926 | ZNF776 | FALSE | Downregulated |
| 0,019701727 | 0,991443926 | ZNF788 | FALSE | Upregulated |
| 0,006040308 | 0,991443926 | ZSWIM8-AS1 | FALSE | Upregulated |
| -0,009896428 | 0,991450719 | PLD2 | FALSE | Downregulated |
| -0,013219168 | 0,991550556 | ACTR10 | FALSE | Downregulated |
| -0,009495701 | 0,991550556 | AKIRIN2 | FALSE | Downregulated |
| 0,008891177 | 0,991550556 | GDF7 | FALSE | Upregulated |
| -0,004293124 | 0,991582089 | HERC1 | FALSE | Downregulated |
| 0,018720963 | 0,991582089 | PIR | FALSE | Upregulated |
| -0,005949346 | 0,991683426 | CARKD | FALSE | Downregulated |
| -0,008933999 | 0,991683426 | CDC27 | FALSE | Downregulated |
| 0,182833504 | 0,991727475 | ACTA1 | FALSE | Upregulated |
| 0,009719474 | 0,991796287 | AC016629.8 | FALSE | Upregulated |
| -0,023842941 | 0,991796287 | ACOXL | FALSE | Downregulated |

| | | | | |
|---|---|---|---|---|
| -0,008362273 | 0,991796287 | ARPP19 | FALSE | Downregulated |
| -0,010368181 | 0,991796287 | ATG3 | FALSE | Downregulated |
| -0,017472949 | 0,991796287 | BET1 | FALSE | Downregulated |
| 0,009402858 | 0,991796287 | CIPC | FALSE | Upregulated |
| 0,020321573 | 0,991796287 | FAM60A | FALSE | Upregulated |
| 0,013827173 | 0,991796287 | GARNL3 | FALSE | Upregulated |
| 0,017526115 | 0,991796287 | LAGE3 | FALSE | Upregulated |
| -0,017299699 | 0,991796287 | LOC101927000 | FALSE | Downregulated |
| -0,018196858 | 0,991796287 | NT5C3A | FALSE | Downregulated |
| 0,013812481 | 0,991796287 | RECK | FALSE | Upregulated |
| 0,007388603 | 0,991796287 | SPATS2L | FALSE | Upregulated |
| -0,005814222 | 0,991796287 | WTAP | FALSE | Downregulated |
| 0,007829184 | 0,991796287 | ZNF784 | FALSE | Upregulated |
| -0,014635779 | 0,991801815 | LOC101927806 | FALSE | Downregulated |
| -0,023728786 | 0,991842916 | SLITRK3 | FALSE | Downregulated |
| 0,153244399 | 0,99184699 | DES | FALSE | Upregulated |
| 0,011622363 | 0,99193692 | CINP | FALSE | Upregulated |
| 0,008831349 | 0,991970453 | PPRC1 | FALSE | Upregulated |
| 0,006865407 | 0,992062808 | TRIM4 | FALSE | Upregulated |
| 0,015855568 | 0,992131707 | GOLGA6C | FALSE | Upregulated |
| 0,006804672 | 0,992131707 | LARP4 | FALSE | Upregulated |
| 0,024882904 | 0,992131707 | LOC102724795 | FALSE | Upregulated |
| -0,029432184 | 0,992166714 | LRRC17 | FALSE | Downregulated |
| -0,017019361 | 0,992166714 | VWA8-AS1 | FALSE | Downregulated |
| 0,011329175 | 0,992231066 | LOC100287166 | FALSE | Upregulated |
| 0,017470789 | 0,992258415 | TWSG1 | FALSE | Upregulated |
| -0,025842197 | 0,992342373 | HLA-DQA2 | FALSE | Downregulated |
| 0,024575039 | 0,992408318 | PIGP | FALSE | Upregulated |
| 0,015289684 | 0,992592863 | DNTTIP1 | FALSE | Upregulated |
| -0,005445443 | 0,992808635 | TRPM7 | FALSE | Downregulated |
| -0,011222797 | 0,992808635 | ZNF707 | FALSE | Downregulated |
| -0,028719675 | 0,992812685 | MEX3B | FALSE | Downregulated |
| 0,018976835 | 0,992874801 | ARPC4-TTLL3 | FALSE | Upregulated |
| 0,009454235 | 0,992959849 | RP1-256G22.2 | FALSE | Upregulated |
| -0,007759262 | 0,99298399 | CLPB | FALSE | Downregulated |
| 0,031483614 | 0,99298399 | GPC3 | FALSE | Upregulated |
| -0,045137661 | 0,993033193 | MOB4 | FALSE | Downregulated |
| 0,016063853 | 0,993101207 | DOK3 | FALSE | Upregulated |
| 0,020065564 | 0,99326979 | IZUMO4 | FALSE | Upregulated |
| 0,009724994 | 0,99326979 | NFXL1 | FALSE | Upregulated |
| 0,012231456 | 0,993331098 | LINC00870 | FALSE | Upregulated |
| 0,151781681 | 0,993383503 | XIRP2 | FALSE | Upregulated |
| -0,018194209 | 0,993430292 | ALG10 | FALSE | Downregulated |
| 0,006385717 | 0,993430292 | BRSK2 | FALSE | Upregulated |
| -0,00510377 | 0,993430292 | CHDH | FALSE | Downregulated |
| -0,004524598 | 0,993430292 | HYI | FALSE | Downregulated |
| 0,010248325 | 0,993430292 | LINC00482 | FALSE | Upregulated |
| -0,0065043 | 0,993430292 | LRRC41 | FALSE | Downregulated |
| 0,013000869 | 0,993430292 | MSANTD4 | FALSE | Upregulated |
| 0,009713823 | 0,993430292 | RGS5 | FALSE | Upregulated |
| 0,00933725 | 0,993430292 | ZNF326 | FALSE | Upregulated |

| | | | | |
|---|---|---|---|---|
| 0,003550729 | 0,993465201 | KIAA0100 | FALSE | Upregulated |
| -0,007093509 | 0,993696039 | LSM3 | FALSE | Downregulated |
| -0,010360411 | 0,993696039 | PRDM11 | FALSE | Downregulated |
| -0,014834246 | 0,993781024 | ANKHD1-EIF4EBP3 | FALSE | Downregulated |
| 0,017460193 | 0,993781024 | ZMYND15 | FALSE | Upregulated |
| -0,007263862 | 0,993864452 | LRRC8D | FALSE | Downregulated |
| 0,004840063 | 0,993864452 | TPP1 | FALSE | Upregulated |
| -0,008412382 | 0,993864452 | TRMT1 | FALSE | Downregulated |
| -0,009046682 | 0,994079706 | GOLGA8A | FALSE | Downregulated |
| 0,004271692 | 0,994195863 | ABLIM1 | FALSE | Upregulated |
| -0,019036621 | 0,994195863 | ADAM21P1 | FALSE | Downregulated |
| -0,019462528 | 0,994195863 | ANKRD20A9P | FALSE | Downregulated |
| -0,016167763 | 0,994195863 | APOLD1 | FALSE | Downregulated |
| -0,012840373 | 0,994195863 | AS3MT | FALSE | Downregulated |
| -0,004487508 | 0,994195863 | BID | FALSE | Downregulated |
| -0,008454415 | 0,994195863 | BOK-AS1 | FALSE | Downregulated |
| -0,004291335 | 0,994195863 | CCDC42B | FALSE | Downregulated |
| -0,011017092 | 0,994195863 | CLEC10A | FALSE | Downregulated |
| 0,007248582 | 0,994195863 | COPS2 | FALSE | Upregulated |
| -0,020765957 | 0,994195863 | CXorf57 | FALSE | Downregulated |
| -0,004158572 | 0,994195863 | EIF4E2 | FALSE | Downregulated |
| -0,00826039 | 0,994195863 | ELMO3 | FALSE | Downregulated |
| 0,006144814 | 0,994195863 | FAM46A | FALSE | Upregulated |
| -0,007342103 | 0,994195863 | IGBP1P1 | FALSE | Downregulated |
| -0,006624326 | 0,994195863 | KIAA1715 | FALSE | Downregulated |
| -0,008026714 | 0,994195863 | KIFAP3 | FALSE | Downregulated |
| -0,01555025 | 0,994195863 | LINC01011 | FALSE | Downregulated |
| -0,010797316 | 0,994195863 | LOC101928356 | FALSE | Downregulated |
| 0,004584497 | 0,994195863 | LOC101929385 | FALSE | Upregulated |
| -0,018573148 | 0,994195863 | LOC101929643 | FALSE | Downregulated |
| 0,04962264 | 0,994195863 | LOC102723943 | FALSE | Upregulated |
| -0,004491067 | 0,994195863 | MAP3K15 | FALSE | Downregulated |
| 0,005581777 | 0,994195863 | MAPK1IP1L | FALSE | Upregulated |
| 0,007169693 | 0,994195863 | MARCO | FALSE | Upregulated |
| -0,01980033 | 0,994195863 | MGC32805 | FALSE | Downregulated |
| -0,016971752 | 0,994195863 | MTNR1B | FALSE | Downregulated |
| 0,135217847 | 0,994195863 | MYH7 | FALSE | Upregulated |
| 0,008464144 | 0,994195863 | NAPG | FALSE | Upregulated |
| 0,003688177 | 0,994195863 | PEA15 | FALSE | Upregulated |
| -0,015145612 | 0,994195863 | PGBD4 | FALSE | Downregulated |
| 0,008858837 | 0,994195863 | PIGL | FALSE | Upregulated |
| 0,003564476 | 0,994195863 | PNN | FALSE | Upregulated |
| 0,01263571 | 0,994195863 | PODXL2 | FALSE | Upregulated |
| -0,010135357 | 0,994195863 | RAD51 | FALSE | Downregulated |
| -0,008035729 | 0,994195863 | RAP2A | FALSE | Downregulated |
| -0,015789665 | 0,994195863 | SMIM8 | FALSE | Downregulated |
| 0,003662714 | 0,994195863 | STIM1 | FALSE | Upregulated |
| -0,016803907 | 0,994195863 | STPG1 | FALSE | Downregulated |
| 0,007243001 | 0,994195863 | TBCK | FALSE | Upregulated |
| 0,005322251 | 0,994195863 | TICAM1 | FALSE | Upregulated |

| | | | | |
|---|---|---|---|---|
| -0,022076508 | 0,994195863 | TNFRSF19 | FALSE | Downregulated |
| -0,010976624 | 0,994195863 | TTC39B | FALSE | Downregulated |
| 0,003741909 | 0,994195863 | USH2A | FALSE | Upregulated |
| 0,007494287 | 0,994195863 | VDAC3 | FALSE | Upregulated |
| -0,005760629 | 0,994195863 | VNN1 | FALSE | Downregulated |
| 0,011972254 | 0,994195863 | XG | FALSE | Upregulated |
| 0,017488091 | 0,994195863 | ZNF738 | FALSE | Upregulated |
| 0,008077356 | 0,994266239 | KNOP1 | FALSE | Upregulated |
| 0,004331375 | 0,994276784 | PRPF38A | FALSE | Upregulated |
| -0,010478291 | 0,994382296 | ABHD11 | FALSE | Downregulated |
| -0,029565881 | 0,994382296 | ARL17B | FALSE | Downregulated |
| 0,017616432 | 0,994382296 | CD83 | FALSE | Upregulated |
| -0,003637292 | 0,994382296 | DCP2 | FALSE | Downregulated |
| 0,008194624 | 0,994382296 | KANSL2 | FALSE | Upregulated |
| 0,015550986 | 0,994382296 | LINC00535 | FALSE | Upregulated |
| 0,01241682 | 0,994382296 | LOC101060179 | FALSE | Upregulated |
| -0,004142364 | 0,994382296 | PPP4C | FALSE | Downregulated |
| -0,008215484 | 0,994382296 | WHAMMP1 | FALSE | Downregulated |
| -0,013431612 | 0,994399129 | LOC102724807 | FALSE | Downregulated |
| 0,006870055 | 0,994498811 | AATF | FALSE | Upregulated |
| -0,012696199 | 0,994498811 | AC091878.1 | FALSE | Downregulated |
| -0,005877532 | 0,994498811 | ADRB2 | FALSE | Downregulated |
| 0,011787661 | 0,994498811 | C9orf117 | FALSE | Upregulated |
| -0,002999226 | 0,994498811 | CTNNB1 | FALSE | Downregulated |
| -0,00497487 | 0,994498811 | HS1BP3 | FALSE | Downregulated |
| 0,017396057 | 0,994498811 | LHX6 | FALSE | Upregulated |
| -0,019863672 | 0,994498811 | LOC101927886 | FALSE | Downregulated |
| 0,018585452 | 0,994498811 | LOC653160 | FALSE | Upregulated |
| -0,006735419 | 0,994498811 | SRSF7 | FALSE | Downregulated |
| 0,00652677 | 0,994498811 | TUBG2 | FALSE | Upregulated |
| 0,003748094 | 0,994994258 | NCKAP1 | FALSE | Upregulated |
| 0,004986088 | 0,995108169 | C11orf30 | FALSE | Upregulated |
| 0,016090684 | 0,995108169 | MIPEPP3 | FALSE | Upregulated |
| 0,007837292 | 0,995108169 | SUSD3 | FALSE | Upregulated |
| -0,013518635 | 0,995108169 | ZNF880 | FALSE | Downregulated |
| 0,005561838 | 0,995139789 | EMC4 | FALSE | Upregulated |
| -0,012557028 | 0,995191908 | GOLGA7B | FALSE | Downregulated |
| -0,004330389 | 0,995277968 | ABCD1 | FALSE | Downregulated |
| 0,008499729 | 0,995277968 | ATP6V0E2-AS1 | FALSE | Upregulated |
| 0,005613838 | 0,995277968 | FKRP | FALSE | Upregulated |
| 0,006787027 | 0,995277968 | FLJ41481 | FALSE | Upregulated |
| -0,005368177 | 0,995277968 | GRAP | FALSE | Downregulated |
| -0,004943348 | 0,995277968 | HTATSF1 | FALSE | Downregulated |
| -0,008030138 | 0,995277968 | LOC101927002 | FALSE | Downregulated |
| -0,007185779 | 0,995277968 | LOC101927187 | FALSE | Downregulated |
| -0,004331647 | 0,995277968 | PMM1 | FALSE | Downregulated |
| 0,005389523 | 0,995277968 | SF3B4 | FALSE | Upregulated |
| -0,004740249 | 0,995277968 | SGMS1 | FALSE | Downregulated |
| -0,009115343 | 0,995277968 | SLX4IP | FALSE | Downregulated |
| -0,011455215 | 0,995277968 | SNORA53 | FALSE | Downregulated |
| -0,040801401 | 0,995277968 | TMEM256-PLSCR3 | FALSE | Downregulated |

| | | | | |
|---|---|---|---|---|
| -0,003727436 | 0,995277968 | TP73-AS1 | FALSE | Downregulated |
| -0,018596947 | 0,995277968 | TRPM2-AS | FALSE | Downregulated |
| -0,002714258 | 0,995277968 | U2AF2 | FALSE | Downregulated |
| -0,002795255 | 0,995277968 | UBE4A | FALSE | Downregulated |
| 0,013573752 | 0,99556257 | VRK1 | FALSE | Upregulated |
| -0,016054156 | 0,995591267 | CATSPERG | FALSE | Downregulated |
| -0,004574131 | 0,995713906 | HSD3BP4 | FALSE | Downregulated |
| -0,003278913 | 0,995713906 | RAB22A | FALSE | Downregulated |
| -0,009107273 | 0,995877544 | LOC101927858 | FALSE | Downregulated |
| 0,006706338 | 0,996048022 | LOC285638 | FALSE | Upregulated |
| 0,004746228 | 0,996355181 | POLK | FALSE | Upregulated |
| -0,003490767 | 0,996536206 | PCSK1 | FALSE | Downregulated |
| 0,009764462 | 0,996841809 | RGS2 | FALSE | Upregulated |
| -0,007981196 | 0,996841809 | SLC6A16 | FALSE | Downregulated |
| 0,005683242 | 0,996855495 | FZD8 | FALSE | Upregulated |
| 0,00515198 | 0,996855495 | METTL2B | FALSE | Upregulated |
| 0,003104429 | 0,996855495 | MMP24-AS1 | FALSE | Upregulated |
| -0,008114166 | 0,996855495 | SOX17 | FALSE | Downregulated |
| -0,003648209 | 0,996907035 | AFAP1L1 | FALSE | Downregulated |
| 0,002600136 | 0,996907035 | BDNF | FALSE | Upregulated |
| 0,001854666 | 0,996907035 | BMPR2 | FALSE | Upregulated |
| -0,008355977 | 0,996907035 | KATNBL1 | FALSE | Downregulated |
| -0,008828822 | 0,996907035 | LOC101927842 | FALSE | Downregulated |
| -0,001869893 | 0,996907035 | PPP6R1 | FALSE | Downregulated |
| -0,002484622 | 0,996907035 | SPIN1 | FALSE | Downregulated |
| -0,007742609 | 0,997119313 | SAYSD1 | FALSE | Downregulated |
| 0,008472881 | 0,997119313 | SH3GL1P1 | FALSE | Upregulated |
| 0,003336315 | 0,997119313 | UBE2G1 | FALSE | Upregulated |
| 0,003626905 | 0,997199732 | PTPRU | FALSE | Upregulated |
| -0,007461744 | 0,997273017 | ALKBH1 | FALSE | Downregulated |
| -0,004470562 | 0,997273017 | C6orf48 | FALSE | Downregulated |
| 0,003544992 | 0,997273017 | IGIP | FALSE | Upregulated |
| 0,003936737 | 0,997273017 | LOC101927841 | FALSE | Upregulated |
| -0,005314138 | 0,997273017 | PSMG2 | FALSE | Downregulated |
| 0,003272364 | 0,997273017 | SCAMP1 | FALSE | Upregulated |
| 0,004158536 | 0,997273017 | TMEM62 | FALSE | Upregulated |
| 0,002396141 | 0,997273017 | TXNDC17 | FALSE | Upregulated |
| -0,007723992 | 0,997294099 | TPX2 | FALSE | Downregulated |
| 0,004176485 | 0,997705366 | C5orf51 | FALSE | Upregulated |
| -0,001719029 | 0,997705366 | CCDC47 | FALSE | Downregulated |
| -0,001790099 | 0,997705366 | EDEM3 | FALSE | Downregulated |
| -0,002026756 | 0,997705366 | EPRS | FALSE | Downregulated |
| 0,002672366 | 0,997705366 | GPR108 | FALSE | Upregulated |
| -0,001775787 | 0,997705366 | KIAA1033 | FALSE | Downregulated |
| -0,002370608 | 0,997705366 | LACTB | FALSE | Downregulated |
| 0,008787831 | 0,997705366 | LOC100506286 | FALSE | Upregulated |
| -0,031174675 | 0,997705366 | NME1-NME2 | FALSE | Downregulated |
| -0,00196737 | 0,997705366 | RAP2C | FALSE | Downregulated |
| -0,002672615 | 0,997705366 | RIMKLA | FALSE | Downregulated |
| -0,004929337 | 0,997705366 | TCF21 | FALSE | Downregulated |
| 0,005456978 | 0,997705366 | TXNL4B | FALSE | Upregulated |

| | | | | |
|---|---|---|---|---|
| -0,00274066 | 0,997710188 | PPWD1 | FALSE | Downregulated |
| -0,002944896 | 0,997859798 | UGT2B11 | FALSE | Downregulated |
| 0,004014229 | 0,997923707 | LOC729887 | FALSE | Upregulated |
| -0,003435801 | 0,997936488 | LOC101928967 | FALSE | Downregulated |
| -0,006050056 | 0,997936488 | LOC101929959 | FALSE | Downregulated |
| -0,002619249 | 0,997936488 | LOC283922 | FALSE | Downregulated |
| -0,005299716 | 0,997936488 | OVCA2 | FALSE | Downregulated |
| -0,003855399 | 0,997936488 | RABL3 | FALSE | Downregulated |
| 0,012545385 | 0,997936488 | RIMBP2 | FALSE | Upregulated |
| 0,007246233 | 0,997936488 | TM2D1 | FALSE | Upregulated |
| 0,00171788 | 0,997936488 | ZNF282 | FALSE | Upregulated |
| -0,001379886 | 0,998232439 | YTHDF1 | FALSE | Downregulated |
| 0,005058463 | 0,998252356 | AC009473.1 | FALSE | Upregulated |
| -0,001867769 | 0,998252356 | CPNE1 | FALSE | Downregulated |
| -0,002472144 | 0,998252356 | TCEAL3 | FALSE | Downregulated |
| -0,002202215 | 0,998252356 | TMEM179B | FALSE | Downregulated |
| 0,002383227 | 0,998277829 | TFEB | FALSE | Upregulated |
| -0,003072303 | 0,998393299 | SP4 | FALSE | Downregulated |
| -0,001620777 | 0,998404379 | APPBP2 | FALSE | Downregulated |
| 0,001772907 | 0,998404379 | DEAF1 | FALSE | Upregulated |
| -0,002078801 | 0,998404379 | FNDC8 | FALSE | Downregulated |
| 0,003221201 | 0,998404379 | HIST2H2AB | FALSE | Upregulated |
| 0,002092789 | 0,998404379 | LINC00244 | FALSE | Upregulated |
| -0,003619591 | 0,998404379 | LINC00678 | FALSE | Downregulated |
| -0,001252944 | 0,998404379 | LOC100133091 | FALSE | Downregulated |
| 0,006154926 | 0,998404379 | MS4A14 | FALSE | Upregulated |
| -0,002721892 | 0,998405305 | MOB2 | FALSE | Downregulated |
| 0,001945922 | 0,998458145 | N4BP2L2-IT2 | FALSE | Upregulated |
| 0,00102193 | 0,998525078 | ATRX | FALSE | Upregulated |
| 0,001193505 | 0,998525078 | AVIL | FALSE | Upregulated |
| 0,001912029 | 0,998525078 | CBY1 | FALSE | Upregulated |
| 0,002111955 | 0,998525078 | CLTCL1 | FALSE | Upregulated |
| 0,002921163 | 0,998525078 | DCLK2 | FALSE | Upregulated |
| -0,001423633 | 0,998525078 | DHCR7 | FALSE | Downregulated |
| 0,002073043 | 0,998525078 | INTS7 | FALSE | Upregulated |
| -0,00060128 | 0,998525078 | ITM2B | FALSE | Downregulated |
| 0,001778385 | 0,998525078 | LOC101927445 | FALSE | Upregulated |
| 0,003242503 | 0,998525078 | MPV17 | FALSE | Upregulated |
| 0,00074474 | 0,998525078 | REEP5 | FALSE | Upregulated |
| 0,001015389 | 0,998525078 | RP11-887P2.5 | FALSE | Upregulated |
| -0,003198915 | 0,998525078 | TOX2 | FALSE | Downregulated |
| 0,002025248 | 0,99858819 | C16orf89 | FALSE | Upregulated |
| -0,000807384 | 0,998729249 | CD68 | FALSE | Downregulated |
| -0,002698159 | 0,998729249 | LINC00115 | FALSE | Downregulated |
| -0,000952888 | 0,998729249 | PPP1R12A | FALSE | Downregulated |
| -0,002450989 | 0,99878378 | FMO6P | FALSE | Downregulated |
| 0,002400191 | 0,998790679 | LIPT2 | FALSE | Upregulated |
| -0,000937752 | 0,998790679 | RTN4RL2 | FALSE | Downregulated |
| 0,0007481 | 0,999212808 | TMCO1 | FALSE | Upregulated |
| 0,002193903 | 0,999212808 | XXYLT1 | FALSE | Upregulated |
| 0,000909371 | 0,999382328 | CHRAC1 | FALSE | Upregulated |

| | | | | |
|---|---|---|---|---|
| -0,001781714 | 0,999382328 | SHISA7 | FALSE | Downregulated |
| -0,002991181 | 0,999382328 | TOX | FALSE | Downregulated |
| 0,000556203 | 0,999382328 | TTC38 | FALSE | Upregulated |
| -0,000871176 | 0,999382328 | UBE2E3 | FALSE | Downregulated |
| -0,000440214 | 0,999391284 | GCFC2 | FALSE | Downregulated |
| 0,000878224 | 0,999391284 | RIPPLY1 | FALSE | Upregulated |
| -0,002319006 | 0,999391284 | SENP3-EIF4A1 | FALSE | Downregulated |
| -0,000566713 | 0,999417719 | ZSCAN18 | FALSE | Downregulated |
| -0,000365489 | 0,999614142 | EIF5B | FALSE | Downregulated |
| 0,000827028 | 0,999614142 | LOC100288152 | FALSE | Upregulated |
| 0,000755188 | 0,999614142 | RP11-1007G5.2 | FALSE | Upregulated |
| 0,000278052 | 0,999638473 | LMNA | FALSE | Upregulated |
| -0,000322381 | 0,999731201 | SLC47A2 | FALSE | Downregulated |
| 0,000322101 | 0,99975384 | TMEM180 | FALSE | Upregulated |
| -0,000244242 | 0,99975384 | ZNF335 | FALSE | Downregulated |
| -9,98875E-05 | 0,999972994 | ARHGEF18 | FALSE | Downregulated |
| 0,00014917 | 0,999972994 | XIAP | FALSE | Upregulated |
| 0,000141181 | 0,999973068 | HSPA1A | FALSE | Upregulated |
| 0 | 1 | ATP6V1G2-DDX39B | FALSE | Upregulated |
| -0,000174039 | 1 | CDRT1 | FALSE | Downregulated |
| -0,000136606 | 1 | G6PC3 | FALSE | Downregulated |
| 0 | 1 | H19 | FALSE | Upregulated |
| 0 | 1 | KRT17 | FALSE | Upregulated |
| 0 | 1 | KRT6A | FALSE | Upregulated |
| 6,82381E-06 | 1 | LOC101927910 | FALSE | Upregulated |
| 0 | 1 | TMX2-CTNND1 | FALSE | Upregulated |
| 1,118112852 | | A3GALT2 | | Upregulated |
| | | AA06 | | |
| | | AACSP1 | | |
| | | AADACL3 | | |
| 0,608246941 | | AADACL4 | FALSE | Upregulated |
| | | AARD | | |
| -0,177683088 | | AATK-AS1 | FALSE | Downregulated |
| 0,771187124 | | ABCA12 | FALSE | Upregulated |
| 0,030159423 | | ABCB5 | FALSE | Upregulated |
| -0,222855544 | | ABCC12 | FALSE | Downregulated |
| 0 | | ABCC13 | FALSE | Upregulated |
| 0,665683691 | | ABCC5-AS1 | FALSE | Upregulated |
| 0 | | ABCC8 | FALSE | Upregulated |
| -0,734825342 | | ABCD2 | FALSE | Downregulated |
| 0,07478914 | | ABCG4 | FALSE | Upregulated |
| | | ABHD14A-ACY1 | | |
| 0,588315691 | | ABO | FALSE | Upregulated |
| 0,12747017 | | AC002116.7 | FALSE | Upregulated |
| -0,238131749 | | AC002398.5 | FALSE | Downregulated |
| 0 | | AC002454.1 | FALSE | Upregulated |
| | | AC002463.3 | | |
| -0,218384846 | | AC002511.1 | FALSE | Downregulated |
| | | AC003051.1 | | |
| 0 | | AC003102.3 | FALSE | Upregulated |
| | | AC003958.2 | | |

| | | | |
|---|---|---|---|
| | AC003984.1 | | |
| | AC004603.4 | | |
| 0,597480774 | AC005022.1 | FALSE | Upregulated |
| 0 | AC005062.2 | FALSE | Upregulated |
| | AC005235.1 | | |
| 2,811501618 | AC005358.3 | | Upregulated |
| 0,254423982 | AC005550.4 | FALSE | Upregulated |
| 0,117723908 | AC005592.2 | FALSE | Upregulated |
| | AC006004.1 | | |
| | AC006196.1 | | |
| 0,098822319 | AC006372.4 | FALSE | Upregulated |
| 1,026304245 | AC007040.7 | | Upregulated |
| 1,034842413 | AC007131.1 | | Upregulated |
| 0 | AC007359.6 | FALSE | Upregulated |
| -0,333834456 | AC007365.1 | FALSE | Downregulated |
| 1,292946193 | AC007386.2 | | Upregulated |
| 0 | AC007392.3 | FALSE | Upregulated |
| 2,015881616 | AC007392.4 | | Upregulated |
| | AC007403.2 | | |
| 0 | AC007405.4 | FALSE | Upregulated |
| | AC007463.2 | | |
| 0 | AC007682.1 | FALSE | Upregulated |
| 0,969853863 | AC007740.1 | FALSE | Upregulated |
| 0,415413327 | AC007743.1 | FALSE | Upregulated |
| 0,390077535 | AC007773.2 | FALSE | Upregulated |
| | AC007879.2 | | |
| -0,176619188 | AC007880.1 | FALSE | Downregulated |
| -0,169025238 | AC007966.1 | FALSE | Downregulated |
| | AC008697.1 | | |
| 0,450612395 | AC008703.1 | FALSE | Upregulated |
| 0 | AC009120.4 | FALSE | Upregulated |
| | AC009227.2 | | |
| | AC009518.4 | | |
| 0,048656215 | AC010091.1 | FALSE | Upregulated |
| 0,062821265 | AC010524.2 | FALSE | Upregulated |
| | AC010731.4 | | |
| 0,174662837 | AC010890.1 | FALSE | Upregulated |
| | AC010975.1 | | |
| | AC010987.6 | | |
| | AC011298.2 | | |
| 0,06784886 | AC011518.1 | FALSE | Upregulated |
| -0,143527104 | AC011609.3 | FALSE | Downregulated |
| | AC011718.2 | | |
| | AC012075.2 | | |
| | AC012494.1 | | |
| 1,190423605 | AC012507.3 | | Upregulated |
| 0,84726383 | AC015922.6 | FALSE | Upregulated |
| | AC016995.3 | | |
| 0,986318603 | AC017060.1 | FALSE | Upregulated |
| | AC018464.3 | | |
| 0 | AC018470.4 | FALSE | Upregulated |

| | | | |
|---|---|---|---|
| 0 | AC018799.1 | FALSE | Upregulated |
| 0,963679593 | AC019048.1 | FALSE | Upregulated |
| | AC019118.2 | | |
| | AC022311.1 | | |
| | AC027269.2 | | |
| 0 | AC068831.10 | FALSE | Upregulated |
| 2,088665987 | AC068858.1 | | Upregulated |
| | AC069277.1 | | |
| -0,091472596 | AC072062.1 | FALSE | Downregulated |
| 0 | AC073128.10 | FALSE | Upregulated |
| -0,77023888 | AC073133.1 | FALSE | Downregulated |
| 0,059322817 | AC073283.4 | FALSE | Upregulated |
| 0 | AC073321.3 | FALSE | Upregulated |
| | AC073628.1 | | |
| -0,354431812 | AC074289.1 | FALSE | Downregulated |
| 0 | AC079145.4 | FALSE | Upregulated |
| -0,157159874 | AC079586.1 | FALSE | Downregulated |
| 0,380867842 | AC079610.2 | FALSE | Upregulated |
| | AC079776.3 | | |
| -0,206687471 | AC087859.1 | FALSE | Downregulated |
| 0,305603432 | AC090044.1 | FALSE | Upregulated |
| 0 | AC090505.6 | FALSE | Upregulated |
| -0,056031977 | AC091969.1 | FALSE | Downregulated |
| -0,295767328 | AC093375.1 | FALSE | Downregulated |
| -0,255944134 | AC093391.2 | FALSE | Downregulated |
| 0 | AC093590.1 | FALSE | Upregulated |
| 0,879129185 | AC095067.1 | FALSE | Upregulated |
| -0,057454521 | AC096559.1 | FALSE | Downregulated |
| 0,138127506 | AC097468.4 | FALSE | Upregulated |
| -0,126579952 | AC097500.2 | FALSE | Downregulated |
| | AC098823.3 | | |
| | AC099342.1 | | |
| 0,608246941 | AC104024.1 | FALSE | Upregulated |
| 1,074853582 | AC104653.1 | | Upregulated |
| 1,398744682 | AC104654.1 | | Upregulated |
| 0 | AC104654.2 | FALSE | Upregulated |
| -0,307747965 | AC104777.2 | FALSE | Downregulated |
| | AC104777.4 | | |
| 1,11320998 | AC104820.2 | | Upregulated |
| 0,048287507 | AC105053.3 | FALSE | Upregulated |
| | AC105253.1 | | |
| 0 | AC105339.1 | FALSE | Upregulated |
| -0,377694111 | AC105760.3 | FALSE | Downregulated |
| 0 | AC107070.1 | FALSE | Upregulated |
| -1,076673761 | AC107622.1 | | Downregulated |
| | AC108142.1 | | |
| | AC113607.2 | | |
| | AC116035.1 | | |
| -0,625752788 | AC125421.1 | FALSE | Downregulated |
| 0,617678979 | AC131097.3 | FALSE | Upregulated |
| -0,360054407 | AC136896.1 | FALSE | Downregulated |

| | | | |
|---|---|---|---|
| -0,093354979 | AC159540.1 | FALSE | Downregulated |
| 0,02569507 | AC226118.1 | FALSE | Upregulated |
| -0,312291437 | ACBD7 | FALSE | Downregulated |
| -0,34969677 | ACCSL | FALSE | Downregulated |
| 0,065073665 | ACER1 | FALSE | Upregulated |
| 0,384312479 | ACPP | FALSE | Upregulated |
| 0,450124955 | ACPT | FALSE | Upregulated |
| -0,116684499 | ACR | FALSE | Downregulated |
| 0,397526912 | ACRV1 | FALSE | Upregulated |
| -0,083897522 | ACSM4 | FALSE | Downregulated |
| 0,688251927 | ACTA2-AS1 | FALSE | Upregulated |
| 0,360832372 | ACTC1 | FALSE | Upregulated |
| 1,123383632 | ACTG1P4 | | Upregulated |
| 1,15627051 | ACTG2 | | Upregulated |
| -0,435118795 | ACTL6B | FALSE | Downregulated |
| 0,036767477 | ACTL7B | FALSE | Upregulated |
| | ACTL8 | | |
| 0 | ACTL9 | FALSE | Upregulated |
| | ACTN1-AS1 | | |
| -0,174015385 | ACTR3BP2 | FALSE | Downregulated |
| 0 | ACTR3BP5 | FALSE | Upregulated |
| | ACTRT1 | | |
| | ACTRT2 | | |
| 0,645291912 | ACYP1 | FALSE | Upregulated |
| -1,591292359 | AD000090.2 | | Downregulated |
| | ADAD1 | | |
| 0,620235175 | ADAD2 | FALSE | Upregulated |
| | ADAM18 | | |
| | ADAM2 | | |
| 1,557581543 | ADAM23 | | Upregulated |
| 0,487886613 | ADAM29 | FALSE | Upregulated |
| 0,669756935 | ADAM30 | FALSE | Upregulated |
| 1,116847879 | ADAM32 | | Upregulated |
| 0,039005732 | ADAM3A | FALSE | Upregulated |
| | ADAM5 | | |
| 0,075728143 | ADAM7 | FALSE | Upregulated |
| 1,544816292 | ADAMDEC1 | | Upregulated |
| 1,600044226 | ADAMTS14 | | Upregulated |
| -0,001452899 | ADAMTS18 | FALSE | Downregulated |
| 0 | ADAMTS19 | FALSE | Upregulated |
| | ADAMTS20 | | |
| -0,200889911 | ADAMTS3 | FALSE | Downregulated |
| 0 | ADAMTS7P1 | FALSE | Upregulated |
| 0,944850273 | ADARB2-AS1 | FALSE | Upregulated |
| 0 | ADCY2 | FALSE | Upregulated |
| 0,914768082 | ADCY8 | FALSE | Upregulated |
| 0,796439842 | ADCYAP1 | FALSE | Upregulated |
| 1,231840587 | ADD2 | | Upregulated |
| 0,91880985 | ADD3-AS1 | FALSE | Upregulated |
| | ADGB | | |
| | ADIG | | |

| | | | |
|---|---|---|---|
| 0,688357234 | ADIPOQ | FALSE | Upregulated |
| -0,20237643 | ADORA1 | FALSE | Downregulated |
| 0,203376495 | ADPRHL1 | FALSE | Upregulated |
| 0,396349643 | ADRA1D | FALSE | Upregulated |
| 2,019062433 | ADRA2C | | Upregulated |
| -0,223303852 | ADRB1 | FALSE | Downregulated |
| | ADRB3 | | |
| -0,569545384 | ADTRP | FALSE | Downregulated |
| | AF064858.7 | | |
| -0,088487369 | AFF2 | FALSE | Downregulated |
| 0,523360633 | AFP | FALSE | Upregulated |
| -0,311839939 | AGAP1-IT1 | FALSE | Downregulated |
| 0 | AGBL1 | FALSE | Upregulated |
| | AGBL1-AS1 | | |
| -1,470808257 | AGBL4 | | Downregulated |
| | AGBL4-IT1 | | |
| 0,13256632 | AGBL5-AS1 | FALSE | Upregulated |
| 0,304573134 | AGPAT4-IT1 | FALSE | Upregulated |
| | AGR3 | | |
| | AGTR2 | | |
| -0,424745169 | AHCTF1P1 | FALSE | Downregulated |
| | AHSP | | |
| 0 | AICDA | FALSE | Upregulated |
| 2,558964083 | AIM1L | | Upregulated |
| | AIPL1 | | |
| -0,06089979 | AIRE | FALSE | Downregulated |
| 0,437630684 | AK7 | FALSE | Upregulated |
| 0,031712089 | AK8 | FALSE | Upregulated |
| 0,137936337 | AKAP14 | FALSE | Upregulated |
| | AKAP17A-2 | | |
| 0 | AKAP4 | FALSE | Upregulated |
| 0,485711093 | AKNAD1 | FALSE | Upregulated |
| 0,731711798 | AKR1B15 | FALSE | Upregulated |
| | AL022344.4 | | |
| 0,039005732 | AL109763.2 | FALSE | Upregulated |
| 0,9702217 | AL132709.5 | FALSE | Upregulated |
| -0,682620527 | ALDH3A1 | FALSE | Downregulated |
| -0,892097699 | ALDH3B2 | FALSE | Downregulated |
| 1,779145242 | ALG1L2 | | Upregulated |
| -0,070955797 | ALLC | FALSE | Downregulated |
| -0,079169848 | ALMS1-IT1 | FALSE | Downregulated |
| 1,72911928 | ALOX12B | | Upregulated |
| -0,2414467 | ALOX15 | FALSE | Downregulated |
| 0,971874621 | ALOX15B | FALSE | Upregulated |
| 0,129325714 | ALOX15P1 | FALSE | Upregulated |
| 0,479813092 | ALOX5AP | FALSE | Upregulated |
| 0,582571468 | ALOXE3 | FALSE | Upregulated |
| | ALPI | | |
| -0,115613886 | ALPP | FALSE | Downregulated |
| | ALPPL2 | | |
| 0 | ALX1 | FALSE | Upregulated |

| | | | |
|---|---|---|---|
| 0 | AMBN | FALSE | Upregulated |
| 0,732196563 | AMELX | FALSE | Upregulated |
| | AMELY | | |
| 0 | AMER2 | FALSE | Upregulated |
| | AMER3 | | |
| -0,934888741 | AMH | FALSE | Downregulated |
| 0,301362353 | AMHR2 | FALSE | Upregulated |
| 1,372229245 | AMPD1 | | Upregulated |
| | AMTN | | |
| | AMY1A | | |
| | AMY1B | | |
| | AMY1C | | |
| | AMY2A | | |
| 0 | ANAPC1P1 | FALSE | Upregulated |
| -0,689970151 | ANGPT1 | FALSE | Downregulated |
| 0,581599095 | ANGPT4 | FALSE | Upregulated |
| 0,97618794 | ANGPTL5 | FALSE | Upregulated |
| 0,039540157 | ANHX | FALSE | Upregulated |
| 0,019985931 | ANKFN1 | FALSE | Upregulated |
| 0,532265625 | ANKK1 | FALSE | Upregulated |
| 1,853798574 | ANKRD1 | | Upregulated |
| 1,505746717 | ANKRD18B | | Upregulated |
| 0 | ANKRD18CP | FALSE | Upregulated |
| -0,172379338 | ANKRD18DP | FALSE | Downregulated |
| -0,854013786 | ANKRD2 | FALSE | Downregulated |
| 0,082729154 | ANKRD20A11P | FALSE | Upregulated |
| | ANKRD20A19P | | |
| -0,195351544 | ANKRD20A3 | FALSE | Downregulated |
| -0,379911266 | ANKRD20A4 | FALSE | Downregulated |
| -0,183457651 | ANKRD20A5P | FALSE | Downregulated |
| 0,058930564 | ANKRD20A8P | FALSE | Upregulated |
| | ANKRD26P1 | | |
| 0,273321258 | ANKRD26P3 | FALSE | Upregulated |
| 0,892490695 | ANKRD29 | FALSE | Upregulated |
| | ANKRD30A | | |
| 0 | ANKRD30B | FALSE | Upregulated |
| | ANKRD30BL | | |
| 0,57117889 | ANKRD30BP2 | FALSE | Upregulated |
| | ANKRD30BP3 | | |
| 0,210032009 | ANKRD31 | FALSE | Upregulated |
| 0 | ANKRD33 | FALSE | Upregulated |
| 0,239947299 | ANKRD34B | FALSE | Upregulated |
| | ANKRD34C | | |
| 0,527274061 | ANKRD36BP2 | FALSE | Upregulated |
| 0,130922456 | ANKRD44-IT1 | FALSE | Upregulated |
| -0,10131956 | ANKRD45 | FALSE | Downregulated |
| | ANKRD60 | | |
| 0 | ANKRD62 | FALSE | Upregulated |
| 0,585417161 | ANKRD62P1-PARP4P3 | FALSE | Upregulated |
| 0,339907739 | ANKRD63 | FALSE | Upregulated |
| 0 | ANKRD66 | FALSE | Upregulated |

| | | | |
|---|---|---|---|
| | ANKRD7 | | |
| | ANKUB1 | | |
| 0,846964406 | ANLN | FALSE | Upregulated |
| 1,53937162 | ANO2 | | Upregulated |
| 0,76933811 | ANO4 | FALSE | Upregulated |
| 0 | ANP32C | FALSE | Upregulated |
| | ANP32D | | |
| 0,092296489 | ANTXRL | FALSE | Upregulated |
| 2,623594368 | ANTXRLP1 | | Upregulated |
| 0 | ANXA2P3 | FALSE | Upregulated |
| 1,37210734 | ANXA3 | | Upregulated |
| 0,679243174 | ANXA8L1 | FALSE | Upregulated |
| -0,272271359 | AOX2P | FALSE | Downregulated |
| | AP000289.6 | | |
| -0,424341588 | AP000330.8 | FALSE | Downregulated |
| | AP000345.1 | | |
| | AP000469.2 | | |
| | AP000475.2 | | |
| 1,568516992 | AP000688.8 | | Upregulated |
| -0,117082984 | AP003025.2 | FALSE | Downregulated |
| | AP1B1P1 | | |
| 1,189603037 | AP1M2 | | Upregulated |
| 1,673871949 | AP1S3 | | Upregulated |
| 0 | AP3B2 | FALSE | Upregulated |
| 0 | APCDD1L-AS1 | FALSE | Upregulated |
| | APELA | | |
| 0,174006501 | APLN | FALSE | Upregulated |
| 0,848285487 | APLP1 | FALSE | Upregulated |
| | APOBEC1 | | |
| -0,007220502 | APOBEC3A | FALSE | Downregulated |
| 1,998414872 | APOBEC3B | | Upregulated |
| 0 | APOBEC3B-AS1 | FALSE | Upregulated |
| -0,785374669 | APOBEC3H | FALSE | Downregulated |
| 0,068479342 | APOD | FALSE | Upregulated |
| 0 | APOOP5 | FALSE | Upregulated |
| | AQP10 | | |
| | AQP12A | | |
| | AQP12B | | |
| -0,001452899 | AQP2 | FALSE | Downregulated |
| 2,261521819 | AQP5 | | Upregulated |
| -1,036233634 | AQP7P3 | | Downregulated |
| 0,087836139 | AQP8 | FALSE | Upregulated |
| -0,104398655 | AQPEP | FALSE | Downregulated |
| 1,104569889 | ARC | | Upregulated |
| 0 | AREG | FALSE | Upregulated |
| 1,923425218 | ARGFX | | Upregulated |
| 0,643572601 | ARHGAP11B | FALSE | Upregulated |
| 2,095478682 | ARHGAP19-SLIT1 | | Upregulated |
| -0,038452025 | ARHGAP26-IT1 | FALSE | Downregulated |
| 1,704395443 | ARHGAP28 | | Upregulated |
| | ARHGAP36 | | |

| | | | |
|---|---|---|---|
| 0 | ARHGAP40 | FALSE | Upregulated |
| -0,116585707 | ARHGAP8 | FALSE | Downregulated |
| 0 | ARHGDIG | FALSE | Upregulated |
| 0,687143177 | ARHGEF26-AS1 | FALSE | Upregulated |
| 0 | ARHGEF3-AS1 | FALSE | Upregulated |
| 1,045664789 | ARHGEF38-IT1 | | Upregulated |
| 0,658663119 | ARL14 | FALSE | Upregulated |
| 0 | ARL14EPL | FALSE | Upregulated |
| 0,68485754 | ARL5C | FALSE | Upregulated |
| 0 | ARL9 | FALSE | Upregulated |
| 0,823353192 | ARMC12 | FALSE | Upregulated |
| | ARMC3 | | |
| 1,310301233 | ARMC4 | | Upregulated |
| | ARMC4P1 | | |
| 0,582058638 | ARMS2 | FALSE | Upregulated |
| 0,032020854 | ARRDC5 | FALSE | Upregulated |
| -0,624241645 | ARSF | FALSE | Downregulated |
| | ARSH | | |
| -0,393578227 | ARSI | FALSE | Downregulated |
| 1,878724011 | ART3 | | Upregulated |
| -0,490145707 | ART5 | FALSE | Downregulated |
| -0,141459944 | ARX | FALSE | Downregulated |
| -0,601751138 | ASAH2 | FALSE | Downregulated |
| 0,339502577 | ASAP1-IT1 | FALSE | Upregulated |
| 0,473813682 | ASB10 | FALSE | Upregulated |
| -0,101494514 | ASB11 | FALSE | Downregulated |
| 0,057442013 | ASB17 | FALSE | Upregulated |
| 0,77720106 | ASB18 | FALSE | Upregulated |
| 0 | ASB5 | FALSE | Upregulated |
| 0 | ASCL4 | FALSE | Upregulated |
| 0,70210981 | ASCL5 | FALSE | Upregulated |
| 0 | ASIC2 | FALSE | Upregulated |
| | ASMT | | |
| | ASMT-2 | | |
| | ASMTL-2 | | |
| | ASMTL-AS1-2 | | |
| 0 | ASTL | FALSE | Upregulated |
| 1,319761323 | ASTN1 | | Upregulated |
| | ASZ1 | | |
| 0,700273045 | ATCAY | FALSE | Upregulated |
| | ATOH1 | | |
| -0,055970583 | ATP12A | FALSE | Downregulated |
| 0,859746507 | ATP13A4 | FALSE | Upregulated |
| 0,407449939 | ATP13A5 | FALSE | Upregulated |
| 0,471984849 | ATP1A3 | FALSE | Upregulated |
| 0,175546899 | ATP1A4 | FALSE | Upregulated |
| -1,020349279 | ATP1B4 | | Downregulated |
| -0,124707131 | ATP2B3 | FALSE | Downregulated |
| 0,048956355 | ATP4A | FALSE | Upregulated |
| 0,087836139 | ATP5EP2 | FALSE | Upregulated |
| 1,018046467 | ATP5L2 | | Upregulated |

| | | | |
|---|---|---|---|
| -1,210950155 | ATP6AP1L | | Downregulated |
| | ATP6V0A4 | | |
| 0,644586486 | ATP6V0CP3 | FALSE | Upregulated |
| -0,151013818 | ATP6V0D2 | FALSE | Downregulated |
| 0,210428453 | ATP6V1B1 | FALSE | Upregulated |
| | ATP6V1G3 | | |
| 0,870490769 | ATP8A2 | FALSE | Upregulated |
| 0,363392679 | ATP8B3 | FALSE | Upregulated |
| -0,418634999 | ATP8B5P | FALSE | Downregulated |
| 0 | ATXN3L | FALSE | Upregulated |
| | ATXN8OS | | |
| -0,270622236 | AURKB | FALSE | Downregulated |
| -0,125280919 | AURKC | FALSE | Downregulated |
| | AVP | | |
| 2,101060855 | AVPR1B | | Upregulated |
| 0,072331122 | AWAT2 | FALSE | Upregulated |
| -0,077227676 | AXDND1 | FALSE | Downregulated |
| -0,373352502 | AZU1 | FALSE | Downregulated |
| -0,201876498 | B3GALT1 | FALSE | Downregulated |
| 2,013855566 | B3GALT5 | | Upregulated |
| 0,209407149 | B3GNT3 | FALSE | Upregulated |
| 0,034128514 | B3GNT6 | FALSE | Upregulated |
| 0 | B4GALNT2 | FALSE | Upregulated |
| 0,301850534 | B4GALNT4 | FALSE | Upregulated |
| 0,669756935 | BAALCOS | FALSE | Upregulated |
| 0,029387898 | BAGE2 | FALSE | Upregulated |
| 1,662456477 | BAI2 | | Upregulated |
| | BANCR | | |
| 0,669756935 | BANF2 | FALSE | Upregulated |
| | BARHL2 | | |
| 0,497936904 | BARX1 | FALSE | Upregulated |
| 1,177182509 | BARX2 | | Upregulated |
| 0 | BASP1P1 | FALSE | Upregulated |
| -0,214029303 | BATF3 | FALSE | Downregulated |
| -0,047713447 | BBS12 | FALSE | Downregulated |
| 0 | BCAR4 | FALSE | Upregulated |
| 1,295558413 | BCAS1 | | Upregulated |
| -0,088899496 | BCL2A1 | FALSE | Downregulated |
| 0,570450811 | BCL2L15 | FALSE | Upregulated |
| 1,571482204 | BCL2L2-PABPN1 | | Upregulated |
| 0,783551112 | BCMO1 | FALSE | Upregulated |
| 0,753508021 | BCORP1 | FALSE | Upregulated |
| 0,165476691 | BCRP2 | FALSE | Upregulated |
| -1,037765714 | BCRP3 | | Downregulated |
| 1,169753992 | BCYRN1 | | Upregulated |
| 1,651675542 | BDKRB1 | | Upregulated |
| 0,880612809 | BDKRB2 | FALSE | Upregulated |
| | BEND2 | | |
| -0,456131977 | BEND5 | FALSE | Downregulated |
| 0,777779356 | BEND6 | FALSE | Upregulated |
| 0,104723245 | BEST3 | FALSE | Upregulated |

| | | | |
|---|---|---|---|
| 0,424657371 | BEST4 | FALSE | Upregulated |
| -0,660936445 | BEX1 | FALSE | Downregulated |
| 0,529326338 | BEX2 | FALSE | Upregulated |
| -0,672285914 | BEX4 | FALSE | Downregulated |
| 0,236440052 | BEX5 | FALSE | Upregulated |
| 0,04955067 | BFSP1 | FALSE | Upregulated |
| 1,865959063 | BFSP2 | | Upregulated |
| 0 | BFSP2-AS1 | FALSE | Upregulated |
| | BGLAP | | |
| 0,048656215 | BHLHA9 | FALSE | Upregulated |
| 0,608246941 | BHLHE23 | FALSE | Upregulated |
| 2,229649036 | BHLHE40-AS1 | | Upregulated |
| -0,524782109 | BIN3-IT1 | FALSE | Downregulated |
| 0,609585253 | BIRC5 | FALSE | Upregulated |
| 0,597480774 | BIRC7 | FALSE | Upregulated |
| | BIRC8 | | |
| -0,061971943 | BLACAT1 | FALSE | Downregulated |
| 0 | BLACE | FALSE | Upregulated |
| 1,155811035 | BLID | | Upregulated |
| 0,453846632 | BLM | FALSE | Upregulated |
| -1,380649034 | BLOC1S5-TXNDC5 | | Downregulated |
| | BMP15 | | |
| 1,005320276 | BMP3 | | Upregulated |
| 0 | BMP7 | FALSE | Upregulated |
| -1,216246886 | BMPR1B | | Downregulated |
| -0,233784713 | BNC1 | FALSE | Downregulated |
| 0,463322856 | BOD1L2 | FALSE | Upregulated |
| 1,406685975 | BPESC1 | | Upregulated |
| 0,151324073 | BPI | FALSE | Upregulated |
| | BPIFA1 | | |
| | BPIFA2 | | |
| | BPIFA3 | | |
| | BPIFA4P | | |
| 0,532090765 | BPIFB1 | FALSE | Upregulated |
| -0,002529603 | BPIFB2 | FALSE | Downregulated |
| | BPIFB3 | | |
| | BPIFB4 | | |
| | BPIFB6 | | |
| -0,451469183 | BPIFC | FALSE | Downregulated |
| | BRDT | | |
| 0,085217661 | BREA2 | FALSE | Upregulated |
| 0,314390177 | BRINP1 | FALSE | Upregulated |
| -0,070023259 | BRINP2 | FALSE | Downregulated |
| | BRINP3 | | |
| -0,633225612 | BRS3 | FALSE | Downregulated |
| 0 | BRWD1-AS1 | FALSE | Upregulated |
| 0,463980724 | BSN-AS2 | FALSE | Upregulated |
| 0,001293581 | BSND | FALSE | Upregulated |
| | BSPH1 | | |
| | BSX | | |
| -0,289422207 | BTBD16 | FALSE | Downregulated |

| | | | |
|---|---|---|---|
| 0,931567804 | BTBD8 | FALSE | Upregulated |
| 1,11859242 | BTC | | Upregulated |
| 0 | BTF3P11 | FALSE | Upregulated |
| | BTG4 | | |
| 1,207539246 | BTLA | | Upregulated |
| 0,530030368 | BTNL10 | FALSE | Upregulated |
| 1,062006821 | BTNL2 | | Upregulated |
| -0,926070808 | BTNL8 | FALSE | Downregulated |
| -0,160031898 | BUB1 | FALSE | Downregulated |
| 0,703213409 | BUB1B | FALSE | Upregulated |
| 0,167105872 | BVES | FALSE | Upregulated |
| | BVES-AS1 | | |
| 0 | BX255923.3 | FALSE | Upregulated |
| 0,103908999 | C10orf107 | FALSE | Upregulated |
| -0,61257648 | C10orf111 | FALSE | Downregulated |
| | C10orf113 | | |
| 0 | C10orf115 | FALSE | Upregulated |
| | C10orf120 | | |
| -0,43001229 | C10orf126 | FALSE | Downregulated |
| 1,369210529 | C10orf129 | | Upregulated |
| 0,941063579 | C10orf131 | FALSE | Upregulated |
| -0,265165121 | C10orf35 | FALSE | Downregulated |
| 0,041547119 | C10orf40 | FALSE | Upregulated |
| 0,048656215 | C10orf53 | FALSE | Upregulated |
| -0,371412646 | C10orf67 | FALSE | Downregulated |
| | C10orf71-AS1 | | |
| | C10orf82 | | |
| 1,001547319 | C10orf85 | | Upregulated |
| 0,97704728 | C10orf90 | FALSE | Upregulated |
| 0 | C10orf91 | FALSE | Upregulated |
| | C10orf99 | | |
| | C11orf40 | | |
| | C11orf44 | | |
| | C11orf53 | | |
| 2,350588422 | C11orf63 | | Upregulated |
| -0,14887559 | C11orf65 | FALSE | Downregulated |
| | C11orf70 | | |
| 1,363321584 | C11orf82 | | Upregulated |
| -0,273893438 | C11orf85 | FALSE | Downregulated |
| | C11orf86 | | |
| -0,780636808 | C11orf87 | FALSE | Downregulated |
| -0,00897377 | C11orf88 | FALSE | Downregulated |
| 0 | C11orf91 | FALSE | Upregulated |
| | C11orf94 | | |
| 0,736224514 | C12orf36 | FALSE | Upregulated |
| 0,006988064 | C12orf39 | FALSE | Upregulated |
| -0,435449173 | C12orf40 | FALSE | Downregulated |
| 0,015134022 | C12orf42 | FALSE | Upregulated |
| | C12orf50 | | |
| | C12orf54 | | |
| 1,214960706 | C12orf55 | | Upregulated |

| | | | |
|---|---|---|---|
| -0,757190004 | C12orf56 | FALSE | Downregulated |
| -0,24702647 | C12orf68 | FALSE | Downregulated |
| -0,261325842 | C12orf74 | FALSE | Downregulated |
| | C12orf77 | | |
| -0,169773574 | C12orf79 | FALSE | Downregulated |
| 0 | C12orf80 | FALSE | Upregulated |
| | C13orf45 | | |
| | C14orf177 | | |
| 0,606031026 | C14orf183 | FALSE | Upregulated |
| -0,115927293 | C14orf23 | FALSE | Downregulated |
| 0 | C14orf39 | FALSE | Upregulated |
| 0,064230946 | C14orf64 | FALSE | Upregulated |
| 0,128771935 | C15orf26 | FALSE | Upregulated |
| | C15orf32 | | |
| 0,865714552 | C15orf43 | FALSE | Upregulated |
| 0 | C15orf48 | FALSE | Upregulated |
| 0 | C15orf53 | FALSE | Upregulated |
| 0 | C15orf56 | FALSE | Upregulated |
| -1,129741029 | C15orf59 | | Downregulated |
| -0,41921401 | C15orf65 | FALSE | Downregulated |
| | C16orf11 | | |
| 0,520244182 | C16orf47 | FALSE | Upregulated |
| 0,251940551 | C16orf74 | FALSE | Upregulated |
| -0,436188202 | C16orf78 | FALSE | Downregulated |
| | C16orf82 | | |
| 0,718329206 | C16orf92 | FALSE | Upregulated |
| | C16orf97 | | |
| | C17orf102 | | |
| 0 | C17orf105 | FALSE | Upregulated |
| | C17orf112 | | |
| 0,092736499 | C17orf50 | FALSE | Upregulated |
| 1,649790717 | C17orf64 | | Upregulated |
| 0,036208106 | C17orf74 | FALSE | Upregulated |
| | C17orf77 | | |
| | C17orf98 | | |
| -0,08530186 | C17orf99 | FALSE | Downregulated |
| 0,071623549 | C18orf12 | FALSE | Upregulated |
| 0 | C18orf42 | FALSE | Upregulated |
| 0,850062865 | C18orf54 | FALSE | Upregulated |
| -1,122444588 | C18orf56 | | Downregulated |
| | C18orf61 | | |
| 0,518361158 | C18orf63 | FALSE | Upregulated |
| | C18orf64 | | |
| -0,131619652 | C19orf18 | FALSE | Downregulated |
| 1,214120973 | C19orf33 | | Upregulated |
| 0,12459073 | C19orf38 | FALSE | Upregulated |
| -0,054802128 | C19orf45 | FALSE | Downregulated |
| 0,749561648 | C19orf59 | FALSE | Upregulated |
| 0,682411001 | C19orf69 | FALSE | Upregulated |
| -0,092319168 | C19orf73 | FALSE | Downregulated |
| -0,191807042 | C19orf81 | FALSE | Downregulated |

| | | | |
|---|---|---|---|
| -0,041288106 | C19orf83 | FALSE | Downregulated |
| 0,765474462 | C1orf100 | FALSE | Upregulated |
| 0,510374654 | C1orf101 | FALSE | Upregulated |
| 0,616592716 | C1orf105 | FALSE | Upregulated |
| 0 | C1orf110 | FALSE | Upregulated |
| 1,448157457 | C1orf127 | | Upregulated |
| | C1orf137 | | |
| | C1orf141 | | |
| 0 | C1orf146 | FALSE | Upregulated |
| | C1orf158 | | |
| -1,159332236 | C1orf173 | | Downregulated |
| 0,848736018 | C1orf180 | FALSE | Upregulated |
| -0,18463646 | C1orf186 | FALSE | Downregulated |
| | C1orf189 | | |
| 1,365533197 | C1orf194 | | Upregulated |
| 0,046708943 | C1orf227 | FALSE | Upregulated |
| -0,63897085 | C1orf233 | FALSE | Downregulated |
| 0,027563344 | C1orf234 | FALSE | Upregulated |
| 0 | C1orf61 | FALSE | Upregulated |
| | C1orf64 | | |
| 0,063262378 | C1orf65 | FALSE | Upregulated |
| 0 | C1orf68 | FALSE | Upregulated |
| | C1orf87 | | |
| | C1orf94 | | |
| 0,958231352 | C1QL1 | FALSE | Upregulated |
| -0,087792529 | C1QL2 | FALSE | Downregulated |
| | C1QL4 | | |
| -0,961964477 | C1QTNF2 | FALSE | Downregulated |
| 0,704424435 | C1QTNF4 | FALSE | Upregulated |
| 0 | C1QTNF8 | FALSE | Upregulated |
| 0,249715628 | C1QTNF9 | FALSE | Upregulated |
| 1,407737618 | C1QTNF9B-AS1 | | Upregulated |
| | C20orf141 | | |
| -0,216944163 | C20orf166 | FALSE | Downregulated |
| -0,540523238 | C20orf166-AS1 | FALSE | Downregulated |
| 0 | C20orf181 | FALSE | Upregulated |
| -0,756893054 | C20orf196 | FALSE | Downregulated |
| 0,012343797 | C20orf197 | FALSE | Upregulated |
| -0,508372692 | C20orf202 | FALSE | Downregulated |
| 0,955534004 | C20orf203 | FALSE | Upregulated |
| 0,653329933 | C20orf26 | FALSE | Upregulated |
| | C20orf57 | | |
| | C20orf78 | | |
| 0 | C20orf85 | FALSE | Upregulated |
| 0 | C21orf128 | FALSE | Upregulated |
| 0,115212105 | C21orf15 | FALSE | Upregulated |
| 0,542241926 | C21orf37 | FALSE | Upregulated |
| 0,010023953 | C21orf49 | FALSE | Upregulated |
| | C21orf54 | | |
| 2,074758043 | C21orf88 | | Upregulated |
| | C21orf91-OT1 | | |

| | | | |
|---|---|---|---|
| 0,861705729 | C22orf24 | FALSE | Upregulated |
| 0,177594914 | C22orf42 | FALSE | Upregulated |
| 1,80796468 | C22orf43 | | Upregulated |
| 0,581435022 | C2CD4C | FALSE | Upregulated |
| 1,223626141 | C2orf27A | | Upregulated |
| 0 | C2orf27B | FALSE | Upregulated |
| 1,116413744 | C2orf40 | | Upregulated |
| -0,051645049 | C2orf48 | FALSE | Downregulated |
| 0 | C2orf50 | FALSE | Upregulated |
| 0 | C2orf53 | FALSE | Upregulated |
| 0,857199672 | C2orf57 | FALSE | Upregulated |
| -0,052967525 | C2orf61 | FALSE | Downregulated |
| 0,587730794 | C2orf62 | FALSE | Upregulated |
| 0,984805419 | C2orf66 | FALSE | Upregulated |
| | C2orf70 | | |
| 0,96693199 | C2orf71 | FALSE | Upregulated |
| -0,488037606 | C2orf76 | FALSE | Downregulated |
| -0,072129931 | C2orf78 | FALSE | Downregulated |
| -0,00897377 | C2orf80 | FALSE | Downregulated |
| 0,005026269 | C2orf83 | FALSE | Upregulated |
| 1,169162886 | C2orf91 | | Upregulated |
| 0,26315926 | C3orf14 | FALSE | Upregulated |
| -0,699432488 | C3orf20 | FALSE | Downregulated |
| 0 | C3orf27 | FALSE | Upregulated |
| -0,809048381 | C3orf33 | FALSE | Downregulated |
| 0,186590836 | C3orf36 | FALSE | Upregulated |
| 0,087930286 | C3orf43 | FALSE | Upregulated |
| 1,101093764 | C3orf52 | | Upregulated |
| 0 | C3orf55 | FALSE | Upregulated |
| | C3orf56 | | |
| 0,141641885 | C3orf72 | FALSE | Upregulated |
| 0,59274454 | C3orf79 | FALSE | Upregulated |
| -1,744881421 | C3orf80 | | Downregulated |
| -0,49398773 | C4orf17 | FALSE | Downregulated |
| -0,179650428 | C4orf22 | FALSE | Downregulated |
| -0,000507374 | C4orf26 | FALSE | Downregulated |
| 0,257706725 | C4orf36 | FALSE | Upregulated |
| 0,630368758 | C4orf40 | FALSE | Upregulated |
| 1,837639342 | C4orf45 | | Upregulated |
| 0,467259232 | C4orf46 | FALSE | Upregulated |
| 0 | C4orf47 | FALSE | Upregulated |
| 0,154445106 | C4orf48 | FALSE | Upregulated |
| 0,290192445 | C4orf50 | FALSE | Upregulated |
| | C4orf51 | | |
| -0,874284029 | C5orf30 | FALSE | Downregulated |
| -0,029106533 | C5orf38 | FALSE | Downregulated |
| | C5orf46 | | |
| 0 | C5orf47 | FALSE | Upregulated |
| -0,051749038 | C5orf48 | FALSE | Downregulated |
| 0,837656437 | C5orf52 | FALSE | Upregulated |
| 0,566209924 | C5orf60 | FALSE | Upregulated |

| | | | |
|---|---|---|---|
| 1,838592957 | C5orf64 | | Upregulated |
| | C6orf100 | | |
| | C6orf118 | | |
| | C6orf15 | | |
| 0,726568321 | C6orf163 | FALSE | Upregulated |
| -0,283091759 | C6orf165 | FALSE | Downregulated |
| -0,071932489 | C6orf195 | FALSE | Downregulated |
| 0 | C6orf222 | FALSE | Upregulated |
| -0,556517444 | C6orf223 | FALSE | Downregulated |
| 0,904174903 | C6orf229 | FALSE | Upregulated |
| 0 | C6orf52 | FALSE | Upregulated |
| -1,104222144 | C6orf57 | | Downregulated |
| 0,665316988 | C6orf58 | FALSE | Upregulated |
| 0,958166295 | C6orf7 | FALSE | Upregulated |
| 0,768745371 | C6orf99 | FALSE | Upregulated |
| 0 | C7orf33 | FALSE | Upregulated |
| 1,216593452 | C7orf34 | | Upregulated |
| -0,122043091 | C7orf57 | FALSE | Downregulated |
| -0,10551374 | C7orf61 | FALSE | Downregulated |
| 0,477705478 | C7orf62 | FALSE | Upregulated |
| 0 | C7orf65 | FALSE | Upregulated |
| | C7orf66 | | |
| -0,099217573 | C7orf69 | FALSE | Downregulated |
| | C7orf72 | | |
| 0,554881589 | C7orf76 | FALSE | Upregulated |
| 0 | C8orf12 | FALSE | Upregulated |
| 0 | C8orf22 | FALSE | Upregulated |
| | C8orf34 | | |
| 0,625647963 | C8orf44-SGK3 | FALSE | Upregulated |
| 0,024803348 | C8orf48 | FALSE | Upregulated |
| 0,076658198 | C8orf69 | FALSE | Upregulated |
| -0,096724386 | C8orf74 | FALSE | Downregulated |
| 0,580082963 | C8orf86 | FALSE | Upregulated |
| | C8orf87 | | |
| 0,699188224 | C8orf88 | FALSE | Upregulated |
| 0,89246021 | C9orf106 | FALSE | Upregulated |
| 0 | C9orf135 | FALSE | Upregulated |
| | C9orf135-AS1 | | |
| -0,049155051 | C9orf153 | FALSE | Downregulated |
| 0,012449554 | C9orf170 | FALSE | Upregulated |
| -1,29894301 | C9orf24 | | Downregulated |
| -0,566217986 | C9orf43 | FALSE | Downregulated |
| -0,80463305 | C9orf47 | FALSE | Downregulated |
| 1,899592149 | C9orf50 | | Upregulated |
| 0 | C9orf53 | FALSE | Upregulated |
| -0,44144418 | C9orf57 | FALSE | Downregulated |
| | C9orf62 | | |
| | C9orf92 | | |
| 1,070538422 | CA10 | | Upregulated |
| 0,292194899 | CA12 | FALSE | Upregulated |
| 0,647721709 | CA4 | FALSE | Upregulated |

| | | | |
|---|---|---|---|
| | CA6 | | |
| 0,805327349 | CA7 | FALSE | Upregulated |
| 0,597017317 | CA8 | FALSE | Upregulated |
| 0,196419147 | CA9 | FALSE | Upregulated |
| -0,107369268 | CABP1 | FALSE | Downregulated |
| | CABP2 | | |
| | CABP5 | | |
| 0 | CABS1 | FALSE | Upregulated |
| -0,033818286 | CACNA1B | FALSE | Downregulated |
| 0 | CACNA1C-IT3 | FALSE | Upregulated |
| 1,196211073 | CACNA1E | | Upregulated |
| 0,360173078 | CACNA1F | FALSE | Upregulated |
| 0,586304306 | CACNA1G | FALSE | Upregulated |
| 1,231847437 | CACNA1I | | Upregulated |
| 0,340609445 | CACNA2D3 | FALSE | Upregulated |
| -0,000844217 | CACNA2D3-AS1 | FALSE | Downregulated |
| 1,450213528 | CACNB4 | | Upregulated |
| | CACNG2 | | |
| 0 | CACNG3 | FALSE | Upregulated |
| 0 | CACNG5 | FALSE | Upregulated |
| 0,034819348 | CACNG6 | FALSE | Upregulated |
| | CACNG7 | | |
| -0,351565836 | CACNG8 | FALSE | Downregulated |
| 0,292361091 | CADM2 | FALSE | Upregulated |
| 0 | CADM2-AS2 | FALSE | Upregulated |
| 1,876970176 | CADPS | | Upregulated |
| 0,793309831 | CAGE1 | FALSE | Upregulated |
| 0,015994382 | CALB2 | FALSE | Upregulated |
| 0 | CALCB | FALSE | Upregulated |
| 0 | CALCR | FALSE | Upregulated |
| 0 | CALHM3 | FALSE | Upregulated |
| 0,809488183 | CALML5 | FALSE | Upregulated |
| -0,034054071 | CALML6 | FALSE | Downregulated |
| 0,050688952 | CALR3 | FALSE | Upregulated |
| -0,011929323 | CALY | FALSE | Downregulated |
| 0,029393789 | CAMK1G | FALSE | Upregulated |
| -0,961901991 | CAMK2A | FALSE | Downregulated |
| 0 | CAMKV | FALSE | Upregulated |
| | CAMP | | |
| 0,003701477 | CAND1.11 | FALSE | Upregulated |
| 0,052830966 | CAPN11 | FALSE | Upregulated |
| 0,436722792 | CAPN13 | FALSE | Upregulated |
| 0,363931409 | CAPN14 | FALSE | Upregulated |
| 0,498085028 | CAPN6 | FALSE | Upregulated |
| 0,614975361 | CAPN8 | FALSE | Upregulated |
| -0,101938701 | CAPN8-2 | FALSE | Downregulated |
| 0,005551624 | CAPN9 | FALSE | Upregulated |
| 0 | CAPNS2 | FALSE | Upregulated |
| | CAPSL | | |
| 0 | CAPZA3 | FALSE | Upregulated |
| | CARD17 | | |

| | | | |
|---|---|---|---|
| | CARD18 | | |
| | CARTPT | | |
| | CASC11 | | |
| 0,673892142 | CASC15 | FALSE | Upregulated |
| 1,240070276 | CASC16 | | Upregulated |
| | CASC17 | | |
| 1,413847977 | CASC18 | | Upregulated |
| 0,945858897 | CASC2 | FALSE | Upregulated |
| 0,176787165 | CASC20 | FALSE | Upregulated |
| | CASC6 | | |
| -0,219756881 | CASC8 | FALSE | Downregulated |
| 0 | CASC9 | FALSE | Upregulated |
| 1,011551836 | CASP12 | | Upregulated |
| 0,49588129 | CASP14 | FALSE | Upregulated |
| 0 | CASP5 | FALSE | Upregulated |
| -0,702612084 | CASQ2 | FALSE | Downregulated |
| 0,703023151 | CASR | FALSE | Upregulated |
| 0,028082375 | CATSPER1 | FALSE | Upregulated |
| 0,572319312 | CATSPER2 | FALSE | Upregulated |
| -1,438078376 | CATSPER3 | | Downregulated |
| | CATSPER4 | | |
| 2,250674109 | CATSPERB | | Upregulated |
| | CATSPERD | | |
| 0,207892218 | CAV3 | FALSE | Upregulated |
| -0,011558913 | CBLN1 | FALSE | Downregulated |
| -0,181317841 | CBLN2 | FALSE | Downregulated |
| 1,615420302 | CBR3-AS1 | | Upregulated |
| 0,87193976 | CBX3P2 | FALSE | Upregulated |
| 1,037536513 | CCAT2 | | Upregulated |
| 0,00074495 | CCDC103 | FALSE | Upregulated |
| | CCDC105 | | |
| -0,36583505 | CCDC11 | FALSE | Downregulated |
| 0,588986961 | CCDC110 | FALSE | Upregulated |
| 2,194272586 | CCDC112 | | Upregulated |
| 0,010860768 | CCDC114 | FALSE | Upregulated |
| 0,110367471 | CCDC116 | FALSE | Upregulated |
| 0,415306423 | CCDC129 | FALSE | Upregulated |
| -0,801554641 | CCDC13 | FALSE | Downregulated |
| -0,239140184 | CCDC13-AS1 | FALSE | Downregulated |
| -0,143635721 | CCDC135 | FALSE | Downregulated |
| 0,230249082 | CCDC136 | FALSE | Upregulated |
| -0,412827226 | CCDC138 | FALSE | Downregulated |
| | CCDC140 | | |
| 0,465705151 | CCDC144CP | FALSE | Upregulated |
| 0 | CCDC144NL | FALSE | Upregulated |
| 0,612605564 | CCDC147 | FALSE | Upregulated |
| 0,217893484 | CCDC147-AS1 | FALSE | Upregulated |
| -0,321806469 | CCDC148 | FALSE | Downregulated |
| 1,180954481 | CCDC148-AS1 | | Upregulated |
| 0,728610997 | CCDC151 | FALSE | Upregulated |
| 0,131580646 | CCDC153 | FALSE | Upregulated |

| | | | |
|---|---|---|---|
| 0,220292424 | CCDC154 | FALSE | Upregulated |
| 0,076959828 | CCDC155 | FALSE | Upregulated |
| 0,667862228 | CCDC160 | FALSE | Upregulated |
| -0,088626364 | CCDC162P | FALSE | Downregulated |
| -0,092650542 | CCDC163P | FALSE | Downregulated |
| 0 | CCDC166 | FALSE | Upregulated |
| | CCDC169 | | |
| | CCDC169-SOHLH2 | | |
| | CCDC172 | | |
| -0,107503178 | CCDC175 | FALSE | Downregulated |
| 0,180776777 | CCDC177 | FALSE | Upregulated |
| 0,039005732 | CCDC178 | FALSE | Upregulated |
| -0,83717299 | CCDC179 | FALSE | Downregulated |
| 0 | CCDC182 | FALSE | Upregulated |
| 0 | CCDC19 | FALSE | Upregulated |
| -0,370840636 | CCDC27 | FALSE | Downregulated |
| 0,100310787 | CCDC38 | FALSE | Upregulated |
| 0,981692785 | CCDC41-AS1 | FALSE | Upregulated |
| 0,19423022 | CCDC42 | FALSE | Upregulated |
| -0,836148379 | CCDC58 | FALSE | Downregulated |
| -0,072774583 | CCDC62 | FALSE | Downregulated |
| 0 | CCDC63 | FALSE | Upregulated |
| 2,322798507 | CCDC74A | | Upregulated |
| 0 | CCDC74B | FALSE | Upregulated |
| 0,039005732 | CCDC79 | FALSE | Upregulated |
| 0,655587539 | CCDC81 | FALSE | Upregulated |
| 0,528804664 | CCDC89 | FALSE | Upregulated |
| 1,029590313 | CCDC96 | | Upregulated |
| | CCER1 | | |
| 0,050688952 | CCIN | FALSE | Upregulated |
| 0,637011154 | CCK | FALSE | Upregulated |
| | CCKAR | | |
| 0,50298882 | CCKBR | FALSE | Upregulated |
| | CCL1 | | |
| 0,607016054 | CCL11 | FALSE | Upregulated |
| 0,099396057 | CCL13 | FALSE | Upregulated |
| -0,633112024 | CCL17 | FALSE | Downregulated |
| 0,807315303 | CCL18 | FALSE | Upregulated |
| 0 | CCL20 | FALSE | Upregulated |
| 0 | CCL24 | FALSE | Upregulated |
| 0 | CCL25 | FALSE | Upregulated |
| | CCL26 | | |
| 0,429269333 | CCL27 | FALSE | Upregulated |
| 1,33239485 | CCL28 | | Upregulated |
| 0,746162899 | CCL4 | FALSE | Upregulated |
| 0,667775146 | CCL4L1 | FALSE | Upregulated |
| | CCL7 | | |
| 1,116763397 | CCL8 | | Upregulated |
| 0,132478011 | CCNA1 | FALSE | Upregulated |
| 0,143295065 | CCNB1 | FALSE | Upregulated |
| 1,208168598 | CCNB3 | | Upregulated |

| | | | |
|---|---|---|---|
| 0,566699194 | CCNE1 | FALSE | Upregulated |
| 0,791799536 | CCNJL | FALSE | Upregulated |
| 0 | CCNO | FALSE | Upregulated |
| | CCNYL2 | | |
| 1,885612995 | CCR3 | | Upregulated |
| -0,129235179 | CCR4 | FALSE | Downregulated |
| 1,496154387 | CCR6 | | Upregulated |
| 1,030636081 | CCRL2 | | Upregulated |
| -1,261366573 | CCT6B | | Downregulated |
| 0 | CCT8L2 | FALSE | Upregulated |
| 0,019985931 | CD164L2 | FALSE | Upregulated |
| | CD177 | | |
| 1,463449334 | CD19 | | Upregulated |
| 0 | CD1A | FALSE | Upregulated |
| 0 | CD1B | FALSE | Upregulated |
| 0,645694261 | CD1C | FALSE | Upregulated |
| -0,024263679 | CD1E | FALSE | Downregulated |
| 0 | CD200R1L | FALSE | Upregulated |
| 1,336267987 | CD207 | | Upregulated |
| | CD300LD | | |
| 2,334412272 | CD300LG | | Upregulated |
| -0,045014015 | CD3D | FALSE | Downregulated |
| 1,179785558 | CD69 | | Upregulated |
| 0 | CD70 | FALSE | Upregulated |
| | CD99-2 | | |
| | CD99P1-2 | | |
| 0 | CDC14C | FALSE | Upregulated |
| 0,145922586 | CDC20B | FALSE | Upregulated |
| 0,531475854 | CDC25A | FALSE | Upregulated |
| 1,052213563 | CDC25C | | Upregulated |
| 0 | CDC45 | FALSE | Upregulated |
| 0,014129285 | CDC6 | FALSE | Upregulated |
| 0,09792113 | CDCA2 | FALSE | Upregulated |
| 0,410089394 | CDCA3 | FALSE | Upregulated |
| 0 | CDCA7 | FALSE | Upregulated |
| 0,396638128 | CDCA8 | FALSE | Upregulated |
| 0 | CDCP2 | FALSE | Upregulated |
| | CDH10 | | |
| -0,100944603 | CDH12 | FALSE | Downregulated |
| -0,074879495 | CDH16 | FALSE | Downregulated |
| 0,589774564 | CDH17 | FALSE | Upregulated |
| | CDH18 | | |
| 1,114359023 | CDH22 | | Upregulated |
| -0,180093623 | CDH26 | FALSE | Downregulated |
| -0,618000743 | CDH3 | FALSE | Downregulated |
| | CDH7 | | |
| 0 | CDH8 | FALSE | Upregulated |
| 0 | CDH9 | FALSE | Upregulated |
| 0,742502815 | CDHR4 | FALSE | Upregulated |
| | CDIPT-AS1 | | |
| 0,525975894 | CDK1 | FALSE | Upregulated |

| | | | |
|---|---|---|---|
| 0 | CDK15 | FALSE | Upregulated |
| | CDK5R2 | | |
| 0 | CDKL2 | FALSE | Upregulated |
| 0,193752442 | CDKL3 | FALSE | Upregulated |
| 0,000577308 | CDKL4 | FALSE | Upregulated |
| 1,265476735 | CDKN2A | | Upregulated |
| 1,782705797 | CDKN2B-AS1 | | Upregulated |
| -0,341701042 | CDKN2D | FALSE | Downregulated |
| | CDRT15L2 | | |
| | CDRT15P2 | | |
| | CDRT7 | | |
| | CDRT8 | | |
| -1,67677039 | CDX1 | | Downregulated |
| | CDX4 | | |
| | CEACAM18 | | |
| 0,038794166 | CEACAM20 | FALSE | Upregulated |
| 1,169166814 | CEACAM3 | | Upregulated |
| -0,005336337 | CEACAM4 | FALSE | Downregulated |
| 0,616631163 | CEACAM5 | FALSE | Upregulated |
| 2,497637143 | CEACAM6 | | Upregulated |
| | CEACAM7 | | |
| | CEBPE | | |
| -0,135365639 | CECR3 | FALSE | Downregulated |
| 0,443491641 | CECR7 | FALSE | Upregulated |
| -1,132251617 | CEL | | Downregulated |
| | CELA1 | | |
| -0,080743019 | CELA2A | FALSE | Downregulated |
| 0 | CELA2B | FALSE | Upregulated |
| | CELA3A | | |
| | CELA3B | | |
| -1,201046872 | CELF5 | | Downregulated |
| 0 | CELP | FALSE | Upregulated |
| 0,453035468 | CENPA | FALSE | Upregulated |
| 0,626687785 | CENPE | FALSE | Upregulated |
| -0,275657481 | CENPH | FALSE | Downregulated |
| 1,303204078 | CENPI | | Upregulated |
| 0,919560825 | CENPK | FALSE | Upregulated |
| -0,216765877 | CENPL | FALSE | Downregulated |
| 0,342169152 | CENPM | FALSE | Upregulated |
| 1,043800451 | CENPQ | | Upregulated |
| | CENPVP1 | | |
| | CENPVP2 | | |
| 0,272754063 | CENPW | FALSE | Upregulated |
| 0,029410567 | CEP170P1 | FALSE | Upregulated |
| 0,412057488 | CEP55 | FALSE | Upregulated |
| | CER1 | | |
| 0 | CES1P2 | FALSE | Upregulated |
| | CES5AP1 | | |
| | CETN1 | | |
| -0,7815498 | CETN3 | FALSE | Downregulated |
| 0 | CETN4P | FALSE | Upregulated |

| | | | |
|---|---|---|---|
| | CFC1 | | |
| | CFC1B | | |
| 0 | CFL1P1 | FALSE | Upregulated |
| | CGA | | |
| 0,019184167 | CGB | FALSE | Upregulated |
| | CGB1 | | |
| 0 | CGB2 | FALSE | Upregulated |
| 0,111070327 | CGB5 | FALSE | Upregulated |
| | CGB8 | | |
| 0,51540738 | CH25H | FALSE | Upregulated |
| -0,015677605 | CHAC1 | FALSE | Downregulated |
| 0,711886838 | CHAF1B | FALSE | Upregulated |
| | CHAT | | |
| -0,030315846 | CHD5 | FALSE | Downregulated |
| -0,013862029 | CHDC2 | FALSE | Downregulated |
| | CHEK2P2 | | |
| 0 | CHGB | FALSE | Upregulated |
| 1,490314345 | CHI3L2 | | Upregulated |
| 0 | CHIA | FALSE | Upregulated |
| 0 | CHIAP2 | FALSE | Upregulated |
| 1,008462906 | CHIT1 | | Upregulated |
| -0,247064724 | CHKB-AS1 | FALSE | Downregulated |
| -0,940877022 | CHKB-CPT1B | FALSE | Downregulated |
| 0,759402487 | CHMP1B2P | FALSE | Upregulated |
| -0,046578688 | CHN1 | FALSE | Downregulated |
| 0 | CHODL | FALSE | Upregulated |
| | CHODL-AS1 | | |
| -0,038454387 | CHP2 | FALSE | Downregulated |
| 1,83650865 | CHRDL1 | | Upregulated |
| 2,681706331 | CHRDL2 | | Upregulated |
| 1,750768882 | CHRFAM7A | | Upregulated |
| -0,09793047 | CHRM1 | FALSE | Downregulated |
| 0,712071348 | CHRM2 | FALSE | Upregulated |
| -0,298161158 | CHRM3-AS1 | FALSE | Downregulated |
| 1,787895594 | CHRM3-AS2 | | Upregulated |
| 0,068447037 | CHRM4 | FALSE | Upregulated |
| 1,190954457 | CHRNA1 | | Upregulated |
| 0,557751485 | CHRNA10 | FALSE | Upregulated |
| 0,509166511 | CHRNA3 | FALSE | Upregulated |
| 0,44238191 | CHRNA5 | FALSE | Upregulated |
| -0,189001521 | CHRNA6 | FALSE | Downregulated |
| 0,359213221 | CHRNA7 | FALSE | Upregulated |
| 0,731194966 | CHRNA9 | FALSE | Upregulated |
| | CHRNB3 | | |
| 0 | CHRNB4 | FALSE | Upregulated |
| 0,492935587 | CHST10 | FALSE | Upregulated |
| -0,255266281 | CHST5 | FALSE | Downregulated |
| 0,294590979 | CHST6 | FALSE | Upregulated |
| -0,022202627 | CHST8 | FALSE | Downregulated |
| 0,220401516 | CIB2 | FALSE | Upregulated |
| | CIB3 | | |

| | | | |
|---|---|---|---|
| 0 | CIB4 | FALSE | Upregulated |
| 0,004287099 | CIDEA | FALSE | Upregulated |
| 1,150253247 | CIDEC | | Upregulated |
| 1,584771496 | CIDECP | | Upregulated |
| 0,357727074 | CILP2 | FALSE | Upregulated |
| | CISTR | | |
| 0,068157417 | CITED1 | FALSE | Upregulated |
| 0,972402774 | CKAP2L | FALSE | Upregulated |
| 1,049641057 | CKLF-CMTM1 | | Upregulated |
| -0,002201933 | CKMT1A | FALSE | Downregulated |
| -0,281202289 | CKMT1B | FALSE | Downregulated |
| 0,184905969 | CKS2 | FALSE | Upregulated |
| 0,019241929 | CLC | FALSE | Upregulated |
| 0,007217966 | CLCA1 | FALSE | Upregulated |
| 0 | CLCA2 | FALSE | Upregulated |
| -0,281122554 | CLCA4 | FALSE | Downregulated |
| 0,189811059 | CLCN1 | FALSE | Upregulated |
| 0,169287231 | CLCNKA | FALSE | Upregulated |
| 0 | CLCNKB | FALSE | Upregulated |
| | CLDN10-AS1 | | |
| -0,104078958 | CLDN16 | FALSE | Downregulated |
| | CLDN17 | | |
| 0,479673037 | CLDN19 | FALSE | Upregulated |
| -0,035556561 | CLDN25 | FALSE | Downregulated |
| 1,537918373 | CLDN6 | | Upregulated |
| | CLDN8 | | |
| -0,082393251 | CLEC12B | FALSE | Downregulated |
| 0,721283626 | CLEC17A | FALSE | Upregulated |
| 0,348714889 | CLEC18B | FALSE | Upregulated |
| | CLEC19A | | |
| 0,001375889 | CLEC1A | FALSE | Upregulated |
| 0 | CLEC2A | FALSE | Upregulated |
| | CLEC2L | | |
| 0 | CLEC3A | FALSE | Upregulated |
| 1,175429911 | CLEC4A | | Upregulated |
| 1,125315974 | CLEC4C | | Upregulated |
| 0,721821071 | CLEC4D | FALSE | Upregulated |
| -0,019051137 | CLEC4E | FALSE | Downregulated |
| 0,542002973 | CLEC4F | FALSE | Upregulated |
| -1,068533793 | CLEC4GP1 | | Downregulated |
| 0,579790272 | CLEC6A | FALSE | Upregulated |
| 0,352154985 | CLEC9A | FALSE | Upregulated |
| 1,066427415 | CLECL1 | | Upregulated |
| -0,145021646 | CLGN | FALSE | Downregulated |
| -0,461626976 | CLIC3 | FALSE | Downregulated |
| | CLK2P | | |
| 0,931807582 | CLLU1 | FALSE | Upregulated |
| | CLLU1OS | | |
| -0,794135623 | CLNK | FALSE | Downregulated |
| | CLPS | | |
| | CLPSL1 | | |

| | | | |
|---|---|---|---|
| | CLPSL2 | | |
| -0,256004109 | CLRN1-AS1 | FALSE | Downregulated |
| 0,170287303 | CLRN2 | FALSE | Upregulated |
| 1,642373941 | CLSPN | | Upregulated |
| -0,76736314 | CLUL1 | FALSE | Downregulated |
| 1,209593174 | CLVS2 | | Upregulated |
| 0,738437896 | CMA1 | FALSE | Upregulated |
| 0 | CMTM5 | FALSE | Upregulated |
| | CNBD1 | | |
| 0,243171474 | CNBD2 | FALSE | Upregulated |
| 0 | CNFN | FALSE | Upregulated |
| | CNGA2 | | |
| | CNGA3 | | |
| 0,099706539 | CNGA4 | FALSE | Upregulated |
| | CNGB1 | | |
| -0,032751007 | CNIH3 | FALSE | Downregulated |
| 0,441881935 | CNN1 | FALSE | Upregulated |
| 0 | CNPY1 | FALSE | Upregulated |
| 1,782697708 | CNR1 | | Upregulated |
| 1,095314604 | CNR2 | | Upregulated |
| -0,009736784 | CNTD2 | FALSE | Downregulated |
| 0,141702167 | CNTF | FALSE | Upregulated |
| -1,461140573 | CNTN1 | | Downregulated |
| 0 | CNTN4-AS2 | FALSE | Upregulated |
| 0 | CNTN5 | FALSE | Upregulated |
| -0,187983533 | CNTN6 | FALSE | Downregulated |
| | CNTNAP5 | | |
| 1,250479469 | COL11A1 | | Upregulated |
| -0,33420963 | COL13A1 | FALSE | Downregulated |
| -0,409948275 | COL20A1 | FALSE | Downregulated |
| 1,909359611 | COL22A1 | | Upregulated |
| -0,185286553 | COL24A1 | FALSE | Downregulated |
| 0 | COL2A1 | FALSE | Upregulated |
| 0,377981572 | COL4A2-AS1 | FALSE | Upregulated |
| | COL6A4P1 | | |
| 1,218763799 | COL6A4P2 | | Upregulated |
| 0 | COL6A5 | FALSE | Upregulated |
| 0 | COL9A1 | FALSE | Upregulated |
| 0,584511496 | COLCA2 | FALSE | Upregulated |
| 2,254729093 | COLEC12 | | Upregulated |
| -0,974065358 | COMMD8 | FALSE | Downregulated |
| 1,358047727 | COMP | | Upregulated |
| 0,002844687 | COQ3 | FALSE | Upregulated |
| 0,965301338 | COX4I2 | FALSE | Upregulated |
| -0,961537536 | COX6A2 | FALSE | Downregulated |
| | COX7B2 | | |
| 0,076658198 | COX8C | FALSE | Upregulated |
| 0,988036419 | CPA1 | FALSE | Upregulated |
| | CPA2 | | |
| 0,785791973 | CPA3 | FALSE | Upregulated |
| 0,301973568 | CPA4 | FALSE | Upregulated |

| | | | |
|---|---|---|---|
| -0,121408961 | CPA5 | FALSE | Downregulated |
| | CPA6 | | |
| 0,018395979 | CPB1 | FALSE | Upregulated |
| -0,282749668 | CPEB2-AS1 | FALSE | Downregulated |
| 0 | CPLX3 | FALSE | Upregulated |
| | CPLX4 | | |
| -0,683149943 | CPNE4 | FALSE | Downregulated |
| -0,357441205 | CPNE7 | FALSE | Downregulated |
| 0,224248983 | CPNE9 | FALSE | Upregulated |
| 0 | CPO | FALSE | Upregulated |
| 0,004618298 | CPSF4L | FALSE | Upregulated |
| -0,870057646 | CPT1C | FALSE | Downregulated |
| | CPXCR1 | | |
| 1,354789336 | CPXM1 | | Upregulated |
| 1,122319873 | CR1L | | Upregulated |
| 0,361991637 | CR2 | FALSE | Upregulated |
| | CRABP1 | | |
| 0,331270531 | CRABP2 | FALSE | Upregulated |
| 0 | CRB1 | FALSE | Upregulated |
| | CRCT1 | | |
| 0,131633777 | CREG2 | FALSE | Upregulated |
| | CRHR1 | | |
| 0,518160927 | CRISP1 | FALSE | Upregulated |
| 0,369953466 | CRISP2 | FALSE | Upregulated |
| 0,861127216 | CRISP3 | FALSE | Upregulated |
| -0,766940105 | CRISPLD1 | FALSE | Downregulated |
| | CRLF2 | | |
| | CRLF2-2 | | |
| 0,502986666 | CRNDE | FALSE | Upregulated |
| 0,147409039 | CRNN | FALSE | Upregulated |
| 0 | CRSP8P | FALSE | Upregulated |
| -0,484979272 | CRTAC1 | FALSE | Downregulated |
| -0,287405738 | CRTAM | FALSE | Downregulated |
| | CRYBA2 | | |
| | CRYBA4 | | |
| 0,266796156 | CRYBB1 | FALSE | Upregulated |
| -0,34695377 | CRYBB2 | FALSE | Downregulated |
| 0,201953505 | CRYBB3 | FALSE | Upregulated |
| | CRYGA | | |
| | CRYGB | | |
| | CRYGC | | |
| | CRYGD | | |
| 0,048656215 | CRYGN | FALSE | Upregulated |
| 0,470723624 | CRYGS | FALSE | Upregulated |
| -0,011727444 | CRYM-AS1 | FALSE | Downregulated |
| | CSAG1 | | |
| | CSAG3 | | |
| | CSAG4 | | |
| | CSF2RA-2 | | |
| | CSH1 | | |
| | CSH2 | | |

| | | | |
|---|---|---|---|
| | CSHL1 | | |
| 1,11489921 | CSMD1 | | Upregulated |
| 0,991085411 | CSMD2 | FALSE | Upregulated |
| -0,545613817 | CSMD3 | FALSE | Downregulated |
| | CSN1S1 | | |
| | CSN1S2AP | | |
| | CSN1S2BP | | |
| | CSN2 | | |
| | CSN3 | | |
| 0,167939653 | CSNK1A1L | FALSE | Upregulated |
| 0,84236931 | CSNK2A3 | FALSE | Upregulated |
| -0,065076736 | CSPG5 | FALSE | Downregulated |
| -0,915322359 | CSRP3 | FALSE | Downregulated |
| | CST1 | | |
| 0,705562129 | CST11 | FALSE | Upregulated |
| | CST13P | | |
| | CST2 | | |
| | CST4 | | |
| | CST5 | | |
| -0,023202137 | CST6 | FALSE | Downregulated |
| | CST8 | | |
| | CST9 | | |
| | CST9L | | |
| | CSTF3-AS1 | | |
| | CSTL1 | | |
| | CT45A1 | | |
| | CT45A2 | | |
| | CT45A3 | | |
| | CT45A5 | | |
| | CT45A6 | | |
| | CT47A1 | | |
| | CT47A10 | | |
| | CT47A11 | | |
| | CT47A12 | | |
| | CT47A2 | | |
| | CT47A3 | | |
| | CT47A4 | | |
| | CT47A5 | | |
| 0 | CT47A6 | FALSE | Upregulated |
| 0 | CT47A7 | FALSE | Upregulated |
| | CT47A8 | | |
| | CT47A9 | | |
| -0,057823576 | CT47B1 | FALSE | Downregulated |
| -0,138126573 | CT55 | FALSE | Downregulated |
| -0,470585932 | CT62 | FALSE | Downregulated |
| | CT83 | | |
| | CTAG1A | | |
| | CTAG1B | | |
| 0,608246941 | CTAG2 | FALSE | Upregulated |
| -0,609535589 | CTAGE1 | FALSE | Downregulated |
| -0,06287783 | CTAGE10P | FALSE | Downregulated |

| | | | |
|---|---|---|---|
| -0,140180739 | CTAGE11P | FALSE | Downregulated |
| -0,79142683 | CTAGE6 | FALSE | Downregulated |
| | CTB-12O2.1 | | |
| 2,557778622 | CTB-174D11.1 | | Upregulated |
| -0,292226016 | CTB-178M22.2 | FALSE | Downregulated |
| 0,825561522 | CTC-436P18.1 | FALSE | Upregulated |
| 0,054183774 | CTCFL | FALSE | Upregulated |
| | CTD-2118P12.1 | | |
| 0 | CTD-2127H9.1 | FALSE | Upregulated |
| | CTD-2194D22.4 | | |
| -0,830506842 | CTD-2258A20.5 | FALSE | Downregulated |
| -0,163472826 | CTD-2270F17.1 | FALSE | Downregulated |
| | CTD-2297D10.2 | | |
| | CTD-2337A12.1 | | |
| 0,497080574 | CTD-2350J17.1 | FALSE | Upregulated |
| -1,232186565 | CTF1 | | Downregulated |
| 0,736161667 | CTLA4 | FALSE | Upregulated |
| 0,924834071 | CTNNA2 | FALSE | Upregulated |
| 0,532440074 | CTRB1 | FALSE | Upregulated |
| | CTRB2 | | |
| -0,055970583 | CTRC | FALSE | Downregulated |
| 0,098625748 | CTSE | FALSE | Upregulated |
| | CTSL3P | | |
| | CTSLP8 | | |
| | CTSV | | |
| 0,612371553 | CTXN1 | FALSE | Upregulated |
| 0,161914213 | CTXN2 | FALSE | Upregulated |
| | CTXN3 | | |
| 0,708689618 | CUZD1 | FALSE | Upregulated |
| -0,693133536 | CWH43 | FALSE | Downregulated |
| 0,561266566 | CXADRP3 | FALSE | Upregulated |
| 1,231177875 | CXCL1 | | Upregulated |
| 2,102205335 | CXCL11 | | Upregulated |
| 0,769887499 | CXCL17 | FALSE | Upregulated |
| 0,052590986 | CXCL3 | FALSE | Upregulated |
| 0 | CXCL5 | FALSE | Upregulated |
| 2,830915048 | CXCL6 | | Upregulated |
| 0,048556976 | CXCR1 | FALSE | Upregulated |
| -1,750063522 | CXCR2 | | Downregulated |
| 2,158809707 | CXorf21 | | Upregulated |
| 0,027887956 | CXorf22 | FALSE | Upregulated |
| | CXorf27 | | |
| | CXorf28 | | |
| | CXorf30 | | |
| -0,361005277 | CXorf31 | FALSE | Downregulated |
| | CXorf49 | | |
| | CXorf49B | | |
| | CXorf51A | | |
| | CXorf51B | | |
| 0,150227037 | CXorf58 | FALSE | Upregulated |
| 0,004929336 | CXorf64 | FALSE | Upregulated |

| | | | |
|---|---|---|---|
| 0,089040084 | CXorf65 | FALSE | Upregulated |
| -0,099173502 | CXorf66 | FALSE | Downregulated |
| 0 | CXorf67 | FALSE | Upregulated |
| 1,166107846 | CXXC11 | | Upregulated |
| | CXXC1P1 | | |
| | CYLC1 | | |
| | CYLC2 | | |
| | CYMP | | |
| | CYorf17 | | |
| 2,73571812 | CYP11B1 | | Upregulated |
| 0 | CYP11B2 | FALSE | Upregulated |
| 0 | CYP19A1 | FALSE | Upregulated |
| 0,459236425 | CYP1B1-AS1 | FALSE | Upregulated |
| 0,010990595 | CYP24A1 | FALSE | Upregulated |
| -0,115454141 | CYP26C1 | FALSE | Downregulated |
| 0,184914411 | CYP27C1 | FALSE | Upregulated |
| -0,848393183 | CYP2G1P | FALSE | Downregulated |
| -1,776021381 | CYP46A1 | | Downregulated |
| 0 | CYP4B1 | FALSE | Upregulated |
| 0,159565689 | CYP4F24P | FALSE | Upregulated |
| | CYP4F30P | | |
| 0,74316013 | CYP4F35P | FALSE | Upregulated |
| | CYP4F62P | | |
| | CYP4F8 | | |
| 0,172294172 | CYP4Z1 | FALSE | Upregulated |
| 0,064020781 | CYP4Z2P | FALSE | Upregulated |
| 1,546666745 | CYSLTR1 | | Upregulated |
| 1,787478911 | CYSLTR2 | | Upregulated |
| 0 | CYTL1 | FALSE | Upregulated |
| | D21S2088E | | |
| 0,041547119 | DACH2 | FALSE | Upregulated |
| 1,562507673 | DACT3-AS1 | | Upregulated |
| -0,081552495 | DAND5 | FALSE | Downregulated |
| | DAOA | | |
| | DAOA-AS1 | | |
| | DAPL1 | | |
| 0,87700141 | DARC | FALSE | Upregulated |
| | DAW1 | | |
| -0,014886722 | DAZ1 | FALSE | Downregulated |
| -0,041888993 | DAZ4 | FALSE | Downregulated |
| | DAZL | | |
| -0,010660197 | DBIL5P2 | FALSE | Downregulated |
| -0,705366443 | DBX1 | FALSE | Downregulated |
| 0 | DBX2 | FALSE | Upregulated |
| | DCAF12L1 | | |
| -0,648384131 | DCAF12L2 | FALSE | Downregulated |
| 0,390828834 | DCAF13P3 | FALSE | Upregulated |
| 0,048656215 | DCAF4L2 | FALSE | Upregulated |
| | DCAF8L1 | | |
| | DCAF8L2 | | |
| 1,760665614 | DCC | | Upregulated |

| | | | | |
|---|---|---|---|---|
| 1,578183512 | DCD | Upregulated | | |
| | DCDC1 | | | |
| -0,289516058 | | DCDC2C | FALSE | Downregulated |
| -0,074012693 | | DCLK3 | FALSE | Downregulated |
| 0,008164751 | DCT | FALSE | Upregulated | |
| 0,642633641 | DCTN1-AS1 | FALSE | Upregulated | |
| 0 | DCX | FALSE | Upregulated | |
| 0,701708183 | DDIT4L | FALSE | Upregulated | |
| 0,041631275 | DDN | FALSE | Upregulated | |
| 0,199319128 | DDX11-AS1 | FALSE | Upregulated | |
| -0,440738902 | | DDX11L10 | FALSE | Downregulated |
| -0,343581399 | | DDX25 | FALSE | Downregulated |
| | DDX4 | | | |
| 0,000577308 | DEC1 | FALSE | Upregulated | |
| -0,352853278 | | DEFA1 | FALSE | Downregulated |
| | DEFA10P | | | |
| | DEFA11P | | | |
| -0,352853278 | | DEFA1B | FALSE | Downregulated |
| -0,201970585 | | DEFA3 | FALSE | Downregulated |
| 0 | DEFA4 | FALSE | Upregulated | |
| | DEFA5 | | | |
| | DEFA6 | | | |
| | DEFA8P | | | |
| | DEFA9P | | | |
| | DEFB103A | | | |
| | DEFB103B | | | |
| | DEFB104A | | | |
| | DEFB104B | | | |
| | DEFB105A | | | |
| | DEFB105B | | | |
| | DEFB106A | | | |
| | DEFB106B | | | |
| | DEFB107A | | | |
| | DEFB107B | | | |
| 0,041547119 | DEFB108B | FALSE | Upregulated | |
| 0 | DEFB109P1 | FALSE | Upregulated | |
| 0,942670697 | DEFB109P1B | FALSE | Upregulated | |
| | DEFB110 | | | |
| | DEFB112 | | | |
| | DEFB113 | | | |
| | DEFB114 | | | |
| | DEFB115 | | | |
| | DEFB116 | | | |
| | DEFB118 | | | |
| | DEFB119 | | | |
| | DEFB121 | | | |
| | DEFB122 | | | |
| | DEFB123 | | | |
| | DEFB124 | | | |
| | DEFB125 | | | |
| | DEFB126 | | | |

| | | | |
|---|---|---|---|
| | DEFB127 | | |
| | DEFB128 | | |
| | DEFB129 | | |
| | DEFB130 | | |
| | DEFB131 | | |
| 0,045026042 | DEFB132 | FALSE | Upregulated |
| | DEFB133 | | |
| | DEFB134 | | |
| | DEFB135 | | |
| | DEFB136 | | |
| | DEFB4A | | |
| | DEFB4B | | |
| | DEFT1P | | |
| | DEFT1P2 | | |
| 0,042470539 | DEPDC1 | FALSE | Upregulated |
| 0,157235154 | DEPDC1B | FALSE | Upregulated |
| -1,150627412 | DGCR6 | | Downregulated |
| 0,618068279 | DGKI | FALSE | Upregulated |
| | DGKK | | |
| -0,642358351 | DGUOK-AS1 | FALSE | Downregulated |
| 0,086558559 | DHDH | FALSE | Upregulated |
| 0,42560591 | DHH | FALSE | Upregulated |
| 0 | DHRS7C | FALSE | Upregulated |
| | DHRSX-2 | | |
| 0,588657992 | DIAPH3 | FALSE | Upregulated |
| 0 | DIAPH3-AS1 | FALSE | Upregulated |
| | DIAPH3-AS2 | | |
| 0,680525484 | DIO2 | FALSE | Upregulated |
| 0,839204995 | DIO2-AS1 | FALSE | Upregulated |
| 1,58449064 | DIO3 | | Upregulated |
| -0,341712613 | DIO3OS | FALSE | Downregulated |
| 0 | DIP2A-IT1 | FALSE | Upregulated |
| 0,639904603 | DIRAS1 | FALSE | Upregulated |
| -0,129313035 | DIRAS2 | FALSE | Downregulated |
| 0 | DIRC1 | FALSE | Upregulated |
| 1,543859682 | DIRC3 | | Upregulated |
| 0,26851989 | DISC1FP1 | FALSE | Upregulated |
| 0 | DKFZP434A062 | FALSE | Upregulated |
| -0,721124811 | DKFZP434F142 | FALSE | Downregulated |
| -0,277033428 | DKFZp434J0226 | FALSE | Downregulated |
| | DKFZP434L187 | | |
| | DKFZp434L192 | | |
| 0,450402505 | DKFZp451B082 | FALSE | Upregulated |
| 0,021391672 | DKFZp566F0947 | FALSE | Upregulated |
| | DKFZp686K1684 | | |
| 0,315076281 | DKFZp686L13185 | FALSE | Upregulated |
| 0,671913102 | DKFZp779M0652 | FALSE | Upregulated |
| -0,15811379 | DKK1 | FALSE | Downregulated |
| 0,425797669 | DKK2 | FALSE | Upregulated |
| 0,800854028 | DKK4 | FALSE | Upregulated |
| 1,857636199 | DKKL1 | | Upregulated |

| | | | |
|---|---|---|---|
| 0,00108448 | DLEU7-AS1 | FALSE | Upregulated |
| 1,686464104 | DLG3-AS1 | | Upregulated |
| 1,756578716 | DLGAP1 | | Upregulated |
| 1,131142502 | DLGAP1-AS3 | | Upregulated |
| | DLGAP1-AS4 | | |
| 0,858824927 | DLGAP3 | FALSE | Upregulated |
| 0 | DLGAP5 | FALSE | Upregulated |
| 0,164739241 | DLK1 | FALSE | Upregulated |
| | DLL3 | | |
| -0,15536325 | DLX1 | FALSE | Downregulated |
| | DLX2 | | |
| -0,137641416 | DLX3 | FALSE | Downregulated |
| 0 | DLX4 | FALSE | Upregulated |
| | DLX5 | | |
| | DLX6 | | |
| | DLX6-AS1 | | |
| 1,307176469 | DMBT1 | | Upregulated |
| | DMBT1P1 | | |
| | DMBX1 | | |
| 0,753369831 | DMC1 | FALSE | Upregulated |
| 0,273752446 | DMP1 | FALSE | Upregulated |
| | DMRT1 | | |
| 0,029950463 | DMRT2 | FALSE | Upregulated |
| | DMRT3 | | |
| | DMRTA2 | | |
| | DMRTB1 | | |
| | DMRTC1 | | |
| | DMRTC1B | | |
| | DMRTC2 | | |
| 0,498228063 | DNAAF1 | FALSE | Upregulated |
| 0,499871343 | DNAAF3 | FALSE | Upregulated |
| -1,167515236 | DNAH12 | | Downregulated |
| 0 | DNAH17-AS1 | FALSE | Upregulated |
| 0,590135242 | DNAH8 | FALSE | Upregulated |
| -1,496961194 | DNAI1 | | Downregulated |
| 1,311224866 | DNAI2 | | Upregulated |
| | DNAJA1P5 | | |
| 0,298001141 | DNAJB8 | FALSE | Upregulated |
| | DNAJB8-AS1 | | |
| 0,671322133 | DNAJC27-AS1 | FALSE | Upregulated |
| 0,056528709 | DNAJC5B | FALSE | Upregulated |
| 0 | DNAJC5G | FALSE | Upregulated |
| -0,506173201 | DNAJC6 | FALSE | Downregulated |
| -0,007543259 | DNASE2B | FALSE | Downregulated |
| 0,50800674 | DNER | FALSE | Upregulated |
| 0,169817191 | DNM1P35 | FALSE | Upregulated |
| 0,875855456 | DNM1P46 | FALSE | Upregulated |
| -0,515808798 | DNM1P50 | FALSE | Downregulated |
| -1,565472332 | DNMT3L | | Downregulated |
| 0,087836139 | DNTT | FALSE | Upregulated |
| 0,049548285 | DOC2GP | FALSE | Upregulated |

| | | | |
|---|---|---|---|
| 0,12674641 | DOCK9-AS1 | FALSE | Upregulated |
| 0 | DOK5 | FALSE | Upregulated |
| 0,067855057 | DPCR1 | FALSE | Upregulated |
| 0 | DPEP1 | FALSE | Upregulated |
| -0,101510146 | DPEP3 | FALSE | Downregulated |
| 0,684207201 | DPF1 | FALSE | Upregulated |
| -0,940507683 | DPH6-AS1 | FALSE | Downregulated |
| 0,104051414 | DPP10 | FALSE | Upregulated |
| 0 | DPP10-AS1 | FALSE | Upregulated |
| | DPP10-AS3 | | |
| 0,468731724 | DPP6 | FALSE | Upregulated |
| | DPPA2 | | |
| 0 | DPPA2P3 | FALSE | Upregulated |
| 0 | DPPA3 | FALSE | Upregulated |
| -0,870430579 | DPPA4 | FALSE | Downregulated |
| | DPPA5 | | |
| | DPRX | | |
| 0,026921574 | DPRXP4 | FALSE | Upregulated |
| 1,7352683 | DPY19L2 | | Upregulated |
| -0,863918126 | DPY19L2P1 | FALSE | Downregulated |
| | DPYSL5 | | |
| 0,787305944 | DQX1 | FALSE | Upregulated |
| 0 | DRAXIN | FALSE | Upregulated |
| -0,056788597 | DRC1 | FALSE | Downregulated |
| 0,443012842 | DRD2 | FALSE | Upregulated |
| 0 | DRD3 | FALSE | Upregulated |
| | DRGX | | |
| 0 | DRP2 | FALSE | Upregulated |
| 1,468034529 | DSC3 | | Upregulated |
| 1,224476161 | DSCAM-AS1 | | Upregulated |
| -0,002529603 | DSCAM-IT1 | FALSE | Downregulated |
| | DSCR10 | | |
| | DSCR4 | | |
| | DSCR8 | | |
| 1,554452359 | DSG3 | | Upregulated |
| 0 | DSG4 | FALSE | Upregulated |
| 0,672427203 | DSPP | FALSE | Upregulated |
| -0,182533884 | DSTNP2 | FALSE | Downregulated |
| | DSTNP4 | | |
| 1,906847191 | DTL | | Upregulated |
| 0,997561407 | DUOXA1 | FALSE | Upregulated |
| 0,610731196 | DUOXA2 | FALSE | Upregulated |
| -0,228929729 | DUPD1 | FALSE | Downregulated |
| 0,18356024 | DUSP13 | FALSE | Upregulated |
| 0,574978172 | DUSP15 | FALSE | Upregulated |
| -0,110309228 | DUSP21 | FALSE | Downregulated |
| 0,837366563 | DUSP26 | FALSE | Upregulated |
| -0,237255688 | DUSP27 | FALSE | Downregulated |
| 0,037837864 | DUSP5P1 | FALSE | Upregulated |
| 0,553523287 | DUSP9 | FALSE | Upregulated |
| | DUX2 | | |

| | | | |
|---|---|---|---|
| | DUX4 | | |
| 0 | DUX4L | FALSE | Upregulated |
| | DUX4L2 | | |
| | DUX4L3 | | |
| | DUX4L4 | | |
| | DUX4L5 | | |
| | DUX4L6 | | |
| | DUX4L7 | | |
| | DUXA | | |
| 0 | DYDC1 | FALSE | Upregulated |
| 0,049800891 | DYDC2 | FALSE | Upregulated |
| 0,02352612 | DYNAP | FALSE | Upregulated |
| 0,712675305 | DYNC1I1 | FALSE | Upregulated |
| -0,001946822 | DYNLRB2 | FALSE | Downregulated |
| | DYTN | | |
| 1,087528441 | DYX1C1 | | Upregulated |
| 0 | DYX1C1-CCPG1 | FALSE | Upregulated |
| 0,77747632 | DZANK1 | FALSE | Upregulated |
| -0,80198675 | E2F7 | FALSE | Downregulated |
| 1,680191856 | E2F8 | | Upregulated |
| 2,116268693 | EAF2 | | Upregulated |
| 1,155701454 | EBF2 | | Upregulated |
| | EBLN1 | | |
| 0,31676668 | ECEL1 | FALSE | Upregulated |
| -0,142856985 | ECEL1P2 | FALSE | Downregulated |
| -0,108155414 | ECRP | FALSE | Downregulated |
| 0,010060206 | ECSCR | FALSE | Upregulated |
| -0,060185244 | ECSCR-2 | FALSE | Downregulated |
| 3,217132654 | EDA2R | | Upregulated |
| 0,057083623 | EDARADD | FALSE | Upregulated |
| 0,052380504 | EDDM3A | FALSE | Upregulated |
| 0,251749218 | EDDM3B | FALSE | Upregulated |
| 0,500214232 | EDIL3 | FALSE | Upregulated |
| -0,399040958 | EDN2 | FALSE | Downregulated |
| 0,376993558 | EDN3 | FALSE | Upregulated |
| 0,180358643 | EEF1DP3 | FALSE | Upregulated |
| -0,467622609 | EEF1E1 | FALSE | Downregulated |
| 0,299862242 | EFCAB1 | FALSE | Upregulated |
| 0,441018673 | EFCAB14-AS1 | FALSE | Upregulated |
| | EFCAB3 | | |
| -0,335699656 | EFCAB5 | FALSE | Downregulated |
| | EFCAB8 | | |
| 0 | EFCAB9 | FALSE | Upregulated |
| -0,306385758 | EFHB | FALSE | Downregulated |
| 0,702148165 | EFHC2 | FALSE | Upregulated |
| -0,173262609 | EFNA3 | FALSE | Downregulated |
| -0,168240452 | EFNA5 | FALSE | Downregulated |
| 0 | EFTUD1P1 | FALSE | Upregulated |
| 0,57940799 | EGF | FALSE | Upregulated |
| 1,369337638 | EGFEM1P | | Upregulated |
| 0 | EGFL6 | FALSE | Upregulated |

| | | | |
|---|---|---|---|
| 1,456864568 | EGR3 | | Upregulated |
| 0 | EGR4 | FALSE | Upregulated |
| -1,015307741 | EID2B | | Downregulated |
| 0,35966749 | EID3 | FALSE | Upregulated |
| | EIF3IP1 | | |
| 0,253519479 | EIF4E1B | FALSE | Upregulated |
| 0 | ELANE | FALSE | Upregulated |
| | ELAVL2 | | |
| -0,818277595 | ELAVL3 | FALSE | Downregulated |
| 0 | ELAVL4 | FALSE | Upregulated |
| 0,58353866 | ELF5 | FALSE | Upregulated |
| 1,406295615 | ELFN2 | | Upregulated |
| -0,283951432 | ELMOD1 | FALSE | Downregulated |
| -0,028639907 | ELOVL2-AS1 | FALSE | Downregulated |
| 0 | ELOVL3 | FALSE | Upregulated |
| -1,052165527 | ELOVL4 | | Downregulated |
| -0,184779798 | ELOVL7 | FALSE | Downregulated |
| | ELSPBP1 | | |
| 0 | EMR3 | FALSE | Upregulated |
| 0 | EMX2 | FALSE | Upregulated |
| 0 | EN1 | FALSE | Upregulated |
| 0,112290726 | EN2 | FALSE | Upregulated |
| 0,642273502 | ENO1-AS1 | FALSE | Upregulated |
| 0,196393704 | ENOX1 | FALSE | Upregulated |
| -0,359076625 | ENPP6 | FALSE | Downregulated |
| | ENTHD1 | | |
| 1,31011125 | ENTPD2 | | Upregulated |
| 0,002661768 | ENTPD3-AS1 | FALSE | Upregulated |
| 2,682319758 | EPCAM | | Upregulated |
| -0,368336958 | EPGN | FALSE | Downregulated |
| 1,786497692 | EPHA1-AS1 | | Upregulated |
| 0,670051742 | EPHA10 | FALSE | Upregulated |
| 0 | EPHA5 | FALSE | Upregulated |
| 0,549868681 | EPHA6 | FALSE | Upregulated |
| 0,4596918 | EPHA7 | FALSE | Upregulated |
| | EPHA8 | | |
| 0 | EPHX3 | FALSE | Upregulated |
| | EPHX4 | | |
| -1,162404209 | EPN2-IT1 | | Downregulated |
| -0,980833995 | EPO | FALSE | Downregulated |
| 0 | EPPIN | FALSE | Upregulated |
| 0 | EPPIN-WFDC6 | FALSE | Upregulated |
| 0,560871592 | EPS8L3 | FALSE | Upregulated |
| | EPYC | | |
| | EQTN | | |
| 0,019888931 | ERAS | FALSE | Upregulated |
| -0,195172572 | ERC2 | FALSE | Downregulated |
| 1,350185645 | ERC2-IT1 | | Upregulated |
| 0 | EREG | FALSE | Upregulated |
| -0,081744461 | ERICH1-AS1 | FALSE | Downregulated |
| -0,035975477 | ERICH2 | FALSE | Downregulated |

| | | | |
|---|---|---|---|
| -0,095036631 | ERMN | FALSE | Downregulated |
| 1,63300375 | ERN2 | | Upregulated |
| 2,018660217 | ERP27 | | Upregulated |
| 0 | ERVMER34-1 | FALSE | Upregulated |
| 0,546590264 | ERVV-1 | FALSE | Upregulated |
| 0,0141269 | ERVV-2 | FALSE | Upregulated |
| -0,101006706 | ESCO2 | FALSE | Downregulated |
| 0,159966803 | ESM1 | FALSE | Upregulated |
| 0 | ESRG | FALSE | Upregulated |
| 0,255511601 | ESRP1 | FALSE | Upregulated |
| 1,967481847 | ESRRG | | Upregulated |
| 0 | ESX1 | FALSE | Upregulated |
| -0,138132445 | ETV3L | FALSE | Downregulated |
| 1,267304507 | ETV7 | | Upregulated |
| | EVX1 | | |
| | EVX2 | | |
| 0,034128514 | EXD1 | FALSE | Upregulated |
| 0,009051348 | EXO1 | FALSE | Upregulated |
| 0 | EZR-AS1 | FALSE | Upregulated |
| 0 | FA2H | FALSE | Upregulated |
| | FABP12 | | |
| 0,646656035 | FABP4 | FALSE | Upregulated |
| -0,086825838 | FABP5P3 | FALSE | Downregulated |
| | FABP6 | | |
| 0,012680241 | FABP7 | FALSE | Upregulated |
| | FABP9 | | |
| 1,449770892 | FAHD2CP | | Upregulated |
| 0,236027478 | FAIM2 | FALSE | Upregulated |
| -1,744227997 | FAM106A | | Downregulated |
| 0,060356782 | FAM106B | FALSE | Upregulated |
| 0,344911444 | FAM106CP | FALSE | Upregulated |
| 0,367289932 | FAM110D | FALSE | Upregulated |
| 2,012619224 | FAM127C | | Upregulated |
| 0 | FAM131C | FALSE | Upregulated |
| 1,347259379 | FAM132A | | Upregulated |
| 0 | FAM133A | FALSE | Upregulated |
| 2,259071443 | FAM133CP | | Upregulated |
| 1,202581359 | FAM135B | | Upregulated |
| | FAM138A | | |
| 0,027871319 | FAM138B | FALSE | Upregulated |
| | FAM138C | | |
| 0 | FAM138D | FALSE | Upregulated |
| 0 | FAM138E | FALSE | Upregulated |
| | FAM138F | | |
| 1,200468957 | FAM150A | | Upregulated |
| 0,660987923 | FAM151B | FALSE | Upregulated |
| -0,882867729 | FAM153A | FALSE | Downregulated |
| 1,562435578 | FAM153B | | Upregulated |
| 0,471371554 | FAM153C | FALSE | Upregulated |
| 0,073197161 | FAM154A | FALSE | Upregulated |
| -0,24666738 | FAM155A | FALSE | Downregulated |

| | | | |
|---|---|---|---|
| 0 | FAM155A-IT1 | FALSE | Upregulated |
| 0,492674106 | FAM155B | FALSE | Upregulated |
| 0 | FAM157B | FALSE | Upregulated |
| -0,097580797 | FAM159A | FALSE | Downregulated |
| -0,111394973 | FAM159B | FALSE | Downregulated |
| 0,684051113 | FAM161A | FALSE | Upregulated |
| 0 | FAM162B | FALSE | Upregulated |
| -0,372007401 | FAM163A | FALSE | Downregulated |
| 0,196741476 | FAM167A | FALSE | Upregulated |
| -0,644111833 | FAM170A | FALSE | Downregulated |
| | FAM170B | | |
| -0,116660471 | FAM170B-AS1 | FALSE | Downregulated |
| 1,268915807 | FAM171B | | Upregulated |
| 0 | FAM172BP | FALSE | Upregulated |
| 1,040740258 | FAM177B | | Upregulated |
| 2,614371364 | FAM179A | | Upregulated |
| 0,743159607 | FAM181A | FALSE | Upregulated |
| | FAM181A-AS1 | | |
| 0,019985931 | FAM181B | FALSE | Upregulated |
| 0,372333337 | FAM182A | FALSE | Upregulated |
| 0 | FAM182B | FALSE | Upregulated |
| 0 | FAM183A | FALSE | Upregulated |
| 0,047409049 | FAM183B | FALSE | Upregulated |
| 0,417484258 | FAM183CP | FALSE | Upregulated |
| 0 | FAM187B | FALSE | Upregulated |
| 1,045399246 | FAM189A1 | | Upregulated |
| 0,009364939 | FAM189A2 | FALSE | Upregulated |
| -0,124986448 | FAM194A | FALSE | Downregulated |
| 0,019701585 | FAM194B | FALSE | Upregulated |
| | FAM197Y2 | | |
| | FAM197Y4 | | |
| | FAM197Y5 | | |
| 0,084649888 | FAM19A1 | FALSE | Upregulated |
| -1,637858264 | FAM19A2 | | Downregulated |
| | FAM19A3 | | |
| 0,019985931 | FAM19A4 | FALSE | Upregulated |
| 1,095091663 | FAM201A | | Upregulated |
| -0,191490471 | FAM201B | FALSE | Downregulated |
| -0,104611498 | FAM209A | FALSE | Downregulated |
| | FAM209B | | |
| 0,573174453 | FAM215A | FALSE | Upregulated |
| 0,018764371 | FAM216B | FALSE | Upregulated |
| 0 | FAM218A | FALSE | Upregulated |
| 0 | FAM220BP | FALSE | Upregulated |
| 1,152992983 | FAM223B | | Upregulated |
| | FAM225B | | |
| 0,902067932 | FAM226A | FALSE | Upregulated |
| 0,94899285 | FAM226B | FALSE | Upregulated |
| 0,703773219 | FAM228A | FALSE | Upregulated |
| -0,400134262 | FAM228B | FALSE | Downregulated |
| | FAM230A | | |

| | | | |
|---|---|---|---|
| 0,064844243 | FAM230B | FALSE | Upregulated |
| | FAM230C | | |
| | FAM231B | | |
| 0,339235667 | FAM231C | FALSE | Upregulated |
| 0,098997119 | FAM231D | FALSE | Upregulated |
| 0,669025015 | FAM24A | FALSE | Upregulated |
| 0 | FAM24B | FALSE | Upregulated |
| | FAM24B-CUZD1 | | |
| 0 | FAM25A | FALSE | Upregulated |
| 0 | FAM25B | FALSE | Upregulated |
| | FAM25C | | |
| | FAM25G | | |
| | FAM26D | | |
| -0,013528086 | FAM27B | FALSE | Downregulated |
| 1,484052402 | FAM27C | | Upregulated |
| | FAM27L | | |
| -0,844662245 | FAM3B | FALSE | Downregulated |
| | FAM41AY1 | | |
| 0,912183246 | FAM41C | FALSE | Upregulated |
| 1,476782375 | FAM43B | | Upregulated |
| 0,206483314 | FAM45B | FALSE | Upregulated |
| | FAM46D | | |
| | FAM47A | | |
| | FAM47B | | |
| -0,116946067 | FAM47C | FALSE | Downregulated |
| | FAM57B | | |
| 0,401315203 | FAM66A | FALSE | Upregulated |
| -1,580290445 | FAM66B | | Downregulated |
| 0,071416694 | FAM66C | FALSE | Upregulated |
| 0,158649807 | FAM66D | FALSE | Upregulated |
| | FAM66E | | |
| -1,300847434 | FAM69C | | Downregulated |
| 0 | FAM71B | FALSE | Upregulated |
| 0,932127686 | FAM71D | FALSE | Upregulated |
| 0 | FAM71E2 | FALSE | Upregulated |
| 0 | FAM71F1 | FALSE | Upregulated |
| 1,29966121 | FAM71F2 | | Upregulated |
| 0,364785557 | FAM72A | FALSE | Upregulated |
| -0,399429801 | FAM72B | FALSE | Downregulated |
| 0 | FAM74A1 | FALSE | Upregulated |
| -0,345516358 | FAM74A3 | FALSE | Downregulated |
| -0,099784179 | FAM74A4 | FALSE | Downregulated |
| | FAM74A7 | | |
| 0,637120423 | FAM81A | FALSE | Upregulated |
| -0,646734855 | FAM81B | FALSE | Downregulated |
| -0,472929978 | FAM83A-AS1 | FALSE | Downregulated |
| -0,615934646 | FAM83B | FALSE | Downregulated |
| 1,096182465 | FAM83C-AS1 | | Upregulated |
| -0,608482748 | FAM83E | FALSE | Downregulated |
| 0,745810615 | FAM83H-AS1 | FALSE | Upregulated |
| -0,830994383 | FAM85A | FALSE | Downregulated |

| | | | |
|---|---|---|---|
| 0,24587647 | FAM86B2 | FALSE | Upregulated |
| -0,290741701 | FAM86EP | FALSE | Downregulated |
| 0,608246941 | FAM86KP | FALSE | Upregulated |
| | FAM90A10P | | |
| | FAM90A27P | | |
| 0,884917716 | FAM90A2P | FALSE | Upregulated |
| 0,23059854 | FAM90A7P | FALSE | Upregulated |
| 1,159147046 | FAM92B | | Upregulated |
| 0,04823374 | FAM95A | FALSE | Upregulated |
| -0,252243358 | FAM95B1 | FALSE | Downregulated |
| 0,004618298 | FAM9A | FALSE | Upregulated |
| -0,601063164 | FAM9B | FALSE | Downregulated |
| 0 | FAM9C | FALSE | Upregulated |
| -0,296010811 | FANCB | FALSE | Downregulated |
| 0,240287407 | FANCD2OS | FALSE | Upregulated |
| | FANK1 | | |
| | FANK1-AS1 | | |
| 1,712070295 | FAP | | Upregulated |
| | FAR2P1 | | |
| -1,150405521 | FAR2P2 | | Downregulated |
| 0,831305035 | FAR2P3 | FALSE | Upregulated |
| | FATE1 | | |
| -0,34969677 | FAXC | FALSE | Downregulated |
| 1,241661105 | FBLL1 | | Upregulated |
| 0,355539562 | FBXO15 | FALSE | Upregulated |
| 0,058432515 | FBXO16 | FALSE | Upregulated |
| 1,448371584 | FBXO36 | | Upregulated |
| -0,673067426 | FBXO39 | FALSE | Downregulated |
| -0,035392672 | FBXO43 | FALSE | Downregulated |
| | FBXO47 | | |
| | FBXW12 | | |
| 0,699710393 | FCAR | FALSE | Upregulated |
| 0,056304337 | FCER1A | FALSE | Upregulated |
| 1,071142823 | FCER2 | | Upregulated |
| 1,260099784 | FCGR1C | | Upregulated |
| 0,105201985 | FCRL1 | FALSE | Upregulated |
| 2,023169965 | FCRL2 | | Upregulated |
| 0 | FCRL4 | FALSE | Upregulated |
| -0,319700419 | FCRLA | FALSE | Downregulated |
| 0,560494289 | FCRLB | FALSE | Upregulated |
| | FDCSP | | |
| | FER1L6 | | |
| 0,554782283 | FER1L6-AS1 | FALSE | Upregulated |
| 0,525922388 | FER1L6-AS2 | FALSE | Upregulated |
| 0 | FERD3L | FALSE | Upregulated |
| | FEV | | |
| -0,336268645 | FEZF1 | FALSE | Downregulated |
| 0,742834087 | FEZF1-AS1 | FALSE | Upregulated |
| | FEZF2 | | |
| 0,669025015 | FFAR1 | FALSE | Upregulated |
| -0,080376676 | FFAR2 | FALSE | Downregulated |

| | | | |
|---|---|---|---|
| 0,068555674 | FFAR3 | FALSE | Upregulated |
| 1,404005806 | FGD5P1 | | Upregulated |
| 1,048736443 | FGF10 | | Upregulated |
| -0,045342599 | FGF10-AS1 | FALSE | Downregulated |
| -0,923159107 | FGF12 | FALSE | Downregulated |
| 1,968461743 | FGF13 | | Upregulated |
| 1,84644806 | FGF13-AS1 | | Upregulated |
| -0,860524065 | FGF14-AS1 | FALSE | Downregulated |
| -1,048536396 | FGF14-IT1 | | Downregulated |
| 0 | FGF16 | FALSE | Upregulated |
| 0,648224889 | FGF18 | FALSE | Upregulated |
| 0 | FGF19 | FALSE | Upregulated |
| | FGF20 | | |
| -1,079579812 | FGF21 | | Downregulated |
| -0,498791113 | FGF22 | FALSE | Downregulated |
| -1,156647347 | FGF23 | | Downregulated |
| | FGF3 | | |
| | FGF4 | | |
| -0,223542949 | FGF5 | FALSE | Downregulated |
| 0,052376707 | FGF6 | FALSE | Upregulated |
| 0,076658198 | FGF8 | FALSE | Upregulated |
| 0 | FGF9 | FALSE | Upregulated |
| 0 | FGFBP1 | FALSE | Upregulated |
| 1,556964491 | FGFBP2 | | Upregulated |
| 0,076825603 | FHAD1 | FALSE | Upregulated |
| 1,171847916 | FHL5 | | Upregulated |
| 0,321232383 | FIBCD1 | FALSE | Upregulated |
| 0,360062072 | FIGF | FALSE | Upregulated |
| | FIGLA | | |
| 0,657758048 | FIRRE | FALSE | Upregulated |
| 0,024051535 | FITM1 | FALSE | Upregulated |
| 0,376726497 | FKBP1A-SDCBP2 | FALSE | Upregulated |
| -1,41948502 | FKBP1B | | Downregulated |
| -0,617156303 | FKBP6 | FALSE | Downregulated |
| 0,676252355 | FKBP9L | FALSE | Upregulated |
| 0,59938295 | FKSG29 | FALSE | Upregulated |
| | FLJ16171 | | |
| | FLJ16779 | | |
| 0,492523584 | FLJ20021 | FALSE | Upregulated |
| 0,063316295 | FLJ20444 | FALSE | Upregulated |
| 0,029934468 | FLJ20518 | FALSE | Upregulated |
| | FLJ21408 | | |
| 0 | FLJ22447 | FALSE | Upregulated |
| 0,790342954 | FLJ22763 | FALSE | Upregulated |
| 0,272618312 | FLJ25363 | FALSE | Upregulated |
| 0 | FLJ25715 | FALSE | Upregulated |
| | FLJ25758 | | |
| | FLJ25917 | | |
| | FLJ26245 | | |
| 0 | FLJ26850 | FALSE | Upregulated |
| 0,466283634 | FLJ27255 | FALSE | Upregulated |

| | | | |
|---|---|---|---|
| -0,99884124 | FLJ27354 | FALSE | Downregulated |
| 1,169012168 | FLJ30375 | | Upregulated |
| 0 | FLJ30679 | FALSE | Upregulated |
| 0,60454628 | FLJ31104 | FALSE | Upregulated |
| 0 | FLJ31356 | FALSE | Upregulated |
| | FLJ31662 | | |
| -0,197530725 | FLJ33360 | FALSE | Downregulated |
| | FLJ33534 | | |
| 0,964423624 | FLJ33544 | FALSE | Upregulated |
| -0,55298932 | FLJ34503 | FALSE | Downregulated |
| 0,039220225 | FLJ35934 | FALSE | Upregulated |
| 0,669025015 | FLJ36000 | FALSE | Upregulated |
| -0,506010318 | FLJ37201 | FALSE | Downregulated |
| 0 | FLJ37505 | FALSE | Upregulated |
| -0,54206709 | FLJ38379 | FALSE | Downregulated |
| 0,038287376 | FLJ38723 | FALSE | Upregulated |
| 0 | FLJ39080 | FALSE | Upregulated |
| -1,475827705 | FLJ39632 | | Downregulated |
| 0,151170448 | FLJ40194 | FALSE | Upregulated |
| 0 | FLJ40288 | FALSE | Upregulated |
| 0 | FLJ41200 | FALSE | Upregulated |
| | FLJ41278 | | |
| 0,455400398 | FLJ41309 | FALSE | Upregulated |
| | FLJ41423 | | |
| 0 | FLJ41733 | FALSE | Upregulated |
| 0,679394792 | FLJ41941 | FALSE | Upregulated |
| -0,106162615 | FLJ42102 | FALSE | Downregulated |
| -0,076358527 | FLJ42351 | FALSE | Downregulated |
| | FLJ42842 | | |
| 0 | FLJ42969 | FALSE | Upregulated |
| 0,294647392 | FLJ43315 | FALSE | Upregulated |
| -1,765054592 | FLJ43585 | | Downregulated |
| 1,639057341 | FLJ43681 | | Upregulated |
| 0,701809199 | FLJ43879 | FALSE | Upregulated |
| 3,449819551 | FLJ44006 | | Upregulated |
| -0,117044605 | FLJ44087 | FALSE | Downregulated |
| -0,319505164 | FLJ44313 | FALSE | Downregulated |
| 0 | FLJ44635 | FALSE | Upregulated |
| -0,056907036 | FLJ44790 | FALSE | Downregulated |
| | FLJ44838 | | |
| -0,083064364 | FLJ45079 | FALSE | Downregulated |
| 0,373254007 | FLJ45139 | FALSE | Upregulated |
| 0,217220008 | FLJ45256 | FALSE | Upregulated |
| -0,059175856 | FLJ45743 | FALSE | Downregulated |
| 0,987978511 | FLJ45825 | FALSE | Upregulated |
| | FLJ45964 | | |
| | FLJ45974 | | |
| | FLJ46066 | | |
| -0,064559782 | FLJ46284 | FALSE | Downregulated |
| 1,057606016 | FLT3 | | Upregulated |
| -0,179341901 | FLVCR1-AS1 | FALSE | Downregulated |

| | | | |
|---|---|---|---|
| -0,451406959 | FMO1 | FALSE | Downregulated |
| | FMO9P | | |
| 0 | FMR1NB | FALSE | Upregulated |
| 0,033944548 | FNDC7 | FALSE | Upregulated |
| | FOLR3 | | |
| 0,82167394 | FOLR4 | FALSE | Upregulated |
| 1,117807197 | FOSB | | Upregulated |
| 0 | FOXB1 | FALSE | Upregulated |
| | FOXB2 | | |
| -0,612977145 | FOXC2 | FALSE | Downregulated |
| 0 | FOXD1 | FALSE | Upregulated |
| 0,613018311 | FOXD3 | FALSE | Upregulated |
| -0,788876389 | FOXD4 | FALSE | Downregulated |
| 0,633919627 | FOXD4L1 | FALSE | Upregulated |
| 0,087047623 | FOXD4L3 | FALSE | Upregulated |
| 0 | FOXD4L6 | FALSE | Upregulated |
| 0,468548378 | FOXF2 | FALSE | Upregulated |
| 0 | FOXG1 | FALSE | Upregulated |
| | FOXI1 | | |
| 0,087771113 | FOXI2 | FALSE | Upregulated |
| | FOXI3 | | |
| 1,547514037 | FOXJ1 | | Upregulated |
| 1,541528872 | FOXL1 | | Upregulated |
| -0,003401657 | FOXL2 | FALSE | Downregulated |
| | FOXN1 | | |
| 0,000592325 | FOXR1 | FALSE | Upregulated |
| 0,073122974 | FOXR2 | FALSE | Upregulated |
| 1,470432482 | FOXS1 | | Upregulated |
| -0,585167559 | FPGT-TNNI3K | FALSE | Downregulated |
| 0,799478414 | FPR2 | FALSE | Upregulated |
| -0,044556088 | FRG2B | FALSE | Downregulated |
| 1,076103931 | FRMD1 | | Upregulated |
| 0,029342466 | FRMD6-AS2 | FALSE | Upregulated |
| 1,88829518 | FRMD8P1 | | Upregulated |
| 0 | FRMPD2 | FALSE | Upregulated |
| 0,747384596 | FRMPD2P1 | FALSE | Upregulated |
| 0 | FRMPD4 | FALSE | Upregulated |
| | FRRS1L | | |
| 0,571539722 | FSBP | FALSE | Upregulated |
| 0,034128514 | FSCB | FALSE | Upregulated |
| | FSCN3 | | |
| -0,257800184 | FSHB | FALSE | Downregulated |
| 0,837176148 | FSHR | FALSE | Upregulated |
| -0,174458722 | FSIP1 | FALSE | Downregulated |
| 0,705562129 | FSTL4 | FALSE | Upregulated |
| | FSTL5 | | |
| | FTH1P18 | | |
| -0,381430565 | FTH1P3 | FALSE | Downregulated |
| | FTHL17 | | |
| | FTLP10 | | |
| | FTMT | | |

| | | | |
|---|---|---|---|
| -0,652029279 | FTO-IT1 | FALSE | Downregulated |
| 2,627001452 | FUT5 | | Upregulated |
| 0,813952045 | FUT9 | FALSE | Upregulated |
| | FXYD4 | | |
| 0,397124959 | FXYD6-FXYD2 | FALSE | Upregulated |
| -0,094555195 | FXYD7 | FALSE | Downregulated |
| 0,597480774 | FZD10 | FALSE | Upregulated |
| 0,347629282 | FZD10-AS1 | FALSE | Upregulated |
| 0,752571143 | FZD2 | FALSE | Upregulated |
| 0,723787147 | FZD9 | FALSE | Upregulated |
| 0 | GAB4 | FALSE | Upregulated |
| 1,045105716 | GABBR2 | | Upregulated |
| | GABRA1 | | |
| 0 | GABRA2 | FALSE | Upregulated |
| 0,627101712 | GABRA3 | FALSE | Upregulated |
| 0 | GABRA4 | FALSE | Upregulated |
| | GABRA5 | | |
| | GABRA6 | | |
| 0,989682689 | GABRB1 | FALSE | Upregulated |
| | GABRB2 | | |
| 0 | GABRD | FALSE | Upregulated |
| 0 | GABRG1 | FALSE | Upregulated |
| | GABRG2 | | |
| 0,735274014 | GABRG3 | FALSE | Upregulated |
| | GABRQ | | |
| 0,554132311 | GABRR1 | FALSE | Upregulated |
| -0,303613517 | GABRR2 | FALSE | Downregulated |
| | GABRR3 | | |
| 0,648104729 | GAD1 | FALSE | Upregulated |
| | GAD2 | | |
| -0,38066924 | GADL1 | FALSE | Downregulated |
| | GAGE1 | | |
| | GAGE12C | | |
| | GAGE12D | | |
| | GAGE12E | | |
| | GAGE12G | | |
| | GAGE12H | | |
| | GAGE12I | | |
| | GAGE12J | | |
| | GAGE2A | | |
| | GAGE2C | | |
| | GAGE2E | | |
| 0 | GAL | FALSE | Upregulated |
| 0,071357241 | GAL3ST3 | FALSE | Upregulated |
| 0,45309944 | GALNT12 | FALSE | Upregulated |
| 0,937748323 | GALNT13 | FALSE | Upregulated |
| -1,287274503 | GALNT14 | | Downregulated |
| 0 | GALNT5 | FALSE | Upregulated |
| -0,104848139 | GALNT8 | FALSE | Downregulated |
| 0,062037882 | GALNT9-2 | FALSE | Upregulated |
| | GALNTL5 | | |

| | | | |
|---|---|---|---|
| -0,129741731 | GALNTL6 | FALSE | Downregulated |
| -0,017737585 | GALP | FALSE | Downregulated |
| 0,669756935 | GALR1 | FALSE | Upregulated |
| 0,040463092 | GALR2 | FALSE | Upregulated |
| -0,806872358 | GALR3 | FALSE | Downregulated |
| 1,252999794 | GAP43 | | Upregulated |
| 0,477509384 | GAPT | FALSE | Upregulated |
| -0,306163004 | GAR1 | FALSE | Downregulated |
| 0,708392948 | GAS2L2 | FALSE | Upregulated |
| 0,5570553 | GAS6-AS2 | FALSE | Upregulated |
| 0,076658198 | GAST | FALSE | Upregulated |
| 1,010070169 | GATA3-AS1 | | Upregulated |
| -0,114823843 | GBP1P1 | FALSE | Downregulated |
| -0,580788963 | GBP6 | FALSE | Downregulated |
| 0,894983492 | GBX1 | FALSE | Upregulated |
| 0,125988383 | GBX2 | FALSE | Upregulated |
| 0,608246941 | GCG | FALSE | Upregulated |
| 0 | GCM1 | FALSE | Upregulated |
| -0,521758363 | GCNT3 | FALSE | Downregulated |
| 0 | GCSAML | FALSE | Upregulated |
| 0,746004426 | GCSAML-AS1 | FALSE | Upregulated |
| -0,018285463 | GDF10 | FALSE | Downregulated |
| | GDF3 | | |
| 0,537596555 | GDF5 | FALSE | Upregulated |
| 0,128121092 | GDF9 | FALSE | Upregulated |
| 1,134555721 | GDPD1 | | Upregulated |
| -0,893713259 | GDPD2 | FALSE | Downregulated |
| 0,832166426 | GDPD3 | FALSE | Upregulated |
| | GFI1B | | |
| 1,803314599 | GFPT2 | | Upregulated |
| 0 | GFRA3 | FALSE | Upregulated |
| 0,470164621 | GFRA4 | FALSE | Upregulated |
| 0,834386225 | GFRAL | FALSE | Upregulated |
| | GFY | | |
| 1,129301634 | GGNBP1 | | Upregulated |
| 0,709756191 | GGT2 | FALSE | Upregulated |
| 0 | GGT3P | FALSE | Upregulated |
| 0,383645052 | GGT6 | FALSE | Upregulated |
| 1,399251089 | GGT8P | | Upregulated |
| 0,644251792 | GGTLC1 | FALSE | Upregulated |
| | GH1 | | |
| | GH2 | | |
| | GHRH | | |
| 0 | GHRHR | FALSE | Upregulated |
| -0,017014159 | GHSR | FALSE | Downregulated |
| 0,071318393 | GIF | FALSE | Upregulated |
| | GIMD1 | | |
| -0,156210459 | GINS3 | FALSE | Downregulated |
| | GIP | | |
| 1,234863412 | GIPR | | Upregulated |
| | GJA10 | | |

| | | | |
|---|---|---|---|
| 0,393639649 | GJA3 | FALSE | Upregulated |
| -1,298565038 | GJA5 | | Downregulated |
| 0 | GJA8 | FALSE | Upregulated |
| 0 | GJB6 | FALSE | Upregulated |
| | GJD2 | | |
| 0 | GJD4 | FALSE | Upregulated |
| | GK2 | | |
| | GKN1 | | |
| | GKN2 | | |
| 0,59878823 | GLB1L3 | FALSE | Upregulated |
| -0,399559145 | GLDN | FALSE | Downregulated |
| 0,447990213 | GLIPR1L2 | FALSE | Upregulated |
| 1,419288084 | GLIS1 | | Upregulated |
| 0,955126284 | GLMN | FALSE | Upregulated |
| -0,889022149 | GLOD5 | FALSE | Downregulated |
| -0,269022566 | GLP1R | FALSE | Downregulated |
| -1,033277665 | GLP2R | | Downregulated |
| -0,081358359 | GLRA2 | FALSE | Downregulated |
| 0 | GLRA3 | FALSE | Upregulated |
| 0 | GLRA4 | FALSE | Upregulated |
| 0,360310223 | GLRB | FALSE | Upregulated |
| | GLT6D1 | | |
| | GLYATL2 | | |
| 0,106212194 | GLYATL3 | FALSE | Upregulated |
| | GLYCAM1 | | |
| | GM140 | | |
| 0,478309004 | GMCL1P1 | FALSE | Upregulated |
| 0 | GML | FALSE | Upregulated |
| 0,027563344 | GNAS-AS1 | FALSE | Upregulated |
| | GNAT3 | | |
| 1,345858758 | GNB3 | | Upregulated |
| 0,072462536 | GNG13 | FALSE | Upregulated |
| 0 | GNG4 | FALSE | Upregulated |
| | GNG8 | | |
| | GNGT1 | | |
| 1,414315863 | GNGT2 | | Upregulated |
| 0,184369135 | GNLY | FALSE | Upregulated |
| -0,283451292 | GNRH2 | FALSE | Downregulated |
| 0 | GOLGA2P2Y | FALSE | Upregulated |
| | GOLGA6L1 | | |
| -0,115927293 | GOLGA6L2 | FALSE | Downregulated |
| 0,280392452 | GOLGA6L3 | FALSE | Upregulated |
| 0 | GOLGA6L6 | FALSE | Upregulated |
| 1,205770481 | GOLGA6L7P | | Upregulated |
| | GOLGA8CP | | |
| | GOLGA8DP | | |
| 0 | GOLGA8EP | FALSE | Upregulated |
| 0 | GOLGA8F | FALSE | Upregulated |
| | GOLGA8G | | |
| -1,603868433 | GOLGA8I | | Downregulated |
| -0,33344166 | GOLGA8J | FALSE | Downregulated |

| | | | |
|---|---|---|---|
| | GOT1L1 | | |
| 0,424471069 | GP1BB | FALSE | Upregulated |
| 0 | GP2 | FALSE | Upregulated |
| 0,70511874 | GP9 | FALSE | Upregulated |
| 0 | GPA33 | FALSE | Upregulated |
| 0,977485012 | GPC2 | FALSE | Upregulated |
| -0,319863947 | GPC5-AS1 | FALSE | Downregulated |
| -0,194296413 | GPC6-AS2 | FALSE | Downregulated |
| | GPHB5 | | |
| 0,903808964 | GPR1 | FALSE | Upregulated |
| 0,669025015 | GPR1-AS | FALSE | Upregulated |
| 1,710834797 | GPR101 | | Upregulated |
| 0,597480774 | GPR110 | FALSE | Upregulated |
| 0 | GPR111 | FALSE | Upregulated |
| | GPR112 | | |
| | GPR115 | | |
| 0 | GPR119 | FALSE | Upregulated |
| 0,624593177 | GPR12 | FALSE | Upregulated |
| 0,310147787 | GPR137C | FALSE | Upregulated |
| | GPR139 | | |
| 0,899163113 | GPR141 | FALSE | Upregulated |
| -0,382612094 | GPR142 | FALSE | Downregulated |
| 0,597480774 | GPR144 | FALSE | Upregulated |
| | GPR148 | | |
| 0,158403636 | GPR149 | FALSE | Upregulated |
| 0,480063189 | GPR15 | FALSE | Upregulated |
| 0,845544031 | GPR158 | FALSE | Upregulated |
| -1,278854558 | GPR158-AS1 | | Downregulated |
| 0,319112919 | GPR19 | FALSE | Upregulated |
| 0,49682077 | GPR20 | FALSE | Upregulated |
| 1,470554564 | GPR25 | | Upregulated |
| 1,358256915 | GPR27 | | Upregulated |
| 1,169337336 | GPR3 | | Upregulated |
| 0 | GPR31 | FALSE | Upregulated |
| | GPR32 | | |
| -0,239268539 | GPR33 | FALSE | Downregulated |
| 0,14586273 | GPR35 | FALSE | Upregulated |
| 1,244410402 | GPR37L1 | | Upregulated |
| 0 | GPR50 | FALSE | Upregulated |
| 2,188834393 | GPR55 | | Upregulated |
| | GPR6 | | |
| 0,314086034 | GPR63 | FALSE | Upregulated |
| 0 | GPR78 | FALSE | Upregulated |
| 0,302511183 | GPR83 | FALSE | Upregulated |
| 0,822713636 | GPR85 | FALSE | Upregulated |
| 0,359645428 | GPR87 | FALSE | Upregulated |
| -0,338837521 | GPR97 | FALSE | Downregulated |
| 0,012656317 | GPRC5A | FALSE | Upregulated |
| -0,181479389 | GPRC6A | FALSE | Downregulated |
| -0,485125107 | GPX5 | FALSE | Downregulated |
| | GPX6 | | |

| | | | |
|---|---|---|---|
| -0,896501831 | GRIA1 | FALSE | Downregulated |
| 0,202265176 | GRIA2 | FALSE | Upregulated |
| -0,021330068 | GRID1-AS1 | FALSE | Downregulated |
| | GRID2 | | |
| 1,572309258 | GRID2IP | | Upregulated |
| | GRIK1-AS1 | | |
| -0,251904412 | GRIK1-AS2 | FALSE | Downregulated |
| -0,052650191 | GRIK2 | FALSE | Downregulated |
| 0 | GRIK3 | FALSE | Upregulated |
| 0,021199179 | GRIK4 | FALSE | Upregulated |
| 0,254338938 | GRIK5 | FALSE | Upregulated |
| 2,47711383 | GRIN1 | | Upregulated |
| | GRK1 | | |
| | GRK1-2 | | |
| 0 | GRK7 | FALSE | Upregulated |
| 0,147007189 | GRM1 | FALSE | Upregulated |
| 0,891525728 | GRM2 | FALSE | Upregulated |
| 0,69621897 | GRM4 | FALSE | Upregulated |
| 0,593775133 | GRM5 | FALSE | Upregulated |
| 1,197235815 | GRM5-AS1 | | Upregulated |
| | GRP | | |
| 0 | GRPR | FALSE | Upregulated |
| | GRXCR1 | | |
| | GRXCR2 | | |
| | GS1-122H1.2 | | |
| | GS1-18A18.1 | | |
| 0,947886097 | GS1-204I12.1 | FALSE | Upregulated |
| 0 | GS1-24F4.2 | FALSE | Upregulated |
| 0,057044605 | GS1-259H13.2 | FALSE | Upregulated |
| -0,082333459 | GS1-279B7.1 | FALSE | Downregulated |
| | GS1-600G8.3 | | |
| 0 | GSC | FALSE | Upregulated |
| | GSC2 | | |
| -0,158029492 | GSDMA | FALSE | Downregulated |
| 0 | GSDMC | FALSE | Upregulated |
| -0,033582502 | GSG1L | FALSE | Downregulated |
| 0,45705359 | GSTA3 | FALSE | Upregulated |
| -0,18409821 | GSTA5 | FALSE | Downregulated |
| -0,825482581 | GSTM2P1 | FALSE | Downregulated |
| 0,017925419 | GSTM5 | FALSE | Upregulated |
| | GSTT2 | | |
| 0 | GSTTP1 | FALSE | Upregulated |
| | GSX1 | | |
| | GSX2 | | |
| 0,386980368 | GTF2A1L | FALSE | Upregulated |
| | GTPBP6-2 | | |
| 0,050688952 | GTSCR1 | FALSE | Upregulated |
| -0,018068965 | GTSF1 | FALSE | Downregulated |
| | GTSF1L | | |
| -0,055805981 | GUCA1A | FALSE | Downregulated |
| | GUCA1C | | |

| | | | |
|---|---|---|---|
| -0,170016351 | GUCA2A | FALSE | Downregulated |
| -0,933450132 | GUCA2B | FALSE | Downregulated |
| 1,326384976 | GUCY1B2 | | Upregulated |
| 0,717814116 | GUCY2D | FALSE | Upregulated |
| 0,653779848 | GUCY2EP | FALSE | Upregulated |
| | GUCY2F | | |
| | GUCY2GP | | |
| 1,404456134 | GULP1 | | Upregulated |
| | GUSBP10 | | |
| -0,491737485 | GUSBP5 | FALSE | Downregulated |
| 1,189214122 | GXYLT2 | | Upregulated |
| -0,253147897 | GYG2P1 | FALSE | Downregulated |
| 0,671601434 | GYPA | FALSE | Upregulated |
| 0,034943632 | GYPB | FALSE | Upregulated |
| -0,159746757 | GZMB | FALSE | Downregulated |
| 1,186341845 | GZMH | | Upregulated |
| 0,648152817 | GZMK | FALSE | Upregulated |
| -0,84987817 | GZMM | FALSE | Downregulated |
| -0,321887456 | H1FOO | FALSE | Downregulated |
| | H2BFM | | |
| | H2BFWT | | |
| 0,023863845 | H2BFXP | FALSE | Upregulated |
| 0,417280097 | H3F3C | FALSE | Upregulated |
| | HAND1 | | |
| 0,118783856 | HAND2-AS1 | FALSE | Upregulated |
| -1,044526811 | HAP1 | | Downregulated |
| 0,016268545 | HAPLN1 | FALSE | Upregulated |
| -0,233107359 | HAPLN2 | FALSE | Downregulated |
| 0 | HAR1B | FALSE | Upregulated |
| -0,569279638 | HARBI1 | FALSE | Downregulated |
| 0,737210084 | HAS1 | FALSE | Upregulated |
| 0,947362043 | HAS2 | FALSE | Upregulated |
| -0,125943706 | HAS2-AS1 | FALSE | Downregulated |
| 0,238895704 | HAUS1 | FALSE | Upregulated |
| -0,120918792 | HAVCR1 | FALSE | Downregulated |
| 0,568815088 | HBEGF | FALSE | Upregulated |
| 0 | HBZ | FALSE | Upregulated |
| 0,597731594 | HCAR1 | FALSE | Upregulated |
| 1,115276186 | HCCAT4 | | Upregulated |
| 1,788689869 | HCCAT5 | | Upregulated |
| -0,150140129 | HCG14 | FALSE | Downregulated |
| -0,053184988 | HCG16 | FALSE | Downregulated |
| | HCG17 | | |
| 0 | HCG22 | FALSE | Upregulated |
| 0,004929336 | HCG23 | FALSE | Upregulated |
| 0 | HCN1 | FALSE | Upregulated |
| 1,717465025 | HCN4 | | Upregulated |
| | HCRT | | |
| | HCRTR2 | | |
| 1,517414031 | HCST | | Upregulated |
| 0 | HDAC11-AS1 | FALSE | Upregulated |

| | | | |
|---|---|---|---|
| | HDGFL1 | | |
| 2,15161411 | HECW1 | | Upregulated |
| | HELT | | |
| -0,495275683 | HEMGN | FALSE | Downregulated |
| | HEPACAM2 | | |
| 0,046846073 | HERC2P10 | FALSE | Upregulated |
| 0,680203578 | HERC2P4 | FALSE | Upregulated |
| 0 | HES2 | FALSE | Upregulated |
| 0 | HES3 | FALSE | Upregulated |
| -1,243370702 | HES5 | | Downregulated |
| 0 | HES7 | FALSE | Upregulated |
| 0,263177882 | HESX1 | FALSE | Upregulated |
| 0,587620761 | HEY1 | FALSE | Upregulated |
| 0 | HFM1 | FALSE | Upregulated |
| -0,618912322 | HGC6.3 | FALSE | Downregulated |
| 0,582224807 | HHATL | FALSE | Upregulated |
| 0,038974311 | HHIP-AS1 | FALSE | Upregulated |
| 0,039005732 | HHIPL2 | FALSE | Upregulated |
| | HHLA1 | | |
| -0,255526361 | HHLA2 | FALSE | Downregulated |
| -0,705981535 | HHLA3 | FALSE | Downregulated |
| 1,240070276 | HIF1A-AS1 | | Upregulated |
| 0 | HIGD2B | FALSE | Upregulated |
| 0,078950011 | HIST1H1T | FALSE | Upregulated |
| -0,055338013 | HIST1H2AA | FALSE | Downregulated |
| 0,596154551 | HIST1H2AB | FALSE | Upregulated |
| 1,311569773 | HIST1H2AH | | Upregulated |
| -0,836870605 | HIST1H2AJ | FALSE | Downregulated |
| 0,270168288 | HIST1H2BB | FALSE | Upregulated |
| -0,33576814 | HIST1H2BH | FALSE | Downregulated |
| 1,437452952 | HIST1H2BI | | Upregulated |
| -0,586558363 | HIST1H2BJ | FALSE | Downregulated |
| 0,020071015 | HIST1H2BL | FALSE | Upregulated |
| 1,353497533 | HIST1H2BM | | Upregulated |
| 0,416268606 | HIST1H2BO | FALSE | Upregulated |
| 0,003329517 | HIST1H3A | FALSE | Upregulated |
| 1,494539677 | HIST1H3E | | Upregulated |
| 0,15021217 | HIST1H3F | FALSE | Upregulated |
| -0,227103669 | HIST1H3G | FALSE | Downregulated |
| 0,979019652 | HIST1H3J | FALSE | Upregulated |
| -0,450712562 | HIST1H4A | FALSE | Downregulated |
| -0,261614355 | HIST1H4F | FALSE | Downregulated |
| | HIST1H4G | | |
| 1,891525224 | HIST1H4L | | Upregulated |
| 0,080331977 | HIST2H2BA | FALSE | Upregulated |
| 0,657922523 | HIST2H2BC | FALSE | Upregulated |
| 0,354960692 | HIST3H2A | FALSE | Upregulated |
| 1,911938619 | HIST3H2BB | | Upregulated |
| 1,79885849 | HIST3H3 | | Upregulated |
| 0,710672756 | HLA-DOB | FALSE | Upregulated |
| 0,737068109 | HLA-DPB2 | FALSE | Upregulated |

| | | | |
|---|---|---|---|
| 1,070460564 | HLA-G | | Upregulated |
| -0,58237765 | HLTF-AS1 | FALSE | Downregulated |
| | HMGA1P7 | | |
| -0,225077169 | HMGA2 | FALSE | Downregulated |
| -0,222706706 | HMGB3P1 | FALSE | Downregulated |
| -0,052643786 | HMGB4 | FALSE | Downregulated |
| 0,238878931 | HMGCLL1 | FALSE | Upregulated |
| 0,202269177 | HMGN2P46 | FALSE | Upregulated |
| | HMHB1 | | |
| 0,561702473 | HMMR | FALSE | Upregulated |
| -0,164552746 | HMP19 | FALSE | Downregulated |
| 0 | HMSD | FALSE | Upregulated |
| | HMX1 | | |
| | HMX2 | | |
| | HMX3 | | |
| -0,081343447 | HNRNPA1P33 | FALSE | Downregulated |
| 0,518767415 | HNRNPA3P1 | FALSE | Upregulated |
| 0 | HNRNPCL1 | FALSE | Upregulated |
| | HNRNPCP5 | | |
| | HNRNPKP3 | | |
| -0,11953799 | HOMER1 | FALSE | Downregulated |
| -0,550434287 | HOPX | FALSE | Downregulated |
| -0,06862836 | HORMAD1 | FALSE | Downregulated |
| -0,004397735 | HORMAD2 | FALSE | Downregulated |
| | HOTAIR | | |
| 2,113503414 | HOTAIRM1 | | Upregulated |
| 0 | HOTTIP | FALSE | Upregulated |
| -0,187920184 | HOXA1 | FALSE | Downregulated |
| -0,108898621 | HOXA10 | FALSE | Downregulated |
| 0,308414236 | HOXA10-AS | FALSE | Upregulated |
| | HOXA10-HOXA9 | | |
| | HOXA11 | | |
| | HOXA11-AS | | |
| 0 | HOXA13 | FALSE | Upregulated |
| -0,184111873 | HOXA5 | FALSE | Downregulated |
| 0,309095896 | HOXA6 | FALSE | Upregulated |
| 0,611339383 | HOXA7 | FALSE | Upregulated |
| 0,542123486 | HOXA9 | FALSE | Upregulated |
| | HOXB-AS1 | | |
| 1,691449461 | HOXB-AS3 | | Upregulated |
| | HOXB1 | | |
| | HOXB13 | | |
| -0,112784781 | HOXB5 | FALSE | Downregulated |
| 0,229755598 | HOXB7 | FALSE | Upregulated |
| 0,068951176 | HOXB8 | FALSE | Upregulated |
| 0,669756935 | HOXB9 | FALSE | Upregulated |
| | HOXC-AS1 | | |
| | HOXC-AS2 | | |
| | HOXC-AS3 | | |
| 0,597480774 | HOXC10 | FALSE | Upregulated |
| 0 | HOXC11 | FALSE | Upregulated |

| | | | |
|---|---|---|---|
| | HOXC12 | | |
| 0 | HOXC13 | FALSE | Upregulated |
| | HOXC13-AS | | |
| -0,007236136 | HOXC4 | FALSE | Downregulated |
| | HOXC5 | | |
| 0,740029782 | HOXC6 | FALSE | Upregulated |
| | HOXC8 | | |
| 0 | HOXC9 | FALSE | Upregulated |
| 2,03734594 | HOXD-AS1 | | Upregulated |
| 2,980166962 | HOXD-AS2 | | Upregulated |
| 0 | HOXD1 | FALSE | Upregulated |
| | HOXD10 | | |
| 0,597480774 | HOXD11 | FALSE | Upregulated |
| | HOXD12 | | |
| | HOXD13 | | |
| 1,936837552 | HOXD3 | | Upregulated |
| 0,542914772 | HOXD4 | FALSE | Upregulated |
| 2,865255527 | HOXD8 | | Upregulated |
| 0 | HOXD9 | FALSE | Upregulated |
| | HP08942 | | |
| 0,276261963 | HPCA | FALSE | Upregulated |
| 0,907374667 | HPCAL4 | FALSE | Upregulated |
| 0 | HPSE2 | FALSE | Upregulated |
| -0,001452899 | HPVC1 | FALSE | Downregulated |
| | HPYR1 | | |
| -0,026536793 | HRASLS | FALSE | Downregulated |
| -0,709639656 | HRASLS2 | FALSE | Downregulated |
| -0,295999605 | HRASLS5 | FALSE | Downregulated |
| 1,175234098 | HRCT1 | | Upregulated |
| -0,165701272 | HRH3 | FALSE | Downregulated |
| -0,052041736 | HRH4 | FALSE | Downregulated |
| | HRK | | |
| 0 | HS3ST4 | FALSE | Upregulated |
| 0,581082149 | HS3ST6 | FALSE | Upregulated |
| 0,116218884 | HS6ST2 | FALSE | Upregulated |
| | HS6ST2-AS1 | | |
| | HS6ST3 | | |
| 0,676226335 | HSD17B7P2 | FALSE | Upregulated |
| 0 | HSD52 | FALSE | Upregulated |
| 0 | HSF2BP | FALSE | Upregulated |
| -0,367027591 | HSF5 | FALSE | Downregulated |
| -0,25412008 | HSFX2 | FALSE | Downregulated |
| -0,599904519 | HSFY1 | FALSE | Downregulated |
| | HSFY1P1 | | |
| -0,54989971 | HSFY2 | FALSE | Downregulated |
| -1,023366587 | HSPA1L | | Downregulated |
| 0,872871263 | HSPB2 | FALSE | Upregulated |
| -0,470674759 | HSPB2-C11orf52 | FALSE | Downregulated |
| | HSPB3 | | |
| | HTN1 | | |
| | HTN3 | | |

| | | | |
|---|---|---|---|
| | HTR1A | | |
| | HTR1B | | |
| -0,063006437 | HTR1D | FALSE | Downregulated |
| | HTR1E | | |
| 1,336718821 | HTR1F | | Upregulated |
| -0,049007014 | HTR2A-AS1 | FALSE | Downregulated |
| | HTR3A | | |
| | HTR3B | | |
| | HTR3C | | |
| | HTR3D | | |
| 0 | HTR4 | FALSE | Upregulated |
| | HTR5A | | |
| 0 | HTR5A-AS1 | FALSE | Upregulated |
| -0,316873705 | HTRA4 | FALSE | Downregulated |
| 1,131333962 | HUS1B | | Upregulated |
| 0,027936955 | HYAL4 | FALSE | Upregulated |
| | HYALP1 | | |
| 0,563874735 | HYMAI | FALSE | Upregulated |
| -0,170219809 | IAPP | FALSE | Downregulated |
| 1,58027573 | IBA57-AS1 | | Upregulated |
| 0,766309303 | IBSP | FALSE | Upregulated |
| 2,155059663 | ICAM4 | | Upregulated |
| 1,969463007 | ICAM5 | | Upregulated |
| 1,542792387 | ICOS | | Upregulated |
| -0,475441256 | IDH1-AS1 | FALSE | Downregulated |
| -0,105535139 | IFI27L1 | FALSE | Downregulated |
| -1,01112267 | IFI27L2 | | Downregulated |
| | IFITM4P | | |
| 0,525609033 | IFLTD1 | FALSE | Upregulated |
| | IFNA1 | | |
| 0 | IFNA10 | FALSE | Upregulated |
| | IFNA13 | | |
| | IFNA14 | | |
| 0 | IFNA16 | FALSE | Upregulated |
| | IFNA17 | | |
| | IFNA2 | | |
| 0,0770659 | IFNA21 | FALSE | Upregulated |
| 0,048656215 | IFNA22P | FALSE | Upregulated |
| 0,041547119 | IFNA4 | FALSE | Upregulated |
| | IFNA5 | | |
| | IFNA6 | | |
| 0 | IFNA7 | FALSE | Upregulated |
| | IFNA8 | | |
| 0,772802397 | IFNB1 | FALSE | Upregulated |
| | IFNE | | |
| 0,188635298 | IFNL1 | FALSE | Upregulated |
| | IFNL2 | | |
| | IFNL3 | | |
| | IFNL4 | | |
| -0,263848555 | IFNW1 | FALSE | Downregulated |
| -0,475053256 | IGDCC3 | FALSE | Downregulated |

| | | | |
|---|---|---|---|
| 0,569585689 | IGDCC4 | FALSE | Upregulated |
| -0,249234658 | IGF2BP1 | FALSE | Downregulated |
| 1,907634576 | IGFBPL1 | | Upregulated |
| 0 | IGFL1 | FALSE | Upregulated |
| 0,583044479 | IGFL2 | FALSE | Upregulated |
| -0,471881029 | IGFL3 | FALSE | Downregulated |
| 0,504308699 | IGFL4 | FALSE | Upregulated |
| 0 | IGLL1 | FALSE | Upregulated |
| -0,08016827 | IGLL3P | FALSE | Downregulated |
| 0 | IGSF1 | FALSE | Upregulated |
| -0,578652386 | IGSF11 | FALSE | Downregulated |
| 1,708195728 | IGSF11-AS1 | | Upregulated |
| 0 | IGSF5 | FALSE | Upregulated |
| 0,38316508 | IL10 | FALSE | Upregulated |
| 0,082275127 | IL11 | FALSE | Upregulated |
| 0,126111389 | IL12A | FALSE | Upregulated |
| -0,153635372 | IL12A-AS1 | FALSE | Downregulated |
| -0,262286623 | IL12B | FALSE | Downregulated |
| 0 | IL13 | FALSE | Upregulated |
| 0,885788857 | IL13RA2 | FALSE | Upregulated |
| 0,669756935 | IL17A | FALSE | Upregulated |
| 0,097252145 | IL17B | FALSE | Upregulated |
| 0,550567457 | IL17C | FALSE | Upregulated |
| | IL17F | | |
| -0,585390808 | IL17RD | FALSE | Downregulated |
| 0,661206439 | IL19 | FALSE | Upregulated |
| | IL1F10 | | |
| 0,495456334 | IL1RAPL1 | FALSE | Upregulated |
| 1,199387236 | IL1RL1 | | Upregulated |
| | IL2 | | |
| 0 | IL20 | FALSE | Upregulated |
| 0,07115386 | IL20RA | FALSE | Upregulated |
| 1,012291541 | IL21 | | Upregulated |
| 0 | IL21-AS1 | FALSE | Upregulated |
| 1,036958357 | IL22 | | Upregulated |
| | IL22RA2 | | |
| -0,046212234 | IL23R | FALSE | Downregulated |
| 0 | IL25 | FALSE | Upregulated |
| 1,348678414 | IL26 | | Upregulated |
| 2,325681097 | IL2RA | | Upregulated |
| -0,214185237 | IL31 | FALSE | Downregulated |
| -0,191825742 | IL31RA | FALSE | Downregulated |
| | IL36A | | |
| | IL36B | | |
| -0,00897377 | IL36G | FALSE | Downregulated |
| 0 | IL36RN | FALSE | Upregulated |
| 0 | IL37 | FALSE | Upregulated |
| | IL3RA-2 | | |
| 0,669756935 | IL4 | FALSE | Upregulated |
| 0,160220985 | IL6 | FALSE | Upregulated |
| 1,951421784 | IL8 | | Upregulated |

| | | | |
|---|---|---|---|
| | IL9 | | |
| -0,617793993 | IL9R | FALSE | Downregulated |
| | IL9R-2 | | |
| -1,003159789 | IMPG2 | | Downregulated |
| 0 | INA | FALSE | Upregulated |
| 0 | INHA | FALSE | Upregulated |
| -0,178801651 | INHBA-AS1 | FALSE | Downregulated |
| 2,387103486 | INMT-FAM188B | | Upregulated |
| 0,377300841 | INPP5J | FALSE | Upregulated |
| 0,101288363 | INSC | FALSE | Upregulated |
| 2,04792071 | INSL3 | | Upregulated |
| | INSL4 | | |
| -0,127789149 | INSL5 | FALSE | Downregulated |
| 0,599121438 | INSL6 | FALSE | Upregulated |
| -0,057856841 | INSM1 | FALSE | Downregulated |
| -0,419368786 | INSRR | FALSE | Downregulated |
| 0,710810744 | INTS4L1 | FALSE | Upregulated |
| -0,643174705 | INTS4L2 | FALSE | Downregulated |
| -0,238058769 | INTS6-AS1 | FALSE | Downregulated |
| 0,109786502 | IPO11-LRRC70 | FALSE | Upregulated |
| 1,864815547 | IQCA1 | | Upregulated |
| | IQCA1P1 | | |
| | IQCF1 | | |
| | IQCF2 | | |
| | IQCF3 | | |
| | IQCF4 | | |
| | IQCF5 | | |
| 0 | IQCF5-AS1 | FALSE | Upregulated |
| | IQCF6 | | |
| | IQCJ | | |
| -0,741912774 | IQUB | FALSE | Downregulated |
| 0,021842065 | IRG1 | FALSE | Upregulated |
| 1,020397711 | IRGC | | Upregulated |
| 0,076793636 | IRGM | FALSE | Upregulated |
| 0,041547119 | IRS4 | FALSE | Upregulated |
| 0,597480774 | IRX1 | FALSE | Upregulated |
| 1,001509591 | IRX2 | | Upregulated |
| 0 | IRX4 | FALSE | Upregulated |
| 0,572261675 | IRX5 | FALSE | Upregulated |
| -0,024808033 | IRX6 | FALSE | Downregulated |
| | ISL1 | | |
| 0,762776835 | ISL2 | FALSE | Upregulated |
| -0,62340407 | ISLR2 | FALSE | Downregulated |
| 0 | ISM2 | FALSE | Upregulated |
| 0,500137898 | ISX | FALSE | Upregulated |
| -0,800391077 | ITGA2B | FALSE | Downregulated |
| -0,29047307 | ITGAE | FALSE | Downregulated |
| 1,073275444 | ITGB1BP2 | | Upregulated |
| 1,214330587 | ITGB6 | | Upregulated |
| 0,157161776 | ITLN2 | FALSE | Upregulated |
| 0,458389491 | ITPR1-AS1 | FALSE | Upregulated |

| | | | |
|---|---|---|---|
| -0,307333623 | IVL | FALSE | Downregulated |
| 0,045982731 | IZUMO1 | FALSE | Upregulated |
| | IZUMO2 | | |
| | IZUMO3 | | |
| | JAZF1-AS1 | | |
| 0,995577576 | JPH1 | FALSE | Upregulated |
| -0,775318769 | JPH4 | FALSE | Downregulated |
| | JRKL-AS1 | | |
| 1,975179991 | JSRP1 | | Upregulated |
| | KATNBL1P6 | | |
| -0,205559064 | KAZALD1 | FALSE | Downregulated |
| -0,618545266 | KC6 | FALSE | Downregulated |
| 0,597480774 | KCNA1 | FALSE | Upregulated |
| -0,235228965 | KCNA10 | FALSE | Downregulated |
| 1,521980451 | KCNA2 | | Upregulated |
| | KCNA4 | | |
| 1,982301266 | KCNA5 | | Upregulated |
| 0 | KCNAB1-AS1 | FALSE | Upregulated |
| 0 | KCNAB1-AS2 | FALSE | Upregulated |
| | KCNB2 | | |
| 0 | KCNC4-AS1 | FALSE | Upregulated |
| 0,866352401 | KCND2 | FALSE | Upregulated |
| 0,161594362 | KCND3-IT1 | FALSE | Upregulated |
| 0 | KCNE1L | FALSE | Upregulated |
| 0,866765114 | KCNE4 | FALSE | Upregulated |
| 0 | KCNF1 | FALSE | Upregulated |
| 0,558881836 | KCNG1 | FALSE | Upregulated |
| | KCNG3 | | |
| 0 | KCNG4 | FALSE | Upregulated |
| | KCNH1-IT1 | | |
| 2,325629022 | KCNH2 | | Upregulated |
| | KCNH5 | | |
| 1,657166427 | KCNIP1 | | Upregulated |
| 0 | KCNIP4-IT1 | FALSE | Upregulated |
| 0,510639616 | KCNJ11 | FALSE | Upregulated |
| -0,214683502 | KCNJ12 | FALSE | Downregulated |
| 1,11001956 | KCNJ16 | | Upregulated |
| 0,001493849 | KCNJ18 | FALSE | Upregulated |
| -0,080417528 | KCNJ2-AS1 | FALSE | Downregulated |
| 0,408086942 | KCNJ6 | FALSE | Upregulated |
| 0,449580934 | KCNK15 | FALSE | Upregulated |
| | KCNK18 | | |
| | KCNK2 | | |
| 0 | KCNK4 | FALSE | Upregulated |
| | KCNK9 | | |
| 0,471789958 | KCNMB4 | FALSE | Upregulated |
| 1,193082869 | KCNN1 | | Upregulated |
| 1,477451232 | KCNN4 | | Upregulated |
| -0,269228069 | KCNQ1DN | FALSE | Downregulated |
| | KCNQ2 | | |
| -0,32940482 | KCNQ3 | FALSE | Downregulated |

| | | | |
|---|---|---|---|
| -0,771974518 | KCNQ4 | FALSE | Downregulated |
| 0,498365761 | KCNQ5 | FALSE | Upregulated |
| 0,898106462 | KCNQ5-AS1 | FALSE | Upregulated |
| | KCNQ5-IT1 | | |
| | KCNU1 | | |
| -0,221367562 | KCNV1 | FALSE | Downregulated |
| -0,963087607 | KCP | FALSE | Downregulated |
| -0,723760239 | KCTD14 | FALSE | Downregulated |
| -0,023929158 | KCTD16 | FALSE | Downregulated |
| 0,437091584 | KCTD19 | FALSE | Upregulated |
| 0 | KCTD8 | FALSE | Upregulated |
| 1,653667319 | KEL | | Upregulated |
| | KERA | | |
| -0,569507264 | KHDC1 | FALSE | Downregulated |
| | KHDC1L | | |
| 0 | KHDC3L | FALSE | Upregulated |
| -0,314498492 | KHDRBS2 | FALSE | Downregulated |
| 0,672994949 | KIAA0101 | FALSE | Upregulated |
| | KIAA1024L | | |
| 0,082698893 | KIAA1210 | FALSE | Upregulated |
| -0,063787496 | KIAA1257 | FALSE | Downregulated |
| 2,565187358 | KIAA1377 | | Upregulated |
| 0,964531284 | KIAA1524 | FALSE | Upregulated |
| 0,484706002 | KIAA2022 | FALSE | Upregulated |
| 0,234249067 | KIF14 | FALSE | Upregulated |
| 1,08284319 | KIF15 | | Upregulated |
| 0,916081123 | KIF18A | FALSE | Upregulated |
| 2,158575591 | KIF18B | | Upregulated |
| 0,72990678 | KIF23 | FALSE | Upregulated |
| 0,221472845 | KIF25 | FALSE | Upregulated |
| 0,004929336 | KIF25-AS1 | FALSE | Upregulated |
| 0 | KIF2B | FALSE | Upregulated |
| -0,208303232 | KIF4A | FALSE | Downregulated |
| 0,070345622 | KIF4B | FALSE | Upregulated |
| 0,865809728 | KIF5A | FALSE | Upregulated |
| 0,492088552 | KIF5C | FALSE | Upregulated |
| -0,16932988 | KIR2DL1 | FALSE | Downregulated |
| 0,087576343 | KIR2DL3 | FALSE | Upregulated |
| 0 | KIR2DL4 | FALSE | Upregulated |
| 0,947886097 | KIR2DS4 | FALSE | Upregulated |
| 0,004929336 | KIR3DL1 | FALSE | Upregulated |
| 1,612670879 | KIR3DL2 | | Upregulated |
| | KIR3DL3 | | |
| 0,514938579 | KIR3DX1 | FALSE | Upregulated |
| | KIRREL2 | | |
| | KIRREL3-AS2 | | |
| 0 | KIRREL3-AS3 | FALSE | Upregulated |
| -0,845714399 | KISS1 | FALSE | Downregulated |
| 0 | KISS1R | FALSE | Upregulated |
| 0,390794663 | KL | FALSE | Upregulated |
| 0,154718451 | KLF1 | FALSE | Upregulated |

| | | | |
|---|---|---|---|
| | KLF14 | | |
| 0 | KLF17 | FALSE | Upregulated |
| 0 | KLHL1 | FALSE | Upregulated |
| 1,263563033 | KLHL14 | | Upregulated |
| 0,753067204 | KLHL30 | FALSE | Upregulated |
| -0,118183059 | KLHL34 | FALSE | Downregulated |
| 0,672371582 | KLHL38 | FALSE | Upregulated |
| 0,375494385 | KLHL4 | FALSE | Upregulated |
| 0,575422489 | KLHL40 | FALSE | Upregulated |
| -0,014740827 | KLHL7-AS1 | FALSE | Downregulated |
| 0,034128514 | KLK1 | FALSE | Upregulated |
| 0,057957529 | KLK10 | FALSE | Upregulated |
| 0,701259204 | KLK11 | FALSE | Upregulated |
| 0,476348149 | KLK12 | FALSE | Upregulated |
| | KLK13 | | |
| 0,292335845 | KLK14 | FALSE | Upregulated |
| | KLK15 | | |
| | KLK2 | | |
| | KLK3 | | |
| | KLK4 | | |
| 0,842847381 | KLK5 | FALSE | Upregulated |
| | KLK6 | | |
| | KLK7 | | |
| 0 | KLK8 | FALSE | Upregulated |
| | KLK9 | | |
| | KLKP1 | | |
| 0,585262989 | KLRAP1 | FALSE | Upregulated |
| -0,289587309 | KLRB1 | FALSE | Downregulated |
| 0,721590403 | KLRC1 | FALSE | Upregulated |
| 0,215204501 | KLRC2 | FALSE | Upregulated |
| -0,226546389 | KLRC3 | FALSE | Downregulated |
| -0,352135239 | KLRC4 | FALSE | Downregulated |
| 1,217700561 | KLRC4-KLRK1 | | Upregulated |
| -0,56972221 | KLRF1 | FALSE | Downregulated |
| | KLRF2 | | |
| 2,282060437 | KLRG1 | | Upregulated |
| -0,940803784 | KLRG2 | FALSE | Downregulated |
| | KNCN | | |
| 0 | KPRP | FALSE | Upregulated |
| 0,669756935 | KRBOX1 | FALSE | Upregulated |
| 1,165395542 | KRT12 | | Upregulated |
| | KRT13 | | |
| 1,260164307 | KRT15 | | Upregulated |
| 0,95989879 | KRT16 | FALSE | Upregulated |
| | KRT16P1 | | |
| 0 | KRT16P2 | FALSE | Upregulated |
| | KRT16P3 | | |
| 0 | KRT18P55 | FALSE | Upregulated |
| -0,394589878 | KRT20 | FALSE | Downregulated |
| | KRT24 | | |
| | KRT25 | | |

| | | | |
|---|---|---|---|
| | KRT26 | | |
| 0 | KRT27 | FALSE | Upregulated |
| | KRT28 | | |
| | KRT3 | | |
| 0 | KRT31 | FALSE | Upregulated |
| 0 | KRT32 | FALSE | Upregulated |
| | KRT33A | | |
| | KRT33B | | |
| | KRT34 | | |
| 1,079291341 | KRT35 | | Upregulated |
| 0 | KRT36 | FALSE | Upregulated |
| 0 | KRT37 | FALSE | Upregulated |
| 0,49536133 | KRT38 | FALSE | Upregulated |
| -0,255418185 | KRT39 | FALSE | Downregulated |
| 0,001493849 | KRT4 | FALSE | Upregulated |
| | KRT40 | | |
| | KRT42P | | |
| 0,565446491 | KRT6B | FALSE | Upregulated |
| 0 | KRT6C | FALSE | Upregulated |
| 0 | KRT71 | FALSE | Upregulated |
| | KRT72 | | |
| 0,779066345 | KRT73 | FALSE | Upregulated |
| 1,529401634 | KRT73-AS1 | | Upregulated |
| | KRT74 | | |
| | KRT75 | | |
| | KRT76 | | |
| 1,266125084 | KRT77 | | Upregulated |
| 0 | KRT78 | FALSE | Upregulated |
| | KRT79 | | |
| 1,852567331 | KRT80 | | Upregulated |
| 2,13567075 | KRT81 | | Upregulated |
| 0 | KRT82 | FALSE | Upregulated |
| 0 | KRT83 | FALSE | Upregulated |
| | KRT84 | | |
| 0,771743768 | KRT85 | FALSE | Upregulated |
| 0,385198445 | KRT86 | FALSE | Upregulated |
| | KRT9 | | |
| | KRTAP1-1 | | |
| | KRTAP1-3 | | |
| | KRTAP1-4 | | |
| 0,661206439 | KRTAP1-5 | FALSE | Upregulated |
| | KRTAP10-1 | | |
| 0 | KRTAP10-10 | FALSE | Upregulated |
| -0,142652043 | KRTAP10-11 | FALSE | Downregulated |
| 0 | KRTAP10-12 | FALSE | Upregulated |
| 0,103096489 | KRTAP10-2 | FALSE | Upregulated |
| | KRTAP10-3 | | |
| -0,299463865 | KRTAP10-4 | FALSE | Downregulated |
| | KRTAP10-5 | | |
| -0,206384657 | KRTAP10-6 | FALSE | Downregulated |
| 0 | KRTAP10-7 | FALSE | Upregulated |

| | | | |
|---|---|---|---|
| 0 | KRTAP10-8 | FALSE | Upregulated |
| -0,237008546 | KRTAP10-9 | FALSE | Downregulated |
| | KRTAP11-1 | | |
| | KRTAP12-1 | | |
| | KRTAP12-2 | | |
| | KRTAP12-3 | | |
| | KRTAP12-4 | | |
| | KRTAP13-1 | | |
| | KRTAP13-2 | | |
| | KRTAP13-3 | | |
| | KRTAP13-4 | | |
| | KRTAP15-1 | | |
| | KRTAP16-1 | | |
| | KRTAP17-1 | | |
| | KRTAP19-1 | | |
| | KRTAP19-2 | | |
| | KRTAP19-3 | | |
| | KRTAP19-4 | | |
| | KRTAP19-5 | | |
| | KRTAP19-6 | | |
| | KRTAP19-7 | | |
| -0,01164514 | KRTAP19-8 | FALSE | Downregulated |
| | KRTAP2-1 | | |
| | KRTAP2-2 | | |
| | KRTAP2-3 | | |
| | KRTAP2-4 | | |
| | KRTAP20-1 | | |
| | KRTAP20-2 | | |
| | KRTAP20-3 | | |
| | KRTAP20-4 | | |
| | KRTAP21-1 | | |
| | KRTAP21-2 | | |
| | KRTAP21-3 | | |
| | KRTAP22-1 | | |
| | KRTAP22-2 | | |
| | KRTAP23-1 | | |
| | KRTAP24-1 | | |
| | KRTAP25-1 | | |
| | KRTAP26-1 | | |
| | KRTAP27-1 | | |
| | KRTAP29-1 | | |
| 0 | KRTAP3-1 | FALSE | Upregulated |
| 0 | KRTAP3-2 | FALSE | Upregulated |
| 0 | KRTAP3-3 | FALSE | Upregulated |
| | KRTAP4-1 | | |
| | KRTAP4-11 | | |
| | KRTAP4-12 | | |
| | KRTAP4-2 | | |
| | KRTAP4-3 | | |
| | KRTAP4-4 | | |
| | KRTAP4-5 | | |

| | | | |
|---|---|---|---|
| | KRTAP4-6 | | |
| | KRTAP4-7 | | |
| | KRTAP4-8 | | |
| | KRTAP4-9 | | |
| 0,047090512 | KRTAP5-1 | FALSE | Upregulated |
| 1,225926768 | KRTAP5-10 | | Upregulated |
| -0,201005193 | KRTAP5-11 | FALSE | Downregulated |
| 0,779081541 | KRTAP5-2 | FALSE | Upregulated |
| | KRTAP5-3 | | |
| 0 | KRTAP5-4 | FALSE | Upregulated |
| | KRTAP5-5 | | |
| 0,042210277 | KRTAP5-6 | FALSE | Upregulated |
| 0 | KRTAP5-7 | FALSE | Upregulated |
| 0 | KRTAP5-8 | FALSE | Upregulated |
| 0,336473626 | KRTAP5-9 | FALSE | Upregulated |
| | KRTAP6-1 | | |
| | KRTAP6-2 | | |
| | KRTAP6-3 | | |
| | KRTAP7-1 | | |
| -0,03943459 | KRTAP8-1 | FALSE | Downregulated |
| | KRTAP9-1 | | |
| | KRTAP9-2 | | |
| | KRTAP9-3 | | |
| | KRTAP9-4 | | |
| | KRTAP9-6 | | |
| | KRTAP9-7 | | |
| | KRTAP9-8 | | |
| | KRTAP9-9 | | |
| 1,125597315 | KRTDAP | | Upregulated |
| 0,078181035 | KSR2 | FALSE | Upregulated |
| 0,902567389 | KTN1-AS1 | FALSE | Upregulated |
| 0,791297139 | L1CAM | FALSE | Upregulated |
| -0,43867343 | L1TD1 | FALSE | Downregulated |
| | LACRT | | |
| | LACTBL1 | | |
| -0,00897377 | LAIR2 | FALSE | Downregulated |
| | LALBA | | |
| 2,063232425 | LAMA1 | | Upregulated |
| 0,787531787 | LAMB2P1 | FALSE | Upregulated |
| -0,122571677 | LAMB4 | FALSE | Downregulated |
| 0,175098163 | LAMP3 | FALSE | Upregulated |
| 0,479664945 | LAMP5 | FALSE | Upregulated |
| 0,463187218 | LAMTOR5-AS1 | FALSE | Upregulated |
| 0,05434211 | LANCL3 | FALSE | Upregulated |
| 1,023196346 | LAYN | | Upregulated |
| 0 | LBX1 | FALSE | Upregulated |
| 0 | LBX1-AS1 | FALSE | Upregulated |
| 1,534558799 | LCA5 | | Upregulated |
| 0 | LCE1A | FALSE | Upregulated |
| | LCE1B | | |
| 0 | LCE1C | FALSE | Upregulated |

| | | | |
|---|---|---|---|
| 0 | LCE1D | FALSE | Upregulated |
| | LCE1E | | |
| | LCE1F | | |
| | LCE2A | | |
| 0 | LCE2B | FALSE | Upregulated |
| | LCE2C | | |
| 0,525497584 | LCE2D | FALSE | Upregulated |
| | LCE3A | | |
| | LCE3B | | |
| | LCE3C | | |
| | LCE3D | | |
| | LCE3E | | |
| | LCE4A | | |
| 0 | LCE5A | FALSE | Upregulated |
| | LCE6A | | |
| | LCN1 | | |
| 0,688342387 | LCN10 | FALSE | Upregulated |
| 0,75091619 | LCN15 | FALSE | Upregulated |
| 1,298759828 | LCN2 | | Upregulated |
| 0 | LCN6 | FALSE | Upregulated |
| | LCN8 | | |
| | LCN9 | | |
| 1,118924113 | LCNL1 | | Upregulated |
| 0,179558027 | LDHC | FALSE | Upregulated |
| 1,096512747 | LDLRAD1 | | Upregulated |
| -0,135286668 | LDOC1 | FALSE | Downregulated |
| 1,589639347 | LEF1-AS1 | | Upregulated |
| 1,162550129 | LEFTY2 | | Upregulated |
| | LELP1 | | |
| 0,062391697 | LEMD1 | FALSE | Upregulated |
| 0,669482247 | LEMD1-AS1 | FALSE | Upregulated |
| 0,001138561 | LEP | FALSE | Upregulated |
| | LEUTX | | |
| 0 | LGALS12 | FALSE | Upregulated |
| | LGALS13 | | |
| | LGALS14 | | |
| | LGALS16 | | |
| | LGALS17A | | |
| 0,089820997 | LGALS2 | FALSE | Upregulated |
| 0 | LGALS7 | FALSE | Upregulated |
| 0 | LGALS7B | FALSE | Upregulated |
| -0,213420405 | LGALS9B | FALSE | Downregulated |
| 0,058843558 | LGALS9C | FALSE | Upregulated |
| -0,128674165 | LGI1 | FALSE | Downregulated |
| -0,412925108 | LGI3 | FALSE | Downregulated |
| 0,705548428 | LHB | FALSE | Upregulated |
| 0,027563344 | LHFPL1 | FALSE | Upregulated |
| 0,688847322 | LHFPL3 | FALSE | Upregulated |
| | LHFPL3-AS1 | | |
| 0,515415467 | LHFPL3-AS2 | FALSE | Upregulated |
| 0,087836139 | LHFPL5 | FALSE | Upregulated |

| | | | |
|---|---|---|---|
| | LHX1 | | |
| 0 | LHX3 | FALSE | Upregulated |
| -0,122885028 | LHX5 | FALSE | Downregulated |
| | LHX8 | | |
| 0,689493386 | LILRP2 | FALSE | Upregulated |
| 1,451102212 | LIM2 | | Upregulated |
| 0 | LIN28B | FALSE | Upregulated |
| 0,604581333 | LIN7B | FALSE | Upregulated |
| 0,109684913 | LINC-ROR | FALSE | Upregulated |
| | LINC00028 | | |
| | LINC00029 | | |
| 0 | LINC00032 | FALSE | Upregulated |
| 1,239330991 | LINC00051 | | Upregulated |
| 0 | LINC00052 | FALSE | Upregulated |
| 0,669756935 | LINC00083 | FALSE | Upregulated |
| -0,580673662 | LINC00086 | FALSE | Downregulated |
| | LINC00102-2 | | |
| | LINC00111 | | |
| 0,060169122 | LINC00112 | FALSE | Upregulated |
| 0,681690965 | LINC00113 | FALSE | Upregulated |
| | LINC00114 | | |
| 0,669756935 | LINC00158 | FALSE | Upregulated |
| | LINC00159 | | |
| | LINC00160 | | |
| 0 | LINC00162 | FALSE | Upregulated |
| 1,266732075 | LINC00163 | | Upregulated |
| 0,830430891 | LINC00173 | FALSE | Upregulated |
| 0 | LINC00184 | FALSE | Upregulated |
| -0,13473005 | LINC00189 | FALSE | Downregulated |
| -0,004298224 | LINC00200 | FALSE | Downregulated |
| 0,03967952 | LINC00202-2 | FALSE | Upregulated |
| | LINC00207 | | |
| 0,48896978 | LINC00208 | FALSE | Upregulated |
| | LINC00210 | | |
| -0,092931712 | LINC00222 | FALSE | Downregulated |
| | LINC00226 | | |
| | LINC00229 | | |
| 1,029175656 | LINC00235 | | Upregulated |
| 0,405670017 | LINC00239 | FALSE | Upregulated |
| -0,059611036 | LINC00240 | FALSE | Downregulated |
| 0,489921649 | LINC00243 | FALSE | Upregulated |
| 0,076856326 | LINC00251 | FALSE | Upregulated |
| 0,063001565 | LINC00254 | FALSE | Upregulated |
| 0,30644269 | LINC00260 | FALSE | Upregulated |
| -0,086594713 | LINC00264 | FALSE | Downregulated |
| -0,267171915 | LINC00266-1 | FALSE | Downregulated |
| 0 | LINC00266-3 | FALSE | Upregulated |
| | LINC00269 | | |
| 0,33662206 | LINC00271 | FALSE | Upregulated |
| 0 | LINC00272 | FALSE | Upregulated |
| | LINC00273 | | |

| | | | |
|---|---|---|---|
| | LINC00276 | | |
| 0 | LINC00277 | FALSE | Upregulated |
| 1,286913344 | LINC00278 | | Upregulated |
| | LINC00284 | | |
| | LINC00290 | | |
| | LINC00293 | | |
| 0 | LINC00297 | FALSE | Upregulated |
| 0 | LINC00298 | FALSE | Upregulated |
| -0,311793195 | LINC00299 | FALSE | Downregulated |
| | LINC00301 | | |
| 0 | LINC00303 | FALSE | Upregulated |
| | LINC00304 | | |
| 0 | LINC00305 | FALSE | Upregulated |
| | LINC00307 | | |
| | LINC00308 | | |
| 0,756343669 | LINC00309 | FALSE | Upregulated |
| 0,280016495 | LINC00310 | FALSE | Upregulated |
| 0,499705214 | LINC00312 | FALSE | Upregulated |
| 0,63706753 | LINC00313 | FALSE | Upregulated |
| -0,021556756 | LINC00316 | FALSE | Downregulated |
| | LINC00317 | | |
| | LINC00320 | | |
| | LINC00322 | | |
| | LINC00323 | | |
| | LINC00326 | | |
| 0 | LINC00327 | FALSE | Upregulated |
| -0,00897377 | LINC00330 | FALSE | Downregulated |
| | LINC00331 | | |
| | LINC00333 | | |
| 0 | LINC00336 | FALSE | Upregulated |
| -0,660736953 | LINC00337 | FALSE | Downregulated |
| | LINC00343 | | |
| | LINC00344 | | |
| 1,125283805 | LINC00347 | | Upregulated |
| | LINC00348 | | |
| | LINC00351 | | |
| | LINC00358 | | |
| | LINC00359 | | |
| 0,034943632 | LINC00366 | FALSE | Upregulated |
| | LINC00367 | | |
| -0,00897377 | LINC00371 | FALSE | Downregulated |
| | LINC00378 | | |
| | LINC00379 | | |
| | LINC00380 | | |
| | LINC00381 | | |
| 0,739583884 | LINC00384 | FALSE | Upregulated |
| | LINC00395 | | |
| | LINC00397 | | |
| 0,530061296 | LINC00398 | FALSE | Upregulated |
| | LINC00403 | | |
| | LINC00408 | | |

| | | | |
|---|---|---|---|
| 0 | LINC00410 | FALSE | Upregulated |
| | LINC00417 | | |
| | LINC00421 | | |
| 0,019985931 | LINC00423 | FALSE | Upregulated |
| | LINC00424 | | |
| -0,486798641 | LINC00426 | FALSE | Downregulated |
| | LINC00433 | | |
| 0,567031158 | LINC00441 | FALSE | Upregulated |
| | LINC00442 | | |
| | LINC00443 | | |
| | LINC00445 | | |
| | LINC00446 | | |
| 0 | LINC00452 | FALSE | Upregulated |
| | LINC00454 | | |
| 0 | LINC00457 | FALSE | Upregulated |
| | LINC00458 | | |
| | LINC00460 | | |
| | LINC00461 | | |
| 0 | LINC00462 | FALSE | Upregulated |
| | LINC00466 | | |
| | LINC00469 | | |
| 1,15793261 | LINC00470 | | Upregulated |
| -0,370614672 | LINC00471 | FALSE | Downregulated |
| | LINC00473 | | |
| | LINC00474 | | |
| -0,059179219 | LINC00475 | FALSE | Downregulated |
| 0,518396015 | LINC00477 | FALSE | Upregulated |
| -0,035041161 | LINC00479 | FALSE | Downregulated |
| 0,579440397 | LINC00483 | FALSE | Upregulated |
| 0 | LINC00486 | FALSE | Upregulated |
| -0,258085557 | LINC00487 | FALSE | Downregulated |
| | LINC00488 | | |
| | LINC00489 | | |
| | LINC00491 | | |
| | LINC00492 | | |
| -0,176755271 | LINC00494 | FALSE | Downregulated |
| | LINC00499 | | |
| -0,173959778 | LINC00501 | FALSE | Downregulated |
| | LINC00502 | | |
| | LINC00507 | | |
| | LINC00511 | | |
| 0,809452915 | LINC00515 | FALSE | Upregulated |
| -0,091034966 | LINC00518 | FALSE | Downregulated |
| | LINC00521 | | |
| 0 | LINC00523 | FALSE | Upregulated |
| 0 | LINC00534 | FALSE | Upregulated |
| | LINC00536 | | |
| 0,512271162 | LINC00540 | FALSE | Upregulated |
| | LINC00544 | | |
| | LINC00545 | | |
| -0,070458119 | LINC00547 | FALSE | Downregulated |

| | | | |
|---|---|---|---|
| 0,332168243 | LINC00548 | FALSE | Upregulated |
| | LINC00550 | | |
| | LINC00551 | | |
| 0,192264829 | LINC00552 | FALSE | Upregulated |
| | LINC00554 | | |
| | LINC00558 | | |
| 0,924425046 | LINC00559 | FALSE | Upregulated |
| | LINC00560 | | |
| 1,150742307 | LINC00565 | | Upregulated |
| -0,007021186 | LINC00568 | FALSE | Downregulated |
| 0 | LINC00570 | FALSE | Upregulated |
| 0,308658113 | LINC00571 | FALSE | Upregulated |
| -0,25941793 | LINC00572 | FALSE | Downregulated |
| | LINC00575 | | |
| | LINC00577 | | |
| | LINC00578 | | |
| | LINC00581 | | |
| | LINC00583 | | |
| | LINC00587 | | |
| | LINC00588 | | |
| | LINC00589 | | |
| | LINC00592 | | |
| 0 | LINC00593 | FALSE | Upregulated |
| 0,004929336 | LINC00595 | FALSE | Upregulated |
| -0,131429165 | LINC00597 | FALSE | Downregulated |
| | LINC00599 | | |
| 0,034943632 | LINC00601 | FALSE | Upregulated |
| 2,635027197 | LINC00602 | | Upregulated |
| | LINC00603 | | |
| 0,11862495 | LINC00605 | FALSE | Upregulated |
| | LINC00606 | | |
| 2,2655777 | LINC00607 | | Upregulated |
| | LINC00608 | | |
| | LINC00609 | | |
| | LINC00613 | | |
| | LINC00615 | | |
| | LINC00616 | | |
| 0 | LINC00617 | FALSE | Upregulated |
| | LINC00618 | | |
| | LINC00619 | | |
| | LINC00620 | | |
| -0,960251127 | LINC00622 | FALSE | Downregulated |
| 0,411293129 | LINC00623-2 | FALSE | Upregulated |
| 0,096206777 | LINC00624 | FALSE | Upregulated |
| 0,023115621 | LINC00626 | FALSE | Upregulated |
| 0,020294071 | LINC00628 | FALSE | Upregulated |
| 0 | LINC00629 | FALSE | Upregulated |
| 0 | LINC00632 | FALSE | Upregulated |
| 0 | LINC00633 | FALSE | Upregulated |
| | LINC00634 | | |
| 0 | LINC00635 | FALSE | Upregulated |

| | | | |
|---|---|---|---|
| | LINC00636 | | |
| | LINC00640 | | |
| | LINC00642 | | |
| 0,049000194 | LINC00643 | FALSE | Upregulated |
| | LINC00644 | | |
| 0,49826773 | LINC00645 | FALSE | Upregulated |
| 0,001493849 | LINC00648 | FALSE | Upregulated |
| 0,09299022 | LINC00656 | FALSE | Upregulated |
| -0,017737585 | LINC00658 | FALSE | Downregulated |
| 0,081723142 | LINC00661 | FALSE | Upregulated |
| -0,535475928 | LINC00664 | FALSE | Downregulated |
| | LINC00668 | | |
| | LINC00669 | | |
| 0 | LINC00670 | FALSE | Upregulated |
| 0,116895171 | LINC00672 | FALSE | Upregulated |
| 1,09327279 | LINC00673 | | Upregulated |
| 0,17763398 | LINC00676 | FALSE | Upregulated |
| | LINC00681 | | |
| | LINC00682 | | |
| 0 | LINC00684 | FALSE | Upregulated |
| | LINC00684-2 | | |
| | LINC00685-2 | | |
| | LINC00687 | | |
| -0,112244938 | LINC00689 | FALSE | Downregulated |
| 0 | LINC00690 | FALSE | Upregulated |
| | LINC00692 | | |
| -0,999600712 | LINC00693 | FALSE | Downregulated |
| -0,076931481 | LINC00696 | FALSE | Downregulated |
| | LINC00698 | | |
| -0,00897377 | LINC00700 | FALSE | Downregulated |
| | LINC00701 | | |
| 0 | LINC00702 | FALSE | Upregulated |
| | LINC00703 | | |
| 0,069215674 | LINC00704 | FALSE | Upregulated |
| 0,440356339 | LINC00706 | FALSE | Upregulated |
| 0 | LINC00708 | FALSE | Upregulated |
| | LINC00709 | | |
| 1,074951073 | LINC00710 | | Upregulated |
| | LINC00836 | | |
| -0,134571071 | LINC00837 | FALSE | Downregulated |
| -0,019429105 | LINC00838 | FALSE | Downregulated |
| 0,894273472 | LINC00839 | FALSE | Upregulated |
| | LINC00840 | | |
| | LINC00841 | | |
| 0,65484825 | LINC00842 | FALSE | Upregulated |
| | LINC00845 | | |
| | LINC00850 | | |
| -0,002529603 | LINC00851 | FALSE | Downregulated |
| 0 | LINC00853 | FALSE | Upregulated |
| | LINC00856 | | |
| 1,327340987 | LINC00857 | | Upregulated |

| | | | |
|---|---|---|---|
| | LINC00858 | | |
| -0,238784963 | LINC00862 | FALSE | Downregulated |
| 0,588777084 | LINC00864 | FALSE | Upregulated |
| 0 | LINC00866 | FALSE | Upregulated |
| 0 | LINC00867 | FALSE | Upregulated |
| | LINC00871 | | |
| 0,51227241 | LINC00877 | FALSE | Upregulated |
| | LINC00879 | | |
| -0,189001521 | LINC00882 | FALSE | Downregulated |
| -0,334746595 | LINC00887 | FALSE | Downregulated |
| 0 | LINC00889 | FALSE | Upregulated |
| -0,016805483 | LINC00891 | FALSE | Downregulated |
| 0,941378306 | LINC00892 | FALSE | Upregulated |
| | LINC00895 | | |
| | LINC00898 | | |
| 0,570166837 | LINC00901 | FALSE | Upregulated |
| | LINC00904 | | |
| | LINC00905 | | |
| 0,754656326 | LINC00906 | FALSE | Upregulated |
| -0,00995385 | LINC00908 | FALSE | Downregulated |
| | LINC00911 | | |
| | LINC00917 | | |
| | LINC00919 | | |
| 0,425080818 | LINC00920 | FALSE | Upregulated |
| | LINC00922 | | |
| -0,230808275 | LINC00923 | FALSE | Downregulated |
| 1,451078487 | LINC00924 | | Upregulated |
| -0,267874129 | LINC00925 | FALSE | Downregulated |
| 1,317898665 | LINC00926 | | Upregulated |
| | LINC00927 | | |
| 0,553763092 | LINC00928 | FALSE | Upregulated |
| | LINC00929 | | |
| -0,334437769 | LINC00930 | FALSE | Downregulated |
| 0 | LINC00934 | FALSE | Upregulated |
| 0,442524855 | LINC00935 | FALSE | Upregulated |
| 1,051067331 | LINC00937 | | Upregulated |
| 0,035722597 | LINC00940 | FALSE | Upregulated |
| 0,355956501 | LINC00942 | FALSE | Upregulated |
| | LINC00944 | | |
| 0,512359721 | LINC00945 | FALSE | Upregulated |
| -0,00897377 | LINC00948 | FALSE | Downregulated |
| | LINC00951 | | |
| | LINC00955 | | |
| 0,678429153 | LINC00958 | FALSE | Upregulated |
| 0,039032745 | LINC00964 | FALSE | Upregulated |
| -0,301096306 | LINC00965 | FALSE | Downregulated |
| 0 | LINC00968 | FALSE | Upregulated |
| -1,513924346 | LINC00969 | | Downregulated |
| 0 | LINC00970 | FALSE | Upregulated |
| 0,694424628 | LINC00971 | FALSE | Upregulated |
| | LINC00974 | | |

| | | | |
|---|---|---|---|
| 0,580472435 | LINC00977 | FALSE | Upregulated |
| -0,344336704 | LINC00982 | FALSE | Downregulated |
| 0 | LINC00989 | FALSE | Upregulated |
| | LINC00992 | | |
| | LINC00993 | | |
| | LINC00994 | | |
| 0,044955958 | LINC00996 | FALSE | Upregulated |
| -0,425100531 | LINC01001 | FALSE | Downregulated |
| 0 | LINC01002 | FALSE | Upregulated |
| | LINC01005 | | |
| 0,074493693 | LINC01006 | FALSE | Upregulated |
| 0,386556914 | LINC01007 | FALSE | Upregulated |
| 1,211402421 | LINC01012 | | Upregulated |
| | LINC01013 | | |
| 0,073122974 | LINC01014 | FALSE | Upregulated |
| | LINC01015 | | |
| 0,098245791 | LINC01016 | FALSE | Upregulated |
| | LINC01019 | | |
| | LINC01020 | | |
| 0 | LINC01021 | FALSE | Upregulated |
| -1,034489824 | LINC01023 | | Downregulated |
| 0 | LINC01028 | FALSE | Upregulated |
| | LINC01029 | | |
| -0,330852343 | LINC01030 | FALSE | Downregulated |
| | LINC01056 | | |
| 1,601877959 | LINC01057 | | Upregulated |
| | LINC01060 | | |
| -0,00897377 | LINC01063 | FALSE | Downregulated |
| 0,117080686 | LINC01069 | FALSE | Upregulated |
| 1,783866953 | LINC01070 | | Upregulated |
| 0,336580269 | LINC01080 | FALSE | Upregulated |
| | LINC01081 | | |
| 0,020913352 | LINC01082 | FALSE | Upregulated |
| | LINC01085 | | |
| 1,054457632 | LINC01087 | | Upregulated |
| 0 | LINC01088 | FALSE | Upregulated |
| -0,058549357 | LINC01091 | FALSE | Downregulated |
| 1,129652647 | LINC01094 | | Upregulated |
| | LINC01095 | | |
| 0,608246941 | LINC01096 | FALSE | Upregulated |
| | LINC01097 | | |
| 0,027563344 | LINC01098 | FALSE | Upregulated |
| 1,168459888 | LINC01099 | | Upregulated |
| | LINC01100 | | |
| 0,554942727 | LINC01101 | FALSE | Upregulated |
| 0 | LINC01102 | FALSE | Upregulated |
| | LINC01103 | | |
| 0 | LINC01104 | FALSE | Upregulated |
| | LINC01105 | | |
| | LINC01111 | | |
| | LINC01114 | | |

| | | | |
|---|---|---|---|
| 0 | LINC01115 | FALSE | Upregulated |
| 0 | LINC01116 | FALSE | Upregulated |
| 0,013318773 | LINC01118 | FALSE | Upregulated |
| 0,455946406 | LINC01119 | FALSE | Upregulated |
| | LINC01120 | | |
| 0,051489162 | LINC01121 | FALSE | Upregulated |
| | LINC01122 | | |
| 1,48934264 | LINC01125 | | Upregulated |
| -0,388498699 | LINC01132 | FALSE | Downregulated |
| 1,380730452 | LINC01134 | | Upregulated |
| -0,087066628 | LINC01135 | FALSE | Downregulated |
| | LINC01136 | | |
| 0 | LINC01139 | FALSE | Upregulated |
| 0,606834907 | LINC01140 | FALSE | Upregulated |
| -0,00897377 | LINC01141 | FALSE | Downregulated |
| | LINC01142 | | |
| 0 | LINC01149 | FALSE | Upregulated |
| | LINC01150 | | |
| 0,578963231 | LINC01153 | FALSE | Upregulated |
| | LINC01158 | | |
| 0,573283345 | LINC01160 | FALSE | Upregulated |
| | LINC01163 | | |
| | LINC01164 | | |
| 1,444798409 | LINC01165 | | Upregulated |
| 0,027563344 | LINC01171 | FALSE | Upregulated |
| 0,197929257 | LINC01176 | FALSE | Upregulated |
| -0,001452899 | LINC01179 | FALSE | Downregulated |
| | LINC01180 | | |
| | LINC01182 | | |
| -0,572314304 | LINC01183 | FALSE | Downregulated |
| 0 | LINC01185 | FALSE | Upregulated |
| 0 | LINC01186 | FALSE | Upregulated |
| 0,073148769 | LINC01187 | FALSE | Upregulated |
| 1,462575403 | LINC01189 | | Upregulated |
| 0,16381368 | LINC01191 | FALSE | Upregulated |
| | LINC01192 | | |
| 0 | LINC01193 | FALSE | Upregulated |
| 0 | LINC01194 | FALSE | Upregulated |
| 0,154154637 | LINC01197 | FALSE | Upregulated |
| | LINC01216 | | |
| 0,799918842 | LINGO2 | FALSE | Upregulated |
| | LIPF | | |
| 2,91990302 | LIPI | | Upregulated |
| -0,745488933 | LIPJ | FALSE | Downregulated |
| | LIPK | | |
| 0,436158848 | LIPM | FALSE | Upregulated |
| 0,498756945 | LIPN | FALSE | Upregulated |
| | LL22NC01-81G9.3 | | |
| | LMAN1L | | |
| 0,781562824 | LMCD1-AS1 | FALSE | Upregulated |
| 0 | LMO1 | FALSE | Upregulated |

| | | | |
|---|---|---|---|
| 0 | LMX1A | FALSE | Upregulated |
| 0,460138338 | LMX1B | FALSE | Upregulated |
| | LOC100125918 | | |
| -0,077204098 | LOC100126447 | FALSE | Downregulated |
| -0,254448056 | LOC100127974 | FALSE | Downregulated |
| 0,179730424 | LOC100128002 | FALSE | Upregulated |
| 0 | LOC100128076 | FALSE | Upregulated |
| 0 | LOC100128108 | FALSE | Upregulated |
| -0,02993411 | LOC100128127 | FALSE | Downregulated |
| | LOC100128139 | | |
| 1,236843314 | LOC100128164 | | Upregulated |
| | LOC100128233 | | |
| 0,60253556 | LOC100128288 | FALSE | Upregulated |
| 0,782505214 | LOC100128325 | FALSE | Upregulated |
| 0,502839747 | LOC100128560 | FALSE | Upregulated |
| 0,54867459 | LOC100128568 | FALSE | Upregulated |
| | LOC100128575 | | |
| 0,698582719 | LOC100128593 | FALSE | Upregulated |
| 0,004929336 | LOC100128885 | FALSE | Upregulated |
| 0 | LOC100128905 | FALSE | Upregulated |
| -0,731073832 | LOC100128908 | FALSE | Downregulated |
| 1,656154903 | LOC100128966 | | Upregulated |
| 0,566917048 | LOC100129055 | FALSE | Upregulated |
| -0,290544529 | LOC100129098 | FALSE | Downregulated |
| 0,30753392 | LOC100129138 | FALSE | Upregulated |
| 0 | LOC100129216 | FALSE | Upregulated |
| 0 | LOC100129223 | FALSE | Upregulated |
| 0,465919451 | LOC100129307 | FALSE | Upregulated |
| 0,152098595 | LOC100129316 | FALSE | Upregulated |
| -0,00897377 | LOC100129455 | FALSE | Downregulated |
| 0,829354007 | LOC100129461 | FALSE | Upregulated |
| -0,305026812 | LOC100129476 | FALSE | Downregulated |
| 0,290646485 | LOC100129516 | FALSE | Upregulated |
| | LOC100129520 | | |
| | LOC100129603 | | |
| | LOC100129636 | | |
| -0,448833363 | LOC100129781 | FALSE | Downregulated |
| 0 | LOC100129860 | FALSE | Upregulated |
| 0,051098818 | LOC100129924 | FALSE | Upregulated |
| 0,019985931 | LOC100129935 | FALSE | Upregulated |
| 0 | LOC100130207 | FALSE | Upregulated |
| 0,871954775 | LOC100130298 | FALSE | Upregulated |
| | LOC100130301 | | |
| 0 | LOC100130417 | FALSE | Upregulated |
| 0,087836139 | LOC100130452 | FALSE | Upregulated |
| 0,889618182 | LOC100130458 | FALSE | Upregulated |
| 1,620923534 | LOC100130502 | | Upregulated |
| 0,053948761 | LOC100130520 | FALSE | Upregulated |
| 0,077371757 | LOC100130673 | FALSE | Upregulated |
| 0,338575409 | LOC100130698 | FALSE | Upregulated |
| | LOC100130849 | | |

| | | | |
|---|---|---|---|
| | LOC100130938 | | |
| | LOC100130964 | | |
| 0 | LOC100131138 | FALSE | Upregulated |
| 1,083050648 | LOC100131289 | | Upregulated |
| 0,601256507 | LOC100131315 | FALSE | Upregulated |
| 0 | LOC100131372 | FALSE | Upregulated |
| 0,468573696 | LOC100131472 | FALSE | Upregulated |
| 1,236721247 | LOC100131530 | | Upregulated |
| -1,24708954 | LOC100131532 | | Downregulated |
| | LOC100131635 | | |
| -0,456438547 | LOC100131655 | FALSE | Downregulated |
| 0 | LOC100132004 | FALSE | Upregulated |
| | LOC100132146 | | |
| 0,587455968 | LOC100132154 | FALSE | Upregulated |
| | LOC100132174 | | |
| | LOC100132202 | | |
| 0 | LOC100132354 | FALSE | Upregulated |
| -0,169899365 | LOC100132735 | FALSE | Downregulated |
| -0,016805483 | LOC100132741 | FALSE | Downregulated |
| | LOC100132831 | | |
| 0 | LOC100132874 | FALSE | Upregulated |
| 1,633261141 | LOC100133050 | | Upregulated |
| | LOC100133267 | | |
| -0,110187422 | LOC100133315 | FALSE | Downregulated |
| -0,186598739 | LOC100133669 | FALSE | Downregulated |
| 0,019627125 | LOC100133746 | FALSE | Upregulated |
| -0,754493998 | LOC100133985 | FALSE | Downregulated |
| 0,025268453 | LOC100134317 | FALSE | Upregulated |
| 0,273668423 | LOC100134368 | FALSE | Upregulated |
| -0,169974652 | LOC100134391 | FALSE | Downregulated |
| 1,867027817 | LOC100134868 | | Upregulated |
| -0,605298441 | LOC100188947 | FALSE | Downregulated |
| | LOC100240728 | | |
| 0 | LOC100240734 | FALSE | Upregulated |
| 0,091218734 | LOC100240735 | FALSE | Upregulated |
| -0,247431216 | LOC100270804 | FALSE | Downregulated |
| | LOC100286906 | | |
| -0,284336258 | LOC100287010 | FALSE | Downregulated |
| | LOC100287072 | | |
| 1,104428836 | LOC100287082 | | Upregulated |
| 1,477290535 | LOC100287098 | | Upregulated |
| | LOC100287294 | | |
| -0,098854098 | LOC100287387 | FALSE | Downregulated |
| 0 | LOC100287413 | FALSE | Upregulated |
| | LOC100287592 | | |
| 0,001493849 | LOC100287610 | FALSE | Upregulated |
| 0,067880217 | LOC100287651 | FALSE | Upregulated |
| | LOC100287704 | | |
| | LOC100287728 | | |
| | LOC100287823 | | |
| | LOC100287834 | | |

| | | | |
|---|---|---|---|
| | LOC100287877 | | |
| -0,253530659 | LOC100287944 | FALSE | Downregulated |
| 0 | LOC100288562 | FALSE | Upregulated |
| 0 | LOC100288570 | FALSE | Upregulated |
| 0,080504022 | LOC100288637 | FALSE | Upregulated |
| 0,579745677 | LOC100288748 | FALSE | Upregulated |
| 0,339388954 | LOC100288814 | FALSE | Upregulated |
| 0,141171812 | LOC100288842 | FALSE | Upregulated |
| 0,171751803 | LOC100288911 | FALSE | Upregulated |
| | LOC100288966 | | |
| -0,829144326 | LOC100288974 | FALSE | Downregulated |
| 1,823780877 | LOC100289092 | | Upregulated |
| -0,15050354 | LOC100289098 | FALSE | Downregulated |
| 0,82900717 | LOC100289333 | FALSE | Upregulated |
| 0,034943632 | LOC100289473 | FALSE | Upregulated |
| -0,810344552 | LOC100289495 | FALSE | Downregulated |
| 0 | LOC100289656 | FALSE | Upregulated |
| -0,13497837 | LOC100289673 | FALSE | Downregulated |
| | LOC100293612 | | |
| | LOC100422212 | | |
| -1,153775596 | LOC100422556 | | Downregulated |
| 0,046435231 | LOC100500773 | FALSE | Upregulated |
| 0,688818519 | LOC100505498 | FALSE | Upregulated |
| 0 | LOC100505501 | FALSE | Upregulated |
| | LOC100505504 | | |
| 0 | LOC100505515 | FALSE | Upregulated |
| -0,055970583 | LOC100505534 | FALSE | Downregulated |
| -0,052200366 | LOC100505555 | FALSE | Downregulated |
| 1,214768345 | LOC100505585 | | Upregulated |
| 1,398059705 | LOC100505588 | | Upregulated |
| 1,240070276 | LOC100505609 | | Upregulated |
| 0,425301944 | LOC100505622 | FALSE | Upregulated |
| 0 | LOC100505664 | FALSE | Upregulated |
| 0 | LOC100505685 | FALSE | Upregulated |
| 0,025225456 | LOC100505710 | FALSE | Upregulated |
| | LOC100505711 | | |
| -0,078821002 | LOC100505774 | FALSE | Downregulated |
| | LOC100505784 | | |
| | LOC100505797 | | |
| 0,423271461 | LOC100505798 | FALSE | Upregulated |
| | LOC100505811 | | |
| 0,340613374 | LOC100505812 | FALSE | Upregulated |
| 1,104168012 | LOC100505817 | | Upregulated |
| 0 | LOC100505824 | FALSE | Upregulated |
| | LOC100505835 | | |
| | LOC100505841 | | |
| | LOC100505853 | | |
| | LOC100505874 | | |
| | LOC100505878 | | |
| | LOC100505887 | | |
| -2,172842233 | LOC100505902 | | Downregulated |

| | | | |
|---|---|---|---|
| -0,85920953 | LOC100505912 | FALSE | Downregulated |
| | LOC100505920 | | |
| | LOC100505940 | | |
| | LOC100505948 | | |
| | LOC100505984 | | |
| -0,537890523 | LOC100506014 | FALSE | Downregulated |
| | LOC100506016 | | |
| 1,875424426 | LOC100506022 | | Upregulated |
| 0,21998387 | LOC100506036 | FALSE | Upregulated |
| -0,373146325 | LOC100506047 | FALSE | Downregulated |
| 0,039005732 | LOC100506082 | FALSE | Upregulated |
| | LOC100506098 | | |
| | LOC100506107 | | |
| 0,353087955 | LOC100506157 | FALSE | Upregulated |
| 0 | LOC100506159 | FALSE | Upregulated |
| 0,556485273 | LOC100506188 | FALSE | Upregulated |
| -0,981258578 | LOC100506235 | FALSE | Downregulated |
| | LOC100506236 | | |
| | LOC100506253 | | |
| | LOC100506271 | | |
| | LOC100506274 | | |
| 1,624462916 | LOC100506282 | | Upregulated |
| | LOC100506289 | | |
| | LOC100506317 | | |
| 0 | LOC100506331 | FALSE | Upregulated |
| -0,450711979 | LOC100506351 | FALSE | Downregulated |
| 1,535178922 | LOC100506358 | | Upregulated |
| 0 | LOC100506371 | FALSE | Upregulated |
| | LOC100506403 | | |
| | LOC100506405 | | |
| | LOC100506406 | | |
| -0,451180989 | LOC100506411 | FALSE | Downregulated |
| 0 | LOC100506422 | FALSE | Upregulated |
| | LOC100506470 | | |
| 1,952770898 | LOC100506473 | | Upregulated |
| 0,02924872 | LOC100506474 | FALSE | Upregulated |
| -0,214820576 | LOC100506489 | FALSE | Downregulated |
| -1,231796668 | LOC100506498 | | Downregulated |
| 1,140813552 | LOC100506526 | | Upregulated |
| 0 | LOC100506530 | FALSE | Upregulated |
| | LOC100506532 | | |
| -0,034410891 | LOC100506557 | FALSE | Downregulated |
| 0 | LOC100506558 | FALSE | Upregulated |
| -0,334615786 | LOC100506563 | FALSE | Downregulated |
| | LOC100506664 | | |
| | LOC100506675 | | |
| 0,553665393 | LOC100506688 | FALSE | Upregulated |
| 1,489617394 | LOC100506691 | | Upregulated |
| 1,112847915 | LOC100506713 | | Upregulated |
| -0,51968707 | LOC100506747 | FALSE | Downregulated |
| | LOC100506790 | | |

| | | | |
|---|---|---|---|
| | LOC100506791 | | |
| 0,713088708 | LOC100506797 | FALSE | Upregulated |
| 2,671831718 | LOC100506827 | | Upregulated |
| | LOC100506851 | | |
| | LOC100506858 | | |
| 0,511722423 | LOC100506869 | FALSE | Upregulated |
| 1,161704309 | LOC100506870 | | Upregulated |
| 0 | LOC100506882 | FALSE | Upregulated |
| | LOC100506885 | | |
| 1,938146686 | LOC100506901 | | Upregulated |
| 0,085572821 | LOC100506908 | FALSE | Upregulated |
| 0 | LOC100506929 | FALSE | Upregulated |
| 0 | LOC100506937 | FALSE | Upregulated |
| | LOC100506944 | | |
| -0,216369155 | LOC100506974 | FALSE | Downregulated |
| 0 | LOC100506999 | FALSE | Upregulated |
| -0,047966471 | LOC100507054 | FALSE | Downregulated |
| -1,368335534 | LOC100507103 | | Downregulated |
| | LOC100507165 | | |
| -0,550347871 | LOC100507195 | FALSE | Downregulated |
| | LOC100507205 | | |
| | LOC100507210 | | |
| | LOC100507221 | | |
| | LOC100507236 | | |
| -0,122754744 | LOC100507277 | FALSE | Downregulated |
| 0,588428662 | LOC100507336 | FALSE | Upregulated |
| 0,517352518 | LOC100507351 | FALSE | Upregulated |
| | LOC100507353 | | |
| | LOC100507384 | | |
| 0 | LOC100507387 | FALSE | Upregulated |
| -1,646652092 | LOC100507395 | | Downregulated |
| | LOC100507403 | | |
| | LOC100507420 | | |
| | LOC100507443 | | |
| 0,240433864 | LOC100507460 | FALSE | Upregulated |
| | LOC100507461 | | |
| -0,012802884 | LOC100507462 | FALSE | Downregulated |
| | LOC100507464 | | |
| | LOC100507468 | | |
| 0 | LOC100507480 | FALSE | Upregulated |
| 1,452902547 | LOC100507487 | | Upregulated |
| -0,226597628 | LOC100507513 | FALSE | Downregulated |
| | LOC100507521 | | |
| 0,836216418 | LOC100507534 | FALSE | Upregulated |
| 0,039005732 | LOC100507548 | FALSE | Upregulated |
| 0,748484195 | LOC100507554 | FALSE | Upregulated |
| | LOC100507560 | | |
| 0,585832712 | LOC100507562 | FALSE | Upregulated |
| -0,018730409 | LOC100507600 | FALSE | Downregulated |
| 2,421315348 | LOC100507634 | | Upregulated |
| 1,761643704 | LOC100507642 | | Upregulated |

| | | | |
|---|---|---|---|
| 0,677673744 | LOC100507657 | FALSE | Upregulated |
| 0,696211899 | LOC100507747 | FALSE | Upregulated |
| 0 | LOC100508046 | FALSE | Upregulated |
| 0 | LOC100508631 | FALSE | Upregulated |
| | LOC100509303 | | |
| -0,078249579 | LOC100616530 | FALSE | Downregulated |
| 1,142771976 | LOC100652777 | | Upregulated |
| 0,2183666 | LOC100652807 | FALSE | Upregulated |
| -0,645048177 | LOC100652911 | FALSE | Downregulated |
| -0,278920569 | LOC100652930 | FALSE | Downregulated |
| | LOC100652931 | | |
| 0,293758865 | LOC100653005 | FALSE | Upregulated |
| 1,627628244 | LOC100653133 | | Upregulated |
| 0,730701628 | LOC100996246 | FALSE | Upregulated |
| | LOC100996249 | | |
| | LOC100996266 | | |
| -0,337677212 | LOC100996274 | FALSE | Downregulated |
| -0,184760187 | LOC100996288 | FALSE | Downregulated |
| | LOC100996290 | | |
| -0,16789423 | LOC100996294 | FALSE | Downregulated |
| 0 | LOC100996301 | FALSE | Upregulated |
| -0,114849297 | LOC100996316 | FALSE | Downregulated |
| -0,193002525 | LOC100996335 | FALSE | Downregulated |
| -0,383520569 | LOC100996337 | FALSE | Downregulated |
| | LOC100996338 | | |
| -0,206818001 | LOC100996342 | FALSE | Downregulated |
| 0,70739719 | LOC100996345 | FALSE | Upregulated |
| | LOC100996349 | | |
| | LOC100996365 | | |
| -1,11669738 | LOC100996369 | | Downregulated |
| -0,228700567 | LOC100996379 | FALSE | Downregulated |
| -0,133403106 | LOC100996385 | FALSE | Downregulated |
| -0,22301446 | LOC100996401 | FALSE | Downregulated |
| 1,085344132 | LOC100996404 | | Upregulated |
| | LOC100996414 | | |
| | LOC100996415 | | |
| 0,44592872 | LOC100996425 | FALSE | Upregulated |
| 0 | LOC100996428 | FALSE | Upregulated |
| 1,061641194 | LOC100996455 | | Upregulated |
| 0,937422436 | LOC100996457 | FALSE | Upregulated |
| 0,98123119 | LOC100996465 | FALSE | Upregulated |
| -0,365819332 | LOC100996478 | FALSE | Downregulated |
| 0 | LOC100996482 | FALSE | Upregulated |
| -0,153558247 | LOC100996521 | FALSE | Downregulated |
| -0,09498569 | LOC100996549 | FALSE | Downregulated |
| 0,505100383 | LOC100996557 | FALSE | Upregulated |
| 0 | LOC100996570 | FALSE | Upregulated |
| 0,712652892 | LOC100996598 | FALSE | Upregulated |
| 0,700831534 | LOC100996602 | FALSE | Upregulated |
| 0,066229525 | LOC100996624 | FALSE | Upregulated |
| | LOC100996630 | | |

| | | | |
|---|---|---|---|
| | LOC100996637 | | |
| | LOC100996645 | | |
| 0,720281875 | LOC100996660 | FALSE | Upregulated |
| -0,36137744 | LOC100996662 | FALSE | Downregulated |
| 0 | LOC100996663 | FALSE | Upregulated |
| 0,427738576 | LOC100996664 | FALSE | Upregulated |
| | LOC100996665 | | |
| 0,304426964 | LOC100996668 | FALSE | Upregulated |
| | LOC100996681 | | |
| 0,405048638 | LOC100996695 | FALSE | Upregulated |
| | LOC100996696 | | |
| 0 | LOC100996701 | FALSE | Upregulated |
| 0,841811342 | LOC100996713 | FALSE | Upregulated |
| 1,740959815 | LOC100996721 | | Upregulated |
| 0,086916024 | LOC100996732 | FALSE | Upregulated |
| 0,226969606 | LOC100996735 | FALSE | Upregulated |
| | LOC100996756 | | |
| 2,197453037 | LOC100996760 | | Upregulated |
| | LOC100996770 | | |
| 0 | LOC100996792 | FALSE | Upregulated |
| | LOC100996792-2 | | |
| 0,027563344 | LOC100996808 | FALSE | Upregulated |
| | LOC100996902 | | |
| -0,727060514 | LOC100996909 | FALSE | Downregulated |
| 0,565254063 | LOC101054525 | FALSE | Upregulated |
| | LOC101059914 | | |
| | LOC101059915 | | |
| 0,004273529 | LOC101059948 | FALSE | Upregulated |
| -0,147296433 | LOC101059954 | FALSE | Downregulated |
| | LOC101060004 | | |
| | LOC101060017 | | |
| | LOC101060024 | | |
| 0,000142015 | LOC101060026 | FALSE | Upregulated |
| 1,752647981 | LOC101060038 | | Upregulated |
| 0,94388997 | LOC101060085 | FALSE | Upregulated |
| 0,0061783 | LOC101060097 | FALSE | Upregulated |
| 0 | LOC101060106 | FALSE | Upregulated |
| | LOC101060128 | | |
| -1,046752587 | LOC101060153 | | Downregulated |
| 0 | LOC101060157 | FALSE | Upregulated |
| -0,00897377 | LOC101060183 | FALSE | Downregulated |
| | LOC101060187 | | |
| | LOC101060211 | | |
| | LOC101060233 | | |
| 1,036767546 | LOC101060236 | | Upregulated |
| | LOC101060254 | | |
| | LOC101060264 | | |
| 0 | LOC101060301 | FALSE | Upregulated |
| 1,908995442 | LOC101060321 | | Upregulated |
| 0,465085529 | LOC101060341 | FALSE | Upregulated |
| 0,597480774 | LOC101060363 | FALSE | Upregulated |

| | | | |
|---|---|---|---|
| 0,076658198 | LOC101060391 | FALSE | Upregulated |
| 0 | LOC101060400 | FALSE | Upregulated |
| 0 | LOC101060498 | FALSE | Upregulated |
| 0 | LOC101060532 | FALSE | Upregulated |
| 1,142876935 | LOC101060569 | | Upregulated |
| 0,60349869 | LOC101060626 | FALSE | Upregulated |
| 0,160025956 | LOC101409256 | FALSE | Upregulated |
| 0 | LOC101559451 | FALSE | Upregulated |
| -0,004298224 | LOC101805489 | FALSE | Downregulated |
| 0 | LOC101805491 | FALSE | Upregulated |
| | LOC101926886 | | |
| | LOC101926893 | | |
| -0,527799262 | LOC101926896 | FALSE | Downregulated |
| | LOC101926897 | | |
| 0 | LOC101926899 | FALSE | Upregulated |
| 0 | LOC101926900 | FALSE | Upregulated |
| -0,567723139 | LOC101926902 | FALSE | Downregulated |
| | LOC101926904 | | |
| -0,312059416 | LOC101926905 | FALSE | Downregulated |
| 1,164983528 | LOC101926907 | | Upregulated |
| 1,466696847 | LOC101926909 | | Upregulated |
| | LOC101926912 | | |
| 0,118494715 | LOC101926916 | FALSE | Upregulated |
| -0,172369401 | LOC101926917 | FALSE | Downregulated |
| | LOC101926921 | | |
| 0,994002939 | LOC101926922 | FALSE | Upregulated |
| | LOC101926923 | | |
| | LOC101926929 | | |
| 0,028394446 | LOC101926936 | FALSE | Upregulated |
| -0,050481942 | LOC101926937 | FALSE | Downregulated |
| 0 | LOC101926940 | FALSE | Upregulated |
| 0,919814296 | LOC101926948 | FALSE | Upregulated |
| | LOC101926951 | | |
| | LOC101926952 | | |
| | LOC101926953 | | |
| -0,878290765 | LOC101926956 | FALSE | Downregulated |
| | LOC101926958 | | |
| 0 | LOC101926959 | FALSE | Upregulated |
| -1,447100595 | LOC101926960 | | Downregulated |
| | LOC101926961 | | |
| | LOC101926965 | | |
| | LOC101926966 | | |
| 0 | LOC101926968 | FALSE | Upregulated |
| 1,878592829 | LOC101926969 | | Upregulated |
| | LOC101926970 | | |
| | LOC101926972 | | |
| | LOC101926974 | | |
| | LOC101926975 | | |
| 0,144130654 | LOC101926977 | FALSE | Upregulated |
| | LOC101926982 | | |
| | LOC101926983 | | |

| | | | |
|---|---|---|---|
| 0 | LOC101926989 | FALSE | Upregulated |
| 0 | LOC101926992 | FALSE | Upregulated |
| 1,065886102 | LOC101926997 | | Upregulated |
| | LOC101926998 | | |
| | LOC101926999 | | |
| | LOC101927008 | | |
| 0,51451481 | LOC101927010 | FALSE | Upregulated |
| | LOC101927012 | | |
| | LOC101927013 | | |
| | LOC101927016 | | |
| | LOC101927019 | | |
| | LOC101927022 | | |
| | LOC101927024 | | |
| | LOC101927025 | | |
| -0,170967497 | LOC101927026 | FALSE | Downregulated |
| -0,413887795 | LOC101927029 | FALSE | Downregulated |
| | LOC101927037 | | |
| 0,823176671 | LOC101927038 | FALSE | Upregulated |
| | LOC101927039 | | |
| | LOC101927042 | | |
| | LOC101927046 | | |
| | LOC101927050 | | |
| | LOC101927052 | | |
| 1,177231422 | LOC101927053 | | Upregulated |
| | LOC101927059 | | |
| -0,292280232 | LOC101927063 | FALSE | Downregulated |
| | LOC101927065 | | |
| 0,123760739 | LOC101927070 | FALSE | Upregulated |
| | LOC101927072 | | |
| | LOC101927080 | | |
| 0,372356382 | LOC101927088 | FALSE | Upregulated |
| | LOC101927089 | | |
| | LOC101927091 | | |
| 0,111124171 | LOC101927093 | FALSE | Upregulated |
| | LOC101927094 | | |
| 0 | LOC101927095 | FALSE | Upregulated |
| 1,957047104 | LOC101927104 | | Upregulated |
| | LOC101927108 | | |
| | LOC101927110 | | |
| 0 | LOC101927112 | FALSE | Upregulated |
| -0,849575857 | LOC101927116 | FALSE | Downregulated |
| | LOC101927118 | | |
| -0,842457615 | LOC101927121 | FALSE | Downregulated |
| 1,371488726 | LOC101927122 | | Upregulated |
| -0,303116283 | LOC101927126 | FALSE | Downregulated |
| | LOC101927127 | | |
| | LOC101927129 | | |
| | LOC101927133 | | |
| 0 | LOC101927135 | FALSE | Upregulated |
| | LOC101927137 | | |
| | LOC101927141 | | |

| | | | |
|---|---|---|---|
| -0,168204558 | LOC101927144 | FALSE | Downregulated |
| | LOC101927145 | | |
| | LOC101927147 | | |
| | LOC101927153 | | |
| | LOC101927154 | | |
| 0,588235444 | LOC101927159 | FALSE | Upregulated |
| 0 | LOC101927165 | FALSE | Upregulated |
| 1,382994698 | LOC101927166 | | Upregulated |
| 0 | LOC101927168 | FALSE | Upregulated |
| -0,00897377 | LOC101927169 | FALSE | Downregulated |
| -1,034995805 | LOC101927170 | | Downregulated |
| 0,034128514 | LOC101927172 | FALSE | Upregulated |
| | LOC101927174 | | |
| -0,018423281 | LOC101927179 | FALSE | Downregulated |
| | LOC101927182 | | |
| | LOC101927191 | | |
| 0,374557714 | LOC101927192 | FALSE | Upregulated |
| 0 | LOC101927199 | FALSE | Upregulated |
| | LOC101927200 | | |
| | LOC101927202 | | |
| | LOC101927205 | | |
| | LOC101927209 | | |
| | LOC101927210 | | |
| | LOC101927212 | | |
| | LOC101927213 | | |
| 0 | LOC101927216 | FALSE | Upregulated |
| -0,115927293 | LOC101927225 | FALSE | Downregulated |
| -0,236272124 | LOC101927226 | FALSE | Downregulated |
| 1,228084231 | LOC101927228 | | Upregulated |
| 0,033356717 | LOC101927229 | FALSE | Upregulated |
| 0 | LOC101927232 | FALSE | Upregulated |
| 0,002370413 | LOC101927234 | FALSE | Upregulated |
| 1,370803231 | LOC101927235 | | Upregulated |
| | LOC101927238 | | |
| | LOC101927241 | | |
| 0,736460285 | LOC101927245 | FALSE | Upregulated |
| | LOC101927247 | | |
| 0,035887847 | LOC101927248 | FALSE | Upregulated |
| 1,765054397 | LOC101927250 | | Upregulated |
| 0,371537189 | LOC101927252 | FALSE | Upregulated |
| | LOC101927253 | | |
| 0,698772907 | LOC101927254 | FALSE | Upregulated |
| -1,016588569 | LOC101927259 | | Downregulated |
| 1,11951213 | LOC101927260 | | Upregulated |
| 1,891167043 | LOC101927262 | | Upregulated |
| 1,704638587 | LOC101927263 | | Upregulated |
| -0,848329081 | LOC101927268 | FALSE | Downregulated |
| | LOC101927271 | | |
| | LOC101927277 | | |
| 0 | LOC101927278 | FALSE | Upregulated |
| | LOC101927280 | | |

| | | | |
|---|---|---|---|
| | LOC101927281 | | |
| 2,277179101 | LOC101927283 | | Upregulated |
| | LOC101927284 | | |
| 0 | LOC101927287 | FALSE | Upregulated |
| | LOC101927291 | | |
| 0 | LOC101927293 | FALSE | Upregulated |
| | LOC101927297 | | |
| | LOC101927299 | | |
| 0,534502809 | LOC101927301 | FALSE | Upregulated |
| | LOC101927304 | | |
| | LOC101927306 | | |
| 0 | LOC101927311 | FALSE | Upregulated |
| 0,597480774 | LOC101927318 | FALSE | Upregulated |
| | LOC101927319 | | |
| 0,203392548 | LOC101927320 | FALSE | Upregulated |
| 0 | LOC101927322 | FALSE | Upregulated |
| 0 | LOC101927325 | FALSE | Upregulated |
| -0,126310572 | LOC101927326 | FALSE | Downregulated |
| 0 | LOC101927328 | FALSE | Upregulated |
| | LOC101927329 | | |
| 0,822993936 | LOC101927330 | FALSE | Upregulated |
| -1,147861344 | LOC101927331 | | Downregulated |
| | LOC101927332 | | |
| 0,043729014 | LOC101927333 | FALSE | Upregulated |
| | LOC101927335 | | |
| | LOC101927337 | | |
| 0,087890481 | LOC101927338 | FALSE | Upregulated |
| | LOC101927341 | | |
| | LOC101927344 | | |
| 0 | LOC101927346 | FALSE | Upregulated |
| -0,115927293 | LOC101927348 | FALSE | Downregulated |
| | LOC101927351 | | |
| -0,115927293 | LOC101927353 | FALSE | Downregulated |
| 0,123193073 | LOC101927354 | FALSE | Upregulated |
| 0 | LOC101927357 | FALSE | Upregulated |
| | LOC101927361 | | |
| | LOC101927362 | | |
| | LOC101927363 | | |
| -0,290155017 | LOC101927366 | FALSE | Downregulated |
| 0,661206439 | LOC101927367 | FALSE | Upregulated |
| 0 | LOC101927369 | FALSE | Upregulated |
| 0 | LOC101927375 | FALSE | Upregulated |
| -0,219418189 | LOC101927376 | FALSE | Downregulated |
| | LOC101927383 | | |
| | LOC101927387 | | |
| 0,072407843 | LOC101927388 | FALSE | Upregulated |
| | LOC101927389 | | |
| -0,189001521 | LOC101927392 | FALSE | Downregulated |
| | LOC101927395 | | |
| | LOC101927397 | | |
| | LOC101927398 | | |

| | | | |
|---|---|---|---|
| 1,294367944 | LOC101927400 | | Upregulated |
| 0,341753817 | LOC101927401 | FALSE | Upregulated |
| 0,019985931 | LOC101927403 | FALSE | Upregulated |
| | LOC101927404 | | |
| | LOC101927405 | | |
| | LOC101927409 | | |
| | LOC101927413 | | |
| | LOC101927414 | | |
| -0,107864403 | LOC101927417 | FALSE | Downregulated |
| | LOC101927426 | | |
| | LOC101927429 | | |
| 1,267816593 | LOC101927430 | | Upregulated |
| -0,259057601 | LOC101927432 | FALSE | Downregulated |
| -0,465826144 | LOC101927435 | FALSE | Downregulated |
| | LOC101927436 | | |
| | LOC101927439 | | |
| 0 | LOC101927441 | FALSE | Upregulated |
| -0,065578176 | LOC101927446 | FALSE | Downregulated |
| 0,232162346 | LOC101927447 | FALSE | Upregulated |
| -0,080134345 | LOC101927448 | FALSE | Downregulated |
| | LOC101927452 | | |
| 1,123994684 | LOC101927454 | | Upregulated |
| -0,110143851 | LOC101927458 | FALSE | Downregulated |
| | LOC101927461 | | |
| | LOC101927466 | | |
| | LOC101927467 | | |
| 1,380026792 | LOC101927471 | | Upregulated |
| | LOC101927473 | | |
| | LOC101927480 | | |
| 0,18316667 | LOC101927481 | FALSE | Upregulated |
| | LOC101927483 | | |
| | LOC101927484 | | |
| 0,002866231 | LOC101927486 | FALSE | Upregulated |
| | LOC101927488 | | |
| -0,183381777 | LOC101927490 | FALSE | Downregulated |
| | LOC101927491 | | |
| | LOC101927493 | | |
| | LOC101927494 | | |
| | LOC101927496 | | |
| 1,681752163 | LOC101927499 | | Upregulated |
| 0,51346959 | LOC101927503 | FALSE | Upregulated |
| | LOC101927504 | | |
| 0,020984766 | LOC101927505 | FALSE | Upregulated |
| 0,576586618 | LOC101927508 | FALSE | Upregulated |
| 0 | LOC101927510 | FALSE | Upregulated |
| -0,018408691 | LOC101927511 | FALSE | Downregulated |
| 0,671032953 | LOC101927512 | FALSE | Upregulated |
| | LOC101927518 | | |
| | LOC101927522 | | |
| 0,076658198 | LOC101927525 | FALSE | Upregulated |
| 0,648160182 | LOC101927526 | FALSE | Upregulated |

| | | | |
|---|---|---|---|
| | LOC101927527 | | |
| 0,026700416 | LOC101927531 | FALSE | Upregulated |
| | LOC101927532 | | |
| | LOC101927533 | | |
| | LOC101927534 | | |
| 1,240070276 | LOC101927537 | | Upregulated |
| 0 | LOC101927538 | FALSE | Upregulated |
| 0,012749217 | LOC101927544 | FALSE | Upregulated |
| 1,23593508 | LOC101927546 | | Upregulated |
| | LOC101927548 | | |
| | LOC101927553 | | |
| 0,660982636 | LOC101927555 | FALSE | Upregulated |
| | LOC101927558 | | |
| | LOC101927561 | | |
| 2,064993651 | LOC101927562 | | Upregulated |
| 0 | LOC101927566 | FALSE | Upregulated |
| 0,099617812 | LOC101927567 | FALSE | Upregulated |
| 0,669198441 | LOC101927568 | FALSE | Upregulated |
| -0,710285324 | LOC101927571 | FALSE | Downregulated |
| | LOC101927573 | | |
| -0,139620996 | LOC101927576 | FALSE | Downregulated |
| 0 | LOC101927582 | FALSE | Upregulated |
| | LOC101927584 | | |
| -0,090062818 | LOC101927589 | FALSE | Downregulated |
| 0,754038479 | LOC101927594 | FALSE | Upregulated |
| 0,302043746 | LOC101927600 | FALSE | Upregulated |
| | LOC101927602 | | |
| 0 | LOC101927603 | FALSE | Upregulated |
| | LOC101927605 | | |
| | LOC101927606 | | |
| 0,709972026 | LOC101927611 | FALSE | Upregulated |
| -0,640456821 | LOC101927613 | FALSE | Downregulated |
| 0,537078947 | LOC101927615 | FALSE | Upregulated |
| -0,358791725 | LOC101927617 | FALSE | Downregulated |
| | LOC101927627 | | |
| 0,464770103 | LOC101927628 | FALSE | Upregulated |
| | LOC101927629 | | |
| -0,1386046 | LOC101927632 | FALSE | Downregulated |
| 0 | LOC101927633 | FALSE | Upregulated |
| 0 | LOC101927634 | FALSE | Upregulated |
| -0,126325251 | LOC101927635 | FALSE | Downregulated |
| 0,234127952 | LOC101927646 | FALSE | Upregulated |
| -1,031654716 | LOC101927651 | | Downregulated |
| -0,374281254 | LOC101927652 | FALSE | Downregulated |
| | LOC101927656 | | |
| 0,039005732 | LOC101927660 | FALSE | Upregulated |
| | LOC101927661 | | |
| 0,64221863 | LOC101927662 | FALSE | Upregulated |
| | LOC101927664 | | |
| -0,197443336 | LOC101927665 | FALSE | Downregulated |
| 0,660982636 | LOC101927669 | FALSE | Upregulated |

| | | | |
|---|---|---|---|
| 2,55041504 | LOC101927670 | | Upregulated |
| -0,287752564 | LOC101927674 | FALSE | Downregulated |
| | LOC101927676 | | |
| -0,839355131 | LOC101927680 | FALSE | Downregulated |
| | LOC101927684 | | |
| 1,164945423 | LOC101927686 | | Upregulated |
| | LOC101927687 | | |
| | LOC101927690 | | |
| | LOC101927691 | | |
| | LOC101927695 | | |
| 0,034128514 | LOC101927696 | FALSE | Upregulated |
| 0,010905726 | LOC101927697 | FALSE | Upregulated |
| 0,447503288 | LOC101927702 | FALSE | Upregulated |
| 0,870041512 | LOC101927703 | FALSE | Upregulated |
| | LOC101927704 | | |
| -0,530625997 | LOC101927705 | FALSE | Downregulated |
| 0,547799545 | LOC101927708 | FALSE | Upregulated |
| | LOC101927709 | | |
| 0,626077297 | LOC101927710 | FALSE | Upregulated |
| | LOC101927711 | | |
| 0 | LOC101927712 | FALSE | Upregulated |
| | LOC101927713 | | |
| 0,602864676 | LOC101927720 | FALSE | Upregulated |
| -0,267442638 | LOC101927721 | FALSE | Downregulated |
| 0 | LOC101927722 | FALSE | Upregulated |
| -0,361819243 | LOC101927725 | FALSE | Downregulated |
| | LOC101927729 | | |
| | LOC101927731 | | |
| 0,114728402 | LOC101927736 | FALSE | Upregulated |
| 0,398227005 | LOC101927737 | FALSE | Upregulated |
| | LOC101927738 | | |
| | LOC101927744 | | |
| -0,202094534 | LOC101927748 | FALSE | Downregulated |
| 1,369292829 | LOC101927754 | | Upregulated |
| | LOC101927757 | | |
| | LOC101927760 | | |
| | LOC101927762 | | |
| 0,250623826 | LOC101927763 | FALSE | Upregulated |
| -0,918696895 | LOC101927764 | FALSE | Downregulated |
| -0,560653286 | LOC101927765 | FALSE | Downregulated |
| 0,020152941 | LOC101927773 | FALSE | Upregulated |
| 0 | LOC101927774 | FALSE | Upregulated |
| 0,980694607 | LOC101927775 | FALSE | Upregulated |
| | LOC101927777 | | |
| | LOC101927779 | | |
| 1,384757732 | LOC101927783 | | Upregulated |
| | LOC101927786 | | |
| | LOC101927788 | | |
| -0,557997673 | LOC101927789 | FALSE | Downregulated |
| 0,638975576 | LOC101927792 | FALSE | Upregulated |
| | LOC101927794 | | |

| | | | |
|---|---|---|---|
| | LOC101927798 | | |
| 1,744685198 | LOC101927799 | | Upregulated |
| -0,564435659 | LOC101927800 | FALSE | Downregulated |
| | LOC101927801 | | |
| 0 | LOC101927804 | FALSE | Upregulated |
| | LOC101927805 | | |
| | LOC101927807 | | |
| 0,756620501 | LOC101927810 | FALSE | Upregulated |
| 0 | LOC101927811 | FALSE | Upregulated |
| 0,061332278 | LOC101927814 | FALSE | Upregulated |
| | LOC101927816 | | |
| 0,06183228 | LOC101927820 | FALSE | Upregulated |
| | LOC101927823 | | |
| 0,130252765 | LOC101927824 | FALSE | Upregulated |
| 1,727104084 | LOC101927825 | | Upregulated |
| -0,366545548 | LOC101927826 | FALSE | Downregulated |
| 0,642076523 | LOC101927834 | FALSE | Upregulated |
| | LOC101927835 | | |
| 0,56123892 | LOC101927837 | FALSE | Upregulated |
| | LOC101927844 | | |
| 0 | LOC101927846 | FALSE | Upregulated |
| 0 | LOC101927852 | FALSE | Upregulated |
| 0 | LOC101927853 | FALSE | Upregulated |
| 0,264041923 | LOC101927855 | FALSE | Upregulated |
| | LOC101927856 | | |
| 0,597328857 | LOC101927857 | FALSE | Upregulated |
| -0,023856574 | LOC101927859 | FALSE | Downregulated |
| 0,767664462 | LOC101927863 | FALSE | Upregulated |
| | LOC101927866 | | |
| 0 | LOC101927867 | FALSE | Upregulated |
| -0,201521565 | LOC101927872 | FALSE | Downregulated |
| 0,039005732 | LOC101927874 | FALSE | Upregulated |
| 0 | LOC101927875 | FALSE | Upregulated |
| | LOC101927877 | | |
| | LOC101927878 | | |
| 0 | LOC101927879 | FALSE | Upregulated |
| -0,168643645 | LOC101927880 | FALSE | Downregulated |
| | LOC101927881 | | |
| 0,62595979 | LOC101927882 | FALSE | Upregulated |
| 0,541385612 | LOC101927885 | FALSE | Upregulated |
| 0,65457448 | LOC101927888 | FALSE | Upregulated |
| 0 | LOC101927890 | FALSE | Upregulated |
| | LOC101927892 | | |
| 0 | LOC101927896 | FALSE | Upregulated |
| 1,059655666 | LOC101927902 | | Upregulated |
| | LOC101927904 | | |
| 0 | LOC101927906 | FALSE | Upregulated |
| | LOC101927908 | | |
| | LOC101927913 | | |
| | LOC101927915 | | |
| | LOC101927916 | | |

| | | | |
|---|---|---|---|
| | LOC101927918 | | |
| -0,025424043 | LOC101927919 | FALSE | Downregulated |
| | LOC101927921 | | |
| 0 | LOC101927922 | FALSE | Upregulated |
| 0,039601705 | LOC101927929 | FALSE | Upregulated |
| 0,089661664 | LOC101927931 | FALSE | Upregulated |
| 0,632907472 | LOC101927932 | FALSE | Upregulated |
| -0,127211226 | LOC101927933 | FALSE | Downregulated |
| | LOC101927935 | | |
| | LOC101927936 | | |
| -0,430800066 | LOC101927942 | FALSE | Downregulated |
| 0,255993941 | LOC101927943 | FALSE | Upregulated |
| | LOC101927946 | | |
| 0 | LOC101927949 | FALSE | Upregulated |
| -0,305905482 | LOC101927953 | FALSE | Downregulated |
| | LOC101927956 | | |
| -0,002529603 | LOC101927957 | FALSE | Downregulated |
| 0 | LOC101927958 | FALSE | Upregulated |
| | LOC101927959 | | |
| | LOC101927960 | | |
| | LOC101927961 | | |
| | LOC101927962 | | |
| | LOC101927967 | | |
| 0,459622315 | LOC101927971 | FALSE | Upregulated |
| 0,705562129 | LOC101927973 | FALSE | Upregulated |
| -0,920714179 | LOC101927974 | FALSE | Downregulated |
| 0,608246941 | LOC101927975 | FALSE | Upregulated |
| 0 | LOC101927978 | FALSE | Upregulated |
| | LOC101927982 | | |
| 0 | LOC101927988 | FALSE | Upregulated |
| 0 | LOC101927991 | FALSE | Upregulated |
| 0 | LOC101927993 | FALSE | Upregulated |
| -0,388409208 | LOC101927995 | FALSE | Downregulated |
| | LOC101927998 | | |
| 1,23323058 | LOC101928000 | | Upregulated |
| 0,165687872 | LOC101928004 | FALSE | Upregulated |
| 0,278436493 | LOC101928005 | FALSE | Upregulated |
| -1,75361785 | LOC101928006 | | Downregulated |
| 0 | LOC101928007 | FALSE | Upregulated |
| | LOC101928011 | | |
| | LOC101928014 | | |
| 0,493117071 | LOC101928021 | FALSE | Upregulated |
| | LOC101928026 | | |
| | LOC101928027 | | |
| | LOC101928028 | | |
| | LOC101928031 | | |
| | LOC101928032 | | |
| | LOC101928037 | | |
| 0 | LOC101928039 | FALSE | Upregulated |
| 0,057300436 | LOC101928044 | FALSE | Upregulated |
| | LOC101928046 | | |

| | | | |
|---|---|---|---|
| -0,578545415 | LOC101928048 | FALSE | Downregulated |
| | LOC101928055-2 | | |
| | LOC101928056 | | |
| 0,790404814 | LOC101928062 | FALSE | Upregulated |
| | LOC101928066 | | |
| 0 | LOC101928067 | FALSE | Upregulated |
| | LOC101928070-2 | | |
| 0,979504131 | LOC101928072 | FALSE | Upregulated |
| | LOC101928075 | | |
| -0,470100792 | LOC101928076 | FALSE | Downregulated |
| | LOC101928077 | | |
| 0,669025015 | LOC101928080 | FALSE | Upregulated |
| | LOC101928082 | | |
| | LOC101928084 | | |
| 0,060388786 | LOC101928085 | FALSE | Upregulated |
| 0 | LOC101928087 | FALSE | Upregulated |
| 0 | LOC101928090 | FALSE | Upregulated |
| -0,471055823 | LOC101928092 | FALSE | Downregulated |
| -0,414938386 | LOC101928096 | FALSE | Downregulated |
| 0,496400713 | LOC101928102 | FALSE | Upregulated |
| 1,502281876 | LOC101928105 | | Upregulated |
| | LOC101928108 | | |
| -0,454575759 | LOC101928109 | FALSE | Downregulated |
| 0 | LOC101928110 | FALSE | Upregulated |
| 1,729422538 | LOC101928111 | | Upregulated |
| 0,244819921 | LOC101928113 | FALSE | Upregulated |
| | LOC101928114 | | |
| 0,597480774 | LOC101928115 | FALSE | Upregulated |
| 0 | LOC101928117 | FALSE | Upregulated |
| | LOC101928121 | | |
| | LOC101928124 | | |
| | LOC101928125 | | |
| 0,064924611 | LOC101928126 | FALSE | Upregulated |
| | LOC101928127 | | |
| | LOC101928128 | | |
| | LOC101928129 | | |
| | LOC101928130 | | |
| | LOC101928131 | | |
| | LOC101928132 | | |
| | LOC101928135 | | |
| | LOC101928144 | | |
| | LOC101928146 | | |
| -0,004298224 | LOC101928149 | FALSE | Downregulated |
| | LOC101928153 | | |
| 0,552019472 | LOC101928156 | FALSE | Upregulated |
| | LOC101928157 | | |
| -0,042008937 | LOC101928160 | FALSE | Downregulated |
| | LOC101928162 | | |
| | LOC101928166 | | |
| | LOC101928167 | | |
| | LOC101928169 | | |

| | | | |
|---|---|---|---|
| 0,661206439 | LOC101928171 | FALSE | Upregulated |
| -0,25853991 | LOC101928175 | FALSE | Downregulated |
| | LOC101928176 | | |
| | LOC101928177 | | |
| 0,405328033 | LOC101928178 | FALSE | Upregulated |
| -1,02994426 | LOC101928180 | | Downregulated |
| 0,04504514 | LOC101928186 | FALSE | Upregulated |
| | LOC101928190 | | |
| 2,38709079 | LOC101928191 | | Upregulated |
| 0,263580151 | LOC101928193 | FALSE | Upregulated |
| 0 | LOC101928196 | FALSE | Upregulated |
| 2,025159996 | LOC101928197 | | Upregulated |
| | LOC101928199 | | |
| | LOC101928207 | | |
| 0,034128514 | LOC101928208 | FALSE | Upregulated |
| | LOC101928216 | | |
| | LOC101928217 | | |
| 0 | LOC101928219 | FALSE | Upregulated |
| 1,543641347 | LOC101928220 | | Upregulated |
| | LOC101928225 | | |
| | LOC101928226 | | |
| 0,153126361 | LOC101928228 | FALSE | Upregulated |
| 0,632050826 | LOC101928231 | FALSE | Upregulated |
| -0,685659632 | LOC101928233 | FALSE | Downregulated |
| | LOC101928234 | | |
| 0,737912451 | LOC101928235 | FALSE | Upregulated |
| -0,446752538 | LOC101928237 | FALSE | Downregulated |
| 1,120459318 | LOC101928238 | | Upregulated |
| 0,016581916 | LOC101928239 | FALSE | Upregulated |
| 1,089371897 | LOC101928245 | | Upregulated |
| 0,120608052 | LOC101928246 | FALSE | Upregulated |
| 0,096454085 | LOC101928247 | FALSE | Upregulated |
| 0 | LOC101928248 | FALSE | Upregulated |
| 0,925186726 | LOC101928249 | FALSE | Upregulated |
| | LOC101928250 | | |
| 1,436910641 | LOC101928251 | | Upregulated |
| -1,100395294 | LOC101928253 | | Downregulated |
| 0 | LOC101928255 | FALSE | Upregulated |
| | LOC101928257 | | |
| 0 | LOC101928258 | FALSE | Upregulated |
| -0,857987851 | LOC101928261 | FALSE | Downregulated |
| 1,177302437 | LOC101928262 | | Upregulated |
| 0,947886097 | LOC101928264 | FALSE | Upregulated |
| -0,073257748 | LOC101928269 | FALSE | Downregulated |
| | LOC101928271 | | |
| | LOC101928272 | | |
| | LOC101928273 | | |
| 0,692223634 | LOC101928274 | FALSE | Upregulated |
| | LOC101928275 | | |
| -0,619358284 | LOC101928277 | FALSE | Downregulated |
| | LOC101928278 | | |

| | | | |
|---|---|---|---|
| 1,180126863 | LOC101928279 | | Upregulated |
| | LOC101928282 | | |
| | LOC101928285 | | |
| 0 | LOC101928286 | FALSE | Upregulated |
| 0 | LOC101928288 | FALSE | Upregulated |
| 0 | LOC101928300 | FALSE | Upregulated |
| 0,027563344 | LOC101928306 | FALSE | Upregulated |
| 0,008212946 | LOC101928307 | FALSE | Upregulated |
| | LOC101928309 | | |
| | LOC101928310 | | |
| -0,11763152 | LOC101928311 | FALSE | Downregulated |
| -0,047343088 | LOC101928312 | FALSE | Downregulated |
| 0,065205747 | LOC101928314 | FALSE | Upregulated |
| | LOC101928316 | | |
| 0,171842827 | LOC101928317 | FALSE | Upregulated |
| 0 | LOC101928325 | FALSE | Upregulated |
| 0,204741576 | LOC101928326 | FALSE | Upregulated |
| 0 | LOC101928327 | FALSE | Upregulated |
| | LOC101928329 | | |
| 0,669025015 | LOC101928330 | FALSE | Upregulated |
| 0,211604647 | LOC101928331 | FALSE | Upregulated |
| -0,011217418 | LOC101928338 | FALSE | Downregulated |
| | LOC101928339 | | |
| -0,017711534 | LOC101928343 | FALSE | Downregulated |
| | LOC101928346 | | |
| 0,300894368 | LOC101928347 | FALSE | Upregulated |
| 0 | LOC101928349 | FALSE | Upregulated |
| | LOC101928350 | | |
| 0,464272715 | LOC101928357 | FALSE | Upregulated |
| | LOC101928358 | | |
| -0,472062833 | LOC101928359 | FALSE | Downregulated |
| 0,896073524 | LOC101928360 | FALSE | Upregulated |
| 1,013083344 | LOC101928361 | | Upregulated |
| 0,034943632 | LOC101928362 | FALSE | Upregulated |
| | LOC101928364 | | |
| | LOC101928367 | | |
| | LOC101928368 | | |
| -0,055772032 | LOC101928369 | FALSE | Downregulated |
| -2,017954303 | LOC101928370 | | Downregulated |
| | LOC101928375 | | |
| 0,07916956 | LOC101928377 | FALSE | Upregulated |
| | LOC101928380 | | |
| -0,223460718 | LOC101928382 | FALSE | Downregulated |
| -0,409864097 | LOC101928383 | FALSE | Downregulated |
| | LOC101928385 | | |
| 0 | LOC101928387 | FALSE | Upregulated |
| 0,67752523 | LOC101928388 | FALSE | Upregulated |
| -0,058224512 | LOC101928389 | FALSE | Downregulated |
| | LOC101928391 | | |
| | LOC101928392 | | |
| | LOC101928393 | | |

| | | | |
|---|---|---|---|
| | LOC101928394 | | |
| 0 | LOC101928397 | FALSE | Upregulated |
| 0,333156087 | LOC101928399 | FALSE | Upregulated |
| -0,055970583 | LOC101928401 | FALSE | Downregulated |
| -0,187950981 | LOC101928402 | FALSE | Downregulated |
| 0,324044931 | LOC101928403 | FALSE | Upregulated |
| | LOC101928405 | | |
| | LOC101928406 | | |
| -0,93122783 | LOC101928407 | FALSE | Downregulated |
| 0,466740211 | LOC101928412 | FALSE | Upregulated |
| 0 | LOC101928414 | FALSE | Upregulated |
| 1,062248702 | LOC101928419 | | Upregulated |
| 0,073122974 | LOC101928421 | FALSE | Upregulated |
| | LOC101928422 | | |
| | LOC101928423 | | |
| | LOC101928426 | | |
| 0 | LOC101928427 | FALSE | Upregulated |
| -0,202572298 | LOC101928428 | FALSE | Downregulated |
| | LOC101928430 | | |
| 0 | LOC101928432 | FALSE | Upregulated |
| 1,289395499 | LOC101928434 | | Upregulated |
| | LOC101928436 | | |
| 1,763548505 | LOC101928437 | | Upregulated |
| | LOC101928447 | | |
| | LOC101928448 | | |
| | LOC101928450 | | |
| | LOC101928455 | | |
| | LOC101928457 | | |
| | LOC101928458 | | |
| 0 | LOC101928459 | FALSE | Upregulated |
| -0,534326762 | LOC101928462 | FALSE | Downregulated |
| | LOC101928463 | | |
| 1,309563836 | LOC101928464 | | Upregulated |
| | LOC101928469 | | |
| 0,014712482 | LOC101928472 | FALSE | Upregulated |
| -0,421262694 | LOC101928474 | FALSE | Downregulated |
| | LOC101928475 | | |
| -1,617418086 | LOC101928477 | | Downregulated |
| | LOC101928478 | | |
| | LOC101928481 | | |
| | LOC101928484 | | |
| -0,362643805 | LOC101928487 | FALSE | Downregulated |
| 0,172576653 | LOC101928489 | FALSE | Upregulated |
| -1,1279267 | LOC101928504 | | Downregulated |
| | LOC101928506 | | |
| 0 | LOC101928513 | FALSE | Upregulated |
| -0,1540447 | LOC101928514 | FALSE | Downregulated |
| | LOC101928519 | | |
| 0,034128514 | LOC101928522 | FALSE | Upregulated |
| | LOC101928523 | | |
| 0 | LOC101928526 | FALSE | Upregulated |

| | | | |
|---|---|---|---|
| -0,444899439 | LOC101928529 | FALSE | Downregulated |
| 1,054158892 | LOC101928530 | | Upregulated |
| 0 | LOC101928533 | FALSE | Upregulated |
| | LOC101928535 | | |
| | LOC101928538 | | |
| | LOC101928543 | | |
| -0,001452899 | LOC101928549 | FALSE | Downregulated |
| 0,975362814 | LOC101928551 | FALSE | Upregulated |
| -0,142722773 | LOC101928556 | FALSE | Downregulated |
| -0,741045319 | LOC101928560 | FALSE | Downregulated |
| 0,823231787 | LOC101928562 | FALSE | Upregulated |
| | LOC101928563 | | |
| | LOC101928564 | | |
| | LOC101928566 | | |
| -0,101142365 | LOC101928568 | FALSE | Downregulated |
| | LOC101928569 | | |
| 0 | LOC101928570 | FALSE | Upregulated |
| 0 | LOC101928572 | FALSE | Upregulated |
| | LOC101928573 | | |
| | LOC101928579 | | |
| | LOC101928581 | | |
| | LOC101928582 | | |
| 2,126805767 | LOC101928583 | | Upregulated |
| 1,264893454 | LOC101928584 | | Upregulated |
| | LOC101928586 | | |
| | LOC101928587 | | |
| 1,387432404 | LOC101928589 | | Upregulated |
| 0 | LOC101928591 | FALSE | Upregulated |
| 1,352419694 | LOC101928592 | | Upregulated |
| | LOC101928593 | | |
| -0,512662986 | LOC101928596 | FALSE | Downregulated |
| | LOC101928598 | | |
| 0,325398395 | LOC101928600 | FALSE | Upregulated |
| 0 | LOC101928602 | FALSE | Upregulated |
| -0,161195308 | LOC101928608 | FALSE | Downregulated |
| | LOC101928611 | | |
| -0,246267043 | LOC101928612 | FALSE | Downregulated |
| | LOC101928614 | | |
| | LOC101928617 | | |
| 0,140153121 | LOC101928618 | FALSE | Upregulated |
| | LOC101928620 | | |
| | LOC101928623 | | |
| -0,409778376 | LOC101928624 | FALSE | Downregulated |
| -0,323779267 | LOC101928625 | FALSE | Downregulated |
| | LOC101928629 | | |
| 0,058865037 | LOC101928632 | FALSE | Upregulated |
| 0,148896881 | LOC101928633 | FALSE | Upregulated |
| | LOC101928634 | | |
| | LOC101928636 | | |
| 0 | LOC101928638 | FALSE | Upregulated |
| 0 | LOC101928639 | FALSE | Upregulated |

| | | | |
|---|---|---|---|
| | LOC101928640 | | |
| | LOC101928643 | | |
| -0,966321592 | LOC101928645 | FALSE | Downregulated |
| 1,846069641 | LOC101928647 | | Upregulated |
| -0,06413353 | LOC101928652 | FALSE | Downregulated |
| | LOC101928654 | | |
| -0,664303603 | LOC101928655 | FALSE | Downregulated |
| 1,026433999 | LOC101928659 | | Upregulated |
| | LOC101928662 | | |
| 0,533053073 | LOC101928665 | FALSE | Upregulated |
| | LOC101928666 | | |
| 0,842068843 | LOC101928667 | FALSE | Upregulated |
| -0,765715851 | LOC101928671 | FALSE | Downregulated |
| 1,322114743 | LOC101928683 | | Upregulated |
| 0,027377332 | LOC101928684 | FALSE | Upregulated |
| | LOC101928688 | | |
| 0,238584417 | LOC101928692 | FALSE | Upregulated |
| 0,016260394 | LOC101928695 | FALSE | Upregulated |
| | LOC101928697 | | |
| -0,091553114 | LOC101928701 | FALSE | Downregulated |
| | LOC101928702 | | |
| | LOC101928704 | | |
| -0,013976448 | LOC101928705 | FALSE | Downregulated |
| | LOC101928710 | | |
| | LOC101928711 | | |
| | LOC101928713 | | |
| 1,32608087 | LOC101928718 | | Upregulated |
| | LOC101928721 | | |
| 0,562433805 | LOC101928722 | FALSE | Upregulated |
| 0,51175222 | LOC101928725 | FALSE | Upregulated |
| 1,424698786 | LOC101928727 | | Upregulated |
| 1,100317816 | LOC101928730 | | Upregulated |
| 0,537725177 | LOC101928733 | FALSE | Upregulated |
| | LOC101928735 | | |
| 0,680077947 | LOC101928740 | FALSE | Upregulated |
| 0,062812485 | LOC101928742 | FALSE | Upregulated |
| | LOC101928743 | | |
| | LOC101928747 | | |
| 0 | LOC101928754 | FALSE | Upregulated |
| -0,699393619 | LOC101928756 | FALSE | Downregulated |
| 0,515696588 | LOC101928761 | FALSE | Upregulated |
| | LOC101928764 | | |
| 0,046162758 | LOC101928765 | FALSE | Upregulated |
| -0,055970583 | LOC101928767 | FALSE | Downregulated |
| | LOC101928770 | | |
| -0,146624803 | LOC101928774 | FALSE | Downregulated |
| | LOC101928775 | | |
| 0 | LOC101928779 | FALSE | Upregulated |
| | LOC101928785 | | |
| 1,300508473 | LOC101928786 | | Upregulated |
| 0 | LOC101928787 | FALSE | Upregulated |

| | | | |
|---|---|---|---|
| 0,734684809 | LOC101928792 | FALSE | Upregulated |
| 0 | LOC101928796 | FALSE | Upregulated |
| | LOC101928800 | | |
| 0,560466993 | LOC101928801 | FALSE | Upregulated |
| 1,008703979 | LOC101928803 | | Upregulated |
| 0 | LOC101928804 | FALSE | Upregulated |
| -0,264760596 | LOC101928806 | FALSE | Downregulated |
| 0,77551617 | LOC101928807 | FALSE | Upregulated |
| -0,173959778 | LOC101928808 | FALSE | Downregulated |
| -1,197400558 | LOC101928810 | | Downregulated |
| | LOC101928813 | | |
| 1,31194132 | LOC101928814 | | Upregulated |
| 0,479474246 | LOC101928815 | FALSE | Upregulated |
| | LOC101928818 | | |
| 0,73018175 | LOC101928819 | FALSE | Upregulated |
| 0,661206439 | LOC101928824 | FALSE | Upregulated |
| -0,531087413 | LOC101928825 | FALSE | Downregulated |
| | LOC101928826 | | |
| | LOC101928828 | | |
| 0 | LOC101928829 | FALSE | Upregulated |
| | LOC101928832 | | |
| | LOC101928833 | | |
| | LOC101928836 | | |
| 0,03893095 | LOC101928838 | FALSE | Upregulated |
| | LOC101928839 | | |
| | LOC101928842 | | |
| 0,583590522 | LOC101928845 | FALSE | Upregulated |
| | LOC101928849 | | |
| 0,003697762 | LOC101928850 | FALSE | Upregulated |
| -0,190978669 | LOC101928852 | FALSE | Downregulated |
| | LOC101928856 | | |
| | LOC101928859 | | |
| | LOC101928860 | | |
| | LOC101928863 | | |
| 0,041547119 | LOC101928864 | FALSE | Upregulated |
| | LOC101928866 | | |
| | LOC101928867 | | |
| -0,21880063 | LOC101928868 | FALSE | Downregulated |
| 0,084636682 | LOC101928870 | FALSE | Upregulated |
| 0,624833244 | LOC101928876 | FALSE | Upregulated |
| -0,358081948 | LOC101928880 | FALSE | Downregulated |
| 0,801507571 | LOC101928881 | FALSE | Upregulated |
| -1,102931809 | LOC101928882 | | Downregulated |
| -0,14231155 | LOC101928887 | FALSE | Downregulated |
| 0 | LOC101928890 | FALSE | Upregulated |
| 1,080193763 | LOC101928891 | | Upregulated |
| | LOC101928893 | | |
| | LOC101928898 | | |
| 0 | LOC101928899 | FALSE | Upregulated |
| 0 | LOC101928902 | FALSE | Upregulated |
| | LOC101928905 | | |

| | | | |
|---|---|---|---|
| -0,806589476 | LOC101928906 | FALSE | Downregulated |
| -0,042948389 | LOC101928907 | FALSE | Downregulated |
| | LOC101928910 | | |
| 0 | LOC101928911 | FALSE | Upregulated |
| | LOC101928912 | | |
| 0,267609212 | LOC101928913 | FALSE | Upregulated |
| 0,131316274 | LOC101928915 | FALSE | Upregulated |
| -0,883463765 | LOC101928916 | FALSE | Downregulated |
| 0 | LOC101928918 | FALSE | Upregulated |
| | LOC101928919 | | |
| 0,891123338 | LOC101928921 | FALSE | Upregulated |
| | LOC101928922 | | |
| | LOC101928923 | | |
| 0,024251327 | LOC101928924 | FALSE | Upregulated |
| | LOC101928928 | | |
| 0 | LOC101928929 | FALSE | Upregulated |
| 0 | LOC101928932 | FALSE | Upregulated |
| 0,330638778 | LOC101928933 | FALSE | Upregulated |
| | LOC101928941 | | |
| 0,661206439 | LOC101928948 | FALSE | Upregulated |
| | LOC101928950 | | |
| 0,606233987 | LOC101928955 | FALSE | Upregulated |
| | LOC101928957 | | |
| | LOC101928962 | | |
| 0,339542265 | LOC101928963 | FALSE | Upregulated |
| | LOC101928964 | | |
| -0,133077926 | LOC101928965 | FALSE | Downregulated |
| 0,321963855 | LOC101928968 | FALSE | Upregulated |
| 0,948025245 | LOC101928970 | FALSE | Upregulated |
| | LOC101928972 | | |
| 1,215899272 | LOC101928974 | | Upregulated |
| -0,652420729 | LOC101928976 | FALSE | Downregulated |
| | LOC101928977 | | |
| | LOC101928978 | | |
| 0 | LOC101928981 | FALSE | Upregulated |
| -0,002529603 | LOC101928987 | FALSE | Downregulated |
| -0,095268562 | LOC101928991 | FALSE | Downregulated |
| | LOC101929002 | | |
| | LOC101929003 | | |
| -0,004736509 | LOC101929006 | FALSE | Downregulated |
| -0,039240178 | LOC101929008 | FALSE | Downregulated |
| 1,740370588 | LOC101929011 | | Upregulated |
| | LOC101929014 | | |
| | LOC101929016 | | |
| | LOC101929021 | | |
| | LOC101929022 | | |
| 0 | LOC101929027 | FALSE | Upregulated |
| | LOC101929028 | | |
| | LOC101929031 | | |
| | LOC101929034 | | |
| -0,552873173 | LOC101929035 | FALSE | Downregulated |

| | | | |
|---|---|---|---|
| 0,34277968 | LOC101929036 | FALSE | Upregulated |
| -0,162853293 | LOC101929038 | FALSE | Downregulated |
| | LOC101929043 | | |
| 0 | LOC101929044 | FALSE | Upregulated |
| | LOC101929048 | | |
| | LOC101929050 | | |
| 1,649258233 | LOC101929054 | | Upregulated |
| 0,000577308 | LOC101929055 | FALSE | Upregulated |
| | LOC101929056 | | |
| 0 | LOC101929058 | FALSE | Upregulated |
| 0,035914265 | LOC101929060 | FALSE | Upregulated |
| 0,047293497 | LOC101929067 | FALSE | Upregulated |
| | LOC101929069 | | |
| 0,019985931 | LOC101929077 | FALSE | Upregulated |
| | LOC101929082 | | |
| 0,048656215 | LOC101929085 | FALSE | Upregulated |
| 0,050688952 | LOC101929087 | FALSE | Upregulated |
| | LOC101929088 | | |
| 0,074083951 | LOC101929090 | FALSE | Upregulated |
| 0 | LOC101929094 | FALSE | Upregulated |
| -1,098441939 | LOC101929097 | | Downregulated |
| 0,28614179 | LOC101929098 | FALSE | Upregulated |
| -0,533223087 | LOC101929107 | FALSE | Downregulated |
| | LOC101929108 | | |
| | LOC101929114 | | |
| 0 | LOC101929119 | FALSE | Upregulated |
| | LOC101929121 | | |
| 0,067880217 | LOC101929124 | FALSE | Upregulated |
| -0,133702137 | LOC101929125 | FALSE | Downregulated |
| | LOC101929129 | | |
| | LOC101929131 | | |
| -0,346566868 | LOC101929132 | FALSE | Downregulated |
| | LOC101929134 | | |
| | LOC101929137 | | |
| 1,202024379 | LOC101929138 | | Upregulated |
| 0 | LOC101929142 | FALSE | Upregulated |
| -0,326169999 | LOC101929143 | FALSE | Downregulated |
| | LOC101929144 | | |
| 0,66422122 | LOC101929149 | FALSE | Upregulated |
| -0,010637249 | LOC101929151 | FALSE | Downregulated |
| | LOC101929152 | | |
| 0 | LOC101929153 | FALSE | Upregulated |
| 0,087836139 | LOC101929154 | FALSE | Upregulated |
| 0,669025015 | LOC101929155 | FALSE | Upregulated |
| | LOC101929156 | | |
| | LOC101929157 | | |
| -0,133864172 | LOC101929159 | FALSE | Downregulated |
| | LOC101929161 | | |
| | LOC101929164 | | |
| 0,377197545 | LOC101929167 | FALSE | Upregulated |
| 0 | LOC101929174 | FALSE | Upregulated |

| | | | |
|---|---|---|---|
| -0,043304873 | LOC101929177 | FALSE | Downregulated |
| -0,189058594 | LOC101929178 | FALSE | Downregulated |
| | LOC101929180 | | |
| 0 | LOC101929184 | FALSE | Upregulated |
| | LOC101929187 | | |
| | LOC101929190 | | |
| 0,629566366 | LOC101929191 | FALSE | Upregulated |
| | LOC101929192 | | |
| | LOC101929194 | | |
| 0,183193756 | LOC101929198 | FALSE | Upregulated |
| -0,42014776 | LOC101929200 | FALSE | Downregulated |
| | LOC101929201 | | |
| | LOC101929202 | | |
| -0,291992746 | LOC101929206 | FALSE | Downregulated |
| | LOC101929214 | | |
| | LOC101929216 | | |
| | LOC101929217 | | |
| 0,437951765 | LOC101929220 | FALSE | Upregulated |
| 0,368041357 | LOC101929223 | FALSE | Upregulated |
| | LOC101929228 | | |
| | LOC101929229 | | |
| 1,3016584 | LOC101929230 | | Upregulated |
| | LOC101929232 | | |
| | LOC101929236 | | |
| 1,038010452 | LOC101929241 | | Upregulated |
| -0,737270904 | LOC101929243 | FALSE | Downregulated |
| 2,728805517 | LOC101929248 | | Upregulated |
| 0,304795027 | LOC101929249 | FALSE | Upregulated |
| | LOC101929250 | | |
| -0,839763646 | LOC101929251 | FALSE | Downregulated |
| | LOC101929256 | | |
| | LOC101929257 | | |
| 0 | LOC101929258 | FALSE | Upregulated |
| 0,521258759 | LOC101929259 | FALSE | Upregulated |
| | LOC101929261 | | |
| 0 | LOC101929266 | FALSE | Upregulated |
| | LOC101929267 | | |
| 0,149401382 | LOC101929269 | FALSE | Upregulated |
| | LOC101929278 | | |
| -0,311108783 | LOC101929279 | FALSE | Downregulated |
| -0,616070315 | LOC101929283 | FALSE | Downregulated |
| 1,028540566 | LOC101929285 | | Upregulated |
| | LOC101929286 | | |
| -0,159557613 | LOC101929287 | FALSE | Downregulated |
| 0 | LOC101929288 | FALSE | Upregulated |
| 0,145421741 | LOC101929289 | FALSE | Upregulated |
| | LOC101929298 | | |
| 0,669756935 | LOC101929300 | FALSE | Upregulated |
| | LOC101929301 | | |
| 0 | LOC101929304 | FALSE | Upregulated |
| 0 | LOC101929305 | FALSE | Upregulated |

| | | | |
|---|---|---|---|
| | LOC101929307 | | |
| -0,306978372 | LOC101929313 | FALSE | Downregulated |
| 0 | LOC101929314 | FALSE | Upregulated |
| | LOC101929316 | | |
| 0 | LOC101929319 | FALSE | Upregulated |
| | LOC101929320 | | |
| | LOC101929321 | | |
| 0 | LOC101929322 | FALSE | Upregulated |
| 0 | LOC101929325 | FALSE | Upregulated |
| | LOC101929326 | | |
| -0,006204521 | LOC101929327 | FALSE | Downregulated |
| 0,065074583 | LOC101929341 | FALSE | Upregulated |
| -0,624885944 | LOC101929346 | FALSE | Downregulated |
| | LOC101929348 | | |
| | LOC101929350 | | |
| | LOC101929351 | | |
| | LOC101929355 | | |
| -0,756391392 | LOC101929356 | FALSE | Downregulated |
| | LOC101929358 | | |
| | LOC101929359 | | |
| | LOC101929363 | | |
| 0 | LOC101929367 | FALSE | Upregulated |
| | LOC101929368 | | |
| -0,348549391 | LOC101929371 | FALSE | Downregulated |
| | LOC101929373 | | |
| 0 | LOC101929374 | FALSE | Upregulated |
| 0 | LOC101929375 | FALSE | Upregulated |
| 0 | LOC101929379 | FALSE | Upregulated |
| -0,257029915 | LOC101929381 | FALSE | Downregulated |
| | LOC101929382 | | |
| | LOC101929386 | | |
| | LOC101929388 | | |
| -0,089204153 | LOC101929393 | FALSE | Downregulated |
| | LOC101929398 | | |
| 0 | LOC101929408 | FALSE | Upregulated |
| | LOC101929411 | | |
| 0 | LOC101929414 | FALSE | Upregulated |
| 0,041547119 | LOC101929418 | FALSE | Upregulated |
| | LOC101929419 | | |
| -0,245656948 | LOC101929422 | FALSE | Downregulated |
| -0,15373786 | LOC101929427 | FALSE | Downregulated |
| -0,039013048 | LOC101929431 | FALSE | Downregulated |
| | LOC101929432 | | |
| -0,238219566 | LOC101929434 | FALSE | Downregulated |
| | LOC101929435 | | |
| | LOC101929437 | | |
| -0,333146256 | LOC101929443 | FALSE | Downregulated |
| | LOC101929445 | | |
| | LOC101929446 | | |
| 0,478083631 | LOC101929447 | FALSE | Upregulated |
| | LOC101929453 | | |

| | | | |
|---|---|---|---|
| | LOC101929456 | | |
| 0,113975405 | LOC101929457 | FALSE | Upregulated |
| 0 | LOC101929459 | FALSE | Upregulated |
| 0 | LOC101929460 | FALSE | Upregulated |
| -0,190154769 | LOC101929462 | FALSE | Downregulated |
| -0,520225049 | LOC101929463 | FALSE | Downregulated |
| 1,222793163 | LOC101929465 | | Upregulated |
| -0,627218251 | LOC101929467 | FALSE | Downregulated |
| 0,778241907 | LOC101929475 | FALSE | Upregulated |
| | LOC101929476 | | |
| | LOC101929477 | | |
| | LOC101929478 | | |
| 0 | LOC101929484 | FALSE | Upregulated |
| | LOC101929485 | | |
| | LOC101929489 | | |
| 0 | LOC101929491 | FALSE | Upregulated |
| | LOC101929492 | | |
| | LOC101929493 | | |
| -0,455105185 | LOC101929494 | FALSE | Downregulated |
| | LOC101929495 | | |
| | LOC101929498 | | |
| | LOC101929507 | | |
| | LOC101929510 | | |
| -0,657653824 | LOC101929511 | FALSE | Downregulated |
| | LOC101929514 | | |
| | LOC101929515 | | |
| 0 | LOC101929518 | FALSE | Upregulated |
| 0,216506542 | LOC101929521 | FALSE | Upregulated |
| 0,095716332 | LOC101929528 | FALSE | Upregulated |
| | LOC101929530 | | |
| 0 | LOC101929531 | FALSE | Upregulated |
| 0 | LOC101929534 | FALSE | Upregulated |
| | LOC101929535 | | |
| 1,090933242 | LOC101929536 | | Upregulated |
| 1,213904787 | LOC101929538 | | Upregulated |
| -0,290263925 | LOC101929542 | FALSE | Downregulated |
| | LOC101929544 | | |
| 0,830236448 | LOC101929547 | FALSE | Upregulated |
| 0 | LOC101929550 | FALSE | Upregulated |
| -0,155425874 | LOC101929552 | FALSE | Downregulated |
| -0,335568703 | LOC101929555 | FALSE | Downregulated |
| | LOC101929557 | | |
| 0,407998754 | LOC101929558 | FALSE | Upregulated |
| 0 | LOC101929560 | FALSE | Upregulated |
| -0,326821635 | LOC101929561 | FALSE | Downregulated |
| | LOC101929562 | | |
| | LOC101929563 | | |
| | LOC101929564 | | |
| 1,240070276 | LOC101929566 | | Upregulated |
| | LOC101929575 | | |
| | LOC101929576 | | |

| | | | |
|---|---|---|---|
| -0,45345984 | LOC101929587 | FALSE | Downregulated |
| 1,301021567 | LOC101929590 | | Upregulated |
| | LOC101929593 | | |
| -1,370710397 | LOC101929594 | | Downregulated |
| 0 | LOC101929601 | FALSE | Upregulated |
| 0,195529993 | LOC101929602 | FALSE | Upregulated |
| 0 | LOC101929604 | FALSE | Upregulated |
| 1,069098127 | LOC101929609 | | Upregulated |
| | LOC101929610 | | |
| | LOC101929612 | | |
| -0,091445324 | LOC101929613 | FALSE | Downregulated |
| 0,154741553 | LOC101929614 | FALSE | Upregulated |
| 1,463066782 | LOC101929615 | | Upregulated |
| | LOC101929617 | | |
| | LOC101929618 | | |
| 0,083694752 | LOC101929619 | FALSE | Upregulated |
| | LOC101929620 | | |
| 0,287857476 | LOC101929625 | FALSE | Upregulated |
| 0 | LOC101929627 | FALSE | Upregulated |
| | LOC101929628 | | |
| 0,612488704 | LOC101929634 | FALSE | Upregulated |
| 0,021842814 | LOC101929637 | FALSE | Upregulated |
| 0,44121598 | LOC101929638 | FALSE | Upregulated |
| 0 | LOC101929641 | FALSE | Upregulated |
| | LOC101929644 | | |
| 0,186755661 | LOC101929645 | FALSE | Upregulated |
| | LOC101929646 | | |
| 1,248404362 | LOC101929647 | | Upregulated |
| 0 | LOC101929650 | FALSE | Upregulated |
| | LOC101929654 | | |
| | LOC101929658 | | |
| -0,405069745 | LOC101929661 | FALSE | Downregulated |
| 0,03188151 | LOC101929666 | FALSE | Upregulated |
| | LOC101929674 | | |
| 0 | LOC101929675 | FALSE | Upregulated |
| | LOC101929681 | | |
| 0,572554177 | LOC101929685 | FALSE | Upregulated |
| | LOC101929688 | | |
| | LOC101929690 | | |
| | LOC101929692 | | |
| 0,001678174 | LOC101929696 | FALSE | Upregulated |
| | LOC101929697 | | |
| | LOC101929700 | | |
| | LOC101929702 | | |
| 0,376586624 | LOC101929704 | FALSE | Upregulated |
| -0,530925474 | LOC101929713 | FALSE | Downregulated |
| 0,579114017 | LOC101929724 | FALSE | Upregulated |
| | LOC101929725 | | |
| -0,059237365 | LOC101929726 | FALSE | Downregulated |
| -0,039724636 | LOC101929727 | FALSE | Downregulated |
| -0,645093252 | LOC101929728 | FALSE | Downregulated |

| | | | |
|---|---|---|---|
| | LOC101929733 | | |
| 0 | LOC101929738 | FALSE | Upregulated |
| -0,01148885 | LOC101929740 | FALSE | Downregulated |
| | LOC101929745 | | |
| | LOC101929748 | | |
| | LOC101929750 | | |
| 0 | LOC101929753 | FALSE | Upregulated |
| 0,394737361 | LOC101929754 | FALSE | Upregulated |
| | LOC101929755 | | |
| | LOC101929757 | | |
| | LOC101929759 | | |
| -0,811320552 | LOC101929761 | FALSE | Downregulated |
| 0 | LOC101929762 | FALSE | Upregulated |
| | LOC101929763 | | |
| -0,030720259 | LOC101929766 | FALSE | Downregulated |
| | LOC101929770 | | |
| 0 | LOC101929777 | FALSE | Upregulated |
| -0,339924205 | LOC101929788 | FALSE | Downregulated |
| 1,069682595 | LOC101929798 | | Upregulated |
| 0 | LOC101929800 | FALSE | Upregulated |
| 0,669025015 | LOC101929803 | FALSE | Upregulated |
| | LOC101929806 | | |
| 0,13180328 | LOC101929814 | FALSE | Upregulated |
| | LOC101929815 | | |
| | LOC101929818 | | |
| 0 | LOC101929826 | FALSE | Upregulated |
| | LOC101929831 | | |
| -1,042131907 | LOC101929846 | | Downregulated |
| 0,815248655 | LOC101929860 | FALSE | Upregulated |
| | LOC101929863 | | |
| | LOC101929866 | | |
| 0,396571267 | LOC101929882 | FALSE | Upregulated |
| -0,204532005 | LOC101929887 | FALSE | Downregulated |
| 0,071214531 | LOC101929891 | FALSE | Upregulated |
| -0,090270291 | LOC101929897 | FALSE | Downregulated |
| -0,256975773 | LOC101929918 | FALSE | Downregulated |
| | LOC101929922 | | |
| 0,25746069 | LOC101929946 | FALSE | Upregulated |
| 1,168459888 | LOC101929953 | | Upregulated |
| 0,966356091 | LOC101929964 | FALSE | Upregulated |
| | LOC101929972 | | |
| -0,004298224 | LOC101929974 | FALSE | Downregulated |
| | LOC101929976 | | |
| -0,30623759 | LOC101929977 | FALSE | Downregulated |
| | LOC101929983 | | |
| | LOC101929989 | | |
| 0,947555375 | LOC101929998 | FALSE | Upregulated |
| 0 | LOC101930010 | FALSE | Upregulated |
| 0 | LOC101930026 | FALSE | Upregulated |
| | LOC101930048 | | |
| | LOC101930053 | | |

| | | | |
|---|---|---|---|
| 0 | LOC101930071 | FALSE | Upregulated |
| | LOC101930091 | | |
| | LOC101930098 | | |
| 0,62367386 | LOC101930100 | FALSE | Upregulated |
| | LOC101930109 | | |
| 0,018025242 | LOC101930114 | FALSE | Upregulated |
| -0,172897284 | LOC101930120 | FALSE | Downregulated |
| | LOC101930164 | | |
| 1,885034661 | LOC101930232 | | Upregulated |
| 0,076658198 | LOC101930249 | FALSE | Upregulated |
| | LOC101930259 | | |
| 0 | LOC101930276 | FALSE | Upregulated |
| | LOC101930294 | | |
| -0,045688495 | LOC101930305 | FALSE | Downregulated |
| | LOC101930310 | | |
| 0,340269001 | LOC101930332 | FALSE | Upregulated |
| | LOC101930343 | | |
| 0,146474839 | LOC101930357 | FALSE | Upregulated |
| 0 | LOC101930370 | FALSE | Upregulated |
| | LOC101930384 | | |
| 0 | LOC101930420 | FALSE | Upregulated |
| 0 | LOC101930434 | FALSE | Upregulated |
| -0,518087133 | LOC101930460 | FALSE | Downregulated |
| | LOC101930476 | | |
| | LOC101930484 | | |
| 0 | LOC101930496 | FALSE | Upregulated |
| | LOC101930523 | | |
| | LOC101930531 | | |
| 0,103583191 | LOC101930580 | FALSE | Upregulated |
| | LOC101930593 | | |
| -0,343425381 | LOC101930644 | FALSE | Downregulated |
| | LOC101930665 | | |
| | LOC102467074 | | |
| | LOC102467075 | | |
| | LOC102467079 | | |
| 2,50468443 | LOC102467080 | | Upregulated |
| | LOC102467081 | | |
| -0,439497863 | LOC102467146 | FALSE | Downregulated |
| 0 | LOC102467147 | FALSE | Upregulated |
| | LOC102467212 | | |
| | LOC102467213 | | |
| | LOC102467214 | | |
| 0 | LOC102467216 | FALSE | Upregulated |
| -0,295467241 | LOC102467217 | FALSE | Downregulated |
| | LOC102467223 | | |
| | LOC102467224 | | |
| -0,06420295 | LOC102467225 | FALSE | Downregulated |
| | LOC102467226 | | |
| | LOC102467655 | | |
| | LOC102477328 | | |
| | LOC102503427 | | |

| | | | |
|---|---|---|---|
| | LOC102503429 | | |
| 1,035911545 | LOC102524628 | | Upregulated |
| | LOC102546175 | | |
| | LOC102546227 | | |
| -1,985312369 | LOC102546228 | | Downregulated |
| 0,024123522 | LOC102546229 | FALSE | Upregulated |
| -0,773603267 | LOC102546294 | FALSE | Downregulated |
| 0 | LOC102546298 | FALSE | Upregulated |
| | LOC102546299 | | |
| | LOC102557615 | | |
| | LOC102577424 | | |
| 0,705195276 | LOC102577426 | FALSE | Upregulated |
| | LOC102606466 | | |
| -0,004298224 | LOC102723305 | FALSE | Downregulated |
| -0,077307883 | LOC102723307 | FALSE | Downregulated |
| | LOC102723308 | | |
| 0,589430723 | LOC102723310 | FALSE | Upregulated |
| | LOC102723315 | | |
| 0,050688952 | LOC102723316 | FALSE | Upregulated |
| | LOC102723318 | | |
| 0,673539638 | LOC102723319 | FALSE | Upregulated |
| | LOC102723320 | | |
| | LOC102723321 | | |
| | LOC102723322 | | |
| 0,804695863 | LOC102723323 | FALSE | Upregulated |
| 0,610235076 | LOC102723324 | FALSE | Upregulated |
| | LOC102723325 | | |
| | LOC102723326 | | |
| 0,076498784 | LOC102723327 | FALSE | Upregulated |
| | LOC102723330 | | |
| 0,370004437 | LOC102723332 | FALSE | Upregulated |
| | LOC102723334 | | |
| | LOC102723336 | | |
| 0,095260654 | LOC102723337 | FALSE | Upregulated |
| | LOC102723338 | | |
| | LOC102723339 | | |
| -0,160429062 | LOC102723340 | FALSE | Downregulated |
| | LOC102723341 | | |
| | LOC102723343 | | |
| | LOC102723344 | | |
| | LOC102723345 | | |
| | LOC102723346 | | |
| | LOC102723347 | | |
| 0 | LOC102723348 | FALSE | Upregulated |
| 0 | LOC102723349 | FALSE | Upregulated |
| 0,019985931 | LOC102723350 | FALSE | Upregulated |
| | LOC102723351 | | |
| | LOC102723352 | | |
| -0,263780027 | LOC102723353 | FALSE | Downregulated |
| -0,079728583 | LOC102723354 | FALSE | Downregulated |
| | LOC102723355 | | |

| | | | |
|---|---|---|---|
| 0 | LOC102723357 | FALSE | Upregulated |
| | LOC102723358 | | |
| -0,058312068 | LOC102723359 | FALSE | Downregulated |
| 0,477600315 | LOC102723361 | FALSE | Upregulated |
| | LOC102723362 | | |
| | LOC102723363 | | |
| | LOC102723364 | | |
| -0,417449314 | LOC102723365 | FALSE | Downregulated |
| -0,4035784 | LOC102723366 | FALSE | Downregulated |
| 0,540659404 | LOC102723367 | FALSE | Upregulated |
| | LOC102723368 | | |
| | LOC102723370 | | |
| 0 | LOC102723371 | FALSE | Upregulated |
| | LOC102723372 | | |
| | LOC102723373 | | |
| -0,024107812 | LOC102723374 | FALSE | Downregulated |
| 0,90024217 | LOC102723375 | FALSE | Upregulated |
| | LOC102723376 | | |
| 0,197789756 | LOC102723377 | FALSE | Upregulated |
| | LOC102723378 | | |
| | LOC102723379 | | |
| 1,978233063 | LOC102723380 | | Upregulated |
| | LOC102723384 | | |
| 0 | LOC102723385 | FALSE | Upregulated |
| | LOC102723386 | | |
| | LOC102723388 | | |
| 0,683498456 | LOC102723389 | FALSE | Upregulated |
| | LOC102723390 | | |
| 0,608246941 | LOC102723391 | FALSE | Upregulated |
| 0 | LOC102723392 | FALSE | Upregulated |
| | LOC102723393 | | |
| | LOC102723394 | | |
| | LOC102723395 | | |
| | LOC102723396 | | |
| | LOC102723397 | | |
| 0,418045939 | LOC102723398 | FALSE | Upregulated |
| | LOC102723402 | | |
| | LOC102723403 | | |
| | LOC102723404 | | |
| 2,821430008 | LOC102723407 | | Upregulated |
| -0,04648513 | LOC102723408 | FALSE | Downregulated |
| 0,060329295 | LOC102723409 | FALSE | Upregulated |
| -2,535274043 | LOC102723410 | | Downregulated |
| | LOC102723411 | | |
| 0,567426611 | LOC102723412 | FALSE | Upregulated |
| | LOC102723413 | | |
| | LOC102723414 | | |
| 0,067498176 | LOC102723415 | FALSE | Upregulated |
| | LOC102723417 | | |
| | LOC102723418 | | |
| | LOC102723419 | | |

| | | | |
|---|---|---|---|
| -0,016101783 | LOC102723420 | FALSE | Downregulated |
| 0,003598903 | LOC102723421 | FALSE | Upregulated |
| | LOC102723423 | | |
| | LOC102723426 | | |
| | LOC102723427 | | |
| 0,282655013 | LOC102723428 | FALSE | Upregulated |
| | LOC102723429 | | |
| | LOC102723430 | | |
| | LOC102723431 | | |
| | LOC102723434 | | |
| | LOC102723435 | | |
| | LOC102723436 | | |
| 1,186960448 | LOC102723437 | | Upregulated |
| | LOC102723439 | | |
| | LOC102723440 | | |
| 0,087836139 | LOC102723441 | FALSE | Upregulated |
| | LOC102723442 | | |
| | LOC102723443 | | |
| | LOC102723445 | | |
| 0,464605708 | LOC102723446 | FALSE | Upregulated |
| | LOC102723447 | | |
| | LOC102723448 | | |
| | LOC102723449 | | |
| -0,100573543 | LOC102723450 | FALSE | Downregulated |
| 1,101847017 | LOC102723452 | | Upregulated |
| | LOC102723453 | | |
| | LOC102723454 | | |
| 0 | LOC102723455 | FALSE | Upregulated |
| | LOC102723456 | | |
| | LOC102723458 | | |
| | LOC102723459 | | |
| | LOC102723460 | | |
| | LOC102723463 | | |
| -0,181967548 | LOC102723464 | FALSE | Downregulated |
| -0,200417874 | LOC102723467 | FALSE | Downregulated |
| -0,06413575 | LOC102723468 | FALSE | Downregulated |
| 0,034943632 | LOC102723470 | FALSE | Upregulated |
| | LOC102723471 | | |
| | LOC102723472 | | |
| | LOC102723473 | | |
| | LOC102723474 | | |
| | LOC102723476 | | |
| | LOC102723477 | | |
| | LOC102723479 | | |
| | LOC102723480 | | |
| -0,076792818 | LOC102723481 | FALSE | Downregulated |
| 0 | LOC102723482 | FALSE | Upregulated |
| -0,005165736 | LOC102723483 | FALSE | Downregulated |
| | LOC102723484 | | |
| | LOC102723485 | | |
| | LOC102723486 | | |

| | | | |
|---|---|---|---|
| | LOC102723487 | | |
| 0,589563406 | LOC102723488 | FALSE | Upregulated |
| | LOC102723490 | | |
| | LOC102723491 | | |
| 0 | LOC102723492 | FALSE | Upregulated |
| 1,274906575 | LOC102723493 | | Upregulated |
| | LOC102723494 | | |
| | LOC102723495 | | |
| | LOC102723496 | | |
| -1,33524409 | LOC102723497 | | Downregulated |
| | LOC102723499 | | |
| | LOC102723502 | | |
| | LOC102723503 | | |
| | LOC102723505 | | |
| 0,042050302 | LOC102723507 | FALSE | Upregulated |
| | LOC102723508 | | |
| 0,399383748 | LOC102723510 | FALSE | Upregulated |
| 5,46553E-05 | LOC102723513 | FALSE | Upregulated |
| | LOC102723514 | | |
| | LOC102723515 | | |
| | LOC102723516 | | |
| 1,386647366 | LOC102723517 | | Upregulated |
| | LOC102723518 | | |
| | LOC102723522 | | |
| | LOC102723523 | | |
| | LOC102723524 | | |
| 0 | LOC102723525 | FALSE | Upregulated |
| | LOC102723526 | | |
| | LOC102723527 | | |
| | LOC102723529 | | |
| | LOC102723530 | | |
| | LOC102723532 | | |
| | LOC102723533 | | |
| -1,763121212 | LOC102723534 | | Downregulated |
| | LOC102723536 | | |
| | LOC102723538 | | |
| 0,116577837 | LOC102723540 | FALSE | Upregulated |
| | LOC102723541 | | |
| 1,452768998 | LOC102723545 | | Upregulated |
| 0,001309969 | LOC102723546 | FALSE | Upregulated |
| 0,135716669 | LOC102723547 | FALSE | Upregulated |
| | LOC102723548 | | |
| -0,365287962 | LOC102723549 | FALSE | Downregulated |
| | LOC102723550 | | |
| | LOC102723551 | | |
| -0,002311421 | LOC102723552 | FALSE | Downregulated |
| 0,957758509 | LOC102723553 | FALSE | Upregulated |
| | LOC102723557 | | |
| 0 | LOC102723558 | FALSE | Upregulated |
| -0,364595513 | LOC102723559 | FALSE | Downregulated |
| | LOC102723560 | | |

| | | | |
|---|---|---|---|
| -0,127229664 | LOC102723561 | FALSE | Downregulated |
| | LOC102723562 | | |
| 0,53464797 | LOC102723563 | FALSE | Upregulated |
| 0,427560075 | LOC102723565 | FALSE | Upregulated |
| | LOC102723567 | | |
| | LOC102723568 | | |
| 1,126199447 | LOC102723569 | | Upregulated |
| -0,06049023 | LOC102723570 | FALSE | Downregulated |
| | LOC102723571 | | |
| 0 | LOC102723572 | FALSE | Upregulated |
| -0,024205792 | LOC102723575 | FALSE | Downregulated |
| 0,719672681 | LOC102723576 | FALSE | Upregulated |
| | LOC102723577 | | |
| | LOC102723578 | | |
| 0,89906048 | LOC102723579 | FALSE | Upregulated |
| -0,376727464 | LOC102723587 | FALSE | Downregulated |
| | LOC102723588 | | |
| 0,608246941 | LOC102723590 | FALSE | Upregulated |
| 0,006863373 | LOC102723591 | FALSE | Upregulated |
| | LOC102723593 | | |
| -0,086155806 | LOC102723594 | FALSE | Downregulated |
| | LOC102723596 | | |
| | LOC102723599 | | |
| -0,185149432 | LOC102723602 | FALSE | Downregulated |
| | LOC102723605 | | |
| | LOC102723606 | | |
| 0 | LOC102723607 | FALSE | Upregulated |
| | LOC102723608 | | |
| | LOC102723609 | | |
| -0,083192521 | LOC102723610 | FALSE | Downregulated |
| | LOC102723612 | | |
| 0,318603036 | LOC102723616 | FALSE | Upregulated |
| | LOC102723617 | | |
| -0,002089411 | LOC102723619 | FALSE | Downregulated |
| | LOC102723620 | | |
| | LOC102723621 | | |
| 0,041547119 | LOC102723623 | FALSE | Upregulated |
| | LOC102723624 | | |
| -0,086488968 | LOC102723627 | FALSE | Downregulated |
| | LOC102723628 | | |
| | LOC102723629 | | |
| 0,043670653 | LOC102723630 | FALSE | Upregulated |
| | LOC102723631 | | |
| | LOC102723633 | | |
| 0 | LOC102723635 | FALSE | Upregulated |
| -0,823089547 | LOC102723636 | FALSE | Downregulated |
| | LOC102723637 | | |
| | LOC102723638 | | |
| | LOC102723639 | | |
| -1,223601723 | LOC102723640 | | Downregulated |
| -0,929865479 | LOC102723641 | FALSE | Downregulated |

| | | | |
|---|---|---|---|
| 1,333494076 | LOC102723642 | | Upregulated |
| 0,204249669 | LOC102723643 | FALSE | Upregulated |
| | LOC102723644 | | |
| | LOC102723645 | | |
| | LOC102723646 | | |
| 0,067831501 | LOC102723647 | FALSE | Upregulated |
| -0,91050176 | LOC102723648 | FALSE | Downregulated |
| | LOC102723649 | | |
| | LOC102723650 | | |
| 0,120687012 | LOC102723652 | FALSE | Upregulated |
| | LOC102723653 | | |
| | LOC102723654 | | |
| 0,673310182 | LOC102723655 | FALSE | Upregulated |
| -0,064217755 | LOC102723656 | FALSE | Downregulated |
| | LOC102723657 | | |
| | LOC102723658 | | |
| -0,66595939 | LOC102723659 | FALSE | Downregulated |
| | LOC102723661 | | |
| | LOC102723662 | | |
| 0 | LOC102723663 | FALSE | Upregulated |
| | LOC102723664 | | |
| | LOC102723667 | | |
| | LOC102723669 | | |
| 0,076658198 | LOC102723670 | FALSE | Upregulated |
| -0,073259509 | LOC102723672 | FALSE | Downregulated |
| | LOC102723673 | | |
| | LOC102723674 | | |
| 0,669756935 | LOC102723676 | FALSE | Upregulated |
| -1,150938594 | LOC102723677 | | Downregulated |
| | LOC102723680 | | |
| 0,074989643 | LOC102723682 | FALSE | Upregulated |
| | LOC102723684 | | |
| 0 | LOC102723685 | FALSE | Upregulated |
| | LOC102723686 | | |
| -0,122575717 | LOC102723687 | FALSE | Downregulated |
| -0,183868532 | LOC102723690 | FALSE | Downregulated |
| 1,081905769 | LOC102723691 | | Upregulated |
| -0,123513219 | LOC102723693 | FALSE | Downregulated |
| 0 | LOC102723695 | FALSE | Upregulated |
| 1,215944313 | LOC102723696 | | Upregulated |
| 0,439870423 | LOC102723697 | FALSE | Upregulated |
| | LOC102723698 | | |
| | LOC102723699 | | |
| 0 | LOC102723700 | FALSE | Upregulated |
| | LOC102723702 | | |
| 0,692995391 | LOC102723704 | FALSE | Upregulated |
| | LOC102723705 | | |
| | LOC102723706 | | |
| | LOC102723710 | | |
| | LOC102723712 | | |
| | LOC102723713 | | |

| | | | |
|---|---|---|---|
| | LOC102723714 | | |
| -0,1555044 | LOC102723715 | FALSE | Downregulated |
| 0,579426065 | LOC102723716 | FALSE | Upregulated |
| 1,287451867 | LOC102723717 | | Upregulated |
| 1,454669866 | LOC102723718 | | Upregulated |
| 1,649940707 | LOC102723720 | | Upregulated |
| 0 | LOC102723721 | FALSE | Upregulated |
| | LOC102723723 | | |
| | LOC102723724 | | |
| | LOC102723725 | | |
| | LOC102723727 | | |
| | LOC102723729 | | |
| 0 | LOC102723730 | FALSE | Upregulated |
| | LOC102723731 | | |
| -0,522412636 | LOC102723732 | FALSE | Downregulated |
| 0,899877773 | LOC102723733 | FALSE | Upregulated |
| | LOC102723734 | | |
| | LOC102723736 | | |
| | LOC102723737 | | |
| | LOC102723738 | | |
| 0,654980006 | LOC102723739 | FALSE | Upregulated |
| | LOC102723740 | | |
| | LOC102723742 | | |
| -0,026872299 | LOC102723744 | FALSE | Downregulated |
| | LOC102723745 | | |
| -0,016775302 | LOC102723746 | FALSE | Downregulated |
| | LOC102723749 | | |
| | LOC102723751 | | |
| | LOC102723752 | | |
| | LOC102723754 | | |
| 0,619934301 | LOC102723755 | FALSE | Upregulated |
| -0,974997331 | LOC102723756 | FALSE | Downregulated |
| 0,213119458 | LOC102723757 | FALSE | Upregulated |
| -0,403443648 | LOC102723759 | FALSE | Downregulated |
| | LOC102723760 | | |
| 0,060421595 | LOC102723761 | FALSE | Upregulated |
| | LOC102723763 | | |
| | LOC102723764 | | |
| | LOC102723765 | | |
| | LOC102723768 | | |
| 1,418632075 | LOC102723770 | | Upregulated |
| -0,004298224 | LOC102723771 | FALSE | Downregulated |
| 0,007214246 | LOC102723774 | FALSE | Upregulated |
| 0 | LOC102723775 | FALSE | Upregulated |
| -0,56028627 | LOC102723777 | FALSE | Downregulated |
| | LOC102723778 | | |
| | LOC102723780 | | |
| 0 | LOC102723781 | FALSE | Upregulated |
| | LOC102723783 | | |
| | LOC102723784 | | |
| | LOC102723785 | | |

| | | | |
|---|---|---|---|
| | LOC102723786 | | |
| | LOC102723787 | | |
| | LOC102723789 | | |
| 0,151823721 | LOC102723790 | FALSE | Upregulated |
| 1,265120123 | LOC102723791 | | Upregulated |
| | LOC102723793 | | |
| | LOC102723795 | | |
| | LOC102723796 | | |
| -0,195834301 | LOC102723797 | FALSE | Downregulated |
| | LOC102723800 | | |
| 0,345196987 | LOC102723801 | FALSE | Upregulated |
| | LOC102723802 | | |
| | LOC102723803 | | |
| 1,226556905 | LOC102723805 | | Upregulated |
| | LOC102723807 | | |
| -0,227393897 | LOC102723808 | FALSE | Downregulated |
| | LOC102723810 | | |
| | LOC102723812 | | |
| 1,700533667 | LOC102723813 | | Upregulated |
| -0,458216764 | LOC102723814 | FALSE | Downregulated |
| | LOC102723816 | | |
| 0,811481981 | LOC102723817 | FALSE | Upregulated |
| | LOC102723818 | | |
| | LOC102723819 | | |
| | LOC102723821 | | |
| | LOC102723822 | | |
| | LOC102723823 | | |
| | LOC102723825 | | |
| | LOC102723826 | | |
| 0 | LOC102723828 | FALSE | Upregulated |
| 0,090676974 | LOC102723829 | FALSE | Upregulated |
| | LOC102723830 | | |
| | LOC102723831 | | |
| | LOC102723832 | | |
| | LOC102723833 | | |
| | LOC102723834 | | |
| | LOC102723835 | | |
| -0,293893978 | LOC102723836 | FALSE | Downregulated |
| | LOC102723837 | | |
| | LOC102723838 | | |
| 0 | LOC102723839 | FALSE | Upregulated |
| -0,126685733 | LOC102723840 | FALSE | Downregulated |
| | LOC102723841 | | |
| | LOC102723842 | | |
| 1,015331251 | LOC102723843 | | Upregulated |
| 0 | LOC102723844 | FALSE | Upregulated |
| | LOC102723846 | | |
| | LOC102723847 | | |
| | LOC102723848 | | |
| | LOC102723849 | | |
| | LOC102723850 | | |

| | | | |
|---|---|---|---|
| 0,039005732 | LOC102723854 | FALSE | Upregulated |
| | LOC102723855 | | |
| 0,136472829 | LOC102723856 | FALSE | Upregulated |
| | LOC102723857 | | |
| | LOC102723858 | | |
| -0,308930456 | LOC102723859 | FALSE | Downregulated |
| | LOC102723860 | | |
| 1,278006564 | LOC102723861 | | Upregulated |
| | LOC102723862 | | |
| 0 | LOC102723865 | FALSE | Upregulated |
| -0,305782263 | LOC102723866 | FALSE | Downregulated |
| -0,125728908 | LOC102723867 | FALSE | Downregulated |
| | LOC102723868 | | |
| | LOC102723869 | | |
| 0,279552331 | LOC102723870 | FALSE | Upregulated |
| | LOC102723871 | | |
| -0,966945859 | LOC102723872 | FALSE | Downregulated |
| | LOC102723873 | | |
| | LOC102723875 | | |
| 0,004929336 | LOC102723876 | FALSE | Upregulated |
| | LOC102723878 | | |
| | LOC102723879 | | |
| | LOC102723880 | | |
| | LOC102723881 | | |
| | LOC102723882 | | |
| | LOC102723883 | | |
| -0,766464902 | LOC102723884 | FALSE | Downregulated |
| 0,573831474 | LOC102723885 | FALSE | Upregulated |
| | LOC102723887 | | |
| 0,608246941 | LOC102723888 | FALSE | Upregulated |
| 0,539379722 | LOC102723889 | FALSE | Upregulated |
| | LOC102723890 | | |
| 0,02849032 | LOC102723891 | FALSE | Upregulated |
| 0 | LOC102723892 | FALSE | Upregulated |
| 0,608246941 | LOC102723894 | FALSE | Upregulated |
| | LOC102723895 | | |
| -0,080118378 | LOC102723896 | FALSE | Downregulated |
| | LOC102723898 | | |
| 0,036748944 | LOC102723899 | FALSE | Upregulated |
| | LOC102723900 | | |
| 0,063452594 | LOC102723901 | FALSE | Upregulated |
| 0 | LOC102723902 | FALSE | Upregulated |
| | LOC102723903 | | |
| 0 | LOC102723904 | FALSE | Upregulated |
| 0 | LOC102723905 | FALSE | Upregulated |
| | LOC102723906 | | |
| | LOC102723907 | | |
| | LOC102723909 | | |
| | LOC102723910 | | |
| | LOC102723911 | | |
| | LOC102723912 | | |

| | | | |
|---|---|---|---|
| 0 | LOC102723913 | FALSE | Upregulated |
| 0,000577308 | LOC102723914 | FALSE | Upregulated |
| 0,499635131 | LOC102723915 | FALSE | Upregulated |
| 0 | LOC102723916 | FALSE | Upregulated |
| | LOC102723918 | | |
| 1,659579534 | LOC102723919 | | Upregulated |
| -0,24946932 | LOC102723920 | FALSE | Downregulated |
| | LOC102723921 | | |
| 0 | LOC102723922 | FALSE | Upregulated |
| | LOC102723924 | | |
| 0,117238243 | LOC102723925 | FALSE | Upregulated |
| 1,333288826 | LOC102723926 | | Upregulated |
| | LOC102723928 | | |
| | LOC102723931 | | |
| | LOC102723932 | | |
| | LOC102723933 | | |
| 0 | LOC102723934 | FALSE | Upregulated |
| | LOC102723935 | | |
| | LOC102723936 | | |
| | LOC102723937 | | |
| | LOC102723938 | | |
| | LOC102723939 | | |
| 0 | LOC102723940 | FALSE | Upregulated |
| | LOC102723941 | | |
| -0,463652211 | LOC102723942 | FALSE | Downregulated |
| | LOC102723944 | | |
| 0 | LOC102723946 | FALSE | Upregulated |
| | LOC102723947 | | |
| | LOC102723948 | | |
| -0,445023454 | LOC102723949 | FALSE | Downregulated |
| | LOC102723950 | | |
| | LOC102723951 | | |
| | LOC102723952 | | |
| -2,030619326 | LOC102723953 | | Downregulated |
| 0,340526815 | LOC102723955 | FALSE | Upregulated |
| 1,775089527 | LOC102723956 | | Upregulated |
| | LOC102723957 | | |
| 0,701300886 | LOC102723958 | FALSE | Upregulated |
| 0,687011092 | LOC102723959 | FALSE | Upregulated |
| | LOC102723961 | | |
| | LOC102723962 | | |
| 0 | LOC102723963 | FALSE | Upregulated |
| 0 | LOC102723966 | FALSE | Upregulated |
| | LOC102723967 | | |
| | LOC102723968 | | |
| | LOC102723969 | | |
| | LOC102723970 | | |
| | LOC102723971 | | |
| -0,07374155 | LOC102723973 | FALSE | Downregulated |
| -0,872492988 | LOC102723974 | FALSE | Downregulated |
| -0,381097947 | LOC102723975 | FALSE | Downregulated |

| | | | |
|---|---|---|---|
| | LOC102723976 | | |
| | LOC102723977 | | |
| | LOC102723978 | | |
| | LOC102723979 | | |
| 0,998881662 | LOC102723981 | FALSE | Upregulated |
| 0,742851002 | LOC102723983 | FALSE | Upregulated |
| | LOC102723984 | | |
| 0,076658198 | LOC102723985 | FALSE | Upregulated |
| | LOC102723986 | | |
| | LOC102723987 | | |
| -0,435492063 | LOC102723988 | FALSE | Downregulated |
| 0,004929336 | LOC102723993 | FALSE | Upregulated |
| | LOC102723994 | | |
| 0,543565227 | LOC102723995 | FALSE | Upregulated |
| | LOC102724000 | | |
| | LOC102724001 | | |
| 0,768157716 | LOC102724003 | FALSE | Upregulated |
| | LOC102724004 | | |
| | LOC102724007 | | |
| | LOC102724008 | | |
| | LOC102724011 | | |
| 0 | LOC102724012 | FALSE | Upregulated |
| | LOC102724013 | | |
| | LOC102724014 | | |
| -0,189001521 | LOC102724015 | FALSE | Downregulated |
| -0,003746071 | LOC102724017 | FALSE | Downregulated |
| -0,00897377 | LOC102724018 | FALSE | Downregulated |
| -1,664823745 | LOC102724019 | | Downregulated |
| | LOC102724020 | | |
| | LOC102724021 | | |
| | LOC102724022 | | |
| | LOC102724027 | | |
| -0,018921051 | LOC102724028 | FALSE | Downregulated |
| | LOC102724029 | | |
| 0,79678003 | LOC102724030 | FALSE | Upregulated |
| 0 | LOC102724031 | FALSE | Upregulated |
| | LOC102724032 | | |
| | LOC102724033 | | |
| 0,373496789 | LOC102724034 | FALSE | Upregulated |
| | LOC102724036 | | |
| | LOC102724038 | | |
| | LOC102724039 | | |
| | LOC102724041 | | |
| | LOC102724044 | | |
| | LOC102724045 | | |
| | LOC102724046 | | |
| | LOC102724048 | | |
| | LOC102724050 | | |
| | LOC102724051 | | |
| | LOC102724052 | | |
| | LOC102724053 | | |

| | | | |
|---|---|---|---|
| | LOC102724054 | | |
| | LOC102724055 | | |
| | LOC102724056 | | |
| 0,049958823 | LOC102724057 | FALSE | Upregulated |
| | LOC102724058 | | |
| -0,329062631 | LOC102724059 | FALSE | Downregulated |
| -0,036713102 | LOC102724060 | FALSE | Downregulated |
| | LOC102724062 | | |
| | LOC102724063 | | |
| 0,777898326 | LOC102724065 | FALSE | Upregulated |
| | LOC102724067 | | |
| 0,179772992 | LOC102724068 | FALSE | Upregulated |
| | LOC102724069 | | |
| | LOC102724071 | | |
| -0,469131466 | LOC102724072 | FALSE | Downregulated |
| -0,709704977 | LOC102724073 | FALSE | Downregulated |
| | LOC102724074 | | |
| 0,1586921 | LOC102724075 | FALSE | Upregulated |
| | LOC102724076 | | |
| | LOC102724078 | | |
| 0 | LOC102724079 | FALSE | Upregulated |
| | LOC102724080 | | |
| | LOC102724081 | | |
| | LOC102724082 | | |
| 0,048656215 | LOC102724084 | FALSE | Upregulated |
| | LOC102724085 | | |
| | LOC102724086 | | |
| | LOC102724087 | | |
| | LOC102724088 | | |
| | LOC102724089 | | |
| 0,048656215 | LOC102724090 | FALSE | Upregulated |
| | LOC102724091 | | |
| -0,263192064 | LOC102724092 | FALSE | Downregulated |
| -1,56553234 | LOC102724093 | | Downregulated |
| 0,039005732 | LOC102724094 | FALSE | Upregulated |
| | LOC102724095 | | |
| 0,857011261 | LOC102724096 | FALSE | Upregulated |
| 0 | LOC102724097 | FALSE | Upregulated |
| -0,02168579 | LOC102724098 | FALSE | Downregulated |
| | LOC102724099 | | |
| | LOC102724101 | | |
| | LOC102724102 | | |
| | LOC102724104 | | |
| 0,323484293 | LOC102724106 | FALSE | Upregulated |
| | LOC102724107 | | |
| -0,222530342 | LOC102724108 | FALSE | Downregulated |
| 0,078625993 | LOC102724109 | FALSE | Upregulated |
| | LOC102724111 | | |
| -0,056585813 | LOC102724112 | FALSE | Downregulated |
| | LOC102724113 | | |
| | LOC102724114 | | |

| | | | |
|---|---|---|---|
| | LOC102724115 | | |
| | LOC102724118 | | |
| | LOC102724119 | | |
| -0,980663107 | LOC102724120 | FALSE | Downregulated |
| -0,790994477 | LOC102724121 | FALSE | Downregulated |
| 0,669756935 | LOC102724122 | FALSE | Upregulated |
| | LOC102724123 | | |
| | LOC102724125 | | |
| | LOC102724127 | | |
| | LOC102724128 | | |
| 0 | LOC102724130 | FALSE | Upregulated |
| | LOC102724131 | | |
| -0,079171183 | LOC102724132 | FALSE | Downregulated |
| | LOC102724134 | | |
| | LOC102724136 | | |
| | LOC102724137 | | |
| 0,597480774 | LOC102724138 | FALSE | Upregulated |
| 0,669756935 | LOC102724139 | FALSE | Upregulated |
| 0 | LOC102724140 | FALSE | Upregulated |
| | LOC102724142 | | |
| -0,624636197 | LOC102724143 | FALSE | Downregulated |
| | LOC102724145 | | |
| | LOC102724146 | | |
| | LOC102724148 | | |
| -0,052715095 | LOC102724149 | FALSE | Downregulated |
| | LOC102724150 | | |
| | LOC102724151 | | |
| | LOC102724152 | | |
| 0 | LOC102724154 | FALSE | Upregulated |
| 1,604035081 | LOC102724155 | | Upregulated |
| 0,450323671 | LOC102724156 | FALSE | Upregulated |
| | LOC102724158 | | |
| | LOC102724160 | | |
| | LOC102724161 | | |
| 0 | LOC102724162 | FALSE | Upregulated |
| | LOC102724165 | | |
| | LOC102724166 | | |
| | LOC102724167 | | |
| 1,117622297 | LOC102724168 | | Upregulated |
| -0,27963675 | LOC102724169 | FALSE | Downregulated |
| 0,02582963 | LOC102724170 | FALSE | Upregulated |
| 0 | LOC102724171 | FALSE | Upregulated |
| | LOC102724173 | | |
| | LOC102724174 | | |
| 0 | LOC102724176 | FALSE | Upregulated |
| | LOC102724180 | | |
| | LOC102724182 | | |
| | LOC102724183 | | |
| -1,564855002 | LOC102724184 | | Downregulated |
| | LOC102724185 | | |
| | LOC102724186 | | |

| | | | |
|---|---|---|---|
| | LOC102724188 | | |
| | LOC102724189 | | |
| 0,170724258 | LOC102724190 | FALSE | Upregulated |
| | LOC102724194 | | |
| 1,219567064 | LOC102724195 | | Upregulated |
| 0,626631668 | LOC102724197 | FALSE | Upregulated |
| | LOC102724199 | | |
| -0,374144785 | LOC102724201 | FALSE | Downregulated |
| | LOC102724202 | | |
| | LOC102724203 | | |
| | LOC102724204 | | |
| | LOC102724205 | | |
| | LOC102724207 | | |
| -0,085629784 | LOC102724208 | FALSE | Downregulated |
| 0,465737295 | LOC102724209 | FALSE | Upregulated |
| | LOC102724210 | | |
| | LOC102724211 | | |
| | LOC102724212 | | |
| 0 | LOC102724213 | FALSE | Upregulated |
| | LOC102724214 | | |
| | LOC102724215 | | |
| | LOC102724216 | | |
| | LOC102724218 | | |
| | LOC102724220 | | |
| | LOC102724222 | | |
| | LOC102724224 | | |
| | LOC102724225 | | |
| | LOC102724226 | | |
| | LOC102724227 | | |
| | LOC102724228 | | |
| 0,751440556 | LOC102724229 | FALSE | Upregulated |
| 0 | LOC102724231 | FALSE | Upregulated |
| 0,391344838 | LOC102724232 | FALSE | Upregulated |
| | LOC102724234 | | |
| 2,221553947 | LOC102724237 | | Upregulated |
| | LOC102724239 | | |
| | LOC102724241 | | |
| | LOC102724242 | | |
| 0,622656528 | LOC102724244 | FALSE | Upregulated |
| 0,772076968 | LOC102724245 | FALSE | Upregulated |
| | LOC102724246 | | |
| | LOC102724247 | | |
| -0,46198856 | LOC102724248 | FALSE | Downregulated |
| 0,148317405 | LOC102724249 | FALSE | Upregulated |
| | LOC102724251 | | |
| -0,377930988 | LOC102724252 | FALSE | Downregulated |
| | LOC102724253 | | |
| -0,450904191 | LOC102724256 | FALSE | Downregulated |
| | LOC102724257 | | |
| -0,856298929 | LOC102724259 | FALSE | Downregulated |
| | LOC102724260 | | |

| | | | |
|---|---|---|---|
| 0,429460632 | LOC102724261 | FALSE | Upregulated |
| -1,350783451 | LOC102724263 | | Downregulated |
| | LOC102724264 | | |
| 0,099718299 | LOC102724265 | FALSE | Upregulated |
| | LOC102724266 | | |
| | LOC102724267 | | |
| | LOC102724268 | | |
| | LOC102724270 | | |
| | LOC102724271 | | |
| 0,066130885 | LOC102724272 | FALSE | Upregulated |
| 1,565773311 | LOC102724274 | | Upregulated |
| 0,718944626 | LOC102724275 | FALSE | Upregulated |
| -0,321362385 | LOC102724276 | FALSE | Downregulated |
| | LOC102724277 | | |
| | LOC102724278 | | |
| | LOC102724279 | | |
| | LOC102724280 | | |
| -0,002529603 | LOC102724282 | FALSE | Downregulated |
| -0,042495736 | LOC102724283 | FALSE | Downregulated |
| | LOC102724284 | | |
| 0,073681855 | LOC102724285 | FALSE | Upregulated |
| | LOC102724287 | | |
| | LOC102724288 | | |
| | LOC102724289 | | |
| | LOC102724290 | | |
| | LOC102724294 | | |
| 0,509366965 | LOC102724295 | FALSE | Upregulated |
| 1,410066292 | LOC102724296 | | Upregulated |
| 0 | LOC102724297 | FALSE | Upregulated |
| 0 | LOC102724298 | FALSE | Upregulated |
| | LOC102724299 | | |
| 0 | LOC102724300 | FALSE | Upregulated |
| | LOC102724301 | | |
| | LOC102724302 | | |
| | LOC102724304 | | |
| | LOC102724305 | | |
| -0,121288552 | LOC102724306 | FALSE | Downregulated |
| -0,591078944 | LOC102724307 | FALSE | Downregulated |
| | LOC102724308 | | |
| 0 | LOC102724309 | FALSE | Upregulated |
| | LOC102724311 | | |
| | LOC102724313 | | |
| | LOC102724315 | | |
| | LOC102724317 | | |
| | LOC102724318 | | |
| | LOC102724321 | | |
| 1,384929244 | LOC102724323 | | Upregulated |
| | LOC102724324 | | |
| | LOC102724325 | | |
| | LOC102724326 | | |
| | LOC102724327 | | |

| | | | |
|---|---|---|---|
| -0,068277487 | LOC102724328 | FALSE | Downregulated |
| -0,074895435 | LOC102724329 | FALSE | Downregulated |
| | LOC102724331 | | |
| | LOC102724332 | | |
| | LOC102724333 | | |
| -0,436607183 | LOC102724334 | FALSE | Downregulated |
| | LOC102724337 | | |
| | LOC102724338 | | |
| 0,028485156 | LOC102724339 | FALSE | Upregulated |
| | LOC102724340 | | |
| | LOC102724341 | | |
| -0,203321707 | LOC102724343 | FALSE | Downregulated |
| | LOC102724344 | | |
| | LOC102724346 | | |
| | LOC102724347 | | |
| 0,009923003 | LOC102724349 | FALSE | Upregulated |
| 0 | LOC102724351 | FALSE | Upregulated |
| 1,408942445 | LOC102724352 | | Upregulated |
| | LOC102724353 | | |
| 0,734212819 | LOC102724354 | FALSE | Upregulated |
| | LOC102724355 | | |
| | LOC102724357 | | |
| | LOC102724358 | | |
| | LOC102724359 | | |
| | LOC102724360 | | |
| 0 | LOC102724361 | FALSE | Upregulated |
| | LOC102724362 | | |
| 1,082360453 | LOC102724363 | | Upregulated |
| | LOC102724366 | | |
| | LOC102724367 | | |
| | LOC102724373 | | |
| | LOC102724375 | | |
| 2,116534509 | LOC102724377 | | Upregulated |
| | LOC102724379 | | |
| 0,661206439 | LOC102724380 | FALSE | Upregulated |
| | LOC102724381 | | |
| | LOC102724382 | | |
| 0,505198863 | LOC102724384 | FALSE | Upregulated |
| -0,050200932 | LOC102724385 | FALSE | Downregulated |
| | LOC102724386 | | |
| 1,149604336 | LOC102724389 | | Upregulated |
| 0,0196787 | LOC102724392 | FALSE | Upregulated |
| | LOC102724393 | | |
| -1,206062839 | LOC102724394 | | Downregulated |
| | LOC102724395 | | |
| | LOC102724396 | | |
| 0,728353424 | LOC102724397 | FALSE | Upregulated |
| -0,333752244 | LOC102724399 | FALSE | Downregulated |
| | LOC102724400 | | |
| -0,090496284 | LOC102724402 | FALSE | Downregulated |
| | LOC102724403 | | |

| | | | |
|---|---|---|---|
| 0 | LOC102724404 | FALSE | Upregulated |
| | LOC102724405 | | |
| 0 | LOC102724406 | FALSE | Upregulated |
| | LOC102724407 | | |
| -1,09354529 | LOC102724409 | | Downregulated |
| | LOC102724410 | | |
| 0 | LOC102724411 | FALSE | Upregulated |
| | LOC102724412 | | |
| | LOC102724414 | | |
| | LOC102724415 | | |
| 0 | LOC102724416 | FALSE | Upregulated |
| | LOC102724417 | | |
| | LOC102724418 | | |
| | LOC102724419 | | |
| 1,016790746 | LOC102724420 | | Upregulated |
| | LOC102724421 | | |
| | LOC102724422 | | |
| 0 | LOC102724424 | FALSE | Upregulated |
| | LOC102724425 | | |
| -1,083085713 | LOC102724426 | | Downregulated |
| | LOC102724427 | | |
| | LOC102724429 | | |
| | LOC102724431 | | |
| 0 | LOC102724432 | FALSE | Upregulated |
| 0 | LOC102724433 | FALSE | Upregulated |
| 0 | LOC102724434 | FALSE | Upregulated |
| | LOC102724435 | | |
| 0,520373733 | LOC102724436 | FALSE | Upregulated |
| | LOC102724438 | | |
| | LOC102724439 | | |
| | LOC102724440 | | |
| | LOC102724443 | | |
| 0,228609982 | LOC102724444 | FALSE | Upregulated |
| 0 | LOC102724446 | FALSE | Upregulated |
| 0,542685633 | LOC102724449 | FALSE | Upregulated |
| 0,903968615 | LOC102724450 | FALSE | Upregulated |
| 0,510486514 | LOC102724451 | FALSE | Upregulated |
| | LOC102724452 | | |
| | LOC102724453 | | |
| | LOC102724454 | | |
| | LOC102724456 | | |
| 0,556457948 | LOC102724457 | FALSE | Upregulated |
| | LOC102724458 | | |
| | LOC102724460 | | |
| | LOC102724461 | | |
| -1,596386408 | LOC102724462 | | Downregulated |
| 0 | LOC102724463 | FALSE | Upregulated |
| 0,055721111 | LOC102724464 | FALSE | Upregulated |
| | LOC102724465 | | |
| 0 | LOC102724467 | FALSE | Upregulated |
| | LOC102724469 | | |

| | | | |
|---|---|---|---|
| | LOC102724470 | | |
| | LOC102724471 | | |
| -0,032225332 | LOC102724472 | FALSE | Downregulated |
| | LOC102724473 | | |
| 0,054054106 | LOC102724477 | FALSE | Upregulated |
| | LOC102724478 | | |
| | LOC102724479 | | |
| | LOC102724480 | | |
| 0,608246941 | LOC102724481 | FALSE | Upregulated |
| | LOC102724482 | | |
| | LOC102724485 | | |
| | LOC102724486 | | |
| 1,325899997 | LOC102724487 | | Upregulated |
| 0,608246941 | LOC102724489 | FALSE | Upregulated |
| | LOC102724491 | | |
| | LOC102724492 | | |
| | LOC102724494 | | |
| | LOC102724495 | | |
| | LOC102724496 | | |
| -0,164169113 | LOC102724497 | FALSE | Downregulated |
| | LOC102724498 | | |
| | LOC102724500 | | |
| | LOC102724501 | | |
| 0,037661308 | LOC102724502 | FALSE | Upregulated |
| | LOC102724503 | | |
| | LOC102724505 | | |
| -0,190977614 | LOC102724506 | FALSE | Downregulated |
| 0,138077959 | LOC102724508 | FALSE | Upregulated |
| | LOC102724509 | | |
| | LOC102724510 | | |
| | LOC102724511 | | |
| | LOC102724512 | | |
| 0 | LOC102724513 | FALSE | Upregulated |
| -0,75425598 | LOC102724517 | FALSE | Downregulated |
| -0,291660749 | LOC102724519 | FALSE | Downregulated |
| | LOC102724520 | | |
| | LOC102724521 | | |
| | LOC102724523 | | |
| 0,659585802 | LOC102724524 | FALSE | Upregulated |
| | LOC102724527 | | |
| | LOC102724529 | | |
| 0,621336202 | LOC102724530 | FALSE | Upregulated |
| | LOC102724531 | | |
| -0,58332519 | LOC102724532 | FALSE | Downregulated |
| | LOC102724533 | | |
| | LOC102724534 | | |
| 1,606081669 | LOC102724537 | | Upregulated |
| | LOC102724538 | | |
| 0,074613204 | LOC102724539 | FALSE | Upregulated |
| | LOC102724540 | | |
| -0,603983005 | LOC102724541 | FALSE | Downregulated |

| | | | |
|---|---|---|---|
| | LOC102724542 | | |
| 0,360979786 | LOC102724543 | FALSE | Upregulated |
| | LOC102724544 | | |
| 0 | LOC102724551 | FALSE | Upregulated |
| 0 | LOC102724553 | FALSE | Upregulated |
| 0 | LOC102724555 | FALSE | Upregulated |
| | LOC102724557 | | |
| -0,00897377 | LOC102724558 | FALSE | Downregulated |
| | LOC102724561 | | |
| | LOC102724564 | | |
| | LOC102724565 | | |
| | LOC102724566 | | |
| | LOC102724567 | | |
| 0,044987085 | LOC102724568 | FALSE | Upregulated |
| 0,786185101 | LOC102724569 | FALSE | Upregulated |
| -0,067600543 | LOC102724570 | FALSE | Downregulated |
| 1,387402844 | LOC102724571 | | Upregulated |
| | LOC102724572 | | |
| 0,50766667 | LOC102724573 | FALSE | Upregulated |
| | LOC102724574 | | |
| | LOC102724575 | | |
| | LOC102724576 | | |
| | LOC102724577 | | |
| | LOC102724579 | | |
| -0,257770106 | LOC102724580 | FALSE | Downregulated |
| 2,199851056 | LOC102724581 | | Upregulated |
| | LOC102724582 | | |
| 0,996223401 | LOC102724586 | FALSE | Upregulated |
| | LOC102724588 | | |
| | LOC102724589 | | |
| | LOC102724590 | | |
| | LOC102724591 | | |
| | LOC102724592 | | |
| 0,122754612 | LOC102724593 | FALSE | Upregulated |
| -1,466308106 | LOC102724594 | | Downregulated |
| | LOC102724595 | | |
| 0 | LOC102724596 | FALSE | Upregulated |
| 1,168459888 | LOC102724599 | | Upregulated |
| 0,671791365 | LOC102724604 | FALSE | Upregulated |
| | LOC102724605 | | |
| | LOC102724606 | | |
| -0,226624815 | LOC102724608 | FALSE | Downregulated |
| | LOC102724609 | | |
| 0,132080374 | LOC102724610 | FALSE | Upregulated |
| | LOC102724612 | | |
| | LOC102724613 | | |
| -1,650738034 | LOC102724614 | | Downregulated |
| | LOC102724616 | | |
| | LOC102724617 | | |
| | LOC102724618 | | |
| | LOC102724619 | | |

| | | | |
|---|---|---|---|
| 1,72354773 | LOC102724620 | | Upregulated |
| | LOC102724622 | | |
| | LOC102724623 | | |
| | LOC102724625 | | |
| | LOC102724627 | | |
| | LOC102724628 | | |
| | LOC102724629 | | |
| -0,185416757 | LOC102724630 | FALSE | Downregulated |
| | LOC102724631 | | |
| 0,669756935 | LOC102724632 | FALSE | Upregulated |
| 0 | LOC102724633 | FALSE | Upregulated |
| -0,055772032 | LOC102724636 | FALSE | Downregulated |
| | LOC102724637 | | |
| | LOC102724638 | | |
| | LOC102724640 | | |
| 0,498768847 | LOC102724641 | FALSE | Upregulated |
| 0,087534599 | LOC102724642 | FALSE | Upregulated |
| | LOC102724644 | | |
| -1,452694943 | LOC102724645 | | Downregulated |
| | LOC102724646 | | |
| | LOC102724647 | | |
| | LOC102724648 | | |
| | LOC102724649 | | |
| | LOC102724650 | | |
| | LOC102724651 | | |
| 0 | LOC102724653 | FALSE | Upregulated |
| | LOC102724654 | | |
| -0,426094288 | LOC102724655 | FALSE | Downregulated |
| | LOC102724657 | | |
| 0,429352598 | LOC102724659 | FALSE | Upregulated |
| 0,550979871 | LOC102724660 | FALSE | Upregulated |
| 0 | LOC102724661 | FALSE | Upregulated |
| -2,207654786 | LOC102724662 | | Downregulated |
| 0,902741766 | LOC102724663 | FALSE | Upregulated |
| | LOC102724664 | | |
| | LOC102724665 | | |
| | LOC102724666 | | |
| 0 | LOC102724667 | FALSE | Upregulated |
| 0,590647912 | LOC102724668 | FALSE | Upregulated |
| | LOC102724669 | | |
| | LOC102724670 | | |
| | LOC102724671 | | |
| -0,16785226 | LOC102724675 | FALSE | Downregulated |
| -0,00897377 | LOC102724676 | FALSE | Downregulated |
| | LOC102724678 | | |
| | LOC102724679 | | |
| | LOC102724680 | | |
| 2,344633279 | LOC102724681 | | Upregulated |
| | LOC102724684 | | |
| 0,165824509 | LOC102724685 | FALSE | Upregulated |
| -0,158929593 | LOC102724686 | FALSE | Downregulated |

| | | | |
|---|---|---|---|
| -0,455240242 | LOC102724687 | FALSE | Downregulated |
| | LOC102724688 | | |
| | LOC102724689 | | |
| | LOC102724690 | | |
| | LOC102724692 | | |
| | LOC102724693 | | |
| | LOC102724694 | | |
| -0,54940068 | LOC102724695 | FALSE | Downregulated |
| 0,689799043 | LOC102724697 | FALSE | Upregulated |
| | LOC102724698 | | |
| | LOC102724700 | | |
| 0 | LOC102724702 | FALSE | Upregulated |
| 0,070131628 | LOC102724703 | FALSE | Upregulated |
| | LOC102724704 | | |
| | LOC102724707 | | |
| 0,565825897 | LOC102724708 | FALSE | Upregulated |
| | LOC102724710 | | |
| -0,111329651 | LOC102724711 | FALSE | Downregulated |
| | LOC102724712 | | |
| | LOC102724713 | | |
| 0,642064891 | LOC102724714 | FALSE | Upregulated |
| 0 | LOC102724715 | FALSE | Upregulated |
| | LOC102724716 | | |
| | LOC102724717 | | |
| | LOC102724718 | | |
| 0,4456711 | LOC102724719 | FALSE | Upregulated |
| | LOC102724721 | | |
| | LOC102724722 | | |
| | LOC102724723 | | |
| | LOC102724728 | | |
| | LOC102724729 | | |
| 2,679273517 | LOC102724731 | | Upregulated |
| | LOC102724732 | | |
| | LOC102724734 | | |
| | LOC102724735 | | |
| | LOC102724736 | | |
| -0,921574501 | LOC102724737 | FALSE | Downregulated |
| | LOC102724738 | | |
| | LOC102724739 | | |
| 0,242453561 | LOC102724740 | FALSE | Upregulated |
| | LOC102724741 | | |
| | LOC102724742 | | |
| -0,158363053 | LOC102724743 | FALSE | Downregulated |
| | LOC102724744 | | |
| 0 | LOC102724745 | FALSE | Upregulated |
| 0,805668975 | LOC102724746 | FALSE | Upregulated |
| | LOC102724747 | | |
| 0 | LOC102724749 | FALSE | Upregulated |
| 1,545466228 | LOC102724751 | | Upregulated |
| 0,734831864 | LOC102724752 | FALSE | Upregulated |
| | LOC102724753 | | |

| | | | |
|---|---|---|---|
| | LOC102724755 | | |
| 0 | LOC102724758 | FALSE | Upregulated |
| 0,051407596 | LOC102724759 | FALSE | Upregulated |
| -0,189001521 | LOC102724762 | FALSE | Downregulated |
| | LOC102724763 | | |
| -0,06838183 | LOC102724764 | FALSE | Downregulated |
| 0 | LOC102724765 | FALSE | Upregulated |
| -0,297373158 | LOC102724766 | FALSE | Downregulated |
| | LOC102724767 | | |
| -0,040163916 | LOC102724768 | FALSE | Downregulated |
| | LOC102724771 | | |
| | LOC102724772 | | |
| 0 | LOC102724774 | FALSE | Upregulated |
| 0 | LOC102724775 | FALSE | Upregulated |
| | LOC102724776 | | |
| | LOC102724777 | | |
| 1,336877609 | LOC102724778 | | Upregulated |
| | LOC102724779 | | |
| | LOC102724780 | | |
| 0 | LOC102724781 | FALSE | Upregulated |
| -0,327419339 | LOC102724783 | FALSE | Downregulated |
| | LOC102724785 | | |
| 1,168459888 | LOC102724786 | | Upregulated |
| 0,70843959 | LOC102724792 | FALSE | Upregulated |
| | LOC102724793 | | |
| | LOC102724794 | | |
| | LOC102724796 | | |
| 1,446277118 | LOC102724797 | | Upregulated |
| | LOC102724798 | | |
| | LOC102724799 | | |
| -1,262534371 | LOC102724800 | | Downregulated |
| | LOC102724801 | | |
| 0,386325887 | LOC102724802 | FALSE | Upregulated |
| | LOC102724803 | | |
| 0,237971061 | LOC102724804 | FALSE | Upregulated |
| 0,005745106 | LOC102724805 | FALSE | Upregulated |
| 0,223974264 | LOC102724806 | FALSE | Upregulated |
| | LOC102724809 | | |
| | LOC102724810 | | |
| 0 | LOC102724812 | FALSE | Upregulated |
| | LOC102724815 | | |
| | LOC102724817 | | |
| | LOC102724818 | | |
| | LOC102724819 | | |
| | LOC102724820 | | |
| -0,055970583 | LOC102724821 | FALSE | Downregulated |
| -0,193121751 | LOC102724824 | FALSE | Downregulated |
| | LOC102724825 | | |
| 0 | LOC102724826 | FALSE | Upregulated |
| 0 | LOC102724827 | FALSE | Upregulated |
| | LOC102724829 | | |

| | | | |
|---|---|---|---|
| | LOC102724830 | | |
| 0,540480851 | LOC102724831 | FALSE | Upregulated |
| 0,419264701 | LOC102724832 | FALSE | Upregulated |
| | LOC102724834 | | |
| | LOC102724835 | | |
| | LOC102724836 | | |
| | LOC102724837 | | |
| | LOC102724838 | | |
| | LOC102724839 | | |
| | LOC102724840 | | |
| 2,528316964 | LOC102724842 | | Upregulated |
| | LOC102724844 | | |
| | LOC102724845 | | |
| | LOC102724846 | | |
| | LOC102724847 | | |
| | LOC102724848 | | |
| | LOC102724849 | | |
| | LOC102724854 | | |
| | LOC102724855 | | |
| 0,076658198 | LOC102724856 | FALSE | Upregulated |
| -0,009545041 | LOC102724857 | FALSE | Downregulated |
| | LOC102724858 | | |
| | LOC102724859 | | |
| -0,384868941 | LOC102724860 | FALSE | Downregulated |
| 0,705562129 | LOC102724861 | FALSE | Upregulated |
| -0,781308361 | LOC102724862 | FALSE | Downregulated |
| | LOC102724863 | | |
| 0,763055934 | LOC102724864 | FALSE | Upregulated |
| 0,030075729 | LOC102724865 | FALSE | Upregulated |
| 0,173526178 | LOC102724869 | FALSE | Upregulated |
| 0 | LOC102724870 | FALSE | Upregulated |
| | LOC102724871 | | |
| | LOC102724874 | | |
| | LOC102724875 | | |
| | LOC102724876 | | |
| | LOC102724877 | | |
| | LOC102724878 | | |
| | LOC102724879 | | |
| 0,560593255 | LOC102724880 | FALSE | Upregulated |
| | LOC102724882 | | |
| 0,152953736 | LOC102724883 | FALSE | Upregulated |
| 0,631017592 | LOC102724884 | FALSE | Upregulated |
| | LOC102724885 | | |
| | LOC102724887 | | |
| | LOC102724888 | | |
| | LOC102724890 | | |
| | LOC102724891 | | |
| | LOC102724892 | | |
| -0,138758398 | LOC102724893 | FALSE | Downregulated |
| | LOC102724894 | | |
| | LOC102724896 | | |

| | | | |
|---|---|---|---|
| 0,661206439 | LOC102724897 | FALSE | Upregulated |
| 0 | LOC102724898 | FALSE | Upregulated |
| | LOC102724899 | | |
| | LOC102724900 | | |
| -0,990159181 | LOC102724901 | FALSE | Downregulated |
| | LOC102724902 | | |
| 1,587777654 | LOC102724903 | | Upregulated |
| | LOC102724904 | | |
| | LOC102724905 | | |
| 0,039005732 | LOC102724906 | FALSE | Upregulated |
| 0,749881718 | LOC102724907 | FALSE | Upregulated |
| 1,522725823 | LOC102724908 | | Upregulated |
| | LOC102724910 | | |
| 0,725717469 | LOC102724911 | FALSE | Upregulated |
| | LOC102724912 | | |
| | LOC102724913 | | |
| | LOC102724914 | | |
| | LOC102724915 | | |
| | LOC102724917 | | |
| | LOC102724918 | | |
| 0,076658198 | LOC102724919 | FALSE | Upregulated |
| 0,669756935 | LOC102724923 | FALSE | Upregulated |
| | LOC102724924 | | |
| | LOC102724925 | | |
| -0,301730223 | LOC102724927 | FALSE | Downregulated |
| | LOC102724928 | | |
| | LOC102724929 | | |
| | LOC102724932 | | |
| | LOC102724934 | | |
| 0,059524118 | LOC102724935 | FALSE | Upregulated |
| | LOC102724936 | | |
| | LOC102724939 | | |
| 0,46375456 | LOC102724940 | FALSE | Upregulated |
| | LOC102724942 | | |
| 0 | LOC102724943 | FALSE | Upregulated |
| 0,460522863 | LOC102724944 | FALSE | Upregulated |
| | LOC102724945 | | |
| 0 | LOC102724946 | FALSE | Upregulated |
| 0,217525419 | LOC102724947 | FALSE | Upregulated |
| 0,056834859 | LOC102724948 | FALSE | Upregulated |
| | LOC102724949 | | |
| 0,488727753 | LOC102724950 | FALSE | Upregulated |
| | LOC102724952 | | |
| | LOC102724953 | | |
| | LOC102724954 | | |
| | LOC102724955 | | |
| | LOC102724957 | | |
| | LOC102724958 | | |
| | LOC102724959 | | |
| | LOC102724960 | | |
| | LOC102724963 | | |

| | | | |
|---|---|---|---|
| | LOC102724964 | | |
| | LOC102724966 | | |
| -0,574822193 | LOC102724967 | FALSE | Downregulated |
| | LOC102724968 | | |
| -0,080759468 | LOC102724970 | FALSE | Downregulated |
| 0,584693576 | LOC102724971 | FALSE | Upregulated |
| | LOC102724972 | | |
| -0,474044539 | LOC102724973 | FALSE | Downregulated |
| | LOC128322 | | |
| -0,835811991 | LOC145474 | FALSE | Downregulated |
| 1,67078468 | LOC145845 | | Upregulated |
| | LOC146481 | | |
| | LOC147004 | | |
| 0,938921723 | LOC149134 | FALSE | Upregulated |
| 0 | LOC149684 | FALSE | Upregulated |
| | LOC149950 | | |
| 0 | LOC150935 | FALSE | Upregulated |
| -0,818199395 | LOC151174 | FALSE | Downregulated |
| -0,337786632 | LOC151760 | FALSE | Downregulated |
| -0,302496494 | LOC152225 | FALSE | Downregulated |
| | LOC152586 | | |
| | LOC154872 | | |
| -0,013754319 | LOC158434 | FALSE | Downregulated |
| -1,246315249 | LOC158960 | | Downregulated |
| | LOC1720 | | |
| 0,523929805 | LOC254896 | FALSE | Upregulated |
| | LOC255187 | | |
| -0,231662319 | LOC256880 | FALSE | Downregulated |
| -0,036330689 | LOC257358 | FALSE | Downregulated |
| -0,494862157 | LOC283028 | FALSE | Downregulated |
| | LOC283045 | | |
| 0,266920299 | LOC283070 | FALSE | Upregulated |
| -0,101653809 | LOC283214 | FALSE | Downregulated |
| | LOC283482 | | |
| | LOC283585 | | |
| -0,00897377 | LOC283682 | FALSE | Downregulated |
| -0,077895188 | LOC283683 | FALSE | Downregulated |
| 2,046945604 | LOC283693 | | Upregulated |
| 0 | LOC283854 | FALSE | Upregulated |
| 0,923768843 | LOC284009 | FALSE | Upregulated |
| 0,461079155 | LOC284240 | FALSE | Upregulated |
| -0,196454479 | LOC284344 | FALSE | Downregulated |
| 0,190368488 | LOC284379 | FALSE | Upregulated |
| 0 | LOC284395 | FALSE | Upregulated |
| 0,15162139 | LOC284561 | FALSE | Upregulated |
| -0,475996957 | LOC284578 | FALSE | Downregulated |
| 0,415737245 | LOC284600 | FALSE | Upregulated |
| 0 | LOC284648 | FALSE | Upregulated |
| | LOC284801 | | |
| 1,993885069 | LOC284865 | | Upregulated |
| 2,192141051 | LOC284898 | | Upregulated |

| | | | |
|---|---|---|---|
| | LOC285000 | | |
| 0,055323234 | LOC285043 | FALSE | Upregulated |
| | LOC285692 | | |
| -0,300078755 | LOC285768 | FALSE | Downregulated |
| -0,534913079 | LOC286178 | FALSE | Downregulated |
| 0 | LOC286186 | FALSE | Upregulated |
| 0,039005732 | LOC286189 | FALSE | Upregulated |
| 0,034128514 | LOC286359 | FALSE | Upregulated |
| 0 | LOC339166 | FALSE | Upregulated |
| -0,458999454 | LOC339240 | FALSE | Downregulated |
| | LOC339260 | | |
| 0 | LOC339298 | FALSE | Upregulated |
| 0,054575782 | LOC339539 | FALSE | Upregulated |
| 0,533519241 | LOC339593 | FALSE | Upregulated |
| 2,924682281 | LOC339622 | | Upregulated |
| 0,597480774 | LOC339862 | FALSE | Upregulated |
| | LOC339978 | | |
| -1,146729112 | LOC340073 | | Downregulated |
| | LOC340090 | | |
| 0,696646091 | LOC340113 | FALSE | Upregulated |
| 0,922562491 | LOC340508 | FALSE | Upregulated |
| 0,050143828 | LOC341056 | FALSE | Upregulated |
| 0 | LOC344887 | FALSE | Upregulated |
| 0,887313159 | LOC345051 | FALSE | Upregulated |
| 0 | LOC374890 | FALSE | Upregulated |
| -0,253058816 | LOC387720 | FALSE | Downregulated |
| 0,067880217 | LOC388406 | FALSE | Upregulated |
| 2,353801818 | LOC388436 | | Upregulated |
| 0 | LOC388456 | FALSE | Upregulated |
| 1,16238225 | LOC388499 | | Upregulated |
| | LOC388553 | | |
| -0,414938574 | LOC388780 | FALSE | Downregulated |
| -0,184316361 | LOC388813 | FALSE | Downregulated |
| | LOC389033 | | |
| -0,178642067 | LOC389273 | FALSE | Downregulated |
| 0,34443609 | LOC389641 | FALSE | Upregulated |
| 0,597480774 | LOC389705 | FALSE | Upregulated |
| | LOC389895 | | |
| | LOC390705 | | |
| 0 | LOC390877 | FALSE | Upregulated |
| | LOC390956 | | |
| -0,872747214 | LOC391722 | FALSE | Downregulated |
| -0,079940935 | LOC392232 | FALSE | Downregulated |
| 0,765404232 | LOC392288 | FALSE | Upregulated |
| -0,353239851 | LOC392364 | FALSE | Downregulated |
| | LOC392452 | | |
| -0,161606847 | LOC399815 | FALSE | Downregulated |
| -0,205329595 | LOC400499 | FALSE | Downregulated |
| | LOC400622 | | |
| 0,05942365 | LOC400655 | FALSE | Upregulated |
| -0,350674684 | LOC400685 | FALSE | Downregulated |

| | | | |
|---|---|---|---|
| | LOC400743 | | |
| 1,23789007 | LOC400748 | | Upregulated |
| -0,385920977 | LOC400768 | FALSE | Downregulated |
| -1,099319225 | LOC400863 | | Downregulated |
| -0,090563074 | LOC400891 | FALSE | Downregulated |
| 0,856234947 | LOC400927 | FALSE | Upregulated |
| | LOC400940 | | |
| 0,505580172 | LOC401010 | FALSE | Upregulated |
| 0,678573206 | LOC401098 | FALSE | Upregulated |
| | LOC401177 | | |
| 0 | LOC401242 | FALSE | Upregulated |
| | LOC401312 | | |
| | LOC401324 | | |
| | LOC401357 | | |
| 0 | LOC401442 | FALSE | Upregulated |
| -0,122172769 | LOC401463 | FALSE | Downregulated |
| 1,068345462 | LOC401480 | | Upregulated |
| | LOC401497 | | |
| | LOC401557 | | |
| | LOC402269 | | |
| -0,526059402 | LOC407835 | FALSE | Downregulated |
| | LOC440040 | | |
| -0,510019711 | LOC440243 | FALSE | Downregulated |
| -0,173013616 | LOC440292 | FALSE | Downregulated |
| -0,960200733 | LOC440311 | FALSE | Downregulated |
| 1,101436422 | LOC440313 | | Upregulated |
| | LOC440337 | | |
| 0,94043371 | LOC440461 | FALSE | Upregulated |
| | LOC440518 | | |
| | LOC440602 | | |
| -1,065868911 | LOC440700 | | Downregulated |
| -0,10434982 | LOC440742 | FALSE | Downregulated |
| 0,020467897 | LOC440895 | FALSE | Upregulated |
| | LOC440910 | | |
| 0,849495004 | LOC441052 | FALSE | Upregulated |
| 0,134842458 | LOC441155 | FALSE | Upregulated |
| 0,551285328 | LOC441204 | FALSE | Upregulated |
| 1,65485963 | LOC441455 | | Upregulated |
| 0,374573006 | LOC441493 | FALSE | Upregulated |
| 0 | LOC441601 | FALSE | Upregulated |
| 1,19099262 | LOC441666 | | Upregulated |
| 0 | LOC441728 | FALSE | Upregulated |
| -0,37473345 | LOC441956 | FALSE | Downregulated |
| 0 | LOC442028 | FALSE | Upregulated |
| | LOC442132 | | |
| 0 | LOC494127 | FALSE | Upregulated |
| | LOC497256 | | |
| 0 | LOC550113 | FALSE | Upregulated |
| -0,361828894 | LOC554206 | FALSE | Downregulated |
| 0,719417069 | LOC641515 | FALSE | Upregulated |
| | LOC641746 | | |

| | | | |
|---|---|---|---|
| 0,171045248 | LOC642131 | FALSE | Upregulated |
| -0,01749261 | LOC642236 | FALSE | Downregulated |
| 0 | LOC642249 | FALSE | Upregulated |
| | LOC642366 | | |
| | LOC642643 | | |
| -0,115411262 | LOC642934 | FALSE | Downregulated |
| -0,055970583 | LOC643327 | FALSE | Downregulated |
| -0,102383912 | LOC643387 | FALSE | Downregulated |
| -0,200228976 | LOC643406 | FALSE | Downregulated |
| 0 | LOC643441 | FALSE | Upregulated |
| -0,216240161 | LOC643486 | FALSE | Downregulated |
| | LOC643711 | | |
| -0,241506909 | LOC643733 | FALSE | Downregulated |
| -1,678154809 | LOC643770 | | Downregulated |
| 0 | LOC643802 | FALSE | Upregulated |
| | LOC643923 | | |
| | LOC644070 | | |
| 0 | LOC644086 | FALSE | Upregulated |
| | LOC644145 | | |
| -0,175384607 | LOC644189 | FALSE | Downregulated |
| -0,298196055 | LOC644249 | FALSE | Downregulated |
| 0,548194603 | LOC644554 | FALSE | Upregulated |
| | LOC644669 | | |
| | LOC644717 | | |
| 0 | LOC644919 | FALSE | Upregulated |
| 0,32613371 | LOC644936 | FALSE | Upregulated |
| | LOC645177 | | |
| | LOC645188 | | |
| 0 | LOC645202 | FALSE | Upregulated |
| | LOC645321 | | |
| -0,890824953 | LOC645638 | FALSE | Downregulated |
| -1,4773252 | LOC645752 | | Downregulated |
| -0,015945281 | LOC645769 | FALSE | Downregulated |
| 0,330975107 | LOC646021 | FALSE | Upregulated |
| 0 | LOC646034 | FALSE | Upregulated |
| -0,354870676 | LOC646278 | FALSE | Downregulated |
| | LOC646513 | | |
| | LOC646588 | | |
| -0,39485337 | LOC646670 | FALSE | Downregulated |
| -0,043826348 | LOC646730 | FALSE | Downregulated |
| 0,708067798 | LOC646736 | FALSE | Upregulated |
| | LOC646743 | | |
| | LOC646813 | | |
| | LOC646862 | | |
| 0,909748389 | LOC646903 | FALSE | Upregulated |
| | LOC646938 | | |
| | LOC647264 | | |
| 0 | LOC648691 | FALSE | Upregulated |
| | LOC649330 | | |
| 0,719076969 | LOC649352 | FALSE | Upregulated |
| | LOC650157 | | |

| | | | |
|---|---|---|---|
| 0,726045491 | LOC650226 | FALSE | Upregulated |
| | LOC650293 | | |
| 0 | LOC650368 | FALSE | Upregulated |
| 0,705562129 | LOC653486 | FALSE | Upregulated |
| 0,617707838 | LOC653786 | FALSE | Upregulated |
| 0,243520404 | LOC654342 | FALSE | Upregulated |
| 0,091883761 | LOC723805 | FALSE | Upregulated |
| | LOC727944 | | |
| -0,355777938 | LOC728026 | FALSE | Downregulated |
| | LOC728114 | | |
| 1,144864693 | LOC728158 | | Upregulated |
| 0 | LOC728637 | FALSE | Upregulated |
| -0,336908779 | LOC728715 | FALSE | Downregulated |
| -0,044343502 | LOC728728 | FALSE | Downregulated |
| | LOC728739 | | |
| | LOC728755 | | |
| -0,667364211 | LOC728975 | FALSE | Downregulated |
| -0,468086585 | LOC729041 | FALSE | Downregulated |
| 0,598202128 | LOC729080 | FALSE | Upregulated |
| -0,455260275 | LOC729159 | FALSE | Downregulated |
| | LOC729222 | | |
| 0 | LOC729224 | FALSE | Upregulated |
| 0 | LOC729307 | FALSE | Upregulated |
| | LOC729444 | | |
| | LOC729458 | | |
| 0,326119658 | LOC729468 | FALSE | Upregulated |
| 0 | LOC729506 | FALSE | Upregulated |
| -0,44732329 | LOC729609 | FALSE | Downregulated |
| -0,084026815 | LOC729770 | FALSE | Downregulated |
| 0,608246941 | LOC729800 | FALSE | Upregulated |
| | LOC729815 | | |
| | LOC729911 | | |
| 0 | LOC729966 | FALSE | Upregulated |
| 0,018084743 | LOC730159 | FALSE | Upregulated |
| 0,215288526 | LOC730268 | FALSE | Upregulated |
| 0 | LOC730338 | FALSE | Upregulated |
| 0,207549846 | LOC731157 | FALSE | Upregulated |
| 0,659224562 | LOC731779 | FALSE | Upregulated |
| 0 | LOC732265 | FALSE | Upregulated |
| 0,697736933 | LOC90246 | FALSE | Upregulated |
| 0,616614981 | LOC93432 | FALSE | Upregulated |
| -0,655234281 | LOH12CR2 | FALSE | Downregulated |
| 0,540800482 | LOR | FALSE | Upregulated |
| 0,087085437 | LOXHD1 | FALSE | Upregulated |
| 1,331693158 | LOXL1-AS1 | | Upregulated |
| 0,088361109 | LPAR3 | FALSE | Upregulated |
| -0,027979783 | LPAR4 | FALSE | Downregulated |
| 0,611573165 | LPL | FALSE | Upregulated |
| 0,633418778 | LPO | FALSE | Upregulated |
| -0,286850448 | LPPR4 | FALSE | Downregulated |
| 0,374613874 | LPPR5 | FALSE | Upregulated |

| | | | |
|---|---|---|---|
| -0,388873527 | LRCH2 | FALSE | Downregulated |
| 0 | LRFN2 | FALSE | Upregulated |
| -0,34969677 | LRFN5 | FALSE | Downregulated |
| 1,579296237 | LRGUK | | Upregulated |
| | LRIT1 | | |
| 1,122585155 | LRIT2 | | Upregulated |
| -0,503234122 | LRIT3 | FALSE | Downregulated |
| 1,627620463 | LRRC1 | | Upregulated |
| 0,796861737 | LRRC10 | FALSE | Upregulated |
| -0,467992432 | LRRC10B | FALSE | Downregulated |
| 0,056576497 | LRRC23 | FALSE | Upregulated |
| 0 | LRRC26 | FALSE | Upregulated |
| -0,203801896 | LRRC3-AS1 | FALSE | Downregulated |
| 4,33271E-05 | LRRC30 | FALSE | Upregulated |
| -0,291617454 | LRRC36 | FALSE | Downregulated |
| 0,05947333 | LRRC37A11P | FALSE | Upregulated |
| 0 | LRRC38 | FALSE | Upregulated |
| 0,68080089 | LRRC3B | FALSE | Upregulated |
| 0,22421811 | LRRC3C | FALSE | Upregulated |
| 1,06772873 | LRRC43 | | Upregulated |
| -0,281421152 | LRRC4C | FALSE | Downregulated |
| | LRRC52 | | |
| 0,341254959 | LRRC6 | FALSE | Upregulated |
| 0,005255594 | LRRC63 | FALSE | Upregulated |
| -0,086843917 | LRRC66 | FALSE | Downregulated |
| -0,3023089 | LRRC69 | FALSE | Downregulated |
| 0,267023907 | LRRC70 | FALSE | Upregulated |
| 1,64673218 | LRRC71 | | Upregulated |
| | LRRC72 | | |
| -0,044532498 | LRRC73 | FALSE | Downregulated |
| 0 | LRRD1 | FALSE | Upregulated |
| 0 | LRRIQ1 | FALSE | Upregulated |
| -0,092237265 | LRRIQ3 | FALSE | Downregulated |
| -0,551577105 | LRRIQ4 | FALSE | Downregulated |
| 1,675743215 | LRRN4CL | | Upregulated |
| 1,003183643 | LRRTM1 | | Upregulated |
| 0,566115152 | LRRTM4 | FALSE | Upregulated |
| 0,680588945 | LRTM1 | FALSE | Upregulated |
| -0,0712309 | LRTM2 | FALSE | Downregulated |
| 2,608274981 | LSAMP-AS1 | | Upregulated |
| 0 | LSP1P3 | FALSE | Upregulated |
| 0,337278259 | LTA | FALSE | Upregulated |
| | LUCAT1 | | |
| -0,620621678 | LURAP1 | FALSE | Downregulated |
| 0,086043174 | LUZP2 | FALSE | Upregulated |
| | LUZP4 | | |
| -0,123249576 | LY6D | FALSE | Downregulated |
| -0,05114389 | LY6G5B | FALSE | Downregulated |
| 0,562598898 | LY6G5C | FALSE | Upregulated |
| 0 | LY6G6C | FALSE | Upregulated |
| | LY6G6D | | |

| | | | |
|---|---|---|---|
| | LY6G6E | | |
| | LY6G6F | | |
| 0 | LY6H | FALSE | Upregulated |
| 0,693192897 | LY86-AS1 | FALSE | Upregulated |
| 0,746542693 | LY96 | FALSE | Upregulated |
| -0,39506488 | LYPD2 | FALSE | Downregulated |
| 0,01561478 | LYPD3 | FALSE | Upregulated |
| 0 | LYPD4 | FALSE | Upregulated |
| 1,317672494 | LYPD5 | | Upregulated |
| 0,556715157 | LYPD6 | FALSE | Upregulated |
| -0,477608084 | LYPD6B | FALSE | Downregulated |
| 0,669025015 | LYPD8 | FALSE | Upregulated |
| 0,906741099 | LYZL1 | FALSE | Upregulated |
| 0,050688952 | LYZL2 | FALSE | Upregulated |
| | LYZL4 | | |
| 0,717166439 | LYZL6 | FALSE | Upregulated |
| 0,010604346 | MAATS1 | FALSE | Upregulated |
| -0,095127497 | MACC1-AS1 | FALSE | Downregulated |
| 0 | MACROD2-IT1 | FALSE | Upregulated |
| 0,25339981 | MAD2L1 | FALSE | Upregulated |
| -0,010034835 | MAEL | FALSE | Downregulated |
| -0,188130105 | MAFA | FALSE | Downregulated |
| -0,599549435 | MAFG-AS1 | FALSE | Downregulated |
| -0,081845786 | MAG | FALSE | Downregulated |
| | MAGEA1 | | |
| -0,018346671 | MAGEA10 | FALSE | Downregulated |
| | MAGEA10-MAGEA5 | | |
| | MAGEA11 | | |
| | MAGEA12 | | |
| | MAGEA2 | | |
| | MAGEA2B | | |
| 0 | MAGEA3 | FALSE | Upregulated |
| | MAGEA4 | | |
| 0,760333593 | MAGEA5 | FALSE | Upregulated |
| | MAGEA6 | | |
| | MAGEA8-AS1 | | |
| 0 | MAGEA9 | FALSE | Upregulated |
| 0,669756935 | MAGEA9B | FALSE | Upregulated |
| | MAGEB1 | | |
| | MAGEB10 | | |
| | MAGEB16 | | |
| | MAGEB17 | | |
| | MAGEB18 | | |
| | MAGEB2 | | |
| | MAGEB3 | | |
| | MAGEB4 | | |
| | MAGEB5 | | |
| | MAGEB6 | | |
| | MAGEC1 | | |
| | MAGEC2 | | |
| | MAGEC3 | | |

| | | | |
|---|---|---|---|
| 0,355748166 | MAGED4 | FALSE | Upregulated |
| 0,561434932 | MAGED4B | FALSE | Upregulated |
| -0,069795274 | MAGEE1 | FALSE | Downregulated |
| 0,641904016 | MAGEE2 | FALSE | Upregulated |
| 0,780170283 | MAGEL2 | FALSE | Upregulated |
| | MAGI2-AS2 | | |
| 0 | MAL | FALSE | Upregulated |
| -0,096394018 | MALRD1 | FALSE | Downregulated |
| 0,16422811 | MAMSTR | FALSE | Upregulated |
| -0,122786748 | MAP1LC3B2 | FALSE | Downregulated |
| -0,062552347 | MAP1LC3C | FALSE | Downregulated |
| 0 | MAP2K4P1 | FALSE | Upregulated |
| 0,026382944 | MAP7D2 | FALSE | Upregulated |
| -0,436613321 | MAP9 | FALSE | Downregulated |
| 0,266715333 | MAPK15 | FALSE | Upregulated |
| 0,94891485 | MAPK4 | FALSE | Upregulated |
| 0,052696865 | MAPK8IP2 | FALSE | Upregulated |
| | MAPT-AS1 | | |
| 0,461954044 | MARCH10 | FALSE | Upregulated |
| | MARCH11 | | |
| 0 | MARCH4 | FALSE | Upregulated |
| | MAS1 | | |
| 0 | MAS1L | FALSE | Upregulated |
| 0,84161288 | MATN3 | FALSE | Upregulated |
| 0,695882197 | MATN4 | FALSE | Upregulated |
| 0,305398618 | MB21D2 | FALSE | Upregulated |
| | MBD3L1 | | |
| | MBD3L2 | | |
| 0 | MBD3L3 | FALSE | Upregulated |
| 0 | MBD3L4 | FALSE | Upregulated |
| | MBD3L5 | | |
| | MC2R | | |
| | MC3R | | |
| | MC4R | | |
| | MC5R | | |
| 0,042977681 | MCF2 | FALSE | Upregulated |
| 0,193159114 | MCF2L2 | FALSE | Upregulated |
| | MCHR2 | | |
| | MCHR2-AS1 | | |
| | MCIDAS | | |
| 0,481918042 | MCM10 | FALSE | Upregulated |
| 0,975554221 | MCOLN2 | FALSE | Upregulated |
| 2,479874081 | MDFI | | Upregulated |
| -0,354729193 | MDGA2 | FALSE | Downregulated |
| -0,178840892 | MDH1B | FALSE | Downregulated |
| -1,17235214 | MDP1 | | Downregulated |
| 0 | MDS2 | FALSE | Upregulated |
| | MED15P9 | | |
| 1,720413451 | MEDAG | | Upregulated |
| 0,796768126 | MEF2C-AS1 | FALSE | Upregulated |
| 0 | MEG8 | FALSE | Upregulated |

| | | | |
|---|---|---|---|
| 1,903996421 | MEG9 | | Upregulated |
| 0,766611739 | MEI1 | FALSE | Upregulated |
| -0,28770196 | MEI4 | FALSE | Downregulated |
| | MEIG1 | | |
| | MEIS1-AS3 | | |
| 0,415213995 | MELK | FALSE | Upregulated |
| 0,227220966 | MEOX1 | FALSE | Upregulated |
| 0,028610657 | MEP1A | FALSE | Upregulated |
| -0,777959732 | MESP1 | FALSE | Downregulated |
| 0,92525397 | MESP2 | FALSE | Upregulated |
| | METTL11B | | |
| 1,297490011 | METTL21EP | | Upregulated |
| 1,009571461 | METTL24 | | Upregulated |
| -0,545396332 | MFAP2 | FALSE | Downregulated |
| 0,336769777 | MFAP5 | FALSE | Upregulated |
| -0,193730474 | MFI2-AS1 | FALSE | Downregulated |
| -0,512664796 | MFRP | FALSE | Downregulated |
| -0,21329464 | MGAM | FALSE | Downregulated |
| -0,014782452 | MGARP | FALSE | Downregulated |
| 1,968307974 | MGAT4C | | Upregulated |
| -0,093373931 | MGAT5B | FALSE | Downregulated |
| 0,413266742 | MGC16025 | FALSE | Upregulated |
| 0,45097918 | MGC16142 | FALSE | Upregulated |
| -1,066927556 | MGC27382 | | Downregulated |
| | MGC2889 | | |
| 0,087008956 | MGC34034 | FALSE | Upregulated |
| 0 | MGC34800 | FALSE | Upregulated |
| 0,191100272 | MGC39545 | FALSE | Upregulated |
| | MGC39584 | | |
| 0,758728803 | MGC42157 | FALSE | Upregulated |
| 0 | MGC45800 | FALSE | Upregulated |
| -0,131082119 | MGC45922 | FALSE | Downregulated |
| 0,909468944 | MIA | FALSE | Upregulated |
| -0,087959743 | MIA-RAB4B | FALSE | Downregulated |
| 0 | MID1-2 | FALSE | Upregulated |
| 0,067880217 | MIMT1 | FALSE | Upregulated |
| -1,00631733 | MINOS1-NBL1 | | Downregulated |
| -0,510416949 | MIP | FALSE | Downregulated |
| | MIR1-1 | | |
| | MIR1-2 | | |
| | MIR100 | | |
| | MIR101-1 | | |
| | MIR101-2 | | |
| | MIR103A1 | | |
| | MIR103A2 | | |
| | MIR103B1 | | |
| | MIR103B2 | | |
| | MIR105-1 | | |
| | MIR105-2 | | |
| | MIR106A | | |
| | MIR106B | | |

MIR107
MIR10A
MIR10B
MIR1178
MIR1179
MIR1180
MIR1181
MIR1182
MIR1183
MIR1184-1
MIR1184-2
MIR1184-3
MIR1185-1
MIR1185-2
MIR1193
MIR1197
MIR1199
MIR1200
MIR1202
MIR1203
MIR1204
MIR1205
MIR1206
MIR1207
MIR1208
MIR122
MIR1224
MIR1225
MIR1226
MIR1227
MIR1228
MIR1229
MIR1231
MIR1233-1
MIR1233-2
MIR1236
MIR1237
MIR1238
MIR124-1
MIR124-2
MIR124-3
MIR1243
MIR1244-1
MIR1244-2
MIR1244-3
MIR1245A
MIR1245B
MIR1246
MIR1247
MIR1249
MIR1250

MIR1251
MIR1252
MIR1253
MIR1254-1
MIR1254-2
MIR1255A
MIR1255B1
MIR1255B2
MIR1256
MIR1257
MIR1258
MIR125A
MIR125B1
MIR125B2
MIR126
MIR1260A
MIR1260B
MIR1261
MIR1262
MIR1263
MIR1264
MIR1265
MIR1266
MIR1267
MIR1268A
MIR1268B
MIR1269A
MIR1269B
MIR127
MIR1270-1
MIR1271
MIR1272
MIR1273A
MIR1273C
MIR1273D
MIR1273F
MIR1273G
MIR1273H
MIR1275
MIR1276
MIR1277
MIR1278
MIR1279
MIR128-1
MIR128-2
MIR1281
MIR1282
MIR1283-1
MIR1283-2
MIR1284
MIR1285-1

| | | | |
|---|---|---|---|
| | MIR1285-2 | | |
| | MIR1286 | | |
| | MIR1287 | | |
| | MIR1288 | | |
| | MIR1289-1 | | |
| | MIR1289-2 | | |
| | MIR129-1 | | |
| | MIR129-2 | | |
| | MIR1290 | | |
| | MIR1292 | | |
| | MIR1293 | | |
| | MIR1294 | | |
| | MIR1295A | | |
| | MIR1295B | | |
| | MIR1296 | | |
| | MIR1297 | | |
| | MIR1298 | | |
| | MIR1299 | | |
| | MIR1301 | | |
| | MIR1302-1 | | |
| | MIR1302-10 | | |
| | MIR1302-11 | | |
| | MIR1302-2 | | |
| | MIR1302-3 | | |
| | MIR1302-4 | | |
| 0 | MIR1302-5 | FALSE | Upregulated |
| | MIR1302-6 | | |
| | MIR1302-7 | | |
| | MIR1302-8 | | |
| | MIR1302-9 | | |
| | MIR1303 | | |
| | MIR1304 | | |
| | MIR1305 | | |
| | MIR1306 | | |
| 0 | MIR1307 | FALSE | Upregulated |
| | MIR130A | | |
| | MIR130B | | |
| | MIR132 | | |
| | MIR1321 | | |
| | MIR1322 | | |
| | MIR1323 | | |
| | MIR1324 | | |
| | MIR133A1 | | |
| | MIR133A2 | | |
| | MIR133B | | |
| | MIR134 | | |
| | MIR1343 | | |
| | MIR135A1 | | |
| | MIR135A2 | | |
| | MIR135B | | |
| | MIR136 | | |

| | | | |
|---|---|---|---|
| | MIR137 | | |
| | MIR137HG | | |
| | MIR138-1 | | |
| | MIR138-2 | | |
| | MIR139 | | |
| | MIR140 | | |
| | MIR141 | | |
| | MIR142 | | |
| | MIR143 | | |
| | MIR144 | | |
| | MIR145 | | |
| | MIR1468 | | |
| | MIR1469 | | |
| | MIR146A | | |
| | MIR146B | | |
| | MIR1470 | | |
| | MIR1471 | | |
| | MIR147A | | |
| | MIR147B | | |
| | MIR148A | | |
| | MIR148B | | |
| | MIR149 | | |
| | MIR150 | | |
| | MIR151A | | |
| | MIR151B | | |
| | MIR152 | | |
| | MIR153-1 | | |
| | MIR153-2 | | |
| | MIR1537 | | |
| | MIR1538 | | |
| | MIR1539 | | |
| | MIR154 | | |
| | MIR155 | | |
| | MIR1587 | | |
| | MIR15A | | |
| | MIR15B | | |
| | MIR16-1 | | |
| | MIR16-2 | | |
| | MIR17 | | |
| | MIR181A1 | | |
| 0,669756935 | MIR181A1HG | FALSE | Upregulated |
| 0 | MIR181A2 | FALSE | Upregulated |
| | MIR181B1 | | |
| | MIR181B2 | | |
| | MIR181C | | |
| | MIR181D | | |
| | MIR182 | | |
| | MIR1825 | | |
| | MIR1827 | | |
| | MIR183 | | |
| | MIR184 | | |

| | | | |
|---|---|---|---|
| | MIR185 | | |
| | MIR186 | | |
| | MIR187 | | |
| | MIR188 | | |
| | MIR18A | | |
| | MIR18B | | |
| | MIR1908 | | |
| | MIR1909 | | |
| | MIR190A | | |
| | MIR190B | | |
| | MIR191 | | |
| | MIR1910 | | |
| | MIR1911 | | |
| | MIR1912 | | |
| | MIR1913 | | |
| | MIR1914 | | |
| | MIR1915 | | |
| 0 | MIR192 | FALSE | Upregulated |
| | MIR193A | | |
| | MIR193B | | |
| | MIR194-1 | | |
| | MIR194-2 | | |
| | MIR195 | | |
| | MIR196A1 | | |
| | MIR196A2 | | |
| | MIR196B | | |
| | MIR197 | | |
| | MIR1972-1 | | |
| | MIR1972-2 | | |
| | MIR1973 | | |
| | MIR1976 | | |
| | MIR198 | | |
| | MIR199A1 | | |
| | MIR199A2 | | |
| | MIR199B | | |
| | MIR19A | | |
| | MIR19B1 | | |
| | MIR19B2 | | |
| | MIR200A | | |
| | MIR200B | | |
| | MIR200C | | |
| | MIR202 | | |
| | MIR202HG | | |
| | MIR203 | | |
| | MIR204 | | |
| | MIR205 | | |
| | MIR2052 | | |
| | MIR2053 | | |
| | MIR2054 | | |
| | MIR205HG | | |
| | MIR206 | | |

| | | | |
|---|---|---|---|
| | MIR208A | | |
| | MIR208B | | |
| | MIR20A | | |
| | MIR20B | | |
| | MIR21 | | |
| | MIR210 | | |
| | MIR211 | | |
| | MIR2110 | | |
| | MIR2113 | | |
| | MIR2114 | | |
| | MIR2115 | | |
| | MIR2116 | | |
| | MIR2117 | | |
| | MIR212 | | |
| 0 | MIR214 | FALSE | Upregulated |
| | MIR215 | | |
| | MIR216A | | |
| | MIR216B | | |
| | MIR217 | | |
| | MIR218-1 | | |
| | MIR218-2 | | |
| | MIR219-1 | | |
| | MIR219-2 | | |
| | MIR22 | | |
| | MIR221 | | |
| | MIR222 | | |
| | MIR223 | | |
| | MIR224 | | |
| | MIR2276 | | |
| | MIR2277 | | |
| | MIR2278 | | |
| | MIR2355 | | |
| | MIR2392 | | |
| | MIR23A | | |
| | MIR23B | | |
| | MIR23C | | |
| | MIR24-1 | | |
| | MIR24-2 | | |
| | MIR2467 | | |
| | MIR25 | | |
| | MIR2681 | | |
| | MIR2682 | | |
| | MIR26A1 | | |
| | MIR26A2 | | |
| | MIR26B | | |
| | MIR27A | | |
| | MIR27B | | |
| | MIR28 | | |
| | MIR2861 | | |
| | MIR2909 | | |
| | MIR296 | | |

MIR2964A
MIR297
MIR298
MIR299
MIR29A
MIR29B1
MIR29B2
MIR29C
MIR300
MIR301A
MIR301B
MIR302A
MIR302B
MIR302C
MIR302D
MIR302E
MIR302F
MIR3064
MIR3065
MIR3074
MIR30A
MIR30B
MIR30C1
MIR30C2
MIR30D
MIR30E
MIR31
MIR3115
MIR3116-1
MIR3116-2
MIR3117
MIR3118-1
MIR3118-2
MIR3118-3
MIR3118-4
MIR3118-5
MIR3118-6
MIR3119-1
MIR3119-2
MIR3120
MIR3121
MIR3122
MIR3123
MIR3124
MIR3125
MIR3126
MIR3127
MIR3128
MIR3129
MIR3130-1
MIR3130-2

MIR3131
MIR3132
MIR3133
MIR3134
MIR3135A
MIR3135B
MIR3136
MIR3137
MIR3138
MIR3139
MIR3140
MIR3141
MIR3142
MIR3143
MIR3144
MIR3145
MIR3146
MIR3147
MIR3148
MIR3150A
MIR3150B
MIR3151
MIR3152
MIR3153
MIR3154
MIR3155A
MIR3155B
MIR3156-1
MIR3156-2
MIR3156-3
MIR3157
MIR3158-1
MIR3158-2
MIR3159
MIR3160-1
MIR3160-2
MIR3161
MIR3162
MIR3163
MIR3164
MIR3165
MIR3166
MIR3167
MIR3168
MIR3169
MIR3170
MIR3173
MIR3174
MIR3175
MIR3176
MIR3177

| | | | |
|---|---|---|---|
| | MIR3178 | | |
| | MIR3179-1 | | |
| | MIR3179-2 | | |
| | MIR3179-3 | | |
| | MIR3180-1 | | |
| | MIR3180-2 | | |
| | MIR3180-3 | | |
| | MIR3180-4 | | |
| | MIR3180-5 | | |
| | MIR3182 | | |
| | MIR3183 | | |
| | MIR3184 | | |
| | MIR3185 | | |
| | MIR3186 | | |
| | MIR3187 | | |
| | MIR3188 | | |
| | MIR3189 | | |
| | MIR3190 | | |
| | MIR3191 | | |
| | MIR3192 | | |
| | MIR3193 | | |
| | MIR3194 | | |
| | MIR3196 | | |
| | MIR3197 | | |
| | MIR3198-1 | | |
| | MIR3198-2 | | |
| | MIR3199-1 | | |
| | MIR3199-2 | | |
| 0 | MIR31HG | FALSE | Upregulated |
| | MIR32 | | |
| | MIR3200 | | |
| | MIR3201 | | |
| | MIR3202-1 | | |
| | MIR3202-2 | | |
| | MIR320A | | |
| | MIR320B1 | | |
| | MIR320B2 | | |
| | MIR320C1 | | |
| | MIR320C2 | | |
| | MIR320D1 | | |
| | MIR320D2 | | |
| | MIR320E | | |
| | MIR323A | | |
| | MIR323B | | |
| | MIR324 | | |
| | MIR325 | | |
| | MIR326 | | |
| | MIR328 | | |
| | MIR329-1 | | |
| | MIR329-2 | | |
| | MIR330 | | |

MIR331
MIR335
MIR337
MIR338
MIR339
MIR33A
MIR33B
MIR340
MIR342
MIR345
MIR346
MIR34A
MIR34B
MIR34C
MIR3529
MIR3545
MIR3591
MIR3605
MIR3606
MIR3607
MIR3609
MIR361
MIR3610
MIR3611
MIR3612
MIR3613
MIR3614
MIR3615
MIR3616
MIR3617
MIR3618
MIR3619
MIR362
MIR3620
MIR3621
MIR3622A
MIR3622B
MIR363
MIR3646
MIR3649
MIR3650
MIR3651
MIR3652
MIR3653
MIR3654
MIR3655
MIR3656
MIR3657
MIR3658
MIR3659
MIR365A

| | | | |
|---|---|---|---|
| | MIR365B | | |
| | MIR3660 | | |
| 0 | MIR3661 | FALSE | Upregulated |
| | MIR3662 | | |
| | MIR3663 | | |
| | MIR3664 | | |
| | MIR3665 | | |
| | MIR3666 | | |
| | MIR3667 | | |
| | MIR3668 | | |
| | MIR367 | | |
| | MIR3670-1 | | |
| | MIR3670-2 | | |
| | MIR3671 | | |
| | MIR3672 | | |
| | MIR3674 | | |
| | MIR3675 | | |
| | MIR3677 | | |
| | MIR3678 | | |
| | MIR3679 | | |
| | MIR3680-1 | | |
| | MIR3680-2 | | |
| | MIR3681 | | |
| | MIR3682 | | |
| | MIR3683 | | |
| | MIR3684 | | |
| | MIR3685 | | |
| | MIR3686 | | |
| | MIR3687 | | |
| | MIR3688-1 | | |
| | MIR3688-2 | | |
| | MIR3689A | | |
| | MIR3689B | | |
| | MIR3689C | | |
| | MIR3689D1 | | |
| | MIR3689D2 | | |
| | MIR3689E | | |
| | MIR3689F | | |
| | MIR369 | | |
| | MIR3690 | | |
| | MIR3690-2 | | |
| | MIR3691 | | |
| | MIR3692 | | |
| | MIR370 | | |
| | MIR3713 | | |
| | MIR3714 | | |
| | MIR371A | | |
| | MIR371B | | |
| | MIR372 | | |
| | MIR373 | | |
| | MIR374A | | |

| | | | |
|---|---|---|---|
| | MIR374B | | |
| | MIR374C | | |
| | MIR375 | | |
| | MIR376A1 | | |
| | MIR376A2 | | |
| | MIR376B | | |
| | MIR376C | | |
| | MIR377 | | |
| | MIR378A | | |
| | MIR378B | | |
| | MIR378C | | |
| | MIR378D1 | | |
| | MIR378D2 | | |
| | MIR378E | | |
| | MIR378F | | |
| | MIR378G | | |
| | MIR378H | | |
| | MIR378I | | |
| | MIR378J | | |
| | MIR379 | | |
| | MIR380 | | |
| | MIR381 | | |
| 0 | MIR381HG | FALSE | Upregulated |
| | MIR382 | | |
| | MIR383 | | |
| | MIR384 | | |
| | MIR3907 | | |
| | MIR3908 | | |
| | MIR3909 | | |
| | MIR3910-1 | | |
| | MIR3910-2 | | |
| | MIR3911 | | |
| | MIR3912 | | |
| | MIR3913-1 | | |
| | MIR3913-2 | | |
| | MIR3914-1 | | |
| | MIR3914-2 | | |
| | MIR3915 | | |
| | MIR3916 | | |
| | MIR3917 | | |
| | MIR3918 | | |
| | MIR3919 | | |
| | MIR3920 | | |
| | MIR3921 | | |
| | MIR3922 | | |
| | MIR3923 | | |
| | MIR3924 | | |
| | MIR3925 | | |
| | MIR3926-1 | | |
| | MIR3926-2 | | |
| | MIR3927 | | |

| | | | | |
|---|---|---|---|---|
| | MIR3928 | | | |
| | MIR3929 | | | |
| | MIR3934 | | | |
| | MIR3935 | | | |
| | MIR3936 | | | |
| | MIR3937 | | | |
| | MIR3938 | | | |
| -0,225882925 | | MIR3939 | FALSE | Downregulated |
| | MIR3940 | | | |
| | MIR3941 | | | |
| | MIR3942 | | | |
| | MIR3943 | | | |
| | MIR3944 | | | |
| | MIR3945 | | | |
| | MIR3960 | | | |
| | MIR3972 | | | |
| | MIR3973 | | | |
| | MIR3974 | | | |
| | MIR3975 | | | |
| | MIR3976 | | | |
| | MIR3977 | | | |
| | MIR3978 | | | |
| | MIR409 | | | |
| | MIR410 | | | |
| | MIR411 | | | |
| | MIR412 | | | |
| | MIR421 | | | |
| | MIR422A | | | |
| | MIR423 | | | |
| | MIR424 | | | |
| | MIR425 | | | |
| | MIR4251 | | | |
| | MIR4252 | | | |
| | MIR4253 | | | |
| | MIR4254 | | | |
| | MIR4255 | | | |
| | MIR4256 | | | |
| | MIR4257 | | | |
| | MIR4258 | | | |
| | MIR4260 | | | |
| | MIR4261 | | | |
| | MIR4262 | | | |
| | MIR4263 | | | |
| | MIR4264 | | | |
| | MIR4265 | | | |
| | MIR4266 | | | |
| | MIR4267 | | | |
| | MIR4268 | | | |
| | MIR4269 | | | |
| | MIR4270 | | | |
| | MIR4271 | | | |

MIR4272
MIR4273
MIR4274
MIR4275
MIR4276
MIR4277
MIR4278
MIR4279
MIR4280
MIR4281
MIR4282
MIR4283-1
MIR4283-2
MIR4284
MIR4286
MIR4287
MIR4288
MIR4289
MIR429
MIR4290
MIR4291
MIR4292
MIR4293
MIR4294
MIR4295
MIR4296
MIR4297
MIR4298
MIR4299
MIR4300
MIR4301
MIR4302
MIR4303
MIR4304
MIR4305
MIR4306
MIR4307
MIR4308
MIR4309
MIR431
MIR4310
MIR4311
MIR4312
MIR4313
MIR4314
MIR4315-1
MIR4315-2
MIR4316
MIR4317
MIR4318
MIR4319

MIR432
MIR4320
MIR4321
MIR4322
MIR4323
MIR4324
MIR4325
MIR4326
MIR4327
MIR4328
MIR4329
MIR433
MIR4330
MIR4417
MIR4418
MIR4419A
MIR4419B
MIR4420
MIR4421
MIR4422
MIR4423
MIR4424
MIR4425
MIR4426
MIR4427
MIR4428
MIR4429
MIR4430
MIR4431
MIR4432
MIR4433
MIR4433B
MIR4434
MIR4435-1
MIR4435-2
MIR4436A
MIR4436B1
MIR4436B2
MIR4437
MIR4438
MIR4439
MIR4440
MIR4441
MIR4442
MIR4443
MIR4444-1
MIR4444-2
MIR4445
MIR4446
MIR4447
MIR4448

MIR4449
MIR4450
MIR4451
MIR4452
MIR4453
MIR4454
MIR4455
MIR4456
MIR4457
MIR4458
MIR4459
MIR4460
MIR4461
MIR4462
MIR4463
MIR4464
MIR4465
MIR4466
MIR4467
MIR4468
MIR4469
MIR4470
MIR4471
MIR4472-1
MIR4472-2
MIR4473
MIR4474
MIR4475
MIR4476
MIR4477A
MIR4477B
MIR4478
MIR4479
MIR448
MIR4480
MIR4481
MIR4482-1
MIR4483
MIR4484
MIR4485
MIR4486
MIR4487
MIR4488
MIR4489
MIR4490
MIR4491
MIR4492
MIR4493
MIR4494
MIR4495
MIR4496

| | | | |
|---|---|---|---|
| | MIR4497 | | |
| | MIR4498 | | |
| | MIR4499 | | |
| | MIR449A | | |
| | MIR449B | | |
| | MIR449C | | |
| | MIR4500 | | |
| | MIR4501 | | |
| | MIR4502 | | |
| | MIR4503 | | |
| | MIR4504 | | |
| | MIR4505 | | |
| | MIR4506 | | |
| | MIR4507 | | |
| | MIR4508 | | |
| | MIR4509-1 | | |
| | MIR4509-2 | | |
| | MIR4509-3 | | |
| | MIR450A1 | | |
| | MIR450A2 | | |
| | MIR450B | | |
| | MIR4510 | | |
| | MIR4511 | | |
| | MIR4512 | | |
| | MIR4513 | | |
| | MIR4514 | | |
| | MIR4515 | | |
| | MIR4516 | | |
| | MIR4517 | | |
| | MIR4518 | | |
| | MIR4519 | | |
| | MIR451A | | |
| | MIR451B | | |
| | MIR452 | | |
| | MIR4520A | | |
| | MIR4520B | | |
| | MIR4521 | | |
| | MIR4522 | | |
| | MIR4523 | | |
| | MIR4524A | | |
| -0,79036047 | MIR4524B | FALSE | Downregulated |
| | MIR4525 | | |
| | MIR4526 | | |
| | MIR4527 | | |
| | MIR4529 | | |
| | MIR4530 | | |
| | MIR4531 | | |
| | MIR4532 | | |
| | MIR4533 | | |
| | MIR4534 | | |
| | MIR4535 | | |

MIR4536-1
MIR454
MIR4540
MIR455
MIR4632
MIR4633
MIR4634
MIR4635
MIR4636
MIR4637
MIR4638
MIR4639
MIR4640
MIR4641
MIR4642
MIR4643
MIR4644
MIR4645
MIR4646
MIR4647
MIR4648
MIR4649
MIR4650-1
MIR4650-2
MIR4651
MIR4652
MIR4653
MIR4654
MIR4655
MIR4656
MIR4657
MIR4658
MIR4659A
MIR4659B
MIR4660
MIR4661
MIR4662A
MIR4662B
MIR4663
MIR4664
MIR4665
MIR4666A
MIR4666B
MIR4667
MIR4668
MIR4669
MIR4670
MIR4671
MIR4672
MIR4673
MIR4674

| | | |
|---|---|---|
| | MIR4675 | |
| | MIR4676 | |
| | MIR4677 | |
| | MIR4678 | |
| | MIR4679-1 | |
| | MIR4679-2 | |
| | MIR4680 | |
| | MIR4681 | |
| | MIR4682 | |
| | MIR4683 | |
| | MIR4684 | |
| | MIR4685 | |
| | MIR4686 | |
| | MIR4687 | |
| | MIR4688 | |
| | MIR4689 | |
| | MIR4690 | |
| | MIR4691 | |
| | MIR4692 | |
| | MIR4693 | |
| | MIR4694 | |
| | MIR4695 | |
| | MIR4696 | |
| | MIR4697 | |
| 1,236820205 | MIR4697HG | Upregulated |
| | MIR4698 | |
| | MIR4699 | |
| | MIR4700 | |
| | MIR4701 | |
| | MIR4703 | |
| | MIR4704 | |
| | MIR4705 | |
| | MIR4706 | |
| | MIR4707 | |
| | MIR4708 | |
| | MIR4709 | |
| | MIR4710 | |
| | MIR4711 | |
| | MIR4712 | |
| | MIR4713 | |
| | MIR4714 | |
| | MIR4715 | |
| | MIR4716 | |
| | MIR4717 | |
| | MIR4718 | |
| | MIR4719 | |
| | MIR4720 | |
| | MIR4721 | |
| | MIR4722 | |
| | MIR4723 | |
| | MIR4724 | |

MIR4725
MIR4726
MIR4727
MIR4728
MIR4729
MIR4730
MIR4731
MIR4732
MIR4733
MIR4734
MIR4735
MIR4736
MIR4737
MIR4738
MIR4739
MIR4740
MIR4741
MIR4742
MIR4743
MIR4744
MIR4745
MIR4746
MIR4747
MIR4748
MIR4749
MIR4750
MIR4751
MIR4752
MIR4753
MIR4754
MIR4755
MIR4756
MIR4757
MIR4758
MIR4759
MIR4760
MIR4761
MIR4762
MIR4763
MIR4764
MIR4765
MIR4766
MIR4767
MIR4768
MIR4769
MIR4770
MIR4771-1
MIR4771-2
MIR4772
MIR4773-1
MIR4773-2

MIR4774
MIR4775
MIR4776-1
MIR4776-2
MIR4777
MIR4778
MIR4779
MIR4780
MIR4781
MIR4782
MIR4783
MIR4784
MIR4785
MIR4786
MIR4787
MIR4788
MIR4789
MIR4790
MIR4791
MIR4792
MIR4793
MIR4794
MIR4795
MIR4796
MIR4797
MIR4798
MIR4799
MIR4800
MIR4801
MIR4802
MIR4803
MIR4804
MIR483
MIR484
MIR485
MIR486
MIR486-2
MIR487A
MIR487B
MIR488
MIR489
MIR490
MIR491
MIR492
MIR493
MIR494
MIR495
MIR496
MIR497
MIR498
MIR4999

| | | | |
|---|---|---|---|
| 0,028936087 | MIR499A | FALSE | Upregulated |
| | MIR499B | | |
| | MIR5000 | | |
| | MIR5001 | | |
| | MIR5002 | | |
| | MIR5003 | | |
| | MIR5004 | | |
| | MIR5006 | | |
| | MIR5007 | | |
| | MIR5008 | | |
| | MIR5009 | | |
| | MIR500A | | |
| | MIR500B | | |
| | MIR501 | | |
| | MIR5010 | | |
| | MIR5011 | | |
| | MIR502 | | |
| | MIR503 | | |
| 1,573930851 | MIR503HG | | Upregulated |
| | MIR504 | | |
| | MIR5047 | | |
| | MIR505 | | |
| | MIR506 | | |
| | MIR507 | | |
| | MIR508 | | |
| | MIR5087 | | |
| | MIR5088 | | |
| | MIR5089 | | |
| | MIR509-1 | | |
| | MIR509-2 | | |
| | MIR509-3 | | |
| | MIR5090 | | |
| | MIR5091 | | |
| | MIR5092 | | |
| | MIR5093 | | |
| | MIR5094 | | |
| | MIR5095 | | |
| | MIR5096 | | |
| | MIR510 | | |
| | MIR5100 | | |
| | MIR511-1 | | |
| | MIR512-1 | | |
| | MIR512-2 | | |
| | MIR513A1 | | |
| 0 | MIR513A2 | FALSE | Upregulated |
| | MIR513B | | |
| | MIR513C | | |
| | MIR514A1 | | |
| | MIR514A2 | | |
| | MIR514A3 | | |
| | MIR514B | | |

| | | | |
|---|---|---|---|
| | MIR515-1 | | |
| | MIR515-2 | | |
| | MIR516A1 | | |
| | MIR516A2 | | |
| | MIR516B1 | | |
| | MIR516B2 | | |
| | MIR517A | | |
| | MIR517B | | |
| | MIR517C | | |
| | MIR5186 | | |
| | MIR5187 | | |
| | MIR5188 | | |
| 0 | MIR5189 | FALSE | Upregulated |
| | MIR518A1 | | |
| | MIR518A2 | | |
| | MIR518B | | |
| | MIR518C | | |
| | MIR518D | | |
| | MIR518E | | |
| | MIR518F | | |
| | MIR5190 | | |
| | MIR5191 | | |
| | MIR5192 | | |
| | MIR5193 | | |
| 0 | MIR5194 | FALSE | Upregulated |
| | MIR5196 | | |
| | MIR5197 | | |
| | MIR519A1 | | |
| | MIR519A2 | | |
| | MIR519B | | |
| | MIR519C | | |
| | MIR519D | | |
| | MIR519E | | |
| | MIR520A | | |
| | MIR520B | | |
| | MIR520C | | |
| | MIR520D | | |
| | MIR520E | | |
| | MIR520F | | |
| | MIR520G | | |
| | MIR520H | | |
| | MIR521-1 | | |
| | MIR521-2 | | |
| | MIR522 | | |
| | MIR523 | | |
| | MIR524 | | |
| | MIR525 | | |
| | MIR526A1 | | |
| | MIR526A2 | | |
| | MIR526B | | |
| | MIR527 | | |

MIR539
MIR541
MIR542
MIR543
MIR544A
MIR544B
MIR545
MIR548A1
MIR548A2
MIR548A3
MIR548AA1
MIR548AA2
MIR548AB
MIR548AC
MIR548AD
MIR548AE1
MIR548AE2
MIR548AG1
MIR548AG2
MIR548AH
MIR548AI
MIR548AJ1
MIR548AJ2
MIR548AK
MIR548AL
MIR548AM
MIR548AN
MIR548AO
MIR548AP
MIR548AQ
MIR548AR
MIR548AS
MIR548AT
MIR548AU
MIR548AV
MIR548AW
MIR548AX
MIR548AY
MIR548AZ
MIR548B
MIR548BA
MIR548C
MIR548D1
MIR548D2
MIR548E
MIR548F1
MIR548F2
MIR548F3
MIR548F4
MIR548F5
MIR548G

| | | | |
|---|---|---|---|
| | MIR548H1 | | |
| | MIR548H2 | | |
| | MIR548H3 | | |
| | MIR548H4 | | |
| | MIR548H5 | | |
| 0 | MIR548I1 | FALSE | Upregulated |
| | MIR548I2 | | |
| | MIR548I3 | | |
| | MIR548I4 | | |
| | MIR548J | | |
| | MIR548K | | |
| | MIR548L | | |
| | MIR548M | | |
| | MIR548N | | |
| 0,042861077 | MIR548O | FALSE | Upregulated |
| | MIR548O2 | | |
| | MIR548P | | |
| | MIR548Q | | |
| | MIR548S | | |
| | MIR548T | | |
| | MIR548U | | |
| | MIR548V | | |
| | MIR548W | | |
| | MIR548X | | |
| | MIR548X2 | | |
| | MIR548Y | | |
| | MIR548Z | | |
| | MIR549 | | |
| | MIR550A1 | | |
| | MIR550A2 | | |
| | MIR550A3 | | |
| | MIR550B1 | | |
| | MIR550B2 | | |
| | MIR551A | | |
| | MIR551B | | |
| | MIR552 | | |
| | MIR553 | | |
| | MIR554 | | |
| | MIR555 | | |
| | MIR556 | | |
| | MIR557 | | |
| | MIR5572 | | |
| | MIR5579 | | |
| | MIR558 | | |
| | MIR5580 | | |
| | MIR5581 | | |
| | MIR5582 | | |
| | MIR5583-1 | | |
| | MIR5583-2 | | |
| | MIR5584 | | |
| | MIR5585 | | |

MIR5586
MIR5587
MIR5588
MIR5589
MIR559
MIR5590
MIR5591
MIR561
MIR562
MIR563
MIR564
MIR566
MIR567
MIR568
MIR5680
MIR5681A
MIR5681B
MIR5682
MIR5683
MIR5684
MIR5685
MIR5687
MIR5688
MIR5689
MIR569
MIR5690
MIR5691
MIR5692A1
MIR5692A2
MIR5692B
MIR5692C1
MIR5692C2
MIR5693
MIR5694
MIR5695
MIR5696
MIR5697
MIR5698
MIR5699
MIR570
MIR5700
MIR5701-1
MIR5701-2
MIR5702
MIR5703
MIR5704
MIR5705
MIR5706
MIR5707
MIR5708
MIR571

MIR572
MIR573
MIR5739
MIR574
MIR575
MIR576
MIR577
MIR578
MIR5787
MIR579
MIR580
MIR581
MIR582
MIR583
MIR584
MIR585
MIR586
MIR587
MIR588
MIR589
MIR590
MIR591
MIR592
MIR593
MIR595
MIR596
MIR597
MIR598
MIR599
MIR600
MIR601
MIR602
MIR603
MIR604
MIR605
MIR606
MIR6068
MIR6069
MIR607
MIR6070
MIR6071
MIR6072
MIR6073
MIR6074
MIR6075
MIR6076
MIR6077-2
MIR6078
MIR6079
MIR608
MIR6080

| | | | |
|---|---|---|---|
| | MIR6081 | | |
| | MIR6082 | | |
| | MIR6083 | | |
| | MIR6084 | | |
| | MIR6085 | | |
| | MIR6086 | | |
| | MIR6087 | | |
| | MIR6088 | | |
| | MIR6089-1 | | |
| | MIR6089-1-2 | | |
| | MIR609 | | |
| | MIR6090 | | |
| | MIR610 | | |
| | MIR611 | | |
| 0 | MIR612 | FALSE | Upregulated |
| | MIR6124 | | |
| | MIR6125 | | |
| | MIR6126 | | |
| | MIR6127 | | |
| | MIR6128 | | |
| | MIR6129 | | |
| | MIR613 | | |
| | MIR6130 | | |
| | MIR6131 | | |
| | MIR6132 | | |
| | MIR6133 | | |
| | MIR6134 | | |
| | MIR614 | | |
| | MIR615 | | |
| | MIR616 | | |
| | MIR6165 | | |
| | MIR617 | | |
| | MIR618 | | |
| | MIR619 | | |
| | MIR620 | | |
| | MIR621 | | |
| | MIR622 | | |
| | MIR623 | | |
| | MIR624 | | |
| | MIR625 | | |
| | MIR626 | | |
| | MIR627 | | |
| | MIR628 | | |
| | MIR629 | | |
| | MIR630 | | |
| | MIR631 | | |
| | MIR632 | | |
| | MIR633 | | |
| | MIR634 | | |
| | MIR635 | | |
| | MIR636 | | |

MIR637
MIR638
MIR639
MIR640
MIR641
MIR642A
MIR642B
MIR643
MIR644A
MIR644B
MIR645
MIR646
MIR647
MIR648
MIR649
MIR6499
MIR6500
MIR6501
MIR6502
MIR6503
MIR6504
MIR6505
MIR6506
MIR6507
MIR6508
MIR6509
MIR651
MIR6510
MIR6511A-2
MIR6511B-1
MIR6512
MIR6513
MIR6514
MIR6515
MIR6516
MIR652
MIR653
MIR654
MIR655
MIR656
MIR657
MIR658
MIR659
MIR660
MIR661
MIR662
MIR663A
MIR663B
MIR664
MIR665
MIR668

| | | | |
|---|---|---|---|
| | MIR670 | | |
| 0 | MIR670HG | FALSE | Upregulated |
| | MIR671 | | |
| | MIR6715A | | |
| | MIR6715B | | |
| | MIR6716 | | |
| | MIR6717 | | |
| | MIR6718 | | |
| | MIR6719 | | |
| | MIR6720 | | |
| | MIR6721 | | |
| | MIR6722 | | |
| | MIR6723 | | |
| | MIR6726 | | |
| | MIR6727 | | |
| | MIR6728 | | |
| | MIR6729 | | |
| | MIR6730 | | |
| | MIR6731 | | |
| | MIR6732 | | |
| | MIR6733 | | |
| | MIR6734 | | |
| | MIR6735 | | |
| | MIR6736 | | |
| | MIR6737 | | |
| | MIR6738 | | |
| | MIR6739 | | |
| | MIR6740 | | |
| | MIR6741 | | |
| | MIR6742 | | |
| | MIR6743 | | |
| | MIR6744 | | |
| | MIR6745 | | |
| | MIR6746 | | |
| | MIR6747 | | |
| | MIR6748 | | |
| | MIR6749 | | |
| | MIR675 | | |
| | MIR6750 | | |
| | MIR6751 | | |
| | MIR6752 | | |
| 0,046770501 | MIR6753 | FALSE | Upregulated |
| | MIR6754 | | |
| | MIR6755 | | |
| | MIR6756 | | |
| | MIR6757 | | |
| | MIR6758 | | |
| | MIR6759 | | |
| | MIR676 | | |
| | MIR6760 | | |
| | MIR6761 | | |

MIR6762
MIR6763
MIR6764
MIR6765
MIR6766
MIR6767
MIR6768
MIR6769A
MIR6769B
MIR6770-2
MIR6771
MIR6772
MIR6773
MIR6774
MIR6775
MIR6776
MIR6777
MIR6778
MIR6779
MIR6780A
MIR6780B
MIR6781
MIR6782
MIR6783
MIR6784
MIR6785
MIR6786
MIR6787
MIR6789
MIR6790
MIR6791
MIR6792
MIR6793
MIR6794
MIR6795
MIR6796
MIR6797
MIR6798
MIR6799
MIR6800
MIR6801
MIR6802
MIR6803
MIR6804
MIR6805
MIR6806
MIR6807
MIR6808
MIR6809
MIR6810
MIR6811

MIR6812
MIR6813
MIR6814
MIR6815
MIR6816
MIR6817
MIR6818
MIR6819
MIR6820
MIR6821
MIR6822
MIR6823
MIR6824
MIR6825
MIR6826
MIR6827
MIR6828
MIR6829
MIR6830
MIR6831
MIR6832
MIR6833
MIR6834
MIR6835
MIR6836
MIR6837
MIR6838
MIR6839
MIR6840
MIR6841
MIR6842
MIR6843
MIR6844
MIR6845
MIR6846
MIR6847
MIR6848
MIR6849
MIR6850
MIR6851
MIR6852
MIR6853
MIR6854
MIR6855
MIR6856
MIR6857
MIR6858
MIR6859-1
MIR6859-2
MIR6860
MIR6861

| | | | | |
|---|---|---|---|---|
| | MIR6862-1 | | | |
| | MIR6862-2 | | | |
| | MIR6863 | | | |
| | MIR6864 | | | |
| | MIR6865 | | | |
| | MIR6866 | | | |
| | MIR6867 | | | |
| | MIR6868 | | | |
| | MIR6869 | | | |
| | MIR6870 | | | |
| | MIR6871 | | | |
| | MIR6872 | | | |
| | MIR6873 | | | |
| | MIR6874 | | | |
| | MIR6875 | | | |
| | MIR6876 | | | |
| | MIR6877 | | | |
| | MIR6878 | | | |
| | MIR6879 | | | |
| | MIR6880 | | | |
| | MIR6881 | | | |
| | MIR6882 | | | |
| | MIR6883 | | | |
| | MIR6884 | | | |
| | MIR6885 | | | |
| | MIR6886 | | | |
| | MIR6887 | | | |
| | MIR6888 | | | |
| | MIR6889 | | | |
| | MIR6890 | | | |
| | MIR6891 | | | |
| | MIR6892 | | | |
| | MIR6893 | | | |
| | MIR6894 | | | |
| | MIR6895 | | | |
| | MIR7-1 | | | |
| | MIR7-2 | | | |
| | MIR7-3 | | | |
| -0,360761924 | | MIR7-3HG | FALSE | Downregulated |
| | MIR708 | | | |
| | MIR7106 | | | |
| | MIR7107 | | | |
| | MIR7108 | | | |
| | MIR7109 | | | |
| | MIR711 | | | |
| | MIR7110 | | | |
| | MIR7111 | | | |
| | MIR7112-2 | | | |
| | MIR7113 | | | |
| | MIR7114 | | | |
| | MIR7150 | | | |

MIR7151
MIR7152
MIR7153
MIR7154
MIR7155
MIR7156
MIR7157
MIR7159
MIR7160
MIR7161
MIR7162
MIR718
MIR744
MIR7515
MIR758
MIR759
MIR760
MIR761
MIR762
MIR764
MIR7641-2
MIR765
MIR766
MIR767
MIR769
MIR770
MIR7702
MIR7703
MIR7705
MIR7843
MIR7844
MIR7845
MIR7846
MIR7848
MIR7849
MIR7850
MIR7851
MIR7852
MIR7853
MIR7854
MIR7855
MIR7856
MIR7973-1
MIR7973-2
MIR7974
MIR7975
MIR7976
MIR7977
MIR7978
MIR802
MIR8052

MIR8053
MIR8054
MIR8055
MIR8056
MIR8057
MIR8058
MIR8059
MIR8060
MIR8061
MIR8062
MIR8063
MIR8064
MIR8065
MIR8066
MIR8067
MIR8068
MIR8069
MIR8070
MIR8071-1
MIR8071-2
MIR8072
MIR8073
MIR8074
MIR8075
MIR8076
MIR8077
MIR8078
MIR8079
MIR8080
MIR8081
MIR8082
MIR8083
MIR8084
MIR8085
MIR8086
MIR8087
MIR8088
MIR8089
MIR873
MIR874
MIR875
MIR876
MIR877
MIR885
MIR887
MIR888
MIR889
MIR890
MIR891A
MIR891B
MIR892A

| | | | |
|---|---|---|---|
| | MIR892B | | |
| | MIR892C | | |
| | MIR9-1 | | |
| | MIR9-2 | | |
| | MIR9-3 | | |
| | MIR920 | | |
| | MIR921 | | |
| | MIR922 | | |
| | MIR924 | | |
| | MIR92A1 | | |
| | MIR92A2 | | |
| | MIR92B | | |
| | MIR93 | | |
| | MIR933 | | |
| | MIR934 | | |
| | MIR935 | | |
| | MIR936 | | |
| | MIR937 | | |
| | MIR938 | | |
| | MIR939 | | |
| | MIR940 | | |
| | MIR942 | | |
| | MIR943 | | |
| | MIR944 | | |
| | MIR95 | | |
| | MIR96 | | |
| | MIR98 | | |
| | MIR99A | | |
| | MIR99B | | |
| | MIRLET7A1 | | |
| | MIRLET7A2 | | |
| | MIRLET7A3 | | |
| | MIRLET7B | | |
| | MIRLET7C | | |
| | MIRLET7D | | |
| | MIRLET7E | | |
| | MIRLET7F1 | | |
| | MIRLET7F2 | | |
| | MIRLET7G | | |
| | MIRLET7I | | |
| -0,23433423 | MIS18A | FALSE | Downregulated |
| -0,304905469 | MISP | FALSE | Downregulated |
| 0,346387404 | MKRN3 | FALSE | Upregulated |
| -0,613360537 | MKRN7P | FALSE | Downregulated |
| 0,606730516 | MKRN9P | FALSE | Upregulated |
| -1,039101651 | MLC1 | | Downregulated |
| 0,44499128 | MLF1 | FALSE | Upregulated |
| 0,398522549 | MLIP-IT1 | FALSE | Upregulated |
| | MLN | | |
| -0,338783876 | MLNR | FALSE | Downregulated |
| | MMD2 | | |

MMP1
-0,189001521 MMP10 FALSE Downregulated
1,189042719 MMP11 Upregulated
2,580234106 MMP12 Upregulated
2,907276678 MMP13 Upregulated
0,704541674 MMP16 FALSE Upregulated
0 MMP20 FALSE Upregulated
1,387667309 MMP23A Upregulated
MMP26
-0,360874141 MMP27 FALSE Downregulated
-1,022392536 MMP28 Downregulated
0,367142743 MMP3 FALSE Upregulated
1,244301899 MMP7 Upregulated
0,001493849 MMP8 FALSE Upregulated
-0,185803395 MND1 FALSE Downregulated
0,548847193 MNX1 FALSE Upregulated
MNX1-AS1
-0,021460817 MOBP FALSE Downregulated
-0,378380769 MOG FALSE Downregulated
0 MORC1 FALSE Upregulated
0,571993031 MORN2 FALSE Upregulated
0,549606782 MORN3 FALSE Upregulated
0 MORN5 FALSE Upregulated
MOS
0,202312013 MOV10L1 FALSE Upregulated
0 MPC1L FALSE Upregulated
1,543097873 MPHOSPH6 Upregulated
0,397054929 MPO FALSE Upregulated
-0,39473385 MPP2 FALSE Downregulated
0,815448825 MPP3 FALSE Upregulated
-0,982741043 MPP4 FALSE Downregulated
-0,23521705 MPPED2 FALSE Downregulated
-1,13761216 MRAP2 Downregulated
0,703915073 MRGPRD FALSE Upregulated
0 MRGPRE FALSE Upregulated
MRGPRG
MRGPRG-AS1
MRGPRX1
MRGPRX2
0,102745464 MRGPRX3 FALSE Upregulated
0,694977512 MRGPRX4 FALSE Upregulated
1,985938261 MROH2B Upregulated
0 MROH3P FALSE Upregulated
0,558286723 MROH5 FALSE Upregulated
0,001038729 MRPL42P5 FALSE Upregulated
-0,905289128 MRVI1-AS1 FALSE Downregulated
-0,081019594 MS4A10 FALSE Downregulated
MS4A12
MS4A13
-0,006396454 MS4A15 FALSE Downregulated
0 MS4A18 FALSE Upregulated

| | | | |
|---|---|---|---|
| 0,340256433 | MS4A2 | FALSE | Upregulated |
| 0,000577308 | MS4A3 | FALSE | Upregulated |
| 0,007698686 | MS4A4E | FALSE | Upregulated |
| | MS4A5 | | |
| 0 | MS4A6E | FALSE | Upregulated |
| 0 | MS4A8 | FALSE | Upregulated |
| 0,250488479 | MSANTD1 | FALSE | Upregulated |
| 0,60973033 | MSANTD3-TMEFF1 | FALSE | Upregulated |
| | MSGN1 | | |
| -0,26286396 | MSH4 | FALSE | Downregulated |
| 0,764873889 | MSI1 | FALSE | Upregulated |
| -1,225991345 | MSL3P1 | | Downregulated |
| 0,477039198 | MSLN | FALSE | Upregulated |
| -0,048913396 | MSMB | FALSE | Downregulated |
| -0,96109134 | MSTN | FALSE | Downregulated |
| 0,380029703 | MSX1 | FALSE | Upregulated |
| 0,039005732 | MSX2 | FALSE | Upregulated |
| -0,088193274 | MSX2P1 | FALSE | Downregulated |
| -0,069252445 | MT1A | FALSE | Downregulated |
| -0,44631294 | MT1B | FALSE | Downregulated |
| 0,007091921 | MT1DP | FALSE | Upregulated |
| 0 | MT1HL1 | FALSE | Upregulated |
| 0 | MT1IP | FALSE | Upregulated |
| -0,135057718 | MT1JP | FALSE | Downregulated |
| 0 | MT3 | FALSE | Upregulated |
| | MT4 | | |
| 1,85921473 | MTBP | | Upregulated |
| | MTFR2 | | |
| | MTHFD2P1 | | |
| 2,018537997 | MTL5 | | Upregulated |
| | MTNR1A | | |
| | MTRNR2L1 | | |
| | MTRNR2L10 | | |
| | MTRNR2L2 | | |
| -0,993754104 | MTRNR2L5 | FALSE | Downregulated |
| | MTRNR2L7 | | |
| 0 | MTRNR2L9 | FALSE | Upregulated |
| 1,285954339 | MTVR2 | | Upregulated |
| 1,667825241 | MUC1 | | Upregulated |
| 0,83385629 | MUC16 | FALSE | Upregulated |
| 0 | MUC17 | FALSE | Upregulated |
| 0,513750254 | MUC19 | FALSE | Upregulated |
| 0 | MUC2 | FALSE | Upregulated |
| 0 | MUC21 | FALSE | Upregulated |
| 0,651727757 | MUC22 | FALSE | Upregulated |
| 1,226488144 | MUC5AC | | Upregulated |
| 0,669025015 | MUC7 | FALSE | Upregulated |
| 0 | MUC8 | FALSE | Upregulated |
| | MUCL1 | | |
| 0,960258047 | MURC | FALSE | Upregulated |
| 0 | MYADML | FALSE | Upregulated |

| | | | |
|---|---|---|---|
| 1,931725223 | MYADML2 | | Upregulated |
| 0,79633347 | MYB | FALSE | Upregulated |
| 1,45368317 | MYBPC3 | | Upregulated |
| 1,436176126 | MYBPH | | Upregulated |
| 0,01390731 | MYBPHL | FALSE | Upregulated |
| -0,013313126 | MYCBP2-AS1 | FALSE | Downregulated |
| 1,747379194 | MYCBPAP | | Upregulated |
| 1,357523849 | MYCNOS | | Upregulated |
| 0,088031836 | MYEF2 | FALSE | Upregulated |
| 2,471485586 | MYEOV | | Upregulated |
| | MYF5 | | |
| -0,09019039 | MYF6 | FALSE | Downregulated |
| 0 | MYH13 | FALSE | Upregulated |
| 1,626010541 | MYH15 | | Upregulated |
| 1,78453568 | MYH4 | | Upregulated |
| 0,944540675 | MYH6 | FALSE | Upregulated |
| 0 | MYH8 | FALSE | Upregulated |
| | MYL10 | | |
| 0,06307772 | MYL3 | FALSE | Upregulated |
| 0,81492989 | MYL4 | FALSE | Upregulated |
| | MYL7 | | |
| -0,208033657 | MYO16-AS1 | FALSE | Downregulated |
| -0,042885095 | MYO1A | FALSE | Downregulated |
| 0,197681725 | MYO3A | FALSE | Upregulated |
| 1,115769768 | MYO3B | | Upregulated |
| 0,041338333 | MYOC | FALSE | Upregulated |
| 0,938092515 | MYOCD | FALSE | Upregulated |
| 0 | MYOD1 | FALSE | Upregulated |
| 0,079598255 | MYOG | FALSE | Upregulated |
| -0,066599591 | MYRFL | FALSE | Downregulated |
| 0,681081065 | MYT1 | FALSE | Upregulated |
| 0 | MYT1L | FALSE | Upregulated |
| | MYT1L-AS1 | | |
| -0,333309089 | MYZAP | FALSE | Downregulated |
| 0,291836297 | MZT1 | FALSE | Upregulated |
| 0 | NAA11 | FALSE | Upregulated |
| | NAALADL2-AS3 | | |
| 0 | NACA2 | FALSE | Upregulated |
| 0 | NACAP1 | FALSE | Upregulated |
| 0,391000129 | NAGPA-AS1 | FALSE | Upregulated |
| -0,226738983 | NAMA | FALSE | Downregulated |
| -0,324480969 | NANOG | FALSE | Downregulated |
| | NANOGNB | | |
| -0,033711379 | NANOS2 | FALSE | Downregulated |
| 0,050688952 | NANOS3 | FALSE | Upregulated |
| -0,095668124 | NAP1L2 | FALSE | Downregulated |
| 0,300404684 | NAP1L3 | FALSE | Upregulated |
| 0 | NAP1L6 | FALSE | Upregulated |
| -0,68098639 | NAPA-AS1 | FALSE | Downregulated |
| -1,272777234 | NARR | | Downregulated |
| 0 | NAT16 | FALSE | Upregulated |

| | | | |
|---|---|---|---|
| 0 | NBEAP1 | FALSE | Upregulated |
| | NBPF18P | | |
| -0,76678768 | NBPF22P | FALSE | Downregulated |
| | NBPF23P | | |
| -0,131082119 | NBPF4 | FALSE | Downregulated |
| 0,088218725 | NBPF6 | FALSE | Upregulated |
| 0,1002135 | NBPF7 | FALSE | Upregulated |
| 0,458593949 | NCAN | FALSE | Upregulated |
| 0,381126956 | NCCRP1 | FALSE | Upregulated |
| | NCOR1P1 | | |
| -0,722418398 | NCR1 | FALSE | Downregulated |
| | NCR2 | | |
| -1,374893861 | NCRNA00185 | | Downregulated |
| 0,027563344 | NCRNA00249 | FALSE | Upregulated |
| 0,251517448 | NCRUPAR | FALSE | Upregulated |
| 0 | NDC80 | FALSE | Upregulated |
| | NDFIP2-AS1 | | |
| -0,406969209 | NDNF | FALSE | Downregulated |
| | NDP | | |
| | NDST4 | | |
| 0,406108135 | NDUFAF4P1 | FALSE | Upregulated |
| | NEBL-AS1 | | |
| 0,021977528 | NEDD8-MDP1 | FALSE | Upregulated |
| 0 | NEFL | FALSE | Upregulated |
| 0,119350497 | NEFM | FALSE | Upregulated |
| | NEGR1-IT1 | | |
| -0,070567456 | NEIL3 | FALSE | Downregulated |
| 1,56278623 | NEK2 | | Upregulated |
| -0,109466732 | NEK5 | FALSE | Downregulated |
| | NELL1 | | |
| 2,416094883 | NETO1 | | Upregulated |
| 1,020498588 | NETO2 | | Upregulated |
| | NEU2 | | |
| | NEUROD1 | | |
| | NEUROD4 | | |
| | NEUROG1 | | |
| | NEUROG2 | | |
| | NEUROG3 | | |
| 0,595604803 | NF1P2 | FALSE | Upregulated |
| 0,258440962 | NFE2 | FALSE | Upregulated |
| 1,532475154 | NFE4 | | Upregulated |
| | NGB | | |
| -0,176113186 | NHEG1 | FALSE | Downregulated |
| 0,440606423 | NHLH1 | FALSE | Upregulated |
| 0 | NHLH2 | FALSE | Upregulated |
| -1,551019631 | NHLRC4 | | Downregulated |
| 0,015214724 | NHS-AS1 | FALSE | Upregulated |
| 0 | NKAIN1 | FALSE | Upregulated |
| 1,965501476 | NKAIN4 | | Upregulated |
| -0,303206096 | NKAPL | FALSE | Downregulated |
| | NKX1-2 | | |

| | | | |
|---|---|---|---|
| 0 | NKX2-1 | FALSE | Upregulated |
| | NKX2-1-AS1 | | |
| | NKX2-2 | | |
| 1,169456714 | NKX2-3 | | Upregulated |
| | NKX2-4 | | |
| -0,134088612 | NKX2-5 | FALSE | Downregulated |
| 0 | NKX2-6 | FALSE | Upregulated |
| 0 | NKX2-8 | FALSE | Upregulated |
| 0,497314788 | NKX3-2 | FALSE | Upregulated |
| | NKX6-1 | | |
| 1,642489087 | NKX6-2 | | Upregulated |
| | NKX6-3 | | |
| 0,073105102 | NLGN1 | FALSE | Upregulated |
| 0,300965538 | NLGN4Y-AS1 | FALSE | Upregulated |
| 0,669756935 | NLRP10 | FALSE | Upregulated |
| 0,861345716 | NLRP11 | FALSE | Upregulated |
| 2,108446982 | NLRP12 | | Upregulated |
| | NLRP13 | | |
| 0,821928429 | NLRP2 | FALSE | Upregulated |
| 0 | NLRP4 | FALSE | Upregulated |
| -0,004298224 | NLRP5 | FALSE | Downregulated |
| | NLRP8 | | |
| -0,327242675 | NMB | FALSE | Downregulated |
| | NMBR | | |
| | NME5 | | |
| 0 | NME8 | FALSE | Upregulated |
| -0,27964097 | NMRK2 | FALSE | Downregulated |
| | NMS | | |
| 0 | NMU | FALSE | Upregulated |
| 0 | NMUR2 | FALSE | Upregulated |
| | NOBOX | | |
| -0,35773799 | NOG | FALSE | Downregulated |
| 0,273623208 | NOS1 | FALSE | Upregulated |
| -0,651969495 | NOS2 | FALSE | Downregulated |
| -0,021462513 | NOTO | FALSE | Downregulated |
| 0,187106612 | NOV | FALSE | Upregulated |
| -1,105993794 | NOVA1 | | Downregulated |
| 0,169643662 | NOX1 | FALSE | Upregulated |
| | NOX3 | | |
| 1,382446309 | NOX4 | | Upregulated |
| -0,28788786 | NOX5 | FALSE | Downregulated |
| -0,064191378 | NPAS4 | FALSE | Downregulated |
| 0,395636317 | NPBWR1 | FALSE | Upregulated |
| | NPBWR2 | | |
| | NPFFR1 | | |
| | NPFFR2 | | |
| 0,311782337 | NPHP1 | FALSE | Upregulated |
| | NPHP3-AS1 | | |
| 0 | NPHS1 | FALSE | Upregulated |
| 0,044434384 | NPHS2 | FALSE | Upregulated |
| 2,410983625 | NPIPA3 | | Upregulated |

| | | | |
|---|---|---|---|
| -0,267530285 | NPM2 | FALSE | Downregulated |
| -0,208510035 | NPPA-AS1 | FALSE | Downregulated |
| 0 | NPPB | FALSE | Upregulated |
| | NPPC | | |
| | NPS | | |
| | NPSR1 | | |
| -0,227976756 | NPSR1-AS1 | FALSE | Downregulated |
| 0,420994192 | NPTX1 | FALSE | Upregulated |
| 1,509219334 | NPTX2 | | Upregulated |
| | NPVF | | |
| | NPY | | |
| | NPY2R | | |
| -0,227243408 | NPY5R | FALSE | Downregulated |
| | NR0B1 | | |
| 0,71310767 | NR2E1 | FALSE | Upregulated |
| 0,000577308 | NR5A1 | FALSE | Upregulated |
| 2,101750313 | NRCAM | | Upregulated |
| -0,206567503 | NREP-AS1 | FALSE | Downregulated |
| | NRG1-IT1 | | |
| | NRG1-IT3 | | |
| 0,130103022 | NRN1L | FALSE | Upregulated |
| 0,391222263 | NRXN1 | FALSE | Upregulated |
| -0,091188211 | NRXN3 | FALSE | Downregulated |
| 0,342393609 | NSFP1 | FALSE | Upregulated |
| 0,170443105 | NT5C1A | FALSE | Upregulated |
| -1,512185423 | NT5C1B-RDH14 | | Downregulated |
| -0,73125283 | NT5M | FALSE | Downregulated |
| 0,810397453 | NTM | FALSE | Upregulated |
| | NTM-IT | | |
| 0 | NTNG1 | FALSE | Upregulated |
| 0,935446882 | NTNG2 | FALSE | Upregulated |
| 0 | NTRK1 | FALSE | Upregulated |
| 0,004094583 | NTS | FALSE | Upregulated |
| | NTSR2 | | |
| 0,061186276 | NUDT11 | FALSE | Upregulated |
| 0,559075125 | NUDT4P2 | FALSE | Upregulated |
| 0,533945389 | NUDT9P1 | FALSE | Upregulated |
| 1,261280811 | NUF2 | | Upregulated |
| 0 | NUP210P1 | FALSE | Upregulated |
| -0,724793078 | NUP37 | FALSE | Downregulated |
| 0,835173447 | NUP62CL | FALSE | Upregulated |
| | NUPR1L | | |
| 0,318267341 | NWD1 | FALSE | Upregulated |
| 0 | NXF2 | FALSE | Upregulated |
| 0 | NXF2B | FALSE | Upregulated |
| 0,177355306 | NXF3 | FALSE | Upregulated |
| | NXF4 | | |
| -0,331119661 | NXF5 | FALSE | Downregulated |
| 0,616402069 | NXNL1 | FALSE | Upregulated |
| 0,707458514 | NXNL2 | FALSE | Upregulated |
| 0,879820473 | NXPE1 | FALSE | Upregulated |

| | | | |
|---|---|---|---|
| | NXPE2 | | |
| | NXPH1 | | |
| | NXPH2 | | |
| 0,366256479 | NYAP2 | FALSE | Upregulated |
| | NYX | | |
| | OACYLP | | |
| | OBP2A | | |
| | OBP2B | | |
| | OC90 | | |
| 0,55038243 | OCA2 | FALSE | Upregulated |
| 1,172599647 | OCLM | | Upregulated |
| -0,00897377 | OCM | FALSE | Downregulated |
| 0,027563344 | OCM2 | FALSE | Upregulated |
| | OCSTAMP | | |
| | ODAM | | |
| -0,101166716 | ODF1 | FALSE | Downregulated |
| 0,590021004 | ODF4 | FALSE | Upregulated |
| -0,024808033 | OFCC1 | FALSE | Downregulated |
| 0,702567401 | OLAH | FALSE | Upregulated |
| | OLFM3 | | |
| -0,218755222 | OLFM4 | FALSE | Downregulated |
| 0,709885154 | OLIG1 | FALSE | Upregulated |
| 0,683473083 | OLIG2 | FALSE | Upregulated |
| | OLIG3 | | |
| -0,476084422 | OLR1 | FALSE | Downregulated |
| -0,347151025 | OMP | FALSE | Downregulated |
| 0,608246941 | ONECUT3 | FALSE | Upregulated |
| | OOEP | | |
| | OOSP1 | | |
| | OOSP2 | | |
| | OPALIN | | |
| | OPN1LW | | |
| 0,008253022 | OPN1MW | FALSE | Upregulated |
| | OPN1MW2 | | |
| 0 | OPN4 | FALSE | Upregulated |
| 0,286742061 | OPN5 | FALSE | Upregulated |
| | OPRD1 | | |
| -0,274714335 | OPRK1 | FALSE | Downregulated |
| -0,622397121 | OPRL1 | FALSE | Downregulated |
| 0,073122974 | OPTC | FALSE | Upregulated |
| 0,597480774 | OR10A3 | FALSE | Upregulated |
| 1,920308448 | OR10A4 | | Upregulated |
| | OR10A6 | | |
| | OR10A7 | | |
| | OR10AG1 | | |
| | OR10C1 | | |
| | OR10G4 | | |
| 0 | OR10G7 | FALSE | Upregulated |
| | OR10G8 | | |
| | OR10G9 | | |
| 0 | OR10H1 | FALSE | Upregulated |

| | | | |
|---|---|---|---|
| | OR10H2 | | |
| 0,041547119 | OR10H3 | FALSE | Upregulated |
| | OR10H4 | | |
| 0,076658198 | OR10H5 | FALSE | Upregulated |
| 1,049994213 | OR10J3 | | Upregulated |
| | OR10K1 | | |
| | OR10K2 | | |
| | OR10P1 | | |
| 0 | OR10Q1 | FALSE | Upregulated |
| | OR10R2 | | |
| 0,88929146 | OR10S1 | FALSE | Upregulated |
| | OR10T2 | | |
| -0,140965505 | OR10V1 | FALSE | Downregulated |
| -0,013049104 | OR10W1 | FALSE | Downregulated |
| | OR10X1 | | |
| | OR10Z1 | | |
| 0,651749175 | OR11A1 | FALSE | Upregulated |
| | OR11G2 | | |
| | OR11H1 | | |
| | OR11H12 | | |
| | OR11H2 | | |
| 0,85283006 | OR11H4 | FALSE | Upregulated |
| -0,480288865 | OR11H6 | FALSE | Downregulated |
| 0 | OR11L1 | FALSE | Upregulated |
| | OR12D2 | | |
| | OR12D3 | | |
| | OR13A1 | | |
| 0,035170657 | OR13C2 | FALSE | Upregulated |
| 0 | OR13C3 | FALSE | Upregulated |
| 1,073900912 | OR13C4 | | Upregulated |
| 0 | OR13C5 | FALSE | Upregulated |
| -0,102033188 | OR13C9 | FALSE | Downregulated |
| 0,087035117 | OR13G1 | FALSE | Upregulated |
| 0,292052584 | OR13H1 | FALSE | Upregulated |
| -0,774789466 | OR13J1 | FALSE | Downregulated |
| 0 | OR14A16 | FALSE | Upregulated |
| | OR14C36 | | |
| | OR14I1 | | |
| | OR14J1 | | |
| | OR1A1 | | |
| | OR1A2 | | |
| -0,130269403 | OR1B1 | FALSE | Downregulated |
| 0,532199101 | OR1C1 | FALSE | Upregulated |
| | OR1D2 | | |
| 0,960955046 | OR1D4 | FALSE | Upregulated |
| 0,779034848 | OR1D5 | FALSE | Upregulated |
| 0,675397479 | OR1E1 | FALSE | Upregulated |
| | OR1E2 | | |
| | OR1F1 | | |
| -0,017737585 | OR1F2P | FALSE | Downregulated |
| 0 | OR1G1 | FALSE | Upregulated |

| | | | |
|---|---|---|---|
| | OR1I1 | | |
| -0,056043697 | OR1J1 | FALSE | Downregulated |
| 0,588159512 | OR1J2 | FALSE | Upregulated |
| | OR1J4 | | |
| 0,994939954 | OR1L1 | FALSE | Upregulated |
| 1,42056647 | OR1L3 | | Upregulated |
| 1,441874543 | OR1L8 | | Upregulated |
| | OR1M1 | | |
| 0,170503616 | OR1N1 | FALSE | Upregulated |
| -0,392515761 | OR1N2 | FALSE | Downregulated |
| -0,117514025 | OR1Q1 | FALSE | Downregulated |
| | OR1S1 | | |
| 0 | OR1S2 | FALSE | Upregulated |
| 0,009734566 | OR2A14 | FALSE | Upregulated |
| -0,371391017 | OR2A25 | FALSE | Downregulated |
| 1,356016285 | OR2A42 | | Upregulated |
| 0,618859316 | OR2A5 | FALSE | Upregulated |
| -0,725611873 | OR2AE1 | FALSE | Downregulated |
| 2,543100624 | OR2AG1 | | Upregulated |
| -0,055970583 | OR2AG2 | FALSE | Downregulated |
| 0 | OR2AK2 | FALSE | Upregulated |
| | OR2AP1 | | |
| 0,312513738 | OR2AT4 | FALSE | Upregulated |
| 1,614363582 | OR2B11 | | Upregulated |
| | OR2B3 | | |
| | OR2B6 | | |
| 0,195577862 | OR2C1 | FALSE | Upregulated |
| 0,98554078 | OR2C3 | FALSE | Upregulated |
| | OR2D2 | | |
| 1,06510502 | OR2F1 | | Upregulated |
| 0,715268943 | OR2F2 | FALSE | Upregulated |
| | OR2G2 | | |
| | OR2G3 | | |
| 0 | OR2G6 | FALSE | Upregulated |
| 0,575639011 | OR2H1 | FALSE | Upregulated |
| 0,684419264 | OR2H2 | FALSE | Upregulated |
| -1,379774248 | OR2J2 | | Downregulated |
| | OR2J3 | | |
| | OR2K2 | | |
| | OR2L13 | | |
| 0 | OR2L1P | FALSE | Upregulated |
| | OR2L2 | | |
| | OR2L3 | | |
| | OR2L5 | | |
| | OR2L8 | | |
| | OR2M1P | | |
| | OR2M2 | | |
| | OR2M3 | | |
| | OR2M4 | | |
| | OR2M5 | | |
| | OR2M7 | | |

| | | | |
|---|---|---|---|
| -0,90794168 | OR2S2 | FALSE | Downregulated |
| | OR2T1 | | |
| | OR2T10 | | |
| 0 | OR2T11 | FALSE | Upregulated |
| | OR2T12 | | |
| | OR2T2 | | |
| 0 | OR2T27 | FALSE | Upregulated |
| 0,031420931 | OR2T29 | FALSE | Upregulated |
| 0 | OR2T3 | FALSE | Upregulated |
| | OR2T33 | | |
| | OR2T34 | | |
| | OR2T35 | | |
| | OR2T4 | | |
| 0,703644183 | OR2T5 | FALSE | Upregulated |
| | OR2T6 | | |
| | OR2T8 | | |
| -0,277442297 | OR2V1 | FALSE | Downregulated |
| | OR2V2 | | |
| 0,067880217 | OR2W1 | FALSE | Upregulated |
| | OR2W3 | | |
| 0 | OR2W5 | FALSE | Upregulated |
| | OR2Y1 | | |
| | OR2Z1 | | |
| 0,087836139 | OR3A1 | FALSE | Upregulated |
| 0,062884305 | OR3A2 | FALSE | Upregulated |
| 0,745755883 | OR3A3 | FALSE | Upregulated |
| -0,262412519 | OR3A4P | FALSE | Downregulated |
| | OR4A15 | | |
| | OR4A16 | | |
| | OR4A47 | | |
| | OR4A5 | | |
| | OR4B1 | | |
| | OR4C11 | | |
| | OR4C12 | | |
| | OR4C13 | | |
| | OR4C15 | | |
| | OR4C16 | | |
| | OR4C3 | | |
| | OR4C45 | | |
| | OR4C46 | | |
| | OR4C6 | | |
| 0 | OR4D1 | FALSE | Upregulated |
| | OR4D10 | | |
| | OR4D11 | | |
| 0,782851003 | OR4D2 | FALSE | Upregulated |
| | OR4D5 | | |
| | OR4D6 | | |
| | OR4D9 | | |
| | OR4F13P | | |
| | OR4F15 | | |
| 0 | OR4F16 | FALSE | Upregulated |

| | | | |
|---|---|---|---|
| 0 | OR4F17 | FALSE | Upregulated |
| 0 | OR4F21 | FALSE | Upregulated |
| 0 | OR4F29 | FALSE | Upregulated |
| 0 | OR4F3 | FALSE | Upregulated |
| | OR4F4 | | |
| 0 | OR4F5 | FALSE | Upregulated |
| | OR4F6 | | |
| | OR4K1 | | |
| | OR4K13 | | |
| | OR4K14 | | |
| | OR4K15 | | |
| | OR4K17 | | |
| | OR4K2 | | |
| | OR4K5 | | |
| | OR4L1 | | |
| | OR4M1 | | |
| 0 | OR4M2 | FALSE | Upregulated |
| | OR4N2 | | |
| | OR4N3P | | |
| | OR4N4 | | |
| | OR4N5 | | |
| | OR4P4 | | |
| | OR4Q3 | | |
| | OR4S1 | | |
| | OR4S2 | | |
| | OR4X1 | | |
| | OR4X2 | | |
| | OR51A2 | | |
| | OR51A4 | | |
| | OR51A7 | | |
| -0,286663402 | OR51B2 | FALSE | Downregulated |
| 0 | OR51B4 | FALSE | Upregulated |
| -0,001452899 | OR51B5 | FALSE | Downregulated |
| | OR51D1 | | |
| 0 | OR51E1 | FALSE | Upregulated |
| | OR51E2 | | |
| | OR51F1 | | |
| | OR51F2 | | |
| | OR51G1 | | |
| | OR51G2 | | |
| 0 | OR51I1 | FALSE | Upregulated |
| | OR51L1 | | |
| | OR51S1 | | |
| | OR51T1 | | |
| | OR51V1 | | |
| | OR52A1 | | |
| | OR52A5 | | |
| 0,947886097 | OR52B2 | FALSE | Upregulated |
| -0,315011125 | OR52B4 | FALSE | Downregulated |
| | OR52E2 | | |
| 0 | OR52E4 | FALSE | Upregulated |

| | | | |
|---|---|---|---|
| | OR52E6 | | |
| | OR52E8 | | |
| -0,260399537 | OR52H1 | FALSE | Downregulated |
| -1,340271682 | OR52I2 | | Downregulated |
| | OR52J3 | | |
| 0 | OR52K1 | FALSE | Upregulated |
| 0 | OR52K2 | FALSE | Upregulated |
| | OR52L1 | | |
| 0 | OR52M1 | FALSE | Upregulated |
| 0,077936159 | OR52N1 | FALSE | Upregulated |
| 0 | OR52N2 | FALSE | Upregulated |
| 0,989169839 | OR52N4 | FALSE | Upregulated |
| 0 | OR52N5 | FALSE | Upregulated |
| | OR52R1 | | |
| 0 | OR56A1 | FALSE | Upregulated |
| 0 | OR56A3 | FALSE | Upregulated |
| | OR56A4 | | |
| | OR56A5 | | |
| -0,055967595 | OR56B1 | FALSE | Downregulated |
| | OR5A1 | | |
| 0 | OR5A2 | FALSE | Upregulated |
| | OR5AC2 | | |
| 0,146681389 | OR5AN1 | FALSE | Upregulated |
| | OR5AP2 | | |
| | OR5AR1 | | |
| | OR5AS1 | | |
| | OR5AU1 | | |
| | OR5B12 | | |
| 0 | OR5B17 | FALSE | Upregulated |
| | OR5B2 | | |
| 0 | OR5B21 | FALSE | Upregulated |
| | OR5B3 | | |
| | OR5D13 | | |
| | OR5D14 | | |
| | OR5D16 | | |
| | OR5D18 | | |
| 0 | OR5E1P | FALSE | Upregulated |
| | OR5F1 | | |
| | OR5H1 | | |
| | OR5H14 | | |
| | OR5H15 | | |
| 0 | OR5H2 | FALSE | Upregulated |
| | OR5H6 | | |
| | OR5I1 | | |
| | OR5J2 | | |
| 0 | OR5K1 | FALSE | Upregulated |
| 0,926457851 | OR5K2 | FALSE | Upregulated |
| 0,000577308 | OR5K3 | FALSE | Upregulated |
| | OR5K4 | | |
| | OR5L1 | | |
| | OR5L2 | | |

| | | | |
|---|---|---|---|
| | OR5M1 | | |
| | OR5M10 | | |
| | OR5M11 | | |
| | OR5M3 | | |
| | OR5M8 | | |
| | OR5M9 | | |
| | OR5P2 | | |
| | OR5P3 | | |
| | OR5R1 | | |
| | OR5T1 | | |
| | OR5T2 | | |
| | OR5T3 | | |
| 0 | OR5V1 | FALSE | Upregulated |
| | OR5W2 | | |
| | OR6A2 | | |
| 0,342257692 | OR6B1 | FALSE | Upregulated |
| 0 | OR6B2 | FALSE | Upregulated |
| | OR6B3 | | |
| | OR6C1 | | |
| | OR6C2 | | |
| | OR6C3 | | |
| | OR6C4 | | |
| | OR6C6 | | |
| | OR6C65 | | |
| | OR6C68 | | |
| | OR6C70 | | |
| | OR6C74 | | |
| | OR6C75 | | |
| | OR6C76 | | |
| 0,699751005 | OR6F1 | FALSE | Upregulated |
| 0 | OR6K2 | FALSE | Upregulated |
| | OR6K3 | | |
| 0 | OR6K6 | FALSE | Upregulated |
| 0,209621381 | OR6M1 | FALSE | Upregulated |
| | OR6N1 | | |
| | OR6N2 | | |
| | OR6P1 | | |
| | OR6Q1 | | |
| | OR6S1 | | |
| -0,05157615 | OR6T1 | FALSE | Downregulated |
| -0,155090219 | OR6V1 | FALSE | Downregulated |
| 9,56273E-05 | OR6W1P | FALSE | Upregulated |
| 0,122538635 | OR6X1 | FALSE | Upregulated |
| | OR6Y1 | | |
| 0 | OR7A10 | FALSE | Upregulated |
| | OR7A17 | | |
| | OR7A5 | | |
| 0 | OR7C1 | FALSE | Upregulated |
| | OR7C2 | | |
| 0 | OR7D2 | FALSE | Upregulated |
| 0,479260234 | OR7D4 | FALSE | Upregulated |

1,086711234 OR7E156P Upregulated
OR7E24
0,013936292 OR7E37P FALSE Upregulated
OR7E5P
-0,333871064 OR7E91P FALSE Downregulated
OR7G1
OR7G2
OR7G3
OR8A1
OR8B12
0,669756935 OR8B2 FALSE Upregulated
OR8B3
OR8B4
0 OR8B8 FALSE Upregulated
OR8D1
0,669756935 OR8D2 FALSE Upregulated
OR8D4
OR8G1
OR8G2
OR8G5
OR8H1
OR8H2
OR8H3
OR8I2
OR8J1
OR8J3
OR8K1
OR8K3
OR8K5
OR8S1
OR8U1
OR9A2
0 OR9A4 FALSE Upregulated
OR9G1
OR9G4
-0,119481104 OR9I1 FALSE Downregulated
OR9K2
OR9Q1
OR9Q2
0,379212693 ORC1 FALSE Upregulated
0,131499253 ORC6 FALSE Upregulated
-0,022219706 OSBPL10-AS1 FALSE Downregulated
1,261309933 OSM Upregulated
1,456885912 OSR1 Upregulated
0 OSR2 FALSE Upregulated
-0,111263835 OSTCP2 FALSE Downregulated
OSTM1-AS1
1,217673791 OSTN Upregulated
0 OTOA FALSE Upregulated
-0,356768895 OTOF FALSE Downregulated
OTOG

| | | | |
|---|---|---|---|
| 0 | OTOGL | FALSE | Upregulated |
| | OTOL1 | | |
| | OTOP1 | | |
| | OTOP2 | | |
| -0,457920525 | OTOP3 | FALSE | Downregulated |
| | OTOR | | |
| | OTOS | | |
| | OTP | | |
| 0 | OTUD6A | FALSE | Upregulated |
| | OTX1 | | |
| | OTX2 | | |
| -0,043458808 | OTX2-AS1 | FALSE | Downregulated |
| 1,155519608 | OVCH1 | | Upregulated |
| -0,612864413 | OVCH1-AS1 | FALSE | Downregulated |
| 0,14576601 | OVCH2 | FALSE | Upregulated |
| -0,483828926 | OVOL1 | FALSE | Downregulated |
| 0 | OVOL1-AS1 | FALSE | Upregulated |
| 0,079452622 | OVOL2 | FALSE | Upregulated |
| 0,767647798 | OVOS | FALSE | Upregulated |
| 0 | OVOS2 | FALSE | Upregulated |
| 0 | OXCT1-AS1 | FALSE | Upregulated |
| 1,778555316 | OXCT2 | | Upregulated |
| | OXGR1 | | |
| 0,500808934 | OXTR | FALSE | Upregulated |
| 0,55549919 | P2RX1 | FALSE | Upregulated |
| 0,169140194 | P2RX3 | FALSE | Upregulated |
| -1,48825427 | P2RX5 | | Downregulated |
| | P2RX6P | | |
| 1,213651926 | P2RY10 | | Upregulated |
| 0,892065698 | P2RY12 | FALSE | Upregulated |
| -0,503279432 | P2RY4 | FALSE | Downregulated |
| | P2RY8-2 | | |
| | PABPC1L2A | | |
| | PABPC1L2B | | |
| -0,317514782 | PABPC1P2 | FALSE | Downregulated |
| 0,158944394 | PABPC3 | FALSE | Upregulated |
| 0,314178674 | PABPC4L | FALSE | Upregulated |
| -0,092256922 | PABPC5 | FALSE | Downregulated |
| -0,072785038 | PACRGL | FALSE | Downregulated |
| 0,127347933 | PACSIN1 | FALSE | Upregulated |
| -0,588067951 | PADI1 | FALSE | Downregulated |
| 1,572433173 | PADI2 | | Upregulated |
| | PADI3 | | |
| -0,580839864 | PADI4 | FALSE | Downregulated |
| 0 | PADI6 | FALSE | Upregulated |
| | PAEP | | |
| | PAGE1 | | |
| | PAGE2 | | |
| | PAGE2B | | |
| | PAGE3 | | |
| 0 | PAGE4 | FALSE | Upregulated |

| | | | |
|---|---|---|---|
| -0,004298224 | PAGE5 | FALSE | Downregulated |
| 0,844951587 | PAK3 | FALSE | Upregulated |
| -0,413777732 | PAK7 | FALSE | Downregulated |
| -0,217232138 | PANDAR | FALSE | Downregulated |
| 0,084779715 | PANX3 | FALSE | Upregulated |
| 0 | PAPL | FALSE | Upregulated |
| 0 | PAPOLB | FALSE | Upregulated |
| | PARD3-AS1 | | |
| 1,025666057 | PARPBP | | Upregulated |
| | PASD1 | | |
| 0,464674872 | PATE1 | FALSE | Upregulated |
| | PATE2 | | |
| -0,291467377 | PATE3 | FALSE | Downregulated |
| | PAX1 | | |
| -0,112084432 | PAX2 | FALSE | Downregulated |
| 0 | PAX3 | FALSE | Upregulated |
| | PAX4 | | |
| 0 | PAX7 | FALSE | Upregulated |
| 0,163422301 | PAX9 | FALSE | Upregulated |
| 0,035297072 | PBK | FALSE | Upregulated |
| 1,214208081 | PBX4 | | Upregulated |
| 0,68370997 | PCA3 | FALSE | Upregulated |
| 0,160305866 | PCAT1 | FALSE | Upregulated |
| 0 | PCAT14 | FALSE | Upregulated |
| 0,588690044 | PCAT18 | FALSE | Upregulated |
| 0,572360988 | PCAT4 | FALSE | Upregulated |
| -0,001452899 | PCAT5 | FALSE | Downregulated |
| -0,053295092 | PCAT6 | FALSE | Downregulated |
| 0,906329247 | PCBP2-OT1 | FALSE | Upregulated |
| 0,853847366 | PCBP3 | FALSE | Upregulated |
| -0,072442207 | PCDH10 | FALSE | Downregulated |
| -0,084014365 | PCDH11X | FALSE | Downregulated |
| 0,300286775 | PCDH11Y | FALSE | Upregulated |
| -0,135299144 | PCDH15 | FALSE | Downregulated |
| 1,248647126 | PCDH19 | | Upregulated |
| -0,10936392 | PCDH8 | FALSE | Downregulated |
| 0 | PCDH9-AS2 | FALSE | Upregulated |
| 1,118117796 | PCDH9-AS3 | | Upregulated |
| -0,076512374 | PCDH9-AS4 | FALSE | Downregulated |
| 0,425655636 | PCDP1 | FALSE | Upregulated |
| | PCGEM1 | | |
| | PCP4 | | |
| 0 | PCSK1N | FALSE | Upregulated |
| 1,487587499 | PCSK2 | | Upregulated |
| 0,003686225 | PCYT1B | FALSE | Upregulated |
| -0,64852833 | PDCD2L | FALSE | Downregulated |
| -0,066859753 | PDCD4-AS1 | FALSE | Downregulated |
| | PDCL2 | | |
| -0,374677151 | PDCL3 | FALSE | Downregulated |
| 0,524165655 | PDE1C | FALSE | Upregulated |
| -0,775935994 | PDE6C | FALSE | Downregulated |

| | | | |
|---|---|---|---|
| 0,432296454 | PDE6G | FALSE | Upregulated |
| 1,041361988 | PDE6H | | Upregulated |
| | PDHA2 | | |
| 0,000266784 | PDIA2 | FALSE | Upregulated |
| 0,290328005 | PDILT | FALSE | Upregulated |
| -0,419204424 | PDLIM4 | FALSE | Downregulated |
| 3,256763099 | PDPN | | Upregulated |
| 0,438433923 | PDX1 | FALSE | Upregulated |
| 0 | PDX1-AS1 | FALSE | Upregulated |
| 0 | PDYN | FALSE | Upregulated |
| 0,653275494 | PDZD3 | FALSE | Upregulated |
| 3,544807777 | PDZK1IP1 | | Upregulated |
| 0,068383 | PEBP4 | FALSE | Upregulated |
| | PENK | | |
| 0,187634625 | PER4 | FALSE | Upregulated |
| 0 | PF4 | FALSE | Upregulated |
| | PF4V1 | | |
| -0,823958313 | PFN3 | FALSE | Downregulated |
| -0,179650428 | PGA3 | FALSE | Downregulated |
| 1,799036158 | PGA4 | | Upregulated |
| 1,028701331 | PGA5 | | Upregulated |
| 1,52272145 | PGBD3 | | Upregulated |
| 0,211725021 | PGC | FALSE | Upregulated |
| | PGCP1 | | |
| 2,189346798 | PGK2 | | Upregulated |
| 0,609573608 | PGLYRP3 | FALSE | Upregulated |
| | PGLYRP4 | | |
| 0 | PGM5-AS1 | FALSE | Upregulated |
| 0,498684658 | PGPEP1L | FALSE | Upregulated |
| 0,909648808 | PHACTR3 | FALSE | Upregulated |
| 2,649489323 | PHEX | | Upregulated |
| 0,403084012 | PHEX-AS1 | FALSE | Upregulated |
| 0,076658198 | PHF21B | FALSE | Upregulated |
| 0,142644555 | PHGR1 | FALSE | Upregulated |
| 1,853532279 | PHLDA2 | | Upregulated |
| -0,662962986 | PHOSPHO2-KLHL23 | FALSE | Downregulated |
| | PHOX2B | | |
| 0 | PI15 | FALSE | Upregulated |
| 1,680565976 | PI16 | | Upregulated |
| | PI3 | | |
| 0,024281708 | PIFO | FALSE | Upregulated |
| 1,262311364 | PIH1D2 | | Upregulated |
| | PIH1D3 | | |
| | PIN1P1 | | |
| 0 | PIP | FALSE | Upregulated |
| 0,99648118 | PIP5K1B | FALSE | Upregulated |
| 0,89129883 | PIR-FIGF | FALSE | Upregulated |
| 0 | PISRT1 | FALSE | Upregulated |
| 0 | PITX2 | FALSE | Upregulated |
| 0,050068234 | PITX3 | FALSE | Upregulated |
| -0,247822707 | PIWIL1 | FALSE | Downregulated |

| | | | |
|---|---|---|---|
| | PIWIL3 | | |
| 1,213965742 | PIWIL4 | | Upregulated |
| -0,535610247 | PKD2L1 | FALSE | Downregulated |
| 0,272314441 | PKIB | FALSE | Upregulated |
| 0,624547674 | PKP3 | FALSE | Upregulated |
| 0,388173738 | PLA2G10 | FALSE | Upregulated |
| -0,379019609 | PLA2G1B | FALSE | Downregulated |
| | PLA2G2C | | |
| | PLA2G2E | | |
| 0,907624853 | PLA2G2F | FALSE | Upregulated |
| 0,022866986 | PLA2G3 | FALSE | Upregulated |
| 0 | PLA2G4D | FALSE | Upregulated |
| 0,463715569 | PLA2G4F | FALSE | Upregulated |
| | PLAC1 | | |
| 0,781275177 | PLAC9 | FALSE | Upregulated |
| -1,383793922 | PLAG1 | | Downregulated |
| 1,893443682 | PLCB4 | | Upregulated |
| 2,945053986 | PLCE1-AS1 | | Upregulated |
| -1,750698889 | PLCH1 | | Downregulated |
| 1,31981459 | PLCH1-AS1 | | Upregulated |
| | PLCXD1-2 | | |
| -0,089054121 | PLD5 | FALSE | Downregulated |
| 0,759940927 | PLEKHA8P1 | FALSE | Upregulated |
| 0,028654136 | PLEKHD1 | FALSE | Upregulated |
| 0,658553235 | PLEKHS1 | FALSE | Upregulated |
| 0,571134001 | PLK4 | FALSE | Upregulated |
| 0 | PLK5 | FALSE | Upregulated |
| 1,335251603 | PLP1 | | Upregulated |
| 0 | PLSCR2 | FALSE | Upregulated |
| | PLSCR5 | | |
| 0,159346299 | PMAIP1 | FALSE | Upregulated |
| -0,397398342 | PMCH | FALSE | Downregulated |
| 0,001493849 | PMCHL1 | FALSE | Upregulated |
| 1,395102943 | PMFBP1 | | Upregulated |
| 0,685443617 | PMP2 | FALSE | Upregulated |
| 0,606200847 | PMS2L2 | FALSE | Upregulated |
| -0,148755462 | PMS2P4 | FALSE | Downregulated |
| | PNCK | | |
| -0,092491633 | PNLDC1 | FALSE | Downregulated |
| | PNLIP | | |
| | PNLIPRP1 | | |
| | PNLIPRP2 | | |
| | PNLIPRP3 | | |
| 0,580957834 | PNMA3 | FALSE | Upregulated |
| -0,726185006 | PNMA5 | FALSE | Downregulated |
| 0,243555581 | PNMAL1 | FALSE | Upregulated |
| -1,061746128 | PNMT | | Downregulated |
| | PNPLA1 | | |
| | PNPLA5 | | |
| -0,120938193 | POLN | FALSE | Downregulated |
| 1,91171647 | POLQ | | Upregulated |

| | | | |
|---|---|---|---|
| -0,063686796 | POM121L12 | FALSE | Downregulated |
| | POM121L2 | | |
| | POM121L4P | | |
| -1,065523123 | POMC | | Downregulated |
| 1,862203795 | POPDC2 | | Upregulated |
| 0,046397089 | POPDC3 | FALSE | Upregulated |
| | POTEA | | |
| | POTEB | | |
| | POTEB2 | | |
| | POTEC | | |
| | POTED | | |
| 1,244186912 | POTEE | | Upregulated |
| 0,061245492 | POTEF | FALSE | Upregulated |
| | POTEG | | |
| | POTEH | | |
| 0 | POTEI | FALSE | Upregulated |
| 0,608246941 | POTEJ | FALSE | Upregulated |
| 0 | POTEKP | FALSE | Upregulated |
| 0,209692609 | POTEM | FALSE | Upregulated |
| 1,09593198 | POU1F1 | | Upregulated |
| 0,473524048 | POU2F3 | FALSE | Upregulated |
| -0,312795427 | POU3F1 | FALSE | Downregulated |
| | POU3F3 | | |
| | POU3F4 | | |
| 0,201560057 | POU4F1 | FALSE | Upregulated |
| | POU4F2 | | |
| 0,661206439 | POU4F3 | FALSE | Upregulated |
| 0,687892908 | POU5F1B | FALSE | Upregulated |
| -0,168851877 | POU6F2-AS1 | FALSE | Downregulated |
| 0 | PP12613 | FALSE | Upregulated |
| 0 | PP2672 | FALSE | Upregulated |
| 2,059901844 | PPAP2C | | Upregulated |
| -0,757325666 | PPAPDC1A | FALSE | Downregulated |
| 0,931698393 | PPAPDC3 | FALSE | Upregulated |
| -0,586014997 | PPBP | FALSE | Downregulated |
| -0,16244628 | PPBPP2 | FALSE | Downregulated |
| -0,315504466 | PPEF1 | FALSE | Downregulated |
| | PPEF2 | | |
| -0,06081492 | PPIAL4B | FALSE | Downregulated |
| 0,697474655 | PPIAL4C | FALSE | Upregulated |
| 0,97678313 | PPIAL4D | FALSE | Upregulated |
| | PPIAL4E | | |
| | PPIAL4F | | |
| | PPIAL4G | | |
| 0,667082254 | PPIAP30 | FALSE | Upregulated |
| -0,488328885 | PPM1J | FALSE | Downregulated |
| 0,050025063 | PPP1R14C | FALSE | Upregulated |
| 0,029081059 | PPP1R17 | FALSE | Upregulated |
| 0 | PPP1R2P3 | FALSE | Upregulated |
| | PPP1R2P9 | | |
| -0,034663094 | PPP1R36 | FALSE | Downregulated |

| | | | |
|---|---|---|---|
| 0,576273196 | PPP1R3A | FALSE | Upregulated |
| 0 | PPP1R42 | FALSE | Upregulated |
| 0,936379078 | PPP2R2B | FALSE | Upregulated |
| | PPP2R2B-IT1 | | |
| 0,76132238 | PPP2R2C | FALSE | Upregulated |
| | PPP2R3B-2 | | |
| 1,198887419 | PPP3R2 | | Upregulated |
| 0,781598079 | PPP4R1L | FALSE | Upregulated |
| -0,01378149 | PPP5D1 | FALSE | Downregulated |
| | PPY | | |
| | PPY2 | | |
| | PRAC1 | | |
| | PRAC2 | | |
| | PRAMEF1 | | |
| 0,442556728 | PRAMEF10 | FALSE | Upregulated |
| | PRAMEF11 | | |
| | PRAMEF12 | | |
| | PRAMEF13 | | |
| -0,07374155 | PRAMEF15 | FALSE | Downregulated |
| | PRAMEF17 | | |
| | PRAMEF18 | | |
| | PRAMEF19 | | |
| 0 | PRAMEF2 | FALSE | Upregulated |
| | PRAMEF20 | | |
| | PRAMEF25 | | |
| | PRAMEF26 | | |
| 0 | PRAMEF4 | FALSE | Upregulated |
| | PRAMEF5 | | |
| | PRAMEF6 | | |
| | PRAMEF7 | | |
| | PRAMEF8 | | |
| | PRAMEF9 | | |
| -1,107056839 | PRAMENP | | Downregulated |
| 1,838587983 | PRDM12 | | Upregulated |
| | PRDM13 | | |
| | PRDM14 | | |
| -0,99421245 | PRDM5 | FALSE | Downregulated |
| 0,597480774 | PRDM9 | FALSE | Upregulated |
| | PRG1 | | |
| | PRG3 | | |
| 0,258898043 | PRIMA1 | FALSE | Upregulated |
| -0,00069498 | PRKACG | FALSE | Downregulated |
| 0 | PRKCG | FALSE | Upregulated |
| -1,175341202 | PRKCQ-AS1 | | Downregulated |
| -0,04929643 | PRKG2 | FALSE | Downregulated |
| | PRL | | |
| | PRLH | | |
| | PRLHR | | |
| | PRM1 | | |
| 0,038401465 | PRM2 | FALSE | Upregulated |
| | PRM3 | | |

| | | | |
|---|---|---|---|
| | PRMT8 | | |
| 1,012612997 | PRND | | Upregulated |
| | PRNT | | |
| 0 | PROK1 | FALSE | Upregulated |
| 0,021054878 | PROK2 | FALSE | Upregulated |
| -0,082910291 | PROKR1 | FALSE | Downregulated |
| -0,382697537 | PROKR2 | FALSE | Downregulated |
| 0,661206439 | PROL1 | FALSE | Upregulated |
| 0 | PRPH | FALSE | Upregulated |
| | PRPS1L1 | | |
| 1,119868833 | PRR16 | | Upregulated |
| | PRR20A | | |
| | PRR20B | | |
| | PRR20C | | |
| | PRR20D | | |
| 0 | PRR20E | FALSE | Upregulated |
| | PRR21 | | |
| | PRR23A | | |
| | PRR23B | | |
| | PRR23C | | |
| | PRR23D1 | | |
| | PRR23D2 | | |
| 0,895157967 | PRR25 | FALSE | Upregulated |
| 0 | PRR4 | FALSE | Upregulated |
| 0,814468901 | PRR5-ARHGAP8 | FALSE | Upregulated |
| 0 | PRR9 | FALSE | Upregulated |
| -0,068669872 | PRRG3 | FALSE | Downregulated |
| -1,386777347 | PRRT1 | | Downregulated |
| 0,372594494 | PRRT3-AS1 | FALSE | Upregulated |
| -0,112536207 | PRRT4 | FALSE | Downregulated |
| 0,795162941 | PRRX2 | FALSE | Upregulated |
| -0,394975918 | PRSS16 | FALSE | Downregulated |
| 1,049825626 | PRSS2 | | Upregulated |
| -0,026264688 | PRSS21 | FALSE | Downregulated |
| 1,496509502 | PRSS22 | | Upregulated |
| -0,171514764 | PRSS27 | FALSE | Downregulated |
| -0,010616246 | PRSS3 | FALSE | Downregulated |
| 0,668650025 | PRSS30P | FALSE | Upregulated |
| -0,252505007 | PRSS33 | FALSE | Downregulated |
| 0,147008136 | PRSS35 | FALSE | Upregulated |
| 1,000703382 | PRSS36 | | Upregulated |
| 0,716699383 | PRSS38 | FALSE | Upregulated |
| | PRSS41 | | |
| -0,230726618 | PRSS42 | FALSE | Downregulated |
| 0,884880238 | PRSS44 | FALSE | Upregulated |
| 1,020585726 | PRSS45 | | Upregulated |
| -0,438018248 | PRSS46 | FALSE | Downregulated |
| 0 | PRSS56 | FALSE | Upregulated |
| 0,030936071 | PRSS57 | FALSE | Upregulated |
| 0,159210253 | PRTFDC1 | FALSE | Upregulated |
| 1,687053826 | PRTN3 | | Upregulated |

| | | | |
|---|---|---|---|
| 0,018560247 | PSCA | FALSE | Upregulated |
| 0,877571162 | PSD2 | FALSE | Upregulated |
| -0,470963789 | PSG10P | FALSE | Downregulated |
| 0,548570405 | PSG11 | FALSE | Upregulated |
| 0,141895826 | PSG2 | FALSE | Upregulated |
| -0,669764584 | PSG3 | FALSE | Downregulated |
| -0,098057783 | PSG4 | FALSE | Downregulated |
| -0,371566181 | PSG5 | FALSE | Downregulated |
| -0,389807512 | PSG6 | FALSE | Downregulated |
| -0,121757252 | PSG7 | FALSE | Downregulated |
| -1,25654723 | PSG8 | | Downregulated |
| 1,589254659 | PSG9 | | Upregulated |
| | PSKH2 | | |
| | PSMA8 | | |
| 0,705562129 | PSMB11 | FALSE | Upregulated |
| | PSORS1C1 | | |
| 0,324440766 | PSORS1C2 | FALSE | Upregulated |
| -0,174122915 | PSORS1C3 | FALSE | Downregulated |
| 0 | PTCHD2 | FALSE | Upregulated |
| -0,158930776 | PTCHD3 | FALSE | Downregulated |
| 0,164298787 | PTCRA | FALSE | Upregulated |
| 0 | PTCSC3 | FALSE | Upregulated |
| | PTENP1-AS | | |
| | PTF1A | | |
| -0,046485519 | PTGER1 | FALSE | Downregulated |
| 1,436156433 | PTGER3 | | Upregulated |
| 0,041679534 | PTGER4P2-CDK2AP2P2 | FALSE | Upregulated |
| 2,180922491 | PTGES | | Upregulated |
| | PTGES3L-AARSD1 | | |
| 1,596420231 | PTGFR | | Upregulated |
| 0,909042345 | PTGS2 | FALSE | Upregulated |
| 0,135246338 | PTH | FALSE | Upregulated |
| | PTH2 | | |
| 0,563946489 | PTH2R | FALSE | Upregulated |
| -0,361251989 | PTHLH | FALSE | Downregulated |
| -0,057393283 | PTPLA | FALSE | Downregulated |
| 0,766207977 | PTPN20B | FALSE | Upregulated |
| 0,301935685 | PTPN5 | FALSE | Upregulated |
| 0 | PTPRN | FALSE | Upregulated |
| 1,511845901 | PTPRQ | | Upregulated |
| -1,009547135 | PTPRVP | | Downregulated |
| -0,029160324 | PTPRZ1 | FALSE | Downregulated |
| -0,294878207 | PTTG1 | FALSE | Downregulated |
| 0,503553748 | PTTG2 | FALSE | Upregulated |
| -0,018739516 | PTX3 | FALSE | Downregulated |
| 0,18970391 | PTX4 | FALSE | Upregulated |
| 1,3128721 | PURG | | Upregulated |
| -0,008191535 | PVALB | FALSE | Downregulated |
| 0 | PVRIG2P | FALSE | Upregulated |
| 0 | PVRL3-AS1 | FALSE | Upregulated |
| 0,251554879 | PVRL4 | FALSE | Upregulated |

| | | | |
|---|---|---|---|
| -0,864189435 | PVT1 | FALSE | Downregulated |
| 0,708128549 | PWRN1 | FALSE | Upregulated |
| -0,13315125 | PWRN3 | FALSE | Downregulated |
| 0,293457367 | PXDNL | FALSE | Upregulated |
| | PYDC1 | | |
| | PYDC2 | | |
| -0,669508571 | PYGO1 | FALSE | Downregulated |
| 0,027563344 | PYY | FALSE | Upregulated |
| 0,105110403 | QPCT | FALSE | Upregulated |
| | QRFPR | | |
| -0,082688828 | R3HDML | FALSE | Downregulated |
| 0,720184439 | RAB19 | FALSE | Upregulated |
| 0,162468561 | RAB25 | FALSE | Upregulated |
| -0,060503 | RAB33A | FALSE | Downregulated |
| 0,148956598 | RAB38 | FALSE | Upregulated |
| 0,824066821 | RAB39A | FALSE | Upregulated |
| -1,336240125 | RAB39B | | Downregulated |
| -1,465418648 | RAB3A | | Downregulated |
| -0,513888473 | RAB3C | FALSE | Downregulated |
| 0,702751189 | RAB40AL | FALSE | Upregulated |
| 0,551580751 | RAB44 | FALSE | Upregulated |
| -0,133515551 | RAB6C | FALSE | Downregulated |
| -0,375854706 | RAB6C-AS1 | FALSE | Downregulated |
| 0,902577999 | RAB9B | FALSE | Upregulated |
| 0 | RAB9BP1 | FALSE | Upregulated |
| 0 | RACGAP1P | FALSE | Upregulated |
| | RAD21L1 | | |
| 0,859272985 | RAD51AP1 | FALSE | Upregulated |
| 0,636478465 | RAD51AP2 | FALSE | Upregulated |
| 0,130645374 | RAD51C | FALSE | Upregulated |
| 0,166047866 | RADIL | FALSE | Upregulated |
| -1,085172876 | RAET1G | | Downregulated |
| 0 | RAET1K | FALSE | Upregulated |
| | RAET1L | | |
| -0,169999122 | RALYL | FALSE | Downregulated |
| -0,735137893 | RAPSN | FALSE | Downregulated |
| -0,295819799 | RASD2 | FALSE | Downregulated |
| -1,436736098 | RASGEF1C | | Downregulated |
| 1,173942457 | RASGRF1 | | Upregulated |
| 0,257961625 | RASSF10 | FALSE | Upregulated |
| 1,206475645 | RASSF8-AS1 | | Upregulated |
| | RAX | | |
| | RBAK-RBAKDN | | |
| 0 | RBAKDN | FALSE | Upregulated |
| 0,080739278 | RBBP8NL | FALSE | Upregulated |
| -0,031859907 | RBFADN | FALSE | Downregulated |
| 0,097981992 | RBFOX1 | FALSE | Upregulated |
| -0,00897377 | RBM11 | FALSE | Downregulated |
| 1,220407328 | RBM20 | | Upregulated |
| 0,223112812 | RBM24 | FALSE | Upregulated |
| -1,268179551 | RBM26-AS1 | | Downregulated |

| | | | |
|---|---|---|---|
| 0,070228824 | RBM44 | FALSE | Upregulated |
| 0 | RBM46 | FALSE | Upregulated |
| -0,115173749 | RBMS3-AS3 | FALSE | Downregulated |
| -0,289111252 | RBMXL2 | FALSE | Downregulated |
| | RBMY1A1 | | |
| | RBMY1A3P | | |
| | RBMY1B | | |
| | RBMY1D | | |
| | RBMY1E | | |
| | RBMY1F | | |
| | RBMY1J | | |
| | RBMY3AP | | |
| 0,451390908 | RBP3 | FALSE | Upregulated |
| -1,340508859 | RBP7 | | Downregulated |
| -0,729863187 | RBPJL | FALSE | Downregulated |
| -0,002529603 | RCAN3AS | FALSE | Downregulated |
| -0,126959959 | RCVRN | FALSE | Downregulated |
| -0,97840749 | RD3 | FALSE | Downregulated |
| 0,359200375 | RD3L | FALSE | Upregulated |
| -0,234153435 | RDH8 | FALSE | Downregulated |
| -0,558254806 | RDM1 | FALSE | Downregulated |
| 0,0533316 | REEP2 | FALSE | Upregulated |
| | REG1A | | |
| | REG1B | | |
| | REG1P | | |
| 0 | REG3A | FALSE | Upregulated |
| | REG3G | | |
| -0,519990958 | REG4 | FALSE | Downregulated |
| 0,192066766 | REM1 | FALSE | Upregulated |
| 0,195576625 | REREP3 | FALSE | Upregulated |
| | RERG-AS1 | | |
| 0,902347316 | RERGL | FALSE | Upregulated |
| | RESP18 | | |
| | RETN | | |
| | RETNLB | | |
| 0,606422624 | REXO1L1P | FALSE | Upregulated |
| 0 | REXO1L2P | FALSE | Upregulated |
| -0,563723499 | RFPL2 | FALSE | Downregulated |
| 1,199752781 | RFPL3S | | Upregulated |
| -0,145139395 | RFPL4A | FALSE | Downregulated |
| -0,62275095 | RFPL4AL1 | FALSE | Downregulated |
| | RFPL4B | | |
| | RFX6 | | |
| -0,028098181 | RFX8 | FALSE | Downregulated |
| 0,213883731 | RGAG1 | FALSE | Upregulated |
| 0,802466732 | RGCC | FALSE | Upregulated |
| 0,43467856 | RGMA | FALSE | Upregulated |
| 0,217929341 | RGMB-AS1 | FALSE | Upregulated |
| 0 | RGPD4-AS1 | FALSE | Upregulated |
| | RGR | | |
| 0 | RGS13 | FALSE | Upregulated |

| | | | |
|---|---|---|---|
| 0,628117479 | RGS21 | FALSE | Upregulated |
| | RGS22 | | |
| 0,661206439 | RGS7 | FALSE | Upregulated |
| 0,597480774 | RGS8 | FALSE | Upregulated |
| -0,012810865 | RGS9BP | FALSE | Downregulated |
| -0,667392827 | RGSL1 | FALSE | Downregulated |
| 1,355855723 | RHAG | | Upregulated |
| 0,540376661 | RHBDL3 | FALSE | Upregulated |
| 1,813992006 | RHO | | Upregulated |
| 0 | RHOV | FALSE | Upregulated |
| 0,649901535 | RHOXF1 | FALSE | Upregulated |
| -0,193899219 | RHPN1-AS1 | FALSE | Downregulated |
| 0,440104161 | RIBC2 | FALSE | Upregulated |
| | RIIAD1 | | |
| -0,345921239 | RIMS1 | FALSE | Downregulated |
| -0,116899723 | RIMS2 | FALSE | Downregulated |
| | RIPPLY2 | | |
| 0,638795407 | RIPPLY3 | FALSE | Upregulated |
| -0,744702133 | RIT2 | FALSE | Downregulated |
| 0,327543322 | RLN1 | FALSE | Upregulated |
| 0,028742589 | RLN2 | FALSE | Upregulated |
| 0,060126583 | RLN3 | FALSE | Upregulated |
| 0,397391356 | RMI2 | FALSE | Upregulated |
| 0 | RMST | FALSE | Upregulated |
| | RNA5-8S5 | | |
| | RNA5S1 | | |
| | RNA5S10 | | |
| | RNA5S11 | | |
| | RNA5S12 | | |
| | RNA5S13 | | |
| | RNA5S14 | | |
| | RNA5S15 | | |
| | RNA5S16 | | |
| | RNA5S17 | | |
| | RNA5S2 | | |
| | RNA5S3 | | |
| | RNA5S4 | | |
| | RNA5S5 | | |
| | RNA5S6 | | |
| | RNA5S7 | | |
| 0 | RNA5S8 | FALSE | Upregulated |
| 0,52922938 | RNA5S9 | FALSE | Upregulated |
| | RNASE11 | | |
| | RNASE12 | | |
| 0,338580204 | RNASE2 | FALSE | Upregulated |
| 0,065560115 | RNASE3 | FALSE | Upregulated |
| | RNASE9 | | |
| 0,876944278 | RNASEH2B-AS1 | FALSE | Upregulated |
| 1,013077175 | RNF126P1 | | Upregulated |
| 0,351484181 | RNF133 | FALSE | Upregulated |
| 1,687955121 | RNF144A-AS1 | | Upregulated |

| | | | |
|---|---|---|---|
| 0,780974886 | RNF148 | FALSE | Upregulated |
| -0,53592853 | RNF151 | FALSE | Downregulated |
| -0,225982982 | RNF17 | FALSE | Downregulated |
| 0 | RNF182 | FALSE | Upregulated |
| 0,514953354 | RNF183 | FALSE | Upregulated |
| -0,506451645 | RNF186 | FALSE | Downregulated |
| 0 | RNF212 | FALSE | Upregulated |
| | RNF216-IT1 | | |
| 0 | RNF223 | FALSE | Upregulated |
| -1,357393998 | RNF5P1 | | Downregulated |
| -0,056406317 | RNFT2 | FALSE | Downregulated |
| 0 | RNU105B | FALSE | Upregulated |
| | RNU105C | | |
| -0,854459452 | RNU11 | FALSE | Downregulated |
| 0,3746934 | RNU2-1 | FALSE | Upregulated |
| -0,377247193 | RNU4ATAC | FALSE | Downregulated |
| 0,608246941 | RNU5A-1 | FALSE | Upregulated |
| 0,597480774 | RNU5B-1 | FALSE | Upregulated |
| | RNU5D-1 | | |
| | RNU5E-1 | | |
| | RNU5F-1 | | |
| | RNU6-1 | | |
| | RNU6-10P | | |
| | RNU6-14P | | |
| | RNU6-15P | | |
| | RNU6-16P | | |
| | RNU6-19P | | |
| -0,126647314 | RNU6-2 | FALSE | Downregulated |
| | RNU6-21P | | |
| | RNU6-23P | | |
| | RNU6-26P | | |
| | RNU6-28P | | |
| | RNU6-30P | | |
| | RNU6-31P | | |
| | RNU6-33P | | |
| | RNU6-34P | | |
| | RNU6-35P | | |
| | RNU6-36P | | |
| | RNU6-39P | | |
| | RNU6-42P | | |
| | RNU6-45P | | |
| | RNU6-46P | | |
| | RNU6-48P | | |
| | RNU6-52P | | |
| | RNU6-53P | | |
| | RNU6-55P | | |
| | RNU6-56P | | |
| | RNU6-57P | | |
| | RNU6-58P | | |
| | RNU6-59P | | |
| | RNU6-63P | | |

| | | | |
|---|---|---|---|
| | RNU6-64P | | |
| | RNU6-66P | | |
| | RNU6-67P | | |
| | RNU6-68P | | |
| | RNU6-69P | | |
| | RNU6-7 | | |
| | RNU6-71P | | |
| | RNU6-72P | | |
| | RNU6-75P | | |
| | RNU6-76P | | |
| | RNU6-78P | | |
| | RNU6-79P | | |
| | RNU6-8 | | |
| | RNU6-81P | | |
| | RNU6-82P | | |
| | RNU6-83P | | |
| -0,003571394 | RNU6-9 | FALSE | Downregulated |
| | RNU7-1 | | |
| | RNU86 | | |
| 0,608246941 | RNVU1-10 | FALSE | Upregulated |
| 0,421417791 | RNVU1-14 | FALSE | Upregulated |
| 0 | RNVU1-15 | FALSE | Upregulated |
| | RNVU1-17 | | |
| 0 | RNVU1-19 | FALSE | Upregulated |
| 2,240102178 | RNVU1-3 | | Upregulated |
| 0 | RNVU1-4 | FALSE | Upregulated |
| 0,60159569 | RNVU1-6 | FALSE | Upregulated |
| -1,080536394 | RNY1 | | Downregulated |
| -0,068404365 | RNY3 | FALSE | Downregulated |
| | RNY4 | | |
| | RNY5 | | |
| -0,192299565 | ROCK1P1 | FALSE | Downregulated |
| | ROPN1 | | |
| 0,750950959 | ROPN1B | FALSE | Upregulated |
| 0,381152569 | ROPN1L | FALSE | Upregulated |
| 0,268895542 | RORB | FALSE | Upregulated |
| 0,781761654 | ROS1 | FALSE | Upregulated |
| -0,160334058 | RP1 | FALSE | Downregulated |
| | RP1-104O17.1 | | |
| 0 | RP1-137K24.1 | FALSE | Upregulated |
| 0 | RP1-140J1.1 | FALSE | Upregulated |
| 0 | RP1-167G20.1 | FALSE | Upregulated |
| -0,217829805 | RP1-172N19.2 | FALSE | Downregulated |
| 0 | RP1-207H1.3 | FALSE | Upregulated |
| | RP1-224A6.3 | | |
| 1,294953797 | RP1-257A15.1 | | Upregulated |
| 0 | RP1-272J12.1 | FALSE | Upregulated |
| 0 | RP1-278O22.1 | FALSE | Upregulated |
| 0 | RP1-290I10.5 | FALSE | Upregulated |
| 0,088661369 | RP1-40E16.9 | FALSE | Upregulated |
| | RP1-63G5.5 | | |

| | | | |
|---|---|---|---|
| -0,187024721 | RP1-90L6.2 | FALSE | Downregulated |
| -0,270379443 | RP1-93I3.1 | FALSE | Downregulated |
| 1,612036121 | RP11-1008C21.1 | | Upregulated |
| | RP11-100N20.1 | | |
| -0,257640881 | RP11-1018N14.1 | FALSE | Downregulated |
| | RP11-104D21.3 | | |
| 0 | RP11-1055B8.3 | FALSE | Upregulated |
| 0,381851452 | RP11-1057B8.2 | FALSE | Upregulated |
| | RP11-1079K10.5 | | |
| 0 | RP11-107I14.4 | FALSE | Upregulated |
| 0,019985931 | RP11-107K17.1 | FALSE | Upregulated |
| -0,307713289 | RP11-1080G15.1 | FALSE | Downregulated |
| | RP11-1084I9.1 | | |
| 1,130515448 | RP11-1094H24.4 | | Upregulated |
| | RP11-109J4.1 | | |
| 0,705562129 | RP11-10A14.5 | FALSE | Upregulated |
| | RP11-10L7.1 | | |
| | RP11-10N16.2 | | |
| 0,819407007 | RP11-110H10.2 | FALSE | Upregulated |
| 0,374867897 | RP11-111I12.1 | FALSE | Upregulated |
| -0,00238016 | RP11-1124B17.1 | FALSE | Downregulated |
| -0,532509881 | RP11-112L6.4 | FALSE | Downregulated |
| 0,942371276 | RP11-1134I14.8 | FALSE | Upregulated |
| 0 | RP11-1191J2.2 | FALSE | Upregulated |
| -0,938962811 | RP11-119F19.2 | FALSE | Downregulated |
| 0 | RP11-1223D19.1 | FALSE | Upregulated |
| 0 | RP11-122M14.1 | FALSE | Upregulated |
| | RP11-124K4.1 | | |
| | RP11-125P18.1 | | |
| 0 | RP11-127O4.3 | FALSE | Upregulated |
| | RP11-129J12.2 | | |
| -0,870788631 | RP11-132A1.4 | FALSE | Downregulated |
| | RP11-1338A24.1 | | |
| | RP11-134O21.1 | | |
| 0 | RP11-134P9.3 | FALSE | Upregulated |
| 0,668194022 | RP11-135J2.4 | FALSE | Upregulated |
| 1,156720279 | RP11-138B4.1 | | Upregulated |
| | RP11-13A1.1 | | |
| | RP11-13E5.2 | | |
| | RP11-13P5.2 | | |
| -1,0068E-05 | RP11-140I24.1 | FALSE | Downregulated |
| | RP11-142G7.2 | | |
| | RP11-143A12.3 | | |
| -0,018476542 | RP11-143M1.2 | FALSE | Downregulated |
| | RP11-143M1.3 | | |
| | RP11-146N18.1 | | |
| 0,661206439 | RP11-148L24.1 | FALSE | Upregulated |
| | RP11-14J7.6 | | |
| 0,535916065 | RP11-14N7.2 | FALSE | Upregulated |
| | RP11-150O12.6 | | |
| | RP11-152L7.2 | | |

| | | | |
|---|---|---|---|
| 0,073122974 | RP11-153K16.1 | FALSE | Upregulated |
| 0 | RP11-154D17.1 | FALSE | Upregulated |
| -0,513309175 | RP11-154D6.1 | FALSE | Downregulated |
| | RP11-157D23.1 | | |
| | RP11-157E21.1 | | |
| | RP11-158A8.1 | | |
| | RP11-158D2.2 | | |
| 1,227474507 | RP11-15I11.3 | | Upregulated |
| | RP11-15K3.1 | | |
| -1,223775246 | RP11-160O5.1 | | Downregulated |
| 0,711366696 | RP11-161H23.9 | FALSE | Upregulated |
| -0,551354466 | RP11-163N6.2 | FALSE | Downregulated |
| 0,289500484 | RP11-165H23.1 | FALSE | Upregulated |
| 0,843348191 | RP11-165P7.1 | FALSE | Upregulated |
| | RP11-166A12.1 | | |
| 0,646948212 | RP11-166O4.5 | FALSE | Upregulated |
| 0,423984354 | RP11-167H9.6 | FALSE | Upregulated |
| 0,041547119 | RP11-168E14.1 | FALSE | Upregulated |
| 0,669756935 | RP11-168E17.1 | FALSE | Upregulated |
| 0,269072594 | RP11-168O10.6 | FALSE | Upregulated |
| 0 | RP11-170N11.1 | FALSE | Upregulated |
| | RP11-171A24.2 | | |
| | RP11-174G17.2 | | |
| 0,608246941 | RP11-175K6.1 | FALSE | Upregulated |
| 0 | RP11-178L8.5 | FALSE | Upregulated |
| | RP11-179A16.1 | | |
| | RP11-181F12.1 | | |
| -0,81525795 | RP11-184I16.4 | FALSE | Downregulated |
| -0,627281157 | RP11-188C12.3 | FALSE | Downregulated |
| | RP11-189B4.6 | | |
| | RP11-18F14.1 | | |
| 0,641492265 | RP11-18H21.1 | FALSE | Upregulated |
| 0 | RP11-191L9.4 | FALSE | Upregulated |
| | RP11-191N8.2 | | |
| 0 | RP11-193H5.1 | FALSE | Upregulated |
| | RP11-198M11.2 | | |
| 0,213991357 | RP11-1C1.4 | FALSE | Upregulated |
| -0,222183528 | RP11-208N14.4 | FALSE | Downregulated |
| 0,130384129 | RP11-214C8.2 | FALSE | Upregulated |
| 0 | RP11-214L13.1 | FALSE | Upregulated |
| -0,014983039 | RP11-214L19.1 | FALSE | Downregulated |
| | RP11-215E13.1 | | |
| -0,154496951 | RP11-21L23.3 | FALSE | Downregulated |
| -0,518836106 | RP11-224O19.2 | FALSE | Downregulated |
| 1,407710084 | RP11-227H15.4 | | Upregulated |
| 0 | RP11-22H5.2 | FALSE | Upregulated |
| -0,755457201 | RP11-230B22.1 | FALSE | Downregulated |
| | RP11-230G5.2 | | |
| 1,061056978 | RP11-236J17.5 | | Upregulated |
| | RP11-236P13.1 | | |
| 0,041547119 | RP11-23B15.1 | FALSE | Upregulated |

| | | | |
|---|---|---|---|
| 0 | RP11-243E13.1 | FALSE | Upregulated |
| 0,043406205 | RP11-245M24.1 | FALSE | Upregulated |
| -0,203392405 | RP11-24I21.1 | FALSE | Downregulated |
| 0 | RP11-24P4.1 | FALSE | Upregulated |
| | RP11-251P6.1 | | |
| 0,597480774 | RP11-252C15.1 | FALSE | Upregulated |
| | RP11-256L11.1 | | |
| | RP11-257I8.2 | | |
| | RP11-262K1.1 | | |
| | RP11-265P11.2 | | |
| | RP11-267C16.1 | | |
| 2,815724411 | RP11-26J3.1 | | Upregulated |
| | RP11-271F18.4 | | |
| | RP11-274B18.2 | | |
| 1,53305074 | RP11-276H19.1 | | Upregulated |
| | RP11-27N21.3 | | |
| 0 | RP11-280K24.4 | FALSE | Upregulated |
| | RP11-280O1.2 | | |
| | RP11-286B14.1 | | |
| | RP11-286E11.1 | | |
| -0,259968948 | RP11-290F5.2 | FALSE | Downregulated |
| 0 | RP11-293M10.5 | FALSE | Upregulated |
| -0,070442341 | RP11-294O2.2 | FALSE | Downregulated |
| | RP11-298O21.5 | | |
| 0 | RP11-299H22.1 | FALSE | Upregulated |
| | RP11-299L17.3 | | |
| | RP11-2A4.3 | | |
| -0,485119312 | RP11-305E17.6 | FALSE | Downregulated |
| | RP11-305P22.9 | | |
| 0,041547119 | RP11-307C19.3 | FALSE | Upregulated |
| -0,534933169 | RP11-308N19.4 | FALSE | Downregulated |
| -0,115927293 | RP11-310P5.1 | FALSE | Downregulated |
| | RP11-313C15.1 | | |
| 0,239498204 | RP11-314C16.1 | FALSE | Upregulated |
| 0,672468563 | RP11-317J10.2 | FALSE | Upregulated |
| 1,361976425 | RP11-317M11.1 | | Upregulated |
| | RP11-318I4.1 | | |
| -0,519430754 | RP11-319E12.1 | FALSE | Downregulated |
| | RP11-319E16.1 | | |
| 1,145643056 | RP11-319G6.1 | | Upregulated |
| 0,063667494 | RP11-320G24.1 | FALSE | Upregulated |
| | RP11-324L3.1 | | |
| 0,249819983 | RP11-326F20.5 | FALSE | Upregulated |
| | RP11-327I22.6 | | |
| 0,998529735 | RP11-329J18.2 | FALSE | Upregulated |
| | RP11-32F11.2 | | |
| | RP11-330A16.1 | | |
| -0,048002269 | RP11-333A23.4 | FALSE | Downregulated |
| | RP11-33G16.1 | | |
| 0 | RP11-341A22.2 | FALSE | Upregulated |
| | RP11-343J18.1 | | |

| | | | |
|---|---|---|---|
| 0,929516666 | RP11-344B5.2 | FALSE | Upregulated |
| 1,027949613 | RP11-344E13.3 | | Upregulated |
| | RP11-344G13.1 | | |
| 0,839217369 | RP11-345L23.1 | FALSE | Upregulated |
| | RP11-346D6.6 | | |
| 0,050688952 | RP11-347D21.3 | FALSE | Upregulated |
| -1,99758484 | RP11-34F13.2 | | Downregulated |
| 2,974081751 | RP11-353N14.5 | | Upregulated |
| 0,07687759 | RP11-353N4.5 | FALSE | Upregulated |
| 0,572380361 | RP11-354E11.2 | FALSE | Upregulated |
| | RP11-360P21.2 | | |
| 1,158343344 | RP11-362A1.1 | | Upregulated |
| | RP11-363D24.1 | | |
| | RP11-363G15.2 | | |
| 0,038984786 | RP11-367N14.2 | FALSE | Upregulated |
| | RP11-368I23.2 | | |
| -0,002529603 | RP11-371M22.1 | FALSE | Downregulated |
| -0,040862998 | RP11-378A12.1 | FALSE | Downregulated |
| 0 | RP11-379F4.4 | FALSE | Upregulated |
| 0 | RP11-37B2.1 | FALSE | Upregulated |
| 0,669756935 | RP11-37N22.1 | FALSE | Upregulated |
| | RP11-384J4.1 | | |
| | RP11-386B13.3 | | |
| 0 | RP11-386I8.6 | FALSE | Upregulated |
| | RP11-389K14.3 | | |
| 1,529525193 | RP11-394O9.1 | | Upregulated |
| | RP11-395L14.3 | | |
| 0,811287554 | RP11-397C12.1 | FALSE | Upregulated |
| -0,13125304 | RP11-397C18.2 | FALSE | Downregulated |
| 0,675850581 | RP11-399K21.14 | FALSE | Upregulated |
| | RP11-3B12.2 | | |
| 0 | RP11-3B12.3 | FALSE | Upregulated |
| 0,055530073 | RP11-3P17.4 | FALSE | Upregulated |
| 1,091400271 | RP11-402G3.3 | | Upregulated |
| | RP11-404O13.1 | | |
| -0,001452899 | RP11-406A20.1 | FALSE | Downregulated |
| 1,06119331 | RP11-406A20.4 | | Upregulated |
| | RP11-406A9.2 | | |
| -0,615117705 | RP11-406H4.1 | FALSE | Downregulated |
| 0 | RP11-406O16.1 | FALSE | Upregulated |
| 0,073122974 | RP11-407A16.3 | FALSE | Upregulated |
| | RP11-408H20.2 | | |
| 0 | RP11-40F8.2FALSE | | Upregulated |
| | RP11-413P11.1 | | |
| | RP11-415C15.2 | | |
| 0,463653703 | RP11-415D17.4 | FALSE | Upregulated |
| | RP11-416O18.1 | | |
| 0 | RP11-417E7.2 | FALSE | Upregulated |
| 0,786664948 | RP11-420A23.1 | FALSE | Upregulated |
| -0,318280761 | RP11-423C15.3 | FALSE | Downregulated |
| -0,936216878 | RP11-423H2.3 | FALSE | Downregulated |

| | | | |
|---|---|---|---|
| 0,039005732 | RP11-425I13.3 | FALSE | Upregulated |
| 0,675622584 | RP11-429H9.4 | FALSE | Upregulated |
| 0,166079147 | RP11-430C1.1 | FALSE | Upregulated |
| 0,073122974 | RP11-430H10.1 | FALSE | Upregulated |
| 0 | RP11-433J20.1 | FALSE | Upregulated |
| | RP11-435M3.2 | | |
| 0,274749764 | RP11-436K8.1 | FALSE | Upregulated |
| | RP11-438N16.1 | | |
| -0,730578084 | RP11-439L18.3 | FALSE | Downregulated |
| 0,332109623 | RP11-439L8.3 | FALSE | Upregulated |
| 0,510861848 | RP11-439L8.4 | FALSE | Upregulated |
| -0,002529603 | RP11-440G9.1 | FALSE | Downregulated |
| 1,678610079 | RP11-445H22.3 | | Upregulated |
| -0,043898724 | RP11-446H18.5 | FALSE | Downregulated |
| 0,073122974 | RP11-449D8.1 | FALSE | Upregulated |
| | RP11-451G4.2 | | |
| -0,275851871 | RP11-452H21.4 | FALSE | Downregulated |
| | RP11-452N4.1 | | |
| 1,747913798 | RP11-453E2.2 | | Upregulated |
| 0 | RP11-454C18.2 | FALSE | Upregulated |
| | RP11-454P21.1 | | |
| 0 | RP11-456O19.4 | FALSE | Upregulated |
| 1,080030493 | RP11-45L9.1 | | Upregulated |
| -0,405480352 | RP11-460N16.1 | FALSE | Downregulated |
| 0 | RP11-461F11.1 | FALSE | Upregulated |
| 0 | RP11-461F11.3 | FALSE | Upregulated |
| -0,397247172 | RP11-461O7.1 | FALSE | Downregulated |
| 0,589455582 | RP11-462G2.1 | FALSE | Upregulated |
| 0,073122974 | RP11-462G22.1 | FALSE | Upregulated |
| | RP11-463J7.2 | | |
| 0,520772439 | RP11-465B22.5 | FALSE | Upregulated |
| | RP11-46C24.3 | | |
| -0,543237746 | RP11-474D1.3 | FALSE | Downregulated |
| | RP11-478H13.3 | | |
| | RP11-480C22.1 | | |
| | RP11-482D24.2 | | |
| -0,002347351 | RP11-483E7.1 | FALSE | Downregulated |
| 0,302596001 | RP11-486A14.2 | FALSE | Upregulated |
| | RP11-488I20.3 | | |
| | RP11-489D6.2 | | |
| 0 | RP11-48F14.1 | FALSE | Upregulated |
| 1,215639147 | RP11-490D19.6 | | Upregulated |
| | RP11-493K23.4 | | |
| 0 | RP11-494M8.4 | FALSE | Upregulated |
| | RP11-497K21.1 | | |
| 0,514413109 | RP11-500B12.1 | FALSE | Upregulated |
| | RP11-501G7.1 | | |
| 1,25678501 | RP11-503C24.4 | | Upregulated |
| | RP11-508P1.2 | | |
| -0,585326489 | RP11-509J21.1 | FALSE | Downregulated |
| -0,818963303 | RP11-510D19.1 | FALSE | Downregulated |

| | | | |
|---|---|---|---|
| 0,208992352 | RP11-513O17.2 | FALSE | Upregulated |
| -0,772298287 | RP11-513O17.2-2 | FALSE | Downregulated |
| | RP11-513O17.3 | | |
| 0,266782327 | RP11-514D23.1 | FALSE | Upregulated |
| 0,688720135 | RP11-519M16.1 | FALSE | Upregulated |
| 1,12691923 | RP11-523L1.2 | | Upregulated |
| 0,38170733 | RP11-523L20.2 | FALSE | Upregulated |
| 0 | RP11-526A4.1 | FALSE | Upregulated |
| 0,751271991 | RP11-526P6.1 | FALSE | Upregulated |
| | RP11-527H14.3 | | |
| 0,336890839 | RP11-532N4.2 | FALSE | Upregulated |
| | RP11-536C10.7 | | |
| | RP11-536I6.1 | | |
| 0,110332944 | RP11-539L10.2 | FALSE | Upregulated |
| 0 | RP11-53O19.1 | FALSE | Upregulated |
| 0,001493849 | RP11-540D4.3 | FALSE | Upregulated |
| 0 | RP11-540K16.1 | FALSE | Upregulated |
| | RP11-542B15.1 | | |
| | RP11-542G1.3 | | |
| -0,0296721 | RP11-544M22.1 | FALSE | Downregulated |
| 0,499503618 | RP11-547D23.1 | FALSE | Upregulated |
| | RP11-547I7.2 | | |
| | RP11-550H2.1 | | |
| 0,159036631 | RP11-551M15.1 | FALSE | Upregulated |
| | RP11-553E24.2 | | |
| -0,266556019 | RP11-554A11.6 | FALSE | Downregulated |
| | RP11-554D15.1 | | |
| -0,097621035 | RP11-554F20.1 | FALSE | Downregulated |
| | RP11-555F9.2 | | |
| -0,097622848 | RP11-557H15.4 | FALSE | Downregulated |
| | RP11-561E1.1 | | |
| | RP11-565A3.2 | | |
| -0,023568955 | RP11-567L7.6 | FALSE | Downregulated |
| | RP11-56I23.1 | | |
| 0,516275516 | RP11-572C15.5 | FALSE | Upregulated |
| 0 | RP11-575F12.1 | FALSE | Upregulated |
| 0 | RP11-576D8.4 | FALSE | Upregulated |
| | RP11-576E20.1 | | |
| | RP11-576I22.2 | | |
| -0,045091281 | RP11-579D7.4 | FALSE | Downregulated |
| 0 | RP11-579E24.1 | FALSE | Upregulated |
| 0,622197799 | RP11-584P21.2 | FALSE | Upregulated |
| | RP11-586D19.1 | | |
| 0 | RP11-588G21.2 | FALSE | Upregulated |
| | RP11-588P8.1 | | |
| -0,031788407 | RP11-58E21.3 | FALSE | Downregulated |
| -0,230457162 | RP11-58E21.4 | FALSE | Downregulated |
| 0,427901641 | RP11-593F23.1 | FALSE | Upregulated |
| 0 | RP11-59J16.1 | FALSE | Upregulated |
| 0 | RP11-5P22.3 | FALSE | Upregulated |
| | RP11-600K15.1 | | |

| | | | |
|---|---|---|---|
| 0 | RP11-609L23.1 | FALSE | Upregulated |
| -0,127146923 | RP11-60I3.5 | FALSE | Downregulated |
| 0,98601015 | RP11-614F17.2 | FALSE | Upregulated |
| 0,130051371 | RP11-61J19.3 | FALSE | Upregulated |
| -0,297659542 | RP11-620J15.3 | FALSE | Downregulated |
| 0,250036688 | RP11-624C23.1 | FALSE | Upregulated |
| | RP11-626P14.1 | | |
| 1,303117692 | RP11-629G13.1 | | Upregulated |
| 0 | RP11-632K5.3 | FALSE | Upregulated |
| 0 | RP11-638F5.1 | FALSE | Upregulated |
| -0,241538539 | RP11-63B19.1 | FALSE | Downregulated |
| -0,119072375 | RP11-63P12.6 | FALSE | Downregulated |
| | RP11-646J21.5 | | |
| 0 | RP11-64I17.1 | FALSE | Upregulated |
| | RP11-653G8.2 | | |
| | RP11-65D17.1 | | |
| 0,786536318 | RP11-65J3.1 | FALSE | Upregulated |
| -0,087642203 | RP11-65M17.1 | FALSE | Downregulated |
| | RP11-65M17.3 | | |
| | RP11-664H17.1 | | |
| | RP11-665C14.2 | | |
| 0 | RP11-666I19.2 | FALSE | Upregulated |
| | RP11-669N7.2 | | |
| -0,391066687 | RP11-669N7.3 | FALSE | Downregulated |
| 0,173942518 | RP11-66A2.2 | FALSE | Upregulated |
| 0 | RP11-66B24.1 | FALSE | Upregulated |
| | RP11-673E1.1 | | |
| 0 | RP11-67M1.1 | FALSE | Upregulated |
| 2,399942585 | RP11-682N22.1 | | Upregulated |
| -0,055970583 | RP11-686G23.2 | FALSE | Downregulated |
| | RP11-690C23.2 | | |
| 0,011620842 | RP11-699L21.1 | FALSE | Upregulated |
| | RP11-69H14.6 | | |
| | RP11-69H14.6-2 | | |
| | RP11-700N1.1 | | |
| 0 | RP11-702F3.3 | FALSE | Upregulated |
| | RP11-707A18.1 | | |
| | RP11-707P20.1 | | |
| 0 | RP11-70F11.11 | FALSE | Upregulated |
| | RP11-70F11.8 | | |
| 0,338401557 | RP11-710F7.3 | FALSE | Upregulated |
| | RP11-712C7.1 | | |
| 1,684746562 | RP11-713P17.3 | | Upregulated |
| 0 | RP11-715G15.1 | FALSE | Upregulated |
| 0 | RP11-719J20.1 | FALSE | Upregulated |
| | RP11-71E19.5 | | |
| 0,524531536 | RP11-71G7.1 | FALSE | Upregulated |
| 1,068348782 | RP11-723O4.2 | | Upregulated |
| 0 | RP11-72L22.1 | FALSE | Upregulated |
| | RP11-730N24.1 | | |
| 0,76164688 | RP11-744J10.3 | FALSE | Upregulated |

| | | | |
|---|---|---|---|
| | RP11-745C15.2 | | |
| -0,282995147 | RP11-752D24.3 | FALSE | Downregulated |
| 1,804041407 | RP11-752G15.3 | | Upregulated |
| 1,172643223 | RP11-752G15.6 | | Upregulated |
| 0,033732737 | RP11-756H20.1 | FALSE | Upregulated |
| | RP11-756K15.2 | | |
| 0,310486193 | RP11-75C9.1 | FALSE | Upregulated |
| | RP11-75F3.1 | | |
| | RP11-767I20.1 | | |
| | RP11-76E12.1 | | |
| 0,948760414 | RP11-76N22.2 | FALSE | Upregulated |
| 0,544688284 | RP11-770E5.1 | FALSE | Upregulated |
| -0,188602893 | RP11-770J1.3 | FALSE | Downregulated |
| 0,819904264 | RP11-778O17.4 | FALSE | Upregulated |
| 0,664582726 | RP11-779O18.1 | FALSE | Upregulated |
| -0,401458946 | RP11-789C1.1 | FALSE | Downregulated |
| | RP11-791G22.1 | | |
| | RP11-794P6.1 | | |
| | RP11-794P6.3 | | |
| | RP11-795J1.1 | | |
| 0 | RP11-799P8.1 | FALSE | Upregulated |
| -1,393892266 | RP11-79E3.3 | | Downregulated |
| | RP11-7O14.1 | | |
| 0,009620192 | RP11-806H10.4 | FALSE | Upregulated |
| | RP11-807H7.1 | | |
| | RP11-80F22.15 | | |
| 1,797303237 | RP11-80G7.1 | | Upregulated |
| | RP11-80K21.3 | | |
| 0,557949305 | RP11-814P5.1 | FALSE | Upregulated |
| 0 | RP11-81H14.2 | FALSE | Upregulated |
| | RP11-81H3.2 | | |
| -1,09043449 | RP11-82L2.1 | | Downregulated |
| 0 | RP11-838N2.4 | FALSE | Upregulated |
| 0,072156265 | RP11-83C7.2 | FALSE | Upregulated |
| -0,754255927 | RP11-843A23.1 | FALSE | Downregulated |
| 0,48572693 | RP11-844P9.2 | FALSE | Upregulated |
| | RP11-84A1.3 | | |
| | RP11-84O12.4 | | |
| 0,048656215 | RP11-855O10.2 | FALSE | Upregulated |
| 0,236366227 | RP11-857B24.5 | FALSE | Upregulated |
| | RP11-85G21.2 | | |
| 0,066858979 | RP11-861A13.4 | FALSE | Upregulated |
| 0,050688952 | RP11-87M18.2 | FALSE | Upregulated |
| | RP11-87N24.2 | | |
| -0,127807949 | RP11-885B4.2 | FALSE | Downregulated |
| | RP11-88I21.2 | | |
| | RP11-893F2.6 | | |
| 1,136655517 | RP11-909B2.1 | | Upregulated |
| | RP11-909M7.3 | | |
| | RP11-92G12.3 | | |
| 0 | RP11-93I21.3 | FALSE | Upregulated |

| | | | |
|---|---|---|---|
| | RP11-945C19.1 | | |
| | RP11-946L20.2 | | |
| | RP11-946L20.4 | | |
| 0 | RP11-94H18.1 | FALSE | Upregulated |
| | RP11-955H22.3 | | |
| | RP11-95H3.1 | | |
| 0,978322136 | RP11-95M5.1 | FALSE | Upregulated |
| -0,40250155 | RP11-95P2.1 | FALSE | Downregulated |
| 0 | RP11-960L18.1 | FALSE | Upregulated |
| -0,170423075 | RP11-96P7.1 | FALSE | Downregulated |
| 0 | RP11-973H7.4 | FALSE | Upregulated |
| | RP11-983C2.2 | | |
| 0,669328143 | RP11-98L5.2 | FALSE | Upregulated |
| | RP11-9G1.3 | | |
| -0,359730898 | RP13-210D15.4 | FALSE | Downregulated |
| | RP13-507P19.2 | | |
| 0 | RP13-60M5.2 | FALSE | Upregulated |
| | RP13-614K11.1 | | |
| | RP13-653N12.1 | | |
| | RP13-653N12.2 | | |
| | RP13-895J2.7 | | |
| -0,788139464 | RP1L1 | FALSE | Downregulated |
| -0,080660339 | RP3-322G13.5 | FALSE | Downregulated |
| 0,31899891 | RP3-323P13.2 | FALSE | Upregulated |
| | RP3-326I13.1 | | |
| | RP3-332B22.1 | | |
| 0 | RP3-399L15.3 | FALSE | Upregulated |
| -0,294676268 | RP3-400B16.1 | FALSE | Downregulated |
| | RP3-416J7.4 | | |
| | RP3-417L20.4 | | |
| | RP3-446N13.1 | | |
| 0,675146616 | RP3-473B4.3 | FALSE | Upregulated |
| | RP3-513G18.2 | | |
| 0,863547858 | RP3-522P13.3 | FALSE | Upregulated |
| 0 | RP4-533D7.5 | FALSE | Upregulated |
| 0,194081822 | RP4-562J12.2 | FALSE | Upregulated |
| | RP4-565E6.1 | | |
| | RP4-598G3.1 | | |
| 0,032318042 | RP4-607I7.1 | FALSE | Upregulated |
| -0,318191881 | RP4-613A2.1 | FALSE | Downregulated |
| 0,620855267 | RP4-640H8.2 | FALSE | Upregulated |
| 1,722414967 | RP4-655J12.4 | | Upregulated |
| | RP4-684O24.5 | | |
| 0,034128514 | RP4-694A7.4 | FALSE | Upregulated |
| -0,231890443 | RP4-697P8.3 | FALSE | Downregulated |
| 0,368710838 | RP4-705O1.1 | FALSE | Upregulated |
| 0 | RP4-723E3.1 | FALSE | Upregulated |
| | RP4-738P15.1 | | |
| | RP4-753D10.5 | | |
| 0 | RP4-753M9.1 | FALSE | Upregulated |
| 0,616734111 | RP4-756H11.3 | FALSE | Upregulated |

| | | | |
|---|---|---|---|
| 0 | RP4-760C5.5 | FALSE | Upregulated |
| | RP4-779E11.3 | | |
| 0,509439246 | RP4-781K5.4 | FALSE | Upregulated |
| -0,39824025 | RP4-782L23.1 | FALSE | Downregulated |
| 0,459122372 | RP4-794H19.4 | FALSE | Upregulated |
| 0 | RP5-1004I9.1 | FALSE | Upregulated |
| | RP5-1007G16.1 | | |
| 0,114187276 | RP5-1010E17.2 | FALSE | Upregulated |
| -1,180087561 | RP5-1021I20.1 | | Downregulated |
| | RP5-1022P6.4 | | |
| 0,73270023 | RP5-1024G6.5 | FALSE | Upregulated |
| 0,554642426 | RP5-1043L13.1 | FALSE | Upregulated |
| 1,08149634 | RP5-1043L13.1-2 | | Upregulated |
| 0,092768757 | RP5-1050D4.5 | FALSE | Upregulated |
| 0,019985931 | RP5-1069C8.2 | FALSE | Upregulated |
| | RP5-1070A16.1 | | |
| | RP5-1073O3.2 | | |
| -0,995893687 | RP5-1073O3.7 | FALSE | Downregulated |
| | RP5-1086L22.1 | | |
| 0,652360834 | RP5-1097F14.3 | FALSE | Upregulated |
| -0,528654316 | RP5-1103G7.4 | FALSE | Downregulated |
| -1,026613681 | RP5-1109J22.2 | | Downregulated |
| -0,306803072 | RP5-1112F19.2 | FALSE | Downregulated |
| 0,03087499 | RP5-1119D9.4 | FALSE | Upregulated |
| | RP5-1120P11.1 | | |
| 1,182084045 | RP5-1125A11.1 | | Upregulated |
| 0,399954712 | RP5-1139I1.1 | FALSE | Upregulated |
| -1,434696038 | RP5-1159O4.2 | | Downregulated |
| | RP5-1166F10.1 | | |
| | RP5-828K20.2 | | |
| -0,167117093 | RP5-833A20.1 | FALSE | Downregulated |
| 0 | RP5-857K21.5 | FALSE | Upregulated |
| 0 | RP5-858B6.3 | FALSE | Upregulated |
| | RP5-887A10.1 | | |
| | RP5-896L10.1 | | |
| | RP5-898J17.1 | | |
| 0,270435054 | RP5-899E9.1 | FALSE | Upregulated |
| | RP5-921G16.2 | | |
| | RP5-933E2.1 | | |
| | RP5-944M2.3 | | |
| | RP5-952N6.1 | | |
| -0,018517259 | RP5-968D22.3 | FALSE | Downregulated |
| | RP5-988G17.1 | | |
| 0,170251501 | RPA4 | FALSE | Upregulated |
| 0 | RPE65 | FALSE | Upregulated |
| 0,575236943 | RPEL1 | FALSE | Upregulated |
| 1,219531902 | RPGRIP1 | | Upregulated |
| 0 | RPH3A | FALSE | Upregulated |
| -0,00897377 | RPL10L | FALSE | Downregulated |
| -0,325968265 | RPL13AP20 | FALSE | Downregulated |
| -0,597335152 | RPL13AP3 | FALSE | Downregulated |

| | | | |
|---|---|---|---|
| 0,368638387 | RPL13P5 | FALSE | Upregulated |
| -0,553447076 | RPL23AP32 | FALSE | Downregulated |
| -0,386001634 | RPL23P8 | FALSE | Downregulated |
| 0 | RPL29P2 | FALSE | Upregulated |
| -0,20742055 | RPL34-AS1 | FALSE | Downregulated |
| 0,145680589 | RPL39L | FALSE | Upregulated |
| 1,372521627 | RPL3L | | Upregulated |
| -0,154828482 | RPLP0P2 | FALSE | Downregulated |
| 0,428507732 | RPP40 | FALSE | Upregulated |
| -0,118085778 | RPRM | FALSE | Downregulated |
| -1,14953799 | RPRML | | Downregulated |
| 0 | RPS10-NUDT3 | FALSE | Upregulated |
| 0,587084549 | RPS14P3 | FALSE | Upregulated |
| 0,673419706 | RPS26P11 | FALSE | Upregulated |
| | RPS4Y2 | | |
| -0,118725817 | RPS6KA2-IT1 | FALSE | Downregulated |
| | RPS7P5 | | |
| | RPSAP52 | | |
| -1,411496769 | RPSAP9 | | Downregulated |
| 0 | RPTN | FALSE | Upregulated |
| 0 | RRH | FALSE | Upregulated |
| 0 | RSPH1 | FALSE | Upregulated |
| 0,24314298 | RSPH6A | FALSE | Upregulated |
| -0,230967119 | RSPO1 | FALSE | Downregulated |
| 0,324329077 | RSPO2 | FALSE | Upregulated |
| -0,331534415 | RSPO4 | FALSE | Downregulated |
| -0,868141459 | RSU1P2 | FALSE | Downregulated |
| 0,040572131 | RTBDN | FALSE | Upregulated |
| 0 | RTDR1 | FALSE | Upregulated |
| -1,001202137 | RTEL1-TNFRSF6B | | Downregulated |
| 0,87450665 | RTKN2 | FALSE | Upregulated |
| 0 | RTP1 | FALSE | Upregulated |
| | RTP2 | | |
| 1,650712146 | RUFY4 | | Upregulated |
| -0,814376776 | RXFP1 | FALSE | Downregulated |
| -0,142699562 | RXFP2 | FALSE | Downregulated |
| -0,333463707 | RXFP3 | FALSE | Downregulated |
| -0,585790883 | RXRG | FALSE | Downregulated |
| 0 | S100A12 | FALSE | Upregulated |
| 0 | S100A2 | FALSE | Upregulated |
| 0 | S100A3 | FALSE | Upregulated |
| 0,786614516 | S100A5 | FALSE | Upregulated |
| | S100A7 | | |
| | S100A7A | | |
| | S100A7L2 | | |
| -0,170900965 | S100A8 | FALSE | Downregulated |
| -0,106916098 | S100B | FALSE | Downregulated |
| 0,393289594 | S100G | FALSE | Upregulated |
| 0 | S100Z | FALSE | Upregulated |
| -0,196992415 | S1PR5 | FALSE | Downregulated |
| -0,293948481 | SAA2-SAA4 | FALSE | Downregulated |

| | | | |
|---|---|---|---|
| | SAA3P | | |
| -0,344077445 | SACS-AS1 | FALSE | Downregulated |
| 0 | SAG | FALSE | Upregulated |
| | SAGE1 | | |
| | SALL3 | | |
| 0,497033101 | SAMD12-AS1 | FALSE | Upregulated |
| 0,825566216 | SAMD13 | FALSE | Upregulated |
| 0 | SAMD7 | FALSE | Upregulated |
| | SAMSN1-AS1 | | |
| 0 | SATB2-AS1 | FALSE | Upregulated |
| 1,069357125 | SBF1P1 | | Upregulated |
| -0,01682454 | SBK2 | FALSE | Downregulated |
| 0 | SBK3 | FALSE | Upregulated |
| 0,418185243 | SBSN | FALSE | Upregulated |
| 0,346019296 | SCARNA1 | FALSE | Upregulated |
| -0,002529603 | SCARNA11 | FALSE | Downregulated |
| | SCARNA14 | | |
| | SCARNA15 | | |
| 0,712185461 | SCARNA16 | FALSE | Upregulated |
| -0,092244387 | SCARNA18 | FALSE | Downregulated |
| 0 | SCARNA20 | FALSE | Upregulated |
| 0,100754873 | SCARNA21 | FALSE | Upregulated |
| 0,049231922 | SCARNA22 | FALSE | Upregulated |
| | SCARNA23 | | |
| | SCARNA27 | | |
| -0,559728916 | SCARNA3 | FALSE | Downregulated |
| 0,630225139 | SCARNA4 | FALSE | Upregulated |
| -0,001452899 | SCARNA8 | FALSE | Downregulated |
| 0,126591845 | SCARNA9L | FALSE | Upregulated |
| 1,056972209 | SCEL | | Upregulated |
| 0 | SCG2 | FALSE | Upregulated |
| | SCGB1A1 | | |
| 1,203978557 | SCGB1B2P | | Upregulated |
| 0,669025015 | SCGB1C1 | FALSE | Upregulated |
| | SCGB1D1 | | |
| 0,597480774 | SCGB1D2 | FALSE | Upregulated |
| | SCGB1D4 | | |
| | SCGB2A1 | | |
| | SCGB2A2 | | |
| 0 | SCGB2B2 | FALSE | Upregulated |
| 0,279742761 | SCGB3A1 | FALSE | Upregulated |
| -0,00897377 | SCGB3A2 | FALSE | Downregulated |
| 2,822912982 | SCHIP1 | | Upregulated |
| 0 | SCHLAP1 | FALSE | Upregulated |
| 0,988537128 | SCIN | FALSE | Upregulated |
| -0,307648678 | SCML2 | FALSE | Downregulated |
| | SCN10A | | |
| -0,228067111 | SCN11A | FALSE | Downregulated |
| 0,206758006 | SCN2A | FALSE | Upregulated |
| 0,125497387 | SCN2B | FALSE | Upregulated |
| 1,221595602 | SCN4B | | Upregulated |

| | | | |
|---|---|---|---|
| 1,571394393 | SCN5A | | Upregulated |
| | SCNN1G | | |
| | SCP2D1 | | |
| -0,449404287 | SCRT2 | FALSE | Downregulated |
| 1,829776736 | SCUBE2 | | Upregulated |
| -0,089454244 | SDC4P | FALSE | Downregulated |
| 0 | SDR16C5 | FALSE | Upregulated |
| | SDR16C6P | | |
| -0,594898442 | SDR9C7 | FALSE | Downregulated |
| -1,719070062 | SEC14L5 | | Downregulated |
| -1,01002963 | SEC14L6 | | Downregulated |
| -0,264238692 | SEC24B-AS1 | FALSE | Downregulated |
| | SEL1L2 | | |
| -0,030257859 | SELV | FALSE | Downregulated |
| 0,008993122 | SEMA3A | FALSE | Upregulated |
| -0,567785799 | SEMA5B | FALSE | Downregulated |
| | SEMG1 | | |
| | SEMG2 | | |
| -0,011427407 | SEPT12 | FALSE | Downregulated |
| | SEPT14 | | |
| 0,687855352 | SEPT7P9 | FALSE | Upregulated |
| | SERF1A | | |
| | SERPINA13P | | |
| -0,062934911 | SERPINA9 | FALSE | Downregulated |
| | SERPINB10 | | |
| | SERPINB11 | | |
| 0 | SERPINB12 | FALSE | Upregulated |
| 0 | SERPINB13 | FALSE | Upregulated |
| 0 | SERPINB2 | FALSE | Upregulated |
| | SERPINB3 | | |
| | SERPINB4 | | |
| 0 | SERPINB5 | FALSE | Upregulated |
| | SERPINB7 | | |
| -1,078227555 | SERPINI1 | | Downregulated |
| | SERPINI2 | | |
| -0,146126554 | SERTAD4 | FALSE | Downregulated |
| 2,157647447 | SERTAD4-AS1 | | Upregulated |
| 0 | SERTM1 | FALSE | Upregulated |
| 0,394428248 | SETSIP | FALSE | Upregulated |
| 1,824592208 | SEZ6L | | Upregulated |
| 0,03143979 | SEZ6L2 | FALSE | Upregulated |
| -0,834567467 | SFR1 | FALSE | Downregulated |
| 1,26424767 | SFRP2 | | Upregulated |
| 1,358670703 | SFRP4 | | Upregulated |
| -0,780218038 | SFT2D1 | FALSE | Downregulated |
| 0 | SFTA1P | FALSE | Upregulated |
| 0 | SFTA2 | FALSE | Upregulated |
| | SFTA3 | | |
| -0,229686238 | SFTPA1 | FALSE | Downregulated |
| -0,409651217 | SFTPA2 | FALSE | Downregulated |
| -0,234984403 | SFTPC | FALSE | Downregulated |

| | | | |
|---|---|---|---|
| 0,765293935 | SGCG | FALSE | Upregulated |
| -0,23633433 | SGCZ | FALSE | Downregulated |
| 0,54061093 | SGIP1 | FALSE | Upregulated |
| -1,0755071 | SGOL1-AS1 | | Downregulated |
| 0,027563344 | SH2D4B | FALSE | Upregulated |
| 0,545235497 | SH2D5 | FALSE | Upregulated |
| 0,456174644 | SH2D6 | FALSE | Upregulated |
| 0,541778702 | SH2D7 | FALSE | Upregulated |
| -0,344550019 | SH3BGR | FALSE | Downregulated |
| 0,591240413 | SH3GL2 | FALSE | Upregulated |
| 0,430185313 | SH3GL3 | FALSE | Upregulated |
| -0,867774609 | SHANK1 | FALSE | Downregulated |
| -0,040249788 | SHC4 | FALSE | Downregulated |
| -0,178215584 | SHCBP1 | FALSE | Downregulated |
| | SHCBP1L | | |
| 0,666111402 | SHISA2 | FALSE | Upregulated |
| 1,399912079 | SHISA3 | | Upregulated |
| 0,634313098 | SHISA8 | FALSE | Upregulated |
| 0,027933434 | SHOX | FALSE | Upregulated |
| | SHOX-2 | | |
| | SHOX2 | | |
| | SI | | |
| | SIAH3 | | |
| -0,202228732 | SIGLEC12 | FALSE | Downregulated |
| 1,874633542 | SIGLEC17P | | Upregulated |
| -0,213704601 | SIGLEC6 | FALSE | Downregulated |
| 1,936320249 | SIGLEC8 | | Upregulated |
| 0,658632233 | SIGLECL1 | FALSE | Upregulated |
| 0,543589648 | SIM1 | FALSE | Upregulated |
| 1,528237928 | SIRPG | | Upregulated |
| 0 | SIX1 | FALSE | Upregulated |
| 0,68761263 | SIX2 | FALSE | Upregulated |
| | SIX3 | | |
| 0 | SIX3-AS1 | FALSE | Upregulated |
| -0,933349719 | SIX4 | FALSE | Downregulated |
| -0,004435734 | SIX6 | FALSE | Downregulated |
| 0,720872597 | SKA1 | FALSE | Upregulated |
| 1,648686566 | SKINTL | | Upregulated |
| 1,001966087 | SKOR1 | | Upregulated |
| | SKOR2 | | |
| 0 | SKP1P2 | FALSE | Upregulated |
| -0,049697614 | SLAMF9 | FALSE | Downregulated |
| 0,076281212 | SLC10A2 | FALSE | Upregulated |
| 1,171476927 | SLC10A4 | | Upregulated |
| -0,013957021 | SLC10A6 | FALSE | Downregulated |
| 0,861838397 | SLC12A1 | FALSE | Upregulated |
| 1,81305926 | SLC12A3 | | Upregulated |
| 0 | SLC13A1 | FALSE | Upregulated |
| -1,547703082 | SLC13A2 | | Downregulated |
| -1,660282132 | SLC15A5 | | Downregulated |
| | SLC17A6 | | |

| | | | |
|---|---|---|---|
| 0,65717562 | SLC17A7 | FALSE | Upregulated |
| -0,716834839 | SLC17A8 | FALSE | Downregulated |
| 0 | SLC18A1 | FALSE | Upregulated |
| | SLC18A3 | | |
| | SLC1A6 | | |
| -0,499820212 | SLC22A11 | FALSE | Downregulated |
| 0,74523817 | SLC22A12 | FALSE | Upregulated |
| -1,023160281 | SLC22A13 | | Downregulated |
| 0 | SLC22A14 | FALSE | Upregulated |
| -0,011268819 | SLC22A16 | FALSE | Downregulated |
| 1,043934224 | SLC22A2 | | Upregulated |
| -0,280424341 | SLC22A20 | FALSE | Downregulated |
| -0,418921014 | SLC22A24 | FALSE | Downregulated |
| 0,550715521 | SLC22A31 | FALSE | Upregulated |
| 0,597480774 | SLC22A6 | FALSE | Upregulated |
| | SLC22A8 | | |
| 0,841226891 | SLC24A2 | FALSE | Upregulated |
| 0,769897639 | SLC25A14 | FALSE | Upregulated |
| | SLC25A31 | | |
| | SLC25A3P1 | | |
| 1,019132028 | SLC25A41 | | Upregulated |
| -0,076404905 | SLC25A48 | FALSE | Downregulated |
| | SLC25A51P1 | | |
| 1,273555305 | SLC25A53 | | Upregulated |
| | SLC25A6-2 | | |
| 0,917828155 | SLC26A10 | FALSE | Upregulated |
| 1,11164847 | SLC26A3 | | Upregulated |
| 0,877238415 | SLC26A4 | FALSE | Upregulated |
| 0,171034985 | SLC26A7 | FALSE | Upregulated |
| 1,273806323 | SLC26A9 | | Upregulated |
| -0,017737585 | SLC27A6 | FALSE | Downregulated |
| 0,807465145 | SLC28A3 | FALSE | Upregulated |
| 0 | SLC2A1-AS1 | FALSE | Upregulated |
| 0,629760853 | SLC2A14 | FALSE | Upregulated |
| 0,100063072 | SLC2A5 | FALSE | Upregulated |
| | SLC2A7 | | |
| -0,49434481 | SLC30A2 | FALSE | Downregulated |
| 0 | SLC30A3 | FALSE | Upregulated |
| 0,469689506 | SLC30A8 | FALSE | Upregulated |
| | SLC32A1 | | |
| | SLC35D3 | | |
| 0,990668152 | SLC35E4 | FALSE | Upregulated |
| 2,315409472 | SLC35F2 | | Upregulated |
| 0,783685082 | SLC35F3 | FALSE | Upregulated |
| 0,437629394 | SLC35F4 | FALSE | Upregulated |
| -0,000136166 | SLC35G3 | FALSE | Downregulated |
| | SLC35G4 | | |
| 0,064984529 | SLC35G5 | FALSE | Upregulated |
| -0,138559304 | SLC36A2 | FALSE | Downregulated |
| -0,0236855 | SLC36A3 | FALSE | Downregulated |
| 1,149744764 | SLC38A5 | | Upregulated |

| | | | |
|---|---|---|---|
| | SLC38A8 | | |
| 1,195329714 | SLC39A12 | | Upregulated |
| 0,477769862 | SLC39A2 | FALSE | Upregulated |
| -0,113179417 | SLC44A4 | FALSE | Downregulated |
| 2,168534622 | SLC44A5 | | Upregulated |
| 1,351849741 | SLC45A1 | | Upregulated |
| 0,141753376 | SLC4A1 | FALSE | Upregulated |
| 0,218963628 | SLC4A11 | FALSE | Upregulated |
| 1,289477244 | SLC4A3 | | Upregulated |
| 0,069817553 | SLC52A3 | FALSE | Upregulated |
| -1,097665759 | SLC5A11 | | Downregulated |
| -0,026610131 | SLC5A5 | FALSE | Downregulated |
| -0,000962546 | SLC5A7 | FALSE | Downregulated |
| | SLC5A8 | | |
| -0,00897377 | SLC6A10P | FALSE | Downregulated |
| | SLC6A10PB | | |
| -0,607964721 | SLC6A11 | FALSE | Downregulated |
| | SLC6A14 | | |
| | SLC6A15 | | |
| | SLC6A18 | | |
| -1,010466815 | SLC6A2 | | Downregulated |
| 0,009668202 | SLC6A3 | FALSE | Upregulated |
| | SLC6A5 | | |
| 0 | SLC6A7 | FALSE | Upregulated |
| -0,069651043 | SLC7A11 | FALSE | Downregulated |
| 1,758887001 | SLC7A11-AS1 | | Upregulated |
| 0 | SLC7A13 | FALSE | Upregulated |
| | SLC7A3 | | |
| 1,720156115 | SLC7A5P1 | | Upregulated |
| | SLC8A2 | | |
| -0,161242399 | SLC9A4 | FALSE | Downregulated |
| 0,311228311 | SLC9A7P1 | FALSE | Upregulated |
| 1,879036719 | SLC9C1 | | Upregulated |
| 1,850503621 | SLC9C2 | | Upregulated |
| 0,607795039 | SLCO5A1 | FALSE | Upregulated |
| | SLCO6A1 | | |
| 0 | SLIT2-IT1 | FALSE | Upregulated |
| | SLITRK1 | | |
| -0,037821891 | SLITRK2 | FALSE | Downregulated |
| -0,014925443 | SLITRK4 | FALSE | Downregulated |
| 1,378886569 | SLITRK5 | | Upregulated |
| 0,622512297 | SLITRK6 | FALSE | Upregulated |
| 0,117966154 | SLMO1 | FALSE | Upregulated |
| -0,002285185 | SLN | FALSE | Downregulated |
| -0,629081977 | SLURP1 | FALSE | Downregulated |
| 0,034128514 | SMCO2 | FALSE | Upregulated |
| | SMCP | | |
| -0,027793414 | SMCR9 | FALSE | Downregulated |
| | SMEK3P | | |
| -0,319358412 | SMIM10 | FALSE | Downregulated |
| -0,197839764 | SMIM17 | FALSE | Downregulated |

| | | | |
|---|---|---|---|
| 0,402175275 | SMIM21 | FALSE | Upregulated |
| 0,698241351 | SMIM22 | FALSE | Upregulated |
| -0,34969677 | SMIM23 | FALSE | Downregulated |
| 0,830110316 | SMIM9 | FALSE | Upregulated |
| | SMKR1 | | |
| 1,071723089 | SMPX | | Upregulated |
| | SMR3A | | |
| | SMR3B | | |
| 0,08670835 | SMTNL2 | FALSE | Upregulated |
| -0,974484036 | SMYD1 | FALSE | Downregulated |
| -0,116418995 | SNAI1 | FALSE | Downregulated |
| -0,667086557 | SNAP25-AS1 | FALSE | Downregulated |
| 0 | SNAP91 | FALSE | Upregulated |
| | SNAR-A1 | | |
| | SNAR-A10 | | |
| | SNAR-A11 | | |
| | SNAR-A12 | | |
| | SNAR-A13 | | |
| | SNAR-A14 | | |
| | SNAR-A2 | | |
| | SNAR-A3 | | |
| | SNAR-A4 | | |
| | SNAR-A5 | | |
| | SNAR-A6 | | |
| | SNAR-A7 | | |
| | SNAR-A8 | | |
| | SNAR-A9 | | |
| | SNAR-B1 | | |
| | SNAR-B2 | | |
| | SNAR-C1 | | |
| | SNAR-C2 | | |
| | SNAR-C3 | | |
| 0,001493849 | SNAR-C4 | FALSE | Upregulated |
| | SNAR-C5 | | |
| | SNAR-D | | |
| | SNAR-E | | |
| | SNAR-F | | |
| | SNAR-G1 | | |
| | SNAR-G2 | | |
| | SNAR-H | | |
| | SNAR-I | | |
| 0 | SNCB | FALSE | Upregulated |
| 1,851851382 | SNCG | | Upregulated |
| -0,078253765 | SNHG12 | FALSE | Downregulated |
| 0,17836701 | SNHG15 | FALSE | Upregulated |
| 0,49513049 | SNHG18 | FALSE | Upregulated |
| -0,054677215 | SNHG4 | FALSE | Downregulated |
| -0,120272942 | SNHG9 | FALSE | Downregulated |
| 0,592949859 | SNORA1 | FALSE | Upregulated |
| 0,469894654 | SNORA10 | FALSE | Upregulated |
| 0,104103766 | SNORA11 | FALSE | Upregulated |

| | | | |
|---|---|---|---|
| 0,127363621 | SNORA11B | FALSE | Upregulated |
| | SNORA11C | | |
| | SNORA11D | | |
| | SNORA11E | | |
| 0,456811247 | SNORA12 | FALSE | Upregulated |
| -0,173729462 | SNORA13 | FALSE | Downregulated |
| 1,448932985 | SNORA14A | | Upregulated |
| 0,319494999 | SNORA14B | FALSE | Upregulated |
| -0,223997397 | SNORA15 | FALSE | Downregulated |
| 0 | SNORA16A | FALSE | Upregulated |
| -0,002529603 | SNORA16B | FALSE | Downregulated |
| | SNORA17 | | |
| 0,685282749 | SNORA18 | FALSE | Upregulated |
| | SNORA19 | | |
| -0,508411848 | SNORA20 | FALSE | Downregulated |
| 1,879542616 | SNORA21 | | Upregulated |
| 1,250416916 | SNORA22 | | Upregulated |
| -0,486220038 | SNORA24 | FALSE | Downregulated |
| 0 | SNORA25 | FALSE | Upregulated |
| -0,342989929 | SNORA26 | FALSE | Downregulated |
| 0,697432696 | SNORA27 | FALSE | Upregulated |
| 0,026501848 | SNORA28 | FALSE | Upregulated |
| | SNORA29 | | |
| 0,502928937 | SNORA2A | FALSE | Upregulated |
| | SNORA2B | | |
| -1,099293088 | SNORA3 | | Downregulated |
| 0 | SNORA30 | FALSE | Upregulated |
| -0,251555443 | SNORA31 | FALSE | Downregulated |
| 0 | SNORA32 | FALSE | Upregulated |
| 0 | SNORA33 | FALSE | Upregulated |
| -0,76156127 | SNORA34 | FALSE | Downregulated |
| | SNORA35 | | |
| | SNORA36A | | |
| | SNORA36B | | |
| | SNORA36C | | |
| -0,50636066 | SNORA37 | FALSE | Downregulated |
| 0 | SNORA38 | FALSE | Upregulated |
| -0,037959525 | SNORA38B | FALSE | Downregulated |
| | SNORA39 | | |
| 0,008411063 | SNORA4 | FALSE | Upregulated |
| 0,469313198 | SNORA40 | FALSE | Upregulated |
| 0 | SNORA41 | FALSE | Upregulated |
| 1,104022892 | SNORA42 | | Upregulated |
| -0,104596862 | SNORA43 | FALSE | Downregulated |
| -0,047957604 | SNORA44 | FALSE | Downregulated |
| 0,484852813 | SNORA45 | FALSE | Upregulated |
| -0,045771121 | SNORA46 | FALSE | Downregulated |
| -2,026737745 | SNORA47 | | Downregulated |
| 0,41600283 | SNORA49 | FALSE | Upregulated |
| -0,117498822 | SNORA50 | FALSE | Downregulated |
| | SNORA51 | | |

| | | | |
|---|---|---|---|
| 0,655562622 | SNORA52 | FALSE | Upregulated |
| -0,081220432 | SNORA55 | FALSE | Downregulated |
| | SNORA56 | | |
| 0,446858937 | SNORA58 | FALSE | Upregulated |
| 0 | SNORA59A | FALSE | Upregulated |
| 0 | SNORA59B | FALSE | Upregulated |
| 0,482144481 | SNORA5A | FALSE | Upregulated |
| | SNORA5B | | |
| -0,561265144 | SNORA5C | FALSE | Downregulated |
| -0,012752157 | SNORA6 | FALSE | Downregulated |
| -0,548838483 | SNORA60 | FALSE | Downregulated |
| 0,321593242 | SNORA61 | FALSE | Upregulated |
| | SNORA62 | | |
| -0,377915321 | SNORA64 | FALSE | Downregulated |
| -0,292127081 | SNORA65 | FALSE | Downregulated |
| 0,097638542 | SNORA66 | FALSE | Upregulated |
| 0,432890389 | SNORA67 | FALSE | Upregulated |
| -0,35809567 | SNORA68 | FALSE | Downregulated |
| | SNORA69 | | |
| -0,03641181 | SNORA70 | FALSE | Downregulated |
| | SNORA70B | | |
| | SNORA70C | | |
| | SNORA70D | | |
| | SNORA70E | | |
| | SNORA70F | | |
| -0,446721149 | SNORA70G | FALSE | Downregulated |
| 0,207883769 | SNORA71A | FALSE | Upregulated |
| 1,427506459 | SNORA71B | | Upregulated |
| 0,080407977 | SNORA71C | FALSE | Upregulated |
| -0,146154144 | SNORA71D | FALSE | Downregulated |
| 0,693207765 | SNORA72 | FALSE | Upregulated |
| 1,674989523 | SNORA74A | | Upregulated |
| 0,034128514 | SNORA75 | FALSE | Upregulated |
| -0,935170012 | SNORA76 | FALSE | Downregulated |
| 0,429757255 | SNORA77 | FALSE | Upregulated |
| 0,927671768 | SNORA78 | FALSE | Upregulated |
| | SNORA79 | | |
| -1,785298397 | SNORA8 | | Downregulated |
| -0,148642523 | SNORA80 | FALSE | Downregulated |
| 0 | SNORA80B | FALSE | Upregulated |
| -0,581314591 | SNORA84 | FALSE | Downregulated |
| 0,221036541 | SNORA9 | FALSE | Upregulated |
| | SNORD100 | | |
| | SNORD101 | | |
| | SNORD102 | | |
| | SNORD103A | | |
| | SNORD103B | | |
| | SNORD104 | | |
| | SNORD105 | | |
| | SNORD105B | | |
| | SNORD11 | | |

SNORD110
SNORD111
SNORD111B
SNORD112
SNORD113-1
SNORD113-2
SNORD113-3
SNORD113-4
SNORD113-5
SNORD113-6
SNORD113-7
SNORD113-8
SNORD113-9
SNORD114-1
SNORD114-10
SNORD114-11
SNORD114-12
SNORD114-13
SNORD114-14
SNORD114-15
SNORD114-16
SNORD114-17
SNORD114-18
SNORD114-19
SNORD114-2
SNORD114-20
SNORD114-21
SNORD114-22
SNORD114-23
SNORD114-24
SNORD114-25
SNORD114-26
SNORD114-27
SNORD114-28
SNORD114-29
SNORD114-3
SNORD114-30
SNORD114-31
SNORD114-4
SNORD114-5
SNORD114-6
SNORD114-7
SNORD114-8
SNORD114-9
SNORD117
SNORD119
SNORD11B
SNORD12
SNORD121A
SNORD121B
SNORD123

| | | | |
|---|---|---|---|
| | SNORD124 | | |
| | SNORD125 | | |
| | SNORD126 | | |
| | SNORD127 | | |
| | SNORD12B | | |
| | SNORD12C | | |
| -0,117059024 | SNORD13 | FALSE | Downregulated |
| | SNORD14A | | |
| | SNORD14B | | |
| | SNORD14C | | |
| | SNORD14D | | |
| | SNORD14E | | |
| 0,538258275 | SNORD15A | FALSE | Upregulated |
| | SNORD16 | | |
| | SNORD18A | | |
| | SNORD18B | | |
| | SNORD18C | | |
| | SNORD19 | | |
| | SNORD19B | | |
| | SNORD1A | | |
| | SNORD1B | | |
| | SNORD1C | | |
| | SNORD2 | | |
| | SNORD20 | | |
| | SNORD21 | | |
| 0,232381686 | SNORD23 | FALSE | Upregulated |
| | SNORD24 | | |
| | SNORD25 | | |
| | SNORD26 | | |
| | SNORD27 | | |
| | SNORD28 | | |
| | SNORD29 | | |
| | SNORD30 | | |
| | SNORD31 | | |
| | SNORD32A | | |
| | SNORD32B | | |
| | SNORD33 | | |
| | SNORD34 | | |
| | SNORD35A | | |
| | SNORD35B | | |
| | SNORD36A | | |
| | SNORD36B | | |
| | SNORD36C | | |
| | SNORD37 | | |
| | SNORD38A | | |
| | SNORD38B | | |
| -0,657553406 | SNORD3B-1 | FALSE | Downregulated |
| -0,499837187 | SNORD3D | FALSE | Downregulated |
| | SNORD41 | | |
| | SNORD42A | | |
| | SNORD42B | | |

| | | | |
|---|---|---|---|
| | SNORD43 | | |
| | SNORD44 | | |
| | SNORD45A | | |
| | SNORD45B | | |
| | SNORD45C | | |
| 0 | SNORD46 | FALSE | Upregulated |
| | SNORD47 | | |
| | SNORD48 | | |
| | SNORD49A | | |
| | SNORD49B | | |
| | SNORD4A | | |
| | SNORD4B | | |
| | SNORD5 | | |
| | SNORD50A | | |
| | SNORD50B | | |
| | SNORD51 | | |
| | SNORD52 | | |
| | SNORD53 | | |
| | SNORD54 | | |
| | SNORD55 | | |
| | SNORD56 | | |
| | SNORD56B | | |
| | SNORD57 | | |
| | SNORD58A | | |
| | SNORD58B | | |
| | SNORD58C | | |
| | SNORD59A | | |
| | SNORD59B | | |
| | SNORD6 | | |
| | SNORD60 | | |
| | SNORD61 | | |
| | SNORD62A | | |
| | SNORD62B | | |
| | SNORD63 | | |
| | SNORD65 | | |
| | SNORD66 | | |
| 0,623488178 | SNORD67 | FALSE | Upregulated |
| | SNORD68 | | |
| | SNORD69 | | |
| | SNORD7 | | |
| | SNORD70 | | |
| | SNORD71 | | |
| | SNORD72 | | |
| | SNORD73A | | |
| | SNORD74 | | |
| | SNORD75 | | |
| 0 | SNORD76 | FALSE | Upregulated |
| | SNORD77 | | |
| | SNORD78 | | |
| | SNORD79 | | |
| -0,572237786 | SNORD8 | FALSE | Downregulated |

| | | | |
|---|---|---|---|
| | SNORD80 | | |
| | SNORD81 | | |
| | SNORD82 | | |
| | SNORD83A | | |
| | SNORD83B | | |
| | SNORD84 | | |
| | SNORD85 | | |
| | SNORD86 | | |
| | SNORD87 | | |
| | SNORD88A | | |
| | SNORD88B | | |
| | SNORD88C | | |
| 0,041547119 | SNORD9 | FALSE | Upregulated |
| | SNORD90 | | |
| | SNORD91A | | |
| | SNORD91B | | |
| | SNORD92 | | |
| | SNORD93 | | |
| | SNORD95 | | |
| | SNORD96A | | |
| | SNORD96B | | |
| -0,111000679 | SNORD97 | FALSE | Downregulated |
| | SNORD98 | | |
| | SNORD99 | | |
| 1,39747009 | SNRPD2P2 | Upregulated | |
| 0,266295275 | SNTG1 | FALSE | Upregulated |
| | SNTG2 | | |
| -0,196647619 | SNTN | FALSE | Downregulated |
| 1,103342106 | SNX29P1 | Upregulated | |
| 0,321944268 | SNX31 | FALSE | Upregulated |
| 0,472320158 | SOCS2-AS1 | FALSE | Upregulated |
| 0 | SOHLH1 | FALSE | Upregulated |
| | SOHLH2 | | |
| -0,111453678 | SORCS1 | FALSE | Downregulated |
| 0 | SORCS3 | FALSE | Upregulated |
| | SORCS3-AS1 | | |
| -0,204705918 | SOST | FALSE | Downregulated |
| | SOX1 | | |
| 0 | SOX11 | FALSE | Upregulated |
| 0 | SOX14 | FALSE | Upregulated |
| 0,127124355 | SOX2 | FALSE | Upregulated |
| 0,124531857 | SOX2-OT | FALSE | Upregulated |
| 0,027613641 | SOX21-AS1 | FALSE | Upregulated |
| | SOX3 | | |
| 0,619413494 | SOX30 | FALSE | Upregulated |
| 0,024788959 | SOX8 | FALSE | Upregulated |
| -0,452949333 | SP6 | FALSE | Downregulated |
| 0 | SP8 | FALSE | Upregulated |
| | SP9 | | |
| -0,006673188 | SPA17 | FALSE | Downregulated |
| 0,050688952 | SPACA1 | FALSE | Upregulated |

| | | | |
|---|---|---|---|
| | SPACA3 | | |
| 1,80425168 | SPACA4 | | Upregulated |
| 1,107282227 | SPACA5 | | Upregulated |
| -0,053895118 | SPACA5B | FALSE | Downregulated |
| 2,647728878 | SPACA7 | | Upregulated |
| | SPAG11A | | |
| | SPAG11B | | |
| 0,566321084 | SPAG6 | FALSE | Upregulated |
| | SPAM1 | | |
| | SPANXA1 | | |
| | SPANXA2 | | |
| | SPANXA2-OT1 | | |
| 0,343245384 | SPANXB1 | FALSE | Upregulated |
| | SPANXC | | |
| 0 | SPANXD | FALSE | Upregulated |
| | SPANXN1 | | |
| | SPANXN2 | | |
| | SPANXN3 | | |
| | SPANXN4 | | |
| | SPANXN5 | | |
| | SPATA16 | | |
| 0,067493036 | SPATA17 | FALSE | Upregulated |
| | SPATA19 | | |
| 0,036494884 | SPATA22 | FALSE | Upregulated |
| | SPATA3 | | |
| 0,00721802 | SPATA3-AS1 | FALSE | Upregulated |
| | SPATA31A1 | | |
| | SPATA31A3 | | |
| | SPATA31A5 | | |
| | SPATA31A6 | | |
| | SPATA31A7 | | |
| 0 | SPATA31B1 | FALSE | Upregulated |
| 0 | SPATA31C1 | FALSE | Upregulated |
| 0,239162001 | SPATA31C2 | FALSE | Upregulated |
| | SPATA31D1 | | |
| | SPATA31D3 | | |
| | SPATA31D4 | | |
| 0 | SPATA31D5P | FALSE | Upregulated |
| 0 | SPATA4 | FALSE | Upregulated |
| | SPATA42 | | |
| 0 | SPATA8 | FALSE | Upregulated |
| | SPATA8-AS1 | | |
| 1,505550204 | SPATC1L | | Upregulated |
| -0,002529603 | SPATS1 | FALSE | Downregulated |
| -0,312133415 | SPC24 | FALSE | Downregulated |
| -0,682061777 | SPC25 | FALSE | Downregulated |
| 0,686503512 | SPDEF | FALSE | Upregulated |
| 1,435101336 | SPDYC | | Upregulated |
| 0,076315958 | SPEF1 | FALSE | Upregulated |
| -0,176113186 | SPEM1 | FALSE | Downregulated |
| | SPERT | | |

| | | | |
|---|---|---|---|
| 0,695629535 | SPESP1 | FALSE | Upregulated |
| 0 | SPG20OS | FALSE | Upregulated |
| | SPHAR | | |
| 0 | SPHKAP | FALSE | Upregulated |
| -0,871866427 | SPINK1 | FALSE | Downregulated |
| 0,018162592 | SPINK13 | FALSE | Upregulated |
| 0 | SPINK14 | FALSE | Upregulated |
| 0 | SPINK2 | FALSE | Upregulated |
| | SPINK4 | | |
| 1,043900455 | SPINK5 | | Upregulated |
| -0,206667111 | SPINK6 | FALSE | Downregulated |
| 0,181550643 | SPINK7 | FALSE | Upregulated |
| | SPINK8 | | |
| 1,149220339 | SPINK9 | | Upregulated |
| | SPINT3 | | |
| | SPINT4 | | |
| 0,669025015 | SPO11 | FALSE | Upregulated |
| 1,082436065 | SPOCD1 | | Upregulated |
| 1,247815951 | SPOCK1 | | Upregulated |
| -0,499418471 | SPOCK3 | FALSE | Downregulated |
| | SPPL2C | | |
| 0,49657185 | SPRED3 | FALSE | Upregulated |
| | SPRR1A | | |
| | SPRR1B | | |
| | SPRR2A | | |
| | SPRR2B | | |
| | SPRR2C | | |
| | SPRR2D | | |
| 0 | SPRR2E | FALSE | Upregulated |
| | SPRR2F | | |
| | SPRR2G | | |
| | SPRR3 | | |
| | SPRR4 | | |
| | SPRY3-2 | | |
| 0,616964901 | SPTA1 | FALSE | Upregulated |
| 0 | SPZ1 | FALSE | Upregulated |
| 0,689687477 | SRD5A1P1 | FALSE | Upregulated |
| 1,460889542 | SRRM3 | | Upregulated |
| 0 | SRRM4 | FALSE | Upregulated |
| 0,030223756 | SRY | FALSE | Upregulated |
| | SSMEM1 | | |
| -0,68975753 | SSR4P1 | FALSE | Downregulated |
| | SST | | |
| 0,661246181 | SSTR3 | FALSE | Upregulated |
| | SSTR4 | | |
| -0,665722699 | SSTR5 | FALSE | Downregulated |
| 0 | SSTR5-AS1 | FALSE | Upregulated |
| 0 | SSUH2 | FALSE | Upregulated |
| | SSX1 | | |
| | SSX2 | | |
| | SSX2B | | |

SSX3
SSX4
SSX4B
SSX5
SSX6
SSX7
SSX8
SSX9
-0,362744558 ST13P4 FALSE Downregulated
1,97079304 ST18 Upregulated
0 ST20-MTHFS FALSE Upregulated
0,428238804 ST6GALNAC1 FALSE Upregulated
1,115517142 ST6GALNAC5 Upregulated
-0,001452899 ST7-OT3 FALSE Downregulated
ST8SIA2
-0,060242643 ST8SIA5 FALSE Downregulated
-1,192577544 ST8SIA6 Downregulated
0,088911633 ST8SIA6-AS1 FALSE Upregulated
1,577708332 STAC2 Upregulated
STARD6
STATH
0,427242357 STC1 FALSE Upregulated
0,008165139 STC2 FALSE Upregulated
0 STEAP1B FALSE Upregulated
-0,242963058 STEAP2-AS1 FALSE Downregulated
STH
-0,437139973 STIL FALSE Downregulated
-0,174337547 STK31 FALSE Downregulated
0,464464425 STK33 FALSE Upregulated
0,260116755 STK4-AS1 FALSE Upregulated
1,364244798 STMN2 Upregulated
-0,160765249 STMN4 FALSE Downregulated
0,007508006 STMND1 FALSE Upregulated
STOML3
0,014930436 STON1-GTF2A1L FALSE Upregulated
-0,663911407 STPG2 FALSE Downregulated
0,529358102 STPG2-AS1 FALSE Upregulated
0,407596195 STRA6 FALSE Upregulated
0 STRA8 FALSE Upregulated
1,053888406 STRIP2 Upregulated
-0,731604726 STX18-AS1 FALSE Downregulated
0,579296858 STYK1 FALSE Upregulated
0,177516873 SUGT1P1 FALSE Upregulated
-0,068314018 SUGT1P3 FALSE Downregulated
2,637736483 SULT1A3 Upregulated
0,691064843 SULT1C3 FALSE Upregulated
0,048861806 SULT2B1 FALSE Upregulated
0 SULT4A1 FALSE Upregulated
SULT6B1
0 SUMO1P1 FALSE Upregulated
0,384429398 SUMO4 FALSE Upregulated

| | | | |
|---|---|---|---|
| | SUN3 | | |
| | SUN5 | | |
| 0,113003304 | SUPT20HL1 | FALSE | Upregulated |
| 1,02491309 | SUPT20HL2 | | Upregulated |
| 0,240735872 | SUZ12P1 | FALSE | Upregulated |
| -0,070921818 | SV2B | FALSE | Downregulated |
| 0,028288674 | SV2C | FALSE | Upregulated |
| 1,562008464 | SVILP1 | | Upregulated |
| 0,23026033 | SVOP | FALSE | Upregulated |
| 1,024790319 | SVOPL | | Upregulated |
| -0,345746737 | SYCE1L | FALSE | Downregulated |
| 0,212884739 | SYCE3 | FALSE | Upregulated |
| 0 | SYCN | FALSE | Upregulated |
| | SYCP1 | | |
| 0,9842628 | SYCP2 | FALSE | Upregulated |
| 0 | SYCP2L | FALSE | Upregulated |
| 0,248226709 | SYN2 | FALSE | Upregulated |
| 0,669756935 | SYNDIG1 | FALSE | Upregulated |
| 0,453823514 | SYNDIG1L | FALSE | Upregulated |
| 1,297794996 | SYNGR3 | | Upregulated |
| 0 | SYNGR4 | FALSE | Upregulated |
| 0 | SYNJ2-IT1 | FALSE | Upregulated |
| -1,37396155 | SYNJ2BP-COX16 | | Downregulated |
| 0,360901615 | SYNPR | FALSE | Upregulated |
| | SYNPR-AS1 | | |
| 0 | SYP-AS1 | FALSE | Upregulated |
| -0,710225976 | SYS1-DBNDD2 | FALSE | Downregulated |
| 0,28185588 | SYT10 | FALSE | Upregulated |
| 0,687842945 | SYT13 | FALSE | Upregulated |
| | SYT14 | | |
| -0,258739994 | SYT14L | FALSE | Downregulated |
| | SYT16 | | |
| | SYT3 | | |
| | SYT4 | | |
| 0 | SYT5 | FALSE | Upregulated |
| -0,008901409 | SYT6 | FALSE | Downregulated |
| 1,50665931 | SYT8 | | Upregulated |
| | T | | |
| | TAAR1 | | |
| | TAAR2 | | |
| | TAAR5 | | |
| | TAAR6 | | |
| | TAAR8 | | |
| | TAAR9 | | |
| 0,004929336 | TAC1 | FALSE | Upregulated |
| | TAC3 | | |
| -1,233485185 | TACR3 | | Downregulated |
| 0,247294454 | TAF13 | FALSE | Upregulated |
| -0,485635163 | TAF1L | FALSE | Downregulated |
| 0,452260684 | TAF7L | FALSE | Upregulated |
| | TAGLN3 | | |

| | | | |
|---|---|---|---|
| 0,002178943 | TAL2 | FALSE | Upregulated |
| | TARM1 | | |
| 0,914544325 | TARP | FALSE | Upregulated |
| | TAS1R2 | | |
| | TAS2R1 | | |
| -0,214061636 | TAS2R10 | FALSE | Downregulated |
| | TAS2R16 | | |
| 0,884091765 | TAS2R20 | FALSE | Upregulated |
| 0,382768641 | TAS2R3 | FALSE | Upregulated |
| 1,356410478 | TAS2R31 | | Upregulated |
| -0,273521755 | TAS2R38 | FALSE | Downregulated |
| 0,532536498 | TAS2R4 | FALSE | Upregulated |
| 0,272892486 | TAS2R40 | FALSE | Upregulated |
| | TAS2R41 | | |
| -0,804444067 | TAS2R43 | FALSE | Downregulated |
| 1,393143133 | TAS2R46 | | Upregulated |
| 0,730106854 | TAS2R5 | FALSE | Upregulated |
| 0,858580734 | TAS2R50 | FALSE | Upregulated |
| 1,54832783 | TAS2R60 | | Upregulated |
| 0 | TAS2R7 | FALSE | Upregulated |
| 0,076658198 | TAS2R8 | FALSE | Upregulated |
| 0 | TAS2R9 | FALSE | Upregulated |
| 1,592869021 | TAX1BP3 | | Upregulated |
| 0 | TBATA | FALSE | Upregulated |
| | TBC1D21 | | |
| | TBC1D26 | | |
| 0,381363107 | TBC1D27 | FALSE | Upregulated |
| 0 | TBC1D28 | FALSE | Upregulated |
| 0,862339569 | TBC1D29 | FALSE | Upregulated |
| -0,242534577 | TBC1D3 | FALSE | Downregulated |
| -0,010767837 | TBC1D3B | FALSE | Downregulated |
| | TBC1D3F | | |
| 0 | TBC1D3P1-DHX40P1 | FALSE | Upregulated |
| | TBC1D3P2 | | |
| | TBC1D3P5 | | |
| -1,464683494 | TBL1Y | | Downregulated |
| 0 | TBPL2 | FALSE | Upregulated |
| | TBR1 | | |
| 0,644083396 | TBX18 | FALSE | Upregulated |
| 0 | TBX22 | FALSE | Upregulated |
| 0,502595609 | TBX4 | FALSE | Upregulated |
| 0 | TBX5 | FALSE | Upregulated |
| 0 | TBX5-AS1 | FALSE | Upregulated |
| | TCAM1P | | |
| | TCEAL2 | | |
| | TCEAL5 | | |
| 0 | TCEAL6 | FALSE | Upregulated |
| 0,902573658 | TCEAL7 | FALSE | Upregulated |
| | TCERG1L | | |
| 1,661905182 | TCF15 | | Upregulated |
| 0 | TCF23 | FALSE | Upregulated |

| | | | |
|---|---|---|---|
| -0,941234232 | TCF24 | FALSE | Downregulated |
| 0,024522102 | TCHHL1 | FALSE | Upregulated |
| 0,081722492 | TCL1A | FALSE | Upregulated |
| 1,004126014 | TCL1B | | Upregulated |
| 0 | TCL6 | FALSE | Upregulated |
| 1,082875752 | TCN1 | | Upregulated |
| | TCP10 | | |
| 0 | TCP10L2 | FALSE | Upregulated |
| 0,132045804 | TCP11 | FALSE | Upregulated |
| | TCP11X1 | | |
| | TCP11X2 | | |
| -0,426162523 | TCTE1 | FALSE | Downregulated |
| 0,862642079 | TCTE3 | FALSE | Upregulated |
| 0 | TCTEX1D2 | FALSE | Upregulated |
| -0,00131735 | TDGF1 | FALSE | Downregulated |
| 0 | TDGF1P3 | FALSE | Upregulated |
| 0,661206439 | TDH | FALSE | Upregulated |
| 0,574072129 | TDRD1 | FALSE | Upregulated |
| 0 | TDRD12 | FALSE | Upregulated |
| 0,356127574 | TDRD15 | FALSE | Upregulated |
| | TDRD5 | | |
| 1,295179098 | TDRD9 | | Upregulated |
| | TDRG1 | | |
| 0,597480774 | TECRL | FALSE | Upregulated |
| -1,545654817 | TECTB | | Downregulated |
| -0,056375693 | TEKT1 | FALSE | Downregulated |
| 0,291157985 | TEKT2 | FALSE | Upregulated |
| 0,70900893 | TEKT3 | FALSE | Upregulated |
| | TEKT4 | | |
| -0,918842625 | TEKT5 | FALSE | Downregulated |
| 0 | TEPP | FALSE | Upregulated |
| | TERT | | |
| | TEX101 | | |
| | TEX11 | | |
| 0 | TEX12 | FALSE | Upregulated |
| -0,081465002 | TEX13B | FALSE | Downregulated |
| 0,186276593 | TEX15 | FALSE | Upregulated |
| 0 | TEX19 | FALSE | Upregulated |
| 0,141835285 | TEX21P | FALSE | Upregulated |
| | TEX26 | | |
| | TEX26-AS1 | | |
| 0 | TEX28 | FALSE | Upregulated |
| 0,012315826 | TEX29 | FALSE | Upregulated |
| 0,661206439 | TEX33 | FALSE | Upregulated |
| -0,035293546 | TEX35 | FALSE | Downregulated |
| | TEX36 | | |
| | TEX36-AS1 | | |
| | TEX37 | | |
| -0,138057228 | TEX40 | FALSE | Downregulated |
| -0,418853831 | TEX41 | FALSE | Downregulated |
| -0,001452899 | TFAMP1 | FALSE | Downregulated |

| | | | |
|---|---|---|---|
| 0,445558218 | TFAP2A | FALSE | Upregulated |
| | TFAP2A-AS1 | | |
| 0,97678313 | TFAP2B | FALSE | Upregulated |
| 0,736829301 | TFAP2C | FALSE | Upregulated |
| | TFAP2D | | |
| 0,669756935 | TFDP3 | FALSE | Upregulated |
| -0,060499344 | TFF1 | FALSE | Downregulated |
| 0 | TFF2 | FALSE | Upregulated |
| 1,568928711 | TFF3 | | Upregulated |
| | TGFA-IT1 | | |
| -0,850962649 | TGIF2-C20orf24 | FALSE | Downregulated |
| | TGIF2LX | | |
| | TGIF2LY | | |
| 0,050694383 | TGM3 | FALSE | Upregulated |
| 0 | TGM5 | FALSE | Upregulated |
| -0,146689815 | TGM6 | FALSE | Downregulated |
| | TGM7 | | |
| 0,948333095 | THAP10 | FALSE | Upregulated |
| 0 | THEG | FALSE | Upregulated |
| | THEG5 | | |
| 0 | THEGL | FALSE | Upregulated |
| -0,269174566 | THSD7B | FALSE | Downregulated |
| 1,702824733 | TIFAB | | Upregulated |
| -0,274247649 | TIGD3 | FALSE | Downregulated |
| -0,089166846 | TIGD4 | FALSE | Downregulated |
| 1,099039513 | TIMP4 | | Upregulated |
| | TINAG | | |
| 0,668602714 | TINCR | FALSE | Upregulated |
| | TISP43 | | |
| -0,557014115 | TKTL1 | FALSE | Downregulated |
| | TKTL2 | | |
| 0,86195114 | TLE6 | FALSE | Upregulated |
| 0,662324558 | TLL1 | FALSE | Upregulated |
| -0,075533245 | TLL2 | FALSE | Downregulated |
| 1,396790365 | TLR10 | | Upregulated |
| 0 | TLR8-AS1 | FALSE | Upregulated |
| -2,302264249 | TLX1 | | Downregulated |
| 0,650877532 | TLX2 | FALSE | Upregulated |
| | TLX3 | | |
| 0,734483765 | TM4SF1-AS1 | FALSE | Upregulated |
| 0,753021079 | TM4SF19 | FALSE | Upregulated |
| -0,367511452 | TM4SF19-AS1 | FALSE | Downregulated |
| | TM4SF19-TCTEX1D2 | | |
| 0 | TMC1 | FALSE | Upregulated |
| 1,470398281 | TMC7 | | Upregulated |
| -0,619562983 | TMCC1-AS1 | FALSE | Downregulated |
| -0,514103616 | TMCO2 | FALSE | Downregulated |
| | TMCO5A | | |
| | TMCO5B | | |
| -0,444925431 | TMED11P | FALSE | Downregulated |
| 1,114920903 | TMED7-TICAM2 | | Upregulated |

| | | | |
|---|---|---|---|
| | TMEFF1 | | |
| -0,394908424 | TMEM107 | FALSE | Downregulated |
| 1,246640204 | TMEM110-MUSTN1 | | Upregulated |
| | TMEM114 | | |
| 0,683178608 | TMEM130 | FALSE | Upregulated |
| -0,468413102 | TMEM132B | FALSE | Downregulated |
| 1,300014651 | TMEM132C | | Upregulated |
| 0,547019563 | TMEM144 | FALSE | Upregulated |
| 0 | TMEM145 | FALSE | Upregulated |
| -0,142149123 | TMEM151A | FALSE | Downregulated |
| -0,04811712 | TMEM151B | FALSE | Downregulated |
| 0 | TMEM155 | FALSE | Upregulated |
| 0,801020439 | TMEM156 | FALSE | Upregulated |
| -0,702675803 | TMEM158 | FALSE | Downregulated |
| 0,923130373 | TMEM159 | FALSE | Upregulated |
| -1,934222201 | TMEM161B-AS1 | | Downregulated |
| 0,338734836 | TMEM171 | FALSE | Upregulated |
| | TMEM174 | | |
| 3,049580755 | TMEM178B | | Upregulated |
| -1,084445777 | TMEM182 | | Downregulated |
| 0,388042192 | TMEM190 | FALSE | Upregulated |
| 1,022782596 | TMEM191B | | Upregulated |
| -0,105462418 | TMEM191C | FALSE | Downregulated |
| 0,183814899 | TMEM196 | FALSE | Upregulated |
| 1,95394335 | TMEM200A | | Upregulated |
| 0,324782254 | TMEM202 | FALSE | Upregulated |
| 0,155360957 | TMEM207 | FALSE | Upregulated |
| | TMEM210 | | |
| 0,408716309 | TMEM211 | FALSE | Upregulated |
| 0,044498801 | TMEM212 | FALSE | Upregulated |
| 0 | TMEM213 | FALSE | Upregulated |
| -0,014708095 | TMEM215 | FALSE | Downregulated |
| 0,942546035 | TMEM216 | FALSE | Upregulated |
| 0,782595283 | TMEM225 | FALSE | Upregulated |
| 0 | TMEM229A | FALSE | Upregulated |
| -0,462170514 | TMEM235 | FALSE | Downregulated |
| -0,030911453 | TMEM236 | FALSE | Downregulated |
| | TMEM239 | | |
| 1,329778442 | TMEM240 | | Upregulated |
| 0 | TMEM244 | FALSE | Upregulated |
| 0,010108383 | TMEM247 | FALSE | Upregulated |
| -0,401597584 | TMEM249 | FALSE | Downregulated |
| | TMEM257 | | |
| -0,917401113 | TMEM262 | FALSE | Downregulated |
| -0,309698915 | TMEM30C | FALSE | Downregulated |
| -0,076723412 | TMEM31 | FALSE | Downregulated |
| 0,108240312 | TMEM35 | FALSE | Upregulated |
| 0,597480774 | TMEM40 | FALSE | Upregulated |
| -1,954733843 | TMEM52 | | Downregulated |
| -0,073813648 | TMEM52B | FALSE | Downregulated |
| | TMEM59L | | |

| | | | |
|---|---|---|---|
| 0 | TMEM61 | FALSE | Upregulated |
| 0,060009722 | TMEM63C | FALSE | Upregulated |
| 0,188004752 | TMEM67 | FALSE | Upregulated |
| 1,654129607 | TMEM71 | | Upregulated |
| -0,201352107 | TMEM72-AS1 | FALSE | Downregulated |
| 0,108728615 | TMEM74 | FALSE | Upregulated |
| -0,980555086 | TMEM81 | FALSE | Downregulated |
| 0,418181713 | TMEM88 | FALSE | Upregulated |
| -0,132615305 | TMEM88B | FALSE | Downregulated |
| 0 | TMEM89 | FALSE | Upregulated |
| | TMEM8C | | |
| 0,640716414 | TMEM95 | FALSE | Upregulated |
| | TMIGD1 | | |
| 0 | TMOD4 | FALSE | Upregulated |
| | TMPRSS11B | | |
| | TMPRSS11BNL | | |
| 0,449449468 | TMPRSS11F | FALSE | Upregulated |
| 0,004929336 | TMPRSS11GP | FALSE | Upregulated |
| 0 | TMPRSS12 | FALSE | Upregulated |
| 1,198728809 | TMPRSS13 | | Upregulated |
| 0,686412155 | TMPRSS15 | FALSE | Upregulated |
| 0,756130823 | TMPRSS3 | FALSE | Upregulated |
| | TMPRSS4-AS1 | | |
| -0,480014165 | TMPRSS5 | FALSE | Downregulated |
| | TMPRSS7 | | |
| | TMSB15A | | |
| | TMSB15B | | |
| 1,108029751 | TMSB4Y | | Upregulated |
| 0,579183895 | TNFAIP6 | FALSE | Upregulated |
| 1,451421991 | TNFAIP8L3 | | Upregulated |
| 0,824977024 | TNFRSF13B | FALSE | Upregulated |
| 0,193650154 | TNFRSF13C | FALSE | Upregulated |
| 0,307574943 | TNFRSF18 | FALSE | Upregulated |
| 0,636971153 | TNFRSF4 | FALSE | Upregulated |
| 0,201889693 | TNFRSF8 | FALSE | Upregulated |
| 0,835099263 | TNFSF11 | FALSE | Upregulated |
| 1,678810908 | TNFSF15 | | Upregulated |
| 0 | TNFSF18 | FALSE | Upregulated |
| 0 | TNFSF9 | FALSE | Upregulated |
| 0,731904049 | TNIP3 | FALSE | Upregulated |
| 0,394108604 | TNKS2-AS1 | FALSE | Upregulated |
| 0 | TNNI3 | FALSE | Upregulated |
| | TNNI3K | | |
| 0,498015061 | TNNT2 | FALSE | Upregulated |
| 0 | TNP1 | FALSE | Upregulated |
| 0 | TNP2 | FALSE | Upregulated |
| 0,124815187 | TNR | FALSE | Upregulated |
| 0,704856716 | TOB2P1 | FALSE | Upregulated |
| 0,083498729 | TOLLIP-AS1 | FALSE | Upregulated |
| 0,564170358 | TOMM20L | FALSE | Upregulated |
| | TOP1P2 | | |

| | | | |
|---|---|---|---|
| 0 | TOPAZ1 | FALSE | Upregulated |
| -0,177001621 | TP53AIP1 | FALSE | Downregulated |
| | TP53TG3 | | |
| | TP53TG3B | | |
| | TP53TG3C | | |
| | TP53TG3D | | |
| -0,565066903 | TP63 | FALSE | Downregulated |
| 0,976128622 | TPBGL | FALSE | Upregulated |
| 0,016314734 | TPD52L3 | FALSE | Upregulated |
| 0,629113177 | TPH1 | FALSE | Upregulated |
| | TPH2 | | |
| -0,317862642 | TPI1P2 | FALSE | Downregulated |
| -0,468917547 | TPPP3 | FALSE | Downregulated |
| -0,656809376 | TPRKB | FALSE | Downregulated |
| -0,548496422 | TPRX1 | FALSE | Downregulated |
| -0,002529603 | TPRXL | FALSE | Downregulated |
| 0,559847728 | TPSD1 | FALSE | Upregulated |
| | TPTE | | |
| | TPTE2 | | |
| | TPTE2P2 | | |
| 0,295489189 | TPTE2P3 | FALSE | Upregulated |
| -0,105554203 | TPTE2P6 | FALSE | Downregulated |
| -0,316717893 | TPTEP1 | FALSE | Downregulated |
| -0,333300565 | TRAIP | FALSE | Downregulated |
| 0 | TRAM1L1 | FALSE | Upregulated |
| 0,494663129 | TRAPPC2P1 | FALSE | Upregulated |
| 0,49383299 | TRAPPC3L | FALSE | Upregulated |
| 1,233033613 | TRAT1 | | Upregulated |
| -1,059023005 | TRDN | | Downregulated |
| 0,000638179 | TREM1 | FALSE | Upregulated |
| 0,186242855 | TREM2 | FALSE | Upregulated |
| 0,180922974 | TREML2 | FALSE | Upregulated |
| 0 | TREML3P | FALSE | Upregulated |
| 0,039448723 | TREML4 | FALSE | Upregulated |
| | TREML5P | | |
| 0,006751017 | TRERNA1 | FALSE | Upregulated |
| -0,183110617 | TREX2 | FALSE | Downregulated |
| 0,777208104 | TRG-AS1 | FALSE | Upregulated |
| -0,339587572 | TRH | FALSE | Downregulated |
| 1,463671772 | TRIM31 | | Upregulated |
| 1,027295697 | TRIM36 | | Upregulated |
| | TRIM40 | | |
| -0,157342736 | TRIM42 | FALSE | Downregulated |
| | TRIM43 | | |
| | TRIM43B | | |
| | TRIM48 | | |
| | TRIM49 | | |
| | TRIM49B | | |
| | TRIM49C | | |
| | TRIM49D1 | | |
| | TRIM49D2P | | |

| | | | |
|---|---|---|---|
| -0,745179425 | TRIM50 | FALSE | Downregulated |
| | TRIM51 | | |
| | TRIM51EP | | |
| | TRIM51HP | | |
| | TRIM53AP | | |
| 0,507484123 | TRIM54 | FALSE | Upregulated |
| -1,946880076 | TRIM58 | | Downregulated |
| | TRIM60 | | |
| 0,727957979 | TRIM61 | FALSE | Upregulated |
| 0 | TRIM63 | FALSE | Upregulated |
| | TRIM64 | | |
| | TRIM64B | | |
| | TRIM64C | | |
| | TRIM71 | | |
| 0,988197267 | TRIM72 | FALSE | Upregulated |
| | TRIM77 | | |
| | TRIML1 | | |
| | TRIML2 | | |
| 0,003093665 | TRIP13 | FALSE | Upregulated |
| | TRNA | | |
| | TRNAA-AGC | | |
| | TRNAA-AGC-2 | | |
| | TRNAA-AGC-3 | | |
| | TRNAA1 | | |
| | TRNAA10 | | |
| | TRNAA11 | | |
| | TRNAA12 | | |
| | TRNAA13 | | |
| | TRNAA14 | | |
| | TRNAA15 | | |
| | TRNAA17 | | |
| | TRNAA18 | | |
| | TRNAA19 | | |
| | TRNAA2 | | |
| | TRNAA20 | | |
| | TRNAA20-2 | | |
| | TRNAA20-3 | | |
| | TRNAA23 | | |
| | TRNAA26 | | |
| | TRNAA27 | | |
| | TRNAA28 | | |
| | TRNAA29 | | |
| | TRNAA3 | | |
| | TRNAA30 | | |
| | TRNAA32 | | |
| | TRNAA34 | | |
| | TRNAA36 | | |
| | TRNAA37 | | |
| | TRNAA37-2 | | |
| | TRNAA38 | | |
| | TRNAA39 | | |

TRNAA4
TRNAA40
TRNAA41
TRNAA41-2
TRNAA5
TRNAA6
TRNAA7
TRNAA8
TRNAA9
TRNAC-GCA
TRNAC10
TRNAC11
TRNAC12
TRNAC13
TRNAC14
TRNAC15
TRNAC16
TRNAC18
TRNAC19
TRNAC2
TRNAC20
TRNAC21
TRNAC22
TRNAC23
TRNAC24
TRNAC25
TRNAC26
TRNAC27
TRNAC28
TRNAC29
TRNAC3
TRNAC30
TRNAC4
TRNAC5
TRNAC6
TRNAC7
TRNAC8
TRNAC9
TRNAD10
TRNAD11
TRNAD12
TRNAD13
TRNAD14
TRNAD15
TRNAD16
TRNAD17
TRNAD18
TRNAD5
TRNAD6
TRNAD8
TRNAD9

TRNAE-UUC
TRNAE-UUC-2
TRNAE1
TRNAE11
TRNAE13
TRNAE18
TRNAE19
TRNAE20
TRNAE21
TRNAE22
TRNAE23
TRNAE24
TRNAE26
TRNAE5
TRNAE6
TRNAE7
TRNAF1
TRNAF10
TRNAF11
TRNAF2
TRNAF3
TRNAF4
TRNAF5
TRNAF6
TRNAF7
TRNAF8
TRNAF9
TRNAG-CCC
TRNAG-CCC-2
TRNAG1
TRNAG10
TRNAG11
TRNAG13
TRNAG15
TRNAG16
TRNAG17
TRNAG18
TRNAG19
TRNAG2
TRNAG20
TRNAG21
TRNAG22
TRNAG23
TRNAG24
TRNAG25
TRNAG26
TRNAG27
TRNAG28
TRNAG29
TRNAG3
TRNAG30

TRNAG31
TRNAG4
TRNAG5
TRNAG6
TRNAG7
TRNAG8
TRNAG9
TRNAH1
TRNAH10
TRNAH2
TRNAH3
TRNAH4
TRNAH5
TRNAH6
TRNAH7
TRNAH9
TRNAI-AAU
TRNAI1
TRNAI11
TRNAI12
TRNAI13
TRNAI15
TRNAI17
TRNAI18
TRNAI2
TRNAI20
TRNAI21
TRNAI22
TRNAI23
TRNAI24
TRNAI25
TRNAI25-2
TRNAI26
TRNAI3
TRNAI4
TRNAI5
TRNAI6
TRNAI7
TRNAI9
TRNAK-CUU
TRNAK1
TRNAK11
TRNAK13
TRNAK14
TRNAK15
TRNAK16
TRNAK17
TRNAK19
TRNAK2
TRNAK20
TRNAK21

TRNAK22
TRNAK23
TRNAK26
TRNAK27
TRNAK29
TRNAK3
TRNAK31
TRNAK33
TRNAK34
TRNAK4
TRNAK6
TRNAK8
TRNAK9
TRNAL-CAA
TRNAL1
TRNAL10
TRNAL11
TRNAL12
TRNAL13
TRNAL14
TRNAL15
TRNAL16
TRNAL17
TRNAL19
TRNAL2
TRNAL20
TRNAL22
TRNAL23
TRNAL24
TRNAL26
TRNAL27
TRNAL28
TRNAL29
TRNAL3
TRNAL30
TRNAL32
TRNAL34
TRNAL35
TRNAL36
TRNAL38
TRNAL4
TRNAL5
TRNAL6
TRNAL7
TRNAL8
TRNAM-CAU
TRNAM1
TRNAM11
TRNAM12
TRNAM13
TRNAM14

TRNAM15
TRNAM16
TRNAM16-2
TRNAM17
TRNAM18
TRNAM19
TRNAM2
TRNAM20
TRNAM3
TRNAM4
TRNAM5
TRNAM6
TRNAM8
TRNAM9
TRNAN-GUU
TRNAN-GUU-2
TRNAN-GUU-3
TRNAN-GUU-4
TRNAN-GUU-5
TRNAN-GUU-6
TRNAN-GUU-7
TRNAN1
TRNAN11
TRNAN14
TRNAN15
TRNAN17
TRNAN18
TRNAN19
TRNAN2
TRNAN20
TRNAN22
TRNAN22-2
TRNAN27
TRNAN29
TRNAN3
TRNAN32
TRNAN5
TRNAN5-2
TRNAN7
TRNAN8
TRNAP1
TRNAP10
TRNAP11
TRNAP12
TRNAP13
TRNAP14
TRNAP15
TRNAP16
TRNAP17
TRNAP18
TRNAP19

TRNAP2
TRNAP21
TRNAP3
TRNAP4
TRNAP5
TRNAP6
TRNAP7
TRNAP8
TRNAP9
TRNAQ-CUG
TRNAQ1
TRNAQ10
TRNAQ11
TRNAQ12
TRNAQ13
TRNAQ15
TRNAQ16
TRNAQ17
TRNAQ2
TRNAQ20
TRNAQ21
TRNAQ22
TRNAQ24
TRNAQ25
TRNAQ27
TRNAQ3
TRNAQ32
TRNAQ8
TRNAR1
TRNAR10
TRNAR11
TRNAR12
TRNAR14
TRNAR15
TRNAR16
TRNAR17
TRNAR18
TRNAR19
TRNAR2
TRNAR20
TRNAR21
TRNAR22
TRNAR23
TRNAR24
TRNAR25
TRNAR26
TRNAR27
TRNAR28
TRNAR29
TRNAR3
TRNAR4

TRNAR5
TRNAR6
TRNAR7
TRNAR8
TRNAR9
TRNAS1
TRNAS10
TRNAS11
TRNAS12
TRNAS13
TRNAS14
TRNAS15
TRNAS16
TRNAS17
TRNAS18
TRNAS19
TRNAS2
TRNAS20
TRNAS21
TRNAS22
TRNAS23
TRNAS24
TRNAS25
TRNAS27
TRNAS28
TRNAS3
TRNAS4
TRNAS5
TRNAS6
TRNAS7
TRNAS9
TRNASUP1
TRNAT1
TRNAT10
TRNAT12
TRNAT13
TRNAT14
TRNAT15
TRNAT16
TRNAT17
TRNAT18
TRNAT19
TRNAT2
TRNAT20
TRNAT21
TRNAT22
TRNAT4
TRNAT5
TRNAT6
TRNAT7
TRNAT8

| | | | |
|---|---|---|---|
| | TRNAT9 | | |
| 0 | TRNAU1 | FALSE | Upregulated |
| | TRNAU2 | | |
| | TRNAV-CAC | | |
| | TRNAV-CAC-2 | | |
| | TRNAV-CAC-3 | | |
| | TRNAV1 | | |
| | TRNAV10 | | |
| | TRNAV11 | | |
| | TRNAV12 | | |
| | TRNAV15 | | |
| | TRNAV16 | | |
| | TRNAV17 | | |
| | TRNAV18 | | |
| | TRNAV19 | | |
| | TRNAV2 | | |
| | TRNAV20 | | |
| | TRNAV21 | | |
| | TRNAV22 | | |
| | TRNAV23 | | |
| | TRNAV24 | | |
| | TRNAV25 | | |
| | TRNAV26 | | |
| | TRNAV27 | | |
| | TRNAV28 | | |
| | TRNAV29 | | |
| | TRNAV3 | | |
| | TRNAV31 | | |
| | TRNAV32 | | |
| | TRNAV4 | | |
| | TRNAV5 | | |
| | TRNAV6 | | |
| | TRNAV7 | | |
| | TRNAV8 | | |
| | TRNAV9 | | |
| | TRNAW2 | | |
| | TRNAW3 | | |
| | TRNAW4 | | |
| | TRNAW5 | | |
| | TRNAW6 | | |
| | TRNAW7 | | |
| | TRNAW9 | | |
| | TRNAY1 | | |
| | TRNAY10 | | |
| | TRNAY12 | | |
| | TRNAY13 | | |
| | TRNAY14 | | |
| | TRNAY15 | | |
| | TRNAY2 | | |
| | TRNAY3 | | |
| | TRNAY4 | | |

| | | | |
|---|---|---|---|
| | TRNAY6 | | |
| | TRNAY7 | | |
| | TRNAY8 | | |
| | TRNAY9 | | |
| | TRNC | | |
| | TRND | | |
| | TRNE | | |
| | TRNF | | |
| | TRNG | | |
| | TRNH | | |
| | TRNI | | |
| | TRNK | | |
| | TRNL1 | | |
| | TRNL2 | | |
| | TRNM | | |
| | TRNN | | |
| | TRNP | | |
| | TRNQ | | |
| | TRNR | | |
| | TRNS1 | | |
| | TRNS2 | | |
| | TRNT | | |
| | TRNV | | |
| | TRNW | | |
| | TRNY | | |
| 0,276915715 | TROAP | FALSE | Upregulated |
| -0,155850459 | TRPC2 | FALSE | Downregulated |
| 0,159548771 | TRPC3 | FALSE | Upregulated |
| 0,437510419 | TRPC4 | FALSE | Upregulated |
| 1,106463228 | TRPC6 | | Upregulated |
| 2,65054779 | TRPC7 | | Upregulated |
| -0,474906351 | TRPM6 | FALSE | Downregulated |
| 0,076658198 | TRPV5 | FALSE | Upregulated |
| 0 | TRY2P | FALSE | Upregulated |
| | TSG1 | | |
| -0,550053744 | TSGA10 | FALSE | Downregulated |
| -0,103701885 | TSGA10IP | FALSE | Downregulated |
| 1,545893903 | TSGA13 | | Upregulated |
| -0,081332936 | TSHB | FALSE | Downregulated |
| 0,105698595 | TSKS | FALSE | Upregulated |
| 0,161648244 | TSPAN10 | FALSE | Upregulated |
| 0,301430304 | TSPAN16 | FALSE | Upregulated |
| | TSPAN19 | | |
| 0,053353466 | TSPAN2 | FALSE | Upregulated |
| | TSPY1 | | |
| | TSPY10 | | |
| | TSPY2 | | |
| -0,054509218 | TSPY26P | FALSE | Downregulated |
| | TSPY3 | | |
| | TSPY4 | | |
| | TSPY8 | | |

| | | | |
|---|---|---|---|
| 0,688744583 | TTC23L | FALSE | Upregulated |
| -0,151758888 | TTC24 | FALSE | Downregulated |
| 0,63639078 | TTC25 | FALSE | Upregulated |
| 0,898249493 | TTC26 | FALSE | Upregulated |
| | TTC29 | | |
| 0,316819031 | TTC32 | FALSE | Upregulated |
| -1,842487328 | TTC34 | | Downregulated |
| 0,578064289 | TTC40 | FALSE | Upregulated |
| 0,287114043 | TTC9B | FALSE | Upregulated |
| 0,099889329 | TTK | FALSE | Upregulated |
| -0,102350495 | TTLL1 | FALSE | Downregulated |
| 0,866578279 | TTLL10 | FALSE | Upregulated |
| 2,587304398 | TTLL10-AS1 | | Upregulated |
| 0,49017351 | TTLL6 | FALSE | Upregulated |
| | TTLL8 | | |
| | TTTY1 | | |
| -0,075752292 | TTTY10 | FALSE | Downregulated |
| | TTTY11 | | |
| | TTTY12 | | |
| | TTTY16 | | |
| | TTTY18 | | |
| | TTTY19 | | |
| | TTTY1B | | |
| | TTTY2 | | |
| | TTTY20 | | |
| | TTTY21 | | |
| | TTTY21B | | |
| | TTTY22 | | |
| | TTTY23 | | |
| | TTTY23B | | |
| | TTTY2B | | |
| | TTTY7 | | |
| 0 | TTTY7B | FALSE | Upregulated |
| | TTTY8 | | |
| | TTTY8B | | |
| 0,039005732 | TUBA3C | FALSE | Upregulated |
| 2,178474433 | TUBA4B | | Upregulated |
| | TUBAL3 | | |
| 1,323299194 | TUBB3 | | Upregulated |
| 0,020619335 | TUBB4A | FALSE | Upregulated |
| | TUBB8 | | |
| 0,494364921 | TUBBP5 | FALSE | Upregulated |
| 0,296507408 | TULP1 | FALSE | Upregulated |
| -0,56063022 | TULP2 | FALSE | Downregulated |
| 0,306105915 | TUSC5 | FALSE | Upregulated |
| 2,091205889 | TUSC7 | | Upregulated |
| 0,744812274 | TVP23C-CDRT4 | FALSE | Upregulated |
| 0,895900985 | TWIST2 | FALSE | Upregulated |
| 0,376107528 | TXNDC2 | FALSE | Upregulated |
| 1,126505338 | TXNDC8 | | Upregulated |
| 0 | TXNRD3NB | FALSE | Upregulated |

| | | | |
|---|---|---|---|
| 0 | TYR | FALSE | Upregulated |
| 1,760480393 | TYRP1 | | Upregulated |
| -1,573758985 | UBAC2-AS1 | | Downregulated |
| -0,121266479 | UBE2C | FALSE | Downregulated |
| -0,507467157 | UBE2DNL | FALSE | Downregulated |
| 0,192712702 | UBE2E2-AS1 | FALSE | Upregulated |
| 0 | UBE2MP1 | FALSE | Upregulated |
| 0,001493849 | UBE2NL | FALSE | Upregulated |
| 0 | UBE2Q2L | FALSE | Upregulated |
| 0,067066191 | UBE2Q2P2 | FALSE | Upregulated |
| 0,184874676 | UBE2Q2P3 | FALSE | Upregulated |
| 0,688116061 | UBE2T | FALSE | Upregulated |
| | UBE2U | | |
| 1,212181731 | UBE3D | | Upregulated |
| | UBL4B | | |
| -0,135146778 | UBQLN3 | FALSE | Downregulated |
| 1,425968546 | UBQLNL | | Upregulated |
| | UBTFL1 | | |
| 0,478273451 | UCHL1 | FALSE | Upregulated |
| | UCHL1-AS1 | | |
| | UCMA | | |
| 0,372527821 | UCN | FALSE | Upregulated |
| 0,508866919 | UCN2 | FALSE | Upregulated |
| | UCN3 | | |
| -0,233059293 | UCP1 | FALSE | Downregulated |
| | UGT2A1 | | |
| 0 | UGT2B28 | FALSE | Upregulated |
| 0,171713425 | UGT8 | FALSE | Upregulated |
| | ULBP1 | | |
| 0,042597573 | ULBP2 | FALSE | Upregulated |
| -0,757520004 | ULBP3 | FALSE | Downregulated |
| -0,240256984 | ULK4P3 | FALSE | Downregulated |
| | UMOD | | |
| 1,121135437 | UMODL1 | | Upregulated |
| 1,239201011 | UNC13A | | Upregulated |
| -0,127885288 | UNC13C | FALSE | Downregulated |
| 1,158827948 | UNC45B | | Upregulated |
| | UNC5D | | |
| -0,277600176 | UNC79 | FALSE | Downregulated |
| -0,380216075 | UNC80 | FALSE | Downregulated |
| | UNCX | | |
| 0,420090509 | UNQ6494 | FALSE | Upregulated |
| -0,00897377 | UNQ9370 | FALSE | Downregulated |
| -0,064105517 | UOX | FALSE | Downregulated |
| 0 | UPK1A | FALSE | Upregulated |
| -0,72910816 | UPK1A-AS1 | FALSE | Downregulated |
| 0 | UPK2 | FALSE | Upregulated |
| | UPK3A | | |
| 0,25938223 | UQCRHL | FALSE | Upregulated |
| | URAD | | |
| 0,004618298 | URGCP-MRPS24 | FALSE | Upregulated |

| | | | |
|---|---|---|---|
| 2,251910809 | USH1C | | Upregulated |
| 0,532674779 | USH1G | FALSE | Upregulated |
| -0,352165926 | USP12-AS2 | FALSE | Downregulated |
| 0 | USP17L10 | FALSE | Upregulated |
| 0 | USP17L11 | FALSE | Upregulated |
| | USP17L12 | | |
| | USP17L13 | | |
| | USP17L15 | | |
| | USP17L17 | | |
| | USP17L18 | | |
| | USP17L19 | | |
| 0,671381024 | USP17L1P | FALSE | Upregulated |
| 0 | USP17L20 | FALSE | Upregulated |
| | USP17L21 | | |
| 0,106256579 | USP17L22 | FALSE | Upregulated |
| | USP17L24 | | |
| | USP17L25 | | |
| | USP17L26 | | |
| | USP17L27 | | |
| | USP17L28 | | |
| | USP17L29 | | |
| 1,136679359 | USP17L3 | | Upregulated |
| | USP17L30 | | |
| 1,247869623 | USP17L4 | | Upregulated |
| | USP17L5 | | |
| -0,368949072 | USP17L6P | FALSE | Downregulated |
| 1,207085426 | USP17L8 | | Upregulated |
| | USP17L9P | | |
| -0,718375209 | USP2-AS1 | FALSE | Downregulated |
| | USP26 | | |
| 0,121379454 | USP27X | FALSE | Upregulated |
| 1,532526506 | USP27X-AS1 | | Upregulated |
| | USP29 | | |
| 1,843872876 | USP41 | | Upregulated |
| 1,254113035 | USP46-AS1 | | Upregulated |
| | UTF1 | | |
| 0,294697269 | UTS2 | FALSE | Upregulated |
| -0,091398447 | UTS2R | FALSE | Downregulated |
| | VAMP7-2 | | |
| 1,587374225 | VANGL2 | | Upregulated |
| -1,445794907 | VAT1L | | Downregulated |
| | VAV3-AS1 | | |
| | VAX1 | | |
| 0,642299941 | VAX2 | FALSE | Upregulated |
| -1,018648346 | VCX | | Downregulated |
| 0,696437607 | VCX3A | FALSE | Upregulated |
| -0,570665097 | VCX3B | FALSE | Downregulated |
| | VCY | | |
| -0,149328179 | VCY1B | FALSE | Downregulated |
| 2,296334236 | VENTX | | Upregulated |
| | VENTXP1 | | |

| | | | |
|---|---|---|---|
| 0,047703368 | VENTXP7 | FALSE | Upregulated |
| 0,290821866 | VGF | FALSE | Upregulated |
| 0,087836139 | VGLL1 | FALSE | Upregulated |
| -0,068402078 | VGLL2 | FALSE | Downregulated |
| | VHLL | | |
| -0,949093726 | VIP | FALSE | Downregulated |
| 0 | VIT | FALSE | Upregulated |
| 0,156387072 | VLDLR-AS1 | FALSE | Upregulated |
| 0 | VN1R10P | FALSE | Upregulated |
| 0 | VN1R2 | FALSE | Upregulated |
| | VN1R4 | | |
| 0,548021115 | VN1R5 | FALSE | Upregulated |
| -1,677898958 | VPREB3 | | Downregulated |
| | VRTN | | |
| 0,001249901 | VSIG1 | FALSE | Upregulated |
| 3,17517464 | VSIG2 | | Upregulated |
| 1,946084845 | VSIG8 | | Upregulated |
| | VSTM1 | | |
| | VSTM2A | | |
| | VSTM2B | | |
| 0,05331589 | VSTM5 | FALSE | Upregulated |
| | VSX1 | | |
| -0,04850409 | VSX2 | FALSE | Downregulated |
| 1,599083491 | VTCN1 | | Upregulated |
| | VTRNA1-1 | | |
| | VTRNA1-2 | | |
| | VTRNA1-3 | | |
| | VTRNA2-1 | | |
| 0,951659166 | VWA3A | FALSE | Upregulated |
| 0,267054292 | VWA3B | FALSE | Upregulated |
| 1,346387219 | VWA5B1 | | Upregulated |
| 0,62910548 | VWC2 | FALSE | Upregulated |
| -0,376641928 | VWC2L | FALSE | Downregulated |
| | VWC2L-IT1 | | |
| -0,597054701 | VWDE | FALSE | Downregulated |
| 1,816733417 | WASIR2 | | Upregulated |
| 0,642298481 | WBP11P1 | FALSE | Upregulated |
| 0,898617532 | WBP2NL | FALSE | Upregulated |
| -0,72941465 | WBP5 | FALSE | Downregulated |
| 2,221839892 | WBSCR17 | | Upregulated |
| 0,016283667 | WBSCR28 | FALSE | Upregulated |
| 0,81912958 | WDFY3-AS2 | FALSE | Upregulated |
| -0,188384929 | WDR17 | FALSE | Downregulated |
| | WDR49 | | |
| -0,893419362 | WDR52-AS1 | FALSE | Downregulated |
| -0,183709961 | WDR63 | FALSE | Downregulated |
| -0,805557898 | WDR64 | FALSE | Downregulated |
| -0,264782616 | WDR86-AS1 | FALSE | Downregulated |
| 0,571667097 | WDR87 | FALSE | Upregulated |
| 0,793466517 | WDR96 | FALSE | Upregulated |
| -0,481170653 | WEE2-AS1 | FALSE | Downregulated |

| | | | |
|---|---|---|---|
| | WFDC10A | | |
| | WFDC10B | | |
| | WFDC11 | | |
| 0,06906799 | WFDC12 | FALSE | Upregulated |
| 0,790325166 | WFDC13 | FALSE | Upregulated |
| 0,52232555 | WFDC2 | FALSE | Upregulated |
| | WFDC3 | | |
| 0 | WFDC5 | FALSE | Upregulated |
| | WFDC6 | | |
| | WFDC8 | | |
| | WFDC9 | | |
| 0,418925458 | WFIKKN2 | FALSE | Upregulated |
| | WI2-2373I1.2 | | |
| 0,178297376 | WI2-81516E3.1 | FALSE | Upregulated |
| | WIF1 | | |
| 3,102344971 | WNT1 | | Upregulated |
| 2,374983741 | WNT10A | | Upregulated |
| 1,169205012 | WNT10B | | Upregulated |
| -0,22052917 | WNT2 | FALSE | Downregulated |
| -0,26100752 | WNT3A | FALSE | Downregulated |
| 0,383412094 | WNT6 | FALSE | Upregulated |
| 0 | WNT7A | FALSE | Upregulated |
| 1,85015824 | WNT7B | | Upregulated |
| 0 | WNT8A | FALSE | Upregulated |
| -0,080696683 | WNT9A | FALSE | Downregulated |
| 0 | WNT9B | FALSE | Upregulated |
| 0 | WSCD2 | FALSE | Upregulated |
| 0,824175048 | WT1 | FALSE | Upregulated |
| 1,348012769 | WT1-AS | | Upregulated |
| 0 | WTAPP1 | FALSE | Upregulated |
| 0 | WTH3DI | FALSE | Upregulated |
| 1,671252015 | WWTR1-AS1 | | Upregulated |
| | XAGE-4 | | |
| | XAGE1B | | |
| | XAGE1E | | |
| | XAGE2 | | |
| 0 | XAGE3 | FALSE | Upregulated |
| | XAGE5 | | |
| 1,578047759 | XCL1 | | Upregulated |
| -0,605525658 | XCL2 | FALSE | Downregulated |
| -0,546698165 | XCR1 | FALSE | Downregulated |
| | XGPY2 | | |
| | XIRP2-AS1 | | |
| 0,530656861 | XK | FALSE | Upregulated |
| | XKR3 | | |
| 0,664513546 | XKR4 | FALSE | Upregulated |
| | XKR5 | | |
| 0,047960156 | XKR7 | FALSE | Upregulated |
| 0,046589889 | XKRX | FALSE | Upregulated |
| 0,792449024 | XRCC2 | FALSE | Upregulated |
| 0,642921431 | XRCC4 | FALSE | Upregulated |

| | | | |
|---|---|---|---|
| 0,079370206 | XRCC6P5 | FALSE | Upregulated |
| | XXYLT1-AS1 | | |
| 0 | YBX3P1 | FALSE | Upregulated |
| | YIPF7 | | |
| 0,669025015 | YPLR6490 | FALSE | Upregulated |
| 0,752167963 | YTHDF3-AS1 | FALSE | Upregulated |
| 0 | YWHAEP1 | FALSE | Upregulated |
| 0,252917043 | YWHAEP7 | FALSE | Upregulated |
| | YY1P2 | | |
| -0,120965521 | ZAN | FALSE | Downregulated |
| 1,878992007 | ZAR1 | | Upregulated |
| -0,087413229 | ZAR1L | FALSE | Downregulated |
| | ZBBX | | |
| | ZBED1-2 | | |
| 0,619585613 | ZBED3-AS1 | FALSE | Upregulated |
| | ZBTB8B | | |
| | ZC2HC1B | | |
| -0,645410692 | ZC3HAV1L | FALSE | Downregulated |
| -0,096180009 | ZCCHC12 | FALSE | Downregulated |
| | ZCCHC13 | | |
| 0,265102048 | ZCCHC16 | FALSE | Upregulated |
| | ZCCHC5 | | |
| 1,458970641 | ZCWPW2 | | Upregulated |
| 1,67398535 | ZDHHC13 | | Upregulated |
| 0,167068198 | ZDHHC19 | FALSE | Upregulated |
| -0,312495561 | ZDHHC22 | FALSE | Downregulated |
| -0,288478435 | ZDHHC8P1 | FALSE | Downregulated |
| 0,660627616 | ZEB1-AS1 | FALSE | Upregulated |
| -1,261256686 | ZFP37 | | Downregulated |
| | ZFP42 | | |
| 0 | ZFP57 | FALSE | Upregulated |
| 0,489574927 | ZFP69B | FALSE | Upregulated |
| -0,720996982 | ZFP82 | FALSE | Downregulated |
| -0,01767359 | ZFR2 | FALSE | Downregulated |
| 0,000577308 | ZFX-AS1 | FALSE | Upregulated |
| -0,25635922 | ZG16B | FALSE | Downregulated |
| -1,149201576 | ZHX1-C8ORF76 | | Downregulated |
| | ZIC2 | | |
| | ZIC3 | | |
| 0 | ZIC5 | FALSE | Upregulated |
| 0,669756935 | ZIM3 | FALSE | Upregulated |
| 0,669756935 | ZMAT4 | FALSE | Upregulated |
| -0,606125221 | ZMIZ1-AS1 | FALSE | Downregulated |
| -1,089327729 | ZMYND12 | | Downregulated |
| 0,277733813 | ZNF114 | FALSE | Upregulated |
| 0,51005035 | ZNF157 | FALSE | Upregulated |
| -0,339259611 | ZNF192P1 | FALSE | Downregulated |
| -1,086243682 | ZNF204P | | Downregulated |
| 0,118202939 | ZNF208 | FALSE | Upregulated |
| 0,918692025 | ZNF222 | FALSE | Upregulated |
| 0,600213407 | ZNF280B | FALSE | Upregulated |

| | | | |
|---|---|---|---|
| 0,286746711 | ZNF285 | FALSE | Upregulated |
| 0,191186654 | ZNF286B | FALSE | Upregulated |
| 0 | ZNF295-AS1 | FALSE | Upregulated |
| 0,883859898 | ZNF300P1 | FALSE | Upregulated |
| | ZNF32-AS3 | | |
| 1,480838334 | ZNF321P | | Upregulated |
| -0,996146314 | ZNF33BP1 | FALSE | Downregulated |
| -1,027407686 | ZNF365 | | Downregulated |
| -0,440265117 | ZNF382 | FALSE | Downregulated |
| 0,187518576 | ZNF385D | FALSE | Upregulated |
| 0,119849037 | ZNF454 | FALSE | Upregulated |
| -0,083344562 | ZNF474 | FALSE | Downregulated |
| | ZNF479 | | |
| -0,028438454 | ZNF487 | FALSE | Downregulated |
| 0,311688729 | ZNF488 | FALSE | Upregulated |
| -0,077944283 | ZNF492 | FALSE | Downregulated |
| -1,085235826 | ZNF501 | | Downregulated |
| 0,645411021 | ZNF503-AS1 | FALSE | Upregulated |
| 0,814745763 | ZNF530 | FALSE | Upregulated |
| 0 | ZNF534 | FALSE | Upregulated |
| 0 | ZNF536 | FALSE | Upregulated |
| 1,402505158 | ZNF541 | | Upregulated |
| 0 | ZNF556 | FALSE | Upregulated |
| | ZNF560 | | |
| 0,34368146 | ZNF578 | FALSE | Upregulated |
| 0,516360935 | ZNF610 | FALSE | Upregulated |
| 2,987463112 | ZNF625-ZNF20 | | Upregulated |
| 1,554617187 | ZNF645 | | Upregulated |
| 0 | ZNF663P | FALSE | Upregulated |
| 1,056862033 | ZNF664-FAM101A | | Upregulated |
| -0,75205638 | ZNF665 | FALSE | Downregulated |
| -0,282979222 | ZNF670 | FALSE | Downregulated |
| 0,553418362 | ZNF670-ZNF695 | FALSE | Upregulated |
| -0,072663956 | ZNF674-AS1 | FALSE | Downregulated |
| 0,36173465 | ZNF676 | FALSE | Upregulated |
| 1,515646051 | ZNF677 | | Upregulated |
| | ZNF679 | | |
| 0,985249565 | ZNF681 | FALSE | Upregulated |
| 1,389806183 | ZNF683 | | Upregulated |
| 0,855856009 | ZNF695 | FALSE | Upregulated |
| 0,844472154 | ZNF702P | FALSE | Upregulated |
| | ZNF705A | | |
| | ZNF705B | | |
| | ZNF705D | | |
| -0,603950589 | ZNF705E | FALSE | Downregulated |
| | ZNF705G | | |
| 1,291407805 | ZNF713 | | Upregulated |
| | ZNF716 | | |
| 0,443674023 | ZNF724P | FALSE | Upregulated |
| -0,981445492 | ZNF726 | FALSE | Downregulated |
| 0,477017051 | ZNF727 | FALSE | Upregulated |

| | | | | |
|---|---|---|---|---|
| 0,212604015 | | ZNF728 | FALSE | Upregulated |
| | | ZNF729 | | |
| 0 | | ZNF730 | FALSE | Upregulated |
| | | ZNF733P | | |
| | | ZNF735 | | |
| -2,073531028 | | ZNF774 | | Downregulated |
| 1,268231885 | | ZNF781 | | Upregulated |
| | | ZNF804B | | |
| 0 | | ZNF806 | FALSE | Upregulated |
| 0,59572702 | | ZNF807 | FALSE | Upregulated |
| 1,255452099 | | ZNF812 | | Upregulated |
| 2,206585167 | | ZNF816-ZNF321P | | Upregulated |
| -0,151030449 | | ZNF826P | FALSE | Downregulated |
| -0,008449045 | | ZNF835 | FALSE | Downregulated |
| 1,389161649 | | ZNF85 | | Upregulated |
| -0,461813836 | | ZNF860 | FALSE | Downregulated |
| 0,670070017 | | ZNF883 | FALSE | Upregulated |
| 1,448081224 | | ZNF890P | | Upregulated |
| 0,187585821 | | ZNF90 | FALSE | Upregulated |
| 0,259104992 | | ZNF93 | FALSE | Upregulated |
| | | ZNF98 | | |
| -0,254117225 | | ZNF99 | FALSE | Downregulated |
| | | ZNRF4 | | |
| 0,242292568 | | ZP1 | FALSE | Upregulated |
| | | ZP2 | | |
| -1,231813713 | | ZP3 | | Downregulated |
| | | ZP4 | | |
| | | ZPBP | | |
| | | ZPBP2 | | |
| -0,217466604 | | ZPLD1 | FALSE | Downregulated |
| 1,096722867 | | ZRANB2-AS2 | | Upregulated |
| 0,177416711 | | ZSCAN10 | FALSE | Upregulated |
| -0,134576593 | | ZSCAN23 | FALSE | Downregulated |
| 0 | | ZSCAN4 | FALSE | Upregulated |
| -1,686707598 | | ZSCAN5A | | Downregulated |
| -0,119190583 | | ZSCAN5B | FALSE | Downregulated |
| | | ZSWIM2 | | |

**clinTCMR vs. NHR**

| log2FoldChange | padj | gene_names | significance | direction |
|---|---|---|---|---|
| 4,211163259 | 7,42814E-14 | UBD | TRUE | Upregulated |
| 2,593825535 | 1,23872E-13 | IL32 | TRUE | Upregulated |
| 2,695539432 | 2,06382E-12 | C1orf204 | TRUE | Upregulated |
| -1,196392368 | 2,06382E-12 | ZKSCAN1 | TRUE | Downregulated |
| 1,825184633 | 1,55517E-10 | GLS | TRUE | Upregulated |
| 2,556662501 | 5,33914E-10 | LOC101927848 | TRUE | Upregulated |
| 2,580956698 | 6,23138E-10 | IL21R-AS1 | TRUE | Upregulated |
| 1,889697212 | 2,37913E-09 | AJUBA | TRUE | Upregulated |
| 1,47280772 | 2,37913E-09 | FAM196B | TRUE | Upregulated |
| 1,83420354 | 2,37913E-09 | TAPSAR1 | TRUE | Upregulated |
| -1,257619205 | 9,79791E-09 | ACADSB | TRUE | Downregulated |

| | | | | |
|---|---|---|---|---|
| -1,150905327 | 1,75397E-08 | ALDOB | TRUE | Downregulated |
| -1,639807081 | 3,6314E-08 | BCHE | TRUE | Downregulated |
| 1,07392672 | 4,04472E-08 | FAM168A | TRUE | Upregulated |
| 2,033442374 | 4,13462E-08 | ZBED2 | TRUE | Upregulated |
| 1,656686134 | 6,17481E-08 | LOC101927954 | TRUE | Upregulated |
| 1,892578109 | 6,63595E-08 | SLC4A5 | TRUE | Upregulated |
| -1,48658901 | 1,1027E-07 | HMGCS2 | TRUE | Downregulated |
| 1,345861723 | 1,21449E-07 | LOC100294362 | TRUE | Upregulated |
| 1,708812431 | 1,8617E-07 | STAT1 | TRUE | Upregulated |
| 2,768774993 | 2,0985E-07 | RP11-77C3.3 | TRUE | Upregulated |
| -2,166040916 | 2,32733E-07 | CACNA1H | TRUE | Downregulated |
| -1,04281091 | 2,32733E-07 | HAAO | TRUE | Downregulated |
| 1,077303976 | 2,32733E-07 | MAP7D1 | TRUE | Upregulated |
| 1,247552491 | 2,32733E-07 | TLDC2 | TRUE | Upregulated |
| 2,983211091 | 2,75063E-07 | MKI67 | TRUE | Upregulated |
| 1,795574901 | 3,24799E-07 | FAT1 | TRUE | Upregulated |
| -1,167551617 | 3,24799E-07 | PGLYRP2 | TRUE | Downregulated |
| 1,646890694 | 3,6085E-07 | APOBEC4 | TRUE | Upregulated |
| 1,320120131 | 3,74918E-07 | LIMK2 | TRUE | Upregulated |
| 1,134218849 | 3,76414E-07 | TYMP | TRUE | Upregulated |
| 2,061250767 | 3,83799E-07 | PRRT2 | TRUE | Upregulated |
| 1,153467528 | 5,92997E-07 | NCAPH2 | TRUE | Upregulated |
| -0,694239795 | 1,00528E-06 | HECTD1 | FALSE | Downregulated |
| -0,939254941 | 1,03284E-06 | APOC3 | FALSE | Downregulated |
| 0,847030486 | 1,05524E-06 | RNF213 | FALSE | Upregulated |
| -0,912017463 | 1,08716E-06 | ALAD | FALSE | Downregulated |
| 1,28314347 | 1,3522E-06 | COTL1 | TRUE | Upregulated |
| 4,470220166 | 1,53373E-06 | CXCL10 | TRUE | Upregulated |
| -1,200862984 | 1,66354E-06 | PKLR | TRUE | Downregulated |
| -1,370992971 | 1,66354E-06 | TTR | TRUE | Downregulated |
| 1,687236937 | 1,90788E-06 | IFI30 | TRUE | Upregulated |
| -0,960806396 | 2,09806E-06 | ACAA1 | FALSE | Downregulated |
| 1,894512582 | 2,09806E-06 | IKZF3 | TRUE | Upregulated |
| 1,465791276 | 2,27798E-06 | DBNDD1 | TRUE | Upregulated |
| 1,081195351 | 2,35732E-06 | TAGLN2 | TRUE | Upregulated |
| 1,687261531 | 2,38488E-06 | RP11-401P9.5 | TRUE | Upregulated |
| -0,867310138 | 2,72558E-06 | ABCC9 | FALSE | Downregulated |
| -1,240311096 | 2,72558E-06 | ALB | TRUE | Downregulated |
| 3,475335421 | 2,72558E-06 | CXCL9 | TRUE | Upregulated |
| 1,798139314 | 2,72558E-06 | FRMD5 | TRUE | Upregulated |
| -0,93429724 | 2,72558E-06 | LCAT | FALSE | Downregulated |
| 1,041138957 | 2,72558E-06 | LOC100128653 | TRUE | Upregulated |
| 1,598336358 | 2,72558E-06 | LOC101929732 | TRUE | Upregulated |
| 1,098148325 | 3,10264E-06 | DEXI | TRUE | Upregulated |
| 1,37166706 | 3,57152E-06 | SLC7A6OS | TRUE | Upregulated |
| 1,994178114 | 3,61232E-06 | FSTL3 | TRUE | Upregulated |
| 1,362355521 | 3,61232E-06 | TAP1 | TRUE | Upregulated |
| 2,59741863 | 3,80145E-06 | DTNA | TRUE | Upregulated |
| 1,869664856 | 4,03011E-06 | GPNMB | TRUE | Upregulated |
| 1,281890646 | 4,03011E-06 | VIM-AS1 | TRUE | Upregulated |
| 1,360466126 | 4,07987E-06 | PHOSPHO1 | TRUE | Upregulated |

| | | | | |
|---|---|---|---|---|
| 1,43219369 | 4,17587E-06 | DOCK2 | TRUE | Upregulated |
| 1,86788899 | 4,17587E-06 | RUNX3 | TRUE | Upregulated |
| -0,875829531 | 4,84061E-06 | EPHX2 | FALSE | Downregulated |
| 2,59338267 | 5,3938E-06 | LOC101927085 | TRUE | Upregulated |
| -0,70318239 | 5,9229E-06 | A1CF | FALSE | Downregulated |
| -0,737133407 | 6,63151E-06 | SERPINF2 | FALSE | Downregulated |
| 3,529795132 | 6,74727E-06 | RET | TRUE | Upregulated |
| 0,881459736 | 7,10381E-06 | CD68 | FALSE | Upregulated |
| 1,338216186 | 8,11436E-06 | ZNF333 | TRUE | Upregulated |
| 1,298259031 | 8,81612E-06 | CELF2-AS1 | TRUE | Upregulated |
| 1,489049553 | 9,84843E-06 | MMP25 | TRUE | Upregulated |
| -0,81775379 | 1,04279E-05 | IRS1 | FALSE | Downregulated |
| 1,653382065 | 1,18957E-05 | TAP2 | TRUE | Upregulated |
| -1,108874115 | 1,21152E-05 | ADH6 | TRUE | Downregulated |
| 0,831004353 | 1,25043E-05 | SHB | FALSE | Upregulated |
| 3,403466331 | 1,28277E-05 | HIST1H1B | TRUE | Upregulated |
| 1,371242958 | 1,37415E-05 | RFX5 | TRUE | Upregulated |
| 1,040360252 | 1,48989E-05 | STK10 | TRUE | Upregulated |
| 1,237679069 | 2,12411E-05 | HLA-B | TRUE | Upregulated |
| 1,17702631 | 2,52516E-05 | TMEM120B | TRUE | Upregulated |
| -1,180717827 | 2,69389E-05 | PACSIN3 | TRUE | Downregulated |
| 1,095172415 | 2,78666E-05 | COA1 | TRUE | Upregulated |
| -0,867388935 | 2,78666E-05 | IL6R | FALSE | Downregulated |
| -0,909589232 | 3,0567E-05 | FGFR4 | FALSE | Downregulated |
| 1,710556475 | 3,38325E-05 | HLA-F-AS1 | TRUE | Upregulated |
| -0,661846621 | 3,38325E-05 | RPL32 | FALSE | Downregulated |
| 1,277499154 | 3,39513E-05 | LOC102724536 | TRUE | Upregulated |
| -0,928955719 | 3,59519E-05 | THRB | FALSE | Downregulated |
| 1,659889117 | 3,63816E-05 | COL1A1 | TRUE | Upregulated |
| -0,809255125 | 3,63816E-05 | KHK | FALSE | Downregulated |
| 0,77165698 | 3,63816E-05 | RHCE | FALSE | Upregulated |
| 0,841315016 | 3,75298E-05 | C1orf56 | FALSE | Upregulated |
| 0,854373101 | 4,01484E-05 | PDAP1 | FALSE | Upregulated |
| 1,468284043 | 4,01484E-05 | TMC8 | TRUE | Upregulated |
| 0,733960424 | 4,03114E-05 | NPC1 | FALSE | Upregulated |
| 0,991394097 | 4,11514E-05 | RP6-206I17.2 | FALSE | Upregulated |
| 1,19680843 | 4,23147E-05 | LAPTM5 | TRUE | Upregulated |
| 1,863214827 | 4,47617E-05 | SPOCK2 | TRUE | Upregulated |
| -0,944319665 | 4,54716E-05 | CTSF | FALSE | Downregulated |
| -0,964485329 | 4,65563E-05 | APOM | FALSE | Downregulated |
| -0,89338444 | 4,65563E-05 | SLC6A1 | FALSE | Downregulated |
| 1,575869145 | 4,79352E-05 | C9orf163 | TRUE | Upregulated |
| 1,564463824 | 4,81497E-05 | ENTPD1-AS1 | TRUE | Upregulated |
| 1,274980494 | 4,81497E-05 | RP11-436I24.1 | TRUE | Upregulated |
| 1,161691741 | 5,07783E-05 | TTYH3 | TRUE | Upregulated |
| 2,32888631 | 5,07783E-05 | ZC3H12D | TRUE | Upregulated |
| 0,640199453 | 5,36442E-05 | CRYBG3 | FALSE | Upregulated |
| 2,409362598 | 5,44037E-05 | PRC1-AS1 | TRUE | Upregulated |
| 1,037253013 | 5,44292E-05 | WARS | TRUE | Upregulated |
| -1,032275386 | 5,69807E-05 | CYP2C8 | TRUE | Downregulated |
| 1,760819194 | 5,79033E-05 | GBP5 | TRUE | Upregulated |

| | | | | |
|---|---|---|---|---|
| -1,259832911 | 6,49248E-05 | DGAT2 | TRUE | Downregulated |
| 1,274930906 | 6,49248E-05 | SH3BGRL3 | TRUE | Upregulated |
| 1,099711538 | 6,55049E-05 | PIGF | TRUE | Upregulated |
| 2,276676894 | 6,8724E-05 | FCRL6 | TRUE | Upregulated |
| 1,004848842 | 7,00561E-05 | ABCB4 | TRUE | Upregulated |
| 0,949133291 | 7,00561E-05 | ZYX | FALSE | Upregulated |
| 1,081413927 | 7,18409E-05 | PLEKHO2 | TRUE | Upregulated |
| 1,677321142 | 7,42293E-05 | SMTNL1 | TRUE | Upregulated |
| 0,610638771 | 7,83195E-05 | TMBIM1 | FALSE | Upregulated |
| -0,7044749 | 7,95026E-05 | ACACB | FALSE | Downregulated |
| 0,998593811 | 8,16031E-05 | CD74 | FALSE | Upregulated |
| 1,029435933 | 8,16031E-05 | TMSB10 | TRUE | Upregulated |
| 2,250925777 | 8,50063E-05 | C11orf45 | TRUE | Upregulated |
| 3,426893933 | 8,5464E-05 | SAA2 | TRUE | Upregulated |
| 1,216746252 | 8,55633E-05 | LOC642441 | TRUE | Upregulated |
| 1,139653354 | 8,66501E-05 | LCP1 | TRUE | Upregulated |
| -1,049162973 | 8,70004E-05 | PLEKHA7 | TRUE | Downregulated |
| 0,95866126 | 8,9567E-05 | C19orf71 | FALSE | Upregulated |
| 1,634803355 | 8,99128E-05 | TCTEX1D4 | TRUE | Upregulated |
| 1,161436642 | 9,21549E-05 | HLA-A | TRUE | Upregulated |
| 0,943451463 | 9,21549E-05 | LOC102724814 | FALSE | Upregulated |
| -0,819087255 | 9,23566E-05 | IVD | FALSE | Downregulated |
| 1,197085547 | 9,80335E-05 | CERKL | TRUE | Upregulated |
| 3,234963513 | 9,87096E-05 | CDCP1 | TRUE | Upregulated |
| 0,903297865 | 9,87096E-05 | MXD3 | FALSE | Upregulated |
| -0,824311944 | 9,87096E-05 | SLC27A5 | FALSE | Downregulated |
| 0,657894046 | 9,87096E-05 | VASN | FALSE | Upregulated |
| 1,312784793 | 9,99542E-05 | LSP1 | TRUE | Upregulated |
| -0,571051698 | 0,000101841 | TJP2 | FALSE | Downregulated |
| -0,85230092 | 0,000102945 | SLC4A4 | FALSE | Downregulated |
| 2,143246018 | 0,000105077 | SLAMF7 | TRUE | Upregulated |
| 1,859516006 | 0,000108245 | ITGB2-AS1 | TRUE | Upregulated |
| 3,200204983 | 0,000108245 | SAA1 | TRUE | Upregulated |
| -2,296974485 | 0,000108245 | SLCO4C1 | TRUE | Downregulated |
| 1,329086479 | 0,00010843 | LGALS9 | TRUE | Upregulated |
| 1,4020374 | 0,000117522 | GBP1 | TRUE | Upregulated |
| 1,7269457 | 0,000117868 | AVPR2 | TRUE | Upregulated |
| 1,033763869 | 0,000121256 | SSBP3-AS1 | TRUE | Upregulated |
| -0,743593303 | 0,000121832 | AR | FALSE | Downregulated |
| -0,736862207 | 0,000124846 | APOA1 | FALSE | Downregulated |
| -0,766382042 | 0,00012757 | SLC7A2 | FALSE | Downregulated |
| -0,96398885 | 0,000128212 | SLC22A1 | FALSE | Downregulated |
| 1,811273566 | 0,000137157 | BIRC3 | TRUE | Upregulated |
| 1,183561318 | 0,000137157 | MIR22HG | TRUE | Upregulated |
| -0,942030638 | 0,000144005 | CYP4A11 | FALSE | Downregulated |
| 0,988378655 | 0,000145507 | CENPO | FALSE | Upregulated |
| 2,453994971 | 0,000152193 | LOC101927256 | TRUE | Upregulated |
| -2,243347694 | 0,000158058 | GJB5 | TRUE | Downregulated |
| 1,103710459 | 0,000160265 | C1QC | TRUE | Upregulated |
| 0,999328784 | 0,000160615 | THNSL1 | FALSE | Upregulated |
| 1,667802473 | 0,000161581 | LOC101928152 | TRUE | Upregulated |

| | | | | |
|---|---|---|---|---|
| 2,79201721 | 0,000163693 | CD8A | TRUE | Upregulated |
| -0,720234461 | 0,000170265 | CYP27A1 | FALSE | Downregulated |
| 1,360098363 | 0,000173867 | TDRD6 | TRUE | Upregulated |
| 1,025712934 | 0,000179012 | TUBB1 | TRUE | Upregulated |
| 0,857541094 | 0,000181489 | RP5-1142A6.2 | FALSE | Upregulated |
| 1,32575445 | 0,000182623 | LOC102724153 | TRUE | Upregulated |
| 1,262455722 | 0,000186118 | JUN | TRUE | Upregulated |
| 1,514341147 | 0,000186118 | LOC101928512 | TRUE | Upregulated |
| 0,835496555 | 0,000186369 | LSM4 | FALSE | Upregulated |
| 1,73753936 | 0,000186369 | PRNCR1 | TRUE | Upregulated |
| -0,736727864 | 0,000190566 | PBLD | FALSE | Downregulated |
| 1,665802981 | 0,000192519 | PIN4P1 | TRUE | Upregulated |
| 1,554454144 | 0,000192519 | PSD | TRUE | Upregulated |
| 0,72921892 | 0,000192519 | PYGB | FALSE | Upregulated |
| 2,20013825 | 0,000198198 | BCL11B | TRUE | Upregulated |
| 1,115398087 | 0,000205929 | FMNL1 | TRUE | Upregulated |
| 1,149113902 | 0,000205929 | ITGB2 | TRUE | Upregulated |
| 0,814732337 | 0,000205929 | PYROXD1 | FALSE | Upregulated |
| 1,069596859 | 0,000205929 | TRIM17 | TRUE | Upregulated |
| -0,732054402 | 0,000208834 | GOT2 | FALSE | Downregulated |
| -0,581714856 | 0,000213789 | AKAP9 | FALSE | Downregulated |
| 1,805641029 | 0,000220814 | FOXP3 | TRUE | Upregulated |
| -1,420365827 | 0,000231881 | CYP2D7P | TRUE | Downregulated |
| -0,775793656 | 0,000233039 | LRP6 | FALSE | Downregulated |
| -0,675696529 | 0,000237366 | ALDH5A1 | FALSE | Downregulated |
| 1,093099189 | 0,000237366 | NFKB2 | TRUE | Upregulated |
| 0,69597927 | 0,000237366 | USP24 | FALSE | Upregulated |
| 2,02528301 | 0,000237555 | MARCH3 | TRUE | Upregulated |
| 3,630913696 | 0,000240287 | AC145110.1 | TRUE | Upregulated |
| -0,927402803 | 0,000240287 | ADAMTSL3 | FALSE | Downregulated |
| 1,191400165 | 0,000240287 | PAGR1 | TRUE | Upregulated |
| 2,326893152 | 0,000245863 | IL2RG | TRUE | Upregulated |
| 3,863254029 | 0,000248371 | RP11-93B14.5 | TRUE | Upregulated |
| -1,060606537 | 0,000260271 | GLYAT | TRUE | Downregulated |
| 1,075874383 | 0,000264718 | INPP5D | TRUE | Upregulated |
| 0,803559012 | 0,000277954 | ACTR3 | FALSE | Upregulated |
| -1,361445759 | 0,000286933 | G6PC | TRUE | Downregulated |
| -1,23162965 | 0,000287766 | PPP1R1A | TRUE | Downregulated |
| -0,486240057 | 0,000287766 | SRRM2 | FALSE | Downregulated |
| 1,121982751 | 0,000289922 | NLRC5 | TRUE | Upregulated |
| 0,731574434 | 0,000289922 | RP11-374P20.4 | FALSE | Upregulated |
| -1,036756838 | 0,00029432 | MDN1 | TRUE | Downregulated |
| 0,892424393 | 0,000294519 | ARHGDIB | FALSE | Upregulated |
| 1,933867271 | 0,000295181 | CD6 | TRUE | Upregulated |
| 1,868821198 | 0,000296462 | GAS6-AS1 | TRUE | Upregulated |
| 0,937625361 | 0,000296462 | SAMHD1 | FALSE | Upregulated |
| -0,565171978 | 0,000325216 | RPL31 | FALSE | Downregulated |
| 2,078220319 | 0,000325216 | VCAN | TRUE | Upregulated |
| -0,566035299 | 0,000338429 | F2 | FALSE | Downregulated |
| 1,309901572 | 0,000338429 | FAM115C | TRUE | Upregulated |
| 0,789698703 | 0,000338429 | MSN | FALSE | Upregulated |

| | | | | |
|---|---|---|---|---|
| 0,886534631 | 0,000338429 | ZNF433 | FALSE | Upregulated |
| -1,159897349 | 0,000355682 | GBA3 | TRUE | Downregulated |
| -0,853313415 | 0,000355682 | KDR | FALSE | Downregulated |
| -0,674341212 | 0,000356518 | C16orf58 | FALSE | Downregulated |
| 1,23416044 | 0,000360267 | KCNAB2 | TRUE | Upregulated |
| 1,518270876 | 0,000370046 | KCNJ4 | TRUE | Upregulated |
| -1,092005989 | 0,000373433 | CMBL | TRUE | Downregulated |
| 1,552004638 | 0,000373433 | UHRF1 | TRUE | Upregulated |
| -1,135447303 | 0,000375959 | AKR1D1 | TRUE | Downregulated |
| -0,77354396 | 0,000393603 | C2orf72 | FALSE | Downregulated |
| -0,939640244 | 0,000393623 | HAO2 | FALSE | Downregulated |
| 1,200612667 | 0,00039367 | ICAM1 | TRUE | Upregulated |
| 0,731113409 | 0,00039367 | LOC100128818 | FALSE | Upregulated |
| 1,783110084 | 0,000395017 | MUC12 | TRUE | Upregulated |
| -0,640372658 | 0,00040655 | CPSF1 | FALSE | Downregulated |
| 2,890038111 | 0,00040655 | RRM2 | TRUE | Upregulated |
| -0,810684626 | 0,000410318 | SPTBN2 | FALSE | Downregulated |
| -0,820190684 | 0,000412119 | CES1 | FALSE | Downregulated |
| -0,682587474 | 0,000412119 | ITIH1 | FALSE | Downregulated |
| -0,618482914 | 0,000412119 | MPST | FALSE | Downregulated |
| 1,442896564 | 0,000412119 | VAMP5 | TRUE | Upregulated |
| -0,885486507 | 0,000422864 | SORD | FALSE | Downregulated |
| 0,873681165 | 0,000424276 | IL7 | FALSE | Upregulated |
| 0,767945961 | 0,000438401 | URAHP | FALSE | Upregulated |
| 2,27642192 | 0,000440704 | GPR132 | TRUE | Upregulated |
| 0,935076934 | 0,00045272 | CTSS | FALSE | Upregulated |
| 0,688628612 | 0,00045272 | NUTM2G | FALSE | Upregulated |
| 1,096658046 | 0,000456285 | CCND1 | TRUE | Upregulated |
| 0,913116147 | 0,000464021 | LOC441454 | FALSE | Upregulated |
| 0,676032178 | 0,000489034 | CHKA | FALSE | Upregulated |
| 1,744500822 | 0,000491153 | PSMB9 | TRUE | Upregulated |
| -1,74672337 | 0,000511536 | LOC100507616 | TRUE | Downregulated |
| -1,512326567 | 0,000511536 | RCAN3 | TRUE | Downregulated |
| 0,739940886 | 0,000511536 | STAT5A | FALSE | Upregulated |
| 0,825369274 | 0,000511536 | VASP | FALSE | Upregulated |
| 1,176602221 | 0,000513253 | UBE2L6 | TRUE | Upregulated |
| 1,405790634 | 0,000514418 | THBS2 | TRUE | Upregulated |
| -0,648252969 | 0,000518255 | EPHB4 | FALSE | Downregulated |
| 1,387608349 | 0,000520207 | SLC35C1 | TRUE | Upregulated |
| 0,697505532 | 0,000521419 | ACTB | FALSE | Upregulated |
| 0,78233117 | 0,000521718 | IRF1 | FALSE | Upregulated |
| 0,577597254 | 0,000522549 | JUND | FALSE | Upregulated |
| 1,275243897 | 0,000536106 | COL1A2 | TRUE | Upregulated |
| -0,512833427 | 0,000537177 | CNBP | FALSE | Downregulated |
| 1,063041089 | 0,000551932 | LOC102724466 | TRUE | Upregulated |
| 0,838724828 | 0,000562029 | MRPL37 | FALSE | Upregulated |
| 1,087586275 | 0,000565715 | CPM | TRUE | Upregulated |
| 0,792176946 | 0,000565715 | TMED1 | FALSE | Upregulated |
| -0,549540735 | 0,000565779 | SEMA6A | FALSE | Downregulated |
| 1,026397695 | 0,000568907 | MVP | TRUE | Upregulated |
| 1,157550831 | 0,000568907 | WIPF1 | TRUE | Upregulated |

| | | | | |
|---|---|---|---|---|
| -1,167923621 | 0,000569936 | CNDP1 | TRUE | Downregulated |
| 0,902776947 | 0,000569936 | SAP30L-AS1 | FALSE | Upregulated |
| -0,448072574 | 0,000582357 | CLUH | FALSE | Downregulated |
| 0,792278671 | 0,000582357 | HTRA1 | FALSE | Upregulated |
| 1,636149129 | 0,000582357 | NCS1 | TRUE | Upregulated |
| -0,72469628 | 0,000603831 | ECHS1 | FALSE | Downregulated |
| -0,868726543 | 0,000603831 | GATM | FALSE | Downregulated |
| -0,785995829 | 0,000619121 | CPS1 | FALSE | Downregulated |
| 1,800886313 | 0,000619121 | IL24 | TRUE | Upregulated |
| 1,424254969 | 0,000619121 | RP11-277P12.20 | TRUE | Upregulated |
| -0,785139431 | 0,000641698 | CLCN5 | FALSE | Downregulated |
| 2,813501021 | 0,000649315 | RLTPR | TRUE | Upregulated |
| 2,017922366 | 0,000659144 | PDGFA | TRUE | Upregulated |
| -0,402156461 | 0,00067045 | EIF4B | FALSE | Downregulated |
| 0,762504357 | 0,00067045 | LAMP2 | FALSE | Upregulated |
| 0,401593224 | 0,00067045 | ZNF609 | FALSE | Upregulated |
| -0,952227198 | 0,000677462 | DMD | FALSE | Downregulated |
| 2,007469371 | 0,000677462 | EPSTI1 | TRUE | Upregulated |
| -1,123033679 | 0,000677462 | HSD17B13 | TRUE | Downregulated |
| -0,956968232 | 0,000677462 | SH3PXD2A | FALSE | Downregulated |
| -0,691984433 | 0,000678201 | CYP4F3 | FALSE | Downregulated |
| 0,982920634 | 0,000678201 | SLC51A | FALSE | Upregulated |
| -0,770318048 | 0,000694566 | ZNF395 | FALSE | Downregulated |
| -0,507666906 | 0,000696905 | SHMT2 | FALSE | Downregulated |
| -1,1546146 | 0,000721961 | LPAL2 | TRUE | Downregulated |
| 1,43379711 | 0,00073574 | GLI1 | TRUE | Upregulated |
| 2,056005444 | 0,00073574 | PAQR4 | TRUE | Upregulated |
| -0,826089665 | 0,000739008 | KBTBD11 | FALSE | Downregulated |
| -0,664507664 | 0,00073972 | AMBP | FALSE | Downregulated |
| -0,681770586 | 0,000740877 | ARHGAP29 | FALSE | Downregulated |
| -0,688096516 | 0,000740877 | LDHD | FALSE | Downregulated |
| -1,05121961 | 0,000746873 | SLC30A10 | TRUE | Downregulated |
| 1,120243461 | 0,000751592 | CYBA | TRUE | Upregulated |
| 1,853640896 | 0,000751592 | KCNS1 | TRUE | Upregulated |
| 1,159974583 | 0,000755629 | DXO | TRUE | Upregulated |
| -0,588519014 | 0,000756431 | RPL37A | FALSE | Downregulated |
| 1,098427537 | 0,000760683 | C1QA | TRUE | Upregulated |
| -0,481371493 | 0,000767359 | RPL3 | FALSE | Downregulated |
| 1,011507481 | 0,000767434 | PTPRCAP | TRUE | Upregulated |
| 1,025858214 | 0,000772452 | KRT8 | TRUE | Upregulated |
| 1,263462116 | 0,000776325 | SLA | TRUE | Upregulated |
| -0,794822751 | 0,000781291 | PTH1R | FALSE | Downregulated |
| 1,232541107 | 0,000781885 | TIMP1 | TRUE | Upregulated |
| 2,92960185 | 0,000782133 | LOC102724371 | TRUE | Upregulated |
| 0,679051944 | 0,000793015 | MARCKS | FALSE | Upregulated |
| -1,639050411 | 0,000804199 | SSTR1 | TRUE | Downregulated |
| 1,395581177 | 0,000813216 | APCDD1L | TRUE | Upregulated |
| 2,818134295 | 0,000813216 | BCAT1 | TRUE | Upregulated |
| 1,05164398 | 0,00081404 | CD160 | TRUE | Upregulated |
| 1,448222524 | 0,00081404 | LOC102724141 | TRUE | Upregulated |
| 1,194453311 | 0,00081404 | LOC729683 | TRUE | Upregulated |

| | | | | |
|---|---|---|---|---|
| 1,296203249 | 0,000817717 | C16orf3 | TRUE | Upregulated |
| -0,725672009 | 0,000821814 | APOF | FALSE | Downregulated |
| 0,96207137 | 0,000821814 | PREX1 | FALSE | Upregulated |
| -0,745281994 | 0,000825956 | TCEA3 | FALSE | Downregulated |
| -0,78880645 | 0,00082805 | ACOX2 | FALSE | Downregulated |
| 0,991368353 | 0,00082805 | TMEM41A | FALSE | Upregulated |
| -0,993084198 | 0,000862721 | PCOLCE | FALSE | Downregulated |
| -0,628806693 | 0,000862721 | PROX1 | FALSE | Downregulated |
| 0,976294048 | 0,000889482 | FERMT3 | FALSE | Upregulated |
| 0,592518084 | 0,000891209 | CSNK1D | FALSE | Upregulated |
| -0,459104178 | 0,000891209 | RXRA | FALSE | Downregulated |
| 0,606626061 | 0,000897862 | CIITA | FALSE | Upregulated |
| 1,985470091 | 0,000897862 | ISG15 | TRUE | Upregulated |
| 1,094748324 | 0,000897862 | SCXB | TRUE | Upregulated |
| 0,61176743 | 0,000897902 | ZNF276 | FALSE | Upregulated |
| -0,650265509 | 0,00090035 | MPDZ | FALSE | Downregulated |
| 0,654608437 | 0,000940505 | PHC1 | FALSE | Upregulated |
| -0,850880796 | 0,000952342 | CAT | FALSE | Downregulated |
| -1,104880622 | 0,000952342 | CYP2E1 | TRUE | Downregulated |
| 3,058838631 | 0,000955235 | LOC100131825 | TRUE | Upregulated |
| 1,117327761 | 0,000957558 | CD44 | TRUE | Upregulated |
| -0,781498137 | 0,000965935 | HGFAC | FALSE | Downregulated |
| -0,656671434 | 0,000965935 | SEPP1 | FALSE | Downregulated |
| -0,824637652 | 0,00097563 | DCXR | FALSE | Downregulated |
| 1,429269526 | 0,000980181 | LINC00896 | TRUE | Upregulated |
| -0,775551575 | 0,000980203 | PAH | FALSE | Downregulated |
| -0,704977481 | 0,001004087 | DLC1 | FALSE | Downregulated |
| -1,056603062 | 0,001024557 | FRRS1 | TRUE | Downregulated |
| 3,007270716 | 0,001030667 | LOC100996318 | TRUE | Upregulated |
| 0,785168256 | 0,001037475 | SLC35B2 | FALSE | Upregulated |
| 1,14994169 | 0,001041889 | IKZF1 | TRUE | Upregulated |
| -0,679587123 | 0,001050477 | GLUD1 | FALSE | Downregulated |
| 1,470163528 | 0,001050477 | TNFSF14 | TRUE | Upregulated |
| -0,58332415 | 0,001050539 | AZGP1 | FALSE | Downregulated |
| 0,924850672 | 0,001050539 | MBNL1-AS1 | FALSE | Upregulated |
| 0,662612144 | 0,00105093 | HNRNPAB | FALSE | Upregulated |
| 0,902203355 | 0,001053129 | ENO4 | FALSE | Upregulated |
| 0,652567457 | 0,001062131 | RHOC | FALSE | Upregulated |
| -2,348247758 | 0,001083965 | NCAM2 | TRUE | Downregulated |
| 1,008906613 | 0,00109324 | PTPRC | TRUE | Upregulated |
| 0,855266721 | 0,001101987 | DNMT1 | FALSE | Upregulated |
| 1,058697923 | 0,001106564 | C1QB | TRUE | Upregulated |
| 0,652091574 | 0,001106564 | SZRD1 | FALSE | Upregulated |
| 0,625762017 | 0,001114313 | PIEZO1 | FALSE | Upregulated |
| -0,734920845 | 0,001114313 | POFUT1 | FALSE | Downregulated |
| -0,526828836 | 0,001114313 | SLC9A3R2 | FALSE | Downregulated |
| 1,393562877 | 0,001117934 | FAM212A | TRUE | Upregulated |
| 1,492777764 | 0,001138974 | CORO1A | TRUE | Upregulated |
| -1,315801984 | 0,001144583 | SAMD5 | TRUE | Downregulated |
| -0,843692516 | 0,001175995 | BHMT | FALSE | Downregulated |
| 4,348283081 | 0,001175995 | ESP33 | TRUE | Upregulated |

| | | | | |
|---|---|---|---|---|
| 2,24072391 | 0,00117972 | C6orf147 | TRUE | Upregulated |
| -0,741184323 | 0,001190398 | AMOTL1 | FALSE | Downregulated |
| -1,102807085 | 0,001199067 | CSAD | TRUE | Downregulated |
| -0,640554896 | 0,001209793 | PHKA2 | FALSE | Downregulated |
| -1,327492484 | 0,001211686 | SH3RF2 | TRUE | Downregulated |
| 1,193200519 | 0,001231954 | FCHSD1 | TRUE | Upregulated |
| 0,535388046 | 0,001231954 | TXLNA | FALSE | Upregulated |
| 0,979099917 | 0,001236296 | LOC100506142 | FALSE | Upregulated |
| -0,855722671 | 0,001236296 | PDE2A | FALSE | Downregulated |
| 2,493412592 | 0,001242974 | B3GNT5 | TRUE | Upregulated |
| -1,084876132 | 0,001242974 | CYP4A22 | TRUE | Downregulated |
| -0,70328106 | 0,001242974 | DBH-AS1 | FALSE | Downregulated |
| 1,465778786 | 0,001242974 | LINGO3 | TRUE | Upregulated |
| 0,595186866 | 0,001242974 | YAP1 | FALSE | Upregulated |
| 0,659929294 | 0,001255557 | APOL6 | FALSE | Upregulated |
| 1,146140831 | 0,001277755 | CYTH4 | TRUE | Upregulated |
| 1,715967994 | 0,001283065 | LAIR1 | TRUE | Upregulated |
| 0,679013013 | 0,001283065 | LOC644083 | FALSE | Upregulated |
| -0,686018228 | 0,001307455 | CDO1 | FALSE | Downregulated |
| 0,775514218 | 0,001310121 | LOC101928719 | FALSE | Upregulated |
| 1,676791316 | 0,001321906 | ZAP70 | TRUE | Upregulated |
| 0,68195896 | 0,001322946 | ADPGK-AS1 | FALSE | Upregulated |
| -0,897929321 | 0,001374683 | SNX29P2 | FALSE | Downregulated |
| 1,084316328 | 0,001405788 | C1orf200 | TRUE | Upregulated |
| -1,280044159 | 0,001405788 | SDHAP1 | TRUE | Downregulated |
| -0,49421227 | 0,001405788 | TFR2 | FALSE | Downregulated |
| 0,784069224 | 0,001405788 | TGFB1 | FALSE | Upregulated |
| -0,839061007 | 0,001405788 | VIPR1 | FALSE | Downregulated |
| 1,486374206 | 0,001405788 | XYLT1 | TRUE | Upregulated |
| -0,410061021 | 0,001429301 | EEF2 | FALSE | Downregulated |
| 0,733412636 | 0,001448091 | C12orf61 | FALSE | Upregulated |
| -0,789030246 | 0,0014557 | ASPDH | FALSE | Downregulated |
| 1,216896691 | 0,001462857 | FGD3 | TRUE | Upregulated |
| -0,664507188 | 0,001464185 | DPYD | FALSE | Downregulated |
| -0,59518586 | 0,001464185 | EPB41L5 | FALSE | Downregulated |
| -0,793617437 | 0,001464185 | IYD | FALSE | Downregulated |
| 0,697467576 | 0,001464185 | MINA | FALSE | Upregulated |
| -0,64491582 | 0,001464185 | SDC2 | FALSE | Downregulated |
| -0,480593479 | 0,001464185 | SIGMAR1 | FALSE | Downregulated |
| -0,66347835 | 0,001489853 | ZGPAT | FALSE | Downregulated |
| 0,992273398 | 0,001491757 | CYBB | FALSE | Upregulated |
| 0,73650214 | 0,001493889 | TMSB4X | FALSE | Upregulated |
| -0,46620012 | 0,001502028 | HPN | FALSE | Downregulated |
| -0,522453673 | 0,001502028 | KANK1 | FALSE | Downregulated |
| -0,568768872 | 0,001502028 | PROC | FALSE | Downregulated |
| -1,538194396 | 0,001502028 | TPSG1 | TRUE | Downregulated |
| 0,654234396 | 0,001528769 | CLOCK | FALSE | Upregulated |
| 1,365226794 | 0,001535735 | RP11-356I2.4 | TRUE | Upregulated |
| -0,934759528 | 0,001545905 | LIPC | FALSE | Downregulated |
| -1,268816544 | 0,001550704 | NYNRIN | TRUE | Downregulated |
| 0,97671158 | 0,001583841 | LOC100996740 | FALSE | Upregulated |

| | | | | |
|---|---|---|---|---|
| -0,652969595 | 0,001595457 | AHCY | FALSE | Downregulated |
| 2,317271092 | 0,001596351 | CAPG | TRUE | Upregulated |
| 0,964571654 | 0,001596351 | FCGR3A | FALSE | Upregulated |
| 0,531489501 | 0,001606324 | CTSZ | FALSE | Upregulated |
| 2,818508766 | 0,001606324 | TOP2A | TRUE | Upregulated |
| -0,520340255 | 0,001638899 | ECE1 | FALSE | Downregulated |
| 0,473974864 | 0,001640996 | PFN1 | FALSE | Upregulated |
| -0,65048667 | 0,001647889 | EIF3L | FALSE | Downregulated |
| 1,096878716 | 0,001659977 | ANXA2 | TRUE | Upregulated |
| 0,82733386 | 0,001672721 | TNFAIP2 | FALSE | Upregulated |
| -0,767364356 | 0,001673082 | EPHX1 | FALSE | Downregulated |
| 0,79172064 | 0,001673082 | PLTP | FALSE | Upregulated |
| 0,959760969 | 0,001699548 | SOD2 | FALSE | Upregulated |
| 2,606834005 | 0,001718517 | TNFRSF12A | TRUE | Upregulated |
| 1,0156577 | 0,001719647 | GBP4 | TRUE | Upregulated |
| 1,698456115 | 0,001733537 | TCEA2 | TRUE | Upregulated |
| 1,565300473 | 0,001735172 | MYC | TRUE | Upregulated |
| 1,192447088 | 0,001742136 | SGK1 | TRUE | Upregulated |
| 0,683332848 | 0,001787515 | FBXL2 | FALSE | Upregulated |
| -0,758485702 | 0,001788022 | NR1I2 | FALSE | Downregulated |
| 0,972183954 | 0,001792067 | SUV39H2 | FALSE | Upregulated |
| 1,025570633 | 0,001827333 | SPI1 | TRUE | Upregulated |
| -0,801419188 | 0,001828007 | QDPR | FALSE | Downregulated |
| 1,179695598 | 0,001829242 | IER5 | TRUE | Upregulated |
| -0,814302461 | 0,001844268 | SLCO1B1 | FALSE | Downregulated |
| -1,263720024 | 0,001848792 | PITPNM3 | TRUE | Downregulated |
| 1,443779518 | 0,001855235 | CDKN1A | TRUE | Upregulated |
| 1,158511693 | 0,001855638 | PTGER4 | TRUE | Upregulated |
| 1,705201151 | 0,001900315 | S100A9 | TRUE | Upregulated |
| 1,240386909 | 0,001900799 | RANGRF | TRUE | Upregulated |
| -0,659510416 | 0,001902243 | MARC2 | FALSE | Downregulated |
| 0,900392753 | 0,00190441 | HLA-DRA | FALSE | Upregulated |
| 1,545104455 | 0,001977184 | LOC101927051 | TRUE | Upregulated |
| -0,633549409 | 0,002010149 | USH2A | FALSE | Downregulated |
| 1,758988191 | 0,002019514 | S1PR4 | TRUE | Upregulated |
| 0,868555586 | 0,002019514 | ZC3H12B | FALSE | Upregulated |
| -0,46243321 | 0,002092873 | NFIA | FALSE | Downregulated |
| -0,64145799 | 0,00211718 | AGT | FALSE | Downregulated |
| 1,580960079 | 0,00211718 | BTG2 | TRUE | Upregulated |
| -0,651568782 | 0,00211718 | SMIM14 | FALSE | Downregulated |
| -1,312911995 | 0,002118033 | C14orf132 | TRUE | Downregulated |
| -0,528579271 | 0,002156988 | PGM1 | FALSE | Downregulated |
| -0,563685442 | 0,002165064 | JMJD1C | FALSE | Downregulated |
| 0,949469509 | 0,002165064 | JUNB | FALSE | Upregulated |
| -0,702643146 | 0,002172711 | GLYCTK | FALSE | Downregulated |
| -0,742087583 | 0,002203555 | ENPP1 | FALSE | Downregulated |
| -0,625896288 | 0,002213838 | ALDH2 | FALSE | Downregulated |
| 0,933468522 | 0,002242553 | PIGR | FALSE | Upregulated |
| -0,664158194 | 0,002243769 | MBNL3 | FALSE | Downregulated |
| 1,067803959 | 0,002276109 | TMEM132A | TRUE | Upregulated |
| 0,545073312 | 0,002276109 | ZNFX1 | FALSE | Upregulated |

| | | | | |
|---|---|---|---|---|
| 2,321001683 | 0,002281713 | HIST1H2AG | TRUE | Upregulated |
| 1,945830304 | 0,002281713 | LOC101928465 | TRUE | Upregulated |
| 1,270738475 | 0,002281713 | MICAL1 | TRUE | Upregulated |
| 1,111425405 | 0,002285276 | TLR9 | TRUE | Upregulated |
| 0,845812747 | 0,002295506 | KRT18 | FALSE | Upregulated |
| 2,845329569 | 0,002295506 | SEMA7A | TRUE | Upregulated |
| 1,021371947 | 0,002330577 | ARSG | TRUE | Upregulated |
| 1,512165907 | 0,002346633 | STK17B | TRUE | Upregulated |
| -0,447400626 | 0,002351925 | CTDSP2 | FALSE | Downregulated |
| 2,305386746 | 0,002351925 | GSG2 | TRUE | Upregulated |
| -0,643242045 | 0,002364381 | ATP11C | FALSE | Downregulated |
| -0,390277958 | 0,002364381 | MTSS1 | FALSE | Downregulated |
| 1,418756782 | 0,002368677 | LILRB2 | TRUE | Upregulated |
| -0,410487289 | 0,002374106 | ARHGAP35 | FALSE | Downregulated |
| 0,881507628 | 0,002376599 | ARHGAP4 | FALSE | Upregulated |
| -0,691553171 | 0,002416505 | BDH1 | FALSE | Downregulated |
| -0,573396199 | 0,002416505 | JUP | FALSE | Downregulated |
| -0,650652688 | 0,002421281 | SARDH | FALSE | Downregulated |
| -0,677216188 | 0,002447822 | TMEM176A | FALSE | Downregulated |
| -0,547044648 | 0,002482326 | DPYS | FALSE | Downregulated |
| -0,458047174 | 0,002491356 | RPL29 | FALSE | Downregulated |
| -0,599745441 | 0,002491356 | SERINC1 | FALSE | Downregulated |
| -1,661194425 | 0,002527263 | RP11-475O6.1 | TRUE | Downregulated |
| 0,82662604 | 0,002537114 | HLA-C | FALSE | Upregulated |
| -0,627787169 | 0,002537114 | LIFR | FALSE | Downregulated |
| 0,879021944 | 0,002537114 | TGFBI | FALSE | Upregulated |
| -0,523494475 | 0,002537114 | UBB | FALSE | Downregulated |
| 0,787633839 | 0,002551087 | RP11-197N18.7 | FALSE | Upregulated |
| 1,287452338 | 0,002567327 | SRGN | TRUE | Upregulated |
| 2,061444569 | 0,002578467 | SELL | TRUE | Upregulated |
| 0,532511216 | 0,002591698 | TMEM184A | FALSE | Upregulated |
| 1,356383707 | 0,002636834 | ELF3 | TRUE | Upregulated |
| 0,887119109 | 0,002647477 | ELL3 | FALSE | Upregulated |
| 0,982347594 | 0,002651917 | HLA-DPA1 | FALSE | Upregulated |
| -0,525296231 | 0,002661941 | GLS2 | FALSE | Downregulated |
| 0,515053913 | 0,002680282 | TMEM200B | FALSE | Upregulated |
| 0,722609003 | 0,002680623 | SNX22 | FALSE | Upregulated |
| 0,915075673 | 0,002684113 | AKNA | FALSE | Upregulated |
| 0,514211063 | 0,002684113 | ATG7 | FALSE | Upregulated |
| 1,065916316 | 0,002684113 | ITGA6 | TRUE | Upregulated |
| 1,505429604 | 0,002701087 | MYO1G | TRUE | Upregulated |
| 1,000423496 | 0,002722006 | PPP1R16B | TRUE | Upregulated |
| 1,254943171 | 0,002744213 | LOC729291 | TRUE | Upregulated |
| -0,583977442 | 0,002746659 | ADCY1 | FALSE | Downregulated |
| 0,948777926 | 0,002746659 | ARID5B | FALSE | Upregulated |
| 1,262709056 | 0,002753957 | LYZ | TRUE | Upregulated |
| 0,571875038 | 0,002768892 | PHKG1 | FALSE | Upregulated |
| 1,375690492 | 0,002799686 | C16orf98 | TRUE | Upregulated |
| 1,649292321 | 0,002800842 | HLA-DQA1 | TRUE | Upregulated |
| -1,50734666 | 0,002824781 | RP11-475O6.1-2 | TRUE | Downregulated |
| -1,035366107 | 0,002843397 | DBT | TRUE | Downregulated |

| | | | | |
|---|---|---|---|---|
| 0,840636066 | 0,002856183 | STRADA | FALSE | Upregulated |
| -0,824152742 | 0,002912059 | ALDH1L1 | FALSE | Downregulated |
| -1,498081752 | 0,002912059 | CYP2C19 | TRUE | Downregulated |
| 0,974053382 | 0,002918988 | NLGN3 | FALSE | Upregulated |
| 1,308829414 | 0,002918988 | RUNX1 | TRUE | Upregulated |
| -0,619831121 | 0,002927507 | SLC25A42 | FALSE | Downregulated |
| 1,176350459 | 0,002968025 | TMEM75 | TRUE | Upregulated |
| -0,641932127 | 0,002997897 | TMEM176B | FALSE | Downregulated |
| 0,74027917 | 0,002998491 | COX15 | FALSE | Upregulated |
| -0,781373762 | 0,003064255 | SEPSECS | FALSE | Downregulated |
| -0,600043479 | 0,003073004 | TMEM56 | FALSE | Downregulated |
| 0,715039273 | 0,003074678 | KRR1 | FALSE | Upregulated |
| -0,538489619 | 0,0030762 | SLC2A4RG | FALSE | Downregulated |
| -1,457032685 | 0,0030762 | ST8SIA3 | TRUE | Downregulated |
| -0,609696171 | 0,003098932 | ST13 | FALSE | Downregulated |
| -0,846597263 | 0,003100781 | PEMT | FALSE | Downregulated |
| -0,660107876 | 0,00314673 | SMARCA1 | FALSE | Downregulated |
| -0,572859287 | 0,003167972 | APOC1 | FALSE | Downregulated |
| 0,984540386 | 0,003167972 | RECQL4 | FALSE | Upregulated |
| -0,690445896 | 0,003167972 | SLC27A2 | FALSE | Downregulated |
| -0,466823707 | 0,003167972 | ZHX3 | FALSE | Downregulated |
| 3,549695059 | 0,003263404 | MGC50722 | TRUE | Upregulated |
| 2,378779529 | 0,003263404 | SCML4 | TRUE | Upregulated |
| 0,802262609 | 0,003278226 | SMC4 | FALSE | Upregulated |
| -0,762800976 | 0,003287127 | HLCS | FALSE | Downregulated |
| -0,576506443 | 0,00329239 | COL27A1 | FALSE | Downregulated |
| -1,756076228 | 0,003310053 | GJB4 | TRUE | Downregulated |
| -0,501981707 | 0,003324484 | SPAG9 | FALSE | Downregulated |
| -0,602517944 | 0,00333044 | PDE3B | FALSE | Downregulated |
| 1,796461883 | 0,003345106 | LOC101928123 | TRUE | Upregulated |
| -0,622712377 | 0,003350326 | FPGS | FALSE | Downregulated |
| -0,604300655 | 0,003376752 | MAT1A | FALSE | Downregulated |
| -0,691170398 | 0,003382649 | GCKR | FALSE | Downregulated |
| -0,65351103 | 0,003387562 | KLF12 | FALSE | Downregulated |
| -0,382413687 | 0,003387562 | PCBP2 | FALSE | Downregulated |
| 0,494826237 | 0,003422692 | PTPMT1 | FALSE | Upregulated |
| -0,479610938 | 0,003433232 | EEF1A1 | FALSE | Downregulated |
| 0,911074994 | 0,003450604 | CDC25B | FALSE | Upregulated |
| 1,056898894 | 0,003450604 | EME1 | TRUE | Upregulated |
| 0,749455976 | 0,003450604 | SEMA4D | FALSE | Upregulated |
| 1,147404667 | 0,00352245 | BYSL | TRUE | Upregulated |
| -0,659206435 | 0,003644841 | SHPRH | FALSE | Downregulated |
| -1,541213411 | 0,0036551 | RP11-492A10.1 | TRUE | Downregulated |
| 0,687077306 | 0,003662685 | KLHL29 | FALSE | Upregulated |
| 1,69948722 | 0,003688383 | S100A1 | TRUE | Upregulated |
| 2,083051953 | 0,00373717 | LOC101927255 | TRUE | Upregulated |
| 2,336163151 | 0,003744131 | A2MP1 | TRUE | Upregulated |
| 0,797833376 | 0,003759354 | KLF6 | FALSE | Upregulated |
| 1,075898657 | 0,003767335 | MCM5 | TRUE | Upregulated |
| -0,971964862 | 0,003834746 | PTPRD | FALSE | Downregulated |
| 1,086908421 | 0,003851418 | LRRC20 | TRUE | Upregulated |

| | | | | |
|---|---|---|---|---|
| -0,551264637 | 0,003851418 | SYNE1 | FALSE | Downregulated |
| -0,342963364 | 0,00390929 | TPT1 | FALSE | Downregulated |
| -1,192265962 | 0,003927586 | RANBP3L | TRUE | Downregulated |
| 1,63407848 | 0,00399572 | LOC100129083 | TRUE | Upregulated |
| 0,591542229 | 0,004016999 | C1orf167 | FALSE | Upregulated |
| -0,929183231 | 0,004016999 | FAM214A | FALSE | Downregulated |
| -2,233043099 | 0,004016999 | LHX9 | TRUE | Downregulated |
| 1,941036935 | 0,004016999 | MDK | TRUE | Upregulated |
| 0,542907998 | 0,004016999 | SPATA21 | FALSE | Upregulated |
| -1,667538926 | 0,00409206 | NRARP | TRUE | Downregulated |
| 1,04590375 | 0,00409206 | RASSF1-AS1 | TRUE | Upregulated |
| -1,099987546 | 0,00413492 | SLC45A3 | TRUE | Downregulated |
| 0,977858913 | 0,004209501 | PCNA-AS1 | FALSE | Upregulated |
| 1,166387835 | 0,00422451 | UNC5A | TRUE | Upregulated |
| -1,110535382 | 0,004309431 | LPA | TRUE | Downregulated |
| -0,625463411 | 0,004443386 | ATXN7L3B | FALSE | Downregulated |
| 1,377403874 | 0,004443386 | HLA-DQB1 | TRUE | Upregulated |
| -0,644527882 | 0,004455943 | WDR59 | FALSE | Downregulated |
| -0,772242455 | 0,00445911 | SLC22A25 | FALSE | Downregulated |
| 1,069578506 | 0,004467547 | LRIF1 | TRUE | Upregulated |
| -1,367483945 | 0,00448495 | ADCY10 | TRUE | Downregulated |
| -1,472421932 | 0,00448495 | LOC101928938 | TRUE | Downregulated |
| 0,338629124 | 0,00448495 | MYH9 | FALSE | Upregulated |
| 0,86511151 | 0,00448495 | PPAPDC2 | FALSE | Upregulated |
| -0,678962486 | 0,004517442 | ZNF358 | FALSE | Downregulated |
| 0,794534874 | 0,00453326 | LOC101928291 | FALSE | Upregulated |
| -0,87975747 | 0,004542286 | MLPH | FALSE | Downregulated |
| -0,764936219 | 0,004542286 | ORAI3 | FALSE | Downregulated |
| 0,656497054 | 0,004555688 | ATP8B1 | FALSE | Upregulated |
| 0,549569105 | 0,004558996 | MAB21L2 | FALSE | Upregulated |
| 1,005541316 | 0,004588043 | SUPT4H1 | TRUE | Upregulated |
| 0,390849821 | 0,004620405 | TRIM8 | FALSE | Upregulated |
| 2,380859295 | 0,004625555 | CDSN | TRUE | Upregulated |
| -1,437922662 | 0,004625555 | LRRTM3 | TRUE | Downregulated |
| -0,804628909 | 0,004625555 | PDK4 | FALSE | Downregulated |
| 0,737722496 | 0,004632952 | GSN | FALSE | Upregulated |
| -0,621739169 | 0,00464914 | SNTB1 | FALSE | Downregulated |
| 2,863620699 | 0,004679502 | DUSP2 | TRUE | Upregulated |
| 0,992556151 | 0,004689677 | DOCK8 | FALSE | Upregulated |
| -0,687976417 | 0,004811119 | FAH | FALSE | Downregulated |
| -0,412681155 | 0,004825541 | FURIN | FALSE | Downregulated |
| -0,652848538 | 0,004839586 | HAO1 | FALSE | Downregulated |
| -0,658438604 | 0,004847197 | ANKS1A | FALSE | Downregulated |
| -0,646577718 | 0,004879552 | APOH | FALSE | Downregulated |
| 1,013548132 | 0,004879552 | MYOF | TRUE | Upregulated |
| -0,726889352 | 0,004879552 | PAIP2B | FALSE | Downregulated |
| 0,864935968 | 0,00493516 | COL3A1 | FALSE | Upregulated |
| 0,888410611 | 0,004939772 | CROT | FALSE | Upregulated |
| -0,453228049 | 0,004939772 | MASP1 | FALSE | Downregulated |
| 0,729640638 | 0,004939772 | PTOV1-AS1 | FALSE | Upregulated |
| 0,843409459 | 0,004939772 | RP11-685M7.3 | FALSE | Upregulated |

| | | | | |
|---|---|---|---|---|
| 2,176823696 | 0,005002975 | LOC100507424 | TRUE | Upregulated |
| 0,960276969 | 0,005006064 | LOC101928657 | FALSE | Upregulated |
| 0,653462445 | 0,005011018 | BBS1 | FALSE | Upregulated |
| -0,655247997 | 0,005011018 | SLC39A5 | FALSE | Downregulated |
| -0,672256395 | 0,00503293 | MUT | FALSE | Downregulated |
| 0,751872235 | 0,0050859 | C5orf66 | FALSE | Upregulated |
| -0,569293493 | 0,0050859 | METTL7A | FALSE | Downregulated |
| -0,770907614 | 0,005128494 | ANAPC16 | FALSE | Downregulated |
| -1,035420229 | 0,005173377 | RNF152 | TRUE | Downregulated |
| -0,357811589 | 0,005208 | RPL41 | FALSE | Downregulated |
| -3,421780493 | 0,005216806 | FAM35DP | TRUE | Downregulated |
| -0,724888942 | 0,005216806 | VARS2 | FALSE | Downregulated |
| -0,638219335 | 0,005218643 | COBLL1 | FALSE | Downregulated |
| -1,156901847 | 0,005239395 | PWRN2 | TRUE | Downregulated |
| 1,771358032 | 0,00524481 | CCDC80 | TRUE | Upregulated |
| -0,546455343 | 0,00524481 | UQCRC1 | FALSE | Downregulated |
| 0,414994466 | 0,005302942 | EPB41 | FALSE | Upregulated |
| -0,51191036 | 0,005340897 | AHSG | FALSE | Downregulated |
| -0,549831414 | 0,005480467 | PEX19 | FALSE | Downregulated |
| -0,389630671 | 0,005511995 | BIRC6 | FALSE | Downregulated |
| 1,643318453 | 0,005573849 | IFI6 | TRUE | Upregulated |
| -0,431999075 | 0,005593299 | SLC25A1 | FALSE | Downregulated |
| -0,98786851 | 0,005642829 | GDF7 | FALSE | Downregulated |
| -0,689870952 | 0,005642829 | SEMA5A | FALSE | Downregulated |
| 0,618086799 | 0,005692293 | DNAAF2 | FALSE | Upregulated |
| -0,626112207 | 0,005734833 | LIME1 | FALSE | Downregulated |
| 1,237226008 | 0,005748167 | MYO5A | TRUE | Upregulated |
| 1,057553319 | 0,005771421 | CEACAM16 | TRUE | Upregulated |
| 1,298196158 | 0,005816849 | NFKBIE | TRUE | Upregulated |
| 0,661269805 | 0,005818352 | ABHD16B | FALSE | Upregulated |
| -0,714260883 | 0,005818352 | KLF15 | FALSE | Downregulated |
| 1,971962633 | 0,005848235 | AC011899.9 | TRUE | Upregulated |
| -0,490843346 | 0,005956875 | AQP9 | FALSE | Downregulated |
| 0,91541592 | 0,005971123 | ARL4C | FALSE | Upregulated |
| -0,681317816 | 0,005971533 | ADH1B | FALSE | Downregulated |
| 0,959767438 | 0,005971533 | LOC100287042 | FALSE | Upregulated |
| -0,751584039 | 0,005999809 | CES3 | FALSE | Downregulated |
| -1,477738244 | 0,005999809 | RP11-701P16.5 | TRUE | Downregulated |
| 1,227536237 | 0,006021154 | SIGLEC10 | TRUE | Upregulated |
| 0,755257014 | 0,006082534 | SPON2 | FALSE | Upregulated |
| 0,933107381 | 0,006218236 | LOC388282 | FALSE | Upregulated |
| 1,311470428 | 0,006245546 | MYOM1 | TRUE | Upregulated |
| 1,192997687 | 0,006265842 | NABP1 | TRUE | Upregulated |
| -0,866910267 | 0,006268349 | PKHD1 | FALSE | Downregulated |
| -0,571606717 | 0,006268349 | SEC62 | FALSE | Downregulated |
| 0,940207605 | 0,006304004 | LOC100272216 | FALSE | Upregulated |
| -0,527582685 | 0,006356075 | ABAT | FALSE | Downregulated |
| 1,073871665 | 0,006390039 | MSR1 | TRUE | Upregulated |
| -0,932501159 | 0,006399522 | NDRG1 | FALSE | Downregulated |
| 2,373664971 | 0,00643227 | KLHDC7B | TRUE | Upregulated |
| -1,331630616 | 0,006519337 | GPR180 | TRUE | Downregulated |

| | | | | |
|---|---|---|---|---|
| -0,893490256 | 0,006525729 | SEMA6C | FALSE | Downregulated |
| -0,429703352 | 0,006541585 | EIF4H | FALSE | Downregulated |
| -0,585239331 | 0,006541585 | KLHDC10 | FALSE | Downregulated |
| 2,411057035 | 0,006541585 | PRDM8 | TRUE | Upregulated |
| -0,461330637 | 0,006547391 | STK25 | FALSE | Downregulated |
| 1,719404017 | 0,006552587 | ACER3 | TRUE | Upregulated |
| 0,598835488 | 0,006554462 | TDRP | FALSE | Upregulated |
| 1,207744683 | 0,006642473 | ITGAX | TRUE | Upregulated |
| 0,823544697 | 0,006688501 | FAT2 | FALSE | Upregulated |
| -0,527773528 | 0,006701584 | PLIN5 | FALSE | Downregulated |
| -0,812784504 | 0,006701584 | PON1 | FALSE | Downregulated |
| -0,497332966 | 0,006701584 | ZC3H11A | FALSE | Downregulated |
| -0,591482474 | 0,00673108 | EGFR | FALSE | Downregulated |
| -0,73871572 | 0,006751028 | GCDH | FALSE | Downregulated |
| -0,509588513 | 0,006811701 | ZFHX4 | FALSE | Downregulated |
| -0,910958114 | 0,006856584 | GALK1 | FALSE | Downregulated |
| 0,597413141 | 0,006856584 | LIMD1-AS1 | FALSE | Upregulated |
| -0,477382825 | 0,006866465 | CRAT | FALSE | Downregulated |
| -0,641998787 | 0,006866465 | FXYD1 | FALSE | Downregulated |
| -1,183079584 | 0,006866465 | SLC1A2 | TRUE | Downregulated |
| 1,459873347 | 0,006935407 | JAK3 | TRUE | Upregulated |
| 0,951812396 | 0,007073692 | SERAC1 | FALSE | Upregulated |
| -2,74343488 | 0,007138674 | HYDIN | TRUE | Downregulated |
| 1,029587132 | 0,007140054 | HLA-F | TRUE | Upregulated |
| 0,838546969 | 0,007140054 | LOC730257 | FALSE | Upregulated |
| 1,399553031 | 0,007173692 | HLA-DMB | TRUE | Upregulated |
| -1,79062794 | 0,007173692 | OAT | TRUE | Downregulated |
| 1,362987715 | 0,007176613 | FPR3 | TRUE | Upregulated |
| 1,607960687 | 0,007176613 | IL7R | TRUE | Upregulated |
| 1,599607261 | 0,0071959 | FCN1 | TRUE | Upregulated |
| -0,803786887 | 0,007197098 | EBP | FALSE | Downregulated |
| 1,558111182 | 0,007214057 | SLC9A5 | TRUE | Upregulated |
| 0,690957682 | 0,007239372 | VSIG10 | FALSE | Upregulated |
| -0,484343766 | 0,007253176 | NADK2 | FALSE | Downregulated |
| 1,160344469 | 0,007325122 | PHF19 | TRUE | Upregulated |
| 0,588550123 | 0,007327657 | KDM2B | FALSE | Upregulated |
| -0,447583412 | 0,007338127 | PCYOX1 | FALSE | Downregulated |
| 0,653713185 | 0,007449928 | LOC100133445 | FALSE | Upregulated |
| -0,502167238 | 0,007459633 | PPARA | FALSE | Downregulated |
| 1,550343197 | 0,007462678 | KLHL6 | TRUE | Upregulated |
| 0,973527451 | 0,007462678 | ZNF385A | FALSE | Upregulated |
| 1,628103213 | 0,007547692 | SSBP2 | TRUE | Upregulated |
| 0,980331082 | 0,007575816 | PKM | FALSE | Upregulated |
| 1,019364173 | 0,007581265 | RHBDF2 | TRUE | Upregulated |
| -0,656554345 | 0,007597206 | ARHGAP32 | FALSE | Downregulated |
| -0,441634862 | 0,007597206 | HADHA | FALSE | Downregulated |
| -1,666550929 | 0,007640881 | FRAS1 | TRUE | Downregulated |
| 1,617839445 | 0,007661507 | CCL5 | TRUE | Upregulated |
| 1,439900259 | 0,007673247 | LOC101929698 | TRUE | Upregulated |
| 0,621187047 | 0,007736155 | OGFRL1 | FALSE | Upregulated |
| 1,456603742 | 0,007814331 | KCNA3 | TRUE | Upregulated |

| | | | | |
|---|---|---|---|---|
| -0,673769291 | 0,007845336 | PIK3R4 | FALSE | Downregulated |
| -0,462255152 | 0,007850958 | RNF130 | FALSE | Downregulated |
| 0,4385988 | 0,007850958 | TMEM140 | FALSE | Upregulated |
| -0,588164169 | 0,007880599 | ABCA6 | FALSE | Downregulated |
| 0,678292429 | 0,007880599 | LRRC4 | FALSE | Upregulated |
| 0,968258588 | 0,007880599 | PLEKHG2 | FALSE | Upregulated |
| 0,614633901 | 0,007961001 | PPP1R9B | FALSE | Upregulated |
| -0,581451681 | 0,008013204 | ABHD15 | FALSE | Downregulated |
| 1,549214654 | 0,008013204 | PROB1 | TRUE | Upregulated |
| -0,911424435 | 0,008075911 | AKR1CL1 | FALSE | Downregulated |
| -0,688779423 | 0,008126033 | HOMER2 | FALSE | Downregulated |
| -1,638841723 | 0,008151953 | FGF14-AS2 | TRUE | Downregulated |
| 1,293891303 | 0,008214611 | RP11-47I22.2 | TRUE | Upregulated |
| 1,91506623 | 0,00825934 | HIST2H3A | TRUE | Upregulated |
| 0,977704541 | 0,008281803 | FAM83G | FALSE | Upregulated |
| 2,054042276 | 0,008281803 | ITK | TRUE | Upregulated |
| 0,894794076 | 0,008316331 | YWHAH | FALSE | Upregulated |
| -0,80555048 | 0,008400169 | COLEC10 | FALSE | Downregulated |
| -0,613963764 | 0,008400169 | MTMR4 | FALSE | Downregulated |
| -0,523591697 | 0,008422285 | ZBTB7B | FALSE | Downregulated |
| 1,122046822 | 0,008570105 | PXN-AS1 | TRUE | Upregulated |
| -0,752732122 | 0,008585366 | ELFN1 | FALSE | Downregulated |
| -0,432638115 | 0,008684514 | TSPYL1 | FALSE | Downregulated |
| 0,938360274 | 0,008695357 | LPCAT1 | FALSE | Upregulated |
| 0,87096058 | 0,0087098 | SLA2 | FALSE | Upregulated |
| 0,905756812 | 0,00878418 | AMZ1 | FALSE | Upregulated |
| 1,292238589 | 0,00878418 | CDKN2B | TRUE | Upregulated |
| 0,976661221 | 0,00878418 | CLDN7 | FALSE | Upregulated |
| -0,827845585 | 0,00878418 | PPP1CC | FALSE | Downregulated |
| 1,157839927 | 0,008836938 | CSF2RB | TRUE | Upregulated |
| -0,430664842 | 0,008840137 | BHMT2 | FALSE | Downregulated |
| -0,840856455 | 0,0088686 | GALE | FALSE | Downregulated |
| -0,749531348 | 0,008878466 | UGT2B7 | FALSE | Downregulated |
| -1,470354658 | 0,008892973 | LOC101929265 | TRUE | Downregulated |
| 0,512364307 | 0,008938175 | MPEG1 | FALSE | Upregulated |
| 1,24187741 | 0,008953561 | ATP2A3 | TRUE | Upregulated |
| 1,903451933 | 0,009016167 | ISG20 | TRUE | Upregulated |
| 1,022404687 | 0,009055106 | PLEKHO1 | TRUE | Upregulated |
| -0,988348079 | 0,009151897 | CASC10 | FALSE | Downregulated |
| 1,808296373 | 0,009172988 | ST8SIA4 | TRUE | Upregulated |
| -0,464721151 | 0,009173096 | VTN | FALSE | Downregulated |
| -0,302133333 | 0,009249064 | TNRC6B | FALSE | Downregulated |
| 1,252802989 | 0,009414893 | LOC101929837 | TRUE | Upregulated |
| -0,800959695 | 0,009414893 | RORC | FALSE | Downregulated |
| 1,733519137 | 0,00941979 | ARNTL2 | TRUE | Upregulated |
| 0,714390007 | 0,00941979 | RASSF4 | FALSE | Upregulated |
| 0,706696688 | 0,009447237 | FLJ40448 | FALSE | Upregulated |
| 2,273393995 | 0,009454262 | PAPPA2 | TRUE | Upregulated |
| 1,237265371 | 0,00947072 | FAM111B | TRUE | Upregulated |
| -0,585569986 | 0,009491449 | ZER1 | FALSE | Downregulated |
| 0,804273659 | 0,009587894 | HYOU1 | FALSE | Upregulated |

| | | | | |
|---|---|---|---|---|
| 0,795570184 | 0,009587894 | LCP2 | FALSE | Upregulated |
| -1,085448208 | 0,009606173 | TTLL4 | TRUE | Downregulated |
| 0,659837681 | 0,009644218 | SMARCA5-AS1 | FALSE | Upregulated |
| 1,093389276 | 0,009695169 | DOCK11 | TRUE | Upregulated |
| 2,545416513 | 0,009732526 | LINC00900 | TRUE | Upregulated |
| 1,044371944 | 0,009746724 | TRIM22 | TRUE | Upregulated |
| -1,335013191 | 0,0097693 | LOC400548 | TRUE | Downregulated |
| -0,462192818 | 0,009773327 | SERPIND1 | FALSE | Downregulated |
| -0,929072579 | 0,009873701 | PCK1 | FALSE | Downregulated |
| -0,455247458 | 0,009910519 | IQGAP2 | FALSE | Downregulated |
| -0,847017408 | 0,009942398 | ADH4 | FALSE | Downregulated |
| -0,568436708 | 0,009942398 | IPO5 | FALSE | Downregulated |
| -0,502326707 | 0,010130862 | ACOX1 | FALSE | Downregulated |
| -0,593339056 | 0,010178028 | TMEM110 | FALSE | Downregulated |
| 0,654828305 | 0,01018737 | DAB2IP | FALSE | Upregulated |
| 2,589544791 | 0,01018737 | E2F1 | TRUE | Upregulated |
| 1,327963735 | 0,01018737 | MTRNR2L8 | TRUE | Upregulated |
| 2,273502254 | 0,010219372 | NRSN1 | TRUE | Upregulated |
| -0,484550184 | 0,010239713 | SHANK3 | FALSE | Downregulated |
| -2,010170934 | 0,010257959 | CYP7A1 | TRUE | Downregulated |
| 0,959839938 | 0,010257959 | MCM3 | FALSE | Upregulated |
| -1,100163109 | 0,010267035 | KCNN2 | TRUE | Downregulated |
| -0,608813126 | 0,01028628 | ZCCHC6 | FALSE | Downregulated |
| 0,633585 | 0,010297616 | LOC101928958 | FALSE | Upregulated |
| 0,727117296 | 0,010432014 | LYPD1 | FALSE | Upregulated |
| 0,417679147 | 0,010439892 | AKAP13 | FALSE | Upregulated |
| 0,713863451 | 0,010439892 | VIM | FALSE | Upregulated |
| 0,853673819 | 0,010475429 | HCP5 | FALSE | Upregulated |
| -0,664768279 | 0,010519044 | STAB2 | FALSE | Downregulated |
| -0,622167169 | 0,010575503 | CYP4F2 | FALSE | Downregulated |
| -0,982276718 | 0,010647589 | EXD3 | FALSE | Downregulated |
| 0,638206319 | 0,010647589 | PLOD3 | FALSE | Upregulated |
| 0,771315545 | 0,010713798 | PSMB8 | FALSE | Upregulated |
| -0,626780161 | 0,010724917 | CITED2 | FALSE | Downregulated |
| 0,910438608 | 0,010754983 | FLJ13224 | FALSE | Upregulated |
| 1,858489292 | 0,010796881 | PERM1 | TRUE | Upregulated |
| 0,993583604 | 0,010831223 | RP11-379J5.5 | FALSE | Upregulated |
| 1,244913495 | 0,010930663 | FCGR2A | TRUE | Upregulated |
| 0,705074093 | 0,010930663 | ZNF469 | FALSE | Upregulated |
| 0,650625703 | 0,010935256 | ACTG1 | FALSE | Upregulated |
| 0,384233834 | 0,010935256 | RNF10 | FALSE | Upregulated |
| -0,504850371 | 0,010955419 | APOA2 | FALSE | Downregulated |
| 1,151245121 | 0,01095614 | MX1 | TRUE | Upregulated |
| -1,419456976 | 0,011001779 | ZNF257 | TRUE | Downregulated |
| 0,584346564 | 0,011180473 | RHOBTB2 | FALSE | Upregulated |
| 0,609791724 | 0,011181296 | H2AFY | FALSE | Upregulated |
| -0,644874334 | 0,011181296 | PNPLA7 | FALSE | Downregulated |
| -1,238296357 | 0,011189059 | AQP7P1 | TRUE | Downregulated |
| -0,659459376 | 0,01119209 | SEC16B | FALSE | Downregulated |
| -0,42166437 | 0,011211402 | C8orf82 | FALSE | Downregulated |
| -0,394610739 | 0,011270227 | PLXNB1 | FALSE | Downregulated |

| | | | | |
|---|---|---|---|---|
| 0,58331513 | 0,011281573 | MFSD11 | FALSE | Upregulated |
| -0,45938483 | 0,011317095 | RPS7 | FALSE | Downregulated |
| 0,550684026 | 0,011317095 | UGGT1 | FALSE | Upregulated |
| 1,215567168 | 0,011349055 | DOK1 | TRUE | Upregulated |
| 1,422506786 | 0,011349055 | FMN1 | TRUE | Upregulated |
| 0,795006085 | 0,011349055 | LOC101928552 | FALSE | Upregulated |
| 1,570626651 | 0,011491959 | RASD1 | TRUE | Upregulated |
| 0,812286054 | 0,011572492 | VAMP8 | FALSE | Upregulated |
| -0,532415758 | 0,011578377 | DCAF11 | FALSE | Downregulated |
| -0,672659507 | 0,011589378 | ZNF740 | FALSE | Downregulated |
| -0,489459597 | 0,011622696 | GRHPR | FALSE | Downregulated |
| -0,709850861 | 0,011664905 | ASS1 | FALSE | Downregulated |
| 2,383714176 | 0,011664905 | CENPF | TRUE | Upregulated |
| -0,707976269 | 0,011664905 | GALT | FALSE | Downregulated |
| 1,598864186 | 0,011664905 | TNFAIP8 | TRUE | Upregulated |
| 1,409217246 | 0,011769068 | PRF1 | TRUE | Upregulated |
| -0,650654486 | 0,011840035 | AAMP | FALSE | Downregulated |
| 1,07486036 | 0,011840035 | LOC285762 | TRUE | Upregulated |
| -0,370753645 | 0,011840035 | MASP2 | FALSE | Downregulated |
| 0,512615996 | 0,011866659 | LINC00094 | FALSE | Upregulated |
| -0,913195066 | 0,011949404 | RHEBP2 | FALSE | Downregulated |
| 0,494791275 | 0,012001641 | C12orf76 | FALSE | Upregulated |
| -0,618301562 | 0,012001641 | PTGR1 | FALSE | Downregulated |
| -0,548699447 | 0,012001641 | SCN8A | FALSE | Downregulated |
| 1,360910782 | 0,012029297 | LOC101928173 | TRUE | Upregulated |
| 2,298053751 | 0,012111938 | HELLS | TRUE | Upregulated |
| -0,568059831 | 0,012111938 | KIAA1217 | FALSE | Downregulated |
| 0,559326806 | 0,012111938 | RSBN1L-AS1 | FALSE | Upregulated |
| -1,019734363 | 0,012150108 | ZFP1 | TRUE | Downregulated |
| -0,822089783 | 0,012203147 | HNF1A-AS1 | FALSE | Downregulated |
| 0,799419947 | 0,012203147 | TYROBP | FALSE | Upregulated |
| -0,583521321 | 0,012280182 | DMGDH | FALSE | Downregulated |
| 0,506387779 | 0,012280182 | GRB2 | FALSE | Upregulated |
| 0,545639282 | 0,012280182 | TMEM14E | FALSE | Upregulated |
| 1,009623049 | 0,012304798 | RPA2 | TRUE | Upregulated |
| -0,465012353 | 0,012351478 | VEGFA | FALSE | Downregulated |
| 0,497964014 | 0,0123964 | ITPK1-AS1 | FALSE | Upregulated |
| 2,418695264 | 0,0123964 | KREMEN2 | TRUE | Upregulated |
| -0,474103478 | 0,0123964 | RPL5 | FALSE | Downregulated |
| -0,461931805 | 0,0123964 | STAB1 | FALSE | Downregulated |
| 2,213224657 | 0,012397659 | CD2 | TRUE | Upregulated |
| -0,529868855 | 0,012401238 | HSDL2 | FALSE | Downregulated |
| 0,970402583 | 0,012407208 | FYB | FALSE | Upregulated |
| -0,443686543 | 0,012461148 | RPS5 | FALSE | Downregulated |
| -0,637663637 | 0,012461148 | SLC13A5 | FALSE | Downregulated |
| 1,993908513 | 0,012461148 | SYT11 | TRUE | Upregulated |
| 0,569650689 | 0,012475216 | PAN3-AS1 | FALSE | Upregulated |
| 1,155021327 | 0,012494092 | GINS2 | TRUE | Upregulated |
| 1,088854115 | 0,012494092 | HLA-DOA | TRUE | Upregulated |
| 0,620912103 | 0,012494092 | SCUBE3 | FALSE | Upregulated |
| 0,714492707 | 0,012523095 | AP4B1-AS1 | FALSE | Upregulated |

| | | | | |
|---|---|---|---|---|
| 0,940678287 | 0,012527606 | HIVEP3 | FALSE | Upregulated |
| 0,619393233 | 0,012529027 | FAM86A | FALSE | Upregulated |
| 1,221263186 | 0,012532743 | NUAK2 | TRUE | Upregulated |
| -0,512503435 | 0,012554534 | MAP2K7 | FALSE | Downregulated |
| 2,440416314 | 0,012570351 | HPSE | TRUE | Upregulated |
| -0,92659519 | 0,012623622 | EXPH5 | FALSE | Downregulated |
| -0,634657637 | 0,012623622 | SLC22A3 | FALSE | Downregulated |
| -0,753716193 | 0,012632923 | DAK | FALSE | Downregulated |
| -0,827600598 | 0,012670406 | GSTA1 | FALSE | Downregulated |
| -0,574348616 | 0,012744905 | PDK2 | FALSE | Downregulated |
| -0,534431342 | 0,012759473 | FN1 | FALSE | Downregulated |
| 0,496670659 | 0,012794052 | SBNO2 | FALSE | Upregulated |
| 2,425010492 | 0,012794052 | THEMIS | TRUE | Upregulated |
| -0,768626029 | 0,012794052 | THSD4 | FALSE | Downregulated |
| -1,03313435 | 0,012794052 | UG0898H09 | TRUE | Downregulated |
| 1,584704669 | 0,012874447 | S100A11 | TRUE | Upregulated |
| 0,448864884 | 0,012974678 | GSE1 | FALSE | Upregulated |
| -0,714095618 | 0,012974678 | IGFALS | FALSE | Downregulated |
| 2,860953443 | 0,012974678 | PLA2G2A | TRUE | Upregulated |
| 2,307994458 | 0,012976435 | FCAMR | TRUE | Upregulated |
| -1,445085092 | 0,012985302 | BBOX1 | TRUE | Downregulated |
| -3,781409587 | 0,013010596 | LOC102724010 | TRUE | Downregulated |
| 0,999073251 | 0,013115122 | ABCC5 | FALSE | Upregulated |
| 0,950263631 | 0,013115122 | KLHDC7A | FALSE | Upregulated |
| -0,827220624 | 0,013115122 | MACROD1 | FALSE | Downregulated |
| 1,107293805 | 0,013115122 | UCP2 | TRUE | Upregulated |
| 1,040695853 | 0,013138153 | WHAMMP2 | TRUE | Upregulated |
| 2,185149362 | 0,013168256 | KNDC1 | TRUE | Upregulated |
| 0,471909487 | 0,013168256 | PTMA | FALSE | Upregulated |
| -0,67730024 | 0,013187926 | FAM213A | FALSE | Downregulated |
| -0,502591963 | 0,013258513 | EHHADH | FALSE | Downregulated |
| -0,420227282 | 0,013324388 | DST | FALSE | Downregulated |
| 0,606366511 | 0,013324388 | KIAA1598 | FALSE | Upregulated |
| -0,593790802 | 0,013324388 | RNF217 | FALSE | Downregulated |
| 2,02725128 | 0,013342585 | ADPRH | TRUE | Upregulated |
| 0,877284922 | 0,013342585 | PTPLAD2 | FALSE | Upregulated |
| -0,644641454 | 0,013342585 | SLC25A10 | FALSE | Downregulated |
| -0,82097453 | 0,013356794 | CA2 | FALSE | Downregulated |
| -0,957899118 | 0,013443066 | FAM198A | FALSE | Downregulated |
| 1,237718575 | 0,013490702 | ALK | TRUE | Upregulated |
| 0,64371393 | 0,013587234 | GAS6 | FALSE | Upregulated |
| 0,562723893 | 0,013622197 | SRRM5 | FALSE | Upregulated |
| 1,574413953 | 0,013622197 | ZNF831 | TRUE | Upregulated |
| 2,850618626 | 0,013635806 | AC002480.3 | TRUE | Upregulated |
| 0,940222671 | 0,013635806 | HK3 | FALSE | Upregulated |
| -0,704161642 | 0,013635806 | OXER1 | FALSE | Downregulated |
| 0,934569628 | 0,013693788 | LRRC18 | FALSE | Upregulated |
| -0,674491793 | 0,013693788 | SAT2 | FALSE | Downregulated |
| 0,983095825 | 0,013700341 | NKPD1 | FALSE | Upregulated |
| -0,683248542 | 0,013720422 | LRRC3 | FALSE | Downregulated |
| 1,439268738 | 0,013761539 | SLC29A3 | TRUE | Upregulated |

| | | | | |
|---|---|---|---|---|
| 0,512224779 | 0,013764689 | MAPKAPK2 | FALSE | Upregulated |
| 0,763133288 | 0,013781331 | NIN | FALSE | Upregulated |
| -0,481519507 | 0,013809422 | GNB2L1 | FALSE | Downregulated |
| -0,415814551 | 0,013809442 | ASXL1 | FALSE | Downregulated |
| -0,754516881 | 0,013820389 | CNTLN | FALSE | Downregulated |
| -2,482696451 | 0,013820389 | EPHB1 | TRUE | Downregulated |
| 0,569556918 | 0,013864758 | RAB5C | FALSE | Upregulated |
| 0,496231169 | 0,014015604 | THAP3 | FALSE | Upregulated |
| -0,84162762 | 0,014015604 | ZNRF3 | FALSE | Downregulated |
| 0,560987844 | 0,014160143 | PLEKHG6 | FALSE | Upregulated |
| 0,948640531 | 0,014172579 | ELP5 | FALSE | Upregulated |
| -0,502776788 | 0,014198157 | KLHDC2 | FALSE | Downregulated |
| -0,469621701 | 0,014220111 | BOK | FALSE | Downregulated |
| 1,392312938 | 0,014220111 | CARD11 | TRUE | Upregulated |
| 0,755047882 | 0,014220111 | FAM212B | FALSE | Upregulated |
| 1,067143423 | 0,014220111 | UBE2S | TRUE | Upregulated |
| 0,918296804 | 0,01427102 | KLF2 | FALSE | Upregulated |
| -0,458197368 | 0,01427102 | PHLDB2 | FALSE | Downregulated |
| 0,731731903 | 0,014329262 | C17orf72 | FALSE | Upregulated |
| -0,669330862 | 0,014337524 | PRDX6 | FALSE | Downregulated |
| 0,633993683 | 0,014337524 | RP4-591N18.2 | FALSE | Upregulated |
| -1,088538893 | 0,014338923 | LINC01093 | TRUE | Downregulated |
| 1,248440811 | 0,014400594 | FBLIM1 | TRUE | Upregulated |
| -0,438929308 | 0,014413786 | SERPINA1 | FALSE | Downregulated |
| 0,700731041 | 0,014417257 | GPR82 | FALSE | Upregulated |
| -0,387640905 | 0,014417257 | RPLP0 | FALSE | Downregulated |
| -0,48472287 | 0,014500844 | F11 | FALSE | Downregulated |
| -1,024944973 | 0,014502253 | CFHR4 | TRUE | Downregulated |
| -0,83590209 | 0,014546628 | CNNM2 | FALSE | Downregulated |
| -0,645928883 | 0,014546628 | CYP2J2 | FALSE | Downregulated |
| 2,040413676 | 0,014549582 | GNA15 | TRUE | Upregulated |
| -0,485245496 | 0,014549582 | PHLPP1 | FALSE | Downregulated |
| -1,063467493 | 0,014552628 | ADH1C | TRUE | Downregulated |
| 0,727508081 | 0,014552628 | B4GALT5 | FALSE | Upregulated |
| 1,302006974 | 0,014552628 | PARVG | TRUE | Upregulated |
| -0,824233167 | 0,014552628 | SLCO1B3 | FALSE | Downregulated |
| 0,760857041 | 0,014556181 | ACLY | FALSE | Upregulated |
| 1,192737874 | 0,014556181 | CLEC2D | TRUE | Upregulated |
| -0,48506795 | 0,014556181 | MXI1 | FALSE | Downregulated |
| -1,531028749 | 0,014556181 | NPW | TRUE | Downregulated |
| -0,908311704 | 0,014556181 | SOX5 | FALSE | Downregulated |
| 0,573589246 | 0,014556181 | SRP19 | FALSE | Upregulated |
| 0,741190705 | 0,014556181 | TUBB2A | FALSE | Upregulated |
| 0,558796654 | 0,014556181 | ZBTB11-AS1 | FALSE | Upregulated |
| 0,979600301 | 0,014556181 | ZC3H12A | FALSE | Upregulated |
| 0,850027432 | 0,014562361 | LINC00514 | FALSE | Upregulated |
| -0,531511237 | 0,014757458 | GC | FALSE | Downregulated |
| 0,776596508 | 0,014757458 | LILRB3 | FALSE | Upregulated |
| -0,738261145 | 0,014757458 | LINC01018 | FALSE | Downregulated |
| 1,552159859 | 0,014757458 | PTAFR | TRUE | Upregulated |
| -0,904178555 | 0,01478065 | ARRDC3 | FALSE | Downregulated |

| | | | | |
|---|---|---|---|---|
| -0,80332886 | 0,01478065 | TIA1 | FALSE | Downregulated |
| -0,522767295 | 0,014781948 | CGN | FALSE | Downregulated |
| 1,005787108 | 0,014781948 | LOC388692 | TRUE | Upregulated |
| 0,769130735 | 0,01484276 | C5orf45 | FALSE | Upregulated |
| 1,098736914 | 0,01484276 | MFSD2A | TRUE | Upregulated |
| 0,819165894 | 0,014849997 | SEPT8 | FALSE | Upregulated |
| 0,865165159 | 0,014917744 | DOK6 | FALSE | Upregulated |
| 0,755111013 | 0,014917744 | SH3KBP1 | FALSE | Upregulated |
| 0,731372718 | 0,014956509 | PITPNM1 | FALSE | Upregulated |
| 0,743436231 | 0,015018393 | HTRA2 | FALSE | Upregulated |
| -0,480578759 | 0,01506708 | CPN2 | FALSE | Downregulated |
| -0,528339387 | 0,015082056 | GAMT | FALSE | Downregulated |
| -0,592380247 | 0,015109855 | AGL | FALSE | Downregulated |
| 0,820613607 | 0,015173543 | COL5A1 | FALSE | Upregulated |
| 0,559242594 | 0,015173543 | LOC100505564 | FALSE | Upregulated |
| -0,48016071 | 0,015173543 | PTPRK | FALSE | Downregulated |
| 0,874925514 | 0,015194784 | IFI16 | FALSE | Upregulated |
| 1,094157147 | 0,015218551 | SLBP | TRUE | Upregulated |
| -0,512078881 | 0,015218551 | UPB1 | FALSE | Downregulated |
| -0,717144049 | 0,015356736 | RAI14 | FALSE | Downregulated |
| 0,451586517 | 0,015464988 | LRP10 | FALSE | Upregulated |
| -0,683211596 | 0,015491002 | EIF3E | FALSE | Downregulated |
| 0,691102165 | 0,015491002 | GPR68 | FALSE | Upregulated |
| 0,520009082 | 0,015491002 | YWHAZ | FALSE | Upregulated |
| 0,576034007 | 0,015518475 | CHRNB2 | FALSE | Upregulated |
| 0,809333916 | 0,015518475 | FLVCR2 | FALSE | Upregulated |
| -1,989038436 | 0,015572002 | TRHDE | TRUE | Downregulated |
| 2,802189587 | 0,015627348 | TRIM46 | TRUE | Upregulated |
| 2,090998114 | 0,015709932 | TMEM92 | TRUE | Upregulated |
| 0,586842588 | 0,015787402 | APOL2 | FALSE | Upregulated |
| -0,621033352 | 0,015787402 | GSDMB | FALSE | Downregulated |
| -0,43975245 | 0,015809628 | FZD5 | FALSE | Downregulated |
| 0,818684375 | 0,015809628 | LANCL2 | FALSE | Upregulated |
| 3,084626595 | 0,015810615 | HIST1H2AI | TRUE | Upregulated |
| 0,622473681 | 0,015937027 | LOC100128882 | FALSE | Upregulated |
| 1,086823117 | 0,015937027 | PAQR8 | TRUE | Upregulated |
| 0,516138669 | 0,015948988 | HIP1R | FALSE | Upregulated |
| -0,620577447 | 0,016007684 | FTCD | FALSE | Downregulated |
| -0,717492172 | 0,016079228 | ERI2 | FALSE | Downregulated |
| -0,627580644 | 0,016108311 | MET | FALSE | Downregulated |
| 0,617185644 | 0,016131133 | FLNA | FALSE | Upregulated |
| -0,964364508 | 0,016141671 | SFXN2 | FALSE | Downregulated |
| -0,4561956 | 0,016149539 | SERPINF1 | FALSE | Downregulated |
| -0,785083979 | 0,016176271 | CTNNBL1 | FALSE | Downregulated |
| 2,632739304 | 0,016176271 | RUNX2 | TRUE | Upregulated |
| -0,732379342 | 0,016271922 | PHGDH | FALSE | Downregulated |
| -0,516386753 | 0,016276401 | OGDHL | FALSE | Downregulated |
| -0,405487667 | 0,016422934 | SLC35E2B | FALSE | Downregulated |
| -1,313610419 | 0,016440823 | SCN3A | TRUE | Downregulated |
| 0,900771419 | 0,016440823 | SLFN5 | FALSE | Upregulated |
| 0,570866854 | 0,016485951 | FANCI | FALSE | Upregulated |

| | | | | |
|---|---|---|---|---|
| 0,640171184 | 0,016640575 | C7orf49 | FALSE | Upregulated |
| -0,477771326 | 0,016726411 | GRB10 | FALSE | Downregulated |
| 1,505570704 | 0,016732343 | GPX7 | TRUE | Upregulated |
| -0,826730249 | 0,01684806 | SLC17A2 | FALSE | Downregulated |
| 0,697397678 | 0,0168732 | PACS1 | FALSE | Upregulated |
| 0,45730379 | 0,016907623 | TMOD1 | FALSE | Upregulated |
| 1,484953791 | 0,016934711 | EFEMP1 | TRUE | Upregulated |
| 0,864627195 | 0,016934711 | LYSMD1 | FALSE | Upregulated |
| 1,275263602 | 0,01701174 | LOC102723726 | TRUE | Upregulated |
| 0,483159396 | 0,017209054 | PAN3 | FALSE | Upregulated |
| -0,713335605 | 0,017400162 | TNFSF10 | FALSE | Downregulated |
| -0,54339591 | 0,017409601 | CAPN5 | FALSE | Downregulated |
| -0,864795491 | 0,017415526 | GANC | FALSE | Downregulated |
| 0,979642225 | 0,0174982 | SAMD9L | FALSE | Upregulated |
| -0,703820915 | 0,017551612 | ECSIT | FALSE | Downregulated |
| 0,846397237 | 0,017729948 | DPCD | FALSE | Upregulated |
| 0,68119649 | 0,01773482 | PRPF40B | FALSE | Upregulated |
| 1,938440024 | 0,017785138 | ENAM | TRUE | Upregulated |
| 0,564621102 | 0,01782303 | ODF3B | FALSE | Upregulated |
| -0,620109834 | 0,017836398 | HGD | FALSE | Downregulated |
| -1,494534266 | 0,017846001 | SULT1E1 | TRUE | Downregulated |
| -0,370744464 | 0,017920245 | AKT2 | FALSE | Downregulated |
| -0,998931097 | 0,017920245 | GNMT | FALSE | Downregulated |
| 1,170688562 | 0,017920245 | MBOAT4 | TRUE | Upregulated |
| 0,478490614 | 0,017920245 | SPNS2 | FALSE | Upregulated |
| -0,401063769 | 0,017958919 | EHMT1 | FALSE | Downregulated |
| 0,501323544 | 0,017958919 | SKIV2L2 | FALSE | Upregulated |
| -0,432081046 | 0,017991051 | CDC42BPA | FALSE | Downregulated |
| -0,618888537 | 0,018089263 | FH | FALSE | Downregulated |
| 0,852588204 | 0,018155094 | CDK8 | FALSE | Upregulated |
| 0,620609417 | 0,01824534 | SLC37A2 | FALSE | Upregulated |
| -0,485903102 | 0,018295361 | ALDH8A1 | FALSE | Downregulated |
| 2,088716721 | 0,018534061 | EXTL1 | TRUE | Upregulated |
| -0,767394739 | 0,018534061 | ILVBL | FALSE | Downregulated |
| 1,112859 | 0,018541303 | RASSF2 | TRUE | Upregulated |
| 1,161525217 | 0,018605645 | ALOX5 | TRUE | Upregulated |
| -0,397225664 | 0,018605645 | CNNM3 | FALSE | Downregulated |
| 1,133077227 | 0,018605645 | GAS7 | TRUE | Upregulated |
| -0,450896018 | 0,018605645 | OAF | FALSE | Downregulated |
| -0,491026703 | 0,018646698 | CTAGE5 | FALSE | Downregulated |
| -0,395738851 | 0,018646698 | PPP1R10 | FALSE | Downregulated |
| 0,881235516 | 0,018731503 | NCKAP1L | FALSE | Upregulated |
| -0,345261144 | 0,018788456 | ASGR1 | FALSE | Downregulated |
| -0,771548329 | 0,018788456 | PRLR | FALSE | Downregulated |
| -0,455527093 | 0,018788456 | RAPGEF2 | FALSE | Downregulated |
| 0,576312626 | 0,018792548 | IQCC | FALSE | Upregulated |
| -0,542332725 | 0,018838046 | MYO10 | FALSE | Downregulated |
| 0,53561682 | 0,018839794 | VAMP1 | FALSE | Upregulated |
| -0,484989378 | 0,018881802 | LMAN1 | FALSE | Downregulated |
| -0,394679379 | 0,018881802 | MYO18A | FALSE | Downregulated |
| -0,519024292 | 0,01889758 | DCAF6 | FALSE | Downregulated |

| | | | | |
|---|---|---|---|---|
| -1,672097047 | 0,018901712 | LINC00844 | TRUE | Downregulated |
| 0,610924565 | 0,018901712 | PCGF3 | FALSE | Upregulated |
| 0,523657485 | 0,018901712 | STARD7-AS1 | FALSE | Upregulated |
| 2,270161732 | 0,019098108 | CECR6 | TRUE | Upregulated |
| 0,948818696 | 0,019098108 | SLC44A3 | FALSE | Upregulated |
| -1,163078434 | 0,01917902 | LOC101060351 | TRUE | Downregulated |
| 0,917854219 | 0,019273753 | LOC101928517 | FALSE | Upregulated |
| 0,77855798 | 0,019387377 | EMILIN2 | FALSE | Upregulated |
| 0,683980434 | 0,019387377 | MYL12A | FALSE | Upregulated |
| -1,651570589 | 0,019387377 | TRHDE-AS1 | TRUE | Downregulated |
| -0,559871055 | 0,01939507 | AFMID | FALSE | Downregulated |
| 0,577354881 | 0,01939507 | VWA1 | FALSE | Upregulated |
| 0,861380802 | 0,019402256 | HK1 | FALSE | Upregulated |
| 0,472388805 | 0,019402256 | LRTOMT | FALSE | Upregulated |
| -0,354632321 | 0,019496293 | LRP5 | FALSE | Downregulated |
| 2,204984229 | 0,019621193 | IL12RB1 | TRUE | Upregulated |
| 0,828467282 | 0,019627497 | OR2A1 | FALSE | Upregulated |
| 0,582259978 | 0,019768181 | DSCR3 | FALSE | Upregulated |
| -0,65762389 | 0,019768181 | RDH16 | FALSE | Downregulated |
| -0,622683997 | 0,01980468 | GPR125 | FALSE | Downregulated |
| -0,50968572 | 0,01980935 | ITIH2 | FALSE | Downregulated |
| -0,795358599 | 0,019826807 | RNU4-2 | FALSE | Downregulated |
| -1,268722528 | 0,019951499 | DENND2A | TRUE | Downregulated |
| -0,42378789 | 0,019951499 | RBM47 | FALSE | Downregulated |
| 0,907968577 | 0,019975337 | ADAM9 | FALSE | Upregulated |
| 0,641066125 | 0,019985201 | LOC653513 | FALSE | Upregulated |
| 1,139263574 | 0,02011744 | PLEK | TRUE | Upregulated |
| 0,417171732 | 0,020158832 | GON4L | FALSE | Upregulated |
| 1,870530625 | 0,020220173 | HIST1H1D | TRUE | Upregulated |
| -0,438023471 | 0,020276573 | RPL23 | FALSE | Downregulated |
| 1,481777171 | 0,020285028 | RP11-725G5.2 | TRUE | Upregulated |
| 1,88149368 | 0,020390691 | BHLHE41 | TRUE | Upregulated |
| 0,569147622 | 0,020455064 | C14orf93 | FALSE | Upregulated |
| 1,853750648 | 0,020725154 | HIST1H3D | TRUE | Upregulated |
| 0,597645841 | 0,020739392 | LOC100506603 | FALSE | Upregulated |
| -0,587300297 | 0,020762749 | HADHB | FALSE | Downregulated |
| 0,74283083 | 0,020762749 | IL18BP | FALSE | Upregulated |
| 1,074143448 | 0,020795084 | HIGD1B | TRUE | Upregulated |
| -1,571365852 | 0,020831799 | PMS2 | TRUE | Downregulated |
| -0,665477911 | 0,020965217 | ACVR2B | FALSE | Downregulated |
| -0,410508912 | 0,020965217 | ITIH4 | FALSE | Downregulated |
| 0,755200301 | 0,021066971 | LOC101928268 | FALSE | Upregulated |
| -0,562248161 | 0,021157402 | SLC17A9 | FALSE | Downregulated |
| 2,255977345 | 0,021157957 | SLC24A5 | TRUE | Upregulated |
| -0,330535827 | 0,021234595 | ACADVL | FALSE | Downregulated |
| 0,874956476 | 0,021234595 | OAS3 | FALSE | Upregulated |
| -0,421797271 | 0,021240776 | ARG1 | FALSE | Downregulated |
| 0,865567837 | 0,021359898 | TUBA1A | FALSE | Upregulated |
| 0,614928318 | 0,021383518 | CASKIN1 | FALSE | Upregulated |
| -0,396957215 | 0,02152246 | AGMAT | FALSE | Downregulated |
| -1,107388378 | 0,02152246 | AOC4P | TRUE | Downregulated |

| | | | | |
|---|---|---|---|---|
| 0,561634816 | 0,02152246 | RAD9A | FALSE | Upregulated |
| -0,425167786 | 0,021587634 | APOB | FALSE | Downregulated |
| 0,370122149 | 0,021587634 | BAIAP2L1 | FALSE | Upregulated |
| -0,591484002 | 0,021591329 | PON3 | FALSE | Downregulated |
| -0,572020549 | 0,021591329 | TBX3 | FALSE | Downregulated |
| 2,352820514 | 0,021600353 | CGB7 | TRUE | Upregulated |
| 0,741605277 | 0,021676945 | RAP2B | FALSE | Upregulated |
| -0,501431868 | 0,02171828 | ACADS | FALSE | Downregulated |
| 0,363476501 | 0,02171828 | SPTAN1 | FALSE | Upregulated |
| 0,554769594 | 0,021726029 | C10orf10 | FALSE | Upregulated |
| -0,902521941 | 0,021726029 | CUX2 | FALSE | Downregulated |
| 0,49138093 | 0,021761614 | ATP2A2 | FALSE | Upregulated |
| 0,567443722 | 0,02186895 | SLC35G2 | FALSE | Upregulated |
| 2,083323486 | 0,021930214 | FER1L4 | TRUE | Upregulated |
| 2,133375167 | 0,021938107 | PLA2G7 | TRUE | Upregulated |
| -0,538350984 | 0,021957108 | IDH1 | FALSE | Downregulated |
| -0,477967988 | 0,021958069 | ERBB3 | FALSE | Downregulated |
| 0,541394978 | 0,021967005 | HLA-E | FALSE | Upregulated |
| 0,675627851 | 0,021979352 | APOL3 | FALSE | Upregulated |
| 0,439090398 | 0,022026098 | FAM65A | FALSE | Upregulated |
| 1,340009781 | 0,022076269 | POU5F1 | TRUE | Upregulated |
| 2,302995314 | 0,02208129 | IRF4 | TRUE | Upregulated |
| 0,773081936 | 0,022100081 | PIK3CD | FALSE | Upregulated |
| -0,611006009 | 0,022100254 | BTG1 | FALSE | Downregulated |
| 0,556893646 | 0,022100254 | CARD14 | FALSE | Upregulated |
| 2,110032924 | 0,022100254 | HIST2H4B | TRUE | Upregulated |
| -2,083610347 | 0,022177784 | CXXC4 | TRUE | Downregulated |
| -0,488537869 | 0,022234059 | ERGIC1 | FALSE | Downregulated |
| -0,61041582 | 0,022243683 | NGFR | FALSE | Downregulated |
| 1,293699178 | 0,022334238 | CD48 | TRUE | Upregulated |
| -0,455662535 | 0,022361909 | KDM4B | FALSE | Downregulated |
| 1,150905195 | 0,022414444 | TCF19 | TRUE | Upregulated |
| -0,416844775 | 0,022449022 | AKAP1 | FALSE | Downregulated |
| -0,570909384 | 0,022483273 | DGCR6L | FALSE | Downregulated |
| 0,556444105 | 0,022557887 | CCDC92 | FALSE | Upregulated |
| 1,171295678 | 0,022557887 | TACC3 | TRUE | Upregulated |
| 0,6726894 | 0,02259767 | NFYC-AS1 | FALSE | Upregulated |
| 0,675948234 | 0,022669185 | OPA3 | FALSE | Upregulated |
| 0,457531313 | 0,022669185 | SLC7A6 | FALSE | Upregulated |
| 0,895337908 | 0,022756183 | ACOT6 | FALSE | Upregulated |
| 0,49833533 | 0,022756357 | CNTD1 | FALSE | Upregulated |
| -0,557098835 | 0,022760923 | LGR4 | FALSE | Downregulated |
| -0,355455869 | 0,022813817 | EIF4EBP2 | FALSE | Downregulated |
| -0,576577155 | 0,022825157 | PRDX2 | FALSE | Downregulated |
| 0,749881551 | 0,022825157 | PRR15L | FALSE | Upregulated |
| -0,618135165 | 0,022870682 | ELL2 | FALSE | Downregulated |
| -1,310292108 | 0,022888712 | B3GAT1 | TRUE | Downregulated |
| 1,765323445 | 0,022898506 | CCR5 | TRUE | Upregulated |
| -0,783374312 | 0,022898506 | FUOM | FALSE | Downregulated |
| -0,361103564 | 0,022898506 | RPL14 | FALSE | Downregulated |
| -1,044895845 | 0,02295708 | PPM1K | TRUE | Downregulated |

| | | | | |
|---|---|---|---|---|
| 1,175521536 | 0,022958257 | SLAMF8 | TRUE | Upregulated |
| -0,638334177 | 0,023007299 | ISOC2 | FALSE | Downregulated |
| 0,783366912 | 0,023007299 | LOC101929204 | FALSE | Upregulated |
| -0,392942287 | 0,023007299 | NIPSNAP1 | FALSE | Downregulated |
| -1,799107182 | 0,023175493 | PTPRT | TRUE | Downregulated |
| 2,411439452 | 0,023212453 | CD52 | TRUE | Upregulated |
| -0,498838682 | 0,023228085 | RPS27 | FALSE | Downregulated |
| 0,487384635 | 0,023262861 | ICMT | FALSE | Upregulated |
| -0,592225545 | 0,023299455 | ALDH7A1 | FALSE | Downregulated |
| -0,556605051 | 0,023299455 | IRS2 | FALSE | Downregulated |
| 2,46847665 | 0,023305392 | HIST2H3D | TRUE | Upregulated |
| 0,374826658 | 0,023305392 | MAP4 | FALSE | Upregulated |
| 0,67077287 | 0,023320681 | CTNS | FALSE | Upregulated |
| -0,740392096 | 0,023364136 | HOGA1 | FALSE | Downregulated |
| -0,596769053 | 0,023444645 | LY6E | FALSE | Downregulated |
| 0,327366044 | 0,023444645 | TAPBP | FALSE | Upregulated |
| 1,265317515 | 0,023465327 | LOC101927730 | TRUE | Upregulated |
| -0,603596496 | 0,023507621 | SH3BGRL2 | FALSE | Downregulated |
| 0,60225325 | 0,023546512 | LOC100506922 | FALSE | Upregulated |
| 0,70308356 | 0,02355461 | PARP8 | FALSE | Upregulated |
| -0,437132665 | 0,02372405 | KLC4 | FALSE | Downregulated |
| 0,453395911 | 0,023743078 | CTSD | FALSE | Upregulated |
| 1,205120531 | 0,023743078 | FAM129A | TRUE | Upregulated |
| 1,60078045 | 0,023743078 | PDCD1 | TRUE | Upregulated |
| -2,261215012 | 0,023756247 | DHRS2 | TRUE | Downregulated |
| 0,756039944 | 0,023756247 | IFIH1 | FALSE | Upregulated |
| -0,813281966 | 0,023756247 | LIMCH1 | FALSE | Downregulated |
| -0,485115842 | 0,023756247 | TSPAN9 | FALSE | Downregulated |
| -0,403317836 | 0,023771109 | DIDO1 | FALSE | Downregulated |
| 0,591847137 | 0,023771109 | UBE2Z | FALSE | Upregulated |
| 0,5121323 | 0,023808659 | NME7 | FALSE | Upregulated |
| -0,932543393 | 0,023846266 | ATP6V0B | FALSE | Downregulated |
| -0,630054648 | 0,023968611 | SLC41A2 | FALSE | Downregulated |
| 1,031108789 | 0,023994277 | MIAT | TRUE | Upregulated |
| 0,570763488 | 0,023994277 | NINJ1 | FALSE | Upregulated |
| 0,976955801 | 0,024042031 | PGP | FALSE | Upregulated |
| 0,627138195 | 0,024042031 | SRGAP2-AS1 | FALSE | Upregulated |
| -0,459577668 | 0,024144323 | RPS24 | FALSE | Downregulated |
| 0,391855502 | 0,024226883 | CD163 | FALSE | Upregulated |
| 1,187349048 | 0,024229263 | DSCR9 | TRUE | Upregulated |
| -0,584490246 | 0,024351611 | AHR | FALSE | Downregulated |
| 0,378833752 | 0,024600965 | EMC1 | FALSE | Upregulated |
| 0,414183633 | 0,024606624 | SLC9A3R1 | FALSE | Upregulated |
| 0,931154394 | 0,024659156 | CHST11 | FALSE | Upregulated |
| -0,851331555 | 0,02476081 | GNAI1 | FALSE | Downregulated |
| -0,406002569 | 0,024813258 | ITPR2 | FALSE | Downregulated |
| 0,499672446 | 0,024813258 | KCNQ1OT1 | FALSE | Upregulated |
| -0,485233722 | 0,024869014 | ATG2B | FALSE | Downregulated |
| 1,036209077 | 0,024869014 | LCORL | TRUE | Upregulated |
| -0,367729761 | 0,02505947 | ZFP36L1 | FALSE | Downregulated |
| 0,549078742 | 0,025188122 | IRAK1 | FALSE | Upregulated |

| | | | | |
|---|---|---|---|---|
| -0,481149229 | 0,025217497 | PCK2 | FALSE | Downregulated |
| -1,60248294 | 0,025312621 | GSTA2 | TRUE | Downregulated |
| -0,37422733 | 0,025312621 | HELZ | FALSE | Downregulated |
| -0,599227993 | 0,025312621 | PPAP2B | FALSE | Downregulated |
| -1,324534898 | 0,02539041 | TPPP2 | TRUE | Downregulated |
| -0,619698475 | 0,025417423 | HMGCL | FALSE | Downregulated |
| 1,623902078 | 0,025455045 | LILRB4 | TRUE | Upregulated |
| 0,513784599 | 0,025509743 | PHYKPL | FALSE | Upregulated |
| 0,81436257 | 0,025536725 | LOC101927420 | FALSE | Upregulated |
| -2,166389212 | 0,025566066 | AC007246.3 | TRUE | Downregulated |
| 1,392026768 | 0,025581567 | LOC101927897 | TRUE | Upregulated |
| -0,751920582 | 0,025789614 | BDH2 | FALSE | Downregulated |
| 0,966115069 | 0,025789614 | LILRB1 | FALSE | Upregulated |
| 0,934109651 | 0,025814726 | IQCK | FALSE | Upregulated |
| 0,804076193 | 0,025882414 | RELL2 | FALSE | Upregulated |
| -0,804143034 | 0,025943479 | TMEM120A | FALSE | Downregulated |
| -0,519624759 | 0,025943479 | ZNF275 | FALSE | Downregulated |
| 0,802993062 | 0,026031001 | CEACAM19 | FALSE | Upregulated |
| -0,734923979 | 0,026058426 | ANKRD24 | FALSE | Downregulated |
| -0,582876659 | 0,026071448 | SLC38A4 | FALSE | Downregulated |
| 0,486124345 | 0,026078372 | CARHSP1 | FALSE | Upregulated |
| 0,948476001 | 0,026078372 | SLC8A1 | FALSE | Upregulated |
| -0,46493317 | 0,026078372 | ZCCHC14 | FALSE | Downregulated |
| 1,013088462 | 0,02608853 | AQP4-AS1 | TRUE | Upregulated |
| 1,246964871 | 0,02616895 | MANF | TRUE | Upregulated |
| -0,612234901 | 0,026246619 | STRADB | FALSE | Downregulated |
| 0,548352444 | 0,026468833 | LPGAT1 | FALSE | Upregulated |
| 1,907963117 | 0,026520496 | GPR84 | TRUE | Upregulated |
| 0,833261293 | 0,026550631 | FBXO6 | FALSE | Upregulated |
| -1,625326548 | 0,026550631 | LINC00211 | TRUE | Downregulated |
| 0,530073873 | 0,026703536 | USP35 | FALSE | Upregulated |
| -0,513879958 | 0,026792553 | ALDH6A1 | FALSE | Downregulated |
| -0,50828232 | 0,026876542 | COX6A1 | FALSE | Downregulated |
| 0,430702334 | 0,026949961 | ADCY7 | FALSE | Upregulated |
| -2,241938083 | 0,026979563 | OPCML | TRUE | Downregulated |
| 1,72191267 | 0,027035097 | MEFV | TRUE | Upregulated |
| 0,581838502 | 0,027105508 | HSPA5 | FALSE | Upregulated |
| -0,751188257 | 0,027172138 | RHEB | FALSE | Downregulated |
| -0,32368403 | 0,027244419 | UBR4 | FALSE | Downregulated |
| 0,761789593 | 0,027327015 | TWF1 | FALSE | Upregulated |
| -0,384240625 | 0,027438378 | FAM20C | FALSE | Downregulated |
| 0,500022071 | 0,027438378 | PPP2R5D | FALSE | Upregulated |
| -0,423252063 | 0,027438378 | RPL6 | FALSE | Downregulated |
| 0,711199538 | 0,027705781 | CSF1 | FALSE | Upregulated |
| -0,584952885 | 0,027856989 | FLT4 | FALSE | Downregulated |
| 0,733279453 | 0,02794577 | DOCK3 | FALSE | Upregulated |
| -0,4597683 | 0,027970225 | ADI1 | FALSE | Downregulated |
| 0,538707726 | 0,027993896 | CDC42SE2 | FALSE | Upregulated |
| 1,043026951 | 0,028002294 | SECTM1 | TRUE | Upregulated |
| -0,583778091 | 0,028084053 | NDUFB10 | FALSE | Downregulated |
| -1,674978237 | 0,02811354 | SLC16A10 | TRUE | Downregulated |

| | | | | |
|---|---|---|---|---|
| -0,416222036 | 0,028114242 | PROS1 | FALSE | Downregulated |
| 1,18770922 | 0,028114242 | SLC45A4 | TRUE | Upregulated |
| -2,388010426 | 0,028114242 | SLC6A19 | TRUE | Downregulated |
| 0,759701012 | 0,028114242 | TSPAN3 | FALSE | Upregulated |
| 2,043574853 | 0,028135321 | LOC102724850 | TRUE | Upregulated |
| -0,839536725 | 0,028135321 | PPP1R3C | FALSE | Downregulated |
| 0,436086276 | 0,028175836 | CDC42SE1 | FALSE | Upregulated |
| 1,614468003 | 0,028175836 | SLC1A3 | TRUE | Upregulated |
| 0,458984824 | 0,028175836 | YWHAB | FALSE | Upregulated |
| -0,61295735 | 0,028363668 | ECM1 | FALSE | Downregulated |
| -0,765412695 | 0,028388064 | SLC16A1 | FALSE | Downregulated |
| 0,573083501 | 0,028448469 | ATG16L2 | FALSE | Upregulated |
| -0,620318383 | 0,028448772 | TMEM192 | FALSE | Downregulated |
| 0,819339403 | 0,028448772 | U2AF1 | FALSE | Upregulated |
| 1,057635535 | 0,028479749 | FRMD6 | TRUE | Upregulated |
| 0,825176232 | 0,028481971 | C15orf52 | FALSE | Upregulated |
| -0,33828506 | 0,028503254 | ASGR2 | FALSE | Downregulated |
| -0,477917045 | 0,028503254 | GJB1 | FALSE | Downregulated |
| 0,52428365 | 0,028503254 | TUBB4B | FALSE | Upregulated |
| -0,597343013 | 0,028560341 | PCBD1 | FALSE | Downregulated |
| -0,522465802 | 0,028628424 | ACAA2 | FALSE | Downregulated |
| 0,939102293 | 0,028630936 | NFAM1 | FALSE | Upregulated |
| -2,196371832 | 0,028630936 | RP11-418J17.1 | TRUE | Downregulated |
| 0,674580902 | 0,028635307 | PFKFB3 | FALSE | Upregulated |
| 0,341454446 | 0,028635307 | SEPT9 | FALSE | Upregulated |
| -0,827444931 | 0,028711545 | CFHR1 | FALSE | Downregulated |
| 0,602311941 | 0,028811216 | WNT4 | FALSE | Upregulated |
| 2,863835322 | 0,028822493 | LOC101928163 | TRUE | Upregulated |
| -0,866513914 | 0,028838888 | TLE2 | FALSE | Downregulated |
| 0,755622436 | 0,028964058 | CD97 | FALSE | Upregulated |
| 0,782029923 | 0,028964058 | CSNK1G2-AS1 | FALSE | Upregulated |
| -0,445790544 | 0,029044411 | PLG | FALSE | Downregulated |
| 1,094058108 | 0,029120742 | SLC16A7 | TRUE | Upregulated |
| 0,408799943 | 0,029141852 | CTBP1-AS | FALSE | Upregulated |
| -0,772992438 | 0,029141852 | MMAB | FALSE | Downregulated |
| 2,541212147 | 0,029150667 | LOC101928985 | TRUE | Upregulated |
| 1,597193715 | 0,029154731 | AC007461.2 | TRUE | Upregulated |
| -0,417452043 | 0,029215496 | F7 | FALSE | Downregulated |
| 1,356156376 | 0,029215496 | PSTPIP1 | TRUE | Upregulated |
| -0,542526261 | 0,029226042 | ITGA9 | FALSE | Downregulated |
| 1,274536062 | 0,02926883 | NUP93 | TRUE | Upregulated |
| -0,602629381 | 0,0292776 | SMO | FALSE | Downregulated |
| -0,459956424 | 0,02939425 | VPS51 | FALSE | Downregulated |
| 1,657373221 | 0,029542209 | FCGR3B | TRUE | Upregulated |
| 0,859594339 | 0,029542209 | LOC101928240 | FALSE | Upregulated |
| 0,690588313 | 0,029542209 | NBPF15 | FALSE | Upregulated |
| -0,547586535 | 0,029542209 | SERINC5 | FALSE | Downregulated |
| -0,706400987 | 0,029615764 | MOCOS | FALSE | Downregulated |
| -0,524486826 | 0,029659661 | FMO5 | FALSE | Downregulated |
| 0,446227964 | 0,029659661 | MTOR | FALSE | Upregulated |
| -0,710116842 | 0,029743 | CTDSPL | FALSE | Downregulated |

| | | | | |
|---|---|---|---|---|
| 1,141995644 | 0,029782827 | TRPM2 | TRUE | Upregulated |
| 0,5771142 | 0,029826431 | DARS2 | FALSE | Upregulated |
| 1,795854615 | 0,029826431 | PKMYT1 | TRUE | Upregulated |
| 1,121592466 | 0,02985298 | FAM120C | TRUE | Upregulated |
| 1,170406673 | 0,02985298 | HIST2H2BF | TRUE | Upregulated |
| -0,542930893 | 0,030035659 | AK4 | FALSE | Downregulated |
| -0,435080229 | 0,030035659 | F5 | FALSE | Downregulated |
| 0,720544872 | 0,030035659 | LOC101929714 | FALSE | Upregulated |
| -0,806781116 | 0,030035659 | LYVE1 | FALSE | Downregulated |
| -0,499971961 | 0,030035659 | MLXIPL | FALSE | Downregulated |
| 1,49880194 | 0,030035659 | PSMG3 | TRUE | Upregulated |
| -0,541237631 | 0,030035659 | SHMT1 | FALSE | Downregulated |
| -0,43296001 | 0,030168406 | NUDT4 | FALSE | Downregulated |
| -0,922492121 | 0,030168406 | SRD5A2 | FALSE | Downregulated |
| 0,802682567 | 0,030269793 | LIMK1 | FALSE | Upregulated |
| -0,550021301 | 0,03028516 | HSD17B10 | FALSE | Downregulated |
| 0,680275179 | 0,030450834 | IL10RA | FALSE | Upregulated |
| 1,21519512 | 0,030484095 | IFIT5 | TRUE | Upregulated |
| 0,862289003 | 0,030484095 | LOC100996723 | FALSE | Upregulated |
| 0,435292838 | 0,030550112 | ACTN1 | FALSE | Upregulated |
| -0,59277489 | 0,030550112 | C3P1 | FALSE | Downregulated |
| 0,70571081 | 0,030579668 | GPCPD1 | FALSE | Upregulated |
| 0,922612785 | 0,030579668 | LOC100130075 | FALSE | Upregulated |
| 0,368168399 | 0,030579668 | MCL1 | FALSE | Upregulated |
| 0,490191646 | 0,030599641 | VGLL4 | FALSE | Upregulated |
| 0,523955179 | 0,030609964 | HELZ2 | FALSE | Upregulated |
| -0,530106999 | 0,030609964 | RPS4X | FALSE | Downregulated |
| 1,025537237 | 0,030760197 | HCK | TRUE | Upregulated |
| -0,595255572 | 0,030760197 | PANK1 | FALSE | Downregulated |
| 0,836938485 | 0,030867422 | ELMOD3 | FALSE | Upregulated |
| 0,64439487 | 0,031037098 | ARL8B | FALSE | Upregulated |
| 1,398372068 | 0,031042784 | CD86 | TRUE | Upregulated |
| -1,270647887 | 0,031176995 | EMD | TRUE | Downregulated |
| -0,381958767 | 0,031176995 | NHSL1 | FALSE | Downregulated |
| -0,655385342 | 0,031505957 | ADHFE1 | FALSE | Downregulated |
| -0,794395703 | 0,031581177 | LYRM7 | FALSE | Downregulated |
| -0,433310137 | 0,03178203 | WWP1 | FALSE | Downregulated |
| 2,045166805 | 0,031999604 | PLAUR | TRUE | Upregulated |
| 0,667725008 | 0,032095136 | IFNK | FALSE | Upregulated |
| -0,538099457 | 0,032130414 | BZRAP1 | FALSE | Downregulated |
| 0,516837032 | 0,032178794 | PHACTR1 | FALSE | Upregulated |
| -0,550181326 | 0,032178794 | RELN | FALSE | Downregulated |
| -0,302193082 | 0,032178794 | RPL4 | FALSE | Downregulated |
| 0,44990738 | 0,032206245 | MAN2B1 | FALSE | Upregulated |
| 0,726934633 | 0,032452262 | ARPC1B | FALSE | Upregulated |
| -0,69899753 | 0,032452262 | HTR2A | FALSE | Downregulated |
| -0,523660564 | 0,032452262 | PECR | FALSE | Downregulated |
| 0,455651217 | 0,032555323 | LRRTM2 | FALSE | Upregulated |
| 0,700259279 | 0,032581981 | PLXNA1 | FALSE | Upregulated |
| -0,716072965 | 0,032623509 | GSTZ1 | FALSE | Downregulated |
| -0,326810826 | 0,032623509 | IQSEC1 | FALSE | Downregulated |

| | | | | |
|---|---|---|---|---|
| -0,508456183 | 0,032869556 | TF | FALSE | Downregulated |
| 1,011857897 | 0,03296038 | CXCR2P1 | TRUE | Upregulated |
| 1,288829631 | 0,03296038 | PTGER2 | TRUE | Upregulated |
| 1,399147943 | 0,033000427 | M1AP | TRUE | Upregulated |
| 1,564902117 | 0,033014383 | SERPINB8 | TRUE | Upregulated |
| -1,111787872 | 0,03308302 | ADRA2B | TRUE | Downregulated |
| 0,509572106 | 0,033135015 | HMGXB3 | FALSE | Upregulated |
| -1,150136038 | 0,03316342 | CCL14 | TRUE | Downregulated |
| 0,606665861 | 0,033171991 | FNBP1 | FALSE | Upregulated |
| 0,498662244 | 0,033191804 | C15orf60 | FALSE | Upregulated |
| -0,650328608 | 0,033191804 | NINL | FALSE | Downregulated |
| 1,458570973 | 0,033192351 | RP11-27K13.3 | TRUE | Upregulated |
| -0,513053231 | 0,033331162 | CLIC4 | FALSE | Downregulated |
| 0,593888579 | 0,033421946 | PIK3IP1 | FALSE | Upregulated |
| -0,658500588 | 0,033478512 | IGF1R | FALSE | Downregulated |
| -0,587404044 | 0,03347884 | PHYH | FALSE | Downregulated |
| -0,575066818 | 0,033553429 | ETFDH | FALSE | Downregulated |
| 1,030119069 | 0,033604787 | DOK2 | TRUE | Upregulated |
| -0,404099566 | 0,033604787 | RBP4 | FALSE | Downregulated |
| 0,627527269 | 0,033691652 | ITGAV | FALSE | Upregulated |
| 0,600733255 | 0,033704872 | TNFAIP3 | FALSE | Upregulated |
| 0,555938847 | 0,033729438 | LOC101928793 | FALSE | Upregulated |
| -0,827879861 | 0,033788148 | IL1RAP | FALSE | Downregulated |
| 1,56888119 | 0,033788148 | LOC101929074 | TRUE | Upregulated |
| -1,079138657 | 0,034153254 | CDA | TRUE | Downregulated |
| -0,58529718 | 0,034189936 | IMPDH2 | FALSE | Downregulated |
| 0,560189476 | 0,034189936 | PVR | FALSE | Upregulated |
| -0,572449439 | 0,034240496 | PPFIBP1 | FALSE | Downregulated |
| -0,690681534 | 0,034291955 | SLC23A2 | FALSE | Downregulated |
| 1,077790611 | 0,034331891 | WAS | TRUE | Upregulated |
| -0,498641781 | 0,034379274 | ATP13A3 | FALSE | Downregulated |
| 0,444809192 | 0,034379274 | DALRD3 | FALSE | Upregulated |
| -0,347337762 | 0,034441108 | RETSAT | FALSE | Downregulated |
| 0,639883587 | 0,034508953 | GHRL | FALSE | Upregulated |
| -0,549804124 | 0,034541069 | POLD2 | FALSE | Downregulated |
| -0,56745648 | 0,034541069 | RDH10 | FALSE | Downregulated |
| 1,325789042 | 0,034541069 | ZNF80 | TRUE | Upregulated |
| -0,364464793 | 0,034563817 | PRPF8 | FALSE | Downregulated |
| 0,873321694 | 0,034647979 | IGSF6 | FALSE | Upregulated |
| 1,450662794 | 0,034685114 | AL592528.1 | TRUE | Upregulated |
| 0,765616082 | 0,034756106 | RCSD1 | FALSE | Upregulated |
| 0,698956238 | 0,034790479 | DNMT3A | FALSE | Upregulated |
| -0,547361562 | 0,0348881 | ARHGEF17 | FALSE | Downregulated |
| 0,944329218 | 0,035008133 | KLHL25 | FALSE | Upregulated |
| -0,559619307 | 0,035085103 | ACAD10 | FALSE | Downregulated |
| 0,851156672 | 0,03516476 | C12orf44 | FALSE | Upregulated |
| 0,51497851 | 0,035362947 | CCDC42B | FALSE | Upregulated |
| 1,374870719 | 0,035451564 | RP11-430B1.2 | TRUE | Upregulated |
| 0,977866158 | 0,035458761 | SH3PXD2B | FALSE | Upregulated |
| 0,798818759 | 0,035495801 | EHD4 | FALSE | Upregulated |
| 2,347003204 | 0,035560492 | SH2D2A | TRUE | Upregulated |

| | | | | |
|---|---|---|---|---|
| 2,260994911 | 0,035620381 | RP11-594N15.2 | TRUE | Upregulated |
| 0,650527739 | 0,035783842 | LIPA | FALSE | Upregulated |
| 1,422672251 | 0,03584981 | CD3E | TRUE | Upregulated |
| 1,020978523 | 0,03584981 | ELF4 | TRUE | Upregulated |
| 0,478264371 | 0,03584981 | FMNL2 | FALSE | Upregulated |
| -0,426557342 | 0,03584981 | PIPOX | FALSE | Downregulated |
| -1,322794563 | 0,035859561 | ZNF18 | TRUE | Downregulated |
| 1,698980117 | 0,03602246 | LOC102723401 | TRUE | Upregulated |
| -0,731600337 | 0,036122438 | GCAT | FALSE | Downregulated |
| 0,955696905 | 0,036122438 | GPSM1 | FALSE | Upregulated |
| 0,561623991 | 0,036122438 | RP11-28G8.1 | FALSE | Upregulated |
| 0,438438085 | 0,036159156 | FGL2 | FALSE | Upregulated |
| 0,405339772 | 0,036727215 | PITPNA | FALSE | Upregulated |
| -0,396289337 | 0,036740184 | MARCH6 | FALSE | Downregulated |
| 0,44149427 | 0,036740184 | YPEL1 | FALSE | Upregulated |
| 0,415740513 | 0,036766004 | PARP14 | FALSE | Upregulated |
| -0,539832085 | 0,036821963 | FAM210B | FALSE | Downregulated |
| 1,047395204 | 0,036841232 | TK1 | TRUE | Upregulated |
| 0,507190121 | 0,036870322 | SMA5 | FALSE | Upregulated |
| -0,713770112 | 0,037037408 | AMACR | FALSE | Downregulated |
| 0,613334381 | 0,037037408 | DRAP1 | FALSE | Upregulated |
| 0,509428744 | 0,037148607 | MAFK | FALSE | Upregulated |
| 0,344332699 | 0,03718765 | ARHGDIA | FALSE | Upregulated |
| -0,38616409 | 0,03718765 | PKD1P1 | FALSE | Downregulated |
| 0,380345491 | 0,03725809 | SMARCA4 | FALSE | Upregulated |
| 0,704147433 | 0,03733082 | COL4A2 | FALSE | Upregulated |
| 0,54400918 | 0,037338341 | CROCCP3 | FALSE | Upregulated |
| -0,311524272 | 0,037338341 | EEF1G | FALSE | Downregulated |
| 0,433475036 | 0,037471936 | GNAL | FALSE | Upregulated |
| 0,882816868 | 0,037712509 | CTSC | FALSE | Upregulated |
| -0,405245533 | 0,03771473 | PTPLAD1 | FALSE | Downregulated |
| 1,533494745 | 0,037730471 | LGALS3 | TRUE | Upregulated |
| 0,742647572 | 0,037730471 | THBS1 | FALSE | Upregulated |
| 0,784321297 | 0,037731517 | LOC102724584 | FALSE | Upregulated |
| -0,534630585 | 0,03778211 | EI24 | FALSE | Downregulated |
| -0,527718439 | 0,037837987 | ABCA5 | FALSE | Downregulated |
| -0,858882153 | 0,037837987 | CEP120 | FALSE | Downregulated |
| 1,007138527 | 0,037837987 | GIMAP1 | TRUE | Upregulated |
| 0,513625043 | 0,038013353 | LPAR6 | FALSE | Upregulated |
| 1,066987301 | 0,038013353 | RASAL3 | TRUE | Upregulated |
| -0,365056795 | 0,038315973 | NR2F2 | FALSE | Downregulated |
| -0,549082199 | 0,03841869 | F9 | FALSE | Downregulated |
| -0,811839707 | 0,03841869 | GPAM | FALSE | Downregulated |
| 0,710954325 | 0,03841869 | GPR34 | FALSE | Upregulated |
| 0,866838068 | 0,038512895 | MYL5 | FALSE | Upregulated |
| 0,427141703 | 0,038512895 | SNRK-AS1 | FALSE | Upregulated |
| 1,0246913 | 0,038660853 | CCL21 | TRUE | Upregulated |
| 0,597708443 | 0,038683476 | DCLRE1C | FALSE | Upregulated |
| 0,755162899 | 0,038692647 | CCDC28B | FALSE | Upregulated |
| 0,880044914 | 0,03873598 | TRIM59 | FALSE | Upregulated |
| 1,42214768 | 0,038797367 | ACE | TRUE | Upregulated |

| | | | | |
|---|---|---|---|---|
| 0,589473742 | 0,038797367 | MTF1 | FALSE | Upregulated |
| 1,478609745 | 0,039054245 | USP17L2 | TRUE | Upregulated |
| 0,548797558 | 0,039195649 | HPS3 | FALSE | Upregulated |
| -0,451918726 | 0,039261136 | RPS27A | FALSE | Downregulated |
| -0,832003293 | 0,039370114 | C16orf13 | FALSE | Downregulated |
| -0,65324047 | 0,039552616 | SURF1 | FALSE | Downregulated |
| -0,513942334 | 0,039660781 | ABCC6 | FALSE | Downregulated |
| 1,390315682 | 0,039745822 | ACSL4 | TRUE | Upregulated |
| 1,588604758 | 0,039758242 | SIRPB2 | TRUE | Upregulated |
| -0,558773049 | 0,039780231 | HAL | FALSE | Downregulated |
| 2,261809443 | 0,039780231 | PLK1 | TRUE | Upregulated |
| -0,488762653 | 0,039819258 | ZNF91 | FALSE | Downregulated |
| -1,065623973 | 0,039862939 | LRRC40 | TRUE | Downregulated |
| 1,189017816 | 0,039934607 | AGAP2-AS1 | TRUE | Upregulated |
| 0,678185816 | 0,039940886 | CDH23 | FALSE | Upregulated |
| 0,886764593 | 0,040148742 | TSPEAR | FALSE | Upregulated |
| 1,555139459 | 0,040197145 | CD72 | TRUE | Upregulated |
| 0,572965493 | 0,040208388 | TUBA1B | FALSE | Upregulated |
| -0,678708484 | 0,040329848 | CCNG1 | FALSE | Downregulated |
| 0,741907627 | 0,040397474 | YRDC | FALSE | Upregulated |
| 0,421866385 | 0,040437043 | CARM1 | FALSE | Upregulated |
| 0,813463116 | 0,040453716 | CCND3 | FALSE | Upregulated |
| -0,294441309 | 0,040490301 | RPS14 | FALSE | Downregulated |
| 0,718888022 | 0,0409742 | HLA-DPB1 | FALSE | Upregulated |
| 0,415620087 | 0,041045337 | FDPSP2 | FALSE | Upregulated |
| -0,856714793 | 0,041056691 | LOC730102 | FALSE | Downregulated |
| -0,430239776 | 0,041167867 | RPS10 | FALSE | Downregulated |
| -0,719937094 | 0,041221438 | MME | FALSE | Downregulated |
| 0,757076899 | 0,041238131 | GRK6 | FALSE | Upregulated |
| 0,609962228 | 0,041292368 | RCC2 | FALSE | Upregulated |
| 0,541385875 | 0,041311771 | MCM7 | FALSE | Upregulated |
| -0,472394394 | 0,0414723 | ADH5 | FALSE | Downregulated |
| -0,573357154 | 0,0414723 | SLC25A47 | FALSE | Downregulated |
| 0,447437137 | 0,0414723 | ZACN | FALSE | Upregulated |
| 2,468445397 | 0,041480805 | ACAN | TRUE | Upregulated |
| -1,312690586 | 0,041480805 | C5orf27 | TRUE | Downregulated |
| -0,409583113 | 0,041480805 | PEBP1 | FALSE | Downregulated |
| 0,588099211 | 0,041608375 | AP000688.14 | FALSE | Upregulated |
| 0,532165672 | 0,041608375 | DGKZ | FALSE | Upregulated |
| 0,923644054 | 0,041714869 | GRIP2 | FALSE | Upregulated |
| 0,821116251 | 0,041920818 | EPB41L3 | FALSE | Upregulated |
| -1,854101262 | 0,041959577 | MYOM2 | TRUE | Downregulated |
| 0,539836064 | 0,041997162 | B2M | FALSE | Upregulated |
| 0,735553771 | 0,042200006 | LOC641367 | FALSE | Upregulated |
| 1,068243046 | 0,042200006 | SLPI | TRUE | Upregulated |
| 0,529642045 | 0,042312695 | ATP6V1C2 | FALSE | Upregulated |
| 1,056384928 | 0,042312695 | MPL | TRUE | Upregulated |
| -0,397091856 | 0,042353272 | KIAA2018 | FALSE | Downregulated |
| -0,827955233 | 0,042353313 | BMS1P6 | FALSE | Downregulated |
| 1,835230756 | 0,042353313 | OTUD7A | TRUE | Upregulated |
| -0,475265272 | 0,042353313 | SIK3 | FALSE | Downregulated |

| | | | | |
|---|---|---|---|---|
| 1,855991641 | 0,042390315 | LOC101927734 | TRUE | Upregulated |
| 0,460024615 | 0,042672866 | APC2 | FALSE | Upregulated |
| 0,798046767 | 0,042692297 | LOC101928212 | FALSE | Upregulated |
| 2,282867825 | 0,042703179 | HKDC1 | TRUE | Upregulated |
| -0,598722644 | 0,042703179 | U2SURP | FALSE | Downregulated |
| -0,357551243 | 0,042995743 | EIF5 | FALSE | Downregulated |
| 0,564616406 | 0,042999207 | C1orf122 | FALSE | Upregulated |
| 1,779815993 | 0,042999207 | CAP2 | TRUE | Upregulated |
| -0,4578438 | 0,043115856 | BMP1 | FALSE | Downregulated |
| 0,630760154 | 0,043118094 | RAB6B | FALSE | Upregulated |
| 2,33711536 | 0,043211259 | GRAP2 | TRUE | Upregulated |
| 2,15021622 | 0,043211259 | KCNJ5 | TRUE | Upregulated |
| -1,813183205 | 0,043211259 | NDUFAB1 | TRUE | Downregulated |
| 0,544256822 | 0,043211259 | PARP9 | FALSE | Upregulated |
| 0,904405445 | 0,043211259 | SLC15A3 | FALSE | Upregulated |
| -0,427391549 | 0,043419658 | TNS1 | FALSE | Downregulated |
| -1,020123553 | 0,043465086 | NDUFS6 | TRUE | Downregulated |
| -0,427484579 | 0,043558621 | MAP3K2 | FALSE | Downregulated |
| 0,932362229 | 0,043558621 | PRR7-AS1 | FALSE | Upregulated |
| 1,354704936 | 0,043629635 | ASPM | TRUE | Upregulated |
| -1,142445153 | 0,043716946 | PCOLCE2 | TRUE | Downregulated |
| -0,455367481 | 0,043733959 | FGFR3 | FALSE | Downregulated |
| -0,651555646 | 0,043733959 | NFYA | FALSE | Downregulated |
| -0,62422829 | 0,043733959 | RNF5 | FALSE | Downregulated |
| -0,350406359 | 0,043733959 | RPL9 | FALSE | Downregulated |
| 1,79475709 | 0,044090117 | RP3-437I16.1 | TRUE | Upregulated |
| 1,279922482 | 0,044147857 | MCM2 | TRUE | Upregulated |
| -0,424231326 | 0,044185309 | LDLRAD4 | FALSE | Downregulated |
| -0,56823154 | 0,044204614 | PIK3C2G | FALSE | Downregulated |
| -0,896022822 | 0,044390397 | LOC101928431 | FALSE | Downregulated |
| -1,775982431 | 0,044584302 | KCNJ3 | TRUE | Downregulated |
| 1,498040532 | 0,044584302 | LOC102724652 | TRUE | Upregulated |
| 1,584063171 | 0,044587727 | HVCN1 | TRUE | Upregulated |
| 0,578116715 | 0,044587727 | LOC101928621 | FALSE | Upregulated |
| 0,474910533 | 0,044613294 | M6PR | FALSE | Upregulated |
| 0,717083231 | 0,044613294 | OMD | FALSE | Upregulated |
| 0,47269161 | 0,044728215 | FANCA | FALSE | Upregulated |
| -0,527279487 | 0,044728215 | LPHN2 | FALSE | Downregulated |
| -0,390526358 | 0,044750574 | C1orf43 | FALSE | Downregulated |
| -0,480145846 | 0,04485275 | ACSL1 | FALSE | Downregulated |
| 1,022157366 | 0,04485275 | ADAP1 | TRUE | Upregulated |
| 0,553597353 | 0,045083103 | ALG1 | FALSE | Upregulated |
| 1,699177359 | 0,045083103 | C16orf54 | TRUE | Upregulated |
| 1,02734298 | 0,045279777 | GJB7 | TRUE | Upregulated |
| 0,56410774 | 0,045475951 | DMXL2 | FALSE | Upregulated |
| 0,511612743 | 0,04568013 | SNN | FALSE | Upregulated |
| 1,005632992 | 0,045752877 | NOP16 | TRUE | Upregulated |
| 0,586088679 | 0,04581214 | AC005594.3 | FALSE | Upregulated |
| -1,887794999 | 0,04581214 | ASB9 | TRUE | Downregulated |
| 0,698845102 | 0,04581214 | C7orf43 | FALSE | Upregulated |
| 0,531147463 | 0,04581214 | EHBP1L1 | FALSE | Upregulated |

| | | | | |
|---|---|---|---|---|
| 0,89963726 | 0,04581214 | KIF21B | FALSE | Upregulated |
| -0,311617925 | 0,04581214 | KMT2C | FALSE | Downregulated |
| 1,285950088 | 0,04581214 | LOC101926994 | TRUE | Upregulated |
| -1,712207206 | 0,04581214 | LOC101930375 | TRUE | Downregulated |
| -1,530790464 | 0,04581214 | OSBPL6 | TRUE | Downregulated |
| -1,345546827 | 0,04581214 | VPS37D | TRUE | Downregulated |
| 0,485953961 | 0,045841253 | TMEM181 | FALSE | Upregulated |
| 0,642369013 | 0,045845786 | FOXD2 | FALSE | Upregulated |
| -0,300792046 | 0,045845786 | KMT2D | FALSE | Downregulated |
| -0,709382748 | 0,045845786 | TMEM109 | FALSE | Downregulated |
| 0,707677721 | 0,045845786 | TPM1 | FALSE | Upregulated |
| -0,489886533 | 0,046207088 | CECR2 | FALSE | Downregulated |
| -1,138946914 | 0,046207088 | WNK2 | TRUE | Downregulated |
| 2,352888541 | 0,046210399 | CD79A | TRUE | Upregulated |
| -0,841382954 | 0,046232123 | LSS | FALSE | Downregulated |
| 0,451970053 | 0,046408684 | VAC14 | FALSE | Upregulated |
| -0,371788148 | 0,046877063 | UBE3A | FALSE | Downregulated |
| -0,754818216 | 0,046921954 | FOXP1 | FALSE | Downregulated |
| -0,478889952 | 0,047227565 | ADAMTS13 | FALSE | Downregulated |
| 0,633481603 | 0,047529425 | RAET1E-AS1 | FALSE | Upregulated |
| 1,036230206 | 0,047568554 | NMI | TRUE | Upregulated |
| 0,617635744 | 0,047626413 | DNAH17 | FALSE | Upregulated |
| 0,855091026 | 0,047628233 | KLC3 | FALSE | Upregulated |
| 0,474847249 | 0,047780268 | LOC101929333 | FALSE | Upregulated |
| 0,389012985 | 0,047780268 | TOP2B | FALSE | Upregulated |
| -0,310456602 | 0,04788111 | FOXN3 | FALSE | Downregulated |
| -0,529459914 | 0,04788111 | RHOBTB3 | FALSE | Downregulated |
| -1,413708748 | 0,04788111 | SPATA31E1 | TRUE | Downregulated |
| 0,980346277 | 0,047919865 | NRG1 | FALSE | Upregulated |
| -0,803171621 | 0,048003181 | TBC1D2 | FALSE | Downregulated |
| 0,5525218 | 0,048028331 | ASAH1 | FALSE | Upregulated |
| 0,43165436 | 0,048028331 | FAM160B2 | FALSE | Upregulated |
| 0,722703628 | 0,048065525 | CCDC64 | FALSE | Upregulated |
| 0,828788546 | 0,048301432 | RNFT1 | FALSE | Upregulated |
| 0,822248306 | 0,048301432 | SEL1L3 | FALSE | Upregulated |
| -0,374446157 | 0,048320847 | RPL30 | FALSE | Downregulated |
| -0,271978452 | 0,04838712 | CREBBP | FALSE | Downregulated |
| -0,949113229 | 0,048653622 | CFHR3 | FALSE | Downregulated |
| 0,996913491 | 0,048656419 | ANKRD44 | FALSE | Upregulated |
| 1,129221443 | 0,04870793 | NUSAP1 | TRUE | Upregulated |
| 0,77100776 | 0,04888189 | CHADL | FALSE | Upregulated |
| -1,306467714 | 0,048903346 | BMS1P2 | TRUE | Downregulated |
| 1,042893701 | 0,048904837 | LOC101929188 | TRUE | Upregulated |
| -0,42689891 | 0,04895966 | DAPK1 | FALSE | Downregulated |
| -0,475693152 | 0,04895966 | XDH | FALSE | Downregulated |
| -0,587090277 | 0,049013677 | CHST7 | FALSE | Downregulated |
| -0,656402231 | 0,049098835 | ASPG | FALSE | Downregulated |
| -0,736682867 | 0,049235381 | SDHB | FALSE | Downregulated |
| 1,37517038 | 0,049290625 | NKD2 | TRUE | Upregulated |
| 0,707759738 | 0,049304144 | CCDC88B | FALSE | Upregulated |
| 1,145272144 | 0,049304144 | TTF2 | TRUE | Upregulated |

| | | | | |
|---|---|---|---|---|
| 0,443268923 | 0,049438479 | DGKQ | FALSE | Upregulated |
| 0,67031002 | 0,049438479 | JPH3 | FALSE | Upregulated |
| 0,503407099 | 0,04966904 | LOC101928120 | FALSE | Upregulated |
| 1,577036332 | 0,049675274 | TRNP1 | TRUE | Upregulated |
| 0,586734826 | 0,050065452 | AKAP5 | FALSE | Upregulated |
| 0,744833655 | 0,050065452 | COL4A1 | FALSE | Upregulated |
| -0,685967486 | 0,050065452 | CYP2A6 | FALSE | Downregulated |
| -0,798621629 | 0,050065452 | GJB3 | FALSE | Downregulated |
| 1,45338613 | 0,050070104 | DCK | FALSE | Upregulated |
| -0,524079969 | 0,050074718 | NUDT16 | FALSE | Downregulated |
| 0,413349224 | 0,050074718 | RGL4 | FALSE | Upregulated |
| 0,544087782 | 0,050074718 | STIP1 | FALSE | Upregulated |
| 1,997566039 | 0,050105331 | LTB | FALSE | Upregulated |
| 0,906737766 | 0,050163234 | ZXDB | FALSE | Upregulated |
| -0,543751419 | 0,050212191 | NR1H4 | FALSE | Downregulated |
| -0,496548283 | 0,050278922 | HADH | FALSE | Downregulated |
| -0,454905385 | 0,050307371 | AGXT | FALSE | Downregulated |
| -0,553656659 | 0,050307371 | CPED1 | FALSE | Downregulated |
| 0,443515025 | 0,050307371 | JADE1 | FALSE | Upregulated |
| -0,514709938 | 0,050369542 | RPS21 | FALSE | Downregulated |
| -0,458580452 | 0,050500142 | DCHS1 | FALSE | Downregulated |
| 1,366497227 | 0,05054356 | SLC25A33 | FALSE | Upregulated |
| 0,500186214 | 0,050689992 | SDC3 | FALSE | Upregulated |
| 0,566006064 | 0,050804761 | C19orf12 | FALSE | Upregulated |
| 0,730865086 | 0,050804761 | PRPF38B | FALSE | Upregulated |
| -0,319152379 | 0,050901858 | ORMDL3 | FALSE | Downregulated |
| -2,086874397 | 0,050901858 | PKD1L2 | FALSE | Downregulated |
| 0,676459076 | 0,050917413 | PTPN7 | FALSE | Upregulated |
| 0,414972501 | 0,051109595 | CRLF3 | FALSE | Upregulated |
| -0,682643516 | 0,051204737 | RAB11B-AS1 | FALSE | Downregulated |
| -0,411463266 | 0,051211414 | ACAT1 | FALSE | Downregulated |
| 1,973558025 | 0,051211414 | ATP6V1E2 | FALSE | Upregulated |
| -1,888115444 | 0,051211414 | LOC102723864 | FALSE | Downregulated |
| -0,285785326 | 0,051211414 | RPL19 | FALSE | Downregulated |
| -0,393021341 | 0,051223326 | ECH1 | FALSE | Downregulated |
| 2,175526589 | 0,051312425 | LOC101928292 | FALSE | Upregulated |
| 0,531620446 | 0,051390369 | A1BG-AS1 | FALSE | Upregulated |
| -0,312758027 | 0,051390369 | RPS6 | FALSE | Downregulated |
| 0,969682603 | 0,051390369 | SLC22A18AS | FALSE | Upregulated |
| 0,37407749 | 0,051390369 | TRIM14 | FALSE | Upregulated |
| 0,790907884 | 0,051596386 | LOC101928429 | FALSE | Upregulated |
| 0,445975869 | 0,051596386 | SNX21 | FALSE | Upregulated |
| 1,109594568 | 0,051602729 | PIPSL | FALSE | Upregulated |
| 0,724045606 | 0,051683176 | RP3-476K8.3 | FALSE | Upregulated |
| 1,66274043 | 0,051683176 | WDR62 | FALSE | Upregulated |
| 0,792407806 | 0,051715528 | MAX | FALSE | Upregulated |
| -0,460226356 | 0,051798665 | ABCA8 | FALSE | Downregulated |
| -0,450873418 | 0,051848157 | ST3GAL1 | FALSE | Downregulated |
| -0,963517141 | 0,051876118 | MOGAT2 | FALSE | Downregulated |
| 1,919176099 | 0,051916441 | CA5BP1 | FALSE | Upregulated |
| -1,124191668 | 0,051916441 | COL11A2 | FALSE | Downregulated |

| | | | | |
|---|---|---|---|---|
| -0,650646704 | 0,051916441 | NCKAP5 | FALSE | Downregulated |
| -1,390886182 | 0,052015364 | CCDC87 | FALSE | Downregulated |
| 0,629436889 | 0,052211108 | BOLA3-AS1 | FALSE | Upregulated |
| -0,397493273 | 0,052332098 | ARHGEF10L | FALSE | Downregulated |
| 0,266902261 | 0,052356379 | SQSTM1 | FALSE | Upregulated |
| -0,440728474 | 0,052384333 | UBA52 | FALSE | Downregulated |
| -0,64039024 | 0,052414811 | BPHL | FALSE | Downregulated |
| -0,466960004 | 0,052426488 | F12 | FALSE | Downregulated |
| 0,7933233 | 0,052450726 | RRP1 | FALSE | Upregulated |
| -1,231943767 | 0,052450726 | VIL1 | FALSE | Downregulated |
| -0,834821117 | 0,052602643 | ARHGAP42 | FALSE | Downregulated |
| 1,094562374 | 0,052615109 | CD163L1 | FALSE | Upregulated |
| -0,600615548 | 0,052624329 | ANKRD28 | FALSE | Downregulated |
| 1,153996518 | 0,052681351 | POLD3 | FALSE | Upregulated |
| 0,443650948 | 0,052723577 | PLXNC1 | FALSE | Upregulated |
| 0,319869205 | 0,052906698 | CHD4 | FALSE | Upregulated |
| -0,426923312 | 0,052957001 | PLOD1 | FALSE | Downregulated |
| -0,294409287 | 0,053133239 | LONP2 | FALSE | Downregulated |
| 0,753228633 | 0,053189431 | PRORSD1P | FALSE | Upregulated |
| -0,520336637 | 0,053189431 | ZBTB10 | FALSE | Downregulated |
| 0,802916936 | 0,053193348 | CLUAP1 | FALSE | Upregulated |
| -0,742799483 | 0,053356117 | FRY-AS1 | FALSE | Downregulated |
| 0,853841703 | 0,053356117 | NPEPL1 | FALSE | Upregulated |
| 0,76000175 | 0,053356117 | OGN | FALSE | Upregulated |
| -0,341055724 | 0,053356117 | TOMM70A | FALSE | Downregulated |
| 0,52553256 | 0,053660212 | CHTOP | FALSE | Upregulated |
| 1,041696247 | 0,053660212 | TRIM21 | FALSE | Upregulated |
| -0,354038461 | 0,053733003 | ASH1L | FALSE | Downregulated |
| 2,046871989 | 0,053816939 | RP11-17A4.2 | FALSE | Upregulated |
| 1,756812032 | 0,053856658 | DDX53 | FALSE | Upregulated |
| 1,312201432 | 0,053856658 | MDM1 | FALSE | Upregulated |
| -0,993860422 | 0,053856658 | SUGCT | FALSE | Downregulated |
| 0,519713853 | 0,053883666 | IFIT1 | FALSE | Upregulated |
| 0,592050563 | 0,053915402 | FAM21A | FALSE | Upregulated |
| -0,417109237 | 0,053915402 | TMPRSS6 | FALSE | Downregulated |
| 0,611041942 | 0,053934352 | AKIRIN1 | FALSE | Upregulated |
| 1,499802101 | 0,05396753 | RTN1 | FALSE | Upregulated |
| 0,631737889 | 0,054139149 | FLJ32790 | FALSE | Upregulated |
| 0,786956555 | 0,054139149 | SLC12A5 | FALSE | Upregulated |
| -0,358238528 | 0,054152461 | HECTD4 | FALSE | Downregulated |
| 0,840379003 | 0,054220344 | WDFY4 | FALSE | Upregulated |
| -0,360819997 | 0,054279896 | SMG1 | FALSE | Downregulated |
| 0,877110658 | 0,054285329 | LOC101929668 | FALSE | Upregulated |
| -0,291473284 | 0,054314749 | KMT2A | FALSE | Downregulated |
| 0,426829709 | 0,054314749 | NACC1 | FALSE | Upregulated |
| -1,0664951 | 0,054314749 | WNT5A | FALSE | Downregulated |
| -0,717773633 | 0,054422642 | LIG4 | FALSE | Downregulated |
| 0,875000742 | 0,054469157 | LOC100287808 | FALSE | Upregulated |
| -0,445857088 | 0,054754196 | CNKSR3 | FALSE | Downregulated |
| 0,935559878 | 0,05479825 | MAP1A | FALSE | Upregulated |
| -0,590189102 | 0,054819907 | HSD17B2 | FALSE | Downregulated |

| | | | | |
|---|---|---|---|---|
| 0,647484975 | 0,054819907 | LOXL4 | FALSE | Upregulated |
| -0,752194563 | 0,054819907 | RP11-384K6.6 | FALSE | Downregulated |
| 0,833392226 | 0,054861219 | CDT1 | FALSE | Upregulated |
| 0,834923266 | 0,054993929 | RP13-143G15.3 | FALSE | Upregulated |
| 0,697470256 | 0,055044527 | LRRC37A6P | FALSE | Upregulated |
| -0,477518353 | 0,055061762 | CEP164 | FALSE | Downregulated |
| 0,352555674 | 0,055128234 | QKI | FALSE | Upregulated |
| 0,958328572 | 0,055266103 | PP14571 | FALSE | Upregulated |
| 0,825686791 | 0,055492219 | FAM98B | FALSE | Upregulated |
| -0,441099791 | 0,055492219 | RAPH1 | FALSE | Downregulated |
| 1,07491072 | 0,055505536 | ACSBG1 | FALSE | Upregulated |
| -0,472209225 | 0,055622382 | ACSM2B | FALSE | Downregulated |
| 0,683809078 | 0,055622382 | ADAMTS2 | FALSE | Upregulated |
| 0,748519432 | 0,055622382 | LOC101926887 | FALSE | Upregulated |
| -0,512713421 | 0,055622382 | MKL2 | FALSE | Downregulated |
| 2,403198758 | 0,055622382 | MTMR11 | FALSE | Upregulated |
| -0,651817107 | 0,055622382 | ROBO4 | FALSE | Downregulated |
| 0,718302245 | 0,055622382 | TIPARP-AS1 | FALSE | Upregulated |
| 1,491729786 | 0,055662788 | STAMBPL1 | FALSE | Upregulated |
| -0,444515562 | 0,055719319 | KIAA2026 | FALSE | Downregulated |
| 1,186728586 | 0,055719319 | NCF1 | FALSE | Upregulated |
| -1,106464625 | 0,055747162 | PLIN1 | FALSE | Downregulated |
| 0,72399578 | 0,055792684 | VTI1A | FALSE | Upregulated |
| -0,957855414 | 0,055794508 | SNRNP25 | FALSE | Downregulated |
| 0,4924576 | 0,055834911 | AQP11 | FALSE | Upregulated |
| 0,970813287 | 0,055863653 | SELPLG | FALSE | Upregulated |
| -0,577912872 | 0,056070092 | ZNF281 | FALSE | Downregulated |
| -0,925104421 | 0,056360127 | PREPL | FALSE | Downregulated |
| -0,470390735 | 0,056371152 | CHDH | FALSE | Downregulated |
| -0,998807875 | 0,056371152 | PARD6B | FALSE | Downregulated |
| -0,354996493 | 0,056554441 | FKBP8 | FALSE | Downregulated |
| -0,654422142 | 0,056641818 | FGD5 | FALSE | Downregulated |
| -0,400070279 | 0,056641818 | SPG7 | FALSE | Downregulated |
| -0,510451954 | 0,056641818 | ZSWIM1 | FALSE | Downregulated |
| 0,490386541 | 0,05665111 | LIMD2 | FALSE | Upregulated |
| -0,294697562 | 0,05665111 | RPL15 | FALSE | Downregulated |
| -0,38004018 | 0,056652085 | RPL7 | FALSE | Downregulated |
| 0,596660535 | 0,056782134 | CLDN3 | FALSE | Upregulated |
| -0,650794601 | 0,056815786 | SRCIN1 | FALSE | Downregulated |
| 0,847326507 | 0,056927999 | AMPD3 | FALSE | Upregulated |
| 0,633155574 | 0,057215064 | GBP2 | FALSE | Upregulated |
| 0,650981764 | 0,057496539 | LOC101927732 | FALSE | Upregulated |
| 0,467377819 | 0,057579649 | CAPN2 | FALSE | Upregulated |
| 0,673370278 | 0,057650552 | CYCS | FALSE | Upregulated |
| 0,628055271 | 0,057650552 | NAP1L1 | FALSE | Upregulated |
| 1,719090718 | 0,057650552 | SPIB | FALSE | Upregulated |
| 0,505840234 | 0,057650552 | TIGD6 | FALSE | Upregulated |
| 0,684188845 | 0,05767481 | SLC7A5P2 | FALSE | Upregulated |
| -0,541147579 | 0,05767481 | TINAGL1 | FALSE | Downregulated |
| -0,513913386 | 0,057689312 | AKT1S1 | FALSE | Downregulated |
| -0,669108736 | 0,057750585 | HSD17B8 | FALSE | Downregulated |

| | | | | |
|---|---|---|---|---|
| -0,578158614 | 0,057750585 | LOC100996492 | FALSE | Downregulated |
| -0,510390707 | 0,057750585 | SMOC1 | FALSE | Downregulated |
| -0,489735035 | 0,057750585 | ZBTB40 | FALSE | Downregulated |
| 2,13604229 | 0,057853104 | HIST1H2AL | FALSE | Upregulated |
| 0,430562091 | 0,057919787 | ENTPD7 | FALSE | Upregulated |
| 0,681496523 | 0,057919787 | TSR2 | FALSE | Upregulated |
| 0,631214508 | 0,058025642 | LOC101927839 | FALSE | Upregulated |
| 1,095988767 | 0,058043455 | DRAM1 | FALSE | Upregulated |
| -0,358438232 | 0,058083125 | MAOB | FALSE | Downregulated |
| -0,477186028 | 0,058083125 | RPL17 | FALSE | Downregulated |
| -0,459928448 | 0,05812693 | IVNS1ABP | FALSE | Downregulated |
| 0,659246172 | 0,05812693 | LINC00854 | FALSE | Upregulated |
| -0,24617908 | 0,05812693 | MST1 | FALSE | Downregulated |
| 0,879972555 | 0,05812693 | SLC8A1-AS1 | FALSE | Upregulated |
| -0,661525164 | 0,058156876 | LZTS3 | FALSE | Downregulated |
| -0,698275173 | 0,058266296 | TSPAN33 | FALSE | Downregulated |
| -0,349711719 | 0,058289816 | FGFRL1 | FALSE | Downregulated |
| 2,071976509 | 0,058450629 | ECT2 | FALSE | Upregulated |
| -0,859805831 | 0,05855396 | NAV2-AS4 | FALSE | Downregulated |
| -0,584270723 | 0,058555788 | AUH | FALSE | Downregulated |
| 0,532583762 | 0,058638928 | CCDC97 | FALSE | Upregulated |
| 0,799432304 | 0,058730261 | SRC | FALSE | Upregulated |
| -1,052201551 | 0,058780382 | LOC101927645 | FALSE | Downregulated |
| -0,702799977 | 0,059052615 | B9D1 | FALSE | Downregulated |
| 0,454072757 | 0,059055808 | IDH2 | FALSE | Upregulated |
| 0,997362676 | 0,059191804 | MAP3K14-AS1 | FALSE | Upregulated |
| 0,897092278 | 0,059248428 | PGGT1B | FALSE | Upregulated |
| 1,445626908 | 0,059363647 | TMEM150B | FALSE | Upregulated |
| -1,907368327 | 0,059538986 | TMEM132E | FALSE | Downregulated |
| 0,625133896 | 0,059540958 | XAF1 | FALSE | Upregulated |
| -0,576255139 | 0,059794427 | MARC1 | FALSE | Downregulated |
| -0,460010588 | 0,05990516 | NBPF14 | FALSE | Downregulated |
| 0,496696486 | 0,060029928 | GRN | FALSE | Upregulated |
| -0,703109593 | 0,060148902 | SLC19A2 | FALSE | Downregulated |
| 0,73732599 | 0,060324582 | AEBP1 | FALSE | Upregulated |
| 1,704632965 | 0,060327566 | RP1-102G20.5 | FALSE | Upregulated |
| -0,748442619 | 0,060356676 | GRAMD1C | FALSE | Downregulated |
| 0,438233924 | 0,060581159 | ATP11A-AS1 | FALSE | Upregulated |
| -0,2652385 | 0,060600342 | MPRIP | FALSE | Downregulated |
| 2,182091024 | 0,06086934 | LOC102724585 | FALSE | Upregulated |
| 0,377510094 | 0,06086934 | RARA | FALSE | Upregulated |
| 2,023683356 | 0,060946604 | CD300LF | FALSE | Upregulated |
| -0,576327505 | 0,061071191 | HMCN1 | FALSE | Downregulated |
| 0,59823247 | 0,061071191 | PTPN6 | FALSE | Upregulated |
| 1,201803238 | 0,061229826 | SOCS3 | FALSE | Upregulated |
| 0,468361819 | 0,061232649 | HNRNPUL2 | FALSE | Upregulated |
| 0,722459572 | 0,06126315 | FGD2 | FALSE | Upregulated |
| 0,999139766 | 0,061325169 | SLC38A1 | FALSE | Upregulated |
| -0,439388772 | 0,061436167 | SERPINC1 | FALSE | Downregulated |
| -0,440234554 | 0,061438249 | ACSM2A | FALSE | Downregulated |
| 0,595743238 | 0,061526768 | TTC7A | FALSE | Upregulated |

| | | | | |
|---|---|---|---|---|
| 0,577837919 | 0,06166596 | TSSK4 | FALSE | Upregulated |
| -0,914522067 | 0,061675062 | FUNDC2 | FALSE | Downregulated |
| 1,075930148 | 0,061974379 | ATP6V0E2-AS1 | FALSE | Upregulated |
| 0,610083595 | 0,061974379 | ZDHHC23 | FALSE | Upregulated |
| -1,585529874 | 0,062071329 | TMEM14A | FALSE | Downregulated |
| -0,633096188 | 0,062100783 | AKAP6 | FALSE | Downregulated |
| 0,687192356 | 0,062100783 | RARRES1 | FALSE | Upregulated |
| 2,252534738 | 0,062100783 | TESPA1 | FALSE | Upregulated |
| -0,470007598 | 0,062199185 | ETNK2 | FALSE | Downregulated |
| -0,489470602 | 0,062199185 | HDHD3 | FALSE | Downregulated |
| -1,835114522 | 0,062199185 | LOC102724720 | FALSE | Downregulated |
| 0,730169321 | 0,062199185 | PLEKHF1 | FALSE | Upregulated |
| -1,832846731 | 0,062199185 | TEN1-CDK3 | FALSE | Downregulated |
| 0,338716731 | 0,062281462 | POM121 | FALSE | Upregulated |
| -0,561981423 | 0,062356106 | DSG2 | FALSE | Downregulated |
| -0,772630995 | 0,062356106 | KIF12 | FALSE | Downregulated |
| 0,307386629 | 0,062356106 | SET | FALSE | Upregulated |
| 0,381726251 | 0,062385483 | SMAD3 | FALSE | Upregulated |
| -2,493348753 | 0,062457861 | FAM222A-AS1 | FALSE | Downregulated |
| -0,328971553 | 0,062711705 | ST6GAL1 | FALSE | Downregulated |
| -0,341764565 | 0,062958077 | LRP3 | FALSE | Downregulated |
| 0,319598504 | 0,062958077 | SHISA5 | FALSE | Upregulated |
| -0,834838872 | 0,062958077 | STK36 | FALSE | Downregulated |
| 0,63274186 | 0,062996855 | CHD3 | FALSE | Upregulated |
| -0,399090127 | 0,062996855 | NFIB | FALSE | Downregulated |
| 0,411697425 | 0,062996855 | P2RX5-TAX1BP3 | FALSE | Upregulated |
| 0,861668224 | 0,062996855 | RIC3 | FALSE | Upregulated |
| -1,761826796 | 0,062996855 | TRIM16L | FALSE | Downregulated |
| -0,436729663 | 0,06304287 | SHPK | FALSE | Downregulated |
| 0,49381465 | 0,063407835 | IGFBP7 | FALSE | Upregulated |
| 0,785806475 | 0,063478702 | IFI35 | FALSE | Upregulated |
| 0,678638788 | 0,063698879 | ARIH2OS | FALSE | Upregulated |
| 0,687912693 | 0,063766775 | DPH3P1 | FALSE | Upregulated |
| 0,485226344 | 0,064041818 | PPT1 | FALSE | Upregulated |
| -0,355368831 | 0,064041818 | TENC1 | FALSE | Downregulated |
| 0,406971947 | 0,064074013 | CEMP1 | FALSE | Upregulated |
| 0,637046246 | 0,064131283 | CCDC157 | FALSE | Upregulated |
| 0,525306205 | 0,064190419 | KIAA0930 | FALSE | Upregulated |
| 1,082664477 | 0,064349117 | LOC101928356 | FALSE | Upregulated |
| -0,591005036 | 0,064349117 | SERPINA7 | FALSE | Downregulated |
| -0,478117641 | 0,064395363 | KIF21A | FALSE | Downregulated |
| -0,418368461 | 0,064530278 | CELF1 | FALSE | Downregulated |
| 1,051954416 | 0,064541895 | SLC30A4 | FALSE | Upregulated |
| 0,673495633 | 0,064668472 | C12orf65 | FALSE | Upregulated |
| 0,739786612 | 0,064800053 | TMEM50A | FALSE | Upregulated |
| 0,581857647 | 0,065218026 | ZCWPW1 | FALSE | Upregulated |
| 0,945315432 | 0,065231254 | LOC101929018 | FALSE | Upregulated |
| 2,073215041 | 0,065361985 | AC007750.5 | FALSE | Upregulated |
| -0,365666211 | 0,065361985 | TOLLIP | FALSE | Downregulated |
| -0,673067278 | 0,065487742 | WDR90 | FALSE | Downregulated |
| 0,830966322 | 0,065497512 | LOC100129518 | FALSE | Upregulated |

| | | | | |
|---|---|---|---|---|
| -0,442474393 | 0,065563466 | CHP1 | FALSE | Downregulated |
| 1,112959485 | 0,065563466 | LOC101927675 | FALSE | Upregulated |
| 0,501611262 | 0,065563466 | MICALL1 | FALSE | Upregulated |
| -0,384558618 | 0,065683568 | SFXN5 | FALSE | Downregulated |
| 0,387730959 | 0,065951274 | DLGAP4 | FALSE | Upregulated |
| -0,542327104 | 0,065962895 | ATP7B | FALSE | Downregulated |
| -0,460073388 | 0,066164081 | RPL22 | FALSE | Downregulated |
| -0,284735505 | 0,066164081 | XBP1 | FALSE | Downregulated |
| 0,509334823 | 0,06647116 | CORO1C | FALSE | Upregulated |
| 0,590522305 | 0,06647116 | FAS-AS1 | FALSE | Upregulated |
| 0,595098518 | 0,06647116 | MICALL2 | FALSE | Upregulated |
| -0,354915959 | 0,066524584 | SEPHS2 | FALSE | Downregulated |
| 0,578947835 | 0,066550824 | NAT9 | FALSE | Upregulated |
| -0,557529505 | 0,066572283 | NDUFA4 | FALSE | Downregulated |
| -0,46381964 | 0,066572283 | SHROOM2 | FALSE | Downregulated |
| -0,904254805 | 0,06688606 | ZNF324 | FALSE | Downregulated |
| -1,054496386 | 0,067006916 | CYP1A2 | FALSE | Downregulated |
| -0,3963323 | 0,06704563 | KCTD3 | FALSE | Downregulated |
| 0,554600979 | 0,06706559 | C15orf37 | FALSE | Upregulated |
| -0,648305637 | 0,06706559 | CLDN14 | FALSE | Downregulated |
| -0,372512894 | 0,067079456 | ACAD11 | FALSE | Downregulated |
| -0,421435013 | 0,06745828 | CHRD | FALSE | Downregulated |
| -3,095751225 | 0,06745828 | KIAA1751 | FALSE | Downregulated |
| 0,343306614 | 0,06745828 | PKP4 | FALSE | Upregulated |
| -0,400136247 | 0,06745828 | RPL10A | FALSE | Downregulated |
| 0,412324961 | 0,067614143 | DOPEY1 | FALSE | Upregulated |
| 1,801825433 | 0,067614143 | HCG25 | FALSE | Upregulated |
| 2,360585207 | 0,067614143 | IQGAP3 | FALSE | Upregulated |
| -0,609810714 | 0,067901003 | IKZF5 | FALSE | Downregulated |
| 0,589639086 | 0,068176357 | SATL1 | FALSE | Upregulated |
| 0,986172418 | 0,068182214 | ITGA4 | FALSE | Upregulated |
| 0,777858211 | 0,068208714 | LOC100499194 | FALSE | Upregulated |
| 1,308413855 | 0,068331142 | ZBTB24 | FALSE | Upregulated |
| 1,23474732 | 0,068340106 | CCL19 | FALSE | Upregulated |
| 1,09447863 | 0,068340106 | CMPK2 | FALSE | Upregulated |
| -0,652509535 | 0,068340106 | ERRFI1 | FALSE | Downregulated |
| 0,358537513 | 0,068340106 | PSAP | FALSE | Upregulated |
| 0,546680563 | 0,068583876 | SOX12 | FALSE | Upregulated |
| 0,447547897 | 0,068608857 | JAZF1 | FALSE | Upregulated |
| 0,643291468 | 0,068608905 | NCF2 | FALSE | Upregulated |
| 0,505923646 | 0,06861674 | ANXA6 | FALSE | Upregulated |
| 2,443137975 | 0,068646383 | MMP9 | FALSE | Upregulated |
| -0,375742367 | 0,068851433 | PIK3R1 | FALSE | Downregulated |
| -0,673178481 | 0,069049227 | FGD4 | FALSE | Downregulated |
| -1,614890658 | 0,069049227 | TCEAL8 | FALSE | Downregulated |
| 1,526641042 | 0,069095813 | LOC101928491 | FALSE | Upregulated |
| 1,07583493 | 0,069126705 | LOC102723317 | FALSE | Upregulated |
| -0,362196517 | 0,069262868 | AK3 | FALSE | Downregulated |
| -0,781668782 | 0,069262868 | DEPDC7 | FALSE | Downregulated |
| 1,762013835 | 0,069461608 | BARHL1 | FALSE | Upregulated |
| 0,401191058 | 0,069461608 | CCNL1 | FALSE | Upregulated |

| | | | | |
|---|---|---|---|---|
| -0,705423706 | 0,069461608 | HHEX | FALSE | Downregulated |
| 0,52594855 | 0,06952206 | CLSTN1 | FALSE | Upregulated |
| 1,231177834 | 0,069541684 | SLC2A1 | FALSE | Upregulated |
| 0,469774825 | 0,069671244 | FUCA2 | FALSE | Upregulated |
| 2,152508848 | 0,06973304 | DUSP8 | FALSE | Upregulated |
| 0,449574346 | 0,069994677 | ABR | FALSE | Upregulated |
| -1,096776533 | 0,069994677 | CYP2A7 | FALSE | Downregulated |
| 0,394413182 | 0,069994677 | LRRFIP1 | FALSE | Upregulated |
| -1,154729828 | 0,069994677 | MRGPRF | FALSE | Downregulated |
| 0,537549675 | 0,069994677 | PML | FALSE | Upregulated |
| 0,389541595 | 0,069994677 | TNFRSF10B | FALSE | Upregulated |
| 2,213794179 | 0,0700191 | IGFBP7-AS1 | FALSE | Upregulated |
| -0,369773106 | 0,0700191 | MED13L | FALSE | Downregulated |
| -0,299655908 | 0,070022974 | GLTSCR2 | FALSE | Downregulated |
| -0,71181519 | 0,070022974 | TMEM25 | FALSE | Downregulated |
| -1,33365827 | 0,07006421 | NAALADL2 | FALSE | Downregulated |
| 0,347904512 | 0,070272682 | FMNL3 | FALSE | Upregulated |
| -0,423662854 | 0,070635202 | FXR1 | FALSE | Downregulated |
| 1,166637679 | 0,070635202 | GOLGA2P7 | FALSE | Upregulated |
| 0,441967777 | 0,070635202 | ILK | FALSE | Upregulated |
| -0,508981676 | 0,070728091 | APCS | FALSE | Downregulated |
| 0,610785998 | 0,070813846 | COPG2IT1 | FALSE | Upregulated |
| 0,790162827 | 0,070821859 | LBH | FALSE | Upregulated |
| 0,745832259 | 0,071100306 | CELF3 | FALSE | Upregulated |
| -0,491271714 | 0,071200325 | GPT | FALSE | Downregulated |
| 1,448307057 | 0,071504236 | SLC2A3 | FALSE | Upregulated |
| 0,767428415 | 0,071526539 | LMNB2 | FALSE | Upregulated |
| -1,103066841 | 0,07174886 | ZNF554 | FALSE | Downregulated |
| 0,585859382 | 0,071757744 | C17orf62 | FALSE | Upregulated |
| -0,570669806 | 0,072048433 | MDM4 | FALSE | Downregulated |
| -0,418404312 | 0,072262599 | KLKB1 | FALSE | Downregulated |
| 0,551359797 | 0,072262599 | PTRH2 | FALSE | Upregulated |
| 3,080296764 | 0,072585503 | RIMBP3C | FALSE | Upregulated |
| 0,52901404 | 0,072630724 | ABI1 | FALSE | Upregulated |
| 0,551226037 | 0,072630724 | KIAA0226 | FALSE | Upregulated |
| 1,178518169 | 0,072630724 | STXBP5-AS1 | FALSE | Upregulated |
| 0,679967823 | 0,072715499 | GATS | FALSE | Upregulated |
| -0,699940906 | 0,072735591 | AMER1 | FALSE | Downregulated |
| -0,689393028 | 0,072888781 | SMIM7 | FALSE | Downregulated |
| 1,678680167 | 0,072920168 | LOC101927071 | FALSE | Upregulated |
| 2,812885497 | 0,073187125 | AKR1B10 | FALSE | Upregulated |
| 0,764863553 | 0,073581875 | VSIG4 | FALSE | Upregulated |
| -0,402468325 | 0,073644297 | ACSM3 | FALSE | Downregulated |
| -0,530826804 | 0,073883945 | NPIPB11 | FALSE | Downregulated |
| -0,303516574 | 0,073900205 | UBXN6 | FALSE | Downregulated |
| 2,254151299 | 0,074120909 | GPC3 | FALSE | Upregulated |
| 0,5162797 | 0,074120909 | LRRC24 | FALSE | Upregulated |
| -0,755585451 | 0,074226742 | DRG2 | FALSE | Downregulated |
| 0,532177141 | 0,074226742 | RDH14 | FALSE | Upregulated |
| -0,634706545 | 0,074234483 | CHID1 | FALSE | Downregulated |
| 0,329647608 | 0,074269655 | GATC | FALSE | Upregulated |

| | | | | |
|---|---|---|---|---|
| -0,552380048 | 0,074269655 | PRDX3 | FALSE | Downregulated |
| -0,819097167 | 0,074269655 | TEK | FALSE | Downregulated |
| -1,227303197 | 0,074269655 | TTLL7 | FALSE | Downregulated |
| 0,342165864 | 0,074269655 | ZNRF2 | FALSE | Upregulated |
| 0,68242958 | 0,0743573 | CD5 | FALSE | Upregulated |
| -0,54539712 | 0,074505844 | IL17RC | FALSE | Downregulated |
| -1,17780901 | 0,074505844 | LOC102724175 | FALSE | Downregulated |
| 0,85877599 | 0,074505844 | YBX3 | FALSE | Upregulated |
| 1,514511549 | 0,074512073 | TPX2 | FALSE | Upregulated |
| 0,421443816 | 0,074641977 | B3GALT2 | FALSE | Upregulated |
| -0,59632719 | 0,074659183 | NFKBIZ | FALSE | Downregulated |
| -0,991230596 | 0,074659183 | PMS2CL | FALSE | Downregulated |
| 0,793428413 | 0,074659183 | RABEP2 | FALSE | Upregulated |
| -0,669862827 | 0,074741046 | FBL | FALSE | Downregulated |
| -0,341073815 | 0,074894316 | ADIPOR1 | FALSE | Downregulated |
| 0,860147484 | 0,074974497 | HK2 | FALSE | Upregulated |
| 0,948364191 | 0,0751317 | CHRNA2 | FALSE | Upregulated |
| 0,88525906 | 0,075258001 | TEAD2 | FALSE | Upregulated |
| -0,742238632 | 0,075432064 | NAV2-AS5 | FALSE | Downregulated |
| 3,838312465 | 0,075592383 | TNFSF12-TNFSF13 | FALSE | Upregulated |
| 0,517835349 | 0,075670091 | ARPC5 | FALSE | Upregulated |
| -0,532674726 | 0,075670091 | MOCS2 | FALSE | Downregulated |
| 0,540725867 | 0,075779013 | IFNAR2 | FALSE | Upregulated |
| -0,649379453 | 0,075842137 | METTL7B | FALSE | Downregulated |
| 0,847868574 | 0,075842137 | MMEL1 | FALSE | Upregulated |
| -0,454393301 | 0,075978916 | RPL21 | FALSE | Downregulated |
| -0,690515604 | 0,076013966 | DCUN1D3 | FALSE | Downregulated |
| -0,376215752 | 0,076013966 | PCF11 | FALSE | Downregulated |
| 0,610642702 | 0,076013966 | RRS1 | FALSE | Upregulated |
| 0,608355957 | 0,076222322 | LENG9 | FALSE | Upregulated |
| -0,419125366 | 0,076222322 | OPLAH | FALSE | Downregulated |
| 1,314639634 | 0,076309415 | IGJ | FALSE | Upregulated |
| 0,757097404 | 0,076319365 | DNAJB11 | FALSE | Upregulated |
| -1,19577059 | 0,076319436 | LOC101060376 | FALSE | Downregulated |
| 1,527830835 | 0,076347643 | PART1 | FALSE | Upregulated |
| 1,062795958 | 0,076428057 | TMEM51 | FALSE | Upregulated |
| 1,337291538 | 0,076693585 | LOXL1 | FALSE | Upregulated |
| 0,407354928 | 0,076693585 | TMEM131 | FALSE | Upregulated |
| 0,350446857 | 0,076693585 | VAT1 | FALSE | Upregulated |
| 0,594885647 | 0,076757581 | ARHGEF39 | FALSE | Upregulated |
| 0,365563985 | 0,076843793 | MYO9B | FALSE | Upregulated |
| 0,947757825 | 0,076893424 | KCNG2 | FALSE | Upregulated |
| 0,728005789 | 0,07695409 | ATP13A2 | FALSE | Upregulated |
| 0,974748672 | 0,07695409 | P4HA2-AS1 | FALSE | Upregulated |
| 0,63531379 | 0,077028124 | LOC101928789 | FALSE | Upregulated |
| -0,598802233 | 0,077076197 | REPS1 | FALSE | Downregulated |
| -0,386437704 | 0,077277513 | RMND5A | FALSE | Downregulated |
| 0,723495977 | 0,077562571 | LOC102724026 | FALSE | Upregulated |
| -0,442902538 | 0,077709937 | CXCL12 | FALSE | Downregulated |
| -0,492600769 | 0,077950517 | AKR1C4 | FALSE | Downregulated |
| 0,99115577 | 0,077950517 | C17orf100 | FALSE | Upregulated |

| | | | | |
|---|---|---|---|---|
| -0,276805506 | 0,077950517 | CERS2 | FALSE | Downregulated |
| -1,115625878 | 0,077950517 | HEPACAM | FALSE | Downregulated |
| -0,501995541 | 0,078125238 | NR5A2 | FALSE | Downregulated |
| 0,603977857 | 0,078132809 | TSTD2 | FALSE | Upregulated |
| 0,700764613 | 0,078176379 | ALYREF | FALSE | Upregulated |
| 0,770549967 | 0,078176379 | FABP5 | FALSE | Upregulated |
| 1,363458457 | 0,078176379 | LOC101927963 | FALSE | Upregulated |
| 0,896811053 | 0,078176379 | SNX32 | FALSE | Upregulated |
| 0,3534753 | 0,078176379 | UBN1 | FALSE | Upregulated |
| 1,065462025 | 0,078201792 | LOC102724133 | FALSE | Upregulated |
| -0,672945728 | 0,078201792 | TGFBRAP1 | FALSE | Downregulated |
| 0,575624604 | 0,078226903 | MFSD7 | FALSE | Upregulated |
| -0,549479989 | 0,078226903 | ZNF33B | FALSE | Downregulated |
| -0,676954873 | 0,078351565 | RP11-355N15.1 | FALSE | Downregulated |
| 1,161447791 | 0,078470847 | LOC100506319 | FALSE | Upregulated |
| -0,247130944 | 0,078546355 | FAM8A1 | FALSE | Downregulated |
| 2,029592983 | 0,078546355 | LINC00707 | FALSE | Upregulated |
| 0,452636421 | 0,078546355 | LOC102723600 | FALSE | Upregulated |
| -0,575071227 | 0,078546355 | NUP88 | FALSE | Downregulated |
| 0,410434514 | 0,078546355 | WDR48 | FALSE | Upregulated |
| -0,480533229 | 0,078549498 | LLGL2 | FALSE | Downregulated |
| -0,470504466 | 0,078673585 | RNF185 | FALSE | Downregulated |
| 0,951783001 | 0,078714152 | ABCA3 | FALSE | Upregulated |
| 0,961132445 | 0,078714152 | CHTF18 | FALSE | Upregulated |
| -0,471948493 | 0,078714152 | ONECUT2 | FALSE | Downregulated |
| -0,346856909 | 0,078714152 | ZBED1 | FALSE | Downregulated |
| -0,325357527 | 0,078815497 | PHF3 | FALSE | Downregulated |
| 0,353759998 | 0,078833488 | SUPT7L | FALSE | Upregulated |
| -0,332277435 | 0,07937965 | HAGH | FALSE | Downregulated |
| 0,355817448 | 0,079653293 | SLC12A4 | FALSE | Upregulated |
| -1,310502655 | 0,080112442 | NR2F1-AS1 | FALSE | Downregulated |
| 1,291622394 | 0,080142447 | MITF | FALSE | Upregulated |
| 1,233582807 | 0,080388468 | STMN1 | FALSE | Upregulated |
| -0,457330901 | 0,080416081 | PXDN | FALSE | Downregulated |
| -0,37733975 | 0,080586776 | RPL26 | FALSE | Downregulated |
| 1,441431504 | 0,08066706 | LOC100506454 | FALSE | Upregulated |
| 0,742741508 | 0,080709661 | SLC1A5 | FALSE | Upregulated |
| -0,363929803 | 0,080884729 | REXO1 | FALSE | Downregulated |
| 0,735120643 | 0,081145631 | AGAP2 | FALSE | Upregulated |
| -0,589365374 | 0,081145631 | CLYBL | FALSE | Downregulated |
| 0,46716794 | 0,081145631 | MAPKAPK5-AS1 | FALSE | Upregulated |
| 0,62411708 | 0,081145631 | SLC43A2 | FALSE | Upregulated |
| -0,420737705 | 0,081145631 | STAT5B | FALSE | Downregulated |
| -0,478930152 | 0,081316776 | MAN1C1 | FALSE | Downregulated |
| 1,197206797 | 0,081351697 | CCNA2 | FALSE | Upregulated |
| -0,390531535 | 0,081386355 | PKD1 | FALSE | Downregulated |
| 1,01550948 | 0,081400836 | FLJ38576 | FALSE | Upregulated |
| -0,679549356 | 0,081428265 | AASS | FALSE | Downregulated |
| 0,939149048 | 0,081428265 | LOC101927955 | FALSE | Upregulated |
| -0,677497943 | 0,08151101 | AADAT | FALSE | Downregulated |
| -0,779316954 | 0,08151101 | DIS3L2 | FALSE | Downregulated |

| | | | | |
|---|---|---|---|---|
| -0,968602632 | 0,08164489 | FXN | FALSE | Downregulated |
| 1,682155867 | 0,08164489 | SP140 | FALSE | Upregulated |
| 0,486441228 | 0,081911711 | PVRIG | FALSE | Upregulated |
| -0,359511268 | 0,081926239 | MON2 | FALSE | Downregulated |
| -0,881246822 | 0,082414745 | LOC101928143 | FALSE | Downregulated |
| 0,560787667 | 0,082498156 | AC104134.2 | FALSE | Upregulated |
| -0,780852035 | 0,082622547 | AC007099.1 | FALSE | Downregulated |
| -0,522748734 | 0,082728586 | NSUN6 | FALSE | Downregulated |
| 1,073386614 | 0,082828491 | HPDL | FALSE | Upregulated |
| -0,489411352 | 0,082878705 | C1RL-AS1 | FALSE | Downregulated |
| 1,243928815 | 0,082879986 | LINC00319 | FALSE | Upregulated |
| 0,448736025 | 0,08302431 | CSK | FALSE | Upregulated |
| 0,685165963 | 0,083097473 | HERC6 | FALSE | Upregulated |
| 0,882446759 | 0,08344226 | RP11-536C5.7 | FALSE | Upregulated |
| 0,867277121 | 0,083534168 | CYR61 | FALSE | Upregulated |
| 0,680720393 | 0,083569228 | LOC101927371 | FALSE | Upregulated |
| -0,343038351 | 0,08365724 | TTC3 | FALSE | Downregulated |
| 0,511531729 | 0,083697379 | LOC146880 | FALSE | Upregulated |
| 0,680391475 | 0,083827104 | C9orf66 | FALSE | Upregulated |
| -0,417548507 | 0,083827104 | ECI1 | FALSE | Downregulated |
| 1,430766481 | 0,083907623 | PIK3R6 | FALSE | Upregulated |
| -0,533378914 | 0,083999557 | TTC36 | FALSE | Downregulated |
| 0,418794824 | 0,084055009 | CALR | FALSE | Upregulated |
| -0,488480985 | 0,084055009 | HP | FALSE | Downregulated |
| 0,257570996 | 0,084157329 | EGLN1 | FALSE | Upregulated |
| -0,266366211 | 0,084293331 | MACF1 | FALSE | Downregulated |
| 0,858470498 | 0,084491135 | FAM78A | FALSE | Upregulated |
| -0,457314378 | 0,084509889 | LRRC3DN | FALSE | Downregulated |
| -0,51211238 | 0,084636127 | COLEC11 | FALSE | Downregulated |
| 1,435934251 | 0,084639277 | IKBIP | FALSE | Upregulated |
| 0,937026897 | 0,084873599 | AIF1 | FALSE | Upregulated |
| 0,68562372 | 0,08510695 | RTL1 | FALSE | Upregulated |
| 0,659660729 | 0,085115175 | R3HDM4 | FALSE | Upregulated |
| 1,130006333 | 0,085196993 | DEGS2 | FALSE | Upregulated |
| 0,37882027 | 0,085279704 | ACOT13 | FALSE | Upregulated |
| 0,528008108 | 0,085279704 | RAD9B | FALSE | Upregulated |
| 1,108747505 | 0,085279704 | ZNF107 | FALSE | Upregulated |
| -0,68857451 | 0,085651601 | UBE2N | FALSE | Downregulated |
| -0,791874288 | 0,085774803 | IBA57 | FALSE | Downregulated |
| -0,435485282 | 0,085774803 | NAGLU | FALSE | Downregulated |
| 0,567841176 | 0,085782164 | RNF141 | FALSE | Upregulated |
| 0,650865002 | 0,085782164 | WIPI1 | FALSE | Upregulated |
| -0,443376457 | 0,085891185 | HPX | FALSE | Downregulated |
| -0,490308494 | 0,085891185 | LOC101928716 | FALSE | Downregulated |
| -0,394950805 | 0,085916006 | MAOA | FALSE | Downregulated |
| 0,456256701 | 0,086015127 | RP11-231I16.1 | FALSE | Upregulated |
| -0,538768498 | 0,086174055 | GOLGA2P5 | FALSE | Downregulated |
| -0,562776138 | 0,086174055 | SNX14 | FALSE | Downregulated |
| 1,054577774 | 0,086174055 | TC2N | FALSE | Upregulated |
| -0,48965825 | 0,086230729 | FOXA1 | FALSE | Downregulated |
| -1,97344791 | 0,08661013 | LINC00630 | FALSE | Downregulated |

| | | | | |
|---|---|---|---|---|
| -0,308893971 | 0,086974328 | MAN1A1 | FALSE | Downregulated |
| -0,237197168 | 0,087129652 | SPEN | FALSE | Downregulated |
| -1,765708966 | 0,087486719 | LOC102723589 | FALSE | Downregulated |
| 1,421043525 | 0,087495328 | IPCEF1 | FALSE | Upregulated |
| 0,994927764 | 0,08750237 | FBXO2 | FALSE | Upregulated |
| 0,386968791 | 0,087513003 | CTSA | FALSE | Upregulated |
| 0,558313456 | 0,087513003 | MFSD4 | FALSE | Upregulated |
| 0,373461592 | 0,087640112 | DAZAP1 | FALSE | Upregulated |
| 0,40561585 | 0,087646848 | PDZD2 | FALSE | Upregulated |
| 0,385656996 | 0,087883383 | MPHOSPH8 | FALSE | Upregulated |
| 1,844919409 | 0,08821456 | GTSE1 | FALSE | Upregulated |
| -1,954371596 | 0,08821456 | KLHL13 | FALSE | Downregulated |
| -0,479755879 | 0,08821456 | LOC101929385 | FALSE | Downregulated |
| 0,911895618 | 0,08821456 | NR2C2AP | FALSE | Upregulated |
| 0,975925551 | 0,08821456 | RP11-473M20.7 | FALSE | Upregulated |
| 0,358746156 | 0,088400698 | POLH | FALSE | Upregulated |
| -0,592999968 | 0,088436539 | CYP51A1 | FALSE | Downregulated |
| 0,892186503 | 0,088436539 | LOC101928707 | FALSE | Upregulated |
| 1,547083295 | 0,088460531 | FCGR1A | FALSE | Upregulated |
| -1,420966241 | 0,088491513 | POSTN | FALSE | Downregulated |
| 0,376444556 | 0,088742314 | C19orf44 | FALSE | Upregulated |
| 0,405711441 | 0,088773448 | PANK2 | FALSE | Upregulated |
| -0,48205807 | 0,088773448 | SLC37A4 | FALSE | Downregulated |
| 0,492224547 | 0,088779736 | IKBKB | FALSE | Upregulated |
| 0,660777594 | 0,088860946 | AC016629.8 | FALSE | Upregulated |
| -1,421111335 | 0,088860946 | LRP4 | FALSE | Downregulated |
| 0,716321067 | 0,088860946 | NGFRAP1 | FALSE | Upregulated |
| 0,530237311 | 0,088997854 | RTCB | FALSE | Upregulated |
| -0,403900702 | 0,089574525 | MOCS1 | FALSE | Downregulated |
| -0,359081863 | 0,089598861 | SMLR1 | FALSE | Downregulated |
| -1,515800134 | 0,089661977 | GSTA7P | FALSE | Downregulated |
| 0,876475836 | 0,089719906 | PLEKHG4 | FALSE | Upregulated |
| 0,449214701 | 0,089793589 | PCSK7 | FALSE | Upregulated |
| -0,323310212 | 0,089793589 | STAU1 | FALSE | Downregulated |
| 1,982764647 | 0,090293154 | ODF3L1 | FALSE | Upregulated |
| 0,946351037 | 0,090368883 | KNTC1 | FALSE | Upregulated |
| -0,390634192 | 0,090487243 | ABCC2 | FALSE | Downregulated |
| 0,516760364 | 0,090487243 | TAPBPL | FALSE | Upregulated |
| -1,372432243 | 0,090715086 | LOC101927105 | FALSE | Downregulated |
| 1,801776763 | 0,091049286 | CELF2-AS2 | FALSE | Upregulated |
| -0,273316638 | 0,091049286 | GNE | FALSE | Downregulated |
| 0,445619742 | 0,091049286 | LOC100133091 | FALSE | Upregulated |
| 1,195964497 | 0,091049286 | LOC101927442 | FALSE | Upregulated |
| 0,516688257 | 0,091049286 | LOC102724342 | FALSE | Upregulated |
| 0,627092468 | 0,091049286 | LOC728805 | FALSE | Upregulated |
| -0,895189317 | 0,091049286 | MAFIP | FALSE | Downregulated |
| -0,936210569 | 0,091049286 | RNU4-1 | FALSE | Downregulated |
| 0,547132517 | 0,091049286 | SPRTN | FALSE | Upregulated |
| 0,274402974 | 0,091398689 | CTSB | FALSE | Upregulated |
| 0,468590554 | 0,091398689 | MARCKSL1 | FALSE | Upregulated |
| -0,453919031 | 0,091398689 | MTR | FALSE | Downregulated |

| | | | | |
|---|---|---|---|---|
| -0,321079142 | 0,091398689 | SORL1 | FALSE | Downregulated |
| 1,023318804 | 0,091583325 | LOC100129697 | FALSE | Upregulated |
| -1,722610445 | 0,091890204 | LOC100132813 | FALSE | Downregulated |
| -0,790163187 | 0,092086594 | APLNR | FALSE | Downregulated |
| -0,610606171 | 0,092086594 | RNF125 | FALSE | Downregulated |
| 0,402786657 | 0,09216783 | CERS6-AS1 | FALSE | Upregulated |
| 0,310736446 | 0,09216783 | DTX4 | FALSE | Upregulated |
| 0,697587992 | 0,09236941 | KATNB1 | FALSE | Upregulated |
| -0,92678034 | 0,09236941 | MYO16 | FALSE | Downregulated |
| 0,723316481 | 0,09236941 | RPS27L | FALSE | Upregulated |
| 0,784630179 | 0,092386554 | GRIK1 | FALSE | Upregulated |
| -0,696846037 | 0,09245406 | DMRTA1 | FALSE | Downregulated |
| 0,451080159 | 0,092476557 | KIF24 | FALSE | Upregulated |
| -0,616026018 | 0,09250874 | ADRA1B | FALSE | Downregulated |
| 0,554049814 | 0,092585653 | PPDPF | FALSE | Upregulated |
| 0,588208215 | 0,092813078 | ARL5B | FALSE | Upregulated |
| -2,01006857 | 0,092813078 | LSMEM1 | FALSE | Downregulated |
| -1,705085149 | 0,092813078 | XPNPEP2 | FALSE | Downregulated |
| -0,454927554 | 0,09301703 | SLC35A3 | FALSE | Downregulated |
| -0,50972269 | 0,093046102 | ERN1 | FALSE | Downregulated |
| -0,405744497 | 0,093263076 | DDI2 | FALSE | Downregulated |
| 0,317607741 | 0,093263076 | GTPBP1 | FALSE | Upregulated |
| -0,629037968 | 0,093423475 | AGXT2 | FALSE | Downregulated |
| 0,528986593 | 0,093423475 | LOC102723990 | FALSE | Upregulated |
| -2,172899196 | 0,093577816 | RP11-364P22.1 | FALSE | Downregulated |
| 0,239700306 | 0,093607138 | ACTN4 | FALSE | Upregulated |
| -0,424655159 | 0,093607138 | CYP4F11 | FALSE | Downregulated |
| 0,999757322 | 0,093607138 | XXYLT1-AS2 | FALSE | Upregulated |
| -0,585796752 | 0,093847669 | BCORL1 | FALSE | Downregulated |
| -1,735500545 | 0,093895288 | SLITRK3 | FALSE | Downregulated |
| -0,915364877 | 0,093895288 | ZNF350 | FALSE | Downregulated |
| 0,322490497 | 0,094044686 | VCL | FALSE | Upregulated |
| -0,342423171 | 0,094054708 | ESRP2 | FALSE | Downregulated |
| -2,088561858 | 0,09406557 | COL28A1 | FALSE | Downregulated |
| 0,92755555 | 0,094085861 | LOC101928926 | FALSE | Upregulated |
| -0,417589413 | 0,094151098 | SERPINA5 | FALSE | Downregulated |
| 0,460007969 | 0,094297564 | ASB16 | FALSE | Upregulated |
| 0,44806656 | 0,094297564 | DENND1C | FALSE | Upregulated |
| -0,484946803 | 0,094297564 | GBE1 | FALSE | Downregulated |
| -0,338699997 | 0,094297564 | RPS3A | FALSE | Downregulated |
| 0,812429017 | 0,094297564 | ZFAT-AS1 | FALSE | Upregulated |
| -1,779904648 | 0,094434222 | LOC102546226 | FALSE | Downregulated |
| 1,325440104 | 0,094493672 | SCN3B | FALSE | Upregulated |
| 1,122307309 | 0,094622197 | CERS1 | FALSE | Upregulated |
| 0,771336294 | 0,094642115 | EID2 | FALSE | Upregulated |
| 0,361115682 | 0,094642115 | PIP4K2B | FALSE | Upregulated |
| -0,471951569 | 0,094647365 | UPF2 | FALSE | Downregulated |
| 0,923866744 | 0,094656333 | KLF4 | FALSE | Upregulated |
| -0,339735949 | 0,094656333 | RPL23A | FALSE | Downregulated |
| 0,691615494 | 0,094656333 | SMARCAD1 | FALSE | Upregulated |
| -0,465628742 | 0,094722752 | LARP1B | FALSE | Downregulated |

| | | | | |
|---|---|---|---|---|
| 1,236638995 | 0,094856425 | EMB | FALSE | Upregulated |
| 0,639534412 | 0,094856425 | HCLS1 | FALSE | Upregulated |
| -1,700982229 | 0,094856425 | ZBTB8OS | FALSE | Downregulated |
| -2,325048662 | 0,094862849 | ASB4 | FALSE | Downregulated |
| -0,740486378 | 0,094862849 | BCKDHB | FALSE | Downregulated |
| 0,446296889 | 0,095150136 | PSMC4 | FALSE | Upregulated |
| -0,986967083 | 0,095150136 | VWCE | FALSE | Downregulated |
| -0,728058149 | 0,095404435 | AC005029.1 | FALSE | Downregulated |
| 0,67550211 | 0,095404435 | ARPC3 | FALSE | Upregulated |
| -0,826216525 | 0,095695338 | ADIRF | FALSE | Downregulated |
| -0,415577429 | 0,09589593 | TIMP3 | FALSE | Downregulated |
| 0,481089508 | 0,095930693 | CECR1 | FALSE | Upregulated |
| 0,762634269 | 0,095930693 | GRIP1 | FALSE | Upregulated |
| 0,613661839 | 0,095930693 | PIK3CB | FALSE | Upregulated |
| -0,607307366 | 0,095940042 | OXR1 | FALSE | Downregulated |
| -0,365737138 | 0,096384483 | PTPRB | FALSE | Downregulated |
| -0,261764368 | 0,096384483 | RPL13 | FALSE | Downregulated |
| 0,565604175 | 0,096384483 | VAC14-AS1 | FALSE | Upregulated |
| 0,593414448 | 0,096444574 | HMGA1 | FALSE | Upregulated |
| 0,425564173 | 0,096551608 | ZNF814 | FALSE | Upregulated |
| 0,674568183 | 0,096799961 | RNF24 | FALSE | Upregulated |
| 0,418295358 | 0,096993111 | PKN1 | FALSE | Upregulated |
| 0,356183733 | 0,09706752 | CYP8B1 | FALSE | Upregulated |
| 0,43483922 | 0,097197341 | ARHGEF1 | FALSE | Upregulated |
| -1,643461493 | 0,097197341 | IL1RL2 | FALSE | Downregulated |
| 0,497693149 | 0,097374925 | TNFRSF1B | FALSE | Upregulated |
| -0,782006584 | 0,097518095 | AGAP9 | FALSE | Downregulated |
| 0,375290766 | 0,097533502 | UCKL1-AS1 | FALSE | Upregulated |
| -0,350298429 | 0,097641262 | RPS20 | FALSE | Downregulated |
| 0,486690888 | 0,097643778 | HEIH | FALSE | Upregulated |
| 0,817782823 | 0,097646332 | CELF2 | FALSE | Upregulated |
| -0,321836597 | 0,097868824 | NBR1 | FALSE | Downregulated |
| 1,17310499 | 0,097889002 | OSBPL3 | FALSE | Upregulated |
| 0,402965665 | 0,097917994 | ANKRD36BP1 | FALSE | Upregulated |
| 0,884911502 | 0,098083549 | FHOD3 | FALSE | Upregulated |
| 1,050439524 | 0,098090021 | C17orf97 | FALSE | Upregulated |
| 1,559348106 | 0,098090021 | EMP3 | FALSE | Upregulated |
| 0,331579294 | 0,098090021 | TMEM129 | FALSE | Upregulated |
| 0,37116547 | 0,098217309 | SIPA1L3 | FALSE | Upregulated |
| -0,376352636 | 0,098278825 | EEF1B2 | FALSE | Downregulated |
| 1,59880168 | 0,098278825 | GFI1 | FALSE | Upregulated |
| 0,870626451 | 0,098390451 | LOC102724164 | FALSE | Upregulated |
| 0,752788089 | 0,098483077 | HRAS | FALSE | Upregulated |
| 0,275471663 | 0,098725028 | YBX1 | FALSE | Upregulated |
| 0,438338548 | 0,098762651 | GPX3 | FALSE | Upregulated |
| -0,826262123 | 0,098762651 | LOC730101 | FALSE | Downregulated |
| 0,352873055 | 0,099087623 | SYNC | FALSE | Upregulated |
| -0,92028097 | 0,099155138 | CCL23 | FALSE | Downregulated |
| 1,065627195 | 0,099155138 | RAB31 | FALSE | Upregulated |
| 0,577375941 | 0,099155138 | SIRPA | FALSE | Upregulated |
| -0,915902358 | 0,099315174 | GRB7 | FALSE | Downregulated |

| | | | | |
|---|---|---|---|---|
| 0,487508769 | 0,099370763 | LOC101929240 | FALSE | Upregulated |
| 0,748822411 | 0,099392214 | B3GNT4 | FALSE | Upregulated |
| 1,381104084 | 0,099392214 | DLG1-AS1 | FALSE | Upregulated |
| 0,36345405 | 0,099392214 | OAZ2 | FALSE | Upregulated |
| -1,011240663 | 0,099392214 | ZFAND1 | FALSE | Downregulated |
| 0,774963669 | 0,099495078 | C1orf111 | FALSE | Upregulated |
| 0,643082666 | 0,099504379 | UBALD2 | FALSE | Upregulated |
| 0,588395206 | 0,099537497 | SPRYD3 | FALSE | Upregulated |
| 0,775276136 | 0,099646451 | LOC102724335 | FALSE | Upregulated |
| -1,055094159 | 0,099754821 | POU6F1 | FALSE | Downregulated |
| 0,941972959 | 0,099970874 | ZMAT3 | FALSE | Upregulated |
| 0,438793919 | 0,100457128 | KLHL5 | FALSE | Upregulated |
| -0,360612346 | 0,100483313 | NISCH | FALSE | Downregulated |
| 1,728219382 | 0,100542451 | SPP1 | FALSE | Upregulated |
| 0,723671246 | 0,100614742 | ZDHHC15 | FALSE | Upregulated |
| 0,347705986 | 0,100650473 | CYFIP1 | FALSE | Upregulated |
| -0,440666236 | 0,100857827 | ACBD4 | FALSE | Downregulated |
| 0,994829006 | 0,100857827 | PIK3CG | FALSE | Upregulated |
| 0,761937318 | 0,100857827 | SH3BP1 | FALSE | Upregulated |
| -0,417381398 | 0,101013496 | BAAT | FALSE | Downregulated |
| 0,924354214 | 0,101020395 | PLP2 | FALSE | Upregulated |
| -0,469043925 | 0,101526163 | PTPRS | FALSE | Downregulated |
| 0,333044426 | 0,101540692 | CREB1 | FALSE | Upregulated |
| 0,983781556 | 0,101540692 | SASS6 | FALSE | Upregulated |
| 0,854536582 | 0,101540692 | SLC5A10 | FALSE | Upregulated |
| -0,790516388 | 0,101540692 | ZNF772 | FALSE | Downregulated |
| 0,768510667 | 0,101540692 | ZRANB2-AS1 | FALSE | Upregulated |
| 1,369894521 | 0,101573116 | RSAD2 | FALSE | Upregulated |
| 1,545673962 | 0,101633423 | IGSF3 | FALSE | Upregulated |
| 0,460688451 | 0,101647043 | PKD2L2 | FALSE | Upregulated |
| 1,053164146 | 0,101999552 | AMICA1 | FALSE | Upregulated |
| -0,380661709 | 0,101999552 | MAP2K1 | FALSE | Downregulated |
| 0,412464905 | 0,102086593 | AIRN | FALSE | Upregulated |
| -0,81043502 | 0,102316951 | EBF3 | FALSE | Downregulated |
| 1,725836267 | 0,102316951 | RNF157-AS1 | FALSE | Upregulated |
| 0,873153259 | 0,102443597 | TDP1 | FALSE | Upregulated |
| -0,719051061 | 0,102762632 | KIAA1958 | FALSE | Downregulated |
| 0,430084053 | 0,102907262 | GALNT10 | FALSE | Upregulated |
| 0,630512913 | 0,102907262 | KLHL31 | FALSE | Upregulated |
| 0,471668869 | 0,102907262 | RARB | FALSE | Upregulated |
| -0,407999587 | 0,102907262 | ZNF512B | FALSE | Downregulated |
| 1,348243767 | 0,102920612 | CALML3 | FALSE | Upregulated |
| -0,499440508 | 0,102920612 | HIBADH | FALSE | Downregulated |
| -0,507342814 | 0,102920612 | MRPS18B | FALSE | Downregulated |
| -0,377755567 | 0,102920612 | MTTP | FALSE | Downregulated |
| -0,269296623 | 0,102920612 | RPS15 | FALSE | Downregulated |
| 1,054157677 | 0,103116736 | LOC101928884 | FALSE | Upregulated |
| 0,867293329 | 0,103207201 | EML1 | FALSE | Upregulated |
| 0,541434606 | 0,103443803 | ZNF497 | FALSE | Upregulated |
| 0,667842974 | 0,10358659 | KCTD10 | FALSE | Upregulated |
| -0,345448133 | 0,103588165 | NUCKS1 | FALSE | Downregulated |

| | | | | |
|---|---|---|---|---|
| -0,461170613 | 0,103639347 | SLC2A2 | FALSE | Downregulated |
| -0,432697115 | 0,103647444 | NR1I3 | FALSE | Downregulated |
| -0,659051063 | 0,103647444 | NRK | FALSE | Downregulated |
| 1,303929408 | 0,103647444 | POM121L9P | FALSE | Upregulated |
| -0,580466428 | 0,103647444 | RDH11 | FALSE | Downregulated |
| 0,475424783 | 0,103704168 | GTPBP2 | FALSE | Upregulated |
| 0,585166433 | 0,104215235 | ABCC1 | FALSE | Upregulated |
| -0,292506728 | 0,104215235 | LPP | FALSE | Downregulated |
| -0,480807714 | 0,104215235 | ST3GAL6 | FALSE | Downregulated |
| 0,579427856 | 0,104294207 | CBL | FALSE | Upregulated |
| 0,886576096 | 0,104443824 | CCR1 | FALSE | Upregulated |
| -0,366239226 | 0,104672255 | PPM1A | FALSE | Downregulated |
| 1,369173981 | 0,104690268 | HS3ST1 | FALSE | Upregulated |
| -0,392705447 | 0,104690268 | ST5 | FALSE | Downregulated |
| -0,859013667 | 0,104690268 | YAF2 | FALSE | Downregulated |
| 0,430833944 | 0,104842434 | RARA-AS1 | FALSE | Upregulated |
| 0,648036016 | 0,104953739 | CSNK1G1 | FALSE | Upregulated |
| -1,649789944 | 0,105154932 | AC079117.1 | FALSE | Downregulated |
| 0,405971497 | 0,105154932 | CHD7 | FALSE | Upregulated |
| -0,763950486 | 0,105253469 | S1PR1 | FALSE | Downregulated |
| -0,287046074 | 0,10531481 | C19orf66 | FALSE | Downregulated |
| -0,513366051 | 0,10531481 | ZFYVE1 | FALSE | Downregulated |
| -0,499036002 | 0,105437963 | RNF216 | FALSE | Downregulated |
| 0,316984107 | 0,105443341 | DDX5 | FALSE | Upregulated |
| -0,917918357 | 0,105443341 | NAIF1 | FALSE | Downregulated |
| 0,491163258 | 0,105497227 | GPX2 | FALSE | Upregulated |
| 0,514648476 | 0,105953917 | IDH3G | FALSE | Upregulated |
| -0,502307083 | 0,106175961 | ACADL | FALSE | Downregulated |
| 0,767314978 | 0,106379814 | FLJ32255 | FALSE | Upregulated |
| 1,649955304 | 0,106549425 | EPHB2 | FALSE | Upregulated |
| 2,033157217 | 0,106549425 | RASL12 | FALSE | Upregulated |
| 0,543249085 | 0,106676084 | LGALS3BP | FALSE | Upregulated |
| 0,564086962 | 0,106767525 | COL6A3 | FALSE | Upregulated |
| -0,377242093 | 0,106767525 | FIS1 | FALSE | Downregulated |
| 0,375700713 | 0,106767525 | HIST1H1E | FALSE | Upregulated |
| 0,938139113 | 0,106767525 | RHBDL2 | FALSE | Upregulated |
| 1,748448903 | 0,106767525 | RPL23AP7 | FALSE | Upregulated |
| 0,924742664 | 0,106767525 | TDRD7 | FALSE | Upregulated |
| 1,433581515 | 0,107091864 | PARS2 | FALSE | Upregulated |
| -0,780644031 | 0,107183022 | MAGI2 | FALSE | Downregulated |
| -0,67817364 | 0,107183022 | RTTN | FALSE | Downregulated |
| 0,793544413 | 0,107207759 | PRKCD | FALSE | Upregulated |
| 0,379029864 | 0,10761408 | WSB2 | FALSE | Upregulated |
| 0,48941504 | 0,107618973 | AC009501.4 | FALSE | Upregulated |
| 0,972116742 | 0,107618973 | C1orf216 | FALSE | Upregulated |
| 1,216370162 | 0,107811709 | SEPT1 | FALSE | Upregulated |
| 1,433510306 | 0,108038127 | CD22 | FALSE | Upregulated |
| 0,683771242 | 0,108226114 | MAD2L2 | FALSE | Upregulated |
| -0,972318759 | 0,108306812 | ZNF225 | FALSE | Downregulated |
| 0,600956125 | 0,108661885 | LOC101927681 | FALSE | Upregulated |
| 1,395769998 | 0,108848943 | SEMA4A | FALSE | Upregulated |

| | | | | |
|---|---|---|---|---|
| 0,69626283 | 0,108928937 | FADD | FALSE | Upregulated |
| -1,853330575 | 0,108928937 | FAM71C | FALSE | Downregulated |
| -0,446396334 | 0,108928937 | SMTN | FALSE | Downregulated |
| -1,050337798 | 0,10893273 | MICU3 | FALSE | Downregulated |
| -0,705365856 | 0,10893273 | PPM1H | FALSE | Downregulated |
| 1,191120521 | 0,10893273 | STK39 | FALSE | Upregulated |
| -0,656004975 | 0,10893273 | ZNF446 | FALSE | Downregulated |
| 0,697065402 | 0,108972518 | C2CD2L | FALSE | Upregulated |
| 0,544572689 | 0,108972518 | LOC102060414 | FALSE | Upregulated |
| 0,651235619 | 0,108972518 | NAGK | FALSE | Upregulated |
| -0,937295154 | 0,108972518 | OMA1 | FALSE | Downregulated |
| -0,394308254 | 0,108972518 | UROC1 | FALSE | Downregulated |
| 2,154725535 | 0,109056019 | LOC101927840 | FALSE | Upregulated |
| -0,284847283 | 0,109056019 | RBM33 | FALSE | Downregulated |
| 2,123908094 | 0,109070338 | LOC729162 | FALSE | Upregulated |
| 1,386296678 | 0,109656118 | SERPINE1 | FALSE | Upregulated |
| 1,319252373 | 0,109747545 | LKAAEAR1 | FALSE | Upregulated |
| -0,884731071 | 0,109835835 | CNTFR-AS1 | FALSE | Downregulated |
| -0,609399707 | 0,109896322 | PPP1R12B | FALSE | Downregulated |
| 0,519757681 | 0,110145544 | IQGAP1 | FALSE | Upregulated |
| -0,557177289 | 0,110328582 | APRT | FALSE | Downregulated |
| 0,408531935 | 0,110328582 | FAM149B1 | FALSE | Upregulated |
| 0,531630219 | 0,110328582 | LRFN4 | FALSE | Upregulated |
| -0,339639118 | 0,110369533 | DCAF5 | FALSE | Downregulated |
| 0,411145902 | 0,110369533 | MDM2 | FALSE | Upregulated |
| 0,545828291 | 0,110447345 | IPPK | FALSE | Upregulated |
| -0,511529267 | 0,110447345 | LIX1L | FALSE | Downregulated |
| 0,242748523 | 0,110713071 | TMEM123 | FALSE | Upregulated |
| -0,54875432 | 0,110820002 | ANGEL1 | FALSE | Downregulated |
| -0,527377507 | 0,110820002 | FBXO17 | FALSE | Downregulated |
| 1,193705166 | 0,110820002 | LOC389332 | FALSE | Upregulated |
| 0,620400376 | 0,111083893 | TRAF1 | FALSE | Upregulated |
| 0,385025958 | 0,111083893 | ZNF7 | FALSE | Upregulated |
| 0,895304024 | 0,111125661 | LMNB1 | FALSE | Upregulated |
| -0,254889102 | 0,111183505 | RBL2 | FALSE | Downregulated |
| 0,491963322 | 0,111479011 | RTN4RL2 | FALSE | Upregulated |
| 0,439029788 | 0,111488459 | C22orf23 | FALSE | Upregulated |
| 0,372664543 | 0,111488459 | DTX3L | FALSE | Upregulated |
| -0,852804067 | 0,111557965 | MRPL55 | FALSE | Downregulated |
| 1,02113538 | 0,111595225 | DEF6 | FALSE | Upregulated |
| 0,487482442 | 0,111595225 | OGFOD1 | FALSE | Upregulated |
| 0,859832478 | 0,111595225 | TRIM45 | FALSE | Upregulated |
| -1,908446862 | 0,111944834 | LOC728989 | FALSE | Downregulated |
| -1,034751253 | 0,111957786 | DANCR | FALSE | Downregulated |
| 0,432547634 | 0,112270498 | SPDYE5 | FALSE | Upregulated |
| -0,339703857 | 0,112270498 | TOR1AIP1 | FALSE | Downregulated |
| 0,535040604 | 0,112617656 | RNF207 | FALSE | Upregulated |
| 1,173562559 | 0,112738084 | B3GALT4 | FALSE | Upregulated |
| 0,598561254 | 0,112738084 | FAM198B | FALSE | Upregulated |
| -0,359352993 | 0,112738084 | PHB2 | FALSE | Downregulated |
| 0,421348808 | 0,112871772 | PTPN14 | FALSE | Upregulated |

| | | | | |
|---|---|---|---|---|
| 0,388152599 | 0,112930525 | PSMD7 | FALSE | Upregulated |
| 0,37038186 | 0,112935892 | POU5F1P4 | FALSE | Upregulated |
| 0,482367894 | 0,11303182 | LOC102723520 | FALSE | Upregulated |
| -1,432166005 | 0,113062148 | FREM2 | FALSE | Downregulated |
| -2,350572414 | 0,113062148 | SCN4A | FALSE | Downregulated |
| 0,405985419 | 0,113499288 | LOC100996506 | FALSE | Upregulated |
| -1,203313038 | 0,113664042 | PLEKHH2 | FALSE | Downregulated |
| 0,350704765 | 0,113664042 | RNF19A | FALSE | Upregulated |
| 0,692963305 | 0,113664042 | ZNF841 | FALSE | Upregulated |
| -0,684750282 | 0,113745972 | FAM101B | FALSE | Downregulated |
| 0,722044981 | 0,113745972 | SPSB1 | FALSE | Upregulated |
| -0,357851525 | 0,113877215 | SCAF8 | FALSE | Downregulated |
| 0,720084205 | 0,11393333 | LOC101929786 | FALSE | Upregulated |
| -0,609786833 | 0,11402609 | DUSP10 | FALSE | Downregulated |
| -1,528945522 | 0,11402609 | PHF7 | FALSE | Downregulated |
| -0,423637693 | 0,11402609 | ZNHIT1 | FALSE | Downregulated |
| -0,306380033 | 0,114055484 | SLCO2B1 | FALSE | Downregulated |
| -0,433795142 | 0,11412819 | GAPVD1 | FALSE | Downregulated |
| 0,438626909 | 0,114277069 | C21orf2 | FALSE | Upregulated |
| -0,339928723 | 0,114308995 | BAZ2B | FALSE | Downregulated |
| -0,635942665 | 0,114308995 | RPL34 | FALSE | Downregulated |
| 0,909136944 | 0,114451751 | FGR | FALSE | Upregulated |
| 0,450893711 | 0,114542029 | MAP3K14 | FALSE | Upregulated |
| 1,688988523 | 0,11478029 | CXCR4 | FALSE | Upregulated |
| 0,543242935 | 0,114875858 | C20orf24 | FALSE | Upregulated |
| 1,784876829 | 0,115068593 | NCR3 | FALSE | Upregulated |
| 0,62023508 | 0,115100891 | ARHGAP9 | FALSE | Upregulated |
| -0,602234118 | 0,115100891 | ERCC6 | FALSE | Downregulated |
| 0,699706476 | 0,115100891 | LOC101928059 | FALSE | Upregulated |
| 1,472247294 | 0,115100891 | LOC101929309 | FALSE | Upregulated |
| 0,737199393 | 0,115100891 | PSEN2 | FALSE | Upregulated |
| 0,654882308 | 0,115159354 | WWC1 | FALSE | Upregulated |
| -0,437091542 | 0,115170926 | HEBP2 | FALSE | Downregulated |
| 0,892060024 | 0,115322898 | DDX39A | FALSE | Upregulated |
| -0,573468124 | 0,115322898 | POLM | FALSE | Downregulated |
| -0,37681458 | 0,115565076 | FBXW2 | FALSE | Downregulated |
| 0,731731159 | 0,115622516 | B3GAT3 | FALSE | Upregulated |
| -0,414936688 | 0,115743092 | ADH1A | FALSE | Downregulated |
| 0,5394417 | 0,115743092 | RNF4 | FALSE | Upregulated |
| -0,485154163 | 0,11580211 | ACADM | FALSE | Downregulated |
| -0,95853811 | 0,11580211 | C21orf59 | FALSE | Downregulated |
| -0,882339632 | 0,11580211 | PLGLA | FALSE | Downregulated |
| -0,403521293 | 0,116055759 | CYP2C9 | FALSE | Downregulated |
| -0,303923729 | 0,11617078 | NDST1 | FALSE | Downregulated |
| -1,770531122 | 0,116298303 | EGFLAM-AS4 | FALSE | Downregulated |
| -0,583047904 | 0,116315963 | CDC42EP3 | FALSE | Downregulated |
| -0,58223039 | 0,116326065 | MEST | FALSE | Downregulated |
| 0,72805185 | 0,116351711 | PRKRA | FALSE | Upregulated |
| 1,375819614 | 0,116640216 | LINC01108 | FALSE | Upregulated |
| 0,914167414 | 0,116938707 | SLAIN1 | FALSE | Upregulated |
| 0,418913595 | 0,117171063 | AZI2 | FALSE | Upregulated |

| | | | | |
|---|---|---|---|---|
| 0,659985713 | 0,117177338 | CD84 | FALSE | Upregulated |
| -0,524028481 | 0,117212397 | DIXDC1 | FALSE | Downregulated |
| -1,326607054 | 0,117212397 | EPHB3 | FALSE | Downregulated |
| 0,490419765 | 0,117212397 | LRRC19 | FALSE | Upregulated |
| 1,027284438 | 0,117220032 | FUT8-AS1 | FALSE | Upregulated |
| -0,621783262 | 0,117245262 | GNRHR | FALSE | Downregulated |
| 0,521941226 | 0,117245262 | ZCCHC3 | FALSE | Upregulated |
| -0,322496978 | 0,117364004 | GPRC5C | FALSE | Downregulated |
| 1,900018591 | 0,11738711 | IGSF21 | FALSE | Upregulated |
| -0,408281303 | 0,117418348 | MDC1 | FALSE | Downregulated |
| 0,770359461 | 0,117520831 | CMKLR1 | FALSE | Upregulated |
| 1,361573693 | 0,117623214 | IRX3 | FALSE | Upregulated |
| 1,731277016 | 0,117707189 | TLR8 | FALSE | Upregulated |
| 0,570548648 | 0,117857554 | ANGPTL2 | FALSE | Upregulated |
| -0,572583219 | 0,117857554 | CPNE1 | FALSE | Downregulated |
| -0,555797758 | 0,117857554 | TBX2 | FALSE | Downregulated |
| -0,494021361 | 0,117897043 | N4BP2L1 | FALSE | Downregulated |
| 1,380200149 | 0,117913406 | NCF1B | FALSE | Upregulated |
| 0,604318177 | 0,118068638 | ITGAL | FALSE | Upregulated |
| -0,346107288 | 0,118068638 | RPS25 | FALSE | Downregulated |
| 0,319996179 | 0,118204732 | JMJD8 | FALSE | Upregulated |
| 0,604636274 | 0,118204732 | MARCO | FALSE | Upregulated |
| 0,424922251 | 0,118204732 | MORF4L2 | FALSE | Upregulated |
| 0,432070356 | 0,118312295 | LOC440300 | FALSE | Upregulated |
| -1,383571918 | 0,118312295 | PM20D2 | FALSE | Downregulated |
| -0,594759173 | 0,118312311 | ANK3 | FALSE | Downregulated |
| -0,347615984 | 0,118313751 | AMOTL2 | FALSE | Downregulated |
| 0,503489814 | 0,118823742 | ACTL10 | FALSE | Upregulated |
| 0,586266918 | 0,118929347 | ANKRD19P | FALSE | Upregulated |
| -1,87673226 | 0,118960813 | COL21A1 | FALSE | Downregulated |
| 0,72273754 | 0,11898309 | C21orf67 | FALSE | Upregulated |
| 0,381257281 | 0,11898309 | DENND2D | FALSE | Upregulated |
| -0,468097557 | 0,119058261 | TTC19 | FALSE | Downregulated |
| -0,739002412 | 0,119101732 | ARAP3 | FALSE | Downregulated |
| 0,616901188 | 0,119105597 | USP3-AS1 | FALSE | Upregulated |
| 1,387617655 | 0,119180799 | EDA | FALSE | Upregulated |
| 0,491265986 | 0,119180799 | SERINC2 | FALSE | Upregulated |
| -0,654424375 | 0,119457576 | PORCN | FALSE | Downregulated |
| 1,081391967 | 0,119526896 | LOC388906 | FALSE | Upregulated |
| -0,775071307 | 0,119526896 | MFSD9 | FALSE | Downregulated |
| 0,78973018 | 0,119526896 | SOX15 | FALSE | Upregulated |
| -0,522499102 | 0,119652395 | NRP2 | FALSE | Downregulated |
| 0,962156891 | 0,119675562 | REEP3 | FALSE | Upregulated |
| 1,607722362 | 0,119768551 | RNASE6 | FALSE | Upregulated |
| -0,564152339 | 0,119912406 | PLSCR4 | FALSE | Downregulated |
| 0,511958065 | 0,120058728 | DUSP28 | FALSE | Upregulated |
| -0,747905544 | 0,120144557 | AZGP1P1 | FALSE | Downregulated |
| -0,449029875 | 0,120144557 | SLC40A1 | FALSE | Downregulated |
| 0,308147488 | 0,120144557 | TLN1 | FALSE | Upregulated |
| 1,659704531 | 0,120144557 | UPP2 | FALSE | Upregulated |
| 1,823878863 | 0,120612027 | NEURL3 | FALSE | Upregulated |

| | | | | |
|---|---|---|---|---|
| 0,80354113 | 0,120612027 | RAB8B | FALSE | Upregulated |
| 0,398362194 | 0,120758364 | TTC28-AS1 | FALSE | Upregulated |
| -1,342216885 | 0,120832603 | CA13 | FALSE | Downregulated |
| -0,686165636 | 0,120832603 | PFKM | FALSE | Downregulated |
| -0,326581149 | 0,120920769 | RPS9 | FALSE | Downregulated |
| -0,325660926 | 0,121089819 | GPX4 | FALSE | Downregulated |
| -2,034683704 | 0,121089819 | RP11-118B18.1 | FALSE | Downregulated |
| -0,471408086 | 0,121089819 | TTC37 | FALSE | Downregulated |
| 0,907595055 | 0,121239552 | TRAPPC13 | FALSE | Upregulated |
| 0,683702548 | 0,121324167 | HCG4B | FALSE | Upregulated |
| 0,467535173 | 0,121356259 | CLCC1 | FALSE | Upregulated |
| 0,484303497 | 0,121361963 | DLK2 | FALSE | Upregulated |
| 0,386038705 | 0,121361963 | GALNT11 | FALSE | Upregulated |
| 0,479456022 | 0,121527423 | DNAJC1 | FALSE | Upregulated |
| 0,775225941 | 0,121654553 | GYPC | FALSE | Upregulated |
| 0,546976432 | 0,121879993 | IKBKG | FALSE | Upregulated |
| -0,529174412 | 0,121897577 | DUSP6 | FALSE | Downregulated |
| 0,929140314 | 0,121897577 | GPR39 | FALSE | Upregulated |
| -0,552798929 | 0,121897577 | RNF128 | FALSE | Downregulated |
| -0,885036485 | 0,121935637 | RP11-234K24.3 | FALSE | Downregulated |
| 1,516406067 | 0,12196716 | ITM2A | FALSE | Upregulated |
| 0,447045947 | 0,12196716 | LOC101928862 | FALSE | Upregulated |
| 1,030406311 | 0,122410646 | RP11-540A21.2 | FALSE | Upregulated |
| -0,70162689 | 0,122492684 | GHDC | FALSE | Downregulated |
| 0,861683472 | 0,122492684 | LINC00921 | FALSE | Upregulated |
| 0,462171333 | 0,12277629 | CLTB | FALSE | Upregulated |
| 0,433428267 | 0,123019365 | ESYT3 | FALSE | Upregulated |
| 0,267415991 | 0,123031902 | HCFC1 | FALSE | Upregulated |
| 0,649743592 | 0,123085174 | PRKAG1 | FALSE | Upregulated |
| 0,390688816 | 0,123120779 | ADAMTSL4 | FALSE | Upregulated |
| 1,186198537 | 0,123120779 | LOC102723331 | FALSE | Upregulated |
| 1,585759903 | 0,123120779 | PYHIN1 | FALSE | Upregulated |
| -0,614694153 | 0,123120779 | TSPAN6 | FALSE | Downregulated |
| 1,313880763 | 0,123120779 | ZNF431 | FALSE | Upregulated |
| 0,589859145 | 0,123174691 | ARFGAP3 | FALSE | Upregulated |
| -0,488465384 | 0,123199752 | RTKN | FALSE | Downregulated |
| 0,606507879 | 0,123337598 | GNRH1 | FALSE | Upregulated |
| 0,286064414 | 0,123434608 | ZDHHC5 | FALSE | Upregulated |
| -0,548864742 | 0,123492641 | SULT1A1 | FALSE | Downregulated |
| 0,473979445 | 0,123601779 | LOC101927018 | FALSE | Upregulated |
| -0,303611511 | 0,123601779 | MKLN1 | FALSE | Downregulated |
| 0,553444203 | 0,123800452 | ANKDD1A | FALSE | Upregulated |
| 0,483936194 | 0,123800452 | LPHN1 | FALSE | Upregulated |
| -0,564016944 | 0,123824093 | PLA2G15 | FALSE | Downregulated |
| 0,613881812 | 0,123867964 | CYP21A2 | FALSE | Upregulated |
| 1,872133081 | 0,123867964 | SLC35G1 | FALSE | Upregulated |
| -0,427599325 | 0,123893809 | HSD17B6 | FALSE | Downregulated |
| -0,358271991 | 0,123893809 | SAMD4A | FALSE | Downregulated |
| -0,569405771 | 0,123955926 | CX3CL1 | FALSE | Downregulated |
| 1,617144565 | 0,123984713 | LOC102724428 | FALSE | Upregulated |
| -0,395864945 | 0,124077396 | TNPO2 | FALSE | Downregulated |

| | | | | |
|---|---|---|---|---|
| 1,998708127 | 0,124365085 | GDNF | FALSE | Upregulated |
| 0,69047783 | 0,124365085 | NFATC1 | FALSE | Upregulated |
| 0,493089188 | 0,124486317 | ZSCAN22 | FALSE | Upregulated |
| 0,548088895 | 0,124596524 | MRAP | FALSE | Upregulated |
| 0,350998187 | 0,124603132 | CORO1B | FALSE | Upregulated |
| 1,297520839 | 0,124625493 | NOD2 | FALSE | Upregulated |
| -2,001907955 | 0,124783615 | LOC101927086 | FALSE | Downregulated |
| -2,132813249 | 0,124789459 | SUPT3H | FALSE | Downregulated |
| -1,210890737 | 0,124923441 | PILRB | FALSE | Downregulated |
| 0,560387839 | 0,124923441 | SASH3 | FALSE | Upregulated |
| 1,031702331 | 0,125240845 | E2F3 | FALSE | Upregulated |
| -0,485796397 | 0,125240845 | SIGIRR | FALSE | Downregulated |
| 0,570965021 | 0,125424308 | STX7 | FALSE | Upregulated |
| 0,378108967 | 0,125441959 | SIK2 | FALSE | Upregulated |
| -0,405215706 | 0,125443179 | DNAH1 | FALSE | Downregulated |
| -0,433414928 | 0,125443179 | FLRT3 | FALSE | Downregulated |
| -1,271492153 | 0,125478007 | KCNB1 | FALSE | Downregulated |
| -0,658092782 | 0,125478007 | NBPF10 | FALSE | Downregulated |
| -1,118182376 | 0,125670831 | FOXE1 | FALSE | Downregulated |
| -0,564138589 | 0,125870612 | KCTD21 | FALSE | Downregulated |
| 1,153758203 | 0,125878059 | COL5A3 | FALSE | Upregulated |
| -0,534509975 | 0,12596276 | SESN2 | FALSE | Downregulated |
| 0,774095217 | 0,125990265 | C9orf139 | FALSE | Upregulated |
| -0,643985655 | 0,126106158 | TTLL2 | FALSE | Downregulated |
| 0,629251299 | 0,126229722 | TRAFD1 | FALSE | Upregulated |
| 0,470031607 | 0,126326766 | DND1 | FALSE | Upregulated |
| 0,654372532 | 0,126326766 | GBP3 | FALSE | Upregulated |
| 0,916554647 | 0,126326766 | GRAMD1B | FALSE | Upregulated |
| -0,718603115 | 0,126326766 | INTS8 | FALSE | Downregulated |
| 0,483010123 | 0,126326766 | STK19 | FALSE | Upregulated |
| -0,37593903 | 0,12639055 | FAM193A | FALSE | Downregulated |
| -0,527633868 | 0,126465579 | HHIP | FALSE | Downregulated |
| 0,967478749 | 0,126465579 | LOC102723741 | FALSE | Upregulated |
| -4,686606426 | 0,12671206 | LOC102723943 | FALSE | Downregulated |
| -0,723758133 | 0,12671206 | TMEM254 | FALSE | Downregulated |
| 0,494249608 | 0,126944167 | KCTD12 | FALSE | Upregulated |
| 0,89990481 | 0,127017052 | DPM2 | FALSE | Upregulated |
| -0,765623106 | 0,127017052 | PNP | FALSE | Downregulated |
| 0,67237545 | 0,127023071 | RIBC1 | FALSE | Upregulated |
| -0,975679364 | 0,127145648 | DCSTAMP | FALSE | Downregulated |
| -0,375769892 | 0,127171163 | ARHGEF40 | FALSE | Downregulated |
| 0,360145866 | 0,127219339 | BTBD9 | FALSE | Upregulated |
| 0,365399558 | 0,127338846 | RP11-133O22.6 | FALSE | Upregulated |
| 0,851978876 | 0,127395609 | MCCD1 | FALSE | Upregulated |
| -0,536689247 | 0,127756781 | BIRC2 | FALSE | Downregulated |
| 1,058239138 | 0,127756781 | DOK3 | FALSE | Upregulated |
| 0,842956681 | 0,128077051 | LOC101928816 | FALSE | Upregulated |
| 0,381443221 | 0,128306225 | ZC3H18 | FALSE | Upregulated |
| 0,773600809 | 0,128467021 | GPRIN3 | FALSE | Upregulated |
| 0,886690343 | 0,128486551 | BBC3 | FALSE | Upregulated |
| 0,793224879 | 0,128579021 | FNDC9 | FALSE | Upregulated |

| | | | | |
|---|---|---|---|---|
| 1,264406592 | 0,128579021 | SIT1 | FALSE | Upregulated |
| 1,386341015 | 0,128708166 | EOMES | FALSE | Upregulated |
| 0,616938674 | 0,128708166 | GPR137B | FALSE | Upregulated |
| 0,916615658 | 0,128708166 | KAAG1 | FALSE | Upregulated |
| 0,864338129 | 0,128708166 | RNF39 | FALSE | Upregulated |
| -0,428913213 | 0,12885939 | CPB2 | FALSE | Downregulated |
| -0,378326664 | 0,128928858 | TSHZ1 | FALSE | Downregulated |
| 0,380301525 | 0,128948655 | MED26 | FALSE | Upregulated |
| -0,481133657 | 0,128948655 | NR3C2 | FALSE | Downregulated |
| -1,145882434 | 0,129224921 | C1orf220 | FALSE | Downregulated |
| 1,14384401 | 0,129298328 | LOC729930 | FALSE | Upregulated |
| -0,431641906 | 0,129324292 | SELENBP1 | FALSE | Downregulated |
| 0,891983305 | 0,129324292 | SLC38A9 | FALSE | Upregulated |
| 0,462966047 | 0,129500958 | ATP6V1B2 | FALSE | Upregulated |
| -0,744723674 | 0,129571427 | IFITM10 | FALSE | Downregulated |
| 0,94478413 | 0,129676804 | LOC102724705 | FALSE | Upregulated |
| -1,001033356 | 0,129676804 | RAC3 | FALSE | Downregulated |
| 0,34980771 | 0,129742431 | ANGPT2 | FALSE | Upregulated |
| 2,061706472 | 0,129742431 | DTHD1 | FALSE | Upregulated |
| -0,354924247 | 0,129742431 | TRAP1 | FALSE | Downregulated |
| -0,376905496 | 0,12985857 | ZADH2 | FALSE | Downregulated |
| -0,625147537 | 0,130069026 | GEMIN5 | FALSE | Downregulated |
| -1,447262762 | 0,130113344 | FAM84A | FALSE | Downregulated |
| -1,281372367 | 0,130113344 | PCDH7 | FALSE | Downregulated |
| 1,552520268 | 0,130195464 | ZNF804A | FALSE | Upregulated |
| -0,407251835 | 0,130624534 | AIG1 | FALSE | Downregulated |
| -0,407430685 | 0,130624534 | ALDH9A1 | FALSE | Downregulated |
| 0,417069352 | 0,130624534 | ALPL | FALSE | Upregulated |
| -0,549453 | 0,130624534 | CCS | FALSE | Downregulated |
| -0,878888034 | 0,130624534 | CYP3A43 | FALSE | Downregulated |
| 0,403085453 | 0,130624534 | LOC101930421 | FALSE | Upregulated |
| 0,405940114 | 0,130661047 | ADRM1 | FALSE | Upregulated |
| -0,318048112 | 0,130661047 | ANPEP | FALSE | Downregulated |
| -0,455648382 | 0,130661047 | HFE2 | FALSE | Downregulated |
| -0,28464762 | 0,130661047 | RPS8 | FALSE | Downregulated |
| 0,889322064 | 0,130661704 | SOX9 | FALSE | Upregulated |
| -0,459317616 | 0,130664735 | IGFBP3 | FALSE | Downregulated |
| -0,354719957 | 0,130867452 | C6 | FALSE | Downregulated |
| 0,475564981 | 0,130932559 | OSBPL2 | FALSE | Upregulated |
| -0,411898869 | 0,130946353 | CHD1 | FALSE | Downregulated |
| 0,386102554 | 0,130946353 | GNS | FALSE | Upregulated |
| 1,328047142 | 0,130946353 | KPNA2 | FALSE | Upregulated |
| 0,588889719 | 0,130946353 | MLK7-AS1 | FALSE | Upregulated |
| 0,931813935 | 0,131066297 | IL27RA | FALSE | Upregulated |
| -0,460742035 | 0,131066297 | RPL37 | FALSE | Downregulated |
| -1,062291745 | 0,131183275 | AMN | FALSE | Downregulated |
| 0,8694283 | 0,131183275 | CCDC167 | FALSE | Upregulated |
| -0,400514454 | 0,131183275 | RELL1 | FALSE | Downregulated |
| -1,828194664 | 0,131236959 | MROH2A | FALSE | Downregulated |
| 0,380291726 | 0,131236959 | RP5-886K2.3 | FALSE | Upregulated |
| 0,306954361 | 0,131236959 | TJP1 | FALSE | Upregulated |

| | | | | |
|---|---|---|---|---|
| -0,522944425 | 0,131236959 | ZRANB3 | FALSE | Downregulated |
| 0,947062598 | 0,131633028 | RTN4R | FALSE | Upregulated |
| -0,951948399 | 0,131633028 | STEAP1 | FALSE | Downregulated |
| -0,962678482 | 0,131832819 | TENM2 | FALSE | Downregulated |
| -0,391734612 | 0,131863407 | TMEM63B | FALSE | Downregulated |
| -0,466869978 | 0,132016932 | RASAL2 | FALSE | Downregulated |
| -0,27815141 | 0,132558515 | SLC10A1 | FALSE | Downregulated |
| 0,475282143 | 0,132633131 | STAP2 | FALSE | Upregulated |
| 0,946785337 | 0,132693377 | UBE2E2 | FALSE | Upregulated |
| 0,462456562 | 0,132899298 | DCAF13 | FALSE | Upregulated |
| 0,620248229 | 0,133023677 | DNM1P41 | FALSE | Upregulated |
| 0,708997457 | 0,133255073 | CPVL | FALSE | Upregulated |
| -0,367945238 | 0,133280669 | AKR7A2 | FALSE | Downregulated |
| 0,590272979 | 0,133280669 | C9orf172 | FALSE | Upregulated |
| 0,394353514 | 0,133280669 | ERCC1 | FALSE | Upregulated |
| 0,822686691 | 0,133280669 | RASSF3 | FALSE | Upregulated |
| 0,374576295 | 0,133843729 | LOXL3 | FALSE | Upregulated |
| 0,295826754 | 0,133853755 | ANKH | FALSE | Upregulated |
| -0,53117843 | 0,133853755 | COX7C | FALSE | Downregulated |
| -0,342747059 | 0,133853755 | MED13 | FALSE | Downregulated |
| 0,302633887 | 0,133896318 | RBCK1 | FALSE | Upregulated |
| 0,376228216 | 0,134192136 | PVRL2 | FALSE | Upregulated |
| -0,9381111 | 0,134268948 | ACP6 | FALSE | Downregulated |
| 0,457626919 | 0,13431486 | CACUL1 | FALSE | Upregulated |
| 2,01127925 | 0,13431486 | STXBP5L | FALSE | Upregulated |
| -0,723019069 | 0,134604867 | CYP2B6 | FALSE | Downregulated |
| 0,553345432 | 0,134604867 | IFNGR1 | FALSE | Upregulated |
| -0,78493132 | 0,134604867 | ZNF385B | FALSE | Downregulated |
| 0,569338779 | 0,134669491 | CHRNB1 | FALSE | Upregulated |
| 0,854220281 | 0,134812397 | SLC16A3 | FALSE | Upregulated |
| 0,810800099 | 0,134946816 | ANKEF1 | FALSE | Upregulated |
| 0,926634542 | 0,134967315 | LOC102724597 | FALSE | Upregulated |
| -0,785156204 | 0,135298431 | LINC00478 | FALSE | Downregulated |
| -0,363351289 | 0,13532708 | RUSC2 | FALSE | Downregulated |
| 1,50092244 | 0,135356984 | RPS16P5 | FALSE | Upregulated |
| -0,660264599 | 0,135410585 | MGC12916 | FALSE | Downregulated |
| -0,562669107 | 0,135410585 | ZSCAN18 | FALSE | Downregulated |
| 1,032743552 | 0,135548338 | FAM131B | FALSE | Upregulated |
| -1,733288021 | 0,135548338 | SMOX | FALSE | Downregulated |
| -1,680207106 | 0,135551976 | LOC102724356 | FALSE | Downregulated |
| 0,606330675 | 0,135609694 | TMEM223 | FALSE | Upregulated |
| -0,693436432 | 0,135818244 | ST3GAL3 | FALSE | Downregulated |
| -1,514738112 | 0,135903312 | TP53TG1 | FALSE | Downregulated |
| -0,46863062 | 0,13591041 | PDIA5 | FALSE | Downregulated |
| 0,408178262 | 0,136250596 | EIF4G3 | FALSE | Upregulated |
| -0,577499898 | 0,136311731 | SGK2 | FALSE | Downregulated |
| 0,834538856 | 0,136334328 | RARG | FALSE | Upregulated |
| 0,902035164 | 0,136426069 | LOC101929352 | FALSE | Upregulated |
| 0,913054683 | 0,13647119 | LOC101928373 | FALSE | Upregulated |
| 0,509966891 | 0,136504593 | ASCC3 | FALSE | Upregulated |
| -0,628538969 | 0,136597253 | APOC4 | FALSE | Downregulated |

| | | | | |
|---|---|---|---|---|
| -0,454165094 | 0,136597253 | BTD | FALSE | Downregulated |
| -0,435394583 | 0,136597253 | COX5B | FALSE | Downregulated |
| 0,696769291 | 0,136597253 | LOC101928443 | FALSE | Upregulated |
| 0,635172233 | 0,136597253 | RNF222 | FALSE | Upregulated |
| 0,772110725 | 0,136597253 | ZNF367 | FALSE | Upregulated |
| -0,873541256 | 0,136751922 | CNTNAP4 | FALSE | Downregulated |
| -1,779860909 | 0,136751922 | MUM1L1 | FALSE | Downregulated |
| 0,885683174 | 0,136807652 | PDSS1 | FALSE | Upregulated |
| -1,648807258 | 0,136887436 | FRG1 | FALSE | Downregulated |
| 0,425988653 | 0,13691024 | LOC101927178 | FALSE | Upregulated |
| -0,707237789 | 0,13691024 | LOC644662 | FALSE | Downregulated |
| 0,423621633 | 0,13691024 | TAF9B | FALSE | Upregulated |
| -0,653410248 | 0,137090423 | EVI5 | FALSE | Downregulated |
| 1,003091299 | 0,1377093 | MIR100HG | FALSE | Upregulated |
| 0,326140567 | 0,138913969 | APPL2 | FALSE | Upregulated |
| 0,901595821 | 0,139025753 | C5AR1 | FALSE | Upregulated |
| 0,769534609 | 0,139025753 | GPSM3 | FALSE | Upregulated |
| 1,989345011 | 0,139025753 | TNF | FALSE | Upregulated |
| 0,474751328 | 0,139085837 | TP53BP2 | FALSE | Upregulated |
| 0,358944404 | 0,139125613 | POLR3H | FALSE | Upregulated |
| -0,792478973 | 0,139400039 | ABCA4 | FALSE | Downregulated |
| 1,125147328 | 0,139445426 | COLGALT2 | FALSE | Upregulated |
| 0,489850586 | 0,139445426 | ZNF789 | FALSE | Upregulated |
| 0,640758703 | 0,139462248 | C6orf136 | FALSE | Upregulated |
| 0,750910398 | 0,139467833 | TM4SF20 | FALSE | Upregulated |
| -0,271535049 | 0,139560124 | SERBP1 | FALSE | Downregulated |
| 0,609591844 | 0,139731527 | ANKRD32 | FALSE | Upregulated |
| 1,486762571 | 0,139731527 | LOC102724240 | FALSE | Upregulated |
| 0,674704977 | 0,13976948 | PGM5P2 | FALSE | Upregulated |
| 0,441713998 | 0,139908462 | FBXO22-AS1 | FALSE | Upregulated |
| 0,384038988 | 0,140266096 | LOC100303749 | FALSE | Upregulated |
| 0,797637844 | 0,140348111 | LOC101593348 | FALSE | Upregulated |
| -0,329277999 | 0,140348111 | MBTPS1 | FALSE | Downregulated |
| 0,493661362 | 0,140384419 | INE2 | FALSE | Upregulated |
| 0,490286173 | 0,140384419 | UNC93B1 | FALSE | Upregulated |
| 0,655973652 | 0,140444006 | ARHGEF5 | FALSE | Upregulated |
| 0,79572495 | 0,140444006 | TSSK2 | FALSE | Upregulated |
| -0,375496574 | 0,140786406 | ACY1 | FALSE | Downregulated |
| 0,771753976 | 0,141237091 | LMOD3 | FALSE | Upregulated |
| -1,680961102 | 0,141326944 | OR10J1 | FALSE | Downregulated |
| 0,512623102 | 0,141326944 | SH3RF3-AS1 | FALSE | Upregulated |
| -0,596440324 | 0,141355359 | HCN3 | FALSE | Downregulated |
| -0,419372408 | 0,141588246 | AFM | FALSE | Downregulated |
| -0,245338151 | 0,141588246 | DCAF8 | FALSE | Downregulated |
| 0,446385136 | 0,141588246 | FLJ38668 | FALSE | Upregulated |
| -0,386102413 | 0,141588246 | GRB14 | FALSE | Downregulated |
| 0,458830142 | 0,141588246 | LOC100505767 | FALSE | Upregulated |
| 1,087729018 | 0,141588246 | P2RY8 | FALSE | Upregulated |
| 0,500593249 | 0,1415986 | ABHD1 | FALSE | Upregulated |
| 0,434586118 | 0,14160862 | RBMS1 | FALSE | Upregulated |
| 0,380386948 | 0,14160862 | YARS | FALSE | Upregulated |

| | | | | |
|---|---|---|---|---|
| 1,29753856 | 0,141685081 | TDRKH | FALSE | Upregulated |
| 1,088523948 | 0,141732864 | ATP10B | FALSE | Upregulated |
| -0,276484166 | 0,14202083 | CALM1 | FALSE | Downregulated |
| 0,526098188 | 0,14202083 | KIAA0895L | FALSE | Upregulated |
| 0,662469492 | 0,14202083 | LDLR | FALSE | Upregulated |
| -1,668057042 | 0,14202083 | LOC102724643 | FALSE | Downregulated |
| 0,399588861 | 0,14202083 | MFHAS1 | FALSE | Upregulated |
| 1,252552006 | 0,14202083 | MGC34796 | FALSE | Upregulated |
| -0,330714133 | 0,14202083 | RPL24 | FALSE | Downregulated |
| 0,586615324 | 0,142063173 | FAM50A | FALSE | Upregulated |
| -0,335666461 | 0,142402356 | C8A | FALSE | Downregulated |
| 0,933388319 | 0,142551639 | SH3D21 | FALSE | Upregulated |
| -0,269537092 | 0,142854951 | WHSC1L1 | FALSE | Downregulated |
| 1,33775301 | 0,142855119 | NRXN2 | FALSE | Upregulated |
| 0,768557965 | 0,142998557 | C14orf79 | FALSE | Upregulated |
| -0,45472274 | 0,142998557 | GOLIM4 | FALSE | Downregulated |
| -0,33566154 | 0,143091592 | ADIPOR2 | FALSE | Downregulated |
| 0,450788419 | 0,143091592 | GSR | FALSE | Upregulated |
| 0,455873162 | 0,143186008 | PISD | FALSE | Upregulated |
| 0,385326783 | 0,14319019 | ARHGAP30 | FALSE | Upregulated |
| 0,446309994 | 0,14319019 | UBE2J1 | FALSE | Upregulated |
| 0,396105327 | 0,143329023 | TMEM80 | FALSE | Upregulated |
| -0,372111224 | 0,143602225 | PON2 | FALSE | Downregulated |
| -0,297967386 | 0,143602225 | RPL12 | FALSE | Downregulated |
| 0,350900274 | 0,143956512 | ATP1B1 | FALSE | Upregulated |
| 0,380740859 | 0,143956512 | DBNL | FALSE | Upregulated |
| 0,439902558 | 0,143956512 | OSMR | FALSE | Upregulated |
| 1,402894022 | 0,144018665 | CD96 | FALSE | Upregulated |
| 0,601020203 | 0,144081882 | TIMELESS | FALSE | Upregulated |
| 0,731786396 | 0,144282458 | SLFN14 | FALSE | Upregulated |
| 1,854492006 | 0,144294437 | ST8SIA1 | FALSE | Upregulated |
| 0,667479293 | 0,144294437 | TRIM5 | FALSE | Upregulated |
| -0,525165152 | 0,14435405 | RALGPS2 | FALSE | Downregulated |
| -0,201082124 | 0,14445398 | MLLT6 | FALSE | Downregulated |
| -0,53083557 | 0,144802856 | MSRA | FALSE | Downregulated |
| 0,678191373 | 0,144802856 | RSU1 | FALSE | Upregulated |
| 1,067474298 | 0,144816845 | LOC646626 | FALSE | Upregulated |
| 0,496408358 | 0,144816845 | SLC1A4 | FALSE | Upregulated |
| 0,567925533 | 0,144926651 | CUEDC1 | FALSE | Upregulated |
| 0,491763242 | 0,144926651 | LOC102724552 | FALSE | Upregulated |
| -0,404454421 | 0,145103214 | MLYCD | FALSE | Downregulated |
| 0,91451331 | 0,145103214 | TSC22D1-AS1 | FALSE | Upregulated |
| -1,424763453 | 0,145232856 | GFRA2 | FALSE | Downregulated |
| -0,499513059 | 0,145292015 | STARD13 | FALSE | Downregulated |
| -0,885820568 | 0,145694384 | FCGBP | FALSE | Downregulated |
| -0,894534385 | 0,145848517 | PREX2 | FALSE | Downregulated |
| 0,697622278 | 0,145864753 | ANKRD13D | FALSE | Upregulated |
| 0,8836776 | 0,145864753 | LRRC37A3 | FALSE | Upregulated |
| -0,29288162 | 0,145864753 | NR2F6 | FALSE | Downregulated |
| -0,260302329 | 0,145864753 | RPL35 | FALSE | Downregulated |
| -0,527342935 | 0,145864753 | TAPT1 | FALSE | Downregulated |

| | | | | |
|---|---|---|---|---|
| -0,454442212 | 0,145912758 | MTG2 | FALSE | Downregulated |
| -0,298132875 | 0,146167613 | RARRES2 | FALSE | Downregulated |
| -0,75880158 | 0,146237409 | TMEM143 | FALSE | Downregulated |
| 0,396948921 | 0,146292698 | LOC101929291 | FALSE | Upregulated |
| 1,025634732 | 0,14643469 | PLEKHG4B | FALSE | Upregulated |
| 0,843638718 | 0,146678832 | ITGB1BP1 | FALSE | Upregulated |
| 1,344687393 | 0,146702045 | DNMT3B | FALSE | Upregulated |
| -0,44015579 | 0,146702045 | SCAI | FALSE | Downregulated |
| 0,688320314 | 0,146738471 | NBPF20 | FALSE | Upregulated |
| 0,457440326 | 0,146770616 | PPM1E | FALSE | Upregulated |
| -1,105868773 | 0,146774236 | ATF1 | FALSE | Downregulated |
| 0,432210402 | 0,146868615 | TYW1 | FALSE | Upregulated |
| 0,497436066 | 0,147178426 | ARL8A | FALSE | Upregulated |
| 0,316792615 | 0,147178426 | DPP9 | FALSE | Upregulated |
| -0,283721981 | 0,147282415 | CAMSAP3 | FALSE | Downregulated |
| -0,66426582 | 0,147282415 | GPR116 | FALSE | Downregulated |
| 0,339323768 | 0,147282415 | MMP24 | FALSE | Upregulated |
| -1,568375882 | 0,147282415 | PSTK | FALSE | Downregulated |
| 0,553805369 | 0,147294274 | BTN3A1 | FALSE | Upregulated |
| 0,577943008 | 0,147294274 | LOC102724549 | FALSE | Upregulated |
| -0,448029559 | 0,147754191 | DEPDC5 | FALSE | Downregulated |
| 1,286277662 | 0,147754191 | HEXA-AS1 | FALSE | Upregulated |
| 1,38348577 | 0,147754191 | LOC729739 | FALSE | Upregulated |
| 1,008955248 | 0,147754191 | MCM6 | FALSE | Upregulated |
| -0,995634475 | 0,147754191 | MMRN1 | FALSE | Downregulated |
| 0,436658068 | 0,147754191 | MTA2 | FALSE | Upregulated |
| -0,91714252 | 0,147754191 | NPY1R | FALSE | Downregulated |
| -1,651740085 | 0,147754191 | TMEM133 | FALSE | Downregulated |
| -0,572084962 | 0,147879415 | LOC100132815 | FALSE | Downregulated |
| 1,554448832 | 0,147879415 | SIGLEC5 | FALSE | Upregulated |
| 1,349833015 | 0,148137634 | OPN1SW | FALSE | Upregulated |
| 1,243518542 | 0,148296208 | SLC7A14 | FALSE | Upregulated |
| -0,249550926 | 0,148324794 | CTDSP1 | FALSE | Downregulated |
| 1,572680995 | 0,148324794 | IFNG | FALSE | Upregulated |
| 0,646825551 | 0,148324794 | RAB20 | FALSE | Upregulated |
| 0,392754843 | 0,148777137 | XRCC3 | FALSE | Upregulated |
| 1,100826148 | 0,148946426 | UBXN10 | FALSE | Upregulated |
| 0,444888022 | 0,149132535 | LAP3 | FALSE | Upregulated |
| 0,418551597 | 0,149231776 | CTR9 | FALSE | Upregulated |
| -1,670759403 | 0,149231776 | ENPP3 | FALSE | Downregulated |
| 0,769435941 | 0,149231776 | HMGN4 | FALSE | Upregulated |
| 0,780690031 | 0,149231776 | PHTF2 | FALSE | Upregulated |
| -0,338004773 | 0,149231776 | ZNF33A | FALSE | Downregulated |
| -0,444818392 | 0,149425303 | EPRS | FALSE | Downregulated |
| 0,268545805 | 0,149425303 | LGALS8 | FALSE | Upregulated |
| -0,642532805 | 0,149549361 | ABCC6P1 | FALSE | Downregulated |
| -0,632961829 | 0,149593869 | TMEM219 | FALSE | Downregulated |
| -0,649261743 | 0,14969116 | ANKRD26 | FALSE | Downregulated |
| 0,349706167 | 0,149759495 | PABPN1 | FALSE | Upregulated |
| -0,862752018 | 0,149775019 | DCDC5 | FALSE | Downregulated |
| -0,506610484 | 0,149775019 | SLC1A1 | FALSE | Downregulated |

| | | | | |
|---|---|---|---|---|
| 1,505763538 | 0,150508874 | ADORA3 | FALSE | Upregulated |
| 0,521225807 | 0,150508874 | PCYT1A | FALSE | Upregulated |
| -1,299931955 | 0,150508874 | ZNF253 | FALSE | Downregulated |
| 0,792917518 | 0,150522664 | G6PD | FALSE | Upregulated |
| 0,537283295 | 0,150826693 | DMTN | FALSE | Upregulated |
| -0,313281874 | 0,150826693 | SORBS1 | FALSE | Downregulated |
| -0,855710351 | 0,150833053 | WDR72 | FALSE | Downregulated |
| -0,580758129 | 0,150884252 | LOC101927854 | FALSE | Downregulated |
| 0,350047308 | 0,150884252 | SETD6 | FALSE | Upregulated |
| 0,478708876 | 0,15114208 | GPSM2 | FALSE | Upregulated |
| 0,681654543 | 0,151285901 | NOP2 | FALSE | Upregulated |
| -1,521862211 | 0,151320487 | NEBL | FALSE | Downregulated |
| 0,470024675 | 0,151357209 | INPP5K | FALSE | Upregulated |
| 0,476220673 | 0,151407484 | EGLN2 | FALSE | Upregulated |
| 0,830810934 | 0,151476009 | ITGAM | FALSE | Upregulated |
| 1,48451754 | 0,151476009 | LOC101929741 | FALSE | Upregulated |
| 0,354261057 | 0,151476009 | ZNF584 | FALSE | Upregulated |
| 0,658203859 | 0,151549751 | ARHGEF33 | FALSE | Upregulated |
| -0,499123941 | 0,151549751 | LOC102724607 | FALSE | Downregulated |
| -1,069111293 | 0,15157044 | LOC100288778 | FALSE | Downregulated |
| 0,344647914 | 0,151623487 | PAPLN | FALSE | Upregulated |
| 0,477936015 | 0,151669948 | PGAM2 | FALSE | Upregulated |
| 0,62853095 | 0,151673191 | C7orf50 | FALSE | Upregulated |
| 1,323531418 | 0,15174513 | OR2A7 | FALSE | Upregulated |
| -0,233579735 | 0,151858499 | SEMA4G | FALSE | Downregulated |
| -0,494580412 | 0,151934274 | DNASE1L3 | FALSE | Downregulated |
| -0,456021144 | 0,152423731 | XYLB | FALSE | Downregulated |
| 0,559205597 | 0,152435522 | GJA9-MYCBP | FALSE | Upregulated |
| -0,391799563 | 0,152497947 | GRIN3A | FALSE | Downregulated |
| 0,305036056 | 0,152659844 | CEP170B | FALSE | Upregulated |
| -0,547153069 | 0,152802082 | DSTN | FALSE | Downregulated |
| -0,934012673 | 0,152802082 | RP11-279F6.1 | FALSE | Downregulated |
| -7,551270253 | 0,152870826 | LOC102723399 | FALSE | Downregulated |
| -0,573619911 | 0,153088839 | ETNPPL | FALSE | Downregulated |
| -1,102909138 | 0,153171827 | LOC101929065 | FALSE | Downregulated |
| -0,358606223 | 0,153171827 | SLC25A5 | FALSE | Downregulated |
| 0,299780367 | 0,153335735 | WBSCR16 | FALSE | Upregulated |
| 0,544788587 | 0,153470806 | RCE1 | FALSE | Upregulated |
| 0,332146527 | 0,153540439 | ORAI2 | FALSE | Upregulated |
| 0,586513647 | 0,153540439 | SMS | FALSE | Upregulated |
| -0,349702127 | 0,153540439 | SPRYD4 | FALSE | Downregulated |
| -0,494360425 | 0,153540439 | TTC14 | FALSE | Downregulated |
| -0,478884467 | 0,153647035 | CHST13 | FALSE | Downregulated |
| 1,623242742 | 0,153753541 | LOC101926931 | FALSE | Upregulated |
| 1,299894574 | 0,153850408 | PPM1N | FALSE | Upregulated |
| -0,350638218 | 0,153886546 | HSD17B4 | FALSE | Downregulated |
| -0,346182348 | 0,154084093 | BGN | FALSE | Downregulated |
| 0,536706768 | 0,154104434 | NFKBIB | FALSE | Upregulated |
| -0,479360299 | 0,154320941 | FAM160A2 | FALSE | Downregulated |
| -0,379905741 | 0,154320941 | NCOA6 | FALSE | Downregulated |
| -0,599336519 | 0,154320941 | SLC22A4 | FALSE | Downregulated |

| | | | | |
|---|---|---|---|---|
| 0,360219182 | 0,154323171 | EVL | FALSE | Upregulated |
| 0,446213775 | 0,154412482 | RANGAP1 | FALSE | Upregulated |
| 1,15265133 | 0,154802484 | TBC1D7 | FALSE | Upregulated |
| -0,48727476 | 0,154879961 | KCTD6 | FALSE | Downregulated |
| 0,633741476 | 0,154879961 | TGFB3 | FALSE | Upregulated |
| -0,666542356 | 0,154882139 | KIAA0319 | FALSE | Downregulated |
| 0,336695576 | 0,154991243 | ABLIM1 | FALSE | Upregulated |
| -0,299827977 | 0,154991243 | RALGAPA2 | FALSE | Downregulated |
| 1,217707289 | 0,154991243 | WNT11 | FALSE | Upregulated |
| -0,257996607 | 0,155004734 | EPAS1 | FALSE | Downregulated |
| 0,367147682 | 0,155004734 | LOC100996746 | FALSE | Upregulated |
| 0,663661244 | 0,155004734 | SLC35A2 | FALSE | Upregulated |
| 0,627011148 | 0,155201824 | SMG7-AS1 | FALSE | Upregulated |
| 1,414529573 | 0,155262814 | F13A1 | FALSE | Upregulated |
| -0,526017328 | 0,155262814 | RSF1 | FALSE | Downregulated |
| 0,396007503 | 0,155317517 | FAM175A | FALSE | Upregulated |
| 0,58429805 | 0,155317517 | LOC730668 | FALSE | Upregulated |
| 1,034176984 | 0,155317517 | MEAF6 | FALSE | Upregulated |
| -0,560608719 | 0,155380102 | ARRDC3-AS1 | FALSE | Downregulated |
| -0,617415215 | 0,155621832 | LRP5L | FALSE | Downregulated |
| 0,71444108 | 0,155625247 | LOC100128573 | FALSE | Upregulated |
| -1,032079473 | 0,155733888 | ARID3C | FALSE | Downregulated |
| -0,596657666 | 0,155735588 | CPS1-IT1 | FALSE | Downregulated |
| -0,796777705 | 0,155735588 | LMLN | FALSE | Downregulated |
| 0,515324301 | 0,15582445 | ADCY3 | FALSE | Upregulated |
| 0,57971187 | 0,15582445 | CLN6 | FALSE | Upregulated |
| -0,299377688 | 0,15582445 | DHTKD1 | FALSE | Downregulated |
| -0,551133647 | 0,15582445 | FDPS | FALSE | Downregulated |
| -0,371391054 | 0,15582445 | GCC2 | FALSE | Downregulated |
| 0,434504947 | 0,15582445 | JARID2 | FALSE | Upregulated |
| -0,417585368 | 0,15582445 | PLEKHG3 | FALSE | Downregulated |
| -0,366700617 | 0,15582445 | PNPLA2 | FALSE | Downregulated |
| 0,504834305 | 0,15582445 | RP11-84C10.4 | FALSE | Upregulated |
| 0,382536378 | 0,15582445 | STRN4 | FALSE | Upregulated |
| -0,46571545 | 0,155990252 | SEPHS1 | FALSE | Downregulated |
| -0,388435373 | 0,15609705 | ATRX | FALSE | Downregulated |
| 0,461949312 | 0,156114347 | SIGLEC1 | FALSE | Upregulated |
| 0,947950773 | 0,156125082 | LOC154761 | FALSE | Upregulated |
| 1,108302106 | 0,156148797 | HAVCR2 | FALSE | Upregulated |
| 0,659107085 | 0,156148797 | MAFF | FALSE | Upregulated |
| -0,642216836 | 0,156148797 | SYBU | FALSE | Downregulated |
| 0,828548058 | 0,156197566 | PRKCB | FALSE | Upregulated |
| 0,955526253 | 0,156197566 | RP1-257A7.4 | FALSE | Upregulated |
| -0,43603016 | 0,156280159 | SNX5 | FALSE | Downregulated |
| 0,396194853 | 0,15643072 | PITRM1-AS1 | FALSE | Upregulated |
| 0,536630596 | 0,156436314 | JAK2 | FALSE | Upregulated |
| 0,388837772 | 0,15656523 | MTX1 | FALSE | Upregulated |
| 0,711920576 | 0,156570812 | SMPD5 | FALSE | Upregulated |
| 0,886052034 | 0,156726827 | FLJ42258 | FALSE | Upregulated |
| -0,771999222 | 0,15690976 | CDON | FALSE | Downregulated |
| -1,155747377 | 0,15690976 | FAM64A | FALSE | Downregulated |

| | | | | |
|---|---|---|---|---|
| 1,566880063 | 0,15710091 | ZNF391 | FALSE | Upregulated |
| 1,661851984 | 0,157112056 | PLB1 | FALSE | Upregulated |
| -1,292027774 | 0,157209083 | SALL4 | FALSE | Downregulated |
| 0,571729551 | 0,157343625 | IL15RA | FALSE | Upregulated |
| 1,650678144 | 0,157462987 | F2RL1 | FALSE | Upregulated |
| 1,246146525 | 0,157462987 | PPAN | FALSE | Upregulated |
| -0,489235681 | 0,15773016 | COPS6 | FALSE | Downregulated |
| 1,200865294 | 0,15773016 | LOC101929295 | FALSE | Upregulated |
| -1,110052664 | 0,158004456 | GATM-AS1 | FALSE | Downregulated |
| 1,009233386 | 0,158291742 | HAO2-IT1 | FALSE | Upregulated |
| -0,300809291 | 0,158291742 | WDR81 | FALSE | Downregulated |
| 0,798567745 | 0,158595962 | SLC6A6 | FALSE | Upregulated |
| 0,679642755 | 0,158650943 | PWWP2A | FALSE | Upregulated |
| 1,041381074 | 0,158955067 | CD247 | FALSE | Upregulated |
| -0,764656268 | 0,159164349 | ZC3H12C | FALSE | Downregulated |
| 1,183692033 | 0,15948925 | RALGPS1 | FALSE | Upregulated |
| -0,240617766 | 0,159506777 | EEF1D | FALSE | Downregulated |
| 0,887433551 | 0,159609631 | CD27-AS1 | FALSE | Upregulated |
| 1,554329871 | 0,159609631 | HIST1H2BG | FALSE | Upregulated |
| -0,673289639 | 0,159623309 | CTH | FALSE | Downregulated |
| 0,982961384 | 0,159873757 | ADA | FALSE | Upregulated |
| 0,2395327 | 0,159957441 | ADAR | FALSE | Upregulated |
| 0,486237767 | 0,159957441 | TRAF3 | FALSE | Upregulated |
| -2,042948951 | 0,160053105 | LOC100506737 | FALSE | Downregulated |
| -0,267949429 | 0,160075411 | CHD2 | FALSE | Downregulated |
| 0,660125822 | 0,160140647 | NEDD4L | FALSE | Upregulated |
| 1,221743029 | 0,160247432 | IL4I1 | FALSE | Upregulated |
| 0,646030299 | 0,160247432 | MFAP3 | FALSE | Upregulated |
| 0,38694733 | 0,16049893 | LOC100505736 | FALSE | Upregulated |
| 0,563381636 | 0,160891962 | KCTD9 | FALSE | Upregulated |
| 0,736295281 | 0,160963611 | TBCB | FALSE | Upregulated |
| 0,981957885 | 0,16104332 | FBXL19-AS1 | FALSE | Upregulated |
| 0,827974334 | 0,16104332 | MR1 | FALSE | Upregulated |
| -0,846499636 | 0,161079976 | TMTC1 | FALSE | Downregulated |
| 0,699275801 | 0,161270034 | BASP1 | FALSE | Upregulated |
| 0,537591833 | 0,161533883 | LRRC37A2 | FALSE | Upregulated |
| 0,420947718 | 0,161543945 | LOC100128374 | FALSE | Upregulated |
| 0,700563308 | 0,16205896 | SLC36A1 | FALSE | Upregulated |
| -1,638531974 | 0,162096738 | IHH | FALSE | Downregulated |
| 0,781053848 | 0,162096738 | LOC101060553 | FALSE | Upregulated |
| -0,538895871 | 0,162096738 | SERTAD2 | FALSE | Downregulated |
| 0,337630799 | 0,162123047 | MARK2 | FALSE | Upregulated |
| -0,512214594 | 0,162370112 | PCMTD1 | FALSE | Downregulated |
| -1,061219558 | 0,162370112 | PDE4D | FALSE | Downregulated |
| 0,46167359 | 0,162370112 | UNC13D | FALSE | Upregulated |
| -0,524799078 | 0,162436148 | UROD | FALSE | Downregulated |
| -0,371576987 | 0,162592336 | PCDH1 | FALSE | Downregulated |
| 0,306767283 | 0,162704203 | LASP1 | FALSE | Upregulated |
| 0,474738774 | 0,162704203 | LOC100506990 | FALSE | Upregulated |
| 0,326372678 | 0,162704203 | RFTN1 | FALSE | Upregulated |
| 0,491616228 | 0,162912128 | ANGPTL7 | FALSE | Upregulated |

| | | | | |
|---|---|---|---|---|
| 1,114939225 | 0,162912128 | CCDC170 | FALSE | Upregulated |
| -0,339152251 | 0,162912128 | GPR126 | FALSE | Downregulated |
| 1,165524596 | 0,162912128 | LPCAT2 | FALSE | Upregulated |
| -0,445025763 | 0,162912128 | TRPV1 | FALSE | Downregulated |
| 0,704915662 | 0,163033722 | RARRES3 | FALSE | Upregulated |
| -0,290977822 | 0,163033722 | SLC16A2 | FALSE | Downregulated |
| -0,482733035 | 0,163548874 | ZNF587B | FALSE | Downregulated |
| 0,476948512 | 0,163745196 | LOC101928070 | FALSE | Upregulated |
| 0,552474421 | 0,164088573 | IKZF2 | FALSE | Upregulated |
| -0,678438593 | 0,164088573 | TNFSF12 | FALSE | Downregulated |
| -1,172884549 | 0,164127636 | TATDN3 | FALSE | Downregulated |
| -0,391114552 | 0,164376464 | FRYL | FALSE | Downregulated |
| 0,710580493 | 0,164376464 | TMEM164 | FALSE | Upregulated |
| -0,663693015 | 0,164408955 | ARHGEF28 | FALSE | Downregulated |
| 1,264637385 | 0,164408955 | CRP | FALSE | Upregulated |
| -0,32821502 | 0,164585451 | C6orf89 | FALSE | Downregulated |
| 0,900185921 | 0,164669136 | LOC102724518 | FALSE | Upregulated |
| -0,68683676 | 0,164669136 | TRPM8 | FALSE | Downregulated |
| 0,467695394 | 0,165244002 | ARPC2 | FALSE | Upregulated |
| 1,478625235 | 0,165248367 | BHLHA15 | FALSE | Upregulated |
| -0,503400833 | 0,165248367 | GNAO1 | FALSE | Downregulated |
| -0,470413072 | 0,165248367 | KLF11 | FALSE | Downregulated |
| 1,123871121 | 0,165248367 | SLC25A24 | FALSE | Upregulated |
| -0,570162281 | 0,165449004 | FAM107A | FALSE | Downregulated |
| 1,010842873 | 0,165547516 | CARD9 | FALSE | Upregulated |
| 0,80215732 | 0,165706026 | LOC102723555 | FALSE | Upregulated |
| -0,532541907 | 0,165755752 | DAO | FALSE | Downregulated |
| -0,277081056 | 0,165755752 | GPD1 | FALSE | Downregulated |
| 0,573285404 | 0,165755752 | MDFIC | FALSE | Upregulated |
| -0,250692293 | 0,165755752 | ZZEF1 | FALSE | Downregulated |
| -0,479559703 | 0,165839366 | AQP3 | FALSE | Downregulated |
| 0,80319885 | 0,165839366 | LOC101928154 | FALSE | Upregulated |
| 0,407569015 | 0,165881127 | RAB22A | FALSE | Upregulated |
| -0,442760948 | 0,165891817 | KCNJ8 | FALSE | Downregulated |
| -0,463688501 | 0,165990333 | CD82 | FALSE | Downregulated |
| -0,967852392 | 0,165990333 | DPY19L2P2 | FALSE | Downregulated |
| 0,379604499 | 0,165990333 | FLII | FALSE | Upregulated |
| -1,161791123 | 0,165990333 | POU6F2-AS2 | FALSE | Downregulated |
| 0,572005497 | 0,166286817 | PHF11 | FALSE | Upregulated |
| 0,339496811 | 0,166328891 | DNAJC14 | FALSE | Upregulated |
| 0,279891221 | 0,166328891 | MYO1F | FALSE | Upregulated |
| 1,685226812 | 0,166558033 | LOC102724867 | FALSE | Upregulated |
| 0,516082451 | 0,166638321 | CDK2AP1 | FALSE | Upregulated |
| -0,66018309 | 0,166638321 | ENTPD8 | FALSE | Downregulated |
| 0,634317677 | 0,166638321 | EVA1C | FALSE | Upregulated |
| 0,482434924 | 0,166638321 | FBXW7 | FALSE | Upregulated |
| 2,004353227 | 0,166638321 | FCRL5 | FALSE | Upregulated |
| 0,381742466 | 0,166638321 | FKBP1AP1 | FALSE | Upregulated |
| -0,417983552 | 0,166638321 | FLCN | FALSE | Downregulated |
| 0,41086381 | 0,166638321 | NTMT1 | FALSE | Upregulated |
| -0,286627044 | 0,166638321 | PQLC1 | FALSE | Downregulated |

| | | | | |
|---|---|---|---|---|
| 0,527146357 | 0,166668663 | ARHGAP27 | FALSE | Upregulated |
| 0,783187704 | 0,166692984 | LOC90784 | FALSE | Upregulated |
| -0,520979033 | 0,166799797 | MORN4 | FALSE | Downregulated |
| 0,74196005 | 0,166836452 | CENPN | FALSE | Upregulated |
| -0,878865822 | 0,16688613 | RNU2-2P | FALSE | Downregulated |
| -0,418975887 | 0,166960165 | AHDC1 | FALSE | Downregulated |
| -0,2672523 | 0,166960165 | H3F3B | FALSE | Downregulated |
| -0,533248111 | 0,166960165 | JADE3 | FALSE | Downregulated |
| 0,426107196 | 0,166960165 | MTMR2 | FALSE | Upregulated |
| 0,828765791 | 0,166960165 | PCNXL2 | FALSE | Upregulated |
| -0,41391218 | 0,166960165 | SLC17A4 | FALSE | Downregulated |
| -0,486919918 | 0,167269516 | C1orf168 | FALSE | Downregulated |
| 1,210518524 | 0,167269516 | CD300LB | FALSE | Upregulated |
| 0,444027088 | 0,167269516 | HIF1A-AS2 | FALSE | Upregulated |
| 0,400433122 | 0,167269516 | PSME2 | FALSE | Upregulated |
| 1,355502589 | 0,167327946 | FGF17 | FALSE | Upregulated |
| 0,674588741 | 0,167327946 | ZBTB33 | FALSE | Upregulated |
| -0,914012346 | 0,167403069 | ADORA2B | FALSE | Downregulated |
| 0,552091076 | 0,167403069 | CCNF | FALSE | Upregulated |
| 0,298654609 | 0,167490626 | LOC100379224 | FALSE | Upregulated |
| -1,895094118 | 0,167647888 | ASB3 | FALSE | Downregulated |
| 0,330652574 | 0,167647888 | TCOF1 | FALSE | Upregulated |
| 0,507187009 | 0,168018042 | SERHL | FALSE | Upregulated |
| 0,355888587 | 0,168159563 | ANXA11 | FALSE | Upregulated |
| 0,529212213 | 0,168159563 | ASPN | FALSE | Upregulated |
| -0,563903022 | 0,168175409 | ATP5O | FALSE | Downregulated |
| 0,480859874 | 0,168285505 | B3GNTL1 | FALSE | Upregulated |
| 0,408486486 | 0,168309693 | ARHGAP31 | FALSE | Upregulated |
| -0,621805929 | 0,168309693 | CYP4F12 | FALSE | Downregulated |
| 0,356141096 | 0,168309693 | FLJ10038 | FALSE | Upregulated |
| 0,589076937 | 0,168309693 | TGM2 | FALSE | Upregulated |
| -0,943887991 | 0,168439903 | LGSN | FALSE | Downregulated |
| 0,402283914 | 0,168484557 | ZNF397 | FALSE | Upregulated |
| 0,27220274 | 0,169217729 | WDR1 | FALSE | Upregulated |
| 0,783156211 | 0,16928965 | OAS2 | FALSE | Upregulated |
| 0,428237213 | 0,169402834 | FILIP1L | FALSE | Upregulated |
| 1,299438006 | 0,169405284 | DNAH5 | FALSE | Upregulated |
| -0,287196284 | 0,169429591 | KIAA0195 | FALSE | Downregulated |
| 1,159604413 | 0,1697171 | BTK | FALSE | Upregulated |
| 0,39917405 | 0,16971956 | NSUN5 | FALSE | Upregulated |
| 0,66059693 | 0,169824073 | IDI2-AS1 | FALSE | Upregulated |
| 0,330310976 | 0,169824073 | PRR26 | FALSE | Upregulated |
| -0,329505873 | 0,169856234 | PUM1 | FALSE | Downregulated |
| -0,272145249 | 0,169856234 | SCARNA2 | FALSE | Downregulated |
| 0,543821267 | 0,170084603 | GRAMD1A | FALSE | Upregulated |
| -0,361430601 | 0,170281186 | DBI | FALSE | Downregulated |
| -0,356330424 | 0,170459348 | PPFIBP2 | FALSE | Downregulated |
| -0,339523708 | 0,170587232 | TMEM57 | FALSE | Downregulated |
| 0,560210542 | 0,170654692 | IRF9 | FALSE | Upregulated |
| -1,246293815 | 0,171072996 | ELP4 | FALSE | Downregulated |
| 1,209448491 | 0,171111185 | CDH6 | FALSE | Upregulated |

| | | | | |
|---|---|---|---|---|
| -0,506617533 | 0,171347551 | FCN2 | FALSE | Downregulated |
| 0,427694105 | 0,171548274 | KIAA0556 | FALSE | Upregulated |
| 0,750690154 | 0,171548274 | LOC100128364 | FALSE | Upregulated |
| -0,702910084 | 0,171600722 | LTN1 | FALSE | Downregulated |
| -0,596625769 | 0,171600722 | SCNN1D | FALSE | Downregulated |
| 0,731160271 | 0,171600722 | SPTBN5 | FALSE | Upregulated |
| -1,712309554 | 0,171729799 | ZNF415 | FALSE | Downregulated |
| -1,03194971 | 0,172010328 | ESPNL | FALSE | Downregulated |
| -0,909610267 | 0,172391521 | PNPLA4 | FALSE | Downregulated |
| -0,47982362 | 0,172498045 | FES | FALSE | Downregulated |
| 1,200803218 | 0,172498045 | TTC9 | FALSE | Upregulated |
| 0,352135433 | 0,17250569 | CFL1 | FALSE | Upregulated |
| 0,99517753 | 0,17250569 | LOC101927911 | FALSE | Upregulated |
| -0,396718473 | 0,17250569 | MFAP3L | FALSE | Downregulated |
| 0,359205334 | 0,172656096 | AGBL5 | FALSE | Upregulated |
| -0,312117509 | 0,172656096 | CCDC50 | FALSE | Downregulated |
| -0,435402684 | 0,172656096 | CFHR5 | FALSE | Downregulated |
| -0,394342542 | 0,172656096 | CYP2C18 | FALSE | Downregulated |
| 1,29308335 | 0,172656096 | HIST1H2BN | FALSE | Upregulated |
| -0,557870997 | 0,172656096 | NMRK1 | FALSE | Downregulated |
| -0,548463235 | 0,172656096 | PLA2G16 | FALSE | Downregulated |
| -0,568494237 | 0,172656096 | RPUSD4 | FALSE | Downregulated |
| 0,782530647 | 0,172656096 | TRPV2 | FALSE | Upregulated |
| -0,449591557 | 0,172873084 | OPN3 | FALSE | Downregulated |
| -0,738114513 | 0,172909332 | LOC100289230 | FALSE | Downregulated |
| 0,778658523 | 0,17311372 | LRRC25 | FALSE | Upregulated |
| -0,429820721 | 0,173179654 | QPRT | FALSE | Downregulated |
| -0,485477209 | 0,173181567 | HS3ST3B1 | FALSE | Downregulated |
| -0,611190323 | 0,173555947 | GREM2 | FALSE | Downregulated |
| 0,563229312 | 0,173918967 | DNAJC24 | FALSE | Upregulated |
| 0,376554749 | 0,173958678 | LYRM2 | FALSE | Upregulated |
| -0,533242844 | 0,174379207 | RRNAD1 | FALSE | Downregulated |
| -0,318076089 | 0,174379207 | TSC1 | FALSE | Downregulated |
| -1,567886922 | 0,174423051 | TOB1-AS1 | FALSE | Downregulated |
| -0,261835239 | 0,174471108 | TNKS1BP1 | FALSE | Downregulated |
| -0,799123515 | 0,174795297 | COX18 | FALSE | Downregulated |
| -1,338523939 | 0,174795297 | MEP1B | FALSE | Downregulated |
| 0,25203893 | 0,175063966 | ARHGEF18 | FALSE | Upregulated |
| -0,439861317 | 0,175099941 | UCK1 | FALSE | Downregulated |
| 0,36322221 | 0,175244305 | AAK1 | FALSE | Upregulated |
| -0,371784032 | 0,175266141 | RPLP1 | FALSE | Downregulated |
| -0,380743247 | 0,175439537 | PLA2G12B | FALSE | Downregulated |
| -0,65268172 | 0,175472453 | AMZ2 | FALSE | Downregulated |
| -0,485706678 | 0,175472453 | FNIP2 | FALSE | Downregulated |
| -0,364377517 | 0,175472453 | NXF1 | FALSE | Downregulated |
| -0,672843048 | 0,175514308 | HSD17B14 | FALSE | Downregulated |
| 0,649638558 | 0,175633914 | SLC6A8 | FALSE | Upregulated |
| 0,568665175 | 0,175635312 | PTTG3P | FALSE | Upregulated |
| -0,301444645 | 0,175635312 | TAOK1 | FALSE | Downregulated |
| 0,987582524 | 0,175639605 | LOC100996433 | FALSE | Upregulated |
| -0,886249422 | 0,17571611 | GPR98 | FALSE | Downregulated |

| | | | | |
|---|---|---|---|---|
| 0,282873984 | 0,175992174 | FAM208B | FALSE | Upregulated |
| 0,971912627 | 0,176303134 | NCEH1 | FALSE | Upregulated |
| -0,354299736 | 0,176598128 | LRRC47 | FALSE | Downregulated |
| 0,643365324 | 0,176888256 | BCO2 | FALSE | Upregulated |
| 0,446276375 | 0,176888256 | BRD7P3 | FALSE | Upregulated |
| -0,659326892 | 0,176888256 | GTPBP10 | FALSE | Downregulated |
| 0,808911913 | 0,176888256 | LINC00936 | FALSE | Upregulated |
| -0,730941272 | 0,176888256 | MEIS1 | FALSE | Downregulated |
| 0,425626532 | 0,176888256 | PHPT1 | FALSE | Upregulated |
| -0,826888679 | 0,176888256 | WEE2 | FALSE | Downregulated |
| 0,685780715 | 0,176947411 | SLC1A7 | FALSE | Upregulated |
| -0,585963907 | 0,176968764 | ZNF462 | FALSE | Downregulated |
| -0,518336683 | 0,177116104 | ANKS4B | FALSE | Downregulated |
| -0,981004909 | 0,177116104 | ISPD | FALSE | Downregulated |
| 1,167298656 | 0,177116104 | LINC00665 | FALSE | Upregulated |
| -0,661524866 | 0,177116104 | RNU105A | FALSE | Downregulated |
| -0,976014974 | 0,177116104 | RP11-1096D5.1 | FALSE | Downregulated |
| 0,557928634 | 0,177116104 | TTC33 | FALSE | Upregulated |
| -0,639853454 | 0,177116104 | ZNF532 | FALSE | Downregulated |
| 0,435607535 | 0,177210288 | ETV6 | FALSE | Upregulated |
| -2,171153267 | 0,177419854 | AJAP1 | FALSE | Downregulated |
| -0,744843546 | 0,177419854 | MANSC1 | FALSE | Downregulated |
| -0,386426285 | 0,177458817 | AOX1 | FALSE | Downregulated |
| 0,31190766 | 0,177458817 | LOC101929264 | FALSE | Upregulated |
| -0,307379945 | 0,177458817 | SLC25A13 | FALSE | Downregulated |
| 0,364798222 | 0,178104447 | PSPC1 | FALSE | Upregulated |
| 0,404658858 | 0,179093439 | ZNF79 | FALSE | Upregulated |
| 0,716900493 | 0,179246032 | FAM98A | FALSE | Upregulated |
| 0,656602094 | 0,179406196 | CLDN18 | FALSE | Upregulated |
| 0,559377813 | 0,179490034 | DAPK3 | FALSE | Upregulated |
| -1,712980907 | 0,179490034 | NOL4 | FALSE | Downregulated |
| -0,235557217 | 0,179541485 | CDHR5 | FALSE | Downregulated |
| -0,372679533 | 0,179562056 | APOC2 | FALSE | Downregulated |
| 0,493954907 | 0,179562056 | RIPK4 | FALSE | Upregulated |
| -0,585821284 | 0,17966856 | ARSE | FALSE | Downregulated |
| 0,190671406 | 0,179692861 | MALAT1 | FALSE | Upregulated |
| 0,421737209 | 0,179692861 | NOD1 | FALSE | Upregulated |
| 0,510699924 | 0,179692861 | TP53I3 | FALSE | Upregulated |
| -0,510006168 | 0,179692861 | TTPA | FALSE | Downregulated |
| 0,420110282 | 0,17980646 | ABCB1 | FALSE | Upregulated |
| -0,228873985 | 0,17980646 | BAG6 | FALSE | Downregulated |
| 0,463162845 | 0,17980646 | CABLES2 | FALSE | Upregulated |
| 1,205540456 | 0,17980646 | GNB1L | FALSE | Upregulated |
| 0,759192272 | 0,17980646 | LOC102288414 | FALSE | Upregulated |
| -0,580092441 | 0,17980646 | PARP6 | FALSE | Downregulated |
| 0,560664692 | 0,17980646 | PTGFRN | FALSE | Upregulated |
| 0,749042311 | 0,179819946 | LAMC3 | FALSE | Upregulated |
| 0,886223197 | 0,179819946 | TPRG1-AS2 | FALSE | Upregulated |
| 0,737046112 | 0,180053779 | CRISPLD2 | FALSE | Upregulated |
| -0,41517953 | 0,180255807 | KCND3 | FALSE | Downregulated |
| 1,975576271 | 0,18035946 | KGFLP1 | FALSE | Upregulated |

| | | | | |
|---|---|---|---|---|
| -0,365357088 | 0,18035946 | RPS3 | FALSE | Downregulated |
| 0,567116701 | 0,180570042 | GMPPB | FALSE | Upregulated |
| 0,587867136 | 0,180669158 | LYN | FALSE | Upregulated |
| 0,263136003 | 0,180686189 | GGA2 | FALSE | Upregulated |
| 0,318867365 | 0,180686189 | LOC101927838 | FALSE | Upregulated |
| 0,861708792 | 0,180686189 | PLEKHB2 | FALSE | Upregulated |
| 0,729815518 | 0,180686189 | SAMD9 | FALSE | Upregulated |
| -0,696284597 | 0,180718267 | MRPS10 | FALSE | Downregulated |
| 0,536544266 | 0,180718267 | STARD5 | FALSE | Upregulated |
| 1,282696039 | 0,181062288 | LOC613038 | FALSE | Upregulated |
| -0,459987331 | 0,181257955 | PDXDC2P | FALSE | Downregulated |
| 0,336529374 | 0,181373025 | GZF1 | FALSE | Upregulated |
| 0,349786669 | 0,181580086 | LPIN2 | FALSE | Upregulated |
| -0,510956265 | 0,181635985 | DIP2A | FALSE | Downregulated |
| 1,43728678 | 0,181635985 | RP1-69M21.2 | FALSE | Upregulated |
| -0,255323533 | 0,181635985 | TBC1D9B | FALSE | Downregulated |
| 0,811442371 | 0,181698748 | ZNF575 | FALSE | Upregulated |
| 0,485170875 | 0,181913724 | PRPH2 | FALSE | Upregulated |
| 0,927402303 | 0,182090572 | MIR497HG | FALSE | Upregulated |
| 0,953123401 | 0,182126091 | SLFN13 | FALSE | Upregulated |
| 1,18313214 | 0,18219349 | AC022007.5 | FALSE | Upregulated |
| 0,713898081 | 0,18219349 | C21orf91 | FALSE | Upregulated |
| 0,792328109 | 0,18219349 | C9orf116 | FALSE | Upregulated |
| 0,467693976 | 0,18219349 | SYT15 | FALSE | Upregulated |
| 0,472533176 | 0,182217671 | MYH7B | FALSE | Upregulated |
| 0,797070059 | 0,18253293 | NHS | FALSE | Upregulated |
| 0,582783262 | 0,182641231 | BMF | FALSE | Upregulated |
| 0,565219851 | 0,183177099 | FNDC5 | FALSE | Upregulated |
| 1,79706944 | 0,183479566 | LOC101929824 | FALSE | Upregulated |
| -0,682778091 | 0,18352074 | GSPT2 | FALSE | Downregulated |
| 1,126383115 | 0,18352074 | HAR1A | FALSE | Upregulated |
| 0,856164326 | 0,18352074 | MCAT | FALSE | Upregulated |
| 0,784072745 | 0,18352074 | MTMR3 | FALSE | Upregulated |
| 0,787224622 | 0,183813985 | LOC101928363 | FALSE | Upregulated |
| 0,848448909 | 0,183911937 | DYNLT1 | FALSE | Upregulated |
| 0,927961599 | 0,184643911 | RAX2 | FALSE | Upregulated |
| -1,518916066 | 0,184700132 | LOC101928172 | FALSE | Downregulated |
| 0,382581685 | 0,184700132 | NCAPG | FALSE | Upregulated |
| 0,413461865 | 0,184724956 | AC003665.1 | FALSE | Upregulated |
| 1,417521382 | 0,184757834 | LINC00662 | FALSE | Upregulated |
| -0,383945868 | 0,184774506 | ITGA5 | FALSE | Downregulated |
| -0,397489927 | 0,184774506 | RALGAPA1 | FALSE | Downregulated |
| 0,675670046 | 0,184774506 | UQCRBP1 | FALSE | Upregulated |
| 0,604340547 | 0,184947552 | AMMECR1 | FALSE | Upregulated |
| 0,711410968 | 0,184947552 | HLA-H | FALSE | Upregulated |
| -0,692568627 | 0,184947552 | SLC25A34 | FALSE | Downregulated |
| -0,387315714 | 0,184947552 | SPTY2D1 | FALSE | Downregulated |
| 0,253880868 | 0,185100238 | NEAT1 | FALSE | Upregulated |
| 0,628673995 | 0,185141967 | ELOVL2 | FALSE | Upregulated |
| 0,227023037 | 0,185141967 | NCOR2 | FALSE | Upregulated |
| 0,678357248 | 0,185207654 | CCDC41 | FALSE | Upregulated |

| | | | | |
|---|---|---|---|---|
| -1,077340144 | 0,185207654 | SLC38A11 | FALSE | Downregulated |
| 0,975860615 | 0,185680493 | MKLN1-AS | FALSE | Upregulated |
| -2,225881524 | 0,186069669 | MROH7 | FALSE | Downregulated |
| 0,618289624 | 0,186272283 | PRPS2 | FALSE | Upregulated |
| -0,601035886 | 0,186372082 | C3orf62 | FALSE | Downregulated |
| 2,415567688 | 0,186406644 | ANKRD22 | FALSE | Upregulated |
| 0,349866796 | 0,186406644 | SRR | FALSE | Upregulated |
| 1,071105143 | 0,186732741 | TCEB1 | FALSE | Upregulated |
| 1,581268191 | 0,186800486 | CD28 | FALSE | Upregulated |
| -0,311248298 | 0,1870017 | MPC2 | FALSE | Downregulated |
| 0,496026681 | 0,1870252 | CCDC18 | FALSE | Upregulated |
| 0,517442199 | 0,1870252 | NEDD9 | FALSE | Upregulated |
| 0,578792029 | 0,187100928 | PNMAL2 | FALSE | Upregulated |
| -0,450256311 | 0,18736365 | RAD54L2 | FALSE | Downregulated |
| -0,343388835 | 0,187766955 | MRPL12 | FALSE | Downregulated |
| -0,203805753 | 0,187766955 | SEC14L2 | FALSE | Downregulated |
| 0,533361282 | 0,187929956 | TONSL | FALSE | Upregulated |
| -0,730668105 | 0,187967903 | LAMTOR5 | FALSE | Downregulated |
| -1,592041347 | 0,187967903 | LOC102723583 | FALSE | Downregulated |
| -0,902680251 | 0,188152666 | LOC644794 | FALSE | Downregulated |
| -0,393461272 | 0,188540603 | FAM114A1 | FALSE | Downregulated |
| 0,494233211 | 0,188690991 | GM2A | FALSE | Upregulated |
| -0,45003399 | 0,18919709 | DHRS4-AS1 | FALSE | Downregulated |
| 1,077578291 | 0,18919709 | LOC102724770 | FALSE | Upregulated |
| -0,404175346 | 0,18919709 | RNF170 | FALSE | Downregulated |
| 1,382565899 | 0,18919709 | RP11-372E1.6 | FALSE | Upregulated |
| 1,170880918 | 0,189413872 | LOC101927131 | FALSE | Upregulated |
| 0,750949622 | 0,189604386 | RELB | FALSE | Upregulated |
| -0,573110163 | 0,189746271 | NTHL1 | FALSE | Downregulated |
| -1,710164501 | 0,18989441 | ACN9 | FALSE | Downregulated |
| 0,476561202 | 0,18989441 | LOC100128531 | FALSE | Upregulated |
| 1,050198005 | 0,18989441 | LOC100507507 | FALSE | Upregulated |
| 0,46475232 | 0,18989441 | RAD52 | FALSE | Upregulated |
| -0,707861529 | 0,189970982 | ESAM | FALSE | Downregulated |
| 0,782873145 | 0,190199944 | CMAHP | FALSE | Upregulated |
| 1,127828333 | 0,190230552 | GMPR | FALSE | Upregulated |
| -0,498264793 | 0,190328875 | NEURL2 | FALSE | Downregulated |
| 1,095456312 | 0,190328875 | OGFR-AS1 | FALSE | Upregulated |
| -0,348383726 | 0,190328875 | PANK3 | FALSE | Downregulated |
| -0,365480421 | 0,190328875 | RASSF5 | FALSE | Downregulated |
| 0,737932158 | 0,190328875 | TOMM5 | FALSE | Upregulated |
| -0,394433326 | 0,190364308 | CDK9 | FALSE | Downregulated |
| 1,520851202 | 0,19041264 | HAPLN3 | FALSE | Upregulated |
| 0,27349596 | 0,19041264 | LOC101927556 | FALSE | Upregulated |
| -0,350564221 | 0,190656178 | GYS2 | FALSE | Downregulated |
| 1,065532118 | 0,190953319 | NUTM1 | FALSE | Upregulated |
| 0,361491158 | 0,191025493 | LOC441081 | FALSE | Upregulated |
| -0,611802235 | 0,191025493 | SMDT1 | FALSE | Downregulated |
| 1,763014107 | 0,191075577 | RAB7B | FALSE | Upregulated |
| 0,778163856 | 0,191075577 | TES | FALSE | Upregulated |
| 0,550885396 | 0,191088586 | PLK2 | FALSE | Upregulated |

| | | | | |
|---|---|---|---|---|
| -0,574734956 | 0,191180374 | HLX | FALSE | Downregulated |
| -1,036853972 | 0,19119506 | HSP90B2P | FALSE | Downregulated |
| 0,959594729 | 0,191354225 | ABHD11 | FALSE | Upregulated |
| -0,84685095 | 0,191376966 | MINOS1P1 | FALSE | Downregulated |
| -0,755270408 | 0,19148692 | ZNF329 | FALSE | Downregulated |
| 0,260410117 | 0,191784616 | RRBP1 | FALSE | Upregulated |
| -0,32564323 | 0,191817545 | CYB5A | FALSE | Downregulated |
| 1,327562937 | 0,191817545 | FAM86JP | FALSE | Upregulated |
| -1,963629712 | 0,191817545 | LOC100653233 | FALSE | Downregulated |
| -0,494472855 | 0,191817545 | SDPR | FALSE | Downregulated |
| -0,454340815 | 0,191898664 | RGPD4 | FALSE | Downregulated |
| -0,460269667 | 0,191991361 | POLR1D | FALSE | Downregulated |
| -0,3737811 | 0,192083618 | SYAP1 | FALSE | Downregulated |
| -0,362099275 | 0,192157792 | CLSTN3 | FALSE | Downregulated |
| 0,571585589 | 0,192293107 | LOC101928604 | FALSE | Upregulated |
| 0,475537505 | 0,192293107 | TLE4 | FALSE | Upregulated |
| 0,510994947 | 0,192293107 | TRPM4 | FALSE | Upregulated |
| -1,056609457 | 0,192361332 | GIN1 | FALSE | Downregulated |
| -0,253531401 | 0,192361332 | MAF | FALSE | Downregulated |
| 0,904375602 | 0,192992046 | VSIG10L | FALSE | Upregulated |
| 0,925015914 | 0,193659673 | MBOAT1 | FALSE | Upregulated |
| -0,247947248 | 0,193695899 | RPL8 | FALSE | Downregulated |
| 1,008909623 | 0,193732702 | DGKA | FALSE | Upregulated |
| -0,494490892 | 0,193768648 | CLEC4G | FALSE | Downregulated |
| 0,398277509 | 0,193768648 | DUOX1 | FALSE | Upregulated |
| 0,455280948 | 0,193891903 | LOC101929387 | FALSE | Upregulated |
| -0,244532175 | 0,193891903 | NACA | FALSE | Downregulated |
| 0,613448266 | 0,193891903 | RPS18P9 | FALSE | Upregulated |
| -0,241119883 | 0,193891903 | SETD2 | FALSE | Downregulated |
| -0,482881792 | 0,193951773 | INPP5B | FALSE | Downregulated |
| 0,623894129 | 0,193965929 | TTC13 | FALSE | Upregulated |
| 0,405622497 | 0,194579564 | PPP1R18 | FALSE | Upregulated |
| 0,732032294 | 0,194667961 | BCL2 | FALSE | Upregulated |
| -0,240195031 | 0,194667961 | F10 | FALSE | Downregulated |
| 0,683622941 | 0,194815328 | C21orf58 | FALSE | Upregulated |
| -0,494982309 | 0,195051072 | FRY | FALSE | Downregulated |
| 0,917640999 | 0,195094439 | GALNT6 | FALSE | Upregulated |
| 1,269208731 | 0,195094439 | PRR11 | FALSE | Upregulated |
| 1,153759502 | 0,195197262 | HIST1H2AE | FALSE | Upregulated |
| 0,34113765 | 0,195205727 | MSMP | FALSE | Upregulated |
| 0,730639739 | 0,195332442 | HSF4 | FALSE | Upregulated |
| -1,067959565 | 0,195332442 | TXNL4B | FALSE | Downregulated |
| 0,69036523 | 0,195397264 | TMEM165 | FALSE | Upregulated |
| -0,762755438 | 0,19541689 | CLCN4 | FALSE | Downregulated |
| 0,809723921 | 0,19541689 | MRAS | FALSE | Upregulated |
| -1,080159054 | 0,19541689 | TM6SF2 | FALSE | Downregulated |
| 1,29110822 | 0,195477599 | RHNO1 | FALSE | Upregulated |
| 0,460724243 | 0,195632187 | EFHD2 | FALSE | Upregulated |
| 0,50836257 | 0,195632187 | KCNC3 | FALSE | Upregulated |
| 0,523389511 | 0,195770573 | LOC100126784 | FALSE | Upregulated |
| 0,703750139 | 0,195770573 | NDUFA12 | FALSE | Upregulated |

| | | | | |
|---|---|---|---|---|
| -0,331368037 | 0,195770573 | WBP2 | FALSE | Downregulated |
| 0,821533912 | 0,195853865 | LY9 | FALSE | Upregulated |
| -1,470810835 | 0,195853865 | SLMO2-ATP5E | FALSE | Downregulated |
| 0,416505802 | 0,195953965 | LOC102724761 | FALSE | Upregulated |
| -0,388085695 | 0,19605429 | ABCD3 | FALSE | Downregulated |
| 0,550756891 | 0,196086723 | LINC00954 | FALSE | Upregulated |
| 0,562126449 | 0,196451131 | ALPK1 | FALSE | Upregulated |
| -0,67526426 | 0,196451131 | FAM110B | FALSE | Downregulated |
| -0,979547191 | 0,196451131 | PALM2-AKAP2 | FALSE | Downregulated |
| -1,488187902 | 0,196451131 | PEG10 | FALSE | Downregulated |
| 0,468094391 | 0,196516761 | ARL17A | FALSE | Upregulated |
| 0,43213561 | 0,196547529 | AC137932.6 | FALSE | Upregulated |
| -0,375181286 | 0,196547529 | SENP6 | FALSE | Downregulated |
| -0,311576936 | 0,196547529 | SLC20A2 | FALSE | Downregulated |
| -0,3102445 | 0,196753538 | SOX13 | FALSE | Downregulated |
| 0,454579497 | 0,196832244 | DYNLL1 | FALSE | Upregulated |
| 0,648111478 | 0,196832244 | GHRLOS | FALSE | Upregulated |
| 1,08825569 | 0,196856513 | GPRIN1 | FALSE | Upregulated |
| -0,281805381 | 0,196882367 | RPS17 | FALSE | Downregulated |
| 0,374562757 | 0,196884245 | HSPB1 | FALSE | Upregulated |
| -0,721896833 | 0,196884245 | MTHFSD | FALSE | Downregulated |
| 0,402982738 | 0,196943613 | CEP57 | FALSE | Upregulated |
| 1,769067667 | 0,196943613 | GALNT9 | FALSE | Upregulated |
| 0,406335715 | 0,196943613 | MAEA | FALSE | Upregulated |
| -0,757034975 | 0,196943613 | PIWIL2 | FALSE | Downregulated |
| -0,459658262 | 0,196978759 | FOXA3 | FALSE | Downregulated |
| 0,459019225 | 0,197233293 | IFIT3 | FALSE | Upregulated |
| 1,156816994 | 0,197289584 | KIAA0226L | FALSE | Upregulated |
| 0,622069343 | 0,19731959 | SYK | FALSE | Upregulated |
| -1,23825157 | 0,197571152 | LINC01010 | FALSE | Downregulated |
| 0,251388727 | 0,197571152 | MSH6 | FALSE | Upregulated |
| -0,451928067 | 0,197571152 | SNHG16 | FALSE | Downregulated |
| -0,381421385 | 0,19767862 | STYX | FALSE | Downregulated |
| -1,451815576 | 0,19767862 | TMEM68 | FALSE | Downregulated |
| 0,754180425 | 0,19773324 | CLIP3 | FALSE | Upregulated |
| -0,432477287 | 0,197773773 | ATP5G2 | FALSE | Downregulated |
| 0,676016104 | 0,198029026 | C1orf35 | FALSE | Upregulated |
| -0,165370009 | 0,198264107 | HUWE1 | FALSE | Downregulated |
| 0,555962584 | 0,198358711 | INPP5F | FALSE | Upregulated |
| -0,40549099 | 0,198583798 | ENGASE | FALSE | Downregulated |
| 0,914688709 | 0,198641618 | TACR2 | FALSE | Upregulated |
| 0,304625538 | 0,198929917 | ARG2 | FALSE | Upregulated |
| -0,374464393 | 0,198929917 | TOB1 | FALSE | Downregulated |
| -0,500668713 | 0,199121012 | FAM99A | FALSE | Downregulated |
| 1,565821637 | 0,199198927 | IFNG-AS1 | FALSE | Upregulated |
| 1,620273823 | 0,199198927 | IL1A | FALSE | Upregulated |
| -0,2748105 | 0,199198927 | SCAP | FALSE | Downregulated |
| -0,358146628 | 0,199198927 | TGFBR3 | FALSE | Downregulated |
| 0,985007432 | 0,199650045 | MTOR-AS1 | FALSE | Upregulated |
| 1,048673907 | 0,199703879 | EPS8L1 | FALSE | Upregulated |
| 0,428355871 | 0,199723013 | FLJ32756 | FALSE | Upregulated |

| | | | | |
|---|---|---|---|---|
| 0,479823303 | 0,199723013 | RP11-450K4.1 | FALSE | Upregulated |
| -0,566267503 | 0,199723013 | ZEB1 | FALSE | Downregulated |
| -0,323972989 | 0,199739184 | ASL | FALSE | Downregulated |
| 0,266168628 | 0,199739184 | HNRNPC | FALSE | Upregulated |
| 0,388727717 | 0,200120782 | ERO1LB | FALSE | Upregulated |
| 0,841185961 | 0,200227869 | PLK3 | FALSE | Upregulated |
| -0,344497346 | 0,200227869 | RGN | FALSE | Downregulated |
| 1,310215073 | 0,200300342 | HSPA2 | FALSE | Upregulated |
| 0,471304789 | 0,200402365 | IL16 | FALSE | Upregulated |
| -0,551131032 | 0,200529187 | LOC101927513 | FALSE | Downregulated |
| 0,480763659 | 0,200529187 | WFS1 | FALSE | Upregulated |
| 0,584249224 | 0,200617621 | CBFB | FALSE | Upregulated |
| 0,302793473 | 0,201020427 | CST3 | FALSE | Upregulated |
| 0,444909226 | 0,201020427 | TUBB6 | FALSE | Upregulated |
| 0,268884398 | 0,201020427 | WHSC1 | FALSE | Upregulated |
| 0,378895755 | 0,201043185 | POLR2B | FALSE | Upregulated |
| 0,496759141 | 0,201304131 | RHOD | FALSE | Upregulated |
| 0,322191024 | 0,201382369 | PLEKHJ1 | FALSE | Upregulated |
| 0,526150374 | 0,201382369 | PSME3 | FALSE | Upregulated |
| -0,686634114 | 0,202014713 | LOC101060581 | FALSE | Downregulated |
| -0,560902997 | 0,202014713 | PAQR7 | FALSE | Downregulated |
| -0,548981824 | 0,202278231 | CYHR1 | FALSE | Downregulated |
| 0,440352831 | 0,202466679 | MRS2 | FALSE | Upregulated |
| 0,379402938 | 0,202478913 | MAP1S | FALSE | Upregulated |
| 0,288764024 | 0,202663958 | GGA1 | FALSE | Upregulated |
| -0,214753623 | 0,202663958 | GOLGB1 | FALSE | Downregulated |
| 0,393761313 | 0,202772945 | INPP4A | FALSE | Upregulated |
| 1,26801217 | 0,202772945 | PTCHD1 | FALSE | Upregulated |
| 0,559920808 | 0,202772945 | UBE2W | FALSE | Upregulated |
| 0,779612555 | 0,202920639 | HLA-DMA | FALSE | Upregulated |
| 0,500093496 | 0,202920639 | PIP4K2A | FALSE | Upregulated |
| -0,826104576 | 0,203000993 | BMPER | FALSE | Downregulated |
| 1,302728368 | 0,203000993 | TNFRSF11A | FALSE | Upregulated |
| 0,797533893 | 0,203380184 | LOC101927340 | FALSE | Upregulated |
| -0,443117897 | 0,203668453 | SEC14L4 | FALSE | Downregulated |
| 1,169430799 | 0,20368143 | RHBDL1 | FALSE | Upregulated |
| -0,601894502 | 0,203831513 | PPIP5K2 | FALSE | Downregulated |
| 0,453607115 | 0,203841312 | OSGIN2 | FALSE | Upregulated |
| -0,897243252 | 0,203856652 | AKR7A2P1 | FALSE | Downregulated |
| -0,353944506 | 0,203991962 | FAT4 | FALSE | Downregulated |
| 0,71262236 | 0,204016484 | CHAF1A | FALSE | Upregulated |
| -0,59202283 | 0,204016484 | CHST12 | FALSE | Downregulated |
| 0,897506566 | 0,204016484 | CIT | FALSE | Upregulated |
| 0,661110621 | 0,204016484 | MCM4 | FALSE | Upregulated |
| -0,356059577 | 0,204067305 | CBX7 | FALSE | Downregulated |
| -0,9093164 | 0,204098786 | SLC35B3 | FALSE | Downregulated |
| 0,509036572 | 0,204382026 | RGS3 | FALSE | Upregulated |
| -0,967226864 | 0,204382026 | TXNDC16 | FALSE | Downregulated |
| 0,490656425 | 0,204970606 | NPB | FALSE | Upregulated |
| -0,478953826 | 0,204970606 | NT5DC1 | FALSE | Downregulated |
| -1,637450367 | 0,204970606 | NTRK3 | FALSE | Downregulated |

| | | | | |
|---|---|---|---|---|
| 0,345060033 | 0,204970606 | RPL23AP64 | FALSE | Upregulated |
| -0,245455709 | 0,204970606 | RPL7A | FALSE | Downregulated |
| 0,956349347 | 0,204970606 | SEBOX | FALSE | Upregulated |
| 0,423773614 | 0,204970606 | SUMO1P3 | FALSE | Upregulated |
| 1,440114547 | 0,204970606 | TMEM246 | FALSE | Upregulated |
| -0,329345139 | 0,20522215 | NTN4 | FALSE | Downregulated |
| 0,258232031 | 0,205425558 | HMGN2 | FALSE | Upregulated |
| -0,417032518 | 0,205959887 | PARP3 | FALSE | Downregulated |
| 0,328414969 | 0,206196874 | FBXL17 | FALSE | Upregulated |
| -1,487151106 | 0,206209846 | C1orf228 | FALSE | Downregulated |
| 1,433087406 | 0,206351012 | ARHGAP11A | FALSE | Upregulated |
| -0,534864356 | 0,206368632 | CPOX | FALSE | Downregulated |
| 0,967560686 | 0,206368632 | PRDM1 | FALSE | Upregulated |
| -0,829215522 | 0,206368632 | ZNF778 | FALSE | Downregulated |
| 0,806622147 | 0,206412863 | TLCD2 | FALSE | Upregulated |
| 0,582878305 | 0,206736108 | RDH13 | FALSE | Upregulated |
| -0,318518576 | 0,206736108 | TPRG1L | FALSE | Downregulated |
| 0,452846541 | 0,20688446 | SSBP3 | FALSE | Upregulated |
| 0,417843945 | 0,206922636 | GJD3 | FALSE | Upregulated |
| -0,71161727 | 0,20692395 | ABHD5 | FALSE | Downregulated |
| 1,581487058 | 0,20698983 | NMNAT2 | FALSE | Upregulated |
| 1,356240622 | 0,207203232 | ZNF711 | FALSE | Upregulated |
| 1,459125952 | 0,207786841 | C3orf67 | FALSE | Upregulated |
| 0,479964221 | 0,207786841 | GNA12 | FALSE | Upregulated |
| 0,430774985 | 0,208039493 | IK | FALSE | Upregulated |
| -0,693104013 | 0,208084624 | BOD1 | FALSE | Downregulated |
| -0,381043275 | 0,208084624 | POLR2C | FALSE | Downregulated |
| 0,439539099 | 0,208439802 | LOC100128361 | FALSE | Upregulated |
| -0,266064727 | 0,208553714 | SPATA13 | FALSE | Downregulated |
| 0,569823746 | 0,20884814 | GAPDHS | FALSE | Upregulated |
| -0,647132502 | 0,209075017 | TFEB | FALSE | Downregulated |
| -0,640901835 | 0,209472397 | CBR4 | FALSE | Downregulated |
| 0,380304117 | 0,209472397 | TVP23A | FALSE | Upregulated |
| 1,332529421 | 0,209489981 | AC104655.2 | FALSE | Upregulated |
| 0,788974319 | 0,209489981 | LINC00870 | FALSE | Upregulated |
| -0,476708482 | 0,209765781 | APPBP2 | FALSE | Downregulated |
| -1,228200306 | 0,209765781 | HOXA3 | FALSE | Downregulated |
| -1,081946958 | 0,209765781 | LOC642846 | FALSE | Downregulated |
| 0,298321973 | 0,209765781 | RNASEH2C | FALSE | Upregulated |
| 0,350784432 | 0,210060945 | ZNF562 | FALSE | Upregulated |
| -1,433549533 | 0,210060945 | ZNF773 | FALSE | Downregulated |
| 0,289759732 | 0,210380204 | B4GALT1 | FALSE | Upregulated |
| -0,257085421 | 0,210380204 | NCOA2 | FALSE | Downregulated |
| 0,317874941 | 0,210380204 | PRH2 | FALSE | Upregulated |
| -0,969415131 | 0,210380204 | RNF180 | FALSE | Downregulated |
| 0,409143097 | 0,210380204 | RPL13AP6 | FALSE | Upregulated |
| 0,327911552 | 0,210380204 | STT3B | FALSE | Upregulated |
| -0,350482385 | 0,210412367 | CALCOCO1 | FALSE | Downregulated |
| 0,376212293 | 0,210412367 | ERVFRD-1 | FALSE | Upregulated |
| 0,455665308 | 0,210412367 | FAS | FALSE | Upregulated |
| 0,885068176 | 0,210412367 | LURAP1L | FALSE | Upregulated |

| | | | | |
|---|---|---|---|---|
| 1,084979231 | 0,210468942 | HSPA6 | FALSE | Upregulated |
| 0,714925188 | 0,210468942 | LOC100134822 | FALSE | Upregulated |
| 0,39180203 | 0,210468942 | NUTM2D | FALSE | Upregulated |
| -0,401961536 | 0,210771495 | ABCG8 | FALSE | Downregulated |
| -0,362001158 | 0,210873301 | BTAF1 | FALSE | Downregulated |
| 0,719832446 | 0,210894599 | GOLGA6L5P | FALSE | Upregulated |
| 0,812620628 | 0,210930165 | MAST1 | FALSE | Upregulated |
| 1,036555688 | 0,21093877 | FAIM3 | FALSE | Upregulated |
| -0,680876714 | 0,21093877 | THAP7 | FALSE | Downregulated |
| 1,471064731 | 0,211059898 | MICB | FALSE | Upregulated |
| 0,255047203 | 0,211059898 | MSL1 | FALSE | Upregulated |
| -0,413734067 | 0,211435929 | C11orf54 | FALSE | Downregulated |
| 0,345902419 | 0,211435929 | RNF219-AS1 | FALSE | Upregulated |
| 0,768622557 | 0,211435929 | RP11-173B14.5 | FALSE | Upregulated |
| -0,345540991 | 0,211435929 | ZMYND8 | FALSE | Downregulated |
| -1,223535288 | 0,211678633 | LOC102724423 | FALSE | Downregulated |
| -0,470123683 | 0,211821437 | PMM1 | FALSE | Downregulated |
| 0,837359562 | 0,212235651 | AL022476.2 | FALSE | Upregulated |
| -1,501758882 | 0,212445607 | ADCK1 | FALSE | Downregulated |
| -0,65425289 | 0,212445607 | RNF121 | FALSE | Downregulated |
| 0,508458454 | 0,212445607 | SIGLEC16 | FALSE | Upregulated |
| -0,347934104 | 0,212445607 | TNPO1 | FALSE | Downregulated |
| 1,072100997 | 0,213065464 | MMD | FALSE | Upregulated |
| -0,664793495 | 0,213302111 | ZNF326 | FALSE | Downregulated |
| 0,444739525 | 0,213496858 | LRRN2 | FALSE | Upregulated |
| -0,901275438 | 0,213503385 | KCNE2 | FALSE | Downregulated |
| 0,283310156 | 0,214155653 | DNAJC3 | FALSE | Upregulated |
| -0,772934319 | 0,214155653 | TYW3 | FALSE | Downregulated |
| 0,247976095 | 0,214323649 | MAP3K13 | FALSE | Upregulated |
| -0,31735398 | 0,21445779 | BCKDK | FALSE | Downregulated |
| 0,544363857 | 0,21445779 | LOC100506388 | FALSE | Upregulated |
| 1,32156177 | 0,214694101 | ADAM28 | FALSE | Upregulated |
| -0,97180992 | 0,214694101 | C17orf89 | FALSE | Downregulated |
| 0,317640161 | 0,214717138 | H1F0 | FALSE | Upregulated |
| 0,386423992 | 0,214804925 | METTL12 | FALSE | Upregulated |
| 1,063322162 | 0,214812232 | LINC00997 | FALSE | Upregulated |
| 0,219804929 | 0,215217749 | RBM15B | FALSE | Upregulated |
| 0,96974072 | 0,21522273 | BICD1 | FALSE | Upregulated |
| 0,31836415 | 0,215249522 | PITPNM2 | FALSE | Upregulated |
| -0,724950742 | 0,215303498 | KCNK5 | FALSE | Downregulated |
| 0,383977459 | 0,215303498 | NFRKB | FALSE | Upregulated |
| -0,517278432 | 0,215316707 | NPIPB6 | FALSE | Downregulated |
| -0,317727143 | 0,215345576 | IRF2BP1 | FALSE | Downregulated |
| 0,641223159 | 0,215647811 | RAB36 | FALSE | Upregulated |
| -0,721324146 | 0,215829275 | ZNF843 | FALSE | Downregulated |
| 0,532887875 | 0,215850276 | SEPT6 | FALSE | Upregulated |
| -0,441770357 | 0,2160925 | ERLIN2 | FALSE | Downregulated |
| 0,735369898 | 0,216364958 | RP11-1H15.2 | FALSE | Upregulated |
| 0,682684231 | 0,216410594 | ALPK3 | FALSE | Upregulated |
| -0,314661832 | 0,216410594 | CDS2 | FALSE | Downregulated |
| 0,525299559 | 0,216410594 | HIST2H2BE | FALSE | Upregulated |

| | | | | |
|---|---|---|---|---|
| 0,841162396 | 0,216410594 | LOC286367 | FALSE | Upregulated |
| -0,481404754 | 0,216410594 | RPL36AL | FALSE | Downregulated |
| -0,461846975 | 0,216410594 | SLC39A11 | FALSE | Downregulated |
| -0,338834985 | 0,216410594 | TANC1 | FALSE | Downregulated |
| 0,520046985 | 0,216410594 | TRNAU1AP | FALSE | Upregulated |
| 0,300887639 | 0,21651467 | PIK3AP1 | FALSE | Upregulated |
| -1,994389999 | 0,216564086 | MSH5-SAPCD1 | FALSE | Downregulated |
| 0,736596643 | 0,216598653 | GTF2B | FALSE | Upregulated |
| -0,286448187 | 0,216598653 | HNF4A | FALSE | Downregulated |
| 0,609714866 | 0,216598653 | LPPR3 | FALSE | Upregulated |
| -0,851038789 | 0,216598653 | NARS2 | FALSE | Downregulated |
| 0,452529015 | 0,216598653 | PFDN1 | FALSE | Upregulated |
| -0,361257701 | 0,216598653 | SPHK2 | FALSE | Downregulated |
| -0,637103697 | 0,216788016 | ZNF511 | FALSE | Downregulated |
| 0,33938045 | 0,216933049 | MIDN | FALSE | Upregulated |
| -1,448721654 | 0,217003574 | PRO2852 | FALSE | Downregulated |
| -0,660158597 | 0,217165255 | KIAA1804 | FALSE | Downregulated |
| 0,63107638 | 0,217287324 | ANO1-AS2 | FALSE | Upregulated |
| 0,647731785 | 0,217287324 | LOC102723543 | FALSE | Upregulated |
| 0,548685313 | 0,217771942 | BAG3 | FALSE | Upregulated |
| 0,500562287 | 0,217771942 | CD47 | FALSE | Upregulated |
| 0,458099467 | 0,217771942 | MGC16275 | FALSE | Upregulated |
| -0,398325284 | 0,217771942 | PPID | FALSE | Downregulated |
| 0,31335786 | 0,217846028 | ANKRD6 | FALSE | Upregulated |
| 0,622792806 | 0,217898112 | LOC390937 | FALSE | Upregulated |
| -0,347493111 | 0,217898112 | PRODH2 | FALSE | Downregulated |
| 1,046632195 | 0,217898112 | S1PR2 | FALSE | Upregulated |
| -0,815599102 | 0,218049092 | SCARA3 | FALSE | Downregulated |
| 0,782889433 | 0,218267049 | SLFN11 | FALSE | Upregulated |
| -0,274812067 | 0,218362653 | CPT2 | FALSE | Downregulated |
| 1,256194691 | 0,218368331 | CCR2 | FALSE | Upregulated |
| 0,338276494 | 0,218368331 | FLOT1 | FALSE | Upregulated |
| -0,472643402 | 0,218912855 | PGAP3 | FALSE | Downregulated |
| -0,423578912 | 0,219206161 | USP48 | FALSE | Downregulated |
| -0,456615336 | 0,219343632 | PCSK1 | FALSE | Downregulated |
| -0,554772114 | 0,219564177 | ADAMTS5 | FALSE | Downregulated |
| 0,367750859 | 0,219564177 | NFATC2 | FALSE | Upregulated |
| 0,38574192 | 0,21956901 | LGMN | FALSE | Upregulated |
| -1,099860938 | 0,21956901 | LRRC2-AS1 | FALSE | Downregulated |
| 1,397967331 | 0,219935277 | BCAN | FALSE | Upregulated |
| -1,292473473 | 0,219935277 | LOC102724873 | FALSE | Downregulated |
| -0,363045589 | 0,219935277 | LRRC8A | FALSE | Downregulated |
| 0,44505037 | 0,2200926 | EVA1B | FALSE | Upregulated |
| 0,754449589 | 0,2200926 | LOC100507530 | FALSE | Upregulated |
| 0,801495369 | 0,220137358 | MED4-AS1 | FALSE | Upregulated |
| 0,434168863 | 0,220233915 | NRAP | FALSE | Upregulated |
| 0,604491141 | 0,22047043 | CCDC104 | FALSE | Upregulated |
| -0,319958475 | 0,220790378 | SOWAHC | FALSE | Downregulated |
| -0,273556241 | 0,220790378 | TUBGCP2 | FALSE | Downregulated |
| 0,360975541 | 0,220883559 | GFPT1 | FALSE | Upregulated |
| -0,654641132 | 0,221049126 | RNASE7 | FALSE | Downregulated |

| | | | | |
|---|---|---|---|---|
| -0,866586386 | 0,221049126 | THOC3 | FALSE | Downregulated |
| 0,17456432 | 0,221085985 | HNRNPU | FALSE | Upregulated |
| 0,381526251 | 0,221136232 | DESI1 | FALSE | Upregulated |
| 0,326277999 | 0,221485555 | PSME4 | FALSE | Upregulated |
| -0,47110188 | 0,221618558 | AGMO | FALSE | Downregulated |
| -1,06395016 | 0,221639533 | CTNNA3 | FALSE | Downregulated |
| 0,361393603 | 0,221639533 | UFM1 | FALSE | Upregulated |
| 0,595126962 | 0,22225084 | ABCA7 | FALSE | Upregulated |
| -0,431711382 | 0,22225084 | DCAKD | FALSE | Downregulated |
| -1,041778254 | 0,22225084 | DNM1 | FALSE | Downregulated |
| 0,446949754 | 0,22225084 | LOC101929451 | FALSE | Upregulated |
| 1,35786113 | 0,222277259 | LAX1 | FALSE | Upregulated |
| -0,402034038 | 0,222354429 | FBP1 | FALSE | Downregulated |
| 0,228394012 | 0,222575321 | AHSA2 | FALSE | Upregulated |
| 1,122071061 | 0,222575321 | APOBEC3D | FALSE | Upregulated |
| 0,451434234 | 0,222575321 | BTN3A2 | FALSE | Upregulated |
| 0,508525942 | 0,222575321 | MOK | FALSE | Upregulated |
| 0,49993454 | 0,222575321 | SLC46A2 | FALSE | Upregulated |
| 1,127194917 | 0,222575321 | SPATA24 | FALSE | Upregulated |
| 0,341652864 | 0,222575321 | XPO5 | FALSE | Upregulated |
| -0,349246625 | 0,222575321 | ZNF318 | FALSE | Downregulated |
| -0,89486691 | 0,222869197 | LOC101928668 | FALSE | Downregulated |
| 0,641798507 | 0,222971008 | INCENP | FALSE | Upregulated |
| -0,398599084 | 0,223144532 | ARPC1A | FALSE | Downregulated |
| 0,33871103 | 0,223272145 | ANKRD40 | FALSE | Upregulated |
| 0,357782872 | 0,223461235 | LOC102723666 | FALSE | Upregulated |
| -0,648940539 | 0,223596289 | ARNTL | FALSE | Downregulated |
| -0,445775534 | 0,223596289 | EIF2D | FALSE | Downregulated |
| -0,422773041 | 0,223596289 | NAGS | FALSE | Downregulated |
| -0,732264786 | 0,223629007 | TSPAN7 | FALSE | Downregulated |
| 0,26888091 | 0,223742106 | REEP5 | FALSE | Upregulated |
| 0,917256346 | 0,223899914 | ZNF799 | FALSE | Upregulated |
| -1,113711503 | 0,223927768 | ABCC11 | FALSE | Downregulated |
| 0,94649771 | 0,223927768 | ATP10A | FALSE | Upregulated |
| 0,57338189 | 0,224092255 | ABCF2 | FALSE | Upregulated |
| 0,520354892 | 0,224128137 | TRIM35 | FALSE | Upregulated |
| -0,435249392 | 0,224265619 | USP53 | FALSE | Downregulated |
| -0,378813317 | 0,224426299 | ITCH | FALSE | Downregulated |
| -0,563123291 | 0,224526487 | TTC21B | FALSE | Downregulated |
| -0,848766883 | 0,224654889 | NOVA2 | FALSE | Downregulated |
| -1,74379432 | 0,224712311 | AC006262.5 | FALSE | Downregulated |
| -0,614118986 | 0,224726158 | CNGA1 | FALSE | Downregulated |
| -0,481179091 | 0,224904245 | CENPV | FALSE | Downregulated |
| 1,219884496 | 0,224904245 | SUCNR1 | FALSE | Upregulated |
| 0,343075345 | 0,225181463 | OPTN | FALSE | Upregulated |
| 0,71197131 | 0,225181463 | SLC52A1 | FALSE | Upregulated |
| 0,683010254 | 0,225181463 | ZRSR2 | FALSE | Upregulated |
| 0,945661074 | 0,225181463 | ZSCAN12P1 | FALSE | Upregulated |
| 0,530209586 | 0,225201472 | C10orf95 | FALSE | Upregulated |
| 0,372356684 | 0,225286794 | KIAA0247 | FALSE | Upregulated |
| -0,967738209 | 0,225286794 | PAAF1 | FALSE | Downregulated |

| | | | | |
|---|---|---|---|---|
| 0,322517687 | 0,225324572 | MCM3AP-AS1 | FALSE | Upregulated |
| -0,331774285 | 0,225682429 | FAM63A | FALSE | Downregulated |
| 1,565479324 | 0,225840543 | PIP5KL1 | FALSE | Upregulated |
| 0,792122322 | 0,22586361 | C3AR1 | FALSE | Upregulated |
| 0,415483218 | 0,226156683 | PFN1P2 | FALSE | Upregulated |
| -0,35975118 | 0,226478261 | TAB2 | FALSE | Downregulated |
| -0,913563139 | 0,226592729 | CTTNBP2 | FALSE | Downregulated |
| 0,379323238 | 0,226592729 | TOR1B | FALSE | Upregulated |
| -0,385317799 | 0,226773175 | CFHR2 | FALSE | Downregulated |
| -0,497514542 | 0,227241477 | LDB2 | FALSE | Downregulated |
| 0,421772143 | 0,2273065 | LOC100506860 | FALSE | Upregulated |
| -0,385163564 | 0,227418772 | C6orf47 | FALSE | Downregulated |
| -1,218518012 | 0,22748724 | CYP1A1 | FALSE | Downregulated |
| 1,620454447 | 0,22748724 | TNFRSF17 | FALSE | Upregulated |
| -0,557445634 | 0,227795966 | IL11RA | FALSE | Downregulated |
| -1,015938868 | 0,227795966 | TMEM161B | FALSE | Downregulated |
| 0,476816695 | 0,227802077 | PLN | FALSE | Upregulated |
| -0,770287111 | 0,227802077 | UTS2B | FALSE | Downregulated |
| -0,43520252 | 0,227853836 | RP11-696N14.1 | FALSE | Downregulated |
| 0,29513075 | 0,227853836 | ZNF233 | FALSE | Upregulated |
| 1,400531692 | 0,228229279 | ACRBP | FALSE | Upregulated |
| 0,522418517 | 0,228388096 | SLC35G6 | FALSE | Upregulated |
| -0,47252637 | 0,22847085 | GDF2 | FALSE | Downregulated |
| -0,428526499 | 0,228769876 | ELP2 | FALSE | Downregulated |
| 0,481290206 | 0,228769876 | NAALAD2 | FALSE | Upregulated |
| 0,582155163 | 0,228769876 | SERPINB9 | FALSE | Upregulated |
| 0,721631716 | 0,228769876 | TRIM6 | FALSE | Upregulated |
| -0,640836638 | 0,228769876 | ZNF268 | FALSE | Downregulated |
| 0,604547623 | 0,228832337 | LIPT1 | FALSE | Upregulated |
| -0,376447823 | 0,228872264 | MSL2 | FALSE | Downregulated |
| -1,743364921 | 0,228872264 | SCG5 | FALSE | Downregulated |
| 0,901733188 | 0,229006043 | LUM | FALSE | Upregulated |
| -0,330512281 | 0,229106081 | RFC1 | FALSE | Downregulated |
| -0,47110555 | 0,229183007 | ZNF45 | FALSE | Downregulated |
| 1,156090902 | 0,229185104 | RSPH10B | FALSE | Upregulated |
| 0,28418471 | 0,229356576 | DAG1 | FALSE | Upregulated |
| -0,422710756 | 0,229816881 | LOC102724696 | FALSE | Downregulated |
| 0,387481558 | 0,230174282 | IL17RA | FALSE | Upregulated |
| -0,249352586 | 0,230280932 | RPL18 | FALSE | Downregulated |
| -1,477657426 | 0,230516795 | AGAP11 | FALSE | Downregulated |
| -0,371181437 | 0,230516795 | DNAJC22 | FALSE | Downregulated |
| 0,344469521 | 0,230516795 | HDAC7 | FALSE | Upregulated |
| 0,888794141 | 0,230516795 | PFKP | FALSE | Upregulated |
| -0,22981584 | 0,230516795 | PNPO | FALSE | Downregulated |
| -0,697688832 | 0,230608406 | CCBE1 | FALSE | Downregulated |
| -0,266105477 | 0,230608406 | SORBS2 | FALSE | Downregulated |
| 0,331154156 | 0,231122785 | FCHSD2 | FALSE | Upregulated |
| -0,520640376 | 0,231400293 | PCGF5 | FALSE | Downregulated |
| 0,276064881 | 0,231400293 | PNRC2 | FALSE | Upregulated |
| -1,083979583 | 0,231691624 | MEF2BNB | FALSE | Downregulated |
| 0,830440264 | 0,231754447 | ANXA1 | FALSE | Upregulated |

| | | | | |
|---|---|---|---|---|
| -0,46253251 | 0,231754447 | INSIG2 | FALSE | Downregulated |
| 0,613882566 | 0,232126909 | ATG9B | FALSE | Upregulated |
| 0,925020355 | 0,232126909 | KRT7 | FALSE | Upregulated |
| 0,833716018 | 0,232126909 | LOC101059906 | FALSE | Upregulated |
| -0,51909962 | 0,232126909 | MREG | FALSE | Downregulated |
| 0,401156262 | 0,232126909 | TICAM1 | FALSE | Upregulated |
| -1,114936685 | 0,232126909 | ZNF775 | FALSE | Downregulated |
| 0,342413049 | 0,232256301 | AC012314.8 | FALSE | Upregulated |
| -1,683788979 | 0,232256301 | RIMS4 | FALSE | Downregulated |
| 0,541813966 | 0,232667488 | ANXA5 | FALSE | Upregulated |
| 0,413052569 | 0,232667488 | CFLAR-AS1 | FALSE | Upregulated |
| 0,525952884 | 0,232667488 | GMIP | FALSE | Upregulated |
| 1,607072362 | 0,232667488 | LOC101927886 | FALSE | Upregulated |
| -0,398289771 | 0,232667488 | PRPF19 | FALSE | Downregulated |
| -0,686893224 | 0,232667488 | RAB40B | FALSE | Downregulated |
| 1,338162517 | 0,232667488 | TMEM108 | FALSE | Upregulated |
| 0,405655282 | 0,232861066 | ZMIZ2 | FALSE | Upregulated |
| -0,353819926 | 0,233082821 | C4orf19 | FALSE | Downregulated |
| 0,461802964 | 0,233474827 | RP11-31F15.2 | FALSE | Upregulated |
| -1,311193886 | 0,233596603 | CLEC5A | FALSE | Downregulated |
| 0,538118243 | 0,233860658 | AP1S2 | FALSE | Upregulated |
| -0,29840739 | 0,233860658 | ARVCF | FALSE | Downregulated |
| 0,500956094 | 0,233860658 | MRC2 | FALSE | Upregulated |
| -0,382064271 | 0,233860658 | RP11-713M15.1 | FALSE | Downregulated |
| 0,746815891 | 0,23452607 | FAR1 | FALSE | Upregulated |
| 0,618904757 | 0,23452607 | PIK3R5 | FALSE | Upregulated |
| 0,850659787 | 0,234635017 | CEP78 | FALSE | Upregulated |
| 1,102392189 | 0,234791333 | LOC101929687 | FALSE | Upregulated |
| -0,430086315 | 0,234791333 | UQCR11 | FALSE | Downregulated |
| -0,525144288 | 0,235184022 | ZNF398 | FALSE | Downregulated |
| 0,45612588 | 0,235663075 | CBX1 | FALSE | Upregulated |
| 0,580069063 | 0,235663075 | ETNK1 | FALSE | Upregulated |
| -0,193194934 | 0,235663075 | PPP6R2 | FALSE | Downregulated |
| 0,347389732 | 0,235663075 | RBM12B-AS1 | FALSE | Upregulated |
| -0,362823877 | 0,236096358 | NOTCH4 | FALSE | Downregulated |
| -0,515736199 | 0,236096358 | SCRN2 | FALSE | Downregulated |
| 0,310199832 | 0,236096358 | WDR82 | FALSE | Upregulated |
| 0,336603066 | 0,236233395 | GNPDA1 | FALSE | Upregulated |
| 0,489292414 | 0,236233395 | RNF6 | FALSE | Upregulated |
| 0,289530918 | 0,236277464 | FKBP15 | FALSE | Upregulated |
| 0,294069046 | 0,23643386 | TMEM115 | FALSE | Upregulated |
| -0,382846292 | 0,236633755 | ELMO2 | FALSE | Downregulated |
| 0,258353215 | 0,236782374 | INF2 | FALSE | Upregulated |
| 0,434201983 | 0,236782374 | PARP12 | FALSE | Upregulated |
| -0,279805847 | 0,237257247 | ALDH3A2 | FALSE | Downregulated |
| 0,63464156 | 0,237257247 | LRRC8B | FALSE | Upregulated |
| -0,392063984 | 0,237428094 | LIMS2 | FALSE | Downregulated |
| 1,271805185 | 0,237463653 | BHLHE22 | FALSE | Upregulated |
| 0,772019907 | 0,237497595 | C15orf54 | FALSE | Upregulated |
| 0,916302349 | 0,237739457 | C1orf21 | FALSE | Upregulated |
| 0,306403943 | 0,23775846 | ALCAM | FALSE | Upregulated |

| | | | | |
|---|---|---|---|---|
| -1,582142191 | 0,23775846 | LOC101926901 | FALSE | Downregulated |
| -1,223798176 | 0,23775846 | MECOM | FALSE | Downregulated |
| 0,914672664 | 0,23775846 | RP6-191P20.4 | FALSE | Upregulated |
| 0,61394349 | 0,23775846 | SNHG3 | FALSE | Upregulated |
| -0,377901625 | 0,237788386 | GHR | FALSE | Downregulated |
| 0,372570529 | 0,23793748 | MON1B | FALSE | Upregulated |
| -0,229539152 | 0,238096007 | FCGRT | FALSE | Downregulated |
| 0,925831656 | 0,238096007 | PDGFRL | FALSE | Upregulated |
| 0,818430098 | 0,238324646 | PSMD10 | FALSE | Upregulated |
| -0,657599056 | 0,238324646 | TET1 | FALSE | Downregulated |
| 0,454831923 | 0,23834126 | ALOX12 | FALSE | Upregulated |
| -0,247892934 | 0,238401461 | TMBIM6 | FALSE | Downregulated |
| 0,409141497 | 0,238543881 | FAM184B | FALSE | Upregulated |
| 0,304746291 | 0,23899633 | OGFR | FALSE | Upregulated |
| 1,564567845 | 0,239192855 | HIST1H4I | FALSE | Upregulated |
| -0,287799993 | 0,239213832 | HNF1A | FALSE | Downregulated |
| -0,651962793 | 0,239213832 | PDGFB | FALSE | Downregulated |
| 0,332729587 | 0,239232479 | MYO1D | FALSE | Upregulated |
| -0,47553702 | 0,239232479 | TM7SF2 | FALSE | Downregulated |
| -1,358293643 | 0,239339912 | CAPS2 | FALSE | Downregulated |
| -0,318366132 | 0,239576582 | ANGPTL1 | FALSE | Downregulated |
| 0,348874743 | 0,239580628 | CLDN22 | FALSE | Upregulated |
| 0,358778219 | 0,239580628 | LOC101929545 | FALSE | Upregulated |
| 0,569793908 | 0,239580628 | STAG3 | FALSE | Upregulated |
| -0,557289469 | 0,239581789 | PHF21A | FALSE | Downregulated |
| -0,846076029 | 0,239581789 | ZNF649 | FALSE | Downregulated |
| 0,57714306 | 0,239760112 | C1QL3 | FALSE | Upregulated |
| -0,644164587 | 0,239760112 | FAM117A | FALSE | Downregulated |
| 0,458270084 | 0,239760112 | HAUS6 | FALSE | Upregulated |
| -0,483481838 | 0,239760112 | HDHD2 | FALSE | Downregulated |
| 0,373920327 | 0,239776558 | ZNF529 | FALSE | Upregulated |
| 0,63841822 | 0,239783954 | SRBD1 | FALSE | Upregulated |
| 0,369028646 | 0,239852829 | CHMP4B | FALSE | Upregulated |
| 1,154388449 | 0,240099653 | BAIAP2L2 | FALSE | Upregulated |
| -0,262953434 | 0,240149776 | SORBS3 | FALSE | Downregulated |
| 0,502092708 | 0,240160382 | EIF2AK3 | FALSE | Upregulated |
| -0,418545762 | 0,240175132 | HIBCH | FALSE | Downregulated |
| 0,621402643 | 0,240175132 | PAX8 | FALSE | Upregulated |
| -0,357753847 | 0,240391425 | SLC43A1 | FALSE | Downregulated |
| -0,430777422 | 0,240472472 | DNAJB2 | FALSE | Downregulated |
| -0,968442139 | 0,240476014 | MAMLD1 | FALSE | Downregulated |
| -0,401556915 | 0,240742739 | PROSC | FALSE | Downregulated |
| -0,619488743 | 0,241020751 | LOC102724562 | FALSE | Downregulated |
| 0,47976014 | 0,241155613 | TTL | FALSE | Upregulated |
| 0,309153572 | 0,241292352 | RUVBL1 | FALSE | Upregulated |
| -1,076997605 | 0,241466343 | GPR157 | FALSE | Downregulated |
| 0,386724406 | 0,241539297 | C2CD5 | FALSE | Upregulated |
| 0,49098061 | 0,241539297 | TIMMDC1 | FALSE | Upregulated |
| 0,535882767 | 0,241539297 | TMC6 | FALSE | Upregulated |
| 1,00033767 | 0,241714801 | SPATA32 | FALSE | Upregulated |
| 0,227332877 | 0,241723994 | EIF3B | FALSE | Upregulated |

| | | | | |
|---|---|---|---|---|
| -1,270360681 | 0,241844203 | RP5-1180C10.2 | FALSE | Downregulated |
| 1,206670227 | 0,241844203 | SRP14-AS1 | FALSE | Upregulated |
| -0,37787898 | 0,24213241 | BCAS3 | FALSE | Downregulated |
| -0,496247524 | 0,242329394 | PSAT1 | FALSE | Downregulated |
| 1,321223063 | 0,242525917 | C1orf54 | FALSE | Upregulated |
| 0,321397928 | 0,242796002 | HSP90AA1 | FALSE | Upregulated |
| -1,114295253 | 0,24291724 | KLHL23 | FALSE | Downregulated |
| -0,606965317 | 0,243031732 | NECAB3 | FALSE | Downregulated |
| 1,028336963 | 0,243229711 | LSM2 | FALSE | Upregulated |
| -0,397301344 | 0,243229711 | RGS12 | FALSE | Downregulated |
| -0,563720258 | 0,243229711 | THAP5 | FALSE | Downregulated |
| 1,135884425 | 0,243248758 | LOC101929409 | FALSE | Upregulated |
| 0,675064533 | 0,243447383 | ANO10 | FALSE | Upregulated |
| -0,798628796 | 0,243447383 | HPRT1 | FALSE | Downregulated |
| -0,60422177 | 0,243447383 | ICK | FALSE | Downregulated |
| 0,519937508 | 0,243447383 | KIAA0513 | FALSE | Upregulated |
| 1,075659697 | 0,243447383 | MGAT3 | FALSE | Upregulated |
| 0,452364321 | 0,243447383 | TMPO-AS1 | FALSE | Upregulated |
| -0,338907371 | 0,243447383 | TOMM20 | FALSE | Downregulated |
| -0,308063445 | 0,243879385 | BRPF3 | FALSE | Downregulated |
| 0,384194225 | 0,243879385 | C12orf23 | FALSE | Upregulated |
| -0,437167298 | 0,243879385 | C14orf159 | FALSE | Downregulated |
| 0,792983627 | 0,243886697 | ACOT9 | FALSE | Upregulated |
| -0,369485106 | 0,243886697 | SFXN1 | FALSE | Downregulated |
| -0,45117105 | 0,244048424 | CBS | FALSE | Downregulated |
| 0,449539759 | 0,244048424 | CIB1 | FALSE | Upregulated |
| 0,403493939 | 0,244915214 | CEBPD | FALSE | Upregulated |
| 1,579422704 | 0,244969695 | CASC1 | FALSE | Upregulated |
| -0,482123745 | 0,245052681 | FCN3 | FALSE | Downregulated |
| 0,681693571 | 0,245052681 | NDUFB3 | FALSE | Upregulated |
| -0,734149653 | 0,245052681 | RPUSD3 | FALSE | Downregulated |
| 0,70585675 | 0,245052681 | TBC1D22B | FALSE | Upregulated |
| 1,019569712 | 0,245052681 | TGM4 | FALSE | Upregulated |
| 0,598397271 | 0,245085077 | RNF25 | FALSE | Upregulated |
| 0,594621635 | 0,245085077 | SPATC1 | FALSE | Upregulated |
| 0,625447131 | 0,245117681 | CCHCR1 | FALSE | Upregulated |
| 0,31528153 | 0,245130086 | MIRLET7DHG | FALSE | Upregulated |
| 0,996556971 | 0,245306027 | FANCG | FALSE | Upregulated |
| -1,443093548 | 0,245326177 | CETN2 | FALSE | Downregulated |
| -0,59994614 | 0,245326177 | CYP3A4 | FALSE | Downregulated |
| 0,634508108 | 0,245326177 | LMO3 | FALSE | Upregulated |
| 1,419783284 | 0,245326177 | LOC102724100 | FALSE | Upregulated |
| 0,841772825 | 0,245449705 | HIPK4 | FALSE | Upregulated |
| 0,314416697 | 0,245449705 | ITIH3 | FALSE | Upregulated |
| 0,431621861 | 0,245585599 | FUT2 | FALSE | Upregulated |
| -0,384881965 | 0,245585599 | LOC100507033 | FALSE | Downregulated |
| 0,237685834 | 0,245585599 | RBBP4 | FALSE | Upregulated |
| -0,620101765 | 0,245648786 | RGS5 | FALSE | Downregulated |
| 0,708975208 | 0,245880769 | THOC7-AS1 | FALSE | Upregulated |
| -0,849452735 | 0,245943834 | RBM43 | FALSE | Downregulated |
| 0,585216246 | 0,246095859 | WNK4 | FALSE | Upregulated |

| | | | | |
|---|---|---|---|---|
| 0,40453973 | 0,246217486 | LOC102724965 | FALSE | Upregulated |
| 0,667778233 | 0,24625536 | TMEM253 | FALSE | Upregulated |
| 1,089221331 | 0,246343237 | RBM45 | FALSE | Upregulated |
| -0,716698411 | 0,246388044 | GPATCH1 | FALSE | Downregulated |
| 0,750126802 | 0,246388044 | PTPRE | FALSE | Upregulated |
| 0,322616189 | 0,246456911 | CDIP1 | FALSE | Upregulated |
| 1,161322516 | 0,246456911 | RP11-111E14.1 | FALSE | Upregulated |
| -0,450509571 | 0,246456911 | RSAD1 | FALSE | Downregulated |
| 0,418132364 | 0,246564578 | DNALI1 | FALSE | Upregulated |
| 1,031731911 | 0,246573532 | KIF3A | FALSE | Upregulated |
| -0,371342411 | 0,246573532 | MAP3K4 | FALSE | Downregulated |
| -0,525150253 | 0,246573532 | SNHG7 | FALSE | Downregulated |
| 1,095104491 | 0,246664676 | AC000099.1 | FALSE | Upregulated |
| 0,929926369 | 0,246845585 | GPR114 | FALSE | Upregulated |
| -0,430184027 | 0,246995879 | CNST | FALSE | Downregulated |
| 0,603409144 | 0,247011037 | DPH5 | FALSE | Upregulated |
| -0,335904219 | 0,247011037 | GPHN | FALSE | Downregulated |
| -0,337508392 | 0,247038833 | HSD11B1 | FALSE | Downregulated |
| 0,448627407 | 0,247218463 | SYNGR2 | FALSE | Upregulated |
| 0,372463645 | 0,247259372 | SMCR5 | FALSE | Upregulated |
| 0,359637953 | 0,24742471 | DOC2A | FALSE | Upregulated |
| -1,197205213 | 0,24742471 | FAM200A | FALSE | Downregulated |
| -1,555493001 | 0,247596787 | LOC101927258 | FALSE | Downregulated |
| -1,425277402 | 0,247816458 | LINC00339 | FALSE | Downregulated |
| 0,635384275 | 0,247825537 | MUC4 | FALSE | Upregulated |
| -0,391643432 | 0,247866445 | CLK1 | FALSE | Downregulated |
| -0,703855855 | 0,247866445 | COX14 | FALSE | Downregulated |
| 0,89494515 | 0,247866445 | LOC101928982 | FALSE | Upregulated |
| 0,296171987 | 0,247868943 | LRRC37A4P | FALSE | Upregulated |
| 1,010586207 | 0,248121501 | LOC101929506 | FALSE | Upregulated |
| -0,301410995 | 0,248121501 | PRKAG2 | FALSE | Downregulated |
| 0,307911815 | 0,248121501 | SENP5 | FALSE | Upregulated |
| 0,425546638 | 0,248121501 | VPS37B | FALSE | Upregulated |
| -1,29593305 | 0,248182566 | ADCY10P1 | FALSE | Downregulated |
| 0,330620287 | 0,248182566 | C9orf69 | FALSE | Upregulated |
| 0,275752885 | 0,248182566 | CD59 | FALSE | Upregulated |
| -0,303069137 | 0,248182566 | CDC42EP4 | FALSE | Downregulated |
| -0,183594202 | 0,248182566 | COPA | FALSE | Downregulated |
| 0,456937293 | 0,248182566 | GORASP2 | FALSE | Upregulated |
| 0,553899198 | 0,248182566 | MAP1B | FALSE | Upregulated |
| 1,031643037 | 0,248182566 | SLC28A2 | FALSE | Upregulated |
| 0,397916354 | 0,248182566 | SLC38A2 | FALSE | Upregulated |
| -0,3917064 | 0,248182566 | SRGAP1 | FALSE | Downregulated |
| -0,492800258 | 0,248324516 | LOC102724672 | FALSE | Downregulated |
| -0,453693732 | 0,248342454 | AGTR1 | FALSE | Downregulated |
| -0,859905734 | 0,248342454 | LIFR-AS1 | FALSE | Downregulated |
| 0,95706437 | 0,248342454 | SAMSN1 | FALSE | Upregulated |
| 0,208528308 | 0,248342454 | SLC46A1 | FALSE | Upregulated |
| 0,514044776 | 0,248342454 | TCEB3B | FALSE | Upregulated |
| 0,309829219 | 0,248360192 | MARK3 | FALSE | Upregulated |
| -0,401403218 | 0,248595596 | COQ9 | FALSE | Downregulated |

| | | | | |
|---|---|---|---|---|
| 0,335875392 | 0,248745919 | UBE2L3 | FALSE | Upregulated |
| -1,595093219 | 0,248996089 | C14orf37 | FALSE | Downregulated |
| -0,488620818 | 0,248996089 | HECW2 | FALSE | Downregulated |
| -0,68905804 | 0,24937872 | CNTFR | FALSE | Downregulated |
| 1,084962456 | 0,24937872 | LOC102723604 | FALSE | Upregulated |
| 0,321936609 | 0,249588334 | SYNJ2 | FALSE | Upregulated |
| 0,983182214 | 0,249669174 | TSEN15 | FALSE | Upregulated |
| -0,585517668 | 0,249757086 | LOC102724455 | FALSE | Downregulated |
| -0,728075246 | 0,249978732 | ABHD13 | FALSE | Downregulated |
| 0,462591347 | 0,249978732 | FGL1 | FALSE | Upregulated |
| 0,519215881 | 0,249978732 | MRPS12 | FALSE | Upregulated |
| 1,281893232 | 0,250091886 | LINC00966 | FALSE | Upregulated |
| -0,68871552 | 0,250128032 | FOLH1B | FALSE | Downregulated |
| 0,507396504 | 0,250165856 | DYRK2 | FALSE | Upregulated |
| 1,830772541 | 0,25073306 | LOC101929133 | FALSE | Upregulated |
| 0,747775979 | 0,250762548 | IMPDH1 | FALSE | Upregulated |
| 0,816727508 | 0,250875573 | GRIN2D | FALSE | Upregulated |
| -1,308238874 | 0,250916074 | ALDH1L1-AS1 | FALSE | Downregulated |
| -0,333134637 | 0,251180536 | XAB2 | FALSE | Downregulated |
| -0,45739754 | 0,251340685 | DNAJC19 | FALSE | Downregulated |
| 0,357650796 | 0,251466011 | OXNAD1 | FALSE | Upregulated |
| 0,419428631 | 0,251515246 | RNF139-AS1 | FALSE | Upregulated |
| -0,394901976 | 0,251515246 | SRSF8 | FALSE | Downregulated |
| 0,602448021 | 0,251640685 | LCTL | FALSE | Upregulated |
| 0,352218853 | 0,251640685 | TBL1XR1 | FALSE | Upregulated |
| 1,307875156 | 0,251640685 | UBASH3A | FALSE | Upregulated |
| -0,382912754 | 0,251949969 | MAP7 | FALSE | Downregulated |
| -0,321770068 | 0,251949969 | TPST2 | FALSE | Downregulated |
| -1,001874754 | 0,251949969 | TRMT10A | FALSE | Downregulated |
| 0,362334322 | 0,252022989 | PSMC3IP | FALSE | Upregulated |
| -0,431315045 | 0,252154469 | APOA5 | FALSE | Downregulated |
| 1,046813514 | 0,252292182 | SGCD | FALSE | Upregulated |
| -0,604463971 | 0,252294503 | ABCG2 | FALSE | Downregulated |
| -1,3147594 | 0,252619656 | TSPAN8 | FALSE | Downregulated |
| -0,376553274 | 0,252691054 | ATP2B1 | FALSE | Downregulated |
| 0,55992988 | 0,252691054 | C2orf69 | FALSE | Upregulated |
| 0,331846273 | 0,252691054 | CELSR1 | FALSE | Upregulated |
| -0,406894673 | 0,252691054 | CYP2D6 | FALSE | Downregulated |
| -1,767419463 | 0,252691054 | IL17REL | FALSE | Downregulated |
| -1,143877204 | 0,252691054 | MUC3A | FALSE | Downregulated |
| 0,561408468 | 0,252691054 | PPIL6 | FALSE | Upregulated |
| 0,295133827 | 0,252691054 | PRELID1 | FALSE | Upregulated |
| 0,621329485 | 0,252691054 | RP11-840I19.3 | FALSE | Upregulated |
| -0,69642367 | 0,252691054 | ZDBF2 | FALSE | Downregulated |
| 0,465960197 | 0,252704444 | FYN | FALSE | Upregulated |
| -0,674041358 | 0,252704444 | RNF165 | FALSE | Downregulated |
| 0,32903633 | 0,252704444 | USP5 | FALSE | Upregulated |
| -0,352376507 | 0,252930642 | DHRS1 | FALSE | Downregulated |
| -1,590461505 | 0,252930642 | RP11-778J15.1 | FALSE | Downregulated |
| 0,388905289 | 0,252930642 | RP4-724E16.2 | FALSE | Upregulated |
| -0,437463574 | 0,252930642 | SLC25A38 | FALSE | Downregulated |

| | | | | |
|---|---|---|---|---|
| -0,762023922 | 0,252930642 | TRAPPC6A | FALSE | Downregulated |
| -0,482685451 | 0,252930642 | ZNF503 | FALSE | Downregulated |
| 0,319166717 | 0,253055027 | ATG14 | FALSE | Upregulated |
| 0,361374901 | 0,253087782 | CLEC16A | FALSE | Upregulated |
| 0,460947414 | 0,25326225 | ZMYM6 | FALSE | Upregulated |
| -0,239627365 | 0,253343912 | CLU | FALSE | Downregulated |
| -0,651879557 | 0,253343912 | LOC101928051 | FALSE | Downregulated |
| -0,472395385 | 0,253373466 | RPS13 | FALSE | Downregulated |
| 0,64792115 | 0,253432381 | ARHGAP25 | FALSE | Upregulated |
| 0,528950267 | 0,253710512 | FLI1 | FALSE | Upregulated |
| 0,269222616 | 0,253710512 | KANSL1L | FALSE | Upregulated |
| -1,34999125 | 0,253732685 | EBLN2 | FALSE | Downregulated |
| 0,537541022 | 0,253921192 | CHFR | FALSE | Upregulated |
| 0,64753676 | 0,253921192 | OR10AD1 | FALSE | Upregulated |
| -1,159729995 | 0,253943649 | DSCAML1 | FALSE | Downregulated |
| 0,886682177 | 0,253960299 | AACS | FALSE | Upregulated |
| 0,725839799 | 0,253960299 | GCNT1 | FALSE | Upregulated |
| 0,4896666 | 0,253960299 | HMOX1 | FALSE | Upregulated |
| -0,361606669 | 0,253960299 | PEX5 | FALSE | Downregulated |
| 0,916239497 | 0,253960299 | PYCARD | FALSE | Upregulated |
| -0,341140737 | 0,253960299 | TBL1X | FALSE | Downregulated |
| -0,68474043 | 0,254282175 | LOC101928384 | FALSE | Downregulated |
| 0,664880125 | 0,254367885 | LOC102724077 | FALSE | Upregulated |
| 0,433996278 | 0,254519764 | CDK17 | FALSE | Upregulated |
| 0,584956392 | 0,25458569 | AAGAB | FALSE | Upregulated |
| 0,450159771 | 0,25465263 | LOC102724345 | FALSE | Upregulated |
| 1,415506074 | 0,25465263 | S100A4 | FALSE | Upregulated |
| 0,360071201 | 0,25465263 | SIX5 | FALSE | Upregulated |
| 0,416064426 | 0,254772898 | CASK | FALSE | Upregulated |
| 0,434449078 | 0,254894074 | LSM3 | FALSE | Upregulated |
| -0,437726691 | 0,255082039 | MPND | FALSE | Downregulated |
| -0,268303514 | 0,255416757 | GPBP1L1 | FALSE | Downregulated |
| -0,524389289 | 0,255520939 | ARMCX4 | FALSE | Downregulated |
| 0,411727029 | 0,25557436 | BST2 | FALSE | Upregulated |
| 0,557637772 | 0,25557436 | RIT1 | FALSE | Upregulated |
| 0,611811594 | 0,255829104 | LINC00482 | FALSE | Upregulated |
| -0,733112939 | 0,256024096 | EBPL | FALSE | Downregulated |
| -0,613124626 | 0,256319912 | ELN | FALSE | Downregulated |
| 0,441642705 | 0,25652914 | PCSK9 | FALSE | Upregulated |
| 0,402064508 | 0,256649161 | NUTM2B | FALSE | Upregulated |
| 0,277649892 | 0,257125635 | BRI3 | FALSE | Upregulated |
| -0,706887531 | 0,257125635 | PLEKHF2 | FALSE | Downregulated |
| 1,253464284 | 0,257125635 | RHPN1 | FALSE | Upregulated |
| 0,505851171 | 0,257125635 | SEZ6 | FALSE | Upregulated |
| -1,036287175 | 0,257298948 | C1orf52 | FALSE | Downregulated |
| 0,281328815 | 0,257298948 | ITPR3 | FALSE | Upregulated |
| 1,384998476 | 0,257539952 | RIPK3 | FALSE | Upregulated |
| 0,4047685 | 0,257786774 | GCNT7 | FALSE | Upregulated |
| -0,508869946 | 0,257786774 | MYCL | FALSE | Downregulated |
| -0,537068176 | 0,257786774 | NPR1 | FALSE | Downregulated |
| 0,544972493 | 0,257893665 | MOB1A | FALSE | Upregulated |

| | | | | |
|---|---|---|---|---|
| 0,813352593 | 0,257893665 | PAPSS1 | FALSE | Upregulated |
| 1,375177726 | 0,257893665 | SLC4A8 | FALSE | Upregulated |
| 0,554519345 | 0,258032628 | GVINP1 | FALSE | Upregulated |
| 0,505119736 | 0,258040185 | ATL3 | FALSE | Upregulated |
| 0,769192357 | 0,258550882 | MFI2 | FALSE | Upregulated |
| -0,942180985 | 0,258649712 | OR51M1 | FALSE | Downregulated |
| 0,485457689 | 0,258649712 | RSPH3 | FALSE | Upregulated |
| 1,842064304 | 0,25882783 | AGAP7 | FALSE | Upregulated |
| 0,529248133 | 0,25980712 | RALGDS | FALSE | Upregulated |
| -0,270243341 | 0,259961659 | RC3H1 | FALSE | Downregulated |
| -0,533226 | 0,260146908 | NAAA | FALSE | Downregulated |
| 0,513203565 | 0,260302738 | ADSS | FALSE | Upregulated |
| -0,566402339 | 0,260500321 | MRPL45 | FALSE | Downregulated |
| 0,768251772 | 0,260527783 | RP11-43N5.1 | FALSE | Upregulated |
| 0,844234991 | 0,260842721 | LOC389199 | FALSE | Upregulated |
| -0,350289998 | 0,260852959 | CROCCP2 | FALSE | Downregulated |
| -0,348345505 | 0,260852959 | ENTPD5 | FALSE | Downregulated |
| -0,983461471 | 0,260882449 | ALKBH8 | FALSE | Downregulated |
| -0,46151565 | 0,260912538 | SC5D | FALSE | Downregulated |
| 0,500308948 | 0,260972754 | CITED4 | FALSE | Upregulated |
| 0,504202919 | 0,261226823 | MED8 | FALSE | Upregulated |
| -1,564257462 | 0,261226823 | PCNAP1 | FALSE | Downregulated |
| 0,292348683 | 0,261345752 | LARP4B | FALSE | Upregulated |
| 0,337540019 | 0,261477175 | NUDT13 | FALSE | Upregulated |
| -0,703414026 | 0,261716834 | NBPF13P | FALSE | Downregulated |
| -0,575061044 | 0,261897191 | MSMO1 | FALSE | Downregulated |
| 0,519069411 | 0,26205744 | GRAP | FALSE | Upregulated |
| 0,911147828 | 0,26205744 | PCGF6 | FALSE | Upregulated |
| 2,087092536 | 0,262162745 | LOC102723615 | FALSE | Upregulated |
| -1,375930404 | 0,26216579 | LINC00493 | FALSE | Downregulated |
| 1,412373866 | 0,262216033 | HIST1H2BE | FALSE | Upregulated |
| -0,668730512 | 0,26228841 | LRRC28 | FALSE | Downregulated |
| 1,057936003 | 0,262613258 | LOC102723989 | FALSE | Upregulated |
| -0,946454487 | 0,262952419 | LOC101927559 | FALSE | Downregulated |
| -0,444431003 | 0,262952419 | NEU4 | FALSE | Downregulated |
| -1,365109948 | 0,262952419 | NHLRC1 | FALSE | Downregulated |
| -0,401383083 | 0,262952419 | PCTP | FALSE | Downregulated |
| 0,432423232 | 0,262952419 | RAVER1 | FALSE | Upregulated |
| -0,36781557 | 0,262952419 | SPG21 | FALSE | Downregulated |
| 0,701057754 | 0,262952419 | TP53TG5 | FALSE | Upregulated |
| 0,37555414 | 0,262952419 | ZNF550 | FALSE | Upregulated |
| 1,37522742 | 0,263012215 | DNAJB13 | FALSE | Upregulated |
| 1,06096777 | 0,263057187 | ABCC4 | FALSE | Upregulated |
| -0,572920357 | 0,263057187 | MXRA5 | FALSE | Downregulated |
| 0,377634432 | 0,263057187 | SNCAIP | FALSE | Upregulated |
| -0,465885531 | 0,263057187 | TMEM183A | FALSE | Downregulated |
| 0,458343477 | 0,263388666 | BCL3 | FALSE | Upregulated |
| 0,378347395 | 0,263985317 | ZNF710 | FALSE | Upregulated |
| -0,600375091 | 0,264048368 | LINC00671 | FALSE | Downregulated |
| 0,241689398 | 0,264068036 | CNP | FALSE | Upregulated |
| 0,843912709 | 0,264945948 | C19orf67 | FALSE | Upregulated |

| | | | | |
|---|---|---|---|---|
| -0,42197814 | 0,264949649 | NDUFV2 | FALSE | Downregulated |
| -0,300011185 | 0,26509324 | FGA | FALSE | Downregulated |
| 0,395297153 | 0,265243444 | LOC101927903 | FALSE | Upregulated |
| -0,255586165 | 0,265257881 | CIRBP | FALSE | Downregulated |
| 1,249096986 | 0,265257881 | ZNF667 | FALSE | Upregulated |
| -0,752656947 | 0,265400962 | CES4A | FALSE | Downregulated |
| -0,999101812 | 0,265400962 | PKHD1L1 | FALSE | Downregulated |
| 1,861672702 | 0,265476655 | C10orf55 | FALSE | Upregulated |
| 0,599234948 | 0,265500937 | EGOT | FALSE | Upregulated |
| 0,286834053 | 0,265500937 | LRCH4 | FALSE | Upregulated |
| -0,227174164 | 0,265682294 | ZNF592 | FALSE | Downregulated |
| -0,332832015 | 0,265761399 | DCBLD2 | FALSE | Downregulated |
| 0,197264696 | 0,266085584 | GGCX | FALSE | Upregulated |
| -0,24432905 | 0,266085584 | PNISR | FALSE | Downregulated |
| 0,379522249 | 0,266315587 | ATAD2B | FALSE | Upregulated |
| -0,182913965 | 0,266534477 | RPL18A | FALSE | Downregulated |
| -0,52115219 | 0,266534477 | UQCC1 | FALSE | Downregulated |
| 0,952589531 | 0,266575703 | PCED1B | FALSE | Upregulated |
| -0,749049528 | 0,266596794 | PPP1R3G | FALSE | Downregulated |
| 1,255865513 | 0,266754396 | RHCG | FALSE | Upregulated |
| -0,445411362 | 0,267030992 | MPLKIP | FALSE | Downregulated |
| 0,601012971 | 0,267030992 | RPS6KA1 | FALSE | Upregulated |
| 0,52471805 | 0,267120956 | C19orf68 | FALSE | Upregulated |
| 0,838569122 | 0,267222106 | TFEC | FALSE | Upregulated |
| -1,488465803 | 0,267364118 | EYA2 | FALSE | Downregulated |
| -0,392449595 | 0,267702256 | AKR1C3 | FALSE | Downregulated |
| 0,866541125 | 0,2679785 | UBASH3B | FALSE | Upregulated |
| 0,44352627 | 0,268244897 | LOC101927831 | FALSE | Upregulated |
| 1,14684852 | 0,26834352 | DNAH3 | FALSE | Upregulated |
| 0,608452048 | 0,26834352 | RASGRP2 | FALSE | Upregulated |
| 0,721777472 | 0,268480061 | CDKN2C | FALSE | Upregulated |
| 0,300775881 | 0,268480061 | PINK1-AS | FALSE | Upregulated |
| 0,489093532 | 0,268480061 | POU5F1P3 | FALSE | Upregulated |
| -0,407756136 | 0,268499625 | AMN1 | FALSE | Downregulated |
| 0,354648508 | 0,268499625 | DCTN2 | FALSE | Upregulated |
| -0,400066109 | 0,268499625 | EIF2B5 | FALSE | Downregulated |
| 0,338901035 | 0,268499625 | PIP5K1C | FALSE | Upregulated |
| 0,474932433 | 0,268499625 | SH2D3A | FALSE | Upregulated |
| -0,608647167 | 0,268499625 | TMEM86B | FALSE | Downregulated |
| 0,729953219 | 0,269051105 | C1orf213 | FALSE | Upregulated |
| -0,490050432 | 0,269051105 | LYPLA1 | FALSE | Downregulated |
| -1,159392073 | 0,269115524 | SEPSECS-AS1 | FALSE | Downregulated |
| -0,345236973 | 0,269207957 | TOM1L2 | FALSE | Downregulated |
| 0,43094212 | 0,269249748 | MED19 | FALSE | Upregulated |
| 0,50769077 | 0,269249748 | TNFRSF21 | FALSE | Upregulated |
| 0,795235558 | 0,269627766 | CCDC65 | FALSE | Upregulated |
| 0,644633853 | 0,269700696 | ARID3A | FALSE | Upregulated |
| -0,476922554 | 0,27007297 | DHRS4 | FALSE | Downregulated |
| 0,499683762 | 0,270183784 | ARHGAP44 | FALSE | Upregulated |
| 0,31304461 | 0,270704685 | HEG1 | FALSE | Upregulated |
| 0,616976192 | 0,270784286 | CR1 | FALSE | Upregulated |

| | | | | |
|---|---|---|---|---|
| 0,864424182 | 0,270784286 | H2AFX | FALSE | Upregulated |
| -0,411350802 | 0,270784286 | PLEKHG1 | FALSE | Downregulated |
| -0,387665914 | 0,270918966 | ARID4A | FALSE | Downregulated |
| 0,78339061 | 0,270918966 | CD27 | FALSE | Upregulated |
| -0,874278139 | 0,270918966 | EXTL2 | FALSE | Downregulated |
| 1,258216224 | 0,270918966 | FUT7 | FALSE | Upregulated |
| -0,5104464 | 0,270918966 | LOC101927157 | FALSE | Downregulated |
| 0,403219459 | 0,271140878 | ARRDC2 | FALSE | Upregulated |
| 0,600184399 | 0,271140878 | FAM122B | FALSE | Upregulated |
| 0,839198098 | 0,271140878 | NCF4 | FALSE | Upregulated |
| 0,766019602 | 0,271140878 | UXS1 | FALSE | Upregulated |
| 0,463204787 | 0,271256973 | FBN1 | FALSE | Upregulated |
| 0,316837014 | 0,271393544 | RAC1 | FALSE | Upregulated |
| 1,240413285 | 0,271393544 | SLC7A1 | FALSE | Upregulated |
| 0,813159681 | 0,271393544 | TMED6 | FALSE | Upregulated |
| -0,341474618 | 0,271413804 | DAAM2 | FALSE | Downregulated |
| -0,878502476 | 0,271413804 | LOC102723954 | FALSE | Downregulated |
| -0,54743608 | 0,27160929 | TEX264 | FALSE | Downregulated |
| 0,379972141 | 0,271664446 | TRIM52 | FALSE | Upregulated |
| 1,058526294 | 0,271780594 | BTN1A1 | FALSE | Upregulated |
| -0,442873151 | 0,271797934 | C9orf3 | FALSE | Downregulated |
| 0,36516589 | 0,271797934 | SPON1 | FALSE | Upregulated |
| 0,397815703 | 0,271800954 | B9D2 | FALSE | Upregulated |
| 1,275279736 | 0,271846709 | RHOH | FALSE | Upregulated |
| 0,282601181 | 0,271926301 | POM121C | FALSE | Upregulated |
| 0,561487029 | 0,271999486 | SRGAP3-AS3 | FALSE | Upregulated |
| 0,779628351 | 0,272311001 | INE1 | FALSE | Upregulated |
| 0,339448197 | 0,272311001 | LOC101929533 | FALSE | Upregulated |
| 0,464983309 | 0,272311001 | TAGLN | FALSE | Upregulated |
| -0,299508152 | 0,272528599 | GCLC | FALSE | Downregulated |
| -0,629076785 | 0,272528599 | PARP16 | FALSE | Downregulated |
| -0,335008534 | 0,272646948 | CAPN7 | FALSE | Downregulated |
| -0,470728519 | 0,272646948 | SLC19A3 | FALSE | Downregulated |
| 0,629277709 | 0,273465661 | ASB7 | FALSE | Upregulated |
| -0,365077763 | 0,273543966 | FBXO21 | FALSE | Downregulated |
| 0,318087097 | 0,273543966 | PNPLA6 | FALSE | Upregulated |
| -0,354721778 | 0,273543966 | SHF | FALSE | Downregulated |
| 0,380157376 | 0,273771494 | PIK3R3 | FALSE | Upregulated |
| -0,688388944 | 0,273785998 | HELQ | FALSE | Downregulated |
| 0,642635969 | 0,273879723 | DDHD1 | FALSE | Upregulated |
| -0,515068141 | 0,273900619 | LRIG3 | FALSE | Downregulated |
| -0,441203355 | 0,273900619 | ZMYM3 | FALSE | Downregulated |
| 1,142895165 | 0,27393335 | CFTR | FALSE | Upregulated |
| -0,393627977 | 0,27393335 | MICU1 | FALSE | Downregulated |
| -0,275078714 | 0,27393335 | PGPEP1 | FALSE | Downregulated |
| -0,747992614 | 0,274584075 | ZNF561 | FALSE | Downregulated |
| -0,906548643 | 0,274723985 | LOC101928615 | FALSE | Downregulated |
| 0,808632501 | 0,274723985 | VWA8-AS1 | FALSE | Upregulated |
| -0,276952465 | 0,274908048 | BNIP3 | FALSE | Downregulated |
| 0,410118798 | 0,274908048 | LOC101929747 | FALSE | Upregulated |
| 1,176688256 | 0,275547413 | SLAMF6 | FALSE | Upregulated |

| | | | | |
|---|---|---|---|---|
| 1,134684273 | 0,275561984 | LOC729737 | FALSE | Upregulated |
| 0,27942021 | 0,27584952 | EIF4E2 | FALSE | Upregulated |
| -0,809718688 | 0,275957364 | APOC1P1 | FALSE | Downregulated |
| 0,75232935 | 0,275957364 | LGALS4 | FALSE | Upregulated |
| 0,893263998 | 0,27600618 | KCNJ2 | FALSE | Upregulated |
| 0,274924524 | 0,276245118 | CCDC85C | FALSE | Upregulated |
| 0,652111625 | 0,276384548 | TFPT | FALSE | Upregulated |
| -0,219585356 | 0,276401242 | RPL13A | FALSE | Downregulated |
| 1,260382251 | 0,276658023 | RP11-473M20.14 | FALSE | Upregulated |
| -0,549301121 | 0,276658023 | WDR18 | FALSE | Downregulated |
| 0,270437046 | 0,277015612 | PTPN11 | FALSE | Upregulated |
| 0,472877775 | 0,277134074 | ZNF57 | FALSE | Upregulated |
| 0,237155099 | 0,277277483 | MLEC | FALSE | Upregulated |
| -0,414541406 | 0,277512889 | HEATR5B | FALSE | Downregulated |
| -0,247441609 | 0,277570324 | BCR | FALSE | Downregulated |
| -0,798103799 | 0,27760908 | TRPV3 | FALSE | Downregulated |
| -2,508354946 | 0,277658894 | LINC00520 | FALSE | Downregulated |
| 1,181039414 | 0,277765649 | ANKLE1 | FALSE | Upregulated |
| 0,351687505 | 0,277765649 | ARF4 | FALSE | Upregulated |
| 0,822655272 | 0,277802102 | CKAP2 | FALSE | Upregulated |
| -0,299625803 | 0,277949947 | C5 | FALSE | Downregulated |
| 0,396511576 | 0,277949947 | LRRC59 | FALSE | Upregulated |
| -0,423466925 | 0,278076551 | BRAF | FALSE | Downregulated |
| 0,719965593 | 0,278076551 | CEP152 | FALSE | Upregulated |
| 0,924326256 | 0,27862274 | STARD3NL | FALSE | Upregulated |
| 0,67823684 | 0,278725664 | LOC101060494 | FALSE | Upregulated |
| -0,937939122 | 0,278892325 | LINC01152 | FALSE | Downregulated |
| 0,434587237 | 0,279067931 | RPS6KA4 | FALSE | Upregulated |
| -0,887045338 | 0,279158914 | CYS1 | FALSE | Downregulated |
| -0,214704431 | 0,279158914 | RPL36 | FALSE | Downregulated |
| 1,045255008 | 0,279158914 | SH2D1A | FALSE | Upregulated |
| 1,192529166 | 0,279158914 | WIPF3 | FALSE | Upregulated |
| 0,479016945 | 0,279185592 | ANKRD36 | FALSE | Upregulated |
| -0,421789213 | 0,279508156 | IGFBP5 | FALSE | Downregulated |
| 1,754055866 | 0,27961102 | RIMBP3B | FALSE | Upregulated |
| 0,508516341 | 0,279705353 | LIPE | FALSE | Upregulated |
| 0,565841201 | 0,280138419 | CYTH2 | FALSE | Upregulated |
| -0,193352519 | 0,280138419 | HDLBP | FALSE | Downregulated |
| -0,479436479 | 0,280138419 | LONRF3 | FALSE | Downregulated |
| -0,35899989 | 0,280138419 | MAST4 | FALSE | Downregulated |
| 0,325788138 | 0,280138419 | POLG | FALSE | Upregulated |
| 0,590254228 | 0,280138419 | UQCRH | FALSE | Upregulated |
| 0,597563698 | 0,280188575 | RP11-118E18.4 | FALSE | Upregulated |
| -0,541483398 | 0,280307636 | WASH5P | FALSE | Downregulated |
| 0,414057614 | 0,280757053 | ENOSF1 | FALSE | Upregulated |
| 0,303963092 | 0,280775387 | SPDYE3 | FALSE | Upregulated |
| 0,945870526 | 0,280775387 | USP32P2 | FALSE | Upregulated |
| 0,417324022 | 0,280857968 | NTN3 | FALSE | Upregulated |
| 0,769855561 | 0,280867515 | AMPH | FALSE | Upregulated |
| 0,345171327 | 0,280867515 | MKL1 | FALSE | Upregulated |
| 0,807132484 | 0,281020288 | ZNF232 | FALSE | Upregulated |

| | | | | |
|---|---|---|---|---|
| -0,201694045 | 0,281048299 | RN7SK | FALSE | Downregulated |
| -0,765695526 | 0,281086994 | C9orf142 | FALSE | Downregulated |
| -0,34924528 | 0,281244123 | F13B | FALSE | Downregulated |
| 0,862846769 | 0,281311105 | PM20D1 | FALSE | Upregulated |
| 0,744053175 | 0,281417135 | BIN2 | FALSE | Upregulated |
| 0,77914793 | 0,281417135 | IFI44L | FALSE | Upregulated |
| -0,481406243 | 0,281417135 | PEF1 | FALSE | Downregulated |
| 0,640533185 | 0,281417135 | SARS2 | FALSE | Upregulated |
| 0,313006984 | 0,281662951 | TMEM2 | FALSE | Upregulated |
| -0,340417742 | 0,281878215 | PAN2 | FALSE | Downregulated |
| -2,000239545 | 0,281946308 | FAM47E-STBD1 | FALSE | Downregulated |
| -0,298275596 | 0,281993796 | DDAH1 | FALSE | Downregulated |
| 0,310513293 | 0,282224975 | LOC101929290 | FALSE | Upregulated |
| -0,247333198 | 0,282224975 | POLRMT | FALSE | Downregulated |
| 0,503284485 | 0,28244989 | COL6A2 | FALSE | Upregulated |
| 1,023939437 | 0,282548822 | GDAP1 | FALSE | Upregulated |
| -0,443834902 | 0,282548822 | RPP14 | FALSE | Downregulated |
| -0,530727183 | 0,282743239 | KIF26A | FALSE | Downregulated |
| 0,334917689 | 0,282744634 | SPDYE6 | FALSE | Upregulated |
| 0,359083672 | 0,282823778 | FAM111A | FALSE | Upregulated |
| 0,569603509 | 0,283298423 | PDE4B | FALSE | Upregulated |
| -0,604590952 | 0,283298423 | ZNF343 | FALSE | Downregulated |
| -0,904048627 | 0,283325046 | PGAP1 | FALSE | Downregulated |
| 0,427883988 | 0,283438923 | LOC100506497 | FALSE | Upregulated |
| 1,068664546 | 0,283469903 | GCSAM | FALSE | Upregulated |
| 0,683036881 | 0,283469903 | GGN | FALSE | Upregulated |
| -0,530895977 | 0,283469903 | GTF3C3 | FALSE | Downregulated |
| 0,61603635 | 0,283469903 | JOSD2 | FALSE | Upregulated |
| 0,529412344 | 0,283469903 | WNT2B | FALSE | Upregulated |
| 0,327542374 | 0,28349469 | LOC100505502 | FALSE | Upregulated |
| 0,490369935 | 0,283514673 | SEC22C | FALSE | Upregulated |
| 0,997791524 | 0,283557854 | CHST9 | FALSE | Upregulated |
| 0,325638081 | 0,28361489 | RFFL | FALSE | Upregulated |
| 0,395601646 | 0,283713138 | HIGD1A | FALSE | Upregulated |
| -0,365717615 | 0,283748815 | AKR7A3 | FALSE | Downregulated |
| -0,295851636 | 0,283748815 | BLVRB | FALSE | Downregulated |
| -1,606075538 | 0,283748815 | PROM1 | FALSE | Downregulated |
| -0,401614148 | 0,284015507 | GEMIN4 | FALSE | Downregulated |
| 0,53979017 | 0,285107917 | MOB3A | FALSE | Upregulated |
| -0,283759083 | 0,285208705 | C1RL | FALSE | Downregulated |
| -0,32013265 | 0,28522586 | LUC7L2 | FALSE | Downregulated |
| 0,386247355 | 0,285491002 | FAM227B | FALSE | Upregulated |
| -0,262207578 | 0,28549215 | MATR3 | FALSE | Downregulated |
| -0,368988193 | 0,285604441 | ATF7 | FALSE | Downregulated |
| 1,598949882 | 0,285604441 | CCL2 | FALSE | Upregulated |
| -0,391834387 | 0,285765576 | FNIP1 | FALSE | Downregulated |
| 0,826949437 | 0,286084571 | RP11-480I12.7 | FALSE | Upregulated |
| 0,702837718 | 0,286607418 | CASP9 | FALSE | Upregulated |
| -1,126346825 | 0,286708851 | LOC102724493 | FALSE | Downregulated |
| -0,673047898 | 0,286729971 | DLL4 | FALSE | Downregulated |
| 0,321403477 | 0,286729971 | LOC100132356 | FALSE | Upregulated |

| | | | | |
|---|---|---|---|---|
| 0,546559278 | 0,286729971 | SPN | FALSE | Upregulated |
| 0,328656927 | 0,286790503 | RILPL1 | FALSE | Upregulated |
| 0,65653483 | 0,28686722 | HSPB6 | FALSE | Upregulated |
| -0,211788587 | 0,287021701 | BOD1L1 | FALSE | Downregulated |
| -0,749534573 | 0,287021701 | CARF | FALSE | Downregulated |
| -0,302723382 | 0,287021701 | COASY | FALSE | Downregulated |
| 1,345090001 | 0,287021701 | CORO6 | FALSE | Upregulated |
| 0,661569024 | 0,287021701 | MARVELD1 | FALSE | Upregulated |
| 0,340276626 | 0,287021701 | MTRF1L | FALSE | Upregulated |
| -0,642333185 | 0,287021701 | PRDM10 | FALSE | Downregulated |
| -0,62988452 | 0,287021701 | TMEM8B | FALSE | Downregulated |
| 0,250991235 | 0,287021701 | TRIM26 | FALSE | Upregulated |
| -0,598235805 | 0,287021701 | ZNF254 | FALSE | Downregulated |
| -0,646170447 | 0,287059845 | GLCE | FALSE | Downregulated |
| 0,384135786 | 0,287315004 | NFKB1 | FALSE | Upregulated |
| -0,480170036 | 0,287403075 | AGAP1 | FALSE | Downregulated |
| -0,375667305 | 0,287403075 | COX6B1 | FALSE | Downregulated |
| -1,628241656 | 0,287403075 | DAB1-AS1 | FALSE | Downregulated |
| 0,640056631 | 0,287403075 | DHRS7B | FALSE | Upregulated |
| -0,907318626 | 0,287403075 | JAKMIP2 | FALSE | Downregulated |
| 0,320922366 | 0,287403075 | PRPF4B | FALSE | Upregulated |
| 0,405117989 | 0,287403075 | THEMIS2 | FALSE | Upregulated |
| 1,036422497 | 0,287464365 | KLRK1 | FALSE | Upregulated |
| -0,577271884 | 0,287579983 | ZNF35 | FALSE | Downregulated |
| -0,699426673 | 0,287579983 | ZNF70 | FALSE | Downregulated |
| -0,370450816 | 0,287610961 | EIF6 | FALSE | Downregulated |
| -0,597604155 | 0,287621761 | GPR88 | FALSE | Downregulated |
| -0,646628387 | 0,287621761 | TTC6 | FALSE | Downregulated |
| 0,253630491 | 0,287652894 | ABHD17A | FALSE | Upregulated |
| 0,440106384 | 0,287652894 | ARHGAP26 | FALSE | Upregulated |
| 0,7863041 | 0,287652894 | GINS1 | FALSE | Upregulated |
| -0,221054854 | 0,287652894 | PAPSS2 | FALSE | Downregulated |
| 0,305860421 | 0,287654983 | GPR146 | FALSE | Upregulated |
| 0,860010345 | 0,287729552 | CCND2 | FALSE | Upregulated |
| -0,840781434 | 0,287729552 | OBFC1 | FALSE | Downregulated |
| 0,484303312 | 0,287729552 | RHOB | FALSE | Upregulated |
| -0,41329107 | 0,287927148 | ALKBH7 | FALSE | Downregulated |
| 0,305008035 | 0,287927148 | GOPC | FALSE | Upregulated |
| 0,539224015 | 0,287927148 | LOC100131270 | FALSE | Upregulated |
| 0,341250083 | 0,287927148 | LOC102723342 | FALSE | Upregulated |
| 0,638821928 | 0,287927148 | NRADDP | FALSE | Upregulated |
| -0,597058606 | 0,287927148 | RGPD8 | FALSE | Downregulated |
| 0,426491701 | 0,287927148 | TREX1 | FALSE | Upregulated |
| 0,281154884 | 0,287980035 | AKAP8 | FALSE | Upregulated |
| 0,379077816 | 0,288297224 | ARHGAP17 | FALSE | Upregulated |
| 0,853593203 | 0,288494674 | RRN3P2 | FALSE | Upregulated |
| -0,338883912 | 0,288518278 | DENND5B | FALSE | Downregulated |
| 0,466033127 | 0,288518278 | DKC1 | FALSE | Upregulated |
| 1,361845821 | 0,288518278 | LSAMP | FALSE | Upregulated |
| -0,872668583 | 0,288719884 | C5orf22 | FALSE | Downregulated |
| 0,282470111 | 0,288964798 | ARF6 | FALSE | Upregulated |

| | | | | |
|---|---|---|---|---|
| -0,745618966 | 0,288964798 | MAP2 | FALSE | Downregulated |
| -1,093738814 | 0,289417968 | TMEM18 | FALSE | Downregulated |
| 1,095395054 | 0,289544088 | HIST1H2AK | FALSE | Upregulated |
| -0,373466452 | 0,289626742 | SCARNA17 | FALSE | Downregulated |
| -0,839177719 | 0,289705438 | SRMS | FALSE | Downregulated |
| 0,669658852 | 0,289783509 | C17orf78 | FALSE | Upregulated |
| -0,365614112 | 0,290062589 | BAHD1 | FALSE | Downregulated |
| -0,713031826 | 0,290084853 | TFAP4 | FALSE | Downregulated |
| 0,423105823 | 0,290154791 | ZDHHC7 | FALSE | Upregulated |
| 1,029491147 | 0,290255382 | FAM89A | FALSE | Upregulated |
| 0,40996183 | 0,290301921 | ASB14 | FALSE | Upregulated |
| -0,394884742 | 0,290301921 | GAS2 | FALSE | Downregulated |
| 0,521227155 | 0,290466752 | RAPGEF5 | FALSE | Upregulated |
| 0,885502052 | 0,290511047 | CCL3L3 | FALSE | Upregulated |
| 1,278962693 | 0,290591471 | FBN2 | FALSE | Upregulated |
| -1,435716641 | 0,290782902 | SCAND3 | FALSE | Downregulated |
| 0,36646303 | 0,290785329 | FAM91A1 | FALSE | Upregulated |
| 0,390600257 | 0,290785329 | LOC101929599 | FALSE | Upregulated |
| 0,209057243 | 0,291714277 | GNB1 | FALSE | Upregulated |
| 0,431858503 | 0,291798264 | ARAP2 | FALSE | Upregulated |
| 0,37628579 | 0,291798264 | SNTB2 | FALSE | Upregulated |
| 0,293010659 | 0,291925135 | GPR183 | FALSE | Upregulated |
| -1,372857686 | 0,291925135 | ZNF880 | FALSE | Downregulated |
| -0,326161518 | 0,291960496 | AKR1A1 | FALSE | Downregulated |
| 0,283721184 | 0,292191257 | HEXDC | FALSE | Upregulated |
| -0,346749749 | 0,29257378 | RPL39 | FALSE | Downregulated |
| 1,089157566 | 0,29257378 | ZNF354A | FALSE | Upregulated |
| -0,293155769 | 0,292702904 | RABEP1 | FALSE | Downregulated |
| -0,416609033 | 0,292728333 | ZBTB39 | FALSE | Downregulated |
| -0,302114382 | 0,293238283 | ARL6IP1 | FALSE | Downregulated |
| 0,405641461 | 0,293338692 | APBB1IP | FALSE | Upregulated |
| 1,449901971 | 0,293338692 | HSD3B1 | FALSE | Upregulated |
| -0,823606806 | 0,293338692 | LOC101927382 | FALSE | Downregulated |
| -0,212397859 | 0,293338692 | RPL10 | FALSE | Downregulated |
| -0,450334888 | 0,293380055 | BTG3 | FALSE | Downregulated |
| 0,395405143 | 0,293380055 | FAM87B | FALSE | Upregulated |
| -0,387651772 | 0,293380055 | NDEL1 | FALSE | Downregulated |
| 1,401750558 | 0,293380055 | TPTE2P1 | FALSE | Upregulated |
| 0,435353112 | 0,293380055 | TSTA3 | FALSE | Upregulated |
| -0,307675607 | 0,293380055 | ZNF507 | FALSE | Downregulated |
| -0,475794366 | 0,293422903 | GGT1 | FALSE | Downregulated |
| -0,328580375 | 0,293422903 | SIN3B | FALSE | Downregulated |
| -1,037545391 | 0,293589086 | MOGAT1 | FALSE | Downregulated |
| -0,326827954 | 0,293612096 | SNX27 | FALSE | Downregulated |
| -1,09006463 | 0,293620171 | GTF2IRD2 | FALSE | Downregulated |
| -0,351317936 | 0,293635961 | SNAPC4 | FALSE | Downregulated |
| -1,095125594 | 0,29378005 | GLIS3-AS1 | FALSE | Downregulated |
| -0,346415186 | 0,29378005 | SNX2 | FALSE | Downregulated |
| -0,255562239 | 0,29383528 | HRG | FALSE | Downregulated |
| -0,453950977 | 0,293850117 | DHRS4L2 | FALSE | Downregulated |
| -1,015244302 | 0,293850117 | IL27 | FALSE | Downregulated |

| | | | | |
|---|---|---|---|---|
| 0,679113446 | 0,293850117 | KLC2 | FALSE | Upregulated |
| -0,411864109 | 0,293850117 | NELFB | FALSE | Downregulated |
| 0,530680831 | 0,293936983 | MFSD1 | FALSE | Upregulated |
| -0,446363534 | 0,293946717 | PRMT7 | FALSE | Downregulated |
| -1,25820583 | 0,294368452 | TBC1D32 | FALSE | Downregulated |
| 0,968091436 | 0,294547986 | EXOG | FALSE | Upregulated |
| 0,330749992 | 0,294547986 | PTP4A2 | FALSE | Upregulated |
| -0,345614349 | 0,294995863 | GFRA1 | FALSE | Downregulated |
| 0,505802459 | 0,294995863 | SGOL1 | FALSE | Upregulated |
| -0,575202751 | 0,295216529 | CKMT2-AS1 | FALSE | Downregulated |
| 0,85353327 | 0,295216529 | MX2 | FALSE | Upregulated |
| -0,234559372 | 0,295231316 | CDK10 | FALSE | Downregulated |
| 1,453243566 | 0,295368772 | CMTM1 | FALSE | Upregulated |
| -0,46826242 | 0,295368772 | GYG2 | FALSE | Downregulated |
| 0,797682623 | 0,295859644 | HMGB2 | FALSE | Upregulated |
| 0,323468185 | 0,295859644 | RUSC1 | FALSE | Upregulated |
| -1,436597671 | 0,296231044 | CCDC85A | FALSE | Downregulated |
| -0,473219799 | 0,296420547 | RSPRY1 | FALSE | Downregulated |
| -0,419923027 | 0,296420547 | TTC39C | FALSE | Downregulated |
| 0,34865514 | 0,296683371 | HS6ST1 | FALSE | Upregulated |
| 0,808613873 | 0,296706575 | LOC100289533 | FALSE | Upregulated |
| -0,265646913 | 0,296706575 | UBR3 | FALSE | Downregulated |
| -0,478811611 | 0,296736803 | HNF1B | FALSE | Downregulated |
| -0,302937795 | 0,296833206 | CASKIN2 | FALSE | Downregulated |
| 0,316606156 | 0,296833206 | NRON | FALSE | Upregulated |
| -0,45103123 | 0,296858198 | FAAH | FALSE | Downregulated |
| -0,187640792 | 0,296879691 | CDC42BPB | FALSE | Downregulated |
| 1,02837526 | 0,296879691 | EMR1 | FALSE | Upregulated |
| -0,876979485 | 0,296901276 | C2orf74 | FALSE | Downregulated |
| -0,351899535 | 0,296901276 | MRPL38 | FALSE | Downregulated |
| -1,048685465 | 0,296901276 | PLXNA4 | FALSE | Downregulated |
| -0,984297158 | 0,296901276 | RNF208 | FALSE | Downregulated |
| 0,396441588 | 0,296901276 | SLC35E2 | FALSE | Upregulated |
| 0,767814413 | 0,297020832 | HELB | FALSE | Upregulated |
| -0,37464304 | 0,297020832 | IL12RB2 | FALSE | Downregulated |
| 0,363521277 | 0,297282574 | LINC01004 | FALSE | Upregulated |
| 0,315916769 | 0,297526783 | CTDP1 | FALSE | Upregulated |
| -0,323184805 | 0,297526783 | TBC1D5 | FALSE | Downregulated |
| 0,898574069 | 0,297775928 | CRHR2 | FALSE | Upregulated |
| -0,280405864 | 0,297978311 | SLC38A3 | FALSE | Downregulated |
| -0,191583021 | 0,298138235 | NFE2L1 | FALSE | Downregulated |
| 0,318482837 | 0,298138235 | OMG | FALSE | Upregulated |
| 0,8260788 | 0,298138235 | TMEM251 | FALSE | Upregulated |
| 0,606964577 | 0,298305224 | FAM114A2 | FALSE | Upregulated |
| 0,271124161 | 0,298305224 | LMNA | FALSE | Upregulated |
| -0,323053565 | 0,298305224 | TRAPPC8 | FALSE | Downregulated |
| 0,444912821 | 0,298370967 | TWF2 | FALSE | Upregulated |
| -0,595930491 | 0,298703683 | B3GNT1 | FALSE | Downregulated |
| 0,804385577 | 0,298704721 | SPINT2 | FALSE | Upregulated |
| -0,818014696 | 0,298834501 | ENPP4 | FALSE | Downregulated |
| -0,475148899 | 0,298840088 | NDST3 | FALSE | Downregulated |

| | | | | |
|---|---|---|---|---|
| 0,284714501 | 0,298840088 | TSPAN4 | FALSE | Upregulated |
| 0,930022333 | 0,298891954 | BPGM | FALSE | Upregulated |
| -0,605849067 | 0,299053882 | BLMH | FALSE | Downregulated |
| 1,402996364 | 0,29927848 | LOC102723511 | FALSE | Upregulated |
| 0,644449875 | 0,299503031 | LOC100506985 | FALSE | Upregulated |
| 0,256682229 | 0,299503031 | TRIM13 | FALSE | Upregulated |
| -0,497312737 | 0,299529089 | INTS4 | FALSE | Downregulated |
| -0,294621232 | 0,299559827 | PLS3 | FALSE | Downregulated |
| 0,528079219 | 0,299576691 | MFSD10 | FALSE | Upregulated |
| 0,725027817 | 0,299991066 | CNOT6 | FALSE | Upregulated |
| 0,937673707 | 0,299991066 | NUPL2 | FALSE | Upregulated |
| -1,304680798 | 0,299991066 | VIPR2 | FALSE | Downregulated |
| 1,211303752 | 0,300173902 | LOC102724920 | FALSE | Upregulated |
| 0,59088842 | 0,300279633 | AHNAK2 | FALSE | Upregulated |
| 0,285008746 | 0,300279633 | GNRHR2 | FALSE | Upregulated |
| -0,320991722 | 0,300427775 | FMO3 | FALSE | Downregulated |
| -0,728247049 | 0,300427775 | FUT10 | FALSE | Downregulated |
| 0,365907827 | 0,300427775 | MAP1LC3B | FALSE | Upregulated |
| 0,466175842 | 0,300427775 | RP5-977B1.7 | FALSE | Upregulated |
| 0,412140025 | 0,300451488 | ESR2 | FALSE | Upregulated |
| 0,969035001 | 0,300451488 | KCNK7 | FALSE | Upregulated |
| 0,876154097 | 0,300451488 | LLPH | FALSE | Upregulated |
| -0,217640324 | 0,300451488 | SMARCA2 | FALSE | Downregulated |
| -0,670353166 | 0,300451488 | ZNF334 | FALSE | Downregulated |
| -0,407619077 | 0,300569313 | HRNR | FALSE | Downregulated |
| -0,258672754 | 0,301142038 | ALDH4A1 | FALSE | Downregulated |
| -0,579381155 | 0,301324358 | IFT20 | FALSE | Downregulated |
| 1,296464723 | 0,301329409 | PRR7 | FALSE | Upregulated |
| -0,319658565 | 0,301412866 | CEBPA | FALSE | Downregulated |
| -1,363307931 | 0,301413203 | MGC15885 | FALSE | Downregulated |
| 0,375024006 | 0,301985814 | CD93 | FALSE | Upregulated |
| 0,614395127 | 0,302170684 | PRRG1 | FALSE | Upregulated |
| -0,464608826 | 0,302170684 | SAMD8 | FALSE | Downregulated |
| 0,272503394 | 0,302170684 | SPDYE1 | FALSE | Upregulated |
| 0,218724405 | 0,302170684 | TUBB | FALSE | Upregulated |
| -0,392306819 | 0,302195182 | LPIN3 | FALSE | Downregulated |
| -0,978680561 | 0,302195182 | ZNF229 | FALSE | Downregulated |
| -0,407647479 | 0,3022177 | ENTPD3 | FALSE | Downregulated |
| -0,348615894 | 0,3022177 | PROZ | FALSE | Downregulated |
| 0,724407789 | 0,302379179 | FAM150B | FALSE | Upregulated |
| -0,358170385 | 0,302556899 | STARD8 | FALSE | Downregulated |
| 1,162244714 | 0,302602019 | BBS7 | FALSE | Upregulated |
| 0,277872729 | 0,302607898 | ATP11B | FALSE | Upregulated |
| -0,323864653 | 0,302638223 | CLPX | FALSE | Downregulated |
| 1,196615705 | 0,302757762 | LOC100505625 | FALSE | Upregulated |
| -1,028455622 | 0,302993761 | AP000479.1 | FALSE | Downregulated |
| 0,409340445 | 0,302993761 | DR1 | FALSE | Upregulated |
| 0,366091479 | 0,303447113 | SYNGAP1 | FALSE | Upregulated |
| -0,92736125 | 0,303463765 | ENHO | FALSE | Downregulated |
| 0,230012367 | 0,303493508 | ZNF687 | FALSE | Upregulated |
| -0,233219096 | 0,303590849 | USF2 | FALSE | Downregulated |

| | | | | |
|---|---|---|---|---|
| -0,327762523 | 0,303698649 | FBXO7 | FALSE | Downregulated |
| 1,20912332 | 0,303698649 | MCHR1 | FALSE | Upregulated |
| 0,289560482 | 0,303698649 | TMPRSS9 | FALSE | Upregulated |
| 0,627233604 | 0,303719883 | C17orf59 | FALSE | Upregulated |
| 0,866324159 | 0,303719883 | LOC102724548 | FALSE | Upregulated |
| 0,383343884 | 0,303719883 | PDE12 | FALSE | Upregulated |
| -0,649574717 | 0,303831189 | TIMM10 | FALSE | Downregulated |
| 0,458684684 | 0,304056169 | BTN3A3 | FALSE | Upregulated |
| 0,599410309 | 0,304457449 | RPAP2 | FALSE | Upregulated |
| -0,292676253 | 0,304457449 | WBP1L | FALSE | Downregulated |
| 0,311494514 | 0,304481031 | BTBD18 | FALSE | Upregulated |
| 0,378257202 | 0,304481031 | DNAJA1 | FALSE | Upregulated |
| -0,590546435 | 0,304577172 | CEBPA-AS1 | FALSE | Downregulated |
| 0,578139099 | 0,304598347 | HCFC1R1 | FALSE | Upregulated |
| -0,83468316 | 0,304598347 | TMEM5 | FALSE | Downregulated |
| 0,39072983 | 0,30463349 | C15orf38 | FALSE | Upregulated |
| -0,242959617 | 0,304673552 | ZRANB1 | FALSE | Downregulated |
| 0,319219614 | 0,304786545 | SLC16A4 | FALSE | Upregulated |
| -0,451557881 | 0,304866333 | ZC3H7A | FALSE | Downregulated |
| 0,244051142 | 0,305043842 | STK24 | FALSE | Upregulated |
| 0,283625961 | 0,305144957 | LOC102724129 | FALSE | Upregulated |
| -0,318037753 | 0,305148248 | ZHX1 | FALSE | Downregulated |
| -1,270020132 | 0,305241396 | P2RX6 | FALSE | Downregulated |
| -0,293520563 | 0,305322825 | ZCCHC24 | FALSE | Downregulated |
| 0,941581732 | 0,305742637 | CRLF1 | FALSE | Upregulated |
| -0,706604184 | 0,306177456 | KIAA1009 | FALSE | Downregulated |
| -0,867974438 | 0,306177456 | LOC102724322 | FALSE | Downregulated |
| -0,330761835 | 0,306177456 | SYNE2 | FALSE | Downregulated |
| -1,369506533 | 0,306224292 | LOC441124 | FALSE | Downregulated |
| -1,360546462 | 0,306463825 | FBXO48 | FALSE | Downregulated |
| -0,496442827 | 0,306463825 | TUBE1 | FALSE | Downregulated |
| -0,668051015 | 0,306464451 | COX7B | FALSE | Downregulated |
| -0,525657082 | 0,306464451 | CYGB | FALSE | Downregulated |
| -0,672739797 | 0,306464451 | DNAJC12 | FALSE | Downregulated |
| 0,40034735 | 0,306464451 | ZNF394 | FALSE | Upregulated |
| 0,448815816 | 0,306829535 | HTR6 | FALSE | Upregulated |
| -0,920697765 | 0,306963627 | TGFA | FALSE | Downregulated |
| -0,533622118 | 0,306963627 | ZXDA | FALSE | Downregulated |
| -0,265873496 | 0,307022786 | ZNF638 | FALSE | Downregulated |
| 0,70364381 | 0,307077145 | FAM213B | FALSE | Upregulated |
| 0,363114103 | 0,307077145 | LOC101927385 | FALSE | Upregulated |
| 0,511018686 | 0,307077145 | RND1 | FALSE | Upregulated |
| 0,334997952 | 0,307137265 | CACTIN-AS1 | FALSE | Upregulated |
| -1,136606063 | 0,307289972 | SDCBP2 | FALSE | Downregulated |
| -0,676538151 | 0,307289972 | TMPRSS11D | FALSE | Downregulated |
| -0,279356489 | 0,30737973 | ARIH2 | FALSE | Downregulated |
| -0,386504471 | 0,307381264 | MRPL34 | FALSE | Downregulated |
| -0,275766541 | 0,307739911 | AFG3L2 | FALSE | Downregulated |
| 0,59812325 | 0,307739911 | CDC73 | FALSE | Upregulated |
| -0,694718974 | 0,307809118 | C16orf59 | FALSE | Downregulated |
| -0,335212857 | 0,308194323 | TTC38 | FALSE | Downregulated |

| | | | | |
|---|---|---|---|---|
| -0,373207266 | 0,308210135 | FAHD1 | FALSE | Downregulated |
| 0,352335728 | 0,308319539 | FAM118B | FALSE | Upregulated |
| 0,399212482 | 0,308393075 | SP110 | FALSE | Upregulated |
| 0,352169129 | 0,308711243 | FNDC4 | FALSE | Upregulated |
| 1,027247252 | 0,308711243 | LOC81691 | FALSE | Upregulated |
| -1,252180943 | 0,308794374 | LINC01003 | FALSE | Downregulated |
| 1,559992643 | 0,308880728 | HLA-DQB2 | FALSE | Upregulated |
| 0,301483317 | 0,309219195 | GSN-AS1 | FALSE | Upregulated |
| -0,669904113 | 0,309323614 | AGBL2 | FALSE | Downregulated |
| 0,264415159 | 0,30933332 | CLCN7 | FALSE | Upregulated |
| -0,927391079 | 0,30933332 | PPT2 | FALSE | Downregulated |
| -1,574860854 | 0,309347146 | LOC101927249 | FALSE | Downregulated |
| -0,458439218 | 0,30953273 | POLL | FALSE | Downregulated |
| -1,537638857 | 0,309893151 | EGFLAM-AS2 | FALSE | Downregulated |
| 0,68164557 | 0,310024433 | FAM46C | FALSE | Upregulated |
| 0,34689083 | 0,310078612 | ANKIB1 | FALSE | Upregulated |
| -0,21881537 | 0,310078612 | FAM120A | FALSE | Downregulated |
| 1,09310515 | 0,310078612 | LOC645485 | FALSE | Upregulated |
| 0,369667677 | 0,310078612 | MORC4 | FALSE | Upregulated |
| 0,682102619 | 0,310078612 | PAPPA-AS1 | FALSE | Upregulated |
| -0,173808375 | 0,310078612 | TMED10 | FALSE | Downregulated |
| -0,816315417 | 0,310130672 | ST6GAL2 | FALSE | Downregulated |
| 0,588918718 | 0,310209591 | RRAS | FALSE | Upregulated |
| -0,366084824 | 0,310252896 | HINT2 | FALSE | Downregulated |
| 0,413313589 | 0,310252896 | LOC440386 | FALSE | Upregulated |
| 0,441399189 | 0,310252896 | YPEL4 | FALSE | Upregulated |
| 0,262923067 | 0,310570627 | AARS | FALSE | Upregulated |
| 0,641052211 | 0,310570627 | HES4 | FALSE | Upregulated |
| 0,297649058 | 0,310570627 | STAT3 | FALSE | Upregulated |
| -0,720757671 | 0,3107206 | HNF4A-AS1 | FALSE | Downregulated |
| -0,52879015 | 0,310885401 | C3orf58 | FALSE | Downregulated |
| -0,270126595 | 0,310885401 | HSPG2 | FALSE | Downregulated |
| 0,887199824 | 0,311070236 | FLJ46363 | FALSE | Upregulated |
| 0,551038325 | 0,311070236 | PDE6A | FALSE | Upregulated |
| 0,268147834 | 0,311089269 | GPC6 | FALSE | Upregulated |
| 0,361538365 | 0,311089269 | NELFE | FALSE | Upregulated |
| -0,348947205 | 0,311172545 | PID1 | FALSE | Downregulated |
| -0,545188704 | 0,311345642 | CD36 | FALSE | Downregulated |
| 1,391865948 | 0,311345642 | GAS2L3 | FALSE | Upregulated |
| 0,345984102 | 0,311406094 | SHANK2-AS3 | FALSE | Upregulated |
| 0,806503977 | 0,311406094 | SMARCD3 | FALSE | Upregulated |
| 0,442640921 | 0,311410727 | DOLK | FALSE | Upregulated |
| 0,319737903 | 0,311410727 | MAPT | FALSE | Upregulated |
| -0,331880382 | 0,311430212 | ACMSD | FALSE | Downregulated |
| -0,632748276 | 0,311430212 | RP11-115J16.1 | FALSE | Downregulated |
| 0,610772458 | 0,311838305 | AOAH | FALSE | Upregulated |
| -0,841082957 | 0,311838305 | EXOSC5 | FALSE | Downregulated |
| -0,41857203 | 0,311838305 | GDA | FALSE | Downregulated |
| -0,417875135 | 0,311838305 | PEX14 | FALSE | Downregulated |
| -0,319976185 | 0,311838305 | RNH1 | FALSE | Downregulated |
| 0,63539784 | 0,312004938 | PRAM1 | FALSE | Upregulated |

| | | | | |
|---|---|---|---|---|
| 0,387078099 | 0,312310144 | LOC102723762 | FALSE | Upregulated |
| -0,846098519 | 0,312661553 | SRPX | FALSE | Downregulated |
| -0,351711042 | 0,312661553 | ZFP91 | FALSE | Downregulated |
| 0,352805663 | 0,312933742 | CASP8 | FALSE | Upregulated |
| 0,462170583 | 0,31295394 | SLC7A7 | FALSE | Upregulated |
| -0,513188118 | 0,31295394 | ZNF514 | FALSE | Downregulated |
| -0,803749269 | 0,313018949 | SYTL5 | FALSE | Downregulated |
| 0,348725409 | 0,313073887 | PMS2P1 | FALSE | Upregulated |
| 0,5170196 | 0,313189192 | TAGAP | FALSE | Upregulated |
| 0,783447791 | 0,31322864 | CCNI2 | FALSE | Upregulated |
| -0,345506228 | 0,313562331 | JMJD4 | FALSE | Downregulated |
| -0,390235614 | 0,313689512 | NMD3 | FALSE | Downregulated |
| -0,314419192 | 0,313941758 | MYO9A | FALSE | Downregulated |
| 0,327377896 | 0,313941758 | TAF11 | FALSE | Upregulated |
| -0,335221476 | 0,314007294 | LOC101927190 | FALSE | Downregulated |
| -0,459229026 | 0,31432877 | DARS | FALSE | Downregulated |
| -0,341509032 | 0,31432877 | VPS28 | FALSE | Downregulated |
| -0,374837081 | 0,314730191 | OCRL | FALSE | Downregulated |
| 0,375818562 | 0,314913732 | PRCC | FALSE | Upregulated |
| 0,484675723 | 0,3149817 | DDX60 | FALSE | Upregulated |
| -1,253881759 | 0,3149817 | KDELC1 | FALSE | Downregulated |
| 0,219530359 | 0,3149817 | UBE2D3 | FALSE | Upregulated |
| 0,533226624 | 0,314996962 | NME6 | FALSE | Upregulated |
| 1,112663487 | 0,314996962 | RASGEF1A | FALSE | Upregulated |
| -1,044187274 | 0,31526101 | ESPNP | FALSE | Downregulated |
| -1,584778901 | 0,315542771 | ADCYAP1R1 | FALSE | Downregulated |
| -0,536655168 | 0,315542771 | PIGV | FALSE | Downregulated |
| 0,431701703 | 0,315542771 | RGMB | FALSE | Upregulated |
| -0,651662427 | 0,315945972 | BEND7 | FALSE | Downregulated |
| -0,535484804 | 0,315945972 | C1orf27 | FALSE | Downregulated |
| -0,191072815 | 0,315945972 | IGFBP4 | FALSE | Downregulated |
| 0,914745458 | 0,315945972 | PAQR5 | FALSE | Upregulated |
| -0,501811847 | 0,315945972 | TXNRD3 | FALSE | Downregulated |
| -0,333273965 | 0,316156685 | EXOC3L4 | FALSE | Downregulated |
| -0,516148995 | 0,316156685 | TOMM7 | FALSE | Downregulated |
| -0,34767861 | 0,316177817 | ABCA9 | FALSE | Downregulated |
| -0,328498249 | 0,316288421 | GNL1 | FALSE | Downregulated |
| 0,406076759 | 0,316538426 | ORMDL2 | FALSE | Upregulated |
| -0,483106547 | 0,317044697 | STK3 | FALSE | Downregulated |
| -0,475872859 | 0,317044697 | ZNF623 | FALSE | Downregulated |
| -0,413017892 | 0,317236152 | GIPC3 | FALSE | Downregulated |
| 0,34852554 | 0,317236152 | PTBP3 | FALSE | Upregulated |
| 0,490484696 | 0,317342134 | VPS9D1-AS1 | FALSE | Upregulated |
| 0,418049662 | 0,317480667 | DEAF1 | FALSE | Upregulated |
| 0,405727704 | 0,317558522 | TPM4 | FALSE | Upregulated |
| 0,256779674 | 0,317662712 | ANKRD13A | FALSE | Upregulated |
| -0,750292724 | 0,317662712 | C6orf211 | FALSE | Downregulated |
| 0,364405902 | 0,317662712 | LINC-PINT | FALSE | Upregulated |
| 0,327653559 | 0,317662712 | SCO2 | FALSE | Upregulated |
| -0,328969938 | 0,317920391 | EHHADH-AS1 | FALSE | Downregulated |
| -0,812268911 | 0,317920391 | GSTM3 | FALSE | Downregulated |

| | | | | |
|---|---|---|---|---|
| 0,248999025 | 0,317920391 | PPIB | FALSE | Upregulated |
| -0,977679016 | 0,317951018 | LRRC27 | FALSE | Downregulated |
| -0,451202165 | 0,317951018 | TMEM64 | FALSE | Downregulated |
| -0,131096532 | 0,318031886 | ARHGEF12 | FALSE | Downregulated |
| 0,621260499 | 0,318031886 | ID1 | FALSE | Upregulated |
| 0,318007657 | 0,31816739 | LOC102724559 | FALSE | Upregulated |
| -0,376649555 | 0,31816739 | SLC29A1 | FALSE | Downregulated |
| -0,301265283 | 0,318357098 | TXNRD2 | FALSE | Downregulated |
| -0,900785955 | 0,318538431 | LOC283177 | FALSE | Downregulated |
| 1,086258902 | 0,318556855 | GDPGP1 | FALSE | Upregulated |
| 0,449099219 | 0,318772708 | DDIT3 | FALSE | Upregulated |
| 0,920916812 | 0,319024242 | MS4A4A | FALSE | Upregulated |
| -0,557194814 | 0,319024242 | NKAIN3 | FALSE | Downregulated |
| 0,666204652 | 0,319135962 | CCDC78 | FALSE | Upregulated |
| -0,322534448 | 0,319266025 | ATP2B2 | FALSE | Downregulated |
| 0,646014209 | 0,319266025 | C1QTNF5 | FALSE | Upregulated |
| 0,506515017 | 0,319266025 | DENR | FALSE | Upregulated |
| -0,65324473 | 0,319266025 | LMAN2L | FALSE | Downregulated |
| -0,393629506 | 0,319266025 | RGPD3 | FALSE | Downregulated |
| 0,733307773 | 0,319345374 | WDR35 | FALSE | Upregulated |
| -1,39898456 | 0,319421918 | RP11-695J4.2 | FALSE | Downregulated |
| -0,221869279 | 0,319633751 | REV3L | FALSE | Downregulated |
| 0,443356735 | 0,319758383 | SSH2 | FALSE | Upregulated |
| -1,118798225 | 0,32026118 | KLHDC1 | FALSE | Downregulated |
| 1,299453475 | 0,320424015 | THUMPD2 | FALSE | Upregulated |
| 0,440392 | 0,320435355 | DSERG1 | FALSE | Upregulated |
| -0,508825411 | 0,320501333 | NR0B2 | FALSE | Downregulated |
| -0,434772175 | 0,320581386 | SAA4 | FALSE | Downregulated |
| 0,309611795 | 0,320622937 | SAMD11 | FALSE | Upregulated |
| 0,648385508 | 0,320622937 | UNC119 | FALSE | Upregulated |
| -0,224294411 | 0,320729634 | MYO15B | FALSE | Downregulated |
| 0,871106747 | 0,320750295 | CDCA5 | FALSE | Upregulated |
| -0,565357433 | 0,320785198 | FHL1 | FALSE | Downregulated |
| 0,470627066 | 0,321168826 | COL14A1 | FALSE | Upregulated |
| 0,369871196 | 0,321388095 | LOC100293704 | FALSE | Upregulated |
| 0,370619094 | 0,321658827 | THOC2 | FALSE | Upregulated |
| -0,604428009 | 0,321681891 | FLG | FALSE | Downregulated |
| -0,854267553 | 0,321681891 | LOC101927000 | FALSE | Downregulated |
| -0,200932603 | 0,32173757 | KIAA1161 | FALSE | Downregulated |
| 0,474851061 | 0,32173757 | LOC102724009 | FALSE | Upregulated |
| -0,316742298 | 0,32173757 | SUCLG2 | FALSE | Downregulated |
| 0,445413303 | 0,321742514 | CCDC137 | FALSE | Upregulated |
| 1,049178376 | 0,321840692 | TBX1 | FALSE | Upregulated |
| 0,436596947 | 0,321840692 | TGIF1 | FALSE | Upregulated |
| -0,230260501 | 0,322314064 | ZBTB20 | FALSE | Downregulated |
| -0,315022363 | 0,322396171 | LRRC45 | FALSE | Downregulated |
| 0,195280468 | 0,322690465 | AIM1 | FALSE | Upregulated |
| 0,352421971 | 0,322690465 | CXCL16 | FALSE | Upregulated |
| 0,662022843 | 0,322690465 | RP1-149A16.3 | FALSE | Upregulated |
| -0,410602642 | 0,322690465 | SRP68 | FALSE | Downregulated |
| 0,21316233 | 0,322690465 | TCIRG1 | FALSE | Upregulated |

| | | | | |
|---|---|---|---|---|
| -0,318435516 | 0,322746391 | TXNIP | FALSE | Downregulated |
| -0,395894652 | 0,322835624 | ACSF2 | FALSE | Downregulated |
| -1,484417332 | 0,322835624 | FAM124B | FALSE | Downregulated |
| 1,187034116 | 0,322835624 | RP11-875O11.2 | FALSE | Upregulated |
| 0,31567635 | 0,32285227 | CD151 | FALSE | Upregulated |
| -0,469924715 | 0,32285227 | CSNK2B | FALSE | Downregulated |
| 0,579140187 | 0,32285227 | EPDR1 | FALSE | Upregulated |
| -0,866203359 | 0,32285227 | HEPN1 | FALSE | Downregulated |
| -0,596282681 | 0,32285227 | JAGN1 | FALSE | Downregulated |
| -0,396827643 | 0,32285227 | TIPARP | FALSE | Downregulated |
| 1,625009185 | 0,323261557 | LINC00332 | FALSE | Upregulated |
| 0,301977601 | 0,323658924 | CNOT7 | FALSE | Upregulated |
| 0,436107614 | 0,323658924 | SEC23B | FALSE | Upregulated |
| 0,504821447 | 0,323836552 | MED11 | FALSE | Upregulated |
| -0,312833711 | 0,323882855 | PLD1 | FALSE | Downregulated |
| -0,435357186 | 0,323882855 | SLC33A1 | FALSE | Downregulated |
| 0,224333393 | 0,324114836 | STX16 | FALSE | Upregulated |
| 0,305329307 | 0,324433117 | CCSAP | FALSE | Upregulated |
| -0,683392565 | 0,32443712 | DZIP1 | FALSE | Downregulated |
| -0,383610839 | 0,324470529 | GLRX5 | FALSE | Downregulated |
| 0,614942456 | 0,324470529 | GPRC5B | FALSE | Upregulated |
| 0,442802493 | 0,324731647 | TPTE2P5 | FALSE | Upregulated |
| 0,299161449 | 0,325314381 | CLTA | FALSE | Upregulated |
| -0,358412706 | 0,325314381 | ELF2 | FALSE | Downregulated |
| -0,264580122 | 0,325314381 | MTUS1 | FALSE | Downregulated |
| -0,23766932 | 0,325314381 | TSC2 | FALSE | Downregulated |
| -0,264568917 | 0,325450509 | LMAN2 | FALSE | Downregulated |
| -0,615438175 | 0,325450509 | PRDM11 | FALSE | Downregulated |
| 0,633989798 | 0,325507285 | DTNBP1 | FALSE | Upregulated |
| 0,796763303 | 0,325622164 | INPP4B | FALSE | Upregulated |
| -0,244833872 | 0,325752327 | PABPC4 | FALSE | Downregulated |
| 0,263108964 | 0,325943165 | FNDC3B | FALSE | Upregulated |
| 0,225942157 | 0,326043622 | EIF3G | FALSE | Upregulated |
| 3,185606267 | 0,326043622 | LOC440354 | FALSE | Upregulated |
| 0,957326051 | 0,326043622 | PGR | FALSE | Upregulated |
| -1,182076309 | 0,326234657 | CCDC126 | FALSE | Downregulated |
| 0,414602283 | 0,326235497 | RP11-538P18.2 | FALSE | Upregulated |
| -0,234312174 | 0,326332799 | GUSB | FALSE | Downregulated |
| -0,547012436 | 0,326332799 | IGIP | FALSE | Downregulated |
| -0,274402716 | 0,326494425 | MRPL20 | FALSE | Downregulated |
| -0,438485162 | 0,326494425 | TRMT1L | FALSE | Downregulated |
| -0,306762676 | 0,326527888 | ATP5D | FALSE | Downregulated |
| 0,404929101 | 0,326527888 | ATP6V1C1 | FALSE | Upregulated |
| -0,244992392 | 0,326527888 | C11orf95 | FALSE | Downregulated |
| -0,317156969 | 0,326527888 | CNNM4 | FALSE | Downregulated |
| -0,321901497 | 0,326527888 | EDC4 | FALSE | Downregulated |
| 1,235060906 | 0,326527888 | HIST1H3I | FALSE | Upregulated |
| -0,410272999 | 0,326527888 | KIAA0141 | FALSE | Downregulated |
| 0,390237569 | 0,326527888 | NAA20 | FALSE | Upregulated |
| -0,502425029 | 0,326527888 | NARFL | FALSE | Downregulated |
| 0,4118792 | 0,326527888 | RBFA | FALSE | Upregulated |

| | | | | |
|---|---|---|---|---|
| -0,40857621 | 0,326527888 | RPS29 | FALSE | Downregulated |
| 1,362034569 | 0,326532761 | LOC101928947 | FALSE | Upregulated |
| 1,195894557 | 0,326540976 | LOC101929805 | FALSE | Upregulated |
| -0,711779853 | 0,326715554 | TEX14 | FALSE | Downregulated |
| -0,332671083 | 0,326765777 | CHCHD2 | FALSE | Downregulated |
| 0,509812764 | 0,326808173 | SLC37A1 | FALSE | Upregulated |
| 0,257038402 | 0,326857071 | CD14 | FALSE | Upregulated |
| 0,308379819 | 0,327000145 | WDFY2 | FALSE | Upregulated |
| 0,669317766 | 0,32705143 | HN1 | FALSE | Upregulated |
| -0,854452618 | 0,32705143 | NPIPA1 | FALSE | Downregulated |
| 0,955102519 | 0,327264337 | RP11-426L16.8 | FALSE | Upregulated |
| -0,576330279 | 0,327610383 | ZG16 | FALSE | Downregulated |
| 0,29083426 | 0,327737502 | FAM219B | FALSE | Upregulated |
| 0,366844615 | 0,327879809 | DCP1A | FALSE | Upregulated |
| 0,36388593 | 0,327957799 | ZC3HAV1 | FALSE | Upregulated |
| 0,862254727 | 0,328141166 | RP11-260E18.1 | FALSE | Upregulated |
| 0,609004835 | 0,328200236 | IER5L | FALSE | Upregulated |
| 0,446062083 | 0,32854232 | TPT1-AS1 | FALSE | Upregulated |
| -0,19229814 | 0,328805746 | ZNF106 | FALSE | Downregulated |
| -1,029501974 | 0,328901147 | CTNND2 | FALSE | Downregulated |
| -1,098777417 | 0,328901147 | RAG1 | FALSE | Downregulated |
| -1,171662119 | 0,328941473 | LOC440157 | FALSE | Downregulated |
| 0,459512811 | 0,328941473 | ODF2 | FALSE | Upregulated |
| -0,930612307 | 0,328941473 | ZNF426 | FALSE | Downregulated |
| 0,270977944 | 0,328970332 | RANBP1 | FALSE | Upregulated |
| 0,714230138 | 0,328970332 | UPRT | FALSE | Upregulated |
| -0,893816099 | 0,32914149 | LPXN | FALSE | Downregulated |
| 0,385610846 | 0,329256405 | NSUN5P1 | FALSE | Upregulated |
| -0,336205962 | 0,329256405 | PCMTD2 | FALSE | Downregulated |
| -0,567617187 | 0,329260448 | CRADD | FALSE | Downregulated |
| 0,870763797 | 0,329260448 | CTSK | FALSE | Upregulated |
| -0,336025633 | 0,329260448 | ZNF148 | FALSE | Downregulated |
| -1,354825112 | 0,329586957 | MOSPD1 | FALSE | Downregulated |
| 0,466226718 | 0,329755821 | BCAP29 | FALSE | Upregulated |
| 0,292247152 | 0,329833889 | DMTF1 | FALSE | Upregulated |
| 0,295977204 | 0,329855592 | NBPF9 | FALSE | Upregulated |
| 0,384536859 | 0,329855592 | SCARNA12 | FALSE | Upregulated |
| 0,334913612 | 0,329897124 | ATP2B4 | FALSE | Upregulated |
| 0,356468355 | 0,329897124 | LOC102723863 | FALSE | Upregulated |
| 0,280648349 | 0,329897124 | NANOS1 | FALSE | Upregulated |
| 0,271828773 | 0,329897124 | PURB | FALSE | Upregulated |
| 0,257600326 | 0,329897124 | UBA5 | FALSE | Upregulated |
| -0,381587638 | 0,330031349 | BNIP3L | FALSE | Downregulated |
| 0,487742629 | 0,330031349 | IL3RA | FALSE | Upregulated |
| 0,221120578 | 0,330312401 | ADRBK1 | FALSE | Upregulated |
| 0,62765348 | 0,330312401 | COL4A2-AS2 | FALSE | Upregulated |
| -0,30053615 | 0,330312401 | CRY2 | FALSE | Downregulated |
| -0,242476137 | 0,330326016 | ARHGAP5 | FALSE | Downregulated |
| -0,650677922 | 0,330326016 | PGBD5 | FALSE | Downregulated |
| 0,420561694 | 0,330354885 | C19orf54 | FALSE | Upregulated |
| 0,760558556 | 0,330485051 | RAET1E | FALSE | Upregulated |

| | | | | |
|---|---|---|---|---|
| -0,410432333 | 0,330646104 | BABAM1 | FALSE | Downregulated |
| -0,21377953 | 0,330719158 | CCNL2 | FALSE | Downregulated |
| -0,417270917 | 0,330731494 | CTPS2 | FALSE | Downregulated |
| -1,117618858 | 0,330944602 | FRG1B | FALSE | Downregulated |
| -0,351202826 | 0,330944602 | NDUFA10 | FALSE | Downregulated |
| 0,40840806 | 0,331020789 | TCHP | FALSE | Upregulated |
| 0,590488778 | 0,331400961 | GUCY1A3 | FALSE | Upregulated |
| -1,510396816 | 0,331447559 | CBWD3 | FALSE | Downregulated |
| -0,732292952 | 0,332108067 | COL4A5 | FALSE | Downregulated |
| 0,313540079 | 0,332149973 | FRMD8 | FALSE | Upregulated |
| -0,244031081 | 0,332149973 | RBM6 | FALSE | Downregulated |
| 0,380846242 | 0,332625029 | NDUFA2 | FALSE | Upregulated |
| 0,479229827 | 0,332625029 | SESN1 | FALSE | Upregulated |
| 1,274413607 | 0,332658999 | LOC101927608 | FALSE | Upregulated |
| 0,428580255 | 0,332890773 | TNK1 | FALSE | Upregulated |
| 0,38541061 | 0,333064815 | ADAM22 | FALSE | Upregulated |
| -0,519720261 | 0,333064815 | IMPACT | FALSE | Downregulated |
| 0,320572956 | 0,333083328 | CENPP | FALSE | Upregulated |
| 1,010654406 | 0,333083328 | GPR150 | FALSE | Upregulated |
| 0,619082412 | 0,333083328 | NOXO1 | FALSE | Upregulated |
| -0,358251524 | 0,333135435 | RBP1 | FALSE | Downregulated |
| -0,397725836 | 0,333135435 | SLC27A3 | FALSE | Downregulated |
| 0,908325889 | 0,333148472 | LOC101929360 | FALSE | Upregulated |
| 1,113453152 | 0,333148472 | RP11-458D21.1 | FALSE | Upregulated |
| -0,207828176 | 0,333398312 | KIAA0319L | FALSE | Downregulated |
| 0,337793808 | 0,333477409 | CCDC69 | FALSE | Upregulated |
| -0,932182077 | 0,333477409 | LOC101927986 | FALSE | Downregulated |
| 0,615698568 | 0,333477409 | LOC101929787 | FALSE | Upregulated |
| 0,661348686 | 0,333477409 | NOC3L | FALSE | Upregulated |
| 1,246654143 | 0,333726737 | LINC01133 | FALSE | Upregulated |
| -1,027147387 | 0,333875009 | SEC14L3 | FALSE | Downregulated |
| 0,372334723 | 0,334005097 | MAPRE3 | FALSE | Upregulated |
| -0,324842099 | 0,334005097 | MTMR12 | FALSE | Downregulated |
| 0,284071974 | 0,334133623 | RAB8A | FALSE | Upregulated |
| 0,761826309 | 0,334415614 | RP11-797H7.5 | FALSE | Upregulated |
| 0,667882676 | 0,33453073 | UTP14A | FALSE | Upregulated |
| -0,34833923 | 0,334679541 | THRB-AS1 | FALSE | Downregulated |
| 0,24245237 | 0,33479022 | LOC102723521 | FALSE | Upregulated |
| -0,362300027 | 0,334904021 | PIK3C2B | FALSE | Downregulated |
| -0,47679346 | 0,334904021 | REPS2 | FALSE | Downregulated |
| -0,702089499 | 0,334954937 | MTIF3 | FALSE | Downregulated |
| 0,398285857 | 0,334954937 | PPM1G | FALSE | Upregulated |
| -0,386746164 | 0,334954937 | SUMO1 | FALSE | Downregulated |
| 0,467862489 | 0,33497821 | C2CD4D | FALSE | Upregulated |
| 0,560068366 | 0,335074608 | FLJ20464 | FALSE | Upregulated |
| -0,268518768 | 0,335152279 | EPB41L2 | FALSE | Downregulated |
| -0,387582759 | 0,335212603 | XPNPEP3 | FALSE | Downregulated |
| -0,481431321 | 0,335301388 | CLEC4M | FALSE | Downregulated |
| 0,951283922 | 0,335301388 | FAM105A | FALSE | Upregulated |
| -0,766391086 | 0,335301388 | GRPEL2 | FALSE | Downregulated |
| 0,375386972 | 0,335301388 | ZNF496 | FALSE | Upregulated |

| | | | | |
|---|---|---|---|---|
| -0,390118438 | 0,335333199 | LOC100130872 | FALSE | Downregulated |
| -1,327375005 | 0,335333199 | TEX22 | FALSE | Downregulated |
| -0,677054678 | 0,335435539 | SGCB | FALSE | Downregulated |
| -0,910779273 | 0,335435539 | WBP4 | FALSE | Downregulated |
| 0,269251807 | 0,335553828 | STUB1 | FALSE | Upregulated |
| 0,922593925 | 0,335791457 | ERCC5 | FALSE | Upregulated |
| 0,495429316 | 0,335861604 | ATP8B2 | FALSE | Upregulated |
| 0,851884411 | 0,335861604 | RCAN2 | FALSE | Upregulated |
| 0,268615293 | 0,335975611 | NUMB | FALSE | Upregulated |
| -0,441237955 | 0,33606256 | EMP2 | FALSE | Downregulated |
| -0,640109499 | 0,336174082 | ETV5 | FALSE | Downregulated |
| -0,392763059 | 0,336268291 | HEATR4 | FALSE | Downregulated |
| 1,214399657 | 0,33637201 | AC006272.1 | FALSE | Upregulated |
| -0,315692781 | 0,336409286 | ARHGAP1 | FALSE | Downregulated |
| -0,689586435 | 0,336409286 | EBF4 | FALSE | Downregulated |
| 0,196436763 | 0,336409286 | EIF2AK1 | FALSE | Upregulated |
| 0,188290923 | 0,336409286 | HERC1 | FALSE | Upregulated |
| 1,13306817 | 0,336409286 | KCNMB2 | FALSE | Upregulated |
| 0,799263386 | 0,336409286 | KPNA5 | FALSE | Upregulated |
| 0,298043452 | 0,336409286 | RECQL | FALSE | Upregulated |
| -0,330791521 | 0,336409286 | RP11-778H2.1 | FALSE | Downregulated |
| -1,295806626 | 0,336409286 | SNORA57 | FALSE | Downregulated |
| 0,488890018 | 0,336409286 | TNXB | FALSE | Upregulated |
| -0,89234293 | 0,336437676 | RPL32P3 | FALSE | Downregulated |
| -0,290252918 | 0,336437676 | RPS28 | FALSE | Downregulated |
| 0,216649639 | 0,336520444 | FRMD4A | FALSE | Upregulated |
| 0,259975963 | 0,336582567 | SPRN | FALSE | Upregulated |
| 1,312949043 | 0,33662303 | C13orf35 | FALSE | Upregulated |
| 0,773881225 | 0,336920919 | OASL | FALSE | Upregulated |
| 2,133642795 | 0,336949083 | ASCL1 | FALSE | Upregulated |
| -0,19276329 | 0,33763449 | SNRNP200 | FALSE | Downregulated |
| -0,551178137 | 0,337704266 | PIGN | FALSE | Downregulated |
| 0,310466466 | 0,337708933 | FAM196A | FALSE | Upregulated |
| -0,847150158 | 0,337841304 | CLDN4 | FALSE | Downregulated |
| -0,637602368 | 0,338065754 | ARHGEF15 | FALSE | Downregulated |
| 0,648361072 | 0,338065754 | MAP6D1 | FALSE | Upregulated |
| 0,365631392 | 0,338065754 | RRAS2 | FALSE | Upregulated |
| -0,554868668 | 0,338065754 | THNSL2 | FALSE | Downregulated |
| -0,717010812 | 0,338550562 | BLOC1S5 | FALSE | Downregulated |
| -0,479462734 | 0,338550562 | PVRL3 | FALSE | Downregulated |
| -1,324425736 | 0,338565833 | SH3GL1P1 | FALSE | Downregulated |
| -0,360874763 | 0,338898915 | FAF2 | FALSE | Downregulated |
| -0,274672032 | 0,338898915 | SLC22A7 | FALSE | Downregulated |
| 0,914186924 | 0,338907789 | LINC00485 | FALSE | Upregulated |
| 0,432200279 | 0,338972865 | C10orf25 | FALSE | Upregulated |
| 0,364946242 | 0,339428084 | PCID2 | FALSE | Upregulated |
| 0,386756153 | 0,339428084 | RP11-98I9.4 | FALSE | Upregulated |
| -0,325623808 | 0,339484457 | CGREF1 | FALSE | Downregulated |
| -0,326332288 | 0,339484457 | D2HGDH | FALSE | Downregulated |
| 0,297177705 | 0,339484457 | MIR600HG | FALSE | Upregulated |
| 0,377715645 | 0,340148572 | APPL1 | FALSE | Upregulated |

| | | | | |
|---|---|---|---|---|
| 0,664915895 | 0,340452349 | COL16A1 | FALSE | Upregulated |
| 0,349498427 | 0,340452349 | HIVEP2 | FALSE | Upregulated |
| -0,491895233 | 0,340452349 | PLAGL2 | FALSE | Downregulated |
| -1,473451539 | 0,340452349 | PRTG | FALSE | Downregulated |
| -0,217840062 | 0,340821297 | BRD1 | FALSE | Downregulated |
| 1,159302448 | 0,340932356 | CDH19 | FALSE | Upregulated |
| 0,398719904 | 0,340932356 | GPATCH4 | FALSE | Upregulated |
| 0,274835276 | 0,341101956 | CERS5 | FALSE | Upregulated |
| 0,350444243 | 0,341101956 | LOC101928061 | FALSE | Upregulated |
| 0,306421279 | 0,341101956 | NAB2 | FALSE | Upregulated |
| -0,182692421 | 0,341101956 | RERE | FALSE | Downregulated |
| 0,335309937 | 0,341297496 | LRP2BP | FALSE | Upregulated |
| 1,182096009 | 0,341298679 | SRD5A3-AS1 | FALSE | Upregulated |
| 0,586787244 | 0,34146055 | LOC102724157 | FALSE | Upregulated |
| -0,437390126 | 0,341952848 | CREB3L2 | FALSE | Downregulated |
| -0,459685304 | 0,342086941 | FAM162A | FALSE | Downregulated |
| -0,307567996 | 0,342413962 | CCDC125 | FALSE | Downregulated |
| 0,939147562 | 0,342413962 | TTYH2 | FALSE | Upregulated |
| 0,569289296 | 0,342555576 | GALNT7 | FALSE | Upregulated |
| 0,632349414 | 0,34275474 | EXO5 | FALSE | Upregulated |
| -0,595058368 | 0,34275474 | TMEM82 | FALSE | Downregulated |
| 0,246910335 | 0,342777091 | FAM120AOS | FALSE | Upregulated |
| -0,28491353 | 0,3428584 | LUC7L3 | FALSE | Downregulated |
| -0,195624601 | 0,3428584 | OBSL1 | FALSE | Downregulated |
| -0,544349849 | 0,3428584 | PEX11A | FALSE | Downregulated |
| 0,491868334 | 0,342879735 | LOC80154 | FALSE | Upregulated |
| -0,347096549 | 0,342885952 | RIN2 | FALSE | Downregulated |
| 0,286373849 | 0,342926776 | MARCH1 | FALSE | Upregulated |
| -0,551654214 | 0,343570988 | MVK | FALSE | Downregulated |
| -0,884039979 | 0,343632756 | TMEM125 | FALSE | Downregulated |
| 0,937718364 | 0,343722854 | HSPA12A | FALSE | Upregulated |
| -0,260524411 | 0,343722854 | TECR | FALSE | Downregulated |
| -0,248295046 | 0,343964628 | OIT3 | FALSE | Downregulated |
| 0,279780081 | 0,344139838 | ASF1A | FALSE | Upregulated |
| 0,617456322 | 0,344139838 | C8orf59 | FALSE | Upregulated |
| -0,559476782 | 0,344714722 | CMYA5 | FALSE | Downregulated |
| -1,085936993 | 0,344714722 | HERC2P3 | FALSE | Downregulated |
| -0,293783525 | 0,344714722 | IRGQ | FALSE | Downregulated |
| 0,484766137 | 0,344714722 | NT5C1B | FALSE | Upregulated |
| 0,830773732 | 0,345192074 | ZNF827 | FALSE | Upregulated |
| 0,351500319 | 0,345315965 | PDE4DIP | FALSE | Upregulated |
| 1,367503903 | 0,345410018 | CDH24 | FALSE | Upregulated |
| 0,804928105 | 0,345410018 | LOC100996419 | FALSE | Upregulated |
| -1,148687341 | 0,345472654 | SPRNP1 | FALSE | Downregulated |
| 0,600986491 | 0,345502158 | RP11-162J8.2 | FALSE | Upregulated |
| 1,265501854 | 0,345546202 | CD8B | FALSE | Upregulated |
| -0,334821338 | 0,345546202 | CFL2 | FALSE | Downregulated |
| -0,337861259 | 0,345628521 | ARMC6 | FALSE | Downregulated |
| 0,680556511 | 0,345868587 | SNAP25 | FALSE | Upregulated |
| 0,501825157 | 0,345934603 | CSGALNACT2 | FALSE | Upregulated |
| 0,369870837 | 0,345973824 | PPFIA2 | FALSE | Upregulated |

| | | | | |
|---|---|---|---|---|
| -0,391900789 | 0,346260276 | MRP63 | FALSE | Downregulated |
| 0,381419946 | 0,346260276 | PSMB10 | FALSE | Upregulated |
| -0,372586748 | 0,346260276 | TBC1D13 | FALSE | Downregulated |
| -0,374021246 | 0,346260276 | UHRF1BP1L | FALSE | Downregulated |
| 0,494231511 | 0,346758059 | ARHGEF37 | FALSE | Upregulated |
| 0,259590113 | 0,346758059 | C1orf86 | FALSE | Upregulated |
| 1,041472449 | 0,346758059 | LOC102724881 | FALSE | Upregulated |
| 0,376129918 | 0,347349197 | MCMDC2 | FALSE | Upregulated |
| -1,245516783 | 0,347521919 | FLT3LG | FALSE | Downregulated |
| 0,497867302 | 0,347626258 | CHRNG | FALSE | Upregulated |
| 0,744449366 | 0,347899296 | DGUOK | FALSE | Upregulated |
| 0,997466692 | 0,347899296 | MEPE | FALSE | Upregulated |
| 0,243725782 | 0,348079389 | LOC652276 | FALSE | Upregulated |
| -0,446337553 | 0,348079389 | PALM3 | FALSE | Downregulated |
| -0,942232046 | 0,348079389 | RP3-365O12.2 | FALSE | Downregulated |
| 1,181103103 | 0,348124778 | SGOL2 | FALSE | Upregulated |
| 0,419705025 | 0,348271798 | HPS6 | FALSE | Upregulated |
| -0,651846178 | 0,348271798 | METTL6 | FALSE | Downregulated |
| -1,172663172 | 0,348271798 | ZNF223 | FALSE | Downregulated |
| 0,435269451 | 0,348614872 | TXNL4A | FALSE | Upregulated |
| -0,292082198 | 0,348867968 | SNX33 | FALSE | Downregulated |
| 0,27930412 | 0,348897394 | CHD6 | FALSE | Upregulated |
| -1,238788132 | 0,348897394 | LSM5 | FALSE | Downregulated |
| -0,269508628 | 0,348897394 | SLC9A7 | FALSE | Downregulated |
| -0,545658148 | 0,348897394 | TMTC4 | FALSE | Downregulated |
| 0,494938238 | 0,349057675 | LOC101929401 | FALSE | Upregulated |
| 0,387242645 | 0,349063308 | TAF6 | FALSE | Upregulated |
| 0,397094645 | 0,349083243 | RP11-242D8.1 | FALSE | Upregulated |
| 0,596730588 | 0,349113932 | C1orf195 | FALSE | Upregulated |
| -0,882683518 | 0,349113932 | IGF2BP2 | FALSE | Downregulated |
| 0,713125452 | 0,349113932 | PCNA | FALSE | Upregulated |
| -0,401449878 | 0,349113932 | SYDE2 | FALSE | Downregulated |
| -0,405803074 | 0,349379773 | AADAC | FALSE | Downregulated |
| 0,908215587 | 0,349379773 | LOC101927952 | FALSE | Upregulated |
| 0,309925952 | 0,349742085 | OTUD4 | FALSE | Upregulated |
| 0,211760067 | 0,349742085 | PPP6R1 | FALSE | Upregulated |
| 0,418371759 | 0,349742085 | TRPV4 | FALSE | Upregulated |
| 0,81933998 | 0,349830372 | CCDC37 | FALSE | Upregulated |
| 0,355275953 | 0,349830372 | FGD1 | FALSE | Upregulated |
| -0,183768651 | 0,349830372 | HSPA9 | FALSE | Downregulated |
| 1,036171007 | 0,349830372 | LINC00967 | FALSE | Upregulated |
| 0,416674658 | 0,349830372 | VWA9 | FALSE | Upregulated |
| 0,589992334 | 0,350186526 | MFSD8 | FALSE | Upregulated |
| -1,178626327 | 0,351308359 | C4orf27 | FALSE | Downregulated |
| 0,290745445 | 0,351652974 | CRB2 | FALSE | Upregulated |
| 0,405460477 | 0,351652974 | MAPRE1 | FALSE | Upregulated |
| 0,558301697 | 0,351652974 | TMEM173 | FALSE | Upregulated |
| 0,5004824 | 0,351652974 | TNC | FALSE | Upregulated |
| 1,110392343 | 0,351652974 | ZNF165 | FALSE | Upregulated |
| -0,456298441 | 0,351915615 | PTDSS2 | FALSE | Downregulated |
| -0,72038417 | 0,35192997 | EPB41L4A-AS1 | FALSE | Downregulated |

| | | | | |
|---|---|---|---|---|
| 0,219236242 | 0,35192997 | PPFIA1 | FALSE | Upregulated |
| 0,250478698 | 0,35192997 | SERP1 | FALSE | Upregulated |
| 1,298394251 | 0,35192997 | TCEB3CL | FALSE | Upregulated |
| 0,210417608 | 0,352005118 | UBC | FALSE | Upregulated |
| 0,298154054 | 0,352084389 | EIF5B | FALSE | Upregulated |
| -0,474485931 | 0,35209252 | CYP39A1 | FALSE | Downregulated |
| -0,304501778 | 0,35209252 | HIVEP1 | FALSE | Downregulated |
| -0,326958051 | 0,35209252 | POMT1 | FALSE | Downregulated |
| 0,86524392 | 0,35262975 | CD209 | FALSE | Upregulated |
| 0,753811207 | 0,35262975 | CEP89 | FALSE | Upregulated |
| 0,452170492 | 0,35262975 | PPP1R35 | FALSE | Upregulated |
| 0,685891062 | 0,35262975 | RAC2 | FALSE | Upregulated |
| -0,244960365 | 0,352956766 | ECHDC2 | FALSE | Downregulated |
| -0,211044902 | 0,352956766 | ETFB | FALSE | Downregulated |
| 0,516931374 | 0,352956766 | FAM163B | FALSE | Upregulated |
| -0,284291881 | 0,352956766 | FAM53B | FALSE | Downregulated |
| 0,673780303 | 0,352956766 | LOC101927864 | FALSE | Upregulated |
| -0,519661088 | 0,352956766 | PCGF1 | FALSE | Downregulated |
| -0,286127956 | 0,353003585 | COL6A6 | FALSE | Downregulated |
| -0,312848767 | 0,353003585 | HRSP12 | FALSE | Downregulated |
| 0,602095023 | 0,353003585 | RAB42 | FALSE | Upregulated |
| -0,310666096 | 0,353431383 | PRKAB2 | FALSE | Downregulated |
| 1,052500169 | 0,353431383 | RP4-553F4.6 | FALSE | Upregulated |
| 1,080055769 | 0,353630358 | ADAMTSL4-AS1 | FALSE | Upregulated |
| 1,146676921 | 0,353630358 | LOC101927583 | FALSE | Upregulated |
| 1,133955643 | 0,353701072 | FRMPD1 | FALSE | Upregulated |
| -0,852589782 | 0,353701072 | SH2D1B | FALSE | Downregulated |
| -0,444239089 | 0,353716219 | VPS37A | FALSE | Downregulated |
| -0,391917996 | 0,353732614 | RAB33B | FALSE | Downregulated |
| -0,280080464 | 0,353766254 | KTN1 | FALSE | Downregulated |
| 1,215004979 | 0,353844208 | LRRC49 | FALSE | Upregulated |
| 0,493077449 | 0,353871226 | LOC101059949 | FALSE | Upregulated |
| 0,220233926 | 0,353911629 | CDK5RAP2 | FALSE | Upregulated |
| 0,505135025 | 0,353911629 | THY1 | FALSE | Upregulated |
| 0,358745659 | 0,3539594 | LOC102724383 | FALSE | Upregulated |
| 0,737744633 | 0,354415214 | C17orf58 | FALSE | Upregulated |
| 0,84978613 | 0,354446477 | GMNC | FALSE | Upregulated |
| 0,54093512 | 0,354446477 | PIANP | FALSE | Upregulated |
| 0,30154503 | 0,354446477 | SPTY2D1-AS1 | FALSE | Upregulated |
| 0,235757358 | 0,354623326 | ATG2A | FALSE | Upregulated |
| 0,267807054 | 0,35464718 | MAP3K3 | FALSE | Upregulated |
| 0,403714019 | 0,354691813 | SNRPA | FALSE | Upregulated |
| 0,442321055 | 0,354852599 | LOC101928830 | FALSE | Upregulated |
| 1,139506357 | 0,355057478 | MKX | FALSE | Upregulated |
| 0,593188417 | 0,355057478 | STMN3 | FALSE | Upregulated |
| -1,004412411 | 0,355326424 | ASPA | FALSE | Downregulated |
| 0,550469539 | 0,355597615 | POP4 | FALSE | Upregulated |
| 0,35505732 | 0,355664496 | CERS4 | FALSE | Upregulated |
| -0,420169554 | 0,355664496 | LEAP2 | FALSE | Downregulated |
| -0,432087833 | 0,355664496 | LRRK2 | FALSE | Downregulated |
| 1,158704647 | 0,355757313 | SLC24A4 | FALSE | Upregulated |

| | | | | |
|---|---|---|---|---|
| 0,767663005 | 0,355757313 | VLDLR | FALSE | Upregulated |
| 0,69088237 | 0,355965887 | BRIX1 | FALSE | Upregulated |
| 0,299005391 | 0,355965887 | FAM178A | FALSE | Upregulated |
| 0,680562625 | 0,355965887 | LAMA3 | FALSE | Upregulated |
| 0,308422511 | 0,356414351 | NBN | FALSE | Upregulated |
| 1,204813152 | 0,356414351 | SPIC | FALSE | Upregulated |
| -0,28959005 | 0,356600472 | SLC25A23 | FALSE | Downregulated |
| -0,350779371 | 0,356630221 | LYNX1 | FALSE | Downregulated |
| 0,818327393 | 0,35687541 | DTD1 | FALSE | Upregulated |
| 0,360161697 | 0,356929089 | PRKRIR | FALSE | Upregulated |
| 0,680009377 | 0,356929089 | ST6GALNAC4 | FALSE | Upregulated |
| -1,182716798 | 0,357066774 | LOC100996693 | FALSE | Downregulated |
| 0,577434563 | 0,357100577 | POU2F2 | FALSE | Upregulated |
| 0,306761651 | 0,357131054 | GDI1 | FALSE | Upregulated |
| -0,446910834 | 0,357609229 | POLR2F | FALSE | Downregulated |
| -0,501759027 | 0,357694295 | KIAA1683 | FALSE | Downregulated |
| 0,530221153 | 0,357814658 | FAM222A | FALSE | Upregulated |
| -1,176948266 | 0,358063061 | AC078941.1 | FALSE | Downregulated |
| -0,389895028 | 0,358063061 | DHRS7 | FALSE | Downregulated |
| -0,299089563 | 0,358063061 | EYA3 | FALSE | Downregulated |
| 1,274341489 | 0,358063061 | HIST2H2AA4 | FALSE | Upregulated |
| 1,05810884 | 0,358063061 | LOC102724233 | FALSE | Upregulated |
| 0,487009539 | 0,358063061 | MRPL10 | FALSE | Upregulated |
| 0,362550159 | 0,358110444 | CRIP2 | FALSE | Upregulated |
| -0,394217715 | 0,358110444 | LOC595101 | FALSE | Downregulated |
| 0,24774437 | 0,358189334 | ACTR2 | FALSE | Upregulated |
| -0,426397491 | 0,358189334 | LDLRAP1 | FALSE | Downregulated |
| 0,736202955 | 0,358189334 | PIGU | FALSE | Upregulated |
| 0,372106563 | 0,358189334 | RP5-1033H22.2 | FALSE | Upregulated |
| -0,561494516 | 0,358252976 | MRPL52 | FALSE | Downregulated |
| -0,813883474 | 0,358272042 | ZNF691 | FALSE | Downregulated |
| 0,440447883 | 0,358376869 | FAM117B | FALSE | Upregulated |
| 0,747444027 | 0,358414323 | MAP4K1 | FALSE | Upregulated |
| -0,744107493 | 0,358519639 | DACT1 | FALSE | Downregulated |
| -0,533244277 | 0,358747919 | SEMA6D | FALSE | Downregulated |
| 0,25261589 | 0,358889316 | CP | FALSE | Upregulated |
| 0,33473753 | 0,358889316 | NUTM2A | FALSE | Upregulated |
| 0,278324026 | 0,359012459 | CYLD | FALSE | Upregulated |
| 0,586831837 | 0,359012459 | RGS19 | FALSE | Upregulated |
| 0,336566911 | 0,359688207 | CXCR6 | FALSE | Upregulated |
| -0,230376181 | 0,359740736 | COMT | FALSE | Downregulated |
| 0,403589142 | 0,359834564 | SOAT1 | FALSE | Upregulated |
| 0,53055921 | 0,360007738 | LOC102724490 | FALSE | Upregulated |
| -0,587431685 | 0,360211608 | C2orf42 | FALSE | Downregulated |
| -1,351295379 | 0,360211608 | LOC441528 | FALSE | Downregulated |
| 0,199385774 | 0,360211608 | MYO1C | FALSE | Upregulated |
| -0,527206593 | 0,360211608 | PLXDC1 | FALSE | Downregulated |
| -0,225532374 | 0,360211608 | R3HDM2 | FALSE | Downregulated |
| 0,588786547 | 0,360211608 | TRIL | FALSE | Upregulated |
| 0,760101304 | 0,360211608 | ZNF2 | FALSE | Upregulated |
| 0,342543852 | 0,360339011 | IFFO1 | FALSE | Upregulated |

| | | | | |
|---|---|---|---|---|
| 0,262749871 | 0,360339011 | PLD3 | FALSE | Upregulated |
| 0,526855167 | 0,360530464 | CFDP1 | FALSE | Upregulated |
| 0,408509597 | 0,360530464 | FAM132B | FALSE | Upregulated |
| -0,608493808 | 0,360530464 | NPR3 | FALSE | Downregulated |
| -0,827629464 | 0,360530464 | SCT | FALSE | Downregulated |
| 0,793950802 | 0,360556134 | LOC101927521 | FALSE | Upregulated |
| -0,328499279 | 0,360630496 | COG3 | FALSE | Downregulated |
| -0,508985767 | 0,360630496 | DDX55 | FALSE | Downregulated |
| -0,230338114 | 0,360630496 | TCF20 | FALSE | Downregulated |
| -0,436964045 | 0,360632705 | HEATR5A | FALSE | Downregulated |
| 0,353336676 | 0,360742184 | CLPTM1L | FALSE | Upregulated |
| 0,317938537 | 0,360742184 | SRSF10 | FALSE | Upregulated |
| 1,078118301 | 0,360797641 | POU2AF1 | FALSE | Upregulated |
| -0,679496295 | 0,361230147 | BTNL9 | FALSE | Downregulated |
| -0,274896174 | 0,361421661 | C17orf70 | FALSE | Downregulated |
| 0,376514991 | 0,361441155 | FOXH1 | FALSE | Upregulated |
| 0,231421637 | 0,361484548 | RHOQ | FALSE | Upregulated |
| 0,392366192 | 0,361836085 | RBMXL1 | FALSE | Upregulated |
| 0,820528493 | 0,361906777 | CAMK4 | FALSE | Upregulated |
| -0,331714205 | 0,361906777 | DDOST | FALSE | Downregulated |
| -0,223989083 | 0,361906777 | NFIX | FALSE | Downregulated |
| -0,398590148 | 0,361906777 | UBR1 | FALSE | Downregulated |
| 0,326729446 | 0,361933951 | ATAD5 | FALSE | Upregulated |
| -0,778927422 | 0,361933951 | SNORD94 | FALSE | Downregulated |
| -0,560331576 | 0,362173719 | FAM180A | FALSE | Downregulated |
| -0,712572307 | 0,362221726 | FAM124A | FALSE | Downregulated |
| 1,039347994 | 0,362221726 | KLRD1 | FALSE | Upregulated |
| 1,146596887 | 0,362221726 | LOC101927536 | FALSE | Upregulated |
| 0,717304274 | 0,362500284 | ARHGEF38 | FALSE | Upregulated |
| 0,35785528 | 0,362500284 | CLIC1 | FALSE | Upregulated |
| 0,76478432 | 0,362500284 | CRYBB2P1 | FALSE | Upregulated |
| 0,555349364 | 0,362500284 | LOC101929354 | FALSE | Upregulated |
| 0,849797888 | 0,362500284 | RP11-856M7.1 | FALSE | Upregulated |
| 0,321574956 | 0,362500284 | VPS52 | FALSE | Upregulated |
| 1,265344246 | 0,362536076 | MTRNR2L6 | FALSE | Upregulated |
| 0,286643596 | 0,362733521 | ANKRD61 | FALSE | Upregulated |
| -0,569735067 | 0,362733521 | ATPAF2 | FALSE | Downregulated |
| 0,286400545 | 0,362733521 | CHAD | FALSE | Upregulated |
| -0,779911832 | 0,362733521 | FAM172A | FALSE | Downregulated |
| 0,403702371 | 0,362733521 | LOC101927898 | FALSE | Upregulated |
| 0,839068667 | 0,362733521 | LOC101928352 | FALSE | Upregulated |
| -1,285039587 | 0,362733521 | NPFF | FALSE | Downregulated |
| 0,235190774 | 0,362733521 | PSMD3 | FALSE | Upregulated |
| -0,886531723 | 0,362733521 | ZNF600 | FALSE | Downregulated |
| 0,402073381 | 0,36276595 | CCNH | FALSE | Upregulated |
| -0,421418583 | 0,36299347 | CDC5L | FALSE | Downregulated |
| 0,296033126 | 0,36299347 | LSMEM2 | FALSE | Upregulated |
| 0,798134331 | 0,36299347 | ZNF878 | FALSE | Upregulated |
| 0,706309019 | 0,362999043 | ROM1 | FALSE | Upregulated |
| -0,160502444 | 0,363125 | CSDE1 | FALSE | Downregulated |
| 0,479838246 | 0,363203952 | ARHGEF4 | FALSE | Upregulated |

| | | | | |
|---|---|---|---|---|
| 0,891268958 | 0,363203952 | LILRA4 | FALSE | Upregulated |
| 0,392850189 | 0,363203952 | LOC101927989 | FALSE | Upregulated |
| 0,51468537 | 0,363203952 | OAS1 | FALSE | Upregulated |
| 0,322014692 | 0,363382229 | MTRNR2L3 | FALSE | Upregulated |
| -0,253282586 | 0,363848198 | ASPH | FALSE | Downregulated |
| 0,309716394 | 0,36418743 | ABCB11 | FALSE | Upregulated |
| -0,611028851 | 0,36418743 | CCNB1IP1 | FALSE | Downregulated |
| -0,289394614 | 0,36418743 | MAU2 | FALSE | Downregulated |
| -1,197538221 | 0,36418743 | YBX2 | FALSE | Downregulated |
| -0,385368298 | 0,364323688 | HPCAL1 | FALSE | Downregulated |
| 1,328250717 | 0,364332409 | NPM3 | FALSE | Upregulated |
| -0,351042945 | 0,364332409 | TMEM220 | FALSE | Downregulated |
| -0,846294825 | 0,364357106 | ACVR1C | FALSE | Downregulated |
| -0,272210036 | 0,364357106 | PCCB | FALSE | Downregulated |
| 1,243109216 | 0,364357106 | TLR5 | FALSE | Upregulated |
| -0,655625089 | 0,364519736 | P2RY2 | FALSE | Downregulated |
| -0,366095756 | 0,364712598 | SLC44A1 | FALSE | Downregulated |
| 0,213799276 | 0,364969075 | VPRBP | FALSE | Upregulated |
| -0,216867471 | 0,365187648 | PGRMC2 | FALSE | Downregulated |
| -0,437593128 | 0,365187648 | S100A16 | FALSE | Downregulated |
| -0,32780065 | 0,365249122 | GPT2 | FALSE | Downregulated |
| -1,389289612 | 0,365249122 | LOC101059936 | FALSE | Downregulated |
| 0,183147246 | 0,365249122 | SZT2 | FALSE | Upregulated |
| 1,064376527 | 0,365400217 | ZNF169 | FALSE | Upregulated |
| 0,84538457 | 0,366027849 | KCNC1 | FALSE | Upregulated |
| 1,277065859 | 0,36614075 | LOC100505851 | FALSE | Upregulated |
| -0,423010195 | 0,366308657 | SLC28A1 | FALSE | Downregulated |
| -0,265609652 | 0,366592898 | NDRG2 | FALSE | Downregulated |
| 0,230720271 | 0,366659759 | USP8 | FALSE | Upregulated |
| -0,396185212 | 0,36675153 | AC011343.1 | FALSE | Downregulated |
| 0,268580668 | 0,36675153 | LOC100131496 | FALSE | Upregulated |
| -0,503472455 | 0,36675153 | MARCH5 | FALSE | Downregulated |
| -0,312260902 | 0,366958175 | C4BPB | FALSE | Downregulated |
| -0,46643551 | 0,366974519 | CXCL2 | FALSE | Downregulated |
| -0,424089557 | 0,366974519 | MYNN | FALSE | Downregulated |
| 0,370393363 | 0,366974519 | PECAM1 | FALSE | Upregulated |
| 0,621624015 | 0,366974519 | RP11-692N5.1 | FALSE | Upregulated |
| -1,116674375 | 0,367003153 | LINC01181 | FALSE | Downregulated |
| -0,682462546 | 0,367077199 | LRRC37BP1 | FALSE | Downregulated |
| 0,851886491 | 0,367313846 | WNT16 | FALSE | Upregulated |
| -1,084873101 | 0,367427855 | LOC100506518 | FALSE | Downregulated |
| 0,426376397 | 0,367427855 | TAC4 | FALSE | Upregulated |
| 0,524663386 | 0,367427855 | TTLL9 | FALSE | Upregulated |
| 0,90158088 | 0,367811386 | TMEM229B | FALSE | Upregulated |
| 0,225299088 | 0,367825657 | CCAR2 | FALSE | Upregulated |
| 0,973909156 | 0,367828379 | GSTO2 | FALSE | Upregulated |
| 0,208729118 | 0,367828379 | MAZ | FALSE | Upregulated |
| -0,216837913 | 0,367871529 | RPS23 | FALSE | Downregulated |
| -1,351343404 | 0,36812966 | LOC101928936 | FALSE | Downregulated |
| -0,974611115 | 0,368164548 | GLB1L2 | FALSE | Downregulated |
| 0,257678286 | 0,368177373 | TBC1D10B | FALSE | Upregulated |

| | | | | |
|---|---|---|---|---|
| -0,676554187 | 0,36828787 | TSTD1 | FALSE | Downregulated |
| -0,299823155 | 0,368428565 | TFPI | FALSE | Downregulated |
| -0,365516504 | 0,368529661 | CECR5 | FALSE | Downregulated |
| 0,480065543 | 0,368529661 | TAS1R3 | FALSE | Upregulated |
| 0,317447108 | 0,368551561 | CCDC71L | FALSE | Upregulated |
| 1,04455742 | 0,368562545 | RXFP4 | FALSE | Upregulated |
| -0,348771812 | 0,368570569 | NDUFC2 | FALSE | Downregulated |
| 0,328038437 | 0,368812173 | CIRBP-AS1 | FALSE | Upregulated |
| 0,359587036 | 0,368812173 | DERL3 | FALSE | Upregulated |
| 1,025953736 | 0,368812173 | EMP1 | FALSE | Upregulated |
| 0,210638109 | 0,368812173 | ITGB1 | FALSE | Upregulated |
| -0,586224408 | 0,368812173 | SLC17A3 | FALSE | Downregulated |
| -0,771751219 | 0,368812173 | SNHG5 | FALSE | Downregulated |
| -0,320182134 | 0,368812173 | TCEA1 | FALSE | Downregulated |
| 0,476978138 | 0,368812173 | TRIAP1 | FALSE | Upregulated |
| 0,774103787 | 0,368812173 | ZSCAN9 | FALSE | Upregulated |
| 1,380868832 | 0,368881228 | JAKMIP2-AS1 | FALSE | Upregulated |
| 0,236082601 | 0,368881228 | PPP2R5A | FALSE | Upregulated |
| -0,199973622 | 0,368881228 | ZNF516 | FALSE | Downregulated |
| 0,601881924 | 0,368950186 | CST7 | FALSE | Upregulated |
| 0,659409919 | 0,368950186 | IFI27 | FALSE | Upregulated |
| 0,297617277 | 0,369681566 | LOC100130744 | FALSE | Upregulated |
| -0,281321325 | 0,369781509 | PXMP2 | FALSE | Downregulated |
| 0,293122519 | 0,370001514 | SPDYE2 | FALSE | Upregulated |
| -0,672352224 | 0,370114013 | AOC1 | FALSE | Downregulated |
| -0,880369511 | 0,370376981 | DBF4B | FALSE | Downregulated |
| 0,422464234 | 0,370680296 | PREP | FALSE | Upregulated |
| 0,484443423 | 0,370680296 | TMEM168 | FALSE | Upregulated |
| 0,471277984 | 0,37068602 | CASP3 | FALSE | Upregulated |
| -0,337996331 | 0,37068602 | DPP7 | FALSE | Downregulated |
| 0,769239911 | 0,37068602 | RLBP1 | FALSE | Upregulated |
| 0,292730026 | 0,37068602 | TFE3 | FALSE | Upregulated |
| -0,632641296 | 0,370978316 | MTRF1 | FALSE | Downregulated |
| 0,503166114 | 0,371263606 | SMCO4 | FALSE | Upregulated |
| -1,071190114 | 0,371374726 | PET117 | FALSE | Downregulated |
| -0,405225428 | 0,371765155 | ING5 | FALSE | Downregulated |
| -1,064172302 | 0,372019869 | TTC3P1 | FALSE | Downregulated |
| 0,367637926 | 0,372149725 | PPP1R15A | FALSE | Upregulated |
| -0,257698315 | 0,372243793 | SIRT5 | FALSE | Downregulated |
| -0,411221796 | 0,372340675 | RWDD2B | FALSE | Downregulated |
| 0,85607926 | 0,372451998 | LOC101928019 | FALSE | Upregulated |
| 0,922808831 | 0,372451998 | TNFRSF11B | FALSE | Upregulated |
| 0,572072978 | 0,372581796 | LOC101928482 | FALSE | Upregulated |
| -0,650417097 | 0,372687206 | LOC101929343 | FALSE | Downregulated |
| -1,080569057 | 0,372878552 | NEK10 | FALSE | Downregulated |
| -0,220308946 | 0,372878552 | TUFM | FALSE | Downregulated |
| -0,301211147 | 0,3729549 | SLC25A25 | FALSE | Downregulated |
| 0,326364195 | 0,3729549 | ZSCAN16 | FALSE | Upregulated |
| -0,363137321 | 0,37305494 | SNAPC5 | FALSE | Downregulated |
| 0,361842451 | 0,373128491 | CREB3 | FALSE | Upregulated |
| 0,650845568 | 0,373128491 | LOC101928263 | FALSE | Upregulated |

| | | | | |
|---|---|---|---|---|
| 0,958430115 | 0,373128491 | LOC102724147 | FALSE | Upregulated |
| 0,288517259 | 0,373155162 | GPR22 | FALSE | Upregulated |
| 0,377457397 | 0,373522419 | NOA1 | FALSE | Upregulated |
| 0,36774178 | 0,373640872 | P2RX4 | FALSE | Upregulated |
| 0,392206583 | 0,373640872 | PRKCH | FALSE | Upregulated |
| 0,367676653 | 0,373678194 | ASB9P1 | FALSE | Upregulated |
| 0,586364918 | 0,373678194 | DNAJC2 | FALSE | Upregulated |
| 1,107899346 | 0,373678194 | EFCAB12 | FALSE | Upregulated |
| -0,304725723 | 0,373678194 | PSMD8 | FALSE | Downregulated |
| -1,103213818 | 0,373678194 | RNF122 | FALSE | Downregulated |
| 0,263687335 | 0,373697109 | CRBN | FALSE | Upregulated |
| 0,81785848 | 0,373697109 | MAGOH | FALSE | Upregulated |
| 0,461897299 | 0,373702337 | PPIL4 | FALSE | Upregulated |
| -0,383841418 | 0,374622943 | CD2AP | FALSE | Downregulated |
| -0,446130635 | 0,374675089 | LOC101929633 | FALSE | Downregulated |
| 0,252274217 | 0,374696038 | RABL6 | FALSE | Upregulated |
| 1,008284847 | 0,374752705 | BLVRA | FALSE | Upregulated |
| -0,358217488 | 0,374893948 | FBXO28 | FALSE | Downregulated |
| 0,429627444 | 0,374956497 | LOC166994 | FALSE | Upregulated |
| -0,239477859 | 0,374956497 | MAN2A1 | FALSE | Downregulated |
| -0,338255117 | 0,374956497 | PPIF | FALSE | Downregulated |
| 0,687071677 | 0,374956497 | RAB40A | FALSE | Upregulated |
| -0,374766669 | 0,374956497 | UFC1 | FALSE | Downregulated |
| -0,273798825 | 0,37505129 | RNASE4 | FALSE | Downregulated |
| 0,301884757 | 0,375187023 | ALAS1 | FALSE | Upregulated |
| 0,227557302 | 0,37554921 | TP53I13 | FALSE | Upregulated |
| -0,336263601 | 0,375566101 | ATF7IP2 | FALSE | Downregulated |
| 0,765986358 | 0,375566101 | SLC36A4 | FALSE | Upregulated |
| -0,23763589 | 0,375787638 | KAT2B | FALSE | Downregulated |
| 0,231117601 | 0,375787638 | SMIM5 | FALSE | Upregulated |
| 0,416260848 | 0,375858699 | NCK2 | FALSE | Upregulated |
| 0,380854487 | 0,37589207 | LOC102723851 | FALSE | Upregulated |
| 0,307136172 | 0,37600903 | ZNF646 | FALSE | Upregulated |
| -0,402555761 | 0,376044989 | HOOK2 | FALSE | Downregulated |
| -0,459057971 | 0,376414058 | PCMT1 | FALSE | Downregulated |
| 0,326131526 | 0,376576383 | VPS36 | FALSE | Upregulated |
| -0,482613157 | 0,376775888 | LAD1 | FALSE | Downregulated |
| -0,784585222 | 0,37743041 | WNT5B | FALSE | Downregulated |
| 0,890724929 | 0,377479307 | CHRND | FALSE | Upregulated |
| 0,379155847 | 0,377479307 | DENND3 | FALSE | Upregulated |
| -0,3575721 | 0,377479307 | EIF3K | FALSE | Downregulated |
| 0,242965719 | 0,377479307 | GPR18 | FALSE | Upregulated |
| 0,298597479 | 0,377479307 | MNT | FALSE | Upregulated |
| 0,150936918 | 0,377479307 | POLDIP2 | FALSE | Upregulated |
| 0,632767119 | 0,377479307 | SEMA3E | FALSE | Upregulated |
| -0,465798308 | 0,37753504 | HOOK1 | FALSE | Downregulated |
| -1,385533134 | 0,37753504 | SLC6A17 | FALSE | Downregulated |
| 0,267988536 | 0,377615229 | OTUB2 | FALSE | Upregulated |
| 0,782201158 | 0,37788502 | LINC00957 | FALSE | Upregulated |
| -0,299507982 | 0,378216535 | MAGED1 | FALSE | Downregulated |
| 0,423074971 | 0,37841113 | AC004019.13 | FALSE | Upregulated |

| | | | | |
|---|---|---|---|---|
| 0,404269968 | 0,37841113 | LOC101926967 | FALSE | Upregulated |
| 0,250287186 | 0,37841113 | PSMA7 | FALSE | Upregulated |
| -0,516530871 | 0,37841113 | WDR83OS | FALSE | Downregulated |
| 1,540247788 | 0,378464269 | LOC101927745 | FALSE | Upregulated |
| 0,379728574 | 0,378597178 | TXNRD1 | FALSE | Upregulated |
| -0,415427764 | 0,378793802 | NLRX1 | FALSE | Downregulated |
| -0,309745657 | 0,378840115 | CYB561A3 | FALSE | Downregulated |
| -0,360643464 | 0,378840115 | LINC00538 | FALSE | Downregulated |
| 0,843758166 | 0,378840115 | TMLHE-AS1 | FALSE | Upregulated |
| 0,729519879 | 0,378925052 | CDK7 | FALSE | Upregulated |
| -0,646022602 | 0,378925052 | MVB12A | FALSE | Downregulated |
| -1,205940417 | 0,378925052 | ST6GALNAC3 | FALSE | Downregulated |
| -0,679925564 | 0,378925052 | TMCC3 | FALSE | Downregulated |
| 0,243723441 | 0,379068129 | PPIL2 | FALSE | Upregulated |
| 0,445862332 | 0,379072086 | TTC1 | FALSE | Upregulated |
| 0,753517944 | 0,379140672 | CD180 | FALSE | Upregulated |
| -0,622192913 | 0,379140672 | DDX52 | FALSE | Downregulated |
| -0,208285237 | 0,379140672 | ENG | FALSE | Downregulated |
| 1,11167404 | 0,379140672 | EVPL | FALSE | Upregulated |
| 1,107313235 | 0,379140672 | KPNA7 | FALSE | Upregulated |
| -0,720712703 | 0,379140672 | LNP1 | FALSE | Downregulated |
| -0,300711232 | 0,379140672 | MATN2 | FALSE | Downregulated |
| -0,361913581 | 0,379140672 | NOL9 | FALSE | Downregulated |
| 0,92839035 | 0,379140672 | OLFML2B | FALSE | Upregulated |
| -0,241882495 | 0,379140672 | UBE4A | FALSE | Downregulated |
| -0,232639507 | 0,379140672 | ZFAND3 | FALSE | Downregulated |
| 0,345450656 | 0,379186607 | NAP1L5 | FALSE | Upregulated |
| -0,254480266 | 0,379260504 | CARKD | FALSE | Downregulated |
| -0,426943938 | 0,379260504 | TTLL5 | FALSE | Downregulated |
| 0,223811175 | 0,37939956 | DSP | FALSE | Upregulated |
| 0,284882915 | 0,37939956 | PA2G4 | FALSE | Upregulated |
| -0,695673462 | 0,37939956 | POLI | FALSE | Downregulated |
| 0,328288785 | 0,37939956 | VOPP1 | FALSE | Upregulated |
| -0,442257298 | 0,379400511 | C14orf166 | FALSE | Downregulated |
| -0,245313011 | 0,379400511 | ONECUT1 | FALSE | Downregulated |
| 3,617282335 | 0,379535674 | RNF103-CHMP3 | FALSE | Upregulated |
| 0,363246018 | 0,379820686 | USP2 | FALSE | Upregulated |
| -0,238522566 | 0,379891218 | KANSL3 | FALSE | Downregulated |
| -0,948386014 | 0,379934865 | ARHGAP5-AS1 | FALSE | Downregulated |
| 0,338573871 | 0,379934865 | TRIP6 | FALSE | Upregulated |
| -0,493167708 | 0,380219812 | HLX-AS1 | FALSE | Downregulated |
| 0,955412004 | 0,380219812 | LOC101928409 | FALSE | Upregulated |
| -1,005940256 | 0,380219812 | LOC101928961 | FALSE | Downregulated |
| 0,435818603 | 0,380219812 | RAN | FALSE | Upregulated |
| 0,362962703 | 0,380219812 | UBA6 | FALSE | Upregulated |
| -0,392466118 | 0,380268887 | ANKRD36C | FALSE | Downregulated |
| -0,446445649 | 0,380301584 | ABCB7 | FALSE | Downregulated |
| -0,300590009 | 0,380301584 | FAM3A | FALSE | Downregulated |
| 0,668226768 | 0,380301584 | HID1 | FALSE | Upregulated |
| 1,107853108 | 0,380301584 | LINC00691 | FALSE | Upregulated |
| 0,440373018 | 0,380301584 | LINC00963 | FALSE | Upregulated |

| | | | | |
|---|---|---|---|---|
| 0,270756175 | 0,380301584 | SMPD1 | FALSE | Upregulated |
| -0,306275479 | 0,380301584 | SRD5A1 | FALSE | Downregulated |
| 0,821402507 | 0,380301584 | THBS4 | FALSE | Upregulated |
| -0,792501802 | 0,380376498 | LOC553103 | FALSE | Downregulated |
| 0,49947548 | 0,380376498 | NEFH | FALSE | Upregulated |
| -0,365935772 | 0,380376498 | SLC25A39 | FALSE | Downregulated |
| 0,522509659 | 0,380743592 | LOC202181 | FALSE | Upregulated |
| 0,381002743 | 0,380743592 | RP11-809M12.1 | FALSE | Upregulated |
| -0,405431463 | 0,38075118 | RHOT1 | FALSE | Downregulated |
| 0,379124803 | 0,380763797 | NBEAL2 | FALSE | Upregulated |
| 0,424128677 | 0,381275837 | PPA1 | FALSE | Upregulated |
| -0,49620558 | 0,381303187 | L2HGDH | FALSE | Downregulated |
| -0,265568935 | 0,381367067 | TCF12 | FALSE | Downregulated |
| 0,288146201 | 0,381438487 | SUMO3 | FALSE | Upregulated |
| -0,383565895 | 0,381555191 | KYNU | FALSE | Downregulated |
| 1,082220063 | 0,381596865 | CDC20 | FALSE | Upregulated |
| 0,304205767 | 0,38167519 | PPP1R12A | FALSE | Upregulated |
| -0,257952523 | 0,38188894 | RPSA | FALSE | Downregulated |
| 1,232755532 | 0,381934408 | FAM225A | FALSE | Upregulated |
| -0,374394999 | 0,382158635 | SH3RF1 | FALSE | Downregulated |
| -0,357221356 | 0,382222721 | ZNF518A | FALSE | Downregulated |
| 0,245025547 | 0,382237313 | CFB | FALSE | Upregulated |
| -0,628212103 | 0,382237313 | DPEP2 | FALSE | Downregulated |
| 0,673398319 | 0,383078718 | LOC101928215 | FALSE | Upregulated |
| 0,883244057 | 0,383524797 | LOC102724578 | FALSE | Upregulated |
| -0,417968285 | 0,383524797 | MAMDC4 | FALSE | Downregulated |
| 0,587689502 | 0,383647188 | SPRYD7 | FALSE | Upregulated |
| -0,324141645 | 0,383751358 | DDX41 | FALSE | Downregulated |
| -0,806005696 | 0,383751358 | MEIOB | FALSE | Downregulated |
| -0,248589809 | 0,383751358 | TST | FALSE | Downregulated |
| -0,330589505 | 0,3840781 | SEPT10 | FALSE | Downregulated |
| -0,343553358 | 0,384122118 | FAM205A | FALSE | Downregulated |
| 0,260239314 | 0,384122118 | HIST1H1C | FALSE | Upregulated |
| 0,280430152 | 0,384122118 | HSPA8 | FALSE | Upregulated |
| 0,310788644 | 0,384122118 | MIER3 | FALSE | Upregulated |
| -0,258371956 | 0,384122118 | NUPR1 | FALSE | Downregulated |
| 0,306611564 | 0,384122118 | RP11-178C3.2 | FALSE | Upregulated |
| 0,952611099 | 0,384122118 | SLC29A4 | FALSE | Upregulated |
| 0,786482229 | 0,38413569 | CASC14 | FALSE | Upregulated |
| -0,438119863 | 0,38413569 | FNBP1L | FALSE | Downregulated |
| 1,095519181 | 0,38413569 | LOC100131347 | FALSE | Upregulated |
| 0,468515094 | 0,384208676 | CCDC115 | FALSE | Upregulated |
| 0,460699641 | 0,384208676 | LLGL1 | FALSE | Upregulated |
| -0,422356426 | 0,384268758 | C19orf70 | FALSE | Downregulated |
| 0,684439755 | 0,384268758 | CTU1 | FALSE | Upregulated |
| -0,738325884 | 0,384568564 | BMP2 | FALSE | Downregulated |
| -1,249845531 | 0,384757536 | AGR2 | FALSE | Downregulated |
| -0,48825628 | 0,384757536 | SPNS3 | FALSE | Downregulated |
| -0,649103447 | 0,385115632 | LINC00883 | FALSE | Downregulated |
| 0,350469583 | 0,385192264 | ALDOA | FALSE | Upregulated |
| 0,569169963 | 0,385253916 | FAM102B | FALSE | Upregulated |

| | | | | |
|---|---|---|---|---|
| 0,423136573 | 0,385265525 | EIF2S1 | FALSE | Upregulated |
| 0,348930033 | 0,385265525 | LOC102723806 | FALSE | Upregulated |
| -0,247443282 | 0,385265525 | RIF1 | FALSE | Downregulated |
| 0,731543676 | 0,38540539 | C2orf82 | FALSE | Upregulated |
| -0,346080777 | 0,385423915 | CCDC130 | FALSE | Downregulated |
| 0,356539567 | 0,385647221 | KCNJ10 | FALSE | Upregulated |
| 0,242715399 | 0,385750779 | PODXL | FALSE | Upregulated |
| -0,570193731 | 0,385845861 | ATP5L | FALSE | Downregulated |
| 0,512690637 | 0,385887508 | LINC00612 | FALSE | Upregulated |
| 0,332012487 | 0,385983299 | LOC101929396 | FALSE | Upregulated |
| 0,418612994 | 0,386002244 | TNIK | FALSE | Upregulated |
| 0,608977793 | 0,386057339 | MAPK10 | FALSE | Upregulated |
| -0,33943563 | 0,386112893 | ANXA7 | FALSE | Downregulated |
| 1,332901198 | 0,386229971 | BANK1 | FALSE | Upregulated |
| -0,783744015 | 0,386229971 | MSRB3 | FALSE | Downregulated |
| -0,203615421 | 0,386255968 | ADCK3 | FALSE | Downregulated |
| -0,372568962 | 0,38626873 | CRELD1 | FALSE | Downregulated |
| 1,146122782 | 0,38626873 | IL15 | FALSE | Upregulated |
| -0,456491217 | 0,38626873 | SLC6A13 | FALSE | Downregulated |
| -0,285707408 | 0,386472704 | RPRD2 | FALSE | Downregulated |
| -1,171611735 | 0,386526776 | ZNF572 | FALSE | Downregulated |
| -2,482954121 | 0,386997106 | RP4-669L17.10 | FALSE | Downregulated |
| 0,410704819 | 0,387193734 | SPAG5-AS1 | FALSE | Upregulated |
| 0,504145667 | 0,387213663 | APBB1 | FALSE | Upregulated |
| 0,25716751 | 0,387745678 | LOC100506634 | FALSE | Upregulated |
| 1,094877483 | 0,387779924 | SLC51B | FALSE | Upregulated |
| 0,322415982 | 0,388029059 | MAP3K1 | FALSE | Upregulated |
| -0,384299994 | 0,388029059 | NLGN2 | FALSE | Downregulated |
| 0,569495272 | 0,388029059 | NT5DC2 | FALSE | Upregulated |
| 0,487948301 | 0,388029059 | PPP2R5B | FALSE | Upregulated |
| 0,715239412 | 0,388029059 | SBDSP1 | FALSE | Upregulated |
| 0,940805852 | 0,388417479 | BLK | FALSE | Upregulated |
| 0,422616235 | 0,388417479 | C10orf118 | FALSE | Upregulated |
| -0,170559133 | 0,388417479 | SCARB2 | FALSE | Downregulated |
| 1,29271826 | 0,388417479 | WISP2 | FALSE | Upregulated |
| 0,420080075 | 0,389009742 | ANKRD27 | FALSE | Upregulated |
| 0,229985352 | 0,389009742 | RAB11FIP3 | FALSE | Upregulated |
| 0,403123429 | 0,389009742 | SLC5A6 | FALSE | Upregulated |
| 0,248660397 | 0,389009742 | ZNF750 | FALSE | Upregulated |
| -0,598155927 | 0,389081256 | C8orf46 | FALSE | Downregulated |
| 0,424578372 | 0,389081256 | EMG1 | FALSE | Upregulated |
| -0,842168987 | 0,389081256 | PARD6A | FALSE | Downregulated |
| -0,545826447 | 0,389294525 | POMP | FALSE | Downregulated |
| 0,435414111 | 0,38948756 | TAPT1-AS1 | FALSE | Upregulated |
| -0,22995679 | 0,389630518 | RALBP1 | FALSE | Downregulated |
| -0,399177906 | 0,389884336 | ISG20L2 | FALSE | Downregulated |
| 0,554968627 | 0,38989226 | CSRP2 | FALSE | Upregulated |
| 0,322230387 | 0,39027593 | ABHD16A | FALSE | Upregulated |
| 0,268169927 | 0,39027593 | DOCK5 | FALSE | Upregulated |
| 0,539411932 | 0,39027593 | OSBPL5 | FALSE | Upregulated |
| -0,368099237 | 0,390306317 | DDX39B | FALSE | Downregulated |

| | | | | |
|---|---|---|---|---|
| 0,456765385 | 0,390306317 | KLLN | FALSE | Upregulated |
| -0,648955375 | 0,390559166 | MEX3A | FALSE | Downregulated |
| -0,344888683 | 0,390851296 | BCAR3 | FALSE | Downregulated |
| 0,250404291 | 0,390851296 | ELK3 | FALSE | Upregulated |
| 0,369364017 | 0,390851296 | RQCD1 | FALSE | Upregulated |
| 0,6965111 | 0,391071467 | KCNE3 | FALSE | Upregulated |
| -0,813565192 | 0,391261872 | FREM1 | FALSE | Downregulated |
| 0,389453476 | 0,391261872 | P2RX7 | FALSE | Upregulated |
| -0,553933629 | 0,391536511 | DDT | FALSE | Downregulated |
| 0,635107921 | 0,391576341 | IL18 | FALSE | Upregulated |
| 0,318226496 | 0,391955238 | ZBTB46 | FALSE | Upregulated |
| -0,395148626 | 0,392244886 | ADK | FALSE | Downregulated |
| -0,337947537 | 0,392244886 | BACH1 | FALSE | Downregulated |
| 0,266547008 | 0,392244886 | EZR | FALSE | Upregulated |
| -0,226462957 | 0,392244886 | KANSL1 | FALSE | Downregulated |
| 0,925580588 | 0,392244886 | LCK | FALSE | Upregulated |
| 0,516059701 | 0,392244886 | LOC729451 | FALSE | Upregulated |
| -0,456506498 | 0,392244886 | NCALD | FALSE | Downregulated |
| 0,437598786 | 0,392244886 | SIPA1L2 | FALSE | Upregulated |
| 0,361627836 | 0,392244886 | SNRPD2 | FALSE | Upregulated |
| 0,37809853 | 0,392244886 | TARBP1 | FALSE | Upregulated |
| 0,707536516 | 0,392244886 | ZNF320 | FALSE | Upregulated |
| -0,196134646 | 0,392910252 | ROCK2 | FALSE | Downregulated |
| 0,300585894 | 0,392931919 | C11orf49 | FALSE | Upregulated |
| -2,188233345 | 0,39293458 | CCZ1B | FALSE | Downregulated |
| 0,95199853 | 0,39293458 | KIF2C | FALSE | Upregulated |
| -0,403316119 | 0,39298082 | KALRN | FALSE | Downregulated |
| -0,833021115 | 0,39301077 | GNN | FALSE | Downregulated |
| 0,504017523 | 0,393269402 | C9orf91 | FALSE | Upregulated |
| 0,578373583 | 0,393269402 | DHX35 | FALSE | Upregulated |
| -0,374828039 | 0,393269402 | PTPN4 | FALSE | Downregulated |
| 0,273761971 | 0,393294735 | GDAP1L1 | FALSE | Upregulated |
| 0,340886923 | 0,393547531 | MCTP1 | FALSE | Upregulated |
| 0,544800033 | 0,393856415 | KIRREL3 | FALSE | Upregulated |
| -1,254437158 | 0,39390412 | CCDC60 | FALSE | Downregulated |
| -0,275676883 | 0,394219484 | ATE1 | FALSE | Downregulated |
| -1,101557824 | 0,394219484 | LOC100287497 | FALSE | Downregulated |
| 0,393854383 | 0,394219484 | LOC730183 | FALSE | Upregulated |
| 0,419309206 | 0,394312019 | UBE2B | FALSE | Upregulated |
| 0,527347244 | 0,394420789 | SNPH | FALSE | Upregulated |
| -0,614147892 | 0,394761284 | CHRNA4 | FALSE | Downregulated |
| 0,663625258 | 0,394810075 | ZNF582-AS1 | FALSE | Upregulated |
| 0,586463969 | 0,395382412 | ASUN | FALSE | Upregulated |
| 0,464924841 | 0,395382412 | DDAH2 | FALSE | Upregulated |
| -0,3217534 | 0,395416146 | ZFYVE9 | FALSE | Downregulated |
| 0,485411059 | 0,395496963 | LOC102723998 | FALSE | Upregulated |
| 0,42214452 | 0,395723064 | PODNL1 | FALSE | Upregulated |
| -0,857957449 | 0,395766776 | TSPAN32 | FALSE | Downregulated |
| -0,22890859 | 0,395872695 | ESRRA | FALSE | Downregulated |
| 0,6019352 | 0,395872695 | HULC | FALSE | Upregulated |
| -0,389115545 | 0,395872695 | NPC1L1 | FALSE | Downregulated |

| | | | | |
|---|---|---|---|---|
| -0,963449954 | 0,395872695 | TMPRSS11A | FALSE | Downregulated |
| 0,311278652 | 0,395880037 | NOP56 | FALSE | Upregulated |
| 0,627589743 | 0,396067269 | IRAK2 | FALSE | Upregulated |
| -0,348303202 | 0,396067269 | PIK3CA | FALSE | Downregulated |
| 0,431819132 | 0,396080444 | PAFAH1B2 | FALSE | Upregulated |
| 0,989455074 | 0,396109656 | CDCA4 | FALSE | Upregulated |
| -0,602760359 | 0,396109656 | KLHL26 | FALSE | Downregulated |
| -0,338659083 | 0,396109656 | METRN | FALSE | Downregulated |
| 0,377269192 | 0,396287049 | STK16 | FALSE | Upregulated |
| -0,434006027 | 0,396313309 | NR2C1 | FALSE | Downregulated |
| -0,841593663 | 0,396360176 | RP11-15A1.3 | FALSE | Downregulated |
| -0,17631355 | 0,396481194 | MIA3 | FALSE | Downregulated |
| 0,335690051 | 0,396517348 | AGO2 | FALSE | Upregulated |
| -0,239738614 | 0,396596029 | PLEKHA6 | FALSE | Downregulated |
| 0,210926471 | 0,396662771 | HAS3 | FALSE | Upregulated |
| -0,224253111 | 0,396662771 | MAML3 | FALSE | Downregulated |
| 0,656020669 | 0,396778887 | PTCH2 | FALSE | Upregulated |
| 0,644890202 | 0,396778887 | SCARNA9 | FALSE | Upregulated |
| -0,591952448 | 0,396778887 | ZNF287 | FALSE | Downregulated |
| 0,319695543 | 0,396874712 | CCDC73 | FALSE | Upregulated |
| -0,243679965 | 0,396931942 | ZXDC | FALSE | Downregulated |
| -0,727372358 | 0,397172354 | ARHGAP19 | FALSE | Downregulated |
| 1,252705562 | 0,397570941 | PXT1 | FALSE | Upregulated |
| 0,246499612 | 0,397570941 | ZNF544 | FALSE | Upregulated |
| -0,445683853 | 0,398102568 | CYP7B1 | FALSE | Downregulated |
| 0,35655526 | 0,398190254 | AP5B1 | FALSE | Upregulated |
| -0,268896805 | 0,398613918 | EIF1 | FALSE | Downregulated |
| 0,285578405 | 0,398613918 | FAM21C | FALSE | Upregulated |
| -1,28379384 | 0,398613918 | LOC101929608 | FALSE | Downregulated |
| 1,130596901 | 0,3986567 | MATN1 | FALSE | Upregulated |
| -0,307238373 | 0,3986567 | RPS15A | FALSE | Downregulated |
| 0,534358148 | 0,3986567 | ZNF385C | FALSE | Upregulated |
| 0,343194838 | 0,398657694 | DDX19A | FALSE | Upregulated |
| 0,332640475 | 0,398751383 | CDK11B | FALSE | Upregulated |
| -0,265947544 | 0,398751383 | CFI | FALSE | Downregulated |
| 0,640046027 | 0,398751383 | MMP2 | FALSE | Upregulated |
| 0,415074331 | 0,398751383 | NAMPT | FALSE | Upregulated |
| -0,304687705 | 0,398751383 | SMAD2 | FALSE | Downregulated |
| 0,223985061 | 0,398751383 | TATDN2 | FALSE | Upregulated |
| 0,432252166 | 0,399450703 | NFKBIL1 | FALSE | Upregulated |
| 0,596501895 | 0,399453065 | RP11-365O16.6 | FALSE | Upregulated |
| 0,877923306 | 0,399453065 | SYP | FALSE | Upregulated |
| -0,328354814 | 0,399464297 | CSNK1E | FALSE | Downregulated |
| -1,112307092 | 0,399530479 | LINC00087 | FALSE | Downregulated |
| -0,238364049 | 0,399530479 | NRIP1 | FALSE | Downregulated |
| -0,385431112 | 0,399692307 | MAPK14 | FALSE | Downregulated |
| 0,361782303 | 0,399692307 | RP11-4F22.2 | FALSE | Upregulated |
| -0,135929508 | 0,399752962 | SUN2 | FALSE | Downregulated |
| 0,362636951 | 0,400290858 | LRRC37A | FALSE | Upregulated |
| -0,24480296 | 0,400465271 | PTK2 | FALSE | Downregulated |
| 0,6108228 | 0,400523175 | ATE1-AS1 | FALSE | Upregulated |

| | | | | |
|---|---|---|---|---|
| -0,320213111 | 0,400523175 | TBC1D4 | FALSE | Downregulated |
| -0,25158124 | 0,400836716 | HINT1 | FALSE | Downregulated |
| 0,385094809 | 0,400836716 | RAB5A | FALSE | Upregulated |
| -0,451934559 | 0,400884273 | LOC389834 | FALSE | Downregulated |
| 0,360226569 | 0,400902065 | ARHGEF6 | FALSE | Upregulated |
| -0,463545038 | 0,400996142 | SLC7A8 | FALSE | Downregulated |
| -0,212556482 | 0,401645039 | ARFGAP2 | FALSE | Downregulated |
| 0,472138804 | 0,401645039 | LYRM5 | FALSE | Upregulated |
| 0,285135076 | 0,401671135 | ADPGK | FALSE | Upregulated |
| -0,344553413 | 0,40186313 | HPGD | FALSE | Downregulated |
| -0,311116484 | 0,401868844 | ARSD | FALSE | Downregulated |
| 0,345824622 | 0,401868844 | FAM177A1 | FALSE | Upregulated |
| 0,58226659 | 0,401868844 | LOC100289455 | FALSE | Upregulated |
| 0,781923427 | 0,401868844 | NCF1C | FALSE | Upregulated |
| 0,674870235 | 0,401868844 | RFX3 | FALSE | Upregulated |
| -0,36742484 | 0,401911439 | SYNPO2 | FALSE | Downregulated |
| 0,766199565 | 0,401964062 | NFATC4 | FALSE | Upregulated |
| 0,447519543 | 0,402157401 | CALCRL | FALSE | Upregulated |
| -0,200426514 | 0,402212308 | HABP2 | FALSE | Downregulated |
| -0,246649583 | 0,402446846 | DMXL1 | FALSE | Downregulated |
| 0,643352531 | 0,402446846 | FAM166A | FALSE | Upregulated |
| -0,302462582 | 0,402482546 | NDUFB9 | FALSE | Downregulated |
| -0,297357743 | 0,402482546 | UBE2K | FALSE | Downregulated |
| 0,572360061 | 0,402532602 | ENPP2 | FALSE | Upregulated |
| 0,446898988 | 0,40260334 | METTL23 | FALSE | Upregulated |
| 0,257931079 | 0,402613663 | CKAP4 | FALSE | Upregulated |
| -0,67236265 | 0,402613663 | IGFBP1 | FALSE | Downregulated |
| 0,206858074 | 0,402613663 | PTGES3 | FALSE | Upregulated |
| -0,288186122 | 0,402613663 | SLC5A3 | FALSE | Downregulated |
| -0,323275792 | 0,402613663 | UACA | FALSE | Downregulated |
| -0,374697195 | 0,402761997 | FGFR1 | FALSE | Downregulated |
| -0,494774736 | 0,40278334 | MRPL46 | FALSE | Downregulated |
| -0,28869656 | 0,40278334 | MTFR1L | FALSE | Downregulated |
| 0,429318338 | 0,402835271 | LOC115110 | FALSE | Upregulated |
| -0,30705619 | 0,402854284 | DDX56 | FALSE | Downregulated |
| 0,2547723 | 0,402854284 | LYL1 | FALSE | Upregulated |
| -0,389657767 | 0,402854284 | PLEKHA1 | FALSE | Downregulated |
| -0,3717019 | 0,402854284 | STAG3L2 | FALSE | Downregulated |
| 0,557480425 | 0,402854284 | TIPRL | FALSE | Upregulated |
| -0,921059235 | 0,402898744 | CYP17A1 | FALSE | Downregulated |
| -0,327892633 | 0,402898744 | PDHA1 | FALSE | Downregulated |
| -0,4290374 | 0,402898744 | SLC22A18 | FALSE | Downregulated |
| -0,428531114 | 0,403014661 | CREBRF | FALSE | Downregulated |
| -1,207487102 | 0,403014661 | DSCAM | FALSE | Downregulated |
| 0,420963527 | 0,403014661 | MANEAL | FALSE | Upregulated |
| -0,376501059 | 0,403014661 | TMEM87A | FALSE | Downregulated |
| 0,34035256 | 0,403483594 | CDIPT | FALSE | Upregulated |
| -0,382286546 | 0,403497406 | CARD10 | FALSE | Downregulated |
| 0,681212448 | 0,403574088 | C5orf56 | FALSE | Upregulated |
| 0,950896877 | 0,40418734 | APOBEC3G | FALSE | Upregulated |
| 0,473220297 | 0,40418734 | MYLK-AS1 | FALSE | Upregulated |

| | | | | |
|---|---|---|---|---|
| -1,625733896 | 0,40418734 | OR2A4 | FALSE | Downregulated |
| -0,387381224 | 0,40418734 | TLN2 | FALSE | Downregulated |
| -0,732348904 | 0,404280972 | ANKRA2 | FALSE | Downregulated |
| -1,328183381 | 0,404280972 | C6orf141 | FALSE | Downregulated |
| -0,313314647 | 0,40443935 | ATL2 | FALSE | Downregulated |
| 0,548516579 | 0,40443935 | PMEL | FALSE | Upregulated |
| 0,502454946 | 0,40443935 | TRIM47 | FALSE | Upregulated |
| 0,440900803 | 0,404906582 | ODC1 | FALSE | Upregulated |
| 0,1767975 | 0,405038014 | SETD1B | FALSE | Upregulated |
| 0,488330592 | 0,405523477 | FBXL18 | FALSE | Upregulated |
| 0,583719573 | 0,405536505 | ELOVL1 | FALSE | Upregulated |
| 0,32352414 | 0,405536505 | TLE3 | FALSE | Upregulated |
| -0,380221373 | 0,405883083 | KCNS2 | FALSE | Downregulated |
| 0,503079669 | 0,405883083 | RNF115 | FALSE | Upregulated |
| -0,410516596 | 0,405898941 | ACAP2 | FALSE | Downregulated |
| -0,419946313 | 0,405926566 | FUT6 | FALSE | Downregulated |
| 1,46573957 | 0,405926566 | LOC102723678 | FALSE | Upregulated |
| 1,013452365 | 0,405989525 | KRT222 | FALSE | Upregulated |
| 0,247731704 | 0,406103325 | CAP1 | FALSE | Upregulated |
| -0,602527242 | 0,406103325 | GSTP1 | FALSE | Downregulated |
| 0,349037304 | 0,406172174 | ALX3 | FALSE | Upregulated |
| 0,417240611 | 0,406172174 | ATP1A1OS | FALSE | Upregulated |
| 0,401358494 | 0,406172174 | LDOC1L | FALSE | Upregulated |
| -0,380714487 | 0,406172174 | LOC100506730 | FALSE | Downregulated |
| 1,182329561 | 0,406172174 | NUDT10 | FALSE | Upregulated |
| -0,518390208 | 0,406172174 | PGM5 | FALSE | Downregulated |
| -0,321020584 | 0,406172174 | SLC25A46 | FALSE | Downregulated |
| 0,321542256 | 0,406172174 | TYW5 | FALSE | Upregulated |
| -0,443877209 | 0,406172174 | UROS | FALSE | Downregulated |
| 0,263339053 | 0,406172174 | WDR26 | FALSE | Upregulated |
| -1,562955209 | 0,406172174 | ZIM2 | FALSE | Downregulated |
| 0,467641466 | 0,40617428 | HPS5 | FALSE | Upregulated |
| -0,945202085 | 0,406283672 | AC009264.1 | FALSE | Downregulated |
| -0,245731364 | 0,406283672 | APH1A | FALSE | Downregulated |
| 0,792044604 | 0,406283672 | CA5B | FALSE | Upregulated |
| -0,201116115 | 0,406283672 | CD81 | FALSE | Downregulated |
| -0,324305678 | 0,406283672 | CDKN1B | FALSE | Downregulated |
| -0,433437089 | 0,406283672 | GLT1D1 | FALSE | Downregulated |
| -0,872697651 | 0,406283672 | KANK4 | FALSE | Downregulated |
| 0,903805511 | 0,406283672 | LOC100287632 | FALSE | Upregulated |
| -0,549391231 | 0,406283672 | PRKG1 | FALSE | Downregulated |
| 0,455942222 | 0,406283672 | TMCO3 | FALSE | Upregulated |
| -0,711131226 | 0,406283672 | ZNF273 | FALSE | Downregulated |
| 0,309115224 | 0,406460716 | UTP18 | FALSE | Upregulated |
| -0,329059989 | 0,406870389 | CRLS1 | FALSE | Downregulated |
| 0,994828424 | 0,406870389 | MILR1 | FALSE | Upregulated |
| -0,357875133 | 0,407105272 | PRMT1 | FALSE | Downregulated |
| 0,940462737 | 0,407600797 | CD83 | FALSE | Upregulated |
| 0,684235404 | 0,407600797 | LPAR5 | FALSE | Upregulated |
| 1,203039062 | 0,407600797 | TMPRSS4 | FALSE | Upregulated |
| 0,233598547 | 0,407600797 | ZMIZ1 | FALSE | Upregulated |

| | | | | |
|---|---|---|---|---|
| 0,995520553 | 0,407655386 | C19orf84 | FALSE | Upregulated |
| 0,487877549 | 0,407835291 | PLSCR1 | FALSE | Upregulated |
| 0,565266401 | 0,407835291 | RNF8 | FALSE | Upregulated |
| 0,314537537 | 0,407850745 | PROSER2-AS1 | FALSE | Upregulated |
| -0,31686394 | 0,408070261 | ARF5 | FALSE | Downregulated |
| 0,281844318 | 0,408070261 | ASH1L-AS1 | FALSE | Upregulated |
| 0,636952674 | 0,408070261 | GIMAP7 | FALSE | Upregulated |
| -0,261639756 | 0,408395819 | SERPINA6 | FALSE | Downregulated |
| 1,186646085 | 0,408608406 | C5orf58 | FALSE | Upregulated |
| -0,981081898 | 0,408608406 | TIAF1 | FALSE | Downregulated |
| 0,510682925 | 0,408777115 | ASPHD1 | FALSE | Upregulated |
| -0,391597696 | 0,408777115 | SIVA1 | FALSE | Downregulated |
| 0,740342581 | 0,408777115 | SLMO2 | FALSE | Upregulated |
| -0,258491871 | 0,408936456 | RPL38 | FALSE | Downregulated |
| -0,203826346 | 0,409006582 | FAM168B | FALSE | Downregulated |
| 0,588246058 | 0,409033394 | PLEKHA8 | FALSE | Upregulated |
| 0,413911523 | 0,409033394 | PYCR2 | FALSE | Upregulated |
| 0,267738504 | 0,409033394 | SHE | FALSE | Upregulated |
| -0,485058053 | 0,409033394 | SSPO | FALSE | Downregulated |
| 1,087653701 | 0,409033394 | TMEM154 | FALSE | Upregulated |
| -0,524455212 | 0,409033394 | ZFP14 | FALSE | Downregulated |
| -1,091003653 | 0,409164858 | C15orf38-AP3S2 | FALSE | Downregulated |
| 0,765419241 | 0,409276028 | OSCP1 | FALSE | Upregulated |
| 0,879964532 | 0,40938262 | ARHGAP22 | FALSE | Upregulated |
| 0,225641659 | 0,409476601 | YWHAG | FALSE | Upregulated |
| -0,261216901 | 0,409922013 | MIB1 | FALSE | Downregulated |
| -1,175116306 | 0,410362988 | CHIC1 | FALSE | Downregulated |
| -0,307756041 | 0,410362988 | SMPDL3A | FALSE | Downregulated |
| 0,246293587 | 0,410373814 | CDC37 | FALSE | Upregulated |
| 0,268502383 | 0,410373814 | CLDN20 | FALSE | Upregulated |
| 0,226553902 | 0,410373814 | COMMD5 | FALSE | Upregulated |
| -0,561124793 | 0,410671184 | NUDT12 | FALSE | Downregulated |
| -1,096203318 | 0,410671184 | SEMA4F | FALSE | Downregulated |
| -0,626320714 | 0,410724134 | GARNL3 | FALSE | Downregulated |
| -0,511500849 | 0,410724134 | NDUFB4 | FALSE | Downregulated |
| -0,297019219 | 0,410724134 | XRN1 | FALSE | Downregulated |
| 0,705738222 | 0,411060303 | C20orf144 | FALSE | Upregulated |
| 0,959853338 | 0,411060303 | CACNA1C-AS1 | FALSE | Upregulated |
| 0,482699979 | 0,411060303 | MBLAC1 | FALSE | Upregulated |
| -0,255544804 | 0,411310512 | LOC101928451 | FALSE | Downregulated |
| 0,264199122 | 0,411310512 | PTPN21 | FALSE | Upregulated |
| -0,887449162 | 0,411310512 | TMEM117 | FALSE | Downregulated |
| -0,215830006 | 0,411310512 | WWC2 | FALSE | Downregulated |
| 0,251983406 | 0,411401817 | ELAC2 | FALSE | Upregulated |
| 0,40037443 | 0,411401817 | PRSS8 | FALSE | Upregulated |
| 0,255193052 | 0,411442985 | HNRNPH3 | FALSE | Upregulated |
| -0,874901725 | 0,411442985 | LOC101060179 | FALSE | Downregulated |
| 0,458561648 | 0,411550315 | CCDC121 | FALSE | Upregulated |
| -0,34500799 | 0,411550315 | SLC25A11 | FALSE | Downregulated |
| -0,136726985 | 0,411550315 | ZSWIM8 | FALSE | Downregulated |
| -0,201454667 | 0,411854116 | FNBP4 | FALSE | Downregulated |

| | | | | |
|---|---|---|---|---|
| 2,332175961 | 0,411854116 | GTF2H2 | FALSE | Upregulated |
| 0,352844279 | 0,411854116 | SURF6 | FALSE | Upregulated |
| -0,664183689 | 0,411872863 | LOC101928574 | FALSE | Downregulated |
| -0,324373565 | 0,412117113 | ADRA1A | FALSE | Downregulated |
| -0,282957149 | 0,412270168 | WDR65 | FALSE | Downregulated |
| -0,338927057 | 0,412284758 | PSMC2 | FALSE | Downregulated |
| 0,750848412 | 0,412350536 | DLEU7 | FALSE | Upregulated |
| 0,350255085 | 0,412350536 | LAS1L | FALSE | Upregulated |
| 0,287637121 | 0,412866915 | ATG13 | FALSE | Upregulated |
| 0,453333034 | 0,412866915 | CLDN23 | FALSE | Upregulated |
| 0,921339848 | 0,412866915 | LOC101928054 | FALSE | Upregulated |
| 0,267172162 | 0,413107865 | RASGRP1 | FALSE | Upregulated |
| -0,799291057 | 0,41332258 | PLSCR3 | FALSE | Downregulated |
| 0,296091652 | 0,413415877 | SETD3 | FALSE | Upregulated |
| 0,549370138 | 0,413422158 | APBB3 | FALSE | Upregulated |
| -0,999911874 | 0,413422158 | COL7A1 | FALSE | Downregulated |
| 1,133266788 | 0,413665111 | CREB5 | FALSE | Upregulated |
| 0,225948839 | 0,413665111 | GREB1L | FALSE | Upregulated |
| 0,358891666 | 0,413845214 | ADAM15 | FALSE | Upregulated |
| 0,375857207 | 0,413845214 | ANKS1B | FALSE | Upregulated |
| 0,252010584 | 0,413845214 | ARFGAP1 | FALSE | Upregulated |
| 0,19311637 | 0,413845214 | EDF1 | FALSE | Upregulated |
| 0,3204769 | 0,413845214 | LOC100996873 | FALSE | Upregulated |
| -0,31319468 | 0,413845214 | NDUFB11 | FALSE | Downregulated |
| -0,368842024 | 0,413845214 | TARS2 | FALSE | Downregulated |
| 0,243388672 | 0,414119209 | COL4A4 | FALSE | Upregulated |
| -1,149536088 | 0,414167363 | C8orf44 | FALSE | Downregulated |
| -0,299616333 | 0,414303108 | TK2 | FALSE | Downregulated |
| 1,169272589 | 0,414514347 | LOC388210 | FALSE | Upregulated |
| 0,530319752 | 0,414948056 | STXBP1 | FALSE | Upregulated |
| 0,224161264 | 0,414996823 | COPG1 | FALSE | Upregulated |
| 0,266236843 | 0,415408924 | CDK16 | FALSE | Upregulated |
| 0,403514853 | 0,415408924 | FAM110C | FALSE | Upregulated |
| -0,261898451 | 0,415408924 | IGSF8 | FALSE | Downregulated |
| -0,395615285 | 0,415408924 | STX1B | FALSE | Downregulated |
| 0,302119276 | 0,415408924 | VASH2 | FALSE | Upregulated |
| 0,192044872 | 0,415409008 | SLC25A29 | FALSE | Upregulated |
| -0,98246919 | 0,415758353 | OVGP1 | FALSE | Downregulated |
| -0,802457795 | 0,415977123 | FOXF1 | FALSE | Downregulated |
| 0,368715335 | 0,415977123 | HSPH1 | FALSE | Upregulated |
| -1,269442964 | 0,416418454 | TMEM233 | FALSE | Downregulated |
| -0,437525435 | 0,416551812 | BOC | FALSE | Downregulated |
| -0,221222926 | 0,416551812 | CCNI | FALSE | Downregulated |
| -0,748149183 | 0,416551812 | PINX1 | FALSE | Downregulated |
| 0,403739939 | 0,416730637 | EXOC2 | FALSE | Upregulated |
| -0,482105354 | 0,416730637 | SLC17A1 | FALSE | Downregulated |
| 0,447052435 | 0,416730637 | TMA7 | FALSE | Upregulated |
| 0,322490399 | 0,416764422 | RAP2C-AS1 | FALSE | Upregulated |
| 0,238965213 | 0,416764422 | TACC1 | FALSE | Upregulated |
| -0,517371194 | 0,417083577 | APTX | FALSE | Downregulated |
| 0,208942834 | 0,417231871 | PGM3 | FALSE | Upregulated |

| | | | | |
|---|---|---|---|---|
| -0,70488839 | 0,417279453 | HCAR3 | FALSE | Downregulated |
| -0,256047794 | 0,417279453 | JTB | FALSE | Downregulated |
| 0,312232435 | 0,417279453 | SNX8 | FALSE | Upregulated |
| 0,29227912 | 0,417409411 | YTHDF2 | FALSE | Upregulated |
| 0,208377066 | 0,417448704 | FLJ42627 | FALSE | Upregulated |
| -0,288202655 | 0,417697047 | GP1BA | FALSE | Downregulated |
| -0,942471051 | 0,417844938 | LINC00939 | FALSE | Downregulated |
| -0,723159852 | 0,418056213 | ALG10B | FALSE | Downregulated |
| -0,435836567 | 0,418056213 | PMCHL2 | FALSE | Downregulated |
| -0,627560379 | 0,418172456 | ING3 | FALSE | Downregulated |
| -0,230764345 | 0,418172456 | SOD1 | FALSE | Downregulated |
| -0,348812443 | 0,418324934 | FAM211A-AS1 | FALSE | Downregulated |
| 0,399671868 | 0,418525268 | KCTD4 | FALSE | Upregulated |
| -0,643617514 | 0,41859475 | WASH2P | FALSE | Downregulated |
| 0,36870178 | 0,418627994 | LOH12CR1 | FALSE | Upregulated |
| -0,209073825 | 0,418688036 | WDTC1 | FALSE | Downregulated |
| 0,256253604 | 0,418778772 | CALM2 | FALSE | Upregulated |
| 0,571954774 | 0,418778772 | LOC100507191 | FALSE | Upregulated |
| -0,205220243 | 0,418778772 | MTHFD1 | FALSE | Downregulated |
| 0,270426464 | 0,418812524 | BCDIN3D-AS1 | FALSE | Upregulated |
| -0,235705803 | 0,418812524 | DIP2B | FALSE | Downregulated |
| -0,412141461 | 0,419007277 | LOC102723972 | FALSE | Downregulated |
| -0,242049206 | 0,419017474 | RAB17 | FALSE | Downregulated |
| 0,735887682 | 0,419017474 | TNFRSF10A | FALSE | Upregulated |
| 0,24909726 | 0,419031578 | CADM1 | FALSE | Upregulated |
| -1,313521384 | 0,419092587 | LIMS3-LOC440895 | FALSE | Downregulated |
| 0,380047776 | 0,419092587 | LOX | FALSE | Upregulated |
| -0,263093134 | 0,419200632 | INTS3 | FALSE | Downregulated |
| 0,76137694 | 0,419207607 | LOC100996574 | FALSE | Upregulated |
| 0,776635391 | 0,419275149 | EXOSC9 | FALSE | Upregulated |
| -0,774529723 | 0,419275149 | UCHL3 | FALSE | Downregulated |
| 0,362389016 | 0,419318028 | BMP6 | FALSE | Upregulated |
| 0,504752138 | 0,419318028 | KDELC2 | FALSE | Upregulated |
| 0,222292303 | 0,419318028 | LOC100128494 | FALSE | Upregulated |
| -0,212521755 | 0,419318028 | PXN | FALSE | Downregulated |
| 0,404583006 | 0,419318028 | TRIM65 | FALSE | Upregulated |
| 0,99518466 | 0,419493056 | FPR1 | FALSE | Upregulated |
| 0,479212086 | 0,419538315 | LOC100287934 | FALSE | Upregulated |
| 0,374239841 | 0,419538315 | NEDD8 | FALSE | Upregulated |
| -0,411758562 | 0,42039559 | ALS2CR11 | FALSE | Downregulated |
| -0,187988535 | 0,42039559 | ANKRD17 | FALSE | Downregulated |
| 0,892258848 | 0,42039559 | FAM27E2 | FALSE | Upregulated |
| 0,257954308 | 0,42039559 | GLIS2 | FALSE | Upregulated |
| -0,386108343 | 0,42039559 | MKNK1 | FALSE | Downregulated |
| 1,082846404 | 0,42039559 | PRIM2 | FALSE | Upregulated |
| 0,440184746 | 0,42039559 | PTPN2 | FALSE | Upregulated |
| -0,294063734 | 0,420638779 | FAM35A | FALSE | Downregulated |
| 0,210616313 | 0,420638779 | GBF1 | FALSE | Upregulated |
| 0,526890417 | 0,420638779 | LOC102724709 | FALSE | Upregulated |
| 0,32345871 | 0,420727191 | TPM3 | FALSE | Upregulated |
| 0,206279651 | 0,421149021 | TPPP | FALSE | Upregulated |

| | | | | |
|---|---|---|---|---|
| 0,281287342 | 0,422089989 | FBXO33 | FALSE | Upregulated |
| 0,817310843 | 0,422089989 | SIGLEC14 | FALSE | Upregulated |
| -0,479363568 | 0,422341671 | PHYHD1 | FALSE | Downregulated |
| -0,712466861 | 0,42274681 | USMG5 | FALSE | Downregulated |
| -0,558300227 | 0,42275927 | PRKXP1 | FALSE | Downregulated |
| -0,249031744 | 0,422816615 | ADCY5 | FALSE | Downregulated |
| -0,503099049 | 0,422825515 | TFRC | FALSE | Downregulated |
| -0,43737951 | 0,422861241 | KCNC4 | FALSE | Downregulated |
| 0,325276164 | 0,423226703 | PLAC4 | FALSE | Upregulated |
| 0,735837599 | 0,423555345 | NKX3-1 | FALSE | Upregulated |
| -0,824412046 | 0,423679636 | CTAGE9 | FALSE | Downregulated |
| 0,584627723 | 0,423679636 | LOC101927227 | FALSE | Upregulated |
| 0,81051056 | 0,423865017 | NBL1 | FALSE | Upregulated |
| -0,259976087 | 0,424004614 | C2orf88 | FALSE | Downregulated |
| -0,364056826 | 0,424117819 | TRAPPC5 | FALSE | Downregulated |
| 1,072509011 | 0,424368426 | EPN3 | FALSE | Upregulated |
| 1,588348123 | 0,424368426 | GTF2IP1 | FALSE | Upregulated |
| 0,616647788 | 0,424368426 | LOC399900 | FALSE | Upregulated |
| 0,685687793 | 0,424368426 | MFGE8 | FALSE | Upregulated |
| -0,489464434 | 0,424368426 | PAQR6 | FALSE | Downregulated |
| -0,159810029 | 0,424368426 | PRRC2C | FALSE | Downregulated |
| 0,142842994 | 0,424368426 | SPTBN1 | FALSE | Upregulated |
| 0,535982752 | 0,424368426 | STAM | FALSE | Upregulated |
| 0,252255931 | 0,424479084 | RICTOR | FALSE | Upregulated |
| -0,623070207 | 0,424540202 | MCEE | FALSE | Downregulated |
| -0,390163604 | 0,424671024 | BSN | FALSE | Downregulated |
| -0,65162365 | 0,42480245 | AQP7 | FALSE | Downregulated |
| 0,269032457 | 0,42480245 | DNPEP | FALSE | Upregulated |
| -0,537836314 | 0,42480245 | GID4 | FALSE | Downregulated |
| 0,987619294 | 0,42480245 | LOC101927069 | FALSE | Upregulated |
| 0,246896538 | 0,42480245 | TMED10P1 | FALSE | Upregulated |
| 0,415124041 | 0,424871861 | AK5 | FALSE | Upregulated |
| 0,497749754 | 0,424970846 | ZNF833P | FALSE | Upregulated |
| -0,210391785 | 0,42513684 | ATP5A1 | FALSE | Downregulated |
| 0,340900287 | 0,42513684 | IFNGR2 | FALSE | Upregulated |
| -0,352241945 | 0,42513684 | ITGB7 | FALSE | Downregulated |
| -0,488512492 | 0,42513684 | PLEKHA3 | FALSE | Downregulated |
| 0,329768142 | 0,42513684 | RAP1B | FALSE | Upregulated |
| 0,571140664 | 0,42513684 | RP11-996F15.2 | FALSE | Upregulated |
| 0,290595814 | 0,42513684 | RPS6KB1 | FALSE | Upregulated |
| 0,365632837 | 0,42513684 | SNIP1 | FALSE | Upregulated |
| 0,228483996 | 0,425216927 | LOC101927469 | FALSE | Upregulated |
| 0,22768227 | 0,425216927 | TRAM1 | FALSE | Upregulated |
| 0,944691539 | 0,425232134 | LOC100630918 | FALSE | Upregulated |
| -0,40965394 | 0,425286267 | ADAMTS9 | FALSE | Downregulated |
| -1,136603132 | 0,425286267 | CTSLP2 | FALSE | Downregulated |
| 0,416570355 | 0,425354689 | C10orf76 | FALSE | Upregulated |
| -0,403871735 | 0,425354689 | C17orf80 | FALSE | Downregulated |
| 0,450180189 | 0,425354689 | IFITM1 | FALSE | Upregulated |
| 0,91113489 | 0,425354689 | LOC388820 | FALSE | Upregulated |
| -0,936815197 | 0,425354689 | MRPL1 | FALSE | Downregulated |

| | | | | |
|---|---|---|---|---|
| 0,244475444 | 0,425354689 | SMG6 | FALSE | Upregulated |
| -1,011319703 | 0,425354689 | ULK4P2 | FALSE | Downregulated |
| 0,499373398 | 0,425372232 | LINC00472 | FALSE | Upregulated |
| 1,025945339 | 0,425372232 | LOC101927767 | FALSE | Upregulated |
| -0,870046746 | 0,425440308 | TAL1 | FALSE | Downregulated |
| 0,198369977 | 0,425951105 | CNDP2 | FALSE | Upregulated |
| 1,159126257 | 0,426038781 | CREB3L1 | FALSE | Upregulated |
| -1,158683745 | 0,426284285 | KCNK17 | FALSE | Downregulated |
| -0,264753039 | 0,426284285 | VARS | FALSE | Downregulated |
| -0,26207017 | 0,426386756 | NOP14 | FALSE | Downregulated |
| -0,818952412 | 0,426452985 | LOC101926894 | FALSE | Downregulated |
| 0,33425446 | 0,426452985 | TPRN | FALSE | Upregulated |
| -0,381634336 | 0,426598603 | YPEL2 | FALSE | Downregulated |
| 0,253734081 | 0,426685256 | DDX58 | FALSE | Upregulated |
| -0,247757958 | 0,426685256 | RPS6KA3 | FALSE | Downregulated |
| 0,853670686 | 0,426738448 | FAM90A25P | FALSE | Upregulated |
| 0,357758807 | 0,426959792 | PELI3 | FALSE | Upregulated |
| 0,480937611 | 0,427177831 | LOC101927870 | FALSE | Upregulated |
| 0,331478865 | 0,427609811 | ATAD3A | FALSE | Upregulated |
| -0,539378825 | 0,427609811 | METTL2B | FALSE | Downregulated |
| -0,25998067 | 0,427825094 | ARID5A | FALSE | Downregulated |
| 0,263143185 | 0,427832313 | FUBP1 | FALSE | Upregulated |
| -0,987415545 | 0,427832313 | LOC102724598 | FALSE | Downregulated |
| 0,251037157 | 0,427832313 | SUPT16H | FALSE | Upregulated |
| -0,453295557 | 0,427842648 | AMOT | FALSE | Downregulated |
| -0,281114019 | 0,427842648 | ATP5C1 | FALSE | Downregulated |
| 0,759312281 | 0,427842648 | GSKIP | FALSE | Upregulated |
| 1,169235006 | 0,427842648 | MT1L | FALSE | Upregulated |
| 0,551211267 | 0,427842648 | SLX1A | FALSE | Upregulated |
| -0,633698425 | 0,428061549 | MOB2 | FALSE | Downregulated |
| -0,566852506 | 0,428061549 | RPL13AP5 | FALSE | Downregulated |
| 1,096129852 | 0,428061549 | TMEM191A | FALSE | Upregulated |
| -0,295073717 | 0,428061549 | TRIB1 | FALSE | Downregulated |
| -1,880492429 | 0,428108121 | LOC102724159 | FALSE | Downregulated |
| 0,346130723 | 0,428108121 | RP11-864J10.4 | FALSE | Upregulated |
| -0,523896762 | 0,428396718 | DISP1 | FALSE | Downregulated |
| 0,313003656 | 0,428396718 | LOC100506504 | FALSE | Upregulated |
| -0,306713027 | 0,428396718 | PEG3 | FALSE | Downregulated |
| 0,331933681 | 0,428396718 | PLCB2 | FALSE | Upregulated |
| 0,593561502 | 0,428396718 | STAMBP | FALSE | Upregulated |
| -0,489899969 | 0,42908155 | PCDH9 | FALSE | Downregulated |
| 0,200672318 | 0,42911811 | NUP50 | FALSE | Upregulated |
| 0,715080082 | 0,42913911 | ADAMTSL5 | FALSE | Upregulated |
| -1,261672861 | 0,429208848 | KIAA0087 | FALSE | Downregulated |
| -1,177644085 | 0,429208848 | LOC344967 | FALSE | Downregulated |
| -0,507172177 | 0,429227124 | GTPBP4 | FALSE | Downregulated |
| 0,331409821 | 0,429453608 | LILRB5 | FALSE | Upregulated |
| -0,268195054 | 0,429478533 | FBXO38 | FALSE | Downregulated |
| -0,459171008 | 0,429478533 | SMARCAL1 | FALSE | Downregulated |
| 0,216877393 | 0,429496008 | NNAT | FALSE | Upregulated |
| 0,210755503 | 0,429605121 | HYI | FALSE | Upregulated |

| | | | | |
|---|---|---|---|---|
| 0,264559882 | 0,429633313 | MXRA7 | FALSE | Upregulated |
| 0,500347412 | 0,429702266 | LOC101927028 | FALSE | Upregulated |
| -0,894548457 | 0,42981282 | BMP10 | FALSE | Downregulated |
| 0,640424286 | 0,42981282 | MAPK11 | FALSE | Upregulated |
| -0,565503675 | 0,42981282 | SFRP1 | FALSE | Downregulated |
| -0,365382723 | 0,429930657 | PTCH1 | FALSE | Downregulated |
| 0,275263848 | 0,429930657 | SEMA4B | FALSE | Upregulated |
| 0,529947632 | 0,429997941 | CASP6 | FALSE | Upregulated |
| -0,187130448 | 0,430277803 | CLASP1 | FALSE | Downregulated |
| -0,610557203 | 0,430333229 | BOLA1 | FALSE | Downregulated |
| -0,382403133 | 0,431202575 | HNRNPUL2-BSCL2 | FALSE | Downregulated |
| -0,502949834 | 0,431202575 | TCTN3 | FALSE | Downregulated |
| 1,004019358 | 0,431202575 | ZNF92 | FALSE | Upregulated |
| -0,307648018 | 0,431324282 | IDH3B | FALSE | Downregulated |
| -0,185126969 | 0,431337984 | MDH2 | FALSE | Downregulated |
| 0,470137602 | 0,431463674 | MID2 | FALSE | Upregulated |
| -0,808938854 | 0,431574922 | PPP1R1C | FALSE | Downregulated |
| 0,221098554 | 0,432192075 | TSR1 | FALSE | Upregulated |
| 0,584624876 | 0,43228365 | BLOC1S3 | FALSE | Upregulated |
| -0,407452534 | 0,43228365 | FRAT1 | FALSE | Downregulated |
| -0,596115508 | 0,43228365 | IQCJ-SCHIP1 | FALSE | Downregulated |
| 0,251888904 | 0,43228365 | JOSD1 | FALSE | Upregulated |
| -0,275666282 | 0,43228365 | PLXNA2 | FALSE | Downregulated |
| -0,265324406 | 0,43228365 | URB1 | FALSE | Downregulated |
| -0,407711015 | 0,432365677 | MIOS | FALSE | Downregulated |
| 0,426780905 | 0,432433119 | RNF166 | FALSE | Upregulated |
| 0,192481612 | 0,432606206 | PAFAH1B1 | FALSE | Upregulated |
| 0,22843724 | 0,432761668 | IL17RB | FALSE | Upregulated |
| -0,411138773 | 0,432920862 | FKBP7 | FALSE | Downregulated |
| 0,479708044 | 0,432920862 | METTL15 | FALSE | Upregulated |
| 0,362202621 | 0,432920862 | PRRT3 | FALSE | Upregulated |
| 0,293378053 | 0,432966231 | ENTPD4 | FALSE | Upregulated |
| 0,808756011 | 0,432966231 | LOC100130705 | FALSE | Upregulated |
| -0,239939086 | 0,432966231 | TRPM7 | FALSE | Downregulated |
| 0,987884894 | 0,433287947 | AC005264.2 | FALSE | Upregulated |
| 0,216078845 | 0,433377223 | PPP1CB | FALSE | Upregulated |
| 0,261662137 | 0,433417062 | DAB2 | FALSE | Upregulated |
| 1,100274902 | 0,433515141 | LOC728752 | FALSE | Upregulated |
| -1,990861363 | 0,433552181 | RIMBP2 | FALSE | Downregulated |
| 0,208975824 | 0,433962845 | BTBD6 | FALSE | Upregulated |
| 0,541284031 | 0,433962845 | LOC101928244 | FALSE | Upregulated |
| -0,236645303 | 0,433962845 | PALLD | FALSE | Downregulated |
| -1,200526653 | 0,433962845 | SLC7A4 | FALSE | Downregulated |
| -0,931168475 | 0,433963359 | CRHR1-IT1 | FALSE | Downregulated |
| -0,378787866 | 0,433969354 | ACP1 | FALSE | Downregulated |
| -1,005698339 | 0,433969354 | DRD1 | FALSE | Downregulated |
| 0,716999182 | 0,434074611 | KCNH7 | FALSE | Upregulated |
| 0,900636206 | 0,434074611 | RNASEH2A | FALSE | Upregulated |
| -0,26838825 | 0,434789784 | CLASRP | FALSE | Downregulated |
| -0,241668706 | 0,434836147 | TTC31 | FALSE | Downregulated |
| 0,78826042 | 0,434856366 | BRE-AS1 | FALSE | Upregulated |

| | | | | |
|---|---|---|---|---|
| 0,301050272 | 0,434856366 | TSC22D4 | FALSE | Upregulated |
| -0,984011537 | 0,435006182 | AC091878.1 | FALSE | Downregulated |
| -0,569433223 | 0,435006182 | CBWD5 | FALSE | Downregulated |
| 0,526508402 | 0,435006182 | TLCD1 | FALSE | Upregulated |
| -0,503237995 | 0,435170681 | IL17RE | FALSE | Downregulated |
| -0,415410621 | 0,435203729 | LOC102723406 | FALSE | Downregulated |
| -0,373835056 | 0,435383467 | MFSD3 | FALSE | Downregulated |
| 0,227952013 | 0,435511426 | KDM4A | FALSE | Upregulated |
| 0,453684404 | 0,435540563 | RIPPLY1 | FALSE | Upregulated |
| -0,281387098 | 0,435699096 | ELMO1 | FALSE | Downregulated |
| -0,375231688 | 0,435699096 | UBE2Q1 | FALSE | Downregulated |
| 0,353190885 | 0,435980242 | AKIRIN2 | FALSE | Upregulated |
| 0,506166532 | 0,435980242 | CELSR3 | FALSE | Upregulated |
| 0,85382311 | 0,435980242 | HCG11 | FALSE | Upregulated |
| 0,446454325 | 0,435980242 | NDE1 | FALSE | Upregulated |
| -0,312865915 | 0,435980242 | TANGO6 | FALSE | Downregulated |
| -0,362865602 | 0,436187842 | NEU1 | FALSE | Downregulated |
| -0,597251226 | 0,43640062 | BMS1P4 | FALSE | Downregulated |
| -0,269689247 | 0,436524006 | N4BP2 | FALSE | Downregulated |
| -1,340019952 | 0,436541477 | KLHDC8A | FALSE | Downregulated |
| 0,213309839 | 0,436619535 | GPR124 | FALSE | Upregulated |
| -0,326951175 | 0,436619535 | GRAMD4 | FALSE | Downregulated |
| 0,644733831 | 0,436619535 | GTPBP8 | FALSE | Upregulated |
| 0,309675395 | 0,436619535 | SERPINB6 | FALSE | Upregulated |
| 0,285126905 | 0,436619535 | TPI1P3 | FALSE | Upregulated |
| -0,223923754 | 0,436695961 | ASMTL | FALSE | Downregulated |
| -0,612130686 | 0,436738481 | MIR17HG | FALSE | Downregulated |
| -0,258026312 | 0,436784694 | RAD21 | FALSE | Downregulated |
| 0,226021067 | 0,436812098 | NAV1 | FALSE | Upregulated |
| -0,36264943 | 0,436929709 | ZNF580 | FALSE | Downregulated |
| 0,184499833 | 0,436992961 | MAT2A | FALSE | Upregulated |
| -0,486687917 | 0,437026475 | GPR61 | FALSE | Downregulated |
| -0,796457809 | 0,437058807 | GSTM2 | FALSE | Downregulated |
| -1,203793719 | 0,437058807 | LOC101929574 | FALSE | Downregulated |
| 0,929030939 | 0,437058807 | LOC102723311 | FALSE | Upregulated |
| -0,457939934 | 0,437058807 | SPRY1 | FALSE | Downregulated |
| -0,205522684 | 0,437105464 | SAR1B | FALSE | Downregulated |
| -0,206546591 | 0,437230954 | AGFG2 | FALSE | Downregulated |
| 0,30960654 | 0,437307821 | NOMO1 | FALSE | Upregulated |
| -0,244858743 | 0,437307821 | SRSF11 | FALSE | Downregulated |
| 1,132257975 | 0,437508433 | FAM171A2 | FALSE | Upregulated |
| 0,579998529 | 0,437607323 | NUDT17 | FALSE | Upregulated |
| -0,579386184 | 0,43801966 | GEMIN8 | FALSE | Downregulated |
| -0,34010298 | 0,438101198 | TP53 | FALSE | Downregulated |
| 0,381345976 | 0,438154299 | USP13 | FALSE | Upregulated |
| -0,25428098 | 0,438519708 | VWA8 | FALSE | Downregulated |
| 0,256403277 | 0,438783455 | ITPKC | FALSE | Upregulated |
| 0,367981925 | 0,439002367 | R3HDM1 | FALSE | Upregulated |
| -1,146006083 | 0,439170144 | LIPH | FALSE | Downregulated |
| 0,221793596 | 0,439332844 | RHOF | FALSE | Upregulated |
| -0,375950672 | 0,439418143 | EDNRB | FALSE | Downregulated |

| | | | | |
|---|---|---|---|---|
| -0,56034201 | 0,439418143 | LRFN3 | FALSE | Downregulated |
| 0,819047867 | 0,439537881 | HIST2H3C | FALSE | Upregulated |
| 0,635140176 | 0,439537881 | IKBKE | FALSE | Upregulated |
| 0,591576825 | 0,439537881 | MSC | FALSE | Upregulated |
| -1,240923454 | 0,439537881 | RP11-180I22.2 | FALSE | Downregulated |
| 0,80525085 | 0,439537881 | RTP4 | FALSE | Upregulated |
| -0,148838905 | 0,439537881 | SF1 | FALSE | Downregulated |
| -0,296002564 | 0,439537881 | SFSWAP | FALSE | Downregulated |
| -0,333900307 | 0,439537881 | SLC25A43 | FALSE | Downregulated |
| 0,801965513 | 0,439537881 | VILL | FALSE | Upregulated |
| -0,959226306 | 0,439638467 | LOC102724601 | FALSE | Downregulated |
| -0,382748909 | 0,439662391 | AP5Z1 | FALSE | Downregulated |
| -0,220565493 | 0,439662391 | ATP5B | FALSE | Downregulated |
| -0,45449688 | 0,439906866 | HSBP1L1 | FALSE | Downregulated |
| -0,317821241 | 0,439906866 | SEMA4C | FALSE | Downregulated |
| 0,338517869 | 0,43998685 | ASAP2 | FALSE | Upregulated |
| 0,320496603 | 0,43998685 | COA5 | FALSE | Upregulated |
| 0,391663642 | 0,43998685 | FOSL2 | FALSE | Upregulated |
| 0,762483253 | 0,43998685 | PLXNB3 | FALSE | Upregulated |
| -0,252443673 | 0,43998685 | PRKAA1 | FALSE | Downregulated |
| 0,217608207 | 0,43998685 | RASA1 | FALSE | Upregulated |
| 0,319694663 | 0,440145755 | FBXL22 | FALSE | Upregulated |
| 0,284613813 | 0,440366064 | ESR1 | FALSE | Upregulated |
| 0,644007202 | 0,440366064 | SETD9 | FALSE | Upregulated |
| 0,320492185 | 0,440784561 | IFITM2 | FALSE | Upregulated |
| 0,365102488 | 0,44079036 | FABP3 | FALSE | Upregulated |
| -0,286386721 | 0,44079036 | SLC23A1 | FALSE | Downregulated |
| -0,230673331 | 0,441050548 | DDX54 | FALSE | Downregulated |
| -0,283595674 | 0,441050548 | RNF13 | FALSE | Downregulated |
| 0,286095106 | 0,441272014 | FANCL | FALSE | Upregulated |
| 0,27478999 | 0,441272014 | RGL1 | FALSE | Upregulated |
| -1,59165357 | 0,441462008 | CRYAB | FALSE | Downregulated |
| -0,754339357 | 0,441462008 | LILRA1 | FALSE | Downregulated |
| -0,287865248 | 0,441746316 | C11orf31 | FALSE | Downregulated |
| 0,330425669 | 0,441880804 | NQO1 | FALSE | Upregulated |
| -0,309178983 | 0,442063424 | RITA1 | FALSE | Downregulated |
| -1,268854515 | 0,442213338 | RP11-295M18.2 | FALSE | Downregulated |
| -0,358332703 | 0,442237879 | GLRX | FALSE | Downregulated |
| -0,380791892 | 0,442237879 | HUS1 | FALSE | Downregulated |
| 0,384632274 | 0,44230636 | CLCF1 | FALSE | Upregulated |
| 0,382257646 | 0,442451286 | RP11-15A1.7 | FALSE | Upregulated |
| 0,762033981 | 0,442478523 | LOC285819 | FALSE | Upregulated |
| -0,358456183 | 0,442813813 | L3MBTL4 | FALSE | Downregulated |
| 0,276605884 | 0,442863163 | C9orf37 | FALSE | Upregulated |
| 0,71382342 | 0,442912179 | RP11-64D24.4 | FALSE | Upregulated |
| 0,293189142 | 0,442956562 | PIGB | FALSE | Upregulated |
| -0,79727192 | 0,443044997 | HRC | FALSE | Downregulated |
| 0,831130798 | 0,443044997 | LOC100132891 | FALSE | Upregulated |
| 0,334844977 | 0,443044997 | MAP3K5 | FALSE | Upregulated |
| 1,173353581 | 0,443044997 | ZBP1 | FALSE | Upregulated |
| -0,197009968 | 0,443104611 | NDUFV1 | FALSE | Downregulated |

| | | | | |
|---|---|---|---|---|
| -0,227929052 | 0,443104611 | ZNF862 | FALSE | Downregulated |
| -0,840951228 | 0,443337569 | LOC102723845 | FALSE | Downregulated |
| -0,240055553 | 0,443480884 | GNPTG | FALSE | Downregulated |
| 0,578691753 | 0,443480884 | MED17 | FALSE | Upregulated |
| -0,526972769 | 0,443509324 | TMEM208 | FALSE | Downregulated |
| -0,800698628 | 0,443716934 | AP4M1 | FALSE | Downregulated |
| -0,862813649 | 0,443716934 | LOC101927858 | FALSE | Downregulated |
| 0,284788861 | 0,444430305 | CABP7 | FALSE | Upregulated |
| 0,597069065 | 0,444430305 | SFTPB | FALSE | Upregulated |
| 0,565508086 | 0,44500131 | ADM2 | FALSE | Upregulated |
| -0,27219214 | 0,44500131 | LIPG | FALSE | Downregulated |
| 0,330725267 | 0,44500131 | LOC101929046 | FALSE | Upregulated |
| -0,276859609 | 0,44500131 | NDUFB7 | FALSE | Downregulated |
| 0,632274518 | 0,44500131 | TOX2 | FALSE | Upregulated |
| -0,202318839 | 0,445360925 | ERP44 | FALSE | Downregulated |
| -1,108003957 | 0,445360925 | SMIM1 | FALSE | Downregulated |
| 0,963450788 | 0,445420425 | LINC00654 | FALSE | Upregulated |
| -0,794816027 | 0,445420425 | LOC102724040 | FALSE | Downregulated |
| -0,332633017 | 0,445475588 | ZNF770 | FALSE | Downregulated |
| -1,40355434 | 0,445509839 | LOC101930033 | FALSE | Downregulated |
| -0,28868697 | 0,445509839 | PKN2 | FALSE | Downregulated |
| 1,164120904 | 0,44568801 | LOC100996535 | FALSE | Upregulated |
| -0,352097267 | 0,44568801 | TIE1 | FALSE | Downregulated |
| 0,387723914 | 0,445822775 | ADRBK2 | FALSE | Upregulated |
| -0,271651094 | 0,445822775 | CACNB2 | FALSE | Downregulated |
| -0,288184596 | 0,445822775 | INTS10 | FALSE | Downregulated |
| 0,861629145 | 0,44593293 | SIRPB1 | FALSE | Upregulated |
| -0,520355869 | 0,445946869 | LSM7 | FALSE | Downregulated |
| 1,237523137 | 0,446023253 | GATSL2 | FALSE | Upregulated |
| -0,741919648 | 0,446079219 | E2F6 | FALSE | Downregulated |
| -0,315698303 | 0,446297735 | KRIT1 | FALSE | Downregulated |
| -0,227245331 | 0,446653007 | CYC1 | FALSE | Downregulated |
| -0,860910228 | 0,446684416 | TMEM256 | FALSE | Downregulated |
| 0,406162845 | 0,446711288 | IMPA2 | FALSE | Upregulated |
| -0,907765203 | 0,446868626 | EYA4 | FALSE | Downregulated |
| 0,277917838 | 0,447198383 | SRF | FALSE | Upregulated |
| -1,109202796 | 0,447340302 | ADAMTS6 | FALSE | Downregulated |
| 0,293477005 | 0,447796482 | PTPN12 | FALSE | Upregulated |
| -0,533211765 | 0,447910939 | ZFAT | FALSE | Downregulated |
| 0,489475213 | 0,448367705 | ZBED5 | FALSE | Upregulated |
| -0,329744915 | 0,448915243 | PFAS | FALSE | Downregulated |
| -0,260012747 | 0,448957589 | TAF1 | FALSE | Downregulated |
| 0,588286695 | 0,449072959 | NCAPG2 | FALSE | Upregulated |
| 0,435036628 | 0,449072959 | PTCD1 | FALSE | Upregulated |
| 0,302879634 | 0,449372894 | CTRL | FALSE | Upregulated |
| 0,836807623 | 0,449536956 | CACNA2D1 | FALSE | Upregulated |
| 0,314617713 | 0,449536956 | CNN2 | FALSE | Upregulated |
| 0,918476075 | 0,449536956 | GATA1 | FALSE | Upregulated |
| 0,275754375 | 0,449536956 | MFSD6 | FALSE | Upregulated |
| -0,322722861 | 0,449536956 | SPRED2 | FALSE | Downregulated |
| -0,730459574 | 0,449536956 | XKR9 | FALSE | Downregulated |

| | | | | |
|---|---|---|---|---|
| -0,648842314 | 0,449536956 | ZSWIM3 | FALSE | Downregulated |
| 0,799505101 | 0,449605978 | LOC101929937 | FALSE | Upregulated |
| 0,309358062 | 0,449716635 | WWC3 | FALSE | Upregulated |
| -0,374364672 | 0,449777553 | LOC100506100 | FALSE | Downregulated |
| 1,052536798 | 0,449909386 | DBF4 | FALSE | Upregulated |
| -0,398554265 | 0,450031595 | BIVM | FALSE | Downregulated |
| 0,329926709 | 0,450031595 | KPNA3 | FALSE | Upregulated |
| -0,464785082 | 0,450031595 | NDUFS4 | FALSE | Downregulated |
| -0,162884832 | 0,450031595 | SETD7 | FALSE | Downregulated |
| 0,173317777 | 0,450031595 | SLC22A23 | FALSE | Upregulated |
| -0,224402542 | 0,450031595 | TBC1D14 | FALSE | Downregulated |
| 0,220138178 | 0,450346297 | GLTSCR1 | FALSE | Upregulated |
| 0,396762438 | 0,450506456 | ERAP2 | FALSE | Upregulated |
| -0,240467046 | 0,450812037 | RPN2 | FALSE | Downregulated |
| -0,955352146 | 0,450959257 | MIR4500HG | FALSE | Downregulated |
| -0,666493808 | 0,450966366 | TCF21 | FALSE | Downregulated |
| 0,442876821 | 0,451007258 | GRIN2C | FALSE | Upregulated |
| 0,697289469 | 0,451222798 | GDF1 | FALSE | Upregulated |
| 0,58517072 | 0,451401988 | CD55 | FALSE | Upregulated |
| -0,394536648 | 0,451401988 | PRADC1 | FALSE | Downregulated |
| 0,818559748 | 0,451678176 | LOC101927756 | FALSE | Upregulated |
| -0,516661848 | 0,451678176 | MUSTN1 | FALSE | Downregulated |
| -0,411565096 | 0,451763104 | PEX2 | FALSE | Downregulated |
| 0,263459634 | 0,45178779 | HMGN1 | FALSE | Upregulated |
| 0,76260769 | 0,45178779 | RP11-680F20.6 | FALSE | Upregulated |
| 0,398741663 | 0,45178779 | USB1 | FALSE | Upregulated |
| 0,944060347 | 0,452057723 | MFSD2B | FALSE | Upregulated |
| 0,215705436 | 0,45207449 | HEATR1 | FALSE | Upregulated |
| 0,32062835 | 0,452218497 | VMP1 | FALSE | Upregulated |
| -0,237118873 | 0,452472556 | DENND4A | FALSE | Downregulated |
| 0,477836982 | 0,452884707 | AFAP1 | FALSE | Upregulated |
| 0,594789353 | 0,452963433 | CDYL2 | FALSE | Upregulated |
| -0,318543621 | 0,453058267 | SLC35F5 | FALSE | Downregulated |
| 0,452121487 | 0,453146823 | FBXO40 | FALSE | Upregulated |
| -0,512090261 | 0,453146823 | FTSJ2 | FALSE | Downregulated |
| -1,01636212 | 0,453621705 | C9orf85 | FALSE | Downregulated |
| 0,170087497 | 0,453621705 | VPS13C | FALSE | Upregulated |
| -0,243884046 | 0,453642902 | LARGE | FALSE | Downregulated |
| 0,21926991 | 0,454055937 | CAPN12 | FALSE | Upregulated |
| -0,693766622 | 0,454055937 | DCP1B | FALSE | Downregulated |
| -0,228458508 | 0,454055937 | F11-AS1 | FALSE | Downregulated |
| 0,204245229 | 0,454055937 | IFITM3 | FALSE | Upregulated |
| -0,505423758 | 0,454055937 | MYH3 | FALSE | Downregulated |
| 0,172915915 | 0,454055937 | NPLOC4 | FALSE | Upregulated |
| -0,290898034 | 0,454111619 | SOS2 | FALSE | Downregulated |
| 0,996572348 | 0,454148125 | PFKFB4 | FALSE | Upregulated |
| -1,170999974 | 0,454211025 | BDNF-AS | FALSE | Downregulated |
| 0,305795744 | 0,454211025 | BECN1 | FALSE | Upregulated |
| 0,227339865 | 0,454211025 | HSP90AB1 | FALSE | Upregulated |
| -1,102518412 | 0,454211025 | LOC100128398 | FALSE | Downregulated |
| 0,425116126 | 0,454211025 | LOC441179 | FALSE | Upregulated |

| | | | | |
|---|---|---|---|---|
| 0,322083013 | 0,454211025 | MFN1 | FALSE | Upregulated |
| -0,498369042 | 0,454211025 | PDE3A | FALSE | Downregulated |
| -0,303797448 | 0,454394355 | DOCK6 | FALSE | Downregulated |
| 0,297390564 | 0,454827505 | FGGY | FALSE | Upregulated |
| -0,240997741 | 0,454827505 | TMUB1 | FALSE | Downregulated |
| -0,841657471 | 0,455223174 | SPTSSB | FALSE | Downregulated |
| 0,486488581 | 0,45524836 | TUBG1 | FALSE | Upregulated |
| -0,41304498 | 0,455465585 | TRAPPC4 | FALSE | Downregulated |
| 0,595542646 | 0,455675752 | ATRIP | FALSE | Upregulated |
| -0,326607021 | 0,455743417 | HIC2 | FALSE | Downregulated |
| 1,16492438 | 0,455743417 | TCHH | FALSE | Upregulated |
| -0,641452559 | 0,455770446 | ARHGAP15 | FALSE | Downregulated |
| -0,447401786 | 0,455770446 | EMBP1 | FALSE | Downregulated |
| 0,266927722 | 0,455770446 | ITGA11 | FALSE | Upregulated |
| 0,227988458 | 0,455770446 | ZFPM1 | FALSE | Upregulated |
| -1,013886367 | 0,456016499 | EMCN | FALSE | Downregulated |
| -0,299215296 | 0,456447784 | CLN8 | FALSE | Downregulated |
| -1,45549087 | 0,456447784 | ITLN1 | FALSE | Downregulated |
| -0,451475603 | 0,456447784 | NKAIN2 | FALSE | Downregulated |
| -0,171135957 | 0,456447784 | PSMB7 | FALSE | Downregulated |
| -0,204838565 | 0,456447784 | USP47 | FALSE | Downregulated |
| 0,208163497 | 0,456447784 | YWHAQ | FALSE | Upregulated |
| 0,246710072 | 0,456535723 | CSF1R | FALSE | Upregulated |
| 0,403465938 | 0,456535723 | DNAJC9 | FALSE | Upregulated |
| 0,247168687 | 0,456535723 | FLJ31945 | FALSE | Upregulated |
| 0,42030582 | 0,456535723 | RAP1GAP | FALSE | Upregulated |
| -0,74978714 | 0,456535723 | UTP11L | FALSE | Downregulated |
| -0,888708449 | 0,456535723 | ZFHX4-AS1 | FALSE | Downregulated |
| 0,415812116 | 0,456932294 | CDH4 | FALSE | Upregulated |
| 0,192274423 | 0,456990537 | SLC25A6 | FALSE | Upregulated |
| -0,569356279 | 0,457200312 | HSD17B3 | FALSE | Downregulated |
| 0,486854802 | 0,457473264 | HIGD1C | FALSE | Upregulated |
| 0,236691898 | 0,457664248 | AMIGO3 | FALSE | Upregulated |
| 0,390573848 | 0,457664248 | COL8A2 | FALSE | Upregulated |
| 0,280438647 | 0,457664248 | DAGLB | FALSE | Upregulated |
| 0,434635366 | 0,457664248 | FZD6 | FALSE | Upregulated |
| -0,186129716 | 0,457664248 | HNRNPF | FALSE | Downregulated |
| 0,366159539 | 0,457750155 | YPEL3 | FALSE | Upregulated |
| 0,579619532 | 0,457764184 | SLC25A19 | FALSE | Upregulated |
| 0,457855109 | 0,457899315 | HMGB3 | FALSE | Upregulated |
| 1,783787816 | 0,457977437 | DUOX2 | FALSE | Upregulated |
| -0,92520234 | 0,457977437 | PGBD1 | FALSE | Downregulated |
| -0,697930781 | 0,457983313 | DCUN1D5 | FALSE | Downregulated |
| 0,378304589 | 0,457983313 | OSTF1 | FALSE | Upregulated |
| 0,22402226 | 0,457983313 | PDIA3 | FALSE | Upregulated |
| -0,694359633 | 0,457983313 | YBEY | FALSE | Downregulated |
| 0,439681772 | 0,458112827 | LOC100132731 | FALSE | Upregulated |
| -0,225083157 | 0,458112827 | SAP30L | FALSE | Downregulated |
| -0,253314565 | 0,458201869 | STRN3 | FALSE | Downregulated |
| -0,5186434 | 0,458420724 | CDHR2 | FALSE | Downregulated |
| -0,439571198 | 0,458420724 | COX7A2L | FALSE | Downregulated |

| | | | | |
|---|---|---|---|---|
| -0,305474298 | 0,458420724 | DIAPH2 | FALSE | Downregulated |
| 0,617819185 | 0,458420724 | ITGBL1 | FALSE | Upregulated |
| -0,30730971 | 0,458420724 | PPP2R2A | FALSE | Downregulated |
| -0,427443737 | 0,458420724 | PSMB1 | FALSE | Downregulated |
| -1,052915467 | 0,458576381 | NEUROD6 | FALSE | Downregulated |
| -0,24769605 | 0,458626622 | ANG | FALSE | Downregulated |
| 0,66391341 | 0,458626622 | LOC101929573 | FALSE | Upregulated |
| 0,566466707 | 0,458846458 | SNRPF | FALSE | Upregulated |
| -0,223797029 | 0,458866646 | AKAP12 | FALSE | Downregulated |
| 0,244511438 | 0,458866646 | CD37 | FALSE | Upregulated |
| 0,626791026 | 0,458866646 | COPZ2 | FALSE | Upregulated |
| -0,418066115 | 0,458866646 | FXYD6 | FALSE | Downregulated |
| -0,807677183 | 0,458866646 | IFT43 | FALSE | Downregulated |
| 0,598962247 | 0,458866646 | KIF20A | FALSE | Upregulated |
| 0,449945248 | 0,458866646 | LOC101928318 | FALSE | Upregulated |
| 0,435588139 | 0,458866646 | MBLAC2 | FALSE | Upregulated |
| 0,756427894 | 0,458866646 | PDZD7 | FALSE | Upregulated |
| -0,481423001 | 0,458866646 | PIGL | FALSE | Downregulated |
| 1,059467222 | 0,458866646 | POLR2J3 | FALSE | Upregulated |
| 0,602514708 | 0,458866646 | RRAGC | FALSE | Upregulated |
| 0,340279126 | 0,458866646 | TNIP2 | FALSE | Upregulated |
| -0,874614954 | 0,458866646 | USP32P1 | FALSE | Downregulated |
| 0,210039251 | 0,458904599 | AGPAT2 | FALSE | Upregulated |
| 0,771339093 | 0,458939016 | C10orf128 | FALSE | Upregulated |
| 0,653507176 | 0,458939016 | C14orf119 | FALSE | Upregulated |
| -0,187298455 | 0,458939016 | FETUB | FALSE | Downregulated |
| 0,555790509 | 0,458939016 | GPR65 | FALSE | Upregulated |
| 0,308121854 | 0,458939016 | PPP4R4 | FALSE | Upregulated |
| 0,453875011 | 0,459001934 | LOC101928206 | FALSE | Upregulated |
| -0,31165607 | 0,459057697 | SCAPER | FALSE | Downregulated |
| 0,36832369 | 0,459057697 | THAP9 | FALSE | Upregulated |
| 0,540953754 | 0,459454416 | CSTB | FALSE | Upregulated |
| 0,431851202 | 0,459910229 | EIF2B3 | FALSE | Upregulated |
| 0,271670241 | 0,459910229 | FAM222B | FALSE | Upregulated |
| 0,717283485 | 0,459910229 | METTL21C | FALSE | Upregulated |
| 0,481821413 | 0,459910229 | PASK | FALSE | Upregulated |
| 0,435681794 | 0,460185272 | FAM65B | FALSE | Upregulated |
| -0,423611037 | 0,46020532 | CNIH1 | FALSE | Downregulated |
| 0,857167516 | 0,460361444 | C8orf76 | FALSE | Upregulated |
| -0,221051222 | 0,460361444 | CHD9 | FALSE | Downregulated |
| 0,555507653 | 0,460361444 | LOC100996643 | FALSE | Upregulated |
| -0,581152929 | 0,460361444 | LOC101927187 | FALSE | Downregulated |
| 0,264995301 | 0,460361444 | LOC101927393 | FALSE | Upregulated |
| -0,354442427 | 0,460361444 | TOP3B | FALSE | Downregulated |
| 0,34021764 | 0,460361444 | VEZT | FALSE | Upregulated |
| 0,307673299 | 0,460412305 | LINC00476 | FALSE | Upregulated |
| -1,174743292 | 0,460412305 | LOC102724348 | FALSE | Downregulated |
| -0,681863467 | 0,460412305 | UNC93A | FALSE | Downregulated |
| 1,037831242 | 0,460429979 | PATL2 | FALSE | Upregulated |
| 0,559217213 | 0,460582871 | LONRF2 | FALSE | Upregulated |
| -1,182683793 | 0,460582871 | SEMA3C | FALSE | Downregulated |

| | | | | |
|---|---|---|---|---|
| 0,272710005 | 0,46073895 | LAT2 | FALSE | Upregulated |
| 0,67185722 | 0,46073895 | LOC101928473 | FALSE | Upregulated |
| 0,547780459 | 0,46073895 | PARVB | FALSE | Upregulated |
| 0,463573299 | 0,46076562 | PPP1R13L | FALSE | Upregulated |
| 0,578552616 | 0,460913769 | C12orf49 | FALSE | Upregulated |
| -0,92051743 | 0,461383854 | CDNF | FALSE | Downregulated |
| -0,67329473 | 0,461383854 | KIF6 | FALSE | Downregulated |
| 0,573762868 | 0,461469162 | RWDD4 | FALSE | Upregulated |
| 0,215779256 | 0,461615814 | EXT1 | FALSE | Upregulated |
| 1,048176986 | 0,461736417 | CD101 | FALSE | Upregulated |
| -0,35353209 | 0,461736417 | ENC1 | FALSE | Downregulated |
| -0,20059412 | 0,461736417 | RPL11 | FALSE | Downregulated |
| 0,363679602 | 0,461736417 | TMEM45B | FALSE | Upregulated |
| 0,270218098 | 0,461771293 | CAPZB | FALSE | Upregulated |
| 0,363743014 | 0,461790686 | C10orf12 | FALSE | Upregulated |
| -0,293608765 | 0,461790686 | LTBP1 | FALSE | Downregulated |
| -1,84431192 | 0,461805319 | MYOM3 | FALSE | Downregulated |
| -0,212091196 | 0,461805319 | ZNF451 | FALSE | Downregulated |
| -0,258306868 | 0,461813404 | AP2S1 | FALSE | Downregulated |
| 0,437593179 | 0,461872172 | CFD | FALSE | Upregulated |
| 0,218692562 | 0,462095398 | EVI2B | FALSE | Upregulated |
| -0,622095655 | 0,462186537 | ASXL3 | FALSE | Downregulated |
| -0,349560357 | 0,462186537 | LETMD1 | FALSE | Downregulated |
| 1,023885427 | 0,462442947 | FAM157A | FALSE | Upregulated |
| 0,3511064 | 0,462459195 | ABHD12 | FALSE | Upregulated |
| 0,371263721 | 0,46262938 | IGSF23 | FALSE | Upregulated |
| 0,260499091 | 0,462785243 | TEP1 | FALSE | Upregulated |
| -0,210605474 | 0,462824944 | COX4I1 | FALSE | Downregulated |
| 0,298352254 | 0,462824944 | MRPS5 | FALSE | Upregulated |
| -0,365973131 | 0,462873064 | AMY2B | FALSE | Downregulated |
| 0,284958317 | 0,463126136 | DENND5A | FALSE | Upregulated |
| 0,274808928 | 0,46326735 | SDCBP | FALSE | Upregulated |
| 0,869934458 | 0,46326735 | SLC18B1 | FALSE | Upregulated |
| 0,349200049 | 0,46326735 | SPSB2 | FALSE | Upregulated |
| -0,334955047 | 0,463316986 | HES1 | FALSE | Downregulated |
| 0,566052372 | 0,463316986 | LEPROTL1 | FALSE | Upregulated |
| -0,318706628 | 0,463316986 | LTBP4 | FALSE | Downregulated |
| 0,730754553 | 0,463615292 | FAM110A | FALSE | Upregulated |
| -0,459192565 | 0,463615292 | MCUR1 | FALSE | Downregulated |
| 0,609805033 | 0,463761539 | DCST1 | FALSE | Upregulated |
| -0,54272016 | 0,463800279 | C14orf80 | FALSE | Downregulated |
| 0,844938877 | 0,463800561 | LOC101929959 | FALSE | Upregulated |
| 0,161708285 | 0,464025088 | SSR1 | FALSE | Upregulated |
| 0,24227296 | 0,464242168 | TARBP2 | FALSE | Upregulated |
| -0,788552549 | 0,464384793 | RSG1 | FALSE | Downregulated |
| 0,44574268 | 0,464384793 | ZBTB22 | FALSE | Upregulated |
| -0,420818686 | 0,464455064 | LAMA4 | FALSE | Downregulated |
| 0,209313523 | 0,464513704 | SMG7 | FALSE | Upregulated |
| -0,176451761 | 0,464629617 | SELO | FALSE | Downregulated |
| 0,424950152 | 0,464761391 | LOC100130992 | FALSE | Upregulated |
| 0,467847542 | 0,464853072 | RCBTB1 | FALSE | Upregulated |

| | | | | |
|---|---|---|---|---|
| -0,746516771 | 0,464905069 | APOPT1 | FALSE | Downregulated |
| 0,358614108 | 0,464905069 | PLIN3 | FALSE | Upregulated |
| 0,401137888 | 0,465271097 | LOC101927124 | FALSE | Upregulated |
| -0,196916881 | 0,465304383 | C1orf115 | FALSE | Downregulated |
| -0,309639597 | 0,465304383 | CYP2R1 | FALSE | Downregulated |
| 0,857164112 | 0,465304383 | FITM2 | FALSE | Upregulated |
| -0,436125855 | 0,465304383 | LOC101929522 | FALSE | Downregulated |
| -0,247012008 | 0,465304383 | PRPF3 | FALSE | Downregulated |
| 0,679935477 | 0,465304383 | RNF175 | FALSE | Upregulated |
| -0,187866336 | 0,465304383 | SIN3A | FALSE | Downregulated |
| 0,577811447 | 0,465304383 | ZNF174 | FALSE | Upregulated |
| -0,659649401 | 0,465461276 | ANKMY2 | FALSE | Downregulated |
| 0,513953547 | 0,465461276 | FICD | FALSE | Upregulated |
| -0,106287434 | 0,465461276 | PLEC | FALSE | Downregulated |
| 0,317285857 | 0,465461276 | SLC48A1 | FALSE | Upregulated |
| 0,679754075 | 0,465473262 | APBA2 | FALSE | Upregulated |
| 0,194254867 | 0,465473262 | SLC26A1 | FALSE | Upregulated |
| -0,430948044 | 0,465481748 | CYB5R1 | FALSE | Downregulated |
| 0,670174062 | 0,465481748 | INSM2 | FALSE | Upregulated |
| 0,544052473 | 0,465481748 | SHQ1 | FALSE | Upregulated |
| 0,34897872 | 0,465577826 | NARF | FALSE | Upregulated |
| 0,425934133 | 0,465577826 | SLIT1 | FALSE | Upregulated |
| -0,348436089 | 0,465806121 | EXOC6B | FALSE | Downregulated |
| 0,335834171 | 0,465806121 | LOC101929005 | FALSE | Upregulated |
| 0,401744405 | 0,465806121 | S100A10 | FALSE | Upregulated |
| 0,469586939 | 0,465809378 | CBY1 | FALSE | Upregulated |
| -0,442902312 | 0,465881791 | SHFM1 | FALSE | Downregulated |
| 0,314001522 | 0,466129578 | C2CD3 | FALSE | Upregulated |
| -0,351268272 | 0,466129578 | LINC01061 | FALSE | Downregulated |
| -0,154123732 | 0,466340444 | RPS2 | FALSE | Downregulated |
| -0,403578268 | 0,466837226 | LOC100506175 | FALSE | Downregulated |
| 0,90004567 | 0,46729036 | ANKRD65 | FALSE | Upregulated |
| -0,837012696 | 0,467417615 | RTN4RL1 | FALSE | Downregulated |
| -0,405053098 | 0,467546695 | ANKRD42 | FALSE | Downregulated |
| -0,415415145 | 0,467546695 | MRE11A | FALSE | Downregulated |
| -0,368363828 | 0,467701118 | CUTA | FALSE | Downregulated |
| -0,274471605 | 0,467701118 | FERMT2 | FALSE | Downregulated |
| -0,310188255 | 0,467701118 | ZNF280D | FALSE | Downregulated |
| 0,58764517 | 0,467827456 | HACE1 | FALSE | Upregulated |
| 0,26107121 | 0,467903605 | AGFG1 | FALSE | Upregulated |
| -0,746152056 | 0,467903605 | NRROS | FALSE | Downregulated |
| -0,206374156 | 0,46827274 | ATF4 | FALSE | Downregulated |
| 0,356282278 | 0,46827274 | CLDN24 | FALSE | Upregulated |
| -0,192419835 | 0,46827274 | DVL3 | FALSE | Downregulated |
| -0,687325966 | 0,46827274 | GRHL2 | FALSE | Downregulated |
| -0,226717052 | 0,46827274 | KPNA6 | FALSE | Downregulated |
| -0,497228894 | 0,46827274 | TIMM17B | FALSE | Downregulated |
| -0,753514037 | 0,46827274 | ZNF311 | FALSE | Downregulated |
| -0,552967559 | 0,46827274 | ZNF324B | FALSE | Downregulated |
| -0,166164612 | 0,468521333 | PDE8A | FALSE | Downregulated |
| 0,249046358 | 0,468630787 | TLK2 | FALSE | Upregulated |

| | | | | |
|---|---|---|---|---|
| -0,369823463 | 0,468694358 | TMTC3 | FALSE | Downregulated |
| -1,054455057 | 0,468766398 | MICALCL | FALSE | Downregulated |
| -0,421062552 | 0,468766398 | TRABD2B | FALSE | Downregulated |
| -0,959332861 | 0,469143515 | ZNF468 | FALSE | Downregulated |
| 0,248686132 | 0,469463192 | HSPA4 | FALSE | Upregulated |
| 0,340137599 | 0,469463192 | JDP2 | FALSE | Upregulated |
| -0,378755737 | 0,469463192 | STARD13-AS | FALSE | Downregulated |
| 0,213664465 | 0,46977309 | RBM5-AS1 | FALSE | Upregulated |
| -0,218370712 | 0,469956521 | FAM107B | FALSE | Downregulated |
| 0,210486236 | 0,470035499 | RRN3P3 | FALSE | Upregulated |
| 0,855873826 | 0,470285411 | FOXC1 | FALSE | Upregulated |
| 0,923674066 | 0,470285411 | HDX | FALSE | Upregulated |
| 0,597768257 | 0,470285411 | RP4-777D9.2 | FALSE | Upregulated |
| 0,303771402 | 0,470331085 | ARMCX3 | FALSE | Upregulated |
| -0,196250458 | 0,470331085 | C1R-2 | FALSE | Downregulated |
| -0,156920534 | 0,470331085 | DIAPH1 | FALSE | Downregulated |
| 0,331627118 | 0,470331085 | DONSON | FALSE | Upregulated |
| -0,20271942 | 0,470331085 | HDAC5 | FALSE | Downregulated |
| -0,828499712 | 0,470331085 | KRTCAP3 | FALSE | Downregulated |
| -0,244539116 | 0,470694212 | STOML2 | FALSE | Downregulated |
| -0,27010479 | 0,470849317 | SCARF1 | FALSE | Downregulated |
| 0,63559262 | 0,471212419 | CBX8 | FALSE | Upregulated |
| 0,380493957 | 0,471212419 | CDR2 | FALSE | Upregulated |
| -0,44851555 | 0,471212419 | FBXL12 | FALSE | Downregulated |
| -0,333017358 | 0,471212419 | GRAMD3 | FALSE | Downregulated |
| -0,399704907 | 0,471212419 | YPEL5 | FALSE | Downregulated |
| -0,86642732 | 0,471516736 | SALL2 | FALSE | Downregulated |
| 1,353890714 | 0,471525452 | GIMAP1-GIMAP5 | FALSE | Upregulated |
| 0,439761044 | 0,471645298 | LOC102724372 | FALSE | Upregulated |
| -0,358431002 | 0,471645298 | TMEM69 | FALSE | Downregulated |
| 0,244235536 | 0,471934093 | EVI2A | FALSE | Upregulated |
| -0,964034211 | 0,471997063 | RNASEH1-AS1 | FALSE | Downregulated |
| -0,614336452 | 0,47252176 | CYP2B7P | FALSE | Downregulated |
| -0,70351863 | 0,47252176 | FAM167B | FALSE | Downregulated |
| -0,380507482 | 0,472596927 | DENND5B-AS1 | FALSE | Downregulated |
| 1,079319127 | 0,472794254 | ARL6 | FALSE | Upregulated |
| -0,638393603 | 0,472794254 | LOC100130876 | FALSE | Downregulated |
| 0,712740838 | 0,473269148 | RP11-611D20.2 | FALSE | Upregulated |
| 0,354666023 | 0,473364775 | INMT | FALSE | Upregulated |
| -0,538441931 | 0,473516656 | LOC101929195 | FALSE | Downregulated |
| 0,528226419 | 0,473516656 | TMEM255A | FALSE | Upregulated |
| -0,629497776 | 0,473564701 | MKS1 | FALSE | Downregulated |
| 0,209242233 | 0,473815578 | ASB6 | FALSE | Upregulated |
| -0,360110226 | 0,473815578 | CPNE8 | FALSE | Downregulated |
| 0,323953452 | 0,473834014 | ALKBH3-AS1 | FALSE | Upregulated |
| -0,31682503 | 0,474069156 | CCDC14 | FALSE | Downregulated |
| -0,206178361 | 0,474069156 | VASH1 | FALSE | Downregulated |
| -0,217294713 | 0,474108505 | PCCA | FALSE | Downregulated |
| 0,657390669 | 0,47438982 | C9orf117 | FALSE | Upregulated |
| -0,302815084 | 0,47438982 | CACNA1D | FALSE | Downregulated |
| 0,408971967 | 0,47438982 | COA7 | FALSE | Upregulated |

| | | | | |
|---|---|---|---|---|
| 0,665530449 | 0,47438982 | LOC101928025 | FALSE | Upregulated |
| 0,290743326 | 0,47438982 | LOC102723328 | FALSE | Upregulated |
| 1,041128095 | 0,47438982 | SCN1A | FALSE | Upregulated |
| 0,766288114 | 0,47438982 | TYRO3P | FALSE | Upregulated |
| 0,939026711 | 0,474804044 | SAAL1 | FALSE | Upregulated |
| 0,213581387 | 0,474877906 | ANGPTL6 | FALSE | Upregulated |
| -0,286826422 | 0,474877906 | HPS4 | FALSE | Downregulated |
| 0,211741669 | 0,474945289 | C3orf17 | FALSE | Upregulated |
| 0,169673455 | 0,474945289 | SH2B3 | FALSE | Upregulated |
| -0,853583068 | 0,474945289 | TMPRSS11E | FALSE | Downregulated |
| 0,624806664 | 0,475249581 | RNF150 | FALSE | Upregulated |
| -0,385054111 | 0,475469954 | LOC101927761 | FALSE | Downregulated |
| 0,458164846 | 0,475548017 | TMEM194A | FALSE | Upregulated |
| -0,221391083 | 0,475574312 | DLG1 | FALSE | Downregulated |
| -0,994389514 | 0,475574312 | LOC101928895 | FALSE | Downregulated |
| 1,052262546 | 0,475574312 | RP11-87E22.2 | FALSE | Upregulated |
| 0,644373554 | 0,475574312 | SYTL3 | FALSE | Upregulated |
| 0,626196009 | 0,475943588 | MADCAM1 | FALSE | Upregulated |
| -0,1004061 | 0,475944357 | MAVS | FALSE | Downregulated |
| -0,802203552 | 0,475979689 | AKR1E2 | FALSE | Downregulated |
| 0,242371247 | 0,475979689 | PACSIN2 | FALSE | Upregulated |
| -0,428300806 | 0,476016983 | ZNF524 | FALSE | Downregulated |
| 0,394742293 | 0,476030638 | MEGF6 | FALSE | Upregulated |
| -0,265297322 | 0,476030638 | PDDC1 | FALSE | Downregulated |
| 0,439290593 | 0,476440029 | EFTUD1 | FALSE | Upregulated |
| 0,50054737 | 0,476440029 | SFMBT2 | FALSE | Upregulated |
| 0,762382505 | 0,476440029 | SLCO2A1 | FALSE | Upregulated |
| 0,792240099 | 0,476681508 | HIST1H4K | FALSE | Upregulated |
| -0,564454425 | 0,476681508 | NDRG3 | FALSE | Downregulated |
| -0,314017867 | 0,476691207 | LOC102724281 | FALSE | Downregulated |
| 0,403193097 | 0,476792386 | ZNF319 | FALSE | Upregulated |
| -0,771200595 | 0,476853942 | CD34 | FALSE | Downregulated |
| -0,431462911 | 0,477312965 | ZNF680 | FALSE | Downregulated |
| -0,378219739 | 0,477313481 | MAP2K5 | FALSE | Downregulated |
| -0,77250976 | 0,477313481 | SEMA3B | FALSE | Downregulated |
| -0,548383561 | 0,477313481 | SOWAHA | FALSE | Downregulated |
| 0,487991338 | 0,477463731 | NIPSNAP3A | FALSE | Upregulated |
| -1,60763612 | 0,477620393 | TPSAB1 | FALSE | Downregulated |
| 0,508951252 | 0,477804656 | RP11-432I5.1 | FALSE | Upregulated |
| 0,312077338 | 0,478197267 | ANAPC13 | FALSE | Upregulated |
| 0,516861026 | 0,478197267 | SCPEP1 | FALSE | Upregulated |
| -0,540140304 | 0,47844924 | ZNF418 | FALSE | Downregulated |
| -0,321477085 | 0,478614444 | ZNF780A | FALSE | Downregulated |
| 0,483419919 | 0,478762498 | POLG2 | FALSE | Upregulated |
| -0,216887111 | 0,478940067 | ERGIC3 | FALSE | Downregulated |
| -0,224398406 | 0,478947319 | C8B | FALSE | Downregulated |
| 0,184851337 | 0,478947319 | DIO1 | FALSE | Upregulated |
| 0,33465528 | 0,478947319 | PQLC3 | FALSE | Upregulated |
| 0,902462797 | 0,478947319 | RP11-829H16.3 | FALSE | Upregulated |
| 0,711783429 | 0,478947319 | S100A6 | FALSE | Upregulated |
| -0,460780114 | 0,478947319 | SLC22A10 | FALSE | Downregulated |

| | | | | |
|---|---|---|---|---|
| -0,522618509 | 0,478947319 | YIF1B | FALSE | Downregulated |
| 0,210503974 | 0,479044621 | CD46 | FALSE | Upregulated |
| 0,261286699 | 0,479179277 | GABPB1 | FALSE | Upregulated |
| 0,27847736 | 0,479179277 | ICA1L | FALSE | Upregulated |
| 0,37659812 | 0,479179277 | RASGRF2 | FALSE | Upregulated |
| -0,29551004 | 0,479179277 | TMED4 | FALSE | Downregulated |
| 0,566948024 | 0,479179277 | ZNF443 | FALSE | Upregulated |
| -0,250896145 | 0,479318208 | PIAS1 | FALSE | Downregulated |
| -0,685911343 | 0,479430708 | NHEJ1 | FALSE | Downregulated |
| -0,397963864 | 0,479471625 | SLC16A13 | FALSE | Downregulated |
| 0,213909881 | 0,479616407 | SMARCA5 | FALSE | Upregulated |
| 0,286162583 | 0,479784325 | EIF2AK2 | FALSE | Upregulated |
| 0,332951047 | 0,480078514 | TOMM34 | FALSE | Upregulated |
| 0,72561888 | 0,480155962 | CEP135 | FALSE | Upregulated |
| 0,29516652 | 0,480155962 | FAM169A | FALSE | Upregulated |
| -0,65374636 | 0,480155962 | FASTKD1 | FALSE | Downregulated |
| 0,275942984 | 0,480259917 | JAM3 | FALSE | Upregulated |
| -0,169542005 | 0,480493245 | TMEM248 | FALSE | Downregulated |
| -0,288617578 | 0,480707826 | C4A | FALSE | Downregulated |
| 0,321359191 | 0,480790339 | ACOT8 | FALSE | Upregulated |
| 0,193839686 | 0,480790339 | FKBP9 | FALSE | Upregulated |
| 0,765031533 | 0,480790339 | LOC101930275 | FALSE | Upregulated |
| -0,382300565 | 0,480790339 | LTB4R | FALSE | Downregulated |
| 0,42041014 | 0,480790339 | RP11-54O15.3 | FALSE | Upregulated |
| -0,261832802 | 0,480790339 | TGFBR3L | FALSE | Downregulated |
| -0,769736902 | 0,480801376 | COX17 | FALSE | Downregulated |
| 0,732633194 | 0,480819284 | SLC9B1 | FALSE | Upregulated |
| 0,303260226 | 0,480839651 | LOC101929040 | FALSE | Upregulated |
| 0,58269722 | 0,480890451 | C16orf93 | FALSE | Upregulated |
| 0,270893085 | 0,480972503 | NUP62 | FALSE | Upregulated |
| -0,306051805 | 0,481192252 | ABCD4 | FALSE | Downregulated |
| 0,476686496 | 0,481192252 | SYT1 | FALSE | Upregulated |
| 0,1808636 | 0,481202754 | CLIP1 | FALSE | Upregulated |
| 0,717509583 | 0,481202754 | DBR1 | FALSE | Upregulated |
| -0,604194977 | 0,481202754 | DDO | FALSE | Downregulated |
| 0,289845955 | 0,481202754 | ECM2 | FALSE | Upregulated |
| -0,71528131 | 0,481202754 | FAM86B3P | FALSE | Downregulated |
| 0,401594456 | 0,481202754 | SAMD1 | FALSE | Upregulated |
| 0,245885061 | 0,481202754 | SBF2-AS1 | FALSE | Upregulated |
| 0,620990425 | 0,481202754 | SLC9A6 | FALSE | Upregulated |
| -0,248192046 | 0,481202754 | TPP2 | FALSE | Downregulated |
| -0,367399777 | 0,481202754 | ZCCHC7 | FALSE | Downregulated |
| 0,340853183 | 0,481319526 | B3GAT2 | FALSE | Upregulated |
| 0,40869184 | 0,481713189 | GIMAP4 | FALSE | Upregulated |
| 0,279281005 | 0,481713189 | WBP11 | FALSE | Upregulated |
| 0,170272006 | 0,481803194 | FAM208A | FALSE | Upregulated |
| -0,210179245 | 0,482041517 | CXXC5 | FALSE | Downregulated |
| -1,012091805 | 0,482041517 | DLEU2L | FALSE | Downregulated |
| 0,501308753 | 0,482041517 | KY | FALSE | Upregulated |
| -0,404417609 | 0,482041517 | SMG9 | FALSE | Downregulated |
| -0,193583488 | 0,482051849 | LINC01000 | FALSE | Downregulated |

| | | | | |
|---|---|---|---|---|
| -0,278670761 | 0,482052159 | GATA4 | FALSE | Downregulated |
| -0,335580372 | 0,48206861 | CCDC90B | FALSE | Downregulated |
| -0,371087431 | 0,482256638 | OR13D1 | FALSE | Downregulated |
| -0,144567009 | 0,482256638 | RPS18 | FALSE | Downregulated |
| -0,176265797 | 0,482256638 | SHANK2 | FALSE | Downregulated |
| 0,273857907 | 0,482256638 | ZNF227 | FALSE | Upregulated |
| 0,218181947 | 0,482407474 | UBE2D4 | FALSE | Upregulated |
| 0,206427708 | 0,482646167 | RABGAP1L | FALSE | Upregulated |
| -1,220524979 | 0,482710156 | LINC00890 | FALSE | Downregulated |
| -0,391926904 | 0,482710156 | SYT12 | FALSE | Downregulated |
| 0,718822513 | 0,482991961 | TBX21 | FALSE | Upregulated |
| 0,478372566 | 0,483026408 | LOC101928967 | FALSE | Upregulated |
| 0,414163029 | 0,483177919 | HSPB9 | FALSE | Upregulated |
| -0,398389395 | 0,483374222 | SEC22A | FALSE | Downregulated |
| -0,229923746 | 0,483374222 | SYT9 | FALSE | Downregulated |
| -0,179549937 | 0,484050398 | C6orf106 | FALSE | Downregulated |
| -0,228302446 | 0,484050398 | INO80D | FALSE | Downregulated |
| 0,287306915 | 0,484050398 | PSMD1 | FALSE | Upregulated |
| 0,303701439 | 0,484050398 | RAB18 | FALSE | Upregulated |
| 0,661994809 | 0,484050398 | RNF219 | FALSE | Upregulated |
| -0,783964671 | 0,484101329 | LOC102724938 | FALSE | Downregulated |
| 0,577613235 | 0,484162471 | AC159540.1-2 | FALSE | Upregulated |
| 0,478332075 | 0,484162471 | AKAP7 | FALSE | Upregulated |
| -0,506775386 | 0,484162471 | CDKL5 | FALSE | Downregulated |
| -0,196126459 | 0,484162471 | CES2 | FALSE | Downregulated |
| 0,426410611 | 0,484162471 | COQ7 | FALSE | Upregulated |
| -0,259194753 | 0,484162471 | DDRGK1 | FALSE | Downregulated |
| -0,543484712 | 0,484162471 | GLUD2 | FALSE | Downregulated |
| -0,282786044 | 0,484162471 | LAMA5 | FALSE | Downregulated |
| -0,488713309 | 0,484162471 | LINC00161 | FALSE | Downregulated |
| -0,555305189 | 0,484162471 | LOC101927610 | FALSE | Downregulated |
| -0,6333891 | 0,484162471 | LOC102723405 | FALSE | Downregulated |
| -0,416921202 | 0,484162471 | LOC102724921 | FALSE | Downregulated |
| -0,35896915 | 0,484162471 | NHLRC3 | FALSE | Downregulated |
| -0,28982857 | 0,484162471 | PCIF1 | FALSE | Downregulated |
| 0,383343626 | 0,484162471 | PPP1R8 | FALSE | Upregulated |
| 0,240299006 | 0,484162471 | RAVER2 | FALSE | Upregulated |
| 0,200133677 | 0,484162471 | RHOA | FALSE | Upregulated |
| 0,713477528 | 0,484162471 | RP11-264C15.2 | FALSE | Upregulated |
| -0,264431354 | 0,484162471 | SEC24B | FALSE | Downregulated |
| 0,241704709 | 0,484162471 | SLC24A1 | FALSE | Upregulated |
| 0,293569874 | 0,484162471 | TIAL1 | FALSE | Upregulated |
| 0,336121958 | 0,484181625 | ITPKB | FALSE | Upregulated |
| -0,161387634 | 0,484290913 | TCEB3 | FALSE | Downregulated |
| 0,432465983 | 0,484575716 | KIAA1045 | FALSE | Upregulated |
| 0,443652776 | 0,48499602 | CCR7 | FALSE | Upregulated |
| 0,337715807 | 0,485067677 | BANF1 | FALSE | Upregulated |
| -0,212223794 | 0,485067677 | CUL5 | FALSE | Downregulated |
| -0,337690047 | 0,485067677 | DDC | FALSE | Downregulated |
| 0,447518695 | 0,485067677 | LOC101929670 | FALSE | Upregulated |
| 0,480366184 | 0,485067677 | LOC93622 | FALSE | Upregulated |

| | | | | |
|---|---|---|---|---|
| -0,217014621 | 0,485067677 | TMEM30A | FALSE | Downregulated |
| -0,251481493 | 0,485067677 | UGT2B4 | FALSE | Downregulated |
| -0,394289131 | 0,485067677 | ZNF490 | FALSE | Downregulated |
| -0,358827097 | 0,485179755 | KBTBD7 | FALSE | Downregulated |
| 0,254930822 | 0,485186854 | HN1L | FALSE | Upregulated |
| -0,478857138 | 0,485186854 | PPIC | FALSE | Downregulated |
| -0,366644604 | 0,485285383 | ALDOC | FALSE | Downregulated |
| -0,31201976 | 0,485314133 | RPS12 | FALSE | Downregulated |
| -0,229807774 | 0,485368704 | KLHDC3 | FALSE | Downregulated |
| 0,719623992 | 0,48538074 | LOC101929790 | FALSE | Upregulated |
| -0,298213356 | 0,48538074 | MST1L | FALSE | Downregulated |
| -0,395734225 | 0,48538074 | NDUFB2 | FALSE | Downregulated |
| 0,738937264 | 0,48538074 | VEGFC | FALSE | Upregulated |
| 0,264629054 | 0,485795711 | SETMAR | FALSE | Upregulated |
| 0,269842338 | 0,485830308 | CCNK | FALSE | Upregulated |
| 0,2951447 | 0,485830308 | HINT3 | FALSE | Upregulated |
| 0,293474664 | 0,486049189 | PLCH2 | FALSE | Upregulated |
| -0,636777807 | 0,486214135 | GGPS1 | FALSE | Downregulated |
| 0,361421967 | 0,486214135 | RP3-325F22.3 | FALSE | Upregulated |
| 0,866634605 | 0,48627891 | LOC101929656 | FALSE | Upregulated |
| 0,213018817 | 0,486355433 | SDR39U1 | FALSE | Upregulated |
| -0,172348752 | 0,486753099 | PARD3 | FALSE | Downregulated |
| 0,212828544 | 0,486823635 | DHDDS | FALSE | Upregulated |
| -0,350259477 | 0,487163673 | ZNF189 | FALSE | Downregulated |
| 0,260552548 | 0,487360188 | PPM1M | FALSE | Upregulated |
| 0,320858686 | 0,487360188 | ZNF791 | FALSE | Upregulated |
| -0,235789587 | 0,487800742 | CTIF | FALSE | Downregulated |
| 0,379765621 | 0,487800742 | DOCK10 | FALSE | Upregulated |
| 0,516653551 | 0,487800742 | FAM53B-AS1 | FALSE | Upregulated |
| 0,707632125 | 0,487800742 | FAM60A | FALSE | Upregulated |
| 0,180110651 | 0,487800742 | GLTPD2 | FALSE | Upregulated |
| 0,465828895 | 0,488293232 | RIC8B | FALSE | Upregulated |
| -0,332287535 | 0,488341385 | KIAA0753 | FALSE | Downregulated |
| 0,292581908 | 0,488341385 | LOC400553 | FALSE | Upregulated |
| 0,960420692 | 0,488341385 | RIN1 | FALSE | Upregulated |
| 0,589160975 | 0,488359518 | KCTD1 | FALSE | Upregulated |
| -0,194538596 | 0,488359518 | TRMT2A | FALSE | Downregulated |
| -0,29004932 | 0,488465661 | PANX1 | FALSE | Downregulated |
| -0,234152104 | 0,488503828 | BCLAF1 | FALSE | Downregulated |
| 0,37281439 | 0,488503828 | MIF4GD | FALSE | Upregulated |
| -0,215599671 | 0,488591701 | BAHCC1 | FALSE | Downregulated |
| 0,980073016 | 0,488634626 | ANKRD13B | FALSE | Upregulated |
| -0,302127939 | 0,488753722 | DCN | FALSE | Downregulated |
| 0,176872019 | 0,489000199 | FBXO11 | FALSE | Upregulated |
| 0,678720282 | 0,489000199 | LINS | FALSE | Upregulated |
| 0,879019954 | 0,489096782 | LOC100132249 | FALSE | Upregulated |
| -0,927086146 | 0,489353774 | KPTN | FALSE | Downregulated |
| -0,460415812 | 0,489457404 | LINC00574 | FALSE | Downregulated |
| -0,244536297 | 0,489457404 | PURA | FALSE | Downregulated |
| 0,869569964 | 0,489492082 | ZMYND10 | FALSE | Upregulated |
| -0,320351718 | 0,490049038 | UBXN7 | FALSE | Downregulated |

| | | | | |
|---|---|---|---|---|
| -0,258349222 | 0,490060103 | VPS9D1 | FALSE | Downregulated |
| 0,257999321 | 0,490084713 | SLC39A13 | FALSE | Upregulated |
| 0,363534383 | 0,490152433 | ADAM8 | FALSE | Upregulated |
| 0,294829136 | 0,490152433 | GNAI3 | FALSE | Upregulated |
| -0,413659852 | 0,490152433 | SLC25A26 | FALSE | Downregulated |
| 0,610796015 | 0,490345099 | ARSJ | FALSE | Upregulated |
| 0,205838247 | 0,490355226 | NCOA3 | FALSE | Upregulated |
| 0,759970114 | 0,490677729 | ARPC4-TTLL3 | FALSE | Upregulated |
| -0,457764803 | 0,490720308 | FBXO3 | FALSE | Downregulated |
| 0,663429215 | 0,490816908 | DCDC2 | FALSE | Upregulated |
| 0,709151078 | 0,490816908 | PTGES3L | FALSE | Upregulated |
| -0,336500564 | 0,490872401 | CAPN3 | FALSE | Downregulated |
| 0,429272681 | 0,490872401 | CLEC2B | FALSE | Upregulated |
| 0,226711835 | 0,490882003 | AXIN1 | FALSE | Upregulated |
| -0,223487555 | 0,491059169 | GTF2IRD2B | FALSE | Downregulated |
| 0,191962755 | 0,491059169 | PDXK | FALSE | Upregulated |
| 0,728750037 | 0,491080436 | LOC101927125 | FALSE | Upregulated |
| 0,524005654 | 0,491291867 | TM4SF1 | FALSE | Upregulated |
| -0,659990148 | 0,491291867 | ZNF366 | FALSE | Downregulated |
| 0,535261585 | 0,49133376 | ADM5 | FALSE | Upregulated |
| -0,659087282 | 0,491446922 | ZNF573 | FALSE | Downregulated |
| -0,31521502 | 0,491527411 | AARS2 | FALSE | Downregulated |
| -0,122208259 | 0,491591358 | ZC3H7B | FALSE | Downregulated |
| -0,196175899 | 0,491692746 | EIF4A2 | FALSE | Downregulated |
| -0,275627382 | 0,491692746 | MESTIT1 | FALSE | Downregulated |
| 0,860993204 | 0,491725625 | WDR38 | FALSE | Upregulated |
| 0,262749844 | 0,491780052 | ZNHIT3 | FALSE | Upregulated |
| 0,199546987 | 0,491788008 | KIF5B | FALSE | Upregulated |
| 0,219744262 | 0,491788008 | LOC286190 | FALSE | Upregulated |
| 0,662634619 | 0,491788008 | ZEB2-AS1 | FALSE | Upregulated |
| -0,420272323 | 0,492149715 | LAMTOR3 | FALSE | Downregulated |
| -0,191632818 | 0,492149715 | NPEPPS | FALSE | Downregulated |
| -0,172391392 | 0,492149715 | SCYL1 | FALSE | Downregulated |
| -0,23688926 | 0,492304693 | PTPN3 | FALSE | Downregulated |
| 0,769607101 | 0,492791971 | RASEF | FALSE | Upregulated |
| 0,811805367 | 0,492874575 | CLDND2 | FALSE | Upregulated |
| -0,708034027 | 0,492893537 | ZNF485 | FALSE | Downregulated |
| -0,196478904 | 0,49301184 | CLDN1 | FALSE | Downregulated |
| -0,377475128 | 0,49301184 | MRPS30 | FALSE | Downregulated |
| -0,829931262 | 0,493115937 | TMEM38A | FALSE | Downregulated |
| 0,520034256 | 0,493288468 | FRMD3 | FALSE | Upregulated |
| 1,187404174 | 0,493288468 | NELL2 | FALSE | Upregulated |
| -0,751681514 | 0,49337571 | POLE2 | FALSE | Downregulated |
| -0,705309364 | 0,49337571 | SLC16A5 | FALSE | Downregulated |
| 0,365482001 | 0,49337571 | ZSWIM4 | FALSE | Upregulated |
| -0,58209929 | 0,493462597 | SLC7A9 | FALSE | Downregulated |
| 1,011444624 | 0,493791855 | PLD4 | FALSE | Upregulated |
| 0,350886939 | 0,493980659 | C14orf1 | FALSE | Upregulated |
| 0,287662359 | 0,493980659 | FGFR2 | FALSE | Upregulated |
| 0,26577217 | 0,493980659 | GK3P | FALSE | Upregulated |
| -0,458060162 | 0,493980659 | SFRP5 | FALSE | Downregulated |

| | | | | |
|---|---|---|---|---|
| 0,366289329 | 0,493980659 | YARS2 | FALSE | Upregulated |
| 0,189754582 | 0,494000847 | CMPK1 | FALSE | Upregulated |
| 0,423763152 | 0,494118951 | DDX50 | FALSE | Upregulated |
| -0,876681329 | 0,494353721 | LEFTY1 | FALSE | Downregulated |
| 0,250975469 | 0,494630254 | BRAT1 | FALSE | Upregulated |
| -0,234504811 | 0,49474997 | SCAND1 | FALSE | Downregulated |
| 0,70239294 | 0,494804517 | NUTM2F | FALSE | Upregulated |
| 0,745162845 | 0,494804517 | RGS7BP | FALSE | Upregulated |
| -0,306354337 | 0,495072562 | LOC102724016 | FALSE | Downregulated |
| -0,201652564 | 0,495296283 | LTBP3 | FALSE | Downregulated |
| -0,256167445 | 0,495398654 | DNAJC16 | FALSE | Downregulated |
| 0,509047243 | 0,495483892 | HAGHL | FALSE | Upregulated |
| -0,22123889 | 0,495483892 | LYST | FALSE | Downregulated |
| 1,075255146 | 0,495483892 | RUNX1-IT1 | FALSE | Upregulated |
| -3,85101918 | 0,495483892 | ZNF559-ZNF177 | FALSE | Downregulated |
| 0,654573735 | 0,495489961 | RNF2 | FALSE | Upregulated |
| 0,308843367 | 0,495630903 | GK | FALSE | Upregulated |
| -1,105539227 | 0,495672608 | HILPDA | FALSE | Downregulated |
| -0,433999363 | 0,495734587 | HMGXB4 | FALSE | Downregulated |
| -0,249842819 | 0,495988115 | HIATL1 | FALSE | Downregulated |
| -0,203937572 | 0,496020428 | FBXL3 | FALSE | Downregulated |
| 0,279827047 | 0,496020428 | MTMR1 | FALSE | Upregulated |
| 0,601520257 | 0,496020428 | TRMT61B | FALSE | Upregulated |
| -0,331895853 | 0,496020428 | ZNF780B | FALSE | Downregulated |
| -0,255373341 | 0,496135849 | SLC39A9 | FALSE | Downregulated |
| 0,489932106 | 0,496278399 | MED18 | FALSE | Upregulated |
| -0,207988448 | 0,496278399 | SLTM | FALSE | Downregulated |
| 0,480951605 | 0,496278399 | SRXN1 | FALSE | Upregulated |
| 0,20712098 | 0,496338453 | SPARC | FALSE | Upregulated |
| -0,350941908 | 0,496365817 | DMAP1 | FALSE | Downregulated |
| 0,201672597 | 0,496365817 | VAV2 | FALSE | Upregulated |
| -0,859163824 | 0,49637326 | C7orf13 | FALSE | Downregulated |
| -0,264841219 | 0,49637548 | YIPF3 | FALSE | Downregulated |
| -0,243219533 | 0,496398063 | ATMIN | FALSE | Downregulated |
| -0,675987716 | 0,496398063 | CYP3A7 | FALSE | Downregulated |
| -0,757262142 | 0,496398063 | FAT3 | FALSE | Downregulated |
| -0,18909137 | 0,496586259 | DYRK1A | FALSE | Downregulated |
| 0,366121602 | 0,496586259 | ZBTB42 | FALSE | Upregulated |
| 0,401164433 | 0,496910474 | SNX29 | FALSE | Upregulated |
| 0,14910864 | 0,49697035 | SP1 | FALSE | Upregulated |
| -0,638924953 | 0,496981154 | TNN | FALSE | Downregulated |
| -0,807062397 | 0,497002102 | CEP57L1 | FALSE | Downregulated |
| 0,335914726 | 0,497224774 | DRG1 | FALSE | Upregulated |
| 0,43621628 | 0,497322776 | ELAC1 | FALSE | Upregulated |
| 0,439568041 | 0,497332703 | ADAM19 | FALSE | Upregulated |
| 0,661293394 | 0,497379097 | TNFSF13B | FALSE | Upregulated |
| 0,262525919 | 0,497741605 | NSFL1C | FALSE | Upregulated |
| -0,335567668 | 0,497756646 | YIPF4 | FALSE | Downregulated |
| -1,467721039 | 0,497760941 | BIVM-ERCC5 | FALSE | Downregulated |
| -0,219640601 | 0,497760941 | NFAT5 | FALSE | Downregulated |
| 0,520422749 | 0,497760941 | PRKCQ | FALSE | Upregulated |

| | | | | |
|---|---|---|---|---|
| 0,69170025 | 0,497835233 | C14orf182 | FALSE | Upregulated |
| -0,657775538 | 0,497847047 | LOC102724514 | FALSE | Downregulated |
| 0,960575978 | 0,497847047 | PLCL1 | FALSE | Upregulated |
| 0,322029428 | 0,497847047 | SNRNP35 | FALSE | Upregulated |
| -0,590195651 | 0,497887149 | MIEF2 | FALSE | Downregulated |
| 0,561789164 | 0,497887149 | PPIL1 | FALSE | Upregulated |
| -0,798092692 | 0,497887149 | PPP1R14A | FALSE | Downregulated |
| -0,252612068 | 0,497887149 | RPL35A | FALSE | Downregulated |
| -0,235598399 | 0,497887149 | TM9SF4 | FALSE | Downregulated |
| -0,376100448 | 0,497887149 | UBIAD1 | FALSE | Downregulated |
| 1,082625117 | 0,498088263 | C1orf106 | FALSE | Upregulated |
| -0,269842227 | 0,498090049 | LSG1 | FALSE | Downregulated |
| -0,521335174 | 0,49820617 | GPRASP2 | FALSE | Downregulated |
| 0,762344147 | 0,49820617 | IFITM5 | FALSE | Upregulated |
| 0,245669288 | 0,49820617 | KCTD21-AS1 | FALSE | Upregulated |
| -0,808157325 | 0,49820617 | RP11-513G11.3 | FALSE | Downregulated |
| -0,417839183 | 0,498233224 | SUPV3L1 | FALSE | Downregulated |
| -0,649231732 | 0,498272839 | ALKBH2 | FALSE | Downregulated |
| -0,187987005 | 0,498272839 | BAZ1B | FALSE | Downregulated |
| 0,503272559 | 0,498272839 | LOC101927201 | FALSE | Upregulated |
| 0,416296171 | 0,498272839 | PRUNE2 | FALSE | Upregulated |
| 0,185224273 | 0,498414699 | TNFRSF1A | FALSE | Upregulated |
| 0,328202306 | 0,498458536 | TMEM9B | FALSE | Upregulated |
| -0,376374246 | 0,498534862 | LINC00910 | FALSE | Downregulated |
| 0,269820223 | 0,498534862 | MARS | FALSE | Upregulated |
| -0,719814061 | 0,498614229 | KIAA1328 | FALSE | Downregulated |
| -0,206509602 | 0,499302192 | BSDC1 | FALSE | Downregulated |
| -0,413503037 | 0,499302192 | C8orf4 | FALSE | Downregulated |
| 0,251648862 | 0,499302192 | C9orf114 | FALSE | Upregulated |
| -0,223341435 | 0,499302192 | DAAM1 | FALSE | Downregulated |
| -0,179329935 | 0,499302192 | GCGR | FALSE | Downregulated |
| 0,203037459 | 0,499302192 | HMHA1 | FALSE | Upregulated |
| -0,376217013 | 0,499302192 | IGBP1 | FALSE | Downregulated |
| 0,312119777 | 0,499302192 | LHFPL4 | FALSE | Upregulated |
| -0,223018347 | 0,499302192 | QARS | FALSE | Downregulated |
| 1,087479647 | 0,499302192 | TRPM2-AS | FALSE | Upregulated |
| -0,59448699 | 0,499302192 | USP21 | FALSE | Downregulated |
| -0,324010011 | 0,499302192 | VTA1 | FALSE | Downregulated |
| 0,335190015 | 0,499324883 | LOC606724 | FALSE | Upregulated |
| -0,208703491 | 0,499726244 | SSU72 | FALSE | Downregulated |
| -0,20322337 | 0,500006534 | THEM6 | FALSE | Downregulated |
| 0,385606926 | 0,500026319 | C1QTNF7 | FALSE | Upregulated |
| 0,926515293 | 0,500026319 | STK32B | FALSE | Upregulated |
| -0,333632085 | 0,500623574 | SULT1A2 | FALSE | Downregulated |
| -0,185046302 | 0,500736957 | CS | FALSE | Downregulated |
| 0,231427785 | 0,500736957 | DNAJC7 | FALSE | Upregulated |
| 0,727266219 | 0,500736957 | LOC100506472 | FALSE | Upregulated |
| -1,043594471 | 0,500736957 | STON1 | FALSE | Downregulated |
| 0,422557123 | 0,500736957 | TRERF1 | FALSE | Upregulated |
| 0,367760974 | 0,500793667 | ATP6V0A2 | FALSE | Upregulated |
| 0,35912341 | 0,500793667 | DCBLD1 | FALSE | Upregulated |

| | | | | |
|---|---|---|---|---|
| 0,418153055 | 0,500793667 | HIAT1 | FALSE | Upregulated |
| -0,525194715 | 0,500793667 | LOC101929512 | FALSE | Downregulated |
| -0,414623355 | 0,500986535 | BROX | FALSE | Downregulated |
| 0,82656747 | 0,501157681 | SLC16A12 | FALSE | Upregulated |
| 0,888815565 | 0,501157681 | TMEM38B | FALSE | Upregulated |
| 0,487738648 | 0,501354773 | EML2 | FALSE | Upregulated |
| -0,286608963 | 0,501354773 | JAM2 | FALSE | Downregulated |
| 0,460330028 | 0,501707051 | C1orf198 | FALSE | Upregulated |
| 0,477519773 | 0,501707051 | LOC101927479 | FALSE | Upregulated |
| 0,613902753 | 0,501707051 | TSPEAR-AS1 | FALSE | Upregulated |
| -0,578301176 | 0,501949229 | GAS5 | FALSE | Downregulated |
| -0,645291759 | 0,502276289 | RHBG | FALSE | Downregulated |
| -0,235029714 | 0,502300293 | PRG4 | FALSE | Downregulated |
| -0,31001475 | 0,502300293 | PRRC1 | FALSE | Downregulated |
| 1,106501907 | 0,502317881 | FAM129C | FALSE | Upregulated |
| 0,629211193 | 0,502517652 | ALG1L | FALSE | Upregulated |
| 0,40521738 | 0,502557165 | SLC30A6 | FALSE | Upregulated |
| -0,324293862 | 0,502767247 | ZNF197 | FALSE | Downregulated |
| -0,365271216 | 0,502927801 | SYS1 | FALSE | Downregulated |
| -0,355105986 | 0,503019877 | RCL1 | FALSE | Downregulated |
| -0,22779275 | 0,503019877 | SLC35E1 | FALSE | Downregulated |
| 0,730504553 | 0,503019877 | TNS4 | FALSE | Upregulated |
| -0,225243276 | 0,503118218 | MIER1 | FALSE | Downregulated |
| 0,436121116 | 0,503139031 | VAV1 | FALSE | Upregulated |
| 0,265713004 | 0,503341198 | ARHGEF2 | FALSE | Upregulated |
| 0,721949295 | 0,503341198 | CLDN9 | FALSE | Upregulated |
| -0,3704231 | 0,503341198 | FASTKD2 | FALSE | Downregulated |
| 0,386951799 | 0,503341198 | ZNF821 | FALSE | Upregulated |
| -0,172224477 | 0,503370703 | DUS1L | FALSE | Downregulated |
| -0,33500437 | 0,503370703 | NUDT5 | FALSE | Downregulated |
| 0,190774762 | 0,503370703 | PEX10 | FALSE | Upregulated |
| 1,180196178 | 0,503370703 | RP11-290F20.3 | FALSE | Upregulated |
| -0,207119997 | 0,503430779 | NFYC | FALSE | Downregulated |
| 0,639296929 | 0,503739839 | BHLHB9 | FALSE | Upregulated |
| 0,220899291 | 0,503739839 | CEP68 | FALSE | Upregulated |
| 0,398236285 | 0,503739839 | CRMP1 | FALSE | Upregulated |
| -1,04703658 | 0,503739839 | FAM3D | FALSE | Downregulated |
| 0,200010274 | 0,503739839 | GALNT2 | FALSE | Upregulated |
| 0,290654416 | 0,503739839 | GBA | FALSE | Upregulated |
| -0,569279033 | 0,503739839 | HEY2 | FALSE | Downregulated |
| 0,46929859 | 0,503739839 | LOC102724784 | FALSE | Upregulated |
| -0,511704774 | 0,503739839 | PPIE | FALSE | Downregulated |
| -0,261773643 | 0,503739839 | RNA45S5 | FALSE | Downregulated |
| 0,268832937 | 0,503739839 | TCF7L2 | FALSE | Upregulated |
| 0,259355401 | 0,503739839 | TKT | FALSE | Upregulated |
| 0,248934469 | 0,503789272 | HCG18 | FALSE | Upregulated |
| -0,703747108 | 0,503789272 | TREH | FALSE | Downregulated |
| -0,21117168 | 0,503790452 | GPS1 | FALSE | Downregulated |
| -0,288842209 | 0,503790452 | NAA25 | FALSE | Downregulated |
| 0,279423593 | 0,503790452 | VAMP2 | FALSE | Upregulated |
| 1,354683907 | 0,5037964 | TSNAX-DISC1 | FALSE | Upregulated |

| | | | | |
|---|---|---|---|---|
| 0,436490409 | 0,504051288 | DIRAS3 | FALSE | Upregulated |
| 0,395257498 | 0,504051288 | FIGNL2 | FALSE | Upregulated |
| 0,371144508 | 0,504051288 | MYO5C | FALSE | Upregulated |
| -0,53970839 | 0,504124797 | PEX6 | FALSE | Downregulated |
| 0,424621801 | 0,504124797 | SLC2A11 | FALSE | Upregulated |
| 0,448243285 | 0,504134731 | LOC101929012 | FALSE | Upregulated |
| -0,146699834 | 0,504134731 | MAN2C1 | FALSE | Downregulated |
| -0,322807602 | 0,504134731 | SDHAP2 | FALSE | Downregulated |
| -0,555312105 | 0,504134731 | ZNF34 | FALSE | Downregulated |
| 0,170598304 | 0,504297783 | KMO | FALSE | Upregulated |
| 0,900506459 | 0,504309351 | LOC101928854 | FALSE | Upregulated |
| -0,900022676 | 0,504337913 | APCDD1 | FALSE | Downregulated |
| -0,257161132 | 0,504337913 | FAM149A | FALSE | Downregulated |
| -0,412185181 | 0,504337913 | TM2D3 | FALSE | Downregulated |
| 0,253607454 | 0,504380872 | ZNF12 | FALSE | Upregulated |
| -0,397945141 | 0,504631644 | N4BP2L2-IT2 | FALSE | Downregulated |
| 0,656313554 | 0,504748592 | GPR56 | FALSE | Upregulated |
| -0,224116491 | 0,504780495 | CCDC152 | FALSE | Downregulated |
| -0,252101044 | 0,504798365 | PDXP | FALSE | Downregulated |
| -0,269318493 | 0,504801367 | WDR5B | FALSE | Downregulated |
| 0,215325475 | 0,504914314 | LOC100288123 | FALSE | Upregulated |
| -0,255978984 | 0,504978917 | RCN3 | FALSE | Downregulated |
| 0,533094189 | 0,505313494 | CSTF2 | FALSE | Upregulated |
| -0,765032817 | 0,505313494 | EHF | FALSE | Downregulated |
| 0,285325832 | 0,505313494 | GTF2A1 | FALSE | Upregulated |
| 0,766708586 | 0,505313494 | PLCZ1 | FALSE | Upregulated |
| 0,740023625 | 0,505313494 | PTRH1 | FALSE | Upregulated |
| 0,317914048 | 0,505313494 | SNRPB | FALSE | Upregulated |
| 0,560792774 | 0,505313494 | SPATA7 | FALSE | Upregulated |
| 0,190252405 | 0,505361462 | FGD5-AS1 | FALSE | Upregulated |
| -0,606356192 | 0,505719483 | LOC102723542 | FALSE | Downregulated |
| 0,296199698 | 0,505868179 | REEP1 | FALSE | Upregulated |
| 0,364530252 | 0,506221095 | LOC101927044 | FALSE | Upregulated |
| -0,22124103 | 0,506675217 | NDUFS1 | FALSE | Downregulated |
| 0,796079235 | 0,506675217 | RP4-544H6.2 | FALSE | Upregulated |
| 0,259829687 | 0,506675217 | TRAF3IP2 | FALSE | Upregulated |
| 1,455656154 | 0,506972407 | C18orf32 | FALSE | Upregulated |
| -0,284368525 | 0,507194506 | AMDHD1 | FALSE | Downregulated |
| 0,260841909 | 0,507194506 | GFOD1 | FALSE | Upregulated |
| 0,368726964 | 0,507194506 | KLHDC4 | FALSE | Upregulated |
| 0,153375046 | 0,507194506 | MAPK8IP3 | FALSE | Upregulated |
| -0,177902166 | 0,507194506 | MAST3 | FALSE | Downregulated |
| 0,186652114 | 0,507194506 | RP11-367J11.2 | FALSE | Upregulated |
| -0,235569364 | 0,507194506 | ZNF445 | FALSE | Downregulated |
| -0,377870972 | 0,507203488 | LOC101928798 | FALSE | Downregulated |
| -0,229980073 | 0,507292221 | IBTK | FALSE | Downregulated |
| -1,038385731 | 0,507345949 | ESRRB | FALSE | Downregulated |
| 1,051703685 | 0,507361962 | MS4A1 | FALSE | Upregulated |
| 0,182190858 | 0,507410852 | AIDA | FALSE | Upregulated |
| -1,000987715 | 0,507443886 | LOC101929621 | FALSE | Downregulated |
| 0,326362962 | 0,507516962 | MFAP4 | FALSE | Upregulated |

| | | | | |
|---|---|---|---|---|
| -0,269069391 | 0,507534742 | ACSM5 | FALSE | Downregulated |
| -0,605775759 | 0,507534742 | MIPOL1 | FALSE | Downregulated |
| 0,956958202 | 0,507534742 | OXT | FALSE | Upregulated |
| -0,338082518 | 0,507534742 | ZFAND2B | FALSE | Downregulated |
| 0,512354163 | 0,507708051 | PRSS54 | FALSE | Upregulated |
| 0,628580907 | 0,507708051 | TEX38 | FALSE | Upregulated |
| -0,228714913 | 0,507823857 | ADD3 | FALSE | Downregulated |
| -0,927067884 | 0,507842069 | CADM2-AS1 | FALSE | Downregulated |
| 0,231126997 | 0,507842069 | NCBP2 | FALSE | Upregulated |
| 0,42109639 | 0,507842069 | SNX20 | FALSE | Upregulated |
| 0,609448025 | 0,507905816 | SCLT1 | FALSE | Upregulated |
| -0,210937286 | 0,508161627 | TMX4 | FALSE | Downregulated |
| -0,242714581 | 0,508223047 | NEMF | FALSE | Downregulated |
| 0,491746719 | 0,508458022 | FBLN7 | FALSE | Upregulated |
| 0,661934783 | 0,508623924 | NFU1 | FALSE | Upregulated |
| 0,31613961 | 0,508814999 | AMMECR1L | FALSE | Upregulated |
| -0,170918992 | 0,508871123 | CYP4V2 | FALSE | Downregulated |
| -0,549200316 | 0,508947619 | DOPEY2 | FALSE | Downregulated |
| -0,694703017 | 0,508947619 | RWDD3 | FALSE | Downregulated |
| 0,394593235 | 0,509451226 | ZNF32 | FALSE | Upregulated |
| 0,494819558 | 0,509831421 | HSCB | FALSE | Upregulated |
| 0,165530657 | 0,509977175 | DUSP16 | FALSE | Upregulated |
| 0,816090172 | 0,509977175 | GPR151 | FALSE | Upregulated |
| 0,191495419 | 0,510087298 | AKAP17A | FALSE | Upregulated |
| 0,435894822 | 0,510430515 | RNGTT | FALSE | Upregulated |
| -0,803274771 | 0,510807678 | CBWD1 | FALSE | Downregulated |
| 1,14980126 | 0,510807678 | PAX5 | FALSE | Upregulated |
| 0,161347478 | 0,510807678 | UBQLN1 | FALSE | Upregulated |
| 0,67970822 | 0,510839513 | ANKRD49 | FALSE | Upregulated |
| -0,213809516 | 0,510839513 | USO1 | FALSE | Downregulated |
| -0,312817069 | 0,510949846 | OTUD1 | FALSE | Downregulated |
| -0,255670129 | 0,510949846 | RSL1D1 | FALSE | Downregulated |
| 0,211186165 | 0,511022035 | COL6A1 | FALSE | Upregulated |
| 0,504150933 | 0,511082651 | CLEC10A | FALSE | Upregulated |
| 0,50091925 | 0,511082651 | LOC102724782 | FALSE | Upregulated |
| -0,273642121 | 0,511082651 | NDUFV3 | FALSE | Downregulated |
| -0,197671889 | 0,511082651 | SLK | FALSE | Downregulated |
| 0,267057122 | 0,511082651 | TESK1 | FALSE | Upregulated |
| -0,345465574 | 0,511493602 | ATP5S | FALSE | Downregulated |
| 0,38151009 | 0,511994981 | CHRAC1 | FALSE | Upregulated |
| 0,129435767 | 0,511994981 | CIDEB | FALSE | Upregulated |
| 0,34279658 | 0,511994981 | LOC100996813 | FALSE | Upregulated |
| 1,11394884 | 0,511994981 | LOC102724124 | FALSE | Upregulated |
| -0,203681054 | 0,511994981 | LRPPRC | FALSE | Downregulated |
| 0,376834531 | 0,511994981 | ZBTB21 | FALSE | Upregulated |
| -0,493668167 | 0,51243349 | ABHD10 | FALSE | Downregulated |
| -0,21171105 | 0,51243349 | FAM115A | FALSE | Downregulated |
| 0,410535782 | 0,51243349 | HNRNPA1P10 | FALSE | Upregulated |
| 0,410344148 | 0,51243349 | MAP3K6 | FALSE | Upregulated |
| 0,28983389 | 0,512857263 | PSMD13 | FALSE | Upregulated |
| 0,351495171 | 0,513066633 | ZBTB47 | FALSE | Upregulated |

| | | | | |
|---|---|---|---|---|
| -0,981667892 | 0,513103695 | ZNF547 | FALSE | Downregulated |
| 0,251975245 | 0,513395951 | TSPYL4 | FALSE | Upregulated |
| -0,435204717 | 0,513534255 | DAB1 | FALSE | Downregulated |
| 0,605581585 | 0,513534255 | IFI44 | FALSE | Upregulated |
| 0,372452353 | 0,513534255 | USP39 | FALSE | Upregulated |
| -0,141522108 | 0,51357538 | GLG1 | FALSE | Downregulated |
| 0,778852875 | 0,51357538 | ZCCHC10 | FALSE | Upregulated |
| -0,734579682 | 0,513581085 | LOC728407 | FALSE | Downregulated |
| 0,961029389 | 0,513581085 | PDLIM3 | FALSE | Upregulated |
| 0,356254461 | 0,513581085 | VCAM1 | FALSE | Upregulated |
| 0,488409398 | 0,513876568 | RP11-127L20.3 | FALSE | Upregulated |
| 0,404757981 | 0,513883135 | LOC101929636 | FALSE | Upregulated |
| 0,305575649 | 0,513883135 | LOC102723422 | FALSE | Upregulated |
| -0,245211356 | 0,513883135 | PDLIM2 | FALSE | Downregulated |
| 0,263813837 | 0,513883135 | RBM17 | FALSE | Upregulated |
| 0,314212025 | 0,514014496 | WDR36 | FALSE | Upregulated |
| 0,137770686 | 0,514067418 | UBTF | FALSE | Upregulated |
| 0,408415279 | 0,514187143 | TFAP2E | FALSE | Upregulated |
| 0,526905973 | 0,514211554 | BATF2 | FALSE | Upregulated |
| 0,26713958 | 0,514211554 | LTBP2 | FALSE | Upregulated |
| -0,173648257 | 0,514211554 | PAPOLA | FALSE | Downregulated |
| -0,276076995 | 0,514211554 | PEX13 | FALSE | Downregulated |
| 0,19469433 | 0,514355014 | STAT2 | FALSE | Upregulated |
| -0,485723675 | 0,514561797 | LZTFL1 | FALSE | Downregulated |
| -0,333017343 | 0,514561797 | MMAA | FALSE | Downregulated |
| -0,290809536 | 0,514561797 | RNA28S5 | FALSE | Downregulated |
| -0,558248365 | 0,514561797 | STAG3L5P | FALSE | Downregulated |
| 0,234218527 | 0,514561797 | TNPO3 | FALSE | Upregulated |
| 0,261764792 | 0,514655384 | ZNF654 | FALSE | Upregulated |
| 0,178503431 | 0,514659007 | DAZAP2 | FALSE | Upregulated |
| 0,226154741 | 0,514659007 | PPP2CB | FALSE | Upregulated |
| 0,380982286 | 0,514869814 | OLFM2 | FALSE | Upregulated |
| 0,785722032 | 0,514893316 | C8orf31 | FALSE | Upregulated |
| -0,353587831 | 0,514893316 | IGFBP2 | FALSE | Downregulated |
| 1,081368078 | 0,514893316 | LOC101927751 | FALSE | Upregulated |
| 0,299326739 | 0,514893316 | MED31 | FALSE | Upregulated |
| 0,206231492 | 0,514941307 | ELK4 | FALSE | Upregulated |
| 0,652589085 | 0,515000039 | PDZD11 | FALSE | Upregulated |
| -0,328092889 | 0,515071027 | JRK | FALSE | Downregulated |
| 0,638738074 | 0,515071027 | LOC101928875 | FALSE | Upregulated |
| 0,268980404 | 0,515071027 | TNFRSF6B | FALSE | Upregulated |
| -0,187764678 | 0,515202716 | FBXO31 | FALSE | Downregulated |
| 0,236421563 | 0,515546408 | DEF8 | FALSE | Upregulated |
| -0,381251948 | 0,515703834 | NUP155 | FALSE | Downregulated |
| -0,426499757 | 0,515703834 | SCRT1 | FALSE | Downregulated |
| -0,611981815 | 0,51570905 | C19orf52 | FALSE | Downregulated |
| -0,727548434 | 0,515820371 | NDUFAF6 | FALSE | Downregulated |
| -0,340833457 | 0,516208299 | ACAD9 | FALSE | Downregulated |
| 0,275777544 | 0,516208299 | FAM161B | FALSE | Upregulated |
| -0,226719245 | 0,516208299 | IL6ST | FALSE | Downregulated |
| 0,318287432 | 0,516208299 | KLHL8 | FALSE | Upregulated |

| | | | | |
|---|---|---|---|---|
| -0,909061556 | 0,516208299 | LOC101927138 | FALSE | Downregulated |
| 0,189084696 | 0,516208299 | MKNK2 | FALSE | Upregulated |
| -0,770907805 | 0,516208299 | NPNT | FALSE | Downregulated |
| -0,389845957 | 0,516208299 | PRICKLE2-AS3 | FALSE | Downregulated |
| 0,763531606 | 0,516208299 | RP4-639F20.1 | FALSE | Upregulated |
| 0,197907217 | 0,516208299 | SRA1 | FALSE | Upregulated |
| -0,250155396 | 0,516208299 | TMEM51-AS1 | FALSE | Downregulated |
| -0,199705977 | 0,516208299 | TRIM27 | FALSE | Downregulated |
| -0,755625403 | 0,516466592 | SPTBN4 | FALSE | Downregulated |
| 0,233503761 | 0,516544716 | SCIMP | FALSE | Upregulated |
| 0,357012722 | 0,516785476 | C12orf29 | FALSE | Upregulated |
| 0,943275581 | 0,516785476 | LOC100996517 | FALSE | Upregulated |
| 0,223099703 | 0,516785476 | MKNK1-AS1 | FALSE | Upregulated |
| -0,327294929 | 0,516876465 | TBCA | FALSE | Downregulated |
| -0,253588332 | 0,51692294 | F8 | FALSE | Downregulated |
| -0,526703336 | 0,51706287 | RRP7B | FALSE | Downregulated |
| 0,71500809 | 0,517277736 | HRH2 | FALSE | Upregulated |
| -0,201594777 | 0,517277736 | INHBC | FALSE | Downregulated |
| -0,294196264 | 0,517353188 | CSPG4 | FALSE | Downregulated |
| 0,454177704 | 0,517353188 | DNASE1L2 | FALSE | Upregulated |
| 0,564659935 | 0,517353188 | RPS6KA2-AS1 | FALSE | Upregulated |
| 0,349353615 | 0,517353188 | TOPORS-AS1 | FALSE | Upregulated |
| 0,764940929 | 0,517353188 | TSSK1B | FALSE | Upregulated |
| 0,499009549 | 0,517353188 | ZNF732 | FALSE | Upregulated |
| 0,201277694 | 0,51779908 | NEXN | FALSE | Upregulated |
| -0,628553399 | 0,51779908 | PSPH | FALSE | Downregulated |
| -0,442693984 | 0,517911841 | SLC10A5 | FALSE | Downregulated |
| 0,550769585 | 0,518189918 | KIAA1211 | FALSE | Upregulated |
| 0,753330706 | 0,518261268 | HIST1H4H | FALSE | Upregulated |
| -0,394140227 | 0,518595287 | UQCRB | FALSE | Downregulated |
| -0,228087948 | 0,518844827 | ATXN2 | FALSE | Downregulated |
| -0,24061016 | 0,519162491 | MSRB1 | FALSE | Downregulated |
| 0,82908309 | 0,519162491 | SLC26A5 | FALSE | Upregulated |
| 0,353794592 | 0,519162491 | SMAP1 | FALSE | Upregulated |
| -0,999198529 | 0,519403935 | GRIA4 | FALSE | Downregulated |
| 0,30628792 | 0,519450549 | SIDT1 | FALSE | Upregulated |
| -0,296997154 | 0,519502979 | HSD17B11 | FALSE | Downregulated |
| 0,33783301 | 0,519756772 | GNL2 | FALSE | Upregulated |
| -0,491276513 | 0,519771882 | SLC41A3 | FALSE | Downregulated |
| 0,419311771 | 0,519891854 | SNX12 | FALSE | Upregulated |
| -0,549871646 | 0,520134751 | MRPS6 | FALSE | Downregulated |
| 0,406985396 | 0,520157935 | DDR1 | FALSE | Upregulated |
| 0,810846318 | 0,520157935 | MT1H | FALSE | Upregulated |
| -0,210085994 | 0,520157935 | PHB | FALSE | Downregulated |
| 0,6156689 | 0,520196001 | ETV2 | FALSE | Upregulated |
| -0,1454743 | 0,520196001 | FAU | FALSE | Downregulated |
| -0,510417431 | 0,520196001 | S100A13 | FALSE | Downregulated |
| 0,234198365 | 0,5202076 | UCKL1 | FALSE | Upregulated |
| 0,33703921 | 0,52030249 | LOC284889 | FALSE | Upregulated |
| 0,383133271 | 0,52030249 | NIPAL4 | FALSE | Upregulated |
| 0,261837004 | 0,52030249 | PGAM1P5 | FALSE | Upregulated |

| | | | | |
|---|---|---|---|---|
| -0,649000897 | 0,52030249 | ZFP3 | FALSE | Downregulated |
| -0,24083408 | 0,520594703 | SSRP1 | FALSE | Downregulated |
| 0,410162236 | 0,520594703 | YIPF5 | FALSE | Upregulated |
| -0,18477376 | 0,520890891 | WDFY3 | FALSE | Downregulated |
| 0,18829495 | 0,521716676 | SMG1P1 | FALSE | Upregulated |
| 0,487863181 | 0,522247646 | OR2A9P | FALSE | Upregulated |
| 0,808837459 | 0,52242758 | FAM86HP | FALSE | Upregulated |
| -0,455036975 | 0,522723973 | LOC101929384 | FALSE | Downregulated |
| -0,388664819 | 0,522778346 | PHF5A | FALSE | Downregulated |
| -0,326581366 | 0,522778648 | PARD6G-AS1 | FALSE | Downregulated |
| 0,717682867 | 0,522778648 | TRIM10 | FALSE | Upregulated |
| 0,189912587 | 0,522840342 | DECR2 | FALSE | Upregulated |
| -0,482923781 | 0,522871595 | MRPL42 | FALSE | Downregulated |
| -0,19558542 | 0,523013184 | AGO1 | FALSE | Downregulated |
| 0,241043874 | 0,523013184 | AGPAT6 | FALSE | Upregulated |
| 0,3864951 | 0,523013184 | PWWP2B | FALSE | Upregulated |
| 0,220203417 | 0,523013184 | RAB14 | FALSE | Upregulated |
| -0,250106397 | 0,523013184 | TIAM1 | FALSE | Downregulated |
| -0,320644616 | 0,523013184 | ZFAND6 | FALSE | Downregulated |
| -0,314821596 | 0,523142559 | ITFG2 | FALSE | Downregulated |
| 0,347560472 | 0,523232095 | ADAMTS10 | FALSE | Upregulated |
| 0,717564865 | 0,523232095 | ANKRD55 | FALSE | Upregulated |
| -0,472613405 | 0,523232095 | BRI3BP | FALSE | Downregulated |
| -0,118575973 | 0,523232095 | COL18A1 | FALSE | Downregulated |
| -1,036370696 | 0,523232095 | COL18A1-AS2 | FALSE | Downregulated |
| -0,24232329 | 0,523232095 | CPNE3 | FALSE | Downregulated |
| -0,321120844 | 0,523232095 | LEPR | FALSE | Downregulated |
| 0,192292162 | 0,523232095 | MCTS1 | FALSE | Upregulated |
| 0,284745356 | 0,523232095 | PSEN1 | FALSE | Upregulated |
| -0,177633234 | 0,523232095 | RORA | FALSE | Downregulated |
| -0,2779827 | 0,523232095 | SAP30BP | FALSE | Downregulated |
| -0,986089044 | 0,523232095 | SLC4A9 | FALSE | Downregulated |
| 0,282944082 | 0,523232095 | UCHL5 | FALSE | Upregulated |
| 0,168647002 | 0,523232095 | UHMK1 | FALSE | Upregulated |
| -0,364076 | 0,523232095 | USP37 | FALSE | Downregulated |
| 0,336883778 | 0,523232095 | VPS8 | FALSE | Upregulated |
| -0,34806526 | 0,523232095 | ZNF720 | FALSE | Downregulated |
| 0,197015472 | 0,523280183 | ITGB5 | FALSE | Upregulated |
| -0,328885463 | 0,523299258 | RAPGEF4 | FALSE | Downregulated |
| 0,321955233 | 0,523419725 | IL10RB | FALSE | Upregulated |
| -0,21760113 | 0,523502623 | SYNM | FALSE | Downregulated |
| -0,158862799 | 0,523545963 | C1R | FALSE | Downregulated |
| 0,212497915 | 0,523545963 | ERBB2 | FALSE | Upregulated |
| -0,097505544 | 0,523545963 | LINC00657 | FALSE | Downregulated |
| 0,651593213 | 0,523557335 | BOLL | FALSE | Upregulated |
| 0,967690181 | 0,523557335 | CGRRF1 | FALSE | Upregulated |
| 0,269026059 | 0,523557335 | RFC5 | FALSE | Upregulated |
| -0,572140814 | 0,523941245 | USP18 | FALSE | Downregulated |
| 0,351033966 | 0,524111476 | TRAPPC3 | FALSE | Upregulated |
| -0,300556369 | 0,524322812 | COQ4 | FALSE | Downregulated |
| -1,022595282 | 0,524335024 | CBLN4 | FALSE | Downregulated |

| | | | | |
|---|---|---|---|---|
| -0,522789568 | 0,524335024 | MANSC4 | FALSE | Downregulated |
| 0,662518309 | 0,524335024 | SPRY3 | FALSE | Upregulated |
| -0,205821779 | 0,524335024 | TMEM97 | FALSE | Downregulated |
| 0,205684271 | 0,524442807 | ERMP1 | FALSE | Upregulated |
| 0,550321043 | 0,524442807 | GINS4 | FALSE | Upregulated |
| 0,345175911 | 0,524442807 | RRM2B | FALSE | Upregulated |
| 0,506436364 | 0,52450809 | SLC52A2 | FALSE | Upregulated |
| 0,417538779 | 0,524602697 | LOC100996761 | FALSE | Upregulated |
| 0,886404947 | 0,524602697 | PARM1 | FALSE | Upregulated |
| -0,332065338 | 0,52465353 | HSD3BP4 | FALSE | Downregulated |
| 1,10830733 | 0,524726882 | CYP3A7-CYP3AP1 | FALSE | Upregulated |
| -0,470696473 | 0,524726882 | DTWD2 | FALSE | Downregulated |
| -0,29311795 | 0,524726882 | GYS1 | FALSE | Downregulated |
| -0,366323042 | 0,524726882 | MBTPS2 | FALSE | Downregulated |
| -0,163814542 | 0,525120587 | SCAF11 | FALSE | Downregulated |
| 0,809057747 | 0,525168557 | TNFSF4 | FALSE | Upregulated |
| -0,664774471 | 0,525587559 | LOC100287175 | FALSE | Downregulated |
| -0,962407129 | 0,525587559 | RP11-342C23.4 | FALSE | Downregulated |
| -0,825794102 | 0,525769764 | DYRK3 | FALSE | Downregulated |
| 0,238269574 | 0,525775207 | CD63 | FALSE | Upregulated |
| 0,40334642 | 0,525775207 | PANO | FALSE | Upregulated |
| 0,632132899 | 0,525805193 | SPHK1 | FALSE | Upregulated |
| -0,159982342 | 0,525932746 | MCM3AP | FALSE | Downregulated |
| -0,123097897 | 0,526198663 | C6orf62 | FALSE | Downregulated |
| -0,264072905 | 0,526591581 | BET1L | FALSE | Downregulated |
| 0,332294998 | 0,526674268 | MCRS1 | FALSE | Upregulated |
| 0,807433776 | 0,526674268 | TIGD1 | FALSE | Upregulated |
| 0,803317995 | 0,526774139 | LILRA2 | FALSE | Upregulated |
| -0,951169861 | 0,526844401 | TPSB2 | FALSE | Downregulated |
| -0,263444404 | 0,527002472 | CLPP | FALSE | Downregulated |
| 0,325721672 | 0,527038513 | TMEM104 | FALSE | Upregulated |
| 0,317679775 | 0,527082523 | AATK | FALSE | Upregulated |
| 0,14086012 | 0,527082523 | KMT2B | FALSE | Upregulated |
| 0,61354536 | 0,527141122 | BSPRY | FALSE | Upregulated |
| -0,296029422 | 0,527141122 | ZNF302 | FALSE | Downregulated |
| 0,574256559 | 0,527352105 | SRI | FALSE | Upregulated |
| 0,448144462 | 0,527357871 | AC020571.3 | FALSE | Upregulated |
| -0,402012978 | 0,527357871 | C19orf48 | FALSE | Downregulated |
| 0,233415752 | 0,527357871 | GOLGA5 | FALSE | Upregulated |
| 0,759139218 | 0,527357871 | HSPB7 | FALSE | Upregulated |
| -0,164264267 | 0,527357871 | MAML1 | FALSE | Downregulated |
| 0,52979508 | 0,527357871 | RAE1 | FALSE | Upregulated |
| 0,224358893 | 0,527357871 | SLC35A1 | FALSE | Upregulated |
| 0,429414857 | 0,527357871 | USP44 | FALSE | Upregulated |
| -0,144734136 | 0,52743784 | KIF1B | FALSE | Downregulated |
| 1,059415925 | 0,52753985 | FAM101A | FALSE | Upregulated |
| -0,247142468 | 0,527580387 | ORM2 | FALSE | Downregulated |
| 0,107136289 | 0,52794472 | WNK1 | FALSE | Upregulated |
| 0,917217019 | 0,528069175 | NALCN | FALSE | Upregulated |
| 1,044658489 | 0,528237954 | TACSTD2 | FALSE | Upregulated |
| -0,335042233 | 0,528292585 | AFAP1L1 | FALSE | Downregulated |

| | | | | |
|---|---|---|---|---|
| 0,840527041 | 0,528292585 | C12orf75 | FALSE | Upregulated |
| 0,137895958 | 0,528292585 | MORF4L1 | FALSE | Upregulated |
| 0,287862458 | 0,528292585 | ZFAND5 | FALSE | Upregulated |
| -0,190850751 | 0,528371646 | CYB5R3 | FALSE | Downregulated |
| -0,159082807 | 0,528371646 | DENND4C | FALSE | Downregulated |
| -1,037513043 | 0,528400314 | LOC102723383 | FALSE | Downregulated |
| -0,572769655 | 0,528469523 | FAM173B | FALSE | Downregulated |
| 0,299352938 | 0,528804858 | AHCYL2 | FALSE | Upregulated |
| -0,528254483 | 0,528804858 | INHBA | FALSE | Downregulated |
| -0,985324711 | 0,528804858 | LRRC7 | FALSE | Downregulated |
| -0,602690929 | 0,528804858 | NIPAL1 | FALSE | Downregulated |
| 0,31811675 | 0,528804858 | PPAT | FALSE | Upregulated |
| 0,289748904 | 0,528804858 | STRIP1 | FALSE | Upregulated |
| -0,561942673 | 0,528852807 | ZNF891 | FALSE | Downregulated |
| 0,842349358 | 0,528876825 | LSM6 | FALSE | Upregulated |
| 0,211883722 | 0,528905623 | DHRS12 | FALSE | Upregulated |
| 0,386910854 | 0,529010124 | FAM214B | FALSE | Upregulated |
| 0,285183277 | 0,529016669 | ANKRD50 | FALSE | Upregulated |
| -0,189338626 | 0,529016669 | DHCR24 | FALSE | Downregulated |
| 0,46841579 | 0,529016669 | DPYSL3 | FALSE | Upregulated |
| 0,67304746 | 0,529016669 | GEM | FALSE | Upregulated |
| -0,32256901 | 0,529016669 | MRPL23-AS1 | FALSE | Downregulated |
| 0,326793129 | 0,529016669 | PTPRU | FALSE | Upregulated |
| 0,40821815 | 0,529016669 | SFTPD | FALSE | Upregulated |
| 0,401178528 | 0,529016669 | SOX4 | FALSE | Upregulated |
| -0,421101336 | 0,529016669 | SYPL2 | FALSE | Downregulated |
| -0,264592592 | 0,529016669 | YIF1A | FALSE | Downregulated |
| -0,240569153 | 0,529016669 | ZZZ3 | FALSE | Downregulated |
| -0,345388708 | 0,529145055 | SOCS7 | FALSE | Downregulated |
| -0,878469345 | 0,529163973 | LOC100233156 | FALSE | Downregulated |
| -0,769274859 | 0,529349768 | MAP2K6 | FALSE | Downregulated |
| -0,732036883 | 0,529349768 | SAYSD1 | FALSE | Downregulated |
| 0,26784046 | 0,529349768 | TBC1D9 | FALSE | Upregulated |
| -0,197695027 | 0,529349768 | XRCC5 | FALSE | Downregulated |
| 0,252683877 | 0,529349768 | YEATS2 | FALSE | Upregulated |
| 0,20642627 | 0,529482671 | GOLM1 | FALSE | Upregulated |
| 0,685160335 | 0,529505605 | LOC101927623 | FALSE | Upregulated |
| 0,317554711 | 0,529505605 | NSMAF | FALSE | Upregulated |
| 0,164262782 | 0,529505605 | SETX | FALSE | Upregulated |
| 0,651384227 | 0,529505605 | ZNF300 | FALSE | Upregulated |
| -0,227999149 | 0,529611717 | SOWAHB | FALSE | Downregulated |
| -0,497230469 | 0,529705125 | PAXIP1-AS1 | FALSE | Downregulated |
| -0,48061507 | 0,530702605 | MTFMT | FALSE | Downregulated |
| 0,753437755 | 0,530702605 | RP11-158M2.5 | FALSE | Upregulated |
| -0,649244754 | 0,530740284 | RP5-1074L1.1 | FALSE | Downregulated |
| 0,31737684 | 0,530919826 | CSF3R | FALSE | Upregulated |
| -0,239861341 | 0,530919826 | SETBP1 | FALSE | Downregulated |
| 0,531343853 | 0,531091559 | LFNG | FALSE | Upregulated |
| -0,828068977 | 0,531091559 | ZFP92 | FALSE | Downregulated |
| 0,56172909 | 0,531271739 | ARL14EP | FALSE | Upregulated |
| -0,685597511 | 0,531271739 | OPHN1 | FALSE | Downregulated |

| | | | | |
|---|---|---|---|---|
| 0,363825509 | 0,531271739 | POLD1 | FALSE | Upregulated |
| -0,624886447 | 0,531271739 | SCARA5 | FALSE | Downregulated |
| -0,613171648 | 0,531271739 | SLC25A17 | FALSE | Downregulated |
| -0,16863665 | 0,531271739 | TMEM66 | FALSE | Downregulated |
| -0,867734611 | 0,531395307 | EFNB3 | FALSE | Downregulated |
| -0,288905995 | 0,53152207 | CAPRIN2 | FALSE | Downregulated |
| 0,299785863 | 0,53152207 | GLOD4 | FALSE | Upregulated |
| -0,447910026 | 0,53152207 | LINC00861 | FALSE | Downregulated |
| -2,816322012 | 0,53152207 | SLX1B-SULT1A4 | FALSE | Downregulated |
| 0,492665703 | 0,531545017 | TRIQK | FALSE | Upregulated |
| 0,63094105 | 0,531739375 | RENBP | FALSE | Upregulated |
| -0,523543749 | 0,531961237 | DPF3 | FALSE | Downregulated |
| 0,265819041 | 0,53196726 | ATP6AP2 | FALSE | Upregulated |
| -0,248851758 | 0,532365977 | AKR1C1 | FALSE | Downregulated |
| 0,229166174 | 0,532365977 | DNAJB12 | FALSE | Upregulated |
| 0,189611314 | 0,532365977 | SPSB3 | FALSE | Upregulated |
| -0,441080718 | 0,532449635 | ZNF429 | FALSE | Downregulated |
| 0,826715453 | 0,532456706 | CAMKMT | FALSE | Upregulated |
| -0,274341091 | 0,532516996 | FSCN1 | FALSE | Downregulated |
| -0,27316461 | 0,532516996 | GFM1 | FALSE | Downregulated |
| 0,932983583 | 0,532641354 | FCRL3 | FALSE | Upregulated |
| -0,953224994 | 0,532645463 | LINC00659 | FALSE | Downregulated |
| 0,260183186 | 0,532666638 | CYB5D2 | FALSE | Upregulated |
| 0,440237872 | 0,532681668 | CD300A | FALSE | Upregulated |
| -0,194189626 | 0,532681668 | CPEB4 | FALSE | Downregulated |
| -0,29846799 | 0,532681668 | INTS5 | FALSE | Downregulated |
| -0,397624223 | 0,532681668 | LOC101927727 | FALSE | Downregulated |
| -0,608885172 | 0,532681668 | NCMAP | FALSE | Downregulated |
| 0,21124949 | 0,532681668 | PAG1 | FALSE | Upregulated |
| 0,613875024 | 0,532681668 | SAPCD1 | FALSE | Upregulated |
| -0,229787153 | 0,532681668 | TSHR | FALSE | Downregulated |
| -0,310957542 | 0,532681668 | WDR19 | FALSE | Downregulated |
| 0,318342686 | 0,532742711 | BTN2A1 | FALSE | Upregulated |
| -0,426144007 | 0,533135271 | STEAP2 | FALSE | Downregulated |
| -0,563753962 | 0,533151206 | BZW2 | FALSE | Downregulated |
| -0,348593888 | 0,533151206 | ERCC3 | FALSE | Downregulated |
| 0,275897434 | 0,533151206 | GPATCH11 | FALSE | Upregulated |
| 0,159531844 | 0,533151206 | TNRC6C | FALSE | Upregulated |
| -0,61839511 | 0,533247289 | FAM53A | FALSE | Downregulated |
| 0,443966842 | 0,533247289 | RTN4IP1 | FALSE | Upregulated |
| -0,24548733 | 0,533460792 | FAM160A1 | FALSE | Downregulated |
| -0,346991775 | 0,533502058 | PPWD1 | FALSE | Downregulated |
| 0,177281812 | 0,533560132 | HDAC6 | FALSE | Upregulated |
| -0,791426363 | 0,533592003 | IL1RAPL2 | FALSE | Downregulated |
| 0,396325144 | 0,533592003 | LOC101927151 | FALSE | Upregulated |
| 0,244122072 | 0,533650768 | GTF2F1 | FALSE | Upregulated |
| -0,199308371 | 0,533650768 | MGST1 | FALSE | Downregulated |
| 0,228683993 | 0,533758453 | LOC102724177 | FALSE | Upregulated |
| 0,188177653 | 0,533764429 | HSD17B1 | FALSE | Upregulated |
| 0,801115016 | 0,533764429 | LOC100130987 | FALSE | Upregulated |
| -1,011586403 | 0,533764429 | LOC101929819 | FALSE | Downregulated |

| | | | | |
|---|---|---|---|---|
| -0,658617129 | 0,533764429 | LOC389607 | FALSE | Downregulated |
| -0,399818881 | 0,533992601 | POLR3GL | FALSE | Downregulated |
| 0,185786438 | 0,534030128 | ERBB2IP | FALSE | Upregulated |
| -0,195507045 | 0,534333861 | HIP1 | FALSE | Downregulated |
| -0,391773978 | 0,534424749 | MT1E | FALSE | Downregulated |
| 0,703739459 | 0,534772477 | LOC554223 | FALSE | Upregulated |
| -0,77304894 | 0,534772477 | ZNF112 | FALSE | Downregulated |
| 0,327168682 | 0,534999626 | SRPRB | FALSE | Upregulated |
| 0,238249984 | 0,535181624 | DNAJB7 | FALSE | Upregulated |
| 0,279044552 | 0,535181624 | MARCH2 | FALSE | Upregulated |
| -0,204780876 | 0,53523504 | IFNAR1 | FALSE | Downregulated |
| -0,294105532 | 0,53523504 | MRPS24 | FALSE | Downregulated |
| 0,250012104 | 0,53523504 | PELP1 | FALSE | Upregulated |
| -0,241430718 | 0,53523504 | TMEM184C | FALSE | Downregulated |
| -0,528086504 | 0,53523504 | TRIT1 | FALSE | Downregulated |
| -0,292476068 | 0,535263501 | MTIF2 | FALSE | Downregulated |
| -0,335131462 | 0,535730899 | ZFYVE28 | FALSE | Downregulated |
| 0,399467434 | 0,535821084 | PROSER1 | FALSE | Upregulated |
| 0,393677985 | 0,535969859 | GP6 | FALSE | Upregulated |
| -0,323918796 | 0,535969859 | LOC101927437 | FALSE | Downregulated |
| -0,446082775 | 0,535984589 | COL26A1 | FALSE | Downregulated |
| -0,59415195 | 0,536005073 | APLF | FALSE | Downregulated |
| -0,445243338 | 0,536005073 | HOMEZ | FALSE | Downregulated |
| -0,236795219 | 0,536005073 | LOC101929340 | FALSE | Downregulated |
| -0,479302168 | 0,536780399 | HAPLN4 | FALSE | Downregulated |
| -0,440532798 | 0,536804769 | EPB41L4A-AS2 | FALSE | Downregulated |
| 1,216438432 | 0,536861976 | FAM72D | FALSE | Upregulated |
| 0,251574759 | 0,5369947 | LOC101928717 | FALSE | Upregulated |
| -0,283823231 | 0,5369947 | MAP4K3 | FALSE | Downregulated |
| -0,534926979 | 0,537166015 | DNAH9 | FALSE | Downregulated |
| 0,218602088 | 0,53720453 | C9orf84 | FALSE | Upregulated |
| 1,01483062 | 0,5372173 | C1orf51 | FALSE | Upregulated |
| 0,545883846 | 0,537307072 | HCG26 | FALSE | Upregulated |
| -0,307914289 | 0,537701232 | DLEC1 | FALSE | Downregulated |
| -0,744571392 | 0,537701232 | PITX1 | FALSE | Downregulated |
| -0,331638749 | 0,537945204 | TIGD2 | FALSE | Downregulated |
| 0,187159877 | 0,538035251 | GPI | FALSE | Upregulated |
| 0,573806687 | 0,538195112 | LINC00167 | FALSE | Upregulated |
| -1,144628207 | 0,538298964 | BMS1P17 | FALSE | Downregulated |
| 0,742490548 | 0,538314431 | CSF2 | FALSE | Upregulated |
| 0,935227025 | 0,538314431 | EIF1AY | FALSE | Upregulated |
| 0,367452583 | 0,538314431 | LOC101927445 | FALSE | Upregulated |
| -0,642554827 | 0,538596154 | FAM229A | FALSE | Downregulated |
| -0,311764191 | 0,538655823 | TMEM141 | FALSE | Downregulated |
| -0,382462765 | 0,538960142 | LOC401286 | FALSE | Downregulated |
| 0,669074198 | 0,539099934 | LOC392196 | FALSE | Upregulated |
| 0,64814767 | 0,539516909 | LOC101926895 | FALSE | Upregulated |
| 0,230200181 | 0,539565178 | CPPED1 | FALSE | Upregulated |
| -0,577387861 | 0,5396244 | ADAMTSL1 | FALSE | Downregulated |
| -1,021822328 | 0,539726095 | LOC101929165 | FALSE | Downregulated |
| -0,496337677 | 0,539895926 | MRO | FALSE | Downregulated |

| | | | | |
|---|---|---|---|---|
| -0,428660497 | 0,539895926 | THAP2 | FALSE | Downregulated |
| 0,938580456 | 0,539940319 | CLCA3P | FALSE | Upregulated |
| -0,417482358 | 0,539940319 | FLJ44342 | FALSE | Downregulated |
| -0,639411755 | 0,539940319 | LOC102724699 | FALSE | Downregulated |
| 0,50962367 | 0,539940319 | NOP10 | FALSE | Upregulated |
| -0,568401606 | 0,539940319 | RBM48 | FALSE | Downregulated |
| -0,243313966 | 0,539940319 | TP73-AS1 | FALSE | Downregulated |
| -0,168004538 | 0,540064185 | ABL1 | FALSE | Downregulated |
| -0,27923091 | 0,54011162 | RP11-103C16.2 | FALSE | Downregulated |
| 0,245978694 | 0,540359923 | FLOT2 | FALSE | Upregulated |
| 0,600150787 | 0,540489905 | AC022173.2 | FALSE | Upregulated |
| 0,837484861 | 0,540489905 | CUBN | FALSE | Upregulated |
| -0,371575291 | 0,540489905 | EFCAB4A | FALSE | Downregulated |
| 0,425967278 | 0,540489905 | GTPBP3 | FALSE | Upregulated |
| 0,315497687 | 0,540489905 | HSPA14 | FALSE | Upregulated |
| 0,624170319 | 0,540489905 | PTPDC1 | FALSE | Upregulated |
| 0,657662155 | 0,540489905 | SLC39A4 | FALSE | Upregulated |
| 0,762963344 | 0,540489905 | SNHG10 | FALSE | Upregulated |
| -0,305803566 | 0,54052918 | MIR210HG | FALSE | Downregulated |
| -0,2648134 | 0,54052918 | SULF2 | FALSE | Downregulated |
| -0,194789831 | 0,540748105 | CALCOCO2 | FALSE | Downregulated |
| 0,347922021 | 0,54104668 | DYNC1LI1 | FALSE | Upregulated |
| 1,074606993 | 0,541078718 | AANAT | FALSE | Upregulated |
| 0,246263773 | 0,54126748 | UBFD1 | FALSE | Upregulated |
| 0,532765121 | 0,541580606 | DUSP12 | FALSE | Upregulated |
| 0,349747258 | 0,541580606 | METTL16 | FALSE | Upregulated |
| 0,225589434 | 0,541913979 | ARGFXP2 | FALSE | Upregulated |
| 0,443464392 | 0,541913979 | HSPA13 | FALSE | Upregulated |
| -0,713230587 | 0,542168458 | FAM92A1P2 | FALSE | Downregulated |
| 0,619864416 | 0,542438375 | NKAPP1 | FALSE | Upregulated |
| 0,403769396 | 0,542506384 | FAM127A | FALSE | Upregulated |
| -0,297030139 | 0,54282294 | GBP7 | FALSE | Downregulated |
| -0,675802955 | 0,54283137 | AOC2 | FALSE | Downregulated |
| 0,630113608 | 0,54283137 | MTHFD1L | FALSE | Upregulated |
| 0,155058433 | 0,54298402 | P4HB | FALSE | Upregulated |
| 0,30773963 | 0,54298402 | RAP1A | FALSE | Upregulated |
| 0,247999475 | 0,54298402 | SIRT3 | FALSE | Upregulated |
| 0,390113307 | 0,543247218 | UBXN11 | FALSE | Upregulated |
| 0,337975455 | 0,543266423 | SGPP1 | FALSE | Upregulated |
| 0,361373974 | 0,543342434 | VPS53 | FALSE | Upregulated |
| 0,412727478 | 0,5433725 | RNF145 | FALSE | Upregulated |
| 0,739236199 | 0,543497467 | INTU | FALSE | Upregulated |
| 0,404266402 | 0,543664915 | LOC100996876 | FALSE | Upregulated |
| -0,183415275 | 0,543664915 | RAB13 | FALSE | Downregulated |
| 0,526370325 | 0,543887048 | FAAH2 | FALSE | Upregulated |
| 0,4593488 | 0,544001696 | VPS33B | FALSE | Upregulated |
| -0,374094286 | 0,544167961 | LOC102724187 | FALSE | Downregulated |
| -0,426873407 | 0,544167961 | ZKSCAN2 | FALSE | Downregulated |
| -0,498609119 | 0,544351091 | RPUSD2 | FALSE | Downregulated |
| 0,136628089 | 0,544413926 | H1FX | FALSE | Upregulated |
| -0,397457284 | 0,544413926 | THOC1 | FALSE | Downregulated |

| | | | | |
|---|---|---|---|---|
| -0,187887401 | 0,544413926 | TRIM33 | FALSE | Downregulated |
| -0,220926518 | 0,544413926 | UGT2B15 | FALSE | Downregulated |
| 0,18580059 | 0,544413926 | WDR73 | FALSE | Upregulated |
| 0,491874464 | 0,544484977 | CASP1 | FALSE | Upregulated |
| 0,132606703 | 0,544484977 | DNM2 | FALSE | Upregulated |
| 0,431512134 | 0,544484977 | PDIA3P1 | FALSE | Upregulated |
| 0,324639519 | 0,544484977 | RBFOX3 | FALSE | Upregulated |
| -0,659265445 | 0,544484977 | SULT1B1 | FALSE | Downregulated |
| 0,346597365 | 0,544484977 | TNFRSF25 | FALSE | Upregulated |
| 0,219306029 | 0,544538169 | RAB6A | FALSE | Upregulated |
| -0,32960344 | 0,544538169 | ZNF628 | FALSE | Downregulated |
| 0,305672844 | 0,544793437 | DPP3 | FALSE | Upregulated |
| 0,919998516 | 0,544822734 | CLEC18A | FALSE | Upregulated |
| -0,73523378 | 0,544822734 | LOC102724002 | FALSE | Downregulated |
| 0,11719108 | 0,54483125 | SFPQ | FALSE | Upregulated |
| 0,241054707 | 0,544834057 | NCKAP5L | FALSE | Upregulated |
| 0,218354831 | 0,544834057 | PDE7A | FALSE | Upregulated |
| 0,182570054 | 0,544868684 | RNF220 | FALSE | Upregulated |
| -0,463669367 | 0,544930948 | CCDC144B | FALSE | Downregulated |
| 0,354251851 | 0,544930948 | MT2A | FALSE | Upregulated |
| -0,703478292 | 0,545055218 | ASAH2B | FALSE | Downregulated |
| 0,85186318 | 0,545492595 | B4GALNT3 | FALSE | Upregulated |
| -0,398732729 | 0,545647773 | LOC102724350 | FALSE | Downregulated |
| 0,75756005 | 0,545775672 | C1QTNF3 | FALSE | Upregulated |
| 0,313392038 | 0,545775672 | C22orf15 | FALSE | Upregulated |
| -0,303548891 | 0,546040043 | ZNF404 | FALSE | Downregulated |
| 0,706535338 | 0,546364823 | MAGOHB | FALSE | Upregulated |
| -0,200956635 | 0,546364823 | PDPR | FALSE | Downregulated |
| -0,465678039 | 0,546460446 | PMS2P5 | FALSE | Downregulated |
| 0,256099377 | 0,546460446 | PP2D1 | FALSE | Upregulated |
| -0,16843711 | 0,546460446 | PSD4 | FALSE | Downregulated |
| -0,244797496 | 0,546460446 | SLC35B4 | FALSE | Downregulated |
| -0,618012251 | 0,546624572 | ACTR6 | FALSE | Downregulated |
| -0,303741277 | 0,546624572 | APOL4 | FALSE | Downregulated |
| -2,031202881 | 0,546624572 | C21orf140 | FALSE | Downregulated |
| 0,194090839 | 0,546624572 | CRNKL1 | FALSE | Upregulated |
| -0,327097287 | 0,546624572 | ESD | FALSE | Downregulated |
| 0,209686662 | 0,546624572 | FAM211A | FALSE | Upregulated |
| 0,1761815 | 0,546624572 | GATAD2A | FALSE | Upregulated |
| 0,308573666 | 0,546624572 | IQCG | FALSE | Upregulated |
| 0,524657751 | 0,546624572 | LOC101928485 | FALSE | Upregulated |
| -0,304628532 | 0,546624572 | NUP133 | FALSE | Downregulated |
| -0,474008576 | 0,546624572 | PIGM | FALSE | Downregulated |
| 0,478606021 | 0,546624572 | PJA1 | FALSE | Upregulated |
| 0,180403382 | 0,546624572 | PMPCB | FALSE | Upregulated |
| -0,393754443 | 0,546624572 | RBMS3 | FALSE | Downregulated |
| 0,365552847 | 0,546624572 | SLC25A28 | FALSE | Upregulated |
| -0,205319708 | 0,546624572 | SNRNP70 | FALSE | Downregulated |
| -0,203175505 | 0,546624572 | SUN1 | FALSE | Downregulated |
| -0,286471278 | 0,546624572 | TAF2 | FALSE | Downregulated |
| 0,338795627 | 0,546624572 | ZNF620 | FALSE | Upregulated |

| | | | | |
|---|---|---|---|---|
| -0,67613142 | 0,546659471 | AGPAT9 | FALSE | Downregulated |
| -0,604477744 | 0,547435474 | ATP1B2 | FALSE | Downregulated |
| -0,220110604 | 0,547626552 | PI4K2B | FALSE | Downregulated |
| 0,135625526 | 0,547742808 | MYL6 | FALSE | Upregulated |
| 0,422772226 | 0,547742808 | SLC26A11 | FALSE | Upregulated |
| 0,392923257 | 0,547775937 | CHMP2B | FALSE | Upregulated |
| 0,235801121 | 0,547889158 | OSGEPL1 | FALSE | Upregulated |
| 0,547791653 | 0,548380017 | APOBEC3F | FALSE | Upregulated |
| 0,35701905 | 0,548380017 | MANEA | FALSE | Upregulated |
| -0,23031967 | 0,548388855 | TSHZ2 | FALSE | Downregulated |
| 0,631428794 | 0,548437803 | CTSW | FALSE | Upregulated |
| -0,48043379 | 0,548761214 | CD1D | FALSE | Downregulated |
| -0,365804929 | 0,548761214 | DOLPP1 | FALSE | Downregulated |
| -0,52077638 | 0,548761214 | LOC100126584 | FALSE | Downregulated |
| -0,682783301 | 0,548761214 | MAGEA8 | FALSE | Downregulated |
| 0,370137464 | 0,548761214 | SLC26A2 | FALSE | Upregulated |
| 0,293577993 | 0,548956346 | YKT6 | FALSE | Upregulated |
| -0,232749342 | 0,548970011 | MYL9 | FALSE | Downregulated |
| -0,359440525 | 0,548970011 | TCEAL3 | FALSE | Downregulated |
| -0,183917544 | 0,549096294 | HNRNPL | FALSE | Downregulated |
| -0,781706096 | 0,549512191 | SMIM11 | FALSE | Downregulated |
| 0,860438692 | 0,549521931 | DAPP1 | FALSE | Upregulated |
| 0,322018182 | 0,549521931 | EIF2B2 | FALSE | Upregulated |
| 0,206723226 | 0,549521931 | FAM45A | FALSE | Upregulated |
| -0,208675195 | 0,549521931 | KRAS | FALSE | Downregulated |
| 0,713216637 | 0,549521931 | LOC101928760 | FALSE | Upregulated |
| 0,208175011 | 0,549521931 | LOC101929211 | FALSE | Upregulated |
| 0,759095231 | 0,549521931 | NAT8L | FALSE | Upregulated |
| 0,279644591 | 0,549521931 | OSBPL11 | FALSE | Upregulated |
| -0,380366628 | 0,549521931 | THYN1 | FALSE | Downregulated |
| 0,458871799 | 0,549967845 | FAM86B1 | FALSE | Upregulated |
| -0,800662689 | 0,550153528 | RP11-328K4.1 | FALSE | Downregulated |
| -0,185926686 | 0,550191572 | NNT | FALSE | Downregulated |
| 0,208804731 | 0,550477127 | HMOX2 | FALSE | Upregulated |
| -0,156321097 | 0,550477127 | SLC31A1 | FALSE | Downregulated |
| -0,320336968 | 0,550505007 | ADARB1 | FALSE | Downregulated |
| -0,418387406 | 0,550665357 | AASDHPPT | FALSE | Downregulated |
| 0,263937452 | 0,550665357 | SEC24A | FALSE | Upregulated |
| -0,141856322 | 0,550742515 | BAZ2A | FALSE | Downregulated |
| -0,200192456 | 0,550742515 | BDP1 | FALSE | Downregulated |
| 0,763920948 | 0,550742515 | FAM47E | FALSE | Upregulated |
| -0,168254168 | 0,550742515 | IARS2 | FALSE | Downregulated |
| 0,255189952 | 0,550742515 | KMT2E-AS1 | FALSE | Upregulated |
| 0,799717021 | 0,550742515 | LIMS3 | FALSE | Upregulated |
| -0,596233544 | 0,550742515 | PHYHIP | FALSE | Downregulated |
| 0,693011235 | 0,550742515 | RP11-298I3.1 | FALSE | Upregulated |
| -0,562472907 | 0,550913456 | CNTN4 | FALSE | Downregulated |
| -0,21950474 | 0,550913456 | CYP3A5 | FALSE | Downregulated |
| -0,761605374 | 0,550913456 | FBP2 | FALSE | Downregulated |
| 0,17352883 | 0,550913456 | GPER1 | FALSE | Upregulated |
| -0,609564565 | 0,550913456 | HHAT | FALSE | Downregulated |

| | | | | |
|---|---|---|---|---|
| 0,870183478 | 0,550913456 | LOC101929643 | FALSE | Upregulated |
| 0,537136541 | 0,550913456 | LOC102723980 | FALSE | Upregulated |
| 0,308299061 | 0,550913456 | LOC102724043 | FALSE | Upregulated |
| 0,534373883 | 0,550913456 | PRICKLE1 | FALSE | Upregulated |
| -0,245804 | 0,550913456 | VKORC1L1 | FALSE | Downregulated |
| -0,160301922 | 0,550913456 | XPO1 | FALSE | Downregulated |
| 0,143670704 | 0,550959483 | YY1 | FALSE | Upregulated |
| -0,511246187 | 0,550963169 | GTDC1 | FALSE | Downregulated |
| -0,639788476 | 0,55101129 | LOC102724314 | FALSE | Downregulated |
| -0,299838718 | 0,551031134 | CCL16 | FALSE | Downregulated |
| -0,160675423 | 0,551031134 | GLTPD1 | FALSE | Downregulated |
| 0,478160417 | 0,551031134 | MRPL17 | FALSE | Upregulated |
| -0,204034774 | 0,551031134 | PYGL | FALSE | Downregulated |
| 0,502443162 | 0,551171309 | KLHL18 | FALSE | Upregulated |
| -0,279983541 | 0,551282864 | MGMT | FALSE | Downregulated |
| -0,377822498 | 0,551282864 | TMEM62 | FALSE | Downregulated |
| -0,166064777 | 0,551340454 | SLC39A14 | FALSE | Downregulated |
| 0,263143288 | 0,551377703 | LOC101927266 | FALSE | Upregulated |
| 0,753719087 | 0,551635423 | HIST2H4A | FALSE | Upregulated |
| 0,311835057 | 0,551797504 | GAS5-AS1 | FALSE | Upregulated |
| 0,317940519 | 0,551797504 | LOC101928576 | FALSE | Upregulated |
| -0,249522272 | 0,551797504 | RPS26 | FALSE | Downregulated |
| 0,161790656 | 0,552014678 | POLR2J4 | FALSE | Upregulated |
| -0,257614444 | 0,552014678 | UBL4A | FALSE | Downregulated |
| 0,387757741 | 0,552054463 | ATP5J2 | FALSE | Upregulated |
| 1,066726633 | 0,552166427 | REM2 | FALSE | Upregulated |
| -1,135824797 | 0,552422411 | CA3 | FALSE | Downregulated |
| -0,57430883 | 0,552749492 | FAM229B | FALSE | Downregulated |
| 0,338007774 | 0,552771652 | LOC100506621 | FALSE | Upregulated |
| -0,213278677 | 0,552771652 | SH3BP2 | FALSE | Downregulated |
| -0,254287711 | 0,552771652 | YY1AP1 | FALSE | Downregulated |
| -0,437840275 | 0,552814679 | ISLR | FALSE | Downregulated |
| -0,554753241 | 0,552838966 | HS3ST3A1 | FALSE | Downregulated |
| -0,204932711 | 0,552840169 | MARCH7 | FALSE | Downregulated |
| -0,27485717 | 0,553221134 | A2M | FALSE | Downregulated |
| -0,173013068 | 0,553221134 | DOCK4 | FALSE | Downregulated |
| -0,278692341 | 0,553249676 | ATF5 | FALSE | Downregulated |
| 0,345071286 | 0,553249676 | LOC100132781 | FALSE | Upregulated |
| -0,356772622 | 0,553436066 | POLK | FALSE | Downregulated |
| -0,438192611 | 0,553659265 | CMTR2 | FALSE | Downregulated |
| 0,365451146 | 0,553659265 | XPR1 | FALSE | Upregulated |
| -0,753199394 | 0,553700612 | AK6 | FALSE | Downregulated |
| 0,621013684 | 0,553700612 | HSH2D | FALSE | Upregulated |
| 0,655413108 | 0,553700612 | LOC102724933 | FALSE | Upregulated |
| -0,151503186 | 0,553700612 | SF3B2 | FALSE | Downregulated |
| 0,55868174 | 0,553700612 | TRIM15 | FALSE | Upregulated |
| 0,428417005 | 0,553791597 | LOC101929425 | FALSE | Upregulated |
| -1,193780372 | 0,553791597 | MYPN | FALSE | Downregulated |
| -0,516037791 | 0,553791597 | PSMG2 | FALSE | Downregulated |
| -0,300023123 | 0,553791597 | RCOR3 | FALSE | Downregulated |
| 0,989106795 | 0,553791597 | RGS16 | FALSE | Upregulated |

| | | | | |
|---|---|---|---|---|
| -0,238113062 | 0,553791597 | SMAD6 | FALSE | Downregulated |
| -0,181755935 | 0,553791597 | TOR1AIP2 | FALSE | Downregulated |
| -0,274496617 | 0,553795032 | GLTSCR1L | FALSE | Downregulated |
| 0,717260628 | 0,553983327 | ARL2 | FALSE | Upregulated |
| -0,194961807 | 0,553983327 | COX11 | FALSE | Downregulated |
| -0,222413659 | 0,553983327 | EFR3A | FALSE | Downregulated |
| 0,610292729 | 0,553983327 | TPRG1 | FALSE | Upregulated |
| -0,240136779 | 0,553983327 | ZNF146 | FALSE | Downregulated |
| -0,439888105 | 0,553986091 | ZNF484 | FALSE | Downregulated |
| -0,243930235 | 0,5540929 | PRPSAP1 | FALSE | Downregulated |
| 0,328499275 | 0,55417647 | PSMD5-AS1 | FALSE | Upregulated |
| 0,217385524 | 0,554254024 | IAH1 | FALSE | Upregulated |
| 0,198834079 | 0,554280124 | SDCBP2-AS1 | FALSE | Upregulated |
| 0,482349392 | 0,554280124 | WRB | FALSE | Upregulated |
| 0,21843605 | 0,554463652 | LOC102723929 | FALSE | Upregulated |
| -0,374827527 | 0,554880507 | COPS7B | FALSE | Downregulated |
| 0,338332693 | 0,554885838 | ADAM21 | FALSE | Upregulated |
| -1,150704675 | 0,55491456 | FMN2 | FALSE | Downregulated |
| 0,737076283 | 0,55491456 | NMUR1 | FALSE | Upregulated |
| 0,168230847 | 0,554947747 | ENO1 | FALSE | Upregulated |
| 0,923397889 | 0,555078235 | CKLF | FALSE | Upregulated |
| -0,323780896 | 0,555166848 | KIAA1919 | FALSE | Downregulated |
| -0,887560543 | 0,555166848 | LOC101927372 | FALSE | Downregulated |
| 0,190482062 | 0,555166848 | LOC101928853 | FALSE | Upregulated |
| -0,399542001 | 0,555166848 | SHROOM4 | FALSE | Downregulated |
| -0,363982416 | 0,555183459 | PBX1 | FALSE | Downregulated |
| -0,758152666 | 0,555183459 | PKIA | FALSE | Downregulated |
| -0,182261297 | 0,555293189 | GALM | FALSE | Downregulated |
| -0,320636649 | 0,555440979 | TFAM | FALSE | Downregulated |
| 0,28985338 | 0,555462473 | MED25 | FALSE | Upregulated |
| -0,2645046 | 0,555462473 | NMRAL1 | FALSE | Downregulated |
| -0,267244461 | 0,555766623 | FAM13B | FALSE | Downregulated |
| -0,274537878 | 0,555794485 | ABL2 | FALSE | Downregulated |
| -0,241466245 | 0,555852233 | C4B | FALSE | Downregulated |
| -0,373265022 | 0,555852233 | FAM185A | FALSE | Downregulated |
| -2,154133005 | 0,555852233 | LOC102724828 | FALSE | Downregulated |
| 0,452258011 | 0,555852233 | SNRNP40 | FALSE | Upregulated |
| 0,392236915 | 0,556121701 | ZNF707 | FALSE | Upregulated |
| 0,34205687 | 0,556223888 | LOC100506731 | FALSE | Upregulated |
| 0,587998412 | 0,556255442 | CLEC11A | FALSE | Upregulated |
| 0,716999025 | 0,556300321 | BRSK1 | FALSE | Upregulated |
| 0,47363377 | 0,556300321 | MIPEP | FALSE | Upregulated |
| 0,453867377 | 0,556300321 | PPP1R3F | FALSE | Upregulated |
| -0,392264871 | 0,556300321 | SH3PXD2A-AS1 | FALSE | Downregulated |
| 0,229261569 | 0,556300321 | TRO | FALSE | Upregulated |
| -0,459406384 | 0,556366036 | DNAJC15 | FALSE | Downregulated |
| -0,954347286 | 0,556366036 | KIF17 | FALSE | Downregulated |
| 0,221194884 | 0,556366036 | MYL12B | FALSE | Upregulated |
| -0,164740108 | 0,556366036 | NCBP1 | FALSE | Downregulated |
| -0,229048435 | 0,556366036 | VEGFB | FALSE | Downregulated |
| -0,160834074 | 0,556366883 | MLF2 | FALSE | Downregulated |

| | | | | |
|---|---|---|---|---|
| -0,330244764 | 0,556399188 | AFF3 | FALSE | Downregulated |
| -0,277241374 | 0,556519668 | PCNXL4 | FALSE | Downregulated |
| 0,694356068 | 0,556607547 | CACNB1 | FALSE | Upregulated |
| 0,265092288 | 0,556879895 | IST1 | FALSE | Upregulated |
| 0,309105823 | 0,556938826 | TRIP10 | FALSE | Upregulated |
| 0,303174259 | 0,557167637 | MLST8 | FALSE | Upregulated |
| -0,250292785 | 0,557264316 | PPP4R2 | FALSE | Downregulated |
| -0,366611537 | 0,5572785 | GOLGA8A | FALSE | Downregulated |
| -0,665715657 | 0,5572785 | LRP2 | FALSE | Downregulated |
| 0,478671567 | 0,5572785 | TRIM34 | FALSE | Upregulated |
| -0,302814804 | 0,557559659 | CCSER2 | FALSE | Downregulated |
| -0,87614849 | 0,557559659 | OR2A2 | FALSE | Downregulated |
| -0,440187736 | 0,557559659 | ZNF304 | FALSE | Downregulated |
| -0,155482322 | 0,557598611 | BRD2 | FALSE | Downregulated |
| 0,328023635 | 0,557598611 | CYSTM1 | FALSE | Upregulated |
| -0,288389025 | 0,557598611 | FRAT2 | FALSE | Downregulated |
| 0,446337888 | 0,557598611 | GMDS | FALSE | Upregulated |
| 0,459113273 | 0,557598611 | LAMB3 | FALSE | Upregulated |
| 0,688081041 | 0,557598611 | LOC101927183 | FALSE | Upregulated |
| 0,168609389 | 0,557598611 | PAQR3 | FALSE | Upregulated |
| -0,494822024 | 0,557598611 | RECK | FALSE | Downregulated |
| -0,200093726 | 0,558119243 | CNPY2 | FALSE | Downregulated |
| -0,974801092 | 0,558119243 | KIAA1324L | FALSE | Downregulated |
| -0,41023348 | 0,558252071 | ACOT1 | FALSE | Downregulated |
| 0,259252299 | 0,558463013 | RAB35 | FALSE | Upregulated |
| -0,716473857 | 0,558485482 | FAM127B | FALSE | Downregulated |
| 0,324746602 | 0,558485482 | PPARD | FALSE | Upregulated |
| 0,252982207 | 0,558485482 | VPS41 | FALSE | Upregulated |
| 0,44028942 | 0,558539263 | NKRF | FALSE | Upregulated |
| 0,425409505 | 0,558597823 | GNG2 | FALSE | Upregulated |
| 0,408722266 | 0,5594625 | FABP2 | FALSE | Upregulated |
| 0,32340667 | 0,559542025 | EMC3-AS1 | FALSE | Upregulated |
| 0,41449634 | 0,559542025 | H2AFY2 | FALSE | Upregulated |
| 0,330041594 | 0,559542025 | LOC642648 | FALSE | Upregulated |
| -0,181783796 | 0,559542025 | SLC35D1 | FALSE | Downregulated |
| 0,166815455 | 0,559726277 | PTPRJ | FALSE | Upregulated |
| 0,176058267 | 0,559817911 | ZSWIM8-AS1 | FALSE | Upregulated |
| -0,391132533 | 0,559857502 | LOC101929328 | FALSE | Downregulated |
| -0,877617129 | 0,559998968 | LOC102723799 | FALSE | Downregulated |
| -0,298387285 | 0,559998968 | MAMDC2 | FALSE | Downregulated |
| -0,441809421 | 0,559998968 | RPAP3 | FALSE | Downregulated |
| -0,391090177 | 0,559998968 | TMEM45A | FALSE | Downregulated |
| 0,284480001 | 0,559998968 | TRA2A | FALSE | Upregulated |
| 0,731991241 | 0,560084482 | CNTNAP3 | FALSE | Upregulated |
| -0,210819337 | 0,560084482 | MSH3 | FALSE | Downregulated |
| 0,273513356 | 0,560558201 | PITHD1 | FALSE | Upregulated |
| -0,424139312 | 0,560654137 | CDHR3 | FALSE | Downregulated |
| 0,451970217 | 0,560672541 | BTN2A2 | FALSE | Upregulated |
| 0,271289941 | 0,560672541 | CCNE2 | FALSE | Upregulated |
| -0,21838713 | 0,560672541 | CD4 | FALSE | Downregulated |
| -0,226044592 | 0,560672541 | EEF2K | FALSE | Downregulated |

| | | | | |
|---|---|---|---|---|
| 0,45347967 | 0,560672541 | RP11-1072A3.2 | FALSE | Upregulated |
| -0,257116447 | 0,560784417 | BRF1 | FALSE | Downregulated |
| -0,422359098 | 0,560784417 | ELMOD2 | FALSE | Downregulated |
| -0,566734009 | 0,560784417 | LINC01089 | FALSE | Downregulated |
| -0,350720294 | 0,560784417 | NUDT22 | FALSE | Downregulated |
| -1,074092327 | 0,560784417 | SENP3 | FALSE | Downregulated |
| -0,605911358 | 0,560784417 | VAV3 | FALSE | Downregulated |
| -0,746756095 | 0,560784417 | ZNF442 | FALSE | Downregulated |
| 0,206256846 | 0,561369114 | LOC101929426 | FALSE | Upregulated |
| 0,295490539 | 0,561477786 | LBP | FALSE | Upregulated |
| -0,225396441 | 0,561566886 | BAG1 | FALSE | Downregulated |
| -0,302741114 | 0,561654536 | FOXO4 | FALSE | Downregulated |
| 0,292475405 | 0,562008511 | CCDC124 | FALSE | Upregulated |
| -0,319522872 | 0,562089648 | NSMCE4A | FALSE | Downregulated |
| 0,256406132 | 0,562089648 | RNF19B | FALSE | Upregulated |
| 0,325130984 | 0,562089648 | TBC1D10A | FALSE | Upregulated |
| 0,208392888 | 0,562104926 | AVL9 | FALSE | Upregulated |
| 0,187272884 | 0,562104926 | NRBP2 | FALSE | Upregulated |
| 0,194913517 | 0,562129081 | RUFY3 | FALSE | Upregulated |
| 0,37903305 | 0,562129081 | TUBA3D | FALSE | Upregulated |
| -0,426579814 | 0,562144031 | AGPAT5 | FALSE | Downregulated |
| 0,572419502 | 0,562144031 | LOC101929185 | FALSE | Upregulated |
| 0,308663186 | 0,562144031 | MBL2 | FALSE | Upregulated |
| 0,711838932 | 0,562191754 | SSPN | FALSE | Upregulated |
| -0,667442581 | 0,562318059 | TRPC5OS | FALSE | Downregulated |
| 0,746381412 | 0,562369829 | CASS4 | FALSE | Upregulated |
| 0,199331464 | 0,562369829 | MED29 | FALSE | Upregulated |
| -0,208438593 | 0,562369829 | PRDX5 | FALSE | Downregulated |
| 0,541754778 | 0,562369829 | SLC8A3 | FALSE | Upregulated |
| 0,254899281 | 0,562369829 | ZNF384 | FALSE | Upregulated |
| -0,256654921 | 0,562430546 | BRD8 | FALSE | Downregulated |
| -0,249171784 | 0,562526881 | C2CD2 | FALSE | Downregulated |
| -0,842332616 | 0,562799622 | SULT1C4 | FALSE | Downregulated |
| 0,42523844 | 0,562924331 | LOC100268168 | FALSE | Upregulated |
| 0,216591943 | 0,562963335 | INTS6 | FALSE | Upregulated |
| -0,368766817 | 0,563128223 | NPIPB9 | FALSE | Downregulated |
| -0,381514644 | 0,563206416 | PPA2 | FALSE | Downregulated |
| 0,279320239 | 0,563206416 | TBC1D20 | FALSE | Upregulated |
| -0,178492712 | 0,563292086 | MRPL43 | FALSE | Downregulated |
| -0,102057646 | 0,563312202 | PLXNB2 | FALSE | Downregulated |
| 0,199577859 | 0,563312202 | SLC9A8 | FALSE | Upregulated |
| -0,244718693 | 0,563472104 | RAB40C | FALSE | Downregulated |
| 0,433301854 | 0,563493914 | ARHGAP39 | FALSE | Upregulated |
| 0,14875385 | 0,563903689 | LOC100507577 | FALSE | Upregulated |
| 0,163488372 | 0,564015921 | CAPRIN1 | FALSE | Upregulated |
| -0,595369319 | 0,564015921 | MED10 | FALSE | Downregulated |
| -0,239950379 | 0,564031458 | FAM69A | FALSE | Downregulated |
| -0,245851338 | 0,564031458 | MTHFS | FALSE | Downregulated |
| 0,476556083 | 0,564031458 | SIGLEC15 | FALSE | Upregulated |
| -0,267021214 | 0,564031458 | TRMT44 | FALSE | Downregulated |
| 0,422249192 | 0,564068954 | EFR3B | FALSE | Upregulated |

| | | | | |
|---|---|---|---|---|
| 0,305377083 | 0,564068954 | KIF22 | FALSE | Upregulated |
| 0,265618848 | 0,564068954 | LDHAL6B | FALSE | Upregulated |
| 0,438586944 | 0,564068954 | SCNN1A | FALSE | Upregulated |
| -0,478869911 | 0,564138791 | CHST14 | FALSE | Downregulated |
| -0,493469391 | 0,564138791 | NUDT14 | FALSE | Downregulated |
| 0,941680649 | 0,564184138 | BOLA2B | FALSE | Upregulated |
| 0,253587307 | 0,564353859 | TEAD3 | FALSE | Upregulated |
| -0,302950929 | 0,565322268 | SLC6A1-AS1 | FALSE | Downregulated |
| 0,313052318 | 0,565535389 | IPO4 | FALSE | Upregulated |
| 0,225250969 | 0,565614855 | HEXB | FALSE | Upregulated |
| -0,302486228 | 0,565721889 | GIPC2 | FALSE | Downregulated |
| 0,279389027 | 0,565910707 | CCT2 | FALSE | Upregulated |
| 0,771729747 | 0,565992593 | DDX11L5 | FALSE | Upregulated |
| 0,331017275 | 0,565992593 | NCDN | FALSE | Upregulated |
| -0,6028366 | 0,565992593 | SLX4IP | FALSE | Downregulated |
| -0,189017739 | 0,565997701 | CYTH1 | FALSE | Downregulated |
| 0,365654618 | 0,566140383 | FTSJ1 | FALSE | Upregulated |
| -0,168201936 | 0,566140383 | GTF2I | FALSE | Downregulated |
| 0,538931068 | 0,566140383 | HM13-AS1 | FALSE | Upregulated |
| -0,171403956 | 0,566140383 | RTEL1 | FALSE | Downregulated |
| 0,198540287 | 0,566140383 | TMPO | FALSE | Upregulated |
| -0,381652383 | 0,566140383 | ZNF155 | FALSE | Downregulated |
| -0,205880514 | 0,566252462 | CEP95 | FALSE | Downregulated |
| -0,193681076 | 0,566252462 | PATL1 | FALSE | Downregulated |
| -0,259193158 | 0,566252462 | SUMF1 | FALSE | Downregulated |
| 0,291705012 | 0,566252462 | ZNF317 | FALSE | Upregulated |
| -0,395503456 | 0,566712639 | CELF4 | FALSE | Downregulated |
| -0,246389126 | 0,566850863 | GSK3A | FALSE | Downregulated |
| 0,511484491 | 0,566850863 | RBPMS-AS1 | FALSE | Upregulated |
| 0,286481227 | 0,566879633 | CBLB | FALSE | Upregulated |
| 0,283245112 | 0,566879633 | CHMP6 | FALSE | Upregulated |
| -0,340510205 | 0,566879633 | DNAJC25 | FALSE | Downregulated |
| -0,236989843 | 0,566879633 | EPS15L1 | FALSE | Downregulated |
| -0,585967022 | 0,566879633 | LOC100132352 | FALSE | Downregulated |
| -0,418694708 | 0,566879633 | NME3 | FALSE | Downregulated |
| -0,467763994 | 0,566879633 | PPHLN1 | FALSE | Downregulated |
| -0,300606151 | 0,566879633 | SPRY2 | FALSE | Downregulated |
| -0,56481855 | 0,566915459 | SULT1C2 | FALSE | Downregulated |
| 0,283491598 | 0,566964815 | DSC2 | FALSE | Upregulated |
| 0,341383227 | 0,56726788 | BCL2L12 | FALSE | Upregulated |
| -0,609058118 | 0,56726788 | CCDC108 | FALSE | Downregulated |
| -0,264086054 | 0,56726788 | FLJ38717 | FALSE | Downregulated |
| -0,303597176 | 0,56726788 | LOC100506548 | FALSE | Downregulated |
| -0,245692734 | 0,56726788 | MBTD1 | FALSE | Downregulated |
| -0,255306948 | 0,56726788 | NCAPD3 | FALSE | Downregulated |
| -0,445565113 | 0,56726788 | PIBF1 | FALSE | Downregulated |
| -0,369678977 | 0,56726788 | SNORD17 | FALSE | Downregulated |
| 0,308715983 | 0,56726788 | SUZ12 | FALSE | Upregulated |
| 0,74639443 | 0,56726788 | TBX6 | FALSE | Upregulated |
| -0,315417489 | 0,56726788 | ZNF500 | FALSE | Downregulated |
| 0,776157513 | 0,567398537 | C12orf71 | FALSE | Upregulated |

| | | | | |
|---|---|---|---|---|
| 0,249540285 | 0,567398537 | GFM2 | FALSE | Upregulated |
| 0,621503127 | 0,567398537 | LOC100128079 | FALSE | Upregulated |
| 0,883571533 | 0,567398537 | LOC102723767 | FALSE | Upregulated |
| -0,233392122 | 0,567398537 | NAPRT1 | FALSE | Downregulated |
| -0,474702678 | 0,567398537 | PDZD4 | FALSE | Downregulated |
| -0,260962269 | 0,567398537 | SDHD | FALSE | Downregulated |
| -0,114303616 | 0,567398537 | SETD5 | FALSE | Downregulated |
| -0,166338232 | 0,567398537 | SH3D19 | FALSE | Downregulated |
| -0,18591827 | 0,567398537 | TP53INP2 | FALSE | Downregulated |
| 0,226542601 | 0,567398537 | TUBA4A | FALSE | Upregulated |
| 0,853349119 | 0,567398537 | VDR | FALSE | Upregulated |
| -0,212375248 | 0,567398537 | VPS54 | FALSE | Downregulated |
| 0,186196292 | 0,56741865 | DMPK | FALSE | Upregulated |
| 0,69782436 | 0,56741865 | FLJ37035 | FALSE | Upregulated |
| -0,153227136 | 0,56741865 | ZNF316 | FALSE | Downregulated |
| -0,162989867 | 0,567470233 | ARAF | FALSE | Downregulated |
| -0,516382605 | 0,567470233 | FAM76A | FALSE | Downregulated |
| -3,352488103 | 0,567470233 | LOC102723595 | FALSE | Downregulated |
| -0,82275265 | 0,567470233 | NOXRED1 | FALSE | Downregulated |
| -0,231264105 | 0,567470233 | PALM2 | FALSE | Downregulated |
| -0,33319986 | 0,567765274 | MGC27345 | FALSE | Downregulated |
| 0,362396917 | 0,568205118 | TIMM22 | FALSE | Upregulated |
| 0,575095734 | 0,568492175 | LOC102724528 | FALSE | Upregulated |
| -0,152982536 | 0,568503176 | AMFR | FALSE | Downregulated |
| -0,194294073 | 0,568503176 | BTF3 | FALSE | Downregulated |
| 0,730585215 | 0,568503176 | LOC100506281 | FALSE | Upregulated |
| 0,26314111 | 0,568503176 | MBOAT7 | FALSE | Upregulated |
| -0,331039353 | 0,568503176 | MCPH1 | FALSE | Downregulated |
| 0,776471912 | 0,568503176 | SPAG17 | FALSE | Upregulated |
| 0,446926504 | 0,568732934 | CD244 | FALSE | Upregulated |
| 0,2863751 | 0,568732934 | PPIP5K1 | FALSE | Upregulated |
| 0,501507819 | 0,568732934 | TRIM69 | FALSE | Upregulated |
| -0,523954693 | 0,569084551 | HIST1H4B | FALSE | Downregulated |
| -0,178238003 | 0,569084551 | SLC47A1 | FALSE | Downregulated |
| -0,398686601 | 0,56908468 | DYNLT3 | FALSE | Downregulated |
| 0,348256992 | 0,56908468 | FARS2 | FALSE | Upregulated |
| 0,233483362 | 0,569129036 | LOC101929017 | FALSE | Upregulated |
| 0,221369063 | 0,569252613 | PITPNB | FALSE | Upregulated |
| 0,984837043 | 0,56938437 | ATP6V1G2 | FALSE | Upregulated |
| -0,504938902 | 0,56953233 | ITGA3 | FALSE | Downregulated |
| 0,401924534 | 0,569703978 | GEMIN8P4 | FALSE | Upregulated |
| -0,395207992 | 0,569703978 | IL17D | FALSE | Downregulated |
| -0,295392001 | 0,569703978 | MAPK8 | FALSE | Downregulated |
| 0,238381079 | 0,569703978 | TMEM39A | FALSE | Upregulated |
| -0,362076796 | 0,569859955 | CHEK1 | FALSE | Downregulated |
| -0,405006659 | 0,569874843 | CD40 | FALSE | Downregulated |
| -0,468139151 | 0,569874843 | GRIN2A | FALSE | Downregulated |
| -0,400676598 | 0,569874843 | KIF13A | FALSE | Downregulated |
| -0,36339087 | 0,569874843 | PRUNE | FALSE | Downregulated |
| -0,243877933 | 0,569874843 | TEF | FALSE | Downregulated |
| 0,24828941 | 0,569874843 | WIPF2 | FALSE | Upregulated |

| | | | | |
|---|---|---|---|---|
| 0,322103212 | 0,569874843 | ZNF30 | FALSE | Upregulated |
| -0,213740125 | 0,569912673 | NHLRC2 | FALSE | Downregulated |
| -0,177951741 | 0,569912673 | PGRMC1 | FALSE | Downregulated |
| -0,176535638 | 0,569953618 | PMM2 | FALSE | Downregulated |
| 0,777782983 | 0,570308549 | PLAU | FALSE | Upregulated |
| -1,029602308 | 0,570485978 | LOC102723794 | FALSE | Downregulated |
| -0,304995084 | 0,57087541 | GADD45A | FALSE | Downregulated |
| -0,195597599 | 0,571134115 | OPA1 | FALSE | Downregulated |
| -0,195860433 | 0,571719393 | AKTIP | FALSE | Downregulated |
| -0,158455059 | 0,571719393 | ASCC2 | FALSE | Downregulated |
| 0,181904419 | 0,571719393 | ATP11A | FALSE | Upregulated |
| -0,247615783 | 0,571719393 | ATP5F1 | FALSE | Downregulated |
| 0,182435223 | 0,571719393 | C11orf57 | FALSE | Upregulated |
| 0,612223538 | 0,571719393 | DAPK2 | FALSE | Upregulated |
| -0,283510281 | 0,571719393 | HSD17B12 | FALSE | Downregulated |
| 0,6003829 | 0,571719393 | KIAA1549L | FALSE | Upregulated |
| 0,580783568 | 0,571719393 | NKG7 | FALSE | Upregulated |
| -0,393489861 | 0,571719393 | NOXA1 | FALSE | Downregulated |
| -0,496352907 | 0,571719393 | RDH5 | FALSE | Downregulated |
| -0,941800335 | 0,571719393 | RP11-421P23.1 | FALSE | Downregulated |
| -0,311221676 | 0,571719393 | SAPCD2 | FALSE | Downregulated |
| -0,67129275 | 0,571719393 | SLC22A17 | FALSE | Downregulated |
| -0,650872748 | 0,571719393 | TCEANC | FALSE | Downregulated |
| -0,269365998 | 0,57192783 | ZC3H6 | FALSE | Downregulated |
| 0,279663611 | 0,572036973 | KLHL42 | FALSE | Upregulated |
| 0,154054863 | 0,572054688 | ANKRD52 | FALSE | Upregulated |
| -0,228068579 | 0,572054688 | ATXN10 | FALSE | Downregulated |
| -0,336049847 | 0,572089199 | C17orf107 | FALSE | Downregulated |
| -0,638821368 | 0,572089199 | CHMP4C | FALSE | Downregulated |
| 1,521666607 | 0,572089199 | LOC284933 | FALSE | Upregulated |
| 0,310776768 | 0,572089199 | SRGAP2 | FALSE | Upregulated |
| 0,269379329 | 0,57218323 | LOC100506258 | FALSE | Upregulated |
| 0,416716024 | 0,57218323 | PDE6B | FALSE | Upregulated |
| -0,15946893 | 0,57218323 | PGAM1 | FALSE | Downregulated |
| 0,475281061 | 0,57218323 | RAD54L | FALSE | Upregulated |
| -0,329608425 | 0,572529544 | BTBD10 | FALSE | Downregulated |
| 0,216366133 | 0,572529544 | GGACT | FALSE | Upregulated |
| 0,245273713 | 0,572529544 | SNORA73A | FALSE | Upregulated |
| -0,387398428 | 0,572529544 | TMEM203 | FALSE | Downregulated |
| 0,578941517 | 0,57282541 | BTBD19 | FALSE | Upregulated |
| 0,36997307 | 0,572839792 | NDUFA4L2 | FALSE | Upregulated |
| 0,211151927 | 0,572878292 | AIMP1 | FALSE | Upregulated |
| 0,600356414 | 0,572878292 | CNIH2 | FALSE | Upregulated |
| 0,391449369 | 0,572878292 | LOC102723708 | FALSE | Upregulated |
| -0,407245501 | 0,572984557 | LAPTM4B | FALSE | Downregulated |
| -0,166698112 | 0,572984557 | MCFD2 | FALSE | Downregulated |
| -0,689749375 | 0,572984557 | MTHFD2L | FALSE | Downregulated |
| 0,454936898 | 0,572984557 | RCCD1 | FALSE | Upregulated |
| 0,309454683 | 0,572984557 | SCG3 | FALSE | Upregulated |
| -0,88045186 | 0,572984557 | WFDC1 | FALSE | Downregulated |
| -0,751841242 | 0,573106223 | IFT52 | FALSE | Downregulated |

| | | | | |
|---|---|---|---|---|
| -0,171495419 | 0,573106223 | RPLP2 | FALSE | Downregulated |
| -0,168114446 | 0,573106223 | VCPIP1 | FALSE | Downregulated |
| -0,5233953 | 0,573240509 | KIAA1875 | FALSE | Downregulated |
| -0,307581743 | 0,573563174 | BCL2L14 | FALSE | Downregulated |
| 0,27386414 | 0,573563174 | CMC2 | FALSE | Upregulated |
| 0,422031802 | 0,573563174 | FLJ39095 | FALSE | Upregulated |
| 0,327829643 | 0,573563174 | KLF16 | FALSE | Upregulated |
| -0,208469196 | 0,573563174 | RFNG | FALSE | Downregulated |
| -0,172578317 | 0,573563174 | SYNRG | FALSE | Downregulated |
| 0,22453849 | 0,573981446 | GPN1 | FALSE | Upregulated |
| -1,008240365 | 0,573981446 | LINC00907 | FALSE | Downregulated |
| 0,718904793 | 0,57399122 | U2AF1L4 | FALSE | Upregulated |
| 0,41708348 | 0,574453135 | ACSBG2 | FALSE | Upregulated |
| -0,453075854 | 0,574453135 | AGBL3 | FALSE | Downregulated |
| 0,525639805 | 0,574453135 | COL9A2 | FALSE | Upregulated |
| -0,265885834 | 0,574453135 | DHPS | FALSE | Downregulated |
| -0,427349044 | 0,574453135 | DUSP5 | FALSE | Downregulated |
| -0,187001153 | 0,574453135 | EFCAB14 | FALSE | Downregulated |
| -0,167813426 | 0,574453135 | FOXK2 | FALSE | Downregulated |
| 0,484940963 | 0,574453135 | GMFG | FALSE | Upregulated |
| 0,202469433 | 0,574453135 | GMPR2 | FALSE | Upregulated |
| 0,396504014 | 0,574453135 | GNAT2 | FALSE | Upregulated |
| -0,173777142 | 0,574453135 | HNRNPDL | FALSE | Downregulated |
| -0,160403081 | 0,574453135 | HYAL1 | FALSE | Downregulated |
| 0,234823507 | 0,574453135 | ISOC1 | FALSE | Upregulated |
| 0,322560802 | 0,574453135 | KCNV2 | FALSE | Upregulated |
| 1,540754052 | 0,574453135 | LOC101060389 | FALSE | Upregulated |
| 0,347389377 | 0,574453135 | MRPL35 | FALSE | Upregulated |
| 0,248673427 | 0,574453135 | PTPN9 | FALSE | Upregulated |
| -0,377196349 | 0,574453135 | RCHY1 | FALSE | Downregulated |
| -0,167059769 | 0,574453135 | SEC63 | FALSE | Downregulated |
| 0,288038229 | 0,574453135 | SLC22A5 | FALSE | Upregulated |
| -0,531024181 | 0,574453135 | SWI5 | FALSE | Downregulated |
| 0,845117813 | 0,574453135 | TRIM52-AS1 | FALSE | Upregulated |
| 0,2149127 | 0,574453135 | YIPF2 | FALSE | Upregulated |
| 0,488196391 | 0,574946943 | GYG1 | FALSE | Upregulated |
| 0,769870637 | 0,575012487 | FBXW9 | FALSE | Upregulated |
| -1,28759525 | 0,575012487 | HSD3B2 | FALSE | Downregulated |
| -0,629400209 | 0,575608262 | LOC100272217 | FALSE | Downregulated |
| -0,253480609 | 0,575658895 | ARMC10 | FALSE | Downregulated |
| -0,423164068 | 0,575658895 | TRADD | FALSE | Downregulated |
| -0,135245116 | 0,57568317 | NKTR | FALSE | Downregulated |
| -0,261784667 | 0,575753694 | SYNE3 | FALSE | Downregulated |
| -0,177812447 | 0,575869931 | EIF3H | FALSE | Downregulated |
| -0,26624914 | 0,575869931 | EPN2 | FALSE | Downregulated |
| -0,244400887 | 0,575869931 | USP33 | FALSE | Downregulated |
| 0,185009952 | 0,575892295 | LOC101928323 | FALSE | Upregulated |
| -0,533798411 | 0,575892295 | PFDN6 | FALSE | Downregulated |
| 0,299668349 | 0,575892295 | SREBF1 | FALSE | Upregulated |
| -0,401133809 | 0,575892295 | ZNF792 | FALSE | Downregulated |
| -0,295480113 | 0,576065786 | LMBRD1 | FALSE | Downregulated |

| | | | | |
|---|---|---|---|---|
| 0,230223996 | 0,576065786 | MMP24-AS1 | FALSE | Upregulated |
| 0,768635269 | 0,576080434 | ATP2C2 | FALSE | Upregulated |
| -0,523979545 | 0,576080434 | GOLGA6C | FALSE | Downregulated |
| 0,477401244 | 0,576080434 | RINL | FALSE | Upregulated |
| 0,222082953 | 0,576228929 | SUMO2 | FALSE | Upregulated |
| 0,494373305 | 0,576342407 | CPLX2 | FALSE | Upregulated |
| 0,503445182 | 0,576422408 | ALG5 | FALSE | Upregulated |
| -0,143931903 | 0,576446541 | AFF1 | FALSE | Downregulated |
| -0,492137516 | 0,576446541 | MYCT1 | FALSE | Downregulated |
| -0,318837092 | 0,576446541 | ZNF697 | FALSE | Downregulated |
| -0,744779591 | 0,576526133 | AMZ2P1 | FALSE | Downregulated |
| -0,153662386 | 0,576754418 | DGCR2 | FALSE | Downregulated |
| 0,224606939 | 0,576754418 | NAT8 | FALSE | Upregulated |
| -0,243448084 | 0,576909704 | SNX30 | FALSE | Downregulated |
| 0,19778018 | 0,577073722 | USP45 | FALSE | Upregulated |
| -0,394697502 | 0,577179014 | TSC22D3 | FALSE | Downregulated |
| 0,190547563 | 0,577473284 | GPX1 | FALSE | Upregulated |
| -0,385542692 | 0,577634712 | GRAMD2 | FALSE | Downregulated |
| -0,757623483 | 0,577634712 | LOC79999 | FALSE | Downregulated |
| -0,191419988 | 0,577634712 | MRPL19 | FALSE | Downregulated |
| -0,352440703 | 0,577634712 | SLC2A8 | FALSE | Downregulated |
| -0,40117831 | 0,577752657 | LOC100129722 | FALSE | Downregulated |
| -0,242335329 | 0,577908612 | PDZD8 | FALSE | Downregulated |
| -0,250553825 | 0,577908612 | SLC25A16 | FALSE | Downregulated |
| 0,291758155 | 0,577908612 | TIGD7 | FALSE | Upregulated |
| 0,251260189 | 0,577965003 | LOC102724611 | FALSE | Upregulated |
| 0,339223399 | 0,578068396 | CCDC106 | FALSE | Upregulated |
| 0,224772978 | 0,578068396 | EMC3 | FALSE | Upregulated |
| 0,372620579 | 0,578068396 | LOC102724733 | FALSE | Upregulated |
| 0,978210713 | 0,578239412 | SPAG4 | FALSE | Upregulated |
| -0,202902759 | 0,578370802 | ZNF655 | FALSE | Downregulated |
| 0,463652655 | 0,578814675 | DGKH | FALSE | Upregulated |
| 0,259228348 | 0,578814675 | ENTHD2 | FALSE | Upregulated |
| 0,181868842 | 0,578814675 | SNW1 | FALSE | Upregulated |
| 0,293905525 | 0,578928841 | CABYR | FALSE | Upregulated |
| 0,498198972 | 0,578928841 | PDP1 | FALSE | Upregulated |
| -0,278110674 | 0,578928841 | UMPS | FALSE | Downregulated |
| 0,35518916 | 0,579083798 | TMPPE | FALSE | Upregulated |
| -0,216567503 | 0,579084671 | EIF1AX | FALSE | Downregulated |
| 0,276610551 | 0,579084671 | PTPRH | FALSE | Upregulated |
| -0,18106189 | 0,579084671 | TM7SF3 | FALSE | Downregulated |
| 0,290563063 | 0,579099891 | STRA13 | FALSE | Upregulated |
| -0,173731724 | 0,579189527 | RTN4 | FALSE | Downregulated |
| 0,38171088 | 0,579285219 | VPS37C | FALSE | Upregulated |
| -0,443182683 | 0,579285219 | ZFP90 | FALSE | Downregulated |
| -0,843465145 | 0,57942405 | SPACA6P | FALSE | Downregulated |
| -0,261092238 | 0,579452413 | ARPP19 | FALSE | Downregulated |
| 0,498193288 | 0,579452413 | LOC101927313 | FALSE | Upregulated |
| 0,178726298 | 0,579629191 | KLF13 | FALSE | Upregulated |
| 0,181553549 | 0,579773932 | APOL1 | FALSE | Upregulated |
| -0,360310965 | 0,579773932 | DUSP1 | FALSE | Downregulated |

| | | | | |
|---|---|---|---|---|
| -0,576404133 | 0,579773932 | EDN1 | FALSE | Downregulated |
| 0,668694075 | 0,579773932 | ISM1-AS1 | FALSE | Upregulated |
| 0,496448046 | 0,579773932 | LOC100996763 | FALSE | Upregulated |
| 0,183305128 | 0,579773932 | NIPA2 | FALSE | Upregulated |
| 0,251432059 | 0,579773932 | TMEM180 | FALSE | Upregulated |
| -0,581762503 | 0,579773932 | TMEM42 | FALSE | Downregulated |
| -0,647488501 | 0,579859398 | FLJ36777 | FALSE | Downregulated |
| -0,319975223 | 0,579965483 | PLEKHM3 | FALSE | Downregulated |
| -0,120581544 | 0,579985069 | HMBOX1 | FALSE | Downregulated |
| -0,331949887 | 0,579985069 | PCBP4 | FALSE | Downregulated |
| 0,569380619 | 0,580344558 | TIMD4 | FALSE | Upregulated |
| -0,734791635 | 0,580344558 | ZNF658B | FALSE | Downregulated |
| 0,185851507 | 0,580358009 | DOCK1 | FALSE | Upregulated |
| 0,503121988 | 0,580358009 | FOLR1 | FALSE | Upregulated |
| 0,227012204 | 0,580358009 | LOC101928892 | FALSE | Upregulated |
| 0,942801254 | 0,580358009 | MSH5 | FALSE | Upregulated |
| 0,201580318 | 0,58040709 | ATP13A1 | FALSE | Upregulated |
| -0,782421217 | 0,58040709 | KCNJ14 | FALSE | Downregulated |
| -0,311517488 | 0,58040709 | RARS | FALSE | Downregulated |
| 0,259725114 | 0,58040709 | TTBK1 | FALSE | Upregulated |
| 0,192350514 | 0,580520465 | AGK | FALSE | Upregulated |
| 0,283086131 | 0,580521312 | ATAD2 | FALSE | Upregulated |
| 0,949651262 | 0,580521312 | C11orf48 | FALSE | Upregulated |
| 0,176031761 | 0,580521312 | CCT5 | FALSE | Upregulated |
| 0,296797253 | 0,580521312 | FBXO41 | FALSE | Upregulated |
| 0,521279377 | 0,580521312 | LOC100506302 | FALSE | Upregulated |
| 0,227586629 | 0,580521312 | LOC153684 | FALSE | Upregulated |
| -0,19592884 | 0,580566403 | TMEM205 | FALSE | Downregulated |
| 0,643870509 | 0,58060486 | ARMC2-AS1 | FALSE | Upregulated |
| -0,212217477 | 0,58060486 | CHPT1 | FALSE | Downregulated |
| -0,279820413 | 0,58060486 | FAM63B | FALSE | Downregulated |
| -0,1458293 | 0,58060486 | WASF2 | FALSE | Downregulated |
| -0,145603075 | 0,580647435 | KAT6A | FALSE | Downregulated |
| -0,166353897 | 0,581228758 | DYSF | FALSE | Downregulated |
| 0,487385331 | 0,581423713 | CD7 | FALSE | Upregulated |
| -0,626735394 | 0,581423713 | LINC00998 | FALSE | Downregulated |
| 0,236580991 | 0,581423713 | NPTN | FALSE | Upregulated |
| -0,257425542 | 0,581575158 | FRS2 | FALSE | Downregulated |
| 0,186624102 | 0,581575158 | GNB4 | FALSE | Upregulated |
| 0,480260606 | 0,581575158 | TMEM126B | FALSE | Upregulated |
| -0,121326927 | 0,581802141 | BPTF | FALSE | Downregulated |
| 0,465744733 | 0,581802141 | CHCHD7 | FALSE | Upregulated |
| 0,464628988 | 0,581802141 | LXN | FALSE | Upregulated |
| 0,286096768 | 0,582309812 | SLC27A4 | FALSE | Upregulated |
| -0,270026169 | 0,582563556 | FOXN2 | FALSE | Downregulated |
| -0,359503021 | 0,582563556 | STOX2 | FALSE | Downregulated |
| -0,187163339 | 0,582563556 | ZNF444 | FALSE | Downregulated |
| -0,297459122 | 0,582563556 | ZNF467 | FALSE | Downregulated |
| -0,61863743 | 0,58256739 | ORC5 | FALSE | Downregulated |
| 0,397872652 | 0,582701286 | PLA2G4C | FALSE | Upregulated |
| -0,171815145 | 0,582701286 | STK40 | FALSE | Downregulated |

| | | | | |
|---|---|---|---|---|
| 0,264447872 | 0,582705797 | CDPF1 | FALSE | Upregulated |
| -0,49992127 | 0,582705797 | KDM4E | FALSE | Downregulated |
| -0,144003234 | 0,582705797 | SNX1 | FALSE | Downregulated |
| -0,209330574 | 0,582705797 | USP31 | FALSE | Downregulated |
| -0,301041294 | 0,582705797 | ZNF264 | FALSE | Downregulated |
| 0,235075194 | 0,582988975 | KIFC3 | FALSE | Upregulated |
| -0,43338152 | 0,582988975 | LINC00886 | FALSE | Downregulated |
| 0,217667579 | 0,583051963 | GCLM | FALSE | Upregulated |
| -0,254761984 | 0,583536013 | HMG20A | FALSE | Downregulated |
| 0,228537776 | 0,583551234 | RAB12 | FALSE | Upregulated |
| 0,231628325 | 0,584063269 | SRP54 | FALSE | Upregulated |
| 0,58485619 | 0,584286797 | LOC100507599 | FALSE | Upregulated |
| 0,26451022 | 0,584886347 | PHF20L1 | FALSE | Upregulated |
| -0,291869848 | 0,584889364 | ANTXR2 | FALSE | Downregulated |
| 1,006639476 | 0,584889364 | FAM169B | FALSE | Upregulated |
| 0,367188474 | 0,584889364 | FBXL21 | FALSE | Upregulated |
| -0,530987926 | 0,584889364 | GYPE | FALSE | Downregulated |
| -0,311046238 | 0,584889364 | HDAC8 | FALSE | Downregulated |
| -0,47625464 | 0,584889364 | LOC101928714 | FALSE | Downregulated |
| 0,507711649 | 0,584889364 | LOC102723506 | FALSE | Upregulated |
| 0,433373648 | 0,584889364 | LOC102724316 | FALSE | Upregulated |
| 2,320523791 | 0,584889364 | LOC102724624 | FALSE | Upregulated |
| 0,19904415 | 0,584889364 | PCDH12 | FALSE | Upregulated |
| 0,162103068 | 0,584889364 | SEPN1 | FALSE | Upregulated |
| -0,654592836 | 0,584889364 | TMEM9B-AS1 | FALSE | Downregulated |
| 0,151226099 | 0,584889364 | XYLT2 | FALSE | Upregulated |
| -0,185744674 | 0,58504757 | EGFR-AS1 | FALSE | Downregulated |
| -0,271255006 | 0,585100179 | RASGEF1B | FALSE | Downregulated |
| -0,273800691 | 0,585175536 | LOC101927514 | FALSE | Downregulated |
| 0,573588526 | 0,585175536 | RBMS3-AS2 | FALSE | Upregulated |
| 0,166588931 | 0,585237772 | EPS8L2 | FALSE | Upregulated |
| -0,141674901 | 0,585237772 | TMEM63A | FALSE | Downregulated |
| 0,317427941 | 0,585589412 | ERH | FALSE | Upregulated |
| 0,177562641 | 0,585618827 | NFE2L3 | FALSE | Upregulated |
| -0,400768374 | 0,585692602 | ENO3 | FALSE | Downregulated |
| 0,15740626 | 0,585776251 | CSNK1G2 | FALSE | Upregulated |
| 0,867974969 | 0,586030842 | BEND4 | FALSE | Upregulated |
| 0,244225827 | 0,586030842 | LARS2 | FALSE | Upregulated |
| 0,577952377 | 0,586045431 | SLC7A10 | FALSE | Upregulated |
| 0,656258065 | 0,586065148 | CDH11 | FALSE | Upregulated |
| -0,293742338 | 0,586065148 | DCAF16 | FALSE | Downregulated |
| 0,155320871 | 0,586065148 | RN7SL2 | FALSE | Upregulated |
| -0,408271327 | 0,586065148 | RP11-307E17.8 | FALSE | Downregulated |
| 0,271339441 | 0,586065148 | SLC35F6 | FALSE | Upregulated |
| 0,288857567 | 0,586065148 | TMEM185B | FALSE | Upregulated |
| 0,937606595 | 0,586098932 | GPR75-ASB3 | FALSE | Upregulated |
| -0,217332011 | 0,586259551 | PHLPP2 | FALSE | Downregulated |
| 0,142262774 | 0,586259551 | SLC12A7 | FALSE | Upregulated |
| -0,377085218 | 0,586259551 | YOD1 | FALSE | Downregulated |
| -0,358769997 | 0,586316746 | SIGLEC11 | FALSE | Downregulated |
| -0,161855774 | 0,586463793 | ACBD5 | FALSE | Downregulated |

| | | | | |
|---|---|---|---|---|
| -0,379184573 | 0,586463793 | AP5M1 | FALSE | Downregulated |
| 0,213576667 | 0,586463793 | BCL2L1 | FALSE | Upregulated |
| 0,385310079 | 0,586463793 | BCL2L10 | FALSE | Upregulated |
| -0,18488756 | 0,586463793 | ETFA | FALSE | Downregulated |
| -0,328151135 | 0,586463793 | GLIS3 | FALSE | Downregulated |
| 0,647845441 | 0,586463793 | LOC100506379 | FALSE | Upregulated |
| -0,349003083 | 0,586463793 | NFXL1 | FALSE | Downregulated |
| -0,196311274 | 0,586463793 | SPIN1 | FALSE | Downregulated |
| 0,243558097 | 0,586463793 | WSB1 | FALSE | Upregulated |
| 0,314691939 | 0,586463793 | ZBTB32 | FALSE | Upregulated |
| -0,571824259 | 0,586539336 | LOC101927759 | FALSE | Downregulated |
| 0,17896023 | 0,586539336 | MAP2K3 | FALSE | Upregulated |
| -0,492753044 | 0,586539336 | PLA2R1 | FALSE | Downregulated |
| 0,270960262 | 0,586581583 | AIMP2 | FALSE | Upregulated |
| -0,358188931 | 0,586747105 | AGAP4 | FALSE | Downregulated |
| 0,270518903 | 0,586747105 | TRIB3 | FALSE | Upregulated |
| 0,347312376 | 0,586747105 | WDR70 | FALSE | Upregulated |
| -0,155342844 | 0,586819713 | LZTR1 | FALSE | Downregulated |
| 0,253417422 | 0,587091815 | CAND2 | FALSE | Upregulated |
| 0,185482042 | 0,58709254 | JADE2 | FALSE | Upregulated |
| 0,159907271 | 0,587274157 | CACFD1 | FALSE | Upregulated |
| 0,194580946 | 0,587274157 | NAA30 | FALSE | Upregulated |
| 0,148030215 | 0,587274157 | SRSF2 | FALSE | Upregulated |
| 0,166106265 | 0,587334095 | VCP | FALSE | Upregulated |
| -0,595006802 | 0,587367736 | SPAG16 | FALSE | Downregulated |
| 0,368211111 | 0,587413571 | RFTN2 | FALSE | Upregulated |
| -0,430261771 | 0,587521519 | CHMP3 | FALSE | Downregulated |
| -0,144209676 | 0,587521519 | GANAB | FALSE | Downregulated |
| -0,465661746 | 0,587521808 | INTS12 | FALSE | Downregulated |
| 0,238142785 | 0,587521808 | RP1-232P20.1 | FALSE | Upregulated |
| 0,571697584 | 0,587590251 | ALKBH3 | FALSE | Upregulated |
| -0,418369237 | 0,587590251 | CNTN3 | FALSE | Downregulated |
| -0,404729467 | 0,587590251 | NECAB1 | FALSE | Downregulated |
| -0,136326194 | 0,587640782 | MAN2A2 | FALSE | Downregulated |
| 0,250656059 | 0,587791164 | UNC45A | FALSE | Upregulated |
| 0,354491906 | 0,588319885 | FER1L5 | FALSE | Upregulated |
| -0,149871624 | 0,588319885 | PLXND1 | FALSE | Downregulated |
| 0,18523387 | 0,588319885 | RAB15 | FALSE | Upregulated |
| 0,994446431 | 0,588343019 | DLGAP1-AS5 | FALSE | Upregulated |
| -0,194172041 | 0,588343019 | KIAA0754 | FALSE | Downregulated |
| 0,214926183 | 0,588343019 | RMDN3 | FALSE | Upregulated |
| -0,25936318 | 0,588343019 | SAP18 | FALSE | Downregulated |
| 0,129437371 | 0,588424289 | ZBTB7A | FALSE | Upregulated |
| -0,466553199 | 0,588551296 | NEGR1 | FALSE | Downregulated |
| 0,940548638 | 0,588579676 | HRH1 | FALSE | Upregulated |
| 0,241307973 | 0,588819185 | MUC6 | FALSE | Upregulated |
| -0,277469553 | 0,588909904 | COQ5 | FALSE | Downregulated |
| 0,171210114 | 0,588909904 | GABPB1-AS1 | FALSE | Upregulated |
| -0,228353522 | 0,588909904 | GLDC | FALSE | Downregulated |
| 0,598476431 | 0,588909904 | LOC100505771 | FALSE | Upregulated |
| 0,296107727 | 0,588909904 | LOC101928188 | FALSE | Upregulated |

| | | | | |
|---|---|---|---|---|
| 0,194455386 | 0,588909904 | LOC101928524 | FALSE | Upregulated |
| -0,134594646 | 0,588909904 | MAGI3 | FALSE | Downregulated |
| -0,288802374 | 0,588909904 | MRPL14 | FALSE | Downregulated |
| 0,283729312 | 0,588909904 | MRPS18A | FALSE | Upregulated |
| 0,181513761 | 0,588909904 | RAB26 | FALSE | Upregulated |
| -0,703789301 | 0,589182542 | AC007163.3 | FALSE | Downregulated |
| 0,633634052 | 0,589774778 | C16orf46 | FALSE | Upregulated |
| -0,302331916 | 0,589869569 | MRPS35 | FALSE | Downregulated |
| 0,375869265 | 0,589898917 | TRAF3IP1 | FALSE | Upregulated |
| -0,284023487 | 0,590033421 | F2R | FALSE | Downregulated |
| -0,425370498 | 0,590048067 | KATNA1 | FALSE | Downregulated |
| 0,337139585 | 0,590102026 | FOLR2 | FALSE | Upregulated |
| 0,109202795 | 0,590105138 | LENG8 | FALSE | Upregulated |
| 0,365207764 | 0,590137574 | CIR1 | FALSE | Upregulated |
| -0,176335669 | 0,590137574 | PTMS | FALSE | Downregulated |
| -0,270116542 | 0,590149596 | CSNK1G3 | FALSE | Downregulated |
| 0,298986383 | 0,590149596 | SLFNL1 | FALSE | Upregulated |
| 0,251675687 | 0,590157385 | KLF10 | FALSE | Upregulated |
| -0,513781761 | 0,590157385 | LACE1 | FALSE | Downregulated |
| 0,154333483 | 0,590157385 | MAFB | FALSE | Upregulated |
| 0,206117896 | 0,590157385 | NLRC3 | FALSE | Upregulated |
| -0,228301293 | 0,590213811 | GPR17 | FALSE | Downregulated |
| -0,292380984 | 0,590451052 | LOC102723722 | FALSE | Downregulated |
| -0,441364026 | 0,590493687 | FMO2 | FALSE | Downregulated |
| -0,123820003 | 0,590515311 | PLIN4 | FALSE | Downregulated |
| 0,347704732 | 0,590515311 | RFWD3 | FALSE | Upregulated |
| 0,211639206 | 0,590515311 | STX4 | FALSE | Upregulated |
| 0,270749548 | 0,590556683 | TMEM101 | FALSE | Upregulated |
| 0,117562921 | 0,590916827 | ITM2B | FALSE | Upregulated |
| -1,948712847 | 0,590941713 | GOLGA8K | FALSE | Downregulated |
| 0,871997048 | 0,591258051 | CXorf57 | FALSE | Upregulated |
| -0,854084296 | 0,591474209 | HSD11B1L | FALSE | Downregulated |
| -0,147138609 | 0,591501733 | NFE2L2 | FALSE | Downregulated |
| 0,181444941 | 0,591501733 | PGAM5 | FALSE | Upregulated |
| -0,259657316 | 0,591943996 | NDUFA11 | FALSE | Downregulated |
| 0,402673895 | 0,591970876 | LOC284412 | FALSE | Upregulated |
| 0,517661643 | 0,591992932 | LOC102724808 | FALSE | Upregulated |
| -0,413933063 | 0,591992932 | OSER1 | FALSE | Downregulated |
| 0,360555605 | 0,592193017 | CASP4 | FALSE | Upregulated |
| -0,345416971 | 0,592193017 | FN3KRP | FALSE | Downregulated |
| -0,254653595 | 0,592193017 | LRRC8E | FALSE | Downregulated |
| 0,410685105 | 0,592193017 | NDNL2 | FALSE | Upregulated |
| 0,148506021 | 0,592281725 | SLC5A2 | FALSE | Upregulated |
| 0,610170805 | 0,592382977 | MNS1 | FALSE | Upregulated |
| -0,172323446 | 0,592382977 | SMG5 | FALSE | Downregulated |
| -0,644620844 | 0,592446725 | DNAH14 | FALSE | Downregulated |
| -0,365920013 | 0,592670616 | CAMK2B | FALSE | Downregulated |
| 0,204624572 | 0,593274397 | TELO2 | FALSE | Upregulated |
| -0,236916772 | 0,593384961 | COL4A3BP | FALSE | Downregulated |
| 0,278218475 | 0,593424878 | ZNHIT2 | FALSE | Upregulated |
| 0,891903388 | 0,593523491 | C1QTNF9B | FALSE | Upregulated |

| | | | | |
|---|---|---|---|---|
| -0,772255967 | 0,593595174 | TLX1NB | FALSE | Downregulated |
| 0,173437954 | 0,593597956 | CRK | FALSE | Upregulated |
| 0,571771703 | 0,59361283 | OXLD1 | FALSE | Upregulated |
| -0,177150018 | 0,593672454 | HP1BP3 | FALSE | Downregulated |
| -0,182800042 | 0,594015716 | MED23 | FALSE | Downregulated |
| -0,271544569 | 0,594075803 | LOC642852 | FALSE | Downregulated |
| -0,314842399 | 0,594368315 | CCDC101 | FALSE | Downregulated |
| 0,49268703 | 0,594368315 | GOLT1B | FALSE | Upregulated |
| 0,753532038 | 0,594368315 | TCTN2 | FALSE | Upregulated |
| 0,336722248 | 0,594423501 | ANKRD13C | FALSE | Upregulated |
| 5,19458836 | 0,594510672 | ATP6V1G2-DDX39B | FALSE | Upregulated |
| 0,309505187 | 0,59465696 | RP11-478P10.1 | FALSE | Upregulated |
| -0,308120439 | 0,59475605 | ARL3 | FALSE | Downregulated |
| 2,048566771 | 0,59475605 | LOC101060216 | FALSE | Upregulated |
| -0,710050915 | 0,59475605 | TMEM136 | FALSE | Downregulated |
| -0,147089872 | 0,595073263 | GAK | FALSE | Downregulated |
| -0,197747231 | 0,595187333 | ABI2 | FALSE | Downregulated |
| -0,502805605 | 0,595187333 | ART1 | FALSE | Downregulated |
| -0,540439914 | 0,595187333 | CRYZL1 | FALSE | Downregulated |
| 0,415049555 | 0,595187333 | DTD2 | FALSE | Upregulated |
| 0,222589954 | 0,595187333 | N4BP1 | FALSE | Upregulated |
| -0,668329177 | 0,595187333 | SLC2A4 | FALSE | Downregulated |
| -0,575801942 | 0,595187333 | TUBD1 | FALSE | Downregulated |
| -0,65868111 | 0,595295476 | SEPT3 | FALSE | Downregulated |
| 0,299894693 | 0,595295476 | TBCK | FALSE | Upregulated |
| 0,640074411 | 0,595481778 | NEUROD2 | FALSE | Upregulated |
| 0,400375816 | 0,59565209 | LOC101928038 | FALSE | Upregulated |
| -0,173579432 | 0,59565209 | PBX2 | FALSE | Downregulated |
| -0,382114581 | 0,595765457 | COPRS | FALSE | Downregulated |
| -0,55731724 | 0,595765457 | RP11-290F20.1 | FALSE | Downregulated |
| -0,255872574 | 0,595765457 | WWTR1 | FALSE | Downregulated |
| -0,366179299 | 0,595766509 | LOC101929272 | FALSE | Downregulated |
| -0,157378735 | 0,595798218 | PACS2 | FALSE | Downregulated |
| 0,608466933 | 0,595976376 | RP11-44F21.3 | FALSE | Upregulated |
| -0,23244098 | 0,595986491 | METTL13 | FALSE | Downregulated |
| -0,279636214 | 0,59606619 | DPY19L4 | FALSE | Downregulated |
| 0,279062194 | 0,59606619 | DUSP14 | FALSE | Upregulated |
| -0,678417333 | 0,59606619 | KCNJ15 | FALSE | Downregulated |
| 0,707325726 | 0,59606619 | SHISA4 | FALSE | Upregulated |
| 0,155925379 | 0,596092899 | GPC1 | FALSE | Upregulated |
| -0,181459313 | 0,596116775 | GTPBP6 | FALSE | Downregulated |
| -0,408203307 | 0,596116775 | SERGEF | FALSE | Downregulated |
| 0,419649095 | 0,596508917 | BACH2 | FALSE | Upregulated |
| -0,342579183 | 0,596508917 | DPH1 | FALSE | Downregulated |
| 0,167856384 | 0,596560174 | C9orf131 | FALSE | Upregulated |
| 0,170687192 | 0,596560174 | LSM14B | FALSE | Upregulated |
| -1,036667046 | 0,596714923 | PLA2G4B | FALSE | Downregulated |
| 0,281251335 | 0,596783318 | DKFZP434H168 | FALSE | Upregulated |
| 0,191208643 | 0,596923253 | FOXN3-AS2 | FALSE | Upregulated |
| 0,403227663 | 0,596923253 | RRP12 | FALSE | Upregulated |
| -0,319826769 | 0,596923253 | UBL5 | FALSE | Downregulated |

| | | | | |
|---|---|---|---|---|
| -0,470563139 | 0,597074833 | LOC220729 | FALSE | Downregulated |
| 0,25038494 | 0,597135461 | FUT11 | FALSE | Upregulated |
| -0,414587874 | 0,597135461 | HINFP | FALSE | Downregulated |
| -0,550099701 | 0,597149486 | LOC101927160 | FALSE | Downregulated |
| 0,51079576 | 0,597184505 | ARSK | FALSE | Upregulated |
| -0,178675084 | 0,597892333 | CLINT1 | FALSE | Downregulated |
| 0,699121834 | 0,597892333 | ZNF763 | FALSE | Upregulated |
| -0,276946861 | 0,597921639 | FSTL1 | FALSE | Downregulated |
| 0,692859228 | 0,598039057 | LOC101930452 | FALSE | Upregulated |
| -0,322670726 | 0,598039057 | SYNJ1 | FALSE | Downregulated |
| 0,231060357 | 0,598197841 | BTBD1 | FALSE | Upregulated |
| 0,183803716 | 0,598197841 | CCT4 | FALSE | Upregulated |
| -0,88445697 | 0,598197841 | GCK | FALSE | Downregulated |
| 0,231988004 | 0,598197841 | RAB24 | FALSE | Upregulated |
| 0,191085862 | 0,598197841 | SP2 | FALSE | Upregulated |
| 0,13074363 | 0,598197841 | STAT6 | FALSE | Upregulated |
| -0,313035189 | 0,598221617 | NOP58 | FALSE | Downregulated |
| 0,774253807 | 0,598307179 | KCNH4 | FALSE | Upregulated |
| 0,254370032 | 0,598307179 | UBE2D2 | FALSE | Upregulated |
| -0,594049405 | 0,598307179 | ZNF416 | FALSE | Downregulated |
| 0,505915989 | 0,59840032 | LOC101927030 | FALSE | Upregulated |
| 0,156341325 | 0,59840032 | MYRIP | FALSE | Upregulated |
| 0,806544428 | 0,59840032 | PWP2 | FALSE | Upregulated |
| 0,143918367 | 0,59840032 | SERINC4 | FALSE | Upregulated |
| 0,45233075 | 0,598470516 | HDAC9 | FALSE | Upregulated |
| 0,361924928 | 0,598470516 | HHIPL1 | FALSE | Upregulated |
| 0,67121226 | 0,598470516 | LOC102723683 | FALSE | Upregulated |
| 0,134042896 | 0,598470516 | SEC16A | FALSE | Upregulated |
| 0,177412468 | 0,598470516 | SLC18A2 | FALSE | Upregulated |
| -0,220813786 | 0,598470516 | TCEB2 | FALSE | Downregulated |
| 0,800455568 | 0,59858573 | EFCAB10 | FALSE | Upregulated |
| -0,16485752 | 0,599183053 | CUX1 | FALSE | Downregulated |
| -0,218291512 | 0,599297111 | ADAMTSL2 | FALSE | Downregulated |
| -0,231320024 | 0,599297111 | C22orf29 | FALSE | Downregulated |
| -0,49457188 | 0,599297111 | KIAA1841 | FALSE | Downregulated |
| -0,169268188 | 0,599297111 | MMRN2 | FALSE | Downregulated |
| 0,384418485 | 0,599297111 | PPM1D | FALSE | Upregulated |
| 0,385246301 | 0,599297111 | STX19 | FALSE | Upregulated |
| 0,154306429 | 0,599297111 | TULP4 | FALSE | Upregulated |
| 0,249095663 | 0,599548889 | PAXBP1-AS1 | FALSE | Upregulated |
| 0,245492933 | 0,599549255 | CAPZA1 | FALSE | Upregulated |
| -0,296212076 | 0,599549255 | CTNNBIP1 | FALSE | Downregulated |
| 0,392707804 | 0,599590947 | RP11-434D9.1 | FALSE | Upregulated |
| -0,580114335 | 0,599887333 | EIF3CL | FALSE | Downregulated |
| -0,275156585 | 0,599887333 | SLIT3 | FALSE | Downregulated |
| 0,21794412 | 0,599887333 | ZBTB8A | FALSE | Upregulated |
| 0,17932611 | 0,599979776 | DHX9 | FALSE | Upregulated |
| -0,388431262 | 0,599979776 | EAPP | FALSE | Downregulated |
| 0,320365752 | 0,5999995 | RGS20 | FALSE | Upregulated |
| -0,218593631 | 0,600076115 | GUF1 | FALSE | Downregulated |
| -0,270374773 | 0,600285562 | C11orf30 | FALSE | Downregulated |

| | | | | |
|---|---|---|---|---|
| -0,257017839 | 0,600285562 | PPARGC1A | FALSE | Downregulated |
| 0,764541513 | 0,600510962 | CHI3L1 | FALSE | Upregulated |
| 0,129041543 | 0,600872525 | SCRIB | FALSE | Upregulated |
| -0,229345534 | 0,600948657 | ANAPC5 | FALSE | Downregulated |
| -0,383289519 | 0,600967757 | TMEM65 | FALSE | Downregulated |
| 0,645244998 | 0,601222375 | IL34 | FALSE | Upregulated |
| 0,583972188 | 0,601222375 | OR7E12P | FALSE | Upregulated |
| 0,199453407 | 0,601222375 | RBM3 | FALSE | Upregulated |
| -0,226787843 | 0,601222375 | RGS14 | FALSE | Downregulated |
| -0,195362292 | 0,601921551 | RP11-143J12.2 | FALSE | Downregulated |
| 0,422083921 | 0,60213485 | CLRN3 | FALSE | Upregulated |
| 0,211801189 | 0,602142645 | SLC25A5-AS1 | FALSE | Upregulated |
| -0,178271247 | 0,602216564 | POLE | FALSE | Downregulated |
| -0,118895754 | 0,602298475 | RPS11 | FALSE | Downregulated |
| -0,449595281 | 0,602343697 | CXorf23 | FALSE | Downregulated |
| -0,266286332 | 0,602343697 | GPR182 | FALSE | Downregulated |
| 0,157484573 | 0,602343697 | SH3GL1 | FALSE | Upregulated |
| -0,135413379 | 0,602355239 | ABCC3 | FALSE | Downregulated |
| -0,188679813 | 0,602355239 | CCNYL1 | FALSE | Downregulated |
| 0,525223934 | 0,602355239 | ZNF286A | FALSE | Upregulated |
| 0,302231904 | 0,60253929 | DDX27 | FALSE | Upregulated |
| 0,644544023 | 0,60253929 | FANCD2 | FALSE | Upregulated |
| -0,471397295 | 0,60253929 | OR52D1 | FALSE | Downregulated |
| -0,279551188 | 0,60253929 | R3HCC1 | FALSE | Downregulated |
| -0,145405773 | 0,602543872 | ARGLU1 | FALSE | Downregulated |
| -0,159477257 | 0,602543872 | TUG1 | FALSE | Downregulated |
| -0,596256562 | 0,602543872 | WBP1 | FALSE | Downregulated |
| 0,415745555 | 0,602603478 | KIAA1279 | FALSE | Upregulated |
| 0,620965175 | 0,602629607 | CCL3 | FALSE | Upregulated |
| -0,846336343 | 0,602629607 | STPG1 | FALSE | Downregulated |
| 0,307029603 | 0,602636986 | LOC101929722 | FALSE | Upregulated |
| -0,407433955 | 0,602636986 | LOC102723908 | FALSE | Downregulated |
| -0,445788899 | 0,602636986 | RSPO3 | FALSE | Downregulated |
| 0,737670572 | 0,602636986 | SBK1 | FALSE | Upregulated |
| -0,292087786 | 0,602773841 | ESCO1 | FALSE | Downregulated |
| -0,502536368 | 0,602795024 | LOC101927321 | FALSE | Downregulated |
| -0,205773396 | 0,602821132 | FOXRED2 | FALSE | Downregulated |
| 0,524168814 | 0,603011373 | DLG3 | FALSE | Upregulated |
| 0,392748878 | 0,603011373 | DOK7 | FALSE | Upregulated |
| 0,291417481 | 0,603011373 | PLET1 | FALSE | Upregulated |
| -0,395761534 | 0,603011373 | UBE2V2 | FALSE | Downregulated |
| 0,396412662 | 0,603032808 | SETD4 | FALSE | Upregulated |
| -0,768995432 | 0,603110328 | LOC644961 | FALSE | Downregulated |
| 0,680726968 | 0,603123545 | RP11-638L3.1 | FALSE | Upregulated |
| -0,603637167 | 0,603575648 | COX6B2 | FALSE | Downregulated |
| -0,196620335 | 0,603575648 | GSK3B | FALSE | Downregulated |
| 0,344883359 | 0,603575648 | NNMT | FALSE | Upregulated |
| -0,462188564 | 0,603749166 | OR5AK2 | FALSE | Downregulated |
| -0,234357123 | 0,603749166 | TTC23 | FALSE | Downregulated |
| 0,301102317 | 0,603749166 | ZNF785 | FALSE | Upregulated |
| 0,21754152 | 0,603769986 | BEND3P3 | FALSE | Upregulated |

| | | | | |
|---|---|---|---|---|
| 0,345701653 | 0,604043842 | SLC25A51 | FALSE | Upregulated |
| -0,333598028 | 0,604043842 | SVIP | FALSE | Downregulated |
| -0,496077106 | 0,604329951 | DCTN6 | FALSE | Downregulated |
| 0,186240313 | 0,60476941 | MKRN2 | FALSE | Upregulated |
| 0,221411173 | 0,60476941 | PRKD2 | FALSE | Upregulated |
| 0,380185886 | 0,60476941 | ZNF251 | FALSE | Upregulated |
| 0,343105549 | 0,604773743 | LOC102723369 | FALSE | Upregulated |
| -0,665411724 | 0,604773743 | TMEM198 | FALSE | Downregulated |
| -0,574256094 | 0,604802861 | KIAA1549 | FALSE | Downregulated |
| 0,382074023 | 0,604802861 | SYTL1 | FALSE | Upregulated |
| 0,775484531 | 0,605046715 | KCNA6 | FALSE | Upregulated |
| -0,164322528 | 0,605046715 | PHC2 | FALSE | Downregulated |
| -0,846311757 | 0,605145602 | FLJ45513 | FALSE | Downregulated |
| 0,211608382 | 0,605145602 | RALGAPB | FALSE | Upregulated |
| 0,736059469 | 0,605216455 | KCND1 | FALSE | Upregulated |
| -0,220393456 | 0,605365606 | FRMD4B | FALSE | Downregulated |
| 0,153382854 | 0,605365606 | GNA11 | FALSE | Upregulated |
| -0,128442326 | 0,605365606 | HERC2 | FALSE | Downregulated |
| 0,243696745 | 0,605365606 | PYCRL | FALSE | Upregulated |
| -0,288978211 | 0,605365606 | SEP15 | FALSE | Downregulated |
| 0,75297361 | 0,605504553 | KLHL41 | FALSE | Upregulated |
| 0,206274328 | 0,605518804 | HTATSF1P2 | FALSE | Upregulated |
| -0,164272689 | 0,605623062 | ERVK13-1 | FALSE | Downregulated |
| 0,233375681 | 0,605623062 | GCH1 | FALSE | Upregulated |
| -0,15761564 | 0,60572818 | CMTM6 | FALSE | Downregulated |
| 0,197880898 | 0,60572818 | MCMBP | FALSE | Upregulated |
| 0,1621199 | 0,60572818 | MEF2D | FALSE | Upregulated |
| 0,603593133 | 0,60572818 | MOGAT3 | FALSE | Upregulated |
| -0,140147055 | 0,60572818 | NONO | FALSE | Downregulated |
| -0,271758351 | 0,60572818 | NT5C3B | FALSE | Downregulated |
| 0,402169338 | 0,60572818 | RP11-35G9.3 | FALSE | Upregulated |
| -0,215508217 | 0,605890966 | DNAJC4 | FALSE | Downregulated |
| -0,234496779 | 0,605890966 | NID2 | FALSE | Downregulated |
| 0,65472391 | 0,605907781 | FZD3 | FALSE | Upregulated |
| 0,334469721 | 0,605907781 | RABGGTA | FALSE | Upregulated |
| -0,137496121 | 0,605907781 | SMEK2 | FALSE | Downregulated |
| -0,209741904 | 0,605907781 | USP10 | FALSE | Downregulated |
| 0,705010562 | 0,606110326 | RPS15AP10 | FALSE | Upregulated |
| -0,542618873 | 0,606110326 | SUSD5 | FALSE | Downregulated |
| -0,216779667 | 0,606144141 | NDUFA13 | FALSE | Downregulated |
| -0,517923104 | 0,606144141 | TRIM74 | FALSE | Downregulated |
| -0,15901105 | 0,606145258 | AP1M1 | FALSE | Downregulated |
| 0,272979383 | 0,606145258 | C1QBP | FALSE | Upregulated |
| -0,351966546 | 0,606145258 | HS2ST1 | FALSE | Downregulated |
| 0,557838547 | 0,606145258 | KIF7 | FALSE | Upregulated |
| -0,554346775 | 0,606145258 | LAGE3 | FALSE | Downregulated |
| 0,345627007 | 0,606145258 | MGME1 | FALSE | Upregulated |
| -0,597006919 | 0,606211199 | C11orf52 | FALSE | Downregulated |
| 0,267097429 | 0,606211199 | CALHM2 | FALSE | Upregulated |
| -0,225324294 | 0,606211199 | IRF6 | FALSE | Downregulated |
| 0,172624177 | 0,606211199 | PLCG1 | FALSE | Upregulated |

| | | | | |
|---|---|---|---|---|
| -0,320277513 | 0,606211199 | PPP3CB | FALSE | Downregulated |
| 0,493121393 | 0,606211199 | RCOR2 | FALSE | Upregulated |
| 0,266163826 | 0,606211199 | SNX10 | FALSE | Upregulated |
| 0,443007458 | 0,606211199 | SPTSSA | FALSE | Upregulated |
| -0,332496748 | 0,606211199 | TRIM3 | FALSE | Downregulated |
| -0,723982744 | 0,606211199 | ZMYM6NB | FALSE | Downregulated |
| 0,726282439 | 0,606227203 | NALCN-AS1 | FALSE | Upregulated |
| 0,576576859 | 0,606441149 | FKBP5 | FALSE | Upregulated |
| -0,181818131 | 0,606441149 | NAV2 | FALSE | Downregulated |
| 0,225515381 | 0,606441149 | SMAD9 | FALSE | Upregulated |
| -0,186434541 | 0,606538306 | IREB2 | FALSE | Downregulated |
| -0,180289195 | 0,606611193 | PDCD6IP | FALSE | Downregulated |
| 0,223058406 | 0,606700819 | CDK2AP2 | FALSE | Upregulated |
| -0,185816261 | 0,606700819 | CPSF3L | FALSE | Downregulated |
| 0,353830049 | 0,606700819 | GIMAP2 | FALSE | Upregulated |
| 0,251359994 | 0,606700819 | KCNJ9 | FALSE | Upregulated |
| -0,389270745 | 0,606700819 | PRPF4 | FALSE | Downregulated |
| 0,190370357 | 0,606851619 | RUFY2 | FALSE | Upregulated |
| 0,362271988 | 0,606951889 | RAD51 | FALSE | Upregulated |
| 0,367049821 | 0,607003481 | CENPBD1 | FALSE | Upregulated |
| -0,789620946 | 0,607143378 | TBC1D19 | FALSE | Downregulated |
| -0,376410754 | 0,607151485 | AMIGO1 | FALSE | Downregulated |
| 0,555430934 | 0,607151485 | N4BP3 | FALSE | Upregulated |
| -0,117260626 | 0,60737146 | APP | FALSE | Downregulated |
| 0,280551578 | 0,607378323 | RASA3 | FALSE | Upregulated |
| 0,117225845 | 0,607430354 | HBP1 | FALSE | Upregulated |
| 0,541155858 | 0,607430354 | IQCB1 | FALSE | Upregulated |
| 0,250467337 | 0,60748021 | MYH11 | FALSE | Upregulated |
| 0,128222199 | 0,607831124 | ATXN2L | FALSE | Upregulated |
| 0,185505817 | 0,607831124 | DPYSL2 | FALSE | Upregulated |
| 0,335629273 | 0,607933858 | FUT8 | FALSE | Upregulated |
| 0,308787524 | 0,607933858 | SUPT20H | FALSE | Upregulated |
| 0,588568221 | 0,607965228 | HIST4H4 | FALSE | Upregulated |
| 0,159597062 | 0,608066991 | ZNF337 | FALSE | Upregulated |
| 0,608439081 | 0,608116129 | TSPAN5 | FALSE | Upregulated |
| 0,277892365 | 0,608597273 | SLX4 | FALSE | Upregulated |
| -0,299132149 | 0,608654333 | GALK2 | FALSE | Downregulated |
| -1,330777775 | 0,608834138 | JMJD7-PLA2G4B | FALSE | Downregulated |
| -0,269461996 | 0,60897661 | C2orf43 | FALSE | Downregulated |
| -0,24293374 | 0,60897661 | PIAS4 | FALSE | Downregulated |
| -0,317332 | 0,60897661 | TMEM53 | FALSE | Downregulated |
| -0,522782766 | 0,609302473 | ATRNL1 | FALSE | Downregulated |
| -0,141807869 | 0,609302473 | BCKDHA | FALSE | Downregulated |
| -0,907822802 | 0,609302473 | GATA2 | FALSE | Downregulated |
| 0,642089647 | 0,609302473 | HIST1H3H | FALSE | Upregulated |
| -0,422588995 | 0,609302473 | LGALSL | FALSE | Downregulated |
| -0,248983559 | 0,609302473 | NCKIPSD | FALSE | Downregulated |
| -0,583880729 | 0,609302473 | OSBPL7 | FALSE | Downregulated |
| 0,494592798 | 0,609302473 | PTGDS | FALSE | Upregulated |
| 0,526787712 | 0,609302473 | REC8 | FALSE | Upregulated |
| -0,274208308 | 0,609302473 | STRBP | FALSE | Downregulated |

| | | | | |
|---|---|---|---|---|
| -0,184669902 | 0,609302473 | TAB3 | FALSE | Downregulated |
| -0,13990892 | 0,609302473 | TRAK1 | FALSE | Downregulated |
| -0,380738148 | 0,609302473 | TRAM2-AS1 | FALSE | Downregulated |
| -0,165832776 | 0,609437533 | NR1H3 | FALSE | Downregulated |
| -0,920539443 | 0,609516149 | KLHL35 | FALSE | Downregulated |
| 0,161940439 | 0,609566086 | ZMYM4 | FALSE | Upregulated |
| 0,198510702 | 0,609947648 | FBXL15 | FALSE | Upregulated |
| -0,620698688 | 0,609947648 | HDDC2 | FALSE | Downregulated |
| 0,246084444 | 0,609947648 | LOC100996255 | FALSE | Upregulated |
| -0,282332378 | 0,609947648 | RAB37 | FALSE | Downregulated |
| 0,536098048 | 0,609947648 | SAP30 | FALSE | Upregulated |
| 0,169985941 | 0,609947648 | SNRPD3 | FALSE | Upregulated |
| 0,19064842 | 0,609947648 | ST7-AS1 | FALSE | Upregulated |
| 0,610157635 | 0,609970744 | BEND3 | FALSE | Upregulated |
| 0,178618108 | 0,609970744 | CAB39 | FALSE | Upregulated |
| 0,185084025 | 0,609970744 | LOC100289511 | FALSE | Upregulated |
| -0,283509368 | 0,609970744 | MUC20 | FALSE | Downregulated |
| 0,210756686 | 0,609970744 | THADA | FALSE | Upregulated |
| -0,192989089 | 0,610085309 | INPP5E | FALSE | Downregulated |
| -0,476289353 | 0,610085309 | LYSMD4 | FALSE | Downregulated |
| -0,149858921 | 0,610335177 | KANK2 | FALSE | Downregulated |
| 0,239361012 | 0,610404587 | LARGE-AS1 | FALSE | Upregulated |
| 0,267354327 | 0,610404587 | ZNF668 | FALSE | Upregulated |
| 0,522831021 | 0,61044062 | LOC101927529 | FALSE | Upregulated |
| -0,147657093 | 0,610522249 | CDC34 | FALSE | Downregulated |
| -0,196975156 | 0,610522249 | NET1 | FALSE | Downregulated |
| -0,477239052 | 0,610775717 | LOC100507547 | FALSE | Downregulated |
| -0,367685508 | 0,610779025 | MYO15A | FALSE | Downregulated |
| -0,233116345 | 0,611140862 | CEBPZ | FALSE | Downregulated |
| 0,177755433 | 0,611140862 | COG1 | FALSE | Upregulated |
| -0,741091984 | 0,611140862 | LOC102723779 | FALSE | Downregulated |
| 0,591050529 | 0,611140862 | LOC729040 | FALSE | Upregulated |
| 0,131646865 | 0,611140862 | TNS3 | FALSE | Upregulated |
| -0,107486437 | 0,611140862 | USP9X | FALSE | Downregulated |
| 0,477683726 | 0,611245913 | LOC101059976 | FALSE | Upregulated |
| 0,613644835 | 0,611303274 | DISC1 | FALSE | Upregulated |
| 0,792527683 | 0,61135344 | UGT3A2 | FALSE | Upregulated |
| -0,166840106 | 0,61135344 | USP40 | FALSE | Downregulated |
| 0,422951202 | 0,611865085 | C16orf52 | FALSE | Upregulated |
| 0,203924832 | 0,611929598 | WDFY1 | FALSE | Upregulated |
| -0,569672379 | 0,612299125 | C11orf73 | FALSE | Downregulated |
| 0,411294563 | 0,612299125 | DLL1 | FALSE | Upregulated |
| 0,223028588 | 0,612515727 | AC004893.11 | FALSE | Upregulated |
| -1,151231254 | 0,612861817 | SYCE1 | FALSE | Downregulated |
| -0,159456064 | 0,613112485 | KDM5C | FALSE | Downregulated |
| -0,807332998 | 0,613144348 | FLJ46552 | FALSE | Downregulated |
| 0,40939872 | 0,613294623 | RHBDF1 | FALSE | Upregulated |
| -0,272416166 | 0,613471802 | MTF2 | FALSE | Downregulated |
| -0,154500403 | 0,613471802 | NFATC3 | FALSE | Downregulated |
| 0,166223424 | 0,613481892 | B3GNT9 | FALSE | Upregulated |
| 0,340309636 | 0,613486544 | C12orf73 | FALSE | Upregulated |

| | | | | |
|---|---|---|---|---|
| -0,607237328 | 0,613486544 | KCNK16 | FALSE | Downregulated |
| -0,312081771 | 0,613495644 | TDO2 | FALSE | Downregulated |
| -0,301687669 | 0,613511803 | SHH | FALSE | Downregulated |
| 0,228312248 | 0,613610349 | CDYL | FALSE | Upregulated |
| 0,77451201 | 0,613802353 | DLG5-AS1 | FALSE | Upregulated |
| -0,154287041 | 0,613802353 | HSPD1 | FALSE | Downregulated |
| 0,336684931 | 0,613802353 | LOC101928069 | FALSE | Upregulated |
| 0,534279227 | 0,613931937 | MT1G | FALSE | Upregulated |
| 0,787469403 | 0,614422865 | ANKRD20A9P | FALSE | Upregulated |
| 0,609992731 | 0,614422865 | CACNG1 | FALSE | Upregulated |
| -0,225546101 | 0,614422865 | FAHD2A | FALSE | Downregulated |
| 0,317896986 | 0,614444535 | LOC101929072 | FALSE | Upregulated |
| 1,065626538 | 0,614444535 | LY75-CD302 | FALSE | Upregulated |
| 0,209188567 | 0,614444535 | PSME1 | FALSE | Upregulated |
| 0,552264865 | 0,614444535 | VWA5B2 | FALSE | Upregulated |
| -0,743239115 | 0,614451755 | GALNT3 | FALSE | Downregulated |
| -0,273388577 | 0,614451755 | GPR108 | FALSE | Downregulated |
| -0,173525572 | 0,614614211 | CTDSPL2 | FALSE | Downregulated |
| -0,325998957 | 0,614787931 | CCDC94 | FALSE | Downregulated |
| -0,701168395 | 0,614860302 | FAM154B | FALSE | Downregulated |
| -0,24833153 | 0,615165451 | ACBD6 | FALSE | Downregulated |
| 0,267620652 | 0,615165451 | GNB5 | FALSE | Upregulated |
| 0,286714526 | 0,615169204 | SLC20A1 | FALSE | Upregulated |
| -0,765690464 | 0,615169204 | SNORA53 | FALSE | Downregulated |
| 0,570899069 | 0,615202787 | RAD51-AS1 | FALSE | Upregulated |
| 0,266879765 | 0,615202787 | SCYL3 | FALSE | Upregulated |
| -0,415435649 | 0,615202787 | TERF1 | FALSE | Downregulated |
| 0,41669659 | 0,615202787 | WDR31 | FALSE | Upregulated |
| 0,184552425 | 0,615202787 | ZC2HC1C | FALSE | Upregulated |
| 0,638727537 | 0,615411777 | LOC101928214 | FALSE | Upregulated |
| 0,2308619 | 0,615411777 | RAB1A | FALSE | Upregulated |
| 0,318626005 | 0,615411777 | SCFD1 | FALSE | Upregulated |
| -0,473655331 | 0,615411777 | ZNF844 | FALSE | Downregulated |
| 0,128838243 | 0,615470924 | DDX6 | FALSE | Upregulated |
| 0,094197424 | 0,615470924 | KIAA1147 | FALSE | Upregulated |
| -0,559646499 | 0,615470924 | LOC101926934 | FALSE | Downregulated |
| -0,160945012 | 0,615486344 | NOTCH2 | FALSE | Downregulated |
| -0,245615013 | 0,615676578 | ALMS1 | FALSE | Downregulated |
| 0,357174996 | 0,615676578 | ATOH8 | FALSE | Upregulated |
| -0,536912269 | 0,615676578 | CMC4 | FALSE | Downregulated |
| -0,466797733 | 0,615676578 | CNTNAP3B | FALSE | Downregulated |
| 0,135616957 | 0,615676578 | COG5 | FALSE | Upregulated |
| -0,283576444 | 0,615676578 | DPY19L1 | FALSE | Downregulated |
| -0,219893004 | 0,615676578 | GCC1 | FALSE | Downregulated |
| -0,254390037 | 0,615676578 | KIAA1429 | FALSE | Downregulated |
| 0,317631154 | 0,615676578 | KIF26B | FALSE | Upregulated |
| 0,175896053 | 0,615676578 | LOC100288162 | FALSE | Upregulated |
| -0,12378916 | 0,615676578 | MYO1B | FALSE | Downregulated |
| -0,782396925 | 0,615676578 | NIM1K | FALSE | Downregulated |
| 0,835328662 | 0,615676578 | NRGN | FALSE | Upregulated |
| 0,113608022 | 0,615676578 | PDPK1 | FALSE | Upregulated |

| | | | | |
|---|---|---|---|---|
| 0,132209365 | 0,615676578 | PTCD3 | FALSE | Upregulated |
| 0,26165392 | 0,615676578 | RAB4A | FALSE | Upregulated |
| 0,457722363 | 0,615676578 | RFX2 | FALSE | Upregulated |
| -0,258668881 | 0,615676578 | RSBN1L | FALSE | Downregulated |
| 0,551241622 | 0,615676578 | YEATS4 | FALSE | Upregulated |
| -0,130887059 | 0,615717751 | PICALM | FALSE | Downregulated |
| -0,450290421 | 0,615885546 | LOC100288102 | FALSE | Downregulated |
| 0,216078045 | 0,615929478 | CDRT15P1 | FALSE | Upregulated |
| 0,21375004 | 0,61620168 | SRSF3 | FALSE | Upregulated |
| -0,43930034 | 0,616341029 | CXorf56 | FALSE | Downregulated |
| -0,254986508 | 0,616565789 | SPR | FALSE | Downregulated |
| -0,390779616 | 0,61662886 | MEG3 | FALSE | Downregulated |
| -0,142378053 | 0,61710221 | USP19 | FALSE | Downregulated |
| 0,319273586 | 0,617241007 | JARID2-AS1 | FALSE | Upregulated |
| -0,386726064 | 0,617263084 | GDPD5 | FALSE | Downregulated |
| 0,120342597 | 0,617263084 | SDF4 | FALSE | Upregulated |
| 0,979871575 | 0,617351102 | PMF1-BGLAP | FALSE | Upregulated |
| 0,588232357 | 0,617498232 | FCHO1 | FALSE | Upregulated |
| -0,16840923 | 0,617498232 | INPPL1 | FALSE | Downregulated |
| -0,525007695 | 0,61754927 | AP4B1 | FALSE | Downregulated |
| -0,542386813 | 0,61754927 | C1QTNF1-AS1 | FALSE | Downregulated |
| -0,462660369 | 0,61754927 | ETHE1 | FALSE | Downregulated |
| 0,52174011 | 0,61754927 | LOC101926889 | FALSE | Upregulated |
| 0,771647893 | 0,61754927 | LOC339192 | FALSE | Upregulated |
| 0,401027143 | 0,61754927 | LPP-AS2 | FALSE | Upregulated |
| -0,26337574 | 0,61754927 | PANX2 | FALSE | Downregulated |
| -0,449086638 | 0,61754927 | PRPF18 | FALSE | Downregulated |
| -0,15383153 | 0,61754927 | RPS16 | FALSE | Downregulated |
| 0,20856625 | 0,61754927 | SPATA20 | FALSE | Upregulated |
| -0,921661314 | 0,617592564 | AC009299.3 | FALSE | Downregulated |
| -0,229071156 | 0,617854925 | ASPSCR1 | FALSE | Downregulated |
| 0,376549018 | 0,617854925 | GTF3C6 | FALSE | Upregulated |
| 0,311265286 | 0,617854925 | TMEM74B | FALSE | Upregulated |
| 0,334329062 | 0,617914654 | LOC101927793 | FALSE | Upregulated |
| 0,748683699 | 0,61795257 | C20orf96 | FALSE | Upregulated |
| -0,387122639 | 0,61795257 | ZNF761 | FALSE | Downregulated |
| 0,896943665 | 0,61811923 | SENP3-EIF4A1 | FALSE | Upregulated |
| 0,173737 | 0,618405376 | CLK3 | FALSE | Upregulated |
| 0,40924061 | 0,618437627 | FBLN2 | FALSE | Upregulated |
| -0,462596016 | 0,618585542 | GCA | FALSE | Downregulated |
| -0,415369424 | 0,618585542 | RP11-1007G5.2 | FALSE | Downregulated |
| -0,367122738 | 0,618629376 | BRF2 | FALSE | Downregulated |
| 0,290736617 | 0,618632967 | MCAM | FALSE | Upregulated |
| 0,313818225 | 0,61869995 | DDX10 | FALSE | Upregulated |
| 0,365887505 | 0,61869995 | DISP2 | FALSE | Upregulated |
| -0,186436205 | 0,61869995 | EPS8 | FALSE | Downregulated |
| 0,330544806 | 0,61869995 | GPBAR1 | FALSE | Upregulated |
| -0,472548623 | 0,61869995 | TEC | FALSE | Downregulated |
| -0,340394372 | 0,61869995 | TRAF3IP3 | FALSE | Downregulated |
| 0,187199102 | 0,618786202 | JMJD1C-AS1 | FALSE | Upregulated |
| 0,487299057 | 0,618786202 | LRCOL1 | FALSE | Upregulated |

| | | | | |
|---|---|---|---|---|
| -2,215961987 | 0,619345784 | TMEM256-PLSCR3 | FALSE | Downregulated |
| 0,183292722 | 0,61941926 | LOC101928874 | FALSE | Upregulated |
| -0,283303723 | 0,61941926 | SYCE2 | FALSE | Downregulated |
| -0,140901878 | 0,619550536 | LMTK2 | FALSE | Downregulated |
| 0,416951667 | 0,619550536 | SIGLEC9 | FALSE | Upregulated |
| -0,196666174 | 0,619642624 | CNOT6L | FALSE | Downregulated |
| 0,129501283 | 0,619642624 | SMURF1 | FALSE | Upregulated |
| 0,428208417 | 0,619647056 | ZNF613 | FALSE | Upregulated |
| 0,199172793 | 0,619828398 | DMWD | FALSE | Upregulated |
| -0,244611464 | 0,619828398 | LOC101927673 | FALSE | Downregulated |
| -0,297591205 | 0,619828398 | ZNF614 | FALSE | Downregulated |
| -0,288724078 | 0,620037849 | FIGN | FALSE | Downregulated |
| 0,508492999 | 0,620114707 | UBE2Q2P1 | FALSE | Upregulated |
| -0,252015642 | 0,620121541 | FUZ | FALSE | Downregulated |
| 0,205366767 | 0,620121541 | RPS6KL1 | FALSE | Upregulated |
| -0,695050754 | 0,620121541 | ZFY | FALSE | Downregulated |
| -0,429416342 | 0,620121541 | ZNF559 | FALSE | Downregulated |
| 0,253959912 | 0,620157683 | TACR1 | FALSE | Upregulated |
| 0,600200003 | 0,620250626 | ATF3 | FALSE | Upregulated |
| -0,338836313 | 0,620288425 | LOC101927741 | FALSE | Downregulated |
| -0,271225938 | 0,620429686 | FBXO32 | FALSE | Downregulated |
| 0,163349343 | 0,620508573 | SYT2 | FALSE | Upregulated |
| 0,117610998 | 0,620610108 | UHRF1BP1 | FALSE | Upregulated |
| 0,49241212 | 0,620644512 | ZNF611 | FALSE | Upregulated |
| 0,442963875 | 0,620662455 | ZNF783 | FALSE | Upregulated |
| 0,225054406 | 0,620801586 | PLEKHH1 | FALSE | Upregulated |
| 0,439096838 | 0,620885511 | TWIST1 | FALSE | Upregulated |
| 0,300198273 | 0,620903743 | LOC646719 | FALSE | Upregulated |
| -0,838670067 | 0,620977629 | RAG2 | FALSE | Downregulated |
| -0,418847493 | 0,620977629 | SF3B14 | FALSE | Downregulated |
| 0,31980405 | 0,620977629 | SLCO3A1 | FALSE | Upregulated |
| 0,28773257 | 0,620977629 | SUCO | FALSE | Upregulated |
| -0,327471317 | 0,620977629 | TSEN34 | FALSE | Downregulated |
| 0,560638562 | 0,620998399 | CYP4F22 | FALSE | Upregulated |
| 0,228617145 | 0,620998399 | HIRIP3 | FALSE | Upregulated |
| 0,179753432 | 0,620998399 | LOC102723489 | FALSE | Upregulated |
| -0,213723137 | 0,620998399 | STK38 | FALSE | Downregulated |
| 0,241461515 | 0,620998399 | ZFYVE21 | FALSE | Upregulated |
| 0,241192688 | 0,621036893 | MPP1 | FALSE | Upregulated |
| -0,225211962 | 0,621160013 | ACAT2 | FALSE | Downregulated |
| -0,221313283 | 0,621517049 | ATP6V1A | FALSE | Downregulated |
| 0,515707075 | 0,621517049 | DOHH | FALSE | Upregulated |
| 0,389232219 | 0,621517049 | FBXO45 | FALSE | Upregulated |
| 0,271386769 | 0,621517049 | GOLT1A | FALSE | Upregulated |
| 0,20064636 | 0,621517049 | KDM4A-AS1 | FALSE | Upregulated |
| -0,212993621 | 0,621517049 | KIAA0922 | FALSE | Downregulated |
| 0,98190672 | 0,621517049 | LAMC2 | FALSE | Upregulated |
| 0,494413673 | 0,621517049 | LINC00649 | FALSE | Upregulated |
| -0,810887781 | 0,621517049 | LOC101927595 | FALSE | Downregulated |
| -0,419932149 | 0,621517049 | LOC102723694 | FALSE | Downregulated |
| -0,704110646 | 0,621517049 | LOC283731 | FALSE | Downregulated |

| | | | | |
|---|---|---|---|---|
| -0,310270265 | 0,621517049 | MAP10 | FALSE | Downregulated |
| 0,592674274 | 0,621517049 | MBIP | FALSE | Upregulated |
| -0,302795279 | 0,621517049 | PRELP | FALSE | Downregulated |
| -1,24780832 | 0,621517049 | PYGM | FALSE | Downregulated |
| 0,290696958 | 0,621517049 | RARS2 | FALSE | Upregulated |
| 0,929030703 | 0,621517049 | STAC | FALSE | Upregulated |
| -0,511312119 | 0,621517049 | TMEM98 | FALSE | Downregulated |
| 0,189770902 | 0,621517049 | ZEB2 | FALSE | Upregulated |
| 0,181756669 | 0,621536759 | GCHFR | FALSE | Upregulated |
| -0,113722174 | 0,621677922 | OS9 | FALSE | Downregulated |
| -0,225458679 | 0,621782314 | GOSR2 | FALSE | Downregulated |
| -0,41614726 | 0,621865141 | ABHD17B | FALSE | Downregulated |
| 0,180883309 | 0,621865141 | BCL9 | FALSE | Upregulated |
| 0,284908593 | 0,621865141 | C19orf35 | FALSE | Upregulated |
| -0,55826709 | 0,621865141 | CHCHD1 | FALSE | Downregulated |
| 0,262382358 | 0,621873251 | PPP4R1 | FALSE | Upregulated |
| -0,274290752 | 0,621994048 | MPHOSPH9 | FALSE | Downregulated |
| 0,395643683 | 0,622213264 | NRSN2 | FALSE | Upregulated |
| -0,282827402 | 0,622269228 | RNR1 | FALSE | Downregulated |
| 0,186433784 | 0,622274517 | UAP1 | FALSE | Upregulated |
| -0,327212061 | 0,622300392 | OSER1-AS1 | FALSE | Downregulated |
| 0,241651454 | 0,622420888 | C11orf83 | FALSE | Upregulated |
| 0,226463907 | 0,622525118 | FCHO2 | FALSE | Upregulated |
| -0,610210487 | 0,622709324 | ACRC | FALSE | Downregulated |
| 0,119430018 | 0,622709324 | ARFGEF2 | FALSE | Upregulated |
| 0,193402174 | 0,622709324 | ST6GALNAC6 | FALSE | Upregulated |
| -0,175753062 | 0,622719676 | PIK3C2A | FALSE | Downregulated |
| 0,767448909 | 0,622744459 | CPNE5 | FALSE | Upregulated |
| 0,259222203 | 0,622744459 | LOC101927345 | FALSE | Upregulated |
| 0,446107346 | 0,622744459 | RYR2 | FALSE | Upregulated |
| 0,181793172 | 0,622863441 | LINC00667 | FALSE | Upregulated |
| -0,422911641 | 0,6229318 | MOSPD3 | FALSE | Downregulated |
| 0,612106691 | 0,622937173 | LINC00933 | FALSE | Upregulated |
| -0,207144872 | 0,622937173 | LRCH1 | FALSE | Downregulated |
| 0,2234612 | 0,622937173 | XPOT | FALSE | Upregulated |
| -0,233381541 | 0,623013075 | MMADHC | FALSE | Downregulated |
| 0,674540835 | 0,623289824 | PDE10A | FALSE | Upregulated |
| -0,773843239 | 0,623384429 | ASPRV1 | FALSE | Downregulated |
| -0,446624081 | 0,623640919 | AAMDC | FALSE | Downregulated |
| 0,177956833 | 0,623640919 | GSDMD | FALSE | Upregulated |
| -0,279778283 | 0,623640919 | HIST1H4C | FALSE | Downregulated |
| -0,218646696 | 0,623640919 | LINC00174 | FALSE | Downregulated |
| -0,208555218 | 0,623640919 | RCAN1 | FALSE | Downregulated |
| 0,457266645 | 0,623805462 | OTUD6B | FALSE | Upregulated |
| 0,452889403 | 0,623808326 | F2RL3 | FALSE | Upregulated |
| -0,621068333 | 0,623808326 | LOC284825 | FALSE | Downregulated |
| -0,55195263 | 0,623808326 | ZNF527 | FALSE | Downregulated |
| 0,556443367 | 0,623815547 | ABRACL | FALSE | Upregulated |
| -0,122136667 | 0,623815547 | CDC14B | FALSE | Downregulated |
| 0,181003019 | 0,623815547 | GIGYF2 | FALSE | Upregulated |
| 0,238705852 | 0,623815547 | NME2 | FALSE | Upregulated |

| | | | | |
|---|---|---|---|---|
| -0,220139444 | 0,623815547 | PABPC1L | FALSE | Downregulated |
| -0,525107434 | 0,623815547 | PYCR1 | FALSE | Downregulated |
| 0,406132631 | 0,623827235 | UGGT2 | FALSE | Upregulated |
| 0,578920544 | 0,623871326 | C6orf226 | FALSE | Upregulated |
| -0,452018193 | 0,623924168 | CCDC59 | FALSE | Downregulated |
| 0,207965353 | 0,624049726 | SLC46A3 | FALSE | Upregulated |
| 0,222531534 | 0,624082871 | ENDOU | FALSE | Upregulated |
| 0,293926328 | 0,624112535 | ALKBH4 | FALSE | Upregulated |
| -0,095692189 | 0,624180786 | LRP1 | FALSE | Downregulated |
| -0,64273998 | 0,62421369 | LOC389831 | FALSE | Downregulated |
| -0,576583406 | 0,624321666 | EDNRA | FALSE | Downregulated |
| 0,212701781 | 0,624321666 | KCNAB1 | FALSE | Upregulated |
| -0,25081259 | 0,624321666 | SUFU | FALSE | Downregulated |
| 0,326807296 | 0,624321666 | WAC-AS1 | FALSE | Upregulated |
| -0,480879443 | 0,624546474 | GNG12-AS1 | FALSE | Downregulated |
| -0,64901978 | 0,624715829 | LOC101930307 | FALSE | Downregulated |
| -0,230899118 | 0,624715829 | SLC9B2 | FALSE | Downregulated |
| 0,341231387 | 0,624715829 | SUSD1 | FALSE | Upregulated |
| 0,144366281 | 0,624753628 | COX20 | FALSE | Upregulated |
| 0,450111873 | 0,624966239 | PDRG1 | FALSE | Upregulated |
| 0,328601865 | 0,625100676 | LOC102723809 | FALSE | Upregulated |
| 0,173425764 | 0,625100676 | MESDC1 | FALSE | Upregulated |
| -0,177621698 | 0,625100676 | PPP5C | FALSE | Downregulated |
| 0,231161234 | 0,625100676 | SPOP | FALSE | Upregulated |
| 0,312090728 | 0,625100676 | TSTD3 | FALSE | Upregulated |
| 0,175966093 | 0,625100676 | UNKL | FALSE | Upregulated |
| -0,168392191 | 0,62515995 | EIF3I | FALSE | Downregulated |
| -0,114214977 | 0,62515995 | MYCBP2 | FALSE | Downregulated |
| 0,205252195 | 0,62515995 | SOGA1 | FALSE | Upregulated |
| 0,206452159 | 0,625271612 | BRPF1 | FALSE | Upregulated |
| -0,428973797 | 0,625271612 | DTWD1 | FALSE | Downregulated |
| 0,138951392 | 0,625540829 | CALM3 | FALSE | Upregulated |
| 0,395116235 | 0,625540829 | CNRIP1 | FALSE | Upregulated |
| -0,744750564 | 0,625540829 | EFCAB6-AS1 | FALSE | Downregulated |
| -0,638796095 | 0,625636473 | ZNF682 | FALSE | Downregulated |
| -0,36584454 | 0,625856815 | FKTN | FALSE | Downregulated |
| -0,118828085 | 0,625856815 | INTS1 | FALSE | Downregulated |
| -0,47597458 | 0,625856815 | LOC102724206 | FALSE | Downregulated |
| 0,703302799 | 0,625856815 | PABPN1L | FALSE | Upregulated |
| 0,158154314 | 0,625856815 | UBE3C | FALSE | Upregulated |
| 0,230960864 | 0,625856815 | WDR20 | FALSE | Upregulated |
| 0,212032281 | 0,626109245 | TYMS | FALSE | Upregulated |
| -0,115242609 | 0,626155306 | PBRM1 | FALSE | Downregulated |
| -0,484992355 | 0,62632741 | LOC101927163 | FALSE | Downregulated |
| -0,124228746 | 0,626328972 | RNF123 | FALSE | Downregulated |
| -0,37961184 | 0,626478127 | SNHG8 | FALSE | Downregulated |
| 0,336959682 | 0,626506185 | LOC101928408 | FALSE | Upregulated |
| -0,258712138 | 0,626506185 | LOC102723804 | FALSE | Downregulated |
| -0,24068855 | 0,626596916 | CEP290 | FALSE | Downregulated |
| 0,328974344 | 0,626596916 | FAM122A | FALSE | Upregulated |
| -0,145960076 | 0,626684344 | CTNND1 | FALSE | Downregulated |

| | | | | |
|---|---|---|---|---|
| 0,45002885 | 0,626684344 | GAREML | FALSE | Upregulated |
| 0,363228771 | 0,626684344 | NRIP3 | FALSE | Upregulated |
| 0,222082314 | 0,626684344 | PREB | FALSE | Upregulated |
| -0,324466025 | 0,626684344 | TMEM55B | FALSE | Downregulated |
| 0,641625149 | 0,626752141 | SMIM3 | FALSE | Upregulated |
| 0,544746407 | 0,626841991 | GLIPR2 | FALSE | Upregulated |
| -0,257199177 | 0,62712236 | BLOC1S1 | FALSE | Downregulated |
| -0,425680423 | 0,62712236 | LOC100129550 | FALSE | Downregulated |
| 0,202059039 | 0,627209747 | ALDH1L2 | FALSE | Upregulated |
| -0,467920976 | 0,627209747 | COX16 | FALSE | Downregulated |
| 0,274870532 | 0,627209747 | SPOPL | FALSE | Upregulated |
| 0,749888478 | 0,627772231 | CACNA1C-AS4 | FALSE | Upregulated |
| 0,23689639 | 0,627947839 | LOC102724006 | FALSE | Upregulated |
| -0,360873253 | 0,627947839 | MRPL53 | FALSE | Downregulated |
| 0,411422607 | 0,628018968 | AKR1B1 | FALSE | Upregulated |
| -0,101466858 | 0,628096694 | REPIN1 | FALSE | Downregulated |
| -0,174300946 | 0,628172507 | PTPRA | FALSE | Downregulated |
| -0,445057278 | 0,628215945 | ZNF528 | FALSE | Downregulated |
| 0,465550107 | 0,628218605 | ZNF678 | FALSE | Upregulated |
| 0,192170857 | 0,62841627 | ZFP36 | FALSE | Upregulated |
| -0,234118647 | 0,628540936 | BRD7 | FALSE | Downregulated |
| 0,33180484 | 0,628540936 | GPRC5D | FALSE | Upregulated |
| 0,216294131 | 0,628540936 | IFIT2 | FALSE | Upregulated |
| 0,339495642 | 0,62861952 | ATP6V1F | FALSE | Upregulated |
| -0,513826224 | 0,628665674 | THG1L | FALSE | Downregulated |
| -0,382672965 | 0,628785056 | ZSCAN26 | FALSE | Downregulated |
| -0,190235777 | 0,628990299 | IPO8 | FALSE | Downregulated |
| 0,168701898 | 0,62902461 | EIF2S2 | FALSE | Upregulated |
| -0,215253364 | 0,629167178 | TMX3 | FALSE | Downregulated |
| 0,260129846 | 0,629183922 | ANKRD37 | FALSE | Upregulated |
| -0,29274472 | 0,629183922 | DTX2P1-UPK3BP1-PMS2P11 | FALSE | Downregulated |
| 0,248523645 | 0,629183922 | FBXO10 | FALSE | Upregulated |
| -0,41895205 | 0,629183922 | FLG2 | FALSE | Downregulated |
| -0,145531514 | 0,629183922 | FUBP3 | FALSE | Downregulated |
| 0,177751344 | 0,629183922 | IL2RB | FALSE | Upregulated |
| -0,772579757 | 0,629183922 | LOC100507258 | FALSE | Downregulated |
| 0,09708927 | 0,629183922 | PCBP1 | FALSE | Upregulated |
| 0,399819223 | 0,629183922 | POLR3D | FALSE | Upregulated |
| -0,275088505 | 0,629183922 | UBAC1 | FALSE | Downregulated |
| 0,792561166 | 0,629396945 | AC007551.3 | FALSE | Upregulated |
| 0,273995938 | 0,629650994 | GNG5 | FALSE | Upregulated |
| 0,300730474 | 0,629906613 | CCDC86 | FALSE | Upregulated |
| -0,472779913 | 0,629906613 | PTGS1 | FALSE | Downregulated |
| 0,157994168 | 0,630010026 | AXL | FALSE | Upregulated |
| 0,2824231 | 0,630010026 | EVI5L | FALSE | Upregulated |
| 0,170413126 | 0,630010026 | LIG3 | FALSE | Upregulated |
| 0,152860981 | 0,630021978 | CRAMP1L | FALSE | Upregulated |
| 0,231513902 | 0,630083084 | SYTL4 | FALSE | Upregulated |
| 0,482659878 | 0,630205568 | LRR1 | FALSE | Upregulated |
| 0,742319728 | 0,630299866 | RBMXL3 | FALSE | Upregulated |

| | | | | |
|---|---|---|---|---|
| -0,570197012 | 0,630299866 | TECTA | FALSE | Downregulated |
| -1,354059014 | 0,630344226 | LOC102723475 | FALSE | Downregulated |
| 0,564445592 | 0,630532518 | CAMK2N2 | FALSE | Upregulated |
| 0,241005001 | 0,630781907 | CALB1 | FALSE | Upregulated |
| 0,557924183 | 0,630949702 | RRP9 | FALSE | Upregulated |
| -0,363661493 | 0,631058502 | AC091729.9 | FALSE | Downregulated |
| 0,590511566 | 0,631058502 | CDH20 | FALSE | Upregulated |
| 0,219956989 | 0,631058502 | DDX21 | FALSE | Upregulated |
| -1,927143407 | 0,631058502 | LOC102724843 | FALSE | Downregulated |
| -0,291005001 | 0,631058502 | TMEM167B | FALSE | Downregulated |
| 0,520600263 | 0,631058502 | ZCCHC18 | FALSE | Upregulated |
| -0,121799099 | 0,631061108 | CENPB | FALSE | Downregulated |
| 0,185680358 | 0,631061108 | MARCH8 | FALSE | Upregulated |
| 0,275093491 | 0,631112549 | COPG2 | FALSE | Upregulated |
| -0,173655811 | 0,631112549 | HIF1AN | FALSE | Downregulated |
| 0,61981487 | 0,631112549 | N6AMT1 | FALSE | Upregulated |
| 0,902119772 | 0,631112549 | PPFIA4 | FALSE | Upregulated |
| 0,121473061 | 0,631112549 | SRSF1 | FALSE | Upregulated |
| -0,235398979 | 0,631746726 | CDK14 | FALSE | Downregulated |
| 0,249614596 | 0,631747249 | CCDC88C | FALSE | Upregulated |
| -0,221419589 | 0,631747249 | DNASE2 | FALSE | Downregulated |
| -0,616384064 | 0,631747249 | FADS6 | FALSE | Downregulated |
| 0,191155509 | 0,631747249 | ITPR1 | FALSE | Upregulated |
| 0,326175311 | 0,631747249 | TMEM194B | FALSE | Upregulated |
| -0,411392615 | 0,631747249 | ZNF568 | FALSE | Downregulated |
| -0,350082947 | 0,631751601 | ZNF212 | FALSE | Downregulated |
| -0,187848868 | 0,632095562 | BAIAP2 | FALSE | Downregulated |
| 0,161250789 | 0,632559588 | IRF7 | FALSE | Upregulated |
| -0,136296264 | 0,632601319 | RBM39 | FALSE | Downregulated |
| -0,325135844 | 0,632601319 | SPATA5L1 | FALSE | Downregulated |
| 0,224679267 | 0,632762097 | AHSA1 | FALSE | Upregulated |
| -0,13110397 | 0,632762097 | MEF2A | FALSE | Downregulated |
| 0,35023614 | 0,632762097 | SLC9A1 | FALSE | Upregulated |
| 0,269420496 | 0,632930931 | LOC101928068 | FALSE | Upregulated |
| -0,278909231 | 0,633269886 | NEK1 | FALSE | Downregulated |
| -0,492069762 | 0,633517802 | ZNF765 | FALSE | Downregulated |
| 0,220319515 | 0,634135936 | FAM173A | FALSE | Upregulated |
| -0,175769952 | 0,634142187 | DCTD | FALSE | Downregulated |
| 0,130977973 | 0,634142187 | HGS | FALSE | Upregulated |
| -0,130629766 | 0,634142187 | KLF3 | FALSE | Downregulated |
| -0,497811445 | 0,634142187 | LOC102723335 | FALSE | Downregulated |
| -0,448169987 | 0,634142187 | LRAT | FALSE | Downregulated |
| -0,796144999 | 0,634142187 | NUTM2A-AS1 | FALSE | Downregulated |
| 0,351837333 | 0,634142187 | TDRD10 | FALSE | Upregulated |
| 0,153253986 | 0,634142187 | TSC22D2 | FALSE | Upregulated |
| -0,244149298 | 0,634142187 | UBE2I | FALSE | Downregulated |
| -0,111782331 | 0,634142187 | UPF1 | FALSE | Downregulated |
| -0,959815225 | 0,634325967 | DCHS2 | FALSE | Downregulated |
| -0,476681562 | 0,634413564 | FEZ1 | FALSE | Downregulated |
| 0,24259101 | 0,634533074 | TRPS1 | FALSE | Upregulated |
| -0,616218668 | 0,634816857 | ZDHHC1 | FALSE | Downregulated |

| | | | | |
|---|---|---|---|---|
| -0,135361117 | 0,634961196 | SERF2 | FALSE | Downregulated |
| -0,215717916 | 0,635208555 | PRDX4 | FALSE | Downregulated |
| 0,137446204 | 0,635275207 | CANX | FALSE | Upregulated |
| -0,342141195 | 0,635275207 | CWC15 | FALSE | Downregulated |
| -0,338540377 | 0,635339529 | CDC23 | FALSE | Downregulated |
| 0,303456522 | 0,635339529 | CHMP4A | FALSE | Upregulated |
| -0,271761977 | 0,635339529 | TSKU | FALSE | Downregulated |
| -0,759409774 | 0,635373507 | CES5A | FALSE | Downregulated |
| 0,621382395 | 0,635499522 | CXorf36 | FALSE | Upregulated |
| -0,60632586 | 0,635633778 | CALCA | FALSE | Downregulated |
| -1,186917018 | 0,635633778 | MYL1 | FALSE | Downregulated |
| 0,312506092 | 0,63571716 | GREB1 | FALSE | Upregulated |
| 0,529617672 | 0,635788549 | LOC102724550 | FALSE | Upregulated |
| 0,280012983 | 0,635828793 | FIZ1 | FALSE | Upregulated |
| -0,269546193 | 0,635828793 | PROX1-AS1 | FALSE | Downregulated |
| 0,279569558 | 0,635828793 | STX12 | FALSE | Upregulated |
| -0,177121873 | 0,63585502 | POLDIP3 | FALSE | Downregulated |
| -0,385569598 | 0,63585502 | ZNF548 | FALSE | Downregulated |
| -0,304493332 | 0,63585502 | ZNF771 | FALSE | Downregulated |
| -0,346347375 | 0,636098179 | COL17A1 | FALSE | Downregulated |
| -0,199521229 | 0,636098179 | FRK | FALSE | Downregulated |
| -0,162285598 | 0,636098179 | GLYATL1 | FALSE | Downregulated |
| 0,745088729 | 0,636098179 | LOC100507656 | FALSE | Upregulated |
| -0,303876987 | 0,636098179 | LOC101928098 | FALSE | Downregulated |
| -0,605762268 | 0,636098179 | MTERFD1 | FALSE | Downregulated |
| 0,282620422 | 0,636098179 | PLAA | FALSE | Upregulated |
| 0,201954196 | 0,636098179 | USP42 | FALSE | Upregulated |
| 0,320274116 | 0,636098179 | ZNF260 | FALSE | Upregulated |
| 0,312421834 | 0,636153281 | SPDYE7P | FALSE | Upregulated |
| 0,407704525 | 0,636251336 | DBN1 | FALSE | Upregulated |
| -0,388273568 | 0,636251336 | TRPT1 | FALSE | Downregulated |
| -0,473283802 | 0,636383287 | EXOC3L1 | FALSE | Downregulated |
| -0,221576612 | 0,636494574 | DHX36 | FALSE | Downregulated |
| -0,383109396 | 0,636699288 | ACSM1 | FALSE | Downregulated |
| -0,225157846 | 0,636737781 | SCYL2 | FALSE | Downregulated |
| 0,236367638 | 0,636860657 | RHOG | FALSE | Upregulated |
| -0,367712768 | 0,636929427 | AQP1 | FALSE | Downregulated |
| 0,270090086 | 0,636929427 | CDC123 | FALSE | Upregulated |
| -0,193265374 | 0,636929427 | FABP1 | FALSE | Downregulated |
| 0,134631946 | 0,637089995 | ABCA2 | FALSE | Upregulated |
| 0,412448926 | 0,637089995 | BCAS2 | FALSE | Upregulated |
| -0,416588283 | 0,637089995 | FAM49A | FALSE | Downregulated |
| 0,345614196 | 0,637089995 | HERC5 | FALSE | Upregulated |
| 0,267214599 | 0,637089995 | HMGCR | FALSE | Upregulated |
| -0,239257122 | 0,637089995 | IRF5 | FALSE | Downregulated |
| -0,235306049 | 0,637089995 | KLHL20 | FALSE | Downregulated |
| 0,195773857 | 0,637089995 | L3MBTL2 | FALSE | Upregulated |
| -0,246080688 | 0,637089995 | MRPL54 | FALSE | Downregulated |
| 0,276612251 | 0,637089995 | MS4A7 | FALSE | Upregulated |
| -0,305724987 | 0,637089995 | PDE1B | FALSE | Downregulated |
| -0,282836377 | 0,637089995 | PHF13 | FALSE | Downregulated |

| | | | | |
|---|---|---|---|---|
| 0,22553963 | 0,637089995 | POLR3A | FALSE | Upregulated |
| -0,648856825 | 0,637089995 | PRB3 | FALSE | Downregulated |
| -0,122716563 | 0,637089995 | RBP5 | FALSE | Downregulated |
| -0,306806388 | 0,637089995 | SIRT4 | FALSE | Downregulated |
| -0,243124538 | 0,637089995 | SKIDA1 | FALSE | Downregulated |
| -0,255428479 | 0,637089995 | TANC2 | FALSE | Downregulated |
| -0,15940611 | 0,637089995 | TOMM40L | FALSE | Downregulated |
| 0,154899429 | 0,637089995 | WASL | FALSE | Upregulated |
| 0,212697267 | 0,637089995 | ZNF571-AS1 | FALSE | Upregulated |
| -0,33262422 | 0,637272674 | CLDN5 | FALSE | Downregulated |
| -0,248637643 | 0,637308493 | SLC22A9 | FALSE | Downregulated |
| 0,187148887 | 0,637473403 | CTAGE7P | FALSE | Upregulated |
| 0,162978013 | 0,637473403 | KRI1 | FALSE | Upregulated |
| 0,466823287 | 0,63752869 | GALNT16 | FALSE | Upregulated |
| -0,261861427 | 0,637852982 | LYRM1 | FALSE | Downregulated |
| 1,072497051 | 0,638022223 | EIF4EBP3 | FALSE | Upregulated |
| -0,625202691 | 0,638022223 | KCNK12 | FALSE | Downregulated |
| -0,508126055 | 0,638022223 | UBAP1L | FALSE | Downregulated |
| -0,260760703 | 0,638144342 | FZD1 | FALSE | Downregulated |
| -0,650774973 | 0,638155591 | KIF1A | FALSE | Downregulated |
| 0,381513244 | 0,638289988 | ATP1B3 | FALSE | Upregulated |
| -0,108583809 | 0,638289988 | TUBGCP6 | FALSE | Downregulated |
| -0,678743888 | 0,638408016 | ADH7 | FALSE | Downregulated |
| 0,204327603 | 0,63862222 | LOC101929781 | FALSE | Upregulated |
| 0,168348947 | 0,63862222 | PGK1 | FALSE | Upregulated |
| -0,573103181 | 0,63862222 | SHD | FALSE | Downregulated |
| 0,217883781 | 0,638717376 | ARHGAP23 | FALSE | Upregulated |
| 0,192283361 | 0,638717376 | BCCIP | FALSE | Upregulated |
| -0,112867168 | 0,638717376 | FLNB | FALSE | Downregulated |
| 0,470222493 | 0,638717376 | PIR | FALSE | Upregulated |
| 0,311304192 | 0,638879231 | MAPK7 | FALSE | Upregulated |
| 0,294398132 | 0,638879231 | ME3 | FALSE | Upregulated |
| 0,627678734 | 0,638879231 | PAFAH1B3 | FALSE | Upregulated |
| -0,403718346 | 0,638893407 | CELF6 | FALSE | Downregulated |
| 0,35079027 | 0,639124987 | IGFLR1 | FALSE | Upregulated |
| 0,157537183 | 0,639460695 | TRAF4 | FALSE | Upregulated |
| 1,102302892 | 0,639462334 | ZFP91-CNTF | FALSE | Upregulated |
| -0,276861502 | 0,639695878 | C9orf64 | FALSE | Downregulated |
| -0,15947647 | 0,639695878 | IER2 | FALSE | Downregulated |
| 0,27435851 | 0,639695878 | LOC102724262 | FALSE | Upregulated |
| -0,248773192 | 0,639695878 | RYK | FALSE | Downregulated |
| 0,208851079 | 0,639729858 | PRR13 | FALSE | Upregulated |
| -0,751108132 | 0,639792824 | C7orf55 | FALSE | Downregulated |
| -0,148597289 | 0,639830631 | SLC25A3 | FALSE | Downregulated |
| -0,239197925 | 0,640295869 | IMP4 | FALSE | Downregulated |
| 0,697789901 | 0,640391137 | COL15A1 | FALSE | Upregulated |
| 0,263727898 | 0,640391137 | SCTR | FALSE | Upregulated |
| -0,260638394 | 0,640576613 | ARSA | FALSE | Downregulated |
| -0,182228966 | 0,640576613 | ATP6V0E2 | FALSE | Downregulated |
| 0,301181665 | 0,640576613 | CEP72 | FALSE | Upregulated |
| 1,277258992 | 0,640576613 | LOC102724504 | FALSE | Upregulated |

| | | | | |
|---|---|---|---|---|
| 0,399095881 | 0,640576613 | METTL5 | FALSE | Upregulated |
| -0,119341442 | 0,640576613 | NEK9 | FALSE | Downregulated |
| 0,185232192 | 0,640576613 | RGAG4 | FALSE | Upregulated |
| -0,564186835 | 0,640576613 | ZNF836 | FALSE | Downregulated |
| -0,20675778 | 0,640681392 | AGO3 | FALSE | Downregulated |
| -0,283984815 | 0,640681392 | C1QTNF6 | FALSE | Downregulated |
| 0,503346856 | 0,640681392 | UPK1B | FALSE | Upregulated |
| 0,373849881 | 0,640701439 | IRAK3 | FALSE | Upregulated |
| 0,300984775 | 0,64081546 | PRKCE | FALSE | Upregulated |
| -0,28107331 | 0,640820536 | SPRY4 | FALSE | Downregulated |
| -0,138448776 | 0,640956695 | ABTB2 | FALSE | Downregulated |
| 0,25124294 | 0,640956695 | AVPI1 | FALSE | Upregulated |
| -0,31703693 | 0,640956695 | BRCA1 | FALSE | Downregulated |
| -0,452715781 | 0,640956695 | ITIH5 | FALSE | Downregulated |
| 0,169835858 | 0,640956695 | PPP4C | FALSE | Upregulated |
| -0,278365698 | 0,640956695 | TSFM | FALSE | Downregulated |
| -0,146223459 | 0,640956695 | ZBTB44 | FALSE | Downregulated |
| 0,381225837 | 0,641219921 | LOC143666 | FALSE | Upregulated |
| -0,184279192 | 0,641301892 | UQCRC2 | FALSE | Downregulated |
| 0,161970994 | 0,641343655 | CRIM1 | FALSE | Upregulated |
| -0,163967145 | 0,641343655 | EFNB1 | FALSE | Downregulated |
| 0,316512669 | 0,641343655 | ENAH | FALSE | Upregulated |
| 0,143816778 | 0,641343655 | GDF11 | FALSE | Upregulated |
| 0,743655492 | 0,641343655 | KIAA1211L | FALSE | Upregulated |
| -0,319423598 | 0,641343655 | THEM4 | FALSE | Downregulated |
| 0,189574361 | 0,641412567 | MAGI1-AS1 | FALSE | Upregulated |
| 0,182651465 | 0,641488216 | CEBPB | FALSE | Upregulated |
| -0,292490115 | 0,641488216 | DEDD2 | FALSE | Downregulated |
| -0,53285584 | 0,641488216 | ZNF71 | FALSE | Downregulated |
| -0,544490068 | 0,6417545 | AC000124.1 | FALSE | Downregulated |
| -0,367090784 | 0,6417545 | CPAMD8 | FALSE | Downregulated |
| 0,300142546 | 0,6417545 | ETV3 | FALSE | Upregulated |
| -0,252944243 | 0,6417545 | GOLPH3L | FALSE | Downregulated |
| -0,194835588 | 0,6417545 | GTF3A | FALSE | Downregulated |
| 0,677420995 | 0,6417545 | LOC101927060 | FALSE | Upregulated |
| -0,443222546 | 0,6417545 | MROH6 | FALSE | Downregulated |
| -0,357891103 | 0,6417545 | MRPL11 | FALSE | Downregulated |
| 0,31058479 | 0,6417545 | NDUFAF7 | FALSE | Upregulated |
| -0,108853813 | 0,6417545 | PFKL | FALSE | Downregulated |
| -0,300540671 | 0,6417545 | SETDB2 | FALSE | Downregulated |
| 0,334606931 | 0,6417545 | ST14 | FALSE | Upregulated |
| 0,182092358 | 0,6417545 | SUV39H1 | FALSE | Upregulated |
| 0,171989617 | 0,6417545 | TUBGCP4 | FALSE | Upregulated |
| 0,389382019 | 0,6417545 | VPS33A | FALSE | Upregulated |
| -0,721722382 | 0,6417545 | ZNF425 | FALSE | Downregulated |
| 0,118954774 | 0,641973933 | EIF3A | FALSE | Upregulated |
| -0,298867445 | 0,641973933 | MRPL9 | FALSE | Downregulated |
| -0,362402893 | 0,642075237 | IDO2 | FALSE | Downregulated |
| 0,647804039 | 0,642122547 | WDHD1 | FALSE | Upregulated |
| 0,170071575 | 0,642122547 | ZNF517 | FALSE | Upregulated |
| 0,199837341 | 0,642242499 | HIST1H2AC | FALSE | Upregulated |

| | | | | |
|---|---|---|---|---|
| -0,15621774 | 0,642306351 | FGB | FALSE | Downregulated |
| 0,204581454 | 0,642556238 | ANO8 | FALSE | Upregulated |
| -0,258684705 | 0,642798209 | POLR1E | FALSE | Downregulated |
| -0,857947323 | 0,642922027 | LOC102723466 | FALSE | Downregulated |
| -0,508770255 | 0,643018544 | C19orf57 | FALSE | Downregulated |
| 0,194428081 | 0,64309079 | MAK16 | FALSE | Upregulated |
| -0,160617487 | 0,64309079 | UBL3 | FALSE | Downregulated |
| -0,315152177 | 0,643191781 | DSG1 | FALSE | Downregulated |
| -0,361582599 | 0,643494142 | LCLAT1 | FALSE | Downregulated |
| -0,109906329 | 0,643562609 | SURF4 | FALSE | Downregulated |
| -0,455857119 | 0,643562609 | TIMM9 | FALSE | Downregulated |
| 0,714537973 | 0,643562609 | ZNF430 | FALSE | Upregulated |
| -0,236346057 | 0,643562609 | ZNF641 | FALSE | Downregulated |
| 0,43153656 | 0,643596022 | BCL10 | FALSE | Upregulated |
| -0,291008928 | 0,643777036 | ANAPC11 | FALSE | Downregulated |
| 0,175973491 | 0,643777036 | LRRC61 | FALSE | Upregulated |
| 0,197265723 | 0,643777036 | LRRK1 | FALSE | Upregulated |
| 0,505765283 | 0,643777036 | USP17L7 | FALSE | Upregulated |
| 0,432849436 | 0,643928777 | CYB5R4 | FALSE | Upregulated |
| -0,20684979 | 0,643928777 | PHF10 | FALSE | Downregulated |
| -0,842887703 | 0,644034226 | AC074138.3 | FALSE | Downregulated |
| 0,521772882 | 0,644063203 | C14orf178 | FALSE | Upregulated |
| 0,239644714 | 0,644201167 | BIN1 | FALSE | Upregulated |
| 0,785305108 | 0,64432517 | LOC440434 | FALSE | Upregulated |
| -0,330576886 | 0,644353204 | KCNIP3 | FALSE | Downregulated |
| 0,206294705 | 0,644752533 | CALU | FALSE | Upregulated |
| 0,22816835 | 0,64486009 | DNAJC8 | FALSE | Upregulated |
| 0,254952945 | 0,645017803 | NCOA7 | FALSE | Upregulated |
| 0,223117908 | 0,645047252 | IDUA | FALSE | Upregulated |
| -0,159199482 | 0,645047252 | OTUD7B | FALSE | Downregulated |
| -0,167246305 | 0,645392825 | EEA1 | FALSE | Downregulated |
| 0,386817083 | 0,645483091 | LOC101928699 | FALSE | Upregulated |
| 0,706828797 | 0,645483091 | LOC102724192 | FALSE | Upregulated |
| -0,693733358 | 0,645700187 | LOC285389 | FALSE | Downregulated |
| 0,11167209 | 0,645717452 | AP3D1 | FALSE | Upregulated |
| -0,626626573 | 0,646152411 | AC009473.1 | FALSE | Downregulated |
| 0,599505825 | 0,646152411 | LOC101928182 | FALSE | Upregulated |
| -0,228041419 | 0,646152411 | OCIAD1 | FALSE | Downregulated |
| 0,194518254 | 0,646152411 | PGD | FALSE | Upregulated |
| 0,200127266 | 0,646209313 | NAIP | FALSE | Upregulated |
| 0,613258891 | 0,646242955 | RAB4B | FALSE | Upregulated |
| -0,335866153 | 0,646242955 | RP1-293L8.2 | FALSE | Downregulated |
| -0,511072788 | 0,646278549 | FRA10AC1 | FALSE | Downregulated |
| -0,722379089 | 0,646278549 | LOC100499484-C9ORF174 | FALSE | Downregulated |
| 0,317393911 | 0,646278549 | NIP7 | FALSE | Upregulated |
| 0,175387887 | 0,646278549 | NRP1 | FALSE | Upregulated |
| 0,698859563 | 0,646278549 | OPRM1 | FALSE | Upregulated |
| -0,276868781 | 0,646278549 | SMNDC1 | FALSE | Downregulated |
| -0,193118342 | 0,646278549 | SSR4 | FALSE | Downregulated |
| 0,146177478 | 0,646278549 | TP53INP1 | FALSE | Upregulated |

| | | | | |
|---|---|---|---|---|
| 0,432964485 | 0,646295726 | METTL18 | FALSE | Upregulated |
| -1,691353135 | 0,646344513 | GUSBP9 | FALSE | Downregulated |
| -0,247155958 | 0,646454182 | APBA3 | FALSE | Downregulated |
| -0,134522457 | 0,646503848 | MEGF8 | FALSE | Downregulated |
| -0,234476053 | 0,647087955 | SGK3 | FALSE | Downregulated |
| 0,310984738 | 0,647214514 | ACSS1 | FALSE | Upregulated |
| -0,260180303 | 0,647214514 | MTM1 | FALSE | Downregulated |
| 0,172184941 | 0,647585344 | ELF1 | FALSE | Upregulated |
| -0,450512335 | 0,647585344 | ZNF585A | FALSE | Downregulated |
| -0,353522535 | 0,647658804 | TMEM160 | FALSE | Downregulated |
| -0,191974874 | 0,647668731 | AGPAT3 | FALSE | Downregulated |
| -0,134008381 | 0,647668731 | DDB1 | FALSE | Downregulated |
| -0,242722254 | 0,647668731 | FCF1 | FALSE | Downregulated |
| 0,161046338 | 0,647668731 | SCAND2P | FALSE | Upregulated |
| 0,284646394 | 0,647668731 | ZNF540 | FALSE | Upregulated |
| 0,453976759 | 0,647678238 | PTGIR | FALSE | Upregulated |
| 0,721551013 | 0,647687915 | ASB2 | FALSE | Upregulated |
| 0,388484904 | 0,647687915 | CD248 | FALSE | Upregulated |
| -0,575664233 | 0,647687915 | MTMR8 | FALSE | Downregulated |
| 0,724973853 | 0,648088326 | NAPSA | FALSE | Upregulated |
| 0,232186088 | 0,648119022 | SLC38A7 | FALSE | Upregulated |
| 0,452658249 | 0,648119022 | SPECC1 | FALSE | Upregulated |
| -0,328758797 | 0,648119022 | ZFAND2A | FALSE | Downregulated |
| 0,136158512 | 0,648767664 | POLR3E | FALSE | Upregulated |
| -0,168591547 | 0,649251738 | FGG | FALSE | Downregulated |
| 0,570145238 | 0,649251738 | GDF15 | FALSE | Upregulated |
| -0,324889739 | 0,649294006 | PAMR1 | FALSE | Downregulated |
| -0,482071863 | 0,649294006 | WDR92 | FALSE | Downregulated |
| -0,122605351 | 0,649407107 | SRPR | FALSE | Downregulated |
| 0,137049535 | 0,649589417 | ROCK1 | FALSE | Upregulated |
| -0,105290575 | 0,649834596 | KHSRP | FALSE | Downregulated |
| 0,30263305 | 0,650008995 | LPPR1 | FALSE | Upregulated |
| -0,167486237 | 0,650452907 | FMR1 | FALSE | Downregulated |
| 0,494504039 | 0,650778908 | LOC101928729 | FALSE | Upregulated |
| 0,246743319 | 0,65081284 | PROCA1 | FALSE | Upregulated |
| -0,445247975 | 0,651076773 | PRKD1 | FALSE | Downregulated |
| -0,272039511 | 0,65111688 | ARL4D | FALSE | Downregulated |
| 0,145129661 | 0,651198 | PARP1 | FALSE | Upregulated |
| 0,166238814 | 0,651364239 | HDGFRP2 | FALSE | Upregulated |
| 0,167769525 | 0,651364239 | PAF1 | FALSE | Upregulated |
| 0,262413046 | 0,651364239 | SHKBP1 | FALSE | Upregulated |
| 0,12220858 | 0,651364239 | TMED5 | FALSE | Upregulated |
| 0,140170603 | 0,651387903 | AP1B1 | FALSE | Upregulated |
| -0,295977229 | 0,651387903 | SCARNA13 | FALSE | Downregulated |
| 0,125055766 | 0,651834378 | FKBP4 | FALSE | Upregulated |
| -0,261339104 | 0,651834378 | OST4 | FALSE | Downregulated |
| -0,251940348 | 0,652077765 | CSNK2A2 | FALSE | Downregulated |
| 0,228987953 | 0,652311541 | ZNF205 | FALSE | Upregulated |
| -0,291140863 | 0,65238788 | TMEM44-AS1 | FALSE | Downregulated |
| 0,700442468 | 0,652435794 | LOC101929225 | FALSE | Upregulated |
| 0,129013315 | 0,652459813 | AGRN | FALSE | Upregulated |

| | | | | |
|---|---|---|---|---|
| -0,1858355 | 0,652459813 | YTHDC1 | FALSE | Downregulated |
| -0,181092388 | 0,652562887 | GGNBP2 | FALSE | Downregulated |
| 0,683165892 | 0,652562887 | LPAR1 | FALSE | Upregulated |
| 0,847629017 | 0,652683437 | LOC101929444 | FALSE | Upregulated |
| -0,451057434 | 0,653293185 | SCGN | FALSE | Downregulated |
| 0,235303378 | 0,653571226 | TBCE | FALSE | Upregulated |
| -0,129527032 | 0,653580179 | DHRS3 | FALSE | Downregulated |
| 0,316750435 | 0,653580179 | PRCD | FALSE | Upregulated |
| 0,297292319 | 0,653580179 | SAMM50 | FALSE | Upregulated |
| -0,367362573 | 0,653580179 | UGT2A3 | FALSE | Downregulated |
| -0,705160951 | 0,653631367 | LOC100130899 | FALSE | Downregulated |
| 0,594602528 | 0,653631367 | ZNF185 | FALSE | Upregulated |
| -0,316797493 | 0,653653218 | HKR1 | FALSE | Downregulated |
| -0,258253714 | 0,653687491 | NFS1 | FALSE | Downregulated |
| -0,295875186 | 0,653687491 | UNG | FALSE | Downregulated |
| 0,315421058 | 0,653690866 | STBD1 | FALSE | Upregulated |
| -0,18425796 | 0,653725598 | DYNLL2 | FALSE | Downregulated |
| 0,145878884 | 0,653726296 | LOC101927377 | FALSE | Upregulated |
| -0,34146842 | 0,653922435 | LOC101929263 | FALSE | Downregulated |
| -0,340269165 | 0,653922435 | LOC102723772 | FALSE | Downregulated |
| -0,171160018 | 0,653922435 | NDUFB8 | FALSE | Downregulated |
| -0,299636639 | 0,653922435 | RP5-1029K10.2 | FALSE | Downregulated |
| -0,164577379 | 0,653922435 | ZDHHC14 | FALSE | Downregulated |
| 0,375893396 | 0,653973017 | MTPN | FALSE | Upregulated |
| 0,192904689 | 0,654034454 | GPR89A | FALSE | Upregulated |
| -0,663191237 | 0,654034454 | GRK4 | FALSE | Downregulated |
| -0,208170401 | 0,654034454 | KAT7 | FALSE | Downregulated |
| -0,345330532 | 0,654034454 | RP11-7F17.5 | FALSE | Downregulated |
| 0,215149381 | 0,654034454 | WDR91 | FALSE | Upregulated |
| -0,135020614 | 0,654090262 | LOC148696 | FALSE | Downregulated |
| 0,262930252 | 0,654096176 | ZFP62 | FALSE | Upregulated |
| -0,256823784 | 0,654133582 | LOC101927644 | FALSE | Downregulated |
| -0,249544281 | 0,654225432 | MRPL16 | FALSE | Downregulated |
| -0,668105612 | 0,65440535 | MALL | FALSE | Downregulated |
| 0,687416133 | 0,654449448 | MGC39372 | FALSE | Upregulated |
| 0,357576384 | 0,654449448 | MRPL21 | FALSE | Upregulated |
| -0,736779355 | 0,654767024 | RP11-806K15.1 | FALSE | Downregulated |
| 0,220401917 | 0,655122369 | SLC2A6 | FALSE | Upregulated |
| -0,794992631 | 0,655122369 | SMC5-AS1 | FALSE | Downregulated |
| 0,41739567 | 0,655238787 | AGA | FALSE | Upregulated |
| -0,310786114 | 0,655238787 | PLS1 | FALSE | Downregulated |
| 0,321249035 | 0,655238787 | SLC12A2 | FALSE | Upregulated |
| 0,48354175 | 0,655332005 | ARL4A | FALSE | Upregulated |
| -0,872196449 | 0,655332005 | CALN1 | FALSE | Downregulated |
| -0,191367269 | 0,655384361 | ZNF236 | FALSE | Downregulated |
| 0,542317123 | 0,655422307 | CD226 | FALSE | Upregulated |
| 0,339840933 | 0,655422307 | GNG3 | FALSE | Upregulated |
| -0,385363137 | 0,655570768 | C5orf51 | FALSE | Downregulated |
| 0,179992973 | 0,65578085 | ARFRP1 | FALSE | Upregulated |
| 0,785488028 | 0,655802784 | DNAJC25-GNG10 | FALSE | Upregulated |
| -0,2475417 | 0,655821928 | IGHMBP2 | FALSE | Downregulated |

| | | | | |
|---|---|---|---|---|
| 0,288957921 | 0,656020543 | LOC102724730 | FALSE | Upregulated |
| -0,202101644 | 0,656084972 | CYP20A1 | FALSE | Downregulated |
| 0,533127688 | 0,656084972 | DOCK9-AS2 | FALSE | Upregulated |
| -0,182745335 | 0,656084972 | EIF3F | FALSE | Downregulated |
| -0,428745637 | 0,656084972 | MED12L | FALSE | Downregulated |
| -0,355573886 | 0,656084972 | PRKG1-AS1 | FALSE | Downregulated |
| 0,244037445 | 0,656084972 | PTGES2 | FALSE | Upregulated |
| 0,104727819 | 0,656109577 | MRFAP1 | FALSE | Upregulated |
| 0,249744523 | 0,656109577 | SDE2 | FALSE | Upregulated |
| -0,149524748 | 0,656284981 | F2RL2 | FALSE | Downregulated |
| -0,408855844 | 0,656407914 | DNAH10 | FALSE | Downregulated |
| 0,101933062 | 0,656407914 | RBPMS | FALSE | Upregulated |
| -0,444725514 | 0,656426566 | C7orf73 | FALSE | Downregulated |
| -0,137605829 | 0,656483402 | HECTD3 | FALSE | Downregulated |
| 0,580295007 | 0,656745188 | BCAS4 | FALSE | Upregulated |
| 0,172528673 | 0,656745188 | CNTRL | FALSE | Upregulated |
| -0,202790958 | 0,656805293 | RPS19BP1 | FALSE | Downregulated |
| 0,14917145 | 0,656840549 | PSIP1 | FALSE | Upregulated |
| -0,168067353 | 0,656901645 | DDX23 | FALSE | Downregulated |
| -0,163910587 | 0,65693643 | C16orf70 | FALSE | Downregulated |
| 0,219276614 | 0,656953682 | CREB3L3 | FALSE | Upregulated |
| 0,533971853 | 0,656953682 | CXADRP2 | FALSE | Upregulated |
| -0,383979728 | 0,656953682 | HIST1H1A | FALSE | Downregulated |
| 0,128648054 | 0,657007786 | OTUD5 | FALSE | Upregulated |
| -0,226055041 | 0,657049102 | UBQLN2 | FALSE | Downregulated |
| -0,57402811 | 0,657144255 | EXTL3-AS1 | FALSE | Downregulated |
| 0,446536699 | 0,657144255 | GADD45G | FALSE | Upregulated |
| -0,35348953 | 0,657144255 | LINC00843 | FALSE | Downregulated |
| -0,29087293 | 0,657144255 | RNF214 | FALSE | Downregulated |
| 0,365373404 | 0,657169488 | ARHGEF34P | FALSE | Upregulated |
| -0,197555644 | 0,657333101 | CPB2-AS1 | FALSE | Downregulated |
| 0,386660221 | 0,657440833 | BACE2 | FALSE | Upregulated |
| -0,108188408 | 0,657486806 | KIAA2013 | FALSE | Downregulated |
| 0,326549726 | 0,657501758 | LOC729652 | FALSE | Upregulated |
| 0,495945177 | 0,657585837 | C15orf41 | FALSE | Upregulated |
| 0,310238722 | 0,657585837 | EMC8 | FALSE | Upregulated |
| 0,525788674 | 0,657585837 | FXYD3 | FALSE | Upregulated |
| 0,198796223 | 0,657585837 | UBXN1 | FALSE | Upregulated |
| 0,468621929 | 0,657703805 | KLHDC9 | FALSE | Upregulated |
| -0,434737054 | 0,657703805 | KLHL17 | FALSE | Downregulated |
| 0,69328113 | 0,657806903 | LOC101929336 | FALSE | Upregulated |
| 0,200068891 | 0,657806903 | ZAK | FALSE | Upregulated |
| -0,6287174 | 0,657851761 | DENND6B | FALSE | Downregulated |
| -0,344251048 | 0,657851761 | PGM2 | FALSE | Downregulated |
| 0,890229929 | 0,657978813 | CBWD6 | FALSE | Upregulated |
| 0,393044716 | 0,657978813 | METRNL | FALSE | Upregulated |
| -0,268642076 | 0,657978813 | VSNL1 | FALSE | Downregulated |
| -0,205394505 | 0,65828719 | NBAS | FALSE | Downregulated |
| 0,171588996 | 0,65846491 | MEN1 | FALSE | Upregulated |
| 0,11326254 | 0,658584742 | HTT | FALSE | Upregulated |
| -0,147554909 | 0,658631003 | PAPD7 | FALSE | Downregulated |

| | | | | |
|---|---|---|---|---|
| 0,298246736 | 0,65881414 | LOC101927940 | FALSE | Upregulated |
| 0,44206886 | 0,659021155 | TEX30 | FALSE | Upregulated |
| 0,455682967 | 0,659129573 | MATK | FALSE | Upregulated |
| -0,205646853 | 0,659207159 | C8G | FALSE | Downregulated |
| -0,344388687 | 0,659319766 | CHML | FALSE | Downregulated |
| 0,522575295 | 0,659319766 | FOSL1 | FALSE | Upregulated |
| 0,352871993 | 0,659319766 | WFIKKN1 | FALSE | Upregulated |
| -0,545667396 | 0,659456632 | LIMS3L | FALSE | Downregulated |
| 0,127986131 | 0,659463907 | CDC42EP1 | FALSE | Upregulated |
| 0,175707661 | 0,659630548 | BCAP31 | FALSE | Upregulated |
| 0,338047818 | 0,659630548 | RAB32 | FALSE | Upregulated |
| 0,479516209 | 0,659630548 | TLDC1 | FALSE | Upregulated |
| 0,145031891 | 0,659647322 | GNPTAB | FALSE | Upregulated |
| -0,314150812 | 0,659647322 | SEC31B | FALSE | Downregulated |
| 0,539428951 | 0,659896846 | UFSP1 | FALSE | Upregulated |
| -0,20306283 | 0,659996914 | PSMA5 | FALSE | Downregulated |
| -0,226377623 | 0,660482847 | TMEM59 | FALSE | Downregulated |
| -0,175874142 | 0,66084264 | CEND1 | FALSE | Downregulated |
| -0,254058226 | 0,66084264 | NLRP9 | FALSE | Downregulated |
| -0,170442962 | 0,66084264 | NR1H2 | FALSE | Downregulated |
| 0,300152192 | 0,661067991 | TMEM87B | FALSE | Upregulated |
| 0,166176314 | 0,66171447 | LPPR2 | FALSE | Upregulated |
| 0,514695421 | 0,662006456 | LOC101927579 | FALSE | Upregulated |
| 0,295902446 | 0,662006456 | METTL9 | FALSE | Upregulated |
| 0,344251094 | 0,662050785 | LOC102724238 | FALSE | Upregulated |
| 0,225770735 | 0,662099807 | CLNS1A | FALSE | Upregulated |
| -0,191370594 | 0,662099807 | MAPK6 | FALSE | Downregulated |
| 0,465180559 | 0,662099807 | RP11-85F14.5 | FALSE | Upregulated |
| -0,223394595 | 0,662326082 | PHF14 | FALSE | Downregulated |
| -0,25657642 | 0,662372796 | NES | FALSE | Downregulated |
| 0,217378929 | 0,662459001 | ARL5A | FALSE | Upregulated |
| 0,905149226 | 0,662576975 | LOC101929583 | FALSE | Upregulated |
| 0,316930239 | 0,662576975 | MSL3 | FALSE | Upregulated |
| -0,278810681 | 0,662576975 | RYBP | FALSE | Downregulated |
| -0,197778113 | 0,662923027 | TNNI1 | FALSE | Downregulated |
| -0,218801569 | 0,662962163 | NPAT | FALSE | Downregulated |
| 0,312740245 | 0,662962163 | UBOX5 | FALSE | Upregulated |
| 0,201268666 | 0,663072876 | ERCC6L2 | FALSE | Upregulated |
| 0,255499725 | 0,663130438 | EXOC1 | FALSE | Upregulated |
| 0,356635174 | 0,663130438 | GGTA1P | FALSE | Upregulated |
| 0,374689596 | 0,663271686 | GSG1 | FALSE | Upregulated |
| 0,158621299 | 0,663271686 | LOC100129361 | FALSE | Upregulated |
| -0,17902933 | 0,663271686 | PATZ1 | FALSE | Downregulated |
| -0,299817251 | 0,663271686 | ST3GAL5 | FALSE | Downregulated |
| 0,229147549 | 0,663271686 | ZNF41 | FALSE | Upregulated |
| 0,451997488 | 0,663405056 | H3F3AP4 | FALSE | Upregulated |
| -0,443349627 | 0,663405056 | LOC102723782 | FALSE | Downregulated |
| 0,288584212 | 0,663405056 | SKA2 | FALSE | Upregulated |
| -0,243688892 | 0,663405056 | SLC39A8 | FALSE | Downregulated |
| -0,478611284 | 0,663479291 | MN1 | FALSE | Downregulated |
| 0,406206538 | 0,663500314 | SDK2 | FALSE | Upregulated |

| | | | | |
|---|---|---|---|---|
| 0,13514845 | 0,663500314 | SH3BP5-AS1 | FALSE | Upregulated |
| 0,270118108 | 0,663500314 | TMEM243 | FALSE | Upregulated |
| 0,368848155 | 0,663515165 | SWSAP1 | FALSE | Upregulated |
| 0,199499298 | 0,663602555 | TMX2 | FALSE | Upregulated |
| -0,568597636 | 0,663770169 | CMSS1 | FALSE | Downregulated |
| 0,553987308 | 0,663770169 | LOC102723827 | FALSE | Upregulated |
| -0,231283225 | 0,663770169 | TTBK2 | FALSE | Downregulated |
| -0,270269087 | 0,663803794 | C11orf96 | FALSE | Downregulated |
| 0,294291785 | 0,663929106 | KCNMB3 | FALSE | Upregulated |
| -0,349855404 | 0,66410688 | NDFIP2 | FALSE | Downregulated |
| -0,498672883 | 0,664127375 | GPR89B | FALSE | Downregulated |
| 0,17354991 | 0,664127375 | KIAA1551 | FALSE | Upregulated |
| 0,247508535 | 0,664127375 | P4HTM | FALSE | Upregulated |
| 0,207403178 | 0,664127375 | SNX3 | FALSE | Upregulated |
| -0,32330498 | 0,664348585 | LACTB2 | FALSE | Downregulated |
| 0,161538142 | 0,664348585 | MEX3D | FALSE | Upregulated |
| 0,087567317 | 0,664348585 | PINK1 | FALSE | Upregulated |
| -0,150391616 | 0,664405617 | CHCHD10 | FALSE | Downregulated |
| -0,157484992 | 0,664486679 | RAD50 | FALSE | Downregulated |
| -0,548705684 | 0,664659933 | ZNF43 | FALSE | Downregulated |
| -0,123271702 | 0,665043867 | CPSF7 | FALSE | Downregulated |
| -0,242763643 | 0,665067675 | FBXO9 | FALSE | Downregulated |
| 0,217317102 | 0,665067675 | GATA6 | FALSE | Upregulated |
| -0,16379384 | 0,665067675 | RECQL5 | FALSE | Downregulated |
| 0,662688494 | 0,665696753 | RNASEK-C17orf49 | FALSE | Upregulated |
| 0,176175919 | 0,665696753 | SPIDR | FALSE | Upregulated |
| 0,17070429 | 0,665781232 | HSPA1A | FALSE | Upregulated |
| 0,157551391 | 0,665884171 | EIF3J | FALSE | Upregulated |
| 0,264905107 | 0,666189975 | TUBG2 | FALSE | Upregulated |
| -0,187630778 | 0,66629325 | GLUD1P3 | FALSE | Downregulated |
| -0,19108353 | 0,666364614 | THBS3 | FALSE | Downregulated |
| 0,414428092 | 0,667282522 | FAM49B | FALSE | Upregulated |
| 0,418568971 | 0,667282522 | LOC101928230 | FALSE | Upregulated |
| -0,352557953 | 0,667282522 | SLC13A3 | FALSE | Downregulated |
| 0,235307843 | 0,667328522 | E2F5 | FALSE | Upregulated |
| -0,119764902 | 0,667328522 | TENM1 | FALSE | Downregulated |
| 0,590863104 | 0,667392098 | C16orf90 | FALSE | Upregulated |
| -0,22153363 | 0,66749059 | RSBN1 | FALSE | Downregulated |
| 0,464360857 | 0,667640411 | FAM26F | FALSE | Upregulated |
| 0,178828562 | 0,667660407 | PSMD6-AS2 | FALSE | Upregulated |
| -0,428613146 | 0,667690286 | TTC18 | FALSE | Downregulated |
| 0,419516899 | 0,667690286 | TWISTNB | FALSE | Upregulated |
| 0,226112715 | 0,667846905 | DDX28 | FALSE | Upregulated |
| 0,302466245 | 0,667846905 | HMCN2 | FALSE | Upregulated |
| -0,171244159 | 0,667992299 | AMPD2 | FALSE | Downregulated |
| 0,496368098 | 0,668164204 | LOC100101148 | FALSE | Upregulated |
| -0,340462954 | 0,668257478 | LOC102723877 | FALSE | Downregulated |
| -0,216199712 | 0,668257478 | MAT2B | FALSE | Downregulated |
| 0,531748747 | 0,668257478 | RP11-381O7.3 | FALSE | Upregulated |
| 0,224899757 | 0,668257478 | SLU7 | FALSE | Upregulated |
| 0,244072395 | 0,668257478 | TBCEL | FALSE | Upregulated |

| | | | | |
|---|---|---|---|---|
| 0,141474394 | 0,6683105 | SLC39A1 | FALSE | Upregulated |
| -0,342039215 | 0,6683105 | TARSL2 | FALSE | Downregulated |
| -0,172958369 | 0,668317215 | CUL9 | FALSE | Downregulated |
| -0,725936536 | 0,668578548 | KLF5 | FALSE | Downregulated |
| -0,170334289 | 0,668611907 | PFDN5 | FALSE | Downregulated |
| -0,15884841 | 0,668714643 | BCAT2 | FALSE | Downregulated |
| 0,250544102 | 0,668714643 | CXCR5 | FALSE | Upregulated |
| -0,45834363 | 0,669112085 | ANKHD1-EIF4EBP3 | FALSE | Downregulated |
| -0,287608039 | 0,669112085 | ATP10D | FALSE | Downregulated |
| -0,237094227 | 0,669112085 | FAM200B | FALSE | Downregulated |
| 0,122875672 | 0,669112085 | HNRNPA2B1 | FALSE | Upregulated |
| 0,332907781 | 0,669112085 | NEXN-AS1 | FALSE | Upregulated |
| 0,659785159 | 0,669112085 | PRRX1 | FALSE | Upregulated |
| 0,262359456 | 0,669112085 | QSOX2 | FALSE | Upregulated |
| 0,195432966 | 0,669112085 | RP11-266L9.2 | FALSE | Upregulated |
| -0,488310778 | 0,669112085 | TMEM27 | FALSE | Downregulated |
| 0,195953276 | 0,669276125 | CCDC9 | FALSE | Upregulated |
| 0,403607912 | 0,669458029 | DAD1 | FALSE | Upregulated |
| -0,687754409 | 0,669458029 | GRM7 | FALSE | Downregulated |
| 0,445609913 | 0,669458029 | STX11 | FALSE | Upregulated |
| -0,301391328 | 0,669810996 | ANKRD16 | FALSE | Downregulated |
| -0,231256624 | 0,669882877 | DYNC2LI1 | FALSE | Downregulated |
| -0,74145358 | 0,670376078 | LOC101928738 | FALSE | Downregulated |
| 0,299700897 | 0,670449291 | ATP6V0E1 | FALSE | Upregulated |
| 0,198252024 | 0,670449291 | CBX6 | FALSE | Upregulated |
| 0,400264435 | 0,670449291 | PHLDA3 | FALSE | Upregulated |
| 0,207476954 | 0,670457599 | DCAF12 | FALSE | Upregulated |
| 0,292260939 | 0,670457599 | DYNC2H1 | FALSE | Upregulated |
| 0,402328606 | 0,670489371 | MOCS3 | FALSE | Upregulated |
| 0,171965453 | 0,670710081 | TARS | FALSE | Upregulated |
| -0,187361673 | 0,670780453 | APOBEC2 | FALSE | Downregulated |
| -0,27584488 | 0,670780453 | LOC101928502 | FALSE | Downregulated |
| 0,447410896 | 0,670796233 | LOC439951 | FALSE | Upregulated |
| -0,30710981 | 0,671158525 | AP5S1 | FALSE | Downregulated |
| 0,210084175 | 0,6711757 | TMEM167A | FALSE | Upregulated |
| -0,7679906 | 0,671179362 | FAM71A | FALSE | Downregulated |
| -0,199875032 | 0,671449882 | CLCN6 | FALSE | Downregulated |
| 0,657224618 | 0,671781568 | CPZ | FALSE | Upregulated |
| -0,185897071 | 0,671796837 | CPQ | FALSE | Downregulated |
| 0,184796259 | 0,671796837 | LOC646652 | FALSE | Upregulated |
| -0,195344299 | 0,671796837 | PAPD5 | FALSE | Downregulated |
| 0,142309564 | 0,672013995 | GOLPH3 | FALSE | Upregulated |
| 0,529611009 | 0,672013995 | PPP1R27 | FALSE | Upregulated |
| 0,595873274 | 0,672141024 | DDX26B | FALSE | Upregulated |
| 0,216947347 | 0,672141024 | UGT3A1 | FALSE | Upregulated |
| -0,29375938 | 0,672185151 | C9orf16 | FALSE | Downregulated |
| 0,250853254 | 0,672185151 | RAB27A | FALSE | Upregulated |
| -0,622934321 | 0,6722003 | EFS | FALSE | Downregulated |
| -0,51085531 | 0,6722003 | G6PC2 | FALSE | Downregulated |
| -0,473458003 | 0,6722003 | RPL21P44 | FALSE | Downregulated |
| -0,180329291 | 0,672270218 | SPATS2L | FALSE | Downregulated |

| | | | | |
|---|---|---|---|---|
| 0,767605821 | 0,67236032 | DNAJC18 | FALSE | Upregulated |
| -0,34028154 | 0,672435431 | SCN1B | FALSE | Downregulated |
| -0,201003132 | 0,672561794 | TP53I11 | FALSE | Downregulated |
| 0,714764651 | 0,672783201 | C2orf81 | FALSE | Upregulated |
| 0,382465121 | 0,672783201 | CHST2 | FALSE | Upregulated |
| -1,109751083 | 0,672783201 | GOLGA8T | FALSE | Downregulated |
| -0,898223496 | 0,672783201 | UGT2B17 | FALSE | Downregulated |
| 0,281276821 | 0,672783201 | ZNF81 | FALSE | Upregulated |
| -0,240211425 | 0,672855356 | BMS1P5 | FALSE | Downregulated |
| -0,109313984 | 0,672855356 | IRF2BP2 | FALSE | Downregulated |
| 0,447066468 | 0,67290009 | RASAL2-AS1 | FALSE | Upregulated |
| -0,30078832 | 0,673080588 | RRM1 | FALSE | Downregulated |
| -0,809433858 | 0,673091056 | TESC | FALSE | Downregulated |
| 0,325682637 | 0,67329725 | SLC7A5 | FALSE | Upregulated |
| 0,241543618 | 0,673305035 | SNX11 | FALSE | Upregulated |
| 0,25567626 | 0,67336721 | LOC102467073 | FALSE | Upregulated |
| -0,531432808 | 0,673452021 | BOLA3 | FALSE | Downregulated |
| -0,11970829 | 0,673452021 | MGAT2 | FALSE | Downregulated |
| -0,573598478 | 0,673662457 | FAM104B | FALSE | Downregulated |
| -0,23081512 | 0,673687606 | ABHD8 | FALSE | Downregulated |
| 0,284586114 | 0,67375852 | LOC100507346 | FALSE | Upregulated |
| 0,131552077 | 0,673808399 | TARDBP | FALSE | Upregulated |
| -0,190215511 | 0,673808399 | XPO7 | FALSE | Downregulated |
| 0,164912694 | 0,673811558 | MAP3K15 | FALSE | Upregulated |
| 0,238972718 | 0,673968247 | CD53 | FALSE | Upregulated |
| -0,353812512 | 0,673968247 | CRY1 | FALSE | Downregulated |
| -0,654935401 | 0,673968247 | DBNDD2 | FALSE | Downregulated |
| 0,520030652 | 0,673968247 | HCG27 | FALSE | Upregulated |
| -0,235493943 | 0,673968247 | ZNF175 | FALSE | Downregulated |
| -0,172525161 | 0,674008902 | RBM26 | FALSE | Downregulated |
| -0,238218357 | 0,67429028 | MRPL28 | FALSE | Downregulated |
| -0,172140055 | 0,67429028 | NUAK1 | FALSE | Downregulated |
| -0,318634304 | 0,674444232 | CCDC159 | FALSE | Downregulated |
| -0,174009546 | 0,674444232 | CGGBP1 | FALSE | Downregulated |
| 0,164887372 | 0,674444232 | DNAJA3 | FALSE | Upregulated |
| 0,173919152 | 0,674444232 | FUCA1 | FALSE | Upregulated |
| 0,39084682 | 0,674444232 | VCPKMT | FALSE | Upregulated |
| 0,166072589 | 0,674449436 | HARS | FALSE | Upregulated |
| 0,394123302 | 0,674710241 | FEN1 | FALSE | Upregulated |
| -0,240031562 | 0,674735541 | MRPS2 | FALSE | Downregulated |
| -0,334405347 | 0,674735541 | SH3YL1 | FALSE | Downregulated |
| -0,495061301 | 0,675006891 | ABLIM2 | FALSE | Downregulated |
| -0,775136247 | 0,675006891 | APOL5 | FALSE | Downregulated |
| 0,312354629 | 0,675006891 | C16orf86 | FALSE | Upregulated |
| 0,458254799 | 0,675006891 | CCDC122 | FALSE | Upregulated |
| 0,219072002 | 0,675006891 | GNG12 | FALSE | Upregulated |
| -0,243064305 | 0,675006891 | GPATCH2L | FALSE | Downregulated |
| -0,384261825 | 0,675006891 | KRBA1 | FALSE | Downregulated |
| -0,150862041 | 0,675006891 | SEC24D | FALSE | Downregulated |
| -0,141360138 | 0,675006891 | SH3BP5 | FALSE | Downregulated |
| -0,349584909 | 0,675006891 | VPS72 | FALSE | Downregulated |

| | | | | |
|---|---|---|---|---|
| -0,686182441 | 0,675006891 | ZNF14 | FALSE | Downregulated |
| 0,159787775 | 0,675006891 | ZNF850 | FALSE | Upregulated |
| 0,474321225 | 0,675175207 | DNAJC28 | FALSE | Upregulated |
| -0,28697195 | 0,675425477 | AC011747.7 | FALSE | Downregulated |
| 0,119738595 | 0,675425477 | CDV3 | FALSE | Upregulated |
| 0,182478101 | 0,675425477 | MASTL | FALSE | Upregulated |
| -0,221176399 | 0,675425477 | MICAL2 | FALSE | Downregulated |
| 0,335520153 | 0,675517668 | OIP5 | FALSE | Upregulated |
| 0,360527447 | 0,675682462 | RP11-106M7.1 | FALSE | Upregulated |
| 0,276733526 | 0,675946951 | DGCR9 | FALSE | Upregulated |
| -0,151587251 | 0,675946951 | PEAK1 | FALSE | Downregulated |
| -0,370659042 | 0,675946951 | POLA2 | FALSE | Downregulated |
| -0,347802436 | 0,675946951 | QRSL1 | FALSE | Downregulated |
| -1,693756896 | 0,675946951 | RAB4B-EGLN2 | FALSE | Downregulated |
| -0,185566439 | 0,675946951 | USP30 | FALSE | Downregulated |
| -0,119024259 | 0,675950327 | GDI2 | FALSE | Downregulated |
| 0,289659988 | 0,675950327 | NOS1AP | FALSE | Upregulated |
| -0,167536432 | 0,676055606 | ACTR1B | FALSE | Downregulated |
| -0,165002964 | 0,676158043 | NEURL1B | FALSE | Downregulated |
| -0,117353469 | 0,676230358 | BMPR2 | FALSE | Downregulated |
| 0,309033207 | 0,676230358 | LOC100129637 | FALSE | Upregulated |
| -0,651003025 | 0,67630255 | NANP | FALSE | Downregulated |
| 0,360469815 | 0,676422808 | LOC101929051 | FALSE | Upregulated |
| 0,493713287 | 0,676577613 | RP4-669L17.10-2 | FALSE | Upregulated |
| 0,34788848 | 0,676674473 | ERICH1 | FALSE | Upregulated |
| -0,167284606 | 0,676674473 | NEK4 | FALSE | Downregulated |
| 1,299344701 | 0,67679673 | ISY1-RAB43 | FALSE | Upregulated |
| 0,288948379 | 0,677153038 | MLKL | FALSE | Upregulated |
| 0,263858493 | 0,677186756 | CLN3 | FALSE | Upregulated |
| -0,181505834 | 0,677270583 | ID2 | FALSE | Downregulated |
| 0,272266032 | 0,677381728 | CHCHD3 | FALSE | Upregulated |
| -0,687332392 | 0,677381728 | LOC102723766 | FALSE | Downregulated |
| 0,698694137 | 0,677446001 | LOC102724476 | FALSE | Upregulated |
| -0,15281594 | 0,677457784 | HMG20B | FALSE | Downregulated |
| -0,321987772 | 0,677553247 | ZNF594 | FALSE | Downregulated |
| -0,100034596 | 0,677593563 | GOLGA4 | FALSE | Downregulated |
| -0,162931646 | 0,677754343 | EPOR | FALSE | Downregulated |
| 0,20547766 | 0,677799878 | COL5A2 | FALSE | Upregulated |
| -0,199904339 | 0,677881438 | TRAPPC10 | FALSE | Downregulated |
| 0,187630224 | 0,67796843 | CEPT1 | FALSE | Upregulated |
| 0,367416789 | 0,67796843 | ERV3-1 | FALSE | Upregulated |
| -0,378656015 | 0,67796843 | ZNF737 | FALSE | Downregulated |
| -0,788926797 | 0,678045808 | LOC102724196 | FALSE | Downregulated |
| 0,310456265 | 0,678045808 | LOC285095 | FALSE | Upregulated |
| -0,284032962 | 0,678045808 | PPP1R21 | FALSE | Downregulated |
| 0,216988284 | 0,678093412 | ZNF503-AS2 | FALSE | Upregulated |
| 0,603223481 | 0,678157111 | HMGN5 | FALSE | Upregulated |
| 0,435879447 | 0,678157111 | LOC400043 | FALSE | Upregulated |
| -0,212197615 | 0,678343958 | FBXO25 | FALSE | Downregulated |
| 0,376339628 | 0,6783506 | AKIP1 | FALSE | Upregulated |
| -0,174482563 | 0,6783506 | CPEB2 | FALSE | Downregulated |

| | | | | |
|---|---|---|---|---|
| -0,162761503 | 0,6783506 | KDM4C | FALSE | Downregulated |
| 0,204805546 | 0,6783506 | LHX4 | FALSE | Upregulated |
| -0,119547401 | 0,6783506 | PDCD4 | FALSE | Downregulated |
| 0,192033453 | 0,6783506 | PHLDB1 | FALSE | Upregulated |
| -0,392477778 | 0,678514896 | RP11-372E1.4 | FALSE | Downregulated |
| 0,322642954 | 0,678921948 | AEN | FALSE | Upregulated |
| -0,147285246 | 0,678921948 | CTSO | FALSE | Downregulated |
| 0,726469189 | 0,678921948 | GOLGA8S | FALSE | Upregulated |
| -0,250066635 | 0,678921948 | MAML2 | FALSE | Downregulated |
| 0,434326933 | 0,678921948 | OR2A1-AS1 | FALSE | Upregulated |
| -0,660151796 | 0,678921948 | PDGFD | FALSE | Downregulated |
| 0,369961264 | 0,678921948 | SLC39A10 | FALSE | Upregulated |
| 0,619864857 | 0,678921948 | SPINT1 | FALSE | Upregulated |
| 0,194741081 | 0,678921948 | TCF3 | FALSE | Upregulated |
| -0,217983641 | 0,679224696 | KLHL36 | FALSE | Downregulated |
| 0,721139128 | 0,679229384 | EFCAB11 | FALSE | Upregulated |
| -0,638985538 | 0,679229384 | LINC00638 | FALSE | Downregulated |
| 0,167397059 | 0,679229384 | ZBTB12 | FALSE | Upregulated |
| 0,235884194 | 0,67985466 | NFASC | FALSE | Upregulated |
| 0,4239666 | 0,680299733 | COL4A6 | FALSE | Upregulated |
| -0,111082088 | 0,680299733 | CSNK1A1 | FALSE | Downregulated |
| -0,200353997 | 0,680299733 | FEM1A | FALSE | Downregulated |
| -0,19187778 | 0,680299733 | FOXA2 | FALSE | Downregulated |
| 1,39628048 | 0,680299733 | LOC102724200 | FALSE | Upregulated |
| -0,226046605 | 0,680299733 | MTO1 | FALSE | Downregulated |
| -0,291812505 | 0,680299733 | XKR8 | FALSE | Downregulated |
| -0,175439811 | 0,680299733 | XPO4 | FALSE | Downregulated |
| -0,14694519 | 0,680591773 | EXOC3 | FALSE | Downregulated |
| 0,253842823 | 0,680591773 | RELT | FALSE | Upregulated |
| 0,25910789 | 0,680591773 | SLC39A3 | FALSE | Upregulated |
| 0,279177417 | 0,680847647 | MVD | FALSE | Upregulated |
| -0,218888279 | 0,680847647 | PDCD7 | FALSE | Downregulated |
| -0,30644839 | 0,681013928 | BBS9 | FALSE | Downregulated |
| 0,172763519 | 0,681013928 | UBAC2 | FALSE | Upregulated |
| 0,146085942 | 0,681054657 | CALML4 | FALSE | Upregulated |
| -0,258621344 | 0,681054657 | IPO9 | FALSE | Downregulated |
| 0,139268115 | 0,681054657 | SLC31A2 | FALSE | Upregulated |
| -0,081424323 | 0,681137072 | ARID1A | FALSE | Downregulated |
| 0,159292904 | 0,681137072 | ZFX | FALSE | Upregulated |
| 0,28184074 | 0,681168719 | DCTN5 | FALSE | Upregulated |
| 0,316669063 | 0,681168719 | NSDHL | FALSE | Upregulated |
| 0,626150107 | 0,681168719 | SNORA7B | FALSE | Upregulated |
| 0,449364591 | 0,681168719 | UBE2D1 | FALSE | Upregulated |
| -0,18636608 | 0,681299713 | SERINC3 | FALSE | Downregulated |
| 0,162290831 | 0,681417637 | FSD2 | FALSE | Upregulated |
| -0,190025689 | 0,681417637 | PMVK | FALSE | Downregulated |
| -0,139780409 | 0,681417637 | RNF11 | FALSE | Downregulated |
| -0,287163964 | 0,681731551 | C5orf15 | FALSE | Downregulated |
| -0,157238613 | 0,681731551 | EIF2A | FALSE | Downregulated |
| 0,371191123 | 0,681925523 | ADM | FALSE | Upregulated |
| -0,549199278 | 0,681925523 | UXT-AS1 | FALSE | Downregulated |

| | | | | |
|---|---|---|---|---|
| 0,170735521 | 0,682136854 | RAPGEF3 | FALSE | Upregulated |
| -0,478548129 | 0,682242227 | CDC42EP2 | FALSE | Downregulated |
| 0,197689731 | 0,682242227 | CPT1B | FALSE | Upregulated |
| 0,400671546 | 0,682242227 | KANSL1-AS1 | FALSE | Upregulated |
| 0,233557249 | 0,682242227 | LDHAL6A | FALSE | Upregulated |
| 0,154512445 | 0,682242227 | NUP188 | FALSE | Upregulated |
| 0,223968078 | 0,682242227 | OGFOD3 | FALSE | Upregulated |
| 0,143282321 | 0,682242227 | PDCD11 | FALSE | Upregulated |
| 0,383972317 | 0,68224864 | TRAF5 | FALSE | Upregulated |
| -0,419229079 | 0,68224864 | ZNF221 | FALSE | Downregulated |
| -0,205281935 | 0,682249563 | CRHBP | FALSE | Downregulated |
| 0,613226012 | 0,682249563 | LOC102724135 | FALSE | Upregulated |
| 0,368156141 | 0,682396583 | LOC101928609 | FALSE | Upregulated |
| 0,340410989 | 0,682579627 | DKK3 | FALSE | Upregulated |
| 0,148054987 | 0,682579627 | EHD3 | FALSE | Upregulated |
| 0,097763258 | 0,682579627 | KHNYN | FALSE | Upregulated |
| -0,150702286 | 0,682579627 | LETM1 | FALSE | Downregulated |
| 0,317569796 | 0,682579627 | LOC100129148 | FALSE | Upregulated |
| 0,146219872 | 0,682579627 | UBR2 | FALSE | Upregulated |
| 0,221986573 | 0,682850704 | CKS1B | FALSE | Upregulated |
| -0,375718725 | 0,682850704 | LOC101927542 | FALSE | Downregulated |
| 0,471491155 | 0,683031524 | TSNAXIP1 | FALSE | Upregulated |
| 0,370874632 | 0,683062693 | PIGS | FALSE | Upregulated |
| 0,309794478 | 0,683239693 | TSNAX | FALSE | Upregulated |
| 0,221663543 | 0,68327279 | LOC102724748 | FALSE | Upregulated |
| -0,140772853 | 0,683710549 | AP4S1 | FALSE | Downregulated |
| -0,153255153 | 0,683710549 | CSPP1 | FALSE | Downregulated |
| -0,233974702 | 0,683710549 | EIF2B4 | FALSE | Downregulated |
| 0,172834912 | 0,683710549 | HERC3 | FALSE | Upregulated |
| -0,249381318 | 0,683710549 | SATB2 | FALSE | Downregulated |
| -0,723133406 | 0,683732654 | TBX20 | FALSE | Downregulated |
| 0,537754146 | 0,683784878 | LOC101929482 | FALSE | Upregulated |
| 0,671882298 | 0,683848412 | TEKT4P2 | FALSE | Upregulated |
| -0,248233867 | 0,683848412 | TMEM47 | FALSE | Downregulated |
| 0,172763017 | 0,683990123 | THEM5 | FALSE | Upregulated |
| -0,192854025 | 0,684073825 | LOC100271836 | FALSE | Downregulated |
| 0,565432645 | 0,684073825 | ZNF230 | FALSE | Upregulated |
| 0,23608931 | 0,684170397 | NOM1 | FALSE | Upregulated |
| -0,187938762 | 0,684563492 | ANKZF1 | FALSE | Downregulated |
| 0,327748528 | 0,684833933 | C5orf34 | FALSE | Upregulated |
| 0,201201037 | 0,685175994 | CCAT1 | FALSE | Upregulated |
| 0,401610406 | 0,685175994 | CEP97 | FALSE | Upregulated |
| 0,255679501 | 0,685175994 | LRRC48 | FALSE | Upregulated |
| -0,325218076 | 0,685175994 | RAB30 | FALSE | Downregulated |
| -0,675122359 | 0,685175994 | RP11-527H14.2 | FALSE | Downregulated |
| 0,152525931 | 0,685175994 | SHANK2-AS1 | FALSE | Upregulated |
| 0,304220015 | 0,685300377 | FAM205B | FALSE | Upregulated |
| 0,224678294 | 0,685300377 | LITAF | FALSE | Upregulated |
| 0,582830518 | 0,685300377 | MED27 | FALSE | Upregulated |
| 0,15555125 | 0,685300377 | NECAP2 | FALSE | Upregulated |
| 0,502939805 | 0,685300377 | NLRP3 | FALSE | Upregulated |

| | | | | |
|---|---|---|---|---|
| -0,375494368 | 0,685300377 | RASL11B | FALSE | Downregulated |
| -0,134535501 | 0,685300377 | RPL7L1 | FALSE | Downregulated |
| -0,296835503 | 0,685334233 | CYB561D2 | FALSE | Downregulated |
| -0,352787936 | 0,685334233 | RUNX1T1 | FALSE | Downregulated |
| -0,216308837 | 0,685357334 | CLPB | FALSE | Downregulated |
| -0,213261631 | 0,685357334 | HYAL2 | FALSE | Downregulated |
| -0,405475667 | 0,685357334 | MBL1P | FALSE | Downregulated |
| 0,109996158 | 0,685357334 | PPL | FALSE | Upregulated |
| 0,17082746 | 0,685357334 | TOR1A | FALSE | Upregulated |
| -0,167724242 | 0,685824697 | FKBP2 | FALSE | Downregulated |
| 0,254509747 | 0,685852221 | ZNF549 | FALSE | Upregulated |
| -0,409335757 | 0,686039068 | ALG11 | FALSE | Downregulated |
| 0,207389089 | 0,686039068 | GNA13 | FALSE | Upregulated |
| 0,379807647 | 0,686039068 | NOC4L | FALSE | Upregulated |
| 0,361506979 | 0,686039068 | ZNF846 | FALSE | Upregulated |
| -0,108952784 | 0,686438581 | BCL9L | FALSE | Downregulated |
| 0,223514401 | 0,686438581 | BTRC | FALSE | Upregulated |
| -0,151007367 | 0,686438581 | CD164 | FALSE | Downregulated |
| 0,158894968 | 0,686438581 | HTR2B | FALSE | Upregulated |
| -0,333270351 | 0,686438581 | MTA3 | FALSE | Downregulated |
| -0,362942327 | 0,686438581 | RBBP8 | FALSE | Downregulated |
| 0,210918201 | 0,686438581 | TOPBP1 | FALSE | Upregulated |
| 0,107593872 | 0,686438581 | TRIO | FALSE | Upregulated |
| -0,176704184 | 0,686581652 | DECR1 | FALSE | Downregulated |
| -0,217023236 | 0,686581652 | USP11 | FALSE | Downregulated |
| -0,201213223 | 0,686581652 | ZNF224 | FALSE | Downregulated |
| -0,174245956 | 0,686588312 | TECPR2 | FALSE | Downregulated |
| -0,193278643 | 0,686645088 | LOC100996573 | FALSE | Downregulated |
| 0,108763094 | 0,686645088 | SYNPO | FALSE | Upregulated |
| -0,416526084 | 0,686668617 | CDRT1 | FALSE | Downregulated |
| 0,248067703 | 0,686668617 | HLA-J | FALSE | Upregulated |
| 0,717950114 | 0,686668617 | LOC102723444 | FALSE | Upregulated |
| 0,715061213 | 0,686668617 | MZB1 | FALSE | Upregulated |
| -0,15831462 | 0,686668617 | ZNF407 | FALSE | Downregulated |
| 0,438285941 | 0,686724624 | STX8 | FALSE | Upregulated |
| 0,233946222 | 0,686800038 | DPY19L2P3 | FALSE | Upregulated |
| -0,286323444 | 0,687269633 | GINM1 | FALSE | Downregulated |
| -0,618159139 | 0,687364156 | GOLGA8R | FALSE | Downregulated |
| 0,436263804 | 0,687364156 | NR2E3 | FALSE | Upregulated |
| 0,176582018 | 0,687364156 | RNF113B | FALSE | Upregulated |
| 0,400413566 | 0,68743889 | IMMP2L | FALSE | Upregulated |
| -0,333155652 | 0,687651143 | MIR155HG | FALSE | Downregulated |
| 0,575668406 | 0,687651143 | RP11-219G17.4 | FALSE | Upregulated |
| -0,293933233 | 0,687697013 | MRPL15 | FALSE | Downregulated |
| 0,586500701 | 0,687706896 | LOC100653515 | FALSE | Upregulated |
| 0,174076483 | 0,687706896 | MRPS14 | FALSE | Upregulated |
| -0,395042166 | 0,687722641 | DAGLA | FALSE | Downregulated |
| 0,177893487 | 0,687722641 | NF2 | FALSE | Upregulated |
| 0,365556526 | 0,687722641 | RINT1 | FALSE | Upregulated |
| -0,154119865 | 0,687722641 | SATB1 | FALSE | Downregulated |
| -0,150096131 | 0,687741845 | POGK | FALSE | Downregulated |

| | | | | |
|---|---|---|---|---|
| 0,351097259 | 0,687882116 | ANKRD53 | FALSE | Upregulated |
| -0,550237223 | 0,687882116 | LOC101928398 | FALSE | Downregulated |
| -0,352075471 | 0,687882116 | METTL20 | FALSE | Downregulated |
| -0,194290306 | 0,687882116 | PAFAH2 | FALSE | Downregulated |
| -0,253297022 | 0,687971563 | MAPK13 | FALSE | Downregulated |
| -0,237736151 | 0,687971563 | SDHAF1 | FALSE | Downregulated |
| -0,761046587 | 0,687971563 | TFCP2L1 | FALSE | Downregulated |
| -0,386565627 | 0,688005024 | LZIC | FALSE | Downregulated |
| 0,306190839 | 0,688218755 | C6orf25 | FALSE | Upregulated |
| -0,211898356 | 0,688218755 | GSTM4 | FALSE | Downregulated |
| 0,294358146 | 0,688218755 | SFXN3 | FALSE | Upregulated |
| -0,167440449 | 0,68842405 | BAIAP3 | FALSE | Downregulated |
| -0,169565786 | 0,68842405 | CPN1 | FALSE | Downregulated |
| 0,340742836 | 0,68842405 | LUZP6 | FALSE | Upregulated |
| 0,126786584 | 0,68842405 | RBMS2 | FALSE | Upregulated |
| -0,281238161 | 0,68842405 | ZNF782 | FALSE | Downregulated |
| -0,154581548 | 0,688532537 | ABHD14B | FALSE | Downregulated |
| 0,518718479 | 0,688532537 | AP000472.2 | FALSE | Upregulated |
| -0,153420841 | 0,688532537 | ATG9A | FALSE | Downregulated |
| -0,28367776 | 0,688532537 | CASP10 | FALSE | Downregulated |
| 0,316601354 | 0,688532537 | EMR2 | FALSE | Upregulated |
| 0,235283864 | 0,688532537 | TMEM184B | FALSE | Upregulated |
| 0,237764583 | 0,688532537 | TRIM7 | FALSE | Upregulated |
| 0,402974624 | 0,688532537 | UNC50 | FALSE | Upregulated |
| -0,704474558 | 0,688532537 | ZNF280C | FALSE | Downregulated |
| 0,412633559 | 0,688730827 | ANAPC4 | FALSE | Upregulated |
| 0,136978226 | 0,688730827 | ARMC5 | FALSE | Upregulated |
| 0,249818833 | 0,688730827 | C19orf25 | FALSE | Upregulated |
| -0,645332689 | 0,688730827 | CYP26B1 | FALSE | Downregulated |
| 0,435822894 | 0,688730827 | FOXN4 | FALSE | Upregulated |
| -0,242250581 | 0,688730827 | HAUS4 | FALSE | Downregulated |
| -0,322260345 | 0,688730827 | KCNIP2 | FALSE | Downregulated |
| -0,180837268 | 0,688730827 | LOC101927339 | FALSE | Downregulated |
| -0,503662923 | 0,688730827 | LOC101928055 | FALSE | Downregulated |
| 0,141956293 | 0,688730827 | LRP11 | FALSE | Upregulated |
| -0,236167516 | 0,688730827 | LRWD1 | FALSE | Downregulated |
| 0,199780602 | 0,688730827 | MMP14 | FALSE | Upregulated |
| 0,183940769 | 0,688730827 | PRC1 | FALSE | Upregulated |
| 0,302002921 | 0,688730827 | PRR18 | FALSE | Upregulated |
| 0,175274905 | 0,688730827 | RABGAP1 | FALSE | Upregulated |
| 0,278158721 | 0,688730827 | RABGGTB | FALSE | Upregulated |
| 0,232411714 | 0,688730827 | SRPK1 | FALSE | Upregulated |
| -0,298553901 | 0,688730827 | TATDN1 | FALSE | Downregulated |
| -0,149365747 | 0,688730827 | ZNF219 | FALSE | Downregulated |
| -0,169436418 | 0,688955144 | RFWD2 | FALSE | Downregulated |
| -0,521080436 | 0,689050146 | RGPD5 | FALSE | Downregulated |
| -0,322464687 | 0,689059839 | WASH7P | FALSE | Downregulated |
| 0,416380223 | 0,689255704 | RP1-27K12.2 | FALSE | Upregulated |
| 0,418601268 | 0,689279415 | COA6 | FALSE | Upregulated |
| 0,160409303 | 0,689313927 | ERCC6L | FALSE | Upregulated |
| 0,359435602 | 0,689313927 | VBP1 | FALSE | Upregulated |

| | | | | |
|---|---|---|---|---|
| -0,611684452 | 0,689445091 | KTI12 | FALSE | Downregulated |
| -0,564791053 | 0,689734246 | LOC102724554 | FALSE | Downregulated |
| -0,189178168 | 0,689769978 | LECT2 | FALSE | Downregulated |
| -0,537925336 | 0,689893794 | ZNF816 | FALSE | Downregulated |
| -0,161692053 | 0,689959249 | BCAM | FALSE | Downregulated |
| 0,245083783 | 0,69004263 | LOC101927185 | FALSE | Upregulated |
| -0,564222945 | 0,690183479 | LOC101926935 | FALSE | Downregulated |
| -0,483476336 | 0,690187759 | OR5AK4P | FALSE | Downregulated |
| 0,73099932 | 0,690305276 | LOC100996716 | FALSE | Upregulated |
| 0,184292487 | 0,690305276 | SMURF2 | FALSE | Upregulated |
| 0,244112703 | 0,690305276 | TMEM106A | FALSE | Upregulated |
| -0,234050027 | 0,690317569 | DHODH | FALSE | Downregulated |
| -0,229992468 | 0,690317569 | KLHL22 | FALSE | Downregulated |
| 0,365422182 | 0,690317569 | TLR6 | FALSE | Upregulated |
| 0,276411054 | 0,690566167 | DPAGT1 | FALSE | Upregulated |
| -0,160743067 | 0,690566167 | RALY | FALSE | Downregulated |
| 0,196189249 | 0,690566167 | RPP38 | FALSE | Upregulated |
| 0,231372785 | 0,690566167 | SPDYA | FALSE | Upregulated |
| -0,269361219 | 0,690566167 | TSN | FALSE | Downregulated |
| 0,494076237 | 0,690806047 | LOC101928706 | FALSE | Upregulated |
| 0,385580126 | 0,69087093 | C11orf80 | FALSE | Upregulated |
| -0,258929307 | 0,691018017 | PTAR1 | FALSE | Downregulated |
| -0,243006833 | 0,691115645 | COPS5 | FALSE | Downregulated |
| 0,575326996 | 0,691141207 | ABHD17C | FALSE | Upregulated |
| -0,268417393 | 0,691141207 | ANKRD34A | FALSE | Downregulated |
| 0,564395108 | 0,691141207 | APOBEC3C | FALSE | Upregulated |
| 0,135684336 | 0,691141207 | CD3EAP | FALSE | Upregulated |
| 0,53538974 | 0,691141207 | FAM227A | FALSE | Upregulated |
| -0,526053623 | 0,691141207 | LOC257396 | FALSE | Downregulated |
| 0,482942117 | 0,691141207 | LOC374443 | FALSE | Upregulated |
| -0,375170309 | 0,691141207 | MED28 | FALSE | Downregulated |
| 0,187734423 | 0,691141207 | NDOR1 | FALSE | Upregulated |
| -0,640478325 | 0,691141207 | PPP1R1B | FALSE | Downregulated |
| 0,171372349 | 0,691141207 | SNAPC3 | FALSE | Upregulated |
| 0,178183541 | 0,691149925 | CNPPD1 | FALSE | Upregulated |
| -0,153565425 | 0,691149925 | KAT6B | FALSE | Downregulated |
| 0,254833134 | 0,691149925 | NSA2 | FALSE | Upregulated |
| -0,128434669 | 0,691149925 | SETD1A | FALSE | Downregulated |
| 0,421541618 | 0,691272148 | EXOSC8 | FALSE | Upregulated |
| 0,150363814 | 0,691295037 | EDEM1 | FALSE | Upregulated |
| 0,127620813 | 0,691447158 | DNAJC5 | FALSE | Upregulated |
| -0,211844271 | 0,691948469 | IMPA1 | FALSE | Downregulated |
| -0,197790294 | 0,692530746 | ACOX3 | FALSE | Downregulated |
| 0,361019572 | 0,692530746 | ANO5 | FALSE | Upregulated |
| -0,219989346 | 0,692530746 | CADPS2 | FALSE | Downregulated |
| 0,144445357 | 0,692530746 | PTK2B | FALSE | Upregulated |
| 0,196407859 | 0,692717161 | SIM2 | FALSE | Upregulated |
| 0,329326947 | 0,692857336 | WDR61 | FALSE | Upregulated |
| -0,998900929 | 0,693272824 | LOC102723360 | FALSE | Downregulated |
| -0,41013433 | 0,693402339 | DERL2 | FALSE | Downregulated |
| 0,152702696 | 0,693724589 | CBR1 | FALSE | Upregulated |

| | | | | |
|---|---|---|---|---|
| -0,502571988 | 0,693724589 | IGF2 | FALSE | Downregulated |
| 0,330783696 | 0,693724589 | OSTM1 | FALSE | Upregulated |
| 0,240759887 | 0,693724589 | PNO1 | FALSE | Upregulated |
| 0,140383181 | 0,693724589 | SLC39A7 | FALSE | Upregulated |
| -0,237051756 | 0,693724589 | SSBP1 | FALSE | Downregulated |
| -0,367426871 | 0,693724589 | TMEM189 | FALSE | Downregulated |
| 0,327444457 | 0,693724589 | ZNF432 | FALSE | Upregulated |
| 0,447886498 | 0,693725432 | ANAPC10 | FALSE | Upregulated |
| 0,343134008 | 0,693725432 | DBIL5P | FALSE | Upregulated |
| 0,247132068 | 0,693725432 | DYNLRB1 | FALSE | Upregulated |
| -0,214957994 | 0,693725432 | NOB1 | FALSE | Downregulated |
| -0,279690429 | 0,693725432 | NT5DC3 | FALSE | Downregulated |
| 0,124298247 | 0,693725432 | RABGEF1 | FALSE | Upregulated |
| -0,552136326 | 0,693788696 | RP11-101E14.2 | FALSE | Downregulated |
| 0,406716919 | 0,69379031 | TMIGD2 | FALSE | Upregulated |
| 0,562874043 | 0,693803428 | LILRA5 | FALSE | Upregulated |
| -0,781373463 | 0,693829213 | BMS1P18 | FALSE | Downregulated |
| 0,423926744 | 0,694105331 | PRKY | FALSE | Upregulated |
| 0,192361357 | 0,694220477 | ELOVL5 | FALSE | Upregulated |
| -0,117321165 | 0,694220477 | SRRT | FALSE | Downregulated |
| 0,331180501 | 0,694220477 | TUBA1C | FALSE | Upregulated |
| 0,181239215 | 0,694588485 | RP11-219B17.1 | FALSE | Upregulated |
| -0,123013053 | 0,694636277 | ITGA1 | FALSE | Downregulated |
| -0,281628264 | 0,694677153 | SLC15A1 | FALSE | Downregulated |
| 0,313426239 | 0,694733821 | DNAJB4 | FALSE | Upregulated |
| -0,308470715 | 0,69484459 | GTF2H4 | FALSE | Downregulated |
| -0,328087505 | 0,694870586 | WHAMMP1 | FALSE | Downregulated |
| 0,168169314 | 0,694921868 | FLJ37453 | FALSE | Upregulated |
| -0,226492148 | 0,694942212 | ZNF330 | FALSE | Downregulated |
| -0,232183548 | 0,695009423 | ST7 | FALSE | Downregulated |
| -0,656082724 | 0,695079794 | FLJ44511 | FALSE | Downregulated |
| 0,24583464 | 0,695079794 | GABPB2 | FALSE | Upregulated |
| -0,136342965 | 0,695079794 | MUM1 | FALSE | Downregulated |
| -0,191429091 | 0,695079794 | NODAL | FALSE | Downregulated |
| 0,822597669 | 0,695079794 | OR2A20P | FALSE | Upregulated |
| 0,19447758 | 0,695079794 | SSBP4 | FALSE | Upregulated |
| -0,339355753 | 0,695079794 | ZSWIM5 | FALSE | Downregulated |
| -0,176600884 | 0,695185374 | ANAPC2 | FALSE | Downregulated |
| 0,284651663 | 0,695185374 | EVC | FALSE | Upregulated |
| -0,188706417 | 0,695185374 | GLE1 | FALSE | Downregulated |
| -0,227627628 | 0,695185374 | NDUFS5 | FALSE | Downregulated |
| 0,306137496 | 0,695185374 | SH3GL1P2 | FALSE | Upregulated |
| 0,409832938 | 0,695185374 | WDPCP | FALSE | Upregulated |
| 0,612456227 | 0,69519115 | ROR1 | FALSE | Upregulated |
| 0,18556656 | 0,695194966 | C11orf42 | FALSE | Upregulated |
| 0,673421201 | 0,695194966 | IRAK1BP1 | FALSE | Upregulated |
| 0,279644367 | 0,695226084 | LOC100507071 | FALSE | Upregulated |
| 0,152519207 | 0,695307326 | TRMT10B | FALSE | Upregulated |
| -0,118191196 | 0,695348778 | VDAC1 | FALSE | Downregulated |
| 0,160173881 | 0,695401216 | LYPLA2 | FALSE | Upregulated |
| -0,158668083 | 0,695529095 | MPI | FALSE | Downregulated |

| | | | | |
|---|---|---|---|---|
| -0,113717391 | 0,695551499 | SAT1 | FALSE | Downregulated |
| 0,347649188 | 0,695814709 | P2RY13 | FALSE | Upregulated |
| 0,670090782 | 0,695833462 | ARHGAP26-AS1 | FALSE | Upregulated |
| 0,169827531 | 0,695833462 | C15orf27 | FALSE | Upregulated |
| 0,135869035 | 0,695833462 | XPO6 | FALSE | Upregulated |
| 0,226281402 | 0,696070344 | DYM | FALSE | Upregulated |
| 0,659468762 | 0,696070344 | FRZB | FALSE | Upregulated |
| -0,333421265 | 0,696070344 | GPR133 | FALSE | Downregulated |
| -1,053781377 | 0,696070344 | GSTM1 | FALSE | Downregulated |
| 0,234995731 | 0,696070344 | LIMS1 | FALSE | Upregulated |
| 0,116613298 | 0,696070344 | MBNL1 | FALSE | Upregulated |
| -0,259008648 | 0,696070344 | TNFRSF10D | FALSE | Downregulated |
| -0,257239964 | 0,696070344 | ZNF408 | FALSE | Downregulated |
| -0,309800252 | 0,696602324 | ACYP2 | FALSE | Downregulated |
| -0,14012563 | 0,696602324 | ALS2 | FALSE | Downregulated |
| -0,205534741 | 0,696602324 | CDC27 | FALSE | Downregulated |
| -0,533163787 | 0,696602324 | LOC101060632 | FALSE | Downregulated |
| 0,328859455 | 0,696602324 | MOB3B | FALSE | Upregulated |
| -0,357049351 | 0,696648402 | C9orf72 | FALSE | Downregulated |
| -0,131924366 | 0,696648402 | GHITM | FALSE | Downregulated |
| 0,38909847 | 0,696648402 | HCG9 | FALSE | Upregulated |
| -0,225817906 | 0,696648402 | PIGG | FALSE | Downregulated |
| 0,136727249 | 0,696648402 | SART3 | FALSE | Upregulated |
| 0,209634272 | 0,69665737 | SH3BP4 | FALSE | Upregulated |
| 0,18190086 | 0,696657392 | SERHL2 | FALSE | Upregulated |
| -0,214335299 | 0,696657392 | TCF4 | FALSE | Downregulated |
| -0,223820501 | 0,697431917 | NBPF12 | FALSE | Downregulated |
| 0,198334607 | 0,697431917 | PIRT | FALSE | Upregulated |
| -0,487094214 | 0,697533302 | SMPD2 | FALSE | Downregulated |
| -0,357936865 | 0,697562978 | DGKG | FALSE | Downregulated |
| -0,3628876 | 0,697562978 | FAM78B | FALSE | Downregulated |
| 0,377963866 | 0,697562978 | KLHL7 | FALSE | Upregulated |
| -0,307737832 | 0,697562978 | LOC101927181 | FALSE | Downregulated |
| -0,222074166 | 0,697562978 | LOC101928101 | FALSE | Downregulated |
| 0,167327251 | 0,697562978 | PPP3R1 | FALSE | Upregulated |
| -0,1698414 | 0,697654757 | ATP9B | FALSE | Downregulated |
| -0,144685386 | 0,697654757 | LINC00674 | FALSE | Downregulated |
| -0,797353327 | 0,697654757 | OR10A2 | FALSE | Downregulated |
| -0,373811889 | 0,697654757 | PPOX | FALSE | Downregulated |
| -0,234379859 | 0,697654757 | ZNF721 | FALSE | Downregulated |
| -0,123700981 | 0,697733127 | APMAP | FALSE | Downregulated |
| -0,309229267 | 0,697733127 | POP7 | FALSE | Downregulated |
| 0,1717649 | 0,69775668 | MAPKAP1 | FALSE | Upregulated |
| -0,375729383 | 0,697779237 | LOC101927206 | FALSE | Downregulated |
| 0,214697411 | 0,697913033 | CSTF3 | FALSE | Upregulated |
| 0,492259688 | 0,697913033 | DSCC1 | FALSE | Upregulated |
| 0,99045899 | 0,697913033 | HNRNPH2 | FALSE | Upregulated |
| -0,138215897 | 0,697913033 | INVS | FALSE | Downregulated |
| 0,246818533 | 0,697913033 | MUC5B | FALSE | Upregulated |
| 0,490155961 | 0,697913033 | ODF3 | FALSE | Upregulated |
| 0,25822075 | 0,697913033 | POMT2 | FALSE | Upregulated |

| | | | | |
|---|---|---|---|---|
| 0,134411181 | 0,697913033 | PRDM2 | FALSE | Upregulated |
| -0,131503164 | 0,697913033 | PRR14L | FALSE | Downregulated |
| 0,468929779 | 0,697913033 | RNF157 | FALSE | Upregulated |
| 0,366461935 | 0,697913033 | TXNDC9 | FALSE | Upregulated |
| 0,124763797 | 0,697913033 | ZBTB11 | FALSE | Upregulated |
| -0,600863019 | 0,698020622 | LINC00893 | FALSE | Downregulated |
| 0,10084449 | 0,698037885 | CGNL1 | FALSE | Upregulated |
| 0,172436493 | 0,698037885 | MAPK1IP1L | FALSE | Upregulated |
| -0,263977646 | 0,698135743 | MANBA | FALSE | Downregulated |
| 0,293123656 | 0,698135743 | PPME1 | FALSE | Upregulated |
| 0,25728729 | 0,698135743 | STON2 | FALSE | Upregulated |
| 0,267493927 | 0,698135743 | TIGIT | FALSE | Upregulated |
| -1,248015584 | 0,698365735 | PRH1 | FALSE | Downregulated |
| 0,177842042 | 0,69863362 | ELAVL1 | FALSE | Upregulated |
| 0,136185566 | 0,69863362 | HDAC4 | FALSE | Upregulated |
| 0,149209724 | 0,69863362 | KBTBD4 | FALSE | Upregulated |
| 0,47220973 | 0,69863362 | LAG3 | FALSE | Upregulated |
| -0,44303295 | 0,69863362 | MMP21 | FALSE | Downregulated |
| -0,192283357 | 0,69863362 | MRPL4 | FALSE | Downregulated |
| 0,22263069 | 0,69863362 | NUPL1 | FALSE | Upregulated |
| 0,307964385 | 0,69863362 | PHYHIPL | FALSE | Upregulated |
| 0,526541443 | 0,698682848 | CA11 | FALSE | Upregulated |
| 0,550282039 | 0,698682848 | LINC00535 | FALSE | Upregulated |
| -0,254070667 | 0,698682848 | NEU3 | FALSE | Downregulated |
| -0,295849076 | 0,698682848 | RASSF1 | FALSE | Downregulated |
| -0,121724987 | 0,69872634 | ACSL5 | FALSE | Downregulated |
| -0,081550223 | 0,698729016 | EPN1 | FALSE | Downregulated |
| -0,137895925 | 0,698864034 | AURKAIP1 | FALSE | Downregulated |
| -0,118750844 | 0,698864034 | OSBPL9 | FALSE | Downregulated |
| -0,177692341 | 0,698966759 | MLLT10 | FALSE | Downregulated |
| 0,163040603 | 0,69945029 | PDZD9 | FALSE | Upregulated |
| -0,162731972 | 0,699534102 | DLD | FALSE | Downregulated |
| 0,201121931 | 0,699534102 | FANCF | FALSE | Upregulated |
| 0,329429931 | 0,699534102 | FGF14 | FALSE | Upregulated |
| -0,7972457 | 0,699534102 | GRM3 | FALSE | Downregulated |
| -0,145210895 | 0,699534102 | INADL | FALSE | Downregulated |
| -0,15888439 | 0,699539398 | C1QTNF1 | FALSE | Downregulated |
| -0,334730435 | 0,699539398 | GCSHP3 | FALSE | Downregulated |
| 0,595407121 | 0,699539398 | HSPE1-MOB4 | FALSE | Upregulated |
| 0,324106554 | 0,699539398 | LOC100499489 | FALSE | Upregulated |
| -0,140257866 | 0,699539398 | SECISBP2L | FALSE | Downregulated |
| -0,90035656 | 0,699539398 | TMEM56-RWDD3 | FALSE | Downregulated |
| 0,216381655 | 0,699651767 | AFAP1-AS1 | FALSE | Upregulated |
| 0,371131112 | 0,699651767 | ASF1B | FALSE | Upregulated |
| 0,254470184 | 0,699651767 | ESF1 | FALSE | Upregulated |
| -0,218794001 | 0,699651767 | MAGEF1 | FALSE | Downregulated |
| -0,256223824 | 0,699651767 | NAA35 | FALSE | Downregulated |
| -0,161663386 | 0,699651767 | SALL1 | FALSE | Downregulated |
| 0,150925066 | 0,699651767 | SRP14 | FALSE | Upregulated |
| -0,488496089 | 0,699668854 | PRELID2 | FALSE | Downregulated |
| 0,726946473 | 0,699772827 | CLIC6 | FALSE | Upregulated |

| | | | | |
|---|---|---|---|---|
| 0,209285876 | 0,699772827 | UPF3A | FALSE | Upregulated |
| -0,927743976 | 0,699833679 | AC004053.1 | FALSE | Downregulated |
| -0,221936413 | 0,699833679 | ALG2 | FALSE | Downregulated |
| -0,1309797 | 0,699833679 | BCL2L13 | FALSE | Downregulated |
| -0,447804424 | 0,699833679 | DACH1 | FALSE | Downregulated |
| -0,254592239 | 0,699833679 | EXOSC2 | FALSE | Downregulated |
| 0,290908324 | 0,699833679 | GGCT | FALSE | Upregulated |
| -0,648808175 | 0,699833679 | KCNH6 | FALSE | Downregulated |
| -0,138253449 | 0,699833679 | N4BP2L2 | FALSE | Downregulated |
| -0,16974465 | 0,699833679 | NUP210 | FALSE | Downregulated |
| -0,171172415 | 0,699833679 | RP11-57H12.3 | FALSE | Downregulated |
| 0,262685674 | 0,699833679 | SH2D3C | FALSE | Upregulated |
| -0,253640109 | 0,699833679 | SOX6 | FALSE | Downregulated |
| -0,187960083 | 0,699833679 | TADA1 | FALSE | Downregulated |
| -0,184710301 | 0,699833679 | TTLL12 | FALSE | Downregulated |
| -0,139026937 | 0,699952425 | GOLGA1 | FALSE | Downregulated |
| 0,546801997 | 0,699981847 | GAL3ST1 | FALSE | Upregulated |
| 0,683554299 | 0,699981847 | LOC100131257 | FALSE | Upregulated |
| 0,497959067 | 0,700033163 | FMR1-AS1 | FALSE | Upregulated |
| -0,340383144 | 0,700033163 | LOC102724974 | FALSE | Downregulated |
| -0,153916988 | 0,700033163 | PTPRM | FALSE | Downregulated |
| -0,344208091 | 0,700033163 | RAPGEFL1 | FALSE | Downregulated |
| 0,180937794 | 0,700033163 | ZDHHC20 | FALSE | Upregulated |
| 0,16903889 | 0,700103811 | ADORA2A | FALSE | Upregulated |
| 0,167121752 | 0,700196954 | LDLRAD2 | FALSE | Upregulated |
| -0,593456478 | 0,700497612 | SLC5A12 | FALSE | Downregulated |
| -0,278035611 | 0,700592782 | SF3B5 | FALSE | Downregulated |
| -0,209107758 | 0,700929856 | DERA | FALSE | Downregulated |
| -0,369436332 | 0,700929856 | LOC101928997 | FALSE | Downregulated |
| 0,300856134 | 0,700930388 | PTK6 | FALSE | Upregulated |
| 0,140286822 | 0,700954451 | ZNF672 | FALSE | Upregulated |
| 0,140048579 | 0,70096324 | IRF3 | FALSE | Upregulated |
| 0,241779895 | 0,70096324 | KIAA1614 | FALSE | Upregulated |
| -0,444900013 | 0,700997338 | JMJD7 | FALSE | Downregulated |
| 0,163708462 | 0,701159416 | ATP5SL | FALSE | Upregulated |
| 0,19359286 | 0,701166696 | C18orf25 | FALSE | Upregulated |
| -0,088484773 | 0,701166696 | NEK6 | FALSE | Downregulated |
| 0,2025147 | 0,701497859 | ZNF44 | FALSE | Upregulated |
| 0,209116313 | 0,701645814 | LAMA2 | FALSE | Upregulated |
| 0,115466116 | 0,701645814 | PNN | FALSE | Upregulated |
| -0,146533623 | 0,701645814 | RBM5 | FALSE | Downregulated |
| 0,163996356 | 0,701707218 | ZSCAN32 | FALSE | Upregulated |
| 0,353450918 | 0,701737637 | AC010226.4 | FALSE | Upregulated |
| -0,501910356 | 0,701737637 | CCDC183 | FALSE | Downregulated |
| -0,26801287 | 0,701737637 | COA3 | FALSE | Downregulated |
| 0,171442715 | 0,701737637 | DIS3 | FALSE | Upregulated |
| 0,257259419 | 0,701737637 | GGT7 | FALSE | Upregulated |
| 0,368050703 | 0,701737637 | SRGAP2C | FALSE | Upregulated |
| -0,4323658 | 0,701779041 | COQ2 | FALSE | Downregulated |
| 0,258974432 | 0,701779041 | COX19 | FALSE | Upregulated |
| 0,556462436 | 0,701779041 | LOC102724841 | FALSE | Upregulated |

| | | | | |
|---|---|---|---|---|
| 0,364310225 | 0,701779041 | MAB21L1 | FALSE | Upregulated |
| -0,119799596 | 0,701779041 | MICAL3 | FALSE | Downregulated |
| -0,385547832 | 0,702115691 | C1orf174 | FALSE | Downregulated |
| -0,089127231 | 0,702115691 | CTBP1 | FALSE | Downregulated |
| 0,331610922 | 0,702115691 | ZNF708 | FALSE | Upregulated |
| 0,15860821 | 0,702171187 | PLEKHM1 | FALSE | Upregulated |
| 0,600348337 | 0,70219145 | ABCA17P | FALSE | Upregulated |
| -1,224085097 | 0,70219145 | MOB4 | FALSE | Downregulated |
| -0,350951799 | 0,70219145 | ODF2L | FALSE | Downregulated |
| 0,259284447 | 0,70219145 | PSMA4 | FALSE | Upregulated |
| -0,169903943 | 0,702323987 | IP6K1 | FALSE | Downregulated |
| -0,264844023 | 0,702468889 | DHRSX | FALSE | Downregulated |
| -0,272936387 | 0,702582674 | CCDC91 | FALSE | Downregulated |
| -0,062364491 | 0,702590349 | PRRC2B | FALSE | Downregulated |
| 0,612507235 | 0,702658877 | AC018742.1 | FALSE | Upregulated |
| -0,199165488 | 0,702658877 | FAM136A | FALSE | Downregulated |
| 0,290006849 | 0,702658877 | RAD51B | FALSE | Upregulated |
| -0,188188179 | 0,702772259 | ING1 | FALSE | Downregulated |
| 0,510211286 | 0,702818414 | LINC00960 | FALSE | Upregulated |
| -0,400113398 | 0,702834643 | C17orf96 | FALSE | Downregulated |
| 0,204236959 | 0,702834643 | LRRC2 | FALSE | Upregulated |
| 0,260631694 | 0,702834643 | RPS2P32 | FALSE | Upregulated |
| 0,282343745 | 0,702834643 | SLC6A4 | FALSE | Upregulated |
| 0,265385434 | 0,702834643 | TSPAN17 | FALSE | Upregulated |
| -0,483898174 | 0,70284203 | ITGAD | FALSE | Downregulated |
| 0,133482481 | 0,702852147 | API5 | FALSE | Upregulated |
| 0,172206946 | 0,702852147 | NEO1 | FALSE | Upregulated |
| 0,611951871 | 0,702852147 | OR2B2 | FALSE | Upregulated |
| 0,534343927 | 0,702937662 | LOC101929007 | FALSE | Upregulated |
| 0,52004325 | 0,703016688 | FNDC1 | FALSE | Upregulated |
| 0,517272965 | 0,703143635 | FGFBP3 | FALSE | Upregulated |
| -0,581002056 | 0,703143635 | RP11-554A11.8 | FALSE | Downregulated |
| 0,133546774 | 0,703183514 | CCRN4L | FALSE | Upregulated |
| 0,165128781 | 0,703263706 | SUV420H1 | FALSE | Upregulated |
| 0,340598797 | 0,703295974 | CREM | FALSE | Upregulated |
| 0,334606695 | 0,703379504 | VIMP | FALSE | Upregulated |
| 0,324778959 | 0,703676185 | C1orf210 | FALSE | Upregulated |
| -0,252324186 | 0,703676185 | NAA16 | FALSE | Downregulated |
| 0,225329338 | 0,703676185 | RSL24D1 | FALSE | Upregulated |
| -0,289091278 | 0,703778129 | LRRC16A | FALSE | Downregulated |
| 0,5029695 | 0,703778129 | OR7E2P | FALSE | Upregulated |
| -0,563766611 | 0,703778129 | PROM2 | FALSE | Downregulated |
| 0,147907206 | 0,703901026 | H2AFV | FALSE | Upregulated |
| 0,143814384 | 0,703901026 | SMARCD2 | FALSE | Upregulated |
| 0,121012283 | 0,703901026 | TRAPPC12 | FALSE | Upregulated |
| -0,150040438 | 0,703959615 | LEMD3 | FALSE | Downregulated |
| -0,213876556 | 0,703959615 | REXO4 | FALSE | Downregulated |
| -0,655443737 | 0,70437452 | C20orf62 | FALSE | Downregulated |
| -0,173258948 | 0,704503681 | GOT1 | FALSE | Downregulated |
| 0,297070221 | 0,704503681 | RAB2B | FALSE | Upregulated |
| -0,235584033 | 0,704503681 | SLC4A1AP | FALSE | Downregulated |

| | | | | |
|---|---|---|---|---|
| -0,236955661 | 0,704503681 | WDR3 | FALSE | Downregulated |
| 0,226250315 | 0,704602943 | NCOA5 | FALSE | Upregulated |
| -0,192200624 | 0,704602943 | SLC30A9 | FALSE | Downregulated |
| -0,14545018 | 0,704639927 | FOXJ3 | FALSE | Downregulated |
| 0,095615582 | 0,704639927 | GNB2 | FALSE | Upregulated |
| -0,163789925 | 0,704639927 | GPRASP1 | FALSE | Downregulated |
| -0,1379144 | 0,704639927 | NR2F1 | FALSE | Downregulated |
| 0,33329758 | 0,704639927 | S1PR3 | FALSE | Upregulated |
| -0,421161787 | 0,704639927 | SNORD22 | FALSE | Downregulated |
| 0,169838629 | 0,704709242 | PARD3B | FALSE | Upregulated |
| -0,678667102 | 0,704774061 | PTGES2-AS1 | FALSE | Downregulated |
| -0,134477925 | 0,704774061 | ZCCHC11 | FALSE | Downregulated |
| 0,162247607 | 0,705043355 | GRPEL1 | FALSE | Upregulated |
| -0,556370454 | 0,705043355 | LOC101929433 | FALSE | Downregulated |
| 0,199754859 | 0,705108843 | SERPINH1 | FALSE | Upregulated |
| -0,327076473 | 0,705108843 | TTC12 | FALSE | Downregulated |
| 0,231302419 | 0,705113044 | ALOX12P2 | FALSE | Upregulated |
| 0,334925884 | 0,705125493 | KBTBD12 | FALSE | Upregulated |
| 0,40429815 | 0,705125493 | MTFP1 | FALSE | Upregulated |
| -0,423662617 | 0,705125493 | PELI2 | FALSE | Downregulated |
| 0,148953508 | 0,705125493 | PROSER2 | FALSE | Upregulated |
| -0,20687254 | 0,705125493 | RNASE13 | FALSE | Downregulated |
| -0,384017375 | 0,705333085 | LOC100505570 | FALSE | Downregulated |
| 0,348594554 | 0,705360839 | CYP11A1 | FALSE | Upregulated |
| -0,554315584 | 0,705360839 | LOC728323 | FALSE | Downregulated |
| -0,101473569 | 0,705360839 | PJA2 | FALSE | Downregulated |
| -0,131837054 | 0,705360839 | RNF187 | FALSE | Downregulated |
| -0,285815454 | 0,705360839 | RP2 | FALSE | Downregulated |
| 0,120953904 | 0,705360839 | ZCCHC2 | FALSE | Upregulated |
| -0,352197489 | 0,705360839 | ZNF10 | FALSE | Downregulated |
| 0,337255544 | 0,705623868 | DDI1 | FALSE | Upregulated |
| 0,704607897 | 0,705623868 | SRGAP2B | FALSE | Upregulated |
| 0,119453559 | 0,70573264 | GORASP1 | FALSE | Upregulated |
| 0,189542497 | 0,70573264 | SDCCAG8 | FALSE | Upregulated |
| -0,165628976 | 0,705858495 | CYP2U1 | FALSE | Downregulated |
| -0,285297611 | 0,705858495 | GMPPA | FALSE | Downregulated |
| -0,318203621 | 0,705858495 | LIAS | FALSE | Downregulated |
| 0,421693662 | 0,705858495 | LOC101928820 | FALSE | Upregulated |
| -0,287275899 | 0,705858495 | MTMR9LP | FALSE | Downregulated |
| -0,072526918 | 0,705858495 | NUMA1 | FALSE | Downregulated |
| -0,660906143 | 0,705858495 | RRN3P1 | FALSE | Downregulated |
| 0,365020526 | 0,705867658 | NOSTRIN | FALSE | Upregulated |
| 0,172840887 | 0,706023238 | RCOR1 | FALSE | Upregulated |
| 0,134923216 | 0,706210955 | ASCC1 | FALSE | Upregulated |
| -0,187025595 | 0,706210955 | GRSF1 | FALSE | Downregulated |
| 0,145677465 | 0,706210955 | SGSM3 | FALSE | Upregulated |
| -0,224308437 | 0,706210955 | SPP2 | FALSE | Downregulated |
| 0,19954458 | 0,706284724 | SMAD1-AS2 | FALSE | Upregulated |
| -0,234239726 | 0,706360348 | MINPP1 | FALSE | Downregulated |
| 0,15402713 | 0,706360348 | NHP2 | FALSE | Upregulated |
| -0,199055092 | 0,706360348 | RAB11FIP5 | FALSE | Downregulated |

| | | | | |
|---|---|---|---|---|
| 0,458269596 | 0,706375103 | CLSTN2 | FALSE | Upregulated |
| 0,298594947 | 0,706684381 | NAA38 | FALSE | Upregulated |
| 0,288235385 | 0,706843584 | TMEM60 | FALSE | Upregulated |
| -0,410286472 | 0,706896689 | C1orf109 | FALSE | Downregulated |
| 0,278550715 | 0,706899836 | NPL | FALSE | Upregulated |
| -0,207142987 | 0,706899836 | STAG3L3 | FALSE | Downregulated |
| 0,149316821 | 0,707030447 | POU2F1 | FALSE | Upregulated |
| -0,181709759 | 0,707030447 | VDAC2 | FALSE | Downregulated |
| 0,098383037 | 0,707092756 | ADCY9 | FALSE | Upregulated |
| -0,29141536 | 0,707092756 | LOC646762 | FALSE | Downregulated |
| 0,300619918 | 0,707189449 | RPP25 | FALSE | Upregulated |
| -0,383370395 | 0,707416301 | BET1 | FALSE | Downregulated |
| -0,14059224 | 0,707421347 | KLHL24 | FALSE | Downregulated |
| 0,309518748 | 0,707561486 | BRE | FALSE | Upregulated |
| 0,213568729 | 0,707561486 | CDC16 | FALSE | Upregulated |
| 0,583972885 | 0,707561486 | LOC101929480 | FALSE | Upregulated |
| -0,333127366 | 0,707561486 | LOC102723701 | FALSE | Downregulated |
| 0,125909877 | 0,707561486 | LRIG1 | FALSE | Upregulated |
| 0,557888645 | 0,707561486 | MROH9 | FALSE | Upregulated |
| 0,185443245 | 0,707561486 | RFX7 | FALSE | Upregulated |
| 0,37337107 | 0,707561486 | RIOK1 | FALSE | Upregulated |
| 0,175309825 | 0,707561486 | RPS6KA2 | FALSE | Upregulated |
| 0,174319435 | 0,707561486 | TM4SF4 | FALSE | Upregulated |
| -0,673293504 | 0,707599022 | SLC45A2 | FALSE | Downregulated |
| -0,55418973 | 0,707599022 | ZNF502 | FALSE | Downregulated |
| -0,250096637 | 0,707842728 | TMEM170B | FALSE | Downregulated |
| 0,669162998 | 0,707874514 | LOC101929042 | FALSE | Upregulated |
| -0,247728715 | 0,707878875 | MTX3 | FALSE | Downregulated |
| -0,11546852 | 0,707878875 | SMAD4 | FALSE | Downregulated |
| -0,370054584 | 0,707895957 | COMMD1 | FALSE | Downregulated |
| 0,177009619 | 0,707895957 | CPSF4 | FALSE | Upregulated |
| -0,186517616 | 0,707895957 | KXD1 | FALSE | Downregulated |
| 0,109084352 | 0,707895957 | LIMD1 | FALSE | Upregulated |
| -0,73467255 | 0,707895957 | LOC101929796 | FALSE | Downregulated |
| -0,501677484 | 0,707895957 | NBPF3 | FALSE | Downregulated |
| 0,235255077 | 0,707895957 | RALB | FALSE | Upregulated |
| 0,227451374 | 0,707895957 | TSSC4 | FALSE | Upregulated |
| 0,37122682 | 0,707922552 | HCCS | FALSE | Upregulated |
| -0,110744704 | 0,70813129 | NMT1 | FALSE | Downregulated |
| -0,252592393 | 0,708307386 | CELSR2 | FALSE | Downregulated |
| -0,317808208 | 0,708457599 | MLLT10P1 | FALSE | Downregulated |
| 0,311071138 | 0,7086703 | HOMER3 | FALSE | Upregulated |
| 0,838076908 | 0,7086703 | SEPT5-GP1BB | FALSE | Upregulated |
| -0,607901634 | 0,7086703 | SNORA23 | FALSE | Downregulated |
| 0,350810676 | 0,708807063 | C11orf84 | FALSE | Upregulated |
| -0,125574572 | 0,708901048 | FEM1B | FALSE | Downregulated |
| -0,140096487 | 0,708901048 | PITRM1 | FALSE | Downregulated |
| 0,145007746 | 0,708901048 | RAB10 | FALSE | Upregulated |
| 0,421375751 | 0,708901048 | ZNRD1 | FALSE | Upregulated |
| 0,580310329 | 0,708909573 | LOC101060195 | FALSE | Upregulated |
| -0,502043125 | 0,708909573 | LOC101928351 | FALSE | Downregulated |

| | | | | |
|---|---|---|---|---|
| 0,627294052 | 0,709006665 | ANXA2R | FALSE | Upregulated |
| -0,145583206 | 0,709059047 | EGFL7 | FALSE | Downregulated |
| -0,526581297 | 0,709059047 | ZNRD1-AS1 | FALSE | Downregulated |
| 0,442114027 | 0,709137213 | DGCR10 | FALSE | Upregulated |
| 0,308358194 | 0,709311583 | FCGR2B | FALSE | Upregulated |
| -0,218557177 | 0,709863292 | EIF4EBP1 | FALSE | Downregulated |
| -0,25466927 | 0,70987985 | ZNF558 | FALSE | Downregulated |
| 0,139094073 | 0,710255563 | WDR45B | FALSE | Upregulated |
| -0,313670972 | 0,71038288 | PZP | FALSE | Downregulated |
| 0,398282815 | 0,710548358 | IL21R | FALSE | Upregulated |
| -0,63787105 | 0,710591499 | LOC283887 | FALSE | Downregulated |
| -0,107775711 | 0,710591499 | MGEA5 | FALSE | Downregulated |
| 0,513560785 | 0,710684365 | BARD1 | FALSE | Upregulated |
| -0,298385644 | 0,710684365 | CSTF1 | FALSE | Downregulated |
| 0,203785481 | 0,710684365 | DNAJA4 | FALSE | Upregulated |
| 0,077603173 | 0,710684365 | FAXDC2 | FALSE | Upregulated |
| 0,331787016 | 0,710684365 | LOC102724788 | FALSE | Upregulated |
| 0,649722231 | 0,710684365 | MCOLN3 | FALSE | Upregulated |
| 0,224051879 | 0,710684365 | NOL10 | FALSE | Upregulated |
| 0,345252129 | 0,710684365 | RSPH4A | FALSE | Upregulated |
| 0,176229152 | 0,710684365 | SLC16A8 | FALSE | Upregulated |
| 0,328054095 | 0,711121323 | ZNF235 | FALSE | Upregulated |
| -0,647624956 | 0,711339208 | LINC01124 | FALSE | Downregulated |
| 0,522261201 | 0,711491843 | TRPM1 | FALSE | Upregulated |
| -0,275360433 | 0,711614277 | TBPL1 | FALSE | Downregulated |
| -0,170637507 | 0,711986728 | METTL3 | FALSE | Downregulated |
| 0,236686909 | 0,712024899 | IQCE | FALSE | Upregulated |
| -0,153706772 | 0,712024899 | ZNF671 | FALSE | Downregulated |
| 0,129062851 | 0,712115641 | DNM1L | FALSE | Upregulated |
| -0,166128731 | 0,712639974 | PHACTR4 | FALSE | Downregulated |
| 0,237047647 | 0,712961374 | STX6 | FALSE | Upregulated |
| 0,158103156 | 0,712988454 | LOC101928817 | FALSE | Upregulated |
| 0,145348891 | 0,712988454 | PAK2 | FALSE | Upregulated |
| -0,112735345 | 0,713143677 | HEXIM2 | FALSE | Downregulated |
| 0,220459872 | 0,713329515 | DHX58 | FALSE | Upregulated |
| 0,173300822 | 0,713353753 | B3GNT8 | FALSE | Upregulated |
| -0,217787675 | 0,713353753 | MAP2K4 | FALSE | Downregulated |
| 0,161854692 | 0,713353753 | STARD9 | FALSE | Upregulated |
| 0,194027827 | 0,713353753 | TXNDC15 | FALSE | Upregulated |
| 0,110515162 | 0,713503822 | RNPEPL1 | FALSE | Upregulated |
| 0,145003734 | 0,713734543 | TMEM8A | FALSE | Upregulated |
| -0,181979312 | 0,713966555 | HDAC10 | FALSE | Downregulated |
| 0,226732375 | 0,713966555 | LOC101930171 | FALSE | Upregulated |
| -0,461477646 | 0,713966555 | NEDD1 | FALSE | Downregulated |
| -0,388355407 | 0,713966555 | POLB | FALSE | Downregulated |
| 0,35543877 | 0,713966555 | SERTAD1 | FALSE | Upregulated |
| -0,262932883 | 0,713966555 | ZNF250 | FALSE | Downregulated |
| -0,340722563 | 0,713966555 | ZW10 | FALSE | Downregulated |
| 0,164395489 | 0,714193097 | SMAD5 | FALSE | Upregulated |
| 0,156887445 | 0,714233852 | KGFLP2 | FALSE | Upregulated |
| -0,248802439 | 0,714233852 | POMGNT2 | FALSE | Downregulated |

| | | | | |
|---|---|---|---|---|
| -0,522451047 | 0,714395501 | ANKDD1B | FALSE | Downregulated |
| -0,468728523 | 0,714395501 | ATAD3C | FALSE | Downregulated |
| 0,214819217 | 0,714395501 | CHSY1 | FALSE | Upregulated |
| -0,110595211 | 0,714395501 | COBL | FALSE | Downregulated |
| -0,178948291 | 0,714395501 | DDX18 | FALSE | Downregulated |
| 0,27065055 | 0,714395501 | DTNB | FALSE | Upregulated |
| 0,352356667 | 0,714395501 | IFT27 | FALSE | Upregulated |
| -1,980320115 | 0,714395501 | LOC102724219 | FALSE | Downregulated |
| 0,372966026 | 0,714395501 | LOC102724976 | FALSE | Upregulated |
| -0,235391231 | 0,714395501 | MALT1 | FALSE | Downregulated |
| -0,398017121 | 0,714395501 | STK11IP | FALSE | Downregulated |
| 0,222581873 | 0,714395501 | SYT7 | FALSE | Upregulated |
| -0,604469646 | 0,714395501 | TENM3 | FALSE | Downregulated |
| -0,665448142 | 0,714426869 | ZNF239 | FALSE | Downregulated |
| -0,125845116 | 0,71461029 | SLC25A22 | FALSE | Downregulated |
| -0,432729805 | 0,714997066 | ANKRD35 | FALSE | Downregulated |
| 0,149270453 | 0,714997066 | DNAJB5 | FALSE | Upregulated |
| -0,241017482 | 0,714997066 | LOC101929173 | FALSE | Downregulated |
| 0,261233846 | 0,714997066 | OSGEP | FALSE | Upregulated |
| -0,171055352 | 0,715261987 | HEXA | FALSE | Downregulated |
| 0,242516891 | 0,71539509 | SWAP70 | FALSE | Upregulated |
| 0,342777928 | 0,715419153 | ARV1 | FALSE | Upregulated |
| -0,118401114 | 0,715487057 | CTSH | FALSE | Downregulated |
| -0,167854597 | 0,715522528 | KLHL21 | FALSE | Downregulated |
| 0,37280358 | 0,715522528 | MMP23B | FALSE | Upregulated |
| 0,286096477 | 0,715564924 | PTPLB | FALSE | Upregulated |
| 0,243227558 | 0,715903959 | PSMA3 | FALSE | Upregulated |
| -0,159173317 | 0,716005724 | EXOSC6 | FALSE | Downregulated |
| 0,405460537 | 0,716106157 | MMS22L | FALSE | Upregulated |
| 0,16111907 | 0,716304327 | TTI2 | FALSE | Upregulated |
| 0,193518963 | 0,716490409 | TIRAP | FALSE | Upregulated |
| 0,588218688 | 0,716573774 | LOC100505938 | FALSE | Upregulated |
| 0,719969954 | 0,716573774 | LOC102031319 | FALSE | Upregulated |
| 0,255907041 | 0,716629762 | DIMT1 | FALSE | Upregulated |
| -0,268689449 | 0,716725258 | PWP1 | FALSE | Downregulated |
| 0,194870709 | 0,716877859 | ACOT4 | FALSE | Upregulated |
| 0,2848963 | 0,716877859 | AK1 | FALSE | Upregulated |
| -0,068354485 | 0,716877859 | HIPK2 | FALSE | Downregulated |
| 0,289752832 | 0,716877859 | POMZP3 | FALSE | Upregulated |
| 0,177214605 | 0,716877859 | SH3GLB2 | FALSE | Upregulated |
| -0,097281017 | 0,716877859 | STARD10 | FALSE | Downregulated |
| 0,232989248 | 0,716877859 | UBALD1 | FALSE | Upregulated |
| 0,299663621 | 0,716999813 | RAP1GAP2 | FALSE | Upregulated |
| -0,184092454 | 0,717101705 | MRPL51 | FALSE | Downregulated |
| 0,344660517 | 0,717155662 | NEK8 | FALSE | Upregulated |
| 0,293901648 | 0,717226448 | LOC101929479 | FALSE | Upregulated |
| 0,166676796 | 0,717294868 | ABHD12B | FALSE | Upregulated |
| 0,177778334 | 0,717294868 | ZNF692 | FALSE | Upregulated |
| -0,449406144 | 0,717320713 | CTC-558O2.1 | FALSE | Downregulated |
| 0,311328529 | 0,717320713 | INCA1 | FALSE | Upregulated |
| 0,160081051 | 0,717320713 | WRNIP1 | FALSE | Upregulated |

| | | | | |
|---|---|---|---|---|
| -0,420301842 | 0,717560611 | ACSL6 | FALSE | Downregulated |
| 0,169573414 | 0,717560611 | HIC1 | FALSE | Upregulated |
| 0,383083457 | 0,717560611 | RAD18 | FALSE | Upregulated |
| -0,130812573 | 0,717560611 | RTN3 | FALSE | Downregulated |
| 0,203054039 | 0,717560611 | SAE1 | FALSE | Upregulated |
| -0,195637054 | 0,717651565 | CMTM8 | FALSE | Downregulated |
| -0,272042293 | 0,717651565 | DPH2 | FALSE | Downregulated |
| -0,15900445 | 0,717651565 | HLF | FALSE | Downregulated |
| 0,424570702 | 0,717651565 | XXYLT1 | FALSE | Upregulated |
| -0,18685974 | 0,717732942 | STX3 | FALSE | Downregulated |
| 0,214726637 | 0,718198203 | ARL6IP5 | FALSE | Upregulated |
| -0,169429327 | 0,718198203 | GALNT1 | FALSE | Downregulated |
| -0,693460495 | 0,718198203 | NR2F2-AS1 | FALSE | Downregulated |
| 0,357807249 | 0,718198203 | RP11-333E1.1 | FALSE | Upregulated |
| 0,191791688 | 0,71826769 | ATAT1 | FALSE | Upregulated |
| 0,554386191 | 0,718482858 | ABCC6P2 | FALSE | Upregulated |
| 0,423473293 | 0,718997484 | LOC102723306 | FALSE | Upregulated |
| -0,513103333 | 0,719017049 | LOC101927032 | FALSE | Downregulated |
| -0,681438398 | 0,719192022 | ASCL3 | FALSE | Downregulated |
| 0,26526569 | 0,719192022 | CACNB3 | FALSE | Upregulated |
| -0,312870287 | 0,719192022 | CTNNAL1 | FALSE | Downregulated |
| 0,3380679 | 0,719192022 | LOC101927002 | FALSE | Upregulated |
| 0,367644464 | 0,719192022 | LRMP | FALSE | Upregulated |
| -0,20264135 | 0,719192022 | NOSIP | FALSE | Downregulated |
| -0,122482364 | 0,719192022 | NUP214 | FALSE | Downregulated |
| -0,505869319 | 0,719192022 | TCEAL1 | FALSE | Downregulated |
| -0,220013062 | 0,719526543 | CA5A | FALSE | Downregulated |
| 0,293686379 | 0,719526543 | LOC100506734 | FALSE | Upregulated |
| 0,180248971 | 0,719526543 | RBPJ | FALSE | Upregulated |
| 0,243270731 | 0,719634495 | AGAP5 | FALSE | Upregulated |
| -0,191763509 | 0,719638802 | PLEKHH3 | FALSE | Downregulated |
| -0,249545224 | 0,719719191 | HDDC3 | FALSE | Downregulated |
| -0,326134203 | 0,719818992 | ZFAS1 | FALSE | Downregulated |
| 0,214070919 | 0,7199851 | ACAP1 | FALSE | Upregulated |
| 0,343848537 | 0,720025745 | C6orf1 | FALSE | Upregulated |
| 0,602882341 | 0,720025745 | CD300E | FALSE | Upregulated |
| 0,229385794 | 0,720025745 | FBXO34 | FALSE | Upregulated |
| 0,297115482 | 0,720025745 | FLJ41481 | FALSE | Upregulated |
| -0,2634423 | 0,720025745 | HDHD1 | FALSE | Downregulated |
| 0,199510173 | 0,720025745 | HP09025 | FALSE | Upregulated |
| 0,284789605 | 0,720025745 | IFT80 | FALSE | Upregulated |
| 0,152503224 | 0,720025745 | KBTBD2 | FALSE | Upregulated |
| 0,131441103 | 0,720025745 | KIAA1033 | FALSE | Upregulated |
| 0,452280916 | 0,720025745 | LDLRAD3 | FALSE | Upregulated |
| -0,450685121 | 0,720025745 | LEPREL2 | FALSE | Downregulated |
| -0,516203249 | 0,720025745 | LOC102723893 | FALSE | Downregulated |
| 0,528595622 | 0,720025745 | LTC4S | FALSE | Upregulated |
| -0,3304571 | 0,720025745 | NUCB2 | FALSE | Downregulated |
| -0,175215059 | 0,720025745 | QSER1 | FALSE | Downregulated |
| 0,158931441 | 0,720025745 | SMAD5-AS1 | FALSE | Upregulated |
| -0,110587985 | 0,720025745 | TOM1L1 | FALSE | Downregulated |

| | | | | |
|---|---|---|---|---|
| 0,135108058 | 0,720056508 | THPO | FALSE | Upregulated |
| 0,148756282 | 0,720140081 | KARS | FALSE | Upregulated |
| 0,304103576 | 0,720528824 | ANKS6 | FALSE | Upregulated |
| 0,206888992 | 0,720780174 | CPEB3 | FALSE | Upregulated |
| -0,289425028 | 0,720783129 | NOL11 | FALSE | Downregulated |
| -0,224640446 | 0,720783129 | TMCO4 | FALSE | Downregulated |
| -0,138354964 | 0,720975196 | SBNO1 | FALSE | Downregulated |
| -0,30347122 | 0,720976723 | NLE1 | FALSE | Downregulated |
| -0,296865857 | 0,720976723 | SHBG | FALSE | Downregulated |
| 0,128792649 | 0,721014554 | RHOU | FALSE | Upregulated |
| -0,263735307 | 0,721113105 | ABCA10 | FALSE | Downregulated |
| 0,174489284 | 0,721113105 | IPO13 | FALSE | Upregulated |
| 0,17228731 | 0,721113105 | ITGA7 | FALSE | Upregulated |
| -0,283332998 | 0,721331294 | MRPL23 | FALSE | Downregulated |
| 0,234128371 | 0,721492705 | GMPS | FALSE | Upregulated |
| 0,290470898 | 0,72169316 | BRMS1 | FALSE | Upregulated |
| 0,245536101 | 0,721703397 | AZI1 | FALSE | Upregulated |
| 0,077874183 | 0,721703397 | CD302 | FALSE | Upregulated |
| 0,145075354 | 0,721703397 | CEP170 | FALSE | Upregulated |
| -0,190730969 | 0,721703397 | COCH | FALSE | Downregulated |
| -0,234972482 | 0,721703397 | DYRK1B | FALSE | Downregulated |
| -0,236179958 | 0,721703397 | GEN1 | FALSE | Downregulated |
| -0,349312351 | 0,721703397 | GMCL1 | FALSE | Downregulated |
| 0,13155724 | 0,721703397 | IL4R | FALSE | Upregulated |
| -0,199474397 | 0,721703397 | KLHL12 | FALSE | Downregulated |
| 0,113196863 | 0,721703397 | KPNA1 | FALSE | Upregulated |
| 0,183613289 | 0,721703397 | LEO1 | FALSE | Upregulated |
| -0,480209831 | 0,721703397 | LOC340074 | FALSE | Downregulated |
| 0,16193501 | 0,721703397 | LRRC39 | FALSE | Upregulated |
| 0,379275584 | 0,721703397 | PSMG1 | FALSE | Upregulated |
| 0,56053612 | 0,721703397 | PTGDR | FALSE | Upregulated |
| 0,119604706 | 0,721703397 | RAI1 | FALSE | Upregulated |
| -0,127075809 | 0,721703397 | SEC23A | FALSE | Downregulated |
| -0,607177012 | 0,72170512 | ACE2 | FALSE | Downregulated |
| 0,555059138 | 0,72170512 | C20orf195 | FALSE | Upregulated |
| 0,165280401 | 0,721731677 | GNPDA2 | FALSE | Upregulated |
| 0,501317178 | 0,721731677 | LOC102723581 | FALSE | Upregulated |
| 0,226273463 | 0,722001933 | LSM8 | FALSE | Upregulated |
| -0,752652344 | 0,722001933 | SNCA | FALSE | Downregulated |
| -0,217700702 | 0,722001933 | SPTLC1 | FALSE | Downregulated |
| -0,363336215 | 0,722128366 | ZKSCAN7 | FALSE | Downregulated |
| -0,375236921 | 0,722188087 | MYL6B | FALSE | Downregulated |
| 0,220482488 | 0,722231619 | SPAG5 | FALSE | Upregulated |
| -0,14326572 | 0,722267244 | LAMB1 | FALSE | Downregulated |
| 0,11258563 | 0,722374562 | NACC2 | FALSE | Upregulated |
| -0,319486805 | 0,722687941 | LOC101928433 | FALSE | Downregulated |
| 0,518164343 | 0,722687941 | ZUFSP | FALSE | Upregulated |
| -0,24066263 | 0,722751785 | SESTD1 | FALSE | Downregulated |
| -0,649633457 | 0,722824157 | RP11-182I10.3 | FALSE | Downregulated |
| 0,188272974 | 0,722824157 | ZNF132 | FALSE | Upregulated |
| 0,095902702 | 0,722859408 | SPG11 | FALSE | Upregulated |

| | | | | |
|---|---|---|---|---|
| -0,166220267 | 0,722897737 | KDELR1 | FALSE | Downregulated |
| 0,411938813 | 0,722897737 | LMTK3 | FALSE | Upregulated |
| -0,355175015 | 0,722897737 | LOC102723997 | FALSE | Downregulated |
| 0,214090347 | 0,722897737 | NRAS | FALSE | Upregulated |
| -0,536889711 | 0,72294822 | EIF3J-AS1 | FALSE | Downregulated |
| -0,194346606 | 0,72294822 | HMCES | FALSE | Downregulated |
| -0,22894746 | 0,72294822 | IFRD1 | FALSE | Downregulated |
| -0,598803499 | 0,72294822 | LOC102724292 | FALSE | Downregulated |
| -0,212047965 | 0,72294822 | RLF | FALSE | Downregulated |
| -0,1203665 | 0,72294822 | STAG1 | FALSE | Downregulated |
| 0,131621856 | 0,72298825 | PALMD | FALSE | Upregulated |
| 0,638122118 | 0,72298825 | ROR2 | FALSE | Upregulated |
| -0,257283074 | 0,72298825 | TUSC2 | FALSE | Downregulated |
| 0,532655411 | 0,723352568 | LOC101929112 | FALSE | Upregulated |
| 0,206696516 | 0,723428843 | ABCC10 | FALSE | Upregulated |
| -0,661100654 | 0,723598556 | C9orf129 | FALSE | Downregulated |
| 0,332353024 | 0,723598556 | LOC101927402 | FALSE | Upregulated |
| -0,336002442 | 0,723822492 | ADCY4 | FALSE | Downregulated |
| -0,282338146 | 0,723833008 | UBXN2B | FALSE | Downregulated |
| -0,199326163 | 0,724297315 | AIP | FALSE | Downregulated |
| -0,158162725 | 0,724297315 | KIN | FALSE | Downregulated |
| -0,202249991 | 0,724297315 | MBD4 | FALSE | Downregulated |
| 0,360768945 | 0,724303237 | GRHL1 | FALSE | Upregulated |
| -0,120053116 | 0,724303237 | NTN1 | FALSE | Downregulated |
| -0,460949901 | 0,724303237 | SUSD2 | FALSE | Downregulated |
| 0,329602143 | 0,724395258 | STAT4 | FALSE | Upregulated |
| 0,494287384 | 0,724452791 | KCNH8 | FALSE | Upregulated |
| -0,150122059 | 0,724467017 | FZR1 | FALSE | Downregulated |
| 0,150834177 | 0,724467017 | ZNF557 | FALSE | Upregulated |
| 0,172401065 | 0,724763319 | LOC102723535 | FALSE | Upregulated |
| 0,11019924 | 0,724763319 | USP54 | FALSE | Upregulated |
| -0,368849593 | 0,724787214 | LOC101928734 | FALSE | Downregulated |
| -0,240778443 | 0,724872837 | DIABLO | FALSE | Downregulated |
| -0,214188762 | 0,724872837 | RPRD1A | FALSE | Downregulated |
| -0,205360837 | 0,724893487 | DHX37 | FALSE | Downregulated |
| -0,207639185 | 0,724893487 | PSMA2 | FALSE | Downregulated |
| 0,525810832 | 0,724970386 | FLVCR1 | FALSE | Upregulated |
| 0,184686367 | 0,724970386 | LOC101928658 | FALSE | Upregulated |
| -0,163217411 | 0,724970386 | NADSYN1 | FALSE | Downregulated |
| -0,091269788 | 0,724970386 | TPCN1 | FALSE | Downregulated |
| 0,327696505 | 0,725036928 | PRR24 | FALSE | Upregulated |
| -0,337152612 | 0,72513812 | NUDT2 | FALSE | Downregulated |
| -0,123163535 | 0,72513812 | PEPD | FALSE | Downregulated |
| -0,361445175 | 0,725266151 | DCAF4L1 | FALSE | Downregulated |
| 0,187966305 | 0,725266151 | DNM3 | FALSE | Upregulated |
| -0,38188761 | 0,725266151 | EGR1 | FALSE | Downregulated |
| -0,153904216 | 0,725266151 | ITFG3 | FALSE | Downregulated |
| -2,089234214 | 0,725266151 | LOC102724956 | FALSE | Downregulated |
| -0,459902669 | 0,725266151 | LOC389765 | FALSE | Downregulated |
| 0,363619804 | 0,725266151 | NXPH4 | FALSE | Upregulated |
| 0,134262826 | 0,725266151 | SLC29A2 | FALSE | Upregulated |

| | | | | |
|---|---|---|---|---|
| 0,278255877 | 0,725266151 | STAU2-AS1 | FALSE | Upregulated |
| -0,10946771 | 0,725359942 | MIEF1 | FALSE | Downregulated |
| 0,130798849 | 0,725359942 | PDS5A | FALSE | Upregulated |
| -0,165677931 | 0,725359942 | ZBTB20-AS1 | FALSE | Downregulated |
| -0,451140373 | 0,725377881 | A4GNT | FALSE | Downregulated |
| -0,681089787 | 0,725377881 | NPIPB8 | FALSE | Downregulated |
| -0,178345538 | 0,725377881 | TCERG1 | FALSE | Downregulated |
| 0,272616649 | 0,725431192 | C8orf47 | FALSE | Upregulated |
| -0,204684038 | 0,725431192 | MRPS7 | FALSE | Downregulated |
| 0,293812747 | 0,725431192 | ZNF486 | FALSE | Upregulated |
| 0,219061338 | 0,725465594 | SLC2A9 | FALSE | Upregulated |
| -0,443173024 | 0,72547159 | CCDC127 | FALSE | Downregulated |
| -0,529322859 | 0,725509675 | GBGT1 | FALSE | Downregulated |
| -0,106220132 | 0,725509675 | KNG1 | FALSE | Downregulated |
| -0,17142584 | 0,725547599 | C19orf24 | FALSE | Downregulated |
| 0,344208597 | 0,725547599 | LOC101928689 | FALSE | Upregulated |
| 0,268678558 | 0,725903165 | PRMT6 | FALSE | Upregulated |
| 0,915970428 | 0,725904544 | CORO7-PAM16 | FALSE | Upregulated |
| -0,667718857 | 0,725904544 | HUNK | FALSE | Downregulated |
| -0,243112687 | 0,726341815 | EFNB2 | FALSE | Downregulated |
| 0,387635273 | 0,72640472 | BMS1P20 | FALSE | Upregulated |
| -0,350122647 | 0,72654283 | NABP2 | FALSE | Downregulated |
| -0,318114942 | 0,72654283 | SCOC | FALSE | Downregulated |
| -0,527913535 | 0,726720619 | ZNF607 | FALSE | Downregulated |
| 0,211004266 | 0,726940201 | NDUFA5 | FALSE | Upregulated |
| 0,129611012 | 0,726986545 | ALDH1A1 | FALSE | Upregulated |
| -0,181026129 | 0,726986545 | COPS7A | FALSE | Downregulated |
| -0,266247632 | 0,726986545 | FGF11 | FALSE | Downregulated |
| -0,114434933 | 0,726986545 | FZD4 | FALSE | Downregulated |
| -0,142205947 | 0,726986545 | GID8 | FALSE | Downregulated |
| 0,259745951 | 0,726986545 | KLHL11 | FALSE | Upregulated |
| 0,314243877 | 0,726986545 | LINC00282 | FALSE | Upregulated |
| 0,166265927 | 0,726986545 | LOC101929751 | FALSE | Upregulated |
| -0,331709681 | 0,726986545 | LOC102723886 | FALSE | Downregulated |
| -0,402002321 | 0,726986545 | NIFK | FALSE | Downregulated |
| -0,454513312 | 0,726986545 | NUDT4P1 | FALSE | Downregulated |
| -0,176333834 | 0,726986545 | OIP5-AS1 | FALSE | Downregulated |
| -0,306309686 | 0,726986545 | PCDH18 | FALSE | Downregulated |
| -0,118565317 | 0,726986545 | TFIP11 | FALSE | Downregulated |
| 0,089869574 | 0,726986545 | TNIP1 | FALSE | Upregulated |
| -0,188199665 | 0,726986545 | ZNF83 | FALSE | Downregulated |
| -0,182576127 | 0,726998305 | AGAP6 | FALSE | Downregulated |
| 0,291307073 | 0,727256501 | LRRC8C | FALSE | Upregulated |
| -0,210571843 | 0,727256501 | PPM1B | FALSE | Downregulated |
| -0,181205876 | 0,727256501 | PPP1R9A | FALSE | Downregulated |
| -0,224983324 | 0,727256501 | RHOBTB1 | FALSE | Downregulated |
| 0,137081403 | 0,727256501 | SNAP29 | FALSE | Upregulated |
| 0,150443613 | 0,727511886 | CAPS | FALSE | Upregulated |
| 0,322128205 | 0,727511886 | LOC101060212 | FALSE | Upregulated |
| 0,474537741 | 0,727511886 | UST | FALSE | Upregulated |
| 0,356230773 | 0,727513975 | LOC101928745 | FALSE | Upregulated |

| | | | | |
|---|---|---|---|---|
| -0,193863302 | 0,727544269 | SCO1 | FALSE | Downregulated |
| 0,174245232 | 0,727583008 | SEC23IP | FALSE | Upregulated |
| 0,084944257 | 0,727609417 | DYNC1H1 | FALSE | Upregulated |
| -0,242416158 | 0,727689193 | MARCH9 | FALSE | Downregulated |
| 0,399697853 | 0,727893319 | LOC101928284 | FALSE | Upregulated |
| 0,227872946 | 0,727893319 | TMEM139 | FALSE | Upregulated |
| -0,339583809 | 0,727895835 | CDADC1 | FALSE | Downregulated |
| -0,207688992 | 0,727895835 | GDAP2 | FALSE | Downregulated |
| -0,297010774 | 0,727895835 | WASF3 | FALSE | Downregulated |
| 0,242959313 | 0,728065505 | NIPAL2 | FALSE | Upregulated |
| -0,271837303 | 0,728098256 | LINC00341 | FALSE | Downregulated |
| 0,094361697 | 0,728302713 | AP1G1 | FALSE | Upregulated |
| -0,12258843 | 0,728302713 | FTL | FALSE | Downregulated |
| 0,742925111 | 0,728302713 | LOC100630923 | FALSE | Upregulated |
| -0,494695193 | 0,728302713 | LOC102724603 | FALSE | Downregulated |
| 0,334120315 | 0,728441117 | CDH13 | FALSE | Upregulated |
| 0,318415418 | 0,728582984 | C1orf226 | FALSE | Upregulated |
| -0,13578258 | 0,728658902 | APBB2 | FALSE | Downregulated |
| -0,183672606 | 0,728658902 | CUEDC2 | FALSE | Downregulated |
| 0,273621208 | 0,728658902 | DTYMK | FALSE | Upregulated |
| -0,173137566 | 0,728658902 | FNTA | FALSE | Downregulated |
| -0,30200585 | 0,728658902 | PA2G4P4 | FALSE | Downregulated |
| 0,123897398 | 0,728658902 | POLD4 | FALSE | Upregulated |
| 0,198307666 | 0,728658902 | SLC19A1 | FALSE | Upregulated |
| 0,092427247 | 0,728705337 | TRIM25 | FALSE | Upregulated |
| 0,217300237 | 0,728722651 | POLR3F | FALSE | Upregulated |
| 0,119634456 | 0,728735378 | HNRNPH1 | FALSE | Upregulated |
| 0,207142109 | 0,728735378 | KCTD17 | FALSE | Upregulated |
| 0,180200192 | 0,728735378 | LUC7L | FALSE | Upregulated |
| 0,226096929 | 0,728735378 | WWC2-AS2 | FALSE | Upregulated |
| 0,43991113 | 0,729007151 | LOC100506859 | FALSE | Upregulated |
| -0,592114795 | 0,729032638 | SNORD89 | FALSE | Downregulated |
| -0,275074893 | 0,729088831 | RAMP3 | FALSE | Downregulated |
| -0,088344048 | 0,729088831 | SSFA2 | FALSE | Downregulated |
| -0,309544559 | 0,729159429 | ENDOV | FALSE | Downregulated |
| -0,139152653 | 0,729188278 | ERLIN1 | FALSE | Downregulated |
| -0,11745108 | 0,729442098 | CCDC88A | FALSE | Downregulated |
| 0,67600383 | 0,729442098 | MOXD1 | FALSE | Upregulated |
| -0,32551051 | 0,729442098 | SAMD12 | FALSE | Downregulated |
| -0,24474126 | 0,729802063 | C1orf123 | FALSE | Downregulated |
| -0,134926651 | 0,729802063 | CCDC93 | FALSE | Downregulated |
| 0,392463657 | 0,729802063 | TRIM16 | FALSE | Upregulated |
| -0,411740917 | 0,72983164 | NDN | FALSE | Downregulated |
| 0,142176549 | 0,729864009 | ZNF217 | FALSE | Upregulated |
| 0,226042331 | 0,730143546 | RWDD2A | FALSE | Upregulated |
| -0,140994964 | 0,730187784 | CCDC176 | FALSE | Downregulated |
| -0,186987658 | 0,730237035 | SPIRE1 | FALSE | Downregulated |
| -0,16922946 | 0,730237035 | TBRG1 | FALSE | Downregulated |
| 0,363610994 | 0,730427979 | FUT3 | FALSE | Upregulated |
| -0,53021837 | 0,730553312 | EHMT1-IT1 | FALSE | Downregulated |
| -0,187638276 | 0,730624477 | HPR | FALSE | Downregulated |

| | | | | |
|---|---|---|---|---|
| 0,307681282 | 0,730635709 | LOC729887 | FALSE | Upregulated |
| 0,503615504 | 0,730663172 | SCAMP5 | FALSE | Upregulated |
| -0,620211904 | 0,730941662 | LOC101930567 | FALSE | Downregulated |
| 0,189046889 | 0,730941662 | LOC645967 | FALSE | Upregulated |
| -0,159118902 | 0,730941662 | ZFP41 | FALSE | Downregulated |
| 0,321733272 | 0,73101276 | RP11-266K4.9 | FALSE | Upregulated |
| 0,363027111 | 0,731276207 | C14orf2 | FALSE | Upregulated |
| -0,215027565 | 0,731276207 | ORMDL1 | FALSE | Downregulated |
| 0,203155486 | 0,731315931 | LOC101927161 | FALSE | Upregulated |
| 0,309315576 | 0,731315931 | TMEM91 | FALSE | Upregulated |
| -0,226780512 | 0,73188439 | GUCY1A2 | FALSE | Downregulated |
| -0,226438449 | 0,731984196 | CCDC61 | FALSE | Downregulated |
| -0,229060705 | 0,732010325 | LOC150776 | FALSE | Downregulated |
| -0,437421253 | 0,732010325 | RAD54B | FALSE | Downregulated |
| 0,211518662 | 0,732010325 | SERPINB1 | FALSE | Upregulated |
| -0,120029135 | 0,732010325 | SMAP2 | FALSE | Downregulated |
| -0,178510819 | 0,732010325 | SMC6 | FALSE | Downregulated |
| 0,377493804 | 0,732164834 | C9orf9 | FALSE | Upregulated |
| 0,256095224 | 0,732180798 | ABHD4 | FALSE | Upregulated |
| -0,368490907 | 0,732180798 | ARR3 | FALSE | Downregulated |
| -0,191079704 | 0,732180798 | GSAP | FALSE | Downregulated |
| 0,276292265 | 0,732180798 | IQCH-AS1 | FALSE | Upregulated |
| 0,567535929 | 0,732180798 | KRT19 | FALSE | Upregulated |
| -0,260650737 | 0,732180798 | NAT2 | FALSE | Downregulated |
| 0,243722471 | 0,732180798 | NIF3L1 | FALSE | Upregulated |
| -0,126230806 | 0,732180798 | RP5-1071N3.1 | FALSE | Downregulated |
| -0,119132598 | 0,732180798 | RPPH1 | FALSE | Downregulated |
| 0,248839071 | 0,732180798 | ZNF766 | FALSE | Upregulated |
| -0,124797955 | 0,732183787 | CCNY | FALSE | Downregulated |
| -0,589309147 | 0,732183787 | LOC339874 | FALSE | Downregulated |
| -0,213388478 | 0,732183787 | MTRR | FALSE | Downregulated |
| 0,508173237 | 0,732183787 | POU5F2 | FALSE | Upregulated |
| -0,292058572 | 0,73225893 | RP3-395M20.9 | FALSE | Downregulated |
| -0,32696218 | 0,73225893 | TNFSF8 | FALSE | Downregulated |
| 0,21032505 | 0,732262268 | MYO7B | FALSE | Upregulated |
| 0,136969872 | 0,732347705 | PAWR | FALSE | Upregulated |
| 0,275021509 | 0,732347705 | TRIP4 | FALSE | Upregulated |
| -0,106041718 | 0,732444386 | ARHGAP21 | FALSE | Downregulated |
| -0,219340251 | 0,732444386 | GABARAPL2 | FALSE | Downregulated |
| -0,200983557 | 0,732444386 | PTPRG-AS1 | FALSE | Downregulated |
| 0,246772321 | 0,732534707 | C2orf54 | FALSE | Upregulated |
| 0,357784271 | 0,732534707 | FAM71E1 | FALSE | Upregulated |
| -0,233913726 | 0,732534707 | NFIL3 | FALSE | Downregulated |
| 0,102076324 | 0,732534707 | STIM1 | FALSE | Upregulated |
| -0,494301588 | 0,732540369 | HTR3E | FALSE | Downregulated |
| -0,235102231 | 0,732540369 | TRMT11 | FALSE | Downregulated |
| -0,262414902 | 0,732585416 | ADSL | FALSE | Downregulated |
| 0,114269054 | 0,732685888 | PPAPDC1B | FALSE | Upregulated |
| 0,210850716 | 0,732844033 | NDUFAF3 | FALSE | Upregulated |
| 0,187833435 | 0,732881412 | LOC642361 | FALSE | Upregulated |
| 0,398498865 | 0,733304533 | CASC5 | FALSE | Upregulated |

| | | | | |
|---|---|---|---|---|
| -0,271689767 | 0,733304533 | NAT1 | FALSE | Downregulated |
| -0,200698019 | 0,733544262 | ATOX1 | FALSE | Downregulated |
| 0,357946008 | 0,733544262 | CCR10 | FALSE | Upregulated |
| -0,20381943 | 0,733544262 | CXCL14 | FALSE | Downregulated |
| 0,130206213 | 0,733574972 | MYO7A | FALSE | Upregulated |
| 0,20354906 | 0,733590349 | CEP63 | FALSE | Upregulated |
| 0,310488731 | 0,733590349 | LIN37 | FALSE | Upregulated |
| -0,215866222 | 0,733590349 | PIDD | FALSE | Downregulated |
| -0,15481597 | 0,733590349 | UGT2B10 | FALSE | Downregulated |
| 0,498783494 | 0,733727775 | LINC00294 | FALSE | Upregulated |
| -0,533241007 | 0,733727775 | ZNF396 | FALSE | Downregulated |
| -0,330268839 | 0,733909578 | CYB5RL | FALSE | Downregulated |
| 0,471149344 | 0,733909778 | LOC101928969 | FALSE | Upregulated |
| 0,199218626 | 0,733909778 | PLD2 | FALSE | Upregulated |
| 0,137567053 | 0,733909778 | SEC61A2 | FALSE | Upregulated |
| 0,251293022 | 0,734205589 | CD320 | FALSE | Upregulated |
| -0,305550962 | 0,734205589 | HTR7P1 | FALSE | Downregulated |
| 0,278836037 | 0,734205589 | POLR3C | FALSE | Upregulated |
| -0,222921865 | 0,734205589 | PRKAR1B | FALSE | Downregulated |
| 0,096265986 | 0,734205589 | TYK2 | FALSE | Upregulated |
| -0,20578067 | 0,73423508 | ANGEL2 | FALSE | Downregulated |
| 1,539133103 | 0,73423508 | RNA18S5 | FALSE | Upregulated |
| 0,110530186 | 0,734411136 | RN7SL1 | FALSE | Upregulated |
| -0,26888913 | 0,734442135 | PHKG2 | FALSE | Downregulated |
| 0,178208527 | 0,734562669 | FUS | FALSE | Upregulated |
| 0,295471932 | 0,734562669 | LINC00641 | FALSE | Upregulated |
| -0,481068575 | 0,734562669 | TMEM187 | FALSE | Downregulated |
| 0,291187174 | 0,734699778 | LOC101929441 | FALSE | Upregulated |
| 0,251868568 | 0,734797265 | BCL6B | FALSE | Upregulated |
| -0,107551256 | 0,734797265 | CC2D1B | FALSE | Downregulated |
| 0,308167389 | 0,734797265 | FAM76B | FALSE | Upregulated |
| 0,848083338 | 0,734797265 | KCNE1 | FALSE | Upregulated |
| 0,195523068 | 0,734797265 | LSM12 | FALSE | Upregulated |
| 0,269005578 | 0,734894823 | IL10RB-AS1 | FALSE | Upregulated |
| -0,238730749 | 0,734894823 | TMEM135 | FALSE | Downregulated |
| 0,120649536 | 0,734970174 | ATF6B | FALSE | Upregulated |
| -0,154285502 | 0,734970174 | HGSNAT | FALSE | Downregulated |
| -0,132152593 | 0,735109301 | CAMK2N1 | FALSE | Downregulated |
| 0,318369829 | 0,735109301 | LCT | FALSE | Upregulated |
| -0,607784555 | 0,735109301 | LOC102723500 | FALSE | Downregulated |
| -0,579952178 | 0,735186651 | GAL3ST2 | FALSE | Downregulated |
| 0,258058189 | 0,735417878 | FFAR4 | FALSE | Upregulated |
| 0,30385114 | 0,735417878 | LOC102723798 | FALSE | Upregulated |
| 0,100397206 | 0,735417878 | TMEM127 | FALSE | Upregulated |
| 0,511367105 | 0,735471683 | RNU6ATAC | FALSE | Upregulated |
| -0,218688407 | 0,735493923 | HOXB3 | FALSE | Downregulated |
| -0,122102211 | 0,735709512 | TTC17 | FALSE | Downregulated |
| 0,178362016 | 0,735779914 | LOC101928917 | FALSE | Upregulated |
| 0,237665183 | 0,73595564 | GOLGA7 | FALSE | Upregulated |
| -0,44103616 | 0,736077598 | C10orf62 | FALSE | Downregulated |
| 0,114175651 | 0,736077598 | RMRP | FALSE | Upregulated |

| | | | | |
|---|---|---|---|---|
| -0,100089881 | 0,736136487 | NFX1 | FALSE | Downregulated |
| -0,280046897 | 0,736173363 | ADCK5 | FALSE | Downregulated |
| 0,124893016 | 0,736173363 | AKAP8L | FALSE | Upregulated |
| 0,204518297 | 0,736173363 | CTB-113P19.1 | FALSE | Upregulated |
| 0,165944693 | 0,736227568 | CHPF2 | FALSE | Upregulated |
| 0,303225188 | 0,736227568 | FLJ46836 | FALSE | Upregulated |
| 0,177917459 | 0,736320216 | CRTC2 | FALSE | Upregulated |
| -0,197161219 | 0,736378432 | ABLIM3 | FALSE | Downregulated |
| 0,169790463 | 0,736523842 | PEX5L | FALSE | Upregulated |
| 0,117964556 | 0,736523842 | RANBP9 | FALSE | Upregulated |
| 0,29761317 | 0,736523842 | SARNP | FALSE | Upregulated |
| 0,188077905 | 0,736523842 | TRMU | FALSE | Upregulated |
| 0,147065608 | 0,736604973 | MYD88 | FALSE | Upregulated |
| -0,248025151 | 0,736668764 | CACNA1A | FALSE | Downregulated |
| 0,290841341 | 0,736668764 | KCTD13 | FALSE | Upregulated |
| -0,112511277 | 0,736668764 | MCCC2 | FALSE | Downregulated |
| 0,57667284 | 0,736668764 | RALGAPA1P | FALSE | Upregulated |
| -0,242257626 | 0,736668764 | SEMA6B | FALSE | Downregulated |
| -0,130738465 | 0,736668764 | SNAP47 | FALSE | Downregulated |
| 0,323469843 | 0,736668764 | SRPK3 | FALSE | Upregulated |
| 0,2179111 | 0,736768802 | DFNB31 | FALSE | Upregulated |
| -0,549749793 | 0,736768802 | IL1R2 | FALSE | Downregulated |
| 0,485298761 | 0,736768802 | ITGB8 | FALSE | Upregulated |
| -0,500189107 | 0,736768802 | MAN1B1-AS1 | FALSE | Downregulated |
| 0,235939346 | 0,736768802 | RGS17 | FALSE | Upregulated |
| 0,316394676 | 0,736912901 | CCDC102A | FALSE | Upregulated |
| 0,369604682 | 0,736912901 | LST1 | FALSE | Upregulated |
| -0,161106148 | 0,736912901 | NUDT21 | FALSE | Downregulated |
| 0,539407508 | 0,736912901 | RGS6 | FALSE | Upregulated |
| 0,539009705 | 0,736912901 | TBC1D31 | FALSE | Upregulated |
| -0,185371412 | 0,737242053 | THUMPD1 | FALSE | Downregulated |
| 0,102242136 | 0,737267855 | RP11-793H13.3 | FALSE | Upregulated |
| 0,214245208 | 0,737267855 | UFSP2 | FALSE | Upregulated |
| -0,369263938 | 0,737435537 | MALSU1 | FALSE | Downregulated |
| -0,070631394 | 0,737614759 | MLLT4 | FALSE | Downregulated |
| 0,201653379 | 0,737744531 | SLC25A30 | FALSE | Upregulated |
| -0,149463186 | 0,737792928 | CTCF | FALSE | Downregulated |
| 0,484450707 | 0,737792928 | SPATA13-AS1 | FALSE | Upregulated |
| 0,105744793 | 0,738079912 | TRAF7 | FALSE | Upregulated |
| 0,149978448 | 0,73831645 | ACSF3 | FALSE | Upregulated |
| 0,232348531 | 0,738344775 | SCAMP3 | FALSE | Upregulated |
| -0,396796569 | 0,738550057 | C17orf67 | FALSE | Downregulated |
| -0,500991755 | 0,738550057 | HAUS8 | FALSE | Downregulated |
| -0,690646949 | 0,738550057 | RPL17-C18orf32 | FALSE | Downregulated |
| 0,22311125 | 0,739038263 | EBAG9 | FALSE | Upregulated |
| -0,328490436 | 0,739038263 | G6PC3 | FALSE | Downregulated |
| 0,121817684 | 0,739180888 | BRD3 | FALSE | Upregulated |
| 0,276111752 | 0,739180888 | DCTN3 | FALSE | Upregulated |
| 0,308957086 | 0,739180888 | LOC286437 | FALSE | Upregulated |
| 0,236344773 | 0,739180888 | TRAF2 | FALSE | Upregulated |
| 0,191351917 | 0,739198517 | LARS2-AS1 | FALSE | Upregulated |

| | | | | |
|---|---|---|---|---|
| 0,367870077 | 0,739198517 | LINC01123 | FALSE | Upregulated |
| -0,095788504 | 0,739198517 | NPIPB4 | FALSE | Downregulated |
| -0,198647274 | 0,739198517 | SNX13 | FALSE | Downregulated |
| 0,08027864 | 0,739198517 | TBC1D8 | FALSE | Upregulated |
| 0,333217172 | 0,739288217 | LOC102724757 | FALSE | Upregulated |
| 0,516616369 | 0,739322065 | C20orf173 | FALSE | Upregulated |
| 0,18765302 | 0,739787936 | RAB11FIP4 | FALSE | Upregulated |
| -0,144918197 | 0,739824379 | EFCAB13 | FALSE | Downregulated |
| -2,951743515 | 0,739824379 | GJA9 | FALSE | Downregulated |
| 0,227738545 | 0,739824379 | JMJD6 | FALSE | Upregulated |
| 0,157607676 | 0,739824379 | PRNP | FALSE | Upregulated |
| -0,33126324 | 0,739824379 | RP11-24J23.2 | FALSE | Downregulated |
| 0,12110925 | 0,740273566 | TIMP2 | FALSE | Upregulated |
| -0,469911094 | 0,740374757 | PCBP1-AS1 | FALSE | Downregulated |
| 0,210356008 | 0,740459581 | HBS1L | FALSE | Upregulated |
| -0,213364276 | 0,74054974 | ERCC2 | FALSE | Downregulated |
| -0,315800644 | 0,740576464 | ARMC1 | FALSE | Downregulated |
| 0,113649605 | 0,740775215 | GTF2IRD1P1 | FALSE | Upregulated |
| 0,29153725 | 0,740775215 | RIPK2 | FALSE | Upregulated |
| 0,156587441 | 0,740775215 | ZNF784 | FALSE | Upregulated |
| 0,210096977 | 0,741087208 | HMGN3-AS1 | FALSE | Upregulated |
| -0,320466526 | 0,741242181 | FAM13C | FALSE | Downregulated |
| -0,148532855 | 0,741242181 | KCNIP4 | FALSE | Downregulated |
| 0,240144365 | 0,741242181 | KRCC1 | FALSE | Upregulated |
| 0,406469626 | 0,741242181 | LOC100130539 | FALSE | Upregulated |
| -0,077674522 | 0,741242181 | TRIP12 | FALSE | Downregulated |
| -0,220215338 | 0,741242181 | UGT2B11 | FALSE | Downregulated |
| 0,361590015 | 0,741305317 | CCDC51 | FALSE | Upregulated |
| 0,402577315 | 0,741531163 | FSCN2 | FALSE | Upregulated |
| 0,144373961 | 0,741531163 | LEMD2 | FALSE | Upregulated |
| -0,409054541 | 0,741531163 | LOC100506023 | FALSE | Downregulated |
| -0,14479804 | 0,741531163 | NEURL4 | FALSE | Downregulated |
| 0,344378847 | 0,741703446 | LOC101241902 | FALSE | Upregulated |
| 0,255163059 | 0,741792141 | CCNG2 | FALSE | Upregulated |
| -0,135253235 | 0,741818172 | LARS | FALSE | Downregulated |
| 0,430732014 | 0,741818172 | LOC102723996 | FALSE | Upregulated |
| -0,177419271 | 0,741818172 | OXA1L | FALSE | Downregulated |
| -0,22251931 | 0,741818172 | ZNF133 | FALSE | Downregulated |
| 0,397564525 | 0,741850116 | SLC25A21 | FALSE | Upregulated |
| -0,337110419 | 0,74186345 | PSMG4 | FALSE | Downregulated |
| -0,111605948 | 0,742061547 | HEXIM1 | FALSE | Downregulated |
| -0,292083052 | 0,742108058 | C6orf48 | FALSE | Downregulated |
| 0,143752071 | 0,742108058 | MSTO2P | FALSE | Upregulated |
| 0,424010898 | 0,742108058 | OR52I1 | FALSE | Upregulated |
| 0,135834505 | 0,742108058 | PSMF1 | FALSE | Upregulated |
| -0,104594565 | 0,742108058 | RAB43 | FALSE | Downregulated |
| -0,131749327 | 0,742120655 | BCAR1 | FALSE | Downregulated |
| -0,49936493 | 0,742120655 | LOC102723651 | FALSE | Downregulated |
| -0,219367937 | 0,742120655 | MACROD2-AS1 | FALSE | Downregulated |
| 0,306205813 | 0,742120941 | SPATS2 | FALSE | Upregulated |
| -0,109555377 | 0,742446431 | HMGB1 | FALSE | Downregulated |

| | | | | |
|---|---|---|---|---|
| 0,238765264 | 0,742674184 | LCA5L | FALSE | Upregulated |
| 0,191790547 | 0,742732434 | IFFO2 | FALSE | Upregulated |
| -0,606655515 | 0,74278581 | GPR171 | FALSE | Downregulated |
| 0,246971854 | 0,742875097 | NUBP1 | FALSE | Upregulated |
| -0,236276031 | 0,742976507 | BBS2 | FALSE | Downregulated |
| -0,273782961 | 0,742976507 | OLA1 | FALSE | Downregulated |
| 0,268953869 | 0,742976507 | POP5 | FALSE | Upregulated |
| -0,5294547 | 0,742984486 | LOC102724387 | FALSE | Downregulated |
| -0,074140891 | 0,742984486 | USP34 | FALSE | Downregulated |
| 0,198195897 | 0,743025522 | PUS1 | FALSE | Upregulated |
| 0,26864561 | 0,743025522 | TRIM39 | FALSE | Upregulated |
| -0,189190726 | 0,743068563 | ANKRD54 | FALSE | Downregulated |
| -0,281949352 | 0,743068563 | CES1P1 | FALSE | Downregulated |
| -0,154352328 | 0,743068563 | G3BP2 | FALSE | Downregulated |
| 0,641789573 | 0,743068563 | IGSF9B | FALSE | Upregulated |
| 0,144820949 | 0,743068563 | NEIL1 | FALSE | Upregulated |
| -0,200550436 | 0,743068563 | TTI1 | FALSE | Downregulated |
| -0,332716441 | 0,743068563 | UBXN2A | FALSE | Downregulated |
| -0,094120237 | 0,743068563 | WDR33 | FALSE | Downregulated |
| 0,20755336 | 0,743191705 | ADAT1 | FALSE | Upregulated |
| -1,148716934 | 0,743191705 | C7orf55-LUC7L2 | FALSE | Downregulated |
| -0,147253147 | 0,743191705 | CAMK1 | FALSE | Downregulated |
| 0,315291992 | 0,743191705 | CCDC70 | FALSE | Upregulated |
| 0,148506289 | 0,743191705 | ICAM3 | FALSE | Upregulated |
| 0,663203246 | 0,743191705 | LOC101929689 | FALSE | Upregulated |
| -0,397576462 | 0,743191705 | LOC102723544 | FALSE | Downregulated |
| -0,173492759 | 0,743191705 | MTG1 | FALSE | Downregulated |
| 0,20492223 | 0,743191705 | NSF | FALSE | Upregulated |
| 0,289967744 | 0,743191705 | RNF215 | FALSE | Upregulated |
| -0,141324487 | 0,743191705 | SEC14L1 | FALSE | Downregulated |
| -0,163165719 | 0,743191705 | SLFNL1-AS1 | FALSE | Downregulated |
| -0,139703392 | 0,743191705 | THOP1 | FALSE | Downregulated |
| 0,222965404 | 0,743196081 | MTMR9 | FALSE | Upregulated |
| 0,237886356 | 0,743196081 | NAGPA | FALSE | Upregulated |
| -0,089119348 | 0,743196081 | PTP4A1 | FALSE | Downregulated |
| -0,29821323 | 0,743196081 | TMEM41B | FALSE | Downregulated |
| 0,255150578 | 0,743196081 | UBA3 | FALSE | Upregulated |
| -0,526453663 | 0,743347619 | C9orf169 | FALSE | Downregulated |
| -0,383834047 | 0,743347619 | ZNF626 | FALSE | Downregulated |
| -0,135528872 | 0,743377472 | PTRF | FALSE | Downregulated |
| -0,193135374 | 0,743683657 | C1GALT1 | FALSE | Downregulated |
| -0,137829589 | 0,743683657 | ECI2 | FALSE | Downregulated |
| -0,831818413 | 0,743683657 | IL20RB | FALSE | Downregulated |
| -0,173213998 | 0,743683657 | KLHL2 | FALSE | Downregulated |
| 0,330847522 | 0,743683657 | MRPL36 | FALSE | Upregulated |
| -0,199642544 | 0,743683657 | POLR2H | FALSE | Downregulated |
| -0,234941054 | 0,743683657 | SCLY | FALSE | Downregulated |
| -0,276403099 | 0,743683657 | TBXA2R | FALSE | Downregulated |
| -0,378333461 | 0,74379223 | LOC102724163 | FALSE | Downregulated |
| -0,241466643 | 0,74379223 | NECAP1 | FALSE | Downregulated |
| -0,36648594 | 0,74379223 | RP11-761I4.3 | FALSE | Downregulated |

| | | | | |
|---|---|---|---|---|
| 0,301727013 | 0,743812712 | SHISA9 | FALSE | Upregulated |
| -0,204413525 | 0,743918366 | LINC00244 | FALSE | Downregulated |
| 0,169416665 | 0,743918366 | ORM1 | FALSE | Upregulated |
| 0,121555806 | 0,743961961 | PHF12 | FALSE | Upregulated |
| 0,128510492 | 0,744119955 | ATP9A | FALSE | Upregulated |
| 0,459149019 | 0,744119955 | HCG8 | FALSE | Upregulated |
| 0,190268478 | 0,744119955 | MID1IP1 | FALSE | Upregulated |
| 0,459972925 | 0,744119955 | MTX2 | FALSE | Upregulated |
| 0,143249173 | 0,744197719 | CYP1B1 | FALSE | Upregulated |
| -0,155102788 | 0,744234973 | C19orf60 | FALSE | Downregulated |
| -0,314656365 | 0,744234973 | SNRNP27 | FALSE | Downregulated |
| 0,128202269 | 0,744300075 | FBXL5 | FALSE | Upregulated |
| -0,137225178 | 0,744425402 | RNF167 | FALSE | Downregulated |
| -0,115776874 | 0,744587575 | ARRDC1 | FALSE | Downregulated |
| -0,214962423 | 0,744587575 | PDK3 | FALSE | Downregulated |
| -0,469093779 | 0,744728899 | CYP4X1 | FALSE | Downregulated |
| 0,085712522 | 0,744728899 | EWSR1 | FALSE | Upregulated |
| -0,142052984 | 0,744728899 | GTF3C5 | FALSE | Downregulated |
| 0,137545057 | 0,744728899 | LOC101928660 | FALSE | Upregulated |
| -0,524190026 | 0,744728899 | NPHP3-ACAD11 | FALSE | Downregulated |
| 0,179019672 | 0,744728899 | RTFDC1 | FALSE | Upregulated |
| -0,06508636 | 0,744829434 | TNRC18 | FALSE | Downregulated |
| 0,203780668 | 0,744892578 | CAMSAP2 | FALSE | Upregulated |
| -0,145629154 | 0,744892578 | GPBP1 | FALSE | Downregulated |
| -0,511257184 | 0,744892578 | ZWILCH | FALSE | Downregulated |
| -0,247859402 | 0,744902383 | ADAMTS7 | FALSE | Downregulated |
| -0,531698259 | 0,744902383 | BTNL3 | FALSE | Downregulated |
| -0,152930462 | 0,744902383 | FAM84B | FALSE | Downregulated |
| 0,369552484 | 0,744902383 | NXPE3 | FALSE | Upregulated |
| -0,269170747 | 0,744902383 | OBSCN | FALSE | Downregulated |
| -0,142162306 | 0,744902383 | PRICKLE2-AS1 | FALSE | Downregulated |
| 0,220765776 | 0,744902383 | PRMT5 | FALSE | Upregulated |
| -0,130998333 | 0,744902383 | PSMC5 | FALSE | Downregulated |
| -0,2441557 | 0,744902383 | PVRL1 | FALSE | Downregulated |
| -0,200815824 | 0,744902383 | TACO1 | FALSE | Downregulated |
| -0,164189903 | 0,744902383 | UFL1 | FALSE | Downregulated |
| -0,384346266 | 0,74496466 | BMP4 | FALSE | Downregulated |
| 0,29274882 | 0,74496466 | PRH1-PRR4 | FALSE | Upregulated |
| -0,199043831 | 0,745065382 | ARHGEF9 | FALSE | Downregulated |
| 0,310501912 | 0,745065382 | CLRN1 | FALSE | Upregulated |
| -0,152426351 | 0,745065382 | LRRC58 | FALSE | Downregulated |
| 0,270083151 | 0,745067902 | DKFZP586I1420 | FALSE | Upregulated |
| -0,289073817 | 0,745278201 | LOC102724023 | FALSE | Downregulated |
| 0,161272258 | 0,745278201 | PSMD11 | FALSE | Upregulated |
| -0,115749416 | 0,745278201 | XRCC6 | FALSE | Downregulated |
| 0,191040336 | 0,745559018 | CCDC66 | FALSE | Upregulated |
| -0,438587732 | 0,745559018 | FOXD2-AS1 | FALSE | Downregulated |
| 0,200100606 | 0,745559018 | RNF34 | FALSE | Upregulated |
| 0,421197752 | 0,745559018 | SMYD3 | FALSE | Upregulated |
| 0,132526491 | 0,745559018 | TECPR1 | FALSE | Upregulated |
| 0,102328333 | 0,745560332 | SMPD4 | FALSE | Upregulated |

| | | | | |
|---|---|---|---|---|
| 0,235606071 | 0,745560332 | TSPO2 | FALSE | Upregulated |
| 0,510219097 | 0,745578471 | P2RY6 | FALSE | Upregulated |
| 0,238665809 | 0,746430833 | TM2D2 | FALSE | Upregulated |
| 0,200878525 | 0,746441235 | LOC101929140 | FALSE | Upregulated |
| 0,276576112 | 0,746441235 | LOC102723382 | FALSE | Upregulated |
| 0,224465689 | 0,746503384 | ANAPC7 | FALSE | Upregulated |
| -0,252729612 | 0,746539142 | ERMARD | FALSE | Downregulated |
| -0,274556894 | 0,746539142 | PNMA6A | FALSE | Downregulated |
| -0,327137551 | 0,746540046 | LOC100287896 | FALSE | Downregulated |
| 0,117671656 | 0,746631961 | ZNF552 | FALSE | Upregulated |
| 0,474041379 | 0,746750047 | DEAR | FALSE | Upregulated |
| -0,277378252 | 0,746750047 | PDE11A | FALSE | Downregulated |
| 0,49962869 | 0,746750047 | SERPINA12 | FALSE | Upregulated |
| 0,208612325 | 0,746889724 | WNT3 | FALSE | Upregulated |
| -0,579262526 | 0,747134444 | TMEM232 | FALSE | Downregulated |
| -0,215431686 | 0,747259856 | LAMTOR2 | FALSE | Downregulated |
| 0,229924204 | 0,747340978 | CLTCL1 | FALSE | Upregulated |
| -0,30297439 | 0,747340978 | LOC101928139 | FALSE | Downregulated |
| -0,085358697 | 0,747415842 | KMT2E | FALSE | Downregulated |
| -0,225271979 | 0,747645496 | ADAMTS16 | FALSE | Downregulated |
| -0,200244758 | 0,747645496 | FKBP10 | FALSE | Downregulated |
| 0,183762891 | 0,747645496 | WDR52 | FALSE | Upregulated |
| -0,299213728 | 0,747688977 | AASDH | FALSE | Downregulated |
| 0,512379422 | 0,747688977 | PHOX2A | FALSE | Upregulated |
| 0,132931424 | 0,747899567 | IGF2-AS | FALSE | Upregulated |
| -0,199967766 | 0,748081424 | ZMAT1 | FALSE | Downregulated |
| -0,411354373 | 0,748163106 | GOLGA6A | FALSE | Downregulated |
| 0,276977365 | 0,748272179 | IGSF22 | FALSE | Upregulated |
| -0,202568933 | 0,748272179 | TPD52L1 | FALSE | Downregulated |
| -0,351916522 | 0,748299082 | SNORA63 | FALSE | Downregulated |
| -0,163796302 | 0,748465689 | RMND1 | FALSE | Downregulated |
| -0,368669373 | 0,748547699 | ARL10 | FALSE | Downregulated |
| 0,28062366 | 0,748547699 | GPD2 | FALSE | Upregulated |
| 0,522825432 | 0,748547699 | MGC32805 | FALSE | Upregulated |
| -0,23302183 | 0,748547699 | ROMO1 | FALSE | Downregulated |
| 0,167993083 | 0,748699785 | EID1 | FALSE | Upregulated |
| 0,131447959 | 0,749107996 | RILP | FALSE | Upregulated |
| 0,229419788 | 0,749157876 | CCDC158 | FALSE | Upregulated |
| 0,217210291 | 0,749210943 | ACTL6A | FALSE | Upregulated |
| -0,312069288 | 0,749210943 | AVPR1A | FALSE | Downregulated |
| -0,331903931 | 0,749210943 | DACT2 | FALSE | Downregulated |
| -0,476929533 | 0,749210943 | GJC1 | FALSE | Downregulated |
| 0,215079381 | 0,749210943 | PSMC6 | FALSE | Upregulated |
| -0,188530219 | 0,749210943 | RUSC1-AS1 | FALSE | Downregulated |
| -0,288598441 | 0,749210943 | TMEM39B | FALSE | Downregulated |
| 0,330514657 | 0,749272929 | HSPA7 | FALSE | Upregulated |
| -0,272501509 | 0,749343057 | PTER | FALSE | Downregulated |
| 0,174593747 | 0,749556152 | LOC102724724 | FALSE | Upregulated |
| 0,464217106 | 0,749699309 | ASCL2 | FALSE | Upregulated |
| -0,087949808 | 0,749699309 | ATM | FALSE | Downregulated |
| -0,094592541 | 0,749699309 | C1S | FALSE | Downregulated |

| | | | | |
|---|---|---|---|---|
| 0,111689726 | 0,749699309 | CCDC47 | FALSE | Upregulated |
| -0,189861052 | 0,749699309 | EMX1 | FALSE | Downregulated |
| 0,106969395 | 0,749699309 | FAM105B | FALSE | Upregulated |
| 0,440621678 | 0,749699309 | FAM188B | FALSE | Upregulated |
| -0,122891894 | 0,749699309 | FAM20B | FALSE | Downregulated |
| -0,191543656 | 0,749699309 | GCNT2 | FALSE | Downregulated |
| -0,110795903 | 0,749699309 | KLB | FALSE | Downregulated |
| -0,596189486 | 0,749699309 | LOC102724037 | FALSE | Downregulated |
| -0,22932707 | 0,749699309 | LOC102724852 | FALSE | Downregulated |
| 0,130833404 | 0,749699309 | LRRC14 | FALSE | Upregulated |
| -0,143036022 | 0,749699309 | MRPS34 | FALSE | Downregulated |
| -0,112911688 | 0,749699309 | MTERFD2 | FALSE | Downregulated |
| 0,181604077 | 0,749699309 | PAQR9 | FALSE | Upregulated |
| -0,279782809 | 0,749699309 | PCDH17 | FALSE | Downregulated |
| -0,194910797 | 0,749699309 | PDSS2 | FALSE | Downregulated |
| -0,35041706 | 0,749699309 | RP11-379B18.5 | FALSE | Downregulated |
| 0,142472591 | 0,749699309 | SIAH2 | FALSE | Upregulated |
| 0,229552861 | 0,749699309 | SLC35B1 | FALSE | Upregulated |
| -0,157382201 | 0,749699309 | SMCHD1 | FALSE | Downregulated |
| 0,153381491 | 0,749699309 | TIAM2 | FALSE | Upregulated |
| -0,179333296 | 0,749699309 | TUBGCP3 | FALSE | Downregulated |
| 0,251954068 | 0,749699309 | WBSCR22 | FALSE | Upregulated |
| -0,171015989 | 0,749699309 | ZNF606 | FALSE | Downregulated |
| 0,133803641 | 0,749779944 | AC093690.1 | FALSE | Upregulated |
| 0,304010789 | 0,749779944 | SLC26A6 | FALSE | Upregulated |
| -0,101327266 | 0,749820413 | COL10A1 | FALSE | Downregulated |
| 0,638998307 | 0,749854835 | BNIPL | FALSE | Upregulated |
| -0,279022559 | 0,749854835 | ELP6 | FALSE | Downregulated |
| -0,463259522 | 0,749854835 | MTNR1B | FALSE | Downregulated |
| 0,104673014 | 0,750324364 | TRMT112 | FALSE | Upregulated |
| -0,19363526 | 0,750466181 | ZNF75D | FALSE | Downregulated |
| -0,135565404 | 0,750494031 | CAD | FALSE | Downregulated |
| 0,460895304 | 0,750494031 | RFPL3 | FALSE | Upregulated |
| -0,151204922 | 0,750494031 | RNF144B | FALSE | Downregulated |
| 0,207574445 | 0,750494031 | ZNF786 | FALSE | Upregulated |
| -0,182533943 | 0,750731365 | SUGP1 | FALSE | Downregulated |
| -0,216261834 | 0,750828386 | FAM73A | FALSE | Downregulated |
| -0,431901335 | 0,750828386 | KCNK1 | FALSE | Downregulated |
| -0,286575175 | 0,750828386 | THOC7 | FALSE | Downregulated |
| -0,105944885 | 0,750828386 | UBA1 | FALSE | Downregulated |
| -0,170046119 | 0,750828386 | ZNF84 | FALSE | Downregulated |
| -0,128998085 | 0,750828386 | ZYG11B | FALSE | Downregulated |
| -0,159580329 | 0,750858771 | CEP192 | FALSE | Downregulated |
| -0,739203255 | 0,750858771 | NDUFA6-AS1 | FALSE | Downregulated |
| 0,106027872 | 0,750858771 | SCARB1 | FALSE | Upregulated |
| -0,141891635 | 0,750858771 | SERPINA11 | FALSE | Downregulated |
| -0,154913905 | 0,750858771 | VNN1 | FALSE | Downregulated |
| 0,419075729 | 0,75134758 | KLF8 | FALSE | Upregulated |
| 0,503086281 | 0,751479418 | RAMP2 | FALSE | Upregulated |
| 0,23011514 | 0,751546151 | BBS5 | FALSE | Upregulated |
| 0,283196766 | 0,751678417 | HSPA12B | FALSE | Upregulated |

| | | | | |
|---|---|---|---|---|
| 0,161880669 | 0,751678417 | LOC646127 | FALSE | Upregulated |
| -0,590477325 | 0,751678417 | UPK3BL | FALSE | Downregulated |
| -0,101285159 | 0,751795404 | CD99 | FALSE | Downregulated |
| 0,088596858 | 0,751851965 | ARAP1 | FALSE | Upregulated |
| -0,209475803 | 0,751922848 | FBXO42 | FALSE | Downregulated |
| 0,272220528 | 0,75222305 | HSD11B2 | FALSE | Upregulated |
| -0,181192482 | 0,75222305 | VAMP3 | FALSE | Downregulated |
| -0,249419761 | 0,752380681 | ZBTB25 | FALSE | Downregulated |
| 0,093462496 | 0,752497809 | SUPT5H | FALSE | Upregulated |
| 0,143002263 | 0,752497809 | TEN1 | FALSE | Upregulated |
| -0,129963037 | 0,752497809 | TMEM43 | FALSE | Downregulated |
| 0,218162766 | 0,752584984 | ZNF585B | FALSE | Upregulated |
| -0,175212845 | 0,752740797 | CCT8 | FALSE | Downregulated |
| 0,258857517 | 0,752740797 | CLEC14A | FALSE | Upregulated |
| 0,625359147 | 0,752740797 | FRG2C | FALSE | Upregulated |
| 0,165705676 | 0,752740797 | NCSTN | FALSE | Upregulated |
| 0,096577349 | 0,752740797 | SAFB | FALSE | Upregulated |
| 1,493563674 | 0,752740797 | SERF2-C15ORF63 | FALSE | Upregulated |
| 0,337602848 | 0,752740797 | TWSG1 | FALSE | Upregulated |
| -0,109838097 | 0,752740797 | YME1L1 | FALSE | Downregulated |
| -0,194600721 | 0,753022049 | CTBS | FALSE | Downregulated |
| -0,253369338 | 0,753022049 | KCNMA1 | FALSE | Downregulated |
| 1,336873325 | 0,753067102 | F8A2 | FALSE | Upregulated |
| -0,310597353 | 0,753067102 | GMFB | FALSE | Downregulated |
| 0,299959073 | 0,753067102 | KIAA1244 | FALSE | Upregulated |
| 0,324928239 | 0,753067102 | OXSM | FALSE | Upregulated |
| -0,376746509 | 0,753067102 | TMEM199 | FALSE | Downregulated |
| 0,442487884 | 0,753067102 | TRDMT1 | FALSE | Upregulated |
| 0,501633987 | 0,753067102 | WDR78 | FALSE | Upregulated |
| -0,144228849 | 0,753067102 | XPA | FALSE | Downregulated |
| -0,297883132 | 0,753067102 | ZKSCAN3 | FALSE | Downregulated |
| -0,194164103 | 0,753067102 | ZNF22 | FALSE | Downregulated |
| 0,301055074 | 0,753077791 | KCTD5 | FALSE | Upregulated |
| -0,194921615 | 0,75308455 | ABTB1 | FALSE | Downregulated |
| 0,193021639 | 0,75308455 | NOL3 | FALSE | Upregulated |
| -0,140218615 | 0,75308455 | SREBF2 | FALSE | Downregulated |
| 0,243235078 | 0,75308455 | TRIM67 | FALSE | Upregulated |
| 0,074564847 | 0,75308455 | VPS13B | FALSE | Upregulated |
| 0,115824428 | 0,753113343 | TPP1 | FALSE | Upregulated |
| -0,128009437 | 0,75334767 | LOC100507520 | FALSE | Downregulated |
| 0,155380514 | 0,75334767 | SMAD7 | FALSE | Upregulated |
| 0,089870951 | 0,753429828 | CREBZF | FALSE | Upregulated |
| -0,208613039 | 0,753604767 | DPH7 | FALSE | Downregulated |
| 0,112438459 | 0,753604767 | ENTPD1 | FALSE | Upregulated |
| -0,449879898 | 0,753604767 | RP11-598F7.4 | FALSE | Downregulated |
| -0,206092412 | 0,753604767 | SAP130 | FALSE | Downregulated |
| -0,226660148 | 0,753734091 | LINC00909 | FALSE | Downregulated |
| -0,330120027 | 0,753734091 | SLC25A52 | FALSE | Downregulated |
| -0,412225107 | 0,753828476 | IMMP1L | FALSE | Downregulated |
| 0,144625022 | 0,753828476 | KIAA1468 | FALSE | Upregulated |
| 0,428536715 | 0,754188517 | ROPN1L-AS1 | FALSE | Upregulated |

| | | | | |
|---|---|---|---|---|
| -0,228659204 | 0,754326614 | GPN2 | FALSE | Downregulated |
| -0,335850991 | 0,754326614 | RP11-96H19.1 | FALSE | Downregulated |
| -0,208100572 | 0,754326614 | SCML1 | FALSE | Downregulated |
| -0,156086261 | 0,754326614 | THSD7A | FALSE | Downregulated |
| 0,644636965 | 0,754341484 | P2RY14 | FALSE | Upregulated |
| -0,153177591 | 0,754395741 | HAX1 | FALSE | Downregulated |
| 0,231011256 | 0,754416049 | ARFIP1 | FALSE | Upregulated |
| -0,275774509 | 0,754416049 | CCDC107 | FALSE | Downregulated |
| -0,514628498 | 0,754416049 | CORO2B | FALSE | Downregulated |
| 0,266355555 | 0,754416049 | DCAF17 | FALSE | Upregulated |
| -0,080169052 | 0,754416049 | KIAA1109 | FALSE | Downregulated |
| 0,081009349 | 0,754416049 | LOC100190986 | FALSE | Upregulated |
| -0,434302222 | 0,754416049 | LOC101927355 | FALSE | Downregulated |
| 0,277800846 | 0,754416049 | LOC102723665 | FALSE | Upregulated |
| -0,099927229 | 0,754416049 | PBXIP1 | FALSE | Downregulated |
| 0,284489499 | 0,754416049 | PCSK5 | FALSE | Upregulated |
| -0,088311517 | 0,754416049 | RIN3 | FALSE | Downregulated |
| 0,158794515 | 0,754416049 | SCRN3 | FALSE | Upregulated |
| -0,087313535 | 0,754416049 | SEC31A | FALSE | Downregulated |
| 0,107784444 | 0,754416049 | ZHX2 | FALSE | Upregulated |
| 0,279675222 | 0,754514761 | TAF9 | FALSE | Upregulated |
| -0,534105381 | 0,754618509 | LOC101927733 | FALSE | Downregulated |
| 0,473963361 | 0,754716611 | FAHD2B | FALSE | Upregulated |
| -0,173953454 | 0,754736924 | DSE | FALSE | Downregulated |
| -0,191830693 | 0,754736924 | KLHL9 | FALSE | Downregulated |
| 0,143196305 | 0,754736924 | SLC47A2 | FALSE | Upregulated |
| -0,284638031 | 0,754736924 | TOX3 | FALSE | Downregulated |
| 0,130668005 | 0,754783385 | LOC644090 | FALSE | Upregulated |
| 0,101739795 | 0,754783385 | TADA3 | FALSE | Upregulated |
| -0,603154501 | 0,75522298 | LOC102723461 | FALSE | Downregulated |
| 0,296856111 | 0,75522298 | SOX7 | FALSE | Upregulated |
| -0,216304059 | 0,755315278 | LOC102724634 | FALSE | Downregulated |
| -0,505717964 | 0,755343296 | ANO3 | FALSE | Downregulated |
| 0,144962884 | 0,755343296 | HABP4 | FALSE | Upregulated |
| -0,227513484 | 0,755343296 | KIFAP3 | FALSE | Downregulated |
| -0,3571896 | 0,755343296 | PRB1 | FALSE | Downregulated |
| -0,120394802 | 0,755343296 | SARM1 | FALSE | Downregulated |
| -0,124897815 | 0,755343296 | UBE2O | FALSE | Downregulated |
| 0,407413527 | 0,755389438 | FLJ42393 | FALSE | Upregulated |
| 0,122331277 | 0,755389438 | KCNRG | FALSE | Upregulated |
| 0,120096987 | 0,755389438 | ZNF335 | FALSE | Upregulated |
| -0,47384275 | 0,75539404 | LOC100996497 | FALSE | Downregulated |
| 0,447894796 | 0,75539404 | RFXAP | FALSE | Upregulated |
| 0,106684553 | 0,75539404 | TBCD | FALSE | Upregulated |
| -0,364019082 | 0,75539404 | ZNF852 | FALSE | Downregulated |
| -0,324379027 | 0,75572708 | EFCAB4B | FALSE | Downregulated |
| -0,260102406 | 0,755730729 | BPNT1 | FALSE | Downregulated |
| 0,231294837 | 0,755730729 | LOC100129203 | FALSE | Upregulated |
| 0,660806522 | 0,755741219 | C4orf6 | FALSE | Upregulated |
| 0,25413878 | 0,755935265 | RP11-203B9.4 | FALSE | Upregulated |
| 0,331128858 | 0,756020669 | PODXL2 | FALSE | Upregulated |

| | | | | |
|---|---|---|---|---|
| 0,259569614 | 0,756109865 | LOC101928466 | FALSE | Upregulated |
| 0,497841332 | 0,756109865 | LOC102724319 | FALSE | Upregulated |
| 0,371490203 | 0,756285637 | FMO6P | FALSE | Upregulated |
| -0,521944784 | 0,756434253 | C22orf31 | FALSE | Downregulated |
| 0,748762766 | 0,756475084 | TMEM183B | FALSE | Upregulated |
| 0,123523723 | 0,756692143 | DCAF7 | FALSE | Upregulated |
| -0,749664031 | 0,756692143 | FCGR2C | FALSE | Downregulated |
| -0,389938396 | 0,756692143 | LRRC37B | FALSE | Downregulated |
| -0,11540506 | 0,756692143 | PTPN18 | FALSE | Downregulated |
| -0,372329506 | 0,756907782 | OAZ3 | FALSE | Downregulated |
| 0,165023933 | 0,757156103 | NANS | FALSE | Upregulated |
| -0,164270815 | 0,757312185 | ZDHHC9 | FALSE | Downregulated |
| -0,090641201 | 0,757407151 | EIF2S3 | FALSE | Downregulated |
| 0,157700325 | 0,757464208 | NUP205 | FALSE | Upregulated |
| 0,962139718 | 0,757464208 | RAB3B | FALSE | Upregulated |
| 0,243266375 | 0,757485424 | DIEXF | FALSE | Upregulated |
| -0,57235469 | 0,757485424 | LPCAT4 | FALSE | Downregulated |
| 0,214955047 | 0,757485424 | NUP107 | FALSE | Upregulated |
| -0,278775662 | 0,757485424 | PRX | FALSE | Downregulated |
| 0,231704338 | 0,757485424 | SMIM13 | FALSE | Upregulated |
| 0,377049735 | 0,757553479 | NUDT1 | FALSE | Upregulated |
| -0,260994425 | 0,757855356 | ICT1 | FALSE | Downregulated |
| 0,190647267 | 0,757855356 | SMAD1 | FALSE | Upregulated |
| -0,2435751 | 0,757993619 | NDUFB6 | FALSE | Downregulated |
| 0,236407382 | 0,757993619 | QTRTD1 | FALSE | Upregulated |
| 0,17907301 | 0,758109265 | H2AFJ | FALSE | Upregulated |
| -0,49943064 | 0,758278356 | OR56B4 | FALSE | Downregulated |
| 0,322799343 | 0,758278356 | SNRPE | FALSE | Upregulated |
| -0,155941745 | 0,758283096 | E2F4 | FALSE | Downregulated |
| 0,178061104 | 0,758500131 | MGAT5 | FALSE | Upregulated |
| -0,549943412 | 0,758731876 | LOC102724889 | FALSE | Downregulated |
| 0,132480017 | 0,75873969 | RNPEP | FALSE | Upregulated |
| 0,123868413 | 0,758789483 | RXRB | FALSE | Upregulated |
| -0,177955297 | 0,758789483 | TMUB2 | FALSE | Downregulated |
| 0,127756591 | 0,758805698 | C22orf26 | FALSE | Upregulated |
| 0,116706318 | 0,758805698 | ZSCAN30 | FALSE | Upregulated |
| 0,345311625 | 0,758835132 | PLCD3 | FALSE | Upregulated |
| -0,089838351 | 0,758875989 | TET3 | FALSE | Downregulated |
| -0,271721894 | 0,758905174 | CASD1 | FALSE | Downregulated |
| -0,161859834 | 0,758905174 | MERTK | FALSE | Downregulated |
| 0,181470965 | 0,758905174 | NKIRAS2 | FALSE | Upregulated |
| -0,226589693 | 0,758905174 | OLFML3 | FALSE | Downregulated |
| 0,288599354 | 0,758905174 | SENP7 | FALSE | Upregulated |
| -0,144917751 | 0,758905174 | TP53BP1 | FALSE | Downregulated |
| 0,377127564 | 0,758967695 | ARMCX2 | FALSE | Upregulated |
| 0,22567097 | 0,759008489 | CDCA7L | FALSE | Upregulated |
| -0,123831257 | 0,759008489 | HDGF | FALSE | Downregulated |
| 0,111864017 | 0,759008489 | SART1 | FALSE | Upregulated |
| -0,207795757 | 0,759008489 | THRSP | FALSE | Downregulated |
| 0,195260156 | 0,759023011 | GDPD4 | FALSE | Upregulated |
| 0,162342034 | 0,759023011 | ZBTB43 | FALSE | Upregulated |

| | | | | |
|---|---|---|---|---|
| 0,181210565 | 0,759023011 | ZNF576 | FALSE | Upregulated |
| 0,472740855 | 0,759023011 | ZNF625 | FALSE | Upregulated |
| -0,14800589 | 0,759128398 | CHST15 | FALSE | Downregulated |
| 0,507332448 | 0,759128398 | SOSTDC1 | FALSE | Upregulated |
| 0,20400702 | 0,759145997 | TMEM134 | FALSE | Upregulated |
| -0,252844024 | 0,759151555 | ZNF226 | FALSE | Downregulated |
| -0,415080899 | 0,75918979 | APH1B | FALSE | Downregulated |
| 0,164558548 | 0,75918979 | CYTH3 | FALSE | Upregulated |
| -0,186492168 | 0,75918979 | HTATSF1 | FALSE | Downregulated |
| -0,25542421 | 0,75918979 | PARL | FALSE | Downregulated |
| 0,162063442 | 0,75918979 | SLC3A2 | FALSE | Upregulated |
| 0,245131267 | 0,75918979 | ZNF706 | FALSE | Upregulated |
| -0,066705459 | 0,759227782 | KIAA1671 | FALSE | Downregulated |
| -0,207626841 | 0,759227782 | LOC101927790 | FALSE | Downregulated |
| -0,149370268 | 0,759227782 | PHC3 | FALSE | Downregulated |
| 0,153487494 | 0,759227782 | PIEZO2 | FALSE | Upregulated |
| 0,096428227 | 0,759227782 | RIC8A | FALSE | Upregulated |
| -0,22149577 | 0,759227782 | SKP2 | FALSE | Downregulated |
| -1,731199624 | 0,759227782 | TMEM189-UBE2V1 | FALSE | Downregulated |
| -0,148339131 | 0,759227782 | UGDH | FALSE | Downregulated |
| 0,318473885 | 0,759404405 | LOC101928984 | FALSE | Upregulated |
| -0,243513782 | 0,759404405 | SPCS1 | FALSE | Downregulated |
| 0,072703597 | 0,759611932 | KIF13B | FALSE | Upregulated |
| 0,358895528 | 0,759733385 | GSTCD | FALSE | Upregulated |
| -0,500941055 | 0,759835601 | CCDC67 | FALSE | Downregulated |
| 0,173856986 | 0,759835601 | POM121L10P | FALSE | Upregulated |
| 0,156994689 | 0,760339672 | RBBP7 | FALSE | Upregulated |
| -0,338218927 | 0,760388808 | SLX1B | FALSE | Downregulated |
| 0,214784267 | 0,760629137 | FKBP14 | FALSE | Upregulated |
| 0,220004571 | 0,760629137 | OSBPL10 | FALSE | Upregulated |
| -0,13586952 | 0,760629137 | PSMB4 | FALSE | Downregulated |
| 0,495596048 | 0,760629137 | SCUBE1 | FALSE | Upregulated |
| 0,231762687 | 0,760629137 | WHAMMP3 | FALSE | Upregulated |
| -0,143864361 | 0,760743441 | C6orf201 | FALSE | Downregulated |
| 0,256931864 | 0,760762997 | CCDC132 | FALSE | Upregulated |
| 0,127206305 | 0,761005779 | ZNF362 | FALSE | Upregulated |
| -0,302280843 | 0,761125031 | NUDT18 | FALSE | Downregulated |
| 0,37884104 | 0,761125031 | PFN4 | FALSE | Upregulated |
| 0,365810662 | 0,761356733 | BACE1-AS | FALSE | Upregulated |
| 0,122154058 | 0,761356733 | CRTC3 | FALSE | Upregulated |
| -0,08395095 | 0,761356733 | PDZK1 | FALSE | Downregulated |
| 0,267042565 | 0,761356733 | PRKX | FALSE | Upregulated |
| 0,156809725 | 0,76144959 | CNTN2 | FALSE | Upregulated |
| -0,334775285 | 0,76144959 | TMEM217 | FALSE | Downregulated |
| 0,282781926 | 0,76144959 | ZNF555 | FALSE | Upregulated |
| 0,116836352 | 0,761518496 | EDEM3 | FALSE | Upregulated |
| -0,210071928 | 0,761518496 | FBXL19 | FALSE | Downregulated |
| 0,520286985 | 0,761518496 | FLJ22184 | FALSE | Upregulated |
| 0,281678077 | 0,761518496 | GOLGA6L9 | FALSE | Upregulated |
| 0,166869435 | 0,761518496 | PLA2G6 | FALSE | Upregulated |
| 0,254597509 | 0,761518496 | PLEKHG5 | FALSE | Upregulated |

| | | | | |
|---|---|---|---|---|
| -0,476598734 | 0,761518496 | SEMA3G | FALSE | Downregulated |
| 0,304960606 | 0,761518496 | SOD3 | FALSE | Upregulated |
| -0,282441663 | 0,761518496 | TMEM132D | FALSE | Downregulated |
| 0,181662831 | 0,761518496 | TXNDC12 | FALSE | Upregulated |
| 0,097666068 | 0,761518496 | ZBED4 | FALSE | Upregulated |
| 0,234298136 | 0,761534808 | C12orf10 | FALSE | Upregulated |
| -0,146062037 | 0,761534808 | CCDC117 | FALSE | Downregulated |
| -0,223561957 | 0,761534808 | ID3 | FALSE | Downregulated |
| -0,185622576 | 0,761534808 | WDR34 | FALSE | Downregulated |
| -0,161772693 | 0,761584451 | DYNC1I2 | FALSE | Downregulated |
| -0,439061634 | 0,761618 | ALG10 | FALSE | Downregulated |
| 0,478947748 | 0,761618 | CDK20 | FALSE | Upregulated |
| 0,186435985 | 0,761618 | LOXL2 | FALSE | Upregulated |
| 0,124045176 | 0,761618 | PRKCZ | FALSE | Upregulated |
| -0,085165972 | 0,761618 | RANBP2 | FALSE | Downregulated |
| 0,248311127 | 0,761618 | RNASE1 | FALSE | Upregulated |
| -0,137113173 | 0,761618 | SKP1 | FALSE | Downregulated |
| -0,383273272 | 0,761656769 | CCDC150 | FALSE | Downregulated |
| 0,106111093 | 0,761888785 | RDX | FALSE | Upregulated |
| 0,347003882 | 0,761994952 | ASIC1 | FALSE | Upregulated |
| -0,230859142 | 0,76216018 | GPS2 | FALSE | Downregulated |
| 0,109452145 | 0,762194927 | ITPK1 | FALSE | Upregulated |
| 0,175458657 | 0,76231914 | FAM89B | FALSE | Upregulated |
| -0,101322555 | 0,76231914 | RAB1B | FALSE | Downregulated |
| 0,435136232 | 0,762525999 | BNIP1 | FALSE | Upregulated |
| 0,3720883 | 0,762939881 | ME1 | FALSE | Upregulated |
| -0,501869012 | 0,762939881 | OR51B6 | FALSE | Downregulated |
| -0,081956407 | 0,763002779 | MTMR10 | FALSE | Downregulated |
| 0,155606993 | 0,763133318 | SFMBT1 | FALSE | Upregulated |
| 0,107070701 | 0,763350347 | HSD3B7 | FALSE | Upregulated |
| 0,640168404 | 0,763350347 | HTT-AS | FALSE | Upregulated |
| -0,107963176 | 0,763350347 | PI4KA | FALSE | Downregulated |
| 0,203316765 | 0,763350347 | ZBTB41 | FALSE | Upregulated |
| 0,425998062 | 0,764139631 | METTL21B | FALSE | Upregulated |
| 0,249411171 | 0,764263366 | FAM204A | FALSE | Upregulated |
| -0,352440332 | 0,764575069 | BRMS1L | FALSE | Downregulated |
| 0,226676267 | 0,764575069 | RASGRP4 | FALSE | Upregulated |
| -0,243295429 | 0,764836532 | ABRA | FALSE | Downregulated |
| -0,195710146 | 0,765052846 | EPC2 | FALSE | Downregulated |
| 0,464992818 | 0,765052846 | HOXA4 | FALSE | Upregulated |
| 0,542432135 | 0,765052846 | VRK1 | FALSE | Upregulated |
| -0,3495344 | 0,765052846 | ZNF788 | FALSE | Downregulated |
| -0,195023872 | 0,765148896 | CARD8 | FALSE | Downregulated |
| -0,24885671 | 0,765148896 | MRPL45P2 | FALSE | Downregulated |
| -0,19447546 | 0,765148896 | RHBDD3 | FALSE | Downregulated |
| -0,192441338 | 0,765161918 | POLR2J | FALSE | Downregulated |
| -0,140743853 | 0,76628981 | RPS6KB2 | FALSE | Downregulated |
| -0,121528949 | 0,76630403 | CPT1A | FALSE | Downregulated |
| -0,1073176 | 0,76630403 | SARS | FALSE | Downregulated |
| 0,107367469 | 0,76630403 | SKIV2L | FALSE | Upregulated |
| 0,095419196 | 0,766376187 | C6orf120 | FALSE | Upregulated |

| | | | | |
|---|---|---|---|---|
| 0,50975069 | 0,766395858 | DOC2B | FALSE | Upregulated |
| -0,220339835 | 0,766395858 | EXD2 | FALSE | Downregulated |
| -0,233479553 | 0,766395858 | TUBA3FP | FALSE | Downregulated |
| 0,344836682 | 0,766584142 | ACHE | FALSE | Upregulated |
| 0,159449715 | 0,766584142 | RMND5B | FALSE | Upregulated |
| 0,267501942 | 0,766799591 | RP11-466A17.1 | FALSE | Upregulated |
| 0,3996363 | 0,767110493 | MC1R | FALSE | Upregulated |
| 0,238917444 | 0,767199096 | FASN | FALSE | Upregulated |
| 0,147384598 | 0,767199096 | MEX3C | FALSE | Upregulated |
| 0,295457623 | 0,767846321 | LOC101928641 | FALSE | Upregulated |
| 0,627462956 | 0,767846321 | NDUFC2-KCTD14 | FALSE | Upregulated |
| 0,365971895 | 0,767846321 | TTC5 | FALSE | Upregulated |
| 0,200333344 | 0,767872972 | EMC9 | FALSE | Upregulated |
| -0,230036567 | 0,767872972 | FAM50B | FALSE | Downregulated |
| -0,221103881 | 0,767872972 | TTLL11 | FALSE | Downregulated |
| -0,187911899 | 0,767973574 | PCP4L1 | FALSE | Downregulated |
| -0,171065845 | 0,768229071 | ANO9 | FALSE | Downregulated |
| -0,156660458 | 0,768246609 | CCT6P3 | FALSE | Downregulated |
| 0,324534945 | 0,768262616 | ZC3H8 | FALSE | Upregulated |
| -0,212541868 | 0,768285967 | CEP85 | FALSE | Downregulated |
| -0,353600851 | 0,768333119 | RILPL2 | FALSE | Downregulated |
| 0,278833796 | 0,768366017 | ARL6IP6 | FALSE | Upregulated |
| -0,567067161 | 0,768366017 | BTN2A3P | FALSE | Downregulated |
| 0,125193732 | 0,768366017 | FDFT1 | FALSE | Upregulated |
| -0,295504562 | 0,768366017 | LENEP | FALSE | Downregulated |
| -0,45457764 | 0,768366017 | MED7 | FALSE | Downregulated |
| 0,103065446 | 0,768366017 | MGA | FALSE | Upregulated |
| -0,273241363 | 0,768366017 | PLA1A | FALSE | Downregulated |
| 0,266753215 | 0,768366017 | RNF113A | FALSE | Upregulated |
| 0,355680492 | 0,768366017 | RUNDC3A | FALSE | Upregulated |
| -0,276371166 | 0,768366017 | TAF1D | FALSE | Downregulated |
| -0,187633787 | 0,768366017 | TPD52 | FALSE | Downregulated |
| -0,175760952 | 0,768366017 | ZC3H3 | FALSE | Downregulated |
| -0,240503134 | 0,768366017 | ZNF248 | FALSE | Downregulated |
| -0,096874518 | 0,768393036 | KIDINS220 | FALSE | Downregulated |
| 0,209377156 | 0,768537144 | CCR9 | FALSE | Upregulated |
| -0,41192856 | 0,768589272 | ACPL2 | FALSE | Downregulated |
| -0,169648752 | 0,768589272 | DCUN1D4 | FALSE | Downregulated |
| -0,175025703 | 0,768589272 | FILIP1 | FALSE | Downregulated |
| -0,311143338 | 0,768589272 | MAGEH1 | FALSE | Downregulated |
| 0,345985257 | 0,768589272 | SV2A | FALSE | Upregulated |
| 0,099894443 | 0,768589272 | TRIM41 | FALSE | Upregulated |
| -0,120304247 | 0,768589272 | ZDHHC18 | FALSE | Downregulated |
| 0,087463316 | 0,768863398 | FBXW5 | FALSE | Upregulated |
| -0,281844196 | 0,76893275 | SLC35A5 | FALSE | Downregulated |
| 0,114912158 | 0,76893275 | USP3 | FALSE | Upregulated |
| 0,105595653 | 0,76893275 | YWHAE | FALSE | Upregulated |
| 0,114969225 | 0,768962164 | MORC2-AS1 | FALSE | Upregulated |
| -0,206694051 | 0,768962164 | PTGR2 | FALSE | Downregulated |
| 0,320630142 | 0,768965442 | IER3 | FALSE | Upregulated |
| 0,18715485 | 0,768986691 | ALDH18A1 | FALSE | Upregulated |

| | | | | |
|---|---|---|---|---|
| -0,326840042 | 0,769071039 | KIAA1456 | FALSE | Downregulated |
| 0,110737242 | 0,769071039 | MRC1 | FALSE | Upregulated |
| 0,125019895 | 0,769230414 | COLGALT1 | FALSE | Upregulated |
| 0,245182242 | 0,769383943 | LINC01043 | FALSE | Upregulated |
| -0,259339762 | 0,769562084 | TM4SF5 | FALSE | Downregulated |
| -0,191974359 | 0,769562084 | ZNF749 | FALSE | Downregulated |
| 0,212768558 | 0,769583216 | MEMO1 | FALSE | Upregulated |
| 0,130686577 | 0,769599538 | SMARCD1 | FALSE | Upregulated |
| -0,121775716 | 0,769721116 | PPP1R26 | FALSE | Downregulated |
| -0,529960961 | 0,769853493 | CD109 | FALSE | Downregulated |
| -0,444585198 | 0,769853493 | CD99P1 | FALSE | Downregulated |
| 0,450158993 | 0,769853493 | PHOSPHO2 | FALSE | Upregulated |
| 0,388180084 | 0,769853493 | SPEG | FALSE | Upregulated |
| 0,505613042 | 0,770203859 | FHL3 | FALSE | Upregulated |
| 0,140446796 | 0,770203859 | LOC285074 | FALSE | Upregulated |
| 0,331854614 | 0,770203859 | VN1R1 | FALSE | Upregulated |
| 0,20640482 | 0,77024962 | IFT46 | FALSE | Upregulated |
| -0,184586696 | 0,77024962 | IQCH | FALSE | Downregulated |
| -0,084679061 | 0,77024962 | JAK1 | FALSE | Downregulated |
| 0,167977258 | 0,77024962 | LOC100506374 | FALSE | Upregulated |
| -0,189734576 | 0,77024962 | OSBPL1A | FALSE | Downregulated |
| 0,219117652 | 0,77024962 | PDCD1LG2 | FALSE | Upregulated |
| -0,115565739 | 0,77024962 | PNKD | FALSE | Downregulated |
| -0,182667257 | 0,77024962 | PTGDR2 | FALSE | Downregulated |
| 0,117951276 | 0,77024962 | ST3GAL2 | FALSE | Upregulated |
| -0,17556968 | 0,770339792 | FAM221B | FALSE | Downregulated |
| 0,172235309 | 0,770339792 | MTMR14 | FALSE | Upregulated |
| -0,218119274 | 0,770339792 | SMAGP | FALSE | Downregulated |
| -0,23309981 | 0,770376865 | POMK | FALSE | Downregulated |
| 0,139445667 | 0,770376865 | UNC119B | FALSE | Upregulated |
| 0,212573257 | 0,77071328 | ACCS | FALSE | Upregulated |
| 0,182999472 | 0,77071328 | ATP6V1E1 | FALSE | Upregulated |
| -0,104005707 | 0,77071328 | DICER1 | FALSE | Downregulated |
| -0,187621165 | 0,77071328 | FOLH1 | FALSE | Downregulated |
| 0,428085434 | 0,77071328 | LIN52 | FALSE | Upregulated |
| 0,326708367 | 0,77071328 | LOC101927895 | FALSE | Upregulated |
| 0,250600487 | 0,77071328 | NCAM1 | FALSE | Upregulated |
| 0,38307182 | 0,77071328 | OVCA2 | FALSE | Upregulated |
| 0,091656985 | 0,77071328 | PNRC1 | FALSE | Upregulated |
| -0,242872262 | 0,77071328 | RCBTB2 | FALSE | Downregulated |
| -0,201490496 | 0,77071328 | RNF135 | FALSE | Downregulated |
| 0,325921423 | 0,77071328 | SLC6A16 | FALSE | Upregulated |
| -0,197232917 | 0,77071328 | SORCS2 | FALSE | Downregulated |
| 0,098453367 | 0,77071328 | TRIM56 | FALSE | Upregulated |
| 0,118805608 | 0,77071328 | TSSK3 | FALSE | Upregulated |
| 2,193379685 | 0,77071328 | UBE2F-SCLY | FALSE | Upregulated |
| -0,107793251 | 0,77071328 | USP32 | FALSE | Downregulated |
| 0,177953082 | 0,771123869 | NLRP1 | FALSE | Upregulated |
| -0,54621072 | 0,771123869 | TMC4 | FALSE | Downregulated |
| 0,339365967 | 0,771261338 | TMEM26 | FALSE | Upregulated |
| -0,100888417 | 0,77145742 | HPD | FALSE | Downregulated |

| | | | | |
|---|---|---|---|---|
| 0,475356319 | 0,771468639 | C16orf91 | FALSE | Upregulated |
| 0,243693553 | 0,771468639 | IL1RN | FALSE | Upregulated |
| -0,23512505 | 0,771468639 | RTCA | FALSE | Downregulated |
| 0,120125785 | 0,771800505 | GNL3L | FALSE | Upregulated |
| -0,112060912 | 0,77181391 | ARHGEF7 | FALSE | Downregulated |
| 0,159735589 | 0,77181391 | GIPC1 | FALSE | Upregulated |
| -0,085411076 | 0,77181391 | KIAA0430 | FALSE | Downregulated |
| -0,296251322 | 0,77181391 | KRBA2 | FALSE | Downregulated |
| 0,537509399 | 0,77181391 | LOC440173 | FALSE | Upregulated |
| -0,224717946 | 0,77181391 | REN | FALSE | Downregulated |
| -0,176201338 | 0,77181391 | SDF2 | FALSE | Downregulated |
| 0,088253179 | 0,771919796 | APEH | FALSE | Upregulated |
| 0,135294923 | 0,771919796 | FTSJ3 | FALSE | Upregulated |
| -0,108096349 | 0,771919796 | NMT2 | FALSE | Downregulated |
| -0,1109173 | 0,771919796 | ORC4 | FALSE | Downregulated |
| 0,107888673 | 0,771919796 | SLC27A1 | FALSE | Upregulated |
| -0,174169598 | 0,771924857 | AC144449.1 | FALSE | Downregulated |
| -0,195128646 | 0,771924857 | BTF3L4 | FALSE | Downregulated |
| 0,501355666 | 0,771924857 | LOC100133920 | FALSE | Upregulated |
| 0,306700677 | 0,771924857 | MGC70870 | FALSE | Upregulated |
| 0,175095915 | 0,771924857 | RBBP5 | FALSE | Upregulated |
| 0,2084361 | 0,771924857 | STS | FALSE | Upregulated |
| -0,481974179 | 0,771924857 | VSTM2L | FALSE | Downregulated |
| 0,103331195 | 0,771961102 | CREG1 | FALSE | Upregulated |
| 0,220475285 | 0,771973654 | IFT172 | FALSE | Upregulated |
| -0,624879971 | 0,771973654 | LOC101929141 | FALSE | Downregulated |
| -0,511036095 | 0,772080192 | POU6F2 | FALSE | Downregulated |
| -0,117395117 | 0,772333684 | PHF8 | FALSE | Downregulated |
| 0,11451646 | 0,772608609 | FXR2 | FALSE | Upregulated |
| -0,201494148 | 0,772608609 | SRSF7 | FALSE | Downregulated |
| -0,167709326 | 0,772981528 | SWT1 | FALSE | Downregulated |
| -0,421532821 | 0,772984229 | GATA6-AS1 | FALSE | Downregulated |
| -0,416748302 | 0,772984229 | GSTT2B | FALSE | Downregulated |
| 0,083349502 | 0,772984229 | TAOK2 | FALSE | Upregulated |
| 0,244977369 | 0,773062459 | TFB1M | FALSE | Upregulated |
| -0,187861311 | 0,773100532 | MAPK9 | FALSE | Downregulated |
| 0,161092245 | 0,773192801 | ZNF410 | FALSE | Upregulated |
| -0,436207931 | 0,773458767 | LOC100652901 | FALSE | Downregulated |
| -0,126726319 | 0,773458767 | PSMB2 | FALSE | Downregulated |
| 0,19146747 | 0,773458767 | VPS35 | FALSE | Upregulated |
| 0,291896171 | 0,773458767 | WDR54 | FALSE | Upregulated |
| 0,471405919 | 0,773492315 | WRAP53 | FALSE | Upregulated |
| 0,087860226 | 0,773578186 | AK2 | FALSE | Upregulated |
| 0,101301176 | 0,773578186 | FGD6 | FALSE | Upregulated |
| -0,45562796 | 0,773578186 | GOLGA7B | FALSE | Downregulated |
| -0,210091815 | 0,773578186 | HARS2 | FALSE | Downregulated |
| 0,18770814 | 0,773578186 | ING4 | FALSE | Upregulated |
| -0,119468385 | 0,773578186 | NCKAP1 | FALSE | Downregulated |
| -0,122972232 | 0,773578186 | SHROOM1 | FALSE | Downregulated |
| 0,206225835 | 0,773578186 | SP140L | FALSE | Upregulated |
| -0,131905806 | 0,773658727 | KLF9 | FALSE | Downregulated |

| | | | | |
|---|---|---|---|---|
| 0,146320712 | 0,773658727 | LACTB | FALSE | Upregulated |
| -0,400511261 | 0,773658727 | LOC101928195 | FALSE | Downregulated |
| -0,338509139 | 0,773658727 | LOC102724791 | FALSE | Downregulated |
| 0,1825255 | 0,773658727 | TPGS1 | FALSE | Upregulated |
| 0,241631262 | 0,773658727 | ZNF346 | FALSE | Upregulated |
| -0,140150927 | 0,773917656 | CHN2 | FALSE | Downregulated |
| -0,133237389 | 0,774131003 | EHMT2 | FALSE | Downregulated |
| -0,165377958 | 0,774268689 | PKIG | FALSE | Downregulated |
| 0,231088682 | 0,774268689 | RP11-279D17.2 | FALSE | Upregulated |
| -0,181535704 | 0,774268689 | SPICE1 | FALSE | Downregulated |
| 0,234636699 | 0,774268689 | ZDHHC12 | FALSE | Upregulated |
| -0,425757452 | 0,774890556 | LOC102723611 | FALSE | Downregulated |
| 0,203234003 | 0,775034173 | RP11-622A1.2 | FALSE | Upregulated |
| 0,312520222 | 0,77533417 | LOC441242 | FALSE | Upregulated |
| 0,407422625 | 0,77533417 | STAG3L1 | FALSE | Upregulated |
| -0,289328906 | 0,77533448 | FAM131A | FALSE | Downregulated |
| -0,09953113 | 0,77533448 | PCYT2 | FALSE | Downregulated |
| 0,110930059 | 0,77543872 | BEST1 | FALSE | Upregulated |
| 0,309223712 | 0,77543872 | HDGFRP3 | FALSE | Upregulated |
| 0,264006981 | 0,77543872 | LECT1 | FALSE | Upregulated |
| 0,099148282 | 0,77543872 | MFN2 | FALSE | Upregulated |
| 0,181141547 | 0,775489821 | IDI2 | FALSE | Upregulated |
| 0,309207535 | 0,775489821 | NME1 | FALSE | Upregulated |
| 0,102548328 | 0,775631322 | CMTR1 | FALSE | Upregulated |
| 0,114056681 | 0,775644615 | ANXA4 | FALSE | Upregulated |
| -0,163986319 | 0,775644615 | RIMKLA | FALSE | Downregulated |
| 0,190147416 | 0,775644615 | RP11-14D22.5 | FALSE | Upregulated |
| 0,204725029 | 0,775644615 | TTC30B | FALSE | Upregulated |
| -0,189711862 | 0,775662844 | ECD | FALSE | Downregulated |
| -0,228330675 | 0,775824106 | SLC35E3 | FALSE | Downregulated |
| 0,12020891 | 0,776257091 | GLUL | FALSE | Upregulated |
| 0,180450982 | 0,776260207 | ATAD3B | FALSE | Upregulated |
| -0,376859443 | 0,776286349 | ZCCHC4 | FALSE | Downregulated |
| -0,166952617 | 0,776420928 | PLBD2 | FALSE | Downregulated |
| -0,180494023 | 0,776447791 | KDM5B | FALSE | Downregulated |
| 0,226586425 | 0,776447791 | MRPL27 | FALSE | Upregulated |
| 0,341959 | 0,776447791 | PGM2L1 | FALSE | Upregulated |
| 0,276439754 | 0,776499911 | MED21 | FALSE | Upregulated |
| -0,351807421 | 0,776499911 | PDE6D | FALSE | Downregulated |
| 0,315387422 | 0,776499911 | WTIP | FALSE | Upregulated |
| -0,436106753 | 0,776758454 | RAPGEF4-AS1 | FALSE | Downregulated |
| -0,51165432 | 0,776789893 | TAS2R13 | FALSE | Downregulated |
| 0,770193325 | 0,77692072 | GOLGA6L4 | FALSE | Upregulated |
| 0,322575712 | 0,77692072 | LIN28A | FALSE | Upregulated |
| -0,230002356 | 0,77692072 | LINC00894 | FALSE | Downregulated |
| -0,262493841 | 0,77692072 | MTUS2 | FALSE | Downregulated |
| 0,146072966 | 0,77692072 | PLCE1 | FALSE | Upregulated |
| 0,426338813 | 0,77692072 | POC5 | FALSE | Upregulated |
| 0,402304703 | 0,77692072 | SLC38A6 | FALSE | Upregulated |
| -0,516047088 | 0,77692072 | ZNF876P | FALSE | Downregulated |
| -0,187712181 | 0,776962921 | NARG2 | FALSE | Downregulated |

| | | | | |
|---|---|---|---|---|
| 0,153269105 | 0,777070459 | ATP5J | FALSE | Upregulated |
| 0,399312555 | 0,777070459 | LOC101927098 | FALSE | Upregulated |
| 0,255948103 | 0,777115028 | ARHGEF16 | FALSE | Upregulated |
| 0,06748048 | 0,777115028 | IGF2R | FALSE | Upregulated |
| 0,385717582 | 0,777115028 | PDZRN4 | FALSE | Upregulated |
| -0,074744948 | 0,777115028 | PRR12 | FALSE | Downregulated |
| 0,296732635 | 0,777132129 | LOC101928505 | FALSE | Upregulated |
| 0,423900309 | 0,777175735 | LOC102723425 | FALSE | Upregulated |
| -0,358697127 | 0,77721227 | COMMD10 | FALSE | Downregulated |
| -0,152040577 | 0,77721227 | IP6K2 | FALSE | Downregulated |
| 0,299344719 | 0,77721227 | ST20 | FALSE | Upregulated |
| 0,103730158 | 0,77746805 | GGA3 | FALSE | Upregulated |
| 0,16524498 | 0,77746805 | UBOX5-AS1 | FALSE | Upregulated |
| 0,138614249 | 0,77755336 | PPP2CA | FALSE | Upregulated |
| -0,079386965 | 0,777736073 | ESYT1 | FALSE | Downregulated |
| -0,078311276 | 0,777736073 | MTSS1L | FALSE | Downregulated |
| -0,135331627 | 0,777736073 | SMC3 | FALSE | Downregulated |
| 0,096968543 | 0,777736073 | TMEM30B | FALSE | Upregulated |
| 0,209593066 | 0,777736073 | UBE2F | FALSE | Upregulated |
| 0,10251328 | 0,777736073 | ZNF140 | FALSE | Upregulated |
| -0,216426334 | 0,7778269 | ATP6V1H | FALSE | Downregulated |
| -0,148073543 | 0,7778269 | DBP | FALSE | Downregulated |
| 0,129817583 | 0,7778269 | PFKFB1 | FALSE | Upregulated |
| 0,159598511 | 0,778127272 | TIMM23 | FALSE | Upregulated |
| 0,515314294 | 0,778131346 | DCLK1 | FALSE | Upregulated |
| 0,141986616 | 0,778166438 | DHX32 | FALSE | Upregulated |
| -0,151160041 | 0,778166438 | LOC101928081 | FALSE | Downregulated |
| 0,301391347 | 0,778166438 | TOR2A | FALSE | Upregulated |
| -0,080525539 | 0,778222603 | FNDC3A | FALSE | Downregulated |
| -0,095455992 | 0,778268194 | DGKD | FALSE | Downregulated |
| 0,254584296 | 0,778302009 | FDXR | FALSE | Upregulated |
| -0,854707367 | 0,778452335 | STX16-NPEPL1 | FALSE | Downregulated |
| 0,140278446 | 0,778568505 | MAK | FALSE | Upregulated |
| 0,247348178 | 0,778568505 | NOMO2 | FALSE | Upregulated |
| 0,140099149 | 0,778568505 | PIP5K1A | FALSE | Upregulated |
| -0,105497592 | 0,778568505 | PPP2R1A | FALSE | Downregulated |
| 0,140324104 | 0,778568505 | TIGD5 | FALSE | Upregulated |
| 0,221676795 | 0,778766211 | CAV2 | FALSE | Upregulated |
| -0,487041091 | 0,778766211 | LINC00950 | FALSE | Downregulated |
| -0,392067941 | 0,778766211 | ZNF214 | FALSE | Downregulated |
| 0,517249389 | 0,778796854 | FLJ45872 | FALSE | Upregulated |
| 0,221026356 | 0,778796854 | LAMTOR4 | FALSE | Upregulated |
| -0,11451891 | 0,778796854 | XIAP | FALSE | Downregulated |
| -0,101030271 | 0,778842281 | EIF2AK4 | FALSE | Downregulated |
| 0,149841673 | 0,778903658 | ACTR8 | FALSE | Upregulated |
| 0,111843101 | 0,778903658 | FLRT1 | FALSE | Upregulated |
| -0,102477928 | 0,779067988 | NDFIP1 | FALSE | Downregulated |
| 0,281793953 | 0,779067988 | PPARG | FALSE | Upregulated |
| 0,25073706 | 0,779105333 | C10orf11 | FALSE | Upregulated |
| -0,244268206 | 0,779364901 | NCK1 | FALSE | Downregulated |
| 0,11769844 | 0,779364901 | VAPB | FALSE | Upregulated |

| | | | | |
|---|---|---|---|---|
| 0,128076827 | 0,779667791 | NUDC | FALSE | Upregulated |
| 0,141768439 | 0,779667791 | THAP4 | FALSE | Upregulated |
| 0,386838343 | 0,779675873 | KIAA1024 | FALSE | Upregulated |
| -0,515590163 | 0,779675873 | LOC101927279 | FALSE | Downregulated |
| 0,40943124 | 0,779675873 | LOC149351 | FALSE | Upregulated |
| 0,165700764 | 0,779773946 | C19orf26 | FALSE | Upregulated |
| 0,34123401 | 0,779945583 | CILP | FALSE | Upregulated |
| 0,413905086 | 0,779945583 | LOC101929410 | FALSE | Upregulated |
| 0,158006617 | 0,779945583 | SOX10 | FALSE | Upregulated |
| -0,367157179 | 0,77998276 | HCN2 | FALSE | Downregulated |
| -0,085178164 | 0,77998276 | RBMX | FALSE | Downregulated |
| -0,138317639 | 0,77998276 | ZBED3 | FALSE | Downregulated |
| -0,317852445 | 0,780018227 | AC147651.4 | FALSE | Downregulated |
| -0,087624985 | 0,780018227 | FAM134A | FALSE | Downregulated |
| -0,17891784 | 0,780018227 | HBCBP | FALSE | Downregulated |
| 0,195098172 | 0,780018227 | MTPAP | FALSE | Upregulated |
| -0,603470305 | 0,7800271 | LOC102724235 | FALSE | Downregulated |
| 0,330338157 | 0,780199926 | NCR3LG1 | FALSE | Upregulated |
| 0,254863173 | 0,780199926 | SRD5A3 | FALSE | Upregulated |
| 0,1834063 | 0,780199926 | WARS2 | FALSE | Upregulated |
| -0,135912036 | 0,780335916 | SPNS1 | FALSE | Downregulated |
| -0,253216895 | 0,780390883 | CHORDC1 | FALSE | Downregulated |
| -0,132571158 | 0,780494336 | C2orf68 | FALSE | Downregulated |
| 0,15037728 | 0,78055712 | SPAG1 | FALSE | Upregulated |
| 0,113665864 | 0,780701885 | FAM104A | FALSE | Upregulated |
| -0,428916791 | 0,780701885 | FLJ38122 | FALSE | Downregulated |
| 0,161171317 | 0,780701885 | PER1 | FALSE | Upregulated |
| 0,17676515 | 0,780701885 | UBXN8 | FALSE | Upregulated |
| 0,135710395 | 0,780742462 | ACP5 | FALSE | Upregulated |
| -0,187144557 | 0,780772224 | CCDC3 | FALSE | Downregulated |
| -0,329368379 | 0,780772224 | HAT1 | FALSE | Downregulated |
| -0,145292563 | 0,7807726 | SDCCAG3 | FALSE | Downregulated |
| -0,094268621 | 0,781038547 | CFH | FALSE | Downregulated |
| -0,132882737 | 0,781038547 | NICN1 | FALSE | Downregulated |
| -0,159027749 | 0,781038547 | NPDC1 | FALSE | Downregulated |
| -0,199637736 | 0,781038547 | PIP5K1P1 | FALSE | Downregulated |
| 0,134024337 | 0,781038547 | PPP1R3E | FALSE | Upregulated |
| 0,360385391 | 0,781038547 | PTPRO | FALSE | Upregulated |
| 0,280985894 | 0,781038547 | SLIT2 | FALSE | Upregulated |
| -0,122130014 | 0,781101362 | GPAA1 | FALSE | Downregulated |
| -1,084340162 | 0,78121766 | ARL2-SNX15 | FALSE | Downregulated |
| -0,562464873 | 0,781514097 | LOC101927934 | FALSE | Downregulated |
| 0,116313118 | 0,781758977 | TERF2IP | FALSE | Upregulated |
| 0,188874067 | 0,781877458 | ACVRL1 | FALSE | Upregulated |
| 0,167153833 | 0,781877458 | HDAC3 | FALSE | Upregulated |
| -0,235955999 | 0,781877458 | LOC100506083 | FALSE | Downregulated |
| 0,319152647 | 0,781877458 | NPPA | FALSE | Upregulated |
| -0,338314636 | 0,781877458 | RP11-899L11.1 | FALSE | Downregulated |
| 0,496250101 | 0,781903999 | PPP1R32 | FALSE | Upregulated |
| 0,13011388 | 0,781913322 | TMED9 | FALSE | Upregulated |
| 0,186999865 | 0,782002693 | AIFM2 | FALSE | Upregulated |

| | | | | |
|---|---|---|---|---|
| 0,214917395 | 0,782072225 | NPIPA5 | FALSE | Upregulated |
| -0,177105279 | 0,782244128 | MAP3K9 | FALSE | Downregulated |
| 0,124737142 | 0,782325533 | NPHP3 | FALSE | Upregulated |
| -0,217290353 | 0,782330429 | ADAMTS8 | FALSE | Downregulated |
| -0,358094732 | 0,782330429 | FLJ12825 | FALSE | Downregulated |
| -0,132052892 | 0,782330429 | TRUB2 | FALSE | Downregulated |
| 0,598665831 | 0,782380015 | LOC653653 | FALSE | Upregulated |
| -0,189185014 | 0,782533057 | LOC101928783 | FALSE | Downregulated |
| -0,130281506 | 0,782533057 | PAIP2 | FALSE | Downregulated |
| -0,30609894 | 0,782533057 | SNORA48 | FALSE | Downregulated |
| -0,183373187 | 0,782533057 | UBA2 | FALSE | Downregulated |
| -0,225356821 | 0,782825056 | MYADM | FALSE | Downregulated |
| 0,290474804 | 0,782825056 | VNN3 | FALSE | Upregulated |
| 0,159581835 | 0,782915772 | AP1AR | FALSE | Upregulated |
| 0,396267329 | 0,782915772 | ZBTB26 | FALSE | Upregulated |
| -0,120735579 | 0,783285812 | ZDHHC8 | FALSE | Downregulated |
| 0,4055573 | 0,783319459 | GATA3 | FALSE | Upregulated |
| -0,140643611 | 0,783373281 | RP11-290F20.2 | FALSE | Downregulated |
| 0,338622657 | 0,783405342 | CERS3 | FALSE | Upregulated |
| 0,179327636 | 0,783405342 | INSIG1 | FALSE | Upregulated |
| -0,126707919 | 0,783405342 | PHF20 | FALSE | Downregulated |
| 0,2808598 | 0,783405342 | RFT1 | FALSE | Upregulated |
| 0,268134001 | 0,783405342 | UBL7 | FALSE | Upregulated |
| -0,096766455 | 0,783405342 | WAPAL | FALSE | Downregulated |
| -0,122918091 | 0,783424027 | PSMD5 | FALSE | Downregulated |
| -0,475716904 | 0,783618155 | RASA4CP | FALSE | Downregulated |
| 0,361337286 | 0,783632173 | LINGO4 | FALSE | Upregulated |
| 0,432131385 | 0,783632173 | LOC101930129 | FALSE | Upregulated |
| 0,095924752 | 0,783632173 | MTCH1 | FALSE | Upregulated |
| -0,163350281 | 0,783632173 | PCNP | FALSE | Downregulated |
| -0,089007145 | 0,783686691 | FAM193B | FALSE | Downregulated |
| -0,233804953 | 0,783686691 | RBP2 | FALSE | Downregulated |
| -1,316713418 | 0,783789021 | BLOC1S1-RDH5 | FALSE | Downregulated |
| -0,06201006 | 0,783789021 | BRD4 | FALSE | Downregulated |
| 0,452569262 | 0,783789021 | EPHA4 | FALSE | Upregulated |
| -0,146072183 | 0,783789021 | FAM211B | FALSE | Downregulated |
| -0,394325061 | 0,783789021 | ITGA10 | FALSE | Downregulated |
| -0,319893343 | 0,783789021 | KIF19 | FALSE | Downregulated |
| 0,250013014 | 0,783789021 | LGI4 | FALSE | Upregulated |
| 0,16794916 | 0,783789021 | MCF2L | FALSE | Upregulated |
| -0,140946726 | 0,783789021 | PSMB3 | FALSE | Downregulated |
| -0,129515281 | 0,783789021 | RNF43 | FALSE | Downregulated |
| -0,184990918 | 0,783789021 | SNORD3A | FALSE | Downregulated |
| -0,190050173 | 0,783789021 | SVEP1 | FALSE | Downregulated |
| -0,413228039 | 0,783789021 | WDYHV1 | FALSE | Downregulated |
| 0,162922684 | 0,783914317 | LNX1 | FALSE | Upregulated |
| -0,138753823 | 0,784125617 | AKR7L | FALSE | Downregulated |
| -0,116277153 | 0,784125617 | FGF2 | FALSE | Downregulated |
| 0,161636055 | 0,784125617 | LOC101929986 | FALSE | Upregulated |
| 0,176208114 | 0,784125617 | SDAD1 | FALSE | Upregulated |
| -0,424410801 | 0,784125617 | TMEM116 | FALSE | Downregulated |

| | | | | |
|---|---|---|---|---|
| -0,321652326 | 0,784125617 | ZNF818P | FALSE | Downregulated |
| 0,356951625 | 0,784140354 | PARK2 | FALSE | Upregulated |
| 0,234203264 | 0,784386369 | TRPA1 | FALSE | Upregulated |
| 0,219424455 | 0,784596905 | FAM126A | FALSE | Upregulated |
| -0,128323693 | 0,784596905 | UBE2J2 | FALSE | Downregulated |
| -0,207387473 | 0,784654159 | EIF4E3 | FALSE | Downregulated |
| 0,144516828 | 0,784685478 | EMC10 | FALSE | Upregulated |
| 0,21604273 | 0,784685478 | LOC100287852 | FALSE | Upregulated |
| -0,114388325 | 0,784685478 | SERPINA4 | FALSE | Downregulated |
| 0,278678729 | 0,784756884 | PI4KAP1 | FALSE | Upregulated |
| 0,149789426 | 0,784941938 | ADAM1A | FALSE | Upregulated |
| -0,277070737 | 0,785170905 | LOC285638 | FALSE | Downregulated |
| 0,289355341 | 0,785299105 | ZNF20 | FALSE | Upregulated |
| 0,259049959 | 0,785330182 | RNASEH1 | FALSE | Upregulated |
| 0,36261709 | 0,785621345 | CYTIP | FALSE | Upregulated |
| -0,27437127 | 0,78565066 | COLQ | FALSE | Downregulated |
| -0,165094977 | 0,785671828 | ZNF660 | FALSE | Downregulated |
| 0,295031177 | 0,785716288 | BAK1 | FALSE | Upregulated |
| -0,134179542 | 0,785716288 | MAPK1 | FALSE | Downregulated |
| 0,13127245 | 0,785716288 | SH3BP5L | FALSE | Upregulated |
| 0,229479178 | 0,785779339 | ZC3HC1 | FALSE | Upregulated |
| 0,334564468 | 0,786015974 | RGS11 | FALSE | Upregulated |
| 0,104624092 | 0,786015974 | SIAE | FALSE | Upregulated |
| -0,255944361 | 0,786125016 | RPL31P11 | FALSE | Downregulated |
| 0,330803162 | 0,78627002 | ICAM2 | FALSE | Upregulated |
| -0,239066811 | 0,786574406 | SLC2A13 | FALSE | Downregulated |
| -0,110243081 | 0,786698747 | NF1 | FALSE | Downregulated |
| -0,229771499 | 0,786698747 | STL | FALSE | Downregulated |
| -0,395162982 | 0,786856746 | CDKN3 | FALSE | Downregulated |
| 0,4846021 | 0,786856746 | EIF5AL1 | FALSE | Upregulated |
| 0,277206036 | 0,786856746 | EOGT | FALSE | Upregulated |
| -0,433400509 | 0,786856746 | HAMP | FALSE | Downregulated |
| -0,172675948 | 0,786856746 | NOS3 | FALSE | Downregulated |
| 0,248870839 | 0,786856746 | UTP6 | FALSE | Upregulated |
| 0,109879959 | 0,786874185 | LINC00242 | FALSE | Upregulated |
| -0,295337733 | 0,787012711 | GRIA3 | FALSE | Downregulated |
| -0,198977545 | 0,787012711 | TSPYL2 | FALSE | Downregulated |
| 0,28001002 | 0,787022498 | BLOC1S2 | FALSE | Upregulated |
| -0,248977646 | 0,787022498 | MCF2L-AS1 | FALSE | Downregulated |
| 0,088088543 | 0,787022498 | MYO1E | FALSE | Upregulated |
| -0,193746073 | 0,787022498 | NENF | FALSE | Downregulated |
| 0,237323309 | 0,787022498 | TMOD2 | FALSE | Upregulated |
| 0,137191677 | 0,787022498 | WDR37 | FALSE | Upregulated |
| 0,120956884 | 0,787106519 | C14orf164 | FALSE | Upregulated |
| -0,132129153 | 0,787106519 | CARS2 | FALSE | Downregulated |
| 0,260623843 | 0,787106519 | PIGH | FALSE | Upregulated |
| 0,168442907 | 0,787106519 | VDAC3 | FALSE | Upregulated |
| -0,130869295 | 0,787175621 | AGGF1 | FALSE | Downregulated |
| 0,293738469 | 0,787175621 | C14orf142 | FALSE | Upregulated |
| 0,108201654 | 0,787175621 | DROSHA | FALSE | Upregulated |
| 0,274791133 | 0,787175621 | GEMIN2 | FALSE | Upregulated |

| | | | | |
|---|---|---|---|---|
| 0,431937081 | 0,787175621 | LOC100652736 | FALSE | Upregulated |
| 0,161236119 | 0,787175621 | LOC101927685 | FALSE | Upregulated |
| 0,089560077 | 0,787175621 | LOC102724445 | FALSE | Upregulated |
| -0,269797259 | 0,787175621 | PPM1L | FALSE | Downregulated |
| 0,101522541 | 0,787175621 | TXNDC11 | FALSE | Upregulated |
| 0,213639089 | 0,787233121 | KCTD15 | FALSE | Upregulated |
| 0,105860307 | 0,78737999 | HM13 | FALSE | Upregulated |
| 0,424224941 | 0,787423597 | LINC01137 | FALSE | Upregulated |
| -0,258546282 | 0,787499602 | ZNF565 | FALSE | Downregulated |
| 0,137911492 | 0,787532021 | DCUN1D1 | FALSE | Upregulated |
| 0,087323273 | 0,787532021 | SNRK | FALSE | Upregulated |
| -0,329750637 | 0,787532021 | SOGA3 | FALSE | Downregulated |
| 0,120165417 | 0,787532021 | TXNDC17 | FALSE | Upregulated |
| -0,187063638 | 0,787741592 | BCDIN3D | FALSE | Downregulated |
| -0,157369338 | 0,787741592 | DPP8 | FALSE | Downregulated |
| -0,338796235 | 0,787741592 | LINC00663 | FALSE | Downregulated |
| 0,157624053 | 0,7878572 | EMILIN3 | FALSE | Upregulated |
| -0,318332961 | 0,7878572 | TMED3 | FALSE | Downregulated |
| -0,253759424 | 0,788081172 | FZD8 | FALSE | Downregulated |
| 0,197993514 | 0,788205331 | ACTA2 | FALSE | Upregulated |
| 0,134155579 | 0,788205331 | AP3B1 | FALSE | Upregulated |
| -0,177995425 | 0,788205331 | C17orf47 | FALSE | Downregulated |
| 0,132493552 | 0,788205331 | ENTPD6 | FALSE | Upregulated |
| 0,391957608 | 0,788205331 | LOC101927842 | FALSE | Upregulated |
| -0,092387331 | 0,788205331 | NPM1 | FALSE | Downregulated |
| 0,444058529 | 0,788205331 | TRABD2A | FALSE | Upregulated |
| -0,170701877 | 0,78832675 | CDK19 | FALSE | Downregulated |
| -0,075340362 | 0,78832675 | KDM2A | FALSE | Downregulated |
| 0,146891684 | 0,78832675 | SAMD10 | FALSE | Upregulated |
| -0,101059829 | 0,788663455 | CDH5 | FALSE | Downregulated |
| 0,108863402 | 0,788663455 | DHX16 | FALSE | Upregulated |
| 0,452816044 | 0,788663455 | MRPS17 | FALSE | Upregulated |
| 0,136667124 | 0,789091722 | RING1 | FALSE | Upregulated |
| -0,234256983 | 0,789217889 | ALG8 | FALSE | Downregulated |
| 0,100271566 | 0,789217889 | DNHD1 | FALSE | Upregulated |
| -0,420365231 | 0,789217889 | MPP7 | FALSE | Downregulated |
| -0,180724996 | 0,789217889 | NRDE2 | FALSE | Downregulated |
| -0,260879676 | 0,789217889 | ZNF569 | FALSE | Downregulated |
| -0,194805041 | 0,789217889 | ZNF621 | FALSE | Downregulated |
| 0,249262581 | 0,789276753 | RPL26L1 | FALSE | Upregulated |
| -0,173822966 | 0,789335419 | ABCB6 | FALSE | Downregulated |
| -0,079590897 | 0,789362663 | SNED1 | FALSE | Downregulated |
| 0,13539839 | 0,789699347 | ASB8 | FALSE | Upregulated |
| -0,235528333 | 0,789699347 | CTU2 | FALSE | Downregulated |
| -0,10186258 | 0,789699347 | SNX17 | FALSE | Downregulated |
| -0,235653486 | 0,789699347 | TMEM11 | FALSE | Downregulated |
| 0,114642501 | 0,789763683 | C1orf63 | FALSE | Upregulated |
| 0,14574405 | 0,789763683 | C22orf39 | FALSE | Upregulated |
| -0,117497345 | 0,790099608 | ZNF154 | FALSE | Downregulated |
| 0,40413517 | 0,79024013 | FAM83D | FALSE | Upregulated |
| -0,16697227 | 0,790260621 | DPP4 | FALSE | Downregulated |

| | | | | |
|---|---|---|---|---|
| 0,274787711 | 0,790260621 | HSDL1 | FALSE | Upregulated |
| -0,107791864 | 0,790260621 | NASP | FALSE | Downregulated |
| -0,393653808 | 0,790419334 | LOC101927693 | FALSE | Downregulated |
| 0,221037795 | 0,790595641 | NVL | FALSE | Upregulated |
| 0,244042791 | 0,790595641 | SLC10A7 | FALSE | Upregulated |
| 0,140694412 | 0,790808474 | HAVCR1P1 | FALSE | Upregulated |
| 0,359485268 | 0,790808474 | LOC102723709 | FALSE | Upregulated |
| 0,254533261 | 0,790808474 | NEK3 | FALSE | Upregulated |
| -0,153239471 | 0,790808474 | NLRC4 | FALSE | Downregulated |
| 0,341355422 | 0,790808474 | PRRG2 | FALSE | Upregulated |
| 0,16940253 | 0,791215106 | DUSP22 | FALSE | Upregulated |
| -0,238194701 | 0,791215106 | FLYWCH2 | FALSE | Downregulated |
| -0,095004093 | 0,791215106 | TRANK1 | FALSE | Downregulated |
| -0,228693322 | 0,791278089 | MIER2 | FALSE | Downregulated |
| 0,203188488 | 0,791394839 | C9orf41 | FALSE | Upregulated |
| -0,368857606 | 0,791403854 | TMEM258 | FALSE | Downregulated |
| 0,071492543 | 0,791589508 | HNRNPA1 | FALSE | Upregulated |
| -0,447355161 | 0,791753537 | LOC100996717 | FALSE | Downregulated |
| -0,215341196 | 0,791777697 | ARHGEF10 | FALSE | Downregulated |
| 0,402763403 | 0,791777697 | FAM83A | FALSE | Upregulated |
| 0,115581503 | 0,791777697 | IWS1 | FALSE | Upregulated |
| -0,308130751 | 0,791777697 | LTV1 | FALSE | Downregulated |
| 0,250530828 | 0,791777697 | SLC23A3 | FALSE | Upregulated |
| 0,154078592 | 0,791831899 | TTN | FALSE | Upregulated |
| -0,566036765 | 0,791963976 | ANKRD20A12P | FALSE | Downregulated |
| -0,516272213 | 0,791963976 | BST1 | FALSE | Downregulated |
| 0,2667623 | 0,791963976 | C9orf156 | FALSE | Upregulated |
| 0,220397342 | 0,791963976 | CCDC68 | FALSE | Upregulated |
| 0,685961867 | 0,791963976 | CDK3 | FALSE | Upregulated |
| -0,136341262 | 0,791963976 | COG7 | FALSE | Downregulated |
| 0,218880836 | 0,791963976 | DKFZP434K028 | FALSE | Upregulated |
| -0,452794265 | 0,791963976 | HERC2P7 | FALSE | Downregulated |
| 0,121269034 | 0,791963976 | LOC101929109 | FALSE | Upregulated |
| 0,235702585 | 0,791963976 | PDE4A | FALSE | Upregulated |
| -0,086008573 | 0,791963976 | POLR2E | FALSE | Downregulated |
| 0,277663134 | 0,791963976 | SLC10A3 | FALSE | Upregulated |
| 0,157885299 | 0,791963976 | SLC44A2 | FALSE | Upregulated |
| 0,120432352 | 0,791963976 | SMCR8 | FALSE | Upregulated |
| -0,308809283 | 0,791963976 | SYNPO2L | FALSE | Downregulated |
| 0,113114784 | 0,791963976 | TAF6L | FALSE | Upregulated |
| -0,234029006 | 0,791963976 | ZNF471 | FALSE | Downregulated |
| -0,200706569 | 0,791963976 | ZNF69 | FALSE | Downregulated |
| -0,434703487 | 0,792293393 | RPL13AP17 | FALSE | Downregulated |
| 0,323195128 | 0,792465541 | LINC00115 | FALSE | Upregulated |
| 0,148842105 | 0,792465541 | TPGS2 | FALSE | Upregulated |
| -0,165113538 | 0,792567914 | LRSAM1 | FALSE | Downregulated |
| 0,145672381 | 0,792567914 | TJAP1 | FALSE | Upregulated |
| 0,434894144 | 0,792663523 | NFKBID | FALSE | Upregulated |
| -0,107092408 | 0,792663523 | WDR11 | FALSE | Downregulated |
| -0,150422956 | 0,79271824 | DHX34 | FALSE | Downregulated |
| 0,208466796 | 0,79271824 | LINC00176 | FALSE | Upregulated |

| | | | | |
|---|---|---|---|---|
| 0,366945974 | 0,79271824 | LOC100499484 | FALSE | Upregulated |
| -0,146830897 | 0,79271824 | SSR2 | FALSE | Downregulated |
| -0,083872177 | 0,79271824 | TNFAIP8L1 | FALSE | Downregulated |
| -0,328827933 | 0,792767371 | C12orf45 | FALSE | Downregulated |
| 0,176234955 | 0,792767371 | CCT6A | FALSE | Upregulated |
| -0,457850137 | 0,792767371 | CNGB3 | FALSE | Downregulated |
| -0,401836386 | 0,792767371 | CSRNP3 | FALSE | Downregulated |
| 0,416504827 | 0,792767371 | DKFZP434I0714 | FALSE | Upregulated |
| 0,127888758 | 0,792767371 | FAM186B | FALSE | Upregulated |
| 0,477151087 | 0,792767371 | KLHL33 | FALSE | Upregulated |
| -0,145988212 | 0,792767371 | NGRN | FALSE | Downregulated |
| -0,087394125 | 0,792767371 | POGZ | FALSE | Downregulated |
| -0,350869846 | 0,792767371 | RPIA | FALSE | Downregulated |
| 0,207794331 | 0,792767371 | SACS | FALSE | Upregulated |
| -0,347994267 | 0,792767371 | SAMD3 | FALSE | Downregulated |
| 0,551284253 | 0,792767371 | SLC34A2 | FALSE | Upregulated |
| -0,296353552 | 0,792767371 | SNAI3-AS1 | FALSE | Downregulated |
| -0,396158277 | 0,792767371 | TENM4 | FALSE | Downregulated |
| -0,110658718 | 0,792767371 | TSR3 | FALSE | Downregulated |
| 0,129622752 | 0,792817413 | TCF7L1 | FALSE | Upregulated |
| -0,444805501 | 0,792848283 | AL121656.5 | FALSE | Downregulated |
| -0,127001056 | 0,792848283 | ETS2 | FALSE | Downregulated |
| -0,322188348 | 0,793062842 | DACT3 | FALSE | Downregulated |
| 0,269920913 | 0,793062842 | LOC102724833 | FALSE | Upregulated |
| -0,277990885 | 0,793062842 | MRM1 | FALSE | Downregulated |
| -0,288047768 | 0,793101497 | LINC00678 | FALSE | Downregulated |
| 0,242527327 | 0,793175391 | RP11-346I3.4 | FALSE | Upregulated |
| -0,614690181 | 0,793303257 | SLC26A4-AS1 | FALSE | Downregulated |
| 0,157932274 | 0,793644801 | FYTTD1 | FALSE | Upregulated |
| 0,202809468 | 0,793651254 | RP11-387D10.3 | FALSE | Upregulated |
| -0,098306809 | 0,793675204 | RPN1 | FALSE | Downregulated |
| 0,169872609 | 0,793730812 | RTP3 | FALSE | Upregulated |
| -0,464789008 | 0,793991839 | CCDC171 | FALSE | Downregulated |
| -0,10060722 | 0,794159225 | TLE1 | FALSE | Downregulated |
| -0,447304508 | 0,79423342 | COLCA1 | FALSE | Downregulated |
| 0,138888916 | 0,79423342 | LRRN4 | FALSE | Upregulated |
| 0,517653164 | 0,79423342 | UBXN10-AS1 | FALSE | Upregulated |
| 0,135204086 | 0,794244001 | TUB | FALSE | Upregulated |
| 0,458014895 | 0,79435645 | RAMP2-AS1 | FALSE | Upregulated |
| 0,597757545 | 0,79435645 | TCEB3CL2 | FALSE | Upregulated |
| -0,22890677 | 0,79435645 | ZNF74 | FALSE | Downregulated |
| -0,127885861 | 0,794509429 | AGO4 | FALSE | Downregulated |
| 0,159415504 | 0,794509429 | CLK2 | FALSE | Upregulated |
| 0,188859738 | 0,794509429 | CSRP2BP | FALSE | Upregulated |
| -0,124947188 | 0,794509429 | PLA2G12A | FALSE | Downregulated |
| -0,082007837 | 0,794604166 | DEDD | FALSE | Downregulated |
| 0,258264519 | 0,794604166 | PIM2 | FALSE | Upregulated |
| 0,163505616 | 0,794656694 | EFHC1 | FALSE | Upregulated |
| 0,372022736 | 0,794728531 | DYRK4 | FALSE | Upregulated |
| 0,150093453 | 0,794856319 | DDHD2 | FALSE | Upregulated |
| -0,386595938 | 0,795110187 | LOC102724336 | FALSE | Downregulated |

| | | | | |
|---|---|---|---|---|
| 0,12185862 | 0,795110187 | PRKAB1 | FALSE | Upregulated |
| -0,545716956 | 0,795334117 | AC016738.3 | FALSE | Downregulated |
| -0,156923713 | 0,795476644 | C16orf71 | FALSE | Downregulated |
| -0,138515511 | 0,795535753 | ASB1 | FALSE | Downregulated |
| -0,093159381 | 0,795535753 | NSD1 | FALSE | Downregulated |
| -0,239041483 | 0,79579249 | ITGB4 | FALSE | Downregulated |
| 0,252905853 | 0,79579249 | P2RX2 | FALSE | Upregulated |
| -0,402853335 | 0,795816328 | PACRG | FALSE | Downregulated |
| -0,393834194 | 0,795933753 | AKR1C6P | FALSE | Downregulated |
| 0,198663763 | 0,795933753 | CTHRC1 | FALSE | Upregulated |
| -0,366998567 | 0,795933753 | FAM86DP | FALSE | Downregulated |
| 0,398090291 | 0,795933753 | FERMT1 | FALSE | Upregulated |
| -0,222407914 | 0,796197201 | MIS12 | FALSE | Downregulated |
| 0,10073003 | 0,796279189 | AGPAT1 | FALSE | Upregulated |
| -0,209523434 | 0,796279189 | PARP15 | FALSE | Downregulated |
| 0,231540696 | 0,796286132 | CDK11A | FALSE | Upregulated |
| -0,289857134 | 0,796286132 | LOC440570 | FALSE | Downregulated |
| -0,252816943 | 0,796286132 | MGST3 | FALSE | Downregulated |
| -0,095657334 | 0,796304097 | STAG3L5P-PVRIG2P-PILRB | FALSE | Downregulated |
| 0,17005026 | 0,796483221 | LOC101929270 | FALSE | Upregulated |
| 0,249295531 | 0,796483221 | P4HA2 | FALSE | Upregulated |
| 0,095553217 | 0,796483221 | PRPF40A | FALSE | Upregulated |
| 0,180177757 | 0,796483221 | RFK | FALSE | Upregulated |
| -0,219701209 | 0,796483221 | SLC11A1 | FALSE | Downregulated |
| -0,403043532 | 0,796483221 | SRRM2-AS1 | FALSE | Downregulated |
| 0,189093056 | 0,796539058 | ZNF417 | FALSE | Upregulated |
| 0,144864547 | 0,796690345 | LOC101929330 | FALSE | Upregulated |
| 0,231646358 | 0,796720723 | CHAMP1 | FALSE | Upregulated |
| -0,077352035 | 0,796720723 | SYMPK | FALSE | Downregulated |
| 0,136747027 | 0,796773924 | CROCC | FALSE | Upregulated |
| 0,152473741 | 0,797019585 | POLR3G | FALSE | Upregulated |
| -0,094558083 | 0,797090095 | CDH2 | FALSE | Downregulated |
| -0,113665579 | 0,797148882 | BDNF | FALSE | Downregulated |
| 0,234751057 | 0,797148882 | KCTD11 | FALSE | Upregulated |
| -0,085319202 | 0,79720139 | GAA | FALSE | Downregulated |
| -0,324828701 | 0,79720139 | MTERF | FALSE | Downregulated |
| -0,112615589 | 0,797623354 | BMP8B | FALSE | Downregulated |
| -0,170341081 | 0,797623354 | CWC25 | FALSE | Downregulated |
| 0,479868052 | 0,797623354 | LOC101929145 | FALSE | Upregulated |
| 0,175401151 | 0,797623354 | NSMCE1 | FALSE | Upregulated |
| 0,330039317 | 0,797812577 | FOXQ1 | FALSE | Upregulated |
| -0,26515228 | 0,797812577 | LRRC16B | FALSE | Downregulated |
| 0,14997874 | 0,797812577 | LRRC8D | FALSE | Upregulated |
| 0,406690045 | 0,797812577 | MEIS3 | FALSE | Upregulated |
| -0,27141705 | 0,797812577 | MRPL22 | FALSE | Downregulated |
| -0,379094862 | 0,797858329 | PRDM16 | FALSE | Downregulated |
| 0,170711081 | 0,797858329 | ZC3H10 | FALSE | Upregulated |
| -0,093332061 | 0,798096476 | CDK5RAP3 | FALSE | Downregulated |
| -1,460848777 | 0,798168709 | MB | FALSE | Downregulated |
| 0,146250956 | 0,798303459 | PIKFYVE | FALSE | Upregulated |

| | | | | |
|---|---|---|---|---|
| -0,124321824 | 0,798704053 | CHUK | FALSE | Downregulated |
| 0,228384089 | 0,798704053 | DRAM2 | FALSE | Upregulated |
| -0,354508588 | 0,798704053 | GRK5-IT1 | FALSE | Downregulated |
| -0,275138472 | 0,798704053 | NUDT19 | FALSE | Downregulated |
| 0,122374009 | 0,798704053 | STXBP2 | FALSE | Upregulated |
| -0,098731894 | 0,798780552 | FDX1 | FALSE | Downregulated |
| 0,458682973 | 0,798972123 | LOC100129215 | FALSE | Upregulated |
| -0,310114969 | 0,798972123 | LOC155060 | FALSE | Downregulated |
| -0,064485468 | 0,798972123 | VPS13D | FALSE | Downregulated |
| -0,082374447 | 0,799048649 | EIF5A | FALSE | Downregulated |
| -0,142861941 | 0,799063808 | WLS | FALSE | Downregulated |
| 0,444810915 | 0,799113747 | C9orf171 | FALSE | Upregulated |
| 0,272457753 | 0,799113747 | DNTTIP1 | FALSE | Upregulated |
| 0,403428784 | 0,799113747 | ELMO1-AS1 | FALSE | Upregulated |
| -0,147017608 | 0,799113747 | SCN9A | FALSE | Downregulated |
| 0,222124188 | 0,799113747 | SPATA41 | FALSE | Upregulated |
| -0,160022349 | 0,799120228 | MRPS15 | FALSE | Downregulated |
| 0,259356854 | 0,799120228 | TMEM170A | FALSE | Upregulated |
| -0,14260835 | 0,799173759 | ANKRD10 | FALSE | Downregulated |
| 0,096128664 | 0,799173759 | HSP90B1 | FALSE | Upregulated |
| -0,2006704 | 0,799173759 | PRSS53 | FALSE | Downregulated |
| 0,238765205 | 0,799234786 | MTRNR2L4 | FALSE | Upregulated |
| -0,220687979 | 0,799408365 | RMDN2 | FALSE | Downregulated |
| 0,105288169 | 0,799448536 | BMP2K | FALSE | Upregulated |
| 0,079783744 | 0,799448536 | HNRNPU-AS1 | FALSE | Upregulated |
| -0,195846792 | 0,799448536 | NLRP6 | FALSE | Downregulated |
| -0,096474621 | 0,799448536 | UGDH-AS1 | FALSE | Downregulated |
| -0,136185733 | 0,799448536 | UVSSA | FALSE | Downregulated |
| -0,208343424 | 0,799525014 | RASIP1 | FALSE | Downregulated |
| 0,122692022 | 0,799599891 | SNF8 | FALSE | Upregulated |
| 0,167357916 | 0,799599891 | YIPF1 | FALSE | Upregulated |
| 0,391907128 | 0,799778429 | TMTC2 | FALSE | Upregulated |
| -0,376907977 | 0,799871744 | ADAM11 | FALSE | Downregulated |
| 0,128070552 | 0,799871744 | MAFG | FALSE | Upregulated |
| -0,292203345 | 0,79988356 | ZDHHC11B | FALSE | Downregulated |
| -0,130245048 | 0,799930236 | ILF2 | FALSE | Downregulated |
| 0,532580349 | 0,800097016 | LINC00582 | FALSE | Upregulated |
| 0,468802582 | 0,800097016 | TMEM119 | FALSE | Upregulated |
| -0,353012673 | 0,800097016 | UBXN7-AS1 | FALSE | Downregulated |
| 0,282924102 | 0,800173664 | NIPSNAP3B | FALSE | Upregulated |
| 0,4544549 | 0,800173664 | PLA2G5 | FALSE | Upregulated |
| -0,095268757 | 0,800173664 | SAMD4B | FALSE | Downregulated |
| 0,19387966 | 0,800173664 | TICRR | FALSE | Upregulated |
| 0,153454375 | 0,800233957 | C14orf105 | FALSE | Upregulated |
| 0,178884731 | 0,800233957 | CISD2 | FALSE | Upregulated |
| -0,294792932 | 0,800233957 | DPM3 | FALSE | Downregulated |
| -0,119994833 | 0,800233957 | FLJ13197 | FALSE | Downregulated |
| -0,153434132 | 0,800488783 | ANKS3 | FALSE | Downregulated |
| -0,237760822 | 0,800488783 | APIP | FALSE | Downregulated |
| 0,357222259 | 0,800488783 | C21orf119 | FALSE | Upregulated |
| -0,433932914 | 0,800488783 | CCM2L | FALSE | Downregulated |

| | | | | |
|---|---|---|---|---|
| 0,1922149 | 0,800488783 | DISC2 | FALSE | Upregulated |
| -0,127101351 | 0,800488783 | DLG5 | FALSE | Downregulated |
| 0,193233107 | 0,800488783 | KNOP1 | FALSE | Upregulated |
| -0,293471878 | 0,800488783 | LOC101927752 | FALSE | Downregulated |
| 0,391837566 | 0,800488783 | LOC101929092 | FALSE | Upregulated |
| 0,378963958 | 0,800488783 | LOC102724851 | FALSE | Upregulated |
| 0,334750822 | 0,800488783 | LOC648987 | FALSE | Upregulated |
| 0,210043761 | 0,800488783 | LRRC15 | FALSE | Upregulated |
| -0,233508847 | 0,800488783 | MSH2 | FALSE | Downregulated |
| -0,163125944 | 0,800488783 | PALM | FALSE | Downregulated |
| 0,287711596 | 0,800488783 | PBDC1 | FALSE | Upregulated |
| 0,344043317 | 0,800488783 | RDH12 | FALSE | Upregulated |
| -0,177242701 | 0,800488783 | SAV1 | FALSE | Downregulated |
| -0,09686181 | 0,800488783 | SUOX | FALSE | Downregulated |
| -1,466843479 | 0,800488783 | TNNC1 | FALSE | Downregulated |
| 0,076424854 | 0,800488783 | USP7 | FALSE | Upregulated |
| 0,159806112 | 0,800488783 | ZNF518B | FALSE | Upregulated |
| -0,294581159 | 0,800574283 | LSM11 | FALSE | Downregulated |
| 0,27094617 | 0,800574283 | METTL2A | FALSE | Upregulated |
| 0,165436402 | 0,800574283 | WDR74 | FALSE | Upregulated |
| 0,12593701 | 0,80061133 | MRPS16 | FALSE | Upregulated |
| -0,198172516 | 0,800997888 | NOTUM | FALSE | Downregulated |
| -0,111117831 | 0,800997888 | OTC | FALSE | Downregulated |
| -0,251344918 | 0,800997888 | SNORA81 | FALSE | Downregulated |
| -0,195565911 | 0,801027396 | TBX15 | FALSE | Downregulated |
| -0,24702484 | 0,801109817 | SP4 | FALSE | Downregulated |
| 0,131109982 | 0,801365831 | RNF169 | FALSE | Upregulated |
| 0,248758752 | 0,80141708 | DET1 | FALSE | Upregulated |
| 0,30130628 | 0,80141708 | KRT10 | FALSE | Upregulated |
| -0,139442953 | 0,80141708 | LOC100287467 | FALSE | Downregulated |
| -0,215155518 | 0,80141708 | SDS | FALSE | Downregulated |
| 0,249950374 | 0,80141708 | WNT8B | FALSE | Upregulated |
| 0,072280458 | 0,801678652 | NCL | FALSE | Upregulated |
| -0,231013902 | 0,80181879 | LMBR1L | FALSE | Downregulated |
| -0,142558246 | 0,801991293 | PKD2 | FALSE | Downregulated |
| 0,130232546 | 0,802064085 | HERC4 | FALSE | Upregulated |
| -0,157676771 | 0,802225093 | ZNF579 | FALSE | Downregulated |
| 0,148437377 | 0,802619425 | DTX1 | FALSE | Upregulated |
| -0,203405145 | 0,802619425 | FOCAD | FALSE | Downregulated |
| -0,348134236 | 0,802651315 | PUS7 | FALSE | Downregulated |
| 0,100095259 | 0,802683856 | MRPS26 | FALSE | Upregulated |
| -0,112939365 | 0,802683856 | PARK7 | FALSE | Downregulated |
| -0,17879818 | 0,802683856 | TUBGCP5 | FALSE | Downregulated |
| -0,142534841 | 0,802797555 | RABEPK | FALSE | Downregulated |
| 0,207366824 | 0,80287136 | ADAMTS1 | FALSE | Upregulated |
| 0,147495617 | 0,80287136 | LINC00675 | FALSE | Upregulated |
| 0,165653223 | 0,802891772 | MECP2 | FALSE | Upregulated |
| 0,114720543 | 0,802903321 | ATP6V0C | FALSE | Upregulated |
| -0,135194792 | 0,80320824 | ALS2CL | FALSE | Downregulated |
| 0,412925276 | 0,803238658 | LOC101928673 | FALSE | Upregulated |
| 0,14499133 | 0,803437307 | KCNH1 | FALSE | Upregulated |

| | | | | |
|---|---|---|---|---|
| -0,233475189 | 0,803606397 | GTF2IRD1 | FALSE | Downregulated |
| -0,176013914 | 0,803606397 | INIP | FALSE | Downregulated |
| -0,12724224 | 0,803606397 | RNMT | FALSE | Downregulated |
| 0,185355953 | 0,803606397 | TMEM19 | FALSE | Upregulated |
| -0,11967925 | 0,803706673 | HDAC1 | FALSE | Downregulated |
| 0,18843688 | 0,803863265 | CHST3 | FALSE | Upregulated |
| 0,418739454 | 0,803863265 | ITPKA | FALSE | Upregulated |
| -0,148508842 | 0,804038748 | NEK11 | FALSE | Downregulated |
| 0,399159917 | 0,80431824 | LZTS1-AS1 | FALSE | Upregulated |
| -0,099994157 | 0,804320285 | PIK3R2 | FALSE | Downregulated |
| -0,171146098 | 0,804320285 | SELT | FALSE | Downregulated |
| -0,120532833 | 0,804331611 | C9 | FALSE | Downregulated |
| -0,265068007 | 0,804370773 | BLOC1S4 | FALSE | Downregulated |
| -0,126694561 | 0,804370773 | NBPF11 | FALSE | Downregulated |
| 0,136177489 | 0,804370773 | PDE8B | FALSE | Upregulated |
| -0,082414867 | 0,804370773 | SDHC | FALSE | Downregulated |
| -0,147631164 | 0,804519816 | LOC102724250 | FALSE | Downregulated |
| -0,191094515 | 0,80459608 | MT1X | FALSE | Downregulated |
| -0,08802555 | 0,804646439 | NCLN | FALSE | Downregulated |
| 0,120685997 | 0,804661034 | CCDC142 | FALSE | Upregulated |
| -0,49199077 | 0,804661034 | LOC101928030 | FALSE | Downregulated |
| -0,179943845 | 0,80473305 | LOC101927894 | FALSE | Downregulated |
| 0,667969548 | 0,804849903 | FAM133DP | FALSE | Upregulated |
| 0,105636996 | 0,804849903 | KPNA4 | FALSE | Upregulated |
| -0,183603185 | 0,804849903 | RP11-383C5.3 | FALSE | Downregulated |
| -0,296788901 | 0,804849903 | SLED1 | FALSE | Downregulated |
| 0,124117703 | 0,804849903 | TNRC6C-AS1 | FALSE | Upregulated |
| 0,344271591 | 0,804849903 | WISP1 | FALSE | Upregulated |
| -0,271481756 | 0,804875545 | MPZL2 | FALSE | Downregulated |
| -0,445933783 | 0,804875545 | MYLK4 | FALSE | Downregulated |
| -0,484895913 | 0,804904427 | ZIK1 | FALSE | Downregulated |
| -0,083061307 | 0,804920446 | TNRC6A | FALSE | Downregulated |
| 0,092184951 | 0,804951866 | PLEKHM2 | FALSE | Upregulated |
| 0,323445115 | 0,804951866 | TMEM220-AS1 | FALSE | Upregulated |
| -0,125422615 | 0,805002618 | EIF2B1 | FALSE | Downregulated |
| 0,134634493 | 0,805002618 | IGF2BP3 | FALSE | Upregulated |
| -0,117349468 | 0,805002618 | MROH1 | FALSE | Downregulated |
| 0,181829459 | 0,805348826 | VPS25 | FALSE | Upregulated |
| 0,15035907 | 0,805401533 | LARP7 | FALSE | Upregulated |
| -0,095590375 | 0,805618684 | YTHDF1 | FALSE | Downregulated |
| 0,087358853 | 0,805649647 | EIF4A1 | FALSE | Upregulated |
| -0,351308803 | 0,805649647 | ZNF596 | FALSE | Downregulated |
| -0,074701125 | 0,805788516 | TEX2 | FALSE | Downregulated |
| 0,114645331 | 0,805792963 | DOCK4-AS1 | FALSE | Upregulated |
| 0,127476714 | 0,805792963 | ZNF768 | FALSE | Upregulated |
| 0,146646896 | 0,80583617 | BLNK | FALSE | Upregulated |
| 0,059499673 | 0,80583617 | CTNNA1 | FALSE | Upregulated |
| 0,330019064 | 0,80583617 | MLLT3 | FALSE | Upregulated |
| -0,142363093 | 0,80583617 | TRIM23 | FALSE | Downregulated |
| -0,184472763 | 0,806479052 | LOC101927841 | FALSE | Downregulated |
| -0,514776696 | 0,806479052 | LOC101929442 | FALSE | Downregulated |

| | | | | |
|---|---|---|---|---|
| -0,089076806 | 0,806479052 | PMPCA | FALSE | Downregulated |
| -0,291488342 | 0,806479052 | PRAF2 | FALSE | Downregulated |
| 0,134405193 | 0,806479052 | TCEAL4 | FALSE | Upregulated |
| 0,396610607 | 0,806479052 | ZNF100 | FALSE | Upregulated |
| -0,072305071 | 0,807016317 | PRKACA | FALSE | Downregulated |
| 0,335494764 | 0,807091645 | FJX1 | FALSE | Upregulated |
| 0,323844966 | 0,807161548 | ABCA11P | FALSE | Upregulated |
| -1,297916507 | 0,807707271 | ACTN2 | FALSE | Downregulated |
| -0,237458778 | 0,807784322 | ANKAR | FALSE | Downregulated |
| 0,477549156 | 0,807784322 | LOC102724312 | FALSE | Upregulated |
| -0,192683343 | 0,807812511 | ATL1 | FALSE | Downregulated |
| -0,132648415 | 0,807812511 | FBXL16 | FALSE | Downregulated |
| -0,566367006 | 0,807812511 | FLNC | FALSE | Downregulated |
| -0,147932138 | 0,807812511 | GARS | FALSE | Downregulated |
| 0,260097055 | 0,807812511 | PSTPIP2 | FALSE | Upregulated |
| -0,420557282 | 0,807812511 | RMDN2-AS1 | FALSE | Downregulated |
| 0,254906791 | 0,807812511 | SHISA6 | FALSE | Upregulated |
| 1,180218185 | 0,807812511 | SLX1A-SULT1A3 | FALSE | Upregulated |
| -0,142440017 | 0,807812511 | ZNF266 | FALSE | Downregulated |
| -0,194892412 | 0,807812511 | ZNF605 | FALSE | Downregulated |
| 0,229405577 | 0,807812511 | ZSWIM7 | FALSE | Upregulated |
| -0,10586182 | 0,807958294 | GET4 | FALSE | Downregulated |
| -0,13083759 | 0,808012583 | TIMM44 | FALSE | Downregulated |
| -0,114441249 | 0,808092187 | OTUB1 | FALSE | Downregulated |
| 0,225792309 | 0,808269439 | PDLIM7 | FALSE | Upregulated |
| -0,212046445 | 0,808286097 | EED | FALSE | Downregulated |
| -0,186345698 | 0,808286097 | IFIT1B | FALSE | Downregulated |
| 0,408803324 | 0,808286097 | MIOX | FALSE | Upregulated |
| -0,293401957 | 0,808286097 | PLAC8 | FALSE | Downregulated |
| 0,18654117 | 0,808286097 | RAP2A | FALSE | Upregulated |
| 0,128606786 | 0,808286097 | TGFBR1 | FALSE | Upregulated |
| -0,085834921 | 0,808286097 | TOM1 | FALSE | Downregulated |
| -0,224414951 | 0,808286097 | ZNF441 | FALSE | Downregulated |
| -0,219101264 | 0,808286097 | ZNHIT6 | FALSE | Downregulated |
| 0,099200639 | 0,80835137 | BCOR | FALSE | Upregulated |
| 0,116877075 | 0,80835137 | UBXN4 | FALSE | Upregulated |
| 0,179787247 | 0,808382576 | LOC101926943 | FALSE | Upregulated |
| -0,249639791 | 0,80848785 | USP43 | FALSE | Downregulated |
| 0,201021095 | 0,80848785 | ZBTB34 | FALSE | Upregulated |
| 0,15507675 | 0,808500751 | LOC388849 | FALSE | Upregulated |
| -0,092658015 | 0,80859426 | GATAD2B | FALSE | Downregulated |
| 0,169505423 | 0,808736708 | APOOL | FALSE | Upregulated |
| -0,282383856 | 0,808736708 | CCDC7 | FALSE | Downregulated |
| -0,090163091 | 0,808736708 | DCTN1 | FALSE | Downregulated |
| 0,110721544 | 0,808736708 | EPT1 | FALSE | Upregulated |
| 0,111544361 | 0,808736708 | FAM69B | FALSE | Upregulated |
| -0,591572036 | 0,808736708 | GNG10 | FALSE | Downregulated |
| -0,267034148 | 0,808736708 | KIAA1107 | FALSE | Downregulated |
| -0,221706084 | 0,808736708 | LOC100507131 | FALSE | Downregulated |
| 0,522289011 | 0,808736708 | LOC101928553 | FALSE | Upregulated |
| -0,158789491 | 0,808736708 | LOC285593 | FALSE | Downregulated |

| | | | | |
|---|---|---|---|---|
| -0,08422508 | 0,808736708 | RAD23A | FALSE | Downregulated |
| 0,42367642 | 0,808736708 | SLAMF1 | FALSE | Upregulated |
| -0,144460684 | 0,808736708 | TMEM37 | FALSE | Downregulated |
| 0,172693837 | 0,808878539 | TMC3 | FALSE | Upregulated |
| 0,141380736 | 0,808924142 | ADAP2 | FALSE | Upregulated |
| -0,284613713 | 0,808924142 | ANKRD39 | FALSE | Downregulated |
| 0,173045222 | 0,808924142 | BAX | FALSE | Upregulated |
| 0,398780533 | 0,808924142 | C9orf40 | FALSE | Upregulated |
| 0,121555106 | 0,808924142 | C9orf78 | FALSE | Upregulated |
| 0,227961003 | 0,808924142 | CSNK1A1P1 | FALSE | Upregulated |
| -0,250035125 | 0,808924142 | FAM174B | FALSE | Downregulated |
| 0,413941235 | 0,808924142 | KCNJ1 | FALSE | Upregulated |
| 0,171620803 | 0,808924142 | LOC100652768 | FALSE | Upregulated |
| 0,204884144 | 0,808924142 | LRRC4B | FALSE | Upregulated |
| 0,134856822 | 0,808924142 | MARVELD3 | FALSE | Upregulated |
| 0,078634519 | 0,808924142 | SMEK1 | FALSE | Upregulated |
| 0,155850722 | 0,808924142 | TSG101 | FALSE | Upregulated |
| 0,069696263 | 0,809001114 | ARF1 | FALSE | Upregulated |
| -0,195473335 | 0,809001114 | GLUD1P7 | FALSE | Downregulated |
| -0,118318015 | 0,809001114 | MED24 | FALSE | Downregulated |
| 0,102197913 | 0,809001114 | PPAP2A | FALSE | Upregulated |
| 0,246036604 | 0,809001114 | RBL1 | FALSE | Upregulated |
| 0,114576223 | 0,809001114 | TESK2 | FALSE | Upregulated |
| -0,214412486 | 0,809040914 | ENDOD1 | FALSE | Downregulated |
| -0,205773095 | 0,809380097 | MPP6 | FALSE | Downregulated |
| 0,253778314 | 0,809466975 | SLC26A8 | FALSE | Upregulated |
| 0,13508105 | 0,80952204 | RYR3 | FALSE | Upregulated |
| -0,346940578 | 0,809560838 | CDC42EP5 | FALSE | Downregulated |
| 0,291403545 | 0,810048956 | AGTRAP | FALSE | Upregulated |
| 0,185142463 | 0,810249221 | USP28 | FALSE | Upregulated |
| 0,372177353 | 0,810364759 | ARL11 | FALSE | Upregulated |
| -0,317299135 | 0,810364759 | RPA3-AS1 | FALSE | Downregulated |
| -0,350320241 | 0,810364759 | ZBTB6 | FALSE | Downregulated |
| 0,187566524 | 0,810394101 | ITPRIP | FALSE | Upregulated |
| -0,15248382 | 0,810782348 | CRIPAK | FALSE | Downregulated |
| 0,292234279 | 0,810782348 | POP1 | FALSE | Upregulated |
| -0,091948925 | 0,810968715 | FSIP2 | FALSE | Downregulated |
| 0,388968952 | 0,811123373 | C2CD4B | FALSE | Upregulated |
| 0,406511391 | 0,811162654 | A2ML1 | FALSE | Upregulated |
| -0,121576203 | 0,811162654 | CTDNEP1 | FALSE | Downregulated |
| -0,276939779 | 0,811162654 | FAM98C | FALSE | Downregulated |
| 0,075536933 | 0,811162654 | KIAA1432 | FALSE | Upregulated |
| -0,308022124 | 0,811162654 | PRR22 | FALSE | Downregulated |
| 0,068998456 | 0,811162654 | UTRN | FALSE | Upregulated |
| -0,096250508 | 0,811162654 | ZNF787 | FALSE | Downregulated |
| 0,19781965 | 0,81142551 | ARHGEF35 | FALSE | Upregulated |
| -0,311557513 | 0,811459074 | COMMD6 | FALSE | Downregulated |
| 0,175983915 | 0,811459074 | SMIM18 | FALSE | Upregulated |
| 0,11843679 | 0,811496813 | NDUFA6 | FALSE | Upregulated |
| -0,153544339 | 0,811646012 | RHPN2 | FALSE | Downregulated |
| -0,31734686 | 0,811646012 | SLC12A8 | FALSE | Downregulated |

| | | | | |
|---|---|---|---|---|
| 0,401153875 | 0,811813703 | ASIC3 | FALSE | Upregulated |
| 0,277876633 | 0,811813703 | COMMD2 | FALSE | Upregulated |
| -0,182758528 | 0,811962682 | ANKRD9 | FALSE | Downregulated |
| 0,130661952 | 0,811962682 | RNASET2 | FALSE | Upregulated |
| 0,196437686 | 0,812095447 | TMEM242 | FALSE | Upregulated |
| 0,15102005 | 0,812166155 | ZNF510 | FALSE | Upregulated |
| -0,149786162 | 0,812233287 | SSNA1 | FALSE | Downregulated |
| -0,103894328 | 0,812298737 | DLST | FALSE | Downregulated |
| -0,181129149 | 0,812298737 | DUSP23 | FALSE | Downregulated |
| -0,376103448 | 0,812298737 | RAB30-AS1 | FALSE | Downregulated |
| 0,134690025 | 0,812298737 | RNF114 | FALSE | Upregulated |
| -0,40549224 | 0,812540415 | HPGDS | FALSE | Downregulated |
| -0,253429586 | 0,81255658 | EIF1B | FALSE | Downregulated |
| -0,227291733 | 0,81265618 | GTF2H5 | FALSE | Downregulated |
| 0,130361318 | 0,812705678 | PARN | FALSE | Upregulated |
| 0,40586378 | 0,812711133 | ATG4C | FALSE | Upregulated |
| 0,122626759 | 0,812711133 | FARSA | FALSE | Upregulated |
| -0,168420766 | 0,812844056 | CDK18 | FALSE | Downregulated |
| 0,209265809 | 0,812872055 | DICER1-AS1 | FALSE | Upregulated |
| 0,135498423 | 0,812872055 | LOC102724975 | FALSE | Upregulated |
| 0,14207079 | 0,812872055 | LRG1 | FALSE | Upregulated |
| -0,079247084 | 0,812930395 | EXOC7 | FALSE | Downregulated |
| -0,115600901 | 0,813069774 | ATG4B | FALSE | Downregulated |
| 0,440224422 | 0,813088892 | C3orf30 | FALSE | Upregulated |
| 0,27098756 | 0,813088892 | SDR42E1 | FALSE | Upregulated |
| 0,129262017 | 0,813092174 | NSMF | FALSE | Upregulated |
| 0,114792415 | 0,813092174 | SNAP23 | FALSE | Upregulated |
| -0,094638082 | 0,813311529 | EZH1 | FALSE | Downregulated |
| -0,513091963 | 0,813311529 | MEX3B | FALSE | Downregulated |
| 0,137640146 | 0,813457234 | P4HA1 | FALSE | Upregulated |
| -0,332377652 | 0,813457234 | ZNF583 | FALSE | Downregulated |
| 0,167383331 | 0,813494124 | GIMAP6 | FALSE | Upregulated |
| -0,261649611 | 0,813494124 | KBTBD6 | FALSE | Downregulated |
| 0,103659736 | 0,813494124 | LOC101927647 | FALSE | Upregulated |
| -0,189343532 | 0,813532395 | CEP44 | FALSE | Downregulated |
| -0,059732651 | 0,813723893 | MYH10 | FALSE | Downregulated |
| 0,137767012 | 0,813910117 | CNOT8 | FALSE | Upregulated |
| 0,13632522 | 0,813910117 | PES1 | FALSE | Upregulated |
| -0,794656001 | 0,813910117 | TCAP | FALSE | Downregulated |
| 0,134371911 | 0,813910117 | THOC5 | FALSE | Upregulated |
| 0,136298964 | 0,813910117 | TMEM214 | FALSE | Upregulated |
| -0,121578728 | 0,814225759 | KDM3A | FALSE | Downregulated |
| 0,325891645 | 0,814225759 | TNRC18P1 | FALSE | Upregulated |
| -0,082583678 | 0,814279926 | SYVN1 | FALSE | Downregulated |
| -0,065801967 | 0,814365682 | RPL27A | FALSE | Downregulated |
| 0,156852528 | 0,814368569 | EPX | FALSE | Upregulated |
| -0,101493287 | 0,814368569 | ZSCAN2 | FALSE | Downregulated |
| 0,084622544 | 0,814400827 | LOC100505658 | FALSE | Upregulated |
| -0,12679253 | 0,814479552 | FN3K | FALSE | Downregulated |
| -0,20327998 | 0,81463991 | ARMCX5 | FALSE | Downregulated |
| -0,416302583 | 0,81463991 | C14orf166B | FALSE | Downregulated |

| | | | | |
|---|---|---|---|---|
| 0,344886306 | 0,81463991 | C1orf162 | FALSE | Upregulated |
| 0,266271855 | 0,81463991 | CDC14A | FALSE | Upregulated |
| -0,151497029 | 0,81463991 | CWF19L2 | FALSE | Downregulated |
| 0,115786951 | 0,81463991 | DERL1 | FALSE | Upregulated |
| 0,192359262 | 0,81463991 | FAM188A | FALSE | Upregulated |
| 0,272377221 | 0,81463991 | FAM221A | FALSE | Upregulated |
| 0,173863784 | 0,81463991 | LONRF1 | FALSE | Upregulated |
| 0,312378999 | 0,81463991 | RBMX2 | FALSE | Upregulated |
| -0,398392958 | 0,814769441 | LOC100289026 | FALSE | Downregulated |
| -0,142169803 | 0,814769441 | LOC101928185 | FALSE | Downregulated |
| 0,177943686 | 0,814866248 | ALX4 | FALSE | Upregulated |
| -0,090741816 | 0,814866248 | CIZ1 | FALSE | Downregulated |
| 0,166802386 | 0,814866248 | DENND6A | FALSE | Upregulated |
| 0,31645288 | 0,814866248 | NXT1 | FALSE | Upregulated |
| 0,20471533 | 0,814866248 | ORAOV1 | FALSE | Upregulated |
| 0,133115702 | 0,814866248 | RRP7A | FALSE | Upregulated |
| 0,078297552 | 0,815107456 | ATXN7L3 | FALSE | Upregulated |
| 0,174777827 | 0,815107456 | NPR2 | FALSE | Upregulated |
| 0,244512423 | 0,815107456 | PPFIA3 | FALSE | Upregulated |
| 0,115558704 | 0,815107456 | TMEM175 | FALSE | Upregulated |
| -0,236580305 | 0,815123965 | GBAS | FALSE | Downregulated |
| 0,533811196 | 0,815172013 | PRKRIP1 | FALSE | Upregulated |
| -0,120594174 | 0,815311619 | CDC40 | FALSE | Downregulated |
| -0,11475804 | 0,815311619 | MED14 | FALSE | Downregulated |
| -0,116940806 | 0,815417833 | CRTC1 | FALSE | Downregulated |
| -0,430143535 | 0,815495953 | RP11-575L7.4 | FALSE | Downregulated |
| 0,125078011 | 0,815562848 | LYSMD3 | FALSE | Upregulated |
| 0,163108276 | 0,815562848 | PTGIS | FALSE | Upregulated |
| -0,11694115 | 0,815834121 | FBXW11 | FALSE | Downregulated |
| 0,148511493 | 0,816039315 | NIPA1 | FALSE | Upregulated |
| 0,132178604 | 0,816039315 | ZSWIM6 | FALSE | Upregulated |
| 0,413362063 | 0,81621429 | ARPP21 | FALSE | Upregulated |
| 0,104810896 | 0,81621429 | C11orf24 | FALSE | Upregulated |
| 0,161048407 | 0,81621429 | CYB561 | FALSE | Upregulated |
| -0,202628825 | 0,81621429 | DEGS1 | FALSE | Downregulated |
| -0,155235868 | 0,81621429 | FAM203A | FALSE | Downregulated |
| -0,264240396 | 0,81621429 | HIST1H2APS1 | FALSE | Downregulated |
| -0,351955099 | 0,81621429 | KCNK10 | FALSE | Downregulated |
| -0,27208766 | 0,81621429 | ST3GAL6-AS1 | FALSE | Downregulated |
| 0,115582729 | 0,81621429 | ZFP69 | FALSE | Upregulated |
| -0,12965642 | 0,816564233 | COX8A | FALSE | Downregulated |
| 0,109338123 | 0,816564233 | PLCL2 | FALSE | Upregulated |
| -0,152780668 | 0,816586088 | METTL14 | FALSE | Downregulated |
| -0,123297374 | 0,816600777 | C3orf22 | FALSE | Downregulated |
| -0,698892809 | 0,816737213 | SKA3 | FALSE | Downregulated |
| -0,139683763 | 0,816809265 | CENPC | FALSE | Downregulated |
| -0,15075867 | 0,816809265 | PDE7B | FALSE | Downregulated |
| -0,145709358 | 0,816840557 | GJA4 | FALSE | Downregulated |
| -0,080770321 | 0,816840557 | ZFYVE16 | FALSE | Downregulated |
| -0,103313368 | 0,816904078 | ASH2L | FALSE | Downregulated |
| 0,157589751 | 0,817063463 | FGFR1OP2 | FALSE | Upregulated |

| | | | | |
|---|---|---|---|---|
| -0,119277098 | 0,81731568 | LOC728024 | FALSE | Downregulated |
| -0,242485782 | 0,817324185 | HIATL2 | FALSE | Downregulated |
| 0,249080473 | 0,817675183 | EPHA3 | FALSE | Upregulated |
| 0,203998192 | 0,817752092 | RBM38 | FALSE | Upregulated |
| -0,121178913 | 0,817828479 | HERPUD2 | FALSE | Downregulated |
| 0,297671014 | 0,817912648 | ICA1 | FALSE | Upregulated |
| 0,13021902 | 0,817922748 | PRKAR2A | FALSE | Upregulated |
| 0,505955847 | 0,817967431 | DDX11L9 | FALSE | Upregulated |
| 0,090942474 | 0,817967431 | DNAJB1 | FALSE | Upregulated |
| -0,142637323 | 0,817967431 | HCFC2 | FALSE | Downregulated |
| -0,128613867 | 0,817967431 | KIAA1656 | FALSE | Downregulated |
| 0,061596293 | 0,817967431 | MAST2 | FALSE | Upregulated |
| 0,153869345 | 0,817967431 | NPC2 | FALSE | Upregulated |
| 0,178579189 | 0,817967431 | PIAS3 | FALSE | Upregulated |
| -0,088702198 | 0,817967431 | USP49 | FALSE | Downregulated |
| -0,260884076 | 0,818354959 | WDR47 | FALSE | Downregulated |
| -0,629423865 | 0,818728488 | INO80B | FALSE | Downregulated |
| 0,68930512 | 0,818781933 | CCDC180 | FALSE | Upregulated |
| -0,170025107 | 0,818781933 | CREB3L4 | FALSE | Downregulated |
| -0,136602924 | 0,818877519 | FER | FALSE | Downregulated |
| -0,267373728 | 0,818877519 | GPR162 | FALSE | Downregulated |
| 0,172948809 | 0,818877519 | LOC728613 | FALSE | Upregulated |
| 0,083313469 | 0,818877519 | LRRFIP2 | FALSE | Upregulated |
| 0,133761611 | 0,818877519 | MAPKAPK3 | FALSE | Upregulated |
| 0,28856612 | 0,818877519 | MIR3648 | FALSE | Upregulated |
| 0,17917418 | 0,818877519 | PKDCC | FALSE | Upregulated |
| -0,09320035 | 0,818877519 | PPP2R2D | FALSE | Downregulated |
| -0,233016696 | 0,818877519 | RGPD2 | FALSE | Downregulated |
| 0,138894219 | 0,818877519 | RHBDD2 | FALSE | Upregulated |
| -0,871494756 | 0,818877519 | RHOXF2B | FALSE | Downregulated |
| 0,07930323 | 0,818877519 | SEC13 | FALSE | Upregulated |
| -0,061839418 | 0,819092775 | ALKBH5 | FALSE | Downregulated |
| 0,143578444 | 0,819092775 | CAMK1D | FALSE | Upregulated |
| 0,116568547 | 0,819092775 | GATAD1 | FALSE | Upregulated |
| 0,096995307 | 0,819092775 | HERC2P2 | FALSE | Upregulated |
| -0,34657105 | 0,819092775 | LOC101927520 | FALSE | Downregulated |
| 0,157315669 | 0,819092775 | LOC283922 | FALSE | Upregulated |
| 0,209127744 | 0,819092775 | MYOZ3 | FALSE | Upregulated |
| 0,132158051 | 0,819092775 | PRKAR2A-AS1 | FALSE | Upregulated |
| 0,045554161 | 0,819092775 | PRRC2A | FALSE | Upregulated |
| -0,128547129 | 0,819092775 | SNX4 | FALSE | Downregulated |
| 0,124042413 | 0,819092775 | SSH3 | FALSE | Upregulated |
| -0,052527908 | 0,819213803 | EP300 | FALSE | Downregulated |
| -0,612518297 | 0,81943459 | C21orf33 | FALSE | Downregulated |
| 0,230436717 | 0,81943459 | EZH2 | FALSE | Upregulated |
| -0,076938019 | 0,81943459 | PCNT | FALSE | Downregulated |
| -0,082824231 | 0,81943459 | TNKS | FALSE | Downregulated |
| -0,129680962 | 0,819472283 | ELP3 | FALSE | Downregulated |
| -0,237753439 | 0,819620575 | OARD1 | FALSE | Downregulated |
| 0,120583837 | 0,819768283 | LOC101928728 | FALSE | Upregulated |
| -0,251341176 | 0,819768283 | TTC39A | FALSE | Downregulated |

| | | | | |
|---|---|---|---|---|
| -0,284841399 | 0,820158343 | CRH | FALSE | Downregulated |
| -0,241486861 | 0,820158343 | CTD-3080P12.3 | FALSE | Downregulated |
| 0,25567042 | 0,820228902 | PLEKHB1 | FALSE | Upregulated |
| 0,355924391 | 0,820556678 | POC1B-GALNT4 | FALSE | Upregulated |
| 0,197156648 | 0,820556678 | ST3GAL4-AS1 | FALSE | Upregulated |
| -0,194894285 | 0,820556678 | TMEM261 | FALSE | Downregulated |
| -0,136986109 | 0,820590768 | SLC25A27 | FALSE | Downregulated |
| -0,185871689 | 0,820591959 | ANP32A-IT1 | FALSE | Downregulated |
| -0,427962051 | 0,820591959 | C6orf164 | FALSE | Downregulated |
| 0,161412967 | 0,820648283 | GGH | FALSE | Upregulated |
| 0,286779637 | 0,820648283 | PKD1L3 | FALSE | Upregulated |
| 0,143609682 | 0,820680271 | C7 | FALSE | Upregulated |
| -0,250699829 | 0,820805318 | ZNF25 | FALSE | Downregulated |
| 0,094303015 | 0,820925572 | CNOT11 | FALSE | Upregulated |
| 0,208662 | 0,820925572 | EFNA2 | FALSE | Upregulated |
| 0,127116728 | 0,820925572 | PLAT | FALSE | Upregulated |
| 0,086651365 | 0,821041113 | PTTG1IP | FALSE | Upregulated |
| 0,234704678 | 0,821285258 | GPR153 | FALSE | Upregulated |
| 0,24324963 | 0,821285258 | HOXA2 | FALSE | Upregulated |
| -0,227361144 | 0,821285258 | LOC101927806 | FALSE | Downregulated |
| 0,305760307 | 0,821285258 | LOC101928558 | FALSE | Upregulated |
| 0,131723272 | 0,821285258 | SMYD4 | FALSE | Upregulated |
| -0,151175477 | 0,82135237 | AREL1 | FALSE | Downregulated |
| -0,193066253 | 0,82135237 | EP400NL | FALSE | Downregulated |
| 0,225045479 | 0,82135237 | GRASP | FALSE | Upregulated |
| 0,27688849 | 0,82135237 | LINC00526 | FALSE | Upregulated |
| 0,113268497 | 0,82135237 | SGTA | FALSE | Upregulated |
| -0,225832286 | 0,821422613 | AC037193.1 | FALSE | Downregulated |
| -0,097317432 | 0,821611637 | DNMBP-AS1 | FALSE | Downregulated |
| 0,260733979 | 0,821611637 | VWA5A | FALSE | Upregulated |
| -0,134506831 | 0,821611637 | ZYG11A | FALSE | Downregulated |
| -0,331622431 | 0,821821865 | GOLGA6B | FALSE | Downregulated |
| -0,1326168 | 0,821821865 | IGF1 | FALSE | Downregulated |
| 0,363360552 | 0,821878833 | ANK1 | FALSE | Upregulated |
| 0,091987695 | 0,82219459 | POLR1A | FALSE | Upregulated |
| 0,302993758 | 0,822269285 | KATNBL1 | FALSE | Upregulated |
| 0,132377875 | 0,822401479 | MAD1L1 | FALSE | Upregulated |
| -0,068254101 | 0,822401479 | MCC | FALSE | Downregulated |
| 0,077173702 | 0,822497673 | SHROOM3 | FALSE | Upregulated |
| -0,134608283 | 0,822915645 | PDHB | FALSE | Downregulated |
| -0,087426039 | 0,822915645 | STK35 | FALSE | Downregulated |
| 0,14784681 | 0,823309645 | ENPP7 | FALSE | Upregulated |
| 0,199672148 | 0,823309645 | FARSB | FALSE | Upregulated |
| -0,298928086 | 0,823309645 | GLI3 | FALSE | Downregulated |
| -0,112556652 | 0,823404023 | ALDH16A1 | FALSE | Downregulated |
| 0,240575748 | 0,823404023 | C1D | FALSE | Upregulated |
| -0,09165942 | 0,823404023 | CKAP5 | FALSE | Downregulated |
| 0,082653512 | 0,823404023 | ESYT2 | FALSE | Upregulated |
| 0,067279101 | 0,823404023 | KDM5A | FALSE | Upregulated |
| 0,659300789 | 0,823404023 | LOC728554 | FALSE | Upregulated |
| -0,189829982 | 0,823404023 | PPCS | FALSE | Downregulated |

| | | | | |
|---|---|---|---|---|
| -0,144072268 | 0,823404023 | SBSPON | FALSE | Downregulated |
| -0,225883053 | 0,823404023 | SREK1IP1 | FALSE | Downregulated |
| -0,224181661 | 0,823404023 | TAS1R1 | FALSE | Downregulated |
| -0,235726197 | 0,823539157 | DENND1B | FALSE | Downregulated |
| -0,116091171 | 0,82366807 | FAM73B | FALSE | Downregulated |
| 0,246354299 | 0,82366807 | NDUFB2-AS1 | FALSE | Upregulated |
| 0,103289151 | 0,823679383 | SULT2A1 | FALSE | Upregulated |
| -0,185037089 | 0,823733651 | HAND2 | FALSE | Downregulated |
| 0,072110027 | 0,823800829 | HNRNPM | FALSE | Upregulated |
| 0,19184972 | 0,823800829 | PIP4K2C | FALSE | Upregulated |
| 0,179803543 | 0,82407116 | LOC100506114 | FALSE | Upregulated |
| 0,213355443 | 0,824109706 | CHGA | FALSE | Upregulated |
| 0,226838473 | 0,824369945 | ZNF8 | FALSE | Upregulated |
| 0,123888729 | 0,824385736 | GPR160 | FALSE | Upregulated |
| 0,15472843 | 0,824385736 | HIST1H2BK | FALSE | Upregulated |
| 0,182346332 | 0,824385736 | IDE | FALSE | Upregulated |
| 0,316932117 | 0,824385736 | LOC91450 | FALSE | Upregulated |
| -0,136473953 | 0,824419408 | ARSB | FALSE | Downregulated |
| 0,133822394 | 0,824520602 | TROVE2 | FALSE | Upregulated |
| -0,164696846 | 0,824610261 | COPS3 | FALSE | Downregulated |
| -0,10426236 | 0,824610261 | EPB41L4B | FALSE | Downregulated |
| -0,411683076 | 0,824610261 | GPC4 | FALSE | Downregulated |
| 0,150205806 | 0,824610261 | IGBP1P1 | FALSE | Upregulated |
| 0,116277783 | 0,824610261 | LATS2 | FALSE | Upregulated |
| 0,121666744 | 0,824610261 | LIG1 | FALSE | Upregulated |
| -0,161189417 | 0,824610261 | LOC150051 | FALSE | Downregulated |
| 0,163425937 | 0,824610261 | NUDT6 | FALSE | Upregulated |
| -0,088096062 | 0,824610261 | PAK4 | FALSE | Downregulated |
| -0,112784812 | 0,824610261 | PUS3 | FALSE | Downregulated |
| -0,219104201 | 0,824610261 | RALA | FALSE | Downregulated |
| -0,115809328 | 0,824610261 | TAZ | FALSE | Downregulated |
| -0,245796631 | 0,824610261 | TSPYL5 | FALSE | Downregulated |
| 0,077618778 | 0,824610261 | ZNF207 | FALSE | Upregulated |
| 0,310239495 | 0,824787174 | LOC101927045 | FALSE | Upregulated |
| 0,258258727 | 0,824815225 | SH2B2 | FALSE | Upregulated |
| -0,096619048 | 0,825223155 | AIFM1 | FALSE | Downregulated |
| 0,168323964 | 0,825223155 | CYB5B | FALSE | Upregulated |
| 0,122468712 | 0,825308872 | ZNF746 | FALSE | Upregulated |
| 0,150569298 | 0,825340147 | ACSS3 | FALSE | Upregulated |
| -0,250644648 | 0,825391402 | SIGLEC7 | FALSE | Downregulated |
| 0,082334114 | 0,825574024 | PPP6R3 | FALSE | Upregulated |
| -0,128678047 | 0,825656772 | LAMTOR1 | FALSE | Downregulated |
| 0,439814982 | 0,825656772 | RFC4 | FALSE | Upregulated |
| -0,197013154 | 0,825656772 | SUGT1 | FALSE | Downregulated |
| 0,464470802 | 0,825803011 | HIST1H2BF | FALSE | Upregulated |
| -0,166918325 | 0,825803011 | PAM | FALSE | Downregulated |
| 0,177666713 | 0,826047405 | TBXAS1 | FALSE | Upregulated |
| -0,095030619 | 0,826061648 | WNK3 | FALSE | Downregulated |
| 0,135414041 | 0,826250281 | FAM46A | FALSE | Upregulated |
| -0,172841852 | 0,826392888 | GNG7 | FALSE | Downregulated |
| -0,078939408 | 0,826392888 | KHDRBS1 | FALSE | Downregulated |

| | | | | |
|---|---|---|---|---|
| -0,321666538 | 0,826392888 | LOC100506538 | FALSE | Downregulated |
| 0,097131957 | 0,82651371 | MLXIP | FALSE | Upregulated |
| 0,140682241 | 0,82651371 | RP11-300J18.1 | FALSE | Upregulated |
| -0,441896985 | 0,82651371 | TXLNB | FALSE | Downregulated |
| -0,102251889 | 0,826588191 | CPSF6 | FALSE | Downregulated |
| 0,16189505 | 0,826588191 | FNDC8 | FALSE | Upregulated |
| -0,26095555 | 0,826771929 | CHCHD5 | FALSE | Downregulated |
| 0,252288519 | 0,826771929 | CNIH4 | FALSE | Upregulated |
| -0,339447766 | 0,826771929 | LINGO1 | FALSE | Downregulated |
| 0,4107182 | 0,826771929 | METTL4 | FALSE | Upregulated |
| -0,206211197 | 0,826771929 | NECAB2 | FALSE | Downregulated |
| -0,20764346 | 0,826771929 | PTRHD1 | FALSE | Downregulated |
| -0,254572082 | 0,826771929 | RP11-267N12.1 | FALSE | Downregulated |
| -0,2155873 | 0,826771929 | SP5 | FALSE | Downregulated |
| -0,128011907 | 0,826771929 | TMEM106B | FALSE | Downregulated |
| -0,131626605 | 0,826771929 | UHRF2 | FALSE | Downregulated |
| 0,123956501 | 0,826771929 | ZNF526 | FALSE | Upregulated |
| -0,186948375 | 0,826856783 | THAP8 | FALSE | Downregulated |
| 0,098709603 | 0,827004719 | ARCN1 | FALSE | Upregulated |
| 0,188612251 | 0,827004719 | ZNF616 | FALSE | Upregulated |
| 0,245996315 | 0,827008203 | DSN1 | FALSE | Upregulated |
| 0,218458297 | 0,827008203 | ZNF577 | FALSE | Upregulated |
| 0,096818114 | 0,827132984 | CHPF | FALSE | Upregulated |
| -0,074540468 | 0,827132984 | IL13RA1 | FALSE | Downregulated |
| -0,175929166 | 0,827280439 | ENOPH1 | FALSE | Downregulated |
| 0,326947888 | 0,827393804 | ZNF709 | FALSE | Upregulated |
| 0,455469873 | 0,827490608 | KIAA1239 | FALSE | Upregulated |
| -0,161212165 | 0,827490608 | TDP2 | FALSE | Downregulated |
| 0,089769286 | 0,827490608 | TSPYL6 | FALSE | Upregulated |
| 0,22904642 | 0,827492903 | LAT | FALSE | Upregulated |
| -0,431309599 | 0,827577292 | AADACL2 | FALSE | Downregulated |
| -0,092247997 | 0,827577292 | PLCB3 | FALSE | Downregulated |
| 0,291495247 | 0,827577292 | RAD51D | FALSE | Upregulated |
| -0,209610968 | 0,827577292 | RP5-1057J7.6 | FALSE | Downregulated |
| 0,121770452 | 0,827577292 | ZNF292 | FALSE | Upregulated |
| -0,476997501 | 0,827652877 | PBOV1 | FALSE | Downregulated |
| 0,238029621 | 0,827652877 | ZMYM1 | FALSE | Upregulated |
| -0,166385136 | 0,827853897 | ANP32E | FALSE | Downregulated |
| -0,141326022 | 0,827959287 | ADNP2 | FALSE | Downregulated |
| -0,058266858 | 0,827959287 | ATXN1 | FALSE | Downregulated |
| 0,415431288 | 0,827994459 | SCN7A | FALSE | Upregulated |
| 0,1503396 | 0,827994459 | TBK1 | FALSE | Upregulated |
| -0,255717356 | 0,828067988 | TMEM185A | FALSE | Downregulated |
| 0,821289141 | 0,828116941 | RASA4B | FALSE | Upregulated |
| -0,076905446 | 0,828219311 | NT5E | FALSE | Downregulated |
| 0,151272588 | 0,828239508 | FBXL14 | FALSE | Upregulated |
| 0,126282201 | 0,828239508 | GLIPR1L1 | FALSE | Upregulated |
| -0,253912437 | 0,828239508 | LINC01107 | FALSE | Downregulated |
| -0,074589448 | 0,828239508 | SLC3A1 | FALSE | Downregulated |
| -0,156348607 | 0,82840333 | DPF2 | FALSE | Downregulated |
| -0,306643284 | 0,82840333 | FBXO4 | FALSE | Downregulated |

| | | | | |
|---|---|---|---|---|
| -0,416744055 | 0,82840333 | LOC100996350 | FALSE | Downregulated |
| -0,16924169 | 0,828415719 | EXOC3L2 | FALSE | Downregulated |
| -0,130226282 | 0,828615512 | MTAP | FALSE | Downregulated |
| 0,310428089 | 0,828645633 | C11orf16 | FALSE | Upregulated |
| 0,124799507 | 0,828645633 | CRIP3 | FALSE | Upregulated |
| -0,621526515 | 0,828645633 | H2AFB2 | FALSE | Downregulated |
| -0,264976988 | 0,828645633 | KRTCAP2 | FALSE | Downregulated |
| -0,206918072 | 0,828645633 | LINC00092 | FALSE | Downregulated |
| 0,370984153 | 0,828645633 | POM121L8P | FALSE | Upregulated |
| 0,088989078 | 0,828645633 | RPL27 | FALSE | Upregulated |
| 0,197127829 | 0,828645633 | SELK | FALSE | Upregulated |
| 0,329716478 | 0,828645633 | UBL7-AS1 | FALSE | Upregulated |
| 0,193102005 | 0,828645633 | ZNF480 | FALSE | Upregulated |
| 0,084797253 | 0,828696245 | DHX38 | FALSE | Upregulated |
| -0,187498181 | 0,828696245 | SKAP1 | FALSE | Downregulated |
| -0,080852397 | 0,828743428 | AHCTF1 | FALSE | Downregulated |
| -0,117738334 | 0,828787521 | TACC2 | FALSE | Downregulated |
| 0,377614322 | 0,828889371 | DUSP4 | FALSE | Upregulated |
| -0,095230256 | 0,829060892 | HEBP1 | FALSE | Downregulated |
| 0,086721358 | 0,829060892 | KDELR2 | FALSE | Upregulated |
| -0,224645101 | 0,829060892 | NUBPL | FALSE | Downregulated |
| 0,15511897 | 0,829272196 | ZNRF2P1 | FALSE | Upregulated |
| 0,193350913 | 0,829308434 | C19orf82 | FALSE | Upregulated |
| 0,199439274 | 0,829375879 | USP46 | FALSE | Upregulated |
| 0,338536778 | 0,829670034 | ZNF267 | FALSE | Upregulated |
| 0,100734654 | 0,829680445 | MYO19 | FALSE | Upregulated |
| -0,158245652 | 0,829745338 | HNF4G | FALSE | Downregulated |
| 0,092868809 | 0,829745338 | KDM1B | FALSE | Upregulated |
| -0,139379213 | 0,829745338 | NLK | FALSE | Downregulated |
| 0,085160942 | 0,829745338 | OLFML1 | FALSE | Upregulated |
| -0,155325403 | 0,829745338 | SIMC1 | FALSE | Downregulated |
| -0,194636751 | 0,829745338 | STX2 | FALSE | Downregulated |
| 0,108841532 | 0,829745338 | TSPAN31 | FALSE | Upregulated |
| -0,179873764 | 0,829745338 | ZMPSTE24 | FALSE | Downregulated |
| -0,389285323 | 0,830056614 | IGFBP6 | FALSE | Downregulated |
| 0,379806041 | 0,830056614 | TGM1 | FALSE | Upregulated |
| -0,20577144 | 0,830066279 | SMC2 | FALSE | Downregulated |
| 0,093303377 | 0,83024412 | PRPF38A | FALSE | Upregulated |
| -0,242974717 | 0,83024412 | RNF216P1 | FALSE | Downregulated |
| 0,197290162 | 0,83024412 | STXBP3 | FALSE | Upregulated |
| 0,280978906 | 0,830423282 | KIF20B | FALSE | Upregulated |
| 0,071723457 | 0,830423282 | SEC61A1 | FALSE | Upregulated |
| 0,120699059 | 0,830449258 | GOLGA8M | FALSE | Upregulated |
| -0,075500114 | 0,830449258 | OGDH | FALSE | Downregulated |
| -0,302457528 | 0,830503323 | WDR66 | FALSE | Downregulated |
| 0,186766941 | 0,83060867 | ALG3 | FALSE | Upregulated |
| 0,293264954 | 0,83060867 | EMR4P | FALSE | Upregulated |
| 0,263593207 | 0,83060867 | LOC101928136 | FALSE | Upregulated |
| 0,150483086 | 0,83060867 | WDR41 | FALSE | Upregulated |
| -0,338815653 | 0,830713954 | ACD | FALSE | Downregulated |
| -0,091482281 | 0,831009526 | MCCC1 | FALSE | Downregulated |

| | | | | |
|---|---|---|---|---|
| 0,114205027 | 0,831022201 | DCTN4 | FALSE | Upregulated |
| -0,222183062 | 0,831099487 | LEPREL1 | FALSE | Downregulated |
| -0,207565303 | 0,831113275 | TMEM179 | FALSE | Downregulated |
| 0,104525845 | 0,831123491 | OGG1 | FALSE | Upregulated |
| 0,096236327 | 0,831142491 | ABI3BP | FALSE | Upregulated |
| -0,264773247 | 0,831142491 | C3orf35 | FALSE | Downregulated |
| -0,199041997 | 0,831142491 | C3orf49 | FALSE | Downregulated |
| -0,178361529 | 0,831142491 | CINP | FALSE | Downregulated |
| -0,07529972 | 0,831142491 | DVL1 | FALSE | Downregulated |
| -0,078531997 | 0,831142491 | F11R | FALSE | Downregulated |
| 0,103294408 | 0,831142491 | FBRS | FALSE | Upregulated |
| -0,187663761 | 0,831142491 | GABBR1 | FALSE | Downregulated |
| 0,071590533 | 0,831142491 | H3F3A | FALSE | Upregulated |
| -0,300681853 | 0,831142491 | HILS1 | FALSE | Downregulated |
| -0,224777977 | 0,831142491 | MPPED1 | FALSE | Downregulated |
| 0,109843863 | 0,831142491 | PGAP2 | FALSE | Upregulated |
| -0,073692766 | 0,831142491 | TCF25 | FALSE | Downregulated |
| 0,071854425 | 0,831142491 | U2AF2 | FALSE | Upregulated |
| 0,122389062 | 0,831196876 | ABCB10 | FALSE | Upregulated |
| 0,086034258 | 0,831212414 | CSRNP2 | FALSE | Upregulated |
| -0,180071945 | 0,831327833 | BUD13 | FALSE | Downregulated |
| 0,303046596 | 0,831327833 | DLGAP1-AS1 | FALSE | Upregulated |
| 0,135747416 | 0,831327833 | FAM178B | FALSE | Upregulated |
| 0,140719885 | 0,831785726 | AHI1 | FALSE | Upregulated |
| -0,160207169 | 0,831785726 | ASNSD1 | FALSE | Downregulated |
| -0,152110846 | 0,831785726 | KLHL15 | FALSE | Downregulated |
| 0,116631325 | 0,831785726 | LOC100996741 | FALSE | Upregulated |
| 0,092239509 | 0,831785726 | SH3GLB1 | FALSE | Upregulated |
| 0,123097229 | 0,831785726 | SLC30A5 | FALSE | Upregulated |
| -0,174467157 | 0,831900045 | GLRA1 | FALSE | Downregulated |
| 0,40596515 | 0,832112853 | TG | FALSE | Upregulated |
| -0,169056118 | 0,832138039 | ADAL | FALSE | Downregulated |
| -0,110869617 | 0,832138039 | CTB-118N6.3 | FALSE | Downregulated |
| 0,209136751 | 0,832138039 | HIST2H2AB | FALSE | Upregulated |
| -0,116406245 | 0,832138039 | SRSF12 | FALSE | Downregulated |
| -0,089646377 | 0,832138039 | TRAPPC9 | FALSE | Downregulated |
| -0,181001025 | 0,832155296 | C8orf33 | FALSE | Downregulated |
| 0,361445232 | 0,832155296 | LOC102723501 | FALSE | Upregulated |
| 0,424289889 | 0,832155296 | RP11-318M2.2 | FALSE | Upregulated |
| -0,058285854 | 0,832282789 | TRRAP | FALSE | Downregulated |
| -0,110296793 | 0,832337236 | NFATC2IP | FALSE | Downregulated |
| -0,624175447 | 0,832337236 | NPIPB3 | FALSE | Downregulated |
| 0,151399147 | 0,832659741 | FBXL6 | FALSE | Upregulated |
| -0,218117127 | 0,832659741 | FOPNL | FALSE | Downregulated |
| 0,14397718 | 0,832659741 | LOC101928355 | FALSE | Upregulated |
| 0,093524827 | 0,832659741 | PI4K2A | FALSE | Upregulated |
| 0,202511817 | 0,832659741 | RP3-400N23.6 | FALSE | Upregulated |
| -0,235633963 | 0,832686283 | MYO1H | FALSE | Downregulated |
| -0,156784603 | 0,832725141 | LPP-AS1 | FALSE | Downregulated |
| 0,090152733 | 0,832725141 | SH2B1 | FALSE | Upregulated |
| -0,168979457 | 0,832963515 | MON1A | FALSE | Downregulated |

| | | | | |
|---|---|---|---|---|
| 0,133119234 | 0,832963515 | NKIRAS1 | FALSE | Upregulated |
| -0,399669635 | 0,832963515 | SLC24A3 | FALSE | Downregulated |
| 0,295147544 | 0,832963515 | VPS13A-AS1 | FALSE | Upregulated |
| 0,211969792 | 0,833157143 | ZNF571 | FALSE | Upregulated |
| 0,122933001 | 0,833285785 | ASTE1 | FALSE | Upregulated |
| 0,337527403 | 0,833285785 | SLC6A9 | FALSE | Upregulated |
| 0,080169262 | 0,83333829 | NGEF | FALSE | Upregulated |
| -0,226855205 | 0,833391967 | DENND2C | FALSE | Downregulated |
| -0,38562856 | 0,833468509 | FSD1L | FALSE | Downregulated |
| 0,074631788 | 0,833468509 | KCTD20 | FALSE | Upregulated |
| -0,081435616 | 0,833468509 | KEAP1 | FALSE | Downregulated |
| 0,081857506 | 0,833468509 | TXNDC5 | FALSE | Upregulated |
| 0,266331432 | 0,83383966 | DEPDC4 | FALSE | Upregulated |
| -0,362967378 | 0,83383966 | DHRS13 | FALSE | Downregulated |
| -0,29899759 | 0,83383966 | EMX2OS | FALSE | Downregulated |
| 0,193894394 | 0,83383966 | FAM151A | FALSE | Upregulated |
| -0,162778267 | 0,83383966 | VPS29 | FALSE | Downregulated |
| -0,21755197 | 0,834100574 | ILKAP | FALSE | Downregulated |
| 0,113680473 | 0,834100574 | IRF2 | FALSE | Upregulated |
| 0,110503271 | 0,834100574 | ST7-AS2 | FALSE | Upregulated |
| -0,226415546 | 0,834231901 | C16orf80 | FALSE | Downregulated |
| -0,239472856 | 0,834231901 | WDR25 | FALSE | Downregulated |
| -0,24010966 | 0,834256726 | MRPL13 | FALSE | Downregulated |
| 0,154407136 | 0,83438111 | S100PBP | FALSE | Upregulated |
| -0,137384614 | 0,83457062 | RGP1 | FALSE | Downregulated |
| -0,092291182 | 0,834638567 | ACAP3 | FALSE | Downregulated |
| 0,270563556 | 0,834638567 | BMI1 | FALSE | Upregulated |
| -0,300537996 | 0,834638567 | FAM83C | FALSE | Downregulated |
| 0,091309764 | 0,834638567 | USP4 | FALSE | Upregulated |
| -0,662485426 | 0,834651726 | ACTN3 | FALSE | Downregulated |
| -0,159022293 | 0,834651726 | AKAP10 | FALSE | Downregulated |
| 0,341617424 | 0,834651726 | LOC646214 | FALSE | Upregulated |
| -0,151948321 | 0,834651726 | SDSL | FALSE | Downregulated |
| 0,169284357 | 0,834651726 | TTLL3 | FALSE | Upregulated |
| 0,087803174 | 0,834771414 | CASC3 | FALSE | Upregulated |
| 0,225952957 | 0,834771414 | ZNF177 | FALSE | Upregulated |
| 0,323849185 | 0,835004092 | RAB34 | FALSE | Upregulated |
| 0,174298728 | 0,8350561 | SLC16A11 | FALSE | Upregulated |
| 0,049884598 | 0,835552499 | SLC38A10 | FALSE | Upregulated |
| -0,081289366 | 0,835552499 | SPTB | FALSE | Downregulated |
| -0,130518186 | 0,835623983 | FGFR1OP | FALSE | Downregulated |
| -0,10509223 | 0,835637672 | AZIN1 | FALSE | Downregulated |
| 0,508449253 | 0,835637672 | LOC101927055 | FALSE | Upregulated |
| 0,213917168 | 0,835760316 | CCDC149 | FALSE | Upregulated |
| 0,314195712 | 0,835760316 | SLC15A2 | FALSE | Upregulated |
| 0,235494829 | 0,835760316 | TASP1 | FALSE | Upregulated |
| 0,11167062 | 0,835781278 | VRK3 | FALSE | Upregulated |
| -0,107308728 | 0,835807572 | ABCB8 | FALSE | Downregulated |
| 0,104154359 | 0,835807572 | C16orf62 | FALSE | Upregulated |
| -0,497840625 | 0,835807572 | IGFN1 | FALSE | Downregulated |
| 0,228346206 | 0,835807572 | LINC00685 | FALSE | Upregulated |

| | | | | |
|---|---|---|---|---|
| 0,066865424 | 0,835807572 | MBD3 | FALSE | Upregulated |
| 0,066575264 | 0,835807572 | PEA15 | FALSE | Upregulated |
| 0,132078125 | 0,835807572 | RP11-760N9.1 | FALSE | Upregulated |
| 0,21690142 | 0,835807572 | STXBP5 | FALSE | Upregulated |
| -0,168474838 | 0,835807572 | TSPO | FALSE | Downregulated |
| -0,316439213 | 0,836021253 | LOC100128006 | FALSE | Downregulated |
| -0,094275634 | 0,836021253 | PSMD4 | FALSE | Downregulated |
| 0,08111407 | 0,836027454 | CCT3 | FALSE | Upregulated |
| -0,256396307 | 0,836027454 | FMOD | FALSE | Downregulated |
| -0,079582276 | 0,836027454 | XPC | FALSE | Downregulated |
| -0,122953741 | 0,836103475 | ACAD8 | FALSE | Downregulated |
| -0,424633848 | 0,836103475 | CLSTN2-AS1 | FALSE | Downregulated |
| 0,508584122 | 0,836103475 | FRMPD3 | FALSE | Upregulated |
| -0,302191423 | 0,836103475 | LOC102724807 | FALSE | Downregulated |
| 0,08832407 | 0,836149698 | HECA | FALSE | Upregulated |
| -0,273614801 | 0,836192933 | LOC101060060 | FALSE | Downregulated |
| 0,345231941 | 0,836192933 | LOC102724293 | FALSE | Upregulated |
| 0,113016451 | 0,836192933 | TMEM138 | FALSE | Upregulated |
| -0,077432075 | 0,836192933 | TMEM245 | FALSE | Downregulated |
| 0,080753782 | 0,836203131 | SLC2A12 | FALSE | Upregulated |
| -0,413020841 | 0,836275822 | LINC00035 | FALSE | Downregulated |
| -2,441377461 | 0,836559332 | RNU1-3 | FALSE | Downregulated |
| 0,211308103 | 0,83660507 | ZDHHC24 | FALSE | Upregulated |
| 0,305865474 | 0,836732047 | CCDC113 | FALSE | Upregulated |
| -0,132671388 | 0,836909319 | CLMP | FALSE | Downregulated |
| 0,093506682 | 0,836912112 | TMEM79 | FALSE | Upregulated |
| -0,101072228 | 0,836912112 | ZBTB45 | FALSE | Downregulated |
| -0,303243436 | 0,836956249 | LINC00637 | FALSE | Downregulated |
| -0,083721168 | 0,836956249 | RANBP10 | FALSE | Downregulated |
| 0,122823941 | 0,836956249 | TMEM206 | FALSE | Upregulated |
| 0,139561001 | 0,836961362 | NR4A3 | FALSE | Upregulated |
| 0,316623432 | 0,836961362 | SPATA12 | FALSE | Upregulated |
| 0,190341293 | 0,837115636 | UAP1L1 | FALSE | Upregulated |
| 0,08123859 | 0,837137838 | NR2C2 | FALSE | Upregulated |
| 0,088157175 | 0,837356234 | TAF4 | FALSE | Upregulated |
| -0,149907171 | 0,837356234 | ZNF436 | FALSE | Downregulated |
| -0,138824297 | 0,837570894 | BNIP2 | FALSE | Downregulated |
| -0,103468564 | 0,837624286 | AKAP11 | FALSE | Downregulated |
| 0,24573973 | 0,837624286 | ALKBH6 | FALSE | Upregulated |
| 0,496163873 | 0,837624286 | LOC101928221 | FALSE | Upregulated |
| 0,164599147 | 0,837624286 | UQCC2 | FALSE | Upregulated |
| 0,075998666 | 0,83768928 | CC2D1A | FALSE | Upregulated |
| 0,204100952 | 0,837978635 | LOC102723622 | FALSE | Upregulated |
| 0,232758049 | 0,838063297 | NSUN5P2 | FALSE | Upregulated |
| 0,102242943 | 0,838063297 | RNF31 | FALSE | Upregulated |
| 0,323002242 | 0,838063297 | RP11-278H7.1 | FALSE | Upregulated |
| 0,235137253 | 0,838063297 | RPL23AP53 | FALSE | Upregulated |
| 0,139134835 | 0,838063297 | ZBED6CL | FALSE | Upregulated |
| 0,290035992 | 0,838063297 | ZNF853 | FALSE | Upregulated |
| -0,317406511 | 0,838092972 | APITD1 | FALSE | Downregulated |
| 0,099667771 | 0,838092972 | CASZ1 | FALSE | Upregulated |

| | | | | |
|---|---|---|---|---|
| -0,090445603 | 0,838092972 | SEMA3F | FALSE | Downregulated |
| -0,15279915 | 0,838092972 | ZNF688 | FALSE | Downregulated |
| -0,161039604 | 0,838094926 | C12orf57 | FALSE | Downregulated |
| -0,221481454 | 0,838132586 | ADPRHL2 | FALSE | Downregulated |
| 0,254237532 | 0,838132586 | ZC4H2 | FALSE | Upregulated |
| 0,355638033 | 0,838313756 | RP11-115H18.1 | FALSE | Upregulated |
| 0,112991965 | 0,838313756 | SPATA1 | FALSE | Upregulated |
| 0,069049748 | 0,838330094 | CABIN1 | FALSE | Upregulated |
| -0,076567131 | 0,838330094 | CBX5 | FALSE | Downregulated |
| -0,071182234 | 0,838330094 | GPATCH8 | FALSE | Downregulated |
| 0,104731084 | 0,838330094 | LOC102723853 | FALSE | Upregulated |
| 0,092554588 | 0,838330094 | RAP2C | FALSE | Upregulated |
| 0,457547878 | 0,838330094 | TOX | FALSE | Upregulated |
| -0,174257507 | 0,838330094 | WDR77 | FALSE | Downregulated |
| -0,118234419 | 0,838330094 | ZNF76 | FALSE | Downregulated |
| 0,313496071 | 0,838330094 | ZSCAN1 | FALSE | Upregulated |
| -0,110420369 | 0,838578277 | ACVR2B-AS1 | FALSE | Downregulated |
| -0,05430143 | 0,838578277 | AES | FALSE | Downregulated |
| -0,208505509 | 0,838578277 | AIF1L | FALSE | Downregulated |
| -0,312709294 | 0,838578277 | CCDC8 | FALSE | Downregulated |
| -0,219486083 | 0,838578277 | DZIP3 | FALSE | Downregulated |
| -0,080846024 | 0,838578277 | ERC1 | FALSE | Downregulated |
| 0,424130512 | 0,838578277 | FLJ46026 | FALSE | Upregulated |
| -0,055126821 | 0,838578277 | GIGYF1 | FALSE | Downregulated |
| -0,18455023 | 0,838578277 | IQSEC3 | FALSE | Downregulated |
| -0,30539182 | 0,838578277 | KITLG | FALSE | Downregulated |
| -0,150180177 | 0,838578277 | KLHL3 | FALSE | Downregulated |
| -0,350936941 | 0,838578277 | LOC100129534 | FALSE | Downregulated |
| -0,090004521 | 0,838578277 | LOC100506710 | FALSE | Downregulated |
| 0,128417689 | 0,838578277 | MAP3K10 | FALSE | Upregulated |
| 0,408899628 | 0,838578277 | MUSK | FALSE | Upregulated |
| 0,126090936 | 0,838578277 | TSEN54 | FALSE | Upregulated |
| -0,054578815 | 0,838578277 | TTPAL | FALSE | Downregulated |
| 0,144991002 | 0,838578277 | UGCG | FALSE | Upregulated |
| 0,139159616 | 0,838578277 | ZNF37BP | FALSE | Upregulated |
| 0,277154922 | 0,838578277 | ZNF525 | FALSE | Upregulated |
| -0,122603577 | 0,838674887 | BMPR1A | FALSE | Downregulated |
| -0,397414903 | 0,838674887 | LINC00202-1 | FALSE | Downregulated |
| 0,329255554 | 0,838691391 | MIXL1 | FALSE | Upregulated |
| -0,107740004 | 0,838691391 | NIT2 | FALSE | Downregulated |
| 0,109205561 | 0,838691391 | PDIA6 | FALSE | Upregulated |
| -0,211655334 | 0,839189529 | C7orf31 | FALSE | Downregulated |
| -0,42015312 | 0,839189529 | RPL36A-HNRNPH2 | FALSE | Downregulated |
| -0,07375031 | 0,839189529 | SIPA1 | FALSE | Downregulated |
| 0,297955144 | 0,839189529 | TMEM105 | FALSE | Upregulated |
| 0,103578341 | 0,839219366 | SEC61B | FALSE | Upregulated |
| 0,175659123 | 0,839237504 | EPB41L1 | FALSE | Upregulated |
| -0,373187525 | 0,839265435 | ABCA13 | FALSE | Downregulated |
| 0,356996225 | 0,839465472 | LOC100093631 | FALSE | Upregulated |
| -0,128062812 | 0,839465472 | SLC17A5 | FALSE | Downregulated |
| 0,102780197 | 0,839465472 | TOMM40 | FALSE | Upregulated |

| | | | | |
|---|---|---|---|---|
| 0,368211844 | 0,839474341 | ATP2A1 | FALSE | Upregulated |
| -0,065002343 | 0,839765096 | CAPN15 | FALSE | Downregulated |
| 0,081237199 | 0,839765096 | DNAJC11 | FALSE | Upregulated |
| -0,153185835 | 0,839878923 | CDKN2AIPNL | FALSE | Downregulated |
| 0,169077173 | 0,839878923 | ST6GALNAC2 | FALSE | Upregulated |
| -0,065001969 | 0,839878923 | UBR5 | FALSE | Downregulated |
| -0,107264334 | 0,839910293 | AC007038.7 | FALSE | Downregulated |
| 0,171145164 | 0,840193353 | C7orf71 | FALSE | Upregulated |
| 0,163884327 | 0,840210472 | TSPAN12 | FALSE | Upregulated |
| -0,08194995 | 0,840324806 | C4BPA | FALSE | Downregulated |
| -0,086086068 | 0,840482511 | CLCN3 | FALSE | Downregulated |
| -0,118896341 | 0,840741838 | PQLC2 | FALSE | Downregulated |
| -0,149020177 | 0,840765453 | LINC00342 | FALSE | Downregulated |
| -0,412285844 | 0,840765453 | LINC00961 | FALSE | Downregulated |
| -0,097432167 | 0,840765453 | ZNF805 | FALSE | Downregulated |
| 0,187226994 | 0,840915993 | ETV4 | FALSE | Upregulated |
| 0,104533009 | 0,841001322 | LOC727978 | FALSE | Upregulated |
| -0,305914489 | 0,841001322 | ULK4P1 | FALSE | Downregulated |
| -0,145729312 | 0,841162152 | CLEC3B | FALSE | Downregulated |
| 0,115692511 | 0,841162152 | EXOSC3 | FALSE | Upregulated |
| 0,08156956 | 0,841162152 | MTMR7 | FALSE | Upregulated |
| -0,158821206 | 0,841162152 | TRPM3 | FALSE | Downregulated |
| -0,085263464 | 0,841209948 | ANO1 | FALSE | Downregulated |
| -0,095198002 | 0,841312118 | ALG12 | FALSE | Downregulated |
| -0,125881996 | 0,841413588 | DNAJB6 | FALSE | Downregulated |
| 0,287475249 | 0,841493221 | LRRCC1 | FALSE | Upregulated |
| -0,1607474 | 0,841493221 | PLEK2 | FALSE | Downregulated |
| -0,089095457 | 0,841493221 | PUS10 | FALSE | Downregulated |
| -0,124335229 | 0,841503786 | BMS1 | FALSE | Downregulated |
| -0,196826851 | 0,841503786 | DPT | FALSE | Downregulated |
| -0,305891622 | 0,841503786 | LY86 | FALSE | Downregulated |
| -0,124705342 | 0,841619002 | CSE1L | FALSE | Downregulated |
| -0,194194615 | 0,841619002 | DDX11L2 | FALSE | Downregulated |
| -0,070277803 | 0,841619002 | FAM134C | FALSE | Downregulated |
| -0,223344184 | 0,841619002 | FBF1 | FALSE | Downregulated |
| 0,104856473 | 0,841619002 | JPX | FALSE | Upregulated |
| 0,395501054 | 0,841619002 | LOC100506161 | FALSE | Upregulated |
| 0,101604613 | 0,841619002 | LOC102723451 | FALSE | Upregulated |
| -0,111973865 | 0,841619002 | MESDC2 | FALSE | Downregulated |
| 0,161709753 | 0,841619002 | NUDT9 | FALSE | Upregulated |
| -0,087582386 | 0,841619002 | SCCPDH | FALSE | Downregulated |
| -0,105952358 | 0,841619002 | SIAH1 | FALSE | Downregulated |
| 0,273429229 | 0,841619002 | SLC5A4 | FALSE | Upregulated |
| 0,070882288 | 0,841619002 | STEAP3 | FALSE | Upregulated |
| 0,182716239 | 0,841619002 | THBD | FALSE | Upregulated |
| 0,167830511 | 0,841713365 | LOC102724243 | FALSE | Upregulated |
| 0,154664995 | 0,841730745 | AKAP3 | FALSE | Upregulated |
| 0,208931959 | 0,841730745 | CHST4 | FALSE | Upregulated |
| -0,094128155 | 0,841730745 | CRTAP | FALSE | Downregulated |
| -0,226439767 | 0,841730745 | OR13F1 | FALSE | Downregulated |
| -0,090619076 | 0,841862701 | SGK494 | FALSE | Downregulated |

| | | | | |
|---|---|---|---|---|
| -0,246584888 | 0,841873134 | AS3MT | FALSE | Downregulated |
| 0,105829194 | 0,841873134 | LOC102724273 | FALSE | Upregulated |
| 0,168004252 | 0,842048035 | USE1 | FALSE | Upregulated |
| 0,380994321 | 0,842136285 | AC016910.1 | FALSE | Upregulated |
| -0,14405534 | 0,842161309 | C11orf35 | FALSE | Downregulated |
| -0,360890249 | 0,842161309 | SEC14L1P1 | FALSE | Downregulated |
| -0,062935352 | 0,842161309 | TMED2 | FALSE | Downregulated |
| 0,109579987 | 0,842184591 | BCL7B | FALSE | Upregulated |
| -0,063231608 | 0,842184591 | CHD8 | FALSE | Downregulated |
| 0,127677658 | 0,842184591 | PARG | FALSE | Upregulated |
| 0,140550584 | 0,842365939 | ZNF252P | FALSE | Upregulated |
| 0,135559383 | 0,842374586 | RNLS | FALSE | Upregulated |
| 0,259543107 | 0,842374586 | TMEM102 | FALSE | Upregulated |
| -0,186452391 | 0,842524326 | MPV17L | FALSE | Downregulated |
| 0,150550439 | 0,842545945 | LOC100996571 | FALSE | Upregulated |
| 0,243742316 | 0,842749958 | SSSCA1-AS1 | FALSE | Upregulated |
| 0,393853773 | 0,843002554 | LINC00506 | FALSE | Upregulated |
| -0,094824868 | 0,843002554 | PTPN1 | FALSE | Downregulated |
| -0,225264582 | 0,843002554 | STOML1 | FALSE | Downregulated |
| -0,187354466 | 0,843238056 | CBR3 | FALSE | Downregulated |
| 0,429734518 | 0,843575141 | TMEM54 | FALSE | Upregulated |
| -0,111328296 | 0,843697388 | LIN7C | FALSE | Downregulated |
| -0,192868029 | 0,843739989 | ANKRD36B | FALSE | Downregulated |
| -0,081396336 | 0,843739989 | CACYBP | FALSE | Downregulated |
| -0,164103426 | 0,843739989 | EDNRB-AS1 | FALSE | Downregulated |
| 0,257028936 | 0,843739989 | LOC729870 | FALSE | Upregulated |
| -0,118169312 | 0,843739989 | MORC3 | FALSE | Downregulated |
| -0,082644859 | 0,843739989 | MXD4 | FALSE | Downregulated |
| -0,204238698 | 0,843739989 | NKAP | FALSE | Downregulated |
| -0,210071623 | 0,843739989 | NTPCR | FALSE | Downregulated |
| -0,103021973 | 0,843739989 | SLC6A12 | FALSE | Downregulated |
| 0,177443761 | 0,843739989 | TMEM209 | FALSE | Upregulated |
| 0,221665429 | 0,844073378 | SPATA18 | FALSE | Upregulated |
| -0,123222348 | 0,844617131 | IDS | FALSE | Downregulated |
| 0,253024079 | 0,844841646 | AGRP | FALSE | Upregulated |
| -0,257654059 | 0,844841646 | GALNT15 | FALSE | Downregulated |
| 0,113882942 | 0,844841646 | GLCCI1 | FALSE | Upregulated |
| 0,305796856 | 0,844841646 | LINC00563 | FALSE | Upregulated |
| 0,120913893 | 0,844841646 | LOC102723824 | FALSE | Upregulated |
| -0,154360729 | 0,844841646 | LOC729173 | FALSE | Downregulated |
| -0,141919845 | 0,844841646 | MKKS | FALSE | Downregulated |
| -0,124972023 | 0,844841646 | RP11-85B7.2 | FALSE | Downregulated |
| 0,15822106 | 0,844884636 | LYPLAL1 | FALSE | Upregulated |
| -0,144097337 | 0,845149159 | FREM3 | FALSE | Downregulated |
| -0,42758555 | 0,845149159 | LHCGR | FALSE | Downregulated |
| -0,17583467 | 0,845149159 | LOC101928741 | FALSE | Downregulated |
| -0,394604527 | 0,845437541 | CHCHD6 | FALSE | Downregulated |
| -0,303398489 | 0,845437541 | RP3-510O8.4 | FALSE | Downregulated |
| 0,388074104 | 0,845484129 | ZNF888 | FALSE | Upregulated |
| 0,129418695 | 0,845711047 | ST7L | FALSE | Upregulated |
| 0,182022369 | 0,845781569 | ANK2 | FALSE | Upregulated |

| | | | | |
|---|---|---|---|---|
| -0,201858275 | 0,845781569 | CEP112 | FALSE | Downregulated |
| 0,071330972 | 0,845781569 | POR | FALSE | Upregulated |
| -0,114777749 | 0,845794874 | NBPF26 | FALSE | Downregulated |
| -0,161059 | 0,845993077 | TEDDM1 | FALSE | Downregulated |
| 0,33022072 | 0,845993077 | TPK1 | FALSE | Upregulated |
| -0,125120528 | 0,845993077 | ZMYND19 | FALSE | Downregulated |
| -0,128558926 | 0,846398455 | C19orf53 | FALSE | Downregulated |
| -0,044268405 | 0,846398455 | HNRNPUL1 | FALSE | Downregulated |
| 0,103401801 | 0,846462635 | ARPC4 | FALSE | Upregulated |
| 0,1221643 | 0,846462635 | CNNM1 | FALSE | Upregulated |
| -0,074121871 | 0,846462635 | KAT2A | FALSE | Downregulated |
| -0,223721112 | 0,846462635 | MRVI1 | FALSE | Downregulated |
| 0,321142494 | 0,846462635 | NPIPA2 | FALSE | Upregulated |
| 0,197671484 | 0,846462635 | NPRL2 | FALSE | Upregulated |
| 0,183532818 | 0,846462635 | PDC | FALSE | Upregulated |
| 0,120569854 | 0,846462635 | PICK1 | FALSE | Upregulated |
| 0,069185148 | 0,846462635 | PRPF31 | FALSE | Upregulated |
| 0,36657238 | 0,846462635 | PTN | FALSE | Upregulated |
| -0,751482389 | 0,846462635 | RNU1-2 | FALSE | Downregulated |
| -0,125632857 | 0,846462635 | RPH3AL | FALSE | Downregulated |
| 0,143973961 | 0,846462635 | S100A14 | FALSE | Upregulated |
| -0,085033433 | 0,846462635 | SSR3 | FALSE | Downregulated |
| 0,097927137 | 0,846462635 | TRNT1 | FALSE | Upregulated |
| -0,269782413 | 0,846462635 | TUFT1 | FALSE | Downregulated |
| -0,115729286 | 0,846510422 | AFTPH | FALSE | Downregulated |
| -0,130556853 | 0,846510422 | C9orf96 | FALSE | Downregulated |
| -0,085532328 | 0,846510422 | MADD | FALSE | Downregulated |
| 0,079831414 | 0,846510422 | SIRT1 | FALSE | Upregulated |
| -0,300972322 | 0,846510422 | SLC9A2 | FALSE | Downregulated |
| 0,224159095 | 0,846510422 | TPM3P9 | FALSE | Upregulated |
| -0,115565204 | 0,846668088 | ABHD6 | FALSE | Downregulated |
| 0,057210862 | 0,846668088 | TNFAIP1 | FALSE | Upregulated |
| 0,521995277 | 0,846668088 | WDR11-AS1 | FALSE | Upregulated |
| -0,115876409 | 0,846727283 | RGL3 | FALSE | Downregulated |
| 0,1721636 | 0,846741173 | GEMIN7 | FALSE | Upregulated |
| -0,144514323 | 0,847034206 | AATF | FALSE | Downregulated |
| -0,226306708 | 0,847034206 | CCDC53 | FALSE | Downregulated |
| 0,121228205 | 0,847034206 | DOCK9 | FALSE | Upregulated |
| -0,077287939 | 0,847034206 | ULK1 | FALSE | Downregulated |
| 0,126663968 | 0,847034206 | XPNPEP1 | FALSE | Upregulated |
| 0,334929582 | 0,847100457 | APOO | FALSE | Upregulated |
| 0,081975381 | 0,847100457 | MEGF11 | FALSE | Upregulated |
| 0,151976007 | 0,847287809 | FLJ30403 | FALSE | Upregulated |
| -0,155605029 | 0,847439024 | CDK2 | FALSE | Downregulated |
| -0,122104863 | 0,847521621 | METAP1 | FALSE | Downregulated |
| -0,076182458 | 0,847521621 | MTA1 | FALSE | Downregulated |
| 0,278613068 | 0,847594549 | LOC644656 | FALSE | Upregulated |
| -0,085852619 | 0,847673435 | MLLT1 | FALSE | Downregulated |
| -0,090129851 | 0,847771389 | RNF146 | FALSE | Downregulated |
| 0,143173232 | 0,847785223 | CACNA2D2 | FALSE | Upregulated |
| 0,129093611 | 0,847785223 | MRPL40 | FALSE | Upregulated |

| | | | | |
|---|---|---|---|---|
| 0,123034007 | 0,847785223 | PHLDB3 | FALSE | Upregulated |
| 0,080472181 | 0,847785223 | QRICH1 | FALSE | Upregulated |
| -0,322962213 | 0,848093692 | CDK5 | FALSE | Downregulated |
| -0,073971811 | 0,848093692 | TRIM24 | FALSE | Downregulated |
| 0,100757869 | 0,848093692 | WDR46 | FALSE | Upregulated |
| -0,12681453 | 0,848093692 | ZNF160 | FALSE | Downregulated |
| 0,089812689 | 0,848114289 | SLC30A7 | FALSE | Upregulated |
| -0,383791284 | 0,848332857 | H2AFB3 | FALSE | Downregulated |
| -0,420844975 | 0,848342174 | DKFZp667F0711 | FALSE | Downregulated |
| 0,060234879 | 0,848389776 | SDC1 | FALSE | Upregulated |
| 0,077272977 | 0,848401572 | PCNX | FALSE | Upregulated |
| 0,20328722 | 0,848612919 | MINOS1 | FALSE | Upregulated |
| -0,06297426 | 0,848672057 | MBD6 | FALSE | Downregulated |
| -0,28624384 | 0,848839614 | GNAZ | FALSE | Downregulated |
| 0,172913685 | 0,848856867 | ZNF662 | FALSE | Upregulated |
| 0,090767194 | 0,849176185 | EIF4ENIF1 | FALSE | Upregulated |
| -0,28435927 | 0,849176185 | MPZL3 | FALSE | Downregulated |
| 0,148013876 | 0,849176185 | NKD1 | FALSE | Upregulated |
| -0,166896266 | 0,849176185 | WWOX | FALSE | Downregulated |
| 0,330565132 | 0,849216192 | MPZ | FALSE | Upregulated |
| 0,110380272 | 0,849216192 | SPIN2A | FALSE | Upregulated |
| 0,132758985 | 0,849430255 | LOC100131395 | FALSE | Upregulated |
| 0,295321768 | 0,849430255 | LOC100505942 | FALSE | Upregulated |
| 0,119417474 | 0,849430255 | MAP3K7 | FALSE | Upregulated |
| 0,164909306 | 0,849430255 | MEIS1-AS2 | FALSE | Upregulated |
| 0,322051584 | 0,849430255 | TUBA3E | FALSE | Upregulated |
| -0,133522549 | 0,849449769 | DYNLL1-AS1 | FALSE | Downregulated |
| -1,809090478 | 0,849541033 | RAD51L3-RFFL | FALSE | Downregulated |
| 0,200568904 | 0,849541033 | SNX24 | FALSE | Upregulated |
| -0,087093475 | 0,84968815 | GGT5 | FALSE | Downregulated |
| 0,264821338 | 0,849705762 | CHSY3 | FALSE | Upregulated |
| -0,120181543 | 0,849713173 | CASP8AP2 | FALSE | Downregulated |
| 0,118429539 | 0,849718853 | A2M-AS1 | FALSE | Upregulated |
| 0,512884298 | 0,849718853 | AOAH-IT1 | FALSE | Upregulated |
| 0,316032629 | 0,849718853 | CYP2S1 | FALSE | Upregulated |
| -0,318724108 | 0,849718853 | GPR26 | FALSE | Downregulated |
| 0,096624573 | 0,849718853 | KDELR3 | FALSE | Upregulated |
| 0,276071132 | 0,849718853 | LOC101929015 | FALSE | Upregulated |
| 0,326581364 | 0,849718853 | MACROD2 | FALSE | Upregulated |
| -0,063722886 | 0,849718853 | NOTCH1 | FALSE | Downregulated |
| 0,30945575 | 0,849718853 | RHOJ | FALSE | Upregulated |
| 0,061598667 | 0,849718853 | TCF7 | FALSE | Upregulated |
| 0,128894786 | 0,849718853 | TIFA | FALSE | Upregulated |
| -0,155180216 | 0,849719198 | LOC101927719 | FALSE | Downregulated |
| -0,166268699 | 0,849719198 | LOC286083 | FALSE | Downregulated |
| -0,107200531 | 0,849719198 | POFUT2 | FALSE | Downregulated |
| 0,121706564 | 0,849843681 | NTSR1 | FALSE | Upregulated |
| 0,188251499 | 0,850108904 | GPR37 | FALSE | Upregulated |
| -0,1921236 | 0,850226143 | CDRT15 | FALSE | Downregulated |
| 0,245032815 | 0,850318964 | MRPS28 | FALSE | Upregulated |
| -0,047938277 | 0,850432336 | CDK13 | FALSE | Downregulated |

| | | | | |
|---|---|---|---|---|
| 0,134629455 | 0,850514923 | VAMP7 | FALSE | Upregulated |
| 0,040080228 | 0,850626456 | LARP1 | FALSE | Upregulated |
| 0,099597627 | 0,850626456 | LOC102724144 | FALSE | Upregulated |
| 0,13859902 | 0,850626456 | ME2 | FALSE | Upregulated |
| 0,242794175 | 0,850626456 | ZNF283 | FALSE | Upregulated |
| -0,095570214 | 0,850688273 | MEIS2 | FALSE | Downregulated |
| 0,413883055 | 0,850984327 | FAM87A | FALSE | Upregulated |
| 0,099211029 | 0,851060376 | BRSK2 | FALSE | Upregulated |
| -0,185259911 | 0,851060376 | SP7 | FALSE | Downregulated |
| -0,191576778 | 0,851060376 | UBTD1 | FALSE | Downregulated |
| -0,167353424 | 0,851060376 | UCA1 | FALSE | Downregulated |
| -0,084514687 | 0,851060376 | ZBTB38 | FALSE | Downregulated |
| 0,349379474 | 0,851060376 | ZNF284 | FALSE | Upregulated |
| -0,589139672 | 0,85117491 | RNU1-27P | FALSE | Downregulated |
| -0,136134915 | 0,85117491 | SF3B4 | FALSE | Downregulated |
| -0,30101172 | 0,851191576 | ADAM20P1 | FALSE | Downregulated |
| 0,402393748 | 0,851363129 | LOC102724621 | FALSE | Upregulated |
| 0,1948797 | 0,851466974 | RPF2 | FALSE | Upregulated |
| 0,378846406 | 0,851776419 | CHL1 | FALSE | Upregulated |
| 0,138245691 | 0,851776419 | FAM86C1 | FALSE | Upregulated |
| -0,143720734 | 0,851776419 | MAL2 | FALSE | Downregulated |
| 0,175738745 | 0,851827457 | LOC101927296 | FALSE | Upregulated |
| -0,208874043 | 0,851827457 | MFNG | FALSE | Downregulated |
| -0,156922572 | 0,851827457 | RABL5 | FALSE | Downregulated |
| 0,188041378 | 0,851827457 | RBM7 | FALSE | Upregulated |
| 0,086190413 | 0,851827457 | RGL2 | FALSE | Upregulated |
| -0,29430543 | 0,852114931 | DDX3Y | FALSE | Downregulated |
| -0,179763719 | 0,852114931 | FAM175B | FALSE | Downregulated |
| 0,134487488 | 0,852273811 | IPP | FALSE | Upregulated |
| 0,252590647 | 0,852273811 | LRRC29 | FALSE | Upregulated |
| -0,115418559 | 0,852360629 | SH2D4A | FALSE | Downregulated |
| 0,142825467 | 0,852360629 | ZCRB1 | FALSE | Upregulated |
| 0,231885294 | 0,852501876 | TTC30A | FALSE | Upregulated |
| 0,202423022 | 0,852619428 | RIOK2 | FALSE | Upregulated |
| 0,172167438 | 0,852687235 | KATNAL1 | FALSE | Upregulated |
| 0,074359367 | 0,852687235 | MYRF | FALSE | Upregulated |
| -0,191489294 | 0,852755506 | CAV1 | FALSE | Downregulated |
| -0,113181294 | 0,852802719 | MRI1 | FALSE | Downregulated |
| 0,162494639 | 0,852802719 | STX18 | FALSE | Upregulated |
| -0,090107619 | 0,852938491 | NAA60 | FALSE | Downregulated |
| 0,413018149 | 0,853063298 | MAB21L3 | FALSE | Upregulated |
| -0,114147083 | 0,853065496 | ANAPC1 | FALSE | Downregulated |
| -0,220114996 | 0,853065496 | DCTPP1 | FALSE | Downregulated |
| -0,321130123 | 0,853065496 | NOMO3 | FALSE | Downregulated |
| 0,119329941 | 0,853065496 | NUDT16L1 | FALSE | Upregulated |
| 0,147653991 | 0,853065496 | SMIM12 | FALSE | Upregulated |
| 0,168028849 | 0,853065496 | SPATA2 | FALSE | Upregulated |
| -0,171199201 | 0,853154191 | DGKE | FALSE | Downregulated |
| 0,186594902 | 0,853154191 | RNASEL | FALSE | Upregulated |
| -0,117010978 | 0,853154191 | TMEM50B | FALSE | Downregulated |
| -0,200760899 | 0,853261235 | BAMBI | FALSE | Downregulated |

| | | | | |
|---|---|---|---|---|
| -0,193910386 | 0,853261235 | CCL22 | FALSE | Downregulated |
| 0,118308618 | 0,853261235 | FAM171A1 | FALSE | Upregulated |
| 0,12369935 | 0,853261235 | PIK3C3 | FALSE | Upregulated |
| 0,120141539 | 0,853261235 | ZC3H15 | FALSE | Upregulated |
| -0,260544214 | 0,853654099 | LEPREL4 | FALSE | Downregulated |
| 0,305880669 | 0,853784949 | SLC9A9-AS1 | FALSE | Upregulated |
| 0,119785038 | 0,853878563 | GPANK1 | FALSE | Upregulated |
| 0,126308014 | 0,853878563 | TGIF2 | FALSE | Upregulated |
| 0,136111851 | 0,853943311 | LGALS8-AS1 | FALSE | Upregulated |
| 0,101035952 | 0,854039431 | CFP | FALSE | Upregulated |
| -0,12622599 | 0,854039431 | FTX | FALSE | Downregulated |
| -0,129706253 | 0,854039431 | SPG20 | FALSE | Downregulated |
| -0,338559344 | 0,854130298 | C18orf21 | FALSE | Downregulated |
| -0,13274518 | 0,854130298 | DDR2 | FALSE | Downregulated |
| 0,383678378 | 0,854130298 | LOC102723603 | FALSE | Upregulated |
| 0,240802855 | 0,854130298 | LOC102724255 | FALSE | Upregulated |
| -0,109731993 | 0,854130298 | PLEKHA5 | FALSE | Downregulated |
| -0,095414632 | 0,854130298 | PLGLB2 | FALSE | Downregulated |
| -0,114184855 | 0,854130298 | SHOC2 | FALSE | Downregulated |
| -0,235684758 | 0,854130298 | WISP3 | FALSE | Downregulated |
| -0,108098629 | 0,854130298 | ZNF829 | FALSE | Downregulated |
| 0,115236375 | 0,85451561 | ARHGAP33 | FALSE | Upregulated |
| 0,118368149 | 0,8546501 | MRPS11 | FALSE | Upregulated |
| 0,120161622 | 0,8546501 | PBX3 | FALSE | Upregulated |
| 0,107886155 | 0,854735787 | CTPS1 | FALSE | Upregulated |
| -0,137352028 | 0,854735787 | MRPS22 | FALSE | Downregulated |
| -0,18713878 | 0,854735787 | SNORD10 | FALSE | Downregulated |
| -0,125156962 | 0,854735787 | UVRAG | FALSE | Downregulated |
| -0,225507766 | 0,854772348 | ARHGEF25 | FALSE | Downregulated |
| 0,163587481 | 0,854772348 | CEP76 | FALSE | Upregulated |
| 0,073004414 | 0,854772348 | DCP2 | FALSE | Upregulated |
| 0,242162361 | 0,854772348 | KIF3C | FALSE | Upregulated |
| 0,376770519 | 0,854772348 | LOC102723874 | FALSE | Upregulated |
| -1,314757827 | 0,854772348 | MYL2 | FALSE | Downregulated |
| 0,117648918 | 0,854772348 | NUP85 | FALSE | Upregulated |
| 0,091578306 | 0,854772348 | RANBP3 | FALSE | Upregulated |
| -0,249117854 | 0,854772348 | RP11-118E18.2 | FALSE | Downregulated |
| -0,146761465 | 0,854772348 | SOGA2 | FALSE | Downregulated |
| -0,093830064 | 0,854772348 | USP25 | FALSE | Downregulated |
| -0,278626044 | 0,854772348 | ZFP2 | FALSE | Downregulated |
| 0,077206583 | 0,854772348 | ZFYVE26 | FALSE | Upregulated |
| 0,067360465 | 0,854875807 | CAMTA2 | FALSE | Upregulated |
| 0,140758946 | 0,854936487 | OR52W1 | FALSE | Upregulated |
| -0,301232861 | 0,854936487 | PTP4A3 | FALSE | Downregulated |
| -0,107571145 | 0,855104154 | FAM57A | FALSE | Downregulated |
| 0,10898869 | 0,855257319 | HLA-L | FALSE | Upregulated |
| -0,095180104 | 0,855257319 | LAPTM4A | FALSE | Downregulated |
| -0,044015742 | 0,855257319 | SRCAP | FALSE | Downregulated |
| -0,141011494 | 0,855257319 | ZNF574 | FALSE | Downregulated |
| 0,165826479 | 0,85545062 | AP4E1 | FALSE | Upregulated |
| 0,094570511 | 0,85545062 | PAIP1 | FALSE | Upregulated |

| | | | | |
|---|---|---|---|---|
| 0,118153504 | 0,856087803 | LOC102724790 | FALSE | Upregulated |
| -0,350137442 | 0,856113876 | CD38 | FALSE | Downregulated |
| -0,066210722 | 0,856168791 | TET2 | FALSE | Downregulated |
| 0,098719428 | 0,856169829 | LRRC41 | FALSE | Upregulated |
| 0,174324428 | 0,856295406 | ENO2 | FALSE | Upregulated |
| 0,118274607 | 0,856295406 | SRM | FALSE | Upregulated |
| -0,237053729 | 0,856310427 | CDC42BPG | FALSE | Downregulated |
| 0,20930621 | 0,856310427 | MRPS33 | FALSE | Upregulated |
| 0,256026113 | 0,856338653 | ZNF648 | FALSE | Upregulated |
| 0,267981462 | 0,856551035 | NTF3 | FALSE | Upregulated |
| 0,122503263 | 0,856725609 | LOC102723769 | FALSE | Upregulated |
| 0,054914753 | 0,856725609 | LONP1 | FALSE | Upregulated |
| -0,106362616 | 0,85676073 | MTFR1 | FALSE | Downregulated |
| 0,107333934 | 0,85676073 | NME9 | FALSE | Upregulated |
| -0,216341448 | 0,856819142 | GAB3 | FALSE | Downregulated |
| -0,131118396 | 0,857113219 | COQ10A | FALSE | Downregulated |
| -0,108553766 | 0,857113219 | HS1BP3 | FALSE | Downregulated |
| 0,261433751 | 0,857146262 | LOC729083 | FALSE | Upregulated |
| 0,127741237 | 0,857146262 | UBE2E3 | FALSE | Upregulated |
| 0,171072192 | 0,857215185 | DGCR5 | FALSE | Upregulated |
| -0,118084606 | 0,857215185 | NDUFS8 | FALSE | Downregulated |
| 0,218617984 | 0,857215185 | PDZRN3 | FALSE | Upregulated |
| 0,141336386 | 0,857215185 | THAP9-AS1 | FALSE | Upregulated |
| 0,072476553 | 0,857347093 | ARL6IP4 | FALSE | Upregulated |
| -0,097288807 | 0,857347093 | CEBPG | FALSE | Downregulated |
| -0,059204571 | 0,857347093 | DDX42 | FALSE | Downregulated |
| 0,105205659 | 0,857347093 | PRCP | FALSE | Upregulated |
| 0,148423421 | 0,857347093 | TBC1D8B | FALSE | Upregulated |
| -0,051941055 | 0,857530817 | APLP2 | FALSE | Downregulated |
| 0,062657667 | 0,857554421 | GBA2 | FALSE | Upregulated |
| 0,075100071 | 0,857554421 | ITSN1 | FALSE | Upregulated |
| -0,203356715 | 0,857554421 | LINC01024 | FALSE | Downregulated |
| 0,159436878 | 0,857554421 | LOC100506596 | FALSE | Upregulated |
| -0,215341992 | 0,857554421 | LOC92249 | FALSE | Downregulated |
| 0,130481966 | 0,857554421 | TRMT1 | FALSE | Upregulated |
| 0,314437455 | 0,857775637 | LOC101929571 | FALSE | Upregulated |
| 0,070489376 | 0,857775637 | MBNL2 | FALSE | Upregulated |
| 0,215906144 | 0,857775637 | PPP1R2 | FALSE | Upregulated |
| -0,059890793 | 0,857775637 | PRPF6 | FALSE | Downregulated |
| -0,920487518 | 0,85783944 | LOC102724516 | FALSE | Downregulated |
| -0,157015033 | 0,857976475 | ATP7A | FALSE | Downregulated |
| 0,171927006 | 0,857976475 | RP11-506O24.1 | FALSE | Upregulated |
| -0,144641668 | 0,857976475 | RSC1A1 | FALSE | Downregulated |
| 0,132895316 | 0,858081842 | RAB11FIP2 | FALSE | Upregulated |
| -0,194324357 | 0,858165794 | LOC102724061 | FALSE | Downregulated |
| 0,547542652 | 0,858399491 | CHURC1-FNTB | FALSE | Upregulated |
| -0,133396589 | 0,858627561 | SOBP | FALSE | Downregulated |
| 0,332170371 | 0,858652242 | PIGP | FALSE | Upregulated |
| 0,086516628 | 0,859065191 | IFT140 | FALSE | Upregulated |
| 0,204040554 | 0,859065191 | TMEM234 | FALSE | Upregulated |
| -0,060597876 | 0,859259959 | MYH14 | FALSE | Downregulated |

| | | | | |
|---|---|---|---|---|
| 0,071456518 | 0,859259959 | NRIP2 | FALSE | Upregulated |
| -0,062859143 | 0,859259959 | PI4KB | FALSE | Downregulated |
| -0,094727822 | 0,859260247 | FBXO46 | FALSE | Downregulated |
| 0,174123797 | 0,859260247 | LOC101927180 | FALSE | Upregulated |
| 0,101084784 | 0,859260247 | PIGT | FALSE | Upregulated |
| -0,091495346 | 0,859906453 | ACOT2 | FALSE | Downregulated |
| 0,244741002 | 0,859912027 | LIX1 | FALSE | Upregulated |
| 0,078828905 | 0,860019754 | EPB41L4A | FALSE | Upregulated |
| 0,118755194 | 0,860019754 | TFCP2 | FALSE | Upregulated |
| 0,206407627 | 0,860180553 | DGCR11 | FALSE | Upregulated |
| -0,115933414 | 0,860180553 | LMBR1 | FALSE | Downregulated |
| 0,368700566 | 0,860180553 | LRRC14B | FALSE | Upregulated |
| 0,082010674 | 0,860207794 | SMU1 | FALSE | Upregulated |
| 0,249127431 | 0,860210443 | ADAMTS4 | FALSE | Upregulated |
| -0,115174142 | 0,860210443 | SPAST | FALSE | Downregulated |
| 0,139055817 | 0,860223056 | ARRDC4 | FALSE | Upregulated |
| 0,22303357 | 0,860267385 | DRD4 | FALSE | Upregulated |
| -0,176405497 | 0,860288145 | LETM2 | FALSE | Downregulated |
| -0,095808834 | 0,860309099 | CSGALNACT1 | FALSE | Downregulated |
| 0,151499939 | 0,860309099 | FBXO30 | FALSE | Upregulated |
| -0,102366459 | 0,860324009 | TRAPPC2 | FALSE | Downregulated |
| 0,144299027 | 0,860493549 | CHAC2 | FALSE | Upregulated |
| 0,127967092 | 0,860618029 | ALDH1L1-AS2 | FALSE | Upregulated |
| -0,095506554 | 0,860751292 | PNPLA3 | FALSE | Downregulated |
| -0,056394382 | 0,860812884 | BAP1 | FALSE | Downregulated |
| 0,121057992 | 0,860812884 | STEAP4 | FALSE | Upregulated |
| 0,314220934 | 0,860812884 | TRPV6 | FALSE | Upregulated |
| 0,267557897 | 0,861015814 | ADAT3 | FALSE | Upregulated |
| 0,093911054 | 0,861163244 | CDC26 | FALSE | Upregulated |
| -0,309950537 | 0,861261507 | FTCDNL1 | FALSE | Downregulated |
| -0,062831624 | 0,861261507 | RAF1 | FALSE | Downregulated |
| -0,457067836 | 0,861346702 | LOC102724369 | FALSE | Downregulated |
| -0,101011249 | 0,861477894 | LSM14A | FALSE | Downregulated |
| -0,114237515 | 0,861546882 | PARVA | FALSE | Downregulated |
| 0,189865312 | 0,861546882 | PDK1 | FALSE | Upregulated |
| 0,202700716 | 0,861546882 | RCN2 | FALSE | Upregulated |
| 0,326419873 | 0,861674425 | LOC102723960 | FALSE | Upregulated |
| 0,262438693 | 0,861981258 | KCNIP2-AS1 | FALSE | Upregulated |
| 0,20341384 | 0,861981258 | LINC00987 | FALSE | Upregulated |
| -0,061787859 | 0,861981258 | NAPA | FALSE | Downregulated |
| -0,180348446 | 0,861981258 | QPCTL | FALSE | Downregulated |
| -0,15853697 | 0,862026985 | ACY3 | FALSE | Downregulated |
| -0,249311344 | 0,862026985 | LZTS1 | FALSE | Downregulated |
| -0,296328979 | 0,862026985 | RPS4Y1 | FALSE | Downregulated |
| 0,08887343 | 0,862026985 | SEC22B | FALSE | Upregulated |
| -0,234110744 | 0,862161961 | MRPS23 | FALSE | Downregulated |
| -0,054251742 | 0,862161961 | NFIC | FALSE | Downregulated |
| 0,198613131 | 0,862183433 | LOC101928158 | FALSE | Upregulated |
| -0,175451628 | 0,862183433 | TSEN2 | FALSE | Downregulated |
| -0,110814048 | 0,862277591 | PEX16 | FALSE | Downregulated |
| 0,172457236 | 0,862377585 | KCNH3 | FALSE | Upregulated |

| | | | | |
|---|---|---|---|---|
| -0,415759138 | 0,862377585 | SLC5A1 | FALSE | Downregulated |
| -0,059270748 | 0,862377585 | SRSF5 | FALSE | Downregulated |
| -0,103584826 | 0,862377585 | YIPF6 | FALSE | Downregulated |
| 0,169423576 | 0,86237788 | PDIK1L | FALSE | Upregulated |
| 0,204214259 | 0,862479818 | ALPK2 | FALSE | Upregulated |
| -0,094525632 | 0,862479818 | NAP1L4 | FALSE | Downregulated |
| -0,201849538 | 0,862499713 | COL23A1 | FALSE | Downregulated |
| 0,082100283 | 0,862499713 | LOC101928445 | FALSE | Upregulated |
| -0,17843344 | 0,862499713 | LOC102723457 | FALSE | Downregulated |
| 0,314118701 | 0,862499713 | ROBO2 | FALSE | Upregulated |
| -0,313061843 | 0,862499713 | YJEFN3 | FALSE | Downregulated |
| 0,071448608 | 0,862499713 | ZSCAN29 | FALSE | Upregulated |
| 0,075366927 | 0,862536098 | PPP2R3A | FALSE | Upregulated |
| 0,197477694 | 0,862659557 | CLEC7A | FALSE | Upregulated |
| 0,121882302 | 0,862659557 | DUT | FALSE | Upregulated |
| -0,115942586 | 0,862659557 | GAREM | FALSE | Downregulated |
| 0,094476162 | 0,862659557 | LOC100288846 | FALSE | Upregulated |
| -0,259395373 | 0,862659557 | LOC101927073 | FALSE | Downregulated |
| -0,31790447 | 0,862659557 | LOC101928083 | FALSE | Downregulated |
| -0,224764499 | 0,862659557 | LOC101929673 | FALSE | Downregulated |
| 0,126618594 | 0,862659557 | MFAP1 | FALSE | Upregulated |
| -0,057920483 | 0,862659557 | PPP2R1B | FALSE | Downregulated |
| -0,058474358 | 0,862659557 | REEP6 | FALSE | Downregulated |
| -0,142353585 | 0,862659557 | RNF181 | FALSE | Downregulated |
| -0,077711557 | 0,862659557 | ZNF644 | FALSE | Downregulated |
| -0,303728421 | 0,863185217 | KBTBD3 | FALSE | Downregulated |
| 0,177007773 | 0,863350195 | MED6 | FALSE | Upregulated |
| -0,080325639 | 0,863365542 | C11orf58 | FALSE | Downregulated |
| -0,134931368 | 0,863365542 | NDUFS3 | FALSE | Downregulated |
| 0,078239093 | 0,863386301 | DNAJC21 | FALSE | Upregulated |
| -0,136700718 | 0,863410591 | ZNF767 | FALSE | Downregulated |
| 0,065883369 | 0,863638935 | BSG | FALSE | Upregulated |
| 0,077168478 | 0,863638935 | CEACAM1 | FALSE | Upregulated |
| -0,104476291 | 0,863638935 | MPPE1 | FALSE | Downregulated |
| 0,139068821 | 0,863699236 | C19orf47 | FALSE | Upregulated |
| 0,323435641 | 0,863699236 | CDS1 | FALSE | Upregulated |
| 0,33998982 | 0,863748968 | XKR6 | FALSE | Upregulated |
| 0,068025604 | 0,863751332 | ERAP1 | FALSE | Upregulated |
| -0,216107064 | 0,863751332 | EXOSC1 | FALSE | Downregulated |
| 0,045511998 | 0,863751332 | KIF1C | FALSE | Upregulated |
| -0,14738431 | 0,863751332 | RCC1 | FALSE | Downregulated |
| 0,090181294 | 0,863986305 | TP73 | FALSE | Upregulated |
| 0,143044972 | 0,863997386 | MOB1B | FALSE | Upregulated |
| 0,077313242 | 0,864320882 | SMPD3 | FALSE | Upregulated |
| 0,055329125 | 0,864417737 | ATN1 | FALSE | Upregulated |
| 0,092153903 | 0,864417737 | BAG4 | FALSE | Upregulated |
| -0,113906745 | 0,864572857 | ATP6V1D | FALSE | Downregulated |
| 0,083015441 | 0,864572857 | PPP2R4 | FALSE | Upregulated |
| 0,1711452 | 0,864572857 | RRP15 | FALSE | Upregulated |
| 0,24546814 | 0,864572857 | TM6SF1 | FALSE | Upregulated |
| -0,14910662 | 0,864572857 | ZNF615 | FALSE | Downregulated |

| | | | | |
|---|---|---|---|---|
| -0,232806143 | 0,864723482 | BRIP1 | FALSE | Downregulated |
| 0,166616218 | 0,864723482 | ERCC4 | FALSE | Upregulated |
| -0,100292445 | 0,865074815 | ZRANB2 | FALSE | Downregulated |
| 0,098756977 | 0,865094613 | PRICKLE4 | FALSE | Upregulated |
| -0,265415143 | 0,865121361 | DNAL4 | FALSE | Downregulated |
| 0,083077617 | 0,865324825 | LHFPL2 | FALSE | Upregulated |
| 0,268087292 | 0,865365538 | TMEFF2 | FALSE | Upregulated |
| 0,111843353 | 0,865630596 | BZRAP1-AS1 | FALSE | Upregulated |
| -0,11263304 | 0,866190869 | CHRNE | FALSE | Downregulated |
| 0,068763441 | 0,866190869 | EDC3 | FALSE | Upregulated |
| -0,097013986 | 0,866190869 | METAP2 | FALSE | Downregulated |
| 0,313711842 | 0,866190869 | NAALADL1 | FALSE | Upregulated |
| 0,261444082 | 0,866190869 | ZNF77 | FALSE | Upregulated |
| 0,084575967 | 0,866215588 | CHMP2A | FALSE | Upregulated |
| -0,104277741 | 0,866297529 | NRG4 | FALSE | Downregulated |
| -0,110680472 | 0,866521608 | CHD1L | FALSE | Downregulated |
| -0,138485337 | 0,866521608 | DHX40 | FALSE | Downregulated |
| -0,350035506 | 0,866521608 | TSLP | FALSE | Downregulated |
| 0,692005421 | 0,866588072 | SCNM1 | FALSE | Upregulated |
| 0,116097449 | 0,866634764 | CNOT4 | FALSE | Upregulated |
| 0,122742119 | 0,866634764 | COL12A1 | FALSE | Upregulated |
| -0,071752276 | 0,866634764 | CSRP1 | FALSE | Downregulated |
| -0,269998759 | 0,866634764 | GMDS-AS1 | FALSE | Downregulated |
| -0,075511646 | 0,866634764 | UBE4B | FALSE | Downregulated |
| -0,34867844 | 0,866751789 | NLGN4Y | FALSE | Downregulated |
| 0,172865005 | 0,866869285 | SYNGR1 | FALSE | Upregulated |
| -0,339866257 | 0,866877664 | MTUS2-AS1 | FALSE | Downregulated |
| -0,152164833 | 0,86702484 | AP1S1 | FALSE | Downregulated |
| 0,254326189 | 0,867179267 | C9orf152 | FALSE | Upregulated |
| 0,192253082 | 0,867179267 | IL23A | FALSE | Upregulated |
| -0,174662925 | 0,867179267 | SSTR2 | FALSE | Downregulated |
| 0,216817023 | 0,867179267 | TAMM41 | FALSE | Upregulated |
| -0,095579467 | 0,867179267 | VKORC1 | FALSE | Downregulated |
| 0,191653871 | 0,867179267 | VMO1 | FALSE | Upregulated |
| -0,17726284 | 0,867205295 | CORT | FALSE | Downregulated |
| 0,219551691 | 0,867209792 | RBX1 | FALSE | Upregulated |
| 0,261955176 | 0,867268098 | RASL10B | FALSE | Upregulated |
| 0,055685804 | 0,867364511 | MMACHC | FALSE | Upregulated |
| -0,066156468 | 0,867364511 | SP100 | FALSE | Downregulated |
| -0,067197259 | 0,867375553 | CEP350 | FALSE | Downregulated |
| -0,105698338 | 0,867386304 | RNF41 | FALSE | Downregulated |
| -0,170505611 | 0,867825962 | CWF19L1 | FALSE | Downregulated |
| -0,172035776 | 0,867825962 | MAPT-IT1 | FALSE | Downregulated |
| 0,175937285 | 0,867825962 | NDUFC1 | FALSE | Upregulated |
| -0,162384012 | 0,867833224 | HIST1H2BA | FALSE | Downregulated |
| -0,110832891 | 0,867853638 | ACKR2 | FALSE | Downregulated |
| -0,061277701 | 0,867853638 | AGAP3 | FALSE | Downregulated |
| -0,058211443 | 0,867853638 | AMT | FALSE | Downregulated |
| -0,258464539 | 0,867853638 | C14orf28 | FALSE | Downregulated |
| 0,200960746 | 0,867853638 | PDCD10 | FALSE | Upregulated |
| 0,200371299 | 0,867853638 | POLR2K | FALSE | Upregulated |

| | | | | |
|---|---|---|---|---|
| 0,07799923 | 0,867853638 | PRSS12 | FALSE | Upregulated |
| -0,10311829 | 0,867853638 | PTDSS1 | FALSE | Downregulated |
| 0,069506462 | 0,867853638 | STK17A | FALSE | Upregulated |
| 0,280280401 | 0,867853638 | SYN3 | FALSE | Upregulated |
| -0,096910196 | 0,867853638 | TPCN2 | FALSE | Downregulated |
| 0,073831645 | 0,867853638 | URGCP | FALSE | Upregulated |
| -0,087688121 | 0,867853638 | ZNF542 | FALSE | Downregulated |
| 0,091486578 | 0,868055067 | TMEM33 | FALSE | Upregulated |
| -0,114727412 | 0,868172826 | ABI3 | FALSE | Downregulated |
| 0,285681237 | 0,868172826 | FAM83F | FALSE | Upregulated |
| -0,097249304 | 0,868172826 | FASTK | FALSE | Downregulated |
| -0,247174083 | 0,868172826 | LCMT1 | FALSE | Downregulated |
| -0,130825351 | 0,868172826 | ZDHHC17 | FALSE | Downregulated |
| -0,176907809 | 0,868172826 | ZNF543 | FALSE | Downregulated |
| 0,198332858 | 0,868408903 | LOC101928381 | FALSE | Upregulated |
| -0,176020879 | 0,868408903 | MRPL39 | FALSE | Downregulated |
| -0,143141093 | 0,868408903 | SOCS5 | FALSE | Downregulated |
| 0,300506076 | 0,868419447 | KCNN3 | FALSE | Upregulated |
| -0,129704871 | 0,868491804 | TOPORS | FALSE | Downregulated |
| -0,082892991 | 0,868517224 | LOC646976 | FALSE | Downregulated |
| -0,150561318 | 0,868993643 | CAMLG | FALSE | Downregulated |
| -0,142287947 | 0,869079465 | MAP4K2 | FALSE | Downregulated |
| -0,107077684 | 0,86943533 | BFAR | FALSE | Downregulated |
| 0,090897875 | 0,86943533 | SPATA2L | FALSE | Upregulated |
| 0,11008022 | 0,86943533 | TBL3 | FALSE | Upregulated |
| -0,142667576 | 0,869445163 | CDKAL1 | FALSE | Downregulated |
| 0,078612041 | 0,869445163 | DHX8 | FALSE | Upregulated |
| 0,091548807 | 0,869445163 | LOC101927208 | FALSE | Upregulated |
| -0,134446327 | 0,869445163 | ZNF277 | FALSE | Downregulated |
| -0,063221391 | 0,869478101 | UGP2 | FALSE | Downregulated |
| 0,118600033 | 0,869481552 | ARRB1 | FALSE | Upregulated |
| 0,240756943 | 0,869481552 | ARTN | FALSE | Upregulated |
| -0,217932556 | 0,869481552 | FAM217A | FALSE | Downregulated |
| -0,092775895 | 0,869481552 | PRR3 | FALSE | Downregulated |
| 0,091383552 | 0,869481552 | TFG | FALSE | Upregulated |
| 0,145932858 | 0,869481552 | WRN | FALSE | Upregulated |
| 0,169762857 | 0,869481552 | ZNF331 | FALSE | Upregulated |
| -0,219550555 | 0,869578351 | C3orf18 | FALSE | Downregulated |
| -0,185450313 | 0,869578351 | ENKD1 | FALSE | Downregulated |
| -0,135688974 | 0,869578351 | GLRX3 | FALSE | Downregulated |
| -0,135521768 | 0,869578351 | GTF2H2B | FALSE | Downregulated |
| 0,078612995 | 0,869578351 | MRS2P2 | FALSE | Upregulated |
| -0,09923728 | 0,869578351 | SF3A3 | FALSE | Downregulated |
| 0,05977829 | 0,869609007 | C15orf62 | FALSE | Upregulated |
| -0,365160365 | 0,869609007 | CLIC5 | FALSE | Downregulated |
| -0,211369786 | 0,869609007 | DPY19L1P1 | FALSE | Downregulated |
| 0,065513433 | 0,869609007 | G3BP1 | FALSE | Upregulated |
| 0,095605336 | 0,869910099 | CCNT1 | FALSE | Upregulated |
| -0,127216056 | 0,869951185 | FAM96A | FALSE | Downregulated |
| 0,08100994 | 0,870129446 | IDNK | FALSE | Upregulated |
| -0,163391761 | 0,870129446 | TP53RK | FALSE | Downregulated |

| | | | | |
|---|---|---|---|---|
| 0,135738839 | 0,870129446 | UQCR10 | FALSE | Upregulated |
| 0,106115242 | 0,87025831 | NDUFA1 | FALSE | Upregulated |
| 0,081822021 | 0,870490988 | ADAM17 | FALSE | Upregulated |
| -0,148405246 | 0,870511119 | C1orf159 | FALSE | Downregulated |
| 0,097139793 | 0,870754225 | FLYWCH1 | FALSE | Upregulated |
| -0,237991657 | 0,870763901 | C7orf63 | FALSE | Downregulated |
| -0,173383567 | 0,870763901 | JRKL | FALSE | Downregulated |
| -0,085905193 | 0,870897312 | IKBKAP | FALSE | Downregulated |
| -0,084730649 | 0,870961565 | FAM92A1 | FALSE | Downregulated |
| -0,096454977 | 0,871114799 | ATPAF1 | FALSE | Downregulated |
| -0,20369755 | 0,871114799 | CLDN10 | FALSE | Downregulated |
| 0,176074001 | 0,871114799 | DUS4L | FALSE | Upregulated |
| -0,170745676 | 0,871114799 | GMEB1 | FALSE | Downregulated |
| 0,143882616 | 0,871114799 | SRGAP3 | FALSE | Upregulated |
| 0,104324531 | 0,871114799 | TAF5 | FALSE | Upregulated |
| -0,145145157 | 0,871114799 | TMEM14C | FALSE | Downregulated |
| 0,104038343 | 0,871114799 | TRAF3IP2-AS1 | FALSE | Upregulated |
| 0,252660627 | 0,871229894 | CNKSR2 | FALSE | Upregulated |
| 0,104819448 | 0,871421876 | ATG16L1 | FALSE | Upregulated |
| 0,08936491 | 0,871421876 | LOC100507217 | FALSE | Upregulated |
| 0,096255518 | 0,871421876 | TMEM44 | FALSE | Upregulated |
| 0,110965195 | 0,871459162 | ALDH3B1 | FALSE | Upregulated |
| 0,140726927 | 0,871459162 | DGCR14 | FALSE | Upregulated |
| -0,229072674 | 0,871459162 | PTPRR | FALSE | Downregulated |
| -0,088022641 | 0,871500951 | RASSF7 | FALSE | Downregulated |
| -0,14861147 | 0,871582563 | DHX57 | FALSE | Downregulated |
| 0,168743724 | 0,871651977 | RP11-521B24.3 | FALSE | Upregulated |
| 0,196293493 | 0,871651977 | TMEM218 | FALSE | Upregulated |
| -0,200121211 | 0,871793873 | SMIM20 | FALSE | Downregulated |
| 0,336889688 | 0,871797461 | RP11-503N18.1 | FALSE | Upregulated |
| 0,22940301 | 0,872210558 | LEF1 | FALSE | Upregulated |
| 0,118775008 | 0,872210558 | TMPRSS2 | FALSE | Upregulated |
| -0,092730505 | 0,872210558 | ZFYVE19 | FALSE | Downregulated |
| -0,271873614 | 0,872233533 | GTF2E1 | FALSE | Downregulated |
| -0,102142384 | 0,872280122 | DHX29 | FALSE | Downregulated |
| -0,302014293 | 0,872280122 | METAP1D | FALSE | Downregulated |
| -0,116740306 | 0,872280122 | TDRD3 | FALSE | Downregulated |
| 0,070637534 | 0,872378205 | AGPAT4 | FALSE | Upregulated |
| -0,147886023 | 0,872451175 | AAR2 | FALSE | Downregulated |
| 0,07504055 | 0,872451175 | UNK | FALSE | Upregulated |
| 0,226296813 | 0,872468347 | C12orf5 | FALSE | Upregulated |
| -0,129967404 | 0,872468347 | LRIG2 | FALSE | Downregulated |
| -0,205146813 | 0,872468347 | ZNF597 | FALSE | Downregulated |
| 0,140459822 | 0,872657748 | TOMM6 | FALSE | Upregulated |
| -0,108334473 | 0,873029152 | NUDT15 | FALSE | Downregulated |
| 0,070704337 | 0,873134546 | LBX2 | FALSE | Upregulated |
| 0,125419813 | 0,873184625 | GNL3 | FALSE | Upregulated |
| -0,238305755 | 0,873184625 | IFT57 | FALSE | Downregulated |
| -0,145453338 | 0,873184625 | RRP8 | FALSE | Downregulated |
| 0,095080199 | 0,873819454 | NELFA | FALSE | Upregulated |
| 0,054503238 | 0,873963877 | XRRA1 | FALSE | Upregulated |

| | | | | |
|---|---|---|---|---|
| 0,097884023 | 0,874322878 | ABCD1 | FALSE | Upregulated |
| 0,994079202 | 0,874326385 | ADIPOQ-AS1 | FALSE | Upregulated |
| 0,104515924 | 0,874326385 | ATP6V0D1 | FALSE | Upregulated |
| 0,115569517 | 0,874326385 | CCDC39 | FALSE | Upregulated |
| -0,0973363 | 0,874326385 | CDC37L1 | FALSE | Downregulated |
| 0,253043076 | 0,874326385 | DCLRE1B | FALSE | Upregulated |
| 0,112052387 | 0,874326385 | DNTTIP2 | FALSE | Upregulated |
| -0,067136522 | 0,874326385 | ERF | FALSE | Downregulated |
| -0,266893841 | 0,874326385 | LOC100652824 | FALSE | Downregulated |
| 0,163316877 | 0,874326385 | STX1A | FALSE | Upregulated |
| -0,065123902 | 0,874326446 | RMDN1 | FALSE | Downregulated |
| 0,131126544 | 0,874386325 | CORIN | FALSE | Upregulated |
| -0,132795766 | 0,874386325 | HSPBP1 | FALSE | Downregulated |
| 0,260345186 | 0,874386325 | LOC101448202 | FALSE | Upregulated |
| 0,101468912 | 0,874386325 | SBDS | FALSE | Upregulated |
| -0,168069217 | 0,874693419 | MID1 | FALSE | Downregulated |
| -1,307041995 | 0,874693419 | TNNT1 | FALSE | Downregulated |
| -0,08442014 | 0,874752236 | ACSL3 | FALSE | Downregulated |
| 0,089355812 | 0,874752236 | BHLHE40 | FALSE | Upregulated |
| -0,187056427 | 0,874752236 | GAN | FALSE | Downregulated |
| -0,180586736 | 0,874752236 | GLIDR | FALSE | Downregulated |
| 0,09026076 | 0,874752236 | GTF3C2 | FALSE | Upregulated |
| -0,148041098 | 0,874752236 | GUSBP1 | FALSE | Downregulated |
| 0,181622237 | 0,874752236 | LRRC55 | FALSE | Upregulated |
| 0,092470108 | 0,874752236 | PER2 | FALSE | Upregulated |
| 0,096587683 | 0,874752236 | PLEKHA2 | FALSE | Upregulated |
| -0,11671521 | 0,874752236 | POLR2D | FALSE | Downregulated |
| -0,068857885 | 0,874752236 | UNC13B | FALSE | Downregulated |
| 0,072500291 | 0,874783436 | INO80 | FALSE | Upregulated |
| -0,085434302 | 0,874783436 | TCAIM | FALSE | Downregulated |
| -0,115903623 | 0,874870996 | ATP5E | FALSE | Downregulated |
| -0,16857223 | 0,874870996 | CRX | FALSE | Downregulated |
| -0,250850729 | 0,874870996 | KIAA1467 | FALSE | Downregulated |
| 0,134053381 | 0,874870996 | PRSS23 | FALSE | Upregulated |
| -0,205330459 | 0,874870996 | USHBP1 | FALSE | Downregulated |
| 0,138756102 | 0,8749705 | ZNF32-AS1 | FALSE | Upregulated |
| 0,266595892 | 0,875211808 | LOC101927861 | FALSE | Upregulated |
| -0,12103752 | 0,875211808 | PLK1S1 | FALSE | Downregulated |
| -0,118924631 | 0,875211808 | PSMG3-AS1 | FALSE | Downregulated |
| 0,062005187 | 0,875211808 | SUMF2 | FALSE | Upregulated |
| 0,140107698 | 0,875316573 | ZBTB2 | FALSE | Upregulated |
| -0,070721998 | 0,875412417 | NAGA | FALSE | Downregulated |
| -0,086759746 | 0,87557453 | EPHA1 | FALSE | Downregulated |
| 0,207438325 | 0,87557453 | EPHB6 | FALSE | Upregulated |
| 0,071387932 | 0,875652465 | PSMC1 | FALSE | Upregulated |
| -0,054295655 | 0,875754063 | KDM3B | FALSE | Downregulated |
| 0,265564041 | 0,875754063 | LOC100131626 | FALSE | Upregulated |
| 0,095486727 | 0,875754063 | ZDHHC16 | FALSE | Upregulated |
| -0,071311833 | 0,875777934 | WDR6 | FALSE | Downregulated |
| -0,104451059 | 0,875878711 | BCL7C | FALSE | Downregulated |
| -0,140731132 | 0,875878711 | CXorf40B | FALSE | Downregulated |

| | | | | |
|---|---|---|---|---|
| -0,18257277 | 0,875878711 | ETV1 | FALSE | Downregulated |
| 0,114892527 | 0,875878711 | GPKOW | FALSE | Upregulated |
| 0,27793323 | 0,875878711 | NPAS1 | FALSE | Upregulated |
| -0,142935217 | 0,875878711 | ZNF141 | FALSE | Downregulated |
| 0,149304387 | 0,875933818 | C2orf44 | FALSE | Upregulated |
| 0,196655133 | 0,875933818 | CCDC40 | FALSE | Upregulated |
| 0,076260744 | 0,875933818 | NOP9 | FALSE | Upregulated |
| 0,056013293 | 0,876071329 | GLYR1 | FALSE | Upregulated |
| -0,138934819 | 0,876071329 | TXNL1 | FALSE | Downregulated |
| 0,143413141 | 0,876197688 | PRPSAP2 | FALSE | Upregulated |
| 0,089319735 | 0,876197688 | USP14 | FALSE | Upregulated |
| -0,164964399 | 0,876580882 | ALG14 | FALSE | Downregulated |
| -0,085226365 | 0,876580882 | LOC101928690 | FALSE | Downregulated |
| 0,054383094 | 0,87683451 | PTOV1 | FALSE | Upregulated |
| -0,091675461 | 0,876870233 | ARF3 | FALSE | Downregulated |
| 0,072903239 | 0,876870233 | FECH | FALSE | Upregulated |
| 0,254656316 | 0,876870233 | LOC100505761 | FALSE | Upregulated |
| -0,069150913 | 0,876870233 | VHL | FALSE | Downregulated |
| -0,129172273 | 0,876995487 | EHD2 | FALSE | Downregulated |
| 0,160865821 | 0,876995487 | LOC101928439 | FALSE | Upregulated |
| -0,177980855 | 0,876995487 | URB2 | FALSE | Downregulated |
| -0,122436459 | 0,877016373 | DUS2 | FALSE | Downregulated |
| -0,064737563 | 0,877016373 | ELMSAN1 | FALSE | Downregulated |
| 0,535458393 | 0,877103129 | RGPD6 | FALSE | Upregulated |
| 0,100006716 | 0,877542667 | PPRC1 | FALSE | Upregulated |
| 0,102527002 | 0,877542667 | SETD8 | FALSE | Upregulated |
| 0,145706133 | 0,877542667 | SMC1B | FALSE | Upregulated |
| -0,165538704 | 0,877542667 | THOC6 | FALSE | Downregulated |
| -0,104439473 | 0,877575718 | PLIN2 | FALSE | Downregulated |
| -0,147795916 | 0,877582384 | KIF27 | FALSE | Downregulated |
| -0,102383503 | 0,877817243 | LNPEP | FALSE | Downregulated |
| 0,163415384 | 0,877967165 | HERC2P9 | FALSE | Upregulated |
| -0,088448505 | 0,878054589 | LOC101928415 | FALSE | Downregulated |
| 0,160462377 | 0,878148389 | CISD1 | FALSE | Upregulated |
| 0,110955832 | 0,878178826 | RNASE10 | FALSE | Upregulated |
| -0,12444131 | 0,878246381 | CCDC120 | FALSE | Downregulated |
| 0,212723906 | 0,878246381 | CHCHD4 | FALSE | Upregulated |
| -0,109022622 | 0,878246381 | CMAS | FALSE | Downregulated |
| 0,497320819 | 0,878246381 | KIAA0408 | FALSE | Upregulated |
| 0,230302901 | 0,878246381 | TTC16 | FALSE | Upregulated |
| -0,219559262 | 0,878246381 | UTP23 | FALSE | Downregulated |
| 0,114158824 | 0,878252495 | ZNF512 | FALSE | Upregulated |
| -0,24766245 | 0,87832083 | ZGLP1 | FALSE | Downregulated |
| -0,173610008 | 0,878412309 | METTL21A | FALSE | Downregulated |
| -0,071914618 | 0,878412309 | THUMPD3 | FALSE | Downregulated |
| -0,152181593 | 0,87848174 | LMOD2 | FALSE | Downregulated |
| 0,211826523 | 0,878568951 | ZMYND15 | FALSE | Upregulated |
| 0,080578226 | 0,87877071 | PDE5A | FALSE | Upregulated |
| -0,220942187 | 0,87877071 | PRKCDBP | FALSE | Downregulated |
| 0,124968115 | 0,878804922 | TDG | FALSE | Upregulated |
| -0,328450219 | 0,878839465 | LOC102724691 | FALSE | Downregulated |

| | | | | |
|---|---|---|---|---|
| 0,102190007 | 0,878839465 | ZNF689 | FALSE | Upregulated |
| 0,090981458 | 0,87907349 | TRIM38 | FALSE | Upregulated |
| -0,12206846 | 0,87907349 | USP12 | FALSE | Downregulated |
| -0,181098821 | 0,879114599 | KIAA1652 | FALSE | Downregulated |
| -0,074481293 | 0,879114599 | YES1 | FALSE | Downregulated |
| -0,123524863 | 0,879209605 | L3MBTL3 | FALSE | Downregulated |
| -0,079870516 | 0,879390764 | FAM102A | FALSE | Downregulated |
| 0,186346752 | 0,879390764 | MB21D1 | FALSE | Upregulated |
| 0,122885199 | 0,879390764 | MRPL30 | FALSE | Upregulated |
| 0,169511529 | 0,879547506 | PDF | FALSE | Upregulated |
| -0,20120462 | 0,879547506 | TGDS | FALSE | Downregulated |
| -0,129437973 | 0,87965773 | ARL13B | FALSE | Downregulated |
| -0,112158591 | 0,879666952 | CAPZA2 | FALSE | Downregulated |
| 0,185649151 | 0,879666952 | RPS10P7 | FALSE | Upregulated |
| 0,081055992 | 0,879666952 | USP1 | FALSE | Upregulated |
| -0,172138112 | 0,87973295 | ING2 | FALSE | Downregulated |
| 0,219448238 | 0,879736354 | RP11-75N4.2 | FALSE | Upregulated |
| -0,204691962 | 0,879802729 | SPATA9 | FALSE | Downregulated |
| -0,097583105 | 0,879920042 | NEK7 | FALSE | Downregulated |
| -0,103322564 | 0,879920042 | TRMT61A | FALSE | Downregulated |
| -0,088907518 | 0,88014627 | CLASP2 | FALSE | Downregulated |
| -0,129274642 | 0,88014627 | SLC39A6 | FALSE | Downregulated |
| -0,219348408 | 0,880267523 | OR51I2 | FALSE | Downregulated |
| -0,178183778 | 0,880291733 | PLGLB1 | FALSE | Downregulated |
| 0,136742397 | 0,88047299 | ACTR10 | FALSE | Upregulated |
| 0,130556435 | 0,88047299 | LOC101927578 | FALSE | Upregulated |
| 0,292464524 | 0,88047299 | LOC101929578 | FALSE | Upregulated |
| -0,277839152 | 0,88047299 | LOC284454 | FALSE | Downregulated |
| 0,111806083 | 0,88047299 | PCED1B-AS1 | FALSE | Upregulated |
| 0,148011003 | 0,88047299 | TSSC1 | FALSE | Upregulated |
| -0,100422126 | 0,880690469 | PER3 | FALSE | Downregulated |
| 0,188598515 | 0,880771429 | WDR4 | FALSE | Upregulated |
| 0,064039554 | 0,880908411 | DYNC1LI2 | FALSE | Upregulated |
| 0,267039743 | 0,880995971 | RP11-379K22.2 | FALSE | Upregulated |
| 0,088807602 | 0,881033206 | HYLS1 | FALSE | Upregulated |
| -0,149653279 | 0,88106879 | AFAP1L2 | FALSE | Downregulated |
| -0,095558276 | 0,881105667 | POC1B | FALSE | Downregulated |
| 0,103961283 | 0,881105667 | STX5 | FALSE | Upregulated |
| 0,20287262 | 0,881187769 | GRM6 | FALSE | Upregulated |
| 0,05007283 | 0,881204596 | PTPN23 | FALSE | Upregulated |
| -0,116463474 | 0,881263814 | C20orf194 | FALSE | Downregulated |
| 0,09987562 | 0,881263814 | COX5A | FALSE | Upregulated |
| 0,070208153 | 0,881263814 | DCAF10 | FALSE | Upregulated |
| 0,132231619 | 0,881263814 | HECTD2 | FALSE | Upregulated |
| 0,061695337 | 0,881263814 | ITPRIPL2 | FALSE | Upregulated |
| 0,360368429 | 0,881263814 | LOC101928557 | FALSE | Upregulated |
| 0,274267522 | 0,881263814 | LYAR | FALSE | Upregulated |
| -0,140494164 | 0,881263814 | PLBD1 | FALSE | Downregulated |
| -0,084456066 | 0,881263814 | PLCG2 | FALSE | Downregulated |
| -0,104359849 | 0,881263814 | RNR2 | FALSE | Downregulated |
| 0,291805879 | 0,881263814 | RP11-261C10.3 | FALSE | Upregulated |

| | | | | |
|---|---|---|---|---|
| 0,305535277 | 0,881263814 | SNX16 | FALSE | Upregulated |
| 0,167933724 | 0,881263814 | TGFB1I1 | FALSE | Upregulated |
| 0,216433352 | 0,881263814 | TSSK6 | FALSE | Upregulated |
| -0,07956851 | 0,881263814 | WDR88 | FALSE | Downregulated |
| -0,100081245 | 0,881263814 | ZNF652 | FALSE | Downregulated |
| -0,171515497 | 0,881355557 | ACTR5 | FALSE | Downregulated |
| 0,16323937 | 0,881355557 | APEX2 | FALSE | Upregulated |
| -0,130151494 | 0,881355557 | CHM | FALSE | Downregulated |
| 0,078782948 | 0,881355557 | CHRM5 | FALSE | Upregulated |
| 0,111321961 | 0,881355557 | EIF4A3 | FALSE | Upregulated |
| -0,308625832 | 0,881355557 | FGF1 | FALSE | Downregulated |
| 0,058236571 | 0,881355557 | KCNJ13 | FALSE | Upregulated |
| -0,15398795 | 0,881355557 | LIN7A | FALSE | Downregulated |
| -0,208906375 | 0,881355557 | LINC00938 | FALSE | Downregulated |
| -0,116420921 | 0,881355557 | LOC101927809 | FALSE | Downregulated |
| 0,058613525 | 0,881355557 | LTBR | FALSE | Upregulated |
| -0,051992058 | 0,881355557 | RAB11B | FALSE | Downregulated |
| -0,137493638 | 0,881355557 | SLC16A6 | FALSE | Downregulated |
| 0,094880487 | 0,881355557 | SNAI3 | FALSE | Upregulated |
| 0,105014181 | 0,881355557 | TOR4A | FALSE | Upregulated |
| -0,137686795 | 0,881355557 | ZIC4 | FALSE | Downregulated |
| 0,096508459 | 0,881428023 | SPATA6L | FALSE | Upregulated |
| 0,062188287 | 0,881498026 | AP1G2 | FALSE | Upregulated |
| -0,279744056 | 0,881572489 | C5orf55 | FALSE | Downregulated |
| 0,150073992 | 0,881677426 | SUSD3 | FALSE | Upregulated |
| -0,118426355 | 0,881796714 | GNPAT | FALSE | Downregulated |
| -0,106831478 | 0,881954627 | DNPH1 | FALSE | Downregulated |
| -0,078269994 | 0,881954627 | TRIM4 | FALSE | Downregulated |
| 0,067265742 | 0,882020206 | ETS1 | FALSE | Upregulated |
| 0,066652758 | 0,882197979 | C17orf103 | FALSE | Upregulated |
| -0,250378878 | 0,882197979 | LOC102724368 | FALSE | Downregulated |
| 0,335463356 | 0,882203228 | ATP4B | FALSE | Upregulated |
| 0,08306877 | 0,882203228 | TBC1D10C | FALSE | Upregulated |
| -0,172205231 | 0,882215936 | SLC41A1 | FALSE | Downregulated |
| 0,149035819 | 0,882296317 | PIGZ | FALSE | Upregulated |
| -0,15587603 | 0,882349635 | UPP1 | FALSE | Downregulated |
| 0,095050048 | 0,882525836 | TBC1D25 | FALSE | Upregulated |
| -0,073705287 | 0,882611556 | FOXM1 | FALSE | Downregulated |
| -0,056710874 | 0,882650156 | SND1 | FALSE | Downregulated |
| -0,11931006 | 0,882650156 | SNRPG | FALSE | Downregulated |
| -0,26114818 | 0,882650156 | TMEM255B | FALSE | Downregulated |
| 0,219283311 | 0,882684349 | CAB39L | FALSE | Upregulated |
| -0,060821013 | 0,882684349 | DAP | FALSE | Downregulated |
| -0,097923132 | 0,882684349 | NEIL2 | FALSE | Downregulated |
| 0,203375168 | 0,882684349 | P2RY1 | FALSE | Upregulated |
| 0,282334252 | 0,882716738 | AC007163.6 | FALSE | Upregulated |
| 0,163778852 | 0,88273189 | FAM216A | FALSE | Upregulated |
| 0,079161254 | 0,882838497 | PP7080 | FALSE | Upregulated |
| -0,062313022 | 0,882838497 | SRRM1 | FALSE | Downregulated |
| 0,202225489 | 0,882838497 | ZNF461 | FALSE | Upregulated |
| -0,158815082 | 0,882994202 | BLZF1 | FALSE | Downregulated |

| | | | | |
|---|---|---|---|---|
| -0,274878814 | 0,882994202 | KIAA1324 | FALSE | Downregulated |
| 0,403741368 | 0,883074646 | INO80B-WBP1 | FALSE | Upregulated |
| -0,242553898 | 0,883074646 | POC1A | FALSE | Downregulated |
| -0,103161912 | 0,883074646 | USP38 | FALSE | Downregulated |
| 0,133839625 | 0,883074646 | ZNF563 | FALSE | Upregulated |
| 0,089978872 | 0,883262801 | TMOD3 | FALSE | Upregulated |
| 0,086125312 | 0,883262801 | ZFP28 | FALSE | Upregulated |
| 0,115537154 | 0,883295004 | CYB5R2 | FALSE | Upregulated |
| 0,257416337 | 0,883295004 | RGS2 | FALSE | Upregulated |
| -0,194906854 | 0,883295004 | SULF1 | FALSE | Downregulated |
| -0,058975865 | 0,883360903 | ABCA1 | FALSE | Downregulated |
| -0,177157653 | 0,88351391 | C11orf1 | FALSE | Downregulated |
| 0,113115901 | 0,88351391 | DCAF15 | FALSE | Upregulated |
| -0,116817894 | 0,88351391 | DNAJC30 | FALSE | Downregulated |
| 0,204032851 | 0,88351391 | NUP35 | FALSE | Upregulated |
| -0,170970766 | 0,88351391 | ZNF653 | FALSE | Downregulated |
| 0,073901375 | 0,883916553 | RAB23 | FALSE | Upregulated |
| -0,053497349 | 0,88418432 | MAP2K2 | FALSE | Downregulated |
| 0,210139875 | 0,884435846 | TAF1B | FALSE | Upregulated |
| 0,290467733 | 0,884520244 | TMIE | FALSE | Upregulated |
| 0,11626066 | 0,884717706 | CTAGE8 | FALSE | Upregulated |
| -0,102411194 | 0,884717706 | SLC25A37 | FALSE | Downregulated |
| -0,056769273 | 0,884822281 | SS18L1 | FALSE | Downregulated |
| 0,073363871 | 0,884836725 | WWP2 | FALSE | Upregulated |
| 0,050020579 | 0,884960755 | ILF3 | FALSE | Upregulated |
| -0,142753473 | 0,884960755 | PI4KAP2 | FALSE | Downregulated |
| -0,134364332 | 0,885139361 | ELMO3 | FALSE | Downregulated |
| -0,099984153 | 0,885227196 | ATP5I | FALSE | Downregulated |
| -0,083019193 | 0,885227196 | DLG2 | FALSE | Downregulated |
| 0,25489041 | 0,885227196 | GS1-251I9.4 | FALSE | Upregulated |
| 0,293438259 | 0,885227196 | HOXB6 | FALSE | Upregulated |
| -0,085459217 | 0,885227196 | MAP1LC3A | FALSE | Downregulated |
| -0,108471791 | 0,885227196 | PAPD4 | FALSE | Downregulated |
| 0,228583336 | 0,885227196 | RP1-234P15.4 | FALSE | Upregulated |
| -0,056890049 | 0,885227196 | SERPING1 | FALSE | Downregulated |
| 0,124344042 | 0,885227196 | SH3BGRL | FALSE | Upregulated |
| -0,078620218 | 0,885227196 | TSC22D1 | FALSE | Downregulated |
| 0,063645326 | 0,885227196 | UBP1 | FALSE | Upregulated |
| 0,101981367 | 0,885227196 | WDR45 | FALSE | Upregulated |
| -0,125756027 | 0,885276496 | HGF | FALSE | Downregulated |
| 0,394879894 | 0,885339424 | P2RY11 | FALSE | Upregulated |
| 0,243382063 | 0,885339424 | TMEM169 | FALSE | Upregulated |
| 0,281126853 | 0,885412713 | CD58 | FALSE | Upregulated |
| 0,05978653 | 0,885412713 | NT5C2 | FALSE | Upregulated |
| -0,071253423 | 0,885412713 | ZNF142 | FALSE | Downregulated |
| 0,085821508 | 0,885510677 | EEPD1 | FALSE | Upregulated |
| 0,062993012 | 0,88595047 | VPS39 | FALSE | Upregulated |
| 0,041405056 | 0,886038152 | INSR | FALSE | Upregulated |
| -0,226816294 | 0,886075386 | MT1M | FALSE | Downregulated |
| -0,091399492 | 0,886075386 | PLOD2 | FALSE | Downregulated |
| -0,09694123 | 0,886075386 | TERF2 | FALSE | Downregulated |

| | | | | |
|---|---|---|---|---|
| -0,115047373 | 0,886147962 | C1orf177 | FALSE | Downregulated |
| -0,077187969 | 0,886147962 | LOC101926920 | FALSE | Downregulated |
| 0,061864302 | 0,886227529 | PHACTR2 | FALSE | Upregulated |
| 0,204692975 | 0,886377664 | ARMCX6 | FALSE | Upregulated |
| -0,130340461 | 0,886477806 | ABHD3 | FALSE | Downregulated |
| 0,215944136 | 0,886477806 | CERCAM | FALSE | Upregulated |
| -0,068493249 | 0,886477806 | CHTF8 | FALSE | Downregulated |
| 0,042866084 | 0,886477806 | FAM13A | FALSE | Upregulated |
| 0,087387646 | 0,886477806 | FAM195B | FALSE | Upregulated |
| 0,106812421 | 0,886477806 | LOC100996273 | FALSE | Upregulated |
| -0,127801531 | 0,886477806 | LOC102723773 | FALSE | Downregulated |
| 0,176899135 | 0,886477806 | LOC102724488 | FALSE | Upregulated |
| -0,220406644 | 0,886477806 | LOC642423 | FALSE | Downregulated |
| -0,193932285 | 0,886477806 | MARS2 | FALSE | Downregulated |
| 0,064001643 | 0,886477806 | MMP15 | FALSE | Upregulated |
| -0,161758233 | 0,886477806 | MORN1 | FALSE | Downregulated |
| 0,235628947 | 0,886477806 | PPIL3 | FALSE | Upregulated |
| -0,111409218 | 0,886477806 | R3HCC1L | FALSE | Downregulated |
| -0,128084557 | 0,886477806 | RABAC1 | FALSE | Downregulated |
| 0,17328361 | 0,886477806 | REEP4 | FALSE | Upregulated |
| 0,119892192 | 0,886477806 | RP1-1J6.2 | FALSE | Upregulated |
| 0,105101302 | 0,886477806 | RP1-60O19.1 | FALSE | Upregulated |
| 0,066953562 | 0,886477806 | SLAIN2 | FALSE | Upregulated |
| -0,049508518 | 0,886477806 | SP3 | FALSE | Downregulated |
| 0,077073405 | 0,886477806 | SPPL2A | FALSE | Upregulated |
| 0,183839376 | 0,886505564 | B3GNT7 | FALSE | Upregulated |
| -0,188257444 | 0,886505564 | LOC101927663 | FALSE | Downregulated |
| 0,135811162 | 0,886505564 | SPACA6P-AS | FALSE | Upregulated |
| 0,150101913 | 0,886510214 | ZNF131 | FALSE | Upregulated |
| -0,122338745 | 0,886641721 | LRRC57 | FALSE | Downregulated |
| 0,156143448 | 0,886641721 | ZNF701 | FALSE | Upregulated |
| -0,081448769 | 0,886669301 | AGPS | FALSE | Downregulated |
| -0,091062123 | 0,886683775 | KCMF1 | FALSE | Downregulated |
| -0,078856731 | 0,886728813 | TAF1C | FALSE | Downregulated |
| 0,172292334 | 0,886900231 | LOC101928678 | FALSE | Upregulated |
| 0,145182801 | 0,886905919 | TMC5 | FALSE | Upregulated |
| 0,056720773 | 0,886925719 | MTDH | FALSE | Upregulated |
| 0,106287282 | 0,887206236 | FAM103A1 | FALSE | Upregulated |
| -0,132374111 | 0,887206236 | NRF1 | FALSE | Downregulated |
| -0,160924152 | 0,887206236 | PCBD2 | FALSE | Downregulated |
| -0,326813223 | 0,887222915 | STAC3 | FALSE | Downregulated |
| -0,221473501 | 0,887290372 | CX3CR1 | FALSE | Downregulated |
| 0,157525393 | 0,887319786 | CARD6 | FALSE | Upregulated |
| 0,064443299 | 0,887319786 | LUZP1 | FALSE | Upregulated |
| 0,076605182 | 0,887319786 | RNF139 | FALSE | Upregulated |
| 0,161333852 | 0,88746501 | PNMA2 | FALSE | Upregulated |
| -0,115503145 | 0,887478624 | ABT1 | FALSE | Downregulated |
| 0,108133151 | 0,887478624 | AC010127.3 | FALSE | Upregulated |
| 0,064586462 | 0,887478624 | FAM13A-AS1 | FALSE | Upregulated |
| -0,057557319 | 0,887478624 | MBD1 | FALSE | Downregulated |
| 0,106555175 | 0,887478624 | RB1 | FALSE | Upregulated |

| | | | | |
|---|---|---|---|---|
| -0,075996811 | 0,887478624 | TBC1D17 | FALSE | Downregulated |
| 0,061657544 | 0,887722392 | EML5 | FALSE | Upregulated |
| 0,054294458 | 0,887722392 | KDM6B | FALSE | Upregulated |
| 0,264669392 | 0,887722392 | LOC100133182 | FALSE | Upregulated |
| -0,206819976 | 0,887722392 | MAD2L1BP | FALSE | Downregulated |
| 0,07095368 | 0,887722392 | MRPL49 | FALSE | Upregulated |
| -0,13736425 | 0,887722392 | MRPS31P5 | FALSE | Downregulated |
| 0,065797877 | 0,887722392 | PDIA4 | FALSE | Upregulated |
| -0,051558977 | 0,887722392 | REV1 | FALSE | Downregulated |
| 0,069929801 | 0,887722392 | SLC8B1 | FALSE | Upregulated |
| 0,081453299 | 0,88777667 | FLT1 | FALSE | Upregulated |
| 0,069947404 | 0,88777667 | GSTO1 | FALSE | Upregulated |
| -0,373567707 | 0,88777667 | LOC101929950 | FALSE | Downregulated |
| 0,17658428 | 0,88777667 | LOC102724587 | FALSE | Upregulated |
| 0,096239542 | 0,887820326 | ATIC | FALSE | Upregulated |
| 0,098442886 | 0,887820326 | PAXIP1-AS2 | FALSE | Upregulated |
| 0,148067158 | 0,887820326 | RAB3D | FALSE | Upregulated |
| 0,104634387 | 0,887867376 | G0S2 | FALSE | Upregulated |
| 0,048310785 | 0,887867376 | GRINA | FALSE | Upregulated |
| -0,134987768 | 0,887867376 | KRBOX4 | FALSE | Downregulated |
| 1,267040265 | 0,88788905 | LOC102723728 | FALSE | Upregulated |
| -0,129149815 | 0,887949427 | BUB3 | FALSE | Downregulated |
| 0,136496883 | 0,888070286 | ZNF440 | FALSE | Upregulated |
| 0,101443933 | 0,888141319 | FKBP3 | FALSE | Upregulated |
| 0,180185217 | 0,888141319 | LMCD1 | FALSE | Upregulated |
| -0,141142075 | 0,888141319 | PDE4C | FALSE | Downregulated |
| 0,097548868 | 0,888188207 | LOC102723992 | FALSE | Upregulated |
| 0,226136394 | 0,888223526 | PGBD4 | FALSE | Upregulated |
| -0,045672929 | 0,888223526 | SKI | FALSE | Downregulated |
| 0,101464057 | 0,888223526 | TRAPPC6B | FALSE | Upregulated |
| -0,143071279 | 0,888235107 | VEPH1 | FALSE | Downregulated |
| 0,103744471 | 0,888287509 | NIPAL3 | FALSE | Upregulated |
| -0,192337349 | 0,888294425 | ZNF17 | FALSE | Downregulated |
| -0,068712658 | 0,888347404 | POMGNT1 | FALSE | Downregulated |
| -0,292554525 | 0,888549197 | LOC728819 | FALSE | Downregulated |
| 0,157137443 | 0,888549197 | PYROXD2 | FALSE | Upregulated |
| 0,150643163 | 0,888631938 | PUSL1 | FALSE | Upregulated |
| 0,137866758 | 0,888641477 | SLC37A3 | FALSE | Upregulated |
| 0,080905048 | 0,888838445 | KAT8 | FALSE | Upregulated |
| 0,124185148 | 0,888838445 | LDHB | FALSE | Upregulated |
| -0,057983513 | 0,888838445 | RAB5B | FALSE | Downregulated |
| -0,125258521 | 0,888838445 | SLC25A35 | FALSE | Downregulated |
| -0,068980067 | 0,888862474 | LMF2 | FALSE | Downregulated |
| 0,105849913 | 0,888895746 | FBXO22 | FALSE | Upregulated |
| -0,113108351 | 0,888952265 | HSPA4L | FALSE | Downregulated |
| -0,071928389 | 0,888975417 | COG4 | FALSE | Downregulated |
| 0,160526962 | 0,888975417 | RP11-1078H9.5 | FALSE | Upregulated |
| 0,243052189 | 0,889053475 | RFC3 | FALSE | Upregulated |
| 0,085166482 | 0,889059892 | FAM199X | FALSE | Upregulated |
| 0,218414587 | 0,889059892 | MIR4458HG | FALSE | Upregulated |
| 0,248889148 | 0,889059892 | OR5C1 | FALSE | Upregulated |

| | | | | |
|---|---|---|---|---|
| -0,046521027 | 0,889059892 | SEC24C | FALSE | Downregulated |
| -0,317292104 | 0,889377604 | ALAS2 | FALSE | Downregulated |
| -0,055517062 | 0,889377604 | ANXA9 | FALSE | Downregulated |
| -0,191079132 | 0,889377604 | HDC | FALSE | Downregulated |
| -0,057800855 | 0,889422044 | ABCG5 | FALSE | Downregulated |
| 0,147485733 | 0,889454882 | CAAP1 | FALSE | Upregulated |
| -0,408850676 | 0,889454882 | SFN | FALSE | Downregulated |
| -0,057206494 | 0,889462538 | CRYL1 | FALSE | Downregulated |
| -0,089912615 | 0,889462538 | EPM2AIP1 | FALSE | Downregulated |
| 0,115241956 | 0,889565158 | LOC100507472 | FALSE | Upregulated |
| -0,150576617 | 0,889565158 | LSM10 | FALSE | Downregulated |
| 0,113784025 | 0,889565158 | MLLT11 | FALSE | Upregulated |
| 0,146475578 | 0,889565158 | NIFK-AS1 | FALSE | Upregulated |
| -0,309243261 | 0,889682896 | KRTAP5-AS1 | FALSE | Downregulated |
| -0,276879264 | 0,88969442 | TUSC3 | FALSE | Downregulated |
| 0,296874824 | 0,889773997 | TNMD | FALSE | Upregulated |
| -0,087739866 | 0,889789352 | EIF3M | FALSE | Downregulated |
| -0,273918933 | 0,889809378 | SMPDL3B | FALSE | Downregulated |
| 0,264204535 | 0,889827021 | LINC01146 | FALSE | Upregulated |
| -0,27243815 | 0,88986456 | ALMS1P | FALSE | Downregulated |
| -0,117129699 | 0,88986456 | BRICD5 | FALSE | Downregulated |
| 0,127000241 | 0,88986456 | MRPS31 | FALSE | Upregulated |
| -0,096489395 | 0,88986456 | NBPF19 | FALSE | Downregulated |
| -0,115458916 | 0,88986456 | ZNF619 | FALSE | Downregulated |
| 0,183519224 | 0,889918005 | LOC101928334 | FALSE | Upregulated |
| 0,207795244 | 0,889918005 | PKNOX2 | FALSE | Upregulated |
| 0,052991439 | 0,889918005 | SEL1L | FALSE | Upregulated |
| 0,048135343 | 0,890113873 | PAICS | FALSE | Upregulated |
| 0,136762056 | 0,890140061 | SOCS4 | FALSE | Upregulated |
| 0,11877105 | 0,890267424 | ZNF473 | FALSE | Upregulated |
| -0,065050496 | 0,890647867 | AP2A2 | FALSE | Downregulated |
| -0,216476265 | 0,890647867 | ZNF793 | FALSE | Downregulated |
| -0,104860368 | 0,890715419 | HSBP1 | FALSE | Downregulated |
| -0,069463435 | 0,890715419 | IPO7 | FALSE | Downregulated |
| 0,124261577 | 0,890715419 | ZCCHC17 | FALSE | Upregulated |
| -0,231647743 | 0,891051622 | MIR143HG | FALSE | Downregulated |
| 0,10642943 | 0,891159852 | RBM28 | FALSE | Upregulated |
| 0,103324514 | 0,891288779 | KIAA1462 | FALSE | Upregulated |
| -0,243533637 | 0,891415098 | GPHA2 | FALSE | Downregulated |
| -0,126745147 | 0,891422668 | NAPSB | FALSE | Downregulated |
| 0,079586205 | 0,891516634 | GADD45GIP1 | FALSE | Upregulated |
| -0,089809989 | 0,89156113 | COPB1 | FALSE | Downregulated |
| -0,131312191 | 0,89156113 | HIST1H2BD | FALSE | Downregulated |
| 0,156247707 | 0,89156113 | MUTYH | FALSE | Upregulated |
| 0,20419929 | 0,891815582 | LOC102724448 | FALSE | Upregulated |
| -0,203660459 | 0,891815582 | LRP4-AS1 | FALSE | Downregulated |
| -0,093107212 | 0,891939278 | ANKMY1 | FALSE | Downregulated |
| 0,051426484 | 0,892265463 | FARP2 | FALSE | Upregulated |
| -0,06678143 | 0,892404664 | AUNIP | FALSE | Downregulated |
| 0,17052543 | 0,892404664 | PTS | FALSE | Upregulated |
| -0,125643537 | 0,8924469 | AGER | FALSE | Downregulated |

| | | | | |
|---|---|---|---|---|
| 0,085765551 | 0,8924469 | ANP32A | FALSE | Upregulated |
| 0,163146314 | 0,8924469 | C10orf2 | FALSE | Upregulated |
| -0,056967324 | 0,8924469 | EHBP1 | FALSE | Downregulated |
| 0,100655782 | 0,8924469 | ERAL1 | FALSE | Upregulated |
| 0,16041341 | 0,8924469 | JAG2 | FALSE | Upregulated |
| 0,094962136 | 0,8924469 | KLF7 | FALSE | Upregulated |
| 0,143294423 | 0,8924469 | LINC00102 | FALSE | Upregulated |
| -0,25088514 | 0,8924469 | LOC100289561 | FALSE | Downregulated |
| 0,08708258 | 0,8924469 | NSG1 | FALSE | Upregulated |
| 0,157541581 | 0,8924469 | RASA4 | FALSE | Upregulated |
| 0,073809559 | 0,8924469 | RBM27 | FALSE | Upregulated |
| 0,055333611 | 0,8924469 | SLC30A1 | FALSE | Upregulated |
| -0,060116061 | 0,8924469 | SOS1 | FALSE | Downregulated |
| 0,058833198 | 0,8924469 | TSPAN14 | FALSE | Upregulated |
| -0,1635833 | 0,8924469 | UBA6-AS1 | FALSE | Downregulated |
| -0,105025411 | 0,8924469 | UQCRFS1 | FALSE | Downregulated |
| 0,091798788 | 0,8924469 | USP6 | FALSE | Upregulated |
| -0,056373442 | 0,8924469 | YTHDF3 | FALSE | Downregulated |
| 0,102776641 | 0,8924469 | ZNF213 | FALSE | Upregulated |
| -0,087811256 | 0,892449321 | GLO1 | FALSE | Downregulated |
| 0,164143615 | 0,892449321 | TAS2R42 | FALSE | Upregulated |
| -0,047095272 | 0,892490887 | HIPK3 | FALSE | Downregulated |
| 0,068707734 | 0,89253439 | SUCLG1 | FALSE | Upregulated |
| -0,131892976 | 0,89273017 | NUDT8 | FALSE | Downregulated |
| 0,226484499 | 0,89273017 | YAE1D1 | FALSE | Upregulated |
| -0,060652646 | 0,892913061 | RBM8A | FALSE | Downregulated |
| 0,195735603 | 0,892975519 | TADA2A | FALSE | Upregulated |
| -0,085796321 | 0,893107972 | ISCU | FALSE | Downregulated |
| 0,166020164 | 0,893459824 | POLR2G | FALSE | Upregulated |
| 0,077151105 | 0,893536755 | CERS6 | FALSE | Upregulated |
| -0,070575938 | 0,893536755 | DHX15 | FALSE | Downregulated |
| 0,200023111 | 0,893536755 | LOC101929070 | FALSE | Upregulated |
| 0,140779611 | 0,893779136 | DNAH6 | FALSE | Upregulated |
| 0,078509912 | 0,894215329 | CRCP | FALSE | Upregulated |
| -0,206702689 | 0,894215329 | SNHG11 | FALSE | Downregulated |
| 0,150031024 | 0,894609062 | SNRPA1 | FALSE | Upregulated |
| 0,055738651 | 0,894677496 | C20orf112 | FALSE | Upregulated |
| -0,104565859 | 0,894677496 | SYNE4 | FALSE | Downregulated |
| -0,118284399 | 0,894677496 | SYTL2 | FALSE | Downregulated |
| -0,20725852 | 0,8947283 | KDM5D | FALSE | Downregulated |
| 0,097424531 | 0,894739786 | CCDC25 | FALSE | Upregulated |
| -0,210151177 | 0,894739786 | PLCXD2 | FALSE | Downregulated |
| -0,142605194 | 0,894776649 | SNRPB2 | FALSE | Downregulated |
| 0,161499614 | 0,894851576 | ZNF345 | FALSE | Upregulated |
| -0,085663209 | 0,89493029 | HEMK1 | FALSE | Downregulated |
| 0,262304632 | 0,895084113 | AQP4 | FALSE | Upregulated |
| -0,057804962 | 0,895084113 | BAZ1A | FALSE | Downregulated |
| -0,076191645 | 0,895084113 | KATNAL2 | FALSE | Downregulated |
| -0,148885402 | 0,895090929 | LOC101929078 | FALSE | Downregulated |
| 0,11001491 | 0,895124525 | NOL12 | FALSE | Upregulated |
| -0,045235771 | 0,895281238 | ACO1 | FALSE | Downregulated |

| | | | | |
|---|---|---|---|---|
| 0,086171958 | 0,895281238 | BRWD3 | FALSE | Upregulated |
| 0,1649782 | 0,895281238 | DNA2 | FALSE | Upregulated |
| 0,076836669 | 0,89530396 | DNASE1 | FALSE | Upregulated |
| -0,063128504 | 0,89530396 | NBPF8 | FALSE | Downregulated |
| 0,112941039 | 0,89530396 | PFKFB2 | FALSE | Upregulated |
| 0,243974254 | 0,895505882 | LARP6 | FALSE | Upregulated |
| 0,045664398 | 0,895598909 | BTBD2 | FALSE | Upregulated |
| 0,203132426 | 0,895598909 | CHST1 | FALSE | Upregulated |
| -0,139547327 | 0,895598909 | HEPHL1 | FALSE | Downregulated |
| 0,159820169 | 0,895598909 | HSPBAP1 | FALSE | Upregulated |
| 0,069093282 | 0,895598909 | LOC101928897 | FALSE | Upregulated |
| -0,217929202 | 0,895598909 | LOC101929519 | FALSE | Downregulated |
| -0,226688172 | 0,895598909 | POLR3K | FALSE | Downregulated |
| 0,100060705 | 0,895598909 | SPDYE2B | FALSE | Upregulated |
| 0,121512841 | 0,895598909 | TMEM201 | FALSE | Upregulated |
| -0,166768107 | 0,895598909 | TTC8 | FALSE | Downregulated |
| -0,110490709 | 0,895598909 | ZMAT2 | FALSE | Downregulated |
| -0,106832888 | 0,895646939 | CXorf38 | FALSE | Downregulated |
| 0,083323164 | 0,895646939 | EPHA2 | FALSE | Upregulated |
| 0,100656605 | 0,895646939 | KIF16B | FALSE | Upregulated |
| 0,104178623 | 0,895646939 | LOC100131564 | FALSE | Upregulated |
| 0,068445366 | 0,895646939 | RRP1B | FALSE | Upregulated |
| -0,033686884 | 0,895646939 | ZFHX3 | FALSE | Downregulated |
| 0,120385421 | 0,895646939 | ZKSCAN4 | FALSE | Upregulated |
| 0,080900039 | 0,895708129 | OFD1 | FALSE | Upregulated |
| 0,103814353 | 0,895708129 | OSTC | FALSE | Upregulated |
| 0,056113372 | 0,895708129 | PUF60 | FALSE | Upregulated |
| -0,058784812 | 0,895782941 | NUGGC | FALSE | Downregulated |
| -0,043127704 | 0,895782941 | PUM2 | FALSE | Downregulated |
| 0,117455684 | 0,895877224 | CNOT10 | FALSE | Upregulated |
| -0,289618666 | 0,895904947 | ZNF813 | FALSE | Downregulated |
| -0,255544018 | 0,895985767 | MFSD6L | FALSE | Downregulated |
| -0,048582619 | 0,896009802 | C2 | FALSE | Downregulated |
| 0,32575058 | 0,896009802 | NPTXR | FALSE | Upregulated |
| 0,125013038 | 0,896079891 | ATP5G1 | FALSE | Upregulated |
| 0,23985127 | 0,896079891 | EFCAB6 | FALSE | Upregulated |
| -0,176710044 | 0,896079891 | FGF7 | FALSE | Downregulated |
| -0,080238913 | 0,896079891 | FLAD1 | FALSE | Downregulated |
| 0,301645597 | 0,896079891 | LINC00346 | FALSE | Upregulated |
| -0,074761634 | 0,896079891 | MLX | FALSE | Downregulated |
| 0,094095591 | 0,896079891 | MRPL3 | FALSE | Upregulated |
| -0,101719236 | 0,896079891 | NSUN4 | FALSE | Downregulated |
| -0,2392766 | 0,896079891 | SNHG1 | FALSE | Downregulated |
| 0,13549187 | 0,896111478 | LOC101929130 | FALSE | Upregulated |
| 0,09508866 | 0,896135667 | BLOC1S6 | FALSE | Upregulated |
| -0,078619411 | 0,896135667 | BRCA2 | FALSE | Downregulated |
| 0,059538119 | 0,896135667 | ENPEP | FALSE | Upregulated |
| -0,067300168 | 0,896135667 | MRPL2 | FALSE | Downregulated |
| 0,100881299 | 0,896135667 | NGLY1 | FALSE | Upregulated |
| -0,064437527 | 0,896135667 | SERPINA3 | FALSE | Downregulated |
| -0,084855674 | 0,896135667 | SH3TC1 | FALSE | Downregulated |

| | | | | |
|---|---|---|---|---|
| 0,256039946 | 0,896135667 | SPIN4 | FALSE | Upregulated |
| 0,038941248 | 0,896135667 | STK11 | FALSE | Upregulated |
| -0,067441693 | 0,896171541 | CAMK2D | FALSE | Downregulated |
| 0,186452046 | 0,89634176 | ERBB4 | FALSE | Upregulated |
| -0,133763494 | 0,896457334 | C12orf66 | FALSE | Downregulated |
| -0,086840598 | 0,896457334 | CPSF2 | FALSE | Downregulated |
| -0,057633649 | 0,896457334 | DGCR8 | FALSE | Downregulated |
| 0,16126327 | 0,896457334 | IFT81 | FALSE | Upregulated |
| 0,270196716 | 0,896457334 | SEMA3D | FALSE | Upregulated |
| 0,053468457 | 0,896476669 | STOM | FALSE | Upregulated |
| 0,07816805 | 0,896655562 | DNASE1L1 | FALSE | Upregulated |
| 0,286387163 | 0,897134628 | BSCL2 | FALSE | Upregulated |
| 0,130366157 | 0,897134628 | RFC2 | FALSE | Upregulated |
| 0,089080361 | 0,897161054 | PLEKHM1P | FALSE | Upregulated |
| -0,229995959 | 0,897161054 | SELP | FALSE | Downregulated |
| 0,155055648 | 0,897161054 | SURF2 | FALSE | Upregulated |
| 0,081131131 | 0,897161054 | TRPC5 | FALSE | Upregulated |
| 0,117827273 | 0,897304564 | ARHGAP24 | FALSE | Upregulated |
| 0,353534927 | 0,897321252 | PRSS55 | FALSE | Upregulated |
| 0,253871376 | 0,897426398 | TBX10 | FALSE | Upregulated |
| 0,092122821 | 0,89754951 | MAPK8IP1 | FALSE | Upregulated |
| -0,109847191 | 0,89754951 | METTL17 | FALSE | Downregulated |
| 0,05699925 | 0,89754951 | RBM12 | FALSE | Upregulated |
| -0,049885034 | 0,89759207 | SRPK2 | FALSE | Downregulated |
| 0,603885079 | 0,89759207 | TTN-AS1 | FALSE | Upregulated |
| 0,149356537 | 0,897866748 | MYLIP | FALSE | Upregulated |
| -0,243448311 | 0,897871242 | CCDC54 | FALSE | Downregulated |
| 0,242044083 | 0,897963911 | LOC101928768 | FALSE | Upregulated |
| 0,050178854 | 0,898013037 | FAM65C | FALSE | Upregulated |
| -0,838578534 | 0,898079911 | PRODH | FALSE | Downregulated |
| 0,080707428 | 0,898079911 | SYDE1 | FALSE | Upregulated |
| 0,098526062 | 0,898211802 | CACNA2D4 | FALSE | Upregulated |
| -0,069450023 | 0,898211802 | ERLEC1 | FALSE | Downregulated |
| 0,242046964 | 0,898211802 | METTL1 | FALSE | Upregulated |
| 0,103998013 | 0,898211802 | MPHOSPH10 | FALSE | Upregulated |
| -0,064983311 | 0,898211802 | SAMD14 | FALSE | Downregulated |
| 0,054619691 | 0,898211802 | TMED7 | FALSE | Upregulated |
| -0,1158993 | 0,898237007 | ISCA1 | FALSE | Downregulated |
| -0,213950591 | 0,898387403 | ARL15 | FALSE | Downregulated |
| 0,067152245 | 0,898387403 | CNOT2 | FALSE | Upregulated |
| -0,202274782 | 0,898429483 | MAP3K7CL | FALSE | Downregulated |
| 0,252823061 | 0,898463091 | LOC728769 | FALSE | Upregulated |
| -0,176385297 | 0,898787639 | DHRS4L1 | FALSE | Downregulated |
| -0,115775859 | 0,898801037 | C3orf83 | FALSE | Downregulated |
| -0,060644637 | 0,898801037 | LMO7 | FALSE | Downregulated |
| -0,106620854 | 0,898801037 | MFF | FALSE | Downregulated |
| -0,046241143 | 0,898801037 | PC | FALSE | Downregulated |
| 0,093475949 | 0,898926479 | RFXANK | FALSE | Upregulated |
| 0,27102042 | 0,899047836 | OXCT1 | FALSE | Upregulated |
| 0,10512365 | 0,899134658 | IRF8 | FALSE | Upregulated |
| 0,096585022 | 0,899134658 | MRPS21 | FALSE | Upregulated |

| | | | | |
|---|---|---|---|---|
| -0,230308316 | 0,899155123 | ACOXL | FALSE | Downregulated |
| -0,095798194 | 0,899155123 | ZNF48 | FALSE | Downregulated |
| 0,252403413 | 0,899158471 | CCDC24 | FALSE | Upregulated |
| 0,125911107 | 0,899264302 | EIF1AD | FALSE | Upregulated |
| -0,313926117 | 0,899264302 | SRGAP2D | FALSE | Downregulated |
| 0,195810977 | 0,899265881 | SAMD15 | FALSE | Upregulated |
| 0,144709601 | 0,899506436 | ISM1 | FALSE | Upregulated |
| -0,1063526 | 0,899655694 | ADCK4 | FALSE | Downregulated |
| 0,147985042 | 0,899655694 | CBWD2 | FALSE | Upregulated |
| 0,219044315 | 0,899655694 | LOC100652758 | FALSE | Upregulated |
| -0,034603225 | 0,899655694 | TMEM259 | FALSE | Downregulated |
| 0,168162799 | 0,899697672 | MSANTD3 | FALSE | Upregulated |
| -0,046092181 | 0,899862791 | STARD7 | FALSE | Downregulated |
| -0,219365076 | 0,899862791 | ZNF135 | FALSE | Downregulated |
| -0,270107508 | 0,899915819 | C5orf49 | FALSE | Downregulated |
| -0,178570874 | 0,899915819 | GYLTL1B | FALSE | Downregulated |
| 0,236325297 | 0,899915819 | LOC101927818 | FALSE | Upregulated |
| 0,069429136 | 0,899915819 | SIKE1 | FALSE | Upregulated |
| 0,271603565 | 0,899915819 | ZBTB7C | FALSE | Upregulated |
| -0,072754972 | 0,900124163 | SLMAP | FALSE | Downregulated |
| -0,15118687 | 0,900173486 | ZNF718 | FALSE | Downregulated |
| -0,26843086 | 0,900179295 | LOC100287015 | FALSE | Downregulated |
| -0,10244099 | 0,900179295 | ZNF700 | FALSE | Downregulated |
| 0,084888539 | 0,900527936 | CHKB | FALSE | Upregulated |
| -0,130665278 | 0,900732799 | COMMD3 | FALSE | Downregulated |
| -0,06870912 | 0,900939414 | CEBPZ-AS1 | FALSE | Downregulated |
| -0,040819066 | 0,900939414 | ERP29 | FALSE | Downregulated |
| -0,072673633 | 0,901000307 | CUL4B | FALSE | Downregulated |
| 0,065357336 | 0,901025265 | CBX4 | FALSE | Upregulated |
| 0,100906552 | 0,901025265 | HTATIP2 | FALSE | Upregulated |
| -0,132052151 | 0,901025265 | NDUFB1 | FALSE | Downregulated |
| -0,044128187 | 0,901025265 | SRSF4 | FALSE | Downregulated |
| 0,161019862 | 0,901097242 | LCN12 | FALSE | Upregulated |
| 0,19452417 | 0,901097242 | MST4 | FALSE | Upregulated |
| -0,235579998 | 0,901097242 | PACRG-AS1 | FALSE | Downregulated |
| -0,056056766 | 0,901097242 | ZBTB4 | FALSE | Downregulated |
| 0,138891856 | 0,901145674 | FAM156B | FALSE | Upregulated |
| -0,058115856 | 0,901145674 | STAG2 | FALSE | Downregulated |
| -0,082008105 | 0,901207493 | USP16 | FALSE | Downregulated |
| -0,068088758 | 0,901352237 | APOA1BP | FALSE | Downregulated |
| 0,119105571 | 0,901352237 | CASP2 | FALSE | Upregulated |
| -0,147579641 | 0,901352237 | CDR1 | FALSE | Downregulated |
| -0,095337018 | 0,901352237 | DHFRL1 | FALSE | Downregulated |
| 0,121614543 | 0,901352237 | GTF2H3 | FALSE | Upregulated |
| -0,195391372 | 0,901352237 | LOC102723703 | FALSE | Downregulated |
| 0,110713035 | 0,901352237 | SPIN3 | FALSE | Upregulated |
| 0,113630383 | 0,901352237 | ZFPL1 | FALSE | Upregulated |
| 0,235736602 | 0,901352237 | ZNF215 | FALSE | Upregulated |
| -0,2701401 | 0,901420991 | RPGRIP1L | FALSE | Downregulated |
| 0,154407804 | 0,901420991 | SLC25A36 | FALSE | Upregulated |
| 0,055258115 | 0,901448737 | CUL7 | FALSE | Upregulated |

| | | | | |
|---|---|---|---|---|
| 0,136478331 | 0,901448737 | GPN3 | FALSE | Upregulated |
| -0,112851365 | 0,901448737 | INHBB | FALSE | Downregulated |
| 0,103537158 | 0,901448737 | LOC101928998 | FALSE | Upregulated |
| 0,155644048 | 0,901448737 | TMEM70 | FALSE | Upregulated |
| 0,084574608 | 0,901672325 | DFNA5 | FALSE | Upregulated |
| -0,090806384 | 0,901683953 | QRICH2 | FALSE | Downregulated |
| 0,139943809 | 0,901825559 | SYN1 | FALSE | Upregulated |
| 0,051422289 | 0,901851849 | AHCYL1 | FALSE | Upregulated |
| 0,078062785 | 0,901851849 | CCDC57 | FALSE | Upregulated |
| 0,090496156 | 0,901851849 | CHMP5 | FALSE | Upregulated |
| 0,055228967 | 0,901851849 | CNN3 | FALSE | Upregulated |
| -0,05336879 | 0,901851849 | EPS15 | FALSE | Downregulated |
| -0,255747578 | 0,901851849 | GJC2 | FALSE | Downregulated |
| -0,094188427 | 0,901851849 | INHBE | FALSE | Downregulated |
| -0,249086882 | 0,901851849 | LINC00528 | FALSE | Downregulated |
| -0,135987329 | 0,901851849 | LMOD1 | FALSE | Downregulated |
| 0,137806438 | 0,901851849 | LYSMD2 | FALSE | Upregulated |
| -0,078121315 | 0,901851849 | MST1P2 | FALSE | Downregulated |
| -0,218121773 | 0,901851849 | NEB | FALSE | Downregulated |
| -0,07776355 | 0,901851849 | PPP1R3B | FALSE | Downregulated |
| 0,110501787 | 0,901851849 | TIMM17A | FALSE | Upregulated |
| 0,096641183 | 0,901851849 | ZNF420 | FALSE | Upregulated |
| 0,080947661 | 0,901883147 | PEX12 | FALSE | Upregulated |
| 0,089474376 | 0,901920046 | HPS1 | FALSE | Upregulated |
| 0,152382627 | 0,901920046 | RPS6KA5 | FALSE | Upregulated |
| 0,079747285 | 0,902054782 | EML6 | FALSE | Upregulated |
| -0,167412162 | 0,902094147 | CAMTA1 | FALSE | Downregulated |
| 0,251709115 | 0,902094147 | MGC57346 | FALSE | Upregulated |
| -0,157785279 | 0,902258055 | LINC00884 | FALSE | Downregulated |
| 0,227913119 | 0,902328266 | NAT14 | FALSE | Upregulated |
| 0,263315582 | 0,902328266 | RFESD | FALSE | Upregulated |
| 0,214657875 | 0,902377536 | CASQ1 | FALSE | Upregulated |
| -0,142065697 | 0,902377536 | ZC2HC1A | FALSE | Downregulated |
| -0,060186593 | 0,902396817 | INPP5A | FALSE | Downregulated |
| -0,246542661 | 0,902401094 | ADAMTS9-AS2 | FALSE | Downregulated |
| 0,069096286 | 0,902430643 | GAB2 | FALSE | Upregulated |
| 0,258720813 | 0,902563537 | ADC | FALSE | Upregulated |
| -0,224279744 | 0,902563537 | CNEP1R1 | FALSE | Downregulated |
| 0,084243044 | 0,902563537 | KIAA1407 | FALSE | Upregulated |
| 0,069366001 | 0,902563537 | LHPP | FALSE | Upregulated |
| -0,142036284 | 0,902563537 | PATE4 | FALSE | Downregulated |
| -0,09011272 | 0,902576372 | SLC12A9 | FALSE | Downregulated |
| 0,196239415 | 0,902743466 | AAED1 | FALSE | Upregulated |
| -0,137132758 | 0,902869018 | LHX2 | FALSE | Downregulated |
| 0,248546602 | 0,902906601 | BMP5 | FALSE | Upregulated |
| 0,122259341 | 0,902906601 | CCNDBP1 | FALSE | Upregulated |
| 0,211789011 | 0,902906601 | DPYSL4 | FALSE | Upregulated |
| 0,152100927 | 0,902906601 | LOC100506060 | FALSE | Upregulated |
| -0,41263649 | 0,902906601 | LY75 | FALSE | Downregulated |
| 0,143980402 | 0,902906601 | MAGI1-IT1 | FALSE | Upregulated |
| 0,040428089 | 0,902906601 | MYLK | FALSE | Upregulated |

| | | | | |
|---|---|---|---|---|
| 0,088620564 | 0,902906601 | PPP2R3B | FALSE | Upregulated |
| -0,059132434 | 0,902937741 | ANGPTL4 | FALSE | Downregulated |
| -0,262421096 | 0,902937741 | TM2D1 | FALSE | Downregulated |
| 0,095070859 | 0,903066306 | UNC5CL | FALSE | Upregulated |
| -0,300766985 | 0,903259096 | LOC101929117 | FALSE | Downregulated |
| 0,192541858 | 0,903390302 | GPC6-AS1 | FALSE | Upregulated |
| 0,1752524 | 0,903390302 | RMI1 | FALSE | Upregulated |
| -0,097984276 | 0,903511684 | DPYD-AS2 | FALSE | Downregulated |
| 0,049770968 | 0,903629817 | RBM10 | FALSE | Upregulated |
| 0,050897553 | 0,903722631 | DDX3X | FALSE | Upregulated |
| -0,094050993 | 0,903722631 | DDX60L | FALSE | Downregulated |
| -0,117163765 | 0,903722631 | FADS2 | FALSE | Downregulated |
| 0,077586188 | 0,903722631 | LOC101927057 | FALSE | Upregulated |
| -0,134239275 | 0,903722631 | PUS7L | FALSE | Downregulated |
| -0,065315951 | 0,903746521 | COG8 | FALSE | Downregulated |
| 0,182059466 | 0,903896846 | CA1 | FALSE | Upregulated |
| 0,085579311 | 0,903896846 | CLIP2 | FALSE | Upregulated |
| 0,093470222 | 0,903896846 | CTD-2201E9.1 | FALSE | Upregulated |
| -0,24896558 | 0,903896846 | KCNS3 | FALSE | Downregulated |
| -0,184445361 | 0,903896846 | PAK1IP1 | FALSE | Downregulated |
| 0,07872498 | 0,903896846 | QSOX1 | FALSE | Upregulated |
| -0,132948279 | 0,903896846 | TBCCD1 | FALSE | Downregulated |
| 0,058990036 | 0,903896846 | TOP1 | FALSE | Upregulated |
| -0,084202299 | 0,903997149 | EFCAB7 | FALSE | Downregulated |
| 0,182646048 | 0,904109098 | AC024592.9 | FALSE | Upregulated |
| -0,223345545 | 0,904109098 | LOC102724475 | FALSE | Downregulated |
| -0,065994774 | 0,904109098 | MYO6 | FALSE | Downregulated |
| -0,089624977 | 0,904109098 | NEURL1 | FALSE | Downregulated |
| -0,087936271 | 0,904109098 | NRM | FALSE | Downregulated |
| 0,094320208 | 0,904109098 | SLC25A32 | FALSE | Upregulated |
| -0,073621208 | 0,904109098 | ZNF618 | FALSE | Downregulated |
| 0,160700392 | 0,904121879 | PLCXD3 | FALSE | Upregulated |
| -0,182481384 | 0,904121879 | WASF1 | FALSE | Downregulated |
| -0,110967952 | 0,904349067 | ARID3B | FALSE | Downregulated |
| 0,227811848 | 0,904349067 | CATSPER2P1 | FALSE | Upregulated |
| -0,22300103 | 0,904349067 | DFNB59 | FALSE | Downregulated |
| -0,061344785 | 0,904349067 | EPG5 | FALSE | Downregulated |
| 0,057141158 | 0,90439086 | MAGT1 | FALSE | Upregulated |
| 0,118589293 | 0,904393116 | AGTPBP1 | FALSE | Upregulated |
| -0,204591662 | 0,904393116 | CMC1 | FALSE | Downregulated |
| -0,196254735 | 0,904393116 | SHISA7 | FALSE | Downregulated |
| -0,199671249 | 0,904393116 | UPK3B | FALSE | Downregulated |
| 0,13752491 | 0,904438319 | ATXN7L2 | FALSE | Upregulated |
| 0,284513696 | 0,904438319 | LINC00598 | FALSE | Upregulated |
| -0,206394622 | 0,904438319 | LOC101928527 | FALSE | Downregulated |
| -0,19337385 | 0,904438319 | LOC643355 | FALSE | Downregulated |
| -0,064753404 | 0,904592137 | MPC1 | FALSE | Downregulated |
| 0,135880622 | 0,904646805 | CCR8 | FALSE | Upregulated |
| -0,099237048 | 0,904646805 | FST | FALSE | Downregulated |
| 0,037464064 | 0,904646805 | GNAI2 | FALSE | Upregulated |
| -0,051639583 | 0,904646805 | GOLGA3 | FALSE | Downregulated |

| | | | | |
|---|---|---|---|---|
| -0,857215956 | 0,904646805 | LOC102724951 | FALSE | Downregulated |
| -0,167966888 | 0,904646805 | SFT2D2 | FALSE | Downregulated |
| -0,169601933 | 0,904721968 | G2E3 | FALSE | Downregulated |
| 0,066267064 | 0,904721968 | STAR | FALSE | Upregulated |
| -0,108069807 | 0,904745484 | TMEM14B | FALSE | Downregulated |
| 0,069513282 | 0,904970814 | DNAH11 | FALSE | Upregulated |
| -0,076308766 | 0,904970814 | SNTA1 | FALSE | Downregulated |
| -0,088337182 | 0,904970814 | TPRA1 | FALSE | Downregulated |
| 0,332501642 | 0,9050475 | LIF | FALSE | Upregulated |
| 0,0918353 | 0,9050475 | MLLT4-AS1 | FALSE | Upregulated |
| 0,136465168 | 0,905192143 | CD81-AS1 | FALSE | Upregulated |
| 0,208908478 | 0,905192143 | CDRT4 | FALSE | Upregulated |
| -0,197873928 | 0,905192143 | LOC101928498 | FALSE | Downregulated |
| -0,038019976 | 0,905307345 | PABPC1 | FALSE | Downregulated |
| 0,114209457 | 0,905386409 | C1orf172 | FALSE | Upregulated |
| 0,17756906 | 0,905386409 | LOC101928289 | FALSE | Upregulated |
| 0,08694916 | 0,905386409 | RAB21 | FALSE | Upregulated |
| -0,064112827 | 0,905386409 | SLC11A2 | FALSE | Downregulated |
| -0,056927379 | 0,905386409 | SPPL2B | FALSE | Downregulated |
| 0,165662821 | 0,905386409 | TTC22 | FALSE | Upregulated |
| 0,055780338 | 0,905386409 | VRK2 | FALSE | Upregulated |
| -0,076989411 | 0,905391981 | ZNF551 | FALSE | Downregulated |
| -0,216405756 | 0,905426998 | C11orf21 | FALSE | Downregulated |
| -0,08262284 | 0,905426998 | CYFIP2 | FALSE | Downregulated |
| -0,069744867 | 0,905426998 | DDX46 | FALSE | Downregulated |
| -0,061090088 | 0,905426998 | DOK4 | FALSE | Downregulated |
| -0,053073457 | 0,905426998 | MAF1 | FALSE | Downregulated |
| -0,037785976 | 0,905426998 | MAN2B2 | FALSE | Downregulated |
| 0,074716704 | 0,905426998 | PEX11G | FALSE | Upregulated |
| 0,157040024 | 0,905426998 | RPGR | FALSE | Upregulated |
| 0,083025209 | 0,905426998 | TANGO2 | FALSE | Upregulated |
| -0,124334016 | 0,905501771 | RPP25L | FALSE | Downregulated |
| -0,058331767 | 0,905541587 | KANK3 | FALSE | Downregulated |
| -0,0606207 | 0,905620188 | EIF3D | FALSE | Downregulated |
| 0,174977288 | 0,905620188 | SOX17 | FALSE | Upregulated |
| -0,075484321 | 0,9056795 | HSPE1 | FALSE | Downregulated |
| -0,050338147 | 0,9056795 | PCM1 | FALSE | Downregulated |
| -0,21010157 | 0,9056795 | PTK7 | FALSE | Downregulated |
| -0,23383499 | 0,905829635 | C5AR2 | FALSE | Downregulated |
| -0,255838271 | 0,905839488 | LPAR2 | FALSE | Downregulated |
| 0,095052832 | 0,906302362 | ABHD14A | FALSE | Upregulated |
| 0,109202502 | 0,906390221 | DSEL | FALSE | Upregulated |
| -0,057164588 | 0,906489573 | CLMN | FALSE | Downregulated |
| -0,117228011 | 0,906489573 | HEATR3 | FALSE | Downregulated |
| -0,127188699 | 0,906489573 | ZNF595 | FALSE | Downregulated |
| -0,113529592 | 0,906530601 | SGCE | FALSE | Downregulated |
| 0,110761321 | 0,906661299 | MED4 | FALSE | Upregulated |
| -0,070955616 | 0,906661299 | PITPNC1 | FALSE | Downregulated |
| -0,142933466 | 0,906661299 | TRUB1 | FALSE | Downregulated |
| -0,152733617 | 0,90669559 | LOC645513 | FALSE | Downregulated |
| 0,046268357 | 0,906891401 | MIB2 | FALSE | Upregulated |

| | | | | |
|---|---|---|---|---|
| 0,239561608 | 0,906891401 | RP11-626I20.3 | FALSE | Upregulated |
| 0,064294194 | 0,906891401 | TBC1D1 | FALSE | Upregulated |
| -0,14488416 | 0,906997342 | LRRC42 | FALSE | Downregulated |
| -0,070894634 | 0,906997342 | MS4A6A | FALSE | Downregulated |
| 0,053877173 | 0,907047491 | CEP250 | FALSE | Upregulated |
| -0,120879516 | 0,907047491 | LOC102723539 | FALSE | Downregulated |
| 0,056138482 | 0,90709898 | MTCH2 | FALSE | Upregulated |
| -0,095882159 | 0,90709898 | OGFOD2 | FALSE | Downregulated |
| -0,068656854 | 0,907154435 | ALDH1B1 | FALSE | Downregulated |
| 0,074426301 | 0,907154435 | CAMK2G | FALSE | Upregulated |
| -0,126278595 | 0,90733444 | BAALC | FALSE | Downregulated |
| 0,142794934 | 0,90733444 | C16orf87 | FALSE | Upregulated |
| 0,225959336 | 0,90733444 | GABARAPL3 | FALSE | Upregulated |
| 0,10666377 | 0,90733444 | LOC283788 | FALSE | Upregulated |
| -0,107685016 | 0,90733444 | MAPK3 | FALSE | Downregulated |
| -0,115125958 | 0,90733444 | NMNAT1 | FALSE | Downregulated |
| 0,099291127 | 0,90733444 | PRKCI | FALSE | Upregulated |
| 0,059650658 | 0,907351358 | GIT1 | FALSE | Upregulated |
| 0,185428458 | 0,907406963 | A4GALT | FALSE | Upregulated |
| -0,145265444 | 0,907406963 | ASB16-AS1 | FALSE | Downregulated |
| 0,070948005 | 0,907406963 | CEP104 | FALSE | Upregulated |
| -0,111964245 | 0,907406963 | CRB3 | FALSE | Downregulated |
| 0,155092414 | 0,907406963 | METTL10 | FALSE | Upregulated |
| 0,202598398 | 0,907406963 | PMP22 | FALSE | Upregulated |
| -0,14584118 | 0,907406963 | RBM18 | FALSE | Downregulated |
| -0,172888775 | 0,907480715 | PITPNA-AS1 | FALSE | Downregulated |
| -0,118865121 | 0,907490512 | LOC727710 | FALSE | Downregulated |
| 0,083969115 | 0,907502829 | ACOT7 | FALSE | Upregulated |
| 0,075407573 | 0,90770024 | DDX49 | FALSE | Upregulated |
| 0,084250103 | 0,90784142 | PRKAG3 | FALSE | Upregulated |
| -0,256804617 | 0,908202284 | CCDC15 | FALSE | Downregulated |
| 0,097048794 | 0,908202284 | DUSP7 | FALSE | Upregulated |
| -0,258384264 | 0,908202284 | ILDR2 | FALSE | Downregulated |
| 0,088061028 | 0,908202284 | UIMC1 | FALSE | Upregulated |
| 0,050148662 | 0,908202284 | WTAP | FALSE | Upregulated |
| -0,05498274 | 0,908353947 | DNAJC13 | FALSE | Downregulated |
| 0,205627398 | 0,908371275 | IL5 | FALSE | Upregulated |
| 0,332911816 | 0,908742825 | CTAGE15 | FALSE | Upregulated |
| 0,149641726 | 0,908742825 | DSC1 | FALSE | Upregulated |
| -0,045583667 | 0,908742825 | PCSK6 | FALSE | Downregulated |
| -0,182878932 | 0,908742825 | TRPC1 | FALSE | Downregulated |
| 0,074167092 | 0,908742825 | USF1 | FALSE | Upregulated |
| 0,079623972 | 0,908824136 | TMEM150A | FALSE | Upregulated |
| -0,105099184 | 0,908905903 | C2orf47 | FALSE | Downregulated |
| 0,093088326 | 0,909017168 | COX7A2 | FALSE | Upregulated |
| 0,09387402 | 0,909096976 | PLXNA3 | FALSE | Upregulated |
| -0,081832243 | 0,909294481 | LOC389906 | FALSE | Downregulated |
| 0,058642278 | 0,909373063 | BBIP1 | FALSE | Upregulated |
| 0,160704257 | 0,909423164 | RAD21-AS1 | FALSE | Upregulated |
| 0,284181932 | 0,909474381 | RP11-329B9.1 | FALSE | Upregulated |
| 0,107561473 | 0,909474381 | UBR7 | FALSE | Upregulated |

| | | | | |
|---|---|---|---|---|
| 0,085707832 | 0,909474381 | VPS45 | FALSE | Upregulated |
| 0,209348307 | 0,909571309 | KCNMB1 | FALSE | Upregulated |
| -0,126661887 | 0,909638856 | KLHDC8B | FALSE | Downregulated |
| 0,100398635 | 0,909847578 | PELI1 | FALSE | Upregulated |
| 0,074547028 | 0,910078862 | DDX20 | FALSE | Upregulated |
| 0,076383799 | 0,910078862 | LEPRE1 | FALSE | Upregulated |
| 0,112662734 | 0,910078862 | METTL22 | FALSE | Upregulated |
| -0,075818549 | 0,910124703 | MAPKAPK5 | FALSE | Downregulated |
| -0,14311455 | 0,910124703 | ZNF423 | FALSE | Downregulated |
| -0,071651663 | 0,910124703 | ZNRF3-AS1 | FALSE | Downregulated |
| 0,072408302 | 0,910546051 | CC2D2A | FALSE | Upregulated |
| -0,086875575 | 0,910720161 | KIAA1715 | FALSE | Downregulated |
| -0,105159117 | 0,910780734 | LOC145783 | FALSE | Downregulated |
| -0,120470787 | 0,910780734 | NREP | FALSE | Downregulated |
| -0,100249801 | 0,910827548 | CCDC22 | FALSE | Downregulated |
| 0,082828292 | 0,910827548 | LOC101929752 | FALSE | Upregulated |
| 0,278233757 | 0,910827548 | LOC102723329 | FALSE | Upregulated |
| -0,047459758 | 0,911204678 | KDSR | FALSE | Downregulated |
| 0,088741473 | 0,911357742 | RRAGD | FALSE | Upregulated |
| -0,0998095 | 0,911454977 | CLEC1B | FALSE | Downregulated |
| 0,072654208 | 0,911454977 | DDX1 | FALSE | Upregulated |
| -0,118540476 | 0,911454977 | ENY2 | FALSE | Downregulated |
| -0,052671092 | 0,911454977 | GABARAPL1 | FALSE | Downregulated |
| 0,061574155 | 0,911454977 | RP11-392P7.6 | FALSE | Upregulated |
| 0,144561028 | 0,911551568 | LINC00899 | FALSE | Upregulated |
| -0,067652357 | 0,911651374 | TRMT5 | FALSE | Downregulated |
| -0,077659865 | 0,911758197 | SMIM19 | FALSE | Downregulated |
| -0,151552037 | 0,911938738 | ERI1 | FALSE | Downregulated |
| -0,085371731 | 0,912063545 | MAP3K19 | FALSE | Downregulated |
| -0,183566566 | 0,912164414 | LOC101928303 | FALSE | Downregulated |
| 0,06357749 | 0,912185941 | C19orf10 | FALSE | Upregulated |
| -0,056744561 | 0,912185941 | GPR155 | FALSE | Downregulated |
| -0,115237685 | 0,912185941 | HAUS3 | FALSE | Downregulated |
| 0,149353786 | 0,912185941 | MPV17L2 | FALSE | Upregulated |
| 0,065377621 | 0,912185941 | MRTO4 | FALSE | Upregulated |
| -0,043382823 | 0,912185941 | SIPA1L1 | FALSE | Downregulated |
| -0,04891585 | 0,912185941 | THRAP3 | FALSE | Downregulated |
| -0,103583 | 0,912185941 | ZNF26 | FALSE | Downregulated |
| 0,075384424 | 0,912220667 | LOC100129917 | FALSE | Upregulated |
| 0,03544389 | 0,912263658 | ARFGEF1 | FALSE | Upregulated |
| 0,081105983 | 0,912263658 | CASP7 | FALSE | Upregulated |
| 0,119471749 | 0,912263658 | CDKN1C | FALSE | Upregulated |
| 0,119213212 | 0,912263658 | FAM122C | FALSE | Upregulated |
| -0,119310379 | 0,912263658 | FIBP | FALSE | Downregulated |
| 0,111986568 | 0,912263658 | KANSL2 | FALSE | Upregulated |
| -0,284469901 | 0,912263658 | LOC101929667 | FALSE | Downregulated |
| 0,237286685 | 0,912263658 | LOC102723625 | FALSE | Upregulated |
| 0,063019335 | 0,912263658 | NARS | FALSE | Upregulated |
| -0,068628477 | 0,912263658 | PIN1 | FALSE | Downregulated |
| 0,065915373 | 0,912263658 | RNF32 | FALSE | Upregulated |
| -0,065288642 | 0,912263658 | STRAP | FALSE | Downregulated |

| | | | | |
|---|---|---|---|---|
| 0,24505495 | 0,912263658 | TXK | FALSE | Upregulated |
| 0,062091989 | 0,912263658 | URI1 | FALSE | Upregulated |
| 0,217402065 | 0,912263658 | ZNF519 | FALSE | Upregulated |
| 0,084649086 | 0,912308611 | CENPU | FALSE | Upregulated |
| -0,073521939 | 0,912662259 | CARS | FALSE | Downregulated |
| -0,172990735 | 0,912662259 | LOC100132111 | FALSE | Downregulated |
| -0,084653082 | 0,912690521 | PSMD6 | FALSE | Downregulated |
| 0,052203592 | 0,913094497 | B3GALNT2 | FALSE | Upregulated |
| 0,168074997 | 0,913192323 | C1orf74 | FALSE | Upregulated |
| -0,333531828 | 0,913192323 | GUSBP2 | FALSE | Downregulated |
| 0,057945587 | 0,913192323 | NAT10 | FALSE | Upregulated |
| -0,113889985 | 0,913243805 | DLEU2 | FALSE | Downregulated |
| -0,209267775 | 0,913243805 | LOC102724376 | FALSE | Downregulated |
| 0,103955805 | 0,913243805 | NOTCH3 | FALSE | Upregulated |
| 0,067443965 | 0,913243805 | SYCP3 | FALSE | Upregulated |
| 0,062556617 | 0,913369346 | TCTA | FALSE | Upregulated |
| -0,09659086 | 0,913426832 | MIIP | FALSE | Downregulated |
| -0,063609661 | 0,913426832 | NPRL3 | FALSE | Downregulated |
| -0,190500749 | 0,913426832 | RHEBL1 | FALSE | Downregulated |
| -0,10745374 | 0,913426832 | ULK2 | FALSE | Downregulated |
| 0,201821022 | 0,913433693 | ASIC4 | FALSE | Upregulated |
| -0,213163567 | 0,913433693 | CD79B | FALSE | Downregulated |
| -0,223611681 | 0,913433693 | DEFB1 | FALSE | Downregulated |
| 0,067762126 | 0,913433693 | FAM120B | FALSE | Upregulated |
| -0,16358774 | 0,913433693 | FAM212B-AS1 | FALSE | Downregulated |
| 0,100058793 | 0,913433693 | ISPD-AS1 | FALSE | Upregulated |
| -0,244520394 | 0,913433693 | LINC00311 | FALSE | Downregulated |
| 0,110647337 | 0,913433693 | LOC100133331 | FALSE | Upregulated |
| 0,090486593 | 0,913433693 | LOC101928499 | FALSE | Upregulated |
| -0,249465535 | 0,913433693 | MARK2P9 | FALSE | Downregulated |
| 0,0952974 | 0,913433693 | MSANTD2 | FALSE | Upregulated |
| -0,138102387 | 0,913433693 | PKI55 | FALSE | Downregulated |
| -0,131480972 | 0,913433693 | SCRG1 | FALSE | Downregulated |
| 0,082516065 | 0,913433693 | SMYD5 | FALSE | Upregulated |
| -0,055969012 | 0,913433693 | SSH1 | FALSE | Downregulated |
| 0,135031027 | 0,913433693 | SYF2 | FALSE | Upregulated |
| -0,190362004 | 0,913433693 | THSD1 | FALSE | Downregulated |
| -0,104101888 | 0,913433693 | TMEM86A | FALSE | Downregulated |
| 0,083226095 | 0,913433693 | ZFYVE27 | FALSE | Upregulated |
| 0,110240202 | 0,913433693 | ZNF589 | FALSE | Upregulated |
| -0,112877374 | 0,913433693 | ZNF703 | FALSE | Downregulated |
| 0,086019539 | 0,913752919 | ARPC5L | FALSE | Upregulated |
| -0,091525462 | 0,913830986 | KIAA0586 | FALSE | Downregulated |
| 0,072270041 | 0,913910433 | KIAA1430 | FALSE | Upregulated |
| 0,106787778 | 0,913986794 | KIF11 | FALSE | Upregulated |
| 0,167053574 | 0,91439059 | LOC439994 | FALSE | Upregulated |
| -0,058474549 | 0,91439059 | RNF126 | FALSE | Downregulated |
| -0,116181234 | 0,91439059 | SAP25 | FALSE | Downregulated |
| -0,058065863 | 0,914580039 | TADA2B | FALSE | Downregulated |
| 0,049703242 | 0,914673924 | MFSD12 | FALSE | Upregulated |
| 0,12717519 | 0,914730222 | DCDC2B | FALSE | Upregulated |

| | | | | |
|---|---|---|---|---|
| -0,038236521 | 0,915070973 | TOB2 | FALSE | Downregulated |
| -0,053855872 | 0,915100188 | LOC101929844 | FALSE | Downregulated |
| -0,188025668 | 0,91525444 | FOXO3B | FALSE | Downregulated |
| 0,197203909 | 0,915334318 | MED30 | FALSE | Upregulated |
| 0,030730159 | 0,915394561 | FARP1 | FALSE | Upregulated |
| 0,081108856 | 0,915781833 | TRIM55 | FALSE | Upregulated |
| 0,088595162 | 0,915911261 | INPP1 | FALSE | Upregulated |
| 0,199908696 | 0,915960641 | AURKAPS1 | FALSE | Upregulated |
| 0,11144417 | 0,915960641 | GLB1L | FALSE | Upregulated |
| 0,052175716 | 0,915960641 | HERPUD1 | FALSE | Upregulated |
| 0,060320896 | 0,915960641 | LOC102724484 | FALSE | Upregulated |
| -0,066011207 | 0,915960641 | ZBTB17 | FALSE | Downregulated |
| 0,076245903 | 0,91596337 | PRR14 | FALSE | Upregulated |
| -0,130926099 | 0,916075661 | NUDT7 | FALSE | Downregulated |
| -0,059362449 | 0,916075661 | TRAPPC11 | FALSE | Downregulated |
| 0,098562369 | 0,916104199 | ANO7 | FALSE | Upregulated |
| -0,030548844 | 0,916241095 | CALD1 | FALSE | Downregulated |
| 0,101570515 | 0,916241095 | FXYD5 | FALSE | Upregulated |
| 0,090032081 | 0,916241095 | HNRNPLL | FALSE | Upregulated |
| 0,268460978 | 0,916241095 | LOC101927082 | FALSE | Upregulated |
| -0,080545411 | 0,916241095 | PLCD1 | FALSE | Downregulated |
| -0,088360976 | 0,916241095 | PLXDC2 | FALSE | Downregulated |
| -0,203842352 | 0,916241095 | PPIEL | FALSE | Downregulated |
| 0,300477363 | 0,916241095 | RP13-228J13.1 | FALSE | Upregulated |
| -0,138351097 | 0,916241095 | WRAP73 | FALSE | Downregulated |
| -0,133225724 | 0,916241095 | ZNF521 | FALSE | Downregulated |
| 0,336514977 | 0,91629174 | PPAN-P2RY11 | FALSE | Upregulated |
| -0,122309135 | 0,91629174 | PPIH | FALSE | Downregulated |
| 0,081835938 | 0,91629174 | TOE1 | FALSE | Upregulated |
| 0,081335464 | 0,916331768 | BTBD7 | FALSE | Upregulated |
| -0,128187262 | 0,916331768 | LOC440390 | FALSE | Downregulated |
| 0,038772151 | 0,916331768 | OSBP | FALSE | Upregulated |
| 0,071107684 | 0,916331768 | PDS5B | FALSE | Upregulated |
| -0,080413812 | 0,916331768 | QTRT1 | FALSE | Downregulated |
| -0,077387158 | 0,916331768 | RAPGEF6 | FALSE | Downregulated |
| 0,371413475 | 0,916331768 | TCEB3C | FALSE | Upregulated |
| 0,142596758 | 0,91636005 | SPDYE4 | FALSE | Upregulated |
| -0,067182967 | 0,916370416 | GAS2L1 | FALSE | Downregulated |
| -0,193122673 | 0,916370416 | ZNF256 | FALSE | Downregulated |
| 0,07002405 | 0,91641088 | YTHDC2 | FALSE | Upregulated |
| -0,145196526 | 0,916412961 | CYYR1 | FALSE | Downregulated |
| 0,047837658 | 0,916516022 | SETDB1 | FALSE | Upregulated |
| 0,197711327 | 0,916707887 | TMEM241 | FALSE | Upregulated |
| 0,096196122 | 0,916839225 | ATP8A1 | FALSE | Upregulated |
| 0,059946401 | 0,916839225 | BCL6 | FALSE | Upregulated |
| -0,228206612 | 0,916839225 | ECE2 | FALSE | Downregulated |
| -0,138395327 | 0,916839225 | FBXO24 | FALSE | Downregulated |
| 0,126493623 | 0,916839225 | PTCD2 | FALSE | Upregulated |
| -0,059725076 | 0,916839225 | SLC25A20 | FALSE | Downregulated |
| -0,108625941 | 0,916839225 | ZNF347 | FALSE | Downregulated |
| 0,080272892 | 0,916855308 | JMY | FALSE | Upregulated |

| | | | | |
|---|---|---|---|---|
| 0,055210276 | 0,916999181 | PEX26 | FALSE | Upregulated |
| -0,06828089 | 0,917120695 | RPAP1 | FALSE | Downregulated |
| -0,039324846 | 0,917547688 | CIAO1 | FALSE | Downregulated |
| 0,110385078 | 0,917547688 | FADS1 | FALSE | Upregulated |
| -0,156700638 | 0,917547688 | HOXB4 | FALSE | Downregulated |
| 0,101179169 | 0,917547688 | LOC101928281 | FALSE | Upregulated |
| 0,078921237 | 0,917547688 | NXN | FALSE | Upregulated |
| 0,130461916 | 0,917547688 | RASSF8 | FALSE | Upregulated |
| -0,164136369 | 0,917547688 | TM4SF18 | FALSE | Downregulated |
| 0,128071752 | 0,917676321 | ISY1 | FALSE | Upregulated |
| 0,078030105 | 0,917676977 | PRPS1 | FALSE | Upregulated |
| 0,12955907 | 0,917873791 | LOC101060398 | FALSE | Upregulated |
| 0,097358723 | 0,917873791 | VSTM4 | FALSE | Upregulated |
| -0,157953567 | 0,917880333 | MED20 | FALSE | Downregulated |
| 0,120261117 | 0,917992769 | NPHP4 | FALSE | Upregulated |
| 0,235862482 | 0,91802765 | CEACAM22P | FALSE | Upregulated |
| -0,050984393 | 0,91802765 | EAF1 | FALSE | Downregulated |
| -0,04289289 | 0,91802765 | GUCD1 | FALSE | Downregulated |
| -0,046974812 | 0,91802765 | KDM1A | FALSE | Downregulated |
| -0,160537335 | 0,91802765 | LINC01011 | FALSE | Downregulated |
| 0,058776439 | 0,91802765 | LOC102723582 | FALSE | Upregulated |
| 0,080065908 | 0,91802765 | MCOLN1 | FALSE | Upregulated |
| 0,046964408 | 0,91802765 | MGRN1 | FALSE | Upregulated |
| -0,072682637 | 0,91802765 | SPCS2 | FALSE | Downregulated |
| -0,06301535 | 0,91802765 | ZNF586 | FALSE | Downregulated |
| 0,085264917 | 0,918282418 | ITPA | FALSE | Upregulated |
| -0,108621599 | 0,918498644 | DNAJC17 | FALSE | Downregulated |
| -0,036832898 | 0,918498644 | MYO5B | FALSE | Downregulated |
| -0,172612515 | 0,918498644 | ZNF184 | FALSE | Downregulated |
| -0,138469206 | 0,918581498 | MYOT | FALSE | Downregulated |
| 0,09382317 | 0,918611404 | BRWD1-IT2 | FALSE | Upregulated |
| 0,03303884 | 0,918611404 | CHERP | FALSE | Upregulated |
| -0,203791499 | 0,918618268 | LOC102723465 | FALSE | Downregulated |
| -0,105941917 | 0,918618268 | LOC102724254 | FALSE | Downregulated |
| -0,046872055 | 0,918618268 | MZT2B | FALSE | Downregulated |
| -0,184917799 | 0,91866851 | C17orf49 | FALSE | Downregulated |
| -0,108045459 | 0,91866851 | LOC729603 | FALSE | Downregulated |
| 0,047069157 | 0,91866851 | LPCAT3 | FALSE | Upregulated |
| -0,034056844 | 0,91866851 | TRIM44 | FALSE | Downregulated |
| -0,131089935 | 0,918716078 | CKB | FALSE | Downregulated |
| 0,160172383 | 0,918716078 | LOC101928628 | FALSE | Upregulated |
| -0,30384344 | 0,918716078 | SEPT5 | FALSE | Downregulated |
| -0,09825629 | 0,918727013 | ZNF593 | FALSE | Downregulated |
| -0,116476505 | 0,918731981 | EMC2 | FALSE | Downregulated |
| -0,092417484 | 0,918801525 | NUDCD2 | FALSE | Downregulated |
| -0,105795147 | 0,918801525 | WDR12 | FALSE | Downregulated |
| -0,064909643 | 0,918821818 | IMP3 | FALSE | Downregulated |
| 0,109868795 | 0,918861098 | ERGIC2 | FALSE | Upregulated |
| 0,070626467 | 0,918861098 | SFI1 | FALSE | Upregulated |
| -0,187380949 | 0,918937568 | ASB15 | FALSE | Downregulated |
| 0,075496926 | 0,918937568 | CLDN15 | FALSE | Upregulated |

| | | | | |
|---|---|---|---|---|
| 0,0351342 | 0,918937568 | LAMP1 | FALSE | Upregulated |
| -0,140131951 | 0,918937568 | LINC00999 | FALSE | Downregulated |
| 0,091144765 | 0,918937568 | LOC729658 | FALSE | Upregulated |
| -0,131876699 | 0,918937568 | NBR2 | FALSE | Downregulated |
| 0,055000256 | 0,918937568 | NOC2L | FALSE | Upregulated |
| 0,162317054 | 0,918937568 | PIGC | FALSE | Upregulated |
| 0,262422133 | 0,918937568 | PSRC1 | FALSE | Upregulated |
| 0,047901508 | 0,918937568 | SS18 | FALSE | Upregulated |
| -0,115695033 | 0,918937568 | THAP6 | FALSE | Downregulated |
| -0,06804526 | 0,918937568 | ZNF776 | FALSE | Downregulated |
| 0,201576915 | 0,919065032 | ACER2 | FALSE | Upregulated |
| -0,128491288 | 0,919065032 | C6orf132 | FALSE | Downregulated |
| 0,09688949 | 0,919065032 | CCDC109B | FALSE | Upregulated |
| -0,055153036 | 0,919065032 | ELK1 | FALSE | Downregulated |
| 0,048257186 | 0,919065032 | EYS | FALSE | Upregulated |
| -0,185284134 | 0,919065032 | GMNN | FALSE | Downregulated |
| 0,228663548 | 0,919065032 | LINC00941 | FALSE | Upregulated |
| 0,095010528 | 0,919065032 | LOC101926985 | FALSE | Upregulated |
| -0,190051213 | 0,919065032 | LOC102723679 | FALSE | Downregulated |
| 0,234498391 | 0,919065032 | MAMDC2-AS1 | FALSE | Upregulated |
| -0,058362899 | 0,919065032 | NAA15 | FALSE | Downregulated |
| 0,047252256 | 0,919065032 | NDUFS2 | FALSE | Upregulated |
| -0,046044235 | 0,919065032 | NFKBIA | FALSE | Downregulated |
| 0,07951188 | 0,919065032 | NLN | FALSE | Upregulated |
| 0,073164606 | 0,919065032 | NOL7 | FALSE | Upregulated |
| 0,193493623 | 0,919065032 | NRG3 | FALSE | Upregulated |
| -0,080801913 | 0,919065032 | NUDT16P1 | FALSE | Downregulated |
| 0,06665136 | 0,919065032 | OSGIN1 | FALSE | Upregulated |
| 0,13884608 | 0,919065032 | RP11-112H10.4 | FALSE | Upregulated |
| -0,062829727 | 0,919065032 | TMCO1 | FALSE | Downregulated |
| 0,083482695 | 0,919074153 | APAF1 | FALSE | Upregulated |
| -0,196304768 | 0,919074153 | CEACAM8 | FALSE | Downregulated |
| 0,185442386 | 0,919074153 | GCM2 | FALSE | Upregulated |
| -0,073276728 | 0,919074153 | LOC101928555 | FALSE | Downregulated |
| 0,221286509 | 0,919074153 | WDR16 | FALSE | Upregulated |
| -0,095354691 | 0,919103071 | FAM184A | FALSE | Downregulated |
| 0,037655575 | 0,919103071 | SUGP2 | FALSE | Upregulated |
| -0,047431846 | 0,919195044 | CTC1 | FALSE | Downregulated |
| -0,23145293 | 0,919195044 | MACC1 | FALSE | Downregulated |
| -0,050748555 | 0,919419533 | SNX18 | FALSE | Downregulated |
| -0,142339015 | 0,919704218 | C8orf58 | FALSE | Downregulated |
| -0,084319372 | 0,919704218 | CBFA2T3 | FALSE | Downregulated |
| 0,106308568 | 0,919704218 | CENPJ | FALSE | Upregulated |
| -0,140488309 | 0,919704218 | DDX12P | FALSE | Downregulated |
| 0,095643114 | 0,919704218 | ETAA1 | FALSE | Upregulated |
| -0,170430724 | 0,919704218 | LOC101927300 | FALSE | Downregulated |
| 0,047097635 | 0,919704218 | RHOT2 | FALSE | Upregulated |
| 0,11220787 | 0,919704218 | RRAGB | FALSE | Upregulated |
| 0,14694479 | 0,919704218 | TMEM121 | FALSE | Upregulated |
| -0,052202543 | 0,919763247 | ERVW-1 | FALSE | Downregulated |
| 0,163283485 | 0,919823286 | USP9Y | FALSE | Upregulated |

| | | | | |
|---|---|---|---|---|
| -0,061488746 | 0,919900237 | MYSM1 | FALSE | Downregulated |
| 0,169166678 | 0,919979153 | NUFIP1 | FALSE | Upregulated |
| 0,050640312 | 0,920253933 | FKBP1A | FALSE | Upregulated |
| 0,246953279 | 0,920253933 | RGS4 | FALSE | Upregulated |
| 0,173305977 | 0,920253933 | SH3TC2 | FALSE | Upregulated |
| -0,163682533 | 0,920253933 | SS18L2 | FALSE | Downregulated |
| 0,052996423 | 0,920523802 | WDR7 | FALSE | Upregulated |
| 0,041976403 | 0,920525026 | C19orf43 | FALSE | Upregulated |
| -1,233977983 | 0,920525026 | DES | FALSE | Downregulated |
| -0,346022879 | 0,920525026 | FAM156A | FALSE | Downregulated |
| 0,154698515 | 0,920525026 | FRS3 | FALSE | Upregulated |
| -0,16981883 | 0,920525026 | LOC100506571 | FALSE | Downregulated |
| -0,032516789 | 0,920525026 | WAC | FALSE | Downregulated |
| -0,153155081 | 0,920554564 | LOC101929115 | FALSE | Downregulated |
| -0,296277479 | 0,920642378 | LOC102723580 | FALSE | Downregulated |
| -0,104218492 | 0,920800491 | MIR181A2HG | FALSE | Downregulated |
| -0,043494599 | 0,920929026 | RPE | FALSE | Downregulated |
| -0,102412909 | 0,920929026 | SMUG1 | FALSE | Downregulated |
| -0,077356923 | 0,920929026 | THTPA | FALSE | Downregulated |
| 0,109674738 | 0,921032215 | TNFSF13 | FALSE | Upregulated |
| 0,026921128 | 0,921337682 | EIF4G1 | FALSE | Upregulated |
| 0,055165679 | 0,921522812 | ANKLE2 | FALSE | Upregulated |
| 0,095847042 | 0,921522812 | MCU | FALSE | Upregulated |
| -0,063785838 | 0,921522812 | MMS19 | FALSE | Downregulated |
| 0,186109306 | 0,921522812 | NPIPB15 | FALSE | Upregulated |
| -0,253577938 | 0,921522812 | PCDH20 | FALSE | Downregulated |
| -0,095238519 | 0,921522812 | PFDN2 | FALSE | Downregulated |
| -0,126218126 | 0,921522812 | SPAG8 | FALSE | Downregulated |
| 0,091291799 | 0,921567078 | BRCC3 | FALSE | Upregulated |
| -0,132105654 | 0,921567078 | ELTD1 | FALSE | Downregulated |
| -0,105966407 | 0,921567078 | FAM135A | FALSE | Downregulated |
| -0,136289724 | 0,921567078 | KLHL10 | FALSE | Downregulated |
| 0,157850993 | 0,921567078 | PCP2 | FALSE | Upregulated |
| 0,094413314 | 0,921567078 | RP11-41O4.2 | FALSE | Upregulated |
| 0,073389327 | 0,921567078 | UBE2M | FALSE | Upregulated |
| -0,121396698 | 0,921567078 | ZDHHC11 | FALSE | Downregulated |
| 0,117700935 | 0,921643975 | NPAS2 | FALSE | Upregulated |
| -0,032098086 | 0,921654145 | TGOLN2 | FALSE | Downregulated |
| 0,138166535 | 0,921755476 | EGFLAM | FALSE | Upregulated |
| -0,167918418 | 0,921823041 | KNSTRN | FALSE | Downregulated |
| 0,114636453 | 0,921823736 | ARHGAP18 | FALSE | Upregulated |
| -0,253143856 | 0,921823981 | POU3F2 | FALSE | Downregulated |
| -0,383597806 | 0,921824627 | LOC102724560 | FALSE | Downregulated |
| 0,094178124 | 0,921824627 | PARP2 | FALSE | Upregulated |
| 0,060543852 | 0,921824627 | SMC5 | FALSE | Upregulated |
| -0,068646566 | 0,921824627 | SPAG7 | FALSE | Downregulated |
| -0,099782413 | 0,921824627 | SPATA5 | FALSE | Downregulated |
| 0,124306958 | 0,921835827 | IL22RA1 | FALSE | Upregulated |
| 0,200616772 | 0,922195131 | LOC100131107 | FALSE | Upregulated |
| -0,056516607 | 0,922265935 | DLG4 | FALSE | Downregulated |
| -0,176155436 | 0,922288242 | LOC101927198 | FALSE | Downregulated |

| | | | | |
|---|---|---|---|---|
| 0,042134505 | 0,922288242 | ZNF664 | FALSE | Upregulated |
| 0,097675183 | 0,922397511 | AC018705.5 | FALSE | Upregulated |
| -0,041438479 | 0,922397511 | ADCY6 | FALSE | Downregulated |
| 0,107406285 | 0,922397511 | DIRC2 | FALSE | Upregulated |
| 0,07554694 | 0,922397511 | DLAT | FALSE | Upregulated |
| 0,056636178 | 0,922397511 | IARS | FALSE | Upregulated |
| -0,116564876 | 0,922397511 | MRPS9 | FALSE | Downregulated |
| 0,043893958 | 0,922397511 | NSUN2 | FALSE | Upregulated |
| 0,087655566 | 0,922397511 | RUNDC1 | FALSE | Upregulated |
| 0,178216889 | 0,922397511 | SLC16A1-AS1 | FALSE | Upregulated |
| 0,035737522 | 0,922479192 | FOXK1 | FALSE | Upregulated |
| -0,081597888 | 0,922568374 | LOC102724683 | FALSE | Downregulated |
| 0,111030341 | 0,922601004 | EMC6 | FALSE | Upregulated |
| 0,13509262 | 0,922632134 | FBXL8 | FALSE | Upregulated |
| -0,159428769 | 0,922632134 | LIPT2 | FALSE | Downregulated |
| 0,078724368 | 0,922632134 | TRIM37 | FALSE | Upregulated |
| -0,108382258 | 0,922632134 | VANGL1 | FALSE | Downregulated |
| -0,119777024 | 0,922632134 | ZNF136 | FALSE | Downregulated |
| -0,048424749 | 0,922671676 | ANKRD23 | FALSE | Downregulated |
| 0,043270545 | 0,922671676 | C16orf72 | FALSE | Upregulated |
| -0,119657917 | 0,922671676 | CD274 | FALSE | Downregulated |
| -0,093250546 | 0,922671676 | CIAPIN1 | FALSE | Downregulated |
| -0,066956722 | 0,922671676 | LOC102724930 | FALSE | Downregulated |
| -0,227079107 | 0,922671676 | MORF4L2-AS1 | FALSE | Downregulated |
| -0,146140625 | 0,922671676 | NTRK2 | FALSE | Downregulated |
| 0,089485791 | 0,922671676 | SLC25A45 | FALSE | Upregulated |
| -0,089414583 | 0,922687337 | ZNF470 | FALSE | Downregulated |
| -0,047464139 | 0,922734497 | SAFB2 | FALSE | Downregulated |
| 0,122923454 | 0,922785458 | LINC00652 | FALSE | Upregulated |
| 0,158084679 | 0,922798949 | DUSP18 | FALSE | Upregulated |
| 0,078193698 | 0,922798949 | SPTLC3 | FALSE | Upregulated |
| 0,04599805 | 0,923012789 | TRIM66 | FALSE | Upregulated |
| -0,454518546 | 0,923024228 | C1QTNF3-AMACR | FALSE | Downregulated |
| -0,061289065 | 0,923149971 | SIRT2 | FALSE | Downregulated |
| 0,10162013 | 0,923268363 | C15orf40 | FALSE | Upregulated |
| -0,04313365 | 0,92332612 | SDC4 | FALSE | Downregulated |
| -0,050800066 | 0,923355843 | ADNP | FALSE | Downregulated |
| 0,047618026 | 0,923355843 | ARHGEF11 | FALSE | Upregulated |
| -0,094879238 | 0,923355843 | ZNF414 | FALSE | Downregulated |
| 0,046715412 | 0,923682417 | CPD | FALSE | Upregulated |
| 0,158694521 | 0,92372917 | HEPH | FALSE | Upregulated |
| -0,048635184 | 0,923814964 | SCARNA10 | FALSE | Downregulated |
| -0,07128532 | 0,923858552 | INS-IGF2 | FALSE | Downregulated |
| 0,097572357 | 0,923878923 | ERG | FALSE | Upregulated |
| -0,16704934 | 0,923878923 | OR7E14P | FALSE | Downregulated |
| -0,099712144 | 0,923878923 | SACM1L | FALSE | Downregulated |
| -0,089595457 | 0,924056882 | C14orf169 | FALSE | Downregulated |
| 0,048538651 | 0,924056882 | CABLES1 | FALSE | Upregulated |
| 0,062678614 | 0,924056882 | E4F1 | FALSE | Upregulated |
| -0,062137444 | 0,924056882 | FAM134B | FALSE | Downregulated |
| -0,177336794 | 0,924056882 | GABRE | FALSE | Downregulated |

| | | | | |
|---|---|---|---|---|
| 0,091003453 | 0,924056882 | KIRREL | FALSE | Upregulated |
| 0,076138057 | 0,924056882 | MGAT4A | FALSE | Upregulated |
| 0,128926511 | 0,924056882 | WASH3P | FALSE | Upregulated |
| 0,034104097 | 0,924056882 | ZC3H4 | FALSE | Upregulated |
| 0,126235057 | 0,924322121 | NGDN | FALSE | Upregulated |
| -0,084117212 | 0,924440871 | ELOVL6 | FALSE | Downregulated |
| -0,140762404 | 0,924440871 | NAE1 | FALSE | Downregulated |
| -0,076039049 | 0,924440871 | NHP2L1 | FALSE | Downregulated |
| 0,106084119 | 0,924784211 | ZNF296 | FALSE | Upregulated |
| 0,142305913 | 0,925080552 | KIAA1586 | FALSE | Upregulated |
| 0,042592014 | 0,925080552 | SASH1 | FALSE | Upregulated |
| -0,066647848 | 0,925098637 | DAP3 | FALSE | Downregulated |
| -0,058037457 | 0,925098637 | OXSR1 | FALSE | Downregulated |
| -0,060874713 | 0,925098637 | WDR13 | FALSE | Downregulated |
| -0,085363503 | 0,925128408 | RBM22 | FALSE | Downregulated |
| -0,08457922 | 0,925128408 | TGS1 | FALSE | Downregulated |
| -0,084297228 | 0,925272877 | SMYD2 | FALSE | Downregulated |
| 0,068992185 | 0,925312551 | AFG3L1P | FALSE | Upregulated |
| 0,193157835 | 0,925312551 | EBI3 | FALSE | Upregulated |
| 0,075030725 | 0,925312551 | RASGRP3 | FALSE | Upregulated |
| 0,094737665 | 0,925362045 | ZNF121 | FALSE | Upregulated |
| -0,062129256 | 0,925460425 | KDM7A | FALSE | Downregulated |
| -0,189617523 | 0,925550137 | CD33 | FALSE | Downregulated |
| 0,149332261 | 0,925619586 | SRFBP1 | FALSE | Upregulated |
| -0,092530673 | 0,925619586 | ZSCAN12 | FALSE | Downregulated |
| -0,034434167 | 0,925665228 | H6PD | FALSE | Downregulated |
| 0,050594786 | 0,925665228 | LEPROT | FALSE | Upregulated |
| -0,141532275 | 0,925665228 | LOC101927308 | FALSE | Downregulated |
| -0,109673153 | 0,926046734 | LOC102723469 | FALSE | Downregulated |
| -0,031825742 | 0,926224331 | SF3A1 | FALSE | Downregulated |
| 0,051266289 | 0,926465445 | AKT3 | FALSE | Upregulated |
| -0,25064897 | 0,926900924 | LOC101929047 | FALSE | Downregulated |
| -0,089761496 | 0,927046757 | HAUS5 | FALSE | Downregulated |
| 0,090626637 | 0,927046757 | ZNF704 | FALSE | Upregulated |
| 0,05707339 | 0,92707207 | CAMSAP1 | FALSE | Upregulated |
| 0,103617463 | 0,927313363 | DUSP11 | FALSE | Upregulated |
| 0,370991578 | 0,927313363 | FNTB | FALSE | Upregulated |
| 0,193800518 | 0,927313363 | LOC101929057 | FALSE | Upregulated |
| 0,380098247 | 0,927313363 | LOC102724370 | FALSE | Upregulated |
| -0,16958716 | 0,927313363 | LOC102724507 | FALSE | Downregulated |
| -0,061410745 | 0,927313363 | RNF14 | FALSE | Downregulated |
| 0,07592616 | 0,927313363 | RP1-256G22.2 | FALSE | Upregulated |
| 0,069359326 | 0,927367879 | CPNE2 | FALSE | Upregulated |
| -0,113096641 | 0,927367879 | KCNK6 | FALSE | Downregulated |
| -0,07177898 | 0,927447179 | BCL2L11 | FALSE | Downregulated |
| 0,047670798 | 0,927447179 | NUP153 | FALSE | Upregulated |
| -0,206935033 | 0,92745977 | CYP26A1 | FALSE | Downregulated |
| 0,06366891 | 0,92745977 | DNAJA2 | FALSE | Upregulated |
| -0,063505751 | 0,92745977 | GUSBP11 | FALSE | Downregulated |
| -0,075234042 | 0,92745977 | LOC643072 | FALSE | Downregulated |
| 0,12246719 | 0,927477 | CCDC36 | FALSE | Upregulated |

| | | | | |
|---|---|---|---|---|
| -0,166760564 | 0,927477 | LOC101928635 | FALSE | Downregulated |
| 0,033236774 | 0,927557307 | ADD1 | FALSE | Upregulated |
| -0,178987169 | 0,927557307 | LOC728392 | FALSE | Downregulated |
| 0,229876611 | 0,927649 | LOC100996758 | FALSE | Upregulated |
| 0,14212772 | 0,927649 | SELM | FALSE | Upregulated |
| -0,069163005 | 0,927842996 | CCNT2-AS1 | FALSE | Downregulated |
| 0,044672006 | 0,927842996 | CUL4A | FALSE | Upregulated |
| -0,064137488 | 0,927842996 | DPH6 | FALSE | Downregulated |
| -0,099477007 | 0,927842996 | EPPK1 | FALSE | Downregulated |
| -0,042068531 | 0,927842996 | LOC101929469 | FALSE | Downregulated |
| -0,138838969 | 0,927842996 | LRRC34 | FALSE | Downregulated |
| -0,160662132 | 0,927842996 | POLR2M | FALSE | Downregulated |
| 0,170395897 | 0,927857401 | CMTM2 | FALSE | Upregulated |
| -0,074021118 | 0,927857401 | RRAGA | FALSE | Downregulated |
| -0,119744145 | 0,927857401 | TMEM252 | FALSE | Downregulated |
| 0,051447016 | 0,927857401 | UBAP1 | FALSE | Upregulated |
| -0,164872946 | 0,927947887 | FASTKD3 | FALSE | Downregulated |
| 0,062789618 | 0,927947887 | PROCR | FALSE | Upregulated |
| 0,036666929 | 0,928197226 | CRKL | FALSE | Upregulated |
| 0,081499366 | 0,928354435 | MRRF | FALSE | Upregulated |
| 0,102415742 | 0,928486654 | CXCR3 | FALSE | Upregulated |
| -0,270836873 | 0,928486654 | FLJ33581 | FALSE | Downregulated |
| -0,043906683 | 0,928486654 | RBM4 | FALSE | Downregulated |
| -0,113976409 | 0,928486654 | TMA16 | FALSE | Downregulated |
| 0,111996188 | 0,928486654 | WDR89 | FALSE | Upregulated |
| -0,135731057 | 0,928651431 | BAI3 | FALSE | Downregulated |
| 0,107059595 | 0,928929279 | LOC101926918 | FALSE | Upregulated |
| 0,047689325 | 0,929001148 | ATP5G3 | FALSE | Upregulated |
| 0,048639583 | 0,929001148 | BBX | FALSE | Upregulated |
| -0,073969328 | 0,929001148 | CD5L | FALSE | Downregulated |
| 0,111430208 | 0,929001148 | FDX1L | FALSE | Upregulated |
| 0,069852036 | 0,929001148 | FHOD1 | FALSE | Upregulated |
| -0,058904006 | 0,929001148 | MBD2 | FALSE | Downregulated |
| -0,032363959 | 0,929001148 | PTBP1 | FALSE | Downregulated |
| 0,045978479 | 0,929001148 | RNPS1 | FALSE | Upregulated |
| -0,179493578 | 0,929001148 | SEC61G | FALSE | Downregulated |
| 0,217546015 | 0,929001148 | SGCA | FALSE | Upregulated |
| -0,083612747 | 0,929001148 | YDJC | FALSE | Downregulated |
| 0,094209907 | 0,929001148 | ZNF182 | FALSE | Upregulated |
| -0,056403422 | 0,929086082 | BZW1 | FALSE | Downregulated |
| 0,11383237 | 0,929086082 | CEP85L | FALSE | Upregulated |
| 0,044449465 | 0,929086082 | DCST2 | FALSE | Upregulated |
| -0,12898565 | 0,929086082 | LOC101927585 | FALSE | Downregulated |
| 0,234408217 | 0,929086082 | SPDL1 | FALSE | Upregulated |
| 0,085276801 | 0,929086082 | TRIM11 | FALSE | Upregulated |
| 0,090938233 | 0,929086082 | ZNF23 | FALSE | Upregulated |
| 0,111636762 | 0,92912155 | ZNF341 | FALSE | Upregulated |
| -0,316746813 | 0,929127679 | HIST2H2AA3 | FALSE | Downregulated |
| 0,062426311 | 0,929127679 | HIST2H2AC | FALSE | Upregulated |
| 0,085021758 | 0,929345594 | ASPHD2 | FALSE | Upregulated |
| 0,106216828 | 0,929414964 | CCBL2 | FALSE | Upregulated |

| | | | | |
|---|---|---|---|---|
| 0,189480029 | 0,929527127 | ERCC8 | FALSE | Upregulated |
| 0,127254413 | 0,929527127 | TCEANC2 | FALSE | Upregulated |
| 0,156291757 | 0,929575259 | LINC00869 | FALSE | Upregulated |
| 0,102733249 | 0,92968923 | NSRP1 | FALSE | Upregulated |
| 0,067231134 | 0,92968923 | TBC1D16 | FALSE | Upregulated |
| -0,049426482 | 0,929870189 | TOR3A | FALSE | Downregulated |
| -0,039593197 | 0,929951796 | TGFBR2 | FALSE | Downregulated |
| -0,040691445 | 0,929951796 | TNKS2 | FALSE | Downregulated |
| 0,097352722 | 0,930036552 | BTBD3 | FALSE | Upregulated |
| 0,129619754 | 0,930036552 | DNAJC3-AS1 | FALSE | Upregulated |
| 0,106897933 | 0,930036552 | NAA40 | FALSE | Upregulated |
| 0,129924921 | 0,930036552 | RP11-147N17.1 | FALSE | Upregulated |
| 0,272402468 | 0,930163401 | ATP1A2 | FALSE | Upregulated |
| 0,119545344 | 0,930163401 | LILRA6 | FALSE | Upregulated |
| -0,11247192 | 0,930187821 | RASA2 | FALSE | Downregulated |
| 0,11031975 | 0,930189247 | TRMT13 | FALSE | Upregulated |
| -0,046427899 | 0,930229238 | NUDT3 | FALSE | Downregulated |
| -0,055988936 | 0,930441892 | NQO2 | FALSE | Downregulated |
| -0,055885425 | 0,930542216 | AQR | FALSE | Downregulated |
| 0,136470099 | 0,930542216 | CLIP4 | FALSE | Upregulated |
| -0,093168785 | 0,930542216 | L3HYPDH | FALSE | Downregulated |
| -0,131545617 | 0,930542216 | P4HA3 | FALSE | Downregulated |
| -0,072905666 | 0,930542216 | PNKP | FALSE | Downregulated |
| 0,079382774 | 0,930542216 | SSB | FALSE | Upregulated |
| -0,172540849 | 0,930542216 | TTC21A | FALSE | Downregulated |
| -0,049345415 | 0,930542216 | ZBTB18 | FALSE | Downregulated |
| -0,091838352 | 0,930553146 | RAMP1 | FALSE | Downregulated |
| 0,071995879 | 0,930587133 | ARL13A | FALSE | Upregulated |
| 0,072457609 | 0,930587133 | CCDC71 | FALSE | Upregulated |
| 0,10614289 | 0,930587133 | PLD6 | FALSE | Upregulated |
| 0,05781142 | 0,930587133 | RTF1 | FALSE | Upregulated |
| 0,035490493 | 0,930597992 | ABHD2 | FALSE | Upregulated |
| -0,155873986 | 0,930597992 | GPR143 | FALSE | Downregulated |
| -0,059880455 | 0,930597992 | PSMB6 | FALSE | Downregulated |
| 0,089891497 | 0,930597992 | SCD | FALSE | Upregulated |
| 0,12570155 | 0,930597992 | SLC25A12 | FALSE | Upregulated |
| 0,051920011 | 0,930610887 | ABCE1 | FALSE | Upregulated |
| 0,03645895 | 0,930610887 | CREBL2 | FALSE | Upregulated |
| -0,129094386 | 0,930610887 | TREML1 | FALSE | Downregulated |
| 0,187132633 | 0,930617433 | TGFB2 | FALSE | Upregulated |
| 0,071625453 | 0,930635278 | FBXO44 | FALSE | Upregulated |
| 0,100589942 | 0,930635278 | RP11-665G4.1 | FALSE | Upregulated |
| 0,064928092 | 0,930865466 | BCS1L | FALSE | Upregulated |
| -0,084893251 | 0,930865466 | NR6A1 | FALSE | Downregulated |
| 0,17900262 | 0,930865466 | RPL21P28 | FALSE | Upregulated |
| -0,045040103 | 0,930865466 | SFT2D3 | FALSE | Downregulated |
| 0,057510608 | 0,930883894 | C9orf173 | FALSE | Upregulated |
| -0,079777783 | 0,930883894 | CLK4 | FALSE | Downregulated |
| 0,069681281 | 0,930883894 | LOC102724364 | FALSE | Upregulated |
| 0,037538146 | 0,930883894 | NAPEPLD | FALSE | Upregulated |
| -0,086119833 | 0,931028836 | EME2 | FALSE | Downregulated |

| | | | | |
|---|---|---|---|---|
| 0,16815721 | 0,931050143 | EIF3C | FALSE | Upregulated |
| -0,155164211 | 0,931053978 | LOC102659288 | FALSE | Downregulated |
| -0,068688331 | 0,931111835 | CXADR | FALSE | Downregulated |
| 0,067590655 | 0,931159402 | ATP6AP1 | FALSE | Upregulated |
| 0,102407564 | 0,931159402 | C3orf38 | FALSE | Upregulated |
| -0,051979327 | 0,931159402 | CCM2 | FALSE | Downregulated |
| 0,088007962 | 0,931159402 | LHFP | FALSE | Upregulated |
| -0,118497675 | 0,931159402 | PIGK | FALSE | Downregulated |
| 0,107148925 | 0,931159402 | RABL2A | FALSE | Upregulated |
| -0,04616568 | 0,931159402 | REST | FALSE | Downregulated |
| 0,044259264 | 0,931159402 | SKIL | FALSE | Upregulated |
| -0,043604292 | 0,931159402 | ZFC3H1 | FALSE | Downregulated |
| -0,053773319 | 0,931169452 | CLVS1 | FALSE | Downregulated |
| 0,06882158 | 0,931169452 | LMO4 | FALSE | Upregulated |
| -0,118548276 | 0,931169452 | MRPL48 | FALSE | Downregulated |
| -0,127474398 | 0,931169452 | UBLCP1 | FALSE | Downregulated |
| 0,03920421 | 0,931232104 | ZNF839 | FALSE | Upregulated |
| 0,039872259 | 0,931322534 | GCFC2 | FALSE | Upregulated |
| 0,062680103 | 0,93133034 | CMTM4 | FALSE | Upregulated |
| -0,10027582 | 0,93133034 | MRPL44 | FALSE | Downregulated |
| -0,084617686 | 0,93133034 | ORC3 | FALSE | Downregulated |
| -0,054499107 | 0,93133034 | PNPT1 | FALSE | Downregulated |
| -0,04266483 | 0,93133034 | SCP2 | FALSE | Downregulated |
| -0,044095235 | 0,931472112 | APEX1 | FALSE | Downregulated |
| -0,053212258 | 0,931490476 | LOC102724535 | FALSE | Downregulated |
| -0,062182923 | 0,931490476 | LOC102724811 | FALSE | Downregulated |
| 1,024944698 | 0,931490476 | PKP1 | FALSE | Upregulated |
| 0,088012614 | 0,931490476 | PSMD12 | FALSE | Upregulated |
| -0,085783146 | 0,931490476 | RND3 | FALSE | Downregulated |
| -0,123797565 | 0,931490476 | ZFP30 | FALSE | Downregulated |
| -0,051910057 | 0,931490476 | ZNF674 | FALSE | Downregulated |
| -0,124443286 | 0,931555661 | C6orf203 | FALSE | Downregulated |
| 0,128249109 | 0,931555661 | GUCY2C | FALSE | Upregulated |
| -0,081759556 | 0,931628907 | LOC100507412 | FALSE | Downregulated |
| -0,184827468 | 0,931628907 | LOC101928669 | FALSE | Downregulated |
| 0,092545945 | 0,931628907 | LRP12 | FALSE | Upregulated |
| -0,060015806 | 0,931628907 | MAPKBP1 | FALSE | Downregulated |
| 0,131450577 | 0,931652998 | HSF2 | FALSE | Upregulated |
| -0,110944268 | 0,931652998 | UPF3B | FALSE | Downregulated |
| -0,059822615 | 0,931657396 | IFNLR1 | FALSE | Downregulated |
| 0,08603623 | 0,931695062 | KHDRBS3 | FALSE | Upregulated |
| 0,070553169 | 0,93170477 | PDCD6 | FALSE | Upregulated |
| -0,198988201 | 0,931778294 | LGR5 | FALSE | Downregulated |
| 0,17419602 | 0,931982796 | PPT2-EGFL8 | FALSE | Upregulated |
| 0,065765639 | 0,932217114 | ATAD1 | FALSE | Upregulated |
| 0,083616559 | 0,932217114 | LOC100287166 | FALSE | Upregulated |
| 0,101815058 | 0,932217114 | MSANTD4 | FALSE | Upregulated |
| 0,025095478 | 0,932217114 | PDXDC1 | FALSE | Upregulated |
| -0,069612232 | 0,932217114 | PPTC7 | FALSE | Downregulated |
| 0,095938983 | 0,932217114 | RP5-1013A22.2 | FALSE | Upregulated |
| -0,06543084 | 0,932217114 | SYPL1 | FALSE | Downregulated |

| | | | | |
|---|---|---|---|---|
| -0,067283065 | 0,932217114 | TRA2B | FALSE | Downregulated |
| 0,084526802 | 0,932474475 | CUL2 | FALSE | Upregulated |
| -0,057169488 | 0,932474475 | EXOC5 | FALSE | Downregulated |
| 0,084482128 | 0,932474475 | RNF138 | FALSE | Upregulated |
| -0,086507351 | 0,932749914 | ROGDI | FALSE | Downregulated |
| 0,131925051 | 0,933114223 | TMEM128 | FALSE | Upregulated |
| 0,273151777 | 0,933131969 | CCL15-CCL14 | FALSE | Upregulated |
| 0,04769827 | 0,933131969 | LBR | FALSE | Upregulated |
| -0,092853234 | 0,933131969 | SCART1 | FALSE | Downregulated |
| 0,196439387 | 0,933131969 | TAS2R19 | FALSE | Upregulated |
| -0,167059503 | 0,933131969 | TMEM163 | FALSE | Downregulated |
| -0,099704741 | 0,933196403 | GALNT18 | FALSE | Downregulated |
| -0,09204739 | 0,933210353 | TMEM17 | FALSE | Downregulated |
| 0,036288718 | 0,933309668 | ANO6 | FALSE | Upregulated |
| 0,049419472 | 0,933309668 | RBM4B | FALSE | Upregulated |
| 0,041480062 | 0,933417497 | CNOT3 | FALSE | Upregulated |
| 0,088883475 | 0,933417497 | FAM43A | FALSE | Upregulated |
| -0,088707987 | 0,933533779 | AARSD1 | FALSE | Downregulated |
| 0,132027195 | 0,933533779 | NSMCE2 | FALSE | Upregulated |
| -0,033119303 | 0,933632387 | APOE | FALSE | Downregulated |
| -0,043884054 | 0,933632387 | C1orf192 | FALSE | Downregulated |
| -0,038383338 | 0,933632387 | KIAA0355 | FALSE | Downregulated |
| -0,049292448 | 0,933632387 | PTPRG | FALSE | Downregulated |
| -0,094655849 | 0,933692953 | CCDC17 | FALSE | Downregulated |
| 0,09940777 | 0,933692953 | LOC101928058 | FALSE | Upregulated |
| 0,049953866 | 0,933698194 | TAB1 | FALSE | Upregulated |
| -0,07713291 | 0,933703653 | C11orf68 | FALSE | Downregulated |
| 0,134345343 | 0,933703653 | SNX7 | FALSE | Upregulated |
| 0,066534819 | 0,933731341 | LINC01128 | FALSE | Upregulated |
| -0,151221463 | 0,933731341 | MUC15 | FALSE | Downregulated |
| -0,17016634 | 0,933731341 | SERP2 | FALSE | Downregulated |
| -0,031498116 | 0,933763014 | MAP3K11 | FALSE | Downregulated |
| 0,094934138 | 0,933878173 | LOC101928794 | FALSE | Upregulated |
| -0,208287299 | 0,934176194 | LOC100287477 | FALSE | Downregulated |
| 0,039158165 | 0,934176194 | TRIP11 | FALSE | Upregulated |
| -0,089285324 | 0,934218487 | PEX11B | FALSE | Downregulated |
| -0,068614987 | 0,934600132 | TBRG4 | FALSE | Downregulated |
| 0,155917697 | 0,934644959 | CAMKK1 | FALSE | Upregulated |
| 0,079402392 | 0,934644959 | GTF2E2 | FALSE | Upregulated |
| -0,104318496 | 0,934644959 | RNASEH2B | FALSE | Downregulated |
| -0,131893502 | 0,934644959 | TMEM200C | FALSE | Downregulated |
| -0,074855319 | 0,934686491 | FAM206A | FALSE | Downregulated |
| 0,026647039 | 0,934829981 | FAM20A | FALSE | Upregulated |
| 0,053138639 | 0,934829981 | RHBDD1 | FALSE | Upregulated |
| -0,075358892 | 0,93562438 | ATG5 | FALSE | Downregulated |
| 0,10490347 | 0,93562438 | DLGAP2 | FALSE | Upregulated |
| -0,126900672 | 0,93562438 | EFCC1 | FALSE | Downregulated |
| -0,160706971 | 0,93562438 | ITPRIPL1 | FALSE | Downregulated |
| -0,048992735 | 0,935786608 | RUFY1 | FALSE | Downregulated |
| -0,05721411 | 0,935811709 | GTSE1-AS1 | FALSE | Downregulated |
| -0,065562116 | 0,93605838 | AOC3 | FALSE | Downregulated |

| | | | | |
|---|---|---|---|---|
| 0,170437525 | 0,93605838 | LOC102724483 | FALSE | Upregulated |
| -0,039437918 | 0,93605838 | SLC25A15 | FALSE | Downregulated |
| -0,029223798 | 0,936101508 | KIAA0100 | FALSE | Downregulated |
| -0,050412705 | 0,936101508 | TOP3A | FALSE | Downregulated |
| 0,074886103 | 0,936187712 | ATP6V1G1 | FALSE | Upregulated |
| 0,032531665 | 0,936187712 | GTF3C1 | FALSE | Upregulated |
| -0,101100708 | 0,936187712 | RIMKLB | FALSE | Downregulated |
| 0,118310478 | 0,936270579 | LOC102724005 | FALSE | Upregulated |
| -0,075002317 | 0,93634251 | C17orf104 | FALSE | Downregulated |
| 0,127468963 | 0,936429775 | LOC102724626 | FALSE | Upregulated |
| 0,056391549 | 0,936463666 | ALG13 | FALSE | Upregulated |
| -0,043152196 | 0,936463666 | ATF7IP | FALSE | Downregulated |
| 0,172157467 | 0,936463666 | EFNA4 | FALSE | Upregulated |
| -0,109642972 | 0,936463666 | FAM133B | FALSE | Downregulated |
| 0,064948765 | 0,936538868 | XRCC1 | FALSE | Upregulated |
| 0,043573857 | 0,93660792 | AKR1C2 | FALSE | Upregulated |
| 0,090699917 | 0,936803485 | VWA2 | FALSE | Upregulated |
| 0,07251422 | 0,936845101 | AP3S1 | FALSE | Upregulated |
| 0,049366992 | 0,936845101 | HNRNPR | FALSE | Upregulated |
| -0,098358009 | 0,936845101 | MLIP | FALSE | Downregulated |
| 0,028759189 | 0,937035964 | AP2M1 | FALSE | Upregulated |
| -0,052000647 | 0,937035964 | MOB3C | FALSE | Downregulated |
| -0,079380379 | 0,937035964 | STARD4 | FALSE | Downregulated |
| -0,181966961 | 0,937120135 | ANKRD20A2 | FALSE | Downregulated |
| -0,053025853 | 0,937120135 | KIAA0232 | FALSE | Downregulated |
| 0,082913784 | 0,937120135 | RP11-395P17.3 | FALSE | Upregulated |
| -0,070185869 | 0,937130032 | PIGO | FALSE | Downregulated |
| 0,143779497 | 0,937130032 | ZNF599 | FALSE | Upregulated |
| 0,141999876 | 0,937293548 | EIF5A2 | FALSE | Upregulated |
| -0,051610829 | 0,93740233 | CDAN1 | FALSE | Downregulated |
| -0,155561089 | 0,93740233 | GEMIN6 | FALSE | Downregulated |
| 0,047970599 | 0,93740233 | LOC284837 | FALSE | Upregulated |
| -0,061508584 | 0,93740233 | PIM1 | FALSE | Downregulated |
| 0,103337195 | 0,93740233 | RNF144A | FALSE | Upregulated |
| 0,060623147 | 0,93740233 | ZNF608 | FALSE | Upregulated |
| 0,038074316 | 0,937543753 | MMP19 | FALSE | Upregulated |
| 0,072354364 | 0,937543753 | PGAM4 | FALSE | Upregulated |
| 0,141597139 | 0,937543753 | PPP1R3D | FALSE | Upregulated |
| -0,040926557 | 0,937543753 | TUT1 | FALSE | Downregulated |
| 0,066196391 | 0,937543753 | ZNF790 | FALSE | Upregulated |
| 0,049313849 | 0,937607159 | RBM12B | FALSE | Upregulated |
| -0,092130942 | 0,937625648 | CWC22 | FALSE | Downregulated |
| -0,06609913 | 0,937705763 | MOAP1 | FALSE | Downregulated |
| -0,10145552 | 0,937744997 | ANXA10 | FALSE | Downregulated |
| -0,075004701 | 0,937744997 | EDRF1 | FALSE | Downregulated |
| 0,030284815 | 0,937744997 | NUFIP2 | FALSE | Upregulated |
| -0,066772663 | 0,937744997 | PTCHD3P1 | FALSE | Downregulated |
| -0,117081751 | 0,937744997 | RFPL1S | FALSE | Downregulated |
| 0,141883538 | 0,937744997 | RPSAP58 | FALSE | Upregulated |
| -0,126978033 | 0,937744997 | TYRO3 | FALSE | Downregulated |
| -0,142332841 | 0,938052194 | AK9 | FALSE | Downregulated |

| | | | | |
|---|---|---|---|---|
| 0,08910635 | 0,938052194 | HMBS | FALSE | Upregulated |
| -0,108287026 | 0,938062965 | MANBAL | FALSE | Downregulated |
| 0,032209595 | 0,938128128 | RPTOR | FALSE | Upregulated |
| -0,18290383 | 0,938226784 | UTY | FALSE | Downregulated |
| 0,08378976 | 0,938455291 | IGSF9 | FALSE | Upregulated |
| 0,223482126 | 0,938455291 | ILDR1 | FALSE | Upregulated |
| 0,033902642 | 0,938455291 | SEPT11 | FALSE | Upregulated |
| -0,070972392 | 0,938499757 | DAXX | FALSE | Downregulated |
| -0,029695345 | 0,938499757 | DOT1L | FALSE | Downregulated |
| 0,097549663 | 0,938499757 | LSM1 | FALSE | Upregulated |
| -0,053023052 | 0,938499757 | PYGO2 | FALSE | Downregulated |
| 0,120621143 | 0,938499757 | TPM2 | FALSE | Upregulated |
| 0,062020882 | 0,938499757 | TTC3-AS1 | FALSE | Upregulated |
| -0,062544768 | 0,938505067 | B3GALT6 | FALSE | Downregulated |
| -0,162537817 | 0,938505067 | LOC101927999 | FALSE | Downregulated |
| -0,041160902 | 0,938505067 | PDLIM5 | FALSE | Downregulated |
| 0,096799154 | 0,938505067 | TTC39B | FALSE | Upregulated |
| -0,052220582 | 0,938505067 | ZNF428 | FALSE | Downregulated |
| -0,041748111 | 0,938629242 | CUL3 | FALSE | Downregulated |
| -0,076146378 | 0,938629242 | DNAL1 | FALSE | Downregulated |
| -0,146848686 | 0,938722405 | C1orf116 | FALSE | Downregulated |
| -0,127371663 | 0,938722405 | CD40LG | FALSE | Downregulated |
| -0,071044131 | 0,938722405 | COPS2 | FALSE | Downregulated |
| 0,131080171 | 0,938722405 | FBXW10 | FALSE | Upregulated |
| 0,092194665 | 0,938722405 | GRIN2B | FALSE | Upregulated |
| -0,054017697 | 0,938722405 | PEG3-AS1 | FALSE | Downregulated |
| 0,039168429 | 0,938722405 | RELA | FALSE | Upregulated |
| -0,070602609 | 0,938722405 | SRRD | FALSE | Downregulated |
| -0,079324574 | 0,938763718 | CDK5RAP1 | FALSE | Downregulated |
| 0,051191385 | 0,938763718 | CSDC2 | FALSE | Upregulated |
| -0,054629785 | 0,938763718 | EXOSC10 | FALSE | Downregulated |
| -0,039571036 | 0,938763718 | FAM195A | FALSE | Downregulated |
| 0,063490788 | 0,938763718 | KIAA0391 | FALSE | Upregulated |
| -0,13759929 | 0,938763718 | TCP10L | FALSE | Downregulated |
| -0,059460059 | 0,938763718 | THAP11 | FALSE | Downregulated |
| -0,180125387 | 0,938763718 | ZNF879 | FALSE | Downregulated |
| 0,064813272 | 0,938774168 | NUMBL | FALSE | Upregulated |
| -0,035438972 | 0,938774168 | ZNF3 | FALSE | Downregulated |
| -0,106720322 | 0,938799439 | C17orf75 | FALSE | Downregulated |
| 0,061235721 | 0,938799439 | CC2D2B | FALSE | Upregulated |
| -0,064491404 | 0,938799439 | CNTNAP1 | FALSE | Downregulated |
| -0,077616086 | 0,938799439 | H2AFZ | FALSE | Downregulated |
| -0,051124485 | 0,938799439 | MARK4 | FALSE | Downregulated |
| -0,077040431 | 0,938799439 | NDUFA9 | FALSE | Downregulated |
| 0,062911201 | 0,938799439 | NTAN1 | FALSE | Upregulated |
| -0,114685362 | 0,938799439 | OR1L4 | FALSE | Downregulated |
| -0,08751399 | 0,938799439 | OR52B6 | FALSE | Downregulated |
| -0,126765244 | 0,938799439 | SPATA6 | FALSE | Downregulated |
| 0,036430868 | 0,938799439 | SRSF6 | FALSE | Upregulated |
| -0,043680416 | 0,938799439 | TBC1D2B | FALSE | Downregulated |
| -0,046234729 | 0,938822237 | PIM3 | FALSE | Downregulated |

| | | | | |
|---|---|---|---|---|
| 0,023726923 | 0,938854275 | ANKFY1 | FALSE | Upregulated |
| 0,138112852 | 0,938854275 | ARMCX1 | FALSE | Upregulated |
| 0,050295135 | 0,938854275 | BCL2L2 | FALSE | Upregulated |
| -0,173953594 | 0,938854275 | C16orf95 | FALSE | Downregulated |
| 0,132013529 | 0,938854275 | ENKUR | FALSE | Upregulated |
| 0,082163179 | 0,938854275 | FBXO27 | FALSE | Upregulated |
| -0,138527007 | 0,938854275 | ISCA2 | FALSE | Downregulated |
| 0,130225633 | 0,938854275 | LINC01168 | FALSE | Upregulated |
| -0,098876975 | 0,938854275 | LOC102724895 | FALSE | Downregulated |
| -0,09559671 | 0,938854275 | METTL8 | FALSE | Downregulated |
| 0,061471066 | 0,938854275 | MIEN1 | FALSE | Upregulated |
| -0,070870989 | 0,938854275 | SCARF2 | FALSE | Downregulated |
| -0,08623403 | 0,938854275 | SMIM2 | FALSE | Downregulated |
| -0,288596319 | 0,938854275 | TBC1D3H | FALSE | Downregulated |
| -0,041844096 | 0,938854275 | TNK2 | FALSE | Downregulated |
| 0,041719776 | 0,938854275 | TXN2 | FALSE | Upregulated |
| -0,035200953 | 0,938854275 | WIZ | FALSE | Downregulated |
| 0,029300332 | 0,938868507 | A1BG | FALSE | Upregulated |
| 0,052632048 | 0,938868507 | ACBD3 | FALSE | Upregulated |
| 0,024764819 | 0,938868507 | ACO2 | FALSE | Upregulated |
| -0,098533933 | 0,938868507 | BBS10 | FALSE | Downregulated |
| 0,140216803 | 0,938868507 | CCDC64B | FALSE | Upregulated |
| -0,03347034 | 0,938868507 | DENND4B | FALSE | Downregulated |
| -0,031349328 | 0,938868507 | EIF4G2 | FALSE | Downregulated |
| -0,04713137 | 0,938868507 | EPN2-AS1 | FALSE | Downregulated |
| 0,07113237 | 0,938868507 | FADS3 | FALSE | Upregulated |
| -0,090047039 | 0,938868507 | FAM210A | FALSE | Downregulated |
| 0,068164736 | 0,938868507 | FKRP | FALSE | Upregulated |
| -0,089867527 | 0,938868507 | HMGCS1 | FALSE | Downregulated |
| 0,030150146 | 0,938868507 | KPNB1 | FALSE | Upregulated |
| 0,10568519 | 0,938868507 | LOC102723356 | FALSE | Upregulated |
| -0,169674524 | 0,938868507 | LOC102724556 | FALSE | Downregulated |
| 0,028835093 | 0,938868507 | MAGI1 | FALSE | Upregulated |
| 0,116882448 | 0,938868507 | MEIS3P1 | FALSE | Upregulated |
| -0,054839606 | 0,938868507 | NUP160 | FALSE | Downregulated |
| -0,061979091 | 0,938868507 | OCIAD2 | FALSE | Downregulated |
| 0,106480603 | 0,938868507 | OR10J5 | FALSE | Upregulated |
| -0,05286495 | 0,938868507 | PLCXD1 | FALSE | Downregulated |
| 0,175803856 | 0,938868507 | RP11-478K7.2 | FALSE | Upregulated |
| -0,15036388 | 0,938868507 | SDHAP3 | FALSE | Downregulated |
| 0,040403778 | 0,938868507 | TAX1BP1 | FALSE | Upregulated |
| -0,04082837 | 0,938868507 | UBE2G2 | FALSE | Downregulated |
| -0,041423989 | 0,938868507 | UBN2 | FALSE | Downregulated |
| -0,04152282 | 0,938868507 | UQCRQ | FALSE | Downregulated |
| -0,118483292 | 0,938868507 | XG | FALSE | Downregulated |
| -0,07592084 | 0,938868507 | ZBTB3 | FALSE | Downregulated |
| 0,054575137 | 0,938868507 | ZNF483 | FALSE | Upregulated |
| -0,054068748 | 0,938868507 | ZNF764 | FALSE | Downregulated |
| -0,139305567 | 0,938992094 | MEOX2 | FALSE | Downregulated |
| 0,071692631 | 0,939010507 | CTBP1-AS2 | FALSE | Upregulated |
| -0,078625132 | 0,939010507 | GPR45 | FALSE | Downregulated |

| | | | | |
|---|---|---|---|---|
| -0,039356269 | 0,939022834 | CARNS1 | FALSE | Downregulated |
| -0,074746803 | 0,939250475 | AXIN2 | FALSE | Downregulated |
| -0,077757794 | 0,939250475 | FAM118A | FALSE | Downregulated |
| -0,048031083 | 0,939250475 | SGMS2 | FALSE | Downregulated |
| 0,093928856 | 0,939250475 | TVP23C | FALSE | Upregulated |
| -0,088455527 | 0,93943901 | SEH1L | FALSE | Downregulated |
| 0,06862295 | 0,939727397 | UNC5B | FALSE | Upregulated |
| -0,059765834 | 0,939861623 | SGMS1 | FALSE | Downregulated |
| -0,078629742 | 0,939990973 | ATP8B4 | FALSE | Downregulated |
| 0,028338732 | 0,939990973 | CERK | FALSE | Upregulated |
| -0,238456732 | 0,939990973 | GCOM1 | FALSE | Downregulated |
| -0,033780695 | 0,939990973 | PPP1R16A | FALSE | Downregulated |
| 0,041319536 | 0,939990973 | PXDC1 | FALSE | Upregulated |
| 0,093901887 | 0,939990973 | RPA3 | FALSE | Upregulated |
| 0,082484774 | 0,939990973 | SPARCL1 | FALSE | Upregulated |
| 0,098963027 | 0,939990973 | TLR1 | FALSE | Upregulated |
| -0,056031519 | 0,939990973 | TM9SF1 | FALSE | Downregulated |
| -0,052029192 | 0,940031744 | ZBTB37 | FALSE | Downregulated |
| -0,049871386 | 0,940057239 | BRD9 | FALSE | Downregulated |
| -0,082535315 | 0,940057239 | LINC01126 | FALSE | Downregulated |
| 0,048610831 | 0,940141364 | ARL1 | FALSE | Upregulated |
| 0,090578222 | 0,940141364 | COMMD4 | FALSE | Upregulated |
| -0,115736425 | 0,940141364 | GPM6A | FALSE | Downregulated |
| 0,079489784 | 0,940285496 | NBEA | FALSE | Upregulated |
| 0,267872876 | 0,940437201 | C19orf77 | FALSE | Upregulated |
| 0,037823358 | 0,940437201 | KIAA0947 | FALSE | Upregulated |
| 0,13139724 | 0,940437201 | LOC100129473 | FALSE | Upregulated |
| 0,170020757 | 0,940437201 | LOC101928168 | FALSE | Upregulated |
| 0,027180158 | 0,940437201 | PRKDC | FALSE | Upregulated |
| -0,063189188 | 0,940437201 | RUNDC3B | FALSE | Downregulated |
| -0,187536259 | 0,940437201 | SLC35F1 | FALSE | Downregulated |
| 0,045034203 | 0,940437201 | TMED8 | FALSE | Upregulated |
| 0,059316235 | 0,940479245 | TMBIM4 | FALSE | Upregulated |
| 0,062470578 | 0,94058283 | GJB2 | FALSE | Upregulated |
| 0,056778682 | 0,940584734 | PFN2 | FALSE | Upregulated |
| 0,066768729 | 0,940611697 | VPS26A | FALSE | Upregulated |
| 0,106057129 | 0,940737751 | BTBD11 | FALSE | Upregulated |
| -0,237342959 | 0,940737751 | GIMAP5 | FALSE | Downregulated |
| -0,148085115 | 0,940737751 | VWA7 | FALSE | Downregulated |
| 0,031714018 | 0,940763838 | NID1 | FALSE | Upregulated |
| 0,112381189 | 0,94082657 | GPR152 | FALSE | Upregulated |
| 0,084777096 | 0,94082657 | LOC102724217 | FALSE | Upregulated |
| -0,120928977 | 0,94082657 | PP13 | FALSE | Downregulated |
| -0,037589053 | 0,94082657 | WDR24 | FALSE | Downregulated |
| 0,041989261 | 0,94085853 | HEATR2 | FALSE | Upregulated |
| -0,063742351 | 0,940877619 | PPP6C | FALSE | Downregulated |
| -0,025285323 | 0,941011422 | AKT1 | FALSE | Downregulated |
| 0,07201796 | 0,941011422 | CDK6 | FALSE | Upregulated |
| -0,053675995 | 0,941011422 | FOXN3-AS1 | FALSE | Downregulated |
| 0,07089333 | 0,941011422 | RP11-508N12.2 | FALSE | Upregulated |
| 0,073081814 | 0,941011422 | SDF2L1 | FALSE | Upregulated |

| | | | | |
|---|---|---|---|---|
| 0,127130645 | 0,941011422 | STAG3L4 | FALSE | Upregulated |
| -0,065112099 | 0,941011422 | ZNF564 | FALSE | Downregulated |
| 0,050666297 | 0,941037845 | LOC102724103 | FALSE | Upregulated |
| -0,045332201 | 0,94137648 | MPG | FALSE | Downregulated |
| 0,031466091 | 0,941541944 | SRSF9 | FALSE | Upregulated |
| 0,035500883 | 0,941645099 | ARHGEF26 | FALSE | Upregulated |
| 0,060978766 | 0,941645099 | RNF26 | FALSE | Upregulated |
| -0,040255573 | 0,941829379 | XRN2 | FALSE | Downregulated |
| -0,167402328 | 0,941829379 | ZNF491 | FALSE | Downregulated |
| 0,037891696 | 0,94198367 | PPP1R15B | FALSE | Upregulated |
| 0,107601141 | 0,94209429 | BICC1 | FALSE | Upregulated |
| 0,10758439 | 0,94209429 | CYP21A1P | FALSE | Upregulated |
| -0,100172686 | 0,94209429 | GLT8D2 | FALSE | Downregulated |
| 0,144772449 | 0,94209429 | MYBL1 | FALSE | Upregulated |
| -0,060033886 | 0,94209429 | PDGFC | FALSE | Downregulated |
| 0,093354281 | 0,94209429 | RSRC1 | FALSE | Upregulated |
| 0,185158138 | 0,942198575 | C17orf82 | FALSE | Upregulated |
| 0,090105006 | 0,942252227 | EGR2 | FALSE | Upregulated |
| 0,058529817 | 0,942458315 | ECT2L | FALSE | Upregulated |
| -0,037460711 | 0,942458315 | EMILIN1 | FALSE | Downregulated |
| -0,072496417 | 0,942458315 | F8A1 | FALSE | Downregulated |
| -0,055208609 | 0,942458315 | ITFG1 | FALSE | Downregulated |
| 0,032685675 | 0,942458315 | KLC1 | FALSE | Upregulated |
| 0,09971271 | 0,942458315 | LOC101926980 | FALSE | Upregulated |
| 0,237116319 | 0,942458315 | LOC647859 | FALSE | Upregulated |
| 0,084608143 | 0,942458315 | NCBP2-AS2 | FALSE | Upregulated |
| -0,03265575 | 0,942458315 | PARP4 | FALSE | Downregulated |
| -0,066600477 | 0,942458315 | PLEKHG7 | FALSE | Downregulated |
| -0,07312656 | 0,942458315 | ROBO1 | FALSE | Downregulated |
| 0,191914816 | 0,942458315 | SLC4A10 | FALSE | Upregulated |
| 0,138654536 | 0,942458315 | ZNF354B | FALSE | Upregulated |
| 0,097663813 | 0,942584102 | ACOT11 | FALSE | Upregulated |
| -0,026846815 | 0,942584102 | AHNAK | FALSE | Downregulated |
| -0,06487904 | 0,942584102 | CLEC12A | FALSE | Downregulated |
| -0,049735131 | 0,942584102 | COL18A1-AS1 | FALSE | Downregulated |
| 0,089974311 | 0,942584102 | COL8A1 | FALSE | Upregulated |
| 0,079168309 | 0,942584102 | EDRF1-AS1 | FALSE | Upregulated |
| -0,082470139 | 0,942584102 | EXOSC7 | FALSE | Downregulated |
| -0,141304489 | 0,942584102 | LINC00263 | FALSE | Downregulated |
| 0,066116438 | 0,942584102 | PPP2R3C | FALSE | Upregulated |
| -0,048733137 | 0,942584102 | RPRD1B | FALSE | Downregulated |
| 0,056771206 | 0,942584102 | SCAMP1 | FALSE | Upregulated |
| -0,047991556 | 0,942584102 | TMEM204 | FALSE | Downregulated |
| -0,026795465 | 0,942584102 | USP22 | FALSE | Downregulated |
| -0,10187382 | 0,942584102 | USP51 | FALSE | Downregulated |
| -0,061817643 | 0,942584102 | WDR60 | FALSE | Downregulated |
| 0,038888571 | 0,942584102 | ZC3H14 | FALSE | Upregulated |
| 0,16398277 | 0,942608764 | MS4A14 | FALSE | Upregulated |
| -0,177569658 | 0,942608764 | PSG1 | FALSE | Downregulated |
| 0,084225988 | 0,942624828 | ADAMTS12 | FALSE | Upregulated |
| 0,22312268 | 0,942624828 | AP3S2 | FALSE | Upregulated |

| | | | | |
|---|---|---|---|---|
| 0,02723209 | 0,942624828 | CLPTM1 | FALSE | Upregulated |
| 0,15787794 | 0,942624828 | FRMD6-AS1 | FALSE | Upregulated |
| 0,084751757 | 0,942624828 | INTS2 | FALSE | Upregulated |
| -0,142314122 | 0,942624828 | LOC101928561 | FALSE | Downregulated |
| 0,067553894 | 0,942624828 | LOC102723309 | FALSE | Upregulated |
| 0,047528023 | 0,942624828 | LPIN1 | FALSE | Upregulated |
| -0,066999739 | 0,942624828 | MRGBP | FALSE | Downregulated |
| -0,02788532 | 0,942624828 | PRAP1 | FALSE | Downregulated |
| 0,08388824 | 0,942624828 | RABL2B | FALSE | Upregulated |
| 0,119338586 | 0,942624828 | ZNF137P | FALSE | Upregulated |
| -0,119571219 | 0,942640174 | GPD1L | FALSE | Downregulated |
| -0,036581861 | 0,942882924 | FOXO3 | FALSE | Downregulated |
| -0,054096906 | 0,942911856 | IFT88 | FALSE | Downregulated |
| 0,145411433 | 0,942911856 | LOC375196 | FALSE | Upregulated |
| -0,044987307 | 0,942911856 | SLC25A44 | FALSE | Downregulated |
| -0,052965011 | 0,942911856 | ZBTB48 | FALSE | Downregulated |
| 0,047039033 | 0,942919906 | CHMP1B | FALSE | Upregulated |
| 0,046047735 | 0,94297466 | C5orf24 | FALSE | Upregulated |
| 0,037564716 | 0,94297466 | CCPG1 | FALSE | Upregulated |
| -0,051914593 | 0,94297466 | CEP70 | FALSE | Downregulated |
| 0,049260253 | 0,94297466 | DVL2 | FALSE | Upregulated |
| -0,054363729 | 0,94297466 | IFT122 | FALSE | Downregulated |
| 0,195332512 | 0,94297466 | IQCD | FALSE | Upregulated |
| 0,019646626 | 0,94297466 | SCAF1 | FALSE | Upregulated |
| -0,051388038 | 0,94297466 | SGSH | FALSE | Downregulated |
| -0,061798496 | 0,94297466 | VPS16 | FALSE | Downregulated |
| -0,096984561 | 0,94297466 | ZNF639 | FALSE | Downregulated |
| -0,123288429 | 0,94308976 | ACTRT3 | FALSE | Downregulated |
| -0,101937167 | 0,94308976 | B4GALNT1 | FALSE | Downregulated |
| -0,15691 | 0,94308976 | PIF1 | FALSE | Downregulated |
| -0,06352685 | 0,943169985 | DPY19L3 | FALSE | Downregulated |
| -0,052939823 | 0,943169985 | GXYLT1 | FALSE | Downregulated |
| 0,049526992 | 0,943169985 | NDRG4 | FALSE | Upregulated |
| 0,054777901 | 0,943195981 | BEAN1 | FALSE | Upregulated |
| 0,179323898 | 0,943265832 | LOC100130238 | FALSE | Upregulated |
| 0,056875363 | 0,943308159 | GTF2H1 | FALSE | Upregulated |
| -0,086145391 | 0,943308159 | NHSL2 | FALSE | Downregulated |
| -0,056877671 | 0,943318543 | ERO1L | FALSE | Downregulated |
| 0,025059314 | 0,943318543 | GSPT1 | FALSE | Upregulated |
| 0,030042265 | 0,943318543 | TRABD | FALSE | Upregulated |
| 0,094320343 | 0,943371548 | ZMYM5 | FALSE | Upregulated |
| 0,130029369 | 0,943543831 | F3 | FALSE | Upregulated |
| 0,115587415 | 0,943605511 | GLI2 | FALSE | Upregulated |
| -0,019855177 | 0,943605511 | NCOA4 | FALSE | Downregulated |
| 0,036202333 | 0,943605511 | ZNF513 | FALSE | Upregulated |
| 0,044019094 | 0,943608189 | CAPNS1 | FALSE | Upregulated |
| 0,040591239 | 0,94362494 | APOBR | FALSE | Upregulated |
| 0,044607816 | 0,943810293 | MXD1 | FALSE | Upregulated |
| -0,046051667 | 0,943880442 | KIF28P | FALSE | Downregulated |
| 0,046052054 | 0,943880442 | LIN54 | FALSE | Upregulated |
| -0,057583832 | 0,943880442 | SEPT7 | FALSE | Downregulated |

| | | | | |
|---|---|---|---|---|
| 0,032597346 | 0,944112639 | ASXL2 | FALSE | Upregulated |
| -0,038349148 | 0,944112639 | ZNF506 | FALSE | Downregulated |
| -0,045974842 | 0,944246487 | PAPPA | FALSE | Downregulated |
| 0,152947861 | 0,944246487 | RP5-855F14.1 | FALSE | Upregulated |
| -0,1273748 | 0,944353667 | COL9A3 | FALSE | Downregulated |
| -0,046467489 | 0,944353667 | ZC3H13 | FALSE | Downregulated |
| 0,070533712 | 0,944571779 | APOA4 | FALSE | Upregulated |
| -0,064605073 | 0,944571779 | ASNA1 | FALSE | Downregulated |
| 0,136537851 | 0,944571779 | C19orf55 | FALSE | Upregulated |
| -0,109502174 | 0,944571779 | C8orf37 | FALSE | Downregulated |
| 0,037003301 | 0,944571779 | CXXC1 | FALSE | Upregulated |
| -0,111999934 | 0,944571779 | GNG11 | FALSE | Downregulated |
| -0,042313234 | 0,944571779 | GPM6B | FALSE | Downregulated |
| -0,045390772 | 0,944571779 | KIAA1731 | FALSE | Downregulated |
| -0,037126361 | 0,944571779 | LDB1 | FALSE | Downregulated |
| 0,050644705 | 0,944571779 | LINC00847 | FALSE | Upregulated |
| -0,156131763 | 0,944571779 | LOC100506286 | FALSE | Downregulated |
| -0,054732626 | 0,944571779 | LOC101928045 | FALSE | Downregulated |
| -0,125638351 | 0,944571779 | MGP | FALSE | Downregulated |
| 0,043561356 | 0,944571779 | MKRN1 | FALSE | Upregulated |
| 0,047939151 | 0,944571779 | MRFAP1L1 | FALSE | Upregulated |
| -0,03353872 | 0,944571779 | NCOA1 | FALSE | Downregulated |
| -0,078147136 | 0,944571779 | NUTF2 | FALSE | Downregulated |
| -0,061937981 | 0,944571779 | PAXIP1 | FALSE | Downregulated |
| -0,044762769 | 0,944571779 | PCED1A | FALSE | Downregulated |
| 0,137154898 | 0,944571779 | PKD1L1 | FALSE | Upregulated |
| -0,054036181 | 0,944571779 | PKDREJ | FALSE | Downregulated |
| 0,065579806 | 0,944571779 | PNPLA8 | FALSE | Upregulated |
| 0,027125725 | 0,944571779 | PPP1R12C | FALSE | Upregulated |
| -0,131127086 | 0,944571779 | PRICKLE2-AS2 | FALSE | Downregulated |
| -0,110286991 | 0,944571779 | PSENEN | FALSE | Downregulated |
| 0,161921331 | 0,944571779 | RANBP17 | FALSE | Upregulated |
| -0,11226446 | 0,944571779 | RP6-109B7.3 | FALSE | Downregulated |
| -0,161472169 | 0,944571779 | RRAD | FALSE | Downregulated |
| 0,047691999 | 0,944571779 | TOP1MT | FALSE | Upregulated |
| -0,073526368 | 0,944571779 | WDR83 | FALSE | Downregulated |
| -0,087817296 | 0,944571779 | ZNF205-AS1 | FALSE | Downregulated |
| -0,064428691 | 0,944679124 | KSR1 | FALSE | Downregulated |
| -0,033937509 | 0,944782609 | ATRN | FALSE | Downregulated |
| 0,095499497 | 0,944782609 | C12orf4 | FALSE | Upregulated |
| -0,107468852 | 0,944782609 | LOC101929280 | FALSE | Downregulated |
| -0,248867346 | 0,944782609 | LOC644172 | FALSE | Downregulated |
| 0,040457455 | 0,944782609 | NUB1 | FALSE | Upregulated |
| -0,078971904 | 0,944782609 | PTPN13 | FALSE | Downregulated |
| 0,139286209 | 0,944782609 | TMEM178A | FALSE | Upregulated |
| 0,046301297 | 0,944782609 | ZDHHC3 | FALSE | Upregulated |
| -0,069025374 | 0,944885459 | LOC101929691 | FALSE | Downregulated |
| -0,089770285 | 0,945097919 | CWC27 | FALSE | Downregulated |
| -0,642318419 | 0,945097919 | EEF1E1-BLOC1S5 | FALSE | Downregulated |
| -0,149482491 | 0,945097919 | LOC101927103 | FALSE | Downregulated |
| 0,042035199 | 0,945097919 | ZNF800 | FALSE | Upregulated |

| | | | | |
|---|---|---|---|---|
| -0,129171968 | 0,945120035 | GUCA1B | FALSE | Downregulated |
| 0,14544819 | 0,945159644 | CYP2F1 | FALSE | Upregulated |
| 0,103497077 | 0,945163235 | CEP128 | FALSE | Upregulated |
| -0,049291304 | 0,945543705 | GSS | FALSE | Downregulated |
| -0,040196695 | 0,945648948 | ELL | FALSE | Downregulated |
| 0,081793141 | 0,945648948 | NR1D1 | FALSE | Upregulated |
| 0,052221141 | 0,945784601 | REL | FALSE | Upregulated |
| -0,03535611 | 0,94580367 | ACTR1A | FALSE | Downregulated |
| -0,093778816 | 0,94580367 | ATG4A | FALSE | Downregulated |
| 0,074916168 | 0,94580367 | DDX59 | FALSE | Upregulated |
| -0,066764142 | 0,94580367 | LOC101928089 | FALSE | Downregulated |
| -0,032905006 | 0,94580367 | NUDCD3 | FALSE | Downregulated |
| 0,171897463 | 0,94580367 | RP11-141O11.2 | FALSE | Upregulated |
| 0,071829005 | 0,94580367 | ZNF717 | FALSE | Upregulated |
| -0,146493753 | 0,945918276 | SULT1A4 | FALSE | Downregulated |
| -0,029628712 | 0,945995648 | PCNXL3 | FALSE | Downregulated |
| -0,174277413 | 0,946051719 | RP11-883G14.2 | FALSE | Downregulated |
| 0,049819411 | 0,946067975 | ZNF124 | FALSE | Upregulated |
| -0,089773749 | 0,94610432 | C15orf57 | FALSE | Downregulated |
| 0,070855596 | 0,94610432 | C1orf112 | FALSE | Upregulated |
| -0,074849705 | 0,94610432 | CDKN2AIP | FALSE | Downregulated |
| -0,081533271 | 0,94610432 | COG6 | FALSE | Downregulated |
| 0,016229115 | 0,94610432 | DDX17 | FALSE | Upregulated |
| -0,080319329 | 0,94610432 | FPGT | FALSE | Downregulated |
| -0,116222428 | 0,94610432 | GPR176 | FALSE | Downregulated |
| 0,088862265 | 0,94610432 | HAUS7 | FALSE | Upregulated |
| -0,065391921 | 0,94610432 | IDI1 | FALSE | Downregulated |
| -0,036768519 | 0,94610432 | KIAA1191 | FALSE | Downregulated |
| 0,173332396 | 0,94610432 | LOC100505909 | FALSE | Upregulated |
| 0,095576153 | 0,94610432 | LRRC9 | FALSE | Upregulated |
| -0,073783763 | 0,94610432 | MRPL18 | FALSE | Downregulated |
| -0,11264414 | 0,94610432 | OR13C8 | FALSE | Downregulated |
| 0,06418729 | 0,94610432 | PGS1 | FALSE | Upregulated |
| 0,037612945 | 0,94610432 | PPM1F | FALSE | Upregulated |
| 0,082702954 | 0,94610432 | SRL | FALSE | Upregulated |
| 0,038698531 | 0,94610432 | TRIOBP | FALSE | Upregulated |
| 0,057843334 | 0,94610432 | URM1 | FALSE | Upregulated |
| -0,116483572 | 0,94610432 | ZNF383 | FALSE | Downregulated |
| 0,039655819 | 0,946317461 | APC | FALSE | Upregulated |
| -0,040721749 | 0,946317461 | FAF1 | FALSE | Downregulated |
| 0,042812382 | 0,946317461 | INO80E | FALSE | Upregulated |
| 0,162195354 | 0,946317461 | LOC729164 | FALSE | Upregulated |
| -0,060501193 | 0,946317461 | NSUN7 | FALSE | Downregulated |
| 0,131196651 | 0,946317461 | PEX7 | FALSE | Upregulated |
| 0,033880086 | 0,946317461 | RBM14 | FALSE | Upregulated |
| -0,124093106 | 0,946317461 | SNORD118 | FALSE | Downregulated |
| 0,06676258 | 0,946317461 | WIBG | FALSE | Upregulated |
| -0,049066012 | 0,946317461 | ZFYVE20 | FALSE | Downregulated |
| 0,129892375 | 0,946398486 | MITD1 | FALSE | Upregulated |
| -0,087967132 | 0,946541051 | LOC727896 | FALSE | Downregulated |
| 0,051423971 | 0,946541051 | LRCH3 | FALSE | Upregulated |

| | | | | |
|---|---|---|---|---|
| 0,051497801 | 0,946541051 | UBE2G1 | FALSE | Upregulated |
| -0,057438379 | 0,946597967 | CBLL1 | FALSE | Downregulated |
| -0,105448495 | 0,946597967 | GORAB | FALSE | Downregulated |
| -0,032647747 | 0,946597967 | PHIP | FALSE | Downregulated |
| -0,116199041 | 0,946597967 | ZMAT5 | FALSE | Downregulated |
| 0,047611433 | 0,94665013 | PPP1R11 | FALSE | Upregulated |
| 0,075506181 | 0,94665013 | TCN2 | FALSE | Upregulated |
| 0,159794991 | 0,946747224 | ADAM33 | FALSE | Upregulated |
| 0,029472563 | 0,946747224 | PRKD3 | FALSE | Upregulated |
| -0,055817653 | 0,946747224 | STK4 | FALSE | Downregulated |
| 0,078020611 | 0,946756747 | FANCM | FALSE | Upregulated |
| -0,051117197 | 0,947047441 | CACNA1C | FALSE | Downregulated |
| -0,066086841 | 0,947158637 | DDX11 | FALSE | Downregulated |
| 0,041396608 | 0,947158637 | ERMAP | FALSE | Upregulated |
| 0,083969787 | 0,947158637 | POGLUT1 | FALSE | Upregulated |
| 0,046926764 | 0,947158637 | SPECC1L | FALSE | Upregulated |
| 0,056788185 | 0,947158637 | SPRED1 | FALSE | Upregulated |
| 0,134293565 | 0,947158637 | ST7-OT4 | FALSE | Upregulated |
| 0,094466629 | 0,947158637 | WDSUB1 | FALSE | Upregulated |
| -0,040853972 | 0,947258633 | GAB1 | FALSE | Downregulated |
| -0,119339953 | 0,947380282 | ANKRD33B | FALSE | Downregulated |
| -0,031027055 | 0,947380282 | MZF1 | FALSE | Downregulated |
| 0,970824415 | 0,94738375 | RNU1-28P | FALSE | Upregulated |
| -0,058318182 | 0,94738375 | VPS11 | FALSE | Downregulated |
| -0,060528255 | 0,947384159 | SCARNA5 | FALSE | Downregulated |
| -0,054191158 | 0,947431251 | HIGD2A | FALSE | Downregulated |
| 0,127583771 | 0,947798489 | GATSL3 | FALSE | Upregulated |
| 0,046490274 | 0,947798489 | LOC101930137 | FALSE | Upregulated |
| -0,062308282 | 0,947934472 | VIPAS39 | FALSE | Downregulated |
| 0,090371035 | 0,947974254 | KIF9-AS1 | FALSE | Upregulated |
| 0,038738319 | 0,94808938 | USP6NL | FALSE | Upregulated |
| 0,033588019 | 0,94808938 | ZNF629 | FALSE | Upregulated |
| -0,060503515 | 0,948258964 | EPM2A | FALSE | Downregulated |
| 0,059401084 | 0,948262211 | LMF1 | FALSE | Upregulated |
| -0,133036751 | 0,948262211 | LRFN1 | FALSE | Downregulated |
| -0,124188526 | 0,948262211 | ZNF624 | FALSE | Downregulated |
| 0,025203916 | 0,948320094 | RAPGEF1 | FALSE | Upregulated |
| -0,036503098 | 0,948578337 | FAM32A | FALSE | Downregulated |
| -0,037447858 | 0,948707215 | SLCO1A2 | FALSE | Downregulated |
| 0,037704772 | 0,948753705 | RFX1 | FALSE | Upregulated |
| 0,070082238 | 0,948753705 | SLC25A40 | FALSE | Upregulated |
| 0,161545699 | 0,9488048 | HYDIN2 | FALSE | Upregulated |
| 0,046185343 | 0,949011315 | NDUFS7 | FALSE | Upregulated |
| -0,07548093 | 0,949011315 | RAB3IP | FALSE | Downregulated |
| -0,052118795 | 0,949031186 | EMC4 | FALSE | Downregulated |
| 0,065151166 | 0,949031186 | LOC100507373 | FALSE | Upregulated |
| -0,044338939 | 0,94905591 | BRAP | FALSE | Downregulated |
| 0,075435649 | 0,94905591 | C16orf89 | FALSE | Upregulated |
| 0,070683111 | 0,94905591 | DNAJB14 | FALSE | Upregulated |
| 0,08549386 | 0,94905591 | FAM217B | FALSE | Upregulated |
| 0,03636097 | 0,94905591 | TRAK2 | FALSE | Upregulated |

| | | | | |
|---|---|---|---|---|
| -0,153954656 | 0,949113359 | LOC284023 | FALSE | Downregulated |
| -0,033447952 | 0,949174828 | ATF6 | FALSE | Downregulated |
| -0,075526878 | 0,949174828 | C2orf49 | FALSE | Downregulated |
| -0,035596631 | 0,949174828 | HIPK1 | FALSE | Downregulated |
| 0,118289751 | 0,949174828 | HS1BP3-IT1 | FALSE | Upregulated |
| 0,030473604 | 0,949174828 | IMPAD1 | FALSE | Upregulated |
| 0,067540404 | 0,949174828 | PRRG4 | FALSE | Upregulated |
| -0,04460179 | 0,949174828 | PTEN | FALSE | Downregulated |
| -0,064742448 | 0,949174828 | STIM2 | FALSE | Downregulated |
| 0,057165394 | 0,949174828 | TANK | FALSE | Upregulated |
| 0,113748245 | 0,949321978 | C1orf131 | FALSE | Upregulated |
| 0,13400583 | 0,94932507 | HSPB11 | FALSE | Upregulated |
| -0,13211448 | 0,949330704 | SMIM8 | FALSE | Downregulated |
| -0,106466324 | 0,949537687 | PRIM1 | FALSE | Downregulated |
| -0,039273353 | 0,949564705 | GALNS | FALSE | Downregulated |
| -0,120603207 | 0,949564705 | NLRP7 | FALSE | Downregulated |
| -0,078091294 | 0,949581724 | C17orf51 | FALSE | Downregulated |
| 0,048203828 | 0,949581724 | LOC100134040 | FALSE | Upregulated |
| 0,095937201 | 0,949979567 | CCDC102B | FALSE | Upregulated |
| 0,052761015 | 0,949979567 | CD80 | FALSE | Upregulated |
| 0,067368581 | 0,950009109 | MBD5 | FALSE | Upregulated |
| 0,141834593 | 0,95008188 | C2CD4A | FALSE | Upregulated |
| 0,078950787 | 0,95008188 | HEATR6 | FALSE | Upregulated |
| 0,039977336 | 0,95008188 | MTERFD3 | FALSE | Upregulated |
| 0,050210467 | 0,95008188 | SGTB | FALSE | Upregulated |
| 0,027580194 | 0,95008188 | SMC1A | FALSE | Upregulated |
| 0,039571332 | 0,950163965 | LOC102723743 | FALSE | Upregulated |
| 0,027255158 | 0,950163965 | MAP4K4 | FALSE | Upregulated |
| -0,047946752 | 0,950163965 | PDGFRA | FALSE | Downregulated |
| -0,032812506 | 0,950163965 | PSKH1 | FALSE | Downregulated |
| -0,936320263 | 0,950163965 | RNU1-1 | FALSE | Downregulated |
| -0,141344512 | 0,950230253 | PRB4 | FALSE | Downregulated |
| 0,051046558 | 0,950480747 | DCPS | FALSE | Upregulated |
| -0,07295223 | 0,950531895 | TRIM32 | FALSE | Downregulated |
| 0,071218622 | 0,950963409 | SEPT7P2 | FALSE | Upregulated |
| 0,01681255 | 0,950987998 | GNAS | FALSE | Upregulated |
| -0,048399267 | 0,951090284 | FEM1C | FALSE | Downregulated |
| -0,048887109 | 0,951090284 | GPR113 | FALSE | Downregulated |
| 0,099774049 | 0,951090284 | HIST1H4D | FALSE | Upregulated |
| -0,040848975 | 0,951090284 | KIAA1522 | FALSE | Downregulated |
| -0,036606069 | 0,951090284 | RPAIN | FALSE | Downregulated |
| -0,029604462 | 0,951090284 | TTC28 | FALSE | Downregulated |
| 0,047448373 | 0,951268776 | KCNT2 | FALSE | Upregulated |
| 0,071152155 | 0,951306749 | TCP11L2 | FALSE | Upregulated |
| -0,0553561 | 0,951685602 | POLE3 | FALSE | Downregulated |
| 0,036538881 | 0,951854155 | C2orf16 | FALSE | Upregulated |
| -0,052854712 | 0,951854155 | CBY3 | FALSE | Downregulated |
| 0,094957501 | 0,951854155 | CRYBA1 | FALSE | Upregulated |
| 0,120165926 | 0,951854155 | FSD1 | FALSE | Upregulated |
| -0,087965943 | 0,951854155 | LOC101929449 | FALSE | Downregulated |
| 0,117765914 | 0,951854155 | LOC401127 | FALSE | Upregulated |

| | | | | |
|---|---|---|---|---|
| 0,034590324 | 0,951854155 | NME4 | FALSE | Upregulated |
| -0,030662943 | 0,951854155 | PHF2 | FALSE | Downregulated |
| -0,020382949 | 0,951854155 | PRKCSH | FALSE | Downregulated |
| 0,10293886 | 0,951854155 | RAB27B | FALSE | Upregulated |
| -0,097713226 | 0,951854155 | RP11-16E12.2 | FALSE | Downregulated |
| 0,076879811 | 0,951854155 | SMA4 | FALSE | Upregulated |
| 0,037075722 | 0,951854155 | ZNF865 | FALSE | Upregulated |
| 0,097881009 | 0,952049835 | TSPAN1 | FALSE | Upregulated |
| 0,025670301 | 0,952184067 | DUSP3 | FALSE | Upregulated |
| 0,099011507 | 0,952184067 | PIGX | FALSE | Upregulated |
| -0,069291432 | 0,952184539 | PDP2 | FALSE | Downregulated |
| 0,031814031 | 0,95232561 | ARHGAP12 | FALSE | Upregulated |
| -0,076476096 | 0,952595232 | ARHGAP6 | FALSE | Downregulated |
| 0,101029711 | 0,952595232 | C1orf95 | FALSE | Upregulated |
| -0,036381378 | 0,952595232 | CDKL1 | FALSE | Downregulated |
| -0,030886742 | 0,952595232 | LOC101927803 | FALSE | Downregulated |
| 0,060400397 | 0,952595232 | PRDM15 | FALSE | Upregulated |
| 0,03964133 | 0,952595232 | PSMA6 | FALSE | Upregulated |
| -0,036120408 | 0,952613312 | GAS8 | FALSE | Downregulated |
| 0,054014798 | 0,952613312 | GPR137 | FALSE | Upregulated |
| -0,120660153 | 0,952613312 | MRPL50 | FALSE | Downregulated |
| -0,031877873 | 0,952613312 | MTHFR | FALSE | Downregulated |
| -0,088156694 | 0,952613312 | NAF1 | FALSE | Downregulated |
| -0,041446377 | 0,952613312 | UBE3B | FALSE | Downregulated |
| 0,050820395 | 0,952651903 | CCDC82 | FALSE | Upregulated |
| -0,022386862 | 0,952651903 | MINK1 | FALSE | Downregulated |
| -0,076056597 | 0,952651903 | PCSK4 | FALSE | Downregulated |
| -0,040288337 | 0,952651903 | SCARNA7 | FALSE | Downregulated |
| -0,06628015 | 0,952651903 | SEC1P | FALSE | Downregulated |
| 0,075415329 | 0,952660825 | CUTC | FALSE | Upregulated |
| 0,058421493 | 0,952660825 | DFFA | FALSE | Upregulated |
| 0,043778322 | 0,952660825 | ITM2C | FALSE | Upregulated |
| -0,147237065 | 0,952660825 | LRP1B | FALSE | Downregulated |
| 0,043752759 | 0,952660825 | STX10 | FALSE | Upregulated |
| 0,04553353 | 0,952660825 | ULK3 | FALSE | Upregulated |
| 0,045350899 | 0,952714375 | ASAP3 | FALSE | Upregulated |
| 0,056761134 | 0,952714375 | LINC00984 | FALSE | Upregulated |
| -0,045539781 | 0,952717621 | RP11-61I13.3 | FALSE | Downregulated |
| 0,050421237 | 0,952814363 | ATG3 | FALSE | Upregulated |
| 0,030879986 | 0,952814363 | DENND1A | FALSE | Upregulated |
| -0,04141598 | 0,952814363 | KDM6A | FALSE | Downregulated |
| -0,160911408 | 0,952814363 | LINC01144 | FALSE | Downregulated |
| -0,104231614 | 0,952814363 | NUDCD1 | FALSE | Downregulated |
| 0,046218624 | 0,952814363 | PMF1 | FALSE | Upregulated |
| 0,096222576 | 0,952814363 | PTPRN2 | FALSE | Upregulated |
| -0,133044229 | 0,952814363 | THAP1 | FALSE | Downregulated |
| -0,100696571 | 0,952814363 | WSCD1 | FALSE | Downregulated |
| 0,056887982 | 0,952917328 | TXN | FALSE | Upregulated |
| 0,043208575 | 0,953098107 | CYB5D1 | FALSE | Upregulated |
| -0,13762636 | 0,953167122 | HES6 | FALSE | Downregulated |
| 0,119890024 | 0,953167122 | SLIRP | FALSE | Upregulated |

| | | | | |
|---|---|---|---|---|
| 0,044252671 | 0,953442468 | KCTD2 | FALSE | Upregulated |
| -0,029030362 | 0,953442468 | PARP10 | FALSE | Downregulated |
| -0,103126858 | 0,953478076 | CEP19 | FALSE | Downregulated |
| 0,048563124 | 0,953478076 | EFEMP2 | FALSE | Upregulated |
| -0,056364307 | 0,953478076 | GPR75 | FALSE | Downregulated |
| -0,051134784 | 0,953478076 | ZNF259 | FALSE | Downregulated |
| -0,086056696 | 0,953872688 | ZNF438 | FALSE | Downregulated |
| -0,062280301 | 0,954069238 | LOC101927850 | FALSE | Downregulated |
| 0,156591345 | 0,95410693 | FAM35BP | FALSE | Upregulated |
| 0,130600735 | 0,954581798 | SELE | FALSE | Upregulated |
| 0,090623056 | 0,954672969 | GKAP1 | FALSE | Upregulated |
| -0,026149809 | 0,954672969 | IL1R1 | FALSE | Downregulated |
| 0,046810945 | 0,954672969 | OCLN | FALSE | Upregulated |
| 0,055727354 | 0,954729863 | ARHGAP10 | FALSE | Upregulated |
| -0,028268488 | 0,954729863 | DDX24 | FALSE | Downregulated |
| 0,109009818 | 0,954729863 | LINC00539 | FALSE | Upregulated |
| 0,224515928 | 0,954729863 | LOC101928344 | FALSE | Upregulated |
| 0,01814911 | 0,954729863 | PHRF1 | FALSE | Upregulated |
| -0,025326604 | 0,954758986 | GLIPR1 | FALSE | Downregulated |
| -0,138315566 | 0,954758986 | GLRX2 | FALSE | Downregulated |
| 0,026936035 | 0,954787778 | CASC4 | FALSE | Upregulated |
| 0,063986053 | 0,954787778 | COIL | FALSE | Upregulated |
| 0,056451683 | 0,954787778 | NAT8B | FALSE | Upregulated |
| 0,037582573 | 0,954799844 | NUBP2 | FALSE | Upregulated |
| 0,037022802 | 0,954884677 | IFRD2 | FALSE | Upregulated |
| 0,082153348 | 0,954884677 | NT5C3A | FALSE | Upregulated |
| -0,030432694 | 0,954884677 | SUDS3 | FALSE | Downregulated |
| -0,032528104 | 0,955003983 | CDH1 | FALSE | Downregulated |
| -0,063111365 | 0,955003983 | CETP | FALSE | Downregulated |
| -0,079395772 | 0,955003983 | MAGIX | FALSE | Downregulated |
| -0,074862365 | 0,955007828 | H1FX-AS1 | FALSE | Downregulated |
| -0,04045408 | 0,955007828 | NUS1 | FALSE | Downregulated |
| 0,046202608 | 0,955007828 | TOMM22 | FALSE | Upregulated |
| -0,069466335 | 0,955007828 | ULK4 | FALSE | Downregulated |
| -0,041073468 | 0,955013202 | C10orf105 | FALSE | Downregulated |
| -0,060516442 | 0,955013202 | NUP43 | FALSE | Downregulated |
| -0,024426373 | 0,955013202 | SHC1 | FALSE | Downregulated |
| -0,053099365 | 0,95502788 | SIRT7 | FALSE | Downregulated |
| -0,028081555 | 0,955030092 | ACP2 | FALSE | Downregulated |
| 0,092347456 | 0,955030092 | CNTNAP2 | FALSE | Upregulated |
| -0,040874538 | 0,955030092 | GLB1 | FALSE | Downregulated |
| -0,046745037 | 0,955030092 | GRWD1 | FALSE | Downregulated |
| 0,872084021 | 0,955030092 | KRT1 | FALSE | Upregulated |
| 0,035133173 | 0,955030092 | LOC101929079 | FALSE | Upregulated |
| -0,080035386 | 0,955030092 | LRRC56 | FALSE | Downregulated |
| 0,16097074 | 0,955030092 | LYRM9 | FALSE | Upregulated |
| 0,080803087 | 0,955030092 | POT1 | FALSE | Upregulated |
| -0,031942615 | 0,955030092 | STT3A | FALSE | Downregulated |
| -0,05493788 | 0,955030092 | TEFM | FALSE | Downregulated |
| -0,0342274 | 0,955030092 | TM9SF2 | FALSE | Downregulated |
| -0,079139061 | 0,955030092 | ZNF684 | FALSE | Downregulated |

| | | | | |
|---|---|---|---|---|
| 0,046705068 | 0,955176379 | POLR3B | FALSE | Upregulated |
| -0,085986197 | 0,955321509 | IL5RA | FALSE | Downregulated |
| -0,081222089 | 0,955770973 | TEX9 | FALSE | Downregulated |
| 0,048604341 | 0,955797729 | AAAS | FALSE | Upregulated |
| -0,054006089 | 0,955797729 | HIST1H4E | FALSE | Downregulated |
| -0,029455804 | 0,955797729 | MED16 | FALSE | Downregulated |
| -0,065717666 | 0,955797729 | NUP54 | FALSE | Downregulated |
| 0,029340343 | 0,955797729 | SESN3 | FALSE | Upregulated |
| 0,123468886 | 0,955797729 | ZNF714 | FALSE | Upregulated |
| 0,032683963 | 0,955814295 | ASMTL-AS1 | FALSE | Upregulated |
| 0,047241362 | 0,955814295 | SLC2A10 | FALSE | Upregulated |
| 0,074910319 | 0,955858547 | AC058791.1 | FALSE | Upregulated |
| -0,043309684 | 0,955858547 | MIRLET7BHG | FALSE | Downregulated |
| -0,079011961 | 0,955858547 | RND2 | FALSE | Downregulated |
| -0,045347899 | 0,955858547 | WDR43 | FALSE | Downregulated |
| 0,105091784 | 0,955858547 | ZNF736 | FALSE | Upregulated |
| -0,031110828 | 0,955881001 | PSMD2 | FALSE | Downregulated |
| 0,030545914 | 0,956015573 | AMBRA1 | FALSE | Upregulated |
| 0,032578641 | 0,956015573 | PRKCA | FALSE | Upregulated |
| 0,094155044 | 0,956015573 | ZBTB9 | FALSE | Upregulated |
| -0,226274112 | 0,9564534 | SIK1 | FALSE | Downregulated |
| -0,046152477 | 0,956507322 | CENPT | FALSE | Downregulated |
| -0,042372609 | 0,956724065 | ARMC7 | FALSE | Downregulated |
| -0,031669504 | 0,956724065 | BLCAP | FALSE | Downregulated |
| 0,10146801 | 0,956724065 | CCDC23 | FALSE | Upregulated |
| 0,045924742 | 0,956724065 | CTC-338M12.4 | FALSE | Upregulated |
| 0,108364103 | 0,956724065 | YY2 | FALSE | Upregulated |
| -0,071786705 | 0,956747266 | ANAPC15 | FALSE | Downregulated |
| -0,04349674 | 0,956747266 | C17orf85 | FALSE | Downregulated |
| 0,082412304 | 0,956747266 | DCLK2 | FALSE | Upregulated |
| -0,072564055 | 0,956747266 | DDIT4 | FALSE | Downregulated |
| 0,076743056 | 0,956747266 | DFFB | FALSE | Upregulated |
| 0,12220889 | 0,956747266 | FZD7 | FALSE | Upregulated |
| -0,151283309 | 0,956747266 | GABRB3 | FALSE | Downregulated |
| -0,030307088 | 0,956747266 | GPR107 | FALSE | Downregulated |
| -0,077536811 | 0,956747266 | GUCY1B3 | FALSE | Downregulated |
| -0,073000476 | 0,956747266 | HOXA-AS2 | FALSE | Downregulated |
| -0,048955612 | 0,956747266 | KREMEN1 | FALSE | Downregulated |
| 0,049074714 | 0,956747266 | NDUFA3 | FALSE | Upregulated |
| -0,115264577 | 0,956747266 | NINJ2 | FALSE | Downregulated |
| -0,045928377 | 0,956747266 | NRTN | FALSE | Downregulated |
| 0,056889931 | 0,956747266 | PEAR1 | FALSE | Upregulated |
| 0,064134522 | 0,956747266 | PIGY | FALSE | Upregulated |
| -0,074250257 | 0,956747266 | RBM34 | FALSE | Downregulated |
| -0,052790132 | 0,956747266 | RP11-983P16.4 | FALSE | Downregulated |
| 0,051551209 | 0,956747266 | SEC11C | FALSE | Upregulated |
| -0,028785591 | 0,956747266 | SLC4A2 | FALSE | Downregulated |
| -0,123770499 | 0,956747266 | SMN1 | FALSE | Downregulated |
| -0,047148697 | 0,956747266 | TMEM179B | FALSE | Downregulated |
| 0,034493209 | 0,956747266 | TMLHE | FALSE | Upregulated |
| 0,04159681 | 0,956747266 | UBAP2 | FALSE | Upregulated |

| | | | | |
|---|---|---|---|---|
| 0,04041874 | 0,956747266 | ZBTB1 | FALSE | Upregulated |
| -0,048291535 | 0,956747266 | ZFP64 | FALSE | Downregulated |
| -0,033340365 | 0,956747266 | ZNF282 | FALSE | Downregulated |
| 0,076372693 | 0,956747266 | ZNF658 | FALSE | Upregulated |
| 0,093986062 | 0,956828247 | MRPL47 | FALSE | Upregulated |
| 0,096592625 | 0,957182238 | ROBO3 | FALSE | Upregulated |
| 0,104457821 | 0,957257295 | SNX15 | FALSE | Upregulated |
| -0,021026407 | 0,957302949 | SERPINA10 | FALSE | Downregulated |
| 0,082394162 | 0,957358847 | ADAM12 | FALSE | Upregulated |
| 0,02126993 | 0,957358847 | ATXN7 | FALSE | Upregulated |
| 0,054149087 | 0,957358847 | IGLON5 | FALSE | Upregulated |
| 0,043099916 | 0,957358847 | LOC101927451 | FALSE | Upregulated |
| -0,030157851 | 0,957358847 | MEPCE | FALSE | Downregulated |
| 0,056342525 | 0,957358847 | MYPOP | FALSE | Upregulated |
| -0,03937037 | 0,957358847 | PLEKHA4 | FALSE | Downregulated |
| -0,035818963 | 0,957358847 | RBM25 | FALSE | Downregulated |
| -0,029764994 | 0,957358847 | UTP14C | FALSE | Downregulated |
| -0,040614057 | 0,957358847 | ZNF747 | FALSE | Downregulated |
| -0,035266604 | 0,957358847 | ZSCAN21 | FALSE | Downregulated |
| 0,075796277 | 0,957388288 | SMG8 | FALSE | Upregulated |
| 0,093151992 | 0,957458912 | AMIGO2 | FALSE | Upregulated |
| -0,048781896 | 0,957458912 | ATRAID | FALSE | Downregulated |
| -0,030327587 | 0,957458912 | CACTIN | FALSE | Downregulated |
| -0,123819605 | 0,957458912 | ESPL1 | FALSE | Downregulated |
| 0,133148917 | 0,957458912 | FRMD7 | FALSE | Upregulated |
| -0,031691583 | 0,957458912 | GNAQ | FALSE | Downregulated |
| 0,1298818 | 0,957458912 | HLA-DQA2 | FALSE | Upregulated |
| -0,04346514 | 0,957458912 | NPY6R | FALSE | Downregulated |
| 0,035576705 | 0,957458912 | PXK | FALSE | Upregulated |
| -0,052244243 | 0,957458912 | TMEM238 | FALSE | Downregulated |
| -0,037795328 | 0,957458912 | VPS4B | FALSE | Downregulated |
| 0,058121616 | 0,957592646 | C18orf8 | FALSE | Upregulated |
| 0,066323756 | 0,957592646 | SCRN1 | FALSE | Upregulated |
| -0,045100709 | 0,957606829 | AMD1 | FALSE | Downregulated |
| -0,029673986 | 0,957606829 | ARID2 | FALSE | Downregulated |
| -0,04304766 | 0,957606829 | C12orf60 | FALSE | Downregulated |
| -0,075396536 | 0,957606829 | COX10-AS1 | FALSE | Downregulated |
| -0,067925574 | 0,957606829 | LOC100288152 | FALSE | Downregulated |
| -0,044462297 | 0,957606829 | ORAI1 | FALSE | Downregulated |
| 0,065209179 | 0,957606829 | PDCD5 | FALSE | Upregulated |
| -0,060325272 | 0,957606829 | ZNF101 | FALSE | Downregulated |
| -0,022940725 | 0,957649792 | FTH1 | FALSE | Downregulated |
| 0,103376671 | 0,957671104 | WDR53 | FALSE | Upregulated |
| 0,069097407 | 0,95803798 | IL18R1 | FALSE | Upregulated |
| 0,037970613 | 0,95803798 | LOC101929004 | FALSE | Upregulated |
| 0,078056492 | 0,958312852 | C9orf89 | FALSE | Upregulated |
| -0,054497194 | 0,958567414 | GLTP | FALSE | Downregulated |
| -0,038335741 | 0,958567414 | SNX6 | FALSE | Downregulated |
| 0,114216743 | 0,958567414 | TTTY15 | FALSE | Upregulated |
| -0,106326846 | 0,958567414 | ZNF667-AS1 | FALSE | Downregulated |
| -0,051173574 | 0,958657984 | USP50 | FALSE | Downregulated |

| | | | | |
|---|---|---|---|---|
| -0,035374354 | 0,958782947 | NOP14-AS1 | FALSE | Downregulated |
| 0,093698109 | 0,958938547 | ADRA2A | FALSE | Upregulated |
| -0,024767474 | 0,958938547 | ANGPTL3 | FALSE | Downregulated |
| -0,03437718 | 0,958938547 | ARRB2 | FALSE | Downregulated |
| 0,050665902 | 0,958938547 | ATHL1 | FALSE | Upregulated |
| 0,030814135 | 0,958938547 | ATP2C1 | FALSE | Upregulated |
| -0,035638305 | 0,958938547 | C21orf62 | FALSE | Downregulated |
| -0,056913862 | 0,958938547 | CCP110 | FALSE | Downregulated |
| 0,064588533 | 0,958938547 | CRYM | FALSE | Upregulated |
| -0,022513369 | 0,958938547 | CTNNB1 | FALSE | Downregulated |
| 0,066885021 | 0,958938547 | CTTNBP2NL | FALSE | Upregulated |
| -0,035243502 | 0,958938547 | DEK | FALSE | Downregulated |
| 0,088820806 | 0,958938547 | FAM86FP | FALSE | Upregulated |
| 0,062085168 | 0,958938547 | FBLN5 | FALSE | Upregulated |
| 0,029302376 | 0,958938547 | FBXO18 | FALSE | Upregulated |
| -0,03351187 | 0,958938547 | FUK | FALSE | Downregulated |
| 0,12871859 | 0,958938547 | GUSBP3 | FALSE | Upregulated |
| -0,07400783 | 0,958938547 | IFT74 | FALSE | Downregulated |
| 0,024565736 | 0,958938547 | KIF3B | FALSE | Upregulated |
| 0,052215146 | 0,958938547 | LGALS1 | FALSE | Upregulated |
| -0,081726804 | 0,958938547 | LINC00852 | FALSE | Downregulated |
| 0,155865087 | 0,958938547 | LOC100288181 | FALSE | Upregulated |
| -0,136511772 | 0,958938547 | LOC101927699 | FALSE | Downregulated |
| -0,118790879 | 0,958938547 | LOC101929440 | FALSE | Downregulated |
| -0,046660141 | 0,958938547 | LOC101929792 | FALSE | Downregulated |
| -0,023067368 | 0,958938547 | MPP5 | FALSE | Downregulated |
| 0,152005791 | 0,958938547 | MROH7-TTC4 | FALSE | Upregulated |
| -0,051134157 | 0,958938547 | PRKACB | FALSE | Downregulated |
| -0,01374852 | 0,958938547 | SBF1 | FALSE | Downregulated |
| 0,038684347 | 0,958938547 | SCAMP2 | FALSE | Upregulated |
| -0,050963064 | 0,958938547 | SLC50A1 | FALSE | Downregulated |
| 0,031691685 | 0,958938547 | SREK1 | FALSE | Upregulated |
| 0,044771912 | 0,958938547 | ZFHX2 | FALSE | Upregulated |
| -0,046240932 | 0,958938547 | ZNF271 | FALSE | Downregulated |
| -0,049918907 | 0,958938547 | ZSCAN25 | FALSE | Downregulated |
| 0,026958493 | 0,958964001 | ZNF598 | FALSE | Upregulated |
| -0,05384281 | 0,959090057 | MRPL32 | FALSE | Downregulated |
| -0,097260126 | 0,959090057 | TSPAN15 | FALSE | Downregulated |
| -0,029715491 | 0,959194238 | RNF168 | FALSE | Downregulated |
| 0,053756372 | 0,959294182 | CCDC173 | FALSE | Upregulated |
| -0,045693605 | 0,95974954 | MUL1 | FALSE | Downregulated |
| 0,025878399 | 0,959773749 | TAOK3 | FALSE | Upregulated |
| -0,111543611 | 0,959780647 | APOLD1 | FALSE | Downregulated |
| -0,073943996 | 0,959780647 | C14orf180 | FALSE | Downregulated |
| 0,047568557 | 0,959780647 | DSTYK | FALSE | Upregulated |
| 0,049305226 | 0,959780647 | THAP7-AS1 | FALSE | Upregulated |
| -0,069856646 | 0,959780647 | ZNF211 | FALSE | Downregulated |
| -0,08481002 | 0,95986423 | LOC100289097 | FALSE | Downregulated |
| -0,088875736 | 0,95986423 | RP11-307C12.11 | FALSE | Downregulated |
| -0,042525386 | 0,959956665 | ZCCHC8 | FALSE | Downregulated |
| -0,023750912 | 0,95998893 | MAN1A2 | FALSE | Downregulated |

| | | | | |
|---|---|---|---|---|
| 0,072164873 | 0,960236784 | ALG1L9P | FALSE | Upregulated |
| -0,017832882 | 0,960236784 | AP2A1 | FALSE | Downregulated |
| 0,053706257 | 0,960236784 | BIN3 | FALSE | Upregulated |
| 0,087713242 | 0,960236784 | C17orf66 | FALSE | Upregulated |
| 0,056692643 | 0,960236784 | COG2 | FALSE | Upregulated |
| -0,101819342 | 0,960236784 | CTAGE4 | FALSE | Downregulated |
| -0,043603264 | 0,960236784 | EIF1B-AS1 | FALSE | Downregulated |
| -0,160053538 | 0,960236784 | HR | FALSE | Downregulated |
| -0,057121686 | 0,960236784 | KLHL28 | FALSE | Downregulated |
| 0,027219757 | 0,960236784 | LOC102724391 | FALSE | Upregulated |
| -0,057896538 | 0,960236784 | PCOLCE-AS1 | FALSE | Downregulated |
| 0,124258982 | 0,960236784 | RP11-677M14.7 | FALSE | Upregulated |
| 0,0694182 | 0,960236784 | SFXN4 | FALSE | Upregulated |
| -0,111241764 | 0,960236784 | SNHG19 | FALSE | Downregulated |
| 0,047227707 | 0,960236784 | ZNF622 | FALSE | Upregulated |
| -0,062004782 | 0,960358998 | GAS1 | FALSE | Downregulated |
| 0,064562996 | 0,960358998 | PARP11 | FALSE | Upregulated |
| -0,032037056 | 0,960382544 | EFNA1 | FALSE | Downregulated |
| 0,092178492 | 0,960383761 | AIFM3 | FALSE | Upregulated |
| -0,037756287 | 0,960518265 | LOC101928192 | FALSE | Downregulated |
| -0,038713362 | 0,960534919 | GMEB2 | FALSE | Downregulated |
| 0,035690538 | 0,960602446 | DDX51 | FALSE | Upregulated |
| -0,030668642 | 0,960602446 | DGAT1 | FALSE | Downregulated |
| -0,024824525 | 0,960602446 | RNF103 | FALSE | Downregulated |
| -0,075630142 | 0,960610314 | MAPK12 | FALSE | Downregulated |
| -0,035511079 | 0,960620176 | TUSC1 | FALSE | Downregulated |
| -0,039269833 | 0,960828288 | GFER | FALSE | Downregulated |
| -0,02045587 | 0,960828288 | HNRNPA3 | FALSE | Downregulated |
| -0,042771836 | 0,960828288 | VPS18 | FALSE | Downregulated |
| -0,1179083 | 0,960837822 | LINC00885 | FALSE | Downregulated |
| 0,045896348 | 0,960837822 | MIS18BP1 | FALSE | Upregulated |
| -0,044505442 | 0,960852024 | ART4 | FALSE | Downregulated |
| 0,024432006 | 0,960852024 | COPB2 | FALSE | Upregulated |
| -0,025162775 | 0,960852024 | DIP2C | FALSE | Downregulated |
| -0,080736724 | 0,960852024 | LINC01138 | FALSE | Downregulated |
| -0,034563874 | 0,960852024 | PKP2 | FALSE | Downregulated |
| 0,034055969 | 0,960852024 | TMEM230 | FALSE | Upregulated |
| -0,048257258 | 0,960852024 | TMX1 | FALSE | Downregulated |
| 0,032150797 | 0,960893484 | TAF10 | FALSE | Upregulated |
| -0,472541588 | 0,960926387 | LOC102724117 | FALSE | Downregulated |
| 0,046205831 | 0,960983408 | LOC101928927 | FALSE | Upregulated |
| -0,070152481 | 0,960983408 | PET100 | FALSE | Downregulated |
| -0,108588151 | 0,961078003 | CRYAA | FALSE | Downregulated |
| -0,024846537 | 0,961078003 | UBAP2L | FALSE | Downregulated |
| -0,033181028 | 0,961078003 | UEVLD | FALSE | Downregulated |
| 0,030787882 | 0,961096653 | AP3M1 | FALSE | Upregulated |
| -0,070664042 | 0,961096653 | FKBP11 | FALSE | Downregulated |
| -0,058349862 | 0,961096653 | HCRTR1 | FALSE | Downregulated |
| -0,153248772 | 0,961096653 | LOC541473 | FALSE | Downregulated |
| 0,376853476 | 0,961096653 | TNNI2 | FALSE | Upregulated |
| -0,086016328 | 0,961137068 | RP11-261N11.8 | FALSE | Downregulated |

| | | | | |
|---|---|---|---|---|
| -0,056908134 | 0,961222729 | ANP32AP1 | FALSE | Downregulated |
| -0,026887698 | 0,961222729 | ETF1 | FALSE | Downregulated |
| -0,028363579 | 0,961305164 | PHKB | FALSE | Downregulated |
| -0,029385351 | 0,961305164 | TBC1D24 | FALSE | Downregulated |
| -0,080145649 | 0,961305164 | TSPAN13 | FALSE | Downregulated |
| 0,072635603 | 0,961322475 | COMTD1 | FALSE | Upregulated |
| -0,032252593 | 0,961322475 | FBXL13 | FALSE | Downregulated |
| -0,027780193 | 0,961322475 | TALDO1 | FALSE | Downregulated |
| 0,032304034 | 0,961498349 | ASB13 | FALSE | Upregulated |
| 0,036340997 | 0,961498349 | EARS2 | FALSE | Upregulated |
| -0,058485657 | 0,961498349 | FUT4 | FALSE | Downregulated |
| 0,056604145 | 0,961498349 | L3MBTL1 | FALSE | Upregulated |
| 0,113848475 | 0,961498349 | LIN9 | FALSE | Upregulated |
| -0,064323338 | 0,961498349 | LOC100288069 | FALSE | Downregulated |
| 0,064723263 | 0,961498349 | MCM8 | FALSE | Upregulated |
| 0,098121124 | 0,961498349 | MSS51 | FALSE | Upregulated |
| -0,037473442 | 0,961498349 | RBFOX2 | FALSE | Downregulated |
| 0,081774215 | 0,961498349 | RIMS3 | FALSE | Upregulated |
| 0,021937839 | 0,961498349 | SEPT2 | FALSE | Upregulated |
| 0,089953426 | 0,961498349 | ZNF16 | FALSE | Upregulated |
| -0,046966104 | 0,961498349 | ZNF322 | FALSE | Downregulated |
| 0,050813193 | 0,961795966 | HSPA1B | FALSE | Upregulated |
| -0,099460688 | 0,961795966 | LINC01127 | FALSE | Downregulated |
| -0,054762162 | 0,961874804 | ARL16 | FALSE | Downregulated |
| -0,071480513 | 0,961874804 | CLCN2 | FALSE | Downregulated |
| 0,096260648 | 0,961874804 | FAM157C | FALSE | Upregulated |
| 0,027025638 | 0,961874804 | PCGF2 | FALSE | Upregulated |
| 0,04598859 | 0,961909049 | FBXW8 | FALSE | Upregulated |
| -0,046725884 | 0,961945981 | C19orf80 | FALSE | Downregulated |
| -0,064822578 | 0,96202121 | PYURF | FALSE | Downregulated |
| 0,059937833 | 0,962029528 | PODN | FALSE | Upregulated |
| 0,036861176 | 0,962139301 | DNAJB9 | FALSE | Upregulated |
| 0,03000289 | 0,962253457 | ADAM10 | FALSE | Upregulated |
| 0,135307205 | 0,962253457 | CHRM3 | FALSE | Upregulated |
| 0,049522544 | 0,962253457 | SNRNP48 | FALSE | Upregulated |
| 0,103618007 | 0,962452736 | PLA2G4A | FALSE | Upregulated |
| -0,033206656 | 0,962455789 | COA4 | FALSE | Downregulated |
| 0,087709222 | 0,962455789 | DPYD-AS1 | FALSE | Upregulated |
| -0,045921584 | 0,962455789 | FIP1L1 | FALSE | Downregulated |
| 0,037075575 | 0,962455789 | GOLGA8N | FALSE | Upregulated |
| 0,172010884 | 0,962455789 | ITIH6 | FALSE | Upregulated |
| -0,073831234 | 0,962455789 | LOC200772 | FALSE | Downregulated |
| 0,021558645 | 0,962455789 | PPIA | FALSE | Upregulated |
| -0,089902834 | 0,962455789 | RP11-473M20.5 | FALSE | Downregulated |
| -0,128268042 | 0,962455789 | SPIN2B | FALSE | Downregulated |
| -0,029178156 | 0,962455789 | TCP1 | FALSE | Downregulated |
| -0,048415247 | 0,962455789 | UTP3 | FALSE | Downregulated |
| 0,025439462 | 0,962455789 | WIPI2 | FALSE | Upregulated |
| 0,057089882 | 0,962455789 | ZNF566 | FALSE | Upregulated |
| 0,019597389 | 0,962488838 | IMMT | FALSE | Upregulated |
| 0,100559968 | 0,962488838 | LOC728290 | FALSE | Upregulated |

| | | | | |
|---|---|---|---|---|
| 0,031991345 | 0,962488838 | MIA2 | FALSE | Upregulated |
| -0,049085582 | 0,962488838 | NDUFB5 | FALSE | Downregulated |
| -0,037196908 | 0,962488838 | PPP1R7 | FALSE | Downregulated |
| 0,082284854 | 0,962488838 | RASL10A | FALSE | Upregulated |
| -0,026977337 | 0,962488838 | SECISBP2 | FALSE | Downregulated |
| 0,062011917 | 0,962488838 | TMEM147 | FALSE | Upregulated |
| -0,255354164 | 0,962598098 | EEF1A2 | FALSE | Downregulated |
| -0,067178821 | 0,96266625 | FBLN1 | FALSE | Downregulated |
| -0,091060376 | 0,96266625 | FOXO6 | FALSE | Downregulated |
| -0,070725825 | 0,96266625 | LOC101927023 | FALSE | Downregulated |
| -0,029831931 | 0,96266625 | PIGQ | FALSE | Downregulated |
| 0,019381376 | 0,96266625 | VPS13A | FALSE | Upregulated |
| 0,072745437 | 0,962826152 | FCER1G | FALSE | Upregulated |
| 0,031217506 | 0,962826152 | MAGED2 | FALSE | Upregulated |
| 0,022463751 | 0,962826152 | TRAM2 | FALSE | Upregulated |
| -0,084155992 | 0,962826152 | ZNF845 | FALSE | Downregulated |
| -0,039286914 | 0,962873235 | B4GALT7 | FALSE | Downregulated |
| -0,069296516 | 0,963002346 | RAI2 | FALSE | Downregulated |
| -0,097539805 | 0,963002346 | RP5-1027G4.3 | FALSE | Downregulated |
| -0,057732989 | 0,963002346 | TMEM126A | FALSE | Downregulated |
| -0,06753155 | 0,963044458 | PRICKLE3 | FALSE | Downregulated |
| 0,067490517 | 0,963056898 | CTGF | FALSE | Upregulated |
| 0,104378782 | 0,963061516 | AC012309.5 | FALSE | Upregulated |
| -0,038621658 | 0,963061516 | MRPS18C | FALSE | Downregulated |
| 0,043993888 | 0,963061516 | RP11-426A6.5 | FALSE | Upregulated |
| 0,030016095 | 0,963072552 | ITGA2 | FALSE | Upregulated |
| 0,038578524 | 0,963179365 | ELOF1 | FALSE | Upregulated |
| -0,034797418 | 0,963191615 | DHX33 | FALSE | Downregulated |
| -0,124539893 | 0,963191615 | FBN3 | FALSE | Downregulated |
| 0,049413184 | 0,963191615 | SMCO3 | FALSE | Upregulated |
| -0,043334426 | 0,963191615 | ZNF808 | FALSE | Downregulated |
| -0,054844025 | 0,963375724 | FOXP2 | FALSE | Downregulated |
| -0,133483824 | 0,963375724 | HS3ST5 | FALSE | Downregulated |
| 0,038006708 | 0,963375724 | MRPS27 | FALSE | Upregulated |
| -0,094479469 | 0,963375724 | TRMT6 | FALSE | Downregulated |
| 0,05875353 | 0,963388776 | DZIP1L | FALSE | Upregulated |
| 0,02409453 | 0,963388776 | PPP1CA | FALSE | Upregulated |
| 0,111004729 | 0,96360218 | CLHC1 | FALSE | Upregulated |
| 0,027017047 | 0,963706406 | PAXBP1 | FALSE | Upregulated |
| 0,030931383 | 0,963781334 | ATG4D | FALSE | Upregulated |
| -0,093251143 | 0,963781334 | LOC101927899 | FALSE | Downregulated |
| 0,043253159 | 0,963801594 | EFCAB2 | FALSE | Upregulated |
| -0,032413262 | 0,963801594 | GOLGA8B | FALSE | Downregulated |
| 0,052531354 | 0,963801594 | IL33 | FALSE | Upregulated |
| 0,523588524 | 0,963801594 | KRT5 | FALSE | Upregulated |
| -0,033669961 | 0,963801594 | LOC100996724 | FALSE | Downregulated |
| 0,026170594 | 0,963801594 | PDCD2 | FALSE | Upregulated |
| -0,057782232 | 0,963801594 | ZBTB16 | FALSE | Downregulated |
| 0,025324887 | 0,963837187 | CCT7 | FALSE | Upregulated |
| -0,068361185 | 0,963837187 | CDR2L | FALSE | Downregulated |
| -0,021733101 | 0,963837187 | CFLAR | FALSE | Downregulated |

| | | | | |
|---|---|---|---|---|
| 0,113444314 | 0,963837187 | CPNE6 | FALSE | Upregulated |
| -0,058934501 | 0,963837187 | DCLRE1A | FALSE | Downregulated |
| -0,038712592 | 0,963837187 | DDA1 | FALSE | Downregulated |
| -0,102950038 | 0,963837187 | DNAJC27 | FALSE | Downregulated |
| 0,027196441 | 0,963837187 | FAR2 | FALSE | Upregulated |
| -0,100091409 | 0,963837187 | GRM8 | FALSE | Downregulated |
| 0,116477104 | 0,963837187 | MEF2BNB-MEF2B | FALSE | Upregulated |
| 0,067159958 | 0,963837187 | PILRA | FALSE | Upregulated |
| 0,035815864 | 0,963837187 | PRR5 | FALSE | Upregulated |
| 0,05711859 | 0,963837187 | SAC3D1 | FALSE | Upregulated |
| -0,040661172 | 0,963837187 | SCMH1 | FALSE | Downregulated |
| 0,015426572 | 0,963837187 | SMARCC2 | FALSE | Upregulated |
| -0,032739289 | 0,963837187 | VEZF1 | FALSE | Downregulated |
| -0,023306173 | 0,963837187 | ZNF24 | FALSE | Downregulated |
| -0,034345003 | 0,963837187 | ZNF696 | FALSE | Downregulated |
| -0,112816895 | 0,963952546 | DDX11L1 | FALSE | Downregulated |
| 0,050670846 | 0,964027137 | COL4A3 | FALSE | Upregulated |
| -0,056576965 | 0,964139081 | COQ10B | FALSE | Downregulated |
| 0,066098931 | 0,964341489 | AC006145.4 | FALSE | Upregulated |
| 0,034107386 | 0,964388965 | ADPRM | FALSE | Upregulated |
| -0,046751236 | 0,964388965 | CD9 | FALSE | Downregulated |
| 0,040166781 | 0,964388965 | CNTROB | FALSE | Upregulated |
| 0,047002709 | 0,964388965 | CSRNP1 | FALSE | Upregulated |
| 0,032416628 | 0,964388965 | DOCK7 | FALSE | Upregulated |
| 0,054348578 | 0,964388965 | FAM207A | FALSE | Upregulated |
| -0,020852913 | 0,964388965 | GUK1 | FALSE | Downregulated |
| 0,028138997 | 0,964388965 | LOC100505915 | FALSE | Upregulated |
| 0,076057954 | 0,964388965 | MPV17 | FALSE | Upregulated |
| -0,037899967 | 0,964388965 | RANBP6 | FALSE | Downregulated |
| 0,085345441 | 0,964388965 | RNF224 | FALSE | Upregulated |
| 0,029218356 | 0,964388965 | SMARCE1 | FALSE | Upregulated |
| -0,032928125 | 0,964388965 | UBQLN4 | FALSE | Downregulated |
| -0,054760285 | 0,964660023 | NDUFAF2 | FALSE | Downregulated |
| 0,034671799 | 0,96468259 | ACOT12 | FALSE | Upregulated |
| 0,021172369 | 0,96468259 | CISD3 | FALSE | Upregulated |
| -0,031007961 | 0,96475044 | ATR | FALSE | Downregulated |
| -0,033730697 | 0,964977692 | EDEM2 | FALSE | Downregulated |
| 0,071244363 | 0,965024997 | FDXACB1 | FALSE | Upregulated |
| -0,094392725 | 0,965024997 | LYG1 | FALSE | Downregulated |
| -0,029156002 | 0,965110925 | ACACA | FALSE | Downregulated |
| -0,07822347 | 0,965110925 | B3GALTL | FALSE | Downregulated |
| -0,030238304 | 0,965110925 | BORA | FALSE | Downregulated |
| -0,068818366 | 0,965110925 | C19orf40 | FALSE | Downregulated |
| 0,031929722 | 0,965110925 | CKMT2 | FALSE | Upregulated |
| -0,133230999 | 0,965110925 | DGKB | FALSE | Downregulated |
| -0,025419972 | 0,965110925 | EML3 | FALSE | Downregulated |
| -0,015126644 | 0,965110925 | EP400 | FALSE | Downregulated |
| 0,065584248 | 0,965110925 | FAM109B | FALSE | Upregulated |
| 0,116589673 | 0,965110925 | HIF3A | FALSE | Upregulated |
| -0,03406127 | 0,965110925 | HLTF | FALSE | Downregulated |
| -0,045557974 | 0,965110925 | KCTD7 | FALSE | Downregulated |

| | | | | |
|---|---|---|---|---|
| 0,036079069 | 0,965110925 | LATS1 | FALSE | Upregulated |
| 0,099029925 | 0,965110925 | LOC100129940 | FALSE | Upregulated |
| 0,030260385 | 0,965110925 | LOC101927066 | FALSE | Upregulated |
| -0,026659158 | 0,965110925 | LOC101928946 | FALSE | Downregulated |
| -0,052176549 | 0,965110925 | MAGOH2 | FALSE | Downregulated |
| 0,025473262 | 0,965110925 | MBP | FALSE | Upregulated |
| 0,018596501 | 0,965110925 | MED1 | FALSE | Upregulated |
| -0,058640008 | 0,965110925 | MOSPD2 | FALSE | Downregulated |
| 0,048092799 | 0,965110925 | PEX3 | FALSE | Upregulated |
| 0,037116409 | 0,965110925 | PPP2R5C | FALSE | Upregulated |
| 0,119100931 | 0,965110925 | PRKAG2-AS1 | FALSE | Upregulated |
| 0,04481102 | 0,965110925 | RBM41 | FALSE | Upregulated |
| 0,074237878 | 0,965110925 | RPL36A | FALSE | Upregulated |
| 0,029235855 | 0,965110925 | SF3A2 | FALSE | Upregulated |
| 0,083760773 | 0,965110925 | SLC34A3 | FALSE | Upregulated |
| 0,034129575 | 0,965110925 | TFDP1 | FALSE | Upregulated |
| 0,024899454 | 0,965110925 | TRIM62 | FALSE | Upregulated |
| -0,075821457 | 0,965110925 | ZNF449 | FALSE | Downregulated |
| 0,019451524 | 0,965750328 | ADORA2A-AS1 | FALSE | Upregulated |
| -0,080897622 | 0,965750328 | IL3 | FALSE | Downregulated |
| -0,031036633 | 0,965750328 | PHKA2-AS1 | FALSE | Downregulated |
| -0,029711694 | 0,965750328 | SLC35A4 | FALSE | Downregulated |
| 0,021194193 | 0,965920716 | LOC148413 | FALSE | Upregulated |
| -0,066092958 | 0,965950353 | PRDM7 | FALSE | Downregulated |
| 0,02226581 | 0,966011799 | COQ6 | FALSE | Upregulated |
| 0,025682466 | 0,966011799 | FBXW4 | FALSE | Upregulated |
| -0,026472783 | 0,966011799 | HNRNPA0 | FALSE | Downregulated |
| 0,036335105 | 0,966011799 | MSTO1 | FALSE | Upregulated |
| 0,023401128 | 0,966011799 | NELFCD | FALSE | Upregulated |
| -0,037482071 | 0,966011799 | PIAS2 | FALSE | Downregulated |
| -0,038850762 | 0,966011799 | PSMD9 | FALSE | Downregulated |
| -0,026502327 | 0,966011799 | SAR1A | FALSE | Downregulated |
| 0,06438277 | 0,966011799 | SDHAF2 | FALSE | Upregulated |
| 0,033905516 | 0,966011799 | SYT17 | FALSE | Upregulated |
| 0,047737353 | 0,966011799 | TMEM221 | FALSE | Upregulated |
| -0,03858792 | 0,96605552 | ATPIF1 | FALSE | Downregulated |
| 0,0442463 | 0,96605552 | COX6C | FALSE | Upregulated |
| 0,038888778 | 0,96605552 | EMC7 | FALSE | Upregulated |
| -0,033774119 | 0,96605552 | EXOC8 | FALSE | Downregulated |
| 0,062555747 | 0,96605552 | FAM220A | FALSE | Upregulated |
| -0,022220169 | 0,96605552 | FLJ31306 | FALSE | Downregulated |
| -0,041982551 | 0,96605552 | LOC100132832 | FALSE | Downregulated |
| -0,026938074 | 0,96605552 | MARVELD2 | FALSE | Downregulated |
| 0,073721538 | 0,96605552 | MFSD5 | FALSE | Upregulated |
| -0,015526555 | 0,96605552 | MRPS25 | FALSE | Downregulated |
| 0,03925121 | 0,96605552 | PLRG1 | FALSE | Upregulated |
| 0,035017725 | 0,96605552 | RBM23 | FALSE | Upregulated |
| -0,026232269 | 0,966170303 | PHF1 | FALSE | Downregulated |
| 0,042437981 | 0,966217239 | MAP4K5 | FALSE | Upregulated |
| 0,052707975 | 0,966333888 | TTF1 | FALSE | Upregulated |
| 0,070883611 | 0,966337739 | SLC6A20 | FALSE | Upregulated |

| | | | | |
|---|---|---|---|---|
| -0,039092036 | 0,966362936 | ADSSL1 | FALSE | Downregulated |
| 0,054305707 | 0,966362936 | FBXL7 | FALSE | Upregulated |
| 0,079595157 | 0,966437613 | NAV3 | FALSE | Upregulated |
| 0,075889535 | 0,966437613 | NLGN4X | FALSE | Upregulated |
| 0,048374402 | 0,966437613 | NXPH3 | FALSE | Upregulated |
| 0,021440269 | 0,966455248 | CAMKK2 | FALSE | Upregulated |
| -0,030783674 | 0,966626163 | LOC102723750 | FALSE | Downregulated |
| -0,057415897 | 0,966688665 | MTHFD2 | FALSE | Downregulated |
| -0,015735217 | 0,966701619 | RER1 | FALSE | Downregulated |
| 0,027879309 | 0,966801475 | APOC4-APOC2 | FALSE | Upregulated |
| 0,044710668 | 0,966922853 | ACTR3C | FALSE | Upregulated |
| 0,069481152 | 0,966922853 | C1orf50 | FALSE | Upregulated |
| 0,01597054 | 0,966922853 | CAST | FALSE | Upregulated |
| -0,087020638 | 0,966922853 | CBX2 | FALSE | Downregulated |
| 0,097627098 | 0,966922853 | CCDC77 | FALSE | Upregulated |
| -0,029411092 | 0,966922853 | CHMP7 | FALSE | Downregulated |
| 0,034500973 | 0,966922853 | CIPC | FALSE | Upregulated |
| -0,021938953 | 0,966922853 | CLN5 | FALSE | Downregulated |
| 0,010422349 | 0,966922853 | CNOT1 | FALSE | Upregulated |
| -0,023445215 | 0,966922853 | CTBP2 | FALSE | Downregulated |
| 0,021354028 | 0,966922853 | ENDOG | FALSE | Upregulated |
| -0,034627431 | 0,966922853 | FAM109A | FALSE | Downregulated |
| -0,047408129 | 0,966922853 | FAM99B | FALSE | Downregulated |
| 0,055223527 | 0,966922853 | FLRT2 | FALSE | Upregulated |
| 0,02691543 | 0,966922853 | GPR64 | FALSE | Upregulated |
| -0,095065594 | 0,966922853 | GTF2H2C | FALSE | Downregulated |
| -0,036944636 | 0,966922853 | HIST1H2BC | FALSE | Downregulated |
| -0,015697558 | 0,966922853 | HNRNPK | FALSE | Downregulated |
| 0,02514517 | 0,966922853 | HPN-AS1 | FALSE | Upregulated |
| -0,03771198 | 0,966922853 | INTS9 | FALSE | Downregulated |
| -0,049054648 | 0,966922853 | KCNQ1 | FALSE | Downregulated |
| -0,141146487 | 0,966922853 | LOC102724374 | FALSE | Downregulated |
| 0,040977406 | 0,966922853 | LOC102724474 | FALSE | Upregulated |
| -0,053974473 | 0,966922853 | MAP3K8 | FALSE | Downregulated |
| -0,0202103 | 0,966922853 | MGLL | FALSE | Downregulated |
| -0,023658718 | 0,966922853 | MLH3 | FALSE | Downregulated |
| 0,029125632 | 0,966922853 | NAA50 | FALSE | Upregulated |
| 0,014699135 | 0,966922853 | NIPBL | FALSE | Upregulated |
| -0,048688636 | 0,966922853 | NR1D2 | FALSE | Downregulated |
| 0,037863858 | 0,966922853 | PHTF1 | FALSE | Upregulated |
| 0,052586913 | 0,966922853 | PMS2P3 | FALSE | Upregulated |
| -0,0270206 | 0,966922853 | PPP2R5E | FALSE | Downregulated |
| 0,051025777 | 0,966922853 | PRIMPOL | FALSE | Upregulated |
| 0,038306811 | 0,966922853 | PSMA1 | FALSE | Upregulated |
| 0,047705128 | 0,966922853 | PSMD14 | FALSE | Upregulated |
| 0,02907517 | 0,966922853 | RC3H2 | FALSE | Upregulated |
| -0,029930771 | 0,966922853 | RNF38 | FALSE | Downregulated |
| -0,048104467 | 0,966922853 | RRP36 | FALSE | Downregulated |
| 0,037294618 | 0,966922853 | SCFD2 | FALSE | Upregulated |
| 0,020314413 | 0,966922853 | SLC12A6 | FALSE | Upregulated |
| -0,108752514 | 0,966922853 | SLC16A9 | FALSE | Downregulated |

| | | | | |
|---|---|---|---|---|
| -0,066710652 | 0,966922853 | SNORA74B | FALSE | Downregulated |
| -0,033046827 | 0,966922853 | SPPL3 | FALSE | Downregulated |
| -0,033254907 | 0,966922853 | STARD3 | FALSE | Downregulated |
| 0,038319174 | 0,966922853 | TCP11L1 | FALSE | Upregulated |
| -0,0932225 | 0,966922853 | TEAD4 | FALSE | Downregulated |
| 0,035549211 | 0,966922853 | TH | FALSE | Upregulated |
| -0,031099791 | 0,966922853 | TMEM198B | FALSE | Downregulated |
| 0,074864518 | 0,966922853 | TMEM237 | FALSE | Upregulated |
| 0,106312559 | 0,966922853 | VGLL3 | FALSE | Upregulated |
| -0,015552433 | 0,966922853 | VPS4A | FALSE | Downregulated |
| -0,027730657 | 0,966922853 | WDR5 | FALSE | Downregulated |
| -0,023235637 | 0,966922853 | ZBTB5 | FALSE | Downregulated |
| 0,041551577 | 0,966922853 | ZDHHC4 | FALSE | Upregulated |
| -0,025024507 | 0,967013837 | C4orf3 | FALSE | Downregulated |
| 0,046468964 | 0,967089349 | MAP7D3 | FALSE | Upregulated |
| 0,020561766 | 0,967089349 | ZNF460 | FALSE | Upregulated |
| -0,04187672 | 0,967235046 | EPC1 | FALSE | Downregulated |
| 0,092229082 | 0,967338454 | KCNT1 | FALSE | Upregulated |
| -0,0216662 | 0,967414213 | MED12 | FALSE | Downregulated |
| 0,038002273 | 0,967414213 | SEPW1 | FALSE | Upregulated |
| 0,058156463 | 0,967414213 | SQLE | FALSE | Upregulated |
| -0,032055129 | 0,967421327 | CMTM3 | FALSE | Downregulated |
| -0,062482859 | 0,967421327 | MRPL33 | FALSE | Downregulated |
| -0,045337931 | 0,967421327 | NAPB | FALSE | Downregulated |
| -0,029968075 | 0,967421327 | SOX18 | FALSE | Downregulated |
| -0,020142281 | 0,967488247 | SF3B3 | FALSE | Downregulated |
| 0,044950279 | 0,967488247 | ZNF790-AS1 | FALSE | Upregulated |
| -0,082672978 | 0,967591904 | RP11-673C5.2 | FALSE | Downregulated |
| 0,020074183 | 0,967783093 | TRIM28 | FALSE | Upregulated |
| -0,039711485 | 0,967808426 | CBX3 | FALSE | Downregulated |
| -0,0918831 | 0,967826551 | HOXB2 | FALSE | Downregulated |
| 0,072249803 | 0,967826551 | LINC01184 | FALSE | Upregulated |
| 0,028295439 | 0,967826551 | ZNF546 | FALSE | Upregulated |
| -0,098181065 | 0,967989482 | FAM86C2P | FALSE | Downregulated |
| 0,048524604 | 0,968195727 | TMEM186 | FALSE | Upregulated |
| -0,051067787 | 0,968195727 | ZNF143 | FALSE | Downregulated |
| -0,046664262 | 0,96822199 | LOC100126582 | FALSE | Downregulated |
| -0,023371737 | 0,96822199 | NBEAL1 | FALSE | Downregulated |
| 0,115911295 | 0,968235032 | DDX43 | FALSE | Upregulated |
| -0,099582021 | 0,968235032 | IP6K3 | FALSE | Downregulated |
| 0,029663997 | 0,968235032 | PEX1 | FALSE | Upregulated |
| 0,036434218 | 0,968235032 | SNRPC | FALSE | Upregulated |
| -0,127105746 | 0,968235032 | SPECC1L-ADORA2A | FALSE | Downregulated |
| 0,045814372 | 0,968235032 | ZNF627 | FALSE | Upregulated |
| -0,017520608 | 0,968392745 | HOOK3 | FALSE | Downregulated |
| -0,018175352 | 0,968462098 | ARNT | FALSE | Downregulated |
| -0,028565322 | 0,968462098 | FOXO1 | FALSE | Downregulated |
| 0,067414163 | 0,968462098 | LOC101927941 | FALSE | Upregulated |
| 0,076020647 | 0,968618994 | C4orf21 | FALSE | Upregulated |
| 0,080593761 | 0,968618994 | C6orf10 | FALSE | Upregulated |

| | | | | |
|---|---|---|---|---|
| -0,018644949 | 0,968618994 | SDHA | FALSE | Downregulated |
| 0,071592173 | 0,968618994 | SNRPD1 | FALSE | Upregulated |
| 0,025164926 | 0,968677654 | C5orf42 | FALSE | Upregulated |
| -0,078803512 | 0,968677654 | LOC101927407 | FALSE | Downregulated |
| 0,019425154 | 0,968680116 | GRTP1 | FALSE | Upregulated |
| 0,018454554 | 0,96879547 | MEGF9 | FALSE | Upregulated |
| -0,036192227 | 0,968877932 | CPLX1 | FALSE | Downregulated |
| -0,026099788 | 0,968877932 | LCOR | FALSE | Downregulated |
| -0,035046551 | 0,968877932 | LRRN1 | FALSE | Downregulated |
| 0,023482606 | 0,968949809 | MAN1B1 | FALSE | Upregulated |
| 0,042912609 | 0,969045038 | B4GALT6 | FALSE | Upregulated |
| 0,047516178 | 0,969045038 | CCDC183-AS1 | FALSE | Upregulated |
| -0,01745087 | 0,969045038 | GSTK1 | FALSE | Downregulated |
| -0,096648586 | 0,969045038 | LDB3 | FALSE | Downregulated |
| -0,066819956 | 0,969045038 | RFPL1 | FALSE | Downregulated |
| 0,046653805 | 0,969045038 | SUCLA2 | FALSE | Upregulated |
| -0,024335164 | 0,969045038 | TXLNG | FALSE | Downregulated |
| -0,090967603 | 0,969045038 | ZNF570 | FALSE | Downregulated |
| 0,040508409 | 0,969145489 | COX7A1 | FALSE | Upregulated |
| 0,039464482 | 0,969145489 | DESI2 | FALSE | Upregulated |
| -0,074767469 | 0,969145489 | FAM174A | FALSE | Downregulated |
| -0,092333041 | 0,969145489 | FOXD4L4 | FALSE | Downregulated |
| -0,040637718 | 0,969145489 | IQSEC2 | FALSE | Downregulated |
| -0,023572844 | 0,969145489 | PRSS48 | FALSE | Downregulated |
| 0,077225963 | 0,969145489 | RACGAP1 | FALSE | Upregulated |
| -0,021855544 | 0,969145489 | RGPD1 | FALSE | Downregulated |
| -0,061795697 | 0,969145489 | SERPINE2 | FALSE | Downregulated |
| 0,018217214 | 0,969145489 | SNX9 | FALSE | Upregulated |
| 0,026558833 | 0,969145489 | TLR4 | FALSE | Upregulated |
| -0,049404824 | 0,969145489 | ZNF134 | FALSE | Downregulated |
| 0,018446676 | 0,969302362 | PRKAR1A | FALSE | Upregulated |
| -0,032793993 | 0,969326877 | KIAA0196 | FALSE | Downregulated |
| -0,031075302 | 0,969449486 | DDX19B | FALSE | Downregulated |
| 0,021213816 | 0,969449486 | LOC101928042 | FALSE | Upregulated |
| 0,049925298 | 0,969449486 | SHC3 | FALSE | Upregulated |
| -0,071064063 | 0,96950298 | ATG10 | FALSE | Downregulated |
| 0,070781961 | 0,96950298 | EVC2 | FALSE | Upregulated |
| -0,02173224 | 0,96950298 | EXTL3 | FALSE | Downregulated |
| 0,031293637 | 0,96950298 | GNAT1 | FALSE | Upregulated |
| -0,030960205 | 0,96950298 | KIAA0907 | FALSE | Downregulated |
| 0,022528722 | 0,96950298 | PSD3 | FALSE | Upregulated |
| 0,038044015 | 0,96950298 | TRMT2B | FALSE | Upregulated |
| -0,022041299 | 0,96950298 | ZNRF1 | FALSE | Downregulated |
| -0,037822549 | 0,969564131 | C4orf29 | FALSE | Downregulated |
| -0,089417065 | 0,969619086 | LOC102724442 | FALSE | Downregulated |
| 0,118816553 | 0,969922827 | CCL15 | FALSE | Upregulated |
| 0,047477127 | 0,969922827 | CYP2W1 | FALSE | Upregulated |
| -0,043042792 | 0,970072374 | KIAA0895 | FALSE | Downregulated |
| 0,085671444 | 0,970078005 | DUSP19 | FALSE | Upregulated |
| 0,03890999 | 0,970174053 | ZDHHC21 | FALSE | Upregulated |
| 0,029752878 | 0,970184489 | IKZF4 | FALSE | Upregulated |

| | | | | |
|---|---|---|---|---|
| -0,074956977 | 0,970184489 | ODF3L2 | FALSE | Downregulated |
| -0,050156044 | 0,970200361 | CAPN10-AS1 | FALSE | Downregulated |
| 0,086130718 | 0,970287126 | CSF2RA | FALSE | Upregulated |
| 0,059933577 | 0,970433549 | C15orf61 | FALSE | Upregulated |
| -0,029413448 | 0,970433549 | FAM192A | FALSE | Downregulated |
| 0,052913559 | 0,970433549 | LOC102724286 | FALSE | Upregulated |
| 0,023909594 | 0,970433549 | NFYB | FALSE | Upregulated |
| -0,040704797 | 0,970433549 | RABIF | FALSE | Downregulated |
| -0,020802974 | 0,970433549 | RIPK1 | FALSE | Downregulated |
| 0,031886575 | 0,970433549 | SENP8 | FALSE | Upregulated |
| 0,017333161 | 0,970433549 | SIDT2 | FALSE | Upregulated |
| 0,016493241 | 0,970433549 | STX17 | FALSE | Upregulated |
| 0,021332523 | 0,970433549 | TAF15 | FALSE | Upregulated |
| 0,043018617 | 0,970433549 | TICAM2 | FALSE | Upregulated |
| -0,050237096 | 0,970528526 | FASTKD5 | FALSE | Downregulated |
| -0,050185544 | 0,970656148 | EXOC6 | FALSE | Downregulated |
| -0,01069648 | 0,970656148 | NCOR1 | FALSE | Downregulated |
| 0,025804053 | 0,970714328 | LOC102723566 | FALSE | Upregulated |
| 0,04988326 | 0,970714328 | MTCP1 | FALSE | Upregulated |
| -0,039779183 | 0,970822404 | MGST2 | FALSE | Downregulated |
| 0,034745822 | 0,970868974 | ANKRD46 | FALSE | Upregulated |
| 0,081799222 | 0,970868974 | CCDC83 | FALSE | Upregulated |
| 0,192538101 | 0,970868974 | FLG-AS1 | FALSE | Upregulated |
| -0,053693838 | 0,970868974 | HYPK | FALSE | Downregulated |
| 0,015122449 | 0,970868974 | IRF2BPL | FALSE | Upregulated |
| -0,048457218 | 0,970868974 | LOC102723381 | FALSE | Downregulated |
| 0,044596431 | 0,970868974 | LOC493754 | FALSE | Upregulated |
| 0,012077377 | 0,970868974 | MGAT4B | FALSE | Upregulated |
| 0,020081471 | 0,970868974 | NUP98 | FALSE | Upregulated |
| 0,080273265 | 0,970868974 | OLFML2A | FALSE | Upregulated |
| -0,055350278 | 0,970868974 | PDE1A | FALSE | Downregulated |
| -0,054125415 | 0,970868974 | PMEPA1 | FALSE | Downregulated |
| -0,056358192 | 0,970868974 | PPCDC | FALSE | Downregulated |
| -0,04959346 | 0,970868974 | RAB9A | FALSE | Downregulated |
| 0,029387473 | 0,970868974 | RBBP9 | FALSE | Upregulated |
| 0,02962035 | 0,970868974 | RP11-475I24.8 | FALSE | Upregulated |
| 0,023561305 | 0,970868974 | SRP72 | FALSE | Upregulated |
| 0,049381597 | 0,970868974 | UBE2Q2 | FALSE | Upregulated |
| 0,021106083 | 0,970868974 | WDR55 | FALSE | Upregulated |
| 0,024029831 | 0,970868974 | ZNF274 | FALSE | Upregulated |
| 0,069180313 | 0,970868974 | ZSCAN20 | FALSE | Upregulated |
| -0,098237474 | 0,970889406 | TTTY14 | FALSE | Downregulated |
| 0,025932139 | 0,971011049 | BOP1 | FALSE | Upregulated |
| -0,06475143 | 0,971011049 | FAM90A1 | FALSE | Downregulated |
| -0,033437993 | 0,971011049 | LOC102723811 | FALSE | Downregulated |
| -0,088752305 | 0,971037632 | S100P | FALSE | Downregulated |
| 0,026436338 | 0,971037632 | SND1-IT1 | FALSE | Upregulated |
| 0,014261551 | 0,971077773 | ANKRD11 | FALSE | Upregulated |
| -0,034108529 | 0,971077773 | C20orf27 | FALSE | Downregulated |
| 0,081864134 | 0,971077773 | C7orf60 | FALSE | Upregulated |
| -0,058093204 | 0,971077773 | CBLN3 | FALSE | Downregulated |

| | | | | |
|---|---|---|---|---|
| -0,02807646 | 0,971077773 | CLDN12 | FALSE | Downregulated |
| 0,011817189 | 0,971077773 | EML4 | FALSE | Upregulated |
| -0,046989266 | 0,971077773 | EXOSC4 | FALSE | Downregulated |
| 0,082185991 | 0,971077773 | FUT1 | FALSE | Upregulated |
| -0,020578324 | 0,971077773 | FYCO1 | FALSE | Downregulated |
| 0,048977668 | 0,971077773 | GPATCH2 | FALSE | Upregulated |
| -0,067072358 | 0,971077773 | HDAC11 | FALSE | Downregulated |
| 0,010888054 | 0,971077773 | LOC100129034 | FALSE | Upregulated |
| 0,087673045 | 0,971077773 | LOC102724515 | FALSE | Upregulated |
| -0,029376338 | 0,971077773 | MLH1 | FALSE | Downregulated |
| 0,024761236 | 0,971077773 | OSBP2 | FALSE | Upregulated |
| -0,023594195 | 0,971077773 | PPP3CA | FALSE | Downregulated |
| -0,067460071 | 0,971077773 | PTCHD4 | FALSE | Downregulated |
| 0,022731811 | 0,971077773 | RNF149 | FALSE | Upregulated |
| 0,019253981 | 0,971077773 | SCAF4 | FALSE | Upregulated |
| -0,035741066 | 0,971077773 | TAT | FALSE | Downregulated |
| 0,050858896 | 0,971077773 | TTC21B-AS1 | FALSE | Upregulated |
| -0,042732298 | 0,971077773 | TTC27 | FALSE | Downregulated |
| -0,04559635 | 0,971169316 | GLI4 | FALSE | Downregulated |
| -0,082764441 | 0,971181659 | ANKRD18A | FALSE | Downregulated |
| -0,024938742 | 0,971181659 | MIF | FALSE | Downregulated |
| 0,014102451 | 0,971181659 | NIT1 | FALSE | Upregulated |
| -0,039631946 | 0,971181659 | PHKA1 | FALSE | Downregulated |
| 0,059804378 | 0,971181659 | ZSCAN16-AS1 | FALSE | Upregulated |
| -0,024866998 | 0,971258403 | ATXN3 | FALSE | Downregulated |
| 0,032037706 | 0,971258403 | FLJ46906 | FALSE | Upregulated |
| 0,033121852 | 0,971258403 | INS | FALSE | Upregulated |
| 0,029274742 | 0,971258403 | PIH1D1 | FALSE | Upregulated |
| -0,036279416 | 0,971258403 | POLR2I | FALSE | Downregulated |
| -0,024826389 | 0,971258403 | RAB2A | FALSE | Downregulated |
| 0,044502655 | 0,971258403 | RAD17 | FALSE | Upregulated |
| -0,041124649 | 0,971258403 | SLC9A9 | FALSE | Downregulated |
| 0,072487091 | 0,971258403 | TNFAIP8L2-SCNM1 | FALSE | Upregulated |
| -0,018975367 | 0,971276108 | ACIN1 | FALSE | Downregulated |
| 0,04036904 | 0,971276108 | AURKA | FALSE | Upregulated |
| 0,066267008 | 0,971276108 | GPR135 | FALSE | Upregulated |
| 0,053557866 | 0,971276108 | LOC100862671 | FALSE | Upregulated |
| 0,036573565 | 0,971276108 | LOC613037 | FALSE | Upregulated |
| -0,033845411 | 0,971276108 | MSRB2 | FALSE | Downregulated |
| -0,033276546 | 0,971276108 | PPARGC1B | FALSE | Downregulated |
| -0,105252735 | 0,971276108 | TEX13A | FALSE | Downregulated |
| -0,026049931 | 0,971276108 | UBE2V1 | FALSE | Downregulated |
| -0,03079016 | 0,971276108 | USP30-AS1 | FALSE | Downregulated |
| 0,046456567 | 0,971276108 | ZSCAN31 | FALSE | Upregulated |
| -0,038002803 | 0,971290253 | ADCK2 | FALSE | Downregulated |
| 0,040130878 | 0,971290253 | EDAR | FALSE | Upregulated |
| 0,021350281 | 0,971290253 | ITGB3 | FALSE | Upregulated |
| -0,090306315 | 0,971290253 | LOC285556 | FALSE | Downregulated |
| -0,054279711 | 0,971290253 | PRKAR2B | FALSE | Downregulated |
| -0,033706767 | 0,971347264 | LOC728763 | FALSE | Downregulated |
| -0,030442729 | 0,971347264 | SLC35C2 | FALSE | Downregulated |

| | | | | |
|---|---|---|---|---|
| -0,033076428 | 0,971469147 | ABCG1 | FALSE | Downregulated |
| 0,033942878 | 0,971469147 | WDR44 | FALSE | Upregulated |
| -0,118177089 | 0,971508227 | AKAP2 | FALSE | Downregulated |
| -0,190508318 | 0,971508227 | F8A3 | FALSE | Downregulated |
| 0,064710663 | 0,971508227 | HCCAT3 | FALSE | Upregulated |
| -0,051038529 | 0,971508227 | MED9 | FALSE | Downregulated |
| -0,02899935 | 0,971508227 | OCEL1 | FALSE | Downregulated |
| -0,066816279 | 0,971508227 | RP5-968D22.1 | FALSE | Downregulated |
| -0,013570127 | 0,971508227 | SF3B1 | FALSE | Downregulated |
| 0,042804737 | 0,971802428 | ZNRF2P2 | FALSE | Upregulated |
| -0,032378491 | 0,97189754 | ALG9 | FALSE | Downregulated |
| 0,030179235 | 0,971900475 | HFE | FALSE | Upregulated |
| 0,046195866 | 0,971900475 | ZNF180 | FALSE | Upregulated |
| -0,049793116 | 0,971932498 | TRAPPC2L | FALSE | Downregulated |
| -0,064266102 | 0,971978409 | LINC00623 | FALSE | Downregulated |
| 0,067512957 | 0,971978409 | TXLNG2P | FALSE | Upregulated |
| 0,074665458 | 0,971980147 | LOC201651 | FALSE | Upregulated |
| 0,090039778 | 0,972120844 | REP15 | FALSE | Upregulated |
| 0,068206549 | 0,97217088 | HCAR2 | FALSE | Upregulated |
| 0,041701068 | 0,972315818 | CORO2A | FALSE | Upregulated |
| 0,024301662 | 0,972315818 | FBXO8 | FALSE | Upregulated |
| -0,072479844 | 0,972315818 | IZUMO4 | FALSE | Downregulated |
| 0,041462681 | 0,972315818 | LOC101928762 | FALSE | Upregulated |
| -0,020054755 | 0,972315818 | MRPL41 | FALSE | Downregulated |
| -0,030905927 | 0,972315818 | NMNAT3 | FALSE | Downregulated |
| -0,029230101 | 0,972315818 | PAOX | FALSE | Downregulated |
| 0,013472265 | 0,972315818 | SGPL1 | FALSE | Upregulated |
| 0,022819114 | 0,972315818 | USP20 | FALSE | Upregulated |
| 0,02130786 | 0,972380488 | SLC25A18 | FALSE | Upregulated |
| 0,018852273 | 0,972426313 | PPP1R13B | FALSE | Upregulated |
| -0,022478852 | 0,972473413 | C12orf43 | FALSE | Downregulated |
| -0,018699154 | 0,97256451 | ACVR2A | FALSE | Downregulated |
| 0,010215991 | 0,97256451 | CLTC | FALSE | Upregulated |
| 0,225729929 | 0,97256451 | COMMD3-BMI1 | FALSE | Upregulated |
| 0,075454451 | 0,97256451 | FIBIN | FALSE | Upregulated |
| 0,030509681 | 0,97256451 | MEF2C | FALSE | Upregulated |
| -0,096967526 | 0,97256451 | TUBB2B | FALSE | Downregulated |
| 0,018910916 | 0,97256451 | ZKSCAN8 | FALSE | Upregulated |
| -0,010625606 | 0,972719481 | ARID1B | FALSE | Downregulated |
| -0,052656927 | 0,972719481 | CLP1 | FALSE | Downregulated |
| -0,043658655 | 0,972719481 | TFB2M | FALSE | Downregulated |
| -0,076644513 | 0,972770814 | ASIP | FALSE | Downregulated |
| 0,025262735 | 0,972903783 | SMIM15 | FALSE | Upregulated |
| 0,060142731 | 0,97298693 | CACNA1G-AS1 | FALSE | Upregulated |
| 0,058600494 | 0,973053068 | TOP1P1 | FALSE | Upregulated |
| 0,024426476 | 0,973075704 | DHCR7 | FALSE | Upregulated |
| -0,024541907 | 0,973075704 | GNA14 | FALSE | Downregulated |
| 0,029831197 | 0,973075704 | RPS6KC1 | FALSE | Upregulated |
| 0,072796128 | 0,97313044 | ZNF19 | FALSE | Upregulated |
| -0,018927089 | 0,973152625 | RNF44 | FALSE | Downregulated |
| -0,074537574 | 0,973170864 | FAM26E | FALSE | Downregulated |

| | | | | |
|---|---|---|---|---|
| 0,040332135 | 0,973170864 | ZNF117 | FALSE | Upregulated |
| 0,05611676 | 0,973170864 | ZNF419 | FALSE | Upregulated |
| -0,066877686 | 0,97329945 | GJC3 | FALSE | Downregulated |
| 0,030416973 | 0,973336801 | PAK1 | FALSE | Upregulated |
| 0,043369188 | 0,973466268 | ZNF202 | FALSE | Upregulated |
| -0,019792772 | 0,973495498 | AVIL | FALSE | Downregulated |
| 0,036576995 | 0,973495498 | COPS8 | FALSE | Upregulated |
| -0,048961736 | 0,973495498 | FHL2 | FALSE | Downregulated |
| -0,022737414 | 0,973495498 | LOC102723554 | FALSE | Downregulated |
| -0,023142437 | 0,973495498 | MCM9 | FALSE | Downregulated |
| 0,014104635 | 0,973495498 | RAB7A | FALSE | Upregulated |
| -0,02362589 | 0,973495498 | STRN | FALSE | Downregulated |
| 0,063748643 | 0,973495498 | ZNF138 | FALSE | Upregulated |
| -0,072525555 | 0,973542203 | LOC101927009 | FALSE | Downregulated |
| 0,076903804 | 0,973578564 | ICOSLG | FALSE | Upregulated |
| 0,13387046 | 0,973578564 | LOC102723584 | FALSE | Upregulated |
| -0,044651778 | 0,973578564 | PKN3 | FALSE | Downregulated |
| -0,011588127 | 0,973774355 | NUCB1 | FALSE | Downregulated |
| 0,035644941 | 0,973856483 | CABP4 | FALSE | Upregulated |
| 0,04614465 | 0,973856483 | SORT1 | FALSE | Upregulated |
| 0,023586514 | 0,973910344 | CSNK2A1 | FALSE | Upregulated |
| -0,022303368 | 0,973910344 | EHD1 | FALSE | Downregulated |
| -0,026430536 | 0,973910344 | ORC2 | FALSE | Downregulated |
| -0,026221527 | 0,973910344 | TMEM9 | FALSE | Downregulated |
| -0,029868608 | 0,974025268 | CDC42P3 | FALSE | Downregulated |
| -0,013573539 | 0,974050026 | GNPNAT1 | FALSE | Downregulated |
| 0,03648798 | 0,974050026 | MAP6 | FALSE | Upregulated |
| 0,051074824 | 0,974050026 | POLE4 | FALSE | Upregulated |
| 0,016474037 | 0,974050026 | SYNCRIP | FALSE | Upregulated |
| -0,023177329 | 0,974050026 | TIMM8B | FALSE | Downregulated |
| 0,027814528 | 0,974050026 | ZBTB14 | FALSE | Upregulated |
| 0,041077934 | 0,974064898 | LEKR1 | FALSE | Upregulated |
| -0,03064633 | 0,974097391 | CENPBD1P1 | FALSE | Downregulated |
| 0,023519861 | 0,974097391 | PSMB5 | FALSE | Upregulated |
| -0,076759836 | 0,974193532 | LINC00338 | FALSE | Downregulated |
| 0,051452173 | 0,974193532 | PIGA | FALSE | Upregulated |
| -0,044949749 | 0,974193532 | TTC9C | FALSE | Downregulated |
| 0,035751424 | 0,974335053 | CCT6P1 | FALSE | Upregulated |
| 0,021260471 | 0,974341747 | GPR179 | FALSE | Upregulated |
| -0,018301685 | 0,974382027 | BID | FALSE | Downregulated |
| 0,022961208 | 0,974382027 | KIAA1199 | FALSE | Upregulated |
| -0,012650669 | 0,974413074 | RREB1 | FALSE | Downregulated |
| -0,020707932 | 0,974418959 | AC009948.5 | FALSE | Downregulated |
| -0,079515263 | 0,974432518 | ADAM21P1 | FALSE | Downregulated |
| -0,027017688 | 0,974432518 | HACL1 | FALSE | Downregulated |
| -0,012791115 | 0,974467911 | AUTS2 | FALSE | Downregulated |
| -0,236083979 | 0,974602277 | C10orf32-ASMT | FALSE | Downregulated |
| -0,059586102 | 0,974602277 | CLIC2 | FALSE | Downregulated |
| 0,021105941 | 0,974602277 | CNPY4 | FALSE | Upregulated |
| -0,048296665 | 0,974602277 | LOC101927054 | FALSE | Downregulated |
| 0,057250908 | 0,974602277 | SLC9A3 | FALSE | Upregulated |

| | | | | |
|---|---|---|---|---|
| 0,027419238 | 0,974602277 | TBC1D15 | FALSE | Upregulated |
| 0,041809318 | 0,974602277 | TRIM68 | FALSE | Upregulated |
| -0,048391839 | 0,974610286 | DNLZ | FALSE | Downregulated |
| 0,057458989 | 0,974610286 | LOC101929719 | FALSE | Upregulated |
| -0,022376265 | 0,974610286 | MPDU1 | FALSE | Downregulated |
| 0,013769587 | 0,974610286 | NRD1 | FALSE | Upregulated |
| -0,021467158 | 0,974610286 | ZNF263 | FALSE | Downregulated |
| -0,058312459 | 0,974784582 | SPEF2 | FALSE | Downregulated |
| -0,015540389 | 0,974784582 | ZMYM2 | FALSE | Downregulated |
| -0,068335757 | 0,974886457 | C22orf34 | FALSE | Downregulated |
| -0,047611068 | 0,974886457 | FLJ23867 | FALSE | Downregulated |
| -0,016905511 | 0,974886457 | SLC25A4 | FALSE | Downregulated |
| -0,067180371 | 0,974887216 | JHDM1D-AS1 | FALSE | Downregulated |
| 0,038064252 | 0,974887216 | RAD1 | FALSE | Upregulated |
| -0,035000465 | 0,975037777 | RBKS | FALSE | Downregulated |
| -0,087831503 | 0,975196146 | RP11-789C17.3 | FALSE | Downregulated |
| -0,026367088 | 0,975293673 | ACSS2 | FALSE | Downregulated |
| -0,242775486 | 0,975293673 | MYO18B | FALSE | Downregulated |
| -0,028451005 | 0,975323715 | ARFIP2 | FALSE | Downregulated |
| -0,065228257 | 0,975323715 | SLC25A21-AS1 | FALSE | Downregulated |
| 0,036764471 | 0,975363447 | C7orf25 | FALSE | Upregulated |
| 0,062739401 | 0,975363447 | CORO7 | FALSE | Upregulated |
| 0,070726099 | 0,975363447 | GPR52 | FALSE | Upregulated |
| 0,019753313 | 0,975363447 | NOLC1 | FALSE | Upregulated |
| 0,017038642 | 0,975363447 | NOTCH2NL | FALSE | Upregulated |
| 0,024579512 | 0,975363447 | PRKAA2 | FALSE | Upregulated |
| -0,063318605 | 0,975363447 | RP11-637A17.2 | FALSE | Downregulated |
| 0,031220739 | 0,975363447 | SERPINE3 | FALSE | Upregulated |
| 0,039759706 | 0,975363447 | SOCS2 | FALSE | Upregulated |
| 0,041576872 | 0,975363447 | STYXL1 | FALSE | Upregulated |
| 0,035319749 | 0,975363447 | TERC | FALSE | Upregulated |
| 0,020282769 | 0,975363447 | TMF1 | FALSE | Upregulated |
| 0,017010802 | 0,975363447 | TRPC4AP | FALSE | Upregulated |
| -0,025338525 | 0,975372022 | RAB3IL1 | FALSE | Downregulated |
| 0,047128931 | 0,97548176 | CCDC43 | FALSE | Upregulated |
| 0,025619345 | 0,975517824 | B4GALT2 | FALSE | Upregulated |
| 0,01922926 | 0,975547991 | CNPY3 | FALSE | Upregulated |
| -0,019669881 | 0,975547991 | EVA1A | FALSE | Downregulated |
| -0,0138826 | 0,975547991 | ITSN2 | FALSE | Downregulated |
| 0,009330944 | 0,975626458 | LSR | FALSE | Upregulated |
| 0,042339309 | 0,975626458 | SNORA7A | FALSE | Upregulated |
| -0,039902882 | 0,975626458 | TBCC | FALSE | Downregulated |
| -0,047434598 | 0,975864963 | C1orf145 | FALSE | Downregulated |
| 0,066388952 | 0,975864963 | STK32C | FALSE | Upregulated |
| 0,058595983 | 0,975907727 | ITGB3BP | FALSE | Upregulated |
| 0,030892747 | 0,976054795 | SRPX2 | FALSE | Upregulated |
| -0,09611128 | 0,976164294 | RP11-506B6.6 | FALSE | Downregulated |
| -0,041290059 | 0,976176882 | POLR1C | FALSE | Downregulated |
| -0,025257694 | 0,976178214 | ERI3 | FALSE | Downregulated |
| 0,058060702 | 0,976178214 | RS1 | FALSE | Upregulated |
| 0,0324817 | 0,976223019 | ZNF195 | FALSE | Upregulated |

| | | | | |
|---|---|---|---|---|
| -0,075405026 | 0,976249256 | ADAMTS15 | FALSE | Downregulated |
| -0,042796808 | 0,976249256 | CCZ1 | FALSE | Downregulated |
| 0,089285722 | 0,976249256 | CDHR1 | FALSE | Upregulated |
| 0,015280135 | 0,976249256 | EXT2 | FALSE | Upregulated |
| -0,026345289 | 0,976249256 | NRL | FALSE | Downregulated |
| -0,025979689 | 0,976249256 | RNPC3 | FALSE | Downregulated |
| -0,016962408 | 0,976249256 | RP11-887P2.5 | FALSE | Downregulated |
| -0,013987494 | 0,976249256 | UFD1L | FALSE | Downregulated |
| 0,017912157 | 0,976288615 | COPE | FALSE | Upregulated |
| -0,018274725 | 0,976288615 | PPP1R37 | FALSE | Downregulated |
| 0,016911025 | 0,976326342 | DHX30 | FALSE | Upregulated |
| 0,051616531 | 0,976326342 | LOC101927910 | FALSE | Upregulated |
| -0,028423457 | 0,976326342 | ZNF830 | FALSE | Downregulated |
| 0,020277495 | 0,976359257 | C10orf54 | FALSE | Upregulated |
| 0,035638988 | 0,976545467 | CHIC2 | FALSE | Upregulated |
| 0,172478139 | 0,976545467 | MYCBP | FALSE | Upregulated |
| 0,059722641 | 0,976560706 | CLDN11 | FALSE | Upregulated |
| 0,058259439 | 0,976560706 | LY6K | FALSE | Upregulated |
| 0,039089123 | 0,976560706 | PAPOLG | FALSE | Upregulated |
| -0,048265778 | 0,976560706 | SLCO1B7 | FALSE | Downregulated |
| -0,011488006 | 0,976560706 | VPS26B | FALSE | Downregulated |
| -0,038651265 | 0,976617094 | ADAT2 | FALSE | Downregulated |
| -0,092303164 | 0,976617094 | CCDC168 | FALSE | Downregulated |
| 0,444386599 | 0,976617094 | RNU1-4 | FALSE | Upregulated |
| 0,06104442 | 0,976617094 | TRPM5 | FALSE | Upregulated |
| -0,0351405 | 0,976645471 | VWF | FALSE | Downregulated |
| -0,02991961 | 0,976916441 | TBKBP1 | FALSE | Downregulated |
| -0,020004198 | 0,976999351 | CCDC6 | FALSE | Downregulated |
| 0,018910085 | 0,976999351 | GOSR1 | FALSE | Upregulated |
| 0,045008518 | 0,976999351 | GPIHBP1 | FALSE | Upregulated |
| -0,050287875 | 0,976999351 | NDUFAF5 | FALSE | Downregulated |
| -0,018499315 | 0,977113769 | FAM129B | FALSE | Downregulated |
| -0,033857674 | 0,977113769 | GPAT2 | FALSE | Downregulated |
| 0,034096882 | 0,977113769 | LOC101928595 | FALSE | Upregulated |
| 0,059407633 | 0,977113769 | LOC102724126 | FALSE | Upregulated |
| 0,017966628 | 0,977113769 | MICA | FALSE | Upregulated |
| -0,031958899 | 0,977113769 | PSAPL1 | FALSE | Downregulated |
| -0,035271651 | 0,977166103 | AEBP2 | FALSE | Downregulated |
| -0,03019093 | 0,977166103 | EFHD1 | FALSE | Downregulated |
| -0,02952819 | 0,977166103 | ENOX2 | FALSE | Downregulated |
| -0,089487114 | 0,977166103 | GALNT4 | FALSE | Downregulated |
| 0,082581407 | 0,977166103 | SMN2 | FALSE | Upregulated |
| -0,045557377 | 0,977172285 | LOC100506639 | FALSE | Downregulated |
| 0,024455975 | 0,97719409 | ADRB2 | FALSE | Upregulated |
| -0,041379049 | 0,977206675 | LOC102723313 | FALSE | Downregulated |
| -0,014411935 | 0,977206675 | RAB3GAP2 | FALSE | Downregulated |
| -0,07471339 | 0,977344279 | CADM3 | FALSE | Downregulated |
| -0,022303263 | 0,977344279 | CADM4 | FALSE | Downregulated |
| 0,021408716 | 0,977344279 | FAM126B | FALSE | Upregulated |
| -0,053138532 | 0,977344279 | FBXO5 | FALSE | Downregulated |
| -0,045524545 | 0,977344279 | LOC113230 | FALSE | Downregulated |

| | | | | |
|---|---|---|---|---|
| 0,015542551 | 0,977344279 | MLANA | FALSE | Upregulated |
| 0,055562679 | 0,977344279 | SSSCA1 | FALSE | Upregulated |
| -0,024487849 | 0,977344279 | STAM2 | FALSE | Downregulated |
| 0,01866842 | 0,977344279 | UBA7 | FALSE | Upregulated |
| 0,036912631 | 0,977344279 | ZNF32-AS2 | FALSE | Upregulated |
| -0,061492119 | 0,977344279 | ZNF699 | FALSE | Downregulated |
| 0,015388195 | 0,977469142 | CANT1 | FALSE | Upregulated |
| 0,035958674 | 0,977469142 | IPO9-AS1 | FALSE | Upregulated |
| 0,023717168 | 0,977469142 | KCTD18 | FALSE | Upregulated |
| -0,027479324 | 0,977469142 | NAPG | FALSE | Downregulated |
| 0,025213066 | 0,977469142 | NT5C | FALSE | Upregulated |
| 0,027744098 | 0,977565332 | RNMTL1 | FALSE | Upregulated |
| -0,037694562 | 0,977644906 | TVP23B | FALSE | Downregulated |
| 0,017327157 | 0,977775462 | GABPA | FALSE | Upregulated |
| 0,032613004 | 0,977809724 | CPSF3 | FALSE | Upregulated |
| 0,03430222 | 0,977809724 | FAM96B | FALSE | Upregulated |
| -0,017661277 | 0,977809724 | GOLGA8H | FALSE | Downregulated |
| -0,035907882 | 0,977809724 | TRMT12 | FALSE | Downregulated |
| 0,038307351 | 0,977809724 | WASH1 | FALSE | Upregulated |
| -0,043858752 | 0,977809724 | ZNF581 | FALSE | Downregulated |
| -0,01957302 | 0,977926249 | TFDP2 | FALSE | Downregulated |
| -0,050745099 | 0,978053013 | AC109828.1 | FALSE | Downregulated |
| -0,051753884 | 0,978053013 | ALKBH1 | FALSE | Downregulated |
| 0,018124977 | 0,978053013 | AMDHD2 | FALSE | Upregulated |
| 0,041881928 | 0,978053013 | AP3M2 | FALSE | Upregulated |
| -0,026026184 | 0,978053013 | ATF2 | FALSE | Downregulated |
| 0,064408159 | 0,978053013 | CCDC134 | FALSE | Upregulated |
| -0,016136992 | 0,978053013 | CHMP1A | FALSE | Downregulated |
| -0,02339532 | 0,978053013 | COMMD9 | FALSE | Downregulated |
| 0,017514289 | 0,978053013 | EFTUD2 | FALSE | Upregulated |
| 0,073716827 | 0,978053013 | H2AFB1 | FALSE | Upregulated |
| -0,019242607 | 0,978053013 | JAG1 | FALSE | Downregulated |
| 0,034881786 | 0,978053013 | LENG1 | FALSE | Upregulated |
| 0,095787519 | 0,978053013 | LINC01106 | FALSE | Upregulated |
| -0,021851236 | 0,978053013 | LRRC32 | FALSE | Downregulated |
| -0,025792036 | 0,978053013 | MRPL24 | FALSE | Downregulated |
| 0,019095582 | 0,978053013 | MZT2A | FALSE | Upregulated |
| 0,011629025 | 0,978053013 | NADK | FALSE | Upregulated |
| -0,040842799 | 0,978053013 | NDUFAF4 | FALSE | Downregulated |
| -0,040971833 | 0,978053013 | PDCL | FALSE | Downregulated |
| 0,034996108 | 0,978053013 | PHF6 | FALSE | Upregulated |
| 0,030334688 | 0,978053013 | PIN4 | FALSE | Upregulated |
| -0,04554634 | 0,978053013 | PLAGL1 | FALSE | Downregulated |
| 0,025002666 | 0,978053013 | PRDM4 | FALSE | Upregulated |
| -0,034134674 | 0,978053013 | SUV420H2 | FALSE | Downregulated |
| 0,020725832 | 0,978053013 | TTYH1 | FALSE | Upregulated |
| 0,029575176 | 0,978053013 | WDR27 | FALSE | Upregulated |
| 0,047328886 | 0,978053013 | ZNF567 | FALSE | Upregulated |
| 0,018129829 | 0,978055702 | BANP | FALSE | Upregulated |
| 0,029199834 | 0,978084352 | POLA1 | FALSE | Upregulated |
| 0,042699738 | 0,978084352 | ZNF28 | FALSE | Upregulated |

| | | | | |
|---|---|---|---|---|
| -0,013992843 | 0,978220928 | MORC2 | FALSE | Downregulated |
| 0,019815998 | 0,978220928 | PGLS | FALSE | Upregulated |
| -0,018806744 | 0,978220928 | SEC11A | FALSE | Downregulated |
| -0,032871957 | 0,978270167 | IPMK | FALSE | Downregulated |
| -0,057047168 | 0,978270167 | KLHL32 | FALSE | Downregulated |
| 0,061182277 | 0,97836638 | ASAP1-IT2 | FALSE | Upregulated |
| -0,025545188 | 0,97836638 | C10orf32 | FALSE | Downregulated |
| -0,024107726 | 0,97836638 | FMO4 | FALSE | Downregulated |
| -0,012144106 | 0,97836638 | YLPM1 | FALSE | Downregulated |
| 0,025460663 | 0,978392258 | LOC100507156 | FALSE | Upregulated |
| -0,026192291 | 0,978434778 | CBLC | FALSE | Downregulated |
| -0,033982918 | 0,978434778 | DPH3 | FALSE | Downregulated |
| 0,046599272 | 0,978434778 | PRB2 | FALSE | Upregulated |
| -0,023598241 | 0,978434778 | TIMM10B | FALSE | Downregulated |
| 0,019299733 | 0,978463819 | APBA1 | FALSE | Upregulated |
| 0,009861266 | 0,978463819 | ATP1A1 | FALSE | Upregulated |
| 0,029231354 | 0,978463819 | ILF3-AS1 | FALSE | Upregulated |
| 0,046503429 | 0,978463819 | LRRC31 | FALSE | Upregulated |
| -0,028847197 | 0,978623961 | CCBL1 | FALSE | Downregulated |
| 0,020454436 | 0,978623961 | SLC5A9 | FALSE | Upregulated |
| -0,016529199 | 0,978623961 | USP15 | FALSE | Downregulated |
| -0,033956815 | 0,978627533 | RABL3 | FALSE | Downregulated |
| 0,019803394 | 0,978627533 | RBAK | FALSE | Upregulated |
| 0,027808972 | 0,978714058 | NSL1 | FALSE | Upregulated |
| -0,04348283 | 0,97880308 | FOS | FALSE | Downregulated |
| 0,018416202 | 0,979006735 | CCAR1 | FALSE | Upregulated |
| 0,023752389 | 0,979006735 | TBL2 | FALSE | Upregulated |
| -0,102248215 | 0,979200295 | TNXA | FALSE | Downregulated |
| 0,024774986 | 0,979487408 | IER3IP1 | FALSE | Upregulated |
| 0,021364333 | 0,979487408 | MPZL1 | FALSE | Upregulated |
| 0,027349083 | 0,979487408 | RPUSD1 | FALSE | Upregulated |
| 0,011591774 | 0,979487408 | TM9SF3 | FALSE | Upregulated |
| 0,015669516 | 0,979496962 | CRYZ | FALSE | Upregulated |
| -0,022841132 | 0,979537605 | CYB561D1 | FALSE | Downregulated |
| 0,058063918 | 0,979539161 | ANKHD1 | FALSE | Upregulated |
| -0,019149311 | 0,979539161 | FTO | FALSE | Downregulated |
| -0,025132193 | 0,979633803 | CCDC12 | FALSE | Downregulated |
| 0,033768629 | 0,979633803 | DHRS11 | FALSE | Upregulated |
| 0,015570318 | 0,979633803 | FOXP4 | FALSE | Upregulated |
| 0,017167908 | 0,979633803 | FOXRED1 | FALSE | Upregulated |
| -0,028282166 | 0,979633803 | GPR128 | FALSE | Downregulated |
| 0,037656339 | 0,979633803 | HYAL3 | FALSE | Upregulated |
| 0,015067211 | 0,979633803 | KAZN | FALSE | Upregulated |
| 0,012842214 | 0,979633803 | RPL28 | FALSE | Upregulated |
| -0,03527405 | 0,979633803 | SH3RF3 | FALSE | Downregulated |
| 0,023041163 | 0,979633803 | TCFL5 | FALSE | Upregulated |
| 0,01786943 | 0,979633803 | ZBED6 | FALSE | Upregulated |
| -0,016804147 | 0,979667125 | FAM219A | FALSE | Downregulated |
| 0,020515454 | 0,979667125 | GDE1 | FALSE | Upregulated |
| -0,009953466 | 0,979667125 | MOV10 | FALSE | Downregulated |
| 0,043561351 | 0,979667125 | RP1-249H1.4 | FALSE | Upregulated |

| | | | | |
|---|---|---|---|---|
| -0,02729479 | 0,97979437 | LYG2 | FALSE | Downregulated |
| -0,01512256 | 0,979882932 | MVB12B | FALSE | Downregulated |
| 0,037478471 | 0,979968541 | ADAM20 | FALSE | Upregulated |
| -0,027331631 | 0,980019473 | PRSS37 | FALSE | Downregulated |
| -0,376615977 | 0,980422253 | MUC13 | FALSE | Downregulated |
| 0,019224554 | 0,98056895 | EXOC4 | FALSE | Upregulated |
| 0,0162661 | 0,980627117 | CA14 | FALSE | Upregulated |
| -0,014142289 | 0,980732313 | LAMC1 | FALSE | Downregulated |
| -0,03614279 | 0,980732313 | LCMT2 | FALSE | Downregulated |
| 0,018619045 | 0,980732313 | LTB4R2 | FALSE | Upregulated |
| 0,035898621 | 0,980843612 | FAM27E3 | FALSE | Upregulated |
| -0,025655912 | 0,981018713 | FBXL4 | FALSE | Downregulated |
| -0,032784601 | 0,981209221 | NDUFA8 | FALSE | Downregulated |
| -0,020821466 | 0,98135675 | FEZ2 | FALSE | Downregulated |
| -0,013486529 | 0,98135675 | PROX2 | FALSE | Downregulated |
| -0,022393633 | 0,981516368 | C1orf85 | FALSE | Downregulated |
| -0,014480053 | 0,981516368 | ECHDC3 | FALSE | Downregulated |
| 0,072303161 | 0,981516368 | RP11-434C1.1 | FALSE | Upregulated |
| 0,018344643 | 0,981516368 | SPTLC2 | FALSE | Upregulated |
| 0,014901587 | 0,981619881 | TRIM2 | FALSE | Upregulated |
| 0,035791077 | 0,981642136 | CACHD1 | FALSE | Upregulated |
| 0,025251333 | 0,981663112 | UCK2 | FALSE | Upregulated |
| -0,02540605 | 0,981716077 | EIF4E | FALSE | Downregulated |
| -0,016468876 | 0,981716077 | KIF2A | FALSE | Downregulated |
| 0,023864112 | 0,981716077 | MTURN | FALSE | Upregulated |
| -0,362287991 | 0,981716077 | MYBPC1 | FALSE | Downregulated |
| 0,081005246 | 0,981716077 | RNVU1-7 | FALSE | Upregulated |
| 0,012248723 | 0,981747011 | AUP1 | FALSE | Upregulated |
| 0,038800503 | 0,981747011 | CCDC141 | FALSE | Upregulated |
| 0,024254125 | 0,981747011 | PLLP | FALSE | Upregulated |
| -0,016691977 | 0,981747011 | SPIRE2 | FALSE | Downregulated |
| 0,018688654 | 0,981747011 | UBE2A | FALSE | Upregulated |
| 0,027524656 | 0,981747011 | ZNF823 | FALSE | Upregulated |
| 0,053963765 | 0,981786736 | DNAH7 | FALSE | Upregulated |
| -0,026307958 | 0,981977954 | PKNOX1 | FALSE | Downregulated |
| -0,020858513 | 0,982003099 | ADO | FALSE | Downregulated |
| -0,022794355 | 0,982003099 | HDAC2 | FALSE | Downregulated |
| 0,031150966 | 0,982003099 | KCNC2 | FALSE | Upregulated |
| 0,008094073 | 0,982090545 | RPS19 | FALSE | Upregulated |
| -0,018516317 | 0,982174959 | RAB7L1 | FALSE | Downregulated |
| 0,040362615 | 0,982234146 | HNRNPA1L2 | FALSE | Upregulated |
| 0,011485713 | 0,982276573 | BMP8A | FALSE | Upregulated |
| -0,012602049 | 0,982276573 | LDHA | FALSE | Downregulated |
| 0,024089649 | 0,982276573 | TMCO6 | FALSE | Upregulated |
| -0,014220317 | 0,982461728 | ZNF181 | FALSE | Downregulated |
| -0,013773759 | 0,982525857 | ARID4B | FALSE | Downregulated |
| -0,046610233 | 0,982525857 | GJA1 | FALSE | Downregulated |
| 0,035632046 | 0,982525857 | LOC100289361 | FALSE | Upregulated |
| -0,028073055 | 0,982525857 | RNASEK | FALSE | Downregulated |
| 0,011362058 | 0,982530539 | MOGS | FALSE | Upregulated |
| -0,04424832 | 0,982620989 | XIRP1 | FALSE | Downregulated |

| | | | | |
|---|---|---|---|---|
| 0,029351129 | 0,982653575 | C6orf123 | FALSE | Upregulated |
| -0,055908324 | 0,982653575 | CASP16 | FALSE | Downregulated |
| -0,060751387 | 0,982653575 | GREM1 | FALSE | Downregulated |
| -0,021255061 | 0,982653575 | TBC1D12 | FALSE | Downregulated |
| -0,008391128 | 0,982661383 | FAN1 | FALSE | Downregulated |
| -0,020736781 | 0,982757108 | PRPF39 | FALSE | Downregulated |
| 0,020101233 | 0,98279693 | ARMC8 | FALSE | Upregulated |
| -0,022772217 | 0,98279693 | DDX47 | FALSE | Downregulated |
| -0,053837514 | 0,982811031 | AC016582.2 | FALSE | Downregulated |
| -0,016177363 | 0,982811031 | EEFSEC | FALSE | Downregulated |
| -0,008988948 | 0,982811031 | LAMB2 | FALSE | Downregulated |
| 0,009000516 | 0,982811031 | LRBA | FALSE | Upregulated |
| 0,026335951 | 0,982811031 | NNT-AS1 | FALSE | Upregulated |
| 0,038910747 | 0,982811031 | OLFM1 | FALSE | Upregulated |
| 0,022440636 | 0,98294523 | TMEM106C | FALSE | Upregulated |
| -0,015739548 | 0,982991511 | TBC1D22A | FALSE | Downregulated |
| -0,022842231 | 0,983004183 | GPATCH3 | FALSE | Downregulated |
| 0,013028861 | 0,983004183 | KAT5 | FALSE | Upregulated |
| -0,017589376 | 0,983004183 | SEPT4 | FALSE | Downregulated |
| -0,047687895 | 0,98301445 | SLC14A2 | FALSE | Downregulated |
| -0,013083597 | 0,98308636 | POLR2L | FALSE | Downregulated |
| 0,018388586 | 0,98308636 | SMIM6 | FALSE | Upregulated |
| 0,011930612 | 0,983210352 | CMIP | FALSE | Upregulated |
| 0,044849438 | 0,983363626 | DGAT2L6 | FALSE | Upregulated |
| 0,032504071 | 0,983363626 | LOC100505727 | FALSE | Upregulated |
| -0,034003566 | 0,983363626 | PLVAP | FALSE | Downregulated |
| -0,021223481 | 0,983363626 | ZNF354C | FALSE | Downregulated |
| 0,01663474 | 0,983585549 | DNM3OS | FALSE | Upregulated |
| -0,051289413 | 0,983585549 | ID4 | FALSE | Downregulated |
| 0,013864422 | 0,983585549 | SPCS3 | FALSE | Upregulated |
| 0,018780626 | 0,983585549 | TIMM50 | FALSE | Upregulated |
| 0,017616691 | 0,983585549 | ZNF777 | FALSE | Upregulated |
| -0,015421928 | 0,98367063 | FAM160B1 | FALSE | Downregulated |
| -0,036235194 | 0,98367063 | MT1F | FALSE | Downregulated |
| 0,039414929 | 0,98367063 | RP11-81N13.1 | FALSE | Upregulated |
| 0,016017729 | 0,98367063 | RP5-914P20.5 | FALSE | Upregulated |
| 0,046679355 | 0,98367063 | RPL23AP82 | FALSE | Upregulated |
| 0,037815281 | 0,983744807 | ZNF669 | FALSE | Upregulated |
| 0,059032758 | 0,98377733 | RAB41 | FALSE | Upregulated |
| 0,01280745 | 0,98379169 | SLC43A3 | FALSE | Upregulated |
| -0,014452534 | 0,983904554 | CBFA2T2 | FALSE | Downregulated |
| 0,041913458 | 0,98401873 | BNC2 | FALSE | Upregulated |
| 0,011454106 | 0,984162643 | HIRA | FALSE | Upregulated |
| 0,314468457 | 0,98424873 | ATP5J2-PTCD1 | FALSE | Upregulated |
| -0,016207514 | 0,98424873 | C2orf15 | FALSE | Downregulated |
| -0,017899409 | 0,98424873 | CCDC85B | FALSE | Downregulated |
| -0,010625948 | 0,98424873 | CDC42 | FALSE | Downregulated |
| 0,007981708 | 0,98424873 | CTTN | FALSE | Upregulated |
| 0,029017018 | 0,98424873 | DPY30 | FALSE | Upregulated |
| -0,014903603 | 0,98424873 | GLT8D1 | FALSE | Downregulated |
| 0,01134555 | 0,98424873 | LINC00261 | FALSE | Upregulated |

| | | | | |
|---|---|---|---|---|
| -0,031883883 | 0,98424873 | MDGA1 | FALSE | Downregulated |
| -0,011616547 | 0,98424873 | SIL1 | FALSE | Downregulated |
| -0,319902435 | 0,98424873 | SNORD3C | FALSE | Downregulated |
| 0,008164552 | 0,98424873 | SVIL | FALSE | Upregulated |
| -0,019858212 | 0,98424873 | TRAPPC1 | FALSE | Downregulated |
| -0,015839627 | 0,98424873 | UTP20 | FALSE | Downregulated |
| -0,016830337 | 0,984441843 | MED22 | FALSE | Downregulated |
| 0,020597471 | 0,984441843 | NPIPB5 | FALSE | Upregulated |
| 0,013197559 | 0,984483699 | CD276 | FALSE | Upregulated |
| 0,02410681 | 0,984483699 | SENP1 | FALSE | Upregulated |
| 0,031620309 | 0,984617255 | ARMC9 | FALSE | Upregulated |
| 0,012933338 | 0,984743065 | CHURC1 | FALSE | Upregulated |
| -0,015979897 | 0,984743065 | FAM53C | FALSE | Downregulated |
| 0,044570872 | 0,984743065 | RSPH10B2 | FALSE | Upregulated |
| 0,022669369 | 0,984892611 | IDH3A | FALSE | Upregulated |
| -0,014707693 | 0,984957194 | RAB3GAP1 | FALSE | Downregulated |
| -0,261759694 | 0,984957194 | XIST | FALSE | Downregulated |
| -0,014268964 | 0,984997317 | DDTL | FALSE | Downregulated |
| 0,04259606 | 0,985010287 | C10orf88 | FALSE | Upregulated |
| -0,008543391 | 0,985124831 | FAM83H | FALSE | Downregulated |
| -0,008286906 | 0,985185929 | BRWD1 | FALSE | Downregulated |
| -0,007886835 | 0,985185929 | GCN1L1 | FALSE | Downregulated |
| 0,302262678 | 0,985185929 | KRT14 | FALSE | Upregulated |
| 0,037343208 | 0,985185929 | LOC100507670 | FALSE | Upregulated |
| -0,030406267 | 0,985185929 | LOC102724047 | FALSE | Downregulated |
| -0,02173781 | 0,985185929 | LRP8 | FALSE | Downregulated |
| -0,014471775 | 0,985185929 | NCAPD2 | FALSE | Downregulated |
| 0,023057364 | 0,985185929 | NDUFAF1 | FALSE | Upregulated |
| 0,030434438 | 0,985185929 | NUP210L | FALSE | Upregulated |
| 0,012549518 | 0,985185929 | RBBP6 | FALSE | Upregulated |
| -0,008746176 | 0,985185929 | RCN1 | FALSE | Downregulated |
| -0,012444369 | 0,985185929 | SNX19 | FALSE | Downregulated |
| -0,014390528 | 0,985185929 | TBC1D23 | FALSE | Downregulated |
| 0,051806574 | 0,985204435 | PPP1R14D | FALSE | Upregulated |
| -0,012451287 | 0,985204435 | TJP3 | FALSE | Downregulated |
| 0,017751309 | 0,985260089 | IMPG1 | FALSE | Upregulated |
| 0,029277181 | 0,985325703 | DTX3 | FALSE | Upregulated |
| -0,045602885 | 0,985325703 | LOC101928879 | FALSE | Downregulated |
| 0,013641774 | 0,985325703 | LTA4H | FALSE | Upregulated |
| 0,046526537 | 0,985325703 | PDE9A | FALSE | Upregulated |
| -0,009149316 | 0,985325703 | SHC2 | FALSE | Downregulated |
| -0,008770758 | 0,985368327 | CD99L2 | FALSE | Downregulated |
| 0,013038456 | 0,985500745 | FOXJ2 | FALSE | Upregulated |
| 0,016530213 | 0,985500745 | LOC102723735 | FALSE | Upregulated |
| -0,042407502 | 0,985500745 | NTN5 | FALSE | Downregulated |
| 0,033257466 | 0,985500745 | SNAI2 | FALSE | Upregulated |
| -0,014949371 | 0,98558202 | RRN3 | FALSE | Downregulated |
| -0,031987235 | 0,985609616 | MNAT1 | FALSE | Downregulated |
| 0,025274225 | 0,985648418 | MICU2 | FALSE | Upregulated |
| 0,031584593 | 0,985648418 | SRCRB4D | FALSE | Upregulated |
| 0,010749757 | 0,985685184 | ATP6V0A1 | FALSE | Upregulated |

| | | | | |
|---|---|---|---|---|
| 0,016534901 | 0,985685184 | FAM3C | FALSE | Upregulated |
| 0,017904017 | 0,985685184 | GTF3C4 | FALSE | Upregulated |
| 0,019707789 | 0,985685184 | IRAK4 | FALSE | Upregulated |
| -0,006048058 | 0,985685184 | RAD23B | FALSE | Downregulated |
| 0,006786407 | 0,985685184 | RNF40 | FALSE | Upregulated |
| -0,035101089 | 0,985685184 | TRMT10C | FALSE | Downregulated |
| -0,012677297 | 0,98574615 | GCSH | FALSE | Downregulated |
| 0,02341161 | 0,98574615 | HEYL | FALSE | Upregulated |
| -0,025667513 | 0,98574615 | PGBD2 | FALSE | Downregulated |
| 0,051302117 | 0,98574615 | POLR2J2 | FALSE | Upregulated |
| -0,049865584 | 0,98574615 | TAF1A | FALSE | Downregulated |
| 0,029975442 | 0,985763278 | BEGAIN | FALSE | Upregulated |
| 0,007814002 | 0,985763278 | DHFR | FALSE | Upregulated |
| -0,023048228 | 0,985763278 | RHD | FALSE | Downregulated |
| 0,016848921 | 0,985763278 | RNF138P1 | FALSE | Upregulated |
| 0,010941634 | 0,985763278 | VAPA | FALSE | Upregulated |
| 0,012647807 | 0,985902206 | ANP32B | FALSE | Upregulated |
| 0,01917865 | 0,985973447 | TEX10 | FALSE | Upregulated |
| 0,018785088 | 0,986140715 | KIAA0020 | FALSE | Upregulated |
| -0,01400177 | 0,986140715 | TMEM222 | FALSE | Downregulated |
| 0,016932371 | 0,986140715 | TUBA8 | FALSE | Upregulated |
| -0,012284438 | 0,986140715 | ZNF837 | FALSE | Downregulated |
| -0,026178329 | 0,986319796 | CIRH1A | FALSE | Downregulated |
| -0,008797382 | 0,986319796 | ENSA | FALSE | Downregulated |
| 0,012808809 | 0,986319796 | TAF5L | FALSE | Upregulated |
| 0,020750966 | 0,986452929 | ANTXR1 | FALSE | Upregulated |
| 0,038221646 | 0,986584049 | IGSF10 | FALSE | Upregulated |
| 0,032554009 | 0,986602633 | ZBTB49 | FALSE | Upregulated |
| 0,018034541 | 0,986711868 | UNC5C | FALSE | Upregulated |
| -0,017986219 | 0,986743409 | C11orf71 | FALSE | Downregulated |
| -0,009616695 | 0,986861971 | GABARAP | FALSE | Downregulated |
| -0,029800553 | 0,986861971 | LOC100129617 | FALSE | Downregulated |
| 0,011574989 | 0,986988253 | BAD | FALSE | Upregulated |
| -0,012402937 | 0,986988253 | BICD2 | FALSE | Downregulated |
| -0,015635995 | 0,986988253 | BRK1 | FALSE | Downregulated |
| -0,039207337 | 0,986988253 | OR51Q1 | FALSE | Downregulated |
| 0,026786924 | 0,987101346 | FIGNL1 | FALSE | Upregulated |
| 0,020586167 | 0,987101346 | INTS7 | FALSE | Upregulated |
| 0,024018763 | 0,987101346 | LINC00116 | FALSE | Upregulated |
| 0,050509472 | 0,987101346 | LOC102723537 | FALSE | Upregulated |
| 0,011394513 | 0,987101346 | LOC102724499 | FALSE | Upregulated |
| 0,008779213 | 0,987101346 | OGT | FALSE | Upregulated |
| -0,020485577 | 0,987101346 | PAX8-AS1 | FALSE | Downregulated |
| 0,033304795 | 0,987101346 | PGLYRP1 | FALSE | Upregulated |
| 0,015817549 | 0,987101346 | PXMP4 | FALSE | Upregulated |
| 0,026134015 | 0,987114795 | ACTR3B | FALSE | Upregulated |
| 0,005559296 | 0,987114795 | POLR2A | FALSE | Upregulated |
| -0,041057795 | 0,987114795 | SMOC2 | FALSE | Downregulated |
| 0,007837048 | 0,987114795 | TEAD1 | FALSE | Upregulated |
| -0,02169064 | 0,987343922 | RP11-890B15.2 | FALSE | Downregulated |
| -0,013090925 | 0,987343922 | SENP2 | FALSE | Downregulated |

| | | | | |
|---|---|---|---|---|
| -0,0306725 | 0,987385053 | GSTA4 | FALSE | Downregulated |
| 0,013477637 | 0,987482135 | BAG5 | FALSE | Upregulated |
| -0,023585085 | 0,987585246 | CCDC84 | FALSE | Downregulated |
| -0,0269167 | 0,987585246 | CDK5R1 | FALSE | Downregulated |
| 0,036604043 | 0,987585246 | LOC101927119 | FALSE | Upregulated |
| 0,0299063 | 0,987585246 | MNDA | FALSE | Upregulated |
| 0,258463076 | 0,987585246 | MYH1 | FALSE | Upregulated |
| -0,01276255 | 0,987585246 | PDHX | FALSE | Downregulated |
| 0,016199982 | 0,987585246 | PMS1 | FALSE | Upregulated |
| 0,020630236 | 0,987585246 | THUMPD3-AS1 | FALSE | Upregulated |
| -0,055592057 | 0,987684094 | FAM19A5 | FALSE | Downregulated |
| -0,017161118 | 0,987686445 | CLUHP3 | FALSE | Downregulated |
| -0,031532526 | 0,987686445 | KIAA1755 | FALSE | Downregulated |
| -0,032834491 | 0,987699641 | CHEK2 | FALSE | Downregulated |
| -0,031959916 | 0,987699641 | CYCSP52 | FALSE | Downregulated |
| 0,010742325 | 0,987699641 | GRIPAP1 | FALSE | Upregulated |
| 0,011328874 | 0,987699641 | HIF1A | FALSE | Upregulated |
| 0,061581274 | 0,987699641 | TTC4 | FALSE | Upregulated |
| -0,029778159 | 0,98778489 | WDR93 | FALSE | Downregulated |
| 0,018596931 | 0,987989613 | CCDC146 | FALSE | Upregulated |
| -0,010975019 | 0,987989613 | RNF111 | FALSE | Downregulated |
| 0,029061073 | 0,988047397 | MMGT1 | FALSE | Upregulated |
| 0,019558805 | 0,988084568 | AC011747.4 | FALSE | Upregulated |
| -0,025704023 | 0,988084568 | C17orf53 | FALSE | Downregulated |
| -0,01094772 | 0,988084568 | CCNC | FALSE | Downregulated |
| 0,02025208 | 0,988084568 | FIG4 | FALSE | Upregulated |
| -0,017423367 | 0,988084568 | LOC102723758 | FALSE | Downregulated |
| -0,014880828 | 0,988084568 | NAA10 | FALSE | Downregulated |
| 0,021898143 | 0,988084568 | SNAPC2 | FALSE | Upregulated |
| -0,020240825 | 0,988084568 | SPATA25 | FALSE | Downregulated |
| -0,017091381 | 0,988084568 | TBP | FALSE | Downregulated |
| 0,018153865 | 0,988084568 | TMEM161A | FALSE | Upregulated |
| -0,007340737 | 0,988084568 | TPR | FALSE | Downregulated |
| -0,017646732 | 0,988218134 | RNF7 | FALSE | Downregulated |
| -0,014744678 | 0,988321683 | WEE1 | FALSE | Downregulated |
| 0,011947564 | 0,988325482 | BAIAP2-AS1 | FALSE | Upregulated |
| 0,010776905 | 0,988325482 | COPZ1 | FALSE | Upregulated |
| 0,014030142 | 0,988325482 | LYRM4 | FALSE | Upregulated |
| -0,020538998 | 0,988325482 | MAGI2-AS3 | FALSE | Downregulated |
| -0,019836098 | 0,988325482 | REXO2 | FALSE | Downregulated |
| 0,007501386 | 0,988325482 | ZNF587 | FALSE | Upregulated |
| 0,023987047 | 0,988390793 | LOC101929548 | FALSE | Upregulated |
| -0,012626356 | 0,988531973 | B3GNT2 | FALSE | Downregulated |
| -0,021298971 | 0,988531973 | COX10 | FALSE | Downregulated |
| 0,012685537 | 0,988531973 | LNX2 | FALSE | Upregulated |
| 0,019051854 | 0,988531973 | LOC100294145 | FALSE | Upregulated |
| 0,037164861 | 0,988531973 | LOC102723671 | FALSE | Upregulated |
| -0,227859442 | 0,988531973 | LOC102724441 | FALSE | Downregulated |
| -0,036623395 | 0,988531973 | LOC90834 | FALSE | Downregulated |
| -0,02731993 | 0,988531973 | PGF | FALSE | Downregulated |
| 0,016606167 | 0,988531973 | SERTAD3 | FALSE | Upregulated |

| | | | | |
|---|---|---|---|---|
| -0,023473953 | 0,98853622 | SSX2IP | FALSE | Downregulated |
| -0,006383111 | 0,98853622 | UBE2R2 | FALSE | Downregulated |
| 0,030789666 | 0,988782543 | EYA1 | FALSE | Upregulated |
| 0,006504273 | 0,988782543 | MGAT1 | FALSE | Upregulated |
| -0,014346342 | 0,988805782 | LOC102724172 | FALSE | Downregulated |
| -0,006100621 | 0,988805782 | NR3C1 | FALSE | Downregulated |
| -0,127492825 | 0,988805782 | TRIM6-TRIM34 | FALSE | Downregulated |
| 0,025223319 | 0,988882061 | INO80C | FALSE | Upregulated |
| -0,020106929 | 0,988882061 | KIAA0825 | FALSE | Downregulated |
| -0,016675643 | 0,988882061 | PRR5L | FALSE | Downregulated |
| -0,028235905 | 0,988882061 | RAB28 | FALSE | Downregulated |
| -0,007195801 | 0,988882061 | RP11-545I5.3 | FALSE | Downregulated |
| -0,008949148 | 0,988992892 | TNFAIP8L2 | FALSE | Downregulated |
| 0,006774016 | 0,988994982 | ESPN | FALSE | Upregulated |
| -0,016191663 | 0,988994982 | KIF9 | FALSE | Downregulated |
| -0,011944167 | 0,988994982 | LANCL1 | FALSE | Downregulated |
| -0,016907 | 0,988994982 | LOC100130285 | FALSE | Downregulated |
| -0,005399167 | 0,988994982 | LRPAP1 | FALSE | Downregulated |
| 0,007991513 | 0,988994982 | PPP1R14B | FALSE | Upregulated |
| -0,029896431 | 0,988994982 | RYR1 | FALSE | Downregulated |
| 0,015865701 | 0,988994982 | SLC15A4 | FALSE | Upregulated |
| 0,005973437 | 0,988994982 | TPI1 | FALSE | Upregulated |
| 0,025230337 | 0,989022328 | LOC102723788 | FALSE | Upregulated |
| 0,006642605 | 0,989091257 | ATXN1L | FALSE | Upregulated |
| 0,019227819 | 0,989091257 | SDK1 | FALSE | Upregulated |
| -0,023859335 | 0,989091257 | TCTN1 | FALSE | Downregulated |
| -0,019704917 | 0,989091257 | ZNF493 | FALSE | Downregulated |
| 0,007309279 | 0,989104158 | LRRN3 | FALSE | Upregulated |
| -0,011657846 | 0,989104158 | STAU2 | FALSE | Downregulated |
| -0,00862021 | 0,989233162 | ABCF1 | FALSE | Downregulated |
| 0,033547046 | 0,989233162 | HSPB8 | FALSE | Upregulated |
| 0,012763565 | 0,989233162 | PDGFRB | FALSE | Upregulated |
| 0,006803599 | 0,989312516 | CTSL | FALSE | Upregulated |
| -0,030684566 | 0,989312516 | LOC388242 | FALSE | Downregulated |
| -0,011881464 | 0,989312516 | MUS81 | FALSE | Downregulated |
| -0,02550403 | 0,989412121 | ASTN2 | FALSE | Downregulated |
| 0,028414276 | 0,989412121 | GPR21 | FALSE | Upregulated |
| 0,009981033 | 0,989412121 | HMGN3 | FALSE | Upregulated |
| 0,013306339 | 0,989412121 | LOC731656 | FALSE | Upregulated |
| -0,018284347 | 0,989432773 | WDR75 | FALSE | Downregulated |
| 0,025712445 | 0,989462288 | GRHL3 | FALSE | Upregulated |
| 0,024052547 | 0,989462288 | METTL25 | FALSE | Upregulated |
| 0,048599721 | 0,989462288 | PRR15 | FALSE | Upregulated |
| -0,007494798 | 0,989462288 | RB1CC1 | FALSE | Downregulated |
| -0,011952768 | 0,989462288 | ZNF37A | FALSE | Downregulated |
| 0,013125456 | 0,989464128 | NRBF2 | FALSE | Upregulated |
| -0,014090993 | 0,989849446 | LMO2 | FALSE | Downregulated |
| -0,009727299 | 0,989938415 | POLR1B | FALSE | Downregulated |
| 0,011115931 | 0,989938415 | TYSND1 | FALSE | Upregulated |
| 0,010347121 | 0,989958052 | DUS3L | FALSE | Upregulated |
| 0,020194898 | 0,989975585 | C5orf63 | FALSE | Upregulated |

| | | | | |
|---|---|---|---|---|
| 0,017991748 | 0,989975585 | DCAF4 | FALSE | Upregulated |
| 0,012415148 | 0,989975585 | DNAJC9-AS1 | FALSE | Upregulated |
| 0,039062151 | 0,989975585 | ERCC6-PGBD3 | FALSE | Upregulated |
| 0,014268181 | 0,989975585 | GPR62 | FALSE | Upregulated |
| -0,028313178 | 0,989975585 | HTRA3 | FALSE | Downregulated |
| 0,018977742 | 0,989975585 | IPO11 | FALSE | Upregulated |
| -0,017514723 | 0,989975585 | LOC101060181 | FALSE | Downregulated |
| -0,013374679 | 0,989975585 | LOC101927031 | FALSE | Downregulated |
| 0,030439751 | 0,989975585 | LOC102724193 | FALSE | Upregulated |
| 0,017716146 | 0,989975585 | MROH8 | FALSE | Upregulated |
| 0,007753393 | 0,989975585 | MYBBP1A | FALSE | Upregulated |
| 0,021066835 | 0,989975585 | NAT6 | FALSE | Upregulated |
| -0,008608853 | 0,989975585 | RPF1 | FALSE | Downregulated |
| -0,026931582 | 0,989975585 | SEMA3B-AS1 | FALSE | Downregulated |
| 0,015142403 | 0,989975585 | SSC5D | FALSE | Upregulated |
| 0,0090192 | 0,989975585 | TPD52L2 | FALSE | Upregulated |
| -0,031951138 | 0,989975585 | UNC5B-AS1 | FALSE | Downregulated |
| 0,028163619 | 0,989975585 | ZNF439 | FALSE | Upregulated |
| -0,097761688 | 0,989993695 | TNNT3 | FALSE | Downregulated |
| 0,013697894 | 0,989994929 | THRA | FALSE | Upregulated |
| 0,021848612 | 0,990104694 | ZNF200 | FALSE | Upregulated |
| 0,02010754 | 0,990289148 | C3orf65 | FALSE | Upregulated |
| 0,035940337 | 0,990404153 | ACTBL2 | FALSE | Upregulated |
| -0,015638533 | 0,990404153 | STARD4-AS1 | FALSE | Downregulated |
| -0,015633338 | 0,990404153 | TLR3 | FALSE | Downregulated |
| 0,013491771 | 0,990404153 | TMEM177 | FALSE | Upregulated |
| -0,00758484 | 0,990505735 | ACVR1B | FALSE | Downregulated |
| -0,009831768 | 0,990505735 | ANXA2P1 | FALSE | Downregulated |
| -0,017603869 | 0,990505735 | COPS4 | FALSE | Downregulated |
| -0,023511962 | 0,990505735 | LOC101929513 | FALSE | Downregulated |
| 0,017504186 | 0,990505735 | LOC102724872 | FALSE | Upregulated |
| 0,010141072 | 0,990505735 | LZTS2 | FALSE | Upregulated |
| -0,015210888 | 0,990505735 | SIRT6 | FALSE | Downregulated |
| 0,008268049 | 0,990505735 | SLC13A4 | FALSE | Upregulated |
| 0,003950726 | 0,990505735 | SON | FALSE | Upregulated |
| -0,045807155 | 0,990575549 | FOXD4L5 | FALSE | Downregulated |
| 0,008659088 | 0,990672139 | ACVR1 | FALSE | Upregulated |
| 0,025363895 | 0,990672139 | ANXA13 | FALSE | Upregulated |
| -0,010307534 | 0,990672139 | B4GALT3 | FALSE | Downregulated |
| -0,016342243 | 0,990672139 | BAG2 | FALSE | Downregulated |
| -0,010891248 | 0,990672139 | BBS4 | FALSE | Downregulated |
| 0,014160614 | 0,990672139 | GPALPP1 | FALSE | Upregulated |
| 0,027165567 | 0,990672139 | LOC101929500 | FALSE | Upregulated |
| 0,01633608 | 0,990672139 | LOC339803 | FALSE | Upregulated |
| -0,007377942 | 0,990672139 | MXRA8 | FALSE | Downregulated |
| 0,011127562 | 0,990672139 | PLCB1 | FALSE | Upregulated |
| -0,019528408 | 0,990672139 | RP11-97O12.2 | FALSE | Downregulated |
| -0,009233725 | 0,990694285 | HNRNPD | FALSE | Downregulated |
| 0,00984716 | 0,990694285 | ZKSCAN5 | FALSE | Upregulated |
| 0,005723365 | 0,990710706 | AP2B1 | FALSE | Upregulated |
| 0,013331272 | 0,990710706 | CCDC28A | FALSE | Upregulated |

| | | | | |
|---|---|---|---|---|
| 0,014596223 | 0,990710706 | EMID1 | FALSE | Upregulated |
| -0,008949352 | 0,990710706 | GIT2 | FALSE | Downregulated |
| -0,028686901 | 0,990710706 | GLA | FALSE | Downregulated |
| -0,010368619 | 0,990710706 | GOLGA8O | FALSE | Downregulated |
| 0,008098966 | 0,990710706 | LDLRAD4-AS1 | FALSE | Upregulated |
| 0,006568121 | 0,990710706 | LOC646471 | FALSE | Upregulated |
| -0,005818878 | 0,990710706 | SMARCC1 | FALSE | Downregulated |
| 0,00987992 | 0,990858809 | SGPP2 | FALSE | Upregulated |
| -0,013186102 | 0,990858809 | UBTD2 | FALSE | Downregulated |
| 0,008310547 | 0,990886162 | RPA1 | FALSE | Upregulated |
| -0,009526205 | 0,990900529 | TSNARE1 | FALSE | Downregulated |
| -0,01595535 | 0,990918276 | LOC728743 | FALSE | Downregulated |
| 0,01862188 | 0,990976953 | NDUFA7 | FALSE | Upregulated |
| -0,023758846 | 0,990976953 | SERF1B | FALSE | Downregulated |
| -0,105761957 | 0,990976953 | TNNC2 | FALSE | Downregulated |
| 0,012137067 | 0,991001523 | B4GALT4 | FALSE | Upregulated |
| 0,019996463 | 0,991390122 | SLC14A1 | FALSE | Upregulated |
| -0,007673697 | 0,991467156 | ASAP1 | FALSE | Downregulated |
| -0,011558292 | 0,991467156 | AVEN | FALSE | Downregulated |
| -0,00964176 | 0,991467156 | TPST1 | FALSE | Downregulated |
| -0,013022933 | 0,991500789 | VAMP4 | FALSE | Downregulated |
| 0,017988108 | 0,99160097 | ALS2CR12 | FALSE | Upregulated |
| 0,021369418 | 0,99160097 | CCDC144A | FALSE | Upregulated |
| 0,019445912 | 0,99160097 | LINC00943 | FALSE | Upregulated |
| 0,009758086 | 0,99160097 | LOC101929347 | FALSE | Upregulated |
| 0,013458846 | 0,99160097 | MSI2 | FALSE | Upregulated |
| 0,008317274 | 0,99160097 | PARD6G | FALSE | Upregulated |
| 0,006364945 | 0,99160097 | ZFR | FALSE | Upregulated |
| 0,008688367 | 0,991704171 | PLCD4 | FALSE | Upregulated |
| 0,008329274 | 0,991708821 | RNF20 | FALSE | Upregulated |
| 0,00734754 | 0,991798285 | DNMBP | FALSE | Upregulated |
| -0,008076582 | 0,991798285 | SQRDL | FALSE | Downregulated |
| 0,01378424 | 0,991798285 | TLR2 | FALSE | Upregulated |
| -0,019251207 | 0,991897975 | C4orf32 | FALSE | Downregulated |
| 0,005771449 | 0,991897975 | CAPN1 | FALSE | Upregulated |
| 0,010808508 | 0,991897975 | CLYBL-AS2 | FALSE | Upregulated |
| 0,015730328 | 0,991897975 | DNAH2 | FALSE | Upregulated |
| -0,007708081 | 0,991897975 | EBNA1BP2 | FALSE | Downregulated |
| 0,00418717 | 0,991897975 | FBRSL1 | FALSE | Upregulated |
| 0,006596761 | 0,991897975 | GOLGA2 | FALSE | Upregulated |
| -0,010634397 | 0,991897975 | LOC101928170 | FALSE | Downregulated |
| 0,007865171 | 0,991897975 | PQBP1 | FALSE | Upregulated |
| 0,003728306 | 0,991897975 | SUPT6H | FALSE | Upregulated |
| 0,014689242 | 0,991897975 | TIMM21 | FALSE | Upregulated |
| 0,163315447 | 0,991897975 | TSIX | FALSE | Upregulated |
| -0,010039458 | 0,991897975 | ZDHHC2 | FALSE | Downregulated |
| -0,011746048 | 0,991897975 | ZFAND4 | FALSE | Downregulated |
| -0,011330462 | 0,991897975 | ZNF638-IT1 | FALSE | Downregulated |
| -0,011893493 | 0,991977263 | RHOXF2 | FALSE | Downregulated |
| -0,008885299 | 0,992003928 | AFF4 | FALSE | Downregulated |
| -0,004178848 | 0,992003928 | ARIH1 | FALSE | Downregulated |

| | | | | |
|---|---|---|---|---|
| -0,034172844 | 0,992003928 | BOLA2 | FALSE | Downregulated |
| 0,01916564 | 0,992003928 | CD24 | FALSE | Upregulated |
| 0,007483293 | 0,992003928 | CLDN2 | FALSE | Upregulated |
| -0,037128518 | 0,992003928 | CTGLF12P | FALSE | Downregulated |
| 0,007293833 | 0,992003928 | GART | FALSE | Upregulated |
| -0,006008618 | 0,992003928 | LOC101929648 | FALSE | Downregulated |
| -0,018711278 | 0,992003928 | LOC643797 | FALSE | Downregulated |
| 0,022251501 | 0,992003928 | NRN1 | FALSE | Upregulated |
| -0,00816764 | 0,992003928 | PAX6 | FALSE | Downregulated |
| 0,006372431 | 0,992003928 | TEX261 | FALSE | Upregulated |
| -0,007758994 | 0,992003928 | TSPAN18 | FALSE | Downregulated |
| 0,005777908 | 0,992003928 | USP36 | FALSE | Upregulated |
| -0,01518116 | 0,992003928 | UTP15 | FALSE | Downregulated |
| 0,015983601 | 0,992003928 | WDR86 | FALSE | Upregulated |
| -0,015069687 | 0,992073749 | MRPS36 | FALSE | Downregulated |
| 0,034243345 | 0,99219926 | RIMBP3 | FALSE | Upregulated |
| -0,009923593 | 0,992225388 | FAM189B | FALSE | Downregulated |
| -0,007502752 | 0,992351851 | GIMAP8 | FALSE | Downregulated |
| 0,00731912 | 0,992449223 | PET112 | FALSE | Upregulated |
| 0,008992415 | 0,992523723 | GRK5 | FALSE | Upregulated |
| -0,013021374 | 0,992545933 | GTF2A2 | FALSE | Downregulated |
| 0,028695954 | 0,992545933 | RERG | FALSE | Upregulated |
| 0,006622286 | 0,992545933 | TLK1 | FALSE | Upregulated |
| 0,014305516 | 0,992590725 | SLC16A14 | FALSE | Upregulated |
| -0,004933662 | 0,992657673 | PSMC3 | FALSE | Downregulated |
| -0,012201141 | 0,992682877 | ARHGAP20 | FALSE | Downregulated |
| 0,003381919 | 0,992682877 | CDK12 | FALSE | Upregulated |
| 0,005656101 | 0,992682877 | NDST2 | FALSE | Upregulated |
| 0,007265673 | 0,992682877 | RAB11FIP1 | FALSE | Upregulated |
| -0,005100386 | 0,992721427 | CCNT2 | FALSE | Downregulated |
| 0,006346859 | 0,992819485 | FLJ35282 | FALSE | Upregulated |
| 0,00777458 | 0,992819485 | WHAMM | FALSE | Upregulated |
| -0,010717705 | 0,992830132 | ZNF234 | FALSE | Downregulated |
| -0,021826341 | 0,99288771 | KCNAB3 | FALSE | Downregulated |
| 0,019310783 | 0,993000359 | ASIC5 | FALSE | Upregulated |
| -0,011150776 | 0,993151828 | TAF8 | FALSE | Downregulated |
| -0,008546586 | 0,993151828 | ZDHHC6 | FALSE | Downregulated |
| 0,006272723 | 0,993195139 | NEDD4 | FALSE | Upregulated |
| -0,021186746 | 0,993195139 | TPBG | FALSE | Downregulated |
| 0,004150339 | 0,993221555 | TINF2 | FALSE | Upregulated |
| 0,010611653 | 0,993240778 | DCUN1D2 | FALSE | Upregulated |
| 0,017339957 | 0,993245706 | RASAL1 | FALSE | Upregulated |
| 0,003767749 | 0,993269239 | SGSM2 | FALSE | Upregulated |
| 0,00771236 | 0,993269239 | TRIB2 | FALSE | Upregulated |
| 0,020352438 | 0,993320408 | EBF1 | FALSE | Upregulated |
| -0,019116409 | 0,993320408 | LOC102724583 | FALSE | Downregulated |
| 0,012597221 | 0,993354332 | LOC101928510 | FALSE | Upregulated |
| -0,013769296 | 0,993373248 | TAF12 | FALSE | Downregulated |
| -0,011019541 | 0,993428487 | DRD5 | FALSE | Downregulated |
| 0,01882451 | 0,993507511 | CPXM2 | FALSE | Upregulated |
| 0,012547211 | 0,993507511 | TIMM8A | FALSE | Upregulated |

| | | | | |
|---|---|---|---|---|
| 0,006332107 | 0,993507511 | TPMT | FALSE | Upregulated |
| -0,006965974 | 0,99350882 | ISYNA1 | FALSE | Downregulated |
| -0,057991784 | 0,993512393 | MYLPF | FALSE | Downregulated |
| 0,008987962 | 0,993547856 | C7orf26 | FALSE | Upregulated |
| -0,008971034 | 0,993547856 | CRIPT | FALSE | Downregulated |
| -0,011715287 | 0,993547856 | LTK | FALSE | Downregulated |
| 0,012320642 | 0,993547856 | RPP30 | FALSE | Upregulated |
| 0,008806169 | 0,993547856 | SGK223 | FALSE | Upregulated |
| 0,004584447 | 0,99357073 | FAM186A | FALSE | Upregulated |
| 0,006636501 | 0,99357073 | LOC101928588 | FALSE | Upregulated |
| 0,013867901 | 0,993593296 | CCDC181 | FALSE | Upregulated |
| 0,004349767 | 0,993593296 | DBH | FALSE | Upregulated |
| 0,02195874 | 0,993593296 | FAM46B | FALSE | Upregulated |
| 0,010547608 | 0,993593296 | LOC101928548 | FALSE | Upregulated |
| 0,009459455 | 0,993593296 | LOC102723820 | FALSE | Upregulated |
| 0,012574068 | 0,993593296 | PTBP2 | FALSE | Upregulated |
| 0,00648984 | 0,993593296 | SCAMP4 | FALSE | Upregulated |
| 0,013565905 | 0,993593296 | TSHZ3 | FALSE | Upregulated |
| 0,01265725 | 0,993593296 | ZIC1 | FALSE | Upregulated |
| 0,007309946 | 0,993688156 | ZCCHC9 | FALSE | Upregulated |
| 0,02017871 | 0,993743962 | OR1L6 | FALSE | Upregulated |
| 0,017290512 | 0,993751307 | NR4A1 | FALSE | Upregulated |
| -0,018600419 | 0,99376934 | ACTL7A | FALSE | Downregulated |
| -0,008090214 | 0,99376934 | BOK-AS1 | FALSE | Downregulated |
| 0,004912835 | 0,99376934 | C16orf45 | FALSE | Upregulated |
| 0,012133976 | 0,99376934 | CPE | FALSE | Upregulated |
| -0,003424756 | 0,99376934 | DIS3L | FALSE | Downregulated |
| 0,003670396 | 0,99376934 | FANCC | FALSE | Upregulated |
| 0,003234057 | 0,99376934 | GPLD1 | FALSE | Upregulated |
| -0,011603522 | 0,99376934 | JKAMP | FALSE | Downregulated |
| -0,003757478 | 0,99376934 | LOC100130093 | FALSE | Downregulated |
| -0,020865054 | 0,99376934 | LOC101927143 | FALSE | Downregulated |
| -0,004491589 | 0,99376934 | MDH1 | FALSE | Downregulated |
| 0,009389291 | 0,99376934 | OTUD3 | FALSE | Upregulated |
| 0,005590885 | 0,99376934 | PRMT2 | FALSE | Upregulated |
| 0,01276566 | 0,99376934 | PRMT3 | FALSE | Upregulated |
| 0,009461326 | 0,99376934 | RSPH9 | FALSE | Upregulated |
| -0,008782733 | 0,993811065 | ZFPM2 | FALSE | Downregulated |
| 0,006526649 | 0,993889174 | DEPTOR | FALSE | Upregulated |
| -0,013448397 | 0,994013039 | FAM58A | FALSE | Downregulated |
| -0,01475948 | 0,994013039 | TMC2 | FALSE | Downregulated |
| -0,008902112 | 0,994044311 | GALC | FALSE | Downregulated |
| -0,006825011 | 0,994044311 | SUB1 | FALSE | Downregulated |
| 0,003251481 | 0,994044311 | UBE2H | FALSE | Upregulated |
| 0,005212078 | 0,994044311 | VTI1B | FALSE | Upregulated |
| 0,112801505 | 0,994061859 | RNVU1-18 | FALSE | Upregulated |
| 0,008803359 | 0,99422974 | LOC102723597 | FALSE | Upregulated |
| 0,009259667 | 0,994235747 | VMA21 | FALSE | Upregulated |
| 0,003148973 | 0,994238018 | CDK4 | FALSE | Upregulated |
| 0,015971587 | 0,994238018 | LINC00314 | FALSE | Upregulated |
| 0,005863433 | 0,994238018 | PHAX | FALSE | Upregulated |

| | | | | |
|---|---|---|---|---|
| -0,006785964 | 0,994238018 | PRMT9 | FALSE | Downregulated |
| -0,008476286 | 0,994238018 | SNAPIN | FALSE | Downregulated |
| -0,00636204 | 0,994392184 | KIAA1143 | FALSE | Downregulated |
| 0,003811924 | 0,994392184 | MED15 | FALSE | Upregulated |
| 0,015900756 | 0,994392184 | RPL22L1 | FALSE | Upregulated |
| 0,004552384 | 0,994426434 | CD2BP2 | FALSE | Upregulated |
| -0,009180507 | 0,994426434 | DPY19L2P4 | FALSE | Downregulated |
| -0,006212762 | 0,994426434 | LMBRD2 | FALSE | Downregulated |
| 0,005511688 | 0,994426434 | RSRC2 | FALSE | Upregulated |
| -0,003161769 | 0,9945092 | PRDX1 | FALSE | Downregulated |
| 0,004127015 | 0,994539066 | RP11-142C4.6 | FALSE | Upregulated |
| -0,002163496 | 0,994544198 | PTPRF | FALSE | Downregulated |
| 0,015166411 | 0,994606331 | C4orf33 | FALSE | Upregulated |
| -0,012620418 | 0,994606331 | CEP41 | FALSE | Downregulated |
| -0,002956391 | 0,994791542 | DLEU1 | FALSE | Downregulated |
| -0,003093041 | 0,994887184 | ECHDC1 | FALSE | Downregulated |
| 0,002995572 | 0,994887184 | MAP3K12 | FALSE | Upregulated |
| 0,007209385 | 0,994887184 | TYW1B | FALSE | Upregulated |
| -0,006070239 | 0,995015345 | LINC00324 | FALSE | Downregulated |
| 0,004741669 | 0,995130005 | ZNF582 | FALSE | Upregulated |
| 0,006281657 | 0,995163717 | MTMR6 | FALSE | Upregulated |
| -0,013743735 | 0,995214417 | LINC00639 | FALSE | Downregulated |
| 0,00401233 | 0,995214417 | RBM42 | FALSE | Upregulated |
| -0,017530707 | 0,995217383 | LOC100505985 | FALSE | Downregulated |
| 0,022425106 | 0,99528968 | RBM14-RBM4 | FALSE | Upregulated |
| -0,004612325 | 0,995446196 | SHARPIN | FALSE | Downregulated |
| -0,006819667 | 0,995480657 | ARHGEF3 | FALSE | Downregulated |
| -0,004891408 | 0,995480657 | C15orf39 | FALSE | Downregulated |
| -0,005903028 | 0,995480657 | FAM179B | FALSE | Downregulated |
| -0,004197056 | 0,995480657 | HNMT | FALSE | Downregulated |
| 0,009553928 | 0,995480657 | HSD17B7 | FALSE | Upregulated |
| 0,006818766 | 0,995480657 | MECR | FALSE | Upregulated |
| -0,003813968 | 0,995480657 | PERP | FALSE | Downregulated |
| -0,008359707 | 0,995480657 | RASSF6 | FALSE | Downregulated |
| -0,011056268 | 0,995480657 | STXBP4 | FALSE | Downregulated |
| 0,030371042 | 0,995626807 | HIST1H4J | FALSE | Upregulated |
| -0,014917757 | 0,995732725 | LOC100506113 | FALSE | Downregulated |
| -0,00378118 | 0,995796753 | PPIG | FALSE | Downregulated |
| -0,014494364 | 0,996059727 | ASNS | FALSE | Downregulated |
| 0,005962229 | 0,996059727 | LOC100996405 | FALSE | Upregulated |
| -0,002707418 | 0,996059727 | SYNJ2BP | FALSE | Downregulated |
| -0,009800283 | 0,996061491 | SULT1C2P1 | FALSE | Downregulated |
| 0,00793083 | 0,996102844 | ARNT2 | FALSE | Upregulated |
| 0,008590371 | 0,996102844 | ASB12 | FALSE | Upregulated |
| -0,009158465 | 0,996102844 | GTF2F2 | FALSE | Downregulated |
| -0,003458656 | 0,996102844 | RAB11A | FALSE | Downregulated |
| 0,008623504 | 0,996109214 | CISH | FALSE | Upregulated |
| 0,002797481 | 0,996143991 | CUL1 | FALSE | Upregulated |
| 0,005187568 | 0,99620144 | ATXN7L1 | FALSE | Upregulated |
| -0,00401423 | 0,99620144 | FBXL20 | FALSE | Downregulated |
| 0,005802799 | 0,99620144 | SKAP2 | FALSE | Upregulated |

| | | | | |
|---|---|---|---|---|
| -0,002667848 | 0,996221237 | ACKR4 | FALSE | Downregulated |
| 0,004945291 | 0,996221237 | CRELD2 | FALSE | Upregulated |
| 0,004288735 | 0,996221237 | DTX2 | FALSE | Upregulated |
| -0,00520408 | 0,996221237 | GRIN3B | FALSE | Downregulated |
| -0,005657103 | 0,996221237 | LOC101929657 | FALSE | Downregulated |
| -0,077862746 | 0,996221237 | MYH7 | FALSE | Downregulated |
| 0,002749354 | 0,996221237 | RBM15 | FALSE | Upregulated |
| -0,002998694 | 0,996221237 | SOCS6 | FALSE | Downregulated |
| -0,004822444 | 0,996221237 | ST3GAL4 | FALSE | Downregulated |
| 0,002831495 | 0,996236127 | ANKRD12 | FALSE | Upregulated |
| 0,004895209 | 0,996236127 | CAPN10 | FALSE | Upregulated |
| -0,010909445 | 0,996236127 | LOC101928189 | FALSE | Downregulated |
| -0,036045812 | 0,996236127 | MYBPC2 | FALSE | Downregulated |
| 0,006925085 | 0,996236127 | NDC1 | FALSE | Upregulated |
| 0,00531504 | 0,996236127 | NS3BP | FALSE | Upregulated |
| 0,004468729 | 0,996236127 | SCARNA6 | FALSE | Upregulated |
| 0,002753396 | 0,996236127 | TIMM13 | FALSE | Upregulated |
| -0,012119537 | 0,996245509 | FAIM | FALSE | Downregulated |
| 0,002006299 | 0,996253407 | GAPDH | FALSE | Upregulated |
| -0,00262362 | 0,996253407 | MAPRE2 | FALSE | Downregulated |
| 0,010712048 | 0,996253407 | OR1K1 | FALSE | Upregulated |
| 0,004775781 | 0,996253407 | PHLDA1 | FALSE | Upregulated |
| -0,003140271 | 0,996253407 | TRAF6 | FALSE | Downregulated |
| 0,002869584 | 0,996253407 | UBE2E1 | FALSE | Upregulated |
| 0,004046379 | 0,996421093 | ADAMTS17 | FALSE | Upregulated |
| -0,001843866 | 0,996421093 | C3 | FALSE | Downregulated |
| -0,068491714 | 0,996421093 | MYH2 | FALSE | Downregulated |
| 0,002567509 | 0,996421093 | RLIM | FALSE | Upregulated |
| -0,005106659 | 0,996553969 | CXorf40A | FALSE | Downregulated |
| -0,011398501 | 0,996553969 | PCLO | FALSE | Downregulated |
| -0,009970366 | 0,996553969 | RP9 | FALSE | Downregulated |
| 0,008575393 | 0,996553969 | ZNF630 | FALSE | Upregulated |
| -0,003277953 | 0,99656671 | SNX25 | FALSE | Downregulated |
| -0,011107013 | 0,9966591 | H1FNT | FALSE | Downregulated |
| 0,003094783 | 0,996716922 | GFOD2 | FALSE | Upregulated |
| 0,01191755 | 0,996740065 | LINC00880 | FALSE | Upregulated |
| 0,002812858 | 0,996974247 | KIFC2 | FALSE | Upregulated |
| -0,007045311 | 0,996974247 | LOC102723614 | FALSE | Downregulated |
| 0,003325379 | 0,996979305 | C22orf46 | FALSE | Upregulated |
| -0,004576568 | 0,997045935 | GK5 | FALSE | Downregulated |
| -0,002857716 | 0,997045935 | TTC7B | FALSE | Downregulated |
| -0,00419046 | 0,997045935 | UXT | FALSE | Downregulated |
| 0,007029806 | 0,997154765 | AWAT1 | FALSE | Upregulated |
| 0,002127181 | 0,997154765 | CCNJ | FALSE | Upregulated |
| -0,002732002 | 0,997154765 | LIMA1 | FALSE | Downregulated |
| -0,007821183 | 0,997154765 | LOC102723512 | FALSE | Downregulated |
| 0,002864716 | 0,997154765 | LOC401296 | FALSE | Upregulated |
| 0,003023351 | 0,997154765 | LOC401320 | FALSE | Upregulated |
| 0,002809968 | 0,997154765 | PHF23 | FALSE | Upregulated |
| -0,005522257 | 0,997382856 | C1GALT1C1 | FALSE | Downregulated |
| -0,00526883 | 0,997456894 | RBPMS2 | FALSE | Downregulated |

| | | | | |
|---|---|---|---|---|
| 0,003170608 | 0,997486977 | GADD45B | FALSE | Upregulated |
| 0,002893871 | 0,99780598 | CCDC174 | FALSE | Upregulated |
| 0,0029011 | 0,99780598 | DDX31 | FALSE | Upregulated |
| -0,004812429 | 0,99780598 | GP5 | FALSE | Downregulated |
| -0,001381295 | 0,99780598 | KIAA0368 | FALSE | Downregulated |
| -0,004692171 | 0,99780598 | LOC100270746 | FALSE | Downregulated |
| -0,003845647 | 0,99780598 | TMEM260 | FALSE | Downregulated |
| -0,050104134 | 0,99780598 | XIRP2 | FALSE | Downregulated |
| -0,001713529 | 0,997808089 | ABCF3 | FALSE | Downregulated |
| 0,041834293 | 0,997808089 | ACTA1 | FALSE | Upregulated |
| -0,002695309 | 0,997808089 | BCL7A | FALSE | Downregulated |
| -0,001187948 | 0,997808089 | CIC | FALSE | Downregulated |
| -0,001788294 | 0,997808089 | CSTF2T | FALSE | Downregulated |
| 0,003683257 | 0,997808089 | HAUS2 | FALSE | Upregulated |
| 0,001864231 | 0,997808089 | LARP4 | FALSE | Upregulated |
| -0,001625954 | 0,997808089 | LBX2-AS1 | FALSE | Downregulated |
| 0,006740145 | 0,997808089 | LINC00353 | FALSE | Upregulated |
| 0,00597921 | 0,997808089 | LOC101927596 | FALSE | Upregulated |
| -0,004288505 | 0,997808089 | LOC101928424 | FALSE | Downregulated |
| -0,003047639 | 0,997808089 | LOC102723897 | FALSE | Downregulated |
| -0,006032818 | 0,997808089 | LRRC37A5P | FALSE | Downregulated |
| 0,023577919 | 0,997808089 | NPIPA8 | FALSE | Upregulated |
| 0,001905239 | 0,997808089 | NRBP1 | FALSE | Upregulated |
| 0,004008708 | 0,997808089 | OSGEPL1-AS1 | FALSE | Upregulated |
| -0,003749274 | 0,997808089 | PALD1 | FALSE | Downregulated |
| 0,002802744 | 0,997808089 | SNUPN | FALSE | Upregulated |
| 0,004787902 | 0,998022641 | MCTP2 | FALSE | Upregulated |
| 0,002942601 | 0,998022641 | PRICKLE2 | FALSE | Upregulated |
| -0,001158595 | 0,998096045 | LOC101927509 | FALSE | Downregulated |
| 0,0064346 | 0,998298277 | NPAS3 | FALSE | Upregulated |
| 0,001841967 | 0,998298277 | TSPAN11 | FALSE | Upregulated |
| 0,001677378 | 0,9983036 | DNAJC10 | FALSE | Upregulated |
| -0,001590835 | 0,99833832 | NOL6 | FALSE | Downregulated |
| 0,001368145 | 0,99833832 | OSBPL8 | FALSE | Upregulated |
| -0,001053296 | 0,99833832 | ZMYND11 | FALSE | Downregulated |
| 0,002035771 | 0,998389608 | RAP1GDS1 | FALSE | Upregulated |
| -0,001282506 | 0,998389608 | TNFRSF14 | FALSE | Downregulated |
| -0,001929009 | 0,998573088 | LRRC46 | FALSE | Downregulated |
| -0,002133316 | 0,998616805 | KIAA0040 | FALSE | Downregulated |
| 0,00246461 | 0,998630346 | DPM1 | FALSE | Upregulated |
| -0,003540767 | 0,998630346 | RP11-863K10.2 | FALSE | Downregulated |
| 0,001899891 | 0,998668329 | BUD31 | FALSE | Upregulated |
| 0,001021967 | 0,998668329 | CAND1 | FALSE | Upregulated |
| -0,006090389 | 0,998668329 | EGFL8 | FALSE | Downregulated |
| 0,000852981 | 0,998668329 | GPR161 | FALSE | Upregulated |
| -0,00157199 | 0,998668329 | NBPF1 | FALSE | Downregulated |
| 0,000849868 | 0,998668329 | RBM19 | FALSE | Upregulated |
| -0,00158657 | 0,998668329 | SLC35D2 | FALSE | Downregulated |
| -0,001205446 | 0,998668329 | SMARCB1 | FALSE | Downregulated |
| 0,000834132 | 0,998668329 | SRP9 | FALSE | Upregulated |
| 0,001468282 | 0,998668329 | TMCC1 | FALSE | Upregulated |

| | | | | |
|---|---|---|---|---|
| 0,001507429 | 0,998668329 | USPL1 | FALSE | Upregulated |
| -0,001622613 | 0,998715821 | PANK4 | FALSE | Downregulated |
| -0,001798514 | 0,998993346 | STK38L | FALSE | Downregulated |
| 0,001183157 | 0,999020153 | ATG12 | FALSE | Upregulated |
| -0,001849712 | 0,999020153 | COMMD7 | FALSE | Downregulated |
| 0,001618213 | 0,999037875 | PPP3CC | FALSE | Upregulated |
| 0,001854499 | 0,999050394 | PNMA1 | FALSE | Upregulated |
| 0,000469804 | 0,999191314 | BACE1 | FALSE | Upregulated |
| -0,00091803 | 0,999206659 | LOC101928319 | FALSE | Downregulated |
| 0,003175876 | 0,999206659 | LOC101929540 | FALSE | Upregulated |
| 0,001822385 | 0,999206659 | ZNF75A | FALSE | Upregulated |
| -0,000653438 | 0,99942727 | NAB1 | FALSE | Downregulated |
| 0,001051522 | 0,999674166 | ARL2BP | FALSE | Upregulated |
| -0,000704814 | 0,999674166 | ATP5H | FALSE | Downregulated |
| 0,00111464 | 0,999674166 | LACC1 | FALSE | Upregulated |
| -0,000700596 | 0,999674166 | PELO | FALSE | Downregulated |
| -0,000928676 | 0,999681111 | ALDH1A3 | FALSE | Downregulated |
| 0,000747656 | 0,999687863 | VMAC | FALSE | Upregulated |
| -0,001214022 | 0,999804044 | C1orf53 | FALSE | Downregulated |
| -0,0008536 | 0,999804044 | COL25A1 | FALSE | Downregulated |
| -0,000847632 | 0,999804044 | CYBRD1 | FALSE | Downregulated |
| -0,00034067 | 0,999804044 | HSF1 | FALSE | Downregulated |
| -0,000424496 | 0,999804044 | LOC729732 | FALSE | Downregulated |
| -0,000427555 | 0,999804044 | NOL8 | FALSE | Downregulated |
| -0,000271525 | 0,999804044 | OAZ1 | FALSE | Downregulated |
| -0,000424492 | 0,999804044 | PDLIM1 | FALSE | Downregulated |
| 0,00036262 | 0,999804044 | PIGW | FALSE | Upregulated |
| 0,00034616 | 0,999804044 | RIOK3 | FALSE | Upregulated |
| 0,000812305 | 0,999804044 | RWDD1 | FALSE | Upregulated |
| -0,000306279 | 0,999804044 | SBF2 | FALSE | Downregulated |
| -0,001014715 | 0,999804044 | SLC4A7 | FALSE | Downregulated |
| 0,001329196 | 0,999804044 | SPATA33 | FALSE | Upregulated |
| -0,000942479 | 0,999804044 | TULP3 | FALSE | Downregulated |
| -0,000319407 | 0,999819257 | RUVBL2 | FALSE | Downregulated |
| -0,005253087 | 0,999830904 | CKM | FALSE | Downregulated |
| 0,00072291 | 0,999830904 | TMEM254-AS1 | FALSE | Upregulated |
| 0,000280934 | 1 | ALG6 | FALSE | Upregulated |
| -0,000122039 | 1 | CLDND1 | FALSE | Downregulated |
| -0,000181267 | 1 | DDB2 | FALSE | Downregulated |
| 0 | 1 | H19 | FALSE | Upregulated |
| 5,07363E-05 | 1 | KDM8 | FALSE | Upregulated |
| 0 | 1 | KRT17 | FALSE | Upregulated |
| 0 | 1 | KRT2 | FALSE | Upregulated |
| 0 | 1 | KRT6A | FALSE | Upregulated |
| 3,16299E-05 | 1 | LOC102724024 | FALSE | Upregulated |
| 9,73676E-06 | 1 | MEA1 | FALSE | Upregulated |
| -0,000267371 | 1 | MGC72080 | FALSE | Downregulated |
| -0,000368573 | 1 | PALB2 | FALSE | Downregulated |
| 2,62582E-06 | 1 | TAF3 | FALSE | Upregulated |
| 0,000297927 | 1 | TMEM150C | FALSE | Upregulated |
| 0 | 1 | TMX2-CTNND1 | FALSE | Upregulated |

| | | | | |
|---|---|---|---|---|
| -3,49566E-05 | 1 | ZFP36L2 | FALSE | Downregulated |
| 0,593632788 | | A3GALT2 | FALSE | Upregulated |
| | | AA06 | | |
| | | AACSP1 | | |
| | | AADACL3 | | |
| 0 | | AADACL4 | FALSE | Upregulated |
| | | AARD | | |
| 0,226133485 | | AATK-AS1 | FALSE | Upregulated |
| 1,184041233 | | ABCA12 | | Upregulated |
| -0,084143895 | | ABCB5 | FALSE | Downregulated |
| 0,143719692 | | ABCB9 | FALSE | Upregulated |
| -0,063219906 | | ABCC12 | FALSE | Downregulated |
| 0 | | ABCC13 | FALSE | Upregulated |
| -0,148682774 | | ABCC5-AS1 | FALSE | Downregulated |
| 0 | | ABCC8 | FALSE | Upregulated |
| 0,493389768 | | ABCD2 | FALSE | Upregulated |
| -0,365827366 | | ABCG4 | FALSE | Downregulated |
| | | ABHD14A-ACY1 | | |
| 0,177811333 | | ABO | FALSE | Upregulated |
| -0,216990473 | | AC002116.7 | FALSE | Downregulated |
| -0,01152389 | | AC002398.5 | FALSE | Downregulated |
| 0,453099891 | | AC002454.1 | FALSE | Upregulated |
| | | AC002463.3 | | |
| 0,16100274 | | AC002511.1 | FALSE | Upregulated |
| | | AC003051.1 | | |
| 0,390950345 | | AC003102.3 | FALSE | Upregulated |
| | | AC003958.2 | | |
| | | AC003984.1 | | |
| | | AC004603.4 | | |
| -1,521599119 | | AC004862.6 | | Downregulated |
| 0 | | AC005022.1 | FALSE | Upregulated |
| 0 | | AC005062.2 | FALSE | Upregulated |
| | | AC005235.1 | | |
| 0,77108844 | | AC005358.3 | FALSE | Upregulated |
| 0,3541948 | | AC005550.4 | FALSE | Upregulated |
| -0,088191227 | | AC005592.2 | FALSE | Downregulated |
| | | AC006004.1 | | |
| | | AC006196.1 | | |
| -0,059004178 | | AC006372.4 | FALSE | Downregulated |
| -0,096557243 | | AC007040.7 | FALSE | Downregulated |
| 1,677635987 | | AC007131.1 | | Upregulated |
| 0,406787727 | | AC007359.6 | FALSE | Upregulated |
| -0,044742671 | | AC007365.1 | FALSE | Downregulated |
| 0,085182977 | | AC007386.2 | FALSE | Upregulated |
| 0 | | AC007392.3 | FALSE | Upregulated |
| 0,206291025 | | AC007392.4 | FALSE | Upregulated |
| | | AC007403.2 | | |
| 0 | | AC007405.4 | FALSE | Upregulated |
| | | AC007463.2 | | |
| 0,538436058 | | AC007682.1 | FALSE | Upregulated |
| 1,021194812 | | AC007740.1 | | Upregulated |

| | | | |
|---|---|---|---|
| -1,063093726 | AC007743.1 | | Downregulated |
| 0,393688067 | AC007773.2 | FALSE | Upregulated |
| | AC007879.2 | | |
| -0,340115606 | AC007880.1 | FALSE | Downregulated |
| 0,153275352 | AC007966.1 | FALSE | Upregulated |
| | AC008697.1 | | |
| 0,816008347 | AC008703.1 | FALSE | Upregulated |
| 0 | AC009120.4 | FALSE | Upregulated |
| | AC009227.2 | | |
| | AC009518.4 | | |
| 0,020789395 | AC010091.1 | FALSE | Upregulated |
| 0,514981842 | AC010524.2 | FALSE | Upregulated |
| | AC010731.4 | | |
| -0,574677908 | AC010890.1 | FALSE | Downregulated |
| | AC010975.1 | | |
| | AC010987.6 | | |
| | AC011298.2 | | |
| 0,010835277 | AC011518.1 | FALSE | Upregulated |
| 0,666860273 | AC011609.3 | FALSE | Upregulated |
| | AC011718.2 | | |
| 1,02126275 | AC011738.4 | | Upregulated |
| | AC012075.2 | | |
| | AC012494.1 | | |
| 0,784963894 | AC012507.3 | FALSE | Upregulated |
| 0,711780861 | AC015922.6 | FALSE | Upregulated |
| | AC016995.3 | | |
| 0,191018546 | AC017060.1 | FALSE | Upregulated |
| | AC018464.3 | | |
| 0 | AC018470.4 | FALSE | Upregulated |
| 0,948746428 | AC018647.3 | FALSE | Upregulated |
| 0 | AC018799.1 | FALSE | Upregulated |
| 0,384192098 | AC019048.1 | FALSE | Upregulated |
| | AC019118.2 | | |
| | AC022311.1 | | |
| | AC027269.2 | | |
| 0 | AC068831.10 | FALSE | Upregulated |
| -0,37671932 | AC068858.1 | FALSE | Downregulated |
| | AC069277.1 | | |
| -0,098012884 | AC072062.1 | FALSE | Downregulated |
| 0,358002007 | AC073128.10 | FALSE | Upregulated |
| -0,911042289 | AC073133.1 | FALSE | Downregulated |
| -0,08693459 | AC073283.4 | FALSE | Downregulated |
| 0 | AC073321.3 | FALSE | Upregulated |
| | AC073628.1 | | |
| 0,365400799 | AC074286.1 | FALSE | Upregulated |
| -0,507361323 | AC074289.1 | FALSE | Downregulated |
| 0 | AC079145.4 | FALSE | Upregulated |
| -0,045864993 | AC079586.1 | FALSE | Downregulated |
| 0,75458729 | AC079610.2 | FALSE | Upregulated |
| | AC079776.3 | | |
| -0,327467014 | AC087859.1 | FALSE | Downregulated |

| | | | |
|---|---|---|---|
| 0,664780692 | AC090044.1 | FALSE | Upregulated |
| 0 | AC090505.6 | FALSE | Upregulated |
| 0,688355021 | AC091969.1 | FALSE | Upregulated |
| 1,248990079 | AC092295.7 | | Upregulated |
| -0,840041698 | AC093375.1 | FALSE | Downregulated |
| 0,753960039 | AC093391.2 | FALSE | Upregulated |
| 0 | AC093590.1 | FALSE | Upregulated |
| -0,747132712 | AC095067.1 | FALSE | Downregulated |
| 0,043320765 | AC096559.1 | FALSE | Upregulated |
| -0,426505955 | AC097468.4 | FALSE | Downregulated |
| -0,071541939 | AC097500.2 | FALSE | Downregulated |
| -0,212001447 | AC097662.2 | FALSE | Downregulated |
| | AC098823.3 | | |
| | AC099342.1 | | |
| 0 | AC104024.1 | FALSE | Upregulated |
| 0,486377151 | AC104653.1 | FALSE | Upregulated |
| 0,615505805 | AC104654.1 | FALSE | Upregulated |
| 0,330467719 | AC104654.2 | FALSE | Upregulated |
| 0,424826421 | AC104777.2 | FALSE | Upregulated |
| | AC104777.4 | | |
| 0,338617235 | AC104820.2 | FALSE | Upregulated |
| -0,750763672 | AC105053.3 | FALSE | Downregulated |
| | AC105253.1 | | |
| 0,448212425 | AC105339.1 | FALSE | Upregulated |
| -0,57327632 | AC105760.3 | FALSE | Downregulated |
| 0 | AC107070.1 | FALSE | Upregulated |
| -1,378152027 | AC107622.1 | | Downregulated |
| | AC108142.1 | | |
| | AC113607.2 | | |
| | AC116035.1 | | |
| -0,562561664 | AC125421.1 | FALSE | Downregulated |
| 0,506630316 | AC131097.3 | FALSE | Upregulated |
| -0,864162569 | AC136896.1 | FALSE | Downregulated |
| 0,067965549 | AC159540.1 | FALSE | Upregulated |
| 0,306141934 | AC226118.1 | FALSE | Upregulated |
| 0,079634069 | ACBD7 | FALSE | Upregulated |
| -0,082073388 | ACCSL | FALSE | Downregulated |
| 0,48101537 | ACER1 | FALSE | Upregulated |
| 0,005840273 | ACKR3 | FALSE | Upregulated |
| 0,918975837 | ACPP | FALSE | Upregulated |
| 0 | ACPT | FALSE | Upregulated |
| 0,768613555 | ACR | FALSE | Upregulated |
| 0,717701195 | ACRV1 | FALSE | Upregulated |
| 0,904810425 | ACSM4 | FALSE | Upregulated |
| 0,622059094 | ACTA2-AS1 | FALSE | Upregulated |
| -0,358768913 | ACTC1 | FALSE | Downregulated |
| 0,197069033 | ACTG1P4 | FALSE | Upregulated |
| 0,008978576 | ACTG2 | FALSE | Upregulated |
| 0,365625131 | ACTL6B | FALSE | Upregulated |
| 0,021563842 | ACTL7B | FALSE | Upregulated |
| | ACTL8 | | |

| | | | |
|---|---|---|---|
| 0,40942204 | ACTL9 | FALSE | Upregulated |
| | ACTN1-AS1 | | |
| -0,369314903 | ACTR3BP2 | FALSE | Downregulated |
| 0 | ACTR3BP5 | FALSE | Upregulated |
| | ACTRT1 | | |
| | ACTRT2 | | |
| 0,428660274 | ACYP1 | FALSE | Upregulated |
| -0,421540546 | AD000090.2 | FALSE | Downregulated |
| | ADAD1 | | |
| -0,129530543 | ADAD2 | FALSE | Downregulated |
| | ADAM18 | | |
| | ADAM2 | | |
| 0,318834709 | ADAM23 | FALSE | Upregulated |
| 0,092024074 | ADAM29 | FALSE | Upregulated |
| 0 | ADAM30 | FALSE | Upregulated |
| 0,526136307 | ADAM32 | FALSE | Upregulated |
| 0,046678197 | ADAM3A | FALSE | Upregulated |
| | ADAM5 | | |
| 0,211047881 | ADAM7 | FALSE | Upregulated |
| 1,589322539 | ADAMDEC1 | | Upregulated |
| 2,250968996 | ADAMTS14 | | Upregulated |
| 0,070401644 | ADAMTS18 | FALSE | Upregulated |
| 0,390950345 | ADAMTS19 | FALSE | Upregulated |
| | ADAMTS20 | | |
| 0,703681735 | ADAMTS3 | FALSE | Upregulated |
| 0 | ADAMTS7P1 | FALSE | Upregulated |
| -1,304778792 | ADARB2 | | Downregulated |
| 0,554233466 | ADARB2-AS1 | FALSE | Upregulated |
| 0 | ADCY2 | FALSE | Upregulated |
| 0,459640654 | ADCY8 | FALSE | Upregulated |
| 0 | ADCYAP1 | FALSE | Upregulated |
| 0,920433045 | ADD2 | FALSE | Upregulated |
| 0,769392891 | ADD3-AS1 | FALSE | Upregulated |
| | ADGB | | |
| | ADIG | | |
| 0,360694188 | ADIPOQ | FALSE | Upregulated |
| -0,989048575 | ADORA1 | FALSE | Downregulated |
| -0,735273763 | ADPRHL1 | FALSE | Downregulated |
| 1,169890043 | ADRA1D | | Upregulated |
| 0,39049268 | ADRA2C | FALSE | Upregulated |
| -0,927049729 | ADRB1 | FALSE | Downregulated |
| | ADRB3 | | |
| -0,542997591 | ADTRP | FALSE | Downregulated |
| | AF064858.7 | | |
| 0,59419965 | AFF2 | FALSE | Upregulated |
| -0,912951423 | AFP | FALSE | Downregulated |
| -0,233635262 | AGAP1-IT1 | FALSE | Downregulated |
| 0 | AGBL1 | FALSE | Upregulated |
| | AGBL1-AS1 | | |
| -1,696069011 | AGBL4 | | Downregulated |
| | AGBL4-IT1 | | |

| | | | |
|---|---|---|---|
| 0,909706914 | AGBL5-AS1 | FALSE | Upregulated |
| 0,53265747 | AGPAT4-IT1 | FALSE | Upregulated |
| | AGR3 | | |
| | AGTR2 | | |
| -0,402755918 | AHCTF1P1 | FALSE | Downregulated |
| -0,011313464 | AHRR | FALSE | Downregulated |
| | AHSP | | |
| 0 | AICDA | FALSE | Upregulated |
| 1,975952708 | AIM1L | | Upregulated |
| 1,102593391 | AIM2 | | Upregulated |
| | AIPL1 | | |
| 0,785296432 | AIRE | FALSE | Upregulated |
| 0,10618591 | AK7 | FALSE | Upregulated |
| 0,357426913 | AK8 | FALSE | Upregulated |
| 0,423403028 | AKAP14 | FALSE | Upregulated |
| | AKAP17A-2 | | |
| 0 | AKAP4 | FALSE | Upregulated |
| 0,329578028 | AKNAD1 | FALSE | Upregulated |
| 0,476674187 | AKR1B15 | FALSE | Upregulated |
| | AL022344.4 | | |
| 0,046678197 | AL109763.2 | FALSE | Upregulated |
| -0,022320049 | AL132709.5 | FALSE | Downregulated |
| -0,695865891 | ALDH1A2 | FALSE | Downregulated |
| 0,887084733 | ALDH3A1 | FALSE | Upregulated |
| -0,262160821 | ALDH3B2 | FALSE | Downregulated |
| -0,38299803 | ALG1L2 | FALSE | Downregulated |
| -0,265530071 | ALLC | FALSE | Downregulated |
| 0,471623712 | ALMS1-IT1 | FALSE | Upregulated |
| -0,660766809 | ALOX12B | FALSE | Downregulated |
| -0,270332337 | ALOX15 | FALSE | Downregulated |
| 1,289649236 | ALOX15B | | Upregulated |
| -0,08735247 | ALOX15P1 | FALSE | Downregulated |
| 1,117963271 | ALOX5AP | | Upregulated |
| 0 | ALOXE3 | FALSE | Upregulated |
| | ALPI | | |
| -0,045290798 | ALPP | FALSE | Downregulated |
| | ALPPL2 | | |
| 0 | ALX1 | FALSE | Upregulated |
| 0,485868897 | AMBN | FALSE | Upregulated |
| 0 | AMELX | FALSE | Upregulated |
| | AMELY | | |
| 0 | AMER2 | FALSE | Upregulated |
| | AMER3 | | |
| 0,757255657 | AMH | FALSE | Upregulated |
| -0,386864191 | AMHR2 | FALSE | Downregulated |
| 0,391970119 | AMPD1 | FALSE | Upregulated |
| | AMTN | | |
| | AMY1A | | |
| | AMY1B | | |
| | AMY1C | | |
| | AMY2A | | |

| | | | |
|---|---|---|---|
| 0 | ANAPC1P1 | FALSE | Upregulated |
| -0,08859704 | ANGPT1 | FALSE | Downregulated |
| -0,012241586 | ANGPT4 | FALSE | Downregulated |
| 0,602534929 | ANGPTL5 | FALSE | Upregulated |
| 0,029177462 | ANHX | FALSE | Upregulated |
| 0,068128077 | ANKFN1 | FALSE | Upregulated |
| -0,828690697 | ANKK1 | FALSE | Downregulated |
| 0,675673274 | ANKRD1 | FALSE | Upregulated |
| -0,348493724 | ANKRD18B | FALSE | Downregulated |
| 0 | ANKRD18CP | FALSE | Upregulated |
| 0,104949029 | ANKRD18DP | FALSE | Upregulated |
| -0,389296299 | ANKRD2 | FALSE | Downregulated |
| -0,563933492 | ANKRD20A1 | FALSE | Downregulated |
| 0,677906935 | ANKRD20A11P | FALSE | Upregulated |
| | ANKRD20A19P | | |
| 0,007372946 | ANKRD20A3 | FALSE | Upregulated |
| 1,900014111 | ANKRD20A4 | | Upregulated |
| 0,370215998 | ANKRD20A5P | FALSE | Upregulated |
| 0,020960867 | ANKRD20A8P | FALSE | Upregulated |
| | ANKRD26P1 | | |
| -0,277095259 | ANKRD26P3 | FALSE | Downregulated |
| 1,108247668 | ANKRD29 | | Upregulated |
| | ANKRD30A | | |
| 0,567584455 | ANKRD30B | FALSE | Upregulated |
| | ANKRD30BL | | |
| 0 | ANKRD30BP2 | FALSE | Upregulated |
| | ANKRD30BP3 | | |
| 0,099740637 | ANKRD31 | FALSE | Upregulated |
| 0 | ANKRD33 | FALSE | Upregulated |
| -0,120881603 | ANKRD34B | FALSE | Downregulated |
| | ANKRD34C | | |
| 0,030773169 | ANKRD36BP2 | FALSE | Upregulated |
| -0,323128068 | ANKRD44-IT1 | FALSE | Downregulated |
| 0,40426344 | ANKRD45 | FALSE | Upregulated |
| | ANKRD60 | | |
| 0 | ANKRD62 | FALSE | Upregulated |
| 0,946072485 | ANKRD62P1-PARP4P3 | FALSE | Upregulated |
| 0,530417656 | ANKRD63 | FALSE | Upregulated |
| 0,623359442 | ANKRD66 | FALSE | Upregulated |
| | ANKRD7 | | |
| | ANKUB1 | | |
| 1,680210198 | ANLN | | Upregulated |
| 0,296738253 | ANO2 | FALSE | Upregulated |
| 0,240360702 | ANO4 | FALSE | Upregulated |
| 0,38647539 | ANP32C | FALSE | Upregulated |
| | ANP32D | | |
| -0,007661083 | ANTXRL | FALSE | Downregulated |
| 0,476657233 | ANTXRLP1 | FALSE | Upregulated |
| 0 | ANXA2P3 | FALSE | Upregulated |
| 0,26655477 | ANXA3 | FALSE | Upregulated |
| -0,555016204 | ANXA8 | FALSE | Downregulated |

| | | | |
|---|---|---|---|
| 0,641302183 | ANXA8L1 | FALSE | Upregulated |
| 0,314777401 | AOX2P | FALSE | Upregulated |
| | AP000289.6 | | |
| -0,61434031 | AP000330.8 | FALSE | Downregulated |
| | AP000345.1 | | |
| | AP000469.2 | | |
| | AP000475.2 | | |
| 0,380186195 | AP000688.8 | FALSE | Upregulated |
| 1,623052223 | AP001258.4 | | Upregulated |
| -0,106355206 | AP003025.2 | FALSE | Downregulated |
| | AP1B1P1 | | |
| 0,262794268 | AP1M2 | FALSE | Upregulated |
| -0,224200055 | AP1S3 | FALSE | Downregulated |
| 0,486142573 | AP3B2 | FALSE | Upregulated |
| 0,406787727 | APCDD1L-AS1 | FALSE | Upregulated |
| | APELA | | |
| -0,488043307 | APITD1-CORT | FALSE | Downregulated |
| -0,361974018 | APLN | FALSE | Downregulated |
| 0,32614553 | APLP1 | FALSE | Upregulated |
| | APOBEC1 | | |
| 0,951110823 | APOBEC3A | FALSE | Upregulated |
| 2,342338233 | APOBEC3B | | Upregulated |
| 0,362544021 | APOBEC3B-AS1 | FALSE | Upregulated |
| -0,484981354 | APOBEC3H | FALSE | Downregulated |
| 0,165076244 | APOD | FALSE | Upregulated |
| 0 | APOOP5 | FALSE | Upregulated |
| | AQP10 | | |
| | AQP12A | | |
| | AQP12B | | |
| 0,070401644 | AQP2 | FALSE | Upregulated |
| 0,552602958 | AQP5 | FALSE | Upregulated |
| 0,265604008 | AQP6 | FALSE | Upregulated |
| -1,446648599 | AQP7P3 | | Downregulated |
| -0,045460148 | AQP8 | FALSE | Downregulated |
| -0,082794598 | AQPEP | FALSE | Downregulated |
| -0,283655911 | ARC | FALSE | Downregulated |
| 0 | AREG | FALSE | Upregulated |
| 0,528771572 | ARGFX | FALSE | Upregulated |
| 0,729827364 | ARHGAP11B | FALSE | Upregulated |
| 1,228255689 | ARHGAP19-SLIT1 | | Upregulated |
| -0,186123223 | ARHGAP26-IT1 | FALSE | Downregulated |
| 1,095742374 | ARHGAP28 | | Upregulated |
| | ARHGAP36 | | |
| 0 | ARHGAP40 | FALSE | Upregulated |
| -0,635514619 | ARHGAP8 | FALSE | Downregulated |
| 0 | ARHGDIG | FALSE | Upregulated |
| 1,525157426 | ARHGEF19 | | Upregulated |
| -0,028907643 | ARHGEF26-AS1 | FALSE | Downregulated |
| 0 | ARHGEF3-AS1 | FALSE | Upregulated |
| 0 | ARHGEF38-IT1 | FALSE | Upregulated |
| 0,946424396 | ARL14 | FALSE | Upregulated |

| | | | |
|---|---|---|---|
| 0 | ARL14EPL | FALSE | Upregulated |
| -0,67137338 | ARL17B | FALSE | Downregulated |
| 0 | ARL5C | FALSE | Upregulated |
| 0,406787727 | ARL9 | FALSE | Upregulated |
| -1,178573079 | ARMC12 | | Downregulated |
| 0,420469891 | ARMC2 | FALSE | Upregulated |
| | ARMC3 | | |
| -0,043894828 | ARMC4 | FALSE | Downregulated |
| | ARMC4P1 | | |
| -2,030151268 | ARMCX5-GPRASP2 | | Downregulated |
| 0,539612447 | ARMS2 | FALSE | Upregulated |
| 0,524385887 | ARRDC5 | FALSE | Upregulated |
| -0,673300148 | ARSF | FALSE | Downregulated |
| | ARSH | | |
| 0,199570368 | ARSI | FALSE | Upregulated |
| -0,163534735 | ART3 | FALSE | Downregulated |
| 0,141726905 | ART5 | FALSE | Upregulated |
| 0,201800988 | ARX | FALSE | Upregulated |
| -0,440077999 | ASAH2 | FALSE | Downregulated |
| -0,016234776 | ASAP1-IT1 | FALSE | Downregulated |
| 0,114488476 | ASB10 | FALSE | Upregulated |
| 0,641850491 | ASB11 | FALSE | Upregulated |
| 0,012306863 | ASB17 | FALSE | Upregulated |
| 0,207100351 | ASB18 | FALSE | Upregulated |
| 0,346435571 | ASB5 | FALSE | Upregulated |
| 0 | ASCL4 | FALSE | Upregulated |
| 0,885392699 | ASCL5 | FALSE | Upregulated |
| 0 | ASIC2 | FALSE | Upregulated |
| | ASMT | | |
| | ASMT-2 | | |
| | ASMTL-2 | | |
| | ASMTL-AS1-2 | | |
| 1,690830387 | ASRGL1 | | Upregulated |
| 0 | ASTL | FALSE | Upregulated |
| -0,394600244 | ASTN1 | FALSE | Downregulated |
| | ASZ1 | | |
| 0,00905151 | ATCAY | FALSE | Upregulated |
| | ATOH1 | | |
| -1,521755887 | ATOH7 | | Downregulated |
| 0,146690165 | ATP12A | FALSE | Upregulated |
| -0,475812224 | ATP13A4 | FALSE | Downregulated |
| 0,044678282 | ATP13A5 | FALSE | Upregulated |
| 0,486060549 | ATP1A3 | FALSE | Upregulated |
| -0,265353325 | ATP1A4 | FALSE | Downregulated |
| -0,091431256 | ATP1B4 | FALSE | Downregulated |
| -0,22843617 | ATP2B3 | FALSE | Downregulated |
| 0,00048018 | ATP4A | FALSE | Upregulated |
| -0,045460148 | ATP5EP2 | FALSE | Downregulated |
| -0,170397657 | ATP5L2 | FALSE | Downregulated |
| -1,48683113 | ATP6AP1L | | Downregulated |
| | ATP6V0A4 | | |

| | | | |
|---|---|---|---|
| 0,591358694 | ATP6V0CP3 | FALSE | Upregulated |
| -0,072969565 | ATP6V0D2 | FALSE | Downregulated |
| -1,249346595 | ATP6V1B1 | | Downregulated |
| | ATP6V1G3 | | |
| 0,323344695 | ATP8A2 | FALSE | Upregulated |
| 1,712662876 | ATP8B3 | | Upregulated |
| -0,705297715 | ATP8B5P | FALSE | Downregulated |
| 0,356618795 | ATXN3L | FALSE | Upregulated |
| | ATXN8OS | | |
| 1,426046754 | AURKB | | Upregulated |
| -1,038855308 | AURKC | | Downregulated |
| | AVP | | |
| -0,25701796 | AVPR1B | FALSE | Downregulated |
| -0,091846163 | AWAT2 | FALSE | Downregulated |
| 0,421528573 | AXDND1 | FALSE | Upregulated |
| 0,201184564 | AZU1 | FALSE | Upregulated |
| 0,866775686 | B3GALNT1 | FALSE | Upregulated |
| 0,035655664 | B3GALT1 | FALSE | Upregulated |
| -0,332987455 | B3GALT5 | FALSE | Downregulated |
| 0,141925876 | B3GNT3 | FALSE | Upregulated |
| 0,055378136 | B3GNT6 | FALSE | Upregulated |
| 0 | B4GALNT2 | FALSE | Upregulated |
| 1,002382939 | B4GALNT4 | | Upregulated |
| 0 | BAALCOS | FALSE | Upregulated |
| 0,318162266 | BAGE2 | FALSE | Upregulated |
| 0,306755915 | BAI1 | FALSE | Upregulated |
| -0,056671736 | BAI2 | FALSE | Downregulated |
| | BANCR | | |
| 0 | BANF2 | FALSE | Upregulated |
| | BARHL2 | | |
| 0,233118511 | BARX1 | FALSE | Upregulated |
| 0 | BARX2 | FALSE | Upregulated |
| 0,38647539 | BASP1P1 | FALSE | Upregulated |
| 1,916111301 | BATF | | Upregulated |
| 1,413157727 | BATF3 | | Upregulated |
| 1,083586679 | BBS12 | | Upregulated |
| 0 | BCAR4 | FALSE | Upregulated |
| 0,40079134 | BCAS1 | FALSE | Upregulated |
| 0,802665665 | BCL11A | FALSE | Upregulated |
| 0,501909949 | BCL2A1 | FALSE | Upregulated |
| 0,539412575 | BCL2L15 | FALSE | Upregulated |
| -0,34985534 | BCL2L2-PABPN1 | FALSE | Downregulated |
| 0,700335951 | BCMO1 | FALSE | Upregulated |
| 0,774771577 | BCORP1 | FALSE | Upregulated |
| -0,512626122 | BCRP2 | FALSE | Downregulated |
| -0,9442976 | BCRP3 | FALSE | Downregulated |
| 0,028702733 | BCYRN1 | FALSE | Upregulated |
| 1,499665756 | BDKRB1 | | Upregulated |
| 0,823624091 | BDKRB2 | FALSE | Upregulated |
| | BEND2 | | |
| -0,310250315 | BEND5 | FALSE | Downregulated |

| | | | |
|---|---|---|---|
| -0,207225908 | BEND6 | FALSE | Downregulated |
| -0,589455868 | BEST2 | FALSE | Downregulated |
| 0,005422933 | BEST3 | FALSE | Upregulated |
| 1,728764196 | BEST4 | | Upregulated |
| 0,211715194 | BEX1 | FALSE | Upregulated |
| 0,368869352 | BEX2 | FALSE | Upregulated |
| 0,382133797 | BEX4 | FALSE | Upregulated |
| 0,272939013 | BEX5 | FALSE | Upregulated |
| -0,388878898 | BFSP1 | FALSE | Downregulated |
| 0,26973982 | BFSP2 | FALSE | Upregulated |
| 0,453695514 | BFSP2-AS1 | FALSE | Upregulated |
| | BGLAP | | |
| 0,020789395 | BHLHA9 | FALSE | Upregulated |
| 0 | BHLHE23 | FALSE | Upregulated |
| 0 | BHLHE40-AS1 | FALSE | Upregulated |
| 0,449316216 | BIK | FALSE | Upregulated |
| -1,664867602 | BIN3-IT1 | | Downregulated |
| 0,82776473 | BIRC5 | FALSE | Upregulated |
| 0 | BIRC7 | FALSE | Upregulated |
| | BIRC8 | | |
| 0,149296437 | BLACAT1 | FALSE | Upregulated |
| 0,64925946 | BLACE | FALSE | Upregulated |
| 0,069170908 | BLID | FALSE | Upregulated |
| 1,185780035 | BLM | | Upregulated |
| -0,882060979 | BLOC1S5-TXNDC5 | FALSE | Downregulated |
| | BMP15 | | |
| 0,311435677 | BMP3 | FALSE | Upregulated |
| 0 | BMP7 | FALSE | Upregulated |
| -1,61230464 | BMPR1B | | Downregulated |
| 0,713966754 | BMX | FALSE | Upregulated |
| -0,159210223 | BNC1 | FALSE | Downregulated |
| -0,501928351 | BOD1L2 | FALSE | Downregulated |
| 0 | BPESC1 | FALSE | Upregulated |
| 0,085186009 | BPI | FALSE | Upregulated |
| | BPIFA1 | | |
| | BPIFA2 | | |
| | BPIFA3 | | |
| | BPIFA4P | | |
| 0,009020768 | BPIFB1 | FALSE | Upregulated |
| 0,099452879 | BPIFB2 | FALSE | Upregulated |
| | BPIFB3 | | |
| | BPIFB4 | | |
| | BPIFB6 | | |
| 0,265834497 | BPIFC | FALSE | Upregulated |
| | BRDT | | |
| -0,777309945 | BREA2 | FALSE | Downregulated |
| 0,00716563 | BRINP1 | FALSE | Upregulated |
| -0,839909955 | BRINP2 | FALSE | Downregulated |
| | BRINP3 | | |
| 0,478351672 | BRS3 | FALSE | Upregulated |
| 1,014703147 | BRWD1-AS1 | | Upregulated |

| | | | |
|---|---|---|---|
| 0,051672937 | BSN-AS2 | FALSE | Upregulated |
| 0,384758161 | BSND | FALSE | Upregulated |
| | BSPH1 | | |
| | BSX | | |
| -0,628657599 | BTBD16 | FALSE | Downregulated |
| 1,212478771 | BTBD17 | | Upregulated |
| -1,037826076 | BTBD8 | | Downregulated |
| 0,847965963 | BTC | FALSE | Upregulated |
| 0,406787727 | BTF3P11 | FALSE | Upregulated |
| | BTG4 | | |
| 0,697914941 | BTLA | FALSE | Upregulated |
| 0,50012082 | BTNL10 | FALSE | Upregulated |
| -0,295817819 | BTNL2 | FALSE | Downregulated |
| -0,488915988 | BTNL8 | FALSE | Downregulated |
| -0,506558445 | BUB1 | FALSE | Downregulated |
| 1,502666257 | BUB1B | | Upregulated |
| 0,121434675 | BVES | FALSE | Upregulated |
| | BVES-AS1 | | |
| 0 | BX255923.3 | FALSE | Upregulated |
| 0,40555295 | C10orf107 | FALSE | Upregulated |
| 0,192365142 | C10orf111 | FALSE | Upregulated |
| | C10orf113 | | |
| 0 | C10orf115 | FALSE | Upregulated |
| | C10orf120 | | |
| -0,284225066 | C10orf126 | FALSE | Downregulated |
| 0,164212711 | C10orf129 | FALSE | Upregulated |
| -0,07189348 | C10orf131 | FALSE | Downregulated |
| 0,706871549 | C10orf35 | FALSE | Upregulated |
| 0,000977115 | C10orf40 | FALSE | Upregulated |
| 0,020789395 | C10orf53 | FALSE | Upregulated |
| 0,121776204 | C10orf67 | FALSE | Upregulated |
| 0,35924767 | C10orf71 | FALSE | Upregulated |
| | C10orf71-AS1 | | |
| | C10orf82 | | |
| 0,036193894 | C10orf85 | FALSE | Upregulated |
| -0,170391513 | C10orf90 | FALSE | Downregulated |
| 0,648875876 | C10orf91 | FALSE | Upregulated |
| | C10orf99 | | |
| | C11orf40 | | |
| | C11orf44 | | |
| | C11orf53 | | |
| 0,954967069 | C11orf63 | FALSE | Upregulated |
| 0,141639158 | C11orf65 | FALSE | Upregulated |
| | C11orf70 | | |
| 0,1894058 | C11orf74 | FALSE | Upregulated |
| 1,52659589 | C11orf82 | | Upregulated |
| 0,032521485 | C11orf85 | FALSE | Upregulated |
| | C11orf86 | | |
| 0,848117222 | C11orf87 | FALSE | Upregulated |
| 0,100713889 | C11orf88 | FALSE | Upregulated |
| 0,376664839 | C11orf91 | FALSE | Upregulated |

| | | | |
|---|---|---|---|
| | C11orf94 | | |
| 0 | C12orf36 | FALSE | Upregulated |
| 0,302725208 | C12orf39 | FALSE | Upregulated |
| -0,304106049 | C12orf40 | FALSE | Downregulated |
| 0,305281715 | C12orf42 | FALSE | Upregulated |
| | C12orf50 | | |
| | C12orf54 | | |
| 0,35729113 | C12orf55 | FALSE | Upregulated |
| -0,683920788 | C12orf56 | FALSE | Downregulated |
| -0,25371426 | C12orf68 | FALSE | Downregulated |
| 0,280965999 | C12orf74 | FALSE | Upregulated |
| | C12orf77 | | |
| -0,035573082 | C12orf79 | FALSE | Downregulated |
| 0 | C12orf80 | FALSE | Upregulated |
| | C13orf45 | | |
| | C14orf177 | | |
| 0,875038491 | C14orf183 | FALSE | Upregulated |
| -0,093662559 | C14orf23 | FALSE | Downregulated |
| 0 | C14orf39 | FALSE | Upregulated |
| -0,04694985 | C14orf64 | FALSE | Downregulated |
| -0,459273935 | C15orf26 | FALSE | Downregulated |
| | C15orf32 | | |
| 0,466775079 | C15orf43 | FALSE | Upregulated |
| 0 | C15orf48 | FALSE | Upregulated |
| 0 | C15orf53 | FALSE | Upregulated |
| 0 | C15orf56 | FALSE | Upregulated |
| -0,872920248 | C15orf59 | FALSE | Downregulated |
| -0,668320351 | C15orf65 | FALSE | Downregulated |
| | C16orf11 | | |
| 0 | C16orf47 | FALSE | Upregulated |
| -0,174834022 | C16orf74 | FALSE | Downregulated |
| -0,344636409 | C16orf78 | FALSE | Downregulated |
| | C16orf82 | | |
| 0,583356869 | C16orf92 | FALSE | Upregulated |
| -0,650046492 | C16orf96 | FALSE | Downregulated |
| | C16orf97 | | |
| | C17orf102 | | |
| 0 | C17orf105 | FALSE | Upregulated |
| | C17orf112 | | |
| 0,241229988 | C17orf50 | FALSE | Upregulated |
| 0,686022675 | C17orf64 | FALSE | Upregulated |
| 0,246298777 | C17orf74 | FALSE | Upregulated |
| | C17orf77 | | |
| | C17orf98 | | |
| 0,156446097 | C17orf99 | FALSE | Upregulated |
| 0,038938467 | C18orf12 | FALSE | Upregulated |
| 0 | C18orf42 | FALSE | Upregulated |
| 0,100873573 | C18orf54 | FALSE | Upregulated |
| 1,714349417 | C18orf56 | | Upregulated |
| | C18orf61 | | |
| -0,085048472 | C18orf63 | FALSE | Downregulated |

| | | | |
|---|---|---|---|
| | C18orf64 | | |
| -0,463402831 | C19orf18 | FALSE | Downregulated |
| 0,314110952 | C19orf33 | FALSE | Upregulated |
| 0,009914605 | C19orf38 | FALSE | Upregulated |
| -0,212688465 | C19orf45 | FALSE | Downregulated |
| 0,592034168 | C19orf59 | FALSE | Upregulated |
| -0,501468014 | C19orf69 | FALSE | Downregulated |
| 0,587529068 | C19orf73 | FALSE | Upregulated |
| 0,171977856 | C19orf81 | FALSE | Upregulated |
| 0,101805999 | C19orf83 | FALSE | Upregulated |
| 0 | C1orf100 | FALSE | Upregulated |
| 0,049368133 | C1orf101 | FALSE | Upregulated |
| 1,037202108 | C1orf105 | | Upregulated |
| 1,192132139 | C1orf110 | | Upregulated |
| 0,198648994 | C1orf127 | FALSE | Upregulated |
| | C1orf137 | | |
| -1,400988888 | C1orf140 | | Downregulated |
| | C1orf141 | | |
| 0 | C1orf146 | FALSE | Upregulated |
| | C1orf158 | | |
| -0,080112937 | C1orf173 | FALSE | Downregulated |
| 0,151611645 | C1orf180 | FALSE | Upregulated |
| -0,493825697 | C1orf185 | FALSE | Downregulated |
| -0,402176435 | C1orf186 | FALSE | Downregulated |
| | C1orf189 | | |
| 0,148771569 | C1orf194 | FALSE | Upregulated |
| 0,26579746 | C1orf227 | FALSE | Upregulated |
| 1,179809905 | C1orf229 | | Upregulated |
| 1,327361907 | C1orf233 | | Upregulated |
| 0,048276727 | C1orf234 | FALSE | Upregulated |
| 0 | C1orf61 | FALSE | Upregulated |
| | C1orf64 | | |
| 0,481086388 | C1orf65 | FALSE | Upregulated |
| 0 | C1orf68 | FALSE | Upregulated |
| | C1orf87 | | |
| | C1orf94 | | |
| 0,607227277 | C1QL1 | FALSE | Upregulated |
| 0,123248774 | C1QL2 | FALSE | Upregulated |
| | C1QL4 | | |
| -0,160127912 | C1QTNF2 | FALSE | Downregulated |
| 1,19466136 | C1QTNF4 | | Upregulated |
| 0 | C1QTNF8 | FALSE | Upregulated |
| 0,850047581 | C1QTNF9 | FALSE | Upregulated |
| -0,803848358 | C1QTNF9B-AS1 | FALSE | Downregulated |
| | C20orf141 | | |
| -0,218103073 | C20orf166 | FALSE | Downregulated |
| 0,071529586 | C20orf166-AS1 | FALSE | Upregulated |
| 0,460528582 | C20orf181 | FALSE | Upregulated |
| 0,462375706 | C20orf196 | FALSE | Upregulated |
| 0,623688187 | C20orf197 | FALSE | Upregulated |
| 0,138461527 | C20orf202 | FALSE | Upregulated |

| | | | |
|---|---|---|---|
| 0,363009252 | C20orf203 | FALSE | Upregulated |
| 0 | C20orf26 | FALSE | Upregulated |
| | C20orf57 | | |
| | C20orf78 | | |
| 0 | C20orf85 | FALSE | Upregulated |
| 0,416459879 | C21orf128 | FALSE | Upregulated |
| -0,117060314 | C21orf15 | FALSE | Downregulated |
| 0,005107265 | C21orf37 | FALSE | Upregulated |
| 0,501757352 | C21orf49 | FALSE | Upregulated |
| | C21orf54 | | |
| 0,263791679 | C21orf88 | FALSE | Upregulated |
| -0,026731683 | C21orf90 | FALSE | Downregulated |
| | C21orf91-OT1 | | |
| 0,364041279 | C22orf24 | FALSE | Upregulated |
| -0,331504539 | C22orf42 | FALSE | Downregulated |
| 0,609214579 | C22orf43 | FALSE | Upregulated |
| 0,20303467 | C2CD4C | FALSE | Upregulated |
| -0,400152566 | C2orf27A | FALSE | Downregulated |
| 0 | C2orf27B | FALSE | Upregulated |
| 0,3303547 | C2orf40 | FALSE | Upregulated |
| 0,21651875 | C2orf48 | FALSE | Upregulated |
| 0 | C2orf50 | FALSE | Upregulated |
| 0,465237344 | C2orf53 | FALSE | Upregulated |
| 0,45617281 | C2orf57 | FALSE | Upregulated |
| 0,23285437 | C2orf61 | FALSE | Upregulated |
| 0 | C2orf62 | FALSE | Upregulated |
| 0 | C2orf66 | FALSE | Upregulated |
| | C2orf70 | | |
| 2,344356 | C2orf71 | | Upregulated |
| 0,370718861 | C2orf73 | FALSE | Upregulated |
| -0,110320355 | C2orf76 | FALSE | Downregulated |
| 0,554710298 | C2orf78 | FALSE | Upregulated |
| 0,100713889 | C2orf80 | FALSE | Upregulated |
| 0,035090141 | C2orf83 | FALSE | Upregulated |
| 0,265633693 | C2orf91 | FALSE | Upregulated |
| 0,170785421 | C3orf14 | FALSE | Upregulated |
| -0,088379243 | C3orf20 | FALSE | Downregulated |
| 0 | C3orf27 | FALSE | Upregulated |
| -0,830585205 | C3orf33 | FALSE | Downregulated |
| -0,231083014 | C3orf36 | FALSE | Downregulated |
| -0,795479354 | C3orf43 | FALSE | Downregulated |
| 1,449014093 | C3orf52 | | Upregulated |
| 0 | C3orf55 | FALSE | Upregulated |
| | C3orf56 | | |
| -1,222441465 | C3orf70 | | Downregulated |
| -0,08223701 | C3orf72 | FALSE | Downregulated |
| 0 | C3orf79 | FALSE | Upregulated |
| -0,364144348 | C3orf80 | FALSE | Downregulated |
| 0,652701864 | C3orf84 | FALSE | Upregulated |
| -0,402732143 | C4orf17 | FALSE | Downregulated |
| -0,034062844 | C4orf22 | FALSE | Downregulated |

| | | | |
|---|---|---|---|
| 0,328473495 | C4orf26 | FALSE | Upregulated |
| -0,383811389 | C4orf36 | FALSE | Downregulated |
| 1,249006088 | C4orf40 | | Upregulated |
| -0,68902824 | C4orf45 | FALSE | Downregulated |
| 0,558103654 | C4orf46 | FALSE | Upregulated |
| 0,68724913 | C4orf47 | FALSE | Upregulated |
| 1,924179064 | C4orf48 | | Upregulated |
| -0,450010058 | C4orf50 | FALSE | Downregulated |
| | C4orf51 | | |
| 1,694916023 | C5orf20 | | Upregulated |
| 1,753706106 | C5orf28 | | Upregulated |
| -0,033327359 | C5orf30 | FALSE | Downregulated |
| 0,120816263 | C5orf38 | FALSE | Upregulated |
| | C5orf46 | | |
| 0 | C5orf47 | FALSE | Upregulated |
| 0,136785908 | C5orf48 | FALSE | Upregulated |
| 0,112680632 | C5orf52 | FALSE | Upregulated |
| 1,226178586 | C5orf54 | | Upregulated |
| 0,417538527 | C5orf60 | FALSE | Upregulated |
| 0,13568641 | C5orf64 | FALSE | Upregulated |
| | C6orf100 | | |
| | C6orf118 | | |
| | C6orf15 | | |
| 0,965543066 | C6orf163 | FALSE | Upregulated |
| 0,011182354 | C6orf165 | FALSE | Upregulated |
| -0,116682619 | C6orf195 | FALSE | Downregulated |
| 0,660083095 | C6orf222 | FALSE | Upregulated |
| -0,143297461 | C6orf223 | FALSE | Downregulated |
| -0,564394881 | C6orf229 | FALSE | Downregulated |
| 0 | C6orf52 | FALSE | Upregulated |
| 0,191610677 | C6orf57 | FALSE | Upregulated |
| 0,429166696 | C6orf58 | FALSE | Upregulated |
| 0,545837784 | C6orf7 | FALSE | Upregulated |
| -0,180633716 | C6orf99 | FALSE | Downregulated |
| 0,578426401 | C7orf33 | FALSE | Upregulated |
| 1,533866596 | C7orf34 | | Upregulated |
| -0,28942601 | C7orf57 | FALSE | Downregulated |
| 0,124221433 | C7orf61 | FALSE | Upregulated |
| 0,082383625 | C7orf62 | FALSE | Upregulated |
| 0 | C7orf65 | FALSE | Upregulated |
| | C7orf66 | | |
| 0,87033635 | C7orf69 | FALSE | Upregulated |
| | C7orf72 | | |
| -0,998239473 | C7orf76 | FALSE | Downregulated |
| 0,390950345 | C8orf12 | FALSE | Upregulated |
| 0 | C8orf22 | FALSE | Upregulated |
| | C8orf34 | | |
| -0,383444619 | C8orf44-SGK3 | FALSE | Downregulated |
| 0,164104066 | C8orf48 | FALSE | Upregulated |
| 0,404174652 | C8orf49 | FALSE | Upregulated |
| -0,016422875 | C8orf69 | FALSE | Downregulated |

| | | | |
|---|---|---|---|
| 0,117568759 | C8orf74 | FALSE | Upregulated |
| 0 | C8orf86 | FALSE | Upregulated |
| | C8orf87 | | |
| 0,369064525 | C8orf88 | FALSE | Upregulated |
| 1,008798773 | C9orf106 | | Upregulated |
| 0 | C9orf135 | FALSE | Upregulated |
| | C9orf135-AS1 | | |
| 0,195913201 | C9orf153 | FALSE | Upregulated |
| -0,208482806 | C9orf170 | FALSE | Downregulated |
| 1,009276662 | C9orf24 | | Upregulated |
| 0,344807854 | C9orf43 | FALSE | Upregulated |
| -0,826355754 | C9orf47 | FALSE | Downregulated |
| 0,962434638 | C9orf50 | FALSE | Upregulated |
| 0,472855489 | C9orf53 | FALSE | Upregulated |
| -0,065407283 | C9orf57 | FALSE | Downregulated |
| | C9orf62 | | |
| | C9orf92 | | |
| 0,873244891 | CA10 | FALSE | Upregulated |
| 0,337111678 | CA12 | FALSE | Upregulated |
| 0,43048677 | CA4 | FALSE | Upregulated |
| | CA6 | | |
| 0,79371823 | CA7 | FALSE | Upregulated |
| 0,480635035 | CA8 | FALSE | Upregulated |
| -0,353431844 | CA9 | FALSE | Downregulated |
| -0,080414653 | CABP1 | FALSE | Downregulated |
| | CABP2 | | |
| | CABP5 | | |
| 0 | CABS1 | FALSE | Upregulated |
| 0,192901972 | CACNA1B | FALSE | Upregulated |
| 0 | CACNA1C-IT3 | FALSE | Upregulated |
| 0,780418341 | CACNA1E | FALSE | Upregulated |
| -0,031054503 | CACNA1F | FALSE | Downregulated |
| 1,039316994 | CACNA1G | | Upregulated |
| 0,520478888 | CACNA1I | FALSE | Upregulated |
| -0,280601307 | CACNA1S | FALSE | Downregulated |
| 0,088413284 | CACNA2D3 | FALSE | Upregulated |
| 0,089465978 | CACNA2D3-AS1 | FALSE | Upregulated |
| 0,439994782 | CACNB4 | FALSE | Upregulated |
| | CACNG2 | | |
| 0 | CACNG3 | FALSE | Upregulated |
| -0,936298782 | CACNG4 | FALSE | Downregulated |
| 0,38647539 | CACNG5 | FALSE | Upregulated |
| 0,320886704 | CACNG6 | FALSE | Upregulated |
| | CACNG7 | | |
| -0,306052959 | CACNG8 | FALSE | Downregulated |
| -0,437814483 | CADM2 | FALSE | Downregulated |
| 0,439397163 | CADM2-AS2 | FALSE | Upregulated |
| -0,980335487 | CADPS | FALSE | Downregulated |
| 0,919604528 | CAGE1 | FALSE | Upregulated |
| 0,886321742 | CAHM | FALSE | Upregulated |
| -0,227976517 | CALB2 | FALSE | Downregulated |

| | | | |
|---|---|---|---|
| 0 | CALCB | FALSE | Upregulated |
| 0 | CALCR | FALSE | Upregulated |
| -0,213271499 | CALHM1 | FALSE | Downregulated |
| 0 | CALHM3 | FALSE | Upregulated |
| 0 | CALML5 | FALSE | Upregulated |
| -0,081271964 | CALML6 | FALSE | Downregulated |
| 0,01196447 | CALR3 | FALSE | Upregulated |
| 0,120694397 | CALY | FALSE | Upregulated |
| 0,356238178 | CAMK1G | FALSE | Upregulated |
| -0,245729754 | CAMK2A | FALSE | Downregulated |
| 0,390950345 | CAMKV | FALSE | Upregulated |
| | CAMP | | |
| 0,079031134 | CAND1.11 | FALSE | Upregulated |
| 0,276340821 | CAPN11 | FALSE | Upregulated |
| 0,375966824 | CAPN13 | FALSE | Upregulated |
| 0,101644342 | CAPN14 | FALSE | Upregulated |
| -0,373056748 | CAPN6 | FALSE | Downregulated |
| 0,459092302 | CAPN8 | FALSE | Upregulated |
| -0,140978463 | CAPN8-2 | FALSE | Downregulated |
| 0,500829697 | CAPN9 | FALSE | Upregulated |
| 0 | CAPNS2 | FALSE | Upregulated |
| | CAPSL | | |
| 0,454416666 | CAPZA3 | FALSE | Upregulated |
| -0,02566398 | CARD16 | FALSE | Downregulated |
| | CARD17 | | |
| | CARD18 | | |
| | CARTPT | | |
| | CASC11 | | |
| 0,09231727 | CASC15 | FALSE | Upregulated |
| 0 | CASC16 | FALSE | Upregulated |
| | CASC17 | | |
| -0,303260158 | CASC18 | FALSE | Downregulated |
| -0,374691898 | CASC2 | FALSE | Downregulated |
| -0,158784957 | CASC20 | FALSE | Downregulated |
| | CASC6 | | |
| 0,497037102 | CASC8 | FALSE | Upregulated |
| 0,66841044 | CASC9 | FALSE | Upregulated |
| 0 | CASP12 | FALSE | Upregulated |
| 0 | CASP14 | FALSE | Upregulated |
| 0,893372355 | CASP5 | FALSE | Upregulated |
| -0,511610002 | CASQ2 | FALSE | Downregulated |
| -0,259816935 | CASR | FALSE | Downregulated |
| 0,042261831 | CATSPER1 | FALSE | Upregulated |
| 0,161010712 | CATSPER2 | FALSE | Upregulated |
| 0,134889346 | CATSPER3 | FALSE | Upregulated |
| | CATSPER4 | | |
| 0,276243883 | CATSPERB | FALSE | Upregulated |
| | CATSPERD | | |
| -0,171707569 | CATSPERG | FALSE | Downregulated |
| -1,139068851 | CAV3 | | Downregulated |
| -0,018497756 | CBLN1 | FALSE | Downregulated |

| | | | |
|---|---|---|---|
| 0,006138245 | CBLN2 | FALSE | Upregulated |
| 1,238916201 | CBR3-AS1 | | Upregulated |
| -0,129758761 | CBX3P2 | FALSE | Downregulated |
| 0,613657183 | CCAT2 | FALSE | Upregulated |
| 0,318291854 | CCDC103 | FALSE | Upregulated |
| | CCDC105 | | |
| -0,025572308 | CCDC11 | FALSE | Downregulated |
| 1,0929434 | CCDC110 | | Upregulated |
| 0,537543595 | CCDC112 | FALSE | Upregulated |
| 0,162273381 | CCDC114 | FALSE | Upregulated |
| 1,412585968 | CCDC116 | | Upregulated |
| -0,199257786 | CCDC129 | FALSE | Downregulated |
| 0,011707374 | CCDC13 | FALSE | Upregulated |
| 0,699699207 | CCDC13-AS1 | FALSE | Upregulated |
| 0,354595348 | CCDC135 | FALSE | Upregulated |
| 0,129413864 | CCDC136 | FALSE | Upregulated |
| -0,840370018 | CCDC138 | FALSE | Downregulated |
| | CCDC140 | | |
| 0,799222581 | CCDC144CP | FALSE | Upregulated |
| 0 | CCDC144NL | FALSE | Upregulated |
| -0,064225807 | CCDC147 | FALSE | Downregulated |
| -0,858735763 | CCDC147-AS1 | FALSE | Downregulated |
| 0,580380563 | CCDC148 | FALSE | Upregulated |
| 0 | CCDC148-AS1 | FALSE | Upregulated |
| 0,602505424 | CCDC151 | FALSE | Upregulated |
| 0,727681382 | CCDC153 | FALSE | Upregulated |
| 1,061300401 | CCDC154 | | Upregulated |
| 0,212507595 | CCDC155 | FALSE | Upregulated |
| 0,424472607 | CCDC160 | FALSE | Upregulated |
| 0,506892705 | CCDC162P | FALSE | Upregulated |
| -0,863385626 | CCDC163P | FALSE | Downregulated |
| 0,328577881 | CCDC166 | FALSE | Upregulated |
| | CCDC169 | | |
| | CCDC169-SOHLH2 | | |
| | CCDC172 | | |
| 0,398857134 | CCDC175 | FALSE | Upregulated |
| -0,002028357 | CCDC177 | FALSE | Downregulated |
| 0,046678197 | CCDC178 | FALSE | Upregulated |
| 0,236347415 | CCDC179 | FALSE | Upregulated |
| 0,474856595 | CCDC182 | FALSE | Upregulated |
| 0 | CCDC19 | FALSE | Upregulated |
| -0,080809746 | CCDC27 | FALSE | Downregulated |
| -0,896733108 | CCDC30 | FALSE | Downregulated |
| 1,951600171 | CCDC33 | | Upregulated |
| 0,653749771 | CCDC34 | FALSE | Upregulated |
| 0,200331798 | CCDC38 | FALSE | Upregulated |
| 0,298165602 | CCDC41-AS1 | FALSE | Upregulated |
| 0,912694179 | CCDC42 | FALSE | Upregulated |
| 0,591608423 | CCDC58 | FALSE | Upregulated |
| 0,073299551 | CCDC62 | FALSE | Upregulated |
| 0,493832612 | CCDC63 | FALSE | Upregulated |

| | | | |
|---|---|---|---|
| 0,561125785 | CCDC74A | FALSE | Upregulated |
| 0,954734417 | CCDC74B | FALSE | Upregulated |
| 0,046678197 | CCDC79 | FALSE | Upregulated |
| 0,16327958 | CCDC81 | FALSE | Upregulated |
| 0,105959515 | CCDC89 | FALSE | Upregulated |
| 0,39732916 | CCDC96 | FALSE | Upregulated |
| | CCER1 | | |
| 1,446387472 | CCER2 | | Upregulated |
| 0,01196447 | CCIN | FALSE | Upregulated |
| 0 | CCK | FALSE | Upregulated |
| | CCKAR | | |
| 0,986637442 | CCKBR | FALSE | Upregulated |
| | CCL1 | | |
| 0,528030348 | CCL11 | FALSE | Upregulated |
| -0,067539962 | CCL13 | FALSE | Downregulated |
| 0,450672596 | CCL17 | FALSE | Upregulated |
| 0,396026609 | CCL18 | FALSE | Upregulated |
| 0,474955637 | CCL20 | FALSE | Upregulated |
| 0,595143106 | CCL24 | FALSE | Upregulated |
| 0,621407122 | CCL25 | FALSE | Upregulated |
| | CCL26 | | |
| -0,303884952 | CCL27 | FALSE | Downregulated |
| 0,570075592 | CCL28 | FALSE | Upregulated |
| 1,352468513 | CCL4 | | Upregulated |
| 0 | CCL4L1 | FALSE | Upregulated |
| | CCL7 | | |
| 0,750869211 | CCL8 | FALSE | Upregulated |
| -0,095539013 | CCNA1 | FALSE | Downregulated |
| 2,667635597 | CCNB1 | | Upregulated |
| 0,949274696 | CCNB2 | FALSE | Upregulated |
| -0,289693185 | CCNB3 | FALSE | Downregulated |
| 0,87126609 | CCNE1 | FALSE | Upregulated |
| -1,204814281 | CCNJL | | Downregulated |
| 0 | CCNO | FALSE | Upregulated |
| | CCNYL2 | | |
| 1,618761124 | CCR3 | | Upregulated |
| 0,088719882 | CCR4 | FALSE | Upregulated |
| 0,487744309 | CCR6 | FALSE | Upregulated |
| 1,333330833 | CCRL2 | | Upregulated |
| -0,423843089 | CCSER1 | FALSE | Downregulated |
| -0,349719193 | CCT6B | FALSE | Downregulated |
| 0 | CCT8L2 | FALSE | Upregulated |
| 0,068128077 | CD164L2 | FALSE | Upregulated |
| | CD177 | | |
| 1,470254051 | CD19 | | Upregulated |
| 0,386818922 | CD1A | FALSE | Upregulated |
| 0 | CD1B | FALSE | Upregulated |
| -1,096765317 | CD1C | | Downregulated |
| -0,134059965 | CD1E | FALSE | Downregulated |
| 0,548437969 | CD200 | FALSE | Upregulated |
| 0,689888397 | CD200R1 | FALSE | Upregulated |

0,38713585 CD200R1L FALSE Upregulated
0,332687977 CD207 FALSE Upregulated
0,84516977 CD300C FALSE Upregulated
CD300LD
0,893219449 CD300LG FALSE Upregulated
0,936520834 CD3D FALSE Upregulated
1,5001713 CD3G Upregulated
0,816517129 CD69 FALSE Upregulated
0 CD70 FALSE Upregulated
CD99-2
CD99P1-2
0 CDC14C FALSE Upregulated
0,861398311 CDC20B FALSE Upregulated
1,335725723 CDC25A Upregulated
1,399690821 CDC25C Upregulated
1,908648015 CDC45 Upregulated
0,42922031 CDC6 FALSE Upregulated
0,751231053 CDC7 FALSE Upregulated
1,128483478 CDCA2 Upregulated
0,106353184 CDCA3 FALSE Upregulated
0 CDCA7 FALSE Upregulated
1,542199781 CDCA8 Upregulated
0 CDCP2 FALSE Upregulated
CDH10
0,580819283 CDH12 FALSE Upregulated
-1,013990123 CDH15 Downregulated
1,051831025 CDH16 Upregulated
-0,472405457 CDH17 FALSE Downregulated
CDH18
0 CDH22 FALSE Upregulated
-0,463687412 CDH26 FALSE Downregulated
-0,134476161 CDH3 FALSE Downregulated
CDH7
0,447639589 CDH8 FALSE Upregulated
0 CDH9 FALSE Upregulated
0 CDHR4 FALSE Upregulated
CDIPT-AS1
1,008849178 CDK1 Upregulated
0,280669967 CDK15 FALSE Upregulated
CDK5R2
0,579381879 CDKL2 FALSE Upregulated
-0,511000288 CDKL3 FALSE Downregulated
0,084976497 CDKL4 FALSE Upregulated
0,887196729 CDKN2A FALSE Upregulated
0,373540435 CDKN2B-AS1 FALSE Upregulated
0,312678272 CDKN2D FALSE Upregulated
CDRT15L2
CDRT15P2
CDRT7
CDRT8
-0,250197963 CDX1 FALSE Downregulated

| | | | |
|---|---|---|---|
| 0,066078334 | CDX2 | FALSE | Upregulated |
| | CDX4 | | |
| | CEACAM18 | | |
| 1,056007238 | CEACAM20 | | Upregulated |
| 1,282729468 | CEACAM21 | | Upregulated |
| 0,596927213 | CEACAM3 | FALSE | Upregulated |
| 1,309805179 | CEACAM4 | | Upregulated |
| 0,004961237 | CEACAM5 | FALSE | Upregulated |
| 0,627919288 | CEACAM6 | FALSE | Upregulated |
| | CEACAM7 | | |
| | CEBPE | | |
| 0,253508347 | CECR3 | FALSE | Upregulated |
| 0,615216167 | CECR5-AS1 | FALSE | Upregulated |
| -0,359736821 | CECR7 | FALSE | Downregulated |
| -0,629573738 | CEL | FALSE | Downregulated |
| | CELA1 | | |
| -0,158175166 | CELA2A | FALSE | Downregulated |
| 0,301684148 | CELA2B | FALSE | Upregulated |
| | CELA3A | | |
| | CELA3B | | |
| -0,406169983 | CELF5 | FALSE | Downregulated |
| 1,240861412 | CELP | | Upregulated |
| 1,838225453 | CENPA | | Upregulated |
| 1,143534986 | CENPE | | Upregulated |
| 0,451061891 | CENPH | FALSE | Upregulated |
| 0,83472297 | CENPI | FALSE | Upregulated |
| 1,145542343 | CENPK | | Upregulated |
| 1,499134787 | CENPL | | Upregulated |
| 1,414037167 | CENPM | | Upregulated |
| -0,007818019 | CENPQ | FALSE | Downregulated |
| | CENPVP1 | | |
| | CENPVP2 | | |
| 0,875661584 | CENPW | FALSE | Upregulated |
| 0,04069224 | CEP170P1 | FALSE | Upregulated |
| 1,234141846 | CEP55 | | Upregulated |
| | CER1 | | |
| 0 | CES1P2 | FALSE | Upregulated |
| | CES5AP1 | | |
| | CETN1 | | |
| 0,692417913 | CETN3 | FALSE | Upregulated |
| 0 | CETN4P | FALSE | Upregulated |
| | CFC1 | | |
| | CFC1B | | |
| 0 | CFL1P1 | FALSE | Upregulated |
| | CGA | | |
| 0,037986104 | CGB | FALSE | Upregulated |
| | CGB1 | | |
| 0 | CGB2 | FALSE | Upregulated |
| 0,18515478 | CGB5 | FALSE | Upregulated |
| | CGB8 | | |
| 0,396784648 | CH25H | FALSE | Upregulated |

| | | | |
|---|---|---|---|
| -0,485025606 | CHAC1 | FALSE | Downregulated |
| 1,191447762 | CHAF1B | | Upregulated |
| | CHAT | | |
| 0,106588172 | CHD5 | FALSE | Upregulated |
| 0,097466287 | CHDC2 | FALSE | Upregulated |
| | CHEK2P2 | | |
| 0 | CHGB | FALSE | Upregulated |
| 0,35536735 | CHI3L2 | FALSE | Upregulated |
| 0 | CHIA | FALSE | Upregulated |
| 0 | CHIAP2 | FALSE | Upregulated |
| 0,923730744 | CHIT1 | FALSE | Upregulated |
| 0,202309212 | CHKB-AS1 | FALSE | Upregulated |
| 1,170222062 | CHKB-CPT1B | | Upregulated |
| 0,247539279 | CHMP1B2P | FALSE | Upregulated |
| 0,683033798 | CHN1 | FALSE | Upregulated |
| 0 | CHODL | FALSE | Upregulated |
| | CHODL-AS1 | | |
| 0,130175184 | CHP2 | FALSE | Upregulated |
| -1,332871403 | CHRDL1 | | Downregulated |
| 1,400361328 | CHRDL2 | | Upregulated |
| -1,290751056 | CHRFAM7A | | Downregulated |
| -0,641284152 | CHRM1 | FALSE | Downregulated |
| 0,109808653 | CHRM2 | FALSE | Upregulated |
| 0,335305022 | CHRM3-AS1 | FALSE | Upregulated |
| -0,997256937 | CHRM3-AS2 | FALSE | Downregulated |
| -0,003414792 | CHRM4 | FALSE | Downregulated |
| 0 | CHRNA1 | FALSE | Upregulated |
| -0,576093031 | CHRNA10 | FALSE | Downregulated |
| 1,652415166 | CHRNA3 | | Upregulated |
| 0,107919349 | CHRNA5 | FALSE | Upregulated |
| 0,011211767 | CHRNA6 | FALSE | Upregulated |
| 0,447252983 | CHRNA7 | FALSE | Upregulated |
| 0,307006748 | CHRNA9 | FALSE | Upregulated |
| | CHRNB3 | | |
| 0 | CHRNB4 | FALSE | Upregulated |
| -0,03276895 | CHST10 | FALSE | Downregulated |
| -0,873754048 | CHST5 | FALSE | Downregulated |
| 0,478201489 | CHST6 | FALSE | Upregulated |
| -0,222561359 | CHST8 | FALSE | Downregulated |
| -0,157419388 | CIB2 | FALSE | Downregulated |
| | CIB3 | | |
| 0 | CIB4 | FALSE | Upregulated |
| 0,332887923 | CIDEA | FALSE | Upregulated |
| 0,822221062 | CIDEC | FALSE | Upregulated |
| 0,633386987 | CIDECP | FALSE | Upregulated |
| 2,218375592 | CILP2 | | Upregulated |
| | CISTR | | |
| 0,019407401 | CITED1 | FALSE | Upregulated |
| 1,509985586 | CKAP2L | | Upregulated |
| 0,14027195 | CKLF-CMTM1 | FALSE | Upregulated |
| 0,065768359 | CKMT1A | FALSE | Upregulated |

| | | | |
|---|---|---|---|
| 0,155868025 | CKMT1B | FALSE | Upregulated |
| -0,502585883 | CKS2 | FALSE | Downregulated |
| 0,099291718 | CLC | FALSE | Upregulated |
| 0,348426097 | CLCA1 | FALSE | Upregulated |
| 0 | CLCA2 | FALSE | Upregulated |
| 1,480393792 | CLCA4 | | Upregulated |
| -0,133527332 | CLCN1 | FALSE | Downregulated |
| 0,150714305 | CLCNKA | FALSE | Upregulated |
| 0,744247047 | CLCNKB | FALSE | Upregulated |
| | CLDN10-AS1 | | |
| 0,111828508 | CLDN16 | FALSE | Upregulated |
| | CLDN17 | | |
| 0,596378665 | CLDN19 | FALSE | Upregulated |
| -0,238674566 | CLDN25 | FALSE | Downregulated |
| 0,771602163 | CLDN6 | FALSE | Upregulated |
| | CLDN8 | | |
| 0,71600811 | CLEC12B | FALSE | Upregulated |
| 0,1634289 | CLEC17A | FALSE | Upregulated |
| -0,276623766 | CLEC18B | FALSE | Downregulated |
| -0,64449067 | CLEC18C | FALSE | Downregulated |
| | CLEC19A | | |
| 0,250179583 | CLEC1A | FALSE | Upregulated |
| 0,381808184 | CLEC2A | FALSE | Upregulated |
| | CLEC2L | | |
| 0 | CLEC3A | FALSE | Upregulated |
| -0,384148488 | CLEC4A | FALSE | Downregulated |
| 0,489429145 | CLEC4C | FALSE | Upregulated |
| 0,353391266 | CLEC4D | FALSE | Upregulated |
| 0,909007095 | CLEC4E | FALSE | Upregulated |
| -0,751472264 | CLEC4F | FALSE | Downregulated |
| -0,606813933 | CLEC4GP1 | FALSE | Downregulated |
| 0 | CLEC6A | FALSE | Upregulated |
| -0,153867899 | CLEC9A | FALSE | Downregulated |
| 1,836435131 | CLECL1 | | Upregulated |
| 1,042996939 | CLGN | | Upregulated |
| 0,217917715 | CLIC3 | FALSE | Upregulated |
| | CLK2P | | |
| 0,784877637 | CLLU1 | FALSE | Upregulated |
| | CLLU1OS | | |
| -0,981693792 | CLNK | FALSE | Downregulated |
| | CLPS | | |
| | CLPSL1 | | |
| | CLPSL2 | | |
| -0,112889542 | CLRN1-AS1 | FALSE | Downregulated |
| 0,176210568 | CLRN2 | FALSE | Upregulated |
| 2,527318907 | CLSPN | | Upregulated |
| 0,563549077 | CLUL1 | FALSE | Upregulated |
| 0,110199541 | CLVS2 | FALSE | Upregulated |
| 0,427078919 | CMA1 | FALSE | Upregulated |
| 0 | CMTM5 | FALSE | Upregulated |
| 0,770925166 | CMTM7 | FALSE | Upregulated |

| | | | |
|---|---|---|---|
| | CNBD1 | | |
| 0,186525306 | CNBD2 | FALSE | Upregulated |
| 0 | CNFN | FALSE | Upregulated |
| | CNGA2 | | |
| | CNGA3 | | |
| 1,280026079 | CNGA4 | | Upregulated |
| | CNGB1 | | |
| 0,036429414 | CNIH3 | FALSE | Upregulated |
| -0,039368796 | CNKSR1 | FALSE | Downregulated |
| -0,093423203 | CNN1 | FALSE | Downregulated |
| 0,469226316 | CNPY1 | FALSE | Upregulated |
| 0 | CNR1 | FALSE | Upregulated |
| 1,184965712 | CNR2 | | Upregulated |
| 0,3692251 | CNTD2 | FALSE | Upregulated |
| -0,121633312 | CNTF | FALSE | Downregulated |
| -1,824356188 | CNTN1 | | Downregulated |
| 0 | CNTN4-AS2 | FALSE | Upregulated |
| 0,402112793 | CNTN5 | FALSE | Upregulated |
| 0,00311518 | CNTN6 | FALSE | Upregulated |
| | CNTNAP5 | | |
| 0,626865773 | COL11A1 | FALSE | Upregulated |
| 0,492599187 | COL13A1 | FALSE | Upregulated |
| 0,776254917 | COL19A1 | FALSE | Upregulated |
| 0,728901259 | COL20A1 | FALSE | Upregulated |
| 0,343816707 | COL22A1 | FALSE | Upregulated |
| 0,179271996 | COL24A1 | FALSE | Upregulated |
| 0,569239807 | COL2A1 | FALSE | Upregulated |
| 0,511498076 | COL4A2-AS1 | FALSE | Upregulated |
| | COL6A4P1 | | |
| 0,541634058 | COL6A4P2 | FALSE | Upregulated |
| 0,435169528 | COL6A5 | FALSE | Upregulated |
| 0,721351799 | COL9A1 | FALSE | Upregulated |
| 0,426374242 | COLCA2 | FALSE | Upregulated |
| 0,54725115 | COLEC12 | FALSE | Upregulated |
| 0,492950853 | COMMD8 | FALSE | Upregulated |
| 2,076672083 | COMP | | Upregulated |
| 0,359630446 | COQ3 | FALSE | Upregulated |
| -0,197556728 | COX4I2 | FALSE | Downregulated |
| -0,984435519 | COX6A2 | FALSE | Downregulated |
| | COX7B2 | | |
| -0,016422875 | COX8C | FALSE | Downregulated |
| 0,041588349 | CPA1 | FALSE | Upregulated |
| | CPA2 | | |
| 0,598729959 | CPA3 | FALSE | Upregulated |
| 0,265721399 | CPA4 | FALSE | Upregulated |
| -0,112713765 | CPA5 | FALSE | Downregulated |
| | CPA6 | | |
| 0,039609977 | CPB1 | FALSE | Upregulated |
| 0,309440876 | CPEB1 | FALSE | Upregulated |
| -0,139519348 | CPEB2-AS1 | FALSE | Downregulated |
| 0,5142626 | CPLX3 | FALSE | Upregulated |

| | | | |
|---|---|---|---|
| | CPLX4 | | |
| -0,323834485 | CPNE4 | FALSE | Downregulated |
| 0,67677176 | CPNE7 | FALSE | Upregulated |
| -0,579380225 | CPNE9 | FALSE | Downregulated |
| 0,387012887 | CPO | FALSE | Upregulated |
| -0,281136308 | CPSF4L | FALSE | Downregulated |
| -0,751515411 | CPT1C | FALSE | Downregulated |
| | CPXCR1 | | |
| 1,821791917 | CPXM1 | | Upregulated |
| -0,08476837 | CR1L | FALSE | Downregulated |
| 0,697201195 | CR2 | FALSE | Upregulated |
| | CRABP1 | | |
| 0,372111698 | CRABP2 | FALSE | Upregulated |
| 0,536094979 | CRB1 | FALSE | Upregulated |
| | CRCT1 | | |
| -0,1823741 | CREG2 | FALSE | Downregulated |
| | CRHR1 | | |
| -0,153531719 | CRIP1 | FALSE | Downregulated |
| 0,044267336 | CRISP1 | FALSE | Upregulated |
| 0,134499244 | CRISP2 | FALSE | Upregulated |
| 0,015617861 | CRISP3 | FALSE | Upregulated |
| -0,81158103 | CRISPLD1 | FALSE | Downregulated |
| | CRLF2 | | |
| | CRLF2-2 | | |
| 0,269777382 | CRNDE | FALSE | Upregulated |
| 0,070748857 | CRNN | FALSE | Upregulated |
| 0 | CRSP8P | FALSE | Upregulated |
| 0,5461063 | CRTAC1 | FALSE | Upregulated |
| 1,391111769 | CRTAM | | Upregulated |
| | CRYBA2 | | |
| | CRYBA4 | | |
| -0,785000105 | CRYBB1 | FALSE | Downregulated |
| 0,125865767 | CRYBB2 | FALSE | Upregulated |
| -0,494732993 | CRYBB3 | FALSE | Downregulated |
| | CRYGA | | |
| | CRYGB | | |
| | CRYGC | | |
| | CRYGD | | |
| 0,020789395 | CRYGN | FALSE | Upregulated |
| -0,146389461 | CRYGS | FALSE | Downregulated |
| -0,256520684 | CRYM-AS1 | FALSE | Downregulated |
| | CSAG1 | | |
| | CSAG3 | | |
| | CSAG4 | | |
| | CSF2RA-2 | | |
| 0,920786376 | CSF3 | FALSE | Upregulated |
| | CSH1 | | |
| | CSH2 | | |
| | CSHL1 | | |
| -0,161314915 | CSMD1 | FALSE | Downregulated |
| -0,706303227 | CSMD2 | FALSE | Downregulated |

| | | | |
|---|---|---|---|
| -0,533924082 | CSMD3 | FALSE | Downregulated |
| | CSN1S1 | | |
| | CSN1S2AP | | |
| | CSN1S2BP | | |
| | CSN2 | | |
| | CSN3 | | |
| -0,16988644 | CSNK1A1L | FALSE | Downregulated |
| 0,421742046 | CSNK2A3 | FALSE | Upregulated |
| 0,811409203 | CSPG5 | FALSE | Upregulated |
| -0,775376218 | CSRP3 | FALSE | Downregulated |
| | CST1 | | |
| 0 | CST11 | FALSE | Upregulated |
| | CST13P | | |
| | CST2 | | |
| | CST4 | | |
| | CST5 | | |
| 0,133478735 | CST6 | FALSE | Upregulated |
| | CST8 | | |
| | CST9 | | |
| | CST9L | | |
| 0,762223462 | CSTA | FALSE | Upregulated |
| | CSTF3-AS1 | | |
| | CSTL1 | | |
| | CT45A1 | | |
| | CT45A2 | | |
| | CT45A3 | | |
| | CT45A5 | | |
| | CT45A6 | | |
| | CT47A1 | | |
| | CT47A10 | | |
| | CT47A11 | | |
| | CT47A12 | | |
| | CT47A2 | | |
| | CT47A3 | | |
| | CT47A4 | | |
| | CT47A5 | | |
| 0,591684679 | CT47A6 | FALSE | Upregulated |
| 0 | CT47A7 | FALSE | Upregulated |
| | CT47A8 | | |
| | CT47A9 | | |
| -0,356038839 | CT47B1 | FALSE | Downregulated |
| -0,867055402 | CT55 | FALSE | Downregulated |
| -0,518339736 | CT62 | FALSE | Downregulated |
| | CT83 | | |
| | CTAG1A | | |
| | CTAG1B | | |
| 0 | CTAG2 | FALSE | Upregulated |
| 0,092075149 | CTAGE1 | FALSE | Upregulated |
| 0,355805897 | CTAGE10P | FALSE | Upregulated |
| -0,378594167 | CTAGE11P | FALSE | Downregulated |
| 1,835515982 | CTAGE6 | | Upregulated |

| | | | |
|---|---|---|---|
| | CTB-12O2.1 | | |
| 0,070626087 | CTB-174D11.1 | FALSE | Upregulated |
| 0,23052653 | CTB-178M22.2 | FALSE | Upregulated |
| 0,081120613 | CTC-436P18.1 | FALSE | Upregulated |
| 0,161546121 | CTCFL | FALSE | Upregulated |
| | CTD-2118P12.1 | | |
| 0 | CTD-2127H9.1 | FALSE | Upregulated |
| | CTD-2194D22.4 | | |
| -1,324372598 | CTD-2201I18.1 | | Downregulated |
| -0,234115407 | CTD-2258A20.5 | FALSE | Downregulated |
| -0,134289565 | CTD-2270F17.1 | FALSE | Downregulated |
| | CTD-2297D10.2 | | |
| | CTD-2337A12.1 | | |
| -0,002902526 | CTD-2350J17.1 | FALSE | Downregulated |
| -0,267556575 | CTF1 | FALSE | Downregulated |
| 1,331166041 | CTLA4 | | Upregulated |
| 0,119918239 | CTNNA2 | FALSE | Upregulated |
| 0,440588535 | CTRB1 | FALSE | Upregulated |
| | CTRB2 | | |
| 0,146690165 | CTRC | FALSE | Upregulated |
| 0,696782748 | CTSE | FALSE | Upregulated |
| -0,73384514 | CTSG | FALSE | Downregulated |
| | CTSL3P | | |
| | CTSLP8 | | |
| | CTSV | | |
| 0,485995477 | CTXN1 | FALSE | Upregulated |
| 0,14225089 | CTXN2 | FALSE | Upregulated |
| | CTXN3 | | |
| 0,348583885 | CUZD1 | FALSE | Upregulated |
| -0,310081006 | CWH43 | FALSE | Downregulated |
| 0,168318765 | CXADRP3 | FALSE | Upregulated |
| 0,794446918 | CXCL1 | FALSE | Upregulated |
| 1,796366992 | CXCL11 | | Upregulated |
| -0,133065901 | CXCL13 | FALSE | Downregulated |
| -0,93792432 | CXCL17 | FALSE | Downregulated |
| 0,700676986 | CXCL3 | FALSE | Upregulated |
| 1,266689304 | CXCL5 | | Upregulated |
| 0,514220976 | CXCL6 | FALSE | Upregulated |
| -0,652908795 | CXCR1 | FALSE | Downregulated |
| -0,900058504 | CXCR2 | FALSE | Downregulated |
| 1,39601219 | CXorf21 | | Upregulated |
| 0,295424179 | CXorf22 | FALSE | Upregulated |
| | CXorf27 | | |
| | CXorf28 | | |
| | CXorf30 | | |
| 0,210474061 | CXorf31 | FALSE | Upregulated |
| | CXorf49 | | |
| | CXorf49B | | |
| | CXorf51A | | |
| | CXorf51B | | |
| 0,310933109 | CXorf58 | FALSE | Upregulated |

| | | | |
|---|---|---|---|
| 0,047839331 | CXorf64 | FALSE | Upregulated |
| -0,055189187 | CXorf65 | FALSE | Downregulated |
| 0,772615389 | CXorf66 | FALSE | Upregulated |
| 0 | CXorf67 | FALSE | Upregulated |
| 0,616922096 | CXXC11 | FALSE | Upregulated |
| | CXXC1P1 | | |
| | CYLC1 | | |
| | CYLC2 | | |
| | CYMP | | |
| | CYorf17 | | |
| 2,59816203 | CYP11B1 | | Upregulated |
| 0,402112793 | CYP11B2 | FALSE | Upregulated |
| 0 | CYP19A1 | FALSE | Upregulated |
| 0,638050686 | CYP1B1-AS1 | FALSE | Upregulated |
| 0,098678714 | CYP24A1 | FALSE | Upregulated |
| 0,055896655 | CYP26C1 | FALSE | Upregulated |
| 0,20801728 | CYP27B1 | FALSE | Upregulated |
| -0,411875426 | CYP27C1 | FALSE | Downregulated |
| -0,11387228 | CYP2A13 | FALSE | Downregulated |
| -0,343495205 | CYP2G1P | FALSE | Downregulated |
| -1,059816818 | CYP46A1 | | Downregulated |
| 0,41050045 | CYP4B1 | FALSE | Upregulated |
| 0,394607018 | CYP4F24P | FALSE | Upregulated |
| | CYP4F30P | | |
| 0,440883232 | CYP4F35P | FALSE | Upregulated |
| | CYP4F62P | | |
| | CYP4F8 | | |
| 0,059002368 | CYP4Z1 | FALSE | Upregulated |
| 0,290684077 | CYP4Z2P | FALSE | Upregulated |
| 1,755173244 | CYSLTR1 | | Upregulated |
| 1,546422594 | CYSLTR2 | | Upregulated |
| 0,448212425 | CYTL1 | FALSE | Upregulated |
| | D21S2088E | | |
| 0,000977115 | DACH2 | FALSE | Upregulated |
| 1,137586054 | DACT3-AS1 | | Upregulated |
| -0,125729769 | DAND5 | FALSE | Downregulated |
| | DAOA | | |
| | DAOA-AS1 | | |
| | DAPL1 | | |
| 0,779035161 | DARC | FALSE | Upregulated |
| | DAW1 | | |
| 1,497409451 | DAZ1 | | Upregulated |
| 0,645891263 | DAZ4 | FALSE | Upregulated |
| | DAZL | | |
| 0,11079169 | DBIL5P2 | FALSE | Upregulated |
| 1,435609634 | DBX1 | | Upregulated |
| 0 | DBX2 | FALSE | Upregulated |
| | DCAF12L1 | | |
| -0,181972966 | DCAF12L2 | FALSE | Downregulated |
| 0,000835676 | DCAF13P3 | FALSE | Upregulated |
| 0,020789395 | DCAF4L2 | FALSE | Upregulated |

| | | | |
|---|---|---|---|
| | DCAF8L1 | | |
| | DCAF8L2 | | |
| -0,391559549 | DCC | FALSE | Downregulated |
| 0 | DCD | FALSE | Upregulated |
| | DCDC1 | | |
| -0,229588308 | DCDC2C | FALSE | Downregulated |
| -0,287764473 | DCLK3 | FALSE | Downregulated |
| -0,730074753 | DCT | FALSE | Downregulated |
| 0 | DCTN1-AS1 | FALSE | Upregulated |
| 0,773058652 | DCX | FALSE | Upregulated |
| 0,361064773 | DDIT4L | FALSE | Upregulated |
| -0,31068926 | DDN | FALSE | Downregulated |
| 0,008765599 | DDX11-AS1 | FALSE | Upregulated |
| 1,106252924 | DDX11L10 | | Upregulated |
| 0,197522658 | DDX25 | FALSE | Upregulated |
| | DDX4 | | |
| 0,084976497 | DEC1 | FALSE | Upregulated |
| 0,605452767 | DEFA1 | FALSE | Upregulated |
| | DEFA10P | | |
| | DEFA11P | | |
| 0,605452767 | DEFA1B | FALSE | Upregulated |
| 0,041226344 | DEFA3 | FALSE | Upregulated |
| 0 | DEFA4 | FALSE | Upregulated |
| | DEFA5 | | |
| | DEFA6 | | |
| | DEFA8P | | |
| | DEFA9P | | |
| | DEFB103A | | |
| | DEFB103B | | |
| | DEFB104A | | |
| | DEFB104B | | |
| | DEFB105A | | |
| | DEFB105B | | |
| | DEFB106A | | |
| | DEFB106B | | |
| | DEFB107A | | |
| | DEFB107B | | |
| 0,000977115 | DEFB108B | FALSE | Upregulated |
| 0 | DEFB109P1 | FALSE | Upregulated |
| 0,33250392 | DEFB109P1B | FALSE | Upregulated |
| | DEFB110 | | |
| | DEFB112 | | |
| | DEFB113 | | |
| | DEFB114 | | |
| | DEFB115 | | |
| | DEFB116 | | |
| | DEFB118 | | |
| | DEFB119 | | |
| | DEFB121 | | |
| | DEFB122 | | |
| | DEFB123 | | |

| | | | |
|---|---|---|---|
| | DEFB124 | | |
| | DEFB125 | | |
| | DEFB126 | | |
| | DEFB127 | | |
| | DEFB128 | | |
| | DEFB129 | | |
| | DEFB130 | | |
| | DEFB131 | | |
| 0,265504515 | DEFB132 | FALSE | Upregulated |
| | DEFB133 | | |
| | DEFB134 | | |
| | DEFB135 | | |
| | DEFB136 | | |
| | DEFB4A | | |
| | DEFB4B | | |
| | DEFT1P | | |
| | DEFT1P2 | | |
| 1,549762331 | DEPDC1 | | Upregulated |
| 0,517103579 | DEPDC1B | FALSE | Upregulated |
| -1,43800444 | DGCR6 | | Downregulated |
| 1,007758981 | DGKI | | Upregulated |
| | DGKK | | |
| -0,293260916 | DGUOK-AS1 | FALSE | Downregulated |
| 0,415048848 | DHDH | FALSE | Upregulated |
| -0,203490716 | DHH | FALSE | Downregulated |
| 0 | DHRS7C | FALSE | Upregulated |
| 0,880436729 | DHRS9 | FALSE | Upregulated |
| | DHRSX-2 | | |
| 2,027979103 | DIAPH3 | | Upregulated |
| 0,3729134 | DIAPH3-AS1 | FALSE | Upregulated |
| | DIAPH3-AS2 | | |
| 0 | DIO2 | FALSE | Upregulated |
| 0,924440965 | DIO2-AS1 | FALSE | Upregulated |
| -0,100822658 | DIO3 | FALSE | Downregulated |
| -0,839681846 | DIO3OS | FALSE | Downregulated |
| 0,387012887 | DIP2A-IT1 | FALSE | Upregulated |
| 0,637124991 | DIRAS1 | FALSE | Upregulated |
| 0,663743676 | DIRAS2 | FALSE | Upregulated |
| 0 | DIRC1 | FALSE | Upregulated |
| -0,346223579 | DIRC3 | FALSE | Downregulated |
| 0,558226505 | DISC1FP1 | FALSE | Upregulated |
| 1,122854238 | DKFZP434A062 | | Upregulated |
| 0,019357528 | DKFZP434F142 | FALSE | Upregulated |
| -0,221799377 | DKFZp434J0226 | FALSE | Downregulated |
| | DKFZP434L187 | | |
| | DKFZp434L192 | | |
| 1,12125936 | DKFZp451B082 | | Upregulated |
| 0,797234041 | DKFZp566F0947 | FALSE | Upregulated |
| | DKFZp686K1684 | | |
| 0,539031043 | DKFZp686L13185 | FALSE | Upregulated |
| 0,31037223 | DKFZp779M0652 | FALSE | Upregulated |

| | | | |
|---|---|---|---|
| 0,284111029 | DKK1 | FALSE | Upregulated |
| 0,417252082 | DKK2 | FALSE | Upregulated |
| 1,312826097 | DKK4 | | Upregulated |
| 0,143436297 | DKKL1 | FALSE | Upregulated |
| 0,045603021 | DLEU7-AS1 | FALSE | Upregulated |
| -0,602535142 | DLG3-AS1 | FALSE | Downregulated |
| -0,830200867 | DLGAP1 | FALSE | Downregulated |
| 0,653507792 | DLGAP1-AS3 | FALSE | Upregulated |
| | DLGAP1-AS4 | | |
| -0,493249118 | DLGAP3 | FALSE | Downregulated |
| 1,578881479 | DLGAP5 | | Upregulated |
| -0,106766 | DLK1 | FALSE | Downregulated |
| | DLL3 | | |
| -0,044012589 | DLX1 | FALSE | Downregulated |
| | DLX2 | | |
| 0,239281757 | DLX3 | FALSE | Upregulated |
| 0 | DLX4 | FALSE | Upregulated |
| | DLX5 | | |
| | DLX6 | | |
| | DLX6-AS1 | | |
| -0,32079097 | DMBT1 | FALSE | Downregulated |
| | DMBT1P1 | | |
| | DMBX1 | | |
| 0,839729769 | DMC1 | FALSE | Upregulated |
| -0,038902623 | DMKN | FALSE | Downregulated |
| 0,584087795 | DMP1 | FALSE | Upregulated |
| | DMRT1 | | |
| 0,073103429 | DMRT2 | FALSE | Upregulated |
| | DMRT3 | | |
| | DMRTA2 | | |
| | DMRTB1 | | |
| | DMRTC1 | | |
| | DMRTC1B | | |
| | DMRTC2 | | |
| 0,501259948 | DNAAF1 | FALSE | Upregulated |
| -0,034966979 | DNAAF3 | FALSE | Downregulated |
| -1,465669466 | DNAH12 | | Downregulated |
| 0 | DNAH17-AS1 | FALSE | Upregulated |
| 0 | DNAH8 | FALSE | Upregulated |
| -1,459072401 | DNAI1 | | Downregulated |
| 0,49156737 | DNAI2 | FALSE | Upregulated |
| | DNAJA1P5 | | |
| -0,351017771 | DNAJB8 | FALSE | Downregulated |
| | DNAJB8-AS1 | | |
| 0,030632837 | DNAJC27-AS1 | FALSE | Upregulated |
| 0,634798023 | DNAJC5B | FALSE | Upregulated |
| 0 | DNAJC5G | FALSE | Upregulated |
| 0,333224588 | DNAJC6 | FALSE | Upregulated |
| 0,298959597 | DNASE2B | FALSE | Upregulated |
| 0,615843206 | DNER | FALSE | Upregulated |
| 1,21074186 | DNM1P35 | | Upregulated |

| | | | |
|---|---|---|---|
| -0,306193827 | DNM1P46 | FALSE | Downregulated |
| -0,334298676 | DNM1P50 | FALSE | Downregulated |
| -2,154905758 | DNMT3L | | Downregulated |
| -0,045460148 | DNTT | FALSE | Downregulated |
| -0,010267802 | DOC2GP | FALSE | Downregulated |
| -0,568760675 | DOCK9-AS1 | FALSE | Downregulated |
| 0,573447433 | DOK5 | FALSE | Upregulated |
| -0,862999712 | DPCR1 | FALSE | Downregulated |
| 0,400765505 | DPEP1 | FALSE | Upregulated |
| 0,328784773 | DPEP3 | FALSE | Upregulated |
| 0,738545124 | DPF1 | FALSE | Upregulated |
| -0,21604683 | DPH6-AS1 | FALSE | Downregulated |
| -0,606612181 | DPP10 | FALSE | Downregulated |
| 0 | DPP10-AS1 | FALSE | Upregulated |
| | DPP10-AS3 | | |
| 0,10010064 | DPP6 | FALSE | Upregulated |
| | DPPA2 | | |
| 0,925610555 | DPPA2P3 | FALSE | Upregulated |
| 0,330334158 | DPPA3 | FALSE | Upregulated |
| -0,478680455 | DPPA4 | FALSE | Downregulated |
| | DPPA5 | | |
| | DPRX | | |
| 1,464838403 | DPRXP4 | | Upregulated |
| -0,486284315 | DPY19L2 | FALSE | Downregulated |
| 0,311925576 | DPY19L2P1 | FALSE | Upregulated |
| | DPYSL5 | | |
| 0,338467867 | DQX1 | FALSE | Upregulated |
| 0,62342437 | DRAXIN | FALSE | Upregulated |
| 0,13893015 | DRC1 | FALSE | Upregulated |
| 0,048324477 | DRD2 | FALSE | Upregulated |
| 0 | DRD3 | FALSE | Upregulated |
| | DRGX | | |
| 0 | DRP2 | FALSE | Upregulated |
| 0,494571317 | DSC3 | FALSE | Upregulated |
| -1,258008699 | DSCAM-AS1 | | Downregulated |
| 0,099452879 | DSCAM-IT1 | FALSE | Upregulated |
| | DSCR10 | | |
| | DSCR4 | | |
| | DSCR8 | | |
| 1,518730074 | DSG3 | | Upregulated |
| 0 | DSG4 | FALSE | Upregulated |
| 0,976297564 | DSPP | FALSE | Upregulated |
| 0,538019959 | DSTNP2 | FALSE | Upregulated |
| | DSTNP4 | | |
| 1,095018446 | DTL | | Upregulated |
| 1,091807806 | DUOXA1 | | Upregulated |
| 1,408838035 | DUOXA2 | | Upregulated |
| 1,656819633 | DUPD1 | | Upregulated |
| -0,199605959 | DUSP13 | FALSE | Downregulated |
| -0,13831876 | DUSP15 | FALSE | Downregulated |
| -0,065859994 | DUSP21 | FALSE | Downregulated |

| | | | |
|---|---|---|---|
| 0,130507466 | DUSP26 | FALSE | Upregulated |
| -0,179696775 | DUSP27 | FALSE | Downregulated |
| 0,516739951 | DUSP5P1 | FALSE | Upregulated |
| 0,034168903 | DUSP9 | FALSE | Upregulated |
| | DUX2 | | |
| | DUX4 | | |
| 0 | DUX4L | FALSE | Upregulated |
| | DUX4L2 | | |
| | DUX4L3 | | |
| | DUX4L4 | | |
| | DUX4L5 | | |
| | DUX4L6 | | |
| | DUX4L7 | | |
| | DUXA | | |
| 0,75365014 | DYDC1 | FALSE | Upregulated |
| 0,038016313 | DYDC2 | FALSE | Upregulated |
| 0,807686007 | DYNAP | FALSE | Upregulated |
| 0,43891296 | DYNC1I1 | FALSE | Upregulated |
| 0,082364975 | DYNLRB2 | FALSE | Upregulated |
| | DYTN | | |
| 0,123636948 | DYX1C1 | FALSE | Upregulated |
| 0 | DYX1C1-CCPG1 | FALSE | Upregulated |
| -0,184836473 | DZANK1 | FALSE | Downregulated |
| 2,306255633 | E2F2 | | Upregulated |
| 0,067949278 | E2F7 | FALSE | Upregulated |
| 1,923468375 | E2F8 | | Upregulated |
| -0,379313116 | EAF2 | FALSE | Downregulated |
| 1,070936107 | EBF2 | | Upregulated |
| | EBLN1 | | |
| -0,406686463 | ECEL1 | FALSE | Downregulated |
| -0,057039413 | ECEL1P2 | FALSE | Downregulated |
| 0,675153655 | ECRP | FALSE | Upregulated |
| -0,261591624 | ECSCR | FALSE | Downregulated |
| 0,103586473 | ECSCR-2 | FALSE | Upregulated |
| 0,797365729 | EDA2R | FALSE | Upregulated |
| -0,015330384 | EDARADD | FALSE | Downregulated |
| 0,187271544 | EDDM3A | FALSE | Upregulated |
| 0,186054115 | EDDM3B | FALSE | Upregulated |
| 0,824323232 | EDIL3 | FALSE | Upregulated |
| 0,257197025 | EDN2 | FALSE | Upregulated |
| 0,022812395 | EDN3 | FALSE | Upregulated |
| -0,551238528 | EEF1DP3 | FALSE | Downregulated |
| -0,091072591 | EEF1E1 | FALSE | Downregulated |
| 0,07469156 | EFCAB1 | FALSE | Upregulated |
| -0,097404008 | EFCAB14-AS1 | FALSE | Downregulated |
| | EFCAB3 | | |
| 0,383761433 | EFCAB5 | FALSE | Upregulated |
| | EFCAB8 | | |
| 0 | EFCAB9 | FALSE | Upregulated |
| 0,119250059 | EFHB | FALSE | Upregulated |
| 0 | EFHC2 | FALSE | Upregulated |

| | | | |
|---|---|---|---|
| 0,749046933 | EFNA3 | FALSE | Upregulated |
| 0,479138066 | EFNA5 | FALSE | Upregulated |
| 0 | EFTUD1P1 | FALSE | Upregulated |
| -0,138887168 | EGF | FALSE | Downregulated |
| -0,095719535 | EGFEM1P | FALSE | Downregulated |
| 0,448212425 | EGFL6 | FALSE | Upregulated |
| -0,188364904 | EGLN3 | FALSE | Downregulated |
| 1,573707373 | EGR3 | | Upregulated |
| 0,569526539 | EGR4 | FALSE | Upregulated |
| -0,146267082 | EID2B | FALSE | Downregulated |
| 0,355464672 | EID3 | FALSE | Upregulated |
| | EIF3IP1 | | |
| -0,020829128 | EIF4E1B | FALSE | Downregulated |
| 0,618886382 | ELANE | FALSE | Upregulated |
| | ELAVL2 | | |
| 0,565297977 | ELAVL3 | FALSE | Upregulated |
| 0,402591809 | ELAVL4 | FALSE | Upregulated |
| 0,244668085 | ELF5 | FALSE | Upregulated |
| 0,737281708 | ELFN2 | FALSE | Upregulated |
| 0,350663318 | ELMOD1 | FALSE | Upregulated |
| -0,224996982 | ELOVL2-AS1 | FALSE | Downregulated |
| 0,330203447 | ELOVL3 | FALSE | Upregulated |
| -0,793003402 | ELOVL4 | FALSE | Downregulated |
| 0,303060421 | ELOVL7 | FALSE | Upregulated |
| | ELSPBP1 | | |
| 1,202112563 | EMR3 | | Upregulated |
| 0,593766607 | EMX2 | FALSE | Upregulated |
| 0 | EN1 | FALSE | Upregulated |
| -0,046746105 | EN2 | FALSE | Downregulated |
| 0 | ENO1-AS1 | FALSE | Upregulated |
| -0,152085962 | ENOX1 | FALSE | Downregulated |
| 1,62010014 | ENPP5 | | Upregulated |
| -0,17894239 | ENPP6 | FALSE | Downregulated |
| | ENTHD1 | | |
| 0,420954221 | ENTPD2 | FALSE | Upregulated |
| -0,042926163 | ENTPD3-AS1 | FALSE | Downregulated |
| -0,287702081 | EPB42 | FALSE | Downregulated |
| 0,685933066 | EPCAM | FALSE | Upregulated |
| 0,09760077 | EPGN | FALSE | Upregulated |
| 1,042562137 | EPHA1-AS1 | | Upregulated |
| 0,47198245 | EPHA10 | FALSE | Upregulated |
| 0 | EPHA5 | FALSE | Upregulated |
| 0,286792782 | EPHA6 | FALSE | Upregulated |
| 0 | EPHA7 | FALSE | Upregulated |
| | EPHA8 | | |
| 0 | EPHX3 | FALSE | Upregulated |
| | EPHX4 | | |
| 0,311395901 | EPN2-IT1 | FALSE | Upregulated |
| -1,185777967 | EPO | | Downregulated |
| 0,363219515 | EPPIN | FALSE | Upregulated |
| 0,769845493 | EPPIN-WFDC6 | FALSE | Upregulated |

| | | | |
|---|---|---|---|
| 0,255427272 | EPS8L3 | FALSE | Upregulated |
| | EPYC | | |
| | EQTN | | |
| 0,068398161 | ERAS | FALSE | Upregulated |
| -0,321855015 | ERC2 | FALSE | Downregulated |
| 0,092961682 | ERC2-IT1 | FALSE | Upregulated |
| 0 | EREG | FALSE | Upregulated |
| 0,112630021 | ERI3-IT1 | FALSE | Upregulated |
| -0,141027546 | ERICH1-AS1 | FALSE | Downregulated |
| -0,347115469 | ERICH2 | FALSE | Downregulated |
| 0,167324789 | ERMN | FALSE | Upregulated |
| -0,627910137 | ERN2 | FALSE | Downregulated |
| 0,082729146 | ERP27 | FALSE | Upregulated |
| 0 | ERVMER34-1 | FALSE | Upregulated |
| 1,553065159 | ERVV-1 | | Upregulated |
| -0,280105311 | ERVV-2 | FALSE | Downregulated |
| 1,085880879 | ESCO2 | | Upregulated |
| -0,50250673 | ESM1 | FALSE | Downregulated |
| 1,073061357 | ESRG | | Upregulated |
| 0,60108469 | ESRP1 | FALSE | Upregulated |
| -0,044266149 | ESRRG | FALSE | Downregulated |
| 0 | ESX1 | FALSE | Upregulated |
| -0,401280695 | ETV3L | FALSE | Downregulated |
| 1,836225111 | ETV7 | | Upregulated |
| -1,61046355 | EVPLL | | Downregulated |
| | EVX1 | | |
| | EVX2 | | |
| 0,055378136 | EXD1 | FALSE | Upregulated |
| 1,623531175 | EXO1 | | Upregulated |
| 0,426437581 | EZR-AS1 | FALSE | Upregulated |
| 0 | FA2H | FALSE | Upregulated |
| | FABP12 | | |
| 0,428315704 | FABP4 | FALSE | Upregulated |
| 0,599059237 | FABP5P3 | FALSE | Upregulated |
| | FABP6 | | |
| -0,591375528 | FABP7 | FALSE | Downregulated |
| | FABP9 | | |
| -0,015395554 | FAHD2CP | FALSE | Downregulated |
| 0,050176392 | FAIM2 | FALSE | Upregulated |
| -0,207783797 | FAM106A | FALSE | Downregulated |
| 0,807537171 | FAM106B | FALSE | Upregulated |
| -0,569211865 | FAM106CP | FALSE | Downregulated |
| 0,651471774 | FAM110D | FALSE | Upregulated |
| 1,536125493 | FAM127C | | Upregulated |
| 0 | FAM131C | FALSE | Upregulated |
| 0,149503795 | FAM132A | FALSE | Upregulated |
| 0,387012887 | FAM133A | FALSE | Upregulated |
| -0,359846873 | FAM133CP | FALSE | Downregulated |
| -0,229829673 | FAM135B | FALSE | Downregulated |
| | FAM138A | | |
| 0,049492144 | FAM138B | FALSE | Upregulated |

| | | | |
|---|---|---|---|
| | FAM138C | | |
| 0,474555919 | FAM138D | FALSE | Upregulated |
| 0 | FAM138E | FALSE | Upregulated |
| | FAM138F | | |
| 0 | FAM150A | FALSE | Upregulated |
| 1,729653302 | FAM151B | | Upregulated |
| -0,027928909 | FAM153A | FALSE | Downregulated |
| 0,672480903 | FAM153B | FALSE | Upregulated |
| -0,079145066 | FAM153C | FALSE | Downregulated |
| 0,238868474 | FAM154A | FALSE | Upregulated |
| 0,360417106 | FAM155A | FALSE | Upregulated |
| 0 | FAM155A-IT1 | FALSE | Upregulated |
| 1,028839835 | FAM155B | | Upregulated |
| 0,146159005 | FAM157B | FALSE | Upregulated |
| 0,850627739 | FAM159A | FALSE | Upregulated |
| -0,137021932 | FAM159B | FALSE | Downregulated |
| -0,589035953 | FAM161A | FALSE | Downregulated |
| 0,36855666 | FAM162B | FALSE | Upregulated |
| 0,150451706 | FAM163A | FALSE | Upregulated |
| 0,812423469 | FAM166B | FALSE | Upregulated |
| 1,35314329 | FAM167A | | Upregulated |
| -0,022808615 | FAM170A | FALSE | Downregulated |
| | FAM170B | | |
| -0,408175595 | FAM170B-AS1 | FALSE | Downregulated |
| -0,078131673 | FAM171B | FALSE | Downregulated |
| 0 | FAM172BP | FALSE | Upregulated |
| 0,244186856 | FAM177B | FALSE | Upregulated |
| 1,133392532 | FAM179A | | Upregulated |
| -0,190471849 | FAM180B | FALSE | Downregulated |
| 0,369693616 | FAM181A | FALSE | Upregulated |
| | FAM181A-AS1 | | |
| 0,068128077 | FAM181B | FALSE | Upregulated |
| -0,019487826 | FAM182A | FALSE | Downregulated |
| 0,704323738 | FAM182B | FALSE | Upregulated |
| 0 | FAM183A | FALSE | Upregulated |
| 0,250187067 | FAM183B | FALSE | Upregulated |
| 0,140564669 | FAM183CP | FALSE | Upregulated |
| 0,454038235 | FAM187B | FALSE | Upregulated |
| 0,492539551 | FAM189A1 | FALSE | Upregulated |
| -0,411647606 | FAM189A2 | FALSE | Downregulated |
| -0,37444562 | FAM194A | FALSE | Downregulated |
| 0,531432132 | FAM194B | FALSE | Upregulated |
| | FAM197Y2 | | |
| | FAM197Y4 | | |
| | FAM197Y5 | | |
| -0,114283075 | FAM19A1 | FALSE | Downregulated |
| -1,024422958 | FAM19A2 | | Downregulated |
| | FAM19A3 | | |
| 0,068128077 | FAM19A4 | FALSE | Upregulated |
| 0,181621986 | FAM201A | FALSE | Upregulated |
| 0,648546614 | FAM201B | FALSE | Upregulated |

| | | | |
|---|---|---|---|
| 0,143255598 | FAM209A | FALSE | Upregulated |
| | FAM209B | | |
| 0,195189426 | FAM215A | FALSE | Upregulated |
| 0,546715595 | FAM216B | FALSE | Upregulated |
| 0 | FAM218A | FALSE | Upregulated |
| 0,514277319 | FAM220BP | FALSE | Upregulated |
| 0,684747803 | FAM223B | FALSE | Upregulated |
| | FAM225B | | |
| 0 | FAM226A | FALSE | Upregulated |
| 0 | FAM226B | FALSE | Upregulated |
| 1,425870937 | FAM228A | | Upregulated |
| -0,075807991 | FAM228B | FALSE | Downregulated |
| | FAM230A | | |
| 0,223985806 | FAM230B | FALSE | Upregulated |
| | FAM230C | | |
| -0,74578559 | FAM231A | FALSE | Downregulated |
| | FAM231B | | |
| -0,01485634 | FAM231C | FALSE | Downregulated |
| 0,689250758 | FAM231D | FALSE | Upregulated |
| 0 | FAM24A | FALSE | Upregulated |
| 0,615083886 | FAM24B | FALSE | Upregulated |
| | FAM24B-CUZD1 | | |
| 0 | FAM25A | FALSE | Upregulated |
| 0 | FAM25B | FALSE | Upregulated |
| | FAM25C | | |
| | FAM25G | | |
| | FAM26D | | |
| -0,634156645 | FAM27B | FALSE | Downregulated |
| -0,008754004 | FAM27C | FALSE | Downregulated |
| | FAM27L | | |
| -0,432218253 | FAM3B | FALSE | Downregulated |
| | FAM41AY1 | | |
| 0,734125312 | FAM41C | FALSE | Upregulated |
| 1,845772309 | FAM43B | | Upregulated |
| 0,516183051 | FAM45B | FALSE | Upregulated |
| | FAM46D | | |
| | FAM47A | | |
| | FAM47B | | |
| -0,066703911 | FAM47C | FALSE | Downregulated |
| | FAM57B | | |
| -0,051911575 | FAM66A | FALSE | Downregulated |
| 0,839671936 | FAM66B | FALSE | Upregulated |
| -0,431487795 | FAM66C | FALSE | Downregulated |
| 0,124735371 | FAM66D | FALSE | Upregulated |
| | FAM66E | | |
| -0,311396666 | FAM69C | FALSE | Downregulated |
| 0 | FAM71B | FALSE | Upregulated |
| -0,122551042 | FAM71D | FALSE | Downregulated |
| 0 | FAM71E2 | FALSE | Upregulated |
| 0,595652528 | FAM71F1 | FALSE | Upregulated |
| 0,407026734 | FAM71F2 | FALSE | Upregulated |

| | | | |
|---|---|---|---|
| -0,506870889 | FAM72A | FALSE | Downregulated |
| 0,140866698 | FAM72B | FALSE | Upregulated |
| -0,487802165 | FAM72C | FALSE | Downregulated |
| 0,47738212 | FAM74A1 | FALSE | Upregulated |
| -0,125597896 | FAM74A3 | FALSE | Downregulated |
| -0,098238713 | FAM74A4 | FALSE | Downregulated |
| | FAM74A7 | | |
| 0 | FAM81A | FALSE | Upregulated |
| -1,007123255 | FAM81B | | Downregulated |
| -0,975582852 | FAM83A-AS1 | FALSE | Downregulated |
| 0,408084869 | FAM83B | FALSE | Upregulated |
| 0 | FAM83C-AS1 | FALSE | Upregulated |
| 0,488329061 | FAM83E | FALSE | Upregulated |
| 0,241353042 | FAM83H-AS1 | FALSE | Upregulated |
| 1,500109197 | FAM85A | | Upregulated |
| 0,172874535 | FAM86B2 | FALSE | Upregulated |
| -0,246537119 | FAM86EP | FALSE | Downregulated |
| 0 | FAM86KP | FALSE | Upregulated |
| | FAM90A10P | | |
| | FAM90A27P | | |
| -0,217557331 | FAM90A2P | FALSE | Downregulated |
| -1,251451053 | FAM90A7P | | Downregulated |
| 0 | FAM92B | FALSE | Upregulated |
| 0,248587991 | FAM95A | FALSE | Upregulated |
| -0,362031382 | FAM95B1 | FALSE | Downregulated |
| 0,132832363 | FAM95C | FALSE | Upregulated |
| -0,281136308 | FAM9A | FALSE | Downregulated |
| -0,864009391 | FAM9B | FALSE | Downregulated |
| 0 | FAM9C | FALSE | Upregulated |
| 0,643737851 | FANCB | FALSE | Upregulated |
| 0,22244292 | FANCD2OS | FALSE | Upregulated |
| -0,633212411 | FANCE | FALSE | Downregulated |
| | FANK1 | | |
| | FANK1-AS1 | | |
| 1,33011675 | FAP | | Upregulated |
| | FAR2P1 | | |
| -0,452901723 | FAR2P2 | FALSE | Downregulated |
| 0,089385366 | FAR2P3 | FALSE | Upregulated |
| 1,928110295 | FASLG | | Upregulated |
| | FATE1 | | |
| -0,082073388 | FAXC | FALSE | Downregulated |
| 0,296136231 | FBLL1 | FALSE | Upregulated |
| -0,44702946 | FBXO15 | FALSE | Downregulated |
| 0,708657996 | FBXO16 | FALSE | Upregulated |
| 0,316272893 | FBXO36 | FALSE | Upregulated |
| 0,162206848 | FBXO39 | FALSE | Upregulated |
| 0,322366034 | FBXO43 | FALSE | Upregulated |
| | FBXO47 | | |
| | FBXW12 | | |
| -0,021194983 | FBXW4P1 | FALSE | Downregulated |
| 0,899079435 | FCAR | FALSE | Upregulated |

| | | | |
|---|---|---|---|
| -1,349008254 | FCER1A | | Downregulated |
| 0,112761561 | FCER2 | FALSE | Upregulated |
| 2,379030633 | FCGR1B | | Upregulated |
| 0,949090617 | FCGR1C | FALSE | Upregulated |
| 0,353157364 | FCRL1 | FALSE | Upregulated |
| 0,15291445 | FCRL2 | FALSE | Upregulated |
| 0 | FCRL4 | FALSE | Upregulated |
| 0,576442888 | FCRLA | FALSE | Upregulated |
| -0,055578054 | FCRLB | FALSE | Downregulated |
| | FDCSP | | |
| 0,390431532 | FENDRR | FALSE | Upregulated |
| | FER1L6 | | |
| 0 | FER1L6-AS1 | FALSE | Upregulated |
| 0,024432587 | FER1L6-AS2 | FALSE | Upregulated |
| 0,284148579 | FERD3L | FALSE | Upregulated |
| | FEV | | |
| -0,264844068 | FEZF1 | FALSE | Downregulated |
| 0 | FEZF1-AS1 | FALSE | Upregulated |
| | FEZF2 | | |
| 0 | FFAR1 | FALSE | Upregulated |
| 0,339005763 | FFAR2 | FALSE | Upregulated |
| -0,003832824 | FFAR3 | FALSE | Downregulated |
| 0,078048296 | FGD5P1 | FALSE | Upregulated |
| 0,321071485 | FGF10 | FALSE | Upregulated |
| 0,137394601 | FGF10-AS1 | FALSE | Upregulated |
| -0,562338406 | FGF12 | FALSE | Downregulated |
| 1,140473351 | FGF13 | | Upregulated |
| -0,138845591 | FGF13-AS1 | FALSE | Downregulated |
| 0,626465379 | FGF14-AS1 | FALSE | Upregulated |
| -0,38341162 | FGF14-IT1 | FALSE | Downregulated |
| 0,453099891 | FGF16 | FALSE | Upregulated |
| 0,954693417 | FGF18 | FALSE | Upregulated |
| 0,6218627 | FGF19 | FALSE | Upregulated |
| | FGF20 | | |
| 0,309001832 | FGF21 | FALSE | Upregulated |
| 0,92671219 | FGF22 | FALSE | Upregulated |
| -0,631468283 | FGF23 | FALSE | Downregulated |
| | FGF3 | | |
| | FGF4 | | |
| 0,206058355 | FGF5 | FALSE | Upregulated |
| 0,02221731 | FGF6 | FALSE | Upregulated |
| -0,016422875 | FGF8 | FALSE | Downregulated |
| 0 | FGF9 | FALSE | Upregulated |
| 0 | FGFBP1 | FALSE | Upregulated |
| 0,869515468 | FGFBP2 | FALSE | Upregulated |
| -0,48970934 | FHAD1 | FALSE | Downregulated |
| -0,201511331 | FHDC1 | FALSE | Downregulated |
| 0,203353189 | FHIT | FALSE | Upregulated |
| -0,533452178 | FHL5 | FALSE | Downregulated |
| -0,003907576 | FIBCD1 | FALSE | Downregulated |
| 0,138627036 | FIGF | FALSE | Upregulated |

| | | | |
|---|---|---|---|
| | FIGLA | | |
| -0,151148436 | FIRRE | FALSE | Downregulated |
| -1,173632862 | FITM1 | | Downregulated |
| 0,293197359 | FKBP1A-SDCBP2 | FALSE | Upregulated |
| -0,166806026 | FKBP1B | FALSE | Downregulated |
| -0,174501398 | FKBP6 | FALSE | Downregulated |
| -0,135034483 | FKBP9L | FALSE | Downregulated |
| -1,228002344 | FKBPL | | Downregulated |
| -0,233394035 | FKSG29 | FALSE | Downregulated |
| | FLJ16171 | | |
| | FLJ16779 | | |
| 0,786550634 | FLJ20021 | FALSE | Upregulated |
| 0,323134797 | FLJ20444 | FALSE | Upregulated |
| 0,295047946 | FLJ20518 | FALSE | Upregulated |
| -0,623392572 | FLJ20712 | FALSE | Downregulated |
| | FLJ21408 | | |
| 0 | FLJ22447 | FALSE | Upregulated |
| -1,262310871 | FLJ22763 | | Downregulated |
| 0,26352608 | FLJ25363 | FALSE | Upregulated |
| 0,579257685 | FLJ25715 | FALSE | Upregulated |
| | FLJ25758 | | |
| | FLJ25917 | | |
| | FLJ26245 | | |
| 0,582991481 | FLJ26850 | FALSE | Upregulated |
| 0,273156147 | FLJ27255 | FALSE | Upregulated |
| 1,194195236 | FLJ27354 | | Upregulated |
| 0,557423545 | FLJ30375 | FALSE | Upregulated |
| 0,583809761 | FLJ30679 | FALSE | Upregulated |
| 0,437291381 | FLJ31104 | FALSE | Upregulated |
| 0 | FLJ31356 | FALSE | Upregulated |
| | FLJ31662 | | |
| -1,24438805 | FLJ33360 | | Downregulated |
| | FLJ33534 | | |
| 0,643242593 | FLJ33544 | FALSE | Upregulated |
| -0,730724141 | FLJ34503 | FALSE | Downregulated |
| 0,825787108 | FLJ35934 | FALSE | Upregulated |
| 0 | FLJ36000 | FALSE | Upregulated |
| -0,995192915 | FLJ37201 | FALSE | Downregulated |
| 0,448212425 | FLJ37505 | FALSE | Upregulated |
| -0,50410537 | FLJ38379 | FALSE | Downregulated |
| -1,212810099 | FLJ38723 | | Downregulated |
| 0 | FLJ39080 | FALSE | Upregulated |
| -1,476566208 | FLJ39632 | | Downregulated |
| -0,15575464 | FLJ40194 | FALSE | Downregulated |
| 0,345410865 | FLJ40288 | FALSE | Upregulated |
| 0 | FLJ41200 | FALSE | Upregulated |
| | FLJ41278 | | |
| 0,273791205 | FLJ41309 | FALSE | Upregulated |
| | FLJ41423 | | |
| 0 | FLJ41733 | FALSE | Upregulated |
| 0,452784315 | FLJ41941 | FALSE | Upregulated |

| | | | |
|---|---|---|---|
| 0,138002674 | FLJ42102 | FALSE | Upregulated |
| -0,133240375 | FLJ42351 | FALSE | Downregulated |
| | FLJ42842 | | |
| 0 | FLJ42969 | FALSE | Upregulated |
| 0,00507025 | FLJ43315 | FALSE | Upregulated |
| -0,592175473 | FLJ43585 | FALSE | Downregulated |
| 0,031689391 | FLJ43681 | FALSE | Upregulated |
| 0 | FLJ43879 | FALSE | Upregulated |
| 0,371086072 | FLJ44006 | FALSE | Upregulated |
| -0,059581248 | FLJ44087 | FALSE | Downregulated |
| -0,86702768 | FLJ44313 | FALSE | Downregulated |
| 0,512094441 | FLJ44635 | FALSE | Upregulated |
| -0,077586755 | FLJ44790 | FALSE | Downregulated |
| | FLJ44838 | | |
| 1,029353061 | FLJ45079 | | Upregulated |
| -0,536302921 | FLJ45139 | FALSE | Downregulated |
| 0,40774821 | FLJ45256 | FALSE | Upregulated |
| 0,079558098 | FLJ45743 | FALSE | Upregulated |
| -0,357932036 | FLJ45825 | FALSE | Downregulated |
| | FLJ45964 | | |
| | FLJ45974 | | |
| | FLJ46066 | | |
| -0,152884811 | FLJ46284 | FALSE | Downregulated |
| 0,239272927 | FLT3 | FALSE | Upregulated |
| 0,191158045 | FLVCR1-AS1 | FALSE | Upregulated |
| -0,211989842 | FMO1 | FALSE | Downregulated |
| | FMO9P | | |
| 0 | FMR1NB | FALSE | Upregulated |
| 0,0059811 | FNDC7 | FALSE | Upregulated |
| | FOLR3 | | |
| 0,656665144 | FOLR4 | FALSE | Upregulated |
| 1,833504597 | FOSB | | Upregulated |
| 0,953022254 | FOXB1 | FALSE | Upregulated |
| | FOXB2 | | |
| -0,418664376 | FOXC2 | FALSE | Downregulated |
| 0 | FOXD1 | FALSE | Upregulated |
| 0 | FOXD3 | FALSE | Upregulated |
| -0,28462558 | FOXD4 | FALSE | Downregulated |
| -0,137087868 | FOXD4L1 | FALSE | Downregulated |
| 0,209172455 | FOXD4L3 | FALSE | Upregulated |
| 0 | FOXD4L6 | FALSE | Upregulated |
| -0,833766309 | FOXE3 | FALSE | Downregulated |
| 0,351230547 | FOXF2 | FALSE | Upregulated |
| 0,448212425 | FOXG1 | FALSE | Upregulated |
| | FOXI1 | | |
| -0,366590103 | FOXI2 | FALSE | Downregulated |
| | FOXI3 | | |
| -0,205831664 | FOXJ1 | FALSE | Downregulated |
| 0,294331789 | FOXL1 | FALSE | Upregulated |
| -0,239253208 | FOXL2 | FALSE | Downregulated |
| | FOXN1 | | |

| | | | |
|---|---|---|---|
| 0,094422424 | FOXR1 | FALSE | Upregulated |
| -0,017135352 | FOXR2 | FALSE | Downregulated |
| 0,466903789 | FOXS1 | FALSE | Upregulated |
| 0,423361895 | FPGT-TNNI3K | FALSE | Upregulated |
| 0,932403525 | FPR2 | FALSE | Upregulated |
| -0,034874615 | FRG2 | FALSE | Downregulated |
| -0,110715806 | FRG2B | FALSE | Downregulated |
| 0,737213956 | FRMD1 | FALSE | Upregulated |
| 0,059382767 | FRMD6-AS2 | FALSE | Upregulated |
| 0,412420007 | FRMD8P1 | FALSE | Upregulated |
| 0,460301712 | FRMPD2 | FALSE | Upregulated |
| -0,043782127 | FRMPD2P1 | FALSE | Downregulated |
| 0 | FRMPD4 | FALSE | Upregulated |
| | FRRS1L | | |
| -1,057313135 | FSBP | | Downregulated |
| 0,055378136 | FSCB | FALSE | Upregulated |
| | FSCN3 | | |
| 0,349564733 | FSHB | FALSE | Upregulated |
| 0 | FSHR | FALSE | Upregulated |
| 0,029189768 | FSIP1 | FALSE | Upregulated |
| 0 | FSTL4 | FALSE | Upregulated |
| | FSTL5 | | |
| | FTH1P18 | | |
| 0,282352318 | FTH1P3 | FALSE | Upregulated |
| | FTHL17 | | |
| | FTLP10 | | |
| | FTMT | | |
| -0,38551335 | FTO-IT1 | FALSE | Downregulated |
| 0,375913519 | FUNDC1 | FALSE | Upregulated |
| 0,894702324 | FUNDC2P2 | FALSE | Upregulated |
| 1,113801258 | FUT5 | | Upregulated |
| -0,280984005 | FUT9 | FALSE | Downregulated |
| -1,314483067 | FXYD2 | | Downregulated |
| | FXYD4 | | |
| -0,034172124 | FXYD6-FXYD2 | FALSE | Downregulated |
| -0,123114942 | FXYD7 | FALSE | Downregulated |
| 0 | FZD10 | FALSE | Upregulated |
| 0,469781679 | FZD10-AS1 | FALSE | Upregulated |
| 0,655212537 | FZD2 | FALSE | Upregulated |
| -0,268205246 | FZD9 | FALSE | Downregulated |
| 0 | GAB4 | FALSE | Upregulated |
| 0,329324616 | GABBR2 | FALSE | Upregulated |
| | GABRA1 | | |
| 0 | GABRA2 | FALSE | Upregulated |
| 0,448724951 | GABRA3 | FALSE | Upregulated |
| 0,350521811 | GABRA4 | FALSE | Upregulated |
| | GABRA5 | | |
| | GABRA6 | | |
| 1,170681006 | GABRB1 | | Upregulated |
| | GABRB2 | | |
| 0,438934441 | GABRD | FALSE | Upregulated |

| | | | |
|---|---|---|---|
| 0 | GABRG1 | FALSE | Upregulated |
| | GABRG2 | | |
| -0,136409683 | GABRG3 | FALSE | Downregulated |
| -0,213357591 | GABRP | FALSE | Downregulated |
| | GABRQ | | |
| 0,25075882 | GABRR1 | FALSE | Upregulated |
| 0,304824471 | GABRR2 | FALSE | Upregulated |
| | GABRR3 | | |
| 1,037454947 | GAD1 | | Upregulated |
| | GAD2 | | |
| -0,467670303 | GADL1 | FALSE | Downregulated |
| | GAGE1 | | |
| | GAGE12C | | |
| | GAGE12D | | |
| | GAGE12E | | |
| | GAGE12G | | |
| | GAGE12H | | |
| | GAGE12I | | |
| | GAGE12J | | |
| | GAGE2A | | |
| | GAGE2C | | |
| | GAGE2E | | |
| 0 | GAL | FALSE | Upregulated |
| 0,221444284 | GAL3ST3 | FALSE | Upregulated |
| 0,204445513 | GAL3ST4 | FALSE | Upregulated |
| 0,411681245 | GALNT12 | FALSE | Upregulated |
| 0 | GALNT13 | FALSE | Upregulated |
| -0,636934736 | GALNT14 | FALSE | Downregulated |
| 0,481637332 | GALNT5 | FALSE | Upregulated |
| 0,146572651 | GALNT8 | FALSE | Upregulated |
| 0,016367963 | GALNT9-2 | FALSE | Upregulated |
| | GALNTL5 | | |
| -0,315239805 | GALNTL6 | FALSE | Downregulated |
| -0,223061762 | GALP | FALSE | Downregulated |
| 0 | GALR1 | FALSE | Upregulated |
| 0,263190331 | GALR2 | FALSE | Upregulated |
| 0,564564902 | GALR3 | FALSE | Upregulated |
| -0,390881377 | GAP43 | FALSE | Downregulated |
| 1,390497241 | GAPT | | Upregulated |
| 0,828746935 | GAR1 | FALSE | Upregulated |
| 1,039722034 | GAS2L2 | | Upregulated |
| 0,891985136 | GAS6-AS2 | FALSE | Upregulated |
| -0,016422875 | GAST | FALSE | Downregulated |
| 0 | GATA3-AS1 | FALSE | Upregulated |
| 1,415641074 | GATA5 | | Upregulated |
| 0,8130083 | GBAP1 | FALSE | Upregulated |
| 0,825165373 | GBP1P1 | FALSE | Upregulated |
| 0,746034156 | GBP6 | FALSE | Upregulated |
| 0,783415868 | GBX1 | FALSE | Upregulated |
| 0,147965921 | GBX2 | FALSE | Upregulated |
| 0 | GCG | FALSE | Upregulated |

| | | | |
|---|---|---|---|
| 0 | GCM1 | FALSE | Upregulated |
| -0,295574817 | GCNT3 | FALSE | Downregulated |
| 0,479743784 | GCNT4 | FALSE | Upregulated |
| 0 | GCSAML | FALSE | Upregulated |
| 0,285560611 | GCSAML-AS1 | FALSE | Upregulated |
| -0,000159195 | GDF10 | FALSE | Downregulated |
| | GDF3 | | |
| -0,002522957 | GDF5 | FALSE | Downregulated |
| -1,383437343 | GDF6 | | Downregulated |
| 0,874798864 | GDF9 | FALSE | Upregulated |
| 0,306330019 | GDNF-AS1 | FALSE | Upregulated |
| 1,027789587 | GDPD1 | | Upregulated |
| 0,495978768 | GDPD2 | FALSE | Upregulated |
| -0,154480919 | GDPD3 | FALSE | Downregulated |
| 0,971603145 | GFAP | FALSE | Upregulated |
| | GFI1B | | |
| 0,594546926 | GFPT2 | FALSE | Upregulated |
| 0 | GFRA3 | FALSE | Upregulated |
| 0,051052741 | GFRA4 | FALSE | Upregulated |
| 0,464439191 | GFRAL | FALSE | Upregulated |
| | GFY | | |
| 0,676732647 | GGNBP1 | FALSE | Upregulated |
| -0,324053444 | GGT2 | FALSE | Downregulated |
| 0 | GGT3P | FALSE | Upregulated |
| 0,058222282 | GGT6 | FALSE | Upregulated |
| 0,429173714 | GGT8P | FALSE | Upregulated |
| 0 | GGTLC1 | FALSE | Upregulated |
| 0,252439545 | GGTLC2 | FALSE | Upregulated |
| | GH1 | | |
| | GH2 | | |
| | GHRH | | |
| 0 | GHRHR | FALSE | Upregulated |
| -0,552991351 | GHSR | FALSE | Downregulated |
| 0,491564866 | GIF | FALSE | Upregulated |
| | GIMD1 | | |
| 0,799543906 | GINS3 | FALSE | Upregulated |
| | GIP | | |
| 0,079585026 | GIPR | FALSE | Upregulated |
| | GJA10 | | |
| -0,075398329 | GJA3 | FALSE | Downregulated |
| 0,567406591 | GJA5 | FALSE | Upregulated |
| 0 | GJA8 | FALSE | Upregulated |
| 0,491599431 | GJB6 | FALSE | Upregulated |
| | GJD2 | | |
| 0 | GJD4 | FALSE | Upregulated |
| | GK2 | | |
| | GKN1 | | |
| | GKN2 | | |
| 0,728703149 | GLB1L3 | FALSE | Upregulated |
| 1,174068962 | GLDN | | Upregulated |
| -0,365648633 | GLIPR1L2 | FALSE | Downregulated |

| | | | |
|---|---|---|---|
| 0,497131609 | GLIS1 | FALSE | Upregulated |
| 0,210002901 | GLMN | FALSE | Upregulated |
| -0,009579265 | GLOD5 | FALSE | Downregulated |
| 0,14121115 | GLP1R | FALSE | Upregulated |
| 0,427929979 | GLP2R | FALSE | Upregulated |
| 0,25533779 | GLRA2 | FALSE | Upregulated |
| 0 | GLRA3 | FALSE | Upregulated |
| 0 | GLRA4 | FALSE | Upregulated |
| -0,127437312 | GLRB | FALSE | Downregulated |
| | GLT6D1 | | |
| | GLYATL2 | | |
| -1,316151731 | GLYATL3 | | Downregulated |
| | GLYCAM1 | | |
| | GM140 | | |
| 0,340920405 | GMCL1P1 | FALSE | Upregulated |
| 0 | GML | FALSE | Upregulated |
| 0,048276727 | GNAS-AS1 | FALSE | Upregulated |
| | GNAT3 | | |
| 1,482046552 | GNB3 | | Upregulated |
| -0,034199672 | GNG13 | FALSE | Downregulated |
| 0,06047791 | GNG4 | FALSE | Upregulated |
| | GNG8 | | |
| | GNGT1 | | |
| 0,59726906 | GNGT2 | FALSE | Upregulated |
| 0,697583673 | GNLY | FALSE | Upregulated |
| 0,570352992 | GNRH2 | FALSE | Upregulated |
| 0 | GOLGA2P2Y | FALSE | Upregulated |
| 0,687443322 | GOLGA6D | FALSE | Upregulated |
| | GOLGA6L1 | | |
| -0,327025408 | GOLGA6L10 | FALSE | Downregulated |
| -0,093662559 | GOLGA6L2 | FALSE | Downregulated |
| -0,149002952 | GOLGA6L3 | FALSE | Downregulated |
| 0 | GOLGA6L6 | FALSE | Upregulated |
| 1,689831736 | GOLGA6L7P | | Upregulated |
| | GOLGA8CP | | |
| | GOLGA8DP | | |
| 0 | GOLGA8EP | FALSE | Upregulated |
| 0 | GOLGA8F | FALSE | Upregulated |
| | GOLGA8G | | |
| 0,643948232 | GOLGA8I | FALSE | Upregulated |
| -0,288975971 | GOLGA8J | FALSE | Downregulated |
| | GOT1L1 | | |
| 0,025091487 | GP1BB | FALSE | Upregulated |
| 0 | GP2 | FALSE | Upregulated |
| 0,330784987 | GP9 | FALSE | Upregulated |
| 0,95757838 | GPA33 | FALSE | Upregulated |
| 0,901373199 | GPC2 | FALSE | Upregulated |
| -1,735280526 | GPC5 | | Downregulated |
| -0,507041722 | GPC5-AS1 | FALSE | Downregulated |
| 0,891394561 | GPC6-AS2 | FALSE | Upregulated |
| | GPHB5 | | |

| | | | |
|---|---|---|---|
| -0,37122612 | GPR1 | FALSE | Downregulated |
| 0 | GPR1-AS | FALSE | Upregulated |
| -0,002828111 | GPR101 | FALSE | Downregulated |
| 0 | GPR110 | FALSE | Upregulated |
| 0 | GPR111 | FALSE | Upregulated |
| | GPR112 | | |
| | GPR115 | | |
| 0,623359442 | GPR119 | FALSE | Upregulated |
| 0 | GPR12 | FALSE | Upregulated |
| 0,370875124 | GPR123 | FALSE | Upregulated |
| 1,364564691 | GPR137C | | Upregulated |
| | GPR139 | | |
| 0,496018884 | GPR141 | FALSE | Upregulated |
| -0,909156869 | GPR142 | FALSE | Downregulated |
| 0 | GPR144 | FALSE | Upregulated |
| | GPR148 | | |
| 0,062410568 | GPR149 | FALSE | Upregulated |
| 0,747869217 | GPR15 | FALSE | Upregulated |
| 0,815509396 | GPR156 | FALSE | Upregulated |
| 0,046336957 | GPR158 | FALSE | Upregulated |
| -0,873798803 | GPR158-AS1 | FALSE | Downregulated |
| -1,213892533 | GPR173 | | Downregulated |
| 2,17921791 | GPR174 | | Upregulated |
| 0,840925378 | GPR19 | FALSE | Upregulated |
| 0,213512911 | GPR20 | FALSE | Upregulated |
| 2,259609321 | GPR25 | | Upregulated |
| 0,31266235 | GPR27 | FALSE | Upregulated |
| -0,485982596 | GPR3 | FALSE | Downregulated |
| 0 | GPR31 | FALSE | Upregulated |
| | GPR32 | | |
| 0,225970377 | GPR33 | FALSE | Upregulated |
| 0,995758712 | GPR35 | FALSE | Upregulated |
| 1,193749891 | GPR37L1 | | Upregulated |
| 0,653913124 | GPR4 | FALSE | Upregulated |
| 0 | GPR50 | FALSE | Upregulated |
| 1,233657553 | GPR55 | | Upregulated |
| | GPR6 | | |
| 0,007534374 | GPR63 | FALSE | Upregulated |
| 0,390950345 | GPR78 | FALSE | Upregulated |
| 0,604883375 | GPR83 | FALSE | Upregulated |
| 0,46679631 | GPR85 | FALSE | Upregulated |
| 0,467271238 | GPR87 | FALSE | Upregulated |
| 0,420091568 | GPR97 | FALSE | Upregulated |
| 0,378216337 | GPRC5A | FALSE | Upregulated |
| -0,01910556 | GPRC6A | FALSE | Downregulated |
| -1,105074618 | GPRIN2 | | Downregulated |
| -0,386655844 | GPX5 | FALSE | Downregulated |
| | GPX6 | | |
| 0,61030232 | GPX8 | FALSE | Upregulated |
| -1,609856736 | GRAPL | | Downregulated |
| 0,568047586 | GRIA1 | FALSE | Upregulated |

| | | | |
|---|---|---|---|
| -0,367753623 | GRIA2 | FALSE | Downregulated |
| -0,376947307 | GRID1 | FALSE | Downregulated |
| -0,221267829 | GRID1-AS1 | FALSE | Downregulated |
| | GRID2 | | |
| 0,95210352 | GRID2IP | FALSE | Upregulated |
| | GRIK1-AS1 | | |
| 0,575805942 | GRIK1-AS2 | FALSE | Upregulated |
| -0,59673032 | GRIK2 | FALSE | Downregulated |
| 0 | GRIK3 | FALSE | Upregulated |
| 0,061604238 | GRIK4 | FALSE | Upregulated |
| 0,768185145 | GRIK5 | FALSE | Upregulated |
| 0,649688444 | GRIN1 | FALSE | Upregulated |
| | GRK1 | | |
| | GRK1-2 | | |
| 0 | GRK7 | FALSE | Upregulated |
| -0,117686075 | GRM1 | FALSE | Downregulated |
| 0,236823198 | GRM2 | FALSE | Upregulated |
| 0 | GRM4 | FALSE | Upregulated |
| 0,838740667 | GRM5 | FALSE | Upregulated |
| -0,031195803 | GRM5-AS1 | FALSE | Downregulated |
| | GRP | | |
| 0,453311518 | GRPR | FALSE | Upregulated |
| | GRXCR1 | | |
| | GRXCR2 | | |
| | GS1-122H1.2 | | |
| | GS1-18A18.1 | | |
| 0 | GS1-204I12.1 | FALSE | Upregulated |
| 0 | GS1-24F4.2 | FALSE | Upregulated |
| 0,263851197 | GS1-259H13.2 | FALSE | Upregulated |
| 0,142243521 | GS1-279B7.1 | FALSE | Upregulated |
| | GS1-600G8.3 | | |
| 0 | GSC | FALSE | Upregulated |
| | GSC2 | | |
| 0,22292287 | GSDMA | FALSE | Upregulated |
| 0 | GSDMC | FALSE | Upregulated |
| 0,121450971 | GSG1L | FALSE | Upregulated |
| 0,148142446 | GSTA3 | FALSE | Upregulated |
| -0,359708941 | GSTA5 | FALSE | Downregulated |
| 0,103883719 | GSTM2P1 | FALSE | Upregulated |
| -1,046601901 | GSTM5 | | Downregulated |
| | GSTT2 | | |
| 0,469226316 | GSTTP1 | FALSE | Upregulated |
| | GSX1 | | |
| | GSX2 | | |
| 0,159143715 | GTF2A1L | FALSE | Upregulated |
| | GTPBP6-2 | | |
| 0,01196447 | GTSCR1 | FALSE | Upregulated |
| 0,062731089 | GTSF1 | FALSE | Upregulated |
| | GTSF1L | | |
| 0,061389277 | GUCA1A | FALSE | Upregulated |
| | GUCA1C | | |

| | | | |
|---|---|---|---|
| -0,076593115 | GUCA2A | FALSE | Downregulated |
| -0,362746959 | GUCA2B | FALSE | Downregulated |
| 0,416114835 | GUCY1B2 | FALSE | Upregulated |
| -0,455233205 | GUCY2D | FALSE | Downregulated |
| -0,229492885 | GUCY2EP | FALSE | Downregulated |
| | GUCY2F | | |
| | GUCY2GP | | |
| -0,262084891 | GULP1 | FALSE | Downregulated |
| | GUSBP10 | | |
| 0,064180021 | GUSBP4 | FALSE | Upregulated |
| -1,639832786 | GUSBP5 | | Downregulated |
| 0,649761178 | GXYLT2 | FALSE | Upregulated |
| -0,265418372 | GYG2P1 | FALSE | Downregulated |
| 0,048202176 | GYPA | FALSE | Upregulated |
| 0,033128344 | GYPB | FALSE | Upregulated |
| 1,711070733 | GZMA | | Upregulated |
| 1,692638118 | GZMB | | Upregulated |
| 0,644338142 | GZMH | FALSE | Upregulated |
| -0,395639493 | GZMK | FALSE | Downregulated |
| 0,966003817 | GZMM | FALSE | Upregulated |
| 0,584664717 | H1FOO | FALSE | Upregulated |
| | H2BFM | | |
| | H2BFWT | | |
| -0,005444838 | H2BFXP | FALSE | Downregulated |
| -0,707750687 | H3F3C | FALSE | Downregulated |
| | HAND1 | | |
| -0,184799526 | HAND2-AS1 | FALSE | Downregulated |
| -1,491350342 | HAP1 | | Downregulated |
| 0,102084982 | HAPLN1 | FALSE | Upregulated |
| -0,273930416 | HAPLN2 | FALSE | Downregulated |
| 0,399795344 | HAR1B | FALSE | Upregulated |
| -0,048804098 | HARBI1 | FALSE | Downregulated |
| 0,935352243 | HAS1 | FALSE | Upregulated |
| 0,608205811 | HAS2 | FALSE | Upregulated |
| 1,082769573 | HAS2-AS1 | | Upregulated |
| 0,243136301 | HAUS1 | FALSE | Upregulated |
| 0,510137217 | HAVCR1 | FALSE | Upregulated |
| 0,590940248 | HBEGF | FALSE | Upregulated |
| 0 | HBZ | FALSE | Upregulated |
| -0,105281799 | HCAR1 | FALSE | Downregulated |
| 0,340762212 | HCCAT4 | FALSE | Upregulated |
| 0 | HCCAT5 | FALSE | Upregulated |
| -0,171409095 | HCG14 | FALSE | Downregulated |
| 0,083856723 | HCG16 | FALSE | Upregulated |
| | HCG17 | | |
| 0 | HCG22 | FALSE | Upregulated |
| 0,047839331 | HCG23 | FALSE | Upregulated |
| 0,442999609 | HCG4 | FALSE | Upregulated |
| 0,387897571 | HCN1 | FALSE | Upregulated |
| 0,825562507 | HCN4 | FALSE | Upregulated |
| | HCRT | | |

| | | | |
|---|---|---|---|
| | HCRTR2 | | |
| 0,53457012 | HCST | FALSE | Upregulated |
| 0 | HDAC11-AS1 | FALSE | Upregulated |
| | HDGFL1 | | |
| 0,349982521 | HECW1 | FALSE | Upregulated |
| | HELT | | |
| 0,144750879 | HEMGN | FALSE | Upregulated |
| -0,772506086 | HENMT1 | FALSE | Downregulated |
| | HEPACAM2 | | |
| -0,083653522 | HERC2P10 | FALSE | Downregulated |
| 0,502997959 | HERC2P4 | FALSE | Upregulated |
| 0,404049146 | HES2 | FALSE | Upregulated |
| 0 | HES3 | FALSE | Upregulated |
| 0,979080423 | HES5 | FALSE | Upregulated |
| 0 | HES7 | FALSE | Upregulated |
| 0,262132668 | HESX1 | FALSE | Upregulated |
| -0,675214976 | HEY1 | FALSE | Downregulated |
| 0,446303474 | HFM1 | FALSE | Upregulated |
| -0,322914799 | HGC6.3 | FALSE | Downregulated |
| -0,027473165 | HHATL | FALSE | Downregulated |
| 0,332527747 | HHIP-AS1 | FALSE | Upregulated |
| 0,046678197 | HHIPL2 | FALSE | Upregulated |
| | HHLA1 | | |
| -0,56390698 | HHLA2 | FALSE | Downregulated |
| -0,70096735 | HHLA3 | FALSE | Downregulated |
| 0 | HIF1A-AS1 | FALSE | Upregulated |
| 0 | HIGD2B | FALSE | Upregulated |
| -0,027829761 | HIST1H1T | FALSE | Downregulated |
| -0,059191005 | HIST1H2AA | FALSE | Downregulated |
| 1,265184881 | HIST1H2AB | | Upregulated |
| 1,24986221 | HIST1H2AD | | Upregulated |
| 2,474270051 | HIST1H2AH | | Upregulated |
| 1,16417016 | HIST1H2AJ | | Upregulated |
| 1,983715022 | HIST1H2AM | | Upregulated |
| 1,021189587 | HIST1H2BB | | Upregulated |
| 1,56097307 | HIST1H2BH | | Upregulated |
| 0,872904937 | HIST1H2BI | FALSE | Upregulated |
| 0,463849111 | HIST1H2BJ | FALSE | Upregulated |
| 0,871142277 | HIST1H2BL | FALSE | Upregulated |
| 1,369954945 | HIST1H2BM | | Upregulated |
| 2,255389124 | HIST1H2BO | | Upregulated |
| 1,96947119 | HIST1H3A | | Upregulated |
| 2,572717695 | HIST1H3B | | Upregulated |
| 0,164086543 | HIST1H3C | FALSE | Upregulated |
| 0,402964025 | HIST1H3E | FALSE | Upregulated |
| 2,355945413 | HIST1H3F | | Upregulated |
| 0,431256166 | HIST1H3G | FALSE | Upregulated |
| 1,823795362 | HIST1H3J | | Upregulated |
| 1,011087667 | HIST1H4A | | Upregulated |
| 1,280712758 | HIST1H4F | | Upregulated |
| | HIST1H4G | | |

| | | | |
|---|---|---|---|
| 0,911631594 | HIST1H4L | FALSE | Upregulated |
| 0,245035521 | HIST2H2BA | FALSE | Upregulated |
| -0,15685042 | HIST2H2BC | FALSE | Downregulated |
| 0,944394906 | HIST3H2A | FALSE | Upregulated |
| 2,14718856 | HIST3H2BB | | Upregulated |
| 1,718277803 | HIST3H3 | | Upregulated |
| 2,062666611 | HJURP | | Upregulated |
| 1,214778298 | HLA-DOB | | Upregulated |
| 0 | HLA-DPB2 | FALSE | Upregulated |
| 0,731723606 | HLA-G | FALSE | Upregulated |
| 0,489393812 | HLTF-AS1 | FALSE | Upregulated |
| | HMGA1P7 | | |
| 0,120731044 | HMGA2 | FALSE | Upregulated |
| 1,007587809 | HMGB3P1 | | Upregulated |
| 0,554829331 | HMGB4 | FALSE | Upregulated |
| 0,17130081 | HMGCLL1 | FALSE | Upregulated |
| -0,185413306 | HMGN2P46 | FALSE | Downregulated |
| | HMHB1 | | |
| 1,22215967 | HMMR | | Upregulated |
| -0,118480417 | HMP19 | FALSE | Downregulated |
| 0 | HMSD | FALSE | Upregulated |
| | HMX1 | | |
| | HMX2 | | |
| | HMX3 | | |
| -0,847050673 | HNRNPA1P33 | FALSE | Downregulated |
| -0,000807366 | HNRNPA3P1 | FALSE | Downregulated |
| 0 | HNRNPCL1 | FALSE | Upregulated |
| | HNRNPCP5 | | |
| | HNRNPKP3 | | |
| -0,728496083 | HOMER1 | FALSE | Downregulated |
| 1,33559717 | HOPX | | Upregulated |
| 0,315966311 | HORMAD1 | FALSE | Upregulated |
| -0,746105503 | HORMAD2 | FALSE | Downregulated |
| | HOTAIR | | |
| 1,195551821 | HOTAIRM1 | | Upregulated |
| 0,390950345 | HOTTIP | FALSE | Upregulated |
| 0,588252785 | HOXA-AS3 | FALSE | Upregulated |
| 0,830077549 | HOXA1 | FALSE | Upregulated |
| 0,724589444 | HOXA10 | FALSE | Upregulated |
| 0,259864409 | HOXA10-AS | FALSE | Upregulated |
| | HOXA10-HOXA9 | | |
| | HOXA11 | | |
| | HOXA11-AS | | |
| 0 | HOXA13 | FALSE | Upregulated |
| -1,180741856 | HOXA5 | | Downregulated |
| -0,336844868 | HOXA6 | FALSE | Downregulated |
| 0 | HOXA7 | FALSE | Upregulated |
| 0 | HOXA9 | FALSE | Upregulated |
| | HOXB-AS1 | | |
| 0,681429053 | HOXB-AS3 | FALSE | Upregulated |
| | HOXB1 | | |

| | | | |
|---|---|---|---|
| | HOXB13 | | |
| -0,357439114 | HOXB5 | FALSE | Downregulated |
| 0,040477135 | HOXB7 | FALSE | Upregulated |
| -0,037835658 | HOXB8 | FALSE | Downregulated |
| 0 | HOXB9 | FALSE | Upregulated |
| | HOXC-AS1 | | |
| | HOXC-AS2 | | |
| | HOXC-AS3 | | |
| 0 | HOXC10 | FALSE | Upregulated |
| 0 | HOXC11 | FALSE | Upregulated |
| | HOXC12 | | |
| 0 | HOXC13 | FALSE | Upregulated |
| | HOXC13-AS | | |
| 0,104684312 | HOXC4 | FALSE | Upregulated |
| | HOXC5 | | |
| 0 | HOXC6 | FALSE | Upregulated |
| | HOXC8 | | |
| 0 | HOXC9 | FALSE | Upregulated |
| -0,010054113 | HOXD-AS1 | FALSE | Downregulated |
| 0,290312159 | HOXD-AS2 | FALSE | Upregulated |
| 0 | HOXD1 | FALSE | Upregulated |
| | HOXD10 | | |
| 0 | HOXD11 | FALSE | Upregulated |
| | HOXD12 | | |
| | HOXD13 | | |
| 0,141649698 | HOXD3 | FALSE | Upregulated |
| -0,062029229 | HOXD4 | FALSE | Downregulated |
| -0,100107743 | HOXD8 | FALSE | Downregulated |
| 0 | HOXD9 | FALSE | Upregulated |
| | HP08942 | | |
| 0,295578805 | HPCA | FALSE | Upregulated |
| 0,210988542 | HPCAL4 | FALSE | Upregulated |
| 0,71319046 | HPSE2 | FALSE | Upregulated |
| 0,070401644 | HPVC1 | FALSE | Upregulated |
| | HPYR1 | | |
| -0,216746548 | HRASLS | FALSE | Downregulated |
| -0,740739382 | HRASLS2 | FALSE | Downregulated |
| -0,095269989 | HRASLS5 | FALSE | Downregulated |
| 0,43234596 | HRCT1 | FALSE | Upregulated |
| 0,534776073 | HRH3 | FALSE | Upregulated |
| 0,043926082 | HRH4 | FALSE | Upregulated |
| | HRK | | |
| 2,250767884 | HS3ST2 | | Upregulated |
| 0 | HS3ST4 | FALSE | Upregulated |
| 0 | HS3ST6 | FALSE | Upregulated |
| 0,916482653 | HS6ST2 | FALSE | Upregulated |
| | HS6ST2-AS1 | | |
| | HS6ST3 | | |
| 0,488797049 | HSD17B7P2 | FALSE | Upregulated |
| 0 | HSD52 | FALSE | Upregulated |
| 0,691289715 | HSF2BP | FALSE | Upregulated |

| | | | |
|---|---|---|---|
| -0,462233016 | HSF5 | FALSE | Downregulated |
| -0,689761609 | HSFX1 | FALSE | Downregulated |
| 0,380597113 | HSFX2 | FALSE | Upregulated |
| -2,576019499 | HSFY1 | | Downregulated |
| | HSFY1P1 | | |
| -0,514628286 | HSFY2 | FALSE | Downregulated |
| 0,915799924 | HSP90AB4P | FALSE | Upregulated |
| -0,439967231 | HSPA1L | FALSE | Downregulated |
| 0,508207887 | HSPB2 | FALSE | Upregulated |
| 0,376120077 | HSPB2-C11orf52 | FALSE | Upregulated |
| | HSPB3 | | |
| | HTN1 | | |
| | HTN3 | | |
| | HTR1A | | |
| | HTR1B | | |
| -0,174197925 | HTR1D | FALSE | Downregulated |
| | HTR1E | | |
| 0,761192721 | HTR1F | FALSE | Upregulated |
| -0,523553258 | HTR2A-AS1 | FALSE | Downregulated |
| -0,972577243 | HTR2C | FALSE | Downregulated |
| | HTR3A | | |
| | HTR3B | | |
| | HTR3C | | |
| | HTR3D | | |
| 0,755753703 | HTR4 | FALSE | Upregulated |
| | HTR5A | | |
| 0 | HTR5A-AS1 | FALSE | Upregulated |
| -0,070755564 | HTR7 | FALSE | Downregulated |
| 0,324154931 | HTRA4 | FALSE | Upregulated |
| -0,099541264 | HUS1B | FALSE | Downregulated |
| 0,056809858 | HYAL4 | FALSE | Upregulated |
| | HYALP1 | | |
| -0,977794936 | HYKK | FALSE | Downregulated |
| 1,452106558 | HYMAI | | Upregulated |
| -0,455054682 | IAPP | FALSE | Downregulated |
| 1,202286877 | IBA57-AS1 | | Upregulated |
| 0,031365483 | IBSP | FALSE | Upregulated |
| 0,130935545 | ICAM4 | FALSE | Upregulated |
| 1,060245027 | ICAM5 | | Upregulated |
| 1,438641106 | ICOS | | Upregulated |
| -0,615144611 | IDH1-AS1 | FALSE | Downregulated |
| 0,556865703 | IDO1 | FALSE | Upregulated |
| -0,090341388 | IFI27L1 | FALSE | Downregulated |
| -0,348669256 | IFI27L2 | FALSE | Downregulated |
| | IFITM4P | | |
| 0,007015577 | IFLTD1 | FALSE | Upregulated |
| | IFNA1 | | |
| 0 | IFNA10 | FALSE | Upregulated |
| | IFNA13 | | |
| | IFNA14 | | |
| 0 | IFNA16 | FALSE | Upregulated |

| | | | |
|---|---|---|---|
| | IFNA17 | | |
| | IFNA2 | | |
| 0,479634233 | IFNA21 | FALSE | Upregulated |
| 0,020789395 | IFNA22P | FALSE | Upregulated |
| 0,000977115 | IFNA4 | FALSE | Upregulated |
| | IFNA5 | | |
| | IFNA6 | | |
| 0 | IFNA7 | FALSE | Upregulated |
| | IFNA8 | | |
| -0,399856971 | IFNB1 | FALSE | Downregulated |
| | IFNE | | |
| -0,221973404 | IFNL1 | FALSE | Downregulated |
| | IFNL2 | | |
| | IFNL3 | | |
| | IFNL4 | | |
| 0,372179381 | IFNW1 | FALSE | Upregulated |
| -0,354270578 | IGDCC3 | FALSE | Downregulated |
| 1,267682505 | IGDCC4 | | Upregulated |
| -0,829014167 | IGF2BP1 | FALSE | Downregulated |
| 0,98204992 | IGFBPL1 | FALSE | Upregulated |
| 0 | IGFL1 | FALSE | Upregulated |
| 0 | IGFL2 | FALSE | Upregulated |
| -0,21713708 | IGFL3 | FALSE | Downregulated |
| 0,561106516 | IGFL4 | FALSE | Upregulated |
| 0,38647539 | IGLL1 | FALSE | Upregulated |
| -0,19510494 | IGLL3P | FALSE | Downregulated |
| 0,355816177 | IGSF1 | FALSE | Upregulated |
| 2,202595999 | IGSF11 | | Upregulated |
| 0,034840507 | IGSF11-AS1 | FALSE | Upregulated |
| 0 | IGSF5 | FALSE | Upregulated |
| 0,390905567 | IL10 | FALSE | Upregulated |
| -0,036774468 | IL11 | FALSE | Downregulated |
| -0,065261816 | IL12A | FALSE | Downregulated |
| -0,01629869 | IL12A-AS1 | FALSE | Downregulated |
| -0,207680125 | IL12B | FALSE | Downregulated |
| 0 | IL13 | FALSE | Upregulated |
| -0,785132217 | IL13RA2 | FALSE | Downregulated |
| 0 | IL17A | FALSE | Upregulated |
| 0,184302529 | IL17B | FALSE | Upregulated |
| -0,022245538 | IL17C | FALSE | Downregulated |
| | IL17F | | |
| -0,881179119 | IL17RD | FALSE | Downregulated |
| 1,047481963 | IL18RAP | | Upregulated |
| 0 | IL19 | FALSE | Upregulated |
| 1,432556776 | IL1B | | Upregulated |
| | IL1F10 | | |
| -0,076205187 | IL1RAPL1 | FALSE | Downregulated |
| 0,386191538 | IL1RL1 | FALSE | Upregulated |
| | IL2 | | |
| 0,355623397 | IL20 | FALSE | Upregulated |
| 0,013795592 | IL20RA | FALSE | Upregulated |

| | | | |
|---|---|---|---|
| 0 | IL21 | FALSE | Upregulated |
| 0,74621294 | IL21-AS1 | FALSE | Upregulated |
| 0 | IL22 | FALSE | Upregulated |
| | IL22RA2 | | |
| 0,7745078 | IL23R | FALSE | Upregulated |
| 0 | IL25 | FALSE | Upregulated |
| 1,603721343 | IL26 | | Upregulated |
| 0,688124025 | IL2RA | FALSE | Upregulated |
| -0,069444444 | IL31 | FALSE | Downregulated |
| -0,06991492 | IL31RA | FALSE | Downregulated |
| | IL36A | | |
| | IL36B | | |
| 0,100713889 | IL36G | FALSE | Upregulated |
| 0 | IL36RN | FALSE | Upregulated |
| 0 | IL37 | FALSE | Upregulated |
| | IL3RA-2 | | |
| 0 | IL4 | FALSE | Upregulated |
| 0,38967398 | IL6 | FALSE | Upregulated |
| 1,01025564 | IL8 | | Upregulated |
| | IL9 | | |
| -0,91791643 | IL9R | FALSE | Downregulated |
| | IL9R-2 | | |
| -0,64709753 | IMPG2 | FALSE | Downregulated |
| 0 | INA | FALSE | Upregulated |
| 0,811473216 | INGX | FALSE | Upregulated |
| 0,990581773 | INHA | FALSE | Upregulated |
| 0,365549998 | INHBA-AS1 | FALSE | Upregulated |
| -0,812132684 | INMT-FAM188B | FALSE | Downregulated |
| 0,305000939 | INPP5J | FALSE | Upregulated |
| 0,023509318 | INSC | FALSE | Upregulated |
| -0,775255255 | INSL3 | FALSE | Downregulated |
| | INSL4 | | |
| -0,128331034 | INSL5 | FALSE | Downregulated |
| 0,300198459 | INSL6 | FALSE | Upregulated |
| -0,14135959 | INSM1 | FALSE | Downregulated |
| -0,64526415 | INSRR | FALSE | Downregulated |
| 0,375508037 | INTS4L1 | FALSE | Upregulated |
| -0,532910727 | INTS4L2 | FALSE | Downregulated |
| 0,82819295 | INTS6-AS1 | FALSE | Upregulated |
| -0,887719168 | IPO11-LRRC70 | FALSE | Downregulated |
| 0,313255157 | IQCA1 | FALSE | Upregulated |
| | IQCA1P1 | | |
| | IQCF1 | | |
| | IQCF2 | | |
| | IQCF3 | | |
| | IQCF4 | | |
| | IQCF5 | | |
| 0 | IQCF5-AS1 | FALSE | Upregulated |
| | IQCF6 | | |
| | IQCJ | | |
| 0,258147671 | IQUB | FALSE | Upregulated |

| | | | |
|---|---|---|---|
| 0,355483568 | IRG1 | FALSE | Upregulated |
| 0 | IRGC | FALSE | Upregulated |
| 0,208647909 | IRGM | FALSE | Upregulated |
| 0,000977115 | IRS4 | FALSE | Upregulated |
| 0 | IRX1 | FALSE | Upregulated |
| 0 | IRX2 | FALSE | Upregulated |
| 0 | IRX4 | FALSE | Upregulated |
| 0 | IRX5 | FALSE | Upregulated |
| -0,224486717 | IRX6 | FALSE | Downregulated |
| | ISL1 | | |
| 0 | ISL2 | FALSE | Upregulated |
| -0,606151439 | ISLR2 | FALSE | Downregulated |
| 0,981253543 | ISM2 | FALSE | Upregulated |
| 0,177886509 | ISX | FALSE | Upregulated |
| 0,09324234 | ITGA2B | FALSE | Upregulated |
| -0,395652075 | ITGA8 | FALSE | Downregulated |
| 0,359527695 | ITGAE | FALSE | Upregulated |
| -0,322551542 | ITGB1BP2 | FALSE | Downregulated |
| 1,160692899 | ITGB6 | | Upregulated |
| 0,108088777 | ITLN2 | FALSE | Upregulated |
| 0,699163315 | ITPKB-IT1 | FALSE | Upregulated |
| -0,581522124 | ITPR1-AS1 | FALSE | Downregulated |
| 1,551657935 | IVL | | Upregulated |
| 0,257038776 | IZUMO1 | FALSE | Upregulated |
| | IZUMO2 | | |
| | IZUMO3 | | |
| 2,087637292 | JAKMIP1 | | Upregulated |
| -1,091201895 | JAKMIP3 | | Downregulated |
| | JAZF1-AS1 | | |
| 1,006983605 | JPH1 | | Upregulated |
| 0,002248811 | JPH2 | FALSE | Upregulated |
| 0,417996115 | JPH4 | FALSE | Upregulated |
| | JRKL-AS1 | | |
| -0,430776928 | JSRP1 | FALSE | Downregulated |
| 0,004235587 | KAL1 | FALSE | Upregulated |
| | KATNBL1P6 | | |
| -0,070296501 | KAZALD1 | FALSE | Downregulated |
| 0,464331737 | KBTBD13 | FALSE | Upregulated |
| -0,780245834 | KBTBD8 | FALSE | Downregulated |
| 0,333732009 | KC6 | FALSE | Upregulated |
| 0 | KCNA1 | FALSE | Upregulated |
| 0,331597433 | KCNA10 | FALSE | Upregulated |
| 0,446595999 | KCNA2 | FALSE | Upregulated |
| | KCNA4 | | |
| 0,967899325 | KCNA5 | FALSE | Upregulated |
| 1,907495635 | KCNA7 | | Upregulated |
| 0 | KCNAB1-AS1 | FALSE | Upregulated |
| 0 | KCNAB1-AS2 | FALSE | Upregulated |
| | KCNB2 | | |
| 0 | KCNC4-AS1 | FALSE | Upregulated |
| 0,619029611 | KCND2 | FALSE | Upregulated |

| | | | |
|---|---|---|---|
| 0,408261566 | KCND3-IT1 | FALSE | Upregulated |
| 0,259615441 | KCNE1L | FALSE | Upregulated |
| 0,528286554 | KCNE4 | FALSE | Upregulated |
| 0 | KCNF1 | FALSE | Upregulated |
| -0,345475781 | KCNG1 | FALSE | Downregulated |
| | KCNG3 | | |
| 0,66265915 | KCNG4 | FALSE | Upregulated |
| | KCNH1-IT1 | | |
| 0,293149657 | KCNH2 | FALSE | Upregulated |
| | KCNH5 | | |
| 0,308528262 | KCNIP1 | FALSE | Upregulated |
| 0,469226316 | KCNIP4-IT1 | FALSE | Upregulated |
| 0,665584541 | KCNJ11 | FALSE | Upregulated |
| -0,24581263 | KCNJ12 | FALSE | Downregulated |
| -0,697737772 | KCNJ16 | FALSE | Downregulated |
| 0,094615407 | KCNJ18 | FALSE | Upregulated |
| 0,166000541 | KCNJ2-AS1 | FALSE | Upregulated |
| 0,323596452 | KCNJ6 | FALSE | Upregulated |
| 0,616408506 | KCNK13 | FALSE | Upregulated |
| -0,235195059 | KCNK15 | FALSE | Downregulated |
| | KCNK18 | | |
| | KCNK2 | | |
| -2,169019509 | KCNK3 | | Downregulated |
| 0 | KCNK4 | FALSE | Upregulated |
| | KCNK9 | | |
| 1,756104205 | KCNMB4 | | Upregulated |
| -0,158815307 | KCNN1 | FALSE | Downregulated |
| 0,564018788 | KCNN4 | FALSE | Upregulated |
| 0,029952855 | KCNQ1DN | FALSE | Upregulated |
| | KCNQ2 | | |
| -0,278822395 | KCNQ3 | FALSE | Downregulated |
| 0,286266228 | KCNQ4 | FALSE | Upregulated |
| 0,818732698 | KCNQ5 | FALSE | Upregulated |
| 1,138138616 | KCNQ5-AS1 | | Upregulated |
| | KCNQ5-IT1 | | |
| | KCNU1 | | |
| -0,306890282 | KCNV1 | FALSE | Downregulated |
| -1,032360094 | KCP | | Downregulated |
| -1,088058021 | KCTD14 | | Downregulated |
| 0,258286674 | KCTD16 | FALSE | Upregulated |
| -0,548305911 | KCTD19 | FALSE | Downregulated |
| 0 | KCTD8 | FALSE | Upregulated |
| 0,480052189 | KDM4D | FALSE | Upregulated |
| 1,403644994 | KEL | | Upregulated |
| | KERA | | |
| 0,355228459 | KHDC1 | FALSE | Upregulated |
| | KHDC1L | | |
| 0,450737483 | KHDC3L | FALSE | Upregulated |
| -0,833801552 | KHDRBS2 | FALSE | Downregulated |
| 0,108192853 | KIAA0101 | FALSE | Upregulated |
| | KIAA1024L | | |

| | | | |
|---|---|---|---|
| -0,015873957 | KIAA1210 | FALSE | Downregulated |
| 0,117102027 | KIAA1257 | FALSE | Upregulated |
| 1,280220794 | KIAA1377 | | Upregulated |
| 0,750023306 | KIAA1524 | FALSE | Upregulated |
| 1,110526648 | KIAA1644 | | Upregulated |
| -0,36599383 | KIAA2022 | FALSE | Downregulated |
| 1,320521317 | KIF14 | | Upregulated |
| 0,184424549 | KIF15 | FALSE | Upregulated |
| 1,468072494 | KIF18A | | Upregulated |
| 3,354449003 | KIF18B | | Upregulated |
| 1,889357216 | KIF23 | | Upregulated |
| -0,119276782 | KIF25 | FALSE | Downregulated |
| 0,047839331 | KIF25-AS1 | FALSE | Upregulated |
| 0 | KIF2B | FALSE | Upregulated |
| 1,454285413 | KIF4A | | Upregulated |
| -0,728317161 | KIF4B | FALSE | Downregulated |
| 0,1318101 | KIF5A | FALSE | Upregulated |
| 0,039514575 | KIF5C | FALSE | Upregulated |
| 1,838065578 | KIFC1 | | Upregulated |
| -0,051703949 | KIR2DL1 | FALSE | Downregulated |
| 0,212823132 | KIR2DL3 | FALSE | Upregulated |
| 0 | KIR2DL4 | FALSE | Upregulated |
| 0 | KIR2DS4 | FALSE | Upregulated |
| 0,047839331 | KIR3DL1 | FALSE | Upregulated |
| 0 | KIR3DL2 | FALSE | Upregulated |
| | KIR3DL3 | | |
| -0,089118313 | KIR3DX1 | FALSE | Downregulated |
| | KIRREL2 | | |
| | KIRREL3-AS2 | | |
| 0 | KIRREL3-AS3 | FALSE | Upregulated |
| -0,628973305 | KISS1 | FALSE | Downregulated |
| 1,04285232 | KISS1R | | Upregulated |
| -1,03682081 | KIT | | Downregulated |
| 0,29716089 | KL | FALSE | Upregulated |
| -0,128073681 | KLF1 | FALSE | Downregulated |
| | KLF14 | | |
| 0 | KLF17 | FALSE | Upregulated |
| 0,435761853 | KLHL1 | FALSE | Upregulated |
| -0,903824738 | KLHL14 | FALSE | Downregulated |
| 0,428833328 | KLHL30 | FALSE | Upregulated |
| 0,546997918 | KLHL34 | FALSE | Upregulated |
| -0,558511034 | KLHL38 | FALSE | Downregulated |
| 0,920762071 | KLHL4 | FALSE | Upregulated |
| 0,492771714 | KLHL40 | FALSE | Upregulated |
| 0,082960342 | KLHL7-AS1 | FALSE | Upregulated |
| 0,055378136 | KLK1 | FALSE | Upregulated |
| 0,283755403 | KLK10 | FALSE | Upregulated |
| 0 | KLK11 | FALSE | Upregulated |
| 0,110187985 | KLK12 | FALSE | Upregulated |
| | KLK13 | | |
| -0,033527764 | KLK14 | FALSE | Downregulated |

| | | | |
|---|---|---|---|
| | KLK15 | | |
| | KLK2 | | |
| | KLK3 | | |
| | KLK4 | | |
| 0 | KLK5 | FALSE | Upregulated |
| | KLK6 | | |
| | KLK7 | | |
| 0 | KLK8 | FALSE | Upregulated |
| | KLK9 | | |
| | KLKP1 | | |
| 0,036316335 | KLRAP1 | FALSE | Upregulated |
| 1,151674915 | KLRB1 | | Upregulated |
| 0,662654732 | KLRC1 | FALSE | Upregulated |
| 0,559508069 | KLRC2 | FALSE | Upregulated |
| -0,233128371 | KLRC3 | FALSE | Downregulated |
| 0,353206904 | KLRC4 | FALSE | Upregulated |
| -0,179461052 | KLRC4-KLRK1 | FALSE | Downregulated |
| -0,097798734 | KLRF1 | FALSE | Downregulated |
| | KLRF2 | | |
| -0,077684751 | KLRG1 | FALSE | Downregulated |
| -1,141337758 | KLRG2 | | Downregulated |
| | KNCN | | |
| 0 | KPRP | FALSE | Upregulated |
| 0 | KRBOX1 | FALSE | Upregulated |
| 0,305373983 | KRT12 | FALSE | Upregulated |
| | KRT13 | | |
| 0,440937483 | KRT15 | FALSE | Upregulated |
| 0 | KRT16 | FALSE | Upregulated |
| | KRT16P1 | | |
| 0,387012887 | KRT16P2 | FALSE | Upregulated |
| | KRT16P3 | | |
| 0,453099891 | KRT18P55 | FALSE | Upregulated |
| 0,30152783 | KRT19P2 | FALSE | Upregulated |
| -0,761029148 | KRT20 | FALSE | Downregulated |
| 1,027578871 | KRT23 | | Upregulated |
| | KRT24 | | |
| | KRT25 | | |
| | KRT26 | | |
| 0,546538923 | KRT27 | FALSE | Upregulated |
| | KRT28 | | |
| | KRT3 | | |
| 0 | KRT31 | FALSE | Upregulated |
| 0 | KRT32 | FALSE | Upregulated |
| | KRT33A | | |
| | KRT33B | | |
| | KRT34 | | |
| 0 | KRT35 | FALSE | Upregulated |
| 0 | KRT36 | FALSE | Upregulated |
| 0 | KRT37 | FALSE | Upregulated |
| 0 | KRT38 | FALSE | Upregulated |
| 0,330623236 | KRT39 | FALSE | Upregulated |

| | | | |
|---|---|---|---|
| 0,094615407 | KRT4 | FALSE | Upregulated |
| | KRT40 | | |
| | KRT42P | | |
| 0,615341206 | KRT6B | FALSE | Upregulated |
| 0 | KRT6C | FALSE | Upregulated |
| 0,387012887 | KRT71 | FALSE | Upregulated |
| | KRT72 | | |
| 0,207122854 | KRT73 | FALSE | Upregulated |
| 0,229404528 | KRT73-AS1 | FALSE | Upregulated |
| | KRT74 | | |
| | KRT75 | | |
| | KRT76 | | |
| 0 | KRT77 | FALSE | Upregulated |
| 0 | KRT78 | FALSE | Upregulated |
| | KRT79 | | |
| -0,06112239 | KRT80 | FALSE | Downregulated |
| 0,739519483 | KRT81 | FALSE | Upregulated |
| 0 | KRT82 | FALSE | Upregulated |
| 0 | KRT83 | FALSE | Upregulated |
| | KRT84 | | |
| 0,350021073 | KRT85 | FALSE | Upregulated |
| 0,001634662 | KRT86 | FALSE | Upregulated |
| 0,840199504 | KRT8P41 | FALSE | Upregulated |
| | KRT9 | | |
| | KRTAP1-1 | | |
| | KRTAP1-3 | | |
| | KRTAP1-4 | | |
| 0 | KRTAP1-5 | FALSE | Upregulated |
| | KRTAP10-1 | | |
| 0,291637382 | KRTAP10-10 | FALSE | Upregulated |
| -0,035781883 | KRTAP10-11 | FALSE | Downregulated |
| 0 | KRTAP10-12 | FALSE | Upregulated |
| 0,181508125 | KRTAP10-2 | FALSE | Upregulated |
| | KRTAP10-3 | | |
| -0,236153243 | KRTAP10-4 | FALSE | Downregulated |
| | KRTAP10-5 | | |
| 0,02839685 | KRTAP10-6 | FALSE | Upregulated |
| 0,584697452 | KRTAP10-7 | FALSE | Upregulated |
| 0,38647539 | KRTAP10-8 | FALSE | Upregulated |
| -0,231904545 | KRTAP10-9 | FALSE | Downregulated |
| | KRTAP11-1 | | |
| | KRTAP12-1 | | |
| | KRTAP12-2 | | |
| | KRTAP12-3 | | |
| | KRTAP12-4 | | |
| | KRTAP13-1 | | |
| | KRTAP13-2 | | |
| | KRTAP13-3 | | |
| | KRTAP13-4 | | |
| | KRTAP15-1 | | |
| | KRTAP16-1 | | |

| | | | |
|---|---|---|---|
| | KRTAP17-1 | | |
| | KRTAP19-1 | | |
| | KRTAP19-2 | | |
| | KRTAP19-3 | | |
| | KRTAP19-4 | | |
| | KRTAP19-5 | | |
| | KRTAP19-6 | | |
| | KRTAP19-7 | | |
| 0,128328249 | KRTAP19-8 | FALSE | Upregulated |
| | KRTAP2-1 | | |
| | KRTAP2-2 | | |
| | KRTAP2-3 | | |
| | KRTAP2-4 | | |
| | KRTAP20-1 | | |
| | KRTAP20-2 | | |
| | KRTAP20-3 | | |
| | KRTAP20-4 | | |
| | KRTAP21-1 | | |
| | KRTAP21-2 | | |
| | KRTAP21-3 | | |
| | KRTAP22-1 | | |
| | KRTAP22-2 | | |
| | KRTAP23-1 | | |
| | KRTAP24-1 | | |
| | KRTAP25-1 | | |
| | KRTAP26-1 | | |
| | KRTAP27-1 | | |
| | KRTAP29-1 | | |
| 0 | KRTAP3-1 | FALSE | Upregulated |
| 0 | KRTAP3-2 | FALSE | Upregulated |
| 0,393935747 | KRTAP3-3 | FALSE | Upregulated |
| | KRTAP4-1 | | |
| | KRTAP4-11 | | |
| | KRTAP4-12 | | |
| | KRTAP4-2 | | |
| | KRTAP4-3 | | |
| | KRTAP4-4 | | |
| | KRTAP4-5 | | |
| | KRTAP4-6 | | |
| | KRTAP4-7 | | |
| | KRTAP4-8 | | |
| | KRTAP4-9 | | |
| 1,163461036 | KRTAP5-1 | | Upregulated |
| 0 | KRTAP5-10 | FALSE | Upregulated |
| 0,028760091 | KRTAP5-11 | FALSE | Upregulated |
| 1,399329186 | KRTAP5-2 | | Upregulated |
| | KRTAP5-3 | | |
| 0 | KRTAP5-4 | FALSE | Upregulated |
| | KRTAP5-5 | | |
| 0,258514524 | KRTAP5-6 | FALSE | Upregulated |
| 1,363272293 | KRTAP5-7 | | Upregulated |

| | | | |
|---|---|---|---|
| 0 | KRTAP5-8 | FALSE | Upregulated |
| -0,120833722 | KRTAP5-9 | FALSE | Downregulated |
| | KRTAP6-1 | | |
| | KRTAP6-2 | | |
| | KRTAP6-3 | | |
| | KRTAP7-1 | | |
| 0,114293466 | KRTAP8-1 | FALSE | Upregulated |
| | KRTAP9-1 | | |
| | KRTAP9-2 | | |
| | KRTAP9-3 | | |
| | KRTAP9-4 | | |
| | KRTAP9-6 | | |
| | KRTAP9-7 | | |
| | KRTAP9-8 | | |
| | KRTAP9-9 | | |
| 0 | KRTDAP | FALSE | Upregulated |
| 1,373896858 | KSR2 | | Upregulated |
| 0,039701882 | KTN1-AS1 | FALSE | Upregulated |
| 0,57389072 | L1CAM | FALSE | Upregulated |
| 0,479149142 | L1TD1 | FALSE | Upregulated |
| | LACRT | | |
| | LACTBL1 | | |
| 0,100713889 | LAIR2 | FALSE | Upregulated |
| | LALBA | | |
| 1,326960404 | LAMA1 | | Upregulated |
| 0,283489842 | LAMB2P1 | FALSE | Upregulated |
| 0,118308307 | LAMB4 | FALSE | Upregulated |
| 1,162223084 | LAMP3 | | Upregulated |
| 0,109498015 | LAMP5 | FALSE | Upregulated |
| -0,827719401 | LAMTOR5-AS1 | FALSE | Downregulated |
| 0,249722131 | LANCL3 | FALSE | Upregulated |
| 0,357327355 | LAYN | FALSE | Upregulated |
| 0,433782989 | LBX1 | FALSE | Upregulated |
| 0 | LBX1-AS1 | FALSE | Upregulated |
| 0,214480529 | LCA5 | FALSE | Upregulated |
| 0 | LCE1A | FALSE | Upregulated |
| | LCE1B | | |
| 0 | LCE1C | FALSE | Upregulated |
| 0 | LCE1D | FALSE | Upregulated |
| | LCE1E | | |
| | LCE1F | | |
| | LCE2A | | |
| 0 | LCE2B | FALSE | Upregulated |
| | LCE2C | | |
| 0 | LCE2D | FALSE | Upregulated |
| | LCE3A | | |
| | LCE3B | | |
| | LCE3C | | |
| | LCE3D | | |
| | LCE3E | | |
| | LCE4A | | |

| | | | |
|---|---|---|---|
| 0 | LCE5A | FALSE | Upregulated |
| | LCE6A | | |
| | LCN1 | | |
| -0,071948597 | LCN10 | FALSE | Downregulated |
| 0,441365315 | LCN15 | FALSE | Upregulated |
| 1,527207267 | LCN2 | | Upregulated |
| 0,406787727 | LCN6 | FALSE | Upregulated |
| | LCN8 | | |
| | LCN9 | | |
| 0,276340979 | LCNL1 | FALSE | Upregulated |
| -0,182863946 | LDHC | FALSE | Downregulated |
| 0,051043006 | LDLRAD1 | FALSE | Upregulated |
| -0,939685983 | LDOC1 | FALSE | Downregulated |
| 1,737470674 | LEF1-AS1 | | Upregulated |
| 0 | LEFTY2 | FALSE | Upregulated |
| | LELP1 | | |
| 0,006521199 | LEMD1 | FALSE | Upregulated |
| 0 | LEMD1-AS1 | FALSE | Upregulated |
| 0,307404653 | LEP | FALSE | Upregulated |
| | LEUTX | | |
| 0,527384605 | LGALS12 | FALSE | Upregulated |
| | LGALS13 | | |
| | LGALS14 | | |
| | LGALS16 | | |
| | LGALS17A | | |
| -0,903569623 | LGALS2 | FALSE | Downregulated |
| 0 | LGALS7 | FALSE | Upregulated |
| 0 | LGALS7B | FALSE | Upregulated |
| 0,437966603 | LGALS9B | FALSE | Upregulated |
| 0,245778845 | LGALS9C | FALSE | Upregulated |
| 0,082544477 | LGI1 | FALSE | Upregulated |
| 1,170780381 | LGI2 | | Upregulated |
| 0,533999912 | LGI3 | FALSE | Upregulated |
| 0,121505888 | LGR6 | FALSE | Upregulated |
| 0,252971461 | LHB | FALSE | Upregulated |
| 0,048276727 | LHFPL1 | FALSE | Upregulated |
| -0,110459992 | LHFPL3 | FALSE | Downregulated |
| | LHFPL3-AS1 | | |
| -0,000395795 | LHFPL3-AS2 | FALSE | Downregulated |
| -0,045460148 | LHFPL5 | FALSE | Downregulated |
| | LHX1 | | |
| 0,453099891 | LHX3 | FALSE | Upregulated |
| -0,054821256 | LHX5 | FALSE | Downregulated |
| -0,747010157 | LHX6 | FALSE | Downregulated |
| | LHX8 | | |
| 0 | LILRP2 | FALSE | Upregulated |
| 0,380774019 | LIM2 | FALSE | Upregulated |
| 0 | LIN28B | FALSE | Upregulated |
| 0,170116166 | LIN7B | FALSE | Upregulated |
| 0,397188812 | LINC-ROR | FALSE | Upregulated |
| | LINC00028 | | |

| | | | |
|---|---|---|---|
| | LINC00029 | | |
| 0,388189839 | LINC00032 | FALSE | Upregulated |
| 0,77374962 | LINC00051 | FALSE | Upregulated |
| 1,655284786 | LINC00052 | | Upregulated |
| 0 | LINC00083 | FALSE | Upregulated |
| -0,177279023 | LINC00086 | FALSE | Downregulated |
| | LINC00102-2 | | |
| | LINC00111 | | |
| 0,216326834 | LINC00112 | FALSE | Upregulated |
| 0,474080644 | LINC00113 | FALSE | Upregulated |
| | LINC00114 | | |
| 1,756199956 | LINC00152 | | Upregulated |
| 0 | LINC00158 | FALSE | Upregulated |
| | LINC00159 | | |
| | LINC00160 | | |
| 0 | LINC00162 | FALSE | Upregulated |
| 0,023327823 | LINC00163 | FALSE | Upregulated |
| 0,607989142 | LINC00173 | FALSE | Upregulated |
| 0 | LINC00184 | FALSE | Upregulated |
| -0,390481236 | LINC00189 | FALSE | Downregulated |
| 0,078076584 | LINC00200 | FALSE | Upregulated |
| 0,271122071 | LINC00202-2 | FALSE | Upregulated |
| | LINC00207 | | |
| 0,172282074 | LINC00208 | FALSE | Upregulated |
| | LINC00210 | | |
| 1,092036268 | LINC00222 | | Upregulated |
| | LINC00226 | | |
| | LINC00229 | | |
| -0,224006491 | LINC00235 | FALSE | Downregulated |
| 0,678035441 | LINC00238 | FALSE | Upregulated |
| -0,136986812 | LINC00239 | FALSE | Downregulated |
| -0,147953542 | LINC00240 | FALSE | Downregulated |
| -0,174641557 | LINC00243 | FALSE | Downregulated |
| 0,212240224 | LINC00251 | FALSE | Upregulated |
| 0,001790571 | LINC00254 | FALSE | Upregulated |
| 1,026750034 | LINC00260 | | Upregulated |
| 0,080855109 | LINC00264 | FALSE | Upregulated |
| -0,314892279 | LINC00265 | FALSE | Downregulated |
| -0,750043683 | LINC00266-1 | FALSE | Downregulated |
| 0,29877405 | LINC00266-3 | FALSE | Upregulated |
| | LINC00269 | | |
| 0,108966091 | LINC00271 | FALSE | Upregulated |
| 0,448212425 | LINC00272 | FALSE | Upregulated |
| | LINC00273 | | |
| | LINC00276 | | |
| 0 | LINC00277 | FALSE | Upregulated |
| 0,707086313 | LINC00278 | FALSE | Upregulated |
| | LINC00284 | | |
| | LINC00290 | | |
| | LINC00293 | | |
| 0 | LINC00297 | FALSE | Upregulated |

| | | | |
|---|---|---|---|
| 0 | LINC00298 | FALSE | Upregulated |
| -0,220544241 | LINC00299 | FALSE | Downregulated |
| | LINC00301 | | |
| 0,390950345 | LINC00303 | FALSE | Upregulated |
| | LINC00304 | | |
| 0 | LINC00305 | FALSE | Upregulated |
| | LINC00307 | | |
| | LINC00308 | | |
| 0,288974728 | LINC00309 | FALSE | Upregulated |
| 0,051548537 | LINC00310 | FALSE | Upregulated |
| -2,032539807 | LINC00312 | | Downregulated |
| 1,311390405 | LINC00313 | | Upregulated |
| 0,331746789 | LINC00316 | FALSE | Upregulated |
| | LINC00317 | | |
| | LINC00320 | | |
| | LINC00322 | | |
| | LINC00323 | | |
| | LINC00326 | | |
| 0 | LINC00327 | FALSE | Upregulated |
| 0,100713889 | LINC00330 | FALSE | Upregulated |
| | LINC00331 | | |
| | LINC00333 | | |
| 0 | LINC00336 | FALSE | Upregulated |
| -0,006598336 | LINC00337 | FALSE | Downregulated |
| | LINC00343 | | |
| | LINC00344 | | |
| -0,073468397 | LINC00347 | FALSE | Downregulated |
| | LINC00348 | | |
| | LINC00351 | | |
| | LINC00358 | | |
| | LINC00359 | | |
| 0,033128344 | LINC00366 | FALSE | Upregulated |
| | LINC00367 | | |
| 0,100713889 | LINC00371 | FALSE | Upregulated |
| | LINC00378 | | |
| | LINC00379 | | |
| | LINC00380 | | |
| | LINC00381 | | |
| 0 | LINC00384 | FALSE | Upregulated |
| | LINC00395 | | |
| | LINC00397 | | |
| -0,32406705 | LINC00398 | FALSE | Downregulated |
| | LINC00403 | | |
| | LINC00408 | | |
| 0 | LINC00410 | FALSE | Upregulated |
| | LINC00417 | | |
| | LINC00421 | | |
| 0,068128077 | LINC00423 | FALSE | Upregulated |
| | LINC00424 | | |
| 1,2243619 | LINC00426 | | Upregulated |
| | LINC00433 | | |

| | | | |
|---|---|---|---|
| 0,233308128 | LINC00441 | FALSE | Upregulated |
| | LINC00442 | | |
| | LINC00443 | | |
| | LINC00445 | | |
| | LINC00446 | | |
| 1,53456453 | LINC00452 | | Upregulated |
| | LINC00454 | | |
| 0 | LINC00457 | FALSE | Upregulated |
| | LINC00458 | | |
| | LINC00460 | | |
| | LINC00461 | | |
| 0 | LINC00462 | FALSE | Upregulated |
| | LINC00466 | | |
| -0,173910785 | LINC00467 | FALSE | Downregulated |
| | LINC00469 | | |
| 0,377897344 | LINC00470 | FALSE | Upregulated |
| 0,558668125 | LINC00471 | FALSE | Upregulated |
| | LINC00473 | | |
| | LINC00474 | | |
| 0,30347676 | LINC00475 | FALSE | Upregulated |
| 0 | LINC00477 | FALSE | Upregulated |
| 0,286337239 | LINC00479 | FALSE | Upregulated |
| -0,044681968 | LINC00483 | FALSE | Downregulated |
| 0 | LINC00486 | FALSE | Upregulated |
| -0,022979945 | LINC00487 | FALSE | Downregulated |
| | LINC00488 | | |
| | LINC00489 | | |
| | LINC00491 | | |
| | LINC00492 | | |
| -0,009346983 | LINC00494 | FALSE | Downregulated |
| | LINC00499 | | |
| -0,049412723 | LINC00501 | FALSE | Downregulated |
| | LINC00502 | | |
| | LINC00507 | | |
| | LINC00511 | | |
| 1,227273928 | LINC00515 | | Upregulated |
| 0,581614473 | LINC00518 | FALSE | Upregulated |
| | LINC00521 | | |
| 0 | LINC00523 | FALSE | Upregulated |
| -1,661080628 | LINC00525 | | Downregulated |
| 0 | LINC00534 | FALSE | Upregulated |
| | LINC00536 | | |
| -0,315234845 | LINC00540 | FALSE | Downregulated |
| | LINC00544 | | |
| | LINC00545 | | |
| 0,201726435 | LINC00547 | FALSE | Upregulated |
| 1,815374404 | LINC00548 | | Upregulated |
| | LINC00550 | | |
| | LINC00551 | | |
| -0,186818216 | LINC00552 | FALSE | Downregulated |
| | LINC00554 | | |

| | | | |
|---|---|---|---|
| | LINC00558 | | |
| 0,701957856 | LINC00559 | FALSE | Upregulated |
| | LINC00560 | | |
| 0,394163909 | LINC00565 | FALSE | Upregulated |
| -0,289683864 | LINC00568 | FALSE | Downregulated |
| 0,504036439 | LINC00570 | FALSE | Upregulated |
| -0,019615572 | LINC00571 | FALSE | Downregulated |
| -0,235601844 | LINC00572 | FALSE | Downregulated |
| | LINC00575 | | |
| | LINC00577 | | |
| | LINC00578 | | |
| | LINC00581 | | |
| | LINC00583 | | |
| | LINC00587 | | |
| | LINC00588 | | |
| | LINC00589 | | |
| | LINC00592 | | |
| 0 | LINC00593 | FALSE | Upregulated |
| 0,047839331 | LINC00595 | FALSE | Upregulated |
| 0,890257814 | LINC00597 | FALSE | Upregulated |
| | LINC00599 | | |
| 0,033128344 | LINC00601 | FALSE | Upregulated |
| 0 | LINC00602 | FALSE | Upregulated |
| | LINC00603 | | |
| 0,100562008 | LINC00605 | FALSE | Upregulated |
| | LINC00606 | | |
| 0,508126497 | LINC00607 | FALSE | Upregulated |
| | LINC00608 | | |
| | LINC00609 | | |
| | LINC00613 | | |
| | LINC00615 | | |
| | LINC00616 | | |
| 0 | LINC00617 | FALSE | Upregulated |
| | LINC00618 | | |
| | LINC00619 | | |
| | LINC00620 | | |
| -0,690120119 | LINC00622 | FALSE | Downregulated |
| 0,820539834 | LINC00623-2 | FALSE | Upregulated |
| 0,224782169 | LINC00624 | FALSE | Upregulated |
| 0,562891403 | LINC00626 | FALSE | Upregulated |
| -0,051522294 | LINC00628 | FALSE | Downregulated |
| 0 | LINC00629 | FALSE | Upregulated |
| 0,493605298 | LINC00632 | FALSE | Upregulated |
| 0 | LINC00633 | FALSE | Upregulated |
| | LINC00634 | | |
| 0 | LINC00635 | FALSE | Upregulated |
| | LINC00636 | | |
| | LINC00640 | | |
| | LINC00642 | | |
| -0,016434257 | LINC00643 | FALSE | Downregulated |
| | LINC00644 | | |

| | | | |
|---|---|---|---|
| 0,228196273 | LINC00645 | FALSE | Upregulated |
| 0,094615407 | LINC00648 | FALSE | Upregulated |
| -0,031013215 | LINC00656 | FALSE | Downregulated |
| -0,223061762 | LINC00658 | FALSE | Downregulated |
| -0,021810926 | LINC00661 | FALSE | Downregulated |
| 0,580614572 | LINC00664 | FALSE | Upregulated |
| | LINC00668 | | |
| | LINC00669 | | |
| 0 | LINC00670 | FALSE | Upregulated |
| -0,028589076 | LINC00672 | FALSE | Downregulated |
| 0,949466511 | LINC00673 | FALSE | Upregulated |
| -0,328955281 | LINC00676 | FALSE | Downregulated |
| | LINC00681 | | |
| | LINC00682 | | |
| 0,388614369 | LINC00684 | FALSE | Upregulated |
| | LINC00684-2 | | |
| | LINC00685-2 | | |
| | LINC00687 | | |
| -0,757111377 | LINC00689 | FALSE | Downregulated |
| 0,406787727 | LINC00690 | FALSE | Upregulated |
| | LINC00692 | | |
| -0,715575558 | LINC00693 | FALSE | Downregulated |
| 0,06135006 | LINC00696 | FALSE | Upregulated |
| | LINC00698 | | |
| 0,100713889 | LINC00700 | FALSE | Upregulated |
| | LINC00701 | | |
| 0 | LINC00702 | FALSE | Upregulated |
| | LINC00703 | | |
| 0,763824994 | LINC00704 | FALSE | Upregulated |
| -0,336974574 | LINC00705 | FALSE | Downregulated |
| 0,036683546 | LINC00706 | FALSE | Upregulated |
| 0 | LINC00708 | FALSE | Upregulated |
| | LINC00709 | | |
| -0,160657373 | LINC00710 | FALSE | Downregulated |
| | LINC00836 | | |
| -0,044093552 | LINC00837 | FALSE | Downregulated |
| 0,157487943 | LINC00838 | FALSE | Upregulated |
| 0,419750001 | LINC00839 | FALSE | Upregulated |
| | LINC00840 | | |
| | LINC00841 | | |
| -0,181442647 | LINC00842 | FALSE | Downregulated |
| | LINC00845 | | |
| | LINC00850 | | |
| 0,099452879 | LINC00851 | FALSE | Upregulated |
| 0 | LINC00853 | FALSE | Upregulated |
| | LINC00856 | | |
| 1,030588769 | LINC00857 | | Upregulated |
| | LINC00858 | | |
| -0,127727684 | LINC00862 | FALSE | Downregulated |
| -0,459046932 | LINC00864 | FALSE | Downregulated |
| -0,578492066 | LINC00865 | FALSE | Downregulated |

| | | | |
|---|---|---|---|
| 0 | LINC00866 | FALSE | Upregulated |
| 0 | LINC00867 | FALSE | Upregulated |
| | LINC00871 | | |
| 0,03509905 | LINC00877 | FALSE | Upregulated |
| | LINC00879 | | |
| 0,179257113 | LINC00881 | FALSE | Upregulated |
| 0,011211767 | LINC00882 | FALSE | Upregulated |
| 0,144876206 | LINC00887 | FALSE | Upregulated |
| 1,078054945 | LINC00888 | | Upregulated |
| 0 | LINC00889 | FALSE | Upregulated |
| 0,080868669 | LINC00891 | FALSE | Upregulated |
| 0,573639285 | LINC00892 | FALSE | Upregulated |
| | LINC00895 | | |
| | LINC00898 | | |
| 0,119766416 | LINC00901 | FALSE | Upregulated |
| | LINC00904 | | |
| | LINC00905 | | |
| 0 | LINC00906 | FALSE | Upregulated |
| 0,066672251 | LINC00908 | FALSE | Upregulated |
| | LINC00911 | | |
| | LINC00917 | | |
| | LINC00919 | | |
| -0,044254879 | LINC00920 | FALSE | Downregulated |
| | LINC00922 | | |
| 0,191844754 | LINC00923 | FALSE | Upregulated |
| 1,578303148 | LINC00924 | | Upregulated |
| -0,63430065 | LINC00925 | FALSE | Downregulated |
| -0,503708019 | LINC00926 | FALSE | Downregulated |
| | LINC00927 | | |
| 0,455206391 | LINC00928 | FALSE | Upregulated |
| | LINC00929 | | |
| -0,110939333 | LINC00930 | FALSE | Downregulated |
| 0 | LINC00934 | FALSE | Upregulated |
| -0,192393293 | LINC00935 | FALSE | Downregulated |
| 0,193223511 | LINC00937 | FALSE | Upregulated |
| -0,964074213 | LINC00940 | FALSE | Downregulated |
| 0,043804698 | LINC00942 | FALSE | Upregulated |
| | LINC00944 | | |
| 0,300764852 | LINC00945 | FALSE | Upregulated |
| 0,100713889 | LINC00948 | FALSE | Upregulated |
| | LINC00951 | | |
| | LINC00955 | | |
| 0,407290161 | LINC00958 | FALSE | Upregulated |
| 0,248925056 | LINC00959 | FALSE | Upregulated |
| 0,076428257 | LINC00964 | FALSE | Upregulated |
| -0,398487069 | LINC00965 | FALSE | Downregulated |
| 0 | LINC00968 | FALSE | Upregulated |
| -0,054696575 | LINC00969 | FALSE | Downregulated |
| 0 | LINC00970 | FALSE | Upregulated |
| -0,01636575 | LINC00971 | FALSE | Downregulated |
| | LINC00974 | | |

| | | | |
|---|---|---|---|
| 0 | LINC00977 | FALSE | Upregulated |
| 0,360103896 | LINC00982 | FALSE | Upregulated |
| 0 | LINC00989 | FALSE | Upregulated |
| | LINC00992 | | |
| | LINC00993 | | |
| | LINC00994 | | |
| 0,964354954 | LINC00996 | FALSE | Upregulated |
| 0,424766175 | LINC01001 | FALSE | Upregulated |
| 0 | LINC01002 | FALSE | Upregulated |
| | LINC01005 | | |
| -1,427589656 | LINC01006 | | Downregulated |
| -1,350020244 | LINC01007 | | Downregulated |
| -0,163991406 | LINC01012 | FALSE | Downregulated |
| | LINC01013 | | |
| -0,017135352 | LINC01014 | FALSE | Downregulated |
| | LINC01015 | | |
| 0,203134476 | LINC01016 | FALSE | Upregulated |
| | LINC01019 | | |
| | LINC01020 | | |
| 0 | LINC01021 | FALSE | Upregulated |
| 1,204396947 | LINC01023 | | Upregulated |
| 0 | LINC01028 | FALSE | Upregulated |
| | LINC01029 | | |
| -0,204554684 | LINC01030 | FALSE | Downregulated |
| | LINC01056 | | |
| -0,148730887 | LINC01057 | FALSE | Downregulated |
| | LINC01060 | | |
| 0,100713889 | LINC01063 | FALSE | Upregulated |
| -0,439980502 | LINC01069 | FALSE | Downregulated |
| -0,488490091 | LINC01070 | FALSE | Downregulated |
| -0,927885241 | LINC01071 | FALSE | Downregulated |
| 1,427667446 | LINC01080 | | Upregulated |
| | LINC01081 | | |
| 0,260482352 | LINC01082 | FALSE | Upregulated |
| | LINC01085 | | |
| 1,1228702 | LINC01087 | | Upregulated |
| 0 | LINC01088 | FALSE | Upregulated |
| 0,451775535 | LINC01091 | FALSE | Upregulated |
| -0,390259212 | LINC01094 | FALSE | Downregulated |
| | LINC01095 | | |
| 0 | LINC01096 | FALSE | Upregulated |
| | LINC01097 | | |
| 0,048276727 | LINC01098 | FALSE | Upregulated |
| 0 | LINC01099 | FALSE | Upregulated |
| | LINC01100 | | |
| 0,603975082 | LINC01101 | FALSE | Upregulated |
| 0,465237344 | LINC01102 | FALSE | Upregulated |
| | LINC01103 | | |
| 0,448212425 | LINC01104 | FALSE | Upregulated |
| | LINC01105 | | |
| | LINC01111 | | |

| | | | |
|---|---|---|---|
| | LINC01114 | | |
| 0 | LINC01115 | FALSE | Upregulated |
| 0,402756206 | LINC01116 | FALSE | Upregulated |
| -0,288421918 | LINC01118 | FALSE | Downregulated |
| 0,138154281 | LINC01119 | FALSE | Upregulated |
| | LINC01120 | | |
| 0,010932239 | LINC01121 | FALSE | Upregulated |
| | LINC01122 | | |
| -0,366298289 | LINC01125 | FALSE | Downregulated |
| -0,186493317 | LINC01132 | FALSE | Downregulated |
| 0,456042235 | LINC01134 | FALSE | Upregulated |
| -0,237043404 | LINC01135 | FALSE | Downregulated |
| | LINC01136 | | |
| 0,448212425 | LINC01139 | FALSE | Upregulated |
| 0,147444475 | LINC01140 | FALSE | Upregulated |
| 0,100713889 | LINC01141 | FALSE | Upregulated |
| | LINC01142 | | |
| 0 | LINC01149 | FALSE | Upregulated |
| | LINC01150 | | |
| -0,401028339 | LINC01153 | FALSE | Downregulated |
| | LINC01158 | | |
| 0,399828724 | LINC01160 | FALSE | Upregulated |
| | LINC01163 | | |
| | LINC01164 | | |
| 0,948584899 | LINC01165 | FALSE | Upregulated |
| 0,048276727 | LINC01171 | FALSE | Upregulated |
| -0,955380594 | LINC01176 | FALSE | Downregulated |
| 0,070401644 | LINC01179 | FALSE | Upregulated |
| | LINC01180 | | |
| | LINC01182 | | |
| 0,508496845 | LINC01183 | FALSE | Upregulated |
| 0,458886499 | LINC01185 | FALSE | Upregulated |
| 0,448212425 | LINC01186 | FALSE | Upregulated |
| 0,009212607 | LINC01187 | FALSE | Upregulated |
| -0,133948481 | LINC01189 | FALSE | Downregulated |
| 0,448647563 | LINC01191 | FALSE | Upregulated |
| | LINC01192 | | |
| 0 | LINC01193 | FALSE | Upregulated |
| 0 | LINC01194 | FALSE | Upregulated |
| 0,433185006 | LINC01197 | FALSE | Upregulated |
| | LINC01216 | | |
| 1,076750576 | LINGO2 | | Upregulated |
| 0,782219138 | LIPE-AS1 | FALSE | Upregulated |
| | LIPF | | |
| -0,410892716 | LIPI | FALSE | Downregulated |
| -0,095746872 | LIPJ | FALSE | Downregulated |
| | LIPK | | |
| 1,737889779 | LIPM | | Upregulated |
| 0,358624888 | LIPN | FALSE | Upregulated |
| | LL22NC01-81G9.3 | | |
| | LMAN1L | | |

| | | | |
|---|---|---|---|
| -0,687795139 | LMCD1-AS1 | FALSE | Downregulated |
| 0 | LMO1 | FALSE | Upregulated |
| 0 | LMX1A | FALSE | Upregulated |
| 0,112060207 | LMX1B | FALSE | Upregulated |
| 0,328331498 | LNX1-AS1 | FALSE | Upregulated |
| 1,212915821 | LNX1-AS2 | | Upregulated |
| | LOC100125918 | | |
| 0,15230243 | LOC100126447 | FALSE | Upregulated |
| 0,340118147 | LOC100127974 | FALSE | Upregulated |
| 0,001597502 | LOC100128002 | FALSE | Upregulated |
| 0 | LOC100128076 | FALSE | Upregulated |
| 0 | LOC100128108 | FALSE | Upregulated |
| 0,135297238 | LOC100128127 | FALSE | Upregulated |
| | LOC100128139 | | |
| -0,106569921 | LOC100128164 | FALSE | Downregulated |
| | LOC100128233 | | |
| 0,470928142 | LOC100128276 | FALSE | Upregulated |
| -0,875471031 | LOC100128288 | FALSE | Downregulated |
| 0 | LOC100128325 | FALSE | Upregulated |
| -0,507635694 | LOC100128560 | FALSE | Downregulated |
| 0,095132412 | LOC100128568 | FALSE | Upregulated |
| | LOC100128575 | | |
| 0,349717391 | LOC100128593 | FALSE | Upregulated |
| 0,047839331 | LOC100128885 | FALSE | Upregulated |
| 0,489639432 | LOC100128905 | FALSE | Upregulated |
| 0,045511312 | LOC100128908 | FALSE | Upregulated |
| 0,040817358 | LOC100128966 | FALSE | Upregulated |
| 0 | LOC100129055 | FALSE | Upregulated |
| 0,059364086 | LOC100129098 | FALSE | Upregulated |
| 0,283972127 | LOC100129138 | FALSE | Upregulated |
| 0,449808511 | LOC100129216 | FALSE | Upregulated |
| 0 | LOC100129223 | FALSE | Upregulated |
| -0,246765556 | LOC100129307 | FALSE | Downregulated |
| 0,641970584 | LOC100129316 | FALSE | Upregulated |
| 0,100713889 | LOC100129455 | FALSE | Upregulated |
| 0,270177977 | LOC100129461 | FALSE | Upregulated |
| 0,544577245 | LOC100129476 | FALSE | Upregulated |
| 0,272578967 | LOC100129516 | FALSE | Upregulated |
| | LOC100129520 | | |
| | LOC100129603 | | |
| | LOC100129636 | | |
| -0,326743372 | LOC100129781 | FALSE | Downregulated |
| 0 | LOC100129860 | FALSE | Upregulated |
| -0,019980256 | LOC100129924 | FALSE | Downregulated |
| 0,068128077 | LOC100129935 | FALSE | Upregulated |
| 0 | LOC100130207 | FALSE | Upregulated |
| 0,208692683 | LOC100130298 | FALSE | Upregulated |
| | LOC100130301 | | |
| 0 | LOC100130417 | FALSE | Upregulated |
| -0,045460148 | LOC100130452 | FALSE | Downregulated |
| -0,058842351 | LOC100130458 | FALSE | Downregulated |

| | | | |
|---|---|---|---|
| 0 | LOC100130502 | FALSE | Upregulated |
| 0,492964015 | LOC100130520 | FALSE | Upregulated |
| -0,009388405 | LOC100130673 | FALSE | Downregulated |
| -0,035694244 | LOC100130698 | FALSE | Downregulated |
| | LOC100130849 | | |
| | LOC100130938 | | |
| | LOC100130964 | | |
| 0 | LOC100131138 | FALSE | Upregulated |
| 0 | LOC100131289 | FALSE | Upregulated |
| -0,294161343 | LOC100131303 | FALSE | Downregulated |
| -0,108094793 | LOC100131315 | FALSE | Downregulated |
| 0,70484267 | LOC100131372 | FALSE | Upregulated |
| 0,11171199 | LOC100131472 | FALSE | Upregulated |
| 0,568706958 | LOC100131530 | FALSE | Upregulated |
| -1,482732902 | LOC100131532 | | Downregulated |
| | LOC100131635 | | |
| 0,076221525 | LOC100131655 | FALSE | Upregulated |
| 0,465845006 | LOC100132004 | FALSE | Upregulated |
| | LOC100132146 | | |
| 0,069188854 | LOC100132154 | FALSE | Upregulated |
| | LOC100132174 | | |
| | LOC100132202 | | |
| 0,59274944 | LOC100132354 | FALSE | Upregulated |
| -0,020263018 | LOC100132735 | FALSE | Downregulated |
| 0,080868669 | LOC100132741 | FALSE | Upregulated |
| | LOC100132831 | | |
| 0 | LOC100132874 | FALSE | Upregulated |
| -0,017382664 | LOC100133050 | FALSE | Downregulated |
| | LOC100133267 | | |
| -0,934894457 | LOC100133315 | FALSE | Downregulated |
| -0,426851377 | LOC100133669 | FALSE | Downregulated |
| 0,301853486 | LOC100133746 | FALSE | Upregulated |
| -0,024722478 | LOC100133985 | FALSE | Downregulated |
| 0,505792733 | LOC100134317 | FALSE | Upregulated |
| -0,937390813 | LOC100134368 | FALSE | Downregulated |
| -0,046140311 | LOC100134391 | FALSE | Downregulated |
| 0,015485442 | LOC100134868 | FALSE | Upregulated |
| -0,14829313 | LOC100188947 | FALSE | Downregulated |
| | LOC100240728 | | |
| 0 | LOC100240734 | FALSE | Upregulated |
| 0,197202605 | LOC100240735 | FALSE | Upregulated |
| 0,583170165 | LOC100270804 | FALSE | Upregulated |
| | LOC100286906 | | |
| -0,339967383 | LOC100287010 | FALSE | Downregulated |
| | LOC100287072 | | |
| -0,455664984 | LOC100287082 | FALSE | Downregulated |
| -0,107121679 | LOC100287098 | FALSE | Downregulated |
| | LOC100287294 | | |
| -0,135607094 | LOC100287387 | FALSE | Downregulated |
| 0 | LOC100287413 | FALSE | Upregulated |
| | LOC100287592 | | |

| | | | |
|---|---|---|---|
| 0,094615407 | LOC100287610 | FALSE | Upregulated |
| -0,010122896 | LOC100287651 | FALSE | Downregulated |
| | LOC100287704 | | |
| | LOC100287728 | | |
| | LOC100287823 | | |
| | LOC100287834 | | |
| | LOC100287877 | | |
| 0,984050421 | LOC100287944 | FALSE | Upregulated |
| 1,324706814 | LOC100288175 | | Upregulated |
| 0 | LOC100288562 | FALSE | Upregulated |
| 0 | LOC100288570 | FALSE | Upregulated |
| 1,268123504 | LOC100288637 | | Upregulated |
| 0 | LOC100288748 | FALSE | Upregulated |
| -0,082962276 | LOC100288814 | FALSE | Downregulated |
| 0,09111031 | LOC100288842 | FALSE | Upregulated |
| -0,074665626 | LOC100288911 | FALSE | Downregulated |
| | LOC100288966 | | |
| -0,654741953 | LOC100288974 | FALSE | Downregulated |
| -0,246737129 | LOC100289092 | FALSE | Downregulated |
| 0,053250412 | LOC100289098 | FALSE | Upregulated |
| 0 | LOC100289333 | FALSE | Upregulated |
| 0,033128344 | LOC100289473 | FALSE | Upregulated |
| 0,366717607 | LOC100289495 | FALSE | Upregulated |
| 0,450737483 | LOC100289656 | FALSE | Upregulated |
| -0,414267818 | LOC100289673 | FALSE | Downregulated |
| | LOC100293612 | | |
| 0,871487948 | LOC100335030 | FALSE | Upregulated |
| | LOC100422212 | | |
| 0,897908626 | LOC100422556 | FALSE | Upregulated |
| 0,029670605 | LOC100500773 | FALSE | Upregulated |
| 0 | LOC100505498 | FALSE | Upregulated |
| 0,201859774 | LOC100505501 | FALSE | Upregulated |
| | LOC100505504 | | |
| 0 | LOC100505515 | FALSE | Upregulated |
| 0,146690165 | LOC100505534 | FALSE | Upregulated |
| 0,614787293 | LOC100505555 | FALSE | Upregulated |
| 1,018478214 | LOC100505585 | | Upregulated |
| -0,76057055 | LOC100505588 | FALSE | Downregulated |
| 0 | LOC100505609 | FALSE | Upregulated |
| 0,395239572 | LOC100505622 | FALSE | Upregulated |
| 0,553037954 | LOC100505650 | FALSE | Upregulated |
| 0 | LOC100505664 | FALSE | Upregulated |
| 0,386400182 | LOC100505685 | FALSE | Upregulated |
| 0,56196501 | LOC100505710 | FALSE | Upregulated |
| | LOC100505711 | | |
| -0,177278143 | LOC100505774 | FALSE | Downregulated |
| | LOC100505784 | | |
| | LOC100505797 | | |
| 0,754799678 | LOC100505798 | FALSE | Upregulated |
| | LOC100505811 | | |
| -0,074744589 | LOC100505812 | FALSE | Downregulated |

| | | | |
|---|---|---|---|
| -0,130877405 | LOC100505817 | FALSE | Downregulated |
| 0 | LOC100505824 | FALSE | Upregulated |
| | LOC100505835 | | |
| | LOC100505841 | | |
| | LOC100505853 | | |
| | LOC100505874 | | |
| | LOC100505878 | | |
| | LOC100505887 | | |
| -0,880813388 | LOC100505902 | FALSE | Downregulated |
| -0,00514207 | LOC100505912 | FALSE | Downregulated |
| | LOC100505920 | | |
| | LOC100505940 | | |
| | LOC100505948 | | |
| | LOC100505984 | | |
| 0,055590857 | LOC100506014 | FALSE | Upregulated |
| | LOC100506016 | | |
| -0,393118032 | LOC100506022 | FALSE | Downregulated |
| -0,89880098 | LOC100506036 | FALSE | Downregulated |
| -0,585839932 | LOC100506047 | FALSE | Downregulated |
| 0,046678197 | LOC100506082 | FALSE | Upregulated |
| | LOC100506098 | | |
| | LOC100506107 | | |
| -0,935763654 | LOC100506124 | FALSE | Downregulated |
| -0,221635138 | LOC100506127 | FALSE | Downregulated |
| -0,129819475 | LOC100506157 | FALSE | Downregulated |
| 0 | LOC100506159 | FALSE | Upregulated |
| 0,543976112 | LOC100506188 | FALSE | Upregulated |
| 0,463046596 | LOC100506235 | FALSE | Upregulated |
| | LOC100506236 | | |
| | LOC100506253 | | |
| | LOC100506271 | | |
| | LOC100506274 | | |
| 0,393002133 | LOC100506282 | FALSE | Upregulated |
| | LOC100506289 | | |
| | LOC100506317 | | |
| 0,50538557 | LOC100506331 | FALSE | Upregulated |
| 1,131096663 | LOC100506351 | | Upregulated |
| -0,835484231 | LOC100506358 | FALSE | Downregulated |
| 0,531870374 | LOC100506371 | FALSE | Upregulated |
| -0,573303089 | LOC100506384 | FALSE | Downregulated |
| | LOC100506403 | | |
| | LOC100506405 | | |
| | LOC100506406 | | |
| -0,316611821 | LOC100506411 | FALSE | Downregulated |
| 0,465237344 | LOC100506422 | FALSE | Upregulated |
| | LOC100506470 | | |
| 0,592767318 | LOC100506473 | FALSE | Upregulated |
| 0,0423658 | LOC100506474 | FALSE | Upregulated |
| -0,471155096 | LOC100506489 | FALSE | Downregulated |
| -1,139748898 | LOC100506498 | | Downregulated |
| 0,635233351 | LOC100506526 | FALSE | Upregulated |

| | | | |
|---|---|---|---|
| 0,448212425 | LOC100506530 | FALSE | Upregulated |
| | LOC100506532 | | |
| 0,102083717 | LOC100506557 | FALSE | Upregulated |
| 0 | LOC100506558 | FALSE | Upregulated |
| 0,150999812 | LOC100506563 | FALSE | Upregulated |
| | LOC100506664 | | |
| | LOC100506675 | | |
| -0,209210154 | LOC100506688 | FALSE | Downregulated |
| -0,619995854 | LOC100506691 | FALSE | Downregulated |
| -0,237282501 | LOC100506713 | FALSE | Downregulated |
| 0,517353039 | LOC100506714 | FALSE | Upregulated |
| -1,56288682 | LOC100506746 | | Downregulated |
| -0,218612789 | LOC100506747 | FALSE | Downregulated |
| -0,669203724 | LOC100506753 | FALSE | Downregulated |
| | LOC100506790 | | |
| | LOC100506791 | | |
| 0,281159454 | LOC100506797 | FALSE | Upregulated |
| -0,102132881 | LOC100506827 | FALSE | Downregulated |
| | LOC100506851 | | |
| | LOC100506858 | | |
| 0,366060677 | LOC100506869 | FALSE | Upregulated |
| -0,38837589 | LOC100506870 | FALSE | Downregulated |
| 0 | LOC100506882 | FALSE | Upregulated |
| | LOC100506885 | | |
| 1,10451807 | LOC100506901 | | Upregulated |
| -0,052830528 | LOC100506908 | FALSE | Downregulated |
| 0 | LOC100506929 | FALSE | Upregulated |
| 0 | LOC100506937 | FALSE | Upregulated |
| | LOC100506944 | | |
| -0,243730142 | LOC100506974 | FALSE | Downregulated |
| 0 | LOC100506999 | FALSE | Upregulated |
| 0,066817203 | LOC100507054 | FALSE | Upregulated |
| 0,760879093 | LOC100507103 | FALSE | Upregulated |
| | LOC100507165 | | |
| 0,02293814 | LOC100507195 | FALSE | Upregulated |
| | LOC100507205 | | |
| | LOC100507210 | | |
| | LOC100507221 | | |
| | LOC100507236 | | |
| 0,064714915 | LOC100507277 | FALSE | Upregulated |
| 0,971753924 | LOC100507316 | FALSE | Upregulated |
| 0,390961325 | LOC100507336 | FALSE | Upregulated |
| 0,021228839 | LOC100507351 | FALSE | Upregulated |
| | LOC100507353 | | |
| | LOC100507384 | | |
| 0 | LOC100507387 | FALSE | Upregulated |
| -1,280911807 | LOC100507395 | | Downregulated |
| | LOC100507403 | | |
| | LOC100507420 | | |
| | LOC100507443 | | |
| 0,718317788 | LOC100507460 | FALSE | Upregulated |

| | | | |
|---|---|---|---|
| | LOC100507461 | | |
| -0,329453638 | LOC100507462 | FALSE | Downregulated |
| | LOC100507464 | | |
| | LOC100507468 | | |
| 0 | LOC100507480 | FALSE | Upregulated |
| 0,382648241 | LOC100507487 | FALSE | Upregulated |
| -0,320699424 | LOC100507513 | FALSE | Downregulated |
| | LOC100507521 | | |
| 1,148680499 | LOC100507534 | | Upregulated |
| 0,046678197 | LOC100507548 | FALSE | Upregulated |
| -0,293935572 | LOC100507554 | FALSE | Downregulated |
| | LOC100507560 | | |
| 0 | LOC100507562 | FALSE | Upregulated |
| 1,661913168 | LOC100507600 | | Upregulated |
| 1,688506853 | LOC100507634 | | Upregulated |
| -0,48244438 | LOC100507642 | FALSE | Downregulated |
| 0,646499101 | LOC100507657 | FALSE | Upregulated |
| 0,033666535 | LOC100507747 | FALSE | Upregulated |
| 0 | LOC100508046 | FALSE | Upregulated |
| 0 | LOC100508631 | FALSE | Upregulated |
| | LOC100509303 | | |
| -1,22044736 | LOC100509620 | | Downregulated |
| 0,938680091 | LOC100510707 | FALSE | Upregulated |
| -0,140813085 | LOC100616530 | FALSE | Downregulated |
| 1,0160353 | LOC100652777 | | Upregulated |
| 1,132638444 | LOC100652807 | | Upregulated |
| -0,282252482 | LOC100652911 | FALSE | Downregulated |
| -0,260290159 | LOC100652930 | FALSE | Downregulated |
| | LOC100652931 | | |
| -0,60207833 | LOC100653005 | FALSE | Downregulated |
| 0 | LOC100653133 | FALSE | Upregulated |
| 0,568109114 | LOC100996246 | FALSE | Upregulated |
| | LOC100996249 | | |
| | LOC100996266 | | |
| -0,012105588 | LOC100996274 | FALSE | Downregulated |
| -0,753922835 | LOC100996288 | FALSE | Downregulated |
| | LOC100996290 | | |
| 0,168303385 | LOC100996294 | FALSE | Upregulated |
| 0 | LOC100996301 | FALSE | Upregulated |
| -0,413062756 | LOC100996316 | FALSE | Downregulated |
| -0,021553912 | LOC100996335 | FALSE | Downregulated |
| 0,596578244 | LOC100996337 | FALSE | Upregulated |
| | LOC100996338 | | |
| -0,551265825 | LOC100996342 | FALSE | Downregulated |
| 0,651449745 | LOC100996345 | FALSE | Upregulated |
| | LOC100996349 | | |
| | LOC100996365 | | |
| 0,616365949 | LOC100996369 | FALSE | Upregulated |
| -0,869378873 | LOC100996379 | FALSE | Downregulated |
| 0,533875346 | LOC100996385 | FALSE | Upregulated |
| 0,255645866 | LOC100996401 | FALSE | Upregulated |

| | | | |
|---|---|---|---|
| 0,121447981 | LOC100996404 | FALSE | Upregulated |
| | LOC100996414 | | |
| | LOC100996415 | | |
| 0,103239985 | LOC100996425 | FALSE | Upregulated |
| 0 | LOC100996428 | FALSE | Upregulated |
| 0,514947092 | LOC100996442 | FALSE | Upregulated |
| 0,534781983 | LOC100996455 | FALSE | Upregulated |
| -0,025423627 | LOC100996457 | FALSE | Downregulated |
| -0,265959086 | LOC100996465 | FALSE | Downregulated |
| 1,423253387 | LOC100996478 | | Upregulated |
| 0 | LOC100996482 | FALSE | Upregulated |
| -0,001172311 | LOC100996521 | FALSE | Downregulated |
| 0,114873054 | LOC100996549 | FALSE | Upregulated |
| 0,53906251 | LOC100996557 | FALSE | Upregulated |
| 0 | LOC100996570 | FALSE | Upregulated |
| 0,138064057 | LOC100996598 | FALSE | Upregulated |
| 1,54195097 | LOC100996602 | | Upregulated |
| 0,266840769 | LOC100996624 | FALSE | Upregulated |
| | LOC100996630 | | |
| 1,173937716 | LOC100996634 | | Upregulated |
| | LOC100996637 | | |
| | LOC100996645 | | |
| 0,025693334 | LOC100996660 | FALSE | Upregulated |
| -0,339520063 | LOC100996662 | FALSE | Downregulated |
| 0,486884909 | LOC100996663 | FALSE | Upregulated |
| 0,43423632 | LOC100996664 | FALSE | Upregulated |
| | LOC100996665 | | |
| -0,006230663 | LOC100996668 | FALSE | Downregulated |
| | LOC100996681 | | |
| 0,428478597 | LOC100996695 | FALSE | Upregulated |
| | LOC100996696 | | |
| 0 | LOC100996701 | FALSE | Upregulated |
| 2,490482766 | LOC100996713 | | Upregulated |
| 0,357918658 | LOC100996721 | FALSE | Upregulated |
| 0,065562112 | LOC100996732 | FALSE | Upregulated |
| 0,80069767 | LOC100996735 | FALSE | Upregulated |
| | LOC100996756 | | |
| -0,133494346 | LOC100996760 | FALSE | Downregulated |
| | LOC100996770 | | |
| 0,390950345 | LOC100996792 | FALSE | Upregulated |
| | LOC100996792-2 | | |
| 0,048276727 | LOC100996808 | FALSE | Upregulated |
| | LOC100996902 | | |
| 0,091996709 | LOC100996909 | FALSE | Upregulated |
| 0,034586977 | LOC101054525 | FALSE | Upregulated |
| | LOC101059914 | | |
| | LOC101059915 | | |
| 0,31689826 | LOC101059948 | FALSE | Upregulated |
| 0,381594945 | LOC101059954 | FALSE | Upregulated |
| | LOC101060004 | | |
| | LOC101060017 | | |

| | | | |
|---|---|---|---|
| | LOC101060024 | | |
| 0,732302063 | LOC101060026 | FALSE | Upregulated |
| 1,618659388 | LOC101060038 | | Upregulated |
| 0,043844642 | LOC101060085 | FALSE | Upregulated |
| 0,314224801 | LOC101060097 | FALSE | Upregulated |
| 0 | LOC101060106 | FALSE | Upregulated |
| | LOC101060128 | | |
| -0,393158412 | LOC101060153 | FALSE | Downregulated |
| 0 | LOC101060157 | FALSE | Upregulated |
| 0,100713889 | LOC101060183 | FALSE | Upregulated |
| | LOC101060187 | | |
| | LOC101060211 | | |
| | LOC101060233 | | |
| 1,047508676 | LOC101060236 | | Upregulated |
| | LOC101060254 | | |
| | LOC101060264 | | |
| 0 | LOC101060301 | FALSE | Upregulated |
| -0,868367339 | LOC101060321 | FALSE | Downregulated |
| -1,389119884 | LOC101060341 | | Downregulated |
| 0 | LOC101060363 | FALSE | Upregulated |
| -0,016422875 | LOC101060391 | FALSE | Downregulated |
| 0,26024305 | LOC101060400 | FALSE | Upregulated |
| 0 | LOC101060498 | FALSE | Upregulated |
| 0 | LOC101060532 | FALSE | Upregulated |
| 0,783730397 | LOC101060569 | FALSE | Upregulated |
| -0,397169342 | LOC101060626 | FALSE | Downregulated |
| 0,41846942 | LOC101409256 | FALSE | Upregulated |
| 0,384082583 | LOC101559451 | FALSE | Upregulated |
| 0,078076584 | LOC101805489 | FALSE | Upregulated |
| 0,416292759 | LOC101805491 | FALSE | Upregulated |
| | LOC101926886 | | |
| | LOC101926893 | | |
| 0,633587867 | LOC101926896 | FALSE | Upregulated |
| | LOC101926897 | | |
| 0 | LOC101926899 | FALSE | Upregulated |
| 0 | LOC101926900 | FALSE | Upregulated |
| -0,088178338 | LOC101926902 | FALSE | Downregulated |
| | LOC101926904 | | |
| -0,148688399 | LOC101926905 | FALSE | Downregulated |
| 1,48409924 | LOC101926907 | | Upregulated |
| 0,521366027 | LOC101926909 | FALSE | Upregulated |
| | LOC101926912 | | |
| 0,287869037 | LOC101926916 | FALSE | Upregulated |
| 0,01644159 | LOC101926917 | FALSE | Upregulated |
| | LOC101926921 | | |
| -0,076761582 | LOC101926922 | FALSE | Downregulated |
| | LOC101926923 | | |
| | LOC101926929 | | |
| 0,028911096 | LOC101926936 | FALSE | Upregulated |
| 0,166441118 | LOC101926937 | FALSE | Upregulated |
| 0 | LOC101926940 | FALSE | Upregulated |

| | | | |
|---|---|---|---|
| 1,607716011 | LOC101926948 | | Upregulated |
| | LOC101926951 | | |
| | LOC101926952 | | |
| | LOC101926953 | | |
| -1,15515851 | LOC101926956 | | Downregulated |
| | LOC101926958 | | |
| 0,542470538 | LOC101926959 | FALSE | Upregulated |
| 0,259662233 | LOC101926960 | FALSE | Upregulated |
| | LOC101926961 | | |
| | LOC101926965 | | |
| | LOC101926966 | | |
| 0 | LOC101926968 | FALSE | Upregulated |
| 0,183004282 | LOC101926969 | FALSE | Upregulated |
| | LOC101926970 | | |
| | LOC101926972 | | |
| | LOC101926974 | | |
| | LOC101926975 | | |
| 0,10910589 | LOC101926977 | FALSE | Upregulated |
| | LOC101926982 | | |
| | LOC101926983 | | |
| 0 | LOC101926989 | FALSE | Upregulated |
| 0 | LOC101926992 | FALSE | Upregulated |
| 0,81537062 | LOC101926997 | FALSE | Upregulated |
| | LOC101926998 | | |
| | LOC101926999 | | |
| | LOC101927008 | | |
| 0,898671176 | LOC101927010 | FALSE | Upregulated |
| -0,677032528 | LOC101927011 | FALSE | Downregulated |
| | LOC101927012 | | |
| | LOC101927013 | | |
| | LOC101927016 | | |
| | LOC101927019 | | |
| | LOC101927022 | | |
| | LOC101927024 | | |
| | LOC101927025 | | |
| 0,149465259 | LOC101927026 | FALSE | Upregulated |
| -0,184656823 | LOC101927029 | FALSE | Downregulated |
| | LOC101927037 | | |
| -1,215693566 | LOC101927038 | | Downregulated |
| | LOC101927039 | | |
| | LOC101927042 | | |
| | LOC101927046 | | |
| | LOC101927050 | | |
| | LOC101927052 | | |
| 0 | LOC101927053 | FALSE | Upregulated |
| | LOC101927059 | | |
| 0,027239538 | LOC101927063 | FALSE | Upregulated |
| | LOC101927065 | | |
| -0,08107785 | LOC101927070 | FALSE | Downregulated |
| | LOC101927072 | | |
| | LOC101927080 | | |

| | | | |
|---|---|---|---|
| -0,82776572 | LOC101927088 | FALSE | Downregulated |
| | LOC101927089 | | |
| | LOC101927091 | | |
| -0,078116701 | LOC101927093 | FALSE | Downregulated |
| | LOC101927094 | | |
| 0 | LOC101927095 | FALSE | Upregulated |
| 0,838733658 | LOC101927104 | FALSE | Upregulated |
| | LOC101927108 | | |
| | LOC101927110 | | |
| 0,416338963 | LOC101927112 | FALSE | Upregulated |
| -0,798650197 | LOC101927116 | FALSE | Downregulated |
| | LOC101927118 | | |
| 0,084247393 | LOC101927121 | FALSE | Upregulated |
| 0,110456049 | LOC101927122 | FALSE | Upregulated |
| -0,80839086 | LOC101927126 | FALSE | Downregulated |
| | LOC101927127 | | |
| | LOC101927129 | | |
| | LOC101927133 | | |
| 0 | LOC101927135 | FALSE | Upregulated |
| | LOC101927137 | | |
| | LOC101927141 | | |
| -0,506108792 | LOC101927144 | FALSE | Downregulated |
| | LOC101927145 | | |
| | LOC101927147 | | |
| | LOC101927153 | | |
| | LOC101927154 | | |
| -0,297144531 | LOC101927159 | FALSE | Downregulated |
| 0,481637332 | LOC101927165 | FALSE | Upregulated |
| 0 | LOC101927166 | FALSE | Upregulated |
| 0 | LOC101927168 | FALSE | Upregulated |
| 0,100713889 | LOC101927169 | FALSE | Upregulated |
| -0,045596956 | LOC101927170 | FALSE | Downregulated |
| 0,055378136 | LOC101927172 | FALSE | Upregulated |
| | LOC101927174 | | |
| 1,303329551 | LOC101927179 | | Upregulated |
| | LOC101927182 | | |
| 0,476133394 | LOC101927188 | FALSE | Upregulated |
| | LOC101927191 | | |
| 0,572387211 | LOC101927192 | FALSE | Upregulated |
| 0 | LOC101927199 | FALSE | Upregulated |
| | LOC101927200 | | |
| | LOC101927202 | | |
| | LOC101927205 | | |
| | LOC101927209 | | |
| | LOC101927210 | | |
| | LOC101927212 | | |
| | LOC101927213 | | |
| 0,596781979 | LOC101927216 | FALSE | Upregulated |
| -0,093662559 | LOC101927225 | FALSE | Downregulated |
| 0,318393058 | LOC101927226 | FALSE | Upregulated |
| 0,608326541 | LOC101927228 | FALSE | Upregulated |

| | | | |
|---|---|---|---|
| 0,037542934 | LOC101927229 | FALSE | Upregulated |
| 0 | LOC101927232 | FALSE | Upregulated |
| 0,102508153 | LOC101927234 | FALSE | Upregulated |
| 0,608141033 | LOC101927235 | FALSE | Upregulated |
| | LOC101927238 | | |
| | LOC101927241 | | |
| 0 | LOC101927245 | FALSE | Upregulated |
| | LOC101927247 | | |
| -0,422950134 | LOC101927248 | FALSE | Downregulated |
| -0,251701115 | LOC101927250 | FALSE | Downregulated |
| 0,512417143 | LOC101927252 | FALSE | Upregulated |
| | LOC101927253 | | |
| 0 | LOC101927254 | FALSE | Upregulated |
| 0,879186142 | LOC101927259 | FALSE | Upregulated |
| 0,10515934 | LOC101927260 | FALSE | Upregulated |
| 0,382590898 | LOC101927262 | FALSE | Upregulated |
| 0,969195493 | LOC101927263 | FALSE | Upregulated |
| 0,517790899 | LOC101927268 | FALSE | Upregulated |
| | LOC101927271 | | |
| | LOC101927277 | | |
| 0,438697884 | LOC101927278 | FALSE | Upregulated |
| | LOC101927280 | | |
| | LOC101927281 | | |
| 0,366883488 | LOC101927283 | FALSE | Upregulated |
| | LOC101927284 | | |
| 0 | LOC101927287 | FALSE | Upregulated |
| | LOC101927291 | | |
| 0 | LOC101927293 | FALSE | Upregulated |
| | LOC101927297 | | |
| | LOC101927299 | | |
| -0,001751729 | LOC101927301 | FALSE | Downregulated |
| | LOC101927304 | | |
| | LOC101927306 | | |
| 0 | LOC101927311 | FALSE | Upregulated |
| 0 | LOC101927318 | FALSE | Upregulated |
| | LOC101927319 | | |
| 0,303871772 | LOC101927320 | FALSE | Upregulated |
| 1,378261162 | LOC101927322 | | Upregulated |
| 0 | LOC101927325 | FALSE | Upregulated |
| -0,381980191 | LOC101927326 | FALSE | Downregulated |
| 0 | LOC101927328 | FALSE | Upregulated |
| | LOC101927329 | | |
| -1,477527259 | LOC101927330 | | Downregulated |
| -0,932184406 | LOC101927331 | FALSE | Downregulated |
| | LOC101927332 | | |
| 0,038422272 | LOC101927333 | FALSE | Upregulated |
| | LOC101927335 | | |
| | LOC101927337 | | |
| 0,171416254 | LOC101927338 | FALSE | Upregulated |
| | LOC101927341 | | |
| | LOC101927344 | | |

| | | | |
|---|---|---|---|
| 0 | LOC101927346 | FALSE | Upregulated |
| -0,093662559 | LOC101927348 | FALSE | Downregulated |
| | LOC101927351 | | |
| -0,093662559 | LOC101927353 | FALSE | Downregulated |
| -0,730878961 | LOC101927354 | FALSE | Downregulated |
| 0 | LOC101927357 | FALSE | Upregulated |
| | LOC101927361 | | |
| | LOC101927362 | | |
| | LOC101927363 | | |
| -0,489265842 | LOC101927366 | FALSE | Downregulated |
| 0 | LOC101927367 | FALSE | Upregulated |
| 0,38647539 | LOC101927369 | FALSE | Upregulated |
| -1,537056634 | LOC101927374 | | Downregulated |
| 0,570144997 | LOC101927375 | FALSE | Upregulated |
| 0,035861011 | LOC101927376 | FALSE | Upregulated |
| | LOC101927383 | | |
| | LOC101927387 | | |
| -0,012093025 | LOC101927388 | FALSE | Downregulated |
| | LOC101927389 | | |
| 0,011211767 | LOC101927392 | FALSE | Upregulated |
| | LOC101927395 | | |
| | LOC101927397 | | |
| | LOC101927398 | | |
| -0,175716851 | LOC101927400 | FALSE | Downregulated |
| 0,448169036 | LOC101927401 | FALSE | Upregulated |
| 0,068128077 | LOC101927403 | FALSE | Upregulated |
| | LOC101927404 | | |
| | LOC101927405 | | |
| | LOC101927409 | | |
| | LOC101927413 | | |
| | LOC101927414 | | |
| 0,879330744 | LOC101927417 | FALSE | Upregulated |
| | LOC101927426 | | |
| | LOC101927429 | | |
| 0,594114703 | LOC101927430 | FALSE | Upregulated |
| -0,296168615 | LOC101927432 | FALSE | Downregulated |
| 0,216583583 | LOC101927435 | FALSE | Upregulated |
| | LOC101927436 | | |
| | LOC101927439 | | |
| 0 | LOC101927441 | FALSE | Upregulated |
| 0,652631022 | LOC101927446 | FALSE | Upregulated |
| 0,289606026 | LOC101927447 | FALSE | Upregulated |
| -0,131181527 | LOC101927448 | FALSE | Downregulated |
| | LOC101927452 | | |
| 0,402509707 | LOC101927454 | FALSE | Upregulated |
| -0,111030623 | LOC101927458 | FALSE | Downregulated |
| | LOC101927461 | | |
| | LOC101927466 | | |
| | LOC101927467 | | |
| 0,329969025 | LOC101927471 | FALSE | Upregulated |
| | LOC101927473 | | |

| | | | |
|---|---|---|---|
| 0,491240154 | LOC101927474 | FALSE | Upregulated |
| | LOC101927480 | | |
| 0,369532281 | LOC101927481 | FALSE | Upregulated |
| | LOC101927483 | | |
| | LOC101927484 | | |
| 0,305247724 | LOC101927486 | FALSE | Upregulated |
| | LOC101927488 | | |
| -1,061983104 | LOC101927490 | | Downregulated |
| | LOC101927491 | | |
| | LOC101927493 | | |
| | LOC101927494 | | |
| | LOC101927496 | | |
| 0,139571958 | LOC101927499 | FALSE | Upregulated |
| 0,03255475 | LOC101927503 | FALSE | Upregulated |
| | LOC101927504 | | |
| 0,277615404 | LOC101927505 | FALSE | Upregulated |
| -0,794774244 | LOC101927506 | FALSE | Downregulated |
| 0,779290787 | LOC101927508 | FALSE | Upregulated |
| 1,031656608 | LOC101927510 | | Upregulated |
| 0,154504817 | LOC101927511 | FALSE | Upregulated |
| 0,25542064 | LOC101927512 | FALSE | Upregulated |
| | LOC101927518 | | |
| | LOC101927522 | | |
| -0,016422875 | LOC101927525 | FALSE | Downregulated |
| 0,381308326 | LOC101927526 | FALSE | Upregulated |
| | LOC101927527 | | |
| 0,069564407 | LOC101927531 | FALSE | Upregulated |
| | LOC101927532 | | |
| | LOC101927533 | | |
| | LOC101927534 | | |
| 0 | LOC101927537 | FALSE | Upregulated |
| 0,426437581 | LOC101927538 | FALSE | Upregulated |
| 0,279794584 | LOC101927544 | FALSE | Upregulated |
| 0,097410515 | LOC101927546 | FALSE | Upregulated |
| | LOC101927548 | | |
| | LOC101927553 | | |
| 0 | LOC101927555 | FALSE | Upregulated |
| | LOC101927558 | | |
| | LOC101927561 | | |
| -0,81275588 | LOC101927562 | FALSE | Downregulated |
| 0 | LOC101927566 | FALSE | Upregulated |
| 0,880722536 | LOC101927567 | FALSE | Upregulated |
| 0 | LOC101927568 | FALSE | Upregulated |
| -0,9050455 | LOC101927571 | FALSE | Downregulated |
| | LOC101927573 | | |
| 0,15101716 | LOC101927576 | FALSE | Upregulated |
| 0 | LOC101927582 | FALSE | Upregulated |
| | LOC101927584 | | |
| 0,290774137 | LOC101927589 | FALSE | Upregulated |
| 0,065962645 | LOC101927594 | FALSE | Upregulated |
| -0,374446946 | LOC101927600 | FALSE | Downregulated |

| | | | |
|---|---|---|---|
| | LOC101927602 | | |
| 0 | LOC101927603 | FALSE | Upregulated |
| | LOC101927605 | | |
| | LOC101927606 | | |
| -0,990731862 | LOC101927609 | FALSE | Downregulated |
| -0,549040581 | LOC101927611 | FALSE | Downregulated |
| -0,434291584 | LOC101927613 | FALSE | Downregulated |
| 0 | LOC101927615 | FALSE | Upregulated |
| -0,975461078 | LOC101927617 | FALSE | Downregulated |
| | LOC101927627 | | |
| 1,336290434 | LOC101927628 | | Upregulated |
| | LOC101927629 | | |
| -0,328922024 | LOC101927632 | FALSE | Downregulated |
| 0,481637332 | LOC101927633 | FALSE | Upregulated |
| 0 | LOC101927634 | FALSE | Upregulated |
| -0,181642703 | LOC101927635 | FALSE | Downregulated |
| -0,17411381 | LOC101927646 | FALSE | Downregulated |
| -0,311979234 | LOC101927651 | FALSE | Downregulated |
| -0,60269172 | LOC101927652 | FALSE | Downregulated |
| | LOC101927656 | | |
| 0,046678197 | LOC101927660 | FALSE | Upregulated |
| | LOC101927661 | | |
| 0,280783262 | LOC101927662 | FALSE | Upregulated |
| | LOC101927664 | | |
| 0,266589749 | LOC101927665 | FALSE | Upregulated |
| 0 | LOC101927669 | FALSE | Upregulated |
| -0,79394535 | LOC101927670 | FALSE | Downregulated |
| -0,180377081 | LOC101927674 | FALSE | Downregulated |
| | LOC101927676 | | |
| -0,484942929 | LOC101927680 | FALSE | Downregulated |
| | LOC101927684 | | |
| 0 | LOC101927686 | FALSE | Upregulated |
| | LOC101927687 | | |
| | LOC101927690 | | |
| | LOC101927691 | | |
| | LOC101927695 | | |
| 0,055378136 | LOC101927696 | FALSE | Upregulated |
| 0,060381005 | LOC101927697 | FALSE | Upregulated |
| 0,406117894 | LOC101927702 | FALSE | Upregulated |
| -0,222941789 | LOC101927703 | FALSE | Downregulated |
| | LOC101927704 | | |
| -0,090805522 | LOC101927705 | FALSE | Downregulated |
| -0,768832574 | LOC101927708 | FALSE | Downregulated |
| | LOC101927709 | | |
| 0 | LOC101927710 | FALSE | Upregulated |
| | LOC101927711 | | |
| 0 | LOC101927712 | FALSE | Upregulated |
| | LOC101927713 | | |
| 0 | LOC101927720 | FALSE | Upregulated |
| 0,372206583 | LOC101927721 | FALSE | Upregulated |
| 0 | LOC101927722 | FALSE | Upregulated |

| | | | |
|---|---|---|---|
| 1,077690527 | LOC101927723 | | Upregulated |
| 0,387582209 | LOC101927725 | FALSE | Upregulated |
| | LOC101927729 | | |
| | LOC101927731 | | |
| 0,366403851 | LOC101927736 | FALSE | Upregulated |
| -0,172787552 | LOC101927737 | FALSE | Downregulated |
| | LOC101927738 | | |
| | LOC101927744 | | |
| 0,257520667 | LOC101927748 | FALSE | Upregulated |
| 0,425769929 | LOC101927754 | FALSE | Upregulated |
| | LOC101927757 | | |
| | LOC101927760 | | |
| | LOC101927762 | | |
| -0,044155334 | LOC101927763 | FALSE | Downregulated |
| -1,519489114 | LOC101927764 | | Downregulated |
| -0,375569081 | LOC101927765 | FALSE | Downregulated |
| 0,433438419 | LOC101927772 | FALSE | Upregulated |
| 0,357037814 | LOC101927773 | FALSE | Upregulated |
| 0 | LOC101927774 | FALSE | Upregulated |
| -0,489807952 | LOC101927775 | FALSE | Downregulated |
| | LOC101927777 | | |
| | LOC101927779 | | |
| 0,258571957 | LOC101927783 | FALSE | Upregulated |
| | LOC101927786 | | |
| | LOC101927788 | | |
| -0,243691618 | LOC101927789 | FALSE | Downregulated |
| 0,322151801 | LOC101927792 | FALSE | Upregulated |
| | LOC101927794 | | |
| | LOC101927798 | | |
| 0,718003842 | LOC101927799 | FALSE | Upregulated |
| 0,303617988 | LOC101927800 | FALSE | Upregulated |
| | LOC101927801 | | |
| 0,406787727 | LOC101927804 | FALSE | Upregulated |
| | LOC101927805 | | |
| | LOC101927807 | | |
| 0,55170453 | LOC101927810 | FALSE | Upregulated |
| 0 | LOC101927811 | FALSE | Upregulated |
| -0,000640574 | LOC101927814 | FALSE | Downregulated |
| | LOC101927816 | | |
| 0,016710144 | LOC101927820 | FALSE | Upregulated |
| | LOC101927823 | | |
| 0,155121689 | LOC101927824 | FALSE | Upregulated |
| 0,796405214 | LOC101927825 | FALSE | Upregulated |
| 0,409688538 | LOC101927826 | FALSE | Upregulated |
| -0,139184475 | LOC101927834 | FALSE | Downregulated |
| | LOC101927835 | | |
| 0,785027219 | LOC101927837 | FALSE | Upregulated |
| | LOC101927844 | | |
| 0 | LOC101927846 | FALSE | Upregulated |
| 0 | LOC101927852 | FALSE | Upregulated |
| 0 | LOC101927853 | FALSE | Upregulated |

| | | | |
|---|---|---|---|
| -0,667676541 | LOC101927855 | FALSE | Downregulated |
| | LOC101927856 | | |
| -0,101310524 | LOC101927857 | FALSE | Downregulated |
| 0,377871117 | LOC101927859 | FALSE | Upregulated |
| 0,117747048 | LOC101927863 | FALSE | Upregulated |
| | LOC101927866 | | |
| 0 | LOC101927867 | FALSE | Upregulated |
| 1,240760101 | LOC101927871 | | Upregulated |
| -0,119269724 | LOC101927872 | FALSE | Downregulated |
| 0,046678197 | LOC101927874 | FALSE | Upregulated |
| 0 | LOC101927875 | FALSE | Upregulated |
| | LOC101927877 | | |
| | LOC101927878 | | |
| 0 | LOC101927879 | FALSE | Upregulated |
| -0,042997165 | LOC101927880 | FALSE | Downregulated |
| | LOC101927881 | | |
| 0,04752935 | LOC101927882 | FALSE | Upregulated |
| -0,322503355 | LOC101927885 | FALSE | Downregulated |
| 0 | LOC101927888 | FALSE | Upregulated |
| 0 | LOC101927890 | FALSE | Upregulated |
| | LOC101927892 | | |
| 0,481637332 | LOC101927896 | FALSE | Upregulated |
| 0,334837082 | LOC101927902 | FALSE | Upregulated |
| | LOC101927904 | | |
| 0,333597385 | LOC101927906 | FALSE | Upregulated |
| | LOC101927908 | | |
| | LOC101927913 | | |
| | LOC101927915 | | |
| | LOC101927916 | | |
| | LOC101927918 | | |
| 0,395165154 | LOC101927919 | FALSE | Upregulated |
| | LOC101927921 | | |
| 0,556380272 | LOC101927922 | FALSE | Upregulated |
| 0,029809716 | LOC101927929 | FALSE | Upregulated |
| -0,051921164 | LOC101927931 | FALSE | Downregulated |
| 0 | LOC101927932 | FALSE | Upregulated |
| -0,13927752 | LOC101927933 | FALSE | Downregulated |
| | LOC101927935 | | |
| | LOC101927936 | | |
| -0,953769625 | LOC101927942 | FALSE | Downregulated |
| 1,016908104 | LOC101927943 | | Upregulated |
| | LOC101927946 | | |
| 0 | LOC101927949 | FALSE | Upregulated |
| 0,367590571 | LOC101927953 | FALSE | Upregulated |
| | LOC101927956 | | |
| 0,099452879 | LOC101927957 | FALSE | Upregulated |
| 0,231858932 | LOC101927958 | FALSE | Upregulated |
| | LOC101927959 | | |
| | LOC101927960 | | |
| | LOC101927961 | | |
| | LOC101927962 | | |

| | | | |
|---|---|---|---|
| | LOC101927967 | | |
| 0,123640864 | LOC101927971 | FALSE | Upregulated |
| 0 | LOC101927973 | FALSE | Upregulated |
| -1,288030653 | LOC101927974 | | Downregulated |
| 0 | LOC101927975 | FALSE | Upregulated |
| 0,387012887 | LOC101927978 | FALSE | Upregulated |
| | LOC101927982 | | |
| 0 | LOC101927988 | FALSE | Upregulated |
| 0 | LOC101927991 | FALSE | Upregulated |
| 0,664255834 | LOC101927993 | FALSE | Upregulated |
| 0,269388226 | LOC101927995 | FALSE | Upregulated |
| | LOC101927998 | | |
| 0,331881071 | LOC101928000 | FALSE | Upregulated |
| 0,06568889 | LOC101928004 | FALSE | Upregulated |
| 0,205850743 | LOC101928005 | FALSE | Upregulated |
| 0,661146488 | LOC101928006 | FALSE | Upregulated |
| 0 | LOC101928007 | FALSE | Upregulated |
| | LOC101928011 | | |
| | LOC101928014 | | |
| -1,097239117 | LOC101928021 | | Downregulated |
| | LOC101928026 | | |
| | LOC101928027 | | |
| | LOC101928028 | | |
| | LOC101928031 | | |
| | LOC101928032 | | |
| | LOC101928037 | | |
| 0,337646995 | LOC101928039 | FALSE | Upregulated |
| -0,893855869 | LOC101928040 | FALSE | Downregulated |
| 0,371230259 | LOC101928044 | FALSE | Upregulated |
| | LOC101928046 | | |
| 0,588422738 | LOC101928047 | FALSE | Upregulated |
| 0,178542089 | LOC101928048 | FALSE | Upregulated |
| 0,597692113 | LOC101928049 | FALSE | Upregulated |
| | LOC101928055-2 | | |
| | LOC101928056 | | |
| 0,680971064 | LOC101928062 | FALSE | Upregulated |
| | LOC101928066 | | |
| 0 | LOC101928067 | FALSE | Upregulated |
| | LOC101928070-2 | | |
| 0,703407598 | LOC101928072 | FALSE | Upregulated |
| | LOC101928075 | | |
| -0,050946007 | LOC101928076 | FALSE | Downregulated |
| | LOC101928077 | | |
| 0 | LOC101928080 | FALSE | Upregulated |
| | LOC101928082 | | |
| | LOC101928084 | | |
| -0,021706002 | LOC101928085 | FALSE | Downregulated |
| 0 | LOC101928087 | FALSE | Upregulated |
| 0 | LOC101928090 | FALSE | Upregulated |
| -0,036662035 | LOC101928092 | FALSE | Downregulated |
| 0,280929801 | LOC101928096 | FALSE | Upregulated |

| | | | |
|---|---|---|---|
| 1,049919098 | LOC101928102 | | Upregulated |
| 1,570476784 | LOC101928105 | | Upregulated |
| | LOC101928108 | | |
| 0,247041127 | LOC101928109 | FALSE | Upregulated |
| 0 | LOC101928110 | FALSE | Upregulated |
| -0,122475778 | LOC101928111 | FALSE | Downregulated |
| 1,313781831 | LOC101928113 | | Upregulated |
| | LOC101928114 | | |
| 0 | LOC101928115 | FALSE | Upregulated |
| 0 | LOC101928117 | FALSE | Upregulated |
| | LOC101928121 | | |
| | LOC101928124 | | |
| | LOC101928125 | | |
| -0,113310343 | LOC101928126 | FALSE | Downregulated |
| | LOC101928127 | | |
| | LOC101928128 | | |
| | LOC101928129 | | |
| | LOC101928130 | | |
| | LOC101928131 | | |
| | LOC101928132 | | |
| | LOC101928135 | | |
| | LOC101928144 | | |
| | LOC101928146 | | |
| 0,078076584 | LOC101928149 | FALSE | Upregulated |
| | LOC101928153 | | |
| 0 | LOC101928156 | FALSE | Upregulated |
| | LOC101928157 | | |
| 0,199495513 | LOC101928160 | FALSE | Upregulated |
| | LOC101928162 | | |
| | LOC101928166 | | |
| | LOC101928167 | | |
| | LOC101928169 | | |
| 0 | LOC101928171 | FALSE | Upregulated |
| 1,026829595 | LOC101928175 | | Upregulated |
| | LOC101928176 | | |
| | LOC101928177 | | |
| -0,129922473 | LOC101928178 | FALSE | Downregulated |
| -1,030685968 | LOC101928180 | | Downregulated |
| -0,013375756 | LOC101928186 | FALSE | Downregulated |
| | LOC101928190 | | |
| -0,038231478 | LOC101928191 | FALSE | Downregulated |
| -0,915981035 | LOC101928193 | FALSE | Downregulated |
| 0,404704938 | LOC101928196 | FALSE | Upregulated |
| 0,655951358 | LOC101928197 | FALSE | Upregulated |
| | LOC101928199 | | |
| | LOC101928207 | | |
| 0,055378136 | LOC101928208 | FALSE | Upregulated |
| | LOC101928216 | | |
| | LOC101928217 | | |
| 0 | LOC101928219 | FALSE | Upregulated |
| 0,452470816 | LOC101928220 | FALSE | Upregulated |

| | | | |
|---|---|---|---|
| | LOC101928225 | | |
| | LOC101928226 | | |
| -0,504168798 | LOC101928228 | FALSE | Downregulated |
| 0,682076449 | LOC101928231 | FALSE | Upregulated |
| -0,345222894 | LOC101928233 | FALSE | Downregulated |
| | LOC101928234 | | |
| 0,303232695 | LOC101928235 | FALSE | Upregulated |
| 1,236859302 | LOC101928236 | | Upregulated |
| 0,384660877 | LOC101928237 | FALSE | Upregulated |
| -0,332342854 | LOC101928238 | FALSE | Downregulated |
| 0,31609178 | LOC101928239 | FALSE | Upregulated |
| 0,76916817 | LOC101928245 | FALSE | Upregulated |
| -0,068065352 | LOC101928246 | FALSE | Downregulated |
| 0,146230128 | LOC101928247 | FALSE | Upregulated |
| 0 | LOC101928248 | FALSE | Upregulated |
| -0,425322697 | LOC101928249 | FALSE | Downregulated |
| | LOC101928250 | | |
| -0,221606007 | LOC101928251 | FALSE | Downregulated |
| -0,936502242 | LOC101928253 | FALSE | Downregulated |
| 0,596257416 | LOC101928255 | FALSE | Upregulated |
| | LOC101928257 | | |
| 0 | LOC101928258 | FALSE | Upregulated |
| -0,063117964 | LOC101928261 | FALSE | Downregulated |
| 0 | LOC101928262 | FALSE | Upregulated |
| 0 | LOC101928264 | FALSE | Upregulated |
| 0,038484245 | LOC101928269 | FALSE | Upregulated |
| | LOC101928271 | | |
| | LOC101928272 | | |
| | LOC101928273 | | |
| 0,790367853 | LOC101928274 | FALSE | Upregulated |
| | LOC101928275 | | |
| -0,1007568 | LOC101928277 | FALSE | Downregulated |
| | LOC101928278 | | |
| 0,586141051 | LOC101928279 | FALSE | Upregulated |
| | LOC101928282 | | |
| | LOC101928285 | | |
| 0 | LOC101928286 | FALSE | Upregulated |
| 0 | LOC101928288 | FALSE | Upregulated |
| 0,448212425 | LOC101928300 | FALSE | Upregulated |
| 0,048276727 | LOC101928306 | FALSE | Upregulated |
| 0,066520504 | LOC101928307 | FALSE | Upregulated |
| | LOC101928309 | | |
| | LOC101928310 | | |
| -0,067802957 | LOC101928311 | FALSE | Downregulated |
| 0,595398569 | LOC101928312 | FALSE | Upregulated |
| -0,019532186 | LOC101928314 | FALSE | Downregulated |
| | LOC101928316 | | |
| 1,286894578 | LOC101928317 | | Upregulated |
| 0 | LOC101928325 | FALSE | Upregulated |
| 0,269102277 | LOC101928326 | FALSE | Upregulated |
| 0 | LOC101928327 | FALSE | Upregulated |

| | | | |
|---|---|---|---|
| | LOC101928329 | | |
| 0 | LOC101928330 | FALSE | Upregulated |
| 0,45906372 | LOC101928331 | FALSE | Upregulated |
| -0,280735714 | LOC101928338 | FALSE | Downregulated |
| | LOC101928339 | | |
| 0,037268864 | LOC101928343 | FALSE | Upregulated |
| | LOC101928346 | | |
| 0,450350708 | LOC101928347 | FALSE | Upregulated |
| 0 | LOC101928349 | FALSE | Upregulated |
| | LOC101928350 | | |
| 0,507793696 | LOC101928357 | FALSE | Upregulated |
| | LOC101928358 | | |
| 0,044126719 | LOC101928359 | FALSE | Upregulated |
| -0,157746712 | LOC101928360 | FALSE | Downregulated |
| -0,472941208 | LOC101928361 | FALSE | Downregulated |
| 0,033128344 | LOC101928362 | FALSE | Upregulated |
| | LOC101928364 | | |
| | LOC101928367 | | |
| | LOC101928368 | | |
| 0,148500228 | LOC101928369 | FALSE | Upregulated |
| -0,414809209 | LOC101928370 | FALSE | Downregulated |
| | LOC101928375 | | |
| 0,258892196 | LOC101928377 | FALSE | Upregulated |
| | LOC101928380 | | |
| -0,759101783 | LOC101928382 | FALSE | Downregulated |
| -0,055430561 | LOC101928383 | FALSE | Downregulated |
| | LOC101928385 | | |
| 0 | LOC101928387 | FALSE | Upregulated |
| 0,698106815 | LOC101928388 | FALSE | Upregulated |
| 0,843954617 | LOC101928389 | FALSE | Upregulated |
| | LOC101928391 | | |
| | LOC101928392 | | |
| | LOC101928393 | | |
| | LOC101928394 | | |
| 0 | LOC101928397 | FALSE | Upregulated |
| 1,168583582 | LOC101928399 | | Upregulated |
| 0,146690165 | LOC101928401 | FALSE | Upregulated |
| -0,060493287 | LOC101928402 | FALSE | Downregulated |
| -0,026568429 | LOC101928403 | FALSE | Downregulated |
| | LOC101928405 | | |
| | LOC101928406 | | |
| -0,129497672 | LOC101928407 | FALSE | Downregulated |
| 0 | LOC101928412 | FALSE | Upregulated |
| 0 | LOC101928414 | FALSE | Upregulated |
| 0,751449464 | LOC101928419 | FALSE | Upregulated |
| -0,017135352 | LOC101928421 | FALSE | Downregulated |
| | LOC101928422 | | |
| | LOC101928423 | | |
| | LOC101928426 | | |
| 0 | LOC101928427 | FALSE | Upregulated |
| -0,079954235 | LOC101928428 | FALSE | Downregulated |

| | | | |
|---|---|---|---|
| | LOC101928430 | | |
| 0 | LOC101928432 | FALSE | Upregulated |
| 1,515005095 | LOC101928434 | | Upregulated |
| | LOC101928436 | | |
| 0,349762851 | LOC101928437 | FALSE | Upregulated |
| | LOC101928447 | | |
| | LOC101928448 | | |
| | LOC101928450 | | |
| | LOC101928455 | | |
| | LOC101928457 | | |
| | LOC101928458 | | |
| 0,42139433 | LOC101928459 | FALSE | Upregulated |
| -0,206086995 | LOC101928462 | FALSE | Downregulated |
| | LOC101928463 | | |
| -0,585405087 | LOC101928464 | FALSE | Downregulated |
| | LOC101928469 | | |
| 0,08832291 | LOC101928472 | FALSE | Upregulated |
| -0,07999261 | LOC101928474 | FALSE | Downregulated |
| | LOC101928475 | | |
| 0,120750007 | LOC101928477 | FALSE | Upregulated |
| | LOC101928478 | | |
| | LOC101928481 | | |
| | LOC101928484 | | |
| -0,351218217 | LOC101928487 | FALSE | Downregulated |
| -0,415698639 | LOC101928489 | FALSE | Downregulated |
| 0,298378831 | LOC101928504 | FALSE | Upregulated |
| | LOC101928506 | | |
| 0 | LOC101928513 | FALSE | Upregulated |
| 0,377265125 | LOC101928514 | FALSE | Upregulated |
| | LOC101928519 | | |
| 0,055378136 | LOC101928522 | FALSE | Upregulated |
| | LOC101928523 | | |
| 0 | LOC101928526 | FALSE | Upregulated |
| 0,046744076 | LOC101928529 | FALSE | Upregulated |
| -0,256007325 | LOC101928530 | FALSE | Downregulated |
| 0,450871336 | LOC101928533 | FALSE | Upregulated |
| | LOC101928535 | | |
| | LOC101928538 | | |
| | LOC101928543 | | |
| 0,070401644 | LOC101928549 | FALSE | Upregulated |
| 0,956281973 | LOC101928551 | FALSE | Upregulated |
| 0,885749967 | LOC101928554 | FALSE | Upregulated |
| 1,353531551 | LOC101928556 | | Upregulated |
| -1,018954523 | LOC101928560 | | Downregulated |
| 0,160596907 | LOC101928562 | FALSE | Upregulated |
| | LOC101928563 | | |
| | LOC101928564 | | |
| | LOC101928566 | | |
| -1,097056617 | LOC101928568 | | Downregulated |
| | LOC101928569 | | |
| 0,401837705 | LOC101928570 | FALSE | Upregulated |

| | | | |
|---|---|---|---|
| -0,28400383 | LOC101928571 | FALSE | Downregulated |
| 0,626917413 | LOC101928572 | FALSE | Upregulated |
| | LOC101928573 | | |
| | LOC101928579 | | |
| | LOC101928581 | | |
| | LOC101928582 | | |
| 0,228881225 | LOC101928583 | FALSE | Upregulated |
| 0,147182423 | LOC101928584 | FALSE | Upregulated |
| | LOC101928586 | | |
| | LOC101928587 | | |
| 0,349784196 | LOC101928589 | FALSE | Upregulated |
| 0 | LOC101928591 | FALSE | Upregulated |
| 0,330827601 | LOC101928592 | FALSE | Upregulated |
| | LOC101928593 | | |
| 0,161043187 | LOC101928596 | FALSE | Upregulated |
| | LOC101928598 | | |
| 0,407193485 | LOC101928600 | FALSE | Upregulated |
| 0,390950345 | LOC101928602 | FALSE | Upregulated |
| -0,09453735 | LOC101928608 | FALSE | Downregulated |
| | LOC101928611 | | |
| 0,001011524 | LOC101928612 | FALSE | Upregulated |
| | LOC101928614 | | |
| | LOC101928617 | | |
| 1,777700275 | LOC101928618 | | Upregulated |
| | LOC101928620 | | |
| | LOC101928623 | | |
| -0,239232856 | LOC101928624 | FALSE | Downregulated |
| 0,298307066 | LOC101928625 | FALSE | Upregulated |
| | LOC101928629 | | |
| 0,021773956 | LOC101928632 | FALSE | Upregulated |
| 0,574947982 | LOC101928633 | FALSE | Upregulated |
| | LOC101928634 | | |
| | LOC101928636 | | |
| 0 | LOC101928638 | FALSE | Upregulated |
| 0 | LOC101928639 | FALSE | Upregulated |
| | LOC101928640 | | |
| | LOC101928643 | | |
| -0,121743676 | LOC101928645 | FALSE | Downregulated |
| 0,375413753 | LOC101928647 | FALSE | Upregulated |
| -0,15372858 | LOC101928652 | FALSE | Downregulated |
| | LOC101928654 | | |
| 1,460810572 | LOC101928655 | | Upregulated |
| 0,125063918 | LOC101928659 | FALSE | Upregulated |
| | LOC101928662 | | |
| 0,700566212 | LOC101928665 | FALSE | Upregulated |
| | LOC101928666 | | |
| -1,014897286 | LOC101928667 | | Downregulated |
| -1,3438684 | LOC101928671 | | Downregulated |
| 1,069242724 | LOC101928676 | | Upregulated |
| -0,16565178 | LOC101928683 | FALSE | Downregulated |
| 0,371149589 | LOC101928684 | FALSE | Upregulated |

| | | | |
|---|---|---|---|
| | LOC101928688 | | |
| -0,286755659 | LOC101928692 | FALSE | Downregulated |
| 0,832759009 | LOC101928695 | FALSE | Upregulated |
| | LOC101928697 | | |
| 0,475681836 | LOC101928701 | FALSE | Upregulated |
| | LOC101928702 | | |
| | LOC101928704 | | |
| 0,330873438 | LOC101928705 | FALSE | Upregulated |
| | LOC101928710 | | |
| | LOC101928711 | | |
| | LOC101928713 | | |
| -0,091603759 | LOC101928718 | FALSE | Downregulated |
| | LOC101928721 | | |
| 1,671774391 | LOC101928722 | | Upregulated |
| 0,256875024 | LOC101928725 | FALSE | Upregulated |
| 0,719188933 | LOC101928727 | FALSE | Upregulated |
| -0,081062865 | LOC101928730 | FALSE | Downregulated |
| 0,212633972 | LOC101928733 | FALSE | Upregulated |
| | LOC101928735 | | |
| 0,058640153 | LOC101928740 | FALSE | Upregulated |
| 0,006569718 | LOC101928742 | FALSE | Upregulated |
| | LOC101928743 | | |
| | LOC101928747 | | |
| 0 | LOC101928754 | FALSE | Upregulated |
| 0,769785599 | LOC101928756 | FALSE | Upregulated |
| 0,725487827 | LOC101928761 | FALSE | Upregulated |
| | LOC101928764 | | |
| 0,019062697 | LOC101928765 | FALSE | Upregulated |
| 0,146690165 | LOC101928767 | FALSE | Upregulated |
| | LOC101928770 | | |
| 0,008411194 | LOC101928774 | FALSE | Upregulated |
| | LOC101928775 | | |
| 0 | LOC101928779 | FALSE | Upregulated |
| | LOC101928785 | | |
| 0 | LOC101928786 | FALSE | Upregulated |
| 0 | LOC101928787 | FALSE | Upregulated |
| 0 | LOC101928792 | FALSE | Upregulated |
| 0,929802544 | LOC101928796 | FALSE | Upregulated |
| | LOC101928800 | | |
| -0,320622884 | LOC101928801 | FALSE | Downregulated |
| -0,973500541 | LOC101928803 | FALSE | Downregulated |
| 0 | LOC101928804 | FALSE | Upregulated |
| -0,606488253 | LOC101928806 | FALSE | Downregulated |
| 0 | LOC101928807 | FALSE | Upregulated |
| -0,049412723 | LOC101928808 | FALSE | Downregulated |
| 0,761622678 | LOC101928810 | FALSE | Upregulated |
| | LOC101928813 | | |
| 0,700110925 | LOC101928814 | FALSE | Upregulated |
| 0,166956144 | LOC101928815 | FALSE | Upregulated |
| | LOC101928818 | | |
| 0,807911573 | LOC101928819 | FALSE | Upregulated |

| | | | |
|---|---|---|---|
| 0 | LOC101928824 | FALSE | Upregulated |
| -0,288549797 | LOC101928825 | FALSE | Downregulated |
| | LOC101928826 | | |
| | LOC101928828 | | |
| 0 | LOC101928829 | FALSE | Upregulated |
| | LOC101928832 | | |
| | LOC101928833 | | |
| | LOC101928836 | | |
| -0,308206531 | LOC101928838 | FALSE | Downregulated |
| | LOC101928839 | | |
| -1,261576277 | LOC101928841 | | Downregulated |
| | LOC101928842 | | |
| 1,472040697 | LOC101928845 | | Upregulated |
| | LOC101928849 | | |
| 0,087116065 | LOC101928850 | FALSE | Upregulated |
| 0,17512632 | LOC101928852 | FALSE | Upregulated |
| | LOC101928856 | | |
| | LOC101928859 | | |
| | LOC101928860 | | |
| | LOC101928863 | | |
| 0,000977115 | LOC101928864 | FALSE | Upregulated |
| | LOC101928866 | | |
| | LOC101928867 | | |
| 0,031444366 | LOC101928868 | FALSE | Upregulated |
| 0,008780918 | LOC101928870 | FALSE | Upregulated |
| 0,271869175 | LOC101928872 | FALSE | Upregulated |
| -0,056531012 | LOC101928876 | FALSE | Downregulated |
| 0,11758063 | LOC101928880 | FALSE | Upregulated |
| -0,10007505 | LOC101928881 | FALSE | Downregulated |
| -0,644645094 | LOC101928882 | FALSE | Downregulated |
| 0,53281018 | LOC101928886 | FALSE | Upregulated |
| -0,42988234 | LOC101928887 | FALSE | Downregulated |
| 0 | LOC101928890 | FALSE | Upregulated |
| 0,18478786 | LOC101928891 | FALSE | Upregulated |
| | LOC101928893 | | |
| | LOC101928898 | | |
| 0,402112793 | LOC101928899 | FALSE | Upregulated |
| 0 | LOC101928902 | FALSE | Upregulated |
| | LOC101928905 | | |
| -0,196923371 | LOC101928906 | FALSE | Downregulated |
| 1,115273945 | LOC101928907 | | Upregulated |
| | LOC101928910 | | |
| 0 | LOC101928911 | FALSE | Upregulated |
| | LOC101928912 | | |
| -0,776069033 | LOC101928913 | FALSE | Downregulated |
| 1,11106994 | LOC101928915 | | Upregulated |
| -0,113530287 | LOC101928916 | FALSE | Downregulated |
| 0 | LOC101928918 | FALSE | Upregulated |
| | LOC101928919 | | |
| 0,41145699 | LOC101928921 | FALSE | Upregulated |
| | LOC101928922 | | |

| | | | |
|---|---|---|---|
| | LOC101928923 | | |
| 0,039629489 | LOC101928924 | FALSE | Upregulated |
| | LOC101928928 | | |
| 0 | LOC101928929 | FALSE | Upregulated |
| 0 | LOC101928932 | FALSE | Upregulated |
| 0,290674103 | LOC101928933 | FALSE | Upregulated |
| | LOC101928941 | | |
| 0 | LOC101928948 | FALSE | Upregulated |
| | LOC101928950 | | |
| 0,257602184 | LOC101928955 | FALSE | Upregulated |
| | LOC101928957 | | |
| | LOC101928962 | | |
| 0,158633196 | LOC101928963 | FALSE | Upregulated |
| | LOC101928964 | | |
| -0,460953322 | LOC101928965 | FALSE | Downregulated |
| 0,789710771 | LOC101928968 | FALSE | Upregulated |
| 0,191985849 | LOC101928970 | FALSE | Upregulated |
| | LOC101928972 | | |
| -1,174472962 | LOC101928974 | | Downregulated |
| -0,423329558 | LOC101928976 | FALSE | Downregulated |
| | LOC101928977 | | |
| | LOC101928978 | | |
| 0 | LOC101928981 | FALSE | Upregulated |
| 0,099452879 | LOC101928987 | FALSE | Upregulated |
| 0,415555839 | LOC101928991 | FALSE | Upregulated |
| -1,101698136 | LOC101928993 | | Downregulated |
| | LOC101929002 | | |
| | LOC101929003 | | |
| 0,084236701 | LOC101929006 | FALSE | Upregulated |
| -0,209506005 | LOC101929008 | FALSE | Downregulated |
| -0,373168277 | LOC101929011 | FALSE | Downregulated |
| | LOC101929014 | | |
| | LOC101929016 | | |
| | LOC101929021 | | |
| | LOC101929022 | | |
| 0,402112793 | LOC101929027 | FALSE | Upregulated |
| | LOC101929028 | | |
| | LOC101929031 | | |
| | LOC101929034 | | |
| -0,005294613 | LOC101929035 | FALSE | Downregulated |
| 0,83923934 | LOC101929036 | FALSE | Upregulated |
| 0,219382241 | LOC101929038 | FALSE | Upregulated |
| -0,422066544 | LOC101929039 | FALSE | Downregulated |
| | LOC101929043 | | |
| 0 | LOC101929044 | FALSE | Upregulated |
| | LOC101929048 | | |
| | LOC101929050 | | |
| 1,838974995 | LOC101929054 | | Upregulated |
| 0,084976497 | LOC101929055 | FALSE | Upregulated |
| | LOC101929056 | | |
| 0 | LOC101929058 | FALSE | Upregulated |

| | | | |
|---|---|---|---|
| 0,19373402 | LOC101929060 | FALSE | Upregulated |
| 0,24167425 | LOC101929067 | FALSE | Upregulated |
| | LOC101929069 | | |
| 0,068128077 | LOC101929077 | FALSE | Upregulated |
| | LOC101929082 | | |
| 0,020789395 | LOC101929085 | FALSE | Upregulated |
| 0,01196447 | LOC101929087 | FALSE | Upregulated |
| | LOC101929088 | | |
| 1,05005471 | LOC101929090 | | Upregulated |
| 0 | LOC101929094 | FALSE | Upregulated |
| -1,720965332 | LOC101929097 | | Downregulated |
| 0,301940391 | LOC101929098 | FALSE | Upregulated |
| 0,115153585 | LOC101929107 | FALSE | Upregulated |
| | LOC101929108 | | |
| | LOC101929114 | | |
| 0 | LOC101929119 | FALSE | Upregulated |
| | LOC101929121 | | |
| -0,010122896 | LOC101929124 | FALSE | Downregulated |
| 0,765555261 | LOC101929125 | FALSE | Upregulated |
| | LOC101929129 | | |
| | LOC101929131 | | |
| 0,466849363 | LOC101929132 | FALSE | Upregulated |
| | LOC101929134 | | |
| | LOC101929137 | | |
| -0,168197512 | LOC101929138 | FALSE | Downregulated |
| 0 | LOC101929142 | FALSE | Upregulated |
| -0,510066061 | LOC101929143 | FALSE | Downregulated |
| | LOC101929144 | | |
| 0 | LOC101929149 | FALSE | Upregulated |
| 0,635319022 | LOC101929151 | FALSE | Upregulated |
| | LOC101929152 | | |
| 0,448212425 | LOC101929153 | FALSE | Upregulated |
| -0,045460148 | LOC101929154 | FALSE | Downregulated |
| 0 | LOC101929155 | FALSE | Upregulated |
| | LOC101929156 | | |
| | LOC101929157 | | |
| -0,085651793 | LOC101929159 | FALSE | Downregulated |
| 0,21216126 | LOC101929160 | FALSE | Upregulated |
| | LOC101929161 | | |
| | LOC101929164 | | |
| 0,182252854 | LOC101929167 | FALSE | Upregulated |
| 0,453099891 | LOC101929174 | FALSE | Upregulated |
| -0,118080133 | LOC101929177 | FALSE | Downregulated |
| 0,980479811 | LOC101929178 | FALSE | Upregulated |
| | LOC101929180 | | |
| 0 | LOC101929184 | FALSE | Upregulated |
| | LOC101929187 | | |
| | LOC101929190 | | |
| 0,134737612 | LOC101929191 | FALSE | Upregulated |
| | LOC101929192 | | |
| | LOC101929194 | | |

| | | | |
|---|---|---|---|
| 0,500642585 | LOC101929198 | FALSE | Upregulated |
| 0,850964121 | LOC101929200 | FALSE | Upregulated |
| | LOC101929201 | | |
| | LOC101929202 | | |
| -0,141402278 | LOC101929206 | FALSE | Downregulated |
| | LOC101929214 | | |
| | LOC101929216 | | |
| | LOC101929217 | | |
| 0,236968756 | LOC101929220 | FALSE | Upregulated |
| -0,462025785 | LOC101929223 | FALSE | Downregulated |
| 1,130944776 | LOC101929227 | | Upregulated |
| | LOC101929228 | | |
| | LOC101929229 | | |
| 0,438834547 | LOC101929230 | FALSE | Upregulated |
| | LOC101929232 | | |
| | LOC101929236 | | |
| -0,374597617 | LOC101929241 | FALSE | Downregulated |
| -0,038745249 | LOC101929243 | FALSE | Downregulated |
| -0,039390294 | LOC101929248 | FALSE | Downregulated |
| -0,246379348 | LOC101929249 | FALSE | Downregulated |
| | LOC101929250 | | |
| 0,291843099 | LOC101929251 | FALSE | Upregulated |
| | LOC101929256 | | |
| | LOC101929257 | | |
| 0 | LOC101929258 | FALSE | Upregulated |
| 1,104347137 | LOC101929259 | | Upregulated |
| | LOC101929261 | | |
| 0 | LOC101929266 | FALSE | Upregulated |
| | LOC101929267 | | |
| -0,097426824 | LOC101929269 | FALSE | Downregulated |
| | LOC101929278 | | |
| 0,330669149 | LOC101929279 | FALSE | Upregulated |
| -0,841440235 | LOC101929283 | FALSE | Downregulated |
| 0,227981602 | LOC101929285 | FALSE | Upregulated |
| | LOC101929286 | | |
| -0,191196079 | LOC101929287 | FALSE | Downregulated |
| 0,475746456 | LOC101929288 | FALSE | Upregulated |
| 0,010008972 | LOC101929289 | FALSE | Upregulated |
| -0,088497722 | LOC101929296 | FALSE | Downregulated |
| | LOC101929298 | | |
| 0 | LOC101929300 | FALSE | Upregulated |
| | LOC101929301 | | |
| 0 | LOC101929304 | FALSE | Upregulated |
| 0 | LOC101929305 | FALSE | Upregulated |
| | LOC101929307 | | |
| -0,595547083 | LOC101929313 | FALSE | Downregulated |
| 0 | LOC101929314 | FALSE | Upregulated |
| | LOC101929316 | | |
| 0 | LOC101929319 | FALSE | Upregulated |
| | LOC101929320 | | |
| | LOC101929321 | | |

| | | | |
|---|---|---|---|
| 0 | LOC101929322 | FALSE | Upregulated |
| 0,418348818 | LOC101929325 | FALSE | Upregulated |
| | LOC101929326 | | |
| 0,335892759 | LOC101929327 | FALSE | Upregulated |
| -0,103288866 | LOC101929341 | FALSE | Downregulated |
| -0,341946601 | LOC101929346 | FALSE | Downregulated |
| | LOC101929348 | | |
| | LOC101929350 | | |
| | LOC101929351 | | |
| | LOC101929355 | | |
| -0,539084912 | LOC101929356 | FALSE | Downregulated |
| | LOC101929358 | | |
| | LOC101929359 | | |
| | LOC101929363 | | |
| 0 | LOC101929367 | FALSE | Upregulated |
| | LOC101929368 | | |
| 1,611916876 | LOC101929371 | | Upregulated |
| | LOC101929373 | | |
| 0,684840547 | LOC101929374 | FALSE | Upregulated |
| 0 | LOC101929375 | FALSE | Upregulated |
| 0,448212425 | LOC101929379 | FALSE | Upregulated |
| 0,184894664 | LOC101929381 | FALSE | Upregulated |
| | LOC101929382 | | |
| | LOC101929386 | | |
| | LOC101929388 | | |
| 0,988245917 | LOC101929393 | FALSE | Upregulated |
| | LOC101929398 | | |
| 0 | LOC101929408 | FALSE | Upregulated |
| | LOC101929411 | | |
| 0 | LOC101929414 | FALSE | Upregulated |
| 0,000977115 | LOC101929418 | FALSE | Upregulated |
| | LOC101929419 | | |
| 1,032069789 | LOC101929422 | | Upregulated |
| -0,802162851 | LOC101929427 | FALSE | Downregulated |
| -0,240143474 | LOC101929431 | FALSE | Downregulated |
| | LOC101929432 | | |
| 0,485330885 | LOC101929434 | FALSE | Upregulated |
| | LOC101929435 | | |
| | LOC101929437 | | |
| -1,132822635 | LOC101929443 | | Downregulated |
| | LOC101929445 | | |
| | LOC101929446 | | |
| 1,164500396 | LOC101929447 | | Upregulated |
| | LOC101929453 | | |
| | LOC101929456 | | |
| 0,420224994 | LOC101929457 | FALSE | Upregulated |
| 0,432670682 | LOC101929459 | FALSE | Upregulated |
| 0,417174874 | LOC101929460 | FALSE | Upregulated |
| -0,259344562 | LOC101929462 | FALSE | Downregulated |
| -0,33750326 | LOC101929463 | FALSE | Downregulated |
| 0,51169317 | LOC101929465 | FALSE | Upregulated |

| | | | |
|---|---|---|---|
| 0,458697222 | LOC101929467 | FALSE | Upregulated |
| 0,629901862 | LOC101929475 | FALSE | Upregulated |
| | LOC101929476 | | |
| | LOC101929477 | | |
| | LOC101929478 | | |
| 0,436450146 | LOC101929484 | FALSE | Upregulated |
| | LOC101929485 | | |
| | LOC101929489 | | |
| -0,205895092 | LOC101929490 | FALSE | Downregulated |
| 0 | LOC101929491 | FALSE | Upregulated |
| | LOC101929492 | | |
| | LOC101929493 | | |
| -0,53508256 | LOC101929494 | FALSE | Downregulated |
| | LOC101929495 | | |
| | LOC101929498 | | |
| | LOC101929507 | | |
| | LOC101929510 | | |
| -0,817442816 | LOC101929511 | FALSE | Downregulated |
| | LOC101929514 | | |
| | LOC101929515 | | |
| 0,334461714 | LOC101929518 | FALSE | Upregulated |
| 0,397470953 | LOC101929521 | FALSE | Upregulated |
| 2,102048362 | LOC101929523 | | Upregulated |
| 0,208631819 | LOC101929528 | FALSE | Upregulated |
| | LOC101929530 | | |
| 0,573063949 | LOC101929531 | FALSE | Upregulated |
| 0 | LOC101929534 | FALSE | Upregulated |
| | LOC101929535 | | |
| 0,475053529 | LOC101929536 | FALSE | Upregulated |
| 0,392211057 | LOC101929538 | FALSE | Upregulated |
| 0,886134975 | LOC101929542 | FALSE | Upregulated |
| | LOC101929544 | | |
| -0,396400231 | LOC101929547 | FALSE | Downregulated |
| 0,390950345 | LOC101929550 | FALSE | Upregulated |
| -1,302837283 | LOC101929552 | | Downregulated |
| 0,456007988 | LOC101929555 | FALSE | Upregulated |
| | LOC101929557 | | |
| 0,257583763 | LOC101929558 | FALSE | Upregulated |
| 0 | LOC101929560 | FALSE | Upregulated |
| 0,531372532 | LOC101929561 | FALSE | Upregulated |
| | LOC101929562 | | |
| | LOC101929563 | | |
| | LOC101929564 | | |
| 0 | LOC101929566 | FALSE | Upregulated |
| | LOC101929575 | | |
| | LOC101929576 | | |
| -0,344033122 | LOC101929580 | FALSE | Downregulated |
| 1,096162668 | LOC101929587 | | Upregulated |
| 0 | LOC101929590 | FALSE | Upregulated |
| | LOC101929593 | | |
| 0,608287046 | LOC101929594 | FALSE | Upregulated |

| | | | |
|---|---|---|---|
| 0 | LOC101929601 | FALSE | Upregulated |
| 0,07357428 | LOC101929602 | FALSE | Upregulated |
| 0,445661805 | LOC101929604 | FALSE | Upregulated |
| -0,23581695 | LOC101929609 | FALSE | Downregulated |
| | LOC101929610 | | |
| | LOC101929612 | | |
| 0,951550858 | LOC101929613 | FALSE | Upregulated |
| -0,188264822 | LOC101929614 | FALSE | Downregulated |
| -0,687602365 | LOC101929615 | FALSE | Downregulated |
| | LOC101929617 | | |
| | LOC101929618 | | |
| -0,334505345 | LOC101929619 | FALSE | Downregulated |
| | LOC101929620 | | |
| 0,307948109 | LOC101929625 | FALSE | Upregulated |
| 0 | LOC101929627 | FALSE | Upregulated |
| | LOC101929628 | | |
| 0,200854292 | LOC101929634 | FALSE | Upregulated |
| 0,100859122 | LOC101929637 | FALSE | Upregulated |
| 0,39768975 | LOC101929638 | FALSE | Upregulated |
| 0,38647539 | LOC101929641 | FALSE | Upregulated |
| | LOC101929644 | | |
| 0,975618017 | LOC101929645 | FALSE | Upregulated |
| | LOC101929646 | | |
| 0,133266244 | LOC101929647 | FALSE | Upregulated |
| 0,603435907 | LOC101929650 | FALSE | Upregulated |
| | LOC101929654 | | |
| | LOC101929658 | | |
| -0,353956412 | LOC101929661 | FALSE | Downregulated |
| 1,019403742 | LOC101929664 | | Upregulated |
| -0,4370808 | LOC101929666 | FALSE | Downregulated |
| | LOC101929674 | | |
| 0 | LOC101929675 | FALSE | Upregulated |
| | LOC101929681 | | |
| 0,797237485 | LOC101929683 | FALSE | Upregulated |
| 0,555718702 | LOC101929685 | FALSE | Upregulated |
| | LOC101929688 | | |
| | LOC101929690 | | |
| | LOC101929692 | | |
| -1,275522951 | LOC101929696 | | Downregulated |
| | LOC101929697 | | |
| | LOC101929700 | | |
| | LOC101929702 | | |
| 0,336449023 | LOC101929704 | FALSE | Upregulated |
| -0,061917153 | LOC101929713 | FALSE | Downregulated |
| -1,872385665 | LOC101929715 | | Downregulated |
| 0,354639565 | LOC101929724 | FALSE | Upregulated |
| | LOC101929725 | | |
| 0,292642911 | LOC101929726 | FALSE | Upregulated |
| 0,135515379 | LOC101929727 | FALSE | Upregulated |
| 0,306047487 | LOC101929728 | FALSE | Upregulated |
| | LOC101929733 | | |

| | | | |
|---|---|---|---|
| 0 | LOC101929738 | FALSE | Upregulated |
| 1,054401675 | LOC101929740 | | Upregulated |
| | LOC101929745 | | |
| | LOC101929748 | | |
| | LOC101929750 | | |
| 0 | LOC101929753 | FALSE | Upregulated |
| 0,328150729 | LOC101929754 | FALSE | Upregulated |
| | LOC101929755 | | |
| | LOC101929757 | | |
| 0,10005252 | LOC101929758 | FALSE | Upregulated |
| | LOC101929759 | | |
| 0,371515547 | LOC101929761 | FALSE | Upregulated |
| 0,583838154 | LOC101929762 | FALSE | Upregulated |
| | LOC101929763 | | |
| 0,142103894 | LOC101929766 | FALSE | Upregulated |
| | LOC101929770 | | |
| 0 | LOC101929777 | FALSE | Upregulated |
| -0,044626022 | LOC101929788 | FALSE | Downregulated |
| -0,162797754 | LOC101929798 | FALSE | Downregulated |
| 0 | LOC101929800 | FALSE | Upregulated |
| 0 | LOC101929803 | FALSE | Upregulated |
| | LOC101929806 | | |
| 1,492328052 | LOC101929814 | | Upregulated |
| | LOC101929815 | | |
| | LOC101929818 | | |
| 0 | LOC101929826 | FALSE | Upregulated |
| | LOC101929831 | | |
| -0,808488121 | LOC101929846 | FALSE | Downregulated |
| 0,452744178 | LOC101929860 | FALSE | Upregulated |
| | LOC101929863 | | |
| | LOC101929866 | | |
| -0,149242914 | LOC101929882 | FALSE | Downregulated |
| 1,393194061 | LOC101929887 | | Upregulated |
| 0,171951332 | LOC101929891 | FALSE | Upregulated |
| -0,447739312 | LOC101929897 | FALSE | Downregulated |
| 0,197677025 | LOC101929901 | FALSE | Upregulated |
| 0,196946248 | LOC101929918 | FALSE | Upregulated |
| | LOC101929922 | | |
| -0,290292076 | LOC101929946 | FALSE | Downregulated |
| 0,456903488 | LOC101929949 | FALSE | Upregulated |
| 0 | LOC101929953 | FALSE | Upregulated |
| -0,304812984 | LOC101929964 | FALSE | Downregulated |
| | LOC101929972 | | |
| 0,078076584 | LOC101929974 | FALSE | Upregulated |
| | LOC101929976 | | |
| -0,463467652 | LOC101929977 | FALSE | Downregulated |
| | LOC101929983 | | |
| | LOC101929989 | | |
| -0,510755639 | LOC101929998 | FALSE | Downregulated |
| 0 | LOC101930010 | FALSE | Upregulated |
| 0 | LOC101930026 | FALSE | Upregulated |

| | | | |
|---|---|---|---|
| | LOC101930048 | | |
| | LOC101930053 | | |
| 0 | LOC101930071 | FALSE | Upregulated |
| | LOC101930091 | | |
| | LOC101930098 | | |
| 0,449974312 | LOC101930100 | FALSE | Upregulated |
| | LOC101930109 | | |
| -0,010291857 | LOC101930114 | FALSE | Downregulated |
| -0,025773226 | LOC101930120 | FALSE | Downregulated |
| | LOC101930164 | | |
| 0 | LOC101930232 | FALSE | Upregulated |
| -0,016422875 | LOC101930249 | FALSE | Downregulated |
| | LOC101930259 | | |
| 0,30134032 | LOC101930276 | FALSE | Upregulated |
| | LOC101930294 | | |
| 0,270942022 | LOC101930305 | FALSE | Upregulated |
| | LOC101930310 | | |
| 0,224284321 | LOC101930332 | FALSE | Upregulated |
| | LOC101930343 | | |
| 0,731580245 | LOC101930357 | FALSE | Upregulated |
| 0,627905519 | LOC101930370 | FALSE | Upregulated |
| | LOC101930384 | | |
| 0,430773263 | LOC101930420 | FALSE | Upregulated |
| 0 | LOC101930434 | FALSE | Upregulated |
| -0,458843072 | LOC101930460 | FALSE | Downregulated |
| | LOC101930476 | | |
| | LOC101930484 | | |
| 0 | LOC101930496 | FALSE | Upregulated |
| | LOC101930523 | | |
| | LOC101930531 | | |
| 0,183137672 | LOC101930580 | FALSE | Upregulated |
| | LOC101930593 | | |
| 0,504877758 | LOC101930644 | FALSE | Upregulated |
| | LOC101930665 | | |
| | LOC102467074 | | |
| | LOC102467075 | | |
| | LOC102467079 | | |
| 0,400680188 | LOC102467080 | FALSE | Upregulated |
| | LOC102467081 | | |
| -0,355843433 | LOC102467146 | FALSE | Downregulated |
| 0 | LOC102467147 | FALSE | Upregulated |
| | LOC102467212 | | |
| | LOC102467213 | | |
| | LOC102467214 | | |
| 0 | LOC102467216 | FALSE | Upregulated |
| 0,215935724 | LOC102467217 | FALSE | Upregulated |
| | LOC102467223 | | |
| | LOC102467224 | | |
| 0,159266912 | LOC102467225 | FALSE | Upregulated |
| | LOC102467226 | | |
| | LOC102467655 | | |

| | | | |
|---|---|---|---|
| | LOC102477328 | | |
| | LOC102503427 | | |
| | LOC102503429 | | |
| -0,513638617 | LOC102524628 | FALSE | Downregulated |
| | LOC102546175 | | |
| | LOC102546227 | | |
| 0,320282505 | LOC102546228 | FALSE | Upregulated |
| 0,021505929 | LOC102546229 | FALSE | Upregulated |
| 0,233672704 | LOC102546294 | FALSE | Upregulated |
| 0 | LOC102546298 | FALSE | Upregulated |
| | LOC102546299 | | |
| | LOC102557615 | | |
| | LOC102577424 | | |
| 0 | LOC102577426 | FALSE | Upregulated |
| 0,165794655 | LOC102606465 | FALSE | Upregulated |
| | LOC102606466 | | |
| 0,078076584 | LOC102723305 | FALSE | Upregulated |
| 0,116482142 | LOC102723307 | FALSE | Upregulated |
| | LOC102723308 | | |
| 0,530157258 | LOC102723310 | FALSE | Upregulated |
| | LOC102723315 | | |
| 0,01196447 | LOC102723316 | FALSE | Upregulated |
| | LOC102723318 | | |
| -0,248462243 | LOC102723319 | FALSE | Downregulated |
| | LOC102723320 | | |
| | LOC102723321 | | |
| | LOC102723322 | | |
| -0,101226159 | LOC102723323 | FALSE | Downregulated |
| 0 | LOC102723324 | FALSE | Upregulated |
| | LOC102723325 | | |
| | LOC102723326 | | |
| -0,026071775 | LOC102723327 | FALSE | Downregulated |
| | LOC102723330 | | |
| -0,090338124 | LOC102723332 | FALSE | Downregulated |
| | LOC102723334 | | |
| | LOC102723336 | | |
| -0,023222185 | LOC102723337 | FALSE | Downregulated |
| | LOC102723338 | | |
| | LOC102723339 | | |
| -0,108708519 | LOC102723340 | FALSE | Downregulated |
| | LOC102723341 | | |
| | LOC102723343 | | |
| | LOC102723344 | | |
| | LOC102723345 | | |
| | LOC102723346 | | |
| | LOC102723347 | | |
| 0 | LOC102723348 | FALSE | Upregulated |
| 0,448212425 | LOC102723349 | FALSE | Upregulated |
| 0,068128077 | LOC102723350 | FALSE | Upregulated |
| | LOC102723351 | | |
| | LOC102723352 | | |

| | | | |
|---|---|---|---|
| 0,11912217 | LOC102723353 | FALSE | Upregulated |
| -0,116415395 | LOC102723354 | FALSE | Downregulated |
| | LOC102723355 | | |
| 0,599347263 | LOC102723357 | FALSE | Upregulated |
| | LOC102723358 | | |
| 1,014786555 | LOC102723359 | | Upregulated |
| 1,651123519 | LOC102723361 | | Upregulated |
| | LOC102723362 | | |
| | LOC102723363 | | |
| | LOC102723364 | | |
| -0,445919519 | LOC102723365 | FALSE | Downregulated |
| 0,203280391 | LOC102723366 | FALSE | Upregulated |
| 0,860502373 | LOC102723367 | FALSE | Upregulated |
| | LOC102723368 | | |
| | LOC102723370 | | |
| 0,529594143 | LOC102723371 | FALSE | Upregulated |
| | LOC102723372 | | |
| | LOC102723373 | | |
| 0,270869774 | LOC102723374 | FALSE | Upregulated |
| 0,314501411 | LOC102723375 | FALSE | Upregulated |
| | LOC102723376 | | |
| -0,202825612 | LOC102723377 | FALSE | Downregulated |
| | LOC102723378 | | |
| | LOC102723379 | | |
| 0,783306272 | LOC102723380 | FALSE | Upregulated |
| | LOC102723384 | | |
| 0 | LOC102723385 | FALSE | Upregulated |
| | LOC102723386 | | |
| | LOC102723388 | | |
| 0,137290392 | LOC102723389 | FALSE | Upregulated |
| | LOC102723390 | | |
| 0 | LOC102723391 | FALSE | Upregulated |
| 0 | LOC102723392 | FALSE | Upregulated |
| | LOC102723393 | | |
| | LOC102723394 | | |
| | LOC102723395 | | |
| | LOC102723396 | | |
| | LOC102723397 | | |
| -0,167597909 | LOC102723398 | FALSE | Downregulated |
| | LOC102723402 | | |
| | LOC102723403 | | |
| | LOC102723404 | | |
| 0,791535284 | LOC102723407 | FALSE | Upregulated |
| 0,031650427 | LOC102723408 | FALSE | Upregulated |
| 0,253622302 | LOC102723409 | FALSE | Upregulated |
| -1,7266145 | LOC102723410 | | Downregulated |
| | LOC102723411 | | |
| 0 | LOC102723412 | FALSE | Upregulated |
| | LOC102723413 | | |
| | LOC102723414 | | |
| -1,253529564 | LOC102723415 | | Downregulated |

| | | | |
|---|---|---|---|
| | LOC102723417 | | |
| | LOC102723418 | | |
| | LOC102723419 | | |
| 0,253974349 | LOC102723420 | FALSE | Upregulated |
| 0,172907575 | LOC102723421 | FALSE | Upregulated |
| | LOC102723423 | | |
| | LOC102723426 | | |
| | LOC102723427 | | |
| -0,124914911 | LOC102723428 | FALSE | Downregulated |
| | LOC102723429 | | |
| | LOC102723430 | | |
| | LOC102723431 | | |
| -0,625254282 | LOC102723433 | FALSE | Downregulated |
| | LOC102723434 | | |
| | LOC102723435 | | |
| | LOC102723436 | | |
| 0,444187748 | LOC102723437 | FALSE | Upregulated |
| | LOC102723439 | | |
| | LOC102723440 | | |
| -0,045460148 | LOC102723441 | FALSE | Downregulated |
| | LOC102723442 | | |
| | LOC102723443 | | |
| | LOC102723445 | | |
| 0,094390613 | LOC102723446 | FALSE | Upregulated |
| | LOC102723447 | | |
| | LOC102723448 | | |
| | LOC102723449 | | |
| -0,848920286 | LOC102723450 | FALSE | Downregulated |
| -0,119338803 | LOC102723452 | FALSE | Downregulated |
| | LOC102723453 | | |
| | LOC102723454 | | |
| 0,439670088 | LOC102723455 | FALSE | Upregulated |
| | LOC102723456 | | |
| | LOC102723458 | | |
| | LOC102723459 | | |
| | LOC102723460 | | |
| | LOC102723463 | | |
| 0,464212101 | LOC102723464 | FALSE | Upregulated |
| -0,320885215 | LOC102723467 | FALSE | Downregulated |
| 0,005843502 | LOC102723468 | FALSE | Upregulated |
| 0,033128344 | LOC102723470 | FALSE | Upregulated |
| | LOC102723471 | | |
| | LOC102723472 | | |
| | LOC102723473 | | |
| | LOC102723474 | | |
| | LOC102723476 | | |
| | LOC102723477 | | |
| | LOC102723479 | | |
| | LOC102723480 | | |
| 0,269418619 | LOC102723481 | FALSE | Upregulated |
| 0 | LOC102723482 | FALSE | Upregulated |

| | | | |
|---|---|---|---|
| 0,092550917 | LOC102723483 | FALSE | Upregulated |
| | LOC102723484 | | |
| | LOC102723485 | | |
| | LOC102723486 | | |
| | LOC102723487 | | |
| 0,468492047 | LOC102723488 | FALSE | Upregulated |
| | LOC102723490 | | |
| | LOC102723491 | | |
| 0 | LOC102723492 | FALSE | Upregulated |
| 1,97130894 | LOC102723493 | | Upregulated |
| | LOC102723494 | | |
| | LOC102723495 | | |
| | LOC102723496 | | |
| 0,226475358 | LOC102723497 | FALSE | Upregulated |
| | LOC102723499 | | |
| | LOC102723502 | | |
| | LOC102723503 | | |
| | LOC102723505 | | |
| 0,290473579 | LOC102723507 | FALSE | Upregulated |
| | LOC102723508 | | |
| 0,170669073 | LOC102723510 | FALSE | Upregulated |
| 0,081618748 | LOC102723513 | FALSE | Upregulated |
| | LOC102723514 | | |
| | LOC102723515 | | |
| | LOC102723516 | | |
| 0,006294609 | LOC102723517 | FALSE | Upregulated |
| | LOC102723518 | | |
| | LOC102723522 | | |
| | LOC102723523 | | |
| | LOC102723524 | | |
| 0 | LOC102723525 | FALSE | Upregulated |
| | LOC102723526 | | |
| | LOC102723527 | | |
| | LOC102723529 | | |
| | LOC102723530 | | |
| | LOC102723532 | | |
| | LOC102723533 | | |
| -0,444383056 | LOC102723534 | FALSE | Downregulated |
| | LOC102723536 | | |
| | LOC102723538 | | |
| -0,153523227 | LOC102723540 | FALSE | Downregulated |
| | LOC102723541 | | |
| 0,105475574 | LOC102723545 | FALSE | Upregulated |
| 0,745890535 | LOC102723546 | FALSE | Upregulated |
| 0,132793124 | LOC102723547 | FALSE | Upregulated |
| | LOC102723548 | | |
| -1,298470955 | LOC102723549 | | Downregulated |
| | LOC102723550 | | |
| | LOC102723551 | | |
| 0,335193947 | LOC102723552 | FALSE | Upregulated |
| -0,490783522 | LOC102723553 | FALSE | Downregulated |

| | | | |
|---|---|---|---|
| | LOC102723557 | | |
| 0,807721791 | LOC102723558 | FALSE | Upregulated |
| 0,326001878 | LOC102723559 | FALSE | Upregulated |
| | LOC102723560 | | |
| -0,09569471 | LOC102723561 | FALSE | Downregulated |
| | LOC102723562 | | |
| 0,517346638 | LOC102723563 | FALSE | Upregulated |
| 0,504379764 | LOC102723564 | FALSE | Upregulated |
| 0,10365882 | LOC102723565 | FALSE | Upregulated |
| | LOC102723567 | | |
| | LOC102723568 | | |
| 0,23393195 | LOC102723569 | FALSE | Upregulated |
| -0,204452305 | LOC102723570 | FALSE | Downregulated |
| | LOC102723571 | | |
| 0,686917838 | LOC102723572 | FALSE | Upregulated |
| 0,094199532 | LOC102723575 | FALSE | Upregulated |
| 0 | LOC102723576 | FALSE | Upregulated |
| | LOC102723577 | | |
| | LOC102723578 | | |
| 0,754557901 | LOC102723579 | FALSE | Upregulated |
| -0,040248954 | LOC102723587 | FALSE | Downregulated |
| | LOC102723588 | | |
| 0 | LOC102723590 | FALSE | Upregulated |
| 0,068980827 | LOC102723591 | FALSE | Upregulated |
| | LOC102723593 | | |
| -0,134847158 | LOC102723594 | FALSE | Downregulated |
| | LOC102723596 | | |
| | LOC102723599 | | |
| -0,009203821 | LOC102723602 | FALSE | Downregulated |
| | LOC102723605 | | |
| | LOC102723606 | | |
| 0 | LOC102723607 | FALSE | Upregulated |
| | LOC102723608 | | |
| | LOC102723609 | | |
| -1,830937873 | LOC102723610 | | Downregulated |
| | LOC102723612 | | |
| -0,575895344 | LOC102723616 | FALSE | Downregulated |
| | LOC102723617 | | |
| -0,180314928 | LOC102723619 | FALSE | Downregulated |
| | LOC102723620 | | |
| | LOC102723621 | | |
| 0,000977115 | LOC102723623 | FALSE | Upregulated |
| | LOC102723624 | | |
| -0,135034152 | LOC102723627 | FALSE | Downregulated |
| | LOC102723628 | | |
| | LOC102723629 | | |
| -0,023935704 | LOC102723630 | FALSE | Downregulated |
| | LOC102723631 | | |
| | LOC102723633 | | |
| 0 | LOC102723635 | FALSE | Upregulated |
| -0,513073238 | LOC102723636 | FALSE | Downregulated |

| | | | |
|---|---|---|---|
| | LOC102723637 | | |
| | LOC102723638 | | |
| | LOC102723639 | | |
| -0,647648914 | LOC102723640 | FALSE | Downregulated |
| -0,92176614 | LOC102723641 | FALSE | Downregulated |
| 0 | LOC102723642 | FALSE | Upregulated |
| -0,297549841 | LOC102723643 | FALSE | Downregulated |
| | LOC102723644 | | |
| | LOC102723645 | | |
| | LOC102723646 | | |
| -0,066647456 | LOC102723647 | FALSE | Downregulated |
| -1,386902624 | LOC102723648 | | Downregulated |
| | LOC102723649 | | |
| | LOC102723650 | | |
| -1,182138864 | LOC102723652 | | Downregulated |
| | LOC102723653 | | |
| | LOC102723654 | | |
| 0,472873733 | LOC102723655 | FALSE | Upregulated |
| 0,15921971 | LOC102723656 | FALSE | Upregulated |
| | LOC102723657 | | |
| | LOC102723658 | | |
| -0,445731034 | LOC102723659 | FALSE | Downregulated |
| | LOC102723661 | | |
| | LOC102723662 | | |
| 0 | LOC102723663 | FALSE | Upregulated |
| | LOC102723664 | | |
| | LOC102723667 | | |
| | LOC102723669 | | |
| -0,016422875 | LOC102723670 | FALSE | Downregulated |
| -0,167018048 | LOC102723672 | FALSE | Downregulated |
| | LOC102723673 | | |
| | LOC102723674 | | |
| 1,158286363 | LOC102723675 | | Upregulated |
| 0 | LOC102723676 | FALSE | Upregulated |
| -0,236656113 | LOC102723677 | FALSE | Downregulated |
| | LOC102723680 | | |
| 0,242342669 | LOC102723682 | FALSE | Upregulated |
| | LOC102723684 | | |
| 0,922956969 | LOC102723685 | FALSE | Upregulated |
| | LOC102723686 | | |
| -0,08369448 | LOC102723687 | FALSE | Downregulated |
| 0,014368547 | LOC102723690 | FALSE | Upregulated |
| -0,352769749 | LOC102723691 | FALSE | Downregulated |
| 0,058121574 | LOC102723692 | FALSE | Upregulated |
| 0,115722174 | LOC102723693 | FALSE | Upregulated |
| 0,351020734 | LOC102723695 | FALSE | Upregulated |
| -0,387124007 | LOC102723696 | FALSE | Downregulated |
| 0,025949278 | LOC102723697 | FALSE | Upregulated |
| | LOC102723698 | | |
| | LOC102723699 | | |
| 0 | LOC102723700 | FALSE | Upregulated |

| | | | |
|---|---|---|---|
| | LOC102723702 | | |
| -0,649534749 | LOC102723704 | FALSE | Downregulated |
| | LOC102723705 | | |
| | LOC102723706 | | |
| | LOC102723710 | | |
| | LOC102723712 | | |
| | LOC102723713 | | |
| | LOC102723714 | | |
| -0,081998664 | LOC102723715 | FALSE | Downregulated |
| -0,016349516 | LOC102723716 | FALSE | Downregulated |
| 0,522673212 | LOC102723717 | FALSE | Upregulated |
| -0,285256493 | LOC102723718 | FALSE | Downregulated |
| 0,18949436 | LOC102723719 | FALSE | Upregulated |
| 0,221502465 | LOC102723720 | FALSE | Upregulated |
| 0 | LOC102723721 | FALSE | Upregulated |
| | LOC102723723 | | |
| | LOC102723724 | | |
| | LOC102723725 | | |
| | LOC102723727 | | |
| | LOC102723729 | | |
| 0,444370831 | LOC102723730 | FALSE | Upregulated |
| | LOC102723731 | | |
| 1,792248319 | LOC102723732 | | Upregulated |
| 0,314732696 | LOC102723733 | FALSE | Upregulated |
| | LOC102723734 | | |
| | LOC102723736 | | |
| | LOC102723737 | | |
| | LOC102723738 | | |
| 0,443828724 | LOC102723739 | FALSE | Upregulated |
| | LOC102723740 | | |
| | LOC102723742 | | |
| 0,147395631 | LOC102723744 | FALSE | Upregulated |
| | LOC102723745 | | |
| 0,360304716 | LOC102723746 | FALSE | Upregulated |
| | LOC102723749 | | |
| | LOC102723751 | | |
| | LOC102723752 | | |
| | LOC102723754 | | |
| 0,454371813 | LOC102723755 | FALSE | Upregulated |
| -0,854900922 | LOC102723756 | FALSE | Downregulated |
| 0,542476731 | LOC102723757 | FALSE | Upregulated |
| -0,109373793 | LOC102723759 | FALSE | Downregulated |
| | LOC102723760 | | |
| 0,203935778 | LOC102723761 | FALSE | Upregulated |
| | LOC102723763 | | |
| | LOC102723764 | | |
| | LOC102723765 | | |
| | LOC102723768 | | |
| 1,243891943 | LOC102723770 | | Upregulated |
| 0,078076584 | LOC102723771 | FALSE | Upregulated |
| 0,069919228 | LOC102723774 | FALSE | Upregulated |

| | | | |
|---|---|---|---|
| 1,035934204 | LOC102723775 | | Upregulated |
| -0,309046216 | LOC102723777 | FALSE | Downregulated |
| | LOC102723778 | | |
| | LOC102723780 | | |
| 0,448212425 | LOC102723781 | FALSE | Upregulated |
| | LOC102723783 | | |
| | LOC102723784 | | |
| | LOC102723785 | | |
| | LOC102723786 | | |
| | LOC102723787 | | |
| | LOC102723789 | | |
| -0,111266856 | LOC102723790 | FALSE | Downregulated |
| 0,569778518 | LOC102723791 | FALSE | Upregulated |
| | LOC102723793 | | |
| | LOC102723795 | | |
| | LOC102723796 | | |
| 0,381069615 | LOC102723797 | FALSE | Upregulated |
| | LOC102723800 | | |
| -0,028588359 | LOC102723801 | FALSE | Downregulated |
| | LOC102723802 | | |
| | LOC102723803 | | |
| 0,71398756 | LOC102723805 | FALSE | Upregulated |
| | LOC102723807 | | |
| 0,20285811 | LOC102723808 | FALSE | Upregulated |
| | LOC102723810 | | |
| | LOC102723812 | | |
| 0,043864383 | LOC102723813 | FALSE | Upregulated |
| 0,189519651 | LOC102723814 | FALSE | Upregulated |
| | LOC102723816 | | |
| 0,681347877 | LOC102723817 | FALSE | Upregulated |
| | LOC102723818 | | |
| | LOC102723819 | | |
| | LOC102723821 | | |
| | LOC102723822 | | |
| | LOC102723823 | | |
| | LOC102723825 | | |
| | LOC102723826 | | |
| 0,392667046 | LOC102723828 | FALSE | Upregulated |
| 0,208450826 | LOC102723829 | FALSE | Upregulated |
| | LOC102723830 | | |
| | LOC102723831 | | |
| | LOC102723832 | | |
| | LOC102723833 | | |
| | LOC102723834 | | |
| | LOC102723835 | | |
| 0,228707221 | LOC102723836 | FALSE | Upregulated |
| | LOC102723837 | | |
| | LOC102723838 | | |
| 0 | LOC102723839 | FALSE | Upregulated |
| -0,171857362 | LOC102723840 | FALSE | Downregulated |
| | LOC102723841 | | |

| | | | |
|---|---|---|---|
| | LOC102723842 | | |
| 0,953953413 | LOC102723843 | FALSE | Upregulated |
| 0,448212425 | LOC102723844 | FALSE | Upregulated |
| | LOC102723846 | | |
| | LOC102723847 | | |
| | LOC102723848 | | |
| | LOC102723849 | | |
| | LOC102723850 | | |
| 0,046678197 | LOC102723854 | FALSE | Upregulated |
| | LOC102723855 | | |
| -0,127634009 | LOC102723856 | FALSE | Downregulated |
| | LOC102723857 | | |
| | LOC102723858 | | |
| 2,181625116 | LOC102723859 | | Upregulated |
| | LOC102723860 | | |
| 0,121515086 | LOC102723861 | FALSE | Upregulated |
| | LOC102723862 | | |
| 0,444271204 | LOC102723865 | FALSE | Upregulated |
| -0,190735019 | LOC102723866 | FALSE | Downregulated |
| 0,898021308 | LOC102723867 | FALSE | Upregulated |
| | LOC102723868 | | |
| | LOC102723869 | | |
| 1,20280153 | LOC102723870 | | Upregulated |
| | LOC102723871 | | |
| -0,572280035 | LOC102723872 | FALSE | Downregulated |
| | LOC102723873 | | |
| | LOC102723875 | | |
| 0,047839331 | LOC102723876 | FALSE | Upregulated |
| | LOC102723878 | | |
| | LOC102723879 | | |
| | LOC102723880 | | |
| | LOC102723881 | | |
| | LOC102723882 | | |
| | LOC102723883 | | |
| -0,190280408 | LOC102723884 | FALSE | Downregulated |
| 1,080720272 | LOC102723885 | | Upregulated |
| | LOC102723887 | | |
| 0 | LOC102723888 | FALSE | Upregulated |
| -0,444028373 | LOC102723889 | FALSE | Downregulated |
| | LOC102723890 | | |
| 1,126591037 | LOC102723891 | | Upregulated |
| 0 | LOC102723892 | FALSE | Upregulated |
| 0 | LOC102723894 | FALSE | Upregulated |
| | LOC102723895 | | |
| 0,434952741 | LOC102723896 | FALSE | Upregulated |
| | LOC102723898 | | |
| 0,020709324 | LOC102723899 | FALSE | Upregulated |
| | LOC102723900 | | |
| -0,205838745 | LOC102723901 | FALSE | Downregulated |
| 0 | LOC102723902 | FALSE | Upregulated |
| | LOC102723903 | | |

| | | | |
|---|---|---|---|
| 0 | LOC102723904 | FALSE | Upregulated |
| 0,643575461 | LOC102723905 | FALSE | Upregulated |
| | LOC102723906 | | |
| | LOC102723907 | | |
| | LOC102723909 | | |
| | LOC102723910 | | |
| | LOC102723911 | | |
| | LOC102723912 | | |
| 0 | LOC102723913 | FALSE | Upregulated |
| 0,084976497 | LOC102723914 | FALSE | Upregulated |
| 0,055871801 | LOC102723915 | FALSE | Upregulated |
| 0 | LOC102723916 | FALSE | Upregulated |
| 1,289497996 | LOC102723917 | | Upregulated |
| | LOC102723918 | | |
| 0,035001641 | LOC102723919 | FALSE | Upregulated |
| 0,228698663 | LOC102723920 | FALSE | Upregulated |
| | LOC102723921 | | |
| 0 | LOC102723922 | FALSE | Upregulated |
| 0,654564644 | LOC102723923 | FALSE | Upregulated |
| | LOC102723924 | | |
| 0,43475858 | LOC102723925 | FALSE | Upregulated |
| -0,454652691 | LOC102723926 | FALSE | Downregulated |
| -0,071161952 | LOC102723927 | FALSE | Downregulated |
| | LOC102723928 | | |
| | LOC102723931 | | |
| | LOC102723932 | | |
| | LOC102723933 | | |
| 0,333941866 | LOC102723934 | FALSE | Upregulated |
| | LOC102723935 | | |
| | LOC102723936 | | |
| | LOC102723937 | | |
| | LOC102723938 | | |
| | LOC102723939 | | |
| 0 | LOC102723940 | FALSE | Upregulated |
| | LOC102723941 | | |
| -0,272434551 | LOC102723942 | FALSE | Downregulated |
| | LOC102723944 | | |
| 0,38647539 | LOC102723946 | FALSE | Upregulated |
| | LOC102723947 | | |
| | LOC102723948 | | |
| -1,267072041 | LOC102723949 | | Downregulated |
| | LOC102723950 | | |
| | LOC102723951 | | |
| | LOC102723952 | | |
| -1,027438858 | LOC102723953 | | Downregulated |
| 0,052447532 | LOC102723955 | FALSE | Upregulated |
| 0,284854014 | LOC102723956 | FALSE | Upregulated |
| | LOC102723957 | | |
| 0,548787992 | LOC102723958 | FALSE | Upregulated |
| 0 | LOC102723959 | FALSE | Upregulated |
| | LOC102723961 | | |

| | | | |
|---|---|---|---|
| | LOC102723962 | | |
| 0,496657307 | LOC102723963 | FALSE | Upregulated |
| -0,645036089 | LOC102723965 | FALSE | Downregulated |
| 0 | LOC102723966 | FALSE | Upregulated |
| | LOC102723967 | | |
| | LOC102723968 | | |
| | LOC102723969 | | |
| | LOC102723970 | | |
| | LOC102723971 | | |
| -0,148637222 | LOC102723973 | FALSE | Downregulated |
| -1,047870941 | LOC102723974 | | Downregulated |
| -0,452885273 | LOC102723975 | FALSE | Downregulated |
| | LOC102723976 | | |
| | LOC102723977 | | |
| | LOC102723978 | | |
| | LOC102723979 | | |
| 0,939675907 | LOC102723981 | FALSE | Upregulated |
| -0,178631953 | LOC102723983 | FALSE | Downregulated |
| | LOC102723984 | | |
| -0,016422875 | LOC102723985 | FALSE | Downregulated |
| | LOC102723986 | | |
| | LOC102723987 | | |
| 0,420155632 | LOC102723988 | FALSE | Upregulated |
| 0,047839331 | LOC102723993 | FALSE | Upregulated |
| | LOC102723994 | | |
| 1,073726718 | LOC102723995 | | Upregulated |
| | LOC102724000 | | |
| | LOC102724001 | | |
| 0,742574852 | LOC102724003 | FALSE | Upregulated |
| | LOC102724004 | | |
| | LOC102724007 | | |
| | LOC102724008 | | |
| | LOC102724011 | | |
| 0 | LOC102724012 | FALSE | Upregulated |
| | LOC102724013 | | |
| | LOC102724014 | | |
| 0,011211767 | LOC102724015 | FALSE | Upregulated |
| 0,31364839 | LOC102724017 | FALSE | Upregulated |
| 0,100713889 | LOC102724018 | FALSE | Upregulated |
| -1,913438142 | LOC102724019 | | Downregulated |
| | LOC102724020 | | |
| | LOC102724021 | | |
| | LOC102724022 | | |
| | LOC102724027 | | |
| 0,386015862 | LOC102724028 | FALSE | Upregulated |
| | LOC102724029 | | |
| -0,365096571 | LOC102724030 | FALSE | Downregulated |
| 0,669595622 | LOC102724031 | FALSE | Upregulated |
| | LOC102724032 | | |
| | LOC102724033 | | |
| 0,552509917 | LOC102724034 | FALSE | Upregulated |

| | | | |
|---|---|---|---|
| | LOC102724036 | | |
| | LOC102724038 | | |
| | LOC102724039 | | |
| | LOC102724041 | | |
| | LOC102724044 | | |
| | LOC102724045 | | |
| | LOC102724046 | | |
| | LOC102724048 | | |
| 0,970885411 | LOC102724049 | FALSE | Upregulated |
| | LOC102724050 | | |
| | LOC102724051 | | |
| | LOC102724052 | | |
| | LOC102724053 | | |
| | LOC102724054 | | |
| | LOC102724055 | | |
| | LOC102724056 | | |
| 0,730149675 | LOC102724057 | FALSE | Upregulated |
| | LOC102724058 | | |
| 0,099414026 | LOC102724059 | FALSE | Upregulated |
| 0,291699957 | LOC102724060 | FALSE | Upregulated |
| | LOC102724062 | | |
| | LOC102724063 | | |
| -0,398669229 | LOC102724064 | FALSE | Downregulated |
| 0 | LOC102724065 | FALSE | Upregulated |
| | LOC102724067 | | |
| -0,0229992 | LOC102724068 | FALSE | Downregulated |
| | LOC102724069 | | |
| | LOC102724071 | | |
| 0,490438862 | LOC102724072 | FALSE | Upregulated |
| 0,579881847 | LOC102724073 | FALSE | Upregulated |
| | LOC102724074 | | |
| 0,391021834 | LOC102724075 | FALSE | Upregulated |
| | LOC102724076 | | |
| | LOC102724078 | | |
| 0,398213488 | LOC102724079 | FALSE | Upregulated |
| | LOC102724080 | | |
| | LOC102724081 | | |
| | LOC102724082 | | |
| 0,020789395 | LOC102724084 | FALSE | Upregulated |
| | LOC102724085 | | |
| | LOC102724086 | | |
| | LOC102724087 | | |
| | LOC102724088 | | |
| | LOC102724089 | | |
| 0,020789395 | LOC102724090 | FALSE | Upregulated |
| | LOC102724091 | | |
| 0,71038366 | LOC102724092 | FALSE | Upregulated |
| -0,279257629 | LOC102724093 | FALSE | Downregulated |
| 0,046678197 | LOC102724094 | FALSE | Upregulated |
| | LOC102724095 | | |
| 0,962258438 | LOC102724096 | FALSE | Upregulated |

| | | | |
|---|---|---|---|
| 0 | LOC102724097 | FALSE | Upregulated |
| -0,237065891 | LOC102724098 | FALSE | Downregulated |
| | LOC102724099 | | |
| | LOC102724101 | | |
| | LOC102724102 | | |
| | LOC102724104 | | |
| -0,971085146 | LOC102724105 | FALSE | Downregulated |
| -0,027923931 | LOC102724106 | FALSE | Downregulated |
| | LOC102724107 | | |
| 0,882648809 | LOC102724108 | FALSE | Upregulated |
| -0,026346587 | LOC102724109 | FALSE | Downregulated |
| | LOC102724111 | | |
| -0,57369258 | LOC102724112 | FALSE | Downregulated |
| | LOC102724113 | | |
| | LOC102724114 | | |
| | LOC102724115 | | |
| | LOC102724118 | | |
| | LOC102724119 | | |
| -0,327595011 | LOC102724120 | FALSE | Downregulated |
| 0,154840479 | LOC102724121 | FALSE | Upregulated |
| 0 | LOC102724122 | FALSE | Upregulated |
| | LOC102724123 | | |
| | LOC102724125 | | |
| | LOC102724127 | | |
| | LOC102724128 | | |
| 0 | LOC102724130 | FALSE | Upregulated |
| | LOC102724131 | | |
| 0,29769581 | LOC102724132 | FALSE | Upregulated |
| | LOC102724134 | | |
| | LOC102724136 | | |
| | LOC102724137 | | |
| 0 | LOC102724138 | FALSE | Upregulated |
| 0 | LOC102724139 | FALSE | Upregulated |
| 0 | LOC102724140 | FALSE | Upregulated |
| | LOC102724142 | | |
| -0,200220488 | LOC102724143 | FALSE | Downregulated |
| | LOC102724145 | | |
| | LOC102724146 | | |
| | LOC102724148 | | |
| 0,129370336 | LOC102724149 | FALSE | Upregulated |
| | LOC102724150 | | |
| | LOC102724151 | | |
| | LOC102724152 | | |
| 0 | LOC102724154 | FALSE | Upregulated |
| 0,394015708 | LOC102724155 | FALSE | Upregulated |
| 0,102875907 | LOC102724156 | FALSE | Upregulated |
| | LOC102724158 | | |
| | LOC102724160 | | |
| | LOC102724161 | | |
| 0,572335353 | LOC102724162 | FALSE | Upregulated |
| | LOC102724165 | | |

| | | | |
|---|---|---|---|
| | LOC102724166 | | |
| | LOC102724167 | | |
| 0,627505582 | LOC102724168 | FALSE | Upregulated |
| 0,026076447 | LOC102724169 | FALSE | Upregulated |
| 0,285548704 | LOC102724170 | FALSE | Upregulated |
| 0,398213488 | LOC102724171 | FALSE | Upregulated |
| | LOC102724173 | | |
| | LOC102724174 | | |
| 0 | LOC102724176 | FALSE | Upregulated |
| | LOC102724180 | | |
| | LOC102724182 | | |
| | LOC102724183 | | |
| -1,189546042 | LOC102724184 | | Downregulated |
| | LOC102724185 | | |
| | LOC102724186 | | |
| | LOC102724188 | | |
| | LOC102724189 | | |
| -0,033062129 | LOC102724190 | FALSE | Downregulated |
| | LOC102724194 | | |
| 0 | LOC102724195 | FALSE | Upregulated |
| 0,389457687 | LOC102724197 | FALSE | Upregulated |
| | LOC102724199 | | |
| 0,088235463 | LOC102724201 | FALSE | Upregulated |
| | LOC102724202 | | |
| | LOC102724203 | | |
| | LOC102724204 | | |
| | LOC102724205 | | |
| | LOC102724207 | | |
| -0,144843098 | LOC102724208 | FALSE | Downregulated |
| 0,321581367 | LOC102724209 | FALSE | Upregulated |
| | LOC102724210 | | |
| | LOC102724211 | | |
| | LOC102724212 | | |
| 0 | LOC102724213 | FALSE | Upregulated |
| | LOC102724214 | | |
| | LOC102724215 | | |
| | LOC102724216 | | |
| | LOC102724218 | | |
| | LOC102724220 | | |
| | LOC102724222 | | |
| | LOC102724224 | | |
| | LOC102724225 | | |
| | LOC102724226 | | |
| | LOC102724227 | | |
| | LOC102724228 | | |
| 0,073403815 | LOC102724229 | FALSE | Upregulated |
| 1,00885435 | LOC102724230 | | Upregulated |
| 0,227145094 | LOC102724231 | FALSE | Upregulated |
| 0,451117039 | LOC102724232 | FALSE | Upregulated |
| | LOC102724234 | | |
| 1,636084593 | LOC102724237 | | Upregulated |

| | | | |
|---|---|---|---|
| | LOC102724239 | | |
| | LOC102724241 | | |
| | LOC102724242 | | |
| -0,303669921 | LOC102724244 | FALSE | Downregulated |
| 0,514930209 | LOC102724245 | FALSE | Upregulated |
| | LOC102724246 | | |
| | LOC102724247 | | |
| 0,408731196 | LOC102724248 | FALSE | Upregulated |
| 0,145726673 | LOC102724249 | FALSE | Upregulated |
| | LOC102724251 | | |
| -1,733363684 | LOC102724252 | | Downregulated |
| | LOC102724253 | | |
| -0,400532563 | LOC102724256 | FALSE | Downregulated |
| | LOC102724257 | | |
| -0,227638475 | LOC102724258 | FALSE | Downregulated |
| 1,445103073 | LOC102724259 | | Upregulated |
| | LOC102724260 | | |
| 0,407886572 | LOC102724261 | FALSE | Upregulated |
| -0,869400333 | LOC102724263 | FALSE | Downregulated |
| | LOC102724264 | | |
| -0,032083618 | LOC102724265 | FALSE | Downregulated |
| | LOC102724266 | | |
| | LOC102724267 | | |
| | LOC102724268 | | |
| | LOC102724270 | | |
| | LOC102724271 | | |
| -0,000173492 | LOC102724272 | FALSE | Downregulated |
| 1,765184015 | LOC102724274 | | Upregulated |
| 0,66375256 | LOC102724275 | FALSE | Upregulated |
| 1,201266313 | LOC102724276 | | Upregulated |
| | LOC102724277 | | |
| | LOC102724278 | | |
| | LOC102724279 | | |
| | LOC102724280 | | |
| 0,099452879 | LOC102724282 | FALSE | Upregulated |
| -0,173880312 | LOC102724283 | FALSE | Downregulated |
| | LOC102724284 | | |
| -0,129000727 | LOC102724285 | FALSE | Downregulated |
| | LOC102724287 | | |
| | LOC102724288 | | |
| | LOC102724289 | | |
| | LOC102724290 | | |
| | LOC102724294 | | |
| -0,29639627 | LOC102724295 | FALSE | Downregulated |
| -0,426357749 | LOC102724296 | FALSE | Downregulated |
| 0 | LOC102724297 | FALSE | Upregulated |
| 0 | LOC102724298 | FALSE | Upregulated |
| | LOC102724299 | | |
| 0,453099891 | LOC102724300 | FALSE | Upregulated |
| | LOC102724301 | | |
| | LOC102724302 | | |

| | | | |
|---|---|---|---|
| | LOC102724304 | | |
| | LOC102724305 | | |
| 1,010362506 | LOC102724306 | | Upregulated |
| 0,320316568 | LOC102724307 | FALSE | Upregulated |
| | LOC102724308 | | |
| 0 | LOC102724309 | FALSE | Upregulated |
| | LOC102724311 | | |
| | LOC102724313 | | |
| | LOC102724315 | | |
| | LOC102724317 | | |
| | LOC102724318 | | |
| | LOC102724321 | | |
| 1,719922231 | LOC102724323 | | Upregulated |
| | LOC102724324 | | |
| | LOC102724325 | | |
| | LOC102724326 | | |
| | LOC102724327 | | |
| 0,150952716 | LOC102724328 | FALSE | Upregulated |
| 0,142089508 | LOC102724329 | FALSE | Upregulated |
| 1,14553016 | LOC102724330 | | Upregulated |
| | LOC102724331 | | |
| | LOC102724332 | | |
| | LOC102724333 | | |
| 1,042043054 | LOC102724334 | | Upregulated |
| | LOC102724337 | | |
| | LOC102724338 | | |
| 0,064850485 | LOC102724339 | FALSE | Upregulated |
| | LOC102724340 | | |
| | LOC102724341 | | |
| 0,742063562 | LOC102724343 | FALSE | Upregulated |
| | LOC102724344 | | |
| | LOC102724346 | | |
| | LOC102724347 | | |
| -0,276096979 | LOC102724349 | FALSE | Downregulated |
| 0 | LOC102724351 | FALSE | Upregulated |
| 0 | LOC102724352 | FALSE | Upregulated |
| | LOC102724353 | | |
| 0,266428315 | LOC102724354 | FALSE | Upregulated |
| | LOC102724355 | | |
| | LOC102724357 | | |
| | LOC102724358 | | |
| | LOC102724359 | | |
| | LOC102724360 | | |
| 0,397241336 | LOC102724361 | FALSE | Upregulated |
| | LOC102724362 | | |
| 0,201311087 | LOC102724363 | FALSE | Upregulated |
| | LOC102724366 | | |
| | LOC102724367 | | |
| | LOC102724373 | | |
| | LOC102724375 | | |
| 0,554073811 | LOC102724377 | FALSE | Upregulated |

| | | | |
|---|---|---|---|
| 0,420594267 | LOC102724378 | FALSE | Upregulated |
| | LOC102724379 | | |
| 0 | LOC102724380 | FALSE | Upregulated |
| | LOC102724381 | | |
| | LOC102724382 | | |
| 0,069389839 | LOC102724384 | FALSE | Upregulated |
| -0,202295959 | LOC102724385 | FALSE | Downregulated |
| | LOC102724386 | | |
| 0,065913471 | LOC102724389 | FALSE | Upregulated |
| 0,331978959 | LOC102724392 | FALSE | Upregulated |
| | LOC102724393 | | |
| 0,495298615 | LOC102724394 | FALSE | Upregulated |
| | LOC102724395 | | |
| | LOC102724396 | | |
| 0,354248997 | LOC102724397 | FALSE | Upregulated |
| -0,237332289 | LOC102724398 | FALSE | Downregulated |
| -0,45342311 | LOC102724399 | FALSE | Downregulated |
| | LOC102724400 | | |
| -0,142647833 | LOC102724402 | FALSE | Downregulated |
| | LOC102724403 | | |
| 0 | LOC102724404 | FALSE | Upregulated |
| | LOC102724405 | | |
| 0,560291623 | LOC102724406 | FALSE | Upregulated |
| | LOC102724407 | | |
| -0,808020603 | LOC102724409 | FALSE | Downregulated |
| | LOC102724410 | | |
| 0 | LOC102724411 | FALSE | Upregulated |
| | LOC102724412 | | |
| 0,68806821 | LOC102724413 | FALSE | Upregulated |
| | LOC102724414 | | |
| | LOC102724415 | | |
| 0,448212425 | LOC102724416 | FALSE | Upregulated |
| | LOC102724417 | | |
| | LOC102724418 | | |
| | LOC102724419 | | |
| 0,253138533 | LOC102724420 | FALSE | Upregulated |
| | LOC102724421 | | |
| | LOC102724422 | | |
| 0,359524145 | LOC102724424 | FALSE | Upregulated |
| | LOC102724425 | | |
| -0,089757783 | LOC102724426 | FALSE | Downregulated |
| | LOC102724427 | | |
| | LOC102724429 | | |
| | LOC102724431 | | |
| 0,390950345 | LOC102724432 | FALSE | Upregulated |
| 0 | LOC102724433 | FALSE | Upregulated |
| 0 | LOC102724434 | FALSE | Upregulated |
| | LOC102724435 | | |
| -0,743804323 | LOC102724436 | FALSE | Downregulated |
| | LOC102724438 | | |
| | LOC102724439 | | |

| | | | |
|---|---|---|---|
| | LOC102724440 | | |
| | LOC102724443 | | |
| 0,52304738 | LOC102724444 | FALSE | Upregulated |
| 0,426437581 | LOC102724446 | FALSE | Upregulated |
| -0,619645897 | LOC102724449 | FALSE | Downregulated |
| 1,271028078 | LOC102724450 | | Upregulated |
| 0,459180725 | LOC102724451 | FALSE | Upregulated |
| | LOC102724452 | | |
| | LOC102724453 | | |
| | LOC102724454 | | |
| | LOC102724456 | | |
| 0,397913108 | LOC102724457 | FALSE | Upregulated |
| | LOC102724458 | | |
| | LOC102724460 | | |
| | LOC102724461 | | |
| -1,00273559 | LOC102724462 | | Downregulated |
| 0,277024005 | LOC102724463 | FALSE | Upregulated |
| -0,069831969 | LOC102724464 | FALSE | Downregulated |
| | LOC102724465 | | |
| 0 | LOC102724467 | FALSE | Upregulated |
| | LOC102724469 | | |
| | LOC102724470 | | |
| | LOC102724471 | | |
| 0,090458955 | LOC102724472 | FALSE | Upregulated |
| | LOC102724473 | | |
| 0,041943384 | LOC102724477 | FALSE | Upregulated |
| | LOC102724478 | | |
| | LOC102724479 | | |
| | LOC102724480 | | |
| 0 | LOC102724481 | FALSE | Upregulated |
| | LOC102724482 | | |
| | LOC102724485 | | |
| | LOC102724486 | | |
| -0,957718793 | LOC102724487 | FALSE | Downregulated |
| 0 | LOC102724489 | FALSE | Upregulated |
| | LOC102724491 | | |
| | LOC102724492 | | |
| | LOC102724494 | | |
| | LOC102724495 | | |
| | LOC102724496 | | |
| 0,477459454 | LOC102724497 | FALSE | Upregulated |
| | LOC102724498 | | |
| | LOC102724500 | | |
| | LOC102724501 | | |
| 0,034473335 | LOC102724502 | FALSE | Upregulated |
| | LOC102724503 | | |
| | LOC102724505 | | |
| 0,199014448 | LOC102724506 | FALSE | Upregulated |
| 1,273356522 | LOC102724508 | | Upregulated |
| | LOC102724509 | | |
| | LOC102724510 | | |

| | | | |
|---|---|---|---|
| | LOC102724511 | | |
| | LOC102724512 | | |
| 0,426437581 | LOC102724513 | FALSE | Upregulated |
| -0,864132794 | LOC102724517 | FALSE | Downregulated |
| 0,072209577 | LOC102724519 | FALSE | Upregulated |
| | LOC102724520 | | |
| | LOC102724521 | | |
| | LOC102724523 | | |
| -0,497016012 | LOC102724524 | FALSE | Downregulated |
| | LOC102724527 | | |
| | LOC102724529 | | |
| -0,117520658 | LOC102724530 | FALSE | Downregulated |
| | LOC102724531 | | |
| -1,185302397 | LOC102724532 | | Downregulated |
| | LOC102724533 | | |
| | LOC102724534 | | |
| 0,157066565 | LOC102724537 | FALSE | Upregulated |
| | LOC102724538 | | |
| 0,206477519 | LOC102724539 | FALSE | Upregulated |
| | LOC102724540 | | |
| -0,185972058 | LOC102724541 | FALSE | Downregulated |
| | LOC102724542 | | |
| 0,528280024 | LOC102724543 | FALSE | Upregulated |
| | LOC102724544 | | |
| 2,350921259 | LOC102724545 | | Upregulated |
| 0 | LOC102724551 | FALSE | Upregulated |
| 0 | LOC102724553 | FALSE | Upregulated |
| 0,436414226 | LOC102724555 | FALSE | Upregulated |
| | LOC102724557 | | |
| 0,100713889 | LOC102724558 | FALSE | Upregulated |
| | LOC102724561 | | |
| | LOC102724564 | | |
| | LOC102724565 | | |
| | LOC102724566 | | |
| | LOC102724567 | | |
| 0,012211947 | LOC102724568 | FALSE | Upregulated |
| -1,053336775 | LOC102724569 | | Downregulated |
| -0,141041393 | LOC102724570 | FALSE | Downregulated |
| 0,805902686 | LOC102724571 | FALSE | Upregulated |
| | LOC102724572 | | |
| 0,046404755 | LOC102724573 | FALSE | Upregulated |
| | LOC102724574 | | |
| | LOC102724575 | | |
| | LOC102724576 | | |
| | LOC102724577 | | |
| | LOC102724579 | | |
| -1,919177395 | LOC102724580 | | Downregulated |
| 0,885412267 | LOC102724581 | FALSE | Upregulated |
| | LOC102724582 | | |
| -0,275294902 | LOC102724586 | FALSE | Downregulated |
| | LOC102724588 | | |

| | | | |
|---|---|---|---|
| | LOC102724589 | | |
| | LOC102724590 | | |
| | LOC102724591 | | |
| | LOC102724592 | | |
| 1,052324972 | LOC102724593 | | Upregulated |
| -2,092581552 | LOC102724594 | | Downregulated |
| | LOC102724595 | | |
| 0 | LOC102724596 | FALSE | Upregulated |
| 0 | LOC102724599 | FALSE | Upregulated |
| 0,640519847 | LOC102724604 | FALSE | Upregulated |
| | LOC102724605 | | |
| | LOC102724606 | | |
| 0,259011551 | LOC102724608 | FALSE | Upregulated |
| | LOC102724609 | | |
| 0,136937676 | LOC102724610 | FALSE | Upregulated |
| | LOC102724612 | | |
| | LOC102724613 | | |
| 0,223714896 | LOC102724614 | FALSE | Upregulated |
| -1,417208086 | LOC102724615 | | Downregulated |
| | LOC102724616 | | |
| | LOC102724617 | | |
| | LOC102724618 | | |
| | LOC102724619 | | |
| 0 | LOC102724620 | FALSE | Upregulated |
| | LOC102724622 | | |
| | LOC102724623 | | |
| | LOC102724625 | | |
| | LOC102724627 | | |
| | LOC102724628 | | |
| | LOC102724629 | | |
| -0,029163266 | LOC102724630 | FALSE | Downregulated |
| | LOC102724631 | | |
| 0 | LOC102724632 | FALSE | Upregulated |
| 0,481637332 | LOC102724633 | FALSE | Upregulated |
| 0,148500228 | LOC102724636 | FALSE | Upregulated |
| | LOC102724637 | | |
| | LOC102724638 | | |
| | LOC102724640 | | |
| 0,057433286 | LOC102724641 | FALSE | Upregulated |
| 0,210333217 | LOC102724642 | FALSE | Upregulated |
| | LOC102724644 | | |
| -1,52634425 | LOC102724645 | | Downregulated |
| | LOC102724646 | | |
| | LOC102724647 | | |
| | LOC102724648 | | |
| | LOC102724649 | | |
| | LOC102724650 | | |
| | LOC102724651 | | |
| 0 | LOC102724653 | FALSE | Upregulated |
| | LOC102724654 | | |
| -0,964518652 | LOC102724655 | FALSE | Downregulated |

| | | | |
|---|---|---|---|
| | LOC102724657 | | |
| 0,47825147 | LOC102724659 | FALSE | Upregulated |
| 0,689444672 | LOC102724660 | FALSE | Upregulated |
| 1,380608288 | LOC102724661 | | Upregulated |
| -0,511808847 | LOC102724662 | FALSE | Downregulated |
| -0,132371374 | LOC102724663 | FALSE | Downregulated |
| | LOC102724664 | | |
| | LOC102724665 | | |
| | LOC102724666 | | |
| 0 | LOC102724667 | FALSE | Upregulated |
| -0,070194979 | LOC102724668 | FALSE | Downregulated |
| | LOC102724669 | | |
| | LOC102724670 | | |
| | LOC102724671 | | |
| 0,330407326 | LOC102724675 | FALSE | Upregulated |
| 0,100713889 | LOC102724676 | FALSE | Upregulated |
| | LOC102724678 | | |
| | LOC102724679 | | |
| | LOC102724680 | | |
| -0,259772288 | LOC102724681 | FALSE | Downregulated |
| -0,976848953 | LOC102724682 | FALSE | Downregulated |
| | LOC102724684 | | |
| -1,21117824 | LOC102724685 | | Downregulated |
| 0,231969019 | LOC102724686 | FALSE | Upregulated |
| -0,314042033 | LOC102724687 | FALSE | Downregulated |
| | LOC102724688 | | |
| | LOC102724689 | | |
| | LOC102724690 | | |
| | LOC102724692 | | |
| | LOC102724693 | | |
| | LOC102724694 | | |
| -0,139858749 | LOC102724695 | FALSE | Downregulated |
| 0,227373709 | LOC102724697 | FALSE | Upregulated |
| | LOC102724698 | | |
| | LOC102724700 | | |
| 0 | LOC102724702 | FALSE | Upregulated |
| 0,001888561 | LOC102724703 | FALSE | Upregulated |
| | LOC102724704 | | |
| | LOC102724707 | | |
| -0,584466749 | LOC102724708 | FALSE | Downregulated |
| | LOC102724710 | | |
| 0,124239177 | LOC102724711 | FALSE | Upregulated |
| | LOC102724712 | | |
| | LOC102724713 | | |
| 0 | LOC102724714 | FALSE | Upregulated |
| 0 | LOC102724715 | FALSE | Upregulated |
| | LOC102724716 | | |
| | LOC102724717 | | |
| | LOC102724718 | | |
| 0,009626889 | LOC102724719 | FALSE | Upregulated |
| | LOC102724721 | | |

| | | | |
|---|---|---|---|
| | LOC102724722 | | |
| | LOC102724723 | | |
| | LOC102724728 | | |
| | LOC102724729 | | |
| -0,210514977 | LOC102724731 | FALSE | Downregulated |
| | LOC102724732 | | |
| | LOC102724734 | | |
| | LOC102724735 | | |
| | LOC102724736 | | |
| 0,084815353 | LOC102724737 | FALSE | Upregulated |
| | LOC102724738 | | |
| | LOC102724739 | | |
| -0,719800457 | LOC102724740 | FALSE | Downregulated |
| | LOC102724741 | | |
| | LOC102724742 | | |
| -0,017453011 | LOC102724743 | FALSE | Downregulated |
| | LOC102724744 | | |
| 0 | LOC102724745 | FALSE | Upregulated |
| 0,413348775 | LOC102724746 | FALSE | Upregulated |
| | LOC102724747 | | |
| 0 | LOC102724749 | FALSE | Upregulated |
| 0,021317681 | LOC102724751 | FALSE | Upregulated |
| -0,23021486 | LOC102724752 | FALSE | Downregulated |
| | LOC102724753 | | |
| 0,168247394 | LOC102724754 | FALSE | Upregulated |
| | LOC102724755 | | |
| 0,787285238 | LOC102724756 | FALSE | Upregulated |
| 0 | LOC102724758 | FALSE | Upregulated |
| 0,514358572 | LOC102724759 | FALSE | Upregulated |
| 0,011211767 | LOC102724762 | FALSE | Upregulated |
| | LOC102724763 | | |
| -0,499925376 | LOC102724764 | FALSE | Downregulated |
| 0,821527439 | LOC102724765 | FALSE | Upregulated |
| 0,526196821 | LOC102724766 | FALSE | Upregulated |
| | LOC102724767 | | |
| 0,149599665 | LOC102724768 | FALSE | Upregulated |
| | LOC102724771 | | |
| | LOC102724772 | | |
| 0 | LOC102724774 | FALSE | Upregulated |
| 0 | LOC102724775 | FALSE | Upregulated |
| | LOC102724776 | | |
| | LOC102724777 | | |
| -0,517311673 | LOC102724778 | FALSE | Downregulated |
| | LOC102724779 | | |
| | LOC102724780 | | |
| 0,387012887 | LOC102724781 | FALSE | Upregulated |
| 0,767329973 | LOC102724783 | FALSE | Upregulated |
| | LOC102724785 | | |
| 0 | LOC102724786 | FALSE | Upregulated |
| 0,256345384 | LOC102724792 | FALSE | Upregulated |
| | LOC102724793 | | |

| | | | |
|---|---|---|---|
| | LOC102724794 | | |
| -0,154164418 | LOC102724795 | FALSE | Downregulated |
| | LOC102724796 | | |
| 0,425499871 | LOC102724797 | FALSE | Upregulated |
| | LOC102724798 | | |
| | LOC102724799 | | |
| 0,294535322 | LOC102724800 | FALSE | Upregulated |
| | LOC102724801 | | |
| 0,513339129 | LOC102724802 | FALSE | Upregulated |
| | LOC102724803 | | |
| -0,308307697 | LOC102724804 | FALSE | Downregulated |
| 0,067587303 | LOC102724805 | FALSE | Upregulated |
| 0,349806604 | LOC102724806 | FALSE | Upregulated |
| | LOC102724809 | | |
| | LOC102724810 | | |
| 0,583809761 | LOC102724812 | FALSE | Upregulated |
| | LOC102724815 | | |
| | LOC102724817 | | |
| | LOC102724818 | | |
| | LOC102724819 | | |
| | LOC102724820 | | |
| 0,146690165 | LOC102724821 | FALSE | Upregulated |
| -0,275908745 | LOC102724824 | FALSE | Downregulated |
| | LOC102724825 | | |
| 0 | LOC102724826 | FALSE | Upregulated |
| 0,706208838 | LOC102724827 | FALSE | Upregulated |
| | LOC102724829 | | |
| | LOC102724830 | | |
| 0,233741859 | LOC102724831 | FALSE | Upregulated |
| 0,00160691 | LOC102724832 | FALSE | Upregulated |
| | LOC102724834 | | |
| | LOC102724835 | | |
| | LOC102724836 | | |
| | LOC102724837 | | |
| | LOC102724838 | | |
| | LOC102724839 | | |
| | LOC102724840 | | |
| 1,294173538 | LOC102724842 | | Upregulated |
| | LOC102724844 | | |
| | LOC102724845 | | |
| | LOC102724846 | | |
| | LOC102724847 | | |
| | LOC102724848 | | |
| | LOC102724849 | | |
| | LOC102724854 | | |
| | LOC102724855 | | |
| -0,016422875 | LOC102724856 | FALSE | Downregulated |
| 0,060579764 | LOC102724857 | FALSE | Upregulated |
| | LOC102724858 | | |
| | LOC102724859 | | |
| -0,233839014 | LOC102724860 | FALSE | Downregulated |

| | | | |
|---|---|---|---|
| 0 | LOC102724861 | FALSE | Upregulated |
| 0,210750242 | LOC102724862 | FALSE | Upregulated |
| | LOC102724863 | | |
| -0,288068341 | LOC102724864 | FALSE | Downregulated |
| 0,041016456 | LOC102724865 | FALSE | Upregulated |
| 0,043214973 | LOC102724869 | FALSE | Upregulated |
| 0 | LOC102724870 | FALSE | Upregulated |
| | LOC102724871 | | |
| | LOC102724874 | | |
| | LOC102724875 | | |
| | LOC102724876 | | |
| | LOC102724877 | | |
| | LOC102724878 | | |
| | LOC102724879 | | |
| -0,146071155 | LOC102724880 | FALSE | Downregulated |
| | LOC102724882 | | |
| -0,432417532 | LOC102724883 | FALSE | Downregulated |
| 1,629671471 | LOC102724884 | | Upregulated |
| | LOC102724885 | | |
| | LOC102724887 | | |
| | LOC102724888 | | |
| | LOC102724890 | | |
| | LOC102724891 | | |
| | LOC102724892 | | |
| -0,155836992 | LOC102724893 | FALSE | Downregulated |
| | LOC102724894 | | |
| | LOC102724896 | | |
| 0 | LOC102724897 | FALSE | Upregulated |
| 0,599446544 | LOC102724898 | FALSE | Upregulated |
| | LOC102724899 | | |
| | LOC102724900 | | |
| -0,594747876 | LOC102724901 | FALSE | Downregulated |
| | LOC102724902 | | |
| 0,258687376 | LOC102724903 | FALSE | Upregulated |
| | LOC102724904 | | |
| | LOC102724905 | | |
| 0,046678197 | LOC102724906 | FALSE | Upregulated |
| 1,43915856 | LOC102724907 | | Upregulated |
| 1,16121716 | LOC102724908 | | Upregulated |
| | LOC102724910 | | |
| 0,354638858 | LOC102724911 | FALSE | Upregulated |
| | LOC102724912 | | |
| | LOC102724913 | | |
| | LOC102724914 | | |
| | LOC102724915 | | |
| 0,249444751 | LOC102724916 | FALSE | Upregulated |
| | LOC102724917 | | |
| | LOC102724918 | | |
| -0,016422875 | LOC102724919 | FALSE | Downregulated |
| 0 | LOC102724923 | FALSE | Upregulated |
| | LOC102724924 | | |

| | | | |
|---|---|---|---|
| | LOC102724925 | | |
| 0,750066567 | LOC102724927 | FALSE | Upregulated |
| | LOC102724928 | | |
| | LOC102724929 | | |
| | LOC102724932 | | |
| | LOC102724934 | | |
| 0,216153162 | LOC102724935 | FALSE | Upregulated |
| | LOC102724936 | | |
| | LOC102724939 | | |
| 0,024537031 | LOC102724940 | FALSE | Upregulated |
| | LOC102724942 | | |
| 0,519426049 | LOC102724943 | FALSE | Upregulated |
| -0,067446426 | LOC102724944 | FALSE | Downregulated |
| | LOC102724945 | | |
| 0 | LOC102724946 | FALSE | Upregulated |
| -0,330089407 | LOC102724947 | FALSE | Downregulated |
| 0,247897015 | LOC102724948 | FALSE | Upregulated |
| | LOC102724949 | | |
| 0,578467735 | LOC102724950 | FALSE | Upregulated |
| | LOC102724952 | | |
| | LOC102724953 | | |
| | LOC102724954 | | |
| | LOC102724955 | | |
| | LOC102724957 | | |
| | LOC102724958 | | |
| | LOC102724959 | | |
| | LOC102724960 | | |
| | LOC102724963 | | |
| | LOC102724964 | | |
| | LOC102724966 | | |
| 0,498213449 | LOC102724967 | FALSE | Upregulated |
| | LOC102724968 | | |
| -0,15914789 | LOC102724970 | FALSE | Downregulated |
| 0,506156656 | LOC102724971 | FALSE | Upregulated |
| | LOC102724972 | | |
| 0,45403248 | LOC102724973 | FALSE | Upregulated |
| | LOC128322 | | |
| 0,135118412 | LOC145474 | FALSE | Upregulated |
| 0 | LOC145845 | FALSE | Upregulated |
| | LOC146481 | | |
| | LOC147004 | | |
| 0,045955031 | LOC149134 | FALSE | Upregulated |
| 0 | LOC149684 | FALSE | Upregulated |
| | LOC149950 | | |
| 0 | LOC150935 | FALSE | Upregulated |
| 0,73432955 | LOC151174 | FALSE | Upregulated |
| -0,238634544 | LOC151760 | FALSE | Downregulated |
| 0,87883721 | LOC152225 | FALSE | Upregulated |
| | LOC152586 | | |
| | LOC154872 | | |
| -1,044615842 | LOC158434 | | Downregulated |

| | | | |
|---|---|---|---|
| -0,593073125 | LOC158960 | FALSE | Downregulated |
| | LOC1720 | | |
| -0,388154694 | LOC254896 | FALSE | Downregulated |
| | LOC255187 | | |
| 0,209108567 | LOC256880 | FALSE | Upregulated |
| 0,018916552 | LOC257358 | FALSE | Upregulated |
| -0,99661586 | LOC283028 | FALSE | Downregulated |
| | LOC283045 | | |
| 0,460783754 | LOC283070 | FALSE | Upregulated |
| 0,111331027 | LOC283214 | FALSE | Upregulated |
| | LOC283482 | | |
| | LOC283585 | | |
| 0,100713889 | LOC283682 | FALSE | Upregulated |
| 0,65784742 | LOC283683 | FALSE | Upregulated |
| 0,424124561 | LOC283693 | FALSE | Upregulated |
| 0 | LOC283854 | FALSE | Upregulated |
| 0,689586393 | LOC284009 | FALSE | Upregulated |
| -0,276828974 | LOC284240 | FALSE | Downregulated |
| 0,139973606 | LOC284344 | FALSE | Upregulated |
| -0,173650669 | LOC284379 | FALSE | Downregulated |
| 0 | LOC284395 | FALSE | Upregulated |
| 0,345105599 | LOC284561 | FALSE | Upregulated |
| 0,489771484 | LOC284578 | FALSE | Upregulated |
| -0,882236668 | LOC284581 | FALSE | Downregulated |
| -0,262474267 | LOC284600 | FALSE | Downregulated |
| 0,719490425 | LOC284648 | FALSE | Upregulated |
| | LOC284801 | | |
| 0,412509968 | LOC284865 | FALSE | Upregulated |
| 0,054547125 | LOC284898 | FALSE | Upregulated |
| | LOC285000 | | |
| 0,512898706 | LOC285043 | FALSE | Upregulated |
| | LOC285692 | | |
| -0,199474511 | LOC285768 | FALSE | Downregulated |
| -0,273178786 | LOC286178 | FALSE | Downregulated |
| 0 | LOC286186 | FALSE | Upregulated |
| 0,046678197 | LOC286189 | FALSE | Upregulated |
| 0,055378136 | LOC286359 | FALSE | Upregulated |
| 0 | LOC339166 | FALSE | Upregulated |
| 0,031418641 | LOC339240 | FALSE | Upregulated |
| | LOC339260 | | |
| 0 | LOC339298 | FALSE | Upregulated |
| 0,042435648 | LOC339539 | FALSE | Upregulated |
| 1,056079553 | LOC339593 | | Upregulated |
| 0,770301824 | LOC339622 | FALSE | Upregulated |
| 0 | LOC339862 | FALSE | Upregulated |
| | LOC339978 | | |
| -0,958304886 | LOC340073 | FALSE | Downregulated |
| | LOC340090 | | |
| -0,209493638 | LOC340113 | FALSE | Downregulated |
| 0,586032367 | LOC340508 | FALSE | Upregulated |
| 0,006448442 | LOC341056 | FALSE | Upregulated |

| | | | |
|---|---|---|---|
| 0,481637332 | LOC344887 | FALSE | Upregulated |
| -0,720783929 | LOC345051 | FALSE | Downregulated |
| 0,406787727 | LOC374890 | FALSE | Upregulated |
| -0,219052433 | LOC387720 | FALSE | Downregulated |
| -0,010122896 | LOC388406 | FALSE | Downregulated |
| 1,201514324 | LOC388436 | | Upregulated |
| 0 | LOC388456 | FALSE | Upregulated |
| 1,117122804 | LOC388499 | | Upregulated |
| | LOC388553 | | |
| -0,157072374 | LOC388780 | FALSE | Downregulated |
| -0,011472086 | LOC388813 | FALSE | Downregulated |
| | LOC389033 | | |
| 0,000292929 | LOC389273 | FALSE | Upregulated |
| -0,637521835 | LOC389602 | FALSE | Downregulated |
| 1,343131394 | LOC389641 | | Upregulated |
| 0 | LOC389705 | FALSE | Upregulated |
| | LOC389895 | | |
| | LOC390705 | | |
| 0 | LOC390877 | FALSE | Upregulated |
| | LOC390956 | | |
| -0,416597077 | LOC391722 | FALSE | Downregulated |
| 0,021140613 | LOC392232 | FALSE | Upregulated |
| 0,292100727 | LOC392288 | FALSE | Upregulated |
| -0,182349845 | LOC392364 | FALSE | Downregulated |
| | LOC392452 | | |
| 0,437058573 | LOC399815 | FALSE | Upregulated |
| 0,805955642 | LOC400499 | FALSE | Upregulated |
| | LOC400622 | | |
| -0,022422417 | LOC400655 | FALSE | Downregulated |
| 0,44784075 | LOC400685 | FALSE | Upregulated |
| | LOC400743 | | |
| -0,370781858 | LOC400748 | FALSE | Downregulated |
| 0,125397098 | LOC400768 | FALSE | Upregulated |
| -1,00344662 | LOC400863 | | Downregulated |
| -0,111924019 | LOC400891 | FALSE | Downregulated |
| 0,441768489 | LOC400927 | FALSE | Upregulated |
| | LOC400940 | | |
| 0,043590164 | LOC401010 | FALSE | Upregulated |
| 0,407214743 | LOC401098 | FALSE | Upregulated |
| | LOC401177 | | |
| 0 | LOC401242 | FALSE | Upregulated |
| | LOC401312 | | |
| | LOC401324 | | |
| | LOC401357 | | |
| 0,638325905 | LOC401442 | FALSE | Upregulated |
| 0,154954169 | LOC401463 | FALSE | Upregulated |
| -0,024403513 | LOC401480 | FALSE | Downregulated |
| | LOC401497 | | |
| | LOC401557 | | |
| -2,916961144 | LOC401589 | | Downregulated |
| | LOC402269 | | |

| | | | |
|---|---|---|---|
| -0,065573436 | LOC407835 | FALSE | Downregulated |
| | LOC440040 | | |
| 0,059638472 | LOC440243 | FALSE | Upregulated |
| -0,005752436 | LOC440292 | FALSE | Downregulated |
| -0,277731019 | LOC440311 | FALSE | Downregulated |
| 0,543267877 | LOC440313 | FALSE | Upregulated |
| | LOC440337 | | |
| 0,03663068 | LOC440461 | FALSE | Upregulated |
| | LOC440518 | | |
| | LOC440602 | | |
| -0,324812531 | LOC440700 | FALSE | Downregulated |
| -0,130258793 | LOC440742 | FALSE | Downregulated |
| -1,113501687 | LOC440895 | | Downregulated |
| | LOC440910 | | |
| -0,634066739 | LOC440934 | FALSE | Downregulated |
| 0,985222755 | LOC441052 | FALSE | Upregulated |
| -1,148336817 | LOC441155 | | Downregulated |
| -0,014135225 | LOC441204 | FALSE | Downregulated |
| 0,153006162 | LOC441455 | FALSE | Upregulated |
| 0,468614724 | LOC441493 | FALSE | Upregulated |
| 0,390950345 | LOC441601 | FALSE | Upregulated |
| -0,561392084 | LOC441666 | FALSE | Downregulated |
| 0 | LOC441728 | FALSE | Upregulated |
| -0,10748866 | LOC441956 | FALSE | Downregulated |
| 0 | LOC442028 | FALSE | Upregulated |
| | LOC442132 | | |
| 0 | LOC494127 | FALSE | Upregulated |
| 0,248500749 | LOC494141 | FALSE | Upregulated |
| | LOC497256 | | |
| 0 | LOC550113 | FALSE | Upregulated |
| 0,787420853 | LOC550643 | FALSE | Upregulated |
| -0,873342245 | LOC554206 | FALSE | Downregulated |
| -0,12818129 | LOC641515 | FALSE | Downregulated |
| | LOC641746 | | |
| 0,320371613 | LOC642131 | FALSE | Upregulated |
| 0,096952174 | LOC642236 | FALSE | Upregulated |
| 0,374151092 | LOC642249 | FALSE | Upregulated |
| | LOC642366 | | |
| | LOC642643 | | |
| -0,123186959 | LOC642934 | FALSE | Downregulated |
| 0,146690165 | LOC643327 | FALSE | Upregulated |
| 0,229111216 | LOC643387 | FALSE | Upregulated |
| -0,688112511 | LOC643406 | FALSE | Downregulated |
| 0 | LOC643441 | FALSE | Upregulated |
| -0,19561308 | LOC643486 | FALSE | Downregulated |
| | LOC643711 | | |
| -0,588845259 | LOC643733 | FALSE | Downregulated |
| 0,061744886 | LOC643770 | FALSE | Upregulated |
| 0 | LOC643802 | FALSE | Upregulated |
| | LOC643923 | | |
| | LOC644070 | | |

| | | | |
|---|---|---|---|
| 0 | LOC644086 | FALSE | Upregulated |
| | LOC644145 | | |
| -0,004678179 | LOC644189 | FALSE | Downregulated |
| -1,270509353 | LOC644249 | | Downregulated |
| 0,813895963 | LOC644554 | FALSE | Upregulated |
| | LOC644669 | | |
| | LOC644717 | | |
| 0 | LOC644919 | FALSE | Upregulated |
| -0,127332581 | LOC644936 | FALSE | Downregulated |
| | LOC645177 | | |
| | LOC645188 | | |
| 0 | LOC645202 | FALSE | Upregulated |
| | LOC645321 | | |
| -0,989713067 | LOC645638 | FALSE | Downregulated |
| -1,056769541 | LOC645752 | | Downregulated |
| 0,459027178 | LOC645769 | FALSE | Upregulated |
| 0,845344192 | LOC646021 | FALSE | Upregulated |
| 0 | LOC646034 | FALSE | Upregulated |
| 0,134430958 | LOC646278 | FALSE | Upregulated |
| | LOC646513 | | |
| | LOC646588 | | |
| 0,492505672 | LOC646670 | FALSE | Upregulated |
| 0,146800464 | LOC646730 | FALSE | Upregulated |
| 0,379567892 | LOC646736 | FALSE | Upregulated |
| | LOC646743 | | |
| | LOC646813 | | |
| | LOC646862 | | |
| 0,061087223 | LOC646903 | FALSE | Upregulated |
| | LOC646938 | | |
| | LOC647264 | | |
| -0,215866233 | LOC648044 | FALSE | Downregulated |
| 0 | LOC648691 | FALSE | Upregulated |
| | LOC649330 | | |
| 1,015933428 | LOC649352 | | Upregulated |
| | LOC650157 | | |
| 0,916122764 | LOC650226 | FALSE | Upregulated |
| | LOC650293 | | |
| 0 | LOC650368 | FALSE | Upregulated |
| -0,108063349 | LOC653160 | FALSE | Downregulated |
| 0 | LOC653486 | FALSE | Upregulated |
| 0,89741048 | LOC653786 | FALSE | Upregulated |
| -0,304196061 | LOC654342 | FALSE | Downregulated |
| -1,103664786 | LOC723805 | | Downregulated |
| | LOC727944 | | |
| -1,338646082 | LOC728026 | | Downregulated |
| | LOC728114 | | |
| 0,843883178 | LOC728158 | FALSE | Upregulated |
| 0 | LOC728637 | FALSE | Upregulated |
| -0,119301617 | LOC728715 | FALSE | Downregulated |
| -0,162259798 | LOC728728 | FALSE | Downregulated |
| | LOC728739 | | |

| | | | |
|---|---|---|---|
| | LOC728755 | | |
| -0,85664601 | LOC728975 | FALSE | Downregulated |
| -0,705430191 | LOC729041 | FALSE | Downregulated |
| 0,82187353 | LOC729080 | FALSE | Upregulated |
| -0,075484322 | LOC729159 | FALSE | Downregulated |
| | LOC729222 | | |
| 0 | LOC729224 | FALSE | Upregulated |
| 0 | LOC729307 | FALSE | Upregulated |
| | LOC729444 | | |
| | LOC729458 | | |
| 0,02574575 | LOC729468 | FALSE | Upregulated |
| 0,469226316 | LOC729506 | FALSE | Upregulated |
| -0,358932154 | LOC729609 | FALSE | Downregulated |
| 0,189583965 | LOC729770 | FALSE | Upregulated |
| 0 | LOC729800 | FALSE | Upregulated |
| | LOC729815 | | |
| | LOC729911 | | |
| 0 | LOC729966 | FALSE | Upregulated |
| 0,281071753 | LOC730159 | FALSE | Upregulated |
| -0,082800075 | LOC730268 | FALSE | Downregulated |
| 0 | LOC730338 | FALSE | Upregulated |
| 0,239407276 | LOC731157 | FALSE | Upregulated |
| 0,37760071 | LOC731779 | FALSE | Upregulated |
| 0,455051771 | LOC732265 | FALSE | Upregulated |
| 0,013393295 | LOC90246 | FALSE | Upregulated |
| 0,721829496 | LOC93432 | FALSE | Upregulated |
| 0,34197773 | LOH12CR2 | FALSE | Upregulated |
| 0 | LOR | FALSE | Upregulated |
| 0,07078209 | LOXHD1 | FALSE | Upregulated |
| 0,549278767 | LOXL1-AS1 | FALSE | Upregulated |
| 0,867082908 | LPAR3 | FALSE | Upregulated |
| 0,397593873 | LPAR4 | FALSE | Upregulated |
| 1,104307736 | LPHN3 | | Upregulated |
| -0,431176227 | LPL | FALSE | Downregulated |
| 0,286795353 | LPO | FALSE | Upregulated |
| 0,037280637 | LPPR4 | FALSE | Upregulated |
| 0,691885611 | LPPR5 | FALSE | Upregulated |
| -0,723244583 | LRCH2 | FALSE | Downregulated |
| 0 | LRFN2 | FALSE | Upregulated |
| -0,082073388 | LRFN5 | FALSE | Downregulated |
| 1,373271726 | LRGUK | | Upregulated |
| | LRIT1 | | |
| 0,7875871 | LRIT2 | FALSE | Upregulated |
| 0,194561131 | LRIT3 | FALSE | Upregulated |
| -0,385864636 | LRRC1 | FALSE | Downregulated |
| -0,022207775 | LRRC10 | FALSE | Downregulated |
| 0,159977258 | LRRC10B | FALSE | Upregulated |
| 0,192994701 | LRRC17 | FALSE | Upregulated |
| 0,273867609 | LRRC23 | FALSE | Upregulated |
| 0,406787727 | LRRC26 | FALSE | Upregulated |
| -0,049729584 | LRRC3-AS1 | FALSE | Downregulated |

| | | | |
|---|---|---|---|
| 0,10640248 | LRRC30 | FALSE | Upregulated |
| -0,199059577 | LRRC36 | FALSE | Downregulated |
| 0,202891845 | LRRC37A11P | FALSE | Upregulated |
| 0 | LRRC38 | FALSE | Upregulated |
| 0 | LRRC3B | FALSE | Upregulated |
| 0,36112554 | LRRC3C | FALSE | Upregulated |
| 0,406995585 | LRRC43 | FALSE | Upregulated |
| -0,823264838 | LRRC4C | FALSE | Downregulated |
| | LRRC52 | | |
| -0,17563083 | LRRC6 | FALSE | Downregulated |
| 1,265207885 | LRRC63 | | Upregulated |
| 0,044238438 | LRRC66 | FALSE | Upregulated |
| 0,005530207 | LRRC69 | FALSE | Upregulated |
| 0,479545559 | LRRC70 | FALSE | Upregulated |
| 0,833465176 | LRRC71 | FALSE | Upregulated |
| | LRRC72 | | |
| 0,459773333 | LRRC73 | FALSE | Upregulated |
| 0 | LRRD1 | FALSE | Upregulated |
| 0,450737483 | LRRIQ1 | FALSE | Upregulated |
| -0,134893338 | LRRIQ3 | FALSE | Downregulated |
| -0,573338299 | LRRIQ4 | FALSE | Downregulated |
| 0,493507559 | LRRN4CL | FALSE | Upregulated |
| 0,081030379 | LRRTM1 | FALSE | Upregulated |
| 0,2150349 | LRRTM4 | FALSE | Upregulated |
| 0,387601104 | LRTM1 | FALSE | Upregulated |
| -0,109753614 | LRTM2 | FALSE | Downregulated |
| 0,328553406 | LSAMP-AS1 | FALSE | Upregulated |
| 0,448212425 | LSP1P3 | FALSE | Upregulated |
| 2,851089343 | LTA | | Upregulated |
| -0,82287339 | LTF | FALSE | Downregulated |
| | LUCAT1 | | |
| -0,594398123 | LURAP1 | FALSE | Downregulated |
| -0,052486792 | LUZP2 | FALSE | Downregulated |
| | LUZP4 | | |
| 0,216583809 | LY6D | FALSE | Upregulated |
| -1,189421893 | LY6G5B | | Downregulated |
| 0,641239547 | LY6G5C | FALSE | Upregulated |
| 0 | LY6G6C | FALSE | Upregulated |
| | LY6G6D | | |
| | LY6G6E | | |
| | LY6G6F | | |
| 0,406787727 | LY6H | FALSE | Upregulated |
| 0 | LY86-AS1 | FALSE | Upregulated |
| -0,794414081 | LY96 | FALSE | Downregulated |
| -0,146351536 | LYPD2 | FALSE | Downregulated |
| 0,020209519 | LYPD3 | FALSE | Upregulated |
| 0 | LYPD4 | FALSE | Upregulated |
| -0,044753103 | LYPD5 | FALSE | Downregulated |
| -0,317398826 | LYPD6 | FALSE | Downregulated |
| 0,235645387 | LYPD6B | FALSE | Upregulated |
| 0 | LYPD8 | FALSE | Upregulated |

| | | | |
|---|---|---|---|
| -0,008479017 | LYZL1 | FALSE | Downregulated |
| 0,01196447 | LYZL2 | FALSE | Upregulated |
| | LYZL4 | | |
| 0,617847823 | LYZL6 | FALSE | Upregulated |
| 0,655204262 | MAATS1 | FALSE | Upregulated |
| -0,144181131 | MACC1-AS1 | FALSE | Downregulated |
| 0,406787727 | MACROD2-IT1 | FALSE | Upregulated |
| 0,771175325 | MAD2L1 | FALSE | Upregulated |
| -0,03878737 | MAEL | FALSE | Downregulated |
| -0,278614093 | MAFA | FALSE | Downregulated |
| 0,124220018 | MAFG-AS1 | FALSE | Upregulated |
| 0,337297572 | MAG | FALSE | Upregulated |
| | MAGEA1 | | |
| 0,564816214 | MAGEA10 | FALSE | Upregulated |
| | MAGEA10-MAGEA5 | | |
| | MAGEA11 | | |
| | MAGEA12 | | |
| | MAGEA2 | | |
| | MAGEA2B | | |
| 0 | MAGEA3 | FALSE | Upregulated |
| | MAGEA4 | | |
| 0,22640051 | MAGEA5 | FALSE | Upregulated |
| | MAGEA6 | | |
| | MAGEA8-AS1 | | |
| 0 | MAGEA9 | FALSE | Upregulated |
| 0 | MAGEA9B | FALSE | Upregulated |
| | MAGEB1 | | |
| | MAGEB10 | | |
| | MAGEB16 | | |
| | MAGEB17 | | |
| | MAGEB18 | | |
| | MAGEB2 | | |
| | MAGEB3 | | |
| | MAGEB4 | | |
| | MAGEB5 | | |
| | MAGEB6 | | |
| | MAGEC1 | | |
| | MAGEC2 | | |
| | MAGEC3 | | |
| 0,270520226 | MAGED4 | FALSE | Upregulated |
| -0,586349742 | MAGED4B | FALSE | Downregulated |
| -0,705550373 | MAGEE1 | FALSE | Downregulated |
| -0,679954469 | MAGEE2 | FALSE | Downregulated |
| 0,699399738 | MAGEL2 | FALSE | Upregulated |
| | MAGI2-AS2 | | |
| 0,538972569 | MAL | FALSE | Upregulated |
| -0,010700793 | MALRD1 | FALSE | Downregulated |
| -0,907543798 | MAMSTR | FALSE | Downregulated |
| 0,063019324 | MANEA-AS1 | FALSE | Upregulated |
| 0,380758732 | MAP1LC3B2 | FALSE | Upregulated |
| -0,325242162 | MAP1LC3C | FALSE | Downregulated |

| | | | |
|---|---|---|---|
| 0,335247729 | MAP2K4P1 | FALSE | Upregulated |
| -0,173669825 | MAP7D2 | FALSE | Downregulated |
| -0,465875261 | MAP9 | FALSE | Downregulated |
| 0,935233567 | MAPK15 | FALSE | Upregulated |
| -1,643595522 | MAPK4 | | Downregulated |
| 0,008022596 | MAPK8IP2 | FALSE | Upregulated |
| | MAPT-AS1 | | |
| 0,206584935 | MARCH10 | FALSE | Upregulated |
| | MARCH11 | | |
| 0,604442847 | MARCH4 | FALSE | Upregulated |
| 0,69911347 | MARK1 | FALSE | Upregulated |
| | MAS1 | | |
| 0 | MAS1L | FALSE | Upregulated |
| 0,838985139 | MATN1-AS1 | FALSE | Upregulated |
| 0,92914276 | MATN3 | FALSE | Upregulated |
| 0 | MATN4 | FALSE | Upregulated |
| 1,385134672 | MB21D2 | | Upregulated |
| | MBD3L1 | | |
| | MBD3L2 | | |
| 0,387012887 | MBD3L3 | FALSE | Upregulated |
| 0,387012887 | MBD3L4 | FALSE | Upregulated |
| | MBD3L5 | | |
| -0,013959494 | MBOAT2 | FALSE | Downregulated |
| | MC2R | | |
| | MC3R | | |
| | MC4R | | |
| | MC5R | | |
| 0,094461153 | MCF2 | FALSE | Upregulated |
| 1,102754158 | MCF2L2 | | Upregulated |
| | MCHR2 | | |
| | MCHR2-AS1 | | |
| | MCIDAS | | |
| 1,907941293 | MCM10 | | Upregulated |
| 1,820841966 | MCOLN2 | | Upregulated |
| 0,757652898 | MDFI | FALSE | Upregulated |
| -0,692912924 | MDGA2 | FALSE | Downregulated |
| -0,32973317 | MDH1B | FALSE | Downregulated |
| -1,262693377 | MDP1 | | Downregulated |
| 0,393594903 | MDS2 | FALSE | Upregulated |
| | MED15P9 | | |
| 0,437571796 | MEDAG | FALSE | Upregulated |
| -2,168902148 | MEF2B | | Downregulated |
| 0 | MEF2C-AS1 | FALSE | Upregulated |
| 0 | MEG8 | FALSE | Upregulated |
| 0,359200513 | MEG9 | FALSE | Upregulated |
| -0,967767258 | MEGF10 | FALSE | Downregulated |
| 0,363772497 | MEI1 | FALSE | Upregulated |
| -0,638852501 | MEI4 | FALSE | Downregulated |
| | MEIG1 | | |
| | MEIS1-AS3 | | |
| 1,577371142 | MELK | | Upregulated |

| | | | |
|---|---|---|---|
| 0,086195408 | MEOX1 | FALSE | Upregulated |
| 0,296089328 | MEP1A | FALSE | Upregulated |
| 0,100840871 | MESP1 | FALSE | Upregulated |
| 0,367173259 | MESP2 | FALSE | Upregulated |
| | METTL11B | | |
| 0,317168616 | METTL21EP | FALSE | Upregulated |
| -0,267381212 | METTL24 | FALSE | Downregulated |
| -0,17586119 | MFAP2 | FALSE | Downregulated |
| 0,191699542 | MFAP5 | FALSE | Upregulated |
| -0,396251809 | MFI2-AS1 | FALSE | Downregulated |
| 0,439179336 | MFRP | FALSE | Upregulated |
| -0,061659792 | MGAM | FALSE | Downregulated |
| 0,1083082 | MGARP | FALSE | Upregulated |
| 0,68064098 | MGAT4C | FALSE | Upregulated |
| 0,452694287 | MGAT5B | FALSE | Upregulated |
| -0,956957183 | MGC16025 | FALSE | Downregulated |
| 0,515031241 | MGC16142 | FALSE | Upregulated |
| -1,103784533 | MGC27382 | | Downregulated |
| | MGC2889 | | |
| -1,155415907 | MGC34034 | | Downregulated |
| 0 | MGC34800 | FALSE | Upregulated |
| -0,854143559 | MGC39545 | FALSE | Downregulated |
| | MGC39584 | | |
| -0,113239553 | MGC42157 | FALSE | Downregulated |
| 0 | MGC45800 | FALSE | Upregulated |
| -0,053959858 | MGC45922 | FALSE | Downregulated |
| 0,348954274 | MIA | FALSE | Upregulated |
| -0,188261782 | MIA-RAB4B | FALSE | Downregulated |
| 0,674478212 | MID1-2 | FALSE | Upregulated |
| -0,010122896 | MIMT1 | FALSE | Downregulated |
| -1,147963249 | MINOS1-NBL1 | | Downregulated |
| -0,619482032 | MIP | FALSE | Downregulated |
| 0,036141165 | MIPEPP3 | FALSE | Upregulated |
| | MIR1-1 | | |
| | MIR1-2 | | |
| | MIR100 | | |
| | MIR101-1 | | |
| | MIR101-2 | | |
| | MIR103A1 | | |
| | MIR103A2 | | |
| | MIR103B1 | | |
| | MIR103B2 | | |
| | MIR105-1 | | |
| | MIR105-2 | | |
| | MIR106A | | |
| | MIR106B | | |
| | MIR107 | | |
| | MIR10A | | |
| | MIR10B | | |
| | MIR1178 | | |
| | MIR1179 | | |

MIR1180
MIR1181
MIR1182
MIR1183
MIR1184-1
MIR1184-2
MIR1184-3
MIR1185-1
MIR1185-2
MIR1193
MIR1197
MIR1199
MIR1200
MIR1202
MIR1203
MIR1204
MIR1205
MIR1206
MIR1207
MIR1208
MIR122
MIR1224
MIR1225
MIR1226
MIR1227
MIR1228
MIR1229
MIR1231
MIR1233-1
MIR1233-2
MIR1236
MIR1237
MIR1238
MIR124-1
MIR124-2
MIR124-3
MIR1243
MIR1244-1
MIR1244-2
MIR1244-3
MIR1245A
MIR1245B
MIR1246
MIR1247
MIR1249
MIR1250
MIR1251
MIR1252
MIR1253
MIR1254-1
MIR1254-2

MIR1255A
MIR1255B1
MIR1255B2
MIR1256
MIR1257
MIR1258
MIR125A
MIR125B1
MIR125B2
MIR126
MIR1260A
MIR1260B
MIR1261
MIR1262
MIR1263
MIR1264
MIR1265
MIR1266
MIR1267
MIR1268A
MIR1268B
MIR1269A
MIR1269B
MIR127
MIR1270-1
MIR1271
MIR1272
MIR1273A
MIR1273C
MIR1273D
MIR1273F
MIR1273G
MIR1273H
MIR1275
MIR1276
MIR1277
MIR1278
MIR1279
MIR128-1
MIR128-2
MIR1281
MIR1282
MIR1283-1
MIR1283-2
MIR1284
MIR1285-1
MIR1285-2
MIR1286
MIR1287
MIR1288
MIR1289-1

| | | | |
|---|---|---|---|
| | MIR1289-2 | | |
| | MIR129-1 | | |
| | MIR129-2 | | |
| | MIR1290 | | |
| | MIR1292 | | |
| | MIR1293 | | |
| | MIR1294 | | |
| | MIR1295A | | |
| | MIR1295B | | |
| | MIR1296 | | |
| | MIR1297 | | |
| | MIR1298 | | |
| | MIR1299 | | |
| | MIR1301 | | |
| | MIR1302-1 | | |
| | MIR1302-10 | | |
| | MIR1302-11 | | |
| | MIR1302-2 | | |
| | MIR1302-3 | | |
| | MIR1302-4 | | |
| 0 | MIR1302-5 | FALSE | Upregulated |
| | MIR1302-6 | | |
| | MIR1302-7 | | |
| | MIR1302-8 | | |
| | MIR1302-9 | | |
| | MIR1303 | | |
| | MIR1304 | | |
| | MIR1305 | | |
| | MIR1306 | | |
| 0 | MIR1307 | FALSE | Upregulated |
| | MIR130A | | |
| | MIR130B | | |
| | MIR132 | | |
| | MIR1321 | | |
| | MIR1322 | | |
| | MIR1323 | | |
| | MIR1324 | | |
| | MIR133A1 | | |
| | MIR133A2 | | |
| | MIR133B | | |
| | MIR134 | | |
| | MIR1343 | | |
| | MIR135A1 | | |
| | MIR135A2 | | |
| | MIR135B | | |
| | MIR136 | | |
| | MIR137 | | |
| | MIR137HG | | |
| | MIR138-1 | | |
| | MIR138-2 | | |
| | MIR139 | | |

| | | | |
|---|---|---|---|
| | MIR140 | | |
| | MIR141 | | |
| | MIR142 | | |
| | MIR143 | | |
| | MIR144 | | |
| | MIR145 | | |
| | MIR1468 | | |
| | MIR1469 | | |
| | MIR146A | | |
| | MIR146B | | |
| | MIR1470 | | |
| | MIR1471 | | |
| | MIR147A | | |
| | MIR147B | | |
| | MIR148A | | |
| | MIR148B | | |
| | MIR149 | | |
| | MIR150 | | |
| | MIR151A | | |
| | MIR151B | | |
| | MIR152 | | |
| | MIR153-1 | | |
| | MIR153-2 | | |
| | MIR1537 | | |
| | MIR1538 | | |
| | MIR1539 | | |
| | MIR154 | | |
| | MIR155 | | |
| | MIR1587 | | |
| | MIR15A | | |
| | MIR15B | | |
| | MIR16-1 | | |
| | MIR16-2 | | |
| | MIR17 | | |
| | MIR181A1 | | |
| 0 | MIR181A1HG | FALSE | Upregulated |
| 0 | MIR181A2 | FALSE | Upregulated |
| | MIR181B1 | | |
| | MIR181B2 | | |
| | MIR181C | | |
| | MIR181D | | |
| | MIR182 | | |
| | MIR1825 | | |
| | MIR1827 | | |
| | MIR183 | | |
| | MIR184 | | |
| | MIR185 | | |
| | MIR186 | | |
| | MIR187 | | |
| | MIR188 | | |
| | MIR18A | | |

| | | | |
|---|---|---|---|
| | MIR18B | | |
| | MIR1908 | | |
| | MIR1909 | | |
| | MIR190A | | |
| | MIR190B | | |
| | MIR191 | | |
| | MIR1910 | | |
| | MIR1911 | | |
| | MIR1912 | | |
| | MIR1913 | | |
| | MIR1914 | | |
| | MIR1915 | | |
| 0 | MIR192 | FALSE | Upregulated |
| | MIR193A | | |
| | MIR193B | | |
| | MIR194-1 | | |
| | MIR194-2 | | |
| | MIR195 | | |
| | MIR196A1 | | |
| | MIR196A2 | | |
| | MIR196B | | |
| | MIR197 | | |
| | MIR1972-1 | | |
| | MIR1972-2 | | |
| | MIR1973 | | |
| | MIR1976 | | |
| | MIR198 | | |
| | MIR199A1 | | |
| | MIR199A2 | | |
| | MIR199B | | |
| | MIR19A | | |
| | MIR19B1 | | |
| | MIR19B2 | | |
| | MIR200A | | |
| | MIR200B | | |
| | MIR200C | | |
| | MIR202 | | |
| | MIR202HG | | |
| | MIR203 | | |
| | MIR204 | | |
| | MIR205 | | |
| | MIR2052 | | |
| | MIR2053 | | |
| | MIR2054 | | |
| | MIR205HG | | |
| | MIR206 | | |
| | MIR208A | | |
| | MIR208B | | |
| | MIR20A | | |
| | MIR20B | | |
| | MIR21 | | |

| | | | |
|---|---|---|---|
| | MIR210 | | |
| | MIR211 | | |
| | MIR2110 | | |
| | MIR2113 | | |
| | MIR2114 | | |
| | MIR2115 | | |
| | MIR2116 | | |
| | MIR2117 | | |
| | MIR212 | | |
| 0 | MIR214 | FALSE | Upregulated |
| | MIR215 | | |
| | MIR216A | | |
| | MIR216B | | |
| | MIR217 | | |
| | MIR218-1 | | |
| | MIR218-2 | | |
| | MIR219-1 | | |
| | MIR219-2 | | |
| | MIR22 | | |
| | MIR221 | | |
| | MIR222 | | |
| | MIR223 | | |
| | MIR224 | | |
| | MIR2276 | | |
| | MIR2277 | | |
| | MIR2278 | | |
| | MIR2355 | | |
| | MIR2392 | | |
| | MIR23A | | |
| | MIR23B | | |
| | MIR23C | | |
| | MIR24-1 | | |
| | MIR24-2 | | |
| | MIR2467 | | |
| | MIR25 | | |
| | MIR2681 | | |
| | MIR2682 | | |
| | MIR26A1 | | |
| | MIR26A2 | | |
| | MIR26B | | |
| | MIR27A | | |
| | MIR27B | | |
| | MIR28 | | |
| | MIR2861 | | |
| | MIR2909 | | |
| | MIR296 | | |
| | MIR2964A | | |
| | MIR297 | | |
| | MIR298 | | |
| | MIR299 | | |
| | MIR29A | | |

MIR29B1
MIR29B2
MIR29C
MIR300
MIR301A
MIR301B
MIR302A
MIR302B
MIR302C
MIR302D
MIR302E
MIR302F
MIR3064
MIR3065
MIR3074
MIR30A
MIR30B
MIR30C1
MIR30C2
MIR30D
MIR30E
MIR31
MIR3115
MIR3116-1
MIR3116-2
MIR3117
MIR3118-1
MIR3118-2
MIR3118-3
MIR3118-4
MIR3118-5
MIR3118-6
MIR3119-1
MIR3119-2
MIR3120
MIR3121
MIR3122
MIR3123
MIR3124
MIR3125
MIR3126
MIR3127
MIR3128
MIR3129
MIR3130-1
MIR3130-2
MIR3131
MIR3132
MIR3133
MIR3134
MIR3135A

MIR3135B
MIR3136
MIR3137
MIR3138
MIR3139
MIR3140
MIR3141
MIR3142
MIR3143
MIR3144
MIR3145
MIR3146
MIR3147
MIR3148
MIR3150A
MIR3150B
MIR3151
MIR3152
MIR3153
MIR3154
MIR3155A
MIR3155B
MIR3156-1
MIR3156-2
MIR3156-3
MIR3157
MIR3158-1
MIR3158-2
MIR3159
MIR3160-1
MIR3160-2
MIR3161
MIR3162
MIR3163
MIR3164
MIR3165
MIR3166
MIR3167
MIR3168
MIR3169
MIR3170
MIR3173
MIR3174
MIR3175
MIR3176
MIR3177
MIR3178
MIR3179-1
MIR3179-2
MIR3179-3
MIR3180-1

| | | | |
|---|---|---|---|
| | MIR3180-2 | | |
| | MIR3180-3 | | |
| | MIR3180-4 | | |
| | MIR3180-5 | | |
| | MIR3182 | | |
| | MIR3183 | | |
| | MIR3184 | | |
| | MIR3185 | | |
| | MIR3186 | | |
| | MIR3187 | | |
| | MIR3188 | | |
| | MIR3189 | | |
| | MIR3190 | | |
| | MIR3191 | | |
| | MIR3192 | | |
| | MIR3193 | | |
| | MIR3194 | | |
| | MIR3196 | | |
| | MIR3197 | | |
| | MIR3198-1 | | |
| | MIR3198-2 | | |
| | MIR3199-1 | | |
| | MIR3199-2 | | |
| 0 | MIR31HG | FALSE | Upregulated |
| | MIR32 | | |
| | MIR3200 | | |
| | MIR3201 | | |
| | MIR3202-1 | | |
| | MIR3202-2 | | |
| | MIR320A | | |
| | MIR320B1 | | |
| | MIR320B2 | | |
| | MIR320C1 | | |
| | MIR320C2 | | |
| | MIR320D1 | | |
| | MIR320D2 | | |
| | MIR320E | | |
| | MIR323A | | |
| | MIR323B | | |
| | MIR324 | | |
| | MIR325 | | |
| | MIR326 | | |
| | MIR328 | | |
| | MIR329-1 | | |
| | MIR329-2 | | |
| | MIR330 | | |
| | MIR331 | | |
| | MIR335 | | |
| | MIR337 | | |
| | MIR338 | | |
| | MIR339 | | |

| | | | |
|---|---|---|---|
| | MIR33A | | |
| | MIR33B | | |
| | MIR340 | | |
| | MIR342 | | |
| | MIR345 | | |
| | MIR346 | | |
| | MIR34A | | |
| | MIR34B | | |
| | MIR34C | | |
| | MIR3529 | | |
| | MIR3545 | | |
| | MIR3591 | | |
| | MIR3605 | | |
| | MIR3606 | | |
| | MIR3607 | | |
| | MIR3609 | | |
| | MIR361 | | |
| | MIR3610 | | |
| | MIR3611 | | |
| | MIR3612 | | |
| | MIR3613 | | |
| | MIR3614 | | |
| | MIR3615 | | |
| | MIR3616 | | |
| | MIR3617 | | |
| | MIR3618 | | |
| | MIR3619 | | |
| | MIR362 | | |
| | MIR3620 | | |
| | MIR3621 | | |
| | MIR3622A | | |
| | MIR3622B | | |
| | MIR363 | | |
| | MIR3646 | | |
| | MIR3649 | | |
| | MIR3650 | | |
| | MIR3651 | | |
| | MIR3652 | | |
| | MIR3653 | | |
| | MIR3654 | | |
| | MIR3655 | | |
| | MIR3656 | | |
| | MIR3657 | | |
| | MIR3658 | | |
| | MIR3659 | | |
| | MIR365A | | |
| | MIR365B | | |
| | MIR3660 | | |
| 0 | MIR3661 | FALSE | Upregulated |
| | MIR3662 | | |
| | MIR3663 | | |

MIR3664
MIR3665
MIR3666
MIR3667
MIR3668
MIR367
MIR3670-1
MIR3670-2
MIR3671
MIR3672
MIR3674
MIR3675
MIR3677
MIR3678
MIR3679
MIR3680-1
MIR3680-2
MIR3681
MIR3682
MIR3683
MIR3684
MIR3685
MIR3686
MIR3687
MIR3688-1
MIR3688-2
MIR3689A
MIR3689B
MIR3689C
MIR3689D1
MIR3689D2
MIR3689E
MIR3689F
MIR369
MIR3690
MIR3690-2
MIR3691
MIR3692
MIR370
MIR3713
MIR3714
MIR371A
MIR371B
MIR372
MIR373
MIR374A
MIR374B
MIR374C
MIR375
MIR376A1
MIR376A2

| | | | |
|---|---|---|---|
| | MIR376B | | |
| | MIR376C | | |
| | MIR377 | | |
| | MIR378A | | |
| | MIR378B | | |
| | MIR378C | | |
| | MIR378D1 | | |
| | MIR378D2 | | |
| | MIR378E | | |
| | MIR378F | | |
| | MIR378G | | |
| | MIR378H | | |
| | MIR378I | | |
| | MIR378J | | |
| | MIR379 | | |
| | MIR380 | | |
| | MIR381 | | |
| 0,448212425 | MIR381HG | FALSE | Upregulated |
| | MIR382 | | |
| | MIR383 | | |
| | MIR384 | | |
| | MIR3907 | | |
| | MIR3908 | | |
| | MIR3909 | | |
| | MIR3910-1 | | |
| | MIR3910-2 | | |
| | MIR3911 | | |
| | MIR3912 | | |
| | MIR3913-1 | | |
| | MIR3913-2 | | |
| | MIR3914-1 | | |
| | MIR3914-2 | | |
| | MIR3915 | | |
| | MIR3916 | | |
| | MIR3917 | | |
| | MIR3918 | | |
| | MIR3919 | | |
| | MIR3920 | | |
| | MIR3921 | | |
| | MIR3922 | | |
| | MIR3923 | | |
| | MIR3924 | | |
| | MIR3925 | | |
| | MIR3926-1 | | |
| | MIR3926-2 | | |
| | MIR3927 | | |
| | MIR3928 | | |
| | MIR3929 | | |
| | MIR3934 | | |
| | MIR3935 | | |
| | MIR3936 | | |

| | | | |
|---|---|---|---|
| | MIR3937 | | |
| | MIR3938 | | |
| 0,085971414 | MIR3939 | FALSE | Upregulated |
| | MIR3940 | | |
| | MIR3941 | | |
| | MIR3942 | | |
| | MIR3943 | | |
| | MIR3944 | | |
| | MIR3945 | | |
| | MIR3960 | | |
| | MIR3972 | | |
| | MIR3973 | | |
| | MIR3974 | | |
| | MIR3975 | | |
| | MIR3976 | | |
| | MIR3977 | | |
| | MIR3978 | | |
| | MIR409 | | |
| | MIR410 | | |
| | MIR411 | | |
| | MIR412 | | |
| | MIR421 | | |
| | MIR422A | | |
| | MIR423 | | |
| | MIR424 | | |
| | MIR425 | | |
| | MIR4251 | | |
| | MIR4252 | | |
| | MIR4253 | | |
| | MIR4254 | | |
| | MIR4255 | | |
| | MIR4256 | | |
| | MIR4257 | | |
| | MIR4258 | | |
| | MIR4260 | | |
| | MIR4261 | | |
| | MIR4262 | | |
| | MIR4263 | | |
| | MIR4264 | | |
| | MIR4265 | | |
| | MIR4266 | | |
| | MIR4267 | | |
| | MIR4268 | | |
| | MIR4269 | | |
| | MIR4270 | | |
| | MIR4271 | | |
| | MIR4272 | | |
| | MIR4273 | | |
| | MIR4274 | | |
| | MIR4275 | | |
| | MIR4276 | | |

MIR4277
MIR4278
MIR4279
MIR4280
MIR4281
MIR4282
MIR4283-1
MIR4283-2
MIR4284
MIR4286
MIR4287
MIR4288
MIR4289
MIR429
MIR4290
MIR4291
MIR4292
MIR4293
MIR4294
MIR4295
MIR4296
MIR4297
MIR4298
MIR4299
MIR4300
MIR4301
MIR4302
MIR4303
MIR4304
MIR4305
MIR4306
MIR4307
MIR4308
MIR4309
MIR431
MIR4310
MIR4311
MIR4312
MIR4313
MIR4314
MIR4315-1
MIR4315-2
MIR4316
MIR4317
MIR4318
MIR4319
MIR432
MIR4320
MIR4321
MIR4322
MIR4323

| | | |
|---|---|---|
| | MIR4324 | |
| | MIR4325 | |
| | MIR4326 | |
| | MIR4327 | |
| | MIR4328 | |
| | MIR4329 | |
| | MIR433 | |
| | MIR4330 | |
| | MIR4417 | |
| | MIR4418 | |
| | MIR4419A | |
| | MIR4419B | |
| | MIR4420 | |
| | MIR4421 | |
| | MIR4422 | |
| | MIR4423 | |
| | MIR4424 | |
| | MIR4425 | |
| | MIR4426 | |
| | MIR4427 | |
| | MIR4428 | |
| | MIR4429 | |
| | MIR4430 | |
| | MIR4431 | |
| | MIR4432 | |
| | MIR4433 | |
| | MIR4433B | |
| | MIR4434 | |
| | MIR4435-1 | |
| 2,099879859 | MIR4435-1HG | Upregulated |
| | MIR4435-2 | |
| | MIR4436A | |
| | MIR4436B1 | |
| | MIR4436B2 | |
| | MIR4437 | |
| | MIR4438 | |
| | MIR4439 | |
| | MIR4440 | |
| | MIR4441 | |
| | MIR4442 | |
| | MIR4443 | |
| | MIR4444-1 | |
| | MIR4444-2 | |
| | MIR4445 | |
| | MIR4446 | |
| | MIR4447 | |
| | MIR4448 | |
| | MIR4449 | |
| | MIR4450 | |
| | MIR4451 | |
| | MIR4452 | |

MIR4453
MIR4454
MIR4455
MIR4456
MIR4457
MIR4458
MIR4459
MIR4460
MIR4461
MIR4462
MIR4463
MIR4464
MIR4465
MIR4466
MIR4467
MIR4468
MIR4469
MIR4470
MIR4471
MIR4472-1
MIR4472-2
MIR4473
MIR4474
MIR4475
MIR4476
MIR4477A
MIR4477B
MIR4478
MIR4479
MIR448
MIR4480
MIR4481
MIR4482-1
MIR4483
MIR4484
MIR4485
MIR4486
MIR4487
MIR4488
MIR4489
MIR4490
MIR4491
MIR4492
MIR4493
MIR4494
MIR4495
MIR4496
MIR4497
MIR4498
MIR4499
MIR449A

| | | | | |
|---|---|---|---|---|
| | MIR449B | | | |
| | MIR449C | | | |
| | MIR4500 | | | |
| | MIR4501 | | | |
| | MIR4502 | | | |
| | MIR4503 | | | |
| | MIR4504 | | | |
| | MIR4505 | | | |
| | MIR4506 | | | |
| | MIR4507 | | | |
| | MIR4508 | | | |
| | MIR4509-1 | | | |
| | MIR4509-2 | | | |
| | MIR4509-3 | | | |
| | MIR450A1 | | | |
| | MIR450A2 | | | |
| | MIR450B | | | |
| | MIR4510 | | | |
| | MIR4511 | | | |
| | MIR4512 | | | |
| | MIR4513 | | | |
| | MIR4514 | | | |
| | MIR4515 | | | |
| | MIR4516 | | | |
| | MIR4517 | | | |
| | MIR4518 | | | |
| | MIR4519 | | | |
| | MIR451A | | | |
| | MIR451B | | | |
| | MIR452 | | | |
| | MIR4520A | | | |
| | MIR4520B | | | |
| | MIR4521 | | | |
| | MIR4522 | | | |
| | MIR4523 | | | |
| | MIR4524A | | | |
| -0,768054534 | | MIR4524B | FALSE | Downregulated |
| | MIR4525 | | | |
| | MIR4526 | | | |
| | MIR4527 | | | |
| | MIR4529 | | | |
| | MIR4530 | | | |
| | MIR4531 | | | |
| | MIR4532 | | | |
| | MIR4533 | | | |
| | MIR4534 | | | |
| | MIR4535 | | | |
| | MIR4536-1 | | | |
| | MIR454 | | | |
| | MIR4540 | | | |
| | MIR455 | | | |

MIR4632
MIR4633
MIR4634
MIR4635
MIR4636
MIR4637
MIR4638
MIR4639
MIR4640
MIR4641
MIR4642
MIR4643
MIR4644
MIR4645
MIR4646
MIR4647
MIR4648
MIR4649
MIR4650-1
MIR4650-2
MIR4651
MIR4652
MIR4653
MIR4654
MIR4655
MIR4656
MIR4657
MIR4658
MIR4659A
MIR4659B
MIR4660
MIR4661
MIR4662A
MIR4662B
MIR4663
MIR4664
MIR4665
MIR4666A
MIR4666B
MIR4667
MIR4668
MIR4669
MIR4670
MIR4671
MIR4672
MIR4673
MIR4674
MIR4675
MIR4676
MIR4677
MIR4678

| | | | | |
|---|---|---|---|---|
| | MIR4679-1 | | | |
| | MIR4679-2 | | | |
| | MIR4680 | | | |
| | MIR4681 | | | |
| | MIR4682 | | | |
| | MIR4683 | | | |
| | MIR4684 | | | |
| | MIR4685 | | | |
| | MIR4686 | | | |
| | MIR4687 | | | |
| | MIR4688 | | | |
| | MIR4689 | | | |
| | MIR4690 | | | |
| | MIR4691 | | | |
| | MIR4692 | | | |
| | MIR4693 | | | |
| | MIR4694 | | | |
| | MIR4695 | | | |
| | MIR4696 | | | |
| | MIR4697 | | | |
| -0,146956074 | | MIR4697HG | FALSE | Downregulated |
| | MIR4698 | | | |
| | MIR4699 | | | |
| | MIR4700 | | | |
| | MIR4701 | | | |
| | MIR4703 | | | |
| | MIR4704 | | | |
| | MIR4705 | | | |
| | MIR4706 | | | |
| | MIR4707 | | | |
| | MIR4708 | | | |
| | MIR4709 | | | |
| | MIR4710 | | | |
| | MIR4711 | | | |
| | MIR4712 | | | |
| | MIR4713 | | | |
| | MIR4714 | | | |
| | MIR4715 | | | |
| | MIR4716 | | | |
| | MIR4717 | | | |
| | MIR4718 | | | |
| | MIR4719 | | | |
| | MIR4720 | | | |
| | MIR4721 | | | |
| | MIR4722 | | | |
| | MIR4723 | | | |
| | MIR4724 | | | |
| | MIR4725 | | | |
| | MIR4726 | | | |
| | MIR4727 | | | |
| | MIR4728 | | | |

MIR4729
MIR4730
MIR4731
MIR4732
MIR4733
MIR4734
MIR4735
MIR4736
MIR4737
MIR4738
MIR4739
MIR4740
MIR4741
MIR4742
MIR4743
MIR4744
MIR4745
MIR4746
MIR4747
MIR4748
MIR4749
MIR4750
MIR4751
MIR4752
MIR4753
MIR4754
MIR4755
MIR4756
MIR4757
MIR4758
MIR4759
MIR4760
MIR4761
MIR4762
MIR4763
MIR4764
MIR4765
MIR4766
MIR4767
MIR4768
MIR4769
MIR4770
MIR4771-1
MIR4771-2
MIR4772
MIR4773-1
MIR4773-2
MIR4774
MIR4775
MIR4776-1
MIR4776-2

| | | | |
|---|---|---|---|
| | MIR4777 | | |
| | MIR4778 | | |
| | MIR4779 | | |
| | MIR4780 | | |
| | MIR4781 | | |
| | MIR4782 | | |
| | MIR4783 | | |
| | MIR4784 | | |
| | MIR4785 | | |
| | MIR4786 | | |
| | MIR4787 | | |
| | MIR4788 | | |
| | MIR4789 | | |
| | MIR4790 | | |
| | MIR4791 | | |
| | MIR4792 | | |
| | MIR4793 | | |
| | MIR4794 | | |
| | MIR4795 | | |
| | MIR4796 | | |
| | MIR4797 | | |
| | MIR4798 | | |
| | MIR4799 | | |
| | MIR4800 | | |
| | MIR4801 | | |
| | MIR4802 | | |
| | MIR4803 | | |
| | MIR4804 | | |
| | MIR483 | | |
| | MIR484 | | |
| | MIR485 | | |
| | MIR486 | | |
| | MIR486-2 | | |
| | MIR487A | | |
| | MIR487B | | |
| | MIR488 | | |
| | MIR489 | | |
| | MIR490 | | |
| | MIR491 | | |
| | MIR492 | | |
| | MIR493 | | |
| | MIR494 | | |
| | MIR495 | | |
| | MIR496 | | |
| | MIR497 | | |
| | MIR498 | | |
| | MIR4999 | | |
| 0,29700963 | MIR499A | FALSE | Upregulated |
| | MIR499B | | |
| | MIR5000 | | |
| | MIR5001 | | |

| | | | |
|---|---|---|---|
| | MIR5002 | | |
| | MIR5003 | | |
| | MIR5004 | | |
| | MIR5006 | | |
| | MIR5007 | | |
| | MIR5008 | | |
| | MIR5009 | | |
| | MIR500A | | |
| | MIR500B | | |
| | MIR501 | | |
| | MIR5010 | | |
| | MIR5011 | | |
| | MIR502 | | |
| | MIR503 | | |
| -0,40500368 | MIR503HG | FALSE | Downregulated |
| | MIR504 | | |
| | MIR5047 | | |
| | MIR505 | | |
| | MIR506 | | |
| | MIR507 | | |
| | MIR508 | | |
| | MIR5087 | | |
| | MIR5088 | | |
| | MIR5089 | | |
| | MIR509-1 | | |
| | MIR509-2 | | |
| | MIR509-3 | | |
| | MIR5090 | | |
| | MIR5091 | | |
| | MIR5092 | | |
| | MIR5093 | | |
| | MIR5094 | | |
| | MIR5095 | | |
| | MIR5096 | | |
| | MIR510 | | |
| | MIR5100 | | |
| | MIR511-1 | | |
| | MIR512-1 | | |
| | MIR512-2 | | |
| | MIR513A1 | | |
| 0 | MIR513A2 | FALSE | Upregulated |
| | MIR513B | | |
| | MIR513C | | |
| | MIR514A1 | | |
| | MIR514A2 | | |
| | MIR514A3 | | |
| | MIR514B | | |
| | MIR515-1 | | |
| | MIR515-2 | | |
| | MIR516A1 | | |
| | MIR516A2 | | |

| | | | |
|---|---|---|---|
| | MIR516B1 | | |
| | MIR516B2 | | |
| | MIR517A | | |
| | MIR517B | | |
| | MIR517C | | |
| | MIR5186 | | |
| | MIR5187 | | |
| | MIR5188 | | |
| 0,390950345 | MIR5189 | FALSE | Upregulated |
| | MIR518A1 | | |
| | MIR518A2 | | |
| | MIR518B | | |
| | MIR518C | | |
| | MIR518D | | |
| | MIR518E | | |
| | MIR518F | | |
| | MIR5190 | | |
| | MIR5191 | | |
| | MIR5192 | | |
| | MIR5193 | | |
| 0 | MIR5194 | FALSE | Upregulated |
| | MIR5196 | | |
| | MIR5197 | | |
| | MIR519A1 | | |
| | MIR519A2 | | |
| | MIR519B | | |
| | MIR519C | | |
| | MIR519D | | |
| | MIR519E | | |
| | MIR520A | | |
| | MIR520B | | |
| | MIR520C | | |
| | MIR520D | | |
| | MIR520E | | |
| | MIR520F | | |
| | MIR520G | | |
| | MIR520H | | |
| | MIR521-1 | | |
| | MIR521-2 | | |
| | MIR522 | | |
| | MIR523 | | |
| | MIR524 | | |
| | MIR525 | | |
| | MIR526A1 | | |
| | MIR526A2 | | |
| | MIR526B | | |
| | MIR527 | | |
| | MIR539 | | |
| | MIR541 | | |
| | MIR542 | | |
| | MIR543 | | |

MIR544A
MIR544B
MIR545
MIR548A1
MIR548A2
MIR548A3
MIR548AA1
MIR548AA2
MIR548AB
MIR548AC
MIR548AD
MIR548AE1
MIR548AE2
MIR548AG1
MIR548AG2
MIR548AH
MIR548AI
MIR548AJ1
MIR548AJ2
MIR548AK
MIR548AL
MIR548AM
MIR548AN
MIR548AO
MIR548AP
MIR548AQ
MIR548AR
MIR548AS
MIR548AT
MIR548AU
MIR548AV
MIR548AW
MIR548AX
MIR548AY
MIR548AZ
MIR548B
MIR548BA
MIR548C
MIR548D1
MIR548D2
MIR548E
MIR548F1
MIR548F2
MIR548F3
MIR548F4
MIR548F5
MIR548G
MIR548H1
MIR548H2
MIR548H3
MIR548H4

| | | | | |
|---|---|---|---|---|
| | MIR548H5 | | | |
| 0 | MIR548I1 | FALSE | Upregulated | |
| | MIR548I2 | | | |
| | MIR548I3 | | | |
| | MIR548I4 | | | |
| | MIR548J | | | |
| | MIR548K | | | |
| | MIR548L | | | |
| | MIR548M | | | |
| | MIR548N | | | |
| -0,012355032 | | MIR548O | FALSE | Downregulated |
| | MIR548O2 | | | |
| | MIR548P | | | |
| | MIR548Q | | | |
| | MIR548S | | | |
| | MIR548T | | | |
| | MIR548U | | | |
| | MIR548V | | | |
| | MIR548W | | | |
| | MIR548X | | | |
| | MIR548X2 | | | |
| | MIR548Y | | | |
| | MIR548Z | | | |
| | MIR549 | | | |
| | MIR550A1 | | | |
| | MIR550A2 | | | |
| | MIR550A3 | | | |
| | MIR550B1 | | | |
| | MIR550B2 | | | |
| | MIR551A | | | |
| | MIR551B | | | |
| | MIR552 | | | |
| | MIR553 | | | |
| | MIR554 | | | |
| | MIR555 | | | |
| | MIR556 | | | |
| | MIR557 | | | |
| | MIR5572 | | | |
| | MIR5579 | | | |
| | MIR558 | | | |
| | MIR5580 | | | |
| | MIR5581 | | | |
| | MIR5582 | | | |
| | MIR5583-1 | | | |
| | MIR5583-2 | | | |
| | MIR5584 | | | |
| | MIR5585 | | | |
| | MIR5586 | | | |
| | MIR5587 | | | |
| | MIR5588 | | | |
| | MIR5589 | | | |

MIR559
MIR5590
MIR5591
MIR561
MIR562
MIR563
MIR564
MIR566
MIR567
MIR568
MIR5680
MIR5681A
MIR5681B
MIR5682
MIR5683
MIR5684
MIR5685
MIR5687
MIR5688
MIR5689
MIR569
MIR5690
MIR5691
MIR5692A1
MIR5692A2
MIR5692B
MIR5692C1
MIR5692C2
MIR5693
MIR5694
MIR5695
MIR5696
MIR5697
MIR5698
MIR5699
MIR570
MIR5700
MIR5701-1
MIR5701-2
MIR5702
MIR5703
MIR5704
MIR5705
MIR5706
MIR5707
MIR5708
MIR571
MIR572
MIR573
MIR5739
MIR574

MIR575
MIR576
MIR577
MIR578
MIR5787
MIR579
MIR580
MIR581
MIR582
MIR583
MIR584
MIR585
MIR586
MIR587
MIR588
MIR589
MIR590
MIR591
MIR592
MIR593
MIR595
MIR596
MIR597
MIR598
MIR599
MIR600
MIR601
MIR602
MIR603
MIR604
MIR605
MIR606
MIR6068
MIR6069
MIR607
MIR6070
MIR6071
MIR6072
MIR6073
MIR6074
MIR6075
MIR6076
MIR6077-2
MIR6078
MIR6079
MIR608
MIR6080
MIR6081
MIR6082
MIR6083
MIR6084

| | | | |
|---|---|---|---|
| | MIR6085 | | |
| | MIR6086 | | |
| | MIR6087 | | |
| | MIR6088 | | |
| | MIR6089-1 | | |
| | MIR6089-1-2 | | |
| | MIR609 | | |
| | MIR6090 | | |
| | MIR610 | | |
| | MIR611 | | |
| 0 | MIR612 | FALSE | Upregulated |
| | MIR6124 | | |
| | MIR6125 | | |
| | MIR6126 | | |
| | MIR6127 | | |
| | MIR6128 | | |
| | MIR6129 | | |
| | MIR613 | | |
| | MIR6130 | | |
| | MIR6131 | | |
| | MIR6132 | | |
| | MIR6133 | | |
| | MIR6134 | | |
| | MIR614 | | |
| | MIR615 | | |
| | MIR616 | | |
| | MIR6165 | | |
| | MIR617 | | |
| | MIR618 | | |
| | MIR619 | | |
| | MIR620 | | |
| | MIR621 | | |
| | MIR622 | | |
| | MIR623 | | |
| | MIR624 | | |
| | MIR625 | | |
| | MIR626 | | |
| | MIR627 | | |
| | MIR628 | | |
| | MIR629 | | |
| | MIR630 | | |
| | MIR631 | | |
| | MIR632 | | |
| | MIR633 | | |
| | MIR634 | | |
| | MIR635 | | |
| | MIR636 | | |
| | MIR637 | | |
| | MIR638 | | |
| | MIR639 | | |
| | MIR640 | | |

| | | | |
|---|---|---|---|
| | MIR641 | | |
| | MIR642A | | |
| | MIR642B | | |
| | MIR643 | | |
| | MIR644A | | |
| | MIR644B | | |
| | MIR645 | | |
| | MIR646 | | |
| | MIR647 | | |
| | MIR648 | | |
| | MIR649 | | |
| | MIR6499 | | |
| | MIR6500 | | |
| | MIR6501 | | |
| | MIR6502 | | |
| | MIR6503 | | |
| | MIR6504 | | |
| | MIR6505 | | |
| | MIR6506 | | |
| | MIR6507 | | |
| | MIR6508 | | |
| | MIR6509 | | |
| | MIR651 | | |
| | MIR6510 | | |
| | MIR6511A-2 | | |
| | MIR6511B-1 | | |
| | MIR6512 | | |
| | MIR6513 | | |
| | MIR6514 | | |
| | MIR6515 | | |
| | MIR6516 | | |
| | MIR652 | | |
| | MIR653 | | |
| | MIR654 | | |
| | MIR655 | | |
| | MIR656 | | |
| | MIR657 | | |
| | MIR658 | | |
| | MIR659 | | |
| | MIR660 | | |
| | MIR661 | | |
| | MIR662 | | |
| | MIR663A | | |
| | MIR663B | | |
| | MIR664 | | |
| | MIR665 | | |
| | MIR668 | | |
| | MIR670 | | |
| 0 | MIR670HG | FALSE | Upregulated |
| | MIR671 | | |
| | MIR6715A | | |

| | | | |
|---|---|---|---|
| | MIR6715B | | |
| | MIR6716 | | |
| | MIR6717 | | |
| | MIR6718 | | |
| | MIR6719 | | |
| | MIR6720 | | |
| | MIR6721 | | |
| | MIR6722 | | |
| | MIR6723 | | |
| | MIR6726 | | |
| | MIR6727 | | |
| | MIR6728 | | |
| | MIR6729 | | |
| | MIR6730 | | |
| | MIR6731 | | |
| | MIR6732 | | |
| | MIR6733 | | |
| | MIR6734 | | |
| | MIR6735 | | |
| | MIR6736 | | |
| | MIR6737 | | |
| | MIR6738 | | |
| | MIR6739 | | |
| | MIR6740 | | |
| | MIR6741 | | |
| | MIR6742 | | |
| | MIR6743 | | |
| | MIR6744 | | |
| | MIR6745 | | |
| | MIR6746 | | |
| | MIR6747 | | |
| | MIR6748 | | |
| | MIR6749 | | |
| | MIR675 | | |
| | MIR6750 | | |
| | MIR6751 | | |
| | MIR6752 | | |
| 0,524932626 | MIR6753 | FALSE | Upregulated |
| | MIR6754 | | |
| | MIR6755 | | |
| | MIR6756 | | |
| | MIR6757 | | |
| | MIR6758 | | |
| | MIR6759 | | |
| | MIR676 | | |
| | MIR6760 | | |
| | MIR6761 | | |
| | MIR6762 | | |
| | MIR6763 | | |
| | MIR6764 | | |
| | MIR6765 | | |

MIR6766
MIR6767
MIR6768
MIR6769A
MIR6769B
MIR6770-2
MIR6771
MIR6772
MIR6773
MIR6774
MIR6775
MIR6776
MIR6777
MIR6778
MIR6779
MIR6780A
MIR6780B
MIR6781
MIR6782
MIR6783
MIR6784
MIR6785
MIR6786
MIR6787
MIR6789
MIR6790
MIR6791
MIR6792
MIR6793
MIR6794
MIR6795
MIR6796
MIR6797
MIR6798
MIR6799
MIR6800
MIR6801
MIR6802
MIR6803
MIR6804
MIR6805
MIR6806
MIR6807
MIR6808
MIR6809
MIR6810
MIR6811
MIR6812
MIR6813
MIR6814
MIR6815

MIR6816
MIR6817
MIR6818
MIR6819
MIR6820
MIR6821
MIR6822
MIR6823
MIR6824
MIR6825
MIR6826
MIR6827
MIR6828
MIR6829
MIR6830
MIR6831
MIR6832
MIR6833
MIR6834
MIR6835
MIR6836
MIR6837
MIR6838
MIR6839
MIR6840
MIR6841
MIR6842
MIR6843
MIR6844
MIR6845
MIR6846
MIR6847
MIR6848
MIR6849
MIR6850
MIR6851
MIR6852
MIR6853
MIR6854
MIR6855
MIR6856
MIR6857
MIR6858
MIR6859-1
MIR6859-2
MIR6860
MIR6861
MIR6862-1
MIR6862-2
MIR6863
MIR6864

| | | | | |
|---|---|---|---|---|
| | MIR6865 | | | |
| | MIR6866 | | | |
| | MIR6867 | | | |
| | MIR6868 | | | |
| | MIR6869 | | | |
| | MIR6870 | | | |
| | MIR6871 | | | |
| | MIR6872 | | | |
| | MIR6873 | | | |
| | MIR6874 | | | |
| | MIR6875 | | | |
| | MIR6876 | | | |
| | MIR6877 | | | |
| | MIR6878 | | | |
| | MIR6879 | | | |
| | MIR6880 | | | |
| | MIR6881 | | | |
| | MIR6882 | | | |
| | MIR6883 | | | |
| | MIR6884 | | | |
| | MIR6885 | | | |
| | MIR6886 | | | |
| | MIR6887 | | | |
| | MIR6888 | | | |
| | MIR6889 | | | |
| | MIR6890 | | | |
| | MIR6891 | | | |
| | MIR6892 | | | |
| | MIR6893 | | | |
| | MIR6894 | | | |
| | MIR6895 | | | |
| | MIR7-1 | | | |
| | MIR7-2 | | | |
| | MIR7-3 | | | |
| -0,456901859 | | MIR7-3HG | FALSE | Downregulated |
| | MIR708 | | | |
| | MIR7106 | | | |
| | MIR7107 | | | |
| | MIR7108 | | | |
| | MIR7109 | | | |
| | MIR711 | | | |
| | MIR7110 | | | |
| | MIR7111 | | | |
| | MIR7112-2 | | | |
| | MIR7113 | | | |
| | MIR7114 | | | |
| | MIR7150 | | | |
| | MIR7151 | | | |
| | MIR7152 | | | |
| | MIR7153 | | | |
| | MIR7154 | | | |

MIR7155
MIR7156
MIR7157
MIR7159
MIR7160
MIR7161
MIR7162
MIR718
MIR744
MIR7515
MIR758
MIR759
MIR760
MIR761
MIR762
MIR764
MIR7641-2
MIR765
MIR766
MIR767
MIR769
MIR770
MIR7702
MIR7703
MIR7705
MIR7843
MIR7844
MIR7845
MIR7846
MIR7848
MIR7849
MIR7850
MIR7851
MIR7852
MIR7853
MIR7854
MIR7855
MIR7856
MIR7973-1
MIR7973-2
MIR7974
MIR7975
MIR7976
MIR7977
MIR7978
MIR802
MIR8052
MIR8053
MIR8054
MIR8055
MIR8056

MIR8057
MIR8058
MIR8059
MIR8060
MIR8061
MIR8062
MIR8063
MIR8064
MIR8065
MIR8066
MIR8067
MIR8068
MIR8069
MIR8070
MIR8071-1
MIR8071-2
MIR8072
MIR8073
MIR8074
MIR8075
MIR8076
MIR8077
MIR8078
MIR8079
MIR8080
MIR8081
MIR8082
MIR8083
MIR8084
MIR8085
MIR8086
MIR8087
MIR8088
MIR8089
MIR873
MIR874
MIR875
MIR876
MIR877
MIR885
MIR887
MIR888
MIR889
MIR890
MIR891A
MIR891B
MIR892A
MIR892B
MIR892C
MIR9-1
MIR9-2

| | | | |
|---|---|---|---|
| | MIR9-3 | | |
| | MIR920 | | |
| | MIR921 | | |
| | MIR922 | | |
| | MIR924 | | |
| | MIR92A1 | | |
| | MIR92A2 | | |
| | MIR92B | | |
| | MIR93 | | |
| | MIR933 | | |
| | MIR934 | | |
| | MIR935 | | |
| | MIR936 | | |
| | MIR937 | | |
| | MIR938 | | |
| | MIR939 | | |
| | MIR940 | | |
| | MIR942 | | |
| | MIR943 | | |
| | MIR944 | | |
| | MIR95 | | |
| | MIR96 | | |
| | MIR98 | | |
| | MIR99A | | |
| | MIR99B | | |
| | MIRLET7A1 | | |
| | MIRLET7A2 | | |
| | MIRLET7A3 | | |
| | MIRLET7B | | |
| | MIRLET7C | | |
| | MIRLET7D | | |
| | MIRLET7E | | |
| | MIRLET7F1 | | |
| | MIRLET7F2 | | |
| | MIRLET7G | | |
| | MIRLET7I | | |
| -1,276139087 | MIS18A | | Downregulated |
| -0,168580574 | MISP | FALSE | Downregulated |
| -0,134625218 | MKRN3 | FALSE | Downregulated |
| -0,860159259 | MKRN7P | FALSE | Downregulated |
| 0,526989961 | MKRN9P | FALSE | Upregulated |
| 0,52785254 | MLC1 | FALSE | Upregulated |
| 0,379621997 | MLF1 | FALSE | Upregulated |
| -0,14351687 | MLIP-IT1 | FALSE | Downregulated |
| | MLN | | |
| 0,353171799 | MLNR | FALSE | Upregulated |
| | MMD2 | | |
| | MMP1 | | |
| 0,011211767 | MMP10 | FALSE | Upregulated |
| 0,053018698 | MMP11 | FALSE | Upregulated |
| 0,906198175 | MMP12 | FALSE | Upregulated |

| | | | |
|---|---|---|---|
| 0,568757548 | MMP13 | FALSE | Upregulated |
| -1,584115743 | MMP16 | | Downregulated |
| 0,53050339 | MMP17 | FALSE | Upregulated |
| 0 | MMP20 | FALSE | Upregulated |
| -0,821630976 | MMP23A | FALSE | Downregulated |
| | MMP26 | | |
| 0,154747657 | MMP27 | FALSE | Upregulated |
| -0,260208238 | MMP28 | FALSE | Downregulated |
| -0,05132957 | MMP3 | FALSE | Downregulated |
| 1,666013697 | MMP7 | | Upregulated |
| 0,094615407 | MMP8 | FALSE | Upregulated |
| 0,269265151 | MND1 | FALSE | Upregulated |
| 0,065771367 | MNX1 | FALSE | Upregulated |
| | MNX1-AS1 | | |
| 0,087446864 | MOBP | FALSE | Upregulated |
| 0,979384798 | MOG | FALSE | Upregulated |
| 0 | MORC1 | FALSE | Upregulated |
| 0,32562122 | MORN2 | FALSE | Upregulated |
| 2,171353903 | MORN3 | | Upregulated |
| 0 | MORN5 | FALSE | Upregulated |
| | MOS | | |
| -0,982527402 | MOV10L1 | FALSE | Downregulated |
| 1,063000049 | MPC1L | | Upregulated |
| -0,354459813 | MPHOSPH6 | FALSE | Downregulated |
| 0,789387447 | MPO | FALSE | Upregulated |
| -0,473257197 | MPP2 | FALSE | Downregulated |
| -0,331872925 | MPP3 | FALSE | Downregulated |
| -0,181370436 | MPP4 | FALSE | Downregulated |
| 0,843637178 | MPPED2 | FALSE | Upregulated |
| -0,647829672 | MRAP2 | FALSE | Downregulated |
| -1,054037937 | MRGPRD | | Downregulated |
| 0,648142267 | MRGPRE | FALSE | Upregulated |
| | MRGPRG | | |
| | MRGPRG-AS1 | | |
| | MRGPRX1 | | |
| | MRGPRX2 | | |
| 0,458156795 | MRGPRX3 | FALSE | Upregulated |
| 0,562919481 | MRGPRX4 | FALSE | Upregulated |
| 0,049555238 | MROH2B | FALSE | Upregulated |
| 1,175219225 | MROH3P | | Upregulated |
| 0,228500348 | MROH5 | FALSE | Upregulated |
| -0,287323565 | MRPL42P5 | FALSE | Downregulated |
| 0,615980518 | MRVI1-AS1 | FALSE | Upregulated |
| 0,135062745 | MS4A10 | FALSE | Upregulated |
| | MS4A12 | | |
| | MS4A13 | | |
| 0,067860353 | MS4A15 | FALSE | Upregulated |
| 0 | MS4A18 | FALSE | Upregulated |
| -0,024257716 | MS4A2 | FALSE | Downregulated |
| 0,084976497 | MS4A3 | FALSE | Upregulated |
| 0,984911688 | MS4A4E | FALSE | Upregulated |

| | | | |
|---|---|---|---|
| | MS4A5 | | |
| 0 | MS4A6E | FALSE | Upregulated |
| 0,481637332 | MS4A8 | FALSE | Upregulated |
| 0,157597626 | MSANTD1 | FALSE | Upregulated |
| 0,446302111 | MSANTD3-TMEFF1 | FALSE | Upregulated |
| | MSGN1 | | |
| 0,0683804 | MSH4 | FALSE | Upregulated |
| 0,108532346 | MSI1 | FALSE | Upregulated |
| 0,019968801 | MSL3P1 | FALSE | Upregulated |
| 0,094942661 | MSLN | FALSE | Upregulated |
| -0,033747407 | MSMB | FALSE | Downregulated |
| -0,194903958 | MST1R | FALSE | Downregulated |
| 0,235761657 | MSTN | FALSE | Upregulated |
| 0,394521883 | MSX1 | FALSE | Upregulated |
| 0,046678197 | MSX2 | FALSE | Upregulated |
| 0,369337071 | MSX2P1 | FALSE | Upregulated |
| 1,095137663 | MT1A | Upregulated | |
| 0,663401605 | MT1B | FALSE | Upregulated |
| 0,632945189 | MT1DP | FALSE | Upregulated |
| 0 | MT1HL1 | FALSE | Upregulated |
| 0 | MT1IP | FALSE | Upregulated |
| 0,153388006 | MT1JP | FALSE | Upregulated |
| 0 | MT3 | FALSE | Upregulated |
| | MT4 | | |
| 1,26247074 | MTBP | Upregulated | |
| | MTFR2 | | |
| | MTHFD2P1 | | |
| 0,300797412 | MTL5 | FALSE | Upregulated |
| | MTNR1A | | |
| | MTRNR2L1 | | |
| | MTRNR2L10 | | |
| | MTRNR2L2 | | |
| -0,986086753 | MTRNR2L5 | FALSE | Downregulated |
| | MTRNR2L7 | | |
| 0 | MTRNR2L9 | FALSE | Upregulated |
| 0,417430176 | MTVR2 | FALSE | Upregulated |
| 0,244098977 | MUC1 | FALSE | Upregulated |
| 0,2815502 | MUC16 | FALSE | Upregulated |
| 0 | MUC17 | FALSE | Upregulated |
| 0,026354217 | MUC19 | FALSE | Upregulated |
| 0,387012887 | MUC2 | FALSE | Upregulated |
| 0 | MUC21 | FALSE | Upregulated |
| -0,990846497 | MUC22 | FALSE | Downregulated |
| 0,715382035 | MUC5AC | FALSE | Upregulated |
| 0 | MUC7 | FALSE | Upregulated |
| 0 | MUC8 | FALSE | Upregulated |
| | MUCL1 | | |
| 0,206565954 | MURC | FALSE | Upregulated |
| 0,682943152 | MYADML | FALSE | Upregulated |
| 0,189559777 | MYADML2 | FALSE | Upregulated |
| 0,616742118 | MYB | FALSE | Upregulated |

| | | | |
|---|---|---|---|
| 1,32298185 | MYBL2 | | Upregulated |
| 0,913959291 | MYBPC3 | FALSE | Upregulated |
| 0,214120785 | MYBPH | FALSE | Upregulated |
| 0,055349761 | MYBPHL | FALSE | Upregulated |
| -0,000270819 | MYCBP2-AS1 | FALSE | Downregulated |
| -0,448697367 | MYCBPAP | FALSE | Downregulated |
| 0,79891634 | MYCN | FALSE | Upregulated |
| -0,094389325 | MYCNOS | FALSE | Downregulated |
| 0,715945062 | MYEF2 | FALSE | Upregulated |
| 0,73501047 | MYEOV | FALSE | Upregulated |
| -0,696200265 | MYEOV2 | FALSE | Downregulated |
| | MYF5 | | |
| -0,097327969 | MYF6 | FALSE | Downregulated |
| 0,365603332 | MYH13 | FALSE | Upregulated |
| -0,279705112 | MYH15 | FALSE | Downregulated |
| -0,593657373 | MYH16 | FALSE | Downregulated |
| 0,518704519 | MYH4 | FALSE | Upregulated |
| 0,763359276 | MYH6 | FALSE | Upregulated |
| 0 | MYH8 | FALSE | Upregulated |
| | MYL10 | | |
| -1,119227591 | MYL3 | | Downregulated |
| 0,182764147 | MYL4 | FALSE | Upregulated |
| | MYL7 | | |
| -0,057359742 | MYLK2 | FALSE | Downregulated |
| 0,044348866 | MYLK3 | FALSE | Upregulated |
| -0,058264289 | MYO16-AS1 | FALSE | Downregulated |
| 0,43833725 | MYO1A | FALSE | Upregulated |
| 0,294801667 | MYO3A | FALSE | Upregulated |
| 0,12025818 | MYO3B | FALSE | Upregulated |
| 0,741551403 | MYOC | FALSE | Upregulated |
| 0,244261577 | MYOCD | FALSE | Upregulated |
| 0,450785219 | MYOD1 | FALSE | Upregulated |
| 0,388072717 | MYOG | FALSE | Upregulated |
| -0,896286994 | MYOZ1 | FALSE | Downregulated |
| 0,207224989 | MYOZ2 | FALSE | Upregulated |
| 0,258873037 | MYRFL | FALSE | Upregulated |
| 0,638483591 | MYT1 | FALSE | Upregulated |
| 0,390950345 | MYT1L | FALSE | Upregulated |
| | MYT1L-AS1 | | |
| -1,196127312 | MYZAP | | Downregulated |
| 0,009047024 | MZT1 | FALSE | Upregulated |
| -1,416375154 | N6AMT2 | | Downregulated |
| 0 | NAA11 | FALSE | Upregulated |
| | NAALADL2-AS3 | | |
| 0 | NACA2 | FALSE | Upregulated |
| -0,726888177 | NACAD | FALSE | Downregulated |
| 0 | NACAP1 | FALSE | Upregulated |
| -0,265543298 | NAGPA-AS1 | FALSE | Downregulated |
| 0,011851082 | NAMA | FALSE | Upregulated |
| -0,180257885 | NANOG | FALSE | Downregulated |
| | NANOGNB | | |

| | | | |
|---|---|---|---|
| 0,041225872 | NANOS2 | FALSE | Upregulated |
| 0,01196447 | NANOS3 | FALSE | Upregulated |
| 0,84138498 | NAP1L2 | FALSE | Upregulated |
| -0,538382791 | NAP1L3 | FALSE | Downregulated |
| 0,481637332 | NAP1L6 | FALSE | Upregulated |
| -0,617882777 | NAPA-AS1 | FALSE | Downregulated |
| 0,585796375 | NARR | FALSE | Upregulated |
| 0,453099891 | NAT16 | FALSE | Upregulated |
| 0,448212425 | NBEAP1 | FALSE | Upregulated |
| | NBPF18P | | |
| 0,014172328 | NBPF22P | FALSE | Upregulated |
| | NBPF23P | | |
| 1,353692419 | NBPF25P | | Upregulated |
| -0,053959858 | NBPF4 | FALSE | Downregulated |
| 0,225977738 | NBPF6 | FALSE | Upregulated |
| -0,058731059 | NBPF7 | FALSE | Downregulated |
| 0,837598079 | NCAM1-AS1 | FALSE | Upregulated |
| 0,364547562 | NCAN | FALSE | Upregulated |
| 1,36497049 | NCAPH | | Upregulated |
| 0,14466454 | NCCRP1 | FALSE | Upregulated |
| | NCOR1P1 | | |
| 0,055639426 | NCR1 | FALSE | Upregulated |
| | NCR2 | | |
| -0,151289092 | NCRNA00185 | FALSE | Downregulated |
| 0,048276727 | NCRNA00249 | FALSE | Upregulated |
| 1,280056424 | NCRUPAR | | Upregulated |
| 1,473572642 | NDC80 | | Upregulated |
| | NDFIP2-AS1 | | |
| 0,300003722 | NDNF | FALSE | Upregulated |
| | NDP | | |
| | NDST4 | | |
| -0,631567278 | NDUFAF4P1 | FALSE | Downregulated |
| | NEBL-AS1 | | |
| 0,245607152 | NEDD8-MDP1 | FALSE | Upregulated |
| 0,451778045 | NEFL | FALSE | Upregulated |
| 0,16805274 | NEFM | FALSE | Upregulated |
| | NEGR1-IT1 | | |
| 0,696859561 | NEIL3 | FALSE | Upregulated |
| 0,562378284 | NEK2 | FALSE | Upregulated |
| 0,692317484 | NEK5 | FALSE | Upregulated |
| | NELL1 | | |
| 0 | NETO1 | FALSE | Upregulated |
| 0,851281058 | NETO2 | FALSE | Upregulated |
| | NEU2 | | |
| | NEUROD1 | | |
| | NEUROD4 | | |
| | NEUROG1 | | |
| | NEUROG2 | | |
| | NEUROG3 | | |
| 0 | NF1P2 | FALSE | Upregulated |
| -0,106122027 | NFE2 | FALSE | Downregulated |

| | | | |
|---|---|---|---|
| 0 | NFE4 | FALSE | Upregulated |
| | NGB | | |
| -0,693922134 | NGF | FALSE | Downregulated |
| 0,008689745 | NHEG1 | FALSE | Upregulated |
| 1,310864601 | NHLH1 | | Upregulated |
| 0 | NHLH2 | FALSE | Upregulated |
| -1,37442553 | NHLRC4 | | Downregulated |
| 0,130635146 | NHS-AS1 | FALSE | Upregulated |
| 0 | NKAIN1 | FALSE | Upregulated |
| 0,338881832 | NKAIN4 | FALSE | Upregulated |
| 0,335840414 | NKAPL | FALSE | Upregulated |
| | NKX1-2 | | |
| 0 | NKX2-1 | FALSE | Upregulated |
| | NKX2-1-AS1 | | |
| | NKX2-2 | | |
| 0 | NKX2-3 | FALSE | Upregulated |
| | NKX2-4 | | |
| 0,353101349 | NKX2-5 | FALSE | Upregulated |
| 0 | NKX2-6 | FALSE | Upregulated |
| 0,450737483 | NKX2-8 | FALSE | Upregulated |
| 0 | NKX3-2 | FALSE | Upregulated |
| | NKX6-1 | | |
| 0,324455058 | NKX6-2 | FALSE | Upregulated |
| | NKX6-3 | | |
| -0,012975624 | NLGN1 | FALSE | Downregulated |
| 0,2811253 | NLGN4Y-AS1 | FALSE | Upregulated |
| 0 | NLRP10 | FALSE | Upregulated |
| -1,355284328 | NLRP11 | | Downregulated |
| -0,01625624 | NLRP12 | FALSE | Downregulated |
| | NLRP13 | | |
| 0,087400384 | NLRP14 | FALSE | Upregulated |
| -0,170891882 | NLRP2 | FALSE | Downregulated |
| 0 | NLRP4 | FALSE | Upregulated |
| 0,078076584 | NLRP5 | FALSE | Upregulated |
| | NLRP8 | | |
| -0,566966241 | NMB | FALSE | Downregulated |
| | NMBR | | |
| -0,392560442 | NME1-NME2 | FALSE | Downregulated |
| | NME5 | | |
| 0 | NME8 | FALSE | Upregulated |
| -0,212167257 | NMRK2 | FALSE | Downregulated |
| | NMS | | |
| 0 | NMU | FALSE | Upregulated |
| 0,705485218 | NMUR2 | FALSE | Upregulated |
| | NOBOX | | |
| -0,130604395 | NOG | FALSE | Downregulated |
| 1,330264839 | NOS1 | | Upregulated |
| -0,724572185 | NOS2 | FALSE | Downregulated |
| 0,17504072 | NOTO | FALSE | Upregulated |
| 0,570612348 | NOV | FALSE | Upregulated |
| 0,008729795 | NOVA1 | FALSE | Upregulated |

| | | | |
|---|---|---|---|
| -0,911282848 | NOX1 | FALSE | Downregulated |
| | NOX3 | | |
| -0,041116898 | NOX4 | FALSE | Downregulated |
| -0,530027705 | NOX5 | FALSE | Downregulated |
| -1,539114651 | NPAP1 | | Downregulated |
| 0,112204508 | NPAS4 | FALSE | Upregulated |
| -0,830908626 | NPBWR1 | FALSE | Downregulated |
| | NPBWR2 | | |
| | NPFFR1 | | |
| | NPFFR2 | | |
| 0,482750998 | NPHP1 | FALSE | Upregulated |
| | NPHP3-AS1 | | |
| 0 | NPHS1 | FALSE | Upregulated |
| -0,666400631 | NPHS2 | FALSE | Downregulated |
| 1,117798057 | NPIPA3 | | Upregulated |
| -1,196093425 | NPIPA7 | | Downregulated |
| 0,581520167 | NPM2 | FALSE | Upregulated |
| 0,139230045 | NPPA-AS1 | FALSE | Upregulated |
| 0,390950345 | NPPB | FALSE | Upregulated |
| | NPPC | | |
| | NPS | | |
| | NPSR1 | | |
| -0,257837977 | NPSR1-AS1 | FALSE | Downregulated |
| 1,671666531 | NPTN-IT1 | | Upregulated |
| 0,349135009 | NPTX1 | FALSE | Upregulated |
| 1,443575841 | NPTX2 | | Upregulated |
| | NPVF | | |
| | NPY | | |
| | NPY2R | | |
| 1,374375617 | NPY4R | | Upregulated |
| -0,225591029 | NPY5R | FALSE | Downregulated |
| | NR0B1 | | |
| 0 | NR2E1 | FALSE | Upregulated |
| 0,151876185 | NR4A2 | FALSE | Upregulated |
| 0,084976497 | NR5A1 | FALSE | Upregulated |
| 0,356417405 | NRCAM | FALSE | Upregulated |
| 0,233113235 | NREP-AS1 | FALSE | Upregulated |
| | NRG1-IT1 | | |
| | NRG1-IT3 | | |
| -0,723907674 | NRG2 | FALSE | Downregulated |
| -0,126237551 | NRN1L | FALSE | Downregulated |
| 0,568273745 | NRXN1 | FALSE | Upregulated |
| -1,84642941 | NRXN3 | | Downregulated |
| 1,268525091 | NSFP1 | | Upregulated |
| -1,121899341 | NSUN3 | | Downregulated |
| -0,271181117 | NT5C1A | FALSE | Downregulated |
| -1,388226405 | NT5C1B-RDH14 | | Downregulated |
| -0,170439096 | NT5M | FALSE | Downregulated |
| 1,407637913 | NTF4 | | Upregulated |
| 1,003760593 | NTM | | Upregulated |
| | NTM-IT | | |

| | | | |
|---|---|---|---|
| 0,465237344 | NTNG1 | FALSE | Upregulated |
| 0,379517742 | NTNG2 | FALSE | Upregulated |
| 0,529718301 | NTRK1 | FALSE | Upregulated |
| 0,094624341 | NTRK3-AS1 | FALSE | Upregulated |
| 0,058513485 | NTS | FALSE | Upregulated |
| | NTSR2 | | |
| 1,007701852 | NUDT11 | | Upregulated |
| -0,213903448 | NUDT4P2 | FALSE | Downregulated |
| 0 | NUDT9P1 | FALSE | Upregulated |
| 0,686934145 | NUF2 | FALSE | Upregulated |
| 0 | NUP210P1 | FALSE | Upregulated |
| -0,296203423 | NUP37 | FALSE | Downregulated |
| 1,019415167 | NUP62CL | | Upregulated |
| | NUPR1L | | |
| -0,009033441 | NWD1 | FALSE | Downregulated |
| 0 | NXF2 | FALSE | Upregulated |
| 0 | NXF2B | FALSE | Upregulated |
| 1,050754735 | NXF3 | | Upregulated |
| | NXF4 | | |
| -0,173772022 | NXF5 | FALSE | Downregulated |
| 0,465815203 | NXNL1 | FALSE | Upregulated |
| -0,172917446 | NXNL2 | FALSE | Downregulated |
| 0,130990349 | NXPE1 | FALSE | Upregulated |
| | NXPE2 | | |
| 1,523335606 | NXPE4 | | Upregulated |
| | NXPH1 | | |
| | NXPH2 | | |
| -0,509886049 | NXT2 | FALSE | Downregulated |
| 0,483466088 | NYAP1 | FALSE | Upregulated |
| 0,447210364 | NYAP2 | FALSE | Upregulated |
| | NYX | | |
| | OACYLP | | |
| | OBP2A | | |
| | OBP2B | | |
| | OC90 | | |
| -0,397736392 | OCA2 | FALSE | Downregulated |
| 1,564897666 | OCLM | | Upregulated |
| 0,100713889 | OCM | FALSE | Upregulated |
| 0,048276727 | OCM2 | FALSE | Upregulated |
| | OCSTAMP | | |
| | ODAM | | |
| -0,123959324 | ODF1 | FALSE | Downregulated |
| 0 | ODF4 | FALSE | Upregulated |
| -0,224486717 | OFCC1 | FALSE | Downregulated |
| 0 | OLAH | FALSE | Upregulated |
| | OLFM3 | | |
| -0,307782012 | OLFM4 | FALSE | Downregulated |
| 0,37098566 | OLIG1 | FALSE | Upregulated |
| -0,195217795 | OLIG2 | FALSE | Downregulated |
| | OLIG3 | | |
| 0,371382633 | OLR1 | FALSE | Upregulated |

| | | | |
|---|---|---|---|
| -0,804887861 | OMP | FALSE | Downregulated |
| 0 | ONECUT3 | FALSE | Upregulated |
| | OOEP | | |
| | OOSP1 | | |
| | OOSP2 | | |
| 1,080858906 | OPA1-AS1 | | Upregulated |
| | OPALIN | | |
| | OPN1LW | | |
| 0,093533232 | OPN1MW | FALSE | Upregulated |
| | OPN1MW2 | | |
| 0 | OPN4 | FALSE | Upregulated |
| 0,243078858 | OPN5 | FALSE | Upregulated |
| | OPRD1 | | |
| -0,195622773 | OPRK1 | FALSE | Downregulated |
| 1,075658397 | OPRL1 | | Upregulated |
| -0,017135352 | OPTC | FALSE | Downregulated |
| 0 | OR10A3 | FALSE | Upregulated |
| 1,564781802 | OR10A4 | | Upregulated |
| -0,634167401 | OR10A5 | FALSE | Downregulated |
| | OR10A6 | | |
| | OR10A7 | | |
| | OR10AG1 | | |
| | OR10C1 | | |
| | OR10G4 | | |
| 0,740528653 | OR10G7 | FALSE | Upregulated |
| | OR10G8 | | |
| | OR10G9 | | |
| 0 | OR10H1 | FALSE | Upregulated |
| | OR10H2 | | |
| 0,000977115 | OR10H3 | FALSE | Upregulated |
| | OR10H4 | | |
| -0,016422875 | OR10H5 | FALSE | Downregulated |
| 0,011195929 | OR10J3 | FALSE | Upregulated |
| | OR10K1 | | |
| | OR10K2 | | |
| | OR10P1 | | |
| 0 | OR10Q1 | FALSE | Upregulated |
| | OR10R2 | | |
| 0,957914693 | OR10S1 | FALSE | Upregulated |
| | OR10T2 | | |
| 0,195022159 | OR10V1 | FALSE | Upregulated |
| 1,754919247 | OR10V2P | | Upregulated |
| 0,372242678 | OR10W1 | FALSE | Upregulated |
| | OR10X1 | | |
| | OR10Z1 | | |
| -0,154016557 | OR11A1 | FALSE | Downregulated |
| | OR11G2 | | |
| | OR11H1 | | |
| | OR11H12 | | |
| | OR11H2 | | |
| -0,461155124 | OR11H4 | FALSE | Downregulated |

| | | | |
|---|---|---|---|
| -0,312094816 | OR11H6 | FALSE | Downregulated |
| 0,634977414 | OR11L1 | FALSE | Upregulated |
| | OR12D2 | | |
| | OR12D3 | | |
| | OR13A1 | | |
| 0,715536309 | OR13C2 | FALSE | Upregulated |
| 0,63829882 | OR13C3 | FALSE | Upregulated |
| 0,157950125 | OR13C4 | FALSE | Upregulated |
| 0,450369977 | OR13C5 | FALSE | Upregulated |
| 0,036522142 | OR13C9 | FALSE | Upregulated |
| -0,397079112 | OR13G1 | FALSE | Downregulated |
| 0,280206877 | OR13H1 | FALSE | Upregulated |
| -0,849078719 | OR13J1 | FALSE | Downregulated |
| 1,124147042 | OR14A16 | | Upregulated |
| | OR14C36 | | |
| | OR14I1 | | |
| | OR14J1 | | |
| | OR1A1 | | |
| | OR1A2 | | |
| -0,228348513 | OR1B1 | FALSE | Downregulated |
| -0,037620036 | OR1C1 | FALSE | Downregulated |
| | OR1D2 | | |
| 0 | OR1D4 | FALSE | Upregulated |
| 0,491535227 | OR1D5 | FALSE | Upregulated |
| 0 | OR1E1 | FALSE | Upregulated |
| | OR1E2 | | |
| | OR1F1 | | |
| -0,223061762 | OR1F2P | FALSE | Downregulated |
| 0 | OR1G1 | FALSE | Upregulated |
| | OR1I1 | | |
| 0,040155671 | OR1J1 | FALSE | Upregulated |
| 0,530658558 | OR1J2 | FALSE | Upregulated |
| | OR1J4 | | |
| 0,243456225 | OR1L1 | FALSE | Upregulated |
| -0,633583114 | OR1L3 | FALSE | Downregulated |
| 0,423943412 | OR1L8 | FALSE | Upregulated |
| | OR1M1 | | |
| 0,452741095 | OR1N1 | FALSE | Upregulated |
| -0,435956873 | OR1N2 | FALSE | Downregulated |
| -0,057355695 | OR1Q1 | FALSE | Downregulated |
| | OR1S1 | | |
| 0 | OR1S2 | FALSE | Upregulated |
| 0,393124818 | OR2A12 | FALSE | Upregulated |
| -0,018256213 | OR2A14 | FALSE | Downregulated |
| -0,159901948 | OR2A25 | FALSE | Downregulated |
| -0,267215694 | OR2A42 | FALSE | Downregulated |
| -0,232458397 | OR2A5 | FALSE | Downregulated |
| -0,44373851 | OR2AE1 | FALSE | Downregulated |
| 0,617890487 | OR2AG1 | FALSE | Upregulated |
| 0,146690165 | OR2AG2 | FALSE | Upregulated |
| 0,390950345 | OR2AK2 | FALSE | Upregulated |

| | | | |
|---|---|---|---|
| | OR2AP1 | | |
| -1,260429282 | OR2AT4 | | Downregulated |
| 1,970766582 | OR2B11 | | Upregulated |
| | OR2B3 | | |
| | OR2B6 | | |
| -0,182311755 | OR2C1 | FALSE | Downregulated |
| -0,067786 | OR2C3 | FALSE | Downregulated |
| | OR2D2 | | |
| 0,707483052 | OR2D3 | FALSE | Upregulated |
| 0,987556125 | OR2F1 | FALSE | Upregulated |
| 0,845906628 | OR2F2 | FALSE | Upregulated |
| | OR2G2 | | |
| | OR2G3 | | |
| 0,635566436 | OR2G6 | FALSE | Upregulated |
| -0,040118071 | OR2H1 | FALSE | Downregulated |
| 0,171839097 | OR2H2 | FALSE | Upregulated |
| -1,32764273 | OR2J2 | | Downregulated |
| | OR2J3 | | |
| | OR2K2 | | |
| | OR2L13 | | |
| 0 | OR2L1P | FALSE | Upregulated |
| | OR2L2 | | |
| | OR2L3 | | |
| | OR2L5 | | |
| | OR2L8 | | |
| | OR2M1P | | |
| | OR2M2 | | |
| | OR2M3 | | |
| | OR2M4 | | |
| | OR2M5 | | |
| | OR2M7 | | |
| 0,299933601 | OR2S2 | FALSE | Upregulated |
| | OR2T1 | | |
| | OR2T10 | | |
| 0,38647539 | OR2T11 | FALSE | Upregulated |
| | OR2T12 | | |
| | OR2T2 | | |
| 0 | OR2T27 | FALSE | Upregulated |
| 0,062794636 | OR2T29 | FALSE | Upregulated |
| 0,387340965 | OR2T3 | FALSE | Upregulated |
| | OR2T33 | | |
| | OR2T34 | | |
| | OR2T35 | | |
| | OR2T4 | | |
| 0 | OR2T5 | FALSE | Upregulated |
| | OR2T6 | | |
| | OR2T8 | | |
| -0,241585319 | OR2V1 | FALSE | Downregulated |
| | OR2V2 | | |
| -0,010122896 | OR2W1 | FALSE | Downregulated |
| | OR2W3 | | |

| | | | |
|---|---|---|---|
| 0,465237344 | OR2W5 | FALSE | Upregulated |
| | OR2Y1 | | |
| | OR2Z1 | | |
| -0,045460148 | OR3A1 | FALSE | Downregulated |
| 0,245944968 | OR3A2 | FALSE | Upregulated |
| -0,060969738 | OR3A3 | FALSE | Downregulated |
| -0,275967019 | OR3A4P | FALSE | Downregulated |
| | OR4A15 | | |
| | OR4A16 | | |
| | OR4A47 | | |
| | OR4A5 | | |
| | OR4B1 | | |
| | OR4C11 | | |
| | OR4C12 | | |
| | OR4C13 | | |
| | OR4C15 | | |
| | OR4C16 | | |
| | OR4C3 | | |
| | OR4C45 | | |
| | OR4C46 | | |
| | OR4C6 | | |
| 0 | OR4D1 | FALSE | Upregulated |
| | OR4D10 | | |
| | OR4D11 | | |
| 0,263723382 | OR4D2 | FALSE | Upregulated |
| | OR4D5 | | |
| | OR4D6 | | |
| | OR4D9 | | |
| | OR4F13P | | |
| | OR4F15 | | |
| 0,396991113 | OR4F16 | FALSE | Upregulated |
| 0 | OR4F17 | FALSE | Upregulated |
| 0,335009167 | OR4F21 | FALSE | Upregulated |
| 0,396991113 | OR4F29 | FALSE | Upregulated |
| 0,396991113 | OR4F3 | FALSE | Upregulated |
| | OR4F4 | | |
| 0 | OR4F5 | FALSE | Upregulated |
| | OR4F6 | | |
| | OR4K1 | | |
| | OR4K13 | | |
| | OR4K14 | | |
| | OR4K15 | | |
| | OR4K17 | | |
| | OR4K2 | | |
| | OR4K5 | | |
| | OR4L1 | | |
| | OR4M1 | | |
| 0 | OR4M2 | FALSE | Upregulated |
| | OR4N2 | | |
| | OR4N3P | | |
| | OR4N4 | | |

| | | | |
|---|---|---|---|
| | OR4N5 | | |
| | OR4P4 | | |
| | OR4Q3 | | |
| | OR4S1 | | |
| | OR4S2 | | |
| | OR4X1 | | |
| | OR4X2 | | |
| | OR51A2 | | |
| | OR51A4 | | |
| | OR51A7 | | |
| -0,196525926 | OR51B2 | FALSE | Downregulated |
| 0 | OR51B4 | FALSE | Upregulated |
| 0,070401644 | OR51B5 | FALSE | Upregulated |
| | OR51D1 | | |
| 0,390950345 | OR51E1 | FALSE | Upregulated |
| | OR51E2 | | |
| | OR51F1 | | |
| | OR51F2 | | |
| | OR51G1 | | |
| | OR51G2 | | |
| 0,403518811 | OR51I1 | FALSE | Upregulated |
| | OR51L1 | | |
| | OR51S1 | | |
| | OR51T1 | | |
| | OR51V1 | | |
| | OR52A1 | | |
| | OR52A5 | | |
| 0 | OR52B2 | FALSE | Upregulated |
| -0,299072686 | OR52B4 | FALSE | Downregulated |
| | OR52E2 | | |
| 0,403369601 | OR52E4 | FALSE | Upregulated |
| | OR52E6 | | |
| | OR52E8 | | |
| -0,20672557 | OR52H1 | FALSE | Downregulated |
| -0,130742102 | OR52I2 | FALSE | Downregulated |
| | OR52J3 | | |
| 0 | OR52K1 | FALSE | Upregulated |
| 0 | OR52K2 | FALSE | Upregulated |
| | OR52L1 | | |
| 0,453099891 | OR52M1 | FALSE | Upregulated |
| 0,176454894 | OR52N1 | FALSE | Upregulated |
| 0,956396979 | OR52N2 | FALSE | Upregulated |
| 0,226680328 | OR52N4 | FALSE | Upregulated |
| 0 | OR52N5 | FALSE | Upregulated |
| | OR52R1 | | |
| 0 | OR56A1 | FALSE | Upregulated |
| 0 | OR56A3 | FALSE | Upregulated |
| | OR56A4 | | |
| | OR56A5 | | |
| 0,631047947 | OR56B1 | FALSE | Upregulated |
| | OR5A1 | | |

| | | | |
|---|---|---|---|
| 0 | OR5A2 | FALSE | Upregulated |
| | OR5AC2 | | |
| -0,110095061 | OR5AN1 | FALSE | Downregulated |
| | OR5AP2 | | |
| | OR5AR1 | | |
| | OR5AS1 | | |
| | OR5AU1 | | |
| | OR5B12 | | |
| 0 | OR5B17 | FALSE | Upregulated |
| | OR5B2 | | |
| 0 | OR5B21 | FALSE | Upregulated |
| | OR5B3 | | |
| | OR5D13 | | |
| | OR5D14 | | |
| | OR5D16 | | |
| | OR5D18 | | |
| 0 | OR5E1P | FALSE | Upregulated |
| | OR5F1 | | |
| | OR5H1 | | |
| | OR5H14 | | |
| | OR5H15 | | |
| 0,423572456 | OR5H2 | FALSE | Upregulated |
| | OR5H6 | | |
| | OR5I1 | | |
| | OR5J2 | | |
| 0,925991586 | OR5K1 | FALSE | Upregulated |
| 0,329802644 | OR5K2 | FALSE | Upregulated |
| 0,084976497 | OR5K3 | FALSE | Upregulated |
| | OR5K4 | | |
| | OR5L1 | | |
| | OR5L2 | | |
| | OR5M1 | | |
| | OR5M10 | | |
| | OR5M11 | | |
| | OR5M3 | | |
| | OR5M8 | | |
| | OR5M9 | | |
| | OR5P2 | | |
| | OR5P3 | | |
| | OR5R1 | | |
| | OR5T1 | | |
| | OR5T2 | | |
| | OR5T3 | | |
| 0,964205252 | OR5V1 | FALSE | Upregulated |
| | OR5W2 | | |
| | OR6A2 | | |
| 0,647574642 | OR6B1 | FALSE | Upregulated |
| 0 | OR6B2 | FALSE | Upregulated |
| | OR6B3 | | |
| | OR6C1 | | |
| | OR6C2 | | |

| | | | |
|---|---|---|---|
| | OR6C3 | | |
| | OR6C4 | | |
| | OR6C6 | | |
| | OR6C65 | | |
| | OR6C68 | | |
| | OR6C70 | | |
| | OR6C74 | | |
| | OR6C75 | | |
| | OR6C76 | | |
| 0,829723883 | OR6F1 | FALSE | Upregulated |
| 0,390950345 | OR6K2 | FALSE | Upregulated |
| | OR6K3 | | |
| 0 | OR6K6 | FALSE | Upregulated |
| -0,225955506 | OR6M1 | FALSE | Downregulated |
| | OR6N1 | | |
| | OR6N2 | | |
| | OR6P1 | | |
| | OR6Q1 | | |
| | OR6S1 | | |
| 0,056294023 | OR6T1 | FALSE | Upregulated |
| 1,646745118 | OR6V1 | | Upregulated |
| 0,343814623 | OR6W1P | FALSE | Upregulated |
| -0,064372499 | OR6X1 | FALSE | Downregulated |
| | OR6Y1 | | |
| 0,469226316 | OR7A10 | FALSE | Upregulated |
| | OR7A17 | | |
| | OR7A5 | | |
| 0 | OR7C1 | FALSE | Upregulated |
| | OR7C2 | | |
| 0,448212425 | OR7D2 | FALSE | Upregulated |
| 0,214617194 | OR7D4 | FALSE | Upregulated |
| 0 | OR7E156P | FALSE | Upregulated |
| | OR7E24 | | |
| 0,312877951 | OR7E37P | FALSE | Upregulated |
| | OR7E5P | | |
| 0,79523711 | OR7E91P | FALSE | Upregulated |
| | OR7G1 | | |
| | OR7G2 | | |
| | OR7G3 | | |
| | OR8A1 | | |
| | OR8B12 | | |
| 0 | OR8B2 | FALSE | Upregulated |
| | OR8B3 | | |
| | OR8B4 | | |
| 0 | OR8B8 | FALSE | Upregulated |
| | OR8D1 | | |
| 0 | OR8D2 | FALSE | Upregulated |
| | OR8D4 | | |
| | OR8G1 | | |
| | OR8G2 | | |
| | OR8G5 | | |

| | | | |
|---|---|---|---|
| | OR8H1 | | |
| | OR8H2 | | |
| | OR8H3 | | |
| | OR8I2 | | |
| | OR8J1 | | |
| | OR8J3 | | |
| | OR8K1 | | |
| | OR8K3 | | |
| | OR8K5 | | |
| | OR8S1 | | |
| | OR8U1 | | |
| | OR9A2 | | |
| 0 | OR9A4 | FALSE | Upregulated |
| | OR9G1 | | |
| | OR9G4 | | |
| 0,500671817 | OR9I1 | FALSE | Upregulated |
| | OR9K2 | | |
| | OR9Q1 | | |
| | OR9Q2 | | |
| -0,271935161 | ORC1 | FALSE | Downregulated |
| 1,364999135 | ORC6 | | Upregulated |
| 0,276491075 | OSBPL10-AS1 | FALSE | Upregulated |
| 0,596989796 | OSCAR | FALSE | Upregulated |
| 0,091368632 | OSM | FALSE | Upregulated |
| 0,600616762 | OSR1 | FALSE | Upregulated |
| 0,588586397 | OSR2 | FALSE | Upregulated |
| -0,201747852 | OSTCP1 | FALSE | Downregulated |
| 0,055112592 | OSTCP2 | FALSE | Upregulated |
| | OSTM1-AS1 | | |
| -0,010602911 | OSTN | FALSE | Downregulated |
| 0 | OTOA | FALSE | Upregulated |
| 1,401913484 | OTOF | | Upregulated |
| | OTOG | | |
| 0,334078767 | OTOGL | FALSE | Upregulated |
| | OTOL1 | | |
| | OTOP1 | | |
| | OTOP2 | | |
| -0,278879694 | OTOP3 | FALSE | Downregulated |
| | OTOR | | |
| | OTOS | | |
| | OTP | | |
| 0 | OTUD6A | FALSE | Upregulated |
| | OTX1 | | |
| | OTX2 | | |
| 0,138147205 | OTX2-AS1 | FALSE | Upregulated |
| -0,471423775 | OVCH1 | FALSE | Downregulated |
| -0,307564732 | OVCH1-AS1 | FALSE | Downregulated |
| -0,124359763 | OVCH2 | FALSE | Downregulated |
| -0,441728592 | OVOL1 | FALSE | Downregulated |
| 0,481637332 | OVOL1-AS1 | FALSE | Upregulated |
| -0,302619998 | OVOL2 | FALSE | Downregulated |

| | | | |
|---|---|---|---|
| 0,1694671 | OVOL3 | FALSE | Upregulated |
| 1,469154969 | OVOS | | Upregulated |
| 0,7201258 | OVOS2 | FALSE | Upregulated |
| 0 | OXCT1-AS1 | FALSE | Upregulated |
| 1,166908201 | OXCT2 | | Upregulated |
| | OXGR1 | | |
| -0,620696603 | OXTR | FALSE | Downregulated |
| -0,613475506 | P2RX1 | FALSE | Downregulated |
| -0,253285476 | P2RX3 | FALSE | Downregulated |
| -0,006487869 | P2RX5 | FALSE | Downregulated |
| | P2RX6P | | |
| 1,882116236 | P2RY10 | | Upregulated |
| 0,257461141 | P2RY12 | FALSE | Upregulated |
| -0,100526176 | P2RY4 | FALSE | Downregulated |
| | P2RY8-2 | | |
| | PABPC1L2A | | |
| | PABPC1L2B | | |
| 0,255071058 | PABPC1P2 | FALSE | Upregulated |
| -0,049842112 | PABPC3 | FALSE | Downregulated |
| 0,046796618 | PABPC4L | FALSE | Upregulated |
| 0,037581429 | PABPC5 | FALSE | Upregulated |
| 0,200636299 | PACRGL | FALSE | Upregulated |
| 0,398723658 | PACSIN1 | FALSE | Upregulated |
| -1,786508505 | PADI1 | | Downregulated |
| 1,819865577 | PADI2 | | Upregulated |
| | PADI3 | | |
| 0,376675171 | PADI4 | FALSE | Upregulated |
| 0,390950345 | PADI6 | FALSE | Upregulated |
| | PAEP | | |
| | PAGE1 | | |
| | PAGE2 | | |
| | PAGE2B | | |
| | PAGE3 | | |
| 0 | PAGE4 | FALSE | Upregulated |
| 0,078076584 | PAGE5 | FALSE | Upregulated |
| -0,6041106 | PAK3 | FALSE | Downregulated |
| 1,027991254 | PAK6 | | Upregulated |
| -0,206400678 | PAK7 | FALSE | Downregulated |
| -0,801243047 | PAM16 | FALSE | Downregulated |
| 0,5119987 | PANDAR | FALSE | Upregulated |
| 0,01071271 | PANX3 | FALSE | Upregulated |
| 0 | PAPL | FALSE | Upregulated |
| 0,594026806 | PAPOLB | FALSE | Upregulated |
| | PARD3-AS1 | | |
| 0,650323776 | PARPBP | FALSE | Upregulated |
| | PASD1 | | |
| 0,709266127 | PATE1 | FALSE | Upregulated |
| | PATE2 | | |
| -0,942169784 | PATE3 | FALSE | Downregulated |
| | PAX1 | | |
| 0,49264775 | PAX2 | FALSE | Upregulated |

| | | | |
|---|---|---|---|
| 0 | PAX3 | FALSE | Upregulated |
| | PAX4 | | |
| 0 | PAX7 | FALSE | Upregulated |
| -0,097797913 | PAX9 | FALSE | Downregulated |
| 0,074503121 | PBK | FALSE | Upregulated |
| 0,23189537 | PBX4 | FALSE | Upregulated |
| 0,477573145 | PCA3 | FALSE | Upregulated |
| -0,297337839 | PCAT1 | FALSE | Downregulated |
| 0,332905271 | PCAT14 | FALSE | Upregulated |
| -0,724296347 | PCAT18 | FALSE | Downregulated |
| -1,019068189 | PCAT19 | | Downregulated |
| 0,394127105 | PCAT4 | FALSE | Upregulated |
| 0,070401644 | PCAT5 | FALSE | Upregulated |
| 0,055253924 | PCAT6 | FALSE | Upregulated |
| 0,975519621 | PCBP2-OT1 | FALSE | Upregulated |
| 1,052590901 | PCBP3 | | Upregulated |
| -0,516836333 | PCCA-AS1 | FALSE | Downregulated |
| -0,187029646 | PCDH10 | FALSE | Downregulated |
| 0,394791577 | PCDH11X | FALSE | Upregulated |
| -0,32206493 | PCDH11Y | FALSE | Downregulated |
| 0,066571983 | PCDH15 | FALSE | Upregulated |
| -0,400312771 | PCDH19 | FALSE | Downregulated |
| -0,086563094 | PCDH8 | FALSE | Downregulated |
| 0 | PCDH9-AS2 | FALSE | Upregulated |
| 0,073536151 | PCDH9-AS3 | FALSE | Upregulated |
| 0,266094032 | PCDH9-AS4 | FALSE | Upregulated |
| 1,050755996 | PCDP1 | | Upregulated |
| | PCGEM1 | | |
| | PCP4 | | |
| 0,816684676 | PCSK1N | FALSE | Upregulated |
| 0,481881975 | PCSK2 | FALSE | Upregulated |
| 1,034651139 | PCYOX1L | | Upregulated |
| -0,017130037 | PCYT1B | FALSE | Downregulated |
| -0,190988893 | PDCD2L | FALSE | Downregulated |
| 0,563105418 | PDCD4-AS1 | FALSE | Upregulated |
| | PDCL2 | | |
| -0,083273574 | PDCL3 | FALSE | Downregulated |
| -0,462508287 | PDCL3P4 | FALSE | Downregulated |
| 0,643787048 | PDE1C | FALSE | Upregulated |
| -1,145756742 | PDE6C | | Downregulated |
| 0,6371801 | PDE6G | FALSE | Upregulated |
| 0,531131662 | PDE6H | FALSE | Upregulated |
| | PDHA2 | | |
| 0,093333093 | PDIA2 | FALSE | Upregulated |
| -0,301926241 | PDILT | FALSE | Downregulated |
| 0,433193398 | PDLIM4 | FALSE | Upregulated |
| 0,684077285 | PDPN | FALSE | Upregulated |
| -0,789632713 | PDX1 | FALSE | Downregulated |
| 0 | PDX1-AS1 | FALSE | Upregulated |
| 0 | PDYN | FALSE | Upregulated |
| -0,795426288 | PDZD3 | FALSE | Downregulated |

| | | | |
|---|---|---|---|
| 2,65382798 | PDZK1IP1 | | Upregulated |
| 0,455398636 | PEBP4 | FALSE | Upregulated |
| | PENK | | |
| 0,09761926 | PER4 | FALSE | Upregulated |
| 0,463425026 | PF4 | FALSE | Upregulated |
| | PF4V1 | | |
| 1,966416725 | PFDN4 | | Upregulated |
| -0,783853013 | PFN3 | FALSE | Downregulated |
| -0,034062844 | PGA3 | FALSE | Downregulated |
| -0,499034524 | PGA4 | FALSE | Downregulated |
| -0,359634016 | PGA5 | FALSE | Downregulated |
| 0 | PGBD3 | FALSE | Upregulated |
| 0,471920853 | PGC | FALSE | Upregulated |
| | PGCP1 | | |
| 0,247256219 | PGK2 | FALSE | Upregulated |
| 0 | PGLYRP3 | FALSE | Upregulated |
| | PGLYRP4 | | |
| 0 | PGM5-AS1 | FALSE | Upregulated |
| 0,177342184 | PGPEP1L | FALSE | Upregulated |
| -0,708487954 | PHACTR3 | FALSE | Downregulated |
| 0,762948464 | PHEX | FALSE | Upregulated |
| 0,947546776 | PHEX-AS1 | FALSE | Upregulated |
| -0,016422875 | PHF21B | FALSE | Downregulated |
| 0,864551104 | PHGR1 | FALSE | Upregulated |
| -0,315722553 | PHLDA2 | FALSE | Downregulated |
| -0,415515828 | PHOSPHO2-KLHL23 | FALSE | Downregulated |
| | PHOX2B | | |
| 0,607447775 | PI15 | FALSE | Upregulated |
| 0,317847272 | PI16 | FALSE | Upregulated |
| | PI3 | | |
| 0,070863998 | PIFO | FALSE | Upregulated |
| 0,656880672 | PIH1D2 | FALSE | Upregulated |
| | PIH1D3 | | |
| | PIN1P1 | | |
| 0,670578066 | PINLYP | FALSE | Upregulated |
| 0 | PIP | FALSE | Upregulated |
| -0,045478829 | PIP5K1B | FALSE | Downregulated |
| -0,881510426 | PIR-FIGF | FALSE | Downregulated |
| 0,402112793 | PISRT1 | FALSE | Upregulated |
| 0 | PITX2 | FALSE | Upregulated |
| 0,028432464 | PITX3 | FALSE | Upregulated |
| 0,32486297 | PIWIL1 | FALSE | Upregulated |
| | PIWIL3 | | |
| 0,322809818 | PIWIL4 | FALSE | Upregulated |
| -0,093350726 | PKD2L1 | FALSE | Downregulated |
| 0,724180235 | PKIB | FALSE | Upregulated |
| 1,296641843 | PKP3 | | Upregulated |
| 0,692127093 | PLA2G10 | FALSE | Upregulated |
| -0,694421708 | PLA2G1B | FALSE | Downregulated |
| | PLA2G2C | | |
| 1,037799206 | PLA2G2D | | Upregulated |

| | | | |
|---|---|---|---|
| | PLA2G2E | | |
| 0,730566968 | PLA2G2F | FALSE | Upregulated |
| 0,043636993 | PLA2G3 | FALSE | Upregulated |
| 0 | PLA2G4D | FALSE | Upregulated |
| 0,554700761 | PLA2G4E | FALSE | Upregulated |
| 0 | PLA2G4F | FALSE | Upregulated |
| | PLAC1 | | |
| -0,411392015 | PLAC8L1 | FALSE | Downregulated |
| 0,353608366 | PLAC9 | FALSE | Upregulated |
| -1,879806077 | PLAG1 | | Downregulated |
| 1,342141376 | PLCB4 | | Upregulated |
| 0,993855549 | PLCE1-AS1 | FALSE | Upregulated |
| -0,939863525 | PLCH1 | FALSE | Downregulated |
| 1,421425224 | PLCH1-AS1 | | Upregulated |
| | PLCXD1-2 | | |
| 0,456066018 | PLD5 | FALSE | Upregulated |
| 0,479722424 | PLEKHA8P1 | FALSE | Upregulated |
| 0,29627825 | PLEKHD1 | FALSE | Upregulated |
| 0,627151295 | PLEKHN1 | FALSE | Upregulated |
| -0,128344214 | PLEKHS1 | FALSE | Downregulated |
| 0,079021366 | PLGRKT | FALSE | Upregulated |
| 0,521520715 | PLK4 | FALSE | Upregulated |
| 0,596290793 | PLK5 | FALSE | Upregulated |
| 1,212799633 | PLP1 | | Upregulated |
| 0,34395989 | PLSCR2 | FALSE | Upregulated |
| | PLSCR5 | | |
| -0,192991491 | PMAIP1 | FALSE | Downregulated |
| 0,844534424 | PMCH | FALSE | Upregulated |
| 0,094615407 | PMCHL1 | FALSE | Upregulated |
| -0,247101941 | PMFBP1 | FALSE | Downregulated |
| 0 | PMP2 | FALSE | Upregulated |
| -0,358502432 | PMS2L2 | FALSE | Downregulated |
| 0,670174088 | PMS2P4 | FALSE | Upregulated |
| | PNCK | | |
| -0,104177108 | PNLDC1 | FALSE | Downregulated |
| | PNLIP | | |
| | PNLIPRP1 | | |
| | PNLIPRP2 | | |
| | PNLIPRP3 | | |
| -0,014594569 | PNMA3 | FALSE | Downregulated |
| 0,067876097 | PNMA5 | FALSE | Upregulated |
| -1,268942584 | PNMAL1 | | Downregulated |
| -1,137645169 | PNMT | | Downregulated |
| 0,155533894 | PNOC | FALSE | Upregulated |
| | PNPLA1 | | |
| | PNPLA5 | | |
| -0,468097733 | POF1B | FALSE | Downregulated |
| -0,380839998 | POLN | FALSE | Downregulated |
| 2,357144585 | POLQ | | Upregulated |
| -0,197997249 | POM121L12 | FALSE | Downregulated |
| | POM121L2 | | |

| | | | |
|---|---|---|---|
| | POM121L4P | | |
| -0,434156551 | POMC | FALSE | Downregulated |
| 0,825896801 | POPDC2 | FALSE | Upregulated |
| -0,701348585 | POPDC3 | FALSE | Downregulated |
| | POTEA | | |
| | POTEB | | |
| | POTEB2 | | |
| | POTEC | | |
| | POTED | | |
| -0,285661369 | POTEE | FALSE | Downregulated |
| 0,021196969 | POTEF | FALSE | Upregulated |
| | POTEG | | |
| | POTEH | | |
| 0,330882068 | POTEI | FALSE | Upregulated |
| 0 | POTEJ | FALSE | Upregulated |
| 0 | POTEKP | FALSE | Upregulated |
| 0,298515972 | POTEM | FALSE | Upregulated |
| 0,909737847 | POU1F1 | FALSE | Upregulated |
| 0,659673757 | POU2F3 | FALSE | Upregulated |
| 1,437325547 | POU3F1 | | Upregulated |
| | POU3F3 | | |
| | POU3F4 | | |
| -0,548578849 | POU4F1 | FALSE | Downregulated |
| | POU4F2 | | |
| 0 | POU4F3 | FALSE | Upregulated |
| 0,869070491 | POU5F1B | FALSE | Upregulated |
| 0,507606822 | POU6F2-AS1 | FALSE | Upregulated |
| 0 | PP12613 | FALSE | Upregulated |
| 0,414973413 | PP2672 | FALSE | Upregulated |
| 0,133724683 | PPAP2C | FALSE | Upregulated |
| 0,180540802 | PPAPDC1A | FALSE | Upregulated |
| 1,167935997 | PPAPDC3 | | Upregulated |
| -0,576538314 | PPBP | FALSE | Downregulated |
| 0,21482791 | PPBPP2 | FALSE | Upregulated |
| -0,448782964 | PPEF1 | FALSE | Downregulated |
| | PPEF2 | | |
| 0,049490576 | PPIAL4B | FALSE | Upregulated |
| 0,019409064 | PPIAL4C | FALSE | Upregulated |
| 0 | PPIAL4D | FALSE | Upregulated |
| | PPIAL4E | | |
| | PPIAL4F | | |
| | PPIAL4G | | |
| -0,127003911 | PPIAP30 | FALSE | Downregulated |
| -0,157077378 | PPM1J | FALSE | Downregulated |
| 0,025337272 | PPP1R14C | FALSE | Upregulated |
| 0,049085716 | PPP1R17 | FALSE | Upregulated |
| -0,912886761 | PPP1R26-AS1 | FALSE | Downregulated |
| 0 | PPP1R2P3 | FALSE | Upregulated |
| | PPP1R2P9 | | |
| -0,214715193 | PPP1R36 | FALSE | Downregulated |
| -0,04603773 | PPP1R3A | FALSE | Downregulated |

| | | | |
|---|---|---|---|
| 0 | PPP1R42 | FALSE | Upregulated |
| 0,76360876 | PPP2R2B | FALSE | Upregulated |
| | PPP2R2B-IT1 | | |
| 0,372878749 | PPP2R2C | FALSE | Upregulated |
| | PPP2R3B-2 | | |
| 0,089078305 | PPP3R2 | FALSE | Upregulated |
| -0,487334711 | PPP4R1L | FALSE | Downregulated |
| -0,700596725 | PPP5D1 | FALSE | Downregulated |
| | PPY | | |
| | PPY2 | | |
| | PRAC1 | | |
| | PRAC2 | | |
| | PRAMEF1 | | |
| 1,363611528 | PRAMEF10 | | Upregulated |
| | PRAMEF11 | | |
| | PRAMEF12 | | |
| | PRAMEF13 | | |
| -0,148637222 | PRAMEF15 | FALSE | Downregulated |
| | PRAMEF17 | | |
| | PRAMEF18 | | |
| | PRAMEF19 | | |
| 0 | PRAMEF2 | FALSE | Upregulated |
| | PRAMEF20 | | |
| | PRAMEF25 | | |
| | PRAMEF26 | | |
| 0 | PRAMEF4 | FALSE | Upregulated |
| | PRAMEF5 | | |
| | PRAMEF6 | | |
| | PRAMEF7 | | |
| | PRAMEF8 | | |
| | PRAMEF9 | | |
| 0,535808176 | PRAMENP | FALSE | Upregulated |
| -0,16265259 | PRDM12 | FALSE | Downregulated |
| | PRDM13 | | |
| | PRDM14 | | |
| -0,539032542 | PRDM5 | FALSE | Downregulated |
| -0,711798502 | PRDM6 | FALSE | Downregulated |
| 0 | PRDM9 | FALSE | Upregulated |
| | PRG1 | | |
| -0,910898211 | PRG2 | FALSE | Downregulated |
| | PRG3 | | |
| -0,481170355 | PRIMA1 | FALSE | Downregulated |
| 0,801908519 | PRKACG | FALSE | Upregulated |
| 0 | PRKCG | FALSE | Upregulated |
| 0,223171254 | PRKCQ-AS1 | FALSE | Upregulated |
| 0,077366797 | PRKG2 | FALSE | Upregulated |
| | PRL | | |
| | PRLH | | |
| | PRLHR | | |
| | PRM1 | | |
| 0,302545561 | PRM2 | FALSE | Upregulated |

| | | | |
|---|---|---|---|
| | PRM3 | | |
| | PRMT8 | | |
| 0,686665621 | PRND | FALSE | Upregulated |
| | PRNT | | |
| 0 | PROK1 | FALSE | Upregulated |
| 0,093232123 | PROK2 | FALSE | Upregulated |
| -0,137653782 | PROKR1 | FALSE | Downregulated |
| -0,067722796 | PROKR2 | FALSE | Downregulated |
| 0 | PROL1 | FALSE | Upregulated |
| -1,13668429 | PROP1 | | Downregulated |
| 0,824376246 | PRPH | FALSE | Upregulated |
| | PRPS1L1 | | |
| 0,124495595 | PRR16 | FALSE | Upregulated |
| 1,437195877 | PRR19 | | Upregulated |
| | PRR20A | | |
| | PRR20B | | |
| | PRR20C | | |
| | PRR20D | | |
| 0 | PRR20E | FALSE | Upregulated |
| | PRR21 | | |
| | PRR23A | | |
| | PRR23B | | |
| | PRR23C | | |
| | PRR23D1 | | |
| | PRR23D2 | | |
| 0,564398649 | PRR25 | FALSE | Upregulated |
| 0,448212425 | PRR4 | FALSE | Upregulated |
| -0,377609275 | PRR5-ARHGAP8 | FALSE | Downregulated |
| 0 | PRR9 | FALSE | Upregulated |
| 0,160849491 | PRRG3 | FALSE | Upregulated |
| -0,479175809 | PRRT1 | FALSE | Downregulated |
| 0,27883575 | PRRT3-AS1 | FALSE | Upregulated |
| 0,280756568 | PRRT4 | FALSE | Upregulated |
| 0 | PRRX2 | FALSE | Upregulated |
| 0,202626599 | PRSS16 | FALSE | Upregulated |
| 0,816838695 | PRSS2 | FALSE | Upregulated |
| 0,132892609 | PRSS21 | FALSE | Upregulated |
| 0,404218506 | PRSS22 | FALSE | Upregulated |
| 1,325132379 | PRSS27 | | Upregulated |
| 0,115033032 | PRSS3 | FALSE | Upregulated |
| 0,231135863 | PRSS30P | FALSE | Upregulated |
| 0,379688888 | PRSS33 | FALSE | Upregulated |
| -0,806568263 | PRSS35 | FALSE | Downregulated |
| -0,47016762 | PRSS36 | FALSE | Downregulated |
| 0,288182347 | PRSS38 | FALSE | Upregulated |
| | PRSS41 | | |
| -0,472520495 | PRSS42 | FALSE | Downregulated |
| 0,549193361 | PRSS44 | FALSE | Upregulated |
| 0,10250785 | PRSS45 | FALSE | Upregulated |
| -0,336510651 | PRSS46 | FALSE | Downregulated |
| -0,67118175 | PRSS50 | FALSE | Downregulated |

| | | | |
|---|---|---|---|
| 0 | PRSS56 | FALSE | Upregulated |
| -0,299630467 | PRSS57 | FALSE | Downregulated |
| 1,400726087 | PRTFDC1 | | Upregulated |
| 0 | PRTN3 | FALSE | Upregulated |
| 0,307155178 | PSCA | FALSE | Upregulated |
| -0,734419404 | PSD2 | FALSE | Downregulated |
| -0,29970387 | PSG10P | FALSE | Downregulated |
| 0,440229147 | PSG11 | FALSE | Upregulated |
| 0,831137763 | PSG2 | FALSE | Upregulated |
| -0,614459958 | PSG3 | FALSE | Downregulated |
| 0,452310953 | PSG4 | FALSE | Upregulated |
| -0,232911962 | PSG5 | FALSE | Downregulated |
| -0,233844428 | PSG6 | FALSE | Downregulated |
| -0,523620068 | PSG7 | FALSE | Downregulated |
| -0,741788019 | PSG8 | FALSE | Downregulated |
| 1,149632153 | PSG9 | | Upregulated |
| 0,529783454 | PSIMCT-1 | FALSE | Upregulated |
| | PSKH2 | | |
| | PSMA8 | | |
| 0 | PSMB11 | FALSE | Upregulated |
| | PSORS1C1 | | |
| -0,054121778 | PSORS1C2 | FALSE | Downregulated |
| -0,753814435 | PSORS1C3 | FALSE | Downregulated |
| -0,76971463 | PSPN | FALSE | Downregulated |
| 0,495272167 | PTCHD2 | FALSE | Upregulated |
| -0,053919153 | PTCHD3 | FALSE | Downregulated |
| 0,196910008 | PTCRA | FALSE | Upregulated |
| 0,469226316 | PTCSC3 | FALSE | Upregulated |
| | PTENP1-AS | | |
| | PTF1A | | |
| 0,757565918 | PTGER1 | FALSE | Upregulated |
| 1,723082963 | PTGER3 | | Upregulated |
| 0,165429418 | PTGER4P2-CDK2AP2P2 | FALSE | Upregulated |
| 0,597123081 | PTGES | FALSE | Upregulated |
| | PTGES3L-AARSD1 | | |
| 0,860304072 | PTGFR | FALSE | Upregulated |
| 0,712483161 | PTGS2 | FALSE | Upregulated |
| -0,765714548 | PTH | FALSE | Downregulated |
| | PTH2 | | |
| 0,382995791 | PTH2R | FALSE | Upregulated |
| 0,261336026 | PTHLH | FALSE | Upregulated |
| 0,152873859 | PTPLA | FALSE | Upregulated |
| 0,649858145 | PTPN20B | FALSE | Upregulated |
| 0,81559344 | PTPN22 | FALSE | Upregulated |
| 0,775604075 | PTPN5 | FALSE | Upregulated |
| 0 | PTPRN | FALSE | Upregulated |
| -0,218604548 | PTPRQ | FALSE | Downregulated |
| 0,381143071 | PTPRVP | FALSE | Upregulated |
| 0,129992828 | PTPRZ1 | FALSE | Upregulated |
| 0,597605941 | PTTG1 | FALSE | Upregulated |
| 0,024387902 | PTTG2 | FALSE | Upregulated |

| | | | |
|---|---|---|---|
| -0,570798307 | PTX3 | FALSE | Downregulated |
| 0,307221631 | PTX4 | FALSE | Upregulated |
| 0,905653182 | PURG | FALSE | Upregulated |
| 0,141569822 | PVALB | FALSE | Upregulated |
| 1,417262843 | PVRIG2P | | Upregulated |
| 0 | PVRL3-AS1 | FALSE | Upregulated |
| 0,763945285 | PVRL4 | FALSE | Upregulated |
| 0,223829749 | PVT1 | FALSE | Upregulated |
| 0 | PWRN1 | FALSE | Upregulated |
| -0,164038696 | PWRN3 | FALSE | Downregulated |
| -1,329372717 | PXDNL | | Downregulated |
| | PYDC1 | | |
| | PYDC2 | | |
| 0,616282567 | PYGO1 | FALSE | Upregulated |
| 0,048276727 | PYY | FALSE | Upregulated |
| -0,199341544 | PYY2 | FALSE | Downregulated |
| 0,938069313 | QPCT | FALSE | Upregulated |
| 0,01215623 | QRFP | FALSE | Upregulated |
| | QRFPR | | |
| 0,018120412 | R3HDML | FALSE | Upregulated |
| 0,150066384 | RAB19 | FALSE | Upregulated |
| -0,520162565 | RAB25 | FALSE | Downregulated |
| 0,134328508 | RAB33A | FALSE | Upregulated |
| -0,577656626 | RAB38 | FALSE | Downregulated |
| -0,26357995 | RAB39A | FALSE | Downregulated |
| 0,344949433 | RAB39B | FALSE | Upregulated |
| 0,241840096 | RAB3A | FALSE | Upregulated |
| -0,25173773 | RAB3C | FALSE | Downregulated |
| 0,131174124 | RAB40AL | FALSE | Upregulated |
| 0,169326778 | RAB44 | FALSE | Upregulated |
| -0,439461244 | RAB6C | FALSE | Downregulated |
| 0,375284626 | RAB6C-AS1 | FALSE | Upregulated |
| 0,14712375 | RAB9B | FALSE | Upregulated |
| 0 | RAB9BP1 | FALSE | Upregulated |
| 0 | RACGAP1P | FALSE | Upregulated |
| | RAD21L1 | | |
| 1,302918749 | RAD51AP1 | | Upregulated |
| -0,150363357 | RAD51AP2 | FALSE | Downregulated |
| -0,650195517 | RAD51C | FALSE | Downregulated |
| 0,033273572 | RADIL | FALSE | Upregulated |
| -1,022962637 | RAET1G | | Downregulated |
| 0 | RAET1K | FALSE | Upregulated |
| | RAET1L | | |
| -0,11368263 | RALYL | FALSE | Downregulated |
| -0,06355641 | RAPSN | FALSE | Downregulated |
| 0,748986395 | RASD2 | FALSE | Upregulated |
| -1,496656942 | RASGEF1C | | Downregulated |
| 1,113577277 | RASGRF1 | | Upregulated |
| -0,950723767 | RASL11A | FALSE | Downregulated |
| 0,067976305 | RASSF10 | FALSE | Upregulated |
| -0,566997728 | RASSF8-AS1 | FALSE | Downregulated |

| | | | |
|---|---|---|---|
| 0,698179302 | RASSF9 | FALSE | Upregulated |
| | RAX | | |
| | RBAK-RBAKDN | | |
| 0,406086649 | RBAKDN | FALSE | Upregulated |
| -0,024711188 | RBBP8NL | FALSE | Downregulated |
| -0,216351445 | RBFADN | FALSE | Downregulated |
| 0,247746115 | RBFOX1 | FALSE | Upregulated |
| 0,100713889 | RBM11 | FALSE | Upregulated |
| -0,081635728 | RBM20 | FALSE | Downregulated |
| 0,222356212 | RBM24 | FALSE | Upregulated |
| -1,182451831 | RBM26-AS1 | | Downregulated |
| 0,744396553 | RBM44 | FALSE | Upregulated |
| 0 | RBM46 | FALSE | Upregulated |
| -0,42158786 | RBMS3-AS3 | FALSE | Downregulated |
| -0,696707319 | RBMXL2 | FALSE | Downregulated |
| | RBMY1A1 | | |
| | RBMY1A3P | | |
| | RBMY1B | | |
| | RBMY1D | | |
| | RBMY1E | | |
| | RBMY1F | | |
| | RBMY1J | | |
| | RBMY3AP | | |
| -0,037066187 | RBP3 | FALSE | Downregulated |
| -1,165618345 | RBP7 | | Downregulated |
| -0,133245522 | RBPJL | FALSE | Downregulated |
| 0,099452879 | RCAN3AS | FALSE | Upregulated |
| 0,138718581 | RCVRN | FALSE | Upregulated |
| -1,283890161 | RD3 | | Downregulated |
| 1,091926846 | RD3L | | Upregulated |
| -0,090565098 | RDH8 | FALSE | Downregulated |
| -0,811417072 | RDM1 | FALSE | Downregulated |
| 0,086000466 | REEP2 | FALSE | Upregulated |
| | REG1A | | |
| | REG1B | | |
| | REG1P | | |
| 0 | REG3A | FALSE | Upregulated |
| | REG3G | | |
| -1,027999634 | REG4 | | Downregulated |
| 0,457886474 | REM1 | FALSE | Upregulated |
| -0,055704272 | REREP3 | FALSE | Downregulated |
| | RERG-AS1 | | |
| 0,659321804 | RERGL | FALSE | Upregulated |
| | RESP18 | | |
| | RETN | | |
| | RETNLB | | |
| 0 | REXO1L1P | FALSE | Upregulated |
| 0,66265915 | REXO1L2P | FALSE | Upregulated |
| -0,700887816 | RFPL2 | FALSE | Downregulated |
| 0,368039171 | RFPL3S | FALSE | Upregulated |
| -0,228662983 | RFPL4A | FALSE | Downregulated |

| | | | |
|---|---|---|---|
| 0,034614116 | RFPL4AL1 | FALSE | Upregulated |
| | RFPL4B | | |
| 2,130369673 | RFX4 | | Upregulated |
| | RFX6 | | |
| -0,205086965 | RFX8 | FALSE | Downregulated |
| -0,752540153 | RGAG1 | FALSE | Downregulated |
| 1,025508477 | RGCC | | Upregulated |
| -0,173391078 | RGMA | FALSE | Downregulated |
| 0,393997074 | RGMB-AS1 | FALSE | Upregulated |
| 0,520151917 | RGPD4-AS1 | FALSE | Upregulated |
| | RGR | | |
| 1,26493704 | RGS1 | | Upregulated |
| 1,147744079 | RGS10 | | Upregulated |
| 0 | RGS13 | FALSE | Upregulated |
| -0,137675573 | RGS18 | FALSE | Downregulated |
| 0,691925266 | RGS21 | FALSE | Upregulated |
| | RGS22 | | |
| 0 | RGS7 | FALSE | Upregulated |
| 0 | RGS8 | FALSE | Upregulated |
| -0,133600684 | RGS9 | FALSE | Downregulated |
| 0,329685147 | RGS9BP | FALSE | Upregulated |
| 0,182405334 | RGSL1 | FALSE | Upregulated |
| 0 | RHAG | FALSE | Upregulated |
| 0,451387982 | RHBDL3 | FALSE | Upregulated |
| 0,93177775 | RHO | FALSE | Upregulated |
| 0 | RHOV | FALSE | Upregulated |
| -0,472944259 | RHOXF1 | FALSE | Downregulated |
| 0,608330413 | RHPN1-AS1 | FALSE | Upregulated |
| 0,054140574 | RIBC2 | FALSE | Upregulated |
| | RIIAD1 | | |
| -0,504518038 | RIMS1 | FALSE | Downregulated |
| -0,926337564 | RIMS2 | FALSE | Downregulated |
| | RIPPLY2 | | |
| 0,44721048 | RIPPLY3 | FALSE | Upregulated |
| -0,492336561 | RIT2 | FALSE | Downregulated |
| -0,127449465 | RLN1 | FALSE | Downregulated |
| 0,041008843 | RLN2 | FALSE | Upregulated |
| 0,24067187 | RLN3 | FALSE | Upregulated |
| 2,385647302 | RMI2 | | Upregulated |
| 0 | RMST | FALSE | Upregulated |
| | RNA5-8S5 | | |
| | RNA5S1 | | |
| | RNA5S10 | | |
| | RNA5S11 | | |
| | RNA5S12 | | |
| | RNA5S13 | | |
| | RNA5S14 | | |
| | RNA5S15 | | |
| | RNA5S16 | | |
| | RNA5S17 | | |
| | RNA5S2 | | |

| | | | |
|---|---|---|---|
| | RNA5S3 | | |
| | RNA5S4 | | |
| | RNA5S5 | | |
| | RNA5S6 | | |
| | RNA5S7 | | |
| 0 | RNA5S8 | FALSE | Upregulated |
| -1,093784214 | RNA5S9 | | Downregulated |
| | RNASE11 | | |
| | RNASE12 | | |
| 1,221294533 | RNASE2 | | Upregulated |
| 0,264480942 | RNASE3 | FALSE | Upregulated |
| 0,117739634 | RNASE8 | FALSE | Upregulated |
| | RNASE9 | | |
| 1,181535488 | RNASEH2B-AS1 | | Upregulated |
| 0,130356863 | RNF112 | FALSE | Upregulated |
| 0,506402691 | RNF126P1 | FALSE | Upregulated |
| 0,356220356 | RNF133 | FALSE | Upregulated |
| 0,757013709 | RNF144A-AS1 | FALSE | Upregulated |
| 0,136116193 | RNF148 | FALSE | Upregulated |
| 0,184101668 | RNF151 | FALSE | Upregulated |
| -0,06248387 | RNF17 | FALSE | Downregulated |
| 0 | RNF182 | FALSE | Upregulated |
| 0,020699669 | RNF183 | FALSE | Upregulated |
| 0,154660303 | RNF186 | FALSE | Upregulated |
| 0,388076022 | RNF212 | FALSE | Upregulated |
| | RNF216-IT1 | | |
| 0,177020051 | RNF223 | FALSE | Upregulated |
| -0,420564375 | RNF5P1 | FALSE | Downregulated |
| -0,509736465 | RNFT2 | FALSE | Downregulated |
| 0 | RNU105B | FALSE | Upregulated |
| | RNU105C | | |
| -0,91344837 | RNU11 | FALSE | Downregulated |
| -0,90720673 | RNU12 | FALSE | Downregulated |
| 0,720810288 | RNU2-1 | FALSE | Upregulated |
| 0,213867534 | RNU4ATAC | FALSE | Upregulated |
| 0 | RNU5A-1 | FALSE | Upregulated |
| 0 | RNU5B-1 | FALSE | Upregulated |
| | RNU5D-1 | | |
| | RNU5E-1 | | |
| | RNU5F-1 | | |
| | RNU6-1 | | |
| | RNU6-10P | | |
| | RNU6-14P | | |
| | RNU6-15P | | |
| | RNU6-16P | | |
| | RNU6-19P | | |
| -0,101200907 | RNU6-2 | FALSE | Downregulated |
| | RNU6-21P | | |
| | RNU6-23P | | |
| | RNU6-26P | | |
| | RNU6-28P | | |

| | | | |
|---|---|---|---|
| | RNU6-30P | | |
| | RNU6-31P | | |
| | RNU6-33P | | |
| | RNU6-34P | | |
| | RNU6-35P | | |
| | RNU6-36P | | |
| | RNU6-39P | | |
| | RNU6-42P | | |
| | RNU6-45P | | |
| | RNU6-46P | | |
| | RNU6-48P | | |
| | RNU6-52P | | |
| | RNU6-53P | | |
| | RNU6-55P | | |
| | RNU6-56P | | |
| | RNU6-57P | | |
| | RNU6-58P | | |
| | RNU6-59P | | |
| | RNU6-63P | | |
| | RNU6-64P | | |
| | RNU6-66P | | |
| | RNU6-67P | | |
| | RNU6-68P | | |
| | RNU6-69P | | |
| | RNU6-7 | | |
| | RNU6-71P | | |
| | RNU6-72P | | |
| | RNU6-75P | | |
| | RNU6-76P | | |
| | RNU6-78P | | |
| | RNU6-79P | | |
| | RNU6-8 | | |
| | RNU6-81P | | |
| | RNU6-82P | | |
| | RNU6-83P | | |
| 0,069142038 | RNU6-9 | FALSE | Upregulated |
| | RNU7-1 | | |
| | RNU86 | | |
| 0 | RNVU1-10 | FALSE | Upregulated |
| 0,387758742 | RNVU1-14 | FALSE | Upregulated |
| 0,387012887 | RNVU1-15 | FALSE | Upregulated |
| | RNVU1-17 | | |
| 0,453099891 | RNVU1-19 | FALSE | Upregulated |
| 0,467369141 | RNVU1-3 | FALSE | Upregulated |
| 0 | RNVU1-4 | FALSE | Upregulated |
| 0 | RNVU1-6 | FALSE | Upregulated |
| -0,780933713 | RNY1 | FALSE | Downregulated |
| 0,19139178 | RNY3 | FALSE | Upregulated |
| | RNY4 | | |
| | RNY5 | | |
| 0,030997953 | ROCK1P1 | FALSE | Upregulated |

| | | | |
|---|---|---|---|
| | ROPN1 | | |
| 0,372721394 | ROPN1B | FALSE | Upregulated |
| 0,303700635 | ROPN1L | FALSE | Upregulated |
| 0,595532146 | RORB | FALSE | Upregulated |
| 1,263980241 | ROS1 | | Upregulated |
| -0,829610844 | RP1 | FALSE | Downregulated |
| | RP1-104O17.1 | | |
| 0 | RP1-137K24.1 | FALSE | Upregulated |
| 0 | RP1-140J1.1 | FALSE | Upregulated |
| 0 | RP1-167G20.1 | FALSE | Upregulated |
| 0,119064107 | RP1-172N19.2 | FALSE | Upregulated |
| 0,88657196 | RP1-207H1.3 | FALSE | Upregulated |
| | RP1-224A6.3 | | |
| 0,46074938 | RP1-257A15.1 | FALSE | Upregulated |
| 0 | RP1-272J12.1 | FALSE | Upregulated |
| 0 | RP1-278O22.1 | FALSE | Upregulated |
| 0,706208838 | RP1-290I10.5 | FALSE | Upregulated |
| -0,022929419 | RP1-40E16.9 | FALSE | Downregulated |
| | RP1-63G5.5 | | |
| -0,043748929 | RP1-90L6.2 | FALSE | Downregulated |
| 0,487411502 | RP1-93I3.1 | FALSE | Upregulated |
| 0,177448562 | RP11-1008C21.1 | FALSE | Upregulated |
| | RP11-100N20.1 | | |
| 0,277717961 | RP11-1018N14.1 | FALSE | Upregulated |
| | RP11-104D21.3 | | |
| 0 | RP11-1055B8.3 | FALSE | Upregulated |
| 1,041526604 | RP11-1057B8.2 | | Upregulated |
| | RP11-1079K10.5 | | |
| 0 | RP11-107I14.4 | FALSE | Upregulated |
| 0,068128077 | RP11-107K17.1 | FALSE | Upregulated |
| 0,406367769 | RP11-1080G15.1 | FALSE | Upregulated |
| | RP11-1084I9.1 | | |
| 0,680673459 | RP11-1094H24.4 | FALSE | Upregulated |
| | RP11-109J4.1 | | |
| 0 | RP11-10A14.5 | FALSE | Upregulated |
| | RP11-10L7.1 | | |
| | RP11-10N16.2 | | |
| 0,138511454 | RP11-110H10.2 | FALSE | Upregulated |
| 0,426806073 | RP11-111I12.1 | FALSE | Upregulated |
| -0,026615206 | RP11-1124B17.1 | FALSE | Downregulated |
| -0,586857391 | RP11-112L6.4 | FALSE | Downregulated |
| 0,686284964 | RP11-1134I14.8 | FALSE | Upregulated |
| 0 | RP11-1191J2.2 | FALSE | Upregulated |
| 0,621192376 | RP11-119F19.2 | FALSE | Upregulated |
| 0,800362633 | RP11-1223D19.1 | FALSE | Upregulated |
| 0 | RP11-122M14.1 | FALSE | Upregulated |
| | RP11-124K4.1 | | |
| | RP11-125P18.1 | | |
| 0,485044108 | RP11-127O4.3 | FALSE | Upregulated |
| | RP11-129J12.2 | | |
| -0,456905502 | RP11-12A2.3 | FALSE | Downregulated |

| | | | |
|---|---|---|---|
| 1,760319562 | RP11-132A1.4 | | Upregulated |
| | RP11-1338A24.1 | | |
| | RP11-134O21.1 | | |
| 0,826876442 | RP11-134P9.3 | FALSE | Upregulated |
| 0,389008972 | RP11-135J2.4 | FALSE | Upregulated |
| 0,624175217 | RP11-138B4.1 | FALSE | Upregulated |
| | RP11-13A1.1 | | |
| | RP11-13E5.2 | | |
| | RP11-13P5.2 | | |
| 0,081836352 | RP11-140I24.1 | FALSE | Upregulated |
| | RP11-142G7.2 | | |
| | RP11-143A12.3 | | |
| 0,124339487 | RP11-143M1.2 | FALSE | Upregulated |
| | RP11-143M1.3 | | |
| | RP11-146N18.1 | | |
| 0 | RP11-148L24.1 | FALSE | Upregulated |
| | RP11-14J7.6 | | |
| 0,814513061 | RP11-14N7.2 | FALSE | Upregulated |
| | RP11-150O12.6 | | |
| | RP11-152L7.2 | | |
| -0,017135352 | RP11-153K16.1 | FALSE | Downregulated |
| 0 | RP11-154D17.1 | FALSE | Upregulated |
| 0,374624708 | RP11-154D6.1 | FALSE | Upregulated |
| | RP11-157D23.1 | | |
| | RP11-157E21.1 | | |
| | RP11-158A8.1 | | |
| | RP11-158D2.2 | | |
| 0,800442624 | RP11-15I11.3 | FALSE | Upregulated |
| | RP11-15K3.1 | | |
| -0,293158673 | RP11-160O5.1 | FALSE | Downregulated |
| 0,569455074 | RP11-161H23.9 | FALSE | Upregulated |
| -0,141027261 | RP11-163N6.2 | FALSE | Downregulated |
| 0,035241679 | RP11-165H23.1 | FALSE | Upregulated |
| 0,889218212 | RP11-165P7.1 | FALSE | Upregulated |
| | RP11-166A12.1 | | |
| 0,564813046 | RP11-166O4.5 | FALSE | Upregulated |
| 0,14101986 | RP11-167H9.6 | FALSE | Upregulated |
| 0,000977115 | RP11-168E14.1 | FALSE | Upregulated |
| 0 | RP11-168E17.1 | FALSE | Upregulated |
| 1,203413868 | RP11-168O10.6 | | Upregulated |
| 0 | RP11-170N11.1 | FALSE | Upregulated |
| | RP11-171A24.2 | | |
| | RP11-174G17.2 | | |
| 0 | RP11-175K6.1 | FALSE | Upregulated |
| 0 | RP11-178L8.5 | FALSE | Upregulated |
| | RP11-179A16.1 | | |
| | RP11-181F12.1 | | |
| -0,765981975 | RP11-184I16.4 | FALSE | Downregulated |
| -0,540034238 | RP11-188C12.3 | FALSE | Downregulated |
| | RP11-189B4.6 | | |
| | RP11-18F14.1 | | |

| | | | |
|---|---|---|---|
| 0,491537534 | RP11-18H21.1 | FALSE | Upregulated |
| 0 | RP11-191L9.4 | FALSE | Upregulated |
| | RP11-191N8.2 | | |
| 0 | RP11-193H5.1 | FALSE | Upregulated |
| | RP11-198M11.2 | | |
| 0,179592153 | RP11-1C1.4 | FALSE | Upregulated |
| -2,040834898 | RP11-205M3.3 | | Downregulated |
| -0,28877873 | RP11-208N14.4 | FALSE | Downregulated |
| -0,333794931 | RP11-214C8.2 | FALSE | Downregulated |
| 0 | RP11-214L13.1 | FALSE | Upregulated |
| 0,583782829 | RP11-214L19.1 | FALSE | Upregulated |
| | RP11-215E13.1 | | |
| -0,035547113 | RP11-21L23.3 | FALSE | Downregulated |
| 0,536032639 | RP11-222A5.1 | FALSE | Upregulated |
| 0,943474159 | RP11-224O19.2 | FALSE | Upregulated |
| 0,92141755 | RP11-227H15.4 | FALSE | Upregulated |
| 0,448809301 | RP11-22H5.2 | FALSE | Upregulated |
| 0,23372779 | RP11-230B22.1 | FALSE | Upregulated |
| | RP11-230G5.2 | | |
| 1,694680377 | RP11-234B24.2 | | Upregulated |
| 0,710924557 | RP11-236J17.5 | FALSE | Upregulated |
| | RP11-236P13.1 | | |
| 0,000977115 | RP11-23B15.1 | FALSE | Upregulated |
| 0 | RP11-243E13.1 | FALSE | Upregulated |
| -0,314771954 | RP11-245M24.1 | FALSE | Downregulated |
| -0,016192915 | RP11-24I21.1 | FALSE | Downregulated |
| 0 | RP11-24P4.1 | FALSE | Upregulated |
| | RP11-251P6.1 | | |
| 0 | RP11-252C15.1 | FALSE | Upregulated |
| | RP11-256L11.1 | | |
| | RP11-257I8.2 | | |
| | RP11-262K1.1 | | |
| | RP11-265P11.2 | | |
| | RP11-267C16.1 | | |
| 0,709254359 | RP11-26J3.1 | FALSE | Upregulated |
| | RP11-271F18.4 | | |
| | RP11-274B18.2 | | |
| -0,029738205 | RP11-276H19.1 | FALSE | Downregulated |
| | RP11-27N21.3 | | |
| 0,556474714 | RP11-280K24.4 | FALSE | Upregulated |
| | RP11-280O1.2 | | |
| | RP11-286B14.1 | | |
| | RP11-286E11.1 | | |
| 0,170748624 | RP11-290F5.2 | FALSE | Upregulated |
| 0 | RP11-293M10.5 | FALSE | Upregulated |
| 0,668488752 | RP11-294O2.2 | FALSE | Upregulated |
| | RP11-298O21.5 | | |
| 0 | RP11-299H22.1 | FALSE | Upregulated |
| | RP11-299L17.3 | | |
| | RP11-2A4.3 | | |
| 2,274109077 | RP11-2O17.2 | | Upregulated |

| | | | |
|---|---|---|---|
| 0,275700672 | RP11-305E17.6 | FALSE | Upregulated |
| | RP11-305P22.9 | | |
| 0,000977115 | RP11-307C19.3 | FALSE | Upregulated |
| -0,208035499 | RP11-308N19.4 | FALSE | Downregulated |
| -0,093662559 | RP11-310P5.1 | FALSE | Downregulated |
| | RP11-313C15.1 | | |
| -0,385878212 | RP11-314C16.1 | FALSE | Downregulated |
| -0,972254904 | RP11-314D7.2 | FALSE | Downregulated |
| -0,091940924 | RP11-317J10.2 | FALSE | Downregulated |
| -0,112927299 | RP11-317M11.1 | FALSE | Downregulated |
| | RP11-318I4.1 | | |
| -0,57943636 | RP11-319E12.1 | FALSE | Downregulated |
| | RP11-319E16.1 | | |
| -0,010929374 | RP11-319G6.1 | FALSE | Downregulated |
| -0,04169029 | RP11-320G24.1 | FALSE | Downregulated |
| | RP11-324L3.1 | | |
| 0,404420545 | RP11-326F20.5 | FALSE | Upregulated |
| | RP11-327I22.6 | | |
| 0,807034374 | RP11-329J18.2 | FALSE | Upregulated |
| | RP11-32F11.2 | | |
| | RP11-330A16.1 | | |
| 0,333642613 | RP11-333A23.4 | FALSE | Upregulated |
| | RP11-33G16.1 | | |
| 0 | RP11-341A22.2 | FALSE | Upregulated |
| | RP11-343J18.1 | | |
| 0,536541952 | RP11-344B5.2 | FALSE | Upregulated |
| 0,462640691 | RP11-344E13.3 | FALSE | Upregulated |
| | RP11-344G13.1 | | |
| 0,892556093 | RP11-345L23.1 | FALSE | Upregulated |
| | RP11-346D6.6 | | |
| 0,01196447 | RP11-347D21.3 | FALSE | Upregulated |
| -1,271402063 | RP11-34F13.2 | | Downregulated |
| 0,577436718 | RP11-353N14.5 | FALSE | Upregulated |
| 0,461553133 | RP11-353N4.5 | FALSE | Upregulated |
| 0,985435554 | RP11-354E11.2 | FALSE | Upregulated |
| | RP11-360P21.2 | | |
| 0,724636085 | RP11-362A1.1 | FALSE | Upregulated |
| 0,90474753 | RP11-362F19.1 | FALSE | Upregulated |
| | RP11-363D24.1 | | |
| | RP11-363G15.2 | | |
| 0,892485082 | RP11-367N14.2 | FALSE | Upregulated |
| | RP11-368I23.2 | | |
| 0,099452879 | RP11-371M22.1 | FALSE | Upregulated |
| 0,16312708 | RP11-378A12.1 | FALSE | Upregulated |
| 0,867952225 | RP11-379F4.4 | FALSE | Upregulated |
| 0,652182388 | RP11-37B2.1 | FALSE | Upregulated |
| 0 | RP11-37N22.1 | FALSE | Upregulated |
| | RP11-384J4.1 | | |
| | RP11-386B13.3 | | |
| 0,546726216 | RP11-386I8.6 | FALSE | Upregulated |
| | RP11-389K14.3 | | |

| | | | |
|---|---|---|---|
| 0 | RP11-394O9.1 | FALSE | Upregulated |
| | RP11-395L14.3 | | |
| -0,786246605 | RP11-397C12.1 | FALSE | Downregulated |
| 0,172675256 | RP11-397C18.2 | FALSE | Upregulated |
| 0,662486676 | RP11-399K21.14 | FALSE | Upregulated |
| | RP11-3B12.2 | | |
| 0 | RP11-3B12.3 | FALSE | Upregulated |
| 0,465926925 | RP11-3P17.4 | FALSE | Upregulated |
| 0,124372589 | RP11-402G3.3 | FALSE | Upregulated |
| 0,623717402 | RP11-402L6.1 | FALSE | Upregulated |
| | RP11-404O13.1 | | |
| 0,070401644 | RP11-406A20.1 | FALSE | Upregulated |
| 0 | RP11-406A20.4 | FALSE | Upregulated |
| | RP11-406A9.2 | | |
| -0,991787847 | RP11-406H4.1 | FALSE | Downregulated |
| 0,399742508 | RP11-406O16.1 | FALSE | Upregulated |
| -0,017135352 | RP11-407A16.3 | FALSE | Downregulated |
| | RP11-408H20.2 | | |
| 0,373094292 | RP11-40F8.2 | FALSE | Upregulated |
| | RP11-413P11.1 | | |
| | RP11-415C15.2 | | |
| -0,373680771 | RP11-415D17.4 | FALSE | Downregulated |
| | RP11-416O18.1 | | |
| 0 | RP11-417E7.2 | FALSE | Upregulated |
| 0,45255205 | RP11-420A23.1 | FALSE | Upregulated |
| -0,183645786 | RP11-423C15.3 | FALSE | Downregulated |
| -1,352996174 | RP11-423H2.3 | | Downregulated |
| 0,046678197 | RP11-425I13.3 | FALSE | Upregulated |
| 0 | RP11-429H9.4 | FALSE | Upregulated |
| 0,073022978 | RP11-430C1.1 | FALSE | Upregulated |
| -0,017135352 | RP11-430H10.1 | FALSE | Downregulated |
| 0,435291647 | RP11-433J20.1 | FALSE | Upregulated |
| | RP11-435M3.2 | | |
| -0,037470309 | RP11-436K8.1 | FALSE | Downregulated |
| | RP11-438N16.1 | | |
| -0,467296237 | RP11-439L18.3 | FALSE | Downregulated |
| -0,041235707 | RP11-439L8.3 | FALSE | Downregulated |
| 0,70351451 | RP11-439L8.4 | FALSE | Upregulated |
| 0,099452879 | RP11-440G9.1 | FALSE | Upregulated |
| 1,698371615 | RP11-445H22.3 | | Upregulated |
| 0,146458703 | RP11-446H18.5 | FALSE | Upregulated |
| -0,017135352 | RP11-449D8.1 | FALSE | Downregulated |
| | RP11-451G4.2 | | |
| -0,204480531 | RP11-452H21.4 | FALSE | Downregulated |
| | RP11-452N4.1 | | |
| 0 | RP11-453E2.2 | FALSE | Upregulated |
| 0 | RP11-454C18.2 | FALSE | Upregulated |
| | RP11-454P21.1 | | |
| 3,305533863 | RP11-456O19.4 | | Upregulated |
| 0,769112976 | RP11-45L9.1 | FALSE | Upregulated |
| 0,293754954 | RP11-460N16.1 | FALSE | Upregulated |

| | | | |
|---|---|---|---|
| 0 | RP11-461F11.1 | FALSE | Upregulated |
| 0 | RP11-461F11.3 | FALSE | Upregulated |
| 1,10127793 | RP11-461O7.1 | | Upregulated |
| 0,9645539 | RP11-462G2.1 | FALSE | Upregulated |
| -0,017135352 | RP11-462G22.1 | FALSE | Downregulated |
| | RP11-463J7.2 | | |
| 0,060233881 | RP11-465B22.5 | FALSE | Upregulated |
| 0,307638558 | RP11-466F5.8 | FALSE | Upregulated |
| | RP11-46C24.3 | | |
| -0,890768998 | RP11-474D1.3 | FALSE | Downregulated |
| 0,972668863 | RP11-475N22.4 | FALSE | Upregulated |
| | RP11-478H13.3 | | |
| | RP11-480C22.1 | | |
| | RP11-482D24.2 | | |
| -0,189568222 | RP11-483E7.1 | FALSE | Downregulated |
| -0,136740525 | RP11-486A14.2 | FALSE | Downregulated |
| 0,179585951 | RP11-486O12.2 | FALSE | Upregulated |
| | RP11-488I20.3 | | |
| | RP11-489D6.2 | | |
| 0 | RP11-48F14.1 | FALSE | Upregulated |
| 0,082169757 | RP11-490D19.6 | FALSE | Upregulated |
| | RP11-493K23.4 | | |
| 0 | RP11-494M8.4 | FALSE | Upregulated |
| | RP11-497K21.1 | | |
| 0,389315727 | RP11-500B12.1 | FALSE | Upregulated |
| | RP11-501G7.1 | | |
| -0,555503458 | RP11-503C24.4 | FALSE | Downregulated |
| | RP11-508P1.2 | | |
| -0,328116171 | RP11-509J21.1 | FALSE | Downregulated |
| -0,654354351 | RP11-510D19.1 | FALSE | Downregulated |
| 0,524766686 | RP11-513O17.2 | FALSE | Upregulated |
| 0,11187309 | RP11-513O17.2-2 | FALSE | Upregulated |
| | RP11-513O17.3 | | |
| -0,189298318 | RP11-514D23.1 | FALSE | Downregulated |
| 0,366354622 | RP11-519M16.1 | FALSE | Upregulated |
| 0,726413699 | RP11-523L1.2 | FALSE | Upregulated |
| 0,226837822 | RP11-523L20.2 | FALSE | Upregulated |
| 0 | RP11-526A4.1 | FALSE | Upregulated |
| 0,259935197 | RP11-526P6.1 | FALSE | Upregulated |
| | RP11-527H14.3 | | |
| 0,439635506 | RP11-532N4.2 | FALSE | Upregulated |
| -0,313170267 | RP11-534N16.1 | FALSE | Downregulated |
| | RP11-536C10.7 | | |
| | RP11-536I6.1 | | |
| -0,347115236 | RP11-539L10.2 | FALSE | Downregulated |
| 0 | RP11-53O19.1 | FALSE | Upregulated |
| 0,094615407 | RP11-540D4.3 | FALSE | Upregulated |
| 0 | RP11-540K16.1 | FALSE | Upregulated |
| | RP11-542B15.1 | | |
| | RP11-542G1.3 | | |
| -0,05181386 | RP11-544M22.1 | FALSE | Downregulated |

| | | | |
|---|---|---|---|
| -0,30550091 | RP11-547D23.1 | FALSE | Downregulated |
| | RP11-547I7.2 | | |
| | RP11-550H2.1 | | |
| -0,022984798 | RP11-551M15.1 | FALSE | Downregulated |
| | RP11-553E24.2 | | |
| 0,145548624 | RP11-554A11.6 | FALSE | Upregulated |
| | RP11-554D15.1 | | |
| -0,103383867 | RP11-554F20.1 | FALSE | Downregulated |
| | RP11-555F9.2 | | |
| -0,34532712 | RP11-557H15.4 | FALSE | Downregulated |
| | RP11-561E1.1 | | |
| -1,017116371 | RP11-563J2.2 | | Downregulated |
| | RP11-565A3.2 | | |
| 0,646475571 | RP11-567L7.6 | FALSE | Upregulated |
| 1,40417203 | RP11-569G13.2 | | Upregulated |
| | RP11-56I23.1 | | |
| 0 | RP11-572C15.5 | FALSE | Upregulated |
| 1,125654616 | RP11-572M11.3 | | Upregulated |
| 0 | RP11-575F12.1 | FALSE | Upregulated |
| 0 | RP11-576D8.4 | FALSE | Upregulated |
| | RP11-576E20.1 | | |
| | RP11-576I22.2 | | |
| 0,138628825 | RP11-579D7.4 | FALSE | Upregulated |
| 0,465237344 | RP11-579E24.1 | FALSE | Upregulated |
| -0,117292076 | RP11-584P21.2 | FALSE | Downregulated |
| | RP11-586D19.1 | | |
| 0 | RP11-588G21.2 | FALSE | Upregulated |
| | RP11-588P8.1 | | |
| 0,869886009 | RP11-58E21.3 | FALSE | Upregulated |
| -0,005390577 | RP11-58E21.4 | FALSE | Downregulated |
| 1,119108476 | RP11-593F23.1 | | Upregulated |
| 0 | RP11-59J16.1 | FALSE | Upregulated |
| 0 | RP11-5P22.3 | FALSE | Upregulated |
| | RP11-600K15.1 | | |
| 0,387978195 | RP11-609L23.1 | FALSE | Upregulated |
| -0,09569753 | RP11-60I3.5 | FALSE | Downregulated |
| 0,0587918 | RP11-614F17.2 | FALSE | Upregulated |
| -0,761700164 | RP11-61J19.3 | FALSE | Downregulated |
| 0,366743863 | RP11-620J15.2 | FALSE | Upregulated |
| -0,284853672 | RP11-620J15.3 | FALSE | Downregulated |
| 0,16273326 | RP11-624C23.1 | FALSE | Upregulated |
| | RP11-626P14.1 | | |
| 0,632862435 | RP11-629G13.1 | FALSE | Upregulated |
| 0,110603211 | RP11-630D6.5 | FALSE | Upregulated |
| 0,441485498 | RP11-632K5.3 | FALSE | Upregulated |
| 0,599522451 | RP11-638F5.1 | FALSE | Upregulated |
| -0,270664024 | RP11-63B19.1 | FALSE | Downregulated |
| -0,114529807 | RP11-63P12.6 | FALSE | Downregulated |
| | RP11-646J21.5 | | |
| 0 | RP11-64I17.1 | FALSE | Upregulated |
| | RP11-653G8.2 | | |

| | | | |
|---|---|---|---|
| | RP11-65D17.1 | | |
| 0,309293937 | RP11-65J3.1 | FALSE | Upregulated |
| 1,123944529 | RP11-65M17.1 | | Upregulated |
| | RP11-65M17.3 | | |
| -0,562684293 | RP11-661P17.1 | FALSE | Downregulated |
| | RP11-664H17.1 | | |
| | RP11-665C14.2 | | |
| 0,362888415 | RP11-666I19.2 | FALSE | Upregulated |
| | RP11-669N7.2 | | |
| -0,324956427 | RP11-669N7.3 | FALSE | Downregulated |
| -0,245018407 | RP11-66A2.2 | FALSE | Downregulated |
| 0 | RP11-66B24.1 | FALSE | Upregulated |
| | RP11-673E1.1 | | |
| 0 | RP11-67M1.1 | FALSE | Upregulated |
| 0,423179916 | RP11-682N22.1 | FALSE | Upregulated |
| 0,146690165 | RP11-686G23.2 | FALSE | Upregulated |
| | RP11-690C23.2 | | |
| 0,283725107 | RP11-699L21.1 | FALSE | Upregulated |
| | RP11-69H14.6 | | |
| | RP11-69H14.6-2 | | |
| | RP11-700N1.1 | | |
| 0 | RP11-702F3.3 | FALSE | Upregulated |
| | RP11-707A18.1 | | |
| | RP11-707P20.1 | | |
| 0 | RP11-70F11.11 | FALSE | Upregulated |
| | RP11-70F11.8 | | |
| 0,509136788 | RP11-710F7.3 | FALSE | Upregulated |
| | RP11-712C7.1 | | |
| -0,017489115 | RP11-713P17.3 | FALSE | Downregulated |
| 0,525673441 | RP11-715G15.1 | FALSE | Upregulated |
| 0,394337678 | RP11-719J20.1 | FALSE | Upregulated |
| | RP11-71E19.5 | | |
| 0,02889597 | RP11-71G7.1 | FALSE | Upregulated |
| 0,782361676 | RP11-723O4.2 | FALSE | Upregulated |
| 0,831936932 | RP11-72L22.1 | FALSE | Upregulated |
| | RP11-730N24.1 | | |
| -1,807387934 | RP11-736K20.6 | | Downregulated |
| 0,622025622 | RP11-744J10.3 | FALSE | Upregulated |
| | RP11-745C15.2 | | |
| 0,103164319 | RP11-752D24.3 | FALSE | Upregulated |
| 0 | RP11-752G15.3 | FALSE | Upregulated |
| -0,430416258 | RP11-752G15.6 | FALSE | Downregulated |
| -0,515943415 | RP11-756H20.1 | FALSE | Downregulated |
| | RP11-756K15.2 | | |
| 0,093740128 | RP11-75C9.1 | FALSE | Upregulated |
| | RP11-75F3.1 | | |
| | RP11-767I20.1 | | |
| | RP11-76E12.1 | | |
| -0,690189843 | RP11-76N22.2 | FALSE | Downregulated |
| 0,449370359 | RP11-770E5.1 | FALSE | Upregulated |
| 0,261482648 | RP11-770J1.3 | FALSE | Upregulated |

| | | | |
|---|---|---|---|
| 0,911694801 | RP11-778O17.4 | FALSE | Upregulated |
| 0,614289298 | RP11-779O18.1 | FALSE | Upregulated |
| -0,58699505 | RP11-789C1.1 | FALSE | Downregulated |
| | RP11-791G22.1 | | |
| | RP11-794P6.1 | | |
| | RP11-794P6.3 | | |
| | RP11-795J1.1 | | |
| 0 | RP11-799P8.1 | FALSE | Upregulated |
| -1,139920023 | RP11-79E3.3 | | Downregulated |
| | RP11-7O14.1 | | |
| 0,010647018 | RP11-806H10.4 | FALSE | Upregulated |
| | RP11-807H7.1 | | |
| | RP11-80F22.15 | | |
| 1,239258427 | RP11-80G7.1 | | Upregulated |
| | RP11-80K21.3 | | |
| 0,173522651 | RP11-814P5.1 | FALSE | Upregulated |
| 0 | RP11-81H14.2 | FALSE | Upregulated |
| | RP11-81H3.2 | | |
| 0,068944527 | RP11-82L2.1 | FALSE | Upregulated |
| 0 | RP11-838N2.4 | FALSE | Upregulated |
| 0,233144672 | RP11-83C7.2 | FALSE | Upregulated |
| -1,352880773 | RP11-843A23.1 | | Downregulated |
| 0,559079574 | RP11-844P9.2 | FALSE | Upregulated |
| | RP11-84A1.3 | | |
| | RP11-84O12.4 | | |
| 0,020789395 | RP11-855O10.2 | FALSE | Upregulated |
| 0,313837853 | RP11-857B24.5 | FALSE | Upregulated |
| | RP11-85G21.2 | | |
| 0,666070953 | RP11-861A13.4 | FALSE | Upregulated |
| 0,01196447 | RP11-87M18.2 | FALSE | Upregulated |
| | RP11-87N24.2 | | |
| -0,095102896 | RP11-885B4.2 | FALSE | Downregulated |
| | RP11-88I21.2 | | |
| | RP11-893F2.6 | | |
| 0,655293473 | RP11-909B2.1 | FALSE | Upregulated |
| | RP11-909M7.3 | | |
| | RP11-92G12.3 | | |
| 0 | RP11-93I21.3 | FALSE | Upregulated |
| | RP11-945C19.1 | | |
| | RP11-946L20.2 | | |
| | RP11-946L20.4 | | |
| 0,426437581 | RP11-94H18.1 | FALSE | Upregulated |
| | RP11-955H22.3 | | |
| | RP11-95H3.1 | | |
| 0,449311415 | RP11-95M5.1 | FALSE | Upregulated |
| -0,41271236 | RP11-95P2.1 | FALSE | Downregulated |
| 0 | RP11-960L18.1 | FALSE | Upregulated |
| -0,064299419 | RP11-96P7.1 | FALSE | Downregulated |
| 0 | RP11-973H7.4 | FALSE | Upregulated |
| | RP11-983C2.2 | | |
| 0 | RP11-98L5.2 | FALSE | Upregulated |

| | | | |
|---|---|---|---|
| | RP11-9G1.3 | | |
| -0,15199761 | RP13-210D15.4 | FALSE | Downregulated |
| | RP13-507P19.2 | | |
| 0 | RP13-60M5.2 | FALSE | Upregulated |
| | RP13-614K11.1 | | |
| | RP13-653N12.1 | | |
| | RP13-653N12.2 | | |
| | RP13-895J2.7 | | |
| -0,522914252 | RP1L1 | FALSE | Downregulated |
| 0,144440244 | RP3-322G13.5 | FALSE | Upregulated |
| -0,191641325 | RP3-323P13.2 | FALSE | Downregulated |
| | RP3-326I13.1 | | |
| | RP3-332B22.1 | | |
| 0 | RP3-399L15.3 | FALSE | Upregulated |
| 0,056335539 | RP3-400B16.1 | FALSE | Upregulated |
| | RP3-416J7.4 | | |
| | RP3-417L20.4 | | |
| | RP3-446N13.1 | | |
| 0,389695919 | RP3-473B4.3 | FALSE | Upregulated |
| | RP3-513G18.2 | | |
| 0,655070946 | RP3-522P13.3 | FALSE | Upregulated |
| 0 | RP4-533D7.5 | FALSE | Upregulated |
| 0,205319103 | RP4-562J12.2 | FALSE | Upregulated |
| | RP4-565E6.1 | | |
| | RP4-598G3.1 | | |
| 0,288640243 | RP4-607I7.1 | FALSE | Upregulated |
| 1,253044559 | RP4-613A2.1 | | Upregulated |
| 0 | RP4-640H8.2 | FALSE | Upregulated |
| 0,274345255 | RP4-655J12.4 | FALSE | Upregulated |
| | RP4-684O24.5 | | |
| 0,055378136 | RP4-694A7.4 | FALSE | Upregulated |
| 0,318364146 | RP4-697P8.3 | FALSE | Upregulated |
| -0,058036121 | RP4-705O1.1 | FALSE | Downregulated |
| 0 | RP4-723E3.1 | FALSE | Upregulated |
| | RP4-738P15.1 | | |
| | RP4-753D10.5 | | |
| 0 | RP4-753M9.1 | FALSE | Upregulated |
| 1,933322247 | RP4-756H11.3 | | Upregulated |
| 0 | RP4-760C5.5 | FALSE | Upregulated |
| | RP4-779E11.3 | | |
| 0,292570017 | RP4-781K5.4 | FALSE | Upregulated |
| -0,18814683 | RP4-782L23.1 | FALSE | Downregulated |
| 0,125157492 | RP4-794H19.4 | FALSE | Upregulated |
| 0 | RP5-1004I9.1 | FALSE | Upregulated |
| | RP5-1007G16.1 | | |
| -0,03899582 | RP5-1010E17.2 | FALSE | Downregulated |
| -0,933936153 | RP5-1021I20.1 | FALSE | Downregulated |
| | RP5-1022P6.4 | | |
| -0,801773631 | RP5-1024G6.5 | FALSE | Downregulated |
| 0,56211961 | RP5-1043L13.1 | FALSE | Upregulated |
| 1,822538023 | RP5-1043L13.1-2 | | Upregulated |

| | | | |
|---|---|---|---|
| -0,194857442 | RP5-1050D4.5 | FALSE | Downregulated |
| 0,068128077 | RP5-1069C8.2 | FALSE | Upregulated |
| | RP5-1070A16.1 | | |
| | RP5-1073O3.2 | | |
| 0,037231474 | RP5-1073O3.7 | FALSE | Upregulated |
| | RP5-1086L22.1 | | |
| 0 | RP5-1097F14.3 | FALSE | Upregulated |
| -0,442769088 | RP5-1103G7.4 | FALSE | Downregulated |
| 0,913732092 | RP5-1109J22.2 | FALSE | Upregulated |
| -0,155120091 | RP5-1112F19.2 | FALSE | Downregulated |
| 0,286908051 | RP5-1119D9.4 | FALSE | Upregulated |
| | RP5-1120P11.1 | | |
| 0,989791679 | RP5-1125A11.1 | FALSE | Upregulated |
| 0,475915078 | RP5-1139I1.1 | FALSE | Upregulated |
| 0,171270674 | RP5-1159O4.2 | FALSE | Upregulated |
| | RP5-1166F10.1 | | |
| | RP5-828K20.2 | | |
| -0,089600426 | RP5-833A20.1 | FALSE | Downregulated |
| 0,40144897 | RP5-857K21.5 | FALSE | Upregulated |
| 0,387012887 | RP5-858B6.3 | FALSE | Upregulated |
| | RP5-887A10.1 | | |
| | RP5-896L10.1 | | |
| | RP5-898J17.1 | | |
| -0,430033995 | RP5-899E9.1 | FALSE | Downregulated |
| | RP5-921G16.2 | | |
| | RP5-933E2.1 | | |
| | RP5-944M2.3 | | |
| | RP5-952N6.1 | | |
| 0,691801792 | RP5-968D22.3 | FALSE | Upregulated |
| | RP5-988G17.1 | | |
| 0,919393972 | RP9P | FALSE | Upregulated |
| 0,273195875 | RPA4 | FALSE | Upregulated |
| 0 | RPE65 | FALSE | Upregulated |
| 0,525217744 | RPEL1 | FALSE | Upregulated |
| 0,517881752 | RPGRIP1 | FALSE | Upregulated |
| 0,410173639 | RPH3A | FALSE | Upregulated |
| 0,100713889 | RPL10L | FALSE | Upregulated |
| -0,079632341 | RPL13AP20 | FALSE | Downregulated |
| -0,740526493 | RPL13AP3 | FALSE | Downregulated |
| 0,109080189 | RPL13P5 | FALSE | Upregulated |
| -1,113338657 | RPL19P12 | | Downregulated |
| 0,371196951 | RPL23AP32 | FALSE | Upregulated |
| 0,176366671 | RPL23P8 | FALSE | Upregulated |
| 0 | RPL29P2 | FALSE | Upregulated |
| 0,348013951 | RPL34-AS1 | FALSE | Upregulated |
| 0,781799566 | RPL39L | FALSE | Upregulated |
| 0,306860599 | RPL3L | FALSE | Upregulated |
| -0,005919442 | RPLP0P2 | FALSE | Downregulated |
| -0,423692508 | RPP21 | FALSE | Downregulated |
| 0,335260906 | RPP40 | FALSE | Upregulated |
| -0,051641041 | RPRM | FALSE | Downregulated |

| | | | |
|---|---|---|---|
| 0,623340876 | RPRML | FALSE | Upregulated |
| 0,711258955 | RPS10-NUDT3 | FALSE | Upregulated |
| 0,165393252 | RPS14P3 | FALSE | Upregulated |
| 0,034082806 | RPS26P11 | FALSE | Upregulated |
| | RPS4Y2 | | |
| -0,076022802 | RPS6KA2-IT1 | FALSE | Downregulated |
| -1,23282922 | RPS6KA6 | | Downregulated |
| | RPS7P5 | | |
| | RPSAP52 | | |
| -0,216251629 | RPSAP9 | FALSE | Downregulated |
| 0,44160309 | RPTN | FALSE | Upregulated |
| 0 | RRH | FALSE | Upregulated |
| 0,426437581 | RSPH1 | FALSE | Upregulated |
| -0,064070425 | RSPH6A | FALSE | Downregulated |
| -0,205943797 | RSPO1 | FALSE | Downregulated |
| 0,440630727 | RSPO2 | FALSE | Upregulated |
| -0,061339497 | RSPO4 | FALSE | Downregulated |
| -1,284210149 | RSU1P2 | | Downregulated |
| 0,00135902 | RTBDN | FALSE | Upregulated |
| 0 | RTDR1 | FALSE | Upregulated |
| -2,299038607 | RTEL1-TNFRSF6B | | Downregulated |
| 1,301456355 | RTKN2 | | Upregulated |
| 0,470497181 | RTN2 | FALSE | Upregulated |
| 0,438038401 | RTP1 | FALSE | Upregulated |
| | RTP2 | | |
| 1,121519356 | RUFY4 | | Upregulated |
| -1,062261132 | RXFP1 | | Downregulated |
| -0,075006126 | RXFP2 | FALSE | Downregulated |
| 0,032916356 | RXFP3 | FALSE | Upregulated |
| -0,19344173 | RXRG | FALSE | Downregulated |
| 0,715983771 | S100A12 | FALSE | Upregulated |
| 0 | S100A2 | FALSE | Upregulated |
| 0,402112793 | S100A3 | FALSE | Upregulated |
| 0,593395278 | S100A5 | FALSE | Upregulated |
| | S100A7 | | |
| | S100A7A | | |
| | S100A7L2 | | |
| 0,750693267 | S100A8 | FALSE | Upregulated |
| -0,464202862 | S100B | FALSE | Downregulated |
| 0,646684671 | S100G | FALSE | Upregulated |
| 0 | S100Z | FALSE | Upregulated |
| 1,055200547 | S1PR5 | | Upregulated |
| 2,326985402 | SAA2-SAA4 | | Upregulated |
| | SAA3P | | |
| 0,065989465 | SACS-AS1 | FALSE | Upregulated |
| 0 | SAG | FALSE | Upregulated |
| | SAGE1 | | |
| | SALL3 | | |
| 0,451586036 | SAMD12-AS1 | FALSE | Upregulated |
| 0,147959174 | SAMD13 | FALSE | Upregulated |
| 0 | SAMD7 | FALSE | Upregulated |

| | | | |
|---|---|---|---|
| | SAMSN1-AS1 | | |
| 0 | SATB2-AS1 | FALSE | Upregulated |
| -0,583522187 | SBF1P1 | FALSE | Downregulated |
| 0,11231694 | SBK2 | FALSE | Upregulated |
| 0,345656761 | SBK3 | FALSE | Upregulated |
| 0,278789727 | SBSN | FALSE | Upregulated |
| 0,561915699 | SCARNA1 | FALSE | Upregulated |
| 0,099452879 | SCARNA11 | FALSE | Upregulated |
| | SCARNA14 | | |
| | SCARNA15 | | |
| -0,388850249 | SCARNA16 | FALSE | Downregulated |
| 0,120569394 | SCARNA18 | FALSE | Upregulated |
| 0 | SCARNA20 | FALSE | Upregulated |
| 0,578834532 | SCARNA21 | FALSE | Upregulated |
| 0,011291549 | SCARNA22 | FALSE | Upregulated |
| | SCARNA23 | | |
| | SCARNA27 | | |
| -0,00408499 | SCARNA3 | FALSE | Downregulated |
| -0,101233586 | SCARNA4 | FALSE | Downregulated |
| 0,070401644 | SCARNA8 | FALSE | Upregulated |
| 0,150780904 | SCARNA9L | FALSE | Upregulated |
| 0,000251685 | SCD5 | FALSE | Upregulated |
| 0,463780701 | SCEL | FALSE | Upregulated |
| 0 | SCG2 | FALSE | Upregulated |
| | SCGB1A1 | | |
| -0,352377504 | SCGB1B2P | FALSE | Downregulated |
| 0 | SCGB1C1 | FALSE | Upregulated |
| | SCGB1D1 | | |
| 0 | SCGB1D2 | FALSE | Upregulated |
| | SCGB1D4 | | |
| | SCGB2A1 | | |
| | SCGB2A2 | | |
| 0,389432927 | SCGB2B2 | FALSE | Upregulated |
| -0,721599266 | SCGB2B3P | FALSE | Downregulated |
| 0,526628949 | SCGB3A1 | FALSE | Upregulated |
| 0,100713889 | SCGB3A2 | FALSE | Upregulated |
| 0,455606144 | SCHIP1 | FALSE | Upregulated |
| 0 | SCHLAP1 | FALSE | Upregulated |
| 0,988184148 | SCIN | FALSE | Upregulated |
| -0,452006623 | SCML2 | FALSE | Downregulated |
| | SCN10A | | |
| -0,971597428 | SCN11A | FALSE | Downregulated |
| -0,01495236 | SCN2A | FALSE | Downregulated |
| -0,056206595 | SCN2B | FALSE | Downregulated |
| -0,361762639 | SCN4B | FALSE | Downregulated |
| 0,816495069 | SCN5A | FALSE | Upregulated |
| 1,451167722 | SCNN1B | | Upregulated |
| | SCNN1G | | |
| | SCP2D1 | | |
| 1,331227354 | SCRT2 | | Upregulated |
| 0,267374366 | SCUBE2 | FALSE | Upregulated |

| | | | |
|---|---|---|---|
| -0,218772198 | SDC4P | FALSE | Downregulated |
| 0 | SDR16C5 | FALSE | Upregulated |
| | SDR16C6P | | |
| -0,230366274 | SDR9C7 | FALSE | Downregulated |
| -0,001797254 | SEC14L5 | FALSE | Downregulated |
| -0,820843433 | SEC14L6 | FALSE | Downregulated |
| 0,395609274 | SEC24B-AS1 | FALSE | Upregulated |
| | SEL1L2 | | |
| 0,115661663 | SELV | FALSE | Upregulated |
| 1,950896165 | SEMA3A | | Upregulated |
| -1,672051231 | SEMA5B | | Downregulated |
| | SEMG1 | | |
| | SEMG2 | | |
| -1,316603948 | SENCR | | Downregulated |
| 0,102025395 | SEPT12 | FALSE | Upregulated |
| | SEPT14 | | |
| -0,61158448 | SEPT7P9 | FALSE | Downregulated |
| | SERF1A | | |
| | SERPINA13P | | |
| -0,033972048 | SERPINA9 | FALSE | Downregulated |
| | SERPINB10 | | |
| | SERPINB11 | | |
| 0 | SERPINB12 | FALSE | Upregulated |
| 0 | SERPINB13 | FALSE | Upregulated |
| 0 | SERPINB2 | FALSE | Upregulated |
| | SERPINB3 | | |
| | SERPINB4 | | |
| 0 | SERPINB5 | FALSE | Upregulated |
| | SERPINB7 | | |
| 0,233730556 | SERPINI1 | FALSE | Upregulated |
| | SERPINI2 | | |
| 0,516768131 | SERTAD4 | FALSE | Upregulated |
| -0,322197346 | SERTAD4-AS1 | FALSE | Downregulated |
| 1,333185533 | SERTM1 | | Upregulated |
| 0,097822813 | SETSIP | FALSE | Upregulated |
| -0,236208959 | SEZ6L | FALSE | Downregulated |
| 1,279940666 | SEZ6L2 | | Upregulated |
| 1,502970192 | SFR1 | | Upregulated |
| 1,10890752 | SFRP2 | | Upregulated |
| 1,924262159 | SFRP4 | | Upregulated |
| -0,583540416 | SFT2D1 | FALSE | Downregulated |
| 0 | SFTA1P | FALSE | Upregulated |
| 0 | SFTA2 | FALSE | Upregulated |
| | SFTA3 | | |
| -0,903595944 | SFTPA1 | FALSE | Downregulated |
| -0,073405745 | SFTPA2 | FALSE | Downregulated |
| -0,332766927 | SFTPC | FALSE | Downregulated |
| 0,011479934 | SGCG | FALSE | Upregulated |
| 0,299495456 | SGCZ | FALSE | Upregulated |
| 0,673558464 | SGIP1 | FALSE | Upregulated |
| 1,550978306 | SGOL1-AS1 | | Upregulated |

| | | | |
|---|---|---|---|
| -1,380864722 | SGSM1 | | Downregulated |
| 0,048276727 | SH2D4B | FALSE | Upregulated |
| 0,449453515 | SH2D5 | FALSE | Upregulated |
| -0,384808767 | SH2D6 | FALSE | Downregulated |
| 0,463353273 | SH2D7 | FALSE | Upregulated |
| 0,70800939 | SH3BGR | FALSE | Upregulated |
| -0,05377805 | SH3GL2 | FALSE | Downregulated |
| -0,161766296 | SH3GL3 | FALSE | Downregulated |
| 0,501113051 | SHANK1 | FALSE | Upregulated |
| 0,305677769 | SHC4 | FALSE | Upregulated |
| 1,240785476 | SHCBP1 | | Upregulated |
| | SHCBP1L | | |
| 0 | SHISA2 | FALSE | Upregulated |
| 1,488951488 | SHISA3 | | Upregulated |
| 0,934580156 | SHISA8 | FALSE | Upregulated |
| 0,295492971 | SHOX | FALSE | Upregulated |
| | SHOX-2 | | |
| | SHOX2 | | |
| | SI | | |
| | SIAH3 | | |
| -0,139301673 | SIGLEC12 | FALSE | Downregulated |
| -0,193297303 | SIGLEC17P | FALSE | Downregulated |
| -0,35182606 | SIGLEC6 | FALSE | Downregulated |
| 2,231384741 | SIGLEC8 | | Upregulated |
| 0,427833744 | SIGLECL1 | FALSE | Upregulated |
| 0,094103637 | SIM1 | FALSE | Upregulated |
| 1,497607401 | SIRPD | | Upregulated |
| 1,082928979 | SIRPG | | Upregulated |
| 0 | SIX1 | FALSE | Upregulated |
| 0,381322373 | SIX2 | FALSE | Upregulated |
| | SIX3 | | |
| 0 | SIX3-AS1 | FALSE | Upregulated |
| -0,707531674 | SIX4 | FALSE | Downregulated |
| 0,103572941 | SIX6 | FALSE | Upregulated |
| 0,682642965 | SKA1 | FALSE | Upregulated |
| 0,021380563 | SKINTL | FALSE | Upregulated |
| 0,073763975 | SKOR1 | FALSE | Upregulated |
| | SKOR2 | | |
| 0,394726433 | SKP1P2 | FALSE | Upregulated |
| -0,186033271 | SLAMF9 | FALSE | Downregulated |
| -0,034800701 | SLC10A2 | FALSE | Downregulated |
| 0 | SLC10A4 | FALSE | Upregulated |
| 0,414710117 | SLC10A6 | FALSE | Upregulated |
| 0,170803615 | SLC12A1 | FALSE | Upregulated |
| 0,247879354 | SLC12A3 | FALSE | Upregulated |
| 0,31315295 | SLC13A1 | FALSE | Upregulated |
| -1,259664749 | SLC13A2 | | Downregulated |
| -2,128150914 | SLC15A5 | | Downregulated |
| | SLC17A6 | | |
| 0,142607741 | SLC17A7 | FALSE | Upregulated |
| 0,329998117 | SLC17A8 | FALSE | Upregulated |

| | | | |
|---|---|---|---|
| 0 | SLC18A1 | FALSE | Upregulated |
| | SLC18A3 | | |
| | SLC1A6 | | |
| 0,094523942 | SLC22A11 | FALSE | Upregulated |
| -0,2309745 | SLC22A12 | FALSE | Downregulated |
| -0,468655192 | SLC22A13 | FALSE | Downregulated |
| 0 | SLC22A14 | FALSE | Upregulated |
| -0,311627656 | SLC22A15 | FALSE | Downregulated |
| 0,089359585 | SLC22A16 | FALSE | Upregulated |
| -0,166355914 | SLC22A2 | FALSE | Downregulated |
| 0,569030162 | SLC22A20 | FALSE | Upregulated |
| -0,578166078 | SLC22A24 | FALSE | Downregulated |
| -0,133728907 | SLC22A31 | FALSE | Downregulated |
| 0 | SLC22A6 | FALSE | Upregulated |
| | SLC22A8 | | |
| 0,851555635 | SLC24A2 | FALSE | Upregulated |
| 0,512811721 | SLC25A14 | FALSE | Upregulated |
| | SLC25A31 | | |
| | SLC25A3P1 | | |
| -0,60861387 | SLC25A41 | FALSE | Downregulated |
| 0,715831015 | SLC25A48 | FALSE | Upregulated |
| | SLC25A51P1 | | |
| 0,731487975 | SLC25A53 | FALSE | Upregulated |
| | SLC25A6-2 | | |
| 0,335816313 | SLC26A10 | FALSE | Upregulated |
| 0,285174874 | SLC26A3 | FALSE | Upregulated |
| 0,071356983 | SLC26A4 | FALSE | Upregulated |
| -0,102077927 | SLC26A7 | FALSE | Downregulated |
| 0,280855437 | SLC26A9 | FALSE | Upregulated |
| -0,223061762 | SLC27A6 | FALSE | Downregulated |
| 1,057235929 | SLC28A3 | | Upregulated |
| 0,406787727 | SLC2A1-AS1 | FALSE | Upregulated |
| 0,950180488 | SLC2A14 | FALSE | Upregulated |
| 0,291698555 | SLC2A5 | FALSE | Upregulated |
| | SLC2A7 | | |
| 0,367838398 | SLC30A2 | FALSE | Upregulated |
| 0 | SLC30A3 | FALSE | Upregulated |
| 0,668391575 | SLC30A8 | FALSE | Upregulated |
| | SLC32A1 | | |
| -1,177687636 | SLC34A1 | | Downregulated |
| | SLC35D3 | | |
| 0,230552348 | SLC35E4 | FALSE | Upregulated |
| 0,40285389 | SLC35F2 | FALSE | Upregulated |
| 0,365702334 | SLC35F3 | FALSE | Upregulated |
| -0,375922996 | SLC35F4 | FALSE | Downregulated |
| 0,085202734 | SLC35G3 | FALSE | Upregulated |
| | SLC35G4 | | |
| 0,002651441 | SLC35G5 | FALSE | Upregulated |
| -0,358979985 | SLC36A2 | FALSE | Downregulated |
| -0,039801987 | SLC36A3 | FALSE | Downregulated |
| -0,387882173 | SLC38A5 | FALSE | Downregulated |

| | | | |
|---|---|---|---|
| | SLC38A8 | | |
| 0 | SLC39A12 | FALSE | Upregulated |
| 0,064820903 | SLC39A2 | FALSE | Upregulated |
| -0,088541117 | SLC44A4 | FALSE | Downregulated |
| -0,300563253 | SLC44A5 | FALSE | Downregulated |
| -0,852770052 | SLC45A1 | FALSE | Downregulated |
| 0,316931537 | SLC4A1 | FALSE | Upregulated |
| 0,380550865 | SLC4A11 | FALSE | Upregulated |
| 0,658990228 | SLC4A3 | FALSE | Upregulated |
| 0,273467881 | SLC52A3 | FALSE | Upregulated |
| -0,038268672 | SLC5A11 | FALSE | Downregulated |
| -0,392655327 | SLC5A5 | FALSE | Downregulated |
| 0,089416766 | SLC5A7 | FALSE | Upregulated |
| | SLC5A8 | | |
| 0,100713889 | SLC6A10P | FALSE | Upregulated |
| | SLC6A10PB | | |
| -0,341801435 | SLC6A11 | FALSE | Downregulated |
| | SLC6A14 | | |
| | SLC6A15 | | |
| | SLC6A18 | | |
| -0,494396338 | SLC6A2 | FALSE | Downregulated |
| 0,094931004 | SLC6A3 | FALSE | Upregulated |
| | SLC6A5 | | |
| 0 | SLC6A7 | FALSE | Upregulated |
| 1,023723975 | SLC7A11 | | Upregulated |
| 0,088462045 | SLC7A11-AS1 | FALSE | Upregulated |
| 0,38647539 | SLC7A13 | FALSE | Upregulated |
| | SLC7A3 | | |
| 1,258291891 | SLC7A5P1 | | Upregulated |
| | SLC8A2 | | |
| 0,712954741 | SLC9A4 | FALSE | Upregulated |
| 0,634592375 | SLC9A7P1 | FALSE | Upregulated |
| 0,836226846 | SLC9C1 | FALSE | Upregulated |
| 0,576311717 | SLC9C2 | FALSE | Upregulated |
| -0,583160285 | SLCO1C1 | FALSE | Downregulated |
| 1,561240599 | SLCO4A1 | | Upregulated |
| 0,743127393 | SLCO5A1 | FALSE | Upregulated |
| | SLCO6A1 | | |
| 0,235141206 | SLFN12 | FALSE | Upregulated |
| 0,523420713 | SLFN12L | FALSE | Upregulated |
| 0,390950345 | SLIT2-IT1 | FALSE | Upregulated |
| | SLITRK1 | | |
| 0,121795341 | SLITRK2 | FALSE | Upregulated |
| 0,417554102 | SLITRK4 | FALSE | Upregulated |
| -0,316647538 | SLITRK5 | FALSE | Downregulated |
| -0,246686276 | SLITRK6 | FALSE | Downregulated |
| 0,042379873 | SLMO1 | FALSE | Upregulated |
| 0,13526597 | SLN | FALSE | Upregulated |
| -0,557895571 | SLURP1 | FALSE | Downregulated |
| 0,055378136 | SMCO2 | FALSE | Upregulated |
| | SMCP | | |

| | | | |
|---|---|---|---|
| -0,245778879 | SMCR9 | FALSE | Downregulated |
| | SMEK3P | | |
| -0,004968665 | SMIM10 | FALSE | Downregulated |
| -0,312448644 | SMIM17 | FALSE | Downregulated |
| -0,791889933 | SMIM2-AS1 | FALSE | Downregulated |
| 0,052005358 | SMIM2-IT1 | FALSE | Upregulated |
| 0,331430975 | SMIM21 | FALSE | Upregulated |
| 0,553730843 | SMIM22 | FALSE | Upregulated |
| -0,082073388 | SMIM23 | FALSE | Downregulated |
| -0,292112384 | SMIM4 | FALSE | Downregulated |
| 0 | SMIM9 | FALSE | Upregulated |
| | SMKR1 | | |
| 0,668388027 | SMPX | FALSE | Upregulated |
| | SMR3A | | |
| | SMR3B | | |
| -0,274354591 | SMTNL2 | FALSE | Downregulated |
| -0,962950048 | SMYD1 | FALSE | Downregulated |
| 0,393163673 | SNAI1 | FALSE | Upregulated |
| 0,545393694 | SNAP25-AS1 | FALSE | Upregulated |
| 0 | SNAP91 | FALSE | Upregulated |
| 0,353099643 | SNAPC1 | FALSE | Upregulated |
| | SNAR-A1 | | |
| | SNAR-A10 | | |
| | SNAR-A11 | | |
| | SNAR-A12 | | |
| | SNAR-A13 | | |
| | SNAR-A14 | | |
| | SNAR-A2 | | |
| | SNAR-A3 | | |
| | SNAR-A4 | | |
| | SNAR-A5 | | |
| | SNAR-A6 | | |
| | SNAR-A7 | | |
| | SNAR-A8 | | |
| | SNAR-A9 | | |
| | SNAR-B1 | | |
| | SNAR-B2 | | |
| | SNAR-C1 | | |
| | SNAR-C2 | | |
| | SNAR-C3 | | |
| 0,094615407 | SNAR-C4 | FALSE | Upregulated |
| | SNAR-C5 | | |
| | SNAR-D | | |
| | SNAR-E | | |
| | SNAR-F | | |
| | SNAR-G1 | | |
| | SNAR-G2 | | |
| | SNAR-H | | |
| | SNAR-I | | |
| 0 | SNCB | FALSE | Upregulated |
| 0,825767508 | SNCG | FALSE | Upregulated |

| | | | |
|---|---|---|---|
| 1,264147139 | SNHG12 | | Upregulated |
| -0,388227041 | SNHG15 | FALSE | Downregulated |
| 1,096529913 | SNHG17 | | Upregulated |
| -0,666378551 | SNHG18 | FALSE | Downregulated |
| 0,100463031 | SNHG4 | FALSE | Upregulated |
| -0,5253098 | SNHG6 | FALSE | Downregulated |
| -0,478610795 | SNHG9 | FALSE | Downregulated |
| 0,104204316 | SNORA1 | FALSE | Upregulated |
| 0,582352915 | SNORA10 | FALSE | Upregulated |
| 0,291734858 | SNORA11 | FALSE | Upregulated |
| -0,110151179 | SNORA11B | FALSE | Downregulated |
| | SNORA11C | | |
| | SNORA11D | | |
| | SNORA11E | | |
| -0,223559423 | SNORA12 | FALSE | Downregulated |
| 0,134326649 | SNORA13 | FALSE | Upregulated |
| 0,082524093 | SNORA14A | FALSE | Upregulated |
| -1,048115489 | SNORA14B | | Downregulated |
| -0,268849582 | SNORA15 | FALSE | Downregulated |
| 0 | SNORA16A | FALSE | Upregulated |
| 0,099452879 | SNORA16B | FALSE | Upregulated |
| | SNORA17 | | |
| 0 | SNORA18 | FALSE | Upregulated |
| | SNORA19 | | |
| -0,925141 | SNORA20 | FALSE | Downregulated |
| -0,364123004 | SNORA21 | FALSE | Downregulated |
| 0 | SNORA22 | FALSE | Upregulated |
| -0,512510156 | SNORA24 | FALSE | Downregulated |
| 0,614009575 | SNORA25 | FALSE | Upregulated |
| 0,133630945 | SNORA26 | FALSE | Upregulated |
| 0 | SNORA27 | FALSE | Upregulated |
| -0,300391559 | SNORA28 | FALSE | Downregulated |
| | SNORA29 | | |
| -0,092715755 | SNORA2A | FALSE | Downregulated |
| | SNORA2B | | |
| -0,284924171 | SNORA3 | FALSE | Downregulated |
| 0 | SNORA30 | FALSE | Upregulated |
| 0,003528888 | SNORA31 | FALSE | Upregulated |
| 0 | SNORA32 | FALSE | Upregulated |
| 0,421409674 | SNORA33 | FALSE | Upregulated |
| 0,060407023 | SNORA34 | FALSE | Upregulated |
| | SNORA35 | | |
| | SNORA36A | | |
| | SNORA36B | | |
| | SNORA36C | | |
| -0,108513056 | SNORA37 | FALSE | Downregulated |
| 0 | SNORA38 | FALSE | Upregulated |
| 0,568385209 | SNORA38B | FALSE | Upregulated |
| | SNORA39 | | |
| 0,073594161 | SNORA4 | FALSE | Upregulated |
| 0 | SNORA40 | FALSE | Upregulated |

| | | | |
|---|---|---|---|
| 0 | SNORA41 | FALSE | Upregulated |
| 0,343043991 | SNORA42 | FALSE | Upregulated |
| -0,446379737 | SNORA43 | FALSE | Downregulated |
| 0,322737784 | SNORA44 | FALSE | Upregulated |
| -0,002794568 | SNORA45 | FALSE | Downregulated |
| 0,474887831 | SNORA46 | FALSE | Upregulated |
| -2,420027571 | SNORA47 | | Downregulated |
| 0,421961471 | SNORA49 | FALSE | Upregulated |
| 0,695435219 | SNORA50 | FALSE | Upregulated |
| | SNORA51 | | |
| 0,524806398 | SNORA52 | FALSE | Upregulated |
| -0,109998737 | SNORA54 | FALSE | Downregulated |
| -0,134803338 | SNORA55 | FALSE | Downregulated |
| | SNORA56 | | |
| -0,151019104 | SNORA58 | FALSE | Downregulated |
| 0 | SNORA59A | FALSE | Upregulated |
| 0 | SNORA59B | FALSE | Upregulated |
| 0,709129614 | SNORA5A | FALSE | Upregulated |
| | SNORA5B | | |
| -0,218905729 | SNORA5C | FALSE | Downregulated |
| 0,274100674 | SNORA6 | FALSE | Upregulated |
| 0,230110021 | SNORA60 | FALSE | Upregulated |
| 0,774666824 | SNORA61 | FALSE | Upregulated |
| | SNORA62 | | |
| -0,296157043 | SNORA64 | FALSE | Downregulated |
| -0,189916657 | SNORA65 | FALSE | Downregulated |
| -0,067320087 | SNORA66 | FALSE | Downregulated |
| 0,39639969 | SNORA67 | FALSE | Upregulated |
| -0,091118088 | SNORA68 | FALSE | Downregulated |
| | SNORA69 | | |
| 0,854841347 | SNORA70 | FALSE | Upregulated |
| | SNORA70B | | |
| | SNORA70C | | |
| | SNORA70D | | |
| | SNORA70E | | |
| | SNORA70F | | |
| 0,038052732 | SNORA70G | FALSE | Upregulated |
| 0,487011378 | SNORA71A | FALSE | Upregulated |
| -0,03822646 | SNORA71B | FALSE | Downregulated |
| 0,226071122 | SNORA71C | FALSE | Upregulated |
| 0,539872037 | SNORA71D | FALSE | Upregulated |
| 0 | SNORA72 | FALSE | Upregulated |
| -0,000318601 | SNORA74A | FALSE | Downregulated |
| 0,055378136 | SNORA75 | FALSE | Upregulated |
| -0,614190716 | SNORA76 | FALSE | Downregulated |
| 0,106266214 | SNORA77 | FALSE | Upregulated |
| -0,202895539 | SNORA78 | FALSE | Downregulated |
| | SNORA79 | | |
| -0,358421311 | SNORA8 | FALSE | Downregulated |
| -0,046771944 | SNORA80 | FALSE | Downregulated |
| 0,857612864 | SNORA80B | FALSE | Upregulated |

| | | | |
|---|---|---|---|
| -0,800310048 | SNORA84 | FALSE | Downregulated |
| 0,26223078 | SNORA9 | FALSE | Upregulated |
| | SNORD100 | | |
| | SNORD101 | | |
| | SNORD102 | | |
| | SNORD103A | | |
| | SNORD103B | | |
| | SNORD104 | | |
| | SNORD105 | | |
| | SNORD105B | | |
| | SNORD11 | | |
| | SNORD110 | | |
| | SNORD111 | | |
| | SNORD111B | | |
| | SNORD112 | | |
| | SNORD113-1 | | |
| | SNORD113-2 | | |
| | SNORD113-3 | | |
| | SNORD113-4 | | |
| | SNORD113-5 | | |
| | SNORD113-6 | | |
| | SNORD113-7 | | |
| | SNORD113-8 | | |
| | SNORD113-9 | | |
| | SNORD114-1 | | |
| | SNORD114-10 | | |
| | SNORD114-11 | | |
| | SNORD114-12 | | |
| | SNORD114-13 | | |
| | SNORD114-14 | | |
| | SNORD114-15 | | |
| | SNORD114-16 | | |
| | SNORD114-17 | | |
| | SNORD114-18 | | |
| | SNORD114-19 | | |
| | SNORD114-2 | | |
| | SNORD114-20 | | |
| | SNORD114-21 | | |
| | SNORD114-22 | | |
| | SNORD114-23 | | |
| | SNORD114-24 | | |
| | SNORD114-25 | | |
| | SNORD114-26 | | |
| | SNORD114-27 | | |
| | SNORD114-28 | | |
| | SNORD114-29 | | |
| | SNORD114-3 | | |
| | SNORD114-30 | | |
| | SNORD114-31 | | |
| | SNORD114-4 | | |
| | SNORD114-5 | | |

| | | | |
|---|---|---|---|
| | SNORD114-6 | | |
| | SNORD114-7 | | |
| | SNORD114-8 | | |
| | SNORD114-9 | | |
| | SNORD117 | | |
| | SNORD119 | | |
| | SNORD11B | | |
| | SNORD12 | | |
| | SNORD121A | | |
| | SNORD121B | | |
| | SNORD123 | | |
| | SNORD124 | | |
| | SNORD125 | | |
| | SNORD126 | | |
| | SNORD127 | | |
| | SNORD12B | | |
| | SNORD12C | | |
| -0,066602077 | SNORD13 | FALSE | Downregulated |
| | SNORD14A | | |
| | SNORD14B | | |
| | SNORD14C | | |
| | SNORD14D | | |
| | SNORD14E | | |
| -0,386655808 | SNORD15A | FALSE | Downregulated |
| -1,185214042 | SNORD15B | | Downregulated |
| | SNORD16 | | |
| | SNORD18A | | |
| | SNORD18B | | |
| | SNORD18C | | |
| | SNORD19 | | |
| | SNORD19B | | |
| | SNORD1A | | |
| | SNORD1B | | |
| | SNORD1C | | |
| | SNORD2 | | |
| | SNORD20 | | |
| | SNORD21 | | |
| 0,06894717 | SNORD23 | FALSE | Upregulated |
| | SNORD24 | | |
| | SNORD25 | | |
| | SNORD26 | | |
| | SNORD27 | | |
| | SNORD28 | | |
| | SNORD29 | | |
| | SNORD30 | | |
| | SNORD31 | | |
| | SNORD32A | | |
| | SNORD32B | | |
| | SNORD33 | | |
| | SNORD34 | | |
| | SNORD35A | | |

SNORD35B
SNORD36A
SNORD36B
SNORD36C
SNORD37
SNORD38A
SNORD38B
-0,062546984 SNORD3B-1 FALSE Downregulated
-0,551692712 SNORD3B-2 FALSE Downregulated
1,288070376 SNORD3D Upregulated
SNORD41
SNORD42A
SNORD42B
SNORD43
SNORD44
SNORD45A
SNORD45B
SNORD45C
0 SNORD46 FALSE Upregulated
SNORD47
SNORD48
SNORD49A
SNORD49B
SNORD4A
SNORD4B
SNORD5
SNORD50A
SNORD50B
SNORD51
SNORD52
SNORD53
SNORD54
SNORD55
SNORD56
SNORD56B
SNORD57
SNORD58A
SNORD58B
SNORD58C
SNORD59A
SNORD59B
SNORD6
SNORD60
SNORD61
SNORD62A
SNORD62B
SNORD63
SNORD65
SNORD66
0,450040107 SNORD67 FALSE Upregulated
SNORD68

| | | | |
|---|---|---|---|
| | SNORD69 | | |
| | SNORD7 | | |
| | SNORD70 | | |
| | SNORD71 | | |
| | SNORD72 | | |
| | SNORD73A | | |
| | SNORD74 | | |
| | SNORD75 | | |
| 0,50516356 | SNORD76 | FALSE | Upregulated |
| | SNORD77 | | |
| | SNORD78 | | |
| | SNORD79 | | |
| -0,574004317 | SNORD8 | FALSE | Downregulated |
| | SNORD80 | | |
| | SNORD81 | | |
| | SNORD82 | | |
| | SNORD83A | | |
| | SNORD83B | | |
| | SNORD84 | | |
| | SNORD85 | | |
| | SNORD86 | | |
| | SNORD87 | | |
| | SNORD88A | | |
| | SNORD88B | | |
| | SNORD88C | | |
| 0,000977115 | SNORD9 | FALSE | Upregulated |
| | SNORD90 | | |
| | SNORD91A | | |
| | SNORD91B | | |
| | SNORD92 | | |
| | SNORD93 | | |
| | SNORD95 | | |
| | SNORD96A | | |
| | SNORD96B | | |
| 0,897418517 | SNORD97 | FALSE | Upregulated |
| | SNORD98 | | |
| | SNORD99 | | |
| -0,290312171 | SNRPD2P2 | FALSE | Downregulated |
| -0,018199063 | SNTG1 | FALSE | Downregulated |
| | SNTG2 | | |
| 0,743193518 | SNTN | FALSE | Upregulated |
| -0,480297332 | SNX29P1 | FALSE | Downregulated |
| -0,135148047 | SNX31 | FALSE | Downregulated |
| -1,283719287 | SOAT2 | | Downregulated |
| 0,998956338 | SOCS1 | FALSE | Upregulated |
| 0,883331125 | SOCS2-AS1 | FALSE | Upregulated |
| 0 | SOHLH1 | FALSE | Upregulated |
| | SOHLH2 | | |
| 0,632989218 | SORCS1 | FALSE | Upregulated |
| 0 | SORCS3 | FALSE | Upregulated |
| | SORCS3-AS1 | | |

| | | | |
|---|---|---|---|
| -0,073744938 | SOST | FALSE | Downregulated |
| 0,765008131 | SOWAHD | FALSE | Upregulated |
| | SOX1 | | |
| 0 | SOX11 | FALSE | Upregulated |
| 0,38647539 | SOX14 | FALSE | Upregulated |
| 0,398482536 | SOX2 | FALSE | Upregulated |
| -0,079911709 | SOX2-OT | FALSE | Downregulated |
| -0,595952279 | SOX21 | FALSE | Downregulated |
| 0,112749892 | SOX21-AS1 | FALSE | Upregulated |
| | SOX3 | | |
| 0,811023499 | SOX30 | FALSE | Upregulated |
| 0,691886704 | SOX8 | FALSE | Upregulated |
| 1,621660728 | SOX9-AS1 | | Upregulated |
| -0,380639387 | SP6 | FALSE | Downregulated |
| 0,387012887 | SP8 | FALSE | Upregulated |
| | SP9 | | |
| 0,345274376 | SPA17 | FALSE | Upregulated |
| 0,01196447 | SPACA1 | FALSE | Upregulated |
| | SPACA3 | | |
| 0,49105908 | SPACA4 | FALSE | Upregulated |
| -0,906927972 | SPACA5 | FALSE | Downregulated |
| -0,436085337 | SPACA5B | FALSE | Downregulated |
| 0,48097419 | SPACA7 | FALSE | Upregulated |
| | SPAG11A | | |
| | SPAG11B | | |
| 0,225736514 | SPAG6 | FALSE | Upregulated |
| | SPAM1 | | |
| | SPANXA1 | | |
| | SPANXA2 | | |
| | SPANXA2-OT1 | | |
| -0,012828367 | SPANXB1 | FALSE | Downregulated |
| | SPANXC | | |
| 0 | SPANXD | FALSE | Upregulated |
| | SPANXN1 | | |
| | SPANXN2 | | |
| | SPANXN3 | | |
| | SPANXN4 | | |
| | SPANXN5 | | |
| | SPATA16 | | |
| 0,215773332 | SPATA17 | FALSE | Upregulated |
| | SPATA19 | | |
| -0,00935474 | SPATA22 | FALSE | Downregulated |
| | SPATA3 | | |
| 1,855894226 | SPATA3-AS1 | | Upregulated |
| | SPATA31A1 | | |
| | SPATA31A3 | | |
| | SPATA31A5 | | |
| | SPATA31A6 | | |
| | SPATA31A7 | | |
| 0,4139846 | SPATA31B1 | FALSE | Upregulated |
| 0,219841565 | SPATA31C1 | FALSE | Upregulated |

| | | | |
|---|---|---|---|
| 0,483869869 | SPATA31C2 | FALSE | Upregulated |
| | SPATA31D1 | | |
| | SPATA31D3 | | |
| | SPATA31D4 | | |
| 0 | SPATA31D5P | FALSE | Upregulated |
| 0,559274463 | SPATA4 | FALSE | Upregulated |
| | SPATA42 | | |
| 1,202607425 | SPATA8 | | Upregulated |
| | SPATA8-AS1 | | |
| 0,679994445 | SPATC1L | FALSE | Upregulated |
| 0,099452879 | SPATS1 | FALSE | Upregulated |
| 0,805128282 | SPC24 | FALSE | Upregulated |
| 1,333643863 | SPC25 | | Upregulated |
| 0,633105911 | SPDEF | FALSE | Upregulated |
| -0,582629796 | SPDYC | FALSE | Downregulated |
| 0,206138097 | SPEF1 | FALSE | Upregulated |
| 0,008689745 | SPEM1 | FALSE | Upregulated |
| | SPERT | | |
| -0,162363267 | SPESP1 | FALSE | Downregulated |
| 0 | SPG20OS | FALSE | Upregulated |
| | SPHAR | | |
| 0 | SPHKAP | FALSE | Upregulated |
| 1,543727412 | SPINK1 | | Upregulated |
| 0,491149431 | SPINK13 | FALSE | Upregulated |
| 0 | SPINK14 | FALSE | Upregulated |
| 0 | SPINK2 | FALSE | Upregulated |
| | SPINK4 | | |
| 0,600105503 | SPINK5 | FALSE | Upregulated |
| -0,36968715 | SPINK6 | FALSE | Downregulated |
| -0,132598605 | SPINK7 | FALSE | Downregulated |
| | SPINK8 | | |
| 0,048628632 | SPINK9 | FALSE | Upregulated |
| | SPINT3 | | |
| | SPINT4 | | |
| 0 | SPO11 | FALSE | Upregulated |
| 0 | SPOCD1 | FALSE | Upregulated |
| 0 | SPOCK1 | FALSE | Upregulated |
| -0,487697478 | SPOCK3 | FALSE | Downregulated |
| | SPPL2C | | |
| -0,474391992 | SPRED3 | FALSE | Downregulated |
| | SPRR1A | | |
| | SPRR1B | | |
| | SPRR2A | | |
| | SPRR2B | | |
| | SPRR2C | | |
| | SPRR2D | | |
| 0 | SPRR2E | FALSE | Upregulated |
| | SPRR2F | | |
| | SPRR2G | | |
| | SPRR3 | | |
| | SPRR4 | | |

SPRY3-2
0,642334946 SPSB4 FALSE Upregulated
-0,193022541 SPTA1 FALSE Downregulated
0,626990484 SPZ1 FALSE Upregulated
0,364337466 SRD5A1P1 FALSE Upregulated
0,454093628 SRRM3 FALSE Upregulated
0 SRRM4 FALSE Upregulated
0,750662552 SRY FALSE Upregulated
SSMEM1
-1,534815933 SSR4P1 Downregulated
SST
0,653586369 SSTR3 FALSE Upregulated
SSTR4
-0,374098246 SSTR5 FALSE Downregulated
0 SSTR5-AS1 FALSE Upregulated
0,532565878 SSUH2 FALSE Upregulated
SSX1
SSX2
SSX2B
SSX3
SSX4
SSX4B
SSX5
SSX6
SSX7
SSX8
SSX9
-0,35887786 ST13P4 FALSE Downregulated
0,654819957 ST18 FALSE Upregulated
0 ST20-MTHFS FALSE Upregulated
0,857915018 ST6GALNAC1 FALSE Upregulated
0 ST6GALNAC5 FALSE Upregulated
0,070401644 ST7-OT3 FALSE Upregulated
ST8SIA2
0,546739037 ST8SIA5 FALSE Upregulated
-0,039018682 ST8SIA6 FALSE Downregulated
0,365968245 ST8SIA6-AS1 FALSE Upregulated
0 STAC2 FALSE Upregulated
0,696709817 STAP1 FALSE Upregulated
STARD6
STATH
0,199568568 STC1 FALSE Upregulated
-0,183489439 STC2 FALSE Downregulated
0,62366828 STEAP1B FALSE Upregulated
-0,298777885 STEAP2-AS1 FALSE Downregulated
STH
1,517506682 STIL Upregulated
0,073279458 STK31 FALSE Upregulated
0,771276989 STK32A FALSE Upregulated
-0,00508458 STK33 FALSE Downregulated
-0,587402519 STK4-AS1 FALSE Downregulated

| | | | |
|---|---|---|---|
| 1,332391601 | STMN2 | | Upregulated |
| -0,252907383 | STMN4 | FALSE | Downregulated |
| 0,33609392 | STMND1 | FALSE | Upregulated |
| | STOML3 | | |
| 0,204107626 | STON1-GTF2A1L | FALSE | Upregulated |
| 0,441034309 | STOX1 | FALSE | Upregulated |
| -0,196442461 | STPG2 | FALSE | Downregulated |
| -0,342216457 | STPG2-AS1 | FALSE | Downregulated |
| 0,334527787 | STRA6 | FALSE | Upregulated |
| 0 | STRA8 | FALSE | Upregulated |
| -2,420519003 | STRC | | Downregulated |
| -0,357967873 | STRIP2 | FALSE | Downregulated |
| -0,28443954 | STX18-AS1 | FALSE | Downregulated |
| 0,568083716 | STXBP6 | FALSE | Upregulated |
| -0,00545075 | STYK1 | FALSE | Downregulated |
| 0,042543811 | SUGT1P1 | FALSE | Upregulated |
| 0,261562334 | SUGT1P3 | FALSE | Upregulated |
| 0 | SULT1A3 | FALSE | Upregulated |
| 0 | SULT1C3 | FALSE | Upregulated |
| 0,553923285 | SULT2B1 | FALSE | Upregulated |
| 0,495757205 | SULT4A1 | FALSE | Upregulated |
| | SULT6B1 | | |
| 0,6702091 | SUMO1P1 | FALSE | Upregulated |
| 0,402136364 | SUMO4 | FALSE | Upregulated |
| | SUN3 | | |
| | SUN5 | | |
| 0,136696838 | SUPT20HL1 | FALSE | Upregulated |
| 0,420809547 | SUPT20HL2 | FALSE | Upregulated |
| -0,271272081 | SUSD4 | FALSE | Downregulated |
| -0,379090327 | SUZ12P1 | FALSE | Downregulated |
| -1,003682433 | SV2B | | Downregulated |
| 0,007291679 | SV2C | FALSE | Upregulated |
| 1,824994556 | SVILP1 | | Upregulated |
| 0,617281565 | SVOP | FALSE | Upregulated |
| 0,056304203 | SVOPL | FALSE | Upregulated |
| 0,024573396 | SYCE1L | FALSE | Upregulated |
| 0,435998371 | SYCE3 | FALSE | Upregulated |
| 0 | SYCN | FALSE | Upregulated |
| | SYCP1 | | |
| 0,896699115 | SYCP2 | FALSE | Upregulated |
| 0 | SYCP2L | FALSE | Upregulated |
| -0,143872207 | SYN2 | FALSE | Downregulated |
| 0 | SYNDIG1 | FALSE | Upregulated |
| 0,169049081 | SYNDIG1L | FALSE | Upregulated |
| 1,308029583 | SYNGR3 | | Upregulated |
| 0,426437581 | SYNGR4 | FALSE | Upregulated |
| 0,853261524 | SYNJ2-IT1 | FALSE | Upregulated |
| -0,691055487 | SYNJ2BP-COX16 | FALSE | Downregulated |
| -0,539015749 | SYNPR | FALSE | Downregulated |
| | SYNPR-AS1 | | |
| 0 | SYP-AS1 | FALSE | Upregulated |

| | | | |
|---|---|---|---|
| 0,314067441 | SYS1-DBNDD2 | FALSE | Upregulated |
| 1,019627605 | SYT10 | | Upregulated |
| 0,024021569 | SYT13 | FALSE | Upregulated |
| | SYT14 | | |
| 0,53527609 | SYT14L | FALSE | Upregulated |
| | SYT16 | | |
| | SYT3 | | |
| | SYT4 | | |
| 0,465237344 | SYT5 | FALSE | Upregulated |
| -0,033154669 | SYT6 | FALSE | Downregulated |
| 0,648100554 | SYT8 | FALSE | Upregulated |
| | T | | |
| | TAAR1 | | |
| | TAAR2 | | |
| -0,520184158 | TAAR3 | FALSE | Downregulated |
| | TAAR5 | | |
| | TAAR6 | | |
| | TAAR8 | | |
| | TAAR9 | | |
| 0,047839331 | TAC1 | FALSE | Upregulated |
| | TAC3 | | |
| 0,087058876 | TACR3 | FALSE | Upregulated |
| 0,867438992 | TAF13 | FALSE | Upregulated |
| -0,557036274 | TAF1L | FALSE | Downregulated |
| 0,776980277 | TAF4B | FALSE | Upregulated |
| 0,335120354 | TAF7L | FALSE | Upregulated |
| | TAGLN3 | | |
| 0,102407508 | TAL2 | FALSE | Upregulated |
| | TARM1 | | |
| 0,426214479 | TARP | FALSE | Upregulated |
| | TAS1R2 | | |
| | TAS2R1 | | |
| -0,357423729 | TAS2R10 | FALSE | Downregulated |
| 0,503342699 | TAS2R14 | FALSE | Upregulated |
| | TAS2R16 | | |
| -0,163456423 | TAS2R20 | FALSE | Downregulated |
| 0,055879918 | TAS2R3 | FALSE | Upregulated |
| -0,784775116 | TAS2R30 | FALSE | Downregulated |
| 2,142101379 | TAS2R31 | | Upregulated |
| 0,341507619 | TAS2R38 | FALSE | Upregulated |
| 0,139018172 | TAS2R39 | FALSE | Upregulated |
| 0,019512538 | TAS2R4 | FALSE | Upregulated |
| 0,272980185 | TAS2R40 | FALSE | Upregulated |
| | TAS2R41 | | |
| -0,447275056 | TAS2R43 | FALSE | Downregulated |
| -0,518554155 | TAS2R46 | FALSE | Downregulated |
| -0,218593169 | TAS2R5 | FALSE | Downregulated |
| 0,896159133 | TAS2R50 | FALSE | Upregulated |
| 0,557927274 | TAS2R60 | FALSE | Upregulated |
| 0 | TAS2R7 | FALSE | Upregulated |
| -0,016422875 | TAS2R8 | FALSE | Downregulated |

| | | | |
|---|---|---|---|
| 0 | TAS2R9 | FALSE | Upregulated |
| 0,02373852 | TAX1BP3 | FALSE | Upregulated |
| 0 | TBATA | FALSE | Upregulated |
| | TBC1D21 | | |
| | TBC1D26 | | |
| 0,051384395 | TBC1D27 | FALSE | Upregulated |
| 0 | TBC1D28 | FALSE | Upregulated |
| 0,285556695 | TBC1D29 | FALSE | Upregulated |
| -0,906643111 | TBC1D3 | FALSE | Downregulated |
| 0,633813651 | TBC1D30 | FALSE | Upregulated |
| 0,080786608 | TBC1D3B | FALSE | Upregulated |
| -1,142847883 | TBC1D3C | | Downregulated |
| | TBC1D3F | | |
| 0,413348881 | TBC1D3P1-DHX40P1 | FALSE | Upregulated |
| | TBC1D3P2 | | |
| | TBC1D3P5 | | |
| -0,11566462 | TBL1Y | FALSE | Downregulated |
| 0 | TBPL2 | FALSE | Upregulated |
| | TBR1 | | |
| 0,062681771 | TBX18 | FALSE | Upregulated |
| -0,055917925 | TBX19 | FALSE | Downregulated |
| 0 | TBX22 | FALSE | Upregulated |
| 0,348825319 | TBX4 | FALSE | Upregulated |
| 0 | TBX5 | FALSE | Upregulated |
| 0,448212425 | TBX5-AS1 | FALSE | Upregulated |
| | TCAM1P | | |
| | TCEAL2 | | |
| | TCEAL5 | | |
| 0,448212425 | TCEAL6 | FALSE | Upregulated |
| -0,108850743 | TCEAL7 | FALSE | Downregulated |
| | TCERG1L | | |
| -0,004464972 | TCF15 | FALSE | Downregulated |
| 0,394197379 | TCF23 | FALSE | Upregulated |
| 0,670923411 | TCF24 | FALSE | Upregulated |
| 0,298300915 | TCHHL1 | FALSE | Upregulated |
| -0,164162354 | TCL1A | FALSE | Downregulated |
| 0,509693822 | TCL1B | FALSE | Upregulated |
| 0,38647539 | TCL6 | FALSE | Upregulated |
| 0,715068599 | TCN1 | FALSE | Upregulated |
| | TCP10 | | |
| 0 | TCP10L2 | FALSE | Upregulated |
| 0,023144677 | TCP11 | FALSE | Upregulated |
| | TCP11X1 | | |
| | TCP11X2 | | |
| -0,682624286 | TCTE1 | FALSE | Downregulated |
| 0,119811432 | TCTE3 | FALSE | Upregulated |
| -0,540450509 | TCTEX1D1 | FALSE | Downregulated |
| 1,307086045 | TCTEX1D2 | | Upregulated |
| -0,462196539 | TDGF1 | FALSE | Downregulated |
| 0 | TDGF1P3 | FALSE | Upregulated |
| 0 | TDH | FALSE | Upregulated |

| | | | |
|---|---|---|---|
| -0,194109032 | TDRD1 | FALSE | Downregulated |
| 0,697944726 | TDRD12 | FALSE | Upregulated |
| 0,712488809 | TDRD15 | FALSE | Upregulated |
| | TDRD5 | | |
| 0,187250845 | TDRD9 | FALSE | Upregulated |
| | TDRG1 | | |
| 0 | TECRL | FALSE | Upregulated |
| -1,305060542 | TECTB | | Downregulated |
| 0,168533443 | TEKT1 | FALSE | Upregulated |
| -0,289382089 | TEKT2 | FALSE | Downregulated |
| 0 | TEKT3 | FALSE | Upregulated |
| | TEKT4 | | |
| -0,354337383 | TEKT5 | FALSE | Downregulated |
| 0 | TEPP | FALSE | Upregulated |
| | TERT | | |
| | TEX101 | | |
| | TEX11 | | |
| 0 | TEX12 | FALSE | Upregulated |
| -0,128453992 | TEX13B | FALSE | Downregulated |
| 0,49712467 | TEX15 | FALSE | Upregulated |
| 0 | TEX19 | FALSE | Upregulated |
| 0,241392858 | TEX21P | FALSE | Upregulated |
| | TEX26 | | |
| | TEX26-AS1 | | |
| 0 | TEX28 | FALSE | Upregulated |
| 0,109100671 | TEX29 | FALSE | Upregulated |
| 0 | TEX33 | FALSE | Upregulated |
| -0,210461804 | TEX35 | FALSE | Downregulated |
| | TEX36 | | |
| | TEX36-AS1 | | |
| | TEX37 | | |
| -0,561362833 | TEX40 | FALSE | Downregulated |
| -0,541515211 | TEX41 | FALSE | Downregulated |
| 0,070401644 | TFAMP1 | FALSE | Upregulated |
| 1,027847739 | TFAP2A | | Upregulated |
| | TFAP2A-AS1 | | |
| 0 | TFAP2B | FALSE | Upregulated |
| -0,026033303 | TFAP2C | FALSE | Downregulated |
| | TFAP2D | | |
| 0 | TFDP3 | FALSE | Upregulated |
| 0,395407221 | TFF1 | FALSE | Upregulated |
| 0,481637332 | TFF2 | FALSE | Upregulated |
| 0,343958765 | TFF3 | FALSE | Upregulated |
| -1,055182817 | TFPI2 | | Downregulated |
| | TGFA-IT1 | | |
| -0,523388245 | TGIF2-C20orf24 | FALSE | Downregulated |
| | TGIF2LX | | |
| | TGIF2LY | | |
| 0,410923983 | TGM3 | FALSE | Upregulated |
| 0 | TGM5 | FALSE | Upregulated |
| -0,046806288 | TGM6 | FALSE | Downregulated |

| | | | |
|---|---|---|---|
| | TGM7 | | |
| 0,292498251 | THAP10 | FALSE | Upregulated |
| 0 | THEG | FALSE | Upregulated |
| | THEG5 | | |
| 0,402112793 | THEGL | FALSE | Upregulated |
| 0,014211447 | THSD7B | FALSE | Upregulated |
| 0,224111044 | TIFAB | FALSE | Upregulated |
| 0,35841945 | TIGD3 | FALSE | Upregulated |
| 0,467096012 | TIGD4 | FALSE | Upregulated |
| 0,599774337 | TIMP4 | FALSE | Upregulated |
| | TINAG | | |
| 0 | TINCR | FALSE | Upregulated |
| 0,302058429 | TIPIN | FALSE | Upregulated |
| | TISP43 | | |
| -0,623861278 | TKTL1 | FALSE | Downregulated |
| | TKTL2 | | |
| 0,930921949 | TLE6 | FALSE | Upregulated |
| -0,272064544 | TLL1 | FALSE | Downregulated |
| 0,106062578 | TLL2 | FALSE | Upregulated |
| 1,633860596 | TLR10 | | Upregulated |
| 1,876700681 | TLR7 | | Upregulated |
| 0 | TLR8-AS1 | FALSE | Upregulated |
| -1,475832375 | TLX1 | | Downregulated |
| 1,112161463 | TLX2 | | Upregulated |
| | TLX3 | | |
| 0,640974526 | TM4SF1-AS1 | FALSE | Upregulated |
| 0,365192982 | TM4SF19 | FALSE | Upregulated |
| -0,001405949 | TM4SF19-AS1 | FALSE | Downregulated |
| | TM4SF19-TCTEX1D2 | | |
| 0 | TMC1 | FALSE | Upregulated |
| 0,807664065 | TMC7 | FALSE | Upregulated |
| 0,33860193 | TMCC1-AS1 | FALSE | Upregulated |
| -0,787794668 | TMCC2 | FALSE | Downregulated |
| 0,848371755 | TMCO2 | FALSE | Upregulated |
| | TMCO5A | | |
| | TMCO5B | | |
| -0,116722451 | TMED11P | FALSE | Downregulated |
| -0,082213221 | TMED7-TICAM2 | FALSE | Downregulated |
| | TMEFF1 | | |
| 0,427110868 | TMEM100 | FALSE | Upregulated |
| 0,112366068 | TMEM107 | FALSE | Upregulated |
| 0,68662968 | TMEM110-MUSTN1 | FALSE | Upregulated |
| | TMEM114 | | |
| 0,724610642 | TMEM130 | FALSE | Upregulated |
| -0,022688237 | TMEM132B | FALSE | Downregulated |
| 0,768523972 | TMEM132C | FALSE | Upregulated |
| 0,301837511 | TMEM144 | FALSE | Upregulated |
| 0,44026487 | TMEM145 | FALSE | Upregulated |
| -0,473063339 | TMEM151A | FALSE | Downregulated |
| 0,119038623 | TMEM151B | FALSE | Upregulated |
| 0 | TMEM155 | FALSE | Upregulated |

| | | | |
|---|---|---|---|
| 1,169156004 | TMEM156 | | Upregulated |
| 1,319862356 | TMEM158 | | Upregulated |
| -0,844814823 | TMEM159 | FALSE | Downregulated |
| -1,021792683 | TMEM161B-AS1 | | Downregulated |
| 0,332655161 | TMEM171 | FALSE | Upregulated |
| | TMEM174 | | |
| 0,695722727 | TMEM178B | FALSE | Upregulated |
| 0,910322977 | TMEM182 | FALSE | Upregulated |
| 0,724487283 | TMEM190 | FALSE | Upregulated |
| 0,865193411 | TMEM191B | FALSE | Upregulated |
| -0,334530121 | TMEM191C | FALSE | Downregulated |
| 0,271392248 | TMEM196 | FALSE | Upregulated |
| 0,486189506 | TMEM200A | FALSE | Upregulated |
| -0,073187995 | TMEM202 | FALSE | Downregulated |
| 0,79357455 | TMEM207 | FALSE | Upregulated |
| | TMEM210 | | |
| -0,597940852 | TMEM211 | FALSE | Downregulated |
| 0,042336372 | TMEM212 | FALSE | Upregulated |
| 0,306924291 | TMEM213 | FALSE | Upregulated |
| 0,446646722 | TMEM215 | FALSE | Upregulated |
| -1,775936885 | TMEM216 | | Downregulated |
| -0,11273365 | TMEM225 | FALSE | Downregulated |
| 0,450737483 | TMEM229A | FALSE | Upregulated |
| 1,215340191 | TMEM231 | | Upregulated |
| 0,403220407 | TMEM235 | FALSE | Upregulated |
| -0,196270774 | TMEM236 | FALSE | Downregulated |
| | TMEM239 | | |
| -0,418643879 | TMEM240 | FALSE | Downregulated |
| 0 | TMEM244 | FALSE | Upregulated |
| 0,091031112 | TMEM247 | FALSE | Upregulated |
| -0,119360923 | TMEM249 | FALSE | Downregulated |
| | TMEM257 | | |
| 0,049083946 | TMEM262 | FALSE | Upregulated |
| 0,012247493 | TMEM30C | FALSE | Upregulated |
| 0,210014895 | TMEM31 | FALSE | Upregulated |
| -0,059315317 | TMEM35 | FALSE | Downregulated |
| 0 | TMEM40 | FALSE | Upregulated |
| -0,318425302 | TMEM52 | FALSE | Downregulated |
| 0,193415935 | TMEM52B | FALSE | Upregulated |
| 0,26000628 | TMEM55A | FALSE | Upregulated |
| | TMEM59L | | |
| 0 | TMEM61 | FALSE | Upregulated |
| 0,603946383 | TMEM63C | FALSE | Upregulated |
| -1,305206836 | TMEM67 | | Downregulated |
| 0,88675012 | TMEM71 | FALSE | Upregulated |
| 0,177425945 | TMEM72 | FALSE | Upregulated |
| 0,797051073 | TMEM72-AS1 | FALSE | Upregulated |
| 0,624563063 | TMEM74 | FALSE | Upregulated |
| 0,493023865 | TMEM81 | FALSE | Upregulated |
| 0,805929296 | TMEM88 | FALSE | Upregulated |
| 0,268927902 | TMEM88B | FALSE | Upregulated |

| | | | |
|---|---|---|---|
| 0,274361641 | TMEM89 | FALSE | Upregulated |
| | TMEM8C | | |
| 0 | TMEM95 | FALSE | Upregulated |
| -0,649766653 | TMEM99 | FALSE | Downregulated |
| | TMIGD1 | | |
| 0,860012913 | TMOD4 | FALSE | Upregulated |
| | TMPRSS11B | | |
| | TMPRSS11BNL | | |
| -0,387371309 | TMPRSS11F | FALSE | Downregulated |
| 0,047839331 | TMPRSS11GP | FALSE | Upregulated |
| 0 | TMPRSS12 | FALSE | Upregulated |
| 0,629548801 | TMPRSS13 | FALSE | Upregulated |
| 0 | TMPRSS15 | FALSE | Upregulated |
| 0,203881273 | TMPRSS3 | FALSE | Upregulated |
| | TMPRSS4-AS1 | | |
| -0,203212135 | TMPRSS5 | FALSE | Downregulated |
| | TMPRSS7 | | |
| | TMSB15A | | |
| | TMSB15B | | |
| 1,598930486 | TMSB4Y | | Upregulated |
| -0,054425816 | TNFAIP6 | FALSE | Downregulated |
| 0,654054239 | TNFAIP8L3 | FALSE | Upregulated |
| 1,898269606 | TNFRSF10C | | Upregulated |
| -0,060705066 | TNFRSF13B | FALSE | Downregulated |
| 0,295092824 | TNFRSF13C | FALSE | Upregulated |
| 0,40719634 | TNFRSF18 | FALSE | Upregulated |
| 0,98613647 | TNFRSF19 | FALSE | Upregulated |
| 0,713558784 | TNFRSF4 | FALSE | Upregulated |
| 0,92407555 | TNFRSF8 | FALSE | Upregulated |
| 1,769780638 | TNFRSF9 | | Upregulated |
| 0,445762264 | TNFSF11 | FALSE | Upregulated |
| 0,0254987 | TNFSF15 | FALSE | Upregulated |
| 0,404338391 | TNFSF18 | FALSE | Upregulated |
| 0,622928752 | TNFSF9 | FALSE | Upregulated |
| 1,375877668 | TNIP3 | | Upregulated |
| 0,490879829 | TNKS2-AS1 | FALSE | Upregulated |
| 0 | TNNI3 | FALSE | Upregulated |
| | TNNI3K | | |
| 0,105187726 | TNNT2 | FALSE | Upregulated |
| 0,291111583 | TNP1 | FALSE | Upregulated |
| 0,576875809 | TNP2 | FALSE | Upregulated |
| 0,481821562 | TNR | FALSE | Upregulated |
| 0,027955006 | TOB2P1 | FALSE | Upregulated |
| -1,646997665 | TOLLIP-AS1 | | Downregulated |
| 0,198144427 | TOMM20L | FALSE | Upregulated |
| | TOP1P2 | | |
| 0,402112793 | TOPAZ1 | FALSE | Upregulated |
| 0,442996005 | TP53AIP1 | FALSE | Upregulated |
| | TP53TG3 | | |
| | TP53TG3B | | |
| | TP53TG3C | | |

| | | | |
|---|---|---|---|
| | TP53TG3D | | |
| -0,230761359 | TP63 | FALSE | Downregulated |
| 0,885062352 | TPBGL | FALSE | Upregulated |
| 0,014848181 | TPD52L3 | FALSE | Upregulated |
| -0,107040638 | TPH1 | FALSE | Downregulated |
| | TPH2 | | |
| 0,379694908 | TPI1P2 | FALSE | Upregulated |
| 1,145622919 | TPO | | Upregulated |
| 0,618291058 | TPPP3 | FALSE | Upregulated |
| -1,663951097 | TPRKB | | Downregulated |
| 0,196228486 | TPRX1 | FALSE | Upregulated |
| 0,099452879 | TPRXL | FALSE | Upregulated |
| 0,26472608 | TPSD1 | FALSE | Upregulated |
| | TPTE | | |
| | TPTE2 | | |
| | TPTE2P2 | | |
| -0,002364448 | TPTE2P3 | FALSE | Downregulated |
| -0,061097802 | TPTE2P6 | FALSE | Downregulated |
| -0,148288645 | TPTEP1 | FALSE | Downregulated |
| 0,65273309 | TRAIP | FALSE | Upregulated |
| 0 | TRAM1L1 | FALSE | Upregulated |
| -0,070043787 | TRAPPC2P1 | FALSE | Downregulated |
| -0,323324982 | TRAPPC3L | FALSE | Downregulated |
| 1,008509456 | TRAT1 | | Upregulated |
| -1,455921077 | TRDN | | Downregulated |
| 0,549665746 | TREM1 | FALSE | Upregulated |
| 1,206324242 | TREM2 | | Upregulated |
| -0,4782326 | TREML2 | FALSE | Downregulated |
| 0 | TREML3P | FALSE | Upregulated |
| -0,477915463 | TREML4 | FALSE | Downregulated |
| | TREML5P | | |
| 0,122127586 | TRERNA1 | FALSE | Upregulated |
| -0,131164819 | TREX2 | FALSE | Downregulated |
| 1,692450488 | TRG-AS1 | | Upregulated |
| -0,552742474 | TRH | FALSE | Downregulated |
| -0,480045838 | TRHR | FALSE | Downregulated |
| -0,57286882 | TRIM29 | FALSE | Downregulated |
| 1,671355399 | TRIM31 | | Upregulated |
| 0,309479103 | TRIM36 | FALSE | Upregulated |
| 0,413785866 | TRIM39-RPP21 | FALSE | Upregulated |
| | TRIM40 | | |
| 0,187660988 | TRIM42 | FALSE | Upregulated |
| | TRIM43 | | |
| | TRIM43B | | |
| | TRIM48 | | |
| | TRIM49 | | |
| | TRIM49B | | |
| | TRIM49C | | |
| | TRIM49D1 | | |
| | TRIM49D2P | | |
| 0,230544904 | TRIM50 | FALSE | Upregulated |

| | | | |
|---|---|---|---|
| | TRIM51 | | |
| | TRIM51EP | | |
| | TRIM51HP | | |
| | TRIM53AP | | |
| 0,107883439 | TRIM54 | FALSE | Upregulated |
| -1,185056699 | TRIM58 | | Downregulated |
| | TRIM60 | | |
| 0,456350028 | TRIM61 | FALSE | Upregulated |
| 0,426796786 | TRIM63 | FALSE | Upregulated |
| | TRIM64 | | |
| | TRIM64B | | |
| | TRIM64C | | |
| | TRIM71 | | |
| 0,048568525 | TRIM72 | FALSE | Upregulated |
| 1,153235442 | TRIM73 | | Upregulated |
| | TRIM77 | | |
| 0,295193301 | TRIM9 | FALSE | Upregulated |
| | TRIML1 | | |
| | TRIML2 | | |
| 0,84168071 | TRIP13 | FALSE | Upregulated |
| | TRNA | | |
| | TRNAA-AGC | | |
| | TRNAA-AGC-2 | | |
| | TRNAA-AGC-3 | | |
| | TRNAA1 | | |
| | TRNAA10 | | |
| | TRNAA11 | | |
| | TRNAA12 | | |
| | TRNAA13 | | |
| | TRNAA14 | | |
| | TRNAA15 | | |
| | TRNAA17 | | |
| | TRNAA18 | | |
| | TRNAA19 | | |
| | TRNAA2 | | |
| | TRNAA20 | | |
| | TRNAA20-2 | | |
| | TRNAA20-3 | | |
| | TRNAA23 | | |
| | TRNAA26 | | |
| | TRNAA27 | | |
| | TRNAA28 | | |
| | TRNAA29 | | |
| | TRNAA3 | | |
| | TRNAA30 | | |
| | TRNAA32 | | |
| | TRNAA34 | | |
| | TRNAA36 | | |
| | TRNAA37 | | |
| | TRNAA37-2 | | |
| | TRNAA38 | | |

TRNAA39
TRNAA4
TRNAA40
TRNAA41
TRNAA41-2
TRNAA5
TRNAA6
TRNAA7
TRNAA8
TRNAA9
TRNAC-GCA
TRNAC10
TRNAC11
TRNAC12
TRNAC13
TRNAC14
TRNAC15
TRNAC16
TRNAC18
TRNAC19
TRNAC2
TRNAC20
TRNAC21
TRNAC22
TRNAC23
TRNAC24
TRNAC25
TRNAC26
TRNAC27
TRNAC28
TRNAC29
TRNAC3
TRNAC30
TRNAC4
TRNAC5
TRNAC6
TRNAC7
TRNAC8
TRNAC9
TRNAD10
TRNAD11
TRNAD12
TRNAD13
TRNAD14
TRNAD15
TRNAD16
TRNAD17
TRNAD18
TRNAD5
TRNAD6
TRNAD8

TRNAD9
TRNAE-UUC
TRNAE-UUC-2
TRNAE1
TRNAE11
TRNAE13
TRNAE18
TRNAE19
TRNAE20
TRNAE21
TRNAE22
TRNAE23
TRNAE24
TRNAE26
TRNAE5
TRNAE6
TRNAE7
TRNAF1
TRNAF10
TRNAF11
TRNAF2
TRNAF3
TRNAF4
TRNAF5
TRNAF6
TRNAF7
TRNAF8
TRNAF9
TRNAG-CCC
TRNAG-CCC-2
TRNAG1
TRNAG10
TRNAG11
TRNAG13
TRNAG15
TRNAG16
TRNAG17
TRNAG18
TRNAG19
TRNAG2
TRNAG20
TRNAG21
TRNAG22
TRNAG23
TRNAG24
TRNAG25
TRNAG26
TRNAG27
TRNAG28
TRNAG29
TRNAG3

TRNAG30
TRNAG31
TRNAG4
TRNAG5
TRNAG6
TRNAG7
TRNAG8
TRNAG9
TRNAH1
TRNAH10
TRNAH2
TRNAH3
TRNAH4
TRNAH5
TRNAH6
TRNAH7
TRNAH9
TRNAI-AAU
TRNAI1
TRNAI11
TRNAI12
TRNAI13
TRNAI15
TRNAI17
TRNAI18
TRNAI2
TRNAI20
TRNAI21
TRNAI22
TRNAI23
TRNAI24
TRNAI25
TRNAI25-2
TRNAI26
TRNAI3
TRNAI4
TRNAI5
TRNAI6
TRNAI7
TRNAI9
TRNAK-CUU
TRNAK1
TRNAK11
TRNAK13
TRNAK14
TRNAK15
TRNAK16
TRNAK17
TRNAK19
TRNAK2
TRNAK20

TRNAK21
TRNAK22
TRNAK23
TRNAK26
TRNAK27
TRNAK29
TRNAK3
TRNAK31
TRNAK33
TRNAK34
TRNAK4
TRNAK6
TRNAK8
TRNAK9
TRNAL-CAA
TRNAL1
TRNAL10
TRNAL11
TRNAL12
TRNAL13
TRNAL14
TRNAL15
TRNAL16
TRNAL17
TRNAL19
TRNAL2
TRNAL20
TRNAL22
TRNAL23
TRNAL24
TRNAL26
TRNAL27
TRNAL28
TRNAL29
TRNAL3
TRNAL30
TRNAL32
TRNAL34
TRNAL35
TRNAL36
TRNAL38
TRNAL4
TRNAL5
TRNAL6
TRNAL7
TRNAL8
TRNAM-CAU
TRNAM1
TRNAM11
TRNAM12
TRNAM13

TRNAM14
TRNAM15
TRNAM16
TRNAM16-2
TRNAM17
TRNAM18
TRNAM19
TRNAM2
TRNAM20
TRNAM3
TRNAM4
TRNAM5
TRNAM6
TRNAM8
TRNAM9
TRNAN-GUU
TRNAN-GUU-2
TRNAN-GUU-3
TRNAN-GUU-4
TRNAN-GUU-5
TRNAN-GUU-6
TRNAN-GUU-7
TRNAN1
TRNAN11
TRNAN14
TRNAN15
TRNAN17
TRNAN18
TRNAN19
TRNAN2
TRNAN20
TRNAN22
TRNAN22-2
TRNAN27
TRNAN29
TRNAN3
TRNAN32
TRNAN5
TRNAN5-2
TRNAN7
TRNAN8
TRNAP1
TRNAP10
TRNAP11
TRNAP12
TRNAP13
TRNAP14
TRNAP15
TRNAP16
TRNAP17
TRNAP18

TRNAP19
TRNAP2
TRNAP21
TRNAP3
TRNAP4
TRNAP5
TRNAP6
TRNAP7
TRNAP8
TRNAP9
TRNAQ-CUG
TRNAQ1
TRNAQ10
TRNAQ11
TRNAQ12
TRNAQ13
TRNAQ15
TRNAQ16
TRNAQ17
TRNAQ2
TRNAQ20
TRNAQ21
TRNAQ22
TRNAQ24
TRNAQ25
TRNAQ27
TRNAQ3
TRNAQ32
TRNAQ8
TRNAR1
TRNAR10
TRNAR11
TRNAR12
TRNAR14
TRNAR15
TRNAR16
TRNAR17
TRNAR18
TRNAR19
TRNAR2
TRNAR20
TRNAR21
TRNAR22
TRNAR23
TRNAR24
TRNAR25
TRNAR26
TRNAR27
TRNAR28
TRNAR29
TRNAR3

TRNAR4
TRNAR5
TRNAR6
TRNAR7
TRNAR8
TRNAR9
TRNAS1
TRNAS10
TRNAS11
TRNAS12
TRNAS13
TRNAS14
TRNAS15
TRNAS16
TRNAS17
TRNAS18
TRNAS19
TRNAS2
TRNAS20
TRNAS21
TRNAS22
TRNAS23
TRNAS24
TRNAS25
TRNAS27
TRNAS28
TRNAS3
TRNAS4
TRNAS5
TRNAS6
TRNAS7
TRNAS9
TRNASUP1
TRNAT1
TRNAT10
TRNAT12
TRNAT13
TRNAT14
TRNAT15
TRNAT16
TRNAT17
TRNAT18
TRNAT19
TRNAT2
TRNAT20
TRNAT21
TRNAT22
TRNAT4
TRNAT5
TRNAT6
TRNAT7

| | | | |
|---|---|---|---|
| | TRNAT8 | | |
| | TRNAT9 | | |
| 0 | TRNAU1 | FALSE | Upregulated |
| | TRNAU2 | | |
| | TRNAV-CAC | | |
| | TRNAV-CAC-2 | | |
| | TRNAV-CAC-3 | | |
| | TRNAV1 | | |
| | TRNAV10 | | |
| | TRNAV11 | | |
| | TRNAV12 | | |
| | TRNAV15 | | |
| | TRNAV16 | | |
| | TRNAV17 | | |
| | TRNAV18 | | |
| | TRNAV19 | | |
| | TRNAV2 | | |
| | TRNAV20 | | |
| | TRNAV21 | | |
| | TRNAV22 | | |
| | TRNAV23 | | |
| | TRNAV24 | | |
| | TRNAV25 | | |
| | TRNAV26 | | |
| | TRNAV27 | | |
| | TRNAV28 | | |
| | TRNAV29 | | |
| | TRNAV3 | | |
| | TRNAV31 | | |
| | TRNAV32 | | |
| | TRNAV4 | | |
| | TRNAV5 | | |
| | TRNAV6 | | |
| | TRNAV7 | | |
| | TRNAV8 | | |
| | TRNAV9 | | |
| | TRNAW2 | | |
| | TRNAW3 | | |
| | TRNAW4 | | |
| | TRNAW5 | | |
| | TRNAW6 | | |
| | TRNAW7 | | |
| | TRNAW9 | | |
| | TRNAY1 | | |
| | TRNAY10 | | |
| | TRNAY12 | | |
| | TRNAY13 | | |
| | TRNAY14 | | |
| | TRNAY15 | | |
| | TRNAY2 | | |
| | TRNAY3 | | |

| | | | |
|---|---|---|---|
| | TRNAY4 | | |
| | TRNAY6 | | |
| | TRNAY7 | | |
| | TRNAY8 | | |
| | TRNAY9 | | |
| | TRNC | | |
| | TRND | | |
| | TRNE | | |
| | TRNF | | |
| | TRNG | | |
| | TRNH | | |
| | TRNI | | |
| | TRNK | | |
| | TRNL1 | | |
| | TRNL2 | | |
| | TRNM | | |
| | TRNN | | |
| | TRNP | | |
| | TRNQ | | |
| | TRNR | | |
| | TRNS1 | | |
| | TRNS2 | | |
| | TRNT | | |
| | TRNV | | |
| | TRNW | | |
| | TRNY | | |
| 0,819426592 | TROAP | FALSE | Upregulated |
| 0,280558821 | TRPC2 | FALSE | Upregulated |
| 0,008277399 | TRPC3 | FALSE | Upregulated |
| 0,2994934 | TRPC4 | FALSE | Upregulated |
| -1,030589358 | TRPC6 | | Downregulated |
| 0,948917737 | TRPC7 | FALSE | Upregulated |
| -0,613510257 | TRPM6 | FALSE | Downregulated |
| -0,016422875 | TRPV5 | FALSE | Downregulated |
| 0 | TRY2P | FALSE | Upregulated |
| 1,102693373 | TSACC | | Upregulated |
| | TSG1 | | |
| 0,474844862 | TSGA10 | FALSE | Upregulated |
| -0,078913265 | TSGA10IP | FALSE | Downregulated |
| 1,162381997 | TSGA13 | | Upregulated |
| -0,131651671 | TSHB | FALSE | Downregulated |
| 0,424198559 | TSKS | FALSE | Upregulated |
| -0,2842982 | TSPAN10 | FALSE | Downregulated |
| 0,487170587 | TSPAN16 | FALSE | Upregulated |
| | TSPAN19 | | |
| 0,072716311 | TSPAN2 | FALSE | Upregulated |
| | TSPY1 | | |
| | TSPY10 | | |
| | TSPY2 | | |
| -1,373131128 | TSPY26P | | Downregulated |
| | TSPY3 | | |

| | | | |
|---|---|---|---|
| | TSPY4 | | |
| | TSPY8 | | |
| 0 | TTC23L | FALSE | Upregulated |
| 0,18460671 | TTC24 | FALSE | Upregulated |
| -0,231460411 | TTC25 | FALSE | Downregulated |
| -0,832734195 | TTC26 | FALSE | Downregulated |
| | TTC29 | | |
| 0,448022829 | TTC32 | FALSE | Upregulated |
| -0,62184267 | TTC34 | FALSE | Downregulated |
| 0,572673782 | TTC40 | FALSE | Upregulated |
| 0,544624643 | TTC9B | FALSE | Upregulated |
| -0,154955703 | TTK | FALSE | Downregulated |
| -0,452995673 | TTLL1 | FALSE | Downregulated |
| 0,239480862 | TTLL10 | FALSE | Upregulated |
| -0,137917867 | TTLL10-AS1 | FALSE | Downregulated |
| 2,152834809 | TTLL13 | | Upregulated |
| 0,323367266 | TTLL6 | FALSE | Upregulated |
| | TTLL8 | | |
| | TTTY1 | | |
| -0,029275514 | TTTY10 | FALSE | Downregulated |
| | TTTY11 | | |
| | TTTY12 | | |
| | TTTY16 | | |
| | TTTY18 | | |
| | TTTY19 | | |
| | TTTY1B | | |
| | TTTY2 | | |
| | TTTY20 | | |
| | TTTY21 | | |
| | TTTY21B | | |
| | TTTY22 | | |
| | TTTY23 | | |
| | TTTY23B | | |
| | TTTY2B | | |
| | TTTY7 | | |
| 0,43812016 | TTTY7B | FALSE | Upregulated |
| | TTTY8 | | |
| | TTTY8B | | |
| 0,046678197 | TUBA3C | FALSE | Upregulated |
| 1,419506274 | TUBA4B | | Upregulated |
| | TUBAL3 | | |
| 0,753862177 | TUBB3 | FALSE | Upregulated |
| 0,435977433 | TUBB4A | FALSE | Upregulated |
| | TUBB8 | | |
| -0,324133881 | TUBBP5 | FALSE | Downregulated |
| -0,356473114 | TULP1 | FALSE | Downregulated |
| -0,376029065 | TULP2 | FALSE | Downregulated |
| -0,097432875 | TUSC5 | FALSE | Downregulated |
| -0,75987482 | TUSC7 | FALSE | Downregulated |
| -1,303340821 | TVP23C-CDRT4 | | Downregulated |
| 0,864381557 | TWIST2 | FALSE | Upregulated |

| | | | |
|---|---|---|---|
| -0,10638868 | TXNDC2 | FALSE | Downregulated |
| -0,28315273 | TXNDC8 | FALSE | Downregulated |
| 0,402490181 | TXNRD3NB | FALSE | Upregulated |
| 0 | TYR | FALSE | Upregulated |
| 0,045506768 | TYRP1 | FALSE | Upregulated |
| 0,004067387 | UBAC2-AS1 | FALSE | Upregulated |
| 2,191007539 | UBE2C | | Upregulated |
| -0,082936757 | UBE2DNL | FALSE | Downregulated |
| 0,286482222 | UBE2E2-AS1 | FALSE | Upregulated |
| 0,644949042 | UBE2MP1 | FALSE | Upregulated |
| 0,094615407 | UBE2NL | FALSE | Upregulated |
| 0,393013293 | UBE2Q2L | FALSE | Upregulated |
| 0,701936141 | UBE2Q2P2 | FALSE | Upregulated |
| 0,643142202 | UBE2Q2P3 | FALSE | Upregulated |
| -0,095815255 | UBE2QL1 | FALSE | Downregulated |
| 0,358272789 | UBE2T | FALSE | Upregulated |
| | UBE2U | | |
| -0,104215127 | UBE3D | FALSE | Downregulated |
| | UBL4B | | |
| 0,249682095 | UBQLN3 | FALSE | Upregulated |
| -0,347736803 | UBQLNL | FALSE | Downregulated |
| | UBTFL1 | | |
| 0,215727349 | UCHL1 | FALSE | Upregulated |
| | UCHL1-AS1 | | |
| | UCMA | | |
| 0,014950913 | UCN | FALSE | Upregulated |
| 0,042934558 | UCN2 | FALSE | Upregulated |
| | UCN3 | | |
| 0,522577565 | UCP1 | FALSE | Upregulated |
| 0,001140215 | UCP3 | FALSE | Upregulated |
| | UGT2A1 | | |
| -1,672624714 | UGT2A2 | | Downregulated |
| 0,368593051 | UGT2B28 | FALSE | Upregulated |
| -0,215313841 | UGT8 | FALSE | Downregulated |
| | ULBP1 | | |
| 0,021513939 | ULBP2 | FALSE | Upregulated |
| -0,200727317 | ULBP3 | FALSE | Downregulated |
| 0,809973368 | ULK4P3 | FALSE | Upregulated |
| | UMOD | | |
| 0,945778122 | UMODL1 | FALSE | Upregulated |
| 0,27970543 | UNC13A | FALSE | Upregulated |
| 0,648827164 | UNC13C | FALSE | Upregulated |
| 0,332414725 | UNC45B | FALSE | Upregulated |
| | UNC5D | | |
| -1,192171524 | UNC79 | | Downregulated |
| -0,189872916 | UNC80 | FALSE | Downregulated |
| | UNCX | | |
| 1,117466243 | UNQ6494 | | Upregulated |
| 0,100713889 | UNQ9370 | FALSE | Upregulated |
| -0,162910989 | UOX | FALSE | Downregulated |
| 0,472566982 | UPK1A | FALSE | Upregulated |

| | | | |
|---|---|---|---|
| 0,64710809 | UPK1A-AS1 | FALSE | Upregulated |
| 0 | UPK2 | FALSE | Upregulated |
| | UPK3A | | |
| -0,398156083 | UQCRHL | FALSE | Downregulated |
| | URAD | | |
| -0,281136308 | URGCP-MRPS24 | FALSE | Downregulated |
| -0,37948057 | USH1C | FALSE | Downregulated |
| 0,809640919 | USH1G | FALSE | Upregulated |
| 0,365126748 | USP12-AS2 | FALSE | Upregulated |
| 0 | USP17L10 | FALSE | Upregulated |
| 0 | USP17L11 | FALSE | Upregulated |
| | USP17L12 | | |
| | USP17L13 | | |
| | USP17L15 | | |
| | USP17L17 | | |
| | USP17L18 | | |
| | USP17L19 | | |
| 1,713872149 | USP17L1P | | Upregulated |
| 0 | USP17L20 | FALSE | Upregulated |
| | USP17L21 | | |
| -0,082246368 | USP17L22 | FALSE | Downregulated |
| | USP17L24 | | |
| | USP17L25 | | |
| | USP17L26 | | |
| | USP17L27 | | |
| | USP17L28 | | |
| | USP17L29 | | |
| 0 | USP17L3 | FALSE | Upregulated |
| | USP17L30 | | |
| 0,23865175 | USP17L4 | FALSE | Upregulated |
| | USP17L5 | | |
| 0,030163892 | USP17L6P | FALSE | Upregulated |
| 0 | USP17L8 | FALSE | Upregulated |
| | USP17L9P | | |
| -0,397316029 | USP2-AS1 | FALSE | Downregulated |
| | USP26 | | |
| -0,714504994 | USP27X | FALSE | Downregulated |
| 0,382506043 | USP27X-AS1 | FALSE | Upregulated |
| | USP29 | | |
| 0,568970846 | USP41 | FALSE | Upregulated |
| 0,712501961 | USP46-AS1 | FALSE | Upregulated |
| 0,054350268 | USP6NL-IT1 | FALSE | Upregulated |
| | UTF1 | | |
| -0,21207693 | UTS2 | FALSE | Downregulated |
| 0,345591965 | UTS2R | FALSE | Upregulated |
| | VAMP7-2 | | |
| 0,192000493 | VANGL2 | FALSE | Upregulated |
| -0,226871167 | VAT1L | FALSE | Downregulated |
| | VAV3-AS1 | | |
| | VAX1 | | |
| 0,191394875 | VAX2 | FALSE | Upregulated |

| | | | |
|---|---|---|---|
| -1,03552075 | VCX | | Downregulated |
| -0,490588789 | VCX2 | FALSE | Downregulated |
| -0,203075222 | VCX3A | FALSE | Downregulated |
| 0,4266765 | VCX3B | FALSE | Upregulated |
| | VCY | | |
| 0,454560422 | VCY1B | FALSE | Upregulated |
| -0,514221786 | VENTX | FALSE | Downregulated |
| | VENTXP1 | | |
| 0,041835055 | VENTXP7 | FALSE | Upregulated |
| 0,78160995 | VGF | FALSE | Upregulated |
| -0,045460148 | VGLL1 | FALSE | Downregulated |
| 0,332745443 | VGLL2 | FALSE | Upregulated |
| | VHLL | | |
| 0,400565877 | VIP | FALSE | Upregulated |
| 0 | VIT | FALSE | Upregulated |
| -0,133265175 | VLDLR-AS1 | FALSE | Downregulated |
| 0 | VN1R10P | FALSE | Upregulated |
| 0,415072059 | VN1R2 | FALSE | Upregulated |
| | VN1R4 | | |
| -0,0483761 | VN1R5 | FALSE | Downregulated |
| 1,991959925 | VNN2 | | Upregulated |
| -0,18510955 | VPREB3 | FALSE | Downregulated |
| | VRTN | | |
| 0,125420808 | VSIG1 | FALSE | Upregulated |
| 0,880673048 | VSIG2 | FALSE | Upregulated |
| 0,685546174 | VSIG8 | FALSE | Upregulated |
| | VSTM1 | | |
| | VSTM2A | | |
| | VSTM2B | | |
| 0,252670599 | VSTM5 | FALSE | Upregulated |
| | VSX1 | | |
| -0,224412458 | VSX2 | FALSE | Downregulated |
| 0,762961488 | VTCN1 | FALSE | Upregulated |
| | VTRNA1-1 | | |
| | VTRNA1-2 | | |
| | VTRNA1-3 | | |
| | VTRNA2-1 | | |
| -0,143750946 | VWA3A | FALSE | Downregulated |
| -0,26378599 | VWA3B | FALSE | Downregulated |
| 0,158571891 | VWA5B1 | FALSE | Upregulated |
| 0,109389692 | VWC2 | FALSE | Upregulated |
| -0,522327192 | VWC2L | FALSE | Downregulated |
| | VWC2L-IT1 | | |
| -0,22471138 | VWDE | FALSE | Downregulated |
| 0,372803365 | WASIR2 | FALSE | Upregulated |
| 0 | WBP11P1 | FALSE | Upregulated |
| -1,148391851 | WBP2NL | | Downregulated |
| 0,482429527 | WBP5 | FALSE | Upregulated |
| 0,412010411 | WBSCR17 | FALSE | Upregulated |
| -1,876186705 | WBSCR27 | | Downregulated |
| 0,519237803 | WBSCR28 | FALSE | Upregulated |

| | | | |
|---|---|---|---|
| 0,218267297 | WDFY3-AS2 | FALSE | Upregulated |
| -1,523414089 | WDR17 | | Downregulated |
| | WDR49 | | |
| 0,579401382 | WDR52-AS1 | FALSE | Upregulated |
| 0,627311528 | WDR63 | FALSE | Upregulated |
| -0,389328563 | WDR64 | FALSE | Downregulated |
| 0,320691615 | WDR76 | FALSE | Upregulated |
| -0,341234517 | WDR86-AS1 | FALSE | Downregulated |
| 0,241682878 | WDR87 | FALSE | Upregulated |
| -0,783123266 | WDR96 | FALSE | Downregulated |
| -1,381802156 | WEE2-AS1 | | Downregulated |
| | WFDC10A | | |
| | WFDC10B | | |
| | WFDC11 | | |
| 0,026961126 | WFDC12 | FALSE | Upregulated |
| 0,655494118 | WFDC13 | FALSE | Upregulated |
| 0,01992285 | WFDC2 | FALSE | Upregulated |
| | WFDC3 | | |
| 0 | WFDC5 | FALSE | Upregulated |
| | WFDC6 | | |
| | WFDC8 | | |
| | WFDC9 | | |
| -0,465013192 | WFIKKN2 | FALSE | Downregulated |
| | WI2-2373I1.2 | | |
| -0,178285009 | WI2-81516E3.1 | FALSE | Downregulated |
| | WIF1 | | |
| 1,552901471 | WNT1 | | Upregulated |
| 0,900720338 | WNT10A | FALSE | Upregulated |
| 0,24647162 | WNT10B | FALSE | Upregulated |
| -0,327434427 | WNT2 | FALSE | Downregulated |
| 0,404701373 | WNT3A | FALSE | Upregulated |
| 0,64567674 | WNT6 | FALSE | Upregulated |
| 0,453099891 | WNT7A | FALSE | Upregulated |
| -0,152544047 | WNT7B | FALSE | Downregulated |
| 0 | WNT8A | FALSE | Upregulated |
| 0,861873751 | WNT9A | FALSE | Upregulated |
| 0 | WNT9B | FALSE | Upregulated |
| 0,322253037 | WSCD2 | FALSE | Upregulated |
| 0,609427288 | WT1 | FALSE | Upregulated |
| 0,216565842 | WT1-AS | FALSE | Upregulated |
| 0,656805963 | WTAPP1 | FALSE | Upregulated |
| 0,497913154 | WTH3DI | FALSE | Upregulated |
| -0,082703101 | WWTR1-AS1 | FALSE | Downregulated |
| | XAGE-4 | | |
| | XAGE1B | | |
| | XAGE1E | | |
| | XAGE2 | | |
| 0,359534878 | XAGE3 | FALSE | Upregulated |
| | XAGE5 | | |
| 0,222298825 | XCL1 | FALSE | Upregulated |
| -0,548537596 | XCL2 | FALSE | Downregulated |

| | | | |
|---|---|---|---|
| -0,122600458 | XCR1 | FALSE | Downregulated |
| | XGPY2 | | |
| | XIRP2-AS1 | | |
| -0,206926835 | XK | FALSE | Downregulated |
| | XKR3 | | |
| 0 | XKR4 | FALSE | Upregulated |
| | XKR5 | | |
| 0,291493325 | XKR7 | FALSE | Upregulated |
| 0,004679247 | XKRX | FALSE | Upregulated |
| 0,100749759 | XRCC2 | FALSE | Upregulated |
| 0,239668646 | XRCC4 | FALSE | Upregulated |
| -0,109065489 | XRCC6BP1 | FALSE | Downregulated |
| 0,437206419 | XRCC6P5 | FALSE | Upregulated |
| | XXYLT1-AS1 | | |
| 0 | YBX3P1 | FALSE | Upregulated |
| | YIPF7 | | |
| 0 | YPLR6490 | FALSE | Upregulated |
| -0,347693006 | YTHDF3-AS1 | FALSE | Downregulated |
| 0 | YWHAEP1 | FALSE | Upregulated |
| 0,021715852 | YWHAEP7 | FALSE | Upregulated |
| | YY1P2 | | |
| -0,418051564 | ZAN | FALSE | Downregulated |
| -0,004340485 | ZAR1 | FALSE | Downregulated |
| 0,230537751 | ZAR1L | FALSE | Upregulated |
| | ZBBX | | |
| | ZBED1-2 | | |
| -0,034669312 | ZBED3-AS1 | FALSE | Downregulated |
| -0,310313885 | ZBED5-AS1 | FALSE | Downregulated |
| | ZBTB8B | | |
| | ZC2HC1B | | |
| 0,823205381 | ZC3HAV1L | FALSE | Upregulated |
| 0,132387945 | ZCCHC12 | FALSE | Upregulated |
| | ZCCHC13 | | |
| 0,216901671 | ZCCHC16 | FALSE | Upregulated |
| | ZCCHC5 | | |
| 0,630931031 | ZCWPW2 | FALSE | Upregulated |
| -1,022333261 | ZDHHC13 | | Downregulated |
| 1,110530746 | ZDHHC19 | | Upregulated |
| 0,388147107 | ZDHHC22 | FALSE | Upregulated |
| -0,11544163 | ZDHHC8P1 | FALSE | Downregulated |
| 1,28279585 | ZEB1-AS1 | | Upregulated |
| -0,379217814 | ZFP37 | FALSE | Downregulated |
| | ZFP42 | | |
| 0,396269993 | ZFP57 | FALSE | Upregulated |
| 0,54265949 | ZFP69B | FALSE | Upregulated |
| -0,467354488 | ZFP82 | FALSE | Downregulated |
| 0,334319699 | ZFR2 | FALSE | Upregulated |
| 0,084976497 | ZFX-AS1 | FALSE | Upregulated |
| 0,57126419 | ZG16B | FALSE | Upregulated |
| -0,703099419 | ZHX1-C8ORF76 | FALSE | Downregulated |
| | ZIC2 | | |

| | | | |
|---|---|---|---|
| | ZIC3 | | |
| 0 | ZIC5 | FALSE | Upregulated |
| 0 | ZIM3 | FALSE | Upregulated |
| 0 | ZMAT4 | FALSE | Upregulated |
| -0,486093804 | ZMIZ1-AS1 | FALSE | Downregulated |
| -1,184968742 | ZMYND12 | | Downregulated |
| -0,032638142 | ZNF114 | FALSE | Downregulated |
| 0 | ZNF157 | FALSE | Upregulated |
| -0,250177886 | ZNF192P1 | FALSE | Downregulated |
| -1,042722924 | ZNF204P | | Downregulated |
| -0,161730751 | ZNF208 | FALSE | Downregulated |
| -0,794958558 | ZNF222 | FALSE | Downregulated |
| -0,251138747 | ZNF252P-AS1 | FALSE | Downregulated |
| 0,50149799 | ZNF280B | FALSE | Upregulated |
| -0,343237509 | ZNF285 | FALSE | Downregulated |
| 0,025130565 | ZNF286B | FALSE | Upregulated |
| 0,337962751 | ZNF295-AS1 | FALSE | Upregulated |
| 0,73178485 | ZNF300P1 | FALSE | Upregulated |
| | ZNF32-AS3 | | |
| -0,047826042 | ZNF321P | FALSE | Downregulated |
| -0,551196259 | ZNF33BP1 | FALSE | Downregulated |
| -0,450572978 | ZNF365 | FALSE | Downregulated |
| -0,674749363 | ZNF382 | FALSE | Downregulated |
| -0,372417547 | ZNF385D | FALSE | Downregulated |
| 0,670848702 | ZNF454 | FALSE | Upregulated |
| 0,426904102 | ZNF474 | FALSE | Upregulated |
| | ZNF479 | | |
| 0,228862569 | ZNF487 | FALSE | Upregulated |
| 0,118310792 | ZNF488 | FALSE | Upregulated |
| 0,095999889 | ZNF492 | FALSE | Upregulated |
| -1,318950692 | ZNF501 | | Downregulated |
| 0,930853312 | ZNF503-AS1 | FALSE | Upregulated |
| -0,485590317 | ZNF530 | FALSE | Downregulated |
| 0 | ZNF534 | FALSE | Upregulated |
| 0,390950345 | ZNF536 | FALSE | Upregulated |
| 0,003525741 | ZNF541 | FALSE | Upregulated |
| 0,426437581 | ZNF556 | FALSE | Upregulated |
| | ZNF560 | | |
| -0,95631092 | ZNF578 | FALSE | Downregulated |
| 0,421474701 | ZNF610 | FALSE | Upregulated |
| 2,0223244 | ZNF625-ZNF20 | | Upregulated |
| 0,467615931 | ZNF645 | FALSE | Upregulated |
| 0 | ZNF663P | FALSE | Upregulated |
| 0,745451293 | ZNF664-FAM101A | FALSE | Upregulated |
| 0,530130722 | ZNF665 | FALSE | Upregulated |
| 0,859282184 | ZNF670 | FALSE | Upregulated |
| 0 | ZNF670-ZNF695 | FALSE | Upregulated |
| -1,076413951 | ZNF674-AS1 | | Downregulated |
| -0,72246717 | ZNF675 | FALSE | Downregulated |
| -0,481359961 | ZNF676 | FALSE | Downregulated |
| 0,178603385 | ZNF677 | FALSE | Upregulated |

| | | | |
|---|---|---|---|
| | ZNF679 | | |
| 0,20208147 | ZNF681 | FALSE | Upregulated |
| 1,379496423 | ZNF683 | | Upregulated |
| 0,299305438 | ZNF695 | FALSE | Upregulated |
| -0,039205626 | ZNF702P | FALSE | Downregulated |
| | ZNF705A | | |
| | ZNF705B | | |
| | ZNF705D | | |
| -0,040153539 | ZNF705E | FALSE | Downregulated |
| | ZNF705G | | |
| -0,662774027 | ZNF713 | FALSE | Downregulated |
| | ZNF716 | | |
| 0,380031964 | ZNF724P | FALSE | Upregulated |
| -0,208576711 | ZNF726 | FALSE | Downregulated |
| -0,574923411 | ZNF727 | FALSE | Downregulated |
| -0,168834789 | ZNF728 | FALSE | Downregulated |
| | ZNF729 | | |
| 0,328448511 | ZNF730 | FALSE | Upregulated |
| | ZNF733P | | |
| | ZNF735 | | |
| -1,623633704 | ZNF738 | | Downregulated |
| 1,076686693 | ZNF774 | | Upregulated |
| 0,28040017 | ZNF781 | FALSE | Upregulated |
| | ZNF804B | | |
| 0,448212425 | ZNF806 | FALSE | Upregulated |
| 0,097246984 | ZNF807 | FALSE | Upregulated |
| 0,384972486 | ZNF812 | FALSE | Upregulated |
| 0,502600007 | ZNF815P | FALSE | Upregulated |
| 0 | ZNF816-ZNF321P | FALSE | Upregulated |
| -0,039228149 | ZNF826P | FALSE | Downregulated |
| -0,802537735 | ZNF835 | FALSE | Downregulated |
| -0,121392504 | ZNF85 | FALSE | Downregulated |
| -0,510331741 | ZNF860 | FALSE | Downregulated |
| -0,199139212 | ZNF883 | FALSE | Downregulated |
| 0,004795656 | ZNF890P | FALSE | Upregulated |
| 1,431532369 | ZNF90 | | Upregulated |
| -0,460326052 | ZNF93 | FALSE | Downregulated |
| | ZNF98 | | |
| -0,251760629 | ZNF99 | FALSE | Downregulated |
| | ZNRF4 | | |
| -0,140148275 | ZP1 | FALSE | Downregulated |
| | ZP2 | | |
| -1,319381307 | ZP3 | | Downregulated |
| | ZP4 | | |
| | ZPBP | | |
| | ZPBP2 | | |
| -0,377500679 | ZPLD1 | FALSE | Downregulated |
| 0,250185448 | ZRANB2-AS2 | FALSE | Upregulated |
| 0,27921206 | ZSCAN10 | FALSE | Upregulated |
| -0,389452881 | ZSCAN23 | FALSE | Downregulated |
| 0 | ZSCAN4 | FALSE | Upregulated |

1,443315062 ZSCAN5A Upregulated
-0,373498977 ZSCAN5B FALSE Downregulated
ZSWIM2
1,596452158 ZWINT Upregulated

**subTCMR vs. NHR**

| log2FoldChange | padj | gene_names | significance | direction |
|---|---|---|---|---|
| 1,545352747 | 0,001111939 | IL21R-AS1 | TRUE | Upregulated |
| 1,930622114 | 0,001111939 | RP11-77C3.3 | TRUE | Upregulated |
| 1,616118263 | 0,001111939 | SPOCK2 | TRUE | Upregulated |
| 2,256326744 | 0,001111939 | UBD | TRUE | Upregulated |
| -0,530668027 | 0,001430015 | RPL37A | FALSE | Downregulated |
| 1,234686497 | 0,001668528 | IL32 | TRUE | Upregulated |
| 1,322854328 | 0,001668528 | ZBED2 | TRUE | Upregulated |
| 0,643389048 | 0,00207781 | C1orf56 | FALSE | Upregulated |
| 1,283296808 | 0,003388214 | LINC00896 | TRUE | Upregulated |
| 1,364858169 | 0,003753416 | FAM111B | TRUE | Upregulated |
| 0,798464739 | 0,003753416 | FAM196B | FALSE | Upregulated |
| 1,268333327 | 0,003753416 | HLA-F-AS1 | TRUE | Upregulated |
| 1,283361932 | 0,004741925 | C1orf204 | TRUE | Upregulated |
| 0,776506431 | 0,004948184 | COA1 | FALSE | Upregulated |
| 0,897517652 | 0,005993821 | NLRC5 | FALSE | Upregulated |
| 2,495608706 | 0,005993821 | SAA2 | TRUE | Upregulated |
| 0,786103224 | 0,006290586 | MBNL1-AS1 | FALSE | Upregulated |
| 0,69323981 | 0,006290586 | RP6-206I17.2 | FALSE | Upregulated |
| 0,580704883 | 0,007305546 | FAM168A | FALSE | Upregulated |
| 0,439698871 | 0,007305546 | TMBIM1 | FALSE | Upregulated |
| 0,98643916 | 0,007648918 | APOBEC4 | FALSE | Upregulated |
| 1,38706732 | 0,007648918 | MTRNR2L8 | TRUE | Upregulated |
| 0,847793585 | 0,0094809 | GLS | FALSE | Upregulated |
| 1,172697679 | 0,012414864 | GLI1 | TRUE | Upregulated |
| 1,845733761 | 0,012414864 | IL2RG | TRUE | Upregulated |
| 2,218693407 | 0,012414864 | SAA1 | TRUE | Upregulated |
| 1,045269965 | 0,012414864 | TMEM75 | TRUE | Upregulated |
| 0,892088628 | 0,014400849 | LOC101927954 | FALSE | Upregulated |
| 0,608738835 | 0,016234585 | ZNF397 | FALSE | Upregulated |
| -0,468530167 | 0,016852433 | EPHB4 | FALSE | Downregulated |
| 1,190388749 | 0,019972303 | RUNX3 | TRUE | Upregulated |
| 0,784219152 | 0,021736015 | CELF2-AS1 | FALSE | Upregulated |
| 0,925832313 | 0,021736015 | HIVEP3 | FALSE | Upregulated |
| 0,804557424 | 0,021736015 | LOC102724536 | FALSE | Upregulated |
| 0,869986221 | 0,021736015 | LYSMD1 | FALSE | Upregulated |
| 0,850655537 | 0,021736015 | TAPSAR1 | FALSE | Upregulated |
| 0,778427193 | 0,02233045 | PRKAA2 | FALSE | Upregulated |
| -0,489204369 | 0,028499016 | AP1M1 | FALSE | Downregulated |
| 1,600635559 | 0,028499016 | BCL11B | TRUE | Upregulated |
| 0,81748622 | 0,028499016 | RP11-436I24.1 | FALSE | Upregulated |
| -0,385380691 | 0,030608115 | F2 | FALSE | Downregulated |
| 1,017784992 | 0,030608115 | LOC285762 | TRUE | Upregulated |
| 2,26269243 | 0,031633267 | ATP2A1 | TRUE | Upregulated |
| 0,813914286 | 0,031633267 | DOCK2 | FALSE | Upregulated |

| | | | | |
|---|---|---|---|---|
| 0,707461303 | 0,03261662 | PIGF | FALSE | Upregulated |
| 0,765258389 | 0,032871713 | CERKL | FALSE | Upregulated |
| 0,619106393 | 0,032871713 | ZNF433 | FALSE | Upregulated |
| 0,395931705 | 0,037168419 | CRYBG3 | FALSE | Upregulated |
| 0,685057676 | 0,037168419 | FAS-AS1 | FALSE | Upregulated |
| 1,721262365 | 0,038011143 | GPR132 | TRUE | Upregulated |
| 0,637457406 | 0,038438181 | SAP30L-AS1 | FALSE | Upregulated |
| 0,91262662 | 0,040653125 | PTGER4 | FALSE | Upregulated |
| -0,632871351 | 0,044435498 | EXOSC6 | FALSE | Downregulated |
| 0,610858122 | 0,045739256 | TLDC2 | FALSE | Upregulated |
| 0,518605685 | 0,04929799 | OGFRL1 | FALSE | Upregulated |
| 0,808009054 | 0,049469344 | ARSG | FALSE | Upregulated |
| 1,031314269 | 0,049493101 | LMLN | TRUE | Upregulated |
| 0,584647515 | 0,04950447 | DEXI | FALSE | Upregulated |
| 0,643772823 | 0,04950447 | LOC100506990 | FALSE | Upregulated |
| 0,988046844 | 0,04950447 | MUC4 | FALSE | Upregulated |
| 0,477914689 | 0,04950447 | PLXNC1 | FALSE | Upregulated |
| 1,035989788 | 0,04950447 | TCTEX1D4 | TRUE | Upregulated |
| 0,659275739 | 0,050266783 | LIMK2 | FALSE | Upregulated |
| -0,973681896 | 0,050266783 | SIGLEC11 | FALSE | Downregulated |
| -0,6758788 | 0,051078399 | FRRS1 | FALSE | Downregulated |
| 1,044913361 | 0,053524849 | FAM212A | FALSE | Upregulated |
| 1,157610228 | 0,054231148 | BIRC3 | FALSE | Upregulated |
| 0,915558861 | 0,054231148 | BYSL | FALSE | Upregulated |
| -0,450777697 | 0,055324285 | AHCY | FALSE | Downregulated |
| 0,464333545 | 0,055324285 | RP11-374P20.4 | FALSE | Upregulated |
| 1,136376098 | 0,055324285 | TRIM21 | FALSE | Upregulated |
| -0,33123979 | 0,056471798 | APP | FALSE | Downregulated |
| 0,780043585 | 0,056471798 | C1orf200 | FALSE | Upregulated |
| -0,41957235 | 0,056471798 | ZKSCAN1 | FALSE | Downregulated |
| -0,623345693 | 0,056576429 | DDX41 | FALSE | Downregulated |
| 1,090992098 | 0,056576429 | LOC101928926 | FALSE | Upregulated |
| -1,324292079 | 0,056576429 | SOWAHA | FALSE | Downregulated |
| 0,848515747 | 0,056813108 | LOC729683 | FALSE | Upregulated |
| 1,094528363 | 0,058887214 | LOC101927848 | FALSE | Upregulated |
| 1,854475979 | 0,059011192 | A2MP1 | FALSE | Upregulated |
| 0,505814274 | 0,059011192 | CD37 | FALSE | Upregulated |
| 1,060593748 | 0,059011192 | CLEC2D | FALSE | Upregulated |
| 0,960694307 | 0,059011192 | RP11-277P12.20 | FALSE | Upregulated |
| -0,472463017 | 0,059011192 | SARS | FALSE | Downregulated |
| 0,45910449 | 0,059086504 | KCNQ1OT1 | FALSE | Upregulated |
| -0,468005673 | 0,059086504 | LRRC47 | FALSE | Downregulated |
| 0,62746036 | 0,059086504 | SPRTN | FALSE | Upregulated |
| 1,184348 | 0,062918133 | MUC12 | FALSE | Upregulated |
| 0,945929259 | 0,063471426 | AQP4-AS1 | FALSE | Upregulated |
| 0,542186163 | 0,063471426 | CENPP | FALSE | Upregulated |
| 1,828931818 | 0,063471426 | CXCL9 | FALSE | Upregulated |
| 0,446200736 | 0,063471426 | LOC101927469 | FALSE | Upregulated |
| -0,833527275 | 0,063471426 | NYNRIN | FALSE | Downregulated |
| -0,310447709 | 0,063471426 | RPL3 | FALSE | Downregulated |
| 0,823182128 | 0,063471426 | TMC8 | FALSE | Upregulated |

| | | | | |
|---|---|---|---|---|
| 0,869697761 | 0,064121122 | FGD3 | FALSE | Upregulated |
| -0,870010284 | 0,065243358 | ADCY10 | FALSE | Downregulated |
| -0,746351894 | 0,065618128 | TBCA | FALSE | Downregulated |
| 0,454069177 | 0,065702489 | DHDDS | FALSE | Upregulated |
| 0,98023902 | 0,066679476 | APCDD1L | FALSE | Upregulated |
| 0,956199999 | 0,066679476 | IKZF3 | FALSE | Upregulated |
| 0,409332707 | 0,066679476 | ITPK1-AS1 | FALSE | Upregulated |
| -0,48895099 | 0,066679476 | KDR | FALSE | Downregulated |
| 0,588563482 | 0,071164903 | UNC13D | FALSE | Upregulated |
| 1,188170415 | 0,072443525 | KCNA3 | FALSE | Upregulated |
| 0,647667784 | 0,072787688 | TRIM17 | FALSE | Upregulated |
| -0,470305425 | 0,072839095 | HAO1 | FALSE | Downregulated |
| -0,289488591 | 0,072993412 | EEF1G | FALSE | Downregulated |
| 1,17870906 | 0,072993412 | ZAP70 | FALSE | Upregulated |
| -0,439197422 | 0,072994426 | ASPH | FALSE | Downregulated |
| -0,385169966 | 0,072994426 | GCGR | FALSE | Downregulated |
| 0,422483611 | 0,0794505 | NUTM2G | FALSE | Upregulated |
| -0,423462481 | 0,080067437 | UBA52 | FALSE | Downregulated |
| 0,463398242 | 0,080622198 | PDAP1 | FALSE | Upregulated |
| 0,546785767 | 0,080622198 | STK10 | FALSE | Upregulated |
| 0,381905913 | 0,080622198 | TMOD1 | FALSE | Upregulated |
| 0,302035566 | 0,086128552 | EPB41 | FALSE | Upregulated |
| 1,044168847 | 0,086128552 | FSTL3 | FALSE | Upregulated |
| 0,831260191 | 0,086128552 | IDI2-AS1 | FALSE | Upregulated |
| 0,405230508 | 0,086128552 | LINC00094 | FALSE | Upregulated |
| 0,620063594 | 0,086128552 | NIN | FALSE | Upregulated |
| 0,424373896 | 0,086128552 | RP11-133O22.6 | FALSE | Upregulated |
| 0,829409113 | 0,086128552 | RP11-401P9.5 | FALSE | Upregulated |
| -0,365307895 | 0,086128552 | RPL5 | FALSE | Downregulated |
| -0,291742652 | 0,086128552 | RPS15 | FALSE | Downregulated |
| 0,661140548 | 0,088164028 | LOC102724466 | FALSE | Upregulated |
| 0,655176509 | 0,088164028 | NFKB2 | FALSE | Upregulated |
| 0,53055725 | 0,089119297 | CCDC97 | FALSE | Upregulated |
| 0,665920759 | 0,089119297 | ITGB2 | FALSE | Upregulated |
| 0,845738978 | 0,089485776 | AMPD3 | FALSE | Upregulated |
| -0,239462668 | 0,089721729 | TPT1 | FALSE | Downregulated |
| 0,629172046 | 0,091349574 | MED19 | FALSE | Upregulated |
| 0,556559542 | 0,094672678 | ICA1L | FALSE | Upregulated |
| 0,777098992 | 0,094778591 | FAM115C | FALSE | Upregulated |
| 0,458186512 | 0,095032576 | FBXL2 | FALSE | Upregulated |
| -0,472204703 | 0,095032576 | MFAP3L | FALSE | Downregulated |
| 0,548923417 | 0,095032576 | RASSF4 | FALSE | Upregulated |
| -1,049151583 | 0,095032576 | RPUSD3 | FALSE | Downregulated |
| -0,993438496 | 0,096818373 | DNAJC12 | FALSE | Downregulated |
| -0,316213764 | 0,096818373 | HADHA | FALSE | Downregulated |
| 0,342137388 | 0,096818373 | MYO1F | FALSE | Upregulated |
| 1,177163543 | 0,096818373 | PTGER2 | FALSE | Upregulated |
| 0,505273907 | 0,096818373 | VASP | FALSE | Upregulated |
| 0,414005123 | 0,098524348 | DNAJC14 | FALSE | Upregulated |
| 2,073891266 | 0,098524348 | IRF4 | FALSE | Upregulated |
| 1,113727211 | 0,098524348 | JAK3 | FALSE | Upregulated |

| | | | | |
|---|---|---|---|---|
| -0,423289467 | 0,098524348 | KIAA1217 | FALSE | Downregulated |
| 0,915089431 | 0,098524348 | SMTNL1 | FALSE | Upregulated |
| 0,398295493 | 0,098524348 | ZNF233 | FALSE | Upregulated |
| -0,361175927 | 0,099127461 | AHSG | FALSE | Downregulated |
| 0,555970925 | 0,099127461 | CFLAR-AS1 | FALSE | Upregulated |
| 0,464110596 | 0,099127461 | IRF1 | FALSE | Upregulated |
| -0,46567473 | 0,099127461 | KBTBD11 | FALSE | Downregulated |
| 0,884972843 | 0,099127461 | LOC101928512 | FALSE | Upregulated |
| 0,448215991 | 0,099127461 | MINA | FALSE | Upregulated |
| 1,473872152 | 0,099127461 | POM121L9P | FALSE | Upregulated |
| 0,934520349 | 0,099127461 | PRRT2 | FALSE | Upregulated |
| -0,358459356 | 0,099127461 | RPS9 | FALSE | Downregulated |
| 1,186854057 | 0,099127461 | SLFN13 | FALSE | Upregulated |
| 0,240791754 | 0,099127461 | ZNF609 | FALSE | Upregulated |
| -0,302220477 | 0,0996998 | FAU | FALSE | Downregulated |
| 0,474081401 | 0,0996998 | RP5-1142A6.2 | FALSE | Upregulated |
| 0,663391909 | 0,101399748 | LRRC37A6P | FALSE | Upregulated |
| 1,169743795 | 0,102316664 | CD6 | FALSE | Upregulated |
| 0,538397934 | 0,103205668 | LOC441454 | FALSE | Upregulated |
| 0,637285649 | 0,103741926 | PHOSPHO1 | FALSE | Upregulated |
| 0,417979887 | 0,106999043 | B3GALT2 | FALSE | Upregulated |
| 1,795137983 | 0,106999043 | FER1L4 | FALSE | Upregulated |
| 0,503052495 | 0,106999043 | OGG1 | FALSE | Upregulated |
| 0,403739494 | 0,106999043 | SPDYE1 | FALSE | Upregulated |
| 0,681666271 | 0,107377373 | CCDC64 | FALSE | Upregulated |
| -0,324396419 | 0,108102949 | RPS5 | FALSE | Downregulated |
| -0,533373958 | 0,110472815 | IGFALS | FALSE | Downregulated |
| -0,461319696 | 0,11071216 | ACADSB | FALSE | Downregulated |
| -0,535368063 | 0,11071216 | CD82 | FALSE | Downregulated |
| 0,603158445 | 0,11071216 | COPG2IT1 | FALSE | Upregulated |
| 0,696666747 | 0,11071216 | FGD2 | FALSE | Upregulated |
| 0,464849242 | 0,11071216 | HTRA1 | FALSE | Upregulated |
| 0,944326494 | 0,11071216 | SNX32 | FALSE | Upregulated |
| 0,637964898 | 0,11071216 | SUV39H2 | FALSE | Upregulated |
| 0,659597604 | 0,11071216 | WIPI1 | FALSE | Upregulated |
| 1,237864505 | 0,111241301 | GPX7 | FALSE | Upregulated |
| 0,36524536 | 0,112480176 | CIITA | FALSE | Upregulated |
| 0,81814733 | 0,11270694 | SLC4A5 | FALSE | Upregulated |
| 0,938516476 | 0,114460405 | RP11-1H15.2 | FALSE | Upregulated |
| 1,062819867 | 0,114460405 | SH3D21 | FALSE | Upregulated |
| 0,838062962 | 0,115138009 | GPSM1 | FALSE | Upregulated |
| -0,525963917 | 0,115138009 | HSD17B2 | FALSE | Downregulated |
| -0,332339237 | 0,115138009 | UBB | FALSE | Downregulated |
| 0,496910192 | 0,11952124 | VSIG10 | FALSE | Upregulated |
| -0,969877112 | 0,121061477 | EMD | FALSE | Downregulated |
| 0,612646279 | 0,121061477 | LOC101928268 | FALSE | Upregulated |
| 0,632312615 | 0,121061477 | LOC642441 | FALSE | Upregulated |
| 0,392989944 | 0,121061477 | PAPLN | FALSE | Upregulated |
| 0,670555906 | 0,121061477 | SCXB | FALSE | Upregulated |
| -0,279925305 | 0,121061477 | TJP2 | FALSE | Downregulated |
| 0,343639528 | 0,121061477 | TRABD | FALSE | Upregulated |

| | | | | |
|---|---|---|---|---|
| 0,422609081 | 0,121061477 | VAMP1 | FALSE | Upregulated |
| -0,292715461 | 0,121061477 | ZFP36L1 | FALSE | Downregulated |
| 1,188679841 | 0,122355958 | IL7R | FALSE | Upregulated |
| 0,71808246 | 0,122355958 | LRRC18 | FALSE | Upregulated |
| 0,832833082 | 0,122355958 | RP11-118E18.4 | FALSE | Upregulated |
| -0,858786299 | 0,125332195 | ADRA2B | FALSE | Downregulated |
| -0,281622162 | 0,125332195 | RETSAT | FALSE | Downregulated |
| 0,939963356 | 0,126075412 | CHRNA2 | FALSE | Upregulated |
| 0,52994285 | 0,126075412 | FAM184B | FALSE | Upregulated |
| 0,446626892 | 0,127990797 | RECQL | FALSE | Upregulated |
| 2,145316248 | 0,128759713 | CXCL10 | FALSE | Upregulated |
| 0,904646444 | 0,128759713 | GBP5 | FALSE | Upregulated |
| 0,420422132 | 0,128759713 | LOC100288123 | FALSE | Upregulated |
| 1,076362261 | 0,130707166 | CARD11 | FALSE | Upregulated |
| -0,304561662 | 0,130707166 | SLC9A3R2 | FALSE | Downregulated |
| 1,262866855 | 0,132222569 | DTNA | FALSE | Upregulated |
| 0,68878263 | 0,132222569 | KCNAB2 | FALSE | Upregulated |
| 0,733473196 | 0,132222569 | RP11-379J5.5 | FALSE | Upregulated |
| 0,743982385 | 0,13267352 | RAP1GAP | FALSE | Upregulated |
| 0,452383494 | 0,133418443 | KLHL29 | FALSE | Upregulated |
| -0,393445672 | 0,136052279 | XPC | FALSE | Downregulated |
| -1,46596995 | 0,136206746 | GTF2IRD2 | FALSE | Downregulated |
| 1,031528431 | 0,136206746 | LOC100129697 | FALSE | Upregulated |
| 1,097744866 | 0,137294834 | LAIR1 | FALSE | Upregulated |
| 0,408580505 | 0,138157668 | ADPGK-AS1 | FALSE | Upregulated |
| 0,593544943 | 0,138157668 | INPP5D | FALSE | Upregulated |
| 1,122467521 | 0,138157668 | SSBP2 | FALSE | Upregulated |
| 0,846802676 | 0,140356542 | C14orf79 | FALSE | Upregulated |
| 0,540913694 | 0,140356542 | ZC3H12B | FALSE | Upregulated |
| -0,240890256 | 0,1408642 | EEF2 | FALSE | Downregulated |
| 0,521240348 | 0,1408642 | LOC100294362 | FALSE | Upregulated |
| 0,440960004 | 0,1408642 | SLC35G2 | FALSE | Upregulated |
| 0,4975392 | 0,141171098 | LOC102723451 | FALSE | Upregulated |
| -0,245789642 | 0,142347042 | RPL19 | FALSE | Downregulated |
| 0,576630842 | 0,142749031 | ALOX12 | FALSE | Upregulated |
| -1,073400025 | 0,142749031 | HTR7P1 | FALSE | Downregulated |
| 0,598193037 | 0,142749031 | LOC102723422 | FALSE | Upregulated |
| 0,615072525 | 0,142749031 | LOC728805 | FALSE | Upregulated |
| -0,441241122 | 0,142749031 | PGLYRP2 | FALSE | Downregulated |
| 0,439092335 | 0,142749031 | RAVER2 | FALSE | Upregulated |
| 0,902790203 | 0,144446959 | LOC101929837 | FALSE | Upregulated |
| -0,887871348 | 0,14565255 | RP11-475O6.1 | FALSE | Downregulated |
| 0,429214705 | 0,145741547 | RARA-AS1 | FALSE | Upregulated |
| 0,461276251 | 0,145786303 | COX15 | FALSE | Upregulated |
| -0,507663102 | 0,14607234 | ALDH1L1 | FALSE | Downregulated |
| 0,388831086 | 0,14607234 | TMEM14E | FALSE | Upregulated |
| 0,830163327 | 0,146880367 | SRGN | FALSE | Upregulated |
| 0,641129299 | 0,148167731 | RP5-977B1.7 | FALSE | Upregulated |
| 0,596379794 | 0,149693586 | PQLC3 | FALSE | Upregulated |
| -0,369069987 | 0,150107778 | EIF3L | FALSE | Downregulated |
| 1,231568427 | 0,150107778 | LINC00957 | FALSE | Upregulated |

| | | | | |
|---|---|---|---|---|
| 0,647239407 | 0,150107778 | PPP1R16B | FALSE | Upregulated |
| -0,282151807 | 0,150107778 | RPL36 | FALSE | Downregulated |
| 0,611471445 | 0,151149564 | TSPAN3 | FALSE | Upregulated |
| 0,453277808 | 0,151204643 | ARHGAP31 | FALSE | Upregulated |
| 1,565831831 | 0,151204643 | CD8A | FALSE | Upregulated |
| 0,486720949 | 0,151204643 | ETV6 | FALSE | Upregulated |
| -0,381689719 | 0,151204643 | HAAO | FALSE | Downregulated |
| 0,40072966 | 0,151204643 | M6PR | FALSE | Upregulated |
| 0,912779672 | 0,151204643 | XYLT1 | FALSE | Upregulated |
| 0,577026232 | 0,151666269 | LOC100506142 | FALSE | Upregulated |
| 1,146735827 | 0,153031932 | CCL5 | FALSE | Upregulated |
| 0,388097453 | 0,153924749 | PPP2R5D | FALSE | Upregulated |
| 1,3106665 | 0,154463451 | RALGPS1 | FALSE | Upregulated |
| 1,034202061 | 0,154562137 | IL24 | FALSE | Upregulated |
| -0,364465946 | 0,154562137 | ST13 | FALSE | Downregulated |
| 0,499903632 | 0,156699666 | INPP5K | FALSE | Upregulated |
| 0,788839598 | 0,156699666 | LOC101929668 | FALSE | Upregulated |
| 0,346180647 | 0,156699666 | MPEG1 | FALSE | Upregulated |
| -0,272233247 | 0,156789639 | AKT1 | FALSE | Downregulated |
| 0,903336256 | 0,157203735 | ALK | FALSE | Upregulated |
| 0,399810461 | 0,157593067 | AGBL5 | FALSE | Upregulated |
| -0,425810328 | 0,157593067 | ATP5D | FALSE | Downregulated |
| 0,33593262 | 0,157593067 | C15orf62 | FALSE | Upregulated |
| 0,395040537 | 0,157593067 | LOC644083 | FALSE | Upregulated |
| 0,740197874 | 0,157593067 | RP13-143G15.3 | FALSE | Upregulated |
| 0,527364419 | 0,157593067 | TUBB1 | FALSE | Upregulated |
| 0,429931518 | 0,157593067 | WDFY2 | FALSE | Upregulated |
| 0,377317514 | 0,157593067 | WDR48 | FALSE | Upregulated |
| -0,50929264 | 0,158014715 | BCHE | FALSE | Downregulated |
| 0,379594483 | 0,158014715 | CLOCK | FALSE | Upregulated |
| 0,586520565 | 0,158014715 | FLVCR2 | FALSE | Upregulated |
| 0,606840006 | 0,158014715 | LOC388282 | FALSE | Upregulated |
| -0,542411558 | 0,158014715 | METAP2 | FALSE | Downregulated |
| -0,264169238 | 0,158014715 | NFIA | FALSE | Downregulated |
| 1,035473743 | 0,158014715 | S100A9 | FALSE | Upregulated |
| 0,34338112 | 0,158014715 | SLC7A6 | FALSE | Upregulated |
| 0,343138625 | 0,158074004 | NPC1 | FALSE | Upregulated |
| -0,362331857 | 0,158621699 | ATF4 | FALSE | Downregulated |
| 0,485958243 | 0,158621699 | DCLRE1C | FALSE | Upregulated |
| 0,401051869 | 0,159443918 | AIRN | FALSE | Upregulated |
| 0,65489162 | 0,159443918 | LOC102724009 | FALSE | Upregulated |
| 0,380954269 | 0,159443918 | OLFML1 | FALSE | Upregulated |
| 1,119649642 | 0,159443918 | PSTPIP1 | FALSE | Upregulated |
| -0,309692162 | 0,159443918 | SERPINF2 | FALSE | Downregulated |
| 0,470569272 | 0,159443918 | TPI1P3 | FALSE | Upregulated |
| 1,315568415 | 0,159691184 | SLC1A3 | FALSE | Upregulated |
| 0,464550773 | 0,15972972 | MTMR2 | FALSE | Upregulated |
| -0,461483333 | 0,15972972 | PKLR | FALSE | Downregulated |
| 0,536833328 | 0,15972972 | RP11-84C10.4 | FALSE | Upregulated |
| 0,372521027 | 0,160012295 | CNTD1 | FALSE | Upregulated |
| -0,433568697 | 0,160012295 | IMPDH2 | FALSE | Downregulated |

| | | | | |
|---|---|---|---|---|
| 1,161289276 | 0,16206594 | OSBPL3 | FALSE | Upregulated |
| -0,74207243 | 0,16206594 | PNP | FALSE | Downregulated |
| 0,458633448 | 0,16206594 | TGFB1 | FALSE | Upregulated |
| -0,475107079 | 0,162164489 | RNF5 | FALSE | Downregulated |
| 0,590456669 | 0,16368885 | CD97 | FALSE | Upregulated |
| 0,663705778 | 0,16368885 | LOC101928212 | FALSE | Upregulated |
| 0,588035357 | 0,16387058 | FAM198B | FALSE | Upregulated |
| 0,607767277 | 0,164710015 | CEACAM19 | FALSE | Upregulated |
| 0,411495424 | 0,164710015 | FANCI | FALSE | Upregulated |
| -0,278854398 | 0,164710015 | MASP1 | FALSE | Downregulated |
| -0,660613695 | 0,164710015 | TFAM | FALSE | Downregulated |
| -0,409222043 | 0,164743907 | APOM | FALSE | Downregulated |
| 1,015771874 | 0,164743907 | GAS6-AS1 | FALSE | Upregulated |
| -0,411837186 | 0,164743907 | SPTBN2 | FALSE | Downregulated |
| 0,614450149 | 0,165120416 | DOK6 | FALSE | Upregulated |
| 0,597340852 | 0,165349911 | C12orf65 | FALSE | Upregulated |
| 0,773008068 | 0,165349911 | ENTPD1-AS1 | FALSE | Upregulated |
| -0,350487224 | 0,165349911 | METTL7A | FALSE | Downregulated |
| -1,072309714 | 0,165349911 | MMRN1 | FALSE | Downregulated |
| 1,375158789 | 0,165349911 | SLC7A14 | FALSE | Upregulated |
| 0,473745805 | 0,1661727 | C5orf66 | FALSE | Upregulated |
| 0,605653934 | 0,1661727 | DKFZP434H168 | FALSE | Upregulated |
| 0,66963789 | 0,1661727 | LOC101929732 | FALSE | Upregulated |
| 1,456268383 | 0,1661727 | LOC102723401 | FALSE | Upregulated |
| 0,72590701 | 0,1661727 | LOC388692 | FALSE | Upregulated |
| 0,353321255 | 0,1661727 | LRRTM2 | FALSE | Upregulated |
| -0,85749026 | 0,1661727 | PRDM11 | FALSE | Downregulated |
| 0,676215703 | 0,1661727 | RAPGEF5 | FALSE | Upregulated |
| 0,786446575 | 0,1661727 | RUNX1 | FALSE | Upregulated |
| 0,35181631 | 0,1661727 | UCKL1-AS1 | FALSE | Upregulated |
| 0,416693331 | 0,167615619 | LIMD2 | FALSE | Upregulated |
| -0,308942286 | 0,1684994 | STAB1 | FALSE | Downregulated |
| 0,348598409 | 0,168601985 | ANGPT2 | FALSE | Upregulated |
| 0,858440051 | 0,169070006 | ALOX5 | FALSE | Upregulated |
| 0,920246529 | 0,169070006 | LOC101927340 | FALSE | Upregulated |
| -0,415597715 | 0,169070006 | PON3 | FALSE | Downregulated |
| -0,310233741 | 0,169395826 | CPSF1 | FALSE | Downregulated |
| -0,331485496 | 0,169587551 | RN7SL2 | FALSE | Downregulated |
| -0,266874813 | 0,171119072 | C8orf82 | FALSE | Downregulated |
| 0,364038669 | 0,171119072 | ENTPD7 | FALSE | Upregulated |
| -0,657120396 | 0,17170687 | CYP2D7P | FALSE | Downregulated |
| 0,783002105 | 0,171965432 | MICAL1 | FALSE | Upregulated |
| 0,570516306 | 0,171965432 | TAP1 | FALSE | Upregulated |
| 0,385022557 | 0,17341863 | SRRM5 | FALSE | Upregulated |
| -0,387904526 | 0,174561657 | ALDOB | FALSE | Downregulated |
| -0,457074171 | 0,174561657 | ATP5G2 | FALSE | Downregulated |
| 1,572029205 | 0,174561657 | B3GNT5 | FALSE | Upregulated |
| 0,620252059 | 0,174561657 | CD44 | FALSE | Upregulated |
| -0,679825214 | 0,174561657 | CYGB | FALSE | Downregulated |
| -0,421123251 | 0,174561657 | SMOC1 | FALSE | Downregulated |
| -0,34498289 | 0,174589254 | ATP11C | FALSE | Downregulated |

| | | | | |
|---|---|---|---|---|
| 1,074597317 | 0,174589254 | FCN1 | FALSE | Upregulated |
| 0,566759776 | 0,174589254 | FMNL1 | FALSE | Upregulated |
| -0,238758918 | 0,174589254 | RPL13 | FALSE | Downregulated |
| 0,549209733 | 0,175517516 | TREX1 | FALSE | Upregulated |
| -0,360448159 | 0,176133947 | ITIH2 | FALSE | Downregulated |
| -0,454483304 | 0,176720855 | BHMT | FALSE | Downregulated |
| 0,813474117 | 0,176720855 | GINS2 | FALSE | Upregulated |
| 0,340358155 | 0,178578427 | DALRD3 | FALSE | Upregulated |
| -0,431805365 | 0,179368003 | MAP3K4 | FALSE | Downregulated |
| 0,334058015 | 0,180965913 | DLGAP4 | FALSE | Upregulated |
| 0,568706646 | 0,181018759 | CD160 | FALSE | Upregulated |
| 0,487334593 | 0,182309761 | GPSM2 | FALSE | Upregulated |
| 1,049338327 | 0,182309761 | KCNS1 | FALSE | Upregulated |
| 0,375364715 | 0,183470242 | CLK3 | FALSE | Upregulated |
| -0,380697256 | 0,183470242 | CTSF | FALSE | Downregulated |
| -0,443665843 | 0,183470242 | EIF3E | FALSE | Downregulated |
| 0,324294865 | 0,183470242 | FDPSP2 | FALSE | Upregulated |
| 1,506738326 | 0,183470242 | GSG2 | FALSE | Upregulated |
| 0,947923877 | 0,183470242 | ITGB2-AS1 | FALSE | Upregulated |
| -0,263051324 | 0,183470242 | MIA3 | FALSE | Downregulated |
| 1,166618405 | 0,183470242 | MKI67 | FALSE | Upregulated |
| -0,252581059 | 0,183470242 | RN7SK | FALSE | Downregulated |
| 0,411669906 | 0,183470242 | RPAIN | FALSE | Upregulated |
| -0,334070513 | 0,183470242 | SYVN1 | FALSE | Downregulated |
| 0,460420505 | 0,183470242 | ZNF550 | FALSE | Upregulated |
| 0,656450292 | 0,183553344 | LOC100506621 | FALSE | Upregulated |
| -0,498563039 | 0,183553344 | MDN1 | FALSE | Downregulated |
| -0,666698928 | 0,183553344 | SHH | FALSE | Downregulated |
| -0,526719444 | 0,184815896 | FAM211A-AS1 | FALSE | Downregulated |
| 0,672971839 | 0,184815896 | FCHSD1 | FALSE | Upregulated |
| 0,341282277 | 0,184815896 | FGD6 | FALSE | Upregulated |
| 0,470405749 | 0,184815896 | FLJ32756 | FALSE | Upregulated |
| -0,31605184 | 0,184815896 | GRHPR | FALSE | Downregulated |
| 1,025393476 | 0,184815896 | IFI6 | FALSE | Upregulated |
| -0,336212579 | 0,184815896 | ITIH1 | FALSE | Downregulated |
| -0,33890845 | 0,184815896 | LDHD | FALSE | Downregulated |
| -0,48336005 | 0,184815896 | PAPD5 | FALSE | Downregulated |
| -0,288818434 | 0,184815896 | RPS7 | FALSE | Downregulated |
| -0,439568992 | 0,184815896 | SLC22A1 | FALSE | Downregulated |
| 0,37686226 | 0,184815896 | SRR | FALSE | Upregulated |
| -1,082306599 | 0,184815896 | SYTL5 | FALSE | Downregulated |
| -0,368549632 | 0,184815896 | THRB | FALSE | Downregulated |
| 0,469847491 | 0,184815896 | WARS | FALSE | Upregulated |
| -0,277588423 | 0,185732996 | EEF1A1 | FALSE | Downregulated |
| -0,33504574 | 0,185732996 | EPHX2 | FALSE | Downregulated |
| 0,506810231 | 0,185732996 | RNF141 | FALSE | Upregulated |
| -0,421208636 | 0,186350335 | NR1H4 | FALSE | Downregulated |
| 0,541762516 | 0,186618187 | LOC101929714 | FALSE | Upregulated |
| -0,269230487 | 0,186618187 | RPL13A | FALSE | Downregulated |
| -0,432122431 | 0,186973179 | CYP4A11 | FALSE | Downregulated |
| 0,447378146 | 0,186973179 | KIAA0556 | FALSE | Upregulated |

| | | | | |
|---|---|---|---|---|
| 0,903840354 | 0,186973179 | LOC101927051 | FALSE | Upregulated |
| -0,393928049 | 0,186973179 | MKL2 | FALSE | Downregulated |
| 1,101307636 | 0,187089799 | COL5A3 | FALSE | Upregulated |
| 1,099936946 | 0,187089799 | LOC101927442 | FALSE | Upregulated |
| -0,409063587 | 0,187296427 | ALDH7A1 | FALSE | Downregulated |
| 0,978897514 | 0,189052585 | ANXA1 | FALSE | Upregulated |
| 0,327314331 | 0,189917604 | RGL4 | FALSE | Upregulated |
| 0,681688711 | 0,189917604 | WDFY4 | FALSE | Upregulated |
| 0,31444101 | 0,190123133 | ADCY7 | FALSE | Upregulated |
| 0,529962455 | 0,190123133 | LOC102724930 | FALSE | Upregulated |
| 0,314139645 | 0,190460132 | GGA1 | FALSE | Upregulated |
| 0,81230272 | 0,190460132 | LOC101929425 | FALSE | Upregulated |
| -0,208361761 | 0,190460132 | RPL41 | FALSE | Downregulated |
| 0,619576479 | 0,193239861 | DNAJC9-AS1 | FALSE | Upregulated |
| -0,431423652 | 0,193239861 | EIF1AX | FALSE | Downregulated |
| 0,539628947 | 0,193239861 | GPATCH4 | FALSE | Upregulated |
| 0,36114112 | 0,193239861 | LOC100128818 | FALSE | Upregulated |
| -0,298818044 | 0,193239861 | RBP4 | FALSE | Downregulated |
| -0,418041455 | 0,193584691 | HOMER2 | FALSE | Downregulated |
| 0,838351992 | 0,193584691 | LOC101927955 | FALSE | Upregulated |
| 0,695159688 | 0,194585182 | HIAT1 | FALSE | Upregulated |
| 0,516491376 | 0,194585182 | KIAA0895L | FALSE | Upregulated |
| 0,512810483 | 0,194585182 | LOC101927839 | FALSE | Upregulated |
| 0,567316266 | 0,194585182 | LOC641367 | FALSE | Upregulated |
| 1,047076843 | 0,194585182 | MARCH3 | FALSE | Upregulated |
| 0,341937213 | 0,194585182 | POU5F1P4 | FALSE | Upregulated |
| -0,217905739 | 0,194585182 | RPL4 | FALSE | Downregulated |
| 0,372140142 | 0,194585182 | SMG6 | FALSE | Upregulated |
| -0,683879868 | 0,194585182 | UQCRB | FALSE | Downregulated |
| -0,231152786 | 0,194585182 | XBP1 | FALSE | Downregulated |
| -0,598888052 | 0,194585182 | ZFYVE28 | FALSE | Downregulated |
| -0,715147318 | 0,194758636 | ADIRF | FALSE | Downregulated |
| 0,363902537 | 0,196451835 | GPR183 | FALSE | Upregulated |
| -0,356334373 | 0,196451835 | IVD | FALSE | Downregulated |
| 0,455363717 | 0,196790923 | ZNF469 | FALSE | Upregulated |
| -1,207707695 | 0,19765317 | PLSCR3 | FALSE | Downregulated |
| 0,341206478 | 0,19765317 | SKIV2L2 | FALSE | Upregulated |
| -0,37993216 | 0,19765317 | TXNRD2 | FALSE | Downregulated |
| 0,443843704 | 0,198008229 | GPX2 | FALSE | Upregulated |
| 0,891791603 | 0,198182018 | ZNF846 | FALSE | Upregulated |
| 0,844947186 | 0,198409518 | RP11-47I22.2 | FALSE | Upregulated |
| 0,363415497 | 0,199168499 | FANCA | FALSE | Upregulated |
| 0,536749292 | 0,199706643 | CHRNB1 | FALSE | Upregulated |
| 0,409192504 | 0,200207967 | CROCCP3 | FALSE | Upregulated |
| 0,556490487 | 0,201205156 | ANKS1B | FALSE | Upregulated |
| 1,041007797 | 0,201596149 | LOC101927030 | FALSE | Upregulated |
| -0,58558679 | 0,201596149 | RSRC2 | FALSE | Downregulated |
| -0,825389395 | 0,201670877 | CYP2C19 | FALSE | Downregulated |
| -0,238991873 | 0,201740544 | SHMT2 | FALSE | Downregulated |
| -0,256716383 | 0,20179858 | OBSL1 | FALSE | Downregulated |
| 0,559978413 | 0,20354472 | HTR6 | FALSE | Upregulated |

| | | | | |
|---|---|---|---|---|
| -0,428454758 | 0,20354472 | MTMR12 | FALSE | Downregulated |
| 0,846697649 | 0,20354472 | RP11-173B14.5 | FALSE | Upregulated |
| 0,458419612 | 0,205027768 | BRD7P3 | FALSE | Upregulated |
| -0,619645457 | 0,205766664 | PVRL3 | FALSE | Downregulated |
| 0,595716487 | 0,20618872 | LOC102724584 | FALSE | Upregulated |
| 0,694798082 | 0,207332573 | LILRB1 | FALSE | Upregulated |
| 1,066425361 | 0,207500443 | ATP10B | FALSE | Upregulated |
| -0,235808158 | 0,207500443 | CTDSP2 | FALSE | Downregulated |
| -0,480998394 | 0,207500443 | CTNNBL1 | FALSE | Downregulated |
| -0,333322714 | 0,207500443 | DPYD | FALSE | Downregulated |
| 0,369843893 | 0,207500443 | FCHSD2 | FALSE | Upregulated |
| -0,665737177 | 0,207500443 | FUNDC2 | FALSE | Downregulated |
| -0,338453895 | 0,207500443 | GAMT | FALSE | Downregulated |
| 0,338365859 | 0,207500443 | ICMT | FALSE | Upregulated |
| 0,407689455 | 0,207500443 | KRR1 | FALSE | Upregulated |
| -0,885304862 | 0,207500443 | MAGOH2 | FALSE | Downregulated |
| -0,481875113 | 0,207500443 | RORC | FALSE | Downregulated |
| 0,714441159 | 0,207500443 | SLC1A7 | FALSE | Upregulated |
| 0,430916493 | 0,207500443 | UNC119B | FALSE | Upregulated |
| -0,305209589 | 0,207500443 | UQCRC1 | FALSE | Downregulated |
| 0,496367598 | 0,207509648 | FAT2 | FALSE | Upregulated |
| -0,316839271 | 0,208617138 | APOC1 | FALSE | Downregulated |
| 0,345609025 | 0,208617138 | EVI2B | FALSE | Upregulated |
| -0,374462269 | 0,208617138 | PIK3R4 | FALSE | Downregulated |
| 0,434078569 | 0,208617138 | RP11-197N18.7 | FALSE | Upregulated |
| -0,282943862 | 0,208617138 | SERPINA1 | FALSE | Downregulated |
| -0,38457521 | 0,208619432 | BCLAF1 | FALSE | Downregulated |
| 0,390635507 | 0,208619432 | FAM175A | FALSE | Upregulated |
| 0,367610505 | 0,208619432 | SCAND2P | FALSE | Upregulated |
| 0,541445157 | 0,208982748 | ADAMTS2 | FALSE | Upregulated |
| 0,729021701 | 0,210701755 | FRMD5 | FALSE | Upregulated |
| -0,236926715 | 0,211009277 | RPS6 | FALSE | Downregulated |
| 1,005054484 | 0,211009277 | STXBP5-AS1 | FALSE | Upregulated |
| 0,513400349 | 0,211171169 | FCGR3A | FALSE | Upregulated |
| 1,337186103 | 0,211171169 | LOC101929309 | FALSE | Upregulated |
| 0,455467605 | 0,211871832 | GPR68 | FALSE | Upregulated |
| 0,426729889 | 0,212402249 | FLJ38668 | FALSE | Upregulated |
| -0,397953614 | 0,212402249 | HLCS | FALSE | Downregulated |
| 0,513023882 | 0,212614564 | CCDC18 | FALSE | Upregulated |
| -0,315747732 | 0,214119515 | ALAD | FALSE | Downregulated |
| -0,376157213 | 0,214181891 | PXMP2 | FALSE | Downregulated |
| 0,381536155 | 0,215517539 | BBS1 | FALSE | Upregulated |
| -0,288676036 | 0,215517539 | C16orf58 | FALSE | Downregulated |
| 0,576208708 | 0,215517539 | DOCK8 | FALSE | Upregulated |
| -0,385761953 | 0,215517539 | ENPP1 | FALSE | Downregulated |
| -0,315359952 | 0,215517539 | FIS1 | FALSE | Downregulated |
| 0,965372956 | 0,215517539 | POU5F1 | FALSE | Upregulated |
| 1,057622896 | 0,215517539 | PTAFR | FALSE | Upregulated |
| -0,614148615 | 0,215836973 | ABI3 | FALSE | Downregulated |
| 0,763847662 | 0,215836973 | AC020571.3 | FALSE | Upregulated |
| -0,459346652 | 0,215836973 | ALB | FALSE | Downregulated |

| | | | | |
|---|---|---|---|---|
| 0,6706946 | 0,215836973 | CRMP1 | FALSE | Upregulated |
| 0,891067221 | 0,215836973 | FAM150B | FALSE | Upregulated |
| 0,321157148 | 0,215836973 | GDF11 | FALSE | Upregulated |
| 0,880801705 | 0,215836973 | LOC102724597 | FALSE | Upregulated |
| 0,338177482 | 0,216566544 | IL17RB | FALSE | Upregulated |
| -0,29768897 | 0,219809963 | GNB2L1 | FALSE | Downregulated |
| 1,033522337 | 0,220212128 | GPR114 | FALSE | Upregulated |
| -0,470194302 | 0,2216318 | NDUFV2 | FALSE | Downregulated |
| -0,195154292 | 0,2216318 | SPEN | FALSE | Downregulated |
| 1,06168463 | 0,222887953 | C11orf45 | FALSE | Upregulated |
| -0,350487868 | 0,222887953 | GYS2 | FALSE | Downregulated |
| 0,690703262 | 0,222887953 | TRERF1 | FALSE | Upregulated |
| -0,236322045 | 0,222980506 | GLTSCR2 | FALSE | Downregulated |
| -0,236052429 | 0,222980506 | IGFBP4 | FALSE | Downregulated |
| 0,459565316 | 0,222980506 | LOC100506860 | FALSE | Upregulated |
| 0,795737252 | 0,222980506 | LOC101927296 | FALSE | Upregulated |
| 0,471650398 | 0,222980506 | MFSD7 | FALSE | Upregulated |
| -0,356920072 | 0,222980506 | ZNF358 | FALSE | Downregulated |
| 0,344705052 | 0,223568402 | ZNF544 | FALSE | Upregulated |
| 0,900089533 | 0,223631333 | LOC101927675 | FALSE | Upregulated |
| -0,361782542 | 0,223631333 | RPS4X | FALSE | Downregulated |
| 0,410783537 | 0,223631333 | SLC37A2 | FALSE | Upregulated |
| 0,70498001 | 0,224597936 | CR1 | FALSE | Upregulated |
| 0,792235446 | 0,224597936 | GJB7 | FALSE | Upregulated |
| 0,62136218 | 0,224597936 | METTL23 | FALSE | Upregulated |
| 0,374206777 | 0,224597936 | PYROXD1 | FALSE | Upregulated |
| 0,828107377 | 0,225187669 | DPM2 | FALSE | Upregulated |
| 0,777688331 | 0,225187669 | LMOD1 | FALSE | Upregulated |
| 0,426319937 | 0,225187669 | OGFOD1 | FALSE | Upregulated |
| -0,754090829 | 0,225187669 | TPPP2 | FALSE | Downregulated |
| 0,396305384 | 0,225485135 | LOC102723929 | FALSE | Upregulated |
| -0,501714723 | 0,225485135 | PTPRD | FALSE | Downregulated |
| 0,666806788 | 0,225485135 | UNC5A | FALSE | Upregulated |
| -0,210230232 | 0,225485307 | ARHGAP35 | FALSE | Downregulated |
| -0,323007994 | 0,225537879 | ABCC2 | FALSE | Downregulated |
| -0,267153203 | 0,225537879 | AKAP1 | FALSE | Downregulated |
| 0,395842858 | 0,225537879 | AKT3 | FALSE | Upregulated |
| -0,299182923 | 0,225537879 | ARGLU1 | FALSE | Downregulated |
| -0,51650851 | 0,225537879 | CCDC90B | FALSE | Downregulated |
| -0,325762503 | 0,225537879 | CYB5A | FALSE | Downregulated |
| -0,237348182 | 0,225537879 | EIF4H | FALSE | Downregulated |
| 0,837965182 | 0,225537879 | FOXP3 | FALSE | Upregulated |
| -0,715247067 | 0,225537879 | GLRX3 | FALSE | Downregulated |
| 0,786314281 | 0,225537879 | NFKBIE | FALSE | Upregulated |
| 0,656269596 | 0,225537879 | TRIM59 | FALSE | Upregulated |
| 0,344318831 | 0,225537879 | VGLL4 | FALSE | Upregulated |
| -0,389958091 | 0,225687814 | EPHX1 | FALSE | Downregulated |
| -0,760926184 | 0,225687814 | NAIF1 | FALSE | Downregulated |
| 0,388438701 | 0,22585705 | LOC100128653 | FALSE | Upregulated |
| 0,309032954 | 0,22585705 | ZNF276 | FALSE | Upregulated |
| 0,998822022 | 0,226758257 | CCL19 | FALSE | Upregulated |

| | | | | |
|---|---|---|---|---|
| -0,308422284 | 0,227107096 | EEF1B2 | FALSE | Downregulated |
| 0,707194983 | 0,227107096 | EVC | FALSE | Upregulated |
| 0,353062085 | 0,227107096 | PLEKHG6 | FALSE | Upregulated |
| -0,327554189 | 0,227107096 | ZGPAT | FALSE | Downregulated |
| 0,904206923 | 0,227206962 | PRF1 | FALSE | Upregulated |
| 1,05017773 | 0,228507101 | M1AP | FALSE | Upregulated |
| -0,320996212 | 0,228652927 | ATXN7L3B | FALSE | Downregulated |
| -0,33209872 | 0,228652927 | GC | FALSE | Downregulated |
| 0,488653723 | 0,228652927 | LCP1 | FALSE | Upregulated |
| 0,368150581 | 0,229347193 | ATP8B1 | FALSE | Upregulated |
| 0,655011177 | 0,229347193 | C9orf163 | FALSE | Upregulated |
| 0,434490569 | 0,229347193 | FAM186B | FALSE | Upregulated |
| -0,303832603 | 0,229347193 | VPS51 | FALSE | Downregulated |
| 0,346258558 | 0,229511221 | COL4A4 | FALSE | Upregulated |
| -0,339977276 | 0,229511221 | MBNL3 | FALSE | Downregulated |
| 0,538870629 | 0,229511221 | RARRES1 | FALSE | Upregulated |
| -0,267822545 | 0,229511221 | RPL30 | FALSE | Downregulated |
| -0,614874652 | 0,229511221 | SH3RF2 | FALSE | Downregulated |
| 0,531292851 | 0,229511221 | TAGLN | FALSE | Upregulated |
| -0,277710261 | 0,229929176 | APOB | FALSE | Downregulated |
| -0,363759949 | 0,229929176 | HSD17B10 | FALSE | Downregulated |
| 0,382255138 | 0,229929176 | LOC101928958 | FALSE | Upregulated |
| -0,234321632 | 0,230442514 | RPL29 | FALSE | Downregulated |
| -0,695071556 | 0,230748471 | GPR88 | FALSE | Downregulated |
| 0,301557453 | 0,230880377 | KPNA1 | FALSE | Upregulated |
| 0,395263874 | 0,230880377 | NAA15 | FALSE | Upregulated |
| -0,399083725 | 0,230880377 | PHYH | FALSE | Downregulated |
| -0,476858016 | 0,231382182 | OXR1 | FALSE | Downregulated |
| 1,078840749 | 0,231923349 | FCRL6 | FALSE | Upregulated |
| -0,354791414 | 0,231923349 | GCKR | FALSE | Downregulated |
| 0,710865752 | 0,231923349 | NABP1 | FALSE | Upregulated |
| -0,433477345 | 0,231923349 | UGT2B7 | FALSE | Downregulated |
| 0,67294681 | 0,232505653 | LOC101928069 | FALSE | Upregulated |
| 0,860344211 | 0,233428503 | IFI27 | FALSE | Upregulated |
| 0,388862275 | 0,233798946 | ERVFRD-1 | FALSE | Upregulated |
| 0,381671713 | 0,233798946 | ESYT3 | FALSE | Upregulated |
| 0,819496318 | 0,233798946 | SLAMF8 | FALSE | Upregulated |
| -0,416204976 | 0,234145338 | RNF216 | FALSE | Downregulated |
| 0,81809386 | 0,234145338 | ZNF732 | FALSE | Upregulated |
| -0,409106503 | 0,234303338 | ARPC1A | FALSE | Downregulated |
| 0,431193128 | 0,234646196 | SYNGAP1 | FALSE | Upregulated |
| 0,593127304 | 0,234845509 | CELF3 | FALSE | Upregulated |
| -0,26375021 | 0,23549796 | CERK | FALSE | Downregulated |
| -0,445101829 | 0,23549796 | DUSP6 | FALSE | Downregulated |
| 0,322126054 | 0,23549796 | GPR18 | FALSE | Upregulated |
| 0,358904704 | 0,23549796 | LOC101930421 | FALSE | Upregulated |
| 0,60734036 | 0,23549796 | LOC102724172 | FALSE | Upregulated |
| 0,573796983 | 0,23549796 | OGN | FALSE | Upregulated |
| -0,349592768 | 0,236192944 | ACSM2B | FALSE | Downregulated |
| -0,312889344 | 0,236717295 | AMBP | FALSE | Downregulated |
| -0,750088887 | 0,236717295 | DLL4 | FALSE | Downregulated |

| | | | | |
|---|---|---|---|---|
| -0,38584112 | 0,236717295 | EDC4 | FALSE | Downregulated |
| 0,988951106 | 0,236717295 | EPSTI1 | FALSE | Upregulated |
| -0,274097572 | 0,236717295 | RPL23A | FALSE | Downregulated |
| 0,928154313 | 0,236717295 | SLC16A14 | FALSE | Upregulated |
| 0,439981034 | 0,236717295 | SSBP3-AS1 | FALSE | Upregulated |
| -0,283478279 | 0,236717295 | TOR1AIP1 | FALSE | Downregulated |
| -0,574650556 | 0,236717295 | TSKU | FALSE | Downregulated |
| 0,40417914 | 0,237141082 | OSMR | FALSE | Upregulated |
| 0,517402364 | 0,237460397 | C7orf43 | FALSE | Upregulated |
| -0,248192371 | 0,237651215 | FAM20C | FALSE | Downregulated |
| 1,124740938 | 0,237651215 | RP11-856M7.1 | FALSE | Upregulated |
| -0,351392303 | 0,238796887 | RPL17 | FALSE | Downregulated |
| 0,267800533 | 0,239223913 | KIAA1432 | FALSE | Upregulated |
| -0,213638914 | 0,240651881 | ASGR1 | FALSE | Downregulated |
| 0,47434894 | 0,240651881 | BOLA3-AS1 | FALSE | Upregulated |
| -0,770173506 | 0,240651881 | C21orf59 | FALSE | Downregulated |
| -0,491726803 | 0,241910771 | GALK1 | FALSE | Downregulated |
| 0,756915251 | 0,242183739 | FHOD3 | FALSE | Upregulated |
| 0,756541505 | 0,243108341 | RP11-665G4.1 | FALSE | Upregulated |
| -0,308610208 | 0,243643386 | FOXK2 | FALSE | Downregulated |
| -0,407127618 | 0,244119831 | COLEC11 | FALSE | Downregulated |
| 0,353232026 | 0,245035317 | LOC101928042 | FALSE | Upregulated |
| -0,329615825 | 0,245035317 | TMEM176B | FALSE | Downregulated |
| 1,461024605 | 0,246021231 | FCAMR | FALSE | Upregulated |
| -0,599211924 | 0,246237478 | TMEM254 | FALSE | Downregulated |
| 0,393803004 | 0,246327705 | RARB | FALSE | Upregulated |
| -0,604372294 | 0,247379966 | CFHR4 | FALSE | Downregulated |
| 0,597596552 | 0,247379966 | FYB | FALSE | Upregulated |
| 0,672913107 | 0,247379966 | MFSD2A | FALSE | Upregulated |
| 0,703619249 | 0,247379966 | MIAT | FALSE | Upregulated |
| 0,830726465 | 0,247379966 | PARVG | FALSE | Upregulated |
| -0,323329741 | 0,247379966 | RPPH1 | FALSE | Downregulated |
| 0,215360977 | 0,247379966 | VPS13B | FALSE | Upregulated |
| -0,442870099 | 0,247553899 | DAK | FALSE | Downregulated |
| -0,690858212 | 0,247553899 | TXNL1 | FALSE | Downregulated |
| -0,348122562 | 0,247937453 | APOH | FALSE | Downregulated |
| -0,63935966 | 0,247937453 | CYP3A4 | FALSE | Downregulated |
| -0,520989522 | 0,247937453 | DBT | FALSE | Downregulated |
| -0,31419884 | 0,247937453 | DDI2 | FALSE | Downregulated |
| -0,353829733 | 0,247937453 | HADHB | FALSE | Downregulated |
| -0,225779256 | 0,247937453 | HECTD1 | FALSE | Downregulated |
| 0,553857457 | 0,247937453 | IFI16 | FALSE | Upregulated |
| 0,499329214 | 0,247937453 | JAK2 | FALSE | Upregulated |
| 0,651002694 | 0,247937453 | LOC101927940 | FALSE | Upregulated |
| 0,377749117 | 0,247937453 | LOC101929545 | FALSE | Upregulated |
| -0,282852554 | 0,247937453 | MAP2K2 | FALSE | Downregulated |
| 0,352945436 | 0,247937453 | MARCH1 | FALSE | Upregulated |
| -0,60319677 | 0,247937453 | MBTPS2 | FALSE | Downregulated |
| 0,40210165 | 0,247937453 | MIER3 | FALSE | Upregulated |
| 0,412614012 | 0,247937453 | MTF1 | FALSE | Upregulated |
| 0,432112425 | 0,247937453 | OSBP2 | FALSE | Upregulated |

| | | | | |
|---|---|---|---|---|
| 0,599865322 | 0,247937453 | PRPS2 | FALSE | Upregulated |
| 0,65427053 | 0,247937453 | PXN-AS1 | FALSE | Upregulated |
| -0,223267515 | 0,247937453 | RPL14 | FALSE | Downregulated |
| -0,27466205 | 0,247937453 | RPL7 | FALSE | Downregulated |
| 0,355810256 | 0,247937453 | SHANK2-AS1 | FALSE | Upregulated |
| -0,325364708 | 0,247937453 | SMG5 | FALSE | Downregulated |
| 0,732700102 | 0,247937453 | TFPT | FALSE | Upregulated |
| 0,459858841 | 0,247937453 | TMEM120B | FALSE | Upregulated |
| -0,305872106 | 0,248473081 | KLHDC10 | FALSE | Downregulated |
| 0,809890602 | 0,248473081 | LOC102723798 | FALSE | Upregulated |
| 0,265258191 | 0,248473081 | XRRA1 | FALSE | Upregulated |
| 0,491890645 | 0,248473081 | ZNF791 | FALSE | Upregulated |
| -0,530360448 | 0,248642016 | AGAP1 | FALSE | Downregulated |
| 0,318199376 | 0,248642016 | AKAP8 | FALSE | Upregulated |
| -0,344691568 | 0,248642016 | BDH1 | FALSE | Downregulated |
| -0,300142812 | 0,248642016 | CCNI | FALSE | Downregulated |
| -0,332779615 | 0,248642016 | COASY | FALSE | Downregulated |
| 0,826048889 | 0,248642016 | LOC102723726 | FALSE | Upregulated |
| -0,424199717 | 0,248642016 | LRRC58 | FALSE | Downregulated |
| 0,895637492 | 0,248642016 | LRRC9 | FALSE | Upregulated |
| 0,297639286 | 0,248642016 | LRTOMT | FALSE | Upregulated |
| 0,524885044 | 0,248642016 | PPAT | FALSE | Upregulated |
| -0,264320031 | 0,248642016 | PRPF6 | FALSE | Downregulated |
| 0,579781963 | 0,248642016 | RASSF1-AS1 | FALSE | Upregulated |
| -0,330582691 | 0,248642016 | TCEA3 | FALSE | Downregulated |
| -0,324314046 | 0,25145298 | ACSM2A | FALSE | Downregulated |
| 0,752607072 | 0,25145298 | MAP3K14-AS1 | FALSE | Upregulated |
| -0,429612853 | 0,251655029 | DCAKD | FALSE | Downregulated |
| 0,405092541 | 0,252207867 | AC009501.4 | FALSE | Upregulated |
| -0,312362928 | 0,252207867 | APOC3 | FALSE | Downregulated |
| 0,539942007 | 0,252207867 | DPCD | FALSE | Upregulated |
| 0,34847672 | 0,252207867 | FAM45A | FALSE | Upregulated |
| 0,694294942 | 0,252207867 | FNDC9 | FALSE | Upregulated |
| -0,331387348 | 0,252207867 | NUGGC | FALSE | Downregulated |
| 0,529776052 | 0,253218204 | NLGN3 | FALSE | Upregulated |
| 0,307732946 | 0,253218204 | PRR26 | FALSE | Upregulated |
| 0,739426847 | 0,253333978 | CORO1A | FALSE | Upregulated |
| -0,433155192 | 0,253373535 | SH3PXD2A | FALSE | Downregulated |
| 0,411460763 | 0,253783224 | BRSK2 | FALSE | Upregulated |
| 0,304370925 | 0,25470992 | RNF220 | FALSE | Upregulated |
| -0,485589075 | 0,254900109 | CNDP1 | FALSE | Downregulated |
| 0,290975684 | 0,255395391 | C12orf76 | FALSE | Upregulated |
| -0,673435798 | 0,255395391 | CACNA1H | FALSE | Downregulated |
| 0,627696376 | 0,255395391 | CLDN18 | FALSE | Upregulated |
| 0,356065338 | 0,255395391 | CRB2 | FALSE | Upregulated |
| 0,639396776 | 0,255395391 | DCTN5 | FALSE | Upregulated |
| -0,261278851 | 0,255395391 | HNRNPC | FALSE | Downregulated |
| 0,455952518 | 0,255395391 | IL10RA | FALSE | Upregulated |
| 1,334012776 | 0,255395391 | KNDC1 | FALSE | Upregulated |
| 0,509040386 | 0,255395391 | LOC101927898 | FALSE | Upregulated |
| 0,480405589 | 0,255395391 | LRRC37A | FALSE | Upregulated |

| | | | | |
|---|---|---|---|---|
| 0,439838734 | 0,255395391 | MFSD4 | FALSE | Upregulated |
| 0,245708467 | 0,255395391 | MSH6 | FALSE | Upregulated |
| 0,465462918 | 0,255395391 | NQO1 | FALSE | Upregulated |
| -0,307701262 | 0,255395391 | OGDHL | FALSE | Downregulated |
| -0,292266274 | 0,255395391 | PPM1A | FALSE | Downregulated |
| -0,437172312 | 0,255395391 | PPP4R2 | FALSE | Downregulated |
| -0,433956945 | 0,255395391 | SEC61B | FALSE | Downregulated |
| 0,675439331 | 0,255395391 | STBD1 | FALSE | Upregulated |
| -0,700599581 | 0,255395391 | ZFAND2A | FALSE | Downregulated |
| -0,314275639 | 0,255799724 | PGRMC1 | FALSE | Downregulated |
| 0,29691839 | 0,255897052 | ANKRD13A | FALSE | Upregulated |
| 0,619623768 | 0,255897052 | C21orf67 | FALSE | Upregulated |
| 0,89986125 | 0,255897052 | CD48 | FALSE | Upregulated |
| -0,330806141 | 0,255897052 | CEP164 | FALSE | Downregulated |
| -0,378106778 | 0,255897052 | GSTO1 | FALSE | Downregulated |
| 0,373425369 | 0,255897052 | IAH1 | FALSE | Upregulated |
| -0,283618243 | 0,255897052 | RPS25 | FALSE | Downregulated |
| 0,637898688 | 0,255897052 | TAP2 | FALSE | Upregulated |
| -0,809847369 | 0,255897052 | VPS72 | FALSE | Downregulated |
| -0,23536402 | 0,256457287 | CALM1 | FALSE | Downregulated |
| 1,283029525 | 0,256457287 | COL7A1 | FALSE | Upregulated |
| 0,56199237 | 0,256457287 | ERAP2 | FALSE | Upregulated |
| 0,294852424 | 0,256457287 | FGL2 | FALSE | Upregulated |
| 0,274592999 | 0,256457287 | LOC100379224 | FALSE | Upregulated |
| 1,102354588 | 0,256457287 | LOC101929015 | FALSE | Upregulated |
| 0,31334601 | 0,256457287 | PRH2 | FALSE | Upregulated |
| 0,30942646 | 0,256457287 | RHOF | FALSE | Upregulated |
| -0,294752467 | 0,256457287 | RN7SL1 | FALSE | Downregulated |
| 0,359205888 | 0,256457287 | RP11-231I16.1 | FALSE | Upregulated |
| -0,225651148 | 0,256457287 | RPLP0 | FALSE | Downregulated |
| -0,549565928 | 0,256457287 | SAMD5 | FALSE | Downregulated |
| 0,526451977 | 0,256457287 | SH3BGRL3 | FALSE | Upregulated |
| -0,355879072 | 0,256457287 | SLC22A3 | FALSE | Downregulated |
| -0,530143012 | 0,256457287 | TBC1D2 | FALSE | Downregulated |
| 0,285692855 | 0,256457287 | TINF2 | FALSE | Upregulated |
| -0,201856357 | 0,256773828 | ACADVL | FALSE | Downregulated |
| 0,832502105 | 0,257202537 | LOC102723317 | FALSE | Upregulated |
| 0,484786363 | 0,257879462 | CYBB | FALSE | Upregulated |
| 0,50428218 | 0,257879462 | IKZF2 | FALSE | Upregulated |
| -0,368193394 | 0,257879462 | NGFR | FALSE | Downregulated |
| -0,961860235 | 0,258240963 | OSBPL6 | FALSE | Downregulated |
| -0,784501408 | 0,258405747 | PNMA6A | FALSE | Downregulated |
| 0,810870735 | 0,258545927 | MYO1G | FALSE | Upregulated |
| -0,261516547 | 0,259079975 | MGAT2 | FALSE | Downregulated |
| 0,774540438 | 0,259597409 | SNORA81 | FALSE | Upregulated |
| 0,539911214 | 0,260015249 | STAT1 | FALSE | Upregulated |
| -0,343451698 | 0,260025903 | FAH | FALSE | Downregulated |
| -0,395067962 | 0,260068367 | RPS15A | FALSE | Downregulated |
| 0,367131049 | 0,260068367 | SPDYE6 | FALSE | Upregulated |
| -0,310213439 | 0,260740402 | AGT | FALSE | Downregulated |
| 0,768820265 | 0,260740402 | C17orf100 | FALSE | Upregulated |

| | | | | |
|---|---|---|---|---|
| -0,305990878 | 0,260740402 | CYP4F3 | FALSE | Downregulated |
| 0,445968237 | 0,260740402 | FOXD2 | FALSE | Upregulated |
| 1,086252189 | 0,260740402 | LOC101927963 | FALSE | Upregulated |
| 0,681981721 | 0,260740402 | LOC102724784 | FALSE | Upregulated |
| -0,298359579 | 0,260740402 | MAOA | FALSE | Downregulated |
| -0,252582718 | 0,260740402 | MARCH6 | FALSE | Downregulated |
| -0,25728337 | 0,260740402 | MPST | FALSE | Downregulated |
| -0,389927249 | 0,260740402 | MTG2 | FALSE | Downregulated |
| 0,542487682 | 0,260740402 | NAGK | FALSE | Upregulated |
| 0,446848365 | 0,260740402 | NBPF15 | FALSE | Upregulated |
| -0,467316453 | 0,260740402 | NMRK1 | FALSE | Downregulated |
| -0,551132548 | 0,260740402 | PDE7B | FALSE | Downregulated |
| 0,441662051 | 0,260740402 | PLAC4 | FALSE | Upregulated |
| 0,449118075 | 0,260740402 | PREX1 | FALSE | Upregulated |
| 0,349233531 | 0,260740402 | RAD9A | FALSE | Upregulated |
| 0,538882526 | 0,260740402 | RFX5 | FALSE | Upregulated |
| -0,245816413 | 0,260740402 | STAU1 | FALSE | Downregulated |
| 0,542085363 | 0,260740402 | SUPT4H1 | FALSE | Upregulated |
| 0,778645645 | 0,260740402 | SYNGR1 | FALSE | Upregulated |
| 0,361477014 | 0,260740402 | TMED1 | FALSE | Upregulated |
| -0,355217917 | 0,260740402 | VARS | FALSE | Downregulated |
| -0,687850206 | 0,261447463 | CXorf40B | FALSE | Downregulated |
| -0,28928172 | 0,261447463 | CYP27A1 | FALSE | Downregulated |
| 0,790574215 | 0,261447463 | ERV3-1 | FALSE | Upregulated |
| 0,755286728 | 0,261447463 | HMGCS1 | FALSE | Upregulated |
| 0,830815961 | 0,261447463 | LOC101928173 | FALSE | Upregulated |
| 0,933508947 | 0,261447463 | LOC388906 | FALSE | Upregulated |
| 0,515845217 | 0,261447463 | TRIP10 | FALSE | Upregulated |
| 0,312876686 | 0,261447463 | TSPAN4 | FALSE | Upregulated |
| -0,25698829 | 0,261447463 | VTN | FALSE | Downregulated |
| 0,413929654 | 0,261783703 | PRPF40B | FALSE | Upregulated |
| -0,395154431 | 0,262885781 | RTKN | FALSE | Downregulated |
| 0,68996714 | 0,262893237 | LMOD3 | FALSE | Upregulated |
| 0,465451909 | 0,263678139 | IL18BP | FALSE | Upregulated |
| 0,277191197 | 0,263678139 | POLH | FALSE | Upregulated |
| 0,368845585 | 0,263814555 | LOC101927178 | FALSE | Upregulated |
| 0,477762331 | 0,263814555 | PTPRC | FALSE | Upregulated |
| -0,434884539 | 0,263814555 | VKORC1 | FALSE | Downregulated |
| -0,358102441 | 0,263825897 | SNX27 | FALSE | Downregulated |
| -0,395276764 | 0,265022159 | C11orf54 | FALSE | Downregulated |
| 0,503802506 | 0,265022159 | GBP4 | FALSE | Upregulated |
| -0,236967284 | 0,265022159 | LRP3 | FALSE | Downregulated |
| -0,710734894 | 0,265022159 | NAPSB | FALSE | Downregulated |
| -0,27061338 | 0,265022159 | NUCKS1 | FALSE | Downregulated |
| -0,250217846 | 0,265022159 | PGM1 | FALSE | Downregulated |
| -0,272854452 | 0,265022159 | PTPRK | FALSE | Downregulated |
| 0,298604652 | 0,265022159 | UBA5 | FALSE | Upregulated |
| -0,598153166 | 0,265022159 | ZNF564 | FALSE | Downregulated |
| -0,597519314 | 0,265384539 | PCGF1 | FALSE | Downregulated |
| -0,48361321 | 0,265384539 | SLC3A2 | FALSE | Downregulated |
| -0,280975959 | 0,266049965 | HRG | FALSE | Downregulated |

| | | | | |
|---|---|---|---|---|
| 0,916153713 | 0,267440526 | AACS | FALSE | Upregulated |
| -0,68384869 | 0,267440526 | CD9 | FALSE | Downregulated |
| 0,319244804 | 0,268740995 | MSMP | FALSE | Upregulated |
| -0,53505189 | 0,268999444 | C17orf80 | FALSE | Downregulated |
| 0,638522573 | 0,268999444 | C8orf47 | FALSE | Upregulated |
| 0,269240746 | 0,268999444 | HAS3 | FALSE | Upregulated |
| -0,154068611 | 0,268999444 | HUWE1 | FALSE | Downregulated |
| 0,962361305 | 0,268999444 | LINC00319 | FALSE | Upregulated |
| 0,651590758 | 0,268999444 | LINC00612 | FALSE | Upregulated |
| 0,556072233 | 0,268999444 | LOC115110 | FALSE | Upregulated |
| 0,391304773 | 0,268999444 | LOC653513 | FALSE | Upregulated |
| -0,248706215 | 0,268999444 | PQLC1 | FALSE | Downregulated |
| -0,376011507 | 0,268999444 | PRDX6 | FALSE | Downregulated |
| 0,448738755 | 0,268999444 | PRPH2 | FALSE | Upregulated |
| -0,684747438 | 0,268999444 | RNU4-1 | FALSE | Downregulated |
| -0,288392366 | 0,268999444 | SEC62 | FALSE | Downregulated |
| -0,463121035 | 0,268999444 | SGSH | FALSE | Downregulated |
| -0,497644249 | 0,268999444 | SIVA1 | FALSE | Downregulated |
| 0,354122033 | 0,268999444 | SLC35A1 | FALSE | Upregulated |
| -0,667889189 | 0,268999444 | SLC35A2 | FALSE | Downregulated |
| -0,462185638 | 0,268999444 | TAF2 | FALSE | Downregulated |
| -0,309081379 | 0,269574807 | ECHS1 | FALSE | Downregulated |
| -0,340784869 | 0,269583881 | EIF1 | FALSE | Downregulated |
| -0,442891097 | 0,269583881 | FARSA | FALSE | Downregulated |
| -0,315278739 | 0,269583881 | FGFR4 | FALSE | Downregulated |
| -0,571607967 | 0,269583881 | HSD17B14 | FALSE | Downregulated |
| 0,585252912 | 0,269583881 | MYL5 | FALSE | Upregulated |
| -0,219100618 | 0,269583881 | RPS2 | FALSE | Downregulated |
| 0,391124901 | 0,269583881 | SRGAP2-AS1 | FALSE | Upregulated |
| -0,353358243 | 0,269583881 | VIPR1 | FALSE | Downregulated |
| 0,451845009 | 0,269583881 | ZNF124 | FALSE | Upregulated |
| 1,006078635 | 0,270665 | ZC3H12D | FALSE | Upregulated |
| -0,495825581 | 0,271397051 | ARSB | FALSE | Downregulated |
| -0,311123371 | 0,271397051 | SERPINC1 | FALSE | Downregulated |
| -0,303470674 | 0,271397051 | SLC25A5 | FALSE | Downregulated |
| 0,342151794 | 0,271751123 | C14orf93 | FALSE | Upregulated |
| -0,462428228 | 0,271751123 | SLCO1B3 | FALSE | Downregulated |
| 0,262653555 | 0,272161064 | VASN | FALSE | Upregulated |
| -0,446156792 | 0,272260917 | TTR | FALSE | Downregulated |
| 0,273007051 | 0,27258629 | RNF213 | FALSE | Upregulated |
| -0,355468028 | 0,272833634 | SAT2 | FALSE | Downregulated |
| 0,81415514 | 0,272971798 | PSMB9 | FALSE | Upregulated |
| 0,463573343 | 0,272971798 | RP11-864J10.4 | FALSE | Upregulated |
| 0,351716905 | 0,273601442 | FAM86A | FALSE | Upregulated |
| 0,300380963 | 0,27446859 | LRRC37A4P | FALSE | Upregulated |
| 0,758329861 | 0,275425655 | C1orf210 | FALSE | Upregulated |
| 0,91733109 | 0,275425655 | S100A1 | FALSE | Upregulated |
| -0,481070229 | 0,276085351 | EXPH5 | FALSE | Downregulated |
| 0,477104641 | 0,276085351 | NAALAD2 | FALSE | Upregulated |
| 0,308502273 | 0,276496276 | MFSD12 | FALSE | Upregulated |
| 0,522003966 | 0,276602608 | MAP3K19 | FALSE | Upregulated |

| | | | | |
|---|---|---|---|---|
| -0,426409494 | 0,277749826 | EIF3K | FALSE | Downregulated |
| -0,18512938 | 0,277749826 | PCBP2 | FALSE | Downregulated |
| -0,344015866 | 0,277749826 | RELN | FALSE | Downregulated |
| -0,360766024 | 0,277749826 | SMO | FALSE | Downregulated |
| -0,304618846 | 0,277938736 | TRIM33 | FALSE | Downregulated |
| -0,544307351 | 0,278696643 | ABCB6 | FALSE | Downregulated |
| -0,507051936 | 0,278872834 | CPEB3 | FALSE | Downregulated |
| -0,263983195 | 0,278872834 | NDUFV1 | FALSE | Downregulated |
| -0,465761968 | 0,279177866 | EIF3M | FALSE | Downregulated |
| -0,284686437 | 0,279177866 | GOT2 | FALSE | Downregulated |
| 0,534951563 | 0,279210945 | KLHDC7A | FALSE | Upregulated |
| -0,34689628 | 0,279210945 | LARP1B | FALSE | Downregulated |
| 0,28474395 | 0,279210945 | MAB21L2 | FALSE | Upregulated |
| 1,093403533 | 0,279210945 | SPEG | FALSE | Upregulated |
| -0,300184524 | 0,279210945 | WBP2 | FALSE | Downregulated |
| 0,392989998 | 0,2792805 | ATG4D | FALSE | Upregulated |
| -0,294809653 | 0,2792805 | CD14 | FALSE | Downregulated |
| 0,40767012 | 0,2792805 | GPR82 | FALSE | Upregulated |
| -0,752926533 | 0,2792805 | HEATR3 | FALSE | Downregulated |
| 0,556831359 | 0,2792805 | LGALS9 | FALSE | Upregulated |
| 0,308672598 | 0,2792805 | LOXL3 | FALSE | Upregulated |
| 0,477136828 | 0,2792805 | NEDD9 | FALSE | Upregulated |
| -0,919833137 | 0,2792805 | PLA2R1 | FALSE | Downregulated |
| -0,335551396 | 0,2792805 | PTGR1 | FALSE | Downregulated |
| 0,344191946 | 0,2792805 | RP11-143J12.2 | FALSE | Upregulated |
| -0,642493312 | 0,2792805 | SEC31B | FALSE | Downregulated |
| 0,927353187 | 0,2792805 | SLAMF7 | FALSE | Upregulated |
| -0,315599008 | 0,2792805 | ZCCHC24 | FALSE | Downregulated |
| -0,234439408 | 0,281071375 | A1CF | FALSE | Downregulated |
| -0,318008417 | 0,281071375 | DMGDH | FALSE | Downregulated |
| 0,465171597 | 0,281071375 | EMILIN2 | FALSE | Upregulated |
| 0,525117499 | 0,281071375 | GRAP | FALSE | Upregulated |
| 0,622050862 | 0,281071375 | KLHL25 | FALSE | Upregulated |
| -0,26385377 | 0,281071375 | REV1 | FALSE | Downregulated |
| 0,705477019 | 0,281071375 | SECTM1 | FALSE | Upregulated |
| 0,498271132 | 0,281071375 | SLC7A6OS | FALSE | Upregulated |
| 0,460584557 | 0,281153961 | LOC100996740 | FALSE | Upregulated |
| -0,207501417 | 0,281153961 | RERE | FALSE | Downregulated |
| 0,56091023 | 0,281153961 | RHBDF2 | FALSE | Upregulated |
| 0,680539885 | 0,281153961 | SIGLEC10 | FALSE | Upregulated |
| 0,304111291 | 0,281153961 | SNX21 | FALSE | Upregulated |
| 0,407719347 | 0,281766807 | SMURF2 | FALSE | Upregulated |
| -0,258363127 | 0,282225308 | GUSB | FALSE | Downregulated |
| 0,458384506 | 0,283234725 | SASH3 | FALSE | Upregulated |
| 0,596266618 | 0,283711148 | GRIP1 | FALSE | Upregulated |
| -0,217278765 | 0,283711148 | SNRNP200 | FALSE | Downregulated |
| 0,248245868 | 0,283777052 | ATG7 | FALSE | Upregulated |
| -0,269594273 | 0,283958282 | PABPC4 | FALSE | Downregulated |
| 0,27206222 | 0,284126322 | APPL2 | FALSE | Upregulated |
| -0,377235149 | 0,284126322 | C11orf31 | FALSE | Downregulated |
| -0,307674181 | 0,284126322 | CHP1 | FALSE | Downregulated |

| | | | | |
|---|---|---|---|---|
| -0,772722678 | 0,284126322 | LOC102724175 | FALSE | Downregulated |
| -0,702612371 | 0,284126322 | ST8SIA3 | FALSE | Downregulated |
| -0,38619056 | 0,284358715 | SURF1 | FALSE | Downregulated |
| -0,238353541 | 0,284361842 | FKBP8 | FALSE | Downregulated |
| -1,125995219 | 0,284361842 | IGF2 | FALSE | Downregulated |
| 0,35581492 | 0,284361842 | SIKE1 | FALSE | Upregulated |
| -0,369624664 | 0,284361842 | VPS28 | FALSE | Downregulated |
| 0,862093242 | 0,284389229 | KLHL6 | FALSE | Upregulated |
| -0,388860372 | 0,284545042 | DDOST | FALSE | Downregulated |
| -0,659773558 | 0,284545042 | FAM118A | FALSE | Downregulated |
| 0,480518746 | 0,284545042 | LARS2-AS1 | FALSE | Upregulated |
| -0,735619373 | 0,28488535 | TFAP4 | FALSE | Downregulated |
| -0,246137092 | 0,287050237 | BRD1 | FALSE | Downregulated |
| -0,227546215 | 0,287050237 | MAF | FALSE | Downregulated |
| -0,35877453 | 0,287050237 | RPL39 | FALSE | Downregulated |
| 0,380573037 | 0,287232891 | LOC102724499 | FALSE | Upregulated |
| -0,216807009 | 0,288634421 | DIAPH1 | FALSE | Downregulated |
| -0,202446881 | 0,28992514 | CTBP1 | FALSE | Downregulated |
| -0,45599318 | 0,28992514 | CYP2A6 | FALSE | Downregulated |
| -0,171716697 | 0,28992514 | SCAF1 | FALSE | Downregulated |
| -0,574781129 | 0,28992514 | SSTR1 | FALSE | Downregulated |
| -0,314214134 | 0,290826551 | TOMM20 | FALSE | Downregulated |
| 0,90962194 | 0,290826551 | ZNF80 | FALSE | Upregulated |
| 0,744503549 | 0,291536521 | ADM2 | FALSE | Upregulated |
| -0,315956505 | 0,291536521 | C8B | FALSE | Downregulated |
| 0,852512661 | 0,291536521 | GRHL1 | FALSE | Upregulated |
| 1,596264454 | 0,291536521 | PLA2G2A | FALSE | Upregulated |
| -0,560030714 | 0,291536521 | SAP130 | FALSE | Downregulated |
| 1,289080638 | 0,291960141 | LOC101927734 | FALSE | Upregulated |
| 0,330845081 | 0,291960141 | LYRM2 | FALSE | Upregulated |
| 0,800464138 | 0,291960141 | SFTPB | FALSE | Upregulated |
| -0,372077947 | 0,292413013 | PRPF19 | FALSE | Downregulated |
| 0,409653613 | 0,294178216 | LOC100128361 | FALSE | Upregulated |
| 0,695491055 | 0,294178216 | RP11-162J8.2 | FALSE | Upregulated |
| 0,724662977 | 0,294193685 | CHGA | FALSE | Upregulated |
| -0,300560986 | 0,294193685 | UROC1 | FALSE | Downregulated |
| 0,38376079 | 0,294634325 | LOC101928892 | FALSE | Upregulated |
| -0,237919464 | 0,294881812 | CLU | FALSE | Downregulated |
| -0,265422424 | 0,294881812 | F5 | FALSE | Downregulated |
| 0,282876701 | 0,294881812 | RHCE | FALSE | Upregulated |
| 0,515009373 | 0,296577224 | AJUBA | FALSE | Upregulated |
| 0,48394713 | 0,296577224 | ARL4C | FALSE | Upregulated |
| -0,49976965 | 0,296577224 | CREB3L2 | FALSE | Downregulated |
| 0,433358199 | 0,296577224 | FAM212B | FALSE | Upregulated |
| 0,614597816 | 0,296577224 | FLJ32255 | FALSE | Upregulated |
| -0,478562273 | 0,296873043 | PPP1R1A | FALSE | Downregulated |
| 0,204620738 | 0,296873043 | SLC46A1 | FALSE | Upregulated |
| 0,759610342 | 0,296987246 | PRSS54 | FALSE | Upregulated |
| 0,43168786 | 0,296987246 | SH3KBP1 | FALSE | Upregulated |
| -0,340610452 | 0,297078688 | TSR3 | FALSE | Downregulated |
| 0,267040278 | 0,297442793 | LOC102724445 | FALSE | Upregulated |

| | | | | |
|---|---|---|---|---|
| 0,49573281 | 0,297442793 | SELT | FALSE | Upregulated |
| 0,523721053 | 0,297679907 | LSP1 | FALSE | Upregulated |
| -0,215437483 | 0,297679907 | NACA | FALSE | Downregulated |
| -0,269178232 | 0,298138587 | FAM134C | FALSE | Downregulated |
| -0,437339642 | 0,298138587 | GBA3 | FALSE | Downregulated |
| -0,584961217 | 0,298237233 | CBX3 | FALSE | Downregulated |
| 0,36814287 | 0,298437392 | PTOV1-AS1 | FALSE | Upregulated |
| 0,414698707 | 0,298596575 | SATL1 | FALSE | Upregulated |
| -0,326726265 | 0,298657858 | ACAD10 | FALSE | Downregulated |
| -0,29077148 | 0,298657858 | FN1 | FALSE | Downregulated |
| -0,441932113 | 0,298657858 | IPO13 | FALSE | Downregulated |
| -0,369305097 | 0,298657858 | PID1 | FALSE | Downregulated |
| 0,512183777 | 0,298657858 | PLEKHF1 | FALSE | Upregulated |
| 0,736173028 | 0,298657858 | RP11-692N5.1 | FALSE | Upregulated |
| -0,470596684 | 0,298657858 | RRN3 | FALSE | Downregulated |
| -0,659312306 | 0,298657858 | SLCO4C1 | FALSE | Downregulated |
| 0,483664125 | 0,298657858 | STARD13-AS | FALSE | Upregulated |
| 0,561523255 | 0,298813455 | ABCC5 | FALSE | Upregulated |
| 1,020850238 | 0,298813455 | AC007461.2 | FALSE | Upregulated |
| 0,571229425 | 0,298813455 | FABP5 | FALSE | Upregulated |
| 0,879780517 | 0,298813455 | LOC101927911 | FALSE | Upregulated |
| 0,327971337 | 0,298813455 | PROX2 | FALSE | Upregulated |
| 0,240023002 | 0,298813455 | PTPMT1 | FALSE | Upregulated |
| -0,693415691 | 0,298813455 | RP11-7F17.5 | FALSE | Downregulated |
| 0,988323961 | 0,298813455 | S1PR2 | FALSE | Upregulated |
| 0,322580227 | 0,298813455 | SMPD3 | FALSE | Upregulated |
| -0,255294586 | 0,298813455 | TUFM | FALSE | Downregulated |
| 0,348307723 | 0,299897592 | PVR | FALSE | Upregulated |
| 0,397023363 | 0,30001689 | LOC101928291 | FALSE | Upregulated |
| 0,370458943 | 0,300743563 | SEMA4D | FALSE | Upregulated |
| -0,805525508 | 0,300912276 | MIR17HG | FALSE | Downregulated |
| -0,180209908 | 0,301785961 | EPN1 | FALSE | Downregulated |
| -0,368404658 | 0,301785961 | ERN1 | FALSE | Downregulated |
| 0,3498874 | 0,301785961 | PKD2L2 | FALSE | Upregulated |
| 0,498259978 | 0,30195419 | TBCK | FALSE | Upregulated |
| 0,423721403 | 0,302235313 | LOC101927057 | FALSE | Upregulated |
| -0,275178168 | 0,302267533 | CTSH | FALSE | Downregulated |
| 0,429723914 | 0,302267533 | HLA-B | FALSE | Upregulated |
| 0,609107187 | 0,302478151 | DOCK11 | FALSE | Upregulated |
| 0,378575842 | 0,302478151 | GK3P | FALSE | Upregulated |
| -0,232005654 | 0,302478151 | HAGH | FALSE | Downregulated |
| 0,40021426 | 0,302478151 | LOC100293704 | FALSE | Upregulated |
| 0,287369791 | 0,302478151 | LOC101927377 | FALSE | Upregulated |
| 0,380913708 | 0,302478151 | ZFP69 | FALSE | Upregulated |
| 0,46591413 | 0,302672747 | TTN | FALSE | Upregulated |
| 0,92356039 | 0,302672747 | ZNF831 | FALSE | Upregulated |
| -0,401622345 | 0,303436812 | C4A | FALSE | Downregulated |
| 0,570611575 | 0,303436812 | HRAS | FALSE | Upregulated |
| 0,443477867 | 0,303436812 | TP53I3 | FALSE | Upregulated |
| -0,2672189 | 0,30469638 | KANK2 | FALSE | Downregulated |
| -0,287691595 | 0,30469638 | NTN4 | FALSE | Downregulated |

| | | | | |
|---|---|---|---|---|
| 0,474643681 | 0,304710787 | C9orf172 | FALSE | Upregulated |
| -0,203730606 | 0,304710787 | HPN | FALSE | Downregulated |
| -0,523535736 | 0,304710787 | IGFBP2 | FALSE | Downregulated |
| 0,825102703 | 0,304710787 | RAX2 | FALSE | Upregulated |
| 0,546461793 | 0,304747245 | ZDHHC15 | FALSE | Upregulated |
| 0,92386727 | 0,305149797 | CD3E | FALSE | Upregulated |
| -0,703998287 | 0,305357833 | NVL | FALSE | Downregulated |
| 0,37795629 | 0,305458272 | TBC1D10C | FALSE | Upregulated |
| 0,315933386 | 0,306964837 | AAK1 | FALSE | Upregulated |
| 0,347377746 | 0,306964837 | SLCO1A2 | FALSE | Upregulated |
| 0,51721481 | 0,306964837 | ZCCHC9 | FALSE | Upregulated |
| -0,237510811 | 0,307033386 | ARG1 | FALSE | Downregulated |
| 1,096032798 | 0,307033386 | C6orf147 | FALSE | Upregulated |
| -0,26231838 | 0,307033386 | PHB2 | FALSE | Downregulated |
| -0,266458236 | 0,307033386 | PLG | FALSE | Downregulated |
| 0,492944738 | 0,307033386 | PTTG3P | FALSE | Upregulated |
| 0,372153121 | 0,307033386 | RPS6KL1 | FALSE | Upregulated |
| 1,553724576 | 0,307033386 | RUNX2 | FALSE | Upregulated |
| 0,343822353 | 0,307033386 | TYMP | FALSE | Upregulated |
| -0,366492163 | 0,307746758 | ADH6 | FALSE | Downregulated |
| -0,200099673 | 0,307746758 | CNBP | FALSE | Downregulated |
| 0,384749166 | 0,307746758 | DNMT1 | FALSE | Upregulated |
| 0,3872803 | 0,308717279 | CENPO | FALSE | Upregulated |
| -0,341533245 | 0,308717279 | FTCD | FALSE | Downregulated |
| 0,450203606 | 0,308717279 | LRRC37A2 | FALSE | Upregulated |
| -0,273076391 | 0,308717279 | MTDH | FALSE | Downregulated |
| 0,478834138 | 0,308717279 | RECQL4 | FALSE | Upregulated |
| 0,811886533 | 0,308717279 | TWISTNB | FALSE | Upregulated |
| 0,413801616 | 0,308717279 | VIM-AS1 | FALSE | Upregulated |
| 0,565540301 | 0,309650993 | SGK1 | FALSE | Upregulated |
| 0,80673728 | 0,30987028 | FMN1 | FALSE | Upregulated |
| 0,350203495 | 0,30987028 | GAS6 | FALSE | Upregulated |
| 0,299369067 | 0,30987028 | MED26 | FALSE | Upregulated |
| -0,234525905 | 0,30987028 | PKD1P1 | FALSE | Downregulated |
| 0,905505569 | 0,30987028 | PM20D1 | FALSE | Upregulated |
| 0,392922301 | 0,310018189 | KLF6 | FALSE | Upregulated |
| 0,349659954 | 0,310426183 | SLC35B2 | FALSE | Upregulated |
| -0,491190313 | 0,311671045 | ARSE | FALSE | Downregulated |
| -0,503034353 | 0,311671045 | ST7 | FALSE | Downregulated |
| -0,35240133 | 0,311964155 | CRTAP | FALSE | Downregulated |
| 0,576789196 | 0,311964155 | FAM163B | FALSE | Upregulated |
| 0,486594303 | 0,311964155 | GAS5-AS1 | FALSE | Upregulated |
| -0,287706451 | 0,311964155 | GLUD1 | FALSE | Downregulated |
| 0,520889424 | 0,311964155 | LOC101928429 | FALSE | Upregulated |
| -0,441654301 | 0,311964155 | SSR2 | FALSE | Downregulated |
| -0,357750439 | 0,311964155 | XYLB | FALSE | Downregulated |
| 0,939836948 | 0,312051456 | MIR3648 | FALSE | Upregulated |
| 0,316695073 | 0,312089808 | GSN-AS1 | FALSE | Upregulated |
| 0,551308731 | 0,312089808 | TAS1R3 | FALSE | Upregulated |
| -0,473159497 | 0,312139546 | CDC5L | FALSE | Downregulated |
| -0,398521337 | 0,312139546 | TMEM220 | FALSE | Downregulated |

| | | | | |
|---|---|---|---|---|
| -0,641367775 | 0,312139546 | ZNF593 | FALSE | Downregulated |
| -0,27938798 | 0,312811691 | IPO5 | FALSE | Downregulated |
| -0,337134221 | 0,313500241 | TINAGL1 | FALSE | Downregulated |
| 0,388679746 | 0,313537737 | ARHGAP17 | FALSE | Upregulated |
| -0,346134087 | 0,313537737 | DCXR | FALSE | Downregulated |
| -0,385364256 | 0,313537737 | KLHL21 | FALSE | Downregulated |
| 0,356667586 | 0,313537737 | STAR | FALSE | Upregulated |
| 0,455151511 | 0,313537737 | STK16 | FALSE | Upregulated |
| 0,307333686 | 0,313817016 | ACKR4 | FALSE | Upregulated |
| -0,198815255 | 0,313817016 | ORMDL3 | FALSE | Downregulated |
| 0,394491096 | 0,314385777 | B9D2 | FALSE | Upregulated |
| -0,280718263 | 0,314780963 | ALDH2 | FALSE | Downregulated |
| 0,527634849 | 0,314780963 | ZBTB32 | FALSE | Upregulated |
| 0,531407999 | 0,314836858 | CASP3 | FALSE | Upregulated |
| -0,32594564 | 0,314836858 | NDUFB10 | FALSE | Downregulated |
| -0,218648112 | 0,314836858 | RPS8 | FALSE | Downregulated |
| 0,300452989 | 0,315109556 | LOC100996746 | FALSE | Upregulated |
| 0,447802247 | 0,315124895 | RCE1 | FALSE | Upregulated |
| 0,416455664 | 0,315769172 | CCAT1 | FALSE | Upregulated |
| -0,202426049 | 0,316458166 | GOLGA4 | FALSE | Downregulated |
| -0,362029378 | 0,316458166 | HAO2 | FALSE | Downregulated |
| -0,226257937 | 0,316458166 | NCL | FALSE | Downregulated |
| -0,398838185 | 0,316458166 | TM4SF4 | FALSE | Downregulated |
| 0,453586248 | 0,316688584 | LANCL2 | FALSE | Upregulated |
| -0,431152009 | 0,317637535 | COQ4 | FALSE | Downregulated |
| 0,318704155 | 0,317637535 | LPAR6 | FALSE | Upregulated |
| 0,364969478 | 0,317637535 | MTRNR2L3 | FALSE | Upregulated |
| 0,34341649 | 0,317637535 | NLRC3 | FALSE | Upregulated |
| -0,50853876 | 0,317637535 | STAM2 | FALSE | Downregulated |
| 0,266187016 | 0,317637535 | THAP3 | FALSE | Upregulated |
| -0,897308809 | 0,317819049 | EGR1 | FALSE | Downregulated |
| 0,443651697 | 0,317819049 | PTPRCAP | FALSE | Upregulated |
| 1,047356973 | 0,317912491 | CRP | FALSE | Upregulated |
| -1,494744418 | 0,317912491 | FGF1 | FALSE | Downregulated |
| -0,832164126 | 0,317912491 | LOC100288778 | FALSE | Downregulated |
| 1,036299477 | 0,317912491 | LOC101929074 | FALSE | Upregulated |
| -0,479182978 | 0,317912491 | MGC12916 | FALSE | Downregulated |
| 0,260841252 | 0,317912491 | RHOQ | FALSE | Upregulated |
| 0,31905468 | 0,317912491 | SHE | FALSE | Upregulated |
| -0,306656589 | 0,318478016 | GAPVD1 | FALSE | Downregulated |
| 0,322798142 | 0,318478016 | LOC100132356 | FALSE | Upregulated |
| -0,673528849 | 0,318785987 | RP11-475O6.1-2 | FALSE | Downregulated |
| -0,229839659 | 0,31940706 | ITIH4 | FALSE | Downregulated |
| 0,859151877 | 0,31940706 | SLC9A5 | FALSE | Upregulated |
| 0,316056491 | 0,319661087 | CHRNB2 | FALSE | Upregulated |
| 0,421954116 | 0,319739185 | ARHGEF6 | FALSE | Upregulated |
| -0,290967363 | 0,319739185 | ARL6IP4 | FALSE | Downregulated |
| -0,481576407 | 0,319739185 | CFHR1 | FALSE | Downregulated |
| -0,334368246 | 0,319739185 | FLCN | FALSE | Downregulated |
| 0,336664024 | 0,319739185 | LOC100506603 | FALSE | Upregulated |
| -0,384151854 | 0,320198061 | PPP1CC | FALSE | Downregulated |

| | | | | |
|---|---|---|---|---|
| 0,45629016 | 0,320383512 | TWF1 | FALSE | Upregulated |
| -0,193806218 | 0,321020981 | EEF1D | FALSE | Downregulated |
| -0,56841934 | 0,322057355 | THAP7 | FALSE | Downregulated |
| -0,424164855 | 0,322130833 | MESDC2 | FALSE | Downregulated |
| -0,347058403 | 0,323026672 | RPL37 | FALSE | Downregulated |
| -0,367830051 | 0,323437014 | AKR1C1 | FALSE | Downregulated |
| 0,40623939 | 0,323481109 | LOC101930137 | FALSE | Upregulated |
| 0,697716255 | 0,323669782 | AVPR2 | FALSE | Upregulated |
| -0,589384623 | 0,324839287 | AOC4P | FALSE | Downregulated |
| -0,286084305 | 0,325089898 | IL6R | FALSE | Downregulated |
| 0,36398582 | 0,325405374 | NOL8 | FALSE | Upregulated |
| -0,441597243 | 0,325448142 | AADAC | FALSE | Downregulated |
| -0,44472243 | 0,325448142 | FRAT2 | FALSE | Downregulated |
| -0,294714847 | 0,325448142 | SLC27A5 | FALSE | Downregulated |
| 0,670723184 | 0,325951136 | LOC100506985 | FALSE | Upregulated |
| 0,30329284 | 0,326339752 | TMED10P1 | FALSE | Upregulated |
| -0,338832018 | 0,326374775 | MRPL38 | FALSE | Downregulated |
| -0,328713196 | 0,32640341 | ADH1B | FALSE | Downregulated |
| 0,339597558 | 0,326884123 | LOC101928862 | FALSE | Upregulated |
| 0,437561567 | 0,327192065 | LINC00854 | FALSE | Upregulated |
| 0,583363975 | 0,327255078 | LOC102723543 | FALSE | Upregulated |
| -0,304758441 | 0,327972268 | ZBTB40 | FALSE | Downregulated |
| -0,603672328 | 0,328068069 | POMP | FALSE | Downregulated |
| 0,296886379 | 0,329057696 | CALML4 | FALSE | Upregulated |
| 0,49030526 | 0,329875799 | INPP5F | FALSE | Upregulated |
| 0,475558142 | 0,329916185 | CYBA | FALSE | Upregulated |
| -0,41124027 | 0,329916185 | SLC30A10 | FALSE | Downregulated |
| 0,716742078 | 0,330951723 | TRPM2 | FALSE | Upregulated |
| -0,712337102 | 0,331768026 | ZNF778 | FALSE | Downregulated |
| 0,393794076 | 0,332260806 | IFIT3 | FALSE | Upregulated |
| 0,734297486 | 0,332260806 | PAPPA-AS1 | FALSE | Upregulated |
| -0,211869471 | 0,332504715 | RPL31 | FALSE | Downregulated |
| -0,271434438 | 0,332504715 | RPLP2 | FALSE | Downregulated |
| 0,445348576 | 0,332504715 | SLA2 | FALSE | Upregulated |
| -0,214636212 | 0,333008379 | LAMP1 | FALSE | Downregulated |
| -0,26589268 | 0,333033495 | ECHDC2 | FALSE | Downregulated |
| 0,824090913 | 0,334477234 | TYRO3 | FALSE | Upregulated |
| -0,305674236 | 0,334855859 | LETM1 | FALSE | Downregulated |
| -0,394319706 | 0,33522454 | CCDC130 | FALSE | Downregulated |
| 0,428129687 | 0,33522454 | LAPTM5 | FALSE | Upregulated |
| 0,710657748 | 0,335425652 | STK17B | FALSE | Upregulated |
| -0,20928682 | 0,335428712 | ASXL1 | FALSE | Downregulated |
| 0,514632656 | 0,335428712 | ZNF841 | FALSE | Upregulated |
| 0,486832076 | 0,335966061 | MYO7B | FALSE | Upregulated |
| 0,405934674 | 0,336045172 | HTRA2 | FALSE | Upregulated |
| -0,227153836 | 0,336045172 | PARD3 | FALSE | Downregulated |
| 0,526377954 | 0,336611568 | TMEM243 | FALSE | Upregulated |
| 0,58795564 | 0,337766441 | SLFN14 | FALSE | Upregulated |
| -0,395513559 | 0,338860272 | SUMF1 | FALSE | Downregulated |
| 0,648311755 | 0,338873465 | FAM120C | FALSE | Upregulated |
| -0,263475708 | 0,339992394 | RPS27 | FALSE | Downregulated |

| | | | | |
|---|---|---|---|---|
| 0,415647817 | 0,340432566 | GLIPR1L1 | FALSE | Upregulated |
| -0,514729459 | 0,34162148 | WDR74 | FALSE | Downregulated |
| 0,352544816 | 0,341927034 | FILIP1L | FALSE | Upregulated |
| -0,172207327 | 0,342582802 | RPS14 | FALSE | Downregulated |
| -0,273344845 | 0,342964372 | BGN | FALSE | Downregulated |
| -0,406204589 | 0,343088257 | ASB1 | FALSE | Downregulated |
| 0,298964606 | 0,343311266 | ATG14 | FALSE | Upregulated |
| 0,681212301 | 0,343311266 | BACH2 | FALSE | Upregulated |
| -0,237164079 | 0,343311266 | F7 | FALSE | Downregulated |
| 0,340654011 | 0,343311266 | LOC100128374 | FALSE | Upregulated |
| 0,651248525 | 0,343311266 | PHYHIPL | FALSE | Upregulated |
| 0,321739127 | 0,343311266 | SCUBE3 | FALSE | Upregulated |
| 0,623261894 | 0,343536436 | EML1 | FALSE | Upregulated |
| 0,461818986 | 0,343536436 | USP3-AS1 | FALSE | Upregulated |
| 0,918759382 | 0,343849828 | ST8SIA4 | FALSE | Upregulated |
| 0,622513242 | 0,344056851 | SNPH | FALSE | Upregulated |
| -0,358537267 | 0,344116026 | SRP72 | FALSE | Downregulated |
| 0,79731491 | 0,344687377 | RPS6KA2-AS1 | FALSE | Upregulated |
| -0,239321218 | 0,34578247 | ANPEP | FALSE | Downregulated |
| -0,225241766 | 0,34578247 | GRINA | FALSE | Downregulated |
| 0,750092052 | 0,34578247 | NUP93 | FALSE | Upregulated |
| 0,405801034 | 0,34578247 | PPAPDC2 | FALSE | Upregulated |
| -0,305522277 | 0,348124966 | APOC2 | FALSE | Downregulated |
| -0,364112986 | 0,348124966 | BET1L | FALSE | Downregulated |
| 0,312712873 | 0,348124966 | BTBD18 | FALSE | Upregulated |
| -0,214715037 | 0,348124966 | CAMSAP3 | FALSE | Downregulated |
| -0,292373186 | 0,348124966 | ETFA | FALSE | Downregulated |
| 0,555818276 | 0,348124966 | HECTD2 | FALSE | Upregulated |
| -0,18304767 | 0,348124966 | RXRA | FALSE | Downregulated |
| 0,575716421 | 0,348124966 | SLC8A1-AS1 | FALSE | Upregulated |
| 0,765446872 | 0,348124966 | SLIT2 | FALSE | Upregulated |
| -0,374386516 | 0,348124966 | USP33 | FALSE | Downregulated |
| 0,320234388 | 0,348561287 | BBIP1 | FALSE | Upregulated |
| 0,228696638 | 0,348561287 | FMNL3 | FALSE | Upregulated |
| -0,180672637 | 0,348561287 | LONP2 | FALSE | Downregulated |
| -0,320204319 | 0,348561287 | RPRD2 | FALSE | Downregulated |
| -0,262797817 | 0,348561287 | SDC2 | FALSE | Downregulated |
| 0,282678629 | 0,348561287 | SLC27A1 | FALSE | Upregulated |
| 0,249004986 | 0,348561287 | SPATA21 | FALSE | Upregulated |
| -0,256037047 | 0,348629325 | PUM1 | FALSE | Downregulated |
| 0,527453662 | 0,348739145 | LOC101059949 | FALSE | Upregulated |
| 0,310463722 | 0,349605787 | CCDC92 | FALSE | Upregulated |
| -0,437852721 | 0,349605787 | CYP4A22 | FALSE | Downregulated |
| 0,289509111 | 0,349858562 | KRI1 | FALSE | Upregulated |
| -0,344717098 | 0,349858562 | QPRT | FALSE | Downregulated |
| 0,75527982 | 0,350472097 | TMEM218 | FALSE | Upregulated |
| 0,319227117 | 0,350822273 | NSUN5 | FALSE | Upregulated |
| 0,446459862 | 0,350822273 | YPEL4 | FALSE | Upregulated |
| 0,478481623 | 0,350946708 | ODF2 | FALSE | Upregulated |
| 0,465117425 | 0,351837787 | ARHGAP9 | FALSE | Upregulated |
| -0,340600934 | 0,351837787 | PARP3 | FALSE | Downregulated |

| | | | | |
|---|---|---|---|---|
| -0,349792922 | 0,351837787 | RSL1D1 | FALSE | Downregulated |
| 0,322824671 | 0,351837787 | SPDYE2 | FALSE | Upregulated |
| 0,225496038 | 0,351837787 | VCL | FALSE | Upregulated |
| -0,268911971 | 0,351837787 | ZER1 | FALSE | Downregulated |
| 0,543822312 | 0,352200622 | USP28 | FALSE | Upregulated |
| 0,551364265 | 0,352410934 | NFAM1 | FALSE | Upregulated |
| 0,632124778 | 0,352720004 | AASDH | FALSE | Upregulated |
| 0,622913422 | 0,352720004 | C16orf89 | FALSE | Upregulated |
| 0,385143439 | 0,352720004 | FAM161B | FALSE | Upregulated |
| -0,38672416 | 0,352720004 | HSD17B8 | FALSE | Downregulated |
| 0,52048451 | 0,352720004 | IFI30 | FALSE | Upregulated |
| -0,286726018 | 0,352720004 | LIME1 | FALSE | Downregulated |
| 0,407484273 | 0,352720004 | MGC16275 | FALSE | Upregulated |
| -0,319773501 | 0,352720004 | ORAI3 | FALSE | Downregulated |
| 0,378656624 | 0,352720004 | PACS1 | FALSE | Upregulated |
| 0,660641208 | 0,352720004 | PLP2 | FALSE | Upregulated |
| -0,266656935 | 0,352720004 | RMRP | FALSE | Downregulated |
| -0,352767987 | 0,352720004 | SEC22B | FALSE | Downregulated |
| -0,274880319 | 0,352720004 | SMARCA1 | FALSE | Downregulated |
| -0,443582956 | 0,352720004 | SULT1A2 | FALSE | Downregulated |
| -0,289149707 | 0,352720004 | TANC1 | FALSE | Downregulated |
| -0,333633972 | 0,354100369 | RNF126 | FALSE | Downregulated |
| 0,228448774 | 0,354495503 | ADRBK1 | FALSE | Upregulated |
| 0,549523121 | 0,354495503 | C16orf3 | FALSE | Upregulated |
| 0,358530722 | 0,354495503 | CDAN1 | FALSE | Upregulated |
| 0,394327577 | 0,354495503 | LHX4 | FALSE | Upregulated |
| -0,607291677 | 0,354495503 | TMEM45A | FALSE | Downregulated |
| -0,159236321 | 0,35537013 | ZC3H7B | FALSE | Downregulated |
| -0,273221932 | 0,357658009 | CYC1 | FALSE | Downregulated |
| 0,316495381 | 0,357734328 | LSM4 | FALSE | Upregulated |
| 0,36423093 | 0,357784792 | CTSS | FALSE | Upregulated |
| 0,266010946 | 0,35832767 | BTBD9 | FALSE | Upregulated |
| 0,49650281 | 0,35832767 | RP11-14D22.5 | FALSE | Upregulated |
| -0,316597196 | 0,35877585 | SELENBP1 | FALSE | Downregulated |
| -0,248936461 | 0,358983863 | APOA2 | FALSE | Downregulated |
| -0,465996838 | 0,358983863 | APOC4 | FALSE | Downregulated |
| -0,576857281 | 0,358983863 | MEIS1 | FALSE | Downregulated |
| 0,36988895 | 0,358983863 | TSSK4 | FALSE | Upregulated |
| 0,392861917 | 0,359242844 | ALX3 | FALSE | Upregulated |
| -0,379833334 | 0,359402032 | COA4 | FALSE | Downregulated |
| -0,263666224 | 0,359402032 | CTSL | FALSE | Downregulated |
| 0,268534642 | 0,359402032 | EDC3 | FALSE | Upregulated |
| 0,761531926 | 0,359402032 | LOC154761 | FALSE | Upregulated |
| 0,67497655 | 0,359402032 | MPL | FALSE | Upregulated |
| -0,476860251 | 0,359402032 | POLR2F | FALSE | Downregulated |
| 0,557073434 | 0,359402032 | SLC2A11 | FALSE | Upregulated |
| 0,455164568 | 0,359681338 | AMZ1 | FALSE | Upregulated |
| 0,331185705 | 0,36146318 | RP4-591N18.2 | FALSE | Upregulated |
| 0,202872602 | 0,36146318 | SHISA5 | FALSE | Upregulated |
| 0,299910069 | 0,36146318 | TMEM80 | FALSE | Upregulated |
| 0,635190965 | 0,361665304 | FAM207A | FALSE | Upregulated |

| | | | | |
|---|---|---|---|---|
| -0,202437154 | 0,362194374 | AGMAT | FALSE | Downregulated |
| -0,239913547 | 0,362194374 | PEBP1 | FALSE | Downregulated |
| -0,406160344 | 0,362194374 | SCAMP2 | FALSE | Downregulated |
| -0,252677116 | 0,362324561 | TRAP1 | FALSE | Downregulated |
| 0,243879466 | 0,362768785 | ZMYND11 | FALSE | Upregulated |
| -0,207189179 | 0,362953178 | RNF130 | FALSE | Downregulated |
| 0,22648036 | 0,362953178 | TMEM200B | FALSE | Upregulated |
| -0,374765725 | 0,363347317 | GLYAT | FALSE | Downregulated |
| 0,338895904 | 0,363347317 | PPM1M | FALSE | Upregulated |
| -0,228580976 | 0,363347317 | RPS17 | FALSE | Downregulated |
| 0,387725267 | 0,363435512 | EVA1B | FALSE | Upregulated |
| 0,280915458 | 0,364061769 | ASCC1 | FALSE | Upregulated |
| 0,21043651 | 0,364170858 | TMEM140 | FALSE | Upregulated |
| -0,528884314 | 0,365545711 | TMTC4 | FALSE | Downregulated |
| -0,233340612 | 0,365691369 | OAF | FALSE | Downregulated |
| 0,415023098 | 0,365937445 | PLN | FALSE | Upregulated |
| 0,321237579 | 0,366438108 | LOC101928660 | FALSE | Upregulated |
| -0,400583364 | 0,366438108 | RPS29 | FALSE | Downregulated |
| -0,276857678 | 0,366682154 | AFMID | FALSE | Downregulated |
| 0,496580228 | 0,366682154 | CEACAM16 | FALSE | Upregulated |
| -0,401034513 | 0,366682154 | HPCAL1 | FALSE | Downregulated |
| 0,373959542 | 0,366682154 | NRAP | FALSE | Upregulated |
| 0,635350072 | 0,366682154 | PIGZ | FALSE | Upregulated |
| -0,286552707 | 0,367119914 | PECR | FALSE | Downregulated |
| -0,178689908 | 0,368115785 | CDC42BPB | FALSE | Downregulated |
| -0,40827446 | 0,368115785 | GSTA1 | FALSE | Downregulated |
| -0,305441615 | 0,368115785 | HGFAC | FALSE | Downregulated |
| 0,899928141 | 0,368115785 | LOC101927085 | FALSE | Upregulated |
| -0,288157684 | 0,368115785 | RNASE4 | FALSE | Downregulated |
| 0,222568664 | 0,368115785 | TCF7 | FALSE | Upregulated |
| 0,333062454 | 0,368260783 | IL7 | FALSE | Upregulated |
| 0,491618682 | 0,368761442 | SUCO | FALSE | Upregulated |
| 0,299888124 | 0,369389353 | HDAC7 | FALSE | Upregulated |
| -0,377228028 | 0,369389353 | ILVBL | FALSE | Downregulated |
| 0,600140313 | 0,369389353 | LOC101060553 | FALSE | Upregulated |
| -0,208311821 | 0,369389353 | PHF3 | FALSE | Downregulated |
| -0,219983429 | 0,369389353 | RPL12 | FALSE | Downregulated |
| -0,287923922 | 0,369389353 | XAB2 | FALSE | Downregulated |
| -0,41261845 | 0,369488136 | AKR1D1 | FALSE | Downregulated |
| 1,084750308 | 0,369488136 | FAM47E | FALSE | Upregulated |
| -0,574754162 | 0,369488136 | PPA2 | FALSE | Downregulated |
| 0,743427938 | 0,369860373 | DEF6 | FALSE | Upregulated |
| 0,317709933 | 0,369860373 | FLNA | FALSE | Upregulated |
| -0,196675762 | 0,369860373 | HELZ | FALSE | Downregulated |
| 0,267509912 | 0,369860373 | HNRNPAB | FALSE | Upregulated |
| 0,333945388 | 0,369979129 | PPP1R18 | FALSE | Upregulated |
| 0,389671108 | 0,370133596 | IL16 | FALSE | Upregulated |
| -0,34305624 | 0,370290137 | ATG4B | FALSE | Downregulated |
| 0,36342613 | 0,370290137 | CTNS | FALSE | Upregulated |
| 0,278168634 | 0,370290137 | ELK3 | FALSE | Upregulated |
| -0,466491261 | 0,370290137 | HIST1H4C | FALSE | Downregulated |

| | | | | |
|---|---|---|---|---|
| -0,25988465 | 0,370290137 | IRF2BP1 | FALSE | Downregulated |
| -0,257040812 | 0,370290137 | MAN2A1 | FALSE | Downregulated |
| 0,315196869 | 0,370290137 | MAPT | FALSE | Upregulated |
| 0,189893742 | 0,370290137 | MRPS25 | FALSE | Upregulated |
| 0,388622029 | 0,370290137 | PLEKHO2 | FALSE | Upregulated |
| -0,288235139 | 0,370290137 | PRODH2 | FALSE | Downregulated |
| 0,29094981 | 0,370290137 | USP35 | FALSE | Upregulated |
| 0,296272188 | 0,370294488 | ERCC1 | FALSE | Upregulated |
| 0,631440584 | 0,370294488 | SPATA25 | FALSE | Upregulated |
| -0,732102535 | 0,370294488 | TRPV3 | FALSE | Downregulated |
| 0,19524837 | 0,370604325 | AHSA2 | FALSE | Upregulated |
| 0,269618516 | 0,371180758 | APC2 | FALSE | Upregulated |
| 0,274054069 | 0,371180758 | C11orf57 | FALSE | Upregulated |
| -0,27166493 | 0,371180758 | CLIC4 | FALSE | Downregulated |
| -0,316115682 | 0,371180758 | HGD | FALSE | Downregulated |
| 0,42046138 | 0,371180758 | LINC00675 | FALSE | Upregulated |
| 0,722745664 | 0,371180758 | LOC92249 | FALSE | Upregulated |
| -0,211056134 | 0,371180758 | RANBP2 | FALSE | Downregulated |
| 0,264736258 | 0,371180758 | SPRN | FALSE | Upregulated |
| 0,370305463 | 0,371389759 | TUBB6 | FALSE | Upregulated |
| -0,512226775 | 0,371549784 | SCARNA6 | FALSE | Downregulated |
| 0,491239426 | 0,371719105 | CLDN7 | FALSE | Upregulated |
| -1,334936605 | 0,372881137 | FOXD4L4 | FALSE | Downregulated |
| 0,35763816 | 0,372881137 | LOC102724144 | FALSE | Upregulated |
| -0,348020511 | 0,372881137 | PPP3CA | FALSE | Downregulated |
| -0,277049194 | 0,37313238 | ALDH6A1 | FALSE | Downregulated |
| -0,53516129 | 0,37313238 | CLN3 | FALSE | Downregulated |
| 0,519117044 | 0,37313238 | LRIF1 | FALSE | Upregulated |
| 0,304642776 | 0,37313238 | OMG | FALSE | Upregulated |
| 0,248326763 | 0,37313238 | SMIM5 | FALSE | Upregulated |
| -0,364898715 | 0,373194732 | COLEC10 | FALSE | Downregulated |
| 0,481879779 | 0,373194732 | MMP25 | FALSE | Upregulated |
| -0,463809383 | 0,373539358 | AMD1 | FALSE | Downregulated |
| 0,363145548 | 0,373652071 | THOC2 | FALSE | Upregulated |
| 0,336301448 | 0,373652071 | WNT4 | FALSE | Upregulated |
| 0,971729248 | 0,37389144 | LOC101927256 | FALSE | Upregulated |
| -0,466867855 | 0,37389144 | NPR1 | FALSE | Downregulated |
| -0,699010699 | 0,37389144 | VIL1 | FALSE | Downregulated |
| 0,514186191 | 0,374016473 | MOB3A | FALSE | Upregulated |
| -0,195035869 | 0,374016473 | SERBP1 | FALSE | Downregulated |
| -0,696535882 | 0,374067562 | FGF14-AS2 | FALSE | Downregulated |
| 0,274809454 | 0,374067562 | NME7 | FALSE | Upregulated |
| -0,237250773 | 0,374067562 | PIPOX | FALSE | Downregulated |
| 0,302073319 | 0,374768453 | SPDYE5 | FALSE | Upregulated |
| -0,2667908 | 0,375285502 | RDX | FALSE | Downregulated |
| -0,193216026 | 0,375366574 | APLP2 | FALSE | Downregulated |
| -0,21531168 | 0,375366574 | SMG1 | FALSE | Downregulated |
| -0,380457177 | 0,375897291 | PAFAH2 | FALSE | Downregulated |
| 0,499808985 | 0,376400569 | LOC100996723 | FALSE | Upregulated |
| -0,448096946 | 0,377893611 | GCNT2 | FALSE | Downregulated |
| 0,429213668 | 0,377988673 | CALHM2 | FALSE | Upregulated |

| | | | | |
|---|---|---|---|---|
| 0,287673818 | 0,377988673 | POLR2C | FALSE | Upregulated |
| 0,825027325 | 0,377988673 | RASAL2-AS1 | FALSE | Upregulated |
| -0,306302491 | 0,378747577 | ASPDH | FALSE | Downregulated |
| 0,630142418 | 0,378747577 | LRRC16B | FALSE | Upregulated |
| -0,211225227 | 0,378747577 | RBM47 | FALSE | Downregulated |
| 0,830160929 | 0,378887464 | LOC101927529 | FALSE | Upregulated |
| 0,632588574 | 0,378887464 | ZSWIM7 | FALSE | Upregulated |
| -0,467772453 | 0,37931823 | PTGDR2 | FALSE | Downregulated |
| 0,316387829 | 0,379411978 | LOC100128882 | FALSE | Upregulated |
| 0,392869252 | 0,379488219 | RRS1 | FALSE | Upregulated |
| -0,358330396 | 0,379794395 | METTL13 | FALSE | Downregulated |
| 0,483843506 | 0,380403337 | CALCRL | FALSE | Upregulated |
| -0,217307539 | 0,380403337 | NADK2 | FALSE | Downregulated |
| -0,295847821 | 0,380591433 | GSDMB | FALSE | Downregulated |
| 0,595122502 | 0,380591433 | PHTF2 | FALSE | Upregulated |
| -0,321593565 | 0,380591433 | TM9SF2 | FALSE | Downregulated |
| 0,426489853 | 0,380591433 | ZNF333 | FALSE | Upregulated |
| -0,259030077 | 0,380591433 | ZNF91 | FALSE | Downregulated |
| -0,341419774 | 0,380691773 | FAF1 | FALSE | Downregulated |
| 0,463645818 | 0,380691773 | LOC100131395 | FALSE | Upregulated |
| 0,318842431 | 0,380691773 | PVRIG | FALSE | Upregulated |
| -0,254505513 | 0,380691773 | UPB1 | FALSE | Downregulated |
| -0,194836088 | 0,381077368 | FCGRT | FALSE | Downregulated |
| 0,233106231 | 0,381337297 | DNAJC3 | FALSE | Upregulated |
| -0,961729817 | 0,38136075 | CERS3 | FALSE | Downregulated |
| 0,300493529 | 0,38270821 | C12orf61 | FALSE | Upregulated |
| 0,336456453 | 0,38270821 | LOC102723520 | FALSE | Upregulated |
| 0,284883706 | 0,38270821 | RAB15 | FALSE | Upregulated |
| -0,453152779 | 0,38270821 | TMEM219 | FALSE | Downregulated |
| 0,264866414 | 0,383210097 | RBM8A | FALSE | Upregulated |
| 0,766309431 | 0,383597648 | ALPK2 | FALSE | Upregulated |
| -0,51263081 | 0,383597648 | ESAM | FALSE | Downregulated |
| 0,399471372 | 0,383597648 | LOC102724345 | FALSE | Upregulated |
| 0,404682796 | 0,383597648 | NBEAL2 | FALSE | Upregulated |
| -0,201460227 | 0,383597648 | SMARCA2 | FALSE | Downregulated |
| -0,240650788 | 0,383597648 | SND1 | FALSE | Downregulated |
| 0,650077087 | 0,383700462 | CCR8 | FALSE | Upregulated |
| 0,275447298 | 0,383700462 | CSRNP2 | FALSE | Upregulated |
| -0,242639737 | 0,383700462 | RPL10A | FALSE | Downregulated |
| 0,568199179 | 0,385025708 | FABP2 | FALSE | Upregulated |
| -0,36091245 | 0,385116388 | BPHL | FALSE | Downregulated |
| 0,422710294 | 0,386055652 | HCP5 | FALSE | Upregulated |
| -0,307870576 | 0,386096812 | ATMIN | FALSE | Downregulated |
| 0,238450374 | 0,386096812 | CDC42SE1 | FALSE | Upregulated |
| -0,255827842 | 0,386096812 | G3BP1 | FALSE | Downregulated |
| 0,539840826 | 0,386096812 | LOC101926985 | FALSE | Upregulated |
| 0,32536201 | 0,386096812 | LOC102724814 | FALSE | Upregulated |
| -0,299425415 | 0,386096812 | NR1I2 | FALSE | Downregulated |
| -0,281274751 | 0,386096812 | SLC17A9 | FALSE | Downregulated |
| -0,389645143 | 0,386096812 | SRP68 | FALSE | Downregulated |
| 0,704197566 | 0,387295061 | BTG2 | FALSE | Upregulated |

| | | | | |
|---|---|---|---|---|
| -0,219435341 | 0,387295061 | RPL23 | FALSE | Downregulated |
| 0,424853219 | 0,387295061 | SLC22A5 | FALSE | Upregulated |
| 0,277661976 | 0,387672737 | AGPAT4 | FALSE | Upregulated |
| 0,829529128 | 0,387672737 | LOC101928123 | FALSE | Upregulated |
| -0,193819043 | 0,387672737 | LRPAP1 | FALSE | Downregulated |
| -0,307904359 | 0,387672737 | MTFR1L | FALSE | Downregulated |
| 0,748846271 | 0,387672737 | ZNF678 | FALSE | Upregulated |
| 0,221409105 | 0,387768343 | CTBP1-AS | FALSE | Upregulated |
| 0,319624652 | 0,387772498 | NPHP3 | FALSE | Upregulated |
| -0,312153629 | 0,387772498 | PSMD8 | FALSE | Downregulated |
| 0,50579156 | 0,38781206 | LOC101929786 | FALSE | Upregulated |
| 0,726095286 | 0,38798434 | BCL10 | FALSE | Upregulated |
| -0,484374575 | 0,38798434 | NOP58 | FALSE | Downregulated |
| 0,271202606 | 0,388386473 | AKTIP | FALSE | Upregulated |
| -0,252151412 | 0,388386473 | ASMTL | FALSE | Downregulated |
| -0,33878474 | 0,388386473 | PLA2G12A | FALSE | Downregulated |
| -0,281753736 | 0,390294006 | PRDX2 | FALSE | Downregulated |
| 0,217437347 | 0,390433955 | ENG | FALSE | Upregulated |
| 0,478869122 | 0,390433955 | LRIG2 | FALSE | Upregulated |
| 0,923208877 | 0,391491475 | PDCD1 | FALSE | Upregulated |
| 0,460690378 | 0,391549342 | GDPD4 | FALSE | Upregulated |
| -0,26488387 | 0,391695823 | CAPN5 | FALSE | Downregulated |
| 0,34707049 | 0,391695823 | PTRH2 | FALSE | Upregulated |
| -0,631850075 | 0,392196605 | TYW1B | FALSE | Downregulated |
| -0,635550778 | 0,392522638 | SULT1E1 | FALSE | Downregulated |
| 0,535244331 | 0,392640395 | THBS2 | FALSE | Upregulated |
| 0,377295245 | 0,393168378 | LOC100128531 | FALSE | Upregulated |
| -0,878994288 | 0,393168378 | MEF2BNB | FALSE | Downregulated |
| -0,690615407 | 0,393503741 | LOC101929130 | FALSE | Downregulated |
| 0,340075298 | 0,393503741 | LOC153684 | FALSE | Upregulated |
| 0,428636141 | 0,393503741 | SENP8 | FALSE | Upregulated |
| 0,768811649 | 0,393503741 | ZNF107 | FALSE | Upregulated |
| -0,233031385 | 0,393620107 | RPL24 | FALSE | Downregulated |
| -0,202205268 | 0,393620107 | SEC31A | FALSE | Downregulated |
| -0,210915803 | 0,394132108 | AUTS2 | FALSE | Downregulated |
| -0,29366074 | 0,394132108 | IBTK | FALSE | Downregulated |
| 0,372298809 | 0,394236852 | ACOT8 | FALSE | Upregulated |
| 0,609796921 | 0,394236852 | ACSBG2 | FALSE | Upregulated |
| 0,55192224 | 0,394236852 | AHNAK2 | FALSE | Upregulated |
| 0,289002889 | 0,394236852 | AZI2 | FALSE | Upregulated |
| -0,320087758 | 0,394236852 | CAT | FALSE | Downregulated |
| -0,399599388 | 0,394236852 | CPOX | FALSE | Downregulated |
| -0,399488474 | 0,394236852 | DNAJB9 | FALSE | Downregulated |
| -0,430248449 | 0,394236852 | EMP2 | FALSE | Downregulated |
| -0,302201911 | 0,394236852 | ETFDH | FALSE | Downregulated |
| 0,213033364 | 0,394236852 | FRMD4A | FALSE | Upregulated |
| 0,279251037 | 0,394236852 | GNB4 | FALSE | Upregulated |
| 0,464649448 | 0,394236852 | HMGN3-AS1 | FALSE | Upregulated |
| -0,443345373 | 0,394236852 | IL1RAP | FALSE | Downregulated |
| -0,18767408 | 0,394236852 | KIAA2013 | FALSE | Downregulated |
| -0,261102622 | 0,394236852 | KLF12 | FALSE | Downregulated |

| | | | | |
|---|---|---|---|---|
| 0,631391466 | 0,394236852 | LOC100507191 | FALSE | Upregulated |
| 0,478751933 | 0,394236852 | LRRC20 | FALSE | Upregulated |
| 0,54748528 | 0,394236852 | LYZ | FALSE | Upregulated |
| 0,239029196 | 0,394236852 | MYRIP | FALSE | Upregulated |
| -0,764825021 | 0,394236852 | NCAM2 | FALSE | Downregulated |
| 0,246555165 | 0,394236852 | NNAT | FALSE | Upregulated |
| 0,460468344 | 0,394236852 | PKM | FALSE | Upregulated |
| 0,532326744 | 0,394236852 | PRUNE2 | FALSE | Upregulated |
| -0,328810672 | 0,394236852 | PTPRG | FALSE | Downregulated |
| 0,350583304 | 0,394236852 | RCC2 | FALSE | Upregulated |
| 0,453551522 | 0,394236852 | RP3-325F22.3 | FALSE | Upregulated |
| 0,504723697 | 0,394236852 | RPA2 | FALSE | Upregulated |
| 0,437410856 | 0,394236852 | RSPH3 | FALSE | Upregulated |
| -0,293021233 | 0,394236852 | SCCPDH | FALSE | Downregulated |
| -0,284643076 | 0,394236852 | SERPINA6 | FALSE | Downregulated |
| -0,304967254 | 0,394236852 | SLC2A2 | FALSE | Downregulated |
| 0,258062067 | 0,394236852 | SLC31A2 | FALSE | Upregulated |
| -0,315618265 | 0,394236852 | SORD | FALSE | Downregulated |
| 0,628387613 | 0,394236852 | SRMS | FALSE | Upregulated |
| 0,264260188 | 0,394236852 | STK17A | FALSE | Upregulated |
| -0,617715088 | 0,394236852 | STK38L | FALSE | Downregulated |
| 0,330766055 | 0,394236852 | TSPAN31 | FALSE | Upregulated |
| 0,221189896 | 0,394236852 | WBSCR16 | FALSE | Upregulated |
| -0,240979189 | 0,394236852 | ZCCHC14 | FALSE | Downregulated |
| -0,525216255 | 0,394236852 | ZDHHC4 | FALSE | Downregulated |
| -0,603758134 | 0,394236852 | ZNF334 | FALSE | Downregulated |
| -0,393827953 | 0,395324108 | SLC16A1 | FALSE | Downregulated |
| 0,463742306 | 0,395517742 | INSIG1 | FALSE | Upregulated |
| -0,520989907 | 0,395517742 | RGS5 | FALSE | Downregulated |
| -0,403187036 | 0,395518219 | NEU1 | FALSE | Downregulated |
| 0,277725037 | 0,395836304 | MDM2 | FALSE | Upregulated |
| -0,219051232 | 0,396770829 | SORBS2 | FALSE | Downregulated |
| 0,444880765 | 0,396906463 | TCEB3B | FALSE | Upregulated |
| -0,290595436 | 0,397096056 | VARS2 | FALSE | Downregulated |
| 0,236649532 | 0,397153168 | GORASP1 | FALSE | Upregulated |
| -0,597011271 | 0,397153168 | TRPT1 | FALSE | Downregulated |
| -0,389742711 | 0,397976604 | KYNU | FALSE | Downregulated |
| 1,022050185 | 0,397976604 | RYR1 | FALSE | Upregulated |
| -0,370801945 | 0,39810733 | HMGCS2 | FALSE | Downregulated |
| 0,512023445 | 0,399137801 | C1orf111 | FALSE | Upregulated |
| 0,313133735 | 0,399137801 | PHF20 | FALSE | Upregulated |
| 0,728329713 | 0,399137801 | RP4-777D9.2 | FALSE | Upregulated |
| 0,37453488 | 0,399191298 | ARHGAP4 | FALSE | Upregulated |
| 0,240696471 | 0,39960344 | ACOT13 | FALSE | Upregulated |
| -0,568502903 | 0,39960344 | C7orf26 | FALSE | Downregulated |
| 0,38401687 | 0,400151573 | ASAP3 | FALSE | Upregulated |
| 0,40393532 | 0,400151573 | BTN3A1 | FALSE | Upregulated |
| -0,29056557 | 0,400151573 | CES1 | FALSE | Downregulated |
| -0,295736823 | 0,400151573 | CPB2 | FALSE | Downregulated |
| 0,883165878 | 0,400151573 | GPRIN1 | FALSE | Upregulated |
| 0,810898909 | 0,400151573 | MX2 | FALSE | Upregulated |

| | | | | |
|---|---|---|---|---|
| -0,443667315 | 0,400151573 | PITPNM3 | FALSE | Downregulated |
| 0,389283835 | 0,400151573 | RBFOX2 | FALSE | Upregulated |
| -0,292153339 | 0,400151573 | RHOBTB3 | FALSE | Downregulated |
| 0,266249007 | 0,400151573 | RP5-886K2.3 | FALSE | Upregulated |
| -0,287777873 | 0,400151573 | SFT2D3 | FALSE | Downregulated |
| -0,253205749 | 0,400151573 | THRAP3 | FALSE | Downregulated |
| -0,472091893 | 0,400151573 | TMEM184B | FALSE | Downregulated |
| -0,476691436 | 0,400151573 | VPS11 | FALSE | Downregulated |
| 0,447618886 | 0,400151573 | WIPF1 | FALSE | Upregulated |
| 0,549282312 | 0,400151573 | ZFAT-AS1 | FALSE | Upregulated |
| 0,246372801 | 0,400151573 | ZMYM2 | FALSE | Upregulated |
| -0,277632083 | 0,40037654 | ACBD4 | FALSE | Downregulated |
| -0,680010032 | 0,40037654 | C6orf211 | FALSE | Downregulated |
| -0,437830677 | 0,40037654 | CENPT | FALSE | Downregulated |
| 0,51625817 | 0,40037654 | LOC100129722 | FALSE | Upregulated |
| -0,252333269 | 0,40037654 | PDK2 | FALSE | Downregulated |
| -0,277092993 | 0,40037654 | RPLP1 | FALSE | Downregulated |
| 0,884790763 | 0,40037654 | ZNF300 | FALSE | Upregulated |
| -0,26647114 | 0,402924028 | IDH1 | FALSE | Downregulated |
| -0,243072791 | 0,403053872 | APOA1 | FALSE | Downregulated |
| 0,24640658 | 0,403053872 | P2RX5-TAX1BP3 | FALSE | Upregulated |
| 0,804161692 | 0,403053872 | S1PR4 | FALSE | Upregulated |
| -0,313609286 | 0,403053872 | SLCO1B1 | FALSE | Downregulated |
| -0,276811669 | 0,403053872 | VHL | FALSE | Downregulated |
| 0,533897784 | 0,403053872 | YBX3 | FALSE | Upregulated |
| -0,267132949 | 0,403053872 | ZNF395 | FALSE | Downregulated |
| 0,415483882 | 0,403101297 | E2F5 | FALSE | Upregulated |
| 0,231155965 | 0,403101297 | ROCK1 | FALSE | Upregulated |
| 0,590869237 | 0,403424811 | C1orf198 | FALSE | Upregulated |
| 0,516490266 | 0,403424811 | COL1A1 | FALSE | Upregulated |
| -0,291545087 | 0,403424811 | COX5B | FALSE | Downregulated |
| -0,536691715 | 0,403424811 | HSPBP1 | FALSE | Downregulated |
| -0,29404479 | 0,403424811 | IYD | FALSE | Downregulated |
| 0,35407297 | 0,403424811 | LOC102723554 | FALSE | Upregulated |
| 0,646314059 | 0,405832271 | PCNXL2 | FALSE | Upregulated |
| -0,688140795 | 0,405923134 | DPM1 | FALSE | Downregulated |
| -0,414449968 | 0,406020324 | LRRK2 | FALSE | Downregulated |
| -0,225496674 | 0,406784403 | ASCC2 | FALSE | Downregulated |
| 0,356856367 | 0,406784403 | ENO4 | FALSE | Upregulated |
| 0,360115264 | 0,406784403 | FCHO2 | FALSE | Upregulated |
| 0,303397334 | 0,406784403 | LOC101928719 | FALSE | Upregulated |
| -0,293507912 | 0,40757138 | ABCD3 | FALSE | Downregulated |
| 0,205815808 | 0,40757138 | ANKH | FALSE | Upregulated |
| -0,208891649 | 0,40757138 | AQP9 | FALSE | Downregulated |
| -0,239652552 | 0,40757138 | C19orf43 | FALSE | Downregulated |
| 0,443129959 | 0,40757138 | COPG2 | FALSE | Upregulated |
| 0,198949803 | 0,40757138 | DTX4 | FALSE | Upregulated |
| 0,596662905 | 0,40757138 | FLJ39095 | FALSE | Upregulated |
| 0,35293713 | 0,40757138 | GCNT7 | FALSE | Upregulated |
| -0,280988843 | 0,40757138 | HSD11B1 | FALSE | Downregulated |
| 0,857932482 | 0,40757138 | LOC100996574 | FALSE | Upregulated |

| | | | | |
|---|---|---|---|---|
| 0,44755467 | 0,40757138 | LOC102467073 | FALSE | Upregulated |
| 0,442923175 | 0,40757138 | MRPS12 | FALSE | Upregulated |
| 0,667969944 | 0,40757138 | MYC | FALSE | Upregulated |
| -0,497760189 | 0,40757138 | NDUFS4 | FALSE | Downregulated |
| 0,49098813 | 0,40757138 | NFATC1 | FALSE | Upregulated |
| 1,144827043 | 0,40757138 | POU2AF1 | FALSE | Upregulated |
| 0,944007398 | 0,40757138 | RTN1 | FALSE | Upregulated |
| -0,249687825 | 0,40757138 | SHPK | FALSE | Downregulated |
| -0,20258786 | 0,40757138 | SLCO2B1 | FALSE | Downregulated |
| 0,391046489 | 0,40757138 | TAF5 | FALSE | Upregulated |
| 0,526879118 | 0,408055769 | LOC102723809 | FALSE | Upregulated |
| 0,512710949 | 0,408380998 | LOC100507071 | FALSE | Upregulated |
| 0,370563634 | 0,40858797 | USP2 | FALSE | Upregulated |
| 0,302377836 | 0,409108628 | LOC102724177 | FALSE | Upregulated |
| -0,229071025 | 0,409418498 | ACAT1 | FALSE | Downregulated |
| 0,349682731 | 0,409418498 | CDK11B | FALSE | Upregulated |
| -0,31167172 | 0,409418498 | NOC2L | FALSE | Downregulated |
| -0,277124011 | 0,409537216 | TBX3 | FALSE | Downregulated |
| 0,25095224 | 0,409578639 | ANKRD6 | FALSE | Upregulated |
| 0,710648398 | 0,409578639 | GTPBP8 | FALSE | Upregulated |
| -0,618516161 | 0,409578639 | RNF145 | FALSE | Downregulated |
| 0,304938164 | 0,409759374 | DOC2A | FALSE | Upregulated |
| 0,474776883 | 0,409759374 | RELT | FALSE | Upregulated |
| 0,608417576 | 0,409902717 | ITGA4 | FALSE | Upregulated |
| 0,193096313 | 0,409902717 | NEAT1 | FALSE | Upregulated |
| -0,296907214 | 0,410079912 | CCDC125 | FALSE | Downregulated |
| 0,240119561 | 0,410079912 | ZNF587 | FALSE | Upregulated |
| -0,666916925 | 0,410188485 | ARID3C | FALSE | Downregulated |
| 0,348375923 | 0,410188485 | C3orf22 | FALSE | Upregulated |
| 0,610506515 | 0,410188485 | CCDC167 | FALSE | Upregulated |
| 0,37134316 | 0,410188485 | FUZ | FALSE | Upregulated |
| 0,357336744 | 0,410188485 | LOC100996741 | FALSE | Upregulated |
| 0,24392034 | 0,410188485 | PHC1 | FALSE | Upregulated |
| -0,299082738 | 0,410188485 | QDPR | FALSE | Downregulated |
| 0,469184935 | 0,410188485 | RP11-622A1.2 | FALSE | Upregulated |
| 0,276762233 | 0,410188485 | TMEM181 | FALSE | Upregulated |
| 0,251810075 | 0,410437426 | C9orf131 | FALSE | Upregulated |
| -0,414449253 | 0,410710843 | PPP1R3C | FALSE | Downregulated |
| 0,232734389 | 0,410710843 | TRIM13 | FALSE | Upregulated |
| 0,317237772 | 0,410814819 | JARID2 | FALSE | Upregulated |
| 0,417088397 | 0,410895487 | NUDT16P1 | FALSE | Upregulated |
| -0,256688308 | 0,410957746 | ACAA1 | FALSE | Downregulated |
| 0,718824732 | 0,410957746 | KIAA1211 | FALSE | Upregulated |
| 0,328506515 | 0,410957746 | LINC01004 | FALSE | Upregulated |
| 0,479687278 | 0,410957746 | LOC101928059 | FALSE | Upregulated |
| -0,243376899 | 0,410957746 | PCK2 | FALSE | Downregulated |
| -0,30979396 | 0,410957746 | PLEKHA7 | FALSE | Downregulated |
| 0,727092111 | 0,410957746 | RAB40A | FALSE | Upregulated |
| -0,173619525 | 0,410957746 | RPS18 | FALSE | Downregulated |
| -0,436764407 | 0,410957746 | UQCC1 | FALSE | Downregulated |
| -0,55683009 | 0,410957746 | ZNF766 | FALSE | Downregulated |

| | | | | |
|---|---|---|---|---|
| 0,60976496 | 0,411250477 | MR1 | FALSE | Upregulated |
| 0,451058568 | 0,411481047 | BMF | FALSE | Upregulated |
| 1,42510723 | 0,411481047 | LOC729162 | FALSE | Upregulated |
| 0,409209775 | 0,411593016 | CCDC88B | FALSE | Upregulated |
| 0,413953715 | 0,412916085 | DOLK | FALSE | Upregulated |
| -0,822155852 | 0,413641597 | CYP3A7 | FALSE | Downregulated |
| -0,739153064 | 0,413711356 | ZNF701 | FALSE | Downregulated |
| -0,242596443 | 0,41457311 | ACADS | FALSE | Downregulated |
| -0,282335195 | 0,41457311 | DDX23 | FALSE | Downregulated |
| 0,197618631 | 0,41457311 | GGA2 | FALSE | Upregulated |
| 0,274106873 | 0,41457311 | LOC101929290 | FALSE | Upregulated |
| -0,257083973 | 0,41457311 | RPL22 | FALSE | Downregulated |
| -0,397925597 | 0,41457311 | SLC33A1 | FALSE | Downregulated |
| 0,41264412 | 0,414896584 | SBSPON | FALSE | Upregulated |
| 0,566420983 | 0,416292539 | AOAH | FALSE | Upregulated |
| -0,390740311 | 0,416292539 | COQ5 | FALSE | Downregulated |
| -0,392745639 | 0,416292539 | DPH6 | FALSE | Downregulated |
| 1,733336344 | 0,416292539 | ESP33 | FALSE | Upregulated |
| 0,366591279 | 0,416292539 | FLJ32790 | FALSE | Upregulated |
| -0,378159959 | 0,416292539 | GNA14 | FALSE | Downregulated |
| -0,629186807 | 0,416292539 | KIAA0895 | FALSE | Downregulated |
| -0,421587895 | 0,416292539 | KLHL42 | FALSE | Downregulated |
| 0,268843796 | 0,416292539 | LOC100131496 | FALSE | Upregulated |
| 0,612821074 | 0,416292539 | LOC101928244 | FALSE | Upregulated |
| 0,583313147 | 0,416292539 | LOC102723908 | FALSE | Upregulated |
| -0,729370552 | 0,416292539 | LOC728743 | FALSE | Downregulated |
| 0,42932562 | 0,416292539 | MDFIC | FALSE | Upregulated |
| -0,369863195 | 0,416292539 | MTFR1 | FALSE | Downregulated |
| 0,364181485 | 0,416292539 | NAA20 | FALSE | Upregulated |
| -0,193591773 | 0,416292539 | PJA2 | FALSE | Downregulated |
| -0,758337997 | 0,416292539 | PNMA2 | FALSE | Downregulated |
| 0,515752119 | 0,416292539 | RARRES3 | FALSE | Upregulated |
| 0,478902445 | 0,416292539 | RNFT1 | FALSE | Upregulated |
| 0,268242705 | 0,416292539 | RPL23AP64 | FALSE | Upregulated |
| -0,317248183 | 0,416292539 | SFSWAP | FALSE | Downregulated |
| -0,425028231 | 0,416292539 | SFXN2 | FALSE | Downregulated |
| 0,50284283 | 0,416292539 | SLA | FALSE | Upregulated |
| -0,578802127 | 0,416292539 | ZNF563 | FALSE | Downregulated |
| -0,479290465 | 0,416410262 | CCDC61 | FALSE | Downregulated |
| -0,290309323 | 0,416410262 | LY6E | FALSE | Downregulated |
| 0,25013546 | 0,416410262 | RBM5-AS1 | FALSE | Upregulated |
| -0,300174711 | 0,416599017 | CPNE3 | FALSE | Downregulated |
| -0,439766535 | 0,416721382 | INTS8 | FALSE | Downregulated |
| -0,26706696 | 0,416721382 | RELA | FALSE | Downregulated |
| -0,618221386 | 0,417052025 | PIBF1 | FALSE | Downregulated |
| -0,631642821 | 0,417470664 | PIGM | FALSE | Downregulated |
| -0,269923438 | 0,417470664 | SLC35A3 | FALSE | Downregulated |
| 0,200245145 | 0,417802129 | HNRNPA3 | FALSE | Upregulated |
| 0,547397207 | 0,417802129 | PRR7-AS1 | FALSE | Upregulated |
| 0,482386462 | 0,417865547 | ABCA7 | FALSE | Upregulated |
| 0,270386438 | 0,417865547 | ABR | FALSE | Upregulated |

| | | | | |
|---|---|---|---|---|
| -0,193327975 | 0,417865547 | ADCY9 | FALSE | Downregulated |
| 0,25552968 | 0,417865547 | ANKRD36BP1 | FALSE | Upregulated |
| -0,186981671 | 0,417865547 | BHMT2 | FALSE | Downregulated |
| 0,286212191 | 0,417865547 | C22orf23 | FALSE | Upregulated |
| 0,386258799 | 0,417865547 | CALB1 | FALSE | Upregulated |
| -0,223029428 | 0,417865547 | CDC42EP1 | FALSE | Downregulated |
| 0,502231552 | 0,417865547 | CFD | FALSE | Upregulated |
| 0,596269027 | 0,417865547 | DTD2 | FALSE | Upregulated |
| -0,49181133 | 0,417865547 | DUS2 | FALSE | Downregulated |
| -0,303009 | 0,417865547 | ECSIT | FALSE | Downregulated |
| 0,256252598 | 0,417865547 | EML5 | FALSE | Upregulated |
| -0,348703921 | 0,417865547 | FAM120B | FALSE | Downregulated |
| -0,45765832 | 0,417865547 | GPKOW | FALSE | Downregulated |
| -0,310688688 | 0,417865547 | MYO6 | FALSE | Downregulated |
| -0,305825771 | 0,417865547 | NDUFA4 | FALSE | Downregulated |
| -0,177438067 | 0,417865547 | OS9 | FALSE | Downregulated |
| 0,362004733 | 0,417865547 | PARP8 | FALSE | Upregulated |
| 0,576620445 | 0,417865547 | PLEK | FALSE | Upregulated |
| -0,285655269 | 0,417865547 | RFNG | FALSE | Downregulated |
| -0,204918276 | 0,417865547 | SCYL1 | FALSE | Downregulated |
| 0,55386186 | 0,417865547 | SLC5A10 | FALSE | Upregulated |
| 0,354091575 | 0,417865547 | STRADA | FALSE | Upregulated |
| 0,526556077 | 0,417865547 | TM4SF20 | FALSE | Upregulated |
| -0,323510475 | 0,417865547 | USP48 | FALSE | Downregulated |
| -0,379893795 | 0,41793657 | RPS13 | FALSE | Downregulated |
| 0,329897133 | 0,41809461 | RAD9B | FALSE | Upregulated |
| 0,429052616 | 0,418534344 | AC010127.3 | FALSE | Upregulated |
| -0,323211489 | 0,418534344 | PPP1R12A | FALSE | Downregulated |
| -0,310938526 | 0,418965543 | ATG13 | FALSE | Downregulated |
| 0,584371372 | 0,419251083 | LINGO3 | FALSE | Upregulated |
| -0,276511031 | 0,419251083 | MAU2 | FALSE | Downregulated |
| -0,556834965 | 0,419270303 | OMA1 | FALSE | Downregulated |
| 0,638298619 | 0,419303532 | LAT | FALSE | Upregulated |
| 0,35769338 | 0,419303532 | LOC646127 | FALSE | Upregulated |
| 0,250273256 | 0,419303532 | PDZD2 | FALSE | Upregulated |
| -0,260819194 | 0,419303532 | RNF217 | FALSE | Downregulated |
| -0,341891884 | 0,419303532 | UBL4A | FALSE | Downregulated |
| 0,603164335 | 0,4203074 | C17orf78 | FALSE | Upregulated |
| -0,35923507 | 0,420699555 | DSTN | FALSE | Downregulated |
| -0,302598985 | 0,420998828 | PDDC1 | FALSE | Downregulated |
| 0,195722825 | 0,420998828 | POM121 | FALSE | Upregulated |
| -0,209306775 | 0,420998828 | SORBS3 | FALSE | Downregulated |
| -0,211575726 | 0,421547228 | EPB41L5 | FALSE | Downregulated |
| -0,19878254 | 0,421547228 | NR2F6 | FALSE | Downregulated |
| 0,507815075 | 0,421547228 | PAX8 | FALSE | Upregulated |
| -0,308987019 | 0,421792791 | PACSIN3 | FALSE | Downregulated |
| -0,42630736 | 0,423659825 | GPAM | FALSE | Downregulated |
| -0,585345523 | 0,424551866 | CDKL5 | FALSE | Downregulated |
| 0,323187169 | 0,424551866 | CIRBP-AS1 | FALSE | Upregulated |
| 0,283620409 | 0,424551866 | LOC102723666 | FALSE | Upregulated |
| 0,372611249 | 0,424551866 | PRR15L | FALSE | Upregulated |

| | | | | |
|---|---|---|---|---|
| 0,246847872 | 0,424551866 | SETDB1 | FALSE | Upregulated |
| 0,558295715 | 0,424812885 | KIRREL3 | FALSE | Upregulated |
| 0,382577771 | 0,424812885 | LHFPL4 | FALSE | Upregulated |
| 0,908098143 | 0,424812885 | PERM1 | FALSE | Upregulated |
| 0,348073997 | 0,424812885 | RNF139-AS1 | FALSE | Upregulated |
| -0,311497031 | 0,424947208 | AKR7A3 | FALSE | Downregulated |
| 0,336981359 | 0,425023891 | GHRL | FALSE | Upregulated |
| 0,485400857 | 0,425023891 | SPATC1 | FALSE | Upregulated |
| 0,399442457 | 0,425706142 | ARID5B | FALSE | Upregulated |
| -0,445094893 | 0,425706142 | FKBP10 | FALSE | Downregulated |
| -0,317409768 | 0,426736004 | COX6B1 | FALSE | Downregulated |
| 0,268171431 | 0,427188494 | CNOT11 | FALSE | Upregulated |
| 0,235135271 | 0,427611688 | LOC100505658 | FALSE | Upregulated |
| -0,332216074 | 0,427611688 | PLA2G15 | FALSE | Downregulated |
| 0,255704258 | 0,427611688 | PLCG1 | FALSE | Upregulated |
| -0,36975482 | 0,427611688 | PRKRIR | FALSE | Downregulated |
| -0,222204375 | 0,427611688 | RBM6 | FALSE | Downregulated |
| -0,296852213 | 0,427611688 | ST3GAL6 | FALSE | Downregulated |
| -0,198826038 | 0,427770107 | OSBP | FALSE | Downregulated |
| -0,22190571 | 0,427770107 | WWC2 | FALSE | Downregulated |
| -0,571727814 | 0,428111405 | CCL14 | FALSE | Downregulated |
| 0,26528403 | 0,428111405 | CCPG1 | FALSE | Upregulated |
| 0,154232884 | 0,428111405 | CD302 | FALSE | Upregulated |
| 0,34899666 | 0,428111405 | COTL1 | FALSE | Upregulated |
| 0,591962374 | 0,428111405 | DOK1 | FALSE | Upregulated |
| -0,727537435 | 0,428111405 | LINC00211 | FALSE | Downregulated |
| 0,271232204 | 0,428111405 | MSN | FALSE | Upregulated |
| 0,33190182 | 0,428111405 | PDE4DIP | FALSE | Upregulated |
| -0,205667037 | 0,428111405 | PROS1 | FALSE | Downregulated |
| -0,22224087 | 0,428111405 | RPS10 | FALSE | Downregulated |
| -0,435809018 | 0,428111405 | SLC22A18 | FALSE | Downregulated |
| 0,467819815 | 0,429097446 | EPB41L3 | FALSE | Upregulated |
| -0,240582303 | 0,429239094 | CYB5R3 | FALSE | Downregulated |
| 0,203525086 | 0,429239094 | DUSP16 | FALSE | Upregulated |
| 0,342243897 | 0,429239094 | LOC101929387 | FALSE | Upregulated |
| -0,511404217 | 0,429239094 | NUDCD2 | FALSE | Downregulated |
| 0,574931975 | 0,429481067 | GAS7 | FALSE | Upregulated |
| -0,231148726 | 0,430688032 | PKD1 | FALSE | Downregulated |
| 0,562227835 | 0,430688032 | PLEKHG4 | FALSE | Upregulated |
| 1,037454928 | 0,430963316 | DDX53 | FALSE | Upregulated |
| 0,221412046 | 0,430963316 | KANSL1L | FALSE | Upregulated |
| 0,314854941 | 0,432472627 | MYSM1 | FALSE | Upregulated |
| 0,282442306 | 0,432472627 | RUFY2 | FALSE | Upregulated |
| -0,366923929 | 0,432472627 | SLC17A2 | FALSE | Downregulated |
| 0,57778123 | 0,432472627 | SLPI | FALSE | Upregulated |
| 0,260796364 | 0,432472627 | TSPYL6 | FALSE | Upregulated |
| 0,571264953 | 0,432569112 | OBSCN | FALSE | Upregulated |
| 0,425609852 | 0,432850458 | C10orf76 | FALSE | Upregulated |
| 0,456747945 | 0,432850458 | FNDC8 | FALSE | Upregulated |
| -0,468442577 | 0,433075573 | DOLPP1 | FALSE | Downregulated |
| -0,424862208 | 0,433075573 | FASTKD2 | FALSE | Downregulated |

| | | | | |
|---|---|---|---|---|
| -0,237119689 | 0,433075573 | KLKB1 | FALSE | Downregulated |
| 0,409400094 | 0,433075573 | RELL2 | FALSE | Upregulated |
| 0,418308613 | 0,433075573 | SEZ6 | FALSE | Upregulated |
| 0,355685002 | 0,433888217 | KCNJ10 | FALSE | Upregulated |
| 0,337719679 | 0,433888217 | LARS2 | FALSE | Upregulated |
| 0,248472117 | 0,4340546 | PURB | FALSE | Upregulated |
| -0,289673402 | 0,434117318 | TFPI | FALSE | Downregulated |
| 0,740840296 | 0,434135176 | LOC101928982 | FALSE | Upregulated |
| -0,468686654 | 0,434135176 | PLRG1 | FALSE | Downregulated |
| 0,448338784 | 0,43470428 | LOC101928240 | FALSE | Upregulated |
| -0,238700677 | 0,435557526 | SDC4 | FALSE | Downregulated |
| -0,484913301 | 0,436191308 | ADH1C | FALSE | Downregulated |
| 0,521108549 | 0,436191308 | C2orf44 | FALSE | Upregulated |
| 0,647206516 | 0,436191308 | GEMIN2 | FALSE | Upregulated |
| 0,479819009 | 0,436191308 | LIPT1 | FALSE | Upregulated |
| -0,378704879 | 0,436191308 | LYVE1 | FALSE | Downregulated |
| 0,197450406 | 0,436191308 | SNED1 | FALSE | Upregulated |
| 0,237177365 | 0,436191308 | ZACN | FALSE | Upregulated |
| 0,391872081 | 0,43619189 | CCNH | FALSE | Upregulated |
| 0,41635888 | 0,43619189 | CLCF1 | FALSE | Upregulated |
| 0,955748246 | 0,43619189 | GCSAM | FALSE | Upregulated |
| -0,23043801 | 0,43619189 | HSPD1 | FALSE | Downregulated |
| 0,371247906 | 0,43619189 | LOC100506497 | FALSE | Upregulated |
| -0,237980733 | 0,43619189 | MAT1A | FALSE | Downregulated |
| -0,582438037 | 0,43619189 | NLE1 | FALSE | Downregulated |
| 0,517641339 | 0,43619189 | PHF19 | FALSE | Upregulated |
| 0,360288588 | 0,43619189 | PIRT | FALSE | Upregulated |
| -0,181986863 | 0,43619189 | RPL10 | FALSE | Downregulated |
| 0,44969614 | 0,43619189 | TLR9 | FALSE | Upregulated |
| 0,256213309 | 0,436255338 | LOC101928323 | FALSE | Upregulated |
| 0,249908535 | 0,437318234 | FMNL2 | FALSE | Upregulated |
| 0,360926064 | 0,43758127 | SYTL4 | FALSE | Upregulated |
| 0,253208257 | 0,43760144 | EYS | FALSE | Upregulated |
| -0,189556296 | 0,43760144 | FNDC3A | FALSE | Downregulated |
| 0,281754594 | 0,43760144 | GPR22 | FALSE | Upregulated |
| 0,259881033 | 0,43760144 | JADE2 | FALSE | Upregulated |
| -0,259168143 | 0,43760144 | KIF21A | FALSE | Downregulated |
| 0,316572946 | 0,43760144 | LRRC19 | FALSE | Upregulated |
| 0,340906688 | 0,43760144 | PPP4R4 | FALSE | Upregulated |
| -0,36624043 | 0,43760144 | PRKAR2A | FALSE | Downregulated |
| -0,253813012 | 0,43760144 | PTH1R | FALSE | Downregulated |
| 0,455686068 | 0,43760144 | RRP1 | FALSE | Upregulated |
| 0,644991228 | 0,43760144 | SASS6 | FALSE | Upregulated |
| -0,463928902 | 0,43760144 | SOX6 | FALSE | Downregulated |
| -0,305585802 | 0,43760144 | URI1 | FALSE | Downregulated |
| -0,271709236 | 0,437875984 | CAPN7 | FALSE | Downregulated |
| 0,465528169 | 0,437875984 | NEDD4L | FALSE | Upregulated |
| 0,261902074 | 0,439040912 | CSPP1 | FALSE | Upregulated |
| -0,178221562 | 0,439040912 | EHMT1 | FALSE | Downregulated |
| 0,4430358 | 0,439040912 | RDH13 | FALSE | Upregulated |
| -0,492465626 | 0,439040912 | SUGCT | FALSE | Downregulated |

| | | | | |
|---|---|---|---|---|
| 0,32446441 | 0,440138065 | LYPD1 | FALSE | Upregulated |
| 0,370031363 | 0,440138065 | MYH11 | FALSE | Upregulated |
| -0,477891149 | 0,440138065 | QSOX2 | FALSE | Downregulated |
| -0,221762572 | 0,440358482 | C1R-2 | FALSE | Downregulated |
| -0,252816532 | 0,440385827 | F12 | FALSE | Downregulated |
| -0,426799224 | 0,440532965 | CD80 | FALSE | Downregulated |
| 0,247351529 | 0,440532965 | GNRHR2 | FALSE | Upregulated |
| -0,25072877 | 0,440532965 | IER2 | FALSE | Downregulated |
| -0,257342566 | 0,440532965 | JTB | FALSE | Downregulated |
| -0,302708004 | 0,440532965 | MARC1 | FALSE | Downregulated |
| -0,222099796 | 0,440532965 | MPDZ | FALSE | Downregulated |
| -0,346935072 | 0,440532965 | PFAS | FALSE | Downregulated |
| 0,445955422 | 0,440532965 | VPS9D1-AS1 | FALSE | Upregulated |
| -0,2454232 | 0,440532965 | XRCC5 | FALSE | Downregulated |
| -0,473449265 | 0,44061601 | CYP2B6 | FALSE | Downregulated |
| -0,24487299 | 0,44061601 | DDX54 | FALSE | Downregulated |
| 0,257733576 | 0,44061601 | DNAAF2 | FALSE | Upregulated |
| 0,386649408 | 0,44061601 | FAM132B | FALSE | Upregulated |
| -0,19335505 | 0,44061601 | IQGAP2 | FALSE | Downregulated |
| -0,329136731 | 0,44061601 | KIAA1522 | FALSE | Downregulated |
| 0,256113802 | 0,44061601 | PAN3-AS1 | FALSE | Upregulated |
| -0,257379864 | 0,44061601 | SLC27A2 | FALSE | Downregulated |
| -0,195741473 | 0,44061601 | SYMPK | FALSE | Downregulated |
| 0,226568066 | 0,44061601 | USP24 | FALSE | Upregulated |
| 0,370862274 | 0,441230639 | ABCC1 | FALSE | Upregulated |
| 0,415468424 | 0,441230639 | ANKRD27 | FALSE | Upregulated |
| -0,160276302 | 0,441230639 | BAG6 | FALSE | Downregulated |
| -0,303794364 | 0,441230639 | CYP2C8 | FALSE | Downregulated |
| 0,645153699 | 0,441230639 | DRAM1 | FALSE | Upregulated |
| -0,242386187 | 0,441230639 | HNRNPH1 | FALSE | Downregulated |
| -0,326788915 | 0,441230639 | NAP1L4 | FALSE | Downregulated |
| -0,150012339 | 0,441230639 | SLC38A10 | FALSE | Downregulated |
| 0,98613845 | 0,441290681 | BHLHE41 | FALSE | Upregulated |
| 0,216263133 | 0,441814796 | COL10A1 | FALSE | Upregulated |
| -0,20298082 | 0,441837597 | SLC2A4RG | FALSE | Downregulated |
| -0,223411764 | 0,441837597 | ZBTB44 | FALSE | Downregulated |
| 0,455532444 | 0,4418543 | IFT46 | FALSE | Upregulated |
| 0,320592469 | 0,442093571 | PFN1P2 | FALSE | Upregulated |
| 0,29924175 | 0,442264332 | AC003665.1 | FALSE | Upregulated |
| 0,550180994 | 0,442264332 | ATP2A3 | FALSE | Upregulated |
| 0,3621529 | 0,442264332 | GPR182 | FALSE | Upregulated |
| -0,621327694 | 0,442264332 | LOC101928433 | FALSE | Downregulated |
| 0,332592257 | 0,442264332 | LOC101929599 | FALSE | Upregulated |
| -0,159066533 | 0,442264332 | MASP2 | FALSE | Downregulated |
| -0,211379417 | 0,442264332 | PIK3R1 | FALSE | Downregulated |
| 0,527452542 | 0,442264332 | SARS2 | FALSE | Upregulated |
| -0,523168983 | 0,442320605 | CCBE1 | FALSE | Downregulated |
| -0,362497931 | 0,442320605 | GRAMD3 | FALSE | Downregulated |
| 0,630479925 | 0,442320605 | ZNF737 | FALSE | Upregulated |
| -0,308333219 | 0,442633328 | ARID4A | FALSE | Downregulated |
| -0,239226442 | 0,443346613 | FAM107B | FALSE | Downregulated |

| | | | | |
|---|---|---|---|---|
| -0,423959626 | 0,44388085 | WDR60 | FALSE | Downregulated |
| 0,608352452 | 0,444194359 | FAM53B-AS1 | FALSE | Upregulated |
| 0,583329599 | 0,444444552 | KBTBD12 | FALSE | Upregulated |
| 0,847614749 | 0,444461066 | CHST9 | FALSE | Upregulated |
| 0,407739902 | 0,444461066 | CNTNAP1 | FALSE | Upregulated |
| -0,269785301 | 0,444461066 | DROSHA | FALSE | Downregulated |
| -0,209848049 | 0,444461066 | GRB10 | FALSE | Downregulated |
| -0,563538078 | 0,444461066 | NUDT8 | FALSE | Downregulated |
| 0,383663968 | 0,444461066 | PRRT3 | FALSE | Upregulated |
| 0,368474125 | 0,444461066 | PTPN7 | FALSE | Upregulated |
| -0,334624263 | 0,444461066 | REPS1 | FALSE | Downregulated |
| 0,622944584 | 0,444461066 | RP11-43N5.1 | FALSE | Upregulated |
| 0,333501631 | 0,444461066 | SND1-IT1 | FALSE | Upregulated |
| 0,553732696 | 0,444461066 | UHRF1 | FALSE | Upregulated |
| -0,324056385 | 0,444579386 | COX7C | FALSE | Downregulated |
| -0,293444854 | 0,444579386 | GATM | FALSE | Downregulated |
| -0,229370561 | 0,444579386 | GJB1 | FALSE | Downregulated |
| 0,805309056 | 0,444579386 | LINC00943 | FALSE | Upregulated |
| 0,45531286 | 0,444579386 | PPIL6 | FALSE | Upregulated |
| 0,517223531 | 0,444579386 | PRMT6 | FALSE | Upregulated |
| 0,604370363 | 0,444618396 | FLJ38576 | FALSE | Upregulated |
| 0,701278344 | 0,444618396 | LOC100129083 | FALSE | Upregulated |
| -0,517942042 | 0,444618396 | ZNF329 | FALSE | Downregulated |
| 0,439718988 | 0,44580711 | ABCF2 | FALSE | Upregulated |
| -0,459856437 | 0,44580711 | C19orf80 | FALSE | Downregulated |
| -2,629172704 | 0,44580711 | C21orf140 | FALSE | Downregulated |
| 0,517508794 | 0,44580711 | CENPN | FALSE | Upregulated |
| -0,35838962 | 0,44580711 | DAO | FALSE | Downregulated |
| 0,356241459 | 0,44580711 | HAUS6 | FALSE | Upregulated |
| -0,332613851 | 0,44580711 | HERPUD2 | FALSE | Downregulated |
| 1,425384901 | 0,44580711 | IGFN1 | FALSE | Upregulated |
| -0,315436238 | 0,44580711 | NAGS | FALSE | Downregulated |
| -0,312388888 | 0,44580711 | PEG3 | FALSE | Downregulated |
| 0,505533446 | 0,44580711 | PRRG1 | FALSE | Upregulated |
| -0,393487021 | 0,44580711 | REXO4 | FALSE | Downregulated |
| 0,612064798 | 0,44580711 | RHBDL2 | FALSE | Upregulated |
| 0,222568658 | 0,44580711 | SEPN1 | FALSE | Upregulated |
| 0,190810654 | 0,44580711 | SH2B3 | FALSE | Upregulated |
| 0,492519735 | 0,44580711 | ZXDB | FALSE | Upregulated |
| 0,518874242 | 0,446715868 | ANO10 | FALSE | Upregulated |
| 0,342247755 | 0,446715868 | APPL1 | FALSE | Upregulated |
| -0,591816855 | 0,446715868 | ARHGEF3 | FALSE | Downregulated |
| 0,568107364 | 0,446715868 | CLIP3 | FALSE | Upregulated |
| 0,356858667 | 0,446715868 | CROT | FALSE | Upregulated |
| 0,238983382 | 0,446715868 | DGKQ | FALSE | Upregulated |
| 0,414561924 | 0,446715868 | DXO | FALSE | Upregulated |
| 0,465293845 | 0,446715868 | FAM98B | FALSE | Upregulated |
| -0,340936688 | 0,446715868 | FN3K | FALSE | Downregulated |
| -0,191582021 | 0,446715868 | FZD5 | FALSE | Downregulated |
| 0,550176072 | 0,446715868 | GK5 | FALSE | Upregulated |
| 0,207432 | 0,446715868 | GLIPR1 | FALSE | Upregulated |

| | | | | |
|---|---|---|---|---|
| 0,256255578 | 0,446715868 | GRIN3A | FALSE | Upregulated |
| -0,244032642 | 0,446715868 | HP1BP3 | FALSE | Downregulated |
| 0,343306597 | 0,446715868 | IGF2BP3 | FALSE | Upregulated |
| 0,578966121 | 0,446715868 | ITGAM | FALSE | Upregulated |
| 0,576785022 | 0,446715868 | KNTC1 | FALSE | Upregulated |
| -0,222226698 | 0,446715868 | LCAT | FALSE | Downregulated |
| -0,216029669 | 0,446715868 | LMAN1 | FALSE | Downregulated |
| 0,498403441 | 0,446715868 | LOC101928408 | FALSE | Upregulated |
| 0,559727609 | 0,446715868 | LOC101928466 | FALSE | Upregulated |
| 0,462779717 | 0,446715868 | LOC441179 | FALSE | Upregulated |
| 0,329650472 | 0,446715868 | LRFN4 | FALSE | Upregulated |
| 0,186116971 | 0,446715868 | MYO5B | FALSE | Upregulated |
| -0,516466247 | 0,446715868 | NDUFB6 | FALSE | Downregulated |
| -0,232963215 | 0,446715868 | OPLAH | FALSE | Downregulated |
| -0,318440616 | 0,446715868 | PMVK | FALSE | Downregulated |
| 0,544927525 | 0,446715868 | PP14571 | FALSE | Upregulated |
| 0,404792886 | 0,446715868 | PRPF38B | FALSE | Upregulated |
| -0,18805806 | 0,446715868 | RIN3 | FALSE | Downregulated |
| 0,246177858 | 0,446715868 | RP5-1071N3.1 | FALSE | Upregulated |
| 0,273203237 | 0,446715868 | SEC61A2 | FALSE | Upregulated |
| 0,658989833 | 0,446715868 | SQLE | FALSE | Upregulated |
| 0,465488338 | 0,446715868 | SRGAP3-AS3 | FALSE | Upregulated |
| -0,175800894 | 0,446715868 | TFR2 | FALSE | Downregulated |
| 0,609055625 | 0,446715868 | TM4SF1 | FALSE | Upregulated |
| 0,610121351 | 0,446715868 | UBE2E2 | FALSE | Upregulated |
| 0,157987609 | 0,446715868 | VPS13D | FALSE | Upregulated |
| 0,675876306 | 0,446715868 | VWA8-AS1 | FALSE | Upregulated |
| -0,197589363 | 0,446715868 | ZFHX4 | FALSE | Downregulated |
| -0,754465214 | 0,448387709 | CTNNA3 | FALSE | Downregulated |
| -0,111024985 | 0,448387709 | MAVS | FALSE | Downregulated |
| -0,164251605 | 0,448387709 | SCARB2 | FALSE | Downregulated |
| 0,763003618 | 0,448387709 | TRPC5OS | FALSE | Upregulated |
| 0,572173244 | 0,448802816 | GGTA1P | FALSE | Upregulated |
| 0,518254621 | 0,449067099 | FAM205B | FALSE | Upregulated |
| -0,215107636 | 0,449123335 | CTAGE5 | FALSE | Downregulated |
| 0,915241059 | 0,449274237 | LOC101928491 | FALSE | Upregulated |
| -0,226743069 | 0,450213255 | FPGS | FALSE | Downregulated |
| 0,211894006 | 0,450213255 | PNRC2 | FALSE | Upregulated |
| -0,327821144 | 0,45067264 | LOC100507217 | FALSE | Downregulated |
| 0,290046781 | 0,45067264 | ZNF12 | FALSE | Upregulated |
| 0,404288557 | 0,450823892 | LOC101927809 | FALSE | Upregulated |
| 1,04459469 | 0,450823892 | LOC102724135 | FALSE | Upregulated |
| 0,387322742 | 0,450823892 | RP11-300J18.1 | FALSE | Upregulated |
| -0,317676137 | 0,450823892 | SLC29A1 | FALSE | Downregulated |
| 0,355894741 | 0,450823892 | TMEM206 | FALSE | Upregulated |
| -0,216579616 | 0,451167303 | ADCY1 | FALSE | Downregulated |
| -0,287024176 | 0,451167303 | ELMO1 | FALSE | Downregulated |
| -0,261376953 | 0,451167303 | IGSF8 | FALSE | Downregulated |
| 0,179063935 | 0,451167303 | LGALS8 | FALSE | Upregulated |
| -0,411635563 | 0,451167303 | TUBE1 | FALSE | Downregulated |
| -0,199039711 | 0,451699381 | CD81 | FALSE | Downregulated |

| | | | | |
|---|---|---|---|---|
| 0,50045554 | 0,451699381 | HHIPL1 | FALSE | Upregulated |
| 0,647057633 | 0,451699381 | RTN4R | FALSE | Upregulated |
| 0,732991149 | 0,451699381 | SLC2A1 | FALSE | Upregulated |
| 0,341194171 | 0,451699381 | THEMIS2 | FALSE | Upregulated |
| -0,218906712 | 0,451699381 | TMEM56 | FALSE | Downregulated |
| -0,204779927 | 0,45206499 | EIF3G | FALSE | Downregulated |
| -0,264125043 | 0,45206499 | FBXO18 | FALSE | Downregulated |
| 0,688268648 | 0,45206499 | HAO2-IT1 | FALSE | Upregulated |
| -0,267420832 | 0,45206499 | KIAA0754 | FALSE | Downregulated |
| 0,568678384 | 0,45206499 | LINC00102 | FALSE | Upregulated |
| -0,344943636 | 0,45206499 | LTA4H | FALSE | Downregulated |
| -0,310990458 | 0,45206499 | MLPH | FALSE | Downregulated |
| -0,199902681 | 0,45206499 | RPS23 | FALSE | Downregulated |
| 0,342637996 | 0,453763295 | BTN3A2 | FALSE | Upregulated |
| 0,437926645 | 0,453905174 | GBP1 | FALSE | Upregulated |
| -0,341818156 | 0,453905174 | GDF2 | FALSE | Downregulated |
| -0,143504417 | 0,453905174 | RPL18A | FALSE | Downregulated |
| -0,115674532 | 0,45398098 | LINC00657 | FALSE | Downregulated |
| 0,330613943 | 0,45398098 | PITPNM1 | FALSE | Upregulated |
| 0,23303351 | 0,45398098 | SETD6 | FALSE | Upregulated |
| -0,493219857 | 0,45398098 | SHROOM4 | FALSE | Downregulated |
| -0,240865338 | 0,45398098 | TM7SF3 | FALSE | Downregulated |
| 0,193330932 | 0,454952261 | SMARCA4 | FALSE | Upregulated |
| -0,364072251 | 0,455624092 | IMP4 | FALSE | Downregulated |
| 0,196254739 | 0,455993234 | FAM208B | FALSE | Upregulated |
| 0,881279304 | 0,455993234 | STAMBPL1 | FALSE | Upregulated |
| 0,626995244 | 0,456024916 | RAE1 | FALSE | Upregulated |
| 0,501829061 | 0,456643119 | MX1 | FALSE | Upregulated |
| 0,556560451 | 0,45711004 | ABCA3 | FALSE | Upregulated |
| 0,498555297 | 0,45711004 | LOC101928281 | FALSE | Upregulated |
| 0,350836042 | 0,45711004 | LOC102724006 | FALSE | Upregulated |
| -0,265702017 | 0,45711004 | MAPRE2 | FALSE | Downregulated |
| -0,465479454 | 0,45711004 | PPM1K | FALSE | Downregulated |
| 0,182053565 | 0,45711004 | SVIL | FALSE | Upregulated |
| 0,422167485 | 0,45711004 | TTC1 | FALSE | Upregulated |
| 0,346165014 | 0,457436084 | FOXH1 | FALSE | Upregulated |
| -0,562352751 | 0,457436084 | KLHL26 | FALSE | Downregulated |
| -0,391228902 | 0,458044385 | RGP1 | FALSE | Downregulated |
| 0,493571829 | 0,458677536 | LOC100128573 | FALSE | Upregulated |
| -0,370856942 | 0,458718066 | GLRX | FALSE | Downregulated |
| -0,195837452 | 0,459458052 | GDI2 | FALSE | Downregulated |
| -0,280653089 | 0,459729645 | ECM1 | FALSE | Downregulated |
| 0,293229132 | 0,459729645 | ERMAP | FALSE | Upregulated |
| 0,433637488 | 0,459729645 | PGAM4 | FALSE | Upregulated |
| -0,174611195 | 0,459729645 | RPL9 | FALSE | Downregulated |
| -0,2881658 | 0,459729645 | UCK1 | FALSE | Downregulated |
| -0,241576624 | 0,459729645 | WBP1L | FALSE | Downregulated |
| 0,291264659 | 0,459729645 | ZSCAN32 | FALSE | Upregulated |
| 0,34931083 | 0,460190976 | ACOT4 | FALSE | Upregulated |
| 0,406431267 | 0,460190976 | FAM83G | FALSE | Upregulated |
| -0,176758181 | 0,460190976 | MYO18A | FALSE | Downregulated |

| | | | | |
|---|---|---|---|---|
| 0,502290032 | 0,460546012 | C7orf71 | FALSE | Upregulated |
| -0,273449905 | 0,460546012 | MBP | FALSE | Downregulated |
| -0,517821915 | 0,461929363 | COL11A2 | FALSE | Downregulated |
| 0,206547702 | 0,461929363 | NCBP1 | FALSE | Upregulated |
| 0,942317859 | 0,461929363 | NEB | FALSE | Upregulated |
| 1,096153894 | 0,461929363 | SMOC2 | FALSE | Upregulated |
| -0,22931285 | 0,461929363 | TECR | FALSE | Downregulated |
| -0,454526312 | 0,461988375 | TRABD2B | FALSE | Downregulated |
| -0,449221635 | 0,462114499 | SNRPG | FALSE | Downregulated |
| -0,208853873 | 0,462415443 | GAA | FALSE | Downregulated |
| 0,229524653 | 0,462577437 | C1orf167 | FALSE | Upregulated |
| 0,248200274 | 0,462577437 | EGFR-AS1 | FALSE | Upregulated |
| 0,426313606 | 0,462577437 | HLA-F | FALSE | Upregulated |
| 0,395011345 | 0,462644114 | GRK6 | FALSE | Upregulated |
| 0,321880226 | 0,462644114 | ZFP28 | FALSE | Upregulated |
| -0,450865611 | 0,462781927 | SNRNP25 | FALSE | Downregulated |
| -0,314307502 | 0,463171168 | PHGDH | FALSE | Downregulated |
| 0,780285473 | 0,463300666 | CASC14 | FALSE | Upregulated |
| -0,20315672 | 0,463417551 | SEC61A1 | FALSE | Downregulated |
| -0,310697052 | 0,463590618 | CEBPG | FALSE | Downregulated |
| 0,348265501 | 0,463993445 | GK | FALSE | Upregulated |
| 0,313347311 | 0,464105171 | FAM205A | FALSE | Upregulated |
| 0,505967514 | 0,464508809 | C16orf59 | FALSE | Upregulated |
| 0,761604136 | 0,464508809 | CDCA5 | FALSE | Upregulated |
| 0,312485373 | 0,46451095 | APOL3 | FALSE | Upregulated |
| 0,304821418 | 0,46451095 | CCDC6 | FALSE | Upregulated |
| -0,276805606 | 0,46451095 | DSG2 | FALSE | Downregulated |
| 0,362862619 | 0,46451095 | KCNH1 | FALSE | Upregulated |
| 0,267644702 | 0,46451095 | NCAPG | FALSE | Upregulated |
| -0,223183672 | 0,46451095 | SMIM14 | FALSE | Downregulated |
| -0,591155352 | 0,46451095 | TENM2 | FALSE | Downregulated |
| 0,578873831 | 0,464779759 | FAR1 | FALSE | Upregulated |
| -0,360309275 | 0,464779759 | FGD4 | FALSE | Downregulated |
| -0,392155291 | 0,465352351 | CSNK2B | FALSE | Downregulated |
| -0,206291509 | 0,466175263 | ACOX1 | FALSE | Downregulated |
| -0,475329889 | 0,466307951 | CEP44 | FALSE | Downregulated |
| -0,224954905 | 0,466307951 | FGFR3 | FALSE | Downregulated |
| 0,221160402 | 0,466307951 | MMP24 | FALSE | Upregulated |
| -0,352083954 | 0,466307951 | NDRG1 | FALSE | Downregulated |
| -0,264020292 | 0,466307951 | PDE2A | FALSE | Downregulated |
| -0,232114056 | 0,466307951 | SIK3 | FALSE | Downregulated |
| -0,287137691 | 0,466307951 | TCEA1 | FALSE | Downregulated |
| -0,405932773 | 0,466307951 | UBAC1 | FALSE | Downregulated |
| 0,229280232 | 0,466307951 | ZNF140 | FALSE | Upregulated |
| -0,448362943 | 0,466307951 | ZNF408 | FALSE | Downregulated |
| 0,249467074 | 0,466307951 | ZNF814 | FALSE | Upregulated |
| 0,284861022 | 0,466307951 | ZYX | FALSE | Upregulated |
| -0,331131137 | 0,466885003 | TBX2 | FALSE | Downregulated |
| -0,172466424 | 0,467852773 | INTS1 | FALSE | Downregulated |
| 0,263455656 | 0,46797508 | IQCC | FALSE | Upregulated |
| -0,425198811 | 0,46797508 | UIMC1 | FALSE | Downregulated |

| | | | | |
|---|---|---|---|---|
| -0,660674525 | 0,468019571 | AMN | FALSE | Downregulated |
| 0,654726451 | 0,468019571 | CCDC65 | FALSE | Upregulated |
| 0,604801353 | 0,468019571 | DDIT4 | FALSE | Upregulated |
| 0,322583358 | 0,468019571 | DNAH17 | FALSE | Upregulated |
| -0,214362598 | 0,468019571 | DOCK4 | FALSE | Downregulated |
| 0,279807943 | 0,468019571 | FBXO33 | FALSE | Upregulated |
| -0,195450197 | 0,468019571 | FNBP4 | FALSE | Downregulated |
| 0,249230092 | 0,468019571 | HMGXB3 | FALSE | Upregulated |
| -0,198084321 | 0,468019571 | HNRNPM | FALSE | Downregulated |
| -0,196346174 | 0,468019571 | ITGB1 | FALSE | Downregulated |
| 0,572008117 | 0,468019571 | LRRC37A3 | FALSE | Upregulated |
| 0,759184179 | 0,468019571 | OR5AK4P | FALSE | Upregulated |
| 0,144370747 | 0,468019571 | PDXDC1 | FALSE | Upregulated |
| 0,296241153 | 0,468019571 | ZNF710 | FALSE | Upregulated |
| -0,388644057 | 0,46840066 | HUS1 | FALSE | Downregulated |
| 0,390841636 | 0,46840066 | NFASC | FALSE | Upregulated |
| 0,563603393 | 0,46840066 | RP11-346I3.4 | FALSE | Upregulated |
| -0,225795122 | 0,468920716 | HSD17B4 | FALSE | Downregulated |
| -0,23881726 | 0,469345279 | XDH | FALSE | Downregulated |
| 0,653943727 | 0,469594315 | ST20 | FALSE | Upregulated |
| -0,321196967 | 0,469621736 | HNF1A-AS1 | FALSE | Downregulated |
| 0,313767504 | 0,469621736 | LOC100996873 | FALSE | Upregulated |
| 0,583395552 | 0,469621736 | LOC101927201 | FALSE | Upregulated |
| -0,414554748 | 0,469621736 | MAPK9 | FALSE | Downregulated |
| -0,443033082 | 0,469621736 | METTL17 | FALSE | Downregulated |
| -0,339825328 | 0,469621736 | PRMT7 | FALSE | Downregulated |
| 0,230511684 | 0,469621736 | PYGB | FALSE | Upregulated |
| 0,313143578 | 0,469621736 | ZDHHC20 | FALSE | Upregulated |
| 0,262187477 | 0,469675978 | CCSAP | FALSE | Upregulated |
| -0,246913545 | 0,469685722 | AGL | FALSE | Downregulated |
| 0,29112046 | 0,469685722 | ARHGDIB | FALSE | Upregulated |
| -0,202806663 | 0,469685722 | CCDC50 | FALSE | Downregulated |
| -0,632615222 | 0,469685722 | HEY2 | FALSE | Downregulated |
| 0,547530751 | 0,469685722 | LILRB2 | FALSE | Upregulated |
| 0,794287154 | 0,469685722 | LILRB4 | FALSE | Upregulated |
| 0,238166738 | 0,469685722 | LOC101927803 | FALSE | Upregulated |
| 0,283970234 | 0,469685722 | LOC101929109 | FALSE | Upregulated |
| 0,441493901 | 0,469685722 | OLFM2 | FALSE | Upregulated |
| 0,281769293 | 0,469685722 | PDZD9 | FALSE | Upregulated |
| -0,215589644 | 0,469685722 | SNX9 | FALSE | Downregulated |
| -0,1957678 | 0,469697471 | FAM134A | FALSE | Downregulated |
| -0,564126971 | 0,469697471 | HNF4A-AS1 | FALSE | Downregulated |
| 0,573725526 | 0,469697471 | LOC101927227 | FALSE | Upregulated |
| -0,186418827 | 0,469697471 | MCM3AP | FALSE | Downregulated |
| 0,267979803 | 0,469697471 | MXD3 | FALSE | Upregulated |
| -0,262177486 | 0,469697471 | RXRB | FALSE | Downregulated |
| 0,301371394 | 0,469697471 | STK19 | FALSE | Upregulated |
| -0,442139364 | 0,469697471 | TNFSF12 | FALSE | Downregulated |
| -0,282525198 | 0,469697471 | U2SURP | FALSE | Downregulated |
| 0,303729493 | 0,469697471 | ZSCAN22 | FALSE | Upregulated |
| -0,314094379 | 0,47039557 | ANKRD24 | FALSE | Downregulated |

| | | | | |
|---|---|---|---|---|
| -0,303433951 | 0,47039557 | GOSR1 | FALSE | Downregulated |
| 0,438922561 | 0,47059572 | CISD2 | FALSE | Upregulated |
| 0,558421946 | 0,470774801 | CDKN2B | FALSE | Upregulated |
| -0,579804008 | 0,470846513 | BMS1P4 | FALSE | Downregulated |
| 0,385300642 | 0,470846513 | DPY19L2P3 | FALSE | Upregulated |
| -0,245700966 | 0,470846513 | GPHN | FALSE | Downregulated |
| -0,482023653 | 0,470846513 | GTPBP4 | FALSE | Downregulated |
| -0,435125024 | 0,470846513 | LAMTOR2 | FALSE | Downregulated |
| 0,559937241 | 0,470846513 | LOC101929441 | FALSE | Upregulated |
| 0,267097482 | 0,470846513 | LOC440300 | FALSE | Upregulated |
| 0,410729079 | 0,470846513 | PSMD5-AS1 | FALSE | Upregulated |
| 0,537109871 | 0,470846513 | RAD1 | FALSE | Upregulated |
| -0,421997756 | 0,470846513 | TANK | FALSE | Downregulated |
| -0,509387397 | 0,47088838 | CYP3A43 | FALSE | Downregulated |
| 0,305814491 | 0,47088838 | HIVEP2 | FALSE | Upregulated |
| 0,302166894 | 0,471069299 | LOC101927385 | FALSE | Upregulated |
| 0,526032653 | 0,471164296 | CDKN1A | FALSE | Upregulated |
| 0,263578381 | 0,471896723 | C9orf84 | FALSE | Upregulated |
| 0,323034534 | 0,472011497 | LOC101929004 | FALSE | Upregulated |
| 0,407218396 | 0,472184027 | ATP13A2 | FALSE | Upregulated |
| -0,328907893 | 0,472184027 | ATPAF1 | FALSE | Downregulated |
| -0,307216491 | 0,472184027 | FAHD2A | FALSE | Downregulated |
| 0,659804362 | 0,472184027 | GP5 | FALSE | Upregulated |
| -0,418552681 | 0,472184027 | LMBRD2 | FALSE | Downregulated |
| -0,338694543 | 0,472184027 | PEX14 | FALSE | Downregulated |
| 0,341920088 | 0,472184027 | TIRAP | FALSE | Upregulated |
| 0,304944147 | 0,472184027 | TP53BP2 | FALSE | Upregulated |
| -0,461144633 | 0,472201252 | SLC1A2 | FALSE | Downregulated |
| 0,489002189 | 0,472217503 | C9orf139 | FALSE | Upregulated |
| 0,263619082 | 0,472257437 | C21orf2 | FALSE | Upregulated |
| 0,347573195 | 0,472257437 | CMTM4 | FALSE | Upregulated |
| 0,304106478 | 0,472257437 | CYB5D2 | FALSE | Upregulated |
| -0,195674051 | 0,472257437 | LIMD1 | FALSE | Downregulated |
| -0,285532567 | 0,472257437 | LOC100507033 | FALSE | Downregulated |
| 0,268671575 | 0,472257437 | LOC101929533 | FALSE | Upregulated |
| 0,324153335 | 0,472257437 | LOC102723806 | FALSE | Upregulated |
| 0,5375682 | 0,472257437 | MCCD1 | FALSE | Upregulated |
| 0,346192427 | 0,472257437 | MIR210HG | FALSE | Upregulated |
| 0,241876516 | 0,472257437 | NIPA2 | FALSE | Upregulated |
| 0,244198377 | 0,472257437 | RUFY3 | FALSE | Upregulated |
| 0,385219089 | 0,472257437 | SLC35G6 | FALSE | Upregulated |
| -0,693737918 | 0,472258007 | C12orf4 | FALSE | Downregulated |
| 0,228103914 | 0,472258007 | CD3EAP | FALSE | Upregulated |
| -0,234012413 | 0,472258007 | CHCHD10 | FALSE | Downregulated |
| -0,382549751 | 0,472258007 | DEPDC7 | FALSE | Downregulated |
| -0,390125456 | 0,472258007 | EVI5 | FALSE | Downregulated |
| -0,272699659 | 0,472258007 | F13B | FALSE | Downregulated |
| 0,185577188 | 0,472258007 | FAM208A | FALSE | Upregulated |
| -0,491505016 | 0,472258007 | HAUS5 | FALSE | Downregulated |
| 0,542831026 | 0,472258007 | HSPB6 | FALSE | Upregulated |
| 0,243647949 | 0,472258007 | JAZF1 | FALSE | Upregulated |

| | | | | |
|---|---|---|---|---|
| 0,708295188 | 0,472258007 | KPNA5 | FALSE | Upregulated |
| 0,177846429 | 0,472258007 | MAP3K13 | FALSE | Upregulated |
| 0,412820441 | 0,472258007 | NGFRAP1 | FALSE | Upregulated |
| -0,410966997 | 0,472258007 | PIDD | FALSE | Downregulated |
| -0,178958792 | 0,472258007 | RPL18 | FALSE | Downregulated |
| -0,256197896 | 0,472258007 | SH3BGRL2 | FALSE | Downregulated |
| 0,607027004 | 0,472258007 | TTF2 | FALSE | Upregulated |
| -0,402028852 | 0,472258007 | UBIAD1 | FALSE | Downregulated |
| -0,432538095 | 0,472258007 | ZNF574 | FALSE | Downregulated |
| 0,209867972 | 0,472259564 | MAP3K12 | FALSE | Upregulated |
| 0,618685545 | 0,472259564 | TC2N | FALSE | Upregulated |
| -0,316814031 | 0,472259564 | TTLL12 | FALSE | Downregulated |
| -0,48846126 | 0,472259564 | YAF2 | FALSE | Downregulated |
| 0,309711113 | 0,472259564 | ZNF789 | FALSE | Upregulated |
| -0,31588453 | 0,472691991 | PIN1 | FALSE | Downregulated |
| -0,262733951 | 0,472937195 | SLC37A4 | FALSE | Downregulated |
| -0,243209539 | 0,473403738 | DNAH1 | FALSE | Downregulated |
| -0,150535671 | 0,473403738 | GNE | FALSE | Downregulated |
| 0,227441387 | 0,473403738 | LRRN3 | FALSE | Upregulated |
| 0,180632679 | 0,473747409 | ATN1 | FALSE | Upregulated |
| -0,300185512 | 0,473747409 | DMD | FALSE | Downregulated |
| -0,382917328 | 0,473747409 | MIOS | FALSE | Downregulated |
| 0,707037615 | 0,473747409 | RCOR2 | FALSE | Upregulated |
| 0,364634389 | 0,473762411 | ATAD1 | FALSE | Upregulated |
| -0,333596023 | 0,473762411 | CA2 | FALSE | Downregulated |
| 0,286045095 | 0,473762411 | CD74 | FALSE | Upregulated |
| -0,57557762 | 0,473762411 | GARNL3 | FALSE | Downregulated |
| 0,524207892 | 0,473762411 | HMGN4 | FALSE | Upregulated |
| 0,367475868 | 0,473762411 | ITGAL | FALSE | Upregulated |
| 0,475574151 | 0,473762411 | LMO3 | FALSE | Upregulated |
| 0,213467635 | 0,473762411 | PIEZO1 | FALSE | Upregulated |
| 0,33931379 | 0,473762411 | PLXNA1 | FALSE | Upregulated |
| 0,738333134 | 0,473762411 | RP11-64D24.4 | FALSE | Upregulated |
| -0,339915899 | 0,473762411 | ZNF747 | FALSE | Downregulated |
| -0,331940419 | 0,473762411 | ZNRF3 | FALSE | Downregulated |
| -0,220495011 | 0,473979413 | ABHD15 | FALSE | Downregulated |
| -0,33346591 | 0,473979413 | TTC7B | FALSE | Downregulated |
| -0,582554204 | 0,474428339 | PHF6 | FALSE | Downregulated |
| -0,237314601 | 0,474552796 | KPNA6 | FALSE | Downregulated |
| 0,559259469 | 0,474552796 | RASAL3 | FALSE | Upregulated |
| 0,294688975 | 0,474552796 | RTN4RL2 | FALSE | Upregulated |
| -0,295186148 | 0,474552796 | SEPT10 | FALSE | Downregulated |
| -0,311930754 | 0,475198194 | SCARNA7 | FALSE | Downregulated |
| 0,314947871 | 0,47546831 | NAP1L5 | FALSE | Upregulated |
| 0,254240655 | 0,475780128 | LOC100505736 | FALSE | Upregulated |
| -0,200364963 | 0,475782387 | ARHGAP5 | FALSE | Downregulated |
| 0,230237695 | 0,475782387 | PANK2 | FALSE | Upregulated |
| 0,379260186 | 0,475782387 | PHF11 | FALSE | Upregulated |
| -0,190765209 | 0,475782387 | POLRMT | FALSE | Downregulated |
| 0,34738459 | 0,475782387 | SPRYD3 | FALSE | Upregulated |
| 0,348836825 | 0,47583652 | ATAT1 | FALSE | Upregulated |

| | | | | |
|---|---|---|---|---|
| 0,25900977 | 0,47583652 | NCBP2 | FALSE | Upregulated |
| 0,205931573 | 0,476099707 | LOC148696 | FALSE | Upregulated |
| 0,366708845 | 0,476734816 | CCNT2-AS1 | FALSE | Upregulated |
| 0,256726901 | 0,476993031 | AQP11 | FALSE | Upregulated |
| 0,410063638 | 0,476993031 | COL1A2 | FALSE | Upregulated |
| 0,3280452 | 0,476993031 | ELL3 | FALSE | Upregulated |
| -0,183114814 | 0,476993031 | ETFB | FALSE | Downregulated |
| -0,368556461 | 0,476993031 | LANCL1 | FALSE | Downregulated |
| 0,341217703 | 0,476993031 | LOC101927732 | FALSE | Upregulated |
| 0,605359472 | 0,476993031 | LOC101929698 | FALSE | Upregulated |
| 0,45863432 | 0,476993031 | LOC390937 | FALSE | Upregulated |
| -0,244302679 | 0,476993031 | LRRC3DN | FALSE | Downregulated |
| 0,340300378 | 0,476993031 | PRKAR2A-AS1 | FALSE | Upregulated |
| 0,49699016 | 0,476993031 | PSD | FALSE | Upregulated |
| 0,243061299 | 0,476993031 | RHOBTB2 | FALSE | Upregulated |
| -0,231479419 | 0,476993031 | RPSA | FALSE | Downregulated |
| 0,398245018 | 0,476993031 | TMEM132A | FALSE | Upregulated |
| 0,264084376 | 0,477504302 | LOC102724559 | FALSE | Upregulated |
| 0,519921876 | 0,477733822 | CCL21 | FALSE | Upregulated |
| 0,343848223 | 0,477733822 | DMTN | FALSE | Upregulated |
| 0,557927765 | 0,477733822 | EGR2 | FALSE | Upregulated |
| 0,17821175 | 0,478389967 | GATC | FALSE | Upregulated |
| 0,233140992 | 0,478389967 | RSBN1L-AS1 | FALSE | Upregulated |
| 0,411605668 | 0,478471181 | CLEC12A | FALSE | Upregulated |
| 0,206375494 | 0,478471181 | LOC102723521 | FALSE | Upregulated |
| -0,203806326 | 0,479093825 | SMC1A | FALSE | Downregulated |
| -0,149963667 | 0,479273398 | DCAF8 | FALSE | Downregulated |
| -0,212199318 | 0,479332618 | SHC2 | FALSE | Downregulated |
| -0,561931155 | 0,479382116 | MTA3 | FALSE | Downregulated |
| 0,2024301 | 0,479382116 | SMAD3 | FALSE | Upregulated |
| 0,366353228 | 0,47995185 | ADRB2 | FALSE | Upregulated |
| -0,318947813 | 0,480145674 | AP3B1 | FALSE | Downregulated |
| -0,433229149 | 0,480145674 | ARNTL | FALSE | Downregulated |
| -0,298501347 | 0,480145674 | C19orf60 | FALSE | Downregulated |
| -0,330482273 | 0,480145674 | CPNE1 | FALSE | Downregulated |
| 0,2058239 | 0,480145674 | SNRK-AS1 | FALSE | Upregulated |
| -0,303658395 | 0,480145674 | TLK1 | FALSE | Downregulated |
| 0,24917762 | 0,480253383 | FKBP1AP1 | FALSE | Upregulated |
| 0,217276771 | 0,480253383 | RASA1 | FALSE | Upregulated |
| -0,234663869 | 0,480396395 | GOLGA1 | FALSE | Downregulated |
| 0,229538088 | 0,480692153 | ANGPTL1 | FALSE | Upregulated |
| 0,318742724 | 0,480692153 | LOC102060414 | FALSE | Upregulated |
| -0,25359363 | 0,480692153 | PAH | FALSE | Downregulated |
| -0,151843448 | 0,480719332 | ACO2 | FALSE | Downregulated |
| -0,136221154 | 0,480719332 | COL18A1 | FALSE | Downregulated |
| 0,292788544 | 0,480719332 | LOC101929017 | FALSE | Upregulated |
| -0,482209148 | 0,480719332 | TBP | FALSE | Downregulated |
| 0,338142059 | 0,480719332 | TMEM41A | FALSE | Upregulated |
| 0,371431657 | 0,480814724 | SPI1 | FALSE | Upregulated |
| 0,376271439 | 0,480949571 | DNAJC24 | FALSE | Upregulated |
| -0,262621793 | 0,480949571 | EIF3D | FALSE | Downregulated |

| | | | | |
|---|---|---|---|---|
| 0,936342838 | 0,480949571 | FLVCR1 | FALSE | Upregulated |
| 0,206642487 | 0,480949571 | PAQR3 | FALSE | Upregulated |
| 0,223388315 | 0,480949571 | PHACTR2 | FALSE | Upregulated |
| -0,213356093 | 0,480949571 | ZNF451 | FALSE | Downregulated |
| -0,232612732 | 0,480949571 | ZNHIT1 | FALSE | Downregulated |
| -0,24480869 | 0,481654539 | ANO1 | FALSE | Downregulated |
| 0,370231265 | 0,481746892 | CSNK1G2-AS1 | FALSE | Upregulated |
| -0,533718376 | 0,481746892 | HERC5 | FALSE | Downregulated |
| 0,667012544 | 0,481746892 | LOC101241902 | FALSE | Upregulated |
| 0,534126864 | 0,481746892 | NR2C2AP | FALSE | Upregulated |
| 0,302620759 | 0,481746892 | OPA3 | FALSE | Upregulated |
| 0,331039134 | 0,481746892 | UBE2D2 | FALSE | Upregulated |
| -0,218169886 | 0,482328798 | FAM193A | FALSE | Downregulated |
| -0,243914714 | 0,482328798 | HPX | FALSE | Downregulated |
| 0,598297869 | 0,48300338 | CMPK2 | FALSE | Upregulated |
| 0,225956353 | 0,48300338 | DNM1L | FALSE | Upregulated |
| -0,229023563 | 0,48300338 | MOGS | FALSE | Downregulated |
| -0,154469586 | 0,484240742 | EIF4EBP2 | FALSE | Downregulated |
| 0,236232321 | 0,484980865 | LOC100505564 | FALSE | Upregulated |
| -0,331440927 | 0,484980865 | NDUFA11 | FALSE | Downregulated |
| -0,232066913 | 0,484980865 | TMEM176A | FALSE | Downregulated |
| -0,155147474 | 0,485383234 | HSPA9 | FALSE | Downregulated |
| 0,381544851 | 0,485620464 | IGSF23 | FALSE | Upregulated |
| -0,234709532 | 0,485620464 | POLE | FALSE | Downregulated |
| 0,290494321 | 0,485650632 | LRRC39 | FALSE | Upregulated |
| -0,225953276 | 0,485837276 | FTL | FALSE | Downregulated |
| 0,27360941 | 0,485837276 | ZNF529 | FALSE | Upregulated |
| -0,227575024 | 0,486066352 | APOF | FALSE | Downregulated |
| 0,151613859 | 0,486066352 | BACE1 | FALSE | Upregulated |
| -0,284909279 | 0,487567709 | GABPA | FALSE | Downregulated |
| 0,460527309 | 0,487567709 | RPS6KA1 | FALSE | Upregulated |
| 0,410597078 | 0,487727705 | SESN1 | FALSE | Upregulated |
| -0,312125727 | 0,487727705 | TTC14 | FALSE | Downregulated |
| -0,229254286 | 0,487971457 | TMX4 | FALSE | Downregulated |
| -0,208745307 | 0,488272441 | AR | FALSE | Downregulated |
| 0,572394184 | 0,488503799 | RP11-473M20.7 | FALSE | Upregulated |
| 0,391957089 | 0,488603417 | AP1S2 | FALSE | Upregulated |
| -0,329113258 | 0,488603417 | SLC19A2 | FALSE | Downregulated |
| -0,271488833 | 0,488677786 | CPSF6 | FALSE | Downregulated |
| 0,234681981 | 0,488677786 | SLC18A2 | FALSE | Upregulated |
| 0,336194832 | 0,48877947 | ANKDD1A | FALSE | Upregulated |
| 0,32235077 | 0,48877947 | C1QC | FALSE | Upregulated |
| -0,454730697 | 0,48877947 | CBLC | FALSE | Downregulated |
| -0,153430162 | 0,48877947 | CDHR5 | FALSE | Downregulated |
| -0,442268875 | 0,48877947 | MRPL46 | FALSE | Downregulated |
| 0,60472275 | 0,48877947 | UXS1 | FALSE | Upregulated |
| -0,218540648 | 0,489687152 | ECI1 | FALSE | Downregulated |
| -0,29535048 | 0,489687152 | NINL | FALSE | Downregulated |
| -0,262100241 | 0,489687152 | RBM26 | FALSE | Downregulated |
| 0,571097984 | 0,490159822 | FAM129A | FALSE | Upregulated |
| 0,399378768 | 0,490159822 | IKZF1 | FALSE | Upregulated |

| | | | | |
|---|---|---|---|---|
| -0,364486363 | 0,491194405 | ALG2 | FALSE | Downregulated |
| 0,299468063 | 0,491194405 | NUTM2B | FALSE | Upregulated |
| 0,255304242 | 0,491194405 | SOCS6 | FALSE | Upregulated |
| -0,405902695 | 0,492304871 | ARHGEF28 | FALSE | Downregulated |
| -0,338851993 | 0,492304871 | ERRFI1 | FALSE | Downregulated |
| -0,249569295 | 0,492304871 | TMPO | FALSE | Downregulated |
| 0,36837586 | 0,49354636 | AFAP1-AS1 | FALSE | Upregulated |
| 0,242771525 | 0,49354636 | AGK | FALSE | Upregulated |
| 0,308825593 | 0,49354636 | ARRDC2 | FALSE | Upregulated |
| 0,323615812 | 0,49354636 | ASB14 | FALSE | Upregulated |
| -0,352263091 | 0,49354636 | GYG2 | FALSE | Downregulated |
| -0,242939046 | 0,49354636 | IRS2 | FALSE | Downregulated |
| 0,444756038 | 0,49354636 | LOC100507346 | FALSE | Upregulated |
| 0,23501443 | 0,49354636 | LOC101929333 | FALSE | Upregulated |
| -0,209749913 | 0,49354636 | MTTP | FALSE | Downregulated |
| 0,320673861 | 0,49354636 | NTN3 | FALSE | Upregulated |
| 0,579761535 | 0,49354636 | SCRG1 | FALSE | Upregulated |
| -0,381023357 | 0,49354636 | TACO1 | FALSE | Downregulated |
| -0,490650983 | 0,49354636 | TIMM17B | FALSE | Downregulated |
| -0,352374416 | 0,49354636 | TLN2 | FALSE | Downregulated |
| -0,510300972 | 0,493614945 | BEND7 | FALSE | Downregulated |
| 0,244252494 | 0,493614945 | XRCC3 | FALSE | Upregulated |
| -0,286546611 | 0,49431541 | ACSL3 | FALSE | Downregulated |
| -0,184764424 | 0,49431541 | BCKDHA | FALSE | Downregulated |
| 0,5174628 | 0,49431541 | FAM151A | FALSE | Upregulated |
| 0,34303848 | 0,49431541 | FYN | FALSE | Upregulated |
| 0,296648877 | 0,49431541 | KIFC2 | FALSE | Upregulated |
| 0,552189622 | 0,49431541 | PDSS1 | FALSE | Upregulated |
| 0,333440519 | 0,49431541 | RPS6KA4 | FALSE | Upregulated |
| 0,383391931 | 0,49431541 | TRIM65 | FALSE | Upregulated |
| 0,307735324 | 0,49431541 | ZNF496 | FALSE | Upregulated |
| -0,278408033 | 0,494558065 | ARMC6 | FALSE | Downregulated |
| 0,576838277 | 0,494558065 | ATP6V0E2-AS1 | FALSE | Upregulated |
| 0,484599195 | 0,494558065 | C21orf91 | FALSE | Upregulated |
| 0,215242836 | 0,494558065 | CHKA | FALSE | Upregulated |
| 0,300523983 | 0,494558065 | CNST | FALSE | Upregulated |
| -0,157714656 | 0,494558065 | F10 | FALSE | Downregulated |
| -0,399225265 | 0,494558065 | FAM3C | FALSE | Downregulated |
| 0,423708995 | 0,494558065 | HSPB9 | FALSE | Upregulated |
| 0,407003296 | 0,494558065 | LDLR | FALSE | Upregulated |
| -0,41125417 | 0,494558065 | LINC01093 | FALSE | Downregulated |
| 0,670852321 | 0,494558065 | LOC101926994 | FALSE | Upregulated |
| -0,411830153 | 0,494558065 | LOC101928783 | FALSE | Downregulated |
| -0,377072573 | 0,494558065 | MTRR | FALSE | Downregulated |
| -0,187880231 | 0,494558065 | NMT1 | FALSE | Downregulated |
| -0,298133819 | 0,494558065 | PCOLCE | FALSE | Downregulated |
| -0,237271606 | 0,494558065 | RPS28 | FALSE | Downregulated |
| -0,252357799 | 0,494558065 | SLC43A1 | FALSE | Downregulated |
| -0,481189944 | 0,494558065 | TIMM10 | FALSE | Downregulated |
| -0,235596579 | 0,495164798 | RPL38 | FALSE | Downregulated |
| 0,421262773 | 0,495169591 | TTC13 | FALSE | Upregulated |

| | | | | |
|---|---|---|---|---|
| -0,213216963 | 0,495679785 | ATP5B | FALSE | Downregulated |
| -0,29552374 | 0,495679785 | DNASE1L3 | FALSE | Downregulated |
| 0,421696492 | 0,495679785 | RGS20 | FALSE | Upregulated |
| -0,440243856 | 0,495679785 | RNF135 | FALSE | Downregulated |
| -0,405074557 | 0,496182436 | ADAT1 | FALSE | Downregulated |
| 0,246866922 | 0,496725745 | SP2 | FALSE | Upregulated |
| -0,384207208 | 0,496833136 | MCOLN1 | FALSE | Downregulated |
| 0,388260208 | 0,497757166 | IMPG1 | FALSE | Upregulated |
| 1,382983441 | 0,497757166 | LOC101927055 | FALSE | Upregulated |
| 0,830357518 | 0,497757166 | LOC101927255 | FALSE | Upregulated |
| 0,275103471 | 0,497757166 | LOC102724484 | FALSE | Upregulated |
| -0,254403484 | 0,497757166 | MET | FALSE | Downregulated |
| 0,308355171 | 0,497757166 | PLCH2 | FALSE | Upregulated |
| -0,167827914 | 0,498022679 | RPL32 | FALSE | Downregulated |
| 0,182107083 | 0,498345521 | GNPNAT1 | FALSE | Upregulated |
| -0,333099694 | 0,498345521 | HES1 | FALSE | Downregulated |
| 0,227935068 | 0,498345521 | MIRLET7DHG | FALSE | Upregulated |
| 0,244087781 | 0,498345521 | SRP19 | FALSE | Upregulated |
| 0,336113783 | 0,498345521 | TARBP1 | FALSE | Upregulated |
| 0,300733863 | 0,498345521 | ZCWPW1 | FALSE | Upregulated |
| -0,337284922 | 0,498345521 | ZNF641 | FALSE | Downregulated |
| 0,204494007 | 0,498562094 | ARG2 | FALSE | Upregulated |
| 0,325075033 | 0,498562094 | MVP | FALSE | Upregulated |
| 0,429249733 | 0,498562094 | RGS17 | FALSE | Upregulated |
| 0,321268494 | 0,498734676 | SPIN2A | FALSE | Upregulated |
| 0,35998295 | 0,498741006 | DUSP14 | FALSE | Upregulated |
| -0,518128635 | 0,498741006 | EAPP | FALSE | Downregulated |
| 0,223609703 | 0,498741006 | NEXN | FALSE | Upregulated |
| -0,250517405 | 0,499191592 | ATP5G3 | FALSE | Downregulated |
| -0,196712024 | 0,499191592 | ATXN1L | FALSE | Downregulated |
| 0,270567592 | 0,499191592 | BCCIP | FALSE | Upregulated |
| -0,211362727 | 0,499191592 | DCAF6 | FALSE | Downregulated |
| 0,713788303 | 0,499191592 | DGKA | FALSE | Upregulated |
| -0,357189248 | 0,499191592 | DMRTA1 | FALSE | Downregulated |
| 0,500216511 | 0,499191592 | DOK2 | FALSE | Upregulated |
| -0,396505086 | 0,499191592 | EIF4A3 | FALSE | Downregulated |
| 0,233184725 | 0,499191592 | FAM149B1 | FALSE | Upregulated |
| -0,636296516 | 0,499191592 | IPO9-AS1 | FALSE | Downregulated |
| -0,398246323 | 0,499191592 | LPA | FALSE | Downregulated |
| -0,140886988 | 0,499191592 | POLR2A | FALSE | Downregulated |
| 0,189910467 | 0,499191592 | POLR2J4 | FALSE | Upregulated |
| 0,295851464 | 0,499191592 | PRC1 | FALSE | Upregulated |
| -0,195485922 | 0,499191592 | RAB43 | FALSE | Downregulated |
| -0,195825226 | 0,499191592 | RMND5A | FALSE | Downregulated |
| 0,518122937 | 0,499191592 | RNF8 | FALSE | Upregulated |
| -0,23392502 | 0,499191592 | RPS3 | FALSE | Downregulated |
| -0,182169207 | 0,499191592 | RPS3A | FALSE | Downregulated |
| -0,142272065 | 0,499191592 | UBR4 | FALSE | Downregulated |
| 0,191198866 | 0,499363788 | FAM65A | FALSE | Upregulated |
| 0,407264719 | 0,499494556 | PPP2R3C | FALSE | Upregulated |
| -0,271219631 | 0,499975557 | OTUB1 | FALSE | Downregulated |

| | | | | |
|---|---|---|---|---|
| -0,417743637 | 0,500521063 | N4BP2L2-IT2 | FALSE | Downregulated |
| 0,634267823 | 0,500521063 | PROB1 | FALSE | Upregulated |
| 0,635984232 | 0,501117915 | HMGB2 | FALSE | Upregulated |
| 0,250990248 | 0,501325547 | PCDH12 | FALSE | Upregulated |
| 0,368189937 | 0,501325547 | ZNF518B | FALSE | Upregulated |
| 0,853117028 | 0,50139487 | MEPE | FALSE | Upregulated |
| 0,549995362 | 0,50139487 | NPAS2 | FALSE | Upregulated |
| -0,235125157 | 0,50181001 | CHD1 | FALSE | Downregulated |
| -0,508549325 | 0,502313554 | AGPAT5 | FALSE | Downregulated |
| -0,607295228 | 0,502313554 | APOC1P1 | FALSE | Downregulated |
| -0,143056661 | 0,502313554 | CERS2 | FALSE | Downregulated |
| -0,265963027 | 0,502313554 | EIF3F | FALSE | Downregulated |
| 0,397076891 | 0,502313554 | HK3 | FALSE | Upregulated |
| -0,260903081 | 0,502313554 | HSPA5 | FALSE | Downregulated |
| 0,259028738 | 0,502313554 | LOC100130744 | FALSE | Upregulated |
| 0,279709207 | 0,502313554 | RPL13AP6 | FALSE | Upregulated |
| 0,426272645 | 0,503667187 | CDT1 | FALSE | Upregulated |
| 0,551457575 | 0,503667187 | FGF14 | FALSE | Upregulated |
| 0,515000757 | 0,503667187 | LOC102723382 | FALSE | Upregulated |
| -0,226748285 | 0,503667187 | TMEM97 | FALSE | Downregulated |
| 0,20389729 | 0,503675879 | CC2D1A | FALSE | Upregulated |
| 0,436677174 | 0,503675879 | ZBTB33 | FALSE | Upregulated |
| -0,233910112 | 0,5049991 | PPP1R15B | FALSE | Downregulated |
| -0,226963504 | 0,505395258 | NDRG2 | FALSE | Downregulated |
| 0,243971742 | 0,505395258 | USP45 | FALSE | Upregulated |
| -0,293355419 | 0,505479939 | C9orf78 | FALSE | Downregulated |
| 0,265872833 | 0,505603778 | CORO1C | FALSE | Upregulated |
| -0,272935576 | 0,505603778 | PALM2 | FALSE | Downregulated |
| -0,255978741 | 0,505603778 | PSMD4 | FALSE | Downregulated |
| -0,242293062 | 0,505603778 | TRPC4AP | FALSE | Downregulated |
| 0,526529623 | 0,505603778 | TTC39B | FALSE | Upregulated |
| 0,31754887 | 0,505968786 | GBP2 | FALSE | Upregulated |
| 0,525391062 | 0,505996658 | WAS | FALSE | Upregulated |
| -0,324707744 | 0,506859889 | AHSA1 | FALSE | Downregulated |
| 0,374302225 | 0,506859889 | CDK8 | FALSE | Upregulated |
| -0,240839283 | 0,506859889 | CPS1 | FALSE | Downregulated |
| 0,609765647 | 0,506859889 | FGF7 | FALSE | Upregulated |
| -0,203925876 | 0,506859889 | KHDRBS1 | FALSE | Downregulated |
| -1,052663831 | 0,506859889 | LOC100499484-C9ORF174 | FALSE | Downregulated |
| 0,363733992 | 0,506859889 | LOC730668 | FALSE | Upregulated |
| -0,526907492 | 0,506859889 | MEG3 | FALSE | Downregulated |
| 0,510174797 | 0,506859889 | MYOZ3 | FALSE | Upregulated |
| 0,534735106 | 0,506859889 | NHS | FALSE | Upregulated |
| 0,251789622 | 0,506859889 | RAB6A | FALSE | Upregulated |
| 0,31841653 | 0,506859889 | SPAG1 | FALSE | Upregulated |
| 0,170952296 | 0,506859889 | TCIRG1 | FALSE | Upregulated |
| -0,22029508 | 0,506859889 | TMEM63B | FALSE | Downregulated |
| -0,216064313 | 0,506859889 | TRIM27 | FALSE | Downregulated |
| 0,286191638 | 0,507025409 | AKAP5 | FALSE | Upregulated |
| -0,448992829 | 0,507025409 | MRPL55 | FALSE | Downregulated |

| | | | | |
|---|---|---|---|---|
| 0,317632415 | 0,507025409 | TTYH3 | FALSE | Upregulated |
| -0,259402952 | 0,507110029 | HMGCL | FALSE | Downregulated |
| -0,570545014 | 0,50711078 | DPY19L2P2 | FALSE | Downregulated |
| 0,195847685 | 0,50711078 | PRKD3 | FALSE | Upregulated |
| -0,334677965 | 0,507884494 | UTP20 | FALSE | Downregulated |
| -0,189019311 | 0,507931672 | ARHGAP21 | FALSE | Downregulated |
| 0,605041853 | 0,507931672 | CD27 | FALSE | Upregulated |
| 0,267479278 | 0,508167298 | NEIL1 | FALSE | Upregulated |
| -0,307616324 | 0,508559617 | LOC100130872 | FALSE | Downregulated |
| 0,293015093 | 0,508559617 | MRS2 | FALSE | Upregulated |
| 0,325126487 | 0,508859505 | FAM53C | FALSE | Upregulated |
| 0,644594059 | 0,508859505 | PLEKHG4B | FALSE | Upregulated |
| 0,525725868 | 0,508859505 | WDR75 | FALSE | Upregulated |
| -0,215081711 | 0,508988235 | ERGIC3 | FALSE | Downregulated |
| 0,330069779 | 0,509596101 | GPCPD1 | FALSE | Upregulated |
| 0,158586807 | 0,509913365 | BAIAP2L1 | FALSE | Upregulated |
| 0,31377586 | 0,509913365 | C5orf45 | FALSE | Upregulated |
| -0,170272755 | 0,509913365 | DIDO1 | FALSE | Downregulated |
| 0,207179988 | 0,509913365 | FAM160B2 | FALSE | Upregulated |
| 0,213654927 | 0,509913365 | GNPTAB | FALSE | Upregulated |
| 0,401521927 | 0,509913365 | MFAP3 | FALSE | Upregulated |
| 0,30368765 | 0,509913365 | MUC6 | FALSE | Upregulated |
| 0,981777755 | 0,509913365 | NRSN1 | FALSE | Upregulated |
| 0,206433812 | 0,509913365 | RABGAP1L | FALSE | Upregulated |
| 0,26311373 | 0,509913365 | TOR1B | FALSE | Upregulated |
| 0,404589681 | 0,510092801 | PNPLA8 | FALSE | Upregulated |
| 0,241811119 | 0,510264805 | CARD14 | FALSE | Upregulated |
| 0,422882259 | 0,510264805 | ETV3 | FALSE | Upregulated |
| 0,510715004 | 0,510264805 | LINC00641 | FALSE | Upregulated |
| -0,579539787 | 0,510264805 | LOC220729 | FALSE | Downregulated |
| -0,178922598 | 0,510264805 | NFX1 | FALSE | Downregulated |
| -0,230233249 | 0,510264805 | ZCCHC6 | FALSE | Downregulated |
| 0,314442083 | 0,510325634 | AKNA | FALSE | Upregulated |
| -0,216182712 | 0,510325634 | APH1A | FALSE | Downregulated |
| -0,236187945 | 0,510325634 | CADM1 | FALSE | Downregulated |
| 0,244208242 | 0,510325634 | CDC42SE2 | FALSE | Upregulated |
| -0,484997106 | 0,510325634 | DDT | FALSE | Downregulated |
| 0,237492195 | 0,510325634 | GLIS2 | FALSE | Upregulated |
| -0,254267858 | 0,510325634 | HRSP12 | FALSE | Downregulated |
| 0,35520024 | 0,510325634 | PAGR1 | FALSE | Upregulated |
| -0,476054279 | 0,510325634 | PCOLCE2 | FALSE | Downregulated |
| 0,267230769 | 0,510325634 | TAGLN2 | FALSE | Upregulated |
| 0,416030392 | 0,510325634 | TMEM74B | FALSE | Upregulated |
| -0,272877559 | 0,510325634 | UBE2J2 | FALSE | Downregulated |
| 0,215852019 | 0,510325634 | ZNF7 | FALSE | Upregulated |
| 0,433428554 | 0,510331955 | DPAGT1 | FALSE | Upregulated |
| 0,26841881 | 0,510331955 | FAM111A | FALSE | Upregulated |
| 0,141904561 | 0,510637984 | TRIM8 | FALSE | Upregulated |
| 0,466323251 | 0,511124135 | GPSM3 | FALSE | Upregulated |
| 0,285302142 | 0,511124135 | LOC101927066 | FALSE | Upregulated |
| 0,683960589 | 0,511124135 | ZNF878 | FALSE | Upregulated |

| | | | | |
|---|---|---|---|---|
| -0,215496635 | 0,511691961 | ADAMTS13 | FALSE | Downregulated |
| 0,272926581 | 0,511691961 | AP000688.14 | FALSE | Upregulated |
| 0,3580179 | 0,511691961 | CD53 | FALSE | Upregulated |
| -0,234771528 | 0,511691961 | CYP4F2 | FALSE | Downregulated |
| 0,549133881 | 0,511691961 | IL27RA | FALSE | Upregulated |
| 0,272442343 | 0,511691961 | LOC101928621 | FALSE | Upregulated |
| -0,210449098 | 0,511691961 | NR1H3 | FALSE | Downregulated |
| 0,392223191 | 0,511691961 | PBX1 | FALSE | Upregulated |
| -0,164659521 | 0,511691961 | STARD10 | FALSE | Downregulated |
| -0,343740606 | 0,511691961 | TMEM64 | FALSE | Downregulated |
| 0,353349434 | 0,511691961 | USP13 | FALSE | Upregulated |
| -0,392103227 | 0,511753302 | ERCC2 | FALSE | Downregulated |
| 0,402448253 | 0,511753302 | GNG7 | FALSE | Upregulated |
| -0,238866638 | 0,511916569 | CTIF | FALSE | Downregulated |
| -0,286330315 | 0,511949908 | CDKN1B | FALSE | Downregulated |
| 0,220050215 | 0,511949908 | NFE2L3 | FALSE | Upregulated |
| 0,241930621 | 0,511949908 | NRON | FALSE | Upregulated |
| 0,501976588 | 0,512300747 | TRIM45 | FALSE | Upregulated |
| -0,389028769 | 0,512430859 | G6PC | FALSE | Downregulated |
| -0,277525169 | 0,512678565 | FAM160A2 | FALSE | Downregulated |
| -0,550880968 | 0,512678565 | GTF2H5 | FALSE | Downregulated |
| 0,456644794 | 0,513135272 | CELF2 | FALSE | Upregulated |
| 0,448923486 | 0,513135272 | LOC100287934 | FALSE | Upregulated |
| -0,416838105 | 0,513135272 | PIGN | FALSE | Downregulated |
| 0,203816778 | 0,513135272 | SP100 | FALSE | Upregulated |
| 0,414000975 | 0,513156632 | ME3 | FALSE | Upregulated |
| 0,421940574 | 0,513357511 | ABHD13 | FALSE | Upregulated |
| 0,329381159 | 0,513639035 | LOC101928555 | FALSE | Upregulated |
| -0,174712851 | 0,513639035 | SERPIND1 | FALSE | Downregulated |
| -0,20457164 | 0,513878049 | AHCTF1 | FALSE | Downregulated |
| 0,433496765 | 0,513878049 | RP11-387D10.3 | FALSE | Upregulated |
| -0,144266692 | 0,514803266 | LRP5 | FALSE | Downregulated |
| -0,356555666 | 0,515025591 | TRIM55 | FALSE | Downregulated |
| 0,177399108 | 0,515233044 | BRWD1 | FALSE | Upregulated |
| 0,425919745 | 0,515233044 | IQCK | FALSE | Upregulated |
| 0,352591106 | 0,515364992 | GNRH1 | FALSE | Upregulated |
| 0,304014841 | 0,515978642 | PSMB8 | FALSE | Upregulated |
| 0,241238589 | 0,516091875 | MIR600HG | FALSE | Upregulated |
| -0,193454056 | 0,516199265 | ALDH8A1 | FALSE | Downregulated |
| -0,892892743 | 0,516199265 | XPNPEP2 | FALSE | Downregulated |
| -0,188991841 | 0,516244676 | ABCC9 | FALSE | Downregulated |
| 0,585574696 | 0,516244676 | AP3M2 | FALSE | Upregulated |
| -0,178457892 | 0,516244676 | APOE | FALSE | Downregulated |
| -0,377396258 | 0,516244676 | CUX2 | FALSE | Downregulated |
| -0,441367199 | 0,516244676 | STARD4 | FALSE | Downregulated |
| 0,564258758 | 0,516411414 | TPRG1-AS2 | FALSE | Upregulated |
| 0,39552474 | 0,516463282 | LOC101927124 | FALSE | Upregulated |
| -0,231349529 | 0,516658681 | HERPUD1 | FALSE | Downregulated |
| -0,314972278 | 0,516658681 | MRPL4 | FALSE | Downregulated |
| -0,157462968 | 0,516658681 | RPL8 | FALSE | Downregulated |
| -0,342599347 | 0,516835533 | KIAA0753 | FALSE | Downregulated |

| | | | | |
|---|---|---|---|---|
| 0,328385593 | 0,517103118 | CDC25B | FALSE | Upregulated |
| 0,24397591 | 0,517103118 | NDST2 | FALSE | Upregulated |
| -0,392711333 | 0,517740649 | APOOL | FALSE | Downregulated |
| 0,434381709 | 0,517740649 | CHMP2B | FALSE | Upregulated |
| -0,259679318 | 0,517740649 | DHX15 | FALSE | Downregulated |
| -0,362896604 | 0,517740649 | DHX37 | FALSE | Downregulated |
| 0,204881671 | 0,517740649 | GREB1L | FALSE | Upregulated |
| 0,278274512 | 0,517740649 | HLA-C | FALSE | Upregulated |
| 0,552508261 | 0,517740649 | LGALS4 | FALSE | Upregulated |
| 0,397273498 | 0,517740649 | VSIG4 | FALSE | Upregulated |
| -0,314789302 | 0,517973764 | SNX4 | FALSE | Downregulated |
| 0,239792064 | 0,51805107 | GDAP1L1 | FALSE | Upregulated |
| -0,484211339 | 0,518094744 | ARHGEF15 | FALSE | Downregulated |
| 0,288801984 | 0,518094744 | DHX32 | FALSE | Upregulated |
| 0,401250562 | 0,518094744 | ETNK1 | FALSE | Upregulated |
| -0,165617545 | 0,518094744 | FAM120A | FALSE | Downregulated |
| 0,392736072 | 0,518094744 | FAT1 | FALSE | Upregulated |
| 0,37482496 | 0,518094744 | HLA-J | FALSE | Upregulated |
| -0,535390553 | 0,518094744 | HPRT1 | FALSE | Downregulated |
| 0,304635226 | 0,518094744 | IFNK | FALSE | Upregulated |
| -0,248933562 | 0,518094744 | ITPKC | FALSE | Downregulated |
| -0,185792142 | 0,518094744 | KLHDC2 | FALSE | Downregulated |
| -0,555648544 | 0,518094744 | MOB2 | FALSE | Downregulated |
| -0,273868001 | 0,518094744 | NARS | FALSE | Downregulated |
| 0,464899844 | 0,518094744 | PI4KAP2 | FALSE | Upregulated |
| -0,228016318 | 0,518094744 | POLD2 | FALSE | Downregulated |
| -0,439693824 | 0,518094744 | PREPL | FALSE | Downregulated |
| 0,506850581 | 0,518094744 | PRKCB | FALSE | Upregulated |
| 0,480932623 | 0,518094744 | PRKG1-AS1 | FALSE | Upregulated |
| 0,190031144 | 0,518094744 | SYNC | FALSE | Upregulated |
| -0,512136302 | 0,518094744 | TRIP4 | FALSE | Downregulated |
| -0,276571826 | 0,518094744 | UQCR11 | FALSE | Downregulated |
| 0,598018629 | 0,518094744 | ZNF582-AS1 | FALSE | Upregulated |
| -0,373114973 | 0,518317596 | CLEC4M | FALSE | Downregulated |
| -0,559190015 | 0,518317596 | CXorf56 | FALSE | Downregulated |
| 0,593559159 | 0,518317596 | HIPK4 | FALSE | Upregulated |
| 0,258299451 | 0,518317596 | ITGA11 | FALSE | Upregulated |
| 0,551921444 | 0,518317596 | LOC102724518 | FALSE | Upregulated |
| 0,725396878 | 0,518317596 | MMD | FALSE | Upregulated |
| -0,38496973 | 0,518317596 | NR0B2 | FALSE | Downregulated |
| -0,224706408 | 0,518317596 | PDCD11 | FALSE | Downregulated |
| -0,282903306 | 0,518317596 | RPS19BP1 | FALSE | Downregulated |
| 0,181933863 | 0,518317596 | SUPT7L | FALSE | Upregulated |
| -0,518990037 | 0,518317596 | TSSC1 | FALSE | Downregulated |
| -0,247830101 | 0,518317596 | TXNIP | FALSE | Downregulated |
| -0,15332936 | 0,518488682 | PABPC1 | FALSE | Downregulated |
| 0,545242159 | 0,518870144 | GRASP | FALSE | Upregulated |
| -0,300304704 | 0,518997216 | PMM1 | FALSE | Downregulated |
| 0,313507795 | 0,519425889 | CHD3 | FALSE | Upregulated |
| 0,244916668 | 0,519425889 | FSD2 | FALSE | Upregulated |
| 0,470899201 | 0,519425889 | LOC101929012 | FALSE | Upregulated |

| | | | | |
|---|---|---|---|---|
| 0,532840936 | 0,519425889 | MAP6D1 | FALSE | Upregulated |
| -0,392237523 | 0,519425889 | PRKAR1B | FALSE | Downregulated |
| 0,446491888 | 0,519425889 | SH3PXD2B | FALSE | Upregulated |
| 0,476238334 | 0,519425889 | UGT2A3 | FALSE | Upregulated |
| -0,179004534 | 0,519800831 | BOK | FALSE | Downregulated |
| 0,475802421 | 0,520059072 | MYOM1 | FALSE | Upregulated |
| 0,28759138 | 0,520059072 | OFD1 | FALSE | Upregulated |
| -0,449819264 | 0,520554762 | PIGL | FALSE | Downregulated |
| -0,470470075 | 0,520649947 | COX18 | FALSE | Downregulated |
| 0,472576355 | 0,520649947 | GALNT7 | FALSE | Upregulated |
| 0,710422603 | 0,520649947 | MTHFD1L | FALSE | Upregulated |
| 0,456339289 | 0,520649947 | PTER | FALSE | Upregulated |
| 0,549247698 | 0,520649947 | ZMYM5 | FALSE | Upregulated |
| -0,503304595 | 0,521515771 | PRKXP1 | FALSE | Downregulated |
| 0,222695553 | 0,521667888 | SPDYE3 | FALSE | Upregulated |
| -0,259788627 | 0,521818157 | SCARF1 | FALSE | Downregulated |
| -0,169022921 | 0,521818157 | TMBIM6 | FALSE | Downregulated |
| 0,286976818 | 0,522017808 | C11orf42 | FALSE | Upregulated |
| -0,394627017 | 0,522017808 | C3orf58 | FALSE | Downregulated |
| 0,199643404 | 0,522017808 | CEMP1 | FALSE | Upregulated |
| 0,351994271 | 0,522017808 | CLDN24 | FALSE | Upregulated |
| 0,541340453 | 0,522017808 | CWC27 | FALSE | Upregulated |
| -0,257971309 | 0,522017808 | EPB41L4B | FALSE | Downregulated |
| -0,215586596 | 0,522017808 | FDX1 | FALSE | Downregulated |
| -0,528214714 | 0,522017808 | HSPA13 | FALSE | Downregulated |
| 0,207552421 | 0,522017808 | LPCAT3 | FALSE | Upregulated |
| -0,184231621 | 0,522017808 | MTHFD1 | FALSE | Downregulated |
| -0,18790307 | 0,522017808 | NFIB | FALSE | Downregulated |
| -0,50966346 | 0,522017808 | OR52B6 | FALSE | Downregulated |
| 0,346790209 | 0,522017808 | PLK2 | FALSE | Upregulated |
| -0,27889699 | 0,522017808 | PML | FALSE | Downregulated |
| 0,438694874 | 0,522017808 | RANGRF | FALSE | Upregulated |
| -0,272027035 | 0,522017808 | SLC25A46 | FALSE | Downregulated |
| 0,421649658 | 0,522017808 | SRPX2 | FALSE | Upregulated |
| 0,21640412 | 0,522017808 | ZNF552 | FALSE | Upregulated |
| 0,1821229 | 0,522114154 | ATXN7L3 | FALSE | Upregulated |
| 0,704138219 | 0,522229085 | STK39 | FALSE | Upregulated |
| 0,257196017 | 0,52234861 | AP3M1 | FALSE | Upregulated |
| 0,207824365 | 0,52234861 | CD46 | FALSE | Upregulated |
| 0,295007097 | 0,52234861 | FGFR2 | FALSE | Upregulated |
| 0,287931097 | 0,52234861 | LOC102724552 | FALSE | Upregulated |
| 0,254005683 | 0,52234861 | MKL1 | FALSE | Upregulated |
| 0,230336055 | 0,52234861 | MLANA | FALSE | Upregulated |
| 0,160349978 | 0,522581233 | VPS13C | FALSE | Upregulated |
| -0,24059366 | 0,522983712 | GPT | FALSE | Downregulated |
| -0,444548685 | 0,522983712 | RP11-115J16.1 | FALSE | Downregulated |
| 0,268911126 | 0,523122632 | LRP2BP | FALSE | Upregulated |
| -0,285663609 | 0,5234246 | APRT | FALSE | Downregulated |
| 0,550875259 | 0,5234246 | C12orf49 | FALSE | Upregulated |
| -0,221191331 | 0,5234246 | CLCN5 | FALSE | Downregulated |
| 0,550570535 | 0,5234246 | FLJ42258 | FALSE | Upregulated |

| | | | | |
|---|---|---|---|---|
| 0,355385922 | 0,5234246 | UBE2L6 | FALSE | Upregulated |
| 0,510697374 | 0,524153152 | SLC35A5 | FALSE | Upregulated |
| 0,420903643 | 0,524153152 | WAC-AS1 | FALSE | Upregulated |
| -0,40314384 | 0,524153152 | ZNF772 | FALSE | Downregulated |
| 0,183340712 | 0,524153152 | ZNFX1 | FALSE | Upregulated |
| 0,259686564 | 0,524174383 | RPF1 | FALSE | Upregulated |
| 0,583139545 | 0,524718395 | RNF216P1 | FALSE | Upregulated |
| -0,187171513 | 0,525157184 | ALDH3A2 | FALSE | Downregulated |
| 0,515215738 | 0,525157184 | CMAHP | FALSE | Upregulated |
| 0,319458565 | 0,525157184 | DFNA5 | FALSE | Upregulated |
| 0,232427929 | 0,525157184 | DMTF1 | FALSE | Upregulated |
| -0,331626738 | 0,525157184 | ENC1 | FALSE | Downregulated |
| 0,413043533 | 0,525157184 | FIGNL2 | FALSE | Upregulated |
| -0,39800168 | 0,525157184 | HTATIP2 | FALSE | Downregulated |
| -0,436561602 | 0,525157184 | ISCA1 | FALSE | Downregulated |
| 0,576129387 | 0,525157184 | LOC101927321 | FALSE | Upregulated |
| 0,384303413 | 0,525157184 | LOC101927894 | FALSE | Upregulated |
| -0,276970488 | 0,525157184 | MACROD1 | FALSE | Downregulated |
| -0,423091294 | 0,525157184 | PARD6B | FALSE | Downregulated |
| 0,283558493 | 0,525157184 | PGAM2 | FALSE | Upregulated |
| 0,269370681 | 0,525157184 | PISD | FALSE | Upregulated |
| -0,244601824 | 0,525157184 | PTRF | FALSE | Downregulated |
| -0,678086116 | 0,525157184 | TPM3P9 | FALSE | Downregulated |
| 0,431717848 | 0,525157184 | ZRANB2-AS1 | FALSE | Upregulated |
| -0,294549588 | 0,525160281 | HPGD | FALSE | Downregulated |
| 0,195896099 | 0,525160281 | RBM19 | FALSE | Upregulated |
| -0,294318476 | 0,525160281 | TIA1 | FALSE | Downregulated |
| -0,147904056 | 0,525160281 | ZZEF1 | FALSE | Downregulated |
| -0,219942188 | 0,525172002 | ACAA2 | FALSE | Downregulated |
| 0,307729474 | 0,525237291 | CAND2 | FALSE | Upregulated |
| 0,300781097 | 0,526360507 | WDR91 | FALSE | Upregulated |
| -0,128745537 | 0,526847461 | CLUH | FALSE | Downregulated |
| -0,24219691 | 0,526847461 | LAMB1 | FALSE | Downregulated |
| -0,205341897 | 0,526847461 | NNT | FALSE | Downregulated |
| -0,274981131 | 0,526847461 | TCEB2 | FALSE | Downregulated |
| 0,334629636 | 0,526999289 | NXN | FALSE | Upregulated |
| -0,231396844 | 0,527205573 | IREB2 | FALSE | Downregulated |
| -0,171456301 | 0,528435047 | RAB11B | FALSE | Downregulated |
| -0,25854543 | 0,528435047 | ZFYVE1 | FALSE | Downregulated |
| -0,335289773 | 0,528622894 | LOC642852 | FALSE | Downregulated |
| -0,195524579 | 0,528622894 | RPS16 | FALSE | Downregulated |
| 0,523969002 | 0,528804817 | FAM46C | FALSE | Upregulated |
| -0,322025711 | 0,528804817 | GEMIN5 | FALSE | Downregulated |
| -0,204957852 | 0,528804817 | MRPL12 | FALSE | Downregulated |
| 0,276240611 | 0,528804817 | MRPS5 | FALSE | Upregulated |
| 0,526393008 | 0,528804817 | PTPRE | FALSE | Upregulated |
| 0,486579255 | 0,528804817 | TLCD1 | FALSE | Upregulated |
| -0,116717336 | 0,528807236 | EIF4B | FALSE | Downregulated |
| 0,296586255 | 0,529245674 | ARL17A | FALSE | Upregulated |
| -0,218335174 | 0,529245674 | CHDH | FALSE | Downregulated |
| 0,469940296 | 0,529271206 | TSSK2 | FALSE | Upregulated |

| | | | | |
|---|---|---|---|---|
| 0,398263352 | 0,529552793 | IGSF6 | FALSE | Upregulated |
| -0,295900914 | 0,529552793 | PKHD1 | FALSE | Downregulated |
| 0,288710548 | 0,529679687 | CDYL | FALSE | Upregulated |
| -0,217654627 | 0,529679687 | CXCL12 | FALSE | Downregulated |
| 0,254240791 | 0,529679687 | DARS2 | FALSE | Upregulated |
| 0,431722756 | 0,529679687 | EGOT | FALSE | Upregulated |
| 0,560494351 | 0,529679687 | HLA-DQA1 | FALSE | Upregulated |
| -0,489663288 | 0,529679687 | ID1 | FALSE | Downregulated |
| 0,545349116 | 0,529679687 | LOC102724133 | FALSE | Upregulated |
| 0,475866421 | 0,529679687 | LOC90784 | FALSE | Upregulated |
| 0,338053913 | 0,529679687 | OR2A1 | FALSE | Upregulated |
| 0,425708757 | 0,529679687 | PLBD1 | FALSE | Upregulated |
| 0,430628188 | 0,529679687 | RIC3 | FALSE | Upregulated |
| -0,419403818 | 0,529679687 | RP5-1029K10.2 | FALSE | Downregulated |
| 0,519284519 | 0,529679687 | TMEM255A | FALSE | Upregulated |
| -0,234233132 | 0,529679687 | WNK3 | FALSE | Downregulated |
| -0,657789818 | 0,530357352 | DNM1 | FALSE | Downregulated |
| -0,192586495 | 0,530357352 | SERPING1 | FALSE | Downregulated |
| 0,618624762 | 0,530679641 | MKLN1-AS | FALSE | Upregulated |
| -0,627089504 | 0,530679641 | MRM1 | FALSE | Downregulated |
| 0,393101243 | 0,530986377 | ZC3H12A | FALSE | Upregulated |
| 0,299576185 | 0,531089462 | AKIRIN1 | FALSE | Upregulated |
| 0,666981259 | 0,531089462 | APBA2 | FALSE | Upregulated |
| 0,224117368 | 0,53109534 | ASF1A | FALSE | Upregulated |
| 0,392957458 | 0,5311301 | ANAPC7 | FALSE | Upregulated |
| -0,275862921 | 0,5311301 | BCORL1 | FALSE | Downregulated |
| 0,541743507 | 0,5311301 | C3AR1 | FALSE | Upregulated |
| 0,391370235 | 0,5311301 | CLUAP1 | FALSE | Upregulated |
| 0,228604074 | 0,5311301 | DDX58 | FALSE | Upregulated |
| -0,173057278 | 0,5311301 | EDF1 | FALSE | Downregulated |
| 0,233572913 | 0,5311301 | EIF4G3 | FALSE | Upregulated |
| -0,446157345 | 0,5311301 | FXN | FALSE | Downregulated |
| -0,459187841 | 0,5311301 | KCNMA1 | FALSE | Downregulated |
| -0,250292852 | 0,5311301 | PCMTD2 | FALSE | Downregulated |
| 0,249632022 | 0,5311301 | PSMC3IP | FALSE | Upregulated |
| -0,317894957 | 0,5311301 | SDHB | FALSE | Downregulated |
| 0,191030623 | 0,5311301 | TARDBP | FALSE | Upregulated |
| -0,62288128 | 0,5311301 | TM6SF2 | FALSE | Downregulated |
| -0,524859748 | 0,5311301 | TSTD1 | FALSE | Downregulated |
| -0,385072415 | 0,5311301 | VAMP7 | FALSE | Downregulated |
| 0,446552454 | 0,5311301 | VAV1 | FALSE | Upregulated |
| 0,485333469 | 0,531131063 | SLC52A1 | FALSE | Upregulated |
| -0,23777428 | 0,531235763 | TAF15 | FALSE | Downregulated |
| 0,361887496 | 0,531235763 | ZNF808 | FALSE | Upregulated |
| 0,600619644 | 0,531311104 | APOBEC3F | FALSE | Upregulated |
| -0,254888794 | 0,531311104 | GOLIM4 | FALSE | Downregulated |
| -0,176054582 | 0,531311104 | NOTCH1 | FALSE | Downregulated |
| 0,46557203 | 0,531675736 | CELSR3 | FALSE | Upregulated |
| 0,33266485 | 0,531675736 | CSF3R | FALSE | Upregulated |
| 0,217761885 | 0,531675736 | DBH | FALSE | Upregulated |
| -0,321544605 | 0,531675736 | GANC | FALSE | Downregulated |

| | | | | |
|---|---|---|---|---|
| 0,411546874 | 0,531675736 | MYLK-AS1 | FALSE | Upregulated |
| 0,361849592 | 0,531675736 | OAS3 | FALSE | Upregulated |
| 0,168962489 | 0,531675736 | PEA15 | FALSE | Upregulated |
| -0,164434508 | 0,531675736 | PICALM | FALSE | Downregulated |
| 0,369926131 | 0,531675736 | PLEKHG2 | FALSE | Upregulated |
| 0,368780324 | 0,531675736 | SERAC1 | FALSE | Upregulated |
| 0,454389163 | 0,531965919 | CYR61 | FALSE | Upregulated |
| -0,527097253 | 0,531965919 | MSH2 | FALSE | Downregulated |
| 0,208791207 | 0,532173114 | FSIP2 | FALSE | Upregulated |
| 0,193770983 | 0,532173114 | YPEL1 | FALSE | Upregulated |
| 0,190652972 | 0,53234483 | GNAL | FALSE | Upregulated |
| 0,263157712 | 0,532805853 | MAPKAP1 | FALSE | Upregulated |
| -0,337817469 | 0,532841692 | OCEL1 | FALSE | Downregulated |
| 0,29805902 | 0,533020928 | IPPK | FALSE | Upregulated |
| -0,164715777 | 0,533628869 | A1BG | FALSE | Downregulated |
| -0,216637723 | 0,533628869 | EFNB1 | FALSE | Downregulated |
| 0,143979333 | 0,533628869 | FAM13A | FALSE | Upregulated |
| 0,260367965 | 0,533628869 | FTSJ3 | FALSE | Upregulated |
| -0,189209202 | 0,533628869 | FUBP3 | FALSE | Downregulated |
| 0,380088687 | 0,533628869 | LOC101928517 | FALSE | Upregulated |
| 0,226357219 | 0,533628869 | LOC102723600 | FALSE | Upregulated |
| 0,336342489 | 0,533628869 | SCYL3 | FALSE | Upregulated |
| -0,350752779 | 0,533628869 | STX5 | FALSE | Downregulated |
| 0,382888863 | 0,533628869 | TIMP1 | FALSE | Upregulated |
| -0,163028755 | 0,533830375 | MOV10 | FALSE | Downregulated |
| 0,239091909 | 0,533987545 | JMJD1C-AS1 | FALSE | Upregulated |
| 0,572909115 | 0,533987545 | TRMT61B | FALSE | Upregulated |
| 0,234742232 | 0,534468132 | LOC100505502 | FALSE | Upregulated |
| 0,472700444 | 0,534596361 | AEN | FALSE | Upregulated |
| 0,264562797 | 0,534596361 | ATAD5 | FALSE | Upregulated |
| 0,345971397 | 0,534596361 | FAM50A | FALSE | Upregulated |
| -0,319802142 | 0,534596361 | HSD17B13 | FALSE | Downregulated |
| 0,627572263 | 0,534596361 | KCNB1 | FALSE | Upregulated |
| 0,225734021 | 0,534596361 | KCNRG | FALSE | Upregulated |
| 0,261718406 | 0,534596361 | LOC100505767 | FALSE | Upregulated |
| 0,310788528 | 0,534596361 | NPB | FALSE | Upregulated |
| -0,198958975 | 0,534596361 | SMARCC1 | FALSE | Downregulated |
| -0,395362949 | 0,534596361 | SRRD | FALSE | Downregulated |
| 0,330195482 | 0,534596361 | STARD5 | FALSE | Upregulated |
| -0,428234228 | 0,534596361 | TMEM203 | FALSE | Downregulated |
| -0,422404735 | 0,534596361 | UTP3 | FALSE | Downregulated |
| 0,243909144 | 0,534813615 | ATG16L2 | FALSE | Upregulated |
| 0,366739203 | 0,534943182 | TMC6 | FALSE | Upregulated |
| -0,335442372 | 0,535698501 | HEATR5A | FALSE | Downregulated |
| 0,156115679 | 0,535792992 | AP1G1 | FALSE | Upregulated |
| 0,258637539 | 0,535792992 | ITGA2 | FALSE | Upregulated |
| -0,218577143 | 0,535831522 | KLHDC3 | FALSE | Downregulated |
| 0,236814036 | 0,535831522 | MAP7D1 | FALSE | Upregulated |
| -0,220292365 | 0,535831522 | RPL27 | FALSE | Downregulated |
| 0,684889204 | 0,535831522 | TCEB1 | FALSE | Upregulated |
| -0,18954077 | 0,536044437 | ACBD5 | FALSE | Downregulated |

| | | | | |
|---|---|---|---|---|
| -0,295910692 | 0,536044437 | FAM214A | FALSE | Downregulated |
| 0,407673585 | 0,536044437 | ISPD-AS1 | FALSE | Upregulated |
| -0,456677407 | 0,536044437 | KIF13A | FALSE | Downregulated |
| 0,257439443 | 0,536044437 | NOD1 | FALSE | Upregulated |
| -0,193425808 | 0,536044437 | USH2A | FALSE | Downregulated |
| -0,437613504 | 0,536044437 | ZFPL1 | FALSE | Downregulated |
| 0,331922854 | 0,536244622 | LOC101926967 | FALSE | Upregulated |
| -0,344559455 | 0,53663163 | GPATCH2L | FALSE | Downregulated |
| -0,254783864 | 0,53663163 | N4BP2L1 | FALSE | Downregulated |
| 0,363844239 | 0,53663163 | ZNF322 | FALSE | Upregulated |
| 0,357882478 | 0,537088849 | MANEAL | FALSE | Upregulated |
| -0,511213323 | 0,537125614 | ERCC4 | FALSE | Downregulated |
| -0,189136216 | 0,537125614 | POLR3E | FALSE | Downregulated |
| 0,192962192 | 0,537125614 | SLC12A6 | FALSE | Upregulated |
| -0,38614675 | 0,53720232 | NSMCE1 | FALSE | Downregulated |
| 0,397024112 | 0,53778006 | HPS5 | FALSE | Upregulated |
| -0,481643138 | 0,53778006 | KIF27 | FALSE | Downregulated |
| -0,330717061 | 0,53778006 | LMBR1 | FALSE | Downregulated |
| 0,226567846 | 0,53778006 | PDCD2 | FALSE | Upregulated |
| -0,528907785 | 0,53778006 | PMS2 | FALSE | Downregulated |
| 0,269426128 | 0,53778006 | PTPN12 | FALSE | Upregulated |
| 0,266333588 | 0,53778006 | PXK | FALSE | Upregulated |
| -0,406241184 | 0,53778006 | STOX2 | FALSE | Downregulated |
| 0,253060926 | 0,537963204 | CACUL1 | FALSE | Upregulated |
| -0,297070404 | 0,538062118 | FCN2 | FALSE | Downregulated |
| 0,229786434 | 0,538062118 | IL2RB | FALSE | Upregulated |
| -0,296825828 | 0,538424394 | CMBL | FALSE | Downregulated |
| 0,349978136 | 0,539079783 | TTC33 | FALSE | Upregulated |
| 0,225593896 | 0,539195976 | C10orf10 | FALSE | Upregulated |
| -0,228919339 | 0,539195976 | CXXC1 | FALSE | Downregulated |
| 0,160803765 | 0,539195976 | LRBA | FALSE | Upregulated |
| 0,214510208 | 0,539195976 | NANOS1 | FALSE | Upregulated |
| -0,257211879 | 0,539195976 | TP73-AS1 | FALSE | Downregulated |
| -0,30588318 | 0,539353508 | PCNXL4 | FALSE | Downregulated |
| -0,517525258 | 0,539819909 | CYP2A7 | FALSE | Downregulated |
| 0,362803513 | 0,539819909 | LIPE | FALSE | Upregulated |
| 0,269434302 | 0,539819909 | SUMO1P3 | FALSE | Upregulated |
| 0,192866726 | 0,539855674 | AGPAT2 | FALSE | Upregulated |
| -0,207006279 | 0,539855674 | ATRN | FALSE | Downregulated |
| -0,638635885 | 0,539855674 | CCDC7 | FALSE | Downregulated |
| -0,193025884 | 0,539855674 | DCAF11 | FALSE | Downregulated |
| -0,211650153 | 0,539855674 | EFCAB14 | FALSE | Downregulated |
| -0,16908019 | 0,539855674 | FAM168B | FALSE | Downregulated |
| 0,293103756 | 0,539855674 | FAM177A1 | FALSE | Upregulated |
| 0,474318714 | 0,539855674 | G6PD | FALSE | Upregulated |
| 0,485206733 | 0,539855674 | HSD11B2 | FALSE | Upregulated |
| 0,482404717 | 0,539855674 | LOC102723555 | FALSE | Upregulated |
| 0,505226469 | 0,539855674 | MYLIP | FALSE | Upregulated |
| 0,425918299 | 0,539855674 | NEFH | FALSE | Upregulated |
| 0,429411141 | 0,539855674 | NME3 | FALSE | Upregulated |
| 0,404783582 | 0,539855674 | PRKRA | FALSE | Upregulated |

| | | | | |
|---|---|---|---|---|
| 0,488517608 | 0,539855674 | RASSF3 | FALSE | Upregulated |
| -0,379281721 | 0,539855674 | SYNJ1 | FALSE | Downregulated |
| 0,265588555 | 0,539855674 | ZBTB46 | FALSE | Upregulated |
| 0,423437291 | 0,539855674 | ZNF704 | FALSE | Upregulated |
| -0,484524979 | 0,539855674 | ZNF706 | FALSE | Downregulated |
| 0,599749031 | 0,539926458 | FUT8-AS1 | FALSE | Upregulated |
| -0,537099538 | 0,539926458 | MTIF3 | FALSE | Downregulated |
| 0,301003501 | 0,539926458 | RNF41 | FALSE | Upregulated |
| 0,3695049 | 0,539926458 | TMEM139 | FALSE | Upregulated |
| 0,641046254 | 0,539926458 | ZSCAN12P1 | FALSE | Upregulated |
| 0,36424738 | 0,54074911 | EME1 | FALSE | Upregulated |
| -0,291553015 | 0,54074911 | TIMM44 | FALSE | Downregulated |
| -0,308628906 | 0,540967382 | INTS5 | FALSE | Downregulated |
| 0,494771761 | 0,540967382 | KIAA1279 | FALSE | Upregulated |
| 0,569097717 | 0,540967382 | MYOT | FALSE | Upregulated |
| -0,209034993 | 0,541520861 | EXOC3 | FALSE | Downregulated |
| 0,232192454 | 0,541798266 | ATP6V1C2 | FALSE | Upregulated |
| -0,202860262 | 0,541798266 | MTA1 | FALSE | Downregulated |
| -0,170062176 | 0,541798266 | PROC | FALSE | Downregulated |
| 0,223609246 | 0,541798266 | PUS10 | FALSE | Upregulated |
| -0,419139588 | 0,541798266 | WNT5A | FALSE | Downregulated |
| -0,225609413 | 0,542059415 | INO80 | FALSE | Downregulated |
| 0,348623686 | 0,543277005 | AEBP1 | FALSE | Upregulated |
| -0,288647652 | 0,543305556 | CLDN3 | FALSE | Downregulated |
| 0,391643975 | 0,544140492 | KLC3 | FALSE | Upregulated |
| 0,571186152 | 0,544547535 | FOLR1 | FALSE | Upregulated |
| -0,124743057 | 0,54469298 | PPP6R2 | FALSE | Downregulated |
| 0,307688135 | 0,544750053 | DENND3 | FALSE | Upregulated |
| -0,209210637 | 0,544750053 | MDC1 | FALSE | Downregulated |
| 0,170002205 | 0,544765951 | TMEM129 | FALSE | Upregulated |
| -0,333706836 | 0,545211349 | CPNE2 | FALSE | Downregulated |
| -0,169612839 | 0,545211349 | MED13L | FALSE | Downregulated |
| 0,254041936 | 0,545517251 | ATP2B4 | FALSE | Upregulated |
| 0,677156653 | 0,545517251 | CHSY3 | FALSE | Upregulated |
| 0,246051586 | 0,545517251 | FBXO22-AS1 | FALSE | Upregulated |
| -0,236073371 | 0,545517251 | HEATR2 | FALSE | Downregulated |
| 0,437887722 | 0,545517251 | LOC101929354 | FALSE | Upregulated |
| 0,407734013 | 0,545517251 | LRRC8B | FALSE | Upregulated |
| 0,442645173 | 0,545517251 | OR10AD1 | FALSE | Upregulated |
| -0,524063965 | 0,545517251 | PPHLN1 | FALSE | Downregulated |
| -0,151213492 | 0,545517251 | RPL7A | FALSE | Downregulated |
| 0,273434607 | 0,545517251 | SULF2 | FALSE | Upregulated |
| 0,240067645 | 0,545517251 | ZNF154 | FALSE | Upregulated |
| -0,25932084 | 0,545784317 | ACOT2 | FALSE | Downregulated |
| 0,540040466 | 0,545784317 | BATF2 | FALSE | Upregulated |
| 0,448956497 | 0,545784317 | LOC102724077 | FALSE | Upregulated |
| 0,415291357 | 0,545917901 | LOC101928318 | FALSE | Upregulated |
| -0,412012668 | 0,545965285 | NDUFB4 | FALSE | Downregulated |
| 0,237570037 | 0,546095145 | FAM196A | FALSE | Upregulated |
| -0,436535063 | 0,547154382 | NUDT9 | FALSE | Downregulated |
| 0,350783322 | 0,548198496 | ALPK1 | FALSE | Upregulated |

| | | | | |
|---|---|---|---|---|
| -0,364215345 | 0,548198496 | KLHL15 | FALSE | Downregulated |
| 0,274242301 | 0,548198496 | MST1L | FALSE | Upregulated |
| 0,447985095 | 0,548198496 | NXPH3 | FALSE | Upregulated |
| -0,244256114 | 0,548198496 | PDLIM1 | FALSE | Downregulated |
| 0,16794007 | 0,548198496 | TJP1 | FALSE | Upregulated |
| 0,146288048 | 0,548198496 | TNIP1 | FALSE | Upregulated |
| 0,273155067 | 0,548811441 | NOMO1 | FALSE | Upregulated |
| 0,34296736 | 0,549580338 | C10orf95 | FALSE | Upregulated |
| -0,239235612 | 0,549580338 | LRRC3 | FALSE | Downregulated |
| -0,169774278 | 0,54959354 | DPYS | FALSE | Downregulated |
| -0,331284173 | 0,54959354 | F2R | FALSE | Downregulated |
| -0,210767896 | 0,54959354 | GTF2IRD2B | FALSE | Downregulated |
| -0,406318369 | 0,54959354 | LOC102724852 | FALSE | Downregulated |
| 0,178213739 | 0,54959354 | SRPK2 | FALSE | Upregulated |
| 0,388599068 | 0,550065958 | GPR61 | FALSE | Upregulated |
| 0,262996942 | 0,550065958 | ZSCAN16 | FALSE | Upregulated |
| -0,170671013 | 0,550124253 | RPS20 | FALSE | Downregulated |
| 0,28962285 | 0,550292731 | COA5 | FALSE | Upregulated |
| 0,37566061 | 0,551208336 | C12orf44 | FALSE | Upregulated |
| 0,201647431 | 0,551208336 | ENTPD1 | FALSE | Upregulated |
| -0,424232136 | 0,551208336 | FBXO3 | FALSE | Downregulated |
| 0,463274869 | 0,551208336 | LOC101928482 | FALSE | Upregulated |
| 0,260608044 | 0,551208336 | LPHN1 | FALSE | Upregulated |
| 0,350479462 | 0,551208336 | LRRC46 | FALSE | Upregulated |
| 0,214017073 | 0,551208336 | LYL1 | FALSE | Upregulated |
| 0,244740225 | 0,551208336 | NUTM2D | FALSE | Upregulated |
| -0,181017352 | 0,551208336 | UBR3 | FALSE | Downregulated |
| 0,281124593 | 0,551208336 | WSB1 | FALSE | Upregulated |
| -0,149617745 | 0,551392944 | MAN1A1 | FALSE | Downregulated |
| 0,331557729 | 0,551430466 | BTN3A3 | FALSE | Upregulated |
| -0,579987499 | 0,551535203 | DFFB | FALSE | Downregulated |
| -0,53907586 | 0,551535203 | DPF3 | FALSE | Downregulated |
| 0,34443517 | 0,551535203 | RB1 | FALSE | Upregulated |
| 0,528858003 | 0,551535203 | SLC25A19 | FALSE | Upregulated |
| -0,178515635 | 0,551549757 | ABAT | FALSE | Downregulated |
| -0,185318197 | 0,551669299 | ERBB3 | FALSE | Downregulated |
| -0,326118928 | 0,551669299 | LEPR | FALSE | Downregulated |
| 0,195331394 | 0,551819947 | ACTB | FALSE | Upregulated |
| 0,31878675 | 0,551819947 | C1QA | FALSE | Upregulated |
| -0,153820251 | 0,551819947 | CTTN | FALSE | Downregulated |
| -0,531757231 | 0,551819947 | MRPS9 | FALSE | Downregulated |
| -0,252353657 | 0,551819947 | RGS12 | FALSE | Downregulated |
| 0,538338861 | 0,551998409 | LIFR-AS1 | FALSE | Upregulated |
| 0,26652351 | 0,551998409 | SNTB2 | FALSE | Upregulated |
| -0,236328575 | 0,552202578 | BAIAP2 | FALSE | Downregulated |
| 0,309810372 | 0,552202578 | DDX28 | FALSE | Upregulated |
| 0,387475327 | 0,552202578 | IDI1 | FALSE | Upregulated |
| -0,333672646 | 0,552202578 | LITAF | FALSE | Downregulated |
| 0,682576051 | 0,552202578 | TMLHE-AS1 | FALSE | Upregulated |
| 0,220384834 | 0,552202578 | UNK | FALSE | Upregulated |
| -0,405982082 | 0,552951012 | MT1E | FALSE | Downregulated |

| | | | | |
|---|---|---|---|---|
| 0,23155297 | 0,553208225 | DEF8 | FALSE | Upregulated |
| 0,489974782 | 0,553674505 | FBLN7 | FALSE | Upregulated |
| -0,176514857 | 0,553807342 | ARHGAP29 | FALSE | Downregulated |
| 0,194165037 | 0,553887177 | HEATR1 | FALSE | Upregulated |
| 0,229204385 | 0,553887177 | SLC16A4 | FALSE | Upregulated |
| 0,317413543 | 0,554070945 | ANKRD19P | FALSE | Upregulated |
| 0,524103101 | 0,554070945 | ASB16-AS1 | FALSE | Upregulated |
| -0,234680189 | 0,554070945 | ATP5C1 | FALSE | Downregulated |
| -0,485200456 | 0,554070945 | B3GAT1 | FALSE | Downregulated |
| 0,50748991 | 0,554070945 | BCAS2 | FALSE | Upregulated |
| 0,315608867 | 0,554070945 | C12orf60 | FALSE | Upregulated |
| -0,475785369 | 0,554070945 | CLEC14A | FALSE | Downregulated |
| -0,200655586 | 0,554070945 | DDAH1 | FALSE | Downregulated |
| 0,406670508 | 0,554070945 | FER1L5 | FALSE | Upregulated |
| -0,202173705 | 0,554070945 | FGA | FALSE | Downregulated |
| 0,389522367 | 0,554070945 | GRAMD2 | FALSE | Upregulated |
| -0,523517947 | 0,554070945 | INTS12 | FALSE | Downregulated |
| 0,363259841 | 0,554070945 | KIAA0586 | FALSE | Upregulated |
| 0,437722589 | 0,554070945 | LOC101927028 | FALSE | Upregulated |
| 0,658742412 | 0,554070945 | LOC101928465 | FALSE | Upregulated |
| -0,541483329 | 0,554070945 | LOC102723954 | FALSE | Downregulated |
| 0,327752608 | 0,554070945 | LOC102724549 | FALSE | Upregulated |
| 0,246681371 | 0,554070945 | MAGI1-AS1 | FALSE | Upregulated |
| 0,213506074 | 0,554070945 | MEGF11 | FALSE | Upregulated |
| 0,206739605 | 0,554070945 | MTMR7 | FALSE | Upregulated |
| 0,312908164 | 0,554070945 | NCF2 | FALSE | Upregulated |
| 0,198535602 | 0,554070945 | NRIP2 | FALSE | Upregulated |
| 0,449306591 | 0,554070945 | PAQR8 | FALSE | Upregulated |
| 0,277450546 | 0,554070945 | PEX1 | FALSE | Upregulated |
| -0,356122102 | 0,554070945 | PIGO | FALSE | Downregulated |
| -0,155702274 | 0,554070945 | ROCK2 | FALSE | Downregulated |
| 0,447883773 | 0,554070945 | SLC22A18AS | FALSE | Upregulated |
| -0,24628512 | 0,554070945 | SMAD2 | FALSE | Downregulated |
| 0,428829274 | 0,554070945 | TMEM194A | FALSE | Upregulated |
| -0,208215616 | 0,554070945 | TNPO1 | FALSE | Downregulated |
| -0,522802851 | 0,554070945 | ZNF577 | FALSE | Downregulated |
| -0,244520762 | 0,554149748 | BCAP31 | FALSE | Downregulated |
| 0,60709434 | 0,55455119 | VCAN | FALSE | Upregulated |
| 0,49428599 | 0,55455119 | ZNF575 | FALSE | Upregulated |
| -0,317169547 | 0,555099191 | CEP120 | FALSE | Downregulated |
| 0,581745666 | 0,555099191 | CERS1 | FALSE | Upregulated |
| 0,560374999 | 0,555099191 | DCST1 | FALSE | Upregulated |
| 0,26869317 | 0,55584183 | ARL8B | FALSE | Upregulated |
| 0,41295537 | 0,55584183 | LYG2 | FALSE | Upregulated |
| -0,226240353 | 0,55584183 | SERPINA4 | FALSE | Downregulated |
| -0,164502819 | 0,556152243 | DCAF5 | FALSE | Downregulated |
| -0,307474915 | 0,556152243 | HPR | FALSE | Downregulated |
| -0,2054305 | 0,556152243 | HSP90AB1 | FALSE | Downregulated |
| 0,210103043 | 0,556152243 | LOC100996506 | FALSE | Upregulated |
| 0,655237798 | 0,556152243 | LOC102724255 | FALSE | Upregulated |
| -0,21758328 | 0,556152243 | NAV2 | FALSE | Downregulated |

| | | | | |
|---|---|---|---|---|
| 0,253829134 | 0,556152243 | PIK3IP1 | FALSE | Upregulated |
| -0,547394072 | 0,556152243 | RNASEH1 | FALSE | Downregulated |
| -0,177030847 | 0,556152243 | SERINC1 | FALSE | Downregulated |
| -0,357397331 | 0,556152243 | UBA2 | FALSE | Downregulated |
| 0,456026571 | 0,556377114 | DISP2 | FALSE | Upregulated |
| -0,921485429 | 0,556377114 | GTF2H2C | FALSE | Downregulated |
| 0,308615848 | 0,556628544 | CDK2AP1 | FALSE | Upregulated |
| -0,252213514 | 0,556934889 | AKAP11 | FALSE | Downregulated |
| 0,275601426 | 0,556934889 | C17orf72 | FALSE | Upregulated |
| 0,180048312 | 0,556934889 | COX20 | FALSE | Upregulated |
| 0,158183451 | 0,556934889 | SMURF1 | FALSE | Upregulated |
| -0,472935102 | 0,557086395 | ZNF480 | FALSE | Downregulated |
| -0,439551221 | 0,557215913 | ADAMTS7 | FALSE | Downregulated |
| 0,232199924 | 0,557215913 | LOC644090 | FALSE | Upregulated |
| 0,306137585 | 0,557215913 | LOC645967 | FALSE | Upregulated |
| 0,187251465 | 0,557215913 | MKNK2 | FALSE | Upregulated |
| -0,287936702 | 0,557215913 | SERTAD2 | FALSE | Downregulated |
| 0,275925903 | 0,557369963 | B3GNTL1 | FALSE | Upregulated |
| 0,231618887 | 0,557369963 | TAF9B | FALSE | Upregulated |
| -0,312286128 | 0,557369963 | ZFAND6 | FALSE | Downregulated |
| -0,22006967 | 0,557463513 | ARHGEF17 | FALSE | Downregulated |
| -0,247138867 | 0,557463513 | LGMN | FALSE | Downregulated |
| 0,314787419 | 0,557463513 | LOC102724790 | FALSE | Upregulated |
| -0,187046863 | 0,557463513 | PDE3B | FALSE | Downregulated |
| -0,26487141 | 0,557463513 | RBM12B | FALSE | Downregulated |
| -0,188585123 | 0,557463513 | SEPP1 | FALSE | Downregulated |
| -0,206732529 | 0,557539608 | COBLL1 | FALSE | Downregulated |
| -0,315709821 | 0,557557446 | APBA3 | FALSE | Downregulated |
| 0,252424474 | 0,557557446 | DAGLB | FALSE | Upregulated |
| -0,27877385 | 0,557557446 | FNIP2 | FALSE | Downregulated |
| 0,55667881 | 0,557557446 | HOXA2 | FALSE | Upregulated |
| -0,309016777 | 0,557557446 | ING5 | FALSE | Downregulated |
| -0,205529851 | 0,557557446 | NCOA6 | FALSE | Downregulated |
| -0,373189999 | 0,557557446 | NECAB3 | FALSE | Downregulated |
| -0,231205255 | 0,557557446 | PANK1 | FALSE | Downregulated |
| -0,258849566 | 0,557557446 | RASSF7 | FALSE | Downregulated |
| 0,349021486 | 0,557557446 | RNF24 | FALSE | Upregulated |
| -0,255555865 | 0,557557446 | TKT | FALSE | Downregulated |
| 0,250415636 | 0,558332462 | LOC101927018 | FALSE | Upregulated |
| -0,230786852 | 0,558433585 | SLC40A1 | FALSE | Downregulated |
| -0,320008749 | 0,558822111 | LOC101928588 | FALSE | Downregulated |
| 0,292591927 | 0,558822111 | MRAP | FALSE | Upregulated |
| -0,269356636 | 0,558987975 | LIPC | FALSE | Downregulated |
| 0,551542078 | 0,559088388 | ALG1L9P | FALSE | Upregulated |
| 0,289308791 | 0,559088388 | HLA-A | FALSE | Upregulated |
| 0,2706037 | 0,559088388 | LOC101929396 | FALSE | Upregulated |
| 0,240157344 | 0,559088388 | RP11-392P7.6 | FALSE | Upregulated |
| -0,206425437 | 0,559088388 | TCF12 | FALSE | Downregulated |
| -0,27718204 | 0,559216364 | HIC1 | FALSE | Downregulated |
| -0,552570592 | 0,55922689 | ASXL3 | FALSE | Downregulated |
| -0,260275671 | 0,55922689 | CNPPD1 | FALSE | Downregulated |

| | | | | |
|---|---|---|---|---|
| 0,337929976 | 0,55922689 | KCTD4 | FALSE | Upregulated |
| 0,527988929 | 0,55922689 | NCMAP | FALSE | Upregulated |
| -0,558543684 | 0,559293002 | CCDC87 | FALSE | Downregulated |
| 0,207508413 | 0,559293002 | EDEM3 | FALSE | Upregulated |
| 0,477056697 | 0,559902203 | ZMAT3 | FALSE | Upregulated |
| -0,223571956 | 0,559902203 | ZNF655 | FALSE | Downregulated |
| 0,17851188 | 0,560190002 | ANXA9 | FALSE | Upregulated |
| -0,344607556 | 0,560190002 | AREL1 | FALSE | Downregulated |
| -0,246550174 | 0,560190002 | DLG4 | FALSE | Downregulated |
| 0,219827112 | 0,560190002 | IRAK1 | FALSE | Upregulated |
| 0,213641922 | 0,560190002 | RICTOR | FALSE | Upregulated |
| 0,240429409 | 0,560190002 | RP11-28G8.1 | FALSE | Upregulated |
| 0,484192899 | 0,560190002 | RP11-466A17.1 | FALSE | Upregulated |
| -0,234675735 | 0,560190002 | SSRP1 | FALSE | Downregulated |
| 0,320586077 | 0,560190002 | TONSL | FALSE | Upregulated |
| -0,474256445 | 0,560294487 | BMPER | FALSE | Downregulated |
| -0,244978743 | 0,560294487 | LOC100996492 | FALSE | Downregulated |
| -0,522892277 | 0,560294487 | MOSPD3 | FALSE | Downregulated |
| 0,284007196 | 0,560294487 | PDE8B | FALSE | Upregulated |
| -0,28463141 | 0,560294487 | RBBP7 | FALSE | Downregulated |
| 0,362624693 | 0,560294487 | TDRD6 | FALSE | Upregulated |
| 0,167739599 | 0,560294487 | TMEM184A | FALSE | Upregulated |
| -0,522936933 | 0,560294487 | TRMT12 | FALSE | Downregulated |
| -0,229289942 | 0,560377021 | ARHGAP1 | FALSE | Downregulated |
| -0,167555419 | 0,56100185 | FTH1 | FALSE | Downregulated |
| 0,604751141 | 0,56100185 | TSHZ3 | FALSE | Upregulated |
| 0,209122163 | 0,561144301 | PPP2R3A | FALSE | Upregulated |
| 0,201434558 | 0,561181014 | DMPK | FALSE | Upregulated |
| -0,166886616 | 0,561181014 | SERPINF1 | FALSE | Downregulated |
| 0,46791025 | 0,561181014 | SLC38A1 | FALSE | Upregulated |
| 0,237961924 | 0,561181014 | TAF11 | FALSE | Upregulated |
| -0,33162518 | 0,561405748 | ATP6AP1 | FALSE | Downregulated |
| -0,294231406 | 0,561515589 | OXA1L | FALSE | Downregulated |
| 0,240164258 | 0,561875227 | C19orf71 | FALSE | Upregulated |
| 0,266834464 | 0,562462476 | NAT9 | FALSE | Upregulated |
| 0,481522542 | 0,562803885 | DPYSL3 | FALSE | Upregulated |
| 0,198773902 | 0,562803885 | SYT2 | FALSE | Upregulated |
| 0,245574386 | 0,563053018 | EPN2-AS1 | FALSE | Upregulated |
| 0,340451887 | 0,563053018 | HK1 | FALSE | Upregulated |
| -0,158189183 | 0,563053018 | LAMB2 | FALSE | Downregulated |
| 0,486938461 | 0,563144002 | MED17 | FALSE | Upregulated |
| -0,29800541 | 0,563144002 | METTL7B | FALSE | Downregulated |
| 0,315651964 | 0,563144002 | TSR2 | FALSE | Upregulated |
| -0,188681788 | 0,563213514 | ZNF275 | FALSE | Downregulated |
| 0,486138721 | 0,563400317 | CNOT6 | FALSE | Upregulated |
| -0,208960882 | 0,563918696 | RPS21 | FALSE | Downregulated |
| 0,434412444 | 0,563918696 | SPTBN5 | FALSE | Upregulated |
| 0,468166288 | 0,564138236 | POLG2 | FALSE | Upregulated |
| -0,197471022 | 0,564234025 | DBH-AS1 | FALSE | Downregulated |
| -0,222267198 | 0,564234025 | FAM114A1 | FALSE | Downregulated |
| -0,216913697 | 0,564234025 | ITGA5 | FALSE | Downregulated |

| | | | | |
|---|---|---|---|---|
| 0,490184142 | 0,564234025 | LOC102723381 | FALSE | Upregulated |
| -0,181196785 | 0,564234025 | MCCC2 | FALSE | Downregulated |
| -0,326519569 | 0,564234025 | NPY6R | FALSE | Downregulated |
| -0,497007543 | 0,564234025 | SF3B14 | FALSE | Downregulated |
| -0,243209477 | 0,564234025 | SLC25A18 | FALSE | Downregulated |
| 0,375612664 | 0,564234025 | TNC | FALSE | Upregulated |
| -0,186058671 | 0,564474076 | AKR7A2 | FALSE | Downregulated |
| -0,436381582 | 0,564474076 | RP11-701P16.5 | FALSE | Downregulated |
| -0,216890922 | 0,565224828 | LPHN2 | FALSE | Downregulated |
| 0,256207653 | 0,565224828 | NELFE | FALSE | Upregulated |
| 0,197243436 | 0,565356887 | SIK2 | FALSE | Upregulated |
| -0,553733057 | 0,56612406 | CYP2B7P | FALSE | Downregulated |
| -0,294577017 | 0,566313989 | RSBN1 | FALSE | Downregulated |
| -0,38691166 | 0,566462108 | NUP43 | FALSE | Downregulated |
| 0,351424818 | 0,566870478 | TUBA8 | FALSE | Upregulated |
| 0,287198829 | 0,567199122 | EXOC8 | FALSE | Upregulated |
| -0,234103596 | 0,56746867 | RPL35A | FALSE | Downregulated |
| 0,292851511 | 0,56829628 | KCNJ9 | FALSE | Upregulated |
| -0,2029394 | 0,569222314 | ZNF444 | FALSE | Downregulated |
| -0,313694661 | 0,569479203 | GAREM | FALSE | Downregulated |
| 0,287697181 | 0,569479203 | IFNGR2 | FALSE | Upregulated |
| 0,195950307 | 0,569479203 | JADE1 | FALSE | Upregulated |
| 0,325573352 | 0,570613826 | CCDC174 | FALSE | Upregulated |
| -0,276018563 | 0,570613826 | PRCC | FALSE | Downregulated |
| -0,20487496 | 0,570613826 | SLC6A1 | FALSE | Downregulated |
| 0,366796708 | 0,570796343 | LOC100507472 | FALSE | Upregulated |
| 0,280226429 | 0,570796343 | RP5-1033H22.2 | FALSE | Upregulated |
| 0,172523453 | 0,570796343 | USP47 | FALSE | Upregulated |
| 0,187558942 | 0,570926517 | C20orf112 | FALSE | Upregulated |
| -0,290703935 | 0,571085254 | BRD7 | FALSE | Downregulated |
| -0,276749991 | 0,571085254 | COX8A | FALSE | Downregulated |
| -0,261303638 | 0,571085254 | NRP2 | FALSE | Downregulated |
| 0,160393763 | 0,571085254 | XPO1 | FALSE | Upregulated |
| -0,357396087 | 0,571109319 | CKMT2-AS1 | FALSE | Downregulated |
| 0,469975737 | 0,571109319 | PRNCR1 | FALSE | Upregulated |
| -0,529903402 | 0,571361561 | MOB3B | FALSE | Downregulated |
| -0,531682291 | 0,571361561 | OBFC1 | FALSE | Downregulated |
| -0,386184963 | 0,571361561 | TMEM86B | FALSE | Downregulated |
| -0,314597764 | 0,571363552 | NOL7 | FALSE | Downregulated |
| -0,171463888 | 0,571961901 | RPS24 | FALSE | Downregulated |
| -0,434774538 | 0,571961901 | STX18 | FALSE | Downregulated |
| -0,196992054 | 0,572016173 | ZMYND8 | FALSE | Downregulated |
| -0,290151702 | 0,572251409 | FAM99A | FALSE | Downregulated |
| 0,458042671 | 0,572251409 | LOC101928152 | FALSE | Upregulated |
| 0,199647151 | 0,572251409 | MCM3AP-AS1 | FALSE | Upregulated |
| -0,23722864 | 0,572251409 | NSUN6 | FALSE | Downregulated |
| -0,186817712 | 0,572251409 | RAB13 | FALSE | Downregulated |
| 0,414740965 | 0,572251409 | RNF115 | FALSE | Upregulated |
| -0,521644793 | 0,572251409 | WASH2P | FALSE | Downregulated |
| 0,398022406 | 0,572251409 | ZNF23 | FALSE | Upregulated |
| 0,486970418 | 0,572251409 | ZNF8 | FALSE | Upregulated |

| | | | | |
|---|---|---|---|---|
| 0,455569571 | 0,572529878 | ARHGAP39 | FALSE | Upregulated |
| -0,183036256 | 0,572733249 | BCKDK | FALSE | Downregulated |
| 0,189025254 | 0,572978897 | LOC101929469 | FALSE | Upregulated |
| 0,213242237 | 0,573068785 | CTBP2 | FALSE | Upregulated |
| -0,282733802 | 0,573131634 | FUOM | FALSE | Downregulated |
| 0,351337859 | 0,573409098 | SGOL1 | FALSE | Upregulated |
| 0,398343895 | 0,573661247 | RNF7 | FALSE | Upregulated |
| 0,446612262 | 0,573765042 | C14orf80 | FALSE | Upregulated |
| -0,170417456 | 0,573765042 | PGAM1 | FALSE | Downregulated |
| -0,478518875 | 0,574029322 | CWC22 | FALSE | Downregulated |
| -0,26638812 | 0,574029322 | MPDU1 | FALSE | Downregulated |
| 0,328899568 | 0,574029322 | NCOA5 | FALSE | Upregulated |
| -0,326495098 | 0,574029322 | ZNF22 | FALSE | Downregulated |
| 0,322271015 | 0,574029322 | ZNF749 | FALSE | Upregulated |
| -0,299471101 | 0,574058977 | GATA6 | FALSE | Downregulated |
| 0,196241378 | 0,574190225 | ATP11A-AS1 | FALSE | Upregulated |
| 0,382329412 | 0,574190225 | B3GNT4 | FALSE | Upregulated |
| -0,182237098 | 0,574190225 | CNKSR3 | FALSE | Downregulated |
| -0,316333883 | 0,574190225 | CRIPAK | FALSE | Downregulated |
| -0,166528569 | 0,574190225 | JUP | FALSE | Downregulated |
| -0,148699193 | 0,574190225 | KANK1 | FALSE | Downregulated |
| 0,414495538 | 0,574190225 | PANO | FALSE | Upregulated |
| -0,310992196 | 0,574190225 | SUGP1 | FALSE | Downregulated |
| -0,319398668 | 0,574190225 | ZNF579 | FALSE | Downregulated |
| 0,206054525 | 0,574460934 | USP3 | FALSE | Upregulated |
| -0,32264146 | 0,574891855 | LRIG3 | FALSE | Downregulated |
| -0,203481831 | 0,574891855 | PIK3R2 | FALSE | Downregulated |
| 0,207962155 | 0,575048819 | COL6A6 | FALSE | Upregulated |
| 0,195235918 | 0,575048819 | DENND2D | FALSE | Upregulated |
| 0,490387154 | 0,575063903 | BMP2 | FALSE | Upregulated |
| 0,262044491 | 0,575063903 | SERHL2 | FALSE | Upregulated |
| 0,269955199 | 0,575270036 | HLA-DRA | FALSE | Upregulated |
| 0,399542188 | 0,575578788 | BCL6B | FALSE | Upregulated |
| -0,17864813 | 0,575578788 | FBXO31 | FALSE | Downregulated |
| 0,17378063 | 0,575578788 | KCTD20 | FALSE | Upregulated |
| 0,443630333 | 0,575578788 | LINC01043 | FALSE | Upregulated |
| 0,218290005 | 0,575578788 | RP11-887P2.5 | FALSE | Upregulated |
| 0,207705342 | 0,575578788 | THPO | FALSE | Upregulated |
| -0,300552761 | 0,575578788 | TRMT1L | FALSE | Downregulated |
| -0,312974732 | 0,575578788 | WDR43 | FALSE | Downregulated |
| 0,670636025 | 0,575578788 | WNT11 | FALSE | Upregulated |
| 0,375728037 | 0,575690447 | LOC101927841 | FALSE | Upregulated |
| -0,270720812 | 0,575880328 | CHMP1B | FALSE | Downregulated |
| 0,540026507 | 0,576033695 | C16orf93 | FALSE | Upregulated |
| -0,20415546 | 0,576033695 | EI24 | FALSE | Downregulated |
| 0,278844681 | 0,576033695 | MCMDC2 | FALSE | Upregulated |
| -0,561494728 | 0,5760924 | PILRA | FALSE | Downregulated |
| -0,334308595 | 0,576760964 | CHMP5 | FALSE | Downregulated |
| -0,308317053 | 0,576760964 | CYP2E1 | FALSE | Downregulated |
| -0,433023422 | 0,576760964 | NBPF13P | FALSE | Downregulated |
| -0,316542985 | 0,576760964 | OST4 | FALSE | Downregulated |

| | | | | |
|---|---|---|---|---|
| -0,442324759 | 0,576760964 | RTCA | FALSE | Downregulated |
| 0,296394381 | 0,576903955 | LOC102723804 | FALSE | Upregulated |
| 0,203197121 | 0,576903955 | LOC286190 | FALSE | Upregulated |
| -0,195342516 | 0,577155516 | ATF6B | FALSE | Downregulated |
| 0,897624373 | 0,577155516 | BCAN | FALSE | Upregulated |
| -0,289510209 | 0,577155516 | GMEB2 | FALSE | Downregulated |
| 0,161122632 | 0,577155516 | HNRNPU-AS1 | FALSE | Upregulated |
| -0,493215854 | 0,577155516 | JRKL | FALSE | Downregulated |
| 0,326702179 | 0,577155516 | LOC101929140 | FALSE | Upregulated |
| -0,383538596 | 0,577155516 | TMEM62 | FALSE | Downregulated |
| -0,153123046 | 0,577155516 | TPI1 | FALSE | Downregulated |
| 0,279705939 | 0,577155516 | ZNF266 | FALSE | Upregulated |
| -0,163113182 | 0,577449769 | AZGP1 | FALSE | Downregulated |
| 0,266986171 | 0,577505641 | PRICKLE4 | FALSE | Upregulated |
| 0,29187314 | 0,577505641 | RP11-538P18.2 | FALSE | Upregulated |
| -0,267560128 | 0,578494125 | SRCIN1 | FALSE | Downregulated |
| 0,419188204 | 0,578527397 | COL4A3 | FALSE | Upregulated |
| -0,464645566 | 0,578527397 | NOMO2 | FALSE | Downregulated |
| -0,40754515 | 0,578527397 | SLC22A10 | FALSE | Downregulated |
| 0,2216272 | 0,578577407 | MRS2P2 | FALSE | Upregulated |
| -0,212175088 | 0,578615799 | AKT1S1 | FALSE | Downregulated |
| 0,398176335 | 0,578615799 | C21orf58 | FALSE | Upregulated |
| -0,196570935 | 0,578615799 | CYP4F11 | FALSE | Downregulated |
| 0,252827365 | 0,578615799 | LOC727978 | FALSE | Upregulated |
| 0,381420975 | 0,578615799 | LRRC28 | FALSE | Upregulated |
| 0,515038836 | 0,578615799 | RINL | FALSE | Upregulated |
| 0,312641624 | 0,578615799 | SLC7A5P2 | FALSE | Upregulated |
| -0,368157238 | 0,578615799 | TMEM143 | FALSE | Downregulated |
| -0,226332128 | 0,578615799 | TTC37 | FALSE | Downregulated |
| -0,275623861 | 0,578634545 | SH3RF1 | FALSE | Downregulated |
| 0,381414706 | 0,578659956 | AAGAB | FALSE | Upregulated |
| 0,498279665 | 0,578659956 | BTN2A2 | FALSE | Upregulated |
| -0,320013033 | 0,578659956 | CXADR | FALSE | Downregulated |
| -0,415901185 | 0,578659956 | DENND2A | FALSE | Downregulated |
| 0,211810601 | 0,578659956 | GCHFR | FALSE | Upregulated |
| 0,303590612 | 0,578659956 | HP09025 | FALSE | Upregulated |
| 0,364158158 | 0,578659956 | LOC102723309 | FALSE | Upregulated |
| 0,267744507 | 0,578659956 | LOC729732 | FALSE | Upregulated |
| 0,364056667 | 0,578659956 | NKPD1 | FALSE | Upregulated |
| -0,417018708 | 0,578659956 | PRDM10 | FALSE | Downregulated |
| 0,279157625 | 0,578659956 | PSMB10 | FALSE | Upregulated |
| 0,216569006 | 0,578659956 | RASGRP1 | FALSE | Upregulated |
| -0,210220374 | 0,578659956 | SEMA5A | FALSE | Downregulated |
| 0,346979908 | 0,578659956 | ZNF385A | FALSE | Upregulated |
| -0,189936397 | 0,578754427 | ABCA6 | FALSE | Downregulated |
| 0,23506792 | 0,578754427 | C11orf49 | FALSE | Upregulated |
| -0,402708647 | 0,578754427 | RIPPLY1 | FALSE | Downregulated |
| 0,178892045 | 0,578754427 | RP11-545I5.3 | FALSE | Upregulated |
| -0,200019264 | 0,578754427 | RPL21 | FALSE | Downregulated |
| 0,688755357 | 0,578754427 | SIGLEC14 | FALSE | Upregulated |
| 0,175977695 | 0,578754427 | TNFRSF10B | FALSE | Upregulated |

| | | | | |
|---|---|---|---|---|
| -0,635600906 | 0,579174743 | TWSG1 | FALSE | Downregulated |
| -0,118613893 | 0,579190964 | CSDE1 | FALSE | Downregulated |
| 0,175730515 | 0,579190964 | KCNJ13 | FALSE | Upregulated |
| -0,176832268 | 0,579190964 | MEGF9 | FALSE | Downregulated |
| -0,507251728 | 0,579190964 | NPW | FALSE | Downregulated |
| 0,455674021 | 0,579255227 | INCA1 | FALSE | Upregulated |
| 0,214812011 | 0,579703832 | LOC101928524 | FALSE | Upregulated |
| -0,217131529 | 0,579703832 | POMGNT1 | FALSE | Downregulated |
| -0,170592245 | 0,579703832 | ZNF33A | FALSE | Downregulated |
| -0,221275147 | 0,579787759 | PSMF1 | FALSE | Downregulated |
| -0,324350957 | 0,580062688 | ANKRD26 | FALSE | Downregulated |
| 0,190268059 | 0,580062688 | CERS6-AS1 | FALSE | Upregulated |
| -0,208378109 | 0,580062688 | DCTD | FALSE | Downregulated |
| 0,151490835 | 0,580062688 | HMGN2 | FALSE | Upregulated |
| 0,351375288 | 0,580062688 | KLF16 | FALSE | Upregulated |
| -0,207091573 | 0,580062688 | PDLIM5 | FALSE | Downregulated |
| -0,292769004 | 0,580062688 | PPP1R12B | FALSE | Downregulated |
| -0,251678306 | 0,580062688 | RBP1 | FALSE | Downregulated |
| -0,445419321 | 0,580062688 | SHBG | FALSE | Downregulated |
| 0,364072967 | 0,580062688 | SPN | FALSE | Upregulated |
| 0,210512434 | 0,580062688 | ZNF517 | FALSE | Upregulated |
| -0,273414854 | 0,580209546 | AMACR | FALSE | Downregulated |
| -0,238860397 | 0,580209546 | ATF7 | FALSE | Downregulated |
| 0,357688063 | 0,580209546 | ATP8B2 | FALSE | Upregulated |
| 0,452650031 | 0,580209546 | C3orf38 | FALSE | Upregulated |
| -0,207965978 | 0,580209546 | CLMN | FALSE | Downregulated |
| -0,254664004 | 0,580209546 | DDX46 | FALSE | Downregulated |
| 0,659486673 | 0,580209546 | FAIM3 | FALSE | Upregulated |
| 0,41010676 | 0,580209546 | FAM98A | FALSE | Upregulated |
| -0,194234792 | 0,580209546 | FGB | FALSE | Downregulated |
| 0,287143091 | 0,580209546 | FGFR1OP | FALSE | Upregulated |
| 0,271812641 | 0,580209546 | FUT2 | FALSE | Upregulated |
| -0,230171424 | 0,580209546 | GCDH | FALSE | Downregulated |
| 0,154505282 | 0,580209546 | GON4L | FALSE | Upregulated |
| -0,202449495 | 0,580209546 | HDHD3 | FALSE | Downregulated |
| 0,224122355 | 0,580209546 | IDNK | FALSE | Upregulated |
| 0,286708688 | 0,580209546 | JUNB | FALSE | Upregulated |
| 0,3651995 | 0,580209546 | LOC100131270 | FALSE | Upregulated |
| 0,324662834 | 0,580209546 | LOC100996813 | FALSE | Upregulated |
| -0,251125385 | 0,580209546 | METRN | FALSE | Downregulated |
| -0,431103229 | 0,580209546 | MYADM | FALSE | Downregulated |
| -0,187104573 | 0,580209546 | NUDCD3 | FALSE | Downregulated |
| 0,478678707 | 0,580209546 | P4HA2 | FALSE | Upregulated |
| 0,508932948 | 0,580209546 | PIM2 | FALSE | Upregulated |
| -0,242563515 | 0,580209546 | PRPSAP1 | FALSE | Downregulated |
| -0,156052757 | 0,580209546 | PTPLAD1 | FALSE | Downregulated |
| 0,174950302 | 0,580209546 | RP11-367J11.2 | FALSE | Upregulated |
| 0,656005217 | 0,580209546 | SAMD15 | FALSE | Upregulated |
| -0,123558169 | 0,580209546 | SF1 | FALSE | Downregulated |
| -0,262930216 | 0,580209546 | SLC25A11 | FALSE | Downregulated |
| 0,537731457 | 0,580209546 | SOCS3 | FALSE | Upregulated |

| | | | | |
|---|---|---|---|---|
| -0,184659368 | 0,580209546 | SOWAHC | FALSE | Downregulated |
| 0,289474853 | 0,580209546 | SOX10 | FALSE | Upregulated |
| 0,409879807 | 0,580209546 | SOX15 | FALSE | Upregulated |
| -0,149138092 | 0,580209546 | TUBB | FALSE | Downregulated |
| 0,401256343 | 0,580209546 | UQCRBP1 | FALSE | Upregulated |
| 0,344569836 | 0,580209546 | ZNF620 | FALSE | Upregulated |
| -0,163389095 | 0,580216131 | ALDH5A1 | FALSE | Downregulated |
| 0,268701528 | 0,580216131 | ALDOA | FALSE | Upregulated |
| 0,228275578 | 0,580216131 | C14orf164 | FALSE | Upregulated |
| -0,201547572 | 0,580216131 | CALCOCO1 | FALSE | Downregulated |
| 0,400970139 | 0,580216131 | CCR7 | FALSE | Upregulated |
| 0,391437603 | 0,580216131 | CDCA7L | FALSE | Upregulated |
| -0,175569883 | 0,580216131 | CES2 | FALSE | Downregulated |
| -0,38944464 | 0,580216131 | CFHR3 | FALSE | Downregulated |
| 0,32822822 | 0,580216131 | CHADL | FALSE | Upregulated |
| -0,326993339 | 0,580216131 | COPS5 | FALSE | Downregulated |
| 0,166494177 | 0,580216131 | CSNK1D | FALSE | Upregulated |
| 0,226854866 | 0,580216131 | DAB2IP | FALSE | Upregulated |
| 0,575692136 | 0,580216131 | EFEMP1 | FALSE | Upregulated |
| -0,164864268 | 0,580216131 | EXOC7 | FALSE | Downregulated |
| 0,263943777 | 0,580216131 | FAM199X | FALSE | Upregulated |
| 0,167856951 | 0,580216131 | FLJ42627 | FALSE | Upregulated |
| -0,325111202 | 0,580216131 | FRAT1 | FALSE | Downregulated |
| -0,180784986 | 0,580216131 | GALM | FALSE | Downregulated |
| 0,56036961 | 0,580216131 | GALNT16 | FALSE | Upregulated |
| -0,218386546 | 0,580216131 | GBE1 | FALSE | Downregulated |
| 0,300655917 | 0,580216131 | GJA4 | FALSE | Upregulated |
| 0,323697654 | 0,580216131 | GNL2 | FALSE | Upregulated |
| 0,430749664 | 0,580216131 | GPAT2 | FALSE | Upregulated |
| -0,611803569 | 0,580216131 | GSTA7P | FALSE | Downregulated |
| -0,220631459 | 0,580216131 | HAL | FALSE | Downregulated |
| 0,55530215 | 0,580216131 | HCG26 | FALSE | Upregulated |
| 0,274691622 | 0,580216131 | LARGE-AS1 | FALSE | Upregulated |
| -0,274578069 | 0,580216131 | LIMCH1 | FALSE | Downregulated |
| 0,427822514 | 0,580216131 | LINC00861 | FALSE | Upregulated |
| 0,37846421 | 0,580216131 | LOC102723998 | FALSE | Upregulated |
| 0,832097385 | 0,580216131 | LOC102724428 | FALSE | Upregulated |
| -0,626355864 | 0,580216131 | LOC642846 | FALSE | Downregulated |
| 0,214695339 | 0,580216131 | MAP3K15 | FALSE | Upregulated |
| -0,24755496 | 0,580216131 | NADSYN1 | FALSE | Downregulated |
| 0,311087587 | 0,580216131 | NIPA1 | FALSE | Upregulated |
| 0,251713351 | 0,580216131 | NUS1 | FALSE | Upregulated |
| -0,184677494 | 0,580216131 | PPFIBP2 | FALSE | Downregulated |
| 0,172244896 | 0,580216131 | PPP1R16A | FALSE | Upregulated |
| -0,357589752 | 0,580216131 | PTAR1 | FALSE | Downregulated |
| -0,15681408 | 0,580216131 | RPL6 | FALSE | Downregulated |
| -0,27307438 | 0,580216131 | RSF1 | FALSE | Downregulated |
| -0,394189388 | 0,580216131 | SH3BGRL | FALSE | Downregulated |
| -0,208918181 | 0,580216131 | SLC25A10 | FALSE | Downregulated |
| -0,267414198 | 0,580216131 | TIPARP | FALSE | Downregulated |
| -0,283823055 | 0,580216131 | TM7SF2 | FALSE | Downregulated |

| | | | | |
|---|---|---|---|---|
| -0,21095109 | 0,580216131 | TM9SF4 | FALSE | Downregulated |
| -0,154866408 | 0,580216131 | TOLLIP | FALSE | Downregulated |
| 0,308656963 | 0,580216131 | TPTE2P5 | FALSE | Upregulated |
| 0,493169105 | 0,580216131 | TRIM34 | FALSE | Upregulated |
| 0,324490993 | 0,580216131 | UBALD2 | FALSE | Upregulated |
| 0,297955046 | 0,580216131 | UBOX5-AS1 | FALSE | Upregulated |
| 0,448297612 | 0,580216131 | UPP1 | FALSE | Upregulated |
| -0,194425997 | 0,580216131 | USO1 | FALSE | Downregulated |
| -0,732493604 | 0,580216131 | USP32P1 | FALSE | Downregulated |
| -0,517346338 | 0,580216131 | VPS33B | FALSE | Downregulated |
| -0,320921825 | 0,580216131 | ZNF467 | FALSE | Downregulated |
| -0,259523804 | 0,580216131 | ZRANB3 | FALSE | Downregulated |
| -0,452826087 | 0,580216131 | ZSCAN26 | FALSE | Downregulated |
| -0,151855476 | 0,580494437 | C1R | FALSE | Downregulated |
| -0,44818342 | 0,58078831 | DUSP5 | FALSE | Downregulated |
| 0,331798589 | 0,580900649 | LOC102724026 | FALSE | Upregulated |
| 0,164830877 | 0,581114979 | HEXIM2 | FALSE | Upregulated |
| -0,13812776 | 0,582428036 | SLC10A1 | FALSE | Downregulated |
| 0,287269645 | 0,582494783 | ANO9 | FALSE | Upregulated |
| -0,146918752 | 0,582906343 | MAGI1 | FALSE | Downregulated |
| 0,567913433 | 0,582906343 | RP11-147N17.1 | FALSE | Upregulated |
| 0,428652062 | 0,583089294 | GOLGA6L5P | FALSE | Upregulated |
| -0,163782663 | 0,583089294 | HOOK3 | FALSE | Downregulated |
| 0,198017314 | 0,583089294 | LOC102724391 | FALSE | Upregulated |
| -0,170655969 | 0,583418207 | APMAP | FALSE | Downregulated |
| -0,30668068 | 0,583551056 | FSTL1 | FALSE | Downregulated |
| -0,259409608 | 0,583551056 | UROD | FALSE | Downregulated |
| -0,365082426 | 0,583551056 | ZNF326 | FALSE | Downregulated |
| -0,136005225 | 0,583678987 | BOD1L1 | FALSE | Downregulated |
| 0,437745216 | 0,583940497 | ADAP1 | FALSE | Upregulated |
| 0,276388597 | 0,583940497 | CSF1 | FALSE | Upregulated |
| -0,167532501 | 0,583940497 | TRIM26 | FALSE | Downregulated |
| -0,536359765 | 0,584568529 | FAM76A | FALSE | Downregulated |
| -0,221697532 | 0,584568529 | ISOC2 | FALSE | Downregulated |
| -0,161817133 | 0,584568529 | KNG1 | FALSE | Downregulated |
| -0,568571016 | 0,584568529 | XXYLT1-AS2 | FALSE | Downregulated |
| -0,159458439 | 0,584620975 | USP19 | FALSE | Downregulated |
| 0,263694736 | 0,584678278 | PFKFB3 | FALSE | Upregulated |
| 0,396707983 | 0,585001691 | CLEC2B | FALSE | Upregulated |
| -0,412303206 | 0,585001691 | FAM222A | FALSE | Downregulated |
| -0,536492784 | 0,585001691 | LACE1 | FALSE | Downregulated |
| 0,26700805 | 0,585001691 | LOC101929451 | FALSE | Upregulated |
| 0,370532947 | 0,585001691 | TRAF2 | FALSE | Upregulated |
| -0,410633776 | 0,585001691 | WDR12 | FALSE | Downregulated |
| 0,19358113 | 0,5853435 | IDH2 | FALSE | Upregulated |
| -0,176036854 | 0,5853435 | KDSR | FALSE | Downregulated |
| -0,179695054 | 0,585521567 | DAP | FALSE | Downregulated |
| 0,161283108 | 0,585521567 | DYRK1A | FALSE | Upregulated |
| 0,193597073 | 0,585521567 | GALNT11 | FALSE | Upregulated |
| -0,275642047 | 0,585521567 | LPAL2 | FALSE | Downregulated |
| -0,140138517 | 0,585521567 | PAPSS2 | FALSE | Downregulated |

| | | | | |
|---|---|---|---|---|
| -0,150202176 | 0,586381638 | AFF1 | FALSE | Downregulated |
| -0,158121782 | 0,586381638 | GAK | FALSE | Downregulated |
| 0,60986862 | 0,586381638 | LOC102724770 | FALSE | Upregulated |
| -0,171153422 | 0,586381638 | PGPEP1 | FALSE | Downregulated |
| 0,182453289 | 0,586381638 | PPIL2 | FALSE | Upregulated |
| 0,236842877 | 0,586381638 | ZNHIT3 | FALSE | Upregulated |
| -0,364120395 | 0,586403998 | MANBA | FALSE | Downregulated |
| -0,191505921 | 0,586403998 | PHB | FALSE | Downregulated |
| 0,447371447 | 0,586505574 | PIN4P1 | FALSE | Upregulated |
| 0,276985617 | 0,586588451 | CADM4 | FALSE | Upregulated |
| -0,294494317 | 0,58735183 | RPL34 | FALSE | Downregulated |
| 0,309322707 | 0,587447303 | TRAF1 | FALSE | Upregulated |
| 0,226613514 | 0,587733973 | ARF4 | FALSE | Upregulated |
| -0,233175786 | 0,587733973 | ATP6V0C | FALSE | Downregulated |
| 0,28255667 | 0,587733973 | RP11-242D8.1 | FALSE | Upregulated |
| 0,454057063 | 0,587800786 | LOC100507530 | FALSE | Upregulated |
| -0,202348971 | 0,587837289 | AMBRA1 | FALSE | Downregulated |
| -0,432968256 | 0,587837289 | LOC102724709 | FALSE | Downregulated |
| 0,534225773 | 0,587837289 | VSIG10L | FALSE | Upregulated |
| 1,003800924 | 0,588066696 | LOC102724621 | FALSE | Upregulated |
| 0,198303845 | 0,588181864 | ANKRD23 | FALSE | Upregulated |
| 0,243465329 | 0,588181864 | ARPC4 | FALSE | Upregulated |
| -0,264728124 | 0,588181864 | ASPG | FALSE | Downregulated |
| 0,197444829 | 0,588181864 | BEST1 | FALSE | Upregulated |
| -0,187753393 | 0,588181864 | RPN1 | FALSE | Downregulated |
| 0,219472709 | 0,588181864 | SUV39H1 | FALSE | Upregulated |
| 0,221631986 | 0,588191503 | PLEKHM1 | FALSE | Upregulated |
| -0,53803691 | 0,589084129 | BLOC1S2 | FALSE | Downregulated |
| -0,261424034 | 0,589189071 | C14orf159 | FALSE | Downregulated |
| -0,319996881 | 0,589189071 | MAP2K5 | FALSE | Downregulated |
| 0,260866607 | 0,589189071 | TRAF3 | FALSE | Upregulated |
| -0,202380391 | 0,589682884 | BTAF1 | FALSE | Downregulated |
| 0,187549594 | 0,589682884 | CCRN4L | FALSE | Upregulated |
| -0,127396561 | 0,589682884 | GIGYF1 | FALSE | Downregulated |
| 0,197254564 | 0,589682884 | PKN1 | FALSE | Upregulated |
| 0,247993255 | 0,589682884 | TTC23 | FALSE | Upregulated |
| 0,247039172 | 0,589682884 | VNN1 | FALSE | Upregulated |
| -0,128078287 | 0,59016695 | KHSRP | FALSE | Downregulated |
| -0,351530267 | 0,591131161 | ATF2 | FALSE | Downregulated |
| 0,399227195 | 0,591131161 | FKTN | FALSE | Upregulated |
| 0,196796416 | 0,591131161 | GPR107 | FALSE | Upregulated |
| -0,182354222 | 0,591131161 | SLC4A4 | FALSE | Downregulated |
| -0,411413528 | 0,591131161 | SRGAP3 | FALSE | Downregulated |
| 0,15756705 | 0,591131161 | UBN1 | FALSE | Upregulated |
| 0,330022341 | 0,591428181 | RNF222 | FALSE | Upregulated |
| -0,30471136 | 0,591428181 | TNIP2 | FALSE | Downregulated |
| -0,18895104 | 0,5915238 | C2orf72 | FALSE | Downregulated |
| 0,378055638 | 0,591587862 | PDE6A | FALSE | Upregulated |
| 0,145368213 | 0,591661824 | SYNPO | FALSE | Upregulated |
| -0,190870195 | 0,59181893 | SLC22A7 | FALSE | Downregulated |
| -0,211257313 | 0,591955939 | CYP2J2 | FALSE | Downregulated |

| | | | | |
|---|---|---|---|---|
| 0,337909455 | 0,592096786 | CEP85 | FALSE | Upregulated |
| -0,402647853 | 0,592096786 | DISP1 | FALSE | Downregulated |
| -0,396302602 | 0,592270741 | DYNLT3 | FALSE | Downregulated |
| 0,212665251 | 0,592270741 | SLC24A1 | FALSE | Upregulated |
| 0,368585631 | 0,592270741 | WDR70 | FALSE | Upregulated |
| -0,35511234 | 0,592938425 | ABCG2 | FALSE | Downregulated |
| -0,180094947 | 0,592938425 | CDO1 | FALSE | Downregulated |
| -0,137995116 | 0,592938425 | H3F3B | FALSE | Downregulated |
| -0,388444124 | 0,592938425 | PMS2CL | FALSE | Downregulated |
| 0,521982751 | 0,592938425 | RAB31 | FALSE | Upregulated |
| -0,193838446 | 0,592991148 | CNPY2 | FALSE | Downregulated |
| -0,245826069 | 0,592991148 | GCC1 | FALSE | Downregulated |
| -0,167206856 | 0,592991148 | IRS1 | FALSE | Downregulated |
| 0,211038272 | 0,592991148 | NUDT13 | FALSE | Upregulated |
| 0,235271597 | 0,592991148 | TNFAIP2 | FALSE | Upregulated |
| 0,246057056 | 0,592991148 | WDR37 | FALSE | Upregulated |
| 0,306955215 | 0,593043435 | ACAP1 | FALSE | Upregulated |
| -0,348647666 | 0,593043435 | ZNF268 | FALSE | Downregulated |
| -0,21217501 | 0,593290156 | LARS | FALSE | Downregulated |
| 0,430466401 | 0,593290156 | LOC101928794 | FALSE | Upregulated |
| 0,272341957 | 0,593831387 | NEIL2 | FALSE | Upregulated |
| 0,286555941 | 0,593884319 | ARAP2 | FALSE | Upregulated |
| -0,15264513 | 0,594034831 | C3 | FALSE | Downregulated |
| 0,261742736 | 0,594034831 | DDX19A | FALSE | Upregulated |
| -0,154274838 | 0,595565568 | GLTPD1 | FALSE | Downregulated |
| -0,086711394 | 0,595882936 | ARHGEF12 | FALSE | Downregulated |
| -0,238426374 | 0,595882936 | IFRD2 | FALSE | Downregulated |
| 0,277696606 | 0,595882936 | IFT88 | FALSE | Upregulated |
| -0,182694266 | 0,595882936 | MGRN1 | FALSE | Downregulated |
| -0,157054586 | 0,595882936 | PPARA | FALSE | Downregulated |
| -0,132096965 | 0,595882936 | RNF40 | FALSE | Downregulated |
| 0,406663925 | 0,595882936 | TMEM11 | FALSE | Upregulated |
| -0,146622123 | 0,596176252 | ABCC3 | FALSE | Downregulated |
| 0,289311353 | 0,596176252 | C1QB | FALSE | Upregulated |
| -0,150544556 | 0,596176252 | ZC3H11A | FALSE | Downregulated |
| -0,321576684 | 0,596394949 | COMMD9 | FALSE | Downregulated |
| 0,263895936 | 0,596394949 | SIM2 | FALSE | Upregulated |
| -0,274071817 | 0,596431415 | TGFBRAP1 | FALSE | Downregulated |
| -0,181498871 | 0,596457875 | RNF187 | FALSE | Downregulated |
| -0,179995188 | 0,596457875 | STAT5B | FALSE | Downregulated |
| 0,256198535 | 0,596682126 | LOC102724383 | FALSE | Upregulated |
| 0,292130811 | 0,596682126 | TOMM34 | FALSE | Upregulated |
| -0,177095011 | 0,596684353 | LMAN2 | FALSE | Downregulated |
| 0,443057044 | 0,596684353 | MGST3 | FALSE | Upregulated |
| 0,233941177 | 0,597130248 | FLJ40448 | FALSE | Upregulated |
| -0,163119904 | 0,597180627 | C6orf89 | FALSE | Downregulated |
| -0,159465724 | 0,597180627 | FAM193B | FALSE | Downregulated |
| -0,235096243 | 0,597180627 | ID2 | FALSE | Downregulated |
| 0,262848767 | 0,597180627 | IST1 | FALSE | Upregulated |
| -0,328049064 | 0,597180627 | NARG2 | FALSE | Downregulated |
| 0,786586364 | 0,597180627 | NEUROD6 | FALSE | Upregulated |

| | | | | |
|---|---|---|---|---|
| 0,166866058 | 0,597180627 | PHKG1 | FALSE | Upregulated |
| 0,150789611 | 0,597180627 | SGSM2 | FALSE | Upregulated |
| 0,398453529 | 0,597180627 | SLC11A1 | FALSE | Upregulated |
| -0,209854741 | 0,597433713 | N4BP2 | FALSE | Downregulated |
| 0,218954307 | 0,597433713 | NCAPH2 | FALSE | Upregulated |
| 0,344635182 | 0,597433713 | RND1 | FALSE | Upregulated |
| 0,232054763 | 0,597513383 | SMAD5-AS1 | FALSE | Upregulated |
| -0,1843064 | 0,597540513 | AGXT | FALSE | Downregulated |
| -0,191157889 | 0,597540513 | ENPEP | FALSE | Downregulated |
| 0,425692807 | 0,597540513 | NRIP3 | FALSE | Upregulated |
| -0,205605016 | 0,597667715 | ANG | FALSE | Downregulated |
| 0,34251223 | 0,597667715 | C17orf104 | FALSE | Upregulated |
| 0,255070501 | 0,597667715 | CACNA1D | FALSE | Upregulated |
| -0,465511658 | 0,597667715 | CAV1 | FALSE | Downregulated |
| 0,388441599 | 0,597667715 | HK2 | FALSE | Upregulated |
| -0,308611713 | 0,597667715 | JMY | FALSE | Downregulated |
| -0,193966033 | 0,597667715 | KTN1 | FALSE | Downregulated |
| 0,283553119 | 0,597667715 | LOC100134040 | FALSE | Upregulated |
| 0,659018531 | 0,597667715 | LOC101927355 | FALSE | Upregulated |
| -0,38633049 | 0,597667715 | LYPLAL1 | FALSE | Downregulated |
| 0,232611272 | 0,597667715 | MAPK1IP1L | FALSE | Upregulated |
| -0,415784328 | 0,597667715 | MRPL45P2 | FALSE | Downregulated |
| -0,2483577 | 0,597667715 | MS4A6A | FALSE | Downregulated |
| -0,500394655 | 0,597667715 | MSANTD4 | FALSE | Downregulated |
| 0,45767634 | 0,597667715 | NAA40 | FALSE | Upregulated |
| -0,462574366 | 0,597667715 | PAXIP1-AS1 | FALSE | Downregulated |
| 0,172580386 | 0,597667715 | RFTN1 | FALSE | Upregulated |
| 0,476805923 | 0,597667715 | STAT4 | FALSE | Upregulated |
| -0,473445344 | 0,597667715 | ZFAS1 | FALSE | Downregulated |
| 0,237690313 | 0,597882523 | HIF1A-AS2 | FALSE | Upregulated |
| -0,190266736 | 0,597997471 | PVRL2 | FALSE | Downregulated |
| -0,170855057 | 0,597997471 | ZCCHC2 | FALSE | Downregulated |
| 0,295718329 | 0,598173472 | CTC-338M12.4 | FALSE | Upregulated |
| 0,506841727 | 0,598173472 | FEN1 | FALSE | Upregulated |
| 0,60045277 | 0,598173472 | IGJ | FALSE | Upregulated |
| -0,266272166 | 0,598173472 | SYDE2 | FALSE | Downregulated |
| 0,182854245 | 0,598173472 | YWHAZ | FALSE | Upregulated |
| -0,169622959 | 0,598479912 | PBLD | FALSE | Downregulated |
| -0,527443227 | 0,598995673 | KBTBD6 | FALSE | Downregulated |
| -0,208343203 | 0,598995673 | NR1H2 | FALSE | Downregulated |
| 0,260145552 | 0,598995673 | RFX7 | FALSE | Upregulated |
| 0,243197999 | 0,598995673 | ST7-AS2 | FALSE | Upregulated |
| 0,210710868 | 0,59954645 | C22orf26 | FALSE | Upregulated |
| -0,190958923 | 0,59954645 | CYP2C9 | FALSE | Downregulated |
| 0,308676227 | 0,59954645 | GATS | FALSE | Upregulated |
| -0,144449018 | 0,59954645 | H6PD | FALSE | Downregulated |
| -0,307937449 | 0,59954645 | HACL1 | FALSE | Downregulated |
| 0,362802153 | 0,59954645 | LOC102724335 | FALSE | Upregulated |
| 0,468276566 | 0,59954645 | LZIC | FALSE | Upregulated |
| -0,219272706 | 0,59954645 | NDUFB9 | FALSE | Downregulated |
| -0,136648135 | 0,59954645 | NIPSNAP1 | FALSE | Downregulated |

| | | | | |
|---|---|---|---|---|
| 0,604787261 | 0,59954645 | PAQR4 | FALSE | Upregulated |
| -0,414582094 | 0,59954645 | PDP2 | FALSE | Downregulated |
| -0,363955474 | 0,59954645 | SMIM12 | FALSE | Downregulated |
| -0,244832185 | 0,59954645 | SPNS1 | FALSE | Downregulated |
| 0,300329591 | 0,59954645 | TIGD7 | FALSE | Upregulated |
| 0,347798616 | 0,59954645 | ZNF32 | FALSE | Upregulated |
| -0,222629045 | 0,599915584 | ACADM | FALSE | Downregulated |
| -0,369197382 | 0,599915584 | VPS25 | FALSE | Downregulated |
| 0,229284272 | 0,600051258 | HEIH | FALSE | Upregulated |
| -0,183816787 | 0,600051258 | TXNDC5 | FALSE | Downregulated |
| -0,306566943 | 0,600884537 | ALDOC | FALSE | Downregulated |
| -0,180551202 | 0,600884537 | ARHGEF40 | FALSE | Downregulated |
| -0,390518571 | 0,600884537 | C8orf33 | FALSE | Downregulated |
| -0,215420501 | 0,600884537 | ELK1 | FALSE | Downregulated |
| 0,410624859 | 0,600884537 | ENO3 | FALSE | Upregulated |
| 0,211823677 | 0,600884537 | FAR2 | FALSE | Upregulated |
| 0,174803008 | 0,600884537 | GPR161 | FALSE | Upregulated |
| 0,398988033 | 0,600884537 | ITGAX | FALSE | Upregulated |
| 0,268477971 | 0,600884537 | LOC101928552 | FALSE | Upregulated |
| 0,257158232 | 0,600884537 | LOC102723762 | FALSE | Upregulated |
| 0,402313968 | 0,600884537 | MAP1A | FALSE | Upregulated |
| -0,344960115 | 0,600884537 | MRPL9 | FALSE | Downregulated |
| 0,182585885 | 0,600884537 | POLD4 | FALSE | Upregulated |
| 0,411831627 | 0,600884537 | RP11-356I2.4 | FALSE | Upregulated |
| 0,205255769 | 0,600884537 | SLC5A3 | FALSE | Upregulated |
| 0,337557139 | 0,600884537 | ZNF540 | FALSE | Upregulated |
| 0,21020011 | 0,601036524 | ASB16 | FALSE | Upregulated |
| 0,219391128 | 0,601036524 | DUOX1 | FALSE | Upregulated |
| -0,178193787 | 0,601036524 | LRRFIP2 | FALSE | Downregulated |
| -0,188976551 | 0,601036524 | MTMR4 | FALSE | Downregulated |
| 0,451774503 | 0,601036524 | TACC3 | FALSE | Upregulated |
| -0,191408812 | 0,601036524 | TF | FALSE | Downregulated |
| 0,404291793 | 0,601252169 | C19orf82 | FALSE | Upregulated |
| -0,156556142 | 0,601322518 | CLIP1 | FALSE | Downregulated |
| -0,138975449 | 0,601732198 | ARF1 | FALSE | Downregulated |
| 0,537522781 | 0,601732198 | ARSJ | FALSE | Upregulated |
| -0,426871363 | 0,601732198 | CPAMD8 | FALSE | Downregulated |
| -0,257233715 | 0,601732198 | GART | FALSE | Downregulated |
| 0,473671405 | 0,601732198 | HACE1 | FALSE | Upregulated |
| -0,468415457 | 0,601732198 | HEPACAM | FALSE | Downregulated |
| -0,480344027 | 0,601732198 | NRARP | FALSE | Downregulated |
| -0,255704254 | 0,601732198 | SLC30A9 | FALSE | Downregulated |
| 0,363097311 | 0,601732198 | TTLL11 | FALSE | Upregulated |
| 0,362624717 | 0,601907901 | TIGIT | FALSE | Upregulated |
| -0,157976855 | 0,601907901 | TOM1L1 | FALSE | Downregulated |
| 0,203607298 | 0,602441776 | ZNF850 | FALSE | Upregulated |
| 0,409222473 | 0,602513708 | RASSF2 | FALSE | Upregulated |
| 0,354440549 | 0,602709012 | LOC101927741 | FALSE | Upregulated |
| 0,17478147 | 0,602709012 | POM121C | FALSE | Upregulated |
| 0,158175651 | 0,603804316 | DPP9 | FALSE | Upregulated |
| 0,432869124 | 0,603804316 | FAM102B | FALSE | Upregulated |

| | | | | |
|---|---|---|---|---|
| -0,470674629 | 0,60391742 | ARHGEF16 | FALSE | Downregulated |
| 0,179816357 | 0,60391742 | BMP8A | FALSE | Upregulated |
| 0,226169858 | 0,60391742 | C15orf27 | FALSE | Upregulated |
| -0,171505404 | 0,60391742 | C6 | FALSE | Downregulated |
| -0,228828368 | 0,60391742 | COPS6 | FALSE | Downregulated |
| 0,309566007 | 0,60391742 | CTD-2201E9.1 | FALSE | Upregulated |
| 0,393301888 | 0,60391742 | CXCR2P1 | FALSE | Upregulated |
| -0,269118719 | 0,60391742 | ERCC6 | FALSE | Downregulated |
| 0,210576587 | 0,60391742 | ERCC6L | FALSE | Upregulated |
| -0,337269667 | 0,60391742 | EXOSC2 | FALSE | Downregulated |
| -0,268644669 | 0,60391742 | FOXN2 | FALSE | Downregulated |
| 0,395504091 | 0,60391742 | GEMIN8P4 | FALSE | Upregulated |
| -0,338134093 | 0,60391742 | GNMT | FALSE | Downregulated |
| 0,406701108 | 0,60391742 | NAA38 | FALSE | Upregulated |
| -0,217628143 | 0,60391742 | PDXP | FALSE | Downregulated |
| 0,284094432 | 0,60391742 | PELI3 | FALSE | Upregulated |
| -0,177121149 | 0,60391742 | PEPD | FALSE | Downregulated |
| -0,185167 | 0,60391742 | TRPM7 | FALSE | Downregulated |
| -0,516403525 | 0,60399975 | COL25A1 | FALSE | Downregulated |
| -0,123051943 | 0,60399975 | PRRC2C | FALSE | Downregulated |
| 0,193223407 | 0,604277019 | OTUB2 | FALSE | Upregulated |
| -0,181652933 | 0,604508516 | POFUT1 | FALSE | Downregulated |
| 0,210580131 | 0,6047482 | HPS3 | FALSE | Upregulated |
| 0,537344865 | 0,6047482 | LOC102724791 | FALSE | Upregulated |
| 0,193362492 | 0,6047482 | SDCBP2-AS1 | FALSE | Upregulated |
| 0,498840287 | 0,6047482 | SHISA6 | FALSE | Upregulated |
| -0,244549309 | 0,604765196 | CDC40 | FALSE | Downregulated |
| -0,16972035 | 0,604765196 | CELF1 | FALSE | Downregulated |
| -0,157919736 | 0,604765196 | CPN2 | FALSE | Downregulated |
| 0,234552529 | 0,604765196 | JAM3 | FALSE | Upregulated |
| 0,258801238 | 0,604765196 | PYCRL | FALSE | Upregulated |
| 0,406612335 | 0,604999944 | PTK6 | FALSE | Upregulated |
| -0,167473142 | 0,604999944 | TUG1 | FALSE | Downregulated |
| 0,134904608 | 0,605015928 | AKAP13 | FALSE | Upregulated |
| 0,484349833 | 0,605015928 | NUSAP1 | FALSE | Upregulated |
| -0,414444903 | 0,605528086 | PARL | FALSE | Downregulated |
| 0,588397459 | 0,605566036 | CCDC170 | FALSE | Upregulated |
| 0,245877244 | 0,606154423 | ERCC6L2 | FALSE | Upregulated |
| -0,300341301 | 0,606481996 | MRPS15 | FALSE | Downregulated |
| 0,29203338 | 0,606506193 | SMIM6 | FALSE | Upregulated |
| -0,165869858 | 0,606542334 | SLC20A2 | FALSE | Downregulated |
| -0,342396896 | 0,606687775 | DIABLO | FALSE | Downregulated |
| 0,357386384 | 0,606687775 | GPATCH3 | FALSE | Upregulated |
| -0,15462018 | 0,60672861 | NAPA | FALSE | Downregulated |
| -0,37258015 | 0,60672861 | PRSS53 | FALSE | Downregulated |
| -0,311688526 | 0,60672861 | TXNRD3 | FALSE | Downregulated |
| 0,261161905 | 0,607704864 | RP5-914P20.5 | FALSE | Upregulated |
| 0,405339447 | 0,607717655 | LAMC3 | FALSE | Upregulated |
| 0,443981642 | 0,607727552 | DKK3 | FALSE | Upregulated |
| 0,197613074 | 0,607727552 | EPB41L4A | FALSE | Upregulated |
| 0,311489721 | 0,607866325 | CXCR5 | FALSE | Upregulated |

| | | | | |
|---|---|---|---|---|
| -0,205900095 | 0,608116986 | SLC13A5 | FALSE | Downregulated |
| -0,186013946 | 0,609881016 | ACSL1 | FALSE | Downregulated |
| -0,16712185 | 0,609881016 | ANGPTL3 | FALSE | Downregulated |
| -0,252481439 | 0,609881016 | DHX36 | FALSE | Downregulated |
| 0,255556241 | 0,609881016 | MICALL2 | FALSE | Upregulated |
| -0,248972497 | 0,609906079 | ATP5H | FALSE | Downregulated |
| 0,398836329 | 0,610010905 | ANKRD44 | FALSE | Upregulated |
| 0,283726799 | 0,610115586 | CBLB | FALSE | Upregulated |
| 0,267602658 | 0,610115586 | CBX1 | FALSE | Upregulated |
| 0,210813806 | 0,610115586 | CLEC16A | FALSE | Upregulated |
| -0,240903654 | 0,610115586 | COL4A3BP | FALSE | Downregulated |
| 0,232269802 | 0,610115586 | LOC102723853 | FALSE | Upregulated |
| -0,251639591 | 0,610115586 | NMRAL1 | FALSE | Downregulated |
| 0,507714844 | 0,610115586 | WDSUB1 | FALSE | Upregulated |
| -0,141195385 | 0,610169576 | MYO1C | FALSE | Downregulated |
| 0,302467942 | 0,610402391 | NOL3 | FALSE | Upregulated |
| -0,267474183 | 0,610419843 | ATF5 | FALSE | Downregulated |
| -0,198964855 | 0,610538458 | PTPN3 | FALSE | Downregulated |
| -0,320603266 | 0,610538458 | WDR18 | FALSE | Downregulated |
| -0,205573621 | 0,610799915 | CHPF | FALSE | Downregulated |
| 0,32493763 | 0,610799915 | DCAF15 | FALSE | Upregulated |
| 0,312048239 | 0,610799915 | LOC101926887 | FALSE | Upregulated |
| 0,607725566 | 0,610799915 | SLC9A3 | FALSE | Upregulated |
| 0,282230726 | 0,610833247 | TACR1 | FALSE | Upregulated |
| 0,677099864 | 0,611115473 | HS3ST1 | FALSE | Upregulated |
| -0,225294389 | 0,611115473 | LIX1L | FALSE | Downregulated |
| 0,689106355 | 0,611115473 | OR7E2P | FALSE | Upregulated |
| 0,465105604 | 0,61118737 | CTD-3080P12.3 | FALSE | Upregulated |
| 0,510952758 | 0,611215966 | DEGS2 | FALSE | Upregulated |
| 0,413772032 | 0,611215966 | FRMD6 | FALSE | Upregulated |
| 0,539943284 | 0,611215966 | MDGA1 | FALSE | Upregulated |
| -0,193935173 | 0,611215966 | PYGL | FALSE | Downregulated |
| 0,142752937 | 0,611215966 | SUGP2 | FALSE | Upregulated |
| -0,113095767 | 0,61130067 | EGLN1 | FALSE | Downregulated |
| -0,296218634 | 0,6113411 | ARHGAP42 | FALSE | Downregulated |
| -0,224462069 | 0,6113411 | BCL6 | FALSE | Downregulated |
| -0,242883396 | 0,6113411 | CRLS1 | FALSE | Downregulated |
| 0,460133017 | 0,6113411 | DSCR9 | FALSE | Upregulated |
| 0,462597748 | 0,6113411 | HLA-DMB | FALSE | Upregulated |
| 0,30425952 | 0,6113411 | ICAM1 | FALSE | Upregulated |
| -0,219354918 | 0,6113411 | IGFBP3 | FALSE | Downregulated |
| -0,365009665 | 0,6113411 | NDUFA9 | FALSE | Downregulated |
| 0,269729252 | 0,6113411 | ORMDL2 | FALSE | Upregulated |
| -0,321989318 | 0,6113411 | RAN | FALSE | Downregulated |
| -0,2904692 | 0,6113411 | SSNA1 | FALSE | Downregulated |
| -0,229557576 | 0,6113411 | TEN1 | FALSE | Downregulated |
| 0,165070893 | 0,6113411 | TMEM30B | FALSE | Upregulated |
| -0,244934514 | 0,611405442 | LPIN1 | FALSE | Downregulated |
| 0,370912572 | 0,611439123 | ABHD4 | FALSE | Upregulated |
| -0,20583376 | 0,611439123 | ADAMTSL3 | FALSE | Downregulated |
| 0,362139595 | 0,611439123 | APBB1 | FALSE | Upregulated |

| | | | | |
|---|---|---|---|---|
| 0,210638785 | 0,611439123 | ARGFXP2 | FALSE | Upregulated |
| -0,142942191 | 0,611439123 | C1orf43 | FALSE | Downregulated |
| 0,260886581 | 0,611439123 | CCNE2 | FALSE | Upregulated |
| -0,177698078 | 0,611439123 | CDH5 | FALSE | Downregulated |
| -0,553889132 | 0,611439123 | COMMD2 | FALSE | Downregulated |
| 0,321082127 | 0,611439123 | CYTH4 | FALSE | Upregulated |
| 0,215878959 | 0,611439123 | DCTN2 | FALSE | Upregulated |
| -0,151321133 | 0,611439123 | ECH1 | FALSE | Downregulated |
| 0,209724855 | 0,611439123 | EHBP1L1 | FALSE | Upregulated |
| 0,338668709 | 0,611439123 | ELOVL2 | FALSE | Upregulated |
| 0,324783648 | 0,611439123 | ERH | FALSE | Upregulated |
| -0,258371092 | 0,611439123 | EXOSC10 | FALSE | Downregulated |
| -0,427226807 | 0,611439123 | FAM135A | FALSE | Downregulated |
| -0,122754997 | 0,611439123 | GLG1 | FALSE | Downregulated |
| -0,324429439 | 0,611439123 | IPO9 | FALSE | Downregulated |
| 0,148853474 | 0,611439123 | JMJD8 | FALSE | Upregulated |
| 0,167377199 | 0,611439123 | KIAA0355 | FALSE | Upregulated |
| -0,39869618 | 0,611439123 | KIF11 | FALSE | Downregulated |
| -0,505253584 | 0,611439123 | KLHL7 | FALSE | Downregulated |
| 0,329106247 | 0,611439123 | LOC100499194 | FALSE | Upregulated |
| 0,530683867 | 0,611439123 | LUM | FALSE | Upregulated |
| 0,346204633 | 0,611439123 | MAFF | FALSE | Upregulated |
| -0,154079636 | 0,611439123 | MBTPS1 | FALSE | Downregulated |
| 0,357547562 | 0,611439123 | NDUFAF7 | FALSE | Upregulated |
| -0,187650333 | 0,611439123 | NFAT5 | FALSE | Downregulated |
| -0,283197831 | 0,611439123 | NTHL1 | FALSE | Downregulated |
| -0,181488851 | 0,611439123 | NUPR1 | FALSE | Downregulated |
| 0,197128619 | 0,611439123 | PCSK7 | FALSE | Upregulated |
| -0,143983415 | 0,611439123 | PHLDB2 | FALSE | Downregulated |
| 0,521508557 | 0,611439123 | PIGU | FALSE | Upregulated |
| -0,276120316 | 0,611439123 | PKIG | FALSE | Downregulated |
| 0,299032228 | 0,611439123 | RASGRP3 | FALSE | Upregulated |
| 0,164152906 | 0,611439123 | RBMS2 | FALSE | Upregulated |
| 0,456813111 | 0,611439123 | RHBDF1 | FALSE | Upregulated |
| -0,288943956 | 0,611439123 | RPH3AL | FALSE | Downregulated |
| 0,351897125 | 0,611439123 | RPS2P32 | FALSE | Upregulated |
| -0,458604004 | 0,611439123 | SAC3D1 | FALSE | Downregulated |
| -0,383039302 | 0,611439123 | SLC10A5 | FALSE | Downregulated |
| 0,309172925 | 0,611439123 | SLFNL1 | FALSE | Upregulated |
| -0,230419592 | 0,611439123 | STRAP | FALSE | Downregulated |
| -0,291450295 | 0,611439123 | TLE2 | FALSE | Downregulated |
| 0,258540742 | 0,611439123 | TLE4 | FALSE | Upregulated |
| -0,091658703 | 0,611439123 | TNRC6B | FALSE | Downregulated |
| -0,164533271 | 0,611439123 | UBA1 | FALSE | Downregulated |
| 0,277851518 | 0,611439123 | USP6 | FALSE | Upregulated |
| -0,237601728 | 0,611439123 | YY1AP1 | FALSE | Downregulated |
| 0,163517071 | 0,611439123 | ZBTB11 | FALSE | Upregulated |
| -0,307765195 | 0,611439123 | ZFP64 | FALSE | Downregulated |
| 0,267091289 | 0,611439123 | ZNF205 | FALSE | Upregulated |
| -0,279143238 | 0,611439123 | ZYG11A | FALSE | Downregulated |
| 0,389383004 | 0,61146944 | CENPJ | FALSE | Upregulated |

| | | | | |
|---|---|---|---|---|
| -0,340516454 | 0,61146944 | SLC2A8 | FALSE | Downregulated |
| -0,114613901 | 0,611927633 | KMT2D | FALSE | Downregulated |
| -0,151132223 | 0,612390355 | CIRBP | FALSE | Downregulated |
| -0,377621207 | 0,612390355 | H2AFZ | FALSE | Downregulated |
| 0,54223115 | 0,612390355 | ISG15 | FALSE | Upregulated |
| -0,353609477 | 0,612697572 | LOC730101 | FALSE | Downregulated |
| 0,523594679 | 0,612697572 | RP11-797H7.5 | FALSE | Upregulated |
| 0,217062953 | 0,613259803 | CEP57 | FALSE | Upregulated |
| -0,149124549 | 0,613259803 | LMTK2 | FALSE | Downregulated |
| -0,402773228 | 0,613275953 | PGBD5 | FALSE | Downregulated |
| 0,375222871 | 0,613279152 | EIF4E | FALSE | Upregulated |
| 0,498369225 | 0,613436584 | AC006145.4 | FALSE | Upregulated |
| 0,311699295 | 0,613974844 | ADAMTS10 | FALSE | Upregulated |
| -0,180902693 | 0,613974844 | C5 | FALSE | Downregulated |
| 0,243248451 | 0,613974844 | CABLES2 | FALSE | Upregulated |
| 0,205597497 | 0,613974844 | CNTRL | FALSE | Upregulated |
| 0,198063558 | 0,613974844 | MTX1 | FALSE | Upregulated |
| 0,252007314 | 0,613974844 | PFDN1 | FALSE | Upregulated |
| 0,344083104 | 0,613974844 | SAMD9L | FALSE | Upregulated |
| -0,151063314 | 0,613974844 | SERF2 | FALSE | Downregulated |
| 0,542066999 | 0,613974844 | SPECC1 | FALSE | Upregulated |
| -0,130776994 | 0,613974844 | TBC1D9B | FALSE | Downregulated |
| 0,431196645 | 0,613974844 | TES | FALSE | Upregulated |
| -0,20137185 | 0,614160507 | FOXA1 | FALSE | Downregulated |
| -0,260205005 | 0,614590281 | ACBD6 | FALSE | Downregulated |
| 0,465780996 | 0,614590281 | ATRIP | FALSE | Upregulated |
| -0,165136695 | 0,614590281 | CLDN1 | FALSE | Downregulated |
| 0,392670108 | 0,614590281 | MAP6 | FALSE | Upregulated |
| -0,22363572 | 0,614590281 | NDUFA13 | FALSE | Downregulated |
| 0,156355389 | 0,614590281 | PGM3 | FALSE | Upregulated |
| -0,157301472 | 0,614590281 | PROX1 | FALSE | Downregulated |
| 0,379046702 | 0,614590281 | RP1-293L8.2 | FALSE | Upregulated |
| -0,238426445 | 0,614590281 | TAZ | FALSE | Downregulated |
| -0,16845667 | 0,614671054 | HPD | FALSE | Downregulated |
| 0,398540492 | 0,614671054 | PDE11A | FALSE | Upregulated |
| 0,336039751 | 0,614671054 | STON2 | FALSE | Upregulated |
| 0,144231383 | 0,615021373 | APEH | FALSE | Upregulated |
| 0,462215796 | 0,615021373 | CLRN1 | FALSE | Upregulated |
| 0,295232868 | 0,615021373 | LOC101927161 | FALSE | Upregulated |
| 0,168127558 | 0,615021373 | MPHOSPH8 | FALSE | Upregulated |
| 0,228889735 | 0,615922743 | ABHD16A | FALSE | Upregulated |
| -0,471527103 | 0,615967468 | EXOSC4 | FALSE | Downregulated |
| 0,320391512 | 0,616025901 | SLC12A5 | FALSE | Upregulated |
| -0,290215312 | 0,616125909 | DYM | FALSE | Downregulated |
| -0,377767727 | 0,616125909 | ZNF615 | FALSE | Downregulated |
| -0,274548429 | 0,616547374 | SCNN1D | FALSE | Downregulated |
| -0,146381458 | 0,616678331 | BCR | FALSE | Downregulated |
| 0,172837425 | 0,616678331 | CARHSP1 | FALSE | Upregulated |
| 0,333683136 | 0,616678331 | CORIN | FALSE | Upregulated |
| 0,164387817 | 0,616678331 | CRLF3 | FALSE | Upregulated |
| -0,154907328 | 0,616678331 | LDLRAD4 | FALSE | Downregulated |

| | | | | |
|---|---|---|---|---|
| 0,395247396 | 0,616678331 | LOC102723877 | FALSE | Upregulated |
| 0,33402983 | 0,616678331 | LOC80154 | FALSE | Upregulated |
| 0,28454775 | 0,616678331 | MED22 | FALSE | Upregulated |
| -0,241748641 | 0,616678331 | PRNP | FALSE | Downregulated |
| 0,368789542 | 0,616678331 | RFTN2 | FALSE | Upregulated |
| -0,333957142 | 0,616678331 | RP11-426A6.5 | FALSE | Downregulated |
| -0,305320735 | 0,616678331 | THAP5 | FALSE | Downregulated |
| 0,45801421 | 0,616678331 | TLR6 | FALSE | Upregulated |
| 0,357128371 | 0,616678331 | TSPAN12 | FALSE | Upregulated |
| 0,109950954 | 0,616678331 | USP34 | FALSE | Upregulated |
| 0,237500817 | 0,616935111 | LINC-PINT | FALSE | Upregulated |
| -0,202207658 | 0,616976664 | STAB2 | FALSE | Downregulated |
| -0,173033939 | 0,617148035 | BRPF3 | FALSE | Downregulated |
| -0,311710942 | 0,617148035 | HIST1H2BC | FALSE | Downregulated |
| 0,412609287 | 0,617459863 | RP11-434D9.1 | FALSE | Upregulated |
| -0,451395992 | 0,617717047 | INHBA | FALSE | Downregulated |
| -0,233820383 | 0,617717047 | SLC35A4 | FALSE | Downregulated |
| -0,161673852 | 0,617774067 | BNIP3 | FALSE | Downregulated |
| -0,350055234 | 0,617774067 | CDK6 | FALSE | Downregulated |
| 0,238190726 | 0,618198865 | NFYC-AS1 | FALSE | Upregulated |
| 0,842575093 | 0,61882631 | LDB3 | FALSE | Upregulated |
| -0,380480854 | 0,61882631 | LOC101928384 | FALSE | Downregulated |
| -0,15984634 | 0,61882631 | MCFD2 | FALSE | Downregulated |
| -0,159500398 | 0,61882631 | RAD23A | FALSE | Downregulated |
| 0,222007217 | 0,61882631 | RDH14 | FALSE | Upregulated |
| -0,160825944 | 0,61882631 | TMEM57 | FALSE | Downregulated |
| 0,430492464 | 0,618851954 | ZNF211 | FALSE | Upregulated |
| -0,43325709 | 0,61898075 | BAALC | FALSE | Downregulated |
| 0,402908127 | 0,619177255 | LOC101928816 | FALSE | Upregulated |
| -0,195851592 | 0,619270154 | FH | FALSE | Downregulated |
| -0,198087123 | 0,619270154 | FMO3 | FALSE | Downregulated |
| -0,173028408 | 0,619270154 | HIP1 | FALSE | Downregulated |
| 0,443666052 | 0,619270154 | LOC102723886 | FALSE | Upregulated |
| -0,326249659 | 0,619270154 | PSMB1 | FALSE | Downregulated |
| -0,199263333 | 0,619270154 | SUCLG2 | FALSE | Downregulated |
| 0,232621708 | 0,619270154 | TMSB10 | FALSE | Upregulated |
| 0,267120984 | 0,619270154 | TSEN54 | FALSE | Upregulated |
| -0,197964066 | 0,619967885 | HDGFRP2 | FALSE | Downregulated |
| -0,3756675 | 0,620068537 | DIEXF | FALSE | Downregulated |
| -0,225831056 | 0,620068537 | IGF1R | FALSE | Downregulated |
| 0,381645863 | 0,620068537 | INTS7 | FALSE | Upregulated |
| 0,173884917 | 0,620284091 | PRDM2 | FALSE | Upregulated |
| -0,314651505 | 0,620284091 | TSSC4 | FALSE | Downregulated |
| 0,317857299 | 0,620284091 | TUBA1A | FALSE | Upregulated |
| 0,343503226 | 0,620284091 | UQCC2 | FALSE | Upregulated |
| 0,486485481 | 0,620325476 | LFNG | FALSE | Upregulated |
| 0,279384329 | 0,620325476 | LNX1 | FALSE | Upregulated |
| 0,192438346 | 0,620325476 | RNF44 | FALSE | Upregulated |
| 0,450863161 | 0,620661724 | PCSK5 | FALSE | Upregulated |
| -0,45744741 | 0,620808997 | MIEF2 | FALSE | Downregulated |
| -0,4006134 | 0,620984615 | ADCK5 | FALSE | Downregulated |

| | | | | |
|---|---|---|---|---|
| 0,400928417 | 0,620984615 | TBC1D22B | FALSE | Upregulated |
| -0,261156753 | 0,62114041 | GSTM4 | FALSE | Downregulated |
| 0,13391684 | 0,62114041 | TRIO | FALSE | Upregulated |
| -0,197594246 | 0,621264992 | AHR | FALSE | Downregulated |
| -0,200372538 | 0,621264992 | AKR1C4 | FALSE | Downregulated |
| -0,415515488 | 0,621264992 | CCDC132 | FALSE | Downregulated |
| 0,23183279 | 0,621264992 | CLASP2 | FALSE | Upregulated |
| 0,376405895 | 0,621264992 | CSF2RB | FALSE | Upregulated |
| 0,407719789 | 0,621264992 | GLI4 | FALSE | Upregulated |
| -0,362550387 | 0,621264992 | HSPA1B | FALSE | Downregulated |
| -0,185454837 | 0,621264992 | ITGA9 | FALSE | Downregulated |
| 0,369967503 | 0,621264992 | KCNJ4 | FALSE | Upregulated |
| -0,231234748 | 0,621264992 | LINC01018 | FALSE | Downregulated |
| -0,255121106 | 0,621264992 | MAT2B | FALSE | Downregulated |
| 0,510356903 | 0,621264992 | MIR100HG | FALSE | Upregulated |
| -0,165935261 | 0,621588019 | CRYL1 | FALSE | Downregulated |
| 0,192883731 | 0,621588019 | TERF2IP | FALSE | Upregulated |
| 0,478667442 | 0,621588019 | TMEM185A | FALSE | Upregulated |
| 0,330970336 | 0,621831507 | AGAP5 | FALSE | Upregulated |
| 0,556135969 | 0,621831507 | TLDC1 | FALSE | Upregulated |
| 0,196816315 | 0,622344402 | CLDN22 | FALSE | Upregulated |
| -0,182232824 | 0,622638611 | NR1I3 | FALSE | Downregulated |
| -0,223979953 | 0,622868718 | ALDH16A1 | FALSE | Downregulated |
| -0,139514331 | 0,623269401 | ZBED1 | FALSE | Downregulated |
| 0,467541554 | 0,623286394 | CEP85L | FALSE | Upregulated |
| -0,244038294 | 0,623286394 | TMEM109 | FALSE | Downregulated |
| -0,309532237 | 0,623590324 | EPC2 | FALSE | Downregulated |
| 0,285054882 | 0,623590324 | ZNF720 | FALSE | Upregulated |
| 0,215175518 | 0,623632372 | KCNIP4 | FALSE | Upregulated |
| 0,331359667 | 0,623639479 | IFI35 | FALSE | Upregulated |
| -0,211537322 | 0,624018993 | STARD8 | FALSE | Downregulated |
| -0,171933186 | 0,624026174 | IVNS1ABP | FALSE | Downregulated |
| -0,197518757 | 0,624311023 | FAM32A | FALSE | Downregulated |
| 0,650190038 | 0,624311023 | RP11-44F21.3 | FALSE | Upregulated |
| 0,245193404 | 0,624311023 | SCRN3 | FALSE | Upregulated |
| -0,290149986 | 0,624325448 | C15orf39 | FALSE | Downregulated |
| -0,112635345 | 0,624325448 | RPL15 | FALSE | Downregulated |
| -0,178960045 | 0,624612496 | GNPTG | FALSE | Downregulated |
| 0,422686331 | 0,624962492 | CHTF18 | FALSE | Upregulated |
| -0,232748862 | 0,624962492 | CTDSPL | FALSE | Downregulated |
| 0,271246972 | 0,624962492 | LOC101929005 | FALSE | Upregulated |
| 0,423585731 | 0,624962492 | RPA3 | FALSE | Upregulated |
| 0,213394071 | 0,624962492 | TCTA | FALSE | Upregulated |
| -0,193765735 | 0,624962492 | TOM1L2 | FALSE | Downregulated |
| -0,146100046 | 0,624962492 | ZBTB20 | FALSE | Downregulated |
| 0,307102783 | 0,624962492 | ZBTB47 | FALSE | Upregulated |
| 0,205012496 | 0,62510558 | OXNAD1 | FALSE | Upregulated |
| 0,306546224 | 0,625284687 | ADRBK2 | FALSE | Upregulated |
| -0,326117292 | 0,625284687 | C20orf194 | FALSE | Downregulated |
| 0,431673187 | 0,625284687 | CCDC78 | FALSE | Upregulated |
| -0,342687332 | 0,625284687 | EBAG9 | FALSE | Downregulated |

| | | | | |
|---|---|---|---|---|
| -0,230185927 | 0,625284687 | ERI2 | FALSE | Downregulated |
| -0,193483349 | 0,625284687 | FGG | FALSE | Downregulated |
| -0,21596319 | 0,625284687 | GATA4 | FALSE | Downregulated |
| -0,185269838 | 0,625284687 | GLYCTK | FALSE | Downregulated |
| -0,171904536 | 0,625284687 | GRB14 | FALSE | Downregulated |
| 0,200366227 | 0,625284687 | IGF2-AS | FALSE | Upregulated |
| 0,220602072 | 0,625284687 | KDELR3 | FALSE | Upregulated |
| 0,37189624 | 0,625284687 | LOC101928741 | FALSE | Upregulated |
| -0,130466795 | 0,625284687 | MYO1B | FALSE | Downregulated |
| -0,128644843 | 0,625284687 | PRPF8 | FALSE | Downregulated |
| -0,265343275 | 0,625284687 | RASGEF1B | FALSE | Downregulated |
| 0,322364066 | 0,625284687 | SLC1A5 | FALSE | Upregulated |
| -0,251907393 | 0,625284687 | SLC27A3 | FALSE | Downregulated |
| 0,236309711 | 0,625284687 | SRSF12 | FALSE | Upregulated |
| 0,375101029 | 0,625284687 | UCP2 | FALSE | Upregulated |
| 0,364032972 | 0,62533045 | FLJ20464 | FALSE | Upregulated |
| 0,449999377 | 0,62533045 | MED4-AS1 | FALSE | Upregulated |
| 0,573434329 | 0,62533045 | NCF1C | FALSE | Upregulated |
| 0,287754643 | 0,625357937 | GOLT1A | FALSE | Upregulated |
| 0,194972123 | 0,625357937 | MKNK1-AS1 | FALSE | Upregulated |
| -0,285236409 | 0,625357937 | PDSS2 | FALSE | Downregulated |
| -0,153527391 | 0,626652755 | MPC2 | FALSE | Downregulated |
| 0,462331612 | 0,626664894 | SPATA18 | FALSE | Upregulated |
| -0,384482239 | 0,626664894 | ZNF18 | FALSE | Downregulated |
| 0,491628204 | 0,627037114 | CCNI2 | FALSE | Upregulated |
| 0,362628308 | 0,627037114 | CDKN2AIP | FALSE | Upregulated |
| -0,495829241 | 0,628170711 | ITGA3 | FALSE | Downregulated |
| 0,377780775 | 0,628341037 | ARID3A | FALSE | Upregulated |
| -0,275512809 | 0,628341037 | ATP5I | FALSE | Downregulated |
| 0,25808903 | 0,628341037 | ATP6V1C1 | FALSE | Upregulated |
| -0,333916328 | 0,628341037 | C11orf68 | FALSE | Downregulated |
| 0,220581328 | 0,628341037 | C2orf68 | FALSE | Upregulated |
| 0,512194917 | 0,628341037 | CARD9 | FALSE | Upregulated |
| -0,269105651 | 0,628341037 | DARS | FALSE | Downregulated |
| -0,358189238 | 0,628341037 | ENOPH1 | FALSE | Downregulated |
| 0,38090937 | 0,628341037 | FAM78A | FALSE | Upregulated |
| -0,097310871 | 0,628341037 | FAM8A1 | FALSE | Downregulated |
| 0,220471682 | 0,628341037 | GAB2 | FALSE | Upregulated |
| -0,294441687 | 0,628341037 | GHDC | FALSE | Downregulated |
| -0,58907564 | 0,628341037 | INO80C | FALSE | Downregulated |
| 0,257210383 | 0,628341037 | KLHL12 | FALSE | Upregulated |
| -0,169444803 | 0,628341037 | LRP6 | FALSE | Downregulated |
| 0,367746839 | 0,628341037 | MIR181A2HG | FALSE | Upregulated |
| 0,290523561 | 0,628341037 | PTPLAD2 | FALSE | Upregulated |
| 0,189451261 | 0,628341037 | SEMA3F | FALSE | Upregulated |
| -0,117831338 | 0,628341037 | SLC25A1 | FALSE | Downregulated |
| 0,162477719 | 0,628341037 | SMG7 | FALSE | Upregulated |
| -0,3158156 | 0,628341037 | SNORD3A | FALSE | Downregulated |
| -0,172251791 | 0,628341037 | UBAP2L | FALSE | Downregulated |
| 0,570126299 | 0,628341037 | USP32P2 | FALSE | Upregulated |
| -0,208890458 | 0,628341037 | VEGFB | FALSE | Downregulated |

| | | | | |
|---|---|---|---|---|
| -0,341825678 | 0,628341037 | ZNF254 | FALSE | Downregulated |
| 0,301670548 | 0,628399373 | TOPORS-AS1 | FALSE | Upregulated |
| 0,425211525 | 0,628447235 | NUDT7 | FALSE | Upregulated |
| -0,357301904 | 0,628447235 | PGM5 | FALSE | Downregulated |
| -0,120096959 | 0,628447235 | RPS11 | FALSE | Downregulated |
| -0,332271826 | 0,628447235 | UNC5B | FALSE | Downregulated |
| -0,293373198 | 0,628448396 | CYP39A1 | FALSE | Downregulated |
| 0,338044251 | 0,6284521 | LYRM5 | FALSE | Upregulated |
| 0,219011111 | 0,62867057 | SIRT3 | FALSE | Upregulated |
| 0,32620365 | 0,628955789 | C2orf69 | FALSE | Upregulated |
| 0,425016835 | 0,628955789 | DLL1 | FALSE | Upregulated |
| 0,199467206 | 0,628955789 | DSCR3 | FALSE | Upregulated |
| -0,242344678 | 0,628955789 | RNF38 | FALSE | Downregulated |
| -0,330476406 | 0,628955789 | ZNF786 | FALSE | Downregulated |
| 0,189228289 | 0,629170048 | TRIM62 | FALSE | Upregulated |
| 0,328047537 | 0,629345815 | PWWP2A | FALSE | Upregulated |
| 1,165557371 | 0,629599925 | CCL15-CCL14 | FALSE | Upregulated |
| -0,1922345 | 0,629599925 | HDGF | FALSE | Downregulated |
| 0,245781713 | 0,629599925 | LOC101929040 | FALSE | Upregulated |
| -0,42862572 | 0,629599925 | MTUS2 | FALSE | Downregulated |
| -0,232377369 | 0,629599925 | PON1 | FALSE | Downregulated |
| 0,214815132 | 0,629599925 | SHANK2-AS3 | FALSE | Upregulated |
| 0,171299291 | 0,629599925 | VPS39 | FALSE | Upregulated |
| 0,545211262 | 0,629616263 | FLJ12825 | FALSE | Upregulated |
| -0,2576638 | 0,629773293 | MAPKAPK5 | FALSE | Downregulated |
| 0,540367331 | 0,629773293 | TPM2 | FALSE | Upregulated |
| 0,290017452 | 0,630973722 | UVRAG | FALSE | Upregulated |
| 0,444784101 | 0,631013375 | LY9 | FALSE | Upregulated |
| 0,317527023 | 0,631029958 | NKAIN3 | FALSE | Upregulated |
| -0,188196681 | 0,631117049 | APBB2 | FALSE | Downregulated |
| 0,280868991 | 0,631117049 | DDIT3 | FALSE | Upregulated |
| -0,154549926 | 0,631117049 | HECTD3 | FALSE | Downregulated |
| 0,181679594 | 0,631117049 | LOC100303749 | FALSE | Upregulated |
| -0,16967703 | 0,631117049 | LRPPRC | FALSE | Downregulated |
| -0,144676637 | 0,631117049 | SHANK3 | FALSE | Downregulated |
| 0,1535549 | 0,631117049 | TP53I13 | FALSE | Upregulated |
| 0,459506512 | 0,631117049 | TSC22D1-AS1 | FALSE | Upregulated |
| -0,324780856 | 0,631117049 | USP12 | FALSE | Downregulated |
| 0,29185963 | 0,631124381 | ZNF252P | FALSE | Upregulated |
| -0,384584769 | 0,63124462 | MTRF1 | FALSE | Downregulated |
| 0,390870053 | 0,63124462 | NCBP2-AS2 | FALSE | Upregulated |
| -0,221470145 | 0,63124462 | PERP | FALSE | Downregulated |
| 0,185111133 | 0,63124462 | PPP1R9B | FALSE | Upregulated |
| 0,406583821 | 0,63124462 | RELB | FALSE | Upregulated |
| -0,25089465 | 0,631252383 | ANKRD10 | FALSE | Downregulated |
| -0,204064855 | 0,631252383 | BTD | FALSE | Downregulated |
| 0,184697861 | 0,631252383 | CEP68 | FALSE | Upregulated |
| -0,464073021 | 0,631252383 | COL8A1 | FALSE | Downregulated |
| -0,250371965 | 0,631252383 | EDEM2 | FALSE | Downregulated |
| -0,216406173 | 0,631252383 | EIF2B5 | FALSE | Downregulated |
| -0,457959154 | 0,631252383 | ESPNL | FALSE | Downregulated |

| | | | | |
|---|---|---|---|---|
| 0,227383948 | 0,631252383 | LDLRAD4-AS1 | FALSE | Upregulated |
| 0,437742918 | 0,631252383 | LOC101928699 | FALSE | Upregulated |
| -0,19970718 | 0,631252383 | MOCS2 | FALSE | Downregulated |
| -0,166544162 | 0,631252383 | NDFIP1 | FALSE | Downregulated |
| 0,116923083 | 0,631252383 | RBM15B | FALSE | Upregulated |
| -0,223766244 | 0,631252383 | SERPINA7 | FALSE | Downregulated |
| -0,238002323 | 0,631252383 | SLC44A1 | FALSE | Downregulated |
| 0,513210263 | 0,631252383 | TFEC | FALSE | Upregulated |
| -0,167842161 | 0,631252383 | TNPO2 | FALSE | Downregulated |
| -0,325344758 | 0,631252383 | WNT3 | FALSE | Downregulated |
| 0,36014945 | 0,631818494 | CAV2 | FALSE | Upregulated |
| -0,265082154 | 0,632128542 | TMEM37 | FALSE | Downregulated |
| -0,139806067 | 0,63238884 | DUS1L | FALSE | Downregulated |
| -0,264118503 | 0,63238884 | MFSD3 | FALSE | Downregulated |
| -0,284242197 | 0,63238884 | MRPS18A | FALSE | Downregulated |
| 0,589543156 | 0,632405731 | ERCC5 | FALSE | Upregulated |
| -0,334557339 | 0,632405731 | GINM1 | FALSE | Downregulated |
| -0,40647444 | 0,632405731 | PIGY | FALSE | Downregulated |
| 0,197420678 | 0,632405731 | RNF13 | FALSE | Upregulated |
| 0,233244236 | 0,632405731 | ZNF571-AS1 | FALSE | Upregulated |
| -0,203127014 | 0,632613676 | CFHR2 | FALSE | Downregulated |
| -0,20079601 | 0,632681137 | ELL2 | FALSE | Downregulated |
| 0,187287935 | 0,632681137 | GALNT10 | FALSE | Upregulated |
| -0,18105914 | 0,632681137 | PXDN | FALSE | Downregulated |
| -0,23602104 | 0,632681137 | RNF125 | FALSE | Downregulated |
| -0,164722107 | 0,632681137 | TSHZ1 | FALSE | Downregulated |
| 0,290343779 | 0,633562785 | ALOX12P2 | FALSE | Upregulated |
| 0,220471596 | 0,633562785 | CASP8 | FALSE | Upregulated |
| 0,294611194 | 0,633562785 | DBNDD1 | FALSE | Upregulated |
| -0,309202421 | 0,633562785 | DNAJC25 | FALSE | Downregulated |
| 0,209555472 | 0,633562785 | DOCK4-AS1 | FALSE | Upregulated |
| 0,182663911 | 0,633562785 | FECH | FALSE | Upregulated |
| 0,139900491 | 0,633562785 | GSE1 | FALSE | Upregulated |
| -0,237831417 | 0,633562785 | HIC2 | FALSE | Downregulated |
| 0,739490593 | 0,633562785 | HIST1H1B | FALSE | Upregulated |
| 0,314510749 | 0,633562785 | IL3RA | FALSE | Upregulated |
| 0,299072439 | 0,633562785 | INIP | FALSE | Upregulated |
| 0,192127963 | 0,633562785 | LOC101928874 | FALSE | Upregulated |
| -0,270111579 | 0,633562785 | MRPL28 | FALSE | Downregulated |
| -0,174966402 | 0,633562785 | PTPN18 | FALSE | Downregulated |
| 0,404178559 | 0,633562785 | RNF39 | FALSE | Upregulated |
| -0,239608738 | 0,633562785 | RNU4-2 | FALSE | Downregulated |
| 0,23251032 | 0,633562785 | RP1-232P20.1 | FALSE | Upregulated |
| 0,52899028 | 0,633562785 | RP6-191P20.4 | FALSE | Upregulated |
| -0,17521415 | 0,633562785 | SART1 | FALSE | Downregulated |
| 0,118622194 | 0,633562785 | SEPT9 | FALSE | Upregulated |
| 0,3378452 | 0,633562785 | SERPINE3 | FALSE | Upregulated |
| -0,196786958 | 0,633562785 | SHROOM1 | FALSE | Downregulated |
| -0,377213941 | 0,633562785 | SLC17A3 | FALSE | Downregulated |
| 0,183107286 | 0,633562785 | SMA5 | FALSE | Upregulated |
| 0,20140632 | 0,633562785 | SMARCA5-AS1 | FALSE | Upregulated |

| | | | | |
|---|---|---|---|---|
| 0,145812249 | 0,633562785 | STX17 | FALSE | Upregulated |
| 0,204000715 | 0,633562785 | TAB1 | FALSE | Upregulated |
| -0,160581303 | 0,633562785 | TGFBR2 | FALSE | Downregulated |
| 0,136836332 | 0,633562785 | TMED5 | FALSE | Upregulated |
| -0,301749638 | 0,633562785 | VPS16 | FALSE | Downregulated |
| -0,228656869 | 0,633562785 | ZBED3 | FALSE | Downregulated |
| -0,25833574 | 0,633869575 | LOC100129917 | FALSE | Downregulated |
| -0,363005566 | 0,633869575 | TMEM208 | FALSE | Downregulated |
| 0,401292084 | 0,633869575 | UNC119 | FALSE | Upregulated |
| 0,234969202 | 0,633957627 | LRRN2 | FALSE | Upregulated |
| -0,295089184 | 0,634070478 | MUL1 | FALSE | Downregulated |
| -0,468622795 | 0,63430336 | ARHGAP19 | FALSE | Downregulated |
| -0,154821544 | 0,634432053 | ADI1 | FALSE | Downregulated |
| 0,356866275 | 0,634432053 | C12orf66 | FALSE | Upregulated |
| 0,557419097 | 0,634432053 | C15orf41 | FALSE | Upregulated |
| -0,393304844 | 0,634432053 | FAM43A | FALSE | Downregulated |
| -0,127597099 | 0,634432053 | IRF2BP2 | FALSE | Downregulated |
| -0,401569755 | 0,634432053 | MVB12A | FALSE | Downregulated |
| -0,185343282 | 0,634432053 | MYO1D | FALSE | Downregulated |
| 0,185350785 | 0,634432053 | PHACTR1 | FALSE | Upregulated |
| -0,18722793 | 0,634432053 | PRDX5 | FALSE | Downregulated |
| -0,203295233 | 0,634432053 | SLC39A9 | FALSE | Downregulated |
| 0,182636048 | 0,634432053 | STAT5A | FALSE | Upregulated |
| -0,192073974 | 0,634432053 | STRADB | FALSE | Downregulated |
| 0,226404744 | 0,634485331 | AC137932.6 | FALSE | Upregulated |
| 0,210806663 | 0,634485331 | CSTF2T | FALSE | Upregulated |
| -0,212125946 | 0,634485331 | DECR1 | FALSE | Downregulated |
| 0,559199183 | 0,634485331 | HAVCR2 | FALSE | Upregulated |
| 0,327331759 | 0,634485331 | MVD | FALSE | Upregulated |
| -0,170481124 | 0,634485331 | PNPLA2 | FALSE | Downregulated |
| 0,351468448 | 0,634485331 | TTLL3 | FALSE | Upregulated |
| -0,121232708 | 0,634866115 | C19orf66 | FALSE | Downregulated |
| 0,364568003 | 0,634866115 | CCNDBP1 | FALSE | Upregulated |
| -0,2143008 | 0,634866115 | LSG1 | FALSE | Downregulated |
| 1,097529456 | 0,634866115 | LY75-CD302 | FALSE | Upregulated |
| 0,220070978 | 0,634866115 | PIGW | FALSE | Upregulated |
| 0,273208601 | 0,634866115 | PRPS1 | FALSE | Upregulated |
| 0,201346392 | 0,634866115 | RP11-142C4.6 | FALSE | Upregulated |
| 0,481090326 | 0,634866115 | RYR2 | FALSE | Upregulated |
| -0,181867473 | 0,634866115 | TRAPPC8 | FALSE | Downregulated |
| 0,359099819 | 0,634866115 | ZNF225 | FALSE | Upregulated |
| 0,311421702 | 0,635397873 | MSMO1 | FALSE | Upregulated |
| 0,291550404 | 0,635419324 | FLJ13224 | FALSE | Upregulated |
| 0,322828415 | 0,635419324 | LONRF1 | FALSE | Upregulated |
| -0,186198808 | 0,635419324 | PSMD2 | FALSE | Downregulated |
| -0,238323818 | 0,635419324 | RAB11B-AS1 | FALSE | Downregulated |
| -0,222169949 | 0,635419324 | UBA7 | FALSE | Downregulated |
| 0,616989814 | 0,635491866 | ISG20 | FALSE | Upregulated |
| 0,295962119 | 0,635588585 | ATP6V0A2 | FALSE | Upregulated |
| -0,421124961 | 0,635815596 | AEBP2 | FALSE | Downregulated |
| -0,190020285 | 0,635859933 | AOX1 | FALSE | Downregulated |

| | | | | |
|---|---|---|---|---|
| 0,460911713 | 0,635859933 | CDYL2 | FALSE | Upregulated |
| 0,396396317 | 0,635859933 | GNG2 | FALSE | Upregulated |
| -0,122545432 | 0,635859933 | KIF1B | FALSE | Downregulated |
| 0,228360257 | 0,635859933 | MTMR1 | FALSE | Upregulated |
| 0,151082084 | 0,635859933 | PTPRJ | FALSE | Upregulated |
| -0,167492102 | 0,635859933 | SEC23A | FALSE | Downregulated |
| -0,217528477 | 0,635859933 | SNHG16 | FALSE | Downregulated |
| -0,13632702 | 0,635859933 | UBR5 | FALSE | Downregulated |
| 0,327845006 | 0,635859933 | XPR1 | FALSE | Upregulated |
| 0,466646061 | 0,635881686 | ACOT11 | FALSE | Upregulated |
| 0,34978614 | 0,635881686 | REN | FALSE | Upregulated |
| -0,253064505 | 0,636013431 | FBXO25 | FALSE | Downregulated |
| -0,377730424 | 0,636454093 | L3HYPDH | FALSE | Downregulated |
| 0,313847489 | 0,636548 | SCARNA13 | FALSE | Upregulated |
| -0,153391772 | 0,636896517 | C8A | FALSE | Downregulated |
| 0,548467818 | 0,637791232 | AL592528.1 | FALSE | Upregulated |
| -0,189534458 | 0,637791232 | MAN1C1 | FALSE | Downregulated |
| 0,431693778 | 0,637791232 | MINOS1 | FALSE | Upregulated |
| -0,363133399 | 0,637791232 | OSBPL10 | FALSE | Downregulated |
| -0,139723925 | 0,637869621 | HNRNPF | FALSE | Downregulated |
| -0,184871629 | 0,637869621 | MTR | FALSE | Downregulated |
| -0,403719001 | 0,637909384 | CD1D | FALSE | Downregulated |
| -0,170725874 | 0,637909384 | CRK | FALSE | Downregulated |
| -0,195211551 | 0,637909384 | ECI2 | FALSE | Downregulated |
| 0,305019898 | 0,637909384 | EPM2A | FALSE | Upregulated |
| -0,288916994 | 0,637909384 | FOLH1 | FALSE | Downregulated |
| -0,233249989 | 0,637909384 | GSTZ1 | FALSE | Downregulated |
| -0,443474466 | 0,637909384 | LINC00936 | FALSE | Downregulated |
| 0,336984734 | 0,637909384 | LOC101927578 | FALSE | Upregulated |
| -0,257263587 | 0,637909384 | PEF1 | FALSE | Downregulated |
| -0,229136953 | 0,637909384 | POLR1D | FALSE | Downregulated |
| -0,332127582 | 0,637909384 | S100A10 | FALSE | Downregulated |
| -0,215382631 | 0,637909384 | SESN2 | FALSE | Downregulated |
| -0,369783469 | 0,637909384 | ZNF251 | FALSE | Downregulated |
| -0,277409851 | 0,637976725 | AFAP1L1 | FALSE | Downregulated |
| -0,442076403 | 0,637976725 | FBXL7 | FALSE | Downregulated |
| -0,258560293 | 0,637976725 | HHEX | FALSE | Downregulated |
| 0,40331976 | 0,637976725 | QPCTL | FALSE | Upregulated |
| 0,338387125 | 0,638086923 | RASGRP4 | FALSE | Upregulated |
| 0,204196953 | 0,638171884 | TOR1A | FALSE | Upregulated |
| -0,259592964 | 0,638581804 | WFS1 | FALSE | Downregulated |
| -0,203671802 | 0,639010027 | ATP7B | FALSE | Downregulated |
| -0,274995431 | 0,639010027 | HNRNPUL2-BSCL2 | FALSE | Downregulated |
| -0,153056905 | 0,639010027 | RPS27A | FALSE | Downregulated |
| 0,14011266 | 0,639010027 | VPRBP | FALSE | Upregulated |
| 0,179688891 | 0,63906804 | FAM219B | FALSE | Upregulated |
| 0,120298695 | 0,639312282 | RER1 | FALSE | Upregulated |
| 0,396772483 | 0,639312926 | LINC00921 | FALSE | Upregulated |
| 0,252449185 | 0,639542527 | SPATA6L | FALSE | Upregulated |
| 0,350381439 | 0,639735321 | EPB41L4A-AS2 | FALSE | Upregulated |
| 0,140331123 | 0,639735321 | FBXO11 | FALSE | Upregulated |

| | | | | |
|---|---|---|---|---|
| 0,187057109 | 0,639735321 | RBM12B-AS1 | FALSE | Upregulated |
| -0,236893499 | 0,639735321 | SEMA6C | FALSE | Downregulated |
| 0,241017288 | 0,63997622 | ACTR8 | FALSE | Upregulated |
| -0,21308836 | 0,63997622 | ASS1 | FALSE | Downregulated |
| -0,345116209 | 0,63997622 | PDGFB | FALSE | Downregulated |
| 0,137134098 | 0,63997622 | PPP2R1B | FALSE | Upregulated |
| -0,447439721 | 0,63997622 | RASL11B | FALSE | Downregulated |
| 0,246616371 | 0,63997622 | SPTLC2 | FALSE | Upregulated |
| -0,205778838 | 0,63997622 | TIAM1 | FALSE | Downregulated |
| -0,365093339 | 0,63997622 | TPSG1 | FALSE | Downregulated |
| -0,298984037 | 0,63997622 | TRPM8 | FALSE | Downregulated |
| -0,15503667 | 0,640494983 | ERGIC1 | FALSE | Downregulated |
| 0,1633453 | 0,640494983 | NCOA3 | FALSE | Upregulated |
| -0,154411752 | 0,640494983 | POR | FALSE | Downregulated |
| -0,213273452 | 0,641208792 | GLUD1P3 | FALSE | Downregulated |
| 0,214850972 | 0,641208792 | PPP1R3E | FALSE | Upregulated |
| 0,461073784 | 0,641208792 | SLC26A8 | FALSE | Upregulated |
| 0,143972028 | 0,641504579 | SLC3A1 | FALSE | Upregulated |
| 0,185498974 | 0,641799996 | BCL9 | FALSE | Upregulated |
| 0,211126167 | 0,641799996 | SDCBP | FALSE | Upregulated |
| 0,22973611 | 0,641799996 | TIMM8B | FALSE | Upregulated |
| 0,162209234 | 0,642195515 | HSD17B1 | FALSE | Upregulated |
| 0,277975941 | 0,642454222 | TPT1-AS1 | FALSE | Upregulated |
| -0,374433944 | 0,642588742 | SLC12A2 | FALSE | Downregulated |
| 0,332320088 | 0,642602464 | PFDN2 | FALSE | Upregulated |
| 0,163840627 | 0,64301805 | C3orf17 | FALSE | Upregulated |
| 0,23589397 | 0,643198996 | ZDHHC23 | FALSE | Upregulated |
| 0,244466506 | 0,643214936 | ASB9P1 | FALSE | Upregulated |
| -0,174596217 | 0,643214936 | DGCR6L | FALSE | Downregulated |
| -0,119372048 | 0,643214936 | EPAS1 | FALSE | Downregulated |
| -0,270092951 | 0,643214936 | HSPH1 | FALSE | Downregulated |
| 0,150416005 | 0,643214936 | NAPEPLD | FALSE | Upregulated |
| -0,237455223 | 0,643214936 | PSMB6 | FALSE | Downregulated |
| -0,210947612 | 0,643214936 | STX4 | FALSE | Downregulated |
| 0,183309373 | 0,643214936 | TNK2 | FALSE | Upregulated |
| -0,200714932 | 0,643214936 | UBAP1 | FALSE | Downregulated |
| 0,159440027 | 0,643214936 | ZNF337 | FALSE | Upregulated |
| 0,114569785 | 0,643438194 | ZNF106 | FALSE | Upregulated |
| 0,270303621 | 0,643584003 | MFAP4 | FALSE | Upregulated |
| -0,354235198 | 0,644091021 | LOC283788 | FALSE | Downregulated |
| 0,449832494 | 0,644091021 | MOCS3 | FALSE | Upregulated |
| -0,128836687 | 0,644165007 | BCL9L | FALSE | Downregulated |
| -0,183873604 | 0,644165007 | KDM4C | FALSE | Downregulated |
| 0,242645572 | 0,644165007 | LILRB3 | FALSE | Upregulated |
| -0,16738871 | 0,644165007 | NBPF14 | FALSE | Downregulated |
| -0,177988799 | 0,644165007 | PHF12 | FALSE | Downregulated |
| 0,1325291 | 0,644165007 | QKI | FALSE | Upregulated |
| 0,112690494 | 0,644165007 | RBPMS | FALSE | Upregulated |
| 0,357639336 | 0,644165007 | UBE2S | FALSE | Upregulated |
| -0,312848169 | 0,644165007 | VPS53 | FALSE | Downregulated |
| 0,204013431 | 0,644326647 | ALG1 | FALSE | Upregulated |

| | | | | |
|---|---|---|---|---|
| 0,114391255 | 0,644326647 | CDK12 | FALSE | Upregulated |
| -0,231744144 | 0,644326647 | HNMT | FALSE | Downregulated |
| 0,234679654 | 0,644326647 | OSGIN2 | FALSE | Upregulated |
| 0,162481198 | 0,644326647 | PODXL | FALSE | Upregulated |
| -0,158600133 | 0,644326647 | RIF1 | FALSE | Downregulated |
| -0,272796307 | 0,644326647 | SEP15 | FALSE | Downregulated |
| -0,148069364 | 0,6444821 | SEL1L | FALSE | Downregulated |
| -0,45769221 | 0,645076055 | UBL7 | FALSE | Downregulated |
| 0,216753356 | 0,645147468 | AKR1C3 | FALSE | Upregulated |
| -0,137051784 | 0,645147468 | STARD7 | FALSE | Downregulated |
| 0,416939358 | 0,645193654 | FBXO2 | FALSE | Upregulated |
| -0,252572906 | 0,645193654 | FCN3 | FALSE | Downregulated |
| -0,473188386 | 0,645193654 | SNORA7A | FALSE | Downregulated |
| 0,261510311 | 0,645772568 | LOC101929204 | FALSE | Upregulated |
| -0,16025531 | 0,645772568 | MRPL43 | FALSE | Downregulated |
| -0,252954818 | 0,645772568 | ZNF503 | FALSE | Downregulated |
| 0,404558047 | 0,645772568 | ZNF891 | FALSE | Upregulated |
| 0,32538323 | 0,645862214 | ARL2BP | FALSE | Upregulated |
| 0,346922025 | 0,645862214 | CCHCR1 | FALSE | Upregulated |
| 0,3267311 | 0,645862214 | IFIT1B | FALSE | Upregulated |
| -0,182954707 | 0,645862214 | LLGL2 | FALSE | Downregulated |
| 0,341348746 | 0,645862214 | NAPB | FALSE | Upregulated |
| -0,173769994 | 0,645862214 | PSKH1 | FALSE | Downregulated |
| 0,152452326 | 0,645862214 | SH3BP5-AS1 | FALSE | Upregulated |
| -0,183524438 | 0,645862214 | TAX1BP1 | FALSE | Downregulated |
| 0,174787295 | 0,645862214 | ZBTB11-AS1 | FALSE | Upregulated |
| 0,459271354 | 0,646066125 | LOC101059906 | FALSE | Upregulated |
| -0,149511444 | 0,646066125 | OAZ1 | FALSE | Downregulated |
| 0,187544386 | 0,646470573 | FAM178A | FALSE | Upregulated |
| -0,216874246 | 0,646470573 | RGS14 | FALSE | Downregulated |
| -0,258217284 | 0,646470573 | SCO1 | FALSE | Downregulated |
| 0,193786479 | 0,646470573 | TMEM184C | FALSE | Upregulated |
| 0,262366914 | 0,646470573 | ZNF660 | FALSE | Upregulated |
| 0,283953038 | 0,646568226 | BTF3L4 | FALSE | Upregulated |
| -0,215338659 | 0,646568226 | FAF2 | FALSE | Downregulated |
| -0,140669544 | 0,646568226 | GPBP1L1 | FALSE | Downregulated |
| 0,217527224 | 0,646568226 | GPR17 | FALSE | Upregulated |
| -0,256333448 | 0,646568226 | LONRF3 | FALSE | Downregulated |
| 0,19028141 | 0,646568226 | PIP5K1C | FALSE | Upregulated |
| -0,254752612 | 0,646568226 | ZNF608 | FALSE | Downregulated |
| 0,49431852 | 0,646845732 | PTGIR | FALSE | Upregulated |
| -0,178890979 | 0,646845732 | SEPSECS | FALSE | Downregulated |
| -0,191136604 | 0,646845732 | USP31 | FALSE | Downregulated |
| -0,267613487 | 0,647232426 | HNF1B | FALSE | Downregulated |
| -0,148981298 | 0,647418884 | EHHADH | FALSE | Downregulated |
| 0,184602969 | 0,647418884 | ERVW-1 | FALSE | Upregulated |
| -0,239921891 | 0,647418884 | MME | FALSE | Downregulated |
| 0,39117894 | 0,647418884 | RP11-890B15.2 | FALSE | Upregulated |
| 0,198305369 | 0,647659012 | CASKIN1 | FALSE | Upregulated |
| 0,175818009 | 0,647684035 | AMIGO3 | FALSE | Upregulated |
| 0,138906232 | 0,647684035 | ANKRD52 | FALSE | Upregulated |

| | | | | |
|---|---|---|---|---|
| 0,280747025 | 0,647684035 | LOC100506388 | FALSE | Upregulated |
| -0,163651149 | 0,647684035 | SKIV2L | FALSE | Downregulated |
| 0,351096253 | 0,647684035 | SOX4 | FALSE | Upregulated |
| -0,230560351 | 0,647959189 | ABCD4 | FALSE | Downregulated |
| 0,395781427 | 0,648128923 | MOSPD2 | FALSE | Upregulated |
| -0,330581368 | 0,648128923 | PELI1 | FALSE | Downregulated |
| 0,213277138 | 0,648235881 | LOC101928690 | FALSE | Upregulated |
| -0,18882185 | 0,648235881 | NHLRC2 | FALSE | Downregulated |
| 0,161381507 | 0,648235881 | OGFR | FALSE | Upregulated |
| -0,172515398 | 0,648235881 | RPN2 | FALSE | Downregulated |
| -0,129893998 | 0,648235881 | SRSF4 | FALSE | Downregulated |
| 0,221830644 | 0,648235881 | TNFRSF6B | FALSE | Upregulated |
| 0,30089013 | 0,648235881 | WWC2-AS2 | FALSE | Upregulated |
| 0,242211407 | 0,64827741 | CCT8 | FALSE | Upregulated |
| 0,42133962 | 0,64827741 | RIOK2 | FALSE | Upregulated |
| 0,173877645 | 0,648285084 | LDB1 | FALSE | Upregulated |
| -0,3841859 | 0,648500714 | PLIN1 | FALSE | Downregulated |
| -0,334434349 | 0,648715892 | AGAP4 | FALSE | Downregulated |
| 0,163472669 | 0,648715892 | CNN3 | FALSE | Upregulated |
| -0,485406754 | 0,648715892 | EFCAB4B | FALSE | Downregulated |
| 0,12563747 | 0,648715892 | EMC1 | FALSE | Upregulated |
| 0,526519919 | 0,648715892 | FMO6P | FALSE | Upregulated |
| -0,140352052 | 0,648715892 | NFIX | FALSE | Downregulated |
| -0,458457812 | 0,648715892 | OAT | FALSE | Downregulated |
| 0,241681682 | 0,648715892 | PAXBP1-AS1 | FALSE | Upregulated |
| -0,113602665 | 0,648715892 | PFKL | FALSE | Downregulated |
| -0,148069221 | 0,648715892 | PRPF31 | FALSE | Downregulated |
| -0,281671765 | 0,648715892 | TFCP2 | FALSE | Downregulated |
| 0,475773481 | 0,648855654 | SBDSP1 | FALSE | Upregulated |
| 0,400837178 | 0,648868218 | LOC101929188 | FALSE | Upregulated |
| 0,295426342 | 0,648868218 | USP50 | FALSE | Upregulated |
| -0,165852505 | 0,648868218 | ZNF512B | FALSE | Downregulated |
| 0,353816676 | 0,649069432 | SLC25A40 | FALSE | Upregulated |
| 0,326948507 | 0,649197973 | FAM122B | FALSE | Upregulated |
| 0,741562917 | 0,649197973 | KLHL41 | FALSE | Upregulated |
| 0,292241118 | 0,649197973 | ZNF622 | FALSE | Upregulated |
| 0,267607404 | 0,649328411 | LOC388849 | FALSE | Upregulated |
| -0,436548827 | 0,649526189 | PYURF | FALSE | Downregulated |
| -0,28984481 | 0,649812343 | MRPL45 | FALSE | Downregulated |
| 0,408102817 | 0,649921357 | RSRC1 | FALSE | Upregulated |
| 0,220288396 | 0,650383935 | KIF2A | FALSE | Upregulated |
| -0,266570411 | 0,650532742 | ARRDC3 | FALSE | Downregulated |
| -0,220138639 | 0,650532742 | RPRD1B | FALSE | Downregulated |
| 0,3590257 | 0,650532742 | TEAD2 | FALSE | Upregulated |
| 0,135522964 | 0,650532742 | WHSC1 | FALSE | Upregulated |
| -0,168051901 | 0,651241441 | SLC4A2 | FALSE | Downregulated |
| -0,218956168 | 0,651287016 | NMD3 | FALSE | Downregulated |
| 0,204272815 | 0,651397348 | KDM4A-AS1 | FALSE | Upregulated |
| -0,294702153 | 0,651413874 | SLC25A32 | FALSE | Downregulated |
| -0,435462885 | 0,651413874 | SNRPA1 | FALSE | Downregulated |
| 0,321234305 | 0,65164375 | SLC44A3 | FALSE | Upregulated |

| | | | | |
|---|---|---|---|---|
| -0,274611295 | 0,651861866 | BANF1 | FALSE | Downregulated |
| 0,360441035 | 0,651861866 | EDRF1-AS1 | FALSE | Upregulated |
| -0,339662369 | 0,651861866 | SCRT1 | FALSE | Downregulated |
| -0,278781444 | 0,651861866 | STK36 | FALSE | Downregulated |
| 0,182826036 | 0,652405518 | ANKRD61 | FALSE | Upregulated |
| 0,391229864 | 0,652624306 | C1orf195 | FALSE | Upregulated |
| -0,137828893 | 0,652624306 | COBL | FALSE | Downregulated |
| 0,359511421 | 0,652913175 | C3orf49 | FALSE | Upregulated |
| 0,32903167 | 0,652913175 | CAMK2B | FALSE | Upregulated |
| 0,204290594 | 0,652913175 | GPN1 | FALSE | Upregulated |
| -0,150431939 | 0,652913175 | HSDL2 | FALSE | Downregulated |
| -0,215386549 | 0,652913175 | NCAPD3 | FALSE | Downregulated |
| 0,23304873 | 0,652913175 | PLCXD1 | FALSE | Upregulated |
| 0,636639989 | 0,652913175 | PTCHD1 | FALSE | Upregulated |
| -0,120652526 | 0,652913175 | PUM2 | FALSE | Downregulated |
| 0,203050816 | 0,652984255 | SRF | FALSE | Upregulated |
| -0,20817212 | 0,653228083 | TBC1D4 | FALSE | Downregulated |
| 0,168252407 | 0,653228083 | TDRP | FALSE | Upregulated |
| 0,190095115 | 0,653228083 | TIGD6 | FALSE | Upregulated |
| -0,252129531 | 0,653793123 | DCUN1D3 | FALSE | Downregulated |
| 0,377921418 | 0,654183565 | GTF3C6 | FALSE | Upregulated |
| 0,358704349 | 0,654183565 | HCK | FALSE | Upregulated |
| 0,154311999 | 0,654183565 | KLB | FALSE | Upregulated |
| 1,910303718 | 0,654183565 | LOC440354 | FALSE | Upregulated |
| -0,164066297 | 0,654183565 | MLXIPL | FALSE | Downregulated |
| -0,233693753 | 0,654183565 | NELFA | FALSE | Downregulated |
| -0,16846544 | 0,654183565 | SNRPD3 | FALSE | Downregulated |
| 0,408901022 | 0,654183565 | TAS1R1 | FALSE | Upregulated |
| 0,108512812 | 0,654248604 | FAM20A | FALSE | Upregulated |
| -0,191717735 | 0,654552693 | C1QTNF1 | FALSE | Downregulated |
| -0,177921228 | 0,654552693 | CABLES1 | FALSE | Downregulated |
| 0,202890864 | 0,654552693 | CD276 | FALSE | Upregulated |
| 0,307633122 | 0,654552693 | LOC100507156 | FALSE | Upregulated |
| 0,159374039 | 0,654552693 | SZRD1 | FALSE | Upregulated |
| -0,152401543 | 0,655173678 | AHCYL1 | FALSE | Downregulated |
| 0,246869732 | 0,655386811 | AMMECR1L | FALSE | Upregulated |
| -0,189641857 | 0,655386811 | CPED1 | FALSE | Downregulated |
| 0,218197251 | 0,655386811 | ITGAV | FALSE | Upregulated |
| -0,224974744 | 0,655618051 | AMDHD1 | FALSE | Downregulated |
| 0,195658743 | 0,655912059 | GP1BA | FALSE | Upregulated |
| 0,424717667 | 0,655912059 | PIK3CG | FALSE | Upregulated |
| 0,278929841 | 0,655912059 | SAV1 | FALSE | Upregulated |
| 0,377595004 | 0,655912059 | SYT1 | FALSE | Upregulated |
| 0,299301439 | 0,65630784 | BAX | FALSE | Upregulated |
| 0,42761748 | 0,65630784 | COX10 | FALSE | Upregulated |
| 0,217069867 | 0,656334051 | ACTL10 | FALSE | Upregulated |
| -0,210623047 | 0,656334051 | HIBCH | FALSE | Downregulated |
| 0,346329499 | 0,656334051 | TBCB | FALSE | Upregulated |
| 0,179711658 | 0,656519107 | RAP2C | FALSE | Upregulated |
| -0,298788054 | 0,656519107 | RRAGD | FALSE | Downregulated |
| 0,263635471 | 0,656519107 | RUSC1-AS1 | FALSE | Upregulated |

| | | | | |
|---|---|---|---|---|
| -0,290358678 | 0,656528113 | CYB561D1 | FALSE | Downregulated |
| -0,394775784 | 0,656528113 | CYP1A2 | FALSE | Downregulated |
| 0,224534158 | 0,656528113 | FAM169A | FALSE | Upregulated |
| 0,245372841 | 0,656528113 | L3MBTL4 | FALSE | Upregulated |
| -0,361882984 | 0,656528113 | MFSD8 | FALSE | Downregulated |
| -0,123471158 | 0,656528113 | NFIC | FALSE | Downregulated |
| -0,156715151 | 0,656528113 | TMEM115 | FALSE | Downregulated |
| -0,281997892 | 0,656528113 | XRCC1 | FALSE | Downregulated |
| -0,145837234 | 0,656528113 | ZBED4 | FALSE | Downregulated |
| -0,278533378 | 0,656604094 | ABCB7 | FALSE | Downregulated |
| 0,194662936 | 0,656604094 | DESI1 | FALSE | Upregulated |
| 0,233441808 | 0,656795099 | PSEN1 | FALSE | Upregulated |
| 0,254398997 | 0,656970697 | SLIT3 | FALSE | Upregulated |
| -0,504550313 | 0,656996103 | GINS4 | FALSE | Downregulated |
| 0,16764018 | 0,656996103 | HIF1AN | FALSE | Upregulated |
| 0,335382194 | 0,656996103 | LOC100287808 | FALSE | Upregulated |
| 0,254798163 | 0,656996103 | PSMA2 | FALSE | Upregulated |
| 0,239084119 | 0,656996103 | RMND5B | FALSE | Upregulated |
| -0,178411186 | 0,657337603 | DHX16 | FALSE | Downregulated |
| 0,278152898 | 0,657337603 | FBXO6 | FALSE | Upregulated |
| -0,197086747 | 0,657337603 | RFWD2 | FALSE | Downregulated |
| -0,30393747 | 0,657337603 | TOPORS | FALSE | Downregulated |
| 0,332958762 | 0,658197786 | NDUFA4L2 | FALSE | Upregulated |
| -0,281665142 | 0,658197786 | NES | FALSE | Downregulated |
| 0,1755058 | 0,65820877 | CYLD | FALSE | Upregulated |
| -0,392333899 | 0,65820877 | NDUFB1 | FALSE | Downregulated |
| 0,186198447 | 0,658472824 | LSMEM2 | FALSE | Upregulated |
| -0,139805422 | 0,658502352 | ARHGEF10L | FALSE | Downregulated |
| 0,216197705 | 0,658502352 | CD93 | FALSE | Upregulated |
| 0,508396269 | 0,659366585 | AC022173.2 | FALSE | Upregulated |
| -0,306543655 | 0,659366585 | NRBF2 | FALSE | Downregulated |
| 0,146929254 | 0,659640265 | HYI | FALSE | Upregulated |
| 0,221323876 | 0,659640265 | RRAS2 | FALSE | Upregulated |
| 0,397685116 | 0,659651287 | BLOC1S3 | FALSE | Upregulated |
| 0,234502515 | 0,659651287 | SLC30A5 | FALSE | Upregulated |
| 0,208657052 | 0,659664455 | RP4-724E16.2 | FALSE | Upregulated |
| 0,380325748 | 0,660318029 | KCTD13 | FALSE | Upregulated |
| 0,44247167 | 0,660318029 | KLHL10 | FALSE | Upregulated |
| 0,344042926 | 0,660318029 | LOC100996876 | FALSE | Upregulated |
| 0,149464639 | 0,661289944 | LOC652276 | FALSE | Upregulated |
| -0,170429579 | 0,661289944 | PALMD | FALSE | Downregulated |
| -0,206385206 | 0,661404304 | AGO3 | FALSE | Downregulated |
| 0,229338121 | 0,661404304 | CIB1 | FALSE | Upregulated |
| 0,150896227 | 0,661404304 | ENSA | FALSE | Upregulated |
| 0,26841022 | 0,661404304 | HBCBP | FALSE | Upregulated |
| -0,202648845 | 0,661404304 | HINT2 | FALSE | Downregulated |
| 0,237019861 | 0,661404304 | LDHAL6B | FALSE | Upregulated |
| 0,356305534 | 0,661404304 | LOC102723708 | FALSE | Upregulated |
| -0,263263333 | 0,661404304 | NHLRC3 | FALSE | Downregulated |
| 0,306786628 | 0,661404304 | PLEKHO1 | FALSE | Upregulated |
| 0,132040837 | 0,661404304 | SBF2 | FALSE | Upregulated |

| | | | | |
|---|---|---|---|---|
| 0,194925178 | 0,661404304 | SLC39A13 | FALSE | Upregulated |
| -0,103553678 | 0,661404304 | SRRM2 | FALSE | Downregulated |
| -0,210065534 | 0,661404304 | TNFSF10 | FALSE | Downregulated |
| -0,239980331 | 0,661699739 | NRG4 | FALSE | Downregulated |
| 0,200069785 | 0,661699739 | ZNF587B | FALSE | Upregulated |
| 0,260232747 | 0,661699739 | ZNHIT2 | FALSE | Upregulated |
| 0,184410783 | 0,661801525 | NUAK1 | FALSE | Upregulated |
| 0,481332495 | 0,661893455 | MYOM2 | FALSE | Upregulated |
| 0,336443836 | 0,662406749 | DIRAS3 | FALSE | Upregulated |
| -0,241831898 | 0,662478276 | RABEPK | FALSE | Downregulated |
| -0,128677753 | 0,662976847 | ADCK3 | FALSE | Downregulated |
| 0,266065959 | 0,662976847 | ARL13A | FALSE | Upregulated |
| 0,203052852 | 0,662976847 | BRPF1 | FALSE | Upregulated |
| 0,167421932 | 0,662976847 | DCST2 | FALSE | Upregulated |
| 0,255518801 | 0,662976847 | DENND5B-AS1 | FALSE | Upregulated |
| -0,429672854 | 0,662976847 | DGKG | FALSE | Downregulated |
| -0,188476691 | 0,662976847 | DLD | FALSE | Downregulated |
| -0,107606388 | 0,662976847 | FURIN | FALSE | Downregulated |
| -0,105408867 | 0,662976847 | HDLBP | FALSE | Downregulated |
| -0,320637984 | 0,662976847 | LOC100289230 | FALSE | Downregulated |
| 0,240115947 | 0,662976847 | LOC440386 | FALSE | Upregulated |
| 0,288263752 | 0,662976847 | LOC642648 | FALSE | Upregulated |
| 0,253112258 | 0,662976847 | MARCO | FALSE | Upregulated |
| -0,369920114 | 0,662976847 | NDFIP2 | FALSE | Downregulated |
| 0,185717165 | 0,662976847 | PPP2R4 | FALSE | Upregulated |
| 0,210318449 | 0,662976847 | PROSER2-AS1 | FALSE | Upregulated |
| 0,335263961 | 0,662976847 | SLC6A4 | FALSE | Upregulated |
| 0,233346345 | 0,662976847 | SP110 | FALSE | Upregulated |
| 0,37885878 | 0,662976847 | STX19 | FALSE | Upregulated |
| 0,161933847 | 0,662976847 | TBC1D10B | FALSE | Upregulated |
| -0,159520802 | 0,662976847 | TRAM1 | FALSE | Downregulated |
| 0,29757021 | 0,662976847 | USP39 | FALSE | Upregulated |
| -0,315113324 | 0,662976847 | WDR83OS | FALSE | Downregulated |
| -0,469915526 | 0,663020314 | EPHB3 | FALSE | Downregulated |
| 0,27492968 | 0,663074386 | C15orf52 | FALSE | Upregulated |
| 0,622422036 | 0,663144305 | HSPB7 | FALSE | Upregulated |
| 0,193435035 | 0,663144305 | RP11-778H2.1 | FALSE | Upregulated |
| -0,12252342 | 0,663144305 | UBE3A | FALSE | Downregulated |
| 0,165088054 | 0,663724146 | ADAMTSL4 | FALSE | Upregulated |
| 0,23587526 | 0,663724146 | HLA-L | FALSE | Upregulated |
| 0,305383668 | 0,663774556 | LOC101927719 | FALSE | Upregulated |
| 0,3185015 | 0,663774556 | SH3BP1 | FALSE | Upregulated |
| 0,656597894 | 0,663983162 | CFTR | FALSE | Upregulated |
| -0,174084129 | 0,663983162 | CUL3 | FALSE | Downregulated |
| 0,194849482 | 0,663983162 | KGFLP2 | FALSE | Upregulated |
| 0,224224703 | 0,663983162 | LOC101927266 | FALSE | Upregulated |
| -0,386285052 | 0,663983162 | TEX14 | FALSE | Downregulated |
| -0,409591689 | 0,66400976 | LOC101928668 | FALSE | Downregulated |
| 0,253557575 | 0,66400976 | SIGLEC16 | FALSE | Upregulated |
| 0,415435207 | 0,66400976 | TCEA2 | FALSE | Upregulated |
| 0,2636151 | 0,664244477 | NLRP1 | FALSE | Upregulated |

| | | | | |
|---|---|---|---|---|
| 0,197738476 | 0,664575247 | CCDC69 | FALSE | Upregulated |
| -0,321376212 | 0,664575247 | COX6C | FALSE | Downregulated |
| 0,153937795 | 0,664575247 | DTX3L | FALSE | Upregulated |
| -0,297831436 | 0,664575247 | FBXL14 | FALSE | Downregulated |
| -0,213738919 | 0,664575247 | HHIP | FALSE | Downregulated |
| 0,358733017 | 0,664575247 | LOC93622 | FALSE | Upregulated |
| -0,112982395 | 0,664575247 | SSFA2 | FALSE | Downregulated |
| 0,349166036 | 0,664575247 | ZNF385C | FALSE | Upregulated |
| 0,343873973 | 0,664803506 | TRIM67 | FALSE | Upregulated |
| 0,181820704 | 0,664938072 | RFX1 | FALSE | Upregulated |
| 0,183632276 | 0,664972441 | NAB2 | FALSE | Upregulated |
| -0,345104412 | 0,664972441 | PPIC | FALSE | Downregulated |
| 0,181787913 | 0,664972441 | PPT1 | FALSE | Upregulated |
| -0,127162611 | 0,664972441 | SF3B2 | FALSE | Downregulated |
| 0,159517651 | 0,664972441 | SLC9A7 | FALSE | Upregulated |
| -0,325242308 | 0,664972441 | UNC5C | FALSE | Downregulated |
| 0,287656376 | 0,665172001 | AK5 | FALSE | Upregulated |
| -0,134525836 | 0,665172001 | ARFGAP2 | FALSE | Downregulated |
| -0,122313668 | 0,665172001 | ASH1L | FALSE | Downregulated |
| 0,141842552 | 0,665172001 | CAMTA2 | FALSE | Upregulated |
| 0,397388861 | 0,665172001 | FAM76B | FALSE | Upregulated |
| 0,284056249 | 0,665172001 | LOC101927790 | FALSE | Upregulated |
| 0,499916563 | 0,665172001 | LOC101928473 | FALSE | Upregulated |
| 0,465691667 | 0,665172001 | LOC389199 | FALSE | Upregulated |
| -0,379329256 | 0,665172001 | NDUFAF1 | FALSE | Downregulated |
| -0,20820535 | 0,665172001 | NDUFV3 | FALSE | Downregulated |
| -0,190088769 | 0,665172001 | NEURL4 | FALSE | Downregulated |
| -0,275298286 | 0,665172001 | RANBP3L | FALSE | Downregulated |
| -0,248289124 | 0,665172001 | RTTN | FALSE | Downregulated |
| 0,204569921 | 0,665172001 | SAMHD1 | FALSE | Upregulated |
| -0,192668346 | 0,665172001 | SLC22A25 | FALSE | Downregulated |
| 0,267603173 | 0,665172001 | SPDYA | FALSE | Upregulated |
| -0,174403792 | 0,665172001 | WDR45B | FALSE | Downregulated |
| 0,189724937 | 0,665172001 | ZNF865 | FALSE | Upregulated |
| 0,183995143 | 0,665433669 | LOC101928793 | FALSE | Upregulated |
| 0,193773872 | 0,665433669 | LOC102723582 | FALSE | Upregulated |
| 0,308147752 | 0,665433669 | NBPF20 | FALSE | Upregulated |
| 0,290317145 | 0,665800141 | KLF2 | FALSE | Upregulated |
| 0,152076947 | 0,665800141 | ZNF460 | FALSE | Upregulated |
| -0,253761501 | 0,665827401 | PIM1 | FALSE | Downregulated |
| 0,40624858 | 0,66597823 | COL16A1 | FALSE | Upregulated |
| -0,175616546 | 0,66597823 | REST | FALSE | Downregulated |
| -0,221220165 | 0,666163246 | EXOC5 | FALSE | Downregulated |
| 0,382999815 | 0,666163246 | LINC00870 | FALSE | Upregulated |
| -0,290340541 | 0,666163246 | POLK | FALSE | Downregulated |
| 0,523698231 | 0,666283776 | USP17L7 | FALSE | Upregulated |
| 0,437951423 | 0,666357494 | KCTD1 | FALSE | Upregulated |
| 0,199007575 | 0,666625296 | CLSTN1 | FALSE | Upregulated |
| 0,279819393 | 0,66718018 | ATP1A1OS | FALSE | Upregulated |
| -0,252786339 | 0,66718018 | B3GNT2 | FALSE | Downregulated |
| -0,187184519 | 0,66718018 | CHCHD2 | FALSE | Downregulated |

| | | | | |
|---|---|---|---|---|
| -0,244974851 | 0,66718018 | DNAJC8 | FALSE | Downregulated |
| 0,160205909 | 0,66718018 | NUMB | FALSE | Upregulated |
| 0,280789042 | 0,66718018 | SORCS2 | FALSE | Upregulated |
| -0,361774201 | 0,667320344 | TERC | FALSE | Downregulated |
| -0,206237328 | 0,667431986 | CLN8 | FALSE | Downregulated |
| -0,285255104 | 0,667431986 | CXCL2 | FALSE | Downregulated |
| -0,292123965 | 0,667431986 | HIST1H2BK | FALSE | Downregulated |
| 0,218303754 | 0,667431986 | LOC102723535 | FALSE | Upregulated |
| 0,230809608 | 0,667431986 | RP11-98I9.4 | FALSE | Upregulated |
| -0,144733761 | 0,667431986 | SH3D19 | FALSE | Downregulated |
| -0,41771496 | 0,667431986 | STX1A | FALSE | Downregulated |
| -0,243461998 | 0,667431986 | SYT17 | FALSE | Downregulated |
| 0,159675583 | 0,667431986 | SYT9 | FALSE | Upregulated |
| -0,203043954 | 0,667431986 | USP1 | FALSE | Downregulated |
| 0,32252545 | 0,667574708 | APOA4 | FALSE | Upregulated |
| -0,188237595 | 0,667574708 | CHPT1 | FALSE | Downregulated |
| 0,116092849 | 0,667574708 | FAN1 | FALSE | Upregulated |
| -0,201187659 | 0,667574708 | LRCH1 | FALSE | Downregulated |
| -0,133239289 | 0,667574708 | MIEF1 | FALSE | Downregulated |
| -0,527778727 | 0,667574708 | RPL36A | FALSE | Downregulated |
| -0,258909186 | 0,667574708 | TIGD2 | FALSE | Downregulated |
| -0,21937198 | 0,667574708 | ZNF280D | FALSE | Downregulated |
| -0,123051066 | 0,667596374 | ANKRD11 | FALSE | Downregulated |
| -0,305333134 | 0,667596374 | BOD1 | FALSE | Downregulated |
| -0,167619223 | 0,667596374 | CD164 | FALSE | Downregulated |
| -0,209155909 | 0,667596374 | CNTLN | FALSE | Downregulated |
| -0,100582678 | 0,667596374 | CTNNA1 | FALSE | Downregulated |
| -0,130974538 | 0,667596374 | DHTKD1 | FALSE | Downregulated |
| 0,248589833 | 0,667596374 | DLEC1 | FALSE | Upregulated |
| 0,231539295 | 0,667596374 | ESR2 | FALSE | Upregulated |
| 0,293277475 | 0,667596374 | FARS2 | FALSE | Upregulated |
| 0,322172964 | 0,667596374 | FBXL21 | FALSE | Upregulated |
| -0,159606042 | 0,667596374 | HINT1 | FALSE | Downregulated |
| 0,288153664 | 0,667596374 | LOC101927044 | FALSE | Upregulated |
| 0,181343544 | 0,667596374 | LOC101927647 | FALSE | Upregulated |
| -0,233432815 | 0,667596374 | NBPF26 | FALSE | Downregulated |
| -0,186660708 | 0,667596374 | OSBPL8 | FALSE | Downregulated |
| 0,307034412 | 0,667596374 | RP11-508N12.2 | FALSE | Upregulated |
| -0,269302638 | 0,667596374 | SYS1 | FALSE | Downregulated |
| 0,247729958 | 0,667596374 | ZNF330 | FALSE | Upregulated |
| 0,233622674 | 0,667596374 | ZNF83 | FALSE | Upregulated |
| 0,240784337 | 0,667861013 | CCND1 | FALSE | Upregulated |
| 0,360219614 | 0,667861013 | ELF4 | FALSE | Upregulated |
| 0,174282187 | 0,667861013 | LPPR2 | FALSE | Upregulated |
| -0,290200407 | 0,667861013 | TMEM53 | FALSE | Downregulated |
| 0,260793723 | 0,667861013 | VTI1A | FALSE | Upregulated |
| -0,280646643 | 0,6680953 | SLC6A13 | FALSE | Downregulated |
| 0,166748116 | 0,668101022 | HLA-E | FALSE | Upregulated |
| -0,109316199 | 0,668101022 | MAN2C1 | FALSE | Downregulated |
| 0,269856657 | 0,668145998 | SPTLC3 | FALSE | Upregulated |
| -0,273668634 | 0,668555484 | ATP5E | FALSE | Downregulated |

| | | | | |
|---|---|---|---|---|
| -0,356146794 | 0,668555484 | MINOS1P1 | FALSE | Downregulated |
| -0,380058071 | 0,668615078 | DDX11L2 | FALSE | Downregulated |
| -0,403397955 | 0,668615078 | GPR37 | FALSE | Downregulated |
| 0,340922512 | 0,66897562 | HSF4 | FALSE | Upregulated |
| -0,2658849 | 0,66897562 | PPM1B | FALSE | Downregulated |
| -0,216596797 | 0,669022694 | SPRED2 | FALSE | Downregulated |
| -0,22222949 | 0,6694338 | AGTR1 | FALSE | Downregulated |
| -0,341740645 | 0,6694338 | ATP8B4 | FALSE | Downregulated |
| 0,319907433 | 0,6694338 | CELF4 | FALSE | Upregulated |
| 0,369249614 | 0,6694338 | DNAH6 | FALSE | Upregulated |
| 0,13835851 | 0,6694338 | ERP44 | FALSE | Upregulated |
| 0,179184566 | 0,6694338 | HTR2B | FALSE | Upregulated |
| 0,17389979 | 0,6694338 | KLHL5 | FALSE | Upregulated |
| 0,185524603 | 0,6694338 | MTRF1L | FALSE | Upregulated |
| 0,212639705 | 0,6694338 | MYL12A | FALSE | Upregulated |
| 0,215656646 | 0,6694338 | PCID2 | FALSE | Upregulated |
| 0,211940383 | 0,6694338 | PP2D1 | FALSE | Upregulated |
| 0,289724288 | 0,6694338 | TGFB3 | FALSE | Upregulated |
| -0,149698934 | 0,6694338 | YME1L1 | FALSE | Downregulated |
| -0,251934365 | 0,6694338 | ZCCHC7 | FALSE | Downregulated |
| -0,219397717 | 0,6694338 | ZNF384 | FALSE | Downregulated |
| 0,220275943 | 0,6694338 | ZNF497 | FALSE | Upregulated |
| -0,349702522 | 0,669508059 | BRF2 | FALSE | Downregulated |
| -0,166616897 | 0,669508059 | CITED2 | FALSE | Downregulated |
| 0,132554352 | 0,669508059 | CREB1 | FALSE | Upregulated |
| 0,233492872 | 0,669508059 | CYTH3 | FALSE | Upregulated |
| 0,298565568 | 0,669508059 | DNAL1 | FALSE | Upregulated |
| -0,250794766 | 0,669508059 | EDNRB | FALSE | Downregulated |
| 0,218553151 | 0,669508059 | FAM136A | FALSE | Upregulated |
| -0,222762971 | 0,669508059 | FOXO1 | FALSE | Downregulated |
| 0,188271506 | 0,669508059 | GGACT | FALSE | Upregulated |
| -0,208680517 | 0,669508059 | HEATR5B | FALSE | Downregulated |
| 0,406000503 | 0,669508059 | HIATL2 | FALSE | Upregulated |
| -0,280182958 | 0,669508059 | HOOK1 | FALSE | Downregulated |
| -0,396873778 | 0,669508059 | HSD17B3 | FALSE | Downregulated |
| 0,483412589 | 0,669508059 | IL21R | FALSE | Upregulated |
| -0,227614649 | 0,669508059 | MGMT | FALSE | Downregulated |
| 0,274546518 | 0,669508059 | PODNL1 | FALSE | Upregulated |
| 0,296687953 | 0,669508059 | RNF166 | FALSE | Upregulated |
| 0,263606126 | 0,669508059 | TRIB2 | FALSE | Upregulated |
| -0,130822344 | 0,669508059 | WDR81 | FALSE | Downregulated |
| -0,16630202 | 0,669706977 | ALDH9A1 | FALSE | Downregulated |
| 0,208546771 | 0,670210015 | AP4B1-AS1 | FALSE | Upregulated |
| -0,234121983 | 0,670210015 | CDC27 | FALSE | Downregulated |
| 0,395402744 | 0,670210015 | CSNK1A1P1 | FALSE | Upregulated |
| 0,167488807 | 0,670210015 | LOC646471 | FALSE | Upregulated |
| -0,176491815 | 0,670210015 | PPFIBP1 | FALSE | Downregulated |
| 0,292201919 | 0,670210015 | PTPN2 | FALSE | Upregulated |
| 0,101265046 | 0,670210015 | TAPBP | FALSE | Upregulated |
| 0,240284262 | 0,670210015 | TRAF3IP2-AS1 | FALSE | Upregulated |
| -0,439124302 | 0,670369855 | LOC101928263 | FALSE | Downregulated |

| | | | | |
|---|---|---|---|---|
| -0,205682645 | 0,670626246 | ACSM5 | FALSE | Downregulated |
| -0,39469907 | 0,670626246 | BOLA1 | FALSE | Downregulated |
| -0,42734888 | 0,670626246 | C5orf27 | FALSE | Downregulated |
| 0,222048615 | 0,670626246 | CC2D2A | FALSE | Upregulated |
| -0,150698505 | 0,670626246 | DHCR24 | FALSE | Downregulated |
| 0,461203206 | 0,670626246 | FAM131B | FALSE | Upregulated |
| -0,205653143 | 0,670626246 | GAS2 | FALSE | Downregulated |
| -0,349669687 | 0,670626246 | LAPTM4B | FALSE | Downregulated |
| 0,149641687 | 0,670626246 | MAPKAPK2 | FALSE | Upregulated |
| -0,170871625 | 0,670626246 | MSH3 | FALSE | Downregulated |
| -0,365788819 | 0,670626246 | STEAP1 | FALSE | Downregulated |
| 0,254101987 | 0,670626246 | STRA13 | FALSE | Upregulated |
| 0,186047841 | 0,67076642 | ERO1LB | FALSE | Upregulated |
| -0,272185576 | 0,67108016 | ARL3 | FALSE | Downregulated |
| 0,228833482 | 0,67108016 | ARL5B | FALSE | Upregulated |
| 0,22088933 | 0,67108016 | CROCC | FALSE | Upregulated |
| 0,142122581 | 0,67108016 | LOC101929264 | FALSE | Upregulated |
| 0,150381272 | 0,67108016 | MEF2D | FALSE | Upregulated |
| -0,333554038 | 0,67108016 | PDE3A | FALSE | Downregulated |
| 0,416937806 | 0,67108016 | RASA4 | FALSE | Upregulated |
| -0,228511429 | 0,671400134 | TMEM106B | FALSE | Downregulated |
| 0,20725182 | 0,671534075 | PAPPA | FALSE | Upregulated |
| 0,449437176 | 0,67157451 | GOLT1B | FALSE | Upregulated |
| 0,162429859 | 0,671823715 | C15orf60 | FALSE | Upregulated |
| 0,235789596 | 0,671926158 | CCDC157 | FALSE | Upregulated |
| -0,137337404 | 0,671926158 | CD99 | FALSE | Downregulated |
| -0,335173635 | 0,671926158 | DCDC5 | FALSE | Downregulated |
| -0,447180158 | 0,671926158 | DERL2 | FALSE | Downregulated |
| 0,377267098 | 0,671926158 | DPT | FALSE | Upregulated |
| 0,212352479 | 0,671926158 | DVL2 | FALSE | Upregulated |
| 0,376440644 | 0,671926158 | GGN | FALSE | Upregulated |
| -0,139424526 | 0,671926158 | GUK1 | FALSE | Downregulated |
| -0,491082057 | 0,671926158 | KIAA1456 | FALSE | Downregulated |
| -0,164062712 | 0,671926158 | LGR4 | FALSE | Downregulated |
| 0,342173261 | 0,671926158 | LOC101929657 | FALSE | Upregulated |
| -0,261796962 | 0,671926158 | MRPS35 | FALSE | Downregulated |
| 0,309972713 | 0,671926158 | MYO15A | FALSE | Upregulated |
| -0,19454911 | 0,671926158 | PCTP | FALSE | Downregulated |
| -0,168906609 | 0,671926158 | PDIA4 | FALSE | Downregulated |
| 0,152190798 | 0,671926158 | POLR3H | FALSE | Upregulated |
| -0,202253853 | 0,671926158 | PSME1 | FALSE | Downregulated |
| -0,212665938 | 0,671926158 | SLC35F5 | FALSE | Downregulated |
| 0,204035251 | 0,671926158 | SLC38A2 | FALSE | Upregulated |
| -0,119969243 | 0,671926158 | SPAG9 | FALSE | Downregulated |
| -0,153519373 | 0,671926158 | ST3GAL1 | FALSE | Downregulated |
| -0,2489573 | 0,671926158 | STARD3 | FALSE | Downregulated |
| 0,221733284 | 0,671926158 | ZDHHC16 | FALSE | Upregulated |
| -0,419808736 | 0,671953325 | LOC102724895 | FALSE | Downregulated |
| -0,152361988 | 0,672048018 | ZXDC | FALSE | Downregulated |
| 0,391788399 | 0,672262639 | DET1 | FALSE | Upregulated |
| 0,48392154 | 0,67253754 | RP11-725G5.2 | FALSE | Upregulated |

| | | | | |
|---|---|---|---|---|
| -0,175828977 | 0,672578709 | CLTA | FALSE | Downregulated |
| -0,67066162 | 0,672578709 | HAMP | FALSE | Downregulated |
| 0,188155626 | 0,672578709 | ITPR1 | FALSE | Upregulated |
| 0,200391708 | 0,672578709 | UBFD1 | FALSE | Upregulated |
| 0,25730384 | 0,672607838 | MRPL40 | FALSE | Upregulated |
| 0,169743832 | 0,672914863 | KRAS | FALSE | Upregulated |
| -0,159199515 | 0,673207063 | HADH | FALSE | Downregulated |
| -0,181954534 | 0,673338353 | APCS | FALSE | Downregulated |
| 0,400673015 | 0,673338353 | FAM217B | FALSE | Upregulated |
| 0,143261074 | 0,673338353 | FANCC | FALSE | Upregulated |
| 0,314153602 | 0,673338353 | FGF11 | FALSE | Upregulated |
| -0,404453407 | 0,673338353 | FOXE1 | FALSE | Downregulated |
| -0,27825782 | 0,673338353 | KIF16B | FALSE | Downregulated |
| -0,389294787 | 0,673338353 | LOC101928058 | FALSE | Downregulated |
| -0,416641005 | 0,673338353 | LOC285638 | FALSE | Downregulated |
| -0,190206624 | 0,673338353 | PIGQ | FALSE | Downregulated |
| 0,573001724 | 0,673338353 | RGS7BP | FALSE | Upregulated |
| 0,179093625 | 0,673338353 | RILPL1 | FALSE | Upregulated |
| 0,45451292 | 0,673338353 | RNF150 | FALSE | Upregulated |
| -0,146319517 | 0,673338353 | SLC25A3 | FALSE | Downregulated |
| 0,189675706 | 0,673338353 | SMCR8 | FALSE | Upregulated |
| -0,40115125 | 0,673338353 | ZNF331 | FALSE | Downregulated |
| -0,2149073 | 0,673592898 | PABPC1L | FALSE | Downregulated |
| 0,154772701 | 0,673592898 | PSIP1 | FALSE | Upregulated |
| -0,192988878 | 0,673608867 | TTC36 | FALSE | Downregulated |
| 0,455260273 | 0,673712108 | C1orf21 | FALSE | Upregulated |
| 0,172422066 | 0,673725844 | TUBGCP4 | FALSE | Upregulated |
| 0,286555955 | 0,674141841 | AGAP2 | FALSE | Upregulated |
| -0,385567601 | 0,674141841 | CHAMP1 | FALSE | Downregulated |
| 0,296454567 | 0,674141841 | TRIM22 | FALSE | Upregulated |
| -0,197762846 | 0,674229218 | RNA45S5 | FALSE | Downregulated |
| 0,288349505 | 0,674343005 | DDX10 | FALSE | Upregulated |
| -0,397817919 | 0,674343005 | SENP7 | FALSE | Downregulated |
| -0,094217359 | 0,674409636 | BRD4 | FALSE | Downregulated |
| 0,13168616 | 0,674715582 | CREBL2 | FALSE | Upregulated |
| 0,305396785 | 0,674715582 | CYTH2 | FALSE | Upregulated |
| -0,180758053 | 0,674715582 | FAM213A | FALSE | Downregulated |
| -0,30843452 | 0,674715582 | HDAC2 | FALSE | Downregulated |
| 0,485847425 | 0,674715582 | LINC01152 | FALSE | Upregulated |
| 0,154503063 | 0,674715582 | NAV1 | FALSE | Upregulated |
| 0,21482447 | 0,674715582 | PCSK1 | FALSE | Upregulated |
| 0,425654256 | 0,674715582 | RRAGC | FALSE | Upregulated |
| 0,218954967 | 0,674715582 | SH3RF3-AS1 | FALSE | Upregulated |
| 0,394945832 | 0,674879612 | HN1 | FALSE | Upregulated |
| -0,213090584 | 0,674972042 | INPP5B | FALSE | Downregulated |
| 0,292702998 | 0,675065297 | ZBTB3 | FALSE | Upregulated |
| 0,406402005 | 0,675183356 | DDI1 | FALSE | Upregulated |
| 0,170051497 | 0,675183356 | FLRT3 | FALSE | Upregulated |
| 0,190322467 | 0,675183356 | GUF1 | FALSE | Upregulated |
| 0,543611379 | 0,675316773 | RP11-473M20.5 | FALSE | Upregulated |
| 0,227508392 | 0,675605273 | ADPRM | FALSE | Upregulated |

| | | | | |
|---|---|---|---|---|
| 0,38712932 | 0,675605273 | CTTNBP2NL | FALSE | Upregulated |
| -0,206491017 | 0,675605273 | DNAJC1 | FALSE | Downregulated |
| 0,267349897 | 0,675605273 | FAM185A | FALSE | Upregulated |
| 0,164207123 | 0,675605273 | FAM186A | FALSE | Upregulated |
| -0,145731382 | 0,675605273 | KIAA2026 | FALSE | Downregulated |
| 0,197778649 | 0,675605273 | NUBP2 | FALSE | Upregulated |
| -0,116381027 | 0,675605273 | PTBP1 | FALSE | Downregulated |
| -0,154026865 | 0,675605273 | SARDH | FALSE | Downregulated |
| -0,160353293 | 0,675605273 | SLC39A5 | FALSE | Downregulated |
| 0,257691394 | 0,675605273 | TIMELESS | FALSE | Upregulated |
| 0,301829789 | 0,675605273 | WNK4 | FALSE | Upregulated |
| 0,090351897 | 0,675669506 | CLTC | FALSE | Upregulated |
| 0,457249235 | 0,675669506 | MAP4K1 | FALSE | Upregulated |
| -0,249787697 | 0,675669506 | NUDT15 | FALSE | Downregulated |
| -0,252554403 | 0,675676847 | ABLIM3 | FALSE | Downregulated |
| 0,228748923 | 0,675676847 | COL4A2 | FALSE | Upregulated |
| -0,187101283 | 0,675676847 | KCNJ8 | FALSE | Downregulated |
| 0,280933386 | 0,675676847 | KCNV2 | FALSE | Upregulated |
| 0,251239419 | 0,675676847 | LDOC1L | FALSE | Upregulated |
| -0,176286375 | 0,675676847 | NEURL1B | FALSE | Downregulated |
| -0,195206146 | 0,675676847 | NUP88 | FALSE | Downregulated |
| 0,173488178 | 0,675676847 | TAF1 | FALSE | Upregulated |
| 0,315880018 | 0,675676847 | TUBA3FP | FALSE | Upregulated |
| -0,221368704 | 0,675676847 | UFC1 | FALSE | Downregulated |
| 0,229210559 | 0,675676847 | WDR19 | FALSE | Upregulated |
| 0,233776247 | 0,675816585 | RP11-61I13.3 | FALSE | Upregulated |
| -0,138074774 | 0,675861673 | KANSL1 | FALSE | Downregulated |
| 0,376941258 | 0,675868017 | EML2 | FALSE | Upregulated |
| -0,182279705 | 0,675960844 | COLGALT1 | FALSE | Downregulated |
| 0,237344915 | 0,675960844 | HLA-DPB1 | FALSE | Upregulated |
| -0,92838767 | 0,676121665 | PLA2G4B | FALSE | Downregulated |
| 0,144242502 | 0,676121665 | RAI1 | FALSE | Upregulated |
| 0,263018622 | 0,676121665 | SPOPL | FALSE | Upregulated |
| 0,240523606 | 0,676225221 | GJA9-MYCBP | FALSE | Upregulated |
| -0,153644897 | 0,676361677 | BLVRB | FALSE | Downregulated |
| 0,237408051 | 0,676361677 | CLMP | FALSE | Upregulated |
| -0,199602779 | 0,676361677 | FAHD1 | FALSE | Downregulated |
| 0,181906874 | 0,676361677 | GDI1 | FALSE | Upregulated |
| 0,329541614 | 0,676361677 | LINC00472 | FALSE | Upregulated |
| 0,405806224 | 0,676361677 | LOC286367 | FALSE | Upregulated |
| 0,323051093 | 0,676361677 | LOC729291 | FALSE | Upregulated |
| 0,20415789 | 0,676361677 | LTBP2 | FALSE | Upregulated |
| -0,102252084 | 0,676361677 | NFE2L1 | FALSE | Downregulated |
| -0,187956533 | 0,676361677 | PROSC | FALSE | Downregulated |
| 0,333843449 | 0,676361677 | PSAPL1 | FALSE | Upregulated |
| -0,233742223 | 0,676361677 | SLC25A27 | FALSE | Downregulated |
| 0,272610354 | 0,676361677 | SLFN5 | FALSE | Upregulated |
| 0,287279821 | 0,676361677 | STAG3 | FALSE | Upregulated |
| 0,211397778 | 0,676361677 | SYT15 | FALSE | Upregulated |
| -0,22199139 | 0,676361677 | WWTR1 | FALSE | Downregulated |
| 0,514181576 | 0,676412562 | SLC25A33 | FALSE | Upregulated |

| | | | | |
|---|---|---|---|---|
| 0,258702113 | 0,67656825 | ATP6V1E1 | FALSE | Upregulated |
| -0,140928977 | 0,67656825 | CFH | FALSE | Downregulated |
| -0,356146526 | 0,67656825 | LOC729603 | FALSE | Downregulated |
| -0,158653345 | 0,67656825 | SLC39A7 | FALSE | Downregulated |
| -0,14852305 | 0,676756956 | MOCS1 | FALSE | Downregulated |
| 0,174102262 | 0,677336845 | ATP13A1 | FALSE | Upregulated |
| 0,246594743 | 0,677336845 | B3GAT2 | FALSE | Upregulated |
| -0,230049055 | 0,677336845 | DSE | FALSE | Downregulated |
| 0,212254776 | 0,677336845 | LOC101928070 | FALSE | Upregulated |
| -0,349213405 | 0,677429642 | BRI3BP | FALSE | Downregulated |
| -0,379508079 | 0,677429642 | PLA2G4C | FALSE | Downregulated |
| -0,215359351 | 0,677771231 | FAM195B | FALSE | Downregulated |
| -0,123611215 | 0,677933663 | AK3 | FALSE | Downregulated |
| 0,233895682 | 0,677933663 | ARHGAP27 | FALSE | Upregulated |
| 0,229737686 | 0,677933663 | LNPEP | FALSE | Upregulated |
| 0,177873029 | 0,677933663 | POLR2B | FALSE | Upregulated |
| -0,148135925 | 0,678252888 | C1RL | FALSE | Downregulated |
| 0,333682314 | 0,678252888 | CCDC41 | FALSE | Upregulated |
| 0,431279533 | 0,67854978 | B4GALNT1 | FALSE | Upregulated |
| 0,152540502 | 0,67854978 | SLC2A12 | FALSE | Upregulated |
| -0,408165869 | 0,67854978 | USP21 | FALSE | Downregulated |
| -0,239200337 | 0,678767396 | FAM101B | FALSE | Downregulated |
| -0,268839739 | 0,678811738 | SPRY4 | FALSE | Downregulated |
| 0,360684955 | 0,678988946 | NCAM1 | FALSE | Upregulated |
| -0,213069297 | 0,678988946 | RPL36AL | FALSE | Downregulated |
| -0,152390707 | 0,679038718 | ACY1 | FALSE | Downregulated |
| 0,24863407 | 0,679038718 | FUT6 | FALSE | Upregulated |
| 0,187120783 | 0,679038718 | HTATSF1P2 | FALSE | Upregulated |
| 0,268871429 | 0,679038718 | ITGA6 | FALSE | Upregulated |
| -0,20894767 | 0,679038718 | MAFG | FALSE | Downregulated |
| 0,331612167 | 0,679038718 | MEIS1-AS2 | FALSE | Upregulated |
| -0,330830477 | 0,679038718 | PLSCR1 | FALSE | Downregulated |
| -0,121333496 | 0,679038718 | SRPR | FALSE | Downregulated |
| 0,144487059 | 0,679038718 | TP53INP1 | FALSE | Upregulated |
| 0,187390347 | 0,679038718 | ZNF674 | FALSE | Upregulated |
| -0,25225717 | 0,679396831 | ACAD9 | FALSE | Downregulated |
| -0,162736496 | 0,679396831 | ADH1A | FALSE | Downregulated |
| 0,229224604 | 0,679396831 | ALKBH3-AS1 | FALSE | Upregulated |
| 0,233439386 | 0,679396831 | ANGPTL2 | FALSE | Upregulated |
| 0,168947142 | 0,679396831 | CSF1R | FALSE | Upregulated |
| 0,48450685 | 0,679396831 | LOC101928875 | FALSE | Upregulated |
| -0,138254302 | 0,679396831 | PPIB | FALSE | Downregulated |
| -0,301960125 | 0,679396831 | RCHY1 | FALSE | Downregulated |
| 0,141137292 | 0,679396831 | RNASEH2C | FALSE | Upregulated |
| 0,296783674 | 0,679396831 | TMEM82 | FALSE | Upregulated |
| 0,257986412 | 0,67974229 | FAM117B | FALSE | Upregulated |
| 0,236893374 | 0,67974229 | MTMR14 | FALSE | Upregulated |
| -0,151757803 | 0,679749612 | NDUFS2 | FALSE | Downregulated |
| -0,23569906 | 0,679882719 | CMTM8 | FALSE | Downregulated |
| -0,252705917 | 0,679882719 | KLF7 | FALSE | Downregulated |
| 0,269078452 | 0,679882719 | LOC100131564 | FALSE | Upregulated |

| | | | | |
|---|---|---|---|---|
| -0,252665815 | 0,679882719 | RLF | FALSE | Downregulated |
| -0,166864257 | 0,679882719 | STT3A | FALSE | Downregulated |
| 0,164890495 | 0,679882719 | URAHP | FALSE | Upregulated |
| 0,450427245 | 0,679882719 | WDR4 | FALSE | Upregulated |
| -0,106512042 | 0,680324678 | PNPO | FALSE | Downregulated |
| 0,274172008 | 0,68044417 | DOCK10 | FALSE | Upregulated |
| -0,310942993 | 0,68044417 | FOLR2 | FALSE | Downregulated |
| -0,223353655 | 0,68044417 | NGRN | FALSE | Downregulated |
| 0,250739879 | 0,680762394 | KCTD17 | FALSE | Upregulated |
| 0,293195954 | 0,680762394 | KIF26B | FALSE | Upregulated |
| 0,118065235 | 0,68082138 | MCC | FALSE | Upregulated |
| -0,294300276 | 0,68082138 | ROMO1 | FALSE | Downregulated |
| 0,185376977 | 0,68082138 | SMCR5 | FALSE | Upregulated |
| 0,122419444 | 0,68082138 | TENM1 | FALSE | Upregulated |
| 0,307819416 | 0,68082138 | ZNF616 | FALSE | Upregulated |
| -0,324062982 | 0,680862001 | CFDP1 | FALSE | Downregulated |
| 0,298157657 | 0,680862001 | ITPRIP | FALSE | Upregulated |
| 0,40759278 | 0,680862001 | KIF6 | FALSE | Upregulated |
| 0,186290537 | 0,680862001 | LOC101929240 | FALSE | Upregulated |
| 0,151345235 | 0,680862001 | LSM14B | FALSE | Upregulated |
| -0,195343384 | 0,680862001 | MICU1 | FALSE | Downregulated |
| 0,544812346 | 0,680862001 | PAQR5 | FALSE | Upregulated |
| 0,222650379 | 0,680862001 | RP11-450K4.1 | FALSE | Upregulated |
| 0,428418139 | 0,680862001 | SOD3 | FALSE | Upregulated |
| -0,387791899 | 0,680862001 | THOC3 | FALSE | Downregulated |
| -0,277991792 | 0,680862001 | TNIK | FALSE | Downregulated |
| 0,33250113 | 0,681040262 | ZNF74 | FALSE | Upregulated |
| 0,187116129 | 0,681118805 | LAT2 | FALSE | Upregulated |
| -0,146543328 | 0,681195168 | ONECUT1 | FALSE | Downregulated |
| -0,209251074 | 0,681195168 | TMEM204 | FALSE | Downregulated |
| -0,122378434 | 0,681262439 | KDM3B | FALSE | Downregulated |
| 0,230400014 | 0,681262439 | SDHAP2 | FALSE | Upregulated |
| 0,236152215 | 0,681385853 | SLC12A9 | FALSE | Upregulated |
| 0,171964114 | 0,681508116 | MARCH8 | FALSE | Upregulated |
| -0,157292571 | 0,681626603 | LOC101928451 | FALSE | Downregulated |
| -0,313182454 | 0,681723907 | PWP1 | FALSE | Downregulated |
| -0,302536293 | 0,682013078 | CSRP2BP | FALSE | Downregulated |
| -0,09003388 | 0,682013078 | LRP1 | FALSE | Downregulated |
| -0,215209413 | 0,682013078 | MMADHC | FALSE | Downregulated |
| -0,20616732 | 0,682013078 | NEDD4 | FALSE | Downregulated |
| -0,27811293 | 0,682013078 | NUP155 | FALSE | Downregulated |
| 0,368188954 | 0,682013078 | PARP11 | FALSE | Upregulated |
| -0,573963796 | 0,682013078 | RP5-968D22.1 | FALSE | Downregulated |
| -0,23210962 | 0,682013078 | VPS37B | FALSE | Downregulated |
| 0,147811886 | 0,682013078 | ZCCHC11 | FALSE | Upregulated |
| 0,43362837 | 0,682250198 | GAREML | FALSE | Upregulated |
| -0,136534307 | 0,682250976 | ESPN | FALSE | Downregulated |
| 0,161530256 | 0,682283779 | ARHGAP30 | FALSE | Upregulated |
| -0,225029402 | 0,68230687 | ABCB10 | FALSE | Downregulated |
| 0,298402938 | 0,68230687 | KIAA0020 | FALSE | Upregulated |
| -0,376598486 | 0,68230687 | THOC6 | FALSE | Downregulated |

| | | | | |
|---|---|---|---|---|
| 0,196563942 | 0,682345382 | INSIG2 | FALSE | Upregulated |
| 0,280463142 | 0,682371747 | ARHGEF4 | FALSE | Upregulated |
| -0,337293038 | 0,682371747 | C12orf10 | FALSE | Downregulated |
| 0,455130756 | 0,682371747 | C2orf82 | FALSE | Upregulated |
| 0,242158962 | 0,682371747 | CD47 | FALSE | Upregulated |
| 0,222171957 | 0,682371747 | CSE1L | FALSE | Upregulated |
| 0,165590637 | 0,682371747 | DCAF10 | FALSE | Upregulated |
| -0,223559625 | 0,682371747 | DNAJA2 | FALSE | Downregulated |
| 0,312465687 | 0,682371747 | GPR62 | FALSE | Upregulated |
| -0,273903002 | 0,682371747 | IBA57 | FALSE | Downregulated |
| 0,464448895 | 0,682371747 | LOC101927308 | FALSE | Upregulated |
| 0,475923731 | 0,682371747 | LOC101928729 | FALSE | Upregulated |
| 0,367379605 | 0,682371747 | RABL3 | FALSE | Upregulated |
| 0,381125488 | 0,682371747 | WDR89 | FALSE | Upregulated |
| -0,190881411 | 0,682439298 | FLJ13197 | FALSE | Downregulated |
| -0,215189124 | 0,68265367 | SULT1A1 | FALSE | Downregulated |
| -0,232058545 | 0,68288803 | ANKRD36C | FALSE | Downregulated |
| 0,179813576 | 0,683031925 | IFT140 | FALSE | Upregulated |
| 0,141108502 | 0,683031925 | SYNJ2BP | FALSE | Upregulated |
| -0,253221668 | 0,683053108 | BMS1P6 | FALSE | Downregulated |
| -0,218515848 | 0,683622501 | AKR1CL1 | FALSE | Downregulated |
| -0,1427608 | 0,683622501 | COX6A1 | FALSE | Downregulated |
| -0,153041239 | 0,683622501 | DNAJC11 | FALSE | Downregulated |
| 0,276672263 | 0,683622501 | DNAJC9 | FALSE | Upregulated |
| 0,203576865 | 0,683622501 | FAM21A | FALSE | Upregulated |
| 0,159234792 | 0,683622501 | GOLM1 | FALSE | Upregulated |
| 0,173012274 | 0,683622501 | IWS1 | FALSE | Upregulated |
| -0,30507196 | 0,683622501 | JAGN1 | FALSE | Downregulated |
| -0,260488364 | 0,683622501 | KCNC4 | FALSE | Downregulated |
| 0,508557422 | 0,683622501 | KIAA1549L | FALSE | Upregulated |
| 0,197457387 | 0,683622501 | LOC101929781 | FALSE | Upregulated |
| 0,248257195 | 0,683622501 | LOX | FALSE | Upregulated |
| -0,210365845 | 0,683622501 | POLR1B | FALSE | Downregulated |
| 0,34628126 | 0,683622501 | SLC16A7 | FALSE | Upregulated |
| -0,161470989 | 0,683622501 | SNX33 | FALSE | Downregulated |
| -0,21437473 | 0,683622501 | TCERG1 | FALSE | Downregulated |
| -0,086952974 | 0,683651887 | CREBBP | FALSE | Downregulated |
| 0,218778893 | 0,683651887 | SLC51A | FALSE | Upregulated |
| -0,182060918 | 0,683666636 | ABCB11 | FALSE | Downregulated |
| 0,279663267 | 0,683666636 | BBS2 | FALSE | Upregulated |
| 0,442127787 | 0,683666636 | LOC102724587 | FALSE | Upregulated |
| -0,234976061 | 0,683666636 | MAPK14 | FALSE | Downregulated |
| -0,202855797 | 0,683666636 | PLSCR4 | FALSE | Downregulated |
| 0,391634128 | 0,683957401 | SLC52A2 | FALSE | Upregulated |
| 0,183294871 | 0,684113836 | MICA | FALSE | Upregulated |
| -0,299621807 | 0,684318247 | LAMTOR3 | FALSE | Downregulated |
| -0,200888613 | 0,684318247 | OSBPL2 | FALSE | Downregulated |
| -0,200933724 | 0,684318247 | RAB2A | FALSE | Downregulated |
| 0,208129497 | 0,684318247 | RNF169 | FALSE | Upregulated |
| 0,218149021 | 0,684318247 | RNF207 | FALSE | Upregulated |
| 0,315771911 | 0,684488547 | C1orf112 | FALSE | Upregulated |

| | | | | |
|---|---|---|---|---|
| 0,370330847 | 0,684488547 | FAM49A | FALSE | Upregulated |
| -0,198876575 | 0,684488547 | GLRX5 | FALSE | Downregulated |
| 0,156472663 | 0,684488547 | LOC100506634 | FALSE | Upregulated |
| -0,168536223 | 0,684488547 | PAXBP1 | FALSE | Downregulated |
| 0,269162439 | 0,684488547 | SLC39A3 | FALSE | Upregulated |
| 0,205158353 | 0,684488547 | THNSL1 | FALSE | Upregulated |
| -0,529346111 | 0,684515986 | GOLGA6A | FALSE | Downregulated |
| 0,467238069 | 0,684515986 | KRT10 | FALSE | Upregulated |
| -0,172142439 | 0,684895257 | PLEKHG3 | FALSE | Downregulated |
| 0,27431768 | 0,684916412 | ANP32E | FALSE | Upregulated |
| 0,158939102 | 0,684916412 | CDIP1 | FALSE | Upregulated |
| 0,347785952 | 0,684916412 | DOK7 | FALSE | Upregulated |
| -0,116718877 | 0,684916412 | DOT1L | FALSE | Downregulated |
| -0,159518072 | 0,684916412 | FRYL | FALSE | Downregulated |
| -0,469560248 | 0,684916412 | GSTA2 | FALSE | Downregulated |
| -0,438671751 | 0,684916412 | ITIH5 | FALSE | Downregulated |
| -0,162256693 | 0,684916412 | KAT6B | FALSE | Downregulated |
| -0,225495753 | 0,684916412 | LDB2 | FALSE | Downregulated |
| 0,448082616 | 0,684916412 | LOC400043 | FALSE | Upregulated |
| -0,290328396 | 0,684916412 | LRRC40 | FALSE | Downregulated |
| -0,372326732 | 0,684916412 | LRRC8C | FALSE | Downregulated |
| 0,240954792 | 0,684916412 | LTB4R | FALSE | Upregulated |
| 0,162454273 | 0,684916412 | MEX3D | FALSE | Upregulated |
| -0,191615134 | 0,684916412 | NDEL1 | FALSE | Downregulated |
| -0,393781653 | 0,684916412 | NOVA2 | FALSE | Downregulated |
| 0,36051184 | 0,684916412 | P4HA2-AS1 | FALSE | Upregulated |
| 0,345926761 | 0,684916412 | PDC | FALSE | Upregulated |
| 0,328767442 | 0,684916412 | RAD51 | FALSE | Upregulated |
| -0,128092708 | 0,684916412 | REEP5 | FALSE | Downregulated |
| 0,347569594 | 0,684916412 | RP11-506O24.1 | FALSE | Upregulated |
| -0,183897694 | 0,684916412 | ZFYVE9 | FALSE | Downregulated |
| 0,219307416 | 0,684916412 | ZNF582 | FALSE | Upregulated |
| 0,130894807 | 0,684928358 | C6orf120 | FALSE | Upregulated |
| 0,141827223 | 0,684928358 | COMMD5 | FALSE | Upregulated |
| 0,182354389 | 0,684928358 | FAM92A1 | FALSE | Upregulated |
| -0,192926909 | 0,684928358 | KDELR1 | FALSE | Downregulated |
| 0,517103517 | 0,684928358 | PRC1-AS1 | FALSE | Upregulated |
| 0,30061497 | 0,684928358 | RABGGTA | FALSE | Upregulated |
| 0,436796694 | 0,684928358 | SPINT2 | FALSE | Upregulated |
| -0,244289319 | 0,684928358 | ZNF160 | FALSE | Downregulated |
| -0,197955767 | 0,6850082 | CHN2 | FALSE | Downregulated |
| -0,306439397 | 0,6850082 | F8A1 | FALSE | Downregulated |
| 0,205808853 | 0,6850082 | LOC102724103 | FALSE | Upregulated |
| -0,300608947 | 0,6850082 | TMEM106C | FALSE | Downregulated |
| -0,11542285 | 0,685060867 | CRAT | FALSE | Downregulated |
| -0,275744066 | 0,685060867 | GUCY1A2 | FALSE | Downregulated |
| 0,17063443 | 0,685060867 | LOC100133445 | FALSE | Upregulated |
| -0,179448166 | 0,685071464 | ABCA9 | FALSE | Downregulated |
| -0,114950489 | 0,685071464 | ADD1 | FALSE | Downregulated |
| 0,245854748 | 0,685071464 | ANKRD36 | FALSE | Upregulated |
| -1,954956977 | 0,685071464 | BLOC1S1-RDH5 | FALSE | Downregulated |

| | | | | |
|---|---|---|---|---|
| -0,32459877 | 0,685071464 | KLHL11 | FALSE | Downregulated |
| -0,223610973 | 0,685071464 | LOC100132815 | FALSE | Downregulated |
| 0,260660089 | 0,685071464 | SLC20A1 | FALSE | Upregulated |
| -0,125107841 | 0,685903646 | FAM83H | FALSE | Downregulated |
| 0,372336124 | 0,685903646 | KLC2 | FALSE | Upregulated |
| 0,2426407 | 0,685903646 | LOC101928657 | FALSE | Upregulated |
| 0,171985231 | 0,685982977 | B2M | FALSE | Upregulated |
| -0,300350644 | 0,685982977 | DSG1 | FALSE | Downregulated |
| 0,374656846 | 0,685982977 | P2RY13 | FALSE | Upregulated |
| 0,211460416 | 0,685982977 | RAP1B | FALSE | Upregulated |
| 0,149335049 | 0,685982977 | STUB1 | FALSE | Upregulated |
| 0,118978354 | 0,685982977 | TTC3 | FALSE | Upregulated |
| -0,232379762 | 0,685982977 | TXNRD1 | FALSE | Downregulated |
| -0,11596281 | 0,686661186 | NBR1 | FALSE | Downregulated |
| -0,10584854 | 0,686714063 | ARFGEF1 | FALSE | Downregulated |
| 0,156783713 | 0,686781169 | MFSD11 | FALSE | Upregulated |
| 0,185363884 | 0,687773908 | MASTL | FALSE | Upregulated |
| 0,184725206 | 0,687779343 | GSN | FALSE | Upregulated |
| 0,158614285 | 0,687795999 | MYO7A | FALSE | Upregulated |
| -0,096575127 | 0,687795999 | NEK6 | FALSE | Downregulated |
| -0,163981102 | 0,687868044 | PHF8 | FALSE | Downregulated |
| -0,218447315 | 0,687868044 | TMEM183A | FALSE | Downregulated |
| 0,192206036 | 0,687868044 | YEATS2 | FALSE | Upregulated |
| 0,088023734 | 0,688157956 | ACTN4 | FALSE | Upregulated |
| 0,371137746 | 0,688320023 | REEP3 | FALSE | Upregulated |
| -0,177684484 | 0,688320023 | ZNF282 | FALSE | Downregulated |
| 0,175814226 | 0,688322413 | GAS8 | FALSE | Upregulated |
| -0,261352003 | 0,689215484 | C14orf132 | FALSE | Downregulated |
| 0,205356659 | 0,689399059 | ARMCX3 | FALSE | Upregulated |
| 0,144523317 | 0,689399059 | RMDN1 | FALSE | Upregulated |
| 0,161270856 | 0,689399059 | SIL1 | FALSE | Upregulated |
| -0,221914801 | 0,689809214 | IRF5 | FALSE | Downregulated |
| 0,299833852 | 0,689809214 | KIRREL | FALSE | Upregulated |
| -0,435849638 | 0,689809214 | TXNL4B | FALSE | Downregulated |
| -0,160624386 | 0,689812665 | SHMT1 | FALSE | Downregulated |
| -0,335168881 | 0,690264216 | KIAA1009 | FALSE | Downregulated |
| -0,257705212 | 0,690295367 | BBS4 | FALSE | Downregulated |
| -0,129893786 | 0,690295367 | C1orf115 | FALSE | Downregulated |
| 0,339532073 | 0,690295367 | LOC100294145 | FALSE | Upregulated |
| -0,245577923 | 0,690295367 | RHPN2 | FALSE | Downregulated |
| -0,188377543 | 0,690295367 | SCARNA17 | FALSE | Downregulated |
| 0,288111971 | 0,690408386 | COX19 | FALSE | Upregulated |
| -0,179094797 | 0,690521375 | SSR4 | FALSE | Downregulated |
| 0,254954878 | 0,690521375 | STK4 | FALSE | Upregulated |
| 0,425951917 | 0,690538322 | AMICA1 | FALSE | Upregulated |
| 0,421551991 | 0,690538322 | LOC101927585 | FALSE | Upregulated |
| 0,326770266 | 0,690538322 | LOC493754 | FALSE | Upregulated |
| 0,161396632 | 0,690538322 | MLH3 | FALSE | Upregulated |
| -0,14849147 | 0,690538322 | NAGLU | FALSE | Downregulated |
| -0,28578531 | 0,690538322 | PARP16 | FALSE | Downregulated |
| -0,144423615 | 0,690538322 | PI4KA | FALSE | Downregulated |

| | | | | |
|---|---|---|---|---|
| -0,193870248 | 0,690538322 | RAD54L2 | FALSE | Downregulated |
| -0,130137792 | 0,690881949 | ATP5A1 | FALSE | Downregulated |
| -0,114754458 | 0,690948421 | GCN1L1 | FALSE | Downregulated |
| 0,299581564 | 0,691034349 | SMIM13 | FALSE | Upregulated |
| 0,265955573 | 0,691105688 | DDX27 | FALSE | Upregulated |
| 0,216337911 | 0,691105688 | SOD2 | FALSE | Upregulated |
| -0,196570124 | 0,691151271 | LYNX1 | FALSE | Downregulated |
| -0,211472529 | 0,691151271 | RAB24 | FALSE | Downregulated |
| -0,366426462 | 0,691256023 | HIST1H1A | FALSE | Downregulated |
| -0,178225236 | 0,691256023 | POMT1 | FALSE | Downregulated |
| 0,286750089 | 0,691527925 | ANKRD13D | FALSE | Upregulated |
| 0,14314737 | 0,691527925 | AP1G2 | FALSE | Upregulated |
| 0,152546253 | 0,691527925 | CERS5 | FALSE | Upregulated |
| 0,141593553 | 0,691527925 | GATAD2B | FALSE | Upregulated |
| -0,160621209 | 0,691527925 | INPP5A | FALSE | Downregulated |
| -0,222788333 | 0,691527925 | ITFG2 | FALSE | Downregulated |
| -0,19808641 | 0,691527925 | KAT7 | FALSE | Downregulated |
| 0,377422044 | 0,691527925 | P2RX2 | FALSE | Upregulated |
| 0,243687747 | 0,691527925 | PCP4L1 | FALSE | Upregulated |
| -0,400085273 | 0,691527925 | RINT1 | FALSE | Downregulated |
| -0,260487237 | 0,691527925 | SMNDC1 | FALSE | Downregulated |
| 0,258305747 | 0,691527925 | ZNF30 | FALSE | Upregulated |
| -0,142967923 | 0,691726383 | SOD1 | FALSE | Downregulated |
| 0,186743932 | 0,691812396 | FNBP1 | FALSE | Upregulated |
| -0,142128123 | 0,691812396 | SRSF6 | FALSE | Downregulated |
| -0,130400454 | 0,691838545 | C11orf95 | FALSE | Downregulated |
| -0,39647142 | 0,691838545 | C6orf123 | FALSE | Downregulated |
| -0,161450141 | 0,691838545 | EGFL7 | FALSE | Downregulated |
| -0,207166774 | 0,691838545 | GLB1 | FALSE | Downregulated |
| -0,29900298 | 0,691838545 | GRB7 | FALSE | Downregulated |
| 0,303727343 | 0,691838545 | LOC101927151 | FALSE | Upregulated |
| 0,28894695 | 0,691838545 | LOC101929691 | FALSE | Upregulated |
| 0,407813141 | 0,691838545 | LOC102723542 | FALSE | Upregulated |
| -0,170882149 | 0,691838545 | MATN2 | FALSE | Downregulated |
| -0,183604046 | 0,691838545 | MED14 | FALSE | Downregulated |
| -0,164125961 | 0,691838545 | NUDT16 | FALSE | Downregulated |
| 0,218949414 | 0,691838545 | PCYT1A | FALSE | Upregulated |
| -0,187220829 | 0,691838545 | RCAN1 | FALSE | Downregulated |
| 0,200539544 | 0,691838545 | RFC5 | FALSE | Upregulated |
| -0,204162423 | 0,691838545 | TRAPPC10 | FALSE | Downregulated |
| 0,204379912 | 0,691838545 | TRPC5 | FALSE | Upregulated |
| 0,200824865 | 0,691838545 | TYW5 | FALSE | Upregulated |
| 0,300684979 | 0,691838545 | ZBTB22 | FALSE | Upregulated |
| 0,104489628 | 0,691915767 | CHD4 | FALSE | Upregulated |
| 0,289357954 | 0,691915767 | DTNB | FALSE | Upregulated |
| -0,1354919 | 0,691915767 | DYSF | FALSE | Downregulated |
| -0,177924839 | 0,691915767 | FBXO21 | FALSE | Downregulated |
| -0,153750204 | 0,691915767 | FMO5 | FALSE | Downregulated |
| 0,189568169 | 0,691915767 | OPN3 | FALSE | Upregulated |
| 0,252945525 | 0,691915767 | SDAD1 | FALSE | Upregulated |
| -0,111538406 | 0,691915767 | SEC24C | FALSE | Downregulated |

| | | | | |
|---|---|---|---|---|
| -0,259655078 | 0,691915767 | SRD5A2 | FALSE | Downregulated |
| -0,172844434 | 0,691915767 | SREK1 | FALSE | Downregulated |
| -0,29446918 | 0,691915767 | TSEN34 | FALSE | Downregulated |
| -0,140879411 | 0,691915767 | ZMYM4 | FALSE | Downregulated |
| 0,177819559 | 0,69201687 | LOC100506922 | FALSE | Upregulated |
| 0,207357792 | 0,69201687 | LOC102724611 | FALSE | Upregulated |
| 0,179941717 | 0,69201687 | TMLHE | FALSE | Upregulated |
| -0,20453996 | 0,6921226 | CLEC4G | FALSE | Downregulated |
| -0,106659679 | 0,6921226 | GNB2 | FALSE | Downregulated |
| -0,122696048 | 0,6921226 | GPX4 | FALSE | Downregulated |
| 0,104816964 | 0,6921226 | GSPT1 | FALSE | Upregulated |
| 0,165351706 | 0,6921226 | KIAA1598 | FALSE | Upregulated |
| -0,192582488 | 0,6921226 | MBTD1 | FALSE | Downregulated |
| 0,186339474 | 0,6921226 | MTERFD3 | FALSE | Upregulated |
| -0,191129471 | 0,6921226 | RBM25 | FALSE | Downregulated |
| 0,210383958 | 0,6921226 | RP11-685M7.3 | FALSE | Upregulated |
| 0,194976704 | 0,6921226 | TIAM2 | FALSE | Upregulated |
| 0,338928842 | 0,6921226 | TRIM6 | FALSE | Upregulated |
| 0,217846283 | 0,6921226 | ZNF132 | FALSE | Upregulated |
| 0,35945814 | 0,692553991 | KY | FALSE | Upregulated |
| 0,190154346 | 0,692553991 | PIK3R3 | FALSE | Upregulated |
| 0,315312186 | 0,692553991 | ZNF471 | FALSE | Upregulated |
| -0,123508667 | 0,692557322 | DUSP3 | FALSE | Downregulated |
| -0,101044508 | 0,692557322 | RPL35 | FALSE | Downregulated |
| 0,389864968 | 0,692557322 | USP51 | FALSE | Upregulated |
| -0,351675105 | 0,69259769 | MICU3 | FALSE | Downregulated |
| -0,452869436 | 0,69259769 | NPIPA1 | FALSE | Downregulated |
| -0,148858221 | 0,692900803 | NRBP2 | FALSE | Downregulated |
| 0,13054872 | 0,692900803 | TPR | FALSE | Upregulated |
| 0,531226443 | 0,693180166 | PPAN | FALSE | Upregulated |
| 0,163514165 | 0,693180166 | VWA1 | FALSE | Upregulated |
| -0,398418162 | 0,69324844 | GPR153 | FALSE | Downregulated |
| -0,105969652 | 0,69324844 | H1FX | FALSE | Downregulated |
| -0,291310563 | 0,69324844 | ZNF250 | FALSE | Downregulated |
| 0,141136605 | 0,693254934 | ASB6 | FALSE | Upregulated |
| 0,333627754 | 0,693254934 | DHRS7B | FALSE | Upregulated |
| -0,486374951 | 0,693532963 | AC000124.1 | FALSE | Downregulated |
| -0,367524445 | 0,694090749 | STXBP3 | FALSE | Downregulated |
| -0,238168695 | 0,694198158 | ATOX1 | FALSE | Downregulated |
| 0,259420007 | 0,694198158 | C19orf35 | FALSE | Upregulated |
| 0,407018626 | 0,694198158 | LOC101927160 | FALSE | Upregulated |
| -0,187037116 | 0,694198158 | MOCOS | FALSE | Downregulated |
| 0,241548319 | 0,694494192 | CCDC28B | FALSE | Upregulated |
| 0,290100277 | 0,694538095 | BRCA1 | FALSE | Upregulated |
| -0,400174759 | 0,694538095 | C14orf180 | FALSE | Downregulated |
| 0,24422025 | 0,694538095 | EFHC1 | FALSE | Upregulated |
| 0,303058491 | 0,694538095 | GABBR1 | FALSE | Upregulated |
| -0,118832944 | 0,694538095 | GANAB | FALSE | Downregulated |
| 0,542008715 | 0,694538095 | HIST2H3C | FALSE | Upregulated |
| 0,197978503 | 0,694538095 | LOC100506504 | FALSE | Upregulated |
| 0,372185306 | 0,694538095 | LOC101928373 | FALSE | Upregulated |

| | | | | |
|---|---|---|---|---|
| -0,272110731 | 0,694538095 | MT2A | FALSE | Downregulated |
| 0,194623342 | 0,694538095 | NFKB1 | FALSE | Upregulated |
| 0,173183206 | 0,694538095 | OXER1 | FALSE | Upregulated |
| -0,232233193 | 0,694538095 | PLBD2 | FALSE | Downregulated |
| -0,15506882 | 0,694538095 | POLR1A | FALSE | Downregulated |
| 0,180947142 | 0,694538095 | PP7080 | FALSE | Upregulated |
| -0,341365655 | 0,694538095 | RAB3IP | FALSE | Downregulated |
| 0,415282663 | 0,694538095 | RP11-112H10.4 | FALSE | Upregulated |
| -0,23814765 | 0,694538095 | S100A16 | FALSE | Downregulated |
| -0,232121439 | 0,694538095 | SLC45A3 | FALSE | Downregulated |
| -0,200913424 | 0,694538095 | SUMO1 | FALSE | Downregulated |
| 0,315548207 | 0,694538095 | TCP11L2 | FALSE | Upregulated |
| 0,400311279 | 0,694538095 | ZNF221 | FALSE | Upregulated |
| -0,181597274 | 0,694613837 | AGPAT3 | FALSE | Downregulated |
| 0,318245002 | 0,694613837 | KDELC2 | FALSE | Upregulated |
| -0,369738136 | 0,694613837 | TTF1 | FALSE | Downregulated |
| 0,191847841 | 0,694613837 | TYMS | FALSE | Upregulated |
| 0,141654418 | 0,694716348 | VAC14 | FALSE | Upregulated |
| -0,146327281 | 0,695104278 | CYTH1 | FALSE | Downregulated |
| 0,271399038 | 0,695304142 | C1QL3 | FALSE | Upregulated |
| -0,317595634 | 0,695304142 | DHX57 | FALSE | Downregulated |
| -0,165307261 | 0,695304142 | F9 | FALSE | Downregulated |
| -0,153393843 | 0,695304142 | MIB1 | FALSE | Downregulated |
| 0,156153754 | 0,695304142 | PSPC1 | FALSE | Upregulated |
| -0,341584735 | 0,695304142 | TMEM209 | FALSE | Downregulated |
| -0,353977122 | 0,695640608 | EMID1 | FALSE | Downregulated |
| -0,17841132 | 0,695646831 | ATP9B | FALSE | Downregulated |
| 0,165560838 | 0,695921944 | MXRA7 | FALSE | Upregulated |
| -0,25659604 | 0,695921944 | NUMBL | FALSE | Downregulated |
| -0,330168898 | 0,696393253 | ACYP2 | FALSE | Downregulated |
| -0,348415005 | 0,696393253 | LINC00886 | FALSE | Downregulated |
| 0,238029166 | 0,696393253 | ZNF37A | FALSE | Upregulated |
| -0,142457796 | 0,696401205 | ABCA8 | FALSE | Downregulated |
| -0,263434916 | 0,696401205 | C11orf96 | FALSE | Downregulated |
| -0,220773681 | 0,696401205 | CARD10 | FALSE | Downregulated |
| 0,262626758 | 0,696401205 | CCND3 | FALSE | Upregulated |
| 0,1785275 | 0,696401205 | DNAJB7 | FALSE | Upregulated |
| 0,251736832 | 0,696401205 | DPY19L3 | FALSE | Upregulated |
| 0,21860122 | 0,696401205 | FERMT3 | FALSE | Upregulated |
| -0,201800714 | 0,696401205 | FNTA | FALSE | Downregulated |
| -0,230374118 | 0,696401205 | INTS4 | FALSE | Downregulated |
| 0,762229897 | 0,696401205 | LINC00900 | FALSE | Upregulated |
| 0,461951581 | 0,696401205 | LOC101927941 | FALSE | Upregulated |
| 0,182640491 | 0,696401205 | PMS2P1 | FALSE | Upregulated |
| -0,344516744 | 0,696401205 | RAC3 | FALSE | Downregulated |
| 0,251059704 | 0,696401205 | SERPINB1 | FALSE | Upregulated |
| 0,3430306 | 0,696401205 | TLR2 | FALSE | Upregulated |
| 0,304796468 | 0,696983667 | PIN4 | FALSE | Upregulated |
| 0,399981674 | 0,697837628 | FAAH2 | FALSE | Upregulated |
| 0,159775633 | 0,69873893 | ZSCAN2 | FALSE | Upregulated |
| -0,205888898 | 0,698897569 | DCN | FALSE | Downregulated |

| | | | | |
|---|---|---|---|---|
| -0,151608375 | 0,698940769 | RASSF5 | FALSE | Downregulated |
| 0,348418834 | 0,698966473 | C9orf116 | FALSE | Upregulated |
| -0,405934155 | 0,699039318 | WASH1 | FALSE | Downregulated |
| -0,231554073 | 0,699077652 | FOXP1 | FALSE | Downregulated |
| 0,206698095 | 0,699464965 | C20orf24 | FALSE | Upregulated |
| -0,149023039 | 0,699566733 | ARL6IP1 | FALSE | Downregulated |
| -0,161945014 | 0,699566733 | CEBPA | FALSE | Downregulated |
| -0,126767326 | 0,699566733 | SLC39A14 | FALSE | Downregulated |
| -0,195295208 | 0,699576183 | CDK2AP2 | FALSE | Downregulated |
| 0,172525538 | 0,699576183 | CPEB2 | FALSE | Upregulated |
| 0,359781469 | 0,699576183 | CRX | FALSE | Upregulated |
| -0,262325867 | 0,699576183 | GSPT2 | FALSE | Downregulated |
| 0,323070519 | 0,699576183 | LOC100128364 | FALSE | Upregulated |
| 0,336334193 | 0,699576183 | LOC101928038 | FALSE | Upregulated |
| -0,325012235 | 0,699576183 | METTL6 | FALSE | Downregulated |
| -0,358435633 | 0,699576183 | MEX3A | FALSE | Downregulated |
| -0,164874193 | 0,699576183 | PFDN5 | FALSE | Downregulated |
| -0,233707068 | 0,699576183 | R3HCC1 | FALSE | Downregulated |
| -0,146524776 | 0,699576183 | SECISBP2L | FALSE | Downregulated |
| 0,206392636 | 0,699576183 | XAF1 | FALSE | Upregulated |
| -0,171432995 | 0,700195249 | HNRNPA0 | FALSE | Downregulated |
| -0,181907765 | 0,700500649 | HGSNAT | FALSE | Downregulated |
| -0,222889796 | 0,700500649 | PMF1 | FALSE | Downregulated |
| 0,161202593 | 0,700500649 | TOR3A | FALSE | Upregulated |
| -0,239385143 | 0,700538734 | ZNF446 | FALSE | Downregulated |
| -0,18567427 | 0,70067216 | CRTC1 | FALSE | Downregulated |
| 0,13167551 | 0,701028101 | SBNO2 | FALSE | Upregulated |
| 0,18533492 | 0,701040214 | ABI1 | FALSE | Upregulated |
| -0,142456624 | 0,701040214 | CMTR1 | FALSE | Downregulated |
| -0,136331168 | 0,701040214 | COPG1 | FALSE | Downregulated |
| -0,159760579 | 0,701040214 | CYP2C18 | FALSE | Downregulated |
| -0,169509651 | 0,701040214 | HFE2 | FALSE | Downregulated |
| -0,126501328 | 0,701040214 | KCTD3 | FALSE | Downregulated |
| 0,300677238 | 0,701040214 | NMNAT1 | FALSE | Upregulated |
| -0,168624187 | 0,701040214 | SUCLG1 | FALSE | Downregulated |
| -0,197710501 | 0,701040214 | TPCN2 | FALSE | Downregulated |
| 0,277214835 | 0,70105732 | PRKCE | FALSE | Upregulated |
| 0,328576468 | 0,701365263 | LOC101928707 | FALSE | Upregulated |
| 0,276718921 | 0,701367089 | PGM5P2 | FALSE | Upregulated |
| 0,191064601 | 0,701367089 | RPS6KA2 | FALSE | Upregulated |
| -0,213467826 | 0,701492873 | ADH4 | FALSE | Downregulated |
| 0,177015649 | 0,701492873 | CHRM5 | FALSE | Upregulated |
| 0,205634347 | 0,701492873 | CRIP2 | FALSE | Upregulated |
| 0,17335116 | 0,701492873 | EFCAB13 | FALSE | Upregulated |
| 0,199044251 | 0,701492873 | FAM87B | FALSE | Upregulated |
| 0,214393923 | 0,701492873 | GOLGA8N | FALSE | Upregulated |
| -0,163089676 | 0,701492873 | LMF2 | FALSE | Downregulated |
| -0,206089677 | 0,701492873 | LRRN4 | FALSE | Downregulated |
| -0,128487984 | 0,701492873 | RPL11 | FALSE | Downregulated |
| 0,340112993 | 0,701492873 | TDP1 | FALSE | Upregulated |
| -0,106600879 | 0,702031668 | ATP1A1 | FALSE | Downregulated |

| | | | | |
|---|---|---|---|---|
| -0,213768898 | 0,702031668 | MRPL54 | FALSE | Downregulated |
| -0,345407516 | 0,702031668 | RPL13AP5 | FALSE | Downregulated |
| 0,172792335 | 0,702549839 | C7orf49 | FALSE | Upregulated |
| -0,155622587 | 0,702632856 | AP2A2 | FALSE | Downregulated |
| -0,16238323 | 0,702632856 | CROCCP2 | FALSE | Downregulated |
| -0,167493266 | 0,703024287 | BCAS3 | FALSE | Downregulated |
| 0,429711539 | 0,703024287 | C3orf65 | FALSE | Upregulated |
| -0,161352169 | 0,70306089 | AFM | FALSE | Downregulated |
| -0,334490654 | 0,70306089 | ANTXR1 | FALSE | Downregulated |
| -0,270033335 | 0,70306089 | CLK4 | FALSE | Downregulated |
| 0,319226425 | 0,70306089 | CRISPLD2 | FALSE | Upregulated |
| -0,170991337 | 0,70306089 | DDRGK1 | FALSE | Downregulated |
| -0,181952668 | 0,70306089 | DPP7 | FALSE | Downregulated |
| -0,20873979 | 0,70306089 | GDA | FALSE | Downregulated |
| -0,336416463 | 0,70306089 | IL1RN | FALSE | Downregulated |
| 0,234400019 | 0,70306089 | IQCG | FALSE | Upregulated |
| 0,10224295 | 0,70306089 | LOC100190986 | FALSE | Upregulated |
| 0,242108588 | 0,70306089 | LOC101928576 | FALSE | Upregulated |
| 0,292649993 | 0,70306089 | LOC102723369 | FALSE | Upregulated |
| 0,234609074 | 0,70306089 | MCRS1 | FALSE | Upregulated |
| -0,312726109 | 0,70306089 | METTL2B | FALSE | Downregulated |
| -0,386106883 | 0,70306089 | ODF2L | FALSE | Downregulated |
| 0,451060844 | 0,70306089 | PDGFRL | FALSE | Upregulated |
| 0,161552984 | 0,70306089 | PITRM1-AS1 | FALSE | Upregulated |
| 0,236359459 | 0,70306089 | RNR1 | FALSE | Upregulated |
| -0,186511994 | 0,70306089 | THSD4 | FALSE | Downregulated |
| -0,177187519 | 0,70306089 | TRIB1 | FALSE | Downregulated |
| -0,392941985 | 0,70306089 | TTC18 | FALSE | Downregulated |
| 0,315423786 | 0,70306089 | UBE2V2 | FALSE | Upregulated |
| -0,137753695 | 0,70306089 | ZNF862 | FALSE | Downregulated |
| -0,207588304 | 0,703156254 | AARS2 | FALSE | Downregulated |
| -0,177703507 | 0,703156254 | AGFG1 | FALSE | Downregulated |
| -0,304597455 | 0,703156254 | COPRS | FALSE | Downregulated |
| -0,204522166 | 0,703156254 | PIAS4 | FALSE | Downregulated |
| 0,109954767 | 0,703156254 | RBBP4 | FALSE | Upregulated |
| 0,142627686 | 0,703156254 | SRA1 | FALSE | Upregulated |
| 0,371598558 | 0,703239555 | C5AR1 | FALSE | Upregulated |
| 0,143178022 | 0,703239555 | CRNKL1 | FALSE | Upregulated |
| -0,121004969 | 0,703239555 | CTNNB1 | FALSE | Downregulated |
| 0,140886751 | 0,703239555 | EPB41L2 | FALSE | Upregulated |
| 0,297303276 | 0,703239555 | GPR128 | FALSE | Upregulated |
| 0,301086644 | 0,703239555 | POP4 | FALSE | Upregulated |
| 0,227820836 | 0,703239555 | PRKAG3 | FALSE | Upregulated |
| -0,200242205 | 0,703239555 | SLC9B2 | FALSE | Downregulated |
| -0,14332251 | 0,703588396 | ATP13A3 | FALSE | Downregulated |
| -0,483794341 | 0,703718147 | AP000479.1 | FALSE | Downregulated |
| 0,126515599 | 0,703718147 | SLC26A1 | FALSE | Upregulated |
| 0,087749819 | 0,704130151 | ANKFY1 | FALSE | Upregulated |
| 0,142885444 | 0,704130151 | FLJ31306 | FALSE | Upregulated |
| 0,252181628 | 0,704130151 | LPCAT1 | FALSE | Upregulated |
| 0,396081545 | 0,704130151 | MAPK11 | FALSE | Upregulated |

| | | | | |
|---|---|---|---|---|
| -0,147186977 | 0,704130151 | RNF185 | FALSE | Downregulated |
| 0,237197761 | 0,704130151 | ZNF514 | FALSE | Upregulated |
| 0,130658439 | 0,704150699 | RNF19A | FALSE | Upregulated |
| 0,283969477 | 0,704402762 | SNHG3 | FALSE | Upregulated |
| 0,341228199 | 0,704702566 | HEPHL1 | FALSE | Upregulated |
| -0,38597258 | 0,70479548 | DACT1 | FALSE | Downregulated |
| 0,138980911 | 0,70479548 | FXR1 | FALSE | Upregulated |
| 0,164094885 | 0,705341979 | DGKZ | FALSE | Upregulated |
| 0,340028833 | 0,70548481 | FCGR2A | FALSE | Upregulated |
| -0,197277717 | 0,70548481 | LOC102724281 | FALSE | Downregulated |
| 0,131154224 | 0,70548481 | SPRYD4 | FALSE | Upregulated |
| -0,235531326 | 0,705609337 | CYP4F12 | FALSE | Downregulated |
| -0,167550921 | 0,705609337 | SLC41A2 | FALSE | Downregulated |
| -0,179169224 | 0,705775731 | FBXO17 | FALSE | Downregulated |
| -0,287766536 | 0,705775731 | SF3B5 | FALSE | Downregulated |
| -0,148269016 | 0,705859887 | AMOTL1 | FALSE | Downregulated |
| 0,266921064 | 0,705859887 | NECAP1 | FALSE | Upregulated |
| 0,07897819 | 0,705880947 | MALAT1 | FALSE | Upregulated |
| -0,140055525 | 0,705880947 | NFYC | FALSE | Downregulated |
| 0,243225182 | 0,705960022 | UNC5CL | FALSE | Upregulated |
| -0,121700706 | 0,706127835 | COMT | FALSE | Downregulated |
| 0,140765436 | 0,706374995 | CTC1 | FALSE | Upregulated |
| -0,402034237 | 0,706374995 | GTF2A2 | FALSE | Downregulated |
| -0,195819295 | 0,706374995 | MRP63 | FALSE | Downregulated |
| 0,157821658 | 0,706440091 | DPYSL2 | FALSE | Upregulated |
| 0,390392217 | 0,706440091 | LOC101927730 | FALSE | Upregulated |
| 0,237458024 | 0,706527674 | ATL3 | FALSE | Upregulated |
| 0,237542018 | 0,706527674 | LOC101927420 | FALSE | Upregulated |
| 0,144056666 | 0,706527674 | SENP5 | FALSE | Upregulated |
| 0,26135787 | 0,706527674 | ZDHHC17 | FALSE | Upregulated |
| -0,223439575 | 0,706606972 | CCT2 | FALSE | Downregulated |
| -0,488537775 | 0,706606972 | GALNT15 | FALSE | Downregulated |
| 0,237943648 | 0,706606972 | LOC100287042 | FALSE | Upregulated |
| -0,185804714 | 0,706606972 | MAPK1 | FALSE | Downregulated |
| -0,631023774 | 0,706606972 | SMN1 | FALSE | Downregulated |
| 0,135420775 | 0,706778382 | LRCH4 | FALSE | Upregulated |
| -0,260757027 | 0,706793705 | BAG3 | FALSE | Downregulated |
| 0,170768956 | 0,707181189 | INPP4A | FALSE | Upregulated |
| -0,171527476 | 0,707218887 | USP10 | FALSE | Downregulated |
| -0,213004781 | 0,707259091 | ATP5O | FALSE | Downregulated |
| -0,192383877 | 0,707706401 | NOL9 | FALSE | Downregulated |
| -0,134577435 | 0,707812569 | DICER1 | FALSE | Downregulated |
| -0,310529104 | 0,707812569 | LACTB2 | FALSE | Downregulated |
| 0,217856114 | 0,707851388 | C16orf71 | FALSE | Upregulated |
| 0,21402682 | 0,708138784 | CHMP6 | FALSE | Upregulated |
| 0,214289361 | 0,708138784 | SIDT1 | FALSE | Upregulated |
| -0,335656683 | 0,708327865 | FAM175B | FALSE | Downregulated |
| 0,271346184 | 0,708579747 | ARRDC4 | FALSE | Upregulated |
| 0,14467391 | 0,708579747 | KDM2B | FALSE | Upregulated |
| -0,198442054 | 0,708579747 | RNF14 | FALSE | Downregulated |
| -0,436043698 | 0,708579747 | SCARA5 | FALSE | Downregulated |

| | | | | |
|---|---|---|---|---|
| 0,312490256 | 0,708658327 | SLC9A9 | FALSE | Upregulated |
| -0,162458815 | 0,708768186 | UGT2B15 | FALSE | Downregulated |
| -0,225924268 | 0,708883281 | CYHR1 | FALSE | Downregulated |
| -0,155115903 | 0,70926367 | ANXA4 | FALSE | Downregulated |
| -0,132464312 | 0,709285047 | GPR126 | FALSE | Downregulated |
| -0,182787914 | 0,709529801 | PPIG | FALSE | Downregulated |
| 0,27917495 | 0,709562801 | FZD6 | FALSE | Upregulated |
| 0,272751751 | 0,709958361 | LOC101060351 | FALSE | Upregulated |
| 0,158304437 | 0,710137402 | ABHD16B | FALSE | Upregulated |
| -0,203389254 | 0,710137402 | ELP3 | FALSE | Downregulated |
| -0,18626986 | 0,710137402 | MARK4 | FALSE | Downregulated |
| -0,226410264 | 0,710137402 | MYNN | FALSE | Downregulated |
| 0,195505773 | 0,710137402 | ZNF692 | FALSE | Upregulated |
| -0,231846871 | 0,710212908 | SMDT1 | FALSE | Downregulated |
| -0,188558391 | 0,710261139 | ENTPD6 | FALSE | Downregulated |
| 0,156581479 | 0,710261139 | LOC101928120 | FALSE | Upregulated |
| 0,200475044 | 0,710545152 | RAB18 | FALSE | Upregulated |
| -0,123238665 | 0,710637708 | PNN | FALSE | Downregulated |
| 0,188643118 | 0,710675089 | LOC102724965 | FALSE | Upregulated |
| -0,139167713 | 0,710836676 | GCLC | FALSE | Downregulated |
| -0,269164238 | 0,710836676 | MRGBP | FALSE | Downregulated |
| -0,248660896 | 0,710836676 | TMCO4 | FALSE | Downregulated |
| 0,191342098 | 0,711055968 | HIGD1A | FALSE | Upregulated |
| -0,347236549 | 0,711101994 | UAP1L1 | FALSE | Downregulated |
| 0,120143197 | 0,711194896 | CDK4 | FALSE | Upregulated |
| -0,154577285 | 0,711230556 | ATP9A | FALSE | Downregulated |
| 0,627280162 | 0,711361197 | LOC100093631 | FALSE | Upregulated |
| 0,126812779 | 0,711589463 | DAG1 | FALSE | Upregulated |
| -0,13521342 | 0,711589463 | HSPG2 | FALSE | Downregulated |
| -0,259498977 | 0,711589463 | MRPL24 | FALSE | Downregulated |
| -0,192366074 | 0,711589463 | PIK3CA | FALSE | Downregulated |
| -0,133039372 | 0,711589463 | STK40 | FALSE | Downregulated |
| -0,121906222 | 0,711589463 | SYNE1 | FALSE | Downregulated |
| 0,183718876 | 0,711609654 | ABHD12B | FALSE | Upregulated |
| 0,232408738 | 0,711609654 | ACSS2 | FALSE | Upregulated |
| 0,281289962 | 0,711609654 | SLCO3A1 | FALSE | Upregulated |
| -0,325786 | 0,711609654 | TCN2 | FALSE | Downregulated |
| 0,147131649 | 0,711609654 | TRAPPC9 | FALSE | Upregulated |
| -0,273658659 | 0,711614413 | HARS2 | FALSE | Downregulated |
| 0,142364913 | 0,711906855 | LIMD1-AS1 | FALSE | Upregulated |
| 0,249875137 | 0,712034943 | ELP5 | FALSE | Upregulated |
| 0,128275931 | 0,712034943 | PAN3 | FALSE | Upregulated |
| 0,28136572 | 0,712034943 | RNF138 | FALSE | Upregulated |
| 0,165912023 | 0,712065981 | PARP9 | FALSE | Upregulated |
| -0,134333955 | 0,712579765 | SF3B3 | FALSE | Downregulated |
| 0,270432977 | 0,712707701 | MEMO1 | FALSE | Upregulated |
| -0,276729169 | 0,712707701 | POLR3GL | FALSE | Downregulated |
| 0,335570419 | 0,712707701 | TRPV2 | FALSE | Upregulated |
| 0,242086568 | 0,712785599 | EPX | FALSE | Upregulated |
| 0,185366861 | 0,712948174 | KIAA0226 | FALSE | Upregulated |
| -0,324780844 | 0,712948174 | LOC101929195 | FALSE | Downregulated |

| | | | | |
|---|---|---|---|---|
| 0,219176688 | 0,712948174 | SLC43A2 | FALSE | Upregulated |
| -0,208888508 | 0,713125222 | EXOC6B | FALSE | Downregulated |
| 0,290065268 | 0,713125222 | MSL3 | FALSE | Upregulated |
| -0,187933765 | 0,713125222 | PRDX4 | FALSE | Downregulated |
| -0,241811385 | 0,713125222 | SYCE2 | FALSE | Downregulated |
| 0,154301303 | 0,713125222 | ZNF506 | FALSE | Upregulated |
| -0,104001324 | 0,713213157 | BAZ2A | FALSE | Downregulated |
| 0,145989552 | 0,713325554 | LUZP1 | FALSE | Upregulated |
| -0,136528609 | 0,713325554 | TST | FALSE | Downregulated |
| 0,285764108 | 0,71389648 | WHAMMP2 | FALSE | Upregulated |
| -0,18987157 | 0,714252239 | CCS | FALSE | Downregulated |
| -0,377663616 | 0,714252239 | FAM174B | FALSE | Downregulated |
| 0,189599622 | 0,714252239 | PDE12 | FALSE | Upregulated |
| -0,306884931 | 0,7143654 | AP1S1 | FALSE | Downregulated |
| -0,322869345 | 0,7143654 | ENY2 | FALSE | Downregulated |
| 0,231814655 | 0,7143654 | LINC00954 | FALSE | Upregulated |
| 0,262725564 | 0,714456735 | ATHL1 | FALSE | Upregulated |
| 0,165913153 | 0,714456735 | CDKL1 | FALSE | Upregulated |
| 0,190508778 | 0,714456735 | CYP2R1 | FALSE | Upregulated |
| -0,241812189 | 0,714456735 | ERCC3 | FALSE | Downregulated |
| 0,352968414 | 0,714456735 | LOC100289455 | FALSE | Upregulated |
| 0,240264057 | 0,714456735 | LOC101928188 | FALSE | Upregulated |
| 0,257383109 | 0,714456735 | MOB1A | FALSE | Upregulated |
| -0,327591963 | 0,714456735 | SERGEF | FALSE | Downregulated |
| -0,323154822 | 0,714456735 | ZFAT | FALSE | Downregulated |
| 0,258507337 | 0,714556608 | LOC101928998 | FALSE | Upregulated |
| 0,654454942 | 0,714560685 | DDX11L1 | FALSE | Upregulated |
| -0,319611487 | 0,714560685 | MTFMT | FALSE | Downregulated |
| 0,49522406 | 0,714649053 | NAV3 | FALSE | Upregulated |
| 0,336193174 | 0,714649053 | ZNF543 | FALSE | Upregulated |
| -0,1428701 | 0,714649514 | FAM115A | FALSE | Downregulated |
| -0,512609585 | 0,714649514 | LINC00623 | FALSE | Downregulated |
| -0,299985313 | 0,714649514 | LMAN2L | FALSE | Downregulated |
| 0,314472768 | 0,714649514 | NHSL2 | FALSE | Upregulated |
| -0,093458287 | 0,714854449 | ASGR2 | FALSE | Downregulated |
| 0,396231029 | 0,714930039 | RP1-257A7.4 | FALSE | Upregulated |
| -0,128876592 | 0,714930039 | SETD1A | FALSE | Downregulated |
| 0,288983538 | 0,715133861 | LRP8 | FALSE | Upregulated |
| -0,309296238 | 0,715218116 | POMK | FALSE | Downregulated |
| -0,282014648 | 0,715424306 | MT1X | FALSE | Downregulated |
| 0,226563006 | 0,715424306 | NBPF12 | FALSE | Upregulated |
| 0,228582723 | 0,715711543 | COL4A1 | FALSE | Upregulated |
| 0,164478355 | 0,7158853 | TARS | FALSE | Upregulated |
| 0,212686509 | 0,716037278 | LOC101928185 | FALSE | Upregulated |
| -0,373249849 | 0,716339434 | CAPN10-AS1 | FALSE | Downregulated |
| -0,177614675 | 0,716339434 | CCDC117 | FALSE | Downregulated |
| 0,535051693 | 0,716339434 | LOC101928182 | FALSE | Upregulated |
| 0,193059088 | 0,716339434 | SLC1A4 | FALSE | Upregulated |
| 0,235134343 | 0,717034882 | CCDC71 | FALSE | Upregulated |
| 0,235365805 | 0,717034882 | SCTR | FALSE | Upregulated |
| 0,358164714 | 0,717231306 | ENKD1 | FALSE | Upregulated |

| | | | | |
|---|---|---|---|---|
| 0,466170585 | 0,717231306 | LOC101928628 | FALSE | Upregulated |
| -0,245116742 | 0,717231306 | NARFL | FALSE | Downregulated |
| 0,177132746 | 0,717231306 | TAPBPL | FALSE | Upregulated |
| 0,215729978 | 0,71724435 | LATS1 | FALSE | Upregulated |
| 0,318163213 | 0,717425992 | C17orf59 | FALSE | Upregulated |
| -0,115123026 | 0,717425992 | SRSF5 | FALSE | Downregulated |
| 0,288935849 | 0,717425992 | TMEM242 | FALSE | Upregulated |
| -0,265436907 | 0,717425992 | ZBTB41 | FALSE | Downregulated |
| 0,355788432 | 0,717458913 | C16orf52 | FALSE | Upregulated |
| -0,116972535 | 0,717458913 | C2 | FALSE | Downregulated |
| 0,149004899 | 0,717592627 | C9orf69 | FALSE | Upregulated |
| 0,186434168 | 0,717840701 | TLE3 | FALSE | Upregulated |
| 0,24015165 | 0,718011456 | LOC101928604 | FALSE | Upregulated |
| -0,181961948 | 0,718487482 | ANXA7 | FALSE | Downregulated |
| 0,132006681 | 0,718502093 | WTAP | FALSE | Upregulated |
| -0,100769059 | 0,718745291 | PDE8A | FALSE | Downregulated |
| 0,117657019 | 0,718745291 | PITPNA | FALSE | Upregulated |
| 0,258150965 | 0,718745291 | PPIL4 | FALSE | Upregulated |
| 0,293257419 | 0,718745291 | TRAF3IP1 | FALSE | Upregulated |
| -0,228932509 | 0,718928098 | KIAA0196 | FALSE | Downregulated |
| 0,254553463 | 0,718928098 | MED11 | FALSE | Upregulated |
| 0,223654584 | 0,718928098 | NEURL1 | FALSE | Upregulated |
| -0,140016024 | 0,718928098 | RGN | FALSE | Downregulated |
| -0,169286902 | 0,718928098 | SEPHS1 | FALSE | Downregulated |
| -0,14850397 | 0,718928098 | SMTN | FALSE | Downregulated |
| -0,218944155 | 0,719040349 | AIP | FALSE | Downregulated |
| -0,17147905 | 0,719040349 | ELFN1 | FALSE | Downregulated |
| 0,216949492 | 0,719244959 | LOC100506100 | FALSE | Upregulated |
| -0,337240921 | 0,719244959 | MLLT10P1 | FALSE | Downregulated |
| -0,13164332 | 0,719244959 | MTUS1 | FALSE | Downregulated |
| -0,176727705 | 0,719353537 | PRKAB1 | FALSE | Downregulated |
| 0,166868252 | 0,719353537 | RAPGEF3 | FALSE | Upregulated |
| -0,298652601 | 0,719519122 | CETP | FALSE | Downregulated |
| -0,177039693 | 0,719519122 | GALE | FALSE | Downregulated |
| -0,196422364 | 0,719690044 | ANKS4B | FALSE | Downregulated |
| -0,133277684 | 0,719690044 | ASL | FALSE | Downregulated |
| -0,132370444 | 0,719690044 | EIF2AK4 | FALSE | Downregulated |
| 0,153613924 | 0,719690044 | FOXN3-AS2 | FALSE | Upregulated |
| -0,177863455 | 0,719690044 | GATAD1 | FALSE | Downregulated |
| -0,293915368 | 0,719690044 | GLRA1 | FALSE | Downregulated |
| -0,342114539 | 0,719690044 | LOC101928139 | FALSE | Downregulated |
| -0,275255955 | 0,719690044 | MTMR6 | FALSE | Downregulated |
| -0,124902309 | 0,719690044 | PACS2 | FALSE | Downregulated |
| 0,307653762 | 0,719690044 | PAX8-AS1 | FALSE | Upregulated |
| -0,208545305 | 0,719690044 | TRAPPC5 | FALSE | Downregulated |
| -0,371854341 | 0,719690044 | USP18 | FALSE | Downregulated |
| -0,285206968 | 0,719832931 | CLCN4 | FALSE | Downregulated |
| -0,141985279 | 0,720077908 | BZRAP1 | FALSE | Downregulated |
| -0,158685995 | 0,720294351 | ANAPC16 | FALSE | Downregulated |
| -0,148372808 | 0,720294351 | NF1 | FALSE | Downregulated |
| -0,173358746 | 0,720294351 | RIN2 | FALSE | Downregulated |

| | | | | |
|---|---|---|---|---|
| 0,323789456 | 0,720294351 | TAF8 | FALSE | Upregulated |
| 0,110489629 | 0,720294351 | TXLNA | FALSE | Upregulated |
| 0,166106632 | 0,720294351 | TYW1 | FALSE | Upregulated |
| -0,112623299 | 0,720294351 | ZNF316 | FALSE | Downregulated |
| 0,325818667 | 0,720294351 | ZNF486 | FALSE | Upregulated |
| -0,304518231 | 0,720401254 | ARHGEF35 | FALSE | Downregulated |
| -0,270451079 | 0,720401254 | NPR2 | FALSE | Downregulated |
| 0,170601664 | 0,720510337 | METTL3 | FALSE | Upregulated |
| -0,208857159 | 0,720710279 | GM2A | FALSE | Downregulated |
| 0,330604474 | 0,720710279 | ZNF565 | FALSE | Upregulated |
| -0,118653429 | 0,721040196 | DDB1 | FALSE | Downregulated |
| 0,241564137 | 0,72108192 | C2CD4D | FALSE | Upregulated |
| -0,199574657 | 0,72117598 | HSD17B11 | FALSE | Downregulated |
| -0,278967532 | 0,721439165 | SLC25A45 | FALSE | Downregulated |
| -0,578600408 | 0,721439165 | SYN3 | FALSE | Downregulated |
| -0,116331793 | 0,721439165 | TNFAIP8L1 | FALSE | Downregulated |
| 0,230835686 | 0,721578499 | AC004019.13 | FALSE | Upregulated |
| -0,135451639 | 0,721578499 | AKAP12 | FALSE | Downregulated |
| -0,400050904 | 0,721578499 | ALG10B | FALSE | Downregulated |
| -0,275934458 | 0,721578499 | APTX | FALSE | Downregulated |
| 0,270356943 | 0,721578499 | ATG9B | FALSE | Upregulated |
| 0,242548763 | 0,721578499 | C12orf29 | FALSE | Upregulated |
| 0,262436523 | 0,721578499 | C3orf83 | FALSE | Upregulated |
| 0,236277765 | 0,721578499 | COL6A2 | FALSE | Upregulated |
| 0,1335311 | 0,721578499 | DCP2 | FALSE | Upregulated |
| -0,214197256 | 0,721578499 | FAM198A | FALSE | Downregulated |
| -0,260455758 | 0,721578499 | GPR180 | FALSE | Downregulated |
| -0,126145589 | 0,721578499 | KHK | FALSE | Downregulated |
| -0,240763314 | 0,721578499 | MGC27345 | FALSE | Downregulated |
| -0,184149745 | 0,721578499 | NELFB | FALSE | Downregulated |
| -0,291892492 | 0,721578499 | PLEKHF2 | FALSE | Downregulated |
| -0,138134098 | 0,721578499 | PLS3 | FALSE | Downregulated |
| 0,116499514 | 0,721578499 | PNRC1 | FALSE | Upregulated |
| 0,163793951 | 0,721578499 | RGL1 | FALSE | Upregulated |
| 0,15154093 | 0,721578499 | RNF170 | FALSE | Upregulated |
| 0,204546934 | 0,721578499 | RP11-4F22.2 | FALSE | Upregulated |
| -0,143823373 | 0,721578499 | SPPL2B | FALSE | Downregulated |
| -0,248835951 | 0,721578499 | TBK1 | FALSE | Downregulated |
| 0,161927716 | 0,721578499 | TEP1 | FALSE | Upregulated |
| 0,474657829 | 0,721578499 | TNS4 | FALSE | Upregulated |
| -0,305741611 | 0,721578499 | TRAPPC6A | FALSE | Downregulated |
| -0,075984688 | 0,721578499 | ZSWIM8 | FALSE | Downregulated |
| 0,190564519 | 0,721679887 | PLEKHH1 | FALSE | Upregulated |
| -0,314918925 | 0,722184132 | DLEU2 | FALSE | Downregulated |
| -0,18974528 | 0,722191723 | KDM7A | FALSE | Downregulated |
| 0,125449262 | 0,722191723 | SPNS2 | FALSE | Upregulated |
| 0,181534727 | 0,722258691 | CCDC73 | FALSE | Upregulated |
| 0,164812673 | 0,722258691 | DMWD | FALSE | Upregulated |
| 0,344052269 | 0,722323753 | MCAT | FALSE | Upregulated |
| 0,446456525 | 0,722334868 | UPK1B | FALSE | Upregulated |
| -0,166371436 | 0,723627606 | BAHD1 | FALSE | Downregulated |

| | | | | |
|---|---|---|---|---|
| 0,126716707 | 0,723627606 | FAM120AOS | FALSE | Upregulated |
| 0,167161533 | 0,723627606 | GTPBP2 | FALSE | Upregulated |
| 0,233015252 | 0,723627606 | LOC101929270 | FALSE | Upregulated |
| -0,142013246 | 0,723627606 | PPP2R2D | FALSE | Downregulated |
| -0,238019181 | 0,723847618 | BCKDHB | FALSE | Downregulated |
| 0,140825149 | 0,723847618 | MRPS26 | FALSE | Upregulated |
| 0,321705824 | 0,724199444 | DNAJC2 | FALSE | Upregulated |
| -0,196345867 | 0,724199444 | ENOSF1 | FALSE | Downregulated |
| 0,159170652 | 0,724199444 | MRPL37 | FALSE | Upregulated |
| 0,076834757 | 0,724199444 | NCOA4 | FALSE | Upregulated |
| 0,206825743 | 0,724199444 | PICK1 | FALSE | Upregulated |
| 0,18849399 | 0,724199444 | RAB6B | FALSE | Upregulated |
| 0,309586768 | 0,724199444 | VEPH1 | FALSE | Upregulated |
| 0,334187163 | 0,724199444 | ZNF205-AS1 | FALSE | Upregulated |
| -0,218016173 | 0,724350865 | GIPC2 | FALSE | Downregulated |
| 0,20933555 | 0,724350865 | NEK11 | FALSE | Upregulated |
| 0,132682788 | 0,724350865 | PRPF40A | FALSE | Upregulated |
| -0,126368346 | 0,724350865 | SCARB1 | FALSE | Downregulated |
| -0,100918868 | 0,724350865 | SETD1B | FALSE | Downregulated |
| -0,098601309 | 0,72435312 | AKAP9 | FALSE | Downregulated |
| 0,169335683 | 0,724513645 | SNAPC3 | FALSE | Upregulated |
| 0,223890159 | 0,724513645 | TMEM45B | FALSE | Upregulated |
| -0,094918412 | 0,72487844 | MRFAP1 | FALSE | Downregulated |
| 0,395546217 | 0,725606422 | SRI | FALSE | Upregulated |
| -0,134993286 | 0,725624346 | GOLPH3 | FALSE | Downregulated |
| -0,165965354 | 0,725717441 | ZNF33B | FALSE | Downregulated |
| -0,192546386 | 0,725829193 | FRS2 | FALSE | Downregulated |
| -0,346001863 | 0,725829193 | LOC100288102 | FALSE | Downregulated |
| 0,177619413 | 0,725839385 | NAIP | FALSE | Upregulated |
| 0,270743608 | 0,726311076 | LOC100132731 | FALSE | Upregulated |
| -0,168785007 | 0,726703489 | ADHFE1 | FALSE | Downregulated |
| 0,193437992 | 0,726703489 | COL3A1 | FALSE | Upregulated |
| 0,37615595 | 0,726703489 | GIMAP7 | FALSE | Upregulated |
| 0,175928149 | 0,726703489 | KIAA1731 | FALSE | Upregulated |
| -0,34346441 | 0,726703489 | LOC101928762 | FALSE | Downregulated |
| 0,153100733 | 0,726703489 | LRRC14 | FALSE | Upregulated |
| -0,250473508 | 0,726703489 | NAA10 | FALSE | Downregulated |
| 0,208855731 | 0,726703489 | NUDT16L1 | FALSE | Upregulated |
| 0,303208055 | 0,726703489 | RIC8B | FALSE | Upregulated |
| 0,154192666 | 0,726703489 | SNN | FALSE | Upregulated |
| -0,14987649 | 0,726703489 | URB1 | FALSE | Downregulated |
| -0,241927582 | 0,726703489 | ZNF420 | FALSE | Downregulated |
| -0,195180418 | 0,726703489 | ZNF746 | FALSE | Downregulated |
| 0,156252304 | 0,727042191 | PTP4A2 | FALSE | Upregulated |
| 0,284752933 | 0,727101839 | GRIP2 | FALSE | Upregulated |
| -0,320864676 | 0,727101839 | SGCB | FALSE | Downregulated |
| 0,132384203 | 0,727122365 | APOL6 | FALSE | Upregulated |
| -0,312561439 | 0,727122365 | CES4A | FALSE | Downregulated |
| -0,180019638 | 0,727122365 | FOXA3 | FALSE | Downregulated |
| 0,335543239 | 0,727122365 | FRMD3 | FALSE | Upregulated |
| 0,295823179 | 0,727122365 | GUCY1A3 | FALSE | Upregulated |

| | | | | |
|---|---|---|---|---|
| -0,177858083 | 0,727122365 | HIST1H2AC | FALSE | Downregulated |
| -0,247692932 | 0,727122365 | IFITM10 | FALSE | Downregulated |
| 0,198251306 | 0,727122365 | KRT8 | FALSE | Upregulated |
| 0,34244783 | 0,727122365 | LINC00282 | FALSE | Upregulated |
| 0,165692047 | 0,727122365 | LOC101926920 | FALSE | Upregulated |
| 0,193293668 | 0,727122365 | LOC400553 | FALSE | Upregulated |
| -0,226246463 | 0,727122365 | NOS3 | FALSE | Downregulated |
| -0,084834338 | 0,727122365 | RBL2 | FALSE | Downregulated |
| 0,096110534 | 0,727373865 | AIM1 | FALSE | Upregulated |
| 0,265563646 | 0,727373865 | JARID2-AS1 | FALSE | Upregulated |
| -0,136584734 | 0,727373865 | RHOT2 | FALSE | Downregulated |
| -0,252516816 | 0,727373865 | TDO2 | FALSE | Downregulated |
| -0,216195307 | 0,727373865 | UBE2N | FALSE | Downregulated |
| 0,491181995 | 0,727387953 | DISC1 | FALSE | Upregulated |
| 0,10664834 | 0,727387953 | VPS13A | FALSE | Upregulated |
| -0,332687444 | 0,727828043 | C5orf51 | FALSE | Downregulated |
| -0,190716554 | 0,728284373 | FAM211B | FALSE | Downregulated |
| 0,155609912 | 0,728284373 | SLC25A25 | FALSE | Upregulated |
| 0,12690964 | 0,728352005 | GRB2 | FALSE | Upregulated |
| -0,335932694 | 0,728372678 | RNF215 | FALSE | Downregulated |
| 0,292864171 | 0,728478938 | ATOH8 | FALSE | Upregulated |
| -0,209774745 | 0,728478938 | GLOD4 | FALSE | Downregulated |
| -0,346385273 | 0,728478938 | SARNP | FALSE | Downregulated |
| 0,175735781 | 0,728478938 | TNFAIP3 | FALSE | Upregulated |
| -0,276841935 | 0,728606368 | AP5S1 | FALSE | Downregulated |
| -0,144988009 | 0,728606368 | MCCC1 | FALSE | Downregulated |
| -0,165329879 | 0,728722928 | FERMT2 | FALSE | Downregulated |
| 0,146733768 | 0,728722928 | GNPDA1 | FALSE | Upregulated |
| 0,210767203 | 0,728722928 | MAPRE1 | FALSE | Upregulated |
| -0,145712728 | 0,728722928 | RIPK1 | FALSE | Downregulated |
| -0,166928056 | 0,729334953 | KBTBD2 | FALSE | Downregulated |
| -0,191220163 | 0,729334953 | MST1P2 | FALSE | Downregulated |
| -0,227249954 | 0,729334953 | NLN | FALSE | Downregulated |
| -0,1839666 | 0,729334953 | TAF5L | FALSE | Downregulated |
| 0,35028086 | 0,729334953 | TGFB1I1 | FALSE | Upregulated |
| 0,247629028 | 0,729645479 | ATP10D | FALSE | Upregulated |
| 0,304289313 | 0,729645479 | DTYMK | FALSE | Upregulated |
| -0,213943822 | 0,729645479 | PLIN2 | FALSE | Downregulated |
| 0,221187041 | 0,729645479 | RFXANK | FALSE | Upregulated |
| 0,214203807 | 0,729645479 | RP11-85B7.2 | FALSE | Upregulated |
| 0,154711157 | 0,729645479 | SBF2-AS1 | FALSE | Upregulated |
| -0,148184273 | 0,730117415 | NDUFB8 | FALSE | Downregulated |
| 0,145352342 | 0,730185592 | NOTCH2NL | FALSE | Upregulated |
| -0,150632855 | 0,730185592 | RALGAPA1 | FALSE | Downregulated |
| 0,3367275 | 0,73029123 | KCNK6 | FALSE | Upregulated |
| 0,125315697 | 0,73029123 | PIK3AP1 | FALSE | Upregulated |
| 0,179507812 | 0,73029123 | SMAD9 | FALSE | Upregulated |
| 0,090094419 | 0,73029123 | TBC1D8 | FALSE | Upregulated |
| 0,337007522 | 0,73029123 | TLCD2 | FALSE | Upregulated |
| -0,191562286 | 0,730350533 | BABAM1 | FALSE | Downregulated |
| 0,358379515 | 0,730350533 | CCZ1 | FALSE | Upregulated |

| | | | | |
|---|---|---|---|---|
| -0,263997873 | 0,730350533 | COL8A2 | FALSE | Downregulated |
| -0,138314 | 0,730350533 | IFNAR1 | FALSE | Downregulated |
| 0,195624656 | 0,730350533 | PARD6G | FALSE | Upregulated |
| 0,172187359 | 0,730350533 | PIGB | FALSE | Upregulated |
| -0,274509346 | 0,730350533 | TBX15 | FALSE | Downregulated |
| 0,269374964 | 0,730428993 | GPBAR1 | FALSE | Upregulated |
| 0,98669627 | 0,730428993 | HNRNPH2 | FALSE | Upregulated |
| 0,241474239 | 0,730428993 | LPP-AS1 | FALSE | Upregulated |
| 0,322263755 | 0,730486651 | ADORA2B | FALSE | Upregulated |
| 0,241562968 | 0,730486651 | CYB5R2 | FALSE | Upregulated |
| 0,510451213 | 0,730486651 | HIST1H3D | FALSE | Upregulated |
| -0,122951412 | 0,731356972 | UGP2 | FALSE | Downregulated |
| -0,195099086 | 0,731548415 | APBA1 | FALSE | Downregulated |
| -0,361015546 | 0,731548415 | MICU2 | FALSE | Downregulated |
| -0,133308162 | 0,731548415 | SNTB1 | FALSE | Downregulated |
| -0,115774981 | 0,73162038 | SCAF8 | FALSE | Downregulated |
| -0,37248466 | 0,731774506 | RNF180 | FALSE | Downregulated |
| 0,228638256 | 0,731787871 | RP11-15A1.7 | FALSE | Upregulated |
| 0,11968572 | 0,731895292 | ACTN1 | FALSE | Upregulated |
| -0,187892827 | 0,731895292 | BECN1 | FALSE | Downregulated |
| -0,190205353 | 0,731895292 | CCDC14 | FALSE | Downregulated |
| 0,143177393 | 0,731895292 | COG1 | FALSE | Upregulated |
| 0,180794122 | 0,731895292 | DUSP28 | FALSE | Upregulated |
| -0,189582744 | 0,731895292 | GRAMD4 | FALSE | Downregulated |
| 0,269543163 | 0,731895292 | INCENP | FALSE | Upregulated |
| -0,319506303 | 0,731895292 | LGSN | FALSE | Downregulated |
| -0,212356817 | 0,731895292 | LYRM1 | FALSE | Downregulated |
| -0,215132748 | 0,731895292 | MTM1 | FALSE | Downregulated |
| -0,769426935 | 0,731895292 | SENP3 | FALSE | Downregulated |
| 0,321517861 | 0,731895292 | SRD5A3 | FALSE | Upregulated |
| -0,303267291 | 0,731895292 | TMEM160 | FALSE | Downregulated |
| -0,101826175 | 0,731895292 | TMEM248 | FALSE | Downregulated |
| 0,231658457 | 0,731895292 | TRNAU1AP | FALSE | Upregulated |
| 0,173274015 | 0,731895292 | ZNF227 | FALSE | Upregulated |
| -0,333882533 | 0,732145613 | METTL20 | FALSE | Downregulated |
| 0,371447688 | 0,732145613 | PI4KAP1 | FALSE | Upregulated |
| 0,170530136 | 0,732189952 | VIM | FALSE | Upregulated |
| -0,321355001 | 0,732410054 | BLZF1 | FALSE | Downregulated |
| -0,214862522 | 0,732545438 | AASS | FALSE | Downregulated |
| 0,174910383 | 0,732545438 | ACP5 | FALSE | Upregulated |
| -0,246158525 | 0,732545438 | LOC150776 | FALSE | Downregulated |
| 0,207139384 | 0,732545438 | NARF | FALSE | Upregulated |
| 0,211661589 | 0,732545438 | TTL | FALSE | Upregulated |
| -0,102084739 | 0,73272001 | NCOA2 | FALSE | Downregulated |
| 0,30582256 | 0,73283153 | ANKEF1 | FALSE | Upregulated |
| 0,125281885 | 0,73283153 | HEXIM1 | FALSE | Upregulated |
| 0,229948866 | 0,73305774 | CACNA2D4 | FALSE | Upregulated |
| 0,209463725 | 0,733247122 | CLIP2 | FALSE | Upregulated |
| -0,29684897 | 0,733568768 | ALG8 | FALSE | Downregulated |
| 0,223951302 | 0,733568768 | FEZ2 | FALSE | Upregulated |
| -0,140379445 | 0,733568768 | GABARAP | FALSE | Downregulated |

| | | | | |
|---|---|---|---|---|
| -0,348104516 | 0,733568768 | HULC | FALSE | Downregulated |
| -0,26951746 | 0,733568768 | LOC101927645 | FALSE | Downregulated |
| 0,231648909 | 0,733568768 | TRMT1 | FALSE | Upregulated |
| 0,208586623 | 0,733568768 | TRPV4 | FALSE | Upregulated |
| -0,193293219 | 0,733568768 | TSPAN33 | FALSE | Downregulated |
| 0,133105701 | 0,733568768 | UGDH-AS1 | FALSE | Upregulated |
| -0,35392216 | 0,733784299 | TRIM68 | FALSE | Downregulated |
| 0,326189091 | 0,733784299 | TVP23C | FALSE | Upregulated |
| 0,233895437 | 0,734028487 | MYOF | FALSE | Upregulated |
| 0,147555931 | 0,734069438 | PTPN21 | FALSE | Upregulated |
| 0,20297947 | 0,734069438 | ZNF41 | FALSE | Upregulated |
| -0,11141954 | 0,734286423 | RAB7A | FALSE | Downregulated |
| 0,256254027 | 0,734310572 | GOLGA7 | FALSE | Upregulated |
| -0,27705736 | 0,734466591 | CWC15 | FALSE | Downregulated |
| 0,16709912 | 0,734466591 | FANCL | FALSE | Upregulated |
| 0,208836739 | 0,734466591 | GNAT1 | FALSE | Upregulated |
| 0,248630769 | 0,734466591 | NUP107 | FALSE | Upregulated |
| 0,206899871 | 0,734466591 | POM121L10P | FALSE | Upregulated |
| 0,14273558 | 0,734466591 | SLC29A2 | FALSE | Upregulated |
| 0,213819494 | 0,734466591 | YRDC | FALSE | Upregulated |
| 0,25074641 | 0,734524753 | DESI2 | FALSE | Upregulated |
| -0,13789984 | 0,734524753 | ITSN1 | FALSE | Downregulated |
| 0,247638403 | 0,734524753 | LOC729658 | FALSE | Upregulated |
| -0,145833127 | 0,734524753 | ONECUT2 | FALSE | Downregulated |
| -0,142415941 | 0,734524753 | ZNF219 | FALSE | Downregulated |
| -0,290187105 | 0,73462539 | ETV5 | FALSE | Downregulated |
| 0,329890781 | 0,73462539 | TCF19 | FALSE | Upregulated |
| -0,240878571 | 0,734681434 | TMEM106A | FALSE | Downregulated |
| -0,323531362 | 0,734931669 | CBWD5 | FALSE | Downregulated |
| 0,147832236 | 0,735056682 | ACTR3 | FALSE | Upregulated |
| -0,155898022 | 0,735056682 | HSPA4 | FALSE | Downregulated |
| 0,390859262 | 0,735056682 | PFKP | FALSE | Upregulated |
| 0,145920765 | 0,735353025 | PRPF38A | FALSE | Upregulated |
| 0,266532635 | 0,735353025 | RABIF | FALSE | Upregulated |
| -0,098784885 | 0,735353025 | TRIM25 | FALSE | Downregulated |
| 0,392158977 | 0,735353025 | TSPEAR-AS1 | FALSE | Upregulated |
| 0,209354743 | 0,735448995 | FAM200B | FALSE | Upregulated |
| -0,231238505 | 0,735495767 | BRK1 | FALSE | Downregulated |
| 0,192649545 | 0,735495767 | BRWD3 | FALSE | Upregulated |
| -0,173252655 | 0,735495767 | CLYBL | FALSE | Downregulated |
| 0,203468808 | 0,735495767 | COL5A1 | FALSE | Upregulated |
| -0,09980579 | 0,735495767 | FGFRL1 | FALSE | Downregulated |
| -0,136339012 | 0,735495767 | HDAC4 | FALSE | Downregulated |
| 0,296511192 | 0,735495767 | KATNA1 | FALSE | Upregulated |
| 0,261566654 | 0,735495767 | LBH | FALSE | Upregulated |
| 0,307455516 | 0,735495767 | LOC284412 | FALSE | Upregulated |
| -0,210128785 | 0,735495767 | METAP1 | FALSE | Downregulated |
| -0,10854887 | 0,735495767 | PC | FALSE | Downregulated |
| 0,227964395 | 0,735495767 | RTL1 | FALSE | Upregulated |
| 0,196265754 | 0,735495767 | SMYD4 | FALSE | Upregulated |
| -0,453665949 | 0,735495767 | WBP1 | FALSE | Downregulated |

| | | | | |
|---|---|---|---|---|
| -0,200982649 | 0,736199185 | HMCES | FALSE | Downregulated |
| 0,191299545 | 0,736199185 | TJAP1 | FALSE | Upregulated |
| 0,309520001 | 0,736208685 | NKRF | FALSE | Upregulated |
| -0,082805666 | 0,736493938 | BIRC6 | FALSE | Downregulated |
| 0,25729723 | 0,736493938 | CHRNG | FALSE | Upregulated |
| 0,187102124 | 0,736493938 | FAS | FALSE | Upregulated |
| -0,100903847 | 0,736493938 | GPD1 | FALSE | Downregulated |
| -0,363753956 | 0,736493938 | IGFBP1 | FALSE | Downregulated |
| -0,24806859 | 0,736493938 | MTMR9 | FALSE | Downregulated |
| -0,152654457 | 0,736493938 | NDUFB7 | FALSE | Downregulated |
| 0,258316598 | 0,736493938 | PSEN2 | FALSE | Upregulated |
| 0,349376271 | 0,736493938 | RAD54L | FALSE | Upregulated |
| -0,22108058 | 0,737038317 | ARL4D | FALSE | Downregulated |
| 0,329056149 | 0,737038317 | BBC3 | FALSE | Upregulated |
| 0,237998124 | 0,737038317 | C19orf25 | FALSE | Upregulated |
| 0,209485696 | 0,737038317 | DNAJA4 | FALSE | Upregulated |
| -0,211791214 | 0,737038317 | MYCL | FALSE | Downregulated |
| 0,106364434 | 0,737038317 | PPP6R1 | FALSE | Upregulated |
| 0,29114502 | 0,737038317 | ZBED5 | FALSE | Upregulated |
| -0,147298003 | 0,737038317 | ZFR | FALSE | Downregulated |
| 0,174506758 | 0,737128846 | ASAP1 | FALSE | Upregulated |
| 0,168900509 | 0,737128846 | ZNF837 | FALSE | Upregulated |
| 0,174954784 | 0,737297288 | ENTPD3 | FALSE | Upregulated |
| -0,197981293 | 0,737354384 | AADAT | FALSE | Downregulated |
| 0,263258546 | 0,737354384 | CCDC173 | FALSE | Upregulated |
| 0,222068344 | 0,737354384 | GMPPB | FALSE | Upregulated |
| -0,419985088 | 0,737354384 | H3F3AP4 | FALSE | Downregulated |
| 0,183468477 | 0,737354384 | HINT3 | FALSE | Upregulated |
| -0,137023547 | 0,737354384 | HSP90AA1 | FALSE | Downregulated |
| -0,167488853 | 0,737354384 | LAPTM4A | FALSE | Downregulated |
| -0,282933878 | 0,737354384 | LOC102723897 | FALSE | Downregulated |
| -0,158226953 | 0,737354384 | MRPS34 | FALSE | Downregulated |
| 0,170734226 | 0,737354384 | NBPF11 | FALSE | Upregulated |
| -0,254372124 | 0,737354384 | NFXL1 | FALSE | Downregulated |
| 0,147926453 | 0,737354384 | PRKCA | FALSE | Upregulated |
| -0,237439849 | 0,737354384 | PSMA1 | FALSE | Downregulated |
| 0,184301379 | 0,737354384 | PTPN6 | FALSE | Upregulated |
| 0,282803775 | 0,737354384 | RAB2B | FALSE | Upregulated |
| -0,11924547 | 0,737354384 | SCN8A | FALSE | Downregulated |
| -0,403777585 | 0,737354384 | SLX1B | FALSE | Downregulated |
| -0,282017795 | 0,737354384 | ZNF594 | FALSE | Downregulated |
| 0,227497642 | 0,737354384 | ZNF700 | FALSE | Upregulated |
| -0,263703099 | 0,737430028 | SERPINB9 | FALSE | Downregulated |
| 0,388207734 | 0,737479092 | ADAM12 | FALSE | Upregulated |
| -0,114596987 | 0,737479092 | MAP4K4 | FALSE | Downregulated |
| 0,163484768 | 0,737479092 | PLTP | FALSE | Upregulated |
| -0,143269778 | 0,737510932 | ACOX2 | FALSE | Downregulated |
| -0,32100701 | 0,737510932 | EMC6 | FALSE | Downregulated |
| -0,264611943 | 0,737792166 | C19orf47 | FALSE | Downregulated |
| 0,220645445 | 0,737925124 | AFF4 | FALSE | Upregulated |
| -0,152498716 | 0,737925124 | EXOC3L4 | FALSE | Downregulated |

| | | | | |
|---|---|---|---|---|
| -0,160045598 | 0,737925124 | FAM102A | FALSE | Downregulated |
| -0,243925748 | 0,737925124 | GLIS3 | FALSE | Downregulated |
| 0,308346442 | 0,737925124 | ITGB1BP1 | FALSE | Upregulated |
| 0,31308682 | 0,737925124 | NOXO1 | FALSE | Upregulated |
| 0,225727116 | 0,737925124 | NRL | FALSE | Upregulated |
| -0,330931059 | 0,737925124 | PDE4A | FALSE | Downregulated |
| -0,24648943 | 0,737925124 | RARS2 | FALSE | Downregulated |
| 0,265266869 | 0,737925124 | TMEM86A | FALSE | Upregulated |
| 0,312883146 | 0,738014814 | GRIN2B | FALSE | Upregulated |
| 0,269946927 | 0,738267324 | DGCR14 | FALSE | Upregulated |
| 0,183387574 | 0,738267324 | FBXL22 | FALSE | Upregulated |
| -0,188299132 | 0,738267324 | GDF7 | FALSE | Downregulated |
| 0,212765375 | 0,738267324 | KLHL31 | FALSE | Upregulated |
| -0,174811734 | 0,738267324 | LINC00174 | FALSE | Downregulated |
| 0,3216522 | 0,738267324 | LOC102724316 | FALSE | Upregulated |
| 0,171849971 | 0,738267324 | MARS | FALSE | Upregulated |
| -0,19552089 | 0,738267324 | NLGN2 | FALSE | Downregulated |
| 0,303345899 | 0,738267324 | NR2F1-AS1 | FALSE | Upregulated |
| -0,237327265 | 0,738267324 | PPP3CB | FALSE | Downregulated |
| 0,284803135 | 0,738267324 | TMEM173 | FALSE | Upregulated |
| 0,291903431 | 0,738336547 | BCL2 | FALSE | Upregulated |
| 0,20879952 | 0,738336547 | LBP | FALSE | Upregulated |
| -0,1586783 | 0,738512856 | PPP1R37 | FALSE | Downregulated |
| 0,22618861 | 0,738512856 | UBE2W | FALSE | Upregulated |
| -0,181461463 | 0,738573195 | SH3GLB2 | FALSE | Downregulated |
| -0,18888468 | 0,738589141 | YTHDC2 | FALSE | Downregulated |
| -0,199093015 | 0,738692729 | PRR5 | FALSE | Downregulated |
| 0,207525838 | 0,738709487 | LOC100272216 | FALSE | Upregulated |
| -0,314771985 | 0,738709487 | RNASEK | FALSE | Downregulated |
| -0,110841402 | 0,738976021 | SFXN5 | FALSE | Downregulated |
| -0,148013362 | 0,739069082 | CELSR1 | FALSE | Downregulated |
| 0,250050061 | 0,739242811 | TAGAP | FALSE | Upregulated |
| -0,190786381 | 0,739329894 | SMIM7 | FALSE | Downregulated |
| 0,148936988 | 0,739461362 | POU2F1 | FALSE | Upregulated |
| -0,268470675 | 0,739523026 | CNOT10 | FALSE | Downregulated |
| -0,152843354 | 0,739523026 | ENGASE | FALSE | Downregulated |
| -0,420382127 | 0,739523026 | KIAA1328 | FALSE | Downregulated |
| -0,269712954 | 0,739523026 | LOC101929072 | FALSE | Downregulated |
| -0,157237712 | 0,739523026 | SLC25A20 | FALSE | Downregulated |
| -0,155893148 | 0,739523026 | SLC25A47 | FALSE | Downregulated |
| -0,141521449 | 0,739523026 | TBC1D5 | FALSE | Downregulated |
| -0,280598359 | 0,739523026 | TMEM261 | FALSE | Downregulated |
| -0,141598861 | 0,739523026 | TUBB4B | FALSE | Downregulated |
| 0,213128743 | 0,740183858 | CD5 | FALSE | Upregulated |
| -0,194832208 | 0,740183858 | CPSF2 | FALSE | Downregulated |
| 0,298345451 | 0,740230722 | LOC100506734 | FALSE | Upregulated |
| 0,211247806 | 0,740307538 | SLC27A4 | FALSE | Upregulated |
| -0,141784934 | 0,740431729 | UBE2O | FALSE | Downregulated |
| -0,095613486 | 0,740431729 | ZNF592 | FALSE | Downregulated |
| -0,285139588 | 0,740803249 | EPB41L1 | FALSE | Downregulated |
| 0,139224596 | 0,740803249 | HEG1 | FALSE | Upregulated |

| | | | | |
|---|---|---|---|---|
| 0,221103769 | 0,740803249 | HMGCR | FALSE | Upregulated |
| 0,342481662 | 0,740803249 | MFI2 | FALSE | Upregulated |
| -0,225172169 | 0,740803249 | ZXDA | FALSE | Downregulated |
| -0,204484602 | 0,741121978 | CADPS2 | FALSE | Downregulated |
| 0,239222526 | 0,741121978 | CDH4 | FALSE | Upregulated |
| -0,289073824 | 0,741121978 | COL26A1 | FALSE | Downregulated |
| -0,208577671 | 0,741121978 | KALRN | FALSE | Downregulated |
| 0,348641228 | 0,741121978 | MAMLD1 | FALSE | Upregulated |
| -0,174109193 | 0,741121978 | NAA50 | FALSE | Downregulated |
| -0,125130083 | 0,741451573 | CDK5RAP3 | FALSE | Downregulated |
| 0,199758879 | 0,741451573 | EIF2B4 | FALSE | Upregulated |
| -0,338910078 | 0,742077587 | SRL | FALSE | Downregulated |
| -0,504942552 | 0,742231002 | HYDIN | FALSE | Downregulated |
| -0,19070782 | 0,742231002 | KCTD21 | FALSE | Downregulated |
| -0,145231435 | 0,742231002 | LEMD3 | FALSE | Downregulated |
| -0,482087564 | 0,742231002 | LOC102723996 | FALSE | Downregulated |
| 0,272338197 | 0,742712434 | LOC100129637 | FALSE | Upregulated |
| -0,134729146 | 0,742712434 | PITRM1 | FALSE | Downregulated |
| 0,108975151 | 0,742712434 | SERINC4 | FALSE | Upregulated |
| 0,370256214 | 0,742774871 | RP11-540A21.2 | FALSE | Upregulated |
| -0,261683757 | 0,742930096 | LOC100507412 | FALSE | Downregulated |
| -0,175150216 | 0,743044026 | VRK3 | FALSE | Downregulated |
| -0,334212811 | 0,743090357 | ATF1 | FALSE | Downregulated |
| 0,158547307 | 0,743090357 | C12orf23 | FALSE | Upregulated |
| -0,276835296 | 0,743090357 | C2orf42 | FALSE | Downregulated |
| 0,270022916 | 0,743090357 | CCNG2 | FALSE | Upregulated |
| 0,10120127 | 0,743090357 | CIAO1 | FALSE | Upregulated |
| 0,169710354 | 0,743090357 | DDX18 | FALSE | Upregulated |
| -0,334077115 | 0,743090357 | G6PC3 | FALSE | Downregulated |
| 0,147406674 | 0,743090357 | GGNBP2 | FALSE | Upregulated |
| -0,111408892 | 0,743090357 | IL13RA1 | FALSE | Downregulated |
| 0,354422878 | 0,743090357 | KIF20A | FALSE | Upregulated |
| 0,265065322 | 0,743090357 | KIF21B | FALSE | Upregulated |
| -0,357916366 | 0,743090357 | LOC101928615 | FALSE | Downregulated |
| 0,348875158 | 0,743090357 | LOC101928641 | FALSE | Upregulated |
| 0,153329901 | 0,743090357 | MAP3K14 | FALSE | Upregulated |
| -0,332190967 | 0,743090357 | NTPCR | FALSE | Downregulated |
| -0,115920038 | 0,743090357 | PLOD1 | FALSE | Downregulated |
| 0,238675903 | 0,743090357 | RP11-383C5.3 | FALSE | Upregulated |
| -0,130816129 | 0,743090357 | RPL28 | FALSE | Downregulated |
| 0,154212702 | 0,743090357 | SCO2 | FALSE | Upregulated |
| 0,445025755 | 0,743090357 | SEPT1 | FALSE | Upregulated |
| 0,326526142 | 0,743090357 | SWSAP1 | FALSE | Upregulated |
| -0,147669333 | 0,743090357 | TRAK2 | FALSE | Downregulated |
| 0,241340372 | 0,743090357 | TRIAP1 | FALSE | Upregulated |
| 0,132646922 | 0,743090357 | TUT1 | FALSE | Upregulated |
| -0,169894828 | 0,743090357 | USP20 | FALSE | Downregulated |
| 0,188068571 | 0,743090357 | VAC14-AS1 | FALSE | Upregulated |
| -0,157606049 | 0,743090357 | YIPF3 | FALSE | Downregulated |
| -0,301234839 | 0,743090357 | ZNHIT6 | FALSE | Downregulated |
| -0,107593042 | 0,743275472 | DGCR2 | FALSE | Downregulated |

| | | | | |
|---|---|---|---|---|
| -0,154640479 | 0,743275472 | EIF4ENIF1 | FALSE | Downregulated |
| -0,209406438 | 0,743275472 | LOC100506548 | FALSE | Downregulated |
| 0,207551545 | 0,743275472 | LOC101928101 | FALSE | Upregulated |
| 0,192133904 | 0,743275472 | SURF6 | FALSE | Upregulated |
| -0,078608404 | 0,743275472 | TMEM259 | FALSE | Downregulated |
| 0,115372141 | 0,743281602 | GPC1 | FALSE | Upregulated |
| -0,16797438 | 0,743281602 | PGD | FALSE | Downregulated |
| -0,129257338 | 0,743317484 | TIMP3 | FALSE | Downregulated |
| -0,126956281 | 0,743478312 | BSDC1 | FALSE | Downregulated |
| 0,254012402 | 0,743478312 | FTSJ1 | FALSE | Upregulated |
| 0,109238348 | 0,743497596 | CEP170B | FALSE | Upregulated |
| 0,15921527 | 0,7435678 | CLCC1 | FALSE | Upregulated |
| 0,17303076 | 0,7435678 | CTRL | FALSE | Upregulated |
| 0,144427159 | 0,7435678 | GIT1 | FALSE | Upregulated |
| 0,241348233 | 0,7435678 | PPP1R21 | FALSE | Upregulated |
| 0,344271654 | 0,743668426 | KDM4E | FALSE | Upregulated |
| -0,360914545 | 0,743732788 | MAGIX | FALSE | Downregulated |
| -0,123859752 | 0,743732788 | PLEKHA6 | FALSE | Downregulated |
| -0,226521949 | 0,743732788 | TIFA | FALSE | Downregulated |
| -0,239678139 | 0,743732788 | TMEM55B | FALSE | Downregulated |
| -0,154259637 | 0,743822641 | KAZN | FALSE | Downregulated |
| -0,175492725 | 0,744292863 | ACVR1 | FALSE | Downregulated |
| -0,176878969 | 0,744292863 | CENPV | FALSE | Downregulated |
| 0,25165057 | 0,744292863 | CUL2 | FALSE | Upregulated |
| -0,275221931 | 0,744292863 | DDX50 | FALSE | Downregulated |
| -0,075020529 | 0,744292863 | EIF4G1 | FALSE | Downregulated |
| 0,373965792 | 0,744292863 | HELB | FALSE | Upregulated |
| -0,151039599 | 0,744292863 | HP | FALSE | Downregulated |
| -0,143894827 | 0,744292863 | RDH10 | FALSE | Downregulated |
| 0,354792673 | 0,744292863 | RPL23AP53 | FALSE | Upregulated |
| 0,124252531 | 0,744292863 | RRN3P3 | FALSE | Upregulated |
| 0,108119926 | 0,744292863 | SAFB | FALSE | Upregulated |
| -0,361547598 | 0,744292863 | ZNF684 | FALSE | Downregulated |
| -0,185325642 | 0,744312254 | RPS12 | FALSE | Downregulated |
| 0,192139039 | 0,744320184 | CDPF1 | FALSE | Upregulated |
| 0,164217422 | 0,744320184 | CLVS1 | FALSE | Upregulated |
| 0,509437768 | 0,744320184 | CORO7 | FALSE | Upregulated |
| 0,271585464 | 0,744320184 | MYO5A | FALSE | Upregulated |
| -0,166360529 | 0,744320184 | PCIF1 | FALSE | Downregulated |
| -0,318859726 | 0,74433339 | TMEM126A | FALSE | Downregulated |
| 1,313850242 | 0,744939194 | EEF1A2 | FALSE | Upregulated |
| -0,194729566 | 0,744939194 | ISG20L2 | FALSE | Downregulated |
| 0,204520604 | 0,744939194 | ITPKB | FALSE | Upregulated |
| 0,275688363 | 0,744939194 | SCD | FALSE | Upregulated |
| -0,285193621 | 0,744939194 | VAMP4 | FALSE | Downregulated |
| 0,357805411 | 0,744939194 | ZNF20 | FALSE | Upregulated |
| -0,246798022 | 0,744939194 | ZNF385B | FALSE | Downregulated |
| 0,176415292 | 0,744959904 | JAG1 | FALSE | Upregulated |
| -0,22505744 | 0,744959904 | PDCD6 | FALSE | Downregulated |
| 0,172446815 | 0,745041175 | BRCA2 | FALSE | Upregulated |
| -0,261598554 | 0,745110958 | BLMH | FALSE | Downregulated |

| | | | | |
|---|---|---|---|---|
| -0,396959834 | 0,745110958 | CYP7A1 | FALSE | Downregulated |
| -0,164048716 | 0,745110958 | DERL1 | FALSE | Downregulated |
| -0,278873019 | 0,745110958 | DGKE | FALSE | Downregulated |
| -0,323308513 | 0,745110958 | ENDOV | FALSE | Downregulated |
| -0,16943674 | 0,745110958 | ERLIN2 | FALSE | Downregulated |
| 0,48232639 | 0,745110958 | FAM90A25P | FALSE | Upregulated |
| 0,141850194 | 0,745110958 | FLJ31945 | FALSE | Upregulated |
| -0,194282696 | 0,745110958 | GBP7 | FALSE | Downregulated |
| -0,165954635 | 0,745110958 | GFOD1 | FALSE | Downregulated |
| 0,104065203 | 0,745110958 | JUND | FALSE | Upregulated |
| -0,107996493 | 0,745110958 | KIAA0368 | FALSE | Downregulated |
| 0,320617811 | 0,745110958 | LENG1 | FALSE | Upregulated |
| -0,177878381 | 0,745110958 | LIPA | FALSE | Downregulated |
| 0,120100749 | 0,745110958 | LOC100128494 | FALSE | Upregulated |
| 0,197305482 | 0,745110958 | LOC401320 | FALSE | Upregulated |
| 0,128022423 | 0,745110958 | NR2C2 | FALSE | Upregulated |
| 0,19978749 | 0,745110958 | PIP4K2A | FALSE | Upregulated |
| -0,328761518 | 0,745110958 | PPP1R3F | FALSE | Downregulated |
| -0,199319277 | 0,745110958 | SCRN2 | FALSE | Downregulated |
| 0,307555172 | 0,745110958 | SLX1A | FALSE | Upregulated |
| 0,249806858 | 0,745110958 | TBKBP1 | FALSE | Upregulated |
| 0,166408413 | 0,745110958 | TICAM1 | FALSE | Upregulated |
| 0,21302891 | 0,745110958 | TOE1 | FALSE | Upregulated |
| -0,117596655 | 0,745110958 | TPRG1L | FALSE | Downregulated |
| 0,167950735 | 0,745110958 | TRIM52 | FALSE | Upregulated |
| 0,331909288 | 0,746040239 | MFNG | FALSE | Upregulated |
| -0,19918728 | 0,746075028 | CCL16 | FALSE | Downregulated |
| 0,441703147 | 0,746075028 | LOC101928352 | FALSE | Upregulated |
| 0,181823541 | 0,746092311 | CDH23 | FALSE | Upregulated |
| 0,26399353 | 0,746530065 | GRIK1 | FALSE | Upregulated |
| 0,111000857 | 0,746530065 | MYO9B | FALSE | Upregulated |
| 0,270083794 | 0,746785351 | IDH3A | FALSE | Upregulated |
| -0,177778974 | 0,746785351 | OIP5-AS1 | FALSE | Downregulated |
| -0,312958953 | 0,746785351 | PCBD2 | FALSE | Downregulated |
| -0,350579304 | 0,746857615 | AKR1B1 | FALSE | Downregulated |
| 0,152767115 | 0,746857615 | CTAGE7P | FALSE | Upregulated |
| -0,129536902 | 0,746857615 | MAP2K3 | FALSE | Downregulated |
| -0,197202704 | 0,746857615 | NOB1 | FALSE | Downregulated |
| 0,21612 | 0,746857615 | PPA1 | FALSE | Upregulated |
| 0,089968653 | 0,746857615 | SP1 | FALSE | Upregulated |
| -0,30428256 | 0,746857615 | ZFP90 | FALSE | Downregulated |
| -0,304483298 | 0,747168552 | CCDC144B | FALSE | Downregulated |
| -0,132625496 | 0,747289001 | RB1CC1 | FALSE | Downregulated |
| -0,137936543 | 0,747380694 | ZNF644 | FALSE | Downregulated |
| 0,155192076 | 0,747553446 | RBM4B | FALSE | Upregulated |
| 0,370628731 | 0,747650429 | LOC100506319 | FALSE | Upregulated |
| 0,135299415 | 0,747729542 | NASP | FALSE | Upregulated |
| 0,306300622 | 0,747745087 | PLEKHA8 | FALSE | Upregulated |
| -0,113002804 | 0,747842023 | TEAD1 | FALSE | Downregulated |
| -0,141004463 | 0,74790356 | BCAR1 | FALSE | Downregulated |
| -0,103578106 | 0,74790356 | CPT2 | FALSE | Downregulated |

| | | | | |
|---|---|---|---|---|
| -0,235292075 | 0,74790356 | GRK5 | FALSE | Downregulated |
| 0,442676036 | 0,74790356 | LGALS3 | FALSE | Upregulated |
| -0,220478734 | 0,74790356 | NSF | FALSE | Downregulated |
| -0,1287528 | 0,74790356 | PAK4 | FALSE | Downregulated |
| 0,155975504 | 0,74790356 | ZNF542 | FALSE | Upregulated |
| -0,115868851 | 0,747924573 | NPM1 | FALSE | Downregulated |
| 0,322273944 | 0,747924573 | PRKCQ | FALSE | Upregulated |
| 0,148260339 | 0,748322089 | AC093690.1 | FALSE | Upregulated |
| 0,171745494 | 0,748322089 | FLJ38717 | FALSE | Upregulated |
| 0,158456587 | 0,748322089 | LDLRAD2 | FALSE | Upregulated |
| 0,103974844 | 0,748322089 | TNRC6C | FALSE | Upregulated |
| 1,834841026 | 0,748391273 | MYBPC2 | FALSE | Upregulated |
| 0,091413889 | 0,748391273 | PFN1 | FALSE | Upregulated |
| -0,096203285 | 0,748428704 | DST | FALSE | Downregulated |
| -0,143477781 | 0,748428704 | ENTPD5 | FALSE | Downregulated |
| 0,489410169 | 0,748428704 | HIST1H1D | FALSE | Upregulated |
| 0,17088798 | 0,748428704 | IQGAP1 | FALSE | Upregulated |
| 0,182792408 | 0,748428704 | LOC102724016 | FALSE | Upregulated |
| 0,230748988 | 0,748428704 | LOC728763 | FALSE | Upregulated |
| 0,341185563 | 0,748428704 | MYO1H | FALSE | Upregulated |
| -0,193943123 | 0,748428704 | POLL | FALSE | Downregulated |
| -0,145287444 | 0,748428704 | TMEM192 | FALSE | Downregulated |
| -0,213858266 | 0,748428704 | ZNF500 | FALSE | Downregulated |
| 0,179538735 | 0,748430191 | ZNF606 | FALSE | Upregulated |
| -0,104129823 | 0,74915009 | C1S | FALSE | Downregulated |
| -0,080827253 | 0,749339883 | MGAT4B | FALSE | Downregulated |
| 0,188275479 | 0,74957516 | MTO1 | FALSE | Upregulated |
| 0,278019417 | 0,749722771 | AURKA | FALSE | Upregulated |
| -0,116518289 | 0,749722771 | CST3 | FALSE | Downregulated |
| -0,128706945 | 0,749722771 | DENND4A | FALSE | Downregulated |
| -0,369203691 | 0,749722771 | EPHA3 | FALSE | Downregulated |
| 0,126987601 | 0,749722771 | GCC2 | FALSE | Upregulated |
| 0,293395579 | 0,749722771 | GTF2B | FALSE | Upregulated |
| 0,198586349 | 0,749722771 | LOC100506374 | FALSE | Upregulated |
| 0,148108916 | 0,749722771 | NHP2 | FALSE | Upregulated |
| 0,314837196 | 0,749722771 | RP1-149A16.3 | FALSE | Upregulated |
| 0,14463466 | 0,749722771 | RUSC1 | FALSE | Upregulated |
| 0,284012102 | 0,749722771 | SLC7A5 | FALSE | Upregulated |
| -0,136261275 | 0,749888132 | CCT7 | FALSE | Downregulated |
| 0,242124607 | 0,749888132 | RIT1 | FALSE | Upregulated |
| -0,174393977 | 0,749972141 | HDAC10 | FALSE | Downregulated |
| -0,205091493 | 0,749972141 | LMBRD1 | FALSE | Downregulated |
| 0,171941426 | 0,749972141 | PIEZO2 | FALSE | Upregulated |
| 0,077094328 | 0,749972141 | SFPQ | FALSE | Upregulated |
| -0,147357485 | 0,749972141 | ZNF362 | FALSE | Downregulated |
| -0,15000794 | 0,750551143 | ACVR2B | FALSE | Downregulated |
| -0,110677537 | 0,750551143 | AMOTL2 | FALSE | Downregulated |
| 0,203770418 | 0,750551143 | B4GALT3 | FALSE | Upregulated |
| -0,118555362 | 0,750551143 | CGN | FALSE | Downregulated |
| -0,13415542 | 0,750551143 | CMIP | FALSE | Downregulated |
| -0,127333284 | 0,750551143 | CTSO | FALSE | Downregulated |

| | | | | |
|---|---|---|---|---|
| -0,301105854 | 0,750551143 | DGCR5 | FALSE | Downregulated |
| -0,510773321 | 0,750551143 | EIF3C | FALSE | Downregulated |
| 0,21382596 | 0,750551143 | ENPP7 | FALSE | Upregulated |
| 0,281479907 | 0,750551143 | LOC102723539 | FALSE | Upregulated |
| 0,168716097 | 0,750551143 | MAK | FALSE | Upregulated |
| -0,108764321 | 0,750551143 | PLIN5 | FALSE | Downregulated |
| -0,270007631 | 0,750551143 | POP7 | FALSE | Downregulated |
| -0,372507132 | 0,750551143 | PTS | FALSE | Downregulated |
| -0,200994223 | 0,750551143 | RHOG | FALSE | Downregulated |
| -0,313444598 | 0,750551143 | S100A13 | FALSE | Downregulated |
| 0,128848899 | 0,750551143 | SESN3 | FALSE | Upregulated |
| 0,11738205 | 0,750551143 | YWHAG | FALSE | Upregulated |
| 0,258578718 | 0,750551143 | ZNF195 | FALSE | Upregulated |
| 0,140439305 | 0,750650412 | CLDN20 | FALSE | Upregulated |
| 0,247232892 | 0,750650412 | FKBP14 | FALSE | Upregulated |
| 0,272209951 | 0,750686883 | AK1 | FALSE | Upregulated |
| 0,190576215 | 0,750772142 | ARHGAP26 | FALSE | Upregulated |
| 0,209544304 | 0,750772142 | CENPU | FALSE | Upregulated |
| 0,272684748 | 0,750772142 | GPRIN3 | FALSE | Upregulated |
| 0,287602961 | 0,750772142 | MRPL21 | FALSE | Upregulated |
| 0,208768093 | 0,750772142 | PLIN3 | FALSE | Upregulated |
| 0,25784369 | 0,750772142 | PPP3CC | FALSE | Upregulated |
| 0,109722531 | 0,750881506 | LASP1 | FALSE | Upregulated |
| 0,123999002 | 0,750937848 | SPTB | FALSE | Upregulated |
| -0,143856749 | 0,750949268 | PCBD1 | FALSE | Downregulated |
| 0,186578423 | 0,751239947 | ATP5J | FALSE | Upregulated |
| 0,26051663 | 0,751239947 | CACNB3 | FALSE | Upregulated |
| 0,173638031 | 0,751239947 | CNTN2 | FALSE | Upregulated |
| 0,3098178 | 0,751239947 | CST7 | FALSE | Upregulated |
| -0,280404521 | 0,751239947 | HIST1H2BD | FALSE | Downregulated |
| -0,212092539 | 0,751239947 | MPZL1 | FALSE | Downregulated |
| 0,114390299 | 0,751239947 | MTOR | FALSE | Upregulated |
| 0,227371061 | 0,751239947 | TMEM238 | FALSE | Upregulated |
| 0,193170357 | 0,751239947 | TOMM22 | FALSE | Upregulated |
| 0,198146595 | 0,75174981 | JPH3 | FALSE | Upregulated |
| 0,349172669 | 0,75174981 | SRGAP2C | FALSE | Upregulated |
| -0,28951139 | 0,752474022 | SLC9A1 | FALSE | Downregulated |
| 0,132157952 | 0,752579119 | PAIP2B | FALSE | Upregulated |
| -0,15547252 | 0,752583381 | RMDN3 | FALSE | Downregulated |
| 0,222442688 | 0,752583381 | TMEM50A | FALSE | Upregulated |
| -0,320063579 | 0,752583381 | ZNF34 | FALSE | Downregulated |
| 0,126890812 | 0,752645131 | F11-AS1 | FALSE | Upregulated |
| 0,22154845 | 0,752645131 | TMPRSS2 | FALSE | Upregulated |
| 0,270480435 | 0,75268785 | LOC100130285 | FALSE | Upregulated |
| -0,199745204 | 0,752692896 | PANX2 | FALSE | Downregulated |
| -0,202575821 | 0,752692896 | SLC39A8 | FALSE | Downregulated |
| 0,260655455 | 0,752881175 | BUB3 | FALSE | Upregulated |
| -0,183699832 | 0,752881175 | COL5A2 | FALSE | Downregulated |
| 0,221291982 | 0,752881175 | EMC3-AS1 | FALSE | Upregulated |
| 0,178558824 | 0,752881175 | EML6 | FALSE | Upregulated |
| 0,156673637 | 0,752881175 | GRIPAP1 | FALSE | Upregulated |

| | | | | |
|---|---|---|---|---|
| -0,162735451 | 0,752881175 | IRF6 | FALSE | Downregulated |
| -0,099148247 | 0,752881175 | ITPR2 | FALSE | Downregulated |
| -0,250913771 | 0,752881175 | MAP4K2 | FALSE | Downregulated |
| -0,322281671 | 0,752881175 | NUDT2 | FALSE | Downregulated |
| -0,378689702 | 0,752881175 | PPT2 | FALSE | Downregulated |
| -0,126196278 | 0,752881175 | PTMS | FALSE | Downregulated |
| -0,118416918 | 0,752881175 | RAB5B | FALSE | Downregulated |
| -0,129260156 | 0,752881175 | SART3 | FALSE | Downregulated |
| 0,65691539 | 0,752881175 | SRGAP2B | FALSE | Upregulated |
| -0,09139849 | 0,752881175 | STK25 | FALSE | Downregulated |
| -0,210638667 | 0,752881175 | TBRG4 | FALSE | Downregulated |
| 0,114519771 | 0,752881175 | TRAM2 | FALSE | Upregulated |
| -0,174311213 | 0,752881175 | ZRANB2 | FALSE | Downregulated |
| -0,296009768 | 0,753036315 | INTS2 | FALSE | Downregulated |
| -0,424868701 | 0,753036315 | KCNJ3 | FALSE | Downregulated |
| 0,101246825 | 0,753036315 | LOC101927556 | FALSE | Upregulated |
| -0,103997983 | 0,753036315 | REXO1 | FALSE | Downregulated |
| 0,098348328 | 0,753114661 | GTPBP1 | FALSE | Upregulated |
| -0,109991744 | 0,753343365 | TRAF7 | FALSE | Downregulated |
| -0,155515841 | 0,753796411 | CHST13 | FALSE | Downregulated |
| 0,17859368 | 0,753796411 | DERL3 | FALSE | Upregulated |
| -0,198845229 | 0,753796411 | KBTBD7 | FALSE | Downregulated |
| 0,137843723 | 0,753796411 | PPAP2A | FALSE | Upregulated |
| -0,286267658 | 0,753796411 | SNHG8 | FALSE | Downregulated |
| -0,092313395 | 0,753796411 | ST6GAL1 | FALSE | Downregulated |
| 0,126467635 | 0,753796411 | UBE2D4 | FALSE | Upregulated |
| 0,194062062 | 0,753968352 | RP11-475I24.8 | FALSE | Upregulated |
| -0,114388859 | 0,753968352 | TATDN2 | FALSE | Downregulated |
| 0,169379418 | 0,754330952 | RC3H2 | FALSE | Upregulated |
| -0,289204416 | 0,754561823 | SELK | FALSE | Downregulated |
| 0,162673276 | 0,754669674 | CEBPD | FALSE | Upregulated |
| 0,29007388 | 0,754669674 | LOC101929787 | FALSE | Upregulated |
| 0,258822341 | 0,754753044 | IGSF9 | FALSE | Upregulated |
| -0,258590178 | 0,754753044 | TBPL1 | FALSE | Downregulated |
| -0,314373371 | 0,754775378 | MCEE | FALSE | Downregulated |
| 0,125603952 | 0,75494258 | ADCY5 | FALSE | Upregulated |
| 0,205280703 | 0,75494258 | RP3-476K8.3 | FALSE | Upregulated |
| 0,392230782 | 0,755408654 | PDGFA | FALSE | Upregulated |
| 0,095104912 | 0,755408654 | SLC22A23 | FALSE | Upregulated |
| 0,38496331 | 0,755505126 | ACSL4 | FALSE | Upregulated |
| 0,189622548 | 0,755505126 | CPM | FALSE | Upregulated |
| 0,410914083 | 0,755505126 | GLB1L2 | FALSE | Upregulated |
| -0,087120265 | 0,755505126 | SURF4 | FALSE | Downregulated |
| 0,21888619 | 0,755505126 | U2AF1 | FALSE | Upregulated |
| -0,128597042 | 0,755773059 | AGPAT1 | FALSE | Downregulated |
| -0,114774409 | 0,755773059 | AP1B1 | FALSE | Downregulated |
| 0,235611597 | 0,755773059 | ARHGEF33 | FALSE | Upregulated |
| -0,130920314 | 0,755773059 | ATRX | FALSE | Downregulated |
| 0,159183056 | 0,755773059 | BEND3P3 | FALSE | Upregulated |
| -0,194334492 | 0,755773059 | BLOC1S1 | FALSE | Downregulated |
| 0,13225847 | 0,755773059 | CAPN2 | FALSE | Upregulated |

| | | | | |
|---|---|---|---|---|
| -0,3008955 | 0,755773059 | CHMP3 | FALSE | Downregulated |
| -0,173016574 | 0,755773059 | COPS7A | FALSE | Downregulated |
| 0,285373334 | 0,755773059 | CRYM | FALSE | Upregulated |
| 0,126005493 | 0,755773059 | CUL4A | FALSE | Upregulated |
| 0,108849197 | 0,755773059 | DLEU1 | FALSE | Upregulated |
| -0,204191709 | 0,755773059 | DPP3 | FALSE | Downregulated |
| 0,384178146 | 0,755773059 | ETV2 | FALSE | Upregulated |
| -0,090353529 | 0,755773059 | EWSR1 | FALSE | Downregulated |
| -0,146094943 | 0,755773059 | F8 | FALSE | Downregulated |
| 0,17455947 | 0,755773059 | FAM173A | FALSE | Upregulated |
| -0,12770412 | 0,755773059 | FBXL17 | FALSE | Downregulated |
| 0,122231683 | 0,755773059 | HIP1R | FALSE | Upregulated |
| 0,18531562 | 0,755773059 | KCNT2 | FALSE | Upregulated |
| 0,241096498 | 0,755773059 | KCTD7 | FALSE | Upregulated |
| 0,412258047 | 0,755773059 | LMTK3 | FALSE | Upregulated |
| -0,227965378 | 0,755773059 | LOC101928499 | FALSE | Downregulated |
| 0,203656333 | 0,755773059 | LOC102723769 | FALSE | Upregulated |
| -0,216880311 | 0,755773059 | MLLT4-AS1 | FALSE | Downregulated |
| 0,328795456 | 0,755773059 | MPZL2 | FALSE | Upregulated |
| -0,216419734 | 0,755773059 | MRPS10 | FALSE | Downregulated |
| -0,09362093 | 0,755773059 | NHSL1 | FALSE | Downregulated |
| -0,164426101 | 0,755773059 | NPTN | FALSE | Downregulated |
| -0,193863383 | 0,755773059 | PHLDB3 | FALSE | Downregulated |
| 0,162076205 | 0,755773059 | PITPNC1 | FALSE | Upregulated |
| -0,183119936 | 0,755773059 | PKD2 | FALSE | Downregulated |
| 0,174580066 | 0,755773059 | PPFIA2 | FALSE | Upregulated |
| 0,149735155 | 0,755773059 | PRICKLE2-AS1 | FALSE | Upregulated |
| 0,257633014 | 0,755773059 | PRPSAP2 | FALSE | Upregulated |
| -0,15726709 | 0,755773059 | RBM42 | FALSE | Downregulated |
| -0,149812841 | 0,755773059 | RCN3 | FALSE | Downregulated |
| -0,262835717 | 0,755773059 | RNF165 | FALSE | Downregulated |
| 0,28483845 | 0,755773059 | RTN4IP1 | FALSE | Upregulated |
| 0,229968431 | 0,755773059 | RWDD2A | FALSE | Upregulated |
| -0,100728718 | 0,755773059 | SCAF11 | FALSE | Downregulated |
| 0,099237591 | 0,755773059 | SLC25A29 | FALSE | Upregulated |
| 0,19975115 | 0,755773059 | SLC46A2 | FALSE | Upregulated |
| -0,20241174 | 0,755773059 | SSH2 | FALSE | Downregulated |
| 0,296722569 | 0,755773059 | SUV420H2 | FALSE | Upregulated |
| 0,181963619 | 0,755773059 | TBC1D25 | FALSE | Upregulated |
| -0,253165814 | 0,755773059 | TSPAN17 | FALSE | Downregulated |
| 0,24856856 | 0,755773059 | TSPO | FALSE | Upregulated |
| 0,203960672 | 0,755773059 | TTLL2 | FALSE | Upregulated |
| 0,188018104 | 0,755773059 | TYSND1 | FALSE | Upregulated |
| -0,250145873 | 0,755773059 | UQCR10 | FALSE | Downregulated |
| 0,093112016 | 0,755773059 | UTRN | FALSE | Upregulated |
| -0,21118633 | 0,755773059 | VDAC3 | FALSE | Downregulated |
| -0,357582554 | 0,755773059 | WNT5B | FALSE | Downregulated |
| 0,146131411 | 0,755773059 | ZBTB12 | FALSE | Upregulated |
| -0,129361119 | 0,755773059 | ZFC3H1 | FALSE | Downregulated |
| -0,275665364 | 0,756039091 | TSNAX | FALSE | Downregulated |
| -0,135681458 | 0,756040195 | LAMP2 | FALSE | Downregulated |

| | | | | |
|---|---|---|---|---|
| -0,111669288 | 0,756040195 | SEC63 | FALSE | Downregulated |
| 0,232157505 | 0,756142688 | DFFA | FALSE | Upregulated |
| 0,250367744 | 0,756142688 | KIAA0040 | FALSE | Upregulated |
| 0,250532297 | 0,756142688 | LINC00482 | FALSE | Upregulated |
| -0,165930174 | 0,756142688 | SAP30BP | FALSE | Downregulated |
| -0,168531835 | 0,756769831 | ASPSCR1 | FALSE | Downregulated |
| 0,137263192 | 0,756769831 | TMSB4X | FALSE | Upregulated |
| 0,140066483 | 0,756970705 | ABI2 | FALSE | Upregulated |
| 0,343261139 | 0,756970705 | AMPH | FALSE | Upregulated |
| -0,300613981 | 0,756970705 | CBR3 | FALSE | Downregulated |
| 0,177793515 | 0,756970705 | CYB5D1 | FALSE | Upregulated |
| -0,155756209 | 0,756970705 | GPT2 | FALSE | Downregulated |
| -0,143438375 | 0,756970705 | HSPA1A | FALSE | Downregulated |
| -0,08105127 | 0,756970705 | IQSEC1 | FALSE | Downregulated |
| -0,260896354 | 0,756970705 | LDHB | FALSE | Downregulated |
| 0,139292399 | 0,756970705 | LEPROT | FALSE | Upregulated |
| 0,123446778 | 0,756970705 | LOC102724129 | FALSE | Upregulated |
| 0,180179892 | 0,756970705 | LOC730257 | FALSE | Upregulated |
| 0,353081666 | 0,756970705 | NCF1 | FALSE | Upregulated |
| -0,154709717 | 0,756970705 | NDUFA10 | FALSE | Downregulated |
| -0,283344717 | 0,756970705 | SH3GL1P2 | FALSE | Downregulated |
| -0,112916175 | 0,756970705 | TRAPPC12 | FALSE | Downregulated |
| -0,346127321 | 0,756970705 | WDR35 | FALSE | Downregulated |
| -0,199169209 | 0,756984596 | ERI3 | FALSE | Downregulated |
| -0,239960326 | 0,756984596 | LOC100507616 | FALSE | Downregulated |
| -0,207127671 | 0,756984596 | PAOX | FALSE | Downregulated |
| -0,375828679 | 0,757283592 | ANAPC4 | FALSE | Downregulated |
| -0,134984596 | 0,757283592 | AXIN1 | FALSE | Downregulated |
| 0,235957854 | 0,757283592 | AZI1 | FALSE | Upregulated |
| 0,294009844 | 0,757283592 | DTWD1 | FALSE | Upregulated |
| 0,124475717 | 0,757283592 | IRF7 | FALSE | Upregulated |
| 0,182028937 | 0,757283592 | LCP2 | FALSE | Upregulated |
| -0,318238875 | 0,757283592 | LOC101928714 | FALSE | Downregulated |
| 0,114610591 | 0,757283592 | LOC148413 | FALSE | Upregulated |
| -0,161670652 | 0,757283592 | MMP24-AS1 | FALSE | Downregulated |
| -0,240676745 | 0,757283592 | NR6A1 | FALSE | Downregulated |
| 0,194719562 | 0,757283592 | RAB5A | FALSE | Upregulated |
| 0,170138953 | 0,757283592 | SLC1A1 | FALSE | Upregulated |
| -0,126104269 | 0,757283592 | TIMP2 | FALSE | Downregulated |
| -0,192226148 | 0,757283592 | TMEM50B | FALSE | Downregulated |
| 0,187533356 | 0,757283592 | WDR36 | FALSE | Upregulated |
| -0,125661005 | 0,757299313 | SRSF11 | FALSE | Downregulated |
| -0,179287939 | 0,757299313 | SSBP4 | FALSE | Downregulated |
| 0,143838949 | 0,757299313 | TVP23A | FALSE | Upregulated |
| 0,269528335 | 0,757636828 | LRRC15 | FALSE | Upregulated |
| -0,119918113 | 0,757665988 | EGFR | FALSE | Downregulated |
| -0,176335165 | 0,757732469 | LAMTOR1 | FALSE | Downregulated |
| -0,106179407 | 0,757828267 | ARAF | FALSE | Downregulated |
| 0,120727568 | 0,757949801 | ITPK1 | FALSE | Upregulated |
| -0,177665567 | 0,758120156 | EID1 | FALSE | Downregulated |
| 0,334396067 | 0,758120156 | SLC25A36 | FALSE | Upregulated |

| | | | | |
|---|---|---|---|---|
| -0,362240649 | 0,758280242 | PTBP2 | FALSE | Downregulated |
| 0,247610464 | 0,758280242 | SOCS5 | FALSE | Upregulated |
| -0,128864235 | 0,758575018 | ACIN1 | FALSE | Downregulated |
| -0,12252101 | 0,758575018 | GPS1 | FALSE | Downregulated |
| -0,223512482 | 0,758792455 | ABCG1 | FALSE | Downregulated |
| 0,15113445 | 0,759200743 | THBS3 | FALSE | Upregulated |
| -0,170987549 | 0,75939767 | APOA5 | FALSE | Downregulated |
| 0,219430765 | 0,75939767 | ARHGEF5 | FALSE | Upregulated |
| 0,212674148 | 0,75939767 | C10orf118 | FALSE | Upregulated |
| 0,181641972 | 0,75939767 | C2orf15 | FALSE | Upregulated |
| 0,127975791 | 0,75939767 | CRIM1 | FALSE | Upregulated |
| 0,128363391 | 0,75939767 | FAM13A-AS1 | FALSE | Upregulated |
| -0,093899952 | 0,75939767 | FBRSL1 | FALSE | Downregulated |
| -0,168215019 | 0,75939767 | GCAT | FALSE | Downregulated |
| -0,178964774 | 0,75939767 | KIAA0141 | FALSE | Downregulated |
| 0,343227663 | 0,75939767 | LOC101929449 | FALSE | Upregulated |
| -1,024621038 | 0,75939767 | LOC102724504 | FALSE | Downregulated |
| 0,134703945 | 0,75939767 | MDH1 | FALSE | Upregulated |
| -0,253808587 | 0,75939767 | MED4 | FALSE | Downregulated |
| -0,132820549 | 0,75939767 | NOP14 | FALSE | Downregulated |
| -0,09900416 | 0,75939767 | REEP6 | FALSE | Downregulated |
| 0,153978797 | 0,75939767 | RNF113B | FALSE | Upregulated |
| 0,220485533 | 0,75939767 | SETDB2 | FALSE | Upregulated |
| 0,49424395 | 0,75939767 | SPATA12 | FALSE | Upregulated |
| -0,270613625 | 0,75939767 | SUGT1 | FALSE | Downregulated |
| -0,084947846 | 0,75939767 | TGOLN2 | FALSE | Downregulated |
| 0,153771811 | 0,75939767 | TTI2 | FALSE | Upregulated |
| -0,257289624 | 0,75939767 | WDR77 | FALSE | Downregulated |
| 0,329538003 | 0,75939767 | ZNF345 | FALSE | Upregulated |
| 0,199481462 | 0,75939767 | ZNF57 | FALSE | Upregulated |
| 0,158587061 | 0,75939767 | ZNF784 | FALSE | Upregulated |
| -0,25743503 | 0,759397712 | NIF3L1 | FALSE | Downregulated |
| 0,109655283 | 0,759397712 | TRMT112 | FALSE | Upregulated |
| 0,233482536 | 0,759452494 | SLC37A1 | FALSE | Upregulated |
| -0,157630474 | 0,759533197 | EBP | FALSE | Downregulated |
| 0,223387535 | 0,759533197 | KIAA1614 | FALSE | Upregulated |
| 0,145494744 | 0,759810015 | ZNF79 | FALSE | Upregulated |
| -0,148458232 | 0,759879563 | FRK | FALSE | Downregulated |
| -0,219435495 | 0,759879563 | SUB1 | FALSE | Downregulated |
| -0,435224647 | 0,759968103 | FAM84A | FALSE | Downregulated |
| -0,196090208 | 0,759968103 | KLHL22 | FALSE | Downregulated |
| 0,249841165 | 0,759987045 | C12orf73 | FALSE | Upregulated |
| -0,180396964 | 0,759987045 | PARN | FALSE | Downregulated |
| -0,195375183 | 0,759987045 | SLC28A1 | FALSE | Downregulated |
| 0,276782154 | 0,760052387 | CORO2A | FALSE | Upregulated |
| -0,166924254 | 0,760052387 | FSCN1 | FALSE | Downregulated |
| 0,149993336 | 0,760052387 | RBBP6 | FALSE | Upregulated |
| -0,12217046 | 0,760052387 | YTHDF1 | FALSE | Downregulated |
| 0,160255031 | 0,760329017 | ASB8 | FALSE | Upregulated |
| 0,10288282 | 0,760329017 | TAOK1 | FALSE | Upregulated |
| 0,156929215 | 0,760332817 | CNPY4 | FALSE | Upregulated |

| | | | | |
|---|---|---|---|---|
| 0,20657649 | 0,760458742 | LOC101928068 | FALSE | Upregulated |
| -0,100551697 | 0,760458742 | MXI1 | FALSE | Downregulated |
| -0,165640387 | 0,760458742 | SDHD | FALSE | Downregulated |
| 0,284526544 | 0,760458742 | SLC6A6 | FALSE | Upregulated |
| 0,277715315 | 0,76052057 | OAS2 | FALSE | Upregulated |
| 0,241041125 | 0,760600018 | SH2D3C | FALSE | Upregulated |
| 0,235700138 | 0,760652344 | SKA2 | FALSE | Upregulated |
| 0,192498105 | 0,760652514 | DTX2 | FALSE | Upregulated |
| 0,319927552 | 0,760652514 | LRRC55 | FALSE | Upregulated |
| -0,19968729 | 0,761038958 | HTATSF1 | FALSE | Downregulated |
| -0,31364032 | 0,761038958 | VPS37D | FALSE | Downregulated |
| 0,313189555 | 0,761041711 | LOC101928678 | FALSE | Upregulated |
| -0,09852197 | 0,761752488 | GPLD1 | FALSE | Downregulated |
| 0,203196537 | 0,761752488 | LOC102723722 | FALSE | Upregulated |
| 0,228568748 | 0,761773245 | GVINP1 | FALSE | Upregulated |
| 0,177303433 | 0,761938164 | RYR3 | FALSE | Upregulated |
| 0,225260387 | 0,762217706 | RSU1 | FALSE | Upregulated |
| -0,232356258 | 0,762217706 | ZMAT2 | FALSE | Downregulated |
| 0,150427772 | 0,762244572 | CCDC142 | FALSE | Upregulated |
| -0,184771624 | 0,762244572 | CSAD | FALSE | Downregulated |
| 0,325747889 | 0,762244572 | DAGLA | FALSE | Upregulated |
| -0,173499302 | 0,762244572 | DDX1 | FALSE | Downregulated |
| -0,180021927 | 0,762244572 | DHX34 | FALSE | Downregulated |
| -0,147678171 | 0,762244572 | GOLGA2P5 | FALSE | Downregulated |
| -0,208095867 | 0,762244572 | HFE | FALSE | Downregulated |
| -0,096119518 | 0,762244572 | IRF2BPL | FALSE | Downregulated |
| 0,147139476 | 0,762244572 | LOC284837 | FALSE | Upregulated |
| -0,117613576 | 0,762244572 | MBNL2 | FALSE | Downregulated |
| -0,069916447 | 0,762244572 | MTSS1 | FALSE | Downregulated |
| -0,122948239 | 0,762244572 | NXF1 | FALSE | Downregulated |
| 0,240380682 | 0,762244572 | OSGEP | FALSE | Upregulated |
| 0,137548033 | 0,762244572 | PDE7A | FALSE | Upregulated |
| -0,148544435 | 0,762244572 | PRDX3 | FALSE | Downregulated |
| -0,209233172 | 0,762244572 | PUS1 | FALSE | Downregulated |
| 0,184798206 | 0,762244572 | REL | FALSE | Upregulated |
| -0,126190651 | 0,762244572 | SCAF4 | FALSE | Downregulated |
| -0,276141242 | 0,762244572 | SCN1B | FALSE | Downregulated |
| 0,250803829 | 0,762244572 | SFXN3 | FALSE | Upregulated |
| -0,195400854 | 0,762244572 | STK3 | FALSE | Downregulated |
| -0,177721268 | 0,762244572 | TMEM198B | FALSE | Downregulated |
| -0,254194095 | 0,762244572 | ZNF260 | FALSE | Downregulated |
| 0,227237798 | 0,762244572 | ZNF843 | FALSE | Upregulated |
| 0,133820539 | 0,762244572 | ZNRF1 | FALSE | Upregulated |
| 0,14691618 | 0,762913516 | ATP5SL | FALSE | Upregulated |
| -0,226362061 | 0,762913516 | BCO2 | FALSE | Downregulated |
| -0,153438201 | 0,762913516 | CAPNS1 | FALSE | Downregulated |
| 0,189608154 | 0,762913516 | CCDC88C | FALSE | Upregulated |
| -0,282712713 | 0,762913516 | CENPBD1 | FALSE | Downregulated |
| -0,094954171 | 0,762913516 | CHD8 | FALSE | Downregulated |
| 0,191514039 | 0,762913516 | CKS1B | FALSE | Upregulated |
| 0,213401052 | 0,762913516 | GAPDHS | FALSE | Upregulated |

| | | | | |
|---|---|---|---|---|
| 0,499498104 | 0,762913516 | GOLGA8R | FALSE | Upregulated |
| -0,127353971 | 0,762913516 | LARGE | FALSE | Downregulated |
| 0,182948934 | 0,762913516 | LOC401296 | FALSE | Upregulated |
| 0,316347393 | 0,762913516 | PAPOLG | FALSE | Upregulated |
| 0,214732024 | 0,762913516 | RP11-478P10.1 | FALSE | Upregulated |
| 0,097297646 | 0,762913516 | SLC12A7 | FALSE | Upregulated |
| -0,189841738 | 0,762913516 | TBC1D16 | FALSE | Downregulated |
| -0,123327588 | 0,762913516 | ZFPM1 | FALSE | Downregulated |
| 0,121147865 | 0,762913516 | ZNF750 | FALSE | Upregulated |
| -0,213550774 | 0,762946125 | AMOT | FALSE | Downregulated |
| 0,093127264 | 0,762946125 | CD163 | FALSE | Upregulated |
| -0,080143355 | 0,762946125 | INSR | FALSE | Downregulated |
| -0,117841465 | 0,762946125 | SRRM1 | FALSE | Downregulated |
| -0,26727878 | 0,763055143 | AARSD1 | FALSE | Downregulated |
| -0,22001837 | 0,763055143 | CCT6A | FALSE | Downregulated |
| -0,152364747 | 0,76311562 | EPS15L1 | FALSE | Downregulated |
| -0,188509395 | 0,763151336 | BMS1 | FALSE | Downregulated |
| 0,203621865 | 0,763151336 | EIF2S1 | FALSE | Upregulated |
| 0,128675748 | 0,76324044 | RAC1 | FALSE | Upregulated |
| -0,30085064 | 0,763351311 | COL4A5 | FALSE | Downregulated |
| 0,132723218 | 0,763351311 | CSK | FALSE | Upregulated |
| -0,178968069 | 0,763351311 | PHF20L1 | FALSE | Downregulated |
| -0,10903619 | 0,763351311 | TMPRSS6 | FALSE | Downregulated |
| 0,1411238 | 0,763719656 | APC | FALSE | Upregulated |
| 0,276712071 | 0,763719656 | RAP1GAP2 | FALSE | Upregulated |
| 0,384275491 | 0,764288796 | ACE | FALSE | Upregulated |
| -0,156140724 | 0,764288796 | PIK3C2B | FALSE | Downregulated |
| 0,102040097 | 0,764288796 | RP11-793H13.3 | FALSE | Upregulated |
| -0,076068502 | 0,764288796 | USP9X | FALSE | Downregulated |
| -0,265018638 | 0,764288796 | ZNF248 | FALSE | Downregulated |
| 0,170919938 | 0,764331943 | LAS1L | FALSE | Upregulated |
| -0,145496033 | 0,764331943 | PDIA5 | FALSE | Downregulated |
| -0,196099939 | 0,764345851 | CASP7 | FALSE | Downregulated |
| 0,155867285 | 0,764345851 | DNAH11 | FALSE | Upregulated |
| -0,276211212 | 0,764345851 | DPEP2 | FALSE | Downregulated |
| -0,14741419 | 0,764345851 | ELF2 | FALSE | Downregulated |
| 0,217993397 | 0,764345851 | FAM73A | FALSE | Upregulated |
| 0,629077824 | 0,764345851 | HKDC1 | FALSE | Upregulated |
| 0,186842577 | 0,764345851 | HLA-DPA1 | FALSE | Upregulated |
| -0,194890806 | 0,764345851 | LINC01128 | FALSE | Downregulated |
| 0,14541305 | 0,764345851 | LOC101928817 | FALSE | Upregulated |
| 0,278520748 | 0,764345851 | MARVELD1 | FALSE | Upregulated |
| 0,190268608 | 0,764345851 | MRPS18C | FALSE | Upregulated |
| -0,228857818 | 0,764345851 | NDUFS3 | FALSE | Downregulated |
| -0,115577737 | 0,764345851 | PUF60 | FALSE | Downregulated |
| 0,100102157 | 0,764345851 | SDHC | FALSE | Upregulated |
| -0,282034835 | 0,764345851 | TTC12 | FALSE | Downregulated |
| 0,248175016 | 0,764345851 | UCA1 | FALSE | Upregulated |
| 0,136973735 | 0,764345851 | ZBTB18 | FALSE | Upregulated |
| -0,145586979 | 0,764487726 | ZNF281 | FALSE | Downregulated |
| 0,154380156 | 0,764502167 | DLK2 | FALSE | Upregulated |

| | | | | |
|---|---|---|---|---|
| -0,217339423 | 0,764502167 | LOC101929722 | FALSE | Downregulated |
| 0,111027813 | 0,764544697 | FKBP15 | FALSE | Upregulated |
| -0,125662509 | 0,764591862 | EEA1 | FALSE | Downregulated |
| -0,114525419 | 0,764828298 | DCTN1 | FALSE | Downregulated |
| 0,281373898 | 0,764828298 | ITGB4 | FALSE | Upregulated |
| -0,09533121 | 0,765028575 | PGRMC2 | FALSE | Downregulated |
| -0,155416698 | 0,765778201 | CSNK1E | FALSE | Downregulated |
| -0,266973798 | 0,765778201 | DTWD2 | FALSE | Downregulated |
| -0,326444185 | 0,765778201 | TLR1 | FALSE | Downregulated |
| 0,109341594 | 0,765807683 | BRI3 | FALSE | Upregulated |
| -0,143474029 | 0,765807683 | CTR9 | FALSE | Downregulated |
| 0,137390513 | 0,765807683 | DNAJB5 | FALSE | Upregulated |
| -0,222399636 | 0,765807683 | IFT20 | FALSE | Downregulated |
| 0,136273419 | 0,765807683 | TELO2 | FALSE | Upregulated |
| -0,101833775 | 0,76581667 | ACACB | FALSE | Downregulated |
| -0,113063667 | 0,766068832 | DCHS1 | FALSE | Downregulated |
| -0,243727765 | 0,766102095 | TEDDM1 | FALSE | Downregulated |
| -0,175551461 | 0,766179614 | KLHL20 | FALSE | Downregulated |
| 0,119793914 | 0,766292831 | CEP250 | FALSE | Upregulated |
| -0,171418664 | 0,766292831 | MTG1 | FALSE | Downregulated |
| -0,235250003 | 0,766292831 | PIP5K1P1 | FALSE | Downregulated |
| -0,136503661 | 0,766398495 | MPI | FALSE | Downregulated |
| 0,167699768 | 0,76672845 | SCARNA12 | FALSE | Upregulated |
| 0,325206672 | 0,766863694 | GNG12-AS1 | FALSE | Upregulated |
| -0,136888488 | 0,766863694 | ITCH | FALSE | Downregulated |
| 0,196849259 | 0,766863694 | LOC101927761 | FALSE | Upregulated |
| 0,278146696 | 0,766863694 | LOC102724974 | FALSE | Upregulated |
| 0,340431754 | 0,766863694 | LOC440570 | FALSE | Upregulated |
| 0,149277979 | 0,766863694 | MRPS14 | FALSE | Upregulated |
| -0,124616677 | 0,766863694 | UTP14C | FALSE | Downregulated |
| -0,166695801 | 0,766863694 | ZNF770 | FALSE | Downregulated |
| -0,092861925 | 0,76688394 | JAK1 | FALSE | Downregulated |
| -0,225326897 | 0,766949569 | CTBP1-AS2 | FALSE | Downregulated |
| -0,115285177 | 0,766949569 | TRIM28 | FALSE | Downregulated |
| -0,182858431 | 0,767177419 | CNOT8 | FALSE | Downregulated |
| 0,309360644 | 0,767177419 | LOC102724061 | FALSE | Upregulated |
| 0,171504494 | 0,767294029 | MORC3 | FALSE | Upregulated |
| -0,192611234 | 0,767351582 | DDA1 | FALSE | Downregulated |
| 0,225298769 | 0,767927646 | ANKRD13C | FALSE | Upregulated |
| 0,145241251 | 0,767927646 | FAM222B | FALSE | Upregulated |
| -0,091230024 | 0,767927646 | HECTD4 | FALSE | Downregulated |
| 0,085863872 | 0,767927646 | KMT2E | FALSE | Upregulated |
| -0,507507514 | 0,767927646 | LOC101927999 | FALSE | Downregulated |
| -0,192612825 | 0,767927646 | MRPL3 | FALSE | Downregulated |
| -0,152534768 | 0,767927646 | MTHFS | FALSE | Downregulated |
| -0,147796943 | 0,767927646 | ORM2 | FALSE | Downregulated |
| -0,112292133 | 0,767927646 | PRKAG2 | FALSE | Downregulated |
| 0,269568201 | 0,767927646 | RAB32 | FALSE | Upregulated |
| -0,163557578 | 0,767927646 | SNAPC5 | FALSE | Downregulated |
| -0,141436318 | 0,767927646 | XPO4 | FALSE | Downregulated |
| -0,226505387 | 0,76804035 | THAP7-AS1 | FALSE | Downregulated |

| | | | | |
|---|---|---|---|---|
| -0,089167046 | 0,76807237 | ZNF516 | FALSE | Downregulated |
| -0,127818549 | 0,76809577 | SLC35E1 | FALSE | Downregulated |
| -0,269743937 | 0,768228015 | AP4E1 | FALSE | Downregulated |
| -0,154000115 | 0,768299255 | ALDH1B1 | FALSE | Downregulated |
| 0,201668526 | 0,768299255 | ARFIP2 | FALSE | Upregulated |
| 0,364545623 | 0,768299255 | BICD1 | FALSE | Upregulated |
| 0,164099745 | 0,768299255 | CLIC1 | FALSE | Upregulated |
| -0,428452382 | 0,768299255 | LOC642423 | FALSE | Downregulated |
| 0,267900516 | 0,768299255 | SCART1 | FALSE | Upregulated |
| 0,223680723 | 0,768299255 | TSPEAR | FALSE | Upregulated |
| 0,164295876 | 0,768507812 | ARPP19 | FALSE | Upregulated |
| -0,120600069 | 0,768507812 | GTPBP6 | FALSE | Downregulated |
| 0,306247795 | 0,768507812 | LOC100507547 | FALSE | Upregulated |
| 0,207801089 | 0,768542666 | HNF4G | FALSE | Upregulated |
| -0,193188734 | 0,768673115 | SYPL1 | FALSE | Downregulated |
| -0,370099113 | 0,768712492 | ANKHD1-EIF4EBP3 | FALSE | Downregulated |
| 0,146339447 | 0,768832926 | UBE2J1 | FALSE | Upregulated |
| -0,175386898 | 0,768930784 | DGAT2 | FALSE | Downregulated |
| -0,133655449 | 0,768930784 | DNAJA3 | FALSE | Downregulated |
| 0,150535072 | 0,768930784 | GJD3 | FALSE | Upregulated |
| 0,060113525 | 0,768930784 | PRRC2A | FALSE | Upregulated |
| 0,130235426 | 0,768930784 | RGPD1 | FALSE | Upregulated |
| -0,196818227 | 0,76898777 | PTDSS2 | FALSE | Downregulated |
| -0,131645725 | 0,76898777 | TPP2 | FALSE | Downregulated |
| 0,115293029 | 0,769468586 | CAPN12 | FALSE | Upregulated |
| -0,330062709 | 0,769468586 | FAM133B | FALSE | Downregulated |
| 0,220175528 | 0,769468586 | FAM189B | FALSE | Upregulated |
| 0,080854875 | 0,769468586 | KIAA1109 | FALSE | Upregulated |
| -0,094771557 | 0,769468586 | KLF3 | FALSE | Downregulated |
| 0,22053309 | 0,769468586 | LOC102724187 | FALSE | Upregulated |
| -0,167916836 | 0,769468586 | MERTK | FALSE | Downregulated |
| 0,206387187 | 0,769468586 | NCKAP1L | FALSE | Upregulated |
| 0,128526053 | 0,769468586 | NUP188 | FALSE | Upregulated |
| -0,14475608 | 0,769468586 | RBM27 | FALSE | Downregulated |
| 0,113919349 | 0,769468586 | ZNF24 | FALSE | Upregulated |
| 0,201706647 | 0,769468586 | ZNF790 | FALSE | Upregulated |
| -0,083613962 | 0,76947891 | TPCN1 | FALSE | Downregulated |
| -0,12938951 | 0,769701307 | POLDIP3 | FALSE | Downregulated |
| 0,138168599 | 0,769745551 | KIAA1551 | FALSE | Upregulated |
| -0,092177815 | 0,769745551 | SELO | FALSE | Downregulated |
| -0,230673403 | 0,769745551 | SSB | FALSE | Downregulated |
| 0,148978331 | 0,769838954 | SF3A2 | FALSE | Upregulated |
| 0,137196721 | 0,769894514 | AIMP1 | FALSE | Upregulated |
| -0,281145444 | 0,769894514 | HINFP | FALSE | Downregulated |
| 0,282133019 | 0,769894514 | MBOAT4 | FALSE | Upregulated |
| -0,313128774 | 0,769894514 | SLC41A1 | FALSE | Downregulated |
| 0,140672832 | 0,769894514 | SMARCD1 | FALSE | Upregulated |
| -0,222067593 | 0,769894514 | TRMT11 | FALSE | Downregulated |
| 0,198307147 | 0,769965557 | INHBE | FALSE | Upregulated |
| -0,096613715 | 0,769965557 | JMJD1C | FALSE | Downregulated |

| | | | | |
|---|---|---|---|---|
| 0,246284164 | 0,769965557 | MAP3K6 | FALSE | Upregulated |
| -0,104184006 | 0,769965557 | MED13 | FALSE | Downregulated |
| -0,115282601 | 0,769965557 | PRELID1 | FALSE | Downregulated |
| 0,169084454 | 0,769965557 | TSTD2 | FALSE | Upregulated |
| 0,116137231 | 0,769965557 | USP49 | FALSE | Upregulated |
| -0,097979452 | 0,769965557 | ZRANB1 | FALSE | Downregulated |
| 0,184295624 | 0,770113525 | LOC101928045 | FALSE | Upregulated |
| -0,153168225 | 0,770296487 | ANGEL1 | FALSE | Downregulated |
| -0,194510171 | 0,770296487 | C19orf54 | FALSE | Downregulated |
| -0,14314297 | 0,770296487 | CFHR5 | FALSE | Downregulated |
| 0,354779351 | 0,770296487 | CNTNAP2 | FALSE | Upregulated |
| 0,196018769 | 0,770296487 | DHX58 | FALSE | Upregulated |
| 0,144082603 | 0,770296487 | GPM6B | FALSE | Upregulated |
| -0,142986198 | 0,770296487 | HMGN1 | FALSE | Downregulated |
| -0,126817869 | 0,770296487 | LRRC45 | FALSE | Downregulated |
| -0,204166246 | 0,770296487 | PHF13 | FALSE | Downregulated |
| -0,135839576 | 0,770296487 | PTPRS | FALSE | Downregulated |
| 0,285820223 | 0,770296487 | RBM7 | FALSE | Upregulated |
| -0,309402823 | 0,770296487 | RNASEH2B | FALSE | Downregulated |
| -0,133575319 | 0,770296487 | SNX5 | FALSE | Downregulated |
| -0,290012233 | 0,770296487 | SPATA41 | FALSE | Downregulated |
| 0,267289211 | 0,770296487 | SYTL1 | FALSE | Upregulated |
| -0,184174643 | 0,770296487 | ZNF398 | FALSE | Downregulated |
| 0,324698903 | 0,770352746 | AL022476.2 | FALSE | Upregulated |
| 0,199324897 | 0,770352746 | ANKRD32 | FALSE | Upregulated |
| -0,118401707 | 0,770352746 | ANKS1A | FALSE | Downregulated |
| -0,278591437 | 0,770352746 | CASD1 | FALSE | Downregulated |
| -0,234218475 | 0,770352746 | CDC23 | FALSE | Downregulated |
| -0,130394422 | 0,770352746 | CRTC3 | FALSE | Downregulated |
| -0,266058034 | 0,770352746 | DANCR | FALSE | Downregulated |
| -0,114890548 | 0,770352746 | EXT1 | FALSE | Downregulated |
| 0,186862806 | 0,770352746 | FTX | FALSE | Upregulated |
| -0,218071938 | 0,770352746 | GGH | FALSE | Downregulated |
| -0,172684261 | 0,770352746 | GLCCI1 | FALSE | Downregulated |
| -0,229439866 | 0,770352746 | GOLGA8A | FALSE | Downregulated |
| -0,267893216 | 0,770352746 | KCNQ1 | FALSE | Downregulated |
| -0,217287872 | 0,770352746 | PPP1R35 | FALSE | Downregulated |
| -0,163096218 | 0,770352746 | PTEN | FALSE | Downregulated |
| -0,225580915 | 0,770352746 | PTGR2 | FALSE | Downregulated |
| -0,258874522 | 0,770352746 | RP11-279F6.1 | FALSE | Downregulated |
| 0,139655462 | 0,770352746 | TXLNG | FALSE | Upregulated |
| 0,132314533 | 0,770352746 | UFM1 | FALSE | Upregulated |
| 0,243730615 | 0,770355415 | TIMM22 | FALSE | Upregulated |
| -0,168704433 | 0,770387655 | HS1BP3 | FALSE | Downregulated |
| 0,247444329 | 0,770387655 | SGCE | FALSE | Upregulated |
| 0,59781928 | 0,770508281 | BOLA2B | FALSE | Upregulated |
| -0,235355765 | 0,770542664 | NIPAL2 | FALSE | Downregulated |
| 0,13673237 | 0,771017715 | NBN | FALSE | Upregulated |
| -0,154784 | 0,771060142 | BRD8 | FALSE | Downregulated |
| -0,259579145 | 0,771060142 | CCNB1IP1 | FALSE | Downregulated |
| -0,10514675 | 0,771072597 | ALDH4A1 | FALSE | Downregulated |

| | | | | |
|---|---|---|---|---|
| 0,233469184 | 0,771072597 | SH3PXD2A-AS1 | FALSE | Upregulated |
| 0,246774931 | 0,771130609 | RPAP2 | FALSE | Upregulated |
| -0,203649329 | 0,771538325 | DRG2 | FALSE | Downregulated |
| -0,125824291 | 0,77180488 | UBE2G2 | FALSE | Downregulated |
| 0,062798266 | 0,771837162 | WNK1 | FALSE | Upregulated |
| -0,259407424 | 0,771874941 | CPSF3 | FALSE | Downregulated |
| 0,2045428 | 0,771874941 | LOC101930171 | FALSE | Upregulated |
| 0,163336288 | 0,771874941 | RAD52 | FALSE | Upregulated |
| 0,203625857 | 0,771885391 | SCG3 | FALSE | Upregulated |
| 0,567106548 | 0,771902909 | LOC101928292 | FALSE | Upregulated |
| -0,256143085 | 0,771902909 | MCM4 | FALSE | Downregulated |
| -0,204773562 | 0,771902909 | NDUFAF3 | FALSE | Downregulated |
| -0,295037648 | 0,771902909 | OR2A9P | FALSE | Downregulated |
| 0,099079383 | 0,771902909 | RIC8A | FALSE | Upregulated |
| -0,096425114 | 0,771902909 | SLC9A3R1 | FALSE | Downregulated |
| -0,144770734 | 0,771902909 | SPPL2A | FALSE | Downregulated |
| -0,133302192 | 0,771902909 | VAPA | FALSE | Downregulated |
| 0,108876677 | 0,771902909 | ZNF839 | FALSE | Upregulated |
| -0,150399729 | 0,771904481 | FES | FALSE | Downregulated |
| -0,296549798 | 0,771916886 | GBAS | FALSE | Downregulated |
| 0,159364203 | 0,772013625 | CAMK2G | FALSE | Upregulated |
| 0,123530824 | 0,772013625 | GPR146 | FALSE | Upregulated |
| -0,264394247 | 0,772013625 | GTPBP3 | FALSE | Downregulated |
| -0,164914094 | 0,772013625 | GYS1 | FALSE | Downregulated |
| -0,15368063 | 0,772013625 | KCTD6 | FALSE | Downregulated |
| 0,189477544 | 0,772013625 | MRC2 | FALSE | Upregulated |
| 0,111105422 | 0,772013625 | MRPL20 | FALSE | Upregulated |
| -0,165488156 | 0,772013625 | NEU4 | FALSE | Downregulated |
| -0,158617659 | 0,772013625 | NID2 | FALSE | Downregulated |
| 0,3071555 | 0,772013625 | POT1 | FALSE | Upregulated |
| -0,237957471 | 0,772013625 | RABEP2 | FALSE | Downregulated |
| 0,133795196 | 0,772013625 | SCIMP | FALSE | Upregulated |
| 0,250937736 | 0,772013625 | SNX20 | FALSE | Upregulated |
| 0,138656311 | 0,772013625 | SPIDR | FALSE | Upregulated |
| -0,17882105 | 0,772013625 | SPPL3 | FALSE | Downregulated |
| 0,16064565 | 0,772013625 | TGFBR1 | FALSE | Upregulated |
| -0,206164728 | 0,772013625 | TIMM10B | FALSE | Downregulated |
| 0,266854885 | 0,772013625 | TRIL | FALSE | Upregulated |
| 0,370807824 | 0,772142732 | NXPE3 | FALSE | Upregulated |
| -0,078805999 | 0,772436492 | AP2A1 | FALSE | Downregulated |
| -0,213119073 | 0,772436492 | CYB561 | FALSE | Downregulated |
| 0,182499036 | 0,772541033 | TPM1 | FALSE | Upregulated |
| -0,089778248 | 0,772560372 | ARAP1 | FALSE | Downregulated |
| 0,273554926 | 0,772737609 | ZNF134 | FALSE | Upregulated |
| -0,134302085 | 0,772859788 | GPR125 | FALSE | Downregulated |
| 1,049327815 | 0,772859788 | ISY1-RAB43 | FALSE | Upregulated |
| 0,200015065 | 0,772859788 | LOC101929633 | FALSE | Upregulated |
| 0,261951746 | 0,773271232 | LOC101928967 | FALSE | Upregulated |
| 0,21768367 | 0,773271232 | TMEM168 | FALSE | Upregulated |
| 0,187707856 | 0,773337964 | ABHD12 | FALSE | Upregulated |
| -0,098394394 | 0,773337964 | PHLPP1 | FALSE | Downregulated |

| | | | | |
|---|---|---|---|---|
| -0,202540497 | 0,773337964 | PTGFRN | FALSE | Downregulated |
| 0,210484034 | 0,773337964 | RFK | FALSE | Upregulated |
| 0,169131829 | 0,773337964 | SNX25 | FALSE | Upregulated |
| 0,220061094 | 0,77353454 | CTSC | FALSE | Upregulated |
| 0,222526362 | 0,77353454 | MVK | FALSE | Upregulated |
| -0,135679924 | 0,77359918 | COG8 | FALSE | Downregulated |
| 0,162075855 | 0,77359918 | COL18A1-AS1 | FALSE | Upregulated |
| -0,08810913 | 0,77359918 | CSNK1A1 | FALSE | Downregulated |
| 0,188843278 | 0,77359918 | KCTD9 | FALSE | Upregulated |
| 0,083355098 | 0,773735544 | ARHGDIA | FALSE | Upregulated |
| 0,109173631 | 0,773735544 | KIF5B | FALSE | Upregulated |
| 0,11615808 | 0,773783867 | EVL | FALSE | Upregulated |
| -0,228013613 | 0,773783867 | LGALS1 | FALSE | Downregulated |
| 0,154827534 | 0,773783867 | RP11-266L9.2 | FALSE | Upregulated |
| -0,117478002 | 0,773783867 | SHROOM2 | FALSE | Downregulated |
| -0,162566073 | 0,773783867 | SLC23A2 | FALSE | Downregulated |
| 0,161006168 | 0,773917675 | ELOVL5 | FALSE | Upregulated |
| -0,185918145 | 0,773993339 | CSNK2A2 | FALSE | Downregulated |
| 0,43853508 | 0,774072216 | EDA | FALSE | Upregulated |
| -0,186773308 | 0,774083967 | ARSA | FALSE | Downregulated |
| 0,306692157 | 0,774083967 | DCTPP1 | FALSE | Upregulated |
| 0,0840889 | 0,774083967 | EIF2AK1 | FALSE | Upregulated |
| -0,255351425 | 0,774083967 | NBEA | FALSE | Downregulated |
| -0,116469549 | 0,774083967 | RHOU | FALSE | Downregulated |
| -0,243533797 | 0,774083967 | UQCRH | FALSE | Downregulated |
| 0,295255972 | 0,774091138 | CDKN2C | FALSE | Upregulated |
| -0,277353809 | 0,774091138 | GAS1 | FALSE | Downregulated |
| -0,206174623 | 0,774093781 | ADAMTS17 | FALSE | Downregulated |
| 0,144934459 | 0,774093781 | ADORA2A | FALSE | Upregulated |
| 0,253052419 | 0,774093781 | AFAP1 | FALSE | Upregulated |
| 0,196518969 | 0,774093781 | CCDC39 | FALSE | Upregulated |
| 0,181512442 | 0,774093781 | CHRNE | FALSE | Upregulated |
| -0,159719709 | 0,774093781 | CX3CL1 | FALSE | Downregulated |
| -0,085748102 | 0,774093781 | ESYT1 | FALSE | Downregulated |
| 0,183779739 | 0,774093781 | FABP3 | FALSE | Upregulated |
| -0,285306566 | 0,774093781 | FZD8 | FALSE | Downregulated |
| -0,250188886 | 0,774093781 | KCNK5 | FALSE | Downregulated |
| 0,065175027 | 0,774093781 | KIAA1147 | FALSE | Upregulated |
| 0,24744548 | 0,774093781 | MAP7D3 | FALSE | Upregulated |
| -0,238993305 | 0,774093781 | MGST2 | FALSE | Downregulated |
| 0,112586098 | 0,774093781 | PITPNM2 | FALSE | Upregulated |
| 0,200736586 | 0,774093781 | PLK1S1 | FALSE | Upregulated |
| 0,199884896 | 0,774093781 | POLR3F | FALSE | Upregulated |
| -0,196314548 | 0,774093781 | REPS2 | FALSE | Downregulated |
| 0,124303759 | 0,774093781 | RNF219-AS1 | FALSE | Upregulated |
| -0,182978359 | 0,774093781 | RP11-355N15.1 | FALSE | Downregulated |
| 0,214695759 | 0,774093781 | RPS18P9 | FALSE | Upregulated |
| 0,145741383 | 0,774093781 | SNX30 | FALSE | Upregulated |
| -0,077467101 | 0,774093781 | STK11 | FALSE | Downregulated |
| 0,132097896 | 0,774093781 | TFE3 | FALSE | Upregulated |
| -0,304005168 | 0,774093781 | ZNF528 | FALSE | Downregulated |

| | | | | |
|---|---|---|---|---|
| -0,111889011 | 0,774547432 | CREG1 | FALSE | Downregulated |
| -0,157469523 | 0,774547432 | DNASE2 | FALSE | Downregulated |
| 0,178925657 | 0,774547432 | LOC101927513 | FALSE | Upregulated |
| 0,180568742 | 0,774547432 | PIK3CD | FALSE | Upregulated |
| 0,173110127 | 0,774547432 | TMEM9 | FALSE | Upregulated |
| -0,090104205 | 0,774614061 | DDX6 | FALSE | Downregulated |
| 0,178885578 | 0,774614061 | POLR3G | FALSE | Upregulated |
| 0,199613559 | 0,774912735 | PRORSD1P | FALSE | Upregulated |
| 0,228227255 | 0,774929812 | DYNC2H1 | FALSE | Upregulated |
| -0,168956036 | 0,774929812 | GLO1 | FALSE | Downregulated |
| 0,232360235 | 0,774929812 | LOC101929328 | FALSE | Upregulated |
| -0,094179284 | 0,774929812 | MGEA5 | FALSE | Downregulated |
| -0,28755393 | 0,774929812 | NUDT14 | FALSE | Downregulated |
| -0,330161472 | 0,774937775 | GATM-AS1 | FALSE | Downregulated |
| 0,135682132 | 0,774937775 | TCP1 | FALSE | Upregulated |
| 0,198266484 | 0,774950283 | ZNF37BP | FALSE | Upregulated |
| -0,147040499 | 0,775162144 | GEMIN4 | FALSE | Downregulated |
| -0,146282518 | 0,775162144 | SERINC3 | FALSE | Downregulated |
| -0,178805458 | 0,775305099 | VPS37A | FALSE | Downregulated |
| 0,127229471 | 0,775566724 | FYCO1 | FALSE | Upregulated |
| -0,130282041 | 0,775566724 | PI4K2A | FALSE | Downregulated |
| 0,176111968 | 0,775615217 | KIAA1407 | FALSE | Upregulated |
| 0,417826915 | 0,775615217 | LOC101929790 | FALSE | Upregulated |
| 0,177072877 | 0,775615217 | ZMYND19 | FALSE | Upregulated |
| -0,221285722 | 0,77591897 | C2orf47 | FALSE | Downregulated |
| -0,191825519 | 0,775983205 | GREM2 | FALSE | Downregulated |
| 0,159286459 | 0,776236545 | DBP | FALSE | Upregulated |
| -0,239188673 | 0,776236545 | EFHD1 | FALSE | Downregulated |
| 0,157115596 | 0,776236545 | UBQLN4 | FALSE | Upregulated |
| 0,154087418 | 0,776236545 | XPOT | FALSE | Upregulated |
| 0,129215355 | 0,776260695 | CEP170 | FALSE | Upregulated |
| -0,133518099 | 0,776260695 | FKBP2 | FALSE | Downregulated |
| -0,246300204 | 0,776260695 | RP11-492A10.1 | FALSE | Downregulated |
| -0,288712447 | 0,776260695 | SNORD10 | FALSE | Downregulated |
| 0,28632507 | 0,776260695 | UGGT2 | FALSE | Upregulated |
| -0,104356301 | 0,776361584 | SDHA | FALSE | Downregulated |
| -0,130244608 | 0,776363613 | PIK3C2G | FALSE | Downregulated |
| -0,141675497 | 0,776731207 | DHRS4-AS1 | FALSE | Downregulated |
| 0,207544029 | 0,776731207 | LOC101928143 | FALSE | Upregulated |
| 0,151504704 | 0,776731207 | LOC102724273 | FALSE | Upregulated |
| -0,062637721 | 0,776731207 | PLXNB2 | FALSE | Downregulated |
| 0,114314107 | 0,776731207 | PSMD7 | FALSE | Upregulated |
| 0,181717434 | 0,776731207 | SH2D3A | FALSE | Upregulated |
| -0,137728601 | 0,776731207 | XRN1 | FALSE | Downregulated |
| 0,117820779 | 0,776731207 | ZNF513 | FALSE | Upregulated |
| -0,18709546 | 0,776940241 | CSTF3 | FALSE | Downregulated |
| 0,255581045 | 0,77774862 | ANK2 | FALSE | Upregulated |
| 0,112444711 | 0,77774862 | AXL | FALSE | Upregulated |
| 0,200524867 | 0,77774862 | CD84 | FALSE | Upregulated |
| -0,314608549 | 0,77774862 | CYS1 | FALSE | Downregulated |
| -0,225153991 | 0,77774862 | DDAH2 | FALSE | Downregulated |

| | | | | |
|---|---|---|---|---|
| 0,172424218 | 0,77774862 | DDX19B | FALSE | Upregulated |
| 0,197738636 | 0,77774862 | DSTYK | FALSE | Upregulated |
| -0,167697874 | 0,77774862 | ITM2C | FALSE | Downregulated |
| 0,148203355 | 0,77774862 | LOC102724342 | FALSE | Upregulated |
| 0,184869598 | 0,77774862 | LOC606724 | FALSE | Upregulated |
| 0,160551283 | 0,77774862 | MCTP1 | FALSE | Upregulated |
| -0,121866854 | 0,77774862 | MED15 | FALSE | Downregulated |
| 0,195439843 | 0,77774862 | NR4A3 | FALSE | Upregulated |
| 0,249244616 | 0,77774862 | NRG1 | FALSE | Upregulated |
| -0,252519214 | 0,77774862 | NUDT12 | FALSE | Downregulated |
| -0,417276002 | 0,77774862 | OVCA2 | FALSE | Downregulated |
| 0,182912971 | 0,77774862 | RNR2 | FALSE | Upregulated |
| -0,206443529 | 0,77774862 | RNU105A | FALSE | Downregulated |
| -0,207099812 | 0,77774862 | RYBP | FALSE | Downregulated |
| 0,120601766 | 0,77774862 | SENP6 | FALSE | Upregulated |
| -0,134886054 | 0,77774862 | SLC43A3 | FALSE | Downregulated |
| 0,31595353 | 0,77774862 | SNAI3-AS1 | FALSE | Upregulated |
| -0,061785612 | 0,77774862 | SUN2 | FALSE | Downregulated |
| 0,182778478 | 0,77774862 | TNK1 | FALSE | Upregulated |
| 0,287756271 | 0,77774862 | VPS33A | FALSE | Upregulated |
| -0,227017241 | 0,77774862 | YOD1 | FALSE | Downregulated |
| -0,182358661 | 0,77774862 | ZMAT1 | FALSE | Downregulated |
| 0,12780443 | 0,777913609 | EVI2A | FALSE | Upregulated |
| 0,188949756 | 0,777975257 | POU5F1P3 | FALSE | Upregulated |
| -0,20021957 | 0,777975257 | ZNF490 | FALSE | Downregulated |
| -0,304987256 | 0,777979742 | HAUS7 | FALSE | Downregulated |
| -0,153518459 | 0,77844539 | ANP32B | FALSE | Downregulated |
| 0,225211593 | 0,77844539 | CCDC121 | FALSE | Upregulated |
| -0,150707677 | 0,77844539 | PPP2CA | FALSE | Downregulated |
| 0,096766377 | 0,77844539 | SLC5A2 | FALSE | Upregulated |
| 0,146621543 | 0,77844539 | WBP11 | FALSE | Upregulated |
| 0,183020155 | 0,778597186 | FAM221B | FALSE | Upregulated |
| 0,226791474 | 0,778597186 | FAM50B | FALSE | Upregulated |
| 0,193048165 | 0,778597186 | GNPAT | FALSE | Upregulated |
| -0,16604774 | 0,778597186 | KLF10 | FALSE | Downregulated |
| 0,180547586 | 0,778597186 | NTAN1 | FALSE | Upregulated |
| -0,265809034 | 0,778597186 | PPP1R3G | FALSE | Downregulated |
| -0,278930301 | 0,778597186 | RAPGEFL1 | FALSE | Downregulated |
| 0,289584587 | 0,778597186 | SHISA9 | FALSE | Upregulated |
| 0,173057749 | 0,778597186 | SMAD1-AS2 | FALSE | Upregulated |
| -0,187011107 | 0,778597186 | SPTLC1 | FALSE | Downregulated |
| -0,146445623 | 0,778597186 | TMEM33 | FALSE | Downregulated |
| 0,114709488 | 0,778597186 | WIPI2 | FALSE | Upregulated |
| -0,152463608 | 0,778597186 | ZDHHC3 | FALSE | Downregulated |
| 0,261396779 | 0,778597186 | ZNF441 | FALSE | Upregulated |
| -0,074387593 | 0,778617971 | MYH10 | FALSE | Downregulated |
| -0,276634318 | 0,778692183 | ARHGAP20 | FALSE | Downregulated |
| 0,181269014 | 0,778692183 | BTRC | FALSE | Upregulated |
| -0,155064873 | 0,778692183 | KDM6A | FALSE | Downregulated |
| 0,085743534 | 0,778692183 | KIAA0195 | FALSE | Upregulated |
| -0,189569505 | 0,778692183 | MRPS30 | FALSE | Downregulated |

| | | | | |
|---|---|---|---|---|
| 0,284180238 | 0,778692183 | PLEKHB2 | FALSE | Upregulated |
| -0,082159216 | 0,778692183 | PPP1R10 | FALSE | Downregulated |
| -0,095732474 | 0,778692183 | SIPA1L1 | FALSE | Downregulated |
| 0,227733557 | 0,778874461 | ADAMTS8 | FALSE | Upregulated |
| 0,11721154 | 0,778874461 | DBNL | FALSE | Upregulated |
| -0,11876214 | 0,778874461 | ETF1 | FALSE | Downregulated |
| 0,124736029 | 0,778874461 | FAM211A | FALSE | Upregulated |
| 0,140524684 | 0,778874461 | GOLGA8H | FALSE | Upregulated |
| 0,152735675 | 0,778874461 | IFFO1 | FALSE | Upregulated |
| 0,178195512 | 0,778874461 | KLHL8 | FALSE | Upregulated |
| -0,591383472 | 0,778874461 | LOC100131257 | FALSE | Downregulated |
| -0,172606053 | 0,778874461 | MPP1 | FALSE | Downregulated |
| -0,153942223 | 0,778874461 | MSRA | FALSE | Downregulated |
| -0,127013057 | 0,778874461 | PPP1R26 | FALSE | Downregulated |
| -0,200367137 | 0,778874461 | SEPW1 | FALSE | Downregulated |
| -0,260771729 | 0,778874461 | SLC2A13 | FALSE | Downregulated |
| 0,193303367 | 0,778874461 | ZNF213 | FALSE | Upregulated |
| -0,129309851 | 0,778879097 | ASMTL-AS1 | FALSE | Downregulated |
| 0,282228215 | 0,778879097 | ATP1B3 | FALSE | Upregulated |
| 0,169178069 | 0,778879097 | GNG12 | FALSE | Upregulated |
| -0,1025371 | 0,778879097 | MAP2K1 | FALSE | Downregulated |
| -0,16513809 | 0,778879097 | MMP14 | FALSE | Downregulated |
| -0,188634702 | 0,778879097 | NAAA | FALSE | Downregulated |
| 0,233859797 | 0,778879097 | TMEM221 | FALSE | Upregulated |
| -0,203790185 | 0,779320378 | IMPACT | FALSE | Downregulated |
| 0,163386965 | 0,77947081 | NPAT | FALSE | Upregulated |
| 0,245523701 | 0,779711059 | ALPK3 | FALSE | Upregulated |
| -0,114184976 | 0,779711059 | CLINT1 | FALSE | Downregulated |
| -0,100069569 | 0,779711059 | CTSD | FALSE | Downregulated |
| -0,230484812 | 0,779711059 | FAM214B | FALSE | Downregulated |
| -0,343929648 | 0,779711059 | FAM221A | FALSE | Downregulated |
| 0,077877792 | 0,779711059 | HBP1 | FALSE | Upregulated |
| -0,319370852 | 0,779711059 | LILRA6 | FALSE | Downregulated |
| -0,25637095 | 0,779711059 | LOC100506596 | FALSE | Downregulated |
| -0,21199468 | 0,779711059 | MIIP | FALSE | Downregulated |
| -0,165609533 | 0,779711059 | MREG | FALSE | Downregulated |
| 0,091956391 | 0,779711059 | PDCD4 | FALSE | Upregulated |
| -0,143646218 | 0,779711059 | RAB40C | FALSE | Downregulated |
| 0,142885964 | 0,779760812 | AC005594.3 | FALSE | Upregulated |
| -0,308815688 | 0,779760812 | ANKRD53 | FALSE | Downregulated |
| 0,107200092 | 0,779760812 | ERF | FALSE | Upregulated |
| -0,181208851 | 0,780020381 | CUTA | FALSE | Downregulated |
| 0,152467736 | 0,780252278 | DHX33 | FALSE | Upregulated |
| -0,226640675 | 0,780753442 | METTL9 | FALSE | Downregulated |
| -0,131698052 | 0,780915761 | GFRA1 | FALSE | Downregulated |
| -0,136727127 | 0,781184297 | IP6K1 | FALSE | Downregulated |
| 0,229489696 | 0,781203323 | FBXL18 | FALSE | Upregulated |
| 0,123736016 | 0,781203323 | SGK494 | FALSE | Upregulated |
| -0,077586011 | 0,781319383 | NUCB1 | FALSE | Downregulated |
| 0,136860454 | 0,781636987 | MFSD6 | FALSE | Upregulated |
| 0,184276334 | 0,78188087 | C9orf96 | FALSE | Upregulated |

| | | | | |
|---|---|---|---|---|
| 0,253748021 | 0,781923077 | RNU2-2P | FALSE | Upregulated |
| 0,19201441 | 0,781923077 | SCMH1 | FALSE | Upregulated |
| 0,125666527 | 0,781941628 | ZC2HC1C | FALSE | Upregulated |
| 0,252071645 | 0,781941628 | ZRSR2 | FALSE | Upregulated |
| 0,122361282 | 0,78205394 | PTPN14 | FALSE | Upregulated |
| -0,14459978 | 0,782109449 | PDHA1 | FALSE | Downregulated |
| 0,231658474 | 0,782311485 | WDR83 | FALSE | Upregulated |
| -0,208220939 | 0,782638819 | SMARCAL1 | FALSE | Downregulated |
| 0,85686531 | 0,782641719 | JMJD7-PLA2G4B | FALSE | Upregulated |
| 0,224742019 | 0,782778088 | DIMT1 | FALSE | Upregulated |
| 0,205162868 | 0,783098879 | PRMT5 | FALSE | Upregulated |
| -0,17824589 | 0,783562279 | FBXO9 | FALSE | Downregulated |
| -0,146762066 | 0,783576269 | DNAJB2 | FALSE | Downregulated |
| 0,314388619 | 0,783576269 | GRHL2 | FALSE | Upregulated |
| 0,19833532 | 0,783576269 | PDK3 | FALSE | Upregulated |
| -0,183483044 | 0,783726738 | ARHGEF9 | FALSE | Downregulated |
| -0,124106597 | 0,783765403 | SSH1 | FALSE | Downregulated |
| -0,167414905 | 0,783871749 | ACOT12 | FALSE | Downregulated |
| 0,230141887 | 0,783871749 | C9orf91 | FALSE | Upregulated |
| -0,09362183 | 0,783871749 | GLYR1 | FALSE | Downregulated |
| -0,096005872 | 0,783871749 | GSTK1 | FALSE | Downregulated |
| -0,3352977 | 0,783871749 | LINC01107 | FALSE | Downregulated |
| 0,078229028 | 0,783871749 | PLIN4 | FALSE | Upregulated |
| -0,073127083 | 0,783871749 | PRKCSH | FALSE | Downregulated |
| 0,10867026 | 0,783871749 | SH3BP5 | FALSE | Upregulated |
| -0,114971815 | 0,783871749 | UBP1 | FALSE | Downregulated |
| 0,134170743 | 0,784141569 | C9orf37 | FALSE | Upregulated |
| -0,200641891 | 0,784748317 | C7orf50 | FALSE | Downregulated |
| -0,422543414 | 0,784748317 | DHRS2 | FALSE | Downregulated |
| 0,285535911 | 0,784748317 | MRPL22 | FALSE | Upregulated |
| -0,143957914 | 0,784748317 | MRPL34 | FALSE | Downregulated |
| 0,814659256 | 0,784748317 | PYGM | FALSE | Upregulated |
| -0,404796782 | 0,784851734 | ANKRD33B | FALSE | Downregulated |
| -0,090811192 | 0,784851734 | RAPGEF2 | FALSE | Downregulated |
| -0,341733597 | 0,784851734 | SH2D1B | FALSE | Downregulated |
| 0,074005021 | 0,784858214 | BPTF | FALSE | Upregulated |
| -0,151630218 | 0,784858214 | CAPRIN2 | FALSE | Downregulated |
| 0,227290337 | 0,784858214 | LOC100996405 | FALSE | Upregulated |
| 0,158103135 | 0,784858214 | PROCR | FALSE | Upregulated |
| 0,083653215 | 0,784858214 | SGPL1 | FALSE | Upregulated |
| 0,155253923 | 0,784913837 | MYH7B | FALSE | Upregulated |
| 0,106965697 | 0,784913837 | SLC47A1 | FALSE | Upregulated |
| -0,179297099 | 0,785331469 | CAMK1D | FALSE | Downregulated |
| -0,088364058 | 0,785331469 | DDX42 | FALSE | Downregulated |
| 0,152347166 | 0,785331469 | DDX51 | FALSE | Upregulated |
| -0,375042512 | 0,785331469 | EIF3CL | FALSE | Downregulated |
| 0,252750386 | 0,785331469 | FAM210A | FALSE | Upregulated |
| -0,103609102 | 0,785331469 | HNRNPDL | FALSE | Downregulated |
| 0,157166491 | 0,785331469 | PLEKHA4 | FALSE | Upregulated |
| 0,14916913 | 0,785331469 | PRR3 | FALSE | Upregulated |
| 0,291658888 | 0,785331469 | SLC45A4 | FALSE | Upregulated |

| | | | | |
|---|---|---|---|---|
| 0,175310654 | 0,785331469 | TWF2 | FALSE | Upregulated |
| -0,148458591 | 0,785331469 | UFL1 | FALSE | Downregulated |
| -0,106092141 | 0,785331469 | WDR59 | FALSE | Downregulated |
| -0,26316525 | 0,785331469 | ZFAND1 | FALSE | Downregulated |
| -0,205640566 | 0,785796178 | HIST1H4E | FALSE | Downregulated |
| -0,229624774 | 0,785796178 | LINC00574 | FALSE | Downregulated |
| -0,267917089 | 0,785796178 | MRPL44 | FALSE | Downregulated |
| -0,155482496 | 0,78580502 | PPP1R3B | FALSE | Downregulated |
| -0,180396077 | 0,78580502 | SLC19A1 | FALSE | Downregulated |
| -0,167803789 | 0,78580502 | SLC22A9 | FALSE | Downregulated |
| -0,124239206 | 0,78580502 | ZBTB10 | FALSE | Downregulated |
| 0,288422619 | 0,78580748 | BIN2 | FALSE | Upregulated |
| 0,13841026 | 0,78580748 | LOC102724761 | FALSE | Upregulated |
| 0,18717272 | 0,78588306 | DUSP22 | FALSE | Upregulated |
| -0,141659725 | 0,78588306 | NF2 | FALSE | Downregulated |
| -0,107563631 | 0,78588306 | SLTM | FALSE | Downregulated |
| 0,407115319 | 0,786014549 | DNAH14 | FALSE | Upregulated |
| 0,14678085 | 0,786014549 | IDH3G | FALSE | Upregulated |
| 0,276944509 | 0,786014549 | LOC102723405 | FALSE | Upregulated |
| 0,297600267 | 0,786014549 | RAC2 | FALSE | Upregulated |
| 0,275362984 | 0,786055816 | DNAH10 | FALSE | Upregulated |
| 0,237975533 | 0,786055816 | IL33 | FALSE | Upregulated |
| 0,108157802 | 0,786055816 | ZMIZ1 | FALSE | Upregulated |
| 0,656273191 | 0,786092152 | FLNC | FALSE | Upregulated |
| -0,191824392 | 0,786187721 | ATAD3B | FALSE | Downregulated |
| 0,154671857 | 0,786187721 | CDIPT | FALSE | Upregulated |
| -0,093526467 | 0,786187721 | KDM4B | FALSE | Downregulated |
| -0,208737605 | 0,786187721 | MFSD9 | FALSE | Downregulated |
| 0,259920485 | 0,786187721 | NECAB2 | FALSE | Upregulated |
| -0,110188445 | 0,786187721 | SYNCRIP | FALSE | Downregulated |
| 0,093070352 | 0,786259702 | MBNL1 | FALSE | Upregulated |
| 0,387460586 | 0,786492301 | ADRA2A | FALSE | Upregulated |
| 0,130070968 | 0,786492301 | ASH1L-AS1 | FALSE | Upregulated |
| -0,122409117 | 0,786492301 | CEP95 | FALSE | Downregulated |
| -0,127959231 | 0,786492301 | CHMP1A | FALSE | Downregulated |
| -0,191263159 | 0,786492301 | COQ10A | FALSE | Downregulated |
| -0,278733391 | 0,786492301 | DRD5 | FALSE | Downregulated |
| 0,209851528 | 0,786492301 | ID3 | FALSE | Upregulated |
| -0,175988481 | 0,786492301 | KIAA1919 | FALSE | Downregulated |
| 0,11699306 | 0,786492301 | MORC2-AS1 | FALSE | Upregulated |
| -0,174917336 | 0,786492301 | NUP85 | FALSE | Downregulated |
| -0,093269645 | 0,786492301 | PLXND1 | FALSE | Downregulated |
| -0,194503097 | 0,786492301 | SEC22C | FALSE | Downregulated |
| 0,165151337 | 0,786503548 | ADAM15 | FALSE | Upregulated |
| -0,095203097 | 0,786503548 | DIO1 | FALSE | Downregulated |
| -0,123908882 | 0,786757839 | HSPA8 | FALSE | Downregulated |
| -0,12691773 | 0,786757839 | PDLIM2 | FALSE | Downregulated |
| -0,169719304 | 0,786757839 | RCOR3 | FALSE | Downregulated |
| -0,290883322 | 0,786757839 | TNFSF8 | FALSE | Downregulated |
| -0,134801994 | 0,786759535 | ZFP91 | FALSE | Downregulated |
| -0,168556991 | 0,786831767 | ATG16L1 | FALSE | Downregulated |

| | | | | |
|---|---|---|---|---|
| -0,175909684 | 0,786831767 | CLDN15 | FALSE | Downregulated |
| -0,197326124 | 0,786831767 | EHD4 | FALSE | Downregulated |
| -0,20486573 | 0,786831767 | TCEAL3 | FALSE | Downregulated |
| 0,267204003 | 0,786831767 | ZNF136 | FALSE | Upregulated |
| -0,18076783 | 0,786832294 | MID1IP1 | FALSE | Downregulated |
| -0,229308869 | 0,786923308 | FOLH1B | FALSE | Downregulated |
| -0,124930167 | 0,786923308 | GPAA1 | FALSE | Downregulated |
| -0,12610485 | 0,786923308 | LAMC1 | FALSE | Downregulated |
| 0,085994201 | 0,786923308 | PAICS | FALSE | Upregulated |
| 0,295856717 | 0,78697867 | METRNL | FALSE | Upregulated |
| -0,163938981 | 0,78697867 | SCN9A | FALSE | Downregulated |
| 0,246318182 | 0,78697867 | TOMM5 | FALSE | Upregulated |
| 0,156275509 | 0,787810848 | LYRM7 | FALSE | Upregulated |
| -0,164796588 | 0,788025064 | FANCF | FALSE | Downregulated |
| -0,279652728 | 0,788025064 | ISPD | FALSE | Downregulated |
| 0,190592881 | 0,788066113 | WDR41 | FALSE | Upregulated |
| -0,10890987 | 0,788190355 | DGCR8 | FALSE | Downregulated |
| 0,27497444 | 0,788279932 | CLEC10A | FALSE | Upregulated |
| -0,214964031 | 0,788279932 | MBLAC2 | FALSE | Downregulated |
| -0,143833032 | 0,788487075 | BANP | FALSE | Downregulated |
| -0,272829578 | 0,78872228 | CCBL2 | FALSE | Downregulated |
| -0,194809566 | 0,78872228 | EMG1 | FALSE | Downregulated |
| -0,163004288 | 0,78872228 | FAAH | FALSE | Downregulated |
| 0,708503 | 0,78872228 | GIMAP5 | FALSE | Upregulated |
| 0,127362077 | 0,78872228 | IFIT1 | FALSE | Upregulated |
| 0,286648714 | 0,78872228 | LOC101927402 | FALSE | Upregulated |
| 0,117998473 | 0,78872228 | NBPF9 | FALSE | Upregulated |
| 0,215274593 | 0,78872228 | PLLP | FALSE | Upregulated |
| -0,156437478 | 0,78872228 | PPP1R11 | FALSE | Downregulated |
| 0,218153358 | 0,78872228 | PRDM15 | FALSE | Upregulated |
| -0,140347164 | 0,78872228 | PUS3 | FALSE | Downregulated |
| -0,169403792 | 0,78872228 | TMEM150A | FALSE | Downregulated |
| -0,193453474 | 0,78872228 | TRAPPC4 | FALSE | Downregulated |
| 0,314834929 | 0,78872228 | WASH3P | FALSE | Upregulated |
| 0,288147481 | 0,788810283 | COL23A1 | FALSE | Upregulated |
| -0,252831148 | 0,789101822 | FAM127A | FALSE | Downregulated |
| 0,393089045 | 0,789101822 | LOC101060632 | FALSE | Upregulated |
| 0,280600354 | 0,78917873 | CNTNAP3B | FALSE | Upregulated |
| 0,144248876 | 0,78917873 | NAA25 | FALSE | Upregulated |
| -0,173291442 | 0,78917873 | SDF2 | FALSE | Downregulated |
| -0,763540442 | 0,789575216 | BIVM-ERCC5 | FALSE | Downregulated |
| 0,14651355 | 0,78960765 | HMG20A | FALSE | Upregulated |
| -0,175672331 | 0,789746762 | CCSER2 | FALSE | Downregulated |
| 0,101484749 | 0,789790233 | ORAI2 | FALSE | Upregulated |
| -0,087358899 | 0,789949909 | SEPHS2 | FALSE | Downregulated |
| 0,29390631 | 0,790146923 | EXOC3L1 | FALSE | Upregulated |
| -0,11817933 | 0,79016089 | CAMK2N1 | FALSE | Downregulated |
| 0,091573632 | 0,79016089 | EIF5A | FALSE | Upregulated |
| 0,249556707 | 0,79016089 | LOC102724164 | FALSE | Upregulated |
| 0,099499731 | 0,79016089 | OAZ2 | FALSE | Upregulated |
| -0,162403484 | 0,79016089 | PPP1R7 | FALSE | Downregulated |

| | | | | |
|---|---|---|---|---|
| 0,289107958 | 0,790286441 | FASTKD1 | FALSE | Upregulated |
| 0,272781138 | 0,790463883 | LIN37 | FALSE | Upregulated |
| 0,100054186 | 0,790608274 | C19orf44 | FALSE | Upregulated |
| 0,22108684 | 0,790608274 | CCBL1 | FALSE | Upregulated |
| -0,115996099 | 0,790608274 | EYA3 | FALSE | Downregulated |
| 0,127587222 | 0,790608274 | MARCKSL1 | FALSE | Upregulated |
| 0,121561595 | 0,790608274 | PROSER2 | FALSE | Upregulated |
| -0,220004394 | 0,790608274 | SPATA5L1 | FALSE | Downregulated |
| 0,157718832 | 0,790816608 | DR1 | FALSE | Upregulated |
| 0,141179341 | 0,790816608 | PGAM1P5 | FALSE | Upregulated |
| -0,179270478 | 0,790816608 | ZNF628 | FALSE | Downregulated |
| -0,238550932 | 0,790845144 | ACY3 | FALSE | Downregulated |
| -0,146601614 | 0,790845144 | BORA | FALSE | Downregulated |
| 0,152415317 | 0,790845144 | HAVCR1P1 | FALSE | Upregulated |
| 0,114694355 | 0,790960999 | IRF3 | FALSE | Upregulated |
| -0,227967559 | 0,790960999 | TMEM14C | FALSE | Downregulated |
| 0,149105813 | 0,791165265 | PPDPF | FALSE | Upregulated |
| -0,214813951 | 0,791262628 | SEMA6B | FALSE | Downregulated |
| -0,229612355 | 0,791262628 | SLC36A1 | FALSE | Downregulated |
| 0,168504891 | 0,791344825 | A2M-AS1 | FALSE | Upregulated |
| -0,147159134 | 0,791344825 | CCNG1 | FALSE | Downregulated |
| 0,167820871 | 0,791344825 | HMGA1 | FALSE | Upregulated |
| -0,108586156 | 0,791344825 | HSP90B1 | FALSE | Downregulated |
| -0,111927076 | 0,791344825 | MRC1 | FALSE | Downregulated |
| 0,439560448 | 0,791344825 | POC1B-GALNT4 | FALSE | Upregulated |
| 0,142370815 | 0,791385109 | AC007038.7 | FALSE | Upregulated |
| 0,203140052 | 0,791385109 | AKAP10 | FALSE | Upregulated |
| 0,089161991 | 0,791385109 | COG5 | FALSE | Upregulated |
| 0,188041171 | 0,791385109 | GPR137 | FALSE | Upregulated |
| 0,250507091 | 0,791385109 | HLA-DQB1 | FALSE | Upregulated |
| 0,112422423 | 0,791385109 | NELFCD | FALSE | Upregulated |
| -0,107590819 | 0,791385109 | PCNX | FALSE | Downregulated |
| -0,295537998 | 0,791385109 | POLI | FALSE | Downregulated |
| 0,183457185 | 0,791385109 | RPS6KC1 | FALSE | Upregulated |
| 0,141492037 | 0,791385109 | TMEM39A | FALSE | Upregulated |
| -0,207963559 | 0,791385109 | ZNF767 | FALSE | Downregulated |
| -0,291502593 | 0,791389161 | CD81-AS1 | FALSE | Downregulated |
| 0,213765659 | 0,791734196 | ACTA2 | FALSE | Upregulated |
| 0,250049668 | 0,791734196 | LOC102724217 | FALSE | Upregulated |
| 0,248659867 | 0,791974181 | RABL2B | FALSE | Upregulated |
| -0,12890098 | 0,792030064 | FAM35A | FALSE | Downregulated |
| -0,194427056 | 0,792254527 | DPH7 | FALSE | Downregulated |
| 0,113908911 | 0,792288152 | ZDHHC18 | FALSE | Upregulated |
| -0,141461694 | 0,792606544 | BCAR3 | FALSE | Downregulated |
| 0,147509621 | 0,792606544 | C9orf173 | FALSE | Upregulated |
| 0,146472738 | 0,792606544 | CBX6 | FALSE | Upregulated |
| 0,245135189 | 0,792606544 | CDHR2 | FALSE | Upregulated |
| -0,167334959 | 0,792606544 | COX5A | FALSE | Downregulated |
| 0,151157031 | 0,792606544 | DYNLL1 | FALSE | Upregulated |
| 0,388922439 | 0,792606544 | HIST2H3A | FALSE | Upregulated |
| 0,284735922 | 0,792606544 | IL18 | FALSE | Upregulated |

| | | | | |
|---|---|---|---|---|
| -0,193591145 | 0,792606544 | LAD1 | FALSE | Downregulated |
| -0,300557042 | 0,792606544 | LINC00884 | FALSE | Downregulated |
| -0,193025941 | 0,792606544 | LOC101928355 | FALSE | Downregulated |
| -0,112516144 | 0,792606544 | MED12 | FALSE | Downregulated |
| -0,327713362 | 0,792606544 | MSRB3 | FALSE | Downregulated |
| 0,154406805 | 0,792606544 | NAP1L1 | FALSE | Upregulated |
| -0,101618241 | 0,792606544 | NRIP1 | FALSE | Downregulated |
| 0,162424534 | 0,792606544 | PIKFYVE | FALSE | Upregulated |
| -0,168383294 | 0,792606544 | PPRC1 | FALSE | Downregulated |
| -0,292890441 | 0,792606544 | RBM34 | FALSE | Downregulated |
| 0,157770614 | 0,792606544 | REEP1 | FALSE | Upregulated |
| -0,105432328 | 0,792606544 | SPSB3 | FALSE | Downregulated |
| -0,247108151 | 0,792606544 | TDRD10 | FALSE | Downregulated |
| -0,11102862 | 0,792606544 | TPP1 | FALSE | Downregulated |
| -0,182097504 | 0,792606544 | TPST1 | FALSE | Downregulated |
| 0,266602338 | 0,792606544 | TVP23B | FALSE | Upregulated |
| 0,298584908 | 0,792606544 | TXNDC9 | FALSE | Upregulated |
| 0,194439024 | 0,792606544 | UG0898H09 | FALSE | Upregulated |
| -0,202129516 | 0,792606544 | UNG | FALSE | Downregulated |
| -0,229374556 | 0,792606544 | ZBTB2 | FALSE | Downregulated |
| 0,174974921 | 0,792991939 | C10orf25 | FALSE | Upregulated |
| 0,165730017 | 0,792991939 | PFN2 | FALSE | Upregulated |
| -0,153455859 | 0,793207192 | SPIRE2 | FALSE | Downregulated |
| 0,171009975 | 0,793314919 | LINC00514 | FALSE | Upregulated |
| -0,146942937 | 0,793314919 | SLC16A8 | FALSE | Downregulated |
| -0,114213046 | 0,793562384 | AK4 | FALSE | Downregulated |
| 0,142995915 | 0,793562384 | AKR7L | FALSE | Upregulated |
| 0,130007995 | 0,793562384 | BRAT1 | FALSE | Upregulated |
| -0,111888476 | 0,793562384 | CBX7 | FALSE | Downregulated |
| 0,183996231 | 0,793562384 | IL15RA | FALSE | Upregulated |
| 0,158475948 | 0,793562384 | LOC100506258 | FALSE | Upregulated |
| 0,170407149 | 0,793562384 | OMD | FALSE | Upregulated |
| 0,271063765 | 0,793562384 | ZNF235 | FALSE | Upregulated |
| 0,198820509 | 0,793628461 | LOC100652768 | FALSE | Upregulated |
| 0,125417007 | 0,793861463 | LRRC4 | FALSE | Upregulated |
| 0,215327089 | 0,793861463 | SLIT1 | FALSE | Upregulated |
| -0,134269213 | 0,793946894 | AZIN1 | FALSE | Downregulated |
| -0,114577325 | 0,793946894 | FLOT1 | FALSE | Downregulated |
| -0,072467497 | 0,793946894 | ITM2B | FALSE | Downregulated |
| 0,114982805 | 0,793946894 | LOC441081 | FALSE | Upregulated |
| -0,218314874 | 0,793946894 | MRPL52 | FALSE | Downregulated |
| -0,085498915 | 0,793946894 | MYO15B | FALSE | Downregulated |
| 0,127472848 | 0,793946894 | RAI14 | FALSE | Upregulated |
| 0,153149186 | 0,793946894 | SMIM15 | FALSE | Upregulated |
| 0,179926971 | 0,793946894 | THAP9 | FALSE | Upregulated |
| 0,09351335 | 0,793946894 | TPPP | FALSE | Upregulated |
| -0,248042752 | 0,793946894 | WNK2 | FALSE | Downregulated |
| -0,273435091 | 0,794162175 | REXO2 | FALSE | Downregulated |
| -0,21221468 | 0,794789385 | B4GALT4 | FALSE | Downregulated |
| -0,332814244 | 0,794789385 | C1orf145 | FALSE | Downregulated |
| -0,093294852 | 0,794789385 | CANX | FALSE | Downregulated |

| | | | | |
|---|---|---|---|---|
| 0,304494283 | 0,794789385 | CCDC36 | FALSE | Upregulated |
| -0,213357086 | 0,794789385 | CEP72 | FALSE | Downregulated |
| -0,24877793 | 0,794789385 | MIR155HG | FALSE | Downregulated |
| -0,094757903 | 0,794789385 | PHKA2 | FALSE | Downregulated |
| -0,14364756 | 0,794789385 | PSMB3 | FALSE | Downregulated |
| 0,132531863 | 0,794828911 | ABCB4 | FALSE | Upregulated |
| -0,237967442 | 0,794828911 | ABRA | FALSE | Downregulated |
| 0,236397506 | 0,794828911 | CCDC17 | FALSE | Upregulated |
| -0,217377723 | 0,794828911 | CYB561D2 | FALSE | Downregulated |
| -0,151119693 | 0,794828911 | MAP4K3 | FALSE | Downregulated |
| 0,143717701 | 0,794828911 | PHLDB1 | FALSE | Upregulated |
| 0,126958954 | 0,794828911 | SIX5 | FALSE | Upregulated |
| 0,124762895 | 0,794828911 | SLC30A7 | FALSE | Upregulated |
| 0,167627996 | 0,794828911 | TBCE | FALSE | Upregulated |
| -0,200715774 | 0,794828911 | YIPF1 | FALSE | Downregulated |
| 0,129134596 | 0,794828911 | ZFP36 | FALSE | Upregulated |
| 0,201104161 | 0,794828911 | ZNF277 | FALSE | Upregulated |
| -0,152825314 | 0,795062555 | FAM160B1 | FALSE | Downregulated |
| -0,166824168 | 0,795157168 | SLC17A5 | FALSE | Downregulated |
| 0,326485962 | 0,795157168 | STX11 | FALSE | Upregulated |
| 0,143500463 | 0,795157168 | SYCP3 | FALSE | Upregulated |
| 0,173750623 | 0,795236622 | GPANK1 | FALSE | Upregulated |
| -0,169179818 | 0,795496396 | DSC2 | FALSE | Downregulated |
| 0,133959942 | 0,79565817 | LOC102723743 | FALSE | Upregulated |
| 0,355956734 | 0,795815597 | ROBO3 | FALSE | Upregulated |
| -0,170024431 | 0,795815597 | ZFP1 | FALSE | Downregulated |
| -0,211054542 | 0,795820199 | IRAK4 | FALSE | Downregulated |
| 0,185821247 | 0,796493239 | ALYREF | FALSE | Upregulated |
| -0,087032739 | 0,796493239 | AMFR | FALSE | Downregulated |
| -0,12191851 | 0,796493239 | CDK16 | FALSE | Downregulated |
| -0,213371253 | 0,796493239 | CDON | FALSE | Downregulated |
| 0,301164901 | 0,796493239 | DACT2 | FALSE | Upregulated |
| -0,118513889 | 0,796493239 | DENND5B | FALSE | Downregulated |
| 0,143883109 | 0,796493239 | FBXO8 | FALSE | Upregulated |
| -0,092548501 | 0,796493239 | GPATCH8 | FALSE | Downregulated |
| 0,118493198 | 0,796493239 | KIF24 | FALSE | Upregulated |
| -0,23524686 | 0,796493239 | LOC100133331 | FALSE | Downregulated |
| 0,169457002 | 0,796493239 | LOC101927685 | FALSE | Upregulated |
| 0,136854274 | 0,796493239 | LOC102724535 | FALSE | Upregulated |
| 0,103906386 | 0,796493239 | MMP19 | FALSE | Upregulated |
| -0,331841985 | 0,796493239 | MN1 | FALSE | Downregulated |
| -0,122016078 | 0,796493239 | NAT10 | FALSE | Downregulated |
| 0,427537481 | 0,796493239 | NPIPB15 | FALSE | Upregulated |
| 0,084299284 | 0,796493239 | PKP4 | FALSE | Upregulated |
| 0,114562913 | 0,796493239 | PNPLA6 | FALSE | Upregulated |
| 0,135347891 | 0,796493239 | PNPT1 | FALSE | Upregulated |
| -0,152399483 | 0,796493239 | PRCP | FALSE | Downregulated |
| -0,236041308 | 0,796493239 | PRPF4 | FALSE | Downregulated |
| 0,164963867 | 0,796493239 | RP11-809M12.1 | FALSE | Upregulated |
| -0,263092638 | 0,796493239 | THOC7 | FALSE | Downregulated |
| -0,08919281 | 0,796493239 | TMEM66 | FALSE | Downregulated |

| | | | | |
|---|---|---|---|---|
| -0,119554776 | 0,796493239 | TNFRSF14 | FALSE | Downregulated |
| -0,168622102 | 0,796493239 | TTLL5 | FALSE | Downregulated |
| -0,13207801 | 0,796493239 | TUBA1B | FALSE | Downregulated |
| -0,182644166 | 0,796493239 | UQCRFS1 | FALSE | Downregulated |
| -0,150197603 | 0,796493239 | YIPF6 | FALSE | Downregulated |
| 0,222607662 | 0,796684403 | CCDC28A | FALSE | Upregulated |
| 0,420136353 | 0,796684403 | LOC388820 | FALSE | Upregulated |
| -0,06532 | 0,796684403 | REPIN1 | FALSE | Downregulated |
| 0,138273897 | 0,796684403 | RNF32 | FALSE | Upregulated |
| -0,245252108 | 0,796684403 | ZNF548 | FALSE | Downregulated |
| -0,191587878 | 0,796711628 | RRM2B | FALSE | Downregulated |
| -0,22733749 | 0,796786026 | AC018705.5 | FALSE | Downregulated |
| -0,188815265 | 0,796786026 | CDC123 | FALSE | Downregulated |
| -0,209826842 | 0,796786026 | CDK2 | FALSE | Downregulated |
| 0,248138544 | 0,796786026 | IER5L | FALSE | Upregulated |
| -0,159440719 | 0,796786026 | KCTD2 | FALSE | Downregulated |
| 0,353650086 | 0,796786026 | LOC102723306 | FALSE | Upregulated |
| 0,157132374 | 0,796885493 | LOXL4 | FALSE | Upregulated |
| 0,234203844 | 0,79691064 | C7orf25 | FALSE | Upregulated |
| -0,206898551 | 0,79691064 | CEBPA-AS1 | FALSE | Downregulated |
| -0,068028293 | 0,79691064 | FOXN3 | FALSE | Downregulated |
| -0,286228589 | 0,79691064 | LOC727896 | FALSE | Downregulated |
| -0,099311844 | 0,79691064 | NUP214 | FALSE | Downregulated |
| -0,266933038 | 0,79691064 | PLA1A | FALSE | Downregulated |
| -0,226727462 | 0,79691064 | ZNF141 | FALSE | Downregulated |
| -0,261917451 | 0,79691064 | ZNF649 | FALSE | Downregulated |
| 0,178673415 | 0,79697317 | HSBP1 | FALSE | Upregulated |
| 0,15868186 | 0,79697317 | NDST3 | FALSE | Upregulated |
| -0,337064889 | 0,79697317 | RPGR | FALSE | Downregulated |
| -0,145116308 | 0,797091781 | ATR | FALSE | Downregulated |
| -0,119142684 | 0,797091781 | DLST | FALSE | Downregulated |
| -0,218212826 | 0,797091781 | FAM180A | FALSE | Downregulated |
| -0,137461943 | 0,797091781 | GOT1 | FALSE | Downregulated |
| -0,130781245 | 0,797091781 | HERC3 | FALSE | Downregulated |
| 0,193776795 | 0,797091781 | NT5C1B | FALSE | Upregulated |
| -0,23510771 | 0,797091781 | TRAM2-AS1 | FALSE | Downregulated |
| -0,199016107 | 0,797091781 | ZNF470 | FALSE | Downregulated |
| 0,26596613 | 0,797201025 | OARD1 | FALSE | Upregulated |
| 0,106134121 | 0,797201025 | PINK1-AS | FALSE | Upregulated |
| -0,106860542 | 0,797309361 | SMARCA5 | FALSE | Downregulated |
| 0,262749836 | 0,797348251 | LINC00844 | FALSE | Upregulated |
| 0,117277509 | 0,797580865 | GNL3L | FALSE | Upregulated |
| -0,131904986 | 0,797701442 | ARSD | FALSE | Downregulated |
| -0,112471862 | 0,797701442 | BCOR | FALSE | Downregulated |
| -0,135517119 | 0,797701442 | PDK4 | FALSE | Downregulated |
| 0,103410502 | 0,797701442 | TSPAN14 | FALSE | Upregulated |
| -0,100078368 | 0,797701442 | VCP | FALSE | Downregulated |
| -0,174731778 | 0,797868258 | RNF152 | FALSE | Downregulated |
| 0,116013098 | 0,797888009 | CCNYL1 | FALSE | Upregulated |
| -0,153366311 | 0,797888009 | MORN4 | FALSE | Downregulated |
| 0,222918722 | 0,798016543 | LOC101929670 | FALSE | Upregulated |

| | | | | |
|---|---|---|---|---|
| -0,099394799 | 0,798185169 | GHITM | FALSE | Downregulated |
| 0,180995821 | 0,798185169 | PLET1 | FALSE | Upregulated |
| -0,169523971 | 0,799237274 | RAB11FIP4 | FALSE | Downregulated |
| 0,247660694 | 0,799237274 | SSTR2 | FALSE | Upregulated |
| -0,084232053 | 0,799336083 | WASF2 | FALSE | Downregulated |
| -0,201790797 | 0,799603481 | FN3KRP | FALSE | Downregulated |
| -0,173274653 | 0,799627369 | ANTXR2 | FALSE | Downregulated |
| -0,128385782 | 0,799627369 | AUH | FALSE | Downregulated |
| -0,09958548 | 0,799627369 | DLC1 | FALSE | Downregulated |
| 0,245500253 | 0,799627369 | ENO2 | FALSE | Upregulated |
| -0,174772062 | 0,799627369 | GDAP2 | FALSE | Downregulated |
| -0,120184546 | 0,799627369 | HSPB1 | FALSE | Downregulated |
| 0,355430712 | 0,799627369 | LOC100101148 | FALSE | Upregulated |
| -0,435508672 | 0,799627369 | LOC100289561 | FALSE | Downregulated |
| -0,27156334 | 0,799627369 | LOC102724782 | FALSE | Downregulated |
| -0,066597756 | 0,799627369 | MACF1 | FALSE | Downregulated |
| 0,217370909 | 0,799627369 | MGME1 | FALSE | Upregulated |
| 0,174706118 | 0,799627369 | NLRP9 | FALSE | Upregulated |
| 0,211077853 | 0,799627369 | NOS1AP | FALSE | Upregulated |
| 0,12964054 | 0,799627369 | PDE5A | FALSE | Upregulated |
| 0,070997583 | 0,799627369 | PDPK1 | FALSE | Upregulated |
| -0,161666117 | 0,799627369 | PITHD1 | FALSE | Downregulated |
| -0,149621661 | 0,799627369 | PTPN4 | FALSE | Downregulated |
| 0,177229802 | 0,799627369 | RNASE10 | FALSE | Upregulated |
| -0,077574554 | 0,799627369 | SKI | FALSE | Downregulated |
| -0,259100364 | 0,799627369 | SMA4 | FALSE | Downregulated |
| 0,277065863 | 0,799627369 | SYF2 | FALSE | Upregulated |
| 0,189878326 | 0,799627369 | TUBG2 | FALSE | Upregulated |
| 0,226116858 | 0,799627369 | VAMP5 | FALSE | Upregulated |
| 0,109399038 | 0,799627369 | ZSCAN30 | FALSE | Upregulated |
| -0,159972737 | 0,799666904 | CASC10 | FALSE | Downregulated |
| 0,148343298 | 0,799666904 | GSS | FALSE | Upregulated |
| 0,352687735 | 0,79983592 | CD209 | FALSE | Upregulated |
| -0,276882814 | 0,79983592 | FAM124A | FALSE | Downregulated |
| 0,162378236 | 0,79983592 | NBPF19 | FALSE | Upregulated |
| -0,143783396 | 0,79983592 | SLC25A39 | FALSE | Downregulated |
| 0,296641471 | 0,79983592 | TRIM69 | FALSE | Upregulated |
| 0,163601441 | 0,799904334 | LOH12CR1 | FALSE | Upregulated |
| -0,239778614 | 0,799904334 | SCARA3 | FALSE | Downregulated |
| 0,151005125 | 0,800028736 | C18orf25 | FALSE | Upregulated |
| 0,331033863 | 0,800028736 | S100A11 | FALSE | Upregulated |
| 0,132823862 | 0,800028736 | ZBTB8A | FALSE | Upregulated |
| -0,136220053 | 0,800217076 | GSK3A | FALSE | Downregulated |
| 0,170034513 | 0,800217076 | PALM | FALSE | Upregulated |
| -0,123496204 | 0,800217076 | POLR2L | FALSE | Downregulated |
| 0,117660859 | 0,800394952 | SNX22 | FALSE | Upregulated |
| 0,196850394 | 0,800951376 | LOC401286 | FALSE | Upregulated |
| 0,327559407 | 0,800951376 | SMARCD3 | FALSE | Upregulated |
| 0,301545731 | 0,801001729 | LOC101928215 | FALSE | Upregulated |
| 0,2280848 | 0,801003383 | KLHL28 | FALSE | Upregulated |
| -0,075791017 | 0,801172272 | PCYOX1 | FALSE | Downregulated |

| | | | | |
|---|---|---|---|---|
| -0,082955912 | 0,801196663 | ILF3 | FALSE | Downregulated |
| -0,191837657 | 0,801409009 | TAF3 | FALSE | Downregulated |
| -0,122724842 | 0,801432128 | SNAPC4 | FALSE | Downregulated |
| 0,233975614 | 0,801632796 | GIMAP1 | FALSE | Upregulated |
| 0,223552854 | 0,801756474 | RHOXF2 | FALSE | Upregulated |
| -0,145135468 | 0,801756474 | RPAP1 | FALSE | Downregulated |
| 0,133541567 | 0,801756474 | ZMIZ2 | FALSE | Upregulated |
| 0,082995289 | 0,80186079 | MAML1 | FALSE | Upregulated |
| 0,116359006 | 0,802050463 | APOL2 | FALSE | Upregulated |
| 0,177583924 | 0,802050463 | FOSL2 | FALSE | Upregulated |
| 0,189413341 | 0,802050463 | RAB20 | FALSE | Upregulated |
| -0,146510459 | 0,802089882 | CDK14 | FALSE | Downregulated |
| 0,131359712 | 0,802196344 | NUP62 | FALSE | Upregulated |
| -0,216005297 | 0,802301339 | ZC3H12C | FALSE | Downregulated |
| 0,1855226 | 0,802306461 | KCTD10 | FALSE | Upregulated |
| 0,174272991 | 0,802370811 | KIF22 | FALSE | Upregulated |
| -0,252919838 | 0,802370811 | MRO | FALSE | Downregulated |
| -0,204950373 | 0,802370811 | PLEK2 | FALSE | Downregulated |
| -0,248372093 | 0,802370811 | PZP | FALSE | Downregulated |
| -0,235633551 | 0,802370811 | WHAMMP1 | FALSE | Downregulated |
| 0,136274824 | 0,802381465 | ZBTB20-AS1 | FALSE | Upregulated |
| 0,356143727 | 0,802575605 | FKBP5 | FALSE | Upregulated |
| 0,248572765 | 0,802575605 | RSPO3 | FALSE | Upregulated |
| 0,156227295 | 0,802674463 | ABHD8 | FALSE | Upregulated |
| -0,252829229 | 0,802674463 | CTTNBP2 | FALSE | Downregulated |
| -0,089638437 | 0,802674463 | PCNXL3 | FALSE | Downregulated |
| -0,12540751 | 0,802955065 | ERLEC1 | FALSE | Downregulated |
| 0,282351202 | 0,802962826 | AC010226.4 | FALSE | Upregulated |
| 0,16301191 | 0,802962826 | C19orf53 | FALSE | Upregulated |
| -0,076377809 | 0,802962826 | EIF5 | FALSE | Downregulated |
| 0,097323241 | 0,802962826 | RHOA | FALSE | Upregulated |
| -0,096822796 | 0,802981566 | BSG | FALSE | Downregulated |
| -0,157046407 | 0,802981566 | C17orf85 | FALSE | Downregulated |
| -0,096396776 | 0,802981566 | COX4I1 | FALSE | Downregulated |
| 0,166703652 | 0,802981566 | PLD2 | FALSE | Upregulated |
| -0,220278349 | 0,802981566 | RBP2 | FALSE | Downregulated |
| 0,188452523 | 0,802981566 | RIBC1 | FALSE | Upregulated |
| 0,176112982 | 0,802983823 | LINC00244 | FALSE | Upregulated |
| -0,123865382 | 0,803015132 | ALAS1 | FALSE | Downregulated |
| 0,115208845 | 0,803015132 | GNL1 | FALSE | Upregulated |
| -0,638487648 | 0,803015132 | MROH7 | FALSE | Downregulated |
| 0,177713889 | 0,803015132 | ZC3H10 | FALSE | Upregulated |
| 0,109680885 | 0,803072324 | MRPL19 | FALSE | Upregulated |
| 0,071416285 | 0,803072324 | PRR12 | FALSE | Upregulated |
| -0,101624766 | 0,803101943 | TAOK3 | FALSE | Downregulated |
| 0,119767315 | 0,803288983 | DNAJB12 | FALSE | Upregulated |
| 0,187998054 | 0,803288983 | SUPT20H | FALSE | Upregulated |
| -0,244304149 | 0,803316707 | STX2 | FALSE | Downregulated |
| 0,221010157 | 0,803316707 | UTS2B | FALSE | Upregulated |
| 0,125170701 | 0,803388193 | TMED8 | FALSE | Upregulated |
| -0,096099883 | 0,803435575 | ARIH2 | FALSE | Downregulated |

| | | | | |
|---|---|---|---|---|
| 0,151274666 | 0,803435575 | MAP3K5 | FALSE | Upregulated |
| 0,179175056 | 0,803435575 | MECP2 | FALSE | Upregulated |
| -0,227396018 | 0,803435575 | RAMP1 | FALSE | Downregulated |
| -0,386654302 | 0,803677775 | RGPD5 | FALSE | Downregulated |
| -0,197358573 | 0,804252959 | TXN | FALSE | Downregulated |
| 0,20831257 | 0,804348386 | C2orf54 | FALSE | Upregulated |
| 0,218277682 | 0,804348386 | TMEM165 | FALSE | Upregulated |
| 0,237601409 | 0,804398515 | C16orf98 | FALSE | Upregulated |
| 0,250891599 | 0,80459049 | AC037193.1 | FALSE | Upregulated |
| -0,083840261 | 0,80459049 | ACAD11 | FALSE | Downregulated |
| -0,182821265 | 0,80459049 | ANXA2 | FALSE | Downregulated |
| 0,200357063 | 0,80459049 | ATP7A | FALSE | Upregulated |
| -0,141067921 | 0,80459049 | BRD9 | FALSE | Downregulated |
| -0,07674589 | 0,80459049 | ECE1 | FALSE | Downregulated |
| 0,216726339 | 0,80459049 | GDPD5 | FALSE | Upregulated |
| -0,268330216 | 0,80459049 | LNP1 | FALSE | Downregulated |
| 0,167703173 | 0,80459049 | NKIRAS2 | FALSE | Upregulated |
| -0,323016719 | 0,80459049 | RP11-15A1.3 | FALSE | Downregulated |
| 0,121984756 | 0,80459049 | SEMA4B | FALSE | Upregulated |
| -0,109030223 | 0,80459049 | SKIL | FALSE | Downregulated |
| 0,199880111 | 0,80459049 | TDG | FALSE | Upregulated |
| -0,073922285 | 0,80459049 | WHSC1L1 | FALSE | Downregulated |
| 0,088154802 | 0,80459049 | XYLT2 | FALSE | Upregulated |
| 0,132536699 | 0,80474983 | CA14 | FALSE | Upregulated |
| -0,208124372 | 0,80474983 | PSMA4 | FALSE | Downregulated |
| -0,121471615 | 0,804895357 | ACADL | FALSE | Downregulated |
| -0,226447021 | 0,804895357 | FTSJ2 | FALSE | Downregulated |
| 0,273746124 | 0,804895357 | NUDT19 | FALSE | Upregulated |
| -0,162420858 | 0,804895357 | RSPRY1 | FALSE | Downregulated |
| -0,082154104 | 0,804895357 | UBC | FALSE | Downregulated |
| -0,153914825 | 0,804895357 | ZSWIM6 | FALSE | Downregulated |
| 0,18196974 | 0,805231713 | MS4A7 | FALSE | Upregulated |
| 0,18268813 | 0,805231713 | PAFAH1B2 | FALSE | Upregulated |
| 0,21966698 | 0,805357893 | CCDC146 | FALSE | Upregulated |
| -0,07345115 | 0,805357893 | KMT2B | FALSE | Downregulated |
| -0,252615849 | 0,805708014 | TUBA1C | FALSE | Downregulated |
| 0,168495573 | 0,805762014 | AC016629.8 | FALSE | Upregulated |
| -0,107186591 | 0,805762014 | ALDH1A1 | FALSE | Downregulated |
| 0,248158772 | 0,805762014 | C1QTNF5 | FALSE | Upregulated |
| 0,167594862 | 0,805762014 | EIF1B-AS1 | FALSE | Upregulated |
| 0,230199417 | 0,805762014 | WASF3 | FALSE | Upregulated |
| 0,086508234 | 0,805799832 | VAT1 | FALSE | Upregulated |
| 0,342014768 | 0,805916364 | LOC102723788 | FALSE | Upregulated |
| 0,093080875 | 0,805916364 | WAPAL | FALSE | Upregulated |
| -0,141448137 | 0,806118555 | C9orf3 | FALSE | Downregulated |
| -0,26567327 | 0,806118555 | HAUS2 | FALSE | Downregulated |
| -0,089756033 | 0,806118555 | PCSK6 | FALSE | Downregulated |
| 0,095850608 | 0,806118555 | SLK | FALSE | Upregulated |
| -0,152824082 | 0,806118555 | TFDP1 | FALSE | Downregulated |
| -0,126030847 | 0,806635791 | EFNA1 | FALSE | Downregulated |
| 0,115736723 | 0,806635791 | SAMD11 | FALSE | Upregulated |

| | | | | |
|---|---|---|---|---|
| -0,095486225 | 0,806635791 | STOM | FALSE | Downregulated |
| -0,094410691 | 0,806635791 | ZNF638 | FALSE | Downregulated |
| 0,151107327 | 0,807099272 | MED31 | FALSE | Upregulated |
| 0,098596966 | 0,807354134 | MCTS1 | FALSE | Upregulated |
| 0,163771028 | 0,807354134 | NRK | FALSE | Upregulated |
| -0,079972565 | 0,807354134 | PNISR | FALSE | Downregulated |
| -0,087174277 | 0,807354134 | SRSF2 | FALSE | Downregulated |
| 0,102435313 | 0,807354134 | STARD7-AS1 | FALSE | Upregulated |
| 0,202543337 | 0,807472668 | ACTR3C | FALSE | Upregulated |
| 0,174516696 | 0,807472668 | TMEM223 | FALSE | Upregulated |
| -0,105590292 | 0,807644847 | MLYCD | FALSE | Downregulated |
| -0,159872605 | 0,807728862 | PARVA | FALSE | Downregulated |
| 0,225014808 | 0,80811371 | SLBP | FALSE | Upregulated |
| 0,109483476 | 0,808374238 | CANT1 | FALSE | Upregulated |
| 0,271076164 | 0,808374238 | DENND1B | FALSE | Upregulated |
| -0,212402354 | 0,808374238 | GMPPA | FALSE | Downregulated |
| -1,062376313 | 0,808374238 | HIST1H4J | FALSE | Downregulated |
| -0,081436055 | 0,808374238 | KLC4 | FALSE | Downregulated |
| 0,148129719 | 0,808374238 | KPNA3 | FALSE | Upregulated |
| 0,22187009 | 0,808374238 | LZTFL1 | FALSE | Upregulated |
| -0,097699007 | 0,808374238 | MARC2 | FALSE | Downregulated |
| -0,087184595 | 0,808374238 | MEGF8 | FALSE | Downregulated |
| 0,197068537 | 0,808374238 | MSR1 | FALSE | Upregulated |
| -0,169043959 | 0,808374238 | NBPF10 | FALSE | Downregulated |
| -0,07858252 | 0,808374238 | NDST1 | FALSE | Downregulated |
| -0,238895255 | 0,808374238 | PA2G4P4 | FALSE | Downregulated |
| -0,165989855 | 0,808374238 | PARG | FALSE | Downregulated |
| -0,292357478 | 0,808374238 | RAET1E | FALSE | Downregulated |
| 0,175246534 | 0,808374238 | RBM28 | FALSE | Upregulated |
| -0,134469186 | 0,808374238 | ROBO4 | FALSE | Downregulated |
| -0,127356597 | 0,808374238 | SMARCE1 | FALSE | Downregulated |
| 0,112698868 | 0,808374238 | SMU1 | FALSE | Upregulated |
| -0,168025964 | 0,808374238 | SPP2 | FALSE | Downregulated |
| -0,099017888 | 0,808374238 | STAG3L5P-PVRIG2P-PILRB | FALSE | Downregulated |
| -0,097873191 | 0,808374238 | USP40 | FALSE | Downregulated |
| 0,137524056 | 0,808374238 | WDR13 | FALSE | Upregulated |
| 0,09325891 | 0,808374238 | YTHDF3 | FALSE | Upregulated |
| -0,143872652 | 0,808374238 | ZC3H6 | FALSE | Downregulated |
| -0,156973346 | 0,808385551 | MRPS27 | FALSE | Downregulated |
| 0,127225532 | 0,808385551 | NICN1 | FALSE | Upregulated |
| 0,248727565 | 0,808385551 | SCPEP1 | FALSE | Upregulated |
| 0,128011357 | 0,808424079 | MAK16 | FALSE | Upregulated |
| -0,209187009 | 0,8084265 | MOB1B | FALSE | Downregulated |
| -0,091421303 | 0,808720065 | TNS1 | FALSE | Downregulated |
| -0,118908428 | 0,808771345 | C6orf47 | FALSE | Downregulated |
| -0,306155172 | 0,808771345 | CAMTA1 | FALSE | Downregulated |
| 0,218394124 | 0,808771345 | COMMD3 | FALSE | Upregulated |
| -0,189878401 | 0,808771345 | CTAGE8 | FALSE | Downregulated |
| 0,249225628 | 0,808771345 | CXorf23 | FALSE | Upregulated |
| -0,107694795 | 0,808771345 | GPX1 | FALSE | Downregulated |

| | | | | |
|---|---|---|---|---|
| -0,199991745 | 0,808771345 | HGF | FALSE | Downregulated |
| -0,07953786 | 0,808771345 | HIPK3 | FALSE | Downregulated |
| 0,221018055 | 0,808771345 | LIAS | FALSE | Upregulated |
| 0,118070422 | 0,808771345 | LOC101927190 | FALSE | Upregulated |
| 0,158031921 | 0,808771345 | LOC642361 | FALSE | Upregulated |
| -0,198295876 | 0,808771345 | NIPAL4 | FALSE | Downregulated |
| -0,110204442 | 0,808771345 | NUDT3 | FALSE | Downregulated |
| 0,323344361 | 0,808771345 | P2RY8 | FALSE | Upregulated |
| 0,155072535 | 0,808771345 | P4HA1 | FALSE | Upregulated |
| 0,122191398 | 0,808771345 | PFKFB1 | FALSE | Upregulated |
| -0,374045089 | 0,808771345 | POLR2M | FALSE | Downregulated |
| 0,200665081 | 0,808771345 | RP11-54O15.3 | FALSE | Upregulated |
| 0,195266001 | 0,808771345 | SMIM18 | FALSE | Upregulated |
| -0,148399402 | 0,808771345 | TCEAL4 | FALSE | Downregulated |
| 0,256787751 | 0,808771345 | TDRD7 | FALSE | Upregulated |
| -0,128611199 | 0,808771345 | USP30 | FALSE | Downregulated |
| -0,28950917 | 0,808873456 | ZMYM1 | FALSE | Downregulated |
| 0,105061886 | 0,808915857 | ERMP1 | FALSE | Upregulated |
| -0,254452149 | 0,808958544 | GMDS | FALSE | Downregulated |
| -0,151701658 | 0,809371519 | FUT11 | FALSE | Downregulated |
| 0,135232886 | 0,809475949 | FAM227B | FALSE | Upregulated |
| 0,129757023 | 0,809475949 | LRRK1 | FALSE | Upregulated |
| -0,151839041 | 0,80958657 | SPR | FALSE | Downregulated |
| 0,095347583 | 0,809623271 | EIF4E2 | FALSE | Upregulated |
| -0,12364382 | 0,809623271 | EIF6 | FALSE | Downregulated |
| 0,094747685 | 0,809623271 | EPS8L2 | FALSE | Upregulated |
| -0,281548296 | 0,809623271 | HS3ST3A1 | FALSE | Downregulated |
| -0,211118355 | 0,809623271 | KCNIP3 | FALSE | Downregulated |
| 0,129965067 | 0,809623271 | NUTM2A | FALSE | Upregulated |
| -0,188595747 | 0,809623271 | PRUNE | FALSE | Downregulated |
| 0,152481 | 0,809623271 | RBBP9 | FALSE | Upregulated |
| -0,133375432 | 0,809623271 | SPATA2L | FALSE | Downregulated |
| -0,144897851 | 0,809623271 | SSH3 | FALSE | Downregulated |
| -0,12006611 | 0,809623271 | TEX261 | FALSE | Downregulated |
| -0,137562596 | 0,809623271 | TFDP2 | FALSE | Downregulated |
| 0,171445743 | 0,809623271 | TRIM7 | FALSE | Upregulated |
| 0,109256434 | 0,809623271 | ZNF562 | FALSE | Upregulated |
| 0,307754168 | 0,809707985 | CCND2 | FALSE | Upregulated |
| -0,113620699 | 0,809707985 | ZFX | FALSE | Downregulated |
| -0,078652346 | 0,80985214 | CDC42BPA | FALSE | Downregulated |
| -0,110974755 | 0,80985214 | HSD17B6 | FALSE | Downregulated |
| -0,390156817 | 0,80985214 | MGP | FALSE | Downregulated |
| 0,120817096 | 0,80985214 | MPG | FALSE | Upregulated |
| -0,149494916 | 0,80985214 | NUDT5 | FALSE | Downregulated |
| -0,121125133 | 0,80985214 | RIOK3 | FALSE | Downregulated |
| 0,372129247 | 0,80985214 | SNX15 | FALSE | Upregulated |
| -0,08411912 | 0,80985214 | STIM1 | FALSE | Downregulated |
| -0,443063201 | 0,810027192 | LOC100133182 | FALSE | Downregulated |
| 0,283950531 | 0,810155168 | ITGBL1 | FALSE | Upregulated |
| 0,164449284 | 0,810269511 | C7 | FALSE | Upregulated |
| 0,230752989 | 0,810269511 | LINC00092 | FALSE | Upregulated |

| | | | | |
|---|---|---|---|---|
| 0,197864535 | 0,810269511 | SLC35C1 | FALSE | Upregulated |
| -0,1061618 | 0,81042728 | APEX1 | FALSE | Downregulated |
| -0,100792474 | 0,81042728 | CDC42 | FALSE | Downregulated |
| 0,13755127 | 0,810445686 | AGO2 | FALSE | Upregulated |
| 0,341166759 | 0,810466245 | USP17L2 | FALSE | Upregulated |
| -0,188418384 | 0,810499922 | ATP6V1G1 | FALSE | Downregulated |
| -0,128359126 | 0,810499922 | FNDC4 | FALSE | Downregulated |
| 0,145487996 | 0,810499922 | PLCB2 | FALSE | Upregulated |
| 0,265445736 | 0,810499922 | TWIST1 | FALSE | Upregulated |
| 0,107429513 | 0,810499922 | ZBTB5 | FALSE | Upregulated |
| -0,228787058 | 0,810610027 | GPR133 | FALSE | Downregulated |
| 0,224478106 | 0,811074137 | UBXN2B | FALSE | Upregulated |
| 0,199711236 | 0,811278926 | LOC100130992 | FALSE | Upregulated |
| -0,256515465 | 0,811278926 | PMS2P3 | FALSE | Downregulated |
| -0,196632602 | 0,811372572 | ABHD5 | FALSE | Downregulated |
| -0,191174994 | 0,811372572 | B3GNT1 | FALSE | Downregulated |
| 0,1204453 | 0,811372572 | CAPS | FALSE | Upregulated |
| 0,189606985 | 0,811372572 | CCDC106 | FALSE | Upregulated |
| -0,168563534 | 0,811372572 | CCDC85B | FALSE | Downregulated |
| -0,189777895 | 0,811372572 | CD36 | FALSE | Downregulated |
| -0,140654191 | 0,811372572 | CEP290 | FALSE | Downregulated |
| 0,41932876 | 0,811372572 | ENAM | FALSE | Upregulated |
| 0,162083828 | 0,811372572 | IFFO2 | FALSE | Upregulated |
| -0,12141545 | 0,811372572 | ITFG3 | FALSE | Downregulated |
| -0,095069031 | 0,811372572 | KEAP1 | FALSE | Downregulated |
| 0,15751466 | 0,811372572 | LOC100126784 | FALSE | Upregulated |
| 0,27568464 | 0,811372572 | LOC101928505 | FALSE | Upregulated |
| 0,151049553 | 0,811372572 | LOC101929046 | FALSE | Upregulated |
| -0,145591806 | 0,811372572 | MMAB | FALSE | Downregulated |
| -0,251148774 | 0,811372572 | NMI | FALSE | Downregulated |
| -0,25487725 | 0,811372572 | PDZD4 | FALSE | Downregulated |
| -0,140060292 | 0,811372572 | PELO | FALSE | Downregulated |
| -0,162821405 | 0,811372572 | PLCD1 | FALSE | Downregulated |
| -0,184838404 | 0,811372572 | PRDM4 | FALSE | Downregulated |
| -0,061047135 | 0,811372572 | SETD5 | FALSE | Downregulated |
| 0,148983692 | 0,811372572 | TMEM214 | FALSE | Upregulated |
| 0,218288643 | 0,811372572 | TNFSF14 | FALSE | Upregulated |
| -0,179859213 | 0,811372572 | TRAPPC3 | FALSE | Downregulated |
| -0,097108306 | 0,811372572 | TSC22D2 | FALSE | Downregulated |
| -0,285794954 | 0,811372572 | TTC8 | FALSE | Downregulated |
| 0,14331481 | 0,811372572 | UBA6 | FALSE | Upregulated |
| 0,08872759 | 0,811372572 | YLPM1 | FALSE | Upregulated |
| 0,115116779 | 0,811374066 | LBX2-AS1 | FALSE | Upregulated |
| -0,307660809 | 0,811467192 | PKHD1L1 | FALSE | Downregulated |
| 0,113734606 | 0,811467192 | RRP1B | FALSE | Upregulated |
| 0,196180883 | 0,811657862 | CCR9 | FALSE | Upregulated |
| 0,200852442 | 0,811657862 | DPH5 | FALSE | Upregulated |
| -0,14392132 | 0,811657862 | EPHA2 | FALSE | Downregulated |
| -0,132771778 | 0,811657862 | FAM219A | FALSE | Downregulated |
| 0,208460854 | 0,811657862 | HLA-DOA | FALSE | Upregulated |
| 0,190605964 | 0,811657862 | NPEPL1 | FALSE | Upregulated |

| | | | | |
|---|---|---|---|---|
| 0,248738108 | 0,811657862 | NPHP4 | FALSE | Upregulated |
| -0,108636794 | 0,811657862 | NUP153 | FALSE | Downregulated |
| 0,187814768 | 0,811657862 | PGS1 | FALSE | Upregulated |
| 0,141738604 | 0,811657862 | POLR3A | FALSE | Upregulated |
| -0,337621904 | 0,811657862 | RP11-261N11.8 | FALSE | Downregulated |
| 0,050771002 | 0,811657862 | SBF1 | FALSE | Upregulated |
| -0,163299597 | 0,811657862 | SGPP2 | FALSE | Downregulated |
| 0,228766683 | 0,811657862 | SRXN1 | FALSE | Upregulated |
| -0,091436001 | 0,811657862 | WIZ | FALSE | Downregulated |
| -0,081176913 | 0,811663828 | NONO | FALSE | Downregulated |
| 0,200663936 | 0,811673964 | UBXN11 | FALSE | Upregulated |
| 0,148718898 | 0,811867989 | SNAI3 | FALSE | Upregulated |
| 0,225639533 | 0,811867989 | TMPRSS11D | FALSE | Upregulated |
| -0,172737224 | 0,811871087 | ARL6IP5 | FALSE | Downregulated |
| 0,152848683 | 0,811871087 | BCL3 | FALSE | Upregulated |
| 0,118601291 | 0,812023529 | ADAM17 | FALSE | Upregulated |
| -0,119279175 | 0,812023529 | ANKLE2 | FALSE | Downregulated |
| 0,203325655 | 0,812023529 | ARHGEF10 | FALSE | Upregulated |
| -0,187422747 | 0,812023529 | ASNA1 | FALSE | Downregulated |
| -0,190744337 | 0,812023529 | BCL2L12 | FALSE | Downregulated |
| -0,148102039 | 0,812023529 | BMPR1A | FALSE | Downregulated |
| -0,162145842 | 0,812023529 | BTBD7 | FALSE | Downregulated |
| -0,442863349 | 0,812023529 | C15orf38-AP3S2 | FALSE | Downregulated |
| 0,103160458 | 0,812023529 | C17orf103 | FALSE | Upregulated |
| -0,115655259 | 0,812023529 | C4BPB | FALSE | Downregulated |
| -0,081459373 | 0,812023529 | C6orf106 | FALSE | Downregulated |
| 0,089589104 | 0,812023529 | CARM1 | FALSE | Upregulated |
| -0,141215869 | 0,812023529 | CARS | FALSE | Downregulated |
| -0,123599047 | 0,812023529 | CASZ1 | FALSE | Downregulated |
| 0,187039364 | 0,812023529 | CCDC115 | FALSE | Upregulated |
| -0,106666002 | 0,812023529 | CFI | FALSE | Downregulated |
| 0,23729718 | 0,812023529 | CXCR3 | FALSE | Upregulated |
| -0,112536257 | 0,812023529 | DAAM2 | FALSE | Downregulated |
| 0,121757225 | 0,812023529 | DMXL2 | FALSE | Upregulated |
| 0,058631009 | 0,812023529 | FAXDC2 | FALSE | Upregulated |
| 0,167294885 | 0,812023529 | FBXO41 | FALSE | Upregulated |
| 0,156007069 | 0,812023529 | GTF2H1 | FALSE | Upregulated |
| 0,371873387 | 0,812023529 | HERC2P3 | FALSE | Upregulated |
| -0,081932407 | 0,812023529 | HGS | FALSE | Downregulated |
| -0,100657665 | 0,812023529 | ITIH3 | FALSE | Downregulated |
| -0,132902878 | 0,812023529 | KAT8 | FALSE | Downregulated |
| -0,087963464 | 0,812023529 | KDM6B | FALSE | Downregulated |
| 0,172526114 | 0,812023529 | KIAA0513 | FALSE | Upregulated |
| 0,145792468 | 0,812023529 | LENG9 | FALSE | Upregulated |
| 0,222126657 | 0,812023529 | LOC100129148 | FALSE | Upregulated |
| -0,226158772 | 0,812023529 | LOC100506083 | FALSE | Downregulated |
| 0,275572117 | 0,812023529 | LOC101928510 | FALSE | Upregulated |
| 0,261741177 | 0,812023529 | LOC102723997 | FALSE | Upregulated |
| 0,19677204 | 0,812023529 | LOC613037 | FALSE | Upregulated |
| -0,200633588 | 0,812023529 | MAPK3 | FALSE | Downregulated |
| -0,174595255 | 0,812023529 | MKKS | FALSE | Downregulated |

| | | | | |
|---|---|---|---|---|
| -0,138506639 | 0,812023529 | MZT2A | FALSE | Downregulated |
| -0,285289994 | 0,812023529 | NAF1 | FALSE | Downregulated |
| -0,264747968 | 0,812023529 | NOC3L | FALSE | Downregulated |
| 0,202781431 | 0,812023529 | PGGT1B | FALSE | Upregulated |
| 0,082452201 | 0,812023529 | PI4KB | FALSE | Upregulated |
| 0,254234717 | 0,812023529 | PIPSL | FALSE | Upregulated |
| 0,178883104 | 0,812023529 | PLEKHG7 | FALSE | Upregulated |
| -0,196413301 | 0,812023529 | PLS1 | FALSE | Downregulated |
| -0,190611845 | 0,812023529 | PMS1 | FALSE | Downregulated |
| -0,107002582 | 0,812023529 | PNPLA7 | FALSE | Downregulated |
| 0,190405701 | 0,812023529 | POLR2I | FALSE | Upregulated |
| 0,286586282 | 0,812023529 | PRICKLE1 | FALSE | Upregulated |
| 0,206377048 | 0,812023529 | PRKCD | FALSE | Upregulated |
| 0,164747316 | 0,812023529 | PRSS8 | FALSE | Upregulated |
| -0,10089083 | 0,812023529 | RABL6 | FALSE | Downregulated |
| 0,184138743 | 0,812023529 | S100A14 | FALSE | Upregulated |
| -0,180086051 | 0,812023529 | SEC11C | FALSE | Downregulated |
| -0,074273696 | 0,812023529 | SF3B1 | FALSE | Downregulated |
| -0,19074833 | 0,812023529 | SLC17A1 | FALSE | Downregulated |
| -0,115999441 | 0,812023529 | SNX29P2 | FALSE | Downregulated |
| 0,162448233 | 0,812023529 | TH | FALSE | Upregulated |
| -0,119669468 | 0,812023529 | TK2 | FALSE | Downregulated |
| -0,167882027 | 0,812023529 | TMEM104 | FALSE | Downregulated |
| -0,244834568 | 0,812023529 | TMOD2 | FALSE | Downregulated |
| 0,088605714 | 0,812023529 | TNKS | FALSE | Upregulated |
| 0,135467975 | 0,812023529 | TRAPPC2 | FALSE | Upregulated |
| 0,110459644 | 0,812023529 | TRMT10B | FALSE | Upregulated |
| -0,097278809 | 0,812023529 | TTC17 | FALSE | Downregulated |
| -0,141156366 | 0,812023529 | TTPA | FALSE | Downregulated |
| -0,234660092 | 0,812023529 | TULP3 | FALSE | Downregulated |
| -0,156202677 | 0,812023529 | VMAC | FALSE | Downregulated |
| -0,156543639 | 0,81207511 | CC2D2B | FALSE | Downregulated |
| 0,134836507 | 0,81207511 | INE2 | FALSE | Upregulated |
| 0,295820719 | 0,812179451 | PLEKHB1 | FALSE | Upregulated |
| 0,143658589 | 0,812334054 | LUC7L | FALSE | Upregulated |
| 0,158741997 | 0,81287542 | ADNP2 | FALSE | Upregulated |
| 0,105244607 | 0,812876475 | RECQL5 | FALSE | Upregulated |
| -0,134766925 | 0,812882832 | TRMT44 | FALSE | Downregulated |
| 0,166745147 | 0,812910712 | ATPIF1 | FALSE | Upregulated |
| 0,199878463 | 0,812910712 | FAM65B | FALSE | Upregulated |
| 0,237324913 | 0,812910712 | ZNF559 | FALSE | Upregulated |
| 0,1668377 | 0,812943335 | EXOC1 | FALSE | Upregulated |
| -0,105805321 | 0,813051573 | ATG9A | FALSE | Downregulated |
| -0,126433609 | 0,813051573 | C4B | FALSE | Downregulated |
| -0,147661296 | 0,813051573 | DDC | FALSE | Downregulated |
| -0,157924692 | 0,813051573 | ELOF1 | FALSE | Downregulated |
| 0,163714778 | 0,813051573 | IQCH | FALSE | Upregulated |
| -0,120526735 | 0,813051573 | PIM3 | FALSE | Downregulated |
| 0,154291056 | 0,813051573 | PPP2R3B | FALSE | Upregulated |
| -0,160753632 | 0,813051573 | PTPRG-AS1 | FALSE | Downregulated |
| 0,088538175 | 0,813051573 | RTEL1 | FALSE | Upregulated |

| | | | | |
|---|---|---|---|---|
| -0,09639115 | 0,813051573 | SHPRH | FALSE | Downregulated |
| -0,181532365 | 0,813051573 | ST3GAL4 | FALSE | Downregulated |
| 0,133283756 | 0,813051573 | ZNF546 | FALSE | Upregulated |
| -0,137221097 | 0,813051573 | ZNF776 | FALSE | Downregulated |
| 0,220690151 | 0,813172418 | CHAF1A | FALSE | Upregulated |
| 0,664730016 | 0,813172418 | RNVU1-7 | FALSE | Upregulated |
| -0,113271261 | 0,81332307 | KLF15 | FALSE | Downregulated |
| -0,446691227 | 0,81358822 | CYP26A1 | FALSE | Downregulated |
| 0,217926197 | 0,813638243 | GCSHP3 | FALSE | Upregulated |
| 0,185642553 | 0,813638243 | OSER1-AS1 | FALSE | Upregulated |
| -0,312099798 | 0,813638243 | PLVAP | FALSE | Downregulated |
| 0,135302201 | 0,813638243 | SYDE1 | FALSE | Upregulated |
| -0,264679887 | 0,813667186 | ANKMY2 | FALSE | Downregulated |
| -0,176423514 | 0,813667186 | APLNR | FALSE | Downregulated |
| 0,072886609 | 0,813667186 | ATM | FALSE | Upregulated |
| -0,129638888 | 0,813667186 | C2CD2 | FALSE | Downregulated |
| -0,132933247 | 0,813667186 | C8G | FALSE | Downregulated |
| 0,119753736 | 0,813667186 | CNPY3 | FALSE | Upregulated |
| -0,093581848 | 0,813667186 | CUX1 | FALSE | Downregulated |
| -0,220080461 | 0,813667186 | GPR98 | FALSE | Downregulated |
| 0,240613352 | 0,813667186 | LOC102723622 | FALSE | Upregulated |
| 0,214766087 | 0,813667186 | LOC102724141 | FALSE | Upregulated |
| 0,185915193 | 0,813667186 | LOC102724350 | FALSE | Upregulated |
| 0,172813595 | 0,813667186 | MFAP1 | FALSE | Upregulated |
| 0,183912186 | 0,813667186 | MSANTD2 | FALSE | Upregulated |
| -0,094160436 | 0,813667186 | MZT2B | FALSE | Downregulated |
| 0,121218973 | 0,813667186 | OSGEPL1 | FALSE | Upregulated |
| -0,126631698 | 0,813667186 | OXSR1 | FALSE | Downregulated |
| -0,098574388 | 0,813667186 | PDIA3 | FALSE | Downregulated |
| -0,168478046 | 0,813667186 | POLR2H | FALSE | Downregulated |
| 0,134503567 | 0,813667186 | RAP2C-AS1 | FALSE | Upregulated |
| -0,078508418 | 0,813667186 | SCAP | FALSE | Downregulated |
| 0,194118944 | 0,813667186 | ZKSCAN4 | FALSE | Upregulated |
| -0,106997708 | 0,813983029 | DCBLD2 | FALSE | Downregulated |
| -0,132238126 | 0,814086072 | HS3ST3B1 | FALSE | Downregulated |
| -0,444380771 | 0,814187752 | FAM35DP | FALSE | Downregulated |
| 0,156118678 | 0,814226339 | MGAT5 | FALSE | Upregulated |
| 0,166344362 | 0,814226339 | NMNAT3 | FALSE | Upregulated |
| -0,283661549 | 0,814226339 | PHLDA3 | FALSE | Downregulated |
| -0,110055913 | 0,814226339 | SEC16B | FALSE | Downregulated |
| 0,292689511 | 0,814226339 | SLC8A3 | FALSE | Upregulated |
| 0,438326342 | 0,814318257 | CDCP1 | FALSE | Upregulated |
| -0,092546397 | 0,814318257 | PMM2 | FALSE | Downregulated |
| -0,222666021 | 0,814318257 | TRADD | FALSE | Downregulated |
| -0,210279168 | 0,814429824 | AMIGO1 | FALSE | Downregulated |
| -0,126859988 | 0,814429824 | EPHA1 | FALSE | Downregulated |
| 0,123982026 | 0,814429824 | FAM69A | FALSE | Upregulated |
| 0,25091086 | 0,814429824 | GSG1 | FALSE | Upregulated |
| 0,118692279 | 0,814429824 | PAIP2 | FALSE | Upregulated |
| -0,077931269 | 0,814429824 | USF2 | FALSE | Downregulated |
| 0,214377589 | 0,81443926 | ELF3 | FALSE | Upregulated |

| | | | | |
|---|---|---|---|---|
| 0,098351376 | 0,81443926 | ELK4 | FALSE | Upregulated |
| 0,153952458 | 0,81443926 | KCNC3 | FALSE | Upregulated |
| -0,142331636 | 0,81443926 | LOC101927673 | FALSE | Downregulated |
| -0,232069899 | 0,81443926 | RRP12 | FALSE | Downregulated |
| -0,057379179 | 0,81443926 | SEC14L2 | FALSE | Downregulated |
| -0,093857402 | 0,814770899 | NFKBIA | FALSE | Downregulated |
| 0,127818513 | 0,814858717 | PIP5K1A | FALSE | Upregulated |
| 0,15854182 | 0,815071642 | ERAL1 | FALSE | Upregulated |
| 0,219651683 | 0,815071642 | FLJ41481 | FALSE | Upregulated |
| 0,226816013 | 0,815071642 | HOMER3 | FALSE | Upregulated |
| -0,143876287 | 0,815071642 | LETMD1 | FALSE | Downregulated |
| -0,095500427 | 0,81516802 | MYO10 | FALSE | Downregulated |
| 0,191145621 | 0,815232842 | CLUHP3 | FALSE | Upregulated |
| -0,167383128 | 0,815232842 | NCALD | FALSE | Downregulated |
| 0,2907823 | 0,815404147 | SYNPO2L | FALSE | Upregulated |
| 0,137128488 | 0,815607578 | WLS | FALSE | Upregulated |
| 0,173362862 | 0,815797808 | ACOT6 | FALSE | Upregulated |
| -0,253232289 | 0,815797808 | LPP-AS2 | FALSE | Downregulated |
| 0,137524841 | 0,815797808 | PSMD13 | FALSE | Upregulated |
| 0,092782808 | 0,815797808 | TRIP11 | FALSE | Upregulated |
| -0,058900933 | 0,815876236 | PCBP1 | FALSE | Downregulated |
| -0,134348113 | 0,815876236 | TBC1D22A | FALSE | Downregulated |
| -0,172707941 | 0,815876236 | UBL5 | FALSE | Downregulated |
| 0,225389216 | 0,815876236 | ZNF117 | FALSE | Upregulated |
| 0,110232908 | 0,816014833 | TBC1D2B | FALSE | Upregulated |
| -0,16923471 | 0,816035033 | PPIP5K2 | FALSE | Downregulated |
| -0,162690493 | 0,816040151 | ARAP3 | FALSE | Downregulated |
| 0,109822241 | 0,816040151 | DAB2 | FALSE | Upregulated |
| -0,145295005 | 0,816040151 | DAP3 | FALSE | Downregulated |
| -0,161894852 | 0,816040151 | FKBP3 | FALSE | Downregulated |
| -0,246690949 | 0,816040151 | LOC101927806 | FALSE | Downregulated |
| -0,117399215 | 0,816040151 | MCM7 | FALSE | Downregulated |
| 0,254702875 | 0,816040151 | MED12L | FALSE | Upregulated |
| -0,113997947 | 0,816040151 | SAR1A | FALSE | Downregulated |
| -0,206368448 | 0,816040151 | TMEM201 | FALSE | Downregulated |
| -0,070015195 | 0,816040151 | TSPYL1 | FALSE | Downregulated |
| -0,113013691 | 0,816040151 | UGT2B4 | FALSE | Downregulated |
| -0,075720597 | 0,816040151 | WDR33 | FALSE | Downregulated |
| -0,093165584 | 0,81604803 | CDH2 | FALSE | Downregulated |
| 0,261069204 | 0,81604803 | MEIOB | FALSE | Upregulated |
| -0,108710587 | 0,81604803 | TMEM43 | FALSE | Downregulated |
| -0,193795397 | 0,816320365 | HS2ST1 | FALSE | Downregulated |
| -0,152132847 | 0,816320365 | POLR3B | FALSE | Downregulated |
| 0,14575231 | 0,816320365 | TYROBP | FALSE | Upregulated |
| -0,122863208 | 0,816725254 | ARRB2 | FALSE | Downregulated |
| 0,21472172 | 0,816725254 | ASPHD1 | FALSE | Upregulated |
| -0,202749625 | 0,816725254 | BUD13 | FALSE | Downregulated |
| -0,188152843 | 0,816725254 | C19orf48 | FALSE | Downregulated |
| -0,228750933 | 0,816725254 | C6orf48 | FALSE | Downregulated |
| 0,189768307 | 0,816725254 | LMO2 | FALSE | Upregulated |
| -0,208548829 | 0,816725254 | LOC202181 | FALSE | Downregulated |

| | | | | |
|---|---|---|---|---|
| 0,256265086 | 0,816725254 | METTL21A | FALSE | Upregulated |
| -0,106602858 | 0,816725254 | PI4K2B | FALSE | Downregulated |
| 0,069489112 | 0,816963502 | UHRF1BP1 | FALSE | Upregulated |
| -0,125778549 | 0,817108265 | BZW1 | FALSE | Downregulated |
| -0,124360604 | 0,817108265 | NUDT21 | FALSE | Downregulated |
| -0,197622066 | 0,817108265 | RABL5 | FALSE | Downregulated |
| 0,08233532 | 0,817176713 | SNRK | FALSE | Upregulated |
| 0,242341286 | 0,817297964 | LECT1 | FALSE | Upregulated |
| -0,157759003 | 0,817297964 | MPHOSPH9 | FALSE | Downregulated |
| -0,261312243 | 0,817297964 | NCAPG2 | FALSE | Downregulated |
| -0,18619645 | 0,817297964 | TM2D3 | FALSE | Downregulated |
| -0,114549746 | 0,817561533 | LARP4 | FALSE | Downregulated |
| 0,193261039 | 0,817592699 | METTL15 | FALSE | Upregulated |
| 0,15413419 | 0,817594537 | CLK2 | FALSE | Upregulated |
| -0,117978526 | 0,817594537 | FABP1 | FALSE | Downregulated |
| -0,08345777 | 0,817594537 | MATR3 | FALSE | Downregulated |
| -0,061432292 | 0,817594537 | TRIP12 | FALSE | Downregulated |
| 0,157099201 | 0,817728333 | DPH3P1 | FALSE | Upregulated |
| -0,141747829 | 0,818094675 | QSOX1 | FALSE | Downregulated |
| 0,254781395 | 0,818094675 | TEX9 | FALSE | Upregulated |
| -0,122907747 | 0,818203314 | ARPC5 | FALSE | Downregulated |
| 0,084371701 | 0,818203314 | CCAR2 | FALSE | Upregulated |
| -0,09672758 | 0,818203314 | CSRP1 | FALSE | Downregulated |
| 0,133931026 | 0,818203314 | KDM8 | FALSE | Upregulated |
| 0,162906186 | 0,818203314 | NIPAL3 | FALSE | Upregulated |
| -0,152391407 | 0,818375274 | AMY2B | FALSE | Downregulated |
| -0,296418929 | 0,818375274 | CPE | FALSE | Downregulated |
| -0,128445243 | 0,818375274 | LOC101927903 | FALSE | Downregulated |
| -0,222631621 | 0,818375274 | NUTF2 | FALSE | Downregulated |
| 0,236798275 | 0,818375274 | RP11-996F15.2 | FALSE | Upregulated |
| 0,143168979 | 0,818375274 | RTFDC1 | FALSE | Upregulated |
| -0,207513349 | 0,818375274 | ST6GALNAC2 | FALSE | Downregulated |
| -0,19553593 | 0,818410324 | LSM7 | FALSE | Downregulated |
| 0,457370687 | 0,818475374 | ANKRD20A9P | FALSE | Upregulated |
| -0,091614954 | 0,818475374 | ATP2A2 | FALSE | Downregulated |
| -0,209934077 | 0,818475374 | DNAJB14 | FALSE | Downregulated |
| 0,221797582 | 0,818475374 | LOC102723597 | FALSE | Upregulated |
| -0,094344626 | 0,818475374 | PHIP | FALSE | Downregulated |
| -0,281435863 | 0,818475374 | SCT | FALSE | Downregulated |
| 0,181059377 | 0,818475374 | SNUPN | FALSE | Upregulated |
| 0,08946653 | 0,818475374 | STAG1 | FALSE | Upregulated |
| -0,198209567 | 0,818475374 | ZNF324 | FALSE | Downregulated |
| -0,107110166 | 0,818483517 | MSL2 | FALSE | Downregulated |
| -0,224306762 | 0,818483517 | OTUD3 | FALSE | Downregulated |
| 0,253900485 | 0,818483517 | PET100 | FALSE | Upregulated |
| -0,23379701 | 0,818483517 | RHD | FALSE | Downregulated |
| -0,0973645 | 0,818483517 | SERPINA5 | FALSE | Downregulated |
| 0,270049951 | 0,818561435 | GRIN2D | FALSE | Upregulated |
| 0,12511908 | 0,818849705 | GNPDA2 | FALSE | Upregulated |
| -0,184207424 | 0,818849705 | SLC25A26 | FALSE | Downregulated |
| 0,227196819 | 0,81897905 | PDE6B | FALSE | Upregulated |

| | | | | |
|---|---|---|---|---|
| 0,135517483 | 0,819133463 | ANGPTL7 | FALSE | Upregulated |
| -0,102778039 | 0,819133463 | CCDC93 | FALSE | Downregulated |
| 0,087360301 | 0,819133463 | ECHDC1 | FALSE | Upregulated |
| 0,248071856 | 0,819133463 | MTCP1 | FALSE | Upregulated |
| 0,216889569 | 0,819133463 | PPM1D | FALSE | Upregulated |
| 0,250606737 | 0,819133463 | RASSF6 | FALSE | Upregulated |
| -0,086178065 | 0,819133463 | SAMD4A | FALSE | Downregulated |
| -0,103132603 | 0,819133463 | SNX18 | FALSE | Downregulated |
| 0,171525243 | 0,819133463 | VPS35 | FALSE | Upregulated |
| 0,212783475 | 0,819133463 | ZNF324B | FALSE | Upregulated |
| 0,138560057 | 0,819133463 | ZNF483 | FALSE | Upregulated |
| 0,311733962 | 0,819290367 | LOC389332 | FALSE | Upregulated |
| 0,153808184 | 0,819290367 | MLST8 | FALSE | Upregulated |
| -0,115975287 | 0,819290367 | TBC1D17 | FALSE | Downregulated |
| 0,086274964 | 0,819527575 | AP2B1 | FALSE | Upregulated |
| 0,099488051 | 0,819527575 | CARNS1 | FALSE | Upregulated |
| -0,242894097 | 0,819719481 | CHST14 | FALSE | Downregulated |
| -0,228401696 | 0,819719481 | RPUSD2 | FALSE | Downregulated |
| -0,17778795 | 0,819719481 | SIMC1 | FALSE | Downregulated |
| 0,150880378 | 0,820233437 | RHEBP2 | FALSE | Upregulated |
| -0,180438686 | 0,820328953 | NCDN | FALSE | Downregulated |
| -0,115448349 | 0,820565718 | SKP1 | FALSE | Downregulated |
| -0,198253612 | 0,820578011 | MANSC1 | FALSE | Downregulated |
| 0,063831464 | 0,820578011 | TTPAL | FALSE | Upregulated |
| 0,167221883 | 0,820578011 | YARS2 | FALSE | Upregulated |
| 0,135511267 | 0,820745999 | DDX20 | FALSE | Upregulated |
| -0,153771607 | 0,820912424 | ACOT7 | FALSE | Downregulated |
| 0,286247944 | 0,820912424 | ARHGEF38 | FALSE | Upregulated |
| -0,136825953 | 0,820912424 | CBS | FALSE | Downregulated |
| 0,121235161 | 0,820912424 | CCDC71L | FALSE | Upregulated |
| 0,211531479 | 0,820912424 | CRY1 | FALSE | Upregulated |
| 0,237213291 | 0,820912424 | DBIL5P | FALSE | Upregulated |
| 0,228273948 | 0,820912424 | FICD | FALSE | Upregulated |
| 0,153573391 | 0,820912424 | FKBP7 | FALSE | Upregulated |
| -0,283807978 | 0,820912424 | GAB3 | FALSE | Downregulated |
| 0,150184436 | 0,820912424 | GNRHR | FALSE | Upregulated |
| -0,103730517 | 0,820912424 | HERC2P2 | FALSE | Downregulated |
| -0,100558348 | 0,820912424 | MUT | FALSE | Downregulated |
| 0,239830707 | 0,820912424 | NCS1 | FALSE | Upregulated |
| -0,161236652 | 0,820912424 | NHP2L1 | FALSE | Downregulated |
| -0,138148663 | 0,820912424 | OTUD1 | FALSE | Downregulated |
| 0,169183292 | 0,820912424 | PCBP4 | FALSE | Upregulated |
| -0,153123427 | 0,820912424 | PCGF5 | FALSE | Downregulated |
| -0,281511088 | 0,820912424 | PDP1 | FALSE | Downregulated |
| 0,138716841 | 0,820912424 | PECAM1 | FALSE | Upregulated |
| -0,180432733 | 0,820912424 | RCAN3 | FALSE | Downregulated |
| -0,097422915 | 0,820912424 | SCP2 | FALSE | Downregulated |
| 0,14655734 | 0,820912424 | SOAT1 | FALSE | Upregulated |
| 0,109131885 | 0,820912424 | TNPO3 | FALSE | Upregulated |
| 0,125882388 | 0,820912424 | TNRC6C-AS1 | FALSE | Upregulated |
| -0,226609848 | 0,821101227 | ZNF561 | FALSE | Downregulated |

| | | | | |
|---|---|---|---|---|
| 0,176604353 | 0,821254324 | ARHGAP10 | FALSE | Upregulated |
| -0,114515382 | 0,821254324 | SIGIRR | FALSE | Downregulated |
| -0,122480166 | 0,821254324 | SLC6A12 | FALSE | Downregulated |
| -0,129866312 | 0,821355921 | BICD2 | FALSE | Downregulated |
| 0,158932409 | 0,821355921 | CBY3 | FALSE | Upregulated |
| 0,179510672 | 0,821355921 | CHCHD3 | FALSE | Upregulated |
| 0,148643092 | 0,821355921 | FBN1 | FALSE | Upregulated |
| -0,196913778 | 0,821355921 | GID4 | FALSE | Downregulated |
| 0,316911771 | 0,821355921 | ITGAD | FALSE | Upregulated |
| -0,133435793 | 0,821355921 | LIG4 | FALSE | Downregulated |
| 0,174420593 | 0,821355921 | LOC100132781 | FALSE | Upregulated |
| 0,137967601 | 0,821355921 | LOC100996255 | FALSE | Upregulated |
| 0,140159124 | 0,821355921 | LOC101927831 | FALSE | Upregulated |
| 0,111367524 | 0,821355921 | LOC102723342 | FALSE | Upregulated |
| -0,095481455 | 0,821355921 | MGST1 | FALSE | Downregulated |
| -0,182846604 | 0,821355921 | RSC1A1 | FALSE | Downregulated |
| 0,207806998 | 0,821355921 | STAU2-AS1 | FALSE | Upregulated |
| 0,110546702 | 0,821355921 | TSHR | FALSE | Upregulated |
| 0,164826057 | 0,821355921 | ZNF785 | FALSE | Upregulated |
| 0,407146263 | 0,821624813 | F13A1 | FALSE | Upregulated |
| -0,121230013 | 0,821624813 | GALNT1 | FALSE | Downregulated |
| -0,086095396 | 0,821624813 | PDXK | FALSE | Downregulated |
| -0,089297719 | 0,821624813 | SAP30L | FALSE | Downregulated |
| 0,086671135 | 0,821624813 | TRIM41 | FALSE | Upregulated |
| 0,170932603 | 0,821699054 | SDSL | FALSE | Upregulated |
| -0,169079775 | 0,821699054 | SYT12 | FALSE | Downregulated |
| 0,242402275 | 0,821709522 | RP11-1078H9.5 | FALSE | Upregulated |
| 0,297459278 | 0,821731798 | SAPCD1 | FALSE | Upregulated |
| -0,146431984 | 0,821775997 | AKIRIN2 | FALSE | Downregulated |
| 0,118970113 | 0,822059277 | FAM118B | FALSE | Upregulated |
| 0,29137997 | 0,822059277 | FUT3 | FALSE | Upregulated |
| 0,212362089 | 0,822059277 | POMZP3 | FALSE | Upregulated |
| -0,244736868 | 0,822249585 | C17orf89 | FALSE | Downregulated |
| 0,169961834 | 0,822249585 | CHFR | FALSE | Upregulated |
| -0,241591149 | 0,822305659 | ILKAP | FALSE | Downregulated |
| 0,10078498 | 0,822305659 | MKRN2 | FALSE | Upregulated |
| -0,205650019 | 0,822305659 | UBOX5 | FALSE | Downregulated |
| 0,139903153 | 0,822419395 | UGT3A1 | FALSE | Upregulated |
| -0,093812458 | 0,822686242 | ATP11A | FALSE | Downregulated |
| 0,233366592 | 0,822686242 | ELOVL1 | FALSE | Upregulated |
| -0,275811884 | 0,822686242 | FAM27E3 | FALSE | Downregulated |
| 0,109320789 | 0,822686242 | FBXL15 | FALSE | Upregulated |
| 0,120753121 | 0,822686242 | MRPS16 | FALSE | Upregulated |
| 0,08206929 | 0,822686242 | NUP50 | FALSE | Upregulated |
| -0,12521519 | 0,822686242 | RGPD4 | FALSE | Downregulated |
| -0,130161988 | 0,822750731 | VEZF1 | FALSE | Downregulated |
| 0,083920857 | 0,822791044 | PIP4K2B | FALSE | Upregulated |
| -0,101699382 | 0,822897132 | INTS3 | FALSE | Downregulated |
| 0,136616187 | 0,822897132 | PCSK9 | FALSE | Upregulated |
| -0,185504443 | 0,822897132 | SCFD1 | FALSE | Downregulated |
| 0,150910832 | 0,823105359 | LOC102723992 | FALSE | Upregulated |

| | | | | |
|---|---|---|---|---|
| -0,06989481 | 0,823121027 | NR2F2 | FALSE | Downregulated |
| -0,119417066 | 0,823143926 | HNRNPR | FALSE | Downregulated |
| -0,068286616 | 0,823198745 | ARFGEF2 | FALSE | Downregulated |
| 0,121866368 | 0,823198745 | CAMSAP1 | FALSE | Upregulated |
| 0,119669032 | 0,823198745 | SGTA | FALSE | Upregulated |
| -0,169382227 | 0,823285347 | COX7A2 | FALSE | Downregulated |
| 0,280601156 | 0,823285347 | PTGDS | FALSE | Upregulated |
| -0,138701799 | 0,823345505 | KCMF1 | FALSE | Downregulated |
| 0,193951815 | 0,823345505 | ZNF627 | FALSE | Upregulated |
| 0,160913791 | 0,823367595 | PCED1B-AS1 | FALSE | Upregulated |
| 0,206249231 | 0,823443381 | LOC100996761 | FALSE | Upregulated |
| 0,129392018 | 0,823557227 | MAPRE3 | FALSE | Upregulated |
| 0,366123142 | 0,823557227 | PART1 | FALSE | Upregulated |
| 0,073940478 | 0,823557227 | WDR1 | FALSE | Upregulated |
| 0,160820242 | 0,823592927 | LOC101928830 | FALSE | Upregulated |
| -0,130971693 | 0,823592927 | UVSSA | FALSE | Downregulated |
| -0,180693556 | 0,823628127 | ADAL | FALSE | Downregulated |
| -0,139524533 | 0,823628127 | ANK3 | FALSE | Downregulated |
| -0,067474114 | 0,823628127 | AP3D1 | FALSE | Downregulated |
| -0,084294455 | 0,823628127 | CTNND1 | FALSE | Downregulated |
| 0,198055983 | 0,823628127 | HOMEZ | FALSE | Upregulated |
| 0,275807923 | 0,823628127 | IKBKE | FALSE | Upregulated |
| 0,123769109 | 0,823628127 | LOC100996573 | FALSE | Upregulated |
| -0,106952694 | 0,823628127 | NR5A2 | FALSE | Downregulated |
| 0,077483006 | 0,823628127 | PCF11 | FALSE | Upregulated |
| -0,144138263 | 0,823628127 | PSAT1 | FALSE | Downregulated |
| -0,152202042 | 0,823628127 | RNF6 | FALSE | Downregulated |
| 0,214603559 | 0,823628127 | TIPRL | FALSE | Upregulated |
| 0,131404826 | 0,823812799 | HSPE1 | FALSE | Upregulated |
| 0,150747755 | 0,823892155 | BCL2L11 | FALSE | Upregulated |
| 0,101663053 | 0,823892155 | BMP2K | FALSE | Upregulated |
| 0,182378528 | 0,823892155 | CHST11 | FALSE | Upregulated |
| -0,119787745 | 0,823892155 | DNMBP | FALSE | Downregulated |
| 0,173291293 | 0,823892155 | EMC9 | FALSE | Upregulated |
| -0,083797965 | 0,823892155 | ESRRA | FALSE | Downregulated |
| -0,208204626 | 0,823892155 | ISLR | FALSE | Downregulated |
| -0,082428347 | 0,823892155 | MAFB | FALSE | Downregulated |
| 0,228082173 | 0,823892155 | RPL31P11 | FALSE | Upregulated |
| -0,227550631 | 0,823892155 | RWDD1 | FALSE | Downregulated |
| -0,134907165 | 0,823892155 | TARS2 | FALSE | Downregulated |
| -0,219840877 | 0,824311313 | CTNNAL1 | FALSE | Downregulated |
| 0,116109787 | 0,824311313 | CUL4B | FALSE | Upregulated |
| 0,087758596 | 0,824311313 | PMPCB | FALSE | Upregulated |
| 0,179072916 | 0,824311313 | PTCD1 | FALSE | Upregulated |
| -0,155286291 | 0,824311313 | SGTB | FALSE | Downregulated |
| -0,183818684 | 0,824381361 | ACTL6A | FALSE | Downregulated |
| 0,101532363 | 0,824381361 | ARHGEF1 | FALSE | Upregulated |
| 0,07657093 | 0,824381361 | DEDD | FALSE | Upregulated |
| -0,145830949 | 0,824381361 | DTX1 | FALSE | Downregulated |
| 0,179410291 | 0,824381361 | LOC101926943 | FALSE | Upregulated |
| -0,171124713 | 0,824381361 | PAK1 | FALSE | Downregulated |

| | | | | |
|---|---|---|---|---|
| -0,188403445 | 0,824381361 | PVRL1 | FALSE | Downregulated |
| -0,201678168 | 0,824381361 | SERTAD3 | FALSE | Downregulated |
| -0,090247671 | 0,824381361 | SLC38A3 | FALSE | Downregulated |
| 0,118094788 | 0,824381361 | SNF8 | FALSE | Upregulated |
| 0,129848304 | 0,824381361 | SQRDL | FALSE | Upregulated |
| 0,099229257 | 0,824381361 | ST3GAL2 | FALSE | Upregulated |
| -0,140779649 | 0,824381361 | USP11 | FALSE | Downregulated |
| 0,127314986 | 0,824381361 | ZSCAN18 | FALSE | Upregulated |
| -0,123572304 | 0,824557037 | SLC39A11 | FALSE | Downregulated |
| 0,343430794 | 0,824605714 | AC000099.1 | FALSE | Upregulated |
| -0,251396635 | 0,824605714 | C7orf31 | FALSE | Downregulated |
| 0,185473845 | 0,824605714 | LOC100287166 | FALSE | Upregulated |
| 0,298046548 | 0,824605714 | PRKY | FALSE | Upregulated |
| -0,10343858 | 0,824605714 | SNAP29 | FALSE | Downregulated |
| 0,174063365 | 0,824605714 | ZBTB21 | FALSE | Upregulated |
| 0,1715278 | 0,824689581 | ALG9 | FALSE | Upregulated |
| -0,155531668 | 0,824689581 | FBXO44 | FALSE | Downregulated |
| 0,115384492 | 0,824689581 | FRMD4B | FALSE | Upregulated |
| -0,051218108 | 0,824689581 | MYH9 | FALSE | Downregulated |
| 0,182478725 | 0,824689581 | NDE1 | FALSE | Upregulated |
| 0,11712468 | 0,824863724 | APOBEC2 | FALSE | Upregulated |
| -0,190202455 | 0,824863724 | CHST3 | FALSE | Downregulated |
| -0,083964736 | 0,824863724 | TOM1 | FALSE | Downregulated |
| -0,183881462 | 0,824863724 | ZNF26 | FALSE | Downregulated |
| 0,239606916 | 0,825130048 | ADPRHL2 | FALSE | Upregulated |
| 0,089881541 | 0,825130048 | ARF6 | FALSE | Upregulated |
| 0,20278096 | 0,825130048 | ZNF367 | FALSE | Upregulated |
| 0,143887655 | 0,825830956 | BLNK | FALSE | Upregulated |
| -0,090152529 | 0,825830956 | CALCOCO2 | FALSE | Downregulated |
| -0,15966147 | 0,825830956 | CHSY1 | FALSE | Downregulated |
| -0,210599133 | 0,825830956 | EXOSC7 | FALSE | Downregulated |
| 0,155844757 | 0,825830956 | GPR137B | FALSE | Upregulated |
| 0,182810439 | 0,825830956 | GRIN2C | FALSE | Upregulated |
| 0,160621564 | 0,825830956 | HERC6 | FALSE | Upregulated |
| -0,161903863 | 0,825830956 | LYRM4 | FALSE | Downregulated |
| -0,215154612 | 0,825830956 | PNPLA4 | FALSE | Downregulated |
| 0,075163559 | 0,825830956 | RPTOR | FALSE | Upregulated |
| -0,111568055 | 0,825830956 | SETBP1 | FALSE | Downregulated |
| 0,078066123 | 0,825830956 | SMAD4 | FALSE | Upregulated |
| -0,269644518 | 0,825830956 | SNORA63 | FALSE | Downregulated |
| -0,131378937 | 0,825830956 | SNX3 | FALSE | Downregulated |
| 0,158238199 | 0,825830956 | SYNE4 | FALSE | Upregulated |
| -0,110212883 | 0,825830956 | TRPV1 | FALSE | Downregulated |
| -0,090047645 | 0,825830956 | YWHAE | FALSE | Downregulated |
| 0,160374711 | 0,825830956 | ZDHHC2 | FALSE | Upregulated |
| 0,138812177 | 0,826145732 | DNM3 | FALSE | Upregulated |
| 0,169057886 | 0,826145732 | EFNB2 | FALSE | Upregulated |
| 0,115607475 | 0,826145732 | FAM91A1 | FALSE | Upregulated |
| -0,149389178 | 0,826145732 | HLX | FALSE | Downregulated |
| -0,173377451 | 0,826145732 | NDUFS6 | FALSE | Downregulated |
| -0,271169614 | 0,826145732 | POLR2K | FALSE | Downregulated |

| | | | | |
|---|---|---|---|---|
| -0,180232135 | 0,826145732 | SNORD17 | FALSE | Downregulated |
| -0,154086223 | 0,826145732 | ZNF721 | FALSE | Downregulated |
| -0,062340478 | 0,826146744 | ALKBH5 | FALSE | Downregulated |
| 0,173200705 | 0,826146744 | CHRAC1 | FALSE | Upregulated |
| 0,143813394 | 0,826146744 | OGFOD3 | FALSE | Upregulated |
| 0,158542755 | 0,826146744 | SYT7 | FALSE | Upregulated |
| 0,077870545 | 0,82617099 | ABCA2 | FALSE | Upregulated |
| -0,089192339 | 0,82617099 | AURKAIP1 | FALSE | Downregulated |
| 0,165075678 | 0,82617099 | INS | FALSE | Upregulated |
| 0,15752264 | 0,826250515 | C1QTNF6 | FALSE | Upregulated |
| -0,175473826 | 0,826250515 | TRAPPC1 | FALSE | Downregulated |
| 0,157835102 | 0,826435693 | ADO | FALSE | Upregulated |
| -0,201559955 | 0,826435693 | ILF3-AS1 | FALSE | Downregulated |
| 0,245149534 | 0,826621162 | CHST2 | FALSE | Upregulated |
| 0,131667765 | 0,826621162 | GPR160 | FALSE | Upregulated |
| 0,471726324 | 0,826621162 | LOC102723603 | FALSE | Upregulated |
| 0,105188053 | 0,826755578 | NFATC2 | FALSE | Upregulated |
| 0,216120882 | 0,826755578 | VWF | FALSE | Upregulated |
| -0,224987773 | 0,827101125 | LOC101927187 | FALSE | Downregulated |
| -0,073899606 | 0,827101125 | RARRES2 | FALSE | Downregulated |
| -0,346646067 | 0,827191182 | TEN1-CDK3 | FALSE | Downregulated |
| 0,126132987 | 0,827381239 | CASK | FALSE | Upregulated |
| 0,195195204 | 0,827381239 | LOC100287852 | FALSE | Upregulated |
| -0,273112403 | 0,827381239 | LOC101929573 | FALSE | Downregulated |
| 0,084052953 | 0,827381239 | SH3GL1 | FALSE | Upregulated |
| -0,195279284 | 0,827381239 | SNX12 | FALSE | Downregulated |
| -0,112384215 | 0,827381239 | SPATS2L | FALSE | Downregulated |
| 0,1301507 | 0,827381239 | TMEM180 | FALSE | Upregulated |
| 0,106246932 | 0,827381239 | TNFAIP8L2 | FALSE | Upregulated |
| 0,098922072 | 0,827486582 | USP5 | FALSE | Upregulated |
| -0,049733249 | 0,827771561 | MST1 | FALSE | Downregulated |
| 0,074251197 | 0,827997711 | SIN3A | FALSE | Upregulated |
| 0,227882937 | 0,828031474 | FMO2 | FALSE | Upregulated |
| -0,081491447 | 0,828031474 | ZNF207 | FALSE | Downregulated |
| -0,17683282 | 0,828347372 | LAMTOR5 | FALSE | Downregulated |
| -0,109457574 | 0,828820496 | ATXN10 | FALSE | Downregulated |
| 0,104952482 | 0,828820496 | CD4 | FALSE | Upregulated |
| -0,19044864 | 0,828820496 | CDK5RAP1 | FALSE | Downregulated |
| -0,152904399 | 0,828820496 | HIST2H2BE | FALSE | Downregulated |
| 0,085211527 | 0,828820496 | PPAPDC1B | FALSE | Upregulated |
| 0,253220672 | 0,828820496 | SORT1 | FALSE | Upregulated |
| 0,174789328 | 0,828858164 | ORC3 | FALSE | Upregulated |
| -0,117137372 | 0,828858164 | UACA | FALSE | Downregulated |
| 0,177648499 | 0,828883571 | CCDC91 | FALSE | Upregulated |
| 0,267168045 | 0,828883571 | CCR10 | FALSE | Upregulated |
| 0,075299781 | 0,828883571 | MGAT1 | FALSE | Upregulated |
| 0,119094563 | 0,828883571 | MLLT10 | FALSE | Upregulated |
| -0,085714782 | 0,828883571 | THEM6 | FALSE | Downregulated |
| 0,132932584 | 0,829126046 | SIRPA | FALSE | Upregulated |
| 0,13565017 | 0,829145257 | IKZF4 | FALSE | Upregulated |
| 0,229186922 | 0,829181034 | SLC38A9 | FALSE | Upregulated |

| | | | | |
|---|---|---|---|---|
| 0,184138407 | 0,829390807 | KIAA1045 | FALSE | Upregulated |
| 0,097813637 | 0,829481878 | HNRNPUL2 | FALSE | Upregulated |
| 0,10367561 | 0,829481878 | RNF146 | FALSE | Upregulated |
| -0,103271533 | 0,829547756 | ACMSD | FALSE | Downregulated |
| 0,150170661 | 0,829547756 | DNAJC30 | FALSE | Upregulated |
| -0,172876826 | 0,829547756 | DUSP23 | FALSE | Downregulated |
| 0,112920002 | 0,829547756 | EFTUD2 | FALSE | Upregulated |
| 0,085013039 | 0,829547756 | KDM5C | FALSE | Upregulated |
| 0,090020278 | 0,829547756 | TP53INP2 | FALSE | Upregulated |
| -0,132179082 | 0,829547756 | VMP1 | FALSE | Downregulated |
| -0,16537813 | 0,829547756 | WARS2 | FALSE | Downregulated |
| -0,147028422 | 0,829548587 | AATK | FALSE | Downregulated |
| 0,10400365 | 0,829548587 | ABI3BP | FALSE | Upregulated |
| 0,208325295 | 0,829548587 | C5orf34 | FALSE | Upregulated |
| -0,059788155 | 0,829548587 | EP400 | FALSE | Downregulated |
| 0,256685518 | 0,829548587 | FAM49B | FALSE | Upregulated |
| -0,07651983 | 0,829548587 | GBA2 | FALSE | Downregulated |
| -0,1449155 | 0,829548587 | H2AFJ | FALSE | Downregulated |
| -0,146847452 | 0,829548587 | IL11RA | FALSE | Downregulated |
| -0,165426362 | 0,829548587 | MUC5B | FALSE | Downregulated |
| -0,114604176 | 0,829548587 | PAIP1 | FALSE | Downregulated |
| -0,069089275 | 0,829548587 | PRKACA | FALSE | Downregulated |
| 0,255133855 | 0,829548587 | RFX2 | FALSE | Upregulated |
| -0,149499931 | 0,829548587 | SNIP1 | FALSE | Downregulated |
| 0,199177439 | 0,829548587 | TMEM253 | FALSE | Upregulated |
| -0,091582967 | 0,829548587 | TTC31 | FALSE | Downregulated |
| -0,151847436 | 0,829548587 | XPNPEP1 | FALSE | Downregulated |
| 0,134448772 | 0,829548587 | ZNF197 | FALSE | Upregulated |
| 0,119667026 | 0,829548868 | JPX | FALSE | Upregulated |
| -0,102706664 | 0,829758775 | MLXIP | FALSE | Downregulated |
| 0,2391711 | 0,829807175 | AVPR1A | FALSE | Upregulated |
| -0,149999621 | 0,829887499 | ANGEL2 | FALSE | Downregulated |
| 0,26200811 | 0,829887499 | CLDN10 | FALSE | Upregulated |
| 0,207663478 | 0,829887499 | MTMR3 | FALSE | Upregulated |
| -0,096572909 | 0,829887499 | SNRNP70 | FALSE | Downregulated |
| 0,264623077 | 0,829994176 | LOC102723335 | FALSE | Upregulated |
| -0,193638597 | 0,830019699 | PEX3 | FALSE | Downregulated |
| 0,224561458 | 0,830019699 | TBCC | FALSE | Upregulated |
| 0,239625015 | 0,830097087 | CD27-AS1 | FALSE | Upregulated |
| 0,08865667 | 0,830097087 | GLTSCR1 | FALSE | Upregulated |
| 0,142371687 | 0,830097087 | GRWD1 | FALSE | Upregulated |
| 0,321808604 | 0,830097087 | NBL1 | FALSE | Upregulated |
| 0,134716892 | 0,830097087 | ZDHHC7 | FALSE | Upregulated |
| -0,216525596 | 0,830148158 | CASP6 | FALSE | Downregulated |
| 0,110671235 | 0,830148158 | ESR1 | FALSE | Upregulated |
| 0,123373368 | 0,830148158 | LOC101928728 | FALSE | Upregulated |
| 0,162794115 | 0,830148158 | LRRC48 | FALSE | Upregulated |
| 0,071414074 | 0,830148158 | TNS3 | FALSE | Upregulated |
| -0,134942049 | 0,830254932 | GSAP | FALSE | Downregulated |
| -0,093633625 | 0,830410067 | ARHGAP32 | FALSE | Downregulated |
| 0,149043884 | 0,830410067 | CYP21A2 | FALSE | Upregulated |

| | | | | |
|---|---|---|---|---|
| 0,127915782 | 0,830410067 | HIRIP3 | FALSE | Upregulated |
| -0,078832515 | 0,830410067 | MAP2K7 | FALSE | Downregulated |
| -0,151705265 | 0,830410067 | WASH5P | FALSE | Downregulated |
| -0,111662406 | 0,830410067 | WDFY1 | FALSE | Downregulated |
| 0,155632444 | 0,830410067 | ZDHHC6 | FALSE | Upregulated |
| 0,141171991 | 0,830427351 | BEAN1 | FALSE | Upregulated |
| -0,118354952 | 0,830427351 | PPP2R2A | FALSE | Downregulated |
| 0,19189007 | 0,830463803 | CDC73 | FALSE | Upregulated |
| 0,144191975 | 0,830463803 | SLC6A1-AS1 | FALSE | Upregulated |
| -0,190415271 | 0,830473896 | KHDRBS3 | FALSE | Downregulated |
| -0,082641333 | 0,830473896 | PSMD3 | FALSE | Downregulated |
| -0,087940705 | 0,830620981 | MARK2 | FALSE | Downregulated |
| -0,075090089 | 0,830620981 | NUDT4 | FALSE | Downregulated |
| -0,07361271 | 0,83072853 | GTF3C1 | FALSE | Downregulated |
| -0,205252126 | 0,83072853 | LRRC4B | FALSE | Downregulated |
| -0,096973217 | 0,83090292 | CCT4 | FALSE | Downregulated |
| -0,121215253 | 0,83090292 | CLCN6 | FALSE | Downregulated |
| 0,088230031 | 0,83090292 | FAM63A | FALSE | Upregulated |
| -0,148105115 | 0,83090292 | ORAI1 | FALSE | Downregulated |
| 0,091014961 | 0,83090292 | TOMM40L | FALSE | Upregulated |
| 0,130573811 | 0,830903084 | ADAM22 | FALSE | Upregulated |
| 0,132209324 | 0,830903084 | ADCY3 | FALSE | Upregulated |
| 0,183465818 | 0,830903084 | BIN3 | FALSE | Upregulated |
| 0,229399787 | 0,830903084 | CD244 | FALSE | Upregulated |
| -0,098400217 | 0,830903084 | CKAP4 | FALSE | Downregulated |
| -0,110535024 | 0,830903084 | CTCF | FALSE | Downregulated |
| -0,116620464 | 0,830903084 | DIS3 | FALSE | Downregulated |
| 0,128076837 | 0,830903084 | FAM57A | FALSE | Upregulated |
| -0,26029642 | 0,830903084 | FEZ1 | FALSE | Downregulated |
| 0,119642298 | 0,830903084 | HIF1A | FALSE | Upregulated |
| 0,084273096 | 0,830903084 | NT5C2 | FALSE | Upregulated |
| 0,087358878 | 0,830903084 | OGT | FALSE | Upregulated |
| -0,093485672 | 0,830903084 | PDS5A | FALSE | Downregulated |
| 0,173888781 | 0,830903084 | RUNDC1 | FALSE | Upregulated |
| 0,058916748 | 0,830903084 | SMARCC2 | FALSE | Upregulated |
| -0,075326851 | 0,830903084 | SPG7 | FALSE | Downregulated |
| 0,245483409 | 0,830903084 | TAS2R42 | FALSE | Upregulated |
| -0,376711906 | 0,830903084 | ZFY | FALSE | Downregulated |
| -0,113835824 | 0,831015261 | ELP2 | FALSE | Downregulated |
| -0,215298154 | 0,831168927 | FPGT | FALSE | Downregulated |
| 0,138793437 | 0,831213079 | HCFC2 | FALSE | Upregulated |
| -0,22192764 | 0,831234468 | PPIH | FALSE | Downregulated |
| 0,157814307 | 0,831499833 | ELMOD3 | FALSE | Upregulated |
| 0,132973492 | 0,83163189 | RP11-31F15.2 | FALSE | Upregulated |
| -0,074598884 | 0,8320401 | TSC2 | FALSE | Downregulated |
| 0,122802538 | 0,8320401 | ZBTB37 | FALSE | Upregulated |
| 0,083696809 | 0,832143247 | AFG3L2 | FALSE | Upregulated |
| 0,090385242 | 0,832143247 | RBM4 | FALSE | Upregulated |
| -0,076313188 | 0,832143247 | UBE2H | FALSE | Downregulated |
| 0,078435081 | 0,832143247 | USP54 | FALSE | Upregulated |
| 0,126277692 | 0,832410015 | GPATCH11 | FALSE | Upregulated |

| | | | | |
|---|---|---|---|---|
| -0,165080439 | 0,832410015 | PRELP | FALSE | Downregulated |
| 0,129388908 | 0,832410015 | SHOC2 | FALSE | Upregulated |
| -0,114391535 | 0,832410015 | SLMAP | FALSE | Downregulated |
| 0,159967561 | 0,832410015 | UFSP2 | FALSE | Upregulated |
| 1,029356014 | 0,832425791 | MYCBP | FALSE | Upregulated |
| -0,14029486 | 0,832639346 | CHD1L | FALSE | Downregulated |
| 0,226779638 | 0,832639346 | GGPS1 | FALSE | Upregulated |
| 0,1504691 | 0,832639346 | PNKP | FALSE | Upregulated |
| -0,073512264 | 0,832639346 | TMEM127 | FALSE | Downregulated |
| -0,229808072 | 0,833007753 | RP11-106M7.1 | FALSE | Downregulated |
| 0,145882669 | 0,833118625 | PROCA1 | FALSE | Upregulated |
| -0,142733881 | 0,833118625 | STRBP | FALSE | Downregulated |
| 0,221687398 | 0,833191786 | LOC101060181 | FALSE | Upregulated |
| 0,133756529 | 0,834014191 | EXOC4 | FALSE | Upregulated |
| 0,18312794 | 0,834014191 | LINC00341 | FALSE | Upregulated |
| 0,110163276 | 0,834014191 | RABGAP1 | FALSE | Upregulated |
| -0,095838761 | 0,834014191 | SATB1 | FALSE | Downregulated |
| 0,145771827 | 0,834016554 | C22orf15 | FALSE | Upregulated |
| -0,113439653 | 0,834089899 | C9 | FALSE | Downregulated |
| 0,207623772 | 0,834089899 | OIP5 | FALSE | Upregulated |
| 0,073791584 | 0,83415092 | KMO | FALSE | Upregulated |
| 0,1288292 | 0,83415092 | PLAT | FALSE | Upregulated |
| -0,058963049 | 0,834694515 | AKT2 | FALSE | Downregulated |
| 0,2175931 | 0,834694515 | ARHGEF34P | FALSE | Upregulated |
| 0,208884175 | 0,834694515 | C10orf11 | FALSE | Upregulated |
| -0,118538112 | 0,834694515 | CECR5 | FALSE | Downregulated |
| -0,090658267 | 0,834694515 | CNOT3 | FALSE | Downregulated |
| -0,111760717 | 0,834694515 | DIXDC1 | FALSE | Downregulated |
| 0,150060805 | 0,834694515 | DNM1P41 | FALSE | Upregulated |
| 0,230938906 | 0,834694515 | DYNLT1 | FALSE | Upregulated |
| -0,098957591 | 0,834694515 | GET4 | FALSE | Downregulated |
| 0,25795079 | 0,834694515 | HSPA7 | FALSE | Upregulated |
| 0,331915135 | 0,834694515 | ISM1-AS1 | FALSE | Upregulated |
| 0,191818793 | 0,834694515 | LOC729451 | FALSE | Upregulated |
| -0,245085429 | 0,834694515 | PRR24 | FALSE | Downregulated |
| 0,102956611 | 0,834694515 | RAB22A | FALSE | Upregulated |
| 0,079258332 | 0,834694515 | SIPA1 | FALSE | Upregulated |
| 0,139368119 | 0,834694515 | SNX10 | FALSE | Upregulated |
| -0,243572394 | 0,834694515 | SRPK3 | FALSE | Downregulated |
| -0,106887131 | 0,834694515 | ST6GALNAC6 | FALSE | Downregulated |
| 0,115667481 | 0,834745123 | C15orf37 | FALSE | Upregulated |
| -0,067144834 | 0,834745123 | MAGI3 | FALSE | Downregulated |
| 0,10211719 | 0,834745123 | PATZ1 | FALSE | Upregulated |
| 0,134084654 | 0,834745123 | PCNP | FALSE | Upregulated |
| 0,15344118 | 0,834779243 | CDC42P3 | FALSE | Upregulated |
| -0,172276303 | 0,834779243 | CHEK1 | FALSE | Downregulated |
| 0,107222008 | 0,834779243 | RBMS1 | FALSE | Upregulated |
| -0,223400956 | 0,834779243 | ZNF202 | FALSE | Downregulated |
| 0,087337094 | 0,834904216 | TRIM66 | FALSE | Upregulated |
| 0,223601294 | 0,834921853 | BAG2 | FALSE | Upregulated |
| 0,750660533 | 0,834921853 | FNTB | FALSE | Upregulated |

| | | | | |
|---|---|---|---|---|
| -0,174230674 | 0,834921853 | LINC00671 | FALSE | Downregulated |
| 0,20905442 | 0,834921853 | THOC7-AS1 | FALSE | Upregulated |
| -0,162733458 | 0,834921853 | TNFRSF10D | FALSE | Downregulated |
| -0,099451267 | 0,834964296 | CTDP1 | FALSE | Downregulated |
| -0,116982959 | 0,834964296 | DNASE1 | FALSE | Downregulated |
| -0,184993745 | 0,834964296 | ELMO3 | FALSE | Downregulated |
| -0,093408299 | 0,834964296 | ETS1 | FALSE | Downregulated |
| 0,21077582 | 0,834964296 | HSCB | FALSE | Upregulated |
| 0,223142763 | 0,834964296 | MAPK10 | FALSE | Upregulated |
| 0,107864301 | 0,834964296 | PSMD6-AS2 | FALSE | Upregulated |
| -0,085763808 | 0,834964296 | SLC39A1 | FALSE | Downregulated |
| -0,162009004 | 0,834999205 | AIFM2 | FALSE | Downregulated |
| 0,171322991 | 0,834999205 | AP5M1 | FALSE | Upregulated |
| 0,072036769 | 0,834999205 | STX16 | FALSE | Upregulated |
| -0,109169377 | 0,835111221 | PPP3R1 | FALSE | Downregulated |
| -0,122389548 | 0,835280075 | CHPF2 | FALSE | Downregulated |
| -0,124097697 | 0,835280075 | CUEDC2 | FALSE | Downregulated |
| -0,21221661 | 0,835280075 | GLUD2 | FALSE | Downregulated |
| 0,152642392 | 0,835280075 | KIAA1462 | FALSE | Upregulated |
| 0,100608954 | 0,835280075 | LRRC61 | FALSE | Upregulated |
| 0,090689964 | 0,835280075 | SIRT1 | FALSE | Upregulated |
| 0,173632461 | 0,835280075 | TMEM19 | FALSE | Upregulated |
| -0,302206455 | 0,835280075 | TMEM200C | FALSE | Downregulated |
| 0,103789958 | 0,835396043 | CYB561A3 | FALSE | Upregulated |
| -0,181303626 | 0,83541429 | LOC102723773 | FALSE | Downregulated |
| -0,164423397 | 0,835483875 | PRPF39 | FALSE | Downregulated |
| -0,154899346 | 0,835705996 | OSBPL1A | FALSE | Downregulated |
| 0,148049428 | 0,836022251 | G0S2 | FALSE | Upregulated |
| 0,136912567 | 0,836022251 | GPR34 | FALSE | Upregulated |
| 0,120098714 | 0,836022251 | TBC1D9 | FALSE | Upregulated |
| 0,134591775 | 0,836306985 | UPF3A | FALSE | Upregulated |
| 0,112543722 | 0,836533767 | ISOC1 | FALSE | Upregulated |
| 0,184927921 | 0,836581928 | ELAC1 | FALSE | Upregulated |
| -0,14075016 | 0,836581928 | HPS6 | FALSE | Downregulated |
| 0,144858482 | 0,836581928 | HSPA14 | FALSE | Upregulated |
| 0,124072968 | 0,836581928 | IP6K2 | FALSE | Upregulated |
| 0,229762826 | 0,836581928 | KCNC2 | FALSE | Upregulated |
| 0,490687179 | 0,836581928 | MROH7-TTC4 | FALSE | Upregulated |
| -0,1476705 | 0,836581928 | TMEM47 | FALSE | Downregulated |
| -0,075938548 | 0,836581928 | ZBTB7B | FALSE | Downregulated |
| -0,080651642 | 0,836933099 | PTPN11 | FALSE | Downregulated |
| -0,154369825 | 0,836933099 | USP37 | FALSE | Downregulated |
| -0,165217233 | 0,836933099 | ZNF782 | FALSE | Downregulated |
| -0,134852427 | 0,837054476 | NUP133 | FALSE | Downregulated |
| -0,230782339 | 0,83735757 | APEX2 | FALSE | Downregulated |
| 0,130363725 | 0,83735757 | ARMC7 | FALSE | Upregulated |
| -0,081568752 | 0,83735757 | FBXL3 | FALSE | Downregulated |
| -0,1376807 | 0,83735757 | GOLPH3L | FALSE | Downregulated |
| 0,190943433 | 0,83735757 | PPME1 | FALSE | Upregulated |
| -0,307978474 | 0,83735757 | RCAN2 | FALSE | Downregulated |
| 0,195403035 | 0,83735757 | SMC1B | FALSE | Upregulated |

| | | | | |
|---|---|---|---|---|
| -0,067413345 | 0,837584301 | NR3C1 | FALSE | Downregulated |
| 0,263023818 | 0,837692125 | DOK3 | FALSE | Upregulated |
| 0,086292029 | 0,837692125 | WDR82 | FALSE | Upregulated |
| 0,15142979 | 0,837728191 | FXYD6 | FALSE | Upregulated |
| -0,079869844 | 0,83794316 | ACSM3 | FALSE | Downregulated |
| 0,172066196 | 0,83794316 | C14orf169 | FALSE | Upregulated |
| 0,295957137 | 0,83794316 | C14orf182 | FALSE | Upregulated |
| -0,201707852 | 0,83794316 | HIST1H2BA | FALSE | Downregulated |
| 0,051385651 | 0,83794316 | LOC100129034 | FALSE | Upregulated |
| 0,116756146 | 0,83794316 | NDOR1 | FALSE | Upregulated |
| 0,171629467 | 0,83794316 | NOP2 | FALSE | Upregulated |
| 0,097705263 | 0,83794316 | PRSS12 | FALSE | Upregulated |
| 0,146626869 | 0,83794316 | PSMG3-AS1 | FALSE | Upregulated |
| 0,089351223 | 0,83794316 | RAB26 | FALSE | Upregulated |
| -0,068798378 | 0,83794316 | SPATA13 | FALSE | Downregulated |
| 0,107471505 | 0,83794316 | SPON1 | FALSE | Upregulated |
| 0,158075579 | 0,83794316 | TNXB | FALSE | Upregulated |
| 0,305448971 | 0,83794316 | USP9Y | FALSE | Upregulated |
| -0,174430492 | 0,83794316 | ZG16 | FALSE | Downregulated |
| -0,09109181 | 0,838214593 | ACTR1A | FALSE | Downregulated |
| 0,087966424 | 0,838359326 | WDR24 | FALSE | Upregulated |
| 0,085467092 | 0,83838309 | ALCAM | FALSE | Upregulated |
| -0,084286045 | 0,83838309 | GPC6 | FALSE | Downregulated |
| 0,139118185 | 0,83838309 | P2RX7 | FALSE | Upregulated |
| 0,159202553 | 0,83838309 | RP11-983P16.4 | FALSE | Upregulated |
| 0,108561846 | 0,83838309 | STXBP2 | FALSE | Upregulated |
| -0,148824902 | 0,83838309 | ZNF48 | FALSE | Downregulated |
| 0,536644612 | 0,838519312 | OR2A20P | FALSE | Upregulated |
| 0,253078589 | 0,838723751 | CACHD1 | FALSE | Upregulated |
| 0,147538839 | 0,838723751 | NRAS | FALSE | Upregulated |
| 0,191957254 | 0,83887816 | ADAMTS1 | FALSE | Upregulated |
| -0,064496098 | 0,838963039 | CENPB | FALSE | Downregulated |
| -0,097678338 | 0,839130289 | AMPD2 | FALSE | Downregulated |
| -0,139082806 | 0,839130289 | NLK | FALSE | Downregulated |
| 0,178162587 | 0,839189683 | LOC102724372 | FALSE | Upregulated |
| -0,125725167 | 0,839220306 | LOC101928192 | FALSE | Downregulated |
| -0,14351685 | 0,839220306 | MIEN1 | FALSE | Downregulated |
| -0,206598945 | 0,839220306 | MPV17L | FALSE | Downregulated |
| 0,215565174 | 0,839220306 | MTRNR2L4 | FALSE | Upregulated |
| -0,158579595 | 0,839220306 | NUDT22 | FALSE | Downregulated |
| 0,367519256 | 0,839220306 | PPT2-EGFL8 | FALSE | Upregulated |
| -0,079491571 | 0,839220306 | RALBP1 | FALSE | Downregulated |
| -0,096460482 | 0,839220306 | SOWAHB | FALSE | Downregulated |
| 0,11905755 | 0,839220306 | ZNF45 | FALSE | Upregulated |
| -0,140525067 | 0,839220306 | ZNF623 | FALSE | Downregulated |
| 0,170143514 | 0,839252004 | ANO1-AS2 | FALSE | Upregulated |
| -0,113236272 | 0,839319089 | TRAF6 | FALSE | Downregulated |
| 0,198189467 | 0,839421543 | USP46 | FALSE | Upregulated |
| -0,072444149 | 0,839439091 | EIF4G2 | FALSE | Downregulated |
| -0,092698416 | 0,839439091 | MESDC1 | FALSE | Downregulated |
| -0,104265112 | 0,839439091 | PSMB4 | FALSE | Downregulated |

| | | | | |
|---|---|---|---|---|
| -0,117608393 | 0,839439091 | SEC24A | FALSE | Downregulated |
| 0,118691374 | 0,839725844 | ABHD1 | FALSE | Upregulated |
| -0,130201668 | 0,839725844 | ADK | FALSE | Downregulated |
| 0,093484743 | 0,839725844 | NBPF8 | FALSE | Upregulated |
| -0,14970641 | 0,839725844 | PTCHD3P1 | FALSE | Downregulated |
| -0,122557187 | 0,839725844 | SEC23IP | FALSE | Downregulated |
| -0,133584761 | 0,839725844 | ZNF652 | FALSE | Downregulated |
| -0,167741565 | 0,839948177 | TET1 | FALSE | Downregulated |
| -0,111524391 | 0,839948177 | TMEM175 | FALSE | Downregulated |
| -0,088815084 | 0,840045983 | ABCA5 | FALSE | Downregulated |
| 0,221111633 | 0,840045983 | FAM13C | FALSE | Upregulated |
| -0,078648138 | 0,840045983 | IFITM3 | FALSE | Downregulated |
| 0,105997517 | 0,840045983 | RALGAPB | FALSE | Upregulated |
| -0,092796695 | 0,840045983 | TAB2 | FALSE | Downregulated |
| 0,168858881 | 0,840045983 | TRPM3 | FALSE | Upregulated |
| -0,266860496 | 0,840045983 | XG | FALSE | Downregulated |
| 0,158503409 | 0,840312966 | NAV2-AS4 | FALSE | Upregulated |
| -0,244296836 | 0,8404412 | LOC100130876 | FALSE | Downregulated |
| -0,248934901 | 0,8404412 | LOC101928381 | FALSE | Downregulated |
| -0,202089543 | 0,8404412 | LOC102724238 | FALSE | Downregulated |
| -0,170902373 | 0,8404412 | SCLY | FALSE | Downregulated |
| 0,108106508 | 0,8404412 | TRNT1 | FALSE | Upregulated |
| -0,106732309 | 0,840698122 | ZNF800 | FALSE | Downregulated |
| -0,153443028 | 0,840717492 | DDR2 | FALSE | Downregulated |
| 0,097747519 | 0,840717492 | EIF5B | FALSE | Upregulated |
| -0,180370266 | 0,840717492 | GLCE | FALSE | Downregulated |
| -0,115950095 | 0,840717492 | MIF | FALSE | Downregulated |
| -0,098729474 | 0,840717492 | MRPL49 | FALSE | Downregulated |
| 0,103594432 | 0,840717492 | UBAC2 | FALSE | Upregulated |
| -0,069910239 | 0,840728431 | HABP2 | FALSE | Downregulated |
| -0,07824116 | 0,840728431 | KANSL3 | FALSE | Downregulated |
| -0,053721723 | 0,840728431 | PTPRF | FALSE | Downregulated |
| -0,116233797 | 0,840728431 | SEC11A | FALSE | Downregulated |
| 0,073351054 | 0,840825526 | FARP2 | FALSE | Upregulated |
| 0,076750747 | 0,840979747 | CSNK1G2 | FALSE | Upregulated |
| 0,187910785 | 0,840979747 | LCORL | FALSE | Upregulated |
| 0,135661031 | 0,840979747 | LSM12 | FALSE | Upregulated |
| 0,082306355 | 0,840979747 | NACC1 | FALSE | Upregulated |
| -0,144110497 | 0,840979747 | PPTC7 | FALSE | Downregulated |
| -0,128476316 | 0,840979747 | TM9SF1 | FALSE | Downregulated |
| 0,153364436 | 0,840979747 | TMEM170B | FALSE | Upregulated |
| -0,081865459 | 0,841184068 | ERBB2IP | FALSE | Downregulated |
| -0,117885549 | 0,84127674 | TADA1 | FALSE | Downregulated |
| -0,105407971 | 0,841316835 | ACSF3 | FALSE | Downregulated |
| 0,064867705 | 0,841732226 | ZDHHC5 | FALSE | Upregulated |
| 0,091984783 | 0,841778269 | PPP1R13B | FALSE | Upregulated |
| -0,147261168 | 0,841778269 | TEK | FALSE | Downregulated |
| 0,140515638 | 0,841826583 | HSD3BP4 | FALSE | Upregulated |
| -0,101109736 | 0,841826583 | PSMD5 | FALSE | Downregulated |
| -0,187723333 | 0,841826583 | TSC22D3 | FALSE | Downregulated |
| 0,081423682 | 0,841847276 | GTF2IRD1P1 | FALSE | Upregulated |

| | | | | |
|---|---|---|---|---|
| -0,222318783 | 0,841968013 | CBWD2 | FALSE | Downregulated |
| -0,245586382 | 0,841968013 | CTGF | FALSE | Downregulated |
| 0,134087993 | 0,841968013 | IFNGR1 | FALSE | Upregulated |
| 0,155976328 | 0,841968013 | LOC731656 | FALSE | Upregulated |
| -0,181578065 | 0,841968013 | MBD5 | FALSE | Downregulated |
| -0,069113284 | 0,842029705 | KIAA2018 | FALSE | Downregulated |
| 0,137831688 | 0,842054058 | SEPT8 | FALSE | Upregulated |
| -0,14840551 | 0,842054058 | URM1 | FALSE | Downregulated |
| -0,172484928 | 0,842065352 | CDKAL1 | FALSE | Downregulated |
| -0,076952457 | 0,842065352 | EIF4A2 | FALSE | Downregulated |
| 0,114107945 | 0,842065352 | FAM13B | FALSE | Upregulated |
| -0,203352798 | 0,842065352 | NEGR1 | FALSE | Downregulated |
| 0,132067458 | 0,842065352 | OCIAD2 | FALSE | Upregulated |
| -0,191167787 | 0,842065352 | PASK | FALSE | Downregulated |
| 0,138064449 | 0,842065352 | TDRD3 | FALSE | Upregulated |
| -0,154047778 | 0,842190279 | L2HGDH | FALSE | Downregulated |
| 0,110625268 | 0,842190279 | PNPLA3 | FALSE | Upregulated |
| -0,117017219 | 0,842623893 | RPP14 | FALSE | Downregulated |
| -0,104907869 | 0,842623893 | ZNF146 | FALSE | Downregulated |
| -0,179508928 | 0,842746429 | USE1 | FALSE | Downregulated |
| 0,121343651 | 0,842787684 | CCT6P3 | FALSE | Upregulated |
| -0,115475598 | 0,84322941 | FAM134B | FALSE | Downregulated |
| -0,149168543 | 0,84322941 | SEC22A | FALSE | Downregulated |
| 0,141876991 | 0,843399616 | PTPRU | FALSE | Upregulated |
| 0,127690571 | 0,843602879 | ANKMY1 | FALSE | Upregulated |
| -0,054680415 | 0,843606259 | IGF2R | FALSE | Downregulated |
| 0,105171126 | 0,843606259 | RPS6KB1 | FALSE | Upregulated |
| 0,282653082 | 0,843623521 | DCDC2 | FALSE | Upregulated |
| 0,124783083 | 0,843623521 | GIMAP8 | FALSE | Upregulated |
| -0,1520371 | 0,843623521 | PHTF1 | FALSE | Downregulated |
| 0,166769849 | 0,843623521 | RP1-1J6.2 | FALSE | Upregulated |
| -0,103738482 | 0,843623521 | TBC1D1 | FALSE | Downregulated |
| 0,115098643 | 0,843623521 | TTC7A | FALSE | Upregulated |
| 0,251686071 | 0,843623521 | ZNF17 | FALSE | Upregulated |
| -0,074824399 | 0,844053458 | TSC1 | FALSE | Downregulated |
| -0,110215466 | 0,84422929 | YIF1A | FALSE | Downregulated |
| 0,094627241 | 0,84426251 | RFFL | FALSE | Upregulated |
| -0,190280631 | 0,844284822 | TSEN2 | FALSE | Downregulated |
| 0,127578885 | 0,844562257 | ANXA2P1 | FALSE | Upregulated |
| -0,210744208 | 0,844562257 | FBXO45 | FALSE | Downregulated |
| -0,084656697 | 0,844562257 | FXYD1 | FALSE | Downregulated |
| 0,15646503 | 0,844562257 | GIMAP6 | FALSE | Upregulated |
| 0,144429069 | 0,844562257 | LOC166994 | FALSE | Upregulated |
| -0,17930851 | 0,844562257 | PARP2 | FALSE | Downregulated |
| -0,165794626 | 0,844562257 | PLEKHA3 | FALSE | Downregulated |
| -0,218244212 | 0,844562257 | SAMD12 | FALSE | Downregulated |
| -0,201636278 | 0,844562257 | STAM | FALSE | Downregulated |
| 0,175062311 | 0,844562257 | TMEM194B | FALSE | Upregulated |
| 0,141792328 | 0,844562257 | ZFHX2 | FALSE | Upregulated |
| -0,246409961 | 0,844614306 | ATRNL1 | FALSE | Downregulated |
| -0,176874771 | 0,844614306 | CRB3 | FALSE | Downregulated |

| | | | | |
|---|---|---|---|---|
| 0,118960157 | 0,844614306 | PEX5L | FALSE | Upregulated |
| -0,089706684 | 0,844614306 | PPP5C | FALSE | Downregulated |
| 0,133009026 | 0,844689158 | MUS81 | FALSE | Upregulated |
| -0,158402713 | 0,844689158 | SVIP | FALSE | Downregulated |
| -0,231275567 | 0,844725351 | TMEM252 | FALSE | Downregulated |
| 0,119577799 | 0,845071617 | ITGB7 | FALSE | Upregulated |
| 0,076190101 | 0,845071617 | KLC1 | FALSE | Upregulated |
| -0,080882792 | 0,845402392 | TOR1AIP2 | FALSE | Downregulated |
| 0,214503395 | 0,845549455 | FBLN2 | FALSE | Upregulated |
| -0,153828071 | 0,845549455 | SEMA6D | FALSE | Downregulated |
| -0,128877003 | 0,845582942 | TOP3B | FALSE | Downregulated |
| 0,147490568 | 0,845778893 | IGBP1P1 | FALSE | Upregulated |
| 0,092222739 | 0,845778893 | KIAA1191 | FALSE | Upregulated |
| 0,223704918 | 0,845778893 | LOC101928884 | FALSE | Upregulated |
| 0,088890215 | 0,845778893 | RAB8A | FALSE | Upregulated |
| -0,068547084 | 0,845778893 | SMLR1 | FALSE | Downregulated |
| 0,119556506 | 0,845778893 | WDR20 | FALSE | Upregulated |
| 0,139680425 | 0,845984146 | LDHAL6A | FALSE | Upregulated |
| 0,095542392 | 0,846236029 | GTF3C5 | FALSE | Upregulated |
| -0,150001883 | 0,846319711 | CTH | FALSE | Downregulated |
| -0,142524972 | 0,846319711 | POLR2J | FALSE | Downregulated |
| -0,188079117 | 0,846426613 | FUT10 | FALSE | Downregulated |
| 0,116087759 | 0,846426613 | GCH1 | FALSE | Upregulated |
| 0,281922048 | 0,846426613 | LOC102724652 | FALSE | Upregulated |
| 0,114427009 | 0,846426613 | RRNAD1 | FALSE | Upregulated |
| 0,080597323 | 0,846426613 | STK35 | FALSE | Upregulated |
| -0,17544555 | 0,846431362 | ACP6 | FALSE | Downregulated |
| 0,107326834 | 0,846591592 | VASH2 | FALSE | Upregulated |
| 0,121756059 | 0,846648064 | THOC5 | FALSE | Upregulated |
| 0,277119939 | 0,846648064 | TNFRSF10A | FALSE | Upregulated |
| 0,138034306 | 0,846684691 | WDR3 | FALSE | Upregulated |
| 0,138450823 | 0,846781446 | RCSD1 | FALSE | Upregulated |
| -0,173706261 | 0,846781446 | TATDN1 | FALSE | Downregulated |
| -0,082204573 | 0,846905062 | DMXL1 | FALSE | Downregulated |
| -0,12308923 | 0,847020872 | BAIAP2-AS1 | FALSE | Downregulated |
| 0,133145231 | 0,847020872 | QRICH2 | FALSE | Upregulated |
| 0,109029238 | 0,847020872 | SOGA1 | FALSE | Upregulated |
| 0,144962471 | 0,847098588 | TCFL5 | FALSE | Upregulated |
| -0,123397888 | 0,847098588 | TSPAN6 | FALSE | Downregulated |
| -0,11962968 | 0,847402082 | CLDN2 | FALSE | Downregulated |
| 0,138184261 | 0,847402082 | TOR4A | FALSE | Upregulated |
| -0,100058838 | 0,847433592 | YTHDC1 | FALSE | Downregulated |
| 0,136939446 | 0,847866464 | BCL2L14 | FALSE | Upregulated |
| -0,065155363 | 0,847866464 | CDK10 | FALSE | Downregulated |
| 0,085003288 | 0,847866464 | NR2F1 | FALSE | Upregulated |
| -0,163129968 | 0,847977402 | CRELD2 | FALSE | Downregulated |
| 0,150161073 | 0,848148805 | ADAM21 | FALSE | Upregulated |
| 0,130411245 | 0,848148805 | APOL4 | FALSE | Upregulated |
| 0,261371409 | 0,848148805 | CCDC80 | FALSE | Upregulated |
| -0,124297922 | 0,848148805 | CD2AP | FALSE | Downregulated |
| -0,103615333 | 0,848148805 | FBP1 | FALSE | Downregulated |

| | | | | |
|---|---|---|---|---|
| -0,075142787 | 0,848148805 | FZD4 | FALSE | Downregulated |
| -0,091119813 | 0,848148805 | H2AFV | FALSE | Downregulated |
| 0,14315358 | 0,848148805 | JUN | FALSE | Upregulated |
| 0,100577833 | 0,848148805 | METTL12 | FALSE | Upregulated |
| -0,311950348 | 0,848148805 | MUC15 | FALSE | Downregulated |
| 0,105763577 | 0,848148805 | PCED1A | FALSE | Upregulated |
| 0,082197917 | 0,848148805 | PON2 | FALSE | Upregulated |
| 0,167409819 | 0,848148805 | SMCO4 | FALSE | Upregulated |
| 0,209426666 | 0,848148805 | SYN1 | FALSE | Upregulated |
| 0,160177061 | 0,848148805 | ZNF417 | FALSE | Upregulated |
| -0,13629854 | 0,848337454 | AC005029.1 | FALSE | Downregulated |
| 0,108643717 | 0,848337454 | TSC22D4 | FALSE | Upregulated |
| -0,077024259 | 0,848372013 | PMPCA | FALSE | Downregulated |
| 0,107087898 | 0,848563992 | HABP4 | FALSE | Upregulated |
| 0,150044264 | 0,848563992 | LOC101928927 | FALSE | Upregulated |
| 0,225722339 | 0,848563992 | SCARNA9 | FALSE | Upregulated |
| -0,198153689 | 0,848563992 | TMEM60 | FALSE | Downregulated |
| 0,064475508 | 0,849097828 | FOXK1 | FALSE | Upregulated |
| 0,125831294 | 0,849147677 | DOCK3 | FALSE | Upregulated |
| 0,081756599 | 0,849465777 | BAZ1A | FALSE | Upregulated |
| 0,106730057 | 0,849465777 | CERS6 | FALSE | Upregulated |
| 0,170244023 | 0,849465777 | TFAP2E | FALSE | Upregulated |
| 0,081866741 | 0,849475981 | CAND1 | FALSE | Upregulated |
| -0,197987646 | 0,849475981 | RP11-203B9.4 | FALSE | Downregulated |
| -0,308020629 | 0,84954151 | CYP1A1 | FALSE | Downregulated |
| -0,10312989 | 0,84954151 | TJP3 | FALSE | Downregulated |
| -0,079214546 | 0,849729127 | TBC1D14 | FALSE | Downregulated |
| 0,131170593 | 0,849756075 | AIMP2 | FALSE | Upregulated |
| 0,24520176 | 0,849756075 | CCDC40 | FALSE | Upregulated |
| -0,073277279 | 0,849756075 | ITSN2 | FALSE | Downregulated |
| 0,223110588 | 0,849756075 | NEK8 | FALSE | Upregulated |
| -0,164781235 | 0,849756075 | XKR8 | FALSE | Downregulated |
| -0,147852956 | 0,849759198 | CDA | FALSE | Downregulated |
| 0,178827272 | 0,849759198 | FADS2 | FALSE | Upregulated |
| -0,069217839 | 0,849759198 | HNRNPA2B1 | FALSE | Downregulated |
| 0,088423722 | 0,849759198 | LOC100288162 | FALSE | Upregulated |
| 0,189054271 | 0,849759198 | PRX | FALSE | Upregulated |
| 0,15651115 | 0,849759198 | SPIN3 | FALSE | Upregulated |
| 0,155254254 | 0,84978644 | KLLN | FALSE | Upregulated |
| -0,078010873 | 0,84978644 | RPL7L1 | FALSE | Downregulated |
| -0,134581638 | 0,849832308 | FAM162A | FALSE | Downregulated |
| -0,261113193 | 0,849915276 | LOC102724163 | FALSE | Downregulated |
| -0,191085457 | 0,849915276 | SEH1L | FALSE | Downregulated |
| -0,129638238 | 0,849915276 | YPEL2 | FALSE | Downregulated |
| -0,142977719 | 0,849941497 | RSL24D1 | FALSE | Downregulated |
| 0,091159225 | 0,849970739 | SDC3 | FALSE | Upregulated |
| -0,19979436 | 0,850065343 | CCDC183-AS1 | FALSE | Downregulated |
| 0,163494366 | 0,850065343 | CHMP4A | FALSE | Upregulated |
| -0,123760606 | 0,850065343 | SNRPD2 | FALSE | Downregulated |
| 0,126848578 | 0,850065343 | UBE2G1 | FALSE | Upregulated |
| -0,091046383 | 0,850065343 | ZNF740 | FALSE | Downregulated |

| | | | | |
|---|---|---|---|---|
| -0,106815732 | 0,850067338 | TMEM230 | FALSE | Downregulated |
| 0,083247751 | 0,850074692 | RABEP1 | FALSE | Upregulated |
| -0,122374158 | 0,850155594 | RRP7A | FALSE | Downregulated |
| 0,130444199 | 0,850303062 | CIPC | FALSE | Upregulated |
| -0,079487898 | 0,850303062 | RTN4 | FALSE | Downregulated |
| -0,192287611 | 0,850523797 | LOC100507131 | FALSE | Downregulated |
| 0,196066691 | 0,850593342 | EFR3B | FALSE | Upregulated |
| 0,195988575 | 0,850593342 | STARD4-AS1 | FALSE | Upregulated |
| 0,050734556 | 0,850776707 | FARP1 | FALSE | Upregulated |
| 0,090521325 | 0,850918097 | NAA30 | FALSE | Upregulated |
| 0,126388573 | 0,851130631 | CACNA1C | FALSE | Upregulated |
| -0,089675389 | 0,851130631 | CLPX | FALSE | Downregulated |
| -0,147321542 | 0,851130631 | CNGA1 | FALSE | Downregulated |
| -0,085900282 | 0,851130631 | DAAM1 | FALSE | Downregulated |
| 0,088068927 | 0,851130631 | DNAJC21 | FALSE | Upregulated |
| -0,12195704 | 0,851130631 | IFT122 | FALSE | Downregulated |
| -0,105930417 | 0,851130631 | MLX | FALSE | Downregulated |
| -0,184510245 | 0,851130631 | MRPL18 | FALSE | Downregulated |
| 0,187282296 | 0,851130631 | NRADDP | FALSE | Upregulated |
| 0,103879576 | 0,851130631 | PHPT1 | FALSE | Upregulated |
| 0,073103123 | 0,851130631 | SIDT2 | FALSE | Upregulated |
| 0,166479203 | 0,851130631 | ZFAND4 | FALSE | Upregulated |
| 0,206822313 | 0,851130631 | ZNF555 | FALSE | Upregulated |
| 0,156078252 | 0,851130631 | ZNF821 | FALSE | Upregulated |
| -0,128190541 | 0,85130865 | FAM46A | FALSE | Downregulated |
| 0,191431971 | 0,85130865 | LOC101593348 | FALSE | Upregulated |
| -0,185501138 | 0,85139521 | DDX52 | FALSE | Downregulated |
| 0,085361402 | 0,851436954 | B3GALNT2 | FALSE | Upregulated |
| -0,14106061 | 0,851436954 | FAM103A1 | FALSE | Downregulated |
| 0,146318994 | 0,851436954 | LOC100506175 | FALSE | Upregulated |
| -0,141890007 | 0,851436954 | POLE3 | FALSE | Downregulated |
| -0,116320359 | 0,851468927 | GLTSCR1L | FALSE | Downregulated |
| -0,106712285 | 0,851549168 | C1orf168 | FALSE | Downregulated |
| -0,11777926 | 0,851549168 | TPRN | FALSE | Downregulated |
| -0,220021164 | 0,851559823 | EPB41L4A-AS1 | FALSE | Downregulated |
| -0,179713539 | 0,851934312 | PEX11B | FALSE | Downregulated |
| -0,093484333 | 0,851949279 | GRN | FALSE | Downregulated |
| -0,06306436 | 0,851959317 | ACO1 | FALSE | Downregulated |
| 0,082980587 | 0,851959317 | PLD3 | FALSE | Upregulated |
| -0,139231795 | 0,851959317 | VPS45 | FALSE | Downregulated |
| -0,078131118 | 0,852022321 | PHC2 | FALSE | Downregulated |
| -0,144251933 | 0,852156834 | CCDC101 | FALSE | Downregulated |
| 0,129178388 | 0,852156834 | LOC101929986 | FALSE | Upregulated |
| -0,071235922 | 0,852156834 | SMPD4 | FALSE | Downregulated |
| 0,11285411 | 0,852156834 | TBC1D23 | FALSE | Upregulated |
| 0,14969584 | 0,852332612 | ZNF619 | FALSE | Upregulated |
| 0,146274915 | 0,852502116 | METTL16 | FALSE | Upregulated |
| -0,045994052 | 0,852657319 | ARID1A | FALSE | Downregulated |
| 0,095640776 | 0,852657319 | C2orf16 | FALSE | Upregulated |
| 0,23400582 | 0,852657319 | DNAH2 | FALSE | Upregulated |
| -0,189904507 | 0,852657319 | ERGIC2 | FALSE | Downregulated |

| | | | | |
|---|---|---|---|---|
| 0,148267512 | 0,852657319 | GABPB2 | FALSE | Upregulated |
| 0,199034374 | 0,852657319 | LEKR1 | FALSE | Upregulated |
| 0,088342846 | 0,852657319 | LINC00242 | FALSE | Upregulated |
| 0,070811365 | 0,852657319 | PARP14 | FALSE | Upregulated |
| -0,067620342 | 0,852657319 | PSAP | FALSE | Downregulated |
| -0,172286078 | 0,852657319 | RABGGTB | FALSE | Downregulated |
| -0,101934506 | 0,852657319 | UBXN4 | FALSE | Downregulated |
| 0,116451997 | 0,852657319 | UCHL5 | FALSE | Upregulated |
| -0,102970782 | 0,852657319 | ZFP41 | FALSE | Downregulated |
| 0,119369163 | 0,852704488 | CRIP3 | FALSE | Upregulated |
| -0,219654682 | 0,852713389 | FDX1L | FALSE | Downregulated |
| 0,119859061 | 0,852796547 | C2orf43 | FALSE | Upregulated |
| -0,143986916 | 0,852796547 | LOC100996273 | FALSE | Downregulated |
| 0,108769015 | 0,852796547 | LOC101928061 | FALSE | Upregulated |
| -0,102366611 | 0,852796547 | TAF10 | FALSE | Downregulated |
| 0,065505105 | 0,852861232 | DDX5 | FALSE | Upregulated |
| 0,101539429 | 0,852861232 | DND1 | FALSE | Upregulated |
| 0,173500301 | 0,852861232 | LINC00176 | FALSE | Upregulated |
| -0,135128232 | 0,852861232 | LOC101927157 | FALSE | Downregulated |
| 0,135743519 | 0,852861232 | USP30-AS1 | FALSE | Upregulated |
| 0,106382097 | 0,852955071 | CRCP | FALSE | Upregulated |
| 0,110019436 | 0,853085064 | LOC102723566 | FALSE | Upregulated |
| 0,118978346 | 0,853278275 | BFAR | FALSE | Upregulated |
| 0,115794072 | 0,853278275 | GTSE1-AS1 | FALSE | Upregulated |
| -0,112617308 | 0,853278275 | PREB | FALSE | Downregulated |
| 0,248331185 | 0,853397299 | DCUN1D5 | FALSE | Upregulated |
| -0,259093555 | 0,853397299 | LHX9 | FALSE | Downregulated |
| -0,181471551 | 0,853397299 | STL | FALSE | Downregulated |
| -0,061955171 | 0,853427369 | SRSF1 | FALSE | Downregulated |
| 0,208775496 | 0,853427369 | ZNF493 | FALSE | Upregulated |
| -0,103356879 | 0,853662652 | FBXW11 | FALSE | Downregulated |
| 0,07630327 | 0,853706828 | AGO1 | FALSE | Upregulated |
| -0,130801094 | 0,853706828 | MRPL23-AS1 | FALSE | Downregulated |
| 0,094133073 | 0,853706828 | PCGF3 | FALSE | Upregulated |
| 0,146720541 | 0,8538321 | ABT1 | FALSE | Upregulated |
| 0,136092054 | 0,8538321 | ALS2CR11 | FALSE | Upregulated |
| 0,111006256 | 0,8538321 | CRHBP | FALSE | Upregulated |
| -0,092904121 | 0,8538321 | DHX8 | FALSE | Downregulated |
| -0,110187801 | 0,8538321 | EEPD1 | FALSE | Downregulated |
| -0,170660917 | 0,8538321 | EP400NL | FALSE | Downregulated |
| 0,147270045 | 0,8538321 | GMPS | FALSE | Upregulated |
| -0,172877861 | 0,8538321 | GTF2H3 | FALSE | Downregulated |
| -0,1659154 | 0,8538321 | HDHD1 | FALSE | Downregulated |
| -0,095163238 | 0,8538321 | HIBADH | FALSE | Downregulated |
| 0,148769089 | 0,8538321 | HNRNPLL | FALSE | Upregulated |
| 0,084189425 | 0,8538321 | IL4R | FALSE | Upregulated |
| 0,250146477 | 0,8538321 | KRT7 | FALSE | Upregulated |
| 0,105338505 | 0,8538321 | LOC101928319 | FALSE | Upregulated |
| -0,093677293 | 0,8538321 | MAGED1 | FALSE | Downregulated |
| 0,079469273 | 0,8538321 | MARCKS | FALSE | Upregulated |
| -0,083752195 | 0,8538321 | NDUFS1 | FALSE | Downregulated |

| | | | | |
|---|---|---|---|---|
| -0,088977834 | 0,8538321 | NOTCH4 | FALSE | Downregulated |
| -0,155258857 | 0,8538321 | PHYHD1 | FALSE | Downregulated |
| -0,091673316 | 0,8538321 | PPM1F | FALSE | Downregulated |
| 0,168505551 | 0,8538321 | PRR5L | FALSE | Upregulated |
| 0,084031919 | 0,8538321 | PTK2B | FALSE | Upregulated |
| -0,07530807 | 0,8538321 | RAB11FIP3 | FALSE | Downregulated |
| -0,127693517 | 0,8538321 | RHBDD2 | FALSE | Downregulated |
| -0,137840697 | 0,8538321 | RNF26 | FALSE | Downregulated |
| 0,164437739 | 0,8538321 | RP11-41O4.2 | FALSE | Upregulated |
| 0,098932504 | 0,8538321 | RUFY1 | FALSE | Upregulated |
| -0,192818559 | 0,8538321 | SENP1 | FALSE | Downregulated |
| 0,16730903 | 0,8538321 | SFTPD | FALSE | Upregulated |
| 0,164697381 | 0,8538321 | SLC15A3 | FALSE | Upregulated |
| -0,08777916 | 0,8538321 | SMARCD2 | FALSE | Downregulated |
| -0,062407698 | 0,8538321 | SORL1 | FALSE | Downregulated |
| 0,133168263 | 0,8538321 | SPG20 | FALSE | Upregulated |
| 0,197946759 | 0,8538321 | TP53TG5 | FALSE | Upregulated |
| 0,137051354 | 0,8538321 | TRIM37 | FALSE | Upregulated |
| 0,126144377 | 0,8538321 | TRMU | FALSE | Upregulated |
| -0,090346456 | 0,8538321 | WDR55 | FALSE | Downregulated |
| 0,131233818 | 0,8538321 | YPEL5 | FALSE | Upregulated |
| 0,161279756 | 0,8538321 | ZNF605 | FALSE | Upregulated |
| -0,169600888 | 0,853848528 | ETAA1 | FALSE | Downregulated |
| 0,163270957 | 0,853848528 | HNRNPA1P10 | FALSE | Upregulated |
| 0,098549492 | 0,853848528 | NPIPB11 | FALSE | Upregulated |
| -0,185702403 | 0,854011339 | ENDOD1 | FALSE | Downregulated |
| 0,103782521 | 0,854177482 | IMP3 | FALSE | Upregulated |
| 0,133421088 | 0,854466263 | C3orf62 | FALSE | Upregulated |
| -0,070416157 | 0,854466263 | PPIA | FALSE | Downregulated |
| -0,076321382 | 0,854484035 | CHRD | FALSE | Downregulated |
| -0,08352783 | 0,854836163 | FAT4 | FALSE | Downregulated |
| 0,100724713 | 0,854836163 | LCOR | FALSE | Upregulated |
| -0,074761058 | 0,854836163 | PCYT2 | FALSE | Downregulated |
| 0,185680122 | 0,854960461 | LOC101929512 | FALSE | Upregulated |
| 0,124196584 | 0,854960461 | TMEM101 | FALSE | Upregulated |
| 0,081476096 | 0,85528995 | PEAK1 | FALSE | Upregulated |
| -0,055522786 | 0,85528995 | SETD7 | FALSE | Downregulated |
| -0,109988691 | 0,85528995 | TACC2 | FALSE | Downregulated |
| 0,138830069 | 0,855293993 | GTF3C3 | FALSE | Upregulated |
| -0,067647573 | 0,855293993 | SORBS1 | FALSE | Downregulated |
| -0,173711039 | 0,855303007 | ACOT1 | FALSE | Downregulated |
| -0,109677912 | 0,855303007 | EBNA1BP2 | FALSE | Downregulated |
| -0,08949661 | 0,855303007 | FGF2 | FALSE | Downregulated |
| 0,22147193 | 0,855303007 | OR10J5 | FALSE | Upregulated |
| 0,075393945 | 0,855303007 | VASH1 | FALSE | Upregulated |
| -0,098365873 | 0,855303007 | ZNF181 | FALSE | Downregulated |
| -0,082729955 | 0,855408907 | AIG1 | FALSE | Downregulated |
| 0,176137653 | 0,855408907 | DDX59 | FALSE | Upregulated |
| -0,14611833 | 0,855408907 | GEN1 | FALSE | Downregulated |
| -0,122127124 | 0,855408907 | GORASP2 | FALSE | Downregulated |
| -0,057288179 | 0,855408907 | MYL6 | FALSE | Downregulated |

| | | | | |
|---|---|---|---|---|
| -0,141752208 | 0,855408907 | SCFD2 | FALSE | Downregulated |
| 0,168883342 | 0,855708118 | ACSS1 | FALSE | Upregulated |
| 0,176194149 | 0,855708118 | CCDC68 | FALSE | Upregulated |
| 0,160742385 | 0,855708118 | KCNMB3 | FALSE | Upregulated |
| -0,142024284 | 0,855708118 | PYCR2 | FALSE | Downregulated |
| 0,198032422 | 0,855708118 | RP11-1096D5.1 | FALSE | Upregulated |
| 0,161165493 | 0,85602253 | LOC100129518 | FALSE | Upregulated |
| -0,108906635 | 0,85602253 | VPS4B | FALSE | Downregulated |
| -0,098048385 | 0,856110526 | CEBPZ-AS1 | FALSE | Downregulated |
| -0,1115792 | 0,856110526 | LYSMD3 | FALSE | Downregulated |
| 0,142483037 | 0,856158391 | FLI1 | FALSE | Upregulated |
| 0,204511363 | 0,856158391 | FLRT2 | FALSE | Upregulated |
| -0,134210987 | 0,856166279 | PAPD4 | FALSE | Downregulated |
| -0,134477911 | 0,856320484 | LOC284889 | FALSE | Downregulated |
| -0,136646939 | 0,856320484 | RPRD1A | FALSE | Downregulated |
| 0,171812593 | 0,856739018 | ZNF790-AS1 | FALSE | Upregulated |
| 0,064010118 | 0,856739659 | FBXW5 | FALSE | Upregulated |
| -0,089405041 | 0,856739659 | SERPINA3 | FALSE | Downregulated |
| -0,145181136 | 0,856753142 | CNIH1 | FALSE | Downregulated |
| 0,243125527 | 0,856778389 | CRYBA1 | FALSE | Upregulated |
| -0,103760837 | 0,8568584 | NIT2 | FALSE | Downregulated |
| 0,222577169 | 0,856963439 | CPLX2 | FALSE | Upregulated |
| -0,213282538 | 0,856963439 | OR5AK2 | FALSE | Downregulated |
| -0,082609334 | 0,856963439 | QARS | FALSE | Downregulated |
| 0,121927638 | 0,857066248 | STX7 | FALSE | Upregulated |
| 0,082105974 | 0,857187168 | BDP1 | FALSE | Upregulated |
| -0,150130492 | 0,857187168 | C8orf4 | FALSE | Downregulated |
| -0,084193857 | 0,857187168 | EAF1 | FALSE | Downregulated |
| 0,10094855 | 0,857187168 | KIAA0922 | FALSE | Upregulated |
| 0,113573331 | 0,857187168 | ZNF394 | FALSE | Upregulated |
| -0,136005211 | 0,857357736 | PXMP4 | FALSE | Downregulated |
| 0,212270045 | 0,857968152 | DGKH | FALSE | Upregulated |
| 0,091090675 | 0,857968152 | VPS54 | FALSE | Upregulated |
| -0,249754263 | 0,858071963 | FOXN4 | FALSE | Downregulated |
| 0,169836551 | 0,858114398 | LOC102724490 | FALSE | Upregulated |
| -0,1438186 | 0,858221147 | ZSWIM4 | FALSE | Downregulated |
| 0,119665324 | 0,858401964 | ZNF175 | FALSE | Upregulated |
| 0,211304633 | 0,85848544 | BACE2 | FALSE | Upregulated |
| -0,19444191 | 0,85848544 | LOC102724023 | FALSE | Downregulated |
| -0,124343851 | 0,858591793 | MTF2 | FALSE | Downregulated |
| -0,105189186 | 0,858984673 | C19orf24 | FALSE | Downregulated |
| 0,114051227 | 0,858984673 | EXOSC3 | FALSE | Upregulated |
| -0,230270223 | 0,858984673 | LOC100505570 | FALSE | Downregulated |
| -0,130781239 | 0,858984673 | NUPL1 | FALSE | Downregulated |
| 0,102007084 | 0,858984673 | PLCG2 | FALSE | Upregulated |
| -0,085311374 | 0,858984673 | SPIN1 | FALSE | Downregulated |
| 0,095094363 | 0,859020787 | LOC101927393 | FALSE | Upregulated |
| 0,097125072 | 0,859020787 | SMC3 | FALSE | Upregulated |
| 0,084535978 | 0,859020787 | SMPD1 | FALSE | Upregulated |
| -0,115365271 | 0,859020787 | UBQLN2 | FALSE | Downregulated |
| 0,139717108 | 0,859020787 | USB1 | FALSE | Upregulated |

| | | | | |
|---|---|---|---|---|
| 0,122647626 | 0,859030407 | RAPGEF6 | FALSE | Upregulated |
| -0,201136332 | 0,859074679 | TOX3 | FALSE | Downregulated |
| -0,143963023 | 0,859216264 | RNF121 | FALSE | Downregulated |
| 0,142474691 | 0,859216264 | SMG7-AS1 | FALSE | Upregulated |
| -0,171648191 | 0,859292997 | ABHD10 | FALSE | Downregulated |
| 0,094231922 | 0,859292997 | CES3 | FALSE | Upregulated |
| 0,101369613 | 0,859292997 | EMC3 | FALSE | Upregulated |
| 0,111884357 | 0,859645884 | HYOU1 | FALSE | Upregulated |
| 0,122594785 | 0,859645884 | LRG1 | FALSE | Upregulated |
| -0,105178691 | 0,859645884 | NFKBIZ | FALSE | Downregulated |
| -0,132777582 | 0,859645884 | TMEM150C | FALSE | Downregulated |
| -0,058950992 | 0,859645884 | ZBTB7A | FALSE | Downregulated |
| -0,078532751 | 0,859645884 | ZNF598 | FALSE | Downregulated |
| 0,137453413 | 0,859714406 | SHKBP1 | FALSE | Upregulated |
| 0,061151291 | 0,859714742 | TEX2 | FALSE | Upregulated |
| -0,76117225 | 0,859776148 | C7orf55-LUC7L2 | FALSE | Downregulated |
| 0,301789968 | 0,859776148 | FAM188B | FALSE | Upregulated |
| -0,230897037 | 0,859776148 | LINC00899 | FALSE | Downregulated |
| -0,140591646 | 0,860062753 | CLEC3B | FALSE | Downregulated |
| -0,125751755 | 0,860062753 | IGHMBP2 | FALSE | Downregulated |
| 0,18295982 | 0,860062753 | LRRC25 | FALSE | Upregulated |
| -0,07509249 | 0,860062753 | NGEF | FALSE | Downregulated |
| 0,112158439 | 0,860062753 | RPP38 | FALSE | Upregulated |
| 0,134645481 | 0,860357334 | ARPC3 | FALSE | Upregulated |
| 0,082652821 | 0,860357334 | CLSTN3 | FALSE | Upregulated |
| 0,126442137 | 0,860357334 | GRAMD1A | FALSE | Upregulated |
| -0,088237457 | 0,860357334 | HIPK1 | FALSE | Downregulated |
| -0,176759617 | 0,860357334 | LOC727710 | FALSE | Downregulated |
| -0,113184179 | 0,860357334 | PGLS | FALSE | Downregulated |
| 0,14266414 | 0,860357334 | RNF214 | FALSE | Upregulated |
| -0,088433882 | 0,860357334 | SHF | FALSE | Downregulated |
| -0,138528562 | 0,860357334 | SMAD1 | FALSE | Downregulated |
| -0,207493422 | 0,860357334 | TEC | FALSE | Downregulated |
| 0,087406874 | 0,860676025 | ADNP | FALSE | Upregulated |
| 0,205855247 | 0,860676025 | MAGI1-IT1 | FALSE | Upregulated |
| 0,126177029 | 0,86076142 | LOC102723972 | FALSE | Upregulated |
| 0,098389058 | 0,861184655 | APOC4-APOC2 | FALSE | Upregulated |
| 0,095437038 | 0,861184655 | GAB1 | FALSE | Upregulated |
| 0,086747527 | 0,861184655 | MFHAS1 | FALSE | Upregulated |
| -0,115913399 | 0,861184655 | PQBP1 | FALSE | Downregulated |
| -0,136557488 | 0,861184655 | UBXN8 | FALSE | Downregulated |
| 0,1123592 | 0,861237802 | ANAPC1 | FALSE | Upregulated |
| -0,105531965 | 0,861347638 | UBXN1 | FALSE | Downregulated |
| -0,111029257 | 0,861536597 | ACSF2 | FALSE | Downregulated |
| 0,143183039 | 0,861536597 | EMBP1 | FALSE | Upregulated |
| 0,091488189 | 0,861536597 | EPT1 | FALSE | Upregulated |
| -0,259469628 | 0,861536597 | GPR89B | FALSE | Downregulated |
| 0,07305576 | 0,861536597 | IMPAD1 | FALSE | Upregulated |
| 0,171651708 | 0,861536597 | LOC285095 | FALSE | Upregulated |
| 0,139968176 | 0,861536597 | TAPT1-AS1 | FALSE | Upregulated |
| -0,13875367 | 0,861536597 | TPGS1 | FALSE | Downregulated |

| | | | | |
|---|---|---|---|---|
| -0,068619129 | 0,861536597 | TRMT2A | FALSE | Downregulated |
| 0,123851741 | 0,861536597 | TRPM4 | FALSE | Upregulated |
| 0,183603002 | 0,861536597 | UBTD1 | FALSE | Upregulated |
| -0,126340231 | 0,861536597 | WHAMM | FALSE | Downregulated |
| 0,070373879 | 0,861536597 | YAP1 | FALSE | Upregulated |
| 0,087207501 | 0,861536597 | ZSWIM1 | FALSE | Upregulated |
| -0,20384346 | 0,861703219 | C1orf220 | FALSE | Downregulated |
| 0,191998203 | 0,861703219 | LMNB1 | FALSE | Upregulated |
| -0,223825059 | 0,861731178 | PCNA | FALSE | Downregulated |
| 0,156793748 | 0,861741305 | NAT2 | FALSE | Upregulated |
| 0,205340408 | 0,861741305 | PDK1 | FALSE | Upregulated |
| -0,105463997 | 0,861741305 | PSMD1 | FALSE | Downregulated |
| -0,112404542 | 0,861741305 | TTC39C | FALSE | Downregulated |
| 0,114566093 | 0,861783222 | TMPO-AS1 | FALSE | Upregulated |
| 0,092757047 | 0,861906543 | NFRKB | FALSE | Upregulated |
| -0,217216834 | 0,862161957 | CCDC84 | FALSE | Downregulated |
| -0,070945536 | 0,862304562 | BTBD6 | FALSE | Downregulated |
| 0,140614774 | 0,862311264 | FAM86C1 | FALSE | Upregulated |
| -0,102881671 | 0,862311264 | FOXA2 | FALSE | Downregulated |
| 0,086227323 | 0,862311264 | LINC00667 | FALSE | Upregulated |
| -0,091235826 | 0,862414741 | KARS | FALSE | Downregulated |
| 0,086392318 | 0,862479937 | SRP14 | FALSE | Upregulated |
| -0,114728746 | 0,862648431 | CREB3 | FALSE | Downregulated |
| 0,189335774 | 0,862648431 | DTNBP1 | FALSE | Upregulated |
| 0,131388106 | 0,862648431 | EMC7 | FALSE | Upregulated |
| -0,290924855 | 0,862648431 | HIST1H4K | FALSE | Downregulated |
| -0,156853523 | 0,862648431 | IL17RE | FALSE | Downregulated |
| -0,167667628 | 0,862648431 | NOL11 | FALSE | Downregulated |
| -0,29426813 | 0,862648431 | PGR | FALSE | Downregulated |
| 0,13988005 | 0,862648431 | SCARF2 | FALSE | Upregulated |
| -0,15396763 | 0,862648431 | SNRNP48 | FALSE | Downregulated |
| -0,256978042 | 0,862648431 | STMN1 | FALSE | Downregulated |
| -0,211239078 | 0,863571306 | BAMBI | FALSE | Downregulated |
| -0,061110308 | 0,863571306 | GLS2 | FALSE | Downregulated |
| -0,173851051 | 0,863571306 | POU6F1 | FALSE | Downregulated |
| -0,193427187 | 0,863582835 | CLRN3 | FALSE | Downregulated |
| -0,198317731 | 0,863656881 | BLOC1S5 | FALSE | Downregulated |
| 0,098431774 | 0,863656881 | DLG2 | FALSE | Upregulated |
| 0,110592268 | 0,863656881 | HIST2H2AC | FALSE | Upregulated |
| 0,07227655 | 0,863656881 | LOC101927838 | FALSE | Upregulated |
| 0,097925563 | 0,863656881 | MESTIT1 | FALSE | Upregulated |
| 0,16518392 | 0,863656881 | RPP25L | FALSE | Upregulated |
| -0,053160415 | 0,863656881 | SETD2 | FALSE | Downregulated |
| -0,084746395 | 0,863656881 | SIN3B | FALSE | Downregulated |
| -0,135829338 | 0,863656881 | ZFP62 | FALSE | Downregulated |
| 0,153312104 | 0,863782168 | PGP | FALSE | Upregulated |
| -0,197896318 | 0,863934774 | ING2 | FALSE | Downregulated |
| 0,20371783 | 0,864054464 | PTGS1 | FALSE | Upregulated |
| -0,12375982 | 0,864054464 | SGMS1 | FALSE | Downregulated |
| 0,081934415 | 0,864054464 | STAT3 | FALSE | Upregulated |
| -0,12113916 | 0,86409936 | DOCK9 | FALSE | Downregulated |

| | | | | |
|---|---|---|---|---|
| -0,145972011 | 0,86413715 | AQP7P1 | FALSE | Downregulated |
| 0,064885304 | 0,86413715 | CABIN1 | FALSE | Upregulated |
| -0,133461002 | 0,86413715 | CYP7B1 | FALSE | Downregulated |
| 0,179163849 | 0,86413715 | GPRASP2 | FALSE | Upregulated |
| -0,076321055 | 0,86413715 | PSMC3 | FALSE | Downregulated |
| 0,106687936 | 0,86413715 | SSBP3 | FALSE | Upregulated |
| -0,127924197 | 0,86413715 | TTC3-AS1 | FALSE | Downregulated |
| 0,165009375 | 0,864339753 | GPNMB | FALSE | Upregulated |
| 0,052416921 | 0,864365814 | MAST2 | FALSE | Upregulated |
| -0,129904617 | 0,864475363 | ABCC6P1 | FALSE | Downregulated |
| -0,071952282 | 0,864618169 | COPB2 | FALSE | Downregulated |
| -0,187491319 | 0,864832474 | THAP6 | FALSE | Downregulated |
| -0,077137277 | 0,865172419 | CCNY | FALSE | Downregulated |
| 0,163909336 | 0,865172812 | WBSCR22 | FALSE | Upregulated |
| -0,057009052 | 0,8653173 | HERC2 | FALSE | Downregulated |
| 0,098993477 | 0,865430713 | ANKZF1 | FALSE | Upregulated |
| 0,136919751 | 0,865430713 | FBXO42 | FALSE | Upregulated |
| 0,09703379 | 0,865430713 | GTF2F1 | FALSE | Upregulated |
| -0,082449917 | 0,865430713 | KANK3 | FALSE | Downregulated |
| -0,153642521 | 0,865430713 | LOC101927727 | FALSE | Downregulated |
| 0,083042168 | 0,865430713 | LOC101929385 | FALSE | Upregulated |
| 0,10327424 | 0,865430713 | MEX3C | FALSE | Upregulated |
| -0,064267533 | 0,865430713 | SETX | FALSE | Downregulated |
| 0,134051287 | 0,865430713 | SLC16A13 | FALSE | Upregulated |
| 0,145571101 | 0,865430713 | TMEM135 | FALSE | Upregulated |
| -0,236546064 | 0,865489562 | BICC1 | FALSE | Downregulated |
| 0,054688624 | 0,865489562 | SLC35E2B | FALSE | Upregulated |
| 0,115655319 | 0,865671266 | C11orf83 | FALSE | Upregulated |
| 0,094858904 | 0,865671266 | RNF139 | FALSE | Upregulated |
| -0,138459958 | 0,865980591 | AC007099.1 | FALSE | Downregulated |
| 0,148418405 | 0,865980591 | CSGALNACT2 | FALSE | Upregulated |
| 0,17919566 | 0,865980591 | DCLRE1A | FALSE | Upregulated |
| -0,115129076 | 0,865980591 | DNAJB6 | FALSE | Downregulated |
| 0,102973073 | 0,865980591 | FBXW7 | FALSE | Upregulated |
| 0,068970409 | 0,865980591 | FKBP9 | FALSE | Upregulated |
| -0,193562288 | 0,865980591 | IPMK | FALSE | Downregulated |
| 0,220345702 | 0,865980591 | JKAMP | FALSE | Upregulated |
| 0,172699355 | 0,865980591 | LOC102724157 | FALSE | Upregulated |
| 0,118199635 | 0,865980591 | NDUFS8 | FALSE | Upregulated |
| -0,089045395 | 0,865980591 | PSME2 | FALSE | Downregulated |
| -0,061764022 | 0,865980591 | SAT1 | FALSE | Downregulated |
| 0,187755393 | 0,865980591 | SLC7A9 | FALSE | Upregulated |
| -0,098292308 | 0,865980591 | WDR5B | FALSE | Downregulated |
| 0,124554649 | 0,865992339 | CDK19 | FALSE | Upregulated |
| 0,100540041 | 0,865992339 | RTCB | FALSE | Upregulated |
| 0,163268625 | 0,866490595 | B4GALT6 | FALSE | Upregulated |
| 0,078563298 | 0,866490595 | COX11 | FALSE | Upregulated |
| 0,152328013 | 0,866490595 | NOXA1 | FALSE | Upregulated |
| 0,110895754 | 0,866490595 | SLC2A6 | FALSE | Upregulated |
| 0,088313778 | 0,866490595 | ZEB2 | FALSE | Upregulated |
| -0,07384948 | 0,866515753 | CARKD | FALSE | Downregulated |

| | | | | |
|---|---|---|---|---|
| 0,127362991 | 0,866515753 | SLX4 | FALSE | Upregulated |
| -0,051644255 | 0,866515753 | UBXN6 | FALSE | Downregulated |
| 0,082474339 | 0,86653833 | ANGPTL4 | FALSE | Upregulated |
| -0,088246067 | 0,86653833 | DOK4 | FALSE | Downregulated |
| -0,118200438 | 0,86653833 | LRRC8D | FALSE | Downregulated |
| -0,090827383 | 0,86653833 | MARVELD2 | FALSE | Downregulated |
| -0,15121877 | 0,866729035 | GPS2 | FALSE | Downregulated |
| -0,080002012 | 0,866729035 | HIST1H1C | FALSE | Downregulated |
| 0,160908311 | 0,866729035 | HMGB3 | FALSE | Upregulated |
| 0,066032249 | 0,866729035 | NPEPPS | FALSE | Upregulated |
| -0,086907755 | 0,866729035 | RNF167 | FALSE | Downregulated |
| -0,098525018 | 0,866729035 | TMCO1 | FALSE | Downregulated |
| -0,087179898 | 0,866766944 | DEPDC5 | FALSE | Downregulated |
| -0,117227404 | 0,866849005 | TPGS2 | FALSE | Downregulated |
| -0,492756233 | 0,866895246 | BOLA2 | FALSE | Downregulated |
| -0,107608759 | 0,866895246 | OSGIN1 | FALSE | Downregulated |
| 0,243447434 | 0,866895246 | SOGA3 | FALSE | Upregulated |
| 0,078277844 | 0,866895246 | ZFYVE26 | FALSE | Upregulated |
| 0,088151879 | 0,867002691 | DYNLL2 | FALSE | Upregulated |
| 0,073612233 | 0,867069 | AP4S1 | FALSE | Upregulated |
| -0,156738381 | 0,867069 | BOK-AS1 | FALSE | Downregulated |
| -0,128301994 | 0,867069 | DNTTIP2 | FALSE | Downregulated |
| -0,083307369 | 0,867069 | ERLIN1 | FALSE | Downregulated |
| -0,194212709 | 0,867069 | IFIT5 | FALSE | Downregulated |
| 0,115889584 | 0,867069 | PLEKHM1P | FALSE | Upregulated |
| -0,084121498 | 0,8671448 | BTG1 | FALSE | Downregulated |
| 0,096193159 | 0,8671448 | GPR64 | FALSE | Upregulated |
| -0,116352157 | 0,8671448 | HOOK2 | FALSE | Downregulated |
| 0,13798647 | 0,867474863 | C19orf68 | FALSE | Upregulated |
| -0,125581828 | 0,86756863 | ALDH3B1 | FALSE | Downregulated |
| 0,11337302 | 0,86756863 | ASAP2 | FALSE | Upregulated |
| 0,091874874 | 0,867697628 | ZNF274 | FALSE | Upregulated |
| -0,071994423 | 0,867741725 | ADH5 | FALSE | Downregulated |
| 0,158493227 | 0,867741725 | PRR18 | FALSE | Upregulated |
| 0,142459212 | 0,867811478 | ALDH1A3 | FALSE | Upregulated |
| -0,079989613 | 0,867811478 | FOXJ3 | FALSE | Downregulated |
| -0,20313814 | 0,867811478 | KCTD5 | FALSE | Downregulated |
| 0,164025539 | 0,867811478 | MRGPRF | FALSE | Upregulated |
| 0,084208015 | 0,867811478 | PAF1 | FALSE | Upregulated |
| 0,117914408 | 0,867899727 | LARP7 | FALSE | Upregulated |
| 0,176464699 | 0,868037695 | EMC2 | FALSE | Upregulated |
| 0,195700817 | 0,868037695 | GMFB | FALSE | Upregulated |
| -0,066145226 | 0,868037695 | IL1R1 | FALSE | Downregulated |
| 0,16322594 | 0,868037695 | KCNIP2 | FALSE | Upregulated |
| -0,117109175 | 0,868037695 | LNX2 | FALSE | Downregulated |
| -0,151990568 | 0,868037695 | LOC101929265 | FALSE | Downregulated |
| -0,097209025 | 0,868037695 | LOC285074 | FALSE | Downregulated |
| 0,106447491 | 0,868037695 | RGPD3 | FALSE | Upregulated |
| 0,196834834 | 0,868037695 | RNF144A | FALSE | Upregulated |
| 0,068668208 | 0,868037695 | UFD1L | FALSE | Upregulated |
| 0,245155975 | 0,868039656 | CCL3L3 | FALSE | Upregulated |

| | | | | |
|---|---|---|---|---|
| -0,204371138 | 0,868039656 | HID1 | FALSE | Downregulated |
| 0,169966521 | 0,868137789 | ARHGAP25 | FALSE | Upregulated |
| 0,084252923 | 0,868137789 | ATP6V0E2 | FALSE | Upregulated |
| -0,077521784 | 0,868137789 | CLN5 | FALSE | Downregulated |
| -0,054062719 | 0,868137789 | FLNB | FALSE | Downregulated |
| 0,097398958 | 0,868137789 | LOC101927339 | FALSE | Upregulated |
| 0,088797235 | 0,868137789 | MCMBP | FALSE | Upregulated |
| 0,101058302 | 0,868137789 | MRFAP1L1 | FALSE | Upregulated |
| -0,128270851 | 0,868137789 | SLC4A1AP | FALSE | Downregulated |
| -0,071845504 | 0,868193271 | PRR14L | FALSE | Downregulated |
| 0,121322825 | 0,868193271 | SPSB2 | FALSE | Upregulated |
| 0,109511795 | 0,868394954 | CREB3L3 | FALSE | Upregulated |
| -0,132489032 | 0,868394954 | LIMS1 | FALSE | Downregulated |
| -0,082012226 | 0,868814409 | AIFM1 | FALSE | Downregulated |
| -0,144979887 | 0,868891397 | ATXN7L1 | FALSE | Downregulated |
| 0,185046752 | 0,868891397 | H1FX-AS1 | FALSE | Upregulated |
| -0,132493288 | 0,868891397 | ICK | FALSE | Downregulated |
| 0,172992139 | 0,868891397 | TMEM186 | FALSE | Upregulated |
| -0,143870506 | 0,869025065 | CRADD | FALSE | Downregulated |
| 0,213259359 | 0,869064669 | ASF1B | FALSE | Upregulated |
| -0,454734561 | 0,869064669 | LOC541473 | FALSE | Downregulated |
| -0,090526258 | 0,869317456 | ADAM10 | FALSE | Downregulated |
| -0,104585565 | 0,869317456 | SLC25A43 | FALSE | Downregulated |
| 0,08317454 | 0,869372315 | CNOT7 | FALSE | Upregulated |
| -0,14892901 | 0,869372315 | TMCO3 | FALSE | Downregulated |
| -0,12889057 | 0,869403336 | GLT1D1 | FALSE | Downregulated |
| -0,108596199 | 0,869490927 | KIAA1430 | FALSE | Downregulated |
| -0,174555337 | 0,869511285 | FCGBP | FALSE | Downregulated |
| -0,117617359 | 0,869511285 | SLC35F6 | FALSE | Downregulated |
| -0,085594408 | 0,869522342 | ARCN1 | FALSE | Downregulated |
| -0,087329452 | 0,869658143 | ATP2B1 | FALSE | Downregulated |
| -0,067355743 | 0,869717007 | GTF2I | FALSE | Downregulated |
| 0,266054179 | 0,86985027 | MDK | FALSE | Upregulated |
| 0,176835432 | 0,86985027 | NOP16 | FALSE | Upregulated |
| -0,068561596 | 0,86985027 | ST5 | FALSE | Downregulated |
| -0,085293661 | 0,869964829 | SLC38A4 | FALSE | Downregulated |
| 0,228516367 | 0,870573476 | PILRB | FALSE | Upregulated |
| -0,108343669 | 0,870573476 | RNF20 | FALSE | Downregulated |
| 0,053902714 | 0,870573476 | SEMA6A | FALSE | Upregulated |
| 0,126699103 | 0,870573476 | ZBED6CL | FALSE | Upregulated |
| 0,163952668 | 0,870605443 | ST3GAL4-AS1 | FALSE | Upregulated |
| 0,102257254 | 0,870605443 | THSD7A | FALSE | Upregulated |
| -0,088582851 | 0,870661386 | DDX56 | FALSE | Downregulated |
| -0,168139062 | 0,870661386 | LINC01126 | FALSE | Downregulated |
| -0,090647269 | 0,870756169 | CD63 | FALSE | Downregulated |
| 0,138200066 | 0,870756169 | DENND6A | FALSE | Upregulated |
| 0,130296282 | 0,870756169 | DPYD-AS2 | FALSE | Upregulated |
| -0,128029037 | 0,870756169 | KIAA1958 | FALSE | Downregulated |
| 0,173667805 | 0,870756169 | LGALSL | FALSE | Upregulated |
| -0,185126598 | 0,870756169 | PRPF18 | FALSE | Downregulated |
| 0,179796028 | 0,870756169 | SHQ1 | FALSE | Upregulated |

| | | | | |
|---|---|---|---|---|
| -0,070808463 | 0,870756169 | SLC7A2 | FALSE | Downregulated |
| -0,201892234 | 0,870756169 | SPAG8 | FALSE | Downregulated |
| 0,094149292 | 0,870756169 | SPATA20 | FALSE | Upregulated |
| 0,054965181 | 0,870756169 | TCEB3 | FALSE | Upregulated |
| 0,092667302 | 0,870756169 | THRB-AS1 | FALSE | Upregulated |
| -0,086284948 | 0,870756169 | UBE2K | FALSE | Downregulated |
| -0,089013806 | 0,870978458 | SEC24B | FALSE | Downregulated |
| -0,15890496 | 0,871243794 | MRPS31 | FALSE | Downregulated |
| -0,11155276 | 0,871375526 | DHX29 | FALSE | Downregulated |
| 0,187030586 | 0,871375526 | ZNF32-AS2 | FALSE | Upregulated |
| 0,053849431 | 0,871521119 | KIAA0100 | FALSE | Upregulated |
| -0,113093056 | 0,871684876 | SH2D4A | FALSE | Downregulated |
| 0,098163736 | 0,87174439 | KIFC3 | FALSE | Upregulated |
| -0,10452389 | 0,871846442 | CYP51A1 | FALSE | Downregulated |
| -0,114928244 | 0,871846442 | STX1B | FALSE | Downregulated |
| -0,131415297 | 0,872048691 | ANKRD42 | FALSE | Downregulated |
| -0,121095823 | 0,872048691 | GADD45A | FALSE | Downregulated |
| 0,100925044 | 0,872048691 | LACTB | FALSE | Upregulated |
| 0,186676786 | 0,872048691 | MRPL17 | FALSE | Upregulated |
| -0,161385201 | 0,872048691 | RPAP3 | FALSE | Downregulated |
| -0,136336401 | 0,872059867 | AZGP1P1 | FALSE | Downregulated |
| 0,092463242 | 0,872059867 | CLTB | FALSE | Upregulated |
| -0,06986226 | 0,872059867 | EIF3H | FALSE | Downregulated |
| 0,156026529 | 0,872059867 | FRAS1 | FALSE | Upregulated |
| 0,116357459 | 0,872059867 | LOC101927437 | FALSE | Upregulated |
| -0,122722367 | 0,872059867 | PIK3C3 | FALSE | Downregulated |
| -0,089724141 | 0,872059867 | SOS2 | FALSE | Downregulated |
| -0,3072376 | 0,872059867 | SPRNP1 | FALSE | Downregulated |
| -0,183073289 | 0,872181402 | RAB42 | FALSE | Downregulated |
| 0,130386089 | 0,872181402 | TSFM | FALSE | Upregulated |
| -0,12116706 | 0,872181402 | TTI1 | FALSE | Downregulated |
| 0,162983835 | 0,872181402 | ZNF440 | FALSE | Upregulated |
| -0,19804905 | 0,872193976 | C16orf80 | FALSE | Downregulated |
| -0,159264379 | 0,872193976 | EMC8 | FALSE | Downregulated |
| 0,162805616 | 0,872193976 | TUBG1 | FALSE | Upregulated |
| -0,122159963 | 0,872220703 | KCTD18 | FALSE | Downregulated |
| -0,043385278 | 0,872259114 | MPRIP | FALSE | Downregulated |
| 0,180393904 | 0,872268901 | SREK1IP1 | FALSE | Upregulated |
| -0,181972319 | 0,872704498 | C1orf213 | FALSE | Downregulated |
| -0,075821202 | 0,872770924 | BTF3 | FALSE | Downregulated |
| -0,098696792 | 0,8728685 | BACH1 | FALSE | Downregulated |
| -0,126028971 | 0,8728685 | CABYR | FALSE | Downregulated |
| 0,128994943 | 0,8728685 | MACROD2-AS1 | FALSE | Upregulated |
| 0,079983709 | 0,873069141 | ATXN2 | FALSE | Upregulated |
| 0,165197789 | 0,873262098 | SAP25 | FALSE | Upregulated |
| 0,237224824 | 0,873665688 | AGRP | FALSE | Upregulated |
| -0,16925169 | 0,873665688 | BBOX1 | FALSE | Downregulated |
| -0,089058443 | 0,873665688 | GPRASP1 | FALSE | Downregulated |
| 0,129442606 | 0,873982784 | DTX2P1-UPK3BP1-PMS2P11 | FALSE | Upregulated |
| -0,120368292 | 0,873982784 | ESD | FALSE | Downregulated |

| | | | | |
|---|---|---|---|---|
| -0,080391712 | 0,873982784 | SPTY2D1 | FALSE | Downregulated |
| 0,054018 | 0,874145022 | CHD2 | FALSE | Upregulated |
| -0,086201586 | 0,874304882 | PARK7 | FALSE | Downregulated |
| -0,059658552 | 0,874440511 | MBD3 | FALSE | Downregulated |
| -0,164763069 | 0,874440511 | VPS37C | FALSE | Downregulated |
| -0,108904623 | 0,874466323 | FGFR1 | FALSE | Downregulated |
| -0,106224723 | 0,874527242 | DUS3L | FALSE | Downregulated |
| -0,517081894 | 0,874597983 | INO80B | FALSE | Downregulated |
| 0,112931476 | 0,874597983 | STAU2 | FALSE | Upregulated |
| 0,076886077 | 0,874610097 | ARFGAP1 | FALSE | Upregulated |
| 0,111183972 | 0,874631242 | IFIH1 | FALSE | Upregulated |
| 0,080201659 | 0,874631242 | KBTBD4 | FALSE | Upregulated |
| -0,201889043 | 0,874631242 | LCT | FALSE | Downregulated |
| 0,083421899 | 0,874631242 | PSD3 | FALSE | Upregulated |
| 0,123806487 | 0,874631242 | RUNDC3B | FALSE | Upregulated |
| 0,083488268 | 0,874631242 | UAP1 | FALSE | Upregulated |
| 0,107208205 | 0,874682888 | RAVER1 | FALSE | Upregulated |
| 0,133387286 | 0,874755955 | CELSR2 | FALSE | Upregulated |
| -0,039665547 | 0,875386576 | HNRNPU | FALSE | Downregulated |
| 0,110100693 | 0,875386576 | MRPL16 | FALSE | Upregulated |
| -0,072469544 | 0,875386576 | MYRF | FALSE | Downregulated |
| -0,575462964 | 0,875556834 | LOC102724159 | FALSE | Downregulated |
| -0,131443193 | 0,875660955 | COX7A2L | FALSE | Downregulated |
| -0,069992566 | 0,875660955 | PCCA | FALSE | Downregulated |
| 0,111920492 | 0,875660955 | SEPT4 | FALSE | Upregulated |
| 0,102474785 | 0,875951665 | IFITM2 | FALSE | Upregulated |
| -0,067819913 | 0,876194846 | CBX5 | FALSE | Downregulated |
| 0,20668791 | 0,876242597 | LOC101927023 | FALSE | Upregulated |
| 0,129594819 | 0,876242597 | LOC101927185 | FALSE | Upregulated |
| 0,039497809 | 0,876242597 | NUMA1 | FALSE | Upregulated |
| 0,087514807 | 0,876242597 | RGAG4 | FALSE | Upregulated |
| 0,058528368 | 0,8763955 | CNP | FALSE | Upregulated |
| -0,096218384 | 0,8763955 | DEK | FALSE | Downregulated |
| 0,100634032 | 0,8763955 | IFNLR1 | FALSE | Upregulated |
| 0,076416091 | 0,8763955 | SLC25A4 | FALSE | Upregulated |
| -0,115006854 | 0,8763955 | TMTC3 | FALSE | Downregulated |
| -0,088724838 | 0,8763955 | ZBTB45 | FALSE | Downregulated |
| 0,289838536 | 0,876404499 | OR10J1 | FALSE | Upregulated |
| 0,197051309 | 0,876506211 | IFI44L | FALSE | Upregulated |
| 0,115657507 | 0,876653576 | AVPI1 | FALSE | Upregulated |
| 0,064575603 | 0,876653576 | B4GALT1 | FALSE | Upregulated |
| -0,10923078 | 0,876653576 | BAG5 | FALSE | Downregulated |
| -0,23672794 | 0,876653576 | FOS | FALSE | Downregulated |
| -0,226874808 | 0,876653576 | HEPN1 | FALSE | Downregulated |
| -0,172994258 | 0,876653576 | KCNE2 | FALSE | Downregulated |
| 0,133625514 | 0,876653576 | LOC100506731 | FALSE | Upregulated |
| -0,292212457 | 0,876653576 | LOC101927861 | FALSE | Downregulated |
| 0,152103315 | 0,876653576 | LOC101929343 | FALSE | Upregulated |
| 0,065507969 | 0,876653576 | LTBR | FALSE | Upregulated |
| 0,108682432 | 0,876653576 | LYPLA1 | FALSE | Upregulated |
| 0,053243587 | 0,876653576 | MTMR10 | FALSE | Upregulated |

| | | | | |
|---|---|---|---|---|
| 0,156439634 | 0,876653576 | MUSTN1 | FALSE | Upregulated |
| -0,05652414 | 0,876653576 | NUFIP2 | FALSE | Downregulated |
| 0,093961633 | 0,876653576 | PELP1 | FALSE | Upregulated |
| 0,106791744 | 0,876653576 | PEX12 | FALSE | Upregulated |
| -0,062995616 | 0,876653576 | POLR2E | FALSE | Downregulated |
| -0,269070807 | 0,876653576 | RP11-381O7.3 | FALSE | Downregulated |
| -0,188744145 | 0,876653576 | SOCS2 | FALSE | Downregulated |
| -0,166421395 | 0,876653576 | TM4SF5 | FALSE | Downregulated |
| 0,148451749 | 0,876693184 | IDE | FALSE | Upregulated |
| 0,121638968 | 0,87670027 | DHODH | FALSE | Upregulated |
| -0,058743124 | 0,87670027 | RALGAPA2 | FALSE | Downregulated |
| -0,086819435 | 0,87670027 | TSHZ2 | FALSE | Downregulated |
| -0,120511593 | 0,876792877 | KDM5B | FALSE | Downregulated |
| 0,184903034 | 0,876792877 | TAF9 | FALSE | Upregulated |
| 0,212381065 | 0,876847311 | WISP3 | FALSE | Upregulated |
| 0,13552252 | 0,876864561 | LOC101928443 | FALSE | Upregulated |
| -0,180082557 | 0,876910196 | TMEM132D | FALSE | Downregulated |
| 0,055512313 | 0,877033918 | PTPN23 | FALSE | Upregulated |
| 0,182471501 | 0,877033918 | RIPK2 | FALSE | Upregulated |
| 0,10454612 | 0,877033918 | SHARPIN | FALSE | Upregulated |
| 0,098219221 | 0,877033918 | VPS41 | FALSE | Upregulated |
| -0,12140288 | 0,877033918 | ZNF264 | FALSE | Downregulated |
| -0,101234423 | 0,877066875 | MTIF2 | FALSE | Downregulated |
| -0,17643721 | 0,877066875 | ZNF143 | FALSE | Downregulated |
| -0,13577217 | 0,877072321 | BRCC3 | FALSE | Downregulated |
| -0,054524302 | 0,877072321 | RBP5 | FALSE | Downregulated |
| 0,182656921 | 0,87720029 | RP11-365O16.6 | FALSE | Upregulated |
| 0,206164253 | 0,877509846 | ARL14EP | FALSE | Upregulated |
| 0,084469681 | 0,877509846 | PRSS48 | FALSE | Upregulated |
| -0,075680555 | 0,87763497 | PDCD6IP | FALSE | Downregulated |
| -0,063698406 | 0,87763497 | SS18L1 | FALSE | Downregulated |
| 0,177848345 | 0,877674061 | FBLIM1 | FALSE | Upregulated |
| 0,12604548 | 0,877674061 | RAP1GDS1 | FALSE | Upregulated |
| 0,052100101 | 0,877674061 | SDF4 | FALSE | Upregulated |
| 0,090315096 | 0,877685206 | ACACA | FALSE | Upregulated |
| -0,149532004 | 0,877685206 | CSTF1 | FALSE | Downregulated |
| -0,097640335 | 0,877685206 | DNAJA1 | FALSE | Downregulated |
| 0,133420849 | 0,877685206 | IRF8 | FALSE | Upregulated |
| 0,101011479 | 0,877685206 | LOC101929747 | FALSE | Upregulated |
| 0,10972309 | 0,877685206 | MMAA | FALSE | Upregulated |
| -0,070496948 | 0,877685206 | RFC1 | FALSE | Downregulated |
| -0,107613859 | 0,877685206 | RWDD2B | FALSE | Downregulated |
| 0,128870421 | 0,877685206 | SMARCAD1 | FALSE | Upregulated |
| -0,050834339 | 0,877685206 | ZHX3 | FALSE | Downregulated |
| 0,095041281 | 0,877744185 | ZFYVE19 | FALSE | Upregulated |
| -0,083517857 | 0,878182495 | D2HGDH | FALSE | Downregulated |
| -0,139422954 | 0,878461516 | DPH2 | FALSE | Downregulated |
| 0,179136216 | 0,878538261 | FBXO24 | FALSE | Upregulated |
| 0,101127623 | 0,878538261 | IMPA1 | FALSE | Upregulated |
| 0,05277737 | 0,878538261 | MAP4 | FALSE | Upregulated |
| 0,074091199 | 0,878538261 | QRICH1 | FALSE | Upregulated |

| | | | | |
|---|---|---|---|---|
| -0,069461766 | 0,878538261 | SSU72 | FALSE | Downregulated |
| 0,120919939 | 0,878562425 | EXOC2 | FALSE | Upregulated |
| -0,05395069 | 0,878562425 | HTT | FALSE | Downregulated |
| -0,071693282 | 0,878608558 | PCDH1 | FALSE | Downregulated |
| -0,144488836 | 0,878632273 | ANO7 | FALSE | Downregulated |
| -0,077949739 | 0,878632273 | CACTIN | FALSE | Downregulated |
| 0,122220333 | 0,878841151 | ZNF512 | FALSE | Upregulated |
| -0,156073975 | 0,878925735 | ERG | FALSE | Downregulated |
| -0,079192918 | 0,878925735 | GSDMD | FALSE | Downregulated |
| -0,100806594 | 0,878925735 | IGFBP5 | FALSE | Downregulated |
| 0,094339216 | 0,878925735 | LOC101928415 | FALSE | Upregulated |
| -0,149201399 | 0,879274016 | ATL1 | FALSE | Downregulated |
| -0,097355585 | 0,879274016 | IK | FALSE | Downregulated |
| -0,20474885 | 0,879348153 | SNHG5 | FALSE | Downregulated |
| 0,07821357 | 0,879621179 | THUMPD3 | FALSE | Upregulated |
| 0,105361117 | 0,879782077 | TSPAN18 | FALSE | Upregulated |
| -0,138318827 | 0,87989083 | ARL13B | FALSE | Downregulated |
| -0,074643312 | 0,87989083 | HEBP2 | FALSE | Downregulated |
| -0,109663917 | 0,87989083 | LEAP2 | FALSE | Downregulated |
| -0,105049162 | 0,87989083 | LOC595101 | FALSE | Downregulated |
| 0,185437615 | 0,87989083 | PEX6 | FALSE | Upregulated |
| 0,153077965 | 0,87989083 | PIP4K2C | FALSE | Upregulated |
| 0,180149212 | 0,87989083 | RP11-1072A3.2 | FALSE | Upregulated |
| -0,124428459 | 0,87989083 | SPRED1 | FALSE | Downregulated |
| 0,091298328 | 0,880014171 | GPR179 | FALSE | Upregulated |
| -0,094506094 | 0,880301321 | HOGA1 | FALSE | Downregulated |
| -0,128566683 | 0,880301321 | LOC389834 | FALSE | Downregulated |
| -0,095833493 | 0,880301321 | PDIA6 | FALSE | Downregulated |
| -0,086941863 | 0,880301321 | TECPR2 | FALSE | Downregulated |
| 0,055444372 | 0,880301321 | TMED2 | FALSE | Upregulated |
| -0,068432614 | 0,880301321 | XRCC6 | FALSE | Downregulated |
| 0,112631081 | 0,88042386 | LOC101929752 | FALSE | Upregulated |
| -0,215360714 | 0,88042386 | MB21D1 | FALSE | Downregulated |
| -0,097587124 | 0,880555577 | IRF2 | FALSE | Downregulated |
| 0,168978399 | 0,880555577 | TMC5 | FALSE | Upregulated |
| 0,170256311 | 0,88061745 | MAP3K8 | FALSE | Upregulated |
| -0,158965658 | 0,88061745 | MRPL42 | FALSE | Downregulated |
| 0,095757616 | 0,880648372 | AC004893.11 | FALSE | Upregulated |
| 0,103416923 | 0,880648372 | DNMT3A | FALSE | Upregulated |
| 0,123602556 | 0,880648372 | ING4 | FALSE | Upregulated |
| -0,128657641 | 0,880925344 | CDR2 | FALSE | Downregulated |
| 0,072276543 | 0,880925344 | EHD3 | FALSE | Upregulated |
| 0,105053685 | 0,881026155 | ACP1 | FALSE | Upregulated |
| -0,132318721 | 0,881026155 | ADAM9 | FALSE | Downregulated |
| -0,145119113 | 0,881026155 | ANP32A-IT1 | FALSE | Downregulated |
| 0,110003244 | 0,881026155 | ATIC | FALSE | Upregulated |
| -0,060981072 | 0,881026155 | CALM3 | FALSE | Downregulated |
| 0,0799958 | 0,881026155 | CCNJ | FALSE | Upregulated |
| -0,207203527 | 0,881026155 | CLCN2 | FALSE | Downregulated |
| 0,115655201 | 0,881026155 | EIF2AK3 | FALSE | Upregulated |
| 0,140844055 | 0,881026155 | ERMARD | FALSE | Upregulated |

| | | | | |
|---|---|---|---|---|
| 0,113387435 | 0,881026155 | FAM178B | FALSE | Upregulated |
| -0,16899304 | 0,881026155 | GALNT18 | FALSE | Downregulated |
| -0,096787864 | 0,881026155 | GOSR2 | FALSE | Downregulated |
| -0,173729965 | 0,881026155 | GPN3 | FALSE | Downregulated |
| -0,133478253 | 0,881026155 | HDDC3 | FALSE | Downregulated |
| 0,19908 | 0,881026155 | KANSL1-AS1 | FALSE | Upregulated |
| 0,148260402 | 0,881026155 | LHFP | FALSE | Upregulated |
| 0,083570664 | 0,881026155 | LHFPL2 | FALSE | Upregulated |
| 0,148179142 | 0,881026155 | LOC100129203 | FALSE | Upregulated |
| -0,838286881 | 0,881026155 | LOC102723943 | FALSE | Downregulated |
| 0,121848012 | 0,881026155 | LOC102724153 | FALSE | Upregulated |
| -0,131497163 | 0,881026155 | LTN1 | FALSE | Downregulated |
| -0,065507811 | 0,881026155 | MGLL | FALSE | Downregulated |
| -0,095718035 | 0,881026155 | MPLKIP | FALSE | Downregulated |
| 0,104335506 | 0,881026155 | NANS | FALSE | Upregulated |
| 0,524821889 | 0,881026155 | NPIPB3 | FALSE | Upregulated |
| 0,177118766 | 0,881026155 | PJA1 | FALSE | Upregulated |
| 0,190207018 | 0,881026155 | RASSF8 | FALSE | Upregulated |
| -0,167876109 | 0,881026155 | RBM43 | FALSE | Downregulated |
| -0,104618124 | 0,881026155 | RNF114 | FALSE | Downregulated |
| -0,060717834 | 0,881026155 | RPL26 | FALSE | Downregulated |
| -0,150646239 | 0,881026155 | SLC15A4 | FALSE | Downregulated |
| 0,119057478 | 0,881026155 | SMS | FALSE | Upregulated |
| -0,185335041 | 0,881026155 | SNX24 | FALSE | Downregulated |
| 0,090326323 | 0,881026155 | TBC1D13 | FALSE | Upregulated |
| 0,113845547 | 0,881026155 | TIMMDC1 | FALSE | Upregulated |
| 0,171913398 | 0,881026155 | TRUB1 | FALSE | Upregulated |
| 0,142692118 | 0,881026155 | TSPO2 | FALSE | Upregulated |
| -0,172160841 | 0,881026155 | TTLL7 | FALSE | Downregulated |
| 0,132026874 | 0,881026155 | WNT2B | FALSE | Upregulated |
| -0,10078985 | 0,881026155 | ZCCHC3 | FALSE | Downregulated |
| 0,080118501 | 0,881026155 | ZNF217 | FALSE | Upregulated |
| -0,099050594 | 0,881113376 | TUBB2A | FALSE | Downregulated |
| 0,10694593 | 0,881114409 | LOC101927854 | FALSE | Upregulated |
| -0,151769883 | 0,881381368 | ASB7 | FALSE | Downregulated |
| 0,093430578 | 0,881381368 | BTBD1 | FALSE | Upregulated |
| -0,051109933 | 0,881381368 | GNB1 | FALSE | Downregulated |
| -0,045517968 | 0,881381368 | KIF13B | FALSE | Downregulated |
| -0,18041989 | 0,881381368 | LOC101928439 | FALSE | Downregulated |
| -0,153050177 | 0,881381368 | LOC101929384 | FALSE | Downregulated |
| 0,250501581 | 0,881381368 | LOC102724720 | FALSE | Upregulated |
| -0,115658668 | 0,881381368 | MIR22HG | FALSE | Downregulated |
| -0,145831841 | 0,881381368 | SLC35E3 | FALSE | Downregulated |
| 0,151294365 | 0,881381368 | ZNF589 | FALSE | Upregulated |
| -0,182636431 | 0,88189148 | HOXA-AS2 | FALSE | Downregulated |
| -0,0676877 | 0,88189148 | TMED7 | FALSE | Downregulated |
| -0,08001703 | 0,882012185 | EFR3A | FALSE | Downregulated |
| 0,075827556 | 0,882185091 | DOCK5 | FALSE | Upregulated |
| 0,215849579 | 0,882266726 | LOC102724005 | FALSE | Upregulated |
| 0,083728958 | 0,882266726 | MAST4 | FALSE | Upregulated |
| -0,054186484 | 0,882266726 | RRBP1 | FALSE | Downregulated |

| | | | | |
|---|---|---|---|---|
| -0,098565241 | 0,882266726 | ZNF302 | FALSE | Downregulated |
| 0,145051682 | 0,882376487 | C18orf8 | FALSE | Upregulated |
| 0,08082086 | 0,882376487 | CFL2 | FALSE | Upregulated |
| 0,161321581 | 0,882376487 | DIRC2 | FALSE | Upregulated |
| -0,073245607 | 0,882376487 | FLJ10038 | FALSE | Downregulated |
| 0,150885694 | 0,882376487 | PGM2 | FALSE | Upregulated |
| -0,218671655 | 0,882376487 | RASD1 | FALSE | Downregulated |
| -0,104242754 | 0,882376487 | THAP11 | FALSE | Downregulated |
| 0,10663908 | 0,882495991 | AC011343.1 | FALSE | Upregulated |
| -0,093814511 | 0,882518742 | ADRA1A | FALSE | Downregulated |
| -0,118321082 | 0,882518742 | KIAA0391 | FALSE | Downregulated |
| 0,166587786 | 0,882518742 | ZNF296 | FALSE | Upregulated |
| 0,093180723 | 0,882652438 | RMND1 | FALSE | Upregulated |
| -0,087234977 | 0,88276476 | ARF5 | FALSE | Downregulated |
| 0,131125972 | 0,88276476 | DENR | FALSE | Upregulated |
| -0,04548272 | 0,88276476 | KMT2C | FALSE | Downregulated |
| -0,107955056 | 0,88276476 | LDLRAP1 | FALSE | Downregulated |
| 0,202931775 | 0,88276476 | ZFP30 | FALSE | Upregulated |
| 0,161794864 | 0,883062419 | EFNA2 | FALSE | Upregulated |
| -0,124043192 | 0,883068603 | DIS3L2 | FALSE | Downregulated |
| -0,079764822 | 0,883068603 | ELF1 | FALSE | Downregulated |
| 0,102917095 | 0,883068603 | RSBN1L | FALSE | Upregulated |
| 0,165425314 | 0,883068603 | TMCC3 | FALSE | Upregulated |
| 0,122699213 | 0,883068603 | ZBTB42 | FALSE | Upregulated |
| -0,093894176 | 0,883208457 | BNIP3L | FALSE | Downregulated |
| 0,131040906 | 0,883335022 | KIFAP3 | FALSE | Upregulated |
| 0,171427206 | 0,883335022 | LOC339803 | FALSE | Upregulated |
| -0,116046261 | 0,883335022 | METTL14 | FALSE | Downregulated |
| 0,170561398 | 0,883335022 | MLIP | FALSE | Upregulated |
| 0,064293414 | 0,883335022 | NFATC3 | FALSE | Upregulated |
| -0,178882684 | 0,883335022 | TMEM189 | FALSE | Downregulated |
| 0,074467202 | 0,883451611 | CAB39 | FALSE | Upregulated |
| -0,070916566 | 0,883451611 | LRP11 | FALSE | Downregulated |
| -0,162757891 | 0,883451611 | SETD4 | FALSE | Downregulated |
| 0,161534493 | 0,88350779 | PCDH18 | FALSE | Upregulated |
| -0,533633894 | 0,883617978 | GSTM1 | FALSE | Downregulated |
| 0,137567656 | 0,883708437 | GHRLOS | FALSE | Upregulated |
| 0,178992153 | 0,883877166 | CARD6 | FALSE | Upregulated |
| -0,100656927 | 0,883877166 | CSNK1G3 | FALSE | Downregulated |
| -0,158667493 | 0,883877166 | GEMIN8 | FALSE | Downregulated |
| -0,172896066 | 0,883877166 | LOC100129550 | FALSE | Downregulated |
| -0,140548453 | 0,883877166 | TMEM8B | FALSE | Downregulated |
| -0,07312112 | 0,883877166 | UBE4B | FALSE | Downregulated |
| -0,095114915 | 0,883877166 | VAMP2 | FALSE | Downregulated |
| -0,154785256 | 0,883877166 | ZNF226 | FALSE | Downregulated |
| 0,159459356 | 0,883877166 | ZNF566 | FALSE | Upregulated |
| 0,142248323 | 0,88408016 | GP6 | FALSE | Upregulated |
| 0,114424628 | 0,88408016 | NCK2 | FALSE | Upregulated |
| 0,054674521 | 0,88408016 | USP7 | FALSE | Upregulated |
| 0,115823824 | 0,88408016 | ZNF271 | FALSE | Upregulated |
| -0,159148945 | 0,884150832 | SPATA2 | FALSE | Downregulated |

| | | | | |
|---|---|---|---|---|
| 0,054098256 | 0,884150832 | TMEM63A | FALSE | Upregulated |
| -0,051409347 | 0,884247499 | ADIPOR1 | FALSE | Downregulated |
| 0,097589033 | 0,88440621 | PLCE1 | FALSE | Upregulated |
| 0,0935 | 0,884455158 | RP11-290F20.2 | FALSE | Upregulated |
| 0,065081573 | 0,884455158 | ZSWIM8-AS1 | FALSE | Upregulated |
| 0,070112765 | 0,884601712 | ZNF318 | FALSE | Upregulated |
| -0,147806995 | 0,884687274 | ARMCX5 | FALSE | Downregulated |
| 0,092366335 | 0,884687274 | LOC102723990 | FALSE | Upregulated |
| -0,10582093 | 0,884687936 | ARMCX4 | FALSE | Downregulated |
| -0,084843871 | 0,884687936 | C16orf45 | FALSE | Downregulated |
| -0,107488968 | 0,884687936 | EFCAB7 | FALSE | Downregulated |
| -0,104268416 | 0,884687936 | IGF1 | FALSE | Downregulated |
| 0,164776552 | 0,884687936 | PATE4 | FALSE | Upregulated |
| -0,177157274 | 0,884687936 | RRP7B | FALSE | Downregulated |
| 0,376012398 | 0,884687936 | SENP3-EIF4A1 | FALSE | Upregulated |
| -0,145677436 | 0,885247764 | PREX2 | FALSE | Downregulated |
| 0,112053012 | 0,885426297 | ATAD2 | FALSE | Upregulated |
| -0,145466684 | 0,885426297 | KIF9 | FALSE | Downregulated |
| 0,073337822 | 0,885476142 | ACVR2A | FALSE | Upregulated |
| -0,0898544 | 0,885476142 | KRIT1 | FALSE | Downregulated |
| -0,046827283 | 0,885476142 | PLXNB1 | FALSE | Downregulated |
| -0,133749655 | 0,885476142 | SAMD1 | FALSE | Downregulated |
| -0,134003917 | 0,885529667 | TMEM14B | FALSE | Downregulated |
| -0,061079515 | 0,885668329 | ZFAND3 | FALSE | Downregulated |
| 0,101797125 | 0,885690955 | PTCH1 | FALSE | Upregulated |
| 0,106703634 | 0,885690955 | SH3BP4 | FALSE | Upregulated |
| -0,080920705 | 0,885690955 | SRSF8 | FALSE | Downregulated |
| 0,072271727 | 0,885703101 | SBNO1 | FALSE | Upregulated |
| -0,702531053 | 0,886466824 | FLG-AS1 | FALSE | Downregulated |
| 0,082895124 | 0,886466824 | GCLM | FALSE | Upregulated |
| 0,09353608 | 0,886522648 | KCTD12 | FALSE | Upregulated |
| -0,200879605 | 0,886522648 | PDE1A | FALSE | Downregulated |
| 0,072359264 | 0,886582701 | RBM15 | FALSE | Upregulated |
| -0,074303414 | 0,886649399 | UBN2 | FALSE | Downregulated |
| -0,098724535 | 0,886699191 | COL6A3 | FALSE | Downregulated |
| 0,091147452 | 0,886699191 | CSDC2 | FALSE | Upregulated |
| -0,098706762 | 0,886699191 | SENP2 | FALSE | Downregulated |
| -0,096962935 | 0,886807861 | PSMB5 | FALSE | Downregulated |
| -0,242272833 | 0,886835547 | LOC101930375 | FALSE | Downregulated |
| 0,065271223 | 0,88685464 | ERAP1 | FALSE | Upregulated |
| 0,073757217 | 0,88685464 | MADD | FALSE | Upregulated |
| -0,17377099 | 0,88685464 | SCNN1A | FALSE | Downregulated |
| 0,104247315 | 0,88685464 | ZNF503-AS2 | FALSE | Upregulated |
| -0,196140727 | 0,886982395 | JAG2 | FALSE | Downregulated |
| -0,066509876 | 0,886982395 | MUM1 | FALSE | Downregulated |
| -0,09992134 | 0,886982395 | NCSTN | FALSE | Downregulated |
| 0,090485073 | 0,88749964 | SIAH1 | FALSE | Upregulated |
| 0,078477476 | 0,88749964 | SLC17A4 | FALSE | Upregulated |
| 0,097353191 | 0,887654388 | AGAP6 | FALSE | Upregulated |
| 0,18077298 | 0,887654388 | CDR1 | FALSE | Upregulated |
| -0,130973295 | 0,887654388 | MANEA | FALSE | Downregulated |

| | | | | |
|---|---|---|---|---|
| -0,069602975 | 0,887654388 | NAGA | FALSE | Downregulated |
| -0,074178358 | 0,887654388 | PSMC5 | FALSE | Downregulated |
| 0,066779045 | 0,887654388 | RHOC | FALSE | Upregulated |
| 0,085301603 | 0,887654388 | TAF1C | FALSE | Upregulated |
| 0,105867531 | 0,887654388 | ZMYM6 | FALSE | Upregulated |
| 0,078705874 | 0,887679286 | A1BG-AS1 | FALSE | Upregulated |
| 0,059766888 | 0,887679286 | ABHD17A | FALSE | Upregulated |
| 0,163689932 | 0,887679286 | ADAT2 | FALSE | Upregulated |
| 0,483725487 | 0,887679286 | AKR1B10 | FALSE | Upregulated |
| 0,106618151 | 0,887679286 | ANKRD54 | FALSE | Upregulated |
| -0,172780007 | 0,887679286 | AQP7 | FALSE | Downregulated |
| -0,071974489 | 0,887679286 | C4orf19 | FALSE | Downregulated |
| -0,081147103 | 0,887679286 | CPPED1 | FALSE | Downregulated |
| -0,107348685 | 0,887679286 | EIF4EBP1 | FALSE | Downregulated |
| 0,095287784 | 0,887679286 | ENDOU | FALSE | Upregulated |
| -0,10106503 | 0,887679286 | FAM126B | FALSE | Downregulated |
| 0,24890508 | 0,887679286 | GEM | FALSE | Upregulated |
| 0,076298028 | 0,887679286 | GMPR2 | FALSE | Upregulated |
| -0,111001165 | 0,887679286 | MOAP1 | FALSE | Downregulated |
| -0,104750621 | 0,887679286 | MRPL14 | FALSE | Downregulated |
| 0,069951905 | 0,887679286 | PIK3C2A | FALSE | Upregulated |
| 0,069733586 | 0,887679286 | PLA2G12B | FALSE | Upregulated |
| -0,18107881 | 0,887679286 | PLEKHH2 | FALSE | Downregulated |
| -0,101995662 | 0,887679286 | PPP1R15A | FALSE | Downregulated |
| 0,157480378 | 0,887679286 | RP5-1013A22.2 | FALSE | Upregulated |
| 0,107579002 | 0,887679286 | SLC22A4 | FALSE | Upregulated |
| -0,129224275 | 0,887679286 | SLC25A34 | FALSE | Downregulated |
| -0,147095154 | 0,887679286 | TMEM161A | FALSE | Downregulated |
| -0,128199137 | 0,887679286 | TRIM3 | FALSE | Downregulated |
| -0,104928268 | 0,887679286 | TROVE2 | FALSE | Downregulated |
| -0,06279054 | 0,887679286 | UBE4A | FALSE | Downregulated |
| -0,153418272 | 0,887679286 | WEE2 | FALSE | Downregulated |
| 0,138538018 | 0,887679286 | ZBTB25 | FALSE | Upregulated |
| 0,181886512 | 0,887703895 | C1orf216 | FALSE | Upregulated |
| 0,154479673 | 0,887728363 | SUCLA2 | FALSE | Upregulated |
| -0,145419097 | 0,887755186 | ZNF304 | FALSE | Downregulated |
| -0,054998257 | 0,887918804 | MON2 | FALSE | Downregulated |
| 0,101282437 | 0,887918804 | RBFA | FALSE | Upregulated |
| -0,080842028 | 0,888128703 | MYO19 | FALSE | Downregulated |
| -0,134581092 | 0,888195898 | AKAP3 | FALSE | Downregulated |
| 0,069496776 | 0,888195898 | FOXO3 | FALSE | Upregulated |
| 0,205386223 | 0,888195898 | INPP4B | FALSE | Upregulated |
| 0,074822636 | 0,888195898 | INPP5E | FALSE | Upregulated |
| 0,071980632 | 0,888195898 | IRGQ | FALSE | Upregulated |
| -0,541666133 | 0,888195898 | LOC102723584 | FALSE | Downregulated |
| -0,059823817 | 0,888195898 | MICAL3 | FALSE | Downregulated |
| 0,062768032 | 0,888195898 | RPE | FALSE | Upregulated |
| 0,091242823 | 0,888195898 | SDCCAG3 | FALSE | Upregulated |
| -0,122174534 | 0,888359869 | C6orf136 | FALSE | Downregulated |
| 0,134130109 | 0,888359869 | C9orf41 | FALSE | Upregulated |
| -0,184159095 | 0,888359869 | KLHL18 | FALSE | Downregulated |

| | | | | |
|---|---|---|---|---|
| 0,185638181 | 0,888359869 | LOC100996763 | FALSE | Upregulated |
| -0,139719046 | 0,888359869 | RCC1 | FALSE | Downregulated |
| 0,175756706 | 0,888492474 | FAM213B | FALSE | Upregulated |
| -0,077123116 | 0,888492474 | IPO8 | FALSE | Downregulated |
| -0,19030392 | 0,888492474 | LOC102724833 | FALSE | Downregulated |
| -0,113594775 | 0,888492474 | PPP6C | FALSE | Downregulated |
| 0,124360601 | 0,888492474 | TRIM5 | FALSE | Upregulated |
| 0,149577427 | 0,888492474 | ULK4 | FALSE | Upregulated |
| 0,1772759 | 0,889065494 | MED21 | FALSE | Upregulated |
| 0,123288455 | 0,889305407 | GPR75 | FALSE | Upregulated |
| -0,143090507 | 0,889305407 | RAMP3 | FALSE | Downregulated |
| 0,146622733 | 0,88941717 | JOSD2 | FALSE | Upregulated |
| -0,116930334 | 0,889425643 | CNTROB | FALSE | Downregulated |
| 0,091567964 | 0,889425643 | DHPS | FALSE | Upregulated |
| -0,123629247 | 0,889425643 | UBALD1 | FALSE | Downregulated |
| -0,071242705 | 0,889426887 | ABCC6 | FALSE | Downregulated |
| 0,058042823 | 0,889426887 | AGFG2 | FALSE | Upregulated |
| -0,04025605 | 0,889426887 | ARID1B | FALSE | Downregulated |
| 0,23516869 | 0,889426887 | ARNTL2 | FALSE | Upregulated |
| -0,087887885 | 0,889426887 | BDH2 | FALSE | Downregulated |
| 0,471849051 | 0,889426887 | C18orf32 | FALSE | Upregulated |
| -0,146713173 | 0,889426887 | C1orf172 | FALSE | Downregulated |
| 0,094181339 | 0,889426887 | C22orf39 | FALSE | Upregulated |
| -0,079164914 | 0,889426887 | C3P1 | FALSE | Downregulated |
| -0,096463847 | 0,889426887 | CAPZA1 | FALSE | Downregulated |
| 0,076207524 | 0,889426887 | CCDC176 | FALSE | Upregulated |
| 0,12043434 | 0,889426887 | CCDC94 | FALSE | Upregulated |
| 0,129475038 | 0,889426887 | CLDN23 | FALSE | Upregulated |
| 0,124896945 | 0,889426887 | CLEC1B | FALSE | Upregulated |
| 0,074826617 | 0,889426887 | COG4 | FALSE | Upregulated |
| 0,069876212 | 0,889426887 | CPSF3L | FALSE | Upregulated |
| 0,068702138 | 0,889426887 | CTDSPL2 | FALSE | Upregulated |
| 0,162663716 | 0,889426887 | DDX39A | FALSE | Upregulated |
| 0,073749236 | 0,889426887 | DENND1C | FALSE | Upregulated |
| -0,085691955 | 0,889426887 | ENTPD4 | FALSE | Downregulated |
| -0,061251026 | 0,889426887 | F11R | FALSE | Downregulated |
| -0,181303137 | 0,889426887 | FKBP11 | FALSE | Downregulated |
| -0,077365845 | 0,889426887 | FOXRED2 | FALSE | Downregulated |
| -0,075994702 | 0,889426887 | GALT | FALSE | Downregulated |
| 0,043406182 | 0,889426887 | GGCX | FALSE | Upregulated |
| 0,087745654 | 0,889426887 | GLE1 | FALSE | Upregulated |
| -0,092534613 | 0,889426887 | HDHD2 | FALSE | Downregulated |
| 0,07396697 | 0,889426887 | HIVEP1 | FALSE | Upregulated |
| -0,060919922 | 0,889426887 | HMGB1 | FALSE | Downregulated |
| 0,129653929 | 0,889426887 | IER5 | FALSE | Upregulated |
| 0,090790532 | 0,889426887 | ING1 | FALSE | Upregulated |
| -0,096543276 | 0,889426887 | JADE3 | FALSE | Downregulated |
| -0,048965031 | 0,889426887 | KDM2A | FALSE | Downregulated |
| 0,085996095 | 0,889426887 | LAMA5 | FALSE | Upregulated |
| 0,160396413 | 0,889426887 | LINC00883 | FALSE | Upregulated |
| 0,149207692 | 0,889426887 | LMBR1L | FALSE | Upregulated |

| | | | | |
|---|---|---|---|---|
| 0,071193711 | 0,889426887 | LOC100506710 | FALSE | Upregulated |
| 0,163256842 | 0,889426887 | LOC101928363 | FALSE | Upregulated |
| 0,16639657 | 0,889426887 | LOC101929051 | FALSE | Upregulated |
| -0,556449688 | 0,889426887 | LOC102724560 | FALSE | Downregulated |
| -0,11686382 | 0,889426887 | LOXL2 | FALSE | Downregulated |
| -0,131776704 | 0,889426887 | LRRC57 | FALSE | Downregulated |
| 0,068623558 | 0,889426887 | MAP3K3 | FALSE | Upregulated |
| 0,07687267 | 0,889426887 | MPC1 | FALSE | Upregulated |
| 0,128220834 | 0,889426887 | NAT8B | FALSE | Upregulated |
| -0,148474004 | 0,889426887 | NDUFA12 | FALSE | Downregulated |
| -0,179111584 | 0,889426887 | NOP10 | FALSE | Downregulated |
| -0,058443898 | 0,889426887 | NT5E | FALSE | Downregulated |
| -0,182765774 | 0,889426887 | OR13F1 | FALSE | Downregulated |
| 0,061124791 | 0,889426887 | PEX10 | FALSE | Upregulated |
| -0,122464099 | 0,889426887 | PPP1R8 | FALSE | Downregulated |
| -0,074174391 | 0,889426887 | RAB10 | FALSE | Downregulated |
| -0,089415102 | 0,889426887 | RALGPS2 | FALSE | Downregulated |
| 0,051955911 | 0,889426887 | RBMX | FALSE | Upregulated |
| -0,073246696 | 0,889426887 | RNH1 | FALSE | Downregulated |
| -0,644373636 | 0,889426887 | RNU1-2 | FALSE | Downregulated |
| 0,153619588 | 0,889426887 | RP11-536C5.7 | FALSE | Upregulated |
| 0,177799045 | 0,889426887 | RWDD4 | FALSE | Upregulated |
| 0,077915417 | 0,889426887 | SCAI | FALSE | Upregulated |
| 0,160694321 | 0,889426887 | SLC16A3 | FALSE | Upregulated |
| -0,082973056 | 0,889426887 | SLC25A5-AS1 | FALSE | Downregulated |
| -0,155714893 | 0,889426887 | SLC37A3 | FALSE | Downregulated |
| 0,181955016 | 0,889426887 | SLC4A7 | FALSE | Upregulated |
| 0,165728421 | 0,889426887 | SLFN11 | FALSE | Upregulated |
| 0,250243659 | 0,889426887 | SLMO2-ATP5E | FALSE | Upregulated |
| -0,134252402 | 0,889426887 | SMAGP | FALSE | Downregulated |
| -0,111897852 | 0,889426887 | SOCS7 | FALSE | Downregulated |
| 0,085465138 | 0,889426887 | SOX12 | FALSE | Upregulated |
| 0,146454263 | 0,889426887 | SPARCL1 | FALSE | Upregulated |
| -0,055210641 | 0,889426887 | SRRT | FALSE | Downregulated |
| -0,102299805 | 0,889426887 | STAG3L3 | FALSE | Downregulated |
| 0,135210129 | 0,889426887 | STEAP2 | FALSE | Upregulated |
| -0,071272624 | 0,889426887 | STOML2 | FALSE | Downregulated |
| -0,110655521 | 0,889426887 | TPD52 | FALSE | Downregulated |
| -0,122350816 | 0,889426887 | TRMT2B | FALSE | Downregulated |
| -0,065078871 | 0,889426887 | UBE3C | FALSE | Downregulated |
| -0,127108183 | 0,889426887 | YDJC | FALSE | Downregulated |
| -0,048044775 | 0,889426887 | ZFP36L2 | FALSE | Downregulated |
| -0,069146695 | 0,889426887 | ZHX1 | FALSE | Downregulated |
| 0,060430328 | 0,889426887 | ZNF664 | FALSE | Upregulated |
| -0,105033999 | 0,889535508 | C19orf70 | FALSE | Downregulated |
| -0,123429111 | 0,889539279 | ASNSD1 | FALSE | Downregulated |
| 0,085030477 | 0,889539279 | CD2BP2 | FALSE | Upregulated |
| -0,084126112 | 0,889539279 | DDB2 | FALSE | Downregulated |
| -0,122722916 | 0,889539279 | KIAA1804 | FALSE | Downregulated |
| 0,088940121 | 0,889539279 | MCM9 | FALSE | Upregulated |
| 0,077090816 | 0,889539279 | RAB23 | FALSE | Upregulated |

| | | | | |
|---|---|---|---|---|
| 0,051003374 | 0,889539279 | REV3L | FALSE | Upregulated |
| 0,112147141 | 0,889539279 | RNLS | FALSE | Upregulated |
| -0,102421368 | 0,889539279 | TAF6 | FALSE | Downregulated |
| 0,17331723 | 0,889539279 | WNT8B | FALSE | Upregulated |
| -0,0380451 | 0,889601492 | COPA | FALSE | Downregulated |
| -0,06851075 | 0,889601492 | SAFB2 | FALSE | Downregulated |
| -0,11627577 | 0,889612243 | LOC644662 | FALSE | Downregulated |
| -0,057205456 | 0,889725293 | ABTB2 | FALSE | Downregulated |
| 0,15008908 | 0,889725293 | FADS1 | FALSE | Upregulated |
| -0,057837292 | 0,889837123 | ANO6 | FALSE | Downregulated |
| 0,086771092 | 0,889837123 | CEPT1 | FALSE | Upregulated |
| -0,122282317 | 0,890034725 | BCL7A | FALSE | Downregulated |
| 0,172557855 | 0,890034725 | KIAA1652 | FALSE | Upregulated |
| 0,065784494 | 0,890034725 | SUDS3 | FALSE | Upregulated |
| -0,062080183 | 0,890034725 | TRANK1 | FALSE | Downregulated |
| -0,041292446 | 0,890056191 | GOLGB1 | FALSE | Downregulated |
| -0,196718788 | 0,890056191 | NIPSNAP3B | FALSE | Downregulated |
| 0,069990077 | 0,890263529 | C1orf192 | FALSE | Upregulated |
| -0,105171502 | 0,890263529 | CWF19L2 | FALSE | Downregulated |
| -0,070518446 | 0,890263529 | FKBP1A | FALSE | Downregulated |
| 0,163216472 | 0,890263529 | GCNT1 | FALSE | Upregulated |
| -0,097514125 | 0,890263529 | MTAP | FALSE | Downregulated |
| -0,05244941 | 0,890533753 | MEF2A | FALSE | Downregulated |
| -0,048571693 | 0,890733255 | LONP1 | FALSE | Downregulated |
| -0,156551318 | 0,890743539 | SUSD3 | FALSE | Downregulated |
| -0,130661085 | 0,890851764 | ATP6V1H | FALSE | Downregulated |
| 0,11146835 | 0,890851764 | ECD | FALSE | Upregulated |
| -0,286183063 | 0,890876356 | ZNF100 | FALSE | Downregulated |
| -0,13657597 | 0,891242081 | GPR45 | FALSE | Downregulated |
| 0,084769444 | 0,891242081 | MOB3C | FALSE | Upregulated |
| 0,104754507 | 0,891242081 | THTPA | FALSE | Upregulated |
| 0,104946965 | 0,891242081 | VWA9 | FALSE | Upregulated |
| 0,081155322 | 0,891255198 | PHACTR4 | FALSE | Upregulated |
| -0,066832578 | 0,89139494 | GABARAPL1 | FALSE | Downregulated |
| 0,109578178 | 0,89139494 | SNX11 | FALSE | Upregulated |
| 0,060414707 | 0,89139494 | TBCD | FALSE | Upregulated |
| -0,082980741 | 0,89139494 | TMX3 | FALSE | Downregulated |
| 0,063935302 | 0,89141432 | PLEKHJ1 | FALSE | Upregulated |
| 0,074258434 | 0,891492923 | BCDIN3D-AS1 | FALSE | Upregulated |
| -0,178793459 | 0,891492923 | CD163L1 | FALSE | Downregulated |
| 0,085092616 | 0,891492923 | CXCL16 | FALSE | Upregulated |
| 0,063470134 | 0,891492923 | DYNC1LI2 | FALSE | Upregulated |
| -0,103681741 | 0,891492923 | FOXO4 | FALSE | Downregulated |
| 0,099053055 | 0,891492923 | FRY-AS1 | FALSE | Upregulated |
| -0,150110898 | 0,891492923 | LOC101928938 | FALSE | Downregulated |
| 0,121986573 | 0,891492923 | MRPL10 | FALSE | Upregulated |
| 0,18838404 | 0,891492923 | PAPSS1 | FALSE | Upregulated |
| -0,060642815 | 0,891528674 | SYNRG | FALSE | Downregulated |
| 0,15602289 | 0,891552006 | RARG | FALSE | Upregulated |
| 0,079476777 | 0,891561735 | TMEM51-AS1 | FALSE | Upregulated |
| 0,055863854 | 0,891739005 | ADIPOR2 | FALSE | Upregulated |

| | | | | |
|---|---|---|---|---|
| 0,069593812 | 0,891739005 | ANKRD40 | FALSE | Upregulated |
| 0,111302685 | 0,891739005 | C17orf47 | FALSE | Upregulated |
| -0,132956846 | 0,891739005 | COQ7 | FALSE | Downregulated |
| 0,110537148 | 0,891739005 | DYRK2 | FALSE | Upregulated |
| -0,120222901 | 0,891739005 | ELOVL6 | FALSE | Downregulated |
| -0,068929987 | 0,891739005 | EXTL3 | FALSE | Downregulated |
| -0,182843354 | 0,891739005 | HYPK | FALSE | Downregulated |
| 0,092445839 | 0,891739005 | LEPRE1 | FALSE | Upregulated |
| 0,066519786 | 0,891739005 | LINC00674 | FALSE | Upregulated |
| 0,104930063 | 0,891739005 | LOC730183 | FALSE | Upregulated |
| 0,083148065 | 0,891739005 | MAP1LC3B | FALSE | Upregulated |
| 0,105628081 | 0,891739005 | MAP3K9 | FALSE | Upregulated |
| 0,116935832 | 0,891739005 | MPHOSPH10 | FALSE | Upregulated |
| -0,118519197 | 0,891739005 | MRE11A | FALSE | Downregulated |
| -0,171797095 | 0,891739005 | POLR1C | FALSE | Downregulated |
| -0,079497756 | 0,891739005 | PPP2R5E | FALSE | Downregulated |
| 0,178659102 | 0,891739005 | PSTPIP2 | FALSE | Upregulated |
| -0,10412523 | 0,891739005 | RAB4A | FALSE | Downregulated |
| -0,097290264 | 0,891739005 | SBDS | FALSE | Downregulated |
| -0,148786365 | 0,891739005 | TERF1 | FALSE | Downregulated |
| 0,132201047 | 0,891739005 | TMPPE | FALSE | Upregulated |
| 0,06982389 | 0,891739005 | XPO5 | FALSE | Upregulated |
| 0,101546835 | 0,891739005 | ZNF462 | FALSE | Upregulated |
| 0,075484312 | 0,891776671 | HARS | FALSE | Upregulated |
| 0,19227363 | 0,891776671 | MFGE8 | FALSE | Upregulated |
| -0,128656695 | 0,891776671 | MTURN | FALSE | Downregulated |
| -0,09335432 | 0,891776671 | RNASET2 | FALSE | Downregulated |
| -0,121434675 | 0,891776671 | TGS1 | FALSE | Downregulated |
| 0,091432321 | 0,891776671 | ZNF76 | FALSE | Upregulated |
| 0,083847684 | 0,891852066 | TPD52L2 | FALSE | Upregulated |
| 0,10909498 | 0,891984345 | OSTF1 | FALSE | Upregulated |
| 0,084140163 | 0,892295198 | SNX8 | FALSE | Upregulated |
| 0,168660933 | 0,892365783 | ARHGAP15 | FALSE | Upregulated |
| -0,061759158 | 0,892365783 | BCL2L13 | FALSE | Downregulated |
| 0,1113141 | 0,892365783 | CBFA2T3 | FALSE | Upregulated |
| -0,07667989 | 0,892365783 | COG3 | FALSE | Downregulated |
| 0,069491371 | 0,892365783 | DHRS12 | FALSE | Upregulated |
| -0,096137449 | 0,892365783 | HNRNPD | FALSE | Downregulated |
| 0,077463247 | 0,892365783 | LOC100505915 | FALSE | Upregulated |
| -0,111019033 | 0,892365783 | MED8 | FALSE | Downregulated |
| -0,128399756 | 0,892365783 | OLFML3 | FALSE | Downregulated |
| -0,096761631 | 0,892365783 | SCAMP4 | FALSE | Downregulated |
| -0,150766118 | 0,892365783 | SLC26A11 | FALSE | Downregulated |
| -0,066484512 | 0,892448024 | CAPN1 | FALSE | Downregulated |
| -0,152369186 | 0,892448024 | DNAJB4 | FALSE | Downregulated |
| 0,083775601 | 0,892448024 | LOC101928717 | FALSE | Upregulated |
| 0,074878387 | 0,892509208 | ECHDC3 | FALSE | Upregulated |
| 0,055835713 | 0,892509208 | HYAL1 | FALSE | Upregulated |
| -0,149439598 | 0,892509208 | LOC101929479 | FALSE | Downregulated |
| 0,089958732 | 0,892509208 | PPP1R9A | FALSE | Upregulated |
| -0,069456963 | 0,892509208 | SECISBP2 | FALSE | Downregulated |

| | | | | |
|---|---|---|---|---|
| 0,137022993 | 0,892509208 | TMEM164 | FALSE | Upregulated |
| -0,072174524 | 0,892509208 | TMEM8A | FALSE | Downregulated |
| -0,100523784 | 0,892509208 | TRPS1 | FALSE | Downregulated |
| 0,101093682 | 0,892574865 | RGS3 | FALSE | Upregulated |
| 0,117689002 | 0,892574865 | STX6 | FALSE | Upregulated |
| -0,05804768 | 0,892813719 | DAPK1 | FALSE | Downregulated |
| -0,093326549 | 0,892963686 | ANP32A | FALSE | Downregulated |
| -0,085090168 | 0,892963686 | BHLHE40 | FALSE | Downregulated |
| 0,106218035 | 0,892963686 | CAPZA2 | FALSE | Upregulated |
| 0,126151965 | 0,892963686 | CCDC104 | FALSE | Upregulated |
| -0,065937454 | 0,892963686 | CLCN3 | FALSE | Downregulated |
| -0,080880951 | 0,892963686 | CLPP | FALSE | Downregulated |
| -0,140014404 | 0,892963686 | COIL | FALSE | Downregulated |
| 0,041853487 | 0,892963686 | CTDSP1 | FALSE | Upregulated |
| -0,072926418 | 0,892963686 | FUCA2 | FALSE | Downregulated |
| 0,073701778 | 0,892963686 | GOLGA5 | FALSE | Upregulated |
| 0,069049936 | 0,892963686 | HECA | FALSE | Upregulated |
| -0,078968247 | 0,892963686 | HEXB | FALSE | Downregulated |
| 0,143664141 | 0,892963686 | HIGD1C | FALSE | Upregulated |
| -0,193305869 | 0,892963686 | LOC101928745 | FALSE | Downregulated |
| 0,103672568 | 0,892963686 | MOK | FALSE | Upregulated |
| 0,147778651 | 0,892963686 | NEXN-AS1 | FALSE | Upregulated |
| -0,048940573 | 0,892963686 | NIT1 | FALSE | Downregulated |
| -0,060380164 | 0,892963686 | NSD1 | FALSE | Downregulated |
| -0,10452514 | 0,892963686 | PPP4R1 | FALSE | Downregulated |
| -0,06192606 | 0,892963686 | RAPH1 | FALSE | Downregulated |
| 0,169837753 | 0,892963686 | RBL1 | FALSE | Upregulated |
| 0,165127448 | 0,892963686 | RP11-24J23.2 | FALSE | Upregulated |
| 0,15081177 | 0,892963686 | RP3-400N23.6 | FALSE | Upregulated |
| 0,137001986 | 0,892963686 | SACS | FALSE | Upregulated |
| 0,176705755 | 0,892963686 | SDHAF2 | FALSE | Upregulated |
| 0,308979603 | 0,892963686 | SPP1 | FALSE | Upregulated |
| -0,091918062 | 0,892963686 | STX3 | FALSE | Downregulated |
| -0,175302503 | 0,892963686 | STYXL1 | FALSE | Downregulated |
| -0,11683109 | 0,892963686 | UGCG | FALSE | Downregulated |
| 0,078864674 | 0,892963686 | VPS52 | FALSE | Upregulated |
| 0,13227518 | 0,892963686 | ZNF32-AS1 | FALSE | Upregulated |
| 0,062374433 | 0,893012185 | ARHGEF26 | FALSE | Upregulated |
| -0,125374012 | 0,893065502 | DDX47 | FALSE | Downregulated |
| 0,087442029 | 0,893065502 | DNASE1L1 | FALSE | Upregulated |
| 0,125821091 | 0,893157788 | SLC6A8 | FALSE | Upregulated |
| -0,080277119 | 0,893267126 | ABCB1 | FALSE | Downregulated |
| -0,124663705 | 0,893267126 | AC011747.7 | FALSE | Downregulated |
| -0,056324749 | 0,893267126 | ADCY6 | FALSE | Downregulated |
| -0,072696349 | 0,893267126 | AHDC1 | FALSE | Downregulated |
| -0,194349656 | 0,893267126 | AS3MT | FALSE | Downregulated |
| -0,101870495 | 0,893267126 | B3GALT6 | FALSE | Downregulated |
| -0,074076536 | 0,893267126 | BAIAP3 | FALSE | Downregulated |
| 0,072185706 | 0,893267126 | BMP8B | FALSE | Upregulated |
| 0,085072424 | 0,893267126 | BRF1 | FALSE | Upregulated |
| -0,089341363 | 0,893267126 | CCDC57 | FALSE | Downregulated |

| | | | | |
|---|---|---|---|---|
| 0,085261752 | 0,893267126 | CEP192 | FALSE | Upregulated |
| -0,043647101 | 0,893267126 | CHERP | FALSE | Downregulated |
| 0,085204223 | 0,893267126 | DCUN1D1 | FALSE | Upregulated |
| -0,053661463 | 0,893267126 | DHRS3 | FALSE | Downregulated |
| -0,060890754 | 0,893267126 | DIP2B | FALSE | Downregulated |
| 0,157094903 | 0,893267126 | DPY19L2P4 | FALSE | Upregulated |
| -0,116916279 | 0,893267126 | EFCAB4A | FALSE | Downregulated |
| 0,133808317 | 0,893267126 | ELMOD2 | FALSE | Upregulated |
| -0,138959738 | 0,893267126 | GPALPP1 | FALSE | Downregulated |
| 0,058406881 | 0,893267126 | GPR124 | FALSE | Upregulated |
| 0,131192565 | 0,893267126 | HMCN2 | FALSE | Upregulated |
| -0,075951849 | 0,893267126 | IL17RC | FALSE | Downregulated |
| 0,100403943 | 0,893267126 | LOC101927345 | FALSE | Upregulated |
| 0,128466254 | 0,893267126 | LOC101929636 | FALSE | Upregulated |
| 0,105170964 | 0,893267126 | LOC102724364 | FALSE | Upregulated |
| -0,055368408 | 0,893267126 | MAN1A2 | FALSE | Downregulated |
| 0,066348683 | 0,893267126 | MTCH2 | FALSE | Upregulated |
| -0,046882153 | 0,893267126 | MTSS1L | FALSE | Downregulated |
| -0,043007064 | 0,893267126 | NIPBL | FALSE | Downregulated |
| -0,123139295 | 0,893267126 | NPIPB9 | FALSE | Downregulated |
| -0,092995886 | 0,893267126 | PHF14 | FALSE | Downregulated |
| -0,139900133 | 0,893267126 | PKNOX1 | FALSE | Downregulated |
| 0,18119809 | 0,893267126 | PPP1R2 | FALSE | Upregulated |
| 0,084123691 | 0,893267126 | PYGO2 | FALSE | Upregulated |
| -0,067423724 | 0,893267126 | RAD21 | FALSE | Downregulated |
| -0,329814049 | 0,893267126 | RPL36A-HNRNPH2 | FALSE | Downregulated |
| -0,13613393 | 0,893267126 | RRP36 | FALSE | Downregulated |
| -0,11240571 | 0,893267126 | SGPP1 | FALSE | Downregulated |
| 0,18158096 | 0,893267126 | SLC25A52 | FALSE | Upregulated |
| -0,149493539 | 0,893267126 | SP7 | FALSE | Downregulated |
| 0,092792607 | 0,893267126 | SYNGR2 | FALSE | Upregulated |
| -0,068821651 | 0,893267126 | SYNJ2 | FALSE | Downregulated |
| 0,14825669 | 0,893267126 | TRPA1 | FALSE | Upregulated |
| 0,088848845 | 0,893267126 | UBR1 | FALSE | Upregulated |
| -0,14736542 | 0,893267126 | VANGL1 | FALSE | Downregulated |
| -0,058547761 | 0,893267126 | ZADH2 | FALSE | Downregulated |
| -0,097906809 | 0,893267126 | ZNF189 | FALSE | Downregulated |
| 0,075628404 | 0,893267126 | ZNF557 | FALSE | Upregulated |
| 0,063702651 | 0,893267126 | ZNF787 | FALSE | Upregulated |
| 0,12959156 | 0,893267126 | ZNF792 | FALSE | Upregulated |
| -0,109736895 | 0,893406549 | IPO4 | FALSE | Downregulated |
| 0,160189854 | 0,893406549 | LOC101060212 | FALSE | Upregulated |
| 0,063200205 | 0,893638599 | CD68 | FALSE | Upregulated |
| -0,117169096 | 0,89395124 | FBXL19 | FALSE | Downregulated |
| -0,090111291 | 0,89395124 | NCKIPSD | FALSE | Downregulated |
| -0,136143571 | 0,894142895 | IL17D | FALSE | Downregulated |
| -0,060983262 | 0,894142895 | KDM1A | FALSE | Downregulated |
| -0,113703801 | 0,894142895 | PNO1 | FALSE | Downregulated |
| 0,064922 | 0,894142895 | ZC3H18 | FALSE | Upregulated |
| 0,073015547 | 0,894232283 | C11orf24 | FALSE | Upregulated |
| 0,054899128 | 0,894232283 | VCPIP1 | FALSE | Upregulated |

| | | | | |
|---|---|---|---|---|
| 0,085244203 | 0,894487883 | FBL | FALSE | Upregulated |
| -0,088372723 | 0,894536003 | ARL8A | FALSE | Downregulated |
| -0,128556509 | 0,894632826 | ARFIP1 | FALSE | Downregulated |
| -0,14386302 | 0,894990582 | AFAP1L2 | FALSE | Downregulated |
| 0,055744896 | 0,894990582 | AIDA | FALSE | Upregulated |
| -0,088918944 | 0,894990582 | DYNC1I2 | FALSE | Downregulated |
| -0,039791341 | 0,894990582 | EML4 | FALSE | Downregulated |
| -0,155823754 | 0,894990582 | GMEB1 | FALSE | Downregulated |
| -0,084023226 | 0,894990582 | HERC4 | FALSE | Downregulated |
| 0,128936021 | 0,894990582 | LOC101928206 | FALSE | Upregulated |
| -0,082915023 | 0,894990582 | MAP1LC3A | FALSE | Downregulated |
| -0,066746437 | 0,894990582 | NAB1 | FALSE | Downregulated |
| -0,130019526 | 0,894990582 | PIK3R5 | FALSE | Downregulated |
| -0,091150703 | 0,894990582 | PLOD2 | FALSE | Downregulated |
| -0,111865478 | 0,894990582 | PMCHL2 | FALSE | Downregulated |
| -0,096049711 | 0,894990582 | THUMPD1 | FALSE | Downregulated |
| -0,064775764 | 0,895201594 | AAMP | FALSE | Downregulated |
| 0,108443865 | 0,895201594 | BCAP29 | FALSE | Upregulated |
| -0,094966223 | 0,895201594 | C16orf13 | FALSE | Downregulated |
| 0,067421639 | 0,895201594 | CALM2 | FALSE | Upregulated |
| 0,081923064 | 0,895201594 | CEBPZ | FALSE | Upregulated |
| -0,083675587 | 0,895201594 | DOCK7 | FALSE | Downregulated |
| 0,064890817 | 0,895201594 | EMILIN1 | FALSE | Upregulated |
| -0,093175982 | 0,895201594 | FAM192A | FALSE | Downregulated |
| -0,115716924 | 0,895201594 | FKRP | FALSE | Downregulated |
| 0,082109743 | 0,895201594 | FLJ35282 | FALSE | Upregulated |
| 0,043252667 | 0,895201594 | HERC1 | FALSE | Upregulated |
| -0,181545849 | 0,895201594 | LINC00999 | FALSE | Downregulated |
| -0,150668222 | 0,895201594 | LOC102723694 | FALSE | Downregulated |
| 0,105421098 | 0,895201594 | LOC102723735 | FALSE | Upregulated |
| -0,110528019 | 0,895201594 | MOGAT2 | FALSE | Downregulated |
| -0,152869426 | 0,895201594 | NDNL2 | FALSE | Downregulated |
| -0,148753544 | 0,895201594 | PPIE | FALSE | Downregulated |
| 0,146804715 | 0,895201594 | SELPLG | FALSE | Upregulated |
| 0,175777992 | 0,895201594 | TRAF5 | FALSE | Upregulated |
| -0,135038948 | 0,895201594 | UBA3 | FALSE | Downregulated |
| 0,062913197 | 0,895342521 | KDELR2 | FALSE | Upregulated |
| 0,08115913 | 0,895374137 | LOC728024 | FALSE | Upregulated |
| -0,084167126 | 0,895374137 | RNF128 | FALSE | Downregulated |
| 0,097546171 | 0,895443419 | EMILIN3 | FALSE | Upregulated |
| 0,073891251 | 0,895471845 | NTMT1 | FALSE | Upregulated |
| -0,142445064 | 0,895558296 | GALC | FALSE | Downregulated |
| 0,15941655 | 0,895605842 | ZSCAN31 | FALSE | Upregulated |
| 0,093575924 | 0,895706901 | LOC102723851 | FALSE | Upregulated |
| 0,075585049 | 0,895706901 | SRGAP1 | FALSE | Upregulated |
| 0,147241177 | 0,895717569 | LOC101060494 | FALSE | Upregulated |
| -0,113992234 | 0,895731389 | THRSP | FALSE | Downregulated |
| 0,094981293 | 0,895933884 | ATP6V1D | FALSE | Upregulated |
| -0,052582258 | 0,895933884 | BAZ2B | FALSE | Downregulated |
| 0,071926617 | 0,895933884 | C1RL-AS1 | FALSE | Upregulated |
| 0,131937336 | 0,895933884 | COL17A1 | FALSE | Upregulated |

| | | | | |
|---|---|---|---|---|
| -0,05449791 | 0,895933884 | DVL1 | FALSE | Downregulated |
| 0,072039354 | 0,895933884 | HPN-AS1 | FALSE | Upregulated |
| -0,062309639 | 0,895933884 | MORC2 | FALSE | Downregulated |
| -0,145154242 | 0,895933884 | POP5 | FALSE | Downregulated |
| -0,09442587 | 0,895933884 | PPM1H | FALSE | Downregulated |
| -0,048221522 | 0,895933884 | SEC16A | FALSE | Downregulated |
| 0,111482058 | 0,895933884 | SMAP1 | FALSE | Upregulated |
| 0,105354382 | 0,896033669 | MAML2 | FALSE | Upregulated |
| 0,084382335 | 0,896033669 | NODAL | FALSE | Upregulated |
| 0,062957353 | 0,896033669 | ORC4 | FALSE | Upregulated |
| -0,095202175 | 0,896058617 | ARHGAP33 | FALSE | Downregulated |
| -0,049921038 | 0,896058617 | DHFR | FALSE | Downregulated |
| -0,080475311 | 0,896058617 | HEXA | FALSE | Downregulated |
| 0,049971417 | 0,896058617 | MBD6 | FALSE | Upregulated |
| 0,068483675 | 0,896058617 | PXDC1 | FALSE | Upregulated |
| -0,052627507 | 0,896400118 | RAF1 | FALSE | Downregulated |
| 0,079980661 | 0,896400118 | SIGLEC1 | FALSE | Upregulated |
| 0,120968218 | 0,89647256 | LOC101929401 | FALSE | Upregulated |
| 0,098312237 | 0,89647256 | NKIRAS1 | FALSE | Upregulated |
| 0,086591032 | 0,89647256 | ZDHHC9 | FALSE | Upregulated |
| 0,127443777 | 0,89647256 | ZNF429 | FALSE | Upregulated |
| -0,109620274 | 0,896632284 | DPF2 | FALSE | Downregulated |
| 0,071555385 | 0,896656243 | C4orf3 | FALSE | Upregulated |
| -0,081488187 | 0,896656243 | SCAPER | FALSE | Downregulated |
| 0,087691743 | 0,896922413 | ZNF829 | FALSE | Upregulated |
| 0,117293223 | 0,896930494 | DFNB31 | FALSE | Upregulated |
| 0,120907919 | 0,897026493 | CIAPIN1 | FALSE | Upregulated |
| 0,090298352 | 0,897026493 | CLDN12 | FALSE | Upregulated |
| 0,058160292 | 0,897026493 | EHBP1 | FALSE | Upregulated |
| 0,090910758 | 0,897026493 | KCNS2 | FALSE | Upregulated |
| -0,054361511 | 0,897026493 | LINC01000 | FALSE | Downregulated |
| 0,175332522 | 0,897026493 | METTL18 | FALSE | Upregulated |
| -0,108698156 | 0,897026493 | PAXIP1 | FALSE | Downregulated |
| 0,035728143 | 0,897026493 | PINK1 | FALSE | Upregulated |
| -0,18066759 | 0,897026493 | PMEPA1 | FALSE | Downregulated |
| -1,836099235 | 0,897026493 | RNU1-1 | FALSE | Downregulated |
| 0,036308144 | 0,897026493 | TMED10 | FALSE | Upregulated |
| 0,152720222 | 0,897026493 | VIMP | FALSE | Upregulated |
| -0,065463079 | 0,897026493 | XIAP | FALSE | Downregulated |
| 0,073217092 | 0,897026493 | YIPF2 | FALSE | Upregulated |
| 0,083650256 | 0,897353287 | JAM2 | FALSE | Upregulated |
| 0,104670073 | 0,897353287 | R3HCC1L | FALSE | Upregulated |
| 0,095929289 | 0,897353287 | SNHG7 | FALSE | Upregulated |
| 0,04814031 | 0,89740017 | MSL1 | FALSE | Upregulated |
| 0,096742107 | 0,897532647 | ERO1L | FALSE | Upregulated |
| 0,145344661 | 0,897567632 | C8orf59 | FALSE | Upregulated |
| 0,125079392 | 0,897567632 | CCDC109B | FALSE | Upregulated |
| 0,07634049 | 0,897894595 | ASAH1 | FALSE | Upregulated |
| -0,141411842 | 0,897894595 | DNAJC15 | FALSE | Downregulated |
| 0,067888296 | 0,897894595 | EPG5 | FALSE | Upregulated |
| 0,635209166 | 0,897894595 | F8A3 | FALSE | Upregulated |

| | | | | |
|---|---|---|---|---|
| 0,102236145 | 0,897894595 | KIAA0319 | FALSE | Upregulated |
| 0,0849642 | 0,897894595 | KIAA1199 | FALSE | Upregulated |
| -0,151367701 | 0,897894595 | LAMB3 | FALSE | Downregulated |
| 0,119548585 | 0,897894595 | PAQR6 | FALSE | Upregulated |
| 0,092779611 | 0,897894595 | RANBP6 | FALSE | Upregulated |
| 0,079693417 | 0,897894595 | RCOR1 | FALSE | Upregulated |
| -0,06942787 | 0,897894595 | SNX2 | FALSE | Downregulated |
| -0,043579432 | 0,898172838 | RPL27A | FALSE | Downregulated |
| 0,162309461 | 0,898456118 | SLC39A10 | FALSE | Upregulated |
| -0,075089145 | 0,898641917 | C5orf42 | FALSE | Downregulated |
| -0,063207216 | 0,898641917 | ETNK2 | FALSE | Downregulated |
| 0,086486301 | 0,898641917 | NSUN5P1 | FALSE | Upregulated |
| -0,097262753 | 0,898846533 | QTRT1 | FALSE | Downregulated |
| -0,068004625 | 0,899082243 | ALPL | FALSE | Downregulated |
| -0,083225227 | 0,899082243 | FEM1A | FALSE | Downregulated |
| -0,089194071 | 0,899082243 | PLEKHA2 | FALSE | Downregulated |
| 0,208610294 | 0,899082243 | PLXNA4 | FALSE | Upregulated |
| -1,809033657 | 0,899082243 | RNU1-3 | FALSE | Downregulated |
| -0,087641342 | 0,899082243 | SF3A3 | FALSE | Downregulated |
| -0,07227368 | 0,899082243 | TRAPPC11 | FALSE | Downregulated |
| 0,079631892 | 0,899082243 | WDR46 | FALSE | Upregulated |
| -0,121666372 | 0,899082243 | ZKSCAN2 | FALSE | Downregulated |
| -0,064314195 | 0,899291965 | HM13 | FALSE | Downregulated |
| -0,093350817 | 0,899291965 | KIAA0907 | FALSE | Downregulated |
| -0,047460365 | 0,899291965 | SDC1 | FALSE | Downregulated |
| -0,101736321 | 0,899291965 | SSBP1 | FALSE | Downregulated |
| 0,092834245 | 0,899291965 | TCHP | FALSE | Upregulated |
| 0,214647673 | 0,899347709 | KDM5D | FALSE | Upregulated |
| -0,123360198 | 0,899353611 | CCDC159 | FALSE | Downregulated |
| -0,158716671 | 0,899353611 | CREM | FALSE | Downregulated |
| 0,092259844 | 0,899353611 | DMAP1 | FALSE | Upregulated |
| -0,138485739 | 0,899353611 | EPDR1 | FALSE | Downregulated |
| 0,398651784 | 0,899353611 | FAM47E-STBD1 | FALSE | Upregulated |
| 0,382907313 | 0,899353611 | GCOM1 | FALSE | Upregulated |
| -0,13130378 | 0,899353611 | LOC102724733 | FALSE | Downregulated |
| -0,144902909 | 0,899353611 | LOC644794 | FALSE | Downregulated |
| 0,119050058 | 0,899353611 | LPPR3 | FALSE | Upregulated |
| -0,086944108 | 0,899353611 | MAPKBP1 | FALSE | Downregulated |
| -0,115474027 | 0,899353611 | NKAIN2 | FALSE | Downregulated |
| 0,036672506 | 0,899353611 | PHRF1 | FALSE | Upregulated |
| -0,064367382 | 0,899353611 | RNPS1 | FALSE | Downregulated |
| 0,075021919 | 0,899353611 | RP11-178C3.2 | FALSE | Upregulated |
| -0,059452224 | 0,899353611 | SIRT5 | FALSE | Downregulated |
| -0,165939899 | 0,899353611 | TCF21 | FALSE | Downregulated |
| -0,124094218 | 0,899353611 | TMCO6 | FALSE | Downregulated |
| 0,178560327 | 0,899353611 | TMED6 | FALSE | Upregulated |
| -0,044529856 | 0,899353611 | TNKS1BP1 | FALSE | Downregulated |
| -0,074045752 | 0,899353611 | TRUB2 | FALSE | Downregulated |
| 0,114755738 | 0,899353611 | TSTD3 | FALSE | Upregulated |
| 0,197031239 | 0,899353611 | UBXN10 | FALSE | Upregulated |
| -0,089084126 | 0,899353611 | ZC3H7A | FALSE | Downregulated |

| | | | | |
|---|---|---|---|---|
| 0,107251172 | 0,899384158 | ADAM8 | FALSE | Upregulated |
| 0,135559501 | 0,899384158 | TRIM39 | FALSE | Upregulated |
| -0,103578786 | 0,899639379 | ART4 | FALSE | Downregulated |
| 0,094959576 | 0,899708465 | CNNM1 | FALSE | Upregulated |
| -0,102889468 | 0,899708465 | PHF21A | FALSE | Downregulated |
| 0,082383226 | 0,899852214 | DCP1A | FALSE | Upregulated |
| 0,110511273 | 0,899852214 | HCG4B | FALSE | Upregulated |
| 0,101900037 | 0,899852214 | NAMPT | FALSE | Upregulated |
| -0,120739037 | 0,899852214 | RAB40B | FALSE | Downregulated |
| 0,058216626 | 0,899876892 | WDR73 | FALSE | Upregulated |
| -0,12185821 | 0,900029934 | CRIPT | FALSE | Downregulated |
| 0,070724973 | 0,900058413 | KCTD21-AS1 | FALSE | Upregulated |
| -0,052332479 | 0,900058413 | KIDINS220 | FALSE | Downregulated |
| 0,093449757 | 0,900058413 | LOC102723824 | FALSE | Upregulated |
| 0,067378943 | 0,900330591 | PTPRM | FALSE | Upregulated |
| 0,174426336 | 0,900334063 | ZIC1 | FALSE | Upregulated |
| 0,070419451 | 0,9004789 | CEBPB | FALSE | Upregulated |
| 0,082475162 | 0,90068165 | AKAP6 | FALSE | Upregulated |
| 0,096467762 | 0,90068165 | RP11-760N9.1 | FALSE | Upregulated |
| -0,077499699 | 0,90075014 | RNMT | FALSE | Downregulated |
| -0,082178932 | 0,900903696 | AGMO | FALSE | Downregulated |
| -0,261656389 | 0,900903696 | NPHP3-ACAD11 | FALSE | Downregulated |
| 0,063627332 | 0,900903696 | SARM1 | FALSE | Upregulated |
| 0,095515021 | 0,900903696 | ZC3H3 | FALSE | Upregulated |
| -0,208877642 | 0,900913544 | C12orf5 | FALSE | Downregulated |
| 0,163534091 | 0,900913544 | DSC1 | FALSE | Upregulated |
| -0,158976654 | 0,900913544 | GPR39 | FALSE | Downregulated |
| 0,049230751 | 0,900913544 | INF2 | FALSE | Upregulated |
| -0,154395425 | 0,900913544 | LOC101928574 | FALSE | Downregulated |
| -0,055337647 | 0,901403053 | BMP1 | FALSE | Downregulated |
| -0,123991217 | 0,901403053 | KNOP1 | FALSE | Downregulated |
| 0,128804722 | 0,901403053 | MIS12 | FALSE | Upregulated |
| 0,100731301 | 0,901403053 | UBE2B | FALSE | Upregulated |
| -0,081446892 | 0,901645981 | PHKA2-AS1 | FALSE | Downregulated |
| -0,092357413 | 0,901647268 | TIMM23 | FALSE | Downregulated |
| 0,081212392 | 0,901759432 | ARPC2 | FALSE | Upregulated |
| -0,085598657 | 0,901759432 | GBA | FALSE | Downregulated |
| -0,126332179 | 0,901828487 | C1orf35 | FALSE | Downregulated |
| -0,117132732 | 0,901828487 | EHD2 | FALSE | Downregulated |
| -0,142113939 | 0,901828487 | ZSWIM5 | FALSE | Downregulated |
| 0,09775841 | 0,901830009 | BSN | FALSE | Upregulated |
| -0,057647137 | 0,901830009 | CAMKK2 | FALSE | Downregulated |
| 0,064403489 | 0,901830009 | CCDC152 | FALSE | Upregulated |
| 0,052656685 | 0,901830009 | CMTM6 | FALSE | Upregulated |
| -0,057556561 | 0,901830009 | CPEB4 | FALSE | Downregulated |
| 0,070496847 | 0,901830009 | GTF3A | FALSE | Upregulated |
| 0,073938737 | 0,901830009 | HDAC1 | FALSE | Upregulated |
| 0,062876174 | 0,901830009 | MIDN | FALSE | Upregulated |
| -0,076186594 | 0,901830009 | MPND | FALSE | Downregulated |
| -0,12112726 | 0,901830009 | MYPOP | FALSE | Downregulated |
| -0,077597779 | 0,901830009 | NEO1 | FALSE | Downregulated |

| | | | | |
|---|---|---|---|---|
| -0,049872443 | 0,901830009 | NRD1 | FALSE | Downregulated |
| -0,168090259 | 0,901830009 | PRKD1 | FALSE | Downregulated |
| -0,148749868 | 0,901830009 | PTCD2 | FALSE | Downregulated |
| -0,127679151 | 0,901830009 | RP11-395P17.3 | FALSE | Downregulated |
| 0,145181223 | 0,901830009 | RRP15 | FALSE | Upregulated |
| 0,105951976 | 0,901830009 | SIRT7 | FALSE | Upregulated |
| 0,077642396 | 0,901830009 | TCF7L2 | FALSE | Upregulated |
| -0,112316304 | 0,902013809 | SUPV3L1 | FALSE | Downregulated |
| -0,059854846 | 0,902013809 | UBR2 | FALSE | Downregulated |
| 0,091294645 | 0,902043097 | C1GALT1 | FALSE | Upregulated |
| 0,064984123 | 0,902150617 | AC012314.8 | FALSE | Upregulated |
| 0,048303697 | 0,902150617 | AK2 | FALSE | Upregulated |
| -0,070035986 | 0,902150617 | ALG12 | FALSE | Downregulated |
| -0,086042892 | 0,902150617 | ALMS1 | FALSE | Downregulated |
| -0,079336014 | 0,902150617 | ANO8 | FALSE | Downregulated |
| 0,125313488 | 0,902150617 | ARL16 | FALSE | Upregulated |
| -0,052328815 | 0,902150617 | BAZ1B | FALSE | Downregulated |
| -0,046783617 | 0,902150617 | CIC | FALSE | Downregulated |
| -0,146786322 | 0,902150617 | CSTB | FALSE | Downregulated |
| -0,055822549 | 0,902150617 | DDX24 | FALSE | Downregulated |
| -0,153275759 | 0,902150617 | DLGAP2 | FALSE | Downregulated |
| 0,041482149 | 0,902150617 | ERP29 | FALSE | Upregulated |
| 0,109604608 | 0,902150617 | FADS3 | FALSE | Upregulated |
| 0,149908462 | 0,902150617 | FASTKD5 | FALSE | Upregulated |
| 0,078653126 | 0,902150617 | FLAD1 | FALSE | Upregulated |
| 0,150473184 | 0,902150617 | KIAA0825 | FALSE | Upregulated |
| -0,037635573 | 0,902150617 | KIF1C | FALSE | Downregulated |
| -0,106873395 | 0,902150617 | LINC00963 | FALSE | Downregulated |
| 0,146180612 | 0,902150617 | LOC101929078 | FALSE | Upregulated |
| -0,089219257 | 0,902150617 | NLRX1 | FALSE | Downregulated |
| 0,083793679 | 0,902150617 | PLEKHH3 | FALSE | Upregulated |
| -0,06658084 | 0,902150617 | PRG4 | FALSE | Downregulated |
| -0,148182308 | 0,902150617 | RNASEL | FALSE | Downregulated |
| -0,040308084 | 0,902150617 | RPS19 | FALSE | Downregulated |
| 0,136960792 | 0,902150617 | RSPH9 | FALSE | Upregulated |
| 0,068968256 | 0,902150617 | SLC11A2 | FALSE | Upregulated |
| -0,060083729 | 0,902150617 | TGFBR3 | FALSE | Downregulated |
| 0,05600431 | 0,902150617 | TSR1 | FALSE | Upregulated |
| 0,077709406 | 0,902150617 | ZBTB17 | FALSE | Upregulated |
| -0,104646861 | 0,902150617 | ZNF133 | FALSE | Downregulated |
| 0,074052597 | 0,902181459 | BAG4 | FALSE | Upregulated |
| 0,046558811 | 0,902181459 | IMMT | FALSE | Upregulated |
| -0,034036343 | 0,902181459 | MLLT4 | FALSE | Downregulated |
| 0,090219664 | 0,902181459 | PAX6 | FALSE | Upregulated |
| -0,125858868 | 0,902181459 | RFC2 | FALSE | Downregulated |
| 0,141789386 | 0,902181459 | ZNF257 | FALSE | Upregulated |
| 0,092543481 | 0,902440103 | LOC102724748 | FALSE | Upregulated |
| -0,110063135 | 0,902440103 | PRICKLE2-AS3 | FALSE | Downregulated |
| -0,09208663 | 0,902590302 | MLH1 | FALSE | Downregulated |
| 0,067774789 | 0,903237638 | FRMD8 | FALSE | Upregulated |
| -0,085286914 | 0,903270891 | ATAD3A | FALSE | Downregulated |

| | | | | |
|---|---|---|---|---|
| -0,070672619 | 0,903270891 | CAMK1 | FALSE | Downregulated |
| 0,076876119 | 0,903270891 | CSGALNACT1 | FALSE | Upregulated |
| -0,064975291 | 0,903270891 | DHRS1 | FALSE | Downregulated |
| -0,155770945 | 0,903270891 | MROH6 | FALSE | Downregulated |
| -0,122897372 | 0,903270891 | MRPL53 | FALSE | Downregulated |
| 0,040734839 | 0,903270891 | MYCBP2 | FALSE | Upregulated |
| 0,060100969 | 0,903270891 | PPP1R14B | FALSE | Upregulated |
| 0,06772259 | 0,903270891 | ZNF671 | FALSE | Upregulated |
| -0,085222507 | 0,903286599 | SH3TC1 | FALSE | Downregulated |
| -0,071127568 | 0,903595366 | GPBP1 | FALSE | Downregulated |
| 0,073787191 | 0,903789758 | AC009948.5 | FALSE | Upregulated |
| 0,150243573 | 0,903789758 | ATE1-AS1 | FALSE | Upregulated |
| -0,082930588 | 0,903789758 | B4GALT5 | FALSE | Downregulated |
| 0,064818502 | 0,903789758 | BID | FALSE | Upregulated |
| -0,124487989 | 0,903789758 | C1orf159 | FALSE | Downregulated |
| 0,063022883 | 0,903789758 | CCDC42B | FALSE | Upregulated |
| 0,043077575 | 0,903789758 | CGNL1 | FALSE | Upregulated |
| 0,074884986 | 0,903789758 | COPZ1 | FALSE | Upregulated |
| -0,106726785 | 0,903789758 | FAM96A | FALSE | Downregulated |
| 0,127789944 | 0,903789758 | IFT80 | FALSE | Upregulated |
| -0,057792395 | 0,903789758 | ITGB5 | FALSE | Downregulated |
| 0,075785232 | 0,903789758 | KATNAL2 | FALSE | Upregulated |
| -0,089807109 | 0,903789758 | NDUFA1 | FALSE | Downregulated |
| 0,179027437 | 0,903789758 | PELI2 | FALSE | Upregulated |
| 0,114177597 | 0,903789758 | PIAS3 | FALSE | Upregulated |
| 0,080955436 | 0,903789758 | RAET1E-AS1 | FALSE | Upregulated |
| -0,096284906 | 0,903789758 | RCL1 | FALSE | Downregulated |
| -0,11990284 | 0,903789758 | RP2 | FALSE | Downregulated |
| -0,051383407 | 0,903789758 | SEPT11 | FALSE | Downregulated |
| 0,089088095 | 0,903789758 | SLC5A9 | FALSE | Upregulated |
| -0,132237524 | 0,903789758 | SPACA6P-AS | FALSE | Downregulated |
| 0,05498439 | 0,903789758 | YWHAB | FALSE | Upregulated |
| 0,087401222 | 0,903968409 | SLC38A7 | FALSE | Upregulated |
| 0,054283483 | 0,904083287 | MGA | FALSE | Upregulated |
| -0,061506603 | 0,904103873 | ZNF672 | FALSE | Downregulated |
| -0,071588586 | 0,90434224 | GRPEL1 | FALSE | Downregulated |
| 0,139776827 | 0,90434224 | HEATR6 | FALSE | Upregulated |
| -0,118695287 | 0,90434224 | PPCS | FALSE | Downregulated |
| 0,071376442 | 0,90434224 | RNF31 | FALSE | Upregulated |
| -0,109425543 | 0,90434224 | SKP2 | FALSE | Downregulated |
| -0,12645057 | 0,90434224 | ZNF121 | FALSE | Downregulated |
| 0,096171555 | 0,90434224 | ZNF510 | FALSE | Upregulated |
| -0,125276647 | 0,904369615 | GYPC | FALSE | Downregulated |
| -0,086028323 | 0,904369615 | P2RX4 | FALSE | Downregulated |
| -0,046530834 | 0,904369615 | P4HB | FALSE | Downregulated |
| -0,070058204 | 0,904386528 | SMAD6 | FALSE | Downregulated |
| -0,358284644 | 0,904386528 | SPECC1L-ADORA2A | FALSE | Downregulated |
| -0,080381403 | 0,904386528 | TOP1MT | FALSE | Downregulated |
| -0,05700233 | 0,904421651 | ERC1 | FALSE | Downregulated |
| -0,150152761 | 0,904421651 | HIST2H2AB | FALSE | Downregulated |

| | | | | |
|---|---|---|---|---|
| -0,070109544 | 0,904421651 | JMJD4 | FALSE | Downregulated |
| 0,085246394 | 0,904421651 | LOC102724811 | FALSE | Upregulated |
| -0,103376642 | 0,904421651 | MAP10 | FALSE | Downregulated |
| -0,114105151 | 0,904421651 | MON1A | FALSE | Downregulated |
| 0,185397259 | 0,904421651 | TPRG1 | FALSE | Upregulated |
| 0,069456373 | 0,904421651 | TTC19 | FALSE | Upregulated |
| -0,112839829 | 0,904421651 | ZNF343 | FALSE | Downregulated |
| 0,144488394 | 0,904702923 | C15orf54 | FALSE | Upregulated |
| -0,14311325 | 0,904774945 | MAGI2-AS3 | FALSE | Downregulated |
| -0,052191438 | 0,904774945 | TULP4 | FALSE | Downregulated |
| -0,064754185 | 0,904960941 | ASH2L | FALSE | Downregulated |
| -0,174957624 | 0,904960941 | C5orf56 | FALSE | Downregulated |
| -0,056870695 | 0,904960941 | CCT3 | FALSE | Downregulated |
| -0,100047878 | 0,904960941 | CWC25 | FALSE | Downregulated |
| 0,122781402 | 0,904960941 | MMEL1 | FALSE | Upregulated |
| -0,166335386 | 0,904960941 | PALM2-AKAP2 | FALSE | Downregulated |
| 0,062123648 | 0,904960941 | PCGF2 | FALSE | Upregulated |
| 0,071632594 | 0,904960941 | SETMAR | FALSE | Upregulated |
| -0,038629764 | 0,904960941 | TUBGCP6 | FALSE | Downregulated |
| -0,114671255 | 0,904960941 | ZNF350 | FALSE | Downregulated |
| -0,12323145 | 0,904960941 | ZNF484 | FALSE | Downregulated |
| -0,061690963 | 0,905007043 | LINC00261 | FALSE | Downregulated |
| -0,103758692 | 0,905057954 | CCDC3 | FALSE | Downregulated |
| 0,145323329 | 0,905057954 | GPR65 | FALSE | Upregulated |
| 0,074960518 | 0,905057954 | NDUFC2 | FALSE | Upregulated |
| -0,042908553 | 0,905057954 | RNF123 | FALSE | Downregulated |
| 0,076327653 | 0,905151107 | EIF2AK2 | FALSE | Upregulated |
| 0,075072838 | 0,905151107 | LIN54 | FALSE | Upregulated |
| 0,112946207 | 0,905151107 | STS | FALSE | Upregulated |
| -0,069535317 | 0,905177971 | APOA1BP | FALSE | Downregulated |
| -0,114205466 | 0,905177971 | SLC15A1 | FALSE | Downregulated |
| -0,073891594 | 0,905266418 | FOXRED1 | FALSE | Downregulated |
| 0,160944915 | 0,905454096 | DCAF4L1 | FALSE | Upregulated |
| -0,123496677 | 0,9055452 | CYBRD1 | FALSE | Downregulated |
| 0,072550685 | 0,9055452 | PTBP3 | FALSE | Upregulated |
| 0,075067929 | 0,905579869 | CPB2-AS1 | FALSE | Upregulated |
| 0,331387115 | 0,905614921 | DDX11L9 | FALSE | Upregulated |
| 0,089985854 | 0,905614921 | KIF12 | FALSE | Upregulated |
| 0,096483026 | 0,905614921 | LRRN1 | FALSE | Upregulated |
| 0,076926929 | 0,905974502 | ACVR2B-AS1 | FALSE | Upregulated |
| 0,122756307 | 0,905974502 | L3MBTL1 | FALSE | Upregulated |
| 0,041451689 | 0,905974502 | MINK1 | FALSE | Upregulated |
| -0,127546783 | 0,905974502 | TMEM260 | FALSE | Downregulated |
| -0,07412683 | 0,906025452 | ATAD2B | FALSE | Downregulated |
| -0,051701588 | 0,906025452 | FBXW2 | FALSE | Downregulated |
| -0,139081863 | 0,906025452 | GPRC5B | FALSE | Downregulated |
| -0,033137222 | 0,906025452 | KIAA1671 | FALSE | Downregulated |
| 0,174830654 | 0,906025452 | LOC101927986 | FALSE | Upregulated |
| -0,089698171 | 0,906025452 | PPP2R5C | FALSE | Downregulated |
| -0,157270176 | 0,906025452 | RFT1 | FALSE | Downregulated |
| -0,098149446 | 0,906025452 | SUZ12 | FALSE | Downregulated |

| | | | | |
|---|---|---|---|---|
| 0,101125212 | 0,906545574 | DYRK1B | FALSE | Upregulated |
| 0,06339905 | 0,906598242 | ATP2C1 | FALSE | Upregulated |
| 0,139258749 | 0,906599546 | NPRL2 | FALSE | Upregulated |
| -0,138823041 | 0,906599546 | SNRPB2 | FALSE | Downregulated |
| 0,115786682 | 0,906599546 | TRAF3IP3 | FALSE | Upregulated |
| 0,136203437 | 0,906600985 | ANKS6 | FALSE | Upregulated |
| 0,111248636 | 0,906600985 | BBS9 | FALSE | Upregulated |
| 0,054265379 | 0,906600985 | DOPEY1 | FALSE | Upregulated |
| -0,112752576 | 0,906600985 | EIF2B3 | FALSE | Downregulated |
| -0,071923529 | 0,906600985 | IL12RB2 | FALSE | Downregulated |
| 0,099070212 | 0,906600985 | IMPA2 | FALSE | Upregulated |
| 0,072030568 | 0,906600985 | LOC146880 | FALSE | Upregulated |
| 0,138254936 | 0,906600985 | MAPT-IT1 | FALSE | Upregulated |
| -0,094579445 | 0,906600985 | NOM1 | FALSE | Downregulated |
| -0,136343165 | 0,906600985 | P2RY2 | FALSE | Downregulated |
| -0,135132788 | 0,906600985 | PCDH17 | FALSE | Downregulated |
| 0,180941632 | 0,906600985 | POU6F2-AS2 | FALSE | Upregulated |
| -0,064952177 | 0,906600985 | PPAP2B | FALSE | Downregulated |
| -0,081128298 | 0,906600985 | RNA28S5 | FALSE | Downregulated |
| -0,070058997 | 0,906600985 | RNF149 | FALSE | Downregulated |
| -0,549723263 | 0,906600985 | RP4-669L17.10 | FALSE | Downregulated |
| -0,072142717 | 0,906600985 | SH3BP5L | FALSE | Downregulated |
| 0,079076854 | 0,906600985 | SPATA1 | FALSE | Upregulated |
| -0,115942789 | 0,906600985 | TTC6 | FALSE | Downregulated |
| 0,069844481 | 0,906600985 | XPA | FALSE | Upregulated |
| -0,108954565 | 0,906600985 | ZDBF2 | FALSE | Downregulated |
| -0,066646698 | 0,906600985 | ZNF148 | FALSE | Downregulated |
| -0,253748525 | 0,906682772 | ANKHD1 | FALSE | Downregulated |
| 0,187489361 | 0,906682772 | DAPK2 | FALSE | Upregulated |
| -0,098099666 | 0,906682772 | FMO4 | FALSE | Downregulated |
| 0,095794917 | 0,906682772 | NCOA7 | FALSE | Upregulated |
| 0,062059721 | 0,906682772 | RNF168 | FALSE | Upregulated |
| 0,086769994 | 0,906682772 | RQCD1 | FALSE | Upregulated |
| 0,081352157 | 0,906799075 | STX10 | FALSE | Upregulated |
| -0,081206153 | 0,906962082 | CRELD1 | FALSE | Downregulated |
| -0,064036707 | 0,907271161 | RGL2 | FALSE | Downregulated |
| -0,093478419 | 0,907327552 | ADAMTS5 | FALSE | Downregulated |
| -0,094571949 | 0,907327552 | ATG3 | FALSE | Downregulated |
| -0,11056819 | 0,907327552 | CAPN10 | FALSE | Downregulated |
| 0,154089971 | 0,907327552 | CCDC102A | FALSE | Upregulated |
| -0,052689029 | 0,907327552 | CDS2 | FALSE | Downregulated |
| 0,06744163 | 0,907327552 | CHTOP | FALSE | Upregulated |
| 0,046282934 | 0,907327552 | CRKL | FALSE | Upregulated |
| -0,067711416 | 0,907327552 | FAM149A | FALSE | Downregulated |
| -0,116744687 | 0,907327552 | FOCAD | FALSE | Downregulated |
| 0,054670843 | 0,907327552 | HMHA1 | FALSE | Upregulated |
| -0,152150693 | 0,907327552 | KIAA1244 | FALSE | Downregulated |
| -0,070501548 | 0,907327552 | L3MBTL2 | FALSE | Downregulated |
| 0,169590749 | 0,907327552 | LOC100996433 | FALSE | Upregulated |
| 0,144213799 | 0,907327552 | LONRF2 | FALSE | Upregulated |
| 0,063299688 | 0,907327552 | MAPKAPK5-AS1 | FALSE | Upregulated |

| | | | | |
|---|---|---|---|---|
| 0,103151401 | 0,907327552 | ME2 | FALSE | Upregulated |
| -0,058673323 | 0,907327552 | MTHFR | FALSE | Downregulated |
| 0,072811316 | 0,907327552 | PLEKHG1 | FALSE | Upregulated |
| -0,098778745 | 0,907327552 | SYBU | FALSE | Downregulated |
| 0,051576925 | 0,907327552 | TFIP11 | FALSE | Upregulated |
| -0,066978603 | 0,907327552 | TRIM4 | FALSE | Downregulated |
| -0,056574163 | 0,907327552 | UNC13B | FALSE | Downregulated |
| -0,062761182 | 0,907327552 | ZNF445 | FALSE | Downregulated |
| 0,073884078 | 0,907327552 | ZNF654 | FALSE | Upregulated |
| 0,106889628 | 0,90733822 | MLLT11 | FALSE | Upregulated |
| 0,1288653 | 0,907430479 | EMR2 | FALSE | Upregulated |
| -0,032782127 | 0,907430479 | EP300 | FALSE | Downregulated |
| -0,104666865 | 0,907430479 | GNL3 | FALSE | Downregulated |
| 0,142753056 | 0,907430479 | HAGHL | FALSE | Upregulated |
| -0,088400344 | 0,907430479 | JDP2 | FALSE | Downregulated |
| 0,163991378 | 0,907430479 | LOC102724976 | FALSE | Upregulated |
| -0,075225082 | 0,907430479 | LZTS3 | FALSE | Downregulated |
| 0,13698715 | 0,907430479 | METTL8 | FALSE | Upregulated |
| -0,145946891 | 0,907430479 | POLR3D | FALSE | Downregulated |
| -0,088995374 | 0,907430479 | PPIP5K1 | FALSE | Downregulated |
| 0,136447307 | 0,907430479 | PRKX | FALSE | Upregulated |
| 0,233442301 | 0,907430479 | SLC5A12 | FALSE | Upregulated |
| -0,076286038 | 0,907430479 | SLFNL1-AS1 | FALSE | Downregulated |
| -0,061269316 | 0,907430479 | TOP1 | FALSE | Downregulated |
| 0,082406262 | 0,907430479 | TTBK2 | FALSE | Upregulated |
| -0,066046485 | 0,907430479 | VPS9D1 | FALSE | Downregulated |
| 0,084487018 | 0,907430479 | ZFYVE21 | FALSE | Upregulated |
| 0,103768425 | 0,907430479 | ZSCAN25 | FALSE | Upregulated |
| -0,114540425 | 0,907464452 | IFT172 | FALSE | Downregulated |
| 0,175598122 | 0,907638005 | PLCXD3 | FALSE | Upregulated |
| 0,123024613 | 0,907642965 | NDUFB3 | FALSE | Upregulated |
| -0,11397484 | 0,907642965 | PLGLA | FALSE | Downregulated |
| -0,072144524 | 0,907642965 | TANGO6 | FALSE | Downregulated |
| -0,149894269 | 0,907642965 | TIMM21 | FALSE | Downregulated |
| 0,082910129 | 0,907793335 | CLNS1A | FALSE | Upregulated |
| 0,060007772 | 0,907793335 | EIF3J | FALSE | Upregulated |
| 0,090040345 | 0,907793335 | EMC4 | FALSE | Upregulated |
| 0,052260294 | 0,907793335 | ITPR3 | FALSE | Upregulated |
| -0,148158722 | 0,907793335 | LOC399900 | FALSE | Downregulated |
| -0,116201947 | 0,907793335 | LOC400548 | FALSE | Downregulated |
| -0,087599015 | 0,907793335 | RAPGEF4 | FALSE | Downregulated |
| -0,139597256 | 0,907793335 | VMA21 | FALSE | Downregulated |
| 0,066187004 | 0,907921152 | EHMT2 | FALSE | Upregulated |
| -0,070670459 | 0,907921152 | MAGED2 | FALSE | Downregulated |
| 0,047991231 | 0,907921152 | POGZ | FALSE | Upregulated |
| -0,081958189 | 0,907921152 | PRRC1 | FALSE | Downregulated |
| -0,451480964 | 0,907921152 | STX16-NPEPL1 | FALSE | Downregulated |
| -0,112233476 | 0,907921152 | TSPAN7 | FALSE | Downregulated |
| -0,094013739 | 0,90809059 | R3HDM4 | FALSE | Downregulated |
| 0,251341452 | 0,908102694 | CHI3L1 | FALSE | Upregulated |
| -0,079686553 | 0,908102694 | PARP12 | FALSE | Downregulated |

| | | | | |
|---|---|---|---|---|
| -0,111696341 | 0,908102694 | PPP2R5B | FALSE | Downregulated |
| -0,087948044 | 0,908217705 | FAM89B | FALSE | Downregulated |
| -0,100328708 | 0,908217705 | MAGI2 | FALSE | Downregulated |
| -0,052429544 | 0,908217705 | MAN2B1 | FALSE | Downregulated |
| -0,070523456 | 0,908217705 | PLGLB2 | FALSE | Downregulated |
| 0,082975948 | 0,908217705 | TTYH1 | FALSE | Upregulated |
| -0,093707535 | 0,908217705 | TXNL4A | FALSE | Downregulated |
| 0,072649133 | 0,908228273 | KXD1 | FALSE | Upregulated |
| 0,05478956 | 0,908228273 | WSB2 | FALSE | Upregulated |
| 0,06020297 | 0,908316345 | FBXO38 | FALSE | Upregulated |
| 0,166814739 | 0,908346935 | FMOD | FALSE | Upregulated |
| 0,107135434 | 0,908346935 | THOC1 | FALSE | Upregulated |
| 0,109812607 | 0,908476988 | SYTL2 | FALSE | Upregulated |
| -0,061828569 | 0,908773916 | KPNA4 | FALSE | Downregulated |
| 0,14651648 | 0,909094851 | C6orf132 | FALSE | Upregulated |
| -1,356081878 | 0,909094851 | GJA9 | FALSE | Downregulated |
| 0,072618285 | 0,909094851 | KIF28P | FALSE | Upregulated |
| -0,05651354 | 0,909177382 | KIAA0947 | FALSE | Downregulated |
| 0,132721086 | 0,909278339 | RNF113A | FALSE | Upregulated |
| -0,070885562 | 0,909389713 | C5orf24 | FALSE | Downregulated |
| -0,153597808 | 0,909445958 | LOC100506060 | FALSE | Downregulated |
| -0,061608798 | 0,909472057 | ACTR1B | FALSE | Downregulated |
| -0,103815701 | 0,909472057 | ARHGEF37 | FALSE | Downregulated |
| -0,042947603 | 0,909472057 | CREBZF | FALSE | Downregulated |
| 0,1097363 | 0,909472057 | LOC102723772 | FALSE | Upregulated |
| -0,101355515 | 0,909472057 | PHF5A | FALSE | Downregulated |
| 0,069213488 | 0,909472057 | TNFRSF1B | FALSE | Upregulated |
| 0,076244516 | 0,909500114 | C17orf62 | FALSE | Upregulated |
| -0,101920878 | 0,909500114 | NAA16 | FALSE | Downregulated |
| 0,08871657 | 0,90963323 | RAB21 | FALSE | Upregulated |
| -0,117811366 | 0,910261605 | ATP5J2 | FALSE | Downregulated |
| 0,075177142 | 0,910528046 | ALS2CL | FALSE | Upregulated |
| -0,105951939 | 0,910528046 | C9orf16 | FALSE | Downregulated |
| -0,101607711 | 0,910528046 | COPS7B | FALSE | Downregulated |
| 0,094608986 | 0,910528046 | FBXO22 | FALSE | Upregulated |
| -0,108430457 | 0,910528046 | GREB1 | FALSE | Downregulated |
| -0,041795746 | 0,910528046 | KDM5A | FALSE | Downregulated |
| -0,040648268 | 0,910528046 | NEK9 | FALSE | Downregulated |
| -0,052914705 | 0,910528046 | NOTCH2 | FALSE | Downregulated |
| -0,416446476 | 0,910528046 | TCEB3C | FALSE | Downregulated |
| -0,081019343 | 0,910528046 | UBAP2 | FALSE | Downregulated |
| 0,152032084 | 0,910528046 | UNC93A | FALSE | Upregulated |
| -0,07444557 | 0,910528046 | VTI1B | FALSE | Downregulated |
| -0,073579767 | 0,910528046 | ZC3H13 | FALSE | Downregulated |
| 0,178459529 | 0,910571422 | CLIP4 | FALSE | Upregulated |
| -0,071285694 | 0,91069259 | PANX1 | FALSE | Downregulated |
| -0,115686344 | 0,910814172 | ZNF414 | FALSE | Downregulated |
| 0,043171737 | 0,910931923 | SNX1 | FALSE | Upregulated |
| 0,099024124 | 0,911076938 | ZNRF2P1 | FALSE | Upregulated |
| 0,108461661 | 0,911207358 | LOC728613 | FALSE | Upregulated |
| -0,096741394 | 0,911377262 | ALDH1L1-AS2 | FALSE | Downregulated |

| | | | | |
|---|---|---|---|---|
| 0,058876146 | 0,911377262 | DCAF7 | FALSE | Upregulated |
| -0,12186003 | 0,911377262 | ZNF101 | FALSE | Downregulated |
| 0,055620694 | 0,911605775 | SNW1 | FALSE | Upregulated |
| 0,097866616 | 0,911840165 | C1orf85 | FALSE | Upregulated |
| -0,058105401 | 0,911849953 | LMO7 | FALSE | Downregulated |
| 0,139138418 | 0,911849953 | SNAPC2 | FALSE | Upregulated |
| -0,067200017 | 0,911849953 | WDR5 | FALSE | Downregulated |
| 0,086902252 | 0,912219691 | FZD1 | FALSE | Upregulated |
| -0,076987775 | 0,912219691 | GNAI3 | FALSE | Downregulated |
| -0,124094392 | 0,912219691 | SLC13A3 | FALSE | Downregulated |
| -0,083289881 | 0,912325048 | MAD1L1 | FALSE | Downregulated |
| -0,05185334 | 0,91233771 | DIP2C | FALSE | Downregulated |
| 0,062702803 | 0,912463321 | AVIL | FALSE | Upregulated |
| -0,131226851 | 0,912463321 | FAM122C | FALSE | Downregulated |
| 0,04028435 | 0,912463321 | MAPK8IP3 | FALSE | Upregulated |
| -0,112989079 | 0,912463321 | NAT1 | FALSE | Downregulated |
| -0,073165708 | 0,912463321 | NT5DC1 | FALSE | Downregulated |
| 0,079019727 | 0,912463321 | RBAK | FALSE | Upregulated |
| 0,12879412 | 0,912463321 | SHC3 | FALSE | Upregulated |
| -0,096560321 | 0,912764895 | COL12A1 | FALSE | Downregulated |
| 0,090830198 | 0,912764895 | DAPK3 | FALSE | Upregulated |
| -0,073585656 | 0,912764895 | GAS2L1 | FALSE | Downregulated |
| -0,090574959 | 0,912764895 | GOLGA8O | FALSE | Downregulated |
| -0,049664448 | 0,912777998 | CFLAR | FALSE | Downregulated |
| -0,076819361 | 0,912777998 | HRNR | FALSE | Downregulated |
| 0,101610609 | 0,912777998 | MFSD1 | FALSE | Upregulated |
| 0,073280595 | 0,912777998 | STRN | FALSE | Upregulated |
| 0,115781335 | 0,912809173 | HLA-H | FALSE | Upregulated |
| 0,0887773 | 0,912968859 | TMEM167B | FALSE | Upregulated |
| 0,142782781 | 0,913156761 | LOC102724286 | FALSE | Upregulated |
| 0,161952385 | 0,913156761 | MMP23B | FALSE | Upregulated |
| 0,078953958 | 0,913156761 | SERHL | FALSE | Upregulated |
| 0,044110272 | 0,913156761 | TRIM14 | FALSE | Upregulated |
| 0,128677105 | 0,913170961 | MED18 | FALSE | Upregulated |
| 0,058226 | 0,913381244 | ATF7IP | FALSE | Upregulated |
| 0,047032111 | 0,913381244 | SRP9 | FALSE | Upregulated |
| 0,147748029 | 0,913438591 | BRIX1 | FALSE | Upregulated |
| 0,03973656 | 0,913567007 | USP22 | FALSE | Upregulated |
| 0,110497689 | 0,913567007 | ZNF717 | FALSE | Upregulated |
| -0,122384253 | 0,913587871 | PIANP | FALSE | Downregulated |
| -0,100185492 | 0,913620468 | AATF | FALSE | Downregulated |
| 0,140495293 | 0,913620468 | CCDC149 | FALSE | Upregulated |
| -0,038454306 | 0,913620468 | HNRNPK | FALSE | Downregulated |
| -0,115660462 | 0,913620468 | PHKA1 | FALSE | Downregulated |
| -0,133474246 | 0,913620468 | UTP6 | FALSE | Downregulated |
| 0,093476822 | 0,913700267 | BOC | FALSE | Upregulated |
| 0,099727561 | 0,913700267 | FAM206A | FALSE | Upregulated |
| 0,116038499 | 0,913700267 | FFAR4 | FALSE | Upregulated |
| 0,053382835 | 0,913700267 | GZF1 | FALSE | Upregulated |
| -0,045070476 | 0,913700267 | MAST3 | FALSE | Downregulated |
| -0,092316431 | 0,913700267 | NPC2 | FALSE | Downregulated |

| | | | | |
|---|---|---|---|---|
| -0,103925616 | 0,913700267 | POLD1 | FALSE | Downregulated |
| -0,076096604 | 0,913700267 | RAB35 | FALSE | Downregulated |
| -0,094883885 | 0,913700267 | VCAM1 | FALSE | Downregulated |
| -0,074989361 | 0,913800575 | LOC101928081 | FALSE | Downregulated |
| 0,079860991 | 0,913871318 | NLRC4 | FALSE | Upregulated |
| 0,054225196 | 0,914103741 | MARK3 | FALSE | Upregulated |
| -0,11980731 | 0,914191212 | ICT1 | FALSE | Downregulated |
| -0,055959187 | 0,914208868 | PTPRA | FALSE | Downregulated |
| -0,03307218 | 0,914208868 | RAD23B | FALSE | Downregulated |
| 0,095108772 | 0,914347982 | ABHD14A | FALSE | Upregulated |
| 0,072462363 | 0,914347982 | ATP6AP2 | FALSE | Upregulated |
| -0,11904446 | 0,914347982 | FAM99B | FALSE | Downregulated |
| 0,119186735 | 0,914347982 | LOC100134822 | FALSE | Upregulated |
| -0,061532295 | 0,914347982 | MAP7 | FALSE | Downregulated |
| 0,093596802 | 0,914347982 | PLEKHM3 | FALSE | Upregulated |
| -0,10761835 | 0,914347982 | RBMS3 | FALSE | Downregulated |
| -0,118606896 | 0,914347982 | SLC41A3 | FALSE | Downregulated |
| -0,089596408 | 0,914347982 | TMEM179B | FALSE | Downregulated |
| 0,059084257 | 0,914347982 | ZNF142 | FALSE | Upregulated |
| -0,052722069 | 0,914445252 | RAB17 | FALSE | Downregulated |
| 0,089902836 | 0,91450491 | NDUFA5 | FALSE | Upregulated |
| -0,058993806 | 0,91450491 | ZBTB38 | FALSE | Downregulated |
| 0,086211146 | 0,914592659 | SLC2A10 | FALSE | Upregulated |
| 0,09357236 | 0,914955169 | CARD8 | FALSE | Upregulated |
| 0,128261611 | 0,915122101 | IRAK3 | FALSE | Upregulated |
| -0,07517198 | 0,915275622 | ABL2 | FALSE | Downregulated |
| 0,127408215 | 0,915275622 | LETM2 | FALSE | Upregulated |
| -0,082733526 | 0,915275622 | LINC01061 | FALSE | Downregulated |
| -0,128230556 | 0,915275622 | MAP2 | FALSE | Downregulated |
| 0,118503619 | 0,915275622 | RAB8B | FALSE | Upregulated |
| 0,083341536 | 0,915275622 | SCAMP1 | FALSE | Upregulated |
| -0,187760279 | 0,915275622 | SGCD | FALSE | Downregulated |
| -0,093556762 | 0,915718799 | DEDD2 | FALSE | Downregulated |
| -0,136407912 | 0,915718799 | FGR | FALSE | Downregulated |
| 0,108970516 | 0,915718799 | KCTD15 | FALSE | Upregulated |
| -0,12758624 | 0,915733746 | BBS10 | FALSE | Downregulated |
| 0,049860051 | 0,916713512 | ABLIM1 | FALSE | Upregulated |
| -0,132709071 | 0,916713512 | CAAP1 | FALSE | Downregulated |
| -0,073664078 | 0,916713512 | CEP70 | FALSE | Downregulated |
| 0,069405468 | 0,916713512 | LOC100288846 | FALSE | Upregulated |
| 0,066823701 | 0,916713512 | NCKAP5L | FALSE | Upregulated |
| -0,060153343 | 0,916713512 | PACSIN2 | FALSE | Downregulated |
| 0,058353985 | 0,916713512 | POLG | FALSE | Upregulated |
| -0,062597785 | 0,916713512 | RNPEP | FALSE | Downregulated |
| -0,099316889 | 0,916713512 | SEL1L3 | FALSE | Downregulated |
| 0,087176259 | 0,916713512 | TAC4 | FALSE | Upregulated |
| 0,058357681 | 0,916713512 | TSSK3 | FALSE | Upregulated |
| -0,131400992 | 0,916802674 | MYCT1 | FALSE | Downregulated |
| -0,095781274 | 0,916980644 | ADCK4 | FALSE | Downregulated |
| 0,080100709 | 0,916980644 | ARPC1B | FALSE | Upregulated |
| -0,091221769 | 0,916980644 | ATRAID | FALSE | Downregulated |

| | | | | |
|---|---|---|---|---|
| 0,171301425 | 0,916980644 | BMI1 | FALSE | Upregulated |
| 0,091813709 | 0,916980644 | C17orf107 | FALSE | Upregulated |
| 0,089892553 | 0,916980644 | DYNLL1-AS1 | FALSE | Upregulated |
| -0,143923091 | 0,916980644 | FAM172A | FALSE | Downregulated |
| -0,079180296 | 0,916980644 | GXYLT1 | FALSE | Downregulated |
| -0,073195776 | 0,916980644 | LSM14A | FALSE | Downregulated |
| -0,095738126 | 0,916980644 | MAL2 | FALSE | Downregulated |
| 0,091325881 | 0,916980644 | NGLY1 | FALSE | Upregulated |
| 0,079110456 | 0,916980644 | NT5C3B | FALSE | Upregulated |
| 0,123439633 | 0,916980644 | PLEKHG5 | FALSE | Upregulated |
| -0,094427182 | 0,916980644 | PPARGC1B | FALSE | Downregulated |
| -0,148798614 | 0,916980644 | RBM48 | FALSE | Downregulated |
| -0,070982417 | 0,916980644 | RING1 | FALSE | Downregulated |
| -0,096880415 | 0,916980644 | SHFM1 | FALSE | Downregulated |
| -0,07356785 | 0,916980644 | SMC4 | FALSE | Downregulated |
| 0,090590144 | 0,917097577 | PLCB1 | FALSE | Upregulated |
| 0,113187817 | 0,917097577 | UBTD2 | FALSE | Upregulated |
| -0,069686759 | 0,917180168 | ALKBH7 | FALSE | Downregulated |
| 0,078350414 | 0,917180168 | BTG3 | FALSE | Upregulated |
| -0,125420085 | 0,917180168 | FAM96B | FALSE | Downregulated |
| -0,143769651 | 0,917180168 | LOC102724322 | FALSE | Downregulated |
| -0,133054164 | 0,917180168 | MED6 | FALSE | Downregulated |
| -0,089475675 | 0,917180168 | MRPS21 | FALSE | Downregulated |
| 0,052233852 | 0,917180168 | PALLD | FALSE | Upregulated |
| 0,094021799 | 0,917180168 | RALGDS | FALSE | Upregulated |
| 0,064329375 | 0,917180168 | SLC13A4 | FALSE | Upregulated |
| -0,043808811 | 0,917180168 | SLC25A13 | FALSE | Downregulated |
| 0,085933038 | 0,917180168 | SPICE1 | FALSE | Upregulated |
| 0,049926524 | 0,917180168 | STAT2 | FALSE | Upregulated |
| 0,118652007 | 0,917180168 | STXBP1 | FALSE | Upregulated |
| -0,069588577 | 0,917180168 | TIGD5 | FALSE | Downregulated |
| 0,115405389 | 0,917180168 | ZC3HC1 | FALSE | Upregulated |
| -0,144362246 | 0,917180168 | ZNF585A | FALSE | Downregulated |
| -0,04556035 | 0,917268528 | MYO1E | FALSE | Downregulated |
| 0,073353106 | 0,917274496 | DHRS7 | FALSE | Upregulated |
| -0,054194422 | 0,917327339 | AKAP8L | FALSE | Downregulated |
| 0,146739609 | 0,917343446 | GYG1 | FALSE | Upregulated |
| -0,182879304 | 0,91744289 | A4GNT | FALSE | Downregulated |
| -0,12190088 | 0,917607555 | CCDC107 | FALSE | Downregulated |
| -0,04606535 | 0,917607555 | COL27A1 | FALSE | Downregulated |
| 0,045732662 | 0,917607555 | GBF1 | FALSE | Upregulated |
| -0,058900532 | 0,917607555 | GFPT1 | FALSE | Downregulated |
| 0,129443788 | 0,917607555 | GRAMD1B | FALSE | Upregulated |
| -0,073857427 | 0,917607555 | GUSBP11 | FALSE | Downregulated |
| -0,066639587 | 0,917607555 | INO80E | FALSE | Downregulated |
| 0,123911598 | 0,917607555 | LSM10 | FALSE | Upregulated |
| -0,071232845 | 0,917607555 | NEURL2 | FALSE | Downregulated |
| -0,102715843 | 0,917607555 | SLC35B1 | FALSE | Downregulated |
| -0,083289054 | 0,917612822 | FAM171A1 | FALSE | Downregulated |
| -0,074303328 | 0,917612822 | ZFYVE20 | FALSE | Downregulated |
| -0,124932661 | 0,917657228 | GTF2IRD1 | FALSE | Downregulated |

| | | | | |
|---|---|---|---|---|
| 0,04254942 | 0,917657228 | PAPOLA | FALSE | Upregulated |
| 0,044916964 | 0,917657228 | RCN1 | FALSE | Upregulated |
| -0,060485341 | 0,917657228 | TTC38 | FALSE | Downregulated |
| -0,11002314 | 0,917657228 | ZNF707 | FALSE | Downregulated |
| 0,063756233 | 0,917776894 | ANKIB1 | FALSE | Upregulated |
| 0,039324939 | 0,917776894 | ATXN2L | FALSE | Upregulated |
| -0,053605039 | 0,917776894 | COL6A1 | FALSE | Downregulated |
| -0,043806838 | 0,917776894 | IARS2 | FALSE | Downregulated |
| 0,094156326 | 0,917776894 | LOC101928502 | FALSE | Upregulated |
| -0,113794803 | 0,917776894 | RCBTB2 | FALSE | Downregulated |
| 0,086415017 | 0,917776894 | TIPARP-AS1 | FALSE | Upregulated |
| 0,141086987 | 0,917776894 | TTC39A | FALSE | Upregulated |
| -0,058148125 | 0,917943058 | MRPL41 | FALSE | Downregulated |
| 0,068017532 | 0,917943058 | WDR90 | FALSE | Upregulated |
| -0,053503177 | 0,917984931 | ANXA11 | FALSE | Downregulated |
| -0,070078823 | 0,917984931 | B3GNT8 | FALSE | Downregulated |
| 0,052074781 | 0,917984931 | CACYBP | FALSE | Upregulated |
| -0,068826219 | 0,917984931 | CALU | FALSE | Downregulated |
| 0,067054979 | 0,917984931 | CYP2D6 | FALSE | Upregulated |
| 0,094767945 | 0,917984931 | DYNC1LI1 | FALSE | Upregulated |
| 0,053702126 | 0,917984931 | FBXL5 | FALSE | Upregulated |
| 0,071443727 | 0,917984931 | FLYWCH1 | FALSE | Upregulated |
| 0,107783351 | 0,917984931 | GTF2H4 | FALSE | Upregulated |
| -0,13006068 | 0,917984931 | HSPA12B | FALSE | Downregulated |
| 0,095709917 | 0,917984931 | LOC101928098 | FALSE | Upregulated |
| 0,068729959 | 0,917984931 | LOC102723863 | FALSE | Upregulated |
| -0,046533572 | 0,917984931 | NCLN | FALSE | Downregulated |
| 0,113622329 | 0,917984931 | NECAB1 | FALSE | Upregulated |
| 0,058598363 | 0,917984931 | NINJ1 | FALSE | Upregulated |
| 0,115332577 | 0,917984931 | ORAOV1 | FALSE | Upregulated |
| -0,06683476 | 0,917984931 | PSMA5 | FALSE | Downregulated |
| 0,104701427 | 0,917984931 | RASSF1 | FALSE | Upregulated |
| -0,10614519 | 0,917984931 | RFWD3 | FALSE | Downregulated |
| -0,120000906 | 0,917984931 | RNASE1 | FALSE | Downregulated |
| 0,067869823 | 0,917984931 | RP11-219B17.1 | FALSE | Upregulated |
| -0,0657613 | 0,917984931 | RPS26 | FALSE | Downregulated |
| -0,04470372 | 0,917984931 | SEPT2 | FALSE | Downregulated |
| -0,137220423 | 0,917984931 | SMG8 | FALSE | Downregulated |
| -0,075435074 | 0,917984931 | SPAG7 | FALSE | Downregulated |
| -0,149956196 | 0,917984931 | SPATA7 | FALSE | Downregulated |
| -0,05106664 | 0,917984931 | XPO6 | FALSE | Downregulated |
| -0,093220254 | 0,917984931 | ZNF436 | FALSE | Downregulated |
| -0,148225852 | 0,917984931 | ZNF597 | FALSE | Downregulated |
| -0,104458346 | 0,918012084 | ENAH | FALSE | Downregulated |
| 0,096485659 | 0,918012084 | GTF2H2B | FALSE | Upregulated |
| -0,106650824 | 0,918012084 | LCA5L | FALSE | Downregulated |
| -0,097513906 | 0,918012084 | LOC100132832 | FALSE | Downregulated |
| 0,056077542 | 0,918012084 | LOC100133091 | FALSE | Upregulated |
| 0,127226101 | 0,918012084 | NDUFC1 | FALSE | Upregulated |
| -0,085903047 | 0,918012084 | RHBDD3 | FALSE | Downregulated |
| 0,115880242 | 0,918095752 | AQP1 | FALSE | Upregulated |

| | | | | |
|---|---|---|---|---|
| 0,123137196 | 0,918095752 | HEYL | FALSE | Upregulated |
| -0,09242911 | 0,918118872 | CXorf38 | FALSE | Downregulated |
| 0,116879898 | 0,918133725 | NREP | FALSE | Upregulated |
| 0,129014559 | 0,918133725 | PUSL1 | FALSE | Upregulated |
| 0,03508096 | 0,91823557 | KHNYN | FALSE | Upregulated |
| 0,133634041 | 0,918275302 | ETHE1 | FALSE | Upregulated |
| 0,119786686 | 0,918442964 | KLHDC8B | FALSE | Upregulated |
| 0,042633028 | 0,918442964 | VDAC1 | FALSE | Upregulated |
| -0,049399098 | 0,918668942 | ARRDC1 | FALSE | Downregulated |
| 0,09700536 | 0,918668942 | LOC101929522 | FALSE | Upregulated |
| 0,052501613 | 0,918706929 | MXD4 | FALSE | Upregulated |
| -0,085106608 | 0,918774693 | AHI1 | FALSE | Downregulated |
| -0,116671567 | 0,918774693 | DCUN1D2 | FALSE | Downregulated |
| 0,081920862 | 0,918774693 | FBXO32 | FALSE | Upregulated |
| 0,043170538 | 0,918774693 | GAPDH | FALSE | Upregulated |
| 0,073268714 | 0,918774693 | GFOD2 | FALSE | Upregulated |
| 0,080986222 | 0,918774693 | NTSR1 | FALSE | Upregulated |
| -0,130643715 | 0,918774693 | RP11-521B24.3 | FALSE | Downregulated |
| -0,053631059 | 0,918774693 | SERINC5 | FALSE | Downregulated |
| 0,088532668 | 0,918774693 | SF3B4 | FALSE | Upregulated |
| 0,134640568 | 0,918774693 | UTP14A | FALSE | Upregulated |
| -0,06772105 | 0,918868412 | RTF1 | FALSE | Downregulated |
| -0,063954882 | 0,918868412 | TMED9 | FALSE | Downregulated |
| 0,087809321 | 0,918882621 | ZNF614 | FALSE | Upregulated |
| -0,050938182 | 0,918921837 | CALR | FALSE | Downregulated |
| 0,055818413 | 0,918921837 | CDH1 | FALSE | Upregulated |
| -0,079931809 | 0,918921837 | GDE1 | FALSE | Downregulated |
| -0,043948671 | 0,918921837 | OSBPL9 | FALSE | Downregulated |
| -0,04715375 | 0,918921837 | PPP1CB | FALSE | Downregulated |
| -0,1064568 | 0,918921837 | ROGDI | FALSE | Downregulated |
| -0,106565269 | 0,918921837 | SFRP5 | FALSE | Downregulated |
| 0,133572036 | 0,918921837 | ST6GAL2 | FALSE | Upregulated |
| -0,127171922 | 0,919018568 | ZNF10 | FALSE | Downregulated |
| 0,068590068 | 0,919080112 | TEAD3 | FALSE | Upregulated |
| -0,094256639 | 0,919114471 | CDKN2AIPNL | FALSE | Downregulated |
| -0,05461017 | 0,91914399 | ARID5A | FALSE | Downregulated |
| -0,067134202 | 0,91920339 | LOC101927208 | FALSE | Downregulated |
| 0,033767504 | 0,919269319 | ADAR | FALSE | Upregulated |
| -0,078260543 | 0,919269319 | CAPN3 | FALSE | Downregulated |
| -0,114881576 | 0,919269319 | ELP6 | FALSE | Downregulated |
| 0,080097873 | 0,919269319 | JRK | FALSE | Upregulated |
| 0,062049183 | 0,919269319 | LHPP | FALSE | Upregulated |
| 0,194144162 | 0,919269319 | LOC389831 | FALSE | Upregulated |
| 0,06127543 | 0,919269319 | MAPK6 | FALSE | Upregulated |
| 0,057312039 | 0,919269319 | SLC8B1 | FALSE | Upregulated |
| 0,054137713 | 0,919269319 | STRN3 | FALSE | Upregulated |
| 0,064673538 | 0,919269319 | TRMT5 | FALSE | Upregulated |
| 0,040869455 | 0,919269319 | ZNRF2 | FALSE | Upregulated |
| -0,094520684 | 0,919367205 | GTF3C4 | FALSE | Downregulated |
| -0,047436482 | 0,919771393 | DECR2 | FALSE | Downregulated |
| -0,103630768 | 0,919874976 | ZNF473 | FALSE | Downregulated |

| | | | | |
|---|---|---|---|---|
| 0,104273411 | 0,919969785 | LOC102723758 | FALSE | Upregulated |
| 0,037995627 | 0,919969785 | YY1 | FALSE | Upregulated |
| 0,042574271 | 0,920026238 | CS | FALSE | Upregulated |
| 0,082981246 | 0,920026238 | TBC1D15 | FALSE | Upregulated |
| -0,057297726 | 0,920026238 | UQCRC2 | FALSE | Downregulated |
| 0,046179808 | 0,920026238 | ZNF3 | FALSE | Upregulated |
| 0,09287063 | 0,920083014 | MCM5 | FALSE | Upregulated |
| -0,158519357 | 0,920155441 | SPAG16 | FALSE | Downregulated |
| 0,039944927 | 0,920229601 | MAT2A | FALSE | Upregulated |
| 0,073887686 | 0,920241111 | AHCYL2 | FALSE | Upregulated |
| -0,122712972 | 0,920241111 | C9orf72 | FALSE | Downregulated |
| -0,102099105 | 0,920241111 | CACNA1A | FALSE | Downregulated |
| 0,080976359 | 0,920241111 | CNOT4 | FALSE | Upregulated |
| -0,10326125 | 0,920241111 | FAM126A | FALSE | Downregulated |
| 0,127970212 | 0,920241111 | FLJ46836 | FALSE | Upregulated |
| -0,047740885 | 0,920241111 | GPI | FALSE | Downregulated |
| 0,173723636 | 0,920241111 | HSPA6 | FALSE | Upregulated |
| 0,124296111 | 0,920241111 | LSM1 | FALSE | Upregulated |
| -0,137734347 | 0,920241111 | NABP2 | FALSE | Downregulated |
| 0,037245023 | 0,920241111 | PTP4A1 | FALSE | Upregulated |
| 0,043807609 | 0,920241111 | RORA | FALSE | Upregulated |
| -0,117008185 | 0,920241111 | SCN3A | FALSE | Downregulated |
| -0,066734312 | 0,920241111 | ZBTB39 | FALSE | Downregulated |
| -0,091281413 | 0,920326548 | MXRA5 | FALSE | Downregulated |
| 0,049651042 | 0,920449067 | API5 | FALSE | Upregulated |
| -0,094157387 | 0,920449067 | IER3IP1 | FALSE | Downregulated |
| 0,07402679 | 0,920449067 | RIPK4 | FALSE | Upregulated |
| -0,078597578 | 0,920464639 | UBE2Q1 | FALSE | Downregulated |
| 0,068719801 | 0,920481153 | GLT8D1 | FALSE | Upregulated |
| 0,07482692 | 0,920568873 | DPY19L4 | FALSE | Upregulated |
| 0,109365156 | 0,920568873 | PTPLB | FALSE | Upregulated |
| 0,069353008 | 0,920568873 | PTPN9 | FALSE | Upregulated |
| 0,084168379 | 0,920568873 | RASA3 | FALSE | Upregulated |
| 0,10102367 | 0,920568873 | SMPD5 | FALSE | Upregulated |
| -0,078674661 | 0,920678756 | GPR113 | FALSE | Downregulated |
| -0,032931839 | 0,920678756 | SIGMAR1 | FALSE | Downregulated |
| 0,132165307 | 0,920678756 | ZNF443 | FALSE | Upregulated |
| -0,078851695 | 0,920763241 | CMAS | FALSE | Downregulated |
| 0,109227064 | 0,920763241 | EDAR | FALSE | Upregulated |
| -0,095787473 | 0,920763241 | ZCRB1 | FALSE | Downregulated |
| 0,063433417 | 0,920979177 | N4BP1 | FALSE | Upregulated |
| 0,101510455 | 0,921063111 | NSL1 | FALSE | Upregulated |
| -0,069709262 | 0,921063111 | ZNF764 | FALSE | Downregulated |
| -0,092336891 | 0,921099517 | AGAP9 | FALSE | Downregulated |
| -0,050703926 | 0,921099517 | HMG20B | FALSE | Downregulated |
| -0,120351113 | 0,921099517 | NSRP1 | FALSE | Downregulated |
| 0,050659248 | 0,921099517 | PHYKPL | FALSE | Upregulated |
| 0,069583205 | 0,921099517 | ZNF292 | FALSE | Upregulated |
| 0,107743696 | 0,921226107 | ANP32AP1 | FALSE | Upregulated |
| -0,177623209 | 0,921226107 | ARPC4-TTLL3 | FALSE | Downregulated |
| -0,065473995 | 0,921226107 | BIRC2 | FALSE | Downregulated |

| | | | | |
|---|---|---|---|---|
| 0,119815495 | 0,921226107 | CISD1 | FALSE | Upregulated |
| -0,07328163 | 0,921226107 | CLPB | FALSE | Downregulated |
| -0,088798598 | 0,921226107 | DDX55 | FALSE | Downregulated |
| -0,083480468 | 0,921226107 | DDX60L | FALSE | Downregulated |
| 0,114491945 | 0,921226107 | FBLN5 | FALSE | Upregulated |
| 0,044859651 | 0,921226107 | HNF1A | FALSE | Upregulated |
| 0,059240207 | 0,921226107 | LEMD2 | FALSE | Upregulated |
| -0,072791733 | 0,921226107 | LIMA1 | FALSE | Downregulated |
| 0,07348757 | 0,921226107 | LOC101928917 | FALSE | Upregulated |
| 0,053619812 | 0,921226107 | LOC101929844 | FALSE | Upregulated |
| -0,114082339 | 0,921226107 | LRRTM3 | FALSE | Downregulated |
| 0,144150147 | 0,921226107 | MBOAT1 | FALSE | Upregulated |
| 0,100430097 | 0,921226107 | MCU | FALSE | Upregulated |
| 0,049292509 | 0,921226107 | MED16 | FALSE | Upregulated |
| 0,110936236 | 0,921226107 | NSDHL | FALSE | Upregulated |
| -0,053507991 | 0,921226107 | SLC9A8 | FALSE | Downregulated |
| -0,102819903 | 0,921226107 | SRBD1 | FALSE | Downregulated |
| -0,130363708 | 0,921226107 | UBA6-AS1 | FALSE | Downregulated |
| 0,077727855 | 0,921226107 | WDR45 | FALSE | Upregulated |
| -0,115698858 | 0,921300604 | NCK1 | FALSE | Downregulated |
| 0,064092236 | 0,921300604 | SMC5 | FALSE | Upregulated |
| 0,038187844 | 0,921300604 | SMEK2 | FALSE | Upregulated |
| 0,313774096 | 0,921445011 | INO80B-WBP1 | FALSE | Upregulated |
| 0,115202938 | 0,921445011 | NT5DC2 | FALSE | Upregulated |
| -0,060338794 | 0,921664193 | PITPNB | FALSE | Downregulated |
| 0,083302461 | 0,921664193 | PREP | FALSE | Upregulated |
| -0,103645886 | 0,921677317 | COX7B | FALSE | Downregulated |
| 0,076310099 | 0,921677317 | LOC101927989 | FALSE | Upregulated |
| -0,125113387 | 0,921872106 | ANXA10 | FALSE | Downregulated |
| 0,056752504 | 0,921872106 | APOBR | FALSE | Upregulated |
| -0,051373391 | 0,921872106 | ATE1 | FALSE | Downregulated |
| -0,113870783 | 0,921872106 | C6orf25 | FALSE | Downregulated |
| -0,182354537 | 0,921872106 | CBWD1 | FALSE | Downregulated |
| 0,125492334 | 0,921872106 | CEP152 | FALSE | Upregulated |
| -0,115409602 | 0,921872106 | COG6 | FALSE | Downregulated |
| -0,093150466 | 0,921872106 | JMJD6 | FALSE | Downregulated |
| -0,075445196 | 0,921872106 | LOC101927451 | FALSE | Downregulated |
| 0,08232754 | 0,921872106 | LOC101927644 | FALSE | Upregulated |
| 0,117493654 | 0,921872106 | LOC102723701 | FALSE | Upregulated |
| -0,141199488 | 0,921872106 | LOC102724488 | FALSE | Downregulated |
| -0,079800322 | 0,921872106 | MCF2L | FALSE | Downregulated |
| 0,14294888 | 0,921872106 | OTUD6B | FALSE | Upregulated |
| -0,109752617 | 0,921872106 | PCOLCE-AS1 | FALSE | Downregulated |
| -0,232325134 | 0,921872106 | PWP2 | FALSE | Downregulated |
| 0,053480342 | 0,921872106 | TARBP2 | FALSE | Upregulated |
| -0,117253552 | 0,921872106 | THBD | FALSE | Downregulated |
| -0,062040486 | 0,921872106 | TUBA4A | FALSE | Downregulated |
| -0,070729741 | 0,921872106 | ZNF580 | FALSE | Downregulated |
| -0,071274965 | 0,922092984 | AFTPH | FALSE | Downregulated |
| -0,059530346 | 0,922092984 | ATL2 | FALSE | Downregulated |
| 0,065687337 | 0,922092984 | BCL7B | FALSE | Upregulated |

| | | | | |
|---|---|---|---|---|
| -0,086724094 | 0,922092984 | COPS3 | FALSE | Downregulated |
| -0,047040615 | 0,922092984 | CP | FALSE | Downregulated |
| -0,050594293 | 0,922092984 | CPD | FALSE | Downregulated |
| 0,066552322 | 0,922092984 | FGD1 | FALSE | Upregulated |
| -0,051866371 | 0,922092984 | FXR2 | FALSE | Downregulated |
| -0,062004303 | 0,922092984 | G3BP2 | FALSE | Downregulated |
| 0,097414919 | 0,922092984 | LPPR1 | FALSE | Upregulated |
| -0,121116939 | 0,922092984 | NIFK-AS1 | FALSE | Downregulated |
| 0,070541325 | 0,922092984 | NSG1 | FALSE | Upregulated |
| 0,042849322 | 0,922092984 | PXN | FALSE | Upregulated |
| 0,10381575 | 0,922092984 | SKAP1 | FALSE | Upregulated |
| -0,092290032 | 0,922092984 | SPRY1 | FALSE | Downregulated |
| -0,087861442 | 0,922092984 | VPS26A | FALSE | Downregulated |
| 0,033331963 | 0,922092984 | YBX1 | FALSE | Upregulated |
| 0,064705899 | 0,922092984 | ZNF518A | FALSE | Upregulated |
| 0,065718507 | 0,922115768 | DNAJC19 | FALSE | Upregulated |
| -0,110811548 | 0,922115768 | QRSL1 | FALSE | Downregulated |
| -0,070843032 | 0,922115768 | SAMD10 | FALSE | Downregulated |
| -0,142726634 | 0,922115768 | SCGN | FALSE | Downregulated |
| 0,107356321 | 0,922115768 | USP44 | FALSE | Upregulated |
| 0,089641108 | 0,922362046 | DNAJB11 | FALSE | Upregulated |
| 0,193410233 | 0,922362046 | FAM157C | FALSE | Upregulated |
| -0,078566353 | 0,922362046 | GNB5 | FALSE | Downregulated |
| 0,052864008 | 0,922362046 | GPX3 | FALSE | Upregulated |
| 0,123041751 | 0,922362046 | GRPEL2 | FALSE | Upregulated |
| 0,135701286 | 0,922362046 | GYPE | FALSE | Upregulated |
| 0,069549926 | 0,922362046 | MBOAT7 | FALSE | Upregulated |
| 0,075953669 | 0,922362046 | RARS | FALSE | Upregulated |
| -0,030056931 | 0,922362046 | SEMA4G | FALSE | Downregulated |
| 0,066415281 | 0,922362046 | TPMT | FALSE | Upregulated |
| -0,119807662 | 0,922538793 | LOC100288152 | FALSE | Downregulated |
| 0,061899136 | 0,922658378 | TRAF3IP2 | FALSE | Upregulated |
| 0,132791564 | 0,923059715 | COX16 | FALSE | Upregulated |
| -0,093392704 | 0,923059715 | MYO16 | FALSE | Downregulated |
| 0,068623879 | 0,923059715 | SPOP | FALSE | Upregulated |
| -0,121507 | 0,92322364 | RABL2A | FALSE | Downregulated |
| -0,087241002 | 0,92322364 | STX12 | FALSE | Downregulated |
| -0,068343443 | 0,92322364 | TMEM44 | FALSE | Downregulated |
| -0,046774727 | 0,923255038 | CFB | FALSE | Downregulated |
| 0,073308758 | 0,923255038 | DHRS4 | FALSE | Upregulated |
| 0,080058316 | 0,923255038 | LOC101928431 | FALSE | Upregulated |
| 0,069311473 | 0,923255038 | SPECC1L | FALSE | Upregulated |
| 0,146279832 | 0,923468032 | LRCOL1 | FALSE | Upregulated |
| 0,067420109 | 0,923698181 | USF1 | FALSE | Upregulated |
| -0,068835619 | 0,923798826 | ABCD1 | FALSE | Downregulated |
| -0,119598537 | 0,923798826 | AKAP7 | FALSE | Downregulated |
| -0,066492332 | 0,923798826 | ARF3 | FALSE | Downregulated |
| -0,055723098 | 0,923798826 | CBX4 | FALSE | Downregulated |
| -0,062870576 | 0,923798826 | CCNC | FALSE | Downregulated |
| 0,175367275 | 0,923798826 | CLSTN2 | FALSE | Upregulated |
| -0,076199103 | 0,923798826 | EMX1 | FALSE | Downregulated |

| | | | | |
|---|---|---|---|---|
| -0,041064011 | 0,923798826 | F11 | FALSE | Downregulated |
| 0,059980738 | 0,923798826 | FAM129B | FALSE | Upregulated |
| 0,108293708 | 0,923798826 | FAM204A | FALSE | Upregulated |
| 0,120515687 | 0,923798826 | FAM78B | FALSE | Upregulated |
| -0,064286382 | 0,923798826 | FASTK | FALSE | Downregulated |
| 0,083977013 | 0,923798826 | FGFR1OP2 | FALSE | Upregulated |
| 0,083892859 | 0,923798826 | FLJ46906 | FALSE | Upregulated |
| 0,054079351 | 0,923798826 | FUBP1 | FALSE | Upregulated |
| -0,054194537 | 0,923798826 | GLUL | FALSE | Downregulated |
| 0,12367389 | 0,923798826 | GOLGA6L9 | FALSE | Upregulated |
| -0,053500839 | 0,923798826 | HMOX2 | FALSE | Downregulated |
| 0,055988357 | 0,923798826 | INTS6 | FALSE | Upregulated |
| 0,04969507 | 0,923798826 | LBX2 | FALSE | Upregulated |
| -0,084375334 | 0,923798826 | LOC102724683 | FALSE | Downregulated |
| 0,118899338 | 0,923798826 | LOC729887 | FALSE | Upregulated |
| -0,065304901 | 0,923798826 | LRRC59 | FALSE | Downregulated |
| 0,07052238 | 0,923798826 | LTB4R2 | FALSE | Upregulated |
| -0,115051062 | 0,923798826 | MCF2L-AS1 | FALSE | Downregulated |
| 0,104154092 | 0,923798826 | MLKL | FALSE | Upregulated |
| -0,10960761 | 0,923798826 | MSI2 | FALSE | Downregulated |
| -0,090749872 | 0,923798826 | MTHFSD | FALSE | Downregulated |
| 0,097834998 | 0,923798826 | NAGPA | FALSE | Upregulated |
| 0,124437336 | 0,923798826 | NRSN2 | FALSE | Upregulated |
| -0,069150743 | 0,923798826 | PLA2G16 | FALSE | Downregulated |
| 0,054670705 | 0,923798826 | PRKAB2 | FALSE | Upregulated |
| 0,050199591 | 0,923798826 | PSMC1 | FALSE | Upregulated |
| 0,046354828 | 0,923798826 | RAB3GAP2 | FALSE | Upregulated |
| -0,083413246 | 0,923798826 | RP1-256G22.2 | FALSE | Downregulated |
| -0,105315046 | 0,923798826 | RP11-307E17.8 | FALSE | Downregulated |
| 0,151728001 | 0,923798826 | TAMM41 | FALSE | Upregulated |
| -0,056788211 | 0,923798826 | TGFBR3L | FALSE | Downregulated |
| 0,062950159 | 0,923798826 | TMX2 | FALSE | Upregulated |
| -0,061532657 | 0,923798826 | TP53BP1 | FALSE | Downregulated |
| -0,155326819 | 0,923798826 | TRHDE | FALSE | Downregulated |
| 0,050409141 | 0,923798826 | WDR6 | FALSE | Upregulated |
| 0,060137569 | 0,923798826 | ZNF586 | FALSE | Upregulated |
| 0,062278896 | 0,923798826 | ZNF618 | FALSE | Upregulated |
| 0,120040946 | 0,923798826 | ZNF708 | FALSE | Upregulated |
| -0,05709258 | 0,923989286 | FAM160A1 | FALSE | Downregulated |
| -0,101270211 | 0,924031337 | EBPL | FALSE | Downregulated |
| -0,126161206 | 0,924031337 | LOC101060376 | FALSE | Downregulated |
| 0,06449394 | 0,92412247 | ECM2 | FALSE | Upregulated |
| -0,068148226 | 0,92412247 | GIT2 | FALSE | Downregulated |
| 0,054429334 | 0,92412247 | NOP9 | FALSE | Upregulated |
| 0,123502845 | 0,92412247 | PIGS | FALSE | Upregulated |
| 0,108839161 | 0,92412247 | RP3-395M20.9 | FALSE | Upregulated |
| 0,071630021 | 0,92412247 | SAMD8 | FALSE | Upregulated |
| -0,187547025 | 0,92412247 | ZSCAN1 | FALSE | Downregulated |
| 0,065153002 | 0,924352229 | CKMT2 | FALSE | Upregulated |
| -0,076526625 | 0,924390636 | BIVM | FALSE | Downregulated |
| 0,051602278 | 0,924390636 | C2orf88 | FALSE | Upregulated |

| | | | | |
|---|---|---|---|---|
| -0,051841515 | 0,924390636 | EZR | FALSE | Downregulated |
| -0,037413153 | 0,924390636 | KAT6A | FALSE | Downregulated |
| 0,109326134 | 0,924390636 | LOC101929018 | FALSE | Upregulated |
| -0,10476323 | 0,924390636 | LRFN3 | FALSE | Downregulated |
| -0,040290736 | 0,924390636 | NISCH | FALSE | Downregulated |
| 0,021689565 | 0,924390636 | PRRC2B | FALSE | Upregulated |
| 0,040946949 | 0,924390636 | TNRC6A | FALSE | Upregulated |
| -0,08892683 | 0,924390636 | ZNF688 | FALSE | Downregulated |
| -0,047796529 | 0,924491461 | PCCB | FALSE | Downregulated |
| -0,17103973 | 0,924627666 | MT1M | FALSE | Downregulated |
| 0,068146738 | 0,924627666 | SDCCAG8 | FALSE | Upregulated |
| 0,065851613 | 0,924685162 | NUP205 | FALSE | Upregulated |
| 0,068183383 | 0,924685162 | PIGR | FALSE | Upregulated |
| -0,100278868 | 0,924790011 | PMS2P5 | FALSE | Downregulated |
| -0,072229961 | 0,924790011 | TOPBP1 | FALSE | Downregulated |
| 0,057434723 | 0,92497545 | GALNS | FALSE | Upregulated |
| -0,067725662 | 0,925113932 | SOX18 | FALSE | Downregulated |
| -0,060249616 | 0,925209729 | SEC14L4 | FALSE | Downregulated |
| 0,113130135 | 0,925229418 | COL4A2-AS2 | FALSE | Upregulated |
| 0,050790642 | 0,925229418 | MAFK | FALSE | Upregulated |
| -0,126718489 | 0,925229418 | MBL1P | FALSE | Downregulated |
| 0,078160715 | 0,925229418 | NBPF1 | FALSE | Upregulated |
| 0,088384984 | 0,925229418 | NDUFB5 | FALSE | Upregulated |
| 0,06567194 | 0,925229418 | NSMF | FALSE | Upregulated |
| 0,043721782 | 0,925229418 | PPP6R3 | FALSE | Upregulated |
| 0,10068843 | 0,925229418 | SEC1P | FALSE | Upregulated |
| 0,125448865 | 0,925293321 | ZNF783 | FALSE | Upregulated |
| 0,136937637 | 0,925312609 | SSX2IP | FALSE | Upregulated |
| -0,096714654 | 0,92532536 | ADSL | FALSE | Downregulated |
| -0,098487944 | 0,92532536 | CD320 | FALSE | Downregulated |
| 0,10421208 | 0,92532536 | DNAJC17 | FALSE | Upregulated |
| 0,057862056 | 0,92532536 | FAM84B | FALSE | Upregulated |
| -0,048350407 | 0,92532536 | INO80D | FALSE | Downregulated |
| -0,074014017 | 0,92532536 | INS-IGF2 | FALSE | Downregulated |
| 0,086562332 | 0,92532536 | NENF | FALSE | Upregulated |
| 0,052343038 | 0,92532536 | PAWR | FALSE | Upregulated |
| -0,077539849 | 0,925485459 | ADAMTS9 | FALSE | Downregulated |
| 0,106590055 | 0,925485459 | CWF19L1 | FALSE | Upregulated |
| 0,081063138 | 0,92562707 | ACSS3 | FALSE | Upregulated |
| 0,060693223 | 0,92562707 | GTF3C2 | FALSE | Upregulated |
| -0,086344221 | 0,92562707 | MARCH9 | FALSE | Downregulated |
| -0,078363629 | 0,92562707 | TBCEL | FALSE | Downregulated |
| -0,072355845 | 0,92562707 | YKT6 | FALSE | Downregulated |
| 0,05187951 | 0,925734865 | BCAM | FALSE | Upregulated |
| 0,040131934 | 0,925734865 | CC2D1B | FALSE | Upregulated |
| -0,100866232 | 0,925734865 | GUSBP1 | FALSE | Downregulated |
| 0,080880841 | 0,925734865 | UBE2E3 | FALSE | Upregulated |
| 0,055501033 | 0,926117737 | CACTIN-AS1 | FALSE | Upregulated |
| 0,048626296 | 0,926117737 | HIRA | FALSE | Upregulated |
| -0,08477854 | 0,926117737 | POMGNT2 | FALSE | Downregulated |
| 0,036450001 | 0,926117737 | TYK2 | FALSE | Upregulated |

| | | | | |
|---|---|---|---|---|
| -0,119375545 | 0,926117737 | ZNF569 | FALSE | Downregulated |
| 0,077330358 | 0,926175083 | FNDC5 | FALSE | Upregulated |
| 0,247975889 | 0,926272626 | CHRM3 | FALSE | Upregulated |
| 0,059580109 | 0,926478168 | VPS36 | FALSE | Upregulated |
| -0,077799999 | 0,926528809 | IGBP1 | FALSE | Downregulated |
| 0,049016468 | 0,926528809 | MYO9A | FALSE | Upregulated |
| 0,070661735 | 0,926739048 | ARRDC3-AS1 | FALSE | Upregulated |
| 0,046913274 | 0,926979689 | DNAJC13 | FALSE | Upregulated |
| -0,076501314 | 0,92703944 | CHST12 | FALSE | Downregulated |
| -0,066708159 | 0,92703944 | CTPS1 | FALSE | Downregulated |
| 0,074402087 | 0,92703944 | ESCO1 | FALSE | Upregulated |
| -0,081473621 | 0,92703944 | KCNN2 | FALSE | Downregulated |
| -0,077752781 | 0,92703944 | OR13D1 | FALSE | Downregulated |
| -0,051096713 | 0,92703944 | PAK2 | FALSE | Downregulated |
| -0,051273663 | 0,92703944 | PEX26 | FALSE | Downregulated |
| -0,039404432 | 0,92703944 | SAR1B | FALSE | Downregulated |
| -0,080639142 | 0,92703944 | VSNL1 | FALSE | Downregulated |
| -0,105788167 | 0,92703944 | VWCE | FALSE | Downregulated |
| 0,09834414 | 0,92703944 | WWOX | FALSE | Upregulated |
| -0,123096485 | 0,92703944 | ZNF639 | FALSE | Downregulated |
| 0,112024813 | 0,927057098 | ZBTB16 | FALSE | Upregulated |
| -0,08942507 | 0,927207434 | ZNF155 | FALSE | Downregulated |
| -0,123217111 | 0,927213184 | AKR7A2P1 | FALSE | Downregulated |
| 0,074164432 | 0,927213184 | C14orf166 | FALSE | Upregulated |
| 0,121580264 | 0,927213184 | C9orf156 | FALSE | Upregulated |
| 0,058623004 | 0,927213184 | CCNK | FALSE | Upregulated |
| 0,096523741 | 0,927213184 | CMKLR1 | FALSE | Upregulated |
| -0,033884225 | 0,927213184 | MYLK | FALSE | Downregulated |
| 0,117308086 | 0,927213184 | NEK3 | FALSE | Upregulated |
| 0,056528854 | 0,927213184 | NFYB | FALSE | Upregulated |
| -0,097783116 | 0,927213184 | POLA1 | FALSE | Downregulated |
| 0,102640637 | 0,927213184 | PUS7L | FALSE | Upregulated |
| -0,100839347 | 0,927213184 | RAD51B | FALSE | Downregulated |
| 0,098443323 | 0,927213184 | RP11-234K24.3 | FALSE | Upregulated |
| 0,059570453 | 0,927213184 | SLC25A38 | FALSE | Upregulated |
| -0,0681141 | 0,927213184 | TPM4 | FALSE | Downregulated |
| -0,12518691 | 0,927213184 | TRHDE-AS1 | FALSE | Downregulated |
| 0,05491428 | 0,927213184 | UBE2E1 | FALSE | Upregulated |
| -0,068044517 | 0,927213184 | VTA1 | FALSE | Downregulated |
| -0,111039792 | 0,927213184 | WFIKKN1 | FALSE | Downregulated |
| 0,086001203 | 0,927213184 | ZNF585B | FALSE | Upregulated |
| 0,065611953 | 0,92732579 | ACLY | FALSE | Upregulated |
| -0,064058178 | 0,92732579 | BRAF | FALSE | Downregulated |
| -0,045544468 | 0,92732579 | C17orf70 | FALSE | Downregulated |
| 0,055336465 | 0,92732579 | C9orf114 | FALSE | Upregulated |
| -0,069794671 | 0,92732579 | DPP8 | FALSE | Downregulated |
| -0,067287823 | 0,92732579 | HEMK1 | FALSE | Downregulated |
| 0,056769998 | 0,92732579 | HPS4 | FALSE | Upregulated |
| 0,177703971 | 0,92732579 | LOC101927842 | FALSE | Upregulated |
| -0,068238708 | 0,92732579 | PES1 | FALSE | Downregulated |
| 0,105315926 | 0,92732579 | PMEL | FALSE | Upregulated |

| | | | | |
|---|---|---|---|---|
| -0,062168986 | 0,92732579 | UHRF1BP1L | FALSE | Downregulated |
| -0,071178464 | 0,92732579 | VAMP8 | FALSE | Downregulated |
| 0,068394116 | 0,927346927 | AFG3L1P | FALSE | Upregulated |
| 0,083371435 | 0,927346927 | CACNA2D2 | FALSE | Upregulated |
| 0,073807304 | 0,927346927 | NRM | FALSE | Upregulated |
| -0,046450887 | 0,927346927 | PDPR | FALSE | Downregulated |
| 0,163806202 | 0,927552581 | SERPINE1 | FALSE | Upregulated |
| 0,090143848 | 0,927552581 | ZNF69 | FALSE | Upregulated |
| -0,061462474 | 0,928116198 | ACOX3 | FALSE | Downregulated |
| 0,112659695 | 0,928116198 | TRMT13 | FALSE | Upregulated |
| 0,068650913 | 0,928211069 | SUFU | FALSE | Upregulated |
| -0,062928501 | 0,928708837 | BOP1 | FALSE | Downregulated |
| -0,097158393 | 0,928740911 | CIR1 | FALSE | Downregulated |
| 0,17562391 | 0,928740911 | DDX3Y | FALSE | Upregulated |
| 0,092471708 | 0,928809499 | TOMM6 | FALSE | Upregulated |
| -0,057078997 | 0,928987965 | UGT2B10 | FALSE | Downregulated |
| -0,121055329 | 0,929149533 | ADM5 | FALSE | Downregulated |
| -0,086969595 | 0,92915009 | TAT | FALSE | Downregulated |
| -0,063065472 | 0,929287454 | PRMT1 | FALSE | Downregulated |
| 0,114211308 | 0,929287454 | RP11-127L20.3 | FALSE | Upregulated |
| -0,078556996 | 0,929308149 | SMG9 | FALSE | Downregulated |
| -0,077887795 | 0,929480251 | DCBLD1 | FALSE | Downregulated |
| 0,021761553 | 0,929480251 | DDX17 | FALSE | Upregulated |
| 0,047261288 | 0,929480251 | PGAM5 | FALSE | Upregulated |
| 0,100146518 | 0,930146165 | HMBS | FALSE | Upregulated |
| 0,034822546 | 0,930247379 | TAOK2 | FALSE | Upregulated |
| -0,295126573 | 0,930410086 | AC004053.1 | FALSE | Downregulated |
| -0,062830787 | 0,930410086 | CCAR1 | FALSE | Downregulated |
| -0,060132371 | 0,930410086 | COG7 | FALSE | Downregulated |
| -0,086149843 | 0,930410086 | LOC101927445 | FALSE | Downregulated |
| 0,12288876 | 0,930410086 | POLR2G | FALSE | Upregulated |
| -0,074118305 | 0,930410086 | TMEM185B | FALSE | Downregulated |
| 0,047032111 | 0,930410086 | YARS | FALSE | Upregulated |
| 0,084460593 | 0,930557937 | ADAMTS16 | FALSE | Upregulated |
| 0,153383364 | 0,930557937 | FAM90A1 | FALSE | Upregulated |
| 0,190309176 | 0,930635624 | NOMO3 | FALSE | Upregulated |
| 0,16958498 | 0,930635624 | TIAF1 | FALSE | Upregulated |
| -0,030594053 | 0,930635624 | UBTF | FALSE | Downregulated |
| -0,060269753 | 0,930912904 | PCMTD1 | FALSE | Downregulated |
| 0,048507122 | 0,931310295 | AVL9 | FALSE | Upregulated |
| 0,056346806 | 0,931310295 | BAD | FALSE | Upregulated |
| 0,101636658 | 0,931310295 | BTBD3 | FALSE | Upregulated |
| -0,071737337 | 0,931310295 | C1QBP | FALSE | Downregulated |
| -0,052635807 | 0,931310295 | CACNB2 | FALSE | Downregulated |
| 0,041077614 | 0,931310295 | ERVK13-1 | FALSE | Upregulated |
| 0,096798907 | 0,931310295 | FAM114A2 | FALSE | Upregulated |
| -0,041052006 | 0,931310295 | FGD5-AS1 | FALSE | Downregulated |
| -0,072023662 | 0,931310295 | FYTTD1 | FALSE | Downregulated |
| -0,074988696 | 0,931310295 | GPR116 | FALSE | Downregulated |
| 0,043903004 | 0,931310295 | HSF1 | FALSE | Upregulated |
| -0,100627115 | 0,931310295 | IDO2 | FALSE | Downregulated |

| | | | | |
|---|---|---|---|---|
| -0,120471932 | 0,931310295 | IPO11 | FALSE | Downregulated |
| 0,105149432 | 0,931310295 | KCTD11 | FALSE | Upregulated |
| -0,05654651 | 0,931310295 | KIAA1468 | FALSE | Downregulated |
| 0,04741482 | 0,931310295 | LDHA | FALSE | Upregulated |
| 0,114760687 | 0,931310295 | LOC101927864 | FALSE | Upregulated |
| -0,083476382 | 0,931310295 | LOC102724474 | FALSE | Downregulated |
| 0,059311381 | 0,931310295 | LOC102724607 | FALSE | Upregulated |
| -0,322658174 | 0,931310295 | LOC644172 | FALSE | Downregulated |
| -0,086658755 | 0,931310295 | LOC729173 | FALSE | Downregulated |
| -0,07117604 | 0,931310295 | MAP2K4 | FALSE | Downregulated |
| 0,071935979 | 0,931310295 | MINPP1 | FALSE | Upregulated |
| 0,03477163 | 0,931310295 | MMACHC | FALSE | Upregulated |
| 0,029660316 | 0,931310295 | NCOR2 | FALSE | Upregulated |
| 0,121230039 | 0,931310295 | NGDN | FALSE | Upregulated |
| 0,089771802 | 0,931310295 | NUBP1 | FALSE | Upregulated |
| 0,156940963 | 0,931310295 | NUDT4P1 | FALSE | Upregulated |
| 0,03932739 | 0,931310295 | PARP4 | FALSE | Upregulated |
| 0,056372309 | 0,931310295 | PDXDC2P | FALSE | Upregulated |
| -0,065333973 | 0,931310295 | PEX16 | FALSE | Downregulated |
| -0,069133852 | 0,931310295 | PHAX | FALSE | Downregulated |
| -0,048545256 | 0,931310295 | PHKB | FALSE | Downregulated |
| 0,091825344 | 0,931310295 | PRCD | FALSE | Upregulated |
| -0,048539311 | 0,931310295 | RAB14 | FALSE | Downregulated |
| 0,087601585 | 0,931310295 | RNF25 | FALSE | Upregulated |
| 0,074561675 | 0,931310295 | RP1-60O19.1 | FALSE | Upregulated |
| -0,182558448 | 0,931310295 | RPS4Y1 | FALSE | Downregulated |
| 0,084133518 | 0,931310295 | SLC25A28 | FALSE | Upregulated |
| -0,25397926 | 0,931310295 | SMN2 | FALSE | Downregulated |
| 0,073226675 | 0,931310295 | ST7L | FALSE | Upregulated |
| 0,27069756 | 0,931310295 | TCEB3CL2 | FALSE | Upregulated |
| -0,086266856 | 0,931310295 | THNSL2 | FALSE | Downregulated |
| -0,104344395 | 0,931310295 | TMEM39B | FALSE | Downregulated |
| -0,054443063 | 0,931310295 | TOP3A | FALSE | Downregulated |
| -0,046454773 | 0,931310295 | WDR11 | FALSE | Downregulated |
| -0,048229012 | 0,931310295 | ZYG11B | FALSE | Downregulated |
| 0,123406104 | 0,931540298 | LINC00987 | FALSE | Upregulated |
| -0,040583263 | 0,931600785 | GPER1 | FALSE | Downregulated |
| 0,07676298 | 0,931709684 | PNMAL2 | FALSE | Upregulated |
| 0,12261952 | 0,931720409 | C11orf84 | FALSE | Upregulated |
| 0,046374372 | 0,931720409 | C16orf70 | FALSE | Upregulated |
| 0,115769993 | 0,931720409 | DUS4L | FALSE | Upregulated |
| 0,119855405 | 0,931720409 | MSC | FALSE | Upregulated |
| -0,054838491 | 0,931720409 | NCKAP5 | FALSE | Downregulated |
| 0,071685439 | 0,931720409 | TRAPPC6B | FALSE | Upregulated |
| 0,067813678 | 0,931720409 | TSNARE1 | FALSE | Upregulated |
| -0,033016274 | 0,931750229 | MAN2A2 | FALSE | Downregulated |
| -0,083628386 | 0,932479269 | COA3 | FALSE | Downregulated |
| 0,097181676 | 0,932479269 | DKFZP434K028 | FALSE | Upregulated |
| -0,090518006 | 0,932479269 | THY1 | FALSE | Downregulated |
| 0,055042152 | 0,932502952 | UEVLD | FALSE | Upregulated |
| -0,116890491 | 0,932694281 | NUBPL | FALSE | Downregulated |

| | | | | |
|---|---|---|---|---|
| -0,062897779 | 0,932982105 | ACAD8 | FALSE | Downregulated |
| -0,095204119 | 0,932982105 | ATP5L | FALSE | Downregulated |
| 0,078439748 | 0,932982105 | BCDIN3D | FALSE | Upregulated |
| 0,038494364 | 0,932982105 | CASC4 | FALSE | Upregulated |
| -0,052934116 | 0,932982105 | CLK1 | FALSE | Downregulated |
| 0,064103787 | 0,932982105 | COCH | FALSE | Upregulated |
| 0,181140828 | 0,932982105 | CRYAA | FALSE | Upregulated |
| 0,068771247 | 0,932982105 | CYCS | FALSE | Upregulated |
| -0,047786426 | 0,932982105 | FBXO7 | FALSE | Downregulated |
| 0,048746959 | 0,932982105 | FBXW4 | FALSE | Upregulated |
| 0,094276471 | 0,932982105 | LCTL | FALSE | Upregulated |
| 0,046674302 | 0,932982105 | LOC100129361 | FALSE | Upregulated |
| 0,06649776 | 0,932982105 | MAP3K7 | FALSE | Upregulated |
| 0,03321086 | 0,932982105 | MDH2 | FALSE | Upregulated |
| 0,11673205 | 0,932982105 | MMP2 | FALSE | Upregulated |
| 0,052369819 | 0,932982105 | MSTO2P | FALSE | Upregulated |
| -0,036263036 | 0,932982105 | NADK | FALSE | Downregulated |
| -0,033158991 | 0,932982105 | NPLOC4 | FALSE | Downregulated |
| -0,07106764 | 0,932982105 | NRTN | FALSE | Downregulated |
| 0,04247691 | 0,932982105 | PABPN1 | FALSE | Upregulated |
| -0,090493751 | 0,932982105 | PSMD12 | FALSE | Downregulated |
| 0,081515877 | 0,932982105 | RNMTL1 | FALSE | Upregulated |
| -0,042736018 | 0,932982105 | RTN3 | FALSE | Downregulated |
| 0,138204144 | 0,932982105 | SLMO2 | FALSE | Upregulated |
| -0,071595407 | 0,932982105 | ST3GAL3 | FALSE | Downregulated |
| -0,113825819 | 0,932982105 | TMA16 | FALSE | Downregulated |
| -0,079536213 | 0,932982105 | TMX1 | FALSE | Downregulated |
| -0,057090727 | 0,932982105 | TNNI1 | FALSE | Downregulated |
| 1,295666273 | 0,932982105 | TSIX | FALSE | Upregulated |
| -0,046587707 | 0,932982105 | TXN2 | FALSE | Downregulated |
| -0,032222137 | 0,932982105 | VPS26B | FALSE | Downregulated |
| -0,109761287 | 0,932982105 | ZDHHC11 | FALSE | Downregulated |
| -0,066894904 | 0,932982105 | ZNF410 | FALSE | Downregulated |
| -0,091386781 | 0,933096251 | ADAM19 | FALSE | Downregulated |
| 0,075774957 | 0,933096251 | ADSS | FALSE | Upregulated |
| -0,039699205 | 0,933096251 | CISD3 | FALSE | Downregulated |
| 0,029636836 | 0,933096251 | GNAI2 | FALSE | Upregulated |
| 0,108915634 | 0,933096251 | MCM8 | FALSE | Upregulated |
| -0,051471948 | 0,933096251 | MICALL1 | FALSE | Downregulated |
| 0,050816488 | 0,933096251 | PLOD3 | FALSE | Upregulated |
| 0,057413627 | 0,933096251 | SPON2 | FALSE | Upregulated |
| -0,093526102 | 0,933096251 | TMTC1 | FALSE | Downregulated |
| 0,047947008 | 0,933197715 | BBX | FALSE | Upregulated |
| 0,057761199 | 0,933197715 | GFM2 | FALSE | Upregulated |
| 0,085992346 | 0,933197715 | KRBOX4 | FALSE | Upregulated |
| -0,048755758 | 0,933200535 | RILP | FALSE | Downregulated |
| -0,100823507 | 0,933218908 | ABHD17B | FALSE | Downregulated |
| 0,05563333 | 0,933218908 | ATF7IP2 | FALSE | Upregulated |
| -0,059720248 | 0,933218908 | MRPL51 | FALSE | Downregulated |
| 0,069767791 | 0,933218908 | NSMCE4A | FALSE | Upregulated |
| 0,053676997 | 0,933218908 | OCRL | FALSE | Upregulated |

| | | | | |
|---|---|---|---|---|
| -0,061580372 | 0,933218908 | PLEKHA5 | FALSE | Downregulated |
| -0,042460995 | 0,933218908 | SUOX | FALSE | Downregulated |
| 0,067638509 | 0,933218908 | THBS1 | FALSE | Upregulated |
| -0,047635348 | 0,933364056 | ATP6V0A1 | FALSE | Downregulated |
| -0,046542333 | 0,93340853 | ADRM1 | FALSE | Downregulated |
| 0,052369783 | 0,93340853 | AGGF1 | FALSE | Upregulated |
| -0,056267387 | 0,93340853 | ARL1 | FALSE | Downregulated |
| -0,322036243 | 0,93340853 | CHURC1-FNTB | FALSE | Downregulated |
| -0,060232667 | 0,93340853 | CNNM2 | FALSE | Downregulated |
| 0,037484597 | 0,93340853 | CYP8B1 | FALSE | Upregulated |
| -0,077974714 | 0,93340853 | DYNLRB1 | FALSE | Downregulated |
| -0,070383212 | 0,93340853 | ENTPD8 | FALSE | Downregulated |
| 0,066155111 | 0,93340853 | GPR108 | FALSE | Upregulated |
| 0,119849755 | 0,93340853 | HPDL | FALSE | Upregulated |
| -0,053269543 | 0,93340853 | LAP3 | FALSE | Downregulated |
| 0,090067193 | 0,93340853 | LOC286083 | FALSE | Upregulated |
| 0,064659684 | 0,93340853 | LRRC2 | FALSE | Upregulated |
| 0,034146094 | 0,93340853 | MED1 | FALSE | Upregulated |
| 0,082001263 | 0,93340853 | MRPL32 | FALSE | Upregulated |
| 0,075962906 | 0,93340853 | OGFOD2 | FALSE | Upregulated |
| -0,067494947 | 0,93340853 | ORC2 | FALSE | Downregulated |
| -0,081137592 | 0,93340853 | PEX11A | FALSE | Downregulated |
| 0,048614667 | 0,93340853 | RALY | FALSE | Upregulated |
| -0,037104953 | 0,93340853 | RC3H1 | FALSE | Downregulated |
| -0,036648513 | 0,93340853 | SLC25A6 | FALSE | Downregulated |
| -0,025155046 | 0,93340853 | SQSTM1 | FALSE | Downregulated |
| 0,116774882 | 0,93340853 | TTC21B-AS1 | FALSE | Upregulated |
| 0,083536889 | 0,93340853 | ZDHHC21 | FALSE | Upregulated |
| 0,055506741 | 0,93340853 | ZNF263 | FALSE | Upregulated |
| 0,060787964 | 0,93340853 | ZNF526 | FALSE | Upregulated |
| 0,076248261 | 0,93340853 | ZNF532 | FALSE | Upregulated |
| -0,085772855 | 0,933508372 | ZMPSTE24 | FALSE | Downregulated |
| -0,055286175 | 0,933561963 | DOCK6 | FALSE | Downregulated |
| -0,050800688 | 0,933616371 | CPQ | FALSE | Downregulated |
| 0,073298457 | 0,933797248 | COL14A1 | FALSE | Upregulated |
| 0,058707245 | 0,933832846 | ASCC3 | FALSE | Upregulated |
| -0,060856279 | 0,933832846 | ZNF696 | FALSE | Downregulated |
| -0,068039648 | 0,933874497 | LAMA2 | FALSE | Downregulated |
| 0,092266322 | 0,933899703 | PRICKLE2 | FALSE | Upregulated |
| 0,043301322 | 0,934023343 | LPIN2 | FALSE | Upregulated |
| -0,100757403 | 0,934023343 | PRAM1 | FALSE | Downregulated |
| -0,071565808 | 0,93406603 | FAM203A | FALSE | Downregulated |
| -0,07362361 | 0,934128955 | HBS1L | FALSE | Downregulated |
| 0,103747473 | 0,934128955 | OSTM1 | FALSE | Upregulated |
| 0,05855203 | 0,934128955 | ZNRF3-AS1 | FALSE | Upregulated |
| 0,046596468 | 0,934305304 | LOC102723489 | FALSE | Upregulated |
| -0,110877426 | 0,934305304 | PGBD2 | FALSE | Downregulated |
| 0,051689062 | 0,934389415 | BCL2L1 | FALSE | Upregulated |
| 0,052834798 | 0,934389415 | C1orf122 | FALSE | Upregulated |
| 0,094013479 | 0,934389415 | GLB1L | FALSE | Upregulated |
| -0,120518041 | 0,934389415 | GOLGA2P7 | FALSE | Downregulated |

| | | | | |
|---|---|---|---|---|
| 0,056300357 | 0,934389415 | LATS2 | FALSE | Upregulated |
| 0,051407379 | 0,934389415 | NDUFB11 | FALSE | Upregulated |
| 0,053601703 | 0,934389415 | NFATC2IP | FALSE | Upregulated |
| 0,045755524 | 0,934389415 | PAG1 | FALSE | Upregulated |
| 0,258829374 | 0,934389415 | RP13-228J13.1 | FALSE | Upregulated |
| 0,04069182 | 0,934389415 | SHB | FALSE | Upregulated |
| 0,093988891 | 0,93442649 | ZNF703 | FALSE | Upregulated |
| -0,055526217 | 0,934478158 | RAB12 | FALSE | Downregulated |
| 0,055351858 | 0,934791772 | KCNAB1 | FALSE | Upregulated |
| -0,066982425 | 0,934856886 | AP5Z1 | FALSE | Downregulated |
| 0,08888555 | 0,934856886 | FBXO40 | FALSE | Upregulated |
| 0,147041459 | 0,934856886 | LOXL1 | FALSE | Upregulated |
| 0,067512763 | 0,934856886 | PSME3 | FALSE | Upregulated |
| -0,105528106 | 0,934864577 | ZKSCAN3 | FALSE | Downregulated |
| -0,045869796 | 0,934895639 | NME4 | FALSE | Downregulated |
| -0,124862703 | 0,934895639 | RFPL1S | FALSE | Downregulated |
| -0,044990562 | 0,934941485 | PRPF3 | FALSE | Downregulated |
| -0,065801097 | 0,934944159 | FILIP1 | FALSE | Downregulated |
| -0,036815082 | 0,935012513 | CAPRIN1 | FALSE | Downregulated |
| -0,046187779 | 0,935015112 | CBR1 | FALSE | Downregulated |
| -0,055461718 | 0,935015112 | LRRC41 | FALSE | Downregulated |
| -0,055867392 | 0,935015112 | SIRT2 | FALSE | Downregulated |
| 0,084933983 | 0,935015112 | SPDYE7P | FALSE | Upregulated |
| 0,074734626 | 0,935015112 | TCP11L1 | FALSE | Upregulated |
| 0,077606256 | 0,935015112 | TMC3 | FALSE | Upregulated |
| -0,089700907 | 0,935015112 | ZDHHC12 | FALSE | Downregulated |
| -0,103754336 | 0,935185688 | LOC101926918 | FALSE | Downregulated |
| 0,042626541 | 0,935185688 | TMPRSS9 | FALSE | Upregulated |
| 0,107409569 | 0,935284065 | SDK1 | FALSE | Upregulated |
| 0,046059851 | 0,935303609 | NCKAP1 | FALSE | Upregulated |
| -0,039570113 | 0,935378197 | CLCN7 | FALSE | Downregulated |
| -0,052422753 | 0,935392219 | ADAMTSL2 | FALSE | Downregulated |
| 0,053750805 | 0,935392219 | SERINC2 | FALSE | Upregulated |
| -0,057783117 | 0,935554544 | ACBD3 | FALSE | Downregulated |
| -0,06785249 | 0,935554544 | SATB2 | FALSE | Downregulated |
| -0,104921676 | 0,935631342 | BMS1P2 | FALSE | Downregulated |
| -0,042471793 | 0,935699301 | LOC101928853 | FALSE | Downregulated |
| -0,044877646 | 0,93571278 | AKR1A1 | FALSE | Downregulated |
| 0,039412789 | 0,93571278 | LMNA | FALSE | Upregulated |
| -0,084345621 | 0,93583084 | ADCK2 | FALSE | Downregulated |
| 0,053938921 | 0,93583084 | CBFA2T2 | FALSE | Upregulated |
| 0,091738146 | 0,93583084 | SAMD9 | FALSE | Upregulated |
| 0,05611396 | 0,93583084 | SERPINB6 | FALSE | Upregulated |
| -0,099271875 | 0,93583084 | TFB2M | FALSE | Downregulated |
| -0,09217549 | 0,93583084 | ZNF70 | FALSE | Downregulated |
| -0,041917222 | 0,935836058 | CECR2 | FALSE | Downregulated |
| 0,095971704 | 0,935842483 | TBCCD1 | FALSE | Upregulated |
| 0,439625189 | 0,935842483 | TTN-AS1 | FALSE | Upregulated |
| -0,06676563 | 0,935953806 | KIAA1715 | FALSE | Downregulated |
| -0,06932528 | 0,935959763 | YPEL3 | FALSE | Downregulated |
| -0,056771902 | 0,935988942 | RSAD1 | FALSE | Downregulated |

| | | | | |
|---|---|---|---|---|
| 0,052263231 | 0,936192959 | PHF10 | FALSE | Upregulated |
| 0,082888037 | 0,936267494 | ABCA4 | FALSE | Upregulated |
| 0,058819042 | 0,936267494 | ARL5A | FALSE | Upregulated |
| 0,041740417 | 0,936267494 | DENND1A | FALSE | Upregulated |
| 0,115688968 | 0,936267494 | ENHO | FALSE | Upregulated |
| 0,05968481 | 0,936267494 | IDUA | FALSE | Upregulated |
| -0,044680831 | 0,936267494 | MEPCE | FALSE | Downregulated |
| 0,045993803 | 0,936267494 | PRKCZ | FALSE | Upregulated |
| -0,074455362 | 0,936267494 | RAB11FIP2 | FALSE | Downregulated |
| -0,088323989 | 0,936267494 | RAB36 | FALSE | Downregulated |
| 0,088219045 | 0,936267494 | RASGRP2 | FALSE | Upregulated |
| -0,116286109 | 0,936267494 | RHBG | FALSE | Downregulated |
| -0,135450664 | 0,936267494 | RP3-365O12.2 | FALSE | Downregulated |
| -0,068337288 | 0,936267494 | SEPT7 | FALSE | Downregulated |
| 0,047490382 | 0,936267494 | WWP2 | FALSE | Upregulated |
| 0,068796873 | 0,936351729 | AFF3 | FALSE | Upregulated |
| 0,080499786 | 0,936470866 | EPC1 | FALSE | Upregulated |
| -0,084338544 | 0,936470866 | FBXL4 | FALSE | Downregulated |
| -0,067731486 | 0,936470866 | RAP1A | FALSE | Downregulated |
| 0,041266971 | 0,936528091 | TXNDC11 | FALSE | Upregulated |
| 0,066923798 | 0,936708711 | MUC20 | FALSE | Upregulated |
| 0,083364226 | 0,936708711 | MYH3 | FALSE | Upregulated |
| -0,0783667 | 0,936708711 | TCTN3 | FALSE | Downregulated |
| -0,049680385 | 0,936711681 | CHMP2A | FALSE | Downregulated |
| -0,094433968 | 0,936711681 | NR1D2 | FALSE | Downregulated |
| -0,074106549 | 0,936711681 | NSUN7 | FALSE | Downregulated |
| -0,060527462 | 0,936711681 | PDHX | FALSE | Downregulated |
| -0,094454212 | 0,936711681 | RAD17 | FALSE | Downregulated |
| -0,066119629 | 0,936711681 | TGIF1 | FALSE | Downregulated |
| -0,056007263 | 0,936711681 | XPNPEP3 | FALSE | Downregulated |
| -0,059105314 | 0,936798143 | ZNF84 | FALSE | Downregulated |
| 0,077564631 | 0,93689911 | PSMA3 | FALSE | Upregulated |
| -0,052825267 | 0,936965306 | ANAPC5 | FALSE | Downregulated |
| 0,06126166 | 0,936965306 | ATG12 | FALSE | Upregulated |
| -0,057266485 | 0,936965306 | C10orf105 | FALSE | Downregulated |
| 0,150641391 | 0,936965306 | CCDC144A | FALSE | Upregulated |
| -0,043868184 | 0,936965306 | CHTF8 | FALSE | Downregulated |
| 0,068137543 | 0,936965306 | CLN6 | FALSE | Upregulated |
| 0,042455024 | 0,936965306 | DHX9 | FALSE | Upregulated |
| 0,067757109 | 0,936965306 | DKC1 | FALSE | Upregulated |
| -0,098477241 | 0,936965306 | EBF4 | FALSE | Downregulated |
| -0,043418077 | 0,936965306 | FAM210B | FALSE | Downregulated |
| 0,052030881 | 0,936965306 | FBXO28 | FALSE | Upregulated |
| -0,032051142 | 0,936965306 | KIAA0430 | FALSE | Downregulated |
| -0,08347872 | 0,936965306 | MID2 | FALSE | Downregulated |
| -0,081990767 | 0,936965306 | PAMR1 | FALSE | Downregulated |
| -0,079813884 | 0,936965306 | SLC39A6 | FALSE | Downregulated |
| 0,080532293 | 0,936965306 | SPSB1 | FALSE | Upregulated |
| -0,06877581 | 0,936965306 | SRPK1 | FALSE | Downregulated |
| -0,207303381 | 0,936965306 | UPK3BL | FALSE | Downregulated |
| 0,089203815 | 0,936965306 | ZNF347 | FALSE | Upregulated |

| | | | | |
|---|---|---|---|---|
| -0,052830821 | 0,937088462 | ATP6V1A | FALSE | Downregulated |
| 0,054164882 | 0,937088462 | LOC101929751 | FALSE | Upregulated |
| 0,315212379 | 0,937088462 | RNU1-27P | FALSE | Upregulated |
| -0,069102865 | 0,937105725 | MCPH1 | FALSE | Downregulated |
| 0,071995161 | 0,937105725 | SNRPC | FALSE | Upregulated |
| 0,064755927 | 0,937157126 | AC144449.1 | FALSE | Upregulated |
| -0,048552396 | 0,937157126 | PKN2 | FALSE | Downregulated |
| 0,098101452 | 0,937157126 | STMN3 | FALSE | Upregulated |
| 0,049140255 | 0,937157126 | TRIM2 | FALSE | Upregulated |
| -0,060223718 | 0,937262565 | NOA1 | FALSE | Downregulated |
| -0,054280612 | 0,937307086 | TBRG1 | FALSE | Downregulated |
| -0,072521248 | 0,937353425 | ANKRD46 | FALSE | Downregulated |
| 0,103221799 | 0,937353425 | PPOX | FALSE | Upregulated |
| 0,059648802 | 0,937353425 | UBE2I | FALSE | Upregulated |
| -0,135157553 | 0,937443912 | CTAGE9 | FALSE | Downregulated |
| 0,076357476 | 0,937443912 | DPH1 | FALSE | Upregulated |
| 0,055967213 | 0,937443912 | NDRG4 | FALSE | Upregulated |
| -0,088853332 | 0,937443912 | PPP1R13L | FALSE | Downregulated |
| 0,073913292 | 0,937443912 | SVEP1 | FALSE | Upregulated |
| -0,046555467 | 0,937443912 | TADA2B | FALSE | Downregulated |
| -0,058092944 | 0,937443912 | UBXN7 | FALSE | Downregulated |
| 0,044468889 | 0,937443912 | VAPB | FALSE | Upregulated |
| -0,091934039 | 0,937499402 | PDIA3P1 | FALSE | Downregulated |
| 0,060579387 | 0,93758758 | FAM63B | FALSE | Upregulated |
| -0,056758952 | 0,937854379 | OCIAD1 | FALSE | Downregulated |
| -0,173467056 | 0,937854379 | OPCML | FALSE | Downregulated |
| 0,071220716 | 0,937854379 | RNF138P1 | FALSE | Upregulated |
| 0,07810725 | 0,937854379 | SNX29 | FALSE | Upregulated |
| -0,050332653 | 0,937854379 | STK38 | FALSE | Downregulated |
| -0,032151067 | 0,937854379 | TLN1 | FALSE | Downregulated |
| 0,083898502 | 0,937854379 | TM2D2 | FALSE | Upregulated |
| -0,055206106 | 0,937870382 | SYNE3 | FALSE | Downregulated |
| 0,080096124 | 0,937978945 | CSRNP1 | FALSE | Upregulated |
| -0,034173279 | 0,938030329 | ADORA2A-AS1 | FALSE | Downregulated |
| -0,078209555 | 0,938030329 | ANKRD9 | FALSE | Downregulated |
| 0,252382905 | 0,938030329 | AP3S2 | FALSE | Upregulated |
| -0,039893221 | 0,938030329 | N4BP2L2 | FALSE | Downregulated |
| -0,059374698 | 0,938030329 | NME2 | FALSE | Downregulated |
| 0,089902791 | 0,938041723 | GNAT2 | FALSE | Upregulated |
| -0,028438117 | 0,938043444 | SZT2 | FALSE | Downregulated |
| 0,119097817 | 0,938062075 | NME1 | FALSE | Upregulated |
| -0,05004172 | 0,938062075 | RAB11A | FALSE | Downregulated |
| 0,046108523 | 0,938308639 | ACTG1 | FALSE | Upregulated |
| -0,088708931 | 0,938308639 | LINC00161 | FALSE | Downregulated |
| 0,045162902 | 0,938602778 | CNOT6L | FALSE | Upregulated |
| 0,058319762 | 0,938602778 | TMEM87A | FALSE | Upregulated |
| -0,057810325 | 0,938684181 | PTDSS1 | FALSE | Downregulated |
| -0,076981788 | 0,938712282 | C4orf29 | FALSE | Downregulated |
| 0,109506347 | 0,93891443 | LOC102724705 | FALSE | Upregulated |
| -0,079264766 | 0,939188984 | RPUSD1 | FALSE | Downregulated |
| -0,040325075 | 0,939355956 | GLYATL1 | FALSE | Downregulated |

| | | | | |
|---|---|---|---|---|
| 0,113897599 | 0,939376624 | FREM1 | FALSE | Upregulated |
| -0,061166489 | 0,939642617 | CPNE8 | FALSE | Downregulated |
| -0,064994336 | 0,939642617 | HIGD2A | FALSE | Downregulated |
| 0,050013238 | 0,939642617 | TSPYL4 | FALSE | Upregulated |
| 0,062396331 | 0,939706712 | LOC101928789 | FALSE | Upregulated |
| 0,111556463 | 0,939706712 | PSMD10 | FALSE | Upregulated |
| 0,090418829 | 0,939706712 | PTPN13 | FALSE | Upregulated |
| -0,081992405 | 0,939706712 | STIM2 | FALSE | Downregulated |
| -0,069090852 | 0,93988104 | RAB27A | FALSE | Downregulated |
| 0,102128259 | 0,939953126 | LOC100499489 | FALSE | Upregulated |
| -0,028167779 | 0,940012562 | HNRNPA1 | FALSE | Downregulated |
| -0,11219607 | 0,940187636 | GRIA3 | FALSE | Downregulated |
| 0,108133085 | 0,940187636 | SPATA31E1 | FALSE | Upregulated |
| 0,107214828 | 0,940325367 | LINC00652 | FALSE | Upregulated |
| -0,088023107 | 0,940638186 | SUSD1 | FALSE | Downregulated |
| -0,080082749 | 0,940644298 | ATPAF2 | FALSE | Downregulated |
| -0,041034705 | 0,940648971 | ATF6 | FALSE | Downregulated |
| 0,108934211 | 0,940648971 | CKB | FALSE | Upregulated |
| 0,125548037 | 0,940648971 | LRP2 | FALSE | Upregulated |
| -0,043964912 | 0,940648971 | PAN2 | FALSE | Downregulated |
| 0,079767264 | 0,940648971 | RASIP1 | FALSE | Upregulated |
| 0,069492625 | 0,940648971 | SLC2A9 | FALSE | Upregulated |
| 0,107777064 | 0,94072218 | EZH2 | FALSE | Upregulated |
| -0,039606176 | 0,941079261 | ARHGEF7 | FALSE | Downregulated |
| 0,045178192 | 0,941136145 | MAN1B1 | FALSE | Upregulated |
| -0,04828603 | 0,941278471 | RDH16 | FALSE | Downregulated |
| -0,072036393 | 0,941278471 | SCARNA5 | FALSE | Downregulated |
| -0,101852486 | 0,941284044 | GCA | FALSE | Downregulated |
| -0,153504115 | 0,941284044 | PEG10 | FALSE | Downregulated |
| -0,059453194 | 0,941284044 | SAA4 | FALSE | Downregulated |
| -0,020986455 | 0,941298943 | GNAS | FALSE | Downregulated |
| -0,095961836 | 0,94129952 | CDRT15 | FALSE | Downregulated |
| 0,08639154 | 0,94129952 | IQCH-AS1 | FALSE | Upregulated |
| -0,090334412 | 0,94129952 | SIRT6 | FALSE | Downregulated |
| 0,047769429 | 0,941316366 | UBE2Z | FALSE | Upregulated |
| 0,062899398 | 0,941341579 | CCDC66 | FALSE | Upregulated |
| 0,114989245 | 0,941341579 | E2F3 | FALSE | Upregulated |
| -0,099427235 | 0,941341579 | LYSMD2 | FALSE | Downregulated |
| 0,028038086 | 0,941341579 | SSR1 | FALSE | Upregulated |
| -0,06379917 | 0,941377658 | FNBP1L | FALSE | Downregulated |
| 0,112441446 | 0,941377658 | RBPMS-AS1 | FALSE | Upregulated |
| 0,048623195 | 0,941489845 | ALDH1L2 | FALSE | Upregulated |
| 0,029605357 | 0,941490215 | CCNL2 | FALSE | Upregulated |
| -0,055781527 | 0,941490215 | DDX49 | FALSE | Downregulated |
| -0,103946876 | 0,941490215 | LOC101927105 | FALSE | Downregulated |
| 0,094276762 | 0,941490215 | SNRNP40 | FALSE | Upregulated |
| -0,048617501 | 0,941491229 | MAEA | FALSE | Downregulated |
| 0,089587291 | 0,941698732 | CELF6 | FALSE | Upregulated |
| 0,043448392 | 0,941698732 | CLASRP | FALSE | Upregulated |
| -0,062213865 | 0,941698732 | RAB3IL1 | FALSE | Downregulated |
| 0,03749091 | 0,941698732 | TMEM30A | FALSE | Upregulated |

| | | | | |
|---|---|---|---|---|
| 0,057898732 | 0,941698732 | ZNF404 | FALSE | Upregulated |
| 0,044857705 | 0,941806544 | UCKL1 | FALSE | Upregulated |
| -0,057674241 | 0,941960708 | HSD17B12 | FALSE | Downregulated |
| -0,099757177 | 0,942116481 | C1orf226 | FALSE | Downregulated |
| -0,089281693 | 0,942233798 | DKFZP586I1420 | FALSE | Downregulated |
| -0,086354736 | 0,942233798 | EED | FALSE | Downregulated |
| 0,050092266 | 0,942233798 | ELAVL1 | FALSE | Upregulated |
| -0,061735592 | 0,942233798 | HCN3 | FALSE | Downregulated |
| 0,044754483 | 0,942233798 | KAT5 | FALSE | Upregulated |
| 0,104657445 | 0,942233798 | LRRC56 | FALSE | Upregulated |
| -0,067460639 | 0,942233798 | MAX | FALSE | Downregulated |
| 0,048625683 | 0,942233798 | SLC47A2 | FALSE | Upregulated |
| -0,060403478 | 0,942233798 | SPAST | FALSE | Downregulated |
| 0,071155622 | 0,942233798 | ZNF558 | FALSE | Upregulated |
| -0,051879222 | 0,942324182 | SMCHD1 | FALSE | Downregulated |
| 0,082096012 | 0,942324182 | TARSL2 | FALSE | Upregulated |
| -0,056772192 | 0,942550341 | AP5B1 | FALSE | Downregulated |
| 0,099113668 | 0,942550341 | COLQ | FALSE | Upregulated |
| -0,089722745 | 0,942550341 | DICER1-AS1 | FALSE | Downregulated |
| -0,083816596 | 0,942550341 | FASN | FALSE | Downregulated |
| 0,070056872 | 0,942550341 | KLHDC4 | FALSE | Upregulated |
| 0,042271559 | 0,942550341 | LPGAT1 | FALSE | Upregulated |
| -0,062747498 | 0,942550341 | LRSAM1 | FALSE | Downregulated |
| 0,049937611 | 0,942550341 | MDM4 | FALSE | Upregulated |
| 0,028434483 | 0,942550341 | NKTR | FALSE | Upregulated |
| -0,036219773 | 0,942550341 | PCM1 | FALSE | Downregulated |
| -0,063409477 | 0,942550341 | PIAS2 | FALSE | Downregulated |
| 0,104964774 | 0,942550341 | PLK3 | FALSE | Upregulated |
| -0,129185336 | 0,942550341 | SEMA3B | FALSE | Downregulated |
| -0,043704606 | 0,942550341 | TAF6L | FALSE | Downregulated |
| -0,075693132 | 0,942550341 | TBC1D8B | FALSE | Downregulated |
| 0,060642936 | 0,942550341 | UBE2M | FALSE | Upregulated |
| 0,041329716 | 0,942550341 | WDR26 | FALSE | Upregulated |
| -0,063951469 | 0,942838392 | C10orf12 | FALSE | Downregulated |
| -0,06295826 | 0,942838392 | C9orf64 | FALSE | Downregulated |
| -0,113272757 | 0,942838392 | DCLK2 | FALSE | Downregulated |
| -0,055572866 | 0,942838392 | HYAL2 | FALSE | Downregulated |
| 0,054489778 | 0,942838392 | MRI1 | FALSE | Upregulated |
| -0,063166355 | 0,942838392 | PEX2 | FALSE | Downregulated |
| 0,077232252 | 0,942838392 | RND3 | FALSE | Upregulated |
| -0,084797109 | 0,942838392 | RRAS | FALSE | Downregulated |
| 0,035355809 | 0,942890192 | SIPA1L3 | FALSE | Upregulated |
| -0,043952373 | 0,942890192 | ZDHHC8 | FALSE | Downregulated |
| -0,082367421 | 0,94300308 | MPP6 | FALSE | Downregulated |
| 0,065794008 | 0,943016986 | ATP6V0B | FALSE | Upregulated |
| 0,063896363 | 0,943092493 | KREMEN1 | FALSE | Upregulated |
| 0,108914703 | 0,943092493 | SLC29A3 | FALSE | Upregulated |
| 0,065958025 | 0,943130654 | WWC1 | FALSE | Upregulated |
| 0,05854352 | 0,943622351 | BMP6 | FALSE | Upregulated |
| -0,045230368 | 0,943622351 | MAP1S | FALSE | Downregulated |
| 0,032324611 | 0,943622351 | MPP5 | FALSE | Upregulated |

| | | | | |
|---|---|---|---|---|
| 0,070490741 | 0,943622351 | SESTD1 | FALSE | Upregulated |
| -0,059138129 | 0,943622351 | TPRA1 | FALSE | Downregulated |
| -0,036248024 | 0,943708479 | NSUN2 | FALSE | Downregulated |
| -0,043936279 | 0,943797378 | CEND1 | FALSE | Downregulated |
| -0,06957303 | 0,943797378 | COPS2 | FALSE | Downregulated |
| 0,061823929 | 0,943797378 | ECT2L | FALSE | Upregulated |
| 0,064088891 | 0,943797378 | FAM110C | FALSE | Upregulated |
| -0,02990216 | 0,943797378 | TENC1 | FALSE | Downregulated |
| -0,050829023 | 0,943804297 | CCDC9 | FALSE | Downregulated |
| -0,042784915 | 0,943804297 | CGGBP1 | FALSE | Downregulated |
| -0,054808746 | 0,943804297 | LEO1 | FALSE | Downregulated |
| -0,06911606 | 0,943804297 | LGALS8-AS1 | FALSE | Downregulated |
| -0,096529552 | 0,943804297 | MPPED1 | FALSE | Downregulated |
| 0,046610476 | 0,943804297 | NR3C2 | FALSE | Upregulated |
| -0,047347728 | 0,943804297 | TCAIM | FALSE | Downregulated |
| 0,112453165 | 0,943938614 | DDX12P | FALSE | Upregulated |
| 0,087773366 | 0,943938614 | DNASE1L2 | FALSE | Upregulated |
| -0,104474699 | 0,943938614 | FBF1 | FALSE | Downregulated |
| 0,062088684 | 0,943938614 | MLK7-AS1 | FALSE | Upregulated |
| 0,032593627 | 0,943938614 | TM9SF3 | FALSE | Upregulated |
| 0,051493855 | 0,943938614 | TUB | FALSE | Upregulated |
| -0,074716476 | 0,944059555 | HKR1 | FALSE | Downregulated |
| -0,044660035 | 0,944059555 | NUB1 | FALSE | Downregulated |
| 1,120921464 | 0,944059555 | RNU1-4 | FALSE | Upregulated |
| 0,057863527 | 0,944059555 | TMEM69 | FALSE | Upregulated |
| -0,087839501 | 0,944059555 | TP53RK | FALSE | Downregulated |
| -0,049278683 | 0,944059555 | USP42 | FALSE | Downregulated |
| 0,064287328 | 0,944132913 | NAV2-AS5 | FALSE | Upregulated |
| -0,107120102 | 0,944141889 | LOC102724757 | FALSE | Downregulated |
| -0,077829038 | 0,944294307 | DDR1 | FALSE | Downregulated |
| -0,116142904 | 0,944294307 | GADD45G | FALSE | Downregulated |
| 0,077315021 | 0,944294307 | PDCD1LG2 | FALSE | Upregulated |
| -0,035125603 | 0,944294307 | TNKS2 | FALSE | Downregulated |
| 0,036597553 | 0,944394729 | ANGPTL6 | FALSE | Upregulated |
| 0,060369648 | 0,944394729 | ANKRD37 | FALSE | Upregulated |
| 0,043731442 | 0,944394729 | ARFRP1 | FALSE | Upregulated |
| -0,142240818 | 0,944394729 | C17orf49 | FALSE | Downregulated |
| -0,103445944 | 0,944394729 | C1orf174 | FALSE | Downregulated |
| 0,052580413 | 0,944394729 | CLDN14 | FALSE | Upregulated |
| -0,087547483 | 0,944394729 | ENPP2 | FALSE | Downregulated |
| -0,080995414 | 0,944394729 | FLG | FALSE | Downregulated |
| 0,061367008 | 0,944394729 | GJB2 | FALSE | Upregulated |
| -0,086110362 | 0,944394729 | IGFLR1 | FALSE | Downregulated |
| -0,041708462 | 0,944394729 | LOC100289511 | FALSE | Downregulated |
| -0,044231754 | 0,944394729 | LOC101928897 | FALSE | Downregulated |
| 0,050910351 | 0,944394729 | LOC102723328 | FALSE | Upregulated |
| 0,074843535 | 0,944394729 | MCUR1 | FALSE | Upregulated |
| 0,069915853 | 0,944394729 | MRRF | FALSE | Upregulated |
| -0,041506716 | 0,944394729 | PLXNA2 | FALSE | Downregulated |
| -0,088910578 | 0,944394729 | POLA2 | FALSE | Downregulated |
| 0,037408998 | 0,944394729 | PSMA7 | FALSE | Upregulated |

| | | | | |
|---|---|---|---|---|
| -0,060608012 | 0,944394729 | RBBP5 | FALSE | Downregulated |
| -0,048066774 | 0,944394729 | RNF111 | FALSE | Downregulated |
| 0,050305815 | 0,944394729 | TCF3 | FALSE | Upregulated |
| 0,036767282 | 0,944394729 | UGGT1 | FALSE | Upregulated |
| 0,03552621 | 0,944394729 | USP32 | FALSE | Upregulated |
| -0,035933987 | 0,944394729 | VAV2 | FALSE | Downregulated |
| -0,082075329 | 0,944394729 | ZNF287 | FALSE | Downregulated |
| 0,086059149 | 0,944394729 | ZNF418 | FALSE | Upregulated |
| -0,050619964 | 0,944394729 | ZNF428 | FALSE | Downregulated |
| -0,03857736 | 0,944789831 | RAD50 | FALSE | Downregulated |
| -0,081113327 | 0,944798271 | CCT6P1 | FALSE | Downregulated |
| -0,037091941 | 0,944798271 | ENDOG | FALSE | Downregulated |
| 0,052697651 | 0,944798271 | LILRB5 | FALSE | Upregulated |
| 0,077482754 | 0,944798271 | SACM1L | FALSE | Upregulated |
| -0,029082791 | 0,944995642 | TET3 | FALSE | Downregulated |
| -0,034518752 | 0,945023854 | ABCG5 | FALSE | Downregulated |
| -0,057701989 | 0,945023854 | MEA1 | FALSE | Downregulated |
| 0,038899592 | 0,945023854 | SLAIN2 | FALSE | Upregulated |
| -0,039578639 | 0,945061517 | VWA8 | FALSE | Downregulated |
| -0,084586472 | 0,945143146 | CD300A | FALSE | Downregulated |
| 0,046326767 | 0,945250837 | C22orf29 | FALSE | Upregulated |
| 0,177670624 | 0,945250837 | GUSBP3 | FALSE | Upregulated |
| -0,105339694 | 0,945250837 | LOC101927313 | FALSE | Downregulated |
| -0,140044902 | 0,945250837 | LOC101928195 | FALSE | Downregulated |
| 0,071529546 | 0,945250837 | LOC102724730 | FALSE | Upregulated |
| -0,057781564 | 0,945250837 | MAGEF1 | FALSE | Downregulated |
| 0,066692898 | 0,945250837 | TMEM44-AS1 | FALSE | Upregulated |
| 0,03652542 | 0,945250837 | ZBTB4 | FALSE | Upregulated |
| 0,029698775 | 0,945400102 | AMT | FALSE | Upregulated |
| 0,053427247 | 0,945400102 | ARHGEF39 | FALSE | Upregulated |
| 0,034294246 | 0,945400102 | CCDC88A | FALSE | Upregulated |
| 0,041296681 | 0,945400102 | DNAJC22 | FALSE | Upregulated |
| 0,087563237 | 0,945400102 | DUSP11 | FALSE | Upregulated |
| 0,063472785 | 0,945400102 | LOC283922 | FALSE | Upregulated |
| 0,066413092 | 0,945400102 | LOC646719 | FALSE | Upregulated |
| 0,067232406 | 0,945400102 | MTPAP | FALSE | Upregulated |
| 0,053313637 | 0,945400102 | PDS5B | FALSE | Upregulated |
| 0,099607879 | 0,945400102 | PLGLB1 | FALSE | Upregulated |
| -0,021702015 | 0,945400102 | POLDIP2 | FALSE | Downregulated |
| -0,085751863 | 0,945400102 | POLR3C | FALSE | Downregulated |
| 0,036820948 | 0,945400102 | SEC24D | FALSE | Upregulated |
| -0,044057965 | 0,945400102 | SNX14 | FALSE | Downregulated |
| 0,035837862 | 0,945400102 | SOS1 | FALSE | Upregulated |
| 0,036815934 | 0,945400102 | STAG2 | FALSE | Upregulated |
| 0,06296538 | 0,945400102 | UROS | FALSE | Upregulated |
| 0,048727535 | 0,945400102 | ZAK | FALSE | Upregulated |
| 0,036300613 | 0,945400102 | ZNF507 | FALSE | Upregulated |
| 0,08813192 | 0,945400102 | ZNF568 | FALSE | Upregulated |
| -0,055613095 | 0,945414359 | SPRY2 | FALSE | Downregulated |
| -0,066992106 | 0,945543041 | PLXNA3 | FALSE | Downregulated |
| 0,059216913 | 0,945543041 | ZFYVE27 | FALSE | Upregulated |

| | | | | |
|---|---|---|---|---|
| 0,082154776 | 0,945591851 | SDS | FALSE | Upregulated |
| -0,025790245 | 0,945626904 | SF3A1 | FALSE | Downregulated |
| -0,041880854 | 0,94563426 | ATP2B2 | FALSE | Downregulated |
| 0,039738416 | 0,945904563 | PPP4C | FALSE | Upregulated |
| 0,034921228 | 0,945985869 | FNDC3B | FALSE | Upregulated |
| -0,03266984 | 0,945985869 | RBM10 | FALSE | Downregulated |
| -0,027802675 | 0,945990784 | EIF3A | FALSE | Downregulated |
| -0,110157344 | 0,945990784 | LPAR5 | FALSE | Downregulated |
| 0,088718279 | 0,945990784 | PODN | FALSE | Upregulated |
| 0,080056069 | 0,94605116 | MRPS31P5 | FALSE | Upregulated |
| 0,069907841 | 0,946151438 | COA7 | FALSE | Upregulated |
| 0,037691042 | 0,946151438 | DOCK1 | FALSE | Upregulated |
| 0,038808669 | 0,946151438 | EPOR | FALSE | Upregulated |
| 0,11373939 | 0,946151438 | IFI44 | FALSE | Upregulated |
| 0,11181864 | 0,946151438 | LOC101927895 | FALSE | Upregulated |
| 0,050140529 | 0,946151438 | SNTA1 | FALSE | Upregulated |
| -0,09890729 | 0,946151438 | TRAPPC2L | FALSE | Downregulated |
| -0,02766414 | 0,946313512 | ANKRD17 | FALSE | Downregulated |
| -0,080562901 | 0,946313512 | BPNT1 | FALSE | Downregulated |
| -0,021842324 | 0,946313512 | C6orf62 | FALSE | Downregulated |
| 0,063297518 | 0,946313512 | CAMSAP2 | FALSE | Upregulated |
| 0,033536818 | 0,946313512 | CCDC85C | FALSE | Upregulated |
| -0,041912469 | 0,946313512 | COPE | FALSE | Downregulated |
| -0,044380146 | 0,946313512 | COQ9 | FALSE | Downregulated |
| -0,140216989 | 0,946313512 | CTAGE4 | FALSE | Downregulated |
| 0,038984182 | 0,946313512 | EIF2A | FALSE | Upregulated |
| -0,06567511 | 0,946313512 | EVI5L | FALSE | Downregulated |
| 0,041618035 | 0,946313512 | FAM21C | FALSE | Upregulated |
| -0,031851824 | 0,946313512 | FAM65C | FALSE | Downregulated |
| -0,046648072 | 0,946313512 | FLT1 | FALSE | Downregulated |
| -0,028872842 | 0,946313512 | GPRC5C | FALSE | Downregulated |
| -0,044228091 | 0,946313512 | IARS | FALSE | Downregulated |
| 0,047209901 | 0,946313512 | IFNAR2 | FALSE | Upregulated |
| 0,040683607 | 0,946313512 | IGFBP7 | FALSE | Upregulated |
| 0,089308294 | 0,946313512 | LOC100288069 | FALSE | Upregulated |
| -0,041309565 | 0,946313512 | LOC101928445 | FALSE | Downregulated |
| 0,237742103 | 0,946313512 | LOC102724374 | FALSE | Upregulated |
| -0,058016474 | 0,946313512 | PCK1 | FALSE | Downregulated |
| -0,055425147 | 0,946313512 | PRR14 | FALSE | Downregulated |
| 0,19773533 | 0,946313512 | RPL17-C18orf32 | FALSE | Upregulated |
| 0,026937748 | 0,946313512 | SERPINA10 | FALSE | Upregulated |
| -0,0637252 | 0,946313512 | TBL2 | FALSE | Downregulated |
| 0,054958212 | 0,946313512 | TCF4 | FALSE | Upregulated |
| -0,087138246 | 0,946313512 | TNFSF13 | FALSE | Downregulated |
| -0,066741292 | 0,946313512 | ZNF830 | FALSE | Downregulated |
| -0,048677701 | 0,94644299 | BST2 | FALSE | Downregulated |
| 0,076139703 | 0,94644299 | CNTFR-AS1 | FALSE | Upregulated |
| -0,058452105 | 0,94644299 | DPY19L1 | FALSE | Downregulated |
| -0,080666698 | 0,94644299 | IQSEC3 | FALSE | Downregulated |
| -0,055641347 | 0,94644299 | MRPS2 | FALSE | Downregulated |
| -0,024974984 | 0,946790731 | TRIM44 | FALSE | Downregulated |

| | | | | |
|---|---|---|---|---|
| -0,042019042 | 0,946813069 | ARID4B | FALSE | Downregulated |
| 0,065495519 | 0,946851422 | CNTFR | FALSE | Upregulated |
| 0,469043121 | 0,946851422 | LOC102724624 | FALSE | Upregulated |
| 0,081474625 | 0,946851422 | NNT-AS1 | FALSE | Upregulated |
| 0,072935259 | 0,946851422 | SMCO3 | FALSE | Upregulated |
| 0,098360848 | 0,946874078 | LOC102723741 | FALSE | Upregulated |
| 0,101220318 | 0,947013126 | LOC102723782 | FALSE | Upregulated |
| -0,066534759 | 0,947167623 | AP3S1 | FALSE | Downregulated |
| -0,036122976 | 0,947167623 | C4BPA | FALSE | Downregulated |
| -0,077918837 | 0,947167623 | FOXP2 | FALSE | Downregulated |
| -0,080433551 | 0,947167623 | HAPLN4 | FALSE | Downregulated |
| 0,061154896 | 0,947167623 | NOL10 | FALSE | Upregulated |
| 0,051528084 | 0,947167623 | RAB11FIP1 | FALSE | Upregulated |
| -0,094797254 | 0,947167623 | RDH5 | FALSE | Downregulated |
| 0,021986051 | 0,947167623 | TMEM123 | FALSE | Upregulated |
| -0,06739721 | 0,947180346 | LOC102724562 | FALSE | Downregulated |
| 0,099170148 | 0,947180346 | SDK2 | FALSE | Upregulated |
| -0,041003237 | 0,947319094 | KCND3 | FALSE | Downregulated |
| -0,033829446 | 0,947319094 | LRIG1 | FALSE | Downregulated |
| -0,056278625 | 0,947448411 | CCDC124 | FALSE | Downregulated |
| 0,04859928 | 0,947448411 | GNAO1 | FALSE | Upregulated |
| 0,076433354 | 0,947448411 | LACC1 | FALSE | Upregulated |
| -0,077370704 | 0,947448411 | ZBTB34 | FALSE | Downregulated |
| -0,065849484 | 0,94751924 | PAM | FALSE | Downregulated |
| 0,052417325 | 0,947604136 | FDPS | FALSE | Upregulated |
| 0,034022522 | 0,947607874 | ELMSAN1 | FALSE | Upregulated |
| -0,0536201 | 0,947607874 | RHOT1 | FALSE | Downregulated |
| 0,076756776 | 0,947610142 | LOC102724254 | FALSE | Upregulated |
| 0,032274853 | 0,947610142 | OIT3 | FALSE | Upregulated |
| -0,081649955 | 0,947818001 | CHIC2 | FALSE | Downregulated |
| -0,08711325 | 0,947818001 | MROH8 | FALSE | Downregulated |
| 0,059012814 | 0,947818001 | TGM2 | FALSE | Upregulated |
| 0,032893262 | 0,9479054 | C16orf72 | FALSE | Upregulated |
| 0,106593341 | 0,947937384 | C5orf63 | FALSE | Upregulated |
| -0,041477149 | 0,94807268 | MRPS18B | FALSE | Downregulated |
| 0,064681133 | 0,948192764 | CBFB | FALSE | Upregulated |
| 0,069447107 | 0,948209902 | RABAC1 | FALSE | Upregulated |
| 0,099000094 | 0,948297347 | C11orf80 | FALSE | Upregulated |
| -0,074502122 | 0,948297347 | CASP4 | FALSE | Downregulated |
| -0,094522994 | 0,948297347 | CHST4 | FALSE | Downregulated |
| -0,070777266 | 0,948297347 | DAB1 | FALSE | Downregulated |
| -0,092502549 | 0,948297347 | DBN1 | FALSE | Downregulated |
| 0,080828711 | 0,948297347 | DCAF17 | FALSE | Upregulated |
| 0,072581812 | 0,948297347 | GPRC5D | FALSE | Upregulated |
| 0,094613157 | 0,948297347 | HDAC9 | FALSE | Upregulated |
| -0,063423663 | 0,948297347 | LAMA4 | FALSE | Downregulated |
| 0,036374828 | 0,948297347 | LUC7L2 | FALSE | Upregulated |
| -0,040289686 | 0,948297347 | LYPLA2 | FALSE | Downregulated |
| -0,040447472 | 0,948297347 | MRPL2 | FALSE | Downregulated |
| 0,038172099 | 0,948297347 | ODF3B | FALSE | Upregulated |
| 0,048897921 | 0,948297347 | PEG3-AS1 | FALSE | Upregulated |

| | | | | |
|---|---|---|---|---|
| 0,048524676 | 0,948297347 | PEX11G | FALSE | Upregulated |
| -0,063561463 | 0,948297347 | PLAA | FALSE | Downregulated |
| 0,239296394 | 0,948297347 | PPAN-P2RY11 | FALSE | Upregulated |
| 0,062987142 | 0,948297347 | PRMT9 | FALSE | Upregulated |
| 0,110191097 | 0,948297347 | PSMG3 | FALSE | Upregulated |
| 0,095139773 | 0,948297347 | PTCH2 | FALSE | Upregulated |
| -0,072067036 | 0,948297347 | RRP8 | FALSE | Downregulated |
| -0,066080162 | 0,948297347 | S100PBP | FALSE | Downregulated |
| -0,034189469 | 0,948297347 | SAMD4B | FALSE | Downregulated |
| -0,275116581 | 0,948297347 | SIK1 | FALSE | Downregulated |
| 0,066037923 | 0,948297347 | SLC8A1 | FALSE | Upregulated |
| 0,026152418 | 0,948297347 | SPG11 | FALSE | Upregulated |
| 0,035154398 | 0,948297347 | SUN1 | FALSE | Upregulated |
| 0,048309235 | 0,948297347 | TGFBI | FALSE | Upregulated |
| -0,061593034 | 0,948297347 | TNFRSF21 | FALSE | Downregulated |
| -0,057427772 | 0,948297347 | TOMM7 | FALSE | Downregulated |
| -0,070421175 | 0,948297347 | TRIM11 | FALSE | Downregulated |
| -0,048246734 | 0,948297347 | WWC3 | FALSE | Downregulated |
| 0,070805832 | 0,948406697 | EDNRB-AS1 | FALSE | Upregulated |
| -0,080491395 | 0,948436127 | C9orf142 | FALSE | Downregulated |
| 0,041587384 | 0,948436127 | FBRS | FALSE | Upregulated |
| -0,053832443 | 0,948436127 | MED25 | FALSE | Downregulated |
| -0,060846542 | 0,948436127 | MTX3 | FALSE | Downregulated |
| 0,053363577 | 0,948436127 | PER3 | FALSE | Upregulated |
| -0,06731963 | 0,948436127 | SAMM50 | FALSE | Downregulated |
| -0,054959837 | 0,948436127 | ZEB1 | FALSE | Downregulated |
| -0,045069123 | 0,948512089 | ATP5F1 | FALSE | Downregulated |
| -0,070789859 | 0,948542996 | FAM122A | FALSE | Downregulated |
| -0,047327643 | 0,948542996 | HMGN3 | FALSE | Downregulated |
| -0,072304445 | 0,948542996 | PRRG4 | FALSE | Downregulated |
| -0,028973948 | 0,948542996 | RBM39 | FALSE | Downregulated |
| 0,066434701 | 0,948542996 | ZFP14 | FALSE | Upregulated |
| -0,029024344 | 0,948616544 | H3F3A | FALSE | Downregulated |
| -0,044656038 | 0,948616544 | NDUFA6 | FALSE | Downregulated |
| 0,071736275 | 0,948896328 | NPIPA5 | FALSE | Upregulated |
| 0,074328087 | 0,949021188 | ZNF761 | FALSE | Upregulated |
| 0,039920726 | 0,949206247 | CAMK2D | FALSE | Upregulated |
| 0,068282376 | 0,949206247 | NME6 | FALSE | Upregulated |
| 0,06909201 | 0,949206247 | PHLDA1 | FALSE | Upregulated |
| -0,098299261 | 0,949228544 | C9orf89 | FALSE | Downregulated |
| 0,088957632 | 0,949504894 | LOC440390 | FALSE | Upregulated |
| -0,03801657 | 0,949504894 | SGSM3 | FALSE | Downregulated |
| 0,042921049 | 0,949560854 | CABP7 | FALSE | Upregulated |
| 0,089291767 | 0,949560854 | ZNF658 | FALSE | Upregulated |
| 0,069190123 | 0,949585845 | SLC25A51 | FALSE | Upregulated |
| 0,86861211 | 0,949585845 | XIST | FALSE | Upregulated |
| -0,076776827 | 0,949698885 | TTC27 | FALSE | Downregulated |
| 0,078680279 | 0,949793293 | AC011747.4 | FALSE | Upregulated |
| -0,092447631 | 0,949793293 | YBEY | FALSE | Downregulated |
| -0,09192172 | 0,949907556 | ISY1 | FALSE | Downregulated |
| 0,033937035 | 0,949947441 | B3GNT9 | FALSE | Upregulated |

| | | | | |
|---|---|---|---|---|
| 0,070218657 | 0,949947441 | CAMLG | FALSE | Upregulated |
| -0,040855942 | 0,949947441 | DNAJC4 | FALSE | Downregulated |
| -0,042426702 | 0,949947441 | GSR | FALSE | Downregulated |
| 0,085233768 | 0,949947441 | KAAG1 | FALSE | Upregulated |
| -0,021113166 | 0,949947441 | KMT2A | FALSE | Downregulated |
| 0,060636078 | 0,949947441 | LOC102724262 | FALSE | Upregulated |
| -0,026270291 | 0,949947441 | RREB1 | FALSE | Downregulated |
| -0,027056083 | 0,949947441 | SLC16A2 | FALSE | Downregulated |
| 0,053578072 | 0,949947441 | SLC44A2 | FALSE | Upregulated |
| -0,055763813 | 0,949947441 | SLC5A6 | FALSE | Downregulated |
| -0,068325003 | 0,949947441 | WIBG | FALSE | Downregulated |
| 0,091094712 | 0,950046172 | GLT8D2 | FALSE | Upregulated |
| -0,037865933 | 0,950046172 | PEX5 | FALSE | Downregulated |
| 0,046907054 | 0,950046172 | RNF19B | FALSE | Upregulated |
| -0,07560216 | 0,950046172 | SMIM2 | FALSE | Downregulated |
| 0,054272089 | 0,950046172 | VPS18 | FALSE | Upregulated |
| 0,06295951 | 0,950046172 | ZNF680 | FALSE | Upregulated |
| -0,052489415 | 0,950046172 | ZNF780B | FALSE | Downregulated |
| -0,078967068 | 0,950108131 | CARF | FALSE | Downregulated |
| -0,033864144 | 0,950108131 | TPST2 | FALSE | Downregulated |
| 0,040438292 | 0,950378892 | LOC101929340 | FALSE | Upregulated |
| 0,040552742 | 0,950454532 | LOC101929079 | FALSE | Upregulated |
| -0,033249129 | 0,950454532 | PPP2R1A | FALSE | Downregulated |
| -0,065051349 | 0,950454532 | TSPYL2 | FALSE | Downregulated |
| 0,076421958 | 0,950737062 | DCSTAMP | FALSE | Upregulated |
| -0,035847411 | 0,95075559 | PNKD | FALSE | Downregulated |
| -0,044103095 | 0,95075559 | PSMA6 | FALSE | Downregulated |
| 0,08748398 | 0,950999321 | LOC101928609 | FALSE | Upregulated |
| -0,237343121 | 0,950999321 | ZFP91-CNTF | FALSE | Downregulated |
| -0,030748094 | 0,951090649 | MBD1 | FALSE | Downregulated |
| 0,088795399 | 0,951111745 | CASP1 | FALSE | Upregulated |
| 0,066590442 | 0,951223453 | DDX11 | FALSE | Upregulated |
| -0,237896916 | 0,951405723 | RBM14-RBM4 | FALSE | Downregulated |
| -0,029187364 | 0,951446588 | ACSL5 | FALSE | Downregulated |
| -0,042516351 | 0,951446588 | C16orf62 | FALSE | Downregulated |
| 0,097117231 | 0,951446588 | CD7 | FALSE | Upregulated |
| -0,030721415 | 0,951446588 | DNAJC5 | FALSE | Downregulated |
| -0,030610489 | 0,951446588 | FAM105B | FALSE | Downregulated |
| -0,039974281 | 0,951446588 | LIMS2 | FALSE | Downregulated |
| 0,03657857 | 0,951446588 | NECAP2 | FALSE | Upregulated |
| -0,081598718 | 0,951446588 | NUAK2 | FALSE | Downregulated |
| 0,052072536 | 0,951446588 | PTPRH | FALSE | Upregulated |
| -0,025059768 | 0,951446588 | SCRIB | FALSE | Downregulated |
| -0,060101297 | 0,951446588 | SIPA1L2 | FALSE | Downregulated |
| 0,044522261 | 0,951446588 | TRIP6 | FALSE | Upregulated |
| 0,048791771 | 0,951446588 | ZBTB48 | FALSE | Upregulated |
| 0,091101494 | 0,951627985 | RFX3 | FALSE | Upregulated |
| -0,034226537 | 0,951647118 | BAAT | FALSE | Downregulated |
| 0,045558107 | 0,951647118 | PIGT | FALSE | Upregulated |
| -0,048855544 | 0,951647118 | SMIM19 | FALSE | Downregulated |
| 0,051813333 | 0,951902328 | ASPN | FALSE | Upregulated |

| | | | | |
|---|---|---|---|---|
| -0,033590279 | 0,951902328 | CASKIN2 | FALSE | Downregulated |
| -0,028542815 | 0,951902328 | CDC34 | FALSE | Downregulated |
| -0,049761039 | 0,951902328 | GIPC1 | FALSE | Downregulated |
| -0,053562086 | 0,951902328 | RHOD | FALSE | Downregulated |
| 0,041514634 | 0,951902328 | RNF144B | FALSE | Upregulated |
| 0,042961737 | 0,951902328 | TLR4 | FALSE | Upregulated |
| -0,028616783 | 0,951937137 | CD99L2 | FALSE | Downregulated |
| 0,035353733 | 0,951937137 | LIG3 | FALSE | Upregulated |
| 0,06541856 | 0,951937137 | PCDH9 | FALSE | Upregulated |
| 0,029284563 | 0,951937137 | RAB1B | FALSE | Upregulated |
| 0,077502522 | 0,951937137 | RP11-432I5.1 | FALSE | Upregulated |
| -0,042662489 | 0,951937137 | STAP2 | FALSE | Downregulated |
| -0,042782027 | 0,951937137 | TAPT1 | FALSE | Downregulated |
| 0,043761167 | 0,951937137 | TP53I11 | FALSE | Upregulated |
| 0,064770344 | 0,952163561 | GTF2E2 | FALSE | Upregulated |
| 0,042346137 | 0,952163561 | ILF2 | FALSE | Upregulated |
| 0,055673092 | 0,952163561 | SLC7A7 | FALSE | Upregulated |
| 0,070032822 | 0,952163561 | TBXA2R | FALSE | Upregulated |
| 0,053141416 | 0,952232642 | DRG1 | FALSE | Upregulated |
| -0,044563181 | 0,952232642 | GOLGA8B | FALSE | Downregulated |
| 0,056859623 | 0,952232642 | LIMK1 | FALSE | Upregulated |
| 0,073006012 | 0,952232642 | LOC102723820 | FALSE | Upregulated |
| -0,03896949 | 0,952232642 | MYL9 | FALSE | Downregulated |
| 0,071731818 | 0,952232642 | OLA1 | FALSE | Upregulated |
| -0,096859725 | 0,952232642 | PRDM7 | FALSE | Downregulated |
| -0,198269935 | 0,952345754 | CCL15 | FALSE | Downregulated |
| 0,080048344 | 0,952461931 | ALG14 | FALSE | Upregulated |
| 0,023602526 | 0,952461931 | ARHGEF18 | FALSE | Upregulated |
| -0,070072635 | 0,952461931 | CES1P1 | FALSE | Downregulated |
| 0,044099384 | 0,952461931 | CPSF4 | FALSE | Upregulated |
| -0,049866271 | 0,952461931 | DCUN1D4 | FALSE | Downregulated |
| -0,039517531 | 0,952461931 | EVA1A | FALSE | Downregulated |
| -0,058993966 | 0,952461931 | FAM110B | FALSE | Downregulated |
| 0,105837908 | 0,952461931 | KIAA1549 | FALSE | Upregulated |
| -0,045241912 | 0,952461931 | KMT2E-AS1 | FALSE | Downregulated |
| -0,045094488 | 0,952461931 | LGALS3BP | FALSE | Downregulated |
| -0,062716315 | 0,952461931 | MAP4K5 | FALSE | Downregulated |
| -0,060450111 | 0,952461931 | MRPL30 | FALSE | Downregulated |
| -0,040303759 | 0,952461931 | NUP210 | FALSE | Downregulated |
| -0,083907574 | 0,952461931 | OR52D1 | FALSE | Downregulated |
| 0,074355447 | 0,952461931 | OSBPL5 | FALSE | Upregulated |
| 0,091924156 | 0,952461931 | PCDH7 | FALSE | Upregulated |
| 0,054987001 | 0,952461931 | PCMT1 | FALSE | Upregulated |
| -0,052440991 | 0,952461931 | PDGFC | FALSE | Downregulated |
| 0,082089546 | 0,952461931 | PDLIM7 | FALSE | Upregulated |
| 0,059860758 | 0,952461931 | PTGIS | FALSE | Upregulated |
| -0,034712906 | 0,952461931 | SCARNA10 | FALSE | Downregulated |
| 0,069192171 | 0,952461931 | SPATA5 | FALSE | Upregulated |
| 0,034539364 | 0,952461931 | SYNM | FALSE | Upregulated |
| 0,082847804 | 0,952461931 | TRAPPC13 | FALSE | Upregulated |
| 0,033927855 | 0,952461931 | URGCP | FALSE | Upregulated |

| | | | | |
|---|---|---|---|---|
| 0,025719104 | 0,95250349 | BMPR2 | FALSE | Upregulated |
| 0,044250558 | 0,95250349 | CLPTM1L | FALSE | Upregulated |
| 0,067435343 | 0,95250349 | CMTR2 | FALSE | Upregulated |
| 0,081035931 | 0,95250349 | LOC101927542 | FALSE | Upregulated |
| -0,03899394 | 0,95250349 | PHLPP2 | FALSE | Downregulated |
| 0,082407843 | 0,95250349 | URB2 | FALSE | Upregulated |
| -0,071693636 | 0,952671936 | HIGD1B | FALSE | Downregulated |
| 0,040972778 | 0,952671936 | IKZF5 | FALSE | Upregulated |
| 0,025597988 | 0,952671936 | KPNB1 | FALSE | Upregulated |
| -0,025838408 | 0,952671936 | SHANK2 | FALSE | Downregulated |
| 0,037778513 | 0,952671936 | WDR65 | FALSE | Upregulated |
| -0,033376153 | 0,952671936 | ZNF629 | FALSE | Downregulated |
| -0,066043033 | 0,952671936 | ZNF638-IT1 | FALSE | Downregulated |
| -0,052499463 | 0,952752844 | ZNF317 | FALSE | Downregulated |
| 0,065884062 | 0,953042823 | CMYA5 | FALSE | Upregulated |
| 0,030362134 | 0,953072954 | MZF1 | FALSE | Upregulated |
| 0,088983936 | 0,95310061 | LOC101926980 | FALSE | Upregulated |
| 0,07165986 | 0,953115404 | CXorf40A | FALSE | Upregulated |
| -0,023086421 | 0,953318371 | AP2M1 | FALSE | Downregulated |
| -0,033048749 | 0,953318371 | CCNT2 | FALSE | Downregulated |
| -0,029410604 | 0,953318371 | DAZAP1 | FALSE | Downregulated |
| 0,066462373 | 0,953318371 | DISC2 | FALSE | Upregulated |
| 0,033911809 | 0,953318371 | GOPC | FALSE | Upregulated |
| 0,055257691 | 0,953318371 | KIAA1143 | FALSE | Upregulated |
| 0,05706833 | 0,953318371 | LOC102724921 | FALSE | Upregulated |
| 0,054085885 | 0,953318371 | POLR2D | FALSE | Upregulated |
| -0,058939733 | 0,953318371 | PSMC6 | FALSE | Downregulated |
| -0,050992251 | 0,953318371 | R3HDM1 | FALSE | Downregulated |
| 0,067837576 | 0,953318371 | WRN | FALSE | Upregulated |
| -0,033823245 | 0,953318371 | ZDHHC14 | FALSE | Downregulated |
| 0,081030438 | 0,953355217 | ACSBG1 | FALSE | Upregulated |
| 0,059299037 | 0,953506301 | DUSP7 | FALSE | Upregulated |
| 0,040420682 | 0,953506301 | TP73 | FALSE | Upregulated |
| -0,119169294 | 0,953630357 | LOC102723709 | FALSE | Downregulated |
| 0,07803534 | 0,953630357 | LOC143666 | FALSE | Upregulated |
| -0,031473611 | 0,953665953 | EXT2 | FALSE | Downregulated |
| -0,029696131 | 0,953665953 | MFN2 | FALSE | Downregulated |
| 0,085469439 | 0,953665953 | PFDN6 | FALSE | Upregulated |
| -0,064540149 | 0,953665953 | UBE2F | FALSE | Downregulated |
| 0,034976561 | 0,95368794 | AUNIP | FALSE | Upregulated |
| -0,037373112 | 0,95368794 | CNOT2 | FALSE | Downregulated |
| 0,233552114 | 0,95368794 | CORO7-PAM16 | FALSE | Upregulated |
| 0,044987188 | 0,95368794 | EPM2AIP1 | FALSE | Upregulated |
| -0,079411941 | 0,95368794 | ERICH1 | FALSE | Downregulated |
| 0,078269973 | 0,95368794 | FAM156B | FALSE | Upregulated |
| 0,035356657 | 0,95368794 | GNAQ | FALSE | Upregulated |
| -0,119439941 | 0,95368794 | GOLGA6B | FALSE | Downregulated |
| -0,038858495 | 0,95368794 | HS6ST1 | FALSE | Downregulated |
| -0,039346777 | 0,95368794 | IDH3B | FALSE | Downregulated |
| 0,068134312 | 0,95368794 | KANSL2 | FALSE | Upregulated |
| -0,029721535 | 0,95368794 | LIFR | FALSE | Downregulated |

| | | | | |
|---|---|---|---|---|
| 0,04779477 | 0,95368794 | LOC102724724 | FALSE | Upregulated |
| -0,076519523 | 0,95368794 | LRRC37BP1 | FALSE | Downregulated |
| 0,050261909 | 0,95368794 | MAP3K10 | FALSE | Upregulated |
| 0,063430565 | 0,95368794 | OAS1 | FALSE | Upregulated |
| 0,05056475 | 0,95368794 | PAQR9 | FALSE | Upregulated |
| -0,022171035 | 0,95368794 | PBRM1 | FALSE | Downregulated |
| -0,059524437 | 0,95368794 | RBM22 | FALSE | Downregulated |
| 0,046254312 | 0,95368794 | RPUSD4 | FALSE | Upregulated |
| 0,048521242 | 0,95368794 | TRA2A | FALSE | Upregulated |
| 0,038192588 | 0,95368794 | ZNF236 | FALSE | Upregulated |
| 0,056295416 | 0,953906922 | FBXO34 | FALSE | Upregulated |
| 0,058293118 | 0,954114585 | CCDC12 | FALSE | Upregulated |
| -0,03729834 | 0,954114585 | ELL | FALSE | Downregulated |
| -0,050847045 | 0,954114585 | FAM109A | FALSE | Downregulated |
| 0,041968617 | 0,954114585 | LPIN3 | FALSE | Upregulated |
| 0,029813653 | 0,954114585 | SLC25A22 | FALSE | Upregulated |
| -0,044660646 | 0,954114585 | ZMYM3 | FALSE | Downregulated |
| -0,051687977 | 0,954199672 | DEPTOR | FALSE | Downregulated |
| 0,037712225 | 0,954199672 | IKBKB | FALSE | Upregulated |
| 0,053824993 | 0,954199672 | NDUFB2 | FALSE | Upregulated |
| -0,03033283 | 0,954199672 | PANK3 | FALSE | Downregulated |
| 0,132004789 | 0,954199672 | TTTY15 | FALSE | Upregulated |
| -0,086583782 | 0,95425081 | MUTYH | FALSE | Downregulated |
| -0,030461172 | 0,95425081 | WASL | FALSE | Downregulated |
| -0,026313431 | 0,954271371 | ABHD2 | FALSE | Downregulated |
| -0,055900046 | 0,954271371 | TBC1D10A | FALSE | Downregulated |
| 0,043563532 | 0,954352104 | ISCU | FALSE | Upregulated |
| 0,050339615 | 0,954352104 | ZNF668 | FALSE | Upregulated |
| 0,059168195 | 0,955086147 | EXD2 | FALSE | Upregulated |
| 0,056019146 | 0,955236435 | FREM3 | FALSE | Upregulated |
| -0,142952254 | 0,955249742 | RNASEK-C17orf49 | FALSE | Downregulated |
| -0,039201258 | 0,955249742 | TMF1 | FALSE | Downregulated |
| -0,058262231 | 0,955301929 | PRKACB | FALSE | Downregulated |
| -0,05905652 | 0,955305188 | RRM1 | FALSE | Downregulated |
| 0,041460376 | 0,955407665 | DRAP1 | FALSE | Upregulated |
| 0,063542586 | 0,955494738 | NRF1 | FALSE | Upregulated |
| 0,063475711 | 0,955568068 | ZCCHC17 | FALSE | Upregulated |
| -0,053459895 | 0,955648858 | SCML1 | FALSE | Downregulated |
| -0,057976643 | 0,955818986 | ANKRD34A | FALSE | Downregulated |
| 0,066987574 | 0,955818986 | LCLAT1 | FALSE | Upregulated |
| -0,058919934 | 0,955818986 | NAPG | FALSE | Downregulated |
| 0,034221778 | 0,955818986 | SH3BP2 | FALSE | Upregulated |
| -0,028166012 | 0,955818986 | TRIM56 | FALSE | Downregulated |
| 0,030057771 | 0,955946088 | ARMC5 | FALSE | Upregulated |
| -0,067605383 | 0,955946088 | DDHD1 | FALSE | Downregulated |
| -0,028674887 | 0,955946088 | DDX3X | FALSE | Downregulated |
| 0,07476053 | 0,955946088 | DHX35 | FALSE | Upregulated |
| -0,036934726 | 0,955946088 | FUK | FALSE | Downregulated |
| -0,052930106 | 0,955946088 | GABARAPL2 | FALSE | Downregulated |
| -0,02166476 | 0,955946088 | HCFC1 | FALSE | Downregulated |
| 0,07045474 | 0,955946088 | IL10RB-AS1 | FALSE | Upregulated |

| | | | | |
|---|---|---|---|---|
| -0,058760862 | 0,955946088 | LOC101928170 | FALSE | Downregulated |
| 0,045539567 | 0,955946088 | LOC102724975 | FALSE | Upregulated |
| -0,040190977 | 0,955946088 | MEST | FALSE | Downregulated |
| -0,029461845 | 0,955946088 | PARP1 | FALSE | Downregulated |
| -0,032713323 | 0,955946088 | RBM5 | FALSE | Downregulated |
| 0,042538995 | 0,955946088 | RP11-103C16.2 | FALSE | Upregulated |
| 0,031150187 | 0,955946088 | TMEM110 | FALSE | Upregulated |
| -0,036398177 | 0,955953529 | FUCA1 | FALSE | Downregulated |
| 0,033955309 | 0,956042428 | MORF4L2 | FALSE | Upregulated |
| 0,0331084 | 0,956042428 | USP36 | FALSE | Upregulated |
| 0,058462259 | 0,956106665 | EXOC3L2 | FALSE | Upregulated |
| -0,03898366 | 0,956198317 | EIF2D | FALSE | Downregulated |
| -0,045966159 | 0,956198317 | FER | FALSE | Downregulated |
| -0,031706601 | 0,956198317 | LYST | FALSE | Downregulated |
| 0,039910202 | 0,956285543 | CXCR6 | FALSE | Upregulated |
| 0,039020576 | 0,956386865 | DENND5A | FALSE | Upregulated |
| -0,061571655 | 0,956386865 | ENOX2 | FALSE | Downregulated |
| 0,070309999 | 0,956386865 | FUT4 | FALSE | Upregulated |
| 0,051709157 | 0,956386865 | GALK2 | FALSE | Upregulated |
| 0,08119181 | 0,956386865 | POLD3 | FALSE | Upregulated |
| -0,070334048 | 0,956386865 | TFB1M | FALSE | Downregulated |
| -0,036156453 | 0,956386865 | USP6NL | FALSE | Downregulated |
| 0,028446468 | 0,956554698 | FEM1B | FALSE | Upregulated |
| 0,055073398 | 0,956558541 | FIZ1 | FALSE | Upregulated |
| 0,065392686 | 0,956558541 | RCBTB1 | FALSE | Upregulated |
| 0,041175054 | 0,956578497 | AC104134.2 | FALSE | Upregulated |
| -0,023048382 | 0,956578497 | BAP1 | FALSE | Downregulated |
| 0,040647274 | 0,956578497 | C10orf54 | FALSE | Upregulated |
| -0,063509454 | 0,956578497 | CASP2 | FALSE | Downregulated |
| -0,052115445 | 0,956578497 | GTPBP10 | FALSE | Downregulated |
| -0,058957395 | 0,956578497 | IFITM1 | FALSE | Downregulated |
| -0,075548125 | 0,956578497 | MANSC4 | FALSE | Downregulated |
| 0,051553897 | 0,956578497 | MGAT4A | FALSE | Upregulated |
| 0,084586292 | 0,956578497 | NIPAL1 | FALSE | Upregulated |
| 0,035220966 | 0,956578497 | RASAL2 | FALSE | Upregulated |
| 0,044021007 | 0,956578497 | SFI1 | FALSE | Upregulated |
| 0,050612582 | 0,956578497 | SLC25A30 | FALSE | Upregulated |
| 0,093304708 | 0,956578497 | TEX38 | FALSE | Upregulated |
| 0,024158927 | 0,956578497 | TRAK1 | FALSE | Upregulated |
| -0,077308501 | 0,956578497 | ZDHHC24 | FALSE | Downregulated |
| 0,040548128 | 0,956578497 | ZNF768 | FALSE | Upregulated |
| -0,060399875 | 0,956780084 | AVEN | FALSE | Downregulated |
| -0,032502804 | 0,956780084 | HEBP1 | FALSE | Downregulated |
| -0,074408495 | 0,956940367 | CDKN1C | FALSE | Downregulated |
| -0,06601298 | 0,956940959 | RNGTT | FALSE | Downregulated |
| 0,064808222 | 0,957125475 | DEGS1 | FALSE | Upregulated |
| 0,041160309 | 0,957125475 | LOC102723750 | FALSE | Upregulated |
| 0,059000891 | 0,957125475 | SCAMP3 | FALSE | Upregulated |
| 0,027191316 | 0,957195071 | CACFD1 | FALSE | Upregulated |
| -0,092150876 | 0,957195071 | P4HA3 | FALSE | Downregulated |
| -0,034893307 | 0,957195071 | TRIOBP | FALSE | Downregulated |

| | | | | |
|---|---|---|---|---|
| 0,036056596 | 0,957197228 | IKBKAP | FALSE | Upregulated |
| -0,082009421 | 0,957362878 | CCDC70 | FALSE | Downregulated |
| 0,030751616 | 0,957362878 | POGK | FALSE | Upregulated |
| 0,051012282 | 0,957362878 | SRGAP2 | FALSE | Upregulated |
| -0,042481481 | 0,957362878 | ZNF551 | FALSE | Downregulated |
| 0,079861738 | 0,957644337 | ARL6IP6 | FALSE | Upregulated |
| -0,040823506 | 0,957644337 | BRAP | FALSE | Downregulated |
| 0,031073281 | 0,957644337 | CRBN | FALSE | Upregulated |
| 0,036864094 | 0,957644337 | CYP2U1 | FALSE | Upregulated |
| -0,050800232 | 0,957644337 | IFRD1 | FALSE | Downregulated |
| -0,073800522 | 0,957644337 | LOC101928548 | FALSE | Downregulated |
| 0,070641678 | 0,957644337 | RPP25 | FALSE | Upregulated |
| -0,031517941 | 0,957835525 | SPHK2 | FALSE | Downregulated |
| -0,043540907 | 0,95786034 | AGXT2 | FALSE | Downregulated |
| 0,035195561 | 0,95786034 | HN1L | FALSE | Upregulated |
| -0,048415457 | 0,95786034 | SRM | FALSE | Downregulated |
| 0,024861027 | 0,958227298 | SMEK1 | FALSE | Upregulated |
| 0,018337841 | 0,958264826 | CDK13 | FALSE | Upregulated |
| 0,038483457 | 0,958264826 | CDRT15P1 | FALSE | Upregulated |
| -0,035453696 | 0,958264826 | CRYZ | FALSE | Downregulated |
| 0,034674773 | 0,958264826 | DGAT1 | FALSE | Upregulated |
| -0,037590154 | 0,958264826 | ETS2 | FALSE | Downregulated |
| -0,036076387 | 0,958264826 | GABPB1 | FALSE | Downregulated |
| -0,036071108 | 0,958264826 | HLF | FALSE | Downregulated |
| 0,052773265 | 0,958264826 | ITPA | FALSE | Upregulated |
| -0,018422621 | 0,958264826 | LENG8 | FALSE | Downregulated |
| -0,042077204 | 0,958264826 | LIG1 | FALSE | Downregulated |
| -0,065696972 | 0,958264826 | MIER2 | FALSE | Downregulated |
| -0,02594813 | 0,958264826 | SEC13 | FALSE | Downregulated |
| -0,052010031 | 0,958264826 | TFEB | FALSE | Downregulated |
| -0,026867751 | 0,958264826 | TSPAN9 | FALSE | Downregulated |
| 0,038546725 | 0,958445061 | TEF | FALSE | Upregulated |
| -0,055969397 | 0,958597273 | LINC00478 | FALSE | Downregulated |
| 0,023417636 | 0,958694775 | CTSZ | FALSE | Upregulated |
| -0,059177233 | 0,958694775 | PWWP2B | FALSE | Downregulated |
| -0,050938242 | 0,958821004 | PIGV | FALSE | Downregulated |
| 0,057059933 | 0,958899527 | NT5DC3 | FALSE | Upregulated |
| 0,045947262 | 0,959103039 | PIH1D1 | FALSE | Upregulated |
| -0,019178616 | 0,959116986 | AES | FALSE | Downregulated |
| -0,083440282 | 0,959229806 | ADAMTSL1 | FALSE | Downregulated |
| 0,075744052 | 0,959229806 | FAM131A | FALSE | Upregulated |
| 0,025922864 | 0,959229806 | GUCD1 | FALSE | Upregulated |
| 0,047395273 | 0,959229806 | RAB37 | FALSE | Upregulated |
| 0,034673457 | 0,959229806 | ZNF805 | FALSE | Upregulated |
| 0,068267666 | 0,959301467 | VWA2 | FALSE | Upregulated |
| 0,034491108 | 0,959622493 | GOLGA2 | FALSE | Upregulated |
| -0,026781506 | 0,959927804 | RUSC2 | FALSE | Downregulated |
| 0,038383513 | 0,959954072 | DCTN4 | FALSE | Upregulated |
| -0,053538915 | 0,959954072 | HSPA4L | FALSE | Downregulated |
| 0,030605484 | 0,959954072 | ILK | FALSE | Upregulated |
| -0,037880788 | 0,959954072 | LECT2 | FALSE | Downregulated |

| | | | | |
|---|---|---|---|---|
| 0,02168673 | 0,959954072 | MCL1 | FALSE | Upregulated |
| 0,062843422 | 0,959954072 | MRPL27 | FALSE | Upregulated |
| -0,055074216 | 0,959954072 | OR52W1 | FALSE | Downregulated |
| 0,052011466 | 0,959954072 | PRKAG1 | FALSE | Upregulated |
| -0,062628179 | 0,959954072 | SFRP1 | FALSE | Downregulated |
| -0,075891493 | 0,959954072 | SFXN4 | FALSE | Downregulated |
| 0,036169628 | 0,959954072 | SMAD5 | FALSE | Upregulated |
| 0,027206597 | 0,959954072 | SMAP2 | FALSE | Upregulated |
| 0,034643958 | 0,959954072 | SUV420H1 | FALSE | Upregulated |
| -0,042179312 | 0,959992142 | LSM3 | FALSE | Downregulated |
| -0,046330293 | 0,960043071 | MAPK8IP1 | FALSE | Downregulated |
| 0,027493437 | 0,960162154 | CCDC47 | FALSE | Upregulated |
| -0,09195673 | 0,960162154 | CISH | FALSE | Downregulated |
| -0,066252051 | 0,960162154 | COMMD7 | FALSE | Downregulated |
| -0,069742327 | 0,960162154 | HES4 | FALSE | Downregulated |
| -0,035544432 | 0,960162154 | ICAM3 | FALSE | Downregulated |
| -0,026282405 | 0,960188985 | ARVCF | FALSE | Downregulated |
| 0,039482463 | 0,960266731 | RDH11 | FALSE | Upregulated |
| 0,088287953 | 0,960266731 | SULF1 | FALSE | Upregulated |
| 0,041061132 | 0,960308009 | LRRC8E | FALSE | Upregulated |
| -0,02699638 | 0,960308009 | RNF11 | FALSE | Downregulated |
| -0,039163537 | 0,960308009 | SNX6 | FALSE | Downregulated |
| -0,044768374 | 0,960308009 | TBL3 | FALSE | Downregulated |
| 0,042666316 | 0,960308009 | ZFAND2B | FALSE | Upregulated |
| -0,048413463 | 0,960475872 | P4HTM | FALSE | Downregulated |
| 0,021446382 | 0,960475872 | ZC3H4 | FALSE | Upregulated |
| 0,050163254 | 0,9604973 | MALT1 | FALSE | Upregulated |
| 0,023776379 | 0,9604973 | SHC1 | FALSE | Upregulated |
| 0,044043614 | 0,960533973 | PFKM | FALSE | Upregulated |
| -0,027044483 | 0,960596826 | ACTR2 | FALSE | Downregulated |
| -0,038708516 | 0,960596826 | ARMC10 | FALSE | Downregulated |
| -0,031467767 | 0,960596826 | DHX30 | FALSE | Downregulated |
| 0,069002773 | 0,960596826 | GTDC1 | FALSE | Upregulated |
| 0,032035156 | 0,960596826 | IPO7 | FALSE | Upregulated |
| 0,065455062 | 0,960596826 | PDCD5 | FALSE | Upregulated |
| 0,040616538 | 0,960596826 | RNF4 | FALSE | Upregulated |
| -0,060170972 | 0,960596826 | ZNF234 | FALSE | Downregulated |
| 0,081679326 | 0,960771464 | MGC70870 | FALSE | Upregulated |
| 0,068187005 | 0,960771464 | ZSWIM3 | FALSE | Upregulated |
| -0,042325536 | 0,960781718 | RAP2B | FALSE | Downregulated |
| -0,069851529 | 0,960781718 | TRIT1 | FALSE | Downregulated |
| -0,043879567 | 0,960786217 | BCS1L | FALSE | Downregulated |
| 0,068263321 | 0,960786217 | CSRP2 | FALSE | Upregulated |
| 0,045935849 | 0,960786217 | CUEDC1 | FALSE | Upregulated |
| 0,026934576 | 0,960786217 | GRTP1 | FALSE | Upregulated |
| -0,047348862 | 0,960786217 | TGIF2 | FALSE | Downregulated |
| -0,023157606 | 0,961029034 | CYP4V2 | FALSE | Downregulated |
| -0,035463513 | 0,961029034 | USP25 | FALSE | Downregulated |
| 0,075639446 | 0,961103447 | USMG5 | FALSE | Upregulated |
| -0,0281989 | 0,961174682 | DHX38 | FALSE | Downregulated |
| -0,050164637 | 0,961186708 | LOC102723811 | FALSE | Downregulated |

| | | | | |
|---|---|---|---|---|
| 0,078345367 | 0,961186708 | SH3RF3 | FALSE | Upregulated |
| -0,069490656 | 0,961215549 | NDRG3 | FALSE | Downregulated |
| 0,03693918 | 0,961215549 | PRKD2 | FALSE | Upregulated |
| 0,035605401 | 0,961215549 | UTP18 | FALSE | Upregulated |
| -0,049211545 | 0,961448384 | ARHGAP44 | FALSE | Downregulated |
| -0,022811188 | 0,961448384 | TUBGCP2 | FALSE | Downregulated |
| 0,03209534 | 0,96147314 | FOXM1 | FALSE | Upregulated |
| -0,068244422 | 0,96147314 | LGI4 | FALSE | Downregulated |
| -0,019500564 | 0,96147314 | RBM33 | FALSE | Downregulated |
| -0,033294915 | 0,96158249 | PEMT | FALSE | Downregulated |
| -0,023634142 | 0,961593056 | AGAP3 | FALSE | Downregulated |
| -0,058851022 | 0,961593056 | AGBL2 | FALSE | Downregulated |
| -0,035626127 | 0,961593056 | ANKRD28 | FALSE | Downregulated |
| -0,052169865 | 0,961593056 | BROX | FALSE | Downregulated |
| -0,045852546 | 0,961593056 | BTN2A1 | FALSE | Downregulated |
| -0,017138583 | 0,961593056 | CALD1 | FALSE | Downregulated |
| -0,050938041 | 0,961593056 | CENPBD1P1 | FALSE | Downregulated |
| -0,036955447 | 0,961593056 | CEP104 | FALSE | Downregulated |
| 0,033842036 | 0,961593056 | CHMP4B | FALSE | Upregulated |
| -0,027699247 | 0,961593056 | DNHD1 | FALSE | Downregulated |
| -0,055903571 | 0,961593056 | EVA1C | FALSE | Downregulated |
| -0,037864027 | 0,961593056 | FBXL16 | FALSE | Downregulated |
| -0,029350331 | 0,961593056 | FLT4 | FALSE | Downregulated |
| -0,081881691 | 0,961593056 | GPR162 | FALSE | Downregulated |
| -0,109569671 | 0,961593056 | GSTO2 | FALSE | Downregulated |
| 0,031170572 | 0,961593056 | H2AFY | FALSE | Upregulated |
| -0,060555513 | 0,961593056 | HCFC1R1 | FALSE | Downregulated |
| 0,049786704 | 0,961593056 | KATNB1 | FALSE | Upregulated |
| -0,052036192 | 0,961593056 | KSR1 | FALSE | Downregulated |
| 0,11486613 | 0,961593056 | LINC00294 | FALSE | Upregulated |
| -0,066514817 | 0,961593056 | LOC100268168 | FALSE | Downregulated |
| 0,065355063 | 0,961593056 | LOC101928595 | FALSE | Upregulated |
| 0,063832235 | 0,961593056 | LOC102288414 | FALSE | Upregulated |
| -0,047664816 | 0,961593056 | LOC102724455 | FALSE | Downregulated |
| -0,028411519 | 0,961593056 | MARCH7 | FALSE | Downregulated |
| 0,066316662 | 0,961593056 | MRPL39 | FALSE | Upregulated |
| 0,043526155 | 0,961593056 | MRPS11 | FALSE | Upregulated |
| 0,050165823 | 0,961593056 | NFKBIL1 | FALSE | Upregulated |
| 0,04088972 | 0,961593056 | NME9 | FALSE | Upregulated |
| 0,054771836 | 0,961593056 | NPIPB5 | FALSE | Upregulated |
| -0,043883133 | 0,961593056 | PARD6G-AS1 | FALSE | Downregulated |
| -0,045526792 | 0,961593056 | PCNA-AS1 | FALSE | Downregulated |
| 0,027554094 | 0,961593056 | PPP1CA | FALSE | Upregulated |
| 0,059291721 | 0,961593056 | PRKG1 | FALSE | Upregulated |
| 0,030741879 | 0,961593056 | PSMB2 | FALSE | Upregulated |
| 0,048040209 | 0,961593056 | RBFOX3 | FALSE | Upregulated |
| -0,055787834 | 0,961593056 | RGPD8 | FALSE | Downregulated |
| 0,027355532 | 0,961593056 | RUVBL1 | FALSE | Upregulated |
| 0,033817332 | 0,961593056 | RUVBL2 | FALSE | Upregulated |
| -0,042363268 | 0,961593056 | SERPINH1 | FALSE | Downregulated |
| 0,03226044 | 0,961593056 | SPTY2D1-AS1 | FALSE | Upregulated |

| | | | | |
|---|---|---|---|---|
| -0,051577333 | 0,961593056 | SSPO | FALSE | Downregulated |
| -0,073002517 | 0,961593056 | STXBP5 | FALSE | Downregulated |
| -0,040442965 | 0,961593056 | TMEM25 | FALSE | Downregulated |
| -0,038650612 | 0,961593056 | TMOD3 | FALSE | Downregulated |
| 0,046206632 | 0,961593056 | TRIM35 | FALSE | Upregulated |
| -0,044206303 | 0,961593056 | USP38 | FALSE | Downregulated |
| 0,034931793 | 0,961593056 | USP53 | FALSE | Upregulated |
| 0,081174116 | 0,961593056 | VNN3 | FALSE | Upregulated |
| -0,069215513 | 0,961593056 | WDR61 | FALSE | Downregulated |
| -0,037824099 | 0,961593056 | ZBED6 | FALSE | Downregulated |
| -0,024345534 | 0,961593056 | ZFYVE16 | FALSE | Downregulated |
| -0,0630924 | 0,961593056 | ZNF823 | FALSE | Downregulated |
| -0,097850113 | 0,961641346 | LOC392196 | FALSE | Downregulated |
| 0,047346378 | 0,961641346 | MAPK13 | FALSE | Upregulated |
| 0,063100423 | 0,961641346 | RP5-1057J7.6 | FALSE | Upregulated |
| -0,034043082 | 0,961641346 | WRNIP1 | FALSE | Downregulated |
| -0,064419527 | 0,962049599 | GNG3 | FALSE | Downregulated |
| -0,054634743 | 0,962135741 | CINP | FALSE | Downregulated |
| 0,043165148 | 0,962135741 | CTBS | FALSE | Upregulated |
| -0,023677909 | 0,962135741 | STK24 | FALSE | Downregulated |
| 0,033091442 | 0,962360007 | MROH1 | FALSE | Upregulated |
| 0,019180776 | 0,962401535 | SET | FALSE | Upregulated |
| -0,05276272 | 0,962446396 | ESF1 | FALSE | Downregulated |
| -0,050223551 | 0,962446396 | MEGF6 | FALSE | Downregulated |
| -0,036416034 | 0,962446396 | POLM | FALSE | Downregulated |
| 0,077811235 | 0,962534387 | C17orf97 | FALSE | Upregulated |
| 0,037895514 | 0,962534387 | LINC00538 | FALSE | Upregulated |
| 0,105435329 | 0,962534387 | LOC729737 | FALSE | Upregulated |
| -0,045537069 | 0,962646871 | S1PR1 | FALSE | Downregulated |
| 0,050449761 | 0,962646871 | SPNS3 | FALSE | Upregulated |
| 0,041195659 | 0,962646871 | TRMT61A | FALSE | Upregulated |
| -0,070257693 | 0,962726145 | CD274 | FALSE | Downregulated |
| -0,075265614 | 0,962726145 | LINC01024 | FALSE | Downregulated |
| -0,049862206 | 0,962726145 | NAA35 | FALSE | Downregulated |
| 0,045166574 | 0,962726145 | TPD52L1 | FALSE | Upregulated |
| -0,022757502 | 0,962753094 | MLF2 | FALSE | Downregulated |
| -0,032973288 | 0,962753094 | SRSF10 | FALSE | Downregulated |
| -0,044181159 | 0,962975787 | IL10RB | FALSE | Downregulated |
| -0,063720993 | 0,963216813 | TMEM41B | FALSE | Downregulated |
| -0,039732989 | 0,96327274 | SC5D | FALSE | Downregulated |
| -0,029285335 | 0,963522016 | MLLT1 | FALSE | Downregulated |
| -0,038695856 | 0,963579276 | A2M | FALSE | Downregulated |
| 0,070679077 | 0,963579276 | DNAH9 | FALSE | Upregulated |
| 0,040804461 | 0,963579276 | DONSON | FALSE | Upregulated |
| 0,023519014 | 0,963579276 | DSP | FALSE | Upregulated |
| -0,03718893 | 0,963579276 | HEATR4 | FALSE | Downregulated |
| 0,042442614 | 0,963579276 | LOC101927681 | FALSE | Upregulated |
| 0,031389041 | 0,963579276 | LOC646976 | FALSE | Upregulated |
| -0,060469081 | 0,963579276 | SEPT7P2 | FALSE | Downregulated |
| -0,026636876 | 0,963579276 | SNX17 | FALSE | Downregulated |
| -0,036405117 | 0,963579276 | TIAL1 | FALSE | Downregulated |

| | | | | |
|---|---|---|---|---|
| -0,028868667 | 0,963579276 | TOB1 | FALSE | Downregulated |
| 0,032426614 | 0,963589947 | KIAA0930 | FALSE | Upregulated |
| 0,041540522 | 0,963666733 | ACKR2 | FALSE | Upregulated |
| -0,056081949 | 0,963666733 | CTHRC1 | FALSE | Downregulated |
| -0,055423335 | 0,963666733 | LOC100506114 | FALSE | Downregulated |
| -0,030902937 | 0,963743646 | CHST7 | FALSE | Downregulated |
| 0,035842413 | 0,963743646 | DNAJC10 | FALSE | Upregulated |
| -0,05373025 | 0,963743646 | EID2 | FALSE | Downregulated |
| 0,043293892 | 0,963743646 | LOC102724043 | FALSE | Upregulated |
| 0,027324986 | 0,963743646 | MED23 | FALSE | Upregulated |
| 0,039342341 | 0,963743646 | OSBPL11 | FALSE | Upregulated |
| 0,038760721 | 0,963750466 | ADAM1A | FALSE | Upregulated |
| 0,046108677 | 0,963750466 | APAF1 | FALSE | Upregulated |
| -0,067153396 | 0,963750466 | GRIN2A | FALSE | Downregulated |
| -0,081103527 | 0,963750466 | HDGFRP3 | FALSE | Downregulated |
| 0,043302453 | 0,963750466 | KIF26A | FALSE | Upregulated |
| -0,025025804 | 0,963750466 | LTBP3 | FALSE | Downregulated |
| 0,042904508 | 0,963750466 | SDHAF1 | FALSE | Upregulated |
| -0,043934492 | 0,963750466 | TIMM50 | FALSE | Downregulated |
| 0,032466415 | 0,963750466 | TSC22D1 | FALSE | Upregulated |
| 0,051281519 | 0,963796149 | ASPHD2 | FALSE | Upregulated |
| 0,040205685 | 0,963796149 | CMTM3 | FALSE | Upregulated |
| 0,068137072 | 0,963796149 | CNRIP1 | FALSE | Upregulated |
| 0,042069901 | 0,963796149 | FIGN | FALSE | Upregulated |
| 0,021919288 | 0,963796149 | GLTPD2 | FALSE | Upregulated |
| 0,039111784 | 0,963796149 | GNA12 | FALSE | Upregulated |
| 0,082513696 | 0,963796149 | LINC01138 | FALSE | Upregulated |
| -0,065204552 | 0,963796149 | LOC100126584 | FALSE | Downregulated |
| 0,039884906 | 0,963796149 | NPC1L1 | FALSE | Upregulated |
| -0,056962656 | 0,963796149 | NUP54 | FALSE | Downregulated |
| -0,046795729 | 0,963796149 | PPARD | FALSE | Downregulated |
| 0,071125161 | 0,963796149 | SNAP25 | FALSE | Upregulated |
| 0,041096132 | 0,963796149 | SNRNP35 | FALSE | Upregulated |
| 0,03906456 | 0,963796149 | SPCS2 | FALSE | Upregulated |
| 0,033736683 | 0,963796149 | SRSF3 | FALSE | Upregulated |
| 0,034554637 | 0,963854312 | CYP20A1 | FALSE | Upregulated |
| -0,038915385 | 0,964140548 | KIAA1429 | FALSE | Downregulated |
| -0,046555059 | 0,964140548 | VPS8 | FALSE | Downregulated |
| -0,062461418 | 0,964240848 | ARMC1 | FALSE | Downregulated |
| -0,044556823 | 0,964240848 | LOC101927371 | FALSE | Downregulated |
| -0,048805321 | 0,964240848 | WDR44 | FALSE | Downregulated |
| 0,037791556 | 0,96433426 | USP16 | FALSE | Upregulated |
| -0,064417756 | 0,964449898 | AIF1 | FALSE | Downregulated |
| 0,02923459 | 0,964449898 | DNAJC7 | FALSE | Upregulated |
| 0,060540135 | 0,964449898 | FLYWCH2 | FALSE | Upregulated |
| 0,086808726 | 0,964449898 | GPR56 | FALSE | Upregulated |
| -0,021676085 | 0,964449898 | R3HDM2 | FALSE | Downregulated |
| 0,062050442 | 0,964449898 | RP11-1007G5.2 | FALSE | Upregulated |
| -0,027168656 | 0,964449898 | TAB3 | FALSE | Downregulated |
| 0,039480702 | 0,964486912 | LOC389906 | FALSE | Upregulated |
| -0,036973001 | 0,964490439 | CSPG4 | FALSE | Downregulated |

| | | | | |
|---|---|---|---|---|
| 0,058783635 | 0,964490439 | FIG4 | FALSE | Upregulated |
| 0,064134371 | 0,964490439 | PRH1-PRR4 | FALSE | Upregulated |
| 0,077523501 | 0,964490439 | SLC36A4 | FALSE | Upregulated |
| 0,076553898 | 0,964561743 | LMCD1 | FALSE | Upregulated |
| 0,029399966 | 0,964561743 | NEMF | FALSE | Upregulated |
| 0,038010791 | 0,964561743 | RAB11FIP5 | FALSE | Upregulated |
| 0,039741137 | 0,964637173 | FBXO10 | FALSE | Upregulated |
| -0,03260404 | 0,964637173 | FBXO46 | FALSE | Downregulated |
| 0,077549539 | 0,964637173 | PDZRN3 | FALSE | Upregulated |
| 0,069088319 | 0,964651873 | SIGLEC15 | FALSE | Upregulated |
| 0,041712865 | 0,964670375 | B4GALT7 | FALSE | Upregulated |
| 0,134716145 | 0,964670375 | DNAJC25-GNG10 | FALSE | Upregulated |
| 0,049967343 | 0,964670375 | FHL1 | FALSE | Upregulated |
| -0,451996224 | 0,964690105 | MYL2 | FALSE | Downregulated |
| 0,069223383 | 0,964842039 | LOC101929352 | FALSE | Upregulated |
| 0,030149331 | 0,964842039 | MON1B | FALSE | Upregulated |
| -0,05186431 | 0,965169902 | ATP8A1 | FALSE | Downregulated |
| -0,024443228 | 0,965169902 | PTK2 | FALSE | Downregulated |
| -0,023994739 | 0,965188254 | AARS | FALSE | Downregulated |
| 0,075990146 | 0,965188254 | AC147651.4 | FALSE | Upregulated |
| 0,029800464 | 0,965188254 | ANXA6 | FALSE | Upregulated |
| 0,073886215 | 0,965188254 | CBLN3 | FALSE | Upregulated |
| 0,030712647 | 0,965188254 | CECR1 | FALSE | Upregulated |
| -0,105319908 | 0,965188254 | MIR4500HG | FALSE | Downregulated |
| -0,030885263 | 0,965188254 | MYD88 | FALSE | Downregulated |
| -0,022026851 | 0,965188254 | NID1 | FALSE | Downregulated |
| -0,067242342 | 0,965188254 | SLAIN1 | FALSE | Downregulated |
| 0,057288669 | 0,965188254 | ST14 | FALSE | Upregulated |
| -0,024225147 | 0,965217236 | ACP2 | FALSE | Downregulated |
| 0,029282731 | 0,965217236 | MXRA8 | FALSE | Upregulated |
| 0,024083563 | 0,965217236 | TTC28 | FALSE | Upregulated |
| 0,166710645 | 0,965329557 | AKAP2 | FALSE | Upregulated |
| 0,037226159 | 0,965329557 | IKBKG | FALSE | Upregulated |
| -0,061545322 | 0,965329557 | RBBP8 | FALSE | Downregulated |
| -0,030706086 | 0,965329557 | SERPINA11 | FALSE | Downregulated |
| 0,054092137 | 0,965329557 | THAP8 | FALSE | Upregulated |
| -0,024786154 | 0,965329557 | UBL3 | FALSE | Downregulated |
| -0,028750861 | 0,965340936 | FAM3A | FALSE | Downregulated |
| 0,037599833 | 0,965340936 | ITFG1 | FALSE | Upregulated |
| -0,030167124 | 0,965340936 | KIAA0247 | FALSE | Downregulated |
| 0,046774598 | 0,965340936 | MAP1B | FALSE | Upregulated |
| 0,027895591 | 0,965340936 | RLIM | FALSE | Upregulated |
| -0,036254359 | 0,965340936 | SGK2 | FALSE | Downregulated |
| 0,040195756 | 0,965340936 | TMEM9B | FALSE | Upregulated |
| 0,041456416 | 0,965340936 | TXNDC12 | FALSE | Upregulated |
| -0,027220762 | 0,965494992 | HNRNPH3 | FALSE | Downregulated |
| -0,033098418 | 0,965494992 | SEMA4C | FALSE | Downregulated |
| 0,019263645 | 0,96564064 | TOB2 | FALSE | Upregulated |
| 0,030738549 | 0,965722073 | CHURC1 | FALSE | Upregulated |
| -0,055014349 | 0,965722073 | MECR | FALSE | Downregulated |
| -0,074577158 | 0,965722073 | OR1L4 | FALSE | Downregulated |

| | | | | |
|---|---|---|---|---|
| 0,014354942 | 0,965722073 | ZFHX3 | FALSE | Upregulated |
| 0,031668523 | 0,965756138 | LOC646652 | FALSE | Upregulated |
| -0,027206874 | 0,965756138 | TAF4 | FALSE | Downregulated |
| 0,057598556 | 0,965799803 | RBM38 | FALSE | Upregulated |
| -0,092829224 | 0,965824556 | CD24 | FALSE | Downregulated |
| 0,058117874 | 0,965824556 | COQ10B | FALSE | Upregulated |
| -0,029493701 | 0,965824556 | SULT2A1 | FALSE | Downregulated |
| -0,049566101 | 0,965874667 | CLYBL-AS2 | FALSE | Downregulated |
| 0,051453518 | 0,965874667 | EME2 | FALSE | Upregulated |
| -0,023257343 | 0,965874667 | RNF103 | FALSE | Downregulated |
| -0,030801262 | 0,965874667 | SIAH2 | FALSE | Downregulated |
| 0,050450261 | 0,965874667 | SLC25A35 | FALSE | Upregulated |
| 0,06572969 | 0,965895672 | GSTM3 | FALSE | Upregulated |
| -0,017665531 | 0,965895672 | KIAA1161 | FALSE | Downregulated |
| 0,057118606 | 0,965895672 | LINC00843 | FALSE | Upregulated |
| -0,065055582 | 0,965895672 | LOC102724872 | FALSE | Downregulated |
| 0,069774265 | 0,965895672 | NSUN5P2 | FALSE | Upregulated |
| 0,034638561 | 0,965895672 | NUP160 | FALSE | Upregulated |
| -0,031843378 | 0,965895672 | PGAP2 | FALSE | Downregulated |
| 0,069539981 | 0,965895672 | SLC10A3 | FALSE | Upregulated |
| -0,027160995 | 0,965895672 | SUPT16H | FALSE | Downregulated |
| 0,028329882 | 0,965895672 | TMEM79 | FALSE | Upregulated |
| 0,02907953 | 0,965895672 | ZZZ3 | FALSE | Upregulated |
| 0,040030004 | 0,966059887 | CEP63 | FALSE | Upregulated |
| -0,03015169 | 0,966059887 | MEIS2 | FALSE | Downregulated |
| -0,072201009 | 0,966059887 | SNORA48 | FALSE | Downregulated |
| -0,035266214 | 0,966059887 | TMEM222 | FALSE | Downregulated |
| 0,024173663 | 0,966136577 | DBI | FALSE | Upregulated |
| 0,028869203 | 0,966136577 | ITGB3 | FALSE | Upregulated |
| -0,024613676 | 0,966152742 | KLF13 | FALSE | Downregulated |
| 0,034200935 | 0,966152742 | SFMBT1 | FALSE | Upregulated |
| 0,011879089 | 0,966231784 | CNOT1 | FALSE | Upregulated |
| -0,026541984 | 0,966231784 | CRY2 | FALSE | Downregulated |
| 0,028094825 | 0,966231784 | FDFT1 | FALSE | Upregulated |
| 0,048243656 | 0,966231784 | GLTP | FALSE | Upregulated |
| 0,045968324 | 0,966231784 | NUDT6 | FALSE | Upregulated |
| -0,021814108 | 0,966231784 | STEAP3 | FALSE | Downregulated |
| 0,032441845 | 0,966231784 | VOPP1 | FALSE | Upregulated |
| -0,032794568 | 0,966310102 | PKP2 | FALSE | Downregulated |
| 0,029782499 | 0,966310102 | WDR88 | FALSE | Upregulated |
| 0,074361393 | 0,966440169 | GUCY2C | FALSE | Upregulated |
| 0,180276308 | 0,966440169 | HIST2H2AA3 | FALSE | Upregulated |
| -0,023813844 | 0,966440169 | LARP4B | FALSE | Downregulated |
| -0,021490079 | 0,966440169 | SHROOM3 | FALSE | Downregulated |
| -0,058803085 | 0,966440169 | TLR3 | FALSE | Downregulated |
| -0,027695624 | 0,966448898 | ANKRD12 | FALSE | Downregulated |
| -0,031627464 | 0,966448898 | AQP3 | FALSE | Downregulated |
| -0,03107747 | 0,966448898 | FLOT2 | FALSE | Downregulated |
| 0,030096463 | 0,966448898 | NOLC1 | FALSE | Upregulated |
| -0,036666217 | 0,966448898 | PDHB | FALSE | Downregulated |
| -0,09504996 | 0,966448898 | STAG3L1 | FALSE | Downregulated |

| | | | | |
|---|---|---|---|---|
| -0,035311732 | 0,966448898 | STAG3L2 | FALSE | Downregulated |
| -0,02133009 | 0,966448898 | TET2 | FALSE | Downregulated |
| -0,051144487 | 0,966448898 | TUBA3D | FALSE | Downregulated |
| 0,045468409 | 0,966490771 | LINC00909 | FALSE | Upregulated |
| -0,028923779 | 0,966490771 | MYL12B | FALSE | Downregulated |
| 0,197451694 | 0,966490771 | RGPD6 | FALSE | Upregulated |
| 0,026749796 | 0,966506951 | ACVR1B | FALSE | Upregulated |
| 0,02308144 | 0,966506951 | ALS2 | FALSE | Upregulated |
| 0,030374639 | 0,966506951 | ARHGEF2 | FALSE | Upregulated |
| 0,039273714 | 0,966506951 | ARIH2OS | FALSE | Upregulated |
| 0,062420838 | 0,966506951 | ART1 | FALSE | Upregulated |
| -0,021833535 | 0,966506951 | ASXL2 | FALSE | Downregulated |
| 0,023499214 | 0,966506951 | AUP1 | FALSE | Upregulated |
| 0,025567464 | 0,966506951 | BCAT2 | FALSE | Upregulated |
| -0,056825459 | 0,966506951 | BRMS1 | FALSE | Downregulated |
| -0,030428041 | 0,966506951 | C6orf201 | FALSE | Downregulated |
| 0,024395545 | 0,966506951 | CAP1 | FALSE | Upregulated |
| -0,030178433 | 0,966506951 | CARS2 | FALSE | Downregulated |
| -0,061322804 | 0,966506951 | CASP9 | FALSE | Downregulated |
| 0,043613723 | 0,966506951 | CCDC120 | FALSE | Upregulated |
| -0,058072024 | 0,966506951 | CCR1 | FALSE | Downregulated |
| -0,057671819 | 0,966506951 | CDK11A | FALSE | Downregulated |
| -0,048102912 | 0,966506951 | CLDND1 | FALSE | Downregulated |
| -0,018000508 | 0,966506951 | CLPTM1 | FALSE | Downregulated |
| -0,033196451 | 0,966506951 | CPT1B | FALSE | Downregulated |
| -0,015645881 | 0,966506951 | CTSB | FALSE | Downregulated |
| 0,042591646 | 0,966506951 | CYSTM1 | FALSE | Upregulated |
| -0,042019658 | 0,966506951 | DDX60 | FALSE | Downregulated |
| 0,040480418 | 0,966506951 | DUT | FALSE | Upregulated |
| -0,047075235 | 0,966506951 | EIF4E3 | FALSE | Downregulated |
| -0,054020338 | 0,966506951 | EPPK1 | FALSE | Downregulated |
| 0,074657014 | 0,966506951 | FREM2 | FALSE | Upregulated |
| 0,052779889 | 0,966506951 | GGCT | FALSE | Upregulated |
| -0,040505759 | 0,966506951 | GNG5 | FALSE | Downregulated |
| 0,035018727 | 0,966506951 | GRAMD1C | FALSE | Upregulated |
| 0,033644449 | 0,966506951 | HTR2A | FALSE | Upregulated |
| 0,05842857 | 0,966506951 | KCNH3 | FALSE | Upregulated |
| 0,037161804 | 0,966506951 | KLHL9 | FALSE | Upregulated |
| 0,031773051 | 0,966506951 | KRT18 | FALSE | Upregulated |
| -0,056881963 | 0,966506951 | LEPREL1 | FALSE | Downregulated |
| -0,038098467 | 0,966506951 | LMO4 | FALSE | Downregulated |
| 0,049368435 | 0,966506951 | LOC100507373 | FALSE | Upregulated |
| -0,0625531 | 0,966506951 | LOC101060398 | FALSE | Downregulated |
| -0,063522276 | 0,966506951 | LOC102723665 | FALSE | Downregulated |
| -0,035308207 | 0,966506951 | LOC102724672 | FALSE | Downregulated |
| 0,040150461 | 0,966506951 | LOC285593 | FALSE | Upregulated |
| -0,017085109 | 0,966506951 | LPP | FALSE | Downregulated |
| -0,064456753 | 0,966506951 | LRAT | FALSE | Downregulated |
| -0,076747382 | 0,966506951 | LRRC27 | FALSE | Downregulated |
| -0,038231396 | 0,966506951 | LRRC32 | FALSE | Downregulated |
| -0,044474973 | 0,966506951 | MAD2L2 | FALSE | Downregulated |

| | | | | |
|---|---|---|---|---|
| -0,041655274 | 0,966506951 | MARCH5 | FALSE | Downregulated |
| 0,039383388 | 0,966506951 | MSTO1 | FALSE | Upregulated |
| -0,041456807 | 0,966506951 | NDUFA3 | FALSE | Downregulated |
| 0,043531217 | 0,966506951 | NEU3 | FALSE | Upregulated |
| 0,049764479 | 0,966506951 | NPR3 | FALSE | Upregulated |
| -0,029486405 | 0,966506951 | NSFL1C | FALSE | Downregulated |
| -0,028982531 | 0,966506951 | NUDC | FALSE | Downregulated |
| 0,082599793 | 0,966506951 | NUTM2F | FALSE | Upregulated |
| -0,037840884 | 0,966506951 | PBX3 | FALSE | Downregulated |
| 0,025137977 | 0,966506951 | PGK1 | FALSE | Upregulated |
| 0,060570487 | 0,966506951 | PIGH | FALSE | Upregulated |
| -0,064450725 | 0,966506951 | PIGK | FALSE | Downregulated |
| -0,052395544 | 0,966506951 | PIWIL2 | FALSE | Downregulated |
| 0,060530331 | 0,966506951 | PNMA1 | FALSE | Upregulated |
| -0,046946538 | 0,966506951 | POMT2 | FALSE | Downregulated |
| -0,032230041 | 0,966506951 | QSER1 | FALSE | Downregulated |
| -0,029502649 | 0,966506951 | RITA1 | FALSE | Downregulated |
| -0,693872605 | 0,966506951 | RNU1-28P | FALSE | Downregulated |
| 0,060565204 | 0,966506951 | RP11-266K4.9 | FALSE | Upregulated |
| -0,058712267 | 0,966506951 | S1PR3 | FALSE | Downregulated |
| -0,038373023 | 0,966506951 | SAE1 | FALSE | Downregulated |
| 0,029259594 | 0,966506951 | SGMS2 | FALSE | Upregulated |
| -0,058425437 | 0,966506951 | SLC23A3 | FALSE | Downregulated |
| 0,032875913 | 0,966506951 | SLC35E2 | FALSE | Upregulated |
| 0,06390839 | 0,966506951 | SLC38A11 | FALSE | Upregulated |
| -0,03313339 | 0,966506951 | SMC6 | FALSE | Downregulated |
| 0,040974614 | 0,966506951 | SPDYE2B | FALSE | Upregulated |
| 0,019947247 | 0,966506951 | SRSF9 | FALSE | Upregulated |
| -0,029826058 | 0,966506951 | STARD13 | FALSE | Downregulated |
| -0,021349598 | 0,966506951 | SUMF2 | FALSE | Downregulated |
| -0,027727497 | 0,966506951 | SYNE2 | FALSE | Downregulated |
| -0,024348609 | 0,966506951 | TACC1 | FALSE | Downregulated |
| -0,025133013 | 0,966506951 | TALDO1 | FALSE | Downregulated |
| 0,027240953 | 0,966506951 | TBC1D24 | FALSE | Upregulated |
| 0,028523214 | 0,966506951 | THEM5 | FALSE | Upregulated |
| 0,06874848 | 0,966506951 | TMEM125 | FALSE | Upregulated |
| -0,044475336 | 0,966506951 | TSN | FALSE | Downregulated |
| 0,045979104 | 0,966506951 | TUSC2 | FALSE | Upregulated |
| 0,027078585 | 0,966506951 | UBE2L3 | FALSE | Upregulated |
| 0,058844243 | 0,966506951 | UBE2Q2 | FALSE | Upregulated |
| -0,023714537 | 0,966506951 | ULK1 | FALSE | Downregulated |
| -0,031583184 | 0,966506951 | VDAC2 | FALSE | Downregulated |
| 0,017082204 | 0,966506951 | VPS4A | FALSE | Upregulated |
| -0,032802195 | 0,966506951 | ZBTB1 | FALSE | Downregulated |
| -0,058280813 | 0,966506951 | ZNF180 | FALSE | Downregulated |
| 0,024462722 | 0,966506951 | ZNF335 | FALSE | Upregulated |
| -0,060137853 | 0,966506951 | ZNF432 | FALSE | Downregulated |
| 0,058729479 | 0,966506951 | ZNF571 | FALSE | Upregulated |
| -0,045200192 | 0,966506951 | ZNF81 | FALSE | Downregulated |
| -0,036853005 | 0,966509435 | PIGG | FALSE | Downregulated |
| -0,071922129 | 0,96651198 | PRDM1 | FALSE | Downregulated |

| | | | | |
|---|---|---|---|---|
| -0,066484987 | 0,96651198 | VMO1 | FALSE | Downregulated |
| 0,023476965 | 0,966683954 | RBM14 | FALSE | Upregulated |
| -0,03668653 | 0,966793429 | OCLN | FALSE | Downregulated |
| -0,043897285 | 0,966990341 | FAM179B | FALSE | Downregulated |
| -0,057180332 | 0,966990341 | LOC101927180 | FALSE | Downregulated |
| 0,021364521 | 0,967099208 | PRKAR1A | FALSE | Upregulated |
| -0,012629557 | 0,967099208 | TNRC18 | FALSE | Downregulated |
| -0,058498657 | 0,9671299 | PYROXD2 | FALSE | Downregulated |
| -0,03683271 | 0,96722896 | AOC3 | FALSE | Downregulated |
| 0,022495366 | 0,96722896 | F2RL2 | FALSE | Upregulated |
| 0,064742388 | 0,967240495 | LOC100270746 | FALSE | Upregulated |
| -0,021872371 | 0,967313118 | GCFC2 | FALSE | Downregulated |
| -0,047327065 | 0,967313118 | GIMAP2 | FALSE | Downregulated |
| -0,035922431 | 0,967313118 | SOX5 | FALSE | Downregulated |
| -0,036053778 | 0,967433645 | POLR1E | FALSE | Downregulated |
| 0,057848058 | 0,967445477 | POGLUT1 | FALSE | Upregulated |
| 0,03169293 | 0,96748316 | E4F1 | FALSE | Upregulated |
| 0,025884573 | 0,967544595 | C1orf63 | FALSE | Upregulated |
| -0,057832265 | 0,967544595 | MED28 | FALSE | Downregulated |
| 0,025744087 | 0,967544595 | NET1 | FALSE | Upregulated |
| -0,033003462 | 0,967544595 | POC1B | FALSE | Downregulated |
| -0,078737821 | 0,967771474 | FOXQ1 | FALSE | Downregulated |
| 0,045787487 | 0,967771474 | MAFIP | FALSE | Upregulated |
| 0,026057207 | 0,967836848 | GSK3B | FALSE | Upregulated |
| 0,017026147 | 0,968069713 | MKLN1 | FALSE | Upregulated |
| -0,018337436 | 0,968374988 | SUPT5H | FALSE | Downregulated |
| 0,038231458 | 0,9685806 | CITED4 | FALSE | Upregulated |
| 0,04317115 | 0,968658486 | CYB5B | FALSE | Upregulated |
| 0,055381268 | 0,968658486 | DDO | FALSE | Upregulated |
| -0,034425228 | 0,968658486 | PDGFRA | FALSE | Downregulated |
| -0,027519116 | 0,968785227 | NAPRT1 | FALSE | Downregulated |
| 0,027143641 | 0,968785227 | PLCL2 | FALSE | Upregulated |
| -0,037742627 | 0,968786193 | PLAGL2 | FALSE | Downregulated |
| 0,025098389 | 0,969030282 | DNPEP | FALSE | Upregulated |
| 0,050144431 | 0,969030282 | PALB2 | FALSE | Upregulated |
| 0,043321515 | 0,969048272 | CPVL | FALSE | Upregulated |
| 0,030201465 | 0,969048272 | GTF2A1 | FALSE | Upregulated |
| 0,0335118 | 0,969048272 | LOC101927514 | FALSE | Upregulated |
| -0,032667378 | 0,969048272 | PDCD7 | FALSE | Downregulated |
| -0,02423499 | 0,969048272 | PLD1 | FALSE | Downregulated |
| -0,053328646 | 0,969048272 | SMC2 | FALSE | Downregulated |
| -0,043067583 | 0,969048272 | ST3GAL5 | FALSE | Downregulated |
| 0,045429101 | 0,969048272 | TBXAS1 | FALSE | Upregulated |
| -0,035979243 | 0,969048272 | TTC21B | FALSE | Downregulated |
| -0,049521151 | 0,96909084 | AC091729.9 | FALSE | Downregulated |
| -0,022551069 | 0,96909084 | APOL1 | FALSE | Downregulated |
| -0,035753014 | 0,96909084 | ATP5S | FALSE | Downregulated |
| -0,028999754 | 0,96909084 | C21orf62 | FALSE | Downregulated |
| -0,017612588 | 0,96909084 | CAPN15 | FALSE | Downregulated |
| 0,063009129 | 0,96909084 | CNIH4 | FALSE | Upregulated |
| 0,017919177 | 0,96909084 | DIS3L | FALSE | Upregulated |

| | | | | |
|---|---|---|---|---|
| 0,040353597 | 0,96909084 | EDRF1 | FALSE | Upregulated |
| -0,045220405 | 0,96909084 | EFTUD1 | FALSE | Downregulated |
| 0,04533905 | 0,96909084 | FUT8 | FALSE | Upregulated |
| -0,021770472 | 0,96909084 | KDM4A | FALSE | Downregulated |
| -0,029572446 | 0,96909084 | KIAA0232 | FALSE | Downregulated |
| 0,057114259 | 0,96909084 | LENEP | FALSE | Upregulated |
| -0,05070472 | 0,96909084 | LHX2 | FALSE | Downregulated |
| 0,037641838 | 0,96909084 | LOC101929792 | FALSE | Upregulated |
| 0,045351782 | 0,96909084 | MAPK7 | FALSE | Upregulated |
| -0,053635509 | 0,96909084 | MID1 | FALSE | Downregulated |
| -0,017464208 | 0,96909084 | MLEC | FALSE | Downregulated |
| 0,034137031 | 0,96909084 | NPIPB6 | FALSE | Upregulated |
| 0,025380659 | 0,96909084 | NRP1 | FALSE | Upregulated |
| 0,025123544 | 0,96909084 | PA2G4 | FALSE | Upregulated |
| -0,056294504 | 0,96909084 | PDF | FALSE | Downregulated |
| 0,017095762 | 0,96909084 | PRAP1 | FALSE | Upregulated |
| -0,04892643 | 0,96909084 | RAB30 | FALSE | Downregulated |
| 0,056869542 | 0,96909084 | RASA2 | FALSE | Upregulated |
| -0,043298848 | 0,96909084 | RNASE7 | FALSE | Downregulated |
| -0,018558723 | 0,96909084 | RNPEPL1 | FALSE | Downregulated |
| -0,047059562 | 0,96909084 | SH3YL1 | FALSE | Downregulated |
| 0,017465118 | 0,96909084 | SLC31A1 | FALSE | Upregulated |
| 0,013299334 | 0,96909084 | SON | FALSE | Upregulated |
| -0,021657386 | 0,96909084 | SPARC | FALSE | Downregulated |
| 0,033028059 | 0,96909084 | SWT1 | FALSE | Upregulated |
| 0,02367295 | 0,96909084 | TMEM2 | FALSE | Upregulated |
| 0,040270649 | 0,96909084 | TNFRSF25 | FALSE | Upregulated |
| -0,048328953 | 0,96909084 | TYW3 | FALSE | Downregulated |
| -0,029339202 | 0,96909084 | UBE3B | FALSE | Downregulated |
| -0,017140097 | 0,96909084 | UBQLN1 | FALSE | Downregulated |
| -0,018381215 | 0,96909084 | WDTC1 | FALSE | Downregulated |
| -0,047365039 | 0,969108823 | WHAMMP3 | FALSE | Downregulated |
| 0,058425765 | 0,969234656 | LYSMD4 | FALSE | Upregulated |
| 0,024277926 | 0,969260417 | LRRC8A | FALSE | Upregulated |
| 0,046236153 | 0,969260417 | NPY1R | FALSE | Upregulated |
| -0,032394089 | 0,969431324 | FBXL20 | FALSE | Downregulated |
| 0,023192639 | 0,969431324 | FLRT1 | FALSE | Upregulated |
| 0,024133185 | 0,969431324 | IL6ST | FALSE | Upregulated |
| 0,037003587 | 0,969431324 | LOC101929347 | FALSE | Upregulated |
| -0,042211269 | 0,969431324 | MRPL15 | FALSE | Downregulated |
| -0,050059975 | 0,969699153 | CHORDC1 | FALSE | Downregulated |
| 0,064499365 | 0,969704584 | PRB1 | FALSE | Upregulated |
| -0,041037374 | 0,969737689 | ALKBH4 | FALSE | Downregulated |
| -0,030718772 | 0,96973932 | ABHD6 | FALSE | Downregulated |
| -0,035758828 | 0,96973932 | ABTB1 | FALSE | Downregulated |
| -0,036889098 | 0,96973932 | CD5L | FALSE | Downregulated |
| -0,022271174 | 0,96973932 | CHD7 | FALSE | Downregulated |
| -0,02111282 | 0,96973932 | H1F0 | FALSE | Downregulated |
| 0,03215517 | 0,96973932 | SPIRE1 | FALSE | Upregulated |
| -0,030468267 | 0,969864051 | NCAPD2 | FALSE | Downregulated |
| 0,038398838 | 0,969896645 | IPP | FALSE | Upregulated |

| | | | | |
|---|---|---|---|---|
| -0,020008353 | 0,969968327 | ESYT2 | FALSE | Downregulated |
| 0,05825028 | 0,970230201 | LOC101928158 | FALSE | Upregulated |
| 0,03407011 | 0,970232028 | PRKCH | FALSE | Upregulated |
| 0,030490671 | 0,970239857 | NDUFA2 | FALSE | Upregulated |
| -0,027583189 | 0,970296526 | AMN1 | FALSE | Downregulated |
| 0,02536752 | 0,970296526 | CGREF1 | FALSE | Upregulated |
| -0,035920756 | 0,970296526 | DHFRL1 | FALSE | Downregulated |
| 0,034158895 | 0,970296526 | DNPH1 | FALSE | Upregulated |
| 0,041609386 | 0,970296526 | EBF3 | FALSE | Upregulated |
| 0,066627022 | 0,970296526 | FAM86B3P | FALSE | Upregulated |
| 0,042702676 | 0,970296526 | LOC101927850 | FALSE | Upregulated |
| -0,016426269 | 0,970296526 | MAOB | FALSE | Downregulated |
| 0,044031158 | 0,970296526 | MBLAC1 | FALSE | Upregulated |
| -0,030495122 | 0,970296526 | MPPE1 | FALSE | Downregulated |
| -0,041644686 | 0,970296526 | PLXDC1 | FALSE | Downregulated |
| 0,03822045 | 0,970296526 | SLC50A1 | FALSE | Upregulated |
| 0,031146186 | 0,970296526 | SYNPO2 | FALSE | Upregulated |
| 0,025875862 | 0,970296526 | YTHDF2 | FALSE | Upregulated |
| -0,046283862 | 0,970296526 | ZNF833P | FALSE | Downregulated |
| 0,01979659 | 0,970368314 | AKAP17A | FALSE | Upregulated |
| 0,040546869 | 0,970469841 | NLRP6 | FALSE | Upregulated |
| 0,023900925 | 0,970602369 | BDNF | FALSE | Upregulated |
| 0,028939169 | 0,970602369 | EEFSEC | FALSE | Upregulated |
| -0,02075485 | 0,970602369 | GGA3 | FALSE | Downregulated |
| 0,054504519 | 0,970602369 | MRPS6 | FALSE | Upregulated |
| 0,024014579 | 0,970602369 | ST7-AS1 | FALSE | Upregulated |
| -0,017010456 | 0,970658622 | FETUB | FALSE | Downregulated |
| -0,041899382 | 0,970661509 | PFKFB2 | FALSE | Downregulated |
| 0,043177307 | 0,970661509 | THAP2 | FALSE | Upregulated |
| -0,027249977 | 0,970676115 | UGDH | FALSE | Downregulated |
| 0,042017167 | 0,970676115 | ZNF212 | FALSE | Upregulated |
| 0,050070701 | 0,970680135 | ZW10 | FALSE | Upregulated |
| 0,038867864 | 0,971195026 | FBXW8 | FALSE | Upregulated |
| -0,03016095 | 0,971195026 | FTO | FALSE | Downregulated |
| -0,03763602 | 0,971198965 | SLC7A8 | FALSE | Downregulated |
| -0,037664518 | 0,971461622 | C10orf32 | FALSE | Downregulated |
| -0,022545672 | 0,971546296 | EDEM1 | FALSE | Downregulated |
| -0,057483016 | 0,97157024 | ALG11 | FALSE | Downregulated |
| 0,040860361 | 0,97157024 | ALX4 | FALSE | Upregulated |
| 0,070915135 | 0,97157024 | RP4-669L17.10-2 | FALSE | Upregulated |
| -0,057875748 | 0,97157024 | SLC16A10 | FALSE | Downregulated |
| -0,037670847 | 0,97157024 | SOBP | FALSE | Downregulated |
| -0,032909002 | 0,97157024 | ZNF777 | FALSE | Downregulated |
| 0,050394385 | 0,97158054 | FDXR | FALSE | Upregulated |
| -0,052340918 | 0,971655299 | NUCB2 | FALSE | Downregulated |
| 0,021136301 | 0,971655299 | TTC28-AS1 | FALSE | Upregulated |
| 0,023178744 | 0,971680714 | PSMC4 | FALSE | Upregulated |
| 0,05436291 | 0,971686689 | LEPROTL1 | FALSE | Upregulated |
| 0,03059467 | 0,971686689 | LOC101929330 | FALSE | Upregulated |
| -0,02824119 | 0,971686689 | SNORA73A | FALSE | Downregulated |
| 0,025502881 | 0,971686689 | SPCS3 | FALSE | Upregulated |

| | | | | |
|---|---|---|---|---|
| 0,04973944 | 0,971709367 | EXO5 | FALSE | Upregulated |
| -0,057053155 | 0,971709367 | ZNF844 | FALSE | Downregulated |
| -0,043647598 | 0,971713125 | ATP5G1 | FALSE | Downregulated |
| 0,055933168 | 0,971713125 | CDH13 | FALSE | Upregulated |
| -0,035832668 | 0,971713125 | RHOBTB1 | FALSE | Downregulated |
| 0,030773235 | 0,971713125 | TUBGCP3 | FALSE | Upregulated |
| -0,03998307 | 0,971724654 | C11orf71 | FALSE | Downregulated |
| -0,018304855 | 0,971724654 | ENO1 | FALSE | Downregulated |
| -0,050671872 | 0,971724654 | STAMBP | FALSE | Downregulated |
| 0,055649066 | 0,971724654 | ZNF177 | FALSE | Upregulated |
| 0,02242124 | 0,971756122 | CNNM4 | FALSE | Upregulated |
| -0,023249553 | 0,971756122 | CUL9 | FALSE | Downregulated |
| -0,047214839 | 0,971756122 | GSTP1 | FALSE | Downregulated |
| -0,027702118 | 0,971756122 | THAP4 | FALSE | Downregulated |
| 0,02806256 | 0,971764607 | CSNK2A1 | FALSE | Upregulated |
| 0,033055419 | 0,971764607 | CXCL14 | FALSE | Upregulated |
| 0,047710992 | 0,971764607 | FCGR2B | FALSE | Upregulated |
| 0,048992034 | 0,971764607 | IGSF22 | FALSE | Upregulated |
| -0,020877737 | 0,971764607 | MMP15 | FALSE | Downregulated |
| 0,053898847 | 0,971764607 | OSGEPL1-AS1 | FALSE | Upregulated |
| 0,020493549 | 0,971764607 | PSME4 | FALSE | Upregulated |
| -0,017799196 | 0,971778097 | ITGA1 | FALSE | Downregulated |
| 0,041709884 | 0,971778097 | TICRR | FALSE | Upregulated |
| 0,031636219 | 0,971798855 | PKDREJ | FALSE | Upregulated |
| 0,022904991 | 0,971823414 | ABCF1 | FALSE | Upregulated |
| -0,020087237 | 0,971823414 | ARHGAP12 | FALSE | Downregulated |
| 0,040983539 | 0,971823414 | ATP6V0E1 | FALSE | Upregulated |
| -0,024863969 | 0,971823414 | ATP6V1B2 | FALSE | Downregulated |
| 0,019610699 | 0,971823414 | C1orf86 | FALSE | Upregulated |
| -0,043597541 | 0,971823414 | CBY1 | FALSE | Downregulated |
| -0,039643791 | 0,971823414 | CHM | FALSE | Downregulated |
| -0,012543559 | 0,971823414 | CIDEB | FALSE | Downregulated |
| 0,453761374 | 0,971823414 | CKM | FALSE | Upregulated |
| -0,017784449 | 0,971823414 | CMPK1 | FALSE | Downregulated |
| -0,039627773 | 0,971823414 | COX14 | FALSE | Downregulated |
| 0,015752146 | 0,971823414 | CPSF7 | FALSE | Upregulated |
| -0,05625597 | 0,971823414 | FAM166A | FALSE | Downregulated |
| -0,053864209 | 0,971823414 | FAM229B | FALSE | Downregulated |
| -0,049353848 | 0,971823414 | FOPNL | FALSE | Downregulated |
| 0,043775908 | 0,971823414 | GEMIN7 | FALSE | Upregulated |
| 0,044190933 | 0,971823414 | H2AFY2 | FALSE | Upregulated |
| -0,023458177 | 0,971823414 | INTS10 | FALSE | Downregulated |
| 0,033306763 | 0,971823414 | IRF9 | FALSE | Upregulated |
| -0,047333082 | 0,971823414 | LIN7A | FALSE | Downregulated |
| -0,051995386 | 0,971823414 | LOC101927002 | FALSE | Downregulated |
| 0,042258127 | 0,971823414 | LOC646762 | FALSE | Upregulated |
| -0,032301063 | 0,971823414 | LRWD1 | FALSE | Downregulated |
| 0,060907602 | 0,971823414 | MCM2 | FALSE | Upregulated |
| 0,038570156 | 0,971823414 | NKD1 | FALSE | Upregulated |
| -0,0290802 | 0,971823414 | PQLC2 | FALSE | Downregulated |
| 0,016851369 | 0,971823414 | PTGES3 | FALSE | Upregulated |

| | | | | |
|---|---|---|---|---|
| -0,031106552 | 0,971823414 | RNASE13 | FALSE | Downregulated |
| 0,024871387 | 0,971823414 | SLC25A44 | FALSE | Upregulated |
| -0,020478144 | 0,971823414 | TLE1 | FALSE | Downregulated |
| 0,057326861 | 0,971823414 | TMEM51 | FALSE | Upregulated |
| -0,030850334 | 0,971823414 | TRIB3 | FALSE | Downregulated |
| -0,079957006 | 0,971823414 | TTC9 | FALSE | Downregulated |
| -0,0271136 | 0,971823414 | TUSC1 | FALSE | Downregulated |
| 0,021855749 | 0,971823414 | USP4 | FALSE | Upregulated |
| -0,027097735 | 0,971823414 | ZC3HAV1 | FALSE | Downregulated |
| 0,032544663 | 0,971867194 | BNIP2 | FALSE | Upregulated |
| 0,047155398 | 0,971867194 | BRE | FALSE | Upregulated |
| 0,033158034 | 0,971867194 | CCDC137 | FALSE | Upregulated |
| 0,021654344 | 0,971867194 | CPT1A | FALSE | Upregulated |
| -0,019949303 | 0,971867194 | CUL1 | FALSE | Downregulated |
| -0,028381653 | 0,971867194 | DERA | FALSE | Downregulated |
| 0,036024319 | 0,971867194 | EFCAB2 | FALSE | Upregulated |
| -0,02607787 | 0,971867194 | EGLN2 | FALSE | Downregulated |
| 0,052890004 | 0,971867194 | GLIDR | FALSE | Upregulated |
| 0,026785794 | 0,971867194 | HYLS1 | FALSE | Upregulated |
| -0,081031641 | 0,971867194 | IGSF10 | FALSE | Downregulated |
| -0,051858541 | 0,971867194 | IL22RA1 | FALSE | Downregulated |
| -0,024465524 | 0,971867194 | KIN | FALSE | Downregulated |
| 0,035594802 | 0,971867194 | L3MBTL3 | FALSE | Upregulated |
| 0,050061634 | 0,971867194 | LOC102724788 | FALSE | Upregulated |
| 0,036285224 | 0,971867194 | MRPL35 | FALSE | Upregulated |
| 0,019530138 | 0,971867194 | NMT2 | FALSE | Upregulated |
| -0,020525669 | 0,971867194 | NUP98 | FALSE | Downregulated |
| 0,022064904 | 0,971867194 | OTC | FALSE | Upregulated |
| 0,018923353 | 0,971867194 | PARP10 | FALSE | Upregulated |
| 0,031274879 | 0,971867194 | SNRPB | FALSE | Upregulated |
| 0,018937175 | 0,971867194 | TCOF1 | FALSE | Upregulated |
| -0,031789156 | 0,971867194 | WDR34 | FALSE | Downregulated |
| 0,031831268 | 0,971878788 | EFEMP2 | FALSE | Upregulated |
| 0,015716044 | 0,971878788 | LOC100507577 | FALSE | Upregulated |
| 0,026899632 | 0,97215291 | CCNT1 | FALSE | Upregulated |
| 0,018035214 | 0,972182276 | MTCH1 | FALSE | Upregulated |
| -0,017997269 | 0,972273945 | KAT2A | FALSE | Downregulated |
| 0,062562521 | 0,972278669 | ERBB4 | FALSE | Upregulated |
| 0,016537911 | 0,972278669 | PCNT | FALSE | Upregulated |
| -0,050266284 | 0,972278669 | PKN3 | FALSE | Downregulated |
| -0,019202757 | 0,972278669 | RANBP1 | FALSE | Downregulated |
| 0,037156388 | 0,972278669 | SLC30A6 | FALSE | Upregulated |
| -0,04752664 | 0,972278669 | SPRYD7 | FALSE | Downregulated |
| 0,024268128 | 0,972278669 | TOMM40 | FALSE | Upregulated |
| -0,040903971 | 0,972278669 | ULK2 | FALSE | Downregulated |
| -0,025811251 | 0,972278669 | USP14 | FALSE | Downregulated |
| -0,028573332 | 0,972314575 | DUSP10 | FALSE | Downregulated |
| 0,014100666 | 0,972314575 | TNFAIP1 | FALSE | Upregulated |
| -0,023142854 | 0,97242178 | CYP1B1 | FALSE | Downregulated |
| 0,023795162 | 0,97242178 | KLF11 | FALSE | Upregulated |
| 0,018345566 | 0,97242178 | SLC35D1 | FALSE | Upregulated |

| | | | | |
|---|---|---|---|---|
| 0,030382803 | 0,972515024 | GARS | FALSE | Upregulated |
| -0,045685478 | 0,972610966 | PRIMPOL | FALSE | Downregulated |
| 0,039846562 | 0,972690966 | FBXO30 | FALSE | Upregulated |
| -0,022059297 | 0,972729032 | CD151 | FALSE | Downregulated |
| -0,505046597 | 0,972729032 | DES | FALSE | Downregulated |
| -0,025178508 | 0,972729032 | GFM1 | FALSE | Downregulated |
| -0,024857246 | 0,972729032 | KDM3A | FALSE | Downregulated |
| 0,026322255 | 0,972729032 | LINC00476 | FALSE | Upregulated |
| 0,035547282 | 0,972729032 | LOC102724634 | FALSE | Upregulated |
| 0,028457075 | 0,972729032 | MIRLET7BHG | FALSE | Upregulated |
| -0,027363008 | 0,972729032 | ORM1 | FALSE | Downregulated |
| -0,018527516 | 0,972729032 | PBX2 | FALSE | Downregulated |
| 0,034270027 | 0,972729032 | RALB | FALSE | Upregulated |
| 0,056716639 | 0,972779229 | DTX3 | FALSE | Upregulated |
| -0,06161249 | 0,972825936 | PM20D2 | FALSE | Downregulated |
| -0,066458653 | 0,972859284 | GLI2 | FALSE | Downregulated |
| 0,028762381 | 0,972859284 | MAPK8 | FALSE | Upregulated |
| -0,027545491 | 0,972859284 | MARCH2 | FALSE | Downregulated |
| -0,034293359 | 0,972859284 | MFF | FALSE | Downregulated |
| 0,023520666 | 0,972859284 | NAT8 | FALSE | Upregulated |
| 0,028999535 | 0,972859284 | RBM23 | FALSE | Upregulated |
| 0,023178138 | 0,972859284 | RNF43 | FALSE | Upregulated |
| 0,036368926 | 0,972859284 | SWAP70 | FALSE | Upregulated |
| -0,038854034 | 0,972859284 | TMEM87B | FALSE | Downregulated |
| -0,022567313 | 0,972859284 | TXNDC17 | FALSE | Downregulated |
| -0,050863149 | 0,972859284 | UPF3B | FALSE | Downregulated |
| 0,018759546 | 0,972859284 | ZSCAN29 | FALSE | Upregulated |
| 0,028318344 | 0,972976107 | CBL | FALSE | Upregulated |
| -0,019313068 | 0,972976107 | KLHL24 | FALSE | Downregulated |
| 0,020342916 | 0,972976107 | TMEM205 | FALSE | Upregulated |
| -0,02847023 | 0,973295489 | CASP8AP2 | FALSE | Downregulated |
| -0,025217139 | 0,973295489 | FGD5 | FALSE | Downregulated |
| 0,075672581 | 0,973295489 | FNDC1 | FALSE | Upregulated |
| -0,02356714 | 0,973295489 | PEX13 | FALSE | Downregulated |
| -0,049020293 | 0,973295489 | RGPD2 | FALSE | Downregulated |
| -0,027222893 | 0,973295489 | TMCC1 | FALSE | Downregulated |
| 0,037927639 | 0,973464514 | LOC101929272 | FALSE | Upregulated |
| 0,02113996 | 0,973640321 | LOC101929648 | FALSE | Upregulated |
| 0,050449608 | 0,973640321 | MYL6B | FALSE | Upregulated |
| 0,019389705 | 0,973640321 | SNAP47 | FALSE | Upregulated |
| 0,052449044 | 0,973716162 | PDCL | FALSE | Upregulated |
| 0,047164663 | 0,973740022 | CCL22 | FALSE | Upregulated |
| 0,01929819 | 0,973740022 | CKAP5 | FALSE | Upregulated |
| -0,048068293 | 0,973740022 | DOPEY2 | FALSE | Downregulated |
| -0,04560858 | 0,973740022 | DZIP1L | FALSE | Downregulated |
| -0,036539866 | 0,973740022 | FLJ30403 | FALSE | Downregulated |
| -0,017805759 | 0,973740022 | GALNT2 | FALSE | Downregulated |
| -0,022803618 | 0,973740022 | KLF9 | FALSE | Downregulated |
| 0,047313906 | 0,973740022 | LOC101927206 | FALSE | Upregulated |
| -0,039911324 | 0,973740022 | LOC729652 | FALSE | Downregulated |
| -0,037546608 | 0,973740022 | NNMT | FALSE | Downregulated |

| | | | | |
|---|---|---|---|---|
| -0,01810544 | 0,973740022 | RBM12 | FALSE | Downregulated |
| -0,03783098 | 0,973740022 | ROBO1 | FALSE | Downregulated |
| 0,048188829 | 0,973740022 | SLC30A4 | FALSE | Upregulated |
| -0,029559257 | 0,973740022 | SNRPA | FALSE | Downregulated |
| 0,046644036 | 0,973740022 | STAG3L5P | FALSE | Upregulated |
| 0,026468434 | 0,973740022 | TMEM167A | FALSE | Upregulated |
| -0,02498179 | 0,973740022 | TPM3 | FALSE | Downregulated |
| 0,051001082 | 0,97376615 | PHYHIP | FALSE | Upregulated |
| -0,022947842 | 0,973828342 | SNCAIP | FALSE | Downregulated |
| 0,036253987 | 0,973862628 | CLTCL1 | FALSE | Upregulated |
| 0,015071052 | 0,973862628 | PEX19 | FALSE | Upregulated |
| -0,024195106 | 0,974218236 | PDZD8 | FALSE | Downregulated |
| 0,025870022 | 0,974218958 | EPN2 | FALSE | Upregulated |
| 0,055002521 | 0,974218958 | LOC102723506 | FALSE | Upregulated |
| -0,035490124 | 0,974218958 | RNF181 | FALSE | Downregulated |
| 0,017613834 | 0,974325753 | BAHCC1 | FALSE | Upregulated |
| 0,031933188 | 0,974325753 | LYN | FALSE | Upregulated |
| -0,020883694 | 0,974325753 | SEC14L1 | FALSE | Downregulated |
| -0,021077269 | 0,974325753 | SRD5A1 | FALSE | Downregulated |
| 0,024836176 | 0,974325753 | TRIM38 | FALSE | Upregulated |
| -0,055006368 | 0,974357542 | LOC100289533 | FALSE | Downregulated |
| 0,027996392 | 0,974357542 | PAQR7 | FALSE | Upregulated |
| -0,013613662 | 0,974367432 | CAST | FALSE | Downregulated |
| 0,021786362 | 0,974434099 | HCG18 | FALSE | Upregulated |
| -0,018374077 | 0,974447113 | CFL1 | FALSE | Downregulated |
| -0,027629394 | 0,974447113 | MRPS7 | FALSE | Downregulated |
| -0,03010491 | 0,974447113 | SEPT6 | FALSE | Downregulated |
| -0,033582943 | 0,974447113 | TEFM | FALSE | Downregulated |
| 0,032651134 | 0,974641805 | HLX-AS1 | FALSE | Upregulated |
| 0,026375218 | 0,97467818 | NPDC1 | FALSE | Upregulated |
| -0,015551816 | 0,974794664 | DENND4B | FALSE | Downregulated |
| 0,107826264 | 0,974794664 | LOC100630923 | FALSE | Upregulated |
| 0,032592327 | 0,974794664 | LOC150051 | FALSE | Upregulated |
| 0,266627283 | 0,974794664 | MB | FALSE | Upregulated |
| -0,020661302 | 0,974794664 | TBL1XR1 | FALSE | Downregulated |
| -0,019263199 | 0,974794664 | TIMM13 | FALSE | Downregulated |
| -0,026359617 | 0,974794664 | ULK3 | FALSE | Downregulated |
| 0,038629761 | 0,974802792 | C15orf40 | FALSE | Upregulated |
| -0,032574017 | 0,974802792 | C1orf177 | FALSE | Downregulated |
| -0,07720127 | 0,974837541 | SULT1A4 | FALSE | Downregulated |
| 0,044024355 | 0,975009323 | KCNG2 | FALSE | Upregulated |
| -0,036882429 | 0,975009323 | NOTCH3 | FALSE | Downregulated |
| -0,046264568 | 0,975009323 | SP4 | FALSE | Downregulated |
| -0,047386197 | 0,975110883 | GPATCH2 | FALSE | Downregulated |
| -0,051635336 | 0,975110883 | MEAF6 | FALSE | Downregulated |
| -0,022156879 | 0,975110883 | VKORC1L1 | FALSE | Downregulated |
| 0,025346234 | 0,975110883 | YIPF4 | FALSE | Upregulated |
| 0,031970882 | 0,975110883 | ZNF549 | FALSE | Upregulated |
| 0,022899694 | 0,9752383 | NQO2 | FALSE | Upregulated |
| 0,043916807 | 0,975416022 | CDADC1 | FALSE | Upregulated |
| 0,023711199 | 0,975416022 | IFIT2 | FALSE | Upregulated |

| | | | | |
|---|---|---|---|---|
| -0,009182252 | 0,975639685 | LARP1 | FALSE | Downregulated |
| 0,036906053 | 0,975661876 | AAR2 | FALSE | Upregulated |
| -0,016472194 | 0,975661876 | CORO1B | FALSE | Downregulated |
| -0,031890992 | 0,975661876 | CREB3L4 | FALSE | Downregulated |
| 0,020193618 | 0,975661876 | ELMO2 | FALSE | Upregulated |
| -0,03145234 | 0,975661876 | FST | FALSE | Downregulated |
| -0,043984235 | 0,975661876 | MORN1 | FALSE | Downregulated |
| 0,044312497 | 0,975661876 | TRIQK | FALSE | Upregulated |
| 0,02971425 | 0,975726417 | MEF2C | FALSE | Upregulated |
| -0,024276868 | 0,975805679 | RAB1A | FALSE | Downregulated |
| -0,019599446 | 0,975805679 | SAMD14 | FALSE | Downregulated |
| 0,046311432 | 0,975820839 | C6orf1 | FALSE | Upregulated |
| 0,019855643 | 0,975820839 | LOC101928716 | FALSE | Upregulated |
| -0,036323721 | 0,975820839 | PROSER1 | FALSE | Downregulated |
| 0,022628263 | 0,975820839 | RBM17 | FALSE | Upregulated |
| -0,036690445 | 0,975820839 | SLC16A6 | FALSE | Downregulated |
| 0,038516839 | 0,975820839 | ZNF182 | FALSE | Upregulated |
| 0,018893026 | 0,975831737 | ACAP3 | FALSE | Upregulated |
| -0,033522447 | 0,975831737 | CASP10 | FALSE | Downregulated |
| 0,032868458 | 0,975831737 | GGT7 | FALSE | Upregulated |
| 0,027250603 | 0,975831737 | HDAC3 | FALSE | Upregulated |
| 0,04405968 | 0,975831737 | ISM1 | FALSE | Upregulated |
| 0,029994525 | 0,975831737 | LOC101928798 | FALSE | Upregulated |
| -0,08721448 | 0,975831737 | LOC101929819 | FALSE | Downregulated |
| 0,04274594 | 0,975831737 | LTK | FALSE | Upregulated |
| -0,015192384 | 0,975831737 | NACC2 | FALSE | Downregulated |
| 0,039016322 | 0,975831737 | NDUFAF2 | FALSE | Upregulated |
| -0,032205845 | 0,975831737 | NSA2 | FALSE | Downregulated |
| -0,014209348 | 0,975831737 | PAFAH1B1 | FALSE | Downregulated |
| -0,029895236 | 0,975831737 | PDGFRB | FALSE | Downregulated |
| -0,025274312 | 0,975831737 | RP11-384K6.6 | FALSE | Downregulated |
| 0,016795575 | 0,975831737 | TMEM131 | FALSE | Upregulated |
| -0,015875051 | 0,975831737 | TRIM24 | FALSE | Downregulated |
| -0,026272151 | 0,975831737 | ZKSCAN5 | FALSE | Downregulated |
| -0,02807459 | 0,975831737 | ZNF576 | FALSE | Downregulated |
| 0,039969198 | 0,975851131 | C17orf51 | FALSE | Upregulated |
| -0,026893561 | 0,975851131 | DCAF16 | FALSE | Downregulated |
| 0,058340927 | 0,975851131 | PRB2 | FALSE | Upregulated |
| 0,024098839 | 0,976036817 | APBB1IP | FALSE | Upregulated |
| 0,03390033 | 0,976036817 | COX7A1 | FALSE | Upregulated |
| 0,015743565 | 0,976036817 | EIF4A1 | FALSE | Upregulated |
| 0,025433154 | 0,976036817 | FGL1 | FALSE | Upregulated |
| -0,019818959 | 0,976036817 | NAA60 | FALSE | Downregulated |
| 0,027058193 | 0,976036817 | PROX1-AS1 | FALSE | Upregulated |
| -0,028484775 | 0,976036817 | SRPRB | FALSE | Downregulated |
| -0,029136101 | 0,976036817 | TSG101 | FALSE | Downregulated |
| 0,020501012 | 0,976036817 | USP15 | FALSE | Upregulated |
| -0,067163028 | 0,976036817 | UST | FALSE | Downregulated |
| -0,038387234 | 0,976036817 | ZNF662 | FALSE | Downregulated |
| 0,031625228 | 0,976080774 | THYN1 | FALSE | Upregulated |
| 0,031693933 | 0,976198358 | AMMECR1 | FALSE | Upregulated |

| | | | | |
|---|---|---|---|---|
| -0,043750693 | 0,976198358 | ARHGAP18 | FALSE | Downregulated |
| -0,027614554 | 0,976198358 | CDC16 | FALSE | Downregulated |
| 0,023584521 | 0,976198358 | CNN2 | FALSE | Upregulated |
| -0,039822345 | 0,976198358 | DZIP1 | FALSE | Downregulated |
| 0,013703318 | 0,976198358 | FKBP4 | FALSE | Upregulated |
| 0,032005278 | 0,976198358 | HMGXB4 | FALSE | Upregulated |
| -0,048561315 | 0,976198358 | LCN12 | FALSE | Downregulated |
| -0,093311967 | 0,976198358 | MSH5-SAPCD1 | FALSE | Downregulated |
| -0,039051531 | 0,976198358 | PTRHD1 | FALSE | Downregulated |
| -0,037092413 | 0,976198358 | RMDN2 | FALSE | Downregulated |
| 0,029518036 | 0,976198358 | SPAG5 | FALSE | Upregulated |
| 0,023988244 | 0,976198358 | TTBK1 | FALSE | Upregulated |
| 0,020727978 | 0,976376236 | NPRL3 | FALSE | Upregulated |
| 0,027175745 | 0,976718911 | SLC35C2 | FALSE | Upregulated |
| -0,031572495 | 0,976859301 | LOC100130075 | FALSE | Downregulated |
| 0,014364091 | 0,977073838 | ARNT | FALSE | Upregulated |
| 0,019346773 | 0,977073838 | FZR1 | FALSE | Upregulated |
| -0,02867923 | 0,977073838 | IDI2 | FALSE | Downregulated |
| -0,014921779 | 0,977073838 | SLC25A42 | FALSE | Downregulated |
| -0,03073023 | 0,977075635 | C12orf57 | FALSE | Downregulated |
| 0,029915455 | 0,977075635 | FIP1L1 | FALSE | Upregulated |
| 0,027364575 | 0,977075635 | INMT | FALSE | Upregulated |
| -0,021832716 | 0,977075635 | NOL6 | FALSE | Downregulated |
| -0,014157588 | 0,977082029 | TCF20 | FALSE | Downregulated |
| -0,01934725 | 0,977142366 | ABCG8 | FALSE | Downregulated |
| -0,01521389 | 0,977142366 | CDC42EP4 | FALSE | Downregulated |
| 0,017997539 | 0,977142366 | CHAD | FALSE | Upregulated |
| 0,021158586 | 0,977142366 | FGGY | FALSE | Upregulated |
| -0,029759627 | 0,977142366 | GBP3 | FALSE | Downregulated |
| -0,032166985 | 0,977142366 | IQSEC2 | FALSE | Downregulated |
| -0,038907338 | 0,977142366 | LOC101928154 | FALSE | Downregulated |
| -0,015134271 | 0,977142366 | LRRFIP1 | FALSE | Downregulated |
| 0,044963026 | 0,977142366 | LURAP1L | FALSE | Upregulated |
| -0,01703401 | 0,977142366 | MAGT1 | FALSE | Downregulated |
| 0,012466912 | 0,977142366 | MAP3K11 | FALSE | Upregulated |
| 0,011362544 | 0,977142366 | MORF4L1 | FALSE | Upregulated |
| 0,030135116 | 0,977142366 | ODC1 | FALSE | Upregulated |
| -0,026293672 | 0,977142366 | SLC35D2 | FALSE | Downregulated |
| -0,027199327 | 0,977142366 | SPAG5-AS1 | FALSE | Downregulated |
| -0,022241805 | 0,977142366 | TP53 | FALSE | Downregulated |
| -0,023415642 | 0,977142366 | UBE2V1 | FALSE | Downregulated |
| -0,043103141 | 0,977142366 | ZKSCAN7 | FALSE | Downregulated |
| 0,017620509 | 0,977228466 | EML3 | FALSE | Upregulated |
| -0,019530193 | 0,977228466 | ZNF646 | FALSE | Downregulated |
| -0,014609995 | 0,977272412 | HDAC5 | FALSE | Downregulated |
| 0,042794845 | 0,977336531 | SWI5 | FALSE | Upregulated |
| 0,026919869 | 0,977401941 | AMZ2 | FALSE | Upregulated |
| 0,028065078 | 0,977491075 | ANXA5 | FALSE | Upregulated |
| -0,029192048 | 0,977491075 | CBR4 | FALSE | Downregulated |
| 0,028591752 | 0,977491075 | CYB5R1 | FALSE | Upregulated |
| 0,044209561 | 0,977491075 | CYYR1 | FALSE | Upregulated |

| | | | | |
|---|---|---|---|---|
| 0,028381102 | 0,977491075 | DDX31 | FALSE | Upregulated |
| 0,021966024 | 0,977491075 | DNALI1 | FALSE | Upregulated |
| 0,038910889 | 0,977491075 | DPH3 | FALSE | Upregulated |
| 0,024282251 | 0,977491075 | DYNC2LI1 | FALSE | Upregulated |
| -0,015882579 | 0,977491075 | EIF3I | FALSE | Downregulated |
| 0,045280954 | 0,977491075 | F2RL3 | FALSE | Upregulated |
| 0,019759337 | 0,977491075 | FLJ37453 | FALSE | Upregulated |
| -0,017683757 | 0,977491075 | GIGYF2 | FALSE | Downregulated |
| 0,036911929 | 0,977491075 | GPD2 | FALSE | Upregulated |
| -0,008289093 | 0,977491075 | HIPK2 | FALSE | Downregulated |
| 0,015221353 | 0,977491075 | HSD3B7 | FALSE | Upregulated |
| 0,027387536 | 0,977491075 | INTS9 | FALSE | Upregulated |
| 0,014179484 | 0,977491075 | KAT2B | FALSE | Upregulated |
| -0,023018076 | 0,977491075 | LOC100287467 | FALSE | Downregulated |
| -0,031539275 | 0,977491075 | MFSD10 | FALSE | Downregulated |
| -0,042685115 | 0,977491075 | MTHFD2 | FALSE | Downregulated |
| -0,026711722 | 0,977491075 | NR2C1 | FALSE | Downregulated |
| -0,042101984 | 0,977491075 | NUDT17 | FALSE | Downregulated |
| 0,015696002 | 0,977491075 | OTUD7B | FALSE | Upregulated |
| 0,021214471 | 0,977491075 | PGAP3 | FALSE | Upregulated |
| 0,025843766 | 0,977491075 | RGMB | FALSE | Upregulated |
| -0,041363902 | 0,977491075 | RP11-372E1.4 | FALSE | Downregulated |
| -0,024088538 | 0,977491075 | SKIDA1 | FALSE | Downregulated |
| -0,013679985 | 0,977491075 | TCF25 | FALSE | Downregulated |
| 0,037096561 | 0,977491075 | WDR72 | FALSE | Upregulated |
| 0,047903013 | 0,977491075 | WDR86 | FALSE | Upregulated |
| -0,017194497 | 0,977502586 | UNKL | FALSE | Downregulated |
| -0,022296746 | 0,977582769 | KLHL2 | FALSE | Downregulated |
| 0,027143345 | 0,977618235 | ARMC8 | FALSE | Upregulated |
| 0,025423622 | 0,977787338 | B4GALT2 | FALSE | Upregulated |
| -0,025063886 | 0,977787338 | GGT1 | FALSE | Downregulated |
| -0,019930473 | 0,977879353 | PPIF | FALSE | Downregulated |
| 0,019146579 | 0,977991602 | TESK2 | FALSE | Upregulated |
| 0,02688092 | 0,978086893 | ARPC5L | FALSE | Upregulated |
| 0,010322261 | 0,978086893 | ATXN1 | FALSE | Upregulated |
| -0,017981495 | 0,978086893 | BAG1 | FALSE | Downregulated |
| 0,015999981 | 0,978086893 | CHD6 | FALSE | Upregulated |
| 0,041610593 | 0,978086893 | HCAR3 | FALSE | Upregulated |
| 0,040036106 | 0,978086893 | HLA-DMA | FALSE | Upregulated |
| 0,048663814 | 0,978086893 | KANK4 | FALSE | Upregulated |
| -0,038656694 | 0,978086893 | LOC100996643 | FALSE | Downregulated |
| -0,068784332 | 0,978086893 | LOC100996717 | FALSE | Downregulated |
| -0,034090189 | 0,978086893 | PHKG2 | FALSE | Downregulated |
| 0,023229068 | 0,978086893 | PPM1G | FALSE | Upregulated |
| 0,020958676 | 0,978086893 | PRR13 | FALSE | Upregulated |
| 0,025178534 | 0,978086893 | SNX13 | FALSE | Upregulated |
| 0,018937493 | 0,978086893 | STIP1 | FALSE | Upregulated |
| -0,030271099 | 0,978086893 | SYPL2 | FALSE | Downregulated |
| 0,023360713 | 0,978086893 | VAMP3 | FALSE | Upregulated |
| -0,013900253 | 0,978332451 | ABCA1 | FALSE | Downregulated |
| 0,025377095 | 0,978332451 | ABCC10 | FALSE | Upregulated |

| | | | | |
|---|---|---|---|---|
| 0,028425474 | 0,978332451 | BUD31 | FALSE | Upregulated |
| 0,014034112 | 0,978332451 | CCNL1 | FALSE | Upregulated |
| 0,031329306 | 0,978332451 | CD40 | FALSE | Upregulated |
| -0,041199947 | 0,978332451 | CSTF2 | FALSE | Downregulated |
| -0,019635274 | 0,978332451 | CTB-118N6.3 | FALSE | Downregulated |
| 0,028272716 | 0,978332451 | FADD | FALSE | Upregulated |
| -0,022207432 | 0,978332451 | GFER | FALSE | Downregulated |
| -0,019585006 | 0,978332451 | GLDC | FALSE | Downregulated |
| -0,034644116 | 0,978332451 | HCRTR1 | FALSE | Downregulated |
| 0,073657149 | 0,978332451 | HIST2H2AA4 | FALSE | Upregulated |
| 0,022929862 | 0,978332451 | KLHL36 | FALSE | Upregulated |
| 0,030382793 | 0,978332451 | LINC00894 | FALSE | Upregulated |
| -0,073093581 | 0,978332451 | LRP1B | FALSE | Downregulated |
| -0,024253725 | 0,978332451 | MAMDC4 | FALSE | Downregulated |
| -0,008062819 | 0,978332451 | MLLT6 | FALSE | Downregulated |
| -0,021804515 | 0,978332451 | NDUFS7 | FALSE | Downregulated |
| 0,038891991 | 0,978332451 | PARVB | FALSE | Upregulated |
| -0,020295742 | 0,978332451 | RHBDD1 | FALSE | Downregulated |
| -0,038567191 | 0,978332451 | RPF2 | FALSE | Downregulated |
| -0,018406779 | 0,978332451 | SMPDL3A | FALSE | Downregulated |
| 0,015779849 | 0,978332451 | SSR3 | FALSE | Upregulated |
| 0,025305762 | 0,978332451 | STEAP4 | FALSE | Upregulated |
| 0,027093734 | 0,978332451 | TBC1D12 | FALSE | Upregulated |
| -0,029077476 | 0,978332451 | THEM4 | FALSE | Downregulated |
| -0,021723007 | 0,978332451 | TIE1 | FALSE | Downregulated |
| 0,012976227 | 0,978332451 | TOP2B | FALSE | Upregulated |
| 0,024216268 | 0,978332451 | WDR52 | FALSE | Upregulated |
| -0,0370015 | 0,978332451 | ZNF25 | FALSE | Downregulated |
| 0,01758258 | 0,978568005 | HIATL1 | FALSE | Upregulated |
| -0,013356415 | 0,978712509 | CD59 | FALSE | Downregulated |
| 0,202124586 | 0,978751123 | ACTN2 | FALSE | Upregulated |
| -0,017770997 | 0,978751123 | AGPAT6 | FALSE | Downregulated |
| 0,101569791 | 0,978751123 | C21orf33 | FALSE | Upregulated |
| 0,036975897 | 0,978751123 | FPR3 | FALSE | Upregulated |
| 0,017298529 | 0,978751123 | LBR | FALSE | Upregulated |
| -0,036650704 | 0,978751123 | LOC101927479 | FALSE | Downregulated |
| -0,012614466 | 0,978751123 | MAP3K2 | FALSE | Downregulated |
| -0,015785734 | 0,978751123 | MIER1 | FALSE | Downregulated |
| -0,033048897 | 0,978751123 | OSER1 | FALSE | Downregulated |
| 0,034056178 | 0,978751123 | PDIK1L | FALSE | Upregulated |
| -0,022259029 | 0,978751123 | PLA2G6 | FALSE | Downregulated |
| -0,018651889 | 0,978751123 | RANGAP1 | FALSE | Downregulated |
| -0,030311003 | 0,978751123 | THRA | FALSE | Downregulated |
| -0,038490706 | 0,978751123 | TMEM91 | FALSE | Downregulated |
| -0,026212756 | 0,978751123 | TUBGCP5 | FALSE | Downregulated |
| -0,031612875 | 0,978751123 | ZNF554 | FALSE | Downregulated |
| -0,021972944 | 0,978799045 | TXNDC15 | FALSE | Downregulated |
| -0,011204733 | 0,978805693 | CLASP1 | FALSE | Downregulated |
| -0,013258023 | 0,978805693 | HDAC6 | FALSE | Downregulated |
| 0,029865079 | 0,978824307 | AGER | FALSE | Upregulated |
| 0,023342279 | 0,978824307 | PIK3CB | FALSE | Upregulated |

| | | | | |
|---|---|---|---|---|
| -0,028749252 | 0,978824307 | TTC30B | FALSE | Downregulated |
| 0,02269215 | 0,978824307 | UBE2A | FALSE | Upregulated |
| -0,010012537 | 0,978824307 | WAC | FALSE | Downregulated |
| -0,026862605 | 0,978824307 | ZNF35 | FALSE | Downregulated |
| 0,021067084 | 0,979061962 | LINC00847 | FALSE | Upregulated |
| 0,013856987 | 0,979065716 | NCOA1 | FALSE | Upregulated |
| 0,013237993 | 0,979065716 | PPP2R5A | FALSE | Upregulated |
| -0,01537259 | 0,979416735 | ERBB2 | FALSE | Downregulated |
| 0,01060258 | 0,97948271 | KIAA0319L | FALSE | Upregulated |
| -0,014594231 | 0,979555438 | BRD3 | FALSE | Downregulated |
| 0,024488267 | 0,979555438 | C9orf66 | FALSE | Upregulated |
| 0,016930695 | 0,979555438 | CPN1 | FALSE | Upregulated |
| 0,018211177 | 0,979555438 | CTDNEP1 | FALSE | Upregulated |
| -0,01987367 | 0,979555438 | DDTL | FALSE | Downregulated |
| -0,105693289 | 0,979555438 | FAM156A | FALSE | Downregulated |
| -0,016133362 | 0,979555438 | GGT5 | FALSE | Downregulated |
| 0,036426389 | 0,979555438 | LOC101927382 | FALSE | Upregulated |
| -0,019033816 | 0,979555438 | MBD2 | FALSE | Downregulated |
| 0,432289725 | 0,979555438 | MYH2 | FALSE | Upregulated |
| -0,029278683 | 0,979555438 | PDE4B | FALSE | Downregulated |
| -0,021540972 | 0,979555438 | UHRF2 | FALSE | Downregulated |
| 0,020147957 | 0,979649801 | EFHD2 | FALSE | Upregulated |
| -0,028363082 | 0,979724024 | PRSS23 | FALSE | Downregulated |
| 0,013899669 | 0,979743498 | ATP11B | FALSE | Upregulated |
| -0,022079698 | 0,979743498 | B9D1 | FALSE | Downregulated |
| -0,026365148 | 0,979743498 | C1orf123 | FALSE | Downregulated |
| -0,053813041 | 0,979743498 | DNAH5 | FALSE | Downregulated |
| 0,037690114 | 0,979743498 | FAM216A | FALSE | Upregulated |
| 0,02557225 | 0,979743498 | HOXB3 | FALSE | Upregulated |
| 0,021643191 | 0,979743498 | LOC102724250 | FALSE | Upregulated |
| 0,017098598 | 0,979743498 | RANBP3 | FALSE | Upregulated |
| -0,049325954 | 0,979743498 | SERPINB8 | FALSE | Downregulated |
| 0,029444561 | 0,979743498 | SLC16A11 | FALSE | Upregulated |
| 0,02948618 | 0,979743498 | SMUG1 | FALSE | Upregulated |
| 0,019457771 | 0,979743498 | ZNF224 | FALSE | Upregulated |
| 0,016367925 | 0,979838361 | TLK2 | FALSE | Upregulated |
| -0,031026431 | 0,980054407 | HAUS3 | FALSE | Downregulated |
| 0,028668623 | 0,980054407 | PKDCC | FALSE | Upregulated |
| -0,042151859 | 0,980054407 | ZNF626 | FALSE | Downregulated |
| -0,029494172 | 0,980383229 | CABP4 | FALSE | Downregulated |
| -0,017716287 | 0,980383229 | NOP56 | FALSE | Downregulated |
| 0,017766395 | 0,980383229 | PARD3B | FALSE | Upregulated |
| -0,033546473 | 0,980383229 | RGS19 | FALSE | Downregulated |
| 0,02011878 | 0,980383229 | TMEM120A | FALSE | Upregulated |
| -0,017286331 | 0,980383229 | ZSCAN21 | FALSE | Downregulated |
| -0,020348862 | 0,98039567 | TMEM141 | FALSE | Downregulated |
| 0,024086292 | 0,980473011 | IGIP | FALSE | Upregulated |
| -0,03332935 | 0,980473011 | RRAGB | FALSE | Downregulated |
| 0,026257441 | 0,980502876 | LLGL1 | FALSE | Upregulated |
| -0,034847242 | 0,980514086 | TMEM147 | FALSE | Downregulated |
| -0,029127852 | 0,980548083 | WDR27 | FALSE | Downregulated |

| | | | | |
|---|---|---|---|---|
| -0,018053075 | 0,980677486 | BCL2L2 | FALSE | Downregulated |
| 0,016639347 | 0,980677486 | CAPZB | FALSE | Upregulated |
| -0,032059344 | 0,980677486 | CYP2W1 | FALSE | Downregulated |
| 0,025183642 | 0,980677486 | DHRSX | FALSE | Upregulated |
| 0,033621499 | 0,980677486 | PCSK4 | FALSE | Upregulated |
| -0,028610479 | 0,980677486 | VSTM4 | FALSE | Downregulated |
| -0,011097012 | 0,980712162 | BRD2 | FALSE | Downregulated |
| -0,014356207 | 0,980746215 | PLCB3 | FALSE | Downregulated |
| 0,017013186 | 0,981032799 | LTBP1 | FALSE | Upregulated |
| -0,027244395 | 0,98141389 | INHBB | FALSE | Downregulated |
| -0,011754299 | 0,98141389 | MIB2 | FALSE | Downregulated |
| 0,033371984 | 0,98141389 | RP11-35G9.3 | FALSE | Upregulated |
| -0,018308783 | 0,98141389 | TESK1 | FALSE | Downregulated |
| 0,01671845 | 0,981535391 | HMCN1 | FALSE | Upregulated |
| -0,030769533 | 0,981535391 | NARS2 | FALSE | Downregulated |
| -0,022285001 | 0,981535391 | SAPCD2 | FALSE | Downregulated |
| -0,013401157 | 0,981535391 | TMUB1 | FALSE | Downregulated |
| -0,014490383 | 0,981565933 | MED29 | FALSE | Downregulated |
| -0,020274405 | 0,981565933 | NOSIP | FALSE | Downregulated |
| -0,012622359 | 0,981582668 | SERP1 | FALSE | Downregulated |
| -0,012852161 | 0,981633418 | CRAMP1L | FALSE | Downregulated |
| -0,029199897 | 0,9818184 | AGTPBP1 | FALSE | Downregulated |
| -0,017130363 | 0,9818184 | DLG5 | FALSE | Downregulated |
| -0,038058266 | 0,9818184 | HIST1H2APS1 | FALSE | Downregulated |
| 0,007419846 | 0,9818184 | HNRNPUL1 | FALSE | Upregulated |
| 0,036763421 | 0,9818184 | HSPBAP1 | FALSE | Upregulated |
| -0,016278089 | 0,9818184 | IL17RA | FALSE | Downregulated |
| 0,021047663 | 0,9818184 | LOC730102 | FALSE | Upregulated |
| -0,023881742 | 0,9818184 | PANK4 | FALSE | Downregulated |
| 0,011752115 | 0,9818184 | PRDX1 | FALSE | Upregulated |
| -0,031869685 | 0,9818184 | RAB3D | FALSE | Downregulated |
| 0,014291472 | 0,9818184 | SH2B1 | FALSE | Upregulated |
| -0,021320058 | 0,9818184 | ZC3H15 | FALSE | Downregulated |
| 0,03690061 | 0,981841172 | AGA | FALSE | Upregulated |
| -0,014417876 | 0,981841172 | C11orf58 | FALSE | Downregulated |
| -0,01148808 | 0,981841172 | DAZAP2 | FALSE | Downregulated |
| -0,033537048 | 0,981841172 | KLF4 | FALSE | Downregulated |
| -0,012286444 | 0,981841172 | NTN1 | FALSE | Downregulated |
| 0,013846 | 0,981841172 | ZKSCAN8 | FALSE | Upregulated |
| 0,025895355 | 0,981898954 | ABHD3 | FALSE | Upregulated |
| 0,014544175 | 0,981898954 | YES1 | FALSE | Upregulated |
| 0,03122944 | 0,981958817 | FANCM | FALSE | Upregulated |
| -0,016130655 | 0,98201449 | HAX1 | FALSE | Downregulated |
| 0,024816684 | 0,982051299 | C5orf15 | FALSE | Upregulated |
| 0,02660309 | 0,982051299 | CCP110 | FALSE | Upregulated |
| 0,020011899 | 0,982051299 | VEZT | FALSE | Upregulated |
| -0,019873399 | 0,982140398 | LRCH3 | FALSE | Downregulated |
| -0,034516452 | 0,982391115 | ACOT9 | FALSE | Downregulated |
| -0,019516629 | 0,982391115 | ADAP2 | FALSE | Downregulated |
| 0,022115193 | 0,982391115 | ANAPC11 | FALSE | Upregulated |
| 0,013332724 | 0,982391115 | ARHGEF11 | FALSE | Upregulated |

| | | | | |
|---|---|---|---|---|
| -0,012316814 | 0,982391115 | ATP1B1 | FALSE | Downregulated |
| 0,013586428 | 0,982391115 | BLCAP | FALSE | Upregulated |
| -0,021623528 | 0,982391115 | C14orf1 | FALSE | Downregulated |
| -0,027330882 | 0,982391115 | CHML | FALSE | Downregulated |
| -0,023809702 | 0,982391115 | DHX40 | FALSE | Downregulated |
| -0,015464586 | 0,982391115 | DNAJC16 | FALSE | Downregulated |
| -0,010657513 | 0,982391115 | DVL3 | FALSE | Downregulated |
| 0,018486681 | 0,982391115 | HECW2 | FALSE | Upregulated |
| 0,05803354 | 0,982391115 | ICOSLG | FALSE | Upregulated |
| -0,011455638 | 0,982391115 | KIF3B | FALSE | Downregulated |
| 0,026087732 | 0,982391115 | KRCC1 | FALSE | Upregulated |
| 0,023452381 | 0,982391115 | LINC00342 | FALSE | Upregulated |
| -0,014494856 | 0,982391115 | LOC101928946 | FALSE | Downregulated |
| -0,05356963 | 0,982391115 | LOC79999 | FALSE | Downregulated |
| 0,050679372 | 0,982391115 | MT1H | FALSE | Upregulated |
| 0,006994646 | 0,982391115 | NCOR1 | FALSE | Upregulated |
| -0,010482264 | 0,982391115 | NFE2L2 | FALSE | Downregulated |
| 0,016884103 | 0,982391115 | PHC3 | FALSE | Upregulated |
| 0,01244782 | 0,982391115 | RANBP9 | FALSE | Upregulated |
| -0,015889914 | 0,982391115 | RP11-57H12.3 | FALSE | Downregulated |
| 0,029985447 | 0,982391115 | RUNX1T1 | FALSE | Upregulated |
| -0,012007834 | 0,982391115 | SMG1P1 | FALSE | Downregulated |
| -0,024922771 | 0,982391115 | SYK | FALSE | Downregulated |
| -0,037497426 | 0,982391115 | TACR2 | FALSE | Downregulated |
| 0,015748677 | 0,982391115 | XPO7 | FALSE | Upregulated |
| -0,013878104 | 0,982391115 | ZNF407 | FALSE | Downregulated |
| 0,020256886 | 0,982408114 | CCNF | FALSE | Upregulated |
| 0,012163954 | 0,982408114 | CIZ1 | FALSE | Upregulated |
| 0,020359581 | 0,982408114 | DCPS | FALSE | Upregulated |
| -0,017305971 | 0,982408114 | EARS2 | FALSE | Downregulated |
| 0,025663192 | 0,982408114 | HELQ | FALSE | Upregulated |
| -0,016401322 | 0,982408114 | LOC102724696 | FALSE | Downregulated |
| -0,009850833 | 0,982408114 | UBE2D3 | FALSE | Downregulated |
| -0,018346814 | 0,982408114 | ZFAND5 | FALSE | Downregulated |
| -0,016514668 | 0,982431379 | CFP | FALSE | Downregulated |
| -0,018371798 | 0,982431379 | NEK7 | FALSE | Downregulated |
| 0,020148867 | 0,982431379 | PAXIP1-AS2 | FALSE | Upregulated |
| -0,01473238 | 0,982455766 | PTPN1 | FALSE | Downregulated |
| -0,010324218 | 0,982599934 | ABL1 | FALSE | Downregulated |
| -0,021267985 | 0,982599934 | ALDH18A1 | FALSE | Downregulated |
| -0,016296541 | 0,982599934 | AMDHD2 | FALSE | Downregulated |
| -0,032165278 | 0,982599934 | DZIP3 | FALSE | Downregulated |
| 0,016678829 | 0,982599934 | ITGA7 | FALSE | Upregulated |
| -0,029088892 | 0,982599934 | LOC102723469 | FALSE | Downregulated |
| -0,016767123 | 0,982599934 | PER2 | FALSE | Downregulated |
| 0,029512515 | 0,982599934 | TMEM179 | FALSE | Upregulated |
| 0,008975165 | 0,982599934 | TOMM70A | FALSE | Upregulated |
| -0,015207365 | 0,982675908 | ABCB8 | FALSE | Downregulated |
| 0,022465288 | 0,982675908 | ACVRL1 | FALSE | Upregulated |
| -0,028571102 | 0,982675908 | AGBL3 | FALSE | Downregulated |
| 0,023741893 | 0,982675908 | AXIN2 | FALSE | Upregulated |

| | | | | |
|---|---|---|---|---|
| 0,015310694 | 0,982675908 | C19orf12 | FALSE | Upregulated |
| 0,030233763 | 0,982675908 | EXOC6 | FALSE | Upregulated |
| -0,02053956 | 0,982675908 | KLHL3 | FALSE | Downregulated |
| 0,02210877 | 0,982675908 | LMF1 | FALSE | Upregulated |
| -0,026303506 | 0,982675908 | LOC101929263 | FALSE | Downregulated |
| 0,034927967 | 0,982675908 | LOC102723980 | FALSE | Upregulated |
| 0,026865206 | 0,982675908 | LRP12 | FALSE | Upregulated |
| 0,021234072 | 0,982675908 | MIS18BP1 | FALSE | Upregulated |
| -0,013626866 | 0,982675908 | PIAS1 | FALSE | Downregulated |
| -0,032055927 | 0,982675908 | RND2 | FALSE | Downregulated |
| -0,016375899 | 0,982675908 | RP11-696N14.1 | FALSE | Downregulated |
| 0,022652406 | 0,982675908 | SRC | FALSE | Upregulated |
| 0,021032223 | 0,982705658 | MSRB2 | FALSE | Upregulated |
| 0,034065358 | 0,982705658 | RSPH4A | FALSE | Upregulated |
| -0,015201333 | 0,982716357 | AQR | FALSE | Downregulated |
| -0,023689748 | 0,982716357 | DUSP1 | FALSE | Downregulated |
| -0,014030584 | 0,982716357 | EEF2K | FALSE | Downregulated |
| -0,008693295 | 0,982716357 | EIF3B | FALSE | Downregulated |
| 0,017372059 | 0,982716357 | ENTHD2 | FALSE | Upregulated |
| -0,022993189 | 0,982716357 | FAM184A | FALSE | Downregulated |
| 0,025568984 | 0,982716357 | LRRC16A | FALSE | Upregulated |
| -0,018150544 | 0,982716357 | RAB7L1 | FALSE | Downregulated |
| 0,035659855 | 0,982716357 | SLC25A12 | FALSE | Upregulated |
| -0,356534462 | 0,982727163 | RNVU1-18 | FALSE | Downregulated |
| 0,029106368 | 0,982992941 | LUZP6 | FALSE | Upregulated |
| 0,019903908 | 0,983004194 | TEX264 | FALSE | Upregulated |
| -0,021726982 | 0,983040829 | CDK18 | FALSE | Downregulated |
| -0,009788083 | 0,983040829 | CDV3 | FALSE | Downregulated |
| 0,017423101 | 0,983040829 | FCF1 | FALSE | Upregulated |
| -0,032067681 | 0,983040829 | LAMA3 | FALSE | Downregulated |
| 0,014550528 | 0,983040829 | THADA | FALSE | Upregulated |
| 0,01616827 | 0,983040829 | ZNF780A | FALSE | Upregulated |
| -0,014826038 | 0,983128627 | ASB13 | FALSE | Downregulated |
| 0,008856076 | 0,983128627 | ATXN7 | FALSE | Upregulated |
| 0,024036941 | 0,983128627 | GPATCH1 | FALSE | Upregulated |
| 0,020799881 | 0,983128627 | LSM8 | FALSE | Upregulated |
| -0,024134292 | 0,983128627 | SLC26A2 | FALSE | Downregulated |
| -0,00556278 | 0,983333875 | PLEC | FALSE | Downregulated |
| -0,225244284 | 0,983426027 | TNNT1 | FALSE | Downregulated |
| 0,016331937 | 0,983440074 | RBPJ | FALSE | Upregulated |
| -0,01307354 | 0,983465263 | SPG21 | FALSE | Downregulated |
| -0,018553128 | 0,98356399 | SMYD5 | FALSE | Downregulated |
| 0,010607999 | 0,98356399 | TMEM245 | FALSE | Upregulated |
| 0,02976098 | 0,98356399 | USP43 | FALSE | Upregulated |
| -0,009603334 | 0,98356399 | WWP1 | FALSE | Downregulated |
| 0,027035748 | 0,983565431 | ACSM1 | FALSE | Upregulated |
| 0,010893712 | 0,983565431 | CTSA | FALSE | Upregulated |
| 0,011988081 | 0,983565431 | ELAC2 | FALSE | Upregulated |
| 0,022218002 | 0,983565431 | LOC101927031 | FALSE | Upregulated |
| -0,011761837 | 0,983565431 | RPS6KA3 | FALSE | Downregulated |
| 0,029444824 | 0,983565431 | ZNF718 | FALSE | Upregulated |

| | | | | |
|---|---|---|---|---|
| -0,023946831 | 0,983581143 | FAM188A | FALSE | Downregulated |
| -0,012074661 | 0,983581143 | FAM53B | FALSE | Downregulated |
| 0,018351982 | 0,983581143 | PORCN | FALSE | Upregulated |
| -0,014503324 | 0,983581143 | SLC46A3 | FALSE | Downregulated |
| 0,014526382 | 0,983581143 | SRP54 | FALSE | Upregulated |
| 0,020812531 | 0,983581143 | ZNF524 | FALSE | Upregulated |
| -0,012391595 | 0,983783793 | JOSD1 | FALSE | Downregulated |
| -0,016019536 | 0,983783793 | PRMT2 | FALSE | Downregulated |
| 0,021791621 | 0,983874274 | ACCS | FALSE | Upregulated |
| 0,022444534 | 0,983874274 | CCDC158 | FALSE | Upregulated |
| 0,020212364 | 0,983874274 | DAXX | FALSE | Upregulated |
| -0,010876036 | 0,983874274 | DLG1 | FALSE | Downregulated |
| -0,018051484 | 0,983874274 | DPP4 | FALSE | Downregulated |
| 0,01106904 | 0,983874274 | EZH1 | FALSE | Upregulated |
| 0,015138755 | 0,983874274 | GADD45GIP1 | FALSE | Upregulated |
| 0,020374971 | 0,983874274 | IQCE | FALSE | Upregulated |
| -0,015803584 | 0,983874274 | KIAA1656 | FALSE | Downregulated |
| -0,017088198 | 0,983874274 | LZTS2 | FALSE | Downregulated |
| 0,014602666 | 0,983874274 | MAP3K1 | FALSE | Upregulated |
| 0,02880662 | 0,983874274 | MIPOL1 | FALSE | Upregulated |
| 0,009215742 | 0,983874274 | MYH14 | FALSE | Upregulated |
| 0,015923052 | 0,983874274 | PHF23 | FALSE | Upregulated |
| -0,014491244 | 0,983874274 | PPM1E | FALSE | Downregulated |
| -0,009567571 | 0,983874274 | PSD4 | FALSE | Downregulated |
| -0,023992704 | 0,983874274 | RBKS | FALSE | Downregulated |
| 0,017501962 | 0,983874274 | SLC25A37 | FALSE | Upregulated |
| -0,03247783 | 0,983874274 | SNRPE | FALSE | Downregulated |
| 0,028221187 | 0,983874274 | SOX9 | FALSE | Upregulated |
| -0,028774626 | 0,983874274 | SPTSSA | FALSE | Downregulated |
| 0,017739067 | 0,983874274 | TANC2 | FALSE | Upregulated |
| -0,025365244 | 0,983874274 | THUMPD3-AS1 | FALSE | Downregulated |
| -0,017838763 | 0,983874274 | USPL1 | FALSE | Downregulated |
| -0,012388345 | 0,983971175 | WDR7 | FALSE | Downregulated |
| -0,022827791 | 0,984240924 | TFRC | FALSE | Downregulated |
| 0,013012372 | 0,984280938 | FOXP4 | FALSE | Upregulated |
| 0,024430278 | 0,984280938 | PSMD14 | FALSE | Upregulated |
| -0,016884218 | 0,984477421 | NFKBIB | FALSE | Downregulated |
| 0,011651857 | 0,984679812 | STRN4 | FALSE | Upregulated |
| 0,021655302 | 0,984802093 | ABCA10 | FALSE | Upregulated |
| -0,018940379 | 0,984802093 | C11orf35 | FALSE | Downregulated |
| 0,023578866 | 0,984802093 | CEP76 | FALSE | Upregulated |
| 0,015204434 | 0,984802093 | CHID1 | FALSE | Upregulated |
| -0,020549103 | 0,984802093 | ELN | FALSE | Downregulated |
| -0,011148305 | 0,984802093 | GNS | FALSE | Downregulated |
| -0,018027986 | 0,984802093 | LOC102723406 | FALSE | Downregulated |
| -0,018953636 | 0,984802093 | NFIL3 | FALSE | Downregulated |
| -0,01040458 | 0,984802093 | PHF2 | FALSE | Downregulated |
| -0,014879323 | 0,984802093 | RAB3GAP1 | FALSE | Downregulated |
| -0,016299846 | 0,984802093 | RIMKLA | FALSE | Downregulated |
| 0,015437731 | 0,984802093 | SLC19A3 | FALSE | Upregulated |
| 0,014163002 | 0,984802093 | SLC25A16 | FALSE | Upregulated |

| | | | | |
|---|---|---|---|---|
| -0,014485133 | 0,984802093 | SMAD7 | FALSE | Downregulated |
| -0,023976794 | 0,984802093 | SOCS4 | FALSE | Downregulated |
| 0,007457813 | 0,984802093 | SPTAN1 | FALSE | Upregulated |
| 0,023647824 | 0,984802093 | SSC5D | FALSE | Upregulated |
| 0,023477468 | 0,984802093 | TICAM2 | FALSE | Upregulated |
| -0,016388774 | 0,984802093 | TSTA3 | FALSE | Downregulated |
| 0,021835224 | 0,984802093 | UBR7 | FALSE | Upregulated |
| -0,008626227 | 0,984802093 | VEGFA | FALSE | Downregulated |
| 0,024148085 | 0,984802093 | ZNF346 | FALSE | Upregulated |
| -0,019861896 | 0,984949449 | CCDC86 | FALSE | Downregulated |
| 0,028227086 | 0,984949449 | CD55 | FALSE | Upregulated |
| -0,015457872 | 0,984949449 | CDC42EP3 | FALSE | Downregulated |
| -0,016689451 | 0,984949449 | CTNNBIP1 | FALSE | Downregulated |
| 0,015862101 | 0,984949449 | ETNPPL | FALSE | Upregulated |
| 0,028862595 | 0,984949449 | FBLN1 | FALSE | Upregulated |
| -0,0201865 | 0,984949449 | LOC101060581 | FALSE | Downregulated |
| 0,019201225 | 0,984949449 | LOC101929173 | FALSE | Upregulated |
| 0,010612309 | 0,984949449 | OPA1 | FALSE | Upregulated |
| 0,01620277 | 0,984949449 | PER1 | FALSE | Upregulated |
| 0,028724647 | 0,984949449 | PPARG | FALSE | Upregulated |
| 0,050843826 | 0,984949449 | TCEB3CL | FALSE | Upregulated |
| -0,016801031 | 0,984949449 | TMBIM4 | FALSE | Downregulated |
| 0,015211616 | 0,984949449 | TSPAN11 | FALSE | Upregulated |
| 0,016348705 | 0,985189906 | BZRAP1-AS1 | FALSE | Upregulated |
| -0,010593494 | 0,985189906 | CDC37 | FALSE | Downregulated |
| 0,01610156 | 0,985189906 | FAM107A | FALSE | Upregulated |
| -0,012428626 | 0,985189906 | SNX19 | FALSE | Downregulated |
| -0,021835643 | 0,985189906 | TMEM65 | FALSE | Downregulated |
| 0,019125064 | 0,985189906 | VPS29 | FALSE | Upregulated |
| 0,024252106 | 0,985198786 | ADAMTS12 | FALSE | Upregulated |
| -0,02035768 | 0,985198786 | BRWD1-IT2 | FALSE | Downregulated |
| -0,016705909 | 0,985198786 | C22orf46 | FALSE | Downregulated |
| -0,01120047 | 0,985198786 | FAM104A | FALSE | Downregulated |
| -0,025073308 | 0,985198786 | FAM86B1 | FALSE | Downregulated |
| 0,012472028 | 0,985198786 | LRRC24 | FALSE | Upregulated |
| -0,01665222 | 0,985198786 | PSMD6 | FALSE | Downregulated |
| 0,00895996 | 0,985198786 | TNFRSF1A | FALSE | Upregulated |
| -0,047849141 | 0,985198786 | UTY | FALSE | Downregulated |
| -0,00691763 | 0,985245261 | UPF1 | FALSE | Downregulated |
| 0,02044787 | 0,985390077 | ARHGAP24 | FALSE | Upregulated |
| -0,028663533 | 0,985390077 | BTBD11 | FALSE | Downregulated |
| 0,017440147 | 0,985390077 | C1orf27 | FALSE | Upregulated |
| -0,021446624 | 0,985390077 | C8orf46 | FALSE | Downregulated |
| -0,016221091 | 0,985390077 | CCDC82 | FALSE | Downregulated |
| -0,009235342 | 0,985390077 | CXXC5 | FALSE | Downregulated |
| -0,016369105 | 0,985390077 | DLAT | FALSE | Downregulated |
| 0,025947662 | 0,985390077 | EIF1B | FALSE | Upregulated |
| -0,022325908 | 0,985390077 | FARSB | FALSE | Downregulated |
| 0,019014028 | 0,985390077 | FBXL12 | FALSE | Upregulated |
| 0,020925194 | 0,985390077 | HAND2 | FALSE | Upregulated |
| -0,009658189 | 0,985390077 | INVS | FALSE | Downregulated |

| | | | | |
|---|---|---|---|---|
| 0,017186369 | 0,985390077 | MRPS22 | FALSE | Upregulated |
| 0,021034067 | 0,985390077 | PARP15 | FALSE | Upregulated |
| 0,022714546 | 0,985390077 | PDE4C | FALSE | Upregulated |
| 0,024819852 | 0,985390077 | PLD6 | FALSE | Upregulated |
| -0,024448046 | 0,985390077 | RIMKLB | FALSE | Downregulated |
| 0,011831061 | 0,985390077 | RP11-713M15.1 | FALSE | Upregulated |
| -0,014112843 | 0,985390077 | SGK3 | FALSE | Downregulated |
| 0,027683018 | 0,985390077 | ST6GALNAC4 | FALSE | Upregulated |
| -0,013107155 | 0,985390077 | TCF7L1 | FALSE | Downregulated |
| -0,028385087 | 0,985390077 | ZNF438 | FALSE | Downregulated |
| -0,026945708 | 0,985530814 | ALG6 | FALSE | Downregulated |
| -0,01687086 | 0,985530814 | LINC00910 | FALSE | Downregulated |
| 0,008276877 | 0,985530814 | LRP10 | FALSE | Upregulated |
| 0,010073153 | 0,985530814 | LUC7L3 | FALSE | Upregulated |
| -0,021035788 | 0,985530814 | METTL22 | FALSE | Downregulated |
| -0,061708114 | 0,985530814 | P2RY11 | FALSE | Downregulated |
| 0,008324472 | 0,985530814 | PBXIP1 | FALSE | Upregulated |
| -0,007423253 | 0,985530814 | SCARNA2 | FALSE | Downregulated |
| -0,027188057 | 0,985537554 | NOSTRIN | FALSE | Downregulated |
| -0,02942979 | 0,985688304 | ZFP3 | FALSE | Downregulated |
| -0,01419533 | 0,985708431 | ANAPC13 | FALSE | Downregulated |
| -0,014975757 | 0,985708431 | AP1AR | FALSE | Downregulated |
| 0,008550529 | 0,985708431 | LOC101927509 | FALSE | Upregulated |
| -0,015499152 | 0,985708431 | PTGES2 | FALSE | Downregulated |
| 0,018577754 | 0,985708431 | SOGA2 | FALSE | Upregulated |
| -0,021695247 | 0,985708431 | TMEM17 | FALSE | Downregulated |
| 0,011964647 | 0,985888254 | SNAP23 | FALSE | Upregulated |
| -0,012613331 | 0,986065293 | FOXJ2 | FALSE | Downregulated |
| 0,017301913 | 0,986065293 | UGT2B11 | FALSE | Upregulated |
| 0,011510281 | 0,986087201 | SREBF2 | FALSE | Upregulated |
| -0,050693694 | 0,98609363 | POLR2J2 | FALSE | Downregulated |
| -0,018551934 | 0,986149675 | TIMM17A | FALSE | Downregulated |
| -0,008326911 | 0,986255346 | SOX13 | FALSE | Downregulated |
| -0,019128531 | 0,986255346 | UCK2 | FALSE | Downregulated |
| 0,016917051 | 0,986280921 | C20orf27 | FALSE | Upregulated |
| -0,00721667 | 0,986299066 | ARIH1 | FALSE | Downregulated |
| -0,016264116 | 0,986299066 | BTBD10 | FALSE | Downregulated |
| 0,011461056 | 0,986299066 | C19orf10 | FALSE | Upregulated |
| -0,009871358 | 0,986299066 | CEACAM1 | FALSE | Downregulated |
| 0,012332112 | 0,986299066 | CHMP7 | FALSE | Upregulated |
| 0,018423925 | 0,986299066 | CLDN5 | FALSE | Upregulated |
| 0,012856451 | 0,986299066 | EMC10 | FALSE | Upregulated |
| 0,007633343 | 0,986299066 | ESRP2 | FALSE | Upregulated |
| 0,008130637 | 0,986299066 | HNF4A | FALSE | Upregulated |
| 0,023549616 | 0,986299066 | MTPN | FALSE | Upregulated |
| 0,015541884 | 0,986299066 | NSMAF | FALSE | Upregulated |
| -0,012922663 | 0,986299066 | POFUT2 | FALSE | Downregulated |
| 0,090094495 | 0,986299066 | RASA4B | FALSE | Upregulated |
| -0,012560287 | 0,986299066 | RPA1 | FALSE | Downregulated |
| -0,012836301 | 0,986299066 | UNC93B1 | FALSE | Downregulated |
| -0,027720966 | 0,986299066 | ZDHHC11B | FALSE | Downregulated |

| | | | | |
|---|---|---|---|---|
| 0,014897837 | 0,986426277 | HCLS1 | FALSE | Upregulated |
| 0,274630165 | 0,986459669 | ACTA1 | FALSE | Upregulated |
| 0,019763192 | 0,986459669 | COG2 | FALSE | Upregulated |
| 0,018252592 | 0,986492593 | ARID3B | FALSE | Upregulated |
| -0,018341611 | 0,986492593 | NOTUM | FALSE | Downregulated |
| -0,010676413 | 0,986492593 | OTUD4 | FALSE | Downregulated |
| -0,013483943 | 0,986492593 | TMEM59 | FALSE | Downregulated |
| -0,022140882 | 0,986577376 | AIF1L | FALSE | Downregulated |
| 0,016458879 | 0,986577376 | ATG5 | FALSE | Upregulated |
| 0,019783636 | 0,986577376 | BCL2L10 | FALSE | Upregulated |
| -0,014364533 | 0,986577376 | C19orf26 | FALSE | Downregulated |
| 0,00792434 | 0,986577376 | CYFIP1 | FALSE | Upregulated |
| -0,022376703 | 0,986577376 | SCRN1 | FALSE | Downregulated |
| -0,012504093 | 0,986582826 | ARHGAP23 | FALSE | Downregulated |
| -0,013991856 | 0,986582826 | ISYNA1 | FALSE | Downregulated |
| 0,014920999 | 0,9866831 | ORMDL1 | FALSE | Upregulated |
| 0,021736516 | 0,9866831 | PALD1 | FALSE | Upregulated |
| -0,014453529 | 0,9866831 | TRA2B | FALSE | Downregulated |
| -0,01450917 | 0,9866831 | ZBTB14 | FALSE | Downregulated |
| 0,013266247 | 0,986685576 | LOC101928658 | FALSE | Upregulated |
| 0,019148965 | 0,986685576 | MTMR9LP | FALSE | Upregulated |
| -0,010019171 | 0,986685576 | TECPR1 | FALSE | Downregulated |
| 0,010787957 | 0,98674782 | ABCE1 | FALSE | Upregulated |
| 0,010420262 | 0,98674782 | ANAPC2 | FALSE | Upregulated |
| -0,009449291 | 0,98674782 | AP2S1 | FALSE | Downregulated |
| 0,013144214 | 0,98674782 | ATP6V0D1 | FALSE | Upregulated |
| -0,007249894 | 0,98674782 | CNDP2 | FALSE | Downregulated |
| -0,021500113 | 0,98674782 | COMMD4 | FALSE | Downregulated |
| 0,012931213 | 0,98674782 | DDHD2 | FALSE | Upregulated |
| -0,011384193 | 0,98674782 | DIAPH2 | FALSE | Downregulated |
| 0,022608577 | 0,98674782 | GAN | FALSE | Upregulated |
| -0,011944967 | 0,98674782 | LIN7C | FALSE | Downregulated |
| -0,012142341 | 0,98674782 | LOC100996724 | FALSE | Downregulated |
| -0,017374522 | 0,98674782 | LOC145783 | FALSE | Downregulated |
| -0,016125381 | 0,98674782 | MCM3 | FALSE | Downregulated |
| -0,015547281 | 0,98674782 | MIF4GD | FALSE | Downregulated |
| -0,027409338 | 0,98674782 | MT1G | FALSE | Downregulated |
| 0,018579764 | 0,98674782 | PRSS37 | FALSE | Upregulated |
| -0,009044947 | 0,98674782 | RAB5C | FALSE | Downregulated |
| 0,007736013 | 0,98674782 | RABGEF1 | FALSE | Upregulated |
| -0,007139042 | 0,98674782 | RBCK1 | FALSE | Downregulated |
| -0,008759922 | 0,98674782 | SDR39U1 | FALSE | Downregulated |
| -0,027453192 | 0,98674782 | SERPINE2 | FALSE | Downregulated |
| -0,0096941 | 0,98674782 | SFXN1 | FALSE | Downregulated |
| -0,017958646 | 0,98674782 | SGK223 | FALSE | Downregulated |
| 0,018953988 | 0,98674782 | SKAP2 | FALSE | Upregulated |
| 0,012840136 | 0,98674782 | SLC48A1 | FALSE | Upregulated |
| 0,007787529 | 0,98674782 | TADA3 | FALSE | Upregulated |
| -0,037158044 | 0,98674782 | UPP2 | FALSE | Downregulated |
| 0,020726701 | 0,98674782 | YIF1B | FALSE | Upregulated |
| 0,014552252 | 0,98674782 | ZNF259 | FALSE | Upregulated |

| | | | | |
|---|---|---|---|---|
| -0,016135616 | 0,987142813 | CCDC22 | FALSE | Downregulated |
| -0,010620447 | 0,987142813 | DCAF13 | FALSE | Downregulated |
| -0,00946996 | 0,987142813 | MYBBP1A | FALSE | Downregulated |
| 0,018065426 | 0,987142813 | NS3BP | FALSE | Upregulated |
| -0,010132749 | 0,987142813 | PPID | FALSE | Downregulated |
| -0,007448927 | 0,987302272 | ATG2A | FALSE | Downregulated |
| -0,011976668 | 0,987302272 | DDX39B | FALSE | Downregulated |
| -0,016287243 | 0,987302272 | DGCR9 | FALSE | Downregulated |
| -0,014969499 | 0,987302272 | LINC00324 | FALSE | Downregulated |
| 0,008108611 | 0,987302272 | MAF1 | FALSE | Upregulated |
| 0,011469873 | 0,987387868 | FRY | FALSE | Upregulated |
| -0,012350714 | 0,987387868 | MICAL2 | FALSE | Downregulated |
| -0,011428604 | 0,987387868 | PSMC2 | FALSE | Downregulated |
| 0,017257953 | 0,987392966 | LAMTOR4 | FALSE | Upregulated |
| -0,010098073 | 0,987392966 | MTA2 | FALSE | Downregulated |
| -0,00955204 | 0,987392966 | SUMO3 | FALSE | Downregulated |
| -0,02112006 | 0,987606998 | NAALADL2 | FALSE | Downregulated |
| -0,021862016 | 0,987729895 | SIGLEC9 | FALSE | Downregulated |
| 0,011459989 | 0,987815767 | RGL3 | FALSE | Upregulated |
| -0,010760633 | 0,987930154 | ADRA1B | FALSE | Downregulated |
| 0,009938043 | 0,987930154 | CHUK | FALSE | Upregulated |
| -0,008842639 | 0,987930154 | GPR155 | FALSE | Downregulated |
| 0,017499348 | 0,987930154 | MRPL11 | FALSE | Upregulated |
| -0,009710167 | 0,987930154 | NRBP1 | FALSE | Downregulated |
| 0,006233985 | 0,987930154 | SP3 | FALSE | Upregulated |
| 0,01023374 | 0,987930154 | TMED4 | FALSE | Upregulated |
| -0,049331106 | 0,987930154 | UGT2B17 | FALSE | Downregulated |
| 0,008603572 | 0,988064281 | ZC3H14 | FALSE | Upregulated |
| -0,013336625 | 0,988064281 | ZNF697 | FALSE | Downregulated |
| 0,00595224 | 0,988201091 | AHNAK | FALSE | Upregulated |
| 0,014204391 | 0,988201091 | INPP1 | FALSE | Upregulated |
| 0,025716245 | 0,988201091 | LOC102724938 | FALSE | Upregulated |
| 0,010372382 | 0,988201091 | TFG | FALSE | Upregulated |
| 0,018980503 | 0,98849862 | CUTC | FALSE | Upregulated |
| -0,012178575 | 0,988638733 | CMC2 | FALSE | Downregulated |
| -0,012796351 | 0,988638733 | FHOD1 | FALSE | Downregulated |
| 0,012200388 | 0,988638733 | HPS1 | FALSE | Upregulated |
| 0,013431113 | 0,988638733 | RNF34 | FALSE | Upregulated |
| -0,017321135 | 0,988638733 | SNAPIN | FALSE | Downregulated |
| 0,013756485 | 0,988708847 | CPLX1 | FALSE | Upregulated |
| -0,018275261 | 0,988708847 | TK1 | FALSE | Downregulated |
| -0,01618597 | 0,988899578 | TRIM47 | FALSE | Downregulated |
| -0,017136352 | 0,988899578 | ZFPM2 | FALSE | Downregulated |
| 0,008619326 | 0,988986553 | MSRB1 | FALSE | Upregulated |
| -0,013100478 | 0,989031337 | RTP3 | FALSE | Downregulated |
| 0,011326519 | 0,989155957 | CENPC | FALSE | Upregulated |
| 0,011206178 | 0,989365672 | PPARGC1A | FALSE | Upregulated |
| -0,007480701 | 0,989414076 | FAM195A | FALSE | Downregulated |
| -0,00757312 | 0,989414076 | KIAA1033 | FALSE | Downregulated |
| 0,015772701 | 0,989414076 | LOC101927181 | FALSE | Upregulated |
| 0,019109005 | 0,989414076 | ZC2HC1A | FALSE | Upregulated |

| | | | | |
|---|---|---|---|---|
| 0,013182835 | 0,990084812 | PLXDC2 | FALSE | Upregulated |
| 0,014394491 | 0,990084812 | SP140L | FALSE | Upregulated |
| -0,018867771 | 0,990535202 | DCDC2B | FALSE | Downregulated |
| -0,009182639 | 0,990535202 | EIF2B1 | FALSE | Downregulated |
| -0,049248541 | 0,990535202 | GTF2IP1 | FALSE | Downregulated |
| -0,218645798 | 0,990535202 | MYH1 | FALSE | Downregulated |
| -0,009290629 | 0,990576789 | AGO4 | FALSE | Downregulated |
| -0,008513864 | 0,990576789 | PROZ | FALSE | Downregulated |
| -0,005238833 | 0,990576789 | PSMB7 | FALSE | Downregulated |
| 0,016960033 | 0,990603216 | ANKAR | FALSE | Upregulated |
| 0,005513432 | 0,990603216 | BTBD2 | FALSE | Upregulated |
| 0,009125823 | 0,990603216 | CDC26 | FALSE | Upregulated |
| 0,010372079 | 0,990603216 | COPB1 | FALSE | Upregulated |
| 0,020602392 | 0,990603216 | ERI1 | FALSE | Upregulated |
| 0,007342467 | 0,990603216 | FAM20B | FALSE | Upregulated |
| -0,01466399 | 0,990603216 | LOC101927793 | FALSE | Downregulated |
| 0,022272818 | 0,990603216 | LOC101927897 | FALSE | Upregulated |
| -0,017557595 | 0,990603216 | LOC102723457 | FALSE | Downregulated |
| -0,009204134 | 0,990603216 | MIA2 | FALSE | Downregulated |
| -0,004881975 | 0,990603216 | TRRAP | FALSE | Downregulated |
| -0,010746035 | 0,990603216 | ZNF44 | FALSE | Downregulated |
| 0,050395914 | 0,990646553 | LOC102723360 | FALSE | Upregulated |
| 0,01240722 | 0,990657696 | PWRN2 | FALSE | Upregulated |
| -0,016006896 | 0,99067856 | BTNL9 | FALSE | Downregulated |
| -0,010317882 | 0,990787563 | ASTE1 | FALSE | Downregulated |
| -0,012717623 | 0,990787563 | TTLL4 | FALSE | Downregulated |
| 0,009780756 | 0,990927722 | CDC37L1 | FALSE | Upregulated |
| -0,008143419 | 0,990927722 | GHR | FALSE | Downregulated |
| 0,014189117 | 0,990963912 | AASDHPPT | FALSE | Upregulated |
| -0,013745224 | 0,990963912 | ANKRD16 | FALSE | Downregulated |
| 0,007495797 | 0,990963912 | CDK9 | FALSE | Upregulated |
| 0,004983866 | 0,990963912 | CNNM3 | FALSE | Upregulated |
| -0,01559754 | 0,990963912 | CTU2 | FALSE | Downregulated |
| 0,008279318 | 0,990963912 | FBXL13 | FALSE | Upregulated |
| -0,009804119 | 0,990963912 | FUS | FALSE | Downregulated |
| -0,009921761 | 0,990963912 | HLTF | FALSE | Downregulated |
| -0,018617797 | 0,990963912 | LXN | FALSE | Downregulated |
| 0,00825687 | 0,990963912 | MED24 | FALSE | Upregulated |
| 0,007598606 | 0,990963912 | MEN1 | FALSE | Upregulated |
| 0,009801272 | 0,990963912 | MFN1 | FALSE | Upregulated |
| -0,011586598 | 0,990963912 | NRDE2 | FALSE | Downregulated |
| 0,00547489 | 0,990963912 | PPFIA1 | FALSE | Upregulated |
| 0,011281134 | 0,990963912 | PSMD9 | FALSE | Upregulated |
| -0,012108497 | 0,990963912 | SIRT4 | FALSE | Downregulated |
| 0,010321539 | 0,990963912 | SLU7 | FALSE | Upregulated |
| 0,008536851 | 0,990963912 | STARD9 | FALSE | Upregulated |
| 0,010496312 | 0,990963912 | TANGO2 | FALSE | Upregulated |
| 0,007065284 | 0,990963912 | VRK2 | FALSE | Upregulated |
| -0,007207158 | 0,990963912 | XRN2 | FALSE | Downregulated |
| 0,014220977 | 0,990963912 | ZIC4 | FALSE | Upregulated |
| -0,005934173 | 0,990993381 | USP8 | FALSE | Downregulated |

| | | | | |
|---|---|---|---|---|
| 0,01004813 | 0,99113867 | BIN1 | FALSE | Upregulated |
| 0,017599386 | 0,99113867 | C17orf75 | FALSE | Upregulated |
| -0,013661958 | 0,99113867 | COPS8 | FALSE | Downregulated |
| 0,009166345 | 0,99113867 | DDX21 | FALSE | Upregulated |
| -0,005762255 | 0,99113867 | DGKD | FALSE | Downregulated |
| -0,013479205 | 0,99113867 | DSEL | FALSE | Downregulated |
| -0,010596378 | 0,99113867 | DSERG1 | FALSE | Downregulated |
| -0,012720428 | 0,99113867 | FLJ44342 | FALSE | Downregulated |
| 0,008236478 | 0,99113867 | FNIP1 | FALSE | Upregulated |
| 0,010477171 | 0,99113867 | GJB3 | FALSE | Upregulated |
| -0,014852471 | 0,99113867 | NIPSNAP3A | FALSE | Downregulated |
| -0,016203178 | 0,99113867 | NR1D1 | FALSE | Downregulated |
| 0,017165532 | 0,99113867 | PDE4D | FALSE | Upregulated |
| 0,010715297 | 0,99113867 | RHOB | FALSE | Upregulated |
| 0,00600794 | 0,99113867 | SLC30A1 | FALSE | Upregulated |
| 0,009692736 | 0,99113867 | TERF2 | FALSE | Upregulated |
| 0,006383541 | 0,991145727 | STT3B | FALSE | Upregulated |
| 0,007458661 | 0,99134991 | ABCF3 | FALSE | Upregulated |
| -0,008523524 | 0,99134991 | APPBP2 | FALSE | Downregulated |
| 0,012641043 | 0,99134991 | CHAC2 | FALSE | Upregulated |
| 0,013621382 | 0,99134991 | CNTNAP4 | FALSE | Upregulated |
| 0,015759771 | 0,99134991 | CORT | FALSE | Upregulated |
| 0,011182738 | 0,99134991 | OSTC | FALSE | Upregulated |
| 0,009201442 | 0,99134991 | PARP6 | FALSE | Upregulated |
| -0,009484448 | 0,99134991 | TMUB2 | FALSE | Downregulated |
| 0,013291507 | 0,99134991 | YIPF5 | FALSE | Upregulated |
| 0,006241794 | 0,991638933 | TRAF4 | FALSE | Upregulated |
| -0,012012968 | 0,991695572 | GIMAP4 | FALSE | Downregulated |
| 0,009961726 | 0,991695572 | HMOX1 | FALSE | Upregulated |
| 0,006461928 | 0,991748226 | ARID2 | FALSE | Upregulated |
| 0,00827424 | 0,991748226 | CERS4 | FALSE | Upregulated |
| -0,008790645 | 0,991748226 | DIP2A | FALSE | Downregulated |
| 0,012922648 | 0,991748226 | TTLL9 | FALSE | Upregulated |
| -0,009490636 | 0,991748226 | ZCCHC8 | FALSE | Downregulated |
| -0,015717758 | 0,992049349 | WRB | FALSE | Downregulated |
| 0,011160952 | 0,992100197 | FAM117A | FALSE | Upregulated |
| -0,015822865 | 0,992100197 | VBP1 | FALSE | Downregulated |
| 0,005672249 | 0,992100197 | ZHX2 | FALSE | Upregulated |
| 0,00631832 | 0,992165995 | CASC3 | FALSE | Upregulated |
| -0,010036525 | 0,992165995 | RNPC3 | FALSE | Downregulated |
| -0,009084657 | 0,992188674 | AMER1 | FALSE | Downregulated |
| -0,00908941 | 0,992188674 | BCL7C | FALSE | Downregulated |
| 0,010868857 | 0,992188674 | C1QTNF7 | FALSE | Upregulated |
| 0,004939172 | 0,992188674 | CDK5RAP2 | FALSE | Upregulated |
| -0,014466993 | 0,992188674 | GJB5 | FALSE | Downregulated |
| 0,010191291 | 0,992188674 | GMIP | FALSE | Upregulated |
| 0,008718079 | 0,992188674 | GNA13 | FALSE | Upregulated |
| 0,006388284 | 0,992188674 | LOC101929291 | FALSE | Upregulated |
| 0,004074612 | 0,992188674 | MAN2B2 | FALSE | Upregulated |
| 0,007451141 | 0,992188674 | MXD1 | FALSE | Upregulated |
| 0,008840779 | 0,992188674 | RYK | FALSE | Upregulated |

| | | | | |
|---|---|---|---|---|
| 0,004402723 | 0,992188674 | STAT6 | FALSE | Upregulated |
| -0,010362519 | 0,992188674 | TDP2 | FALSE | Downregulated |
| 0,013087741 | 0,992188674 | WASH7P | FALSE | Upregulated |
| 0,008499345 | 0,992261385 | TRIM23 | FALSE | Upregulated |
| 0,012899683 | 0,992274694 | DCTN3 | FALSE | Upregulated |
| 0,005649428 | 0,992274694 | INPPL1 | FALSE | Upregulated |
| 0,005600287 | 0,992274694 | PTTG1IP | FALSE | Upregulated |
| -0,010459461 | 0,992274694 | VIPAS39 | FALSE | Downregulated |
| 0,008998592 | 0,992274694 | ZNF75D | FALSE | Upregulated |
| -0,008753333 | 0,992289952 | ANKS3 | FALSE | Downregulated |
| -0,008818057 | 0,992289952 | MBD4 | FALSE | Downregulated |
| -0,011221856 | 0,992289952 | SDF2L1 | FALSE | Downregulated |
| -0,013818935 | 0,992289952 | SFMBT2 | FALSE | Downregulated |
| 0,009989314 | 0,992289952 | ZNF511 | FALSE | Upregulated |
| 0,034578133 | 0,992739552 | GIMAP1-GIMAP5 | FALSE | Upregulated |
| 0,016974267 | 0,992739552 | H2AFX | FALSE | Upregulated |
| -0,014637327 | 0,992739552 | ZNF521 | FALSE | Downregulated |
| -0,008069291 | 0,992794895 | FOXN3-AS1 | FALSE | Downregulated |
| -0,008148941 | 0,992851707 | STRIP1 | FALSE | Downregulated |
| 0,014424823 | 0,992862478 | COX10-AS1 | FALSE | Upregulated |
| -0,006213785 | 0,992862478 | EPRS | FALSE | Downregulated |
| -0,00748135 | 0,992862478 | TMEM138 | FALSE | Downregulated |
| 0,008629921 | 0,993208772 | BLOC1S6 | FALSE | Upregulated |
| 0,017583457 | 0,993208772 | DLEU7 | FALSE | Upregulated |
| 0,004676132 | 0,993208772 | PTPRB | FALSE | Upregulated |
| -0,009590709 | 0,993208772 | TMEM134 | FALSE | Downregulated |
| 0,008319857 | 0,993582878 | ADSSL1 | FALSE | Upregulated |
| 0,00691892 | 0,993582878 | ALG13 | FALSE | Upregulated |
| 0,012237127 | 0,993582878 | APBB3 | FALSE | Upregulated |
| 0,007822586 | 0,993582878 | ARFGAP3 | FALSE | Upregulated |
| -0,012669462 | 0,993582878 | ARHGAP6 | FALSE | Downregulated |
| 0,008572529 | 0,993582878 | ARRB1 | FALSE | Upregulated |
| -0,0098371 | 0,993582878 | BBS5 | FALSE | Downregulated |
| 0,009733159 | 0,993582878 | BRICD5 | FALSE | Upregulated |
| -0,005832626 | 0,993582878 | CAD | FALSE | Downregulated |
| 0,00808537 | 0,993582878 | CCDC25 | FALSE | Upregulated |
| 0,010075238 | 0,993582878 | CCL23 | FALSE | Upregulated |
| 0,006045692 | 0,993582878 | CCM2 | FALSE | Upregulated |
| -0,007353717 | 0,993582878 | CHKB | FALSE | Downregulated |
| 0,005169203 | 0,993582878 | COQ6 | FALSE | Upregulated |
| -0,007349983 | 0,993582878 | CYFIP2 | FALSE | Downregulated |
| -0,005636851 | 0,993582878 | EHHADH-AS1 | FALSE | Downregulated |
| -0,005183165 | 0,993582878 | EIF2S2 | FALSE | Downregulated |
| 0,004624137 | 0,993582878 | GOLGA3 | FALSE | Upregulated |
| -0,006060472 | 0,993582878 | GPR89A | FALSE | Downregulated |
| -0,004738909 | 0,993582878 | HEXDC | FALSE | Downregulated |
| -0,012693315 | 0,993582878 | IRAK2 | FALSE | Downregulated |
| 0,007050684 | 0,993582878 | LOC100271836 | FALSE | Upregulated |
| -0,009824144 | 0,993582878 | LOC101928089 | FALSE | Downregulated |
| -0,009013767 | 0,993582878 | LSS | FALSE | Downregulated |
| 0,004131019 | 0,993582878 | LZTR1 | FALSE | Upregulated |

| | | | | |
|---|---|---|---|---|
| -0,004246365 | 0,993582878 | MAZ | FALSE | Downregulated |
| -0,006859694 | 0,993582878 | MMS19 | FALSE | Downregulated |
| -0,010682596 | 0,993582878 | MRPL23 | FALSE | Downregulated |
| 0,006176864 | 0,993582878 | MRTO4 | FALSE | Upregulated |
| -0,009457024 | 0,993582878 | MYO5C | FALSE | Downregulated |
| -0,015704407 | 0,993582878 | NFATC4 | FALSE | Downregulated |
| 0,006688867 | 0,993582878 | NOP14-AS1 | FALSE | Upregulated |
| 0,009698665 | 0,993582878 | PDE1B | FALSE | Upregulated |
| 0,004651159 | 0,993582878 | PLEKHM2 | FALSE | Upregulated |
| -0,010838188 | 0,993582878 | POU2F2 | FALSE | Downregulated |
| 0,010659076 | 0,993582878 | QTRTD1 | FALSE | Upregulated |
| 0,005204046 | 0,993582878 | RANBP10 | FALSE | Upregulated |
| -0,010639025 | 0,993582878 | RP11-279D17.2 | FALSE | Downregulated |
| -0,008292775 | 0,993582878 | SDE2 | FALSE | Downregulated |
| 0,007736186 | 0,993582878 | SEC23B | FALSE | Upregulated |
| -0,005228547 | 0,993582878 | SH3GLB1 | FALSE | Downregulated |
| -0,010131832 | 0,993582878 | TMEM177 | FALSE | Downregulated |
| 0,005129433 | 0,993582878 | UPF2 | FALSE | Upregulated |
| 0,014505864 | 0,993582878 | ZBTB24 | FALSE | Upregulated |
| 0,010301144 | 0,993582878 | ZSCAN12 | FALSE | Upregulated |
| 0,004928249 | 0,993691384 | ABHD14B | FALSE | Upregulated |
| -0,005956966 | 0,993691384 | AGPS | FALSE | Downregulated |
| -0,008216362 | 0,993691384 | CBLL1 | FALSE | Downregulated |
| 0,003204182 | 0,993691384 | DYNC1H1 | FALSE | Upregulated |
| 0,007561712 | 0,993691384 | HAUS4 | FALSE | Upregulated |
| 0,007728911 | 0,993691384 | HDAC8 | FALSE | Upregulated |
| 0,054084896 | 0,993691384 | LOC102724843 | FALSE | Upregulated |
| -0,012247654 | 0,993691384 | MRAS | FALSE | Downregulated |
| -0,009320774 | 0,993691384 | TMA7 | FALSE | Downregulated |
| -0,008849988 | 0,993691384 | ZNF621 | FALSE | Downregulated |
| 0,010192862 | 0,993790756 | EIF1AD | FALSE | Upregulated |
| -0,006380455 | 0,99386719 | DCAF12 | FALSE | Downregulated |
| -0,006949272 | 0,99386719 | DEAF1 | FALSE | Downregulated |
| 0,004925096 | 0,99386719 | DNMBP-AS1 | FALSE | Upregulated |
| -0,00799948 | 0,99386719 | FBXL6 | FALSE | Downregulated |
| -0,00350354 | 0,993955892 | RAPGEF1 | FALSE | Downregulated |
| -0,004004951 | 0,994034658 | GNA11 | FALSE | Downregulated |
| -0,006297995 | 0,99410238 | SAP18 | FALSE | Downregulated |
| -0,009211814 | 0,994271948 | ACTR10 | FALSE | Downregulated |
| -0,007672544 | 0,994271948 | NT5C | FALSE | Downregulated |
| 0,003450539 | 0,994271948 | PPP1R12C | FALSE | Upregulated |
| 0,008160599 | 0,994271948 | THAP9-AS1 | FALSE | Upregulated |
| -0,004729445 | 0,994271948 | UQCRQ | FALSE | Downregulated |
| -0,010516988 | 0,99433534 | SP5 | FALSE | Downregulated |
| 0,00929076 | 0,99454648 | ATP6V1F | FALSE | Upregulated |
| 0,007757079 | 0,994713351 | SMYD2 | FALSE | Upregulated |
| -0,006648179 | 0,994819339 | ADARB1 | FALSE | Downregulated |
| -0,01025004 | 0,994819339 | HERC2P9 | FALSE | Downregulated |
| 0,01110431 | 0,994819339 | LOC101928356 | FALSE | Upregulated |
| 0,004076848 | 0,994819339 | SLC25A15 | FALSE | Upregulated |
| -0,008381567 | 0,994819339 | TEX10 | FALSE | Downregulated |

| | | | | |
|---|---|---|---|---|
| -0,008528381 | 0,994819339 | UXT | FALSE | Downregulated |
| -0,005655676 | 0,994819339 | WIPF2 | FALSE | Downregulated |
| 0,003426755 | 0,994819339 | ZNF687 | FALSE | Upregulated |
| 0,004764726 | 0,994872153 | ADPGK | FALSE | Upregulated |
| -0,005585817 | 0,994872153 | C15orf38 | FALSE | Downregulated |
| 0,008720906 | 0,994872153 | CDHR3 | FALSE | Upregulated |
| 0,00422537 | 0,994872153 | INADL | FALSE | Upregulated |
| -0,007039726 | 0,994872153 | KIAA1683 | FALSE | Downregulated |
| 0,010353146 | 0,994872153 | PPM1L | FALSE | Upregulated |
| -0,004251742 | 0,994872153 | PRKAA1 | FALSE | Downregulated |
| 0,010352107 | 0,994872153 | RAB9A | FALSE | Upregulated |
| 0,007066572 | 0,994872153 | RASGRF2 | FALSE | Upregulated |
| 0,01539364 | 0,994872153 | SLC29A4 | FALSE | Upregulated |
| -0,005105921 | 0,994872153 | SMARCB1 | FALSE | Downregulated |
| -0,00247148 | 0,994872153 | SPTBN1 | FALSE | Downregulated |
| -0,009007675 | 0,994872153 | TRIM32 | FALSE | Downregulated |
| 0,004997682 | 0,994872153 | TRO | FALSE | Upregulated |
| -0,003819299 | 0,995000213 | MTERFD2 | FALSE | Downregulated |
| -0,00280453 | 0,995040628 | CDC14B | FALSE | Downregulated |
| 0,006546719 | 0,995040628 | CREBRF | FALSE | Upregulated |
| -0,007953721 | 0,995040628 | GLUD1P7 | FALSE | Downregulated |
| 0,007246632 | 0,995040628 | LINC00984 | FALSE | Upregulated |
| -0,005343458 | 0,995040628 | PSMD11 | FALSE | Downregulated |
| 0,004771163 | 0,995040628 | SUMO2 | FALSE | Upregulated |
| 0,002576533 | 0,995040628 | SUPT6H | FALSE | Upregulated |
| -0,003550639 | 0,995040628 | YWHAQ | FALSE | Downregulated |
| 0,006112519 | 0,995164922 | PALM3 | FALSE | Upregulated |
| 0,006603162 | 0,995338157 | TRAFD1 | FALSE | Upregulated |
| 0,007105312 | 0,995456789 | GPN2 | FALSE | Upregulated |
| 0,003964129 | 0,995456789 | TBL1X | FALSE | Upregulated |
| 0,004050723 | 0,995488322 | LOC101929211 | FALSE | Upregulated |
| 0,003706411 | 0,99559849 | DNAJB1 | FALSE | Upregulated |
| 0,008101399 | 0,995633527 | FIBP | FALSE | Upregulated |
| 0,005342066 | 0,995646065 | C11orf30 | FALSE | Upregulated |
| -0,006499404 | 0,995646065 | C2CD2L | FALSE | Downregulated |
| -0,006515 | 0,995646065 | GADD45B | FALSE | Downregulated |
| -0,005044028 | 0,995646065 | RAB33B | FALSE | Downregulated |
| 0,003767137 | 0,995955031 | SCAND1 | FALSE | Upregulated |
| 0,0096092 | 0,996062844 | AGAP2-AS1 | FALSE | Upregulated |
| 0,00650765 | 0,996062844 | HSBP1L1 | FALSE | Upregulated |
| 0,007839476 | 0,996062844 | LOC100126582 | FALSE | Upregulated |
| 0,002981585 | 0,996078986 | ATG2B | FALSE | Upregulated |
| 0,004385916 | 0,996078986 | C2CD5 | FALSE | Upregulated |
| 0,002632368 | 0,996078986 | EIF2S3 | FALSE | Upregulated |
| 0,003962448 | 0,996078986 | MVB12B | FALSE | Upregulated |
| 0,002990984 | 0,996078986 | OGDH | FALSE | Upregulated |
| -0,002508913 | 0,996078986 | PDZK1 | FALSE | Downregulated |
| -0,005421964 | 0,996078986 | PPWD1 | FALSE | Downregulated |
| 0,00570983 | 0,996078986 | RRAGA | FALSE | Upregulated |
| -0,008496125 | 0,996078986 | SLC10A7 | FALSE | Downregulated |
| 0,003573772 | 0,996078986 | SLC25A23 | FALSE | Upregulated |

| | | | | |
|---|---|---|---|---|
| 0,008338168 | 0,996078986 | ZNF131 | FALSE | Upregulated |
| 0,005079914 | 0,996128924 | FEM1C | FALSE | Upregulated |
| 0,003188358 | 0,996128924 | MMRN2 | FALSE | Upregulated |
| 0,003612636 | 0,996128924 | STYX | FALSE | Upregulated |
| -0,007563484 | 0,996185055 | ETV4 | FALSE | Downregulated |
| 0,008415299 | 0,996185055 | MANF | FALSE | Upregulated |
| -0,005131589 | 0,996185055 | NDUFS5 | FALSE | Downregulated |
| 0,003828472 | 0,996185055 | PHF1 | FALSE | Upregulated |
| -0,005157727 | 0,996185055 | PRADC1 | FALSE | Downregulated |
| 0,007718618 | 0,996185055 | RECK | FALSE | Upregulated |
| 0,003912667 | 0,996185055 | SETD3 | FALSE | Upregulated |
| 0,00318777 | 0,996668648 | SIAE | FALSE | Upregulated |
| -0,007123297 | 0,996841723 | AC058791.1 | FALSE | Downregulated |
| -0,003076559 | 0,996841723 | AKR1C2 | FALSE | Downregulated |
| 0,004354314 | 0,996841723 | ANKRD50 | FALSE | Upregulated |
| -0,003612196 | 0,996841723 | ATXN3 | FALSE | Downregulated |
| -0,004747964 | 0,996841723 | B3GAT3 | FALSE | Downregulated |
| -0,005551394 | 0,996841723 | BASP1 | FALSE | Downregulated |
| -0,003159226 | 0,996841723 | C12orf43 | FALSE | Downregulated |
| 0,004180964 | 0,996841723 | C14orf105 | FALSE | Upregulated |
| -0,005183331 | 0,996841723 | C2orf49 | FALSE | Downregulated |
| 0,00351062 | 0,996841723 | CDK17 | FALSE | Upregulated |
| 0,0024942 | 0,996841723 | CHD9 | FALSE | Upregulated |
| -0,00558298 | 0,996841723 | CNTN3 | FALSE | Downregulated |
| 0,004013221 | 0,996841723 | CPS1-IT1 | FALSE | Upregulated |
| -0,004690401 | 0,996841723 | CSNK1G1 | FALSE | Downregulated |
| 0,002908503 | 0,996841723 | CUL5 | FALSE | Upregulated |
| -0,00655232 | 0,996841723 | DCP1B | FALSE | Downregulated |
| -0,004529075 | 0,996841723 | DNM3OS | FALSE | Downregulated |
| -0,002559752 | 0,996841723 | EPS8 | FALSE | Downregulated |
| 0,003278675 | 0,996841723 | FAM69B | FALSE | Upregulated |
| 0,005192385 | 0,996841723 | FBXO27 | FALSE | Upregulated |
| -0,007053323 | 0,996841723 | GAS5 | FALSE | Downregulated |
| 0,002272465 | 0,996841723 | GATAD2A | FALSE | Upregulated |
| -0,007831157 | 0,996841723 | GINS1 | FALSE | Downregulated |
| -0,002794777 | 0,996841723 | HELZ2 | FALSE | Downregulated |
| 0,002252096 | 0,996841723 | HIST1H1E | FALSE | Upregulated |
| 0,002856397 | 0,996841723 | HNRNPL | FALSE | Upregulated |
| 0,002689786 | 0,996841723 | INHBC | FALSE | Upregulated |
| 0,006914251 | 0,996841723 | JAKMIP2 | FALSE | Upregulated |
| -0,006202944 | 0,996841723 | LOC101927870 | FALSE | Downregulated |
| -0,005518353 | 0,996841723 | LOC101928051 | FALSE | Downregulated |
| 0,002850875 | 0,996841723 | LOC101929426 | FALSE | Upregulated |
| -0,006721506 | 0,996841723 | LOC101929548 | FALSE | Downregulated |
| -0,006676796 | 0,996841723 | LOC102723356 | FALSE | Downregulated |
| -0,004060009 | 0,996841723 | LRP5L | FALSE | Downregulated |
| -0,00425699 | 0,996841723 | MBL2 | FALSE | Downregulated |
| -0,003302354 | 0,996841723 | MNT | FALSE | Downregulated |
| 0,003903299 | 0,996841723 | MORC4 | FALSE | Upregulated |
| 0,003132532 | 0,996841723 | NBAS | FALSE | Upregulated |
| -0,002684867 | 0,996841723 | NBEAL1 | FALSE | Downregulated |

| | | | | |
|---|---|---|---|---|
| -0,00443556 | 0,996841723 | NOL12 | FALSE | Downregulated |
| 0,002232335 | 0,996841723 | NPIPB4 | FALSE | Upregulated |
| -0,006012228 | 0,996841723 | NPL | FALSE | Downregulated |
| -0,002875308 | 0,996841723 | OPTN | FALSE | Downregulated |
| 0,005041311 | 0,996841723 | PEAR1 | FALSE | Upregulated |
| -0,004016928 | 0,996841723 | PET112 | FALSE | Downregulated |
| 0,00434747 | 0,996841723 | PRKCI | FALSE | Upregulated |
| 0,002106795 | 0,996841723 | PRKDC | FALSE | Upregulated |
| 0,003267024 | 0,996841723 | PRPF4B | FALSE | Upregulated |
| -0,001884734 | 0,996841723 | PTOV1 | FALSE | Downregulated |
| -0,003023057 | 0,996841723 | PURA | FALSE | Downregulated |
| -0,006056162 | 0,996841723 | RAP2A | FALSE | Downregulated |
| 0,00423382 | 0,996841723 | RBMXL1 | FALSE | Upregulated |
| -0,037102185 | 0,996841723 | RNA18S5 | FALSE | Downregulated |
| 0,001611508 | 0,996841723 | RNF10 | FALSE | Upregulated |
| -0,008604894 | 0,996841723 | RP11-267N12.1 | FALSE | Downregulated |
| -0,003336198 | 0,996841723 | SCYL2 | FALSE | Downregulated |
| 0,004530447 | 0,996841723 | SETD8 | FALSE | Upregulated |
| -0,004770682 | 0,996841723 | SPCS1 | FALSE | Downregulated |
| 0,002970434 | 0,996841723 | SS18 | FALSE | Upregulated |
| -0,002878541 | 0,996841723 | SYAP1 | FALSE | Downregulated |
| 0,004514683 | 0,996841723 | TBC1D20 | FALSE | Upregulated |
| 0,009362232 | 0,996841723 | TXLNG2P | FALSE | Upregulated |
| -0,002198321 | 0,996841723 | U2AF2 | FALSE | Downregulated |
| -0,002412522 | 0,996841723 | WDFY3 | FALSE | Downregulated |
| 0,005509245 | 0,996841723 | ZNF595 | FALSE | Upregulated |
| -0,004552192 | 0,996841723 | ZNF771 | FALSE | Downregulated |
| 0,006030564 | 0,996841723 | ZNRF2P2 | FALSE | Upregulated |
| 0,003119884 | 0,996893606 | MRPS24 | FALSE | Upregulated |
| 0,006131897 | 0,997065158 | LOC101927000 | FALSE | Upregulated |
| 0,005596317 | 0,997065158 | RCCD1 | FALSE | Upregulated |
| 0,005102453 | 0,997311581 | DRAM2 | FALSE | Upregulated |
| 0,003547524 | 0,997311581 | NEK1 | FALSE | Upregulated |
| 0,006059163 | 0,997326799 | CYP21A1P | FALSE | Upregulated |
| 0,002586225 | 0,997326799 | FAM73B | FALSE | Upregulated |
| -0,005443242 | 0,997326799 | FLG2 | FALSE | Downregulated |
| -0,002869199 | 0,997326799 | GRSF1 | FALSE | Downregulated |
| 0,001390833 | 0,997326799 | HMBOX1 | FALSE | Upregulated |
| -0,00222777 | 0,997326799 | KDM1B | FALSE | Downregulated |
| 0,005688316 | 0,997326799 | LOC102723313 | FALSE | Upregulated |
| 0,012235233 | 0,997326799 | PRKRIP1 | FALSE | Upregulated |
| 0,003868328 | 0,997326799 | SRSF7 | FALSE | Upregulated |
| -0,003243402 | 0,997436926 | EXD3 | FALSE | Downregulated |
| 0,003807207 | 0,997455825 | ZNF319 | FALSE | Upregulated |
| 0,002498515 | 0,997503298 | GCSH | FALSE | Upregulated |
| -0,001672554 | 0,997503298 | RARA | FALSE | Downregulated |
| -0,015167332 | 0,997503298 | TNXA | FALSE | Downregulated |
| 0,003585072 | 0,997503298 | ZNF354C | FALSE | Upregulated |
| 0,002996806 | 0,997503298 | ZNF689 | FALSE | Upregulated |
| 0,004023473 | 0,997639873 | RP11-840I19.3 | FALSE | Upregulated |
| -0,004617705 | 0,997787499 | ATXN7L2 | FALSE | Downregulated |

| | | | | |
|---|---|---|---|---|
| -0,002799889 | 0,997787499 | BMS1P5 | FALSE | Downregulated |
| -0,001960995 | 0,997787499 | CYP3A5 | FALSE | Downregulated |
| 0,002313942 | 0,997787499 | EHD1 | FALSE | Upregulated |
| 0,003637293 | 0,997787499 | GRIN3B | FALSE | Upregulated |
| 0,004126244 | 0,997787499 | KATNAL1 | FALSE | Upregulated |
| 0,001022702 | 0,997787499 | LSR | FALSE | Upregulated |
| -0,001563727 | 0,997787499 | MAML3 | FALSE | Downregulated |
| -0,002618613 | 0,997787499 | PLEKHA1 | FALSE | Downregulated |
| 0,002077565 | 0,997787499 | PPP2CB | FALSE | Upregulated |
| 0,00208283 | 0,997787499 | SALL1 | FALSE | Upregulated |
| -0,001642322 | 0,997787499 | SASH1 | FALSE | Downregulated |
| 0,002031353 | 0,997787499 | SLC23A1 | FALSE | Upregulated |
| 0,002455416 | 0,997864504 | ZBTB43 | FALSE | Upregulated |
| 0,004430506 | 0,9978882 | CD248 | FALSE | Upregulated |
| -0,004338121 | 0,9978882 | IMPDH1 | FALSE | Downregulated |
| -0,001697138 | 0,99794563 | FMR1 | FALSE | Downregulated |
| -0,047900772 | 0,99794563 | KRT14 | FALSE | Downregulated |
| 0,001253777 | 0,99794563 | PPL | FALSE | Upregulated |
| 0,004198211 | 0,99794563 | SEMA3E | FALSE | Upregulated |
| -0,001545703 | 0,998048147 | ZNF584 | FALSE | Downregulated |
| 0,002217973 | 0,998060429 | DHRS4L2 | FALSE | Upregulated |
| 0,003782462 | 0,998091273 | CHRNA4 | FALSE | Upregulated |
| 0,003282364 | 0,998124785 | LOC102724243 | FALSE | Upregulated |
| 0,003139648 | 0,998352845 | ALG3 | FALSE | Upregulated |
| 0,001674403 | 0,998352845 | LIPG | FALSE | Upregulated |
| -0,003511927 | 0,998352845 | LOC101927610 | FALSE | Downregulated |
| -0,001957332 | 0,998352845 | RHEB | FALSE | Downregulated |
| 0,043593398 | 0,998352845 | SNORD3C | FALSE | Upregulated |
| 0,002020531 | 0,998352845 | UMPS | FALSE | Upregulated |
| 0,002316664 | 0,998352845 | YWHAH | FALSE | Upregulated |
| 0,003427455 | 0,998392829 | ASUN | FALSE | Upregulated |
| -0,004869284 | 0,998392829 | MUC3A | FALSE | Downregulated |
| -0,003056138 | 0,998420018 | LMOD2 | FALSE | Downregulated |
| 0,003177081 | 0,998436005 | ANKRD36B | FALSE | Upregulated |
| -0,001407258 | 0,998482273 | FLII | FALSE | Downregulated |
| 0,002078507 | 0,998482273 | NEDD8 | FALSE | Upregulated |
| 0,039236507 | 0,998482273 | XIRP2 | FALSE | Upregulated |
| 0,002806415 | 0,998504226 | C16orf86 | FALSE | Upregulated |
| 0,001203788 | 0,998504226 | EPS15 | FALSE | Upregulated |
| 0,00122511 | 0,998504226 | GABPB1-AS1 | FALSE | Upregulated |
| -0,001627169 | 0,998568743 | AAAS | FALSE | Downregulated |
| -0,001644408 | 0,998568743 | ACAT2 | FALSE | Downregulated |
| 0,001164329 | 0,998568743 | ADD3 | FALSE | Upregulated |
| -0,001386154 | 0,998568743 | C2CD3 | FALSE | Downregulated |
| 0,001378381 | 0,998568743 | CTPS2 | FALSE | Upregulated |
| -0,001428484 | 0,998568743 | DHCR7 | FALSE | Downregulated |
| 0,000668696 | 0,998568743 | DNM2 | FALSE | Upregulated |
| -0,002236067 | 0,998568743 | FXYD5 | FALSE | Downregulated |
| -0,001071976 | 0,998568743 | GID8 | FALSE | Downregulated |
| -0,001565202 | 0,998568743 | IDS | FALSE | Downregulated |
| -0,001917828 | 0,998568743 | LMNB2 | FALSE | Downregulated |

| | | | | |
|---|---|---|---|---|
| -0,002170737 | 0,998568743 | LOC100996571 | FALSE | Downregulated |
| 0,00175649 | 0,998568743 | LOC643072 | FALSE | Upregulated |
| 0,001585338 | 0,998568743 | LTBP4 | FALSE | Upregulated |
| 0,002982165 | 0,998568743 | MAB21L1 | FALSE | Upregulated |
| 0,002494871 | 0,998568743 | NIP7 | FALSE | Upregulated |
| 0,001018438 | 0,998568743 | PAPD7 | FALSE | Upregulated |
| 0,001508321 | 0,998568743 | PLCD4 | FALSE | Upregulated |
| 0,002077022 | 0,998568743 | RPS27L | FALSE | Upregulated |
| 0,001368062 | 0,998568743 | THOP1 | FALSE | Upregulated |
| -0,001585914 | 0,998568743 | UNC45A | FALSE | Downregulated |
| -0,001954159 | 0,998568743 | WEE1 | FALSE | Downregulated |
| -0,028581726 | 0,998624648 | MYBPC1 | FALSE | Downregulated |
| -0,000953767 | 0,998661793 | RELL1 | FALSE | Downregulated |
| -0,001524449 | 0,998661793 | SDHAP1 | FALSE | Downregulated |
| 0,001670614 | 0,998927057 | RBM41 | FALSE | Upregulated |
| -0,00118948 | 0,998958698 | GOLGA8M | FALSE | Downregulated |
| 0,001289402 | 0,999010199 | MAMDC2 | FALSE | Upregulated |
| -0,000614168 | 0,999062105 | AGRN | FALSE | Downregulated |
| 0,00176284 | 0,999062105 | AOC1 | FALSE | Upregulated |
| 0,001316048 | 0,999062105 | CTB-113P19.1 | FALSE | Upregulated |
| -0,001150921 | 0,999062105 | GNAI1 | FALSE | Downregulated |
| 0,001070033 | 0,999062105 | MARVELD3 | FALSE | Upregulated |
| 0,000905924 | 0,999062105 | MKRN1 | FALSE | Upregulated |
| -0,000884145 | 0,999062105 | RPS6KB2 | FALSE | Downregulated |
| -0,001889606 | 0,999062105 | SCOC | FALSE | Downregulated |
| 0,000892748 | 0,999062105 | SLC35B4 | FALSE | Upregulated |
| -0,001012827 | 0,999069416 | CA5A | FALSE | Downregulated |
| 0,000785426 | 0,999069416 | CHST15 | FALSE | Upregulated |
| -0,000524263 | 0,999069416 | DENND4C | FALSE | Downregulated |
| -0,000895105 | 0,999069416 | E2F4 | FALSE | Downregulated |
| 0,001537388 | 0,999069416 | NDUFA8 | FALSE | Upregulated |
| 0,000873333 | 0,999069416 | PRLR | FALSE | Upregulated |
| -0,000528758 | 0,999069416 | SLC12A4 | FALSE | Downregulated |
| -0,000395421 | 0,999069416 | SRCAP | FALSE | Downregulated |
| 0,000540357 | 0,999069416 | UHMK1 | FALSE | Upregulated |
| -0,000828893 | 0,999138718 | GIPC3 | FALSE | Downregulated |
| 0,00142029 | 0,999159494 | HIST2H2BF | FALSE | Upregulated |
| -0,000521539 | 0,999160264 | ITPRIPL2 | FALSE | Downregulated |
| 0,000712288 | 0,999281696 | SDPR | FALSE | Upregulated |
| -0,000808035 | 0,999363366 | ACAP2 | FALSE | Downregulated |
| 0,001325949 | 0,999363366 | GOLGA6C | FALSE | Upregulated |
| 0,000373677 | 0,999367947 | UBE2R2 | FALSE | Upregulated |
| 0,000339336 | 0,999542353 | PTCD3 | FALSE | Upregulated |
| 0,000973434 | 0,999674166 | GJB4 | FALSE | Upregulated |
| 0,000861857 | 0,999674166 | IQCJ-SCHIP1 | FALSE | Upregulated |
| 0,001280766 | 0,999674166 | MT1F | FALSE | Upregulated |
| 0,000581353 | 0,999804598 | IGLON5 | FALSE | Upregulated |
| -0,000468046 | 0,999804598 | LOC100506730 | FALSE | Downregulated |
| -0,00033325 | 0,999804598 | NEK4 | FALSE | Downregulated |
| -0,000400608 | 0,999804598 | NFYA | FALSE | Downregulated |
| 0,000409029 | 0,999819157 | MKNK1 | FALSE | Upregulated |

| | | | | |
|---|---|---|---|---|
| 0,000409539 | 0,999819157 | NSUN4 | FALSE | Upregulated |
| 0,000218276 | 0,999819157 | OTUD5 | FALSE | Upregulated |
| -0,000213243 | 0,999819157 | PTMA | FALSE | Downregulated |
| 0,000401933 | 0,999819157 | SREBF1 | FALSE | Upregulated |
| -0,000166361 | 0,999881494 | CCT5 | FALSE | Downregulated |
| 6,41323E-05 | 0,999881494 | CEP350 | FALSE | Upregulated |
| 0,000111982 | 0,999881494 | CRTC2 | FALSE | Upregulated |
| -9,10002E-05 | 0,999881494 | CUL7 | FALSE | Downregulated |
| 0,00025688 | 0,999881494 | EIF2B2 | FALSE | Upregulated |
| 0,000137776 | 0,999881494 | LOC100130093 | FALSE | Upregulated |
| 7,3677E-05 | 0,999881494 | LOC100507520 | FALSE | Upregulated |
| -0,000270402 | 0,999881494 | MAPKAPK3 | FALSE | Downregulated |
| 0,000335373 | 0,999881494 | MCAM | FALSE | Upregulated |
| 0,000437118 | 0,999881494 | MRVI1 | FALSE | Upregulated |
| 0,000174315 | 0,999881494 | NFS1 | FALSE | Upregulated |
| 0,000290294 | 0,999881494 | NPIPA2 | FALSE | Upregulated |
| -9,89042E-05 | 0,999881494 | PATL1 | FALSE | Downregulated |
| 0 | 1 | H19 | FALSE | Upregulated |
| 0 | 1 | KRT1 | FALSE | Upregulated |
| -0,000972306 | 1 | MYH7 | FALSE | Downregulated |
| -2,77011E-05 | 1 | RBM3 | FALSE | Downregulated |
| 0,016577626 | | A2ML1 | FALSE | Upregulated |
| 0,114485338 | | A3GALT2 | FALSE | Upregulated |
| -0,508491652 | | A4GALT | FALSE | Downregulated |
| | | AA06 | | |
| | | AACSP1 | | |
| -0,205847813 | | AADACL2 | FALSE | Downregulated |
| | | AADACL3 | | |
| 0 | | AADACL4 | FALSE | Upregulated |
| -0,229893917 | | AAED1 | FALSE | Downregulated |
| -0,150879999 | | AAMDC | FALSE | Downregulated |
| 0,660329049 | | AANAT | FALSE | Upregulated |
| | | AARD | | |
| -0,483862797 | | AATK-AS1 | FALSE | Downregulated |
| 0,354706153 | | ABCA11P | FALSE | Upregulated |
| 0,976827547 | | ABCA12 | FALSE | Upregulated |
| -0,359073539 | | ABCA13 | FALSE | Downregulated |
| 0,408106295 | | ABCA17P | FALSE | Upregulated |
| -0,171138464 | | ABCB5 | FALSE | Downregulated |
| 0,39312809 | | ABCB9 | FALSE | Upregulated |
| -0,302029586 | | ABCC11 | FALSE | Downregulated |
| -0,005249532 | | ABCC12 | FALSE | Downregulated |
| 0 | | ABCC13 | FALSE | Upregulated |
| 0,500122021 | | ABCC4 | FALSE | Upregulated |
| -0,464425816 | | ABCC5-AS1 | FALSE | Downregulated |
| -0,696334966 | | ABCC6P2 | FALSE | Downregulated |
| 0,274635952 | | ABCC8 | FALSE | Upregulated |
| -0,320693274 | | ABCD2 | FALSE | Downregulated |
| -0,453418329 | | ABCG4 | FALSE | Downregulated |
| 0,077540951 | | ABHD11 | FALSE | Upregulated |
| | | ABHD14A-ACY1 | | |

| | | | |
|---|---|---|---|
| 0,45806829 | ABHD17C | FALSE | Upregulated |
| -0,033315576 | ABLIM2 | FALSE | Downregulated |
| 0,111887206 | ABO | FALSE | Upregulated |
| 0,487980006 | ABRACL | FALSE | Upregulated |
| 0,249987172 | AC002116.7 | FALSE | Upregulated |
| 0,128876749 | AC002398.5 | FALSE | Upregulated |
| 0 | AC002454.1 | FALSE | Upregulated |
| | AC002463.3 | | |
| 1,642056422 | AC002480.3 | | Upregulated |
| 0,146484581 | AC002511.1 | FALSE | Upregulated |
| | AC003051.1 | | |
| 0 | AC003102.3 | FALSE | Upregulated |
| | AC003958.2 | | |
| | AC003984.1 | | |
| | AC004603.4 | | |
| -0,441279649 | AC004862.6 | FALSE | Downregulated |
| 0 | AC005022.1 | FALSE | Upregulated |
| 0,297178435 | AC005062.2 | FALSE | Upregulated |
| | AC005235.1 | | |
| 1,59724085 | AC005264.2 | | Upregulated |
| 0,076136157 | AC005358.3 | FALSE | Upregulated |
| 0,289076049 | AC005550.4 | FALSE | Upregulated |
| 0,125377815 | AC005592.2 | FALSE | Upregulated |
| | AC006004.1 | | |
| | AC006196.1 | | |
| -0,302935006 | AC006262.5 | FALSE | Downregulated |
| -0,285892509 | AC006272.1 | FALSE | Downregulated |
| 0,124510669 | AC006372.4 | FALSE | Upregulated |
| -0,181572993 | AC007040.7 | FALSE | Downregulated |
| 0,896406172 | AC007131.1 | FALSE | Upregulated |
| -0,14324096 | AC007163.3 | FALSE | Downregulated |
| -0,570916296 | AC007163.6 | FALSE | Downregulated |
| -0,318182466 | AC007246.3 | FALSE | Downregulated |
| 0 | AC007359.6 | FALSE | Upregulated |
| 0,02179022 | AC007365.1 | FALSE | Upregulated |
| -0,026920307 | AC007386.2 | FALSE | Downregulated |
| 0,291340714 | AC007392.3 | FALSE | Upregulated |
| 0,420331063 | AC007392.4 | FALSE | Upregulated |
| | AC007403.2 | | |
| 0,277764892 | AC007405.4 | FALSE | Upregulated |
| | AC007463.2 | | |
| 0,638855386 | AC007551.3 | FALSE | Upregulated |
| 0,301095482 | AC007682.1 | FALSE | Upregulated |
| 0,339860169 | AC007740.1 | FALSE | Upregulated |
| -0,773880801 | AC007743.1 | FALSE | Downregulated |
| 1,425643819 | AC007750.5 | | Upregulated |
| -0,172794422 | AC007773.2 | FALSE | Downregulated |
| | AC007879.2 | | |
| -0,318275673 | AC007880.1 | FALSE | Downregulated |
| -0,142328311 | AC007966.1 | FALSE | Downregulated |
| | AC008697.1 | | |

| | | | |
|---|---|---|---|
| 0,345146101 | AC008703.1 | FALSE | Upregulated |
| 0,428437689 | AC009120.4 | FALSE | Upregulated |
| | AC009227.2 | | |
| -0,002236874 | AC009264.1 | FALSE | Downregulated |
| -0,2002865 | AC009299.3 | FALSE | Downregulated |
| -0,749450104 | AC009473.1 | FALSE | Downregulated |
| | AC009518.4 | | |
| -0,01330656 | AC010091.1 | FALSE | Downregulated |
| 1,120150795 | AC010524.2 | | Upregulated |
| | AC010731.4 | | |
| 0,12710841 | AC010890.1 | FALSE | Upregulated |
| | AC010975.1 | | |
| | AC010987.6 | | |
| | AC011298.2 | | |
| 0,839764232 | AC011518.1 | FALSE | Upregulated |
| 0,323601213 | AC011609.3 | FALSE | Upregulated |
| | AC011718.2 | | |
| 0,921440631 | AC011738.4 | FALSE | Upregulated |
| 1,257531613 | AC011899.9 | | Upregulated |
| | AC012075.2 | | |
| -0,064237985 | AC012309.5 | FALSE | Downregulated |
| | AC012494.1 | | |
| -0,024753669 | AC012507.3 | FALSE | Downregulated |
| -0,00440522 | AC015922.6 | FALSE | Downregulated |
| -0,037045014 | AC016582.2 | FALSE | Downregulated |
| 1,177604123 | AC016738.3 | | Upregulated |
| -0,073548283 | AC016910.1 | FALSE | Downregulated |
| | AC016995.3 | | |
| 0,465785683 | AC017060.1 | FALSE | Upregulated |
| | AC018464.3 | | |
| 0,273889386 | AC018470.4 | FALSE | Upregulated |
| 1,333704399 | AC018647.3 | | Upregulated |
| -0,323629584 | AC018742.1 | FALSE | Downregulated |
| 0 | AC018799.1 | FALSE | Upregulated |
| 0,830268176 | AC019048.1 | FALSE | Upregulated |
| | AC019118.2 | | |
| -0,00014923 | AC022007.5 | FALSE | Downregulated |
| | AC022311.1 | | |
| -0,441472363 | AC024592.9 | FALSE | Downregulated |
| | AC027269.2 | | |
| 0 | AC068831.10 | FALSE | Upregulated |
| 0,822411653 | AC068858.1 | FALSE | Upregulated |
| | AC069277.1 | | |
| -0,006433782 | AC072062.1 | FALSE | Downregulated |
| 0,278043438 | AC073128.10 | FALSE | Upregulated |
| -1,113033067 | AC073133.1 | | Downregulated |
| 0,06523155 | AC073283.4 | FALSE | Upregulated |
| 0 | AC073321.3 | FALSE | Upregulated |
| | AC073628.1 | | |
| 0,376227152 | AC074138.3 | FALSE | Upregulated |
| 0,88937119 | AC074286.1 | FALSE | Upregulated |

| | | | |
|---|---|---|---|
| 0,374831862 | AC074289.1 | FALSE | Upregulated |
| 0,622769334 | AC078941.1 | FALSE | Upregulated |
| -1,157181585 | AC079117.1 | | Downregulated |
| 0,405141944 | AC079145.4 | FALSE | Upregulated |
| -0,152464419 | AC079586.1 | FALSE | Downregulated |
| 0,299330153 | AC079610.2 | FALSE | Upregulated |
| | AC079776.3 | | |
| -0,16144964 | AC087859.1 | FALSE | Downregulated |
| 0,150534835 | AC090044.1 | FALSE | Upregulated |
| 0 | AC090505.6 | FALSE | Upregulated |
| 0,301027247 | AC091878.1 | FALSE | Upregulated |
| 0,581019849 | AC091969.1 | FALSE | Upregulated |
| 0,576305718 | AC092295.7 | FALSE | Upregulated |
| -0,21555234 | AC093375.1 | FALSE | Downregulated |
| 0,47210539 | AC093391.2 | FALSE | Upregulated |
| 0,426980121 | AC093590.1 | FALSE | Upregulated |
| -0,795980867 | AC095067.1 | FALSE | Downregulated |
| -0,061941738 | AC096559.1 | FALSE | Downregulated |
| -0,302326538 | AC097468.4 | FALSE | Downregulated |
| -0,000802802 | AC097500.2 | FALSE | Downregulated |
| 0,058042192 | AC097662.2 | FALSE | Upregulated |
| | AC098823.3 | | |
| | AC099342.1 | | |
| 0 | AC104024.1 | FALSE | Upregulated |
| 0 | AC104653.1 | FALSE | Upregulated |
| 0,497479295 | AC104654.1 | FALSE | Upregulated |
| 0 | AC104654.2 | FALSE | Upregulated |
| 0,45755797 | AC104655.2 | FALSE | Upregulated |
| 0,181702747 | AC104777.2 | FALSE | Upregulated |
| | AC104777.4 | | |
| 0,122142423 | AC104820.2 | FALSE | Upregulated |
| -0,781490122 | AC105053.3 | FALSE | Downregulated |
| | AC105253.1 | | |
| 0 | AC105339.1 | FALSE | Upregulated |
| -0,004578537 | AC105760.3 | FALSE | Downregulated |
| 0,430562545 | AC107070.1 | FALSE | Upregulated |
| -1,009373314 | AC107622.1 | | Downregulated |
| | AC108142.1 | | |
| -0,287269233 | AC109828.1 | FALSE | Downregulated |
| | AC113607.2 | | |
| | AC116035.1 | | |
| -0,134179876 | AC125421.1 | FALSE | Downregulated |
| 0 | AC131097.3 | FALSE | Upregulated |
| -0,356469962 | AC136896.1 | FALSE | Downregulated |
| 2,444054233 | AC145110.1 | | Upregulated |
| 0,058528266 | AC159540.1 | FALSE | Upregulated |
| 0,121199827 | AC159540.1-2 | FALSE | Upregulated |
| -0,015353369 | AC226118.1 | FALSE | Downregulated |
| 1,491449265 | ACAN | | Upregulated |
| 0,282455585 | ACBD7 | FALSE | Upregulated |
| -0,275401324 | ACCSL | FALSE | Downregulated |

| | | | |
|---|---|---|---|
| -0,122079819 | ACD | FALSE | Downregulated |
| -0,283030643 | ACE2 | FALSE | Downregulated |
| -0,008605167 | ACER1 | FALSE | Downregulated |
| 0,661190858 | ACER2 | FALSE | Upregulated |
| 0,941749929 | ACER3 | FALSE | Upregulated |
| 0,41238761 | ACHE | FALSE | Upregulated |
| 0,694644374 | ACKR3 | FALSE | Upregulated |
| -0,794594866 | ACN9 | FALSE | Downregulated |
| 0,084712109 | ACOXL | FALSE | Upregulated |
| 0,523351247 | ACPL2 | FALSE | Upregulated |
| -0,247571721 | ACPP | FALSE | Downregulated |
| 0,477241335 | ACPT | FALSE | Upregulated |
| 0,908798414 | ACR | FALSE | Upregulated |
| 1,089435562 | ACRBP | | Upregulated |
| -0,064606359 | ACRC | FALSE | Downregulated |
| -0,471259594 | ACRV1 | FALSE | Downregulated |
| 0,688793027 | ACSL6 | FALSE | Upregulated |
| 0,142066498 | ACSM4 | FALSE | Upregulated |
| 0,413445559 | ACTA2-AS1 | FALSE | Upregulated |
| 0,503247294 | ACTBL2 | FALSE | Upregulated |
| 0,911505875 | ACTC1 | FALSE | Upregulated |
| 0,747349931 | ACTG1P4 | FALSE | Upregulated |
| 0,449965363 | ACTG2 | FALSE | Upregulated |
| 1,266756268 | ACTL6B | | Upregulated |
| 0,597925249 | ACTL7A | FALSE | Upregulated |
| 0,131328282 | ACTL7B | FALSE | Upregulated |
| | ACTL8 | | |
| 0,570628545 | ACTL9 | FALSE | Upregulated |
| | ACTN1-AS1 | | |
| 1,840060458 | ACTN3 | | Upregulated |
| 0,046633664 | ACTR3B | FALSE | Upregulated |
| -0,03294099 | ACTR3BP2 | FALSE | Downregulated |
| 0,416116418 | ACTR3BP5 | FALSE | Upregulated |
| -0,174633156 | ACTR5 | FALSE | Downregulated |
| 0,229518777 | ACTR6 | FALSE | Upregulated |
| | ACTRT1 | | |
| | ACTRT2 | | |
| 0,060944475 | ACTRT3 | FALSE | Upregulated |
| -0,273768507 | ACVR1C | FALSE | Downregulated |
| -0,336045091 | ACYP1 | FALSE | Downregulated |
| -1,225722552 | AD000090.2 | | Downregulated |
| 0,183944276 | ADA | FALSE | Upregulated |
| | ADAD1 | | |
| -0,308751071 | ADAD2 | FALSE | Downregulated |
| -0,011380473 | ADAM11 | FALSE | Downregulated |
| | ADAM18 | | |
| | ADAM2 | | |
| -0,067708769 | ADAM20 | FALSE | Downregulated |
| -0,710320125 | ADAM20P1 | FALSE | Downregulated |
| 0,567822958 | ADAM21P1 | FALSE | Upregulated |
| 0,501923678 | ADAM23 | FALSE | Upregulated |

| | | | |
|---|---|---|---|
| 0,185594489 | ADAM28 | FALSE | Upregulated |
| -0,007086128 | ADAM29 | FALSE | Downregulated |
| 0 | ADAM30 | FALSE | Upregulated |
| 0 | ADAM32 | FALSE | Upregulated |
| 0,248632153 | ADAM33 | FALSE | Upregulated |
| -0,006903421 | ADAM3A | FALSE | Downregulated |
| | ADAM5 | | |
| 0,112683018 | ADAM7 | FALSE | Upregulated |
| 0 | ADAMDEC1 | FALSE | Upregulated |
| 0,842875931 | ADAMTS14 | FALSE | Upregulated |
| 0,489055276 | ADAMTS15 | FALSE | Upregulated |
| 0,00139671 | ADAMTS18 | FALSE | Upregulated |
| 0 | ADAMTS19 | FALSE | Upregulated |
| | ADAMTS20 | | |
| 0,323851097 | ADAMTS3 | FALSE | Upregulated |
| 0,077300072 | ADAMTS4 | FALSE | Upregulated |
| 0,189930048 | ADAMTS6 | FALSE | Upregulated |
| 0,446115305 | ADAMTS7P1 | FALSE | Upregulated |
| 0,388357718 | ADAMTS9-AS2 | FALSE | Upregulated |
| 0,710375033 | ADAMTSL4-AS1 | FALSE | Upregulated |
| 0,469120726 | ADAMTSL5 | FALSE | Upregulated |
| -0,98577008 | ADARB2 | FALSE | Downregulated |
| 0,002944767 | ADARB2-AS1 | FALSE | Upregulated |
| 0,247088032 | ADAT3 | FALSE | Upregulated |
| -0,362172348 | ADC | FALSE | Downregulated |
| -0,509037863 | ADCK1 | FALSE | Downregulated |
| -0,845882797 | ADCY10P1 | FALSE | Downregulated |
| 0,598536919 | ADCY2 | FALSE | Upregulated |
| -0,656719532 | ADCY4 | FALSE | Downregulated |
| 0,482674215 | ADCY8 | FALSE | Upregulated |
| 0,670836485 | ADCYAP1 | FALSE | Upregulated |
| -0,964530752 | ADCYAP1R1 | FALSE | Downregulated |
| -0,041459577 | ADD2 | FALSE | Downregulated |
| -0,125269915 | ADD3-AS1 | FALSE | Downregulated |
| | ADGB | | |
| -0,37305809 | ADH7 | FALSE | Downregulated |
| | ADIG | | |
| 0,679403082 | ADIPOQ | FALSE | Upregulated |
| 1,540966197 | ADIPOQ-AS1 | | Upregulated |
| -0,155061081 | ADM | FALSE | Downregulated |
| -0,151476577 | ADORA1 | FALSE | Downregulated |
| -0,0242468 | ADORA3 | FALSE | Downregulated |
| 1,394976639 | ADPRH | | Upregulated |
| 1,133313574 | ADPRHL1 | | Upregulated |
| 1,29640939 | ADRA1D | | Upregulated |
| 0,154178648 | ADRA2C | FALSE | Upregulated |
| -0,099787711 | ADRB1 | FALSE | Downregulated |
| | ADRB3 | | |
| 0,164083282 | ADTRP | FALSE | Upregulated |
| | AF064858.7 | | |
| -0,106104362 | AFF2 | FALSE | Downregulated |

| | | | |
|---|---|---|---|
| -0,718379963 | AFP | FALSE | Downregulated |
| 0,28873888 | AGAP1-IT1 | FALSE | Upregulated |
| -0,262225286 | AGAP11 | FALSE | Downregulated |
| 0,26772077 | AGAP7 | FALSE | Upregulated |
| 0,552531641 | AGBL1 | FALSE | Upregulated |
| | AGBL1-AS1 | | |
| -1,443177571 | AGBL4 | | Downregulated |
| | AGBL4-IT1 | | |
| 1,522003343 | AGBL5-AS1 | | Upregulated |
| 0,278205259 | AGPAT4-IT1 | FALSE | Upregulated |
| 0,008786178 | AGPAT9 | FALSE | Upregulated |
| -0,250432091 | AGR2 | FALSE | Downregulated |
| | AGR3 | | |
| | AGTR2 | | |
| 0,245820153 | AGTRAP | FALSE | Upregulated |
| -0,140458592 | AHCTF1P1 | FALSE | Downregulated |
| -0,598725251 | AHRR | FALSE | Downregulated |
| | AHSP | | |
| 0,428463356 | AICDA | FALSE | Upregulated |
| -6,96269E-05 | AIFM3 | FALSE | Downregulated |
| 0,593960921 | AIM1L | FALSE | Upregulated |
| 1,375445873 | AIM2 | | Upregulated |
| | AIPL1 | | |
| 0,249344334 | AIRE | FALSE | Upregulated |
| -1,006988489 | AJAP1 | | Downregulated |
| -0,783469455 | AK6 | FALSE | Downregulated |
| 0,668779135 | AK7 | FALSE | Upregulated |
| -0,436575405 | AK8 | FALSE | Downregulated |
| 0,384113637 | AK9 | FALSE | Upregulated |
| 0,275712221 | AKAP14 | FALSE | Upregulated |
| | AKAP17A-2 | | |
| 0,443268062 | AKAP4 | FALSE | Upregulated |
| 0,382027821 | AKIP1 | FALSE | Upregulated |
| -0,012624512 | AKNAD1 | FALSE | Downregulated |
| 0 | AKR1B15 | FALSE | Upregulated |
| -0,55480352 | AKR1C6P | FALSE | Downregulated |
| -0,024751308 | AKR1E2 | FALSE | Downregulated |
| | AL022344.4 | | |
| -0,006903421 | AL109763.2 | FALSE | Downregulated |
| -0,560356154 | AL121656.5 | FALSE | Downregulated |
| -0,088048871 | AL132709.5 | FALSE | Downregulated |
| 0,893344215 | ALAS2 | FALSE | Upregulated |
| 0,517751163 | ALDH1A2 | FALSE | Upregulated |
| -0,497800863 | ALDH1L1-AS1 | FALSE | Downregulated |
| 0,243489822 | ALDH3A1 | FALSE | Upregulated |
| -0,279105416 | ALDH3B2 | FALSE | Downregulated |
| -0,139367928 | ALG10 | FALSE | Downregulated |
| 0,250130831 | ALG1L | FALSE | Upregulated |
| 0,159133448 | ALG1L2 | FALSE | Upregulated |
| 0,036504101 | ALG5 | FALSE | Upregulated |
| -0,299060297 | ALKBH1 | FALSE | Downregulated |

| | | | |
|---|---|---|---|
| -0,928003742 | ALKBH2 | FALSE | Downregulated |
| -0,20073945 | ALKBH3 | FALSE | Downregulated |
| 0,244324574 | ALKBH6 | FALSE | Upregulated |
| -0,628621997 | ALKBH8 | FALSE | Downregulated |
| 0,578803667 | ALLC | FALSE | Upregulated |
| 0,276241806 | ALMS1-IT1 | FALSE | Upregulated |
| 0,693558003 | ALMS1P | FALSE | Upregulated |
| -0,130445822 | ALOX12B | FALSE | Downregulated |
| -0,30094364 | ALOX15 | FALSE | Downregulated |
| 0,163083149 | ALOX15B | FALSE | Upregulated |
| 0,124417075 | ALOX15P1 | FALSE | Upregulated |
| 0,367786436 | ALOX5AP | FALSE | Upregulated |
| 0 | ALOXE3 | FALSE | Upregulated |
| | ALPI | | |
| 0,031587424 | ALPP | FALSE | Upregulated |
| | ALPPL2 | | |
| -0,033319296 | ALS2CR12 | FALSE | Downregulated |
| 0,461070237 | ALX1 | FALSE | Upregulated |
| 0,437073458 | AMBN | FALSE | Upregulated |
| 0,403221758 | AMELX | FALSE | Upregulated |
| | AMELY | | |
| 0 | AMER2 | FALSE | Upregulated |
| | AMER3 | | |
| -0,34986911 | AMH | FALSE | Downregulated |
| 0,052503521 | AMHR2 | FALSE | Upregulated |
| -0,095324162 | AMIGO2 | FALSE | Downregulated |
| 0,43322854 | AMPD1 | FALSE | Upregulated |
| | AMTN | | |
| | AMY1A | | |
| | AMY1B | | |
| | AMY1C | | |
| | AMY2A | | |
| -0,169546456 | AMZ2P1 | FALSE | Downregulated |
| -0,639722396 | ANAPC10 | FALSE | Downregulated |
| -0,359313919 | ANAPC15 | FALSE | Downregulated |
| 0,280928672 | ANAPC1P1 | FALSE | Upregulated |
| 0,121158857 | ANGPT1 | FALSE | Upregulated |
| 0,008036281 | ANGPT4 | FALSE | Upregulated |
| 0,588155289 | ANGPTL5 | FALSE | Upregulated |
| 0,127659122 | ANHX | FALSE | Upregulated |
| 1,054585857 | ANK1 | | Upregulated |
| 0,282563824 | ANKDD1B | FALSE | Upregulated |
| -0,002253536 | ANKFN1 | FALSE | Downregulated |
| -0,064552361 | ANKK1 | FALSE | Downregulated |
| 1,067153201 | ANKLE1 | | Upregulated |
| -0,252204101 | ANKRA2 | FALSE | Downregulated |
| 1,476766268 | ANKRD1 | | Upregulated |
| -0,171147952 | ANKRD13B | FALSE | Downregulated |
| 0,367000457 | ANKRD18A | FALSE | Upregulated |
| -0,149835757 | ANKRD18B | FALSE | Downregulated |
| 0,293326262 | ANKRD18CP | FALSE | Upregulated |

| | | | |
|---|---|---|---|
| 1,707324233 | ANKRD18DP | | Upregulated |
| 0,109471418 | ANKRD2 | FALSE | Upregulated |
| -0,356144228 | ANKRD20A1 | FALSE | Downregulated |
| -0,12778095 | ANKRD20A11P | FALSE | Downregulated |
| 0,253519899 | ANKRD20A12P | FALSE | Upregulated |
| | ANKRD20A19P | | |
| 0,387961117 | ANKRD20A2 | FALSE | Upregulated |
| 1,496377096 | ANKRD20A3 | | Upregulated |
| 0,833885208 | ANKRD20A4 | FALSE | Upregulated |
| -0,129962958 | ANKRD20A5P | FALSE | Downregulated |
| -0,005070114 | ANKRD20A8P | FALSE | Downregulated |
| 2,784144522 | ANKRD22 | | Upregulated |
| | ANKRD26P1 | | |
| 0,087258652 | ANKRD26P3 | FALSE | Upregulated |
| 0,050741043 | ANKRD29 | FALSE | Upregulated |
| | ANKRD30A | | |
| 0,294603606 | ANKRD30B | FALSE | Upregulated |
| | ANKRD30BL | | |
| 0,432195028 | ANKRD30BP2 | FALSE | Upregulated |
| | ANKRD30BP3 | | |
| 0,469254082 | ANKRD31 | FALSE | Upregulated |
| 0 | ANKRD33 | FALSE | Upregulated |
| 0,589984707 | ANKRD34B | FALSE | Upregulated |
| | ANKRD34C | | |
| -0,258518156 | ANKRD35 | FALSE | Downregulated |
| 0,316445149 | ANKRD36BP2 | FALSE | Upregulated |
| -0,157886573 | ANKRD39 | FALSE | Downregulated |
| 0,204393924 | ANKRD44-IT1 | FALSE | Upregulated |
| -0,150751703 | ANKRD45 | FALSE | Downregulated |
| 0,906844635 | ANKRD49 | FALSE | Upregulated |
| 0,824491748 | ANKRD55 | FALSE | Upregulated |
| | ANKRD60 | | |
| 0,293326262 | ANKRD62 | FALSE | Upregulated |
| 0,884100635 | ANKRD62P1-PARP4P3 | FALSE | Upregulated |
| 0,600433961 | ANKRD63 | FALSE | Upregulated |
| -0,271591773 | ANKRD65 | FALSE | Downregulated |
| 0 | ANKRD66 | FALSE | Upregulated |
| | ANKRD7 | | |
| | ANKUB1 | | |
| 1,179842578 | ANLN | | Upregulated |
| 0,248485379 | ANO2 | FALSE | Upregulated |
| -0,050044233 | ANO3 | FALSE | Downregulated |
| 0,233046249 | ANO4 | FALSE | Upregulated |
| -0,190539555 | ANO5 | FALSE | Downregulated |
| 0 | ANP32C | FALSE | Upregulated |
| | ANP32D | | |
| 0,256919506 | ANTXRL | FALSE | Upregulated |
| 0,423658763 | ANTXRLP1 | FALSE | Upregulated |
| -0,373236587 | ANXA13 | FALSE | Downregulated |
| 0,284987956 | ANXA2P3 | FALSE | Upregulated |
| 1,42584503 | ANXA2R | | Upregulated |

| | | | |
|---|---|---|---|
| 0,073637576 | ANXA3 | FALSE | Upregulated |
| -0,05037907 | ANXA8 | FALSE | Downregulated |
| 0,470966108 | ANXA8L1 | FALSE | Upregulated |
| 0,652441256 | AOAH-IT1 | FALSE | Upregulated |
| -0,078585441 | AOC2 | FALSE | Downregulated |
| 0,029256947 | AOX2P | FALSE | Upregulated |
| | AP000289.6 | | |
| 0,770343765 | AP000330.8 | FALSE | Upregulated |
| | AP000345.1 | | |
| | AP000469.2 | | |
| -0,115619084 | AP000472.2 | FALSE | Downregulated |
| | AP000475.2 | | |
| -0,038555569 | AP000688.8 | FALSE | Downregulated |
| 0,902773283 | AP001258.4 | FALSE | Upregulated |
| -0,149115345 | AP003025.2 | FALSE | Downregulated |
| | AP1B1P1 | | |
| 0,04263755 | AP1M2 | FALSE | Upregulated |
| 0,067336033 | AP1S3 | FALSE | Upregulated |
| 0,586026444 | AP3B2 | FALSE | Upregulated |
| 0,125914216 | AP4B1 | FALSE | Upregulated |
| -0,627293031 | AP4M1 | FALSE | Downregulated |
| -0,303393833 | APCDD1 | FALSE | Downregulated |
| 0 | APCDD1L-AS1 | FALSE | Upregulated |
| | APELA | | |
| -0,101942832 | APH1B | FALSE | Downregulated |
| -0,441329227 | APIP | FALSE | Downregulated |
| 0,409983363 | APITD1 | FALSE | Upregulated |
| -0,941550603 | APITD1-CORT | FALSE | Downregulated |
| 0,051773184 | APLF | FALSE | Upregulated |
| -0,54172637 | APLN | FALSE | Downregulated |
| -0,0045635 | APLP1 | FALSE | Downregulated |
| | APOBEC1 | | |
| -0,066561283 | APOBEC3A | FALSE | Downregulated |
| 0,27605107 | APOBEC3B | FALSE | Upregulated |
| 0,428143503 | APOBEC3B-AS1 | FALSE | Upregulated |
| -0,725078843 | APOBEC3C | FALSE | Downregulated |
| 0,652264335 | APOBEC3D | FALSE | Upregulated |
| -0,192635581 | APOBEC3G | FALSE | Downregulated |
| -0,070046245 | APOBEC3H | FALSE | Downregulated |
| -0,294983152 | APOD | FALSE | Downregulated |
| -0,162133284 | APOL5 | FALSE | Downregulated |
| -0,618638797 | APOLD1 | FALSE | Downregulated |
| -1,187437295 | APOO | Downregulated | |
| 0,291340714 | APOOP5 | FALSE | Upregulated |
| -0,044633408 | APOPT1 | FALSE | Downregulated |
| | AQP10 | | |
| | AQP12A | | |
| | AQP12B | | |
| 0,00139671 | AQP2 | FALSE | Upregulated |
| 0,002540653 | AQP4 | FALSE | Upregulated |
| 0 | AQP5 | FALSE | Upregulated |

| | | | |
|---|---|---|---|
| -0,428539211 | AQP6 | FALSE | Downregulated |
| -0,317051951 | AQP7P3 | FALSE | Downregulated |
| -0,015040902 | AQP8 | FALSE | Downregulated |
| -0,140560374 | AQPEP | FALSE | Downregulated |
| 0,33930768 | ARC | FALSE | Upregulated |
| 0 | AREG | FALSE | Upregulated |
| 0,601481818 | ARGFX | FALSE | Upregulated |
| 0,400617163 | ARHGAP11A | FALSE | Upregulated |
| 0,805271037 | ARHGAP11B | FALSE | Upregulated |
| 2,465981811 | ARHGAP19-SLIT1 | | Upregulated |
| 0,408363978 | ARHGAP22 | FALSE | Upregulated |
| 1,066185879 | ARHGAP26-AS1 | | Upregulated |
| -0,018209239 | ARHGAP26-IT1 | FALSE | Downregulated |
| 0,261328953 | ARHGAP28 | FALSE | Upregulated |
| | ARHGAP36 | | |
| 0,273422532 | ARHGAP40 | FALSE | Upregulated |
| -0,167905449 | ARHGAP5-AS1 | FALSE | Downregulated |
| 0,082979445 | ARHGAP8 | FALSE | Upregulated |
| 0 | ARHGDIG | FALSE | Upregulated |
| 1,14409711 | ARHGEF19 | | Upregulated |
| 0,200828976 | ARHGEF25 | FALSE | Upregulated |
| 0,016912388 | ARHGEF26-AS1 | FALSE | Upregulated |
| 0 | ARHGEF3-AS1 | FALSE | Upregulated |
| 0 | ARHGEF38-IT1 | FALSE | Upregulated |
| 0,372725083 | ARL10 | FALSE | Upregulated |
| 1,045032456 | ARL11 | | Upregulated |
| 0 | ARL14 | FALSE | Upregulated |
| 0,272277655 | ARL14EPL | FALSE | Upregulated |
| -0,046642014 | ARL15 | FALSE | Downregulated |
| -2,017453089 | ARL17B | | Downregulated |
| -0,200296252 | ARL2 | FALSE | Downregulated |
| -0,349786849 | ARL2-SNX15 | FALSE | Downregulated |
| 0,379557675 | ARL4A | FALSE | Upregulated |
| 0,566691207 | ARL5C | FALSE | Upregulated |
| 0,498839336 | ARL6 | FALSE | Upregulated |
| 0 | ARL9 | FALSE | Upregulated |
| -0,026248638 | ARMC12 | FALSE | Downregulated |
| -1,011191513 | ARMC2 | | Downregulated |
| -0,847540008 | ARMC2-AS1 | FALSE | Downregulated |
| | ARMC3 | | |
| -0,259339952 | ARMC4 | FALSE | Downregulated |
| | ARMC4P1 | | |
| -0,291287351 | ARMC9 | FALSE | Downregulated |
| 0,931721273 | ARMCX1 | FALSE | Upregulated |
| 0,025637935 | ARMCX2 | FALSE | Upregulated |
| -3,123325297 | ARMCX5-GPRASP2 | | Downregulated |
| 0,032296529 | ARMCX6 | FALSE | Upregulated |
| -0,227339641 | ARMS2 | FALSE | Downregulated |
| -0,118129831 | ARNT2 | FALSE | Downregulated |
| 0,752066719 | ARPP21 | FALSE | Upregulated |
| -0,358361173 | ARR3 | FALSE | Downregulated |

| | | | |
|---|---|---|---|
| 0,291233534 | ARRDC5 | FALSE | Upregulated |
| -0,475613151 | ARSF | FALSE | Downregulated |
| | ARSH | | |
| 0,419420082 | ARSI | FALSE | Upregulated |
| 0,584199486 | ARSK | FALSE | Upregulated |
| 0,740309359 | ART3 | FALSE | Upregulated |
| -0,248046334 | ART5 | FALSE | Downregulated |
| -0,296834627 | ARTN | FALSE | Downregulated |
| 0,083105717 | ARV1 | FALSE | Upregulated |
| -0,138043775 | ARX | FALSE | Downregulated |
| -0,532872148 | ASAH2 | FALSE | Downregulated |
| -0,029872935 | ASAH2B | FALSE | Downregulated |
| -0,167014711 | ASAP1-IT1 | FALSE | Downregulated |
| 0,155305304 | ASAP1-IT2 | FALSE | Upregulated |
| 0,552524761 | ASB10 | FALSE | Upregulated |
| -0,148634105 | ASB11 | FALSE | Downregulated |
| 0,227969239 | ASB12 | FALSE | Upregulated |
| -0,13824646 | ASB15 | FALSE | Downregulated |
| 0,135714138 | ASB17 | FALSE | Upregulated |
| 0,421176596 | ASB18 | FALSE | Upregulated |
| 0,452431457 | ASB2 | FALSE | Upregulated |
| -1,041685291 | ASB3 | | Downregulated |
| 0,348050373 | ASB4 | FALSE | Upregulated |
| 0,668306726 | ASB5 | FALSE | Upregulated |
| -0,263141925 | ASB9 | FALSE | Downregulated |
| 0,28824064 | ASCL1 | FALSE | Upregulated |
| 0,386964976 | ASCL2 | FALSE | Upregulated |
| 0,667757387 | ASCL3 | FALSE | Upregulated |
| 0 | ASCL4 | FALSE | Upregulated |
| 0,15813836 | ASCL5 | FALSE | Upregulated |
| 0,715627349 | ASIC1 | FALSE | Upregulated |
| 0 | ASIC2 | FALSE | Upregulated |
| 0,520220208 | ASIC3 | FALSE | Upregulated |
| -0,184922142 | ASIC4 | FALSE | Downregulated |
| 0,293445983 | ASIC5 | FALSE | Upregulated |
| 0,31487039 | ASIP | FALSE | Upregulated |
| | ASMT | | |
| | ASMT-2 | | |
| | ASMTL-2 | | |
| | ASMTL-AS1-2 | | |
| 0,42715208 | ASNS | FALSE | Upregulated |
| -0,041387596 | ASPA | FALSE | Downregulated |
| 0,441986117 | ASPM | FALSE | Upregulated |
| -0,200981765 | ASPRV1 | FALSE | Downregulated |
| -0,059249808 | ASRGL1 | FALSE | Downregulated |
| 0 | ASTL | FALSE | Upregulated |
| 0,063571722 | ASTN1 | FALSE | Upregulated |
| -0,213277203 | ASTN2 | FALSE | Downregulated |
| | ASZ1 | | |
| -0,211025843 | ATAD3C | FALSE | Downregulated |
| 0,960459578 | ATCAY | FALSE | Upregulated |

| | | | |
|---|---|---|---|
| 0,276327097 | ATF3 | FALSE | Upregulated |
| 0,518379061 | ATG10 | FALSE | Upregulated |
| -0,562876062 | ATG4A | FALSE | Downregulated |
| -0,18296079 | ATG4C | FALSE | Downregulated |
| | ATOH1 | | |
| -0,774126267 | ATOH7 | FALSE | Downregulated |
| 0,176088971 | ATP10A | FALSE | Upregulated |
| 0,018839898 | ATP12A | FALSE | Upregulated |
| -0,745371226 | ATP13A4 | FALSE | Downregulated |
| -0,15556678 | ATP13A5 | FALSE | Downregulated |
| 1,153421762 | ATP1A2 | | Upregulated |
| 0,59037783 | ATP1A3 | FALSE | Upregulated |
| -0,046213247 | ATP1A4 | FALSE | Downregulated |
| 0,057702931 | ATP1B2 | FALSE | Upregulated |
| -0,467094199 | ATP1B4 | FALSE | Downregulated |
| -0,157705194 | ATP2B3 | FALSE | Downregulated |
| 0,212611577 | ATP2C2 | FALSE | Upregulated |
| -0,614760581 | ATP4A | FALSE | Downregulated |
| 0,347582353 | ATP4B | FALSE | Upregulated |
| -0,015040902 | ATP5EP2 | FALSE | Downregulated |
| -3,065431029 | ATP5J2-PTCD1 | | Downregulated |
| 0,617110204 | ATP5L2 | FALSE | Upregulated |
| -0,324185042 | ATP6AP1L | FALSE | Downregulated |
| | ATP6V0A4 | | |
| -0,030411814 | ATP6V0CP3 | FALSE | Downregulated |
| 0,158672209 | ATP6V0D2 | FALSE | Upregulated |
| -0,623624758 | ATP6V1B1 | FALSE | Downregulated |
| 0,610869668 | ATP6V1E2 | FALSE | Upregulated |
| 0,96597791 | ATP6V1G2 | FALSE | Upregulated |
| 0 | ATP6V1G2-DDX39B | FALSE | Upregulated |
| | ATP6V1G3 | | |
| 0,263864424 | ATP8A2 | FALSE | Upregulated |
| 0,011421845 | ATP8B3 | FALSE | Upregulated |
| -0,736653128 | ATP8B5P | FALSE | Downregulated |
| 0,428421287 | ATXN3L | FALSE | Upregulated |
| | ATXN8OS | | |
| -0,59139712 | AURKAPS1 | FALSE | Downregulated |
| -0,156228647 | AURKB | FALSE | Downregulated |
| -0,567407799 | AURKC | FALSE | Downregulated |
| | AVP | | |
| 0,392815798 | AVPR1B | FALSE | Upregulated |
| 0,28862639 | AWAT1 | FALSE | Upregulated |
| 0,997987895 | AWAT2 | FALSE | Upregulated |
| 1,075174897 | AXDND1 | | Upregulated |
| -0,091028489 | AZU1 | FALSE | Downregulated |
| 0,996192294 | B3GALNT1 | FALSE | Upregulated |
| -0,142632443 | B3GALT1 | FALSE | Downregulated |
| 0,943583335 | B3GALT4 | FALSE | Upregulated |
| -0,289280665 | B3GALT5 | FALSE | Downregulated |
| -0,288993469 | B3GALTL | FALSE | Downregulated |
| 0,342189037 | B3GNT3 | FALSE | Upregulated |

| | | | |
|---|---|---|---|
| -0,006848587 | B3GNT6 | FALSE | Downregulated |
| -0,448920987 | B3GNT7 | FALSE | Downregulated |
| 0 | B4GALNT2 | FALSE | Upregulated |
| 0,139689892 | B4GALNT3 | FALSE | Upregulated |
| 0,199376117 | B4GALNT4 | FALSE | Upregulated |
| 0 | BAALCOS | FALSE | Upregulated |
| 0,251260027 | BACE1-AS | FALSE | Upregulated |
| -0,01260029 | BAGE2 | FALSE | Downregulated |
| 1,23201387 | BAI1 | | Upregulated |
| 0,288949676 | BAI2 | FALSE | Upregulated |
| -0,107425943 | BAI3 | FALSE | Downregulated |
| 0,283521929 | BAIAP2L2 | FALSE | Upregulated |
| 0,332886226 | BAK1 | FALSE | Upregulated |
| | BANCR | | |
| 0 | BANF2 | FALSE | Upregulated |
| 1,04013984 | BANK1 | | Upregulated |
| -0,141917398 | BARD1 | FALSE | Downregulated |
| 0,327816954 | BARHL1 | FALSE | Upregulated |
| | BARHL2 | | |
| 0,303577425 | BARX1 | FALSE | Upregulated |
| 0 | BARX2 | FALSE | Upregulated |
| 0 | BASP1P1 | FALSE | Upregulated |
| 0,227792868 | BATF | FALSE | Upregulated |
| 0,280932553 | BATF3 | FALSE | Upregulated |
| 0,277666846 | BBS12 | FALSE | Upregulated |
| -0,283964964 | BBS7 | FALSE | Downregulated |
| 0,280928672 | BCAR4 | FALSE | Upregulated |
| 0,308897671 | BCAS1 | FALSE | Upregulated |
| 0,356096663 | BCAS4 | FALSE | Upregulated |
| 1,040162277 | BCAT1 | | Upregulated |
| 0,675952118 | BCL11A | FALSE | Upregulated |
| 0,241434056 | BCL2A1 | FALSE | Upregulated |
| 0,461962517 | BCL2L15 | FALSE | Upregulated |
| 1,885195743 | BCL2L2-PABPN1 | | Upregulated |
| 0,604103814 | BCMO1 | FALSE | Upregulated |
| 0,249182846 | BCORP1 | FALSE | Upregulated |
| 0,98209451 | BCRP2 | FALSE | Upregulated |
| -0,901280849 | BCRP3 | FALSE | Downregulated |
| 0,484342812 | BCYRN1 | FALSE | Upregulated |
| 0,461082084 | BDKRB1 | FALSE | Upregulated |
| 0,152725287 | BDKRB2 | FALSE | Upregulated |
| 0,853710828 | BDNF-AS | FALSE | Upregulated |
| -0,892359965 | BEGAIN | FALSE | Downregulated |
| | BEND2 | | |
| -0,240434078 | BEND3 | FALSE | Downregulated |
| 0,859506209 | BEND4 | FALSE | Upregulated |
| 0,000350942 | BEND5 | FALSE | Upregulated |
| 0,053602426 | BEND6 | FALSE | Upregulated |
| -0,394949907 | BEST2 | FALSE | Downregulated |
| -0,156608059 | BEST3 | FALSE | Downregulated |
| 1,905904248 | BEST4 | | Upregulated |

| | | | |
|---|---|---|---|
| -0,056049448 | BET1 | FALSE | Downregulated |
| -0,055231376 | BEX1 | FALSE | Downregulated |
| 0,157716651 | BEX2 | FALSE | Upregulated |
| -0,956615768 | BEX4 | FALSE | Downregulated |
| -0,116514342 | BEX5 | FALSE | Downregulated |
| 0,121317909 | BFSP1 | FALSE | Upregulated |
| 0,367810861 | BFSP2 | FALSE | Upregulated |
| 0 | BFSP2-AS1 | FALSE | Upregulated |
| | BGLAP | | |
| -1,512887216 | BHLHA15 | | Downregulated |
| -0,01330656 | BHLHA9 | FALSE | Downregulated |
| -0,289477024 | BHLHB9 | FALSE | Downregulated |
| 1,230304843 | BHLHE22 | | Upregulated |
| 0 | BHLHE23 | FALSE | Upregulated |
| 0 | BHLHE40-AS1 | FALSE | Upregulated |
| 0,226419902 | BIK | FALSE | Upregulated |
| -0,257359093 | BIN3-IT1 | FALSE | Downregulated |
| 0,318619676 | BIRC5 | FALSE | Upregulated |
| 0 | BIRC7 | FALSE | Upregulated |
| | BIRC8 | | |
| 0,295640489 | BLACAT1 | FALSE | Upregulated |
| 0 | BLACE | FALSE | Upregulated |
| -0,060768758 | BLID | FALSE | Downregulated |
| 0,427938768 | BLK | FALSE | Upregulated |
| 0,270876139 | BLM | FALSE | Upregulated |
| -0,723059998 | BLOC1S4 | FALSE | Downregulated |
| -0,09661229 | BLOC1S5-TXNDC5 | FALSE | Downregulated |
| -0,096933194 | BLVRA | FALSE | Downregulated |
| -0,112576252 | BMP10 | FALSE | Downregulated |
| | BMP15 | | |
| 0,334058404 | BMP3 | FALSE | Upregulated |
| 0,083461485 | BMP4 | FALSE | Upregulated |
| -0,223172581 | BMP5 | FALSE | Downregulated |
| 0 | BMP7 | FALSE | Upregulated |
| -0,329913795 | BMPR1B | FALSE | Downregulated |
| 0,039540684 | BMS1P17 | FALSE | Upregulated |
| 0,249515614 | BMS1P18 | FALSE | Upregulated |
| -0,786200907 | BMS1P20 | FALSE | Downregulated |
| 1,116252342 | BMX | | Upregulated |
| 0,759474313 | BNC1 | FALSE | Upregulated |
| -0,005100277 | BNC2 | FALSE | Downregulated |
| -0,494485359 | BNIP1 | FALSE | Downregulated |
| -0,126378134 | BNIPL | FALSE | Downregulated |
| 0,196316304 | BOD1L2 | FALSE | Upregulated |
| -0,312439962 | BOLA3 | FALSE | Downregulated |
| 0,186392874 | BOLL | FALSE | Upregulated |
| 0 | BPESC1 | FALSE | Upregulated |
| 0,405075113 | BPGM | FALSE | Upregulated |
| 0,107427199 | BPI | FALSE | Upregulated |
| | BPIFA1 | | |
| | BPIFA2 | | |

| | | | |
|---|---|---|---|
| | BPIFA3 | | |
| | BPIFA4P | | |
| -0,007553621 | BPIFB1 | FALSE | Downregulated |
| 0,001141803 | BPIFB2 | FALSE | Upregulated |
| | BPIFB3 | | |
| | BPIFB4 | | |
| | BPIFB6 | | |
| -0,229282772 | BPIFC | FALSE | Downregulated |
| | BRDT | | |
| 0,577890894 | BRE-AS1 | FALSE | Upregulated |
| -0,453086581 | BREA2 | FALSE | Downregulated |
| -0,165078227 | BRINP1 | FALSE | Downregulated |
| -0,86309838 | BRINP2 | FALSE | Downregulated |
| | BRINP3 | | |
| -0,441598708 | BRIP1 | FALSE | Downregulated |
| 0,079876325 | BRMS1L | FALSE | Upregulated |
| 0,8558933 | BRS3 | FALSE | Upregulated |
| 0,564187137 | BRSK1 | FALSE | Upregulated |
| 0,297788097 | BRWD1-AS1 | FALSE | Upregulated |
| 1,464199669 | BSCL2 | | Upregulated |
| 0,25790522 | BSN-AS2 | FALSE | Upregulated |
| 0,323819304 | BSND | FALSE | Upregulated |
| | BSPH1 | | |
| 0,192939251 | BSPRY | FALSE | Upregulated |
| 0,118458208 | BST1 | FALSE | Upregulated |
| | BSX | | |
| -0,821756025 | BTBD16 | FALSE | Downregulated |
| 1,306302643 | BTBD17 | | Upregulated |
| 0,04016373 | BTBD19 | FALSE | Upregulated |
| -0,013808431 | BTBD8 | FALSE | Downregulated |
| 0,049049729 | BTC | FALSE | Upregulated |
| 0 | BTF3P11 | FALSE | Upregulated |
| | BTG4 | | |
| 0,624981315 | BTK | FALSE | Upregulated |
| 0,143389414 | BTLA | FALSE | Upregulated |
| 0,608136423 | BTN1A1 | FALSE | Upregulated |
| -0,321227474 | BTN2A3P | FALSE | Downregulated |
| 0,117211309 | BTNL10 | FALSE | Upregulated |
| -0,219928255 | BTNL2 | FALSE | Downregulated |
| -0,435530155 | BTNL3 | FALSE | Downregulated |
| 0,115919231 | BTNL8 | FALSE | Upregulated |
| -0,759981127 | BUB1 | FALSE | Downregulated |
| 0,421374113 | BUB1B | FALSE | Upregulated |
| 0,487045694 | BVES | FALSE | Upregulated |
| | BVES-AS1 | | |
| 0 | BX255923.3 | FALSE | Upregulated |
| -0,078110599 | BZW2 | FALSE | Downregulated |
| 0,295084997 | C10orf107 | FALSE | Upregulated |
| -0,119683026 | C10orf111 | FALSE | Downregulated |
| | C10orf113 | | |
| 0,272277655 | C10orf115 | FALSE | Upregulated |

| | | | |
|---|---|---|---|
| | C10orf120 | | |
| 0,536428784 | C10orf126 | FALSE | Upregulated |
| -0,176721437 | C10orf128 | FALSE | Downregulated |
| -0,072048117 | C10orf129 | FALSE | Downregulated |
| 0,264266513 | C10orf131 | FALSE | Upregulated |
| -0,200769457 | C10orf2 | FALSE | Downregulated |
| -0,70169015 | C10orf32-ASMT | FALSE | Downregulated |
| -0,561390356 | C10orf35 | FALSE | Downregulated |
| -0,001674428 | C10orf40 | FALSE | Downregulated |
| -0,01330656 | C10orf53 | FALSE | Downregulated |
| 0,462913518 | C10orf55 | FALSE | Upregulated |
| -0,076110021 | C10orf62 | FALSE | Downregulated |
| -0,262065247 | C10orf67 | FALSE | Downregulated |
| 1,095277594 | C10orf71 | | Upregulated |
| | C10orf71-AS1 | | |
| | C10orf82 | | |
| -0,003523093 | C10orf85 | FALSE | Downregulated |
| -0,329659761 | C10orf88 | FALSE | Downregulated |
| 0,12094579 | C10orf90 | FALSE | Upregulated |
| 0 | C10orf91 | FALSE | Upregulated |
| | C10orf99 | | |
| -0,615999341 | C11orf1 | FALSE | Downregulated |
| 0,715103902 | C11orf16 | FALSE | Upregulated |
| 0,099568241 | C11orf21 | FALSE | Upregulated |
| | C11orf40 | | |
| | C11orf44 | | |
| 1,076226544 | C11orf48 | | Upregulated |
| -0,425817617 | C11orf52 | FALSE | Downregulated |
| | C11orf53 | | |
| 0,559679053 | C11orf63 | FALSE | Upregulated |
| 0,169160421 | C11orf65 | FALSE | Upregulated |
| | C11orf70 | | |
| -0,289356151 | C11orf73 | FALSE | Downregulated |
| -0,004845519 | C11orf74 | FALSE | Downregulated |
| 0,555463535 | C11orf82 | FALSE | Upregulated |
| 0,575131691 | C11orf85 | FALSE | Upregulated |
| | C11orf86 | | |
| 0,073654805 | C11orf87 | FALSE | Upregulated |
| -0,004525881 | C11orf88 | FALSE | Downregulated |
| 0 | C11orf91 | FALSE | Upregulated |
| | C11orf94 | | |
| 0 | C12orf36 | FALSE | Upregulated |
| 0,292128559 | C12orf39 | FALSE | Upregulated |
| -0,433084496 | C12orf40 | FALSE | Downregulated |
| 0,130465212 | C12orf42 | FALSE | Upregulated |
| 0,096442866 | C12orf45 | FALSE | Upregulated |
| | C12orf50 | | |
| | C12orf54 | | |
| -0,02745599 | C12orf55 | FALSE | Downregulated |
| -0,822370423 | C12orf56 | FALSE | Downregulated |
| -0,00541294 | C12orf68 | FALSE | Downregulated |

| | | | |
|---|---|---|---|
| 1,42689669 | C12orf71 | | Upregulated |
| 0,320762555 | C12orf74 | FALSE | Upregulated |
| -0,061633463 | C12orf75 | FALSE | Downregulated |
| | C12orf77 | | |
| 0,441742724 | C12orf79 | FALSE | Upregulated |
| 0,291340714 | C12orf80 | FALSE | Upregulated |
| 0,516040598 | C13orf35 | FALSE | Upregulated |
| | C13orf45 | | |
| 0,558822125 | C14orf119 | FALSE | Upregulated |
| -0,00690394 | C14orf142 | FALSE | Downregulated |
| 0,168951665 | C14orf166B | FALSE | Upregulated |
| | C14orf177 | | |
| 0,036369932 | C14orf178 | FALSE | Upregulated |
| 0,553558269 | C14orf183 | FALSE | Upregulated |
| -0,0088524 | C14orf2 | FALSE | Downregulated |
| -0,158343621 | C14orf23 | FALSE | Downregulated |
| 0,544602679 | C14orf28 | FALSE | Upregulated |
| -0,215307691 | C14orf37 | FALSE | Downregulated |
| 0,277119458 | C14orf39 | FALSE | Upregulated |
| 0,139204399 | C14orf64 | FALSE | Upregulated |
| -0,175617268 | C15orf26 | FALSE | Downregulated |
| | C15orf32 | | |
| -0,014829034 | C15orf43 | FALSE | Downregulated |
| 0,584464732 | C15orf48 | FALSE | Upregulated |
| 0,271886396 | C15orf53 | FALSE | Upregulated |
| 0 | C15orf56 | FALSE | Upregulated |
| 0,130916864 | C15orf57 | FALSE | Upregulated |
| -1,528734111 | C15orf59 | | Downregulated |
| 0,288408796 | C15orf61 | FALSE | Upregulated |
| -0,48410868 | C15orf65 | FALSE | Downregulated |
| | C16orf11 | | |
| -0,153446317 | C16orf46 | FALSE | Downregulated |
| 0 | C16orf47 | FALSE | Upregulated |
| 0,606819228 | C16orf54 | FALSE | Upregulated |
| 0,497768292 | C16orf74 | FALSE | Upregulated |
| -0,436941552 | C16orf78 | FALSE | Downregulated |
| | C16orf82 | | |
| -0,653598747 | C16orf87 | FALSE | Downregulated |
| 0,278076399 | C16orf90 | FALSE | Upregulated |
| -0,755871067 | C16orf91 | FALSE | Downregulated |
| 0,415289659 | C16orf92 | FALSE | Upregulated |
| -0,045596274 | C16orf95 | FALSE | Downregulated |
| 0,634796584 | C16orf96 | FALSE | Upregulated |
| | C16orf97 | | |
| | C17orf102 | | |
| 0,277764892 | C17orf105 | FALSE | Upregulated |
| | C17orf112 | | |
| -0,021735557 | C17orf50 | FALSE | Downregulated |
| 0,50389055 | C17orf53 | FALSE | Upregulated |
| 0,821617927 | C17orf58 | FALSE | Upregulated |
| 1,006303084 | C17orf64 | | Upregulated |

| | | | |
|---|---|---|---|
| -0,481105539 | C17orf66 | FALSE | Downregulated |
| -0,779681037 | C17orf67 | FALSE | Downregulated |
| 0,150130679 | C17orf74 | FALSE | Upregulated |
| | C17orf77 | | |
| 0,120586831 | C17orf82 | FALSE | Upregulated |
| -0,222091063 | C17orf96 | FALSE | Downregulated |
| | C17orf98 | | |
| 0,313207451 | C17orf99 | FALSE | Upregulated |
| -0,130537581 | C18orf12 | FALSE | Downregulated |
| -0,215278211 | C18orf21 | FALSE | Downregulated |
| 0,444895712 | C18orf42 | FALSE | Upregulated |
| 0,249868182 | C18orf54 | FALSE | Upregulated |
| -0,290366514 | C18orf56 | FALSE | Downregulated |
| | C18orf61 | | |
| 0,958185731 | C18orf63 | FALSE | Upregulated |
| | C18orf64 | | |
| 0,02838064 | C19orf18 | FALSE | Upregulated |
| 0,556546631 | C19orf33 | FALSE | Upregulated |
| -0,233062419 | C19orf38 | FALSE | Downregulated |
| 0,267563656 | C19orf40 | FALSE | Upregulated |
| 0,83210228 | C19orf45 | FALSE | Upregulated |
| -0,206671185 | C19orf52 | FALSE | Downregulated |
| -0,504749224 | C19orf55 | FALSE | Downregulated |
| 0,092871931 | C19orf57 | FALSE | Upregulated |
| -0,31791955 | C19orf59 | FALSE | Downregulated |
| -0,096861596 | C19orf67 | FALSE | Downregulated |
| -0,064734459 | C19orf69 | FALSE | Downregulated |
| -0,156196895 | C19orf73 | FALSE | Downregulated |
| 0,924544595 | C19orf77 | FALSE | Upregulated |
| -0,017864251 | C19orf81 | FALSE | Downregulated |
| 0,578865653 | C19orf83 | FALSE | Upregulated |
| -0,156998262 | C19orf84 | FALSE | Downregulated |
| 0,32353433 | C1D | FALSE | Upregulated |
| 0,227417773 | C1GALT1C1 | FALSE | Upregulated |
| 0,295616338 | C1orf100 | FALSE | Upregulated |
| 0,438953257 | C1orf101 | FALSE | Upregulated |
| 0,233758447 | C1orf105 | FALSE | Upregulated |
| 1,066778811 | C1orf106 | | Upregulated |
| -0,657190873 | C1orf109 | FALSE | Downregulated |
| 0,732342261 | C1orf110 | FALSE | Upregulated |
| -0,518654399 | C1orf116 | FALSE | Downregulated |
| 0,074581206 | C1orf127 | FALSE | Upregulated |
| -0,08330625 | C1orf131 | FALSE | Downregulated |
| | C1orf137 | | |
| -0,017526272 | C1orf140 | FALSE | Downregulated |
| | C1orf141 | | |
| 0 | C1orf146 | FALSE | Upregulated |
| | C1orf158 | | |
| 0,089671708 | C1orf162 | FALSE | Upregulated |
| 0,098515582 | C1orf173 | FALSE | Upregulated |
| 0,142897134 | C1orf180 | FALSE | Upregulated |

| | | | |
|---|---|---|---|
| -0,198047111 | C1orf185 | FALSE | Downregulated |
| 0,413170989 | C1orf186 | FALSE | Upregulated |
| | C1orf189 | | |
| 0,695800608 | C1orf194 | FALSE | Upregulated |
| -0,003443887 | C1orf227 | FALSE | Downregulated |
| -0,367790026 | C1orf228 | FALSE | Downregulated |
| -0,620810348 | C1orf229 | FALSE | Downregulated |
| 0,173902367 | C1orf233 | FALSE | Upregulated |
| -0,009465128 | C1orf234 | FALSE | Downregulated |
| 0,124174296 | C1orf50 | FALSE | Upregulated |
| 1,03671695 | C1orf51 | | Upregulated |
| 0,348666482 | C1orf52 | FALSE | Upregulated |
| -0,369900792 | C1orf53 | FALSE | Downregulated |
| 0,563612891 | C1orf54 | FALSE | Upregulated |
| 0 | C1orf61 | FALSE | Upregulated |
| | C1orf64 | | |
| 0,138867668 | C1orf65 | FALSE | Upregulated |
| 0 | C1orf68 | FALSE | Upregulated |
| 0,964720995 | C1orf74 | FALSE | Upregulated |
| | C1orf87 | | |
| | C1orf94 | | |
| 0,777801429 | C1orf95 | FALSE | Upregulated |
| -0,023741254 | C1QL1 | FALSE | Downregulated |
| -0,008152455 | C1QL2 | FALSE | Downregulated |
| | C1QL4 | | |
| -0,175128388 | C1QTNF1-AS1 | FALSE | Downregulated |
| -0,411163583 | C1QTNF2 | FALSE | Downregulated |
| 0,711938334 | C1QTNF3 | FALSE | Upregulated |
| -1,074146514 | C1QTNF3-AMACR | | Downregulated |
| 0,612413021 | C1QTNF4 | FALSE | Upregulated |
| 0 | C1QTNF8 | FALSE | Upregulated |
| 1,066795985 | C1QTNF9 | | Upregulated |
| 0,850446048 | C1QTNF9B | FALSE | Upregulated |
| 0,257970514 | C1QTNF9B-AS1 | FALSE | Upregulated |
| | C20orf141 | | |
| -0,388302688 | C20orf144 | FALSE | Downregulated |
| 0,806567623 | C20orf166 | FALSE | Upregulated |
| -0,138676142 | C20orf166-AS1 | FALSE | Downregulated |
| 0,347819495 | C20orf173 | FALSE | Upregulated |
| 0,425608064 | C20orf181 | FALSE | Upregulated |
| 0,178037285 | C20orf195 | FALSE | Upregulated |
| -0,131002708 | C20orf196 | FALSE | Downregulated |
| 0,133004165 | C20orf197 | FALSE | Upregulated |
| 0,329127789 | C20orf202 | FALSE | Upregulated |
| 0,157410497 | C20orf203 | FALSE | Upregulated |
| 0,420361146 | C20orf26 | FALSE | Upregulated |
| | C20orf57 | | |
| -0,710740577 | C20orf62 | FALSE | Downregulated |
| | C20orf78 | | |
| 0 | C20orf85 | FALSE | Upregulated |
| 0,273674649 | C20orf96 | FALSE | Upregulated |

| | | | |
|---|---|---|---|
| -0,429482821 | C21orf119 | FALSE | Downregulated |
| 0,266178573 | C21orf128 | FALSE | Upregulated |
| 0,125229948 | C21orf15 | FALSE | Upregulated |
| 0,128712315 | C21orf37 | FALSE | Upregulated |
| 0,040335305 | C21orf49 | FALSE | Upregulated |
| | C21orf54 | | |
| 0,406400264 | C21orf88 | FALSE | Upregulated |
| -0,450614194 | C21orf90 | FALSE | Downregulated |
| | C21orf91-OT1 | | |
| 0,056436988 | C22orf24 | FALSE | Upregulated |
| -0,439035351 | C22orf31 | FALSE | Downregulated |
| 0,38061625 | C22orf34 | FALSE | Upregulated |
| 0,668427838 | C22orf42 | FALSE | Upregulated |
| 0,65555158 | C22orf43 | FALSE | Upregulated |
| 0,258686398 | C2CD4A | FALSE | Upregulated |
| -0,280547951 | C2CD4B | FALSE | Downregulated |
| -0,597198955 | C2CD4C | FALSE | Downregulated |
| -0,060608322 | C2orf27A | FALSE | Downregulated |
| 0 | C2orf27B | FALSE | Upregulated |
| 0,019093442 | C2orf40 | FALSE | Upregulated |
| 0,297685691 | C2orf48 | FALSE | Upregulated |
| 0,291149147 | C2orf50 | FALSE | Upregulated |
| 0 | C2orf53 | FALSE | Upregulated |
| 0,007956344 | C2orf57 | FALSE | Upregulated |
| 0,072025 | C2orf61 | FALSE | Upregulated |
| 0,289289663 | C2orf62 | FALSE | Upregulated |
| 0,56406148 | C2orf66 | FALSE | Upregulated |
| | C2orf70 | | |
| 0,901875161 | C2orf71 | FALSE | Upregulated |
| 0,905646479 | C2orf73 | FALSE | Upregulated |
| -0,631278223 | C2orf74 | FALSE | Downregulated |
| 0,098876477 | C2orf76 | FALSE | Upregulated |
| -0,016120402 | C2orf78 | FALSE | Downregulated |
| -0,004525881 | C2orf80 | FALSE | Downregulated |
| 0,378935534 | C2orf81 | FALSE | Upregulated |
| 0,44836087 | C2orf83 | FALSE | Upregulated |
| 0,244418803 | C2orf91 | FALSE | Upregulated |
| 0,453184695 | C3orf14 | FALSE | Upregulated |
| -0,167886169 | C3orf18 | FALSE | Downregulated |
| -0,284561247 | C3orf20 | FALSE | Downregulated |
| 0,464115025 | C3orf27 | FALSE | Upregulated |
| 0,32520154 | C3orf30 | FALSE | Upregulated |
| -0,412257878 | C3orf33 | FALSE | Downregulated |
| -0,58613754 | C3orf35 | FALSE | Downregulated |
| 0,41104987 | C3orf36 | FALSE | Upregulated |
| 0,394304041 | C3orf43 | FALSE | Upregulated |
| 0,620116739 | C3orf52 | FALSE | Upregulated |
| 0 | C3orf55 | FALSE | Upregulated |
| | C3orf56 | | |
| 1,419160138 | C3orf67 | | Upregulated |
| -0,115352657 | C3orf70 | FALSE | Downregulated |

| | | | |
|---|---|---|---|
| 0,144636958 | C3orf72 | FALSE | Upregulated |
| 0 | C3orf79 | FALSE | Upregulated |
| -0,871361036 | C3orf80 | FALSE | Downregulated |
| 0,344518661 | C3orf84 | FALSE | Upregulated |
| 0,171671505 | C4orf17 | FALSE | Upregulated |
| -0,023897228 | C4orf21 | FALSE | Downregulated |
| -0,13840432 | C4orf22 | FALSE | Downregulated |
| 0,002952063 | C4orf26 | FALSE | Upregulated |
| 0,160683357 | C4orf27 | FALSE | Upregulated |
| -0,807570111 | C4orf32 | FALSE | Downregulated |
| 0,627306335 | C4orf33 | FALSE | Upregulated |
| 0,084310437 | C4orf36 | FALSE | Upregulated |
| 0,55456272 | C4orf40 | FALSE | Upregulated |
| 0,714305144 | C4orf45 | FALSE | Upregulated |
| 0,08738952 | C4orf46 | FALSE | Upregulated |
| 0 | C4orf47 | FALSE | Upregulated |
| 0,430442907 | C4orf48 | FALSE | Upregulated |
| 0,340397219 | C4orf50 | FALSE | Upregulated |
| | C4orf51 | | |
| 0,038235552 | C4orf6 | FALSE | Upregulated |
| 0,104517408 | C5AR2 | FALSE | Upregulated |
| 0,758672539 | C5orf20 | FALSE | Upregulated |
| -0,445713895 | C5orf22 | FALSE | Downregulated |
| -0,114768347 | C5orf28 | FALSE | Downregulated |
| -0,650995413 | C5orf30 | FALSE | Downregulated |
| 0,007176694 | C5orf38 | FALSE | Upregulated |
| | C5orf46 | | |
| 0,28181789 | C5orf47 | FALSE | Upregulated |
| 0,299031549 | C5orf48 | FALSE | Upregulated |
| 0,219788127 | C5orf49 | FALSE | Upregulated |
| 0,153812289 | C5orf52 | FALSE | Upregulated |
| 0,808280607 | C5orf54 | FALSE | Upregulated |
| 0,928979453 | C5orf55 | FALSE | Upregulated |
| 0,975201532 | C5orf58 | FALSE | Upregulated |
| 0,658844502 | C5orf60 | FALSE | Upregulated |
| 0,102001382 | C5orf64 | FALSE | Upregulated |
| 0,285756803 | C6orf10 | FALSE | Upregulated |
| | C6orf100 | | |
| | C6orf118 | | |
| 0,089398486 | C6orf141 | FALSE | Upregulated |
| | C6orf15 | | |
| -0,048082282 | C6orf163 | FALSE | Downregulated |
| -0,55661066 | C6orf164 | FALSE | Downregulated |
| 0,278866095 | C6orf165 | FALSE | Upregulated |
| -0,162149878 | C6orf195 | FALSE | Downregulated |
| -0,646297135 | C6orf203 | FALSE | Downregulated |
| 0 | C6orf222 | FALSE | Upregulated |
| 0,021466064 | C6orf223 | FALSE | Upregulated |
| 0,353386891 | C6orf226 | FALSE | Upregulated |
| -0,06299303 | C6orf229 | FALSE | Downregulated |
| 0 | C6orf52 | FALSE | Upregulated |

| | | | |
|---|---|---|---|
| -0,491229501 | C6orf57 | FALSE | Downregulated |
| 0 | C6orf58 | FALSE | Upregulated |
| 0,74081586 | C6orf7 | FALSE | Upregulated |
| 0,127494885 | C6orf99 | FALSE | Upregulated |
| 0,193596816 | C7orf13 | FALSE | Upregulated |
| 0,562654504 | C7orf33 | FALSE | Upregulated |
| 0,111335017 | C7orf34 | FALSE | Upregulated |
| -0,469416596 | C7orf55 | FALSE | Downregulated |
| -0,199426769 | C7orf57 | FALSE | Downregulated |
| -0,04533092 | C7orf60 | FALSE | Downregulated |
| -0,152606291 | C7orf61 | FALSE | Downregulated |
| 0,146669717 | C7orf62 | FALSE | Upregulated |
| -0,086179849 | C7orf63 | FALSE | Downregulated |
| 0 | C7orf65 | FALSE | Upregulated |
| | C7orf66 | | |
| 0,611634665 | C7orf69 | FALSE | Upregulated |
| | C7orf72 | | |
| 0,105440991 | C7orf73 | FALSE | Upregulated |
| -0,37014899 | C7orf76 | FALSE | Downregulated |
| 0 | C8orf12 | FALSE | Upregulated |
| 0,277119458 | C8orf22 | FALSE | Upregulated |
| -0,334754218 | C8orf31 | FALSE | Downregulated |
| | C8orf34 | | |
| 0,478623434 | C8orf37 | FALSE | Upregulated |
| -0,017172163 | C8orf44 | FALSE | Downregulated |
| 0,036514666 | C8orf44-SGK3 | FALSE | Upregulated |
| -0,136344409 | C8orf48 | FALSE | Downregulated |
| 0,364624557 | C8orf49 | FALSE | Upregulated |
| -0,056176412 | C8orf58 | FALSE | Downregulated |
| -0,018149848 | C8orf69 | FALSE | Downregulated |
| -0,009214617 | C8orf74 | FALSE | Downregulated |
| -0,005114237 | C8orf76 | FALSE | Downregulated |
| 0 | C8orf86 | FALSE | Upregulated |
| | C8orf87 | | |
| 0 | C8orf88 | FALSE | Upregulated |
| 0,436159801 | C9orf106 | FALSE | Upregulated |
| 0,343886021 | C9orf117 | FALSE | Upregulated |
| 0,413796295 | C9orf129 | FALSE | Upregulated |
| 0 | C9orf135 | FALSE | Upregulated |
| | C9orf135-AS1 | | |
| -0,161470652 | C9orf152 | FALSE | Downregulated |
| -0,004133033 | C9orf153 | FALSE | Downregulated |
| -1,062406383 | C9orf169 | | Downregulated |
| -0,196366258 | C9orf170 | FALSE | Downregulated |
| 0,982852036 | C9orf171 | FALSE | Upregulated |
| -0,321353818 | C9orf24 | FALSE | Downregulated |
| -0,230634094 | C9orf40 | FALSE | Downregulated |
| 0,651409192 | C9orf43 | FALSE | Upregulated |
| 0,031476799 | C9orf47 | FALSE | Upregulated |
| 0,875702226 | C9orf50 | FALSE | Upregulated |
| 0,438204542 | C9orf53 | FALSE | Upregulated |

| | | | |
|---|---|---|---|
| 0,227000233 | C9orf57 | FALSE | Upregulated |
| | C9orf62 | | |
| -0,265324916 | C9orf85 | FALSE | Downregulated |
| -0,230951853 | C9orf9 | FALSE | Downregulated |
| | C9orf92 | | |
| 0,274943819 | CA1 | FALSE | Upregulated |
| 0,575800853 | CA10 | FALSE | Upregulated |
| -0,273486894 | CA11 | FALSE | Downregulated |
| -0,237294819 | CA12 | FALSE | Downregulated |
| 0,029419996 | CA13 | FALSE | Upregulated |
| 0,214819376 | CA3 | FALSE | Upregulated |
| 0 | CA4 | FALSE | Upregulated |
| -0,365574648 | CA5B | FALSE | Downregulated |
| 1,016703901 | CA5BP1 | | Upregulated |
| | CA6 | | |
| 1,039073759 | CA7 | | Upregulated |
| 0,308139496 | CA8 | FALSE | Upregulated |
| -0,121684151 | CA9 | FALSE | Downregulated |
| 0,3026975 | CAB39L | FALSE | Upregulated |
| 0,415293907 | CABP1 | FALSE | Upregulated |
| | CABP2 | | |
| | CABP5 | | |
| 0 | CABS1 | FALSE | Upregulated |
| 0,151898511 | CACNA1B | FALSE | Upregulated |
| 0,90689264 | CACNA1C-AS1 | FALSE | Upregulated |
| 0,800791968 | CACNA1C-AS4 | FALSE | Upregulated |
| 0 | CACNA1C-IT3 | FALSE | Upregulated |
| 1,240732991 | CACNA1E | | Upregulated |
| 0,021107112 | CACNA1F | FALSE | Upregulated |
| 0,867004619 | CACNA1G | FALSE | Upregulated |
| 0,211899352 | CACNA1G-AS1 | FALSE | Upregulated |
| 0,954015586 | CACNA1I | FALSE | Upregulated |
| 0,898654502 | CACNA1S | FALSE | Upregulated |
| 1,029115081 | CACNA2D1 | | Upregulated |
| 0,029253589 | CACNA2D3 | FALSE | Upregulated |
| -0,009226926 | CACNA2D3-AS1 | FALSE | Downregulated |
| 0,235430577 | CACNB1 | FALSE | Upregulated |
| -0,140939238 | CACNB4 | FALSE | Downregulated |
| 0,335636186 | CACNG1 | FALSE | Upregulated |
| | CACNG2 | | |
| 0,291340714 | CACNG3 | FALSE | Upregulated |
| 0,04650706 | CACNG4 | FALSE | Upregulated |
| 0 | CACNG5 | FALSE | Upregulated |
| 0,270740663 | CACNG6 | FALSE | Upregulated |
| | CACNG7 | | |
| 0,14458351 | CACNG8 | FALSE | Upregulated |
| -0,388763726 | CADM2 | FALSE | Downregulated |
| -0,472535238 | CADM2-AS1 | FALSE | Downregulated |
| 0 | CADM2-AS2 | FALSE | Upregulated |
| 0,171036965 | CADM3 | FALSE | Upregulated |
| -0,83218325 | CADPS | FALSE | Downregulated |

| | | | |
|---|---|---|---|
| 0,698929323 | CAGE1 | FALSE | Upregulated |
| -0,413598459 | CAHM | FALSE | Downregulated |
| 0,242840641 | CALB2 | FALSE | Upregulated |
| -0,020006306 | CALCA | FALSE | Downregulated |
| 0,310048341 | CALCB | FALSE | Upregulated |
| 0,437155367 | CALCR | FALSE | Upregulated |
| -0,719223912 | CALHM1 | FALSE | Downregulated |
| 0 | CALHM3 | FALSE | Upregulated |
| 0,261675802 | CALML3 | FALSE | Upregulated |
| 0 | CALML5 | FALSE | Upregulated |
| -0,186922088 | CALML6 | FALSE | Downregulated |
| -0,510095821 | CALN1 | FALSE | Downregulated |
| -0,011481061 | CALR3 | FALSE | Downregulated |
| 0,132098867 | CALY | FALSE | Upregulated |
| 0,860165134 | CAMK1G | FALSE | Upregulated |
| 0,402404834 | CAMK2A | FALSE | Upregulated |
| 0,062322807 | CAMK2N2 | FALSE | Upregulated |
| 0,416033067 | CAMK4 | FALSE | Upregulated |
| 0,403803414 | CAMKK1 | FALSE | Upregulated |
| -0,425656602 | CAMKMT | FALSE | Downregulated |
| 0 | CAMKV | FALSE | Upregulated |
| | CAMP | | |
| 0,152849933 | CAND1.11 | FALSE | Upregulated |
| 1,62254348 | CAP2 | Upregulated | |
| 0,206318948 | CAPG | FALSE | Upregulated |
| 0,001249705 | CAPN11 | FALSE | Upregulated |
| 0,166336938 | CAPN13 | FALSE | Upregulated |
| 0,009615426 | CAPN14 | FALSE | Upregulated |
| -0,043800725 | CAPN6 | FALSE | Downregulated |
| 0 | CAPN8 | FALSE | Upregulated |
| -0,150550238 | CAPN8-2 | FALSE | Downregulated |
| 0,299999322 | CAPN9 | FALSE | Upregulated |
| 0,328165839 | CAPNS2 | FALSE | Upregulated |
| -0,387019081 | CAPS2 | FALSE | Downregulated |
| | CAPSL | | |
| 0,425639587 | CAPZA3 | FALSE | Upregulated |
| 0,630427196 | CARD16 | FALSE | Upregulated |
| | CARD17 | | |
| | CARD18 | | |
| | CARTPT | | |
| 0,430297976 | CASC1 | FALSE | Upregulated |
| | CASC11 | | |
| 0,671166864 | CASC15 | FALSE | Upregulated |
| 0 | CASC16 | FALSE | Upregulated |
| | CASC17 | | |
| -0,081286303 | CASC18 | FALSE | Downregulated |
| -0,203771248 | CASC2 | FALSE | Downregulated |
| 0,113020858 | CASC20 | FALSE | Upregulated |
| 0,122535246 | CASC5 | FALSE | Upregulated |
| | CASC6 | | |
| 0,306032473 | CASC8 | FALSE | Upregulated |

| | | | |
|---|---|---|---|
| 0,45265829 | CASC9 | FALSE | Upregulated |
| 0 | CASP12 | FALSE | Upregulated |
| 0 | CASP14 | FALSE | Upregulated |
| -0,345551809 | CASP16 | FALSE | Downregulated |
| 0,447234208 | CASP5 | FALSE | Upregulated |
| 0,215521844 | CASQ1 | FALSE | Upregulated |
| 0,093101912 | CASQ2 | FALSE | Upregulated |
| -0,676452401 | CASR | FALSE | Downregulated |
| 0,302353525 | CASS4 | FALSE | Upregulated |
| -0,018106104 | CATSPER1 | FALSE | Downregulated |
| -0,122695986 | CATSPER2 | FALSE | Downregulated |
| 0,021428474 | CATSPER2P1 | FALSE | Upregulated |
| -0,696577396 | CATSPER3 | FALSE | Downregulated |
| | CATSPER4 | | |
| -0,055167508 | CATSPERB | FALSE | Downregulated |
| | CATSPERD | | |
| 0,915702203 | CATSPERG | FALSE | Upregulated |
| -0,523338174 | CAV3 | FALSE | Downregulated |
| -0,412267802 | CBLN1 | FALSE | Downregulated |
| -0,001175257 | CBLN2 | FALSE | Downregulated |
| 0,01278372 | CBLN4 | FALSE | Upregulated |
| 1,017922484 | CBR3-AS1 | | Upregulated |
| -0,063774468 | CBWD3 | FALSE | Downregulated |
| -0,202485137 | CBWD6 | FALSE | Downregulated |
| 0,277751966 | CBX2 | FALSE | Upregulated |
| 0,008363314 | CBX3P2 | FALSE | Upregulated |
| 0,041155596 | CBX8 | FALSE | Upregulated |
| 0,975930703 | CCAT2 | FALSE | Upregulated |
| 0,191341744 | CCDC102B | FALSE | Upregulated |
| 0,428266394 | CCDC103 | FALSE | Upregulated |
| | CCDC105 | | |
| 0,198383042 | CCDC108 | FALSE | Upregulated |
| 0,309444926 | CCDC11 | FALSE | Upregulated |
| -0,018411243 | CCDC110 | FALSE | Downregulated |
| 0,140943955 | CCDC112 | FALSE | Upregulated |
| 0,145679141 | CCDC113 | FALSE | Upregulated |
| -0,011834672 | CCDC114 | FALSE | Downregulated |
| 0,153037263 | CCDC116 | FALSE | Upregulated |
| 0,290343743 | CCDC122 | FALSE | Upregulated |
| -0,729726808 | CCDC126 | FALSE | Downregulated |
| -0,250737632 | CCDC127 | FALSE | Downregulated |
| 0,412128303 | CCDC129 | FALSE | Upregulated |
| -0,462973257 | CCDC13 | FALSE | Downregulated |
| -0,318984832 | CCDC13-AS1 | FALSE | Downregulated |
| -0,284845128 | CCDC134 | FALSE | Downregulated |
| 0,100275753 | CCDC135 | FALSE | Upregulated |
| 0,274494161 | CCDC136 | FALSE | Upregulated |
| 0,160005902 | CCDC138 | FALSE | Upregulated |
| | CCDC140 | | |
| 0,027052869 | CCDC141 | FALSE | Upregulated |
| 0,881593558 | CCDC144CP | FALSE | Upregulated |

| | | | |
|---|---|---|---|
| 0 | CCDC144NL | FALSE | Upregulated |
| 0,536156406 | CCDC147 | FALSE | Upregulated |
| -0,362631439 | CCDC147-AS1 | FALSE | Downregulated |
| 0,107451037 | CCDC148 | FALSE | Upregulated |
| 0 | CCDC148-AS1 | FALSE | Upregulated |
| 0,156625511 | CCDC15 | FALSE | Upregulated |
| -0,006377737 | CCDC150 | FALSE | Downregulated |
| 0,132189102 | CCDC151 | FALSE | Upregulated |
| 0,361061114 | CCDC153 | FALSE | Upregulated |
| 0,77070669 | CCDC154 | FALSE | Upregulated |
| -0,015218057 | CCDC155 | FALSE | Downregulated |
| 0 | CCDC160 | FALSE | Upregulated |
| 0,737139423 | CCDC162P | FALSE | Upregulated |
| -0,046613386 | CCDC163P | FALSE | Downregulated |
| 0 | CCDC166 | FALSE | Upregulated |
| 1,206640722 | CCDC168 | | Upregulated |
| | CCDC169 | | |
| | CCDC169-SOHLH2 | | |
| -0,242461981 | CCDC171 | FALSE | Downregulated |
| | CCDC172 | | |
| -0,217442322 | CCDC175 | FALSE | Downregulated |
| 0,419583477 | CCDC177 | FALSE | Upregulated |
| -0,006903421 | CCDC178 | FALSE | Downregulated |
| 0,406343208 | CCDC179 | FALSE | Upregulated |
| 1,013891452 | CCDC180 | | Upregulated |
| -0,012639094 | CCDC181 | FALSE | Downregulated |
| 0,292622021 | CCDC182 | FALSE | Upregulated |
| 0,066678195 | CCDC183 | FALSE | Upregulated |
| 0,440459379 | CCDC19 | FALSE | Upregulated |
| 0,44105305 | CCDC23 | FALSE | Upregulated |
| -0,920981112 | CCDC24 | FALSE | Downregulated |
| -0,124452488 | CCDC27 | FALSE | Downregulated |
| 0,319595624 | CCDC30 | FALSE | Upregulated |
| 1,724066398 | CCDC33 | | Upregulated |
| -1,20122024 | CCDC34 | | Downregulated |
| 0,337240759 | CCDC37 | FALSE | Upregulated |
| 0,18498278 | CCDC38 | FALSE | Upregulated |
| 0,714306577 | CCDC41-AS1 | FALSE | Upregulated |
| 0,30313425 | CCDC42 | FALSE | Upregulated |
| 0,089667543 | CCDC43 | FALSE | Upregulated |
| 0,247481878 | CCDC51 | FALSE | Upregulated |
| -0,044751662 | CCDC53 | FALSE | Downregulated |
| 0,170442446 | CCDC54 | FALSE | Upregulated |
| 0,653235394 | CCDC58 | FALSE | Upregulated |
| -0,378014322 | CCDC59 | FALSE | Downregulated |
| 1,0246641 | CCDC60 | | Upregulated |
| -0,267278389 | CCDC62 | FALSE | Downregulated |
| 0,414308511 | CCDC63 | FALSE | Upregulated |
| -0,054563688 | CCDC64B | FALSE | Downregulated |
| -0,173654658 | CCDC67 | FALSE | Downregulated |
| 0,681115578 | CCDC74A | FALSE | Upregulated |

| | | | |
|---|---|---|---|
| 0,563725465 | CCDC74B | FALSE | Upregulated |
| 0,696065837 | CCDC77 | FALSE | Upregulated |
| -0,006903421 | CCDC79 | FALSE | Downregulated |
| 0,145443924 | CCDC8 | FALSE | Upregulated |
| -0,377057187 | CCDC81 | FALSE | Downregulated |
| 0,652796693 | CCDC83 | FALSE | Upregulated |
| -0,848202507 | CCDC85A | FALSE | Downregulated |
| 0,122839212 | CCDC89 | FALSE | Upregulated |
| 0,109105524 | CCDC96 | FALSE | Upregulated |
| | CCER1 | | |
| 1,476026304 | CCER2 | | Upregulated |
| -0,011481061 | CCIN | FALSE | Downregulated |
| 0,282267047 | CCK | FALSE | Upregulated |
| | CCKAR | | |
| 0,029216814 | CCKBR | FALSE | Upregulated |
| | CCL1 | | |
| 0 | CCL11 | FALSE | Upregulated |
| -0,019083029 | CCL13 | FALSE | Downregulated |
| 0,342516111 | CCL17 | FALSE | Upregulated |
| -0,789639032 | CCL18 | FALSE | Downregulated |
| 0,853014657 | CCL2 | FALSE | Upregulated |
| 0,698818315 | CCL20 | FALSE | Upregulated |
| 0,431300858 | CCL24 | FALSE | Upregulated |
| 0,272802081 | CCL25 | FALSE | Upregulated |
| | CCL26 | | |
| -0,04842672 | CCL27 | FALSE | Downregulated |
| 0,708955508 | CCL28 | FALSE | Upregulated |
| 0,324086879 | CCL3 | FALSE | Upregulated |
| 0,779043012 | CCL4 | FALSE | Upregulated |
| 0,555287717 | CCL4L1 | FALSE | Upregulated |
| | CCL7 | | |
| 0,299902724 | CCL8 | FALSE | Upregulated |
| -0,229135396 | CCM2L | FALSE | Downregulated |
| 0,123628404 | CCNA1 | FALSE | Upregulated |
| 0,191348445 | CCNA2 | FALSE | Upregulated |
| 0,240536745 | CCNB1 | FALSE | Upregulated |
| 0,364097522 | CCNB2 | FALSE | Upregulated |
| -0,034689538 | CCNB3 | FALSE | Downregulated |
| 0,006353158 | CCNE1 | FALSE | Upregulated |
| -0,624853516 | CCNJL | FALSE | Downregulated |
| 0 | CCNO | FALSE | Upregulated |
| | CCNYL2 | | |
| 0,883013507 | CCR2 | FALSE | Upregulated |
| 0,589849544 | CCR3 | FALSE | Upregulated |
| 0,586240526 | CCR4 | FALSE | Upregulated |
| 1,004795586 | CCR5 | | Upregulated |
| 1,037914612 | CCR6 | | Upregulated |
| -0,333644758 | CCRL2 | FALSE | Downregulated |
| -0,30327337 | CCSER1 | FALSE | Downregulated |
| -0,688516511 | CCT6B | FALSE | Downregulated |
| 0,41097888 | CCT8L2 | FALSE | Upregulated |

| | | | |
|---|---|---|---|
| -0,875252067 | CCZ1B | FALSE | Downregulated |
| 0,751037728 | CD101 | FALSE | Upregulated |
| -0,581371461 | CD109 | FALSE | Downregulated |
| -0,002253536 | CD164L2 | FALSE | Downregulated |
| | CD177 | | |
| -0,328266271 | CD180 | FALSE | Downregulated |
| 0,351844477 | CD19 | FALSE | Upregulated |
| 0 | CD1A | FALSE | Upregulated |
| 0,406118821 | CD1B | FALSE | Upregulated |
| -0,682540434 | CD1C | FALSE | Downregulated |
| 0,668459229 | CD1E | FALSE | Upregulated |
| 1,10692439 | CD2 | | Upregulated |
| 0,237187519 | CD200 | FALSE | Upregulated |
| -0,43224465 | CD200R1 | FALSE | Downregulated |
| 0 | CD200R1L | FALSE | Upregulated |
| 1,046998908 | CD207 | | Upregulated |
| 1,075755691 | CD22 | | Upregulated |
| 0,006126921 | CD226 | FALSE | Upregulated |
| -0,103456804 | CD247 | FALSE | Downregulated |
| 1,614443482 | CD28 | | Upregulated |
| 0,262843111 | CD300C | FALSE | Upregulated |
| 1,225265531 | CD300E | | Upregulated |
| 0,470410435 | CD300LB | FALSE | Upregulated |
| | CD300LD | | |
| 0,300220511 | CD300LF | FALSE | Upregulated |
| 0,325347661 | CD300LG | FALSE | Upregulated |
| 0,734469488 | CD33 | FALSE | Upregulated |
| -0,803674337 | CD34 | FALSE | Downregulated |
| -0,288533809 | CD38 | FALSE | Downregulated |
| 0,667833298 | CD3D | FALSE | Upregulated |
| 1,651520128 | CD3G | | Upregulated |
| 0,40228142 | CD40LG | FALSE | Upregulated |
| 1,60892863 | CD52 | | Upregulated |
| -0,165872183 | CD58 | FALSE | Downregulated |
| 0,958185478 | CD69 | FALSE | Upregulated |
| 0,273422532 | CD70 | FALSE | Upregulated |
| 0,54085172 | CD72 | FALSE | Upregulated |
| 1,331044655 | CD79A | | Upregulated |
| -0,467683832 | CD79B | FALSE | Downregulated |
| 0,311953115 | CD83 | FALSE | Upregulated |
| 0,124912974 | CD86 | FALSE | Upregulated |
| 0,235934193 | CD8B | FALSE | Upregulated |
| 0,995345221 | CD96 | FALSE | Upregulated |
| | CD99-2 | | |
| -0,952896197 | CD99P1 | FALSE | Downregulated |
| | CD99P1-2 | | |
| 0,152814379 | CDC14A | FALSE | Upregulated |
| 0,273422532 | CDC14C | FALSE | Upregulated |
| -0,771797822 | CDC20 | FALSE | Downregulated |
| 0,553437768 | CDC20B | FALSE | Upregulated |
| 0,046829056 | CDC25A | FALSE | Upregulated |

| | | | |
|---|---|---|---|
| 0,252914877 | CDC25C | FALSE | Upregulated |
| 0,233316614 | CDC42BPG | FALSE | Upregulated |
| 0,127870113 | CDC42EP2 | FALSE | Upregulated |
| -0,406976536 | CDC42EP5 | FALSE | Downregulated |
| 0,865063961 | CDC45 | FALSE | Upregulated |
| 0,020511698 | CDC6 | FALSE | Upregulated |
| 0,813981026 | CDC7 | FALSE | Upregulated |
| -0,276256895 | CDCA2 | FALSE | Downregulated |
| -0,465538042 | CDCA3 | FALSE | Downregulated |
| 1,014980757 | CDCA4 | | Upregulated |
| 1,120836944 | CDCA7 | | Upregulated |
| 0,447882646 | CDCA8 | FALSE | Upregulated |
| 0 | CDCP2 | FALSE | Upregulated |
| | CDH10 | | |
| 0,152672491 | CDH11 | FALSE | Upregulated |
| 0,031345428 | CDH12 | FALSE | Upregulated |
| -0,491821072 | CDH15 | FALSE | Downregulated |
| -0,015514431 | CDH16 | FALSE | Downregulated |
| -0,277469926 | CDH17 | FALSE | Downregulated |
| | CDH18 | | |
| 0,004968943 | CDH19 | FALSE | Upregulated |
| 0,724774935 | CDH20 | FALSE | Upregulated |
| 0,57335429 | CDH22 | FALSE | Upregulated |
| 0,236247937 | CDH24 | FALSE | Upregulated |
| -0,255017633 | CDH26 | FALSE | Downregulated |
| 0,073076937 | CDH3 | FALSE | Upregulated |
| -0,531811907 | CDH6 | FALSE | Downregulated |
| | CDH7 | | |
| 0,420297526 | CDH8 | FALSE | Upregulated |
| 0 | CDH9 | FALSE | Upregulated |
| 1,135703897 | CDHR1 | | Upregulated |
| 0 | CDHR4 | FALSE | Upregulated |
| | CDIPT-AS1 | | |
| -0,001112068 | CDK1 | FALSE | Downregulated |
| 0,323366384 | CDK15 | FALSE | Upregulated |
| -0,007636623 | CDK20 | FALSE | Downregulated |
| -1,078314439 | CDK3 | | Downregulated |
| -0,365979654 | CDK5 | FALSE | Downregulated |
| -0,53131551 | CDK5R1 | FALSE | Downregulated |
| | CDK5R2 | | |
| -0,376295342 | CDK7 | FALSE | Downregulated |
| 0,262855808 | CDKL2 | FALSE | Upregulated |
| -0,029236549 | CDKL3 | FALSE | Downregulated |
| -0,00509535 | CDKL4 | FALSE | Downregulated |
| 0,593170796 | CDKN2A | FALSE | Upregulated |
| 0,498949678 | CDKN2B-AS1 | FALSE | Upregulated |
| -0,010670157 | CDKN2D | FALSE | Downregulated |
| -0,090038064 | CDKN3 | FALSE | Downregulated |
| -0,539397932 | CDNF | FALSE | Downregulated |
| -0,212302346 | CDR2L | FALSE | Downregulated |
| -0,137146603 | CDRT1 | FALSE | Downregulated |

| | | | |
|---|---|---|---|
| | CDRT15L2 | | |
| | CDRT15P2 | | |
| 0,248743505 | CDRT4 | FALSE | Upregulated |
| | CDRT7 | | |
| | CDRT8 | | |
| -0,034433992 | CDS1 | FALSE | Downregulated |
| -0,024475305 | CDSN | FALSE | Downregulated |
| -0,500825781 | CDX1 | FALSE | Downregulated |
| 1,144820042 | CDX2 | | Upregulated |
| | CDX4 | | |
| | CEACAM18 | | |
| 0,70349772 | CEACAM20 | FALSE | Upregulated |
| 0,626960029 | CEACAM21 | FALSE | Upregulated |
| 0,052921775 | CEACAM22P | FALSE | Upregulated |
| 0,730547553 | CEACAM3 | FALSE | Upregulated |
| 0,119070896 | CEACAM4 | FALSE | Upregulated |
| -0,00370352 | CEACAM5 | FALSE | Downregulated |
| 0,27322222 | CEACAM6 | FALSE | Upregulated |
| | CEACAM7 | | |
| -0,501842021 | CEACAM8 | FALSE | Downregulated |
| | CEBPE | | |
| 0,170056764 | CECR3 | FALSE | Upregulated |
| 0,861131753 | CECR5-AS1 | FALSE | Upregulated |
| 1,322794287 | CECR6 | | Upregulated |
| 0,098888842 | CECR7 | FALSE | Upregulated |
| -0,253637748 | CEL | FALSE | Downregulated |
| | CELA1 | | |
| -0,017171324 | CELA2A | FALSE | Downregulated |
| 0,694710962 | CELA2B | FALSE | Upregulated |
| | CELA3A | | |
| | CELA3B | | |
| 1,787732247 | CELF2-AS2 | | Upregulated |
| -0,426212346 | CELF5 | FALSE | Downregulated |
| 0,404183497 | CELP | FALSE | Upregulated |
| 0,424803996 | CENPA | FALSE | Upregulated |
| 0,426851325 | CENPE | FALSE | Upregulated |
| -0,306858598 | CENPF | FALSE | Downregulated |
| -0,247675552 | CENPH | FALSE | Downregulated |
| -0,148049244 | CENPI | FALSE | Downregulated |
| 0,020278171 | CENPK | FALSE | Upregulated |
| -0,489507852 | CENPL | FALSE | Downregulated |
| 0,599360526 | CENPM | FALSE | Upregulated |
| 0,071539871 | CENPQ | FALSE | Upregulated |
| | CENPVP1 | | |
| | CENPVP2 | | |
| 0,703550694 | CENPW | FALSE | Upregulated |
| -0,235770555 | CEP112 | FALSE | Downregulated |
| 0,079510404 | CEP128 | FALSE | Upregulated |
| -0,334804295 | CEP135 | FALSE | Downregulated |
| 0,139301056 | CEP170P1 | FALSE | Upregulated |
| -0,327768342 | CEP19 | FALSE | Downregulated |

| | | | |
|---|---|---|---|
| -0,285896077 | CEP41 | FALSE | Downregulated |
| 0,564605054 | CEP55 | FALSE | Upregulated |
| -0,312848023 | CEP57L1 | FALSE | Downregulated |
| 0,474553366 | CEP78 | FALSE | Upregulated |
| 0,584425649 | CEP89 | FALSE | Upregulated |
| 0,276258154 | CEP97 | FALSE | Upregulated |
| | CER1 | | |
| 0,38164863 | CERCAM | FALSE | Upregulated |
| 0,280928672 | CES1P2 | FALSE | Upregulated |
| -0,377681024 | CES5A | FALSE | Downregulated |
| | CES5AP1 | | |
| | CETN1 | | |
| -1,158130024 | CETN2 | | Downregulated |
| 0,070806729 | CETN3 | FALSE | Upregulated |
| 0 | CETN4P | FALSE | Upregulated |
| | CFC1 | | |
| | CFC1B | | |
| 0,274635952 | CFL1P1 | FALSE | Upregulated |
| | CGA | | |
| 0,038216899 | CGB | FALSE | Upregulated |
| | CGB1 | | |
| 0,284731019 | CGB2 | FALSE | Upregulated |
| 0,134806214 | CGB5 | FALSE | Upregulated |
| 1,261748645 | CGB7 | | Upregulated |
| | CGB8 | | |
| 0,223334284 | CGRRF1 | FALSE | Upregulated |
| 0,99024964 | CH25H | FALSE | Upregulated |
| -0,198549649 | CHAC1 | FALSE | Downregulated |
| 0,40795493 | CHAF1B | FALSE | Upregulated |
| | CHAT | | |
| 0,252038941 | CHCHD1 | FALSE | Upregulated |
| -0,469798859 | CHCHD4 | FALSE | Downregulated |
| 0,557461209 | CHCHD5 | FALSE | Upregulated |
| 0,533533949 | CHCHD6 | FALSE | Upregulated |
| 0,131997964 | CHCHD7 | FALSE | Upregulated |
| 0,429614377 | CHD5 | FALSE | Upregulated |
| 0,141209973 | CHDC2 | FALSE | Upregulated |
| 0,225767933 | CHEK2 | FALSE | Upregulated |
| | CHEK2P2 | | |
| 0 | CHGB | FALSE | Upregulated |
| 0,430014684 | CHI3L2 | FALSE | Upregulated |
| 0,293242527 | CHIA | FALSE | Upregulated |
| 0,272277655 | CHIAP2 | FALSE | Upregulated |
| 0,322454314 | CHIC1 | FALSE | Upregulated |
| 0,365401219 | CHIT1 | FALSE | Upregulated |
| -0,027013655 | CHKB-AS1 | FALSE | Downregulated |
| -1,387629453 | CHKB-CPT1B | | Downregulated |
| -1,141761226 | CHL1 | | Downregulated |
| 0,728184803 | CHMP1B2P | FALSE | Upregulated |
| -0,348926909 | CHMP4C | FALSE | Downregulated |
| -0,020856854 | CHN1 | FALSE | Downregulated |

| | | | |
|---|---|---|---|
| 0,290109866 | CHODL | FALSE | Upregulated |
| | CHODL-AS1 | | |
| 0,009608601 | CHP2 | FALSE | Upregulated |
| -1,094708298 | CHRDL1 | | Downregulated |
| 1,005843155 | CHRDL2 | | Upregulated |
| 0,065595332 | CHRFAM7A | FALSE | Upregulated |
| -0,924790453 | CHRM1 | FALSE | Downregulated |
| 0,621475391 | CHRM2 | FALSE | Upregulated |
| 0,290185236 | CHRM3-AS1 | FALSE | Upregulated |
| 0,869848786 | CHRM3-AS2 | FALSE | Upregulated |
| -0,021259612 | CHRM4 | FALSE | Downregulated |
| 0,259644232 | CHRNA1 | FALSE | Upregulated |
| 0,487804666 | CHRNA10 | FALSE | Upregulated |
| 0,383805234 | CHRNA3 | FALSE | Upregulated |
| 0,214348423 | CHRNA5 | FALSE | Upregulated |
| -0,148465128 | CHRNA6 | FALSE | Downregulated |
| -0,16122224 | CHRNA7 | FALSE | Downregulated |
| -0,182244836 | CHRNA9 | FALSE | Downregulated |
| | CHRNB3 | | |
| 0 | CHRNB4 | FALSE | Upregulated |
| 0,924875754 | CHRND | FALSE | Upregulated |
| 0,20013943 | CHST1 | FALSE | Upregulated |
| -0,13034401 | CHST10 | FALSE | Downregulated |
| -0,011565065 | CHST5 | FALSE | Downregulated |
| -0,298257558 | CHST6 | FALSE | Downregulated |
| -0,016345128 | CHST8 | FALSE | Downregulated |
| -0,062837399 | CIB2 | FALSE | Downregulated |
| | CIB3 | | |
| 0,293242527 | CIB4 | FALSE | Upregulated |
| 0,017296193 | CIDEA | FALSE | Upregulated |
| 0,379656111 | CIDEC | FALSE | Upregulated |
| 0,291478389 | CIDECP | FALSE | Upregulated |
| 0,410706327 | CILP | FALSE | Upregulated |
| 0,563405124 | CILP2 | FALSE | Upregulated |
| 0,237321226 | CIRH1A | FALSE | Upregulated |
| | CISTR | | |
| 0,007914223 | CIT | FALSE | Upregulated |
| 0,006077282 | CITED1 | FALSE | Upregulated |
| 0,018304097 | CKAP2 | FALSE | Upregulated |
| 0,841346245 | CKAP2L | FALSE | Upregulated |
| 0,490814582 | CKLF | FALSE | Upregulated |
| 0,286454283 | CKLF-CMTM1 | FALSE | Upregulated |
| 0,152690177 | CKMT1A | FALSE | Upregulated |
| -0,142904312 | CKMT1B | FALSE | Downregulated |
| 0,027159274 | CKS2 | FALSE | Upregulated |
| 0,005143381 | CLC | FALSE | Upregulated |
| 0,303090353 | CLCA1 | FALSE | Upregulated |
| 0 | CLCA2 | FALSE | Upregulated |
| 0,18392589 | CLCA3P | FALSE | Upregulated |
| -0,128871792 | CLCA4 | FALSE | Downregulated |
| 0,65295768 | CLCN1 | FALSE | Upregulated |

| | | | |
|---|---|---|---|
| 0,04587931 | CLCNKA | FALSE | Upregulated |
| 0 | CLCNKB | FALSE | Upregulated |
| | CLDN10-AS1 | | |
| 0,55578013 | CLDN11 | FALSE | Upregulated |
| -0,19291834 | CLDN16 | FALSE | Downregulated |
| | CLDN17 | | |
| -0,151980786 | CLDN19 | FALSE | Downregulated |
| -0,022877478 | CLDN25 | FALSE | Downregulated |
| -0,33197835 | CLDN4 | FALSE | Downregulated |
| -0,260947901 | CLDN6 | FALSE | Downregulated |
| | CLDN8 | | |
| -0,335107656 | CLDN9 | FALSE | Downregulated |
| 0,968768748 | CLDND2 | FALSE | Upregulated |
| 0,61901609 | CLEC11A | FALSE | Upregulated |
| 0,303733493 | CLEC12B | FALSE | Upregulated |
| 1,255594701 | CLEC17A | | Upregulated |
| 0,635937869 | CLEC18A | FALSE | Upregulated |
| 0,450032502 | CLEC18B | FALSE | Upregulated |
| -0,722625828 | CLEC18C | FALSE | Downregulated |
| | CLEC19A | | |
| 0,154831754 | CLEC1A | FALSE | Upregulated |
| 0 | CLEC2A | FALSE | Upregulated |
| | CLEC2L | | |
| 0,277119458 | CLEC3A | FALSE | Upregulated |
| 0,195976276 | CLEC4A | FALSE | Upregulated |
| 0,083497784 | CLEC4C | FALSE | Upregulated |
| 0,546577418 | CLEC4D | FALSE | Upregulated |
| -0,094848055 | CLEC4E | FALSE | Downregulated |
| -0,748770514 | CLEC4F | FALSE | Downregulated |
| -1,099200416 | CLEC4GP1 | | Downregulated |
| -0,278749103 | CLEC5A | FALSE | Downregulated |
| 0 | CLEC6A | FALSE | Upregulated |
| -0,448749326 | CLEC7A | FALSE | Downregulated |
| -0,122707256 | CLEC9A | FALSE | Downregulated |
| 0,980076823 | CLECL1 | FALSE | Upregulated |
| -0,731094918 | CLGN | FALSE | Downregulated |
| -0,319554737 | CLHC1 | FALSE | Downregulated |
| 0,537117257 | CLIC2 | FALSE | Upregulated |
| -0,681996089 | CLIC3 | FALSE | Downregulated |
| 0,5654367 | CLIC5 | FALSE | Upregulated |
| -0,245628296 | CLIC6 | FALSE | Downregulated |
| | CLK2P | | |
| -0,006734809 | CLLU1 | FALSE | Downregulated |
| | CLLU1OS | | |
| 0,609696761 | CLNK | FALSE | Upregulated |
| -0,106035243 | CLP1 | FALSE | Downregulated |
| | CLPS | | |
| | CLPSL1 | | |
| | CLPSL2 | | |
| 0,080191917 | CLRN1-AS1 | FALSE | Upregulated |
| -0,117124783 | CLRN2 | FALSE | Downregulated |

| | | | |
|---|---|---|---|
| 1,125721383 | CLSPN | | Upregulated |
| -0,027404501 | CLSTN2-AS1 | FALSE | Downregulated |
| 0,625336143 | CLUL1 | FALSE | Upregulated |
| 0,47501698 | CLVS2 | FALSE | Upregulated |
| 0,164893961 | CMA1 | FALSE | Upregulated |
| 0,055141121 | CMC1 | FALSE | Upregulated |
| -0,04396371 | CMC4 | FALSE | Downregulated |
| 0,250326968 | CMSS1 | FALSE | Upregulated |
| 0,837835985 | CMTM1 | FALSE | Upregulated |
| -0,032467342 | CMTM2 | FALSE | Downregulated |
| 0,547692162 | CMTM5 | FALSE | Upregulated |
| 0,210875724 | CMTM7 | FALSE | Upregulated |
| | CNBD1 | | |
| 0,411415187 | CNBD2 | FALSE | Upregulated |
| 0,171936181 | CNEP1R1 | FALSE | Upregulated |
| 0 | CNFN | FALSE | Upregulated |
| | CNGA2 | | |
| | CNGA3 | | |
| -0,48611523 | CNGA4 | FALSE | Downregulated |
| | CNGB1 | | |
| -0,363400182 | CNGB3 | FALSE | Downregulated |
| 1,04307658 | CNIH2 | | Upregulated |
| -0,168481899 | CNIH3 | FALSE | Downregulated |
| -0,022532016 | CNKSR1 | FALSE | Downregulated |
| -0,258484029 | CNKSR2 | FALSE | Downregulated |
| -0,260518153 | CNN1 | FALSE | Downregulated |
| 0 | CNPY1 | FALSE | Upregulated |
| 0,55140223 | CNR1 | FALSE | Upregulated |
| 0,740010012 | CNR2 | FALSE | Upregulated |
| 0,533370129 | CNTD2 | FALSE | Upregulated |
| 0,119439873 | CNTF | FALSE | Upregulated |
| -0,890371776 | CNTN1 | FALSE | Downregulated |
| 0,35660477 | CNTN4 | FALSE | Upregulated |
| 1,020190134 | CNTN4-AS2 | | Upregulated |
| 0 | CNTN5 | FALSE | Upregulated |
| -0,127486457 | CNTN6 | FALSE | Downregulated |
| -0,013306265 | CNTNAP3 | FALSE | Downregulated |
| | CNTNAP5 | | |
| -0,722576631 | COA6 | FALSE | Downregulated |
| 0,24334196 | COL11A1 | FALSE | Upregulated |
| 0,043192659 | COL13A1 | FALSE | Upregulated |
| 0,892109056 | COL15A1 | FALSE | Upregulated |
| -0,141970251 | COL18A1-AS2 | FALSE | Downregulated |
| 1,633805466 | COL19A1 | | Upregulated |
| -0,571369004 | COL20A1 | FALSE | Downregulated |
| -0,679979812 | COL21A1 | FALSE | Downregulated |
| 0,48312133 | COL22A1 | FALSE | Upregulated |
| 0,262888677 | COL24A1 | FALSE | Upregulated |
| 0,079467285 | COL28A1 | FALSE | Upregulated |
| 0,445679212 | COL2A1 | FALSE | Upregulated |
| 0,379786953 | COL4A2-AS1 | FALSE | Upregulated |

| | | | |
|---|---|---|---|
| 0,310409666 | COL4A6 | FALSE | Upregulated |
| | COL6A4P1 | | |
| 0,04387623 | COL6A4P2 | FALSE | Upregulated |
| 0,291391126 | COL6A5 | FALSE | Upregulated |
| 0,449519902 | COL9A1 | FALSE | Upregulated |
| 0,342360709 | COL9A2 | FALSE | Upregulated |
| 0,845390271 | COL9A3 | FALSE | Upregulated |
| -0,952803398 | COLCA1 | FALSE | Downregulated |
| -0,844640132 | COLCA2 | FALSE | Downregulated |
| 0,578712418 | COLEC12 | FALSE | Upregulated |
| 0,742082248 | COLGALT2 | FALSE | Upregulated |
| -0,302360816 | COMMD1 | FALSE | Downregulated |
| 0,6082857 | COMMD10 | FALSE | Upregulated |
| 0,611326261 | COMMD3-BMI1 | FALSE | Upregulated |
| -0,337338805 | COMMD6 | FALSE | Downregulated |
| 0,265170952 | COMMD8 | FALSE | Upregulated |
| 0,247485509 | COMP | FALSE | Upregulated |
| 0,100233676 | COMTD1 | FALSE | Upregulated |
| 0,241822363 | COPS4 | FALSE | Upregulated |
| 0,111949954 | COPZ2 | FALSE | Upregulated |
| 0,021006485 | COQ2 | FALSE | Upregulated |
| -0,111182701 | COQ3 | FALSE | Downregulated |
| 0,315407508 | CORO2B | FALSE | Upregulated |
| 1,272721696 | CORO6 | | Upregulated |
| -0,573229022 | COX17 | FALSE | Downregulated |
| -0,17117495 | COX4I2 | FALSE | Downregulated |
| -0,223093396 | COX6A2 | FALSE | Downregulated |
| -0,21816562 | COX6B2 | FALSE | Downregulated |
| | COX7B2 | | |
| -0,018149848 | COX8C | FALSE | Downregulated |
| 0,279576795 | CPA1 | FALSE | Upregulated |
| | CPA2 | | |
| 0,501955774 | CPA3 | FALSE | Upregulated |
| 0,192185989 | CPA4 | FALSE | Upregulated |
| -0,001838095 | CPA5 | FALSE | Downregulated |
| | CPA6 | | |
| 0,26864001 | CPB1 | FALSE | Upregulated |
| 1,375297124 | CPEB1 | | Upregulated |
| -0,817305708 | CPEB2-AS1 | FALSE | Downregulated |
| 0,292899904 | CPLX3 | FALSE | Upregulated |
| | CPLX4 | | |
| -0,309049341 | CPNE4 | FALSE | Downregulated |
| -0,122803949 | CPNE5 | FALSE | Downregulated |
| -0,281673067 | CPNE6 | FALSE | Downregulated |
| -0,404180648 | CPNE7 | FALSE | Downregulated |
| -0,458987616 | CPNE9 | FALSE | Downregulated |
| 0 | CPO | FALSE | Upregulated |
| -0,169495171 | CPSF4L | FALSE | Downregulated |
| -0,17906522 | CPT1C | FALSE | Downregulated |
| | CPXCR1 | | |
| 0,223877409 | CPXM1 | FALSE | Upregulated |

| | | | |
|---|---|---|---|
| -0,322831187 | CPXM2 | FALSE | Downregulated |
| 0,683506233 | CPZ | FALSE | Upregulated |
| 0,245522452 | CR1L | FALSE | Upregulated |
| 0,175116701 | CR2 | FALSE | Upregulated |
| | CRABP1 | | |
| -0,653547318 | CRABP2 | FALSE | Downregulated |
| 0,439488698 | CRB1 | FALSE | Upregulated |
| | CRCT1 | | |
| 1,107460397 | CREB3L1 | | Upregulated |
| 1,013778985 | CREB5 | | Upregulated |
| 0,33130616 | CREG2 | FALSE | Upregulated |
| -0,323030029 | CRH | FALSE | Downregulated |
| | CRHR1 | | |
| 0,306018682 | CRHR1-IT1 | FALSE | Upregulated |
| 0,43404694 | CRHR2 | FALSE | Upregulated |
| 0,669806372 | CRIP1 | FALSE | Upregulated |
| -0,010130287 | CRISP1 | FALSE | Downregulated |
| -0,166531867 | CRISP2 | FALSE | Downregulated |
| 0,139549707 | CRISP3 | FALSE | Upregulated |
| -1,009030554 | CRISPLD1 | | Downregulated |
| 0,55177674 | CRLF1 | FALSE | Upregulated |
| | CRLF2 | | |
| | CRLF2-2 | | |
| 0,120413435 | CRNDE | FALSE | Upregulated |
| 0,258820331 | CRNN | FALSE | Upregulated |
| 0 | CRSP8P | FALSE | Upregulated |
| 0,315087962 | CRTAC1 | FALSE | Upregulated |
| 0,627624284 | CRTAM | FALSE | Upregulated |
| 0,618662509 | CRYAB | FALSE | Upregulated |
| | CRYBA2 | | |
| | CRYBA4 | | |
| 0,265374564 | CRYBB1 | FALSE | Upregulated |
| -0,294217936 | CRYBB2 | FALSE | Downregulated |
| 0,140675837 | CRYBB2P1 | FALSE | Upregulated |
| -0,277595923 | CRYBB3 | FALSE | Downregulated |
| | CRYGA | | |
| | CRYGB | | |
| | CRYGC | | |
| | CRYGD | | |
| -0,01330656 | CRYGN | FALSE | Downregulated |
| 0,415016824 | CRYGS | FALSE | Upregulated |
| -0,021848506 | CRYM-AS1 | FALSE | Downregulated |
| -0,355643691 | CRYZL1 | FALSE | Downregulated |
| | CSAG1 | | |
| | CSAG3 | | |
| | CSAG4 | | |
| -0,218273399 | CSF2 | FALSE | Downregulated |
| 0,49398021 | CSF2RA | FALSE | Upregulated |
| | CSF2RA-2 | | |
| 0,828055324 | CSF3 | FALSE | Upregulated |
| | CSH1 | | |

| | | | |
|---|---|---|---|
| | CSH2 | | |
| | CSHL1 | | |
| 0,318771777 | CSMD1 | FALSE | Upregulated |
| -0,086364066 | CSMD2 | FALSE | Downregulated |
| -0,240631287 | CSMD3 | FALSE | Downregulated |
| | CSN1S1 | | |
| | CSN1S2AP | | |
| | CSN1S2BP | | |
| | CSN2 | | |
| | CSN3 | | |
| 0,129330176 | CSNK1A1L | FALSE | Upregulated |
| 0,110803923 | CSNK2A3 | FALSE | Upregulated |
| 0,267553199 | CSPG5 | FALSE | Upregulated |
| 0,030426557 | CSRNP3 | FALSE | Upregulated |
| -0,537176048 | CSRP3 | FALSE | Downregulated |
| | CST1 | | |
| 0 | CST11 | FALSE | Upregulated |
| | CST13P | | |
| | CST2 | | |
| | CST4 | | |
| | CST5 | | |
| -0,000404566 | CST6 | FALSE | Downregulated |
| | CST8 | | |
| | CST9 | | |
| | CST9L | | |
| 0,341623339 | CSTA | FALSE | Upregulated |
| | CSTF3-AS1 | | |
| | CSTL1 | | |
| | CT45A1 | | |
| | CT45A2 | | |
| | CT45A3 | | |
| | CT45A5 | | |
| | CT45A6 | | |
| | CT47A1 | | |
| | CT47A10 | | |
| | CT47A11 | | |
| | CT47A12 | | |
| | CT47A2 | | |
| | CT47A3 | | |
| | CT47A4 | | |
| | CT47A5 | | |
| 0 | CT47A6 | FALSE | Upregulated |
| 0,296723345 | CT47A7 | FALSE | Upregulated |
| | CT47A8 | | |
| | CT47A9 | | |
| 0,336655484 | CT47B1 | FALSE | Upregulated |
| -1,315818721 | CT55 | | Downregulated |
| -1,089201005 | CT62 | | Downregulated |
| | CT83 | | |
| | CTAG1A | | |
| | CTAG1B | | |

| | | | |
|---|---|---|---|
| 0 | CTAG2 | FALSE | Upregulated |
| 0,056093729 | CTAGE1 | FALSE | Upregulated |
| 0,124156023 | CTAGE10P | FALSE | Upregulated |
| -0,182297911 | CTAGE11P | FALSE | Downregulated |
| 0,690672341 | CTAGE15 | FALSE | Upregulated |
| 0,646419666 | CTAGE6 | FALSE | Upregulated |
| | CTB-12O2.1 | | |
| 0,417323476 | CTB-174D11.1 | FALSE | Upregulated |
| 0,054859118 | CTB-178M22.2 | FALSE | Upregulated |
| -0,063138345 | CTC-436P18.1 | FALSE | Downregulated |
| 0,283869539 | CTC-558O2.1 | FALSE | Upregulated |
| 0,480329563 | CTCFL | FALSE | Upregulated |
| | CTD-2118P12.1 | | |
| 0 | CTD-2127H9.1 | FALSE | Upregulated |
| | CTD-2194D22.4 | | |
| -0,802993975 | CTD-2201I18.1 | FALSE | Downregulated |
| -0,141946699 | CTD-2258A20.5 | FALSE | Downregulated |
| -0,026275106 | CTD-2270F17.1 | FALSE | Downregulated |
| | CTD-2297D10.2 | | |
| | CTD-2337A12.1 | | |
| 0,134666007 | CTD-2350J17.1 | FALSE | Upregulated |
| -0,353822354 | CTF1 | FALSE | Downregulated |
| 0,223706745 | CTGLF12P | FALSE | Upregulated |
| 0,97949936 | CTLA4 | FALSE | Upregulated |
| 0,005042781 | CTNNA2 | FALSE | Upregulated |
| -0,793767577 | CTNND2 | FALSE | Downregulated |
| 0,147699456 | CTRB1 | FALSE | Upregulated |
| | CTRB2 | | |
| 0,018839898 | CTRC | FALSE | Upregulated |
| -0,03138686 | CTSE | FALSE | Downregulated |
| -0,741317376 | CTSG | FALSE | Downregulated |
| 0,39490937 | CTSK | FALSE | Upregulated |
| | CTSL3P | | |
| 0,382670099 | CTSLP2 | FALSE | Upregulated |
| | CTSLP8 | | |
| | CTSV | | |
| -0,15350309 | CTSW | FALSE | Downregulated |
| -0,558512198 | CTU1 | FALSE | Downregulated |
| 0 | CTXN1 | FALSE | Upregulated |
| 0,905983486 | CTXN2 | FALSE | Upregulated |
| | CTXN3 | | |
| 1,082780927 | CUBN | | Upregulated |
| 0,270408815 | CUZD1 | FALSE | Upregulated |
| 0,702715905 | CWH43 | FALSE | Upregulated |
| 0,199555944 | CX3CR1 | FALSE | Upregulated |
| 0,199154073 | CXADRP2 | FALSE | Upregulated |
| -0,260955433 | CXADRP3 | FALSE | Downregulated |
| 0,180052763 | CXCL1 | FALSE | Upregulated |
| 1,629468334 | CXCL11 | | Upregulated |
| 0,172009168 | CXCL13 | FALSE | Upregulated |
| -0,249879027 | CXCL17 | FALSE | Downregulated |

| | | | |
|---|---|---|---|
| 0,146902076 | CXCL3 | FALSE | Upregulated |
| 0,539424444 | CXCL5 | FALSE | Upregulated |
| 0,551110314 | CXCL6 | FALSE | Upregulated |
| -0,314060526 | CXCR1 | FALSE | Downregulated |
| -0,357276553 | CXCR2 | FALSE | Downregulated |
| 1,050874989 | CXCR4 | | Upregulated |
| 1,04455882 | CXorf21 | | Upregulated |
| -0,003363871 | CXorf22 | FALSE | Downregulated |
| | CXorf27 | | |
| | CXorf28 | | |
| | CXorf30 | | |
| 0,4167458 | CXorf31 | FALSE | Upregulated |
| 0,568844915 | CXorf36 | FALSE | Upregulated |
| | CXorf49 | | |
| | CXorf49B | | |
| | CXorf51A | | |
| | CXorf51B | | |
| 0,778790927 | CXorf57 | FALSE | Upregulated |
| -0,026705133 | CXorf58 | FALSE | Downregulated |
| 0,005298914 | CXorf64 | FALSE | Upregulated |
| 0,511626736 | CXorf65 | FALSE | Upregulated |
| 0,585280559 | CXorf66 | FALSE | Upregulated |
| 0 | CXorf67 | FALSE | Upregulated |
| 0,283268271 | CXXC11 | FALSE | Upregulated |
| | CXXC1P1 | | |
| -0,587664728 | CXXC4 | FALSE | Downregulated |
| 0,522486396 | CYB5R4 | FALSE | Upregulated |
| -0,336761149 | CYB5RL | FALSE | Downregulated |
| 0,459899264 | CYCSP52 | FALSE | Upregulated |
| | CYLC1 | | |
| | CYLC2 | | |
| | CYMP | | |
| | CYorf17 | | |
| -0,179162119 | CYP11A1 | FALSE | Downregulated |
| 1,314189478 | CYP11B1 | | Upregulated |
| 0 | CYP11B2 | FALSE | Upregulated |
| 0,000458965 | CYP17A1 | FALSE | Upregulated |
| 0 | CYP19A1 | FALSE | Upregulated |
| 0,410572789 | CYP1B1-AS1 | FALSE | Upregulated |
| 0,154860969 | CYP24A1 | FALSE | Upregulated |
| 0,064137734 | CYP26B1 | FALSE | Upregulated |
| 0,917387158 | CYP26C1 | FALSE | Upregulated |
| 0,349513211 | CYP27B1 | FALSE | Upregulated |
| 0,017285966 | CYP27C1 | FALSE | Upregulated |
| 1,130567931 | CYP2A13 | | Upregulated |
| 0,467329383 | CYP2F1 | FALSE | Upregulated |
| -0,799921278 | CYP2G1P | FALSE | Downregulated |
| -0,70013398 | CYP2S1 | FALSE | Downregulated |
| -0,57773171 | CYP3A7-CYP3AP1 | FALSE | Downregulated |
| -0,936525948 | CYP46A1 | FALSE | Downregulated |
| 0,424910619 | CYP4B1 | FALSE | Upregulated |

| | | | |
|---|---|---|---|
| 0,192024475 | CYP4F22 | FALSE | Upregulated |
| 0,246288097 | CYP4F24P | FALSE | Upregulated |
| | CYP4F30P | | |
| 0,575339929 | CYP4F35P | FALSE | Upregulated |
| | CYP4F62P | | |
| | CYP4F8 | | |
| 0,31106442 | CYP4X1 | FALSE | Upregulated |
| 0,332587224 | CYP4Z1 | FALSE | Upregulated |
| 0,122568281 | CYP4Z2P | FALSE | Upregulated |
| 0,140804261 | CYSLTR1 | FALSE | Upregulated |
| 1,114498942 | CYSLTR2 | | Upregulated |
| -0,317451499 | CYTIP | FALSE | Downregulated |
| 0 | CYTL1 | FALSE | Upregulated |
| | D21S2088E | | |
| 0,713227697 | DAB1-AS1 | FALSE | Upregulated |
| -0,457156926 | DACH1 | FALSE | Downregulated |
| -0,001674428 | DACH2 | FALSE | Downregulated |
| -0,173465451 | DACT3 | FALSE | Downregulated |
| 0,58538125 | DACT3-AS1 | FALSE | Upregulated |
| 0,101604522 | DAD1 | FALSE | Upregulated |
| -0,163229707 | DAND5 | FALSE | Downregulated |
| | DAOA | | |
| | DAOA-AS1 | | |
| | DAPL1 | | |
| -0,044054289 | DAPP1 | FALSE | Downregulated |
| 0,323939971 | DARC | FALSE | Upregulated |
| | DAW1 | | |
| 0,726371372 | DAZ1 | FALSE | Upregulated |
| 0,158267462 | DAZ4 | FALSE | Upregulated |
| | DAZL | | |
| 0,018822941 | DBF4 | FALSE | Upregulated |
| -0,251555583 | DBF4B | FALSE | Downregulated |
| 0,186627454 | DBIL5P2 | FALSE | Upregulated |
| -0,160729077 | DBNDD2 | FALSE | Downregulated |
| 0,553600355 | DBR1 | FALSE | Upregulated |
| 0,040648641 | DBX1 | FALSE | Upregulated |
| 0 | DBX2 | FALSE | Upregulated |
| | DCAF12L1 | | |
| 0,502430699 | DCAF12L2 | FALSE | Upregulated |
| -0,051012279 | DCAF13P3 | FALSE | Downregulated |
| 0,286349669 | DCAF4 | FALSE | Upregulated |
| -0,01330656 | DCAF4L2 | FALSE | Downregulated |
| | DCAF8L1 | | |
| | DCAF8L2 | | |
| 0,05577461 | DCC | FALSE | Upregulated |
| 0 | DCD | FALSE | Upregulated |
| | DCDC1 | | |
| -0,140424593 | DCDC2C | FALSE | Downregulated |
| -0,272817616 | DCHS2 | FALSE | Downregulated |
| 0,908778393 | DCK | FALSE | Upregulated |
| 0,672360757 | DCLK1 | FALSE | Upregulated |

| | | | |
|---|---|---|---|
| -0,182272407 | DCLK3 | FALSE | Downregulated |
| -0,529100146 | DCLRE1B | FALSE | Downregulated |
| -0,138253661 | DCT | FALSE | Downregulated |
| 0,280596027 | DCTN1-AS1 | FALSE | Upregulated |
| -0,688278489 | DCTN6 | FALSE | Downregulated |
| 0 | DCX | FALSE | Upregulated |
| 0,426905045 | DDIT4L | FALSE | Upregulated |
| 0,801719728 | DDN | FALSE | Upregulated |
| 0,114202843 | DDX11-AS1 | FALSE | Upregulated |
| -0,336814673 | DDX11L10 | FALSE | Downregulated |
| 0,659892444 | DDX11L5 | FALSE | Upregulated |
| 0,185713934 | DDX25 | FALSE | Upregulated |
| -0,187710531 | DDX26B | FALSE | Downregulated |
| | DDX4 | | |
| 0,023880715 | DDX43 | FALSE | Upregulated |
| -0,914433608 | DEAR | FALSE | Downregulated |
| -0,00509535 | DEC1 | FALSE | Downregulated |
| 0,045023792 | DEFA1 | FALSE | Upregulated |
| | DEFA10P | | |
| | DEFA11P | | |
| 0,045023792 | DEFA1B | FALSE | Upregulated |
| 0,01027329 | DEFA3 | FALSE | Upregulated |
| 0 | DEFA4 | FALSE | Upregulated |
| | DEFA5 | | |
| | DEFA6 | | |
| | DEFA8P | | |
| | DEFA9P | | |
| 0,306455355 | DEFB1 | FALSE | Upregulated |
| | DEFB103A | | |
| | DEFB103B | | |
| | DEFB104A | | |
| | DEFB104B | | |
| | DEFB105A | | |
| | DEFB105B | | |
| | DEFB106A | | |
| | DEFB106B | | |
| | DEFB107A | | |
| | DEFB107B | | |
| -0,001674428 | DEFB108B | FALSE | Downregulated |
| 0,300241802 | DEFB109P1 | FALSE | Upregulated |
| 0,178263821 | DEFB109P1B | FALSE | Upregulated |
| | DEFB110 | | |
| | DEFB112 | | |
| | DEFB113 | | |
| | DEFB114 | | |
| | DEFB115 | | |
| | DEFB116 | | |
| | DEFB118 | | |
| | DEFB119 | | |
| | DEFB121 | | |
| | DEFB122 | | |

| | | | |
|---|---|---|---|
| | DEFB123 | | |
| | DEFB124 | | |
| | DEFB125 | | |
| | DEFB126 | | |
| | DEFB127 | | |
| | DEFB128 | | |
| | DEFB129 | | |
| | DEFB130 | | |
| | DEFB131 | | |
| 0,281320309 | DEFB132 | FALSE | Upregulated |
| | DEFB133 | | |
| | DEFB134 | | |
| | DEFB135 | | |
| | DEFB136 | | |
| | DEFB4A | | |
| | DEFB4B | | |
| | DEFT1P | | |
| | DEFT1P2 | | |
| -0,160473376 | DENND2C | FALSE | Downregulated |
| -0,784133389 | DENND6B | FALSE | Downregulated |
| 0,134473643 | DEPDC1 | FALSE | Upregulated |
| 0,148867823 | DEPDC1B | FALSE | Upregulated |
| 0,57610252 | DEPDC4 | FALSE | Upregulated |
| -0,282928404 | DFNB59 | FALSE | Downregulated |
| 0,332649389 | DGAT2L6 | FALSE | Upregulated |
| 0,111621453 | DGCR10 | FALSE | Upregulated |
| 0,356240172 | DGCR11 | FALSE | Upregulated |
| -0,59283983 | DGCR6 | FALSE | Downregulated |
| 0,911503353 | DGKB | FALSE | Upregulated |
| 0,448121335 | DGKI | FALSE | Upregulated |
| | DGKK | | |
| -0,295150308 | DGUOK | FALSE | Downregulated |
| 0,093131021 | DGUOK-AS1 | FALSE | Upregulated |
| 0,117090345 | DHDH | FALSE | Upregulated |
| -0,16175303 | DHH | FALSE | Downregulated |
| -0,874441889 | DHRS11 | FALSE | Downregulated |
| 0,803980121 | DHRS13 | FALSE | Upregulated |
| -0,699653674 | DHRS4L1 | FALSE | Downregulated |
| 0 | DHRS7C | FALSE | Upregulated |
| -0,182540969 | DHRS9 | FALSE | Downregulated |
| | DHRSX-2 | | |
| 0,841750024 | DIAPH3 | FALSE | Upregulated |
| 0,43170026 | DIAPH3-AS1 | FALSE | Upregulated |
| | DIAPH3-AS2 | | |
| 0,271815953 | DIO2 | FALSE | Upregulated |
| 0,291950738 | DIO2-AS1 | FALSE | Upregulated |
| 0,404480507 | DIO3 | FALSE | Upregulated |
| -0,23002204 | DIO3OS | FALSE | Downregulated |
| 0 | DIP2A-IT1 | FALSE | Upregulated |
| 0,073039398 | DIRAS1 | FALSE | Upregulated |
| 0,182935905 | DIRAS2 | FALSE | Upregulated |

| | | | |
|---|---|---|---|
| 0 | DIRC1 | FALSE | Upregulated |
| -0,435594891 | DIRC3 | FALSE | Downregulated |
| -0,158868376 | DISC1FP1 | FALSE | Downregulated |
| 0,417355489 | DKFZP434A062 | FALSE | Upregulated |
| -0,122978709 | DKFZP434F142 | FALSE | Downregulated |
| 0,858411201 | DKFZP434I0714 | FALSE | Upregulated |
| -0,150619088 | DKFZp434J0226 | FALSE | Downregulated |
| | DKFZP434L187 | | |
| | DKFZp434L192 | | |
| 1,152618923 | DKFZp451B082 | | Upregulated |
| 0,290200034 | DKFZp566F0947 | FALSE | Upregulated |
| 0,341735614 | DKFZp667F0711 | FALSE | Upregulated |
| | DKFZp686K1684 | | |
| 0,654956749 | DKFZp686L13185 | FALSE | Upregulated |
| -0,362916921 | DKFZp779M0652 | FALSE | Downregulated |
| 0,038141043 | DKK1 | FALSE | Upregulated |
| 0,139701997 | DKK2 | FALSE | Upregulated |
| 1,277256987 | DKK4 | | Upregulated |
| 0,095313551 | DKKL1 | FALSE | Upregulated |
| 0,108644066 | DLEU2L | FALSE | Upregulated |
| 0,038552906 | DLEU7-AS1 | FALSE | Upregulated |
| 0,096963221 | DLG1-AS1 | FALSE | Upregulated |
| 0,316625809 | DLG3 | FALSE | Upregulated |
| 0,804524788 | DLG3-AS1 | FALSE | Upregulated |
| -0,684838847 | DLG5-AS1 | FALSE | Downregulated |
| 0,280898106 | DLGAP1 | FALSE | Upregulated |
| 0,711427247 | DLGAP1-AS1 | FALSE | Upregulated |
| 0,285754264 | DLGAP1-AS3 | FALSE | Upregulated |
| | DLGAP1-AS4 | | |
| 1,438275038 | DLGAP1-AS5 | | Upregulated |
| -0,455847006 | DLGAP3 | FALSE | Downregulated |
| 0,563092807 | DLGAP5 | FALSE | Upregulated |
| -0,00765293 | DLK1 | FALSE | Downregulated |
| | DLL3 | | |
| -0,141162723 | DLX1 | FALSE | Downregulated |
| | DLX2 | | |
| 0,172531243 | DLX3 | FALSE | Upregulated |
| 0,277764892 | DLX4 | FALSE | Upregulated |
| | DLX5 | | |
| | DLX6 | | |
| | DLX6-AS1 | | |
| -0,193438945 | DMBT1 | FALSE | Downregulated |
| | DMBT1P1 | | |
| | DMBX1 | | |
| -0,124702043 | DMC1 | FALSE | Downregulated |
| -0,480258526 | DMKN | FALSE | Downregulated |
| 0,85783575 | DMP1 | FALSE | Upregulated |
| | DMRT1 | | |
| 0,254645666 | DMRT2 | FALSE | Upregulated |
| | DMRT3 | | |
| | DMRTA2 | | |

| | | | |
|---|---|---|---|
| | DMRTB1 | | |
| | DMRTC1 | | |
| | DMRTC1B | | |
| | DMRTC2 | | |
| -0,083071987 | DNA2 | FALSE | Downregulated |
| 0,05955279 | DNAAF1 | FALSE | Upregulated |
| 0,394631192 | DNAAF3 | FALSE | Upregulated |
| -0,779262977 | DNAH12 | FALSE | Downregulated |
| 0 | DNAH17-AS1 | FALSE | Upregulated |
| 0,428065399 | DNAH3 | FALSE | Upregulated |
| -0,196684335 | DNAH7 | FALSE | Downregulated |
| 0,301917595 | DNAH8 | FALSE | Upregulated |
| -0,677034253 | DNAI1 | FALSE | Downregulated |
| 0,132653039 | DNAI2 | FALSE | Upregulated |
| | DNAJA1P5 | | |
| 0,994619707 | DNAJB13 | FALSE | Upregulated |
| 0,119476107 | DNAJB8 | FALSE | Upregulated |
| | DNAJB8-AS1 | | |
| 0,356236453 | DNAJC18 | FALSE | Upregulated |
| -0,652497936 | DNAJC27 | FALSE | Downregulated |
| -0,417352779 | DNAJC27-AS1 | FALSE | Downregulated |
| 0,361995461 | DNAJC28 | FALSE | Upregulated |
| 0,189437398 | DNAJC3-AS1 | FALSE | Upregulated |
| -0,178247384 | DNAJC5B | FALSE | Downregulated |
| 0,277119458 | DNAJC5G | FALSE | Upregulated |
| 0,28886126 | DNAJC6 | FALSE | Upregulated |
| 0,342424545 | DNAL4 | FALSE | Upregulated |
| 0,00513235 | DNASE2B | FALSE | Upregulated |
| 0,321178765 | DNER | FALSE | Upregulated |
| 0,631043583 | DNLZ | FALSE | Upregulated |
| 0,456391569 | DNM1P35 | FALSE | Upregulated |
| 0,001539327 | DNM1P46 | FALSE | Upregulated |
| -0,603016782 | DNM1P50 | FALSE | Downregulated |
| 0,315273473 | DNMT3B | FALSE | Upregulated |
| -1,479426717 | DNMT3L | | Downregulated |
| -0,015040902 | DNTT | FALSE | Downregulated |
| -0,239741385 | DNTTIP1 | FALSE | Downregulated |
| 0,676543337 | DOC2B | FALSE | Upregulated |
| -0,006454352 | DOC2GP | FALSE | Downregulated |
| -0,187395339 | DOCK9-AS1 | FALSE | Downregulated |
| 0,631519689 | DOCK9-AS2 | FALSE | Upregulated |
| 0,653254861 | DOHH | FALSE | Upregulated |
| 0,744236218 | DOK5 | FALSE | Upregulated |
| 0,721231649 | DPCR1 | FALSE | Upregulated |
| 0,426406837 | DPEP1 | FALSE | Upregulated |
| -0,00094525 | DPEP3 | FALSE | Downregulated |
| -0,613441516 | DPF1 | FALSE | Downregulated |
| -0,534393429 | DPH6-AS1 | FALSE | Downregulated |
| 0,109723018 | DPM3 | FALSE | Upregulated |
| 0,140702435 | DPP10 | FALSE | Upregulated |
| 0 | DPP10-AS1 | FALSE | Upregulated |

| | | | |
|---|---|---|---|
| | DPP10-AS3 | | |
| 0,426389313 | DPP6 | FALSE | Upregulated |
| | DPPA2 | | |
| 0 | DPPA2P3 | FALSE | Upregulated |
| 0 | DPPA3 | FALSE | Upregulated |
| -0,206274191 | DPPA4 | FALSE | Downregulated |
| | DPPA5 | | |
| | DPRX | | |
| 0,114064341 | DPRXP4 | FALSE | Upregulated |
| -0,086163087 | DPY19L1P1 | FALSE | Downregulated |
| 0,069951092 | DPY19L2 | FALSE | Upregulated |
| -0,04969678 | DPY19L2P1 | FALSE | Downregulated |
| -0,025346564 | DPY30 | FALSE | Downregulated |
| 0,336300237 | DPYD-AS1 | FALSE | Upregulated |
| -0,523331087 | DPYSL4 | FALSE | Downregulated |
| | DPYSL5 | | |
| 0,296949131 | DQX1 | FALSE | Upregulated |
| 0 | DRAXIN | FALSE | Upregulated |
| 0,009225615 | DRC1 | FALSE | Upregulated |
| -0,899302839 | DRD1 | FALSE | Downregulated |
| -0,179619681 | DRD2 | FALSE | Downregulated |
| 0,297928284 | DRD3 | FALSE | Upregulated |
| -0,350361932 | DRD4 | FALSE | Downregulated |
| | DRGX | | |
| 0,272268835 | DRP2 | FALSE | Upregulated |
| 0,079119311 | DSC3 | FALSE | Upregulated |
| 0,575577442 | DSCAM | FALSE | Upregulated |
| -0,520762604 | DSCAM-AS1 | FALSE | Downregulated |
| 0,001141803 | DSCAM-IT1 | FALSE | Upregulated |
| -0,662727876 | DSCAML1 | FALSE | Downregulated |
| 0,841257125 | DSCC1 | FALSE | Upregulated |
| | DSCR10 | | |
| | DSCR4 | | |
| | DSCR8 | | |
| 1,119118252 | DSG3 | | Upregulated |
| 0,598219649 | DSG4 | FALSE | Upregulated |
| -0,036928678 | DSN1 | FALSE | Downregulated |
| 0,569140168 | DSPP | FALSE | Upregulated |
| -0,564312491 | DSTNP2 | FALSE | Downregulated |
| | DSTNP4 | | |
| 0,346741627 | DTD1 | FALSE | Upregulated |
| 2,423408904 | DTHD1 | | Upregulated |
| -0,134576987 | DTL | FALSE | Downregulated |
| -0,69072454 | DUOX2 | FALSE | Downregulated |
| -0,003587948 | DUOXA1 | FALSE | Downregulated |
| 0,51107172 | DUOXA2 | FALSE | Upregulated |
| 1,127510821 | DUPD1 | | Upregulated |
| 0,096023001 | DUSP12 | FALSE | Upregulated |
| 0,32924123 | DUSP13 | FALSE | Upregulated |
| 0,234078962 | DUSP15 | FALSE | Upregulated |
| -0,225222155 | DUSP18 | FALSE | Downregulated |

| | | | |
|---|---|---|---|
| -0,492121648 | DUSP19 | FALSE | Downregulated |
| 1,614269101 | DUSP2 | | Upregulated |
| 0,000557131 | DUSP21 | FALSE | Upregulated |
| 0,82426223 | DUSP26 | FALSE | Upregulated |
| 0,385835254 | DUSP27 | FALSE | Upregulated |
| 0,168208922 | DUSP4 | FALSE | Upregulated |
| -0,013426609 | DUSP5P1 | FALSE | Downregulated |
| -0,765800443 | DUSP8 | FALSE | Downregulated |
| 0,825921355 | DUSP9 | FALSE | Upregulated |
| | DUX2 | | |
| | DUX4 | | |
| 0 | DUX4L | FALSE | Upregulated |
| | DUX4L2 | | |
| | DUX4L3 | | |
| | DUX4L4 | | |
| | DUX4L5 | | |
| | DUX4L6 | | |
| | DUX4L7 | | |
| | DUXA | | |
| 0 | DYDC1 | FALSE | Upregulated |
| 0,122598213 | DYDC2 | FALSE | Upregulated |
| -0,012999829 | DYNAP | FALSE | Downregulated |
| 1,004218356 | DYNC1I1 | | Upregulated |
| 0,145965493 | DYNLRB2 | FALSE | Upregulated |
| -0,043871893 | DYRK3 | FALSE | Downregulated |
| -0,621895529 | DYRK4 | FALSE | Downregulated |
| | DYTN | | |
| 0,84668127 | DYX1C1 | FALSE | Upregulated |
| 0 | DYX1C1-CCPG1 | FALSE | Upregulated |
| -0,005531268 | DZANK1 | FALSE | Downregulated |
| 0,690666375 | E2F1 | FALSE | Upregulated |
| -0,379482197 | E2F2 | FALSE | Downregulated |
| -0,769239752 | E2F6 | FALSE | Downregulated |
| 0,168024541 | E2F7 | FALSE | Upregulated |
| 0,506066605 | E2F8 | FALSE | Upregulated |
| 0,101376188 | EAF2 | FALSE | Upregulated |
| 0,547425343 | EBF1 | FALSE | Upregulated |
| 1,113109511 | EBF2 | | Upregulated |
| -0,256766736 | EBI3 | FALSE | Downregulated |
| | EBLN1 | | |
| -0,481448029 | EBLN2 | FALSE | Downregulated |
| -0,588156454 | ECE2 | FALSE | Downregulated |
| 0,102521189 | ECEL1 | FALSE | Upregulated |
| -0,153982968 | ECEL1P2 | FALSE | Downregulated |
| 0,509070067 | ECRP | FALSE | Upregulated |
| -0,18170578 | ECSCR | FALSE | Downregulated |
| 0,68535926 | ECSCR-2 | FALSE | Upregulated |
| 1,309949631 | ECT2 | | Upregulated |
| 0,183968387 | EDA2R | FALSE | Upregulated |
| 0,701273311 | EDARADD | FALSE | Upregulated |
| 0,283719947 | EDDM3A | FALSE | Upregulated |

| | | | |
|---|---|---|---|
| -0,291537706 | EDDM3B | FALSE | Downregulated |
| -0,011568548 | EDIL3 | FALSE | Downregulated |
| 0,210049906 | EDN1 | FALSE | Upregulated |
| 0,68996356 | EDN2 | FALSE | Upregulated |
| -0,056141925 | EDN3 | FALSE | Downregulated |
| 0,265355029 | EDNRA | FALSE | Upregulated |
| -0,334521657 | EEF1DP3 | FALSE | Downregulated |
| 0,684071975 | EEF1E1 | FALSE | Upregulated |
| -0,489810801 | EEF1E1-BLOC1S5 | FALSE | Downregulated |
| -0,41834243 | EFCAB1 | FALSE | Downregulated |
| 0,488783129 | EFCAB10 | FALSE | Upregulated |
| -0,149362298 | EFCAB11 | FALSE | Downregulated |
| 0,427044081 | EFCAB12 | FALSE | Upregulated |
| -0,036489593 | EFCAB14-AS1 | FALSE | Downregulated |
| | EFCAB3 | | |
| 0,443009406 | EFCAB5 | FALSE | Upregulated |
| 0,384644718 | EFCAB6 | FALSE | Upregulated |
| -0,176652508 | EFCAB6-AS1 | FALSE | Downregulated |
| | EFCAB8 | | |
| 0,443268062 | EFCAB9 | FALSE | Upregulated |
| 0,030378758 | EFCC1 | FALSE | Upregulated |
| -0,006722349 | EFHB | FALSE | Downregulated |
| 0,279341771 | EFHC2 | FALSE | Upregulated |
| 1,021445999 | EFNA3 | | Upregulated |
| 0,505896445 | EFNA4 | FALSE | Upregulated |
| 0,930096842 | EFNA5 | FALSE | Upregulated |
| 0,230310781 | EFNB3 | FALSE | Upregulated |
| -0,249752963 | EFS | FALSE | Downregulated |
| 0 | EFTUD1P1 | FALSE | Upregulated |
| 0,114656141 | EGF | FALSE | Upregulated |
| 0,382151366 | EGFEM1P | FALSE | Upregulated |
| 0 | EGFL6 | FALSE | Upregulated |
| -0,770683727 | EGFL8 | FALSE | Downregulated |
| -0,912330355 | EGFLAM | FALSE | Downregulated |
| -0,814587298 | EGFLAM-AS2 | FALSE | Downregulated |
| -0,83942184 | EGFLAM-AS4 | FALSE | Downregulated |
| 0,22493655 | EGLN3 | FALSE | Upregulated |
| 0,234592892 | EGR3 | FALSE | Upregulated |
| 0 | EGR4 | FALSE | Upregulated |
| -0,682861743 | EHF | FALSE | Downregulated |
| 0,393998614 | EHMT1-IT1 | FALSE | Upregulated |
| 0,261435859 | EID2B | FALSE | Upregulated |
| 0,77210096 | EID3 | FALSE | Upregulated |
| 0,107685562 | EIF1AY | FALSE | Upregulated |
| | EIF3IP1 | | |
| 0,025824044 | EIF3J-AS1 | FALSE | Upregulated |
| 0,088811567 | EIF4E1B | FALSE | Upregulated |
| -0,310720569 | EIF4EBP3 | FALSE | Downregulated |
| 0,546574095 | EIF5A2 | FALSE | Upregulated |
| 0,278073363 | EIF5AL1 | FALSE | Upregulated |
| 0,626663579 | ELANE | FALSE | Upregulated |

| | | | |
|---|---|---|---|
| | ELAVL2 | | |
| -0,095402824 | ELAVL3 | FALSE | Downregulated |
| 0,270827676 | ELAVL4 | FALSE | Upregulated |
| 0,766846206 | ELF5 | FALSE | Upregulated |
| 0,473425367 | ELFN2 | FALSE | Upregulated |
| 0,415193114 | ELMO1-AS1 | FALSE | Upregulated |
| 0,024341648 | ELMOD1 | FALSE | Upregulated |
| 0,264930358 | ELOVL2-AS1 | FALSE | Upregulated |
| 0 | ELOVL3 | FALSE | Upregulated |
| 0,453051051 | ELOVL4 | FALSE | Upregulated |
| 0,329012152 | ELOVL7 | FALSE | Upregulated |
| -0,242574076 | ELP4 | FALSE | Downregulated |
| | ELSPBP1 | | |
| -0,032215737 | ELTD1 | FALSE | Downregulated |
| 0,760925517 | EMB | FALSE | Upregulated |
| 0,302307159 | EMCN | FALSE | Upregulated |
| -0,244501505 | EMP1 | FALSE | Downregulated |
| 0,722759569 | EMP3 | FALSE | Upregulated |
| 0,125116847 | EMR1 | FALSE | Upregulated |
| 0,712412802 | EMR3 | FALSE | Upregulated |
| -0,72588645 | EMR4P | FALSE | Downregulated |
| 0 | EMX2 | FALSE | Upregulated |
| 0,680889878 | EMX2OS | FALSE | Upregulated |
| 0 | EN1 | FALSE | Upregulated |
| 0,39417772 | EN2 | FALSE | Upregulated |
| 0,237535678 | ENKUR | FALSE | Upregulated |
| 0 | ENO1-AS1 | FALSE | Upregulated |
| -0,315472811 | ENOX1 | FALSE | Downregulated |
| -0,442612279 | ENPP3 | FALSE | Downregulated |
| -0,688937333 | ENPP4 | FALSE | Downregulated |
| 0,226968911 | ENPP5 | FALSE | Upregulated |
| 0,418187551 | ENPP6 | FALSE | Upregulated |
| | ENTHD1 | | |
| 0,813372816 | ENTPD2 | FALSE | Upregulated |
| -0,557417021 | ENTPD3-AS1 | FALSE | Downregulated |
| -0,150942495 | EOGT | FALSE | Downregulated |
| 1,374321944 | EOMES | | Upregulated |
| -0,179145861 | EPB42 | FALSE | Downregulated |
| 0,534495265 | EPCAM | FALSE | Upregulated |
| -0,308487734 | EPGN | FALSE | Downregulated |
| 1,226528734 | EPHA1-AS1 | | Upregulated |
| 0,769040276 | EPHA10 | FALSE | Upregulated |
| 0,71945532 | EPHA4 | FALSE | Upregulated |
| 0 | EPHA5 | FALSE | Upregulated |
| 0,067676302 | EPHA6 | FALSE | Upregulated |
| 0,579782421 | EPHA7 | FALSE | Upregulated |
| | EPHA8 | | |
| -0,983580424 | EPHB1 | FALSE | Downregulated |
| 1,298840425 | EPHB2 | | Upregulated |
| 0,075711834 | EPHB6 | FALSE | Upregulated |
| 0,588758211 | EPHX3 | FALSE | Upregulated |

| | | | |
|---|---|---|---|
| | EPHX4 | | |
| 0,303254974 | EPN2-IT1 | FALSE | Upregulated |
| 0,006339326 | EPN3 | FALSE | Upregulated |
| -1,194202155 | EPO | | Downregulated |
| 0 | EPPIN | FALSE | Upregulated |
| 0,420021567 | EPPIN-WFDC6 | FALSE | Upregulated |
| 0,563566947 | EPS8L1 | FALSE | Upregulated |
| -0,148246334 | EPS8L3 | FALSE | Downregulated |
| | EPYC | | |
| | EQTN | | |
| -0,005542756 | ERAS | FALSE | Downregulated |
| -0,312625037 | ERC2 | FALSE | Downregulated |
| 0,176333791 | ERC2-IT1 | FALSE | Upregulated |
| 0,829726066 | ERCC6-PGBD3 | FALSE | Upregulated |
| -0,372278139 | ERCC8 | FALSE | Downregulated |
| 0 | EREG | FALSE | Upregulated |
| 1,194139344 | ERI3-IT1 | | Upregulated |
| -0,154185831 | ERICH1-AS1 | FALSE | Downregulated |
| 0,374351127 | ERICH2 | FALSE | Upregulated |
| 0,705446134 | ERMN | FALSE | Upregulated |
| 0,056119283 | ERN2 | FALSE | Upregulated |
| 0,097270708 | ERP27 | FALSE | Upregulated |
| 0 | ERVMER34-1 | FALSE | Upregulated |
| 0,586319544 | ERVV-1 | FALSE | Upregulated |
| -0,179681256 | ERVV-2 | FALSE | Downregulated |
| 0,000203023 | ESCO2 | FALSE | Upregulated |
| -0,030537936 | ESM1 | FALSE | Downregulated |
| -1,324248168 | ESPL1 | | Downregulated |
| -0,94720514 | ESPNP | FALSE | Downregulated |
| 0 | ESRG | FALSE | Upregulated |
| 0,34613275 | ESRP1 | FALSE | Upregulated |
| 0,662026092 | ESRRB | FALSE | Upregulated |
| 1,061904219 | ESRRG | | Upregulated |
| 0,434653506 | ESX1 | FALSE | Upregulated |
| -0,678726464 | ETV1 | FALSE | Downregulated |
| -0,326599829 | ETV3L | FALSE | Downregulated |
| 1,171859004 | ETV7 | | Upregulated |
| 0,881035108 | EVC2 | FALSE | Upregulated |
| 0,370342442 | EVPL | FALSE | Upregulated |
| 0,469567095 | EVPLL | FALSE | Upregulated |
| | EVX1 | | |
| | EVX2 | | |
| -0,006848587 | EXD1 | FALSE | Downregulated |
| 0,570783024 | EXO1 | FALSE | Upregulated |
| 0,773198939 | EXOG | FALSE | Upregulated |
| -1,187092739 | EXOSC1 | | Downregulated |
| -0,304595512 | EXOSC5 | FALSE | Downregulated |
| 0,221975561 | EXOSC8 | FALSE | Upregulated |
| -0,083592663 | EXOSC9 | FALSE | Downregulated |
| -0,068317034 | EXTL1 | FALSE | Downregulated |
| -0,636230911 | EXTL2 | FALSE | Downregulated |

| | | | |
|---|---|---|---|
| 0,119064744 | EXTL3-AS1 | FALSE | Upregulated |
| 0,066312154 | EYA1 | FALSE | Upregulated |
| 0,290938849 | EYA2 | FALSE | Upregulated |
| -0,012382302 | EYA4 | FALSE | Downregulated |
| 0 | EZR-AS1 | FALSE | Upregulated |
| 1,156163376 | F2RL1 | | Upregulated |
| 0,176816996 | F3 | FALSE | Upregulated |
| 0,947133325 | F8A2 | FALSE | Upregulated |
| 0 | FA2H | FALSE | Upregulated |
| | FABP12 | | |
| 0,128480123 | FABP4 | FALSE | Upregulated |
| 0,004204782 | FABP5P3 | FALSE | Upregulated |
| | FABP6 | | |
| -0,415418232 | FABP7 | FALSE | Downregulated |
| | FABP9 | | |
| -0,390381442 | FADS6 | FALSE | Downregulated |
| 0,273719098 | FAHD2B | FALSE | Upregulated |
| 1,095289671 | FAHD2CP | | Upregulated |
| 0,093330185 | FAIM | FALSE | Upregulated |
| 0,333228084 | FAIM2 | FALSE | Upregulated |
| 0,326249438 | FAM101A | FALSE | Upregulated |
| 0,893272504 | FAM104B | FALSE | Upregulated |
| 0,565100505 | FAM105A | FALSE | Upregulated |
| -1,118085337 | FAM106A | | Downregulated |
| -0,279219099 | FAM106B | FALSE | Downregulated |
| -0,5558468 | FAM106CP | FALSE | Downregulated |
| -0,13053786 | FAM109B | FALSE | Downregulated |
| -0,123809546 | FAM110A | FALSE | Downregulated |
| 0,096288433 | FAM110D | FALSE | Upregulated |
| 0,112747576 | FAM124B | FALSE | Upregulated |
| 0,169134647 | FAM127B | FALSE | Upregulated |
| 0,293716733 | FAM127C | FALSE | Upregulated |
| -0,047472545 | FAM129C | FALSE | Downregulated |
| 0,273422532 | FAM131C | FALSE | Upregulated |
| 0,088153293 | FAM132A | FALSE | Upregulated |
| 0 | FAM133A | FALSE | Upregulated |
| 0,0250359 | FAM133CP | FALSE | Upregulated |
| 1,819821894 | FAM133DP | | Upregulated |
| -0,228165195 | FAM135B | FALSE | Downregulated |
| | FAM138A | | |
| -0,00857108 | FAM138B | FALSE | Downregulated |
| | FAM138C | | |
| 0,426187702 | FAM138D | FALSE | Upregulated |
| 0 | FAM138E | FALSE | Upregulated |
| | FAM138F | | |
| 0 | FAM150A | FALSE | Upregulated |
| 0,676065797 | FAM151B | FALSE | Upregulated |
| -0,376044232 | FAM153A | FALSE | Downregulated |
| 0 | FAM153B | FALSE | Upregulated |
| -0,029232273 | FAM153C | FALSE | Downregulated |
| 0,414240606 | FAM154A | FALSE | Upregulated |

| | | | |
|---|---|---|---|
| 0,768654804 | FAM154B | FALSE | Upregulated |
| 0,170128994 | FAM155A | FALSE | Upregulated |
| 0 | FAM155A-IT1 | FALSE | Upregulated |
| 0,142733133 | FAM155B | FALSE | Upregulated |
| 0,459868701 | FAM157A | FALSE | Upregulated |
| 1,217627266 | FAM157B | | Upregulated |
| -0,002165611 | FAM159A | FALSE | Downregulated |
| 0,00160549 | FAM159B | FALSE | Upregulated |
| 0,499134111 | FAM161A | FALSE | Upregulated |
| 0,541823933 | FAM162B | FALSE | Upregulated |
| 0,297308464 | FAM163A | FALSE | Upregulated |
| -0,566993478 | FAM166B | FALSE | Downregulated |
| 0,378208474 | FAM167A | FALSE | Upregulated |
| -0,410460547 | FAM167B | FALSE | Downregulated |
| 0,198491105 | FAM169B | FALSE | Upregulated |
| 0,187925706 | FAM170A | FALSE | Upregulated |
| | FAM170B | | |
| -1,447689372 | FAM170B-AS1 | | Downregulated |
| 0,437848642 | FAM171A2 | FALSE | Upregulated |
| -0,64770429 | FAM171B | FALSE | Downregulated |
| 0,269374097 | FAM172BP | FALSE | Upregulated |
| -0,263095893 | FAM173B | FALSE | Downregulated |
| 0,115539804 | FAM174A | FALSE | Upregulated |
| 0,263922671 | FAM177B | FALSE | Upregulated |
| 0,527739304 | FAM179A | FALSE | Upregulated |
| 0,756696463 | FAM180B | FALSE | Upregulated |
| 0,664602753 | FAM181A | FALSE | Upregulated |
| | FAM181A-AS1 | | |
| -0,002253536 | FAM181B | FALSE | Downregulated |
| -0,488528723 | FAM182A | FALSE | Downregulated |
| 0,370085894 | FAM182B | FALSE | Upregulated |
| 0,425110553 | FAM183A | FALSE | Upregulated |
| -0,00952407 | FAM183B | FALSE | Downregulated |
| 0,348418388 | FAM183CP | FALSE | Upregulated |
| 0 | FAM187B | FALSE | Upregulated |
| -0,471765504 | FAM189A1 | FALSE | Downregulated |
| -0,000145457 | FAM189A2 | FALSE | Downregulated |
| -0,587595076 | FAM194A | FALSE | Downregulated |
| 0,287801306 | FAM194B | FALSE | Upregulated |
| | FAM197Y2 | | |
| | FAM197Y4 | | |
| | FAM197Y5 | | |
| -0,272293481 | FAM19A1 | FALSE | Downregulated |
| -0,97682686 | FAM19A2 | FALSE | Downregulated |
| | FAM19A3 | | |
| -0,002253536 | FAM19A4 | FALSE | Downregulated |
| -0,248552644 | FAM19A5 | FALSE | Downregulated |
| -0,100285207 | FAM200A | FALSE | Downregulated |
| 0,257123438 | FAM201A | FALSE | Upregulated |
| -0,558429416 | FAM201B | FALSE | Downregulated |
| -0,14531021 | FAM209A | FALSE | Downregulated |

| | | | |
|---|---|---|---|
| | FAM209B | | |
| 0,262737944 | FAM212B-AS1 | FALSE | Upregulated |
| -0,005047929 | FAM215A | FALSE | Downregulated |
| 0,00153605 | FAM216B | FALSE | Upregulated |
| 0,427425383 | FAM217A | FALSE | Upregulated |
| 0,978185146 | FAM218A | FALSE | Upregulated |
| -0,526763494 | FAM220A | FALSE | Downregulated |
| 0 | FAM220BP | FALSE | Upregulated |
| -0,371054264 | FAM222A-AS1 | FALSE | Downregulated |
| 0,461360541 | FAM223B | FALSE | Upregulated |
| 0,932563859 | FAM225A | FALSE | Upregulated |
| | FAM225B | | |
| 0,297978352 | FAM226A | FALSE | Upregulated |
| 0 | FAM226B | FALSE | Upregulated |
| 0,644109951 | FAM227A | FALSE | Upregulated |
| 1,100799594 | FAM228A | | Upregulated |
| 0,211363635 | FAM228B | FALSE | Upregulated |
| -0,251359394 | FAM229A | FALSE | Downregulated |
| | FAM230A | | |
| -0,011168865 | FAM230B | FALSE | Downregulated |
| | FAM230C | | |
| -0,080778882 | FAM231A | FALSE | Downregulated |
| | FAM231B | | |
| -0,009085168 | FAM231C | FALSE | Downregulated |
| 0,563258838 | FAM231D | FALSE | Upregulated |
| 0 | FAM24A | FALSE | Upregulated |
| 0,295017524 | FAM24B | FALSE | Upregulated |
| | FAM24B-CUZD1 | | |
| 0 | FAM25A | FALSE | Upregulated |
| 0 | FAM25B | FALSE | Upregulated |
| | FAM25C | | |
| | FAM25G | | |
| | FAM26D | | |
| -0,461249365 | FAM26E | FALSE | Downregulated |
| 0,175870363 | FAM26F | FALSE | Upregulated |
| 0,465375917 | FAM27B | FALSE | Upregulated |
| 0,278362943 | FAM27C | FALSE | Upregulated |
| 1,032674965 | FAM27E2 | | Upregulated |
| | FAM27L | | |
| 0,379798998 | FAM35BP | FALSE | Upregulated |
| -0,721237008 | FAM3B | FALSE | Downregulated |
| -0,472621381 | FAM3D | FALSE | Downregulated |
| | FAM41AY1 | | |
| 0,285642989 | FAM41C | FALSE | Upregulated |
| 0,714996582 | FAM43B | FALSE | Upregulated |
| 0,463470093 | FAM45B | FALSE | Upregulated |
| -0,664430791 | FAM46B | FALSE | Downregulated |
| | FAM46D | | |
| | FAM47A | | |
| | FAM47B | | |
| -0,148513368 | FAM47C | FALSE | Downregulated |

| | | | |
|---|---|---|---|
| 0,339418271 | FAM53A | FALSE | Upregulated |
| | FAM57B | | |
| -0,15038168 | FAM58A | FALSE | Downregulated |
| -0,124767259 | FAM60A | FALSE | Downregulated |
| -0,647920472 | FAM64A | FALSE | Downregulated |
| 0,124470872 | FAM66A | FALSE | Upregulated |
| -0,745774124 | FAM66B | FALSE | Downregulated |
| -0,575052288 | FAM66C | FALSE | Downregulated |
| -0,025301999 | FAM66D | FALSE | Downregulated |
| | FAM66E | | |
| -0,732565942 | FAM69C | FALSE | Downregulated |
| -0,185687317 | FAM71A | FALSE | Downregulated |
| 0 | FAM71B | FALSE | Upregulated |
| 0,156316706 | FAM71C | FALSE | Upregulated |
| 1,168101235 | FAM71D | | Upregulated |
| 0,012663999 | FAM71E1 | FALSE | Upregulated |
| 0 | FAM71E2 | FALSE | Upregulated |
| 0,288020774 | FAM71F1 | FALSE | Upregulated |
| -0,085067374 | FAM71F2 | FALSE | Downregulated |
| 0,533327652 | FAM72A | FALSE | Upregulated |
| -0,003802111 | FAM72B | FALSE | Downregulated |
| 0,026388983 | FAM72C | FALSE | Upregulated |
| 0,899489527 | FAM72D | FALSE | Upregulated |
| 0 | FAM74A1 | FALSE | Upregulated |
| -0,281375954 | FAM74A3 | FALSE | Downregulated |
| -0,007421221 | FAM74A4 | FALSE | Downregulated |
| | FAM74A7 | | |
| 0,423119254 | FAM81A | FALSE | Upregulated |
| -1,324010673 | FAM81B | | Downregulated |
| 0,804281205 | FAM83A | FALSE | Upregulated |
| 0,048586618 | FAM83A-AS1 | FALSE | Upregulated |
| 0,393081959 | FAM83B | FALSE | Upregulated |
| 1,149328994 | FAM83C | | Upregulated |
| 0 | FAM83C-AS1 | FALSE | Upregulated |
| 0,011520088 | FAM83D | FALSE | Upregulated |
| -0,110296503 | FAM83E | FALSE | Downregulated |
| 0,413246373 | FAM83F | FALSE | Upregulated |
| 0,901967679 | FAM83H-AS1 | FALSE | Upregulated |
| 1,144795147 | FAM85A | | Upregulated |
| 0,632429103 | FAM86B2 | FALSE | Upregulated |
| 0,953332884 | FAM86C2P | FALSE | Upregulated |
| 0,578982921 | FAM86DP | FALSE | Upregulated |
| -1,016712279 | FAM86EP | | Downregulated |
| 0,325011993 | FAM86FP | FALSE | Upregulated |
| 0,528121132 | FAM86HP | FALSE | Upregulated |
| 0,708476455 | FAM86JP | FALSE | Upregulated |
| 0 | FAM86KP | FALSE | Upregulated |
| 1,548467162 | FAM87A | | Upregulated |
| 0,471118855 | FAM89A | FALSE | Upregulated |
| | FAM90A10P | | |
| | FAM90A27P | | |

| | | | |
|---|---|---|---|
| 0,295209532 | FAM90A2P | FALSE | Upregulated |
| -0,501591377 | FAM90A7P | FALSE | Downregulated |
| 0,008601379 | FAM92A1P2 | FALSE | Upregulated |
| 0 | FAM92B | FALSE | Upregulated |
| 0,149312596 | FAM95A | FALSE | Upregulated |
| 0,703337879 | FAM95B1 | FALSE | Upregulated |
| -0,173352632 | FAM95C | FALSE | Downregulated |
| -0,049836066 | FAM98C | FALSE | Downregulated |
| -0,169495171 | FAM9A | FALSE | Downregulated |
| -0,76849066 | FAM9B | FALSE | Downregulated |
| 0 | FAM9C | FALSE | Upregulated |
| -0,336960432 | FANCB | FALSE | Downregulated |
| 0,463462845 | FANCD2 | FALSE | Upregulated |
| 0,529932184 | FANCD2OS | FALSE | Upregulated |
| -0,370581501 | FANCE | FALSE | Downregulated |
| 0,210322973 | FANCG | FALSE | Upregulated |
| | FANK1 | | |
| | FANK1-AS1 | | |
| -0,005224117 | FAP | FALSE | Downregulated |
| | FAR2P1 | | |
| -0,398860376 | FAR2P2 | FALSE | Downregulated |
| 0,470171613 | FAR2P3 | FALSE | Upregulated |
| 1,110752902 | FASLG | | Upregulated |
| -0,399481704 | FASTKD3 | FALSE | Downregulated |
| -0,470429491 | FAT3 | FALSE | Downregulated |
| | FATE1 | | |
| -0,275401324 | FAXC | FALSE | Downregulated |
| -0,158145496 | FBLL1 | FALSE | Downregulated |
| 0,290748752 | FBN2 | FALSE | Upregulated |
| 0,325079253 | FBN3 | FALSE | Upregulated |
| 0,214310651 | FBP2 | FALSE | Upregulated |
| 0,298408479 | FBXL19-AS1 | FALSE | Upregulated |
| 0,091200836 | FBXL8 | FALSE | Upregulated |
| -0,322720312 | FBXO15 | FALSE | Downregulated |
| 0,160749154 | FBXO16 | FALSE | Upregulated |
| 0,811411635 | FBXO36 | FALSE | Upregulated |
| 1,626415976 | FBXO39 | | Upregulated |
| -0,227450277 | FBXO4 | FALSE | Downregulated |
| 0,431121228 | FBXO43 | FALSE | Upregulated |
| | FBXO47 | | |
| -0,886450079 | FBXO48 | FALSE | Downregulated |
| -0,490886321 | FBXO5 | FALSE | Downregulated |
| 0,0574466 | FBXW10 | FALSE | Upregulated |
| | FBXW12 | | |
| -0,109139586 | FBXW4P1 | FALSE | Downregulated |
| 0,351200904 | FBXW9 | FALSE | Upregulated |
| 0,266886614 | FCAR | FALSE | Upregulated |
| -0,159607366 | FCER1A | FALSE | Downregulated |
| -0,285933027 | FCER1G | FALSE | Downregulated |
| 0,287656741 | FCER2 | FALSE | Upregulated |
| 1,150947915 | FCGR1A | | Upregulated |

| | | | |
|---|---|---|---|
| 1,056981161 | FCGR1B | | Upregulated |
| 0,95651047 | FCGR1C | FALSE | Upregulated |
| -0,329657978 | FCGR2C | FALSE | Downregulated |
| 1,257809829 | FCGR3B | | Upregulated |
| 0,533452132 | FCHO1 | FALSE | Upregulated |
| 0,280036067 | FCRL1 | FALSE | Upregulated |
| 0,395509863 | FCRL2 | FALSE | Upregulated |
| 1,266084255 | FCRL3 | | Upregulated |
| 0 | FCRL4 | FALSE | Upregulated |
| 1,166226239 | FCRL5 | | Upregulated |
| -0,001298455 | FCRLA | FALSE | Downregulated |
| -0,244066553 | FCRLB | FALSE | Downregulated |
| | FDCSP | | |
| 0,345874215 | FDXACB1 | FALSE | Upregulated |
| 0,172592684 | FENDRR | FALSE | Upregulated |
| | FER1L6 | | |
| 0,741011926 | FER1L6-AS1 | FALSE | Upregulated |
| -0,017308649 | FER1L6-AS2 | FALSE | Downregulated |
| 0,422407744 | FERD3L | FALSE | Upregulated |
| -0,247927622 | FERMT1 | FALSE | Downregulated |
| | FEV | | |
| -0,439235401 | FEZF1 | FALSE | Downregulated |
| 0,554979017 | FEZF1-AS1 | FALSE | Upregulated |
| | FEZF2 | | |
| 0 | FFAR1 | FALSE | Upregulated |
| 1,010412502 | FFAR2 | | Upregulated |
| 0,132243142 | FFAR3 | FALSE | Upregulated |
| 0,149099677 | FGD5P1 | FALSE | Upregulated |
| 0,711171247 | FGF10 | FALSE | Upregulated |
| 0,011828808 | FGF10-AS1 | FALSE | Upregulated |
| -0,492497101 | FGF12 | FALSE | Downregulated |
| 0,425804654 | FGF13 | FALSE | Upregulated |
| -0,487811374 | FGF13-AS1 | FALSE | Downregulated |
| 0,572334205 | FGF14-AS1 | FALSE | Upregulated |
| -0,74333418 | FGF14-IT1 | FALSE | Downregulated |
| 0 | FGF16 | FALSE | Upregulated |
| 1,107772064 | FGF17 | | Upregulated |
| 0,253508575 | FGF18 | FALSE | Upregulated |
| 0 | FGF19 | FALSE | Upregulated |
| | FGF20 | | |
| -0,984970599 | FGF21 | FALSE | Downregulated |
| 0,144893681 | FGF22 | FALSE | Upregulated |
| -0,745467041 | FGF23 | FALSE | Downregulated |
| | FGF3 | | |
| | FGF4 | | |
| 0,013603336 | FGF5 | FALSE | Upregulated |
| -0,010800911 | FGF6 | FALSE | Downregulated |
| -0,018149848 | FGF8 | FALSE | Downregulated |
| 0,287167769 | FGF9 | FALSE | Upregulated |
| 0 | FGFBP1 | FALSE | Upregulated |
| -0,017734654 | FGFBP2 | FALSE | Downregulated |

| | | | |
|---|---|---|---|
| -0,122951017 | FGFBP3 | FALSE | Downregulated |
| -0,166867844 | FHAD1 | FALSE | Downregulated |
| -0,498695183 | FHDC1 | FALSE | Downregulated |
| -0,192415179 | FHIT | FALSE | Downregulated |
| -0,046987061 | FHL2 | FALSE | Downregulated |
| 1,020576182 | FHL3 | | Upregulated |
| -0,120858612 | FHL5 | FALSE | Downregulated |
| 0,009595863 | FIBCD1 | FALSE | Upregulated |
| -0,318933731 | FIBIN | FALSE | Downregulated |
| 0,418438394 | FIGF | FALSE | Upregulated |
| | FIGLA | | |
| 0,41684618 | FIGNL1 | FALSE | Upregulated |
| 0,391106944 | FIRRE | FALSE | Upregulated |
| 0,091462143 | FITM1 | FALSE | Upregulated |
| -0,408371629 | FITM2 | FALSE | Downregulated |
| 0,510474293 | FJX1 | FALSE | Upregulated |
| 0,165991667 | FKBP1A-SDCBP2 | FALSE | Upregulated |
| -0,059514421 | FKBP1B | FALSE | Downregulated |
| 0,00408157 | FKBP6 | FALSE | Upregulated |
| 1,632714499 | FKBP9L | | Upregulated |
| -0,436818311 | FKBPL | FALSE | Downregulated |
| 0,482447159 | FKSG29 | FALSE | Upregulated |
| | FLJ16171 | | |
| | FLJ16779 | | |
| 0,817226351 | FLJ20021 | FALSE | Upregulated |
| 0,000358031 | FLJ20444 | FALSE | Upregulated |
| -0,005486539 | FLJ20518 | FALSE | Downregulated |
| -0,721452446 | FLJ20712 | FALSE | Downregulated |
| | FLJ21408 | | |
| 1,43826879 | FLJ22184 | | Upregulated |
| 0,837119766 | FLJ22447 | FALSE | Upregulated |
| -0,132968024 | FLJ22763 | FALSE | Downregulated |
| -0,442532051 | FLJ23867 | FALSE | Downregulated |
| 0,482394915 | FLJ25363 | FALSE | Upregulated |
| 0,307191428 | FLJ25715 | FALSE | Upregulated |
| | FLJ25758 | | |
| | FLJ25917 | | |
| | FLJ26245 | | |
| 0,415226009 | FLJ26850 | FALSE | Upregulated |
| 0,009677323 | FLJ27255 | FALSE | Upregulated |
| 0,320721557 | FLJ27354 | FALSE | Upregulated |
| 0,565806962 | FLJ30375 | FALSE | Upregulated |
| 0 | FLJ30679 | FALSE | Upregulated |
| 0,132821821 | FLJ31104 | FALSE | Upregulated |
| 0 | FLJ31356 | FALSE | Upregulated |
| | FLJ31662 | | |
| -0,035413855 | FLJ33360 | FALSE | Downregulated |
| | FLJ33534 | | |
| -0,156929476 | FLJ33544 | FALSE | Downregulated |
| 0,482283071 | FLJ33581 | FALSE | Upregulated |
| 0,62289467 | FLJ34503 | FALSE | Upregulated |

| | | | |
|---|---|---|---|
| 0,272355437 | FLJ35934 | FALSE | Upregulated |
| 0 | FLJ36000 | FALSE | Upregulated |
| -0,687389764 | FLJ36777 | FALSE | Downregulated |
| 0,112669202 | FLJ37035 | FALSE | Upregulated |
| -0,277040911 | FLJ37201 | FALSE | Downregulated |
| 0 | FLJ37505 | FALSE | Upregulated |
| 0,31760653 | FLJ38122 | FALSE | Upregulated |
| -0,295014569 | FLJ38379 | FALSE | Downregulated |
| -0,989052045 | FLJ38723 | FALSE | Downregulated |
| 0,271787299 | FLJ39080 | FALSE | Upregulated |
| -0,890721097 | FLJ39632 | FALSE | Downregulated |
| 0,121532474 | FLJ40194 | FALSE | Upregulated |
| 0 | FLJ40288 | FALSE | Upregulated |
| 0,426587325 | FLJ41200 | FALSE | Upregulated |
| | FLJ41278 | | |
| -0,018512747 | FLJ41309 | FALSE | Downregulated |
| | FLJ41423 | | |
| 0 | FLJ41733 | FALSE | Upregulated |
| 0,096073698 | FLJ41941 | FALSE | Upregulated |
| -0,144350802 | FLJ42102 | FALSE | Downregulated |
| -0,162480305 | FLJ42351 | FALSE | Downregulated |
| -0,082163342 | FLJ42393 | FALSE | Downregulated |
| | FLJ42842 | | |
| 0 | FLJ42969 | FALSE | Upregulated |
| -0,132131394 | FLJ43315 | FALSE | Downregulated |
| -0,314301942 | FLJ43585 | FALSE | Downregulated |
| 0,27019767 | FLJ43681 | FALSE | Upregulated |
| 0 | FLJ43879 | FALSE | Upregulated |
| -0,003473094 | FLJ44006 | FALSE | Downregulated |
| -0,432311649 | FLJ44087 | FALSE | Downregulated |
| -0,653974504 | FLJ44313 | FALSE | Downregulated |
| -0,841357391 | FLJ44511 | FALSE | Downregulated |
| 0,556476664 | FLJ44635 | FALSE | Upregulated |
| -0,138536495 | FLJ44790 | FALSE | Downregulated |
| | FLJ44838 | | |
| 0,289455674 | FLJ45079 | FALSE | Upregulated |
| -0,156014369 | FLJ45139 | FALSE | Downregulated |
| 0,005200034 | FLJ45256 | FALSE | Upregulated |
| -0,182330273 | FLJ45513 | FALSE | Downregulated |
| -0,151522101 | FLJ45743 | FALSE | Downregulated |
| 0,11913102 | FLJ45825 | FALSE | Upregulated |
| 0,076020674 | FLJ45872 | FALSE | Upregulated |
| | FLJ45964 | | |
| | FLJ45974 | | |
| 1,068240789 | FLJ46026 | | Upregulated |
| | FLJ46066 | | |
| -0,023445738 | FLJ46284 | FALSE | Downregulated |
| 0,575977546 | FLJ46363 | FALSE | Upregulated |
| -0,460271716 | FLJ46552 | FALSE | Downregulated |
| 1,001739224 | FLT3 | | Upregulated |
| -1,116680515 | FLT3LG | | Downregulated |

| | | | |
|---|---|---|---|
| -0,07171637 | FLVCR1-AS1 | FALSE | Downregulated |
| 0,348263506 | FMN2 | FALSE | Upregulated |
| 0,230585467 | FMO1 | FALSE | Upregulated |
| | FMO9P | | |
| 0,263915078 | FMR1-AS1 | FALSE | Upregulated |
| 0,284987956 | FMR1NB | FALSE | Upregulated |
| 0,142914537 | FNDC7 | FALSE | Upregulated |
| | FOLR3 | | |
| 0,406163295 | FOLR4 | FALSE | Upregulated |
| -0,752555391 | FOSB | FALSE | Downregulated |
| 0,596656299 | FOSL1 | FALSE | Upregulated |
| 0,841600162 | FOXB1 | FALSE | Upregulated |
| | FOXB2 | | |
| 0,212884355 | FOXC1 | FALSE | Upregulated |
| -0,52865188 | FOXC2 | FALSE | Downregulated |
| 0,297538904 | FOXD1 | FALSE | Upregulated |
| -0,505259057 | FOXD2-AS1 | FALSE | Downregulated |
| 0 | FOXD3 | FALSE | Upregulated |
| -0,270504736 | FOXD4 | FALSE | Downregulated |
| 0,114417004 | FOXD4L1 | FALSE | Upregulated |
| 0,413812961 | FOXD4L3 | FALSE | Upregulated |
| 0,486365405 | FOXD4L5 | FALSE | Upregulated |
| 0,423049593 | FOXD4L6 | FALSE | Upregulated |
| 0,093850831 | FOXE3 | FALSE | Upregulated |
| -0,091456641 | FOXF1 | FALSE | Downregulated |
| 0,145036632 | FOXF2 | FALSE | Upregulated |
| 0 | FOXG1 | FALSE | Upregulated |
| | FOXI1 | | |
| -0,43363027 | FOXI2 | FALSE | Downregulated |
| | FOXI3 | | |
| 0,238116163 | FOXJ1 | FALSE | Upregulated |
| 1,022960185 | FOXL1 | | Upregulated |
| -0,037452929 | FOXL2 | FALSE | Downregulated |
| | FOXN1 | | |
| -0,222763455 | FOXO3B | FALSE | Downregulated |
| -0,768096969 | FOXO6 | FALSE | Downregulated |
| 0,138660669 | FOXR1 | FALSE | Upregulated |
| -0,015614825 | FOXR2 | FALSE | Downregulated |
| -0,129892174 | FOXS1 | FALSE | Downregulated |
| -0,167925111 | FPGT-TNNI3K | FALSE | Downregulated |
| 0,414517866 | FPR1 | FALSE | Upregulated |
| 0,816522871 | FPR2 | FALSE | Upregulated |
| -0,966203951 | FRA10AC1 | FALSE | Downregulated |
| -0,622973808 | FRG1 | FALSE | Downregulated |
| -0,111953592 | FRG1B | FALSE | Downregulated |
| -0,831631696 | FRG2 | FALSE | Downregulated |
| 0,605211463 | FRG2B | FALSE | Upregulated |
| 0,817128157 | FRG2C | FALSE | Upregulated |
| 0,399091124 | FRMD1 | FALSE | Upregulated |
| 0,692913623 | FRMD6-AS1 | FALSE | Upregulated |
| -0,008081985 | FRMD6-AS2 | FALSE | Downregulated |

| | | | |
|---|---|---|---|
| 0,918168029 | FRMD7 | FALSE | Upregulated |
| 0,697379337 | FRMD8P1 | FALSE | Upregulated |
| 0,709640313 | FRMPD1 | FALSE | Upregulated |
| 0,280789448 | FRMPD2 | FALSE | Upregulated |
| -0,153247431 | FRMPD2P1 | FALSE | Downregulated |
| -0,923556937 | FRMPD3 | FALSE | Downregulated |
| 0,274060651 | FRMPD4 | FALSE | Upregulated |
| | FRRS1L | | |
| -0,304152107 | FRS3 | FALSE | Downregulated |
| 0,631969583 | FRZB | FALSE | Upregulated |
| 0,611662065 | FSBP | FALSE | Upregulated |
| -0,006848587 | FSCB | FALSE | Downregulated |
| 0,351023263 | FSCN2 | FALSE | Upregulated |
| | FSCN3 | | |
| -0,713448552 | FSD1 | FALSE | Downregulated |
| 0,0467443 | FSD1L | FALSE | Upregulated |
| 0,607541644 | FSHB | FALSE | Upregulated |
| 0,387101371 | FSHR | FALSE | Upregulated |
| 0,090880328 | FSIP1 | FALSE | Upregulated |
| 0 | FSTL4 | FALSE | Upregulated |
| | FSTL5 | | |
| 0,358333731 | FTCDNL1 | FALSE | Upregulated |
| | FTH1P18 | | |
| -0,023438982 | FTH1P3 | FALSE | Downregulated |
| | FTHL17 | | |
| | FTLP10 | | |
| | FTMT | | |
| -0,279923798 | FTO-IT1 | FALSE | Downregulated |
| 0,003850626 | FUNDC1 | FALSE | Upregulated |
| 0,032827584 | FUNDC2P2 | FALSE | Upregulated |
| -0,048250412 | FUT1 | FALSE | Downregulated |
| 0,573524941 | FUT5 | FALSE | Upregulated |
| 0,782310224 | FUT7 | FALSE | Upregulated |
| 1,053834041 | FUT9 | Upregulated | |
| -0,339658523 | FXYD2 | FALSE | Downregulated |
| 0,264628407 | FXYD3 | FALSE | Upregulated |
| | FXYD4 | | |
| -0,004056831 | FXYD6-FXYD2 | FALSE | Downregulated |
| -0,010079356 | FXYD7 | FALSE | Downregulated |
| 0 | FZD10 | FALSE | Upregulated |
| 0,029909091 | FZD10-AS1 | FALSE | Upregulated |
| -0,410686422 | FZD2 | FALSE | Downregulated |
| -0,582074217 | FZD3 | FALSE | Downregulated |
| 0,949203518 | FZD7 | FALSE | Upregulated |
| -0,156928137 | FZD9 | FALSE | Downregulated |
| 0,376400814 | G2E3 | FALSE | Upregulated |
| 0,000868546 | G6PC2 | FALSE | Upregulated |
| 0 | GAB4 | FALSE | Upregulated |
| 0,173384027 | GABARAPL3 | FALSE | Upregulated |
| 0,002827944 | GABBR2 | FALSE | Upregulated |
| | GABRA1 | | |

| | | | |
|---|---|---|---|
| 0,277119458 | GABRA2 | FALSE | Upregulated |
| 0,294278502 | GABRA3 | FALSE | Upregulated |
| 0,40517066 | GABRA4 | FALSE | Upregulated |
| | GABRA5 | | |
| | GABRA6 | | |
| 0,3221015 | GABRB1 | FALSE | Upregulated |
| | GABRB2 | | |
| -0,099542583 | GABRB3 | FALSE | Downregulated |
| 0,437794524 | GABRD | FALSE | Upregulated |
| 0,201064564 | GABRE | FALSE | Upregulated |
| 0,277764892 | GABRG1 | FALSE | Upregulated |
| | GABRG2 | | |
| 0,124396625 | GABRG3 | FALSE | Upregulated |
| -0,167136403 | GABRP | FALSE | Downregulated |
| | GABRQ | | |
| -0,02485822 | GABRR1 | FALSE | Downregulated |
| -0,128454767 | GABRR2 | FALSE | Downregulated |
| | GABRR3 | | |
| 0,727182384 | GAD1 | FALSE | Upregulated |
| | GAD2 | | |
| -0,300748274 | GADL1 | FALSE | Downregulated |
| | GAGE1 | | |
| | GAGE12C | | |
| | GAGE12D | | |
| | GAGE12E | | |
| | GAGE12G | | |
| | GAGE12H | | |
| | GAGE12I | | |
| | GAGE12J | | |
| | GAGE2A | | |
| | GAGE2C | | |
| | GAGE2E | | |
| 0 | GAL | FALSE | Upregulated |
| -0,633403168 | GAL3ST1 | FALSE | Downregulated |
| -0,495458337 | GAL3ST2 | FALSE | Downregulated |
| 0,13477164 | GAL3ST3 | FALSE | Upregulated |
| 0,629210263 | GAL3ST4 | FALSE | Upregulated |
| 0,268201766 | GALNT12 | FALSE | Upregulated |
| 0 | GALNT13 | FALSE | Upregulated |
| 0,764602739 | GALNT14 | FALSE | Upregulated |
| -0,876074815 | GALNT3 | FALSE | Downregulated |
| -0,804649872 | GALNT4 | FALSE | Downregulated |
| 0 | GALNT5 | FALSE | Upregulated |
| 0,323772911 | GALNT6 | FALSE | Upregulated |
| -0,147669431 | GALNT8 | FALSE | Downregulated |
| 1,607181789 | GALNT9 | | Upregulated |
| -0,014221116 | GALNT9-2 | FALSE | Downregulated |
| | GALNTL5 | | |
| 0,268141906 | GALNTL6 | FALSE | Upregulated |
| -0,175713062 | GALP | FALSE | Downregulated |
| 0 | GALR1 | FALSE | Upregulated |

| | | | |
|---|---|---|---|
| 0,279263106 | GALR2 | FALSE | Upregulated |
| 0,064208931 | GALR3 | FALSE | Upregulated |
| 0,476543073 | GAP43 | FALSE | Upregulated |
| 0,130234745 | GAPT | FALSE | Upregulated |
| -0,7277049 | GAR1 | FALSE | Downregulated |
| -0,141412426 | GAS2L2 | FALSE | Downregulated |
| 0,97757161 | GAS2L3 | FALSE | Upregulated |
| -0,246214887 | GAS6-AS2 | FALSE | Downregulated |
| -0,018149848 | GAST | FALSE | Downregulated |
| 0,541080054 | GATA1 | FALSE | Upregulated |
| 0,067458757 | GATA2 | FALSE | Upregulated |
| 0,56644908 | GATA3 | FALSE | Upregulated |
| 0,247386795 | GATA3-AS1 | FALSE | Upregulated |
| 0,911299737 | GATA5 | FALSE | Upregulated |
| -0,02264752 | GATA6-AS1 | FALSE | Downregulated |
| 1,843285049 | GATSL2 | | Upregulated |
| -0,387045717 | GATSL3 | FALSE | Downregulated |
| -1,191797492 | GBAP1 | | Downregulated |
| -0,256335618 | GBGT1 | FALSE | Downregulated |
| 0,400159951 | GBP1P1 | FALSE | Upregulated |
| -0,75905077 | GBP6 | FALSE | Downregulated |
| -0,023419232 | GBX1 | FALSE | Downregulated |
| 0,118953846 | GBX2 | FALSE | Upregulated |
| 0 | GCG | FALSE | Upregulated |
| 0,121727732 | GCK | FALSE | Upregulated |
| 0 | GCM1 | FALSE | Upregulated |
| 0,631851991 | GCM2 | FALSE | Upregulated |
| -0,153890929 | GCNT3 | FALSE | Downregulated |
| -0,473864697 | GCNT4 | FALSE | Downregulated |
| 0 | GCSAML | FALSE | Upregulated |
| 0,270597153 | GCSAML-AS1 | FALSE | Upregulated |
| 0,432059911 | GDAP1 | FALSE | Upregulated |
| -0,050796165 | GDF1 | FALSE | Downregulated |
| -0,177573516 | GDF10 | FALSE | Downregulated |
| -0,238178503 | GDF15 | FALSE | Downregulated |
| | GDF3 | | |
| 0,126606979 | GDF5 | FALSE | Upregulated |
| -0,217904677 | GDF6 | FALSE | Downregulated |
| 1,06685682 | GDF9 | | Upregulated |
| 0,427926302 | GDNF | FALSE | Upregulated |
| 1,174136232 | GDNF-AS1 | | Upregulated |
| 0,453020123 | GDPD1 | FALSE | Upregulated |
| 0,838215868 | GDPD2 | FALSE | Upregulated |
| 0,886312677 | GDPD3 | FALSE | Upregulated |
| 0,098329239 | GDPGP1 | FALSE | Upregulated |
| -0,609673907 | GEMIN6 | FALSE | Downregulated |
| 0,594055056 | GFAP | FALSE | Upregulated |
| 1,1203229 | GFI1 | | Upregulated |
| | GFI1B | | |
| -0,035182796 | GFPT2 | FALSE | Downregulated |
| -1,707849094 | GFRA2 | | Downregulated |

| | | | |
|---|---|---|---|
| 0 | GFRA3 | FALSE | Upregulated |
| 0,012749608 | GFRA4 | FALSE | Upregulated |
| 0,018964661 | GFRAL | FALSE | Upregulated |
| | GFY | | |
| 0,412337995 | GGNBP1 | FALSE | Upregulated |
| -0,179583164 | GGT2 | FALSE | Downregulated |
| 0 | GGT3P | FALSE | Upregulated |
| 0,315668227 | GGT6 | FALSE | Upregulated |
| 0 | GGT8P | FALSE | Upregulated |
| 0 | GGTLC1 | FALSE | Upregulated |
| 1,084234411 | GGTLC2 | | Upregulated |
| | GH1 | | |
| | GH2 | | |
| | GHRH | | |
| 0 | GHRHR | FALSE | Upregulated |
| 0,153198235 | GHSR | FALSE | Upregulated |
| 0,275825549 | GIF | FALSE | Upregulated |
| | GIMD1 | | |
| 0,15450394 | GIN1 | FALSE | Upregulated |
| 0,456614866 | GINS3 | FALSE | Upregulated |
| | GIP | | |
| -0,236356194 | GIPR | FALSE | Downregulated |
| 0,613209847 | GJA1 | FALSE | Upregulated |
| | GJA10 | | |
| 0,555333805 | GJA3 | FALSE | Upregulated |
| -0,497667874 | GJA5 | FALSE | Downregulated |
| 0,295243574 | GJA8 | FALSE | Upregulated |
| 0 | GJB6 | FALSE | Upregulated |
| -0,919155836 | GJC1 | FALSE | Downregulated |
| -0,559016379 | GJC2 | FALSE | Downregulated |
| 0,348945923 | GJC3 | FALSE | Upregulated |
| | GJD2 | | |
| 0 | GJD4 | FALSE | Upregulated |
| | GK2 | | |
| 0,024454908 | GKAP1 | FALSE | Upregulated |
| | GKN1 | | |
| | GKN2 | | |
| 0,442253265 | GLA | FALSE | Upregulated |
| 0,307521572 | GLB1L3 | FALSE | Upregulated |
| 0,738391358 | GLDN | FALSE | Upregulated |
| 0,005726792 | GLI3 | FALSE | Upregulated |
| -0,334431089 | GLIPR1L2 | FALSE | Downregulated |
| 0,267134809 | GLIPR2 | FALSE | Upregulated |
| 0,283437811 | GLIS1 | FALSE | Upregulated |
| -0,196787147 | GLIS3-AS1 | FALSE | Downregulated |
| -0,502535051 | GLMN | FALSE | Downregulated |
| -0,125097608 | GLOD5 | FALSE | Downregulated |
| -0,140715174 | GLP1R | FALSE | Downregulated |
| 0,24202954 | GLP2R | FALSE | Upregulated |
| 0,421261967 | GLRA2 | FALSE | Upregulated |
| 0 | GLRA3 | FALSE | Upregulated |

| | | | |
|---|---|---|---|
| 0,285372545 | GLRA4 | FALSE | Upregulated |
| -0,005005647 | GLRB | FALSE | Downregulated |
| 1,134092338 | GLRX2 | | Upregulated |
| | GLT6D1 | | |
| | GLYATL2 | | |
| 0,239412933 | GLYATL3 | FALSE | Upregulated |
| | GLYCAM1 | | |
| | GM140 | | |
| -0,612576493 | GMCL1 | FALSE | Downregulated |
| -0,004191099 | GMCL1P1 | FALSE | Downregulated |
| 0,505255303 | GMDS-AS1 | FALSE | Upregulated |
| 0,571796061 | GMFG | FALSE | Upregulated |
| 0,538006232 | GML | FALSE | Upregulated |
| -0,059137514 | GMNC | FALSE | Downregulated |
| -0,274220626 | GMNN | FALSE | Downregulated |
| 0,285231862 | GMPR | FALSE | Upregulated |
| 0,706598674 | GNA15 | FALSE | Upregulated |
| -0,009465128 | GNAS-AS1 | FALSE | Downregulated |
| | GNAT3 | | |
| 0,226750327 | GNAZ | FALSE | Upregulated |
| 0,449184411 | GNB1L | FALSE | Upregulated |
| 0,827468535 | GNB3 | FALSE | Upregulated |
| 0,046755381 | GNG10 | FALSE | Upregulated |
| -1,14455682 | GNG11 | | Downregulated |
| 0,283538547 | GNG13 | FALSE | Upregulated |
| 0,380703859 | GNG4 | FALSE | Upregulated |
| | GNG8 | | |
| | GNGT1 | | |
| 1,150566235 | GNGT2 | | Upregulated |
| 0,306892941 | GNLY | FALSE | Upregulated |
| -0,144876487 | GNN | FALSE | Downregulated |
| 0,366318571 | GNRH2 | FALSE | Upregulated |
| 0,285239188 | GOLGA2P2Y | FALSE | Upregulated |
| -1,528469562 | GOLGA6D | | Downregulated |
| | GOLGA6L1 | | |
| 0,503659882 | GOLGA6L10 | FALSE | Upregulated |
| -0,158343621 | GOLGA6L2 | FALSE | Downregulated |
| 0,742542323 | GOLGA6L3 | FALSE | Upregulated |
| 0,51000109 | GOLGA6L4 | FALSE | Upregulated |
| 0 | GOLGA6L6 | FALSE | Upregulated |
| 0,364625982 | GOLGA6L7P | FALSE | Upregulated |
| 0,30667946 | GOLGA7B | FALSE | Upregulated |
| | GOLGA8CP | | |
| | GOLGA8DP | | |
| 0 | GOLGA8EP | FALSE | Upregulated |
| 0 | GOLGA8F | FALSE | Upregulated |
| | GOLGA8G | | |
| -0,841196115 | GOLGA8I | FALSE | Downregulated |
| -0,423045859 | GOLGA8J | FALSE | Downregulated |
| -0,487890293 | GOLGA8K | FALSE | Downregulated |
| 0,875401848 | GOLGA8S | FALSE | Upregulated |

| | | | |
|---|---|---|---|
| -0,796557967 | GOLGA8T | FALSE | Downregulated |
| -0,103601372 | GORAB | FALSE | Downregulated |
| | GOT1L1 | | |
| -0,020898376 | GP1BB | FALSE | Downregulated |
| 0 | GP2 | FALSE | Upregulated |
| 0,268024425 | GP9 | FALSE | Upregulated |
| 0,575069922 | GPA33 | FALSE | Upregulated |
| 0 | GPC2 | FALSE | Upregulated |
| 1,751998087 | GPC3 | | Upregulated |
| -0,279939433 | GPC4 | FALSE | Downregulated |
| -0,719309148 | GPC5 | FALSE | Downregulated |
| -0,463658996 | GPC5-AS1 | FALSE | Downregulated |
| -0,058860673 | GPC6-AS1 | FALSE | Downregulated |
| 0,246341751 | GPC6-AS2 | FALSE | Upregulated |
| -0,202788132 | GPD1L | FALSE | Downregulated |
| -0,460125148 | GPHA2 | FALSE | Downregulated |
| | GPHB5 | | |
| 0,336337139 | GPIHBP1 | FALSE | Upregulated |
| -0,434666594 | GPM6A | FALSE | Downregulated |
| 0,246847156 | GPR1 | FALSE | Upregulated |
| 0 | GPR1-AS | FALSE | Upregulated |
| -0,030381139 | GPR101 | FALSE | Downregulated |
| 0 | GPR110 | FALSE | Upregulated |
| 0 | GPR111 | FALSE | Upregulated |
| | GPR112 | | |
| | GPR115 | | |
| 0 | GPR119 | FALSE | Upregulated |
| 0 | GPR12 | FALSE | Upregulated |
| -1,070251068 | GPR123 | | Downregulated |
| 0,268947471 | GPR135 | FALSE | Upregulated |
| 0,59378855 | GPR137C | FALSE | Upregulated |
| | GPR139 | | |
| 0,929987428 | GPR141 | FALSE | Upregulated |
| 0,207792937 | GPR142 | FALSE | Upregulated |
| -0,424431944 | GPR143 | FALSE | Downregulated |
| 0 | GPR144 | FALSE | Upregulated |
| | GPR148 | | |
| -0,134978682 | GPR149 | FALSE | Downregulated |
| 1,036591756 | GPR15 | | Upregulated |
| 0,56851033 | GPR150 | FALSE | Upregulated |
| -0,322969665 | GPR151 | FALSE | Downregulated |
| 0,185570993 | GPR152 | FALSE | Upregulated |
| 0,575930436 | GPR156 | FALSE | Upregulated |
| 0,123094131 | GPR157 | FALSE | Upregulated |
| -0,224784712 | GPR158 | FALSE | Downregulated |
| -0,354889056 | GPR158-AS1 | FALSE | Downregulated |
| -0,32933812 | GPR171 | FALSE | Downregulated |
| -0,656653668 | GPR173 | FALSE | Downregulated |
| 1,32382246 | GPR174 | | Upregulated |
| -0,079051657 | GPR176 | FALSE | Downregulated |
| 0,464850708 | GPR19 | FALSE | Upregulated |

| | | | |
|---|---|---|---|
| 0,121928927 | GPR20 | FALSE | Upregulated |
| 0,36927663 | GPR21 | FALSE | Upregulated |
| 0,767634957 | GPR25 | FALSE | Upregulated |
| 0,447082254 | GPR26 | FALSE | Upregulated |
| 0,274742921 | GPR27 | FALSE | Upregulated |
| -0,141700675 | GPR3 | FALSE | Downregulated |
| 0,272979989 | GPR31 | FALSE | Upregulated |
| | GPR32 | | |
| 0,134038697 | GPR33 | FALSE | Upregulated |
| 0,376770075 | GPR35 | FALSE | Upregulated |
| 0,855124437 | GPR37L1 | FALSE | Upregulated |
| -0,188955696 | GPR4 | FALSE | Downregulated |
| 0,836167845 | GPR50 | FALSE | Upregulated |
| 0,082347177 | GPR52 | FALSE | Upregulated |
| -0,001973316 | GPR55 | FALSE | Downregulated |
| | GPR6 | | |
| -0,13270436 | GPR63 | FALSE | Downregulated |
| -0,872860628 | GPR75-ASB3 | FALSE | Downregulated |
| 0 | GPR78 | FALSE | Upregulated |
| 0,176569957 | GPR83 | FALSE | Upregulated |
| 0,420427233 | GPR84 | FALSE | Upregulated |
| 0,027884155 | GPR85 | FALSE | Upregulated |
| 0,82144885 | GPR87 | FALSE | Upregulated |
| 0,160480305 | GPR97 | FALSE | Upregulated |
| 0,141585506 | GPRC5A | FALSE | Upregulated |
| -0,142541544 | GPRC6A | FALSE | Downregulated |
| 0,278048401 | GPRIN2 | FALSE | Upregulated |
| -0,313859633 | GPX5 | FALSE | Downregulated |
| | GPX6 | | |
| -0,129021233 | GPX8 | FALSE | Downregulated |
| 1,771162011 | GRAP2 | | Upregulated |
| -0,890156948 | GRAPL | FALSE | Downregulated |
| 0,349025041 | GREM1 | FALSE | Upregulated |
| -0,651207365 | GRHL3 | FALSE | Downregulated |
| -0,927206997 | GRIA1 | FALSE | Downregulated |
| -0,562954103 | GRIA2 | FALSE | Downregulated |
| -0,130349067 | GRIA4 | FALSE | Downregulated |
| 0,523511 | GRID1 | FALSE | Upregulated |
| -0,452936841 | GRID1-AS1 | FALSE | Downregulated |
| | GRID2 | | |
| 0,707920325 | GRID2IP | FALSE | Upregulated |
| | GRIK1-AS1 | | |
| 0,174682212 | GRIK1-AS2 | FALSE | Upregulated |
| -0,039389179 | GRIK2 | FALSE | Downregulated |
| 0,86549168 | GRIK3 | FALSE | Upregulated |
| -0,081805902 | GRIK4 | FALSE | Downregulated |
| 0,748893113 | GRIK5 | FALSE | Upregulated |
| 0,117705792 | GRIN1 | FALSE | Upregulated |
| | GRK1 | | |
| | GRK1-2 | | |
| -0,427718994 | GRK4 | FALSE | Downregulated |

| | | | |
|---|---|---|---|
| 0,055801719 | GRK5-IT1 | FALSE | Upregulated |
| 0 | GRK7 | FALSE | Upregulated |
| 0,157868672 | GRM1 | FALSE | Upregulated |
| 0,587136059 | GRM2 | FALSE | Upregulated |
| 0,109035103 | GRM3 | FALSE | Upregulated |
| 0 | GRM4 | FALSE | Upregulated |
| 0,567457138 | GRM5 | FALSE | Upregulated |
| 0,118719743 | GRM5-AS1 | FALSE | Upregulated |
| 0,129659399 | GRM6 | FALSE | Upregulated |
| 0,468301461 | GRM7 | FALSE | Upregulated |
| -0,104494492 | GRM8 | FALSE | Downregulated |
| | GRP | | |
| 0,284662013 | GRPR | FALSE | Upregulated |
| | GRXCR1 | | |
| | GRXCR2 | | |
| | GS1-122H1.2 | | |
| | GS1-18A18.1 | | |
| 0 | GS1-204I12.1 | FALSE | Upregulated |
| 0 | GS1-24F4.2 | FALSE | Upregulated |
| 0,371123149 | GS1-251I9.4 | FALSE | Upregulated |
| -0,005937314 | GS1-259H13.2 | FALSE | Downregulated |
| -0,119567003 | GS1-279B7.1 | FALSE | Downregulated |
| | GS1-600G8.3 | | |
| 0 | GSC | FALSE | Upregulated |
| | GSC2 | | |
| 0,145418776 | GSDMA | FALSE | Upregulated |
| 0 | GSDMC | FALSE | Upregulated |
| 0,009536804 | GSG1L | FALSE | Upregulated |
| -0,155234188 | GSKIP | FALSE | Downregulated |
| 0,002108976 | GSTA3 | FALSE | Upregulated |
| 0,367683195 | GSTA4 | FALSE | Upregulated |
| -0,170705185 | GSTA5 | FALSE | Downregulated |
| 0,184995447 | GSTCD | FALSE | Upregulated |
| -0,176618435 | GSTM2 | FALSE | Downregulated |
| -0,83931016 | GSTM2P1 | FALSE | Downregulated |
| 0,097010221 | GSTM5 | FALSE | Upregulated |
| | GSTT2 | | |
| -0,026426612 | GSTT2B | FALSE | Downregulated |
| 0 | GSTTP1 | FALSE | Upregulated |
| | GSX1 | | |
| | GSX2 | | |
| -0,022358994 | GTF2A1L | FALSE | Downregulated |
| -0,310850235 | GTF2E1 | FALSE | Downregulated |
| -0,156090683 | GTF2F2 | FALSE | Downregulated |
| 0,719942548 | GTF2H2 | FALSE | Upregulated |
| | GTPBP6-2 | | |
| -0,011481061 | GTSCR1 | FALSE | Downregulated |
| 1,20334203 | GTSE1 | | Upregulated |
| -0,174020604 | GTSF1 | FALSE | Downregulated |
| | GTSF1L | | |
| -0,490433805 | GUCA1A | FALSE | Downregulated |

| | | | |
|---|---|---|---|
| -0,176662645 | GUCA1B | FALSE | Downregulated |
| | GUCA1C | | |
| 0,026179022 | GUCA2A | FALSE | Upregulated |
| -0,402449427 | GUCA2B | FALSE | Downregulated |
| 1,233281697 | GUCY1B2 | | Upregulated |
| -0,793609478 | GUCY1B3 | FALSE | Downregulated |
| -0,142398235 | GUCY2D | FALSE | Downregulated |
| 0,134968739 | GUCY2EP | FALSE | Upregulated |
| | GUCY2F | | |
| | GUCY2GP | | |
| 0,438130622 | GULP1 | FALSE | Upregulated |
| | GUSBP10 | | |
| -1,56636513 | GUSBP2 | | Downregulated |
| 1,391773267 | GUSBP4 | | Upregulated |
| 0,194470992 | GUSBP5 | FALSE | Upregulated |
| -0,562113526 | GUSBP9 | FALSE | Downregulated |
| -0,13846111 | GXYLT2 | FALSE | Downregulated |
| 0,143230172 | GYG2P1 | FALSE | Upregulated |
| -0,10338844 | GYLTL1B | FALSE | Downregulated |
| 0,863424188 | GYPA | FALSE | Upregulated |
| 0,000535661 | GYPB | FALSE | Upregulated |
| 0,636240988 | GZMA | FALSE | Upregulated |
| 0,093974683 | GZMB | FALSE | Upregulated |
| 1,091614166 | GZMH | | Upregulated |
| 0,069781026 | GZMK | FALSE | Upregulated |
| -0,03798482 | GZMM | FALSE | Downregulated |
| 1,120499398 | H1FNT | | Upregulated |
| -0,287772384 | H1FOO | FALSE | Downregulated |
| 0,322566824 | H2AFB1 | FALSE | Upregulated |
| -1,460038656 | H2AFB2 | | Downregulated |
| -0,065573882 | H2AFB3 | FALSE | Downregulated |
| | H2BFM | | |
| | H2BFWT | | |
| 0,007931427 | H2BFXP | FALSE | Upregulated |
| -0,159222125 | H3F3C | FALSE | Downregulated |
| | HAND1 | | |
| -0,259115851 | HAND2-AS1 | FALSE | Downregulated |
| -0,484825429 | HAP1 | FALSE | Downregulated |
| -0,006322129 | HAPLN1 | FALSE | Downregulated |
| -0,001822665 | HAPLN2 | FALSE | Downregulated |
| 1,034189631 | HAPLN3 | | Upregulated |
| 0,734599375 | HAR1A | FALSE | Upregulated |
| 0,283092522 | HAR1B | FALSE | Upregulated |
| 0,966041559 | HARBI1 | FALSE | Upregulated |
| -0,001237159 | HAS1 | FALSE | Downregulated |
| 0,936095565 | HAS2 | FALSE | Upregulated |
| 0,807542838 | HAS2-AS1 | FALSE | Upregulated |
| -0,480444977 | HAT1 | FALSE | Downregulated |
| -0,267587171 | HAUS1 | FALSE | Downregulated |
| -0,152623161 | HAUS8 | FALSE | Downregulated |
| 0,459290636 | HAVCR1 | FALSE | Upregulated |

| | | | |
|---|---|---|---|
| 0,879975426 | HBEGF | FALSE | Upregulated |
| 0,491979732 | HBZ | FALSE | Upregulated |
| -0,607472885 | HCAR1 | FALSE | Downregulated |
| 0,171898854 | HCAR2 | FALSE | Upregulated |
| -0,356449663 | HCCAT3 | FALSE | Downregulated |
| 0,254663908 | HCCAT4 | FALSE | Upregulated |
| 0,721832171 | HCCAT5 | FALSE | Upregulated |
| 0,164525421 | HCCS | FALSE | Upregulated |
| 0,898804713 | HCG11 | FALSE | Upregulated |
| -0,030966505 | HCG14 | FALSE | Downregulated |
| -0,02382056 | HCG16 | FALSE | Downregulated |
| | HCG17 | | |
| 0,277119458 | HCG22 | FALSE | Upregulated |
| 0,005298914 | HCG23 | FALSE | Upregulated |
| 0,850210576 | HCG25 | FALSE | Upregulated |
| 1,123827079 | HCG27 | | Upregulated |
| -0,161608103 | HCG4 | FALSE | Downregulated |
| -0,078688562 | HCG8 | FALSE | Downregulated |
| -0,124672571 | HCG9 | FALSE | Downregulated |
| 0,291534484 | HCN1 | FALSE | Upregulated |
| -0,095025216 | HCN2 | FALSE | Downregulated |
| 1,43992873 | HCN4 | | Upregulated |
| | HCRT | | |
| | HCRTR2 | | |
| 0,083998979 | HCST | FALSE | Upregulated |
| -0,502464663 | HDAC11 | FALSE | Downregulated |
| 0,277833435 | HDAC11-AS1 | FALSE | Upregulated |
| -0,375944623 | HDC | FALSE | Downregulated |
| -0,626320443 | HDDC2 | FALSE | Downregulated |
| | HDGFL1 | | |
| -0,514166478 | HDX | FALSE | Downregulated |
| 0,107176638 | HECW1 | FALSE | Upregulated |
| 0,025724157 | HELLS | FALSE | Upregulated |
| | HELT | | |
| -0,001211241 | HEMGN | FALSE | Downregulated |
| -0,06336297 | HENMT1 | FALSE | Downregulated |
| | HEPACAM2 | | |
| 0,424568163 | HEPH | FALSE | Upregulated |
| -0,175798624 | HERC2P10 | FALSE | Downregulated |
| 0,011328303 | HERC2P4 | FALSE | Upregulated |
| -1,784907317 | HERC2P7 | | Downregulated |
| 0 | HES2 | FALSE | Upregulated |
| 0 | HES3 | FALSE | Upregulated |
| -0,425891564 | HES5 | FALSE | Downregulated |
| -0,214807816 | HES6 | FALSE | Downregulated |
| 0,274635952 | HES7 | FALSE | Upregulated |
| 0,776018722 | HESX1 | FALSE | Upregulated |
| 0,655345322 | HEXA-AS1 | FALSE | Upregulated |
| -1,094086173 | HEY1 | | Downregulated |
| 0,566633487 | HFM1 | FALSE | Upregulated |
| 0,195811242 | HGC6.3 | FALSE | Upregulated |

| | | | |
|---|---|---|---|
| 0,097094684 | HHAT | FALSE | Upregulated |
| 0,398863263 | HHATL | FALSE | Upregulated |
| -0,039071788 | HHIP-AS1 | FALSE | Downregulated |
| -0,006903421 | HHIPL2 | FALSE | Downregulated |
| | HHLA1 | | |
| -0,328156211 | HHLA2 | FALSE | Downregulated |
| 0,136448393 | HHLA3 | FALSE | Upregulated |
| 0 | HIF1A-AS1 | FALSE | Upregulated |
| 0,80126852 | HIF3A | FALSE | Upregulated |
| 0,297178435 | HIGD2B | FALSE | Upregulated |
| -0,855950371 | HILPDA | FALSE | Downregulated |
| 0,78539012 | HILS1 | FALSE | Upregulated |
| -0,013960773 | HIST1H1T | FALSE | Downregulated |
| 0,417613283 | HIST1H2AA | FALSE | Upregulated |
| 0,228278453 | HIST1H2AB | FALSE | Upregulated |
| 0,545796723 | HIST1H2AD | FALSE | Upregulated |
| -0,72533346 | HIST1H2AE | FALSE | Downregulated |
| 0,815679709 | HIST1H2AG | FALSE | Upregulated |
| -0,051664864 | HIST1H2AH | FALSE | Downregulated |
| 0,835242238 | HIST1H2AI | FALSE | Upregulated |
| -0,18670688 | HIST1H2AJ | FALSE | Downregulated |
| 0,18973491 | HIST1H2AK | FALSE | Upregulated |
| -0,490236908 | HIST1H2AL | FALSE | Downregulated |
| 0,063244283 | HIST1H2AM | FALSE | Upregulated |
| 0,112759244 | HIST1H2BB | FALSE | Upregulated |
| 0,859636128 | HIST1H2BE | FALSE | Upregulated |
| -0,451236589 | HIST1H2BF | FALSE | Downregulated |
| 0,667607217 | HIST1H2BG | FALSE | Upregulated |
| -0,348990886 | HIST1H2BH | FALSE | Downregulated |
| -0,326576346 | HIST1H2BI | FALSE | Downregulated |
| -0,131890567 | HIST1H2BJ | FALSE | Downregulated |
| 0,055556946 | HIST1H2BL | FALSE | Upregulated |
| 0,705156384 | HIST1H2BM | FALSE | Upregulated |
| -0,081096308 | HIST1H2BN | FALSE | Downregulated |
| 0,429668866 | HIST1H2BO | FALSE | Upregulated |
| 0,721725048 | HIST1H3A | FALSE | Upregulated |
| -0,603099424 | HIST1H3B | FALSE | Downregulated |
| 0,926187029 | HIST1H3C | FALSE | Upregulated |
| 0,119045971 | HIST1H3E | FALSE | Upregulated |
| 0,697726327 | HIST1H3F | FALSE | Upregulated |
| -0,524478114 | HIST1H3G | FALSE | Downregulated |
| -0,058899363 | HIST1H3H | FALSE | Downregulated |
| 0,259333107 | HIST1H3I | FALSE | Upregulated |
| 0,119767184 | HIST1H3J | FALSE | Upregulated |
| 0,31897014 | HIST1H4A | FALSE | Upregulated |
| -0,252920056 | HIST1H4B | FALSE | Downregulated |
| 0,411555941 | HIST1H4D | FALSE | Upregulated |
| 0,400686095 | HIST1H4F | FALSE | Upregulated |
| | HIST1H4G | | |
| -0,329164702 | HIST1H4H | FALSE | Downregulated |
| 0,605126222 | HIST1H4I | FALSE | Upregulated |

| | | | |
|---|---|---|---|
| 0,36685081 | HIST1H4L | FALSE | Upregulated |
| -0,029729931 | HIST2H2BA | FALSE | Downregulated |
| -0,631764849 | HIST2H2BC | FALSE | Downregulated |
| 0,552417225 | HIST2H3D | FALSE | Upregulated |
| 0,462605067 | HIST2H4A | FALSE | Upregulated |
| 0,803148032 | HIST2H4B | FALSE | Upregulated |
| 0,141025594 | HIST3H2A | FALSE | Upregulated |
| 0,595274815 | HIST3H2BB | FALSE | Upregulated |
| 0,282730609 | HIST3H3 | FALSE | Upregulated |
| 0,467319389 | HIST4H4 | FALSE | Upregulated |
| 0,760075193 | HJURP | FALSE | Upregulated |
| 0,001138372 | HLA-DOB | FALSE | Upregulated |
| 0,559392525 | HLA-DPB2 | FALSE | Upregulated |
| 0,299051581 | HLA-DQA2 | FALSE | Upregulated |
| 1,806121274 | HLA-DQB2 | | Upregulated |
| 0,761331456 | HLA-G | FALSE | Upregulated |
| 0,37943394 | HLTF-AS1 | FALSE | Upregulated |
| 0,148668804 | HM13-AS1 | FALSE | Upregulated |
| | HMGA1P7 | | |
| -0,246550243 | HMGA2 | FALSE | Downregulated |
| 0,743067435 | HMGB3P1 | FALSE | Upregulated |
| 0,134831038 | HMGB4 | FALSE | Upregulated |
| 0,46710069 | HMGCLL1 | FALSE | Upregulated |
| -0,721363766 | HMGN2P46 | FALSE | Downregulated |
| -0,383206756 | HMGN5 | FALSE | Downregulated |
| | HMHB1 | | |
| -0,305878962 | HMMR | FALSE | Downregulated |
| -0,027098078 | HMP19 | FALSE | Downregulated |
| 0,592901165 | HMSD | FALSE | Upregulated |
| | HMX1 | | |
| | HMX2 | | |
| | HMX3 | | |
| 0,595686769 | HNRNPA1L2 | FALSE | Upregulated |
| 0,05518745 | HNRNPA1P33 | FALSE | Upregulated |
| 0,27961062 | HNRNPA3P1 | FALSE | Upregulated |
| 0 | HNRNPCL1 | FALSE | Upregulated |
| | HNRNPCP5 | | |
| | HNRNPKP3 | | |
| -1,402271179 | HOMER1 | | Downregulated |
| 1,01624499 | HOPX | | Upregulated |
| -0,148760664 | HORMAD1 | FALSE | Downregulated |
| -1,200510089 | HORMAD2 | | Downregulated |
| | HOTAIR | | |
| 0,840872496 | HOTAIRM1 | FALSE | Upregulated |
| 0 | HOTTIP | FALSE | Upregulated |
| 0,769982463 | HOXA-AS3 | FALSE | Upregulated |
| -0,168895708 | HOXA1 | FALSE | Downregulated |
| 0,158161207 | HOXA10 | FALSE | Upregulated |
| 0,155215715 | HOXA10-AS | FALSE | Upregulated |
| | HOXA10-HOXA9 | | |
| | HOXA11 | | |

| | | | |
|---|---|---|---|
| | HOXA11-AS | | |
| 0 | HOXA13 | FALSE | Upregulated |
| -0,230550655 | HOXA3 | FALSE | Downregulated |
| -0,060299428 | HOXA4 | FALSE | Downregulated |
| -0,393480546 | HOXA5 | FALSE | Downregulated |
| -0,180433808 | HOXA6 | FALSE | Downregulated |
| 0,427989698 | HOXA7 | FALSE | Upregulated |
| 0 | HOXA9 | FALSE | Upregulated |
| | HOXB-AS1 | | |
| 0,570642185 | HOXB-AS3 | FALSE | Upregulated |
| | HOXB1 | | |
| | HOXB13 | | |
| -0,204979923 | HOXB2 | FALSE | Downregulated |
| 0,077270731 | HOXB4 | FALSE | Upregulated |
| -0,140303884 | HOXB5 | FALSE | Downregulated |
| -0,20677828 | HOXB6 | FALSE | Downregulated |
| -0,154532913 | HOXB7 | FALSE | Downregulated |
| -0,009162334 | HOXB8 | FALSE | Downregulated |
| 0 | HOXB9 | FALSE | Upregulated |
| | HOXC-AS1 | | |
| | HOXC-AS2 | | |
| | HOXC-AS3 | | |
| 0 | HOXC10 | FALSE | Upregulated |
| 0 | HOXC11 | FALSE | Upregulated |
| | HOXC12 | | |
| 0 | HOXC13 | FALSE | Upregulated |
| | HOXC13-AS | | |
| 0,144014046 | HOXC4 | FALSE | Upregulated |
| | HOXC5 | | |
| 0,275337823 | HOXC6 | FALSE | Upregulated |
| | HOXC8 | | |
| 0 | HOXC9 | FALSE | Upregulated |
| 0,28946842 | HOXD-AS1 | FALSE | Upregulated |
| 0,37298162 | HOXD-AS2 | FALSE | Upregulated |
| 0 | HOXD1 | FALSE | Upregulated |
| | HOXD10 | | |
| 0 | HOXD11 | FALSE | Upregulated |
| | HOXD12 | | |
| | HOXD13 | | |
| -0,000467477 | HOXD3 | FALSE | Downregulated |
| 0,262068522 | HOXD4 | FALSE | Upregulated |
| 0,405368737 | HOXD8 | FALSE | Upregulated |
| 0 | HOXD9 | FALSE | Upregulated |
| | HP08942 | | |
| 0,389267327 | HPCA | FALSE | Upregulated |
| 0,649036864 | HPCAL4 | FALSE | Upregulated |
| 0,610548866 | HPGDS | FALSE | Upregulated |
| 1,377977238 | HPSE | Upregulated | |
| 0 | HPSE2 | FALSE | Upregulated |
| 0,00139671 | HPVC1 | FALSE | Upregulated |
| | HPYR1 | | |

| | | | |
|---|---|---|---|
| 0,188523934 | HR | FALSE | Upregulated |
| -0,172279053 | HRASLS | FALSE | Downregulated |
| -0,616343037 | HRASLS2 | FALSE | Downregulated |
| 0,132077442 | HRASLS5 | FALSE | Upregulated |
| 0,023988805 | HRC | FALSE | Upregulated |
| 0,146300369 | HRCT1 | FALSE | Upregulated |
| 0,938476278 | HRH1 | FALSE | Upregulated |
| -0,318044119 | HRH2 | FALSE | Downregulated |
| 0,925107409 | HRH3 | FALSE | Upregulated |
| -0,153593877 | HRH4 | FALSE | Downregulated |
| | HRK | | |
| 0,19859987 | HS1BP3-IT1 | FALSE | Upregulated |
| 0,169846331 | HS3ST2 | FALSE | Upregulated |
| 0,280716753 | HS3ST4 | FALSE | Upregulated |
| 0,659787377 | HS3ST5 | FALSE | Upregulated |
| 0,304857807 | HS3ST6 | FALSE | Upregulated |
| 0,110220923 | HS6ST2 | FALSE | Upregulated |
| | HS6ST2-AS1 | | |
| | HS6ST3 | | |
| -0,598226979 | HSD11B1L | FALSE | Downregulated |
| 0,472286088 | HSD17B7 | FALSE | Upregulated |
| 0,592739539 | HSD17B7P2 | FALSE | Upregulated |
| 0,818541437 | HSD3B1 | FALSE | Upregulated |
| -0,31032143 | HSD3B2 | FALSE | Downregulated |
| 0,277119458 | HSD52 | FALSE | Upregulated |
| -0,121404219 | HSDL1 | FALSE | Downregulated |
| -0,037044724 | HSF2 | FALSE | Downregulated |
| 0,575281014 | HSF2BP | FALSE | Upregulated |
| -0,427796621 | HSF5 | FALSE | Downregulated |
| -1,44524353 | HSFX1 | | Downregulated |
| 0,353087166 | HSFX2 | FALSE | Upregulated |
| -1,647501122 | HSFY1 | | Downregulated |
| | HSFY1P1 | | |
| -0,586924164 | HSFY2 | FALSE | Downregulated |
| 0,249853281 | HSH2D | FALSE | Upregulated |
| 0,346548151 | HSP90AB4P | FALSE | Upregulated |
| -0,063682096 | HSP90B2P | FALSE | Downregulated |
| 0,045054235 | HSPA12A | FALSE | Upregulated |
| -0,637075327 | HSPA1L | FALSE | Downregulated |
| 1,061671193 | HSPA2 | | Upregulated |
| -0,39126517 | HSPB11 | FALSE | Downregulated |
| -0,206476749 | HSPB2 | FALSE | Downregulated |
| 0,02549287 | HSPB2-C11orf52 | FALSE | Upregulated |
| | HSPB3 | | |
| 0,526734416 | HSPB8 | FALSE | Upregulated |
| 0,665277818 | HSPE1-MOB4 | FALSE | Upregulated |
| | HTN1 | | |
| | HTN3 | | |
| | HTR1A | | |
| | HTR1B | | |
| -0,154484467 | HTR1D | FALSE | Downregulated |

| | | | |
|---|---|---|---|
| | HTR1E | | |
| 0,076387306 | HTR1F | FALSE | Upregulated |
| 0,648616528 | HTR2A-AS1 | FALSE | Upregulated |
| 0,263483706 | HTR2C | FALSE | Upregulated |
| | HTR3A | | |
| | HTR3B | | |
| | HTR3C | | |
| | HTR3D | | |
| -0,068055201 | HTR3E | FALSE | Downregulated |
| 0,715047719 | HTR4 | FALSE | Upregulated |
| | HTR5A | | |
| 0,435471857 | HTR5A-AS1 | FALSE | Upregulated |
| 0,304025937 | HTR7 | FALSE | Upregulated |
| -1,0971539 | HTRA3 | | Downregulated |
| 0,219664299 | HTRA4 | FALSE | Upregulated |
| 0,828086293 | HTT-AS | FALSE | Upregulated |
| 0,865973375 | HUNK | FALSE | Upregulated |
| -0,045195214 | HUS1B | FALSE | Downregulated |
| 1,203902286 | HVCN1 | | Upregulated |
| -0,190872849 | HYAL3 | FALSE | Downregulated |
| -0,007085003 | HYAL4 | FALSE | Downregulated |
| | HYALP1 | | |
| -0,296015738 | HYDIN2 | FALSE | Downregulated |
| -0,158701144 | HYKK | FALSE | Downregulated |
| 0,456158751 | HYMAI | FALSE | Upregulated |
| 0,191874101 | IAPP | FALSE | Upregulated |
| 0,260818766 | IBA57-AS1 | FALSE | Upregulated |
| 0,176723186 | IBSP | FALSE | Upregulated |
| -0,194486506 | ICA1 | FALSE | Downregulated |
| -0,149215791 | ICAM2 | FALSE | Downregulated |
| -0,021665043 | ICAM4 | FALSE | Downregulated |
| 0,629514384 | ICAM5 | FALSE | Upregulated |
| 0,846757132 | ICOS | FALSE | Upregulated |
| 0,502803475 | ID4 | FALSE | Upregulated |
| -0,619132819 | IDH1-AS1 | FALSE | Downregulated |
| 1,541663822 | IDO1 | | Upregulated |
| -0,09686558 | IER3 | FALSE | Downregulated |
| -0,278004378 | IFI27L1 | FALSE | Downregulated |
| -0,347648125 | IFI27L2 | FALSE | Downregulated |
| | IFITM4P | | |
| 0,941425204 | IFITM5 | FALSE | Upregulated |
| 0,13116121 | IFLTD1 | FALSE | Upregulated |
| | IFNA1 | | |
| 0 | IFNA10 | FALSE | Upregulated |
| | IFNA13 | | |
| | IFNA14 | | |
| 0,437956466 | IFNA16 | FALSE | Upregulated |
| | IFNA17 | | |
| | IFNA2 | | |
| -0,016172189 | IFNA21 | FALSE | Downregulated |
| -0,01330656 | IFNA22P | FALSE | Downregulated |

| | | | |
|---|---|---|---|
| -0,001674428 | IFNA4 | FALSE | Downregulated |
| | IFNA5 | | |
| | IFNA6 | | |
| 0 | IFNA7 | FALSE | Upregulated |
| | IFNA8 | | |
| -0,579799367 | IFNB1 | FALSE | Downregulated |
| | IFNE | | |
| 0,599456815 | IFNG | FALSE | Upregulated |
| 0,772723597 | IFNG-AS1 | FALSE | Upregulated |
| 0,547759036 | IFNL1 | FALSE | Upregulated |
| | IFNL2 | | |
| | IFNL3 | | |
| | IFNL4 | | |
| 0,608677267 | IFNW1 | FALSE | Upregulated |
| -0,838296957 | IFT27 | FALSE | Downregulated |
| -0,297778539 | IFT43 | FALSE | Downregulated |
| -0,080798989 | IFT52 | FALSE | Downregulated |
| 0,106011106 | IFT57 | FALSE | Upregulated |
| 0,080614988 | IFT74 | FALSE | Upregulated |
| -0,231853632 | IFT81 | FALSE | Downregulated |
| -0,175864578 | IGDCC3 | FALSE | Downregulated |
| 0,265667489 | IGDCC4 | FALSE | Upregulated |
| -0,109456682 | IGF2BP1 | FALSE | Downregulated |
| -0,076268517 | IGF2BP2 | FALSE | Downregulated |
| -0,009350712 | IGFBP6 | FALSE | Downregulated |
| 1,819071067 | IGFBP7-AS1 | | Upregulated |
| 0,312571546 | IGFBPL1 | FALSE | Upregulated |
| 0 | IGFL1 | FALSE | Upregulated |
| 0,325566144 | IGFL2 | FALSE | Upregulated |
| -0,015569658 | IGFL3 | FALSE | Downregulated |
| 0,274494817 | IGFL4 | FALSE | Upregulated |
| 0 | IGLL1 | FALSE | Upregulated |
| 0,033348968 | IGLL3P | FALSE | Upregulated |
| 0,311639588 | IGSF1 | FALSE | Upregulated |
| 0,23072711 | IGSF11 | FALSE | Upregulated |
| 0,692255548 | IGSF11-AS1 | FALSE | Upregulated |
| 1,691625557 | IGSF21 | | Upregulated |
| 0,509351136 | IGSF3 | FALSE | Upregulated |
| 0 | IGSF5 | FALSE | Upregulated |
| 0,886746468 | IGSF9B | FALSE | Upregulated |
| -0,13274488 | IHH | FALSE | Downregulated |
| 0,568228513 | IKBIP | FALSE | Upregulated |
| 0,183644306 | IL10 | FALSE | Upregulated |
| 0,440425204 | IL11 | FALSE | Upregulated |
| -0,018550103 | IL12A | FALSE | Downregulated |
| 0,132878838 | IL12A-AS1 | FALSE | Upregulated |
| 0,133696256 | IL12B | FALSE | Upregulated |
| 1,166503085 | IL12RB1 | | Upregulated |
| 0,435628075 | IL13 | FALSE | Upregulated |
| -0,232225047 | IL13RA2 | FALSE | Downregulated |
| 0,590696456 | IL15 | FALSE | Upregulated |

| | | | |
|---|---|---|---|
| 0 | IL17A | FALSE | Upregulated |
| 0,138167526 | IL17B | FALSE | Upregulated |
| 0,423162917 | IL17C | FALSE | Upregulated |
| | IL17F | | |
| -0,127840432 | IL17RD | FALSE | Downregulated |
| 0,571329795 | IL17REL | FALSE | Upregulated |
| -0,288442344 | IL18R1 | FALSE | Downregulated |
| 1,36448091 | IL18RAP | | Upregulated |
| 0 | IL19 | FALSE | Upregulated |
| 0,531222475 | IL1A | FALSE | Upregulated |
| 1,247853294 | IL1B | | Upregulated |
| | IL1F10 | | |
| -0,676885615 | IL1R2 | FALSE | Downregulated |
| -0,167797872 | IL1RAPL1 | FALSE | Downregulated |
| -0,365557472 | IL1RAPL2 | FALSE | Downregulated |
| 0,323636389 | IL1RL1 | FALSE | Upregulated |
| -1,032345131 | IL1RL2 | | Downregulated |
| | IL2 | | |
| 0,258657557 | IL20 | FALSE | Upregulated |
| 0,517912014 | IL20RA | FALSE | Upregulated |
| 0,040748287 | IL20RB | FALSE | Upregulated |
| 0,463432472 | IL21 | FALSE | Upregulated |
| 0 | IL21-AS1 | FALSE | Upregulated |
| 0,299393637 | IL22 | FALSE | Upregulated |
| | IL22RA2 | | |
| 0,492919227 | IL23A | FALSE | Upregulated |
| 0,157090485 | IL23R | FALSE | Upregulated |
| 0,277119458 | IL25 | FALSE | Upregulated |
| 0,300246382 | IL26 | FALSE | Upregulated |
| -0,719682426 | IL27 | FALSE | Downregulated |
| 0,698149681 | IL2RA | FALSE | Upregulated |
| -0,13457206 | IL3 | FALSE | Downregulated |
| 0,75156536 | IL31 | FALSE | Upregulated |
| 0,355862649 | IL31RA | FALSE | Upregulated |
| -0,244400076 | IL34 | FALSE | Downregulated |
| | IL36A | | |
| | IL36B | | |
| -0,004525881 | IL36G | FALSE | Downregulated |
| 0 | IL36RN | FALSE | Upregulated |
| 0 | IL37 | FALSE | Upregulated |
| | IL3RA-2 | | |
| 0 | IL4 | FALSE | Upregulated |
| 0,429560328 | IL4I1 | FALSE | Upregulated |
| 0,379797004 | IL5 | FALSE | Upregulated |
| -0,399111985 | IL5RA | FALSE | Downregulated |
| 0,676098475 | IL6 | FALSE | Upregulated |
| 0,050189004 | IL8 | FALSE | Upregulated |
| | IL9 | | |
| -0,84892513 | IL9R | FALSE | Downregulated |
| | IL9R-2 | | |
| -0,590974206 | ILDR1 | FALSE | Downregulated |

| | | | |
|---|---|---|---|
| -0,711008656 | ILDR2 | FALSE | Downregulated |
| -0,650737211 | IMMP1L | FALSE | Downregulated |
| -0,32651484 | IMMP2L | FALSE | Downregulated |
| 0,038789333 | IMPG2 | FALSE | Upregulated |
| 0,423980256 | INA | FALSE | Upregulated |
| 0,906127301 | INE1 | FALSE | Upregulated |
| -0,339111676 | ING3 | FALSE | Downregulated |
| 0,207303896 | INGX | FALSE | Upregulated |
| 0,282141347 | INHA | FALSE | Upregulated |
| 0,411713161 | INHBA-AS1 | FALSE | Upregulated |
| 0,08539459 | INMT-FAM188B | FALSE | Upregulated |
| -0,154011267 | INPP5J | FALSE | Downregulated |
| -0,417929025 | INSC | FALSE | Downregulated |
| -0,780771276 | INSL3 | FALSE | Downregulated |
| | INSL4 | | |
| 0,158884609 | INSL5 | FALSE | Upregulated |
| 0,098174718 | INSL6 | FALSE | Upregulated |
| -0,158378854 | INSM1 | FALSE | Downregulated |
| 0,663302782 | INSM2 | FALSE | Upregulated |
| -0,192752411 | INSRR | FALSE | Downregulated |
| 0,045257521 | INTS4L1 | FALSE | Upregulated |
| 0,015133226 | INTS4L2 | FALSE | Upregulated |
| 0,407894166 | INTS6-AS1 | FALSE | Upregulated |
| 0,379654803 | INTU | FALSE | Upregulated |
| 0,423095574 | IP6K3 | FALSE | Upregulated |
| 0,818587625 | IPCEF1 | FALSE | Upregulated |
| -0,296203072 | IPO11-LRRC70 | FALSE | Downregulated |
| 0,139193778 | IQCA1 | FALSE | Upregulated |
| | IQCA1P1 | | |
| 0,114197041 | IQCB1 | FALSE | Upregulated |
| 0,408986314 | IQCD | FALSE | Upregulated |
| | IQCF1 | | |
| | IQCF2 | | |
| | IQCF3 | | |
| | IQCF4 | | |
| | IQCF5 | | |
| 0 | IQCF5-AS1 | FALSE | Upregulated |
| | IQCF6 | | |
| | IQCJ | | |
| 1,34611485 | IQGAP3 | | Upregulated |
| -0,118543138 | IQUB | FALSE | Downregulated |
| 0,253897208 | IRAK1BP1 | FALSE | Upregulated |
| 0,69381537 | IRG1 | FALSE | Upregulated |
| 0,321038117 | IRGC | FALSE | Upregulated |
| -0,013122049 | IRGM | FALSE | Downregulated |
| -0,001674428 | IRS4 | FALSE | Downregulated |
| 0 | IRX1 | FALSE | Upregulated |
| 0 | IRX2 | FALSE | Upregulated |
| 0,284413927 | IRX3 | FALSE | Upregulated |
| 0 | IRX4 | FALSE | Upregulated |
| 0 | IRX5 | FALSE | Upregulated |

| | | | |
|---|---|---|---|
| -0,170643017 | IRX6 | FALSE | Downregulated |
| 0,572047725 | ISCA2 | FALSE | Upregulated |
| | ISL1 | | |
| 0 | ISL2 | FALSE | Upregulated |
| -1,068155203 | ISLR2 | | Downregulated |
| 0 | ISM2 | FALSE | Upregulated |
| 0,348093745 | ISX | FALSE | Upregulated |
| 0,094094353 | ITGA10 | FALSE | Upregulated |
| 0,065990565 | ITGA2B | FALSE | Upregulated |
| -0,691902393 | ITGA8 | FALSE | Downregulated |
| 0,169034606 | ITGAE | FALSE | Upregulated |
| -0,111190188 | ITGB1BP2 | FALSE | Downregulated |
| -0,469064934 | ITGB3BP | FALSE | Downregulated |
| 0,886078705 | ITGB6 | FALSE | Upregulated |
| 0,74977019 | ITGB8 | FALSE | Upregulated |
| 1,280036296 | ITIH6 | | Upregulated |
| 0,634254384 | ITK | FALSE | Upregulated |
| 0,035991614 | ITLN1 | FALSE | Upregulated |
| 0,285957073 | ITLN2 | FALSE | Upregulated |
| 0,277709882 | ITM2A | FALSE | Upregulated |
| 0,079932537 | ITPKA | FALSE | Upregulated |
| -0,062682352 | ITPKB-IT1 | FALSE | Downregulated |
| -0,436433994 | ITPR1-AS1 | FALSE | Downregulated |
| 0,287604069 | ITPRIPL1 | FALSE | Upregulated |
| 0,317643559 | IVL | FALSE | Upregulated |
| -0,022550136 | IZUMO1 | FALSE | Downregulated |
| | IZUMO2 | | |
| | IZUMO3 | | |
| 0,237279131 | IZUMO4 | FALSE | Upregulated |
| 1,948190858 | JAKMIP1 | | Upregulated |
| 0,878095924 | JAKMIP2-AS1 | FALSE | Upregulated |
| -0,715205323 | JAKMIP3 | FALSE | Downregulated |
| | JAZF1-AS1 | | |
| -0,09117196 | JHDM1D-AS1 | FALSE | Downregulated |
| -0,489993817 | JMJD7 | FALSE | Downregulated |
| 0,795412042 | JPH1 | FALSE | Upregulated |
| 1,31145531 | JPH2 | | Upregulated |
| -0,25925751 | JPH4 | FALSE | Downregulated |
| | JRKL-AS1 | | |
| 0,375665576 | JSRP1 | FALSE | Upregulated |
| -0,250256532 | KAL1 | FALSE | Downregulated |
| 0,627068617 | KATNBL1 | FALSE | Upregulated |
| | KATNBL1P6 | | |
| 0,175245772 | KAZALD1 | FALSE | Upregulated |
| 0,150228926 | KBTBD13 | FALSE | Upregulated |
| -0,144773286 | KBTBD3 | FALSE | Downregulated |
| -0,736306002 | KBTBD8 | FALSE | Downregulated |
| 0,296264804 | KC6 | FALSE | Upregulated |
| 0 | KCNA1 | FALSE | Upregulated |
| 0,164111439 | KCNA10 | FALSE | Upregulated |
| -0,013541881 | KCNA2 | FALSE | Downregulated |

| | | | |
|---|---|---|---|
| | KCNA4 | | |
| 0,235171623 | KCNA5 | FALSE | Upregulated |
| 0,835009746 | KCNA6 | FALSE | Upregulated |
| 1,05279546 | KCNA7 | | Upregulated |
| 0,59504892 | KCNAB1-AS1 | FALSE | Upregulated |
| 0,566106101 | KCNAB1-AS2 | FALSE | Upregulated |
| 0,155349964 | KCNAB3 | FALSE | Upregulated |
| | KCNB2 | | |
| 0,956089885 | KCNC1 | FALSE | Upregulated |
| 0,279966523 | KCNC4-AS1 | FALSE | Upregulated |
| 0,496505137 | KCND1 | FALSE | Upregulated |
| 0,189773097 | KCND2 | FALSE | Upregulated |
| 0,02565044 | KCND3-IT1 | FALSE | Upregulated |
| 1,402839075 | KCNE1 | | Upregulated |
| 0 | KCNE1L | FALSE | Upregulated |
| -0,30984139 | KCNE3 | FALSE | Downregulated |
| 0,167902981 | KCNE4 | FALSE | Upregulated |
| 0 | KCNF1 | FALSE | Upregulated |
| 0,290523075 | KCNG1 | FALSE | Upregulated |
| | KCNG3 | | |
| 0 | KCNG4 | FALSE | Upregulated |
| | KCNH1-IT1 | | |
| 0,561056723 | KCNH2 | FALSE | Upregulated |
| 0,499152659 | KCNH4 | FALSE | Upregulated |
| | KCNH5 | | |
| -0,094246977 | KCNH6 | FALSE | Downregulated |
| 0,847467463 | KCNH7 | FALSE | Upregulated |
| -0,144409979 | KCNH8 | FALSE | Downregulated |
| 0,443090009 | KCNIP1 | FALSE | Upregulated |
| 0,197134807 | KCNIP2-AS1 | FALSE | Upregulated |
| 0 | KCNIP4-IT1 | FALSE | Upregulated |
| -0,387844946 | KCNJ1 | FALSE | Downregulated |
| 1,227848713 | KCNJ11 | | Upregulated |
| 0,76097862 | KCNJ12 | FALSE | Upregulated |
| 0,375929485 | KCNJ14 | FALSE | Upregulated |
| 0,648434428 | KCNJ15 | FALSE | Upregulated |
| -0,166493516 | KCNJ16 | FALSE | Downregulated |
| 0,000264809 | KCNJ18 | FALSE | Upregulated |
| 0,237650338 | KCNJ2 | FALSE | Upregulated |
| -0,005475981 | KCNJ2-AS1 | FALSE | Downregulated |
| -0,564941868 | KCNJ5 | FALSE | Downregulated |
| 0,140498104 | KCNJ6 | FALSE | Upregulated |
| 0,453223686 | KCNK1 | FALSE | Upregulated |
| 0,239009842 | KCNK10 | FALSE | Upregulated |
| 0,485618138 | KCNK12 | FALSE | Upregulated |
| 0,169187461 | KCNK13 | FALSE | Upregulated |
| 0,11722918 | KCNK15 | FALSE | Upregulated |
| -0,040095668 | KCNK16 | FALSE | Downregulated |
| -0,355912637 | KCNK17 | FALSE | Downregulated |
| | KCNK18 | | |
| | KCNK2 | | |

| | | | |
|---|---|---|---|
| -1,421989019 | KCNK3 | | Downregulated |
| 0,293326262 | KCNK4 | FALSE | Upregulated |
| 0,917431885 | KCNK7 | FALSE | Upregulated |
| | KCNK9 | | |
| 0,151541558 | KCNMB1 | FALSE | Upregulated |
| 0,910349114 | KCNMB2 | FALSE | Upregulated |
| 0,90810246 | KCNMB4 | FALSE | Upregulated |
| 0,242272293 | KCNN1 | FALSE | Upregulated |
| -0,756640591 | KCNN3 | FALSE | Downregulated |
| 0,811493909 | KCNN4 | FALSE | Upregulated |
| -0,142597797 | KCNQ1DN | FALSE | Downregulated |
| | KCNQ2 | | |
| -0,239589804 | KCNQ3 | FALSE | Downregulated |
| -1,125678281 | KCNQ4 | | Downregulated |
| 0,86592487 | KCNQ5 | FALSE | Upregulated |
| 1,792189039 | KCNQ5-AS1 | | Upregulated |
| | KCNQ5-IT1 | | |
| -0,243426128 | KCNS3 | FALSE | Downregulated |
| 0,146933547 | KCNT1 | FALSE | Upregulated |
| | KCNU1 | | |
| -0,149894289 | KCNV1 | FALSE | Downregulated |
| -0,821470817 | KCP | FALSE | Downregulated |
| -1,449722415 | KCTD14 | | Downregulated |
| 0,221662257 | KCTD16 | FALSE | Upregulated |
| -0,159054816 | KCTD19 | FALSE | Downregulated |
| 0,297178435 | KCTD8 | FALSE | Upregulated |
| -0,442075124 | KDELC1 | FALSE | Downregulated |
| 1,390709459 | KDM4D | | Upregulated |
| 0,295420819 | KEL | FALSE | Upregulated |
| | KERA | | |
| 0,331305283 | KGFLP1 | FALSE | Upregulated |
| -0,402968991 | KHDC1 | FALSE | Downregulated |
| | KHDC1L | | |
| 0 | KHDC3L | FALSE | Upregulated |
| -0,205105506 | KHDRBS2 | FALSE | Downregulated |
| 1,479932015 | KIAA0087 | | Upregulated |
| -0,43889955 | KIAA0101 | FALSE | Downregulated |
| 0,91120582 | KIAA0226L | FALSE | Upregulated |
| 1,522310193 | KIAA0408 | | Upregulated |
| -0,077904003 | KIAA1024 | FALSE | Downregulated |
| | KIAA1024L | | |
| 0,047263404 | KIAA1107 | FALSE | Upregulated |
| 0,119076171 | KIAA1210 | FALSE | Upregulated |
| -0,303441782 | KIAA1211L | FALSE | Downregulated |
| 0,092815286 | KIAA1239 | FALSE | Upregulated |
| 0,158697372 | KIAA1257 | FALSE | Upregulated |
| 0,409623093 | KIAA1324 | FALSE | Upregulated |
| -0,26470366 | KIAA1324L | FALSE | Downregulated |
| 2,049606048 | KIAA1377 | | Upregulated |
| 0,08549257 | KIAA1467 | FALSE | Upregulated |
| -0,06314585 | KIAA1524 | FALSE | Downregulated |

| | | | |
|---|---|---|---|
| -0,345500224 | KIAA1586 | FALSE | Downregulated |
| 0,260244327 | KIAA1644 | FALSE | Upregulated |
| -0,42316747 | KIAA1751 | FALSE | Downregulated |
| 0,918168671 | KIAA1755 | FALSE | Upregulated |
| 0,51926468 | KIAA1841 | FALSE | Upregulated |
| -0,70758692 | KIAA1875 | FALSE | Downregulated |
| 0,478146737 | KIAA2022 | FALSE | Upregulated |
| -0,047461312 | KIF14 | FALSE | Downregulated |
| -0,466261579 | KIF15 | FALSE | Downregulated |
| -0,915585911 | KIF17 | FALSE | Downregulated |
| 0,90426455 | KIF18A | FALSE | Upregulated |
| 0,622191367 | KIF18B | FALSE | Upregulated |
| -0,096843425 | KIF19 | FALSE | Downregulated |
| -0,311025368 | KIF1A | FALSE | Downregulated |
| 0,413972625 | KIF20B | FALSE | Upregulated |
| 1,079694476 | KIF23 | Upregulated | |
| -0,433774987 | KIF25 | FALSE | Downregulated |
| 0,005298914 | KIF25-AS1 | FALSE | Upregulated |
| 0,291340714 | KIF2B | FALSE | Upregulated |
| 0,5887339 | KIF2C | FALSE | Upregulated |
| -0,150267822 | KIF3A | FALSE | Downregulated |
| 0,292745031 | KIF3C | FALSE | Upregulated |
| -0,146817512 | KIF4A | FALSE | Downregulated |
| -0,18022199 | KIF4B | FALSE | Downregulated |
| -0,009877471 | KIF5A | FALSE | Downregulated |
| -0,355917445 | KIF5C | FALSE | Downregulated |
| 0,262313018 | KIF7 | FALSE | Upregulated |
| -0,28548555 | KIF9-AS1 | FALSE | Downregulated |
| 0,789876376 | KIFC1 | FALSE | Upregulated |
| -0,141836667 | KIR2DL1 | FALSE | Downregulated |
| 0,119531094 | KIR2DL3 | FALSE | Upregulated |
| 0 | KIR2DL4 | FALSE | Upregulated |
| 0 | KIR2DS4 | FALSE | Upregulated |
| 0,005298914 | KIR3DL1 | FALSE | Upregulated |
| 0,252392406 | KIR3DL2 | FALSE | Upregulated |
| | KIR3DL3 | | |
| 0,192057565 | KIR3DX1 | FALSE | Upregulated |
| | KIRREL2 | | |
| | KIRREL3-AS2 | | |
| 0,41482555 | KIRREL3-AS3 | FALSE | Upregulated |
| -0,28572042 | KISS1 | FALSE | Downregulated |
| 0,740843246 | KISS1R | FALSE | Upregulated |
| -1,028106595 | KIT | Downregulated | |
| 0,084891 | KITLG | FALSE | Upregulated |
| 0,697856439 | KL | FALSE | Upregulated |
| 0,11987666 | KLF1 | FALSE | Upregulated |
| | KLF14 | | |
| 0 | KLF17 | FALSE | Upregulated |
| 0,479928514 | KLF5 | FALSE | Upregulated |
| 0,253455786 | KLF8 | FALSE | Upregulated |
| -0,455165508 | KLHDC1 | FALSE | Downregulated |

| | | | |
|---|---|---|---|
| 1,764759603 | KLHDC7B | | Upregulated |
| -0,740840032 | KLHDC8A | FALSE | Downregulated |
| 0,181652638 | KLHDC9 | FALSE | Upregulated |
| 0,288213735 | KLHL1 | FALSE | Upregulated |
| -0,038033438 | KLHL13 | FALSE | Downregulated |
| -0,469923653 | KLHL14 | FALSE | Downregulated |
| 0,064952952 | KLHL17 | FALSE | Upregulated |
| 0,204508722 | KLHL23 | FALSE | Upregulated |
| 0,052402745 | KLHL30 | FALSE | Upregulated |
| -0,112592317 | KLHL32 | FALSE | Downregulated |
| 0,495583167 | KLHL33 | FALSE | Upregulated |
| 0,251568999 | KLHL34 | FALSE | Upregulated |
| -0,128506191 | KLHL35 | FALSE | Downregulated |
| 0,313929522 | KLHL38 | FALSE | Upregulated |
| 0,543845019 | KLHL4 | FALSE | Upregulated |
| 0,700686311 | KLHL40 | FALSE | Upregulated |
| 0,146982566 | KLHL7-AS1 | FALSE | Upregulated |
| -0,006848587 | KLK1 | FALSE | Downregulated |
| 0,283161491 | KLK10 | FALSE | Upregulated |
| 0 | KLK11 | FALSE | Upregulated |
| 0,009010674 | KLK12 | FALSE | Upregulated |
| | KLK13 | | |
| -0,170847634 | KLK14 | FALSE | Downregulated |
| | KLK15 | | |
| | KLK2 | | |
| | KLK3 | | |
| | KLK4 | | |
| 0 | KLK5 | FALSE | Upregulated |
| | KLK6 | | |
| | KLK7 | | |
| 0 | KLK8 | FALSE | Upregulated |
| | KLK9 | | |
| | KLKP1 | | |
| -0,087743358 | KLRAP1 | FALSE | Downregulated |
| 0,91780496 | KLRB1 | FALSE | Upregulated |
| 1,066022773 | KLRC1 | | Upregulated |
| 0,037416841 | KLRC2 | FALSE | Upregulated |
| 0,165255953 | KLRC3 | FALSE | Upregulated |
| 0,250638326 | KLRC4 | FALSE | Upregulated |
| -0,580744335 | KLRC4-KLRK1 | FALSE | Downregulated |
| 0,671740135 | KLRD1 | FALSE | Upregulated |
| -0,139524577 | KLRF1 | FALSE | Downregulated |
| | KLRF2 | | |
| 1,180309147 | KLRG1 | | Upregulated |
| -0,668548947 | KLRG2 | FALSE | Downregulated |
| 0,624182533 | KLRK1 | FALSE | Upregulated |
| | KNCN | | |
| -0,640340991 | KNSTRN | FALSE | Downregulated |
| -0,213337195 | KPNA2 | FALSE | Downregulated |
| 0,330109383 | KPNA7 | FALSE | Upregulated |
| 0 | KPRP | FALSE | Upregulated |

| | | | |
|---|---|---|---|
| -0,275325376 | KPTN | FALSE | Downregulated |
| -0,94570761 | KRBA1 | FALSE | Downregulated |
| -0,257224663 | KRBA2 | FALSE | Downregulated |
| 0 | KRBOX1 | FALSE | Upregulated |
| 1,051799312 | KREMEN2 | | Upregulated |
| -0,149176845 | KRT12 | FALSE | Downregulated |
| | KRT13 | | |
| 0,08825974 | KRT15 | FALSE | Upregulated |
| 0 | KRT16 | FALSE | Upregulated |
| | KRT16P1 | | |
| 0 | KRT16P2 | FALSE | Upregulated |
| | KRT16P3 | | |
| 0,425140966 | KRT17 | FALSE | Upregulated |
| 0 | KRT18P55 | FALSE | Upregulated |
| 0,887491471 | KRT19 | FALSE | Upregulated |
| 0,491479911 | KRT19P2 | FALSE | Upregulated |
| 0 | KRT2 | FALSE | Upregulated |
| -0,54137691 | KRT20 | FALSE | Downregulated |
| 0,155319734 | KRT222 | FALSE | Upregulated |
| 0,393034714 | KRT23 | FALSE | Upregulated |
| | KRT24 | | |
| | KRT25 | | |
| | KRT26 | | |
| 0,435024977 | KRT27 | FALSE | Upregulated |
| | KRT28 | | |
| | KRT3 | | |
| 0 | KRT31 | FALSE | Upregulated |
| 0 | KRT32 | FALSE | Upregulated |
| | KRT33A | | |
| | KRT33B | | |
| | KRT34 | | |
| 0 | KRT35 | FALSE | Upregulated |
| 0 | KRT36 | FALSE | Upregulated |
| 0 | KRT37 | FALSE | Upregulated |
| 0 | KRT38 | FALSE | Upregulated |
| 0,15436386 | KRT39 | FALSE | Upregulated |
| 0,000264809 | KRT4 | FALSE | Upregulated |
| | KRT40 | | |
| | KRT42P | | |
| 0,100082 | KRT5 | FALSE | Upregulated |
| 0 | KRT6A | FALSE | Upregulated |
| 0 | KRT6B | FALSE | Upregulated |
| 0 | KRT6C | FALSE | Upregulated |
| 0 | KRT71 | FALSE | Upregulated |
| | KRT72 | | |
| 0,005366493 | KRT73 | FALSE | Upregulated |
| 0,327871496 | KRT73-AS1 | FALSE | Upregulated |
| | KRT74 | | |
| | KRT75 | | |
| | KRT76 | | |
| 0 | KRT77 | FALSE | Upregulated |

| | | | |
|---|---|---|---|
| 0 | KRT78 | FALSE | Upregulated |
| | KRT79 | | |
| 0,400210302 | KRT80 | FALSE | Upregulated |
| -0,188528073 | KRT81 | FALSE | Downregulated |
| 0 | KRT82 | FALSE | Upregulated |
| 0 | KRT83 | FALSE | Upregulated |
| | KRT84 | | |
| 0,105200699 | KRT85 | FALSE | Upregulated |
| -0,366915004 | KRT86 | FALSE | Downregulated |
| -0,087297559 | KRT8P41 | FALSE | Downregulated |
| | KRT9 | | |
| | KRTAP1-1 | | |
| | KRTAP1-3 | | |
| | KRTAP1-4 | | |
| 0 | KRTAP1-5 | FALSE | Upregulated |
| | KRTAP10-1 | | |
| 0 | KRTAP10-10 | FALSE | Upregulated |
| -0,149713684 | KRTAP10-11 | FALSE | Downregulated |
| 0,268052598 | KRTAP10-12 | FALSE | Upregulated |
| -0,019313425 | KRTAP10-2 | FALSE | Downregulated |
| | KRTAP10-3 | | |
| 0,303769328 | KRTAP10-4 | FALSE | Upregulated |
| | KRTAP10-5 | | |
| 0,017141281 | KRTAP10-6 | FALSE | Upregulated |
| 0 | KRTAP10-7 | FALSE | Upregulated |
| 0 | KRTAP10-8 | FALSE | Upregulated |
| -0,011489422 | KRTAP10-9 | FALSE | Downregulated |
| | KRTAP11-1 | | |
| | KRTAP12-1 | | |
| | KRTAP12-2 | | |
| | KRTAP12-3 | | |
| | KRTAP12-4 | | |
| | KRTAP13-1 | | |
| | KRTAP13-2 | | |
| | KRTAP13-3 | | |
| | KRTAP13-4 | | |
| | KRTAP15-1 | | |
| | KRTAP16-1 | | |
| | KRTAP17-1 | | |
| | KRTAP19-1 | | |
| | KRTAP19-2 | | |
| | KRTAP19-3 | | |
| | KRTAP19-4 | | |
| | KRTAP19-5 | | |
| | KRTAP19-6 | | |
| | KRTAP19-7 | | |
| 0,005406653 | KRTAP19-8 | FALSE | Upregulated |
| | KRTAP2-1 | | |
| | KRTAP2-2 | | |
| | KRTAP2-3 | | |
| | KRTAP2-4 | | |

| | | | |
|---|---|---|---|
| | KRTAP20-1 | | |
| | KRTAP20-2 | | |
| | KRTAP20-3 | | |
| | KRTAP20-4 | | |
| | KRTAP21-1 | | |
| | KRTAP21-2 | | |
| | KRTAP21-3 | | |
| | KRTAP22-1 | | |
| | KRTAP22-2 | | |
| | KRTAP23-1 | | |
| | KRTAP24-1 | | |
| | KRTAP25-1 | | |
| | KRTAP26-1 | | |
| | KRTAP27-1 | | |
| | KRTAP29-1 | | |
| 0 | KRTAP3-1 | FALSE | Upregulated |
| 0 | KRTAP3-2 | FALSE | Upregulated |
| 0,272341956 | KRTAP3-3 | FALSE | Upregulated |
| | KRTAP4-1 | | |
| | KRTAP4-11 | | |
| | KRTAP4-12 | | |
| | KRTAP4-2 | | |
| | KRTAP4-3 | | |
| | KRTAP4-4 | | |
| | KRTAP4-5 | | |
| | KRTAP4-6 | | |
| | KRTAP4-7 | | |
| | KRTAP4-8 | | |
| | KRTAP4-9 | | |
| 0,287424956 | KRTAP5-1 | FALSE | Upregulated |
| 0,257619078 | KRTAP5-10 | FALSE | Upregulated |
| 0,004939325 | KRTAP5-11 | FALSE | Upregulated |
| 0,228620561 | KRTAP5-2 | FALSE | Upregulated |
| | KRTAP5-3 | | |
| 0,291340714 | KRTAP5-4 | FALSE | Upregulated |
| | KRTAP5-5 | | |
| -0,010928533 | KRTAP5-6 | FALSE | Downregulated |
| 1,000913785 | KRTAP5-7 | | Upregulated |
| 0,297178435 | KRTAP5-8 | FALSE | Upregulated |
| -0,027583154 | KRTAP5-9 | FALSE | Downregulated |
| -0,737218914 | KRTAP5-AS1 | FALSE | Downregulated |
| | KRTAP6-1 | | |
| | KRTAP6-2 | | |
| | KRTAP6-3 | | |
| | KRTAP7-1 | | |
| 0,157488478 | KRTAP8-1 | FALSE | Upregulated |
| | KRTAP9-1 | | |
| | KRTAP9-2 | | |
| | KRTAP9-3 | | |
| | KRTAP9-4 | | |
| | KRTAP9-6 | | |

| | | | |
|---|---|---|---|
| | KRTAP9-7 | | |
| | KRTAP9-8 | | |
| | KRTAP9-9 | | |
| -0,10173498 | KRTCAP2 | FALSE | Downregulated |
| -0,186420189 | KRTCAP3 | FALSE | Downregulated |
| 0 | KRTDAP | FALSE | Upregulated |
| 0,689132755 | KSR2 | FALSE | Upregulated |
| -0,071364037 | KTI12 | FALSE | Downregulated |
| -0,157240205 | KTN1-AS1 | FALSE | Downregulated |
| 0,789539702 | L1CAM | FALSE | Upregulated |
| 0,168244055 | L1TD1 | FALSE | Upregulated |
| | LACRT | | |
| | LACTBL1 | | |
| 0,0794 | LAG3 | FALSE | Upregulated |
| -0,079395217 | LAGE3 | FALSE | Downregulated |
| -0,004525881 | LAIR2 | FALSE | Downregulated |
| | LALBA | | |
| 0,771891083 | LAMA1 | FALSE | Upregulated |
| 0,736660983 | LAMB2P1 | FALSE | Upregulated |
| 0,459015449 | LAMB4 | FALSE | Upregulated |
| 0,668493328 | LAMC2 | FALSE | Upregulated |
| 1,092422457 | LAMP3 | | Upregulated |
| 0,142101196 | LAMP5 | FALSE | Upregulated |
| -0,30708379 | LAMTOR5-AS1 | FALSE | Downregulated |
| 0,127495752 | LANCL3 | FALSE | Upregulated |
| 0,126826435 | LARP6 | FALSE | Upregulated |
| 1,121443999 | LAX1 | | Upregulated |
| 0,006179703 | LAYN | FALSE | Upregulated |
| 0,42018173 | LBX1 | FALSE | Upregulated |
| 0,277119458 | LBX1-AS1 | FALSE | Upregulated |
| 0,503104812 | LCA5 | FALSE | Upregulated |
| 0 | LCE1A | FALSE | Upregulated |
| | LCE1B | | |
| 0 | LCE1C | FALSE | Upregulated |
| 0 | LCE1D | FALSE | Upregulated |
| | LCE1E | | |
| | LCE1F | | |
| | LCE2A | | |
| 0 | LCE2B | FALSE | Upregulated |
| | LCE2C | | |
| 0 | LCE2D | FALSE | Upregulated |
| | LCE3A | | |
| | LCE3B | | |
| | LCE3C | | |
| | LCE3D | | |
| | LCE3E | | |
| | LCE4A | | |
| 0 | LCE5A | FALSE | Upregulated |
| | LCE6A | | |
| 0,56163374 | LCK | FALSE | Upregulated |
| -0,229542493 | LCMT1 | FALSE | Downregulated |

| | | | |
|---|---|---|---|
| 0,16216896 | LCMT2 | FALSE | Upregulated |
| | LCN1 | | |
| -0,08581094 | LCN10 | FALSE | Downregulated |
| 0,439582982 | LCN15 | FALSE | Upregulated |
| -0,152690229 | LCN2 | FALSE | Downregulated |
| 0 | LCN6 | FALSE | Upregulated |
| | LCN8 | | |
| | LCN9 | | |
| 0,967481536 | LCNL1 | FALSE | Upregulated |
| -0,378585705 | LDHC | FALSE | Downregulated |
| 0,012958663 | LDLRAD1 | FALSE | Upregulated |
| 0,250789129 | LDLRAD3 | FALSE | Upregulated |
| -0,979122228 | LDOC1 | FALSE | Downregulated |
| 0,415317616 | LEF1 | FALSE | Upregulated |
| 0,623553676 | LEF1-AS1 | FALSE | Upregulated |
| -0,22942214 | LEFTY1 | FALSE | Downregulated |
| 0,272894361 | LEFTY2 | FALSE | Upregulated |
| | LELP1 | | |
| -0,011098311 | LEMD1 | FALSE | Downregulated |
| 0 | LEMD1-AS1 | FALSE | Upregulated |
| -0,305056072 | LEP | FALSE | Downregulated |
| 0,131606895 | LEPREL2 | FALSE | Upregulated |
| -0,324360026 | LEPREL4 | FALSE | Downregulated |
| | LEUTX | | |
| 0,297708309 | LGALS12 | FALSE | Upregulated |
| | LGALS13 | | |
| | LGALS14 | | |
| | LGALS16 | | |
| | LGALS17A | | |
| 0,451830932 | LGALS2 | FALSE | Upregulated |
| 0 | LGALS7 | FALSE | Upregulated |
| 0 | LGALS7B | FALSE | Upregulated |
| -0,140527702 | LGALS9B | FALSE | Downregulated |
| 0,276628673 | LGALS9C | FALSE | Upregulated |
| -0,098673866 | LGI1 | FALSE | Downregulated |
| 2,019304394 | LGI2 | | Upregulated |
| 0,650736054 | LGI3 | FALSE | Upregulated |
| -0,1404804 | LGR5 | FALSE | Downregulated |
| 0,811486862 | LGR6 | FALSE | Upregulated |
| 1,011926878 | LHB | | Upregulated |
| 0,269638293 | LHCGR | FALSE | Upregulated |
| -0,009465128 | LHFPL1 | FALSE | Downregulated |
| 0,287580449 | LHFPL3 | FALSE | Upregulated |
| | LHFPL3-AS1 | | |
| 0,290679065 | LHFPL3-AS2 | FALSE | Upregulated |
| -0,015040902 | LHFPL5 | FALSE | Downregulated |
| | LHX1 | | |
| 0 | LHX3 | FALSE | Upregulated |
| -0,001916634 | LHX5 | FALSE | Downregulated |
| -0,484378897 | LHX6 | FALSE | Downregulated |
| | LHX8 | | |

| | | | |
|---|---|---|---|
| -0,693047478 | LIF | FALSE | Downregulated |
| -0,086283635 | LILRA1 | FALSE | Downregulated |
| 0,422001621 | LILRA2 | FALSE | Upregulated |
| -0,492558447 | LILRA4 | FALSE | Downregulated |
| -0,507950953 | LILRA5 | FALSE | Downregulated |
| 0,555169217 | LILRP2 | FALSE | Upregulated |
| 0,416457236 | LIM2 | FALSE | Upregulated |
| 0,243085177 | LIMS3 | FALSE | Upregulated |
| 0,327235043 | LIMS3-LOC440895 | FALSE | Upregulated |
| -0,032882377 | LIMS3L | FALSE | Downregulated |
| 0,489947318 | LIN28A | FALSE | Upregulated |
| 0 | LIN28B | FALSE | Upregulated |
| -0,155254156 | LIN52 | FALSE | Downregulated |
| -1,224405884 | LIN7B | | Downregulated |
| -0,1447601 | LIN9 | FALSE | Downregulated |
| -0,300127308 | LINC-ROR | FALSE | Downregulated |
| | LINC00028 | | |
| | LINC00029 | | |
| 0 | LINC00032 | FALSE | Upregulated |
| 0,273624221 | LINC00035 | FALSE | Upregulated |
| 1,050722124 | LINC00051 | | Upregulated |
| 1,470954412 | LINC00052 | | Upregulated |
| 0 | LINC00083 | FALSE | Upregulated |
| -0,258651339 | LINC00086 | FALSE | Downregulated |
| 0,364121044 | LINC00087 | FALSE | Upregulated |
| | LINC00102-2 | | |
| | LINC00111 | | |
| -0,006682944 | LINC00112 | FALSE | Downregulated |
| 0,327667389 | LINC00113 | FALSE | Upregulated |
| | LINC00114 | | |
| 0,476402658 | LINC00115 | FALSE | Upregulated |
| -0,76773945 | LINC00116 | FALSE | Downregulated |
| -0,684824133 | LINC00152 | FALSE | Downregulated |
| 0 | LINC00158 | FALSE | Upregulated |
| | LINC00159 | | |
| | LINC00160 | | |
| 0 | LINC00162 | FALSE | Upregulated |
| 1,009797469 | LINC00163 | | Upregulated |
| 0,410444235 | LINC00167 | FALSE | Upregulated |
| 0,662138068 | LINC00173 | FALSE | Upregulated |
| 0 | LINC00184 | FALSE | Upregulated |
| -0,133279234 | LINC00189 | FALSE | Downregulated |
| 0,000504523 | LINC00200 | FALSE | Upregulated |
| 0,045834047 | LINC00202-1 | FALSE | Upregulated |
| -0,005984873 | LINC00202-2 | FALSE | Downregulated |
| | LINC00207 | | |
| -0,398211956 | LINC00208 | FALSE | Downregulated |
| | LINC00210 | | |
| 0,026429513 | LINC00222 | FALSE | Upregulated |
| | LINC00226 | | |
| | LINC00229 | | |

| | | | |
|---|---|---|---|
| 0,314204271 | LINC00235 | FALSE | Upregulated |
| 0,784921068 | LINC00238 | FALSE | Upregulated |
| -0,024636467 | LINC00239 | FALSE | Downregulated |
| -0,155068816 | LINC00240 | FALSE | Downregulated |
| -0,224740019 | LINC00243 | FALSE | Downregulated |
| -0,015321927 | LINC00251 | FALSE | Downregulated |
| 0,136335194 | LINC00254 | FALSE | Upregulated |
| 0,568485283 | LINC00260 | FALSE | Upregulated |
| -0,027793361 | LINC00263 | FALSE | Downregulated |
| -0,011369275 | LINC00264 | FALSE | Downregulated |
| -0,18029892 | LINC00265 | FALSE | Downregulated |
| -0,77468653 | LINC00266-1 | FALSE | Downregulated |
| 0,414034817 | LINC00266-3 | FALSE | Upregulated |
| | LINC00269 | | |
| 0,004896266 | LINC00271 | FALSE | Upregulated |
| 0 | LINC00272 | FALSE | Upregulated |
| | LINC00273 | | |
| | LINC00276 | | |
| 0,862789329 | LINC00277 | FALSE | Upregulated |
| 0,676152461 | LINC00278 | FALSE | Upregulated |
| | LINC00284 | | |
| | LINC00290 | | |
| | LINC00293 | | |
| 0 | LINC00297 | FALSE | Upregulated |
| 0 | LINC00298 | FALSE | Upregulated |
| -0,13930464 | LINC00299 | FALSE | Downregulated |
| | LINC00301 | | |
| 0 | LINC00303 | FALSE | Upregulated |
| | LINC00304 | | |
| 0 | LINC00305 | FALSE | Upregulated |
| | LINC00307 | | |
| | LINC00308 | | |
| 0 | LINC00309 | FALSE | Upregulated |
| 0,401542126 | LINC00310 | FALSE | Upregulated |
| -0,802813332 | LINC00311 | FALSE | Downregulated |
| -1,27241192 | LINC00312 | | Downregulated |
| 0 | LINC00313 | FALSE | Upregulated |
| -0,364864271 | LINC00314 | FALSE | Downregulated |
| 0,006980238 | LINC00316 | FALSE | Upregulated |
| | LINC00317 | | |
| | LINC00320 | | |
| | LINC00322 | | |
| | LINC00323 | | |
| | LINC00326 | | |
| 0,277119458 | LINC00327 | FALSE | Upregulated |
| -0,004525881 | LINC00330 | FALSE | Downregulated |
| | LINC00331 | | |
| -0,70775308 | LINC00332 | FALSE | Downregulated |
| | LINC00333 | | |
| 0,290181932 | LINC00336 | FALSE | Upregulated |
| 0,173674703 | LINC00337 | FALSE | Upregulated |

| | | | |
|---|---|---|---|
| -0,31871312 | LINC00338 | FALSE | Downregulated |
| -0,926424588 | LINC00339 | FALSE | Downregulated |
| | LINC00343 | | |
| | LINC00344 | | |
| 0,722596592 | LINC00346 | FALSE | Upregulated |
| 0,100390012 | LINC00347 | FALSE | Upregulated |
| | LINC00348 | | |
| | LINC00351 | | |
| -0,865611154 | LINC00353 | FALSE | Downregulated |
| | LINC00358 | | |
| | LINC00359 | | |
| 0,000535661 | LINC00366 | FALSE | Upregulated |
| | LINC00367 | | |
| -0,004525881 | LINC00371 | FALSE | Downregulated |
| | LINC00378 | | |
| | LINC00379 | | |
| | LINC00380 | | |
| | LINC00381 | | |
| 0,258099012 | LINC00384 | FALSE | Upregulated |
| | LINC00395 | | |
| | LINC00397 | | |
| -0,046480933 | LINC00398 | FALSE | Downregulated |
| | LINC00403 | | |
| | LINC00408 | | |
| 0,277833435 | LINC00410 | FALSE | Upregulated |
| | LINC00417 | | |
| | LINC00421 | | |
| -0,002253536 | LINC00423 | FALSE | Downregulated |
| | LINC00424 | | |
| 0,598675432 | LINC00426 | FALSE | Upregulated |
| | LINC00433 | | |
| -0,03339867 | LINC00441 | FALSE | Downregulated |
| | LINC00442 | | |
| | LINC00443 | | |
| | LINC00445 | | |
| | LINC00446 | | |
| 0,260713346 | LINC00452 | FALSE | Upregulated |
| | LINC00454 | | |
| 0 | LINC00457 | FALSE | Upregulated |
| | LINC00458 | | |
| | LINC00460 | | |
| | LINC00461 | | |
| 0,271886396 | LINC00462 | FALSE | Upregulated |
| | LINC00466 | | |
| 0,037250136 | LINC00467 | FALSE | Upregulated |
| | LINC00469 | | |
| 0,293955122 | LINC00470 | FALSE | Upregulated |
| 0,489212986 | LINC00471 | FALSE | Upregulated |
| | LINC00473 | | |
| | LINC00474 | | |
| -0,016357556 | LINC00475 | FALSE | Downregulated |

| | | | |
|---|---|---|---|
| 0,319222233 | LINC00477 | FALSE | Upregulated |
| -0,006753065 | LINC00479 | FALSE | Downregulated |
| 0,083267889 | LINC00483 | FALSE | Upregulated |
| -0,159244686 | LINC00485 | FALSE | Downregulated |
| 0,277833435 | LINC00486 | FALSE | Upregulated |
| 0,272875084 | LINC00487 | FALSE | Upregulated |
| | LINC00488 | | |
| | LINC00489 | | |
| | LINC00491 | | |
| | LINC00492 | | |
| -0,541557901 | LINC00493 | FALSE | Downregulated |
| 0,308449706 | LINC00494 | FALSE | Upregulated |
| | LINC00499 | | |
| -0,136619946 | LINC00501 | FALSE | Downregulated |
| | LINC00502 | | |
| -0,533356348 | LINC00506 | FALSE | Downregulated |
| | LINC00507 | | |
| | LINC00511 | | |
| 0,804650087 | LINC00515 | FALSE | Upregulated |
| 0,141023274 | LINC00518 | FALSE | Upregulated |
| -1,701544451 | LINC00520 | | Downregulated |
| | LINC00521 | | |
| 0 | LINC00523 | FALSE | Upregulated |
| -0,592099625 | LINC00525 | FALSE | Downregulated |
| 0,266188219 | LINC00526 | FALSE | Upregulated |
| 0,172368192 | LINC00528 | FALSE | Upregulated |
| 0,291340714 | LINC00534 | FALSE | Upregulated |
| 0,395578512 | LINC00535 | FALSE | Upregulated |
| | LINC00536 | | |
| 0,001803163 | LINC00539 | FALSE | Upregulated |
| -0,170951434 | LINC00540 | FALSE | Downregulated |
| | LINC00544 | | |
| | LINC00545 | | |
| 0,300542079 | LINC00547 | FALSE | Upregulated |
| -0,13350662 | LINC00548 | FALSE | Downregulated |
| | LINC00550 | | |
| | LINC00551 | | |
| 0,102169729 | LINC00552 | FALSE | Upregulated |
| | LINC00554 | | |
| | LINC00558 | | |
| -0,331393591 | LINC00559 | FALSE | Downregulated |
| | LINC00560 | | |
| 0,769768055 | LINC00563 | FALSE | Upregulated |
| 0 | LINC00565 | FALSE | Upregulated |
| 0,26275984 | LINC00568 | FALSE | Upregulated |
| 1,207854644 | LINC00570 | | Upregulated |
| -0,13132879 | LINC00571 | FALSE | Downregulated |
| -0,307222115 | LINC00572 | FALSE | Downregulated |
| | LINC00575 | | |
| | LINC00577 | | |
| | LINC00578 | | |

| | | | |
|---|---|---|---|
| | LINC00581 | | |
| 0,28516252 | LINC00582 | FALSE | Upregulated |
| | LINC00583 | | |
| | LINC00587 | | |
| | LINC00588 | | |
| | LINC00589 | | |
| | LINC00592 | | |
| 0,273422532 | LINC00593 | FALSE | Upregulated |
| 0,005298914 | LINC00595 | FALSE | Upregulated |
| 0,965739659 | LINC00597 | FALSE | Upregulated |
| -0,866390585 | LINC00598 | FALSE | Downregulated |
| | LINC00599 | | |
| 0,000535661 | LINC00601 | FALSE | Upregulated |
| 0,278946888 | LINC00602 | FALSE | Upregulated |
| | LINC00603 | | |
| -0,119585398 | LINC00605 | FALSE | Downregulated |
| | LINC00606 | | |
| 0,006226545 | LINC00607 | FALSE | Upregulated |
| | LINC00608 | | |
| | LINC00609 | | |
| | LINC00613 | | |
| | LINC00615 | | |
| | LINC00616 | | |
| 0,274635952 | LINC00617 | FALSE | Upregulated |
| | LINC00618 | | |
| | LINC00619 | | |
| | LINC00620 | | |
| 0,133713742 | LINC00622 | FALSE | Upregulated |
| -0,036677053 | LINC00623-2 | FALSE | Downregulated |
| 0,336315045 | LINC00624 | FALSE | Upregulated |
| 0,288128354 | LINC00626 | FALSE | Upregulated |
| 0,113569352 | LINC00628 | FALSE | Upregulated |
| 0 | LINC00629 | FALSE | Upregulated |
| -0,071717127 | LINC00630 | FALSE | Downregulated |
| 0,432485633 | LINC00632 | FALSE | Upregulated |
| 0,277833435 | LINC00633 | FALSE | Upregulated |
| | LINC00634 | | |
| 0,300241802 | LINC00635 | FALSE | Upregulated |
| | LINC00636 | | |
| 0,574338852 | LINC00637 | FALSE | Upregulated |
| 0,4787073 | LINC00638 | FALSE | Upregulated |
| 0,51582682 | LINC00639 | FALSE | Upregulated |
| | LINC00640 | | |
| | LINC00642 | | |
| 0,291614587 | LINC00643 | FALSE | Upregulated |
| | LINC00644 | | |
| 0,386990434 | LINC00645 | FALSE | Upregulated |
| 0,000264809 | LINC00648 | FALSE | Upregulated |
| -0,318293445 | LINC00649 | FALSE | Downregulated |
| -0,298167659 | LINC00654 | FALSE | Downregulated |
| 0,265122827 | LINC00656 | FALSE | Upregulated |

| | | | |
|---|---|---|---|
| -0,175713062 | LINC00658 | FALSE | Downregulated |
| -0,228730434 | LINC00659 | FALSE | Downregulated |
| 0,127894014 | LINC00661 | FALSE | Upregulated |
| 0,735782905 | LINC00662 | FALSE | Upregulated |
| -0,199736909 | LINC00663 | FALSE | Downregulated |
| -0,081872161 | LINC00664 | FALSE | Downregulated |
| 0,132208644 | LINC00665 | FALSE | Upregulated |
| | LINC00668 | | |
| | LINC00669 | | |
| 0 | LINC00670 | FALSE | Upregulated |
| 0,464243643 | LINC00672 | FALSE | Upregulated |
| 0 | LINC00673 | FALSE | Upregulated |
| 0,650069009 | LINC00676 | FALSE | Upregulated |
| 0,183367424 | LINC00678 | FALSE | Upregulated |
| | LINC00681 | | |
| | LINC00682 | | |
| 0 | LINC00684 | FALSE | Upregulated |
| | LINC00684-2 | | |
| 0,059271661 | LINC00685 | FALSE | Upregulated |
| | LINC00685-2 | | |
| | LINC00687 | | |
| -0,05109928 | LINC00689 | FALSE | Downregulated |
| 0 | LINC00690 | FALSE | Upregulated |
| 0,377433556 | LINC00691 | FALSE | Upregulated |
| | LINC00692 | | |
| -0,912522466 | LINC00693 | FALSE | Downregulated |
| -0,140269318 | LINC00696 | FALSE | Downregulated |
| | LINC00698 | | |
| -0,004525881 | LINC00700 | FALSE | Downregulated |
| | LINC00701 | | |
| 0,293242527 | LINC00702 | FALSE | Upregulated |
| | LINC00703 | | |
| 0,113548436 | LINC00704 | FALSE | Upregulated |
| 0,042372901 | LINC00705 | FALSE | Upregulated |
| 0,561464952 | LINC00706 | FALSE | Upregulated |
| 1,517035333 | LINC00707 | | Upregulated |
| 0 | LINC00708 | FALSE | Upregulated |
| | LINC00709 | | |
| -0,062431993 | LINC00710 | FALSE | Downregulated |
| | LINC00836 | | |
| -0,145176298 | LINC00837 | FALSE | Downregulated |
| 0,00459845 | LINC00838 | FALSE | Upregulated |
| -0,123032118 | LINC00839 | FALSE | Downregulated |
| | LINC00840 | | |
| | LINC00841 | | |
| 0,148106701 | LINC00842 | FALSE | Upregulated |
| | LINC00845 | | |
| | LINC00850 | | |
| 0,001141803 | LINC00851 | FALSE | Upregulated |
| 0,023979727 | LINC00852 | FALSE | Upregulated |
| 0,55324513 | LINC00853 | FALSE | Upregulated |

| | | | |
|---|---|---|---|
| | LINC00856 | | |
| 0,187107305 | LINC00857 | FALSE | Upregulated |
| | LINC00858 | | |
| 0,238445982 | LINC00862 | FALSE | Upregulated |
| 0,326340112 | LINC00864 | FALSE | Upregulated |
| 0,21306234 | LINC00865 | FALSE | Upregulated |
| 0 | LINC00866 | FALSE | Upregulated |
| 0,277119458 | LINC00867 | FALSE | Upregulated |
| 0,060824034 | LINC00869 | FALSE | Upregulated |
| | LINC00871 | | |
| 0,285258172 | LINC00877 | FALSE | Upregulated |
| | LINC00879 | | |
| -1,039584072 | LINC00880 | | Downregulated |
| -0,184649225 | LINC00881 | FALSE | Downregulated |
| -0,148465128 | LINC00882 | FALSE | Downregulated |
| -0,390824807 | LINC00885 | FALSE | Downregulated |
| 0,310843323 | LINC00887 | FALSE | Upregulated |
| -0,767245409 | LINC00888 | FALSE | Downregulated |
| 0 | LINC00889 | FALSE | Upregulated |
| 0,138244854 | LINC00890 | FALSE | Upregulated |
| 0,003711465 | LINC00891 | FALSE | Upregulated |
| 0,305188115 | LINC00892 | FALSE | Upregulated |
| -0,324727552 | LINC00893 | FALSE | Downregulated |
| | LINC00895 | | |
| | LINC00898 | | |
| 1,01487478 | LINC00901 | | Upregulated |
| | LINC00904 | | |
| | LINC00905 | | |
| 0 | LINC00906 | FALSE | Upregulated |
| 0,852391279 | LINC00907 | FALSE | Upregulated |
| 0,15098562 | LINC00908 | FALSE | Upregulated |
| | LINC00911 | | |
| | LINC00917 | | |
| | LINC00919 | | |
| -0,243799842 | LINC00920 | FALSE | Downregulated |
| | LINC00922 | | |
| 0,428511585 | LINC00923 | FALSE | Upregulated |
| 0,607090691 | LINC00924 | FALSE | Upregulated |
| 0,263627103 | LINC00925 | FALSE | Upregulated |
| -0,016214855 | LINC00926 | FALSE | Downregulated |
| | LINC00927 | | |
| 0,00190812 | LINC00928 | FALSE | Upregulated |
| | LINC00929 | | |
| -0,282368772 | LINC00930 | FALSE | Downregulated |
| -0,639389255 | LINC00933 | FALSE | Downregulated |
| 0,277764892 | LINC00934 | FALSE | Upregulated |
| -0,028231434 | LINC00935 | FALSE | Downregulated |
| -0,005208168 | LINC00937 | FALSE | Downregulated |
| 0,095443091 | LINC00938 | FALSE | Upregulated |
| 0,194324225 | LINC00939 | FALSE | Upregulated |
| 0,247222948 | LINC00940 | FALSE | Upregulated |

| | | | |
|---|---|---|---|
| 1,208015419 | LINC00941 | | Upregulated |
| -0,042977333 | LINC00942 | FALSE | Downregulated |
| | LINC00944 | | |
| -0,000490246 | LINC00945 | FALSE | Downregulated |
| -0,004525881 | LINC00948 | FALSE | Downregulated |
| -0,558480237 | LINC00950 | FALSE | Downregulated |
| | LINC00951 | | |
| | LINC00955 | | |
| 0 | LINC00958 | FALSE | Upregulated |
| -0,248915067 | LINC00959 | FALSE | Downregulated |
| 0,269170388 | LINC00960 | FALSE | Upregulated |
| 0,139336791 | LINC00961 | FALSE | Upregulated |
| 0,42584013 | LINC00964 | FALSE | Upregulated |
| -0,520267628 | LINC00965 | FALSE | Downregulated |
| 0,803907967 | LINC00966 | FALSE | Upregulated |
| 0,194658687 | LINC00967 | FALSE | Upregulated |
| 0,26891321 | LINC00968 | FALSE | Upregulated |
| -0,676195225 | LINC00969 | FALSE | Downregulated |
| 0,290109866 | LINC00970 | FALSE | Upregulated |
| -0,15780438 | LINC00971 | FALSE | Downregulated |
| | LINC00974 | | |
| 0,447456621 | LINC00977 | FALSE | Upregulated |
| -0,576223482 | LINC00982 | FALSE | Downregulated |
| 0 | LINC00989 | FALSE | Upregulated |
| | LINC00992 | | |
| | LINC00993 | | |
| | LINC00994 | | |
| 0,561354877 | LINC00996 | FALSE | Upregulated |
| 0,999739804 | LINC00997 | FALSE | Upregulated |
| -1,720863474 | LINC00998 | | Downregulated |
| 0,174455826 | LINC01001 | FALSE | Upregulated |
| 0,284507876 | LINC01002 | FALSE | Upregulated |
| 0,350259179 | LINC01003 | FALSE | Upregulated |
| | LINC01005 | | |
| -0,671481767 | LINC01006 | FALSE | Downregulated |
| 0,796194563 | LINC01007 | FALSE | Upregulated |
| -0,595241226 | LINC01010 | FALSE | Downregulated |
| 0,309299319 | LINC01011 | FALSE | Upregulated |
| 0,141572476 | LINC01012 | FALSE | Upregulated |
| | LINC01013 | | |
| -0,015614825 | LINC01014 | FALSE | Downregulated |
| | LINC01015 | | |
| 0,25765964 | LINC01016 | FALSE | Upregulated |
| | LINC01019 | | |
| | LINC01020 | | |
| 0,433328466 | LINC01021 | FALSE | Upregulated |
| 0,883361616 | LINC01023 | FALSE | Upregulated |
| 0,288474415 | LINC01028 | FALSE | Upregulated |
| | LINC01029 | | |
| 0,018516946 | LINC01030 | FALSE | Upregulated |
| | LINC01056 | | |

| | | | |
|---|---|---|---|
| 0,139274635 | LINC01057 | FALSE | Upregulated |
| | LINC01060 | | |
| -0,004525881 | LINC01063 | FALSE | Downregulated |
| -0,253091837 | LINC01069 | FALSE | Downregulated |
| 1,332618307 | LINC01070 | Upregulated | |
| 0,124039536 | LINC01071 | FALSE | Upregulated |
| 0,436539639 | LINC01080 | FALSE | Upregulated |
| | LINC01081 | | |
| -0,000838345 | LINC01082 | FALSE | Downregulated |
| | LINC01085 | | |
| 0,787104332 | LINC01087 | FALSE | Upregulated |
| 0 | LINC01088 | FALSE | Upregulated |
| -0,587116886 | LINC01089 | FALSE | Downregulated |
| -0,026616627 | LINC01091 | FALSE | Downregulated |
| -0,370633991 | LINC01094 | FALSE | Downregulated |
| | LINC01095 | | |
| 0 | LINC01096 | FALSE | Upregulated |
| | LINC01097 | | |
| -0,009465128 | LINC01098 | FALSE | Downregulated |
| 0 | LINC01099 | FALSE | Upregulated |
| | LINC01100 | | |
| 0,634357644 | LINC01101 | FALSE | Upregulated |
| 0 | LINC01102 | FALSE | Upregulated |
| | LINC01103 | | |
| 0 | LINC01104 | FALSE | Upregulated |
| | LINC01105 | | |
| 0,133565018 | LINC01106 | FALSE | Upregulated |
| 0,68500345 | LINC01108 | FALSE | Upregulated |
| | LINC01111 | | |
| | LINC01114 | | |
| 0,277764892 | LINC01115 | FALSE | Upregulated |
| 0 | LINC01116 | FALSE | Upregulated |
| -0,031748494 | LINC01118 | FALSE | Downregulated |
| 0,160324323 | LINC01119 | FALSE | Upregulated |
| | LINC01120 | | |
| -0,006506989 | LINC01121 | FALSE | Downregulated |
| | LINC01122 | | |
| 0,0791128 | LINC01123 | FALSE | Upregulated |
| -0,325079513 | LINC01124 | FALSE | Downregulated |
| 0,943466456 | LINC01125 | FALSE | Upregulated |
| 0,902445026 | LINC01127 | FALSE | Upregulated |
| 0,162708977 | LINC01132 | FALSE | Upregulated |
| 0,627885433 | LINC01133 | FALSE | Upregulated |
| 0,013163144 | LINC01134 | FALSE | Upregulated |
| 0,525982769 | LINC01135 | FALSE | Upregulated |
| | LINC01136 | | |
| 0,258735643 | LINC01137 | FALSE | Upregulated |
| 0 | LINC01139 | FALSE | Upregulated |
| 0,542282358 | LINC01140 | FALSE | Upregulated |
| -0,004525881 | LINC01141 | FALSE | Downregulated |
| | LINC01142 | | |

| | | | |
|---|---|---|---|
| -0,335316943 | LINC01144 | FALSE | Downregulated |
| 0,638429842 | LINC01146 | FALSE | Upregulated |
| 0 | LINC01149 | FALSE | Upregulated |
| | LINC01150 | | |
| -0,231842884 | LINC01153 | FALSE | Downregulated |
| | LINC01158 | | |
| -0,020023978 | LINC01160 | FALSE | Downregulated |
| | LINC01163 | | |
| | LINC01164 | | |
| 0,415469274 | LINC01165 | FALSE | Upregulated |
| -0,326854802 | LINC01168 | FALSE | Downregulated |
| -0,009465128 | LINC01171 | FALSE | Downregulated |
| 0,072908061 | LINC01176 | FALSE | Upregulated |
| 0,00139671 | LINC01179 | FALSE | Upregulated |
| | LINC01180 | | |
| -0,908863512 | LINC01181 | FALSE | Downregulated |
| | LINC01182 | | |
| 0,189632254 | LINC01183 | FALSE | Upregulated |
| -0,752629457 | LINC01184 | FALSE | Downregulated |
| 0 | LINC01185 | FALSE | Upregulated |
| 0 | LINC01186 | FALSE | Upregulated |
| 0,121828022 | LINC01187 | FALSE | Upregulated |
| -0,046151416 | LINC01189 | FALSE | Downregulated |
| 0,325741425 | LINC01191 | FALSE | Upregulated |
| | LINC01192 | | |
| 0,289235841 | LINC01193 | FALSE | Upregulated |
| 0,277119458 | LINC01194 | FALSE | Upregulated |
| 0,39339115 | LINC01197 | FALSE | Upregulated |
| | LINC01216 | | |
| -0,679961589 | LINGO1 | FALSE | Downregulated |
| -0,326164513 | LINGO2 | FALSE | Downregulated |
| -0,517869521 | LINGO4 | FALSE | Downregulated |
| -0,193350624 | LINS | FALSE | Downregulated |
| 0,500040779 | LIPE-AS1 | FALSE | Upregulated |
| | LIPF | | |
| 0,018761289 | LIPH | FALSE | Upregulated |
| 0,377065114 | LIPI | FALSE | Upregulated |
| -0,599589813 | LIPJ | FALSE | Downregulated |
| | LIPK | | |
| -0,045560959 | LIPM | FALSE | Downregulated |
| 0,007166748 | LIPN | FALSE | Upregulated |
| 0,049900422 | LIPT2 | FALSE | Upregulated |
| 0,059157964 | LIX1 | FALSE | Upregulated |
| 0,714701893 | LKAAEAR1 | FALSE | Upregulated |
| | LL22NC01-81G9.3 | | |
| -0,142070777 | LLPH | FALSE | Downregulated |
| | LMAN1L | | |
| 0,084223604 | LMCD1-AS1 | FALSE | Upregulated |
| 0 | LMO1 | FALSE | Upregulated |
| 0 | LMX1A | FALSE | Upregulated |
| 0,009125516 | LMX1B | FALSE | Upregulated |

| | | | |
|---|---|---|---|
| -0,037757655 | LNX1-AS1 | FALSE | Downregulated |
| 0,305913857 | LNX1-AS2 | FALSE | Upregulated |
| | LOC100125918 | | |
| 0,01648495 | LOC100126447 | FALSE | Upregulated |
| 0,033718007 | LOC100127974 | FALSE | Upregulated |
| 0,633202745 | LOC100128002 | FALSE | Upregulated |
| -0,526191608 | LOC100128006 | FALSE | Downregulated |
| 0 | LOC100128076 | FALSE | Upregulated |
| 0,09678797 | LOC100128079 | FALSE | Upregulated |
| 0 | LOC100128108 | FALSE | Upregulated |
| 0,147470622 | LOC100128127 | FALSE | Upregulated |
| | LOC100128139 | | |
| 0,933476274 | LOC100128164 | FALSE | Upregulated |
| | LOC100128233 | | |
| 0,029601951 | LOC100128276 | FALSE | Upregulated |
| 0,445002624 | LOC100128288 | FALSE | Upregulated |
| 0,385171408 | LOC100128325 | FALSE | Upregulated |
| 0,312137658 | LOC100128398 | FALSE | Upregulated |
| 0,149786071 | LOC100128560 | FALSE | Upregulated |
| -0,432158037 | LOC100128568 | FALSE | Downregulated |
| | LOC100128575 | | |
| 0,716873807 | LOC100128593 | FALSE | Upregulated |
| 0,005298914 | LOC100128885 | FALSE | Upregulated |
| 0,583300082 | LOC100128905 | FALSE | Upregulated |
| -0,08792827 | LOC100128908 | FALSE | Downregulated |
| -0,033913279 | LOC100128966 | FALSE | Downregulated |
| 0,294476532 | LOC100129055 | FALSE | Upregulated |
| 0,140137382 | LOC100129098 | FALSE | Upregulated |
| 0,127902005 | LOC100129138 | FALSE | Upregulated |
| 0,178477338 | LOC100129215 | FALSE | Upregulated |
| 0,414613678 | LOC100129216 | FALSE | Upregulated |
| 0,440211355 | LOC100129223 | FALSE | Upregulated |
| -0,033043381 | LOC100129307 | FALSE | Downregulated |
| -0,327878659 | LOC100129316 | FALSE | Downregulated |
| -0,004525881 | LOC100129455 | FALSE | Downregulated |
| 0,343602283 | LOC100129461 | FALSE | Upregulated |
| -0,771795601 | LOC100129473 | FALSE | Downregulated |
| 0,171687847 | LOC100129476 | FALSE | Upregulated |
| 0,141861991 | LOC100129516 | FALSE | Upregulated |
| | LOC100129520 | | |
| 0,071718138 | LOC100129534 | FALSE | Upregulated |
| | LOC100129603 | | |
| -0,617373051 | LOC100129617 | FALSE | Downregulated |
| | LOC100129636 | | |
| -0,757111052 | LOC100129781 | FALSE | Downregulated |
| 0,545081122 | LOC100129860 | FALSE | Upregulated |
| 0,281911094 | LOC100129924 | FALSE | Upregulated |
| -0,002253536 | LOC100129935 | FALSE | Downregulated |
| -0,345148011 | LOC100129940 | FALSE | Downregulated |
| 0,281984833 | LOC100130207 | FALSE | Upregulated |
| 0,67334009 | LOC100130238 | FALSE | Upregulated |

| | | | |
|---|---|---|---|
| 0,465707814 | LOC100130298 | FALSE | Upregulated |
| | LOC100130301 | | |
| 0,454943503 | LOC100130417 | FALSE | Upregulated |
| -0,015040902 | LOC100130452 | FALSE | Downregulated |
| -0,179979034 | LOC100130458 | FALSE | Downregulated |
| 0 | LOC100130502 | FALSE | Upregulated |
| -0,015055335 | LOC100130520 | FALSE | Downregulated |
| 0,510511888 | LOC100130539 | FALSE | Upregulated |
| -0,024454173 | LOC100130673 | FALSE | Downregulated |
| -0,213311571 | LOC100130698 | FALSE | Downregulated |
| -0,118643295 | LOC100130705 | FALSE | Downregulated |
| | LOC100130849 | | |
| 0,497957047 | LOC100130899 | FALSE | Upregulated |
| | LOC100130938 | | |
| | LOC100130964 | | |
| 0,682960623 | LOC100130987 | FALSE | Upregulated |
| 0,945789792 | LOC100131107 | FALSE | Upregulated |
| 0,670611094 | LOC100131138 | FALSE | Upregulated |
| 0,444945841 | LOC100131289 | FALSE | Upregulated |
| -0,519858051 | LOC100131303 | FALSE | Downregulated |
| 0,429080966 | LOC100131315 | FALSE | Upregulated |
| 0,707094383 | LOC100131347 | FALSE | Upregulated |
| 0,286014522 | LOC100131372 | FALSE | Upregulated |
| 0,004038899 | LOC100131472 | FALSE | Upregulated |
| -0,23621633 | LOC100131530 | FALSE | Downregulated |
| -0,884964848 | LOC100131532 | FALSE | Downregulated |
| 0,413594208 | LOC100131626 | FALSE | Upregulated |
| | LOC100131635 | | |
| -0,657553698 | LOC100131655 | FALSE | Downregulated |
| 1,767762455 | LOC100131825 | | Upregulated |
| 0 | LOC100132004 | FALSE | Upregulated |
| -0,64959458 | LOC100132111 | FALSE | Downregulated |
| | LOC100132146 | | |
| 0,145490944 | LOC100132154 | FALSE | Upregulated |
| | LOC100132174 | | |
| | LOC100132202 | | |
| 0,832983002 | LOC100132249 | FALSE | Upregulated |
| -0,200759359 | LOC100132352 | FALSE | Downregulated |
| 0,540040019 | LOC100132354 | FALSE | Upregulated |
| -0,149604067 | LOC100132735 | FALSE | Downregulated |
| 0,003711465 | LOC100132741 | FALSE | Upregulated |
| -1,111929405 | LOC100132813 | | Downregulated |
| | LOC100132831 | | |
| 0,434759656 | LOC100132874 | FALSE | Upregulated |
| 0,617554751 | LOC100132891 | FALSE | Upregulated |
| 0,069198453 | LOC100133050 | FALSE | Upregulated |
| | LOC100133267 | | |
| 0,319152184 | LOC100133315 | FALSE | Upregulated |
| -0,387777845 | LOC100133669 | FALSE | Downregulated |
| -0,001215276 | LOC100133746 | FALSE | Downregulated |
| 1,088607752 | LOC100133920 | | Upregulated |

| | | | |
|---|---|---|---|
| -0,811863472 | LOC100133985 | FALSE | Downregulated |
| 0,260857161 | LOC100134317 | FALSE | Upregulated |
| 0,017555071 | LOC100134368 | FALSE | Upregulated |
| 0,019772142 | LOC100134391 | FALSE | Upregulated |
| 0,132544124 | LOC100134868 | FALSE | Upregulated |
| 0,141808155 | LOC100188947 | FALSE | Upregulated |
| 0,506395464 | LOC100233156 | FALSE | Upregulated |
| | LOC100240728 | | |
| 0,447071688 | LOC100240734 | FALSE | Upregulated |
| -0,017234027 | LOC100240735 | FALSE | Downregulated |
| 0,337715282 | LOC100270804 | FALSE | Upregulated |
| -0,493075598 | LOC100272217 | FALSE | Downregulated |
| | LOC100286906 | | |
| 0,109350958 | LOC100287010 | FALSE | Upregulated |
| 0,328409337 | LOC100287015 | FALSE | Upregulated |
| | LOC100287072 | | |
| -0,326432928 | LOC100287082 | FALSE | Downregulated |
| 0,566655444 | LOC100287098 | FALSE | Upregulated |
| -0,248506087 | LOC100287175 | FALSE | Downregulated |
| | LOC100287294 | | |
| -0,14349285 | LOC100287387 | FALSE | Downregulated |
| 0 | LOC100287413 | FALSE | Upregulated |
| 0,135606138 | LOC100287477 | FALSE | Upregulated |
| 0,261279668 | LOC100287497 | FALSE | Upregulated |
| | LOC100287592 | | |
| 0,000264809 | LOC100287610 | FALSE | Upregulated |
| 0,492202116 | LOC100287632 | FALSE | Upregulated |
| -0,014724123 | LOC100287651 | FALSE | Downregulated |
| | LOC100287704 | | |
| | LOC100287728 | | |
| | LOC100287823 | | |
| | LOC100287834 | | |
| | LOC100287877 | | |
| -0,560957129 | LOC100287896 | FALSE | Downregulated |
| 0,22187228 | LOC100287944 | FALSE | Upregulated |
| -0,015482207 | LOC100288175 | FALSE | Downregulated |
| -0,335068829 | LOC100288181 | FALSE | Downregulated |
| 0 | LOC100288562 | FALSE | Upregulated |
| 0,272626305 | LOC100288570 | FALSE | Upregulated |
| 0,022876902 | LOC100288637 | FALSE | Upregulated |
| 0 | LOC100288748 | FALSE | Upregulated |
| 0,131734999 | LOC100288814 | FALSE | Upregulated |
| -0,066342136 | LOC100288842 | FALSE | Downregulated |
| -0,287909317 | LOC100288911 | FALSE | Downregulated |
| | LOC100288966 | | |
| -1,235254477 | LOC100288974 | | Downregulated |
| 0,195474033 | LOC100289026 | FALSE | Upregulated |
| 0,068177946 | LOC100289092 | FALSE | Upregulated |
| -0,276682475 | LOC100289097 | FALSE | Downregulated |
| -0,134112818 | LOC100289098 | FALSE | Downregulated |
| 0,546142966 | LOC100289333 | FALSE | Upregulated |

| | | | |
|---|---|---|---|
| -0,163491717 | LOC100289361 | FALSE | Downregulated |
| 0,000535661 | LOC100289473 | FALSE | Upregulated |
| 0,419832298 | LOC100289495 | FALSE | Upregulated |
| 0 | LOC100289656 | FALSE | Upregulated |
| 0,573356176 | LOC100289673 | FALSE | Upregulated |
| | LOC100293612 | | |
| 0,334195568 | LOC100335030 | FALSE | Upregulated |
| | LOC100422212 | | |
| -0,965898421 | LOC100422556 | FALSE | Downregulated |
| 0,214235903 | LOC100499484 | FALSE | Upregulated |
| -0,004118546 | LOC100500773 | FALSE | Downregulated |
| 0 | LOC100505498 | FALSE | Upregulated |
| 0,259323072 | LOC100505501 | FALSE | Upregulated |
| | LOC100505504 | | |
| 0,30722983 | LOC100505515 | FALSE | Upregulated |
| 0,018839898 | LOC100505534 | FALSE | Upregulated |
| 0,890194201 | LOC100505555 | FALSE | Upregulated |
| -0,002473216 | LOC100505585 | FALSE | Downregulated |
| -0,047898296 | LOC100505588 | FALSE | Downregulated |
| 0 | LOC100505609 | FALSE | Upregulated |
| 0,744502934 | LOC100505622 | FALSE | Upregulated |
| 0,25519031 | LOC100505625 | FALSE | Upregulated |
| -0,689731308 | LOC100505650 | FALSE | Downregulated |
| 0,439058315 | LOC100505664 | FALSE | Upregulated |
| 0,286166773 | LOC100505685 | FALSE | Upregulated |
| 0,140166988 | LOC100505710 | FALSE | Upregulated |
| | LOC100505711 | | |
| 0,064145915 | LOC100505727 | FALSE | Upregulated |
| 0,708270418 | LOC100505761 | FALSE | Upregulated |
| -0,469829288 | LOC100505771 | FALSE | Downregulated |
| -0,152565538 | LOC100505774 | FALSE | Downregulated |
| | LOC100505784 | | |
| | LOC100505797 | | |
| 0,080779177 | LOC100505798 | FALSE | Upregulated |
| | LOC100505811 | | |
| -0,743376041 | LOC100505812 | FALSE | Downregulated |
| 0,14200369 | LOC100505817 | FALSE | Upregulated |
| 0,282571604 | LOC100505824 | FALSE | Upregulated |
| | LOC100505835 | | |
| | LOC100505841 | | |
| 0,663837983 | LOC100505851 | FALSE | Upregulated |
| | LOC100505853 | | |
| | LOC100505874 | | |
| | LOC100505878 | | |
| | LOC100505887 | | |
| -1,110259584 | LOC100505902 | | Downregulated |
| -0,077730023 | LOC100505909 | FALSE | Downregulated |
| -0,386337386 | LOC100505912 | FALSE | Downregulated |
| | LOC100505920 | | |
| 0,631776499 | LOC100505938 | FALSE | Upregulated |
| | LOC100505940 | | |

| | | | |
|---|---|---|---|
| 0,309224525 | LOC100505942 | FALSE | Upregulated |
| | LOC100505948 | | |
| | LOC100505984 | | |
| 0,379448318 | LOC100505985 | FALSE | Upregulated |
| -0,321014237 | LOC100506014 | FALSE | Downregulated |
| | LOC100506016 | | |
| 0,190203473 | LOC100506022 | FALSE | Upregulated |
| -0,062493797 | LOC100506023 | FALSE | Downregulated |
| -0,350470405 | LOC100506036 | FALSE | Downregulated |
| -0,310415524 | LOC100506047 | FALSE | Downregulated |
| -0,006903421 | LOC100506082 | FALSE | Downregulated |
| | LOC100506098 | | |
| | LOC100506107 | | |
| 1,203559118 | LOC100506113 | | Upregulated |
| -0,351177545 | LOC100506124 | FALSE | Downregulated |
| 0,191243742 | LOC100506127 | FALSE | Upregulated |
| -0,335775285 | LOC100506157 | FALSE | Downregulated |
| 0,272277655 | LOC100506159 | FALSE | Upregulated |
| 1,200816275 | LOC100506161 | | Upregulated |
| 0,459241435 | LOC100506188 | FALSE | Upregulated |
| 0,685535676 | LOC100506235 | FALSE | Upregulated |
| | LOC100506236 | | |
| | LOC100506253 | | |
| | LOC100506271 | | |
| | LOC100506274 | | |
| 0,60760803 | LOC100506281 | FALSE | Upregulated |
| -0,185299684 | LOC100506282 | FALSE | Downregulated |
| 0,222532169 | LOC100506286 | FALSE | Upregulated |
| | LOC100506289 | | |
| -0,177522826 | LOC100506302 | FALSE | Downregulated |
| | LOC100506317 | | |
| 0,717082118 | LOC100506331 | FALSE | Upregulated |
| -0,119293069 | LOC100506351 | FALSE | Downregulated |
| 0,04596276 | LOC100506358 | FALSE | Upregulated |
| 0 | LOC100506371 | FALSE | Upregulated |
| 0,656994038 | LOC100506379 | FALSE | Upregulated |
| 0,125451431 | LOC100506384 | FALSE | Upregulated |
| | LOC100506403 | | |
| | LOC100506405 | | |
| | LOC100506406 | | |
| -0,080451758 | LOC100506411 | FALSE | Downregulated |
| 0 | LOC100506422 | FALSE | Upregulated |
| 1,234383033 | LOC100506454 | | Upregulated |
| | LOC100506470 | | |
| -0,415611973 | LOC100506472 | FALSE | Downregulated |
| -0,401533297 | LOC100506473 | FALSE | Downregulated |
| -0,00289892 | LOC100506474 | FALSE | Downregulated |
| -0,04657162 | LOC100506489 | FALSE | Downregulated |
| -0,662199691 | LOC100506498 | FALSE | Downregulated |
| -0,609955459 | LOC100506518 | FALSE | Downregulated |
| 0,294971359 | LOC100506526 | FALSE | Upregulated |

| | | | |
|---|---|---|---|
| 0 | LOC100506530 | FALSE | Upregulated |
| | LOC100506532 | | |
| 0,195846079 | LOC100506538 | FALSE | Upregulated |
| 0,009282089 | LOC100506557 | FALSE | Upregulated |
| 0,438880435 | LOC100506558 | FALSE | Upregulated |
| 0,290670908 | LOC100506563 | FALSE | Upregulated |
| 0,140367653 | LOC100506571 | FALSE | Upregulated |
| -0,07147399 | LOC100506639 | FALSE | Downregulated |
| | LOC100506664 | | |
| | LOC100506675 | | |
| 0,120914092 | LOC100506688 | FALSE | Upregulated |
| -0,101304418 | LOC100506691 | FALSE | Downregulated |
| 0,284373016 | LOC100506713 | FALSE | Upregulated |
| 0,05237531 | LOC100506714 | FALSE | Upregulated |
| -0,197794309 | LOC100506737 | FALSE | Downregulated |
| -0,210717482 | LOC100506746 | FALSE | Downregulated |
| -0,550989178 | LOC100506747 | FALSE | Downregulated |
| 0,505068593 | LOC100506753 | FALSE | Upregulated |
| | LOC100506790 | | |
| | LOC100506791 | | |
| 0,409304262 | LOC100506797 | FALSE | Upregulated |
| 0,401163419 | LOC100506827 | FALSE | Upregulated |
| | LOC100506851 | | |
| | LOC100506858 | | |
| -0,358621723 | LOC100506859 | FALSE | Downregulated |
| 0,018449106 | LOC100506869 | FALSE | Upregulated |
| -0,614550515 | LOC100506870 | FALSE | Downregulated |
| 0,284987956 | LOC100506882 | FALSE | Upregulated |
| | LOC100506885 | | |
| 1,887053799 | LOC100506901 | | Upregulated |
| 0,129073625 | LOC100506908 | FALSE | Upregulated |
| 0 | LOC100506929 | FALSE | Upregulated |
| 0 | LOC100506937 | FALSE | Upregulated |
| | LOC100506944 | | |
| 0,637694036 | LOC100506974 | FALSE | Upregulated |
| 0 | LOC100506999 | FALSE | Upregulated |
| 0,266554578 | LOC100507054 | FALSE | Upregulated |
| 0,590535074 | LOC100507103 | FALSE | Upregulated |
| | LOC100507165 | | |
| -0,251723627 | LOC100507195 | FALSE | Downregulated |
| | LOC100507205 | | |
| | LOC100507210 | | |
| | LOC100507221 | | |
| | LOC100507236 | | |
| 0,775797419 | LOC100507258 | FALSE | Upregulated |
| -0,18101615 | LOC100507277 | FALSE | Downregulated |
| -1,045360955 | LOC100507316 | | Downregulated |
| 0,054784744 | LOC100507336 | FALSE | Upregulated |
| 0,145793417 | LOC100507351 | FALSE | Upregulated |
| | LOC100507353 | | |
| | LOC100507384 | | |

| | | | |
|---|---|---|---|
| 0,293242527 | LOC100507387 | FALSE | Upregulated |
| -0,521917227 | LOC100507395 | FALSE | Downregulated |
| | LOC100507403 | | |
| | LOC100507420 | | |
| 0,838101894 | LOC100507424 | FALSE | Upregulated |
| | LOC100507443 | | |
| 0,137680749 | LOC100507460 | FALSE | Upregulated |
| | LOC100507461 | | |
| -0,005017607 | LOC100507462 | FALSE | Downregulated |
| | LOC100507464 | | |
| | LOC100507468 | | |
| 0 | LOC100507480 | FALSE | Upregulated |
| 0,61967807 | LOC100507487 | FALSE | Upregulated |
| -0,033838436 | LOC100507507 | FALSE | Downregulated |
| -0,293690618 | LOC100507513 | FALSE | Downregulated |
| | LOC100507521 | | |
| 0,019395057 | LOC100507534 | FALSE | Upregulated |
| -0,006903421 | LOC100507548 | FALSE | Downregulated |
| 0,133342754 | LOC100507554 | FALSE | Upregulated |
| | LOC100507560 | | |
| 0,432042984 | LOC100507562 | FALSE | Upregulated |
| 0,552695308 | LOC100507599 | FALSE | Upregulated |
| 0,00655966 | LOC100507600 | FALSE | Upregulated |
| 0,405200286 | LOC100507634 | FALSE | Upregulated |
| 0,140933243 | LOC100507642 | FALSE | Upregulated |
| 0,818490316 | LOC100507656 | FALSE | Upregulated |
| 0,27227367 | LOC100507657 | FALSE | Upregulated |
| 0,184625599 | LOC100507670 | FALSE | Upregulated |
| 0,374420052 | LOC100507747 | FALSE | Upregulated |
| 0 | LOC100508046 | FALSE | Upregulated |
| 0,293326262 | LOC100508631 | FALSE | Upregulated |
| | LOC100509303 | | |
| -0,676904099 | LOC100509620 | FALSE | Downregulated |
| 0,379793276 | LOC100510707 | FALSE | Upregulated |
| -0,159016277 | LOC100616530 | FALSE | Downregulated |
| -0,526684148 | LOC100630918 | FALSE | Downregulated |
| -0,193348817 | LOC100652736 | FALSE | Downregulated |
| 0,220290096 | LOC100652758 | FALSE | Upregulated |
| 0,166959055 | LOC100652777 | FALSE | Upregulated |
| -0,129475942 | LOC100652807 | FALSE | Downregulated |
| 0,566567292 | LOC100652824 | FALSE | Upregulated |
| -0,301618309 | LOC100652901 | FALSE | Downregulated |
| 0,274071794 | LOC100652911 | FALSE | Upregulated |
| 0,351986509 | LOC100652930 | FALSE | Upregulated |
| | LOC100652931 | | |
| 0,009680707 | LOC100653005 | FALSE | Upregulated |
| 0,536480351 | LOC100653133 | FALSE | Upregulated |
| -0,377563284 | LOC100653233 | FALSE | Downregulated |
| 0,110416695 | LOC100653515 | FALSE | Upregulated |
| -0,44039614 | LOC100862671 | FALSE | Downregulated |
| -0,577720992 | LOC100996246 | FALSE | Downregulated |

| | | | |
|---|---|---|---|
| | LOC100996249 | | |
| | LOC100996266 | | |
| 0,129726847 | LOC100996274 | FALSE | Upregulated |
| 0,905515486 | LOC100996288 | FALSE | Upregulated |
| | LOC100996290 | | |
| -0,140756304 | LOC100996294 | FALSE | Downregulated |
| 0,315338207 | LOC100996301 | FALSE | Upregulated |
| -0,173612284 | LOC100996316 | FALSE | Downregulated |
| 0,839696522 | LOC100996318 | FALSE | Upregulated |
| 0,007068729 | LOC100996335 | FALSE | Upregulated |
| 0,466837012 | LOC100996337 | FALSE | Upregulated |
| | LOC100996338 | | |
| 0,226364775 | LOC100996342 | FALSE | Upregulated |
| 0,317370137 | LOC100996345 | FALSE | Upregulated |
| | LOC100996349 | | |
| 0,466834573 | LOC100996350 | FALSE | Upregulated |
| | LOC100996365 | | |
| 0,378602154 | LOC100996369 | FALSE | Upregulated |
| -0,528673011 | LOC100996379 | FALSE | Downregulated |
| 0,328877661 | LOC100996385 | FALSE | Upregulated |
| 0,021467334 | LOC100996401 | FALSE | Upregulated |
| -0,01753274 | LOC100996404 | FALSE | Downregulated |
| | LOC100996414 | | |
| | LOC100996415 | | |
| 0,575637282 | LOC100996419 | FALSE | Upregulated |
| 0,158369358 | LOC100996425 | FALSE | Upregulated |
| 0,293326262 | LOC100996428 | FALSE | Upregulated |
| -0,446053752 | LOC100996442 | FALSE | Downregulated |
| 0,469136585 | LOC100996455 | FALSE | Upregulated |
| 1,195278157 | LOC100996457 | | Upregulated |
| 0,099818616 | LOC100996465 | FALSE | Upregulated |
| 0,33145933 | LOC100996478 | FALSE | Upregulated |
| 0 | LOC100996482 | FALSE | Upregulated |
| -0,214935043 | LOC100996497 | FALSE | Downregulated |
| 0,367681321 | LOC100996517 | FALSE | Upregulated |
| 0,000823418 | LOC100996521 | FALSE | Upregulated |
| 1,547380228 | LOC100996535 | | Upregulated |
| -0,1648461 | LOC100996549 | FALSE | Downregulated |
| 0,16216552 | LOC100996557 | FALSE | Upregulated |
| 0 | LOC100996570 | FALSE | Upregulated |
| -0,210512146 | LOC100996598 | FALSE | Downregulated |
| 0,27277909 | LOC100996602 | FALSE | Upregulated |
| 0,683809405 | LOC100996624 | FALSE | Upregulated |
| | LOC100996630 | | |
| 1,306272222 | LOC100996634 | | Upregulated |
| | LOC100996637 | | |
| | LOC100996645 | | |
| 0,39698043 | LOC100996660 | FALSE | Upregulated |
| -0,299625018 | LOC100996662 | FALSE | Downregulated |
| 0,545141292 | LOC100996663 | FALSE | Upregulated |
| 0,014602858 | LOC100996664 | FALSE | Upregulated |

| | | | |
|---|---|---|---|
| | LOC100996665 | | |
| -0,717032001 | LOC100996668 | FALSE | Downregulated |
| | LOC100996681 | | |
| -0,638146505 | LOC100996693 | FALSE | Downregulated |
| 0,308427694 | LOC100996695 | FALSE | Upregulated |
| | LOC100996696 | | |
| 0 | LOC100996701 | FALSE | Upregulated |
| 0,955336461 | LOC100996713 | FALSE | Upregulated |
| 0,619540668 | LOC100996716 | FALSE | Upregulated |
| 1,638876734 | LOC100996721 | | Upregulated |
| 0,030093704 | LOC100996732 | FALSE | Upregulated |
| 0,570442932 | LOC100996735 | FALSE | Upregulated |
| | LOC100996756 | | |
| 0,5068311 | LOC100996758 | FALSE | Upregulated |
| 0,235562379 | LOC100996760 | FALSE | Upregulated |
| | LOC100996770 | | |
| 0 | LOC100996792 | FALSE | Upregulated |
| | LOC100996792-2 | | |
| -0,009465128 | LOC100996808 | FALSE | Downregulated |
| | LOC100996902 | | |
| -0,32508954 | LOC100996909 | FALSE | Downregulated |
| 0,008785567 | LOC101054525 | FALSE | Upregulated |
| | LOC101059914 | | |
| | LOC101059915 | | |
| -0,032880237 | LOC101059936 | FALSE | Downregulated |
| 0,144314145 | LOC101059948 | FALSE | Upregulated |
| -0,33113462 | LOC101059954 | FALSE | Downregulated |
| 0,851133025 | LOC101059976 | FALSE | Upregulated |
| | LOC101060004 | | |
| | LOC101060017 | | |
| | LOC101060024 | | |
| 0,680724821 | LOC101060026 | FALSE | Upregulated |
| 0,286523594 | LOC101060038 | FALSE | Upregulated |
| 0,347709732 | LOC101060060 | FALSE | Upregulated |
| 0,309260602 | LOC101060085 | FALSE | Upregulated |
| -0,003489667 | LOC101060097 | FALSE | Downregulated |
| 0,405141944 | LOC101060106 | FALSE | Upregulated |
| | LOC101060128 | | |
| -0,446474104 | LOC101060153 | FALSE | Downregulated |
| 0 | LOC101060157 | FALSE | Upregulated |
| -0,178870398 | LOC101060179 | FALSE | Downregulated |
| -0,004525881 | LOC101060183 | FALSE | Downregulated |
| | LOC101060187 | | |
| 0,545405294 | LOC101060195 | FALSE | Upregulated |
| | LOC101060211 | | |
| 1,110693951 | LOC101060216 | | Upregulated |
| | LOC101060233 | | |
| 1,215172996 | LOC101060236 | | Upregulated |
| | LOC101060254 | | |
| | LOC101060264 | | |
| 0 | LOC101060301 | FALSE | Upregulated |

| | | | |
|---|---|---|---|
| -0,3165948 | LOC101060321 | FALSE | Downregulated |
| 0,165213766 | LOC101060341 | FALSE | Upregulated |
| 0 | LOC101060363 | FALSE | Upregulated |
| 0,648684722 | LOC101060389 | FALSE | Upregulated |
| -0,018149848 | LOC101060391 | FALSE | Downregulated |
| 0,531280759 | LOC101060400 | FALSE | Upregulated |
| 0 | LOC101060498 | FALSE | Upregulated |
| 0,28181789 | LOC101060532 | FALSE | Upregulated |
| 0,177001047 | LOC101060569 | FALSE | Upregulated |
| -0,307982432 | LOC101060626 | FALSE | Downregulated |
| -0,094697607 | LOC101409256 | FALSE | Downregulated |
| -0,047837046 | LOC101448202 | FALSE | Downregulated |
| 0 | LOC101559451 | FALSE | Upregulated |
| 0,000504523 | LOC101805489 | FALSE | Upregulated |
| 0,285394326 | LOC101805491 | FALSE | Upregulated |
| | LOC101926886 | | |
| 0,907484702 | LOC101926889 | FALSE | Upregulated |
| | LOC101926893 | | |
| -1,145276436 | LOC101926894 | | Downregulated |
| 0,27801981 | LOC101926895 | FALSE | Upregulated |
| 0,669189569 | LOC101926896 | FALSE | Upregulated |
| | LOC101926897 | | |
| 0,291340714 | LOC101926899 | FALSE | Upregulated |
| 0,303479723 | LOC101926900 | FALSE | Upregulated |
| 0,1955622 | LOC101926901 | FALSE | Upregulated |
| -0,538426178 | LOC101926902 | FALSE | Downregulated |
| | LOC101926904 | | |
| -0,276010121 | LOC101926905 | FALSE | Downregulated |
| 1,083416976 | LOC101926907 | | Upregulated |
| 0,266186224 | LOC101926909 | FALSE | Upregulated |
| | LOC101926912 | | |
| 0,114003988 | LOC101926916 | FALSE | Upregulated |
| -0,000731589 | LOC101926917 | FALSE | Downregulated |
| | LOC101926921 | | |
| 0,834120306 | LOC101926922 | FALSE | Upregulated |
| | LOC101926923 | | |
| | LOC101926929 | | |
| 0,247249294 | LOC101926931 | FALSE | Upregulated |
| -0,255291276 | LOC101926934 | FALSE | Downregulated |
| 0,130514398 | LOC101926935 | FALSE | Upregulated |
| 0,396023183 | LOC101926936 | FALSE | Upregulated |
| 0,300299802 | LOC101926937 | FALSE | Upregulated |
| 0 | LOC101926940 | FALSE | Upregulated |
| 0,577977225 | LOC101926948 | FALSE | Upregulated |
| | LOC101926951 | | |
| | LOC101926952 | | |
| | LOC101926953 | | |
| -0,919865337 | LOC101926956 | FALSE | Downregulated |
| | LOC101926958 | | |
| 0 | LOC101926959 | FALSE | Upregulated |
| 0,746031064 | LOC101926960 | FALSE | Upregulated |

| | | | |
|---|---|---|---|
| | LOC101926961 | | |
| | LOC101926965 | | |
| | LOC101926966 | | |
| 0 | LOC101926968 | FALSE | Upregulated |
| 1,121462768 | LOC101926969 | | Upregulated |
| | LOC101926970 | | |
| | LOC101926972 | | |
| | LOC101926974 | | |
| | LOC101926975 | | |
| 0,258602414 | LOC101926977 | FALSE | Upregulated |
| | LOC101926982 | | |
| | LOC101926983 | | |
| 0 | LOC101926989 | FALSE | Upregulated |
| 0,280928672 | LOC101926992 | FALSE | Upregulated |
| -0,141154403 | LOC101926997 | FALSE | Downregulated |
| | LOC101926998 | | |
| | LOC101926999 | | |
| | LOC101927008 | | |
| 0,185507341 | LOC101927009 | FALSE | Upregulated |
| 0,883575074 | LOC101927010 | FALSE | Upregulated |
| -0,093313726 | LOC101927011 | FALSE | Downregulated |
| | LOC101927012 | | |
| | LOC101927013 | | |
| | LOC101927016 | | |
| | LOC101927019 | | |
| | LOC101927022 | | |
| | LOC101927024 | | |
| | LOC101927025 | | |
| -0,048933282 | LOC101927026 | FALSE | Downregulated |
| 1,222359755 | LOC101927029 | | Upregulated |
| 0,300549439 | LOC101927032 | FALSE | Upregulated |
| | LOC101927037 | | |
| -0,174380581 | LOC101927038 | FALSE | Downregulated |
| | LOC101927039 | | |
| | LOC101927042 | | |
| 0,251842554 | LOC101927045 | FALSE | Upregulated |
| | LOC101927046 | | |
| | LOC101927050 | | |
| | LOC101927052 | | |
| 0 | LOC101927053 | FALSE | Upregulated |
| 0,389546194 | LOC101927054 | FALSE | Upregulated |
| | LOC101927059 | | |
| 0,259973192 | LOC101927060 | FALSE | Upregulated |
| 0,000237167 | LOC101927063 | FALSE | Upregulated |
| | LOC101927065 | | |
| 0,725912066 | LOC101927069 | FALSE | Upregulated |
| -0,022695575 | LOC101927070 | FALSE | Downregulated |
| 0,79605923 | LOC101927071 | FALSE | Upregulated |
| | LOC101927072 | | |
| 0,353807228 | LOC101927073 | FALSE | Upregulated |
| | LOC101927080 | | |

| | | | |
|---|---|---|---|
| 1,50682741 | LOC101927082 | | Upregulated |
| -0,264123804 | LOC101927086 | FALSE | Downregulated |
| 0,625402943 | LOC101927088 | FALSE | Upregulated |
| | LOC101927089 | | |
| | LOC101927091 | | |
| 0,124177986 | LOC101927093 | FALSE | Upregulated |
| | LOC101927094 | | |
| 0 | LOC101927095 | FALSE | Upregulated |
| 0,526086217 | LOC101927098 | FALSE | Upregulated |
| 0,926154321 | LOC101927103 | FALSE | Upregulated |
| 0,872239912 | LOC101927104 | FALSE | Upregulated |
| | LOC101927108 | | |
| | LOC101927110 | | |
| 0 | LOC101927112 | FALSE | Upregulated |
| -0,920681644 | LOC101927116 | FALSE | Downregulated |
| | LOC101927118 | | |
| 0,228125477 | LOC101927119 | FALSE | Upregulated |
| -0,396276735 | LOC101927121 | FALSE | Downregulated |
| 0,531195264 | LOC101927122 | FALSE | Upregulated |
| 0,679883169 | LOC101927125 | FALSE | Upregulated |
| -0,49509713 | LOC101927126 | FALSE | Downregulated |
| | LOC101927127 | | |
| | LOC101927129 | | |
| 0,829241641 | LOC101927131 | FALSE | Upregulated |
| | LOC101927133 | | |
| 0,575932082 | LOC101927135 | FALSE | Upregulated |
| | LOC101927137 | | |
| -0,145085608 | LOC101927138 | FALSE | Downregulated |
| | LOC101927141 | | |
| -1,097140193 | LOC101927143 | | Downregulated |
| -0,270930055 | LOC101927144 | FALSE | Downregulated |
| | LOC101927145 | | |
| | LOC101927147 | | |
| | LOC101927153 | | |
| | LOC101927154 | | |
| 1,020431065 | LOC101927159 | | Upregulated |
| 0,227291359 | LOC101927163 | FALSE | Upregulated |
| 0 | LOC101927165 | FALSE | Upregulated |
| 0,361274501 | LOC101927166 | FALSE | Upregulated |
| 0 | LOC101927168 | FALSE | Upregulated |
| -0,004525881 | LOC101927169 | FALSE | Downregulated |
| -0,28749745 | LOC101927170 | FALSE | Downregulated |
| -0,006848587 | LOC101927172 | FALSE | Downregulated |
| | LOC101927174 | | |
| 0,61222794 | LOC101927179 | FALSE | Upregulated |
| | LOC101927182 | | |
| -0,411684933 | LOC101927183 | FALSE | Downregulated |
| 0,946532236 | LOC101927188 | FALSE | Upregulated |
| | LOC101927191 | | |
| 0,030989826 | LOC101927192 | FALSE | Upregulated |
| 0,454390784 | LOC101927198 | FALSE | Upregulated |

| | | | |
|---|---|---|---|
| 0 | LOC101927199 | FALSE | Upregulated |
| | LOC101927200 | | |
| | LOC101927202 | | |
| | LOC101927205 | | |
| | LOC101927209 | | |
| | LOC101927210 | | |
| | LOC101927212 | | |
| | LOC101927213 | | |
| 0,277301071 | LOC101927216 | FALSE | Upregulated |
| -0,158343621 | LOC101927225 | FALSE | Downregulated |
| 0,162110129 | LOC101927226 | FALSE | Upregulated |
| 0,396940688 | LOC101927228 | FALSE | Upregulated |
| -0,00372018 | LOC101927229 | FALSE | Downregulated |
| 0 | LOC101927232 | FALSE | Upregulated |
| 0,139775611 | LOC101927234 | FALSE | Upregulated |
| 0,802938237 | LOC101927235 | FALSE | Upregulated |
| | LOC101927238 | | |
| | LOC101927241 | | |
| 0 | LOC101927245 | FALSE | Upregulated |
| | LOC101927247 | | |
| 0,477460363 | LOC101927248 | FALSE | Upregulated |
| -0,020004397 | LOC101927249 | FALSE | Downregulated |
| -0,399313031 | LOC101927250 | FALSE | Downregulated |
| 0,160457364 | LOC101927252 | FALSE | Upregulated |
| | LOC101927253 | | |
| 0,41796078 | LOC101927254 | FALSE | Upregulated |
| 0,720375977 | LOC101927258 | FALSE | Upregulated |
| 0,335178411 | LOC101927259 | FALSE | Upregulated |
| -0,181677294 | LOC101927260 | FALSE | Downregulated |
| 0,267872763 | LOC101927262 | FALSE | Upregulated |
| 0,499203209 | LOC101927263 | FALSE | Upregulated |
| 0,366186079 | LOC101927268 | FALSE | Upregulated |
| | LOC101927271 | | |
| | LOC101927277 | | |
| 0,432126243 | LOC101927278 | FALSE | Upregulated |
| -0,683677194 | LOC101927279 | FALSE | Downregulated |
| | LOC101927280 | | |
| | LOC101927281 | | |
| -0,215640788 | LOC101927283 | FALSE | Downregulated |
| | LOC101927284 | | |
| 0 | LOC101927287 | FALSE | Upregulated |
| | LOC101927291 | | |
| 0 | LOC101927293 | FALSE | Upregulated |
| | LOC101927297 | | |
| | LOC101927299 | | |
| 0,221268794 | LOC101927300 | FALSE | Upregulated |
| 0,570952607 | LOC101927301 | FALSE | Upregulated |
| | LOC101927304 | | |
| | LOC101927306 | | |
| 0,277119458 | LOC101927311 | FALSE | Upregulated |
| 0 | LOC101927318 | FALSE | Upregulated |

| | | | |
|---|---|---|---|
| | LOC101927319 | | |
| 0,527024197 | LOC101927320 | FALSE | Upregulated |
| 1,512091012 | LOC101927322 | | Upregulated |
| 0 | LOC101927325 | FALSE | Upregulated |
| -0,317175151 | LOC101927326 | FALSE | Downregulated |
| 0,421660595 | LOC101927328 | FALSE | Upregulated |
| | LOC101927329 | | |
| -0,457484485 | LOC101927330 | FALSE | Downregulated |
| -0,296536348 | LOC101927331 | FALSE | Downregulated |
| | LOC101927332 | | |
| 0,276004788 | LOC101927333 | FALSE | Upregulated |
| | LOC101927335 | | |
| | LOC101927337 | | |
| -0,008895443 | LOC101927338 | FALSE | Downregulated |
| | LOC101927341 | | |
| | LOC101927344 | | |
| 0 | LOC101927346 | FALSE | Upregulated |
| -0,158343621 | LOC101927348 | FALSE | Downregulated |
| | LOC101927351 | | |
| -0,158343621 | LOC101927353 | FALSE | Downregulated |
| -0,081110171 | LOC101927354 | FALSE | Downregulated |
| 0,293242527 | LOC101927357 | FALSE | Upregulated |
| | LOC101927361 | | |
| | LOC101927362 | | |
| | LOC101927363 | | |
| 0,412930821 | LOC101927366 | FALSE | Upregulated |
| 0 | LOC101927367 | FALSE | Upregulated |
| 0 | LOC101927369 | FALSE | Upregulated |
| -0,500835562 | LOC101927372 | FALSE | Downregulated |
| 0,276085795 | LOC101927374 | FALSE | Upregulated |
| 1,001667398 | LOC101927375 | | Upregulated |
| 0,150564441 | LOC101927376 | FALSE | Upregulated |
| | LOC101927383 | | |
| | LOC101927387 | | |
| -0,01980817 | LOC101927388 | FALSE | Downregulated |
| | LOC101927389 | | |
| -0,148465128 | LOC101927392 | FALSE | Downregulated |
| | LOC101927395 | | |
| | LOC101927397 | | |
| | LOC101927398 | | |
| 0,808637947 | LOC101927400 | FALSE | Upregulated |
| 0,138255369 | LOC101927401 | FALSE | Upregulated |
| -0,002253536 | LOC101927403 | FALSE | Downregulated |
| | LOC101927404 | | |
| | LOC101927405 | | |
| -0,353757898 | LOC101927407 | FALSE | Downregulated |
| | LOC101927409 | | |
| | LOC101927413 | | |
| | LOC101927414 | | |
| -0,035242766 | LOC101927417 | FALSE | Downregulated |
| | LOC101927426 | | |

| | | | |
|---|---|---|---|
| | LOC101927429 | | |
| 0,774731526 | LOC101927430 | FALSE | Upregulated |
| 0,404720147 | LOC101927432 | FALSE | Upregulated |
| -0,037050381 | LOC101927435 | FALSE | Downregulated |
| | LOC101927436 | | |
| | LOC101927439 | | |
| 0 | LOC101927441 | FALSE | Upregulated |
| 0,067468757 | LOC101927446 | FALSE | Upregulated |
| 0,465251913 | LOC101927447 | FALSE | Upregulated |
| -0,021012718 | LOC101927448 | FALSE | Downregulated |
| | LOC101927452 | | |
| 0,438490037 | LOC101927454 | FALSE | Upregulated |
| -0,015083143 | LOC101927458 | FALSE | Downregulated |
| | LOC101927461 | | |
| | LOC101927466 | | |
| | LOC101927467 | | |
| 0,309780022 | LOC101927471 | FALSE | Upregulated |
| | LOC101927473 | | |
| 1,191500636 | LOC101927474 | | Upregulated |
| | LOC101927480 | | |
| 0,440673999 | LOC101927481 | FALSE | Upregulated |
| | LOC101927483 | | |
| | LOC101927484 | | |
| -0,000730793 | LOC101927486 | FALSE | Downregulated |
| | LOC101927488 | | |
| -0,330191165 | LOC101927490 | FALSE | Downregulated |
| | LOC101927491 | | |
| | LOC101927493 | | |
| | LOC101927494 | | |
| | LOC101927496 | | |
| -0,187503692 | LOC101927499 | FALSE | Downregulated |
| 0,741876451 | LOC101927503 | FALSE | Upregulated |
| | LOC101927504 | | |
| 0,142903709 | LOC101927505 | FALSE | Upregulated |
| -0,468096437 | LOC101927506 | FALSE | Downregulated |
| 0,435797306 | LOC101927508 | FALSE | Upregulated |
| 0 | LOC101927510 | FALSE | Upregulated |
| 0,003509782 | LOC101927511 | FALSE | Upregulated |
| 0,111256518 | LOC101927512 | FALSE | Upregulated |
| | LOC101927518 | | |
| -0,184654946 | LOC101927520 | FALSE | Downregulated |
| 0,509069291 | LOC101927521 | FALSE | Upregulated |
| | LOC101927522 | | |
| -0,018149848 | LOC101927525 | FALSE | Downregulated |
| 0,722491204 | LOC101927526 | FALSE | Upregulated |
| | LOC101927527 | | |
| -0,005653927 | LOC101927531 | FALSE | Downregulated |
| | LOC101927532 | | |
| | LOC101927533 | | |
| | LOC101927534 | | |
| 0,222616674 | LOC101927536 | FALSE | Upregulated |

| | | | |
|---|---|---|---|
| 0 | LOC101927537 | FALSE | Upregulated |
| 0 | LOC101927538 | FALSE | Upregulated |
| -0,001751586 | LOC101927544 | FALSE | Downregulated |
| 0,548188179 | LOC101927546 | FALSE | Upregulated |
| | LOC101927548 | | |
| | LOC101927553 | | |
| 0 | LOC101927555 | FALSE | Upregulated |
| | LOC101927558 | | |
| -0,544714825 | LOC101927559 | FALSE | Downregulated |
| | LOC101927561 | | |
| -0,622050899 | LOC101927562 | FALSE | Downregulated |
| 0 | LOC101927566 | FALSE | Upregulated |
| 0,859886577 | LOC101927567 | FALSE | Upregulated |
| 1,350087397 | LOC101927568 | | Upregulated |
| -0,946687914 | LOC101927571 | FALSE | Downregulated |
| | LOC101927573 | | |
| 0,308240577 | LOC101927576 | FALSE | Upregulated |
| -0,022239343 | LOC101927579 | FALSE | Downregulated |
| 0,274635952 | LOC101927582 | FALSE | Upregulated |
| 1,128932265 | LOC101927583 | | Upregulated |
| | LOC101927584 | | |
| -0,673308323 | LOC101927589 | FALSE | Downregulated |
| 0,25778885 | LOC101927594 | FALSE | Upregulated |
| -0,114277529 | LOC101927595 | FALSE | Downregulated |
| 0,70799641 | LOC101927596 | FALSE | Upregulated |
| 0,037934092 | LOC101927600 | FALSE | Upregulated |
| | LOC101927602 | | |
| 0,288482103 | LOC101927603 | FALSE | Upregulated |
| | LOC101927605 | | |
| | LOC101927606 | | |
| 1,370836313 | LOC101927608 | | Upregulated |
| -0,361730622 | LOC101927609 | FALSE | Downregulated |
| 0,259574757 | LOC101927611 | FALSE | Upregulated |
| 0,324321103 | LOC101927613 | FALSE | Upregulated |
| 0,751489441 | LOC101927615 | FALSE | Upregulated |
| 0,216437704 | LOC101927617 | FALSE | Upregulated |
| 0,402781526 | LOC101927623 | FALSE | Upregulated |
| | LOC101927627 | | |
| 0,075913858 | LOC101927628 | FALSE | Upregulated |
| | LOC101927629 | | |
| 0,355880184 | LOC101927632 | FALSE | Upregulated |
| 0 | LOC101927633 | FALSE | Upregulated |
| 0 | LOC101927634 | FALSE | Upregulated |
| -0,03600178 | LOC101927635 | FALSE | Downregulated |
| -0,080299054 | LOC101927646 | FALSE | Downregulated |
| -0,845596069 | LOC101927651 | FALSE | Downregulated |
| -0,251185123 | LOC101927652 | FALSE | Downregulated |
| | LOC101927656 | | |
| -0,006903421 | LOC101927660 | FALSE | Downregulated |
| | LOC101927661 | | |
| -0,381908359 | LOC101927662 | FALSE | Downregulated |

| | | | |
|---|---|---|---|
| 0,557836309 | LOC101927663 | FALSE | Upregulated |
| | LOC101927664 | | |
| -0,157125757 | LOC101927665 | FALSE | Downregulated |
| 0 | LOC101927669 | FALSE | Upregulated |
| 0,299801288 | LOC101927670 | FALSE | Upregulated |
| -0,289940235 | LOC101927674 | FALSE | Downregulated |
| | LOC101927676 | | |
| -0,256861987 | LOC101927680 | FALSE | Downregulated |
| | LOC101927684 | | |
| 0 | LOC101927686 | FALSE | Upregulated |
| | LOC101927687 | | |
| | LOC101927690 | | |
| | LOC101927691 | | |
| 0,822218268 | LOC101927693 | FALSE | Upregulated |
| | LOC101927695 | | |
| -0,006848587 | LOC101927696 | FALSE | Downregulated |
| -0,001320428 | LOC101927697 | FALSE | Downregulated |
| -0,393067665 | LOC101927699 | FALSE | Downregulated |
| 0,133671337 | LOC101927702 | FALSE | Upregulated |
| -0,570783577 | LOC101927703 | FALSE | Downregulated |
| | LOC101927704 | | |
| -0,168583647 | LOC101927705 | FALSE | Downregulated |
| -0,155394416 | LOC101927708 | FALSE | Downregulated |
| | LOC101927709 | | |
| 0,296754183 | LOC101927710 | FALSE | Upregulated |
| | LOC101927711 | | |
| 0 | LOC101927712 | FALSE | Upregulated |
| | LOC101927713 | | |
| 0,430382497 | LOC101927720 | FALSE | Upregulated |
| 0,175009208 | LOC101927721 | FALSE | Upregulated |
| 0,293326262 | LOC101927722 | FALSE | Upregulated |
| 0,190450677 | LOC101927723 | FALSE | Upregulated |
| -0,25756577 | LOC101927725 | FALSE | Downregulated |
| | LOC101927729 | | |
| | LOC101927731 | | |
| 0,484318865 | LOC101927733 | FALSE | Upregulated |
| 0,272462411 | LOC101927736 | FALSE | Upregulated |
| 0,008754972 | LOC101927737 | FALSE | Upregulated |
| | LOC101927738 | | |
| | LOC101927744 | | |
| 1,26188621 | LOC101927745 | | Upregulated |
| -0,130384421 | LOC101927748 | FALSE | Downregulated |
| 0,94190664 | LOC101927751 | FALSE | Upregulated |
| 0,015982116 | LOC101927752 | FALSE | Upregulated |
| 0,136278449 | LOC101927754 | FALSE | Upregulated |
| 0,75353318 | LOC101927756 | FALSE | Upregulated |
| | LOC101927757 | | |
| -0,153193591 | LOC101927759 | FALSE | Downregulated |
| | LOC101927760 | | |
| | LOC101927762 | | |
| -0,941137576 | LOC101927763 | FALSE | Downregulated |

| | | | |
|---|---|---|---|
| 0,085061822 | LOC101927764 | FALSE | Upregulated |
| 0,745516189 | LOC101927765 | FALSE | Upregulated |
| -0,592326807 | LOC101927767 | FALSE | Downregulated |
| 0,923941405 | LOC101927772 | FALSE | Upregulated |
| 0,172217806 | LOC101927773 | FALSE | Upregulated |
| 0 | LOC101927774 | FALSE | Upregulated |
| -0,336044038 | LOC101927775 | FALSE | Downregulated |
| | LOC101927777 | | |
| | LOC101927779 | | |
| 0,419963414 | LOC101927783 | FALSE | Upregulated |
| | LOC101927786 | | |
| | LOC101927788 | | |
| 0,344988057 | LOC101927789 | FALSE | Upregulated |
| -0,386660374 | LOC101927792 | FALSE | Downregulated |
| | LOC101927794 | | |
| | LOC101927798 | | |
| 0,903286866 | LOC101927799 | FALSE | Upregulated |
| 0,321020877 | LOC101927800 | FALSE | Upregulated |
| | LOC101927801 | | |
| 0 | LOC101927804 | FALSE | Upregulated |
| | LOC101927805 | | |
| | LOC101927807 | | |
| 0,408018455 | LOC101927810 | FALSE | Upregulated |
| 0 | LOC101927811 | FALSE | Upregulated |
| 0,133030207 | LOC101927814 | FALSE | Upregulated |
| | LOC101927816 | | |
| 0,48323834 | LOC101927818 | FALSE | Upregulated |
| -0,014083957 | LOC101927820 | FALSE | Downregulated |
| | LOC101927823 | | |
| 0,123610392 | LOC101927824 | FALSE | Upregulated |
| 0,631835509 | LOC101927825 | FALSE | Upregulated |
| -0,135759753 | LOC101927826 | FALSE | Downregulated |
| -0,037565386 | LOC101927834 | FALSE | Downregulated |
| | LOC101927835 | | |
| -0,489356657 | LOC101927837 | FALSE | Downregulated |
| 1,846686103 | LOC101927840 | | Upregulated |
| | LOC101927844 | | |
| 0,284987956 | LOC101927846 | FALSE | Upregulated |
| 0,445135141 | LOC101927852 | FALSE | Upregulated |
| 0,272626305 | LOC101927853 | FALSE | Upregulated |
| -0,185808683 | LOC101927855 | FALSE | Downregulated |
| | LOC101927856 | | |
| 0,263480715 | LOC101927857 | FALSE | Upregulated |
| 0,250845819 | LOC101927858 | FALSE | Upregulated |
| -0,00297973 | LOC101927859 | FALSE | Downregulated |
| 0,046009919 | LOC101927863 | FALSE | Upregulated |
| | LOC101927866 | | |
| 0 | LOC101927867 | FALSE | Upregulated |
| 1,531268826 | LOC101927871 | | Upregulated |
| -0,798506402 | LOC101927872 | FALSE | Downregulated |
| -0,006903421 | LOC101927874 | FALSE | Downregulated |

| | | | |
|---|---|---|---|
| 0 | LOC101927875 | FALSE | Upregulated |
| | LOC101927877 | | |
| | LOC101927878 | | |
| 0,272626305 | LOC101927879 | FALSE | Upregulated |
| -0,13296674 | LOC101927880 | FALSE | Downregulated |
| | LOC101927881 | | |
| -0,174822249 | LOC101927882 | FALSE | Downregulated |
| -0,176560191 | LOC101927885 | FALSE | Downregulated |
| 1,163550814 | LOC101927886 | | Upregulated |
| 0,298608236 | LOC101927888 | FALSE | Upregulated |
| 0,293242527 | LOC101927890 | FALSE | Upregulated |
| | LOC101927892 | | |
| 0 | LOC101927896 | FALSE | Upregulated |
| 0,312002785 | LOC101927899 | FALSE | Upregulated |
| -0,304285152 | LOC101927902 | FALSE | Downregulated |
| | LOC101927904 | | |
| 0 | LOC101927906 | FALSE | Upregulated |
| | LOC101927908 | | |
| -0,43591757 | LOC101927910 | FALSE | Downregulated |
| | LOC101927913 | | |
| | LOC101927915 | | |
| | LOC101927916 | | |
| | LOC101927918 | | |
| 0,141822178 | LOC101927919 | FALSE | Upregulated |
| | LOC101927921 | | |
| 0 | LOC101927922 | FALSE | Upregulated |
| 0,29270186 | LOC101927929 | FALSE | Upregulated |
| 0,123255145 | LOC101927931 | FALSE | Upregulated |
| 0,299758308 | LOC101927932 | FALSE | Upregulated |
| 0,538360303 | LOC101927933 | FALSE | Upregulated |
| -0,104214008 | LOC101927934 | FALSE | Downregulated |
| | LOC101927935 | | |
| | LOC101927936 | | |
| -0,049945176 | LOC101927942 | FALSE | Downregulated |
| 0,296842071 | LOC101927943 | FALSE | Upregulated |
| | LOC101927946 | | |
| 0,272626305 | LOC101927949 | FALSE | Upregulated |
| 0,223977906 | LOC101927952 | FALSE | Upregulated |
| -0,158936843 | LOC101927953 | FALSE | Downregulated |
| | LOC101927956 | | |
| 0,001141803 | LOC101927957 | FALSE | Upregulated |
| 0,404807106 | LOC101927958 | FALSE | Upregulated |
| | LOC101927959 | | |
| | LOC101927960 | | |
| | LOC101927961 | | |
| | LOC101927962 | | |
| | LOC101927967 | | |
| 0,006179527 | LOC101927971 | FALSE | Upregulated |
| 0 | LOC101927973 | FALSE | Upregulated |
| -0,401499736 | LOC101927974 | FALSE | Downregulated |
| 0 | LOC101927975 | FALSE | Upregulated |

| | | | |
|---|---|---|---|
| 0 | LOC101927978 | FALSE | Upregulated |
| | LOC101927982 | | |
| 0 | LOC101927988 | FALSE | Upregulated |
| 0,280928672 | LOC101927991 | FALSE | Upregulated |
| 0 | LOC101927993 | FALSE | Upregulated |
| -0,457790611 | LOC101927995 | FALSE | Downregulated |
| | LOC101927998 | | |
| 0,27393205 | LOC101928000 | FALSE | Upregulated |
| -0,145318597 | LOC101928004 | FALSE | Downregulated |
| 0,514896432 | LOC101928005 | FALSE | Upregulated |
| -0,138710803 | LOC101928006 | FALSE | Downregulated |
| 0 | LOC101928007 | FALSE | Upregulated |
| | LOC101928011 | | |
| | LOC101928014 | | |
| 0,174661103 | LOC101928019 | FALSE | Upregulated |
| 0,088837979 | LOC101928021 | FALSE | Upregulated |
| 0,438583988 | LOC101928025 | FALSE | Upregulated |
| | LOC101928026 | | |
| | LOC101928027 | | |
| | LOC101928028 | | |
| -0,014556979 | LOC101928030 | FALSE | Downregulated |
| | LOC101928031 | | |
| | LOC101928032 | | |
| | LOC101928037 | | |
| 0 | LOC101928039 | FALSE | Upregulated |
| 0,250314535 | LOC101928040 | FALSE | Upregulated |
| 0,520456753 | LOC101928044 | FALSE | Upregulated |
| | LOC101928046 | | |
| 0,536025097 | LOC101928047 | FALSE | Upregulated |
| -0,458109414 | LOC101928048 | FALSE | Downregulated |
| 0,984995271 | LOC101928049 | FALSE | Upregulated |
| 0,425803194 | LOC101928054 | FALSE | Upregulated |
| 0,209833334 | LOC101928055 | FALSE | Upregulated |
| | LOC101928055-2 | | |
| | LOC101928056 | | |
| 0,538371528 | LOC101928062 | FALSE | Upregulated |
| | LOC101928066 | | |
| 0 | LOC101928067 | FALSE | Upregulated |
| | LOC101928070-2 | | |
| 0,190085433 | LOC101928072 | FALSE | Upregulated |
| | LOC101928075 | | |
| -0,579580683 | LOC101928076 | FALSE | Downregulated |
| | LOC101928077 | | |
| 0 | LOC101928080 | FALSE | Upregulated |
| | LOC101928082 | | |
| -0,487263564 | LOC101928083 | FALSE | Downregulated |
| | LOC101928084 | | |
| 0,585701194 | LOC101928085 | FALSE | Upregulated |
| 0,266049836 | LOC101928087 | FALSE | Upregulated |
| 0 | LOC101928090 | FALSE | Upregulated |
| -0,242353749 | LOC101928092 | FALSE | Downregulated |

| | | | |
|---|---|---|---|
| -0,172818018 | LOC101928096 | FALSE | Downregulated |
| 0,303230855 | LOC101928102 | FALSE | Upregulated |
| 0,214956104 | LOC101928105 | FALSE | Upregulated |
| | LOC101928108 | | |
| 0,127117001 | LOC101928109 | FALSE | Upregulated |
| 0 | LOC101928110 | FALSE | Upregulated |
| -0,30434575 | LOC101928111 | FALSE | Downregulated |
| 0,39821799 | LOC101928113 | FALSE | Upregulated |
| | LOC101928114 | | |
| 0 | LOC101928115 | FALSE | Upregulated |
| 0,53379401 | LOC101928117 | FALSE | Upregulated |
| | LOC101928121 | | |
| | LOC101928124 | | |
| | LOC101928125 | | |
| 0,339096031 | LOC101928126 | FALSE | Upregulated |
| | LOC101928127 | | |
| | LOC101928128 | | |
| | LOC101928129 | | |
| | LOC101928130 | | |
| | LOC101928131 | | |
| | LOC101928132 | | |
| | LOC101928135 | | |
| 0,133373134 | LOC101928136 | FALSE | Upregulated |
| | LOC101928144 | | |
| | LOC101928146 | | |
| 0,000504523 | LOC101928149 | FALSE | Upregulated |
| | LOC101928153 | | |
| 0,31545939 | LOC101928156 | FALSE | Upregulated |
| | LOC101928157 | | |
| 0,141145411 | LOC101928160 | FALSE | Upregulated |
| | LOC101928162 | | |
| 0,923330116 | LOC101928163 | FALSE | Upregulated |
| | LOC101928166 | | |
| | LOC101928167 | | |
| -0,030598495 | LOC101928168 | FALSE | Downregulated |
| | LOC101928169 | | |
| 0 | LOC101928171 | FALSE | Upregulated |
| -0,041189108 | LOC101928172 | FALSE | Downregulated |
| 0,600913467 | LOC101928175 | FALSE | Upregulated |
| | LOC101928176 | | |
| | LOC101928177 | | |
| 0,126732701 | LOC101928178 | FALSE | Upregulated |
| -0,49009306 | LOC101928180 | FALSE | Downregulated |
| 0,15978422 | LOC101928186 | FALSE | Upregulated |
| -0,02258626 | LOC101928189 | FALSE | Downregulated |
| | LOC101928190 | | |
| 1,202137808 | LOC101928191 | | Upregulated |
| 0,371435373 | LOC101928193 | FALSE | Upregulated |
| 0,288417185 | LOC101928196 | FALSE | Upregulated |
| 0,769175654 | LOC101928197 | FALSE | Upregulated |
| | LOC101928199 | | |

| | | | |
|---|---|---|---|
| | LOC101928207 | | |
| -0,006848587 | LOC101928208 | FALSE | Downregulated |
| 0,64503433 | LOC101928214 | FALSE | Upregulated |
| | LOC101928216 | | |
| | LOC101928217 | | |
| 0,43796255 | LOC101928219 | FALSE | Upregulated |
| 0,5521917 | LOC101928220 | FALSE | Upregulated |
| 1,092766779 | LOC101928221 | | Upregulated |
| | LOC101928225 | | |
| | LOC101928226 | | |
| 0,274393502 | LOC101928228 | FALSE | Upregulated |
| 0,061205653 | LOC101928230 | FALSE | Upregulated |
| 0 | LOC101928231 | FALSE | Upregulated |
| -0,247408691 | LOC101928233 | FALSE | Downregulated |
| | LOC101928234 | | |
| -0,29725647 | LOC101928235 | FALSE | Downregulated |
| 0,498183541 | LOC101928236 | FALSE | Upregulated |
| -0,376103394 | LOC101928237 | FALSE | Downregulated |
| 0,340119356 | LOC101928238 | FALSE | Upregulated |
| -0,003426485 | LOC101928239 | FALSE | Downregulated |
| 0,297313667 | LOC101928245 | FALSE | Upregulated |
| 0,118654844 | LOC101928246 | FALSE | Upregulated |
| -0,019175039 | LOC101928247 | FALSE | Downregulated |
| 0 | LOC101928248 | FALSE | Upregulated |
| 0,112931594 | LOC101928249 | FALSE | Upregulated |
| | LOC101928250 | | |
| 0,135614376 | LOC101928251 | FALSE | Upregulated |
| -1,034758021 | LOC101928253 | | Downregulated |
| 0,315249239 | LOC101928255 | FALSE | Upregulated |
| | LOC101928257 | | |
| 0 | LOC101928258 | FALSE | Upregulated |
| 0,098661454 | LOC101928261 | FALSE | Upregulated |
| 0,405941687 | LOC101928262 | FALSE | Upregulated |
| 0 | LOC101928264 | FALSE | Upregulated |
| -0,15392772 | LOC101928269 | FALSE | Downregulated |
| | LOC101928271 | | |
| | LOC101928272 | | |
| | LOC101928273 | | |
| 0,197097142 | LOC101928274 | FALSE | Upregulated |
| | LOC101928275 | | |
| -0,298748942 | LOC101928277 | FALSE | Downregulated |
| | LOC101928278 | | |
| 0,265862193 | LOC101928279 | FALSE | Upregulated |
| | LOC101928282 | | |
| 0,159097727 | LOC101928284 | FALSE | Upregulated |
| | LOC101928285 | | |
| 0 | LOC101928286 | FALSE | Upregulated |
| 0,271886396 | LOC101928288 | FALSE | Upregulated |
| 0,351509943 | LOC101928289 | FALSE | Upregulated |
| 0 | LOC101928300 | FALSE | Upregulated |
| -0,75929169 | LOC101928303 | FALSE | Downregulated |

| | | | |
|---|---|---|---|
| -0,009465128 | LOC101928306 | FALSE | Downregulated |
| 0,587171444 | LOC101928307 | FALSE | Upregulated |
| | LOC101928309 | | |
| | LOC101928310 | | |
| -0,149716866 | LOC101928311 | FALSE | Downregulated |
| 0,724081625 | LOC101928312 | FALSE | Upregulated |
| -0,007714454 | LOC101928314 | FALSE | Downregulated |
| | LOC101928316 | | |
| 1,201188865 | LOC101928317 | | Upregulated |
| 0,290109866 | LOC101928325 | FALSE | Upregulated |
| 0,07835733 | LOC101928326 | FALSE | Upregulated |
| 0,272626305 | LOC101928327 | FALSE | Upregulated |
| | LOC101928329 | | |
| 0 | LOC101928330 | FALSE | Upregulated |
| 0,297958762 | LOC101928331 | FALSE | Upregulated |
| 0,313649936 | LOC101928334 | FALSE | Upregulated |
| -0,111928168 | LOC101928338 | FALSE | Downregulated |
| | LOC101928339 | | |
| -0,113054831 | LOC101928343 | FALSE | Downregulated |
| 0,332683919 | LOC101928344 | FALSE | Upregulated |
| | LOC101928346 | | |
| 0,153244175 | LOC101928347 | FALSE | Upregulated |
| 0,318210677 | LOC101928349 | FALSE | Upregulated |
| | LOC101928350 | | |
| 0,312053857 | LOC101928351 | FALSE | Upregulated |
| 0,528859042 | LOC101928357 | FALSE | Upregulated |
| | LOC101928358 | | |
| 0,338226472 | LOC101928359 | FALSE | Upregulated |
| 0,026122274 | LOC101928360 | FALSE | Upregulated |
| 0,051883707 | LOC101928361 | FALSE | Upregulated |
| 0,000535661 | LOC101928362 | FALSE | Upregulated |
| | LOC101928364 | | |
| | LOC101928367 | | |
| | LOC101928368 | | |
| 0,018607112 | LOC101928369 | FALSE | Upregulated |
| -0,47913288 | LOC101928370 | FALSE | Downregulated |
| | LOC101928375 | | |
| -0,16377886 | LOC101928377 | FALSE | Downregulated |
| | LOC101928380 | | |
| -0,611962575 | LOC101928382 | FALSE | Downregulated |
| -0,108611745 | LOC101928383 | FALSE | Downregulated |
| | LOC101928385 | | |
| 0,293326262 | LOC101928387 | FALSE | Upregulated |
| 0,295885871 | LOC101928388 | FALSE | Upregulated |
| -0,181349047 | LOC101928389 | FALSE | Downregulated |
| | LOC101928391 | | |
| | LOC101928392 | | |
| | LOC101928393 | | |
| | LOC101928394 | | |
| 0,293326262 | LOC101928397 | FALSE | Upregulated |
| 0,484521971 | LOC101928398 | FALSE | Upregulated |

| | | | |
|---|---|---|---|
| 0,434853141 | LOC101928399 | FALSE | Upregulated |
| 0,018839898 | LOC101928401 | FALSE | Upregulated |
| 0,308209304 | LOC101928402 | FALSE | Upregulated |
| -0,535146396 | LOC101928403 | FALSE | Downregulated |
| | LOC101928405 | | |
| | LOC101928406 | | |
| 0,938971532 | LOC101928407 | FALSE | Upregulated |
| 1,057618997 | LOC101928409 | | Upregulated |
| 0 | LOC101928412 | FALSE | Upregulated |
| 0,566771105 | LOC101928414 | FALSE | Upregulated |
| 0,574283617 | LOC101928419 | FALSE | Upregulated |
| -0,015614825 | LOC101928421 | FALSE | Downregulated |
| | LOC101928422 | | |
| | LOC101928423 | | |
| -0,31560142 | LOC101928424 | FALSE | Downregulated |
| | LOC101928426 | | |
| 0 | LOC101928427 | FALSE | Upregulated |
| -0,015225779 | LOC101928428 | FALSE | Downregulated |
| | LOC101928430 | | |
| 0,447071688 | LOC101928432 | FALSE | Upregulated |
| 0,047764937 | LOC101928434 | FALSE | Upregulated |
| | LOC101928436 | | |
| 0,242245192 | LOC101928437 | FALSE | Upregulated |
| | LOC101928447 | | |
| | LOC101928448 | | |
| | LOC101928450 | | |
| | LOC101928455 | | |
| | LOC101928457 | | |
| | LOC101928458 | | |
| 0,431387294 | LOC101928459 | FALSE | Upregulated |
| -0,227635254 | LOC101928462 | FALSE | Downregulated |
| | LOC101928463 | | |
| -0,005380762 | LOC101928464 | FALSE | Downregulated |
| | LOC101928469 | | |
| 0,000845759 | LOC101928472 | FALSE | Upregulated |
| -0,138833536 | LOC101928474 | FALSE | Downregulated |
| | LOC101928475 | | |
| 0,116041491 | LOC101928477 | FALSE | Upregulated |
| | LOC101928478 | | |
| | LOC101928481 | | |
| | LOC101928484 | | |
| -0,190042842 | LOC101928485 | FALSE | Downregulated |
| -0,443676059 | LOC101928487 | FALSE | Downregulated |
| -0,240021375 | LOC101928489 | FALSE | Downregulated |
| -1,142995481 | LOC101928498 | | Downregulated |
| -0,44184501 | LOC101928504 | FALSE | Downregulated |
| | LOC101928506 | | |
| 0,594119494 | LOC101928513 | FALSE | Upregulated |
| 0,67492535 | LOC101928514 | FALSE | Upregulated |
| | LOC101928519 | | |
| -0,006848587 | LOC101928522 | FALSE | Downregulated |

| | | | |
|---|---|---|---|
| | LOC101928523 | | |
| 0,28181789 | LOC101928526 | FALSE | Upregulated |
| -0,341452539 | LOC101928527 | FALSE | Downregulated |
| 0,323203932 | LOC101928529 | FALSE | Upregulated |
| -0,375683041 | LOC101928530 | FALSE | Downregulated |
| 0 | LOC101928533 | FALSE | Upregulated |
| | LOC101928535 | | |
| | LOC101928538 | | |
| | LOC101928543 | | |
| 0,00139671 | LOC101928549 | FALSE | Upregulated |
| 0,466773318 | LOC101928551 | FALSE | Upregulated |
| 0,995365925 | LOC101928553 | FALSE | Upregulated |
| 0,237124356 | LOC101928554 | FALSE | Upregulated |
| 0,956326265 | LOC101928556 | FALSE | Upregulated |
| 0,598332113 | LOC101928557 | FALSE | Upregulated |
| -0,481547644 | LOC101928558 | FALSE | Downregulated |
| 0,213624396 | LOC101928560 | FALSE | Upregulated |
| 0,362519237 | LOC101928561 | FALSE | Upregulated |
| 0,740034462 | LOC101928562 | FALSE | Upregulated |
| | LOC101928563 | | |
| | LOC101928564 | | |
| | LOC101928566 | | |
| 0,254489694 | LOC101928568 | FALSE | Upregulated |
| | LOC101928569 | | |
| 0,281459827 | LOC101928570 | FALSE | Upregulated |
| 0,086391981 | LOC101928571 | FALSE | Upregulated |
| 0,568614766 | LOC101928572 | FALSE | Upregulated |
| | LOC101928573 | | |
| | LOC101928579 | | |
| | LOC101928581 | | |
| | LOC101928582 | | |
| 0,295259937 | LOC101928583 | FALSE | Upregulated |
| -0,077078137 | LOC101928584 | FALSE | Downregulated |
| | LOC101928586 | | |
| | LOC101928587 | | |
| 0,479981647 | LOC101928589 | FALSE | Upregulated |
| 0 | LOC101928591 | FALSE | Upregulated |
| 0,549814631 | LOC101928592 | FALSE | Upregulated |
| | LOC101928593 | | |
| -0,304341495 | LOC101928596 | FALSE | Downregulated |
| | LOC101928598 | | |
| 0,01484109 | LOC101928600 | FALSE | Upregulated |
| 0 | LOC101928602 | FALSE | Upregulated |
| -0,128815538 | LOC101928608 | FALSE | Downregulated |
| | LOC101928611 | | |
| -0,255301491 | LOC101928612 | FALSE | Downregulated |
| | LOC101928614 | | |
| | LOC101928617 | | |
| 0,617913197 | LOC101928618 | FALSE | Upregulated |
| | LOC101928620 | | |
| | LOC101928623 | | |

| | | | |
|---|---|---|---|
| -1,06459017 | LOC101928624 | | Downregulated |
| 0,303078369 | LOC101928625 | FALSE | Upregulated |
| | LOC101928629 | | |
| -0,018618494 | LOC101928632 | FALSE | Downregulated |
| 0,122093338 | LOC101928633 | FALSE | Upregulated |
| | LOC101928634 | | |
| -0,536462717 | LOC101928635 | FALSE | Downregulated |
| | LOC101928636 | | |
| 0,293543259 | LOC101928638 | FALSE | Upregulated |
| 0 | LOC101928639 | FALSE | Upregulated |
| | LOC101928640 | | |
| | LOC101928643 | | |
| -0,432880998 | LOC101928645 | FALSE | Downregulated |
| 1,011145616 | LOC101928647 | | Upregulated |
| -0,161050255 | LOC101928652 | FALSE | Downregulated |
| | LOC101928654 | | |
| 0,178842376 | LOC101928655 | FALSE | Upregulated |
| 0,233353616 | LOC101928659 | FALSE | Upregulated |
| | LOC101928662 | | |
| 0,27368219 | LOC101928665 | FALSE | Upregulated |
| | LOC101928666 | | |
| -0,55431185 | LOC101928667 | FALSE | Downregulated |
| 0,061651742 | LOC101928669 | FALSE | Upregulated |
| -0,659659226 | LOC101928671 | FALSE | Downregulated |
| 1,024184052 | LOC101928673 | | Upregulated |
| 0,212263825 | LOC101928676 | FALSE | Upregulated |
| 0,428581198 | LOC101928683 | FALSE | Upregulated |
| 0,131795029 | LOC101928684 | FALSE | Upregulated |
| | LOC101928688 | | |
| 0,169177753 | LOC101928689 | FALSE | Upregulated |
| -0,006407309 | LOC101928692 | FALSE | Downregulated |
| 0,618716806 | LOC101928695 | FALSE | Upregulated |
| | LOC101928697 | | |
| 0,302532385 | LOC101928701 | FALSE | Upregulated |
| | LOC101928702 | | |
| | LOC101928704 | | |
| 0,437702667 | LOC101928705 | FALSE | Upregulated |
| -0,047806165 | LOC101928706 | FALSE | Downregulated |
| | LOC101928710 | | |
| | LOC101928711 | | |
| | LOC101928713 | | |
| 1,037828911 | LOC101928718 | | Upregulated |
| | LOC101928721 | | |
| 0,593316646 | LOC101928722 | FALSE | Upregulated |
| 0,009876686 | LOC101928725 | FALSE | Upregulated |
| 0 | LOC101928727 | FALSE | Upregulated |
| 0,260942569 | LOC101928730 | FALSE | Upregulated |
| 1,232028275 | LOC101928733 | | Upregulated |
| 0,264011716 | LOC101928734 | FALSE | Upregulated |
| | LOC101928735 | | |
| 0,457473801 | LOC101928738 | FALSE | Upregulated |

| | | | |
|---|---|---|---|
| 0,166020807 | LOC101928740 | FALSE | Upregulated |
| -0,010847412 | LOC101928742 | FALSE | Downregulated |
| | LOC101928743 | | |
| | LOC101928747 | | |
| 0,297178435 | LOC101928754 | FALSE | Upregulated |
| 0,291939173 | LOC101928756 | FALSE | Upregulated |
| 1,232325018 | LOC101928760 | | Upregulated |
| 2,016747451 | LOC101928761 | | Upregulated |
| | LOC101928764 | | |
| -0,009651905 | LOC101928765 | FALSE | Downregulated |
| 0,018839898 | LOC101928767 | FALSE | Upregulated |
| -0,0530396 | LOC101928768 | FALSE | Downregulated |
| | LOC101928770 | | |
| -0,655733181 | LOC101928774 | FALSE | Downregulated |
| | LOC101928775 | | |
| 0 | LOC101928779 | FALSE | Upregulated |
| | LOC101928785 | | |
| 0,247132525 | LOC101928786 | FALSE | Upregulated |
| 0,59675859 | LOC101928787 | FALSE | Upregulated |
| 0 | LOC101928792 | FALSE | Upregulated |
| 0 | LOC101928796 | FALSE | Upregulated |
| | LOC101928800 | | |
| -0,565327671 | LOC101928801 | FALSE | Downregulated |
| -0,48547426 | LOC101928803 | FALSE | Downregulated |
| 0 | LOC101928804 | FALSE | Upregulated |
| -0,488200869 | LOC101928806 | FALSE | Downregulated |
| 0,255904644 | LOC101928807 | FALSE | Upregulated |
| -0,136619946 | LOC101928808 | FALSE | Downregulated |
| 1,083238608 | LOC101928810 | | Upregulated |
| | LOC101928813 | | |
| 1,035337866 | LOC101928814 | | Upregulated |
| 0,032770364 | LOC101928815 | FALSE | Upregulated |
| | LOC101928818 | | |
| 0 | LOC101928819 | FALSE | Upregulated |
| 0,565541386 | LOC101928820 | FALSE | Upregulated |
| 0 | LOC101928824 | FALSE | Upregulated |
| -0,505977276 | LOC101928825 | FALSE | Downregulated |
| | LOC101928826 | | |
| | LOC101928828 | | |
| 0,274576178 | LOC101928829 | FALSE | Upregulated |
| | LOC101928832 | | |
| | LOC101928833 | | |
| | LOC101928836 | | |
| -0,698196277 | LOC101928838 | FALSE | Downregulated |
| | LOC101928839 | | |
| -0,899247864 | LOC101928841 | FALSE | Downregulated |
| | LOC101928842 | | |
| -0,208363135 | LOC101928845 | FALSE | Downregulated |
| | LOC101928849 | | |
| 0,153466334 | LOC101928850 | FALSE | Upregulated |
| -0,167349474 | LOC101928852 | FALSE | Downregulated |

| | | | |
|---|---|---|---|
| 1,110769211 | LOC101928854 | | Upregulated |
| | LOC101928856 | | |
| | LOC101928859 | | |
| | LOC101928860 | | |
| | LOC101928863 | | |
| -0,001674428 | LOC101928864 | FALSE | Downregulated |
| | LOC101928866 | | |
| | LOC101928867 | | |
| 0,317102409 | LOC101928868 | FALSE | Upregulated |
| -0,570515924 | LOC101928870 | FALSE | Downregulated |
| 0,370299123 | LOC101928872 | FALSE | Upregulated |
| 0,251726324 | LOC101928876 | FALSE | Upregulated |
| -0,213552156 | LOC101928879 | FALSE | Downregulated |
| 0,152432289 | LOC101928880 | FALSE | Upregulated |
| 0,576838973 | LOC101928881 | FALSE | Upregulated |
| -0,553255766 | LOC101928882 | FALSE | Downregulated |
| 0,319129918 | LOC101928886 | FALSE | Upregulated |
| -0,15749336 | LOC101928887 | FALSE | Downregulated |
| 0,28181789 | LOC101928890 | FALSE | Upregulated |
| 0,549129556 | LOC101928891 | FALSE | Upregulated |
| | LOC101928893 | | |
| 0,353741722 | LOC101928895 | FALSE | Upregulated |
| | LOC101928898 | | |
| 0 | LOC101928899 | FALSE | Upregulated |
| 0,28181789 | LOC101928902 | FALSE | Upregulated |
| | LOC101928905 | | |
| -0,884183074 | LOC101928906 | FALSE | Downregulated |
| 0,161285685 | LOC101928907 | FALSE | Upregulated |
| | LOC101928910 | | |
| 0,578496533 | LOC101928911 | FALSE | Upregulated |
| | LOC101928912 | | |
| -0,836210352 | LOC101928913 | FALSE | Downregulated |
| 0,76564908 | LOC101928915 | FALSE | Upregulated |
| 0,472147493 | LOC101928916 | FALSE | Upregulated |
| 0,289235841 | LOC101928918 | FALSE | Upregulated |
| | LOC101928919 | | |
| -0,023092029 | LOC101928921 | FALSE | Downregulated |
| | LOC101928922 | | |
| | LOC101928923 | | |
| -0,006257449 | LOC101928924 | FALSE | Downregulated |
| | LOC101928928 | | |
| 0,277764892 | LOC101928929 | FALSE | Upregulated |
| 0,404359425 | LOC101928932 | FALSE | Upregulated |
| 0,105089598 | LOC101928933 | FALSE | Upregulated |
| 0,721897607 | LOC101928936 | FALSE | Upregulated |
| | LOC101928941 | | |
| 0,51780845 | LOC101928947 | FALSE | Upregulated |
| 0 | LOC101928948 | FALSE | Upregulated |
| | LOC101928950 | | |
| 0,233199921 | LOC101928955 | FALSE | Upregulated |
| | LOC101928957 | | |

| | | | |
|---|---|---|---|
| -0,59563529 | LOC101928961 | FALSE | Downregulated |
| | LOC101928962 | | |
| -0,140476228 | LOC101928963 | FALSE | Downregulated |
| | LOC101928964 | | |
| -0,034853333 | LOC101928965 | FALSE | Downregulated |
| 0,323124346 | LOC101928968 | FALSE | Upregulated |
| 0,090535131 | LOC101928969 | FALSE | Upregulated |
| 1,168900562 | LOC101928970 | | Upregulated |
| | LOC101928972 | | |
| 0,108411004 | LOC101928974 | FALSE | Upregulated |
| -0,350249674 | LOC101928976 | FALSE | Downregulated |
| | LOC101928977 | | |
| | LOC101928978 | | |
| 0,442946224 | LOC101928981 | FALSE | Upregulated |
| 0,332984426 | LOC101928984 | FALSE | Upregulated |
| 1,127433106 | LOC101928985 | | Upregulated |
| 0,001141803 | LOC101928987 | FALSE | Upregulated |
| 0,261660608 | LOC101928991 | FALSE | Upregulated |
| -0,181092642 | LOC101928993 | FALSE | Downregulated |
| -0,08863602 | LOC101928997 | FALSE | Downregulated |
| | LOC101929002 | | |
| | LOC101929003 | | |
| 0,719956011 | LOC101929006 | FALSE | Upregulated |
| -0,021021069 | LOC101929007 | FALSE | Downregulated |
| -0,014397653 | LOC101929008 | FALSE | Downregulated |
| 0,854797099 | LOC101929011 | FALSE | Upregulated |
| | LOC101929014 | | |
| | LOC101929016 | | |
| | LOC101929021 | | |
| | LOC101929022 | | |
| 0 | LOC101929027 | FALSE | Upregulated |
| | LOC101929028 | | |
| | LOC101929031 | | |
| | LOC101929034 | | |
| 0,120403089 | LOC101929035 | FALSE | Upregulated |
| 0,291311386 | LOC101929036 | FALSE | Upregulated |
| 0,152191163 | LOC101929038 | FALSE | Upregulated |
| 0,177005243 | LOC101929039 | FALSE | Upregulated |
| 0,133985024 | LOC101929042 | FALSE | Upregulated |
| | LOC101929043 | | |
| 0 | LOC101929044 | FALSE | Upregulated |
| 0,499563574 | LOC101929047 | FALSE | Upregulated |
| | LOC101929048 | | |
| | LOC101929050 | | |
| 1,325140111 | LOC101929054 | | Upregulated |
| -0,00509535 | LOC101929055 | FALSE | Downregulated |
| | LOC101929056 | | |
| -0,442936699 | LOC101929057 | FALSE | Downregulated |
| 0,278669683 | LOC101929058 | FALSE | Upregulated |
| 0,176090055 | LOC101929060 | FALSE | Upregulated |
| -0,430257018 | LOC101929065 | FALSE | Downregulated |

| | | | |
|---|---|---|---|
| 0,143359097 | LOC101929067 | FALSE | Upregulated |
| | LOC101929069 | | |
| 0,289732278 | LOC101929070 | FALSE | Upregulated |
| -0,002253536 | LOC101929077 | FALSE | Downregulated |
| | LOC101929082 | | |
| -0,01330656 | LOC101929085 | FALSE | Downregulated |
| -0,011481061 | LOC101929087 | FALSE | Downregulated |
| | LOC101929088 | | |
| -0,52257787 | LOC101929090 | FALSE | Downregulated |
| 0,009715691 | LOC101929092 | FALSE | Upregulated |
| 0 | LOC101929094 | FALSE | Upregulated |
| -0,901487082 | LOC101929097 | FALSE | Downregulated |
| 0,374587055 | LOC101929098 | FALSE | Upregulated |
| -0,20600681 | LOC101929107 | FALSE | Downregulated |
| | LOC101929108 | | |
| 0,356985713 | LOC101929112 | FALSE | Upregulated |
| | LOC101929114 | | |
| -0,054289628 | LOC101929115 | FALSE | Downregulated |
| -0,17949195 | LOC101929117 | FALSE | Downregulated |
| 0,291340714 | LOC101929119 | FALSE | Upregulated |
| | LOC101929121 | | |
| -0,014724123 | LOC101929124 | FALSE | Downregulated |
| 1,392124534 | LOC101929125 | | Upregulated |
| | LOC101929129 | | |
| | LOC101929131 | | |
| 0,941512347 | LOC101929132 | FALSE | Upregulated |
| 1,440503918 | LOC101929133 | | Upregulated |
| | LOC101929134 | | |
| | LOC101929137 | | |
| 0,711122612 | LOC101929138 | FALSE | Upregulated |
| -1,038043633 | LOC101929141 | | Downregulated |
| 0 | LOC101929142 | FALSE | Upregulated |
| 0,545322859 | LOC101929143 | FALSE | Upregulated |
| | LOC101929144 | | |
| -0,236277306 | LOC101929145 | FALSE | Downregulated |
| 0,421585569 | LOC101929149 | FALSE | Upregulated |
| 0,289078151 | LOC101929151 | FALSE | Upregulated |
| | LOC101929152 | | |
| 0 | LOC101929153 | FALSE | Upregulated |
| -0,015040902 | LOC101929154 | FALSE | Downregulated |
| 0 | LOC101929155 | FALSE | Upregulated |
| | LOC101929156 | | |
| | LOC101929157 | | |
| 0,139741928 | LOC101929159 | FALSE | Upregulated |
| -0,222357594 | LOC101929160 | FALSE | Downregulated |
| | LOC101929161 | | |
| | LOC101929164 | | |
| 0,393112256 | LOC101929165 | FALSE | Upregulated |
| 0,4416558 | LOC101929167 | FALSE | Upregulated |
| 0 | LOC101929174 | FALSE | Upregulated |
| -0,085972597 | LOC101929177 | FALSE | Downregulated |

| | | | |
|---|---|---|---|
| 0,37782653 | LOC101929178 | FALSE | Upregulated |
| | LOC101929180 | | |
| 0 | LOC101929184 | FALSE | Upregulated |
| 0,226081981 | LOC101929185 | FALSE | Upregulated |
| | LOC101929187 | | |
| | LOC101929190 | | |
| 0,011615355 | LOC101929191 | FALSE | Upregulated |
| | LOC101929192 | | |
| | LOC101929194 | | |
| 0,11376788 | LOC101929198 | FALSE | Upregulated |
| 0,069693451 | LOC101929200 | FALSE | Upregulated |
| | LOC101929201 | | |
| | LOC101929202 | | |
| -0,000458989 | LOC101929206 | FALSE | Downregulated |
| | LOC101929214 | | |
| | LOC101929216 | | |
| | LOC101929217 | | |
| 0,717603007 | LOC101929220 | FALSE | Upregulated |
| -0,030899824 | LOC101929223 | FALSE | Downregulated |
| 0,282135885 | LOC101929225 | FALSE | Upregulated |
| -0,919412558 | LOC101929227 | FALSE | Downregulated |
| | LOC101929228 | | |
| | LOC101929229 | | |
| -0,308513875 | LOC101929230 | FALSE | Downregulated |
| | LOC101929232 | | |
| | LOC101929236 | | |
| -0,402083295 | LOC101929241 | FALSE | Downregulated |
| -0,639354456 | LOC101929243 | FALSE | Downregulated |
| -0,140255569 | LOC101929248 | FALSE | Downregulated |
| 0,813343367 | LOC101929249 | FALSE | Upregulated |
| | LOC101929250 | | |
| 0,197228457 | LOC101929251 | FALSE | Upregulated |
| | LOC101929256 | | |
| | LOC101929257 | | |
| 0 | LOC101929258 | FALSE | Upregulated |
| 0,600338228 | LOC101929259 | FALSE | Upregulated |
| | LOC101929261 | | |
| 0 | LOC101929266 | FALSE | Upregulated |
| | LOC101929267 | | |
| 0,542495075 | LOC101929269 | FALSE | Upregulated |
| | LOC101929278 | | |
| 0,684844388 | LOC101929279 | FALSE | Upregulated |
| 0,081158301 | LOC101929280 | FALSE | Upregulated |
| -0,330806378 | LOC101929283 | FALSE | Downregulated |
| -0,622463386 | LOC101929285 | FALSE | Downregulated |
| | LOC101929286 | | |
| -0,382047998 | LOC101929287 | FALSE | Downregulated |
| 0,284419228 | LOC101929288 | FALSE | Upregulated |
| 0,226283985 | LOC101929289 | FALSE | Upregulated |
| 0,766432883 | LOC101929295 | FALSE | Upregulated |
| -0,32205651 | LOC101929296 | FALSE | Downregulated |

| | | | |
|---|---|---|---|
| | LOC101929298 | | |
| 0 | LOC101929300 | FALSE | Upregulated |
| | LOC101929301 | | |
| 1,385066549 | LOC101929304 | | Upregulated |
| 0,272277655 | LOC101929305 | FALSE | Upregulated |
| | LOC101929307 | | |
| -1,175973675 | LOC101929313 | | Downregulated |
| 0 | LOC101929314 | FALSE | Upregulated |
| | LOC101929316 | | |
| 0,285372545 | LOC101929319 | FALSE | Upregulated |
| | LOC101929320 | | |
| | LOC101929321 | | |
| 0 | LOC101929322 | FALSE | Upregulated |
| 0,283626494 | LOC101929325 | FALSE | Upregulated |
| | LOC101929326 | | |
| -0,005401589 | LOC101929327 | FALSE | Downregulated |
| 1,119787436 | LOC101929336 | | Upregulated |
| 0,816764329 | LOC101929341 | FALSE | Upregulated |
| 0,509405639 | LOC101929346 | FALSE | Upregulated |
| | LOC101929348 | | |
| | LOC101929350 | | |
| | LOC101929351 | | |
| | LOC101929355 | | |
| -0,935264797 | LOC101929356 | FALSE | Downregulated |
| | LOC101929358 | | |
| | LOC101929359 | | |
| -0,123696951 | LOC101929360 | FALSE | Downregulated |
| | LOC101929363 | | |
| 0,617568395 | LOC101929367 | FALSE | Upregulated |
| | LOC101929368 | | |
| 0,155492156 | LOC101929371 | FALSE | Upregulated |
| | LOC101929373 | | |
| 0,288833705 | LOC101929374 | FALSE | Upregulated |
| 0 | LOC101929375 | FALSE | Upregulated |
| 0 | LOC101929379 | FALSE | Upregulated |
| 0,264784362 | LOC101929381 | FALSE | Upregulated |
| | LOC101929382 | | |
| | LOC101929386 | | |
| | LOC101929388 | | |
| -0,131938235 | LOC101929393 | FALSE | Downregulated |
| | LOC101929398 | | |
| 0,406635447 | LOC101929408 | FALSE | Upregulated |
| 0,149244831 | LOC101929409 | FALSE | Upregulated |
| -0,81274112 | LOC101929410 | FALSE | Downregulated |
| | LOC101929411 | | |
| 0,293326262 | LOC101929414 | FALSE | Upregulated |
| -0,001674428 | LOC101929418 | FALSE | Downregulated |
| | LOC101929419 | | |
| 0,146899363 | LOC101929422 | FALSE | Upregulated |
| -0,150000974 | LOC101929427 | FALSE | Downregulated |
| 0,268788334 | LOC101929431 | FALSE | Upregulated |

| | | | |
|---|---|---|---|
| | LOC101929432 | | |
| 0,6040779 | LOC101929433 | FALSE | Upregulated |
| -0,425219035 | LOC101929434 | FALSE | Downregulated |
| | LOC101929435 | | |
| | LOC101929437 | | |
| 0,329672248 | LOC101929440 | FALSE | Upregulated |
| -0,176061469 | LOC101929442 | FALSE | Downregulated |
| 0,025306739 | LOC101929443 | FALSE | Upregulated |
| 1,052402078 | LOC101929444 | | Upregulated |
| | LOC101929445 | | |
| | LOC101929446 | | |
| 0,011340145 | LOC101929447 | FALSE | Upregulated |
| | LOC101929453 | | |
| | LOC101929456 | | |
| 0,43136184 | LOC101929457 | FALSE | Upregulated |
| 0,289504779 | LOC101929459 | FALSE | Upregulated |
| 0,271737409 | LOC101929460 | FALSE | Upregulated |
| -0,126973451 | LOC101929462 | FALSE | Downregulated |
| -0,217797111 | LOC101929463 | FALSE | Downregulated |
| 0,44216805 | LOC101929465 | FALSE | Upregulated |
| 0,004536421 | LOC101929467 | FALSE | Upregulated |
| 0,571191327 | LOC101929475 | FALSE | Upregulated |
| | LOC101929476 | | |
| | LOC101929477 | | |
| | LOC101929478 | | |
| -0,07107593 | LOC101929480 | FALSE | Downregulated |
| -0,324789341 | LOC101929482 | FALSE | Downregulated |
| 0 | LOC101929484 | FALSE | Upregulated |
| | LOC101929485 | | |
| | LOC101929489 | | |
| -0,231161027 | LOC101929490 | FALSE | Downregulated |
| 0,284987956 | LOC101929491 | FALSE | Upregulated |
| | LOC101929492 | | |
| | LOC101929493 | | |
| -0,097122947 | LOC101929494 | FALSE | Downregulated |
| | LOC101929495 | | |
| | LOC101929498 | | |
| 0,885405345 | LOC101929500 | FALSE | Upregulated |
| 0,552761732 | LOC101929506 | FALSE | Upregulated |
| | LOC101929507 | | |
| | LOC101929510 | | |
| -0,419575099 | LOC101929511 | FALSE | Downregulated |
| 0,271809546 | LOC101929513 | FALSE | Upregulated |
| | LOC101929514 | | |
| | LOC101929515 | | |
| 0 | LOC101929518 | FALSE | Upregulated |
| -0,087393337 | LOC101929519 | FALSE | Downregulated |
| 0,40330954 | LOC101929521 | FALSE | Upregulated |
| 0,363275729 | LOC101929523 | FALSE | Upregulated |
| 0,276217378 | LOC101929528 | FALSE | Upregulated |
| | LOC101929530 | | |

| | | | |
|---|---|---|---|
| 0,308737698 | LOC101929531 | FALSE | Upregulated |
| 0,293242527 | LOC101929534 | FALSE | Upregulated |
| | LOC101929535 | | |
| 0 | LOC101929536 | FALSE | Upregulated |
| -0,271175399 | LOC101929538 | FALSE | Downregulated |
| -0,792225152 | LOC101929540 | FALSE | Downregulated |
| -0,13030006 | LOC101929542 | FALSE | Downregulated |
| | LOC101929544 | | |
| 0,225065899 | LOC101929547 | FALSE | Upregulated |
| 0 | LOC101929550 | FALSE | Upregulated |
| -0,216844342 | LOC101929552 | FALSE | Downregulated |
| -0,165007652 | LOC101929555 | FALSE | Downregulated |
| | LOC101929557 | | |
| 0,422700259 | LOC101929558 | FALSE | Upregulated |
| 0 | LOC101929560 | FALSE | Upregulated |
| 0,30022777 | LOC101929561 | FALSE | Upregulated |
| | LOC101929562 | | |
| | LOC101929563 | | |
| | LOC101929564 | | |
| 0 | LOC101929566 | FALSE | Upregulated |
| 0,721400953 | LOC101929571 | FALSE | Upregulated |
| -0,394087535 | LOC101929574 | FALSE | Downregulated |
| | LOC101929575 | | |
| | LOC101929576 | | |
| 0,395083287 | LOC101929578 | FALSE | Upregulated |
| -0,126776283 | LOC101929580 | FALSE | Downregulated |
| -0,842106793 | LOC101929583 | FALSE | Downregulated |
| -0,124071034 | LOC101929587 | FALSE | Downregulated |
| 0,533159243 | LOC101929590 | FALSE | Upregulated |
| | LOC101929593 | | |
| 0,668056343 | LOC101929594 | FALSE | Upregulated |
| 0 | LOC101929601 | FALSE | Upregulated |
| -0,149515831 | LOC101929602 | FALSE | Downregulated |
| 0,409967065 | LOC101929604 | FALSE | Upregulated |
| -0,256523533 | LOC101929608 | FALSE | Downregulated |
| 0,435129614 | LOC101929609 | FALSE | Upregulated |
| | LOC101929610 | | |
| | LOC101929612 | | |
| 1,32310013 | LOC101929613 | | Upregulated |
| 0,123637152 | LOC101929614 | FALSE | Upregulated |
| 0,323843546 | LOC101929615 | FALSE | Upregulated |
| | LOC101929617 | | |
| | LOC101929618 | | |
| -0,440242757 | LOC101929619 | FALSE | Downregulated |
| | LOC101929620 | | |
| -0,032480029 | LOC101929621 | FALSE | Downregulated |
| 0,41182066 | LOC101929625 | FALSE | Upregulated |
| 0 | LOC101929627 | FALSE | Upregulated |
| | LOC101929628 | | |
| 0,165039697 | LOC101929634 | FALSE | Upregulated |
| 0,274185108 | LOC101929637 | FALSE | Upregulated |

| | | | |
|---|---|---|---|
| 0,575672192 | LOC101929638 | FALSE | Upregulated |
| 0 | LOC101929641 | FALSE | Upregulated |
| 0,188884928 | LOC101929643 | FALSE | Upregulated |
| | LOC101929644 | | |
| 0,396646981 | LOC101929645 | FALSE | Upregulated |
| | LOC101929646 | | |
| 0,489956281 | LOC101929647 | FALSE | Upregulated |
| 0,450638083 | LOC101929650 | FALSE | Upregulated |
| | LOC101929654 | | |
| 0,468435624 | LOC101929656 | FALSE | Upregulated |
| | LOC101929658 | | |
| 0,289238926 | LOC101929661 | FALSE | Upregulated |
| 0,765042183 | LOC101929664 | FALSE | Upregulated |
| -0,531627908 | LOC101929666 | FALSE | Downregulated |
| 0,775870795 | LOC101929667 | FALSE | Upregulated |
| 0,255498426 | LOC101929673 | FALSE | Upregulated |
| | LOC101929674 | | |
| 0 | LOC101929675 | FALSE | Upregulated |
| | LOC101929681 | | |
| -1,357947092 | LOC101929683 | | Downregulated |
| 0,442795359 | LOC101929685 | FALSE | Upregulated |
| 0,535110724 | LOC101929687 | FALSE | Upregulated |
| | LOC101929688 | | |
| 0,875575817 | LOC101929689 | FALSE | Upregulated |
| | LOC101929690 | | |
| | LOC101929692 | | |
| -1,587939829 | LOC101929696 | | Downregulated |
| | LOC101929697 | | |
| | LOC101929700 | | |
| | LOC101929702 | | |
| 0,157552935 | LOC101929704 | FALSE | Upregulated |
| 0,042498029 | LOC101929713 | FALSE | Upregulated |
| 0,129082067 | LOC101929715 | FALSE | Upregulated |
| 0,106118676 | LOC101929719 | FALSE | Upregulated |
| 0,295610575 | LOC101929724 | FALSE | Upregulated |
| | LOC101929725 | | |
| 0,156211389 | LOC101929726 | FALSE | Upregulated |
| 0,148154173 | LOC101929727 | FALSE | Upregulated |
| -0,609528589 | LOC101929728 | FALSE | Downregulated |
| | LOC101929733 | | |
| 0,293242527 | LOC101929738 | FALSE | Upregulated |
| 0,143927241 | LOC101929740 | FALSE | Upregulated |
| 1,27779142 | LOC101929741 | | Upregulated |
| | LOC101929745 | | |
| | LOC101929748 | | |
| | LOC101929750 | | |
| 0,405490867 | LOC101929753 | FALSE | Upregulated |
| -0,025072958 | LOC101929754 | FALSE | Downregulated |
| | LOC101929755 | | |
| | LOC101929757 | | |
| -0,282155221 | LOC101929758 | FALSE | Downregulated |

| | | | |
|---|---|---|---|
| | LOC101929759 | | |
| -0,399774947 | LOC101929761 | FALSE | Downregulated |
| 0 | LOC101929762 | FALSE | Upregulated |
| | LOC101929763 | | |
| 0,160058532 | LOC101929766 | FALSE | Upregulated |
| | LOC101929770 | | |
| 0,427530075 | LOC101929777 | FALSE | Upregulated |
| -0,321227053 | LOC101929788 | FALSE | Downregulated |
| 0,417618515 | LOC101929796 | FALSE | Upregulated |
| -0,273986773 | LOC101929798 | FALSE | Downregulated |
| 0 | LOC101929800 | FALSE | Upregulated |
| 0 | LOC101929803 | FALSE | Upregulated |
| 0,208431782 | LOC101929805 | FALSE | Upregulated |
| | LOC101929806 | | |
| -0,030288107 | LOC101929814 | FALSE | Downregulated |
| | LOC101929815 | | |
| | LOC101929818 | | |
| 1,607075541 | LOC101929824 | | Upregulated |
| 0,41534535 | LOC101929826 | FALSE | Upregulated |
| | LOC101929831 | | |
| -0,258121889 | LOC101929846 | FALSE | Downregulated |
| -0,731889328 | LOC101929860 | FALSE | Downregulated |
| | LOC101929863 | | |
| | LOC101929866 | | |
| -0,017093013 | LOC101929882 | FALSE | Downregulated |
| 0,709040022 | LOC101929887 | FALSE | Upregulated |
| 0,24618557 | LOC101929891 | FALSE | Upregulated |
| -0,2053457 | LOC101929897 | FALSE | Downregulated |
| 0,276953698 | LOC101929901 | FALSE | Upregulated |
| -0,310738754 | LOC101929918 | FALSE | Downregulated |
| | LOC101929922 | | |
| 0,662865464 | LOC101929937 | FALSE | Upregulated |
| -0,577234298 | LOC101929946 | FALSE | Downregulated |
| 1,411762268 | LOC101929949 | | Upregulated |
| -0,993400296 | LOC101929950 | FALSE | Downregulated |
| 0 | LOC101929953 | FALSE | Upregulated |
| 0,325481494 | LOC101929959 | FALSE | Upregulated |
| -0,29085482 | LOC101929964 | FALSE | Downregulated |
| | LOC101929972 | | |
| 0,000504523 | LOC101929974 | FALSE | Upregulated |
| | LOC101929976 | | |
| 0,000500193 | LOC101929977 | FALSE | Upregulated |
| | LOC101929983 | | |
| | LOC101929989 | | |
| -0,01561674 | LOC101929998 | FALSE | Downregulated |
| 0,42149487 | LOC101930010 | FALSE | Upregulated |
| 0,284308453 | LOC101930026 | FALSE | Upregulated |
| 0,770768981 | LOC101930033 | FALSE | Upregulated |
| | LOC101930048 | | |
| | LOC101930053 | | |
| 0 | LOC101930071 | FALSE | Upregulated |

| | | | |
|---|---|---|---|
| | LOC101930091 | | |
| | LOC101930098 | | |
| 0 | LOC101930100 | FALSE | Upregulated |
| | LOC101930109 | | |
| 0,115519668 | LOC101930114 | FALSE | Upregulated |
| 0,013199952 | LOC101930120 | FALSE | Upregulated |
| 0,046486725 | LOC101930129 | FALSE | Upregulated |
| | LOC101930164 | | |
| 0 | LOC101930232 | FALSE | Upregulated |
| -0,018149848 | LOC101930249 | FALSE | Downregulated |
| | LOC101930259 | | |
| 0,566058288 | LOC101930275 | FALSE | Upregulated |
| 0 | LOC101930276 | FALSE | Upregulated |
| | LOC101930294 | | |
| -0,018235269 | LOC101930305 | FALSE | Downregulated |
| -0,946742207 | LOC101930307 | FALSE | Downregulated |
| | LOC101930310 | | |
| 0,427327564 | LOC101930332 | FALSE | Upregulated |
| | LOC101930343 | | |
| 1,108411159 | LOC101930357 | | Upregulated |
| 0,423417081 | LOC101930370 | FALSE | Upregulated |
| | LOC101930384 | | |
| 0 | LOC101930420 | FALSE | Upregulated |
| 0 | LOC101930434 | FALSE | Upregulated |
| 0,047069579 | LOC101930452 | FALSE | Upregulated |
| 0,270838341 | LOC101930460 | FALSE | Upregulated |
| | LOC101930476 | | |
| | LOC101930484 | | |
| 0 | LOC101930496 | FALSE | Upregulated |
| | LOC101930523 | | |
| | LOC101930531 | | |
| 0,154714158 | LOC101930567 | FALSE | Upregulated |
| -0,018683471 | LOC101930580 | FALSE | Downregulated |
| | LOC101930593 | | |
| -0,160384452 | LOC101930644 | FALSE | Downregulated |
| | LOC101930665 | | |
| 1,689786522 | LOC102031319 | | Upregulated |
| | LOC102467074 | | |
| | LOC102467075 | | |
| | LOC102467079 | | |
| 1,20411219 | LOC102467080 | | Upregulated |
| | LOC102467081 | | |
| -0,426580476 | LOC102467146 | FALSE | Downregulated |
| 0,438153284 | LOC102467147 | FALSE | Upregulated |
| | LOC102467212 | | |
| | LOC102467213 | | |
| | LOC102467214 | | |
| 0,277119458 | LOC102467216 | FALSE | Upregulated |
| -0,189684887 | LOC102467217 | FALSE | Downregulated |
| | LOC102467223 | | |
| | LOC102467224 | | |

| | | | |
|---|---|---|---|
| 0,016809178 | LOC102467225 | FALSE | Upregulated |
| | LOC102467226 | | |
| | LOC102467655 | | |
| | LOC102477328 | | |
| | LOC102503427 | | |
| | LOC102503429 | | |
| -0,322364858 | LOC102524628 | FALSE | Downregulated |
| | LOC102546175 | | |
| 0,03945122 | LOC102546226 | FALSE | Upregulated |
| | LOC102546227 | | |
| -1,128218686 | LOC102546228 | | Downregulated |
| 0,144405216 | LOC102546229 | FALSE | Upregulated |
| 0,598318723 | LOC102546294 | FALSE | Upregulated |
| 0,273422532 | LOC102546298 | FALSE | Upregulated |
| | LOC102546299 | | |
| | LOC102557615 | | |
| | LOC102577424 | | |
| 0 | LOC102577426 | FALSE | Upregulated |
| -0,083813347 | LOC102606465 | FALSE | Downregulated |
| | LOC102606466 | | |
| 0,739930843 | LOC102659288 | FALSE | Upregulated |
| 0,000504523 | LOC102723305 | FALSE | Upregulated |
| 0,404860684 | LOC102723307 | FALSE | Upregulated |
| | LOC102723308 | | |
| 0 | LOC102723310 | FALSE | Upregulated |
| 0,807296236 | LOC102723311 | FALSE | Upregulated |
| | LOC102723315 | | |
| -0,011481061 | LOC102723316 | FALSE | Downregulated |
| | LOC102723318 | | |
| -0,181597877 | LOC102723319 | FALSE | Downregulated |
| | LOC102723320 | | |
| | LOC102723321 | | |
| | LOC102723322 | | |
| 0,263907918 | LOC102723323 | FALSE | Upregulated |
| 0 | LOC102723324 | FALSE | Upregulated |
| | LOC102723325 | | |
| | LOC102723326 | | |
| 0,416638948 | LOC102723327 | FALSE | Upregulated |
| 0,667420868 | LOC102723329 | FALSE | Upregulated |
| | LOC102723330 | | |
| 1,016266112 | LOC102723331 | | Upregulated |
| -0,140953133 | LOC102723332 | FALSE | Downregulated |
| | LOC102723334 | | |
| | LOC102723336 | | |
| -0,013301487 | LOC102723337 | FALSE | Downregulated |
| | LOC102723338 | | |
| | LOC102723339 | | |
| -0,166649266 | LOC102723340 | FALSE | Downregulated |
| | LOC102723341 | | |
| | LOC102723343 | | |
| | LOC102723344 | | |

| | | | |
|---|---|---|---|
| | LOC102723345 | | |
| | LOC102723346 | | |
| | LOC102723347 | | |
| 0 | LOC102723348 | FALSE | Upregulated |
| 0 | LOC102723349 | FALSE | Upregulated |
| -0,002253536 | LOC102723350 | FALSE | Downregulated |
| | LOC102723351 | | |
| | LOC102723352 | | |
| 0,300199083 | LOC102723353 | FALSE | Upregulated |
| -0,161731704 | LOC102723354 | FALSE | Downregulated |
| | LOC102723355 | | |
| 0,269698002 | LOC102723357 | FALSE | Upregulated |
| | LOC102723358 | | |
| 0,836365413 | LOC102723359 | FALSE | Upregulated |
| 0,467585008 | LOC102723361 | FALSE | Upregulated |
| | LOC102723362 | | |
| | LOC102723363 | | |
| | LOC102723364 | | |
| -0,425154926 | LOC102723365 | FALSE | Downregulated |
| -0,198305292 | LOC102723366 | FALSE | Downregulated |
| 0,629745234 | LOC102723367 | FALSE | Upregulated |
| | LOC102723368 | | |
| | LOC102723370 | | |
| 0,304762103 | LOC102723371 | FALSE | Upregulated |
| | LOC102723372 | | |
| | LOC102723373 | | |
| 0,124194032 | LOC102723374 | FALSE | Upregulated |
| 0,283983879 | LOC102723375 | FALSE | Upregulated |
| | LOC102723376 | | |
| 0,262992327 | LOC102723377 | FALSE | Upregulated |
| | LOC102723378 | | |
| | LOC102723379 | | |
| 0,680772234 | LOC102723380 | FALSE | Upregulated |
| -0,349881821 | LOC102723383 | FALSE | Downregulated |
| | LOC102723384 | | |
| 0 | LOC102723385 | FALSE | Upregulated |
| | LOC102723386 | | |
| | LOC102723388 | | |
| -0,310574681 | LOC102723389 | FALSE | Downregulated |
| | LOC102723390 | | |
| 0 | LOC102723391 | FALSE | Upregulated |
| 0,447071688 | LOC102723392 | FALSE | Upregulated |
| | LOC102723393 | | |
| | LOC102723394 | | |
| | LOC102723395 | | |
| | LOC102723396 | | |
| | LOC102723397 | | |
| 0,122846964 | LOC102723398 | FALSE | Upregulated |
| 2,868487708 | LOC102723399 | | Upregulated |
| | LOC102723402 | | |
| | LOC102723403 | | |

| | | | |
|---|---|---|---|
| | LOC102723404 | | |
| 0,161562343 | LOC102723407 | FALSE | Upregulated |
| -0,01316343 | LOC102723408 | FALSE | Downregulated |
| -0,007930109 | LOC102723409 | FALSE | Downregulated |
| -1,470400776 | LOC102723410 | | Downregulated |
| | LOC102723411 | | |
| 0,294320768 | LOC102723412 | FALSE | Upregulated |
| | LOC102723413 | | |
| | LOC102723414 | | |
| -0,745601786 | LOC102723415 | FALSE | Downregulated |
| | LOC102723417 | | |
| | LOC102723418 | | |
| | LOC102723419 | | |
| 0,235449399 | LOC102723420 | FALSE | Upregulated |
| 1,003735405 | LOC102723421 | | Upregulated |
| | LOC102723423 | | |
| 0,441745026 | LOC102723425 | FALSE | Upregulated |
| | LOC102723426 | | |
| | LOC102723427 | | |
| -0,432039791 | LOC102723428 | FALSE | Downregulated |
| | LOC102723429 | | |
| | LOC102723430 | | |
| | LOC102723431 | | |
| 0,78901693 | LOC102723433 | FALSE | Upregulated |
| | LOC102723434 | | |
| | LOC102723435 | | |
| | LOC102723436 | | |
| 0 | LOC102723437 | FALSE | Upregulated |
| | LOC102723439 | | |
| | LOC102723440 | | |
| -0,015040902 | LOC102723441 | FALSE | Downregulated |
| | LOC102723442 | | |
| | LOC102723443 | | |
| 0,600235396 | LOC102723444 | FALSE | Upregulated |
| | LOC102723445 | | |
| 0,009416434 | LOC102723446 | FALSE | Upregulated |
| | LOC102723447 | | |
| | LOC102723448 | | |
| | LOC102723449 | | |
| 0,481927061 | LOC102723450 | FALSE | Upregulated |
| 0,275880911 | LOC102723452 | FALSE | Upregulated |
| | LOC102723453 | | |
| | LOC102723454 | | |
| 0,286939648 | LOC102723455 | FALSE | Upregulated |
| | LOC102723456 | | |
| | LOC102723458 | | |
| | LOC102723459 | | |
| | LOC102723460 | | |
| 1,284694417 | LOC102723461 | | Upregulated |
| | LOC102723463 | | |
| 0,793449256 | LOC102723464 | FALSE | Upregulated |

| | | | |
|---|---|---|---|
| 0,155462379 | LOC102723465 | FALSE | Upregulated |
| -0,671529754 | LOC102723466 | FALSE | Downregulated |
| 0,689319374 | LOC102723467 | FALSE | Upregulated |
| 0,415343158 | LOC102723468 | FALSE | Upregulated |
| 0,000535661 | LOC102723470 | FALSE | Upregulated |
| | LOC102723471 | | |
| | LOC102723472 | | |
| | LOC102723473 | | |
| | LOC102723474 | | |
| -0,581679112 | LOC102723475 | FALSE | Downregulated |
| | LOC102723476 | | |
| | LOC102723477 | | |
| | LOC102723479 | | |
| | LOC102723480 | | |
| -0,049379958 | LOC102723481 | FALSE | Downregulated |
| 0,423237084 | LOC102723482 | FALSE | Upregulated |
| 0,428965416 | LOC102723483 | FALSE | Upregulated |
| | LOC102723484 | | |
| | LOC102723485 | | |
| | LOC102723486 | | |
| | LOC102723487 | | |
| 0,307328229 | LOC102723488 | FALSE | Upregulated |
| | LOC102723490 | | |
| | LOC102723491 | | |
| 0 | LOC102723492 | FALSE | Upregulated |
| 1,435248789 | LOC102723493 | | Upregulated |
| | LOC102723494 | | |
| | LOC102723495 | | |
| | LOC102723496 | | |
| -0,531794044 | LOC102723497 | FALSE | Downregulated |
| | LOC102723499 | | |
| -0,253921295 | LOC102723500 | FALSE | Downregulated |
| -0,099026492 | LOC102723501 | FALSE | Downregulated |
| | LOC102723502 | | |
| | LOC102723503 | | |
| | LOC102723505 | | |
| 0,142147734 | LOC102723507 | FALSE | Upregulated |
| | LOC102723508 | | |
| 0,027741422 | LOC102723510 | FALSE | Upregulated |
| 1,049405374 | LOC102723511 | | Upregulated |
| -0,518763183 | LOC102723512 | FALSE | Downregulated |
| 0,144061787 | LOC102723513 | FALSE | Upregulated |
| | LOC102723514 | | |
| | LOC102723515 | | |
| | LOC102723516 | | |
| 0,545645735 | LOC102723517 | FALSE | Upregulated |
| | LOC102723518 | | |
| | LOC102723522 | | |
| | LOC102723523 | | |
| | LOC102723524 | | |
| 0 | LOC102723525 | FALSE | Upregulated |

| | | | |
|---|---|---|---|
| | LOC102723526 | | |
| | LOC102723527 | | |
| | LOC102723529 | | |
| | LOC102723530 | | |
| | LOC102723532 | | |
| | LOC102723533 | | |
| -0,068802251 | LOC102723534 | FALSE | Downregulated |
| | LOC102723536 | | |
| 1,325195277 | LOC102723537 | | Upregulated |
| | LOC102723538 | | |
| -0,018674369 | LOC102723540 | FALSE | Downregulated |
| | LOC102723541 | | |
| -0,253579801 | LOC102723544 | FALSE | Downregulated |
| 0,261242846 | LOC102723545 | FALSE | Upregulated |
| -0,172815 | LOC102723546 | FALSE | Downregulated |
| 0,117103792 | LOC102723547 | FALSE | Upregulated |
| | LOC102723548 | | |
| -1,245699087 | LOC102723549 | | Downregulated |
| | LOC102723550 | | |
| | LOC102723551 | | |
| 0,295471224 | LOC102723552 | FALSE | Upregulated |
| 0,208148673 | LOC102723553 | FALSE | Upregulated |
| | LOC102723557 | | |
| 0,452690317 | LOC102723558 | FALSE | Upregulated |
| 0,144824001 | LOC102723559 | FALSE | Upregulated |
| | LOC102723560 | | |
| -0,140978214 | LOC102723561 | FALSE | Downregulated |
| | LOC102723562 | | |
| 0,556434823 | LOC102723563 | FALSE | Upregulated |
| -0,403028075 | LOC102723564 | FALSE | Downregulated |
| 0,58811929 | LOC102723565 | FALSE | Upregulated |
| | LOC102723567 | | |
| | LOC102723568 | | |
| 0,596778925 | LOC102723569 | FALSE | Upregulated |
| 1,859747497 | LOC102723570 | | Upregulated |
| | LOC102723571 | | |
| 0 | LOC102723572 | FALSE | Upregulated |
| 0,002192964 | LOC102723575 | FALSE | Upregulated |
| 0,573340707 | LOC102723576 | FALSE | Upregulated |
| | LOC102723577 | | |
| | LOC102723578 | | |
| 0,51075284 | LOC102723579 | FALSE | Upregulated |
| -0,780715241 | LOC102723580 | FALSE | Downregulated |
| 0,212191283 | LOC102723581 | FALSE | Upregulated |
| -0,042225409 | LOC102723583 | FALSE | Downregulated |
| -0,123138763 | LOC102723587 | FALSE | Downregulated |
| | LOC102723588 | | |
| 0,237138229 | LOC102723589 | FALSE | Upregulated |
| 0 | LOC102723590 | FALSE | Upregulated |
| 0,135985779 | LOC102723591 | FALSE | Upregulated |
| | LOC102723593 | | |

| | | | |
|---|---|---|---|
| -0,00036056 | LOC102723594 | FALSE | Downregulated |
| 1,423507778 | LOC102723595 | | Upregulated |
| | LOC102723596 | | |
| | LOC102723599 | | |
| -0,082980816 | LOC102723602 | FALSE | Downregulated |
| 1,494635792 | LOC102723604 | | Upregulated |
| | LOC102723605 | | |
| | LOC102723606 | | |
| 0,542849703 | LOC102723607 | FALSE | Upregulated |
| | LOC102723608 | | |
| | LOC102723609 | | |
| -0,802006484 | LOC102723610 | FALSE | Downregulated |
| 0,061400605 | LOC102723611 | FALSE | Upregulated |
| | LOC102723612 | | |
| 0,502787042 | LOC102723614 | FALSE | Upregulated |
| 1,312644009 | LOC102723615 | | Upregulated |
| -0,094237857 | LOC102723616 | FALSE | Downregulated |
| | LOC102723617 | | |
| -0,136582247 | LOC102723619 | FALSE | Downregulated |
| | LOC102723620 | | |
| | LOC102723621 | | |
| -0,001674428 | LOC102723623 | FALSE | Downregulated |
| | LOC102723624 | | |
| 0,163388047 | LOC102723625 | FALSE | Upregulated |
| -0,153179674 | LOC102723627 | FALSE | Downregulated |
| | LOC102723628 | | |
| | LOC102723629 | | |
| 0,693730834 | LOC102723630 | FALSE | Upregulated |
| | LOC102723631 | | |
| | LOC102723633 | | |
| 0,451076232 | LOC102723635 | FALSE | Upregulated |
| -0,449681625 | LOC102723636 | FALSE | Downregulated |
| | LOC102723637 | | |
| | LOC102723638 | | |
| | LOC102723639 | | |
| -1,318797103 | LOC102723640 | | Downregulated |
| -0,188470041 | LOC102723641 | FALSE | Downregulated |
| 0,542547776 | LOC102723642 | FALSE | Upregulated |
| -0,128033156 | LOC102723643 | FALSE | Downregulated |
| | LOC102723644 | | |
| | LOC102723645 | | |
| | LOC102723646 | | |
| -0,003213111 | LOC102723647 | FALSE | Downregulated |
| -0,983918499 | LOC102723648 | FALSE | Downregulated |
| | LOC102723649 | | |
| | LOC102723650 | | |
| 0,357320701 | LOC102723651 | FALSE | Upregulated |
| 1,22671001 | LOC102723652 | | Upregulated |
| | LOC102723653 | | |
| | LOC102723654 | | |
| -0,213618257 | LOC102723655 | FALSE | Downregulated |

| | | | |
|---|---|---|---|
| 0,016773597 | LOC102723656 | FALSE | Upregulated |
| | LOC102723657 | | |
| | LOC102723658 | | |
| -0,593846568 | LOC102723659 | FALSE | Downregulated |
| | LOC102723661 | | |
| | LOC102723662 | | |
| 0,586208985 | LOC102723663 | FALSE | Upregulated |
| | LOC102723664 | | |
| | LOC102723667 | | |
| | LOC102723669 | | |
| -0,018149848 | LOC102723670 | FALSE | Downregulated |
| -0,084235914 | LOC102723671 | FALSE | Downregulated |
| -0,021275891 | LOC102723672 | FALSE | Downregulated |
| | LOC102723673 | | |
| | LOC102723674 | | |
| 0,822371584 | LOC102723675 | FALSE | Upregulated |
| 0 | LOC102723676 | FALSE | Upregulated |
| -0,204985797 | LOC102723677 | FALSE | Downregulated |
| -0,174580871 | LOC102723678 | FALSE | Downregulated |
| -0,294193834 | LOC102723679 | FALSE | Downregulated |
| | LOC102723680 | | |
| -0,023740011 | LOC102723682 | FALSE | Downregulated |
| 0,369096732 | LOC102723683 | FALSE | Upregulated |
| | LOC102723684 | | |
| 0 | LOC102723685 | FALSE | Upregulated |
| | LOC102723686 | | |
| -0,152478545 | LOC102723687 | FALSE | Downregulated |
| -0,009489398 | LOC102723690 | FALSE | Downregulated |
| -0,029840335 | LOC102723691 | FALSE | Downregulated |
| 0,333200449 | LOC102723692 | FALSE | Upregulated |
| -0,001183301 | LOC102723693 | FALSE | Downregulated |
| 0,264958074 | LOC102723695 | FALSE | Upregulated |
| -0,61262969 | LOC102723696 | FALSE | Downregulated |
| -0,010565857 | LOC102723697 | FALSE | Downregulated |
| | LOC102723698 | | |
| | LOC102723699 | | |
| 0,576104897 | LOC102723700 | FALSE | Upregulated |
| | LOC102723702 | | |
| 0,038238758 | LOC102723703 | FALSE | Upregulated |
| 0,392460782 | LOC102723704 | FALSE | Upregulated |
| | LOC102723705 | | |
| | LOC102723706 | | |
| | LOC102723710 | | |
| | LOC102723712 | | |
| | LOC102723713 | | |
| | LOC102723714 | | |
| -0,000941876 | LOC102723715 | FALSE | Downregulated |
| 0,122214806 | LOC102723716 | FALSE | Upregulated |
| 0,008457559 | LOC102723717 | FALSE | Upregulated |
| -0,273303906 | LOC102723718 | FALSE | Downregulated |
| -1,082563746 | LOC102723719 | | Downregulated |

| | | | |
|---|---|---|---|
| 0,983987355 | LOC102723720 | FALSE | Upregulated |
| 0,242349099 | LOC102723721 | FALSE | Upregulated |
| | LOC102723723 | | |
| | LOC102723724 | | |
| | LOC102723725 | | |
| | LOC102723727 | | |
| 1,076721141 | LOC102723728 | | Upregulated |
| | LOC102723729 | | |
| 1,006670084 | LOC102723730 | | Upregulated |
| | LOC102723731 | | |
| 1,683037491 | LOC102723732 | | Upregulated |
| 0,96170005 | LOC102723733 | FALSE | Upregulated |
| | LOC102723734 | | |
| | LOC102723736 | | |
| | LOC102723737 | | |
| | LOC102723738 | | |
| 0,575909377 | LOC102723739 | FALSE | Upregulated |
| | LOC102723740 | | |
| | LOC102723742 | | |
| -0,004030137 | LOC102723744 | FALSE | Downregulated |
| | LOC102723745 | | |
| 0,007633787 | LOC102723746 | FALSE | Upregulated |
| | LOC102723749 | | |
| | LOC102723751 | | |
| | LOC102723752 | | |
| | LOC102723754 | | |
| 0,238619216 | LOC102723755 | FALSE | Upregulated |
| -1,099439927 | LOC102723756 | | Downregulated |
| 0,225908832 | LOC102723757 | FALSE | Upregulated |
| -0,299584592 | LOC102723759 | FALSE | Downregulated |
| | LOC102723760 | | |
| 0,140386881 | LOC102723761 | FALSE | Upregulated |
| | LOC102723763 | | |
| | LOC102723764 | | |
| | LOC102723765 | | |
| 0,130092209 | LOC102723766 | FALSE | Upregulated |
| -0,262986309 | LOC102723767 | FALSE | Downregulated |
| | LOC102723768 | | |
| -0,580815013 | LOC102723770 | FALSE | Downregulated |
| 0,000504523 | LOC102723771 | FALSE | Upregulated |
| 0,137296842 | LOC102723774 | FALSE | Upregulated |
| 0 | LOC102723775 | FALSE | Upregulated |
| -0,084150885 | LOC102723777 | FALSE | Downregulated |
| | LOC102723778 | | |
| -0,173257877 | LOC102723779 | FALSE | Downregulated |
| | LOC102723780 | | |
| 0 | LOC102723781 | FALSE | Upregulated |
| | LOC102723783 | | |
| | LOC102723784 | | |
| | LOC102723785 | | |
| | LOC102723786 | | |

| | | | |
|---|---|---|---|
| | LOC102723787 | | |
| | LOC102723789 | | |
| -0,029344609 | LOC102723790 | FALSE | Downregulated |
| 0,576497518 | LOC102723791 | FALSE | Upregulated |
| | LOC102723793 | | |
| -0,946122234 | LOC102723794 | FALSE | Downregulated |
| | LOC102723795 | | |
| | LOC102723796 | | |
| 0,91305472 | LOC102723797 | FALSE | Upregulated |
| 0,237140564 | LOC102723799 | FALSE | Upregulated |
| | LOC102723800 | | |
| -0,0107704 | LOC102723801 | FALSE | Downregulated |
| | LOC102723802 | | |
| | LOC102723803 | | |
| 0,904779859 | LOC102723805 | FALSE | Upregulated |
| | LOC102723807 | | |
| 0,119626366 | LOC102723808 | FALSE | Upregulated |
| | LOC102723810 | | |
| | LOC102723812 | | |
| 0,140593622 | LOC102723813 | FALSE | Upregulated |
| 0,039289498 | LOC102723814 | FALSE | Upregulated |
| | LOC102723816 | | |
| 0,179240359 | LOC102723817 | FALSE | Upregulated |
| | LOC102723818 | | |
| | LOC102723819 | | |
| | LOC102723821 | | |
| | LOC102723822 | | |
| | LOC102723823 | | |
| | LOC102723825 | | |
| | LOC102723826 | | |
| -0,748254657 | LOC102723827 | FALSE | Downregulated |
| 0,2781141 | LOC102723828 | FALSE | Upregulated |
| -0,022615687 | LOC102723829 | FALSE | Downregulated |
| | LOC102723830 | | |
| | LOC102723831 | | |
| | LOC102723832 | | |
| | LOC102723833 | | |
| | LOC102723834 | | |
| | LOC102723835 | | |
| -0,196517274 | LOC102723836 | FALSE | Downregulated |
| | LOC102723837 | | |
| | LOC102723838 | | |
| 0,277764892 | LOC102723839 | FALSE | Upregulated |
| -0,02959148 | LOC102723840 | FALSE | Downregulated |
| | LOC102723841 | | |
| | LOC102723842 | | |
| 0,995636139 | LOC102723843 | FALSE | Upregulated |
| 0 | LOC102723844 | FALSE | Upregulated |
| 0,275314501 | LOC102723845 | FALSE | Upregulated |
| | LOC102723846 | | |
| | LOC102723847 | | |

| | | | |
|---|---|---|---|
| | LOC102723848 | | |
| | LOC102723849 | | |
| | LOC102723850 | | |
| -0,006903421 | LOC102723854 | FALSE | Downregulated |
| | LOC102723855 | | |
| -0,283971574 | LOC102723856 | FALSE | Downregulated |
| | LOC102723857 | | |
| | LOC102723858 | | |
| 0,178426101 | LOC102723859 | FALSE | Upregulated |
| | LOC102723860 | | |
| 0,136585728 | LOC102723861 | FALSE | Upregulated |
| | LOC102723862 | | |
| 0,145912032 | LOC102723864 | FALSE | Upregulated |
| 0 | LOC102723865 | FALSE | Upregulated |
| 0,164919604 | LOC102723866 | FALSE | Upregulated |
| -0,01363615 | LOC102723867 | FALSE | Downregulated |
| | LOC102723868 | | |
| | LOC102723869 | | |
| 0,895073077 | LOC102723870 | FALSE | Upregulated |
| | LOC102723871 | | |
| 0,178906824 | LOC102723872 | FALSE | Upregulated |
| | LOC102723873 | | |
| 0,335448743 | LOC102723874 | FALSE | Upregulated |
| | LOC102723875 | | |
| 0,005298914 | LOC102723876 | FALSE | Upregulated |
| | LOC102723878 | | |
| | LOC102723879 | | |
| | LOC102723880 | | |
| | LOC102723881 | | |
| | LOC102723882 | | |
| | LOC102723883 | | |
| -0,330664147 | LOC102723884 | FALSE | Downregulated |
| -0,307628818 | LOC102723885 | FALSE | Downregulated |
| | LOC102723887 | | |
| 0 | LOC102723888 | FALSE | Upregulated |
| 0,872398983 | LOC102723889 | FALSE | Upregulated |
| | LOC102723890 | | |
| 0,052790477 | LOC102723891 | FALSE | Upregulated |
| 0,422238157 | LOC102723892 | FALSE | Upregulated |
| -0,206974937 | LOC102723893 | FALSE | Downregulated |
| 0 | LOC102723894 | FALSE | Upregulated |
| | LOC102723895 | | |
| 0,309012528 | LOC102723896 | FALSE | Upregulated |
| | LOC102723898 | | |
| 0,137498164 | LOC102723899 | FALSE | Upregulated |
| | LOC102723900 | | |
| 0,313493608 | LOC102723901 | FALSE | Upregulated |
| 0,307232554 | LOC102723902 | FALSE | Upregulated |
| | LOC102723903 | | |
| 0,280928672 | LOC102723904 | FALSE | Upregulated |
| 0 | LOC102723905 | FALSE | Upregulated |

| | | | |
|---|---|---|---|
| | LOC102723906 | | |
| | LOC102723907 | | |
| | LOC102723909 | | |
| | LOC102723910 | | |
| | LOC102723911 | | |
| | LOC102723912 | | |
| 0,274582205 | LOC102723913 | FALSE | Upregulated |
| -0,00509535 | LOC102723914 | FALSE | Downregulated |
| 0,010748089 | LOC102723915 | FALSE | Upregulated |
| 0,293242527 | LOC102723916 | FALSE | Upregulated |
| 2,275454844 | LOC102723917 | | Upregulated |
| | LOC102723918 | | |
| -0,281746611 | LOC102723919 | FALSE | Downregulated |
| -0,004519092 | LOC102723920 | FALSE | Downregulated |
| | LOC102723921 | | |
| 0,274635952 | LOC102723922 | FALSE | Upregulated |
| 0,705233943 | LOC102723923 | FALSE | Upregulated |
| | LOC102723924 | | |
| 0,113542482 | LOC102723925 | FALSE | Upregulated |
| -0,841698575 | LOC102723926 | FALSE | Downregulated |
| 0,502288777 | LOC102723927 | FALSE | Upregulated |
| | LOC102723928 | | |
| | LOC102723931 | | |
| | LOC102723932 | | |
| | LOC102723933 | | |
| 0 | LOC102723934 | FALSE | Upregulated |
| | LOC102723935 | | |
| | LOC102723936 | | |
| | LOC102723937 | | |
| | LOC102723938 | | |
| | LOC102723939 | | |
| 0 | LOC102723940 | FALSE | Upregulated |
| | LOC102723941 | | |
| -0,299271535 | LOC102723942 | FALSE | Downregulated |
| | LOC102723944 | | |
| 0 | LOC102723946 | FALSE | Upregulated |
| | LOC102723947 | | |
| | LOC102723948 | | |
| -0,32927514 | LOC102723949 | FALSE | Downregulated |
| | LOC102723950 | | |
| | LOC102723951 | | |
| | LOC102723952 | | |
| -0,892188954 | LOC102723953 | FALSE | Downregulated |
| -0,070916391 | LOC102723955 | FALSE | Downregulated |
| 0,679697872 | LOC102723956 | FALSE | Upregulated |
| | LOC102723957 | | |
| 0,150407336 | LOC102723958 | FALSE | Upregulated |
| 0,287955436 | LOC102723959 | FALSE | Upregulated |
| 0,555651239 | LOC102723960 | FALSE | Upregulated |
| | LOC102723961 | | |
| | LOC102723962 | | |

| | | | |
|---|---|---|---|
| 0 | LOC102723963 | FALSE | Upregulated |
| -0,170671412 | LOC102723965 | FALSE | Downregulated |
| 0,277119458 | LOC102723966 | FALSE | Upregulated |
| | LOC102723967 | | |
| | LOC102723968 | | |
| | LOC102723969 | | |
| | LOC102723970 | | |
| | LOC102723971 | | |
| -0,166026487 | LOC102723973 | FALSE | Downregulated |
| 0,530185499 | LOC102723974 | FALSE | Upregulated |
| -0,312696659 | LOC102723975 | FALSE | Downregulated |
| | LOC102723976 | | |
| | LOC102723977 | | |
| | LOC102723978 | | |
| | LOC102723979 | | |
| 0,291556967 | LOC102723981 | FALSE | Upregulated |
| 0,425103776 | LOC102723983 | FALSE | Upregulated |
| | LOC102723984 | | |
| -0,018149848 | LOC102723985 | FALSE | Downregulated |
| | LOC102723986 | | |
| | LOC102723987 | | |
| 0,001513725 | LOC102723988 | FALSE | Upregulated |
| 0,382738798 | LOC102723989 | FALSE | Upregulated |
| 0,005298914 | LOC102723993 | FALSE | Upregulated |
| | LOC102723994 | | |
| -0,014676578 | LOC102723995 | FALSE | Downregulated |
| | LOC102724000 | | |
| | LOC102724001 | | |
| -0,239015644 | LOC102724002 | FALSE | Downregulated |
| -0,128640305 | LOC102724003 | FALSE | Downregulated |
| | LOC102724004 | | |
| | LOC102724007 | | |
| | LOC102724008 | | |
| -2,35992974 | LOC102724010 | | Downregulated |
| | LOC102724011 | | |
| 0,422443977 | LOC102724012 | FALSE | Upregulated |
| | LOC102724013 | | |
| | LOC102724014 | | |
| -0,148465128 | LOC102724015 | FALSE | Downregulated |
| 0,002278541 | LOC102724017 | FALSE | Upregulated |
| -0,004525881 | LOC102724018 | FALSE | Downregulated |
| -0,717590764 | LOC102724019 | FALSE | Downregulated |
| | LOC102724020 | | |
| | LOC102724021 | | |
| | LOC102724022 | | |
| 0,479717781 | LOC102724024 | FALSE | Upregulated |
| | LOC102724027 | | |
| 0,135529589 | LOC102724028 | FALSE | Upregulated |
| | LOC102724029 | | |
| 0,387939946 | LOC102724030 | FALSE | Upregulated |
| 0 | LOC102724031 | FALSE | Upregulated |

| | | | |
|---|---|---|---|
| | LOC102724032 | | |
| | LOC102724033 | | |
| 0,840092948 | LOC102724034 | FALSE | Upregulated |
| | LOC102724036 | | |
| -0,016389151 | LOC102724037 | FALSE | Downregulated |
| | LOC102724038 | | |
| | LOC102724039 | | |
| -0,200681191 | LOC102724040 | FALSE | Downregulated |
| | LOC102724041 | | |
| | LOC102724044 | | |
| | LOC102724045 | | |
| | LOC102724046 | | |
| 0,142498971 | LOC102724047 | FALSE | Upregulated |
| | LOC102724048 | | |
| 0,374609953 | LOC102724049 | FALSE | Upregulated |
| | LOC102724050 | | |
| | LOC102724051 | | |
| | LOC102724052 | | |
| | LOC102724053 | | |
| | LOC102724054 | | |
| | LOC102724055 | | |
| | LOC102724056 | | |
| 0,29236422 | LOC102724057 | FALSE | Upregulated |
| | LOC102724058 | | |
| 0,158641709 | LOC102724059 | FALSE | Upregulated |
| 0,270009605 | LOC102724060 | FALSE | Upregulated |
| | LOC102724062 | | |
| | LOC102724063 | | |
| -0,464587339 | LOC102724064 | FALSE | Downregulated |
| 0,254961479 | LOC102724065 | FALSE | Upregulated |
| | LOC102724067 | | |
| 0,662112163 | LOC102724068 | FALSE | Upregulated |
| | LOC102724069 | | |
| | LOC102724071 | | |
| 1,019217006 | LOC102724072 | | Upregulated |
| -0,243851105 | LOC102724073 | FALSE | Downregulated |
| | LOC102724074 | | |
| -0,098374975 | LOC102724075 | FALSE | Downregulated |
| | LOC102724076 | | |
| | LOC102724078 | | |
| 0,266315965 | LOC102724079 | FALSE | Upregulated |
| | LOC102724080 | | |
| | LOC102724081 | | |
| | LOC102724082 | | |
| -0,01330656 | LOC102724084 | FALSE | Downregulated |
| | LOC102724085 | | |
| | LOC102724086 | | |
| | LOC102724087 | | |
| | LOC102724088 | | |
| | LOC102724089 | | |
| -0,01330656 | LOC102724090 | FALSE | Downregulated |

| | | | |
|---|---|---|---|
| | LOC102724091 | | |
| 0,179544396 | LOC102724092 | FALSE | Upregulated |
| -0,304960048 | LOC102724093 | FALSE | Downregulated |
| -0,006903421 | LOC102724094 | FALSE | Downregulated |
| | LOC102724095 | | |
| 0,909344173 | LOC102724096 | FALSE | Upregulated |
| 0 | LOC102724097 | FALSE | Upregulated |
| -0,154622615 | LOC102724098 | FALSE | Downregulated |
| | LOC102724099 | | |
| 1,361219552 | LOC102724100 | | Upregulated |
| | LOC102724101 | | |
| | LOC102724102 | | |
| | LOC102724104 | | |
| 0,187238231 | LOC102724105 | FALSE | Upregulated |
| 0,908802684 | LOC102724106 | FALSE | Upregulated |
| | LOC102724107 | | |
| 1,032332141 | LOC102724108 | | Upregulated |
| -0,018442049 | LOC102724109 | FALSE | Downregulated |
| | LOC102724111 | | |
| 0,246109373 | LOC102724112 | FALSE | Upregulated |
| | LOC102724113 | | |
| | LOC102724114 | | |
| | LOC102724115 | | |
| -1,829068384 | LOC102724117 | | Downregulated |
| | LOC102724118 | | |
| | LOC102724119 | | |
| -0,768868175 | LOC102724120 | FALSE | Downregulated |
| 0,541521939 | LOC102724121 | FALSE | Upregulated |
| 0 | LOC102724122 | FALSE | Upregulated |
| | LOC102724123 | | |
| 1,420419301 | LOC102724124 | | Upregulated |
| | LOC102724125 | | |
| 0,350794747 | LOC102724126 | FALSE | Upregulated |
| | LOC102724127 | | |
| | LOC102724128 | | |
| 0,431640268 | LOC102724130 | FALSE | Upregulated |
| | LOC102724131 | | |
| -0,168517806 | LOC102724132 | FALSE | Downregulated |
| | LOC102724134 | | |
| | LOC102724136 | | |
| | LOC102724137 | | |
| 0 | LOC102724138 | FALSE | Upregulated |
| 0 | LOC102724139 | FALSE | Upregulated |
| 0 | LOC102724140 | FALSE | Upregulated |
| | LOC102724142 | | |
| -0,418651007 | LOC102724143 | FALSE | Downregulated |
| | LOC102724145 | | |
| | LOC102724146 | | |
| 0,033451755 | LOC102724147 | FALSE | Upregulated |
| | LOC102724148 | | |
| 0,012062552 | LOC102724149 | FALSE | Upregulated |

| | | | |
|---|---|---|---|
| | LOC102724150 | | |
| | LOC102724151 | | |
| | LOC102724152 | | |
| 0,293326262 | LOC102724154 | FALSE | Upregulated |
| 0,434394258 | LOC102724155 | FALSE | Upregulated |
| 0,01051915 | LOC102724156 | FALSE | Upregulated |
| | LOC102724158 | | |
| | LOC102724160 | | |
| | LOC102724161 | | |
| 0 | LOC102724162 | FALSE | Upregulated |
| | LOC102724165 | | |
| | LOC102724166 | | |
| | LOC102724167 | | |
| 0,761437366 | LOC102724168 | FALSE | Upregulated |
| 0,123914476 | LOC102724169 | FALSE | Upregulated |
| 0,520818897 | LOC102724170 | FALSE | Upregulated |
| 0,266315965 | LOC102724171 | FALSE | Upregulated |
| | LOC102724173 | | |
| | LOC102724174 | | |
| 0,273422532 | LOC102724176 | FALSE | Upregulated |
| | LOC102724180 | | |
| | LOC102724182 | | |
| | LOC102724183 | | |
| -0,391838542 | LOC102724184 | FALSE | Downregulated |
| | LOC102724185 | | |
| | LOC102724186 | | |
| | LOC102724188 | | |
| | LOC102724189 | | |
| 0,012691333 | LOC102724190 | FALSE | Upregulated |
| 0,804753631 | LOC102724192 | FALSE | Upregulated |
| -0,493991331 | LOC102724193 | FALSE | Downregulated |
| | LOC102724194 | | |
| 0,691326429 | LOC102724195 | FALSE | Upregulated |
| 0,780577942 | LOC102724196 | FALSE | Upregulated |
| 1,089849239 | LOC102724197 | | Upregulated |
| | LOC102724199 | | |
| 0,566012746 | LOC102724200 | FALSE | Upregulated |
| -0,098398984 | LOC102724201 | FALSE | Downregulated |
| | LOC102724202 | | |
| | LOC102724203 | | |
| | LOC102724204 | | |
| | LOC102724205 | | |
| -0,095585049 | LOC102724206 | FALSE | Downregulated |
| | LOC102724207 | | |
| -0,150199727 | LOC102724208 | FALSE | Downregulated |
| 0,14927774 | LOC102724209 | FALSE | Upregulated |
| | LOC102724210 | | |
| | LOC102724211 | | |
| | LOC102724212 | | |
| 0 | LOC102724213 | FALSE | Upregulated |
| | LOC102724214 | | |

| | | | |
|---|---|---|---|
| | LOC102724215 | | |
| | LOC102724216 | | |
| | LOC102724218 | | |
| -2,675846115 | LOC102724219 | | Downregulated |
| | LOC102724220 | | |
| | LOC102724222 | | |
| | LOC102724224 | | |
| | LOC102724225 | | |
| | LOC102724226 | | |
| | LOC102724227 | | |
| | LOC102724228 | | |
| 0,608061706 | LOC102724229 | FALSE | Upregulated |
| 1,010904834 | LOC102724230 | | Upregulated |
| 0 | LOC102724231 | FALSE | Upregulated |
| -0,400724049 | LOC102724232 | FALSE | Downregulated |
| 0,731728219 | LOC102724233 | FALSE | Upregulated |
| | LOC102724234 | | |
| 0,526222837 | LOC102724235 | FALSE | Upregulated |
| 1,482716756 | LOC102724237 | | Upregulated |
| | LOC102724239 | | |
| 1,718605713 | LOC102724240 | | Upregulated |
| | LOC102724241 | | |
| | LOC102724242 | | |
| 1,040638426 | LOC102724244 | | Upregulated |
| 0 | LOC102724245 | FALSE | Upregulated |
| | LOC102724246 | | |
| | LOC102724247 | | |
| 0,559803794 | LOC102724248 | FALSE | Upregulated |
| 0,101391275 | LOC102724249 | FALSE | Upregulated |
| | LOC102724251 | | |
| 0,354075638 | LOC102724252 | FALSE | Upregulated |
| | LOC102724253 | | |
| -0,031521782 | LOC102724256 | FALSE | Downregulated |
| | LOC102724257 | | |
| -0,225964989 | LOC102724258 | FALSE | Downregulated |
| -0,114527022 | LOC102724259 | FALSE | Downregulated |
| | LOC102724260 | | |
| 0,010465309 | LOC102724261 | FALSE | Upregulated |
| -1,11510884 | LOC102724263 | | Downregulated |
| | LOC102724264 | | |
| -0,013667189 | LOC102724265 | FALSE | Downregulated |
| | LOC102724266 | | |
| | LOC102724267 | | |
| | LOC102724268 | | |
| | LOC102724270 | | |
| | LOC102724271 | | |
| 0,280082144 | LOC102724272 | FALSE | Upregulated |
| 0,882575709 | LOC102724274 | FALSE | Upregulated |
| 0,503136311 | LOC102724275 | FALSE | Upregulated |
| 0,711582226 | LOC102724276 | FALSE | Upregulated |
| | LOC102724277 | | |

| | | | |
|---|---|---|---|
| | LOC102724278 | | |
| | LOC102724279 | | |
| | LOC102724280 | | |
| 0,001141803 | LOC102724282 | FALSE | Upregulated |
| -0,016630248 | LOC102724283 | FALSE | Downregulated |
| | LOC102724284 | | |
| -0,061847166 | LOC102724285 | FALSE | Downregulated |
| | LOC102724287 | | |
| | LOC102724288 | | |
| | LOC102724289 | | |
| | LOC102724290 | | |
| -0,143426222 | LOC102724292 | FALSE | Downregulated |
| -0,164227161 | LOC102724293 | FALSE | Downregulated |
| | LOC102724294 | | |
| -0,187835935 | LOC102724295 | FALSE | Downregulated |
| -0,427075817 | LOC102724296 | FALSE | Downregulated |
| 0,411019001 | LOC102724297 | FALSE | Upregulated |
| 0 | LOC102724298 | FALSE | Upregulated |
| | LOC102724299 | | |
| 0 | LOC102724300 | FALSE | Upregulated |
| | LOC102724301 | | |
| | LOC102724302 | | |
| | LOC102724304 | | |
| | LOC102724305 | | |
| 0,600699593 | LOC102724306 | FALSE | Upregulated |
| -0,254165245 | LOC102724307 | FALSE | Downregulated |
| | LOC102724308 | | |
| 0,285372545 | LOC102724309 | FALSE | Upregulated |
| | LOC102724311 | | |
| 0,769119369 | LOC102724312 | FALSE | Upregulated |
| | LOC102724313 | | |
| -0,096236994 | LOC102724314 | FALSE | Downregulated |
| | LOC102724315 | | |
| | LOC102724317 | | |
| | LOC102724318 | | |
| 0,411071039 | LOC102724319 | FALSE | Upregulated |
| | LOC102724321 | | |
| 0,860581941 | LOC102724323 | FALSE | Upregulated |
| | LOC102724324 | | |
| | LOC102724325 | | |
| | LOC102724326 | | |
| | LOC102724327 | | |
| 0,761578832 | LOC102724328 | FALSE | Upregulated |
| -0,009053508 | LOC102724329 | FALSE | Downregulated |
| 0,58137459 | LOC102724330 | FALSE | Upregulated |
| | LOC102724331 | | |
| | LOC102724332 | | |
| | LOC102724333 | | |
| 0,276894323 | LOC102724334 | FALSE | Upregulated |
| 0,374105004 | LOC102724336 | FALSE | Upregulated |
| | LOC102724337 | | |

| | | | |
|---|---|---|---|
| | LOC102724338 | | |
| 0,133653855 | LOC102724339 | FALSE | Upregulated |
| | LOC102724340 | | |
| | LOC102724341 | | |
| 0,160020061 | LOC102724343 | FALSE | Upregulated |
| | LOC102724344 | | |
| | LOC102724346 | | |
| | LOC102724347 | | |
| 0,146268218 | LOC102724348 | FALSE | Upregulated |
| 0,110695941 | LOC102724349 | FALSE | Upregulated |
| 0,300241802 | LOC102724351 | FALSE | Upregulated |
| 0 | LOC102724352 | FALSE | Upregulated |
| | LOC102724353 | | |
| 0,558147277 | LOC102724354 | FALSE | Upregulated |
| | LOC102724355 | | |
| -0,983688324 | LOC102724356 | FALSE | Downregulated |
| | LOC102724357 | | |
| | LOC102724358 | | |
| | LOC102724359 | | |
| | LOC102724360 | | |
| 0 | LOC102724361 | FALSE | Upregulated |
| | LOC102724362 | | |
| 0,015512986 | LOC102724363 | FALSE | Upregulated |
| | LOC102724366 | | |
| | LOC102724367 | | |
| -0,319409869 | LOC102724368 | FALSE | Downregulated |
| -0,362768322 | LOC102724369 | FALSE | Downregulated |
| 1,132735822 | LOC102724370 | | Upregulated |
| 1,588930129 | LOC102724371 | | Upregulated |
| | LOC102724373 | | |
| | LOC102724375 | | |
| 0,29331538 | LOC102724376 | FALSE | Upregulated |
| 0,146013126 | LOC102724377 | FALSE | Upregulated |
| 0,11511683 | LOC102724378 | FALSE | Upregulated |
| | LOC102724379 | | |
| 0 | LOC102724380 | FALSE | Upregulated |
| | LOC102724381 | | |
| | LOC102724382 | | |
| 0,153365447 | LOC102724384 | FALSE | Upregulated |
| -0,052587944 | LOC102724385 | FALSE | Downregulated |
| | LOC102724386 | | |
| -0,50616737 | LOC102724387 | FALSE | Downregulated |
| 0,091564311 | LOC102724389 | FALSE | Upregulated |
| 0,003832122 | LOC102724392 | FALSE | Upregulated |
| | LOC102724393 | | |
| -0,367410739 | LOC102724394 | FALSE | Downregulated |
| | LOC102724395 | | |
| | LOC102724396 | | |
| 0,268216551 | LOC102724397 | FALSE | Upregulated |
| -0,836181784 | LOC102724398 | FALSE | Downregulated |
| -0,455017029 | LOC102724399 | FALSE | Downregulated |

| | | | |
|---|---|---|---|
| | LOC102724400 | | |
| -0,147981245 | LOC102724402 | FALSE | Downregulated |
| | LOC102724403 | | |
| 0,573188232 | LOC102724404 | FALSE | Upregulated |
| | LOC102724405 | | |
| 0 | LOC102724406 | FALSE | Upregulated |
| | LOC102724407 | | |
| -0,058741713 | LOC102724409 | FALSE | Downregulated |
| | LOC102724410 | | |
| 0 | LOC102724411 | FALSE | Upregulated |
| | LOC102724412 | | |
| -0,633301681 | LOC102724413 | FALSE | Downregulated |
| | LOC102724414 | | |
| | LOC102724415 | | |
| 0 | LOC102724416 | FALSE | Upregulated |
| | LOC102724417 | | |
| | LOC102724418 | | |
| | LOC102724419 | | |
| 0,697704656 | LOC102724420 | FALSE | Upregulated |
| | LOC102724421 | | |
| | LOC102724422 | | |
| -0,603999289 | LOC102724423 | FALSE | Downregulated |
| 0,400844053 | LOC102724424 | FALSE | Upregulated |
| | LOC102724425 | | |
| -1,220693999 | LOC102724426 | | Downregulated |
| | LOC102724427 | | |
| | LOC102724429 | | |
| | LOC102724431 | | |
| 0 | LOC102724432 | FALSE | Upregulated |
| 0,279397957 | LOC102724433 | FALSE | Upregulated |
| 0 | LOC102724434 | FALSE | Upregulated |
| | LOC102724435 | | |
| -0,014349807 | LOC102724436 | FALSE | Downregulated |
| | LOC102724438 | | |
| | LOC102724439 | | |
| | LOC102724440 | | |
| 3,987990725 | LOC102724441 | | Upregulated |
| 1,188758527 | LOC102724442 | | Upregulated |
| | LOC102724443 | | |
| -0,027283986 | LOC102724444 | FALSE | Downregulated |
| 0 | LOC102724446 | FALSE | Upregulated |
| 0,449896423 | LOC102724448 | FALSE | Upregulated |
| -0,543286899 | LOC102724449 | FALSE | Downregulated |
| 0,218191831 | LOC102724450 | FALSE | Upregulated |
| -1,051222044 | LOC102724451 | | Downregulated |
| | LOC102724452 | | |
| | LOC102724453 | | |
| | LOC102724454 | | |
| | LOC102724456 | | |
| -0,55307588 | LOC102724457 | FALSE | Downregulated |
| | LOC102724458 | | |

| | | | |
|---|---|---|---|
| | LOC102724460 | | |
| | LOC102724461 | | |
| -0,331917982 | LOC102724462 | FALSE | Downregulated |
| 0 | LOC102724463 | FALSE | Upregulated |
| 0,769203285 | LOC102724464 | FALSE | Upregulated |
| | LOC102724465 | | |
| 0,28181789 | LOC102724467 | FALSE | Upregulated |
| | LOC102724469 | | |
| | LOC102724470 | | |
| | LOC102724471 | | |
| 0,008513185 | LOC102724472 | FALSE | Upregulated |
| | LOC102724473 | | |
| -0,109489162 | LOC102724475 | FALSE | Downregulated |
| 0,402509517 | LOC102724476 | FALSE | Upregulated |
| -0,010456707 | LOC102724477 | FALSE | Downregulated |
| | LOC102724478 | | |
| | LOC102724479 | | |
| | LOC102724480 | | |
| 0 | LOC102724481 | FALSE | Upregulated |
| | LOC102724482 | | |
| -0,103377474 | LOC102724483 | FALSE | Downregulated |
| | LOC102724485 | | |
| | LOC102724486 | | |
| -0,504985347 | LOC102724487 | FALSE | Downregulated |
| 0 | LOC102724489 | FALSE | Upregulated |
| | LOC102724491 | | |
| | LOC102724492 | | |
| -0,006131687 | LOC102724493 | FALSE | Downregulated |
| | LOC102724494 | | |
| | LOC102724495 | | |
| | LOC102724496 | | |
| 0,431314023 | LOC102724497 | FALSE | Upregulated |
| | LOC102724498 | | |
| | LOC102724500 | | |
| | LOC102724501 | | |
| 0,273202152 | LOC102724502 | FALSE | Upregulated |
| | LOC102724503 | | |
| | LOC102724505 | | |
| -0,189778708 | LOC102724506 | FALSE | Downregulated |
| -0,192441116 | LOC102724507 | FALSE | Downregulated |
| 1,179430342 | LOC102724508 | | Upregulated |
| | LOC102724509 | | |
| | LOC102724510 | | |
| | LOC102724511 | | |
| | LOC102724512 | | |
| 0 | LOC102724513 | FALSE | Upregulated |
| -0,07076871 | LOC102724514 | FALSE | Downregulated |
| -0,46257702 | LOC102724515 | FALSE | Downregulated |
| 0,306194279 | LOC102724516 | FALSE | Upregulated |
| -0,360727709 | LOC102724517 | FALSE | Downregulated |
| -0,302918722 | LOC102724519 | FALSE | Downregulated |

| | | | |
|---|---|---|---|
| | LOC102724520 | | |
| | LOC102724521 | | |
| | LOC102724523 | | |
| -0,615820231 | LOC102724524 | FALSE | Downregulated |
| | LOC102724527 | | |
| 0,669415132 | LOC102724528 | FALSE | Upregulated |
| | LOC102724529 | | |
| -0,029078411 | LOC102724530 | FALSE | Downregulated |
| | LOC102724531 | | |
| -0,057345553 | LOC102724532 | FALSE | Downregulated |
| | LOC102724533 | | |
| | LOC102724534 | | |
| 0,530064821 | LOC102724537 | FALSE | Upregulated |
| | LOC102724538 | | |
| 0,141283933 | LOC102724539 | FALSE | Upregulated |
| | LOC102724540 | | |
| -0,158628221 | LOC102724541 | FALSE | Downregulated |
| | LOC102724542 | | |
| 0,295720394 | LOC102724543 | FALSE | Upregulated |
| | LOC102724544 | | |
| 0,438176188 | LOC102724545 | FALSE | Upregulated |
| 0,394336523 | LOC102724548 | FALSE | Upregulated |
| -0,148661806 | LOC102724550 | FALSE | Downregulated |
| 0 | LOC102724551 | FALSE | Upregulated |
| 0 | LOC102724553 | FALSE | Upregulated |
| -0,178395561 | LOC102724554 | FALSE | Downregulated |
| 0,423324777 | LOC102724555 | FALSE | Upregulated |
| -0,240151845 | LOC102724556 | FALSE | Downregulated |
| | LOC102724557 | | |
| -0,004525881 | LOC102724558 | FALSE | Downregulated |
| | LOC102724561 | | |
| | LOC102724564 | | |
| | LOC102724565 | | |
| | LOC102724566 | | |
| | LOC102724567 | | |
| 0,127062357 | LOC102724568 | FALSE | Upregulated |
| 0,019793104 | LOC102724569 | FALSE | Upregulated |
| -0,156643593 | LOC102724570 | FALSE | Downregulated |
| -0,169833252 | LOC102724571 | FALSE | Downregulated |
| | LOC102724572 | | |
| 0,139025275 | LOC102724573 | FALSE | Upregulated |
| | LOC102724574 | | |
| | LOC102724575 | | |
| | LOC102724576 | | |
| | LOC102724577 | | |
| 0,411902206 | LOC102724578 | FALSE | Upregulated |
| | LOC102724579 | | |
| -0,662181417 | LOC102724580 | FALSE | Downregulated |
| 0,577791683 | LOC102724581 | FALSE | Upregulated |
| | LOC102724582 | | |
| -0,343154926 | LOC102724583 | FALSE | Downregulated |

| | | | |
|---|---|---|---|
| -0,224089793 | LOC102724585 | FALSE | Downregulated |
| -0,040575503 | LOC102724586 | FALSE | Downregulated |
| | LOC102724588 | | |
| | LOC102724589 | | |
| | LOC102724590 | | |
| | LOC102724591 | | |
| | LOC102724592 | | |
| 0,449270961 | LOC102724593 | FALSE | Upregulated |
| -1,233821495 | LOC102724594 | | Downregulated |
| | LOC102724595 | | |
| 0,285729904 | LOC102724596 | FALSE | Upregulated |
| 0,118347533 | LOC102724598 | FALSE | Upregulated |
| 0 | LOC102724599 | FALSE | Upregulated |
| -0,064776989 | LOC102724601 | FALSE | Downregulated |
| -0,783947916 | LOC102724603 | FALSE | Downregulated |
| -0,023510249 | LOC102724604 | FALSE | Downregulated |
| | LOC102724605 | | |
| | LOC102724606 | | |
| -0,164645642 | LOC102724608 | FALSE | Downregulated |
| | LOC102724609 | | |
| 0,402456424 | LOC102724610 | FALSE | Upregulated |
| | LOC102724612 | | |
| | LOC102724613 | | |
| -0,636363972 | LOC102724614 | FALSE | Downregulated |
| -0,169485574 | LOC102724615 | FALSE | Downregulated |
| | LOC102724616 | | |
| | LOC102724617 | | |
| | LOC102724618 | | |
| | LOC102724619 | | |
| 0 | LOC102724620 | FALSE | Upregulated |
| | LOC102724622 | | |
| | LOC102724623 | | |
| | LOC102724625 | | |
| 0,819554909 | LOC102724626 | FALSE | Upregulated |
| | LOC102724627 | | |
| | LOC102724628 | | |
| | LOC102724629 | | |
| 0,155546522 | LOC102724630 | FALSE | Upregulated |
| | LOC102724631 | | |
| 0 | LOC102724632 | FALSE | Upregulated |
| 0 | LOC102724633 | FALSE | Upregulated |
| 0,018607112 | LOC102724636 | FALSE | Upregulated |
| | LOC102724637 | | |
| | LOC102724638 | | |
| | LOC102724640 | | |
| 0,140202581 | LOC102724641 | FALSE | Upregulated |
| 0,129850086 | LOC102724642 | FALSE | Upregulated |
| -0,194436904 | LOC102724643 | FALSE | Downregulated |
| | LOC102724644 | | |
| -0,376807687 | LOC102724645 | FALSE | Downregulated |
| | LOC102724646 | | |

| | | | |
|---|---|---|---|
| | LOC102724647 | | |
| | LOC102724648 | | |
| | LOC102724649 | | |
| | LOC102724650 | | |
| | LOC102724651 | | |
| 0,277833435 | LOC102724653 | FALSE | Upregulated |
| | LOC102724654 | | |
| -0,860299402 | LOC102724655 | FALSE | Downregulated |
| | LOC102724657 | | |
| 0,435446544 | LOC102724659 | FALSE | Upregulated |
| -0,005031759 | LOC102724660 | FALSE | Downregulated |
| 0 | LOC102724661 | FALSE | Upregulated |
| -0,831157019 | LOC102724662 | FALSE | Downregulated |
| 0,296218597 | LOC102724663 | FALSE | Upregulated |
| | LOC102724664 | | |
| | LOC102724665 | | |
| | LOC102724666 | | |
| 0,273422532 | LOC102724667 | FALSE | Upregulated |
| 0,125176128 | LOC102724668 | FALSE | Upregulated |
| | LOC102724669 | | |
| | LOC102724670 | | |
| | LOC102724671 | | |
| 0,460216695 | LOC102724675 | FALSE | Upregulated |
| -0,004525881 | LOC102724676 | FALSE | Downregulated |
| | LOC102724678 | | |
| | LOC102724679 | | |
| | LOC102724680 | | |
| -0,030541709 | LOC102724681 | FALSE | Downregulated |
| 0,238807503 | LOC102724682 | FALSE | Upregulated |
| | LOC102724684 | | |
| -0,00279278 | LOC102724685 | FALSE | Downregulated |
| 0,004845297 | LOC102724686 | FALSE | Upregulated |
| -0,231070762 | LOC102724687 | FALSE | Downregulated |
| | LOC102724688 | | |
| | LOC102724689 | | |
| | LOC102724690 | | |
| 0,592477025 | LOC102724691 | FALSE | Upregulated |
| | LOC102724692 | | |
| | LOC102724693 | | |
| | LOC102724694 | | |
| -0,139944137 | LOC102724695 | FALSE | Downregulated |
| -0,329843492 | LOC102724697 | FALSE | Downregulated |
| | LOC102724698 | | |
| 0,063584301 | LOC102724699 | FALSE | Upregulated |
| | LOC102724700 | | |
| 0,272277655 | LOC102724702 | FALSE | Upregulated |
| -0,014157948 | LOC102724703 | FALSE | Downregulated |
| | LOC102724704 | | |
| | LOC102724707 | | |
| 0,203918847 | LOC102724708 | FALSE | Upregulated |
| | LOC102724710 | | |

| | | | |
|---|---|---|---|
| -0,000538082 | LOC102724711 | FALSE | Downregulated |
| | LOC102724712 | | |
| | LOC102724713 | | |
| 0 | LOC102724714 | FALSE | Upregulated |
| 0,291340714 | LOC102724715 | FALSE | Upregulated |
| | LOC102724716 | | |
| | LOC102724717 | | |
| | LOC102724718 | | |
| 0,513754433 | LOC102724719 | FALSE | Upregulated |
| | LOC102724721 | | |
| | LOC102724722 | | |
| | LOC102724723 | | |
| | LOC102724728 | | |
| | LOC102724729 | | |
| -0,057764088 | LOC102724731 | FALSE | Downregulated |
| | LOC102724732 | | |
| | LOC102724734 | | |
| | LOC102724735 | | |
| | LOC102724736 | | |
| -0,588561862 | LOC102724737 | FALSE | Downregulated |
| | LOC102724738 | | |
| | LOC102724739 | | |
| 0,321106849 | LOC102724740 | FALSE | Upregulated |
| | LOC102724741 | | |
| | LOC102724742 | | |
| 0,002241872 | LOC102724743 | FALSE | Upregulated |
| | LOC102724744 | | |
| 0 | LOC102724745 | FALSE | Upregulated |
| 1,757459543 | LOC102724746 | | Upregulated |
| | LOC102724747 | | |
| 0,726778993 | LOC102724749 | FALSE | Upregulated |
| 0,265989912 | LOC102724751 | FALSE | Upregulated |
| 0,201295615 | LOC102724752 | FALSE | Upregulated |
| | LOC102724753 | | |
| 0,23418267 | LOC102724754 | FALSE | Upregulated |
| | LOC102724755 | | |
| -0,142594431 | LOC102724756 | FALSE | Downregulated |
| 0 | LOC102724758 | FALSE | Upregulated |
| 0,284168031 | LOC102724759 | FALSE | Upregulated |
| -0,148465128 | LOC102724762 | FALSE | Downregulated |
| | LOC102724763 | | |
| -0,332011495 | LOC102724764 | FALSE | Downregulated |
| 0,720993433 | LOC102724765 | FALSE | Upregulated |
| 0,005477513 | LOC102724766 | FALSE | Upregulated |
| | LOC102724767 | | |
| 0,149801356 | LOC102724768 | FALSE | Upregulated |
| | LOC102724771 | | |
| | LOC102724772 | | |
| 0,280928672 | LOC102724774 | FALSE | Upregulated |
| 0,300241802 | LOC102724775 | FALSE | Upregulated |
| | LOC102724776 | | |

| | | | |
|---|---|---|---|
| | LOC102724777 | | |
| -0,12806895 | LOC102724778 | FALSE | Downregulated |
| | LOC102724779 | | |
| | LOC102724780 | | |
| 0 | LOC102724781 | FALSE | Upregulated |
| 0,354072665 | LOC102724783 | FALSE | Upregulated |
| | LOC102724785 | | |
| 0 | LOC102724786 | FALSE | Upregulated |
| 0,38636847 | LOC102724792 | FALSE | Upregulated |
| | LOC102724793 | | |
| | LOC102724794 | | |
| -0,921089365 | LOC102724795 | FALSE | Downregulated |
| | LOC102724796 | | |
| 0,088803161 | LOC102724797 | FALSE | Upregulated |
| | LOC102724798 | | |
| | LOC102724799 | | |
| -1,027912984 | LOC102724800 | | Downregulated |
| | LOC102724801 | | |
| 0,707447411 | LOC102724802 | FALSE | Upregulated |
| | LOC102724803 | | |
| -0,153478494 | LOC102724804 | FALSE | Downregulated |
| 0,5745575 | LOC102724805 | FALSE | Upregulated |
| -0,01961669 | LOC102724806 | FALSE | Downregulated |
| -0,703560055 | LOC102724807 | FALSE | Downregulated |
| -0,001504888 | LOC102724808 | FALSE | Downregulated |
| | LOC102724809 | | |
| | LOC102724810 | | |
| 0 | LOC102724812 | FALSE | Upregulated |
| | LOC102724815 | | |
| | LOC102724817 | | |
| | LOC102724818 | | |
| | LOC102724819 | | |
| | LOC102724820 | | |
| 0,018839898 | LOC102724821 | FALSE | Upregulated |
| 0,081773135 | LOC102724824 | FALSE | Upregulated |
| | LOC102724825 | | |
| 0 | LOC102724826 | FALSE | Upregulated |
| 0 | LOC102724827 | FALSE | Upregulated |
| -0,784974897 | LOC102724828 | FALSE | Downregulated |
| | LOC102724829 | | |
| | LOC102724830 | | |
| 0,270057135 | LOC102724831 | FALSE | Upregulated |
| 0,536639018 | LOC102724832 | FALSE | Upregulated |
| | LOC102724834 | | |
| | LOC102724835 | | |
| | LOC102724836 | | |
| | LOC102724837 | | |
| | LOC102724838 | | |
| | LOC102724839 | | |
| | LOC102724840 | | |
| 0,092189934 | LOC102724841 | FALSE | Upregulated |

| | | | |
|---|---|---|---|
| 1,38520821 | LOC102724842 | | Upregulated |
| | LOC102724844 | | |
| | LOC102724845 | | |
| | LOC102724846 | | |
| | LOC102724847 | | |
| | LOC102724848 | | |
| | LOC102724849 | | |
| 0,204279741 | LOC102724850 | FALSE | Upregulated |
| 0,226931969 | LOC102724851 | FALSE | Upregulated |
| | LOC102724854 | | |
| | LOC102724855 | | |
| -0,018149848 | LOC102724856 | FALSE | Downregulated |
| 0,006328102 | LOC102724857 | FALSE | Upregulated |
| | LOC102724858 | | |
| | LOC102724859 | | |
| -0,431296211 | LOC102724860 | FALSE | Downregulated |
| 0 | LOC102724861 | FALSE | Upregulated |
| -0,566686872 | LOC102724862 | FALSE | Downregulated |
| | LOC102724863 | | |
| 0,239255342 | LOC102724864 | FALSE | Upregulated |
| 0,133828175 | LOC102724865 | FALSE | Upregulated |
| 0,547182904 | LOC102724867 | FALSE | Upregulated |
| -0,132348974 | LOC102724869 | FALSE | Downregulated |
| 0,285372545 | LOC102724870 | FALSE | Upregulated |
| | LOC102724871 | | |
| -0,096372702 | LOC102724873 | FALSE | Downregulated |
| | LOC102724874 | | |
| | LOC102724875 | | |
| | LOC102724876 | | |
| | LOC102724877 | | |
| | LOC102724878 | | |
| | LOC102724879 | | |
| 0,230574925 | LOC102724880 | FALSE | Upregulated |
| 0,010125178 | LOC102724881 | FALSE | Upregulated |
| | LOC102724882 | | |
| 0,969243443 | LOC102724883 | FALSE | Upregulated |
| 0,900051798 | LOC102724884 | FALSE | Upregulated |
| | LOC102724885 | | |
| | LOC102724887 | | |
| | LOC102724888 | | |
| 0,170831645 | LOC102724889 | FALSE | Upregulated |
| | LOC102724890 | | |
| | LOC102724891 | | |
| | LOC102724892 | | |
| -0,31904471 | LOC102724893 | FALSE | Downregulated |
| | LOC102724894 | | |
| | LOC102724896 | | |
| 0 | LOC102724897 | FALSE | Upregulated |
| 0,743825998 | LOC102724898 | FALSE | Upregulated |
| | LOC102724899 | | |
| | LOC102724900 | | |

| | | | |
|---|---|---|---|
| -0,412876135 | LOC102724901 | FALSE | Downregulated |
| | LOC102724902 | | |
| 0,388414345 | LOC102724903 | FALSE | Upregulated |
| | LOC102724904 | | |
| | LOC102724905 | | |
| -0,006903421 | LOC102724906 | FALSE | Downregulated |
| 0,608225375 | LOC102724907 | FALSE | Upregulated |
| 0,713133028 | LOC102724908 | FALSE | Upregulated |
| | LOC102724910 | | |
| 0 | LOC102724911 | FALSE | Upregulated |
| | LOC102724912 | | |
| | LOC102724913 | | |
| | LOC102724914 | | |
| | LOC102724915 | | |
| -0,249079964 | LOC102724916 | FALSE | Downregulated |
| | LOC102724917 | | |
| | LOC102724918 | | |
| -0,018149848 | LOC102724919 | FALSE | Downregulated |
| 1,045667876 | LOC102724920 | | Upregulated |
| 0 | LOC102724923 | FALSE | Upregulated |
| | LOC102724924 | | |
| | LOC102724925 | | |
| -0,143422155 | LOC102724927 | FALSE | Downregulated |
| | LOC102724928 | | |
| | LOC102724929 | | |
| | LOC102724932 | | |
| 0,169469932 | LOC102724933 | FALSE | Upregulated |
| | LOC102724934 | | |
| 0,27762724 | LOC102724935 | FALSE | Upregulated |
| | LOC102724936 | | |
| | LOC102724939 | | |
| -0,245020818 | LOC102724940 | FALSE | Downregulated |
| | LOC102724942 | | |
| 0,297924766 | LOC102724943 | FALSE | Upregulated |
| 0,169629821 | LOC102724944 | FALSE | Upregulated |
| | LOC102724945 | | |
| 0 | LOC102724946 | FALSE | Upregulated |
| 0,407642546 | LOC102724947 | FALSE | Upregulated |
| 0,13794669 | LOC102724948 | FALSE | Upregulated |
| | LOC102724949 | | |
| 0,006464133 | LOC102724950 | FALSE | Upregulated |
| -2,687285086 | LOC102724951 | | Downregulated |
| | LOC102724952 | | |
| | LOC102724953 | | |
| | LOC102724954 | | |
| | LOC102724955 | | |
| 1,868402414 | LOC102724956 | | Upregulated |
| | LOC102724957 | | |
| | LOC102724958 | | |
| | LOC102724959 | | |
| | LOC102724960 | | |

| | | | |
|---|---|---|---|
| | LOC102724963 | | |
| | LOC102724964 | | |
| | LOC102724966 | | |
| -0,379769159 | LOC102724967 | FALSE | Downregulated |
| | LOC102724968 | | |
| -0,156378515 | LOC102724970 | FALSE | Downregulated |
| -0,232187854 | LOC102724971 | FALSE | Downregulated |
| | LOC102724972 | | |
| -0,137139461 | LOC102724973 | FALSE | Downregulated |
| -0,076491374 | LOC113230 | FALSE | Downregulated |
| | LOC128322 | | |
| 0,488118323 | LOC145474 | FALSE | Upregulated |
| 0,378820425 | LOC145845 | FALSE | Upregulated |
| | LOC146481 | | |
| | LOC147004 | | |
| 0,180028922 | LOC149134 | FALSE | Upregulated |
| 1,014489855 | LOC149351 | | Upregulated |
| 0 | LOC149684 | FALSE | Upregulated |
| | LOC149950 | | |
| 0,293242527 | LOC150935 | FALSE | Upregulated |
| 0,380455157 | LOC151174 | FALSE | Upregulated |
| -0,292613404 | LOC151760 | FALSE | Downregulated |
| 0,173502432 | LOC152225 | FALSE | Upregulated |
| | LOC152586 | | |
| | LOC154872 | | |
| -0,528472105 | LOC155060 | FALSE | Downregulated |
| 0,15661641 | LOC158434 | FALSE | Upregulated |
| -0,316202056 | LOC158960 | FALSE | Downregulated |
| | LOC1720 | | |
| 0,194285118 | LOC200772 | FALSE | Upregulated |
| 0,032017953 | LOC201651 | FALSE | Upregulated |
| -0,677516789 | LOC254896 | FALSE | Downregulated |
| | LOC255187 | | |
| 0,718862624 | LOC256880 | FALSE | Upregulated |
| 0,58054244 | LOC257358 | FALSE | Upregulated |
| -0,281500444 | LOC257396 | FALSE | Downregulated |
| -0,296567742 | LOC283028 | FALSE | Downregulated |
| | LOC283045 | | |
| 0,165378019 | LOC283070 | FALSE | Upregulated |
| -0,469470573 | LOC283177 | FALSE | Downregulated |
| -0,145708153 | LOC283214 | FALSE | Downregulated |
| | LOC283482 | | |
| | LOC283585 | | |
| -0,004525881 | LOC283682 | FALSE | Downregulated |
| 0,016435364 | LOC283683 | FALSE | Upregulated |
| 0,401526616 | LOC283693 | FALSE | Upregulated |
| 0,438371868 | LOC283731 | FALSE | Upregulated |
| 0,437010354 | LOC283854 | FALSE | Upregulated |
| 0,696081518 | LOC283887 | FALSE | Upregulated |
| 0,449538494 | LOC284009 | FALSE | Upregulated |
| 0,130382616 | LOC284023 | FALSE | Upregulated |

| | | | |
|---|---|---|---|
| -0,005300266 | LOC284240 | FALSE | Downregulated |
| 0,349884668 | LOC284344 | FALSE | Upregulated |
| 0,692123706 | LOC284379 | FALSE | Upregulated |
| 0 | LOC284395 | FALSE | Upregulated |
| -0,199767972 | LOC284454 | FALSE | Downregulated |
| 0,249699816 | LOC284561 | FALSE | Upregulated |
| 0,750091675 | LOC284578 | FALSE | Upregulated |
| -0,014595724 | LOC284581 | FALSE | Downregulated |
| -0,172844189 | LOC284600 | FALSE | Downregulated |
| 0 | LOC284648 | FALSE | Upregulated |
| | LOC284801 | | |
| -0,545497984 | LOC284825 | FALSE | Downregulated |
| 0,041632332 | LOC284865 | FALSE | Upregulated |
| -0,144182103 | LOC284898 | FALSE | Downregulated |
| 2,040299508 | LOC284933 | | Upregulated |
| | LOC285000 | | |
| 0,420674548 | LOC285043 | FALSE | Upregulated |
| -0,459066639 | LOC285389 | FALSE | Downregulated |
| 1,132202627 | LOC285556 | | Upregulated |
| | LOC285692 | | |
| -0,022098395 | LOC285768 | FALSE | Downregulated |
| 0,541922044 | LOC285819 | FALSE | Upregulated |
| 0,194042784 | LOC286178 | FALSE | Upregulated |
| 0,277119458 | LOC286186 | FALSE | Upregulated |
| -0,006903421 | LOC286189 | FALSE | Downregulated |
| -0,006848587 | LOC286359 | FALSE | Downregulated |
| -0,496566626 | LOC286437 | FALSE | Downregulated |
| 0,271886396 | LOC339166 | FALSE | Upregulated |
| 0,65539787 | LOC339192 | FALSE | Upregulated |
| -0,482454266 | LOC339240 | FALSE | Downregulated |
| | LOC339260 | | |
| 0 | LOC339298 | FALSE | Upregulated |
| -0,009950398 | LOC339539 | FALSE | Downregulated |
| 0 | LOC339593 | FALSE | Upregulated |
| 0,361490076 | LOC339622 | FALSE | Upregulated |
| 0 | LOC339862 | FALSE | Upregulated |
| 0,785665065 | LOC339874 | FALSE | Upregulated |
| | LOC339978 | | |
| -0,626087808 | LOC340073 | FALSE | Downregulated |
| 0,534578681 | LOC340074 | FALSE | Upregulated |
| | LOC340090 | | |
| 0,107428347 | LOC340113 | FALSE | Upregulated |
| 0 | LOC340508 | FALSE | Upregulated |
| 0,273722389 | LOC341056 | FALSE | Upregulated |
| 0 | LOC344887 | FALSE | Upregulated |
| -0,734039236 | LOC344967 | FALSE | Downregulated |
| -0,414649691 | LOC345051 | FALSE | Downregulated |
| -0,063974576 | LOC374443 | FALSE | Downregulated |
| 0 | LOC374890 | FALSE | Upregulated |
| -0,068677945 | LOC375196 | FALSE | Downregulated |
| -0,136516183 | LOC387720 | FALSE | Downregulated |

| | | | |
|---|---|---|---|
| -0,023655299 | LOC388210 | FALSE | Downregulated |
| 0,134913061 | LOC388242 | FALSE | Upregulated |
| -0,014724123 | LOC388406 | FALSE | Downregulated |
| 2,519474745 | LOC388436 | | Upregulated |
| 0,297178435 | LOC388456 | FALSE | Upregulated |
| 1,121669105 | LOC388499 | | Upregulated |
| | LOC388553 | | |
| -0,139815686 | LOC388780 | FALSE | Downregulated |
| -0,14342438 | LOC388813 | FALSE | Downregulated |
| | LOC389033 | | |
| 0,151269254 | LOC389273 | FALSE | Upregulated |
| 0,07293222 | LOC389602 | FALSE | Upregulated |
| -0,51631413 | LOC389607 | FALSE | Downregulated |
| 0,293563661 | LOC389641 | FALSE | Upregulated |
| 0 | LOC389705 | FALSE | Upregulated |
| -0,031415609 | LOC389765 | FALSE | Downregulated |
| | LOC389895 | | |
| | LOC390705 | | |
| 0 | LOC390877 | FALSE | Upregulated |
| | LOC390956 | | |
| -0,104551176 | LOC391722 | FALSE | Downregulated |
| 0,112521081 | LOC392232 | FALSE | Upregulated |
| 0,272027851 | LOC392288 | FALSE | Upregulated |
| -0,121726386 | LOC392364 | FALSE | Downregulated |
| | LOC392452 | | |
| 0,290688548 | LOC399815 | FALSE | Upregulated |
| 0,086306054 | LOC400499 | FALSE | Upregulated |
| | LOC400622 | | |
| 0,136713725 | LOC400655 | FALSE | Upregulated |
| -0,008787777 | LOC400685 | FALSE | Downregulated |
| | LOC400743 | | |
| 0,123866607 | LOC400748 | FALSE | Upregulated |
| -0,158501962 | LOC400768 | FALSE | Downregulated |
| -0,904718861 | LOC400863 | FALSE | Downregulated |
| -0,011494463 | LOC400891 | FALSE | Downregulated |
| 0,445348632 | LOC400927 | FALSE | Upregulated |
| | LOC400940 | | |
| 0,141281018 | LOC401010 | FALSE | Upregulated |
| 0 | LOC401098 | FALSE | Upregulated |
| -0,045199048 | LOC401127 | FALSE | Downregulated |
| | LOC401177 | | |
| 0,280928672 | LOC401242 | FALSE | Upregulated |
| | LOC401312 | | |
| | LOC401324 | | |
| | LOC401357 | | |
| 0 | LOC401442 | FALSE | Upregulated |
| -0,129942941 | LOC401463 | FALSE | Downregulated |
| -0,063504011 | LOC401480 | FALSE | Downregulated |
| | LOC401497 | | |
| | LOC401557 | | |
| -0,547858945 | LOC401589 | FALSE | Downregulated |

| | | | |
|---|---|---|---|
| | LOC402269 | | |
| -0,168391287 | LOC407835 | FALSE | Downregulated |
| -0,712090997 | LOC439951 | FALSE | Downregulated |
| 0,637992779 | LOC439994 | FALSE | Upregulated |
| | LOC440040 | | |
| -0,430220938 | LOC440157 | FALSE | Downregulated |
| 1,020810941 | LOC440173 | | Upregulated |
| -0,22230704 | LOC440243 | FALSE | Downregulated |
| -0,143352179 | LOC440292 | FALSE | Downregulated |
| -0,794962758 | LOC440311 | FALSE | Downregulated |
| 0,622350737 | LOC440313 | FALSE | Upregulated |
| | LOC440337 | | |
| 0,573799129 | LOC440434 | FALSE | Upregulated |
| -0,061066784 | LOC440461 | FALSE | Downregulated |
| | LOC440518 | | |
| | LOC440602 | | |
| 0,051353081 | LOC440700 | FALSE | Upregulated |
| -0,152805148 | LOC440742 | FALSE | Downregulated |
| 0,357587028 | LOC440895 | FALSE | Upregulated |
| | LOC440910 | | |
| -0,221389993 | LOC440934 | FALSE | Downregulated |
| 0,837699685 | LOC441052 | FALSE | Upregulated |
| -0,077472396 | LOC441124 | FALSE | Downregulated |
| -0,200136757 | LOC441155 | FALSE | Downregulated |
| 0,27959969 | LOC441204 | FALSE | Upregulated |
| 0,18440296 | LOC441242 | FALSE | Upregulated |
| 0,519600778 | LOC441455 | FALSE | Upregulated |
| 0,455715313 | LOC441493 | FALSE | Upregulated |
| -1,334937811 | LOC441528 | | Downregulated |
| 0 | LOC441601 | FALSE | Upregulated |
| -0,25149358 | LOC441666 | FALSE | Downregulated |
| 0,284987956 | LOC441728 | FALSE | Upregulated |
| -0,268975168 | LOC441956 | FALSE | Downregulated |
| 0,407848193 | LOC442028 | FALSE | Upregulated |
| | LOC442132 | | |
| 0,443268062 | LOC494127 | FALSE | Upregulated |
| -0,296105376 | LOC494141 | FALSE | Downregulated |
| | LOC497256 | | |
| 0 | LOC550113 | FALSE | Upregulated |
| 0,464115323 | LOC550643 | FALSE | Upregulated |
| -0,129506483 | LOC553103 | FALSE | Downregulated |
| -0,606101944 | LOC554206 | FALSE | Downregulated |
| 0,086678896 | LOC554223 | FALSE | Upregulated |
| 1,129445073 | LOC613038 | | Upregulated |
| -0,166611983 | LOC641515 | FALSE | Downregulated |
| | LOC641746 | | |
| 0,041975571 | LOC642131 | FALSE | Upregulated |
| 0,149667923 | LOC642236 | FALSE | Upregulated |
| 0 | LOC642249 | FALSE | Upregulated |
| | LOC642366 | | |
| | LOC642643 | | |

| | | | |
|---|---|---|---|
| -0,142777245 | LOC642934 | FALSE | Downregulated |
| 0,018839898 | LOC643327 | FALSE | Upregulated |
| 0,276806401 | LOC643355 | FALSE | Upregulated |
| 0,311821416 | LOC643387 | FALSE | Upregulated |
| -0,459080845 | LOC643406 | FALSE | Downregulated |
| 0 | LOC643441 | FALSE | Upregulated |
| -0,481692479 | LOC643486 | FALSE | Downregulated |
| | LOC643711 | | |
| -0,314145812 | LOC643733 | FALSE | Downregulated |
| -0,33760293 | LOC643770 | FALSE | Downregulated |
| -0,326456936 | LOC643797 | FALSE | Downregulated |
| 0,41482555 | LOC643802 | FALSE | Upregulated |
| | LOC643923 | | |
| | LOC644070 | | |
| 0 | LOC644086 | FALSE | Upregulated |
| | LOC644145 | | |
| -0,136665374 | LOC644189 | FALSE | Downregulated |
| 0,255998845 | LOC644249 | FALSE | Upregulated |
| 0,743066154 | LOC644554 | FALSE | Upregulated |
| -0,358123349 | LOC644656 | FALSE | Downregulated |
| | LOC644669 | | |
| | LOC644717 | | |
| 0 | LOC644919 | FALSE | Upregulated |
| -0,32504683 | LOC644936 | FALSE | Downregulated |
| -0,682695839 | LOC644961 | FALSE | Downregulated |
| | LOC645177 | | |
| | LOC645188 | | |
| 0,300241802 | LOC645202 | FALSE | Upregulated |
| | LOC645321 | | |
| 0,553982606 | LOC645485 | FALSE | Upregulated |
| -0,666948001 | LOC645513 | FALSE | Downregulated |
| -1,150613261 | LOC645638 | | Downregulated |
| -0,544952772 | LOC645752 | FALSE | Downregulated |
| 0,513828101 | LOC645769 | FALSE | Upregulated |
| 0,857023912 | LOC646021 | FALSE | Upregulated |
| 0,287453598 | LOC646034 | FALSE | Upregulated |
| 0,774234717 | LOC646214 | FALSE | Upregulated |
| -0,141252787 | LOC646278 | FALSE | Downregulated |
| | LOC646513 | | |
| | LOC646588 | | |
| -0,226597899 | LOC646626 | FALSE | Downregulated |
| 1,212678237 | LOC646670 | | Upregulated |
| 0,160390816 | LOC646730 | FALSE | Upregulated |
| 0,540237333 | LOC646736 | FALSE | Upregulated |
| | LOC646743 | | |
| | LOC646813 | | |
| | LOC646862 | | |
| -0,021275255 | LOC646903 | FALSE | Downregulated |
| | LOC646938 | | |
| | LOC647264 | | |
| 0,410553113 | LOC647859 | FALSE | Upregulated |

| | | | |
|---|---|---|---|
| 0,384462805 | LOC648044 | FALSE | Upregulated |
| 0 | LOC648691 | FALSE | Upregulated |
| 0,540121036 | LOC648987 | FALSE | Upregulated |
| | LOC649330 | | |
| 0,155519485 | LOC649352 | FALSE | Upregulated |
| | LOC650157 | | |
| 0,303627411 | LOC650226 | FALSE | Upregulated |
| | LOC650293 | | |
| 0 | LOC650368 | FALSE | Upregulated |
| -0,352655165 | LOC653160 | FALSE | Downregulated |
| 0 | LOC653486 | FALSE | Upregulated |
| 0,357988533 | LOC653653 | FALSE | Upregulated |
| 0,271811301 | LOC653786 | FALSE | Upregulated |
| -0,391878853 | LOC654342 | FALSE | Downregulated |
| -0,295709147 | LOC723805 | FALSE | Downregulated |
| | LOC727944 | | |
| -0,240500672 | LOC728026 | FALSE | Downregulated |
| | LOC728114 | | |
| 1,231539813 | LOC728158 | | Upregulated |
| -0,288292721 | LOC728290 | FALSE | Downregulated |
| -0,712262991 | LOC728323 | FALSE | Downregulated |
| 0,082519857 | LOC728392 | FALSE | Upregulated |
| -0,552963679 | LOC728407 | FALSE | Downregulated |
| -0,397888194 | LOC728554 | FALSE | Downregulated |
| 0,432045676 | LOC728637 | FALSE | Upregulated |
| -0,891155734 | LOC728715 | FALSE | Downregulated |
| -0,013241042 | LOC728728 | FALSE | Downregulated |
| | LOC728739 | | |
| -0,551820401 | LOC728752 | FALSE | Downregulated |
| | LOC728755 | | |
| 0,803834291 | LOC728769 | FALSE | Upregulated |
| 1,169050746 | LOC728819 | | Upregulated |
| -0,350854951 | LOC728975 | FALSE | Downregulated |
| -0,021188012 | LOC728989 | FALSE | Downregulated |
| 0,704063488 | LOC729040 | FALSE | Upregulated |
| -0,569782943 | LOC729041 | FALSE | Downregulated |
| 0,597355944 | LOC729080 | FALSE | Upregulated |
| -0,363859456 | LOC729083 | FALSE | Downregulated |
| -0,367036924 | LOC729159 | FALSE | Downregulated |
| 0,113373992 | LOC729164 | FALSE | Upregulated |
| | LOC729222 | | |
| 0,601637389 | LOC729224 | FALSE | Upregulated |
| 0 | LOC729307 | FALSE | Upregulated |
| | LOC729444 | | |
| | LOC729458 | | |
| -0,999041024 | LOC729468 | FALSE | Downregulated |
| 0 | LOC729506 | FALSE | Upregulated |
| -0,256752702 | LOC729609 | FALSE | Downregulated |
| 0,659026301 | LOC729739 | FALSE | Upregulated |
| 0,149412327 | LOC729770 | FALSE | Upregulated |
| 0 | LOC729800 | FALSE | Upregulated |

| | | | |
|---|---|---|---|
| | LOC729815 | | |
| 0,489695919 | LOC729870 | FALSE | Upregulated |
| | LOC729911 | | |
| 0,741914517 | LOC729930 | FALSE | Upregulated |
| 0 | LOC729966 | FALSE | Upregulated |
| -0,002963975 | LOC730159 | FALSE | Downregulated |
| -0,470054853 | LOC730268 | FALSE | Downregulated |
| 0,297178435 | LOC730338 | FALSE | Upregulated |
| -0,300346171 | LOC731157 | FALSE | Downregulated |
| 0,285987685 | LOC731779 | FALSE | Upregulated |
| 0,575074286 | LOC732265 | FALSE | Upregulated |
| 0,042779885 | LOC81691 | FALSE | Upregulated |
| 0,261902018 | LOC90246 | FALSE | Upregulated |
| 0,600334197 | LOC90834 | FALSE | Upregulated |
| 0,280220315 | LOC91450 | FALSE | Upregulated |
| 0 | LOC93432 | FALSE | Upregulated |
| 0,258253715 | LOH12CR2 | FALSE | Upregulated |
| 0 | LOR | FALSE | Upregulated |
| 0,009420174 | LOXHD1 | FALSE | Upregulated |
| 0,694086513 | LOXL1-AS1 | FALSE | Upregulated |
| 0,331498994 | LPAR1 | FALSE | Upregulated |
| 0,674122895 | LPAR2 | FALSE | Upregulated |
| 0,018400286 | LPAR3 | FALSE | Upregulated |
| 0,143482041 | LPAR4 | FALSE | Upregulated |
| 0,655924761 | LPCAT2 | FALSE | Upregulated |
| 0,149719677 | LPCAT4 | FALSE | Upregulated |
| 0,913238591 | LPHN3 | FALSE | Upregulated |
| -0,609079665 | LPL | FALSE | Downregulated |
| 0,048627652 | LPO | FALSE | Upregulated |
| -0,282939489 | LPPR4 | FALSE | Downregulated |
| 0,168428639 | LPPR5 | FALSE | Upregulated |
| 0,12854531 | LPXN | FALSE | Upregulated |
| -0,034735131 | LRCH2 | FALSE | Downregulated |
| 0,140694967 | LRFN1 | FALSE | Upregulated |
| 0 | LRFN2 | FALSE | Upregulated |
| -0,275401324 | LRFN5 | FALSE | Downregulated |
| 0,30850432 | LRGUK | FALSE | Upregulated |
| | LRIT1 | | |
| -0,191170552 | LRIT2 | FALSE | Downregulated |
| -0,44458211 | LRIT3 | FALSE | Downregulated |
| -0,004800456 | LRMP | FALSE | Downregulated |
| -0,464979829 | LRP4 | FALSE | Downregulated |
| -0,197654317 | LRP4-AS1 | FALSE | Downregulated |
| 0,227691959 | LRR1 | FALSE | Upregulated |
| -0,637385474 | LRRC1 | FALSE | Downregulated |
| 0,571254351 | LRRC10 | FALSE | Upregulated |
| -0,431606514 | LRRC10B | FALSE | Downregulated |
| -0,346180845 | LRRC14B | FALSE | Downregulated |
| 0,018244275 | LRRC17 | FALSE | Upregulated |
| 0,194992694 | LRRC2-AS1 | FALSE | Upregulated |
| 0,822348557 | LRRC23 | FALSE | Upregulated |

| | | | |
|---|---|---|---|
| 0 | LRRC26 | FALSE | Upregulated |
| 0,431168091 | LRRC29 | FALSE | Upregulated |
| -0,158821389 | LRRC3-AS1 | FALSE | Downregulated |
| 0,271517363 | LRRC30 | FALSE | Upregulated |
| -0,161067032 | LRRC31 | FALSE | Downregulated |
| -0,114186602 | LRRC34 | FALSE | Downregulated |
| -0,288820769 | LRRC36 | FALSE | Downregulated |
| 0,141200438 | LRRC37A11P | FALSE | Upregulated |
| 0,221887279 | LRRC37A5P | FALSE | Upregulated |
| -0,893764275 | LRRC37B | FALSE | Downregulated |
| 0,298301704 | LRRC38 | FALSE | Upregulated |
| 0,547213979 | LRRC3B | FALSE | Upregulated |
| 0,313595763 | LRRC3C | FALSE | Upregulated |
| -0,128694782 | LRRC42 | FALSE | Downregulated |
| 0,110413417 | LRRC43 | FALSE | Upregulated |
| -0,057553584 | LRRC49 | FALSE | Downregulated |
| -1,216591823 | LRRC4C | | Downregulated |
| | LRRC52 | | |
| -0,547415956 | LRRC6 | FALSE | Downregulated |
| -0,011197196 | LRRC63 | FALSE | Downregulated |
| -0,492739223 | LRRC66 | FALSE | Downregulated |
| -0,143092705 | LRRC69 | FALSE | Downregulated |
| -0,39837977 | LRRC7 | FALSE | Downregulated |
| -0,148184162 | LRRC70 | FALSE | Downregulated |
| 0,771921948 | LRRC71 | FALSE | Upregulated |
| | LRRC72 | | |
| -0,090321066 | LRRC73 | FALSE | Downregulated |
| -0,051933418 | LRRCC1 | FALSE | Downregulated |
| 0 | LRRD1 | FALSE | Upregulated |
| 0 | LRRIQ1 | FALSE | Upregulated |
| -0,158975191 | LRRIQ3 | FALSE | Downregulated |
| -0,19355892 | LRRIQ4 | FALSE | Downregulated |
| 0,109055239 | LRRN4CL | FALSE | Upregulated |
| 0,461650194 | LRRTM1 | FALSE | Upregulated |
| 0,288195097 | LRRTM4 | FALSE | Upregulated |
| 0,574404479 | LRTM1 | FALSE | Upregulated |
| -0,161030532 | LRTM2 | FALSE | Downregulated |
| 1,31805949 | LSAMP | | Upregulated |
| 0,278951017 | LSAMP-AS1 | FALSE | Upregulated |
| -0,039041702 | LSM11 | FALSE | Downregulated |
| -0,303337813 | LSM2 | FALSE | Downregulated |
| -0,709410372 | LSM5 | FALSE | Downregulated |
| -0,521664726 | LSM6 | FALSE | Downregulated |
| -0,538962839 | LSMEM1 | FALSE | Downregulated |
| 0 | LSP1P3 | FALSE | Upregulated |
| 0,453011598 | LST1 | FALSE | Upregulated |
| 0,506385577 | LTA | FALSE | Upregulated |
| 1,79825292 | LTB | | Upregulated |
| -0,310710919 | LTC4S | FALSE | Downregulated |
| -0,841936307 | LTF | FALSE | Downregulated |
| 0,170488145 | LTV1 | FALSE | Upregulated |

| | | | |
|---|---|---|---|
| | LUCAT1 | | |
| -0,463502453 | LURAP1 | FALSE | Downregulated |
| 0,12995662 | LUZP2 | FALSE | Upregulated |
| | LUZP4 | | |
| 0,172566627 | LY6D | FALSE | Upregulated |
| -0,528021359 | LY6G5B | FALSE | Downregulated |
| 0,297490439 | LY6G5C | FALSE | Upregulated |
| 0 | LY6G6C | FALSE | Upregulated |
| | LY6G6D | | |
| | LY6G6E | | |
| | LY6G6F | | |
| 0 | LY6H | FALSE | Upregulated |
| 0,367878738 | LY6K | FALSE | Upregulated |
| 0,049436813 | LY75 | FALSE | Upregulated |
| -0,24298445 | LY86 | FALSE | Downregulated |
| 0,265905879 | LY86-AS1 | FALSE | Upregulated |
| -0,19699687 | LY96 | FALSE | Downregulated |
| -0,353251832 | LYAR | FALSE | Downregulated |
| 0,029723584 | LYG1 | FALSE | Upregulated |
| -0,449981366 | LYPD2 | FALSE | Downregulated |
| 0,041869959 | LYPD3 | FALSE | Upregulated |
| 0,269965519 | LYPD4 | FALSE | Upregulated |
| 0,268527987 | LYPD5 | FALSE | Upregulated |
| -0,068650093 | LYPD6 | FALSE | Downregulated |
| -0,122994482 | LYPD6B | FALSE | Downregulated |
| 0 | LYPD8 | FALSE | Upregulated |
| -0,508719679 | LYRM9 | FALSE | Downregulated |
| 1,136247265 | LYZL1 | Upregulated | |
| -0,011481061 | LYZL2 | FALSE | Downregulated |
| | LYZL4 | | |
| 0,088455781 | LYZL6 | FALSE | Upregulated |
| 0,206771267 | LZTS1 | FALSE | Upregulated |
| 0,581446323 | LZTS1-AS1 | FALSE | Upregulated |
| 1,121944838 | MAATS1 | Upregulated | |
| 0,087833183 | MAB21L3 | FALSE | Upregulated |
| 0,385272461 | MACC1 | FALSE | Upregulated |
| -0,003678263 | MACC1-AS1 | FALSE | Downregulated |
| 0,041010537 | MACROD2 | FALSE | Upregulated |
| 0 | MACROD2-IT1 | FALSE | Upregulated |
| -0,788054239 | MAD2L1 | FALSE | Downregulated |
| -0,672722241 | MAD2L1BP | FALSE | Downregulated |
| 0,823143373 | MADCAM1 | FALSE | Upregulated |
| -0,028865054 | MAEL | FALSE | Downregulated |
| -0,043208212 | MAFA | FALSE | Downregulated |
| 0,330635968 | MAFG-AS1 | FALSE | Upregulated |
| 0,138027869 | MAG | FALSE | Upregulated |
| | MAGEA1 | | |
| 0,012565453 | MAGEA10 | FALSE | Upregulated |
| | MAGEA10-MAGEA5 | | |
| | MAGEA11 | | |
| | MAGEA12 | | |

| | | | |
|---|---|---|---|
| | MAGEA2 | | |
| | MAGEA2B | | |
| 0,288284367 | MAGEA3 | FALSE | Upregulated |
| | MAGEA4 | | |
| -0,022819935 | MAGEA5 | FALSE | Downregulated |
| | MAGEA6 | | |
| -0,069206577 | MAGEA8 | FALSE | Downregulated |
| | MAGEA8-AS1 | | |
| 0 | MAGEA9 | FALSE | Upregulated |
| 0 | MAGEA9B | FALSE | Upregulated |
| | MAGEB1 | | |
| | MAGEB10 | | |
| | MAGEB16 | | |
| | MAGEB17 | | |
| | MAGEB18 | | |
| | MAGEB2 | | |
| | MAGEB3 | | |
| | MAGEB4 | | |
| | MAGEB5 | | |
| | MAGEB6 | | |
| | MAGEC1 | | |
| | MAGEC2 | | |
| | MAGEC3 | | |
| 0,486074479 | MAGED4 | FALSE | Upregulated |
| 0,414356206 | MAGED4B | FALSE | Upregulated |
| 0,293142823 | MAGEE1 | FALSE | Upregulated |
| 0,689407392 | MAGEE2 | FALSE | Upregulated |
| -0,57115698 | MAGEH1 | FALSE | Downregulated |
| -0,013754495 | MAGEL2 | FALSE | Downregulated |
| | MAGI2-AS2 | | |
| 0,160232575 | MAGOH | FALSE | Upregulated |
| -0,371003242 | MAGOHB | FALSE | Downregulated |
| 0,32245329 | MAL | FALSE | Upregulated |
| 0,05027885 | MALL | FALSE | Upregulated |
| 0,402706819 | MALRD1 | FALSE | Upregulated |
| -0,015875119 | MALSU1 | FALSE | Downregulated |
| -0,102362483 | MAMDC2-AS1 | FALSE | Downregulated |
| -0,698769291 | MAMSTR | FALSE | Downregulated |
| -0,90766887 | MAN1B1-AS1 | FALSE | Downregulated |
| -0,681566874 | MANBAL | FALSE | Downregulated |
| 0,635428733 | MANEA-AS1 | FALSE | Upregulated |
| 0,13970478 | MAP1LC3B2 | FALSE | Upregulated |
| -0,470275586 | MAP1LC3C | FALSE | Downregulated |
| 0 | MAP2K4P1 | FALSE | Upregulated |
| -0,14243746 | MAP2K6 | FALSE | Downregulated |
| -0,356925809 | MAP3K7CL | FALSE | Downregulated |
| 0,000161993 | MAP7D2 | FALSE | Upregulated |
| -0,501314395 | MAP9 | FALSE | Downregulated |
| 0,038925962 | MAPK12 | FALSE | Upregulated |
| 0,039997032 | MAPK15 | FALSE | Upregulated |
| 0,289499972 | MAPK4 | FALSE | Upregulated |

| | | | |
|---|---|---|---|
| 0,123390181 | MAPK8IP2 | FALSE | Upregulated |
| | MAPT-AS1 | | |
| 0,506520018 | MARCH10 | FALSE | Upregulated |
| | MARCH11 | | |
| 0 | MARCH4 | FALSE | Upregulated |
| 0,258383404 | MARK1 | FALSE | Upregulated |
| 0,144177969 | MARK2P9 | FALSE | Upregulated |
| -0,152382701 | MARS2 | FALSE | Downregulated |
| | MAS1 | | |
| 0 | MAS1L | FALSE | Upregulated |
| -0,016914195 | MAST1 | FALSE | Downregulated |
| -0,186076942 | MATK | FALSE | Downregulated |
| 1,150907573 | MATN1 | | Upregulated |
| 0,674126225 | MATN1-AS1 | FALSE | Upregulated |
| 1,265915014 | MATN3 | | Upregulated |
| 0 | MATN4 | FALSE | Upregulated |
| -0,308194368 | MB21D2 | FALSE | Downregulated |
| | MBD3L1 | | |
| | MBD3L2 | | |
| 0 | MBD3L3 | FALSE | Upregulated |
| 0 | MBD3L4 | FALSE | Upregulated |
| | MBD3L5 | | |
| 0,595700779 | MBIP | FALSE | Upregulated |
| 0,595012083 | MBOAT2 | FALSE | Upregulated |
| 0,548239105 | MC1R | FALSE | Upregulated |
| | MC2R | | |
| | MC3R | | |
| | MC4R | | |
| | MC5R | | |
| -0,436078845 | MCF2 | FALSE | Downregulated |
| 0,315937883 | MCF2L2 | FALSE | Upregulated |
| 0,696504748 | MCHR1 | FALSE | Upregulated |
| | MCHR2 | | |
| | MCHR2-AS1 | | |
| | MCIDAS | | |
| 0,852792473 | MCM10 | FALSE | Upregulated |
| -0,274846344 | MCM6 | FALSE | Downregulated |
| 1,20933263 | MCOLN2 | | Upregulated |
| 0,942797252 | MCOLN3 | FALSE | Upregulated |
| -0,222083523 | MCTP2 | FALSE | Downregulated |
| 0,692379482 | MDFI | FALSE | Upregulated |
| -0,255242687 | MDGA2 | FALSE | Downregulated |
| 0,33774576 | MDH1B | FALSE | Upregulated |
| 0,13540116 | MDM1 | FALSE | Upregulated |
| -0,478348332 | MDP1 | FALSE | Downregulated |
| 0 | MDS2 | FALSE | Upregulated |
| 0,492820151 | ME1 | FALSE | Upregulated |
| -0,753562973 | MECOM | FALSE | Downregulated |
| 0,078780352 | MED10 | FALSE | Upregulated |
| | MED15P9 | | |
| -0,084301931 | MED20 | FALSE | Downregulated |

| | | | |
|---|---|---|---|
| -0,190395123 | MED27 | FALSE | Downregulated |
| 0,201651397 | MED30 | FALSE | Upregulated |
| 0,016130129 | MED7 | FALSE | Upregulated |
| -0,263641964 | MED9 | FALSE | Downregulated |
| 0,268835309 | MEDAG | FALSE | Upregulated |
| -1,427099711 | MEF2B | | Downregulated |
| 0,955489912 | MEF2BNB-MEF2B | FALSE | Upregulated |
| 1,094204174 | MEF2C-AS1 | | Upregulated |
| 1,162034455 | MEFV | | Upregulated |
| 0 | MEG8 | FALSE | Upregulated |
| 0,65543267 | MEG9 | FALSE | Upregulated |
| -0,282782256 | MEGF10 | FALSE | Downregulated |
| 0,23780893 | MEI1 | FALSE | Upregulated |
| -0,92955574 | MEI4 | FALSE | Downregulated |
| | MEIG1 | | |
| | MEIS1-AS3 | | |
| 0,27144786 | MEIS3 | FALSE | Upregulated |
| 0,154711942 | MEIS3P1 | FALSE | Upregulated |
| 0,048223103 | MELK | FALSE | Upregulated |
| 0,118957215 | MEOX1 | FALSE | Upregulated |
| 0,100562758 | MEOX2 | FALSE | Upregulated |
| -0,013301533 | MEP1A | FALSE | Downregulated |
| -0,12770791 | MEP1B | FALSE | Downregulated |
| 0,673894649 | MESP1 | FALSE | Upregulated |
| 0,013161062 | MESP2 | FALSE | Upregulated |
| -0,495261012 | METAP1D | FALSE | Downregulated |
| -0,028932943 | METTL1 | FALSE | Downregulated |
| 0,208493233 | METTL10 | FALSE | Upregulated |
| | METTL11B | | |
| 0,152169829 | METTL21B | FALSE | Upregulated |
| -0,265422029 | METTL21C | FALSE | Downregulated |
| 0,134113362 | METTL21EP | FALSE | Upregulated |
| 0,541946007 | METTL24 | FALSE | Upregulated |
| -0,110747844 | METTL25 | FALSE | Downregulated |
| 0,436897034 | METTL2A | FALSE | Upregulated |
| 0,210184215 | METTL4 | FALSE | Upregulated |
| -0,13767409 | METTL5 | FALSE | Downregulated |
| -0,440298051 | MEX3B | FALSE | Downregulated |
| -0,282906936 | MFAP2 | FALSE | Downregulated |
| 0,858392039 | MFAP5 | FALSE | Upregulated |
| 0,532935626 | MFI2-AS1 | FALSE | Upregulated |
| 0,007651649 | MFRP | FALSE | Upregulated |
| 0,887993302 | MFSD2B | FALSE | Upregulated |
| -0,169778338 | MFSD5 | FALSE | Downregulated |
| -0,135754785 | MFSD6L | FALSE | Downregulated |
| 0,077982489 | MGAM | FALSE | Upregulated |
| 0,049356895 | MGARP | FALSE | Upregulated |
| 0,789800317 | MGAT3 | FALSE | Upregulated |
| 0,324795199 | MGAT4C | FALSE | Upregulated |
| 0,021524351 | MGAT5B | FALSE | Upregulated |
| -0,443058168 | MGC15885 | FALSE | Downregulated |

| | | | |
|---|---|---|---|
| -0,217212656 | MGC16025 | FALSE | Downregulated |
| -0,291540341 | MGC16142 | FALSE | Downregulated |
| -0,870825588 | MGC27382 | FALSE | Downregulated |
| | MGC2889 | | |
| -0,612734589 | MGC32805 | FALSE | Downregulated |
| -0,930541381 | MGC34034 | FALSE | Downregulated |
| 0,051483871 | MGC34796 | FALSE | Upregulated |
| 0 | MGC34800 | FALSE | Upregulated |
| 0,835632763 | MGC39372 | FALSE | Upregulated |
| -0,489136403 | MGC39545 | FALSE | Downregulated |
| | MGC39584 | | |
| 0,250767873 | MGC42157 | FALSE | Upregulated |
| 0,272626305 | MGC45800 | FALSE | Upregulated |
| -0,143920439 | MGC45922 | FALSE | Downregulated |
| 1,514207083 | MGC50722 | | Upregulated |
| 0,936538668 | MGC57346 | FALSE | Upregulated |
| -0,59732564 | MGC72080 | FALSE | Downregulated |
| 0,037084434 | MIA | FALSE | Upregulated |
| -0,142762222 | MIA-RAB4B | FALSE | Downregulated |
| -0,170479244 | MICALCL | FALSE | Downregulated |
| 0,740179571 | MICB | FALSE | Upregulated |
| 0,460851221 | MID1-2 | FALSE | Upregulated |
| -0,292656911 | MILR1 | FALSE | Downregulated |
| -0,014724123 | MIMT1 | FALSE | Downregulated |
| -0,695424046 | MINOS1-NBL1 | FALSE | Downregulated |
| 0,331710763 | MIOX | FALSE | Upregulated |
| -0,64959475 | MIP | FALSE | Downregulated |
| 0,139401956 | MIPEP | FALSE | Upregulated |
| -0,486883945 | MIPEPP3 | FALSE | Downregulated |
| | MIR1-1 | | |
| | MIR1-2 | | |
| | MIR100 | | |
| | MIR101-1 | | |
| | MIR101-2 | | |
| | MIR103A1 | | |
| | MIR103A2 | | |
| | MIR103B1 | | |
| | MIR103B2 | | |
| | MIR105-1 | | |
| | MIR105-2 | | |
| | MIR106A | | |
| | MIR106B | | |
| | MIR107 | | |
| | MIR10A | | |
| | MIR10B | | |
| | MIR1178 | | |
| | MIR1179 | | |
| | MIR1180 | | |
| | MIR1181 | | |
| | MIR1182 | | |
| | MIR1183 | | |

MIR1184-1
MIR1184-2
MIR1184-3
MIR1185-1
MIR1185-2
MIR1193
MIR1197
MIR1199
MIR1200
MIR1202
MIR1203
MIR1204
MIR1205
MIR1206
MIR1207
MIR1208
MIR122
MIR1224
MIR1225
MIR1226
MIR1227
MIR1228
MIR1229
MIR1231
MIR1233-1
MIR1233-2
MIR1236
MIR1237
MIR1238
MIR124-1
MIR124-2
MIR124-3
MIR1243
MIR1244-1
MIR1244-2
MIR1244-3
MIR1245A
MIR1245B
MIR1246
MIR1247
MIR1249
MIR1250
MIR1251
MIR1252
MIR1253
MIR1254-1
MIR1254-2
MIR1255A
MIR1255B1
MIR1255B2
MIR1256

MIR1257
MIR1258
MIR125A
MIR125B1
MIR125B2
MIR126
MIR1260A
MIR1260B
MIR1261
MIR1262
MIR1263
MIR1264
MIR1265
MIR1266
MIR1267
MIR1268A
MIR1268B
MIR1269A
MIR1269B
MIR127
MIR1270-1
MIR1271
MIR1272
MIR1273A
MIR1273C
MIR1273D
MIR1273F
MIR1273G
MIR1273H
MIR1275
MIR1276
MIR1277
MIR1278
MIR1279
MIR128-1
MIR128-2
MIR1281
MIR1282
MIR1283-1
MIR1283-2
MIR1284
MIR1285-1
MIR1285-2
MIR1286
MIR1287
MIR1288
MIR1289-1
MIR1289-2
MIR129-1
MIR129-2
MIR1290

| | | | |
|---|---|---|---|
| | MIR1292 | | |
| | MIR1293 | | |
| | MIR1294 | | |
| | MIR1295A | | |
| | MIR1295B | | |
| | MIR1296 | | |
| | MIR1297 | | |
| | MIR1298 | | |
| | MIR1299 | | |
| | MIR1301 | | |
| | MIR1302-1 | | |
| | MIR1302-10 | | |
| | MIR1302-11 | | |
| | MIR1302-2 | | |
| | MIR1302-3 | | |
| | MIR1302-4 | | |
| 0 | MIR1302-5 | FALSE | Upregulated |
| | MIR1302-6 | | |
| | MIR1302-7 | | |
| | MIR1302-8 | | |
| | MIR1302-9 | | |
| | MIR1303 | | |
| | MIR1304 | | |
| | MIR1305 | | |
| | MIR1306 | | |
| 0,291340714 | MIR1307 | FALSE | Upregulated |
| | MIR130A | | |
| | MIR130B | | |
| | MIR132 | | |
| | MIR1321 | | |
| | MIR1322 | | |
| | MIR1323 | | |
| | MIR1324 | | |
| | MIR133A1 | | |
| | MIR133A2 | | |
| | MIR133B | | |
| | MIR134 | | |
| | MIR1343 | | |
| | MIR135A1 | | |
| | MIR135A2 | | |
| | MIR135B | | |
| | MIR136 | | |
| | MIR137 | | |
| | MIR137HG | | |
| | MIR138-1 | | |
| | MIR138-2 | | |
| | MIR139 | | |
| | MIR140 | | |
| | MIR141 | | |
| | MIR142 | | |
| | MIR143 | | |

| | | | |
|---|---|---|---|
| -0,398577948 | MIR143HG | FALSE | Downregulated |
| | MIR144 | | |
| | MIR145 | | |
| | MIR1468 | | |
| | MIR1469 | | |
| | MIR146A | | |
| | MIR146B | | |
| | MIR1470 | | |
| | MIR1471 | | |
| | MIR147A | | |
| | MIR147B | | |
| | MIR148A | | |
| | MIR148B | | |
| | MIR149 | | |
| | MIR150 | | |
| | MIR151A | | |
| | MIR151B | | |
| | MIR152 | | |
| | MIR153-1 | | |
| | MIR153-2 | | |
| | MIR1537 | | |
| | MIR1538 | | |
| | MIR1539 | | |
| | MIR154 | | |
| | MIR155 | | |
| | MIR1587 | | |
| | MIR15A | | |
| | MIR15B | | |
| | MIR16-1 | | |
| | MIR16-2 | | |
| | MIR17 | | |
| | MIR181A1 | | |
| 0 | MIR181A1HG | FALSE | Upregulated |
| 0,41482555 | MIR181A2 | FALSE | Upregulated |
| | MIR181B1 | | |
| | MIR181B2 | | |
| | MIR181C | | |
| | MIR181D | | |
| | MIR182 | | |
| | MIR1825 | | |
| | MIR1827 | | |
| | MIR183 | | |
| | MIR184 | | |
| | MIR185 | | |
| | MIR186 | | |
| | MIR187 | | |
| | MIR188 | | |
| | MIR18A | | |
| | MIR18B | | |
| | MIR1908 | | |
| | MIR1909 | | |

| | | | |
|---|---|---|---|
| | MIR190A | | |
| | MIR190B | | |
| | MIR191 | | |
| | MIR1910 | | |
| | MIR1911 | | |
| | MIR1912 | | |
| | MIR1913 | | |
| | MIR1914 | | |
| | MIR1915 | | |
| 0 | MIR192 | FALSE | Upregulated |
| | MIR193A | | |
| | MIR193B | | |
| | MIR194-1 | | |
| | MIR194-2 | | |
| | MIR195 | | |
| | MIR196A1 | | |
| | MIR196A2 | | |
| | MIR196B | | |
| | MIR197 | | |
| | MIR1972-1 | | |
| | MIR1972-2 | | |
| | MIR1973 | | |
| | MIR1976 | | |
| | MIR198 | | |
| | MIR199A1 | | |
| | MIR199A2 | | |
| | MIR199B | | |
| | MIR19A | | |
| | MIR19B1 | | |
| | MIR19B2 | | |
| | MIR200A | | |
| | MIR200B | | |
| | MIR200C | | |
| | MIR202 | | |
| | MIR202HG | | |
| | MIR203 | | |
| | MIR204 | | |
| | MIR205 | | |
| | MIR2052 | | |
| | MIR2053 | | |
| | MIR2054 | | |
| | MIR205HG | | |
| | MIR206 | | |
| | MIR208A | | |
| | MIR208B | | |
| | MIR20A | | |
| | MIR20B | | |
| | MIR21 | | |
| | MIR210 | | |
| | MIR211 | | |
| | MIR2110 | | |

| | | | |
|---|---|---|---|
| | MIR2113 | | |
| | MIR2114 | | |
| | MIR2115 | | |
| | MIR2116 | | |
| | MIR2117 | | |
| | MIR212 | | |
| 0,297178435 | MIR214 | FALSE | Upregulated |
| | MIR215 | | |
| | MIR216A | | |
| | MIR216B | | |
| | MIR217 | | |
| | MIR218-1 | | |
| | MIR218-2 | | |
| | MIR219-1 | | |
| | MIR219-2 | | |
| | MIR22 | | |
| | MIR221 | | |
| | MIR222 | | |
| | MIR223 | | |
| | MIR224 | | |
| | MIR2276 | | |
| | MIR2277 | | |
| | MIR2278 | | |
| | MIR2355 | | |
| | MIR2392 | | |
| | MIR23A | | |
| | MIR23B | | |
| | MIR23C | | |
| | MIR24-1 | | |
| | MIR24-2 | | |
| | MIR2467 | | |
| | MIR25 | | |
| | MIR2681 | | |
| | MIR2682 | | |
| | MIR26A1 | | |
| | MIR26A2 | | |
| | MIR26B | | |
| | MIR27A | | |
| | MIR27B | | |
| | MIR28 | | |
| | MIR2861 | | |
| | MIR2909 | | |
| | MIR296 | | |
| | MIR2964A | | |
| | MIR297 | | |
| | MIR298 | | |
| | MIR299 | | |
| | MIR29A | | |
| | MIR29B1 | | |
| | MIR29B2 | | |
| | MIR29C | | |

MIR300
MIR301A
MIR301B
MIR302A
MIR302B
MIR302C
MIR302D
MIR302E
MIR302F
MIR3064
MIR3065
MIR3074
MIR30A
MIR30B
MIR30C1
MIR30C2
MIR30D
MIR30E
MIR31
MIR3115
MIR3116-1
MIR3116-2
MIR3117
MIR3118-1
MIR3118-2
MIR3118-3
MIR3118-4
MIR3118-5
MIR3118-6
MIR3119-1
MIR3119-2
MIR3120
MIR3121
MIR3122
MIR3123
MIR3124
MIR3125
MIR3126
MIR3127
MIR3128
MIR3129
MIR3130-1
MIR3130-2
MIR3131
MIR3132
MIR3133
MIR3134
MIR3135A
MIR3135B
MIR3136
MIR3137

MIR3138
MIR3139
MIR3140
MIR3141
MIR3142
MIR3143
MIR3144
MIR3145
MIR3146
MIR3147
MIR3148
MIR3150A
MIR3150B
MIR3151
MIR3152
MIR3153
MIR3154
MIR3155A
MIR3155B
MIR3156-1
MIR3156-2
MIR3156-3
MIR3157
MIR3158-1
MIR3158-2
MIR3159
MIR3160-1
MIR3160-2
MIR3161
MIR3162
MIR3163
MIR3164
MIR3165
MIR3166
MIR3167
MIR3168
MIR3169
MIR3170
MIR3173
MIR3174
MIR3175
MIR3176
MIR3177
MIR3178
MIR3179-1
MIR3179-2
MIR3179-3
MIR3180-1
MIR3180-2
MIR3180-3
MIR3180-4

| | | | |
|---|---|---|---|
| | MIR3180-5 | | |
| | MIR3182 | | |
| | MIR3183 | | |
| | MIR3184 | | |
| | MIR3185 | | |
| | MIR3186 | | |
| | MIR3187 | | |
| | MIR3188 | | |
| | MIR3189 | | |
| | MIR3190 | | |
| | MIR3191 | | |
| | MIR3192 | | |
| | MIR3193 | | |
| | MIR3194 | | |
| | MIR3196 | | |
| | MIR3197 | | |
| | MIR3198-1 | | |
| | MIR3198-2 | | |
| | MIR3199-1 | | |
| | MIR3199-2 | | |
| 0,277833435 | MIR31HG | FALSE | Upregulated |
| | MIR32 | | |
| | MIR3200 | | |
| | MIR3201 | | |
| | MIR3202-1 | | |
| | MIR3202-2 | | |
| | MIR320A | | |
| | MIR320B1 | | |
| | MIR320B2 | | |
| | MIR320C1 | | |
| | MIR320C2 | | |
| | MIR320D1 | | |
| | MIR320D2 | | |
| | MIR320E | | |
| | MIR323A | | |
| | MIR323B | | |
| | MIR324 | | |
| | MIR325 | | |
| | MIR326 | | |
| | MIR328 | | |
| | MIR329-1 | | |
| | MIR329-2 | | |
| | MIR330 | | |
| | MIR331 | | |
| | MIR335 | | |
| | MIR337 | | |
| | MIR338 | | |
| | MIR339 | | |
| | MIR33A | | |
| | MIR33B | | |
| | MIR340 | | |

| | | | |
|---|---|---|---|
| | MIR342 | | |
| | MIR345 | | |
| | MIR346 | | |
| | MIR34A | | |
| | MIR34B | | |
| | MIR34C | | |
| | MIR3529 | | |
| | MIR3545 | | |
| | MIR3591 | | |
| | MIR3605 | | |
| | MIR3606 | | |
| | MIR3607 | | |
| | MIR3609 | | |
| | MIR361 | | |
| | MIR3610 | | |
| | MIR3611 | | |
| | MIR3612 | | |
| | MIR3613 | | |
| | MIR3614 | | |
| | MIR3615 | | |
| | MIR3616 | | |
| | MIR3617 | | |
| | MIR3618 | | |
| | MIR3619 | | |
| | MIR362 | | |
| | MIR3620 | | |
| | MIR3621 | | |
| | MIR3622A | | |
| | MIR3622B | | |
| | MIR363 | | |
| | MIR3646 | | |
| | MIR3649 | | |
| | MIR3650 | | |
| | MIR3651 | | |
| | MIR3652 | | |
| | MIR3653 | | |
| | MIR3654 | | |
| | MIR3655 | | |
| | MIR3656 | | |
| | MIR3657 | | |
| | MIR3658 | | |
| | MIR3659 | | |
| | MIR365A | | |
| | MIR365B | | |
| | MIR3660 | | |
| 0,288284367 | MIR3661 | FALSE | Upregulated |
| | MIR3662 | | |
| | MIR3663 | | |
| | MIR3664 | | |
| | MIR3665 | | |
| | MIR3666 | | |

MIR3667
MIR3668
MIR367
MIR3670-1
MIR3670-2
MIR3671
MIR3672
MIR3674
MIR3675
MIR3677
MIR3678
MIR3679
MIR3680-1
MIR3680-2
MIR3681
MIR3682
MIR3683
MIR3684
MIR3685
MIR3686
MIR3687
MIR3688-1
MIR3688-2
MIR3689A
MIR3689B
MIR3689C
MIR3689D1
MIR3689D2
MIR3689E
MIR3689F
MIR369
MIR3690
MIR3690-2
MIR3691
MIR3692
MIR370
MIR3713
MIR3714
MIR371A
MIR371B
MIR372
MIR373
MIR374A
MIR374B
MIR374C
MIR375
MIR376A1
MIR376A2
MIR376B
MIR376C
MIR377

| | | | |
|---|---|---|---|
| | MIR378A | | |
| | MIR378B | | |
| | MIR378C | | |
| | MIR378D1 | | |
| | MIR378D2 | | |
| | MIR378E | | |
| | MIR378F | | |
| | MIR378G | | |
| | MIR378H | | |
| | MIR378I | | |
| | MIR378J | | |
| | MIR379 | | |
| | MIR380 | | |
| | MIR381 | | |
| 0 | MIR381HG | FALSE | Upregulated |
| | MIR382 | | |
| | MIR383 | | |
| | MIR384 | | |
| | MIR3907 | | |
| | MIR3908 | | |
| | MIR3909 | | |
| | MIR3910-1 | | |
| | MIR3910-2 | | |
| | MIR3911 | | |
| | MIR3912 | | |
| | MIR3913-1 | | |
| | MIR3913-2 | | |
| | MIR3914-1 | | |
| | MIR3914-2 | | |
| | MIR3915 | | |
| | MIR3916 | | |
| | MIR3917 | | |
| | MIR3918 | | |
| | MIR3919 | | |
| | MIR3920 | | |
| | MIR3921 | | |
| | MIR3922 | | |
| | MIR3923 | | |
| | MIR3924 | | |
| | MIR3925 | | |
| | MIR3926-1 | | |
| | MIR3926-2 | | |
| | MIR3927 | | |
| | MIR3928 | | |
| | MIR3929 | | |
| | MIR3934 | | |
| | MIR3935 | | |
| | MIR3936 | | |
| | MIR3937 | | |
| | MIR3938 | | |
| 0,007822289 | MIR3939 | FALSE | Upregulated |

MIR3940
MIR3941
MIR3942
MIR3943
MIR3944
MIR3945
MIR3960
MIR3972
MIR3973
MIR3974
MIR3975
MIR3976
MIR3977
MIR3978
MIR409
MIR410
MIR411
MIR412
MIR421
MIR422A
MIR423
MIR424
MIR425
MIR4251
MIR4252
MIR4253
MIR4254
MIR4255
MIR4256
MIR4257
MIR4258
MIR4260
MIR4261
MIR4262
MIR4263
MIR4264
MIR4265
MIR4266
MIR4267
MIR4268
MIR4269
MIR4270
MIR4271
MIR4272
MIR4273
MIR4274
MIR4275
MIR4276
MIR4277
MIR4278
MIR4279

MIR4280
MIR4281
MIR4282
MIR4283-1
MIR4283-2
MIR4284
MIR4286
MIR4287
MIR4288
MIR4289
MIR429
MIR4290
MIR4291
MIR4292
MIR4293
MIR4294
MIR4295
MIR4296
MIR4297
MIR4298
MIR4299
MIR4300
MIR4301
MIR4302
MIR4303
MIR4304
MIR4305
MIR4306
MIR4307
MIR4308
MIR4309
MIR431
MIR4310
MIR4311
MIR4312
MIR4313
MIR4314
MIR4315-1
MIR4315-2
MIR4316
MIR4317
MIR4318
MIR4319
MIR432
MIR4320
MIR4321
MIR4322
MIR4323
MIR4324
MIR4325
MIR4326

| | | | |
|---|---|---|---|
| | MIR4327 | | |
| | MIR4328 | | |
| | MIR4329 | | |
| | MIR433 | | |
| | MIR4330 | | |
| | MIR4417 | | |
| | MIR4418 | | |
| | MIR4419A | | |
| | MIR4419B | | |
| | MIR4420 | | |
| | MIR4421 | | |
| | MIR4422 | | |
| | MIR4423 | | |
| | MIR4424 | | |
| | MIR4425 | | |
| | MIR4426 | | |
| | MIR4427 | | |
| | MIR4428 | | |
| | MIR4429 | | |
| | MIR4430 | | |
| | MIR4431 | | |
| | MIR4432 | | |
| | MIR4433 | | |
| | MIR4433B | | |
| | MIR4434 | | |
| | MIR4435-1 | | |
| 0,999686864 | MIR4435-1HG | FALSE | Upregulated |
| | MIR4435-2 | | |
| | MIR4436A | | |
| | MIR4436B1 | | |
| | MIR4436B2 | | |
| | MIR4437 | | |
| | MIR4438 | | |
| | MIR4439 | | |
| | MIR4440 | | |
| | MIR4441 | | |
| | MIR4442 | | |
| | MIR4443 | | |
| | MIR4444-1 | | |
| | MIR4444-2 | | |
| | MIR4445 | | |
| | MIR4446 | | |
| | MIR4447 | | |
| | MIR4448 | | |
| | MIR4449 | | |
| | MIR4450 | | |
| | MIR4451 | | |
| | MIR4452 | | |
| | MIR4453 | | |
| | MIR4454 | | |
| | MIR4455 | | |

| | | | |
|---|---|---|---|
| | MIR4456 | | |
| | MIR4457 | | |
| | MIR4458 | | |
| 0,57309588 | MIR4458HG | FALSE | Upregulated |
| | MIR4459 | | |
| | MIR4460 | | |
| | MIR4461 | | |
| | MIR4462 | | |
| | MIR4463 | | |
| | MIR4464 | | |
| | MIR4465 | | |
| | MIR4466 | | |
| | MIR4467 | | |
| | MIR4468 | | |
| | MIR4469 | | |
| | MIR4470 | | |
| | MIR4471 | | |
| | MIR4472-1 | | |
| | MIR4472-2 | | |
| | MIR4473 | | |
| | MIR4474 | | |
| | MIR4475 | | |
| | MIR4476 | | |
| | MIR4477A | | |
| | MIR4477B | | |
| | MIR4478 | | |
| | MIR4479 | | |
| | MIR448 | | |
| | MIR4480 | | |
| | MIR4481 | | |
| | MIR4482-1 | | |
| | MIR4483 | | |
| | MIR4484 | | |
| | MIR4485 | | |
| | MIR4486 | | |
| | MIR4487 | | |
| | MIR4488 | | |
| | MIR4489 | | |
| | MIR4490 | | |
| | MIR4491 | | |
| | MIR4492 | | |
| | MIR4493 | | |
| | MIR4494 | | |
| | MIR4495 | | |
| | MIR4496 | | |
| | MIR4497 | | |
| | MIR4498 | | |
| | MIR4499 | | |
| | MIR449A | | |
| | MIR449B | | |
| | MIR449C | | |

| | | | |
|---|---|---|---|
| | MIR4500 | | |
| | MIR4501 | | |
| | MIR4502 | | |
| | MIR4503 | | |
| | MIR4504 | | |
| | MIR4505 | | |
| | MIR4506 | | |
| | MIR4507 | | |
| | MIR4508 | | |
| | MIR4509-1 | | |
| | MIR4509-2 | | |
| | MIR4509-3 | | |
| | MIR450A1 | | |
| | MIR450A2 | | |
| | MIR450B | | |
| | MIR4510 | | |
| | MIR4511 | | |
| | MIR4512 | | |
| | MIR4513 | | |
| | MIR4514 | | |
| | MIR4515 | | |
| | MIR4516 | | |
| | MIR4517 | | |
| | MIR4518 | | |
| | MIR4519 | | |
| | MIR451A | | |
| | MIR451B | | |
| | MIR452 | | |
| | MIR4520A | | |
| | MIR4520B | | |
| | MIR4521 | | |
| | MIR4522 | | |
| | MIR4523 | | |
| | MIR4524A | | |
| 0,383359179 | MIR4524B | FALSE | Upregulated |
| | MIR4525 | | |
| | MIR4526 | | |
| | MIR4527 | | |
| | MIR4529 | | |
| | MIR4530 | | |
| | MIR4531 | | |
| | MIR4532 | | |
| | MIR4533 | | |
| | MIR4534 | | |
| | MIR4535 | | |
| | MIR4536-1 | | |
| | MIR454 | | |
| | MIR4540 | | |
| | MIR455 | | |
| | MIR4632 | | |
| | MIR4633 | | |

MIR4634
MIR4635
MIR4636
MIR4637
MIR4638
MIR4639
MIR4640
MIR4641
MIR4642
MIR4643
MIR4644
MIR4645
MIR4646
MIR4647
MIR4648
MIR4649
MIR4650-1
MIR4650-2
MIR4651
MIR4652
MIR4653
MIR4654
MIR4655
MIR4656
MIR4657
MIR4658
MIR4659A
MIR4659B
MIR4660
MIR4661
MIR4662A
MIR4662B
MIR4663
MIR4664
MIR4665
MIR4666A
MIR4666B
MIR4667
MIR4668
MIR4669
MIR4670
MIR4671
MIR4672
MIR4673
MIR4674
MIR4675
MIR4676
MIR4677
MIR4678
MIR4679-1
MIR4679-2

| | | | |
|---|---|---|---|
| | MIR4680 | | |
| | MIR4681 | | |
| | MIR4682 | | |
| | MIR4683 | | |
| | MIR4684 | | |
| | MIR4685 | | |
| | MIR4686 | | |
| | MIR4687 | | |
| | MIR4688 | | |
| | MIR4689 | | |
| | MIR4690 | | |
| | MIR4691 | | |
| | MIR4692 | | |
| | MIR4693 | | |
| | MIR4694 | | |
| | MIR4695 | | |
| | MIR4696 | | |
| | MIR4697 | | |
| 0,65823327 | MIR4697HG | FALSE | Upregulated |
| | MIR4698 | | |
| | MIR4699 | | |
| | MIR4700 | | |
| | MIR4701 | | |
| | MIR4703 | | |
| | MIR4704 | | |
| | MIR4705 | | |
| | MIR4706 | | |
| | MIR4707 | | |
| | MIR4708 | | |
| | MIR4709 | | |
| | MIR4710 | | |
| | MIR4711 | | |
| | MIR4712 | | |
| | MIR4713 | | |
| | MIR4714 | | |
| | MIR4715 | | |
| | MIR4716 | | |
| | MIR4717 | | |
| | MIR4718 | | |
| | MIR4719 | | |
| | MIR4720 | | |
| | MIR4721 | | |
| | MIR4722 | | |
| | MIR4723 | | |
| | MIR4724 | | |
| | MIR4725 | | |
| | MIR4726 | | |
| | MIR4727 | | |
| | MIR4728 | | |
| | MIR4729 | | |
| | MIR4730 | | |

MIR4731
MIR4732
MIR4733
MIR4734
MIR4735
MIR4736
MIR4737
MIR4738
MIR4739
MIR4740
MIR4741
MIR4742
MIR4743
MIR4744
MIR4745
MIR4746
MIR4747
MIR4748
MIR4749
MIR4750
MIR4751
MIR4752
MIR4753
MIR4754
MIR4755
MIR4756
MIR4757
MIR4758
MIR4759
MIR4760
MIR4761
MIR4762
MIR4763
MIR4764
MIR4765
MIR4766
MIR4767
MIR4768
MIR4769
MIR4770
MIR4771-1
MIR4771-2
MIR4772
MIR4773-1
MIR4773-2
MIR4774
MIR4775
MIR4776-1
MIR4776-2
MIR4777
MIR4778

| | | | | |
|---|---|---|---|---|
| | MIR4779 | | | |
| | MIR4780 | | | |
| | MIR4781 | | | |
| | MIR4782 | | | |
| | MIR4783 | | | |
| | MIR4784 | | | |
| | MIR4785 | | | |
| | MIR4786 | | | |
| | MIR4787 | | | |
| | MIR4788 | | | |
| | MIR4789 | | | |
| | MIR4790 | | | |
| | MIR4791 | | | |
| | MIR4792 | | | |
| | MIR4793 | | | |
| | MIR4794 | | | |
| | MIR4795 | | | |
| | MIR4796 | | | |
| | MIR4797 | | | |
| | MIR4798 | | | |
| | MIR4799 | | | |
| | MIR4800 | | | |
| | MIR4801 | | | |
| | MIR4802 | | | |
| | MIR4803 | | | |
| | MIR4804 | | | |
| | MIR483 | | | |
| | MIR484 | | | |
| | MIR485 | | | |
| | MIR486 | | | |
| | MIR486-2 | | | |
| | MIR487A | | | |
| | MIR487B | | | |
| | MIR488 | | | |
| | MIR489 | | | |
| | MIR490 | | | |
| | MIR491 | | | |
| | MIR492 | | | |
| | MIR493 | | | |
| | MIR494 | | | |
| | MIR495 | | | |
| | MIR496 | | | |
| | MIR497 | | | |
| 0,452468921 | | MIR497HG | FALSE | Upregulated |
| | MIR498 | | | |
| | MIR4999 | | | |
| -0,002638994 | | MIR499A | FALSE | Downregulated |
| | MIR499B | | | |
| | MIR5000 | | | |
| | MIR5001 | | | |
| | MIR5002 | | | |

| | | | |
|---|---|---|---|
| | MIR5003 | | |
| | MIR5004 | | |
| | MIR5006 | | |
| | MIR5007 | | |
| | MIR5008 | | |
| | MIR5009 | | |
| | MIR500A | | |
| | MIR500B | | |
| | MIR501 | | |
| | MIR5010 | | |
| | MIR5011 | | |
| | MIR502 | | |
| | MIR503 | | |
| 0,244168313 | MIR503HG | FALSE | Upregulated |
| | MIR504 | | |
| | MIR5047 | | |
| | MIR505 | | |
| | MIR506 | | |
| | MIR507 | | |
| | MIR508 | | |
| | MIR5087 | | |
| | MIR5088 | | |
| | MIR5089 | | |
| | MIR509-1 | | |
| | MIR509-2 | | |
| | MIR509-3 | | |
| | MIR5090 | | |
| | MIR5091 | | |
| | MIR5092 | | |
| | MIR5093 | | |
| | MIR5094 | | |
| | MIR5095 | | |
| | MIR5096 | | |
| | MIR510 | | |
| | MIR5100 | | |
| | MIR511-1 | | |
| | MIR512-1 | | |
| | MIR512-2 | | |
| | MIR513A1 | | |
| 0 | MIR513A2 | FALSE | Upregulated |
| | MIR513B | | |
| | MIR513C | | |
| | MIR514A1 | | |
| | MIR514A2 | | |
| | MIR514A3 | | |
| | MIR514B | | |
| | MIR515-1 | | |
| | MIR515-2 | | |
| | MIR516A1 | | |
| | MIR516A2 | | |
| | MIR516B1 | | |

| | | | |
|---|---|---|---|
| | MIR516B2 | | |
| | MIR517A | | |
| | MIR517B | | |
| | MIR517C | | |
| | MIR5186 | | |
| | MIR5187 | | |
| | MIR5188 | | |
| 0 | MIR5189 | FALSE | Upregulated |
| | MIR518A1 | | |
| | MIR518A2 | | |
| | MIR518B | | |
| | MIR518C | | |
| | MIR518D | | |
| | MIR518E | | |
| | MIR518F | | |
| | MIR5190 | | |
| | MIR5191 | | |
| | MIR5192 | | |
| | MIR5193 | | |
| 0,405839243 | MIR5194 | FALSE | Upregulated |
| | MIR5196 | | |
| | MIR5197 | | |
| | MIR519A1 | | |
| | MIR519A2 | | |
| | MIR519B | | |
| | MIR519C | | |
| | MIR519D | | |
| | MIR519E | | |
| | MIR520A | | |
| | MIR520B | | |
| | MIR520C | | |
| | MIR520D | | |
| | MIR520E | | |
| | MIR520F | | |
| | MIR520G | | |
| | MIR520H | | |
| | MIR521-1 | | |
| | MIR521-2 | | |
| | MIR522 | | |
| | MIR523 | | |
| | MIR524 | | |
| | MIR525 | | |
| | MIR526A1 | | |
| | MIR526A2 | | |
| | MIR526B | | |
| | MIR527 | | |
| | MIR539 | | |
| | MIR541 | | |
| | MIR542 | | |
| | MIR543 | | |
| | MIR544A | | |

MIR544B
MIR545
MIR548A1
MIR548A2
MIR548A3
MIR548AA1
MIR548AA2
MIR548AB
MIR548AC
MIR548AD
MIR548AE1
MIR548AE2
MIR548AG1
MIR548AG2
MIR548AH
MIR548AI
MIR548AJ1
MIR548AJ2
MIR548AK
MIR548AL
MIR548AM
MIR548AN
MIR548AO
MIR548AP
MIR548AQ
MIR548AR
MIR548AS
MIR548AT
MIR548AU
MIR548AV
MIR548AW
MIR548AX
MIR548AY
MIR548AZ
MIR548B
MIR548BA
MIR548C
MIR548D1
MIR548D2
MIR548E
MIR548F1
MIR548F2
MIR548F3
MIR548F4
MIR548F5
MIR548G
MIR548H1
MIR548H2
MIR548H3
MIR548H4
MIR548H5

| | | | |
|---|---|---|---|
| 0,288284367 | MIR548I1 | FALSE | Upregulated |
| | MIR548I2 | | |
| | MIR548I3 | | |
| | MIR548I4 | | |
| | MIR548J | | |
| | MIR548K | | |
| | MIR548L | | |
| | MIR548M | | |
| | MIR548N | | |
| 0,137088329 | MIR548O | FALSE | Upregulated |
| | MIR548O2 | | |
| | MIR548P | | |
| | MIR548Q | | |
| | MIR548S | | |
| | MIR548T | | |
| | MIR548U | | |
| | MIR548V | | |
| | MIR548W | | |
| | MIR548X | | |
| | MIR548X2 | | |
| | MIR548Y | | |
| | MIR548Z | | |
| | MIR549 | | |
| | MIR550A1 | | |
| | MIR550A2 | | |
| | MIR550A3 | | |
| | MIR550B1 | | |
| | MIR550B2 | | |
| | MIR551A | | |
| | MIR551B | | |
| | MIR552 | | |
| | MIR553 | | |
| | MIR554 | | |
| | MIR555 | | |
| | MIR556 | | |
| | MIR557 | | |
| | MIR5572 | | |
| | MIR5579 | | |
| | MIR558 | | |
| | MIR5580 | | |
| | MIR5581 | | |
| | MIR5582 | | |
| | MIR5583-1 | | |
| | MIR5583-2 | | |
| | MIR5584 | | |
| | MIR5585 | | |
| | MIR5586 | | |
| | MIR5587 | | |
| | MIR5588 | | |
| | MIR5589 | | |
| | MIR559 | | |

MIR5590
MIR5591
MIR561
MIR562
MIR563
MIR564
MIR566
MIR567
MIR568
MIR5680
MIR5681A
MIR5681B
MIR5682
MIR5683
MIR5684
MIR5685
MIR5687
MIR5688
MIR5689
MIR569
MIR5690
MIR5691
MIR5692A1
MIR5692A2
MIR5692B
MIR5692C1
MIR5692C2
MIR5693
MIR5694
MIR5695
MIR5696
MIR5697
MIR5698
MIR5699
MIR570
MIR5700
MIR5701-1
MIR5701-2
MIR5702
MIR5703
MIR5704
MIR5705
MIR5706
MIR5707
MIR5708
MIR571
MIR572
MIR573
MIR5739
MIR574
MIR575

MIR576
MIR577
MIR578
MIR5787
MIR579
MIR580
MIR581
MIR582
MIR583
MIR584
MIR585
MIR586
MIR587
MIR588
MIR589
MIR590
MIR591
MIR592
MIR593
MIR595
MIR596
MIR597
MIR598
MIR599
MIR600
MIR601
MIR602
MIR603
MIR604
MIR605
MIR606
MIR6068
MIR6069
MIR607
MIR6070
MIR6071
MIR6072
MIR6073
MIR6074
MIR6075
MIR6076
MIR6077-2
MIR6078
MIR6079
MIR608
MIR6080
MIR6081
MIR6082
MIR6083
MIR6084
MIR6085

| | | | |
|---|---|---|---|
| | MIR6086 | | |
| | MIR6087 | | |
| | MIR6088 | | |
| | MIR6089-1 | | |
| | MIR6089-1-2 | | |
| | MIR609 | | |
| | MIR6090 | | |
| | MIR610 | | |
| | MIR611 | | |
| 0,272277655 | MIR612 | FALSE | Upregulated |
| | MIR6124 | | |
| | MIR6125 | | |
| | MIR6126 | | |
| | MIR6127 | | |
| | MIR6128 | | |
| | MIR6129 | | |
| | MIR613 | | |
| | MIR6130 | | |
| | MIR6131 | | |
| | MIR6132 | | |
| | MIR6133 | | |
| | MIR6134 | | |
| | MIR614 | | |
| | MIR615 | | |
| | MIR616 | | |
| | MIR6165 | | |
| | MIR617 | | |
| | MIR618 | | |
| | MIR619 | | |
| | MIR620 | | |
| | MIR621 | | |
| | MIR622 | | |
| | MIR623 | | |
| | MIR624 | | |
| | MIR625 | | |
| | MIR626 | | |
| | MIR627 | | |
| | MIR628 | | |
| | MIR629 | | |
| | MIR630 | | |
| | MIR631 | | |
| | MIR632 | | |
| | MIR633 | | |
| | MIR634 | | |
| | MIR635 | | |
| | MIR636 | | |
| | MIR637 | | |
| | MIR638 | | |
| | MIR639 | | |
| | MIR640 | | |
| | MIR641 | | |

| | | | |
|---|---|---|---|
| | MIR642A | | |
| | MIR642B | | |
| | MIR643 | | |
| | MIR644A | | |
| | MIR644B | | |
| | MIR645 | | |
| | MIR646 | | |
| | MIR647 | | |
| | MIR648 | | |
| | MIR649 | | |
| | MIR6499 | | |
| | MIR6500 | | |
| | MIR6501 | | |
| | MIR6502 | | |
| | MIR6503 | | |
| | MIR6504 | | |
| | MIR6505 | | |
| | MIR6506 | | |
| | MIR6507 | | |
| | MIR6508 | | |
| | MIR6509 | | |
| | MIR651 | | |
| | MIR6510 | | |
| | MIR6511A-2 | | |
| | MIR6511B-1 | | |
| | MIR6512 | | |
| | MIR6513 | | |
| | MIR6514 | | |
| | MIR6515 | | |
| | MIR6516 | | |
| | MIR652 | | |
| | MIR653 | | |
| | MIR654 | | |
| | MIR655 | | |
| | MIR656 | | |
| | MIR657 | | |
| | MIR658 | | |
| | MIR659 | | |
| | MIR660 | | |
| | MIR661 | | |
| | MIR662 | | |
| | MIR663A | | |
| | MIR663B | | |
| | MIR664 | | |
| | MIR665 | | |
| | MIR668 | | |
| | MIR670 | | |
| 0,297178435 | MIR670HG | FALSE | Upregulated |
| | MIR671 | | |
| | MIR6715A | | |
| | MIR6715B | | |

| | | | | |
|---|---|---|---|---|
| | MIR6716 | | | |
| | MIR6717 | | | |
| | MIR6718 | | | |
| | MIR6719 | | | |
| | MIR6720 | | | |
| | MIR6721 | | | |
| | MIR6722 | | | |
| | MIR6723 | | | |
| | MIR6726 | | | |
| | MIR6727 | | | |
| | MIR6728 | | | |
| | MIR6729 | | | |
| | MIR6730 | | | |
| | MIR6731 | | | |
| | MIR6732 | | | |
| | MIR6733 | | | |
| | MIR6734 | | | |
| | MIR6735 | | | |
| | MIR6736 | | | |
| | MIR6737 | | | |
| | MIR6738 | | | |
| | MIR6739 | | | |
| | MIR6740 | | | |
| | MIR6741 | | | |
| | MIR6742 | | | |
| | MIR6743 | | | |
| | MIR6744 | | | |
| | MIR6745 | | | |
| | MIR6746 | | | |
| | MIR6747 | | | |
| | MIR6748 | | | |
| | MIR6749 | | | |
| | MIR675 | | | |
| | MIR6750 | | | |
| | MIR6751 | | | |
| | MIR6752 | | | |
| -0,011120974 | | MIR6753 | FALSE | Downregulated |
| | MIR6754 | | | |
| | MIR6755 | | | |
| | MIR6756 | | | |
| | MIR6757 | | | |
| | MIR6758 | | | |
| | MIR6759 | | | |
| | MIR676 | | | |
| | MIR6760 | | | |
| | MIR6761 | | | |
| | MIR6762 | | | |
| | MIR6763 | | | |
| | MIR6764 | | | |
| | MIR6765 | | | |
| | MIR6766 | | | |

MIR6767
MIR6768
MIR6769A
MIR6769B
MIR6770-2
MIR6771
MIR6772
MIR6773
MIR6774
MIR6775
MIR6776
MIR6777
MIR6778
MIR6779
MIR6780A
MIR6780B
MIR6781
MIR6782
MIR6783
MIR6784
MIR6785
MIR6786
MIR6787
MIR6789
MIR6790
MIR6791
MIR6792
MIR6793
MIR6794
MIR6795
MIR6796
MIR6797
MIR6798
MIR6799
MIR6800
MIR6801
MIR6802
MIR6803
MIR6804
MIR6805
MIR6806
MIR6807
MIR6808
MIR6809
MIR6810
MIR6811
MIR6812
MIR6813
MIR6814
MIR6815
MIR6816

MIR6817
MIR6818
MIR6819
MIR6820
MIR6821
MIR6822
MIR6823
MIR6824
MIR6825
MIR6826
MIR6827
MIR6828
MIR6829
MIR6830
MIR6831
MIR6832
MIR6833
MIR6834
MIR6835
MIR6836
MIR6837
MIR6838
MIR6839
MIR6840
MIR6841
MIR6842
MIR6843
MIR6844
MIR6845
MIR6846
MIR6847
MIR6848
MIR6849
MIR6850
MIR6851
MIR6852
MIR6853
MIR6854
MIR6855
MIR6856
MIR6857
MIR6858
MIR6859-1
MIR6859-2
MIR6860
MIR6861
MIR6862-1
MIR6862-2
MIR6863
MIR6864
MIR6865

| | | | | |
|---|---|---|---|---|
| | MIR6866 | | | |
| | MIR6867 | | | |
| | MIR6868 | | | |
| | MIR6869 | | | |
| | MIR6870 | | | |
| | MIR6871 | | | |
| | MIR6872 | | | |
| | MIR6873 | | | |
| | MIR6874 | | | |
| | MIR6875 | | | |
| | MIR6876 | | | |
| | MIR6877 | | | |
| | MIR6878 | | | |
| | MIR6879 | | | |
| | MIR6880 | | | |
| | MIR6881 | | | |
| | MIR6882 | | | |
| | MIR6883 | | | |
| | MIR6884 | | | |
| | MIR6885 | | | |
| | MIR6886 | | | |
| | MIR6887 | | | |
| | MIR6888 | | | |
| | MIR6889 | | | |
| | MIR6890 | | | |
| | MIR6891 | | | |
| | MIR6892 | | | |
| | MIR6893 | | | |
| | MIR6894 | | | |
| | MIR6895 | | | |
| | MIR7-1 | | | |
| | MIR7-2 | | | |
| | MIR7-3 | | | |
| -0,294566193 | | MIR7-3HG | FALSE | Downregulated |
| | MIR708 | | | |
| | MIR7106 | | | |
| | MIR7107 | | | |
| | MIR7108 | | | |
| | MIR7109 | | | |
| | MIR711 | | | |
| | MIR7110 | | | |
| | MIR7111 | | | |
| | MIR7112-2 | | | |
| | MIR7113 | | | |
| | MIR7114 | | | |
| | MIR7150 | | | |
| | MIR7151 | | | |
| | MIR7152 | | | |
| | MIR7153 | | | |
| | MIR7154 | | | |
| | MIR7155 | | | |

MIR7156
MIR7157
MIR7159
MIR7160
MIR7161
MIR7162
MIR718
MIR744
MIR7515
MIR758
MIR759
MIR760
MIR761
MIR762
MIR764
MIR7641-2
MIR765
MIR766
MIR767
MIR769
MIR770
MIR7702
MIR7703
MIR7705
MIR7843
MIR7844
MIR7845
MIR7846
MIR7848
MIR7849
MIR7850
MIR7851
MIR7852
MIR7853
MIR7854
MIR7855
MIR7856
MIR7973-1
MIR7973-2
MIR7974
MIR7975
MIR7976
MIR7977
MIR7978
MIR802
MIR8052
MIR8053
MIR8054
MIR8055
MIR8056
MIR8057

MIR8058
MIR8059
MIR8060
MIR8061
MIR8062
MIR8063
MIR8064
MIR8065
MIR8066
MIR8067
MIR8068
MIR8069
MIR8070
MIR8071-1
MIR8071-2
MIR8072
MIR8073
MIR8074
MIR8075
MIR8076
MIR8077
MIR8078
MIR8079
MIR8080
MIR8081
MIR8082
MIR8083
MIR8084
MIR8085
MIR8086
MIR8087
MIR8088
MIR8089
MIR873
MIR874
MIR875
MIR876
MIR877
MIR885
MIR887
MIR888
MIR889
MIR890
MIR891A
MIR891B
MIR892A
MIR892B
MIR892C
MIR9-1
MIR9-2
MIR9-3

| | | | |
|---|---|---|---|
| | MIR920 | | |
| | MIR921 | | |
| | MIR922 | | |
| | MIR924 | | |
| | MIR92A1 | | |
| | MIR92A2 | | |
| | MIR92B | | |
| | MIR93 | | |
| | MIR933 | | |
| | MIR934 | | |
| | MIR935 | | |
| | MIR936 | | |
| | MIR937 | | |
| | MIR938 | | |
| | MIR939 | | |
| | MIR940 | | |
| | MIR942 | | |
| | MIR943 | | |
| | MIR944 | | |
| | MIR95 | | |
| | MIR96 | | |
| | MIR98 | | |
| | MIR99A | | |
| | MIR99B | | |
| | MIRLET7A1 | | |
| | MIRLET7A2 | | |
| | MIRLET7A3 | | |
| | MIRLET7B | | |
| | MIRLET7C | | |
| | MIRLET7D | | |
| | MIRLET7E | | |
| | MIRLET7F1 | | |
| | MIRLET7F2 | | |
| | MIRLET7G | | |
| | MIRLET7I | | |
| -0,431932103 | MIS18A | FALSE | Downregulated |
| -0,297825903 | MISP | FALSE | Downregulated |
| 0,250336201 | MITD1 | FALSE | Upregulated |
| 0,233969059 | MITF | FALSE | Upregulated |
| -0,06492232 | MIXL1 | FALSE | Downregulated |
| 0,315424621 | MKRN3 | FALSE | Upregulated |
| -0,596379524 | MKRN7P | FALSE | Downregulated |
| 0,294861532 | MKRN9P | FALSE | Upregulated |
| -0,608543342 | MKS1 | FALSE | Downregulated |
| 0,487325297 | MKX | FALSE | Upregulated |
| -0,320242365 | MLC1 | FALSE | Downregulated |
| 0,287199052 | MLF1 | FALSE | Upregulated |
| -0,021688931 | MLIP-IT1 | FALSE | Downregulated |
| 0,023993262 | MLLT3 | FALSE | Upregulated |
| | MLN | | |
| 0,127964277 | MLNR | FALSE | Upregulated |

| | | | |
|---|---|---|---|
| | MMD2 | | |
| 0,354210862 | MMGT1 | FALSE | Upregulated |
| | MMP1 | | |
| -0,148465128 | MMP10 | FALSE | Downregulated |
| -0,011204555 | MMP11 | FALSE | Downregulated |
| 1,143362856 | MMP12 | | Upregulated |
| 0,862219985 | MMP13 | FALSE | Upregulated |
| -1,232019699 | MMP16 | | Downregulated |
| -0,476898137 | MMP17 | FALSE | Downregulated |
| 0 | MMP20 | FALSE | Upregulated |
| -0,093144844 | MMP21 | FALSE | Downregulated |
| -0,198041504 | MMP23A | FALSE | Downregulated |
| | MMP26 | | |
| -0,285187474 | MMP27 | FALSE | Downregulated |
| -0,480591882 | MMP28 | FALSE | Downregulated |
| 0,282941944 | MMP3 | FALSE | Upregulated |
| 0,080781688 | MMP7 | FALSE | Upregulated |
| 0,000264809 | MMP8 | FALSE | Upregulated |
| 1,064283688 | MMP9 | | Upregulated |
| 0,076869387 | MMS22L | FALSE | Upregulated |
| 0,573825921 | MNAT1 | FALSE | Upregulated |
| 0,009842704 | MND1 | FALSE | Upregulated |
| -0,44726742 | MNDA | FALSE | Downregulated |
| 0,064552441 | MNS1 | FALSE | Upregulated |
| 0,077342643 | MNX1 | FALSE | Upregulated |
| | MNX1-AS1 | | |
| 0,027270689 | MOB4 | FALSE | Upregulated |
| -0,014822418 | MOBP | FALSE | Downregulated |
| 0,235576451 | MOG | FALSE | Upregulated |
| 0,097346152 | MOGAT1 | FALSE | Upregulated |
| -0,415405863 | MOGAT3 | FALSE | Downregulated |
| 0,277833435 | MORC1 | FALSE | Upregulated |
| -0,302473572 | MORF4L2-AS1 | FALSE | Downregulated |
| -0,49075564 | MORN2 | FALSE | Downregulated |
| 1,158743723 | MORN3 | | Upregulated |
| 0,289057391 | MORN5 | FALSE | Upregulated |
| | MOS | | |
| -0,32155671 | MOSPD1 | FALSE | Downregulated |
| -0,930547689 | MOV10L1 | FALSE | Downregulated |
| -0,4580108 | MOXD1 | FALSE | Downregulated |
| 0,282164461 | MPC1L | FALSE | Upregulated |
| -0,590532281 | MPHOSPH6 | FALSE | Downregulated |
| 0,609549857 | MPO | FALSE | Upregulated |
| 0,532618447 | MPP2 | FALSE | Upregulated |
| -0,308320008 | MPP3 | FALSE | Downregulated |
| 0,430295184 | MPP4 | FALSE | Upregulated |
| 0,227779914 | MPP7 | FALSE | Upregulated |
| -0,046087652 | MPPED2 | FALSE | Downregulated |
| 0,285964956 | MPV17 | FALSE | Upregulated |
| -0,275364262 | MPV17L2 | FALSE | Downregulated |
| 0,34184398 | MPZ | FALSE | Upregulated |

| | | | |
|---|---|---|---|
| 0,031593554 | MPZL3 | FALSE | Upregulated |
| -0,946671723 | MRAP2 | FALSE | Downregulated |
| -0,727548797 | MRGPRD | FALSE | Downregulated |
| 0,455008338 | MRGPRE | FALSE | Upregulated |
| | MRGPRG | | |
| | MRGPRG-AS1 | | |
| | MRGPRX1 | | |
| | MRGPRX2 | | |
| -1,151993865 | MRGPRX3 | | Downregulated |
| 0,564103575 | MRGPRX4 | FALSE | Upregulated |
| -0,558161141 | MROH2A | FALSE | Downregulated |
| 0,354744715 | MROH2B | FALSE | Upregulated |
| 0,280932347 | MROH3P | FALSE | Upregulated |
| -0,016296357 | MROH5 | FALSE | Downregulated |
| 0,584484355 | MROH9 | FALSE | Upregulated |
| -0,611774141 | MRPL1 | FALSE | Downregulated |
| -0,555724015 | MRPL13 | FALSE | Downregulated |
| 0,090790958 | MRPL33 | FALSE | Upregulated |
| 0,149676119 | MRPL36 | FALSE | Upregulated |
| -0,021616784 | MRPL42P5 | FALSE | Downregulated |
| -0,064362636 | MRPL47 | FALSE | Downregulated |
| 0,171181945 | MRPL48 | FALSE | Upregulated |
| 0,505221066 | MRPL50 | FALSE | Upregulated |
| -0,50120237 | MRPS17 | FALSE | Downregulated |
| 0,282768361 | MRPS23 | FALSE | Upregulated |
| 0,106885985 | MRPS28 | FALSE | Upregulated |
| -0,090005332 | MRPS33 | FALSE | Downregulated |
| -0,328275337 | MRPS36 | FALSE | Downregulated |
| 0,422062709 | MRVI1-AS1 | FALSE | Upregulated |
| 0,101302508 | MS4A1 | FALSE | Upregulated |
| 0,167220707 | MS4A10 | FALSE | Upregulated |
| | MS4A12 | | |
| | MS4A13 | | |
| -0,155611256 | MS4A14 | FALSE | Downregulated |
| -0,010635676 | MS4A15 | FALSE | Downregulated |
| 0,284987956 | MS4A18 | FALSE | Upregulated |
| 0,093770628 | MS4A2 | FALSE | Upregulated |
| -0,00509535 | MS4A3 | FALSE | Downregulated |
| 0,562744406 | MS4A4A | FALSE | Upregulated |
| 1,156529368 | MS4A4E | | Upregulated |
| | MS4A5 | | |
| 0,293242527 | MS4A6E | FALSE | Upregulated |
| 0 | MS4A8 | FALSE | Upregulated |
| 0,362395664 | MSANTD1 | FALSE | Upregulated |
| -1,021876459 | MSANTD3 | | Downregulated |
| 0,116501929 | MSANTD3-TMEFF1 | FALSE | Upregulated |
| | MSGN1 | | |
| -0,030442246 | MSH4 | FALSE | Downregulated |
| 0,538781811 | MSH5 | FALSE | Upregulated |
| -0,256626236 | MSI1 | FALSE | Downregulated |
| -0,964765394 | MSL3P1 | FALSE | Downregulated |

| | | | |
|---|---|---|---|
| 0,148833681 | MSLN | FALSE | Upregulated |
| -0,28269647 | MSMB | FALSE | Downregulated |
| 0,938222043 | MSS51 | FALSE | Upregulated |
| -0,167500238 | MST1R | FALSE | Downregulated |
| 0,619283007 | MST4 | FALSE | Upregulated |
| 0,568960625 | MSTN | FALSE | Upregulated |
| -0,156247267 | MSX1 | FALSE | Downregulated |
| -0,006903421 | MSX2 | FALSE | Downregulated |
| 0,18577459 | MSX2P1 | FALSE | Upregulated |
| 0,980166436 | MT1A | FALSE | Upregulated |
| 1,730265841 | MT1B | | Upregulated |
| 0,340897506 | MT1DP | FALSE | Upregulated |
| 0,289235841 | MT1HL1 | FALSE | Upregulated |
| 0 | MT1IP | FALSE | Upregulated |
| -0,135520095 | MT1JP | FALSE | Downregulated |
| 0,140828578 | MT1L | FALSE | Upregulated |
| 0 | MT3 | FALSE | Upregulated |
| | MT4 | | |
| 1,496669813 | MTBP | | Upregulated |
| -0,193434851 | MTERF | FALSE | Downregulated |
| -0,86025802 | MTERFD1 | FALSE | Downregulated |
| 0,27586933 | MTFP1 | FALSE | Upregulated |
| | MTFR2 | | |
| -0,270131349 | MTHFD2L | FALSE | Downregulated |
| | MTHFD2P1 | | |
| 0,412797667 | MTL5 | FALSE | Upregulated |
| 2,253004947 | MTMR11 | | Upregulated |
| 0,758137133 | MTMR8 | FALSE | Upregulated |
| | MTNR1A | | |
| 0,200318546 | MTNR1B | FALSE | Upregulated |
| 0,580754519 | MTOR-AS1 | FALSE | Upregulated |
| | MTRNR2L1 | | |
| | MTRNR2L10 | | |
| | MTRNR2L2 | | |
| -0,173470795 | MTRNR2L5 | FALSE | Downregulated |
| 1,247693133 | MTRNR2L6 | | Upregulated |
| | MTRNR2L7 | | |
| 0 | MTRNR2L9 | FALSE | Upregulated |
| -0,341703313 | MTUS2-AS1 | FALSE | Downregulated |
| 0,335790284 | MTVR2 | FALSE | Upregulated |
| -0,20438055 | MTX2 | FALSE | Downregulated |
| -0,588074432 | MUC1 | FALSE | Downregulated |
| 0,18383212 | MUC13 | FALSE | Upregulated |
| -0,145321035 | MUC16 | FALSE | Downregulated |
| 0,272275666 | MUC17 | FALSE | Upregulated |
| -0,00780606 | MUC19 | FALSE | Downregulated |
| 0 | MUC2 | FALSE | Upregulated |
| 0 | MUC21 | FALSE | Upregulated |
| 0,824470808 | MUC22 | FALSE | Upregulated |
| 1,388093114 | MUC5AC | | Upregulated |
| 0 | MUC7 | FALSE | Upregulated |

| | | | |
|---|---|---|---|
| 0 | MUC8 | FALSE | Upregulated |
| | MUCL1 | | |
| -1,404619925 | MUM1L1 | | Downregulated |
| 0,420947161 | MURC | FALSE | Upregulated |
| 0,587088747 | MUSK | FALSE | Upregulated |
| 0 | MYADML | FALSE | Upregulated |
| 0,242372597 | MYADML2 | FALSE | Upregulated |
| 0,274818096 | MYB | FALSE | Upregulated |
| 0,40270699 | MYBL1 | FALSE | Upregulated |
| 0,142284307 | MYBL2 | FALSE | Upregulated |
| 0,854783171 | MYBPC3 | FALSE | Upregulated |
| -0,938737645 | MYBPH | FALSE | Downregulated |
| 0,135472683 | MYBPHL | FALSE | Upregulated |
| 0,119693766 | MYCBP2-AS1 | FALSE | Upregulated |
| 0,254236121 | MYCBPAP | FALSE | Upregulated |
| 0,489235565 | MYCN | FALSE | Upregulated |
| 1,100660917 | MYCNOS | | Upregulated |
| -0,618891098 | MYEF2 | FALSE | Downregulated |
| 0,069883298 | MYEOV | FALSE | Upregulated |
| -0,266360124 | MYEOV2 | FALSE | Downregulated |
| | MYF5 | | |
| 0,251660847 | MYF6 | FALSE | Upregulated |
| 0,272090836 | MYH13 | FALSE | Upregulated |
| -0,21708285 | MYH15 | FALSE | Downregulated |
| 0,225527376 | MYH16 | FALSE | Upregulated |
| 0,177632653 | MYH4 | FALSE | Upregulated |
| 0,739217622 | MYH6 | FALSE | Upregulated |
| 0,415944425 | MYH8 | FALSE | Upregulated |
| 0,506660338 | MYL1 | FALSE | Upregulated |
| | MYL10 | | |
| -0,357061535 | MYL3 | FALSE | Downregulated |
| 0,435500765 | MYL4 | FALSE | Upregulated |
| | MYL7 | | |
| 0,798221557 | MYLK2 | FALSE | Upregulated |
| 0,00502982 | MYLK3 | FALSE | Upregulated |
| -0,373578986 | MYLK4 | FALSE | Downregulated |
| 1,695343917 | MYLPF | | Upregulated |
| 0,318325092 | MYO16-AS1 | FALSE | Upregulated |
| 1,147981532 | MYO18B | | Upregulated |
| 0,168893294 | MYO1A | FALSE | Upregulated |
| -0,009036632 | MYO3A | FALSE | Downregulated |
| 0,91854085 | MYO3B | FALSE | Upregulated |
| 0,283850436 | MYOC | FALSE | Upregulated |
| 0,390058801 | MYOCD | FALSE | Upregulated |
| 0,271372711 | MYOD1 | FALSE | Upregulated |
| 0,752084481 | MYOG | FALSE | Upregulated |
| -0,130571425 | MYOM3 | FALSE | Downregulated |
| 1,320134618 | MYOZ1 | | Upregulated |
| 1,605320833 | MYOZ2 | | Upregulated |
| 0,778352048 | MYPN | FALSE | Upregulated |
| 0,629228729 | MYRFL | FALSE | Upregulated |

| | | | |
|---|---|---|---|
| 0 | MYT1 | FALSE | Upregulated |
| 0 | MYT1L | FALSE | Upregulated |
| | MYT1L-AS1 | | |
| 0,737528756 | MYZAP | FALSE | Upregulated |
| 0,525216143 | MZB1 | FALSE | Upregulated |
| -0,299350993 | MZT1 | FALSE | Downregulated |
| 0,262705655 | N4BP3 | FALSE | Upregulated |
| 0,261141915 | N6AMT1 | FALSE | Upregulated |
| -0,408712263 | N6AMT2 | FALSE | Downregulated |
| 0,277119458 | NAA11 | FALSE | Upregulated |
| 0,69952304 | NAALADL1 | FALSE | Upregulated |
| | NAALADL2-AS3 | | |
| 0,293712788 | NACA2 | FALSE | Upregulated |
| -0,422915354 | NACAD | FALSE | Downregulated |
| 0,430927481 | NACAP1 | FALSE | Upregulated |
| 0,034987811 | NAE1 | FALSE | Upregulated |
| 0,083675689 | NAGPA-AS1 | FALSE | Upregulated |
| 0,493187052 | NALCN | FALSE | Upregulated |
| 1,094045155 | NALCN-AS1 | | Upregulated |
| -0,114242024 | NAMA | FALSE | Downregulated |
| -0,305215024 | NANOG | FALSE | Downregulated |
| | NANOGNB | | |
| -0,024037949 | NANOS2 | FALSE | Downregulated |
| -0,011481061 | NANOS3 | FALSE | Downregulated |
| 0,345224636 | NANP | FALSE | Upregulated |
| 0,474864874 | NAP1L2 | FALSE | Upregulated |
| -0,54229715 | NAP1L3 | FALSE | Downregulated |
| 0 | NAP1L6 | FALSE | Upregulated |
| 0,131891218 | NAPA-AS1 | FALSE | Upregulated |
| 0,462855452 | NAPSA | FALSE | Upregulated |
| -0,108762343 | NARR | FALSE | Downregulated |
| -0,136650469 | NAT14 | FALSE | Downregulated |
| 0 | NAT16 | FALSE | Upregulated |
| 0,148879848 | NAT6 | FALSE | Upregulated |
| -0,211263038 | NAT8L | FALSE | Downregulated |
| 0 | NBEAP1 | FALSE | Upregulated |
| | NBPF18P | | |
| -1,138880172 | NBPF22P | | Downregulated |
| | NBPF23P | | |
| 0,509016291 | NBPF25P | FALSE | Upregulated |
| 0,050853493 | NBPF3 | FALSE | Upregulated |
| -0,143920439 | NBPF4 | FALSE | Downregulated |
| -0,01013871 | NBPF6 | FALSE | Downregulated |
| -0,025015387 | NBPF7 | FALSE | Downregulated |
| -0,075111096 | NBR2 | FALSE | Downregulated |
| 0,998075698 | NCAM1-AS1 | FALSE | Upregulated |
| 0,147696772 | NCAN | FALSE | Upregulated |
| -0,019139697 | NCAPH | FALSE | Downregulated |
| 0,191105149 | NCCRP1 | FALSE | Upregulated |
| 0,299072383 | NCEH1 | FALSE | Upregulated |
| 0,789579249 | NCF1B | FALSE | Upregulated |

| | | | |
|---|---|---|---|
| 0,19222505 | NCF4 | FALSE | Upregulated |
| | NCOR1P1 | | |
| -0,98538609 | NCR1 | FALSE | Downregulated |
| | NCR2 | | |
| 0,71367953 | NCR3 | FALSE | Upregulated |
| 0,523804486 | NCR3LG1 | FALSE | Upregulated |
| -0,341989336 | NCRNA00185 | FALSE | Downregulated |
| -0,009465128 | NCRNA00249 | FALSE | Downregulated |
| 0,149196717 | NCRUPAR | FALSE | Upregulated |
| 0,082784888 | NDC1 | FALSE | Upregulated |
| 0,773365891 | NDC80 | FALSE | Upregulated |
| | NDFIP2-AS1 | | |
| 0,206627765 | NDN | FALSE | Upregulated |
| -0,425655687 | NDNF | FALSE | Downregulated |
| | NDP | | |
| | NDST4 | | |
| -0,662661969 | NDUFA6-AS1 | FALSE | Downregulated |
| -0,352071916 | NDUFA7 | FALSE | Downregulated |
| -0,641518134 | NDUFAB1 | FALSE | Downregulated |
| -0,816739184 | NDUFAF4 | FALSE | Downregulated |
| -0,299395829 | NDUFAF4P1 | FALSE | Downregulated |
| 0,433287571 | NDUFAF5 | FALSE | Upregulated |
| 0,038441294 | NDUFAF6 | FALSE | Upregulated |
| 0,605832757 | NDUFB2-AS1 | FALSE | Upregulated |
| -0,260931277 | NDUFC2-KCTD14 | FALSE | Downregulated |
| -0,227508605 | NEBL | FALSE | Downregulated |
| | NEBL-AS1 | | |
| -0,713993552 | NEDD1 | FALSE | Downregulated |
| 0,417811059 | NEDD8-MDP1 | FALSE | Upregulated |
| 0 | NEFL | FALSE | Upregulated |
| 0,723692464 | NEFM | FALSE | Upregulated |
| | NEGR1-IT1 | | |
| 0,557740464 | NEIL3 | FALSE | Upregulated |
| -0,855006601 | NEK10 | FALSE | Downregulated |
| 0,406907307 | NEK2 | FALSE | Upregulated |
| 1,194148624 | NEK5 | | Upregulated |
| | NELL1 | | |
| 1,984199923 | NELL2 | | Upregulated |
| 0 | NETO1 | FALSE | Upregulated |
| -0,02387546 | NETO2 | FALSE | Downregulated |
| | NEU2 | | |
| 0,622686608 | NEURL3 | FALSE | Upregulated |
| | NEUROD1 | | |
| -0,344415972 | NEUROD2 | FALSE | Downregulated |
| | NEUROD4 | | |
| | NEUROG1 | | |
| | NEUROG2 | | |
| | NEUROG3 | | |
| 0,289177209 | NF1P2 | FALSE | Upregulated |
| 0,57111279 | NFE2 | FALSE | Upregulated |
| 0 | NFE4 | FALSE | Upregulated |

| | | | |
|---|---|---|---|
| -0,505289512 | NFKBID | FALSE | Downregulated |
| 0,116451813 | NFU1 | FALSE | Upregulated |
| | NGB | | |
| 0,745113912 | NGF | FALSE | Upregulated |
| -0,137129761 | NHEG1 | FALSE | Downregulated |
| 0,037191857 | NHEJ1 | FALSE | Upregulated |
| 0,316310686 | NHLH1 | FALSE | Upregulated |
| 0,293937211 | NHLH2 | FALSE | Upregulated |
| 0,03826349 | NHLRC1 | FALSE | Upregulated |
| -0,631543268 | NHLRC4 | FALSE | Downregulated |
| 0,128965535 | NHS-AS1 | FALSE | Upregulated |
| -0,459950378 | NIFK | FALSE | Downregulated |
| -0,581432708 | NIM1K | FALSE | Downregulated |
| -0,024434894 | NINJ2 | FALSE | Downregulated |
| 0,277119458 | NKAIN1 | FALSE | Upregulated |
| 0 | NKAIN4 | FALSE | Upregulated |
| -0,626204763 | NKAP | FALSE | Downregulated |
| 0,162803167 | NKAPL | FALSE | Upregulated |
| 0,200874821 | NKAPP1 | FALSE | Upregulated |
| 0,83065464 | NKD2 | FALSE | Upregulated |
| -0,034689711 | NKG7 | FALSE | Downregulated |
| | NKX1-2 | | |
| 0 | NKX2-1 | FALSE | Upregulated |
| | NKX2-1-AS1 | | |
| | NKX2-2 | | |
| 0,269041409 | NKX2-3 | FALSE | Upregulated |
| | NKX2-4 | | |
| 0,433097061 | NKX2-5 | FALSE | Upregulated |
| 0 | NKX2-6 | FALSE | Upregulated |
| 0 | NKX2-8 | FALSE | Upregulated |
| 0,590469156 | NKX3-1 | FALSE | Upregulated |
| 0 | NKX3-2 | FALSE | Upregulated |
| | NKX6-1 | | |
| 0,433966516 | NKX6-2 | FALSE | Upregulated |
| | NKX6-3 | | |
| -0,011328201 | NLGN1 | FALSE | Downregulated |
| -0,309361166 | NLGN4X | FALSE | Downregulated |
| 0,676461221 | NLGN4Y | FALSE | Upregulated |
| 0,727373503 | NLGN4Y-AS1 | FALSE | Upregulated |
| 0 | NLRP10 | FALSE | Upregulated |
| -0,782025685 | NLRP11 | FALSE | Downregulated |
| 1,536879753 | NLRP12 | | Upregulated |
| | NLRP13 | | |
| -0,321270112 | NLRP14 | FALSE | Downregulated |
| 0,468322152 | NLRP2 | FALSE | Upregulated |
| 0,324429624 | NLRP3 | FALSE | Upregulated |
| 0,410243185 | NLRP4 | FALSE | Upregulated |
| 0,000504523 | NLRP5 | FALSE | Upregulated |
| 0,229073597 | NLRP7 | FALSE | Upregulated |
| | NLRP8 | | |
| -0,504326488 | NMB | FALSE | Downregulated |

| | | | |
|---|---|---|---|
| | NMBR | | |
| -0,264467555 | NME1-NME2 | FALSE | Downregulated |
| | NME5 | | |
| 0 | NME8 | FALSE | Upregulated |
| 1,281048914 | NMNAT2 | | Upregulated |
| 0,660583573 | NMRK2 | FALSE | Upregulated |
| | NMS | | |
| 0 | NMU | FALSE | Upregulated |
| 0,479116293 | NMUR1 | FALSE | Upregulated |
| 0 | NMUR2 | FALSE | Upregulated |
| | NOBOX | | |
| -0,347612804 | NOC4L | FALSE | Downregulated |
| 0,288748693 | NOD2 | FALSE | Upregulated |
| -0,276829504 | NOG | FALSE | Downregulated |
| -0,049417804 | NOL4 | FALSE | Downregulated |
| 2,094731692 | NOS1 | | Upregulated |
| -0,396548295 | NOS2 | FALSE | Downregulated |
| -0,01043221 | NOTO | FALSE | Downregulated |
| 0,382357838 | NOV | FALSE | Upregulated |
| -0,344147523 | NOVA1 | FALSE | Downregulated |
| 0,486609968 | NOX1 | FALSE | Upregulated |
| | NOX3 | | |
| -0,00219862 | NOX4 | FALSE | Downregulated |
| -0,470255527 | NOX5 | FALSE | Downregulated |
| 0,091099579 | NOXRED1 | FALSE | Upregulated |
| -0,073330801 | NPAP1 | FALSE | Downregulated |
| 0,628523773 | NPAS1 | FALSE | Upregulated |
| 1,624717174 | NPAS3 | | Upregulated |
| -0,018282339 | NPAS4 | FALSE | Downregulated |
| 0,839372573 | NPBWR1 | FALSE | Upregulated |
| | NPBWR2 | | |
| 0,528890829 | NPFF | FALSE | Upregulated |
| | NPFFR1 | | |
| | NPFFR2 | | |
| 0,900720132 | NPHP1 | FALSE | Upregulated |
| | NPHP3-AS1 | | |
| 0 | NPHS1 | FALSE | Upregulated |
| 0,648937575 | NPHS2 | FALSE | Upregulated |
| 1,320262224 | NPIPA3 | | Upregulated |
| 0,702579875 | NPIPA7 | FALSE | Upregulated |
| 0,819255297 | NPIPA8 | FALSE | Upregulated |
| -1,179138773 | NPIPB8 | | Downregulated |
| -0,415642427 | NPM2 | FALSE | Downregulated |
| 1,227240223 | NPM3 | | Upregulated |
| -0,27140263 | NPNT | FALSE | Downregulated |
| -0,190412819 | NPPA | FALSE | Downregulated |
| 0,19645737 | NPPA-AS1 | FALSE | Upregulated |
| 0 | NPPB | FALSE | Upregulated |
| | NPPC | | |
| | NPS | | |
| | NPSR1 | | |

| | | | |
|---|---|---|---|
| -0,304485961 | NPSR1-AS1 | FALSE | Downregulated |
| 0,407308188 | NPTN-IT1 | FALSE | Upregulated |
| 0,614805498 | NPTX1 | FALSE | Upregulated |
| 0,085348226 | NPTX2 | FALSE | Upregulated |
| 0,226091933 | NPTXR | FALSE | Upregulated |
| | NPVF | | |
| | NPY | | |
| | NPY2R | | |
| 0,397136173 | NPY4R | FALSE | Upregulated |
| 0,120721389 | NPY5R | FALSE | Upregulated |
| | NR0B1 | | |
| 0 | NR2E1 | FALSE | Upregulated |
| 0,42026454 | NR2E3 | FALSE | Upregulated |
| 0,499817848 | NR2F2-AS1 | FALSE | Upregulated |
| -0,130086367 | NR4A1 | FALSE | Downregulated |
| 0,224575923 | NR4A2 | FALSE | Upregulated |
| -0,00509535 | NR5A1 | FALSE | Downregulated |
| 0,540005275 | NRCAM | FALSE | Upregulated |
| 0,158394971 | NREP-AS1 | FALSE | Upregulated |
| | NRG1-IT1 | | |
| | NRG1-IT3 | | |
| 0,22563131 | NRG2 | FALSE | Upregulated |
| 0,105600333 | NRG3 | FALSE | Upregulated |
| 1,227319732 | NRGN | | Upregulated |
| -0,320178712 | NRN1 | FALSE | Downregulated |
| 0,160549594 | NRN1L | FALSE | Upregulated |
| -0,068079431 | NRROS | FALSE | Downregulated |
| 0,140938962 | NRXN1 | FALSE | Upregulated |
| 0,699829035 | NRXN2 | FALSE | Upregulated |
| -1,455319014 | NRXN3 | | Downregulated |
| 1,104385918 | NSFP1 | | Upregulated |
| -0,25746736 | NSMCE2 | FALSE | Downregulated |
| -0,459807782 | NSUN3 | FALSE | Downregulated |
| -0,129944574 | NT5C1A | FALSE | Downregulated |
| -1,051829492 | NT5C1B-RDH14 | | Downregulated |
| -0,225429124 | NT5C3A | FALSE | Downregulated |
| -0,214292654 | NT5M | FALSE | Downregulated |
| 0,415169808 | NTF3 | FALSE | Upregulated |
| 0,761124855 | NTF4 | FALSE | Upregulated |
| 0,384873387 | NTM | FALSE | Upregulated |
| | NTM-IT | | |
| 0,536365331 | NTN5 | FALSE | Upregulated |
| 0 | NTNG1 | FALSE | Upregulated |
| 0,80978657 | NTNG2 | FALSE | Upregulated |
| 0 | NTRK1 | FALSE | Upregulated |
| -0,288860828 | NTRK2 | FALSE | Downregulated |
| -2,269134243 | NTRK3 | | Downregulated |
| 0,407143114 | NTRK3-AS1 | FALSE | Upregulated |
| -0,001600678 | NTS | FALSE | Downregulated |
| | NTSR2 | | |
| 0,208292337 | NUDCD1 | FALSE | Upregulated |

| | | | |
|---|---|---|---|
| -0,107753581 | NUDT1 | FALSE | Downregulated |
| 1,759508264 | NUDT10 | | Upregulated |
| -0,165893912 | NUDT11 | FALSE | Downregulated |
| -0,017483027 | NUDT18 | FALSE | Downregulated |
| -0,669901779 | NUDT4P2 | FALSE | Downregulated |
| 0,603676434 | NUDT9P1 | FALSE | Upregulated |
| 0,692861541 | NUF2 | FALSE | Upregulated |
| 0,008713078 | NUFIP1 | FALSE | Upregulated |
| -0,58461241 | NUP210L | FALSE | Downregulated |
| 0,277764892 | NUP210P1 | FALSE | Upregulated |
| -0,266951735 | NUP35 | FALSE | Downregulated |
| -0,546399354 | NUP37 | FALSE | Downregulated |
| 1,125587006 | NUP62CL | | Upregulated |
| 0,060725277 | NUPL2 | FALSE | Upregulated |
| | NUPR1L | | |
| 0,191456653 | NUTM1 | FALSE | Upregulated |
| 0,498922913 | NUTM2A-AS1 | FALSE | Upregulated |
| 0,014385035 | NWD1 | FALSE | Upregulated |
| 0 | NXF2 | FALSE | Upregulated |
| 0 | NXF2B | FALSE | Upregulated |
| -0,029854804 | NXF3 | FALSE | Downregulated |
| | NXF4 | | |
| -0,273324478 | NXF5 | FALSE | Downregulated |
| 0 | NXNL1 | FALSE | Upregulated |
| 0,147096792 | NXNL2 | FALSE | Upregulated |
| 1,190889678 | NXPE1 | | Upregulated |
| | NXPE2 | | |
| 1,031837432 | NXPE4 | | Upregulated |
| | NXPH1 | | |
| | NXPH2 | | |
| -0,08565732 | NXPH4 | FALSE | Downregulated |
| 0,017177141 | NXT1 | FALSE | Upregulated |
| -0,734031074 | NXT2 | FALSE | Downregulated |
| 0,068649157 | NYAP1 | FALSE | Upregulated |
| 0,022269948 | NYAP2 | FALSE | Upregulated |
| | NYX | | |
| | OACYLP | | |
| 0,307694352 | OASL | FALSE | Upregulated |
| 0,156095774 | OAZ3 | FALSE | Upregulated |
| | OBP2A | | |
| | OBP2B | | |
| | OC90 | | |
| 0,537652986 | OCA2 | FALSE | Upregulated |
| 0,721266674 | OCLM | FALSE | Upregulated |
| -0,004525881 | OCM | FALSE | Downregulated |
| -0,009465128 | OCM2 | FALSE | Downregulated |
| | OCSTAMP | | |
| | ODAM | | |
| -0,138342045 | ODF1 | FALSE | Downregulated |
| -0,061843216 | ODF3 | FALSE | Downregulated |
| 0,976996664 | ODF3L1 | FALSE | Upregulated |

| | | | |
|---|---|---|---|
| -0,067535002 | ODF3L2 | FALSE | Downregulated |
| 0,441645828 | ODF4 | FALSE | Upregulated |
| -0,170643017 | OFCC1 | FALSE | Downregulated |
| 0,872199716 | OGFR-AS1 | FALSE | Upregulated |
| 0,460942322 | OLAH | FALSE | Upregulated |
| 0,640432209 | OLFM1 | FALSE | Upregulated |
| | OLFM3 | | |
| -0,29840341 | OLFM4 | FALSE | Downregulated |
| 0,29195446 | OLFML2A | FALSE | Upregulated |
| -0,352017105 | OLFML2B | FALSE | Downregulated |
| 0,416225316 | OLIG1 | FALSE | Upregulated |
| 0,098881093 | OLIG2 | FALSE | Upregulated |
| | OLIG3 | | |
| -0,569799618 | OLR1 | FALSE | Downregulated |
| -0,230745767 | OMP | FALSE | Downregulated |
| 0 | ONECUT3 | FALSE | Upregulated |
| | OOEP | | |
| | OOSP1 | | |
| | OOSP2 | | |
| 1,333877176 | OPA1-AS1 | | Upregulated |
| | OPALIN | | |
| 0,051179485 | OPHN1 | FALSE | Upregulated |
| | OPN1LW | | |
| 0,146753986 | OPN1MW | FALSE | Upregulated |
| | OPN1MW2 | | |
| 0,272553722 | OPN1SW | FALSE | Upregulated |
| 0 | OPN4 | FALSE | Upregulated |
| 0,001388321 | OPN5 | FALSE | Upregulated |
| | OPRD1 | | |
| -0,151352334 | OPRK1 | FALSE | Downregulated |
| 0,314258633 | OPRL1 | FALSE | Upregulated |
| 0,891252284 | OPRM1 | FALSE | Upregulated |
| -0,015614825 | OPTC | FALSE | Downregulated |
| -1,329485139 | OR10A2 | | Downregulated |
| 0 | OR10A3 | FALSE | Upregulated |
| 0,012856298 | OR10A4 | FALSE | Upregulated |
| 0,171974295 | OR10A5 | FALSE | Upregulated |
| | OR10A6 | | |
| | OR10A7 | | |
| | OR10AG1 | | |
| | OR10C1 | | |
| | OR10G4 | | |
| 0,307698867 | OR10G7 | FALSE | Upregulated |
| | OR10G8 | | |
| | OR10G9 | | |
| 0,297178435 | OR10H1 | FALSE | Upregulated |
| | OR10H2 | | |
| -0,001674428 | OR10H3 | FALSE | Downregulated |
| | OR10H4 | | |
| -0,018149848 | OR10H5 | FALSE | Downregulated |
| -0,022179671 | OR10J3 | FALSE | Downregulated |

| | | | |
|---|---|---|---|
| | OR10K1 | | |
| | OR10K2 | | |
| | OR10P1 | | |
| 0,271886396 | OR10Q1 | FALSE | Upregulated |
| | OR10R2 | | |
| 0,891359484 | OR10S1 | FALSE | Upregulated |
| | OR10T2 | | |
| -0,150828898 | OR10V1 | FALSE | Downregulated |
| 0,366840478 | OR10V2P | FALSE | Upregulated |
| 0,002114818 | OR10W1 | FALSE | Upregulated |
| | OR10X1 | | |
| | OR10Z1 | | |
| -0,039224827 | OR11A1 | FALSE | Downregulated |
| | OR11G2 | | |
| | OR11H1 | | |
| | OR11H12 | | |
| | OR11H2 | | |
| 0,417720626 | OR11H4 | FALSE | Upregulated |
| -0,419977352 | OR11H6 | FALSE | Downregulated |
| 0,445820547 | OR11L1 | FALSE | Upregulated |
| | OR12D2 | | |
| | OR12D3 | | |
| | OR13A1 | | |
| 0,150605571 | OR13C2 | FALSE | Upregulated |
| 0,302800847 | OR13C3 | FALSE | Upregulated |
| 0,425202514 | OR13C4 | FALSE | Upregulated |
| 0,423101892 | OR13C5 | FALSE | Upregulated |
| -0,247451347 | OR13C8 | FALSE | Downregulated |
| 0,009551639 | OR13C9 | FALSE | Upregulated |
| -0,218245436 | OR13G1 | FALSE | Downregulated |
| 0,428079706 | OR13H1 | FALSE | Upregulated |
| -0,62491174 | OR13J1 | FALSE | Downregulated |
| 0,300684786 | OR14A16 | FALSE | Upregulated |
| | OR14C36 | | |
| | OR14I1 | | |
| | OR14J1 | | |
| | OR1A1 | | |
| | OR1A2 | | |
| 0,885784045 | OR1B1 | FALSE | Upregulated |
| 0,088175502 | OR1C1 | FALSE | Upregulated |
| | OR1D2 | | |
| 0 | OR1D4 | FALSE | Upregulated |
| 0 | OR1D5 | FALSE | Upregulated |
| 0,282414516 | OR1E1 | FALSE | Upregulated |
| | OR1E2 | | |
| | OR1F1 | | |
| -0,175713062 | OR1F2P | FALSE | Downregulated |
| 0 | OR1G1 | FALSE | Upregulated |
| | OR1I1 | | |
| 0,097628718 | OR1J1 | FALSE | Upregulated |
| 0 | OR1J2 | FALSE | Upregulated |

| | | | |
|---|---|---|---|
| | OR1J4 | | |
| 0,245392248 | OR1K1 | FALSE | Upregulated |
| -0,145647653 | OR1L1 | FALSE | Downregulated |
| 0,870488141 | OR1L3 | FALSE | Upregulated |
| 0,706985991 | OR1L6 | FALSE | Upregulated |
| 0,254999029 | OR1L8 | FALSE | Upregulated |
| | OR1M1 | | |
| 0,913488214 | OR1N1 | FALSE | Upregulated |
| -0,138110314 | OR1N2 | FALSE | Downregulated |
| -0,048454949 | OR1Q1 | FALSE | Downregulated |
| | OR1S1 | | |
| 0,300241802 | OR1S2 | FALSE | Upregulated |
| -0,856756142 | OR2A1-AS1 | FALSE | Downregulated |
| 0,215426653 | OR2A12 | FALSE | Upregulated |
| 0,348254712 | OR2A14 | FALSE | Upregulated |
| -0,006240395 | OR2A2 | FALSE | Downregulated |
| -0,121281435 | OR2A25 | FALSE | Downregulated |
| 0,48613648 | OR2A4 | FALSE | Upregulated |
| -0,772269878 | OR2A42 | FALSE | Downregulated |
| 0,247115293 | OR2A5 | FALSE | Upregulated |
| 0,010669496 | OR2A7 | FALSE | Upregulated |
| -0,07438595 | OR2AE1 | FALSE | Downregulated |
| 0,383346762 | OR2AG1 | FALSE | Upregulated |
| 0,018839898 | OR2AG2 | FALSE | Upregulated |
| 0 | OR2AK2 | FALSE | Upregulated |
| | OR2AP1 | | |
| -0,70406977 | OR2AT4 | FALSE | Downregulated |
| 1,023282277 | OR2B11 | | Upregulated |
| 0,607844537 | OR2B2 | FALSE | Upregulated |
| | OR2B3 | | |
| | OR2B6 | | |
| 0,671681772 | OR2C1 | FALSE | Upregulated |
| -0,186726496 | OR2C3 | FALSE | Downregulated |
| | OR2D2 | | |
| -0,212439493 | OR2D3 | FALSE | Downregulated |
| 0,428487225 | OR2F1 | FALSE | Upregulated |
| 0,281301282 | OR2F2 | FALSE | Upregulated |
| | OR2G2 | | |
| | OR2G3 | | |
| 0 | OR2G6 | FALSE | Upregulated |
| -0,025847624 | OR2H1 | FALSE | Downregulated |
| -0,189401083 | OR2H2 | FALSE | Downregulated |
| 0,104549027 | OR2J2 | FALSE | Upregulated |
| | OR2J3 | | |
| | OR2K2 | | |
| | OR2L13 | | |
| 0,274635952 | OR2L1P | FALSE | Upregulated |
| | OR2L2 | | |
| | OR2L3 | | |
| | OR2L5 | | |
| | OR2L8 | | |

| | | | |
|---|---|---|---|
| | OR2M1P | | |
| | OR2M2 | | |
| | OR2M3 | | |
| | OR2M4 | | |
| | OR2M5 | | |
| | OR2M7 | | |
| -0,000262672 | OR2S2 | FALSE | Downregulated |
| | OR2T1 | | |
| | OR2T10 | | |
| 0 | OR2T11 | FALSE | Upregulated |
| | OR2T12 | | |
| | OR2T2 | | |
| 0,277833435 | OR2T27 | FALSE | Upregulated |
| 0,13343011 | OR2T29 | FALSE | Upregulated |
| 0 | OR2T3 | FALSE | Upregulated |
| | OR2T33 | | |
| | OR2T34 | | |
| | OR2T35 | | |
| | OR2T4 | | |
| 0,262857102 | OR2T5 | FALSE | Upregulated |
| | OR2T6 | | |
| | OR2T8 | | |
| -0,29346148 | OR2V1 | FALSE | Downregulated |
| | OR2V2 | | |
| -0,014724123 | OR2W1 | FALSE | Downregulated |
| | OR2W3 | | |
| 0 | OR2W5 | FALSE | Upregulated |
| | OR2Y1 | | |
| | OR2Z1 | | |
| -0,015040902 | OR3A1 | FALSE | Downregulated |
| -0,012678267 | OR3A2 | FALSE | Downregulated |
| 0,109783918 | OR3A3 | FALSE | Upregulated |
| -0,289215184 | OR3A4P | FALSE | Downregulated |
| | OR4A15 | | |
| | OR4A16 | | |
| | OR4A47 | | |
| | OR4A5 | | |
| | OR4B1 | | |
| | OR4C11 | | |
| | OR4C12 | | |
| | OR4C13 | | |
| | OR4C15 | | |
| | OR4C16 | | |
| | OR4C3 | | |
| | OR4C45 | | |
| | OR4C46 | | |
| | OR4C6 | | |
| 0 | OR4D1 | FALSE | Upregulated |
| | OR4D10 | | |
| | OR4D11 | | |
| 0 | OR4D2 | FALSE | Upregulated |

| | | | |
|---|---|---|---|
| | OR4D5 | | |
| | OR4D6 | | |
| | OR4D9 | | |
| | OR4F13P | | |
| | OR4F15 | | |
| 0 | OR4F16 | FALSE | Upregulated |
| 0,235453169 | OR4F17 | FALSE | Upregulated |
| 0,270322197 | OR4F21 | FALSE | Upregulated |
| 0 | OR4F29 | FALSE | Upregulated |
| 0 | OR4F3 | FALSE | Upregulated |
| | OR4F4 | | |
| 0,274391281 | OR4F5 | FALSE | Upregulated |
| | OR4F6 | | |
| | OR4K1 | | |
| | OR4K13 | | |
| | OR4K14 | | |
| | OR4K15 | | |
| | OR4K17 | | |
| | OR4K2 | | |
| | OR4K5 | | |
| | OR4L1 | | |
| | OR4M1 | | |
| 0,293326262 | OR4M2 | FALSE | Upregulated |
| | OR4N2 | | |
| | OR4N3P | | |
| | OR4N4 | | |
| | OR4N5 | | |
| | OR4P4 | | |
| | OR4Q3 | | |
| | OR4S1 | | |
| | OR4S2 | | |
| | OR4X1 | | |
| | OR4X2 | | |
| | OR51A2 | | |
| | OR51A4 | | |
| | OR51A7 | | |
| -0,303529671 | OR51B2 | FALSE | Downregulated |
| 0,293326262 | OR51B4 | FALSE | Upregulated |
| 0,00139671 | OR51B5 | FALSE | Upregulated |
| 0,062149515 | OR51B6 | FALSE | Upregulated |
| | OR51D1 | | |
| 0 | OR51E1 | FALSE | Upregulated |
| | OR51E2 | | |
| | OR51F1 | | |
| | OR51F2 | | |
| | OR51G1 | | |
| | OR51G2 | | |
| 0,281313325 | OR51I1 | FALSE | Upregulated |
| 0,242382462 | OR51I2 | FALSE | Upregulated |
| | OR51L1 | | |
| -0,848934301 | OR51M1 | FALSE | Downregulated |

| | | | |
|---|---|---|---|
| -0,480597699 | OR51Q1 | FALSE | Downregulated |
| | OR51S1 | | |
| | OR51T1 | | |
| | OR51V1 | | |
| | OR52A1 | | |
| | OR52A5 | | |
| 0 | OR52B2 | FALSE | Upregulated |
| -0,2516401 | OR52B4 | FALSE | Downregulated |
| | OR52E2 | | |
| 0,266815044 | OR52E4 | FALSE | Upregulated |
| | OR52E6 | | |
| | OR52E8 | | |
| 0,141229643 | OR52H1 | FALSE | Upregulated |
| 0,178512593 | OR52I1 | FALSE | Upregulated |
| -0,994970473 | OR52I2 | FALSE | Downregulated |
| | OR52J3 | | |
| 0 | OR52K1 | FALSE | Upregulated |
| 0 | OR52K2 | FALSE | Upregulated |
| | OR52L1 | | |
| 0 | OR52M1 | FALSE | Upregulated |
| 0,27678039 | OR52N1 | FALSE | Upregulated |
| 0 | OR52N2 | FALSE | Upregulated |
| 0,418336316 | OR52N4 | FALSE | Upregulated |
| 0 | OR52N5 | FALSE | Upregulated |
| | OR52R1 | | |
| 0,277833435 | OR56A1 | FALSE | Upregulated |
| 0,697848333 | OR56A3 | FALSE | Upregulated |
| | OR56A4 | | |
| | OR56A5 | | |
| 0,472824827 | OR56B1 | FALSE | Upregulated |
| 0,387548336 | OR56B4 | FALSE | Upregulated |
| | OR5A1 | | |
| 0,461070237 | OR5A2 | FALSE | Upregulated |
| | OR5AC2 | | |
| 0,257997477 | OR5AN1 | FALSE | Upregulated |
| | OR5AP2 | | |
| | OR5AR1 | | |
| | OR5AS1 | | |
| | OR5AU1 | | |
| | OR5B12 | | |
| 0,291340714 | OR5B17 | FALSE | Upregulated |
| | OR5B2 | | |
| 0,290109866 | OR5B21 | FALSE | Upregulated |
| | OR5B3 | | |
| 0,100616057 | OR5C1 | FALSE | Upregulated |
| | OR5D13 | | |
| | OR5D14 | | |
| | OR5D16 | | |
| | OR5D18 | | |
| 0,320547785 | OR5E1P | FALSE | Upregulated |
| | OR5F1 | | |

| | | | |
|---|---|---|---|
| | OR5H1 | | |
| | OR5H14 | | |
| | OR5H15 | | |
| 0 | OR5H2 | FALSE | Upregulated |
| | OR5H6 | | |
| | OR5I1 | | |
| | OR5J2 | | |
| 0,286067832 | OR5K1 | FALSE | Upregulated |
| -0,121348224 | OR5K2 | FALSE | Downregulated |
| -0,00509535 | OR5K3 | FALSE | Downregulated |
| | OR5K4 | | |
| | OR5L1 | | |
| | OR5L2 | | |
| | OR5M1 | | |
| | OR5M10 | | |
| | OR5M11 | | |
| | OR5M3 | | |
| | OR5M8 | | |
| | OR5M9 | | |
| | OR5P2 | | |
| | OR5P3 | | |
| | OR5R1 | | |
| | OR5T1 | | |
| | OR5T2 | | |
| | OR5T3 | | |
| 0 | OR5V1 | FALSE | Upregulated |
| | OR5W2 | | |
| | OR6A2 | | |
| 0,720836731 | OR6B1 | FALSE | Upregulated |
| 0,297424754 | OR6B2 | FALSE | Upregulated |
| | OR6B3 | | |
| | OR6C1 | | |
| | OR6C2 | | |
| | OR6C3 | | |
| | OR6C4 | | |
| | OR6C6 | | |
| | OR6C65 | | |
| | OR6C68 | | |
| | OR6C70 | | |
| | OR6C74 | | |
| | OR6C75 | | |
| | OR6C76 | | |
| 1,447873065 | OR6F1 | | Upregulated |
| 0 | OR6K2 | FALSE | Upregulated |
| | OR6K3 | | |
| 0 | OR6K6 | FALSE | Upregulated |
| 0,531761286 | OR6M1 | FALSE | Upregulated |
| | OR6N1 | | |
| | OR6N2 | | |
| | OR6P1 | | |
| | OR6Q1 | | |

| | | | |
|---|---|---|---|
| | OR6S1 | | |
| -0,007332004 | OR6T1 | FALSE | Downregulated |
| 0,60951597 | OR6V1 | FALSE | Upregulated |
| -0,002107024 | OR6W1P | FALSE | Downregulated |
| 0,834760791 | OR6X1 | FALSE | Upregulated |
| | OR6Y1 | | |
| 0 | OR7A10 | FALSE | Upregulated |
| | OR7A17 | | |
| | OR7A5 | | |
| 0 | OR7C1 | FALSE | Upregulated |
| | OR7C2 | | |
| 0 | OR7D2 | FALSE | Upregulated |
| -0,181266239 | OR7D4 | FALSE | Downregulated |
| -0,252983326 | OR7E12P | FALSE | Downregulated |
| 0,099934242 | OR7E14P | FALSE | Upregulated |
| 0 | OR7E156P | FALSE | Upregulated |
| | OR7E24 | | |
| 0,423825116 | OR7E37P | FALSE | Upregulated |
| | OR7E5P | | |
| -0,119618736 | OR7E91P | FALSE | Downregulated |
| | OR7G1 | | |
| | OR7G2 | | |
| | OR7G3 | | |
| | OR8A1 | | |
| | OR8B12 | | |
| 0 | OR8B2 | FALSE | Upregulated |
| | OR8B3 | | |
| | OR8B4 | | |
| 0,41482555 | OR8B8 | FALSE | Upregulated |
| | OR8D1 | | |
| 0 | OR8D2 | FALSE | Upregulated |
| | OR8D4 | | |
| | OR8G1 | | |
| | OR8G2 | | |
| | OR8G5 | | |
| | OR8H1 | | |
| | OR8H2 | | |
| | OR8H3 | | |
| | OR8I2 | | |
| | OR8J1 | | |
| | OR8J3 | | |
| | OR8K1 | | |
| | OR8K3 | | |
| | OR8K5 | | |
| | OR8S1 | | |
| | OR8U1 | | |
| | OR9A2 | | |
| 0 | OR9A4 | FALSE | Upregulated |
| | OR9G1 | | |
| | OR9G4 | | |
| 0,595623588 | OR9I1 | FALSE | Upregulated |

| | | | |
|---|---|---|---|
| | OR9K2 | | |
| | OR9Q1 | | |
| | OR9Q2 | | |
| -0,594906014 | ORC1 | FALSE | Downregulated |
| -0,16803694 | ORC5 | FALSE | Downregulated |
| -0,177158074 | ORC6 | FALSE | Downregulated |
| 0,412018967 | OSBPL10-AS1 | FALSE | Upregulated |
| 0,068177674 | OSBPL7 | FALSE | Upregulated |
| 0,402771173 | OSCAR | FALSE | Upregulated |
| -0,067545409 | OSCP1 | FALSE | Downregulated |
| 0,077936549 | OSM | FALSE | Upregulated |
| 0,336655064 | OSR1 | FALSE | Upregulated |
| 0,299817909 | OSR2 | FALSE | Upregulated |
| 0,057200472 | OSTCP1 | FALSE | Upregulated |
| -0,146335101 | OSTCP2 | FALSE | Downregulated |
| | OSTM1-AS1 | | |
| -0,063724589 | OSTN | FALSE | Downregulated |
| 0,737694699 | OTOA | FALSE | Upregulated |
| 0,26654088 | OTOF | FALSE | Upregulated |
| | OTOG | | |
| 0 | OTOGL | FALSE | Upregulated |
| | OTOL1 | | |
| | OTOP1 | | |
| | OTOP2 | | |
| -0,440594498 | OTOP3 | FALSE | Downregulated |
| | OTOR | | |
| | OTOS | | |
| | OTP | | |
| 0 | OTUD6A | FALSE | Upregulated |
| 0,589271444 | OTUD7A | FALSE | Upregulated |
| | OTX1 | | |
| | OTX2 | | |
| 0,151903275 | OTX2-AS1 | FALSE | Upregulated |
| 0,677671127 | OVCH1 | FALSE | Upregulated |
| 0,687135818 | OVCH1-AS1 | FALSE | Upregulated |
| 0,147632422 | OVCH2 | FALSE | Upregulated |
| -0,84962416 | OVGP1 | FALSE | Downregulated |
| -0,704372557 | OVOL1 | FALSE | Downregulated |
| 0 | OVOL1-AS1 | FALSE | Upregulated |
| 0,183942316 | OVOL2 | FALSE | Upregulated |
| -0,400383429 | OVOL3 | FALSE | Downregulated |
| 0,286495597 | OVOS | FALSE | Upregulated |
| 0 | OVOS2 | FALSE | Upregulated |
| 0,258373752 | OXCT1 | FALSE | Upregulated |
| 0,300241802 | OXCT1-AS1 | FALSE | Upregulated |
| 0,804212803 | OXCT2 | FALSE | Upregulated |
| | OXGR1 | | |
| 0,235530664 | OXLD1 | FALSE | Upregulated |
| 0,174558526 | OXSM | FALSE | Upregulated |
| -0,044759782 | OXT | FALSE | Downregulated |
| -0,544241187 | OXTR | FALSE | Downregulated |

| | | | |
|---|---|---|---|
| -0,858083257 | P2RX1 | FALSE | Downregulated |
| 0,163703551 | P2RX3 | FALSE | Upregulated |
| -0,753544186 | P2RX5 | FALSE | Downregulated |
| -1,205815777 | P2RX6 | | Downregulated |
| | P2RX6P | | |
| -0,384774948 | P2RY1 | FALSE | Downregulated |
| 0,95529769 | P2RY10 | FALSE | Upregulated |
| -0,588860519 | P2RY12 | FALSE | Downregulated |
| 0,158620604 | P2RY14 | FALSE | Upregulated |
| -0,157410586 | P2RY4 | FALSE | Downregulated |
| 0,380585685 | P2RY6 | FALSE | Upregulated |
| | P2RY8-2 | | |
| -0,364146288 | PAAF1 | FALSE | Downregulated |
| | PABPC1L2A | | |
| | PABPC1L2B | | |
| -0,453009508 | PABPC1P2 | FALSE | Downregulated |
| -0,161322364 | PABPC3 | FALSE | Downregulated |
| 0,129576449 | PABPC4L | FALSE | Upregulated |
| -0,183682959 | PABPC5 | FALSE | Downregulated |
| 0,497542799 | PABPN1L | FALSE | Upregulated |
| -0,087685826 | PACRG | FALSE | Downregulated |
| 0,210890438 | PACRG-AS1 | FALSE | Upregulated |
| 0,116162334 | PACRGL | FALSE | Upregulated |
| -0,007378234 | PACSIN1 | FALSE | Downregulated |
| -2,043000347 | PADI1 | | Downregulated |
| 0,640580625 | PADI2 | FALSE | Upregulated |
| | PADI3 | | |
| 0,024375104 | PADI4 | FALSE | Upregulated |
| 0 | PADI6 | FALSE | Upregulated |
| | PAEP | | |
| -0,007796837 | PAFAH1B3 | FALSE | Downregulated |
| | PAGE1 | | |
| | PAGE2 | | |
| | PAGE2B | | |
| | PAGE3 | | |
| 0 | PAGE4 | FALSE | Upregulated |
| 0,000504523 | PAGE5 | FALSE | Upregulated |
| -0,568047945 | PAK1IP1 | FALSE | Downregulated |
| 0,500434557 | PAK3 | FALSE | Upregulated |
| 0,768091611 | PAK6 | FALSE | Upregulated |
| -0,317062814 | PAK7 | FALSE | Downregulated |
| -1,155649917 | PAM16 | | Downregulated |
| -0,15564468 | PANDAR | FALSE | Downregulated |
| 0,55857636 | PANX3 | FALSE | Upregulated |
| 0 | PAPL | FALSE | Upregulated |
| 0 | PAPOLB | FALSE | Upregulated |
| 1,018235752 | PAPPA2 | | Upregulated |
| | PARD3-AS1 | | |
| -0,066591194 | PARD6A | FALSE | Downregulated |
| 0,147709294 | PARK2 | FALSE | Upregulated |
| -0,676383239 | PARM1 | FALSE | Downregulated |

| | | | |
|---|---|---|---|
| -0,569728642 | PARPBP | FALSE | Downregulated |
| 0,52514878 | PARS2 | FALSE | Upregulated |
| | PASD1 | | |
| -0,260525553 | PATE1 | FALSE | Downregulated |
| | PATE2 | | |
| -1,362018084 | PATE3 | | Downregulated |
| 0,509374674 | PATL2 | FALSE | Upregulated |
| | PAX1 | | |
| 0,163738024 | PAX2 | FALSE | Upregulated |
| 0,467535788 | PAX3 | FALSE | Upregulated |
| | PAX4 | | |
| 0,46183651 | PAX5 | FALSE | Upregulated |
| 0 | PAX7 | FALSE | Upregulated |
| 0,178194613 | PAX9 | FALSE | Upregulated |
| -0,442669535 | PBDC1 | FALSE | Downregulated |
| 0,11070566 | PBK | FALSE | Upregulated |
| 0,664671186 | PBOV1 | FALSE | Upregulated |
| 0,056806273 | PBX4 | FALSE | Upregulated |
| 1,550706694 | PCA3 | | Upregulated |
| -0,120386414 | PCAT1 | FALSE | Downregulated |
| 0,410239381 | PCAT14 | FALSE | Upregulated |
| 0,853558769 | PCAT18 | FALSE | Upregulated |
| -0,543629089 | PCAT19 | FALSE | Downregulated |
| 0,284008197 | PCAT4 | FALSE | Upregulated |
| 0,00139671 | PCAT5 | FALSE | Upregulated |
| 0,114645915 | PCAT6 | FALSE | Upregulated |
| -0,562589182 | PCBP1-AS1 | FALSE | Downregulated |
| 0,56958149 | PCBP2-OT1 | FALSE | Upregulated |
| 0,400273466 | PCBP3 | FALSE | Upregulated |
| 0,170493671 | PCCA-AS1 | FALSE | Upregulated |
| -0,187683035 | PCDH10 | FALSE | Downregulated |
| 0,323244402 | PCDH11X | FALSE | Upregulated |
| 0,962934315 | PCDH11Y | FALSE | Upregulated |
| 0,764492954 | PCDH15 | FALSE | Upregulated |
| 0,057028342 | PCDH19 | FALSE | Upregulated |
| 0,801020811 | PCDH20 | FALSE | Upregulated |
| -0,155727797 | PCDH8 | FALSE | Downregulated |
| 0 | PCDH9-AS2 | FALSE | Upregulated |
| 0,258888941 | PCDH9-AS3 | FALSE | Upregulated |
| 1,063824407 | PCDH9-AS4 | | Upregulated |
| 0,540190286 | PCDP1 | FALSE | Upregulated |
| 0,02046952 | PCED1B | FALSE | Upregulated |
| | PCGEM1 | | |
| -0,180829782 | PCGF6 | FALSE | Downregulated |
| 0,664673672 | PCLO | FALSE | Upregulated |
| -0,616098229 | PCNAP1 | FALSE | Downregulated |
| 0,149886964 | PCP2 | FALSE | Upregulated |
| | PCP4 | | |
| 0 | PCSK1N | FALSE | Upregulated |
| 0,036838811 | PCSK2 | FALSE | Upregulated |
| 0,478654555 | PCYOX1L | FALSE | Upregulated |

| | | | |
|---|---|---|---|
| 0,305234476 | PCYT1B | FALSE | Upregulated |
| -0,127349728 | PDCD10 | FALSE | Downregulated |
| 0,042528228 | PDCD2L | FALSE | Upregulated |
| -1,016770666 | PDCD4-AS1 | | Downregulated |
| | PDCL2 | | |
| 0,960067853 | PDCL3 | FALSE | Upregulated |
| 0,679601721 | PDCL3P4 | FALSE | Upregulated |
| -0,189348455 | PDE10A | FALSE | Downregulated |
| 0,263077234 | PDE1C | FALSE | Upregulated |
| 0,040563357 | PDE6C | FALSE | Upregulated |
| -0,099295835 | PDE6D | FALSE | Downregulated |
| -0,179778133 | PDE6G | FALSE | Downregulated |
| 0 | PDE6H | FALSE | Upregulated |
| -1,103670924 | PDE9A | | Downregulated |
| -0,21354598 | PDGFD | FALSE | Downregulated |
| | PDHA2 | | |
| 0,148963708 | PDIA2 | FALSE | Upregulated |
| -0,291536868 | PDILT | FALSE | Downregulated |
| 0,875394236 | PDLIM3 | FALSE | Upregulated |
| 0,069244594 | PDLIM4 | FALSE | Upregulated |
| 0,20766658 | PDPN | FALSE | Upregulated |
| -0,096969922 | PDRG1 | FALSE | Downregulated |
| -0,286368559 | PDX1 | FALSE | Downregulated |
| 0,730403311 | PDX1-AS1 | FALSE | Upregulated |
| 0 | PDYN | FALSE | Upregulated |
| -0,090196395 | PDZD11 | FALSE | Downregulated |
| -0,535459905 | PDZD3 | FALSE | Downregulated |
| 0,680643627 | PDZD7 | FALSE | Upregulated |
| 0,546204421 | PDZK1IP1 | FALSE | Upregulated |
| -0,305731601 | PDZRN4 | FALSE | Downregulated |
| 0,233269391 | PEBP4 | FALSE | Upregulated |
| | PENK | | |
| 0,325097589 | PER4 | FALSE | Upregulated |
| -0,116397933 | PET117 | FALSE | Downregulated |
| -1,204894281 | PEX7 | | Downregulated |
| 0,250718253 | PF4 | FALSE | Upregulated |
| | PF4V1 | | |
| 0,472058322 | PFDN4 | FALSE | Upregulated |
| 0,220414702 | PFKFB4 | FALSE | Upregulated |
| -0,509392285 | PFN3 | FALSE | Downregulated |
| 0,680935266 | PFN4 | FALSE | Upregulated |
| -0,13840432 | PGA3 | FALSE | Downregulated |
| -0,451897379 | PGA4 | FALSE | Downregulated |
| 0,744467962 | PGA5 | FALSE | Upregulated |
| 0,309236918 | PGAP1 | FALSE | Upregulated |
| 0,118171008 | PGBD1 | FALSE | Upregulated |
| 0 | PGBD3 | FALSE | Upregulated |
| -0,093920123 | PGBD4 | FALSE | Downregulated |
| 0,548103923 | PGC | FALSE | Upregulated |
| | PGCP1 | | |
| 0,32536494 | PGF | FALSE | Upregulated |

| | | | |
|---|---|---|---|
| 0,126733654 | PGK2 | FALSE | Upregulated |
| 0,055348557 | PGLYRP1 | FALSE | Upregulated |
| 0 | PGLYRP3 | FALSE | Upregulated |
| | PGLYRP4 | | |
| 0,131293312 | PGM2L1 | FALSE | Upregulated |
| 0,272626305 | PGM5-AS1 | FALSE | Upregulated |
| -0,181445647 | PGPEP1L | FALSE | Downregulated |
| -0,581195414 | PHACTR3 | FALSE | Downregulated |
| -0,221532661 | PHEX | FALSE | Downregulated |
| 0,094461128 | PHEX-AS1 | FALSE | Upregulated |
| -0,018149848 | PHF21B | FALSE | Downregulated |
| -0,16719314 | PHF7 | FALSE | Downregulated |
| 0,85502401 | PHGR1 | FALSE | Upregulated |
| -0,029140721 | PHLDA2 | FALSE | Downregulated |
| -0,681821144 | PHOSPHO2 | FALSE | Downregulated |
| -0,744256903 | PHOSPHO2-KLHL23 | FALSE | Downregulated |
| 0,037254067 | PHOX2A | FALSE | Upregulated |
| | PHOX2B | | |
| 0,23602158 | PI15 | FALSE | Upregulated |
| 0,081054266 | PI16 | FALSE | Upregulated |
| | PI3 | | |
| -0,229151456 | PIF1 | FALSE | Downregulated |
| 0,13544129 | PIFO | FALSE | Upregulated |
| -0,19349764 | PIGA | FALSE | Downregulated |
| 0,36380026 | PIGC | FALSE | Upregulated |
| 0,41449789 | PIGP | FALSE | Upregulated |
| 0,129169781 | PIGX | FALSE | Upregulated |
| -0,169446172 | PIH1D2 | FALSE | Downregulated |
| | PIH1D3 | | |
| 0,452534534 | PIK3R6 | FALSE | Upregulated |
| | PIN1P1 | | |
| 0,605346139 | PINLYP | FALSE | Upregulated |
| -0,162738796 | PINX1 | FALSE | Downregulated |
| 0,293242527 | PIP | FALSE | Upregulated |
| 0,398581508 | PIP5K1B | FALSE | Upregulated |
| 0,232607972 | PIP5KL1 | FALSE | Upregulated |
| -0,274641094 | PIR | FALSE | Downregulated |
| 0,470786082 | PIR-FIGF | FALSE | Upregulated |
| 0 | PISRT1 | FALSE | Upregulated |
| 0,116694118 | PITPNA-AS1 | FALSE | Upregulated |
| -0,159349868 | PITX1 | FALSE | Downregulated |
| 0,277119458 | PITX2 | FALSE | Upregulated |
| -0,002904991 | PITX3 | FALSE | Downregulated |
| -0,122236543 | PIWIL1 | FALSE | Downregulated |
| | PIWIL3 | | |
| 0,083419767 | PIWIL4 | FALSE | Upregulated |
| -0,931940399 | PKD1L1 | FALSE | Downregulated |
| -0,164568372 | PKD1L2 | FALSE | Downregulated |
| 0,334446502 | PKD1L3 | FALSE | Upregulated |
| -0,456138134 | PKD2L1 | FALSE | Downregulated |
| -0,244628748 | PKI55 | FALSE | Downregulated |

| | | | |
|---|---|---|---|
| 0,424539429 | PKIA | FALSE | Upregulated |
| 0,078265874 | PKIB | FALSE | Upregulated |
| 0,566310369 | PKMYT1 | FALSE | Upregulated |
| -0,264324683 | PKNOX2 | FALSE | Downregulated |
| 0 | PKP1 | FALSE | Upregulated |
| 0,174146959 | PKP3 | FALSE | Upregulated |
| 0,308063888 | PLA2G10 | FALSE | Upregulated |
| -0,329946168 | PLA2G1B | FALSE | Downregulated |
| | PLA2G2C | | |
| 1,619546883 | PLA2G2D | | Upregulated |
| | PLA2G2E | | |
| 0,572257659 | PLA2G2F | FALSE | Upregulated |
| -0,00852576 | PLA2G3 | FALSE | Downregulated |
| -0,177660024 | PLA2G4A | FALSE | Downregulated |
| 0 | PLA2G4D | FALSE | Upregulated |
| -0,473112727 | PLA2G4E | FALSE | Downregulated |
| 0 | PLA2G4F | FALSE | Upregulated |
| 0,371048555 | PLA2G5 | FALSE | Upregulated |
| 0,319170789 | PLA2G7 | FALSE | Upregulated |
| | PLAC1 | | |
| 0,178516737 | PLAC8 | FALSE | Upregulated |
| -0,07090938 | PLAC8L1 | FALSE | Downregulated |
| 0,263796311 | PLAC9 | FALSE | Upregulated |
| -0,339467021 | PLAG1 | FALSE | Downregulated |
| -0,048796982 | PLAGL1 | FALSE | Downregulated |
| -0,035127068 | PLAU | FALSE | Downregulated |
| 0,761694525 | PLAUR | FALSE | Upregulated |
| 1,065502419 | PLB1 | | Upregulated |
| 0,567492976 | PLCB4 | FALSE | Upregulated |
| 0,563683828 | PLCD3 | FALSE | Upregulated |
| 0,800222593 | PLCE1-AS1 | FALSE | Upregulated |
| -1,37293864 | PLCH1 | | Downregulated |
| 0,964207293 | PLCH1-AS1 | FALSE | Upregulated |
| 1,02303065 | PLCL1 | | Upregulated |
| | PLCXD1-2 | | |
| -0,246684223 | PLCXD2 | FALSE | Downregulated |
| 0,162918539 | PLCZ1 | FALSE | Upregulated |
| 1,1392649 | PLD4 | | Upregulated |
| 0,301683926 | PLD5 | FALSE | Upregulated |
| 0,150546077 | PLEKHA8P1 | FALSE | Upregulated |
| -0,013236655 | PLEKHD1 | FALSE | Downregulated |
| 0,638923408 | PLEKHN1 | FALSE | Upregulated |
| 0,232534575 | PLEKHS1 | FALSE | Upregulated |
| 0,658773382 | PLGRKT | FALSE | Upregulated |
| 0,032161946 | PLK1 | FALSE | Upregulated |
| -0,031923943 | PLK4 | FALSE | Downregulated |
| 0,315221731 | PLK5 | FALSE | Upregulated |
| 0,33685929 | PLP1 | FALSE | Upregulated |
| 0,280305182 | PLSCR2 | FALSE | Upregulated |
| | PLSCR5 | | |
| 0,236628261 | PLXNB3 | FALSE | Upregulated |

| | | | |
|---|---|---|---|
| -0,32785389 | PMAIP1 | FALSE | Downregulated |
| 0,217572888 | PMCH | FALSE | Upregulated |
| 0,000264809 | PMCHL1 | FALSE | Upregulated |
| 1,505221376 | PMF1-BGLAP | | Upregulated |
| -0,381259879 | PMFBP1 | FALSE | Downregulated |
| 0 | PMP2 | FALSE | Upregulated |
| -0,330443638 | PMP22 | FALSE | Downregulated |
| -0,626634647 | PMS2L2 | FALSE | Downregulated |
| -0,794565906 | PMS2P4 | FALSE | Downregulated |
| | PNCK | | |
| -0,018331638 | PNLDC1 | FALSE | Downregulated |
| | PNLIP | | |
| | PNLIPRP1 | | |
| | PNLIPRP2 | | |
| | PNLIPRP3 | | |
| 0,407196149 | PNMA3 | FALSE | Upregulated |
| -0,208070107 | PNMA5 | FALSE | Downregulated |
| -0,062067191 | PNMAL1 | FALSE | Downregulated |
| -0,410745796 | PNMT | FALSE | Downregulated |
| -1,173391788 | PNOC | | Downregulated |
| | PNPLA1 | | |
| | PNPLA5 | | |
| -0,294316014 | POC1A | FALSE | Downregulated |
| 0,489387649 | POC5 | FALSE | Upregulated |
| -0,210121214 | PODXL2 | FALSE | Downregulated |
| -0,912445733 | POF1B | FALSE | Downregulated |
| -1,113554655 | POLB | | Downregulated |
| -0,286698971 | POLE2 | FALSE | Downregulated |
| 0,183516342 | POLE4 | FALSE | Upregulated |
| -0,196387749 | POLN | FALSE | Downregulated |
| 0,686512069 | POLQ | FALSE | Upregulated |
| 0,032626235 | POLR2J3 | FALSE | Upregulated |
| 0,124533825 | POLR3K | FALSE | Upregulated |
| -0,013274547 | POM121L12 | FALSE | Downregulated |
| | POM121L2 | | |
| | POM121L4P | | |
| -0,148828779 | POM121L8P | FALSE | Downregulated |
| 0,400165345 | POMC | FALSE | Upregulated |
| 0,367632319 | POP1 | FALSE | Upregulated |
| 1,496478228 | POPDC2 | | Upregulated |
| 0,512466613 | POPDC3 | FALSE | Upregulated |
| 0,007727351 | POSTN | FALSE | Upregulated |
| | POTEA | | |
| | POTEB | | |
| | POTEB2 | | |
| | POTEC | | |
| | POTED | | |
| 0,076874484 | POTEE | FALSE | Upregulated |
| 0,71030106 | POTEF | FALSE | Upregulated |
| | POTEG | | |
| | POTEH | | |

| | | | |
|---|---|---|---|
| 0 | POTEI | FALSE | Upregulated |
| 0 | POTEJ | FALSE | Upregulated |
| 0,288284367 | POTEKP | FALSE | Upregulated |
| 0,504482876 | POTEM | FALSE | Upregulated |
| 0,53742974 | POU1F1 | FALSE | Upregulated |
| 0,905215364 | POU2F3 | FALSE | Upregulated |
| 0,942980114 | POU3F1 | FALSE | Upregulated |
| 0,792611872 | POU3F2 | FALSE | Upregulated |
| | POU3F3 | | |
| | POU3F4 | | |
| -0,034887306 | POU4F1 | FALSE | Downregulated |
| | POU4F2 | | |
| 0 | POU4F3 | FALSE | Upregulated |
| 0,830542132 | POU5F1B | FALSE | Upregulated |
| 0,689246505 | POU5F2 | FALSE | Upregulated |
| 0,050260053 | POU6F2 | FALSE | Upregulated |
| 0,40996092 | POU6F2-AS1 | FALSE | Upregulated |
| 0,277037085 | PP12613 | FALSE | Upregulated |
| -0,326016032 | PP13 | FALSE | Downregulated |
| 0 | PP2672 | FALSE | Upregulated |
| 0,27242027 | PPAP2C | FALSE | Upregulated |
| -0,072103809 | PPAPDC1A | FALSE | Downregulated |
| 0,391134522 | PPAPDC3 | FALSE | Upregulated |
| -0,152239604 | PPBP | FALSE | Downregulated |
| -0,144285893 | PPBPP2 | FALSE | Downregulated |
| -0,431958325 | PPCDC | FALSE | Downregulated |
| -0,169956571 | PPEF1 | FALSE | Downregulated |
| | PPEF2 | | |
| -0,308971907 | PPFIA3 | FALSE | Downregulated |
| -0,030304467 | PPFIA4 | FALSE | Downregulated |
| -0,018275152 | PPIAL4B | FALSE | Downregulated |
| -0,010905425 | PPIAL4C | FALSE | Downregulated |
| 0 | PPIAL4D | FALSE | Upregulated |
| | PPIAL4E | | |
| | PPIAL4F | | |
| | PPIAL4G | | |
| 0,405741343 | PPIAP30 | FALSE | Upregulated |
| -0,543426889 | PPIEL | FALSE | Downregulated |
| 0,59404318 | PPIL1 | FALSE | Upregulated |
| 0,304613608 | PPIL3 | FALSE | Upregulated |
| -0,590887089 | PPM1J | FALSE | Downregulated |
| 0,435339439 | PPM1N | FALSE | Upregulated |
| -0,374610579 | PPP1R14A | FALSE | Downregulated |
| 0,642845516 | PPP1R14C | FALSE | Upregulated |
| -0,062229756 | PPP1R14D | FALSE | Downregulated |
| 0,277773191 | PPP1R17 | FALSE | Upregulated |
| 0,333386528 | PPP1R1B | FALSE | Upregulated |
| -0,777086652 | PPP1R1C | FALSE | Downregulated |
| -0,421748741 | PPP1R26-AS1 | FALSE | Downregulated |
| -0,355841986 | PPP1R27 | FALSE | Downregulated |
| 0,289753232 | PPP1R2P3 | FALSE | Upregulated |

| | | | |
|---|---|---|---|
| | PPP1R2P9 | | |
| -0,243944121 | PPP1R32 | FALSE | Downregulated |
| -0,167756337 | PPP1R36 | FALSE | Downregulated |
| 1,048784935 | PPP1R3A | | Upregulated |
| 0,452890655 | PPP1R3D | FALSE | Upregulated |
| 0,277119458 | PPP1R42 | FALSE | Upregulated |
| 1,196178229 | PPP2R2B | | Upregulated |
| | PPP2R2B-IT1 | | |
| 0,006980542 | PPP2R2C | FALSE | Upregulated |
| | PPP2R3B-2 | | |
| 0,93711051 | PPP3R2 | FALSE | Upregulated |
| -0,186895142 | PPP4R1L | FALSE | Downregulated |
| 0,180677194 | PPP5D1 | FALSE | Upregulated |
| | PPY | | |
| | PPY2 | | |
| | PRAC1 | | |
| | PRAC2 | | |
| 0,384561969 | PRAF2 | FALSE | Upregulated |
| | PRAMEF1 | | |
| 0,144839008 | PRAMEF10 | FALSE | Upregulated |
| | PRAMEF11 | | |
| | PRAMEF12 | | |
| | PRAMEF13 | | |
| -0,166026487 | PRAMEF15 | FALSE | Downregulated |
| | PRAMEF17 | | |
| | PRAMEF18 | | |
| | PRAMEF19 | | |
| 0,273422532 | PRAMEF2 | FALSE | Upregulated |
| | PRAMEF20 | | |
| | PRAMEF25 | | |
| | PRAMEF26 | | |
| 0,454943503 | PRAMEF4 | FALSE | Upregulated |
| | PRAMEF5 | | |
| | PRAMEF6 | | |
| | PRAMEF7 | | |
| | PRAMEF8 | | |
| | PRAMEF9 | | |
| 0,142529421 | PRAMENP | FALSE | Upregulated |
| -0,154837955 | PRB3 | FALSE | Downregulated |
| 0,183855892 | PRB4 | FALSE | Upregulated |
| 0,23271448 | PRDM12 | FALSE | Upregulated |
| | PRDM13 | | |
| | PRDM14 | | |
| -0,46630851 | PRDM16 | FALSE | Downregulated |
| -0,543304159 | PRDM5 | FALSE | Downregulated |
| 0,125562732 | PRDM6 | FALSE | Upregulated |
| 1,280186768 | PRDM8 | | Upregulated |
| 0 | PRDM9 | FALSE | Upregulated |
| -0,508845413 | PRELID2 | FALSE | Downregulated |
| | PRG1 | | |
| -1,161515306 | PRG2 | | Downregulated |

| | | | |
|---|---|---|---|
| | PRG3 | | |
| 1,430039799 | PRH1 | | Upregulated |
| -0,349187561 | PRICKLE2-AS2 | FALSE | Downregulated |
| 0,028618782 | PRICKLE3 | FALSE | Upregulated |
| 0,112565245 | PRIM1 | FALSE | Upregulated |
| 0,012012906 | PRIM2 | FALSE | Upregulated |
| -0,371471648 | PRIMA1 | FALSE | Downregulated |
| 0,001471298 | PRKACG | FALSE | Upregulated |
| 1,334309002 | PRKAG2-AS1 | | Upregulated |
| -1,264414765 | PRKAR2B | | Downregulated |
| 0,279538796 | PRKCDBP | FALSE | Upregulated |
| 0 | PRKCG | FALSE | Upregulated |
| 0,080805939 | PRKCQ-AS1 | FALSE | Upregulated |
| 0,126744597 | PRKG2 | FALSE | Upregulated |
| | PRL | | |
| | PRLH | | |
| | PRLHR | | |
| | PRM1 | | |
| -0,010174178 | PRM2 | FALSE | Downregulated |
| | PRM3 | | |
| 0,131466579 | PRMT3 | FALSE | Upregulated |
| | PRMT8 | | |
| 0,002608982 | PRND | FALSE | Upregulated |
| | PRNT | | |
| -0,951254974 | PRO2852 | FALSE | Downregulated |
| -1,748897081 | PRODH | | Downregulated |
| 0,593438725 | PROK1 | FALSE | Upregulated |
| -0,006494402 | PROK2 | FALSE | Downregulated |
| 0,133141933 | PROKR1 | FALSE | Upregulated |
| -0,131198911 | PROKR2 | FALSE | Downregulated |
| 0 | PROL1 | FALSE | Upregulated |
| 0,349208205 | PROM1 | FALSE | Upregulated |
| -0,625817509 | PROM2 | FALSE | Downregulated |
| 0,316924804 | PROP1 | FALSE | Upregulated |
| 0,3083613 | PRPH | FALSE | Upregulated |
| | PRPS1L1 | | |
| 0,04024071 | PRR11 | FALSE | Upregulated |
| 2,015234051 | PRR15 | | Upregulated |
| 0,695123192 | PRR16 | FALSE | Upregulated |
| -0,724265217 | PRR19 | FALSE | Downregulated |
| | PRR20A | | |
| | PRR20B | | |
| | PRR20C | | |
| | PRR20D | | |
| 0,591502865 | PRR20E | FALSE | Upregulated |
| | PRR21 | | |
| -0,016618365 | PRR22 | FALSE | Downregulated |
| | PRR23A | | |
| | PRR23B | | |
| | PRR23C | | |
| | PRR23D1 | | |

| | | | |
|---|---|---|---|
| | PRR23D2 | | |
| 0,757482599 | PRR25 | FALSE | Upregulated |
| 0 | PRR4 | FALSE | Upregulated |
| 0,445165368 | PRR5-ARHGAP8 | FALSE | Upregulated |
| 0,27297085 | PRR7 | FALSE | Upregulated |
| 0 | PRR9 | FALSE | Upregulated |
| -0,520825238 | PRRG2 | FALSE | Downregulated |
| 0,025845563 | PRRG3 | FALSE | Upregulated |
| -0,640028134 | PRRT1 | FALSE | Downregulated |
| 0,740248707 | PRRT3-AS1 | FALSE | Upregulated |
| 0,197485081 | PRRT4 | FALSE | Upregulated |
| 0,041019135 | PRRX1 | FALSE | Upregulated |
| 0,281716047 | PRRX2 | FALSE | Upregulated |
| 0,35439564 | PRSS16 | FALSE | Upregulated |
| 0,641882054 | PRSS2 | FALSE | Upregulated |
| 0,003524371 | PRSS21 | FALSE | Upregulated |
| 0,263028712 | PRSS22 | FALSE | Upregulated |
| 0,636362118 | PRSS27 | FALSE | Upregulated |
| 0,272778305 | PRSS3 | FALSE | Upregulated |
| 0,118063207 | PRSS30P | FALSE | Upregulated |
| 0,161070349 | PRSS33 | FALSE | Upregulated |
| -0,508350787 | PRSS35 | FALSE | Downregulated |
| 0,73459343 | PRSS36 | FALSE | Upregulated |
| 0,026801104 | PRSS38 | FALSE | Upregulated |
| | PRSS41 | | |
| -0,829817711 | PRSS42 | FALSE | Downregulated |
| -0,194839941 | PRSS44 | FALSE | Downregulated |
| -0,266779533 | PRSS45 | FALSE | Downregulated |
| -0,429261322 | PRSS46 | FALSE | Downregulated |
| -0,649210819 | PRSS50 | FALSE | Downregulated |
| 0,894632904 | PRSS55 | FALSE | Upregulated |
| 0 | PRSS56 | FALSE | Upregulated |
| -0,184276496 | PRSS57 | FALSE | Downregulated |
| 0,457195239 | PRTFDC1 | FALSE | Upregulated |
| -0,835335841 | PRTG | FALSE | Downregulated |
| 0 | PRTN3 | FALSE | Upregulated |
| -0,0016469 | PSCA | FALSE | Downregulated |
| -0,807665011 | PSD2 | FALSE | Downregulated |
| -0,0149938 | PSENEN | FALSE | Downregulated |
| 0,177577374 | PSG1 | FALSE | Upregulated |
| 0,448872026 | PSG10P | FALSE | Upregulated |
| 0 | PSG11 | FALSE | Upregulated |
| 0,270235333 | PSG2 | FALSE | Upregulated |
| 0,554985995 | PSG3 | FALSE | Upregulated |
| 0,283052731 | PSG4 | FALSE | Upregulated |
| 0,279633574 | PSG5 | FALSE | Upregulated |
| 0,134731877 | PSG6 | FALSE | Upregulated |
| -0,801050282 | PSG7 | FALSE | Downregulated |
| 0,578224103 | PSG8 | FALSE | Upregulated |
| 0,553103593 | PSG9 | FALSE | Upregulated |
| 0,071020344 | PSIMCT-1 | FALSE | Upregulated |

| | | | |
|---|---|---|---|
| | PSKH2 | | |
| | PSMA8 | | |
| 0 | PSMB11 | FALSE | Upregulated |
| 0,259236514 | PSMG1 | FALSE | Upregulated |
| -0,138374118 | PSMG2 | FALSE | Downregulated |
| -0,18297082 | PSMG4 | FALSE | Downregulated |
| | PSORS1C1 | | |
| -0,179357375 | PSORS1C2 | FALSE | Downregulated |
| -0,583212348 | PSORS1C3 | FALSE | Downregulated |
| -0,310119394 | PSPH | FALSE | Downregulated |
| 0,192260022 | PSPN | FALSE | Upregulated |
| 0,068681998 | PSRC1 | FALSE | Upregulated |
| -0,075844883 | PSTK | FALSE | Downregulated |
| 0,287452729 | PTCHD2 | FALSE | Upregulated |
| -0,13535547 | PTCHD3 | FALSE | Downregulated |
| 0,384745841 | PTCHD4 | FALSE | Upregulated |
| -0,707981858 | PTCRA | FALSE | Downregulated |
| 0 | PTCSC3 | FALSE | Upregulated |
| | PTENP1-AS | | |
| | PTF1A | | |
| 0,540332872 | PTGDR | FALSE | Upregulated |
| 0,56520696 | PTGER1 | FALSE | Upregulated |
| -0,072537174 | PTGER3 | FALSE | Downregulated |
| -0,17168968 | PTGER4P2-CDK2AP2P2 | FALSE | Downregulated |
| 0,146226274 | PTGES | FALSE | Upregulated |
| 0,300404929 | PTGES2-AS1 | FALSE | Upregulated |
| 0,553086817 | PTGES3L | FALSE | Upregulated |
| | PTGES3L-AARSD1 | | |
| 0,210320977 | PTGFR | FALSE | Upregulated |
| -0,204521227 | PTGS2 | FALSE | Downregulated |
| -0,012141539 | PTH | FALSE | Downregulated |
| | PTH2 | | |
| 0,493274546 | PTH2R | FALSE | Upregulated |
| 0,041923448 | PTHLH | FALSE | Upregulated |
| 0,703306126 | PTK7 | FALSE | Upregulated |
| -0,224847777 | PTN | FALSE | Downregulated |
| -0,269133632 | PTP4A3 | FALSE | Downregulated |
| -0,135890697 | PTPDC1 | FALSE | Downregulated |
| 0,042989444 | PTPLA | FALSE | Upregulated |
| -0,138382788 | PTPN20B | FALSE | Downregulated |
| -0,078959152 | PTPN22 | FALSE | Downregulated |
| 0,1724022 | PTPN5 | FALSE | Upregulated |
| 0,280928672 | PTPRN | FALSE | Upregulated |
| -0,023076735 | PTPRN2 | FALSE | Downregulated |
| 0,137768144 | PTPRO | FALSE | Upregulated |
| 0,595972781 | PTPRQ | FALSE | Upregulated |
| 0,689103676 | PTPRR | FALSE | Upregulated |
| -0,412417426 | PTPRT | FALSE | Downregulated |
| 0,001180901 | PTPRVP | FALSE | Upregulated |
| 0,575193108 | PTPRZ1 | FALSE | Upregulated |
| 0,227136906 | PTRH1 | FALSE | Upregulated |

| | | | |
|---|---|---|---|
| 0,188725043 | PTTG1 | FALSE | Upregulated |
| 0,005166247 | PTTG2 | FALSE | Upregulated |
| -0,346104661 | PTX3 | FALSE | Downregulated |
| -0,109861864 | PTX4 | FALSE | Downregulated |
| -0,006565342 | PURG | FALSE | Downregulated |
| 0,349448274 | PUS7 | FALSE | Upregulated |
| 0,730355209 | PVALB | FALSE | Upregulated |
| 1,226258631 | PVRIG2P | | Upregulated |
| 0,306619654 | PVRL3-AS1 | FALSE | Upregulated |
| -0,314898939 | PVRL4 | FALSE | Downregulated |
| -0,318120689 | PVT1 | FALSE | Downregulated |
| 0 | PWRN1 | FALSE | Upregulated |
| 0,248778806 | PWRN3 | FALSE | Upregulated |
| -1,089586796 | PXDNL | | Downregulated |
| 0,25299339 | PXT1 | FALSE | Upregulated |
| 0,667172143 | PYCARD | FALSE | Upregulated |
| -0,317706317 | PYCR1 | FALSE | Downregulated |
| | PYDC1 | | |
| | PYDC2 | | |
| -0,112310177 | PYGO1 | FALSE | Downregulated |
| 0,540856109 | PYHIN1 | FALSE | Upregulated |
| -0,009465128 | PYY | FALSE | Downregulated |
| 0,916646377 | PYY2 | FALSE | Upregulated |
| 0,193466618 | QPCT | FALSE | Upregulated |
| -0,767873748 | QRFP | FALSE | Downregulated |
| | QRFPR | | |
| 0,035374686 | R3HDML | FALSE | Upregulated |
| 0,001726029 | RAB19 | FALSE | Upregulated |
| -0,038147173 | RAB25 | FALSE | Downregulated |
| -0,429768634 | RAB27B | FALSE | Downregulated |
| 0,19332647 | RAB28 | FALSE | Upregulated |
| -0,553756066 | RAB30-AS1 | FALSE | Downregulated |
| 0,305494848 | RAB33A | FALSE | Upregulated |
| 0,314912098 | RAB34 | FALSE | Upregulated |
| -0,708665852 | RAB38 | FALSE | Downregulated |
| 0,425125228 | RAB39A | FALSE | Upregulated |
| 0,252679578 | RAB39B | FALSE | Upregulated |
| 0,092460929 | RAB3A | FALSE | Upregulated |
| 0,221511178 | RAB3B | FALSE | Upregulated |
| -0,049553698 | RAB3C | FALSE | Downregulated |
| 0,297723132 | RAB40AL | FALSE | Upregulated |
| -0,00943231 | RAB41 | FALSE | Downregulated |
| 0,561479201 | RAB44 | FALSE | Upregulated |
| -0,332378852 | RAB4B | FALSE | Downregulated |
| 0,821125817 | RAB4B-EGLN2 | FALSE | Upregulated |
| -0,624667246 | RAB6C | FALSE | Downregulated |
| -0,268011461 | RAB6C-AS1 | FALSE | Downregulated |
| 1,224232182 | RAB7B | | Upregulated |
| -0,135336919 | RAB9B | FALSE | Downregulated |
| 0 | RAB9BP1 | FALSE | Upregulated |
| -0,02724448 | RACGAP1 | FALSE | Downregulated |

| | | | |
|---|---|---|---|
| 0 | RACGAP1P | FALSE | Upregulated |
| 0,016011738 | RAD18 | FALSE | Upregulated |
| 0,185981352 | RAD21-AS1 | FALSE | Upregulated |
| | RAD21L1 | | |
| 0,38739908 | RAD51-AS1 | FALSE | Upregulated |
| 0,178904367 | RAD51AP1 | FALSE | Upregulated |
| 0,250515136 | RAD51AP2 | FALSE | Upregulated |
| -0,246994088 | RAD51C | FALSE | Downregulated |
| -0,002029103 | RAD51D | FALSE | Downregulated |
| -1,183872191 | RAD51L3-RFFL | | Downregulated |
| -0,316303554 | RAD54B | FALSE | Downregulated |
| -0,333471204 | RADIL | FALSE | Downregulated |
| -0,37227274 | RAET1G | FALSE | Downregulated |
| 0,277764892 | RAET1K | FALSE | Upregulated |
| | RAET1L | | |
| -0,166639786 | RAG1 | FALSE | Downregulated |
| 0,074245816 | RAG2 | FALSE | Upregulated |
| -0,788911234 | RAI2 | FALSE | Downregulated |
| -0,39072536 | RALA | FALSE | Downregulated |
| 0,470934367 | RALGAPA1P | FALSE | Upregulated |
| -0,05080728 | RALYL | FALSE | Downregulated |
| 0,460299495 | RAMP2 | FALSE | Upregulated |
| 0,954626582 | RAMP2-AS1 | FALSE | Upregulated |
| 0,246280724 | RANBP17 | FALSE | Upregulated |
| 0,39460219 | RAPGEF4-AS1 | FALSE | Upregulated |
| -0,479016437 | RAPSN | FALSE | Downregulated |
| 0,138015046 | RASA4CP | FALSE | Upregulated |
| -0,089020685 | RASAL1 | FALSE | Downregulated |
| 0,728202233 | RASD2 | FALSE | Upregulated |
| -0,470997622 | RASEF | FALSE | Downregulated |
| 0,337643744 | RASGEF1A | FALSE | Upregulated |
| -0,516781458 | RASGEF1C | FALSE | Downregulated |
| 0,650164583 | RASGRF1 | FALSE | Upregulated |
| 0,233637884 | RASL10A | FALSE | Upregulated |
| 0,327246107 | RASL10B | FALSE | Upregulated |
| -0,481585281 | RASL11A | FALSE | Downregulated |
| 1,19131052 | RASL12 | | Upregulated |
| 0,023733638 | RASSF10 | FALSE | Upregulated |
| 0,209178709 | RASSF8-AS1 | FALSE | Upregulated |
| -0,180106288 | RASSF9 | FALSE | Downregulated |
| | RAX | | |
| | RBAK-RBAKDN | | |
| 0 | RBAKDN | FALSE | Upregulated |
| 0,132913408 | RBBP8NL | FALSE | Upregulated |
| -0,172140486 | RBFADN | FALSE | Downregulated |
| 0,464115959 | RBFOX1 | FALSE | Upregulated |
| -0,004525881 | RBM11 | FALSE | Downregulated |
| -0,132748428 | RBM18 | FALSE | Downregulated |
| 0,897753432 | RBM20 | FALSE | Upregulated |
| -0,027509455 | RBM24 | FALSE | Downregulated |
| -0,626997097 | RBM26-AS1 | FALSE | Downregulated |

| | | | |
|---|---|---|---|
| 0,283626875 | RBM44 | FALSE | Upregulated |
| 0,69960154 | RBM45 | FALSE | Upregulated |
| 0,566032492 | RBM46 | FALSE | Upregulated |
| 0,494500305 | RBMS3-AS2 | FALSE | Upregulated |
| -0,045693587 | RBMS3-AS3 | FALSE | Downregulated |
| 0,4309588 | RBMX2 | FALSE | Upregulated |
| 0,001101941 | RBMXL2 | FALSE | Upregulated |
| -0,002601887 | RBMXL3 | FALSE | Downregulated |
| | RBMY1A1 | | |
| | RBMY1A3P | | |
| | RBMY1B | | |
| | RBMY1D | | |
| | RBMY1E | | |
| | RBMY1F | | |
| | RBMY1J | | |
| | RBMY3AP | | |
| -0,281780688 | RBP3 | FALSE | Downregulated |
| -1,234505021 | RBP7 | | Downregulated |
| -0,427693922 | RBPJL | FALSE | Downregulated |
| -0,622406586 | RBPMS2 | FALSE | Downregulated |
| 0,42152646 | RBX1 | FALSE | Upregulated |
| 0,001141803 | RCAN3AS | FALSE | Upregulated |
| 0,090796483 | RCN2 | FALSE | Upregulated |
| 0,318498721 | RCVRN | FALSE | Upregulated |
| -0,588883755 | RD3 | FALSE | Downregulated |
| 0,197094891 | RD3L | FALSE | Upregulated |
| 0,438590987 | RDH12 | FALSE | Upregulated |
| 0,135812987 | RDH8 | FALSE | Upregulated |
| 0,519344935 | RDM1 | FALSE | Upregulated |
| -0,599497273 | REC8 | FALSE | Downregulated |
| -0,202616798 | REEP2 | FALSE | Downregulated |
| 0,091123593 | REEP4 | FALSE | Upregulated |
| | REG1A | | |
| | REG1B | | |
| | REG1P | | |
| 0 | REG3A | FALSE | Upregulated |
| | REG3G | | |
| -0,06583572 | REG4 | FALSE | Downregulated |
| 0,211701277 | REM1 | FALSE | Upregulated |
| 0,686986824 | REM2 | FALSE | Upregulated |
| 0,046221035 | RENBP | FALSE | Upregulated |
| 0,702885213 | REP15 | FALSE | Upregulated |
| -0,680616069 | REREP3 | FALSE | Downregulated |
| 0,360450315 | RERG | FALSE | Upregulated |
| | RERG-AS1 | | |
| 0,735742122 | RERGL | FALSE | Upregulated |
| | RESP18 | | |
| 2,968823084 | RET | | Upregulated |
| | RETN | | |
| | RETNLB | | |
| 0,287063298 | REXO1L1P | FALSE | Upregulated |

| | | | |
|---|---|---|---|
| 0 | REXO1L2P | FALSE | Upregulated |
| 0,469047644 | RFC3 | FALSE | Upregulated |
| 0,426078501 | RFC4 | FALSE | Upregulated |
| 0,739370183 | RFESD | FALSE | Upregulated |
| -0,350492102 | RFPL1 | FALSE | Downregulated |
| 0,160588604 | RFPL2 | FALSE | Upregulated |
| 0,247741083 | RFPL3 | FALSE | Upregulated |
| 0,887702785 | RFPL3S | FALSE | Upregulated |
| 0,460644655 | RFPL4A | FALSE | Upregulated |
| -0,217093102 | RFPL4AL1 | FALSE | Downregulated |
| | RFPL4B | | |
| 1,413438755 | RFX4 | | Upregulated |
| | RFX6 | | |
| 0,128408124 | RFX8 | FALSE | Upregulated |
| 0,380181942 | RFXAP | FALSE | Upregulated |
| -0,562682965 | RGAG1 | FALSE | Downregulated |
| 0,23160382 | RGCC | FALSE | Upregulated |
| -0,074665926 | RGMA | FALSE | Downregulated |
| 1,769461455 | RGMB-AS1 | | Upregulated |
| 0 | RGPD4-AS1 | FALSE | Upregulated |
| | RGR | | |
| 1,095537179 | RGS1 | | Upregulated |
| 0,25017199 | RGS10 | FALSE | Upregulated |
| 0,401664475 | RGS11 | FALSE | Upregulated |
| 0,290109866 | RGS13 | FALSE | Upregulated |
| 0,903048444 | RGS16 | FALSE | Upregulated |
| 0,396179004 | RGS18 | FALSE | Upregulated |
| 0,0444857 | RGS2 | FALSE | Upregulated |
| 0 | RGS21 | FALSE | Upregulated |
| | RGS22 | | |
| -0,268503105 | RGS4 | FALSE | Downregulated |
| 0,476637535 | RGS6 | FALSE | Upregulated |
| 0 | RGS7 | FALSE | Upregulated |
| 0 | RGS8 | FALSE | Upregulated |
| 0,419031762 | RGS9 | FALSE | Upregulated |
| 0,295400949 | RGS9BP | FALSE | Upregulated |
| 0,224236321 | RGSL1 | FALSE | Upregulated |
| 0 | RHAG | FALSE | Upregulated |
| 0,354210983 | RHBDL1 | FALSE | Upregulated |
| 0,878297908 | RHBDL3 | FALSE | Upregulated |
| 0,338739558 | RHCG | FALSE | Upregulated |
| -0,031111794 | RHEBL1 | FALSE | Downregulated |
| -0,046457931 | RHNO1 | FALSE | Downregulated |
| 0,306325279 | RHO | FALSE | Upregulated |
| 0,474263951 | RHOH | FALSE | Upregulated |
| 0,115792124 | RHOJ | FALSE | Upregulated |
| 0 | RHOV | FALSE | Upregulated |
| 0,49486253 | RHOXF1 | FALSE | Upregulated |
| -0,414080984 | RHOXF2B | FALSE | Downregulated |
| 0,83984804 | RHPN1 | FALSE | Upregulated |
| 0,926679029 | RHPN1-AS1 | FALSE | Upregulated |

| | | | |
|---|---|---|---|
| 0,144797762 | RIBC2 | FALSE | Upregulated |
| | RIIAD1 | | |
| -0,373900682 | RILPL2 | FALSE | Downregulated |
| -1,325471372 | RIMBP2 | | Downregulated |
| 0,628411839 | RIMBP3 | FALSE | Upregulated |
| 1,151445933 | RIMBP3B | | Upregulated |
| 0,94549695 | RIMBP3C | FALSE | Upregulated |
| -0,352065949 | RIMS1 | FALSE | Downregulated |
| -0,49390013 | RIMS2 | FALSE | Downregulated |
| -0,59553395 | RIMS3 | FALSE | Downregulated |
| -0,083598136 | RIMS4 | FALSE | Downregulated |
| 0,782246695 | RIN1 | FALSE | Upregulated |
| 0,105613458 | RIOK1 | FALSE | Upregulated |
| 0,905270005 | RIPK3 | FALSE | Upregulated |
| | RIPPLY2 | | |
| 0,721362875 | RIPPLY3 | FALSE | Upregulated |
| -0,233420219 | RIT2 | FALSE | Downregulated |
| 0,230245762 | RLBP1 | FALSE | Upregulated |
| -0,156988955 | RLN1 | FALSE | Downregulated |
| 0,282086417 | RLN2 | FALSE | Upregulated |
| 0,130817146 | RLN3 | FALSE | Upregulated |
| 2,270769358 | RLTPR | | Upregulated |
| -0,21435927 | RMDN2-AS1 | FALSE | Downregulated |
| 0,192451955 | RMI1 | FALSE | Upregulated |
| 0,51778214 | RMI2 | FALSE | Upregulated |
| 0 | RMST | FALSE | Upregulated |
| | RNA5-8S5 | | |
| | RNA5S1 | | |
| | RNA5S10 | | |
| | RNA5S11 | | |
| | RNA5S12 | | |
| | RNA5S13 | | |
| | RNA5S14 | | |
| | RNA5S15 | | |
| | RNA5S16 | | |
| | RNA5S17 | | |
| | RNA5S2 | | |
| | RNA5S3 | | |
| | RNA5S4 | | |
| | RNA5S5 | | |
| | RNA5S6 | | |
| | RNA5S7 | | |
| 0 | RNA5S8 | FALSE | Upregulated |
| -1,207386885 | RNA5S9 | | Downregulated |
| | RNASE11 | | |
| | RNASE12 | | |
| 0,264200922 | RNASE2 | FALSE | Upregulated |
| 0,422091058 | RNASE3 | FALSE | Upregulated |
| 0,355775962 | RNASE6 | FALSE | Upregulated |
| -0,45906994 | RNASE8 | FALSE | Downregulated |
| | RNASE9 | | |

| | | | |
|---|---|---|---|
| -0,37324979 | RNASEH1-AS1 | FALSE | Downregulated |
| 0,307337002 | RNASEH2A | FALSE | Upregulated |
| 1,534002878 | RNASEH2B-AS1 | | Upregulated |
| 0,961833849 | RNF103-CHMP3 | FALSE | Upregulated |
| 0,210499845 | RNF112 | FALSE | Upregulated |
| -0,418062569 | RNF122 | FALSE | Downregulated |
| 0,395994634 | RNF126P1 | FALSE | Upregulated |
| 0,029181907 | RNF133 | FALSE | Upregulated |
| 0,229208781 | RNF144A-AS1 | FALSE | Upregulated |
| -0,187778522 | RNF148 | FALSE | Downregulated |
| -0,01289951 | RNF151 | FALSE | Downregulated |
| 0,552060782 | RNF157 | FALSE | Upregulated |
| 1,070013107 | RNF157-AS1 | | Upregulated |
| 0,14146106 | RNF17 | FALSE | Upregulated |
| 0,078356707 | RNF175 | FALSE | Upregulated |
| 0,454943503 | RNF182 | FALSE | Upregulated |
| -0,002439674 | RNF183 | FALSE | Downregulated |
| -0,351870671 | RNF186 | FALSE | Downregulated |
| 0,006871907 | RNF2 | FALSE | Upregulated |
| -0,735573502 | RNF208 | FALSE | Downregulated |
| 0,9264598 | RNF212 | FALSE | Upregulated |
| | RNF216-IT1 | | |
| 0,520517558 | RNF219 | FALSE | Upregulated |
| 0,259193923 | RNF223 | FALSE | Upregulated |
| 0,105113199 | RNF224 | FALSE | Upregulated |
| 0,461198427 | RNF5P1 | FALSE | Upregulated |
| -0,60805903 | RNFT2 | FALSE | Downregulated |
| 0 | RNU105B | FALSE | Upregulated |
| | RNU105C | | |
| -0,847126038 | RNU11 | FALSE | Downregulated |
| 0,410124573 | RNU12 | FALSE | Upregulated |
| 0,167332563 | RNU2-1 | FALSE | Upregulated |
| -0,119140418 | RNU4ATAC | FALSE | Downregulated |
| 0 | RNU5A-1 | FALSE | Upregulated |
| 0 | RNU5B-1 | FALSE | Upregulated |
| | RNU5D-1 | | |
| | RNU5E-1 | | |
| | RNU5F-1 | | |
| | RNU6-1 | | |
| | RNU6-10P | | |
| | RNU6-14P | | |
| | RNU6-15P | | |
| | RNU6-16P | | |
| | RNU6-19P | | |
| -0,15371215 | RNU6-2 | FALSE | Downregulated |
| | RNU6-21P | | |
| | RNU6-23P | | |
| | RNU6-26P | | |
| | RNU6-28P | | |
| | RNU6-30P | | |
| | RNU6-31P | | |

| | | | |
|---|---|---|---|
| | RNU6-33P | | |
| | RNU6-34P | | |
| | RNU6-35P | | |
| | RNU6-36P | | |
| | RNU6-39P | | |
| | RNU6-42P | | |
| | RNU6-45P | | |
| | RNU6-46P | | |
| | RNU6-48P | | |
| | RNU6-52P | | |
| | RNU6-53P | | |
| | RNU6-55P | | |
| | RNU6-56P | | |
| | RNU6-57P | | |
| | RNU6-58P | | |
| | RNU6-59P | | |
| | RNU6-63P | | |
| | RNU6-64P | | |
| | RNU6-66P | | |
| | RNU6-67P | | |
| | RNU6-68P | | |
| | RNU6-69P | | |
| | RNU6-7 | | |
| | RNU6-71P | | |
| | RNU6-72P | | |
| | RNU6-75P | | |
| | RNU6-76P | | |
| | RNU6-78P | | |
| | RNU6-79P | | |
| | RNU6-8 | | |
| | RNU6-81P | | |
| | RNU6-82P | | |
| | RNU6-83P | | |
| -0,000115243 | RNU6-9 | FALSE | Downregulated |
| 0,35756557 | RNU6ATAC | FALSE | Upregulated |
| | RNU7-1 | | |
| | RNU86 | | |
| 0 | RNVU1-10 | FALSE | Upregulated |
| 0,316000599 | RNVU1-14 | FALSE | Upregulated |
| 0 | RNVU1-15 | FALSE | Upregulated |
| | RNVU1-17 | | |
| 0 | RNVU1-19 | FALSE | Upregulated |
| 0,44686621 | RNVU1-3 | FALSE | Upregulated |
| 0,298871691 | RNVU1-4 | FALSE | Upregulated |
| 0,588277018 | RNVU1-6 | FALSE | Upregulated |
| -0,518732498 | RNY1 | FALSE | Downregulated |
| 0,153186133 | RNY3 | FALSE | Upregulated |
| | RNY4 | | |
| | RNY5 | | |
| 0,422748435 | ROBO2 | FALSE | Upregulated |
| 0,282680929 | ROCK1P1 | FALSE | Upregulated |

0,016475953 ROM1 FALSE Upregulated
ROPN1
-0,511570801 ROPN1B FALSE Downregulated
0,593924306 ROPN1L FALSE Upregulated
0,352473765 ROPN1L-AS1 FALSE Upregulated
0,06909925 ROR1 FALSE Upregulated
-0,102158812 ROR2 FALSE Downregulated
1,049885654 RORB Upregulated
0,89265472 ROS1 FALSE Upregulated
-0,187873523 RP1 FALSE Downregulated
1,098778205 RP1-102G20.5 Upregulated
RP1-104O17.1
0 RP1-137K24.1 FALSE Upregulated
0,281652067 RP1-140J1.1 FALSE Upregulated
0,545475223 RP1-167G20.1 FALSE Upregulated
0,486144626 RP1-172N19.2 FALSE Upregulated
0,600482083 RP1-207H1.3 FALSE Upregulated
RP1-224A6.3
1,152552848 RP1-234P15.4 Upregulated
0,63649986 RP1-249H1.4 FALSE Upregulated
0,176022973 RP1-257A15.1 FALSE Upregulated
0 RP1-272J12.1 FALSE Upregulated
0,28181789 RP1-278O22.1 FALSE Upregulated
-0,141913256 RP1-27K12.2 FALSE Downregulated
0 RP1-290I10.5 FALSE Upregulated
0,280105634 RP1-40E16.9 FALSE Upregulated
RP1-63G5.5
0,833660201 RP1-69M21.2 FALSE Upregulated
-0,006255337 RP1-90L6.2 FALSE Downregulated
0,333186725 RP1-93I3.1 FALSE Upregulated
0,709313466 RP11-1008C21.1 FALSE Upregulated
RP11-100N20.1
0,325320229 RP11-1018N14.1 FALSE Upregulated
-0,306603241 RP11-101E14.2 FALSE Downregulated
RP11-104D21.3
0,293242527 RP11-1055B8.3 FALSE Upregulated
0,383443079 RP11-1057B8.2 FALSE Upregulated
RP11-1079K10.5
0 RP11-107I14.4 FALSE Upregulated
-0,002253536 RP11-107K17.1 FALSE Downregulated
0,915125524 RP11-1080G15.1 FALSE Upregulated
RP11-1084I9.1
0 RP11-1094H24.4 FALSE Upregulated
RP11-109J4.1
0 RP11-10A14.5 FALSE Upregulated
RP11-10L7.1
RP11-10N16.2
-0,011256954 RP11-110H10.2 FALSE Downregulated
0,311264788 RP11-111E14.1 FALSE Upregulated
0,921094884 RP11-111I12.1 FALSE Upregulated
-0,028430776 RP11-1124B17.1 FALSE Downregulated

| | | | |
|---|---|---|---|
| -0,279347828 | RP11-112L6.4 | FALSE | Downregulated |
| 0,32857629 | RP11-1134I14.8 | FALSE | Upregulated |
| -0,003403877 | RP11-115H18.1 | FALSE | Downregulated |
| 0,545037882 | RP11-118B18.1 | FALSE | Upregulated |
| 0,39353557 | RP11-118E18.2 | FALSE | Upregulated |
| 0,293326262 | RP11-1191J2.2 | FALSE | Upregulated |
| -0,507547686 | RP11-119F19.2 | FALSE | Downregulated |
| 0 | RP11-1223D19.1 | FALSE | Upregulated |
| 0,415033641 | RP11-122M14.1 | FALSE | Upregulated |
| | RP11-124K4.1 | | |
| | RP11-125P18.1 | | |
| 0 | RP11-127O4.3 | FALSE | Upregulated |
| | RP11-129J12.2 | | |
| 0,030096083 | RP11-12A2.3 | FALSE | Upregulated |
| 1,165234355 | RP11-132A1.4 | | Upregulated |
| | RP11-1338A24.1 | | |
| | RP11-134O21.1 | | |
| 0 | RP11-134P9.3 | FALSE | Upregulated |
| 0,568196926 | RP11-135J2.4 | FALSE | Upregulated |
| 0,294355549 | RP11-138B4.1 | FALSE | Upregulated |
| | RP11-13A1.1 | | |
| | RP11-13E5.2 | | |
| | RP11-13P5.2 | | |
| 0,143772146 | RP11-140I24.1 | FALSE | Upregulated |
| 0,678554561 | RP11-141O11.2 | FALSE | Upregulated |
| | RP11-142G7.2 | | |
| | RP11-143A12.3 | | |
| -0,00222928 | RP11-143M1.2 | FALSE | Downregulated |
| | RP11-143M1.3 | | |
| | RP11-146N18.1 | | |
| 0 | RP11-148L24.1 | FALSE | Upregulated |
| | RP11-14J7.6 | | |
| 0,277286914 | RP11-14N7.2 | FALSE | Upregulated |
| | RP11-150O12.6 | | |
| | RP11-152L7.2 | | |
| -0,015614825 | RP11-153K16.1 | FALSE | Downregulated |
| 0,28181789 | RP11-154D17.1 | FALSE | Upregulated |
| -0,602569739 | RP11-154D6.1 | FALSE | Downregulated |
| | RP11-157D23.1 | | |
| | RP11-157E21.1 | | |
| | RP11-158A8.1 | | |
| | RP11-158D2.2 | | |
| 0,756645794 | RP11-158M2.5 | FALSE | Upregulated |
| 0,399568572 | RP11-15I11.3 | FALSE | Upregulated |
| | RP11-15K3.1 | | |
| -0,29886975 | RP11-160O5.1 | FALSE | Downregulated |
| 0,689769988 | RP11-161H23.9 | FALSE | Upregulated |
| -0,216803179 | RP11-163N6.2 | FALSE | Downregulated |
| -0,533219454 | RP11-165H23.1 | FALSE | Downregulated |
| 0,237708022 | RP11-165P7.1 | FALSE | Upregulated |
| | RP11-166A12.1 | | |

| | | | |
|---|---|---|---|
| -0,03063553 | RP11-166O4.5 | FALSE | Downregulated |
| 0,164243365 | RP11-167H9.6 | FALSE | Upregulated |
| -0,001674428 | RP11-168E14.1 | FALSE | Downregulated |
| 0 | RP11-168E17.1 | FALSE | Upregulated |
| 0,642852991 | RP11-168O10.6 | FALSE | Upregulated |
| -0,15404221 | RP11-16E12.2 | FALSE | Downregulated |
| 0,290109866 | RP11-170N11.1 | FALSE | Upregulated |
| | RP11-171A24.2 | | |
| | RP11-174G17.2 | | |
| 0 | RP11-175K6.1 | FALSE | Upregulated |
| 0 | RP11-178L8.5 | FALSE | Upregulated |
| | RP11-179A16.1 | | |
| 1,36206337 | RP11-17A4.2 | | Upregulated |
| -0,146017423 | RP11-180I22.2 | FALSE | Downregulated |
| | RP11-181F12.1 | | |
| -0,973393968 | RP11-182I10.3 | FALSE | Downregulated |
| 0,53913616 | RP11-184I16.4 | FALSE | Upregulated |
| -0,186972772 | RP11-188C12.3 | FALSE | Downregulated |
| | RP11-189B4.6 | | |
| | RP11-18F14.1 | | |
| 0,282878546 | RP11-18H21.1 | FALSE | Upregulated |
| 0 | RP11-191L9.4 | FALSE | Upregulated |
| | RP11-191N8.2 | | |
| 0,297178435 | RP11-193H5.1 | FALSE | Upregulated |
| | RP11-198M11.2 | | |
| 0,315889292 | RP11-1C1.4 | FALSE | Upregulated |
| -0,796960069 | RP11-205M3.3 | FALSE | Downregulated |
| -0,161670253 | RP11-208N14.4 | FALSE | Downregulated |
| 0,244860149 | RP11-214C8.2 | FALSE | Upregulated |
| 0 | RP11-214L13.1 | FALSE | Upregulated |
| 0,162673105 | RP11-214L19.1 | FALSE | Upregulated |
| | RP11-215E13.1 | | |
| 0,81482107 | RP11-219G17.4 | FALSE | Upregulated |
| 0,436961738 | RP11-21L23.3 | FALSE | Upregulated |
| 0,559664982 | RP11-222A5.1 | FALSE | Upregulated |
| -0,245771365 | RP11-224O19.2 | FALSE | Downregulated |
| 0,106669364 | RP11-227H15.4 | FALSE | Upregulated |
| 0,433943032 | RP11-22H5.2 | FALSE | Upregulated |
| 0,586623142 | RP11-230B22.1 | FALSE | Upregulated |
| | RP11-230G5.2 | | |
| -0,543380341 | RP11-234B24.2 | FALSE | Downregulated |
| 0,111467343 | RP11-236J17.5 | FALSE | Upregulated |
| | RP11-236P13.1 | | |
| -0,001674428 | RP11-23B15.1 | FALSE | Downregulated |
| 0 | RP11-243E13.1 | FALSE | Upregulated |
| -0,171403242 | RP11-245M24.1 | FALSE | Downregulated |
| 0,447367064 | RP11-24I21.1 | FALSE | Upregulated |
| 0 | RP11-24P4.1 | FALSE | Upregulated |
| | RP11-251P6.1 | | |
| 0 | RP11-252C15.1 | FALSE | Upregulated |
| | RP11-256L11.1 | | |

| | | | |
|---|---|---|---|
| | RP11-257I8.2 | | |
| 0,551876937 | RP11-260E18.1 | FALSE | Upregulated |
| 0,762935329 | RP11-261C10.3 | FALSE | Upregulated |
| | RP11-262K1.1 | | |
| 0,167515107 | RP11-264C15.2 | FALSE | Upregulated |
| | RP11-265P11.2 | | |
| | RP11-267C16.1 | | |
| 0,723767158 | RP11-26J3.1 | FALSE | Upregulated |
| | RP11-271F18.4 | | |
| | RP11-274B18.2 | | |
| 0,250842156 | RP11-276H19.1 | FALSE | Upregulated |
| 0,297237474 | RP11-278H7.1 | FALSE | Upregulated |
| 0,663553443 | RP11-27K13.3 | FALSE | Upregulated |
| | RP11-27N21.3 | | |
| 0 | RP11-280K24.4 | FALSE | Upregulated |
| | RP11-280O1.2 | | |
| | RP11-286B14.1 | | |
| | RP11-286E11.1 | | |
| -0,483561936 | RP11-290F20.1 | FALSE | Downregulated |
| 0,387204635 | RP11-290F20.3 | FALSE | Upregulated |
| 0,742677342 | RP11-290F5.2 | FALSE | Upregulated |
| 0 | RP11-293M10.5 | FALSE | Upregulated |
| 0,008954394 | RP11-294O2.2 | FALSE | Upregulated |
| 0,283658964 | RP11-295M18.2 | FALSE | Upregulated |
| 0,308255697 | RP11-298I3.1 | FALSE | Upregulated |
| | RP11-298O21.5 | | |
| 0 | RP11-299H22.1 | FALSE | Upregulated |
| | RP11-299L17.3 | | |
| | RP11-2A4.3 | | |
| 0,495687872 | RP11-2O17.2 | FALSE | Upregulated |
| 0,459078577 | RP11-305E17.6 | FALSE | Upregulated |
| | RP11-305P22.9 | | |
| -0,019728341 | RP11-307C12.11 | FALSE | Downregulated |
| -0,001674428 | RP11-307C19.3 | FALSE | Downregulated |
| -0,432697215 | RP11-308N19.4 | FALSE | Downregulated |
| -0,158343621 | RP11-310P5.1 | FALSE | Downregulated |
| | RP11-313C15.1 | | |
| -1,221349788 | RP11-314C16.1 | | Downregulated |
| -0,632204044 | RP11-314D7.2 | FALSE | Downregulated |
| 0,842780681 | RP11-317J10.2 | FALSE | Upregulated |
| 0,400600353 | RP11-317M11.1 | FALSE | Upregulated |
| | RP11-318I4.1 | | |
| 0,941196312 | RP11-318M2.2 | FALSE | Upregulated |
| -0,449371889 | RP11-319E12.1 | FALSE | Downregulated |
| | RP11-319E16.1 | | |
| 0,908738687 | RP11-319G6.1 | FALSE | Upregulated |
| 0,140834165 | RP11-320G24.1 | FALSE | Upregulated |
| | RP11-324L3.1 | | |
| -0,278311 | RP11-326F20.5 | FALSE | Downregulated |
| | RP11-327I22.6 | | |
| -1,079946468 | RP11-328K4.1 | | Downregulated |

| | | | |
|---|---|---|---|
| 0,482305127 | RP11-329B9.1 | FALSE | Upregulated |
| 0,463773207 | RP11-329J18.2 | FALSE | Upregulated |
| | RP11-32F11.2 | | |
| | RP11-330A16.1 | | |
| -0,034656718 | RP11-333A23.4 | FALSE | Downregulated |
| -0,043464584 | RP11-333E1.1 | FALSE | Downregulated |
| | RP11-33G16.1 | | |
| 0 | RP11-341A22.2 | FALSE | Upregulated |
| 0,0255141 | RP11-342C23.4 | FALSE | Upregulated |
| | RP11-343J18.1 | | |
| -0,069521337 | RP11-344B5.2 | FALSE | Downregulated |
| 0,289349293 | RP11-344E13.3 | FALSE | Upregulated |
| | RP11-344G13.1 | | |
| -0,005418383 | RP11-345L23.1 | FALSE | Downregulated |
| | RP11-346D6.6 | | |
| -0,011481061 | RP11-347D21.3 | FALSE | Downregulated |
| -1,791177732 | RP11-34F13.2 | | Downregulated |
| 1,327137864 | RP11-353N14.5 | | Upregulated |
| 0,424026594 | RP11-353N4.5 | FALSE | Upregulated |
| 1,023977366 | RP11-354E11.2 | | Upregulated |
| | RP11-360P21.2 | | |
| 0,911193583 | RP11-362A1.1 | FALSE | Upregulated |
| 1,358162247 | RP11-362F19.1 | | Upregulated |
| | RP11-363D24.1 | | |
| | RP11-363G15.2 | | |
| 0,260104589 | RP11-364P22.1 | FALSE | Upregulated |
| 0,126023163 | RP11-367N14.2 | FALSE | Upregulated |
| | RP11-368I23.2 | | |
| 0,001141803 | RP11-371M22.1 | FALSE | Upregulated |
| 0,829753234 | RP11-372E1.6 | FALSE | Upregulated |
| 0,436218789 | RP11-378A12.1 | FALSE | Upregulated |
| -0,381335011 | RP11-379B18.5 | FALSE | Downregulated |
| 0,426798348 | RP11-379F4.4 | FALSE | Upregulated |
| -0,06189308 | RP11-379K22.2 | FALSE | Downregulated |
| 0 | RP11-37B2.1 | FALSE | Upregulated |
| 0 | RP11-37N22.1 | FALSE | Upregulated |
| | RP11-384J4.1 | | |
| | RP11-386B13.3 | | |
| 0,441108464 | RP11-386I8.6 | FALSE | Upregulated |
| | RP11-389K14.3 | | |
| 0,380704344 | RP11-394O9.1 | FALSE | Upregulated |
| | RP11-395L14.3 | | |
| -0,144368585 | RP11-397C12.1 | FALSE | Downregulated |
| 0,135715124 | RP11-397C18.2 | FALSE | Upregulated |
| 0,858946924 | RP11-399K21.14 | FALSE | Upregulated |
| | RP11-3B12.2 | | |
| 0,272973268 | RP11-3B12.3 | FALSE | Upregulated |
| 0,517867929 | RP11-3P17.4 | FALSE | Upregulated |
| -0,074555978 | RP11-402G3.3 | FALSE | Downregulated |
| 0,849131404 | RP11-402L6.1 | FALSE | Upregulated |
| | RP11-404O13.1 | | |

| | | | |
|---|---|---|---|
| 0,00139671 | RP11-406A20.1 | FALSE | Upregulated |
| 0,747020191 | RP11-406A20.4 | FALSE | Upregulated |
| | RP11-406A9.2 | | |
| -0,597034989 | RP11-406H4.1 | FALSE | Downregulated |
| 0 | RP11-406O16.1 | FALSE | Upregulated |
| -0,015614825 | RP11-407A16.3 | FALSE | Downregulated |
| | RP11-408H20.2 | | |
| 0,289336322 | RP11-40F8.2 | FALSE | Upregulated |
| | RP11-413P11.1 | | |
| | RP11-415C15.2 | | |
| 0,237117094 | RP11-415D17.4 | FALSE | Upregulated |
| | RP11-416O18.1 | | |
| 0 | RP11-417E7.2 | FALSE | Upregulated |
| -0,827123812 | RP11-418J17.1 | FALSE | Downregulated |
| 0,435773638 | RP11-420A23.1 | FALSE | Upregulated |
| -0,414896411 | RP11-421P23.1 | FALSE | Downregulated |
| -0,295763848 | RP11-423C15.3 | FALSE | Downregulated |
| -0,803052764 | RP11-423H2.3 | FALSE | Downregulated |
| -0,006903421 | RP11-425I13.3 | FALSE | Downregulated |
| 0,356932356 | RP11-426L16.8 | FALSE | Upregulated |
| 0,282340913 | RP11-429H9.4 | FALSE | Upregulated |
| 0,341145785 | RP11-430B1.2 | FALSE | Upregulated |
| -0,021146283 | RP11-430C1.1 | FALSE | Downregulated |
| -0,015614825 | RP11-430H10.1 | FALSE | Downregulated |
| 0,291262359 | RP11-433J20.1 | FALSE | Upregulated |
| -0,356369944 | RP11-434C1.1 | FALSE | Downregulated |
| | RP11-435M3.2 | | |
| 0,089785983 | RP11-436K8.1 | FALSE | Upregulated |
| | RP11-438N16.1 | | |
| -0,240832478 | RP11-439L18.3 | FALSE | Downregulated |
| 0,003528322 | RP11-439L8.3 | FALSE | Upregulated |
| 0,007803058 | RP11-439L8.4 | FALSE | Upregulated |
| 0,001141803 | RP11-440G9.1 | FALSE | Upregulated |
| 1,415687326 | RP11-445H22.3 | | Upregulated |
| 0,160013747 | RP11-446H18.5 | FALSE | Upregulated |
| -0,015614825 | RP11-449D8.1 | FALSE | Downregulated |
| | RP11-451G4.2 | | |
| 0,844514143 | RP11-452H21.4 | FALSE | Upregulated |
| | RP11-452N4.1 | | |
| 0,27104689 | RP11-453E2.2 | FALSE | Upregulated |
| 0 | RP11-454C18.2 | FALSE | Upregulated |
| | RP11-454P21.1 | | |
| 0 | RP11-456O19.4 | FALSE | Upregulated |
| 0,401551596 | RP11-458D21.1 | FALSE | Upregulated |
| 0,476172313 | RP11-45L9.1 | FALSE | Upregulated |
| 0,657110549 | RP11-460N16.1 | FALSE | Upregulated |
| 0 | RP11-461F11.1 | FALSE | Upregulated |
| 0,306728562 | RP11-461F11.3 | FALSE | Upregulated |
| 1,136762232 | RP11-461O7.1 | | Upregulated |
| 0 | RP11-462G2.1 | FALSE | Upregulated |
| -0,015614825 | RP11-462G22.1 | FALSE | Downregulated |

| | | | |
|---|---|---|---|
| | RP11-463J7.2 | | |
| -0,148749314 | RP11-465B22.5 | FALSE | Downregulated |
| -1,280024942 | RP11-466F5.8 | | Downregulated |
| | RP11-46C24.3 | | |
| 0,602714706 | RP11-473M20.14 | FALSE | Upregulated |
| -0,587866138 | RP11-474D1.3 | FALSE | Downregulated |
| 0,685228457 | RP11-475N22.4 | FALSE | Upregulated |
| | RP11-478H13.3 | | |
| 0,498805718 | RP11-478K7.2 | FALSE | Upregulated |
| | RP11-480C22.1 | | |
| -0,304170872 | RP11-480I12.7 | FALSE | Downregulated |
| | RP11-482D24.2 | | |
| 0,343748548 | RP11-483E7.1 | FALSE | Upregulated |
| -0,305706413 | RP11-486A14.2 | FALSE | Downregulated |
| 0,587765852 | RP11-486O12.2 | FALSE | Upregulated |
| | RP11-488I20.3 | | |
| | RP11-489D6.2 | | |
| 0 | RP11-48F14.1 | FALSE | Upregulated |
| 0,17621051 | RP11-490D19.6 | FALSE | Upregulated |
| | RP11-493K23.4 | | |
| 0,293242527 | RP11-494M8.4 | FALSE | Upregulated |
| | RP11-497K21.1 | | |
| 0,193267572 | RP11-500B12.1 | FALSE | Upregulated |
| | RP11-501G7.1 | | |
| -0,499939428 | RP11-503C24.4 | FALSE | Downregulated |
| -0,221843008 | RP11-503N18.1 | FALSE | Downregulated |
| 0,621790503 | RP11-506B6.6 | FALSE | Upregulated |
| | RP11-508P1.2 | | |
| 0,456362169 | RP11-509J21.1 | FALSE | Upregulated |
| -0,025317717 | RP11-510D19.1 | FALSE | Downregulated |
| 0,08517612 | RP11-513G11.3 | FALSE | Upregulated |
| 0,024971354 | RP11-513O17.2 | FALSE | Upregulated |
| -0,135439512 | RP11-513O17.2-2 | FALSE | Downregulated |
| | RP11-513O17.3 | | |
| -0,583774384 | RP11-514D23.1 | FALSE | Downregulated |
| 0 | RP11-519M16.1 | FALSE | Upregulated |
| -0,391156235 | RP11-523L1.2 | FALSE | Downregulated |
| 0,023324762 | RP11-523L20.2 | FALSE | Upregulated |
| 0,28536712 | RP11-526A4.1 | FALSE | Upregulated |
| 0,408981198 | RP11-526P6.1 | FALSE | Upregulated |
| -0,242454537 | RP11-527H14.2 | FALSE | Downregulated |
| | RP11-527H14.3 | | |
| 1,626224384 | RP11-532N4.2 | | Upregulated |
| -0,665369468 | RP11-534N16.1 | FALSE | Downregulated |
| | RP11-536C10.7 | | |
| | RP11-536I6.1 | | |
| -0,183504666 | RP11-539L10.2 | FALSE | Downregulated |
| 0,284987956 | RP11-53O19.1 | FALSE | Upregulated |
| 0,000264809 | RP11-540D4.3 | FALSE | Upregulated |
| 0,297178435 | RP11-540K16.1 | FALSE | Upregulated |
| | RP11-542B15.1 | | |

| | | | |
|---|---|---|---|
| | RP11-542G1.3 | | |
| 0,539510415 | RP11-544M22.1 | FALSE | Upregulated |
| -0,167450125 | RP11-547D23.1 | FALSE | Downregulated |
| | RP11-547I7.2 | | |
| | RP11-550H2.1 | | |
| 0,173440056 | RP11-551M15.1 | FALSE | Upregulated |
| | RP11-553E24.2 | | |
| -0,103956311 | RP11-554A11.6 | FALSE | Downregulated |
| 0,127492257 | RP11-554A11.8 | FALSE | Upregulated |
| | RP11-554D15.1 | | |
| 0,278093964 | RP11-554F20.1 | FALSE | Upregulated |
| | RP11-555F9.2 | | |
| -0,612483334 | RP11-557H15.4 | FALSE | Downregulated |
| | RP11-561E1.1 | | |
| -1,089804445 | RP11-563J2.2 | | Downregulated |
| | RP11-565A3.2 | | |
| 0,588810802 | RP11-567L7.6 | FALSE | Upregulated |
| 1,777595322 | RP11-569G13.2 | | Upregulated |
| | RP11-56I23.1 | | |
| 0,449479977 | RP11-572C15.5 | FALSE | Upregulated |
| 0,900709318 | RP11-572M11.3 | FALSE | Upregulated |
| 0,293326262 | RP11-575F12.1 | FALSE | Upregulated |
| -0,413534784 | RP11-575L7.4 | FALSE | Downregulated |
| 0,284987956 | RP11-576D8.4 | FALSE | Upregulated |
| | RP11-576E20.1 | | |
| | RP11-576I22.2 | | |
| 0,012698408 | RP11-579D7.4 | FALSE | Upregulated |
| 0 | RP11-579E24.1 | FALSE | Upregulated |
| -0,028882239 | RP11-584P21.2 | FALSE | Downregulated |
| | RP11-586D19.1 | | |
| 0 | RP11-588G21.2 | FALSE | Upregulated |
| | RP11-588P8.1 | | |
| 0,587192508 | RP11-58E21.3 | FALSE | Upregulated |
| 0,129102809 | RP11-58E21.4 | FALSE | Upregulated |
| 0,002891501 | RP11-593F23.1 | FALSE | Upregulated |
| 1,330920503 | RP11-594N15.2 | | Upregulated |
| -0,309428036 | RP11-598F7.4 | FALSE | Downregulated |
| 0,277764892 | RP11-59J16.1 | FALSE | Upregulated |
| 0,45759495 | RP11-5P22.3 | FALSE | Upregulated |
| | RP11-600K15.1 | | |
| 0 | RP11-609L23.1 | FALSE | Upregulated |
| -0,141007843 | RP11-60I3.5 | FALSE | Downregulated |
| 0,019766326 | RP11-611D20.2 | FALSE | Upregulated |
| 0,299674204 | RP11-614F17.2 | FALSE | Upregulated |
| 0,09902053 | RP11-61J19.3 | FALSE | Upregulated |
| 0,58656092 | RP11-620J15.2 | FALSE | Upregulated |
| -0,323593165 | RP11-620J15.3 | FALSE | Downregulated |
| 0,591350038 | RP11-624C23.1 | FALSE | Upregulated |
| 0,144354434 | RP11-626I20.3 | FALSE | Upregulated |
| | RP11-626P14.1 | | |
| 0,280327078 | RP11-629G13.1 | FALSE | Upregulated |

| | | | |
|---|---|---|---|
| 0,297327371 | RP11-630D6.5 | FALSE | Upregulated |
| 0 | RP11-632K5.3 | FALSE | Upregulated |
| 0,133850629 | RP11-637A17.2 | FALSE | Upregulated |
| 0,448434867 | RP11-638F5.1 | FALSE | Upregulated |
| -0,119615791 | RP11-638L3.1 | FALSE | Downregulated |
| -0,144575648 | RP11-63B19.1 | FALSE | Downregulated |
| 0,160459014 | RP11-63P12.6 | FALSE | Upregulated |
| | RP11-646J21.5 | | |
| 0 | RP11-64I17.1 | FALSE | Upregulated |
| | RP11-653G8.2 | | |
| | RP11-65D17.1 | | |
| -0,166784391 | RP11-65J3.1 | FALSE | Downregulated |
| 1,321961417 | RP11-65M17.1 | | Upregulated |
| | RP11-65M17.3 | | |
| 0,158025929 | RP11-661P17.1 | FALSE | Upregulated |
| | RP11-664H17.1 | | |
| | RP11-665C14.2 | | |
| 0,411359293 | RP11-666I19.2 | FALSE | Upregulated |
| | RP11-669N7.2 | | |
| -0,023495361 | RP11-669N7.3 | FALSE | Downregulated |
| 0,155914376 | RP11-66A2.2 | FALSE | Upregulated |
| 0,566807581 | RP11-66B24.1 | FALSE | Upregulated |
| 0,533234901 | RP11-673C5.2 | FALSE | Upregulated |
| | RP11-673E1.1 | | |
| 0,910128102 | RP11-677M14.7 | FALSE | Upregulated |
| 0,277764892 | RP11-67M1.1 | FALSE | Upregulated |
| -0,320642189 | RP11-680F20.6 | FALSE | Downregulated |
| 0,99971659 | RP11-682N22.1 | FALSE | Upregulated |
| 0,018839898 | RP11-686G23.2 | FALSE | Upregulated |
| | RP11-690C23.2 | | |
| -0,002490782 | RP11-695J4.2 | FALSE | Downregulated |
| 0,14332638 | RP11-699L21.1 | FALSE | Upregulated |
| | RP11-69H14.6 | | |
| | RP11-69H14.6-2 | | |
| | RP11-700N1.1 | | |
| 0,274635952 | RP11-702F3.3 | FALSE | Upregulated |
| | RP11-707A18.1 | | |
| | RP11-707P20.1 | | |
| 0 | RP11-70F11.11 | FALSE | Upregulated |
| | RP11-70F11.8 | | |
| -0,725681616 | RP11-710F7.3 | FALSE | Downregulated |
| | RP11-712C7.1 | | |
| -0,158482673 | RP11-713P17.3 | FALSE | Downregulated |
| 0 | RP11-715G15.1 | FALSE | Upregulated |
| 0,266937229 | RP11-719J20.1 | FALSE | Upregulated |
| | RP11-71E19.5 | | |
| -0,018516502 | RP11-71G7.1 | FALSE | Downregulated |
| 0,448221756 | RP11-723O4.2 | FALSE | Upregulated |
| 0,457742365 | RP11-72L22.1 | FALSE | Upregulated |
| | RP11-730N24.1 | | |
| -0,738756144 | RP11-736K20.6 | FALSE | Downregulated |

| | | | |
|---|---|---|---|
| 1,229556226 | RP11-744J10.3 | | Upregulated |
| | RP11-745C15.2 | | |
| -0,101733571 | RP11-752D24.3 | FALSE | Downregulated |
| 0,309364872 | RP11-752G15.3 | FALSE | Upregulated |
| 0,003251737 | RP11-752G15.6 | FALSE | Upregulated |
| -0,592677927 | RP11-756H20.1 | FALSE | Downregulated |
| | RP11-756K15.2 | | |
| -0,00939774 | RP11-75C9.1 | FALSE | Downregulated |
| | RP11-75F3.1 | | |
| 0,052229712 | RP11-75N4.2 | FALSE | Upregulated |
| -0,13701336 | RP11-761I4.3 | FALSE | Downregulated |
| | RP11-767I20.1 | | |
| | RP11-76E12.1 | | |
| -0,463804284 | RP11-76N22.2 | FALSE | Downregulated |
| 0,429211184 | RP11-770E5.1 | FALSE | Upregulated |
| 0,159724398 | RP11-770J1.3 | FALSE | Upregulated |
| 0,571828106 | RP11-778J15.1 | FALSE | Upregulated |
| 1,50727633 | RP11-778O17.4 | | Upregulated |
| 0 | RP11-779O18.1 | FALSE | Upregulated |
| -0,302728927 | RP11-789C1.1 | FALSE | Downregulated |
| 1,182301791 | RP11-789C17.3 | | Upregulated |
| | RP11-791G22.1 | | |
| | RP11-794P6.1 | | |
| | RP11-794P6.3 | | |
| | RP11-795J1.1 | | |
| 0 | RP11-799P8.1 | FALSE | Upregulated |
| -1,069173297 | RP11-79E3.3 | | Downregulated |
| | RP11-7O14.1 | | |
| 0,253282048 | RP11-806H10.4 | FALSE | Upregulated |
| 0,292871412 | RP11-806K15.1 | FALSE | Upregulated |
| | RP11-807H7.1 | | |
| | RP11-80F22.15 | | |
| 1,104830853 | RP11-80G7.1 | | Upregulated |
| | RP11-80K21.3 | | |
| 0,285986212 | RP11-814P5.1 | FALSE | Upregulated |
| 0,579279955 | RP11-81H14.2 | FALSE | Upregulated |
| | RP11-81H3.2 | | |
| -0,052611122 | RP11-81N13.1 | FALSE | Downregulated |
| -0,568017627 | RP11-829H16.3 | FALSE | Downregulated |
| 0,619603196 | RP11-82L2.1 | FALSE | Upregulated |
| 0 | RP11-838N2.4 | FALSE | Upregulated |
| -0,022465308 | RP11-83C7.2 | FALSE | Downregulated |
| -0,405287845 | RP11-843A23.1 | FALSE | Downregulated |
| 0,066386381 | RP11-844P9.2 | FALSE | Upregulated |
| | RP11-84A1.3 | | |
| | RP11-84O12.4 | | |
| -0,01330656 | RP11-855O10.2 | FALSE | Downregulated |
| 0,269933532 | RP11-857B24.5 | FALSE | Upregulated |
| 0,427808452 | RP11-85F14.5 | FALSE | Upregulated |
| | RP11-85G21.2 | | |
| 0,139081471 | RP11-861A13.4 | FALSE | Upregulated |

| | | | |
|---|---|---|---|
| 0,548688153 | RP11-863K10.2 | FALSE | Upregulated |
| -0,366977222 | RP11-875O11.2 | FALSE | Downregulated |
| 1,39446423 | RP11-87E22.2 | | Upregulated |
| -0,011481061 | RP11-87M18.2 | FALSE | Downregulated |
| | RP11-87N24.2 | | |
| 0,628985083 | RP11-883G14.2 | FALSE | Upregulated |
| -0,140832914 | RP11-885B4.2 | FALSE | Downregulated |
| | RP11-88I21.2 | | |
| | RP11-893F2.6 | | |
| -0,010146324 | RP11-899L11.1 | FALSE | Downregulated |
| 0,2842625 | RP11-909B2.1 | FALSE | Upregulated |
| | RP11-909M7.3 | | |
| | RP11-92G12.3 | | |
| 1,760254635 | RP11-93B14.5 | | Upregulated |
| 0,427715419 | RP11-93I21.3 | FALSE | Upregulated |
| | RP11-945C19.1 | | |
| | RP11-946L20.2 | | |
| | RP11-946L20.4 | | |
| 0 | RP11-94H18.1 | FALSE | Upregulated |
| | RP11-955H22.3 | | |
| | RP11-95H3.1 | | |
| 0,434138967 | RP11-95M5.1 | FALSE | Upregulated |
| -0,277193986 | RP11-95P2.1 | FALSE | Downregulated |
| 0 | RP11-960L18.1 | FALSE | Upregulated |
| -1,094596555 | RP11-96H19.1 | | Downregulated |
| -0,133610036 | RP11-96P7.1 | FALSE | Downregulated |
| 0,429963856 | RP11-973H7.4 | FALSE | Upregulated |
| -0,049960614 | RP11-97O12.2 | FALSE | Downregulated |
| | RP11-983C2.2 | | |
| 0 | RP11-98L5.2 | FALSE | Upregulated |
| | RP11-9G1.3 | | |
| -0,240730436 | RP13-210D15.4 | FALSE | Downregulated |
| | RP13-507P19.2 | | |
| 0 | RP13-60M5.2 | FALSE | Upregulated |
| | RP13-614K11.1 | | |
| | RP13-653N12.1 | | |
| | RP13-653N12.2 | | |
| | RP13-895J2.7 | | |
| -0,469864882 | RP1L1 | FALSE | Downregulated |
| 0,156816252 | RP3-322G13.5 | FALSE | Upregulated |
| 0,947416546 | RP3-323P13.2 | FALSE | Upregulated |
| | RP3-326I13.1 | | |
| | RP3-332B22.1 | | |
| 0,285372545 | RP3-399L15.3 | FALSE | Upregulated |
| -0,161804429 | RP3-400B16.1 | FALSE | Downregulated |
| | RP3-416J7.4 | | |
| | RP3-417L20.4 | | |
| 1,262783357 | RP3-437I16.1 | | Upregulated |
| | RP3-446N13.1 | | |
| 0,434324696 | RP3-473B4.3 | FALSE | Upregulated |
| -0,046286016 | RP3-510O8.4 | FALSE | Downregulated |

| | | | |
|---|---|---|---|
| | RP3-513G18.2 | | |
| 0,308839712 | RP3-522P13.3 | FALSE | Upregulated |
| 0,272277655 | RP4-533D7.5 | FALSE | Upregulated |
| 0,460006763 | RP4-544H6.2 | FALSE | Upregulated |
| 0,483371706 | RP4-553F4.6 | FALSE | Upregulated |
| 0,71445602 | RP4-562J12.2 | FALSE | Upregulated |
| | RP4-565E6.1 | | |
| | RP4-598G3.1 | | |
| -0,006101953 | RP4-607I7.1 | FALSE | Downregulated |
| 0,79820799 | RP4-613A2.1 | FALSE | Upregulated |
| 0,442917064 | RP4-639F20.1 | FALSE | Upregulated |
| 0,297039551 | RP4-640H8.2 | FALSE | Upregulated |
| 0,192027847 | RP4-655J12.4 | FALSE | Upregulated |
| | RP4-684O24.5 | | |
| -0,006848587 | RP4-694A7.4 | FALSE | Downregulated |
| 1,26143795 | RP4-697P8.3 | | Upregulated |
| -0,018647511 | RP4-705O1.1 | FALSE | Downregulated |
| 0 | RP4-723E3.1 | FALSE | Upregulated |
| | RP4-738P15.1 | | |
| | RP4-753D10.5 | | |
| 0,285372545 | RP4-753M9.1 | FALSE | Upregulated |
| 0,45839635 | RP4-756H11.3 | FALSE | Upregulated |
| 0,289235841 | RP4-760C5.5 | FALSE | Upregulated |
| | RP4-779E11.3 | | |
| -0,007271989 | RP4-781K5.4 | FALSE | Downregulated |
| -0,284942882 | RP4-782L23.1 | FALSE | Downregulated |
| 0,27391954 | RP4-794H19.4 | FALSE | Upregulated |
| 0,291340714 | RP5-1004I9.1 | FALSE | Upregulated |
| | RP5-1007G16.1 | | |
| 0,127976558 | RP5-1010E17.2 | FALSE | Upregulated |
| -0,843681572 | RP5-1021I20.1 | FALSE | Downregulated |
| | RP5-1022P6.4 | | |
| -0,446772146 | RP5-1024G6.5 | FALSE | Downregulated |
| -0,068318966 | RP5-1027G4.3 | FALSE | Downregulated |
| 0 | RP5-1043L13.1 | FALSE | Upregulated |
| 1,979302846 | RP5-1043L13.1-2 | | Upregulated |
| -0,298719771 | RP5-1050D4.5 | FALSE | Downregulated |
| -0,002253536 | RP5-1069C8.2 | FALSE | Downregulated |
| | RP5-1070A16.1 | | |
| | RP5-1073O3.2 | | |
| -0,481049925 | RP5-1073O3.7 | FALSE | Downregulated |
| -0,179781449 | RP5-1074L1.1 | FALSE | Downregulated |
| | RP5-1086L22.1 | | |
| 0,420175495 | RP5-1097F14.3 | FALSE | Upregulated |
| -0,209361184 | RP5-1103G7.4 | FALSE | Downregulated |
| 1,178142388 | RP5-1109J22.2 | | Upregulated |
| -0,295867158 | RP5-1112F19.2 | FALSE | Downregulated |
| 0,135516 | RP5-1119D9.4 | FALSE | Upregulated |
| | RP5-1120P11.1 | | |
| 0,493858315 | RP5-1125A11.1 | FALSE | Upregulated |
| 0,428592316 | RP5-1139I1.1 | FALSE | Upregulated |

| | | | |
|---|---|---|---|
| -0,611649846 | RP5-1159O4.2 | FALSE | Downregulated |
| | RP5-1166F10.1 | | |
| 0,42540875 | RP5-1180C10.2 | FALSE | Upregulated |
| | RP5-828K20.2 | | |
| 0,194485681 | RP5-833A20.1 | FALSE | Upregulated |
| -0,155009819 | RP5-855F14.1 | FALSE | Downregulated |
| 0,277604347 | RP5-857K21.5 | FALSE | Upregulated |
| 0 | RP5-858B6.3 | FALSE | Upregulated |
| | RP5-887A10.1 | | |
| | RP5-896L10.1 | | |
| | RP5-898J17.1 | | |
| 0,161807908 | RP5-899E9.1 | FALSE | Upregulated |
| | RP5-921G16.2 | | |
| | RP5-933E2.1 | | |
| | RP5-944M2.3 | | |
| | RP5-952N6.1 | | |
| 0,988209872 | RP5-968D22.3 | FALSE | Upregulated |
| | RP5-988G17.1 | | |
| -0,136311623 | RP6-109B7.3 | FALSE | Downregulated |
| -1,410348724 | RP9 | | Downregulated |
| 0,096264199 | RP9P | FALSE | Upregulated |
| 0,076029381 | RPA3-AS1 | FALSE | Upregulated |
| -0,078959486 | RPA4 | FALSE | Downregulated |
| 0,277833435 | RPE65 | FALSE | Upregulated |
| 0 | RPEL1 | FALSE | Upregulated |
| 1,066700537 | RPGRIP1 | | Upregulated |
| 0,436307565 | RPGRIP1L | FALSE | Upregulated |
| 0,276200825 | RPH3A | FALSE | Upregulated |
| 0,290645605 | RPIA | FALSE | Upregulated |
| -0,004525881 | RPL10L | FALSE | Downregulated |
| -0,105707401 | RPL13AP17 | FALSE | Downregulated |
| -0,210849258 | RPL13AP20 | FALSE | Downregulated |
| -0,876576338 | RPL13AP3 | FALSE | Downregulated |
| 0,450423866 | RPL13P5 | FALSE | Upregulated |
| -0,679090916 | RPL19P12 | FALSE | Downregulated |
| -0,511898149 | RPL21P28 | FALSE | Downregulated |
| -0,616008085 | RPL21P44 | FALSE | Downregulated |
| 0,594364637 | RPL22L1 | FALSE | Upregulated |
| 0,094484047 | RPL23AP32 | FALSE | Upregulated |
| 0,297592188 | RPL23AP7 | FALSE | Upregulated |
| -0,067071024 | RPL23AP82 | FALSE | Downregulated |
| 0,164986996 | RPL23P8 | FALSE | Upregulated |
| 0,141704061 | RPL26L1 | FALSE | Upregulated |
| 0 | RPL29P2 | FALSE | Upregulated |
| -0,672065026 | RPL32P3 | FALSE | Downregulated |
| 0,438150824 | RPL34-AS1 | FALSE | Upregulated |
| 0,014272545 | RPL39L | FALSE | Upregulated |
| 0,309630848 | RPL3L | FALSE | Upregulated |
| 0,146674917 | RPLP0P2 | FALSE | Upregulated |
| 0,070533817 | RPP21 | FALSE | Upregulated |
| -0,335846132 | RPP30 | FALSE | Downregulated |

| | | | |
|---|---|---|---|
| 1,487431139 | RPP40 | | Upregulated |
| -0,155180994 | RPRM | FALSE | Downregulated |
| 0,203470606 | RPRML | FALSE | Upregulated |
| 0 | RPS10-NUDT3 | FALSE | Upregulated |
| -0,009885967 | RPS10P7 | FALSE | Downregulated |
| 0,721760492 | RPS14P3 | FALSE | Upregulated |
| -0,788555606 | RPS15AP10 | FALSE | Downregulated |
| 1,531080019 | RPS16P5 | | Upregulated |
| -0,14171202 | RPS26P11 | FALSE | Downregulated |
| | RPS4Y2 | | |
| -0,013376718 | RPS6KA2-IT1 | FALSE | Downregulated |
| 0,02575814 | RPS6KA5 | FALSE | Upregulated |
| -0,679485191 | RPS6KA6 | FALSE | Downregulated |
| | RPS7P5 | | |
| | RPSAP52 | | |
| 0,554295981 | RPSAP58 | FALSE | Upregulated |
| 0,385359361 | RPSAP9 | FALSE | Upregulated |
| 0,283510872 | RPTN | FALSE | Upregulated |
| -0,744120986 | RRAD | FALSE | Downregulated |
| 0 | RRH | FALSE | Upregulated |
| 0,258658035 | RRM2 | FALSE | Upregulated |
| -0,225630521 | RRN3P1 | FALSE | Downregulated |
| 0,185218374 | RRN3P2 | FALSE | Upregulated |
| 0,628913218 | RRP9 | FALSE | Upregulated |
| 0,015176628 | RS1 | FALSE | Upregulated |
| -0,208956608 | RSAD2 | FALSE | Downregulated |
| 0,078338977 | RSG1 | FALSE | Upregulated |
| 0 | RSPH1 | FALSE | Upregulated |
| 1,154145829 | RSPH10B | | Upregulated |
| 0,580286889 | RSPH10B2 | FALSE | Upregulated |
| 0,285639981 | RSPH6A | FALSE | Upregulated |
| 0,269762685 | RSPO1 | FALSE | Upregulated |
| 0,282221053 | RSPO2 | FALSE | Upregulated |
| 0,144163847 | RSPO4 | FALSE | Upregulated |
| 0,080612303 | RSU1P2 | FALSE | Upregulated |
| 0,003992645 | RTBDN | FALSE | Upregulated |
| 0,43150683 | RTDR1 | FALSE | Upregulated |
| -0,24910836 | RTEL1-TNFRSF6B | FALSE | Downregulated |
| 0,333991847 | RTKN2 | FALSE | Upregulated |
| 0,217069295 | RTN2 | FALSE | Upregulated |
| -0,572652114 | RTN4RL1 | FALSE | Downregulated |
| 0,289590908 | RTP1 | FALSE | Upregulated |
| | RTP2 | | |
| -0,293696976 | RTP4 | FALSE | Downregulated |
| 0,270535452 | RUFY4 | FALSE | Upregulated |
| -0,053150469 | RUNDC3A | FALSE | Downregulated |
| 0,762809614 | RUNX1-IT1 | FALSE | Upregulated |
| 0,045089543 | RWDD3 | FALSE | Upregulated |
| -0,053391877 | RXFP1 | FALSE | Downregulated |
| 0,012003502 | RXFP2 | FALSE | Upregulated |
| -0,312123194 | RXFP3 | FALSE | Downregulated |

| | | | |
|---|---|---|---|
| -0,122633387 | RXFP4 | FALSE | Downregulated |
| -0,432579871 | RXRG | FALSE | Downregulated |
| 0 | S100A12 | FALSE | Upregulated |
| 0 | S100A2 | FALSE | Upregulated |
| 0 | S100A3 | FALSE | Upregulated |
| 0,64339108 | S100A4 | FALSE | Upregulated |
| 0,47241678 | S100A5 | FALSE | Upregulated |
| 0,448199481 | S100A6 | FALSE | Upregulated |
| | S100A7 | | |
| | S100A7A | | |
| | S100A7L2 | | |
| 0,737534518 | S100A8 | FALSE | Upregulated |
| -0,332400399 | S100B | FALSE | Downregulated |
| 0,171768134 | S100G | FALSE | Upregulated |
| 0,165664831 | S100P | FALSE | Upregulated |
| 0,290402161 | S100Z | FALSE | Upregulated |
| -0,615863316 | S1PR5 | FALSE | Downregulated |
| 0,899756787 | SAA2-SAA4 | FALSE | Upregulated |
| | SAA3P | | |
| 0,706744267 | SAAL1 | FALSE | Upregulated |
| 0,356118311 | SACS-AS1 | FALSE | Upregulated |
| 0,284987956 | SAG | FALSE | Upregulated |
| | SAGE1 | | |
| -0,433791194 | SALL2 | FALSE | Downregulated |
| | SALL3 | | |
| -0,794120359 | SALL4 | FALSE | Downregulated |
| -0,039909689 | SAMD12-AS1 | FALSE | Downregulated |
| -0,157540923 | SAMD13 | FALSE | Downregulated |
| 0,046639107 | SAMD3 | FALSE | Upregulated |
| 0 | SAMD7 | FALSE | Upregulated |
| -0,161370061 | SAMSN1 | FALSE | Downregulated |
| | SAMSN1-AS1 | | |
| 0,358657416 | SAP30 | FALSE | Upregulated |
| 0 | SATB2-AS1 | FALSE | Upregulated |
| 0,065711067 | SAYSD1 | FALSE | Upregulated |
| -0,304416786 | SBF1P1 | FALSE | Downregulated |
| 0,120203678 | SBK1 | FALSE | Upregulated |
| 0,002328382 | SBK2 | FALSE | Upregulated |
| 0 | SBK3 | FALSE | Upregulated |
| 0,025781019 | SBSN | FALSE | Upregulated |
| -0,018075137 | SCAMP5 | FALSE | Downregulated |
| -0,95613298 | SCAND3 | FALSE | Downregulated |
| 0,03353803 | SCARNA1 | FALSE | Upregulated |
| 0,001141803 | SCARNA11 | FALSE | Upregulated |
| | SCARNA14 | | |
| | SCARNA15 | | |
| 0,164868355 | SCARNA16 | FALSE | Upregulated |
| -0,020427572 | SCARNA18 | FALSE | Downregulated |
| 0,611712534 | SCARNA20 | FALSE | Upregulated |
| 0,042485823 | SCARNA21 | FALSE | Upregulated |
| 0,134954713 | SCARNA22 | FALSE | Upregulated |

| | | | |
|---|---|---|---|
| | SCARNA23 | | |
| | SCARNA27 | | |
| -0,596815818 | SCARNA3 | FALSE | Downregulated |
| 0,543825459 | SCARNA4 | FALSE | Upregulated |
| 0,00139671 | SCARNA8 | FALSE | Upregulated |
| 0,12123727 | SCARNA9L | FALSE | Upregulated |
| -0,454761991 | SCD5 | FALSE | Downregulated |
| 0,101734679 | SCEL | FALSE | Upregulated |
| 0 | SCG2 | FALSE | Upregulated |
| -0,064598131 | SCG5 | FALSE | Downregulated |
| | SCGB1A1 | | |
| -0,30003553 | SCGB1B2P | FALSE | Downregulated |
| 0 | SCGB1C1 | FALSE | Upregulated |
| | SCGB1D1 | | |
| 0 | SCGB1D2 | FALSE | Upregulated |
| | SCGB1D4 | | |
| | SCGB2A1 | | |
| | SCGB2A2 | | |
| 0,290505128 | SCGB2B2 | FALSE | Upregulated |
| 0,179114598 | SCGB2B3P | FALSE | Upregulated |
| -0,143154934 | SCGB3A1 | FALSE | Downregulated |
| -0,004525881 | SCGB3A2 | FALSE | Downregulated |
| 0 | SCHIP1 | FALSE | Upregulated |
| 0,551491419 | SCHLAP1 | FALSE | Upregulated |
| 0,277254331 | SCIN | FALSE | Upregulated |
| -0,504104839 | SCLT1 | FALSE | Downregulated |
| -0,260854777 | SCML2 | FALSE | Downregulated |
| 1,279868152 | SCML4 | | Upregulated |
| | SCN10A | | |
| -0,64189353 | SCN11A | FALSE | Downregulated |
| 0,686352026 | SCN1A | FALSE | Upregulated |
| 1,076672971 | SCN2A | | Upregulated |
| -0,023453053 | SCN2B | FALSE | Downregulated |
| 1,103382276 | SCN3B | | Upregulated |
| -0,269823142 | SCN4A | FALSE | Downregulated |
| -0,262549598 | SCN4B | FALSE | Downregulated |
| 0,713357817 | SCN5A | FALSE | Upregulated |
| -0,119579919 | SCN7A | FALSE | Downregulated |
| -1,018433733 | SCNM1 | | Downregulated |
| 0,283811672 | SCNN1B | FALSE | Upregulated |
| | SCNN1G | | |
| | SCP2D1 | | |
| 0,292388303 | SCRT2 | FALSE | Upregulated |
| 0,248785812 | SCUBE1 | FALSE | Upregulated |
| -0,521911499 | SCUBE2 | FALSE | Downregulated |
| -0,337831963 | SDC4P | FALSE | Downregulated |
| -0,843653725 | SDCBP2 | FALSE | Downregulated |
| 0,087577979 | SDHAP3 | FALSE | Upregulated |
| 0 | SDR16C5 | FALSE | Upregulated |
| | SDR16C6P | | |
| -0,060232431 | SDR42E1 | FALSE | Downregulated |

| | | | |
|---|---|---|---|
| 0,149598053 | SDR9C7 | FALSE | Upregulated |
| -0,269660841 | SEBOX | FALSE | Downregulated |
| -0,061304052 | SEC14L1P1 | FALSE | Downregulated |
| 0,043361187 | SEC14L3 | FALSE | Upregulated |
| -1,013705366 | SEC14L5 | | Downregulated |
| -0,64794875 | SEC14L6 | FALSE | Downregulated |
| 0,209953037 | SEC24B-AS1 | FALSE | Upregulated |
| -0,566891433 | SEC61G | FALSE | Downregulated |
| | SEL1L2 | | |
| 0,420612093 | SELE | FALSE | Upregulated |
| 0,960197728 | SELL | FALSE | Upregulated |
| 0,027919126 | SELM | FALSE | Upregulated |
| -0,232495221 | SELP | FALSE | Downregulated |
| 0,14517521 | SELV | FALSE | Upregulated |
| 0,127375295 | SEMA3A | FALSE | Upregulated |
| 0,807946792 | SEMA3B-AS1 | FALSE | Upregulated |
| -0,378066638 | SEMA3C | FALSE | Downregulated |
| 1,21844752 | SEMA3D | | Upregulated |
| -0,471347733 | SEMA3G | FALSE | Downregulated |
| 1,159602694 | SEMA4A | | Upregulated |
| -0,42425234 | SEMA4F | FALSE | Downregulated |
| -0,918998052 | SEMA5B | FALSE | Downregulated |
| 1,422255009 | SEMA7A | | Upregulated |
| | SEMG1 | | |
| | SEMG2 | | |
| -0,039204526 | SENCR | FALSE | Downregulated |
| -0,021805768 | SEPSECS-AS1 | FALSE | Downregulated |
| 0,148355399 | SEPT12 | FALSE | Upregulated |
| | SEPT14 | | |
| 0,113952293 | SEPT3 | FALSE | Upregulated |
| 0,190681362 | SEPT5 | FALSE | Upregulated |
| 0,130545688 | SEPT5-GP1BB | FALSE | Upregulated |
| -0,32131337 | SEPT7P9 | FALSE | Downregulated |
| | SERF1A | | |
| -0,155441557 | SERF1B | FALSE | Downregulated |
| 1,047928145 | SERF2-C15ORF63 | | Upregulated |
| -0,264125827 | SERP2 | FALSE | Downregulated |
| 0,211071477 | SERPINA12 | FALSE | Upregulated |
| | SERPINA13P | | |
| 0,288805648 | SERPINA9 | FALSE | Upregulated |
| | SERPINB10 | | |
| | SERPINB11 | | |
| 0 | SERPINB12 | FALSE | Upregulated |
| 0 | SERPINB13 | FALSE | Upregulated |
| 0,27174569 | SERPINB2 | FALSE | Upregulated |
| | SERPINB3 | | |
| | SERPINB4 | | |
| 0 | SERPINB5 | FALSE | Upregulated |
| | SERPINB7 | | |
| -0,015892871 | SERPINI1 | FALSE | Downregulated |
| | SERPINI2 | | |

| | | | |
|---|---|---|---|
| 0,044512483 | SERTAD1 | FALSE | Upregulated |
| 0,169829012 | SERTAD4 | FALSE | Upregulated |
| -0,31632382 | SERTAD4-AS1 | FALSE | Downregulated |
| 0,945828921 | SERTM1 | FALSE | Upregulated |
| 0,331987312 | SETD9 | FALSE | Upregulated |
| 0,003182548 | SETSIP | FALSE | Upregulated |
| 0,373483049 | SEZ6L | FALSE | Upregulated |
| -0,130382674 | SEZ6L2 | FALSE | Downregulated |
| 0,167335112 | SFN | FALSE | Upregulated |
| 0,624274412 | SFR1 | FALSE | Upregulated |
| 0,515088713 | SFRP2 | FALSE | Upregulated |
| 0,085369672 | SFRP4 | FALSE | Upregulated |
| -0,092737654 | SFT2D1 | FALSE | Downregulated |
| 0,394972622 | SFT2D2 | FALSE | Upregulated |
| 0 | SFTA1P | FALSE | Upregulated |
| 0 | SFTA2 | FALSE | Upregulated |
| | SFTA3 | | |
| -0,177585051 | SFTPA1 | FALSE | Downregulated |
| -0,247542863 | SFTPA2 | FALSE | Downregulated |
| 0,220401653 | SFTPC | FALSE | Upregulated |
| 1,123681677 | SGCA | | Upregulated |
| 0,148357637 | SGCG | FALSE | Upregulated |
| 0,020847044 | SGCZ | FALSE | Upregulated |
| -0,325582721 | SGIP1 | FALSE | Downregulated |
| -0,422176552 | SGOL1-AS1 | FALSE | Downregulated |
| 1,387094093 | SGOL2 | | Upregulated |
| -0,440507853 | SGSM1 | FALSE | Downregulated |
| -0,318151654 | SH2B2 | FALSE | Downregulated |
| 1,173838137 | SH2D1A | | Upregulated |
| 1,100251589 | SH2D2A | | Upregulated |
| -0,009465128 | SH2D4B | FALSE | Downregulated |
| 0,277319882 | SH2D5 | FALSE | Upregulated |
| -0,628545927 | SH2D6 | FALSE | Downregulated |
| 0,01552207 | SH2D7 | FALSE | Upregulated |
| -0,21534498 | SH3BGR | FALSE | Downregulated |
| -0,681275936 | SH3GL1P1 | FALSE | Downregulated |
| 0,137974415 | SH3GL2 | FALSE | Upregulated |
| 0,117308118 | SH3GL3 | FALSE | Upregulated |
| 0,331730637 | SH3TC2 | FALSE | Upregulated |
| 1,2440881 | SHANK1 | | Upregulated |
| 0,569279171 | SHC4 | FALSE | Upregulated |
| 0,110529566 | SHCBP1 | FALSE | Upregulated |
| | SHCBP1L | | |
| -0,428333269 | SHD | FALSE | Downregulated |
| 0,284175187 | SHISA2 | FALSE | Upregulated |
| 1,009698693 | SHISA3 | | Upregulated |
| 0,84297823 | SHISA4 | FALSE | Upregulated |
| 0,087268994 | SHISA7 | FALSE | Upregulated |
| 0,287454042 | SHISA8 | FALSE | Upregulated |
| -0,003332419 | SHOX | FALSE | Downregulated |
| | SHOX-2 | | |

| | | | |
|---|---|---|---|
| | SHOX2 | | |
| | SI | | |
| | SIAH3 | | |
| -0,18010698 | SIGLEC12 | FALSE | Downregulated |
| 0,666677692 | SIGLEC17P | FALSE | Upregulated |
| 1,194706837 | SIGLEC5 | | Upregulated |
| 0,126396636 | SIGLEC6 | FALSE | Upregulated |
| -0,432961774 | SIGLEC7 | FALSE | Downregulated |
| 0,542966439 | SIGLEC8 | FALSE | Upregulated |
| 0 | SIGLECL1 | FALSE | Upregulated |
| 0,145723252 | SIM1 | FALSE | Upregulated |
| 0,127802114 | SIRPB1 | FALSE | Upregulated |
| 0,801711781 | SIRPB2 | FALSE | Upregulated |
| 1,213847823 | SIRPD | | Upregulated |
| 1,155821533 | SIRPG | | Upregulated |
| 0,183029692 | SIT1 | FALSE | Upregulated |
| 0,552531641 | SIX1 | FALSE | Upregulated |
| 0,269638972 | SIX2 | FALSE | Upregulated |
| | SIX3 | | |
| 0 | SIX3-AS1 | FALSE | Upregulated |
| 0,313530938 | SIX4 | FALSE | Upregulated |
| 0,133540797 | SIX6 | FALSE | Upregulated |
| -0,163084852 | SKA1 | FALSE | Downregulated |
| -1,338937279 | SKA3 | | Downregulated |
| 0,1188537 | SKINTL | FALSE | Upregulated |
| 0,086634246 | SKOR1 | FALSE | Upregulated |
| | SKOR2 | | |
| 0 | SKP1P2 | FALSE | Upregulated |
| 0,450475086 | SLAMF1 | FALSE | Upregulated |
| 1,365035211 | SLAMF6 | | Upregulated |
| 0,265671453 | SLAMF9 | FALSE | Upregulated |
| -0,015149192 | SLC10A2 | FALSE | Downregulated |
| 0 | SLC10A4 | FALSE | Upregulated |
| 0,294353549 | SLC10A6 | FALSE | Upregulated |
| 0,033136787 | SLC12A1 | FALSE | Upregulated |
| 0,395297812 | SLC12A3 | FALSE | Upregulated |
| -0,28026022 | SLC12A8 | FALSE | Downregulated |
| 0,26326675 | SLC13A1 | FALSE | Upregulated |
| -0,748639444 | SLC13A2 | FALSE | Downregulated |
| 0,351547087 | SLC14A1 | FALSE | Upregulated |
| -0,864636542 | SLC14A2 | FALSE | Downregulated |
| 0,001123591 | SLC15A2 | FALSE | Upregulated |
| -0,898801707 | SLC15A5 | FALSE | Downregulated |
| -0,221166925 | SLC16A1-AS1 | FALSE | Downregulated |
| -0,405907881 | SLC16A12 | FALSE | Downregulated |
| 0,416105336 | SLC16A5 | FALSE | Upregulated |
| -0,388070087 | SLC16A9 | FALSE | Downregulated |
| | SLC17A6 | | |
| -0,123591351 | SLC17A7 | FALSE | Downregulated |
| -0,550794526 | SLC17A8 | FALSE | Downregulated |
| 0,271886396 | SLC18A1 | FALSE | Upregulated |

| | | | |
|---|---|---|---|
| | SLC18A3 | | |
| 0,534359863 | SLC18B1 | FALSE | Upregulated |
| | SLC1A6 | | |
| -0,432759723 | SLC22A11 | FALSE | Downregulated |
| 0,104889972 | SLC22A12 | FALSE | Upregulated |
| -0,495076356 | SLC22A13 | FALSE | Downregulated |
| 0 | SLC22A14 | FALSE | Upregulated |
| 0,166023583 | SLC22A15 | FALSE | Upregulated |
| 0,152824518 | SLC22A16 | FALSE | Upregulated |
| -0,324713312 | SLC22A17 | FALSE | Downregulated |
| -0,007696921 | SLC22A2 | FALSE | Downregulated |
| 0,424241875 | SLC22A20 | FALSE | Upregulated |
| -1,030352569 | SLC22A24 | | Downregulated |
| 0,1227196 | SLC22A31 | FALSE | Upregulated |
| 0 | SLC22A6 | FALSE | Upregulated |
| | SLC22A8 | | |
| 0,720140205 | SLC24A2 | FALSE | Upregulated |
| -0,415360719 | SLC24A3 | FALSE | Downregulated |
| 1,137651427 | SLC24A4 | | Upregulated |
| 1,361281145 | SLC24A5 | | Upregulated |
| 0,198326213 | SLC25A14 | FALSE | Upregulated |
| -0,096632413 | SLC25A17 | FALSE | Downregulated |
| -0,120789139 | SLC25A21 | FALSE | Downregulated |
| 0,199668215 | SLC25A21-AS1 | FALSE | Upregulated |
| 0,460438174 | SLC25A24 | FALSE | Upregulated |
| | SLC25A31 | | |
| | SLC25A3P1 | | |
| 0,936097146 | SLC25A41 | FALSE | Upregulated |
| 0,571925016 | SLC25A48 | FALSE | Upregulated |
| | SLC25A51P1 | | |
| 0,521385066 | SLC25A53 | FALSE | Upregulated |
| | SLC25A6-2 | | |
| 0,217963386 | SLC26A10 | FALSE | Upregulated |
| 0,788139735 | SLC26A3 | FALSE | Upregulated |
| -0,076148312 | SLC26A4 | FALSE | Downregulated |
| 0,192063734 | SLC26A4-AS1 | FALSE | Upregulated |
| 0,600621798 | SLC26A5 | FALSE | Upregulated |
| -0,248513473 | SLC26A6 | FALSE | Downregulated |
| 0,286788943 | SLC26A7 | FALSE | Upregulated |
| -0,060442949 | SLC26A9 | FALSE | Downregulated |
| -0,175713062 | SLC27A6 | FALSE | Downregulated |
| 0,73133326 | SLC28A2 | FALSE | Upregulated |
| 0,871836649 | SLC28A3 | FALSE | Upregulated |
| 0 | SLC2A1-AS1 | FALSE | Upregulated |
| -0,481042768 | SLC2A14 | FALSE | Downregulated |
| 0,563653656 | SLC2A3 | FALSE | Upregulated |
| 0,307229727 | SLC2A4 | FALSE | Upregulated |
| 0,177709817 | SLC2A5 | FALSE | Upregulated |
| | SLC2A7 | | |
| 1,24735059 | SLC30A2 | | Upregulated |
| 0 | SLC30A3 | FALSE | Upregulated |

| | | | |
|---|---|---|---|
| 0,474585273 | SLC30A8 | FALSE | Upregulated |
| | SLC32A1 | | |
| 0,124286219 | SLC34A1 | FALSE | Upregulated |
| 0,210554975 | SLC34A2 | FALSE | Upregulated |
| 0,041540553 | SLC34A3 | FALSE | Upregulated |
| 0,048697227 | SLC35B3 | FALSE | Upregulated |
| | SLC35D3 | | |
| -0,435489727 | SLC35E4 | FALSE | Downregulated |
| 0,680622054 | SLC35F1 | FALSE | Upregulated |
| -0,155604884 | SLC35F2 | FALSE | Downregulated |
| 0,010268759 | SLC35F3 | FALSE | Upregulated |
| 0,525394381 | SLC35F4 | FALSE | Upregulated |
| 0,71478243 | SLC35G1 | FALSE | Upregulated |
| 0,278979493 | SLC35G3 | FALSE | Upregulated |
| | SLC35G4 | | |
| 0,263044272 | SLC35G5 | FALSE | Upregulated |
| 0,762129857 | SLC36A2 | FALSE | Upregulated |
| -0,375203172 | SLC36A3 | FALSE | Downregulated |
| -0,421519939 | SLC38A5 | FALSE | Downregulated |
| -0,291045783 | SLC38A6 | FALSE | Downregulated |
| | SLC38A8 | | |
| 0 | SLC39A12 | FALSE | Upregulated |
| 0,416534603 | SLC39A2 | FALSE | Upregulated |
| 0,636664122 | SLC39A4 | FALSE | Upregulated |
| 0,424550319 | SLC44A4 | FALSE | Upregulated |
| -0,336277666 | SLC44A5 | FALSE | Downregulated |
| -0,140525087 | SLC45A1 | FALSE | Downregulated |
| -0,569200936 | SLC45A2 | FALSE | Downregulated |
| 0,329995608 | SLC4A1 | FALSE | Upregulated |
| 0,984126433 | SLC4A10 | FALSE | Upregulated |
| 0,210971414 | SLC4A11 | FALSE | Upregulated |
| 0,811535799 | SLC4A3 | FALSE | Upregulated |
| 0,827618184 | SLC4A8 | FALSE | Upregulated |
| -0,414695983 | SLC4A9 | FALSE | Downregulated |
| 0,235425688 | SLC51B | FALSE | Upregulated |
| 0,107409881 | SLC52A3 | FALSE | Upregulated |
| 1,213892443 | SLC5A1 | | Upregulated |
| 0,198749806 | SLC5A11 | FALSE | Upregulated |
| 0,226544568 | SLC5A4 | FALSE | Upregulated |
| -0,059723358 | SLC5A5 | FALSE | Downregulated |
| -0,00931449 | SLC5A7 | FALSE | Downregulated |
| | SLC5A8 | | |
| -0,004525881 | SLC6A10P | FALSE | Downregulated |
| | SLC6A10PB | | |
| -0,622809504 | SLC6A11 | FALSE | Downregulated |
| | SLC6A14 | | |
| | SLC6A15 | | |
| 0,199111229 | SLC6A16 | FALSE | Upregulated |
| 0,220338448 | SLC6A17 | FALSE | Upregulated |
| | SLC6A18 | | |
| -0,788547388 | SLC6A19 | FALSE | Downregulated |

| | | | |
|---|---|---|---|
| -0,461812389 | SLC6A2 | FALSE | Downregulated |
| -0,014977496 | SLC6A20 | FALSE | Downregulated |
| -0,008980035 | SLC6A3 | FALSE | Downregulated |
| | SLC6A5 | | |
| 0 | SLC6A7 | FALSE | Upregulated |
| 0,310316049 | SLC6A9 | FALSE | Upregulated |
| 0,971987032 | SLC7A1 | FALSE | Upregulated |
| -0,199683197 | SLC7A10 | FALSE | Downregulated |
| 0,228244559 | SLC7A11 | FALSE | Upregulated |
| -0,219856739 | SLC7A11-AS1 | FALSE | Downregulated |
| 0 | SLC7A13 | FALSE | Upregulated |
| | SLC7A3 | | |
| -0,022964415 | SLC7A4 | FALSE | Downregulated |
| -0,06102513 | SLC7A5P1 | FALSE | Downregulated |
| | SLC8A2 | | |
| -0,324707911 | SLC9A2 | FALSE | Downregulated |
| 0,780757608 | SLC9A4 | FALSE | Upregulated |
| 0,494146363 | SLC9A6 | FALSE | Upregulated |
| -0,211490341 | SLC9A7P1 | FALSE | Downregulated |
| 0,259527088 | SLC9A9-AS1 | FALSE | Upregulated |
| -0,238337272 | SLC9B1 | FALSE | Downregulated |
| 1,687223163 | SLC9C1 | | Upregulated |
| 0,701243701 | SLC9C2 | FALSE | Upregulated |
| -0,147800155 | SLCO1B7 | FALSE | Downregulated |
| -0,682102479 | SLCO1C1 | FALSE | Downregulated |
| 0,178223029 | SLCO2A1 | FALSE | Upregulated |
| -0,356398037 | SLCO4A1 | FALSE | Downregulated |
| 1,101539203 | SLCO5A1 | | Upregulated |
| | SLCO6A1 | | |
| -0,252096719 | SLED1 | FALSE | Downregulated |
| -0,502861594 | SLFN12 | FALSE | Downregulated |
| 1,107159922 | SLFN12L | | Upregulated |
| 0,385718794 | SLIRP | FALSE | Upregulated |
| 0 | SLIT2-IT1 | FALSE | Upregulated |
| | SLITRK1 | | |
| 0,466267537 | SLITRK2 | FALSE | Upregulated |
| -0,108672339 | SLITRK3 | FALSE | Downregulated |
| -0,029399242 | SLITRK4 | FALSE | Downregulated |
| 0,254940949 | SLITRK5 | FALSE | Upregulated |
| -0,591109601 | SLITRK6 | FALSE | Downregulated |
| -0,010424476 | SLMO1 | FALSE | Downregulated |
| 0,550026172 | SLN | FALSE | Upregulated |
| -0,465246081 | SLURP1 | FALSE | Downregulated |
| -0,009405637 | SLX1A-SULT1A3 | FALSE | Downregulated |
| -1,080263056 | SLX1B-SULT1A4 | | Downregulated |
| -0,158140334 | SLX4IP | FALSE | Downregulated |
| 0,318792921 | SMC5-AS1 | FALSE | Upregulated |
| -0,006848587 | SMCO2 | FALSE | Downregulated |
| | SMCP | | |
| -0,0148479 | SMCR9 | FALSE | Downregulated |
| | SMEK3P | | |

| | | | |
|---|---|---|---|
| -0,526839098 | SMIM1 | FALSE | Downregulated |
| -0,472401885 | SMIM10 | FALSE | Downregulated |
| -0,634217963 | SMIM11 | FALSE | Downregulated |
| -0,15370376 | SMIM17 | FALSE | Downregulated |
| -0,141497306 | SMIM2-AS1 | FALSE | Downregulated |
| 0,758705358 | SMIM2-IT1 | FALSE | Upregulated |
| -0,628093956 | SMIM20 | FALSE | Downregulated |
| 0,272183678 | SMIM21 | FALSE | Upregulated |
| 0,002828246 | SMIM22 | FALSE | Upregulated |
| -0,275401324 | SMIM23 | FALSE | Downregulated |
| 0,291339596 | SMIM3 | FALSE | Upregulated |
| 0,102867184 | SMIM4 | FALSE | Upregulated |
| 0,220945315 | SMIM8 | FALSE | Upregulated |
| 0 | SMIM9 | FALSE | Upregulated |
| | SMKR1 | | |
| -0,376404109 | SMOX | FALSE | Downregulated |
| 0,046586672 | SMPD2 | FALSE | Upregulated |
| -0,159924288 | SMPDL3B | FALSE | Downregulated |
| 1,000740312 | SMPX | | Upregulated |
| | SMR3A | | |
| | SMR3B | | |
| 0,613604222 | SMTNL2 | FALSE | Upregulated |
| -0,484484791 | SMYD1 | FALSE | Downregulated |
| -0,191532369 | SMYD3 | FALSE | Downregulated |
| 0,318362269 | SNAI1 | FALSE | Upregulated |
| 0,292410882 | SNAI2 | FALSE | Upregulated |
| -0,125914521 | SNAP25-AS1 | FALSE | Downregulated |
| 0 | SNAP91 | FALSE | Upregulated |
| 0,225022584 | SNAPC1 | FALSE | Upregulated |
| | SNAR-A1 | | |
| | SNAR-A10 | | |
| | SNAR-A11 | | |
| | SNAR-A12 | | |
| | SNAR-A13 | | |
| | SNAR-A14 | | |
| | SNAR-A2 | | |
| | SNAR-A3 | | |
| | SNAR-A4 | | |
| | SNAR-A5 | | |
| | SNAR-A6 | | |
| | SNAR-A7 | | |
| | SNAR-A8 | | |
| | SNAR-A9 | | |
| | SNAR-B1 | | |
| | SNAR-B2 | | |
| | SNAR-C1 | | |
| | SNAR-C2 | | |
| | SNAR-C3 | | |
| 0,000264809 | SNAR-C4 | FALSE | Upregulated |
| | SNAR-C5 | | |
| | SNAR-D | | |

| | | | |
|---|---|---|---|
| | SNAR-E | | |
| | SNAR-F | | |
| | SNAR-G1 | | |
| | SNAR-G2 | | |
| | SNAR-H | | |
| | SNAR-I | | |
| 0,235934081 | SNCA | FALSE | Upregulated |
| 0 | SNCB | FALSE | Upregulated |
| 0,807848743 | SNCG | FALSE | Upregulated |
| 0,70726972 | SNHG1 | FALSE | Upregulated |
| -0,482087503 | SNHG10 | FALSE | Downregulated |
| -0,069308702 | SNHG11 | FALSE | Downregulated |
| 1,704243218 | SNHG12 | | Upregulated |
| -0,07655379 | SNHG15 | FALSE | Downregulated |
| -0,338594881 | SNHG17 | FALSE | Downregulated |
| -0,055297668 | SNHG18 | FALSE | Downregulated |
| 0,521888998 | SNHG19 | FALSE | Upregulated |
| -0,03236727 | SNHG4 | FALSE | Downregulated |
| 0,443498515 | SNHG6 | FALSE | Upregulated |
| 0,008690492 | SNHG9 | FALSE | Upregulated |
| -0,276180614 | SNORA1 | FALSE | Downregulated |
| 0,00266258 | SNORA10 | FALSE | Upregulated |
| 0,245497203 | SNORA11 | FALSE | Upregulated |
| 0,402300255 | SNORA11B | FALSE | Upregulated |
| | SNORA11C | | |
| | SNORA11D | | |
| | SNORA11E | | |
| -0,300894405 | SNORA12 | FALSE | Downregulated |
| 0,056757161 | SNORA13 | FALSE | Upregulated |
| -0,04282928 | SNORA14A | FALSE | Downregulated |
| -0,727484985 | SNORA14B | FALSE | Downregulated |
| -0,299345719 | SNORA15 | FALSE | Downregulated |
| 0 | SNORA16A | FALSE | Upregulated |
| 0,001141803 | SNORA16B | FALSE | Upregulated |
| | SNORA17 | | |
| 0,416263104 | SNORA18 | FALSE | Upregulated |
| | SNORA19 | | |
| -0,292394051 | SNORA20 | FALSE | Downregulated |
| 0,874128274 | SNORA21 | FALSE | Upregulated |
| 0,326255124 | SNORA22 | FALSE | Upregulated |
| 0,504596255 | SNORA23 | FALSE | Upregulated |
| -0,362623861 | SNORA24 | FALSE | Downregulated |
| 0 | SNORA25 | FALSE | Upregulated |
| 0,45276524 | SNORA26 | FALSE | Upregulated |
| 0 | SNORA27 | FALSE | Upregulated |
| -0,148366876 | SNORA28 | FALSE | Downregulated |
| | SNORA29 | | |
| -0,286404373 | SNORA2A | FALSE | Downregulated |
| | SNORA2B | | |
| -0,15491222 | SNORA3 | FALSE | Downregulated |
| 0,280928672 | SNORA30 | FALSE | Upregulated |

| | | | |
|---|---|---|---|
| -0,295562409 | SNORA31 | FALSE | Downregulated |
| 0,277764892 | SNORA32 | FALSE | Upregulated |
| 0,284921005 | SNORA33 | FALSE | Upregulated |
| -0,421028184 | SNORA34 | FALSE | Downregulated |
| | SNORA35 | | |
| | SNORA36A | | |
| | SNORA36B | | |
| | SNORA36C | | |
| -0,318929809 | SNORA37 | FALSE | Downregulated |
| 0,435020786 | SNORA38 | FALSE | Upregulated |
| -0,251064819 | SNORA38B | FALSE | Downregulated |
| | SNORA39 | | |
| -0,010019707 | SNORA4 | FALSE | Downregulated |
| 0,324392826 | SNORA40 | FALSE | Upregulated |
| 0,592995606 | SNORA41 | FALSE | Upregulated |
| 0,127236332 | SNORA42 | FALSE | Upregulated |
| -0,533624868 | SNORA43 | FALSE | Downregulated |
| 0,267455483 | SNORA44 | FALSE | Upregulated |
| -0,282942777 | SNORA45 | FALSE | Downregulated |
| -0,448757023 | SNORA46 | FALSE | Downregulated |
| -0,857021942 | SNORA47 | FALSE | Downregulated |
| 0,593379115 | SNORA49 | FALSE | Upregulated |
| 0,173697073 | SNORA50 | FALSE | Upregulated |
| | SNORA51 | | |
| 0,138329681 | SNORA52 | FALSE | Upregulated |
| -0,445661052 | SNORA53 | FALSE | Downregulated |
| -1,618425546 | SNORA54 | | Downregulated |
| 0,130714863 | SNORA55 | FALSE | Upregulated |
| | SNORA56 | | |
| -1,862436042 | SNORA57 | | Downregulated |
| -0,017909019 | SNORA58 | FALSE | Downregulated |
| 0,277902965 | SNORA59A | FALSE | Upregulated |
| 0,277902965 | SNORA59B | FALSE | Upregulated |
| 0,775735582 | SNORA5A | FALSE | Upregulated |
| | SNORA5B | | |
| -0,104508977 | SNORA5C | FALSE | Downregulated |
| 0,440369846 | SNORA6 | FALSE | Upregulated |
| 0,022291406 | SNORA60 | FALSE | Upregulated |
| 0,171623019 | SNORA61 | FALSE | Upregulated |
| | SNORA62 | | |
| 0,986096521 | SNORA64 | FALSE | Upregulated |
| -0,302353036 | SNORA65 | FALSE | Downregulated |
| 0,268318295 | SNORA66 | FALSE | Upregulated |
| 0,158522307 | SNORA67 | FALSE | Upregulated |
| 0,160532784 | SNORA68 | FALSE | Upregulated |
| | SNORA69 | | |
| -0,494718526 | SNORA70 | FALSE | Downregulated |
| | SNORA70B | | |
| | SNORA70C | | |
| | SNORA70D | | |
| | SNORA70E | | |

| | | | |
|---|---|---|---|
| | SNORA70F | | |
| 0,248691534 | SNORA70G | FALSE | Upregulated |
| -0,137096692 | SNORA71A | FALSE | Downregulated |
| 0,31069648 | SNORA71B | FALSE | Upregulated |
| 0,127576938 | SNORA71C | FALSE | Upregulated |
| 0,146144904 | SNORA71D | FALSE | Upregulated |
| 0 | SNORA72 | FALSE | Upregulated |
| -0,257991521 | SNORA74A | FALSE | Downregulated |
| -0,481465958 | SNORA74B | FALSE | Downregulated |
| -0,006848587 | SNORA75 | FALSE | Downregulated |
| 0,145409729 | SNORA76 | FALSE | Upregulated |
| 0,023725602 | SNORA77 | FALSE | Upregulated |
| -0,130229033 | SNORA78 | FALSE | Downregulated |
| | SNORA79 | | |
| 0,247449835 | SNORA7B | FALSE | Upregulated |
| -0,305193501 | SNORA8 | FALSE | Downregulated |
| 0,437273964 | SNORA80 | FALSE | Upregulated |
| 0,423129709 | SNORA80B | FALSE | Upregulated |
| -0,741464379 | SNORA84 | FALSE | Downregulated |
| 0,258896948 | SNORA9 | FALSE | Upregulated |
| | SNORD100 | | |
| | SNORD101 | | |
| | SNORD102 | | |
| | SNORD103A | | |
| | SNORD103B | | |
| | SNORD104 | | |
| | SNORD105 | | |
| | SNORD105B | | |
| | SNORD11 | | |
| | SNORD110 | | |
| | SNORD111 | | |
| | SNORD111B | | |
| | SNORD112 | | |
| | SNORD113-1 | | |
| | SNORD113-2 | | |
| | SNORD113-3 | | |
| | SNORD113-4 | | |
| | SNORD113-5 | | |
| | SNORD113-6 | | |
| | SNORD113-7 | | |
| | SNORD113-8 | | |
| | SNORD113-9 | | |
| | SNORD114-1 | | |
| | SNORD114-10 | | |
| | SNORD114-11 | | |
| | SNORD114-12 | | |
| | SNORD114-13 | | |
| | SNORD114-14 | | |
| | SNORD114-15 | | |
| | SNORD114-16 | | |
| | SNORD114-17 | | |

| | | | |
|---|---|---|---|
| | SNORD114-18 | | |
| | SNORD114-19 | | |
| | SNORD114-2 | | |
| | SNORD114-20 | | |
| | SNORD114-21 | | |
| | SNORD114-22 | | |
| | SNORD114-23 | | |
| | SNORD114-24 | | |
| | SNORD114-25 | | |
| | SNORD114-26 | | |
| | SNORD114-27 | | |
| | SNORD114-28 | | |
| | SNORD114-29 | | |
| | SNORD114-3 | | |
| | SNORD114-30 | | |
| | SNORD114-31 | | |
| | SNORD114-4 | | |
| | SNORD114-5 | | |
| | SNORD114-6 | | |
| | SNORD114-7 | | |
| | SNORD114-8 | | |
| | SNORD114-9 | | |
| | SNORD117 | | |
| 0,02506856 | SNORD118 | FALSE | Upregulated |
| | SNORD119 | | |
| | SNORD11B | | |
| | SNORD12 | | |
| | SNORD121A | | |
| | SNORD121B | | |
| | SNORD123 | | |
| | SNORD124 | | |
| | SNORD125 | | |
| | SNORD126 | | |
| | SNORD127 | | |
| | SNORD12B | | |
| | SNORD12C | | |
| -0,148476667 | SNORD13 | FALSE | Downregulated |
| | SNORD14A | | |
| | SNORD14B | | |
| | SNORD14C | | |
| | SNORD14D | | |
| | SNORD14E | | |
| -0,425986415 | SNORD15A | FALSE | Downregulated |
| -0,666713065 | SNORD15B | FALSE | Downregulated |
| | SNORD16 | | |
| | SNORD18A | | |
| | SNORD18B | | |
| | SNORD18C | | |
| | SNORD19 | | |
| | SNORD19B | | |
| | SNORD1A | | |

| | | | |
|---|---|---|---|
| | SNORD1B | | |
| | SNORD1C | | |
| | SNORD2 | | |
| | SNORD20 | | |
| | SNORD21 | | |
| -0,406168817 | SNORD22 | FALSE | Downregulated |
| -0,130979477 | SNORD23 | FALSE | Downregulated |
| | SNORD24 | | |
| | SNORD25 | | |
| | SNORD26 | | |
| | SNORD27 | | |
| | SNORD28 | | |
| | SNORD29 | | |
| | SNORD30 | | |
| | SNORD31 | | |
| | SNORD32A | | |
| | SNORD32B | | |
| | SNORD33 | | |
| | SNORD34 | | |
| | SNORD35A | | |
| | SNORD35B | | |
| | SNORD36A | | |
| | SNORD36B | | |
| | SNORD36C | | |
| | SNORD37 | | |
| | SNORD38A | | |
| | SNORD38B | | |
| -0,775282308 | SNORD3B-1 | FALSE | Downregulated |
| 0,177165786 | SNORD3B-2 | FALSE | Upregulated |
| -0,621933672 | SNORD3D | FALSE | Downregulated |
| | SNORD41 | | |
| | SNORD42A | | |
| | SNORD42B | | |
| | SNORD43 | | |
| | SNORD44 | | |
| | SNORD45A | | |
| | SNORD45B | | |
| | SNORD45C | | |
| 0,274468217 | SNORD46 | FALSE | Upregulated |
| | SNORD47 | | |
| | SNORD48 | | |
| | SNORD49A | | |
| | SNORD49B | | |
| | SNORD4A | | |
| | SNORD4B | | |
| | SNORD5 | | |
| | SNORD50A | | |
| | SNORD50B | | |
| | SNORD51 | | |
| | SNORD52 | | |
| | SNORD53 | | |

| | | | | |
|---|---|---|---|---|
| | SNORD54 | | | |
| | SNORD55 | | | |
| | SNORD56 | | | |
| | SNORD56B | | | |
| | SNORD57 | | | |
| | SNORD58A | | | |
| | SNORD58B | | | |
| | SNORD58C | | | |
| | SNORD59A | | | |
| | SNORD59B | | | |
| | SNORD6 | | | |
| | SNORD60 | | | |
| | SNORD61 | | | |
| | SNORD62A | | | |
| | SNORD62B | | | |
| | SNORD63 | | | |
| | SNORD65 | | | |
| | SNORD66 | | | |
| 0 | SNORD67 | FALSE | Upregulated | |
| | SNORD68 | | | |
| | SNORD69 | | | |
| | SNORD7 | | | |
| | SNORD70 | | | |
| | SNORD71 | | | |
| | SNORD72 | | | |
| | SNORD73A | | | |
| | SNORD74 | | | |
| | SNORD75 | | | |
| 0 | SNORD76 | FALSE | Upregulated | |
| | SNORD77 | | | |
| | SNORD78 | | | |
| | SNORD79 | | | |
| -0,422891397 | | SNORD8 | FALSE | Downregulated |
| | SNORD80 | | | |
| | SNORD81 | | | |
| | SNORD82 | | | |
| | SNORD83A | | | |
| | SNORD83B | | | |
| | SNORD84 | | | |
| | SNORD85 | | | |
| | SNORD86 | | | |
| | SNORD87 | | | |
| | SNORD88A | | | |
| | SNORD88B | | | |
| | SNORD88C | | | |
| -0,263109468 | | SNORD89 | FALSE | Downregulated |
| -0,001674428 | | SNORD9 | FALSE | Downregulated |
| | SNORD90 | | | |
| | SNORD91A | | | |
| | SNORD91B | | | |
| | SNORD92 | | | |

| | | | |
|---|---|---|---|
| | SNORD93 | | |
| -0,037922079 | SNORD94 | FALSE | Downregulated |
| | SNORD95 | | |
| | SNORD96A | | |
| | SNORD96B | | |
| 1,020470328 | SNORD97 | | Upregulated |
| | SNORD98 | | |
| | SNORD99 | | |
| -0,024247521 | SNRNP27 | FALSE | Downregulated |
| -0,22784134 | SNRPD1 | FALSE | Downregulated |
| 1,369596118 | SNRPD2P2 | | Upregulated |
| -0,176867109 | SNRPF | FALSE | Downregulated |
| -0,124180602 | SNTG1 | FALSE | Downregulated |
| | SNTG2 | | |
| -0,130015768 | SNTN | FALSE | Downregulated |
| 0,818149681 | SNX16 | FALSE | Upregulated |
| 0,803115906 | SNX29P1 | FALSE | Upregulated |
| -0,163680429 | SNX31 | FALSE | Downregulated |
| 0,157691712 | SNX7 | FALSE | Upregulated |
| -1,194448664 | SOAT2 | | Downregulated |
| 0,839134417 | SOCS1 | FALSE | Upregulated |
| 0,300874926 | SOCS2-AS1 | FALSE | Upregulated |
| 0,300241802 | SOHLH1 | FALSE | Upregulated |
| | SOHLH2 | | |
| 0,030186537 | SORCS1 | FALSE | Upregulated |
| 0 | SORCS3 | FALSE | Upregulated |
| | SORCS3-AS1 | | |
| 0,129986624 | SOST | FALSE | Upregulated |
| -0,094906298 | SOSTDC1 | FALSE | Downregulated |
| -0,060248869 | SOWAHD | FALSE | Downregulated |
| | SOX1 | | |
| 0 | SOX11 | FALSE | Upregulated |
| 0 | SOX14 | FALSE | Upregulated |
| -0,940943829 | SOX17 | FALSE | Downregulated |
| 0,536714943 | SOX2 | FALSE | Upregulated |
| 0,120665481 | SOX2-OT | FALSE | Upregulated |
| 0,271243351 | SOX21 | FALSE | Upregulated |
| 0,000186938 | SOX21-AS1 | FALSE | Upregulated |
| | SOX3 | | |
| 0,329302364 | SOX30 | FALSE | Upregulated |
| -0,219213199 | SOX7 | FALSE | Downregulated |
| 0,028164251 | SOX8 | FALSE | Upregulated |
| 0,728057492 | SOX9-AS1 | FALSE | Upregulated |
| 0,502835253 | SP140 | FALSE | Upregulated |
| 0,143036846 | SP6 | FALSE | Upregulated |
| 0 | SP8 | FALSE | Upregulated |
| | SP9 | | |
| 0,04248548 | SPA17 | FALSE | Upregulated |
| -0,011481061 | SPACA1 | FALSE | Downregulated |
| | SPACA3 | | |
| -0,497110121 | SPACA4 | FALSE | Downregulated |

| | | | |
|---|---|---|---|
| 0,140358913 | SPACA5 | FALSE | Upregulated |
| -0,343041936 | SPACA5B | FALSE | Downregulated |
| 0,224255694 | SPACA6P | FALSE | Upregulated |
| 0,760494936 | SPACA7 | FALSE | Upregulated |
| | SPAG11A | | |
| | SPAG11B | | |
| 1,081465509 | SPAG17 | | Upregulated |
| -0,102609997 | SPAG4 | FALSE | Downregulated |
| -0,014948367 | SPAG6 | FALSE | Downregulated |
| | SPAM1 | | |
| | SPANXA1 | | |
| | SPANXA2 | | |
| | SPANXA2-OT1 | | |
| 0,003520835 | SPANXB1 | FALSE | Upregulated |
| | SPANXC | | |
| 0 | SPANXD | FALSE | Upregulated |
| | SPANXN1 | | |
| | SPANXN2 | | |
| | SPANXN3 | | |
| | SPANXN4 | | |
| | SPANXN5 | | |
| 1,079571515 | SPATA13-AS1 | | Upregulated |
| | SPATA16 | | |
| -0,009538492 | SPATA17 | FALSE | Downregulated |
| | SPATA19 | | |
| 0,001118135 | SPATA22 | FALSE | Upregulated |
| 0,436557279 | SPATA24 | FALSE | Upregulated |
| | SPATA3 | | |
| 0,088842351 | SPATA3-AS1 | FALSE | Upregulated |
| | SPATA31A1 | | |
| | SPATA31A3 | | |
| | SPATA31A5 | | |
| | SPATA31A6 | | |
| | SPATA31A7 | | |
| 0 | SPATA31B1 | FALSE | Upregulated |
| 0,244434311 | SPATA31C1 | FALSE | Upregulated |
| 0,468330498 | SPATA31C2 | FALSE | Upregulated |
| | SPATA31D1 | | |
| | SPATA31D3 | | |
| | SPATA31D4 | | |
| 0 | SPATA31D5P | FALSE | Upregulated |
| 0,976802176 | SPATA32 | FALSE | Upregulated |
| -0,402745439 | SPATA33 | FALSE | Downregulated |
| 0,29133661 | SPATA4 | FALSE | Upregulated |
| | SPATA42 | | |
| -0,352468407 | SPATA6 | FALSE | Downregulated |
| 0,536975865 | SPATA8 | FALSE | Upregulated |
| | SPATA8-AS1 | | |
| 0,183782087 | SPATA9 | FALSE | Upregulated |
| 0,344927696 | SPATC1L | FALSE | Upregulated |
| 0,001141803 | SPATS1 | FALSE | Upregulated |

| | | | |
|---|---|---|---|
| 0,096990056 | SPATS2 | FALSE | Upregulated |
| 0,333510036 | SPC24 | FALSE | Upregulated |
| -0,256110126 | SPC25 | FALSE | Downregulated |
| 0,274740643 | SPDEF | FALSE | Upregulated |
| 1,397007124 | SPDL1 | | Upregulated |
| 0,166993232 | SPDYC | FALSE | Upregulated |
| 0,011503654 | SPDYE4 | FALSE | Upregulated |
| 0,286392014 | SPEF1 | FALSE | Upregulated |
| 0,072009995 | SPEF2 | FALSE | Upregulated |
| -0,137129761 | SPEM1 | FALSE | Downregulated |
| | SPERT | | |
| 0,287549886 | SPESP1 | FALSE | Upregulated |
| 0 | SPG20OS | FALSE | Upregulated |
| | SPHAR | | |
| 0,279375282 | SPHK1 | FALSE | Upregulated |
| 0,284987956 | SPHKAP | FALSE | Upregulated |
| 0,725073907 | SPIB | FALSE | Upregulated |
| 0,94121153 | SPIC | FALSE | Upregulated |
| 0,839261157 | SPIN2B | FALSE | Upregulated |
| 0,101851622 | SPIN4 | FALSE | Upregulated |
| -0,085905028 | SPINK1 | FALSE | Downregulated |
| 0,583501642 | SPINK13 | FALSE | Upregulated |
| 0 | SPINK14 | FALSE | Upregulated |
| 0 | SPINK2 | FALSE | Upregulated |
| | SPINK4 | | |
| 0,141020425 | SPINK5 | FALSE | Upregulated |
| 0,60574456 | SPINK6 | FALSE | Upregulated |
| 0,26124614 | SPINK7 | FALSE | Upregulated |
| | SPINK8 | | |
| 0,231170421 | SPINK9 | FALSE | Upregulated |
| 0,737734981 | SPINT1 | FALSE | Upregulated |
| | SPINT3 | | |
| | SPINT4 | | |
| 0 | SPO11 | FALSE | Upregulated |
| 0 | SPOCD1 | FALSE | Upregulated |
| 0,268818918 | SPOCK1 | FALSE | Upregulated |
| -0,234337121 | SPOCK3 | FALSE | Downregulated |
| | SPPL2C | | |
| -0,131287917 | SPRED3 | FALSE | Downregulated |
| | SPRR1A | | |
| | SPRR1B | | |
| | SPRR2A | | |
| | SPRR2B | | |
| | SPRR2C | | |
| | SPRR2D | | |
| 0 | SPRR2E | FALSE | Upregulated |
| | SPRR2F | | |
| | SPRR2G | | |
| | SPRR3 | | |
| | SPRR4 | | |
| 1,358859016 | SPRY3 | | Upregulated |

| | | | |
|---|---|---|---|
| | SPRY3-2 | | |
| -0,869093868 | SPSB4 | FALSE | Downregulated |
| 0,017816394 | SPTA1 | FALSE | Upregulated |
| -0,162383916 | SPTBN4 | FALSE | Downregulated |
| -0,526752629 | SPTSSB | FALSE | Downregulated |
| 0 | SPZ1 | FALSE | Upregulated |
| 0,350088461 | SRCRB4D | FALSE | Upregulated |
| 0,283749894 | SRD5A1P1 | FALSE | Upregulated |
| 0,439444687 | SRD5A3-AS1 | FALSE | Upregulated |
| -0,040265709 | SRFBP1 | FALSE | Downregulated |
| 0,137549897 | SRGAP2D | FALSE | Upregulated |
| 0,597387499 | SRP14-AS1 | FALSE | Upregulated |
| -0,269054434 | SRPX | FALSE | Downregulated |
| -1,116817461 | SRRM2-AS1 | | Downregulated |
| 0,114647919 | SRRM3 | FALSE | Upregulated |
| 0,291340714 | SRRM4 | FALSE | Upregulated |
| -0,010725599 | SRY | FALSE | Downregulated |
| -0,421610895 | SS18L2 | FALSE | Downregulated |
| | SSMEM1 | | |
| 0,201445432 | SSPN | FALSE | Upregulated |
| -0,014046785 | SSR4P1 | FALSE | Downregulated |
| 0,061543669 | SSSCA1 | FALSE | Upregulated |
| 0,573152725 | SSSCA1-AS1 | FALSE | Upregulated |
| | SST | | |
| 0,464094479 | SSTR3 | FALSE | Upregulated |
| | SSTR4 | | |
| -0,42228593 | SSTR5 | FALSE | Downregulated |
| 0,30979538 | SSTR5-AS1 | FALSE | Upregulated |
| 0,285062792 | SSUH2 | FALSE | Upregulated |
| | SSX1 | | |
| | SSX2 | | |
| | SSX2B | | |
| | SSX3 | | |
| | SSX4 | | |
| | SSX4B | | |
| | SSX5 | | |
| | SSX6 | | |
| | SSX7 | | |
| | SSX8 | | |
| | SSX9 | | |
| -0,302213324 | ST13P4 | FALSE | Downregulated |
| 1,154581136 | ST18 | | Upregulated |
| 0,826742176 | ST20-MTHFS | FALSE | Upregulated |
| 0,186981579 | ST3GAL6-AS1 | FALSE | Upregulated |
| 0,678460345 | ST6GALNAC1 | FALSE | Upregulated |
| -1,06192856 | ST6GALNAC3 | | Downregulated |
| 0,71240316 | ST6GALNAC5 | FALSE | Upregulated |
| 0,00139671 | ST7-OT3 | FALSE | Upregulated |
| -0,817614069 | ST7-OT4 | FALSE | Downregulated |
| 0,445369259 | ST8SIA1 | FALSE | Upregulated |
| | ST8SIA2 | | |

| | | | |
|---|---|---|---|
| -0,157456032 | ST8SIA5 | FALSE | Downregulated |
| -0,612187446 | ST8SIA6 | FALSE | Downregulated |
| 0,463079352 | ST8SIA6-AS1 | FALSE | Upregulated |
| 1,249984446 | STAC | | Upregulated |
| 0 | STAC2 | FALSE | Upregulated |
| 0,610468721 | STAC3 | FALSE | Upregulated |
| 0,140206377 | STAG3L4 | FALSE | Upregulated |
| 0,339756146 | STAP1 | FALSE | Upregulated |
| 0,583200372 | STARD3NL | FALSE | Upregulated |
| | STARD6 | | |
| | STATH | | |
| -0,401051635 | STC1 | FALSE | Downregulated |
| -0,313528691 | STC2 | FALSE | Downregulated |
| 0,722608295 | STEAP1B | FALSE | Upregulated |
| -0,003745761 | STEAP2-AS1 | FALSE | Downregulated |
| | STH | | |
| 0,436783795 | STIL | FALSE | Upregulated |
| -0,190108744 | STK11IP | FALSE | Downregulated |
| -0,164873478 | STK31 | FALSE | Downregulated |
| 0,784253432 | STK32A | FALSE | Upregulated |
| 1,088791734 | STK32B | | Upregulated |
| 0,003007923 | STK32C | FALSE | Upregulated |
| 0,250176981 | STK33 | FALSE | Upregulated |
| 0,214059162 | STK4-AS1 | FALSE | Upregulated |
| 0,387098759 | STMN2 | FALSE | Upregulated |
| 0,537949072 | STMN4 | FALSE | Upregulated |
| -0,004290336 | STMND1 | FALSE | Downregulated |
| 0,178045441 | STOML1 | FALSE | Upregulated |
| | STOML3 | | |
| -0,68330274 | STON1 | FALSE | Downregulated |
| 0,081377453 | STON1-GTF2A1L | FALSE | Upregulated |
| 0,306772279 | STOX1 | FALSE | Upregulated |
| -0,284875373 | STPG1 | FALSE | Downregulated |
| 0,494688547 | STPG2 | FALSE | Upregulated |
| 0,399917838 | STPG2-AS1 | FALSE | Upregulated |
| -0,015379263 | STRA6 | FALSE | Downregulated |
| 0,297178435 | STRA8 | FALSE | Upregulated |
| -0,900102141 | STRC | FALSE | Downregulated |
| 0,007420023 | STRIP2 | FALSE | Upregulated |
| 0,092099259 | STX18-AS1 | FALSE | Upregulated |
| -0,102505407 | STX8 | FALSE | Downregulated |
| -0,299170153 | STXBP4 | FALSE | Downregulated |
| -0,962631043 | STXBP5L | FALSE | Downregulated |
| -0,338802528 | STXBP6 | FALSE | Downregulated |
| 0,679663921 | STYK1 | FALSE | Upregulated |
| 0,522878596 | SUCNR1 | FALSE | Upregulated |
| 0,12743372 | SUGT1P1 | FALSE | Upregulated |
| 0,391546002 | SUGT1P3 | FALSE | Upregulated |
| 0 | SULT1A3 | FALSE | Upregulated |
| -0,223649776 | SULT1B1 | FALSE | Downregulated |
| -0,440138267 | SULT1C2 | FALSE | Downregulated |

| | | | |
|---|---|---|---|
| 0,292267771 | SULT1C2P1 | FALSE | Upregulated |
| 0 | SULT1C3 | FALSE | Upregulated |
| -0,043837962 | SULT1C4 | FALSE | Downregulated |
| 0,131500113 | SULT2B1 | FALSE | Upregulated |
| 0,437979753 | SULT4A1 | FALSE | Upregulated |
| | SULT6B1 | | |
| 0 | SUMO1P1 | FALSE | Upregulated |
| -1,394581183 | SUMO4 | | Downregulated |
| | SUN3 | | |
| | SUN5 | | |
| 0,70359644 | SUPT20HL1 | FALSE | Upregulated |
| 1,169257177 | SUPT20HL2 | | Upregulated |
| -0,271018266 | SUPT3H | FALSE | Downregulated |
| 0,158136599 | SURF2 | FALSE | Upregulated |
| -0,16597764 | SUSD2 | FALSE | Downregulated |
| -0,424575889 | SUSD4 | FALSE | Downregulated |
| -0,210071294 | SUSD5 | FALSE | Downregulated |
| 0,113619378 | SUZ12P1 | FALSE | Upregulated |
| -0,022052872 | SV2A | FALSE | Downregulated |
| -0,341622131 | SV2B | FALSE | Downregulated |
| -0,015448814 | SV2C | FALSE | Downregulated |
| 1,193850834 | SVILP1 | | Upregulated |
| 0,16483886 | SVOP | FALSE | Upregulated |
| -0,015973174 | SVOPL | FALSE | Downregulated |
| -0,78873928 | SYCE1 | FALSE | Downregulated |
| -0,023569737 | SYCE1L | FALSE | Downregulated |
| -0,48907731 | SYCE3 | FALSE | Downregulated |
| 0 | SYCN | FALSE | Upregulated |
| | SYCP1 | | |
| 0,763345793 | SYCP2 | FALSE | Upregulated |
| 0,289235841 | SYCP2L | FALSE | Upregulated |
| -0,054558264 | SYN2 | FALSE | Downregulated |
| 0 | SYNDIG1 | FALSE | Upregulated |
| 0,28745854 | SYNDIG1L | FALSE | Upregulated |
| 0,880885956 | SYNGR3 | FALSE | Upregulated |
| 0 | SYNGR4 | FALSE | Upregulated |
| 0,740813178 | SYNJ2-IT1 | FALSE | Upregulated |
| -1,236106987 | SYNJ2BP-COX16 | | Downregulated |
| -0,531082843 | SYNPR | FALSE | Downregulated |
| | SYNPR-AS1 | | |
| -0,218707251 | SYP | FALSE | Downregulated |
| 0,440806366 | SYP-AS1 | FALSE | Upregulated |
| -0,499586589 | SYS1-DBNDD2 | FALSE | Downregulated |
| -0,256807541 | SYT10 | FALSE | Downregulated |
| 1,660740209 | SYT11 | | Upregulated |
| -0,146982809 | SYT13 | FALSE | Downregulated |
| | SYT14 | | |
| 0,049635132 | SYT14L | FALSE | Upregulated |
| | SYT16 | | |
| | SYT3 | | |
| | SYT4 | | |

| | | | |
|---|---|---|---|
| 0 | SYT5 | FALSE | Upregulated |
| 0,248346788 | SYT6 | FALSE | Upregulated |
| 0,017898711 | SYT8 | FALSE | Upregulated |
| 0,507773352 | SYTL3 | FALSE | Upregulated |
| | T | | |
| | TAAR1 | | |
| | TAAR2 | | |
| -0,183163923 | TAAR3 | FALSE | Downregulated |
| | TAAR5 | | |
| | TAAR6 | | |
| | TAAR8 | | |
| | TAAR9 | | |
| 0,005298914 | TAC1 | FALSE | Upregulated |
| | TAC3 | | |
| -0,715359473 | TACR3 | FALSE | Downregulated |
| 0,520280168 | TACSTD2 | FALSE | Upregulated |
| -0,185910153 | TADA2A | FALSE | Downregulated |
| -0,734149218 | TAF12 | FALSE | Downregulated |
| 1,11780281 | TAF13 | | Upregulated |
| 0,13250523 | TAF1A | FALSE | Upregulated |
| 0,075636175 | TAF1B | FALSE | Upregulated |
| -0,559791598 | TAF1D | FALSE | Downregulated |
| -0,323616988 | TAF1L | FALSE | Downregulated |
| -0,932486428 | TAF4B | FALSE | Downregulated |
| 0,155357166 | TAF7L | FALSE | Upregulated |
| | TAGLN3 | | |
| -0,651453591 | TAL1 | FALSE | Downregulated |
| 0,139478558 | TAL2 | FALSE | Upregulated |
| | TARM1 | | |
| -0,016784474 | TARP | FALSE | Downregulated |
| | TAS1R2 | | |
| | TAS2R1 | | |
| 0,054816027 | TAS2R10 | FALSE | Upregulated |
| -0,416363221 | TAS2R13 | FALSE | Downregulated |
| 0,05443578 | TAS2R14 | FALSE | Upregulated |
| | TAS2R16 | | |
| 0,584183235 | TAS2R19 | FALSE | Upregulated |
| -0,398371922 | TAS2R20 | FALSE | Downregulated |
| 1,088033534 | TAS2R3 | | Upregulated |
| 0,220129642 | TAS2R30 | FALSE | Upregulated |
| 0,76801661 | TAS2R31 | FALSE | Upregulated |
| -0,006508655 | TAS2R38 | FALSE | Downregulated |
| -0,479560296 | TAS2R39 | FALSE | Downregulated |
| 0,414934414 | TAS2R4 | FALSE | Upregulated |
| -0,249994592 | TAS2R40 | FALSE | Downregulated |
| | TAS2R41 | | |
| -0,032933408 | TAS2R43 | FALSE | Downregulated |
| -0,581814651 | TAS2R46 | FALSE | Downregulated |
| -0,121774858 | TAS2R5 | FALSE | Downregulated |
| 0,177409765 | TAS2R50 | FALSE | Upregulated |
| -0,041535231 | TAS2R60 | FALSE | Downregulated |

| | | | |
|---|---|---|---|
| 0,552275455 | TAS2R7 | FALSE | Upregulated |
| -0,018149848 | TAS2R8 | FALSE | Downregulated |
| 0 | TAS2R9 | FALSE | Upregulated |
| 0,286348007 | TASP1 | FALSE | Upregulated |
| -0,033664727 | TATDN3 | FALSE | Downregulated |
| 0,095411915 | TAX1BP3 | FALSE | Upregulated |
| 0 | TBATA | FALSE | Upregulated |
| -0,491443113 | TBC1D19 | FALSE | Downregulated |
| | TBC1D21 | | |
| | TBC1D26 | | |
| 0,28805737 | TBC1D27 | FALSE | Upregulated |
| 0,293326262 | TBC1D28 | FALSE | Upregulated |
| 0,328729337 | TBC1D29 | FALSE | Upregulated |
| -0,438599685 | TBC1D3 | FALSE | Downregulated |
| 0,054721054 | TBC1D30 | FALSE | Upregulated |
| -0,114285436 | TBC1D31 | FALSE | Downregulated |
| 0,040547991 | TBC1D32 | FALSE | Upregulated |
| 0,014711513 | TBC1D3B | FALSE | Upregulated |
| -1,68531259 | TBC1D3C | | Downregulated |
| | TBC1D3F | | |
| 0,524300376 | TBC1D3H | FALSE | Upregulated |
| 0 | TBC1D3P1-DHX40P1 | FALSE | Upregulated |
| | TBC1D3P2 | | |
| | TBC1D3P5 | | |
| -0,236885357 | TBC1D7 | FALSE | Downregulated |
| -0,542099307 | TBL1Y | FALSE | Downregulated |
| 0,277833435 | TBPL2 | FALSE | Upregulated |
| | TBR1 | | |
| -0,000950251 | TBX1 | FALSE | Downregulated |
| 0,312932679 | TBX10 | FALSE | Upregulated |
| 0,118836136 | TBX18 | FALSE | Upregulated |
| -0,109839155 | TBX19 | FALSE | Downregulated |
| -0,536854667 | TBX20 | FALSE | Downregulated |
| -0,151939009 | TBX21 | FALSE | Downregulated |
| 0 | TBX22 | FALSE | Upregulated |
| 0,164422926 | TBX4 | FALSE | Upregulated |
| 0,272277655 | TBX5 | FALSE | Upregulated |
| 0 | TBX5-AS1 | FALSE | Upregulated |
| 0,814628005 | TBX6 | FALSE | Upregulated |
| | TCAM1P | | |
| 0,285175489 | TCAP | FALSE | Upregulated |
| 0,019020309 | TCEAL1 | FALSE | Upregulated |
| | TCEAL2 | | |
| | TCEAL5 | | |
| 0 | TCEAL6 | FALSE | Upregulated |
| -0,000121433 | TCEAL7 | FALSE | Downregulated |
| 0,209240485 | TCEAL8 | FALSE | Upregulated |
| -0,394321777 | TCEANC | FALSE | Downregulated |
| 0,080396985 | TCEANC2 | FALSE | Upregulated |
| | TCERG1L | | |
| -0,066330271 | TCF15 | FALSE | Downregulated |

| | | | |
|---|---|---|---|
| 0 | TCF23 | FALSE | Upregulated |
| -0,349174533 | TCF24 | FALSE | Downregulated |
| 0,377613502 | TCHH | FALSE | Upregulated |
| 0,272951823 | TCHHL1 | FALSE | Upregulated |
| 0,387165016 | TCL1A | FALSE | Upregulated |
| 0 | TCL1B | FALSE | Upregulated |
| 0 | TCL6 | FALSE | Upregulated |
| 0 | TCN1 | FALSE | Upregulated |
| | TCP10 | | |
| -0,464727571 | TCP10L | FALSE | Downregulated |
| 0 | TCP10L2 | FALSE | Upregulated |
| 0,007488359 | TCP11 | FALSE | Upregulated |
| | TCP11X1 | | |
| | TCP11X2 | | |
| -0,336916346 | TCTE1 | FALSE | Downregulated |
| -0,201936538 | TCTE3 | FALSE | Downregulated |
| 0,147553239 | TCTEX1D1 | FALSE | Upregulated |
| 0,906095333 | TCTEX1D2 | FALSE | Upregulated |
| -0,37687068 | TCTN1 | FALSE | Downregulated |
| -0,031486082 | TCTN2 | FALSE | Downregulated |
| -0,546132141 | TDGF1 | FALSE | Downregulated |
| 0,279296663 | TDGF1P3 | FALSE | Upregulated |
| 0 | TDH | FALSE | Upregulated |
| 0,703432045 | TDRD1 | FALSE | Upregulated |
| 0 | TDRD12 | FALSE | Upregulated |
| 0,117237813 | TDRD15 | FALSE | Upregulated |
| | TDRD5 | | |
| 1,066870214 | TDRD9 | | Upregulated |
| | TDRG1 | | |
| 0,755412357 | TDRKH | FALSE | Upregulated |
| 0,670926581 | TEAD4 | FALSE | Upregulated |
| 0 | TECRL | FALSE | Upregulated |
| -0,40678694 | TECTA | FALSE | Downregulated |
| -0,85418637 | TECTB | FALSE | Downregulated |
| 0,445570615 | TEKT1 | FALSE | Upregulated |
| -0,032429096 | TEKT2 | FALSE | Downregulated |
| 0,263555872 | TEKT3 | FALSE | Upregulated |
| | TEKT4 | | |
| 0,290862052 | TEKT4P2 | FALSE | Upregulated |
| -0,889465512 | TEKT5 | FALSE | Downregulated |
| -1,499760606 | TENM3 | | Downregulated |
| -0,515678464 | TENM4 | FALSE | Downregulated |
| 0 | TEPP | FALSE | Upregulated |
| | TERT | | |
| -0,920631414 | TESC | FALSE | Downregulated |
| 1,407296137 | TESPA1 | | Upregulated |
| | TEX101 | | |
| | TEX11 | | |
| 0,418177489 | TEX12 | FALSE | Upregulated |
| -0,99351973 | TEX13A | FALSE | Downregulated |
| -0,01732122 | TEX13B | FALSE | Downregulated |

| | | | |
|---|---|---|---|
| 0,025254051 | TEX15 | FALSE | Upregulated |
| 0 | TEX19 | FALSE | Upregulated |
| 0,460327821 | TEX21P | FALSE | Upregulated |
| -0,030110679 | TEX22 | FALSE | Downregulated |
| | TEX26 | | |
| | TEX26-AS1 | | |
| 0,293242527 | TEX28 | FALSE | Upregulated |
| 0,13940991 | TEX29 | FALSE | Upregulated |
| 0,158165459 | TEX30 | FALSE | Upregulated |
| 0 | TEX33 | FALSE | Upregulated |
| -0,168861612 | TEX35 | FALSE | Downregulated |
| | TEX36 | | |
| | TEX36-AS1 | | |
| | TEX37 | | |
| 0,391398517 | TEX40 | FALSE | Upregulated |
| -0,582090944 | TEX41 | FALSE | Downregulated |
| 0,00139671 | TFAMP1 | FALSE | Upregulated |
| 1,058914091 | TFAP2A | | Upregulated |
| | TFAP2A-AS1 | | |
| 0 | TFAP2B | FALSE | Upregulated |
| -0,118893375 | TFAP2C | FALSE | Downregulated |
| | TFAP2D | | |
| -0,513391278 | TFCP2L1 | FALSE | Downregulated |
| 0 | TFDP3 | FALSE | Upregulated |
| 0,011901572 | TFF1 | FALSE | Upregulated |
| 0 | TFF2 | FALSE | Upregulated |
| 0,466086066 | TFF3 | FALSE | Upregulated |
| -1,244779012 | TFPI2 | | Downregulated |
| -0,196397824 | TG | FALSE | Downregulated |
| -0,418462472 | TGDS | FALSE | Downregulated |
| -0,532590814 | TGFA | FALSE | Downregulated |
| | TGFA-IT1 | | |
| 0,24397553 | TGFB2 | FALSE | Upregulated |
| -0,678319056 | TGIF2-C20orf24 | FALSE | Downregulated |
| | TGIF2LX | | |
| | TGIF2LY | | |
| 0,275144381 | TGM1 | FALSE | Upregulated |
| 0,009187949 | TGM3 | FALSE | Upregulated |
| 0,620084998 | TGM4 | FALSE | Upregulated |
| 0,455512512 | TGM5 | FALSE | Upregulated |
| -0,004688595 | TGM6 | FALSE | Downregulated |
| | TGM7 | | |
| 0,092662563 | THAP1 | FALSE | Upregulated |
| 0,983835614 | THAP10 | FALSE | Upregulated |
| 0,658469525 | THBS4 | FALSE | Upregulated |
| 0 | THEG | FALSE | Upregulated |
| | THEG5 | | |
| 0 | THEGL | FALSE | Upregulated |
| 1,853163453 | THEMIS | | Upregulated |
| -0,054246281 | THG1L | FALSE | Downregulated |
| -0,073308144 | THSD1 | FALSE | Downregulated |

| | | | |
|---|---|---|---|
| 0,11355584 | THSD7B | FALSE | Upregulated |
| 0,950560673 | THUMPD2 | FALSE | Upregulated |
| 0,528408739 | TIFAB | FALSE | Upregulated |
| -0,198155145 | TIGD1 | FALSE | Downregulated |
| 0,12443614 | TIGD3 | FALSE | Upregulated |
| 0,80556924 | TIGD4 | FALSE | Upregulated |
| -0,141490068 | TIMD4 | FALSE | Downregulated |
| -0,207589655 | TIMM8A | FALSE | Downregulated |
| -0,531104487 | TIMM9 | FALSE | Downregulated |
| 0,116839868 | TIMP4 | FALSE | Upregulated |
| | TINAG | | |
| 0,43790778 | TINCR | FALSE | Upregulated |
| 0,014980772 | TIPIN | FALSE | Upregulated |
| | TISP43 | | |
| -0,581790989 | TKTL1 | FALSE | Downregulated |
| | TKTL2 | | |
| 1,479598814 | TLE6 | | Upregulated |
| -0,161712465 | TLL1 | FALSE | Downregulated |
| -0,165196348 | TLL2 | FALSE | Downregulated |
| 0,355609112 | TLR10 | FALSE | Upregulated |
| 0,313207728 | TLR5 | FALSE | Upregulated |
| 0,842341233 | TLR7 | FALSE | Upregulated |
| 1,059109156 | TLR8 | | Upregulated |
| 0 | TLR8-AS1 | FALSE | Upregulated |
| -0,337350464 | TLX1 | FALSE | Downregulated |
| 0,020233862 | TLX1NB | FALSE | Upregulated |
| 0,460403919 | TLX2 | FALSE | Upregulated |
| | TLX3 | | |
| -0,346332499 | TM2D1 | FALSE | Downregulated |
| 0,365515895 | TM4SF1-AS1 | FALSE | Upregulated |
| -0,27340822 | TM4SF18 | FALSE | Downregulated |
| 0,284936527 | TM4SF19 | FALSE | Upregulated |
| -0,439516755 | TM4SF19-AS1 | FALSE | Downregulated |
| | TM4SF19-TCTEX1D2 | | |
| 0,501236466 | TM6SF1 | FALSE | Upregulated |
| 0,448393544 | TMC1 | FALSE | Upregulated |
| 0,537217774 | TMC2 | FALSE | Upregulated |
| 0,429473828 | TMC4 | FALSE | Upregulated |
| 0,110196868 | TMC7 | FALSE | Upregulated |
| -0,08427517 | TMCC1-AS1 | FALSE | Downregulated |
| 0,939477956 | TMCC2 | FALSE | Upregulated |
| 0,496210244 | TMCO2 | FALSE | Upregulated |
| | TMCO5A | | |
| | TMCO5B | | |
| -0,264724333 | TMED11P | FALSE | Downregulated |
| -0,717356023 | TMED3 | FALSE | Downregulated |
| -0,065108941 | TMED7-TICAM2 | FALSE | Downregulated |
| | TMEFF1 | | |
| 0,948413279 | TMEFF2 | FALSE | Upregulated |
| 0,337647047 | TMEM100 | FALSE | Upregulated |
| 0,566029301 | TMEM102 | FALSE | Upregulated |

| | | | |
|---|---|---|---|
| -0,256099178 | TMEM105 | FALSE | Downregulated |
| -0,877493254 | TMEM107 | FALSE | Downregulated |
| 0,467751069 | TMEM108 | FALSE | Upregulated |
| 0,723694479 | TMEM110-MUSTN1 | FALSE | Upregulated |
| | TMEM114 | | |
| 0,235811761 | TMEM116 | FALSE | Upregulated |
| 0,164421613 | TMEM117 | FALSE | Upregulated |
| -0,798006903 | TMEM119 | FALSE | Downregulated |
| -0,100814928 | TMEM121 | FALSE | Downregulated |
| 0,03202614 | TMEM126B | FALSE | Upregulated |
| 0,203798691 | TMEM128 | FALSE | Upregulated |
| 0,177537378 | TMEM130 | FALSE | Upregulated |
| 0,002438924 | TMEM132B | FALSE | Upregulated |
| 0,32054005 | TMEM132C | FALSE | Upregulated |
| -1,017878794 | TMEM132E | | Downregulated |
| -0,058651408 | TMEM133 | FALSE | Downregulated |
| 0,158171631 | TMEM136 | FALSE | Upregulated |
| -0,055614222 | TMEM144 | FALSE | Downregulated |
| 0,283293685 | TMEM145 | FALSE | Upregulated |
| -0,335335446 | TMEM14A | FALSE | Downregulated |
| 0,441082115 | TMEM150B | FALSE | Upregulated |
| 0,284415897 | TMEM151A | FALSE | Upregulated |
| 0,072934636 | TMEM151B | FALSE | Upregulated |
| 0,47618102 | TMEM154 | FALSE | Upregulated |
| 0 | TMEM155 | FALSE | Upregulated |
| 1,258299859 | TMEM156 | | Upregulated |
| 0,330148506 | TMEM158 | FALSE | Upregulated |
| -0,109107712 | TMEM159 | FALSE | Downregulated |
| -0,251868044 | TMEM161B | FALSE | Downregulated |
| -1,675147182 | TMEM161B-AS1 | | Downregulated |
| -0,523242366 | TMEM163 | FALSE | Downregulated |
| 0,249175746 | TMEM169 | FALSE | Upregulated |
| -0,147544843 | TMEM170A | FALSE | Downregulated |
| -0,023777462 | TMEM171 | FALSE | Downregulated |
| | TMEM174 | | |
| -0,11385758 | TMEM178A | FALSE | Downregulated |
| 0,278469392 | TMEM178B | FALSE | Upregulated |
| 0,067655167 | TMEM18 | FALSE | Upregulated |
| 0,431807464 | TMEM182 | FALSE | Upregulated |
| 0,811690497 | TMEM183B | FALSE | Upregulated |
| 0,032003827 | TMEM187 | FALSE | Upregulated |
| 0,982778481 | TMEM189-UBE2V1 | FALSE | Upregulated |
| 1,184636743 | TMEM190 | | Upregulated |
| 0,599920752 | TMEM191A | FALSE | Upregulated |
| 0 | TMEM191B | FALSE | Upregulated |
| -0,687294045 | TMEM191C | FALSE | Downregulated |
| 0,838218544 | TMEM196 | FALSE | Upregulated |
| 0,129073112 | TMEM198 | FALSE | Upregulated |
| -0,106363501 | TMEM199 | FALSE | Downregulated |
| 0,823707822 | TMEM200A | FALSE | Upregulated |
| -0,144380317 | TMEM202 | FALSE | Downregulated |

| | | | |
|---|---|---|---|
| 0,458466516 | TMEM207 | FALSE | Upregulated |
| | TMEM210 | | |
| -0,181417356 | TMEM211 | FALSE | Downregulated |
| 0,082016234 | TMEM212 | FALSE | Upregulated |
| 0,665681186 | TMEM213 | FALSE | Upregulated |
| 0,077739607 | TMEM215 | FALSE | Upregulated |
| -0,673183971 | TMEM216 | FALSE | Downregulated |
| -0,239550158 | TMEM217 | FALSE | Downregulated |
| 0,549571745 | TMEM220-AS1 | FALSE | Upregulated |
| 0,494434653 | TMEM225 | FALSE | Upregulated |
| 0 | TMEM229A | FALSE | Upregulated |
| 0,70460645 | TMEM229B | FALSE | Upregulated |
| -0,173284728 | TMEM231 | FALSE | Downregulated |
| 0,2616694 | TMEM232 | FALSE | Upregulated |
| 0,365312123 | TMEM233 | FALSE | Upregulated |
| 0,048983671 | TMEM234 | FALSE | Upregulated |
| -0,248227294 | TMEM235 | FALSE | Downregulated |
| 0,279897213 | TMEM236 | FALSE | Upregulated |
| -0,226244484 | TMEM237 | FALSE | Downregulated |
| | TMEM239 | | |
| 0,494293727 | TMEM240 | FALSE | Upregulated |
| 0,511920331 | TMEM241 | FALSE | Upregulated |
| 0 | TMEM244 | FALSE | Upregulated |
| 0,855110903 | TMEM246 | FALSE | Upregulated |
| 0,271813643 | TMEM247 | FALSE | Upregulated |
| -0,046531603 | TMEM249 | FALSE | Downregulated |
| 0,238548445 | TMEM251 | FALSE | Upregulated |
| -0,143355174 | TMEM254-AS1 | FALSE | Downregulated |
| 0,042939577 | TMEM255B | FALSE | Upregulated |
| 0,037428469 | TMEM256 | FALSE | Upregulated |
| 1,280245964 | TMEM256-PLSCR3 | | Upregulated |
| | TMEM257 | | |
| -0,800283917 | TMEM258 | FALSE | Downregulated |
| 0,162446465 | TMEM26 | FALSE | Upregulated |
| -0,738569215 | TMEM262 | FALSE | Downregulated |
| 0,064513068 | TMEM27 | FALSE | Upregulated |
| 0,684235527 | TMEM30C | FALSE | Upregulated |
| 0,162299727 | TMEM31 | FALSE | Upregulated |
| 0,118076172 | TMEM35 | FALSE | Upregulated |
| -0,26194153 | TMEM38A | FALSE | Downregulated |
| 0,57169538 | TMEM38B | FALSE | Upregulated |
| 0 | TMEM40 | FALSE | Upregulated |
| -0,367080694 | TMEM42 | FALSE | Downregulated |
| -0,10131169 | TMEM5 | FALSE | Downregulated |
| 0,052867145 | TMEM52 | FALSE | Upregulated |
| 0,019383098 | TMEM52B | FALSE | Upregulated |
| -0,50773302 | TMEM54 | FALSE | Downregulated |
| 0,317893526 | TMEM55A | FALSE | Upregulated |
| -0,057779952 | TMEM56-RWDD3 | FALSE | Downregulated |
| | TMEM59L | | |
| 0,285372545 | TMEM61 | FALSE | Upregulated |

| | | | |
|---|---|---|---|
| 0,057601537 | TMEM63C | FALSE | Upregulated |
| -0,958888945 | TMEM67 | FALSE | Downregulated |
| 0,012983444 | TMEM68 | FALSE | Upregulated |
| -0,470301732 | TMEM70 | FALSE | Downregulated |
| 0,228295112 | TMEM71 | FALSE | Upregulated |
| 0,528785912 | TMEM72 | FALSE | Upregulated |
| 0,151520491 | TMEM72-AS1 | FALSE | Upregulated |
| 0,491283069 | TMEM74 | FALSE | Upregulated |
| 0,063020014 | TMEM81 | FALSE | Upregulated |
| 0,504846742 | TMEM88 | FALSE | Upregulated |
| 0,174904778 | TMEM88B | FALSE | Upregulated |
| 0 | TMEM89 | FALSE | Upregulated |
| | TMEM8C | | |
| 1,651951958 | TMEM92 | | Upregulated |
| 0 | TMEM95 | FALSE | Upregulated |
| -0,496619893 | TMEM98 | FALSE | Downregulated |
| -1,395681393 | TMEM99 | | Downregulated |
| -0,240629216 | TMEM9B-AS1 | FALSE | Downregulated |
| -0,006614918 | TMIE | FALSE | Downregulated |
| | TMIGD1 | | |
| 0,347574907 | TMIGD2 | FALSE | Upregulated |
| 0,810900613 | TMOD4 | FALSE | Upregulated |
| 0,212334112 | TMPRSS11A | FALSE | Upregulated |
| | TMPRSS11B | | |
| | TMPRSS11BNL | | |
| 0,102049664 | TMPRSS11E | FALSE | Upregulated |
| 0,077397262 | TMPRSS11F | FALSE | Upregulated |
| 0,005298914 | TMPRSS11GP | FALSE | Upregulated |
| 0 | TMPRSS12 | FALSE | Upregulated |
| 0,40661609 | TMPRSS13 | FALSE | Upregulated |
| 0 | TMPRSS15 | FALSE | Upregulated |
| 0,193863892 | TMPRSS3 | FALSE | Upregulated |
| 0,151106563 | TMPRSS4 | FALSE | Upregulated |
| | TMPRSS4-AS1 | | |
| -0,724450132 | TMPRSS5 | FALSE | Downregulated |
| | TMPRSS7 | | |
| | TMSB15A | | |
| | TMSB15B | | |
| 0,574003908 | TMSB4Y | FALSE | Upregulated |
| 0,572311939 | TMTC2 | FALSE | Upregulated |
| 0 | TMX2-CTNND1 | FALSE | Upregulated |
| 1,456734971 | TNF | | Upregulated |
| 0,283944987 | TNFAIP6 | FALSE | Upregulated |
| 0,808991753 | TNFAIP8 | FALSE | Upregulated |
| 0,097885107 | TNFAIP8L2-SCNM1 | FALSE | Upregulated |
| 0,035636678 | TNFAIP8L3 | FALSE | Upregulated |
| 0,819436702 | TNFRSF10C | FALSE | Upregulated |
| 0,482867035 | TNFRSF11A | FALSE | Upregulated |
| 0,850809251 | TNFRSF11B | FALSE | Upregulated |
| 1,153132147 | TNFRSF12A | | Upregulated |
| 0,013280527 | TNFRSF13B | FALSE | Upregulated |

| | | | |
|---|---|---|---|
| -0,120448579 | TNFRSF13C | FALSE | Downregulated |
| 1,822226842 | TNFRSF17 | | Upregulated |
| 0,151330071 | TNFRSF18 | FALSE | Upregulated |
| 0,817946586 | TNFRSF19 | FALSE | Upregulated |
| 0,5489438 | TNFRSF4 | FALSE | Upregulated |
| 0,148217518 | TNFRSF8 | FALSE | Upregulated |
| 1,374067978 | TNFRSF9 | | Upregulated |
| -0,501098376 | TNFSF11 | FALSE | Downregulated |
| 1,677172717 | TNFSF12-TNFSF13 | | Upregulated |
| -0,093279445 | TNFSF13B | FALSE | Downregulated |
| 0,729056432 | TNFSF15 | FALSE | Upregulated |
| 0,276544896 | TNFSF18 | FALSE | Upregulated |
| -0,34375873 | TNFSF4 | FALSE | Downregulated |
| 0,457977992 | TNFSF9 | FALSE | Upregulated |
| 0,669788064 | TNIP3 | FALSE | Upregulated |
| 0,004342836 | TNKS2-AS1 | FALSE | Upregulated |
| 0,40231215 | TNMD | FALSE | Upregulated |
| -0,268269975 | TNN | FALSE | Downregulated |
| -0,168898343 | TNNC1 | FALSE | Downregulated |
| 2,081493838 | TNNC2 | | Upregulated |
| 2,452439924 | TNNI2 | | Upregulated |
| 0,277119458 | TNNI3 | FALSE | Upregulated |
| | TNNI3K | | |
| 0,000435672 | TNNT2 | FALSE | Upregulated |
| 0,900688878 | TNNT3 | FALSE | Upregulated |
| 0 | TNP1 | FALSE | Upregulated |
| 0 | TNP2 | FALSE | Upregulated |
| -0,435678801 | TNR | FALSE | Downregulated |
| -0,320664985 | TNRC18P1 | FALSE | Downregulated |
| 0,001790719 | TOB1-AS1 | FALSE | Upregulated |
| 0,204861083 | TOB2P1 | FALSE | Upregulated |
| -0,047550522 | TOLLIP-AS1 | FALSE | Downregulated |
| -0,193952307 | TOMM20L | FALSE | Downregulated |
| -0,219672225 | TOP1P1 | FALSE | Downregulated |
| | TOP1P2 | | |
| 0,782392529 | TOP2A | FALSE | Upregulated |
| 0 | TOPAZ1 | FALSE | Upregulated |
| -0,465263647 | TOR2A | FALSE | Downregulated |
| 0,671691549 | TOX | FALSE | Upregulated |
| 0,236958192 | TOX2 | FALSE | Upregulated |
| 0,111488162 | TP53AIP1 | FALSE | Upregulated |
| -0,174929345 | TP53TG1 | FALSE | Downregulated |
| | TP53TG3 | | |
| | TP53TG3B | | |
| | TP53TG3C | | |
| | TP53TG3D | | |
| 0,187740488 | TP63 | FALSE | Upregulated |
| -0,117967158 | TPBG | FALSE | Downregulated |
| 0,732183006 | TPBGL | FALSE | Upregulated |
| 0,086763902 | TPD52L3 | FALSE | Upregulated |
| 0,106337562 | TPH1 | FALSE | Upregulated |

| | | | |
|---|---|---|---|
| | TPH2 | | |
| -0,119305504 | TPI1P2 | FALSE | Downregulated |
| -1,179653936 | TPK1 | | Downregulated |
| 0,010826647 | TPO | FALSE | Upregulated |
| -0,167874072 | TPPP3 | FALSE | Downregulated |
| -0,739181369 | TPRKB | FALSE | Downregulated |
| 0,25222697 | TPRX1 | FALSE | Upregulated |
| 0,001141803 | TPRXL | FALSE | Upregulated |
| -0,804997385 | TPSAB1 | FALSE | Downregulated |
| -0,4661675 | TPSB2 | FALSE | Downregulated |
| 0,115380215 | TPSD1 | FALSE | Upregulated |
| | TPTE | | |
| | TPTE2 | | |
| 0,744456498 | TPTE2P1 | FALSE | Upregulated |
| | TPTE2P2 | | |
| -0,135892616 | TPTE2P3 | FALSE | Downregulated |
| 0,121868589 | TPTE2P6 | FALSE | Upregulated |
| -0,012539817 | TPTEP1 | FALSE | Downregulated |
| 0,304259736 | TPX2 | FALSE | Upregulated |
| -0,066442763 | TRABD2A | FALSE | Downregulated |
| 0,295388503 | TRAIP | FALSE | Upregulated |
| 0 | TRAM1L1 | FALSE | Upregulated |
| -0,319354108 | TRAPPC2P1 | FALSE | Downregulated |
| -0,416267857 | TRAPPC3L | FALSE | Downregulated |
| 0,36845044 | TRAT1 | FALSE | Upregulated |
| -0,628531785 | TRDMT1 | FALSE | Downregulated |
| 0,086721368 | TRDN | FALSE | Upregulated |
| -0,214503258 | TREH | FALSE | Downregulated |
| 0,298318652 | TREM1 | FALSE | Upregulated |
| 0,084273903 | TREM2 | FALSE | Upregulated |
| -0,64144128 | TREML1 | FALSE | Downregulated |
| -0,018938149 | TREML2 | FALSE | Downregulated |
| 0 | TREML3P | FALSE | Upregulated |
| 0,012427543 | TREML4 | FALSE | Upregulated |
| | TREML5P | | |
| 0,562259644 | TRERNA1 | FALSE | Upregulated |
| -0,404882015 | TREX2 | FALSE | Downregulated |
| 0,691997357 | TRG-AS1 | FALSE | Upregulated |
| -0,059557167 | TRH | FALSE | Downregulated |
| -0,35995409 | TRHR | FALSE | Downregulated |
| 0,481387493 | TRIM10 | FALSE | Upregulated |
| 0,446522782 | TRIM15 | FALSE | Upregulated |
| 0,24294026 | TRIM16 | FALSE | Upregulated |
| 0,401744221 | TRIM16L | FALSE | Upregulated |
| -0,464355775 | TRIM29 | FALSE | Downregulated |
| 0,584961913 | TRIM31 | FALSE | Upregulated |
| -0,176348678 | TRIM36 | FALSE | Downregulated |
| 2,078617645 | TRIM39-RPP21 | | Upregulated |
| | TRIM40 | | |
| -0,13691866 | TRIM42 | FALSE | Downregulated |
| | TRIM43 | | |

| | | | |
|---|---|---|---|
| | TRIM43B | | |
| 1,128252103 | TRIM46 | | Upregulated |
| | TRIM48 | | |
| | TRIM49 | | |
| | TRIM49B | | |
| | TRIM49C | | |
| | TRIM49D1 | | |
| | TRIM49D2P | | |
| -0,591197375 | TRIM50 | FALSE | Downregulated |
| | TRIM51 | | |
| | TRIM51EP | | |
| | TRIM51HP | | |
| -0,107974864 | TRIM52-AS1 | FALSE | Downregulated |
| | TRIM53AP | | |
| 0,984770464 | TRIM54 | FALSE | Upregulated |
| -0,840167656 | TRIM58 | FALSE | Downregulated |
| -0,743480429 | TRIM6-TRIM34 | FALSE | Downregulated |
| | TRIM60 | | |
| -0,19224685 | TRIM61 | FALSE | Downregulated |
| 0,413318692 | TRIM63 | FALSE | Upregulated |
| | TRIM64 | | |
| | TRIM64B | | |
| | TRIM64C | | |
| | TRIM71 | | |
| 0,713108094 | TRIM72 | FALSE | Upregulated |
| 0,444033848 | TRIM73 | FALSE | Upregulated |
| -0,308868088 | TRIM74 | FALSE | Downregulated |
| | TRIM77 | | |
| 1,315259134 | TRIM9 | | Upregulated |
| | TRIML1 | | |
| | TRIML2 | | |
| 0,184897055 | TRIP13 | FALSE | Upregulated |
| -0,601494705 | TRMT10A | FALSE | Downregulated |
| 0,539117175 | TRMT10C | FALSE | Upregulated |
| -0,414975972 | TRMT6 | FALSE | Downregulated |
| | TRNA | | |
| | TRNAA-AGC | | |
| | TRNAA-AGC-2 | | |
| | TRNAA-AGC-3 | | |
| | TRNAA1 | | |
| | TRNAA10 | | |
| | TRNAA11 | | |
| | TRNAA12 | | |
| | TRNAA13 | | |
| | TRNAA14 | | |
| | TRNAA15 | | |
| | TRNAA17 | | |
| | TRNAA18 | | |
| | TRNAA19 | | |
| | TRNAA2 | | |
| | TRNAA20 | | |

TRNAA20-2
TRNAA20-3
TRNAA23
TRNAA26
TRNAA27
TRNAA28
TRNAA29
TRNAA3
TRNAA30
TRNAA32
TRNAA34
TRNAA36
TRNAA37
TRNAA37-2
TRNAA38
TRNAA39
TRNAA4
TRNAA40
TRNAA41
TRNAA41-2
TRNAA5
TRNAA6
TRNAA7
TRNAA8
TRNAA9
TRNAC-GCA
TRNAC10
TRNAC11
TRNAC12
TRNAC13
TRNAC14
TRNAC15
TRNAC16
TRNAC18
TRNAC19
TRNAC2
TRNAC20
TRNAC21
TRNAC22
TRNAC23
TRNAC24
TRNAC25
TRNAC26
TRNAC27
TRNAC28
TRNAC29
TRNAC3
TRNAC30
TRNAC4
TRNAC5
TRNAC6

TRNAC7
TRNAC8
TRNAC9
TRNAD10
TRNAD11
TRNAD12
TRNAD13
TRNAD14
TRNAD15
TRNAD16
TRNAD17
TRNAD18
TRNAD5
TRNAD6
TRNAD8
TRNAD9
TRNAE-UUC
TRNAE-UUC-2
TRNAE1
TRNAE11
TRNAE13
TRNAE18
TRNAE19
TRNAE20
TRNAE21
TRNAE22
TRNAE23
TRNAE24
TRNAE26
TRNAE5
TRNAE6
TRNAE7
TRNAF1
TRNAF10
TRNAF11
TRNAF2
TRNAF3
TRNAF4
TRNAF5
TRNAF6
TRNAF7
TRNAF8
TRNAF9
TRNAG-CCC
TRNAG-CCC-2
TRNAG1
TRNAG10
TRNAG11
TRNAG13
TRNAG15
TRNAG16

TRNAG17
TRNAG18
TRNAG19
TRNAG2
TRNAG20
TRNAG21
TRNAG22
TRNAG23
TRNAG24
TRNAG25
TRNAG26
TRNAG27
TRNAG28
TRNAG29
TRNAG3
TRNAG30
TRNAG31
TRNAG4
TRNAG5
TRNAG6
TRNAG7
TRNAG8
TRNAG9
TRNAH1
TRNAH10
TRNAH2
TRNAH3
TRNAH4
TRNAH5
TRNAH6
TRNAH7
TRNAH9
TRNAI-AAU
TRNAI1
TRNAI11
TRNAI12
TRNAI13
TRNAI15
TRNAI17
TRNAI18
TRNAI2
TRNAI20
TRNAI21
TRNAI22
TRNAI23
TRNAI24
TRNAI25
TRNAI25-2
TRNAI26
TRNAI3
TRNAI4

TRNAI5
TRNAI6
TRNAI7
TRNAI9
TRNAK-CUU
TRNAK1
TRNAK11
TRNAK13
TRNAK14
TRNAK15
TRNAK16
TRNAK17
TRNAK19
TRNAK2
TRNAK20
TRNAK21
TRNAK22
TRNAK23
TRNAK26
TRNAK27
TRNAK29
TRNAK3
TRNAK31
TRNAK33
TRNAK34
TRNAK4
TRNAK6
TRNAK8
TRNAK9
TRNAL-CAA
TRNAL1
TRNAL10
TRNAL11
TRNAL12
TRNAL13
TRNAL14
TRNAL15
TRNAL16
TRNAL17
TRNAL19
TRNAL2
TRNAL20
TRNAL22
TRNAL23
TRNAL24
TRNAL26
TRNAL27
TRNAL28
TRNAL29
TRNAL3
TRNAL30

TRNAL32
TRNAL34
TRNAL35
TRNAL36
TRNAL38
TRNAL4
TRNAL5
TRNAL6
TRNAL7
TRNAL8
TRNAM-CAU
TRNAM1
TRNAM11
TRNAM12
TRNAM13
TRNAM14
TRNAM15
TRNAM16
TRNAM16-2
TRNAM17
TRNAM18
TRNAM19
TRNAM2
TRNAM20
TRNAM3
TRNAM4
TRNAM5
TRNAM6
TRNAM8
TRNAM9
TRNAN-GUU
TRNAN-GUU-2
TRNAN-GUU-3
TRNAN-GUU-4
TRNAN-GUU-5
TRNAN-GUU-6
TRNAN-GUU-7
TRNAN1
TRNAN11
TRNAN14
TRNAN15
TRNAN17
TRNAN18
TRNAN19
TRNAN2
TRNAN20
TRNAN22
TRNAN22-2
TRNAN27
TRNAN29
TRNAN3

TRNAN32
TRNAN5
TRNAN5-2
TRNAN7
TRNAN8
TRNAP1
TRNAP10
TRNAP11
TRNAP12
TRNAP13
TRNAP14
TRNAP15
TRNAP16
TRNAP17
TRNAP18
TRNAP19
TRNAP2
TRNAP21
TRNAP3
TRNAP4
TRNAP5
TRNAP6
TRNAP7
TRNAP8
TRNAP9
TRNAQ-CUG
TRNAQ1
TRNAQ10
TRNAQ11
TRNAQ12
TRNAQ13
TRNAQ15
TRNAQ16
TRNAQ17
TRNAQ2
TRNAQ20
TRNAQ21
TRNAQ22
TRNAQ24
TRNAQ25
TRNAQ27
TRNAQ3
TRNAQ32
TRNAQ8
TRNAR1
TRNAR10
TRNAR11
TRNAR12
TRNAR14
TRNAR15
TRNAR16

TRNAR17
TRNAR18
TRNAR19
TRNAR2
TRNAR20
TRNAR21
TRNAR22
TRNAR23
TRNAR24
TRNAR25
TRNAR26
TRNAR27
TRNAR28
TRNAR29
TRNAR3
TRNAR4
TRNAR5
TRNAR6
TRNAR7
TRNAR8
TRNAR9
TRNAS1
TRNAS10
TRNAS11
TRNAS12
TRNAS13
TRNAS14
TRNAS15
TRNAS16
TRNAS17
TRNAS18
TRNAS19
TRNAS2
TRNAS20
TRNAS21
TRNAS22
TRNAS23
TRNAS24
TRNAS25
TRNAS27
TRNAS28
TRNAS3
TRNAS4
TRNAS5
TRNAS6
TRNAS7
TRNAS9
TRNASUP1
TRNAT1
TRNAT10
TRNAT12

| | | | |
|---|---|---|---|
| | TRNAT13 | | |
| | TRNAT14 | | |
| | TRNAT15 | | |
| | TRNAT16 | | |
| | TRNAT17 | | |
| | TRNAT18 | | |
| | TRNAT19 | | |
| | TRNAT2 | | |
| | TRNAT20 | | |
| | TRNAT21 | | |
| | TRNAT22 | | |
| | TRNAT4 | | |
| | TRNAT5 | | |
| | TRNAT6 | | |
| | TRNAT7 | | |
| | TRNAT8 | | |
| | TRNAT9 | | |
| 0,28181789 | TRNAU1 | FALSE | Upregulated |
| | TRNAU2 | | |
| | TRNAV-CAC | | |
| | TRNAV-CAC-2 | | |
| | TRNAV-CAC-3 | | |
| | TRNAV1 | | |
| | TRNAV10 | | |
| | TRNAV11 | | |
| | TRNAV12 | | |
| | TRNAV15 | | |
| | TRNAV16 | | |
| | TRNAV17 | | |
| | TRNAV18 | | |
| | TRNAV19 | | |
| | TRNAV2 | | |
| | TRNAV20 | | |
| | TRNAV21 | | |
| | TRNAV22 | | |
| | TRNAV23 | | |
| | TRNAV24 | | |
| | TRNAV25 | | |
| | TRNAV26 | | |
| | TRNAV27 | | |
| | TRNAV28 | | |
| | TRNAV29 | | |
| | TRNAV3 | | |
| | TRNAV31 | | |
| | TRNAV32 | | |
| | TRNAV4 | | |
| | TRNAV5 | | |
| | TRNAV6 | | |
| | TRNAV7 | | |
| | TRNAV8 | | |
| | TRNAV9 | | |

| | | | |
|---|---|---|---|
| | TRNAW2 | | |
| | TRNAW3 | | |
| | TRNAW4 | | |
| | TRNAW5 | | |
| | TRNAW6 | | |
| | TRNAW7 | | |
| | TRNAW9 | | |
| | TRNAY1 | | |
| | TRNAY10 | | |
| | TRNAY12 | | |
| | TRNAY13 | | |
| | TRNAY14 | | |
| | TRNAY15 | | |
| | TRNAY2 | | |
| | TRNAY3 | | |
| | TRNAY4 | | |
| | TRNAY6 | | |
| | TRNAY7 | | |
| | TRNAY8 | | |
| | TRNAY9 | | |
| | TRNC | | |
| | TRND | | |
| | TRNE | | |
| | TRNF | | |
| | TRNG | | |
| | TRNH | | |
| | TRNI | | |
| | TRNK | | |
| | TRNL1 | | |
| | TRNL2 | | |
| | TRNM | | |
| | TRNN | | |
| | TRNP | | |
| -0,171704369 | TRNP1 | FALSE | Downregulated |
| | TRNQ | | |
| | TRNR | | |
| | TRNS1 | | |
| | TRNS2 | | |
| | TRNT | | |
| | TRNV | | |
| | TRNW | | |
| | TRNY | | |
| -0,102814773 | TROAP | FALSE | Downregulated |
| -0,143771481 | TRPC1 | FALSE | Downregulated |
| 0,747255051 | TRPC2 | FALSE | Upregulated |
| -0,207706868 | TRPC3 | FALSE | Downregulated |
| 0,568057969 | TRPC4 | FALSE | Upregulated |
| 1,492950281 | TRPC6 | | Upregulated |
| 0,362548085 | TRPC7 | FALSE | Upregulated |
| -0,856459484 | TRPM1 | FALSE | Downregulated |
| 0,742071436 | TRPM2-AS | FALSE | Upregulated |

| | | | |
|---|---|---|---|
| 0,796999006 | TRPM5 | FALSE | Upregulated |
| 0,347722239 | TRPM6 | FALSE | Upregulated |
| -0,018149848 | TRPV5 | FALSE | Downregulated |
| 0,259848934 | TRPV6 | FALSE | Upregulated |
| 0,294408303 | TRY2P | FALSE | Upregulated |
| 0,303797644 | TSACC | FALSE | Upregulated |
| 0,53016486 | TSEN15 | FALSE | Upregulated |
| | TSG1 | | |
| -0,425002485 | TSGA10 | FALSE | Downregulated |
| -0,147360392 | TSGA10IP | FALSE | Downregulated |
| 1,093012853 | TSGA13 | | Upregulated |
| -0,157808177 | TSHB | FALSE | Downregulated |
| 1,148336429 | TSKS | | Upregulated |
| 0,174496729 | TSLP | FALSE | Upregulated |
| 0,364621276 | TSNAX-DISC1 | FALSE | Upregulated |
| 0,019369144 | TSNAXIP1 | FALSE | Upregulated |
| -0,002963726 | TSPAN1 | FALSE | Downregulated |
| -0,428815044 | TSPAN10 | FALSE | Downregulated |
| 0,456027339 | TSPAN13 | FALSE | Upregulated |
| -0,397591188 | TSPAN15 | FALSE | Downregulated |
| 0,494340871 | TSPAN16 | FALSE | Upregulated |
| | TSPAN19 | | |
| 0,132489862 | TSPAN2 | FALSE | Upregulated |
| 0,351759592 | TSPAN32 | FALSE | Upregulated |
| 0,417224895 | TSPAN5 | FALSE | Upregulated |
| 0,094538761 | TSPAN8 | FALSE | Upregulated |
| | TSPY1 | | |
| | TSPY10 | | |
| | TSPY2 | | |
| -0,222476899 | TSPY26P | FALSE | Downregulated |
| | TSPY3 | | |
| | TSPY4 | | |
| | TSPY8 | | |
| -0,318967849 | TSPYL5 | FALSE | Downregulated |
| 0,673383455 | TSSK1B | FALSE | Upregulated |
| 0,34912646 | TSSK6 | FALSE | Upregulated |
| -0,469638537 | TTC16 | FALSE | Downregulated |
| -0,549042268 | TTC21A | FALSE | Downregulated |
| -0,141045703 | TTC22 | FALSE | Downregulated |
| 0,287383229 | TTC23L | FALSE | Upregulated |
| 0,441254775 | TTC24 | FALSE | Upregulated |
| 0,537249982 | TTC25 | FALSE | Upregulated |
| -0,299298913 | TTC26 | FALSE | Downregulated |
| | TTC29 | | |
| -0,000772155 | TTC30A | FALSE | Downregulated |
| 0,12438815 | TTC32 | FALSE | Upregulated |
| -0,183906079 | TTC34 | FALSE | Downregulated |
| -0,285739068 | TTC3P1 | FALSE | Downregulated |
| -0,397620558 | TTC4 | FALSE | Downregulated |
| 0,011406539 | TTC40 | FALSE | Upregulated |
| 0,101954511 | TTC5 | FALSE | Upregulated |

| | | | |
|---|---|---|---|
| 0,286797129 | TTC9B | FALSE | Upregulated |
| -0,069504033 | TTC9C | FALSE | Downregulated |
| -0,420501991 | TTK | FALSE | Downregulated |
| 0,687876295 | TTLL1 | FALSE | Upregulated |
| 0,012534082 | TTLL10 | FALSE | Upregulated |
| 0,809777206 | TTLL10-AS1 | FALSE | Upregulated |
| 1,076466038 | TTLL13 | | Upregulated |
| -0,007614149 | TTLL6 | FALSE | Downregulated |
| | TTLL8 | | |
| | TTTY1 | | |
| 0,259849586 | TTTY10 | FALSE | Upregulated |
| | TTTY11 | | |
| | TTTY12 | | |
| -0,294871648 | TTTY14 | FALSE | Downregulated |
| | TTTY16 | | |
| | TTTY18 | | |
| | TTTY19 | | |
| | TTTY1B | | |
| | TTTY2 | | |
| | TTTY20 | | |
| | TTTY21 | | |
| | TTTY21B | | |
| | TTTY22 | | |
| | TTTY23 | | |
| | TTTY23B | | |
| | TTTY2B | | |
| | TTTY7 | | |
| 0,289507097 | TTTY7B | FALSE | Upregulated |
| | TTTY8 | | |
| | TTTY8B | | |
| -0,381194558 | TTYH2 | FALSE | Downregulated |
| -0,006903421 | TUBA3C | FALSE | Downregulated |
| 0,173601545 | TUBA3E | FALSE | Upregulated |
| 0,202988797 | TUBA4B | FALSE | Upregulated |
| | TUBAL3 | | |
| 0,63046572 | TUBB2B | FALSE | Upregulated |
| 0,000207301 | TUBB3 | FALSE | Upregulated |
| -0,185685531 | TUBB4A | FALSE | Downregulated |
| | TUBB8 | | |
| 0,125395671 | TUBBP5 | FALSE | Upregulated |
| -0,672386422 | TUBD1 | FALSE | Downregulated |
| -0,94190531 | TUFT1 | FALSE | Downregulated |
| 0,667443443 | TULP1 | FALSE | Upregulated |
| 0,456367566 | TULP2 | FALSE | Upregulated |
| -0,358779232 | TUSC3 | FALSE | Downregulated |
| 0,225946229 | TUSC5 | FALSE | Upregulated |
| 0,111444994 | TUSC7 | FALSE | Upregulated |
| 0,313912525 | TVP23C-CDRT4 | FALSE | Upregulated |
| 0,99922765 | TWIST2 | FALSE | Upregulated |
| -0,563224377 | TXK | FALSE | Downregulated |
| 1,254253708 | TXLNB | | Upregulated |

| | | | |
|---|---|---|---|
| -0,741295792 | TXNDC16 | FALSE | Downregulated |
| -0,004668834 | TXNDC2 | FALSE | Downregulated |
| -0,072620363 | TXNDC8 | FALSE | Downregulated |
| 0 | TXNRD3NB | FALSE | Upregulated |
| 0 | TYR | FALSE | Upregulated |
| -0,019818849 | TYRO3P | FALSE | Downregulated |
| 0,149598452 | TYRP1 | FALSE | Upregulated |
| 0,90413208 | U2AF1L4 | FALSE | Upregulated |
| -0,617975798 | UBAC2-AS1 | FALSE | Downregulated |
| -0,648605222 | UBAP1L | FALSE | Downregulated |
| 0,848660185 | UBASH3A | FALSE | Upregulated |
| 0,010378761 | UBASH3B | FALSE | Upregulated |
| 0,205401301 | UBE2C | FALSE | Upregulated |
| 0,20121243 | UBE2D1 | FALSE | Upregulated |
| -0,583564252 | UBE2DNL | FALSE | Downregulated |
| -0,037582481 | UBE2E2-AS1 | FALSE | Downregulated |
| 0,522639712 | UBE2F-SCLY | FALSE | Upregulated |
| 0 | UBE2MP1 | FALSE | Upregulated |
| 0,000264809 | UBE2NL | FALSE | Upregulated |
| 0,273940847 | UBE2Q2L | FALSE | Upregulated |
| 0,596668752 | UBE2Q2P1 | FALSE | Upregulated |
| 0,274369622 | UBE2Q2P2 | FALSE | Upregulated |
| -0,33438924 | UBE2Q2P3 | FALSE | Downregulated |
| 0,644062521 | UBE2QL1 | FALSE | Upregulated |
| 0,84757824 | UBE2T | FALSE | Upregulated |
| | UBE2U | | |
| -0,535782675 | UBE3D | FALSE | Downregulated |
| | UBL4B | | |
| -0,627547551 | UBL7-AS1 | FALSE | Downregulated |
| -0,416964284 | UBLCP1 | FALSE | Downregulated |
| -0,044621584 | UBQLN3 | FALSE | Downregulated |
| -0,458904236 | UBQLNL | FALSE | Downregulated |
| | UBTFL1 | | |
| 0,230045202 | UBXN10-AS1 | FALSE | Upregulated |
| -0,027287633 | UBXN2A | FALSE | Downregulated |
| 0,559696896 | UBXN7-AS1 | FALSE | Upregulated |
| 0,563418946 | UCHL1 | FALSE | Upregulated |
| | UCHL1-AS1 | | |
| -0,370485116 | UCHL3 | FALSE | Downregulated |
| | UCMA | | |
| 0,987007149 | UCN | FALSE | Upregulated |
| 0,280077111 | UCN2 | FALSE | Upregulated |
| | UCN3 | | |
| 0,36032317 | UCP1 | FALSE | Upregulated |
| 0,777770895 | UCP3 | FALSE | Upregulated |
| 0,244385395 | UFSP1 | FALSE | Upregulated |
| | UGT2A1 | | |
| -0,714488916 | UGT2A2 | FALSE | Downregulated |
| 0,398410848 | UGT2B28 | FALSE | Upregulated |
| 0,404235824 | UGT3A2 | FALSE | Upregulated |
| 0,553238087 | UGT8 | FALSE | Upregulated |

| | | | |
|---|---|---|---|
| | ULBP1 | | |
| -0,004571254 | ULBP2 | FALSE | Downregulated |
| -0,517110289 | ULBP3 | FALSE | Downregulated |
| 0,180687325 | ULK4P1 | FALSE | Upregulated |
| -0,008315178 | ULK4P2 | FALSE | Downregulated |
| 0,434480502 | ULK4P3 | FALSE | Upregulated |
| | UMOD | | |
| 0 | UMODL1 | FALSE | Upregulated |
| -0,725597915 | UNC13A | FALSE | Downregulated |
| 1,230410365 | UNC13C | | Upregulated |
| 0,88278118 | UNC45B | FALSE | Upregulated |
| 0,339556618 | UNC50 | FALSE | Upregulated |
| -0,23028341 | UNC5B-AS1 | FALSE | Downregulated |
| | UNC5D | | |
| -0,152020315 | UNC79 | FALSE | Downregulated |
| -0,53663031 | UNC80 | FALSE | Downregulated |
| | UNCX | | |
| 0,269239968 | UNQ6494 | FALSE | Upregulated |
| -0,004525881 | UNQ9370 | FALSE | Downregulated |
| -0,01642357 | UOX | FALSE | Downregulated |
| 0 | UPK1A | FALSE | Upregulated |
| 0,043286098 | UPK1A-AS1 | FALSE | Upregulated |
| 0 | UPK2 | FALSE | Upregulated |
| | UPK3A | | |
| 0,122232541 | UPK3B | FALSE | Upregulated |
| 0,19562332 | UPRT | FALSE | Upregulated |
| -0,443099837 | UQCRHL | FALSE | Downregulated |
| | URAD | | |
| -0,169495171 | URGCP-MRPS24 | FALSE | Downregulated |
| -0,121838616 | USH1C | FALSE | Downregulated |
| 1,004658348 | USH1G | | Upregulated |
| -0,272520286 | USHBP1 | FALSE | Downregulated |
| 0,306403154 | USP12-AS2 | FALSE | Upregulated |
| 0,291340714 | USP17L10 | FALSE | Upregulated |
| 0 | USP17L11 | FALSE | Upregulated |
| | USP17L12 | | |
| | USP17L13 | | |
| | USP17L15 | | |
| | USP17L17 | | |
| | USP17L18 | | |
| | USP17L19 | | |
| 0,419216975 | USP17L1P | FALSE | Upregulated |
| 0 | USP17L20 | FALSE | Upregulated |
| | USP17L21 | | |
| 0,269779132 | USP17L22 | FALSE | Upregulated |
| | USP17L24 | | |
| | USP17L25 | | |
| | USP17L26 | | |
| | USP17L27 | | |
| | USP17L28 | | |
| | USP17L29 | | |

| | | | |
|---|---|---|---|
| 0,436161692 | USP17L3 | FALSE | Upregulated |
| | USP17L30 | | |
| 0,283711413 | USP17L4 | FALSE | Upregulated |
| | USP17L5 | | |
| -0,170416555 | USP17L6P | FALSE | Downregulated |
| 0 | USP17L8 | FALSE | Upregulated |
| | USP17L9P | | |
| -0,265944784 | USP2-AS1 | FALSE | Downregulated |
| | USP26 | | |
| -0,271381554 | USP27X | FALSE | Downregulated |
| -0,008196563 | USP27X-AS1 | FALSE | Downregulated |
| | USP29 | | |
| 0,982285029 | USP41 | FALSE | Upregulated |
| 0,379732657 | USP46-AS1 | FALSE | Upregulated |
| -0,435241863 | USP6NL-IT1 | FALSE | Downregulated |
| | UTF1 | | |
| -0,083004018 | UTP11L | FALSE | Downregulated |
| -0,249233912 | UTP15 | FALSE | Downregulated |
| 0,099033684 | UTP23 | FALSE | Upregulated |
| -0,590647259 | UTS2 | FALSE | Downregulated |
| -0,075114489 | UTS2R | FALSE | Downregulated |
| 0,281759529 | UXT-AS1 | FALSE | Upregulated |
| | VAMP7-2 | | |
| 0,325574495 | VANGL2 | FALSE | Upregulated |
| -1,04521439 | VAT1L | | Downregulated |
| 0,211871069 | VAV3 | FALSE | Upregulated |
| | VAV3-AS1 | | |
| | VAX1 | | |
| 0,09261312 | VAX2 | FALSE | Upregulated |
| 0,337680985 | VCPKMT | FALSE | Upregulated |
| -0,968397818 | VCX | FALSE | Downregulated |
| -0,105130501 | VCX2 | FALSE | Downregulated |
| 0,073328888 | VCX3A | FALSE | Upregulated |
| 0,891458144 | VCX3B | FALSE | Upregulated |
| | VCY | | |
| 0,003224405 | VCY1B | FALSE | Upregulated |
| 0,801718714 | VDR | FALSE | Upregulated |
| 1,09266696 | VEGFC | | Upregulated |
| 1,32361505 | VENTX | | Upregulated |
| | VENTXP1 | | |
| -0,017672535 | VENTXP7 | FALSE | Downregulated |
| 0,177830082 | VGF | FALSE | Upregulated |
| -0,015040902 | VGLL1 | FALSE | Downregulated |
| 0,304282266 | VGLL2 | FALSE | Upregulated |
| -0,344084251 | VGLL3 | FALSE | Downregulated |
| | VHLL | | |
| 0,136581954 | VILL | FALSE | Upregulated |
| 0,36525638 | VIP | FALSE | Upregulated |
| -0,705225919 | VIPR2 | FALSE | Downregulated |
| 0,277119458 | VIT | FALSE | Upregulated |
| -0,284132163 | VLDLR | FALSE | Downregulated |

| | | | |
|---|---|---|---|
| -0,023221189 | VLDLR-AS1 | FALSE | Downregulated |
| 0,327359988 | VN1R1 | FALSE | Upregulated |
| 0 | VN1R10P | FALSE | Upregulated |
| 0 | VN1R2 | FALSE | Upregulated |
| | VN1R4 | | |
| 0,006667059 | VN1R5 | FALSE | Upregulated |
| 0,684142363 | VNN2 | FALSE | Upregulated |
| -1,320656419 | VPREB3 | | Downregulated |
| 0,372889364 | VPS13A-AS1 | FALSE | Upregulated |
| 0,443719849 | VRK1 | FALSE | Upregulated |
| | VRTN | | |
| 1,036695802 | VSIG1 | | Upregulated |
| 0,675790926 | VSIG2 | FALSE | Upregulated |
| 0,852802458 | VSIG8 | FALSE | Upregulated |
| | VSTM1 | | |
| | VSTM2A | | |
| | VSTM2B | | |
| 0,254513202 | VSTM2L | FALSE | Upregulated |
| 0,271671151 | VSTM5 | FALSE | Upregulated |
| | VSX1 | | |
| -0,037178009 | VSX2 | FALSE | Downregulated |
| -0,078901254 | VTCN1 | FALSE | Downregulated |
| | VTRNA1-1 | | |
| | VTRNA1-2 | | |
| | VTRNA1-3 | | |
| | VTRNA2-1 | | |
| -0,106848515 | VWA3A | FALSE | Downregulated |
| -0,162428479 | VWA3B | FALSE | Downregulated |
| -0,244762572 | VWA5A | FALSE | Downregulated |
| 0 | VWA5B1 | FALSE | Upregulated |
| -0,07199643 | VWA5B2 | FALSE | Downregulated |
| -0,358001222 | VWA7 | FALSE | Downregulated |
| -0,030183467 | VWC2 | FALSE | Downregulated |
| -0,336890465 | VWC2L | FALSE | Downregulated |
| | VWC2L-IT1 | | |
| -0,563165204 | VWDE | FALSE | Downregulated |
| -0,586919015 | WASF1 | FALSE | Downregulated |
| 0,290407067 | WASIR2 | FALSE | Upregulated |
| 0,433378118 | WBP11P1 | FALSE | Upregulated |
| -0,399766883 | WBP2NL | FALSE | Downregulated |
| -0,212982893 | WBP4 | FALSE | Downregulated |
| -0,462243105 | WBP5 | FALSE | Downregulated |
| 0,42480668 | WBSCR17 | FALSE | Upregulated |
| -0,777239073 | WBSCR27 | FALSE | Downregulated |
| 0,14831639 | WBSCR28 | FALSE | Upregulated |
| 1,150731138 | WDFY3-AS2 | | Upregulated |
| 0,996707056 | WDHD1 | FALSE | Upregulated |
| -0,165173092 | WDPCP | FALSE | Downregulated |
| 0,428112419 | WDR11-AS1 | FALSE | Upregulated |
| 0,415223916 | WDR16 | FALSE | Upregulated |
| -0,183460362 | WDR17 | FALSE | Downregulated |

| | | | |
|---|---|---|---|
| -0,33135265 | WDR25 | FALSE | Downregulated |
| -0,2236585 | WDR31 | FALSE | Downregulated |
| 0,4932289 | WDR38 | FALSE | Upregulated |
| -0,440770267 | WDR47 | FALSE | Downregulated |
| | WDR49 | | |
| -0,2499294 | WDR52-AS1 | FALSE | Downregulated |
| 0,710570179 | WDR53 | FALSE | Upregulated |
| 0,563860378 | WDR54 | FALSE | Upregulated |
| 0,568155221 | WDR62 | FALSE | Upregulated |
| 0,349003276 | WDR63 | FALSE | Upregulated |
| -0,257842606 | WDR64 | FALSE | Downregulated |
| -0,657105544 | WDR66 | FALSE | Downregulated |
| -0,208804488 | WDR76 | FALSE | Downregulated |
| 0,279511297 | WDR78 | FALSE | Upregulated |
| -0,169124147 | WDR86-AS1 | FALSE | Downregulated |
| 0,440741428 | WDR87 | FALSE | Upregulated |
| -0,75835541 | WDR92 | FALSE | Downregulated |
| -0,352129244 | WDR93 | FALSE | Downregulated |
| -0,158595168 | WDR96 | FALSE | Downregulated |
| 0,027183403 | WDYHV1 | FALSE | Upregulated |
| -0,365109302 | WEE2-AS1 | FALSE | Downregulated |
| 0,520990821 | WFDC1 | FALSE | Upregulated |
| | WFDC10A | | |
| | WFDC10B | | |
| | WFDC11 | | |
| 0,126459901 | WFDC12 | FALSE | Upregulated |
| 0,6222394 | WFDC13 | FALSE | Upregulated |
| -0,016194846 | WFDC2 | FALSE | Downregulated |
| | WFDC3 | | |
| 0 | WFDC5 | FALSE | Upregulated |
| | WFDC6 | | |
| | WFDC8 | | |
| | WFDC9 | | |
| 0,374563721 | WFIKKN2 | FALSE | Upregulated |
| | WI2-2373I1.2 | | |
| -0,285866371 | WI2-81516E3.1 | FALSE | Downregulated |
| | WIF1 | | |
| 0,563965223 | WIPF3 | FALSE | Upregulated |
| -0,14984977 | WISP1 | FALSE | Downregulated |
| 1,683477165 | WISP2 | | Upregulated |
| 0,842685872 | WNT1 | FALSE | Upregulated |
| 0,552315155 | WNT10A | FALSE | Upregulated |
| 0,91996857 | WNT10B | FALSE | Upregulated |
| 0,26021043 | WNT16 | FALSE | Upregulated |
| 0,362350915 | WNT2 | FALSE | Upregulated |
| 0,059556897 | WNT3A | FALSE | Upregulated |
| 0,088227696 | WNT6 | FALSE | Upregulated |
| 0 | WNT7A | FALSE | Upregulated |
| -0,423725693 | WNT7B | FALSE | Downregulated |
| 0,585734984 | WNT8A | FALSE | Upregulated |
| 0,017398716 | WNT9A | FALSE | Upregulated |

| | | | |
|---|---|---|---|
| 0 | WNT9B | FALSE | Upregulated |
| 0,009524952 | WRAP53 | FALSE | Upregulated |
| -0,355773783 | WRAP73 | FALSE | Downregulated |
| -0,048264192 | WSCD1 | FALSE | Downregulated |
| 0,405731887 | WSCD2 | FALSE | Upregulated |
| -0,315096729 | WT1 | FALSE | Downregulated |
| -0,005222527 | WT1-AS | FALSE | Downregulated |
| 0,419409904 | WTAPP1 | FALSE | Upregulated |
| 0 | WTH3DI | FALSE | Upregulated |
| 0,512685202 | WTIP | FALSE | Upregulated |
| -0,439564893 | WWTR1-AS1 | FALSE | Downregulated |
| | XAGE-4 | | |
| | XAGE1B | | |
| | XAGE1E | | |
| | XAGE2 | | |
| 0,44021769 | XAGE3 | FALSE | Upregulated |
| | XAGE5 | | |
| -0,186702013 | XCL1 | FALSE | Downregulated |
| -0,192260141 | XCL2 | FALSE | Downregulated |
| 0,450574006 | XCR1 | FALSE | Upregulated |
| | XGPY2 | | |
| -0,633580134 | XIRP1 | FALSE | Downregulated |
| | XIRP2-AS1 | | |
| 0,905662611 | XK | FALSE | Upregulated |
| | XKR3 | | |
| 0,274626966 | XKR4 | FALSE | Upregulated |
| | XKR5 | | |
| -0,648120243 | XKR6 | FALSE | Downregulated |
| 0,268422684 | XKR7 | FALSE | Upregulated |
| -0,473158255 | XKR9 | FALSE | Downregulated |
| 0,134831831 | XKRX | FALSE | Upregulated |
| 0,130136324 | XRCC2 | FALSE | Upregulated |
| -0,40583221 | XRCC4 | FALSE | Downregulated |
| -0,624417544 | XRCC6BP1 | FALSE | Downregulated |
| 0,126136534 | XRCC6P5 | FALSE | Upregulated |
| 0,010093572 | XXYLT1 | FALSE | Upregulated |
| | XXYLT1-AS1 | | |
| 0,486395438 | YAE1D1 | FALSE | Upregulated |
| -0,117724157 | YBX2 | FALSE | Downregulated |
| 0,41482555 | YBX3P1 | FALSE | Upregulated |
| 0,117497632 | YEATS4 | FALSE | Upregulated |
| | YIPF7 | | |
| 0,85180965 | YJEFN3 | FALSE | Upregulated |
| 0 | YPLR6490 | FALSE | Upregulated |
| 0,720956707 | YTHDF3-AS1 | FALSE | Upregulated |
| 0 | YWHAEP1 | FALSE | Upregulated |
| 0,02465988 | YWHAEP7 | FALSE | Upregulated |
| | YY1P2 | | |
| 0,752570238 | YY2 | FALSE | Upregulated |
| -0,03757721 | ZAN | FALSE | Downregulated |
| 0,418273129 | ZAR1 | FALSE | Upregulated |

| | | | |
|---|---|---|---|
| 0,457878838 | ZAR1L | FALSE | Upregulated |
| | ZBBX | | |
| | ZBED1-2 | | |
| -0,151705915 | ZBED3-AS1 | FALSE | Downregulated |
| 0,122173408 | ZBED5-AS1 | FALSE | Upregulated |
| 0,721170924 | ZBP1 | FALSE | Upregulated |
| -0,029397366 | ZBTB26 | FALSE | Downregulated |
| 0,420542085 | ZBTB49 | FALSE | Upregulated |
| -0,267727873 | ZBTB6 | FALSE | Downregulated |
| -1,293821502 | ZBTB7C | | Downregulated |
| | ZBTB8B | | |
| -0,627815061 | ZBTB8OS | FALSE | Downregulated |
| -0,385340711 | ZBTB9 | FALSE | Downregulated |
| | ZC2HC1B | | |
| 0,49571956 | ZC3H8 | FALSE | Upregulated |
| 0,117102482 | ZC3HAV1L | FALSE | Upregulated |
| -0,12779104 | ZC4H2 | FALSE | Downregulated |
| -0,18435203 | ZCCHC10 | FALSE | Downregulated |
| -0,006413853 | ZCCHC12 | FALSE | Downregulated |
| | ZCCHC13 | | |
| 1,317320188 | ZCCHC16 | | Upregulated |
| 0,200222954 | ZCCHC18 | FALSE | Upregulated |
| 0,261089307 | ZCCHC4 | FALSE | Upregulated |
| | ZCCHC5 | | |
| 0,083824257 | ZCWPW2 | FALSE | Upregulated |
| 0,102481557 | ZDHHC1 | FALSE | Upregulated |
| 0,338594996 | ZDHHC13 | FALSE | Upregulated |
| 0,436360449 | ZDHHC19 | FALSE | Upregulated |
| 0,571926609 | ZDHHC22 | FALSE | Upregulated |
| -0,150111394 | ZDHHC8P1 | FALSE | Downregulated |
| 0,128516802 | ZEB1-AS1 | FALSE | Upregulated |
| 0,238535386 | ZEB2-AS1 | FALSE | Upregulated |
| 0,284344543 | ZFHX4-AS1 | FALSE | Upregulated |
| 0,146266116 | ZFP2 | FALSE | Upregulated |
| -1,415963409 | ZFP37 | | Downregulated |
| | ZFP42 | | |
| 0,421729574 | ZFP57 | FALSE | Upregulated |
| 0,416314087 | ZFP69B | FALSE | Upregulated |
| -0,233602832 | ZFP82 | FALSE | Downregulated |
| 0,380409654 | ZFP92 | FALSE | Upregulated |
| 0,149791622 | ZFR2 | FALSE | Upregulated |
| -0,00509535 | ZFX-AS1 | FALSE | Downregulated |
| 0,323029884 | ZG16B | FALSE | Upregulated |
| -0,088683786 | ZGLP1 | FALSE | Downregulated |
| 0,005956873 | ZHX1-C8ORF76 | FALSE | Upregulated |
| | ZIC2 | | |
| | ZIC3 | | |
| 0,280928672 | ZIC5 | FALSE | Upregulated |
| 0,433197587 | ZIK1 | FALSE | Upregulated |
| -0,690551623 | ZIM2 | FALSE | Downregulated |
| 0 | ZIM3 | FALSE | Upregulated |

| | | | |
|---|---|---|---|
| 0 | ZMAT4 | FALSE | Upregulated |
| 0,282781125 | ZMAT5 | FALSE | Upregulated |
| 0,006223013 | ZMIZ1-AS1 | FALSE | Upregulated |
| -0,627129927 | ZMYM6NB | FALSE | Downregulated |
| -0,321568113 | ZMYND10 | FALSE | Downregulated |
| -1,243965294 | ZMYND12 | | Downregulated |
| -0,332225534 | ZMYND15 | FALSE | Downregulated |
| -0,09913665 | ZNF112 | FALSE | Downregulated |
| -0,130296581 | ZNF114 | FALSE | Downregulated |
| 0,130850795 | ZNF135 | FALSE | Upregulated |
| 0,365060786 | ZNF137P | FALSE | Upregulated |
| 0,029078486 | ZNF138 | FALSE | Upregulated |
| 0,063074795 | ZNF14 | FALSE | Upregulated |
| 0,306061429 | ZNF157 | FALSE | Upregulated |
| 0,702098409 | ZNF16 | FALSE | Upregulated |
| 0,829411781 | ZNF165 | FALSE | Upregulated |
| 0,796487402 | ZNF169 | FALSE | Upregulated |
| -0,112995672 | ZNF174 | FALSE | Downregulated |
| 0,232674385 | ZNF184 | FALSE | Upregulated |
| 0,360452664 | ZNF185 | FALSE | Upregulated |
| 0,121034989 | ZNF19 | FALSE | Upregulated |
| -0,149075069 | ZNF192P1 | FALSE | Downregulated |
| 0,083313388 | ZNF2 | FALSE | Upregulated |
| 0,432752008 | ZNF200 | FALSE | Upregulated |
| -0,347920124 | ZNF204P | FALSE | Downregulated |
| -0,703890969 | ZNF208 | FALSE | Downregulated |
| -0,576500535 | ZNF214 | FALSE | Downregulated |
| 0,592329161 | ZNF215 | FALSE | Upregulated |
| 0,8326281 | ZNF222 | FALSE | Upregulated |
| -0,058557417 | ZNF223 | FALSE | Downregulated |
| -0,728138216 | ZNF229 | FALSE | Downregulated |
| 0,36733822 | ZNF230 | FALSE | Upregulated |
| 0,468866249 | ZNF232 | FALSE | Upregulated |
| -0,404299155 | ZNF239 | FALSE | Downregulated |
| 0,372267452 | ZNF252P-AS1 | FALSE | Upregulated |
| -0,60568145 | ZNF253 | FALSE | Downregulated |
| 0,408999294 | ZNF256 | FALSE | Upregulated |
| 0,636687599 | ZNF267 | FALSE | Upregulated |
| -0,727400577 | ZNF273 | FALSE | Downregulated |
| 0,589478174 | ZNF28 | FALSE | Upregulated |
| 0,43066349 | ZNF280B | FALSE | Upregulated |
| 0,474687564 | ZNF280C | FALSE | Upregulated |
| -0,547841092 | ZNF283 | FALSE | Downregulated |
| 0,797302245 | ZNF284 | FALSE | Upregulated |
| -0,453213369 | ZNF285 | FALSE | Downregulated |
| 0,190240007 | ZNF286A | FALSE | Upregulated |
| -0,105971298 | ZNF286B | FALSE | Downregulated |
| 0,406251013 | ZNF295-AS1 | FALSE | Upregulated |
| 0,629242271 | ZNF300P1 | FALSE | Upregulated |
| -0,510744419 | ZNF311 | FALSE | Downregulated |
| | ZNF32-AS3 | | |

| | | | |
|---|---|---|---|
| 0,656423442 | ZNF320 | FALSE | Upregulated |
| 0,222385933 | ZNF321P | FALSE | Upregulated |
| -1,262194747 | ZNF33BP1 | | Downregulated |
| -0,525107475 | ZNF341 | FALSE | Downregulated |
| 1,053467878 | ZNF354A | | Upregulated |
| 0,443481707 | ZNF354B | FALSE | Upregulated |
| -0,155702035 | ZNF365 | FALSE | Downregulated |
| 0,088397135 | ZNF366 | FALSE | Upregulated |
| -0,591591158 | ZNF382 | FALSE | Downregulated |
| 0,255372345 | ZNF383 | FALSE | Upregulated |
| -0,477789708 | ZNF385D | FALSE | Downregulated |
| 0,150593592 | ZNF391 | FALSE | Upregulated |
| 0,119534784 | ZNF396 | FALSE | Upregulated |
| -1,064032295 | ZNF415 | | Downregulated |
| -0,284224704 | ZNF416 | FALSE | Downregulated |
| 0,134040663 | ZNF419 | FALSE | Upregulated |
| -0,418007625 | ZNF423 | FALSE | Downregulated |
| 0,291789897 | ZNF425 | FALSE | Upregulated |
| -0,124696561 | ZNF426 | FALSE | Downregulated |
| -0,097748701 | ZNF43 | FALSE | Downregulated |
| 0,479424277 | ZNF430 | FALSE | Upregulated |
| 0,078707682 | ZNF431 | FALSE | Upregulated |
| 0,016827749 | ZNF439 | FALSE | Upregulated |
| -0,10639777 | ZNF442 | FALSE | Downregulated |
| -0,359988841 | ZNF449 | FALSE | Downregulated |
| 1,343487404 | ZNF454 | | Upregulated |
| -0,203873142 | ZNF461 | FALSE | Downregulated |
| 0,325956177 | ZNF468 | FALSE | Upregulated |
| 0,304313259 | ZNF474 | FALSE | Upregulated |
| | ZNF479 | | |
| -0,118954109 | ZNF485 | FALSE | Downregulated |
| 0,333528855 | ZNF487 | FALSE | Upregulated |
| -0,167379065 | ZNF488 | FALSE | Downregulated |
| 0,018513418 | ZNF491 | FALSE | Upregulated |
| 0,003691769 | ZNF492 | FALSE | Upregulated |
| -0,541712119 | ZNF501 | FALSE | Downregulated |
| 0,087837664 | ZNF502 | FALSE | Upregulated |
| 0,745889332 | ZNF503-AS1 | FALSE | Upregulated |
| 0,508528566 | ZNF519 | FALSE | Upregulated |
| 0,210705799 | ZNF525 | FALSE | Upregulated |
| -0,140941395 | ZNF527 | FALSE | Downregulated |
| 0,24298652 | ZNF530 | FALSE | Upregulated |
| 0,300241802 | ZNF534 | FALSE | Upregulated |
| 0 | ZNF536 | FALSE | Upregulated |
| 0,181511441 | ZNF541 | FALSE | Upregulated |
| 0,347155182 | ZNF547 | FALSE | Upregulated |
| 0 | ZNF556 | FALSE | Upregulated |
| -1,488051096 | ZNF559-ZNF177 | | Downregulated |
| | ZNF560 | | |
| 0,369681923 | ZNF567 | FALSE | Upregulated |
| -1,10085019 | ZNF570 | | Downregulated |

| | | | |
|---|---|---|---|
| 0,328724427 | ZNF572 | FALSE | Upregulated |
| -0,228196368 | ZNF573 | FALSE | Downregulated |
| -1,232138014 | ZNF578 | | Downregulated |
| 0,09074572 | ZNF581 | FALSE | Upregulated |
| -0,858627131 | ZNF583 | FALSE | Downregulated |
| 0,014060068 | ZNF596 | FALSE | Upregulated |
| 0,505330724 | ZNF599 | FALSE | Upregulated |
| 0,171732142 | ZNF600 | FALSE | Upregulated |
| 0,287921181 | ZNF607 | FALSE | Upregulated |
| -0,264074291 | ZNF610 | FALSE | Downregulated |
| 0,513116977 | ZNF611 | FALSE | Upregulated |
| 0,236072053 | ZNF613 | FALSE | Upregulated |
| -0,520043013 | ZNF624 | FALSE | Downregulated |
| -0,087610527 | ZNF625 | FALSE | Downregulated |
| 0,502326332 | ZNF625-ZNF20 | FALSE | Upregulated |
| 0,012448252 | ZNF630 | FALSE | Upregulated |
| 0,282123124 | ZNF645 | FALSE | Upregulated |
| 0,080355575 | ZNF648 | FALSE | Upregulated |
| -0,090531072 | ZNF653 | FALSE | Downregulated |
| -0,713841635 | ZNF658B | FALSE | Downregulated |
| 0,416253503 | ZNF663P | FALSE | Upregulated |
| 0,909264127 | ZNF664-FAM101A | FALSE | Upregulated |
| -0,435856204 | ZNF665 | FALSE | Downregulated |
| 0,973224169 | ZNF667 | FALSE | Upregulated |
| -1,328140972 | ZNF667-AS1 | | Downregulated |
| -0,578267793 | ZNF669 | FALSE | Downregulated |
| 0,007120011 | ZNF670 | FALSE | Upregulated |
| 0,450228244 | ZNF670-ZNF695 | FALSE | Upregulated |
| -0,093145704 | ZNF674-AS1 | FALSE | Downregulated |
| -0,095721881 | ZNF675 | FALSE | Downregulated |
| -0,208489927 | ZNF676 | FALSE | Downregulated |
| 0,30119132 | ZNF677 | FALSE | Upregulated |
| | ZNF679 | | |
| 0,429986614 | ZNF681 | FALSE | Upregulated |
| 0,405540763 | ZNF682 | FALSE | Upregulated |
| 0,645011065 | ZNF683 | FALSE | Upregulated |
| -0,853355941 | ZNF691 | FALSE | Downregulated |
| 0 | ZNF695 | FALSE | Upregulated |
| 0,800843145 | ZNF699 | FALSE | Upregulated |
| 1,046039048 | ZNF702P | | Upregulated |
| | ZNF705A | | |
| | ZNF705B | | |
| | ZNF705D | | |
| -0,59846125 | ZNF705E | FALSE | Downregulated |
| | ZNF705G | | |
| 0,34620147 | ZNF709 | FALSE | Upregulated |
| 0,415770659 | ZNF71 | FALSE | Upregulated |
| 1,411004416 | ZNF711 | | Upregulated |
| 0,594665031 | ZNF713 | FALSE | Upregulated |
| 0,174466224 | ZNF714 | FALSE | Upregulated |
| | ZNF716 | | |

| | | | |
|---|---|---|---|
| 0,146827808 | ZNF724P | FALSE | Upregulated |
| -0,823497215 | ZNF726 | FALSE | Downregulated |
| -0,811770558 | ZNF727 | FALSE | Downregulated |
| 0,154141453 | ZNF728 | FALSE | Upregulated |
| | ZNF729 | | |
| 0 | ZNF730 | FALSE | Upregulated |
| | ZNF733P | | |
| | ZNF735 | | |
| 0,30084271 | ZNF736 | FALSE | Upregulated |
| -0,552624379 | ZNF738 | FALSE | Downregulated |
| -0,273406296 | ZNF75A | FALSE | Downregulated |
| 0,604233502 | ZNF763 | FALSE | Upregulated |
| -0,537799267 | ZNF765 | FALSE | Downregulated |
| 0,123585528 | ZNF77 | FALSE | Upregulated |
| 0,612978379 | ZNF773 | FALSE | Upregulated |
| 0,162707713 | ZNF774 | FALSE | Upregulated |
| -0,296330468 | ZNF775 | FALSE | Downregulated |
| -0,276332869 | ZNF781 | FALSE | Downregulated |
| 0,500644073 | ZNF788 | FALSE | Upregulated |
| 1,019026842 | ZNF793 | | Upregulated |
| 0,525713151 | ZNF799 | FALSE | Upregulated |
| 0,460403404 | ZNF804A | FALSE | Upregulated |
| | ZNF804B | | |
| 0 | ZNF806 | FALSE | Upregulated |
| -0,058771786 | ZNF807 | FALSE | Downregulated |
| 1,128914581 | ZNF812 | | Upregulated |
| 0,110707023 | ZNF813 | FALSE | Upregulated |
| 0,670122249 | ZNF815P | FALSE | Upregulated |
| 0,274812988 | ZNF816 | FALSE | Upregulated |
| 0,308962609 | ZNF816-ZNF321P | FALSE | Upregulated |
| 0,069904821 | ZNF818P | FALSE | Upregulated |
| -0,135205692 | ZNF826P | FALSE | Downregulated |
| 0,877945984 | ZNF827 | FALSE | Upregulated |
| -0,892242055 | ZNF835 | FALSE | Downregulated |
| -0,793059684 | ZNF836 | FALSE | Downregulated |
| 0,457174647 | ZNF845 | FALSE | Upregulated |
| 0,209839724 | ZNF85 | FALSE | Upregulated |
| 0,127646366 | ZNF852 | FALSE | Upregulated |
| 0,154827687 | ZNF853 | FALSE | Upregulated |
| 0,012715321 | ZNF860 | FALSE | Upregulated |
| 0,126593023 | ZNF876P | FALSE | Upregulated |
| -0,304357895 | ZNF879 | FALSE | Downregulated |
| 0,125335201 | ZNF880 | FALSE | Upregulated |
| -0,184859766 | ZNF883 | FALSE | Downregulated |
| -0,050938538 | ZNF888 | FALSE | Downregulated |
| -0,122367156 | ZNF890P | FALSE | Downregulated |
| 0,049570363 | ZNF90 | FALSE | Upregulated |
| 1,068964718 | ZNF92 | | Upregulated |
| 0,403956883 | ZNF93 | FALSE | Upregulated |
| | ZNF98 | | |
| -0,001772849 | ZNF99 | FALSE | Downregulated |

| | | | |
|---|---|---|---|
| 0,369351839 | ZNRD1 | FALSE | Upregulated |
| 0,208811541 | ZNRD1-AS1 | FALSE | Upregulated |
| | ZNRF4 | | |
| 0,122283409 | ZP1 | FALSE | Upregulated |
| | ZP2 | | |
| 1,047078182 | ZP3 | | Upregulated |
| | ZP4 | | |
| | ZPBP | | |
| | ZPBP2 | | |
| -0,283850016 | ZPLD1 | FALSE | Downregulated |
| 0,276967504 | ZRANB2-AS2 | FALSE | Upregulated |
| 0,421370431 | ZSCAN10 | FALSE | Upregulated |
| -0,019864769 | ZSCAN16-AS1 | FALSE | Downregulated |
| -0,303868318 | ZSCAN20 | FALSE | Downregulated |
| -0,274845578 | ZSCAN23 | FALSE | Downregulated |
| 0,291220603 | ZSCAN4 | FALSE | Upregulated |
| 0,405919339 | ZSCAN5A | FALSE | Upregulated |
| -0,507961765 | ZSCAN5B | FALSE | Downregulated |
| -0,300545865 | ZSCAN9 | FALSE | Downregulated |
| | ZSWIM2 | | |
| -0,45536077 | ZUFSP | FALSE | Downregulated |
| 0,142853873 | ZWILCH | FALSE | Upregulated |
| 0,088907475 | ZWINT | FALSE | Upregulated |

**Adjustment of p-value by false discovery rate.**

**Suppl. Table 9: Pathway enrichment of differentially expressed genes in the validation cohort**

**cABMR upregulated**

| q-value | Pathway name | Pathway ID | | | q-value | Pathway name | Pathway ID |
|---|---|---|---|---|---|---|---|
| 1.1678358811 | regulation of le | GO:1903037 | | | 9.4533823298 | Extracellular m | R-HSA-1474244 |
| 2.0818144897 | positive regula | GO:0022409 | | | 6.3927759380 | Cell adhesion n | path:hsa04514 |
| 1.1503799177 | regulation of ly | GO:0051249 | | | 2.1505394230 | Integrin cell su | R-HSA-216083 |
| 6.9128607528 | positive regula | GO:0002696 | | | 4.1061535572 | ECM-receptor i | path:hsa04512 |
| 1.3168453305 | positive regula | GO:0050867 | | | 4.1061535572 | Viral myocardi | path:hsa05416 |
| 8.4324623344 | hemopoiesis | GO:0030097 | | | 0.0001207868 | Focal adhesion | path:hsa04510 |
| 1.2118512111 | regulation of le | GO:1902105 | | | 0.0001671278 | Immune Syster | R-HSA-168256 |
| 8.5881662895 | cellular respon | GO:0071345 | | | 0.0005741855 | RUNX3 regulat | R-HSA-895191 |
| 1.5401202798 | regulation of le | GO:0070663 | | | 0.0006005593 | Allograft reject | path:hsa05330 |
| 5.2656246308 | positive regula | GO:1903708 | | | 0.0006373330 | RUNX3 regulat | R-HSA-895193 |
| 8.0260862804 | positive regula | GO:0050778 | | | 0.0010357842 | Graft-versus-ho | path:hsa05332 |
| 0.0001025818 | positive regula | GO:0001819 | | | 0.0010357842 | Toxoplasmosis | path:hsa05145 |
| 0.0001354776 | ion transmemb | GO:0034220 | | | 0.0010357842 | Epstein-Barr vi | path:hsa05169 |
| 0.0002370102 | regulation of a | GO:0050854 | | | 0.0010825212 | Type I diabetes | path:hsa04940 |
| 0.0002521193 | membrane raft | GO:0031580 | | | 0.0011796964 | Collagen chain | R-HSA-894821 |
| 0.0004644215 | negative regula | GO:0050866 | | | 0.0024571965 | TNFR2 non-car | R-HSA-566854 |
| 0.0006173580 | negative regula | GO:0050777 | | | 0.0024571965 | RUNX3 Regulat | R-HSA-894927 |
| 0.0006254821 | negative regula | GO:0002695 | | | 0.0024571965 | Th17 cell differ | path:hsa04659 |
| 0.0007678738 | regulation of in | GO:0032663 | | | 0.0024571965 | Phagosome - H | path:hsa04145 |
| 0.0009496431 | positive regula | GO:0051272 | | | 0.0028832825 | Primary immu | path:hsa05340 |
| 0.0014048005 | positive regula | GO:0008284 | | | 0.0030349188 | Collagen forma | R-HSA-147429 |
| 0.0021441791 | negative regula | GO:0001818 | | | 0.0030349188 | Signaling by PD | R-HSA-186797 |
| 0.0021441791 | collagen-activa | GO:0038063 | | | 0.0030349188 | Small cell lung | path:hsa05222 |
| 0.0036675809 | regulation of a | GO:0045765 | | | 0.0030349188 | Th1 and Th2 ce | path:hsa04658 |
| 0.0042833482 | regulation of n | GO:0043313 | | | 0.0035022894 | RUNX1 regulat | R-HSA-893924 |
| 0.0046176651 | antigen recept | GO:0050851 | | | 0.0037654707 | Interferon gam | R-HSA-877300 |
| 0.0046591032 | positive regula | GO:0045597 | | | 0.0037654707 | Leishmaniasis - | path:hsa05140 |
| 0.0046591032 | regulation of tr | GO:0034762 | | | 0.0042137273 | Hematopoietic | path:hsa04640 |
| 0.0046591032 | cell morphoger | GO:0000904 | | | 0.0051057293 | Amoebiasis - H | path:hsa05146 |
| 0.0051050320 | negative regula | GO:0002698 | | | 0.0058337463 | ABC transporte | path:hsa02010 |
| 0.0069908453 | regulation of le | GO:0043300 | | | 0.0071011753 | Intestinal imm | path:hsa04672 |
| 0.0072151589 | organic acid tra | GO:1903825 | | | 0.0071011753 | Tuberculosis - | path:hsa05152 |
| 0.0083711571 | plasma membr | GO:0044856 | | | 0.0075212218 | Cytokine Signa | R-HSA-128021 |
| 0.0089910837 | regulation of io | GO:0043269 | | | 0.0077620876 | Collagen biosy | R-HSA-165081 |
| 0.0092709787 | regulation of va | GO:1901342 | | | 0.0081345671 | PI3K-Akt signal | path:hsa04151 |
| 0.0098702322 | interferon-gam | GO:0060333 | | | 0.0081345671 | Scavenging by | R-HSA-300048 |
| 0.0098702322 | negative regula | GO:2000181 | | | 0.0087271321 | Hypertrophic c | path:hsa05410 |
| 0.0113067705 | positive regula | GO:0010912 | | | 0.0098957165 | Neutrophil deg | R-HSA-679869 |
| 0.0114398851 | negative regula | GO:0045596 | | | 0.0098957165 | Autoimmune t | path:hsa05320 |
| 0.0126898316 | regulation of io | GO:0032412 | | | 0.0110844071 | Transcriptiona | R-HSA-887816 |
| 0.0127976467 | response to int | GO:0034341 | | | 0.0120204369 | RUNX1 and FO | R-HSA-887733 |
| 0.0133818747 | positive regula | GO:0010811 | | | 0.0123846706 | Pathways in ca | path:hsa05200 |
| 0.0140589300 | regulation of n | GO:1902563 | | | 0.0133270814 | ECM proteogly | R-HSA-300017 |
| 0.0148740737 | regulation of T- | GO:0045622 | | | 0.0138313767 | Arrhythmogen | path:hsa05412 |
| 0.0148740737 | neutrophil mig | GO:1990266 | | | 0.0139695544 | RUNX2 regulat | R-HSA-894133 |
| 0.0151452247 | anion transpor | GO:0006820 | | | 0.0139695544 | AGE-RAGE signa | path:hsa04933 |
| 0.0159503491 | negative regula | GO:1901343 | | | 0.0139695544 | Antigen proces | path:hsa04612 |
| 0.0163515024 | regulation of p | GO:2000361 | | | 0.0190709878 | Binding and Up | R-HSA-217378 |
| 0.0163515024 | vesicle fusion | GO:0006906 | | | 0.0204119406 | Human papillo | path:hsa05165 |
| 0.0163515024 | neutrophil che | GO:0030593 | | | 0.0229503166 | Semaphorin in | R-HSA-373755 |
| 0.0173165581 | amino acid tra | GO:0006865 | | | 0.0264022657 | Developmenta | R-HSA-126673 |
| 0.0180396174 | T-helper 1 type | GO:0042088 | | | 0.0283886750 | Leukocyte tran | path:hsa04670 |
| 0.0184129822 | T cell mediated | GO:0002456 | | | 0.0285528544 | Assembly of co | R-HSA-202209 |
| 0.0184129822 | cellular respon | GO:0071230 | | | 0.0295797217 | Nervous systen | R-HSA-967510 |
| 0.0185415319 | glycosaminogl | GO:0030203 | | | 0.0324479764 | Central carbon | path:hsa05230 |
| 0.0188625660 | neutrophil me | GO:0002446 | | | 0.0324479764 | Transcriptiona | R-HSA-887815 |
| 0.0188625660 | keratan sulfate | GO:0042340 | | | 0.0333373528 | Staphylococcu | path:hsa05150 |
| 0.0190933235 | actin polymeri | GO:0008154 | | | 0.0333373528 | Innate Immune | R-HSA-168249 |
| 0.0190933235 | neuron project | GO:0031175 | | | 0.0333373528 | Dilated cardior | path:hsa05414 |
| 0.0190933235 | organic anion t | GO:0015711 | | | 0.0333373528 | Axon guidance | R-HSA-422475 |
| 0.0204267451 | organelle mem | GO:0090174 | | | 0.0333373528 | RUNX1 regulat | R-HSA-893924 |
| 0.0205765629 | regulation of ly | GO:2000401 | | | 0.0333373528 | RUNX1 regulat | R-HSA-893596 |
| 0.0212977873 | positive regula | GO:0002699 | | | 0.0347242833 | Cytokine-cytok | path:hsa04060 |
| 0.0214515234 | T cell chemota | GO:0010818 | | | 0.0384415185 | Regulation of I | R-HSA-381426 |
| 0.0227969482 | positive regula | GO:0002687 | | | 0.0384466237 | Regulation of R | R-HSA-893459 |
| 0.0263641091 | aminoglycan ca | GO:0006026 | | | 0.0441641431 | Elastic fibre for | R-HSA-156694 |
| 0.0304960722 | regulation of ty | GO:0002828 | | | 0.0441641431 | RUNX1 regulat | R-HSA-893925 |
| 0.0307361122 | regulation of m | GO:0071675 | | | 0.0441641431 | RUNX1 regulat | R-HSA-893924 |
| 0.0336351777 | lipid phosphor | GO:0046834 | | | 0.0441641431 | RUNX1 regulat | R-HSA-893198 |
| 0.0345187708 | negative regula | GO:0022408 | | | 0.0441641431 | Protein digesti | path:hsa04974 |
| 0.0345187708 | regulation of ep | GO:0030856 | | | 0.0441641431 | Degradation of | R-HSA-147422 |
| 0.0345187708 | regulation of le | GO:0002703 | | | 0.0447881352 | Hedgehog signa | path:hsa04340 |
| 0.0352879881 | osteoclast deve | GO:0036035 | | | 0.0452337295 | T cell receptor | path:hsa04660 |
| 0.0353608968 | post-embryoni | GO:0048050 | | | 0.0466265681 | Other semapho | R-HSA-416700 |
| 0.0353608968 | regulation of a | GO:0002580 | | | 0.0483191973 | Human T-cell le | path:hsa05166 |
| 0.0396294349 | negative regula | GO:0051271 | | | 0.0497025161 | Post-translatio | R-HSA-895727 |
| 0.0400151372 | antigen proces | GO:0019885 | | | 0.0497025161 | JAK-STAT signal | path:hsa04630 |
| 0.0400973282 | regulation of c | GO:0060284 | | | 0.0497025161 | Interferon Sign | R-HSA-913531 |
| 0.0407169083 | regulation of c | GO:0002691 | | | 0.0584219014 | Complement a | path:hsa04610 |
| 0.0407169083 | leukocyte apop | GO:0071887 | | | | | |
| 0.0413998873 | positive regula | GO:0031349 | | | | | |
| 0.0444154174 | renal vesicle m | GO:0072077 | | | | | |
| 0.0456344074 | lacrimal gland | GO:0032808 | | | | | |
| 0.0456344074 | regulation of T | GO:2001188 | | | | | |
| 0.0456344074 | antigen proces | GO:0002476 | | | | | |
| 0.0456344074 | glutathione tra | GO:0034635 | | | | | |
| 0.0462229496 | organic acid tra | GO:0015849 | | | | | |
| 0.0462623843 | regulation of cy | GO:0001959 | | | | | |
| 0.0478684629 | response to ma | GO:0032026 | | | | | |
| 0.0487994199 | metanephros d | GO:0001656 | | | | | |
| 0.0519711355 | negative regula | GO:0051129 | | | | | |
| 0.0519711355 | regulation of in | GO:0045088 | | | | | |
| 0.0519711355 | regulation of T | GO:0001914 | | | | | |
| 0.0519711355 | odontogenesis | GO:0042475 | | | | | |
| 0.0519711355 | myeloid dendr | GO:0043011 | | | | | |
| 0.0522532997 | regulation of p | GO:0043067 | | | | | |
| 0.0543596936 | visual system d | GO:0150063 | | | | | |
| 0.0555190668 | positive regula | GO:0010942 | | | | | |
| 0.0573151793 | adrenal gland c | GO:0030325 | | | | | |
| 0.0590634110 | aminoglycan b | GO:0006023 | | | | | |
| 0.0593767038 | myeloid leuko | GO:0002573 | | | | | |
| 0.0628889224 | negative regula | GO:0001911 | | | | | |
| 0.0628889224 | chemokine-me | GO:0070098 | | | | | |

#### cABMR downregulated

| q-value | Pathway name | Pathway ID | | q-value | Pathway name | Pathway ID |
|---|---|---|---|---|---|---|
| 0.0004058040 | carboxylic acid | GO:0046394 | | 2.53839273102e-05 | Metallothione | R-HSA-566123 |
| 0.0004058040 | response to zin | GO:0010043 | | 3.45944474342e-05 | Response to m | R-HSA-566052 |
| 0.0004812922 | carboxylic acid | GO:0019752 | | 0.00017049668257 | Metabolism | R-HSA-143072 |
| 0.0007315193 | very-low-densi | GO:0034372 | | 0.00486510845736 | Mineral absorp | path:hsa04978 |
| 0.0007969364 | acylglycerol m | GO:0006639 | | 0.0151801130248 | Metabolism of | R-HSA-240850 |
| 0.0009817383 | response to co | GO:0046688 | | 0.0388737514713 | Plasma lipopro | R-HSA-896389 |
| 0.0013697530 | regulation of li | GO:0060191 | | 0.0388737514713 | Plasma lipopro | R-HSA-174824 |
| 0.0013886930 | fatty acid biosy | GO:0006633 | | 0.0388737514713 | Steroid hormo | path:hsa00140 |
| 0.0026016260 | response to cad | GO:0046686 | | 0.0388737514713 | Glycolysis / Glu | path:hsa00010 |
| 0.0026016260 | cellular respon | GO:0071248 | | 0.0388737514713 | Insulin signalin | path:hsa04910 |
| 0.0039806047 | glucose metab | GO:0006006 | | 0.0388737514713 | Complement c | R-HSA-166658 |
| 0.0053271913 | lipid homeosta | GO:0055088 | | 0.0388737514713 | Histidine meta | path:hsa00340 |
| 0.0053271913 | protein-lipid c | GO:0065005 | | 0.041371766525 | PPAR signaling | path:hsa03320 |
| 0.0053271913 | protein-lipid c | GO:0034368 | | 0.0431794305166 | Non-alcoholic | path:hsa04932 |
| 0.0059992123 | response to ery | GO:0036017 | | 0.0431794305166 | Plasma lipopro | R-HSA-896389 |
| 0.0059992123 | regulation of tr | GO:0090207 | | 0.0492087803725 | Complement a | path:hsa04610 |
| 0.0062006169 | hexose biosynt | GO:0019319 | | | | |
| 0.0071841952 | regulation of ca | GO:0043255 | | | | |
| 0.0096116522 | regulation of sy | GO:0003044 | | | | |
| 0.0099835617 | neutral lipid ca | GO:0046461 | | | | |
| 0.0099835617 | steroid biosynt | GO:0006694 | | | | |
| 0.0099835617 | positive regula | GO:0003084 | | | | |
| 0.0111032641 | regulation of li | GO:0046890 | | | | |
| 0.0117857884 | cellular cation | GO:0030003 | | | | |
| 0.0140539879 | ion homeostas | GO:0050801 | | | | |
| 0.0180407960 | acute inflamma | GO:0002526 | | | | |
| 0.0183826561 | ribonucleotide | GO:0009259 | | | | |
| 0.0203708225 | regulation of c | GO:0010675 | | | | |
| 0.0217153860 | positive regula | GO:0062013 | | | | |
| 0.0217153860 | glycerol metab | GO:0006071 | | | | |
| 0.0217153860 | glycerolipid ca | GO:0046503 | | | | |
| 0.0217153860 | entry into host | GO:0044409 | | | | |
| 0.0217153860 | entry into cell | GO:0051806 | | | | |
| 0.0217153860 | glycogen metal | GO:0005977 | | | | |
| 0.0217153860 | lipid transport | GO:0006869 | | | | |
| 0.0217153860 | cellular glucan | GO:0006073 | | | | |
| 0.0217153860 | glucan metabo | GO:0044042 | | | | |
| 0.0222349017 | coenzyme bios | GO:0009108 | | | | |
| 0.0222419548 | organic hydrox | GO:0015850 | | | | |
| 0.0222419548 | establishment | GO:0051905 | | | | |
| 0.0248116743 | steroid catabol | GO:0006706 | | | | |
| 0.0248116743 | pigment granu | GO:0051875 | | | | |
| 0.0289361789 | regulation of c | GO:0010565 | | | | |
| 0.0289361789 | cellular respon | GO:0071549 | | | | |
| 0.0289361789 | carboxylic acid | GO:0046395 | | | | |
| 0.0370976292 | ribonucleoside | GO:0009119 | | | | |
| 0.0418273039 | nucleotide me | GO:0009117 | | | | |
| 0.0423939866 | regulation of ca | GO:0006109 | | | | |

#### clinTCMR upregulated

| q-value | Pathway name | Pathway ID | | | q-value | Pathway name | Pathway ID |
|---|---|---|---|---|---|---|---|
| 6.5987721278 | cellular respon | GO:0071345 | | | 1.1379577583 | Immune Syster | R-HSA-168256 |
| 1.2587286341 | positive regula | GO:0022409 | | | 7.2861799546 | Cytokine Signa | R-HSA-1280215 |
| 1.1776313757 | regulation of le | GO:1903037 | | | 5.8032980613 | Interferon Sign | R-HSA-913531 |
| 1.6708683692 | hemopoiesis | GO:0030097 | | | 6.0935143855 | Epstein-Barr vi | path:hsa05169 |
| 2.3552612530 | response to int | GO:0034341 | | | 1.2367169313 | Interferon gam | R-HSA-877300 |
| 3.7570411280 | regulation of ly | GO:0051249 | | | 1.2474657052 | Primary immu | path:hsa05340 |
| 4.2097812557 | positive regula | GO:0050867 | | | 1.6194365629 | Innate Immune | R-HSA-168249 |
| 8.0944311091 | positive regula | GO:0002696 | | | 1.1573128607 | Cell adhesion n | path:hsa04514 |
| 1.2062278003 | regulation of le | GO:1902105 | | | 1.2751896385 | Neutrophil deg | R-HSA-6798695 |
| 1.7502260232 | positive regula | GO:0050778 | | | 1.3274434458 | Viral myocardi | path:hsa05416 |
| 3.4465426965 | interferon-gam | GO:0060333 | | | 2.7488871392 | Th17 cell differ | path:hsa04659 |
| 4.6003547159 | regulation of le | GO:0070663 | | | 7.1962589969 | Phagosome - H | path:hsa04145 |
| 6.6613346705 | neutrophil me | GO:0002446 | | | 1.3101410190 | Human T-cell le | path:hsa05166 |
| 2.7130498631 | positive regula | GO:0001819 | | | 1.8577909360 | Allograft reject | path:hsa05330 |
| 5.5573743079 | regulation of d | GO:0031347 | | | 1.9386399386 | Th1 and Th2 ce | path:hsa04658 |
| 6.3639299145 | negative regula | GO:0050777 | | | 2.2234531743 | Antigen proces | path:hsa04612 |
| 8.1746763117 | positive regula | GO:1903708 | | | 2.2234531743 | Staphylococcu | path:hsa05150 |
| 1.8966185540 | T cell mediatec | GO:0002456 | | | 4.7270461549 | Graft-versus-ho | path:hsa05332 |
| 2.3441717143 | regulation of ir | GO:0032663 | | | 5.5612165530 | Type I diabetes | path:hsa04940 |
| 2.5345520433 | negative regula | GO:0050866 | | | 2.4340806986 | Adaptive Immu | R-HSA-1280218 |
| 3.7888553081 | negative regula | GO:0002695 | | | 2.7855980747 | Autoimmune t | path:hsa05320 |
| 4.6969082380 | antigen recept | GO:0050851 | | | 5.4880240670 | Interferon alph | R-HSA-909733 |
| 9.0528918877 | negative regula | GO:0031348 | | | 5.9721364013 | TCR signaling | R-HSA-202403 |
| 2.2498070191 | negative regula | GO:0001818 | | | 8.3220636571 | Leishmaniasis - | path:hsa05140 |
| 2.4782492725 | regulation of p | GO:0043067 | | | 0.0001604459 | Inflammatory b | path:hsa05321 |
| 2.9996909037 | regulation of T | GO:0002664 | | | 0.0002180386 | Mitotic G1 pha | R-HSA-453279 |
| 3.7667731542 | regulation of ir | GO:0050727 | | | 0.0004528394 | Generation of s | R-HSA-202433 |
| 4.0361231711 | negative regula | GO:0001911 | | | 0.0005227324 | G1/S Transition | R-HSA-69206 |
| 4.1643428143 | dendritic cell d | GO:0097028 | | | 0.0006755833 | Hematopoietic | path:hsa04640 |
| 5.0098770319 | positive regula | GO:0008284 | | | 0.0008034147 | Immunoregula | R-HSA-198933 |
| 5.0646775873 | positive regula | GO:0002699 | | | 0.0010555126 | T cell receptor | path:hsa04660 |
| 6.0730773945 | regulation of ir | GO:0032649 | | | 0.0016140829 | Tuberculosis - | path:hsa05152 |
| 6.2606339251 | regulation of ly | GO:0002911 | | | 0.0018857828 | Antigen Presen | R-HSA-983170 |
| 6.7686573278 | regulation of ir | GO:0045088 | | | 0.0019638294 | Rheumatoid ar | path:hsa05323 |
| 7.2977683874 | negative regula | GO:0022408 | | | 0.0022660993 | JAK-STAT signal | path:hsa04630 |
| 9.6250646090 | negative regula | GO:2000524 | | | 0.0023720014 | Antigen proces | R-HSA-1236975 |
| 0.0001178141 | type I interfero | GO:0060337 | | | 0.0023720014 | Interleukin-2 fa | R-HSA-451927 |
| 0.0001271659 | positive regula | GO:0031349 | | | 0.0028795479 | Kaposi sarcoma | path:hsa05167 |
| 0.0001370435 | actin polymeri | GO:0008154 | | | 0.0028795479 | Transcriptiona | R-HSA-961622 |
| 0.0001414805 | antigen proces | GO:0019885 | | | 0.0034028556 | B cell receptor | path:hsa04662 |
| 0.0001797432 | leukocyte adhe | GO:0061756 | | | 0.0035827883 | Chagas disease | path:hsa05142 |
| 0.0001807093 | response to typ | GO:0034340 | | | 0.0057645552 | RUNX3 regulat | R-HSA-895193 |
| 0.0002064206 | negative regula | GO:0002698 | | | 0.0057645552 | RUNX3 regulat | R-HSA-895143 |
| 0.0002229865 | regulation of a | GO:0050854 | | | 0.0060140242 | Systemic lupus | path:hsa05322 |
| 0.0003349620 | regulation of le | GO:0002703 | | | 0.0062727902 | PD-L1 expressi | path:hsa05235 |
| 0.0004000051 | positive regula | GO:0002645 | | | 0.0063946552 | Toxoplasmosis | path:hsa05145 |
| 0.0006984525 | regulation of T | GO:0001914 | | | 0.0077891756 | Interleukin-9 si | R-HSA-898594 |
| 0.0007080886 | regulation of a | GO:0002819 | | | 0.0083915389 | Pathways in ca | path:hsa05200 |
| 0.0007907940 | leukocyte aggr | GO:0070486 | | | 0.0096810726 | Human cytome | path:hsa05163 |
| 0.0009601643 | myeloid leuko | GO:0002573 | | | 0.0102989281 | Formyl peptide | R-HSA-444473 |
| 0.0009601643 | neutrophil mig | GO:1990266 | | | 0.0117669473 | Viral protein in | path:hsa04061 |
| 0.0010021665 | regulation of ir | GO:0032655 | | | 0.0117669473 | Cell cycle - Hon | path:hsa04110 |
| 0.0010639853 | antigen proces | GO:0042590 | | | 0.0126653895 | RUNX2 regulat | R-HSA-894133 |
| 0.0011085495 | neutrophil che | GO:0030593 | | | 0.0126653895 | RUNX3 regulat | R-HSA-895191 |
| 0.0012103099 | regulation of T | GO:2001188 | | | 0.0131647918 | Osteoclast diffe | path:hsa04380 |
| 0.0014093240 | regulation of c | GO:0002718 | | | 0.0138999824 | NOD-like recep | path:hsa04621 |
| 0.0014093240 | negative regula | GO:0048525 | | | 0.0144185198 | Bladder cancer | path:hsa05219 |
| 0.0014232732 | regulation of ir | GO:0032675 | | | 0.0146883134 | Signaling by Int | R-HSA-449147 |
| 0.0015423704 | negative regula | GO:0050858 | | | 0.0152877370 | Unwinding of D | R-HSA-176974 |
| 0.0019157138 | membrane raft | GO:0031580 | | | 0.0152877370 | Binding and Up | R-HSA-217378 |
| 0.0020725971 | regulation of ir | GO:0032673 | | | 0.0185588755 | Interleukin-3, I | R-HSA-512988 |
| 0.0021767573 | regulation of v | GO:0050792 | | | 0.0188256285 | Endosomal/Va | R-HSA-123697 |
| 0.0021767573 | positive regula | GO:0031334 | | | 0.0188256285 | Interleukin rec | R-HSA-912526 |
| 0.0022014914 | regulation of m | GO:0071675 | | | 0.0193965021 | Chemokine sig | path:hsa04062 |
| 0.0027494609 | interleukin-21 | GO:0038114 | | | 0.0209428105 | Translocation | R-HSA-202430 |
| 0.0027494609 | response to int | GO:0098756 | | | 0.0222479697 | Advanced glyc | R-HSA-879415 |
| 0.0028336483 | interleukin-10 | GO:0140105 | | | 0.0226872401 | Intestinal imm | path:hsa04672 |
| 0.0028336483 | positive regula | GO:0010942 | | | 0.0241252737 | Small cell lung | path:hsa05222 |
| 0.0028336483 | regulation of c | GO:0002691 | | | 0.0251938414 | Asthma - Homo | path:hsa05310 |
| 0.0028336483 | regulation of p | GO:0002700 | | | 0.0251938414 | ISG15 antiviral | R-HSA-116940 |
| 0.0032081731 | response to int | GO:0070670 | | | 0.0253742247 | Interleukin-4 a | R-HSA-678580 |
| 0.0036294914 | positive regula | GO:0001912 | | | 0.0253742247 | Transcriptiona | R-HSA-887815 |
| 0.0036294914 | regulation of ir | GO:0032653 | | | 0.0265035798 | Cytokine-cytok | path:hsa04060 |
| 0.0036294914 | interleukin-9-r | GO:0038113 | | | 0.0269206413 | Phosphorylatio | R-HSA-202427 |
| 0.0036359237 | negative regula | GO:1903707 | | | 0.0273166292 | Non-small cell | path:hsa05223 |
| 0.0038099905 | positive regula | GO:0033630 | | | 0.0273166292 | Fc gamma R-m | path:hsa04666 |
| 0.0039417241 | leukocyte apop | GO:0071887 | | | 0.0273166292 | Polo-like kinas | R-HSA-156711 |
| 0.0040966410 | G1/S transition | GO:0000082 | | | 0.0273166292 | Transcriptiona | R-HSA-887816 |
| 0.0042130195 | regulation of a | GO:0008064 | | | 0.0273166292 | RUNX3 regulat | R-HSA-894185 |
| 0.0042130195 | regulation of T | GO:0045622 | | | 0.0273166292 | TFAP2 (AP-2) fa | R-HSA-886691 |
| 0.0045614323 | positive regula | GO:0032103 | | | 0.0273166292 | PD-1 signaling | R-HSA-389948 |
| 0.0045614323 | response to int | GO:0071104 | | | 0.0273166292 | DNA strand elo | R-HSA-69190 |
| 0.0045614323 | regulation of n | GO:1902563 | | | 0.0285220530 | Downstream T | R-HSA-202424 |
| 0.0049982447 | positive regula | GO:0009967 | | | 0.0308148846 | G1/S-Specific T | R-HSA-69205 |
| 0.0051744746 | regulation of o | GO:0033043 | | | 0.0308148846 | TNF receptor s | R-HSA-567659 |
| 0.0051982337 | ion homeostas | GO:0050801 | | | 0.0332024361 | Costimulation | R-HSA-388841 |
| 0.0052038917 | cellular cation | GO:0030003 | | | 0.0351754765 | NF-kappa B sig | path:hsa04064 |
| 0.0052645682 | regulation of m | GO:0045637 | | | 0.0355665973 | Natural killer c | path:hsa04650 |
| 0.0055364766 | eosinophil mig | GO:0072677 | | | 0.0356577657 | S Phase | R-HSA-69242 |
| 0.0061920055 | positive regula | GO:0002687 | | | 0.0356577657 | Toll-like recept | path:hsa04620 |
| 0.0064461110 | negative regula | GO:0032102 | | | 0.0356577657 | RUNX1 regulat | R-HSA-893924 |
| 0.0070124613 | phosphorylatio | GO:0016310 | | | 0.0356577657 | RUNX1 regulat | R-HSA-893925 |
| 0.0070124613 | positive regula | GO:0051272 | | | 0.0381005479 | Cell Cycle, Mito | R-HSA-69278 |
| 0.0070311285 | cellular respon | GO:0071396 | | | 0.0381005479 | Other semapho | R-HSA-416700 |
| 0.0070311285 | regulation of n | GO:0042269 | | | 0.0381005479 | Scavenging by | R-HSA-300048 |
| 0.0082073885 | regulation of tu | GO:1903555 | | | 0.0391044885 | Thyroid cancer | path:hsa05216 |
| 0.0083107495 | pattern recogn | GO:0002221 | | | 0.0391044885 | Antiviral mech | R-HSA-116941 |
| 0.0088464422 | regulation of p | GO:0071801 | | | 0.0396774164 | Extracellular m | R-HSA-147424 |
| 0.0091215706 | regulation of c | GO:1900015 | | | 0.0439340165 | MHC class II an | R-HSA-213229 |
| 0.0092563122 | positive regula | GO:0031325 | | | 0.0452978160 | Scavenging by | R-HSA-300047 |
| 0.0099507986 | positive regula | GO:0010647 | | | 0.0453639977 | Complement a | path:hsa04610 |
| 0.0100010238 | regulation of a | GO:0032956 | | | 0.0453639977 | Measles - Hom | path:hsa05162 |
| 0.0105709357 | response to int | GO:0070672 | | | 0.0483853653 | Chemokine rec | R-HSA-380108 |
| 0.0136612226 | regulation of p | GO:0051246 | | | 0.0498405140 | Human papillo | path:hsa05165 |
| 0.0136612226 | negative regula | GO:0060548 | | | | | |
| 0.0136612226 | positive regula | GO:0010604 | | | | | |
| 0.0143347712 | positive regula | GO:0051092 | | | | | |
| 0.0143347712 | vesicle fusion | GO:0006906 | | | | | |
| 0.0146330701 | regulation of p | GO:0032271 | | | | | |
| 0.0146330701 | positive regula | GO:0045597 | | | | | |
| 0.0146539639 | negative regula | GO:0051129 | | | | | |
| 0.0172970826 | podosome asse | GO:0071800 | | | | | |
| 0.0172970826 | regulation of ir | GO:0033623 | | | | | |
| 0.0180419151 | positive regula | GO:0051173 | | | | | |
| 0.0184384010 | DNA recombin | GO:0006310 | | | | | |
| 0.0189868167 | positive regula | GO:0051345 | | | | | |
| 0.0190424096 | organelle mem | GO:0090174 | | | | | |
| 0.0190424096 | regulation of p | GO:0051174 | | | | | |
| 0.0201427098 | negative regula | GO:0010648 | | | | | |
| 0.0205623873 | viral life cycle | GO:0019058 | | | | | |
| 0.0208261415 | regulation of c | GO:0032268 | | | | | |
| 0.0216083142 | response to tu | GO:0034612 | | | | | |
| 0.0216992411 | positive regula | GO:0051130 | | | | | |
| 0.0216992411 | ERK1 and ERK2 | GO:0070371 | | | | | |
| 0.0216992411 | chronic inflam | GO:0002544 | | | | | |
| 0.0218814721 | antigen proces | GO:0002476 | | | | | |
| 0.0218814721 | regulation of n | GO:0043313 | | | | | |
| 0.0230084319 | regulation of ir | GO:1902531 | | | | | |
| 0.0240390044 | myeloid dendr | GO:0043011 | | | | | |
| 0.0240390044 | eosinophil che | GO:0048245 | | | | | |
| 0.0240390044 | regulation of ir | GO:0032674 | | | | | |
| 0.0242263614 | negative regula | GO:1903035 | | | | | |
| 0.0242263614 | chemokine-me | GO:0070098 | | | | | |
| 0.0244728886 | G1 DNA damag | GO:0044783 | | | | | |
| 0.0246729489 | regulation of re | GO:1903034 | | | | | |
| 0.0264370318 | regulation of le | GO:0002688 | | | | | |
| 0.0264649729 | alpha-beta T ce | GO:0002363 | | | | | |
| 0.0272402076 | positive regula | GO:0010811 | | | | | |
| 0.0272663022 | negative regula | GO:0009968 | | | | | |
| 0.0272663022 | maintenance o | GO:0010216 | | | | | |
| 0.0272663022 | interleukin-4-r | GO:0035771 | | | | | |
| 0.0287744431 | regulation of c | GO:0022604 | | | | | |
| 0.0287744431 | CD4-positive o | GO:0043369 | | | | | |
| 0.0287744431 | negative regula | GO:0051346 | | | | | |
| 0.0294632067 | regulation of v | GO:1901342 | | | | | |
| 0.0308727220 | regulation of a | GO:0045765 | | | | | |
| 0.0315022592 | regulation of li | GO:0060191 | | | | | |
| 0.0315022592 | regulation of ty | GO:0032479 | | | | | |
| 0.0326218997 | viral genome re | GO:0019079 | | | | | |
| 0.0340580064 | response to ch | GO:1990868 | | | | | |
| 0.0368064663 | regulation of le | GO:0043300 | | | | | |
| 0.0383724810 | negative regula | GO:0008285 | | | | | |
| 0.0383724810 | mitotic DNA da | GO:0044773 | | | | | |
| 0.0383724810 | calcium ion tra | GO:0097553 | | | | | |
| 0.0383724810 | integrin activa | GO:0033622 | | | | | |

#### clinTCMR downregulated

| q-value | Pathway name | Pathway ID | | | q-value | Pathway name | Pathway ID |
|---|---|---|---|---|---|---|---|
| 1.3815662695 | carboxylic acic | GO:0019752 | | | 2.8677777934 | Metabolism | R-HSA-143072 |
| 2.0653631557 | terpenoid meta | GO:0006721 | | | 4.7795969563 | Biological oxid | R-HSA-211859 |
| 0.0001993396 | carboxylic acic | GO:0046394 | | | 6.4862607818 | Phase I - Functi | R-HSA-211945 |
| 0.0002986701 | lipid hydroxyla | GO:0002933 | | | 0.0001155222 | Drug metaboli | path:hsa00982 |
| 0.0051099309 | branched-chai | GO:0009083 | | | 0.0001575532 | Metabolism of | R-HSA-556833 |
| 0.0051099309 | ethanol metab | GO:0006067 | | | 0.0002081599 | Chemical carci | path:hsa05204 |
| 0.0051099309 | exogenous dru | GO:0042738 | | | 0.0003912779 | Cytochrome P4 | R-HSA-211897 |
| 0.0051972538 | arachidonic ac | GO:0019369 | | | 0.0004888028 | CYP2E1 reactic | R-HSA-211999 |
| 0.0057713897 | steroid catabo | GO:0006706 | | | 0.0016521058 | Bile acid and bi | R-HSA-194068 |
| 0.0103757617 | carboxylic acic | GO:0046395 | | | 0.0027419215 | Branched-chai | R-HSA-70895 |
| 0.0135583057 | bile acid biosyı | GO:0006699 | | | 0.0027419215 | Xenobiotics | R-HSA-211981 |
| 0.0148934194 | nucleotide me | GO:0009117 | | | 0.0046814419 | Steroid hormo | path:hsa00140 |
| 0.0148934194 | thyroid hormo | GO:0070327 | | | 0.0046814419 | Biosynthesis of | R-HSA-902730 |
| 0.0149492671 | cholesterol cat | GO:0006707 | | | 0.0051654768 | SLC-mediated t | R-HSA-425407 |
| 0.0149492671 | sterol catabol | GO:0016127 | | | 0.0051654768 | Linoleic acid m | path:hsa00591 |
| 0.0149492671 | fatty acid biosy | GO:0006633 | | | 0.0051654768 | Glycolysis / Glu | path:hsa00010 |
| 0.0201859392 | alpha-amino ac | GO:1901606 | | | 0.0052057543 | Synthesis of bil | R-HSA-192105 |
| 0.0234612161 | ribonucleotide | GO:0009259 | | | 0.0059318119 | Synthesis of ep | R-HSA-214267 |
| 0.0241350278 | steroid biosynt | GO:0006694 | | | 0.0059318119 | Biosynthesis of | R-HSA-901868 |
| 0.0249655541 | purine nucleot | GO:0006163 | | | 0.0059579860 | Metabolism of | R-HSA-71291 |
| 0.0249655541 | unsaturated fa | GO:0033559 | | | 0.0063197162 | Metabolism of | path:hsa00980 |
| 0.0260827554 | long-chain fatt | GO:0001676 | | | 0.0065605391 | Synthesis of (16 | R-HSA-214281 |
| 0.0391407821 | nucleoside bisp | GO:0033865 | | | 0.0095115303 | Synthesis of bil | R-HSA-193807 |
| 0.0391407821 | purine nucleos | GO:0034032 | | | 0.0100569250 | Fatty acid degr | path:hsa00071 |
| 0.0429147715 | cholesterol me | GO:0008203 | | | 0.0100569250 | Ethanol oxidat | R-HSA-71384 |
| 0.0429147715 | positive regula | GO:0045834 | | | 0.0100569250 | Transport of or | R-HSA-879518 |
| | | | | | 0.0121505395 | Pyruvate metal | path:hsa00620 |
| | | | | | 0.0124431568 | Valine, leucine | path:hsa00280 |
| | | | | | 0.0176083760 | Biosynthesis of | R-HSA-901867 |
| | | | | | 0.0176083760 | Primary bile ac | path:hsa00120 |
| | | | | | 0.0181248842 | Serotonergic sy | path:hsa04726 |
| | | | | | 0.0199688050 | Arachidonic ac | R-HSA-214275 |
| | | | | | 0.0199688050 | Biosynthesis of | R-HSA-901867 |
| | | | | | 0.0201464034 | Arachidonic ac | path:hsa00590 |
| | | | | | 0.0208372672 | Synthesis of bil | R-HSA-193368 |
| | | | | | 0.0211532734 | Metabolism of | R-HSA-895732 |
| | | | | | 0.0211532734 | GnRH secretion | path:hsa04929 |
| | | | | | 0.0243472425 | Retinol metabo | path:hsa00830 |

#### subTCMR upregulated

| q-value | Pathway name | Pathway ID | | q-value | Pathway name | Pathway ID |
|---|---|---|---|---|---|---|
| 0.0508222381 | regulation of e | GO:0050678 | | 0.000268214 | Scavenging by | R-HSA-300047 |
| 0.0508222381 | epidermal cell | GO:0009913 | | 0.000286988 | Formyl peptide | R-HSA-444473 |
| 0.1097396327 | cellular respon | GO:0071345 | | 0.000386211 | Advanced glyco | R-HSA-879415 |
| | | | | 0.002706544 | Binding and Up | R-HSA-217378 |
| | | | | 0.002760795 | G alpha (q) sigr | R-HSA-416476 |
| | | | | 0.007353117 | Class A/1 (Rhoc | R-HSA-373076 |
| | | | | 0.007353117 | Th1 and Th2 ce | path:hsa04658 |
| | | | | 0.015479488 | GPCR ligand bi | R-HSA-500792 |
| | | | | 0.016360455 | Signal Transdu | R-HSA-162582 |

**subTCMR downregulated**

None

**Suppl. Table 10: Unique differentially expressed genes per phenotype**
Adjustment of p-value by false discovery rate.

**Unique cABMR upregulated**

| gene name | log2fold change | padj |
|---|---|---|
| SLC7A14 | 4,22083472 | 4,81871E-08 |
| FGF14 | 2,93556785 | 1,77857E-07 |
| PAPPA-AS1 | 2,74508047 | 1,09512E-06 |
| SLC1A7 | 2,21217984 | 1,41733E-06 |
| AEBP1 | 1,83944162 | 9,07596E-06 |
| LOC100506990 | 1,34250223 | 2,44586E-05 |
| KCNQ1OT1 | 1,03130617 | 3,7778E-05 |
| ZNF433 | 1,20049366 | 5,0477E-05 |
| SAP30L-AS1 | 1,26635264 | 5,72062E-05 |
| MBNL1-AS1 | 1,36411436 | 6,64463E-05 |
| LOC441454 | 1,24130754 | 7,17608E-05 |
| NLGN3 | 1,44929787 | 0,000139676 |
| PVRIG | 1,12004047 | 0,000154854 |
| C10orf10 | 1,0286446 | 0,000169846 |
| LOC100506142 | 1,36234488 | 0,000169846 |
| KLHL29 | 1,02260617 | 0,000221542 |
| RP11-44F21.3 | 2,95728822 | 0,000221542 |
| IGJ | 2,72083331 | 0,000223468 |
| PCNXL2 | 2,05048054 | 0,000236944 |
| GCSAM | 3,10897689 | 0,00024271 |
| GSN | 1,10933998 | 0,000253429 |
| CD84 | 1,54716932 | 0,000292848 |
| KCTD12 | 1,17243224 | 0,000292848 |
| DDI1 | 2,23601819 | 0,00029536 |
| PLAC4 | 1,28301559 | 0,000299252 |
| LRRC18 | 1,49905957 | 0,000321242 |
| PRPF40B | 1,16989553 | 0,000333532 |
| RP6-206I17.2 | 1,08437144 | 0,000386073 |
| TSPAN3 | 1,38403983 | 0,000417815 |
| KRR1 | 1,01221255 | 0,000462896 |
| LAMA2 | 1,3570129 | 0,000466038 |
| LOC102724584 | 1,44611716 | 0,000466038 |
| AC007750.5 | 3,72998583 | 0,00046815 |
| KAAG1 | 2,15165002 | 0,00047886 |
| LINC00691 | 3,26131342 | 0,000494155 |
| CASC14 | 2,5727184 | 0,000611455 |
| CLDN18 | 1,6176357 | 0,000665355 |
| THY1 | 1,56472767 | 0,000739254 |
| ATP8B2 | 1,49354973 | 0,000760984 |
| LOC100289455 | 1,98311712 | 0,000790574 |
| LYSMD1 | 1,37801026 | 0,000791871 |
| PLEKHG4 | 1,78658808 | 0,000812961 |
| COL15A1 | 3,33332017 | 0,000828615 |
| C1orf198 | 1,79161138 | 0,000846284 |
| HMGN4 | 1,66410974 | 0,000846284 |

**Unique cABMR downregulated**

| gene name | log2fold change | padj |
|---|---|---|
| APOC3 | -1,0260515 | 3,15526E-05 |
| PTH1R | -1,2979095 | 3,39131E-05 |
| AVPR1A | -2,9907034 | 0,00010418 |
| CBS | -1,4486147 | 0,000139676 |
| PCDH1 | -1,0653993 | 0,000196695 |
| PPP1R3C | -1,5720208 | 0,000345922 |
| HAO2 | -1,1491007 | 0,000615493 |
| MOCOS | -1,3777136 | 0,000687318 |
| MLPH | -1,2497064 | 0,000731724 |
| CFHR3 | -1,8781383 | 0,000741847 |
| CD82 | -1,3382492 | 0,001057926 |
| HP | -1,0097892 | 0,001067611 |
| HAL | -1,0311858 | 0,001212695 |
| ATP5D | -1,0198892 | 0,00136474 |
| APCS | -1,0060151 | 0,001505014 |
| ECSIT | -1,170779 | 0,00163824 |
| THSD4 | -1,18217 | 0,001711866 |
| ORM1 | -1,2086131 | 0,001753649 |
| ATF5 | -1,2211826 | 0,002121802 |
| MT2A | -1,5870293 | 0,002353067 |
| ADRA1A | -1,0987472 | 0,002495111 |
| MT1E | -1,759747 | 0,00258218 |
| LINC01018 | -1,1006144 | 0,003257886 |
| SH3PXD2A | -1,0646675 | 0,003319852 |
| LIPC | -1,0965879 | 0,00361461 |
| MME | -1,2259485 | 0,003761997 |
| CFHR1 | -1,3082297 | 0,003770057 |
| LYRM1 | -1,5471964 | 0,005280242 |
| C11orf31 | -1,0795825 | 0,005460912 |
| ASPG | -1,0976118 | 0,00547018 |
| ERRFI1 | -1,1428253 | 0,00547018 |
| RHEBP2 | -1,2294892 | 0,005942151 |
| CLEC4M | -1,4520827 | 0,006373766 |
| DNAJC12 | -2,7543149 | 0,007274592 |
| MT1X | -1,573931 | 0,00817191 |
| APOA5 | -1,0259224 | 0,008404614 |
| LINC00671 | -1,4753314 | 0,008404614 |
| OCEL1 | -1,3932408 | 0,008825408 |
| NDUFB4 | -1,702235 | 0,008934361 |
| SLC19A2 | -1,2434628 | 0,010714224 |
| PCK1 | -1,1422283 | 0,011045487 |
| ANKRD24 | -1,0837427 | 0,012787438 |
| UQCRB | -1,3835806 | 0,013473128 |
| HAPLN4 | -1,8049452 | 0,014318188 |
| RHEB | -1,0440108 | 0,014895525 |

**Unique clinTCMR upregulated**

| gene name | log2fold change | padj |
|---|---|---|
| AJUBA | 1,88969721 | 2,37913E-09 |
| FAM168A | 1,07392672 | 4,04472E-08 |
| MAP7D1 | 1,07730398 | 2,32733E-07 |
| MKI67 | 2,98321109 | 2,75063E-07 |
| TYMP | 1,13421885 | 3,76414E-07 |
| NCAPH2 | 1,15346753 | 5,92997E-07 |
| COTL1 | 1,28314347 | 1,3522E-06 |
| IFI30 | 1,68723694 | 1,90788E-06 |
| TAGLN2 | 1,08119535 | 2,35732E-06 |
| RET | 3,52979513 | 6,74727E-06 |
| ZNF333 | 1,33821619 | 8,11436E-06 |
| HIST1H1B | 3,40346633 | 1,28277E-05 |
| RFX5 | 1,37124296 | 1,37415E-05 |
| HLA-B | 1,23767907 | 2,12411E-05 |
| LAPTM5 | 1,19680843 | 4,23147E-05 |
| C9orf163 | 1,57586914 | 4,79352E-05 |
| TTYH3 | 1,16169174 | 5,07783E-05 |
| ZC3H12D | 2,32888631 | 5,07783E-05 |
| PRC1-AS1 | 2,4093626 | 5,44037E-05 |
| WARS | 1,03725301 | 5,44292E-05 |
| SH3BGRL3 | 1,27493091 | 6,49248E-05 |
| ABCB4 | 1,00484884 | 7,00561E-05 |
| PLEKHO2 | 1,08141393 | 7,18409E-05 |
| SMTNL1 | 1,67732114 | 7,42293E-05 |
| TMSB10 | 1,02943593 | 8,16031E-05 |
| SAA2 | 3,42689393 | 8,5464E-05 |
| LCP1 | 1,13965335 | 8,66501E-05 |
| HLA-A | 1,16143664 | 9,21549E-05 |
| ITGB2-AS1 | 1,85951601 | 0,000108245 |
| SAA1 | 3,20020498 | 0,000108245 |
| LGALS9 | 1,32908648 | 0,00010843 |
| GBP1 | 1,4020374 | 0,000117522 |
| SSBP3-AS1 | 1,03376387 | 0,000121256 |
| MIR22HG | 1,18356132 | 0,000137157 |
| C1QC | 1,10371046 | 0,000160265 |
| CD8A | 2,79201721 | 0,000163693 |
| TDRD6 | 1,36009836 | 0,000173867 |
| TUBB1 | 1,02571293 | 0,000179012 |
| LOC102724153 | 1,32575445 | 0,000182623 |
| JUN | 1,26245572 | 0,000186118 |
| PRNCR1 | 1,73753936 | 0,000186369 |
| PSD | 1,55445414 | 0,000192519 |
| FOXP3 | 1,80564103 | 0,000220814 |
| PAGR1 | 1,19140017 | 0,000240287 |
| IL2RG | 2,32689315 | 0,000245863 |

**Unique clinTCMR downregulated**

| gene name | log2fold change | padj |
|---|---|---|
| ZKSCAN1 | -1,1963924 | 2,06382E-12 |
| ACADSB | -1,2576192 | 9,79791E-09 |
| BCHE | -1,6398071 | 3,6314E-08 |
| HMGCS2 | -1,486589 | 1,1027E-07 |
| HAAO | -1,0428109 | 2,32733E-07 |
| PGLYRP2 | -1,1675516 | 3,24799E-07 |
| TTR | -1,370993 | 1,66354E-06 |
| ALB | -1,2403111 | 2,72558E-06 |
| ADH6 | -1,1088741 | 1,21152E-05 |
| CYP2C8 | -1,0322754 | 5,69807E-05 |
| PLEKHA7 | -1,049163 | 8,70004E-05 |
| SLCO4C1 | -2,2969745 | 0,000108245 |
| CYP2D7P | -1,4203658 | 0,000231881 |
| GLYAT | -1,0606065 | 0,000260271 |
| MDN1 | -1,0367568 | 0,00029432 |
| GBA3 | -1,1598973 | 0,000355682 |
| CMBL | -1,092006 | 0,000373433 |
| LOC100507616 | -1,7467234 | 0,000511536 |
| RCAN3 | -1,5123266 | 0,000511536 |
| HSD17B13 | -1,1230337 | 0,000677462 |
| LPAL2 | -1,1546146 | 0,000721961 |
| FRRS1 | -1,0566031 | 0,001024557 |
| SAMD5 | -1,315802 | 0,001144583 |
| CSAD | -1,1028071 | 0,001199067 |
| SH3RF2 | -1,3274925 | 0,001211686 |
| CYP4A22 | -1,0848761 | 0,001242974 |
| SDHAP1 | -1,2800442 | 0,001405788 |
| TPSG1 | -1,5381944 | 0,001502028 |
| PITPNM3 | -1,26372 | 0,001848792 |
| C14orf132 | -1,312912 | 0,002118033 |
| RP11-475O6.1 | -1,6611944 | 0,002527263 |
| RP11-475O6.1-2 | -1,5073467 | 0,002824781 |
| DBT | -1,0353661 | 0,002843397 |
| CYP2C19 | -1,4980818 | 0,002912059 |
| ST8SIA3 | -1,4570327 | 0,0030762 |
| RP11-492A10.1 | -1,5412134 | 0,0036551 |
| RANBP3L | -1,192266 | 0,003927586 |
| LHX9 | -2,2330431 | 0,004016999 |
| NRARP | -1,6675389 | 0,00409206 |
| SLC45A3 | -1,0999875 | 0,00413492 |
| LPA | -1,1105354 | 0,004309431 |
| ADCY10 | -1,3674839 | 0,00448495 |
| LOC101928938 | -1,4724219 | 0,00448495 |
| LRRTM3 | -1,4379227 | 0,004625555 |

#### Unique subTCMR upregulated
None

#### Unique subTCMR downregulated
None

#### Suppl. Table 11: Pathway enrichment of unique pathways within phenotypes cABMR upregulated

| q-value | Pathway name | Pathway ID | | q-value | Pathway name | Pathway ID |
|---|---|---|---|---|---|---|
| 0.000285119194864 | cell morphogene | GO:0000904 | | 2.06236184028e | Extracellular matrix organization | R-HSA-1474244 |
| 0.00120378760213 | organic acid tran | GO:1903825 | | 2.69981741616e | Integrin cell surface interactions | R-HSA-216083 |
| 0.00242259335622 | regulation of leuk | GO:1903037 | | 0.001943241717 | ECM-receptor interaction - Homo sapiens (hum | path:hsa04512 |
| 0.00325879886425 | organic anion tra | GO:0015711 | | 0.005094284869 | Focal adhesion - Homo sapiens (human) | path:hsa04510 |
| 0.00412749453656 | neuron projectio | GO:0031175 | | 0.015656167299 | Collagen chain trimerization | R-HSA-8948216 |
| 0.00412749453656 | positive regulatio | GO:0022409 | | 0.027494663072 | Developmental Biology | R-HSA-1266738 |
| 0.00528504988426 | ion transmembra | GO:0034220 | | 0.027867224364 | Signaling by PDGF | R-HSA-186797 |
| 0.0065134620799 | regulation of ion | GO:0032412 | | 0.027867224364 | Leukocyte transendothelial migration - Homo s | path:hsa04670 |
| 0.00870633304702 | amino acid trans | GO:0006865 | | 0.036872900823 | Hypertrophic cardiomyopathy - Homo sapiens | path:hsa05410 |
| 0.00870633304702 | organic acid tran | GO:0015849 | | 0.036872900823 | Collagen formation | R-HSA-1474290 |
| 0.00998586981008 | positive regulatio | GO:0002696 | | 0.039836101060 | Nervous system development | R-HSA-9675108 |
| 0.00998586981008 | positive regulatio | GO:0051272 | | 0.041435572739 | RUNX3 Regulates Immune Response and Cell M | R-HSA-8949275 |
| 0.00998586981008 | regulation of tran | GO:0034762 | | 0.042033299356 | Collagen biosynthesis and modifying enzymes | R-HSA-1650814 |
| 0.00998586981008 | anion transport | GO:0006820 | | 0.044983214837 | Amoebiasis - Homo sapiens (human) | path:hsa05146 |
| 0.00998586981008 | regulation of cell | GO:0060284 | | 0.051061912390 | Axon guidance | R-HSA-422475 |
| 0.00998586981008 | positive regulatio | GO:0050867 | | 0.052152976264 | ABC transporters - Homo sapiens (human) | path:hsa02010 |
| 0.00998586981008 | regulation of lym | GO:0051249 | | 0.052841274507 | Assembly of collagen fibrils and other multime | R-HSA-2022090 |
| 0.00998586981008 | generation of neu | GO:0048699 | | 0.052841274507 | Cell adhesion molecules - Homo sapiens (huma | path:hsa04514 |
| 0.010484393962 | hemopoiesis | GO:0030097 | | 0.052841274507 | Arrhythmogenic right ventricular cardiomyopa | path:hsa05412 |
| 0.010484393962 | collagen-activate | GO:0038063 | | 0.052841274507 | Toxoplasmosis - Homo sapiens (human) | path:hsa05145 |
| 0.0127982301703 | regulation of ion | GO:0043269 | | 0.058387319449 | TNFR2 non-canonical NF-kB pathway | R-HSA-5668541 |
| 0.0187189271889 | gliogenesis | GO:0042063 | | 0.061664656846 | RUNX2 regulates genes involved in cell migratio | R-HSA-8941332 |
| 0.0201435329127 | regulation of leuk | GO:1902105 | | 0.066839508594 | Complement and coagulation cascades - Homo | path:hsa04610 |
| 0.0210550354206 | negative regulati | GO:0001818 | | 0.066970080945 | ECM proteoglycans | R-HSA-3000178 |
| 0.0249710798271 | regulation of anti | GO:0050854 | | 0.066970080945 | TNFs bind their physiological receptors | R-HSA-5669034 |
| 0.0260644631891 | cellular response | GO:0071345 | | 0.066970080945 | PI3K-Akt signaling pathway - Homo sapiens (hu | path:hsa04151 |
| 0.0324698808341 | negative regulati | GO:0045596 | | 0.066970080945 | Trafficking of AMPA receptors | R-HSA-399719 |
| 0.0324698808341 | T-helper 1 type in | GO:0042088 | | 0.066970080945 | Glutamate binding, activation of AMPA recepto | R-HSA-399721 |
| 0.0374521833396 | lipid phosphoryla | GO:0046834 | | 0.066970080945 | ABC-family proteins mediated transport | R-HSA-382556 |
| 0.0374521833396 | regulation of nerv | GO:0051960 | | 0.066970080945 | Small cell lung cancer - Homo sapiens (human) | path:hsa05222 |
| 0.0374521833396 | lacrimal gland de | GO:0032808 | | 0.066970080945 | RUNX1 and FOXP3 control the development of | R-HSA-8877330 |
| 0.0374521833396 | glutathione trans | GO:0034635 | | 0.066970080945 | Activation of SMO | R-HSA-5635838 |
| 0.0428033400946 | epithelial cell apo | GO:1904019 | | 0.072951631441 | Dilated cardiomyopathy - Homo sapiens (huma | path:hsa05414 |
| 0.0428033400946 | positive regulatio | GO:0010634 | | 0.072951631441 | Amino acid transport across the plasma memb | R-HSA-352230 |
| 0.0428033400946 | positive regulatio | GO:0045597 | | 0.083501385809 | Semaphorin interactions | R-HSA-373755 |
| 0.0458051011639 | negative regulati | GO:2000181 | | | | |
| 0.0458051011639 | negative regulati | GO:1901380 | | | | |
| 0.0458051011639 | negative regulati | GO:0051961 | | | | |
| 0.0475290699376 | antigen receptor- | GO:0050851 | | | | |
| 0.0479244644684 | negative regulati | GO:0051129 | | | | |
| 0.0479244644684 | positive regulatio | GO:0050778 | | | | |
| 0.0498819988935 | positive regulatio | GO:1903708 | | | | |
| 0.0563670620724 | T cell chemotaxis | GO:0010818 | | | | |
| 0.0623350033368 | negative regulati | GO:1901343 | | | | |
| 0.0623350033368 | mesenchymal ste | GO:0072497 | | | | |
| 0.0664648133344 | negative regulati | GO:0051271 | | | | |
| 0.0664648133344 | regulation of type | GO:0002828 | | | | |
| 0.0664648133344 | female gonad dev | GO:0008585 | | | | |
| 0.0664648133344 | anterograde tran | GO:0098916 | | | | |
| 0.0664648133344 | nucleobase catab | GO:0046113 | | | | |
| 0.0664648133344 | negative regulati | GO:1903011 | | | | |
| 0.0664648133344 | visual system dev | GO:0150063 | | | | |
| 0.0664648133344 | negative regulati | GO:0045833 | | | | |
| 0.0664648133344 | modified amino a | GO:0072337 | | | | |
| 0.069985383017 | cellular response | GO:0071230 | | | | |
| 0.0700693232767 | erythrocyte devel | GO:0048821 | | | | |
| 0.0759944824756 | positive regulatio | GO:0002687 | | | | |
| 0.0759944824756 | cardiac muscle ce | GO:0086003 | | | | |
| 0.0759944824756 | regulation of lym | GO:2000401 | | | | |
| 0.0759944824756 | regulation of cell | GO:0002691 | | | | |
| 0.0759944824756 | regulation of glut | GO:1900449 | | | | |
| 0.0759944824756 | vitamin D biosyn | GO:0042368 | | | | |
| 0.0759944824756 | regulation of vita | GO:0030656 | | | | |
| 0.0759944824756 | negative regulati | GO:0032351 | | | | |
| 0.0759944824756 | keratan sulfate ca | GO:0042340 | | | | |
| 0.0759944824756 | oligopeptide tran | GO:0035672 | | | | |

#### Unique cABMR downregulated

| q-value | Pathway name | Pathway ID | | q-value | Pathway name | Pathway ID |
|---|---|---|---|---|---|---|
| 0.0005410384 | very-low-densi | GO:0034372 | | 4.8431362153 | Metallothionei | R-HSA-5661231 |
| 0.0005410384 | response to co | GO:0046688 | | 6.6241258271 | Response to m | R-HSA-5660526 |
| 0.0005410384 | regulation of li | GO:0060191 | | 0.0005604854 | Metabolism | R-HSA-1430728 |
| 0.0011097681 | response to zin | GO:0010043 | | 0.0009815043 | Mineral absorp | path:hsa04978 |
| 0.0011570561 | response to ca | GO:0046686 | | 0.0059084903 | Metabolism of | R-HSA-2408508 |
| 0.0029310141 | fatty acid biosy | GO:0006633 | | 0.0143833722 | Plasma lipopro | R-HSA-174824 |
| 0.0029310141 | protein-lipid c | GO:0065005 | | 0.0156491565 | Plasma lipopro | R-HSA-8963898 |
| 0.0029310141 | protein-lipid c | GO:0034368 | | 0.0197176790 | PPAR signaling | path:hsa03320 |
| 0.0044842755 | response to ery | GO:0036017 | | 0.0241800378 | Plasma lipopro | R-HSA-8963899 |
| 0.0051556395 | carboxylic acic | GO:0019752 | | 0.0241800378 | Metabolism of | R-HSA-196854 |
| 0.0052724063 | regulation of sy | GO:0003044 | | 0.0241800378 | Respiratory ele | R-HSA-611105 |
| 0.0053804172 | neutral lipid ca | GO:0046461 | | 0.0429931973 | Respiratory ele | R-HSA-163200 |
| 0.0053804172 | carboxylic acic | GO:0046394 | | 0.0434241275 | Glycine, serine | path:hsa00260 |
| 0.0067432826 | positive regula | GO:0003084 | | 0.0434241275 | AMPK signaling | path:hsa04152 |
| 0.0077885860 | acylglycerol m | GO:0006639 | | | | |
| 0.0097830745 | lipid homeosta | GO:0055088 | | | | |
| 0.0100418990 | organic hydrox | GO:0015850 | | | | |
| 0.0100418990 | entry into host | GO:0044409 | | | | |
| 0.0100418990 | entry into cell | GO:0051806 | | | | |
| 0.0121358683 | glycerolipid ca | GO:0046503 | | | | |
| 0.0183531293 | establishment | GO:0051905 | | | | |
| 0.0183531293 | cellular respon | GO:0071248 | | | | |
| 0.0188592451 | regulation of li | GO:0046890 | | | | |
| 0.0188592451 | pigment granu | GO:0051875 | | | | |
| 0.0190834127 | regulation of c | GO:0043255 | | | | |
| 0.0199155379 | cellular cation | GO:0030003 | | | | |
| 0.0202582041 | cellular respon | GO:0071549 | | | | |
| 0.0202582041 | ribonucleoside | GO:0009119 | | | | |
| 0.0250711510 | acute inflamm | GO:0002526 | | | | |
| 0.0250711510 | lipid transport | GO:0006869 | | | | |
| 0.0293002428 | regulation of tr | GO:0090207 | | | | |
| 0.0456121947 | regulation of c | GO:0010675 | | | | |
| 0.0485952844 | positive regula | GO:0062013 | | | | |
| 0.0487382813 | ion homeostas | GO:0050801 | | | | |

**Unique clinTCMR upregulated**

| q-value | Pathway name | Pathway ID | | q-value | Pathway name | Pathway ID |
|---|---|---|---|---|---|---|
| 4.88340888096e-09 | cellular respon | GO:0071345 | | 2.0520684235 | Immune Syster | R-HSA-168256 |
| 4.88340888096e-09 | response to int | GO:0034341 | | 3.4921859285 | Interferon Sign | R-HSA-913531 |
| 2.20627151982e-07 | positive regula | GO:0050778 | | 5.8505561622 | Cytokine Signa | R-HSA-1280215 |
| 2.20627151982e-07 | neutrophil me | GO:0002446 | | 8.0842495929 | Innate Immune | R-HSA-168249 |
| 2.00302548216e-06 | positive regula | GO:0022409 | | 3.0019158584 | Neutrophil deg | R-HSA-6798695 |
| 2.31876741428e-06 | interferon-gam | GO:0060333 | | 2.0680558545 | Interferon gam | R-HSA-877300 |
| 5.12314444984e-06 | regulation of T | GO:0002664 | | 7.8684881955 | Epstein-Barr vi | path:hsa05169 |
| 5.1594782451e-06 | regulation of ir | GO:0050727 | | 0.0001253624 | Staphylococcu | path:hsa05150 |
| 6.23170481053e-06 | regulation of ir | GO:0032649 | | 0.0001982883 | G1/S Transitior | R-HSA-69206 |
| 6.23170481053e-06 | regulation of d | GO:0031347 | | 0.0001982883 | Chagas disease | path:hsa05142 |
| 7.47355828218e-06 | regulation of le | GO:1903037 | | 0.0002116705 | Mitotic G1 pha | R-HSA-453279 |
| 1.20868679873e-05 | negative regula | GO:0031348 | | 0.0006378962 | Human T-cell le | path:hsa05166 |
| 1.25300251312e-05 | regulation of ly | GO:0002911 | | 0.0009372165 | Interferon alph | R-HSA-909733 |
| 1.25300251312e-05 | hemopoiesis | GO:0030097 | | 0.0014657734 | Antigen proces | path:hsa04612 |
| 3.18800891029e-05 | negative regula | GO:2000524 | | 0.0014927236 | Th17 cell differ | path:hsa04659 |
| 5.04602615329e-05 | antigen recept | GO:0050851 | | 0.0015102586 | B cell receptor | path:hsa04662 |
| 8.8142271708e-05 | positive regula | GO:0002645 | | 0.0018554564 | Immunoregula | R-HSA-198933 |
| 9.45175951428e-05 | regulation of o | GO:0033043 | | 0.0018554564 | Adaptive Immu | R-HSA-1280218 |
| 0.00014711230923 | regulation of le | GO:1902105 | | 0.0018554564 | Primary immu | path:hsa05340 |
| 0.000156862673927 | negative regula | GO:0001818 | | 0.0022659548 | Phagosome - H | path:hsa04145 |
| 0.000157848139556 | positive regula | GO:0050867 | | 0.0027681985 | Th1 and Th2 ce | path:hsa04658 |
| 0.00017813112017 | regulation of ir | GO:0032675 | | 0.0027681985 | Inflammatory b | path:hsa05321 |
| 0.000259350124814 | T cell mediatec | GO:0002456 | | 0.0028441830 | Generation of s | R-HSA-202433 |
| 0.000282196349999 | positive regula | GO:0001819 | | 0.0034940092 | Formyl peptide | R-HSA-444473 |
| 0.000380782125529 | positive regula | GO:0002696 | | 0.0044565693 | TCR signaling | R-HSA-202403 |
| 0.000527456389065 | regulation of ir | GO:0032655 | | 0.0044565693 | Cell Cycle, Mitc | R-HSA-69278 |
| 0.000585886581381 | regulation of le | GO:0070663 | | 0.0047540056 | Transcriptiona | R-HSA-9616222 |
| 0.000588565894883 | negative regula | GO:0032102 | | 0.0051411064 | Leishmaniasis - | path:hsa05140 |
| 0.000865060906489 | positive regula | GO:0033630 | | 0.0052044268 | Unwinding of D | R-HSA-176974 |
| 0.00125754760563 | leukocyte adhe | GO:0061756 | | 0.0083506528 | DNA strand elo | R-HSA-69190 |
| 0.00127500691775 | regulation of ly | GO:0051249 | | 0.0089929597 | Kaposi sarcoma | path:hsa05167 |
| 0.00134233567682 | positive regula | GO:0002699 | | 0.0101407246 | Viral myocardi | path:hsa05416 |
| 0.00139333790445 | positive regula | GO:0032103 | | 0.0126189749 | Osteoclast diffe | path:hsa04380 |
| 0.00139333790445 | interleukin-10- | GO:0140105 | | 0.0127296096 | Allograft reject | path:hsa05330 |
| 0.00142306959513 | regulation of vi | GO:0050792 | | 0.0127296096 | G1/S-Specific T | R-HSA-69205 |
| 0.00144597955418 | negative regula | GO:0048525 | | 0.0155032316 | Systemic lupus | path:hsa05322 |
| 0.00155546074194 | dendritic cell c | GO:0097028 | | 0.0155032316 | TFAP2 (AP-2) fa | R-HSA-8866911 |
| 0.00208725628155 | negative regula | GO:0050866 | | 0.0155032316 | Interleukin-2 fa | R-HSA-451927 |
| 0.00208725628155 | negative regula | GO:0050777 | | 0.0165773983 | Bladder cancer | path:hsa05219 |
| 0.00233500536725 | negative regula | GO:0050858 | | 0.0176921800 | Graft-versus-ho | path:hsa05332 |
| 0.0023879127824 | regulation of cy | GO:0002718 | | 0.0180446038 | S Phase | R-HSA-69242 |
| 0.0023879127824 | positive regula | GO:1903708 | | 0.0180446038 | Type I diabetes | path:hsa04940 |
| 0.00319400016381 | negative regula | GO:1903035 | | 0.0180446038 | Cell Cycle | R-HSA-1640170 |
| 0.00322341404163 | regulation of ir | GO:0032663 | | 0.0191328430 | Interleukin-3, I | R-HSA-512988 |
| 0.0033411726914 | viral genome re | GO:0019079 | | 0.0219598117 | Cell adhesion n | path:hsa04514 |
| 0.00364465826331 | regulation of p | GO:0043067 | | 0.0219598117 | Synthesis of DN | R-HSA-69239 |
| 0.00366759796983 | regulation of c | GO:0002691 | | 0.0258625384 | Scavenging by ( | R-HSA-3000471 |
| 0.00366759796983 | ERK1 and ERK2 | GO:0070371 | | 0.0275048997 | DNA Replicatio | R-HSA-69306 |
| 0.00376560814731 | type I interfero | GO:0060337 | | 0.0275048997 | Peptide hormo | R-HSA-2980736 |
| 0.00435582018888 | negative regula | GO:0002695 | | 0.0304713417 | Autoimmune t | path:hsa05320 |
| 0.00443298860613 | negative regula | GO:0022408 | | 0.0304713417 | Downstream TC | R-HSA-202424 |
| 0.00484644536627 | regulation of re | GO:1903034 | | 0.0315299121 | Complement a | path:hsa04610 |
| 0.00484644536627 | response to typ | GO:0034340 | | 0.0329953299 | Interleukin rec | R-HSA-912526 |
| 0.0051319340544 | negative regula | GO:0010648 | | 0.0333367289 | Cell cycle - Hor | path:hsa04110 |
| 0.00568628683072 | regulation of ir | GO:1902531 | | 0.0352873953 | Translocation ( | R-HSA-202430 |
| 0.00608263880654 | positive regula | GO:0010942 | | 0.0409416754 | Rheumatoid ar | path:hsa05323 |
| 0.0062788298712 | regulation of p | GO:0002700 | | 0.0414351244 | ISG15 antiviral | R-HSA-1169408 |
| 0.0062788298712 | negative regula | GO:0051129 | | 0.0428939543 | Phosphorylatio | R-HSA-202427 |
| 0.00675441097811 | viral life cycle | GO:0019058 | | 0.0428939543 | Termination of | R-HSA-5656169 |
| 0.00675441097811 | chronic inflam | GO:0002544 | | 0.0428939543 | Fc gamma R-m | path:hsa04666 |
| 0.00675441097811 | positive regula | GO:0031349 | | 0.0428939543 | Yersinia infecti | path:hsa05135 |
| 0.00676007764889 | regulation of p | GO:0051246 | | 0.0428939543 | Tuberculosis - I | path:hsa05152 |
| 0.00687178909081 | regulation of a | GO:0032956 | | 0.0428939543 | PD-1 signaling | R-HSA-389948 |
| 0.00711590616123 | positive regula | GO:0009967 | | 0.0428939543 | Human cytome | path:hsa05163 |
| 0.00711590616123 | negative regula | GO:0009968 | | 0.0428939543 | Hematopoietic | path:hsa04640 |
| 0.00715858545378 | regulation of n | GO:0042269 | | 0.0428939543 | NOD-like recep | path:hsa04621 |
| 0.00715858545378 | regulation of ir | GO:0032674 | | 0.0428939543 | Pyrimidine salv | R-HSA-73614 |
| 0.00734488137169 | DNA biosynthe | GO:0071897 | | 0.0428939543 | Free fatty acids | R-HSA-400451 |
| 0.00739141327038 | positive regula | GO:0010647 | | 0.0428939543 | Dissolution of I | R-HSA-75205 |
| 0.00751420570836 | myeloid leuko | GO:0002573 | | 0.0428939543 | The role of GTS | R-HSA-8852276 |
| 0.00751420570836 | DNA recombin | GO:0006310 | | 0.0428939543 | Synthesis, secr | R-HSA-422085 |
| 0.00770134779911 | regulation of fi | GO:0048145 | | 0.0428939543 | Activation of tl | R-HSA-68962 |
| 0.00839751642838 | antigen proces | GO:0042590 | | 0.0444126339 | Viral protein ir | path:hsa04061 |
| 0.00839751642838 | regulation of le | GO:0002703 | | 0.0449670685 | Coronavirus di | path:hsa05171 |
| 0.00851027563959 | negative regula | GO:0001911 | | 0.0482436328 | WNT5:FZD7-m | R-HSA-9673324 |
| 0.0103768110551 | actin polymeri | GO:0008154 | | 0.0482436328 | Killing mechan | R-HSA-9664420 |
| 0.0105272832757 | regulation of a | GO:0002819 | | 0.0494329109 | T cell receptor | path:hsa04660 |
| 0.0105948164487 | eosinophil mig | GO:0072677 | | | | |
| 0.0108382189515 | regulation of c | GO:0032268 | | | | |
| 0.0134715388177 | positive regula | GO:0001912 | | | | |
| 0.0134715388177 | regulation of ir | GO:0032653 | | | | |
| 0.0142181311829 | ruffle organizat | GO:0031529 | | | | |
| 0.0154982399572 | positive regula | GO:0031334 | | | | |
| 0.0162169790345 | regulation of ir | GO:0045088 | | | | |
| 0.0162349317467 | regulation of a | GO:0008064 | | | | |
| 0.0162349317467 | positive regula | GO:0002687 | | | | |
| 0.0184846688824 | phosphorylatio | GO:0016310 | | | | |
| 0.0197925503218 | regulation of ir | GO:0032673 | | | | |
| 0.0201037817953 | regulation of a | GO:0050854 | | | | |
| 0.0208298095248 | neutrophil che | GO:0030593 | | | | |
| 0.0209629731684 | regulation of c | GO:0022604 | | | | |
| 0.0240047834014 | regulation of m | GO:0045637 | | | | |
| 0.0240047834014 | negative regula | GO:1903707 | | | | |
| 0.0247552335191 | positive regula | GO:0008284 | | | | |
| 0.0252643864573 | regulation of c | GO:0033632 | | | | |
| 0.0252643864573 | pyrimidine nuc | GO:0043097 | | | | |
| 0.0252643864573 | mononuclear c | GO:1903131 | | | | |
| 0.0252643864573 | epithelial cell a | GO:1904019 | | | | |
| 0.0290383036759 | positive regula | GO:0048524 | | | | |
| 0.031027571383 | regulation of T | GO:0001914 | | | | |
| 0.032399139402 | cellular cation | GO:0030003 | | | | |
| 0.032399139402 | positive regula | GO:0090184 | | | | |
| 0.032399139402 | positive regula | GO:0031325 | | | | |
| 0.032399139402 | leukocyte aggr | GO:0070486 | | | | |
| 0.032399139402 | regulation of p | GO:0071801 | | | | |
| 0.032399139402 | acid secretion | GO:0046717 | | | | |
| 0.0332213691583 | regulation of le | GO:0002688 | | | | |
| 0.0340060357784 | DNA replicatio | GO:0006260 | | | | |
| 0.0358279510866 | regulation of p | GO:0032271 | | | | |
| 0.0358279510866 | response to int | GO:0070672 | | | | |
| 0.0358279510866 | regulation of c | GO:1901722 | | | | |
| 0.0359786935658 | negative regula | GO:0002698 | | | | |
| 0.0359786935658 | neutrophil mig | GO:1990266 | | | | |
| 0.0360113063325 | ion homeostas | GO:0050801 | | | | |
| 0.0375638131962 | regulation of p | GO:0051174 | | | | |
| 0.0392944482584 | macrophage cy | GO:0010934 | | | | |

##### Unique clinTCMR downregulated

| q-value | Pathway name | Pathway ID | | q-value | Pathway name | Pathway ID |
|---|---|---|---|---|---|---|
| 0.0004410372 | carboxylic acid | GO:0019752 | | 6.0522370177 | Metabolism | R-HSA-143072 |
| 0.0006160801 | terpenoid meta | GO:0006721 | | 2.5472412354 | Biological oxid | R-HSA-211859 |
| 0.0026301467 | branched-chai | GO:0009083 | | 1.1830624000 | Phase I - Functi | R-HSA-211945 |
| 0.0026301467 | ethanol metab | GO:0006067 | | 0.0003927997 | Drug metaboli | path:hsa00982 |
| 0.0089213366 | lipid hydroxyla | GO:0002933 | | 0.0005916099 | Metabolism of | R-HSA-556833 |
| 0.0089213366 | carboxylic acid | GO:0046394 | | 0.0006000015 | Chemical carci | path:hsa05204 |
| 0.0119636828 | thyroid hormo | GO:0070327 | | 0.0015421734 | Branched-chai | R-HSA-70895 |
| 0.0119636828 | alpha-amino ac | GO:1901606 | | 0.0015556689 | Cytochrome P4 | R-HSA-211897 |
| 0.0120536199 | carboxylic acid | GO:0046395 | | 0.0058922511 | Synthesis of ep | R-HSA-214267 |
| 0.0210229890 | arachidonic ac | GO:0019369 | | 0.0066560915 | Synthesis of (16 | R-HSA-214281 |
| 0.0390500010 | exogenous dru | GO:0042738 | | 0.0066560915 | SLC-mediated t | R-HSA-425407 |
| 0.0390500010 | fatty acid biosy | GO:0006633 | | 0.0066560915 | Metabolism of | R-HSA-71291 |
| 0.0447171428 | steroid catabol | GO:0006706 | | 0.0066560915 | Fatty acid degr | path:hsa00071 |
| 0.0640193041 | nucleotide me | GO:0009117 | | 0.0066560915 | Bile acid and bi | R-HSA-194068 |
| 0.0655661987 | protein-lipid c | GO:0034368 | | 0.0068827200 | CYP2E1 reactic | R-HSA-211999 |
| 0.0724262011 | bile acid biosy | GO:0006699 | | 0.0071376299 | Ethanol oxidat | R-HSA-71384 |
| | | | | 0.0071376299 | Transport of or | R-HSA-879518 |
| | | | | 0.0071376299 | Valine, leucine | path:hsa00280 |
| | | | | 0.0128633368 | Arachidonic ac | R-HSA-214275 |
| | | | | 0.0174350997 | Retinol metabo | path:hsa00830 |
| | | | | 0.0182218400 | Xenobiotics | R-HSA-211981 |
| | | | | 0.0216609529 | Metabolism of | path:hsa00980 |
| | | | | 0.0253696941 | Plasma lipopro | R-HSA-896389 |
| | | | | 0.0301037110 | Linoleic acid m | path:hsa00591 |
| | | | | 0.0308724088 | Synthesis of bil | R-HSA-192105 |
| | | | | 0.0311411118 | Fatty acid meta | R-HSA-897886 |
| | | | | 0.0312448347 | Glyoxylate met | R-HSA-389661 |
| | | | | 0.0312448347 | Amino acid tra | R-HSA-352230 |

##### Suppl. Table 12: Shared differentially expressed genes of cABMR and clinTCMR

Adjustment of p-value by false discovery rate.

##### Shared cABMR and clinTCMR upregulated

| gene name | log2fold change cABMR | padj cABMR | log2fold change clinTCMR | padj clinTCMR |
|---|---|---|---|---|
| GLS | 2,015723806 | 4,81871E-08 | 1,825184633 | 1,55517E-10 |
| C1orf204 | 2,713994837 | 6,60574E-08 | 2,695539432 | 2,06382E-12 |
| KCNA3 | 2,928053701 | 1,77857E-07 | 1,456603742 | 0,007814331 |
| UBD | 3,746335609 | 1,77857E-07 | 4,211163259 | 7,42814E-14 |
| APOBEC4 | 1,921310999 | 1,34775E-06 | 1,646890694 | 3,6085E-07 |
| COA1 | 1,532955726 | 1,34775E-06 | 1,095172415 | 2,78666E-05 |
| DEXI | 1,401015699 | 1,34775E-06 | 1,098148325 | 3,10264E-06 |
| LOC102723401 | 3,956344279 | 1,40505E-06 | 1,698980117 | 0,03602246 |
| NRSN1 | 4,470939161 | 1,98722E-06 | 2,273502254 | 0,010219372 |
| RP11-77C3.3 | 3,144271898 | 2,02652E-06 | 2,768774993 | 2,0985E-07 |
| ARSG | 1,725384899 | 2,63182E-06 | 1,021371947 | 0,002330577 |
| FAM196B | 1,4789798 | 3,36553E-06 | 1,47280772 | 2,37913E-09 |

| | | | | |
|---|---|---|---|---|
| LOC102724536 | 1,725133873 | 3,36553E-06 | 1,277499154 | 3,39513E-05 |
| PHOSPHO1 | 1,700135898 | 3,36553E-06 | 1,360466126 | 4,07987E-06 |
| ZBED2 | 2,146447248 | 3,848E-06 | 2,033442374 | 4,13462E-08 |
| CELF2-AS1 | 1,654153897 | 3,95917E-06 | 1,298259031 | 8,81612E-06 |
| FAT1 | 2,018137379 | 4,44881E-06 | 1,795574901 | 3,24799E-07 |
| HLA-F-AS1 | 2,2089382 | 7,83497E-06 | 1,710556475 | 3,38325E-05 |
| IRF4 | 4,394940508 | 9,07596E-06 | 2,302995314 | 0,02208129 |
| TMEM75 | 1,943556929 | 9,13518E-06 | 1,176350459 | 0,002968025 |
| TAPSAR1 | 1,736729846 | 1,60326E-05 | 1,83420354 | 2,37913E-09 |
| MUC12 | 2,535535868 | 1,68827E-05 | 1,783110084 | 0,000395017 |
| LIMK2 | 1,403794159 | 2,47164E-05 | 1,320120131 | 3,74918E-07 |
| TLDC2 | 1,310437541 | 2,47164E-05 | 1,247552491 | 2,32733E-07 |
| IL32 | 2,002708374 | 2,6018E-05 | 2,593825535 | 1,23872E-13 |
| LOC101928152 | 2,216441366 | 2,6018E-05 | 1,667802473 | 0,000161581 |
| LOC101927954 | 1,634551248 | 3,17772E-05 | 1,656686134 | 6,17481E-08 |
| AQP4-AS1 | 2,007767544 | 3,74818E-05 | 1,013088462 | 0,02608853 |
| EFEMP1 | 2,786666092 | 3,7778E-05 | 1,484953791 | 0,016934711 |
| CXCL10 | 4,590354477 | 4,26497E-05 | 4,470220166 | 1,53373E-06 |
| PIGF | 1,357242447 | 4,626E-05 | 1,099711538 | 6,55049E-05 |
| COL1A1 | 2,025052754 | 5,18423E-05 | 1,659889117 | 3,63816E-05 |
| FAM115C | 1,799899495 | 5,72062E-05 | 1,309901572 | 0,000338429 |
| VIM-AS1 | 1,427232915 | 6,09991E-05 | 1,281890646 | 4,03011E-06 |
| BCAT1 | 3,662833608 | 7,17608E-05 | 2,818134295 | 0,000813216 |
| JAK3 | 2,346984977 | 0,00010418 | 1,459873347 | 0,006935407 |
| LINC00900 | 4,149665906 | 0,00010418 | 2,545416513 | 0,009732526 |
| LOC101928173 | 2,273532745 | 0,00010418 | 1,360910782 | 0,012029297 |
| SPOCK2 | 2,216993085 | 0,00010418 | 1,863214827 | 4,47617E-05 |
| DTNA | 2,655007156 | 0,000135651 | 2,59741863 | 3,80145E-06 |
| ITGA6 | 1,550884374 | 0,000135651 | 1,065916316 | 0,002684113 |
| CXCL9 | 3,586416692 | 0,000136978 | 3,475335421 | 2,72558E-06 |
| COL1A2 | 1,698719435 | 0,000169846 | 1,275243897 | 0,000536106 |
| LOC101927734 | 3,668994054 | 0,000171879 | 1,855991641 | 0,042390315 |
| SLC7A6OS | 1,408416765 | 0,000171879 | 1,37166706 | 3,57152E-06 |
| IL21R-AS1 | 2,086313471 | 0,000183056 | 2,580956698 | 6,23138E-10 |
| LOC100294362 | 1,245563464 | 0,000189564 | 1,345861723 | 1,21449E-07 |
| APCDD1L | 1,805948181 | 0,000189659 | 1,395581177 | 0,000813216 |
| STAT1 | 1,584458033 | 0,00021334 | 1,708812431 | 1,8617E-07 |
| BYSL | 1,670188885 | 0,000215093 | 1,147404667 | 0,00352245 |
| RP11-436I24.1 | 1,44749902 | 0,000215093 | 1,274980494 | 4,81497E-05 |
| FMNL1 | 1,350227918 | 0,000221542 | 1,115398087 | 0,000205929 |
| LOC101927085 | 2,6240642 | 0,000221542 | 2,59338267 | 5,3938E-06 |
| FSTL3 | 1,999701312 | 0,000223015 | 1,994178114 | 3,61232E-06 |
| RUNX3 | 1,876823074 | 0,000236944 | 1,86788899 | 4,17587E-06 |
| SLC4A5 | 1,670903583 | 0,000257895 | 1,892578109 | 6,63595E-08 |
| PRDM8 | 3,413040942 | 0,000274417 | 2,411057035 | 0,006541585 |
| GSG2 | 3,114597943 | 0,000283051 | 2,305386746 | 0,002351925 |
| TMC8 | 1,602152425 | 0,000351153 | 1,468284043 | 4,01484E-05 |

| | | | | |
|---|---|---|---|---|
| LOC100128653 | 1,027849486 | 0,000352635 | 1,041138957 | 2,72558E-06 |
| TMEM120B | 1,260920634 | 0,000360821 | 1,17702631 | 2,52516E-05 |
| ATP2A3 | 1,900618274 | 0,000386073 | 1,24187741 | 0,008953561 |
| LOC101927848 | 1,969624851 | 0,000395353 | 2,556662501 | 5,33914E-10 |
| BCL11B | 2,524919442 | 0,000397849 | 2,20013825 | 0,000198198 |
| C11orf45 | 2,516354379 | 0,000417815 | 2,250925777 | 8,50063E-05 |
| B3GNT5 | 3,073743399 | 0,000469374 | 2,493412592 | 0,001242974 |
| TRIM17 | 1,246738667 | 0,000469374 | 1,069596859 | 0,000205929 |
| CCND1 | 1,350368242 | 0,00047886 | 1,096658046 | 0,000456285 |
| AC007461.2 | 2,678289188 | 0,000529295 | 1,597193715 | 0,029154731 |
| RP11-401P9.5 | 1,595284424 | 0,000532369 | 1,687261531 | 2,38488E-06 |
| BHLHE41 | 2,790126363 | 0,000598201 | 1,88149368 | 0,020390691 |
| XYLT1 | 1,930991769 | 0,000598201 | 1,486374206 | 0,001405788 |
| THBS2 | 1,707086545 | 0,000611455 | 1,405790634 | 0,000514418 |
| GRAP2 | 3,999215077 | 0,00066364 | 2,33711536 | 0,043211259 |
| LYZ | 1,7249586 | 0,000731724 | 1,262709056 | 0,002753957 |
| LOC101928123 | 2,421010648 | 0,000790574 | 1,796461883 | 0,003345106 |
| AVPR2 | 1,832276263 | 0,000807288 | 1,7269457 | 0,000117868 |
| CD48 | 1,99881676 | 0,000828779 | 1,293699178 | 0,022334238 |
| ALK | 1,882852354 | 0,000896516 | 1,237718575 | 0,013490702 |
| STK10 | 1,00936074 | 0,000965709 | 1,040360252 | 1,48989E-05 |
| DOCK2 | 1,325623459 | 0,001057926 | 1,43219369 | 4,17587E-06 |
| CERKL | 1,278070229 | 0,001062372 | 1,197085547 | 9,80335E-05 |
| TMEM132A | 1,370092463 | 0,001119339 | 1,067803959 | 0,002276109 |
| ENTPD1-AS1 | 1,592628407 | 0,001163472 | 1,564463824 | 4,81497E-05 |
| ZNF80 | 2,186425562 | 0,00136474 | 1,325789042 | 0,034541069 |
| CD6 | 2,026609664 | 0,001660242 | 1,933867271 | 0,000295181 |
| ANXA2 | 1,340150496 | 0,001782887 | 1,096878716 | 0,001659977 |
| STK17B | 1,805353409 | 0,00184127 | 1,512165907 | 0,002346633 |
| LOC642441 | 1,233904389 | 0,001897558 | 1,216746252 | 8,55633E-05 |
| SRGN | 1,544516736 | 0,002006952 | 1,287452338 | 0,002567327 |
| SCML4 | 2,776493306 | 0,002206418 | 2,378779529 | 0,003263404 |
| LOC101929732 | 1,384522357 | 0,002234858 | 1,598336358 | 2,72558E-06 |
| DOCK11 | 1,470898497 | 0,002259388 | 1,093389276 | 0,009695169 |
| LINGO3 | 1,737989475 | 0,002266994 | 1,465778786 | 0,001242974 |
| TNFRSF12A | 3,091600536 | 0,002290885 | 2,606834005 | 0,001718517 |
| TCTEX1D4 | 1,617982717 | 0,002435195 | 1,634803355 | 8,99128E-05 |
| MIAT | 1,573597245 | 0,002446323 | 1,031108789 | 0,023994277 |
| PRRT2 | 1,653000471 | 0,002808164 | 2,061250767 | 3,83799E-07 |
| PTPRC | 1,15263336 | 0,003238219 | 1,008906613 | 0,00109324 |
| CDKN2B | 1,680715352 | 0,003319852 | 1,292238589 | 0,00878418 |
| C1orf200 | 1,25204513 | 0,003378269 | 1,084316328 | 0,001405788 |
| ENAM | 2,60066559 | 0,003401495 | 1,938440024 | 0,017785138 |
| LOC102724466 | 1,159079184 | 0,003425513 | 1,063041089 | 0,000551932 |
| DOK1 | 1,575590969 | 0,003542415 | 1,215567168 | 0,011349055 |
| BIRC3 | 1,789868583 | 0,00357376 | 1,811273566 | 0,000137157 |
| IKZF3 | 1,548801889 | 0,003610328 | 1,894512582 | 2,09806E-06 |

| | | | | |
|---|---|---|---|---|
| HLA-DOA | 1,471257023 | 0,003705605 | 1,088854115 | 0,012494092 |
| ZAP70 | 1,848051503 | 0,003705605 | 1,676791316 | 0,001321906 |
| AC002480.3 | 3,848089693 | 0,003761997 | 2,850618626 | 0,013635806 |
| MICAL1 | 1,481800353 | 0,003785234 | 1,270738475 | 0,002281713 |
| RP11-277P12.20 | 1,537134758 | 0,004081151 | 1,424254969 | 0,000619121 |
| ACAN | 3,642765147 | 0,004166834 | 2,468445397 | 0,041480805 |
| SLAMF7 | 1,983944526 | 0,004637094 | 2,143246018 | 0,000105077 |
| TAP1 | 1,126213484 | 0,004946203 | 1,362355521 | 3,61232E-06 |
| CCDC80 | 2,161840422 | 0,005067003 | 1,771358032 | 0,00524481 |
| NFKB2 | 1,069736167 | 0,005280242 | 1,093099189 | 0,000237366 |
| VCAN | 2,036286785 | 0,005597984 | 2,078220319 | 0,000325216 |
| MTRNR2L8 | 1,681303592 | 0,006317444 | 1,327963735 | 0,01018737 |
| PIN4P1 | 1,556888894 | 0,006392205 | 1,665802981 | 0,000192519 |
| C6orf147 | 2,260292941 | 0,006787332 | 2,24072391 | 0,00117972 |
| KCNS1 | 1,845324855 | 0,006907773 | 1,853640896 | 0,000751592 |
| LOC101927255 | 2,354755242 | 0,006907773 | 2,083051953 | 0,00373717 |
| LOC101927256 | 2,244864721 | 0,006958953 | 2,453994971 | 0,000152193 |
| C16orf54 | 2,326182576 | 0,007362006 | 1,699177359 | 0,045083103 |
| NLRC5 | 1,085390211 | 0,00749908 | 1,121982751 | 0,000289922 |
| PAQR8 | 1,423647223 | 0,007548149 | 1,086823117 | 0,015937027 |
| FRMD5 | 1,392069325 | 0,007670825 | 1,798139314 | 2,72558E-06 |
| PRF1 | 1,748350278 | 0,007670825 | 1,409217246 | 0,011769068 |
| SLC9A5 | 1,872581225 | 0,008243573 | 1,558111182 | 0,007214057 |
| HLA-F | 1,265797937 | 0,008257124 | 1,029587132 | 0,007140054 |
| FCHSD1 | 1,240997376 | 0,008793124 | 1,193200519 | 0,001231954 |
| PPP1R16B | 1,088697588 | 0,008793124 | 1,000423496 | 0,002722006 |
| TIMP1 | 1,247157278 | 0,009105799 | 1,232541107 | 0,000781885 |
| CAP2 | 2,561958614 | 0,00922475 | 1,779815993 | 0,042999207 |
| PKMYT1 | 2,327912218 | 0,00980662 | 1,795854615 | 0,029826431 |
| GPNMB | 1,429326709 | 0,010123211 | 1,869664856 | 4,03011E-06 |
| AC145110.1 | 3,168929148 | 0,010140231 | 3,630913696 | 0,000240287 |
| IL7R | 1,820154403 | 0,010444506 | 1,607960687 | 0,007176613 |
| FER1L4 | 2,541351959 | 0,010607128 | 2,083323486 | 0,021930214 |
| PDCD1 | 2,148654132 | 0,010607128 | 1,60078045 | 0,023743078 |
| LOC100996318 | 2,925465325 | 0,010610884 | 3,007270716 | 0,001030667 |
| GLI1 | 1,411677872 | 0,010714224 | 1,43379711 | 0,00073574 |
| BTG2 | 1,605395562 | 0,011208642 | 1,580960079 | 0,00211718 |
| PSTPIP1 | 1,784348668 | 0,011574345 | 1,356156376 | 0,029215496 |
| SLA | 1,268057169 | 0,011692796 | 1,263462116 | 0,000776325 |
| IL24 | 1,742847286 | 0,011698992 | 1,800886313 | 0,000619121 |
| MMP25 | 1,166620231 | 0,01188502 | 1,489049553 | 9,84843E-06 |
| FAM212A | 1,379290301 | 0,011949416 | 1,393562877 | 0,001117934 |
| FAM111B | 1,37807503 | 0,014162096 | 1,237265371 | 0,00947072 |
| ITGB2 | 1,012059493 | 0,01426327 | 1,149113902 | 0,000205929 |
| LSP1 | 1,107398536 | 0,014647563 | 1,312784793 | 9,99542E-05 |
| LOC729683 | 1,139572551 | 0,015005278 | 1,194453311 | 0,00081404 |
| FCRL6 | 1,829862446 | 0,015086435 | 2,276676894 | 6,8724E-05 |

| | | | | |
|---|---|---|---|---|
| MYO1G | 1,508335064 | 0,01537771 | 1,505429604 | 0,002701087 |
| DBNDD1 | 1,058968923 | 0,015803229 | 1,465791276 | 2,27798E-06 |
| SCXB | 1,00483034 | 0,016720027 | 1,094748324 | 0,000897862 |
| GAS7 | 1,391679464 | 0,017522227 | 1,133077227 | 0,018605645 |
| GPR132 | 1,968256022 | 0,017758574 | 2,27642192 | 0,000440704 |
| CLEC2D | 1,393011766 | 0,017946933 | 1,192737874 | 0,014556181 |
| POU5F1 | 1,605082049 | 0,018504767 | 1,340009781 | 0,022076269 |
| MYOF | 1,065671227 | 0,018878471 | 1,013548132 | 0,004879552 |
| LOC285762 | 1,239002685 | 0,018880797 | 1,07486036 | 0,011840035 |
| LOC101928512 | 1,288304122 | 0,018928233 | 1,514341147 | 0,000186118 |
| FRMD6 | 1,363054834 | 0,019798025 | 1,057635535 | 0,028479749 |
| TAP2 | 1,222913142 | 0,020348677 | 1,653382065 | 1,18957E-05 |
| GBP5 | 1,386434228 | 0,020967167 | 1,760819194 | 5,79033E-05 |
| PTAFR | 1,752522509 | 0,02155624 | 1,552159859 | 0,014757458 |
| TRIM22 | 1,162844311 | 0,022360386 | 1,044371944 | 0,009746724 |
| S1PR4 | 1,700908808 | 0,022972413 | 1,758988191 | 0,002019514 |
| PERM1 | 1,986712687 | 0,023651325 | 1,858489292 | 0,010796881 |
| ARNTL2 | 1,830118738 | 0,024785005 | 1,733519137 | 0,00941979 |
| HKDC1 | 2,905984945 | 0,025119442 | 2,282867825 | 0,042703179 |
| RHBDF2 | 1,094604224 | 0,02579982 | 1,019364173 | 0,007581265 |
| LOC101927051 | 1,469541127 | 0,026214961 | 1,545104455 | 0,001977184 |
| SSBP2 | 1,692678396 | 0,026698388 | 1,628103213 | 0,007547692 |
| LOC388692 | 1,122865222 | 0,026835765 | 1,005787108 | 0,014781948 |
| CDCP1 | 2,524586037 | 0,027192616 | 3,234963513 | 9,87096E-05 |
| LOC102724371 | 2,581616969 | 0,027764574 | 2,92960185 | 0,000782133 |
| RP11-47I22.2 | 1,33893085 | 0,028119325 | 1,293891303 | 0,008214611 |
| CD2 | 2,318847073 | 0,028655216 | 2,213224657 | 0,012397659 |
| FMN1 | 1,549069677 | 0,029777804 | 1,422506786 | 0,011349055 |
| IKZF1 | 1,011874227 | 0,030143421 | 1,14994169 | 0,001041889 |
| KCNJ4 | 1,252782488 | 0,031195791 | 1,518270876 | 0,000370046 |
| PROB1 | 1,592433726 | 0,031268816 | 1,549214654 | 0,008013204 |
| UHRF1 | 1,287676139 | 0,032730726 | 1,552004638 | 0,000373433 |
| LRIF1 | 1,07523133 | 0,032898399 | 1,069578506 | 0,004467547 |
| SLAMF8 | 1,370716064 | 0,033088594 | 1,175521536 | 0,022958257 |
| CARD11 | 1,466766355 | 0,035086378 | 1,392312938 | 0,014220111 |
| RASSF1-AS1 | 1,026588844 | 0,035381837 | 1,04590375 | 0,00409206 |
| LGALS3 | 1,891124984 | 0,037888098 | 1,533494745 | 0,037730471 |
| PTGER2 | 1,534421658 | 0,037888098 | 1,288829631 | 0,03296038 |
| MPL | 1,30314176 | 0,038001062 | 1,056384928 | 0,042312695 |
| S100A1 | 1,581930412 | 0,038001062 | 1,69948722 | 0,003688383 |
| RUNX1 | 1,185372047 | 0,038250829 | 1,308829414 | 0,002918988 |
| FAM129A | 1,384103774 | 0,040091266 | 1,205120531 | 0,023743078 |
| FBLIM1 | 1,351361335 | 0,040093804 | 1,248440811 | 0,014400594 |
| HLA-DMB | 1,365697208 | 0,040095319 | 1,399553031 | 0,007173692 |
| RASAL3 | 1,311846733 | 0,040147251 | 1,066987301 | 0,038013353 |
| LOC102723726 | 1,388878655 | 0,041675121 | 1,275263602 | 0,01701174 |
| CAPG | 1,931728198 | 0,043828611 | 2,317271092 | 0,001596351 |

| | | | | |
|---|---|---|---|---|
| RP11-356I2.4 | 1,180889662 | 0,043882584 | 1,365226794 | 0,001535735 |
| MARCH3 | 1,542323827 | 0,045324996 | 2,02528301 | 0,000237555 |
| THEMIS | 2,442746132 | 0,045541931 | 2,425010492 | 0,012794052 |
| CD86 | 1,569737691 | 0,045938697 | 1,398372068 | 0,031042784 |
| ELF4 | 1,165072292 | 0,046680707 | 1,020978523 | 0,03584981 |
| FCN1 | 1,585089673 | 0,046889646 | 1,599607261 | 0,0071959 |
| CORO1A | 1,250958992 | 0,04758834 | 1,492777764 | 0,001138974 |
| MGC50722 | 3,206702407 | 0,049131405 | 3,549695059 | 0,003263404 |
| UCP2 | 1,154978213 | 0,049502327 | 1,107293805 | 0,013115122 |
| HIGD1B | 1,146506762 | 0,049807397 | 1,074143448 | 0,020795084 |

#### Shared cABMR and clinTCMR downregulated

| gene name | log2fold change cABMR | padj cABMR | log2fold change clinTCMR | padj clinTCMR |
|---|---|---|---|---|
| G6PC | -1,819358544 | 9,99001E-05 | -1,361445759 | 0,000286933 |
| ALDOB | -1,038783164 | 0,000112488 | -1,150905327 | 1,75397E-08 |
| PPP1R1A | -1,657580547 | 0,000112488 | -1,23162965 | 0,000287766 |
| GJB5 | -2,88955532 | 0,000436864 | -2,243347694 | 0,000158058 |
| PACSIN3 | -1,288487671 | 0,000510315 | -1,180717827 | 2,69389E-05 |
| CNDP1 | -1,385616863 | 0,001057926 | -1,167923621 | 0,000569936 |
| PKLR | -1,046645047 | 0,001395254 | -1,200862984 | 1,66354E-06 |
| CACNA1H | -1,71259881 | 0,002372783 | -2,166040916 | 2,32733E-07 |
| SLC30A10 | -1,140495814 | 0,004057295 | -1,05121961 | 0,000746873 |
| KCNN2 | -1,580212924 | 0,004173021 | -1,100163109 | 0,010267035 |
| NCAM2 | -2,664171003 | 0,004874893 | -2,348247758 | 0,001083965 |
| GJB4 | -2,122297848 | 0,006203321 | -1,756076228 | 0,003310053 |
| CCL14 | -1,820088461 | 0,007371192 | -1,150136038 | 0,03316342 |
| AOC4P | -1,616059464 | 0,007865033 | -1,107388378 | 0,02152246 |
| TTLL4 | -1,343438384 | 0,01028266 | -1,085448208 | 0,009606173 |
| RNF152 | -1,219670675 | 0,010720631 | -1,035420229 | 0,005173377 |
| DGAT2 | -1,044709988 | 0,012594035 | -1,259832911 | 6,49248E-05 |
| CYP2E1 | -1,065095557 | 0,016201874 | -1,104880622 | 0,000952342 |
| AKR1D1 | -1,015477108 | 0,01708689 | -1,135447303 | 0,000375959 |
| NYNRIN | -1,233065332 | 0,017483548 | -1,268816544 | 0,001550704 |
| LINC01093 | -1,366498204 | 0,01774418 | -1,088538893 | 0,014338923 |
| TPPP2 | -1,926974722 | 0,022932463 | -1,324534898 | 0,02539041 |
| BBOX1 | -1,638144291 | 0,024673021 | -1,445085092 | 0,012985302 |
| RP11-701P16.5 | -1,721226256 | 0,025964132 | -1,477738244 | 0,005999809 |
| SSTR1 | -1,342055646 | 0,030327575 | -1,639050411 | 0,000804199 |
| CFHR4 | -1,150306948 | 0,038057955 | -1,024944973 | 0,014502253 |
| C5orf27 | -1,711290757 | 0,039947942 | -1,312690586 | 0,041480805 |

#### Suppl. Table 13: Pathway enrichment of shared DEGs of cABMR and clinTCMR
#### Common upregulated pathways of cABMR and clinTCMR

| q-value | Pathway nam | Pathway ID |
|---|---|---|
| 4.099699352 | regulation of l | GO:0051249 |
| 5.536131249 | positive regul | GO:0022409 |
| 5.536131249 | regulation of l | GO:1903037 |
| 3.386652207 | positive regul | GO:0002696 |
| 4.570377189 | positive regul | GO:0050867 |
| 2.116083961 | hemopoiesis | GO:0030097 |
| 2.116083961 | regulation of l | GO:1902105 |
| 6.182800642 | cellular respo | GO:0071345 |
| 1.528611105 | regulation of l | GO:0070663 |
| 0.000324087 | positive regul | GO:0001819 |
| 0.000451789 | membrane raf | GO:0031580 |
| 0.000645176 | negative regul | GO:0050777 |
| 0.000645176 | positive regul | GO:1903708 |
| 0.000788205 | positive regul | GO:0050778 |
| 0.001417411 | positive regul | GO:0008284 |
| 0.001417411 | negative regul | GO:0002695 |
| 0.001417411 | interferon-gar | GO:0060333 |
| 0.001518380 | regulation of i | GO:0032663 |
| 0.002295708 | negative regul | GO:0050866 |
| 0.002509523 | response to in | GO:0034341 |
| 0.004089760 | regulation of i | GO:0045088 |
| 0.004146168 | antigen proce | GO:0019885 |
| 0.005696092 | negative regul | GO:0002698 |
| 0.008096375 | negative regul | GO:0001911 |
| 0.008096375 | regulation of p | GO:0043067 |
| 0.009987263 | regulation of T | GO:2001188 |
| 0.009987263 | antigen proce | GO:0002476 |
| 0.009987263 | regulation of r | GO:0043313 |
| 0.013630288 | vesicle fusion | GO:0006906 |
| 0.014830517 | T cell mediate | GO:0002456 |
| 0.014830517 | regulation of a | GO:0050854 |
| 0.014830517 | actin polymer | GO:0008154 |
| 0.015123286 | interleukin-21 | GO:0038114 |
| 0.015123286 | response to in | GO:0098756 |
| 0.015123286 | organelle mer | GO:0090174 |
| 0.015472219 | liver regenera | GO:0097421 |
| 0.016957509 | positive regul | GO:0010912 |
| 0.016957509 | interleukin-9- | GO:0038113 |
| 0.016957509 | protein comp | GO:0051259 |
| 0.017436219 | regulation of a | GO:0045765 |
| 0.018425181 | positive regul | GO:0031349 |
| 0.018682632 | response to in | GO:0070670 |
| 0.018682632 | regulation of r | GO:1902563 |
| 0.018682632 | response to in | GO:0071104 |
| 0.018828425 | negative regul | GO:0022408 |
| 0.019421745 | neutrophil mi | GO:1990266 |
| 0.020934751 | regulation of T | GO:0045622 |
| 0.020934751 | regulation of T | GO:0001914 |
| 0.023869465 | peripheral ner | GO:0048934 |
| 0.023869465 | peripheral ner | GO:0048935 |
| 0.026972792 | regulation of v | GO:1901342 |
| 0.026987168 | leukocyte agg | GO:0070486 |
| 0.027386665 | regulation of e | GO:0050678 |
| 0.032011109 | regulation of l | GO:0043300 |
| 0.032909860 | regulation of l | GO:0002703 |
| 0.032925647 | neutrophil me | GO:0002446 |
| 0.033800898 | dendritic cell | GO:0097028 |
| 0.033800898 | type I interfer | GO:0060337 |
| 0.034965388 | regulation of r | GO:0071675 |
| 0.039884242 | positive regul | GO:0002699 |
| 0.039884242 | response to ty | GO:0034340 |
| 0.041201803 | leukocyte apo | GO:0071887 |
| 0.041201803 | positive regul | GO:0051092 |
| 0.041201803 | renal vesicle n | GO:0072077 |
| 0.041201803 | neutrophil ch | GO:0030593 |
| 0.041201803 | regulation of c | GO:0001959 |
| 0.041201803 | ion transmem | GO:0034220 |
| 0.046842409 | regulation of c | GO:0031347 |

| q-value | Pathway nam | Pathway ID |
|---|---|---|
| 2.387074885 | Immune Syste | R-HSA-168256 |
| 5.094441098 | Cell adhesion | path:hsa0451 |
| 3.484367290 | Primary immu | path:hsa0534 |
| 3.484367290 | Viral myocard | path:hsa0541 |
| 4.530129803 | Epstein-Barr v | path:hsa0516 |
| 7.282532020 | Cytokine Sign | R-HSA-128021 |
| 0.000467057 | Allograft rejec | path:hsa0533 |
| 0.000472548 | Interferon gar | R-HSA-877300 |
| 0.000643537 | Graft-versus-h | path:hsa0533 |
| 0.000643537 | Type I diabete | path:hsa0494 |
| 0.000643537 | Th17 cell diffe | path:hsa0465 |
| 0.000643537 | RUNX3 regula | R-HSA-895193 |
| 0.000726884 | Phagosome - I | path:hsa0414 |
| 0.001260616 | Autoimmune | path:hsa0532 |
| 0.001964206 | Th1 and Th2 c | path:hsa0465 |
| 0.003237329 | RUNX3 regula | R-HSA-895191 |
| 0.006059455 | Interferon Sig | R-HSA-913531 |
| 0.006474473 | Antigen proce | path:hsa0461 |
| 0.006474473 | Human T-cell | path:hsa0516 |
| 0.008285928 | Extracellular r | R-HSA-147424 |
| 0.011580585 | Antigen Prese | R-HSA-983170 |
| 0.011580585 | RUNX1 regula | R-HSA-893924 |
| 0.014801963 | Hematopoieti | path:hsa0464 |
| 0.016350327 | Pathways in c | path:hsa0520 |
| 0.017727859 | T cell recepto | path:hsa0466 |
| 0.018092769 | RUNX3 regula | R-HSA-895143 |
| 0.018605227 | Post-translati | R-HSA-895727 |
| 0.020448572 | JAK-STAT signa | path:hsa0463 |
| 0.020448572 | Interleukin-9 | R-HSA-898594 |
| 0.020448572 | Toxoplasmosi | path:hsa0514 |
| 0.020927200 | Adaptive Imm | R-HSA-128021 |
| 0.023366099 | Innate Immur | R-HSA-168249 |
| 0.023740344 | Transcription | R-HSA-887816 |
| 0.023740344 | TCR signaling | R-HSA-202403 |
| 0.026152216 | Leishmaniasis | path:hsa0514 |
| 0.026152216 | Regulation of | R-HSA-381426 |
| 0.026152216 | Tuberculosis - | path:hsa0515 |
| 0.029533531 | Antigen proce | R-HSA-123697 |
| 0.037941004 | ER-Phagosom | R-HSA-123697 |
| 0.039308896 | ECM-receptor | path:hsa0451 |
| 0.039394459 | PD-L1 express | path:hsa0523 |
| 0.039394459 | Focal adhesio | path:hsa0451 |
| 0.039394459 | Interleukin-4 | R-HSA-678580 |
| 0.039394459 | Intestinal imn | path:hsa0467 |
| 0.039394459 | Small cell lung | path:hsa0522 |
| 0.039394459 | Rheumatoid a | path:hsa0532 |
| 0.039394459 | Neutrophil de | R-HSA-679869 |
| 0.040459647 | Staphylococc | path:hsa0515 |
| 0.040459647 | Transcription | R-HSA-887815 |
| 0.041840226 | TNFR2 non-ca | R-HSA-566854 |
| 0.041840226 | Regulation of | R-HSA-893459 |
| 0.041840226 | TNF receptor s | R-HSA-567659 |
| 0.045386324 | Interleukin-20 | R-HSA-885469 |
| 0.045386324 | AGE-RAGE sigr | path:hsa0493 |

**Shared downregulated pathways of cABMR and clinTCMR**

| q-value | Pathway name | Pathway ID | | q-value | Pathway name | Pathway ID |
|---|---|---|---|---|---|---|
| 0.0335826683 | glucose metab | GO:0006006 | | 0.0139053035 | Steroid hormo | path:hsa0014C |
| 0.0335826683 | carboxylic acic | GO:0046394 | | 0.0139053035 | GnRH secretior | path:hsa04929 |
| 0.0335826683 | hexose catabol | GO:0019320 | | 0.0139053035 | Glycolysis / Glu | path:hsa0001C |
| 0.0599683964 | hexose biosynt | GO:0019319 | | 0.0139053035 | Glycolysis | R-HSA-70171 |
| 0.0599683964 | carboxylic acic | GO:0019752 | | 0.0178970354 | Glucose metab | R-HSA-70326 |
| 0.0599683964 | regulation of ic | GO:0043269 | | | | |
| 0.0599683964 | glycolytic proc | GO:0006096 | | | | |
| 0.0625433701 | terpenoid meta | GO:0006721 | | | | |

**Suppl. Table 14. Characteristics of cDR samples vs controls (NHR)**

Continuous variables reported as median (range), categorical variables as number (frequency). P-values calculated with Kruskal Wallis for continuous variables and Fisher test for categorical variables. A p-value < 0.05 is considered significant.

| Suppl. Table 14 | NHR | CDR | p |
|---|---|---|---|
| sample number | 6 | 4 | |
| female sex [n (%)] | 2 (33.3) | 1 (25.0) | 1.00 |
| age [years] (median (range)) | 47.50 [25.00 - 65.00] | 39.50 [20.00 - 60.00] | 0.39 |
| time after liver transplantation [months] (median (range)) | 51.00 [19.00 - 65.00] | 44.50 [12.00 - 62.00] | 0.75 |
| AST [xULN] (median (range)) | 0.58 [0.23 - 0.77] | 3.36 [2.20 - 12.45] | 0.01 |
| ALT [xULN] (median (range)) | 0.39 [0.31 - 0.41] | 2.32 [1.27 - 16.26] | 0.01 |
| AP [xULN] (median (range)) | 0.55 [0.31 - 1.64] | 2.04 [1.66 - 3.83] | 0.01 |
| GGT [xULN] (median (range)) | 0.29 [0.20 - 0.64] | 4.98 [2.98 - 10.00] | 0.01 |
| bilirubin [xULN] (median (range)) | 0.26 [0.19 - 1.00] | 12.89 [9.35 - 20.71] | 0.01 |
| mHAI (median (range)) | 1.00 [0.00 - 1.00] | 1.50 [0.00 - 4.00] | 0.25 |
| Ishak Fibrosis (median (range)) | 0.00 [0.00 - 1.00] | 2.00 [0.00 - 4.00] | 0.05 |
| LAF score (median (range))* | 0.00 [0.00 - 1.00] | 2.50 [1.00 - 4.00] | 0.13 |
| RAI (median (range)) | 0.50 [0.00 - 1.00] | 4.00 [2.00 - 6.00] | 0.01 |
| TAC [n (%)] | 5 (83.3) | 3 (75.0) | 1.00 |
| CsA [n (%)] | 0 (0) | 0 (0) | NA |
| Everolimus [n (%)] | 1 (16.7) | 0 (0.0) | 1.00 |
| Prednisolone [n (%)] | 2 (33.3) | 3 (75.0) | 0.52 |
| MMF [n (%)] | 5 (83.3) | 4 (100.0) | 1.00 |
| Azathioprine [n (%)] | 0 (0) | 0 (0) | NA |
| *Venturi LAF score available in 5/10 biopsies (NHR 3/6; CDR 2/4) | | | |

**Suppl. Table 15: Diffentially expressed genes of chronic ductopenic rejection compared to controls**

Adjustment of p-value by false discovery rate.

| log2FoldChange | padj | gene_names | significance | direction |
|---|---|---|---|---|
| -2,449162222 | 6,35782E-21 | ALB | TRUE | Downregulated |
| -2,986361483 | 5,39489E-16 | HMGCS2 | TRUE | Downregulated |
| -2,498165867 | 6,99849E-16 | TTR | TRUE | Downregulated |
| 3,988420709 | 7,98632E-16 | FAT1 | TRUE | Upregulated |
| -1,88938605 | 7,12884E-13 | ALDOB | TRUE | Downregulated |
| 2,830027766 | 7,0182E-12 | CCND1 | TRUE | Upregulated |
| -3,697021427 | 3,03925E-11 | GSTA1 | TRUE | Downregulated |
| -1,832613439 | 3,14659E-11 | CES1 | TRUE | Downregulated |
| -3,010904375 | 4,70713E-11 | CYP2C8 | TRUE | Downregulated |
| -2,037548961 | 1,03417E-10 | ZKSCAN1 | TRUE | Downregulated |
| 3,010270513 | 2,04166E-10 | VTI1A | TRUE | Upregulated |
| -2,339052337 | 4,02833E-10 | ADH6 | TRUE | Downregulated |
| -1,91739067 | 4,26085E-10 | ADH1B | TRUE | Downregulated |
| -2,567793722 | 4,76427E-10 | BHMT | TRUE | Downregulated |

| | | | | |
|---|---|---|---|---|
| -1,865969273 | 7,23523E-10 | APOH | TRUE | Downregulated |
| -1,973502876 | 7,23523E-10 | DCXR | TRUE | Downregulated |
| -3,208332659 | 7,23523E-10 | LRRTM3 | TRUE | Downregulated |
| -2,561981227 | 7,5168E-10 | ACADSB | TRUE | Downregulated |
| -2,27223042 | 9,6783E-10 | CYP4A11 | TRUE | Downregulated |
| -1,890421631 | 1,34608E-09 | HPX | TRUE | Downregulated |
| -2,578671185 | 1,90881E-09 | HSD17B6 | TRUE | Downregulated |
| -2,713184016 | 2,55313E-09 | CACNA1H | TRUE | Downregulated |
| -1,785513275 | 5,0304E-09 | APOA1 | TRUE | Downregulated |
| -2,481966339 | 5,73093E-09 | ADH1C | TRUE | Downregulated |
| -1,977165076 | 5,85935E-09 | ACAA1 | TRUE | Downregulated |
| -2,097588608 | 7,48553E-09 | ADH4 | TRUE | Downregulated |
| -2,318053486 | 9,10197E-09 | HP | TRUE | Downregulated |
| -2,031179982 | 9,21907E-09 | SORD | TRUE | Downregulated |
| -4,282425118 | 9,29288E-09 | GSTA2 | TRUE | Downregulated |
| -1,656043978 | 1,36626E-08 | MAT1A | TRUE | Downregulated |
| -1,531548431 | 1,56033E-08 | APOC3 | TRUE | Downregulated |
| -1,783819758 | 2,01388E-08 | AMBP | TRUE | Downregulated |
| -2,402324561 | 2,19634E-08 | ALDH1L1 | TRUE | Downregulated |
| -2,128747782 | 2,3659E-08 | PCK1 | TRUE | Downregulated |
| -4,475484551 | 2,58116E-08 | GNMT | TRUE | Downregulated |
| -3,550495413 | 4,54962E-08 | HAO2 | TRUE | Downregulated |
| -2,02260385 | 6,65962E-08 | TF | TRUE | Downregulated |
| -1,77473617 | 7,14758E-08 | RARRES2 | TRUE | Downregulated |
| -2,886665358 | 8,91502E-08 | RDH16 | TRUE | Downregulated |
| -2,666011567 | 1,4276E-07 | AKR1D1 | TRUE | Downregulated |
| -1,832593115 | 1,4878E-07 | ALDH2 | TRUE | Downregulated |
| -2,140808774 | 1,50004E-07 | ALAD | TRUE | Downregulated |
| -1,641446135 | 1,62853E-07 | GLUD1 | TRUE | Downregulated |
| -3,302058616 | 1,84046E-07 | RHEBP2 | TRUE | Downregulated |
| 2,823426336 | 2,53906E-07 | APOBEC4 | TRUE | Upregulated |
| -1,740261626 | 2,6862E-07 | AOX1 | TRUE | Downregulated |
| -2,063150991 | 3,48401E-07 | SLC22A1 | TRUE | Downregulated |
| 2,532548605 | 4,64381E-07 | CPM | TRUE | Upregulated |
| -1,634014131 | 5,16256E-07 | CCDC152 | TRUE | Downregulated |
| -2,513231742 | 7,01342E-07 | ZNF446 | TRUE | Downregulated |
| 2,129986217 | 7,09062E-07 | NEDD4L | TRUE | Upregulated |
| -1,681537556 | 9,77503E-07 | CYP2C9 | TRUE | Downregulated |
| -1,475945952 | 1,11095E-06 | APOC1 | TRUE | Downregulated |
| 2,345582608 | 1,21144E-06 | SZRD1 | TRUE | Upregulated |
| -2,984775708 | 1,21165E-06 | PPP1R1A | TRUE | Downregulated |
| -1,458944946 | 1,21165E-06 | SERPINF2 | TRUE | Downregulated |
| -1,759863884 | 1,37824E-06 | GATM | TRUE | Downregulated |
| -1,966984723 | 1,41299E-06 | CAT | TRUE | Downregulated |
| -2,23471432 | 1,41299E-06 | DMGDH | TRUE | Downregulated |
| -1,844579224 | 1,41299E-06 | EGFR | TRUE | Downregulated |
| 1,897330319 | 1,41299E-06 | GLS | TRUE | Upregulated |
| -1,673221345 | 1,54463E-06 | ACACB | TRUE | Downregulated |
| -1,992115166 | 1,54463E-06 | ORM1 | TRUE | Downregulated |
| -2,615513461 | 2,00531E-06 | MRPL23-AS1 | TRUE | Downregulated |
| -2,705530425 | 2,50016E-06 | CMBL | TRUE | Downregulated |

| | | | | |
|---|---|---|---|---|
| -2,414756075 | 2,50016E-06 | HAAO | TRUE | Downregulated |
| -1,854692369 | 2,70604E-06 | ASS1 | TRUE | Downregulated |
| -2,208783624 | 3,14418E-06 | ORM2 | TRUE | Downregulated |
| -2,542244259 | 3,36156E-06 | B9D1 | TRUE | Downregulated |
| -1,467382802 | 3,61447E-06 | C1R | TRUE | Downregulated |
| -2,89669882 | 3,70265E-06 | CYP2C19 | TRUE | Downregulated |
| 3,372691964 | 3,71107E-06 | LYZ | TRUE | Upregulated |
| -1,746481742 | 3,71107E-06 | SERPINA5 | TRUE | Downregulated |
| -1,714312535 | 3,73017E-06 | ACSL1 | TRUE | Downregulated |
| 3,995675355 | 4,1657E-06 | LINC00319 | TRUE | Upregulated |
| -1,719279994 | 5,80488E-06 | ADH1A | TRUE | Downregulated |
| -1,489793573 | 6,00837E-06 | PLG | TRUE | Downregulated |
| -1,496618793 | 6,29493E-06 | C6 | TRUE | Downregulated |
| -2,345604961 | 8,32152E-06 | PGLYRP2 | TRUE | Downregulated |
| -1,270815475 | 1,13795E-05 | SEPP1 | TRUE | Downregulated |
| -1,395900846 | 1,15938E-05 | AZGP1 | TRUE | Downregulated |
| -1,697485303 | 1,20091E-05 | GC | TRUE | Downregulated |
| -1,958172061 | 1,24838E-05 | COBLL1 | TRUE | Downregulated |
| -2,434038953 | 1,43656E-05 | GLYAT | TRUE | Downregulated |
| 1,85866891 | 1,63213E-05 | COL1A1 | TRUE | Upregulated |
| -1,853140169 | 1,82951E-05 | ADRA1A | TRUE | Downregulated |
| 2,853665521 | 2,27855E-05 | PRRT2 | TRUE | Upregulated |
| 2,387361961 | 2,27855E-05 | TNFAIP2 | TRUE | Upregulated |
| 2,573188444 | 2,30592E-05 | TCEA2 | TRUE | Upregulated |
| -1,760304682 | 2,33255E-05 | CCDC176 | TRUE | Downregulated |
| -1,901501909 | 2,69728E-05 | APCS | TRUE | Downregulated |
| -2,211304004 | 2,92244E-05 | ZSCAN25 | TRUE | Downregulated |
| 2,648552982 | 3,07902E-05 | C1orf204 | TRUE | Upregulated |
| -1,421270372 | 3,34536E-05 | SLC27A5 | TRUE | Downregulated |
| -1,546826619 | 3,41685E-05 | C1RL-AS1 | TRUE | Downregulated |
| -1,598485527 | 3,46982E-05 | ADI1 | TRUE | Downregulated |
| -2,969297332 | 3,46982E-05 | PHGDH | TRUE | Downregulated |
| -2,312662974 | 3,6796E-05 | AMOTL1 | TRUE | Downregulated |
| -1,501762489 | 3,90467E-05 | DPYS | TRUE | Downregulated |
| -2,519862246 | 3,90467E-05 | ERI2 | TRUE | Downregulated |
| -1,833083167 | 4,42817E-05 | FABP1 | TRUE | Downregulated |
| -2,674882791 | 4,82387E-05 | ACOT11 | TRUE | Downregulated |
| -2,445642355 | 5,1455E-05 | IL1RAP | TRUE | Downregulated |
| -2,028373763 | 5,34777E-05 | SLCO1B1 | TRUE | Downregulated |
| -1,306372418 | 5,43486E-05 | PCK2 | TRUE | Downregulated |
| -1,687279676 | 5,81972E-05 | CUX2 | TRUE | Downregulated |
| -1,927478163 | 6,20403E-05 | APOA5 | TRUE | Downregulated |
| -1,381152514 | 6,24481E-05 | CLU | TRUE | Downregulated |
| -2,02764181 | 7,24343E-05 | PDK4 | TRUE | Downregulated |
| -2,284697781 | 7,48235E-05 | CYP2B6 | TRUE | Downregulated |
| -2,26155438 | 7,98204E-05 | DGAT2 | TRUE | Downregulated |
| -1,664686395 | 7,99607E-05 | IL6R | TRUE | Downregulated |
| -2,109605429 | 8,46988E-05 | CYP2A6 | TRUE | Downregulated |
| -1,711505908 | 9,74421E-05 | MARC2 | TRUE | Downregulated |
| -1,644144991 | 9,93032E-05 | SERPINC1 | TRUE | Downregulated |
| -1,520002866 | 9,9442E-05 | SLC38A4 | TRUE | Downregulated |

| | | | | |
|---|---|---|---|---|
| 2,493336228 | 9,9442E-05 | TPM1 | TRUE | Upregulated |
| -1,91313696 | 0,000102355 | MUT | TRUE | Downregulated |
| -1,78802531 | 0,00010905 | HPR | TRUE | Downregulated |
| 2,025085957 | 0,000112694 | SHB | TRUE | Upregulated |
| -3,583347145 | 0,000113059 | SCN3A | TRUE | Downregulated |
| -1,743210919 | 0,000122432 | ACAT1 | TRUE | Downregulated |
| -1,714389843 | 0,000127446 | LRRC8A | TRUE | Downregulated |
| -1,612787649 | 0,000137249 | ECHS1 | TRUE | Downregulated |
| -2,377187242 | 0,000144787 | HSD17B13 | TRUE | Downregulated |
| -2,257506507 | 0,000144787 | UG0898H09 | TRUE | Downregulated |
| -1,748632126 | 0,000147247 | PKLR | TRUE | Downregulated |
| 2,97281729 | 0,000153724 | PRNCR1 | TRUE | Upregulated |
| 1,896808211 | 0,000170524 | ABCB4 | TRUE | Upregulated |
| -1,790264457 | 0,000176488 | ACADS | TRUE | Downregulated |
| -2,296467944 | 0,000181388 | DHODH | TRUE | Downregulated |
| 2,235004837 | 0,000205224 | TDRD6 | TRUE | Upregulated |
| 1,576123647 | 0,000229544 | C1orf56 | TRUE | Upregulated |
| -3,659404705 | 0,000247821 | CFHR4 | TRUE | Downregulated |
| 2,507689471 | 0,000248789 | JUN | TRUE | Upregulated |
| -1,771986502 | 0,000249599 | KHK | TRUE | Downregulated |
| -1,661984303 | 0,000249599 | ZGPAT | TRUE | Downregulated |
| -1,699417748 | 0,000298167 | C1R-2 | TRUE | Downregulated |
| -1,553308157 | 0,000300689 | LIME1 | TRUE | Downregulated |
| -2,057319514 | 0,000302877 | RGPD3 | TRUE | Downregulated |
| -1,576736832 | 0,000326353 | F9 | TRUE | Downregulated |
| 2,004085499 | 0,000335223 | COL1A2 | TRUE | Upregulated |
| -2,056121235 | 0,000335223 | ZNF324 | TRUE | Downregulated |
| -3,011361223 | 0,000336404 | MME | TRUE | Downregulated |
| 2,395187389 | 0,000336404 | SLC35C1 | TRUE | Upregulated |
| -1,469387747 | 0,000364974 | EPHX1 | TRUE | Downregulated |
| -1,401944409 | 0,00041215 | PIK3R1 | TRUE | Downregulated |
| 1,646398023 | 0,000412339 | COL5A1 | TRUE | Upregulated |
| -1,570213577 | 0,000412339 | UGT2B7 | TRUE | Downregulated |
| -1,849690275 | 0,000437139 | CDO1 | TRUE | Downregulated |
| -2,143121806 | 0,000437139 | PEX6 | TRUE | Downregulated |
| 1,731530823 | 0,000472127 | MFSD2A | TRUE | Upregulated |
| 1,534999345 | 0,000497929 | CROT | TRUE | Upregulated |
| -1,315058254 | 0,000497929 | F12 | TRUE | Downregulated |
| -1,404183202 | 0,000498544 | GRHPR | TRUE | Downregulated |
| 2,478161244 | 0,000513486 | KCNJ4 | TRUE | Upregulated |
| -2,187081165 | 0,000544218 | RUSC1-AS1 | TRUE | Downregulated |
| -1,626258769 | 0,000564375 | ST8SIA3 | TRUE | Downregulated |
| -1,429656157 | 0,000619187 | FAM53B | TRUE | Downregulated |
| 1,537019597 | 0,000621854 | LOC100272216 | TRUE | Upregulated |
| -1,396457112 | 0,00064899 | ITIH1 | TRUE | Downregulated |
| -1,164947216 | 0,00064899 | SARM1 | TRUE | Downregulated |
| -1,142422643 | 0,00064899 | TFR2 | TRUE | Downregulated |
| 2,614249059 | 0,000669064 | FBLIM1 | TRUE | Upregulated |
| -1,505484446 | 0,000693089 | CGREF1 | TRUE | Downregulated |
| -1,418162193 | 0,000693792 | AKR1C3 | TRUE | Downregulated |
| -1,924559966 | 0,000693792 | CYP1A2 | TRUE | Downregulated |

| | | | | |
|---|---|---|---|---|
| 2,658352142 | 0,00070549 | FOXP3 | TRUE | Upregulated |
| -1,760069272 | 0,000721136 | METTL21A | TRUE | Downregulated |
| -2,506324236 | 0,000734621 | LOC102723811 | TRUE | Downregulated |
| -1,739553074 | 0,000753956 | AASS | TRUE | Downregulated |
| -2,110132843 | 0,000772223 | HMGCS1 | TRUE | Downregulated |
| -1,162952834 | 0,000845351 | SERPINA1 | TRUE | Downregulated |
| -1,341688438 | 0,000885299 | ACAA2 | TRUE | Downregulated |
| -1,705482408 | 0,00088743 | CYB5A | TRUE | Downregulated |
| -1,790469931 | 0,000898148 | CYP3A4 | TRUE | Downregulated |
| -1,540978959 | 0,000898148 | FMO5 | TRUE | Downregulated |
| -2,086830848 | 0,000993282 | DRG2 | TRUE | Downregulated |
| -1,28172519 | 0,001064351 | ACSM5 | TRUE | Downregulated |
| -2,077474352 | 0,001064351 | SC5D | TRUE | Downregulated |
| -1,53559742 | 0,001109579 | RP11-696N14.1 | TRUE | Downregulated |
| 2,500619732 | 0,001141329 | ENAH | TRUE | Upregulated |
| -1,385616888 | 0,001159162 | SERPINA3 | TRUE | Downregulated |
| -1,209142001 | 0,001167342 | SARDH | TRUE | Downregulated |
| 1,398521781 | 0,001173734 | C10orf10 | TRUE | Upregulated |
| 1,90874604 | 0,001174731 | TGFBI | TRUE | Upregulated |
| 2,037381055 | 0,001174731 | TTYH3 | TRUE | Upregulated |
| 1,523567004 | 0,001174768 | LOC100128653 | TRUE | Upregulated |
| -1,50067168 | 0,001198302 | SCP2 | TRUE | Downregulated |
| -1,297072491 | 0,001262437 | IQGAP2 | TRUE | Downregulated |
| -1,633922421 | 0,001321477 | HAO1 | TRUE | Downregulated |
| 1,828064466 | 0,001368978 | ZYX | TRUE | Upregulated |
| -1,530221017 | 0,001376204 | ACOX2 | TRUE | Downregulated |
| -1,57144386 | 0,001399248 | SDC2 | TRUE | Downregulated |
| 1,852343123 | 0,001400243 | SYTL4 | TRUE | Upregulated |
| -1,963532278 | 0,001484067 | PTGR1 | TRUE | Downregulated |
| -2,752086288 | 0,001506503 | TMC3 | TRUE | Downregulated |
| 1,258519971 | 0,00152244 | GPX3 | TRUE | Upregulated |
| 2,41129522 | 0,00152575 | KAAG1 | TRUE | Upregulated |
| -2,093259571 | 0,001537783 | ARRDC3 | TRUE | Downregulated |
| -1,315912871 | 0,001631456 | LOC102724535 | TRUE | Downregulated |
| -1,714349339 | 0,001666143 | AC011343.1 | TRUE | Downregulated |
| -1,679860505 | 0,00168044 | FRMD8 | TRUE | Downregulated |
| 1,429345367 | 0,00168044 | HRNR | TRUE | Upregulated |
| -1,486077068 | 0,001819578 | EPHX2 | TRUE | Downregulated |
| -1,408863881 | 0,001897653 | AR | TRUE | Downregulated |
| -1,516059198 | 0,001939525 | ASPG | TRUE | Downregulated |
| -1,519061617 | 0,002012505 | PBLD | TRUE | Downregulated |
| -1,158603284 | 0,002012505 | PEBP1 | TRUE | Downregulated |
| 1,310318818 | 0,002067865 | VIM-AS1 | TRUE | Upregulated |
| -1,609402229 | 0,002110662 | ZCCHC6 | TRUE | Downregulated |
| -1,197064832 | 0,002278722 | SERPINF1 | TRUE | Downregulated |
| -1,385839071 | 0,002278722 | TPR | TRUE | Downregulated |
| -1,126244278 | 0,002288622 | NOP9 | TRUE | Downregulated |
| -1,725204447 | 0,00253076 | SH3PXD2A | TRUE | Downregulated |
| -1,818841025 | 0,002578578 | ALDH7A1 | TRUE | Downregulated |
| 1,75180814 | 0,002594948 | LOC102724153 | TRUE | Upregulated |
| -1,863207755 | 0,00276412 | ABHD15 | TRUE | Downregulated |

| | | | | |
|---|---|---|---|---|
| 2,810217772 | 0,002789155 | LOC101928363 | TRUE | Upregulated |
| 1,103921279 | 0,002806118 | RNF213 | TRUE | Upregulated |
| 2,732207682 | 0,002808305 | SERPINE1 | TRUE | Upregulated |
| -1,660866658 | 0,002824299 | F13B | TRUE | Downregulated |
| 1,922188241 | 0,002824299 | FAM83G | TRUE | Upregulated |
| 1,291348796 | 0,003035103 | INF2 | TRUE | Upregulated |
| 2,232620123 | 0,003053234 | LOC101928240 | TRUE | Upregulated |
| -1,363495506 | 0,003168545 | PGM1 | TRUE | Downregulated |
| -1,443443442 | 0,003282498 | C9 | TRUE | Downregulated |
| -2,182791832 | 0,003471619 | SMPDL3A | TRUE | Downregulated |
| -1,526820374 | 0,00361744 | SLC39A5 | TRUE | Downregulated |
| -1,41249234 | 0,003639312 | FGA | TRUE | Downregulated |
| 1,489286045 | 0,003660879 | HMCN2 | TRUE | Upregulated |
| -1,013572294 | 0,003686406 | SERPIND1 | TRUE | Downregulated |
| -1,09965501 | 0,003835142 | CPS1 | TRUE | Downregulated |
| -1,338144652 | 0,003846181 | AKAP9 | TRUE | Downregulated |
| -1,76990187 | 0,003905964 | HTR2A | TRUE | Downregulated |
| -1,195216321 | 0,003981306 | MDN1 | TRUE | Downregulated |
| -1,521397066 | 0,00416827 | ERRFI1 | TRUE | Downregulated |
| 1,793048501 | 0,00423652 | LOXL4 | TRUE | Upregulated |
| 1,725697949 | 0,004296973 | NBPF15 | TRUE | Upregulated |
| -2,131845133 | 0,00435613 | AKR7A2P1 | TRUE | Downregulated |
| 1,505125548 | 0,004381093 | TAPSAR1 | TRUE | Upregulated |
| -2,039208415 | 0,004433304 | LIPC | TRUE | Downregulated |
| 1,466633366 | 0,004433304 | PPAPDC2 | TRUE | Upregulated |
| -1,283812644 | 0,004528923 | C1S | TRUE | Downregulated |
| -1,926468412 | 0,004628992 | PXMP2 | TRUE | Downregulated |
| 1,766982313 | 0,004732056 | LOC100996813 | TRUE | Upregulated |
| -1,114730971 | 0,004855749 | C3P1 | TRUE | Downregulated |
| -1,826183271 | 0,004855749 | LINC01018 | TRUE | Downregulated |
| -1,092912938 | 0,004982485 | IGF2-AS | TRUE | Downregulated |
| -1,421724875 | 0,005115284 | PRG4 | TRUE | Downregulated |
| -1,491408148 | 0,00523939 | RORC | TRUE | Downregulated |
| -1,542784994 | 0,005345705 | HOMER2 | TRUE | Downregulated |
| 1,77195077 | 0,005345705 | SERINC2 | TRUE | Upregulated |
| -1,680298441 | 0,005631009 | SMIM14 | TRUE | Downregulated |
| -1,278009553 | 0,005651553 | F2 | TRUE | Downregulated |
| -2,451435897 | 0,005754844 | GBA3 | TRUE | Downregulated |
| -0,957766542 | 0,005757212 | AGXT | FALSE | Downregulated |
| -1,48590145 | 0,005775296 | FGG | TRUE | Downregulated |
| -1,36369907 | 0,005775296 | GAMT | TRUE | Downregulated |
| -1,109108025 | 0,005789603 | TSHR | TRUE | Downregulated |
| -1,905041523 | 0,005803193 | CYP4A22 | TRUE | Downregulated |
| -1,07911211 | 0,006164937 | VTN | TRUE | Downregulated |
| -0,960726556 | 0,006284799 | AGT | FALSE | Downregulated |
| -1,308855564 | 0,006284799 | ETNK2 | TRUE | Downregulated |
| 1,370351649 | 0,006370769 | C5orf45 | TRUE | Upregulated |
| -1,138633069 | 0,00640222 | ALDH6A1 | TRUE | Downregulated |
| 1,188643645 | 0,006465976 | GSE1 | TRUE | Upregulated |
| -1,182541638 | 0,006468002 | CYP27A1 | TRUE | Downregulated |
| -1,019642767 | 0,006548518 | A1CF | TRUE | Downregulated |

| | | | | |
|---|---|---|---|---|
| 1,537504697 | 0,006548518 | LPGAT1 | TRUE | Upregulated |
| -1,570289725 | 0,006754644 | SMOC1 | TRUE | Downregulated |
| -1,566833835 | 0,006762729 | CDC42EP3 | TRUE | Downregulated |
| 2,159587029 | 0,007267117 | ARPC1B | TRUE | Upregulated |
| -1,728311731 | 0,007422903 | GCDH | TRUE | Downregulated |
| -1,343013869 | 0,007539833 | PTMS | TRUE | Downregulated |
| 1,551453391 | 0,007555707 | KRT8 | TRUE | Upregulated |
| 1,421957096 | 0,007574409 | COA1 | TRUE | Upregulated |
| 1,315930324 | 0,0083291 | RP6-206I17.2 | TRUE | Upregulated |
| -2,148362235 | 0,008378457 | KDR | TRUE | Downregulated |
| 1,599299793 | 0,008610828 | ELP5 | TRUE | Upregulated |
| -1,731082068 | 0,008610828 | HEATR4 | TRUE | Downregulated |
| -1,599449587 | 0,008610828 | TCEA3 | TRUE | Downregulated |
| -1,198085518 | 0,008615474 | BCAT2 | TRUE | Downregulated |
| 1,133507436 | 0,008615474 | PIEZO1 | TRUE | Upregulated |
| -1,829121994 | 0,008621602 | NUPR1 | TRUE | Downregulated |
| -2,463051313 | 0,008787458 | LOC101928798 | TRUE | Downregulated |
| -2,118273127 | 0,008791301 | MGMT | TRUE | Downregulated |
| -1,448908524 | 0,008836118 | ECH1 | TRUE | Downregulated |
| 1,869592892 | 0,009206099 | CHST15 | TRUE | Upregulated |
| 1,609171155 | 0,009218005 | FLJ40448 | TRUE | Upregulated |
| -1,361577723 | 0,009242103 | ECHDC2 | TRUE | Downregulated |
| 1,482830156 | 0,009318531 | CTNS | TRUE | Upregulated |
| -1,315233831 | 0,009353193 | APOF | TRUE | Downregulated |
| -1,65837945 | 0,009795039 | PON3 | TRUE | Downregulated |
| -0,967489066 | 0,0101998 | RBP4 | FALSE | Downregulated |
| -2,235501516 | 0,010346099 | GJB3 | TRUE | Downregulated |
| -1,700966061 | 0,010582095 | WDR72 | TRUE | Downregulated |
| 1,406831878 | 0,010736998 | PYGB | TRUE | Upregulated |
| -1,184162016 | 0,010974392 | UGT2B4 | TRUE | Downregulated |
| 1,776730152 | 0,011120525 | PAGR1 | TRUE | Upregulated |
| -1,18971627 | 0,011430656 | IVD | TRUE | Downregulated |
| 1,099348512 | 0,011430656 | TMEM184A | TRUE | Upregulated |
| 1,881131272 | 0,011582669 | MYOM1 | TRUE | Upregulated |
| 1,804055271 | 0,011833189 | NBEAL2 | TRUE | Upregulated |
| -1,4396649 | 0,011897335 | CES3 | TRUE | Downregulated |
| -1,307348766 | 0,012126275 | C4B | TRUE | Downregulated |
| 2,000189192 | 0,012320913 | AJUBA | TRUE | Upregulated |
| -1,365519873 | 0,01253494 | STAB2 | TRUE | Downregulated |
| 1,856767028 | 0,01256466 | OMD | TRUE | Upregulated |
| 1,480967936 | 0,012848858 | SAP30L-AS1 | TRUE | Upregulated |
| -1,281969669 | 0,012920967 | JMJD4 | TRUE | Downregulated |
| -2,387827923 | 0,013084905 | STARD4-AS1 | TRUE | Downregulated |
| -2,123342843 | 0,013271369 | TRPM3 | TRUE | Downregulated |
| -1,515657284 | 0,013486373 | ARRDC3-AS1 | TRUE | Downregulated |
| 2,099225879 | 0,013486373 | IL32 | TRUE | Upregulated |
| 2,381932231 | 0,013580645 | TCTEX1D4 | TRUE | Upregulated |
| -1,406061486 | 0,013583984 | CYP4F2 | TRUE | Downregulated |
| -1,075723459 | 0,013910533 | C8A | TRUE | Downregulated |
| 0,9805156 | 0,013910533 | COX15 | FALSE | Upregulated |
| -1,076356836 | 0,013910533 | FN1 | TRUE | Downregulated |

| | | | | |
|---|---|---|---|---|
| 2,170128482 | 0,014085158 | ALPK3 | TRUE | Upregulated |
| -1,361313263 | 0,014091408 | GPR125 | TRUE | Downregulated |
| -1,150510807 | 0,014136392 | TOMM40L | TRUE | Downregulated |
| 1,237968732 | 0,014250505 | FLNA | TRUE | Upregulated |
| -1,616825808 | 0,014250505 | QDPR | TRUE | Downregulated |
| 1,409646044 | 0,014292903 | VIM | TRUE | Upregulated |
| -1,604288248 | 0,014300577 | RP11-355N15.1 | TRUE | Downregulated |
| 1,846547518 | 0,01465482 | PTPRU | TRUE | Upregulated |
| -1,082933423 | 0,014665739 | ACSM2A | TRUE | Downregulated |
| -1,481088327 | 0,014759067 | ACSF2 | TRUE | Downregulated |
| -1,041546494 | 0,015025845 | ACSM2B | TRUE | Downregulated |
| -1,705838123 | 0,015309569 | ACADM | TRUE | Downregulated |
| -1,853882819 | 0,015511802 | NRK | TRUE | Downregulated |
| 1,475371404 | 0,015921625 | LOC100271836 | TRUE | Upregulated |
| -1,336111443 | 0,016079318 | LDHD | TRUE | Downregulated |
| -1,465673471 | 0,016127698 | RNR2 | TRUE | Downregulated |
| 1,894999614 | 0,016136548 | SLC51A | TRUE | Upregulated |
| -0,924719464 | 0,016838167 | HRG | FALSE | Downregulated |
| -1,298720418 | 0,016838167 | HSD11B1 | TRUE | Downregulated |
| -1,358688101 | 0,017170907 | HCN3 | TRUE | Downregulated |
| -1,177022555 | 0,017811035 | SLC2A2 | TRUE | Downregulated |
| 1,742946507 | 0,018131349 | KLF6 | TRUE | Upregulated |
| -1,196948441 | 0,018150926 | MAOB | TRUE | Downregulated |
| -1,245276265 | 0,018612824 | SOD1 | TRUE | Downregulated |
| -1,722502442 | 0,01864191 | FDPS | TRUE | Downregulated |
| -0,962259615 | 0,018732947 | RPL3 | FALSE | Downregulated |
| -1,313699798 | 0,018732947 | TCP1 | TRUE | Downregulated |
| -1,255072943 | 0,018732947 | UBB | TRUE | Downregulated |
| 1,925975401 | 0,018754937 | PLXNA1 | TRUE | Upregulated |
| -3,295016603 | 0,018820902 | LOC101930375 | TRUE | Downregulated |
| 2,710272121 | 0,018821881 | ACSL4 | TRUE | Upregulated |
| 2,246979814 | 0,018935538 | GPRIN3 | TRUE | Upregulated |
| 1,129466794 | 0,018935538 | MTOR | TRUE | Upregulated |
| -1,300734236 | 0,018973357 | UGDH-AS1 | TRUE | Downregulated |
| -1,574188953 | 0,019236293 | G6PC | TRUE | Downregulated |
| -1,111231907 | 0,019573171 | F11-AS1 | TRUE | Downregulated |
| -1,321779193 | 0,019667558 | IGFALS | TRUE | Downregulated |
| -1,02345003 | 0,019667558 | MTHFD1 | TRUE | Downregulated |
| -1,535351339 | 0,019758673 | B4GALT6 | TRUE | Downregulated |
| -1,465083639 | 0,019882009 | NPIPB11 | TRUE | Downregulated |
| -0,814784775 | 0,019895858 | ABAT | FALSE | Downregulated |
| -1,286795277 | 0,020014041 | SERPINA4 | TRUE | Downregulated |
| 1,417577119 | 0,020228516 | COL4A1 | TRUE | Upregulated |
| -1,535340137 | 0,020332974 | GCKR | TRUE | Downregulated |
| 0,907617245 | 0,020642184 | SLC26A1 | FALSE | Upregulated |
| 0,9546438 | 0,02080245 | LOC100996506 | FALSE | Upregulated |
| 1,88353671 | 0,021361339 | SLC7A6OS | TRUE | Upregulated |
| -2,278137622 | 0,021535068 | CYP2E1 | TRUE | Downregulated |
| 2,346168996 | 0,021535068 | FRMD5 | TRUE | Upregulated |
| -1,319615484 | 0,021579123 | ABCC9 | TRUE | Downregulated |
| 1,824093579 | 0,021579123 | RALGDS | TRUE | Upregulated |

| | | | | |
|---|---|---|---|---|
| -1,139979498 | 0,021692399 | GOT2 | TRUE | Downregulated |
| -1,181492184 | 0,021835652 | F11 | TRUE | Downregulated |
| 1,131945694 | 0,021951773 | ABCA2 | TRUE | Upregulated |
| -1,700231844 | 0,022729632 | CITED2 | TRUE | Downregulated |
| -1,181577173 | 0,022781856 | FGB | TRUE | Downregulated |
| 1,190088523 | 0,022781856 | TYMP | TRUE | Upregulated |
| -1,441236752 | 0,023392624 | PRDX6 | TRUE | Downregulated |
| -1,006596138 | 0,02363727 | CNNM3 | TRUE | Downregulated |
| -1,168487611 | 0,02363727 | CPB2-AS1 | TRUE | Downregulated |
| 1,112003386 | 0,023844515 | RP5-1142A6.2 | TRUE | Upregulated |
| -1,188546339 | 0,023844515 | TMEM176B | TRUE | Downregulated |
| -1,182648385 | 0,024427648 | SLC16A2 | TRUE | Downregulated |
| -1,958889311 | 0,024538394 | EI24 | TRUE | Downregulated |
| -1,051854155 | 0,025101498 | EHHADH | TRUE | Downregulated |
| -1,353230798 | 0,025101498 | KLF9 | TRUE | Downregulated |
| -1,342370402 | 0,025110464 | LCAT | TRUE | Downregulated |
| 2,017368017 | 0,026501486 | WWC1 | TRUE | Upregulated |
| -0,905114178 | 0,026949297 | EIF4B | FALSE | Downregulated |
| -1,641119885 | 0,027149637 | GPHN | TRUE | Downregulated |
| 1,282493329 | 0,027149637 | LOC101929333 | TRUE | Upregulated |
| -1,135778583 | 0,027508522 | PLIN5 | TRUE | Downregulated |
| -1,30765624 | 0,02753632 | MRPL43 | TRUE | Downregulated |
| 1,142289391 | 0,027647752 | GJA9-MYCBP | TRUE | Upregulated |
| -1,718844669 | 0,027717358 | TTC36 | TRUE | Downregulated |
| 1,565950683 | 0,027899233 | CD160 | TRUE | Upregulated |
| -1,078859596 | 0,028610314 | C8B | TRUE | Downregulated |
| 1,113892244 | 0,028610314 | DOCK5 | TRUE | Upregulated |
| -1,577458746 | 0,028610314 | LOC102724607 | TRUE | Downregulated |
| -1,129069752 | 0,028713244 | ECE1 | TRUE | Downregulated |
| 1,394855097 | 0,028713244 | LOC101929573 | TRUE | Upregulated |
| -1,142150935 | 0,029874547 | HGD | TRUE | Downregulated |
| -1,336878455 | 0,030210957 | PRLR | TRUE | Downregulated |
| -0,920190771 | 0,030506335 | ITIH2 | FALSE | Downregulated |
| -1,755281594 | 0,030692133 | CA2 | TRUE | Downregulated |
| 1,004628817 | 0,031031703 | MDM2 | TRUE | Upregulated |
| -1,031200144 | 0,032298574 | AFM | TRUE | Downregulated |
| -1,235823138 | 0,032377371 | ETFB | TRUE | Downregulated |
| 1,444774197 | 0,032435962 | BCL2L1 | TRUE | Upregulated |
| 1,377413068 | 0,032435962 | CNNM1 | TRUE | Upregulated |
| -1,916603163 | 0,032435962 | CNTFR-AS1 | TRUE | Downregulated |
| -1,041889956 | 0,032435962 | FXR1 | TRUE | Downregulated |
| -0,83196982 | 0,032435962 | ITPR2 | FALSE | Downregulated |
| -2,734384428 | 0,032435962 | LOC101929787 | TRUE | Downregulated |
| 1,348316708 | 0,033061295 | MAFK | TRUE | Upregulated |
| 0,964974796 | 0,033214177 | LOC100294362 | FALSE | Upregulated |
| -2,291602661 | 0,033431089 | LOC150051 | TRUE | Downregulated |
| 1,535846459 | 0,033431089 | ZEB2 | TRUE | Upregulated |
| -1,218421831 | 0,033715903 | ST13 | TRUE | Downregulated |
| -1,652315935 | 0,034636338 | SLC1A2 | TRUE | Downregulated |
| 2,080139847 | 0,034669173 | DBNDD1 | TRUE | Upregulated |
| 1,68169378 | 0,034669173 | FLG2 | TRUE | Upregulated |

| | | | | |
|---|---|---|---|---|
| -1,946688675 | 0,034669173 | SH3BGRL2 | TRUE | Downregulated |
| -1,053868642 | 0,034692801 | CFH | TRUE | Downregulated |
| -2,28080841 | 0,035151712 | GPR88 | TRUE | Downregulated |
| 1,04414756 | 0,035151712 | NCAPH2 | TRUE | Upregulated |
| -2,096336266 | 0,035249388 | PTPRD | TRUE | Downregulated |
| 2,025973616 | 0,035249388 | RUNX1 | TRUE | Upregulated |
| -1,759709506 | 0,035249388 | SGMS2 | TRUE | Downregulated |
| -0,901200871 | 0,035249388 | SNTB1 | FALSE | Downregulated |
| -1,010438889 | 0,036050727 | HAGH | TRUE | Downregulated |
| -1,283425107 | 0,037095563 | FAM109A | TRUE | Downregulated |
| 2,102017804 | 0,03739403 | TNFSF14 | TRUE | Upregulated |
| 1,110369997 | 0,03777422 | KIAA1551 | TRUE | Upregulated |
| 1,148866928 | 0,037794905 | FAT2 | TRUE | Upregulated |
| 2,07806238 | 0,038315368 | LOC102724077 | TRUE | Upregulated |
| -1,298362912 | 0,038333022 | RP5-1071N3.1 | TRUE | Downregulated |
| -1,172522471 | 0,038802127 | THSD7A | TRUE | Downregulated |
| -0,936971143 | 0,039158634 | SHMT1 | FALSE | Downregulated |
| -0,927817752 | 0,039227139 | DHTKD1 | FALSE | Downregulated |
| -1,149244474 | 0,039227139 | NUCKS1 | TRUE | Downregulated |
| -1,030051637 | 0,039546291 | PCYOX1 | TRUE | Downregulated |
| -0,93154781 | 0,039626087 | SLC9A3R2 | FALSE | Downregulated |
| -1,886079126 | 0,039917046 | ACADL | TRUE | Downregulated |
| 1,087869865 | 0,039917046 | LOC100128882 | TRUE | Upregulated |
| -1,445466079 | 0,040146225 | CFHR5 | TRUE | Downregulated |
| 0,875816113 | 0,040456336 | SFPQ | FALSE | Upregulated |
| 1,450585836 | 0,040506465 | ADAMTS2 | TRUE | Upregulated |
| -0,751774069 | 0,040674502 | ITIH4 | FALSE | Downregulated |
| -1,253681251 | 0,040747125 | KCTD3 | TRUE | Downregulated |
| 1,434399882 | 0,040747125 | KRT18 | TRUE | Upregulated |
| 1,361917806 | 0,040747125 | PIK3IP1 | TRUE | Upregulated |
| -1,165904232 | 0,041846265 | NPAT | TRUE | Downregulated |
| 1,62587081 | 0,042090002 | DXO | TRUE | Upregulated |
| 1,51006306 | 0,042190476 | C19orf12 | TRUE | Upregulated |
| -1,244384617 | 0,042190476 | PON1 | TRUE | Downregulated |
| -1,077082871 | 0,042918939 | MTTP | TRUE | Downregulated |
| -2,413809732 | 0,043799639 | ADAMTS8 | TRUE | Downregulated |
| -0,797538079 | 0,044024658 | APOE | FALSE | Downregulated |
| 1,705900934 | 0,044024658 | SPON2 | TRUE | Upregulated |
| -1,195851308 | 0,044177989 | HADHA | TRUE | Downregulated |
| -1,000390456 | 0,044177989 | SLC7A2 | TRUE | Downregulated |
| 1,748845472 | 0,044215726 | CITED4 | TRUE | Upregulated |
| 1,849122988 | 0,044215726 | MFAP4 | TRUE | Upregulated |
| -1,201400373 | 0,044215726 | WHAMM | TRUE | Downregulated |
| -1,012386661 | 0,04426863 | DAAM2 | TRUE | Downregulated |
| 1,673293972 | 0,044314325 | PLOD3 | TRUE | Upregulated |
| 1,350423681 | 0,044525735 | LOC102724536 | TRUE | Upregulated |
| 1,206177982 | 0,045195033 | TMEM2 | TRUE | Upregulated |
| -1,521989431 | 0,045496659 | CFHR2 | TRUE | Downregulated |
| -2,229249802 | 0,045962978 | PIWIL2 | TRUE | Downregulated |
| 1,620375995 | 0,046177413 | TNC | TRUE | Upregulated |
| -2,086591611 | 0,046254246 | SRD5A2 | TRUE | Downregulated |

| | | | | |
|---|---|---|---|---|
| -1,463866643 | 0,046368096 | BTD | TRUE | Downregulated |
| 1,088700788 | 0,047246728 | LASP1 | TRUE | Upregulated |
| 1,633277316 | 0,048186366 | C14orf105 | TRUE | Upregulated |
| -1,091677392 | 0,048186366 | GNE | TRUE | Downregulated |
| -1,18912659 | 0,048607139 | TBX3 | TRUE | Downregulated |
| 1,737817318 | 0,048813392 | MEGF6 | TRUE | Upregulated |
| 1,296152405 | 0,050130965 | LOC102724342 | FALSE | Upregulated |
| 1,661380763 | 0,050134816 | SRPX2 | FALSE | Upregulated |
| -2,270421442 | 0,050774154 | ACKR2 | FALSE | Downregulated |
| -1,707575326 | 0,05116723 | FAHD2A | FALSE | Downregulated |
| 0,841883013 | 0,05116723 | NEAT1 | FALSE | Upregulated |
| -1,190955561 | 0,052306602 | ENPP1 | FALSE | Downregulated |
| 1,295072858 | 0,052306602 | LOC100126784 | FALSE | Upregulated |
| -1,386683596 | 0,052532519 | HRSP12 | FALSE | Downregulated |
| -1,657580293 | 0,052532519 | KCND3 | FALSE | Downregulated |
| 1,513558193 | 0,05273072 | SIRPA | FALSE | Upregulated |
| 1,413008484 | 0,052788676 | TNXB | FALSE | Upregulated |
| 1,166680537 | 0,053103287 | LAMP2 | FALSE | Upregulated |
| -1,344611465 | 0,054037545 | USH2A | FALSE | Downregulated |
| 1,823647783 | 0,054719904 | LOC102724584 | FALSE | Upregulated |
| 0,823554314 | 0,0548164 | CCNL1 | FALSE | Upregulated |
| 1,75201481 | 0,0548164 | LOC285095 | FALSE | Upregulated |
| -0,984459446 | 0,055091558 | ANGPTL3 | FALSE | Downregulated |
| -0,958658391 | 0,055091558 | FTCD | FALSE | Downregulated |
| -0,913566027 | 0,056134699 | DBH | FALSE | Downregulated |
| -1,359629542 | 0,056513065 | LGR4 | FALSE | Downregulated |
| -0,993418286 | 0,057074915 | NHSL1 | FALSE | Downregulated |
| -1,539968901 | 0,058415335 | SLIT3 | FALSE | Downregulated |
| -1,010594938 | 0,05869695 | C5 | FALSE | Downregulated |
| 1,739257828 | 0,05869695 | PLEKHO2 | FALSE | Upregulated |
| -1,090125104 | 0,058793017 | AMOTL2 | FALSE | Downregulated |
| 0,917712975 | 0,059846605 | SLC12A7 | FALSE | Upregulated |
| -1,323285609 | 0,0602064 | CD4 | FALSE | Downregulated |
| 1,308613485 | 0,060829737 | ZMIZ2 | FALSE | Upregulated |
| -0,959496042 | 0,060839012 | TJP2 | FALSE | Downregulated |
| -1,765430384 | 0,060841837 | FAM200B | FALSE | Downregulated |
| 1,322066431 | 0,060968574 | KIAA1598 | FALSE | Upregulated |
| -1,148586323 | 0,061087981 | KLKB1 | FALSE | Downregulated |
| -1,587300901 | 0,061087981 | TSPEAR | FALSE | Downregulated |
| -2,092016797 | 0,061212254 | SH3RF2 | FALSE | Downregulated |
| -0,823761131 | 0,061458051 | BAAT | FALSE | Downregulated |
| -1,81618656 | 0,061799331 | LOC730101 | FALSE | Downregulated |
| -1,061495381 | 0,062015163 | CFI | FALSE | Downregulated |
| -1,598120987 | 0,062040231 | C12orf60 | FALSE | Downregulated |
| -1,052400813 | 0,062213702 | DNAJC16 | FALSE | Downregulated |
| 0,974368117 | 0,062374063 | PVRIG | FALSE | Upregulated |
| 1,967452417 | 0,062812481 | ABCC5 | FALSE | Upregulated |
| -0,865915107 | 0,062812481 | DCAF11 | FALSE | Downregulated |
| 1,109255463 | 0,062879061 | C5orf66 | FALSE | Upregulated |
| -1,008468317 | 0,062879061 | TMEM56 | FALSE | Downregulated |
| 1,063260325 | 0,062954721 | FAM168A | FALSE | Upregulated |

| | | | | |
|---|---|---|---|---|
| -0,848987427 | 0,063181024 | ST3GAL1 | FALSE | Downregulated |
| 1,414648035 | 0,063181024 | STAT1 | FALSE | Upregulated |
| 1,578747968 | 0,06424171 | SLC46A2 | FALSE | Upregulated |
| -1,174679689 | 0,06484449 | PSMB7 | FALSE | Downregulated |
| 0,924326231 | 0,064932359 | RRN3P3 | FALSE | Upregulated |
| -0,927346631 | 0,065327413 | BHMT2 | FALSE | Downregulated |
| 1,399883882 | 0,065570636 | RP5-977B1.7 | FALSE | Upregulated |
| -0,861225909 | 0,066063451 | ANPEP | FALSE | Downregulated |
| 0,90917404 | 0,066063451 | CEP170B | FALSE | Upregulated |
| 0,96468231 | 0,066179882 | LOC101929240 | FALSE | Upregulated |
| -0,972545779 | 0,06626121 | FBP1 | FALSE | Downregulated |
| 1,171558167 | 0,06626121 | KIFC3 | FALSE | Upregulated |
| 1,126989982 | 0,066288159 | CSNK1D | FALSE | Upregulated |
| -1,468185422 | 0,066299628 | TDO2 | FALSE | Downregulated |
| -1,088804166 | 0,06747298 | CPB2 | FALSE | Downregulated |
| -1,13997516 | 0,06844932 | CAPN5 | FALSE | Downregulated |
| 1,251151843 | 0,068458362 | CD151 | FALSE | Upregulated |
| -1,282184843 | 0,069273584 | EHHADH-AS1 | FALSE | Downregulated |
| -1,597971909 | 0,069273584 | LOC728024 | FALSE | Downregulated |
| -1,340216382 | 0,069866142 | TTLL4 | FALSE | Downregulated |
| -0,934838047 | 0,07008583 | CSDE1 | FALSE | Downregulated |
| -1,20122851 | 0,070468479 | IGF1 | FALSE | Downregulated |
| 1,703713658 | 0,070477173 | LOC101928212 | FALSE | Upregulated |
| -1,484736285 | 0,070785865 | FAH | FALSE | Downregulated |
| 0,759032602 | 0,071107287 | AKAP13 | FALSE | Upregulated |
| -1,204916889 | 0,071387073 | SLC27A2 | FALSE | Downregulated |
| -0,981331222 | 0,071882376 | NIPSNAP1 | FALSE | Downregulated |
| -1,192149731 | 0,072019187 | SELENBP1 | FALSE | Downregulated |
| -0,973125366 | 0,072125924 | C1RL | FALSE | Downregulated |
| 2,083051158 | 0,072125924 | ICAM1 | FALSE | Upregulated |
| -1,019606869 | 0,072125924 | MAF | FALSE | Downregulated |
| -1,317605496 | 0,072125924 | PGRMC2 | FALSE | Downregulated |
| -1,437817471 | 0,072125924 | SYCE2 | FALSE | Downregulated |
| -2,213117766 | 0,072501818 | UQCRB | FALSE | Downregulated |
| 1,38447153 | 0,072844006 | RANGAP1 | FALSE | Upregulated |
| -1,060947184 | 0,073868777 | IDH1 | FALSE | Downregulated |
| -0,972171058 | 0,073868777 | PROC | FALSE | Downregulated |
| -1,406226069 | 0,073921849 | FXYD1 | FALSE | Downregulated |
| -1,023612219 | 0,073921849 | MTERFD2 | FALSE | Downregulated |
| -0,744150502 | 0,073921849 | RXRA | FALSE | Downregulated |
| -1,259922652 | 0,074408145 | PHYH | FALSE | Downregulated |
| -2,249381471 | 0,074757027 | LPA | FALSE | Downregulated |
| -1,134694255 | 0,075594985 | CRAT | FALSE | Downregulated |
| -1,567576535 | 0,075842398 | HADH | FALSE | Downregulated |
| 1,099476854 | 0,076235246 | TNFRSF10B | FALSE | Upregulated |
| -1,075085824 | 0,07646142 | PGRMC1 | FALSE | Downregulated |
| -1,188970817 | 0,076485894 | CYP51A1 | FALSE | Downregulated |
| -1,524471945 | 0,076588341 | GALK1 | FALSE | Downregulated |
| -0,692366805 | 0,0766617 | AHSG | FALSE | Downregulated |
| -0,939108897 | 0,076957535 | C4BPA | FALSE | Downregulated |
| 1,32198478 | 0,076957535 | LOC100506142 | FALSE | Upregulated |

| | | | | |
|---|---|---|---|---|
| 1,312548564 | 0,076957535 | MFHAS1 | FALSE | Upregulated |
| -1,08968518 | 0,076982577 | AKR1C4 | FALSE | Downregulated |
| 1,68391281 | 0,077147339 | LRRC20 | FALSE | Upregulated |
| 1,695836026 | 0,077595804 | LAPTM5 | FALSE | Upregulated |
| 1,251207704 | 0,077717106 | VWA1 | FALSE | Upregulated |
| -1,027242736 | 0,077820589 | OPN3 | FALSE | Downregulated |
| 0,851803992 | 0,077820589 | PDZD2 | FALSE | Upregulated |
| 1,066185396 | 0,078201921 | COL4A2 | FALSE | Upregulated |
| -1,540257284 | 0,078201921 | PSAT1 | FALSE | Downregulated |
| -0,849222175 | 0,078325637 | ATF5 | FALSE | Downregulated |
| 1,28624156 | 0,078325637 | DMXL2 | FALSE | Upregulated |
| -0,765115031 | 0,078325637 | HGFAC | FALSE | Downregulated |
| 0,779560258 | 0,078467011 | LOC101929545 | FALSE | Upregulated |
| 0,928553325 | 0,07944778 | CRYBG3 | FALSE | Upregulated |
| -1,510535638 | 0,079558804 | EBP | FALSE | Downregulated |
| -1,453405949 | 0,080560224 | NPIPB6 | FALSE | Downregulated |
| 1,355344399 | 0,080560224 | PISD | FALSE | Upregulated |
| -1,934565931 | 0,080560224 | SLC22A10 | FALSE | Downregulated |
| 1,331889383 | 0,080560224 | ZFYVE26 | FALSE | Upregulated |
| 1,746405631 | 0,080992437 | OGN | FALSE | Upregulated |
| -1,565189184 | 0,081069449 | APOBR | FALSE | Downregulated |
| -1,728222345 | 0,081069449 | PPP1R3C | FALSE | Downregulated |
| -0,710771378 | 0,081323406 | FURIN | FALSE | Downregulated |
| -1,181719422 | 0,081444908 | ZBTB10 | FALSE | Downregulated |
| -1,250466237 | 0,081890101 | FADS2 | FALSE | Downregulated |
| 1,729891853 | 0,081890101 | ITGA6 | FALSE | Upregulated |
| 1,170723489 | 0,082406509 | ITGB5 | FALSE | Upregulated |
| 1,387726928 | 0,082406509 | PTPN7 | FALSE | Upregulated |
| 1,330652774 | 0,082876828 | GNRH1 | FALSE | Upregulated |
| 0,704204974 | 0,082951191 | CTSB | FALSE | Upregulated |
| 1,244124849 | 0,082951191 | RP4-591N18.2 | FALSE | Upregulated |
| -1,560002548 | 0,083080377 | CYP2J2 | FALSE | Downregulated |
| -0,816480428 | 0,084210802 | OIT3 | FALSE | Downregulated |
| 1,308490032 | 0,084505262 | EHBP1L1 | FALSE | Upregulated |
| -0,865472394 | 0,084954834 | ESRP2 | FALSE | Downregulated |
| 2,164931096 | 0,085222371 | FLG | FALSE | Upregulated |
| -0,804520573 | 0,085324889 | GLYCTK | FALSE | Downregulated |
| -1,90716464 | 0,085324889 | LPAL2 | FALSE | Downregulated |
| 0,722568341 | 0,085324889 | NPIPB4 | FALSE | Upregulated |
| -1,109091324 | 0,085324889 | PPID | FALSE | Downregulated |
| -1,211503809 | 0,085324889 | RPN1 | FALSE | Downregulated |
| -0,813956501 | 0,08615465 | FMO3 | FALSE | Downregulated |
| -0,899635163 | 0,08637205 | TMEM110 | FALSE | Downregulated |
| 1,150947612 | 0,086479104 | CPEB4 | FALSE | Upregulated |
| -0,843284462 | 0,086479104 | SLC5A2 | FALSE | Downregulated |
| -1,632430291 | 0,086641853 | DNASE1L3 | FALSE | Downregulated |
| 1,829870541 | 0,087792214 | KCNAB2 | FALSE | Upregulated |
| -1,121904672 | 0,089525881 | RGPD4 | FALSE | Downregulated |
| 1,398062955 | 0,090343257 | LOC102724335 | FALSE | Upregulated |
| -1,039910714 | 0,090364332 | ARG1 | FALSE | Downregulated |
| 0,829987963 | 0,090364332 | CTSD | FALSE | Upregulated |

| | | | | |
|---|---|---|---|---|
| -1,798327298 | 0,090472215 | NDUFB11 | FALSE | Downregulated |
| -0,61042387 | 0,091460019 | RPPH1 | FALSE | Downregulated |
| -1,087145486 | 0,091591333 | STBD1 | FALSE | Downregulated |
| 0,952977607 | 0,092001736 | CD59 | FALSE | Upregulated |
| -0,801956658 | 0,092001736 | TPT1 | FALSE | Downregulated |
| -1,483409663 | 0,092547607 | FERMT2 | FALSE | Downregulated |
| 1,048005735 | 0,092547607 | GRAMD4 | FALSE | Upregulated |
| -0,939992623 | 0,092547607 | SLC2A4RG | FALSE | Downregulated |
| -1,084731239 | 0,092547607 | ZNF483 | FALSE | Downregulated |
| -1,67335746 | 0,092868915 | CXCL2 | FALSE | Downregulated |
| -0,946280342 | 0,092868915 | KNG1 | FALSE | Downregulated |
| 0,708226434 | 0,095210597 | MALAT1 | FALSE | Upregulated |
| -1,404504395 | 0,095210597 | MT1E | FALSE | Downregulated |
| -1,783752712 | 0,095823124 | HOGA1 | FALSE | Downregulated |
| -1,981146146 | 0,095823124 | HS2ST1 | FALSE | Downregulated |
| -0,833132359 | 0,095823124 | TTC28 | FALSE | Downregulated |
| 1,081624658 | 0,095843996 | PHOSPHO1 | FALSE | Upregulated |
| -1,059727654 | 0,097385704 | PIRT | FALSE | Downregulated |
| -1,849387608 | 0,097827671 | SLC17A2 | FALSE | Downregulated |
| 1,887624323 | 0,098296902 | ZMAT3 | FALSE | Upregulated |
| -1,615455501 | 0,098369737 | PTH1R | FALSE | Downregulated |
| 0,907228667 | 0,098527132 | SEC61A1 | FALSE | Upregulated |
| -1,214501019 | 0,099599898 | ACVR2B-AS1 | FALSE | Downregulated |
| -2,166557854 | 0,099599898 | POU6F2-AS2 | FALSE | Downregulated |
| -1,416373932 | 0,100033354 | C4BPB | FALSE | Downregulated |
| 0,977980143 | 0,100878757 | HTR2B | FALSE | Upregulated |
| 1,056207182 | 0,101088645 | KLHL29 | FALSE | Upregulated |
| -0,842882227 | 0,10135142 | NFATC3 | FALSE | Downregulated |
| -1,842420684 | 0,103476236 | MLIP | FALSE | Downregulated |
| 1,047872356 | 0,103572475 | LOC653513 | FALSE | Upregulated |
| -1,158275275 | 0,103793341 | SHMT2 | FALSE | Downregulated |
| -1,164595174 | 0,103793341 | ZBTB16 | FALSE | Downregulated |
| 1,699109905 | 0,103904505 | SLC27A4 | FALSE | Upregulated |
| -0,861569996 | 0,103996859 | BDH1 | FALSE | Downregulated |
| 1,584544171 | 0,104750646 | SMAD7 | FALSE | Upregulated |
| -1,173596473 | 0,10501822 | MAOA | FALSE | Downregulated |
| 1,286443332 | 0,10501822 | RECQL4 | FALSE | Upregulated |
| -0,919031986 | 0,10501822 | UGT2B10 | FALSE | Downregulated |
| 0,869529325 | 0,105091164 | ERBB2 | FALSE | Upregulated |
| -0,801311776 | 0,105199685 | ADCY1 | FALSE | Downregulated |
| -1,442715253 | 0,105199685 | REN | FALSE | Downregulated |
| -1,43303558 | 0,10564112 | THRSP | FALSE | Downregulated |
| -1,718815264 | 0,106721438 | KCNIP3 | FALSE | Downregulated |
| -1,582934408 | 0,106811549 | NDUFC2 | FALSE | Downregulated |
| -1,603427367 | 0,107781607 | LOC101927157 | FALSE | Downregulated |
| -1,378535211 | 0,107928374 | MSMO1 | FALSE | Downregulated |
| -1,029922441 | 0,10870883 | C4A | FALSE | Downregulated |
| 1,100010857 | 0,10870883 | CCDC97 | FALSE | Upregulated |
| 1,906744095 | 0,10870883 | TNFSF8 | FALSE | Upregulated |
| 1,587946962 | 0,108728256 | DOCK2 | FALSE | Upregulated |
| 0,81645847 | 0,108728256 | LOC101928120 | FALSE | Upregulated |

| | | | | |
|---|---|---|---|---|
| 1,044754885 | 0,108728256 | PDAP1 | FALSE | Upregulated |
| -1,12162362 | 0,109831519 | ANG | FALSE | Downregulated |
| -1,173879058 | 0,109831519 | NUDT4 | FALSE | Downregulated |
| -1,323056133 | 0,110317804 | TEF | FALSE | Downregulated |
| -0,844983786 | 0,1111783 | EIF4EBP2 | FALSE | Downregulated |
| -0,873907509 | 0,111185876 | EPB41L2 | FALSE | Downregulated |
| -0,993068761 | 0,112430065 | FNIP2 | FALSE | Downregulated |
| -0,857802572 | 0,112430065 | IRS1 | FALSE | Downregulated |
| 1,293000223 | 0,112430065 | NINJ1 | FALSE | Upregulated |
| 1,079861283 | 0,112430065 | SLC38A2 | FALSE | Upregulated |
| 1,190428835 | 0,112430065 | TRIM17 | FALSE | Upregulated |
| -1,003919339 | 0,113059248 | GLTSCR2 | FALSE | Downregulated |
| -0,969400257 | 0,113460181 | RPE | FALSE | Downregulated |
| -1,200349095 | 0,113460181 | SLC37A4 | FALSE | Downregulated |
| -0,759544579 | 0,11488463 | CYP4F3 | FALSE | Downregulated |
| -0,972492758 | 0,114945025 | ASGR1 | FALSE | Downregulated |
| -1,910901348 | 0,11496657 | NAV2-AS4 | FALSE | Downregulated |
| -0,977668822 | 0,115745269 | CEBPB | FALSE | Downregulated |
| -1,218325919 | 0,115745269 | RP11-478P10.1 | FALSE | Downregulated |
| -1,472540131 | 0,11677101 | ACBD5 | FALSE | Downregulated |
| 0,940756648 | 0,11677101 | ATP11A | FALSE | Upregulated |
| 1,347494048 | 0,117692562 | STAG3 | FALSE | Upregulated |
| 1,09973127 | 0,118289679 | FAS | FALSE | Upregulated |
| -1,256451127 | 0,118318169 | BLVRB | FALSE | Downregulated |
| -1,033006182 | 0,119461734 | LOC644090 | FALSE | Downregulated |
| 1,340857683 | 0,119496647 | PPDPF | FALSE | Upregulated |
| 1,018528089 | 0,119496647 | SIPA1L3 | FALSE | Upregulated |
| 1,410145392 | 0,120063155 | ATAD2B | FALSE | Upregulated |
| -1,424678611 | 0,120063155 | ERMAP | FALSE | Downregulated |
| 0,783735245 | 0,120070831 | SLC47A1 | FALSE | Upregulated |
| -0,955144133 | 0,120421397 | TAT | FALSE | Downregulated |
| -1,403190618 | 0,120894962 | ATPIF1 | FALSE | Downregulated |
| 0,959912573 | 0,120894962 | FNDC3B | FALSE | Upregulated |
| -0,68480769 | 0,120894962 | RPL13 | FALSE | Downregulated |
| 1,346536918 | 0,120894962 | SLC35G2 | FALSE | Upregulated |
| -0,981864075 | 0,12144348 | RGN | FALSE | Downregulated |
| -1,682549802 | 0,122004089 | LINC01152 | FALSE | Downregulated |
| -0,600945169 | 0,122004089 | LOC102724129 | FALSE | Downregulated |
| -1,03281417 | 0,12324713 | RNASE4 | FALSE | Downregulated |
| 1,582102801 | 0,123683088 | BYSL | FALSE | Upregulated |
| -1,038883556 | 0,123683088 | LOC100507033 | FALSE | Downregulated |
| -0,791696131 | 0,123683088 | SERPING1 | FALSE | Downregulated |
| -1,640542204 | 0,124659981 | SYBU | FALSE | Downregulated |
| -0,704644201 | 0,124694268 | PAH | FALSE | Downregulated |
| -1,148444589 | 0,126183306 | MAP2K1 | FALSE | Downregulated |
| -0,876849114 | 0,12651916 | BSN | FALSE | Downregulated |
| -1,577493886 | 0,126584353 | GIPC2 | FALSE | Downregulated |
| 0,825514478 | 0,130055064 | TLDC2 | FALSE | Upregulated |
| 0,84239154 | 0,130618649 | TDRP | FALSE | Upregulated |
| -0,85585117 | 0,130851503 | RPL29 | FALSE | Downregulated |
| 0,959028625 | 0,130851503 | SNCAIP | FALSE | Upregulated |

| | | | | |
|---|---|---|---|---|
| -0,863801998 | 0,131109517 | NR1I2 | FALSE | Downregulated |
| -1,43119513 | 0,132520892 | TTPA | FALSE | Downregulated |
| -0,95797728 | 0,133219549 | RPL23 | FALSE | Downregulated |
| -1,576552325 | 0,134176087 | TSNARE1 | FALSE | Downregulated |
| -0,819674285 | 0,134320894 | GRIN3A | FALSE | Downregulated |
| -1,055402862 | 0,134320894 | RP11-432I5.1 | FALSE | Downregulated |
| -0,827433172 | 0,135112287 | INS-IGF2 | FALSE | Downregulated |
| -1,712503974 | 0,135112287 | RP1-293L8.2 | FALSE | Downregulated |
| 0,897515151 | 0,135912731 | LOC101928793 | FALSE | Upregulated |
| -0,949655876 | 0,136290302 | HSD17B4 | FALSE | Downregulated |
| -1,559070857 | 0,136401662 | TM7SF2 | FALSE | Downregulated |
| -1,106008238 | 0,136401662 | TNFSF10 | FALSE | Downregulated |
| -1,867152947 | 0,136477076 | TIGD7 | FALSE | Downregulated |
| 1,110326738 | 0,137019382 | ANGPTL7 | FALSE | Upregulated |
| -1,253158327 | 0,137416735 | NAV2-AS5 | FALSE | Downregulated |
| -1,475870418 | 0,138388364 | FAHD1 | FALSE | Downregulated |
| 1,262638853 | 0,139080005 | AEBP1 | FALSE | Upregulated |
| -1,006764681 | 0,139085878 | EPB41L5 | FALSE | Downregulated |
| 0,850817608 | 0,13968845 | DTX4 | FALSE | Upregulated |
| -0,837777809 | 0,13968845 | NFIA | FALSE | Downregulated |
| 1,221390032 | 0,139693757 | LOC400553 | FALSE | Upregulated |
| -0,692507487 | 0,140764073 | LOC646471 | FALSE | Downregulated |
| -0,902231385 | 0,141621908 | PPP1R26 | FALSE | Downregulated |
| 1,092919291 | 0,14219704 | PLTP | FALSE | Upregulated |
| -1,315505078 | 0,142617896 | FUOM | FALSE | Downregulated |
| 1,06080375 | 0,142617896 | URAHP | FALSE | Upregulated |
| -0,656979211 | 0,144242142 | F7 | FALSE | Downregulated |
| -0,777014148 | 0,144573573 | AGFG2 | FALSE | Downregulated |
| -0,803842238 | 0,144625829 | ASGR2 | FALSE | Downregulated |
| 0,942233574 | 0,14520513 | SPTAN1 | FALSE | Upregulated |
| -1,413015051 | 0,148481865 | GSTZ1 | FALSE | Downregulated |
| 0,983214182 | 0,148481865 | MMP15 | FALSE | Upregulated |
| 0,957014516 | 0,148481865 | TACC1 | FALSE | Upregulated |
| -0,749329435 | 0,149454816 | UGP2 | FALSE | Downregulated |
| 1,380779983 | 0,149473742 | TAGLN2 | FALSE | Upregulated |
| 1,111636583 | 0,150539028 | CLPTM1L | FALSE | Upregulated |
| -1,614193377 | 0,150748951 | ACSM1 | FALSE | Downregulated |
| -1,041178254 | 0,150748951 | DCAF6 | FALSE | Downregulated |
| 1,488971722 | 0,150748951 | FCHSD1 | FALSE | Upregulated |
| -0,739039293 | 0,150748951 | PPL | FALSE | Downregulated |
| 0,965955755 | 0,150748951 | SHROOM1 | FALSE | Upregulated |
| 1,362841486 | 0,150748951 | TAF11 | FALSE | Upregulated |
| -0,824041187 | 0,1517478 | MPDZ | FALSE | Downregulated |
| -1,108493566 | 0,151781639 | ACBD4 | FALSE | Downregulated |
| 1,069410146 | 0,152394185 | TULP4 | FALSE | Upregulated |
| 1,499880269 | 0,153091346 | MRC2 | FALSE | Upregulated |
| 1,063161555 | 0,153947664 | LOC728805 | FALSE | Upregulated |
| -0,734215196 | 0,154031745 | PIPOX | FALSE | Downregulated |
| 1,761377156 | 0,154073317 | MYO5A | FALSE | Upregulated |
| 1,850856538 | 0,154894191 | LOC101928783 | FALSE | Upregulated |
| -0,951587466 | 0,154894191 | RAD54L2 | FALSE | Downregulated |

| | | | | |
|---|---|---|---|---|
| -1,286763577 | 0,154918533 | SERPINA7 | FALSE | Downregulated |
| -0,920978595 | 0,156293587 | MTDH | FALSE | Downregulated |
| 1,026912645 | 0,157037751 | CDH23 | FALSE | Upregulated |
| 1,216459635 | 0,157037751 | MIRLET7BHG | FALSE | Upregulated |
| -1,251785317 | 0,157164988 | SLC30A10 | FALSE | Downregulated |
| 1,693324773 | 0,158260961 | ARHGAP4 | FALSE | Upregulated |
| -0,846076698 | 0,158450379 | SDHA | FALSE | Downregulated |
| 1,377704151 | 0,158648433 | PZP | FALSE | Upregulated |
| -1,57353485 | 0,161417348 | LOC102723877 | FALSE | Downregulated |
| 1,007975544 | 0,161561176 | FMNL2 | FALSE | Upregulated |
| -1,300491655 | 0,161561176 | GANC | FALSE | Downregulated |
| -0,807122482 | 0,161702578 | MTMR4 | FALSE | Downregulated |
| 0,820131641 | 0,162292065 | LOC101929599 | FALSE | Upregulated |
| 0,708802241 | 0,162292065 | MGAT4B | FALSE | Upregulated |
| -0,912616555 | 0,163062874 | GPAM | FALSE | Downregulated |
| -0,872681648 | 0,163719824 | RPL18 | FALSE | Downregulated |
| -1,329577148 | 0,164072743 | HADHB | FALSE | Downregulated |
| -1,127696513 | 0,164072743 | LSS | FALSE | Downregulated |
| -0,747230373 | 0,164072743 | UPB1 | FALSE | Downregulated |
| -0,641327946 | 0,164361301 | APOB | FALSE | Downregulated |
| 1,299437284 | 0,164361301 | ASCC3 | FALSE | Upregulated |
| 1,127525449 | 0,164361301 | GAS6 | FALSE | Upregulated |
| -1,419512223 | 0,165323581 | PDLIM2 | FALSE | Downregulated |
| 0,981218393 | 0,165602786 | CD74 | FALSE | Upregulated |
| -1,173132106 | 0,165855193 | SULT1A1 | FALSE | Downregulated |
| 1,239258833 | 0,16609513 | AGAP6 | FALSE | Upregulated |
| -0,707757518 | 0,16609513 | PRAP1 | FALSE | Downregulated |
| 1,010647267 | 0,167976569 | TGM2 | FALSE | Upregulated |
| 0,746188345 | 0,168437464 | ITGA7 | FALSE | Upregulated |
| -1,16273321 | 0,16848464 | HFE2 | FALSE | Downregulated |
| -0,705483726 | 0,170899774 | HECTD1 | FALSE | Downregulated |
| -1,174836561 | 0,171359487 | LOC102724975 | FALSE | Downregulated |
| 0,856269595 | 0,172236877 | ABLIM1 | FALSE | Upregulated |
| -1,398951531 | 0,172365168 | DAO | FALSE | Downregulated |
| 1,260060389 | 0,172365168 | GPR82 | FALSE | Upregulated |
| -1,606683825 | 0,172365168 | UBE2N | FALSE | Downregulated |
| -1,386708382 | 0,172611988 | ATP5D | FALSE | Downregulated |
| -0,742937717 | 0,172611988 | SPTBN2 | FALSE | Downregulated |
| 1,26872744 | 0,172611988 | TGFB1 | FALSE | Upregulated |
| 1,165283965 | 0,172826272 | CORO1C | FALSE | Upregulated |
| -0,765209891 | 0,172826272 | LOC102723422 | FALSE | Downregulated |
| 0,965384981 | 0,173492079 | SPRTN | FALSE | Upregulated |
| 1,279260275 | 0,174084651 | LOC101927954 | FALSE | Upregulated |
| 0,981140569 | 0,175015356 | PYROXD1 | FALSE | Upregulated |
| -1,426132841 | 0,175053404 | JADE3 | FALSE | Downregulated |
| -0,931945411 | 0,175680975 | TMED2 | FALSE | Downregulated |
| 0,866020142 | 0,176398146 | LOC100506603 | FALSE | Upregulated |
| -1,249685514 | 0,176913243 | MIA2 | FALSE | Downregulated |
| -0,776018822 | 0,177137052 | SORL1 | FALSE | Downregulated |
| 0,792397078 | 0,177233513 | CLOCK | FALSE | Upregulated |
| 1,261964444 | 0,177513817 | LOC100996643 | FALSE | Upregulated |

| | | | | |
|---|---|---|---|---|
| -0,79688025 | 0,177750287 | ABCA6 | FALSE | Downregulated |
| -0,835563333 | 0,178007774 | ALDH5A1 | FALSE | Downregulated |
| -0,869800743 | 0,178047473 | FLJ35282 | FALSE | Downregulated |
| -0,844803684 | 0,178538537 | NR2F2 | FALSE | Downregulated |
| 1,447762587 | 0,178538537 | RAB11FIP1 | FALSE | Upregulated |
| -0,947811019 | 0,17939007 | CNBP | FALSE | Downregulated |
| -0,78076884 | 0,17971406 | TSPYL1 | FALSE | Downregulated |
| -0,862495897 | 0,180338232 | WDR65 | FALSE | Downregulated |
| -0,803381995 | 0,180722779 | SULT2A1 | FALSE | Downregulated |
| 1,078452625 | 0,180722779 | THBS1 | FALSE | Upregulated |
| 1,413708178 | 0,180722779 | YEATS2 | FALSE | Upregulated |
| 1,202690594 | 0,181365679 | CELF2-AS1 | FALSE | Upregulated |
| -0,98578735 | 0,184129381 | SLCO1B3 | FALSE | Downregulated |
| -1,378937149 | 0,184236915 | ALS2CR11 | FALSE | Downregulated |
| 1,494444502 | 0,184311568 | ENTPD4 | FALSE | Upregulated |
| 1,125865941 | 0,184373137 | AVL9 | FALSE | Upregulated |
| 0,636869134 | 0,184687411 | CYP8B1 | FALSE | Upregulated |
| 0,919242439 | 0,185498689 | RAB43 | FALSE | Upregulated |
| 0,827049943 | 0,185644283 | C19orf71 | FALSE | Upregulated |
| 0,786260143 | 0,185912682 | SMG1P1 | FALSE | Upregulated |
| -0,931011077 | 0,186309007 | AGL | FALSE | Downregulated |
| 1,424043031 | 0,186778696 | ELOVL2 | FALSE | Upregulated |
| -0,84718234 | 0,186778696 | SCAI | FALSE | Downregulated |
| 0,674216722 | 0,188212427 | TCIRG1 | FALSE | Upregulated |
| 1,042242963 | 0,188667024 | SIGLEC1 | FALSE | Upregulated |
| 1,14529596 | 0,188939531 | KIAA0754 | FALSE | Upregulated |
| -0,675840709 | 0,188977216 | ENG | FALSE | Downregulated |
| -1,070694243 | 0,190554749 | TOR1AIP1 | FALSE | Downregulated |
| 0,697015023 | 0,19293901 | LOC441081 | FALSE | Upregulated |
| 0,887196614 | 0,19293901 | SH3RF3-AS1 | FALSE | Upregulated |
| -1,11482038 | 0,19293901 | TUSC1 | FALSE | Downregulated |
| 1,09830815 | 0,193916005 | ACSS3 | FALSE | Upregulated |
| -1,369576417 | 0,194042693 | AC018705.5 | FALSE | Downregulated |
| 0,944736548 | 0,194042693 | FBXL2 | FALSE | Upregulated |
| -1,092347341 | 0,194042693 | MRPL2 | FALSE | Downregulated |
| -1,339077208 | 0,194653663 | SAFB2 | FALSE | Downregulated |
| -1,182773004 | 0,194756231 | CTSF | FALSE | Downregulated |
| -0,935760608 | 0,195238776 | ACSL5 | FALSE | Downregulated |
| -0,747772309 | 0,195238776 | C2orf88 | FALSE | Downregulated |
| 0,999183153 | 0,195238776 | MECP2 | FALSE | Upregulated |
| 1,341969939 | 0,195238776 | STARD3 | FALSE | Upregulated |
| -0,993469231 | 0,195343484 | PCSK1 | FALSE | Downregulated |
| -0,826613152 | 0,196240559 | PAPSS2 | FALSE | Downregulated |
| -0,964350836 | 0,196586978 | SUCLG2 | FALSE | Downregulated |
| 1,259858475 | 0,196925098 | TMEM75 | FALSE | Upregulated |
| 0,984215968 | 0,19776269 | TRIM26 | FALSE | Upregulated |
| 1,110070909 | 0,197822844 | FAM196B | FALSE | Upregulated |
| 1,079708509 | 0,198310347 | CHD7 | FALSE | Upregulated |
| -0,738579685 | 0,198310347 | COX11 | FALSE | Downregulated |
| -0,774177804 | 0,198310347 | SLC31A1 | FALSE | Downregulated |
| 0,777965407 | 0,198310347 | SMA5 | FALSE | Upregulated |

| | | | | |
|---|---|---|---|---|
| 0,877710829 | 0,198366991 | DNAJC5 | FALSE | Upregulated |
| -1,176538316 | 0,198419464 | KDM8 | FALSE | Downregulated |
| -1,490068962 | 0,198419464 | NDRG1 | FALSE | Downregulated |
| -1,247612311 | 0,198904204 | ZNF518B | FALSE | Downregulated |
| 0,799118439 | 0,200793003 | OR2A1 | FALSE | Upregulated |
| 0,902594659 | 0,200959285 | LOC102723535 | FALSE | Upregulated |
| 1,074043188 | 0,200959285 | MLXIP | FALSE | Upregulated |
| 0,859373156 | 0,200959285 | TIGD6 | FALSE | Upregulated |
| -0,830504315 | 0,20135554 | LRRC37A2 | FALSE | Downregulated |
| -1,298451267 | 0,201673622 | SCART1 | FALSE | Downregulated |
| 1,200505983 | 0,201759905 | ITGB2 | FALSE | Upregulated |
| 1,260558327 | 0,201759905 | NCKAP5L | FALSE | Upregulated |
| 0,889415217 | 0,201759905 | TSSK4 | FALSE | Upregulated |
| -0,818707682 | 0,202257946 | LMAN1 | FALSE | Downregulated |
| -0,88717501 | 0,202257946 | PROS1 | FALSE | Downregulated |
| -1,420287541 | 0,202288769 | PARK7 | FALSE | Downregulated |
| -1,213026728 | 0,203584254 | AKAP12 | FALSE | Downregulated |
| 0,855560319 | 0,204589981 | NPC1 | FALSE | Upregulated |
| -1,395085162 | 0,205240788 | ACVR2B | FALSE | Downregulated |
| -0,649212622 | 0,205522462 | LOC148696 | FALSE | Downregulated |
| 0,910812293 | 0,205522462 | SAMHD1 | FALSE | Upregulated |
| 1,384908835 | 0,206471464 | PLCB2 | FALSE | Upregulated |
| -1,300386869 | 0,208956724 | MARC1 | FALSE | Downregulated |
| -0,760786017 | 0,210228148 | CPN2 | FALSE | Downregulated |
| -0,661952954 | 0,210307131 | LONP2 | FALSE | Downregulated |
| -0,934392001 | 0,210609632 | GYS2 | FALSE | Downregulated |
| -1,31987506 | 0,211855705 | RHEB | FALSE | Downregulated |
| 0,993927369 | 0,212567956 | SPIN2A | FALSE | Upregulated |
| 0,851987486 | 0,212890622 | HELZ2 | FALSE | Upregulated |
| 1,131936445 | 0,213368529 | LIPA | FALSE | Upregulated |
| 0,906511617 | 0,214933753 | GPX1 | FALSE | Upregulated |
| 0,880559181 | 0,215639766 | SBNO2 | FALSE | Upregulated |
| 1,074312854 | 0,215864151 | PACS2 | FALSE | Upregulated |
| -0,861425973 | 0,21609286 | JMJD1C | FALSE | Downregulated |
| 0,998788055 | 0,216484754 | DEXI | FALSE | Upregulated |
| -1,561576789 | 0,217525583 | FAM76B | FALSE | Downregulated |
| 1,368083236 | 0,217753208 | LOC101929732 | FALSE | Upregulated |
| 1,19404078 | 0,217753208 | PIP5K1C | FALSE | Upregulated |
| 1,314796722 | 0,21791693 | TMC8 | FALSE | Upregulated |
| -1,490233999 | 0,218609912 | LOC101929752 | FALSE | Downregulated |
| 0,809934229 | 0,219938301 | CD68 | FALSE | Upregulated |
| -0,824135615 | 0,220585788 | FAM186A | FALSE | Downregulated |
| -0,981181868 | 0,22129566 | RPL37A | FALSE | Downregulated |
| -0,545420555 | 0,221465126 | IGFBP4 | FALSE | Downregulated |
| 1,197214725 | 0,221465126 | INPP4A | FALSE | Upregulated |
| -1,049848161 | 0,221465126 | LDLRAD4-AS1 | FALSE | Downregulated |
| 1,072923054 | 0,221465126 | RP11-436I24.1 | FALSE | Upregulated |
| -0,962375267 | 0,221465126 | RPL26 | FALSE | Downregulated |
| -1,261284199 | 0,221465126 | TTLL2 | FALSE | Downregulated |
| -1,300017865 | 0,221934812 | FH | FALSE | Downregulated |
| 0,699581644 | 0,22339283 | CTSZ | FALSE | Upregulated |

| | | | | |
|---|---|---|---|---|
| 1,210413309 | 0,224007643 | CDC42SE1 | FALSE | Upregulated |
| -0,905766659 | 0,224007643 | QPRT | FALSE | Downregulated |
| -0,854318759 | 0,224197983 | RPS23 | FALSE | Downregulated |
| -0,812984962 | 0,224422368 | ZSWIM1 | FALSE | Downregulated |
| 1,044891278 | 0,224518068 | CAPN2 | FALSE | Upregulated |
| 0,703061604 | 0,225015809 | C7orf49 | FALSE | Upregulated |
| -1,3111805 | 0,225603824 | EMX1 | FALSE | Downregulated |
| -0,989230069 | 0,225603824 | RALGPS2 | FALSE | Downregulated |
| 1,308466407 | 0,225603824 | THBS2 | FALSE | Upregulated |
| 0,939776827 | 0,225827589 | COL6A3 | FALSE | Upregulated |
| 1,124981539 | 0,226606865 | KCTD9 | FALSE | Upregulated |
| -0,860055327 | 0,228825995 | COL6A6 | FALSE | Downregulated |
| -1,341231211 | 0,229179532 | NOP14 | FALSE | Downregulated |
| -1,125148224 | 0,229179532 | POMT1 | FALSE | Downregulated |
| 0,858136148 | 0,230079857 | AGRN | FALSE | Upregulated |
| -1,443137009 | 0,230079857 | DHRS4 | FALSE | Downregulated |
| 1,392267083 | 0,230079857 | NLRC5 | FALSE | Upregulated |
| -1,086698886 | 0,230079857 | SLC41A2 | FALSE | Downregulated |
| 1,165785017 | 0,230079857 | SOD2 | FALSE | Upregulated |
| 1,212437736 | 0,230089258 | LOC101927989 | FALSE | Upregulated |
| -1,024633861 | 0,230620431 | BAIAP2 | FALSE | Downregulated |
| 1,131256694 | 0,230620431 | LOC101929722 | FALSE | Upregulated |
| 0,822268304 | 0,231000994 | TMEM120B | FALSE | Upregulated |
| 1,38845557 | 0,231154174 | IQCK | FALSE | Upregulated |
| 0,853989274 | 0,231219332 | GAB2 | FALSE | Upregulated |
| 1,173879442 | 0,231977901 | PITPNM1 | FALSE | Upregulated |
| 1,449402985 | 0,232966543 | CTR9 | FALSE | Upregulated |
| -0,783613222 | 0,232966543 | EGFR-AS1 | FALSE | Downregulated |
| 0,601110733 | 0,232966543 | GLUL | FALSE | Upregulated |
| 0,829417938 | 0,232966543 | PLXND1 | FALSE | Upregulated |
| 0,984961409 | 0,232966543 | SRA1 | FALSE | Upregulated |
| -0,748241302 | 0,233694617 | CYP4V2 | FALSE | Downregulated |
| -0,733115763 | 0,233694617 | SERPINA10 | FALSE | Downregulated |
| -1,064958475 | 0,233882026 | GOLIM4 | FALSE | Downregulated |
| -0,967500588 | 0,233882026 | QSER1 | FALSE | Downregulated |
| 1,233586659 | 0,239842671 | PTPRH | FALSE | Upregulated |
| -1,404567041 | 0,240341399 | BRCA1 | FALSE | Downregulated |
| -1,27328159 | 0,240341399 | CD5L | FALSE | Downregulated |
| 1,500535343 | 0,240341399 | TRIM22 | FALSE | Upregulated |
| -0,804280485 | 0,242135015 | WWP1 | FALSE | Downregulated |
| -1,529784089 | 0,242135015 | ZNF444 | FALSE | Downregulated |
| 0,993771154 | 0,242670809 | C2orf54 | FALSE | Upregulated |
| 1,302752205 | 0,243510798 | AXIN1 | FALSE | Upregulated |
| 0,70006647 | 0,243510798 | CCNL2 | FALSE | Upregulated |
| 0,71794997 | 0,243510798 | CXCL12 | FALSE | Upregulated |
| -0,8591262 | 0,243510798 | MYO1B | FALSE | Downregulated |
| 1,380218077 | 0,243510798 | PTGIS | FALSE | Upregulated |
| -0,976498518 | 0,244202292 | DBT | FALSE | Downregulated |
| 0,942001684 | 0,244637262 | CENPP | FALSE | Upregulated |
| -1,429550934 | 0,2447515 | CLEC4M | FALSE | Downregulated |
| -1,088100571 | 0,245349782 | GNAT1 | FALSE | Downregulated |

| | | | | |
|---|---|---|---|---|
| -0,743964777 | 0,2456385 | PTPRS | FALSE | Downregulated |
| -0,743798218 | 0,245639404 | CEBPA | FALSE | Downregulated |
| 1,04155368 | 0,24604213 | FLVCR2 | FALSE | Upregulated |
| -0,730178732 | 0,24604213 | MBNL3 | FALSE | Downregulated |
| 0,764671921 | 0,24604213 | ZNFX1 | FALSE | Upregulated |
| 0,820249283 | 0,246216428 | ACTG1 | FALSE | Upregulated |
| 1,138920372 | 0,246310372 | CXCL16 | FALSE | Upregulated |
| 0,726102185 | 0,246911821 | HERC1 | FALSE | Upregulated |
| 1,514985679 | 0,247553401 | LOC101929479 | FALSE | Upregulated |
| 1,14143971 | 0,247589543 | ECM2 | FALSE | Upregulated |
| 0,960532242 | 0,247925306 | ANKRD13A | FALSE | Upregulated |
| -1,121248788 | 0,247925306 | ATP11C | FALSE | Downregulated |
| -1,62074087 | 0,247925306 | CISH | FALSE | Downregulated |
| 1,217818201 | 0,247925306 | CLDN2 | FALSE | Upregulated |
| -0,841278703 | 0,247925306 | EIF3L | FALSE | Downregulated |
| -0,706836309 | 0,248252478 | LOC101927803 | FALSE | Downregulated |
| 1,159053711 | 0,248871217 | CDC25B | FALSE | Upregulated |
| 1,113696837 | 0,248871217 | DNALI1 | FALSE | Upregulated |
| 0,762546501 | 0,249521414 | HNRNPAB | FALSE | Upregulated |
| -0,744095228 | 0,249813438 | ADPRM | FALSE | Downregulated |
| 0,740942919 | 0,25054777 | HERC2P2 | FALSE | Upregulated |
| -0,818272848 | 0,251367795 | NADK | FALSE | Downregulated |
| -0,67625306 | 0,251526804 | RBL2 | FALSE | Downregulated |
| -0,570071724 | 0,251627345 | APOA2 | FALSE | Downregulated |
| -0,590366297 | 0,251627345 | C19orf66 | FALSE | Downregulated |
| 0,990218988 | 0,251627345 | TRIOBP | FALSE | Upregulated |
| -1,019256381 | 0,253174772 | BTG1 | FALSE | Downregulated |
| 1,060088281 | 0,254149605 | IPPK | FALSE | Upregulated |
| -0,833199065 | 0,254907803 | SLC9A7 | FALSE | Downregulated |
| 0,959671473 | 0,255164551 | LOC440300 | FALSE | Upregulated |
| -1,167545533 | 0,255164551 | PLXDC1 | FALSE | Downregulated |
| -0,83153132 | 0,255164551 | SIGMAR1 | FALSE | Downregulated |
| -1,197837885 | 0,255169389 | NME4 | FALSE | Downregulated |
| -0,691647915 | 0,255347952 | LOC729732 | FALSE | Downregulated |
| -1,172983366 | 0,256142963 | SAT2 | FALSE | Downregulated |
| -0,695069718 | 0,256477555 | INHBC | FALSE | Downregulated |
| -1,247574305 | 0,256502182 | CHN2 | FALSE | Downregulated |
| -1,255608025 | 0,258370751 | ADK | FALSE | Downregulated |
| 0,882938722 | 0,259145732 | ACTL10 | FALSE | Upregulated |
| 0,731194811 | 0,259145732 | SMG6 | FALSE | Upregulated |
| 1,273104771 | 0,260075555 | DERL3 | FALSE | Upregulated |
| 0,967730462 | 0,260313948 | HP09025 | FALSE | Upregulated |
| -0,81951278 | 0,260581788 | MPST | FALSE | Downregulated |
| 0,784308636 | 0,260581788 | MYO15B | FALSE | Upregulated |
| 0,811586793 | 0,261814411 | ZSCAN22 | FALSE | Upregulated |
| -0,817999759 | 0,264733812 | RPL11 | FALSE | Downregulated |
| 0,982611373 | 0,264733812 | YWHAZ | FALSE | Upregulated |
| 0,703116276 | 0,264778713 | ARHGEF40 | FALSE | Upregulated |
| -0,837765507 | 0,264778713 | CALM1 | FALSE | Downregulated |
| -0,949905783 | 0,265859065 | AKR1C1 | FALSE | Downregulated |
| 1,604257568 | 0,266505749 | IFI30 | FALSE | Upregulated |

| | | | | |
|---|---|---|---|---|
| 1,139008968 | 0,266505749 | ZNF142 | FALSE | Upregulated |
| -0,858127513 | 0,266557158 | KIAA1217 | FALSE | Downregulated |
| -0,962933243 | 0,266594205 | EMILIN3 | FALSE | Downregulated |
| 0,676095279 | 0,266594205 | GGA2 | FALSE | Upregulated |
| -1,041952393 | 0,267323899 | EIF2A | FALSE | Downregulated |
| -1,010419344 | 0,26890222 | FAM107A | FALSE | Downregulated |
| 1,15335588 | 0,26890222 | PACS1 | FALSE | Upregulated |
| 1,241679076 | 0,269197605 | RASGRP3 | FALSE | Upregulated |
| -1,226821641 | 0,26921112 | BNIP3 | FALSE | Downregulated |
| -1,440280399 | 0,26921112 | CLYBL | FALSE | Downregulated |
| -0,66297098 | 0,26921112 | LOC101929469 | FALSE | Downregulated |
| -1,149284863 | 0,26921112 | OXER1 | FALSE | Downregulated |
| 0,834045603 | 0,271102989 | ACTN1 | FALSE | Upregulated |
| -1,039008564 | 0,273093335 | IARS2 | FALSE | Downregulated |
| -0,834421338 | 0,273678404 | C6orf201 | FALSE | Downregulated |
| -1,523186779 | 0,273678404 | ELP2 | FALSE | Downregulated |
| 0,55257225 | 0,273678404 | NUTM2G | FALSE | Upregulated |
| -0,784809892 | 0,273678404 | RPL9 | FALSE | Downregulated |
| 0,753524692 | 0,274262903 | C21orf2 | FALSE | Upregulated |
| -1,147419479 | 0,274262903 | KCNS2 | FALSE | Downregulated |
| 1,279455967 | 0,274262903 | SASH3 | FALSE | Upregulated |
| -0,771329306 | 0,274397019 | ZBED4 | FALSE | Downregulated |
| -0,842931368 | 0,275052186 | SERINC5 | FALSE | Downregulated |
| 0,596641588 | 0,275286462 | SIK2 | FALSE | Upregulated |
| -0,598367078 | 0,276128883 | PPARA | FALSE | Downregulated |
| 0,711790658 | 0,277579464 | BRI3 | FALSE | Upregulated |
| 1,357319084 | 0,277610868 | RCC2 | FALSE | Upregulated |
| -0,989372686 | 0,27827481 | LARP1B | FALSE | Downregulated |
| -1,105093658 | 0,27827481 | NYNRIN | FALSE | Downregulated |
| -0,800783729 | 0,27827481 | PHLDB2 | FALSE | Downregulated |
| 1,169951168 | 0,278961872 | DAB2 | FALSE | Upregulated |
| 1,293221525 | 0,278961872 | DUSP1 | FALSE | Upregulated |
| -0,956657754 | 0,278961872 | TTC31 | FALSE | Downregulated |
| -0,729512894 | 0,278997956 | IL16 | FALSE | Downregulated |
| -0,918535208 | 0,279614463 | ADCY5 | FALSE | Downregulated |
| -0,90016497 | 0,281052879 | NAGS | FALSE | Downregulated |
| 1,319181974 | 0,28235237 | SLC7A7 | FALSE | Upregulated |
| 0,598489687 | 0,282416611 | MAPK8IP3 | FALSE | Upregulated |
| -0,714828501 | 0,282916366 | GHR | FALSE | Downregulated |
| 1,290059169 | 0,282916366 | SLC38A7 | FALSE | Upregulated |
| 0,704502849 | 0,282916366 | TRIO | FALSE | Upregulated |
| 0,676113074 | 0,283471094 | CEMP1 | FALSE | Upregulated |
| -1,200973652 | 0,285023271 | ALDH1L1-AS2 | FALSE | Downregulated |
| -0,763257496 | 0,285023271 | SLC10A1 | FALSE | Downregulated |
| -0,72757183 | 0,287001651 | TMEM176A | FALSE | Downregulated |
| -0,755558705 | 0,28783605 | RPS20 | FALSE | Downregulated |
| 0,601564928 | 0,288614496 | XRCC3 | FALSE | Upregulated |
| 1,13431744 | 0,289572781 | EMC3-AS1 | FALSE | Upregulated |
| 0,759837988 | 0,289572781 | SMCR5 | FALSE | Upregulated |
| -1,124866535 | 0,290037001 | SAFB | FALSE | Downregulated |
| 1,035120334 | 0,290273459 | SIN3B | FALSE | Upregulated |

| | | | | |
|---|---|---|---|---|
| 0,676931076 | 0,290459844 | ATG7 | FALSE | Upregulated |
| 1,212707561 | 0,290459844 | C15orf52 | FALSE | Upregulated |
| 0,756567889 | 0,290459844 | GRN | FALSE | Upregulated |
| -1,158241711 | 0,290459844 | PROCR | FALSE | Downregulated |
| -0,976888176 | 0,29135737 | FGF2 | FALSE | Downregulated |
| -0,575615797 | 0,29135737 | RPL19 | FALSE | Downregulated |
| 1,141686553 | 0,291930752 | KDM3A | FALSE | Upregulated |
| -0,712894734 | 0,292553278 | C3 | FALSE | Downregulated |
| 1,167364329 | 0,293119099 | EIF4G3 | FALSE | Upregulated |
| 0,852418563 | 0,293119099 | GREB1L | FALSE | Upregulated |
| -1,10496338 | 0,293119099 | NMD3 | FALSE | Downregulated |
| -1,233669475 | 0,294118542 | GBP7 | FALSE | Downregulated |
| 0,870798659 | 0,294118542 | GDA | FALSE | Upregulated |
| -0,782788949 | 0,294118542 | GPT | FALSE | Downregulated |
| -0,99595155 | 0,294118542 | PRDX3 | FALSE | Downregulated |
| -1,090058889 | 0,294150934 | ASB13 | FALSE | Downregulated |
| -1,04221675 | 0,294150934 | SLC35A3 | FALSE | Downregulated |
| -0,981820004 | 0,294649583 | USP15 | FALSE | Downregulated |
| 0,760570708 | 0,294649583 | VPS13C | FALSE | Upregulated |
| -0,806208961 | 0,29653811 | YY1 | FALSE | Downregulated |
| 0,757453992 | 0,296662284 | COL4A4 | FALSE | Upregulated |
| -1,343993895 | 0,296662284 | FCN3 | FALSE | Downregulated |
| 0,79112918 | 0,296807833 | SH3D19 | FALSE | Upregulated |
| -1,15984647 | 0,296851159 | RNF128 | FALSE | Downregulated |
| -0,739965841 | 0,296851159 | TTC19 | FALSE | Downregulated |
| 1,130628819 | 0,297026242 | ANGPTL2 | FALSE | Upregulated |
| 1,079510246 | 0,297189523 | CSF1 | FALSE | Upregulated |
| 0,840713498 | 0,29731221 | NFASC | FALSE | Upregulated |
| -1,089041421 | 0,29829134 | RPL22 | FALSE | Downregulated |
| -0,619405527 | 0,298936752 | CLUH | FALSE | Downregulated |
| -0,60718755 | 0,298936752 | LOC102723554 | FALSE | Downregulated |
| -0,975904435 | 0,300595669 | SLC22A3 | FALSE | Downregulated |
| -0,615819527 | 0,302948417 | CCNYL1 | FALSE | Downregulated |
| 0,874688033 | 0,302948417 | CDC42EP1 | FALSE | Upregulated |
| 0,777831279 | 0,302948417 | COL18A1-AS1 | FALSE | Upregulated |
| -0,836446922 | 0,302948417 | CYP4F11 | FALSE | Downregulated |
| 1,218383132 | 0,302948417 | FGD2 | FALSE | Upregulated |
| -1,110122494 | 0,302948417 | FOXP1 | FALSE | Downregulated |
| -0,837197694 | 0,302948417 | GCLC | FALSE | Downregulated |
| -0,804300757 | 0,302948417 | IYD | FALSE | Downregulated |
| -0,842588303 | 0,302948417 | MET | FALSE | Downregulated |
| 0,861916789 | 0,302948417 | OBSCN | FALSE | Upregulated |
| -0,95443496 | 0,302948417 | RNF152 | FALSE | Downregulated |
| 1,070713918 | 0,303156731 | CDC42SE2 | FALSE | Upregulated |
| -1,164516624 | 0,303156731 | MFAP3L | FALSE | Downregulated |
| 0,711040773 | 0,303156731 | MGLL | FALSE | Upregulated |
| 1,061122609 | 0,303156731 | RBM3 | FALSE | Upregulated |
| 0,874033659 | 0,304419125 | FGFR2 | FALSE | Upregulated |
| -1,133884751 | 0,304419125 | LOC100506258 | FALSE | Downregulated |
| -0,872078159 | 0,304419125 | PEG3-AS1 | FALSE | Downregulated |
| -1,0657432 | 0,304678201 | SKIDA1 | FALSE | Downregulated |

| | | | | |
|---|---|---|---|---|
| -1,270789178 | 0,304802166 | SPNS3 | FALSE | Downregulated |
| 0,945872193 | 0,305195553 | LHFPL2 | FALSE | Upregulated |
| 0,915824864 | 0,305353678 | LIMK2 | FALSE | Upregulated |
| -1,017301418 | 0,305477209 | ETFDH | FALSE | Downregulated |
| -0,650269623 | 0,305477209 | GP1BA | FALSE | Downregulated |
| -0,606111019 | 0,305477209 | HDLBP | FALSE | Downregulated |
| -1,2203967 | 0,305477209 | POLD2 | FALSE | Downregulated |
| -0,732598723 | 0,305523237 | ACSM3 | FALSE | Downregulated |
| 0,70192497 | 0,305523237 | ACTB | FALSE | Upregulated |
| 0,762466549 | 0,305523237 | HS6ST1 | FALSE | Upregulated |
| 1,159506594 | 0,305523237 | RP3-476K8.3 | FALSE | Upregulated |
| -0,704847931 | 0,305531171 | KIN | FALSE | Downregulated |
| 0,645949247 | 0,305848849 | HDAC6 | FALSE | Upregulated |
| 1,121093219 | 0,30635823 | KIAA0930 | FALSE | Upregulated |
| 0,945896128 | 0,30981486 | LINC00514 | FALSE | Upregulated |
| 1,154860184 | 0,310184993 | LOC101928830 | FALSE | Upregulated |
| 1,113248958 | 0,310327181 | ZFP36 | FALSE | Upregulated |
| 0,887670275 | 0,311605301 | CIITA | FALSE | Upregulated |
| 1,264800839 | 0,311660077 | ALK | FALSE | Upregulated |
| 1,246681321 | 0,311660077 | IFNGR2 | FALSE | Upregulated |
| -1,359491253 | 0,311660077 | MAGI2-AS3 | FALSE | Downregulated |
| 1,189012551 | 0,311660077 | QSOX1 | FALSE | Upregulated |
| 0,792271008 | 0,311660077 | YAP1 | FALSE | Upregulated |
| 0,889933808 | 0,314016437 | TMEM129 | FALSE | Upregulated |
| -1,104514036 | 0,314997344 | HPGD | FALSE | Downregulated |
| 0,658376916 | 0,314997344 | SPNS2 | FALSE | Upregulated |
| -0,635964466 | 0,316303007 | GPX4 | FALSE | Downregulated |
| 1,195811193 | 0,316927383 | GPATCH2L | FALSE | Upregulated |
| 0,638676291 | 0,317358993 | PRH2 | FALSE | Upregulated |
| -0,630769177 | 0,318185937 | RPS9 | FALSE | Downregulated |
| -0,759159589 | 0,318400192 | GPR126 | FALSE | Downregulated |
| -1,165027178 | 0,319369631 | PAIP2B | FALSE | Downregulated |
| 1,084821126 | 0,32000764 | KLHL31 | FALSE | Upregulated |
| 0,729661956 | 0,32000764 | MARCKS | FALSE | Upregulated |
| -1,067040276 | 0,320185918 | DHRS4-AS1 | FALSE | Downregulated |
| 1,123561119 | 0,320273296 | RELL2 | FALSE | Upregulated |
| -0,660181029 | 0,321799862 | DAK | FALSE | Downregulated |
| 0,615730693 | 0,321799862 | NKTR | FALSE | Upregulated |
| 1,288876646 | 0,321799862 | PKM | FALSE | Upregulated |
| 0,795404665 | 0,323803105 | RICTOR | FALSE | Upregulated |
| -1,029741282 | 0,324157341 | CNPY3 | FALSE | Downregulated |
| 0,980044459 | 0,324157341 | LINC00854 | FALSE | Upregulated |
| 1,057804877 | 0,324157341 | LITAF | FALSE | Upregulated |
| -0,630368692 | 0,324157341 | MAN1A1 | FALSE | Downregulated |
| 0,907449938 | 0,324157341 | OPA3 | FALSE | Upregulated |
| -1,054453285 | 0,324157341 | PID1 | FALSE | Downregulated |
| -1,334817665 | 0,324157341 | REPS1 | FALSE | Downregulated |
| -0,501397132 | 0,324157341 | RP11-793H13.3 | FALSE | Downregulated |
| -0,720334913 | 0,324311555 | FAM13A-AS1 | FALSE | Downregulated |
| -0,762212974 | 0,324311555 | GSK3B | FALSE | Downregulated |
| 0,871476846 | 0,324311555 | ZNF469 | FALSE | Upregulated |

| | | | | |
|---|---|---|---|---|
| -1,217399409 | 0,324940507 | PALM3 | FALSE | Downregulated |
| -0,821951249 | 0,326059527 | PHF8 | FALSE | Downregulated |
| 1,153660396 | 0,327129214 | GPR34 | FALSE | Upregulated |
| 0,534601347 | 0,32811107 | FLJ42627 | FALSE | Upregulated |
| -0,778951373 | 0,32811107 | RPL8 | FALSE | Downregulated |
| 1,075886052 | 0,32919366 | ACLY | FALSE | Upregulated |
| 1,011159314 | 0,32919366 | IQGAP1 | FALSE | Upregulated |
| 1,085278788 | 0,32919366 | TMSB10 | FALSE | Upregulated |
| -0,992641697 | 0,329975433 | ECT2L | FALSE | Downregulated |
| -0,870360439 | 0,331532574 | UQCRC1 | FALSE | Downregulated |
| -1,188019545 | 0,331979941 | RCL1 | FALSE | Downregulated |
| -0,759211684 | 0,332982715 | CEND1 | FALSE | Downregulated |
| 0,831925072 | 0,334199301 | IL4R | FALSE | Upregulated |
| 0,84153173 | 0,33420541 | ATP11B | FALSE | Upregulated |
| 1,219107902 | 0,33420541 | PCED1B-AS1 | FALSE | Upregulated |
| -0,80199729 | 0,33420541 | SMARCA2 | FALSE | Downregulated |
| -0,846197445 | 0,33420541 | TOB1 | FALSE | Downregulated |
| -1,057935512 | 0,33420541 | VNN1 | FALSE | Downregulated |
| 1,042776926 | 0,334680496 | MAPK1 | FALSE | Upregulated |
| 0,991393107 | 0,336140335 | ENO4 | FALSE | Upregulated |
| -0,674740488 | 0,336398369 | C2orf72 | FALSE | Downregulated |
| 1,032734747 | 0,336398369 | CNOT7 | FALSE | Upregulated |
| 1,348844168 | 0,336398369 | NFKB2 | FALSE | Upregulated |
| 0,592271101 | 0,336398369 | PCSK7 | FALSE | Upregulated |
| 0,805084673 | 0,336398369 | PTTG1IP | FALSE | Upregulated |
| -1,013522084 | 0,336398369 | SEPSECS | FALSE | Downregulated |
| 1,152806706 | 0,336398369 | VPS37B | FALSE | Upregulated |
| 1,010491844 | 0,336398369 | WDR91 | FALSE | Upregulated |
| -0,864105207 | 0,336465129 | SEC63 | FALSE | Downregulated |
| -0,907838085 | 0,337976479 | NR3C2 | FALSE | Downregulated |
| -0,875096208 | 0,338581733 | FGL1 | FALSE | Downregulated |
| 0,985345466 | 0,338581733 | MVP | FALSE | Upregulated |
| 1,011430924 | 0,338581733 | ZCCHC9 | FALSE | Upregulated |
| 0,848817749 | 0,338634241 | CIRBP-AS1 | FALSE | Upregulated |
| 1,14730416 | 0,338634241 | CRIP2 | FALSE | Upregulated |
| 0,91777187 | 0,338634241 | SLC48A1 | FALSE | Upregulated |
| -0,969156762 | 0,33909067 | AFMID | FALSE | Downregulated |
| 1,306432288 | 0,339125497 | LOC101928152 | FALSE | Upregulated |
| 0,65520093 | 0,339125497 | RHOBTB2 | FALSE | Upregulated |
| 0,821683019 | 0,33999995 | LOC101927393 | FALSE | Upregulated |
| 1,100638846 | 0,34084825 | KIAA0556 | FALSE | Upregulated |
| -0,775270905 | 0,34084825 | RP11-778H2.1 | FALSE | Downregulated |
| 1,248699246 | 0,341752667 | LIMK1 | FALSE | Upregulated |
| -1,279293408 | 0,341752667 | OCIAD1 | FALSE | Downregulated |
| -0,633422321 | 0,342432248 | APOL1 | FALSE | Downregulated |
| -0,592542867 | 0,342432248 | STARD10 | FALSE | Downregulated |
| -1,041281536 | 0,342533819 | RNASE13 | FALSE | Downregulated |
| -0,951481618 | 0,343870541 | ZNF181 | FALSE | Downregulated |
| 1,112159355 | 0,344107405 | DENND5A | FALSE | Upregulated |
| -0,890375075 | 0,344107405 | SLC25A5 | FALSE | Downregulated |
| 1,273362065 | 0,344882537 | GRIN2C | FALSE | Upregulated |

| | | | | |
|---|---|---|---|---|
| -1,107675963 | 0,345133215 | GOLGA2P5 | FALSE | Downregulated |
| -1,410682462 | 0,345133215 | LECT2 | FALSE | Downregulated |
| 0,686330052 | 0,345133215 | SHC1 | FALSE | Upregulated |
| -0,62185331 | 0,345357481 | F5 | FALSE | Downregulated |
| -1,075328861 | 0,345392099 | ZNHIT1 | FALSE | Downregulated |
| -0,920691196 | 0,345783212 | C16orf58 | FALSE | Downregulated |
| -0,894299718 | 0,345783212 | RNF130 | FALSE | Downregulated |
| -0,702809731 | 0,345783212 | SEPHS2 | FALSE | Downregulated |
| -0,938608376 | 0,348148924 | GRIN2A | FALSE | Downregulated |
| -0,891299113 | 0,348148924 | POGK | FALSE | Downregulated |
| -0,508845458 | 0,348158187 | CDHR5 | FALSE | Downregulated |
| -0,742781264 | 0,348158187 | FKBP4 | FALSE | Downregulated |
| -0,966639096 | 0,348158187 | SLC4A4 | FALSE | Downregulated |
| -0,879721868 | 0,348300694 | PCBD1 | FALSE | Downregulated |
| -0,842981027 | 0,348425416 | TGFBR3L | FALSE | Downregulated |
| 0,578000334 | 0,348681992 | ADAMTSL4 | FALSE | Upregulated |
| -0,894897813 | 0,348681992 | ATF7 | FALSE | Downregulated |
| 0,976818651 | 0,348681992 | FNDC5 | FALSE | Upregulated |
| 1,211538357 | 0,348693785 | FHOD1 | FALSE | Upregulated |
| 1,035422498 | 0,348693785 | FLYWCH1 | FALSE | Upregulated |
| 0,909209399 | 0,348693785 | FOXD2 | FALSE | Upregulated |
| -0,867324022 | 0,348693785 | KLC4 | FALSE | Downregulated |
| -0,634832066 | 0,348693785 | RP11-713M15.1 | FALSE | Downregulated |
| 0,707921446 | 0,348693785 | SLC4A2 | FALSE | Upregulated |
| 0,620194305 | 0,350031614 | CHRNB2 | FALSE | Upregulated |
| -0,869684094 | 0,35006371 | EMP2 | FALSE | Downregulated |
| 0,60385638 | 0,350695927 | ABCC3 | FALSE | Upregulated |
| 0,831918896 | 0,35112291 | MBNL1-AS1 | FALSE | Upregulated |
| -0,622795149 | 0,351501267 | LINC00261 | FALSE | Downregulated |
| 1,186418222 | 0,352367101 | FAM171A1 | FALSE | Upregulated |
| 0,935145782 | 0,352367101 | NEURL1B | FALSE | Upregulated |
| -1,326475993 | 0,352367101 | SH3PXD2A-AS1 | FALSE | Downregulated |
| -0,506942836 | 0,352367101 | TMBIM6 | FALSE | Downregulated |
| 1,000577666 | 0,353027409 | KCNH1 | FALSE | Upregulated |
| -0,857023051 | 0,353214007 | CTAGE5 | FALSE | Downregulated |
| 0,988299134 | 0,353812315 | ASTE1 | FALSE | Upregulated |
| 1,033979378 | 0,353812315 | DNMT3A | FALSE | Upregulated |
| 0,871086734 | 0,353812315 | LEPR | FALSE | Upregulated |
| 0,638791369 | 0,353812315 | MRPL37 | FALSE | Upregulated |
| 1,056787826 | 0,355492127 | ANKRD32 | FALSE | Upregulated |
| 0,658071024 | 0,355492127 | APPL2 | FALSE | Upregulated |
| -0,792048207 | 0,355492127 | H1FX | FALSE | Downregulated |
| 1,099670009 | 0,355492127 | KIAA0247 | FALSE | Upregulated |
| -0,629438461 | 0,355492127 | METTL7A | FALSE | Downregulated |
| 1,256704933 | 0,355492127 | NEB | FALSE | Upregulated |
| 0,7542536 | 0,355492127 | SLA2 | FALSE | Upregulated |
| 0,744640224 | 0,355492127 | VGLL4 | FALSE | Upregulated |
| 0,88040655 | 0,356394118 | EVA1B | FALSE | Upregulated |
| -1,414943825 | 0,356555156 | RNR1 | FALSE | Downregulated |
| 0,808082114 | 0,358361327 | ENOSF1 | FALSE | Upregulated |
| 0,759483608 | 0,358361327 | LOC101927266 | FALSE | Upregulated |

| | | | | |
|---|---|---|---|---|
| -0,97291151 | 0,358639169 | MXI1 | FALSE | Downregulated |
| 0,890823909 | 0,358639169 | RBM19 | FALSE | Upregulated |
| -0,926443359 | 0,358828547 | ECI2 | FALSE | Downregulated |
| 1,050368264 | 0,358828547 | FMNL1 | FALSE | Upregulated |
| 1,109663019 | 0,35891311 | ASAP1 | FALSE | Upregulated |
| -0,60669094 | 0,35891311 | EEF1D | FALSE | Downregulated |
| 1,132482736 | 0,35891311 | FBXO10 | FALSE | Upregulated |
| -0,705726814 | 0,35891311 | HSPD1 | FALSE | Downregulated |
| 1,194557282 | 0,35891311 | IGFBP1 | FALSE | Upregulated |
| 0,984295921 | 0,35891311 | NSMF | FALSE | Upregulated |
| 1,13427652 | 0,35891311 | PARP12 | FALSE | Upregulated |
| -0,54320676 | 0,35891311 | ST6GAL1 | FALSE | Downregulated |
| 0,637142852 | 0,35891311 | STAT6 | FALSE | Upregulated |
| 0,965150755 | 0,35891311 | TTC7A | FALSE | Upregulated |
| 0,890069389 | 0,361693114 | LOC102723582 | FALSE | Upregulated |
| 0,949619324 | 0,361910455 | LOC102724724 | FALSE | Upregulated |
| -0,782363333 | 0,361910455 | MORN4 | FALSE | Downregulated |
| -0,55751715 | 0,362214283 | SYNE1 | FALSE | Downregulated |
| -0,779767192 | 0,362749895 | ANKRD28 | FALSE | Downregulated |
| -0,688958285 | 0,363801295 | GPD1 | FALSE | Downregulated |
| -1,449001233 | 0,364830353 | LOC286083 | FALSE | Downregulated |
| -0,730105812 | 0,366075772 | ADCK3 | FALSE | Downregulated |
| 0,705256481 | 0,366075772 | EMILIN2 | FALSE | Upregulated |
| -0,652386969 | 0,366075772 | SORBS1 | FALSE | Downregulated |
| 0,944314543 | 0,366779553 | ATG13 | FALSE | Upregulated |
| -0,563734282 | 0,366779553 | FLRT1 | FALSE | Downregulated |
| -0,718486713 | 0,366779553 | GLYATL1 | FALSE | Downregulated |
| 1,000721096 | 0,366779553 | HLA-DRA | FALSE | Upregulated |
| -0,691938504 | 0,366779553 | SERPINA6 | FALSE | Downregulated |
| -0,604485006 | 0,366779553 | SLC6A1 | FALSE | Downregulated |
| 0,81891906 | 0,366779553 | SNX19 | FALSE | Upregulated |
| -0,931368435 | 0,366779553 | TH | FALSE | Downregulated |
| 1,108042153 | 0,367173891 | PREX1 | FALSE | Upregulated |
| -0,940854651 | 0,367173891 | SMO | FALSE | Downregulated |
| -0,627010664 | 0,367245447 | CALML4 | FALSE | Downregulated |
| -0,733940205 | 0,367245447 | MGST1 | FALSE | Downregulated |
| -0,737792178 | 0,367747058 | GFRA1 | FALSE | Downregulated |
| 1,015923748 | 0,368202159 | PARP8 | FALSE | Upregulated |
| 1,022778612 | 0,36842101 | LOC101928192 | FALSE | Upregulated |
| -1,065596492 | 0,36842101 | SLC22A4 | FALSE | Downregulated |
| -0,883714495 | 0,36842101 | XYLB | FALSE | Downregulated |
| 0,825406169 | 0,368726274 | FAM65A | FALSE | Upregulated |
| 1,197784699 | 0,369078773 | MYL5 | FALSE | Upregulated |
| 0,493917043 | 0,370073987 | PHC1 | FALSE | Upregulated |
| -0,889464721 | 0,370634345 | HNRNPM | FALSE | Downregulated |
| -0,587460725 | 0,370634345 | RP5-886K2.3 | FALSE | Downregulated |
| 0,673779182 | 0,370634345 | TYK2 | FALSE | Upregulated |
| -0,874157426 | 0,370871388 | FAM213A | FALSE | Downregulated |
| 1,129494802 | 0,370871388 | FOSL2 | FALSE | Upregulated |
| -1,262196613 | 0,373304181 | GPR182 | FALSE | Downregulated |
| 0,896111754 | 0,373620876 | CAP1 | FALSE | Upregulated |

| | | | | |
|---|---|---|---|---|
| -1,182037058 | 0,373620876 | RCOR3 | FALSE | Downregulated |
| -0,565453751 | 0,373620876 | RGPD1 | FALSE | Downregulated |
| -0,673893811 | 0,373620876 | SLC20A2 | FALSE | Downregulated |
| -0,561864463 | 0,373627637 | HNF1A | FALSE | Downregulated |
| -0,904616295 | 0,373627637 | LOC101927854 | FALSE | Downregulated |
| 0,660915975 | 0,375337182 | NFAT5 | FALSE | Upregulated |
| -0,690690018 | 0,375337182 | RPL30 | FALSE | Downregulated |
| -0,662665949 | 0,375337182 | SIAH1 | FALSE | Downregulated |
| -1,076472478 | 0,375337182 | TRNT1 | FALSE | Downregulated |
| -1,011743348 | 0,375337182 | ZNF264 | FALSE | Downregulated |
| 0,97344324 | 0,375936868 | CEP192 | FALSE | Upregulated |
| -0,610445538 | 0,376104816 | RORA | FALSE | Downregulated |
| -0,907034248 | 0,376269249 | COX6A1 | FALSE | Downregulated |
| 0,683597066 | 0,376470488 | DCAF10 | FALSE | Upregulated |
| 0,995534024 | 0,376708696 | THRA | FALSE | Upregulated |
| -0,74967127 | 0,377827322 | ECHDC1 | FALSE | Downregulated |
| -1,025773256 | 0,378332837 | EBF3 | FALSE | Downregulated |
| -0,884336924 | 0,378332837 | FOXA1 | FALSE | Downregulated |
| -0,89235275 | 0,378332837 | RPL24 | FALSE | Downregulated |
| -0,9562341 | 0,378332837 | ZHX1 | FALSE | Downregulated |
| 0,828828973 | 0,378371832 | LOC101929426 | FALSE | Upregulated |
| -0,82008954 | 0,378452 | PKHD1 | FALSE | Downregulated |
| -0,686736209 | 0,378452 | PLA2G12B | FALSE | Downregulated |
| 0,946608455 | 0,378452 | TUBB2A | FALSE | Upregulated |
| -0,747127073 | 0,379207015 | RPL35 | FALSE | Downregulated |
| -1,113399251 | 0,379443115 | OIP5-AS1 | FALSE | Downregulated |
| -0,657446967 | 0,380047499 | CIRBP | FALSE | Downregulated |
| 0,906417401 | 0,380189976 | NEDD9 | FALSE | Upregulated |
| 0,46727711 | 0,380390269 | PHKG1 | FALSE | Upregulated |
| -0,552945789 | 0,380555386 | OGDHL | FALSE | Downregulated |
| -0,669620171 | 0,381147025 | SEMA3F | FALSE | Downregulated |
| -1,048325256 | 0,382000199 | CYP1B1 | FALSE | Downregulated |
| 0,916974953 | 0,382000199 | DAB2IP | FALSE | Upregulated |
| -0,733142859 | 0,385100838 | CLDN22 | FALSE | Downregulated |
| 0,488586852 | 0,385100838 | LENG8 | FALSE | Upregulated |
| -0,848932997 | 0,385100838 | NCL | FALSE | Downregulated |
| -0,908526128 | 0,386194652 | AKR1A1 | FALSE | Downregulated |
| 1,170860079 | 0,387052117 | EPHA2 | FALSE | Upregulated |
| -0,673614719 | 0,387052117 | RABEP1 | FALSE | Downregulated |
| -1,119729465 | 0,387089183 | AMN1 | FALSE | Downregulated |
| 0,953568522 | 0,387089183 | DGKZ | FALSE | Upregulated |
| 0,690159594 | 0,387089183 | MAST2 | FALSE | Upregulated |
| 0,629043421 | 0,387089183 | SET | FALSE | Upregulated |
| 0,699747389 | 0,387627079 | ITGA11 | FALSE | Upregulated |
| 0,611469377 | 0,388228155 | ZZEF1 | FALSE | Upregulated |
| 0,728073483 | 0,38831659 | LOC101929396 | FALSE | Upregulated |
| 0,97814504 | 0,38831659 | PLEKHM1P | FALSE | Upregulated |
| 0,777451313 | 0,38831659 | SLC25A25 | FALSE | Upregulated |
| -0,808196037 | 0,388704943 | ARHGEF26 | FALSE | Downregulated |
| -1,122437461 | 0,388704943 | GAS2 | FALSE | Downregulated |
| -0,875705374 | 0,388704943 | KCTD6 | FALSE | Downregulated |

| | | | | |
|---|---|---|---|---|
| 1,25282763 | 0,388704943 | NRG1 | FALSE | Upregulated |
| 0,789456314 | 0,388704943 | SLC17A4 | FALSE | Upregulated |
| -1,097754467 | 0,389592212 | FAM114A1 | FALSE | Downregulated |
| -0,57609103 | 0,389592212 | HABP2 | FALSE | Downregulated |
| 1,05683469 | 0,390838514 | LOC101929522 | FALSE | Upregulated |
| 1,061672382 | 0,391727017 | ENTPD6 | FALSE | Upregulated |
| -0,669120812 | 0,392074727 | CARNS1 | FALSE | Downregulated |
| 0,723346192 | 0,392074727 | HMGXB3 | FALSE | Upregulated |
| -0,595106787 | 0,392074727 | LBX2 | FALSE | Downregulated |
| -1,120857417 | 0,392306543 | PIK3R4 | FALSE | Downregulated |
| -1,093178434 | 0,394592768 | MARCO | FALSE | Downregulated |
| -0,925862593 | 0,395640131 | HAVCR1P1 | FALSE | Downregulated |
| -1,110763825 | 0,396081395 | CASC3 | FALSE | Downregulated |
| -0,88206322 | 0,396081395 | CBR1 | FALSE | Downregulated |
| 0,498857657 | 0,396081395 | FDPSP2 | FALSE | Upregulated |
| -0,826455845 | 0,396081395 | MT2A | FALSE | Downregulated |
| 0,942002986 | 0,396173988 | BMP6 | FALSE | Upregulated |
| -0,614127494 | 0,396173988 | RPS14 | FALSE | Downregulated |
| -0,661275875 | 0,396173988 | ZNF517 | FALSE | Downregulated |
| 0,796575272 | 0,396835567 | XAF1 | FALSE | Upregulated |
| 0,840648713 | 0,39745294 | HLA-A | FALSE | Upregulated |
| 0,905826534 | 0,39745294 | VPS9D1-AS1 | FALSE | Upregulated |
| -0,823194082 | 0,397677269 | FGD4 | FALSE | Downregulated |
| -1,067232527 | 0,397677269 | HSD17B11 | FALSE | Downregulated |
| -0,605873624 | 0,397677269 | LOC284889 | FALSE | Downregulated |
| -0,872975995 | 0,397677269 | PANK1 | FALSE | Downregulated |
| 0,978698275 | 0,397742625 | TFRC | FALSE | Upregulated |
| -0,776146976 | 0,398959212 | ESR1 | FALSE | Downregulated |
| -0,827194304 | 0,401506817 | LPIN3 | FALSE | Downregulated |
| 1,028768092 | 0,401771533 | SGSH | FALSE | Upregulated |
| -0,918597752 | 0,402072131 | PPFIBP1 | FALSE | Downregulated |
| -0,850851453 | 0,402072131 | SCARNA17 | FALSE | Downregulated |
| -0,888639869 | 0,402072131 | XRCC5 | FALSE | Downregulated |
| -1,212760959 | 0,402764254 | USP37 | FALSE | Downregulated |
| 0,66154259 | 0,402800215 | DPP9 | FALSE | Upregulated |
| 0,85631816 | 0,402999116 | C1QTNF1 | FALSE | Upregulated |
| 1,035495977 | 0,402999116 | NLGN3 | FALSE | Upregulated |
| 1,129079463 | 0,404012279 | ADAMTS10 | FALSE | Upregulated |
| -0,959648557 | 0,405502906 | BTF3 | FALSE | Downregulated |
| -0,620363743 | 0,405502906 | XDH | FALSE | Downregulated |
| 0,813497324 | 0,405758651 | CHKA | FALSE | Upregulated |
| 1,012585501 | 0,40780482 | GDI1 | FALSE | Upregulated |
| 0,769133404 | 0,407881379 | B3GALNT2 | FALSE | Upregulated |
| 0,900470704 | 0,407881379 | COL5A2 | FALSE | Upregulated |
| 0,638430158 | 0,407881379 | RARA | FALSE | Upregulated |
| -1,129228361 | 0,408109216 | SOX5 | FALSE | Downregulated |
| 0,715747408 | 0,408130552 | CHAD | FALSE | Upregulated |
| -1,00099287 | 0,408221182 | FCN2 | FALSE | Downregulated |
| 0,556800813 | 0,408626737 | SEPT9 | FALSE | Upregulated |
| 1,155403495 | 0,408708912 | FERMT3 | FALSE | Upregulated |
| 1,11273911 | 0,408790724 | NCF2 | FALSE | Upregulated |

| | | | | |
|---|---|---|---|---|
| -0,747756282 | 0,411255061 | HSDL2 | FALSE | Downregulated |
| 0,848167404 | 0,411262425 | LOC100128374 | FALSE | Upregulated |
| 0,840891598 | 0,411754236 | C17orf62 | FALSE | Upregulated |
| 1,244581553 | 0,411764826 | LAMC3 | FALSE | Upregulated |
| 1,038941074 | 0,411764826 | LINC00954 | FALSE | Upregulated |
| -0,492766373 | 0,411764826 | LOC100129034 | FALSE | Downregulated |
| 0,890738014 | 0,411764826 | RASSF4 | FALSE | Upregulated |
| 0,95174581 | 0,41315077 | KRR1 | FALSE | Upregulated |
| -1,174297406 | 0,413463134 | CDH4 | FALSE | Downregulated |
| -0,942097411 | 0,413463134 | IL17RC | FALSE | Downregulated |
| -0,783261132 | 0,413463134 | NR1I3 | FALSE | Downregulated |
| -0,947332888 | 0,413463134 | RDH10 | FALSE | Downregulated |
| -0,851657737 | 0,413463134 | ZNF839 | FALSE | Downregulated |
| 0,901984355 | 0,414079739 | HLA-DPA1 | FALSE | Upregulated |
| -0,490202956 | 0,414415454 | CIDEB | FALSE | Downregulated |
| 1,070704999 | 0,415831414 | INPP5F | FALSE | Upregulated |
| 1,050724845 | 0,416771799 | SEMA4C | FALSE | Upregulated |
| -0,970507704 | 0,416920487 | OR13D1 | FALSE | Downregulated |
| -0,50314113 | 0,416920487 | VEGFA | FALSE | Downregulated |
| -0,539391876 | 0,417238401 | FAN1 | FALSE | Downregulated |
| 0,780978497 | 0,417238401 | FAS-AS1 | FALSE | Upregulated |
| 0,795969265 | 0,417238401 | FBXO18 | FALSE | Upregulated |
| 0,660552934 | 0,417238401 | SHPK | FALSE | Upregulated |
| 0,942832205 | 0,418018224 | FAM111A | FALSE | Upregulated |
| 0,759787084 | 0,418018224 | FLII | FALSE | Upregulated |
| -0,830389133 | 0,418018224 | MPC2 | FALSE | Downregulated |
| 1,03960306 | 0,418018224 | NBPF1 | FALSE | Upregulated |
| 0,815105678 | 0,418465529 | CRNKL1 | FALSE | Upregulated |
| -0,700118756 | 0,418669082 | CFB | FALSE | Downregulated |
| 0,573303211 | 0,419069971 | LRRC4 | FALSE | Upregulated |
| 1,389618359 | 0,4198479 | DCUN1D5 | FALSE | Upregulated |
| -1,182504279 | 0,4198479 | DUSP10 | FALSE | Downregulated |
| -1,219684222 | 0,421089361 | DNAJC7 | FALSE | Downregulated |
| 1,074133728 | 0,421089361 | ELMOD3 | FALSE | Upregulated |
| 0,828958974 | 0,421200171 | ARFGAP1 | FALSE | Upregulated |
| -0,777370267 | 0,421200171 | ITGA2 | FALSE | Downregulated |
| -0,623888136 | 0,421200171 | KLF15 | FALSE | Downregulated |
| -0,684955149 | 0,421200171 | SEC62 | FALSE | Downregulated |
| -0,929470949 | 0,42207803 | PRDX2 | FALSE | Downregulated |
| 0,622826328 | 0,422481971 | GON4L | FALSE | Upregulated |
| 0,625851343 | 0,422489162 | RP11-197N18.7 | FALSE | Upregulated |
| -0,667732986 | 0,422489162 | RP11-85B7.2 | FALSE | Downregulated |
| 0,861312298 | 0,422541138 | AFAP1-AS1 | FALSE | Upregulated |
| 0,745737882 | 0,422541138 | COL3A1 | FALSE | Upregulated |
| -0,987981145 | 0,422541138 | DNAJA1 | FALSE | Downregulated |
| -0,966917235 | 0,422541138 | ERLIN2 | FALSE | Downregulated |
| -0,640449407 | 0,422541138 | GJB1 | FALSE | Downregulated |
| 1,050142225 | 0,422541138 | NDOR1 | FALSE | Upregulated |
| -0,975599053 | 0,422541138 | YWHAQ | FALSE | Downregulated |
| -0,49520968 | 0,423249423 | ARHGAP35 | FALSE | Downregulated |
| 1,001350604 | 0,423249423 | CDK16 | FALSE | Upregulated |

| | | | | |
|---|---|---|---|---|
| -1,063610292 | 0,423575017 | GALT | FALSE | Downregulated |
| -0,596112695 | 0,423575017 | HELZ | FALSE | Downregulated |
| -0,680081529 | 0,423669511 | BTBD6 | FALSE | Downregulated |
| 0,621015373 | 0,423669511 | CRLF3 | FALSE | Upregulated |
| -0,862829996 | 0,423669511 | DDB2 | FALSE | Downregulated |
| 0,85169457 | 0,423669511 | LOC102723328 | FALSE | Upregulated |
| -0,632747887 | 0,423669511 | RPL7A | FALSE | Downregulated |
| 0,505857905 | 0,423669511 | SPDYE3 | FALSE | Upregulated |
| 0,501556414 | 0,424301389 | DUSP16 | FALSE | Upregulated |
| 0,865176304 | 0,424430371 | NICN1 | FALSE | Upregulated |
| 0,805995571 | 0,424430371 | TRAF3IP2-AS1 | FALSE | Upregulated |
| -0,897419554 | 0,424549618 | KTN1 | FALSE | Downregulated |
| 1,017397269 | 0,424549618 | NIN | FALSE | Upregulated |
| 1,219112836 | 0,424549618 | TBC1D12 | FALSE | Upregulated |
| 0,783974929 | 0,424549618 | ZNF433 | FALSE | Upregulated |
| 0,665548312 | 0,424706729 | STX16 | FALSE | Upregulated |
| -0,69709869 | 0,425234409 | PHB2 | FALSE | Downregulated |
| 1,007783168 | 0,425880091 | GK | FALSE | Upregulated |
| -0,709830924 | 0,426678694 | ANKRD23 | FALSE | Downregulated |
| -0,56974484 | 0,426678694 | VPS4A | FALSE | Downregulated |
| -1,100459121 | 0,4272929 | LOC101928875 | FALSE | Downregulated |
| 1,268801169 | 0,428626692 | CCDC88B | FALSE | Upregulated |
| 0,63680431 | 0,429387148 | MAZ | FALSE | Upregulated |
| 1,055088243 | 0,429486736 | MAFG | FALSE | Upregulated |
| 0,58552078 | 0,431501067 | LOC101929290 | FALSE | Upregulated |
| -0,755238359 | 0,431501067 | SLC25A22 | FALSE | Downregulated |
| -1,063963884 | 0,431501067 | UGDH | FALSE | Downregulated |
| -0,961253274 | 0,431503208 | KCNJ8 | FALSE | Downregulated |
| -0,961604655 | 0,431503208 | MAN1A2 | FALSE | Downregulated |
| 0,989181412 | 0,431503208 | VASP | FALSE | Upregulated |
| -0,934019653 | 0,431574252 | AIG1 | FALSE | Downregulated |
| -0,828465396 | 0,431574252 | C11orf54 | FALSE | Downregulated |
| 0,67729849 | 0,431574252 | MPEG1 | FALSE | Upregulated |
| 0,603924686 | 0,431574252 | SREBF1 | FALSE | Upregulated |
| 0,692340262 | 0,431574252 | SSBP3-AS1 | FALSE | Upregulated |
| 0,968831935 | 0,431574252 | ZNF124 | FALSE | Upregulated |
| 0,761105416 | 0,431686377 | ACP5 | FALSE | Upregulated |
| -0,796522454 | 0,43219749 | EPB41L4B | FALSE | Downregulated |
| 0,461076761 | 0,43219749 | THAP3 | FALSE | Upregulated |
| -1,184220804 | 0,436807892 | APOA4 | FALSE | Downregulated |
| 0,715818086 | 0,436807892 | CYFIP1 | FALSE | Upregulated |
| -0,887667657 | 0,436807892 | EIF3E | FALSE | Downregulated |
| -0,692497256 | 0,436807892 | KCTD21 | FALSE | Downregulated |
| -0,88624903 | 0,436807892 | MAN1C1 | FALSE | Downregulated |
| -0,854232547 | 0,436807892 | NNMT | FALSE | Downregulated |
| 0,693079669 | 0,43697736 | ABLIM3 | FALSE | Upregulated |
| -0,562881121 | 0,43697736 | ARG2 | FALSE | Downregulated |
| 0,801731822 | 0,43697736 | DPYSL2 | FALSE | Upregulated |
| 0,93990985 | 0,43697736 | ENPP7 | FALSE | Upregulated |
| -0,54963247 | 0,43697736 | GLS2 | FALSE | Downregulated |
| -0,994338742 | 0,43697736 | HNRNPH3 | FALSE | Downregulated |

| | | | | |
|---|---|---|---|---|
| 0,804797986 | 0,43697736 | MFSD7 | FALSE | Upregulated |
| -0,744096356 | 0,43697736 | NACA | FALSE | Downregulated |
| -0,77958985 | 0,43697736 | RPL5 | FALSE | Downregulated |
| -0,670770618 | 0,43697736 | SMPD3 | FALSE | Downregulated |
| 0,943430585 | 0,437206541 | ZNF618 | FALSE | Upregulated |
| -1,096225177 | 0,437643511 | CLASRP | FALSE | Downregulated |
| -1,046753413 | 0,437643511 | ILVBL | FALSE | Downregulated |
| 1,117764166 | 0,438455148 | CD84 | FALSE | Upregulated |
| -0,856984058 | 0,439174128 | CUL3 | FALSE | Downregulated |
| 0,67839159 | 0,439174128 | ZMIZ1 | FALSE | Upregulated |
| 0,550480536 | 0,439466565 | ERVFRD-1 | FALSE | Upregulated |
| 0,930642057 | 0,439466565 | LOC388282 | FALSE | Upregulated |
| 0,885924968 | 0,439554949 | LOC730257 | FALSE | Upregulated |
| -0,558276575 | 0,439554949 | SLC13A5 | FALSE | Downregulated |
| 0,997351877 | 0,439597433 | ITGAV | FALSE | Upregulated |
| 0,724398902 | 0,440344668 | IAH1 | FALSE | Upregulated |
| -0,817212837 | 0,440765749 | LOC101928443 | FALSE | Downregulated |
| -0,798945478 | 0,441914817 | ARAF | FALSE | Downregulated |
| 1,106589604 | 0,441933581 | TPM4 | FALSE | Upregulated |
| 0,627567709 | 0,442006767 | ARHGEF18 | FALSE | Upregulated |
| 0,45756932 | 0,442006767 | UCKL1-AS1 | FALSE | Upregulated |
| 0,868624312 | 0,442108644 | MYH11 | FALSE | Upregulated |
| 0,665265086 | 0,442379986 | MAPT | FALSE | Upregulated |
| -0,634217416 | 0,442456949 | ZNRF1 | FALSE | Downregulated |
| 0,750253866 | 0,442647917 | POLG | FALSE | Upregulated |
| -0,997239362 | 0,442873378 | TIAL1 | FALSE | Downregulated |
| 0,658115984 | 0,443179816 | CDIP1 | FALSE | Upregulated |
| 1,071763028 | 0,443179816 | LOC101928373 | FALSE | Upregulated |
| -0,494075047 | 0,443179816 | XBP1 | FALSE | Downregulated |
| 0,609357026 | 0,443179816 | ZNF276 | FALSE | Upregulated |
| 1,149569751 | 0,443841665 | DOPEY2 | FALSE | Upregulated |
| -0,91050229 | 0,444207557 | LOC102724177 | FALSE | Downregulated |
| -1,029359058 | 0,444736097 | MAP7 | FALSE | Downregulated |
| -1,030545345 | 0,444736097 | UTY | FALSE | Downregulated |
| 0,80435644 | 0,444854509 | LOC100134040 | FALSE | Upregulated |
| -0,732840264 | 0,444854509 | PHLPP1 | FALSE | Downregulated |
| -0,938468095 | 0,444854509 | ZNF57 | FALSE | Downregulated |
| 0,805875104 | 0,445224181 | ARHGEF39 | FALSE | Upregulated |
| 0,863262241 | 0,445224181 | ARRDC2 | FALSE | Upregulated |
| 1,027252386 | 0,445224181 | MICALL2 | FALSE | Upregulated |
| -1,288219872 | 0,445224181 | RCAN3 | FALSE | Downregulated |
| -0,885696199 | 0,445383197 | C2CD4D | FALSE | Downregulated |
| -0,397813108 | 0,445383197 | EEF1A1 | FALSE | Downregulated |
| -0,557830248 | 0,44573361 | ALDH8A1 | FALSE | Downregulated |
| 0,857458428 | 0,44573361 | ATG16L2 | FALSE | Upregulated |
| 0,781422468 | 0,44573361 | CADM1 | FALSE | Upregulated |
| -0,524832862 | 0,44573361 | DIDO1 | FALSE | Downregulated |
| -0,832071714 | 0,44573361 | GBE1 | FALSE | Downregulated |
| -0,576483947 | 0,44573361 | TCEB3 | FALSE | Downregulated |
| 0,754267862 | 0,445996996 | DCHS1 | FALSE | Upregulated |
| 0,900870833 | 0,447003093 | TMEM132A | FALSE | Upregulated |

| | | | | |
|---|---|---|---|---|
| -0,882411222 | 0,447914785 | CDC42EP4 | FALSE | Downregulated |
| -1,618456868 | 0,447914785 | CHRNA4 | FALSE | Downregulated |
| 0,837296736 | 0,447914785 | LINC01004 | FALSE | Upregulated |
| -0,506962867 | 0,449585982 | ACADVL | FALSE | Downregulated |
| 0,571389073 | 0,449585982 | LINC01000 | FALSE | Upregulated |
| -0,501966444 | 0,449585982 | MYO5B | FALSE | Downregulated |
| -0,7145894 | 0,449585982 | PTPRK | FALSE | Downregulated |
| 0,605561268 | 0,449585982 | TMBIM1 | FALSE | Upregulated |
| -0,42512801 | 0,449820409 | EEF2 | FALSE | Downregulated |
| -0,634239559 | 0,450540717 | SMURF1 | FALSE | Downregulated |
| 0,649639604 | 0,450540717 | SOX12 | FALSE | Upregulated |
| 0,533412506 | 0,450540717 | SPDYE6 | FALSE | Upregulated |
| 1,071502859 | 0,450540717 | ZC3H12A | FALSE | Upregulated |
| -1,09228191 | 0,450540717 | ZDHHC18 | FALSE | Downregulated |
| -0,594503549 | 0,450692438 | FOXN3 | FALSE | Downregulated |
| -0,658997957 | 0,450692438 | GDI2 | FALSE | Downregulated |
| 0,818557652 | 0,450692438 | PIGR | FALSE | Upregulated |
| 0,483671944 | 0,45081309 | ARHGDIA | FALSE | Upregulated |
| -0,771416003 | 0,45081309 | MZT2B | FALSE | Downregulated |
| -0,92221673 | 0,451027157 | HBCBP | FALSE | Downregulated |
| 0,895618883 | 0,451027157 | TEP1 | FALSE | Upregulated |
| -0,637757156 | 0,451027157 | TOB2 | FALSE | Downregulated |
| -0,67752494 | 0,452518084 | CELF1 | FALSE | Downregulated |
| 0,627964974 | 0,452518084 | ZFPM1 | FALSE | Upregulated |
| -0,525241208 | 0,452906743 | PLEKHG3 | FALSE | Downregulated |
| 0,59249217 | 0,45296858 | F11R | FALSE | Upregulated |
| 1,016574895 | 0,453197456 | SLC5A10 | FALSE | Upregulated |
| -0,960523114 | 0,453420983 | DOLK | FALSE | Downregulated |
| 0,790745076 | 0,453420983 | JUNB | FALSE | Upregulated |
| 0,646619237 | 0,453709645 | ADCY7 | FALSE | Upregulated |
| 1,04381576 | 0,453709645 | FLJ13224 | FALSE | Upregulated |
| 0,951273169 | 0,453709645 | LAMA2 | FALSE | Upregulated |
| -0,952475271 | 0,453709645 | LOC101928101 | FALSE | Downregulated |
| 1,183260202 | 0,453709645 | LOC101928884 | FALSE | Upregulated |
| 0,851695019 | 0,454156913 | ITPKC | FALSE | Upregulated |
| 0,499976832 | 0,454156913 | TGOLN2 | FALSE | Upregulated |
| -0,514556249 | 0,454650505 | PPP2R1B | FALSE | Downregulated |
| 0,627533019 | 0,454655911 | TRIM14 | FALSE | Upregulated |
| -0,534851739 | 0,455102496 | HSD17B1 | FALSE | Downregulated |
| 0,970167019 | 0,45593271 | SNTB2 | FALSE | Upregulated |
| -0,718344943 | 0,456486932 | HSD3B7 | FALSE | Downregulated |
| -1,036262539 | 0,456660703 | PPIG | FALSE | Downregulated |
| -0,499198052 | 0,456660703 | SEMA4G | FALSE | Downregulated |
| 0,658943259 | 0,458519404 | ANXA6 | FALSE | Upregulated |
| -0,711783702 | 0,458587873 | C8orf82 | FALSE | Downregulated |
| 0,988666272 | 0,458587873 | FAM212B | FALSE | Upregulated |
| -0,676761591 | 0,458778897 | SALL1 | FALSE | Downregulated |
| -0,683650734 | 0,458778897 | ZNF24 | FALSE | Downregulated |
| 0,968095865 | 0,459430007 | CPPED1 | FALSE | Upregulated |
| -0,718640638 | 0,459477589 | FETUB | FALSE | Downregulated |
| -1,043715631 | 0,459963154 | IRF6 | FALSE | Downregulated |

| | | | | |
|---|---|---|---|---|
| 0,796553875 | 0,461155653 | OPTN | FALSE | Upregulated |
| 0,608308536 | 0,462051904 | CD2BP2 | FALSE | Upregulated |
| 0,89466592 | 0,46230965 | PPFIA2 | FALSE | Upregulated |
| 0,879617122 | 0,46230965 | SVEP1 | FALSE | Upregulated |
| -1,079625175 | 0,463184052 | FOXA3 | FALSE | Downregulated |
| 1,023953866 | 0,463184052 | MAPK1IP1L | FALSE | Upregulated |
| -0,976296542 | 0,463184052 | PSMC1 | FALSE | Downregulated |
| 0,582836973 | 0,463184052 | SCUBE3 | FALSE | Upregulated |
| 0,582661041 | 0,463184052 | TRIM8 | FALSE | Upregulated |
| 0,680924161 | 0,463913503 | ARHGEF11 | FALSE | Upregulated |
| -0,554733194 | 0,464420535 | CBS | FALSE | Downregulated |
| 0,747839543 | 0,464420535 | POMGNT1 | FALSE | Upregulated |
| -0,461709929 | 0,464467886 | ACOX1 | FALSE | Downregulated |
| 0,919971287 | 0,464467886 | ANKRD50 | FALSE | Upregulated |
| -0,828700439 | 0,464467886 | FAM3A | FALSE | Downregulated |
| 0,830668276 | 0,464467886 | FLJ32790 | FALSE | Upregulated |
| -0,621043888 | 0,464467886 | HSP90B1 | FALSE | Downregulated |
| 0,605908278 | 0,464467886 | SETX | FALSE | Upregulated |
| -0,639900276 | 0,464467886 | SORBS3 | FALSE | Downregulated |
| -0,819758252 | 0,464467886 | STYX | FALSE | Downregulated |
| -0,849163169 | 0,464937525 | KAT5 | FALSE | Downregulated |
| -0,709160083 | 0,465343821 | TP53I13 | FALSE | Downregulated |
| 0,532411761 | 0,465433681 | EP400 | FALSE | Upregulated |
| 0,739717436 | 0,465433681 | PPT1 | FALSE | Upregulated |
| 0,50968688 | 0,465433681 | VPS13A | FALSE | Upregulated |
| -0,997207991 | 0,467548466 | NDUFV3 | FALSE | Downregulated |
| -0,629459005 | 0,468395705 | MLF2 | FALSE | Downregulated |
| -0,740617585 | 0,468545537 | AHDC1 | FALSE | Downregulated |
| -0,898631934 | 0,468894371 | ARF3 | FALSE | Downregulated |
| -0,571933324 | 0,470264254 | ANGPTL1 | FALSE | Downregulated |
| -1,008536149 | 0,470264254 | TADA2B | FALSE | Downregulated |
| 0,64109719 | 0,470457611 | POLE | FALSE | Upregulated |
| 1,105934873 | 0,470457611 | SUPT4H1 | FALSE | Upregulated |
| 0,892757602 | 0,475520869 | RP1-232P20.1 | FALSE | Upregulated |
| -1,092278838 | 0,475854516 | RPS29 | FALSE | Downregulated |
| -0,878970206 | 0,475857837 | BCKDHB | FALSE | Downregulated |
| -0,7049232 | 0,476197582 | SLC25A13 | FALSE | Downregulated |
| 1,097733937 | 0,476197582 | TIMP1 | FALSE | Upregulated |
| 0,878704134 | 0,476453425 | CLSTN1 | FALSE | Upregulated |
| 0,604024959 | 0,476453425 | FAM208A | FALSE | Upregulated |
| 0,661699318 | 0,477535161 | CERK | FALSE | Upregulated |
| 0,541647472 | 0,477620985 | DOT1L | FALSE | Upregulated |
| 1,460045476 | 0,477620985 | NTSR1 | FALSE | Upregulated |
| 0,442274836 | 0,477620985 | RGL4 | FALSE | Upregulated |
| 0,786160793 | 0,477620985 | VSIG10 | FALSE | Upregulated |
| -0,774040094 | 0,478353639 | PTK2B | FALSE | Downregulated |
| 0,887499463 | 0,479023618 | FCGR3A | FALSE | Upregulated |
| 0,727429287 | 0,479023618 | LOC100996741 | FALSE | Upregulated |
| -0,663485846 | 0,47953159 | MIER1 | FALSE | Downregulated |
| 0,517624919 | 0,480059191 | MYO1F | FALSE | Upregulated |
| 0,808946999 | 0,480095213 | EXTL3 | FALSE | Upregulated |

| | | | | |
|---|---|---|---|---|
| -0,684007626 | 0,480095213 | PECR | FALSE | Downregulated |
| -0,525775022 | 0,480216276 | RAPH1 | FALSE | Downregulated |
| -0,678106718 | 0,480300267 | RBM6 | FALSE | Downregulated |
| 0,504008038 | 0,480462865 | FTL | FALSE | Upregulated |
| 1,097388887 | 0,480622521 | ATR | FALSE | Upregulated |
| 0,644224556 | 0,480622521 | LOC100133445 | FALSE | Upregulated |
| 0,891962226 | 0,480622521 | STIP1 | FALSE | Upregulated |
| 0,912832609 | 0,481600646 | OGFOD1 | FALSE | Upregulated |
| 0,765972318 | 0,481600646 | SLC35B2 | FALSE | Upregulated |
| -0,912153956 | 0,482165029 | CSAD | FALSE | Downregulated |
| 0,935888741 | 0,482165029 | RPRD1A | FALSE | Upregulated |
| -1,003461986 | 0,48236264 | C1orf226 | FALSE | Downregulated |
| -0,461902709 | 0,48236264 | RPL41 | FALSE | Downregulated |
| -0,701794576 | 0,48236264 | SPATS2L | FALSE | Downregulated |
| 1,016156066 | 0,48236264 | TMEM74B | FALSE | Upregulated |
| 0,902711234 | 0,48236264 | ZNF160 | FALSE | Upregulated |
| -1,003809174 | 0,48240131 | NDRG4 | FALSE | Downregulated |
| 0,94593405 | 0,482639022 | AGAP9 | FALSE | Upregulated |
| -0,618877503 | 0,482639022 | ALKBH5 | FALSE | Downregulated |
| 0,784851482 | 0,482639022 | ALS2CL | FALSE | Upregulated |
| 1,146834301 | 0,482639022 | C16orf3 | FALSE | Upregulated |
| -0,598580017 | 0,482639022 | DBH-AS1 | FALSE | Downregulated |
| -0,773256386 | 0,482639022 | DCAF7 | FALSE | Downregulated |
| 0,969304026 | 0,482639022 | DNAJC10 | FALSE | Upregulated |
| 0,630631164 | 0,482639022 | EPB41 | FALSE | Upregulated |
| -0,643037292 | 0,482639022 | FAU | FALSE | Downregulated |
| -0,761513766 | 0,482639022 | FPGS | FALSE | Downregulated |
| -0,667340714 | 0,482639022 | GCC2 | FALSE | Downregulated |
| -0,416310097 | 0,482639022 | HNF4A | FALSE | Downregulated |
| -0,915064298 | 0,482639022 | ISOC1 | FALSE | Downregulated |
| 0,705784882 | 0,482639022 | KIF28P | FALSE | Upregulated |
| -0,814472527 | 0,482639022 | LIPG | FALSE | Downregulated |
| 0,684921527 | 0,482639022 | LPHN1 | FALSE | Upregulated |
| -0,635701307 | 0,482639022 | MAP2K7 | FALSE | Downregulated |
| 0,71802474 | 0,482639022 | MXRA7 | FALSE | Upregulated |
| -0,785028593 | 0,482639022 | NODAL | FALSE | Downregulated |
| -1,486647236 | 0,482639022 | PARP15 | FALSE | Downregulated |
| -0,424097456 | 0,482639022 | PCBP2 | FALSE | Downregulated |
| -0,656860492 | 0,482639022 | PRDX1 | FALSE | Downregulated |
| 0,698917403 | 0,482639022 | PVRL2 | FALSE | Upregulated |
| -0,52617487 | 0,482639022 | SNORD3A | FALSE | Downregulated |
| 0,848124416 | 0,483170193 | TPD52L2 | FALSE | Upregulated |
| 0,89278864 | 0,483544404 | TOR1B | FALSE | Upregulated |
| 0,675815979 | 0,484170206 | PGAM5 | FALSE | Upregulated |
| -0,722365275 | 0,485241833 | RP11-392P7.6 | FALSE | Downregulated |
| -0,482231846 | 0,485321362 | KANK1 | FALSE | Downregulated |
| -1,026000979 | 0,48562834 | MARCKSL1 | FALSE | Downregulated |
| -0,584219395 | 0,485757745 | RELL1 | FALSE | Downregulated |
| 0,527661362 | 0,485757745 | VASN | FALSE | Upregulated |
| -0,721917222 | 0,486173704 | CTSO | FALSE | Downregulated |
| -0,728597405 | 0,486173704 | KLHDC2 | FALSE | Downregulated |

| | | | | |
|---|---|---|---|---|
| 0,590528037 | 0,486173704 | LOC101928719 | FALSE | Upregulated |
| 0,607877769 | 0,486173704 | OMG | FALSE | Upregulated |
| 0,692655509 | 0,486418277 | AAK1 | FALSE | Upregulated |
| 0,555181688 | 0,486418277 | DOCK4-AS1 | FALSE | Upregulated |
| -0,620594731 | 0,486891638 | ZNF629 | FALSE | Downregulated |
| -0,801425122 | 0,487364144 | ZNF281 | FALSE | Downregulated |
| 0,766868457 | 0,488406296 | POLR3E | FALSE | Upregulated |
| -0,570691874 | 0,489830471 | AHCY | FALSE | Downregulated |
| -0,890474072 | 0,489830471 | BPHL | FALSE | Downregulated |
| -1,015921355 | 0,489830471 | MMAB | FALSE | Downregulated |
| -0,690231956 | 0,489830471 | RMND5A | FALSE | Downregulated |
| 0,666403253 | 0,491450591 | FOXH1 | FALSE | Upregulated |
| -0,640845036 | 0,491886721 | BCDIN3D-AS1 | FALSE | Downregulated |
| 0,507365855 | 0,49203753 | MIRLET7DHG | FALSE | Upregulated |
| 0,597579076 | 0,492203798 | PLD3 | FALSE | Upregulated |
| 0,776580894 | 0,493083292 | XPO6 | FALSE | Upregulated |
| -0,775392091 | 0,493385264 | PLCXD1 | FALSE | Downregulated |
| -0,619621607 | 0,493751806 | SLC25A1 | FALSE | Downregulated |
| 0,658258434 | 0,494677149 | PRPF40B | FALSE | Upregulated |
| -0,766196507 | 0,495645933 | BRPF3 | FALSE | Downregulated |
| 0,956889831 | 0,495662053 | BMS1P6 | FALSE | Upregulated |
| -1,124871062 | 0,495999844 | ARSE | FALSE | Downregulated |
| -0,758386879 | 0,497141659 | LINC00667 | FALSE | Downregulated |
| -0,921710494 | 0,497141659 | PDIA5 | FALSE | Downregulated |
| -0,651468185 | 0,499143364 | KDM4B | FALSE | Downregulated |
| 0,985943361 | 0,499143364 | PPP2CB | FALSE | Upregulated |
| 0,582193958 | 0,499170772 | SETMAR | FALSE | Upregulated |
| -0,882853393 | 0,499332846 | ZZZ3 | FALSE | Downregulated |
| 0,821681087 | 0,499724366 | ATP2B4 | FALSE | Upregulated |
| 0,647067128 | 0,499724366 | CCNT2 | FALSE | Upregulated |
| 0,868454715 | 0,499724366 | CSK | FALSE | Upregulated |
| 0,557937352 | 0,499724366 | ERMP1 | FALSE | Upregulated |
| 0,566835173 | 0,499724366 | LOC101928817 | FALSE | Upregulated |
| -0,527417092 | 0,499724366 | OS9 | FALSE | Downregulated |
| -0,766491067 | 0,50044646 | ID2 | FALSE | Downregulated |
| 0,654342942 | 0,500547804 | GNS | FALSE | Upregulated |
| 0,864963623 | 0,500616348 | GFOD1 | FALSE | Upregulated |
| 0,841952745 | 0,500897855 | IL17RA | FALSE | Upregulated |
| -0,790964601 | 0,501840285 | HEBP1 | FALSE | Downregulated |
| -0,899769196 | 0,501840285 | PROSC | FALSE | Downregulated |
| 0,838073155 | 0,501840285 | THADA | FALSE | Upregulated |
| -0,569590295 | 0,501997313 | HPN-AS1 | FALSE | Downregulated |
| -0,89225777 | 0,503288374 | LOC102724730 | FALSE | Downregulated |
| -0,98813728 | 0,503555698 | PRPSAP1 | FALSE | Downregulated |
| 0,461899382 | 0,503695991 | SPTBN1 | FALSE | Upregulated |
| -0,554055754 | 0,504540646 | ERP44 | FALSE | Downregulated |
| -0,58460384 | 0,504540646 | RP11-57H12.3 | FALSE | Downregulated |
| 0,846644541 | 0,504694282 | NBPF11 | FALSE | Upregulated |
| 0,473707322 | 0,504872596 | APOL6 | FALSE | Upregulated |
| 0,602273801 | 0,504872596 | B3GALT2 | FALSE | Upregulated |
| 0,517405374 | 0,505273434 | TBC1D8 | FALSE | Upregulated |

| | | | | |
|---|---|---|---|---|
| 0,879099049 | 0,506214227 | FAM169A | FALSE | Upregulated |
| 0,750493619 | 0,506421463 | LOC100128361 | FALSE | Upregulated |
| 0,718301286 | 0,506961438 | GLTSCR1 | FALSE | Upregulated |
| 0,758780445 | 0,506961438 | SYT15 | FALSE | Upregulated |
| -0,498457716 | 0,507712224 | SORBS2 | FALSE | Downregulated |
| -0,701361823 | 0,507952232 | JAM3 | FALSE | Downregulated |
| 0,700520547 | 0,507952232 | NACC1 | FALSE | Upregulated |
| 0,748941386 | 0,507952232 | PI4K2A | FALSE | Upregulated |
| 0,771990108 | 0,508108566 | L3MBTL2 | FALSE | Upregulated |
| 0,497605404 | 0,50857789 | FAM160B2 | FALSE | Upregulated |
| -0,998325393 | 0,50857789 | PRPF3 | FALSE | Downregulated |
| 0,560838692 | 0,50857789 | SLC9A3R1 | FALSE | Upregulated |
| 0,676122775 | 0,509684997 | RHOC | FALSE | Upregulated |
| -1,122640679 | 0,510640201 | SLC16A1 | FALSE | Downregulated |
| -0,735508892 | 0,510658386 | PLEKHA7 | FALSE | Downregulated |
| 0,687219026 | 0,510658386 | SMG7 | FALSE | Upregulated |
| -0,63891191 | 0,511110177 | BOD1L1 | FALSE | Downregulated |
| -0,451408176 | 0,511110177 | C2 | FALSE | Downregulated |
| 0,472123986 | 0,511110177 | LRRN3 | FALSE | Upregulated |
| 0,927121705 | 0,511110177 | MAP1S | FALSE | Upregulated |
| -0,418732211 | 0,511110177 | PINK1-AS | FALSE | Downregulated |
| -0,891299324 | 0,512275652 | MESTIT1 | FALSE | Downregulated |
| 0,736080593 | 0,512275652 | MKL1 | FALSE | Upregulated |
| -0,990851604 | 0,512275652 | OSBPL2 | FALSE | Downregulated |
| -0,657913488 | 0,512275652 | RIMKLA | FALSE | Downregulated |
| -0,777253857 | 0,513307571 | APOM | FALSE | Downregulated |
| -0,680771605 | 0,513307571 | CPEB2 | FALSE | Downregulated |
| 0,769195813 | 0,513307571 | HMOX2 | FALSE | Upregulated |
| 0,50563307 | 0,513307571 | RP11-374P20.4 | FALSE | Upregulated |
| 0,763960597 | 0,513433718 | CDK5RAP2 | FALSE | Upregulated |
| -0,453564622 | 0,513673771 | GUCD1 | FALSE | Downregulated |
| -0,466525694 | 0,514315848 | TARDBP | FALSE | Downregulated |
| -0,635823728 | 0,515117597 | AK3 | FALSE | Downregulated |
| 0,610585184 | 0,515117597 | GOLGA8A | FALSE | Upregulated |
| 0,68311657 | 0,515117597 | LRRFIP1 | FALSE | Upregulated |
| -0,898607979 | 0,515117597 | RNF185 | FALSE | Downregulated |
| 0,723653959 | 0,515117597 | URB1 | FALSE | Upregulated |
| -0,819547266 | 0,515118249 | DGCR6L | FALSE | Downregulated |
| -0,417589445 | 0,515118249 | HPN | FALSE | Downregulated |
| 0,798667216 | 0,515118249 | RNF44 | FALSE | Upregulated |
| 0,668262907 | 0,515118249 | ZC3H3 | FALSE | Upregulated |
| 0,8772038 | 0,515491636 | CCNF | FALSE | Upregulated |
| -1,02645683 | 0,515491636 | HAX1 | FALSE | Downregulated |
| -0,811703769 | 0,515491636 | LOC102724023 | FALSE | Downregulated |
| -1,015488923 | 0,515700379 | DDRGK1 | FALSE | Downregulated |
| -0,856239202 | 0,515700379 | GCSH | FALSE | Downregulated |
| -0,423603299 | 0,515700379 | LOC100128494 | FALSE | Downregulated |
| 0,524924407 | 0,515700379 | RRBP1 | FALSE | Upregulated |
| 0,588496332 | 0,515914711 | C1orf21 | FALSE | Upregulated |
| -0,933585328 | 0,515914711 | ZNF621 | FALSE | Downregulated |
| -0,826398707 | 0,5171315 | P4HA1 | FALSE | Downregulated |

| | | | | |
|---|---|---|---|---|
| -0,635699382 | 0,517829752 | ATF4 | FALSE | Downregulated |
| -0,991164507 | 0,518938356 | NOS1AP | FALSE | Downregulated |
| -0,778004258 | 0,518938356 | RFNG | FALSE | Downregulated |
| 0,986590545 | 0,518938356 | ST3GAL4-AS1 | FALSE | Upregulated |
| -0,6710808 | 0,519277281 | ZNF140 | FALSE | Downregulated |
| -0,7532076 | 0,520181981 | CNOT11 | FALSE | Downregulated |
| -0,719478214 | 0,520181981 | RP11-475I24.8 | FALSE | Downregulated |
| -0,686386002 | 0,520508987 | HMGCL | FALSE | Downregulated |
| -0,693403701 | 0,522411954 | ZNF837 | FALSE | Downregulated |
| -0,413078465 | 0,523981176 | PCSK6 | FALSE | Downregulated |
| 0,907563493 | 0,524269976 | ITGAL | FALSE | Upregulated |
| -0,712854666 | 0,524269976 | PDZD8 | FALSE | Downregulated |
| -0,71045671 | 0,524505064 | MARCH6 | FALSE | Downregulated |
| 0,952525639 | 0,52456494 | LOC101928381 | FALSE | Upregulated |
| 0,746203154 | 0,524828438 | SENP5 | FALSE | Upregulated |
| 0,437807281 | 0,524828438 | STEAP3 | FALSE | Upregulated |
| -0,799095122 | 0,524833805 | ABCC6P1 | FALSE | Downregulated |
| 0,548980319 | 0,525197425 | SHROOM3 | FALSE | Upregulated |
| 0,535893365 | 0,525762182 | TRO | FALSE | Upregulated |
| 1,038260789 | 0,526769209 | C1orf200 | FALSE | Upregulated |
| -0,544371447 | 0,526769209 | KLB | FALSE | Downregulated |
| -0,539677446 | 0,526769209 | RPS17 | FALSE | Downregulated |
| 0,901698353 | 0,526789169 | NUS1 | FALSE | Upregulated |
| -0,671099698 | 0,526789169 | TMEM97 | FALSE | Downregulated |
| 0,824270456 | 0,527102775 | FAM198B | FALSE | Upregulated |
| 0,704249087 | 0,527102775 | TACC2 | FALSE | Upregulated |
| -0,82892947 | 0,527460594 | SLTM | FALSE | Downregulated |
| -0,768396052 | 0,527658499 | C11orf58 | FALSE | Downregulated |
| 0,577644535 | 0,527658499 | LOC100130744 | FALSE | Upregulated |
| 0,977053022 | 0,527663086 | TMC6 | FALSE | Upregulated |
| 0,791629432 | 0,527834561 | LPPR1 | FALSE | Upregulated |
| -0,917927855 | 0,528289601 | SEMA6C | FALSE | Downregulated |
| -0,449514811 | 0,528289601 | VPRBP | FALSE | Downregulated |
| 0,860230421 | 0,528783033 | TREX1 | FALSE | Upregulated |
| -0,604927309 | 0,53171373 | PDCD4 | FALSE | Downregulated |
| 0,662563389 | 0,532181465 | AXL | FALSE | Upregulated |
| -0,469411837 | 0,532181465 | CERS2 | FALSE | Downregulated |
| 0,597041685 | 0,532181465 | DAG1 | FALSE | Upregulated |
| -0,719488466 | 0,532181465 | LLGL2 | FALSE | Downregulated |
| -0,91564997 | 0,532538639 | DNAJC21 | FALSE | Downregulated |
| 0,463848853 | 0,532538639 | MCL1 | FALSE | Upregulated |
| 0,668056031 | 0,532538639 | SMARCA4 | FALSE | Upregulated |
| -0,775628257 | 0,533506531 | RPS27 | FALSE | Downregulated |
| 0,777008106 | 0,53522544 | GTPBP2 | FALSE | Upregulated |
| 0,646625205 | 0,53522544 | LOC644083 | FALSE | Upregulated |
| -0,611718462 | 0,535473037 | ANO1 | FALSE | Downregulated |
| 0,793674734 | 0,535473037 | B4GALT7 | FALSE | Upregulated |
| -0,378617253 | 0,535473037 | DCST2 | FALSE | Downregulated |
| -0,776134571 | 0,535473037 | ETNPPL | FALSE | Downregulated |
| 0,723463809 | 0,5357271 | ILK | FALSE | Upregulated |
| -0,705863668 | 0,5357271 | SCIMP | FALSE | Downregulated |

| | | | | |
|---|---|---|---|---|
| -0,524365089 | 0,535913869 | NRAP | FALSE | Downregulated |
| 0,832289982 | 0,536014137 | INPP5K | FALSE | Upregulated |
| 0,503748322 | 0,538079854 | DNAH1 | FALSE | Upregulated |
| -0,763670606 | 0,538094796 | RPS15A | FALSE | Downregulated |
| -0,837947204 | 0,538303097 | BCAS3 | FALSE | Downregulated |
| -0,807454232 | 0,538303097 | CLIC4 | FALSE | Downregulated |
| 0,71468868 | 0,538303097 | H2AFY | FALSE | Upregulated |
| -1,024680163 | 0,538303097 | PLEKHA5 | FALSE | Downregulated |
| 0,663334913 | 0,538303097 | SLC30A7 | FALSE | Upregulated |
| -0,656210399 | 0,538452085 | ARL6IP1 | FALSE | Downregulated |
| -0,524600126 | 0,538452085 | NISCH | FALSE | Downregulated |
| 0,577081743 | 0,539067697 | PGAM2 | FALSE | Upregulated |
| 0,787529532 | 0,539173985 | ZNF451 | FALSE | Upregulated |
| -0,842815472 | 0,54007627 | N4BP2L1 | FALSE | Downregulated |
| 0,608686669 | 0,541626056 | CENPO | FALSE | Upregulated |
| 0,789400364 | 0,541626056 | DOCK1 | FALSE | Upregulated |
| 1,106410986 | 0,541626056 | LRRC59 | FALSE | Upregulated |
| -0,425387332 | 0,541626056 | MAML3 | FALSE | Downregulated |
| 0,572109339 | 0,541626056 | PCNX | FALSE | Upregulated |
| 0,801717382 | 0,542023482 | LOC101927839 | FALSE | Upregulated |
| -0,671055424 | 0,542023482 | PSMD3 | FALSE | Downregulated |
| -0,76460086 | 0,542023482 | SLC25A5-AS1 | FALSE | Downregulated |
| 0,637120037 | 0,542302728 | AP2A1 | FALSE | Upregulated |
| 0,519905486 | 0,542321752 | RHCE | FALSE | Upregulated |
| 0,762627742 | 0,542423707 | MED15 | FALSE | Upregulated |
| -0,526459362 | 0,542423707 | TTC3 | FALSE | Downregulated |
| -1,010414084 | 0,542448065 | FCGBP | FALSE | Downregulated |
| -0,495248817 | 0,542448065 | GLIPR1 | FALSE | Downregulated |
| -0,641649643 | 0,542448065 | MCFD2 | FALSE | Downregulated |
| 0,544290326 | 0,542448065 | MYO1C | FALSE | Upregulated |
| 0,677428466 | 0,542791084 | INPPL1 | FALSE | Upregulated |
| 0,502633419 | 0,543736434 | MTMR7 | FALSE | Upregulated |
| 0,359903023 | 0,543888198 | TNRC18 | FALSE | Upregulated |
| -0,503423182 | 0,544046627 | A1BG-AS1 | FALSE | Downregulated |
| -0,812901674 | 0,544046627 | C1orf168 | FALSE | Downregulated |
| -0,705750037 | 0,544046627 | DBI | FALSE | Downregulated |
| 0,882950966 | 0,544046627 | EIF2B2 | FALSE | Upregulated |
| -0,515947531 | 0,544508376 | KRAS | FALSE | Downregulated |
| -0,484944 | 0,544508376 | SEMA6A | FALSE | Downregulated |
| 0,651370676 | 0,545881511 | ARHGAP21 | FALSE | Upregulated |
| 0,753624686 | 0,546098002 | NR2F1 | FALSE | Upregulated |
| 0,579885213 | 0,546311446 | ATP2B2 | FALSE | Upregulated |
| 0,819346614 | 0,546311446 | CHD3 | FALSE | Upregulated |
| 0,535680299 | 0,546311446 | HBP1 | FALSE | Upregulated |
| 0,535154444 | 0,546311446 | ICMT | FALSE | Upregulated |
| 0,823054454 | 0,546311446 | LOC101928657 | FALSE | Upregulated |
| 0,886431632 | 0,546311446 | MORC4 | FALSE | Upregulated |
| 0,64855359 | 0,546311446 | NUMB | FALSE | Upregulated |
| 0,440855277 | 0,546311446 | RBPMS | FALSE | Upregulated |
| -0,502465274 | 0,546311446 | REEP6 | FALSE | Downregulated |
| 0,830855319 | 0,546311446 | RP11-14D22.5 | FALSE | Upregulated |

| | | | | |
|---|---|---|---|---|
| -0,671834657 | 0,546311446 | SDS | FALSE | Downregulated |
| -0,951413253 | 0,546311446 | SLC25A43 | FALSE | Downregulated |
| 0,996506741 | 0,546311446 | SLC2A10 | FALSE | Upregulated |
| 0,801246287 | 0,546311446 | TRIP6 | FALSE | Upregulated |
| 0,638114616 | 0,546311446 | UNC13B | FALSE | Upregulated |
| -0,670687012 | 0,54634794 | ENO3 | FALSE | Downregulated |
| 0,609209772 | 0,54634794 | STAT2 | FALSE | Upregulated |
| 0,505328109 | 0,546497335 | FAM196A | FALSE | Upregulated |
| -0,51964555 | 0,546497335 | SERBP1 | FALSE | Downregulated |
| 0,56825932 | 0,54662512 | GBA2 | FALSE | Upregulated |
| -0,862435782 | 0,546934998 | PPM1B | FALSE | Downregulated |
| 0,782583904 | 0,547315454 | TBC1D22A | FALSE | Upregulated |
| -0,908493322 | 0,547511071 | SULT1A2 | FALSE | Downregulated |
| 0,764078736 | 0,547511071 | TGFBR1 | FALSE | Upregulated |
| -0,67753951 | 0,548519044 | ALDH9A1 | FALSE | Downregulated |
| -0,579038407 | 0,548519044 | NR2F6 | FALSE | Downregulated |
| 0,539669939 | 0,548532119 | JUND | FALSE | Upregulated |
| 0,510844932 | 0,548532119 | NAV2 | FALSE | Upregulated |
| 0,729277666 | 0,549690873 | RBM12 | FALSE | Upregulated |
| 0,561853512 | 0,549702006 | WBSCR16 | FALSE | Upregulated |
| -0,841242056 | 0,549950344 | ACMSD | FALSE | Downregulated |
| -0,729608335 | 0,549950344 | AKAP6 | FALSE | Downregulated |
| -0,590289057 | 0,549950344 | RPS6 | FALSE | Downregulated |
| 0,823348777 | 0,55002012 | YTHDC2 | FALSE | Upregulated |
| -0,625617502 | 0,550871749 | LOC102723451 | FALSE | Downregulated |
| 0,666933798 | 0,552196394 | LOC100505915 | FALSE | Upregulated |
| -0,696239639 | 0,552196394 | TBL1X | FALSE | Downregulated |
| -1,123433879 | 0,552449604 | PACSIN3 | FALSE | Downregulated |
| 0,576909583 | 0,553288487 | TPTE2P5 | FALSE | Upregulated |
| -0,767565613 | 0,553604977 | AC007099.1 | FALSE | Downregulated |
| 0,780626788 | 0,553604977 | ZNF263 | FALSE | Upregulated |
| 0,733098997 | 0,553698902 | LOC80154 | FALSE | Upregulated |
| 0,720878068 | 0,553807746 | CTSS | FALSE | Upregulated |
| 0,544084914 | 0,554106459 | ENTPD7 | FALSE | Upregulated |
| 0,483846035 | 0,554106459 | LRRTM2 | FALSE | Upregulated |
| -0,511945915 | 0,554106459 | STX17 | FALSE | Downregulated |
| 0,582373319 | 0,555376822 | PHC3 | FALSE | Upregulated |
| 0,751446572 | 0,556679881 | LOC101928268 | FALSE | Upregulated |
| 0,862256975 | 0,557063271 | ASAH1 | FALSE | Upregulated |
| 0,80911656 | 0,557063271 | YRDC | FALSE | Upregulated |
| -1,015194487 | 0,559097721 | POC1B | FALSE | Downregulated |
| 0,569341756 | 0,560059894 | TET3 | FALSE | Upregulated |
| 0,733504098 | 0,560489413 | ATP6V0A1 | FALSE | Upregulated |
| 0,521343717 | 0,560489413 | SCARB1 | FALSE | Upregulated |
| -0,546355999 | 0,560500899 | AP000688.14 | FALSE | Downregulated |
| 0,572401179 | 0,560500899 | AP1B1 | FALSE | Upregulated |
| -0,703909787 | 0,560500899 | GNAO1 | FALSE | Downregulated |
| 0,561587225 | 0,560500899 | HAL | FALSE | Upregulated |
| -0,512369261 | 0,560500899 | KLF13 | FALSE | Downregulated |
| -0,874477101 | 0,560500899 | PSMD8 | FALSE | Downregulated |
| -0,590326198 | 0,560500899 | SLC35D1 | FALSE | Downregulated |

| | | | | |
|---|---|---|---|---|
| -0,633356191 | 0,560969378 | FLRT3 | FALSE | Downregulated |
| -0,435406806 | 0,561217011 | PC | FALSE | Downregulated |
| -0,992344714 | 0,561418802 | DLG2 | FALSE | Downregulated |
| -0,570112308 | 0,561966672 | RPS28 | FALSE | Downregulated |
| -0,669270866 | 0,562801929 | CPED1 | FALSE | Downregulated |
| 0,836341507 | 0,563243165 | HIGD1A | FALSE | Upregulated |
| -0,369466438 | 0,563610247 | RN7SL2 | FALSE | Downregulated |
| 0,737926113 | 0,564254059 | LOC101927732 | FALSE | Upregulated |
| 0,572336912 | 0,56529727 | KIAA0368 | FALSE | Upregulated |
| -0,453046687 | 0,565429727 | RPS18 | FALSE | Downregulated |
| 0,784779932 | 0,565591005 | POLR1A | FALSE | Upregulated |
| -0,567006046 | 0,565591005 | TNNI1 | FALSE | Downregulated |
| 0,590756118 | 0,566016545 | MBNL2 | FALSE | Upregulated |
| -0,454564735 | 0,566016545 | PROX1 | FALSE | Downregulated |
| -0,665493238 | 0,566016545 | RANBP1 | FALSE | Downregulated |
| -0,723553275 | 0,566367948 | MAGED1 | FALSE | Downregulated |
| 0,570424926 | 0,566643011 | EIF4A1 | FALSE | Upregulated |
| 0,738695334 | 0,566643011 | RBM15 | FALSE | Upregulated |
| -0,801691377 | 0,566643011 | TRIM2 | FALSE | Downregulated |
| -0,858131934 | 0,567255105 | SAA4 | FALSE | Downregulated |
| -0,390558044 | 0,568760593 | HEXIM1 | FALSE | Downregulated |
| -0,658011716 | 0,568760593 | MRPL20 | FALSE | Downregulated |
| 0,557810322 | 0,568971071 | BCAR1 | FALSE | Upregulated |
| 0,537705807 | 0,569122015 | GOLGA3 | FALSE | Upregulated |
| 0,63377233 | 0,571156782 | VPS39 | FALSE | Upregulated |
| 0,571915178 | 0,571708169 | ATG2A | FALSE | Upregulated |
| 0,664862169 | 0,571708169 | MSN | FALSE | Upregulated |
| 1,002183412 | 0,571708169 | RP11-384K6.6 | FALSE | Upregulated |
| 0,709511219 | 0,571708169 | SH3TC1 | FALSE | Upregulated |
| 0,687077357 | 0,573275949 | MADD | FALSE | Upregulated |
| 0,564865386 | 0,57425606 | AC003665.1 | FALSE | Upregulated |
| 0,852308667 | 0,57425606 | GLIS3 | FALSE | Upregulated |
| 0,782228642 | 0,57425606 | PCNA-AS1 | FALSE | Upregulated |
| 0,578893352 | 0,57425606 | TJP1 | FALSE | Upregulated |
| -0,504743099 | 0,575618064 | MDH2 | FALSE | Downregulated |
| -0,561757538 | 0,576053772 | AGMAT | FALSE | Downregulated |
| 0,460470095 | 0,576733557 | FAM83H | FALSE | Upregulated |
| -0,620636518 | 0,57715031 | LPCAT3 | FALSE | Downregulated |
| 0,519213178 | 0,57715031 | NSD1 | FALSE | Upregulated |
| 0,607091 | 0,577218194 | DNAJC22 | FALSE | Upregulated |
| -0,840385271 | 0,577218194 | GCHFR | FALSE | Downregulated |
| 0,907717673 | 0,577218194 | LOC729451 | FALSE | Upregulated |
| -0,749234364 | 0,577218194 | RTN3 | FALSE | Downregulated |
| 0,762236621 | 0,577584836 | ARHGEF1 | FALSE | Upregulated |
| -0,749162708 | 0,577584836 | NDUFS2 | FALSE | Downregulated |
| -0,795796222 | 0,577613815 | SRD5A1 | FALSE | Downregulated |
| -0,786633882 | 0,57788607 | COX5B | FALSE | Downregulated |
| 0,446648708 | 0,57788607 | ITPK1-AS1 | FALSE | Upregulated |
| -0,832612794 | 0,57788607 | LOC388849 | FALSE | Downregulated |
| 0,667595539 | 0,57788607 | SRSF4 | FALSE | Upregulated |
| -0,506642909 | 0,578623431 | CEBPD | FALSE | Downregulated |

| | | | | |
|---|---|---|---|---|
| 0,618096929 | 0,578623431 | FGL2 | FALSE | Upregulated |
| -0,897700346 | 0,578623431 | GRAMD1C | FALSE | Downregulated |
| 0,467812169 | 0,578623431 | LRTOMT | FALSE | Upregulated |
| -0,803662215 | 0,580615963 | UBL3 | FALSE | Downregulated |
| 0,883318418 | 0,58110573 | LOC101926887 | FALSE | Upregulated |
| 0,654955234 | 0,582858136 | ABHD17A | FALSE | Upregulated |
| -0,372162464 | 0,584203512 | DST | FALSE | Downregulated |
| 0,486805634 | 0,584203512 | KMT2B | FALSE | Upregulated |
| 0,47293063 | 0,584203512 | MYO9B | FALSE | Upregulated |
| 0,940597672 | 0,584203512 | TYROBP | FALSE | Upregulated |
| -0,77470669 | 0,584560431 | CCT5 | FALSE | Downregulated |
| -0,717932594 | 0,584560431 | UBE2E1 | FALSE | Downregulated |
| -0,635483156 | 0,586783615 | ATF7IP2 | FALSE | Downregulated |
| -0,65147879 | 0,587134734 | BCKDHA | FALSE | Downregulated |
| 0,547745472 | 0,587134734 | SLC23A2 | FALSE | Upregulated |
| 0,804149195 | 0,587134734 | TNFAIP3 | FALSE | Upregulated |
| -0,650230827 | 0,588076728 | FAM221B | FALSE | Downregulated |
| -0,581849847 | 0,588076728 | GCFC2 | FALSE | Downregulated |
| -0,658177658 | 0,588076728 | PPIF | FALSE | Downregulated |
| 0,623597815 | 0,588076728 | RNF19A | FALSE | Upregulated |
| 0,56600285 | 0,588076728 | SERHL2 | FALSE | Upregulated |
| -0,792779078 | 0,588549506 | PER2 | FALSE | Downregulated |
| 0,594205667 | 0,588728326 | FAR2 | FALSE | Upregulated |
| -0,680228581 | 0,589419584 | CNNM4 | FALSE | Downregulated |
| 0,663710748 | 0,589419584 | DLK2 | FALSE | Upregulated |
| 0,910119446 | 0,589419584 | PCYT1A | FALSE | Upregulated |
| 0,827978898 | 0,589419584 | PIGF | FALSE | Upregulated |
| 0,706984905 | 0,589419584 | RTN4RL2 | FALSE | Upregulated |
| -0,635465346 | 0,590492007 | ACY1 | FALSE | Downregulated |
| -0,560804842 | 0,590531427 | PHF3 | FALSE | Downregulated |
| -0,868086531 | 0,590531427 | TALDO1 | FALSE | Downregulated |
| -0,701731722 | 0,590557278 | CHPT1 | FALSE | Downregulated |
| -0,671691077 | 0,590557278 | SCMH1 | FALSE | Downregulated |
| 0,517196743 | 0,59100039 | PFN1P2 | FALSE | Upregulated |
| 0,501520355 | 0,591234233 | GPR183 | FALSE | Upregulated |
| -0,73146654 | 0,591761405 | FOXA2 | FALSE | Downregulated |
| 0,842729545 | 0,591761405 | KCTD4 | FALSE | Upregulated |
| -0,761630162 | 0,591761405 | LDHAL6B | FALSE | Downregulated |
| 0,675420835 | 0,591761405 | LOC101929451 | FALSE | Upregulated |
| -1,097555055 | 0,591761405 | MANSC1 | FALSE | Downregulated |
| -0,7943538 | 0,591761405 | OTUB1 | FALSE | Downregulated |
| 0,41291983 | 0,591761405 | POLH | FALSE | Upregulated |
| 0,410817374 | 0,591761405 | SEC16A | FALSE | Upregulated |
| -0,687407809 | 0,591761405 | YWHAE | FALSE | Downregulated |
| 0,551759415 | 0,592008539 | ADRBK1 | FALSE | Upregulated |
| 0,523989281 | 0,592008539 | LZTR1 | FALSE | Upregulated |
| 0,794199959 | 0,592201356 | LOC100131395 | FALSE | Upregulated |
| -0,539080728 | 0,592275101 | APOBEC2 | FALSE | Downregulated |
| 0,883210937 | 0,592389151 | TIMM23 | FALSE | Upregulated |
| 0,555482719 | 0,592458902 | ACAP3 | FALSE | Upregulated |
| 0,486818948 | 0,592458902 | B4GALT1 | FALSE | Upregulated |

| | | | | |
|---|---|---|---|---|
| -0,498994219 | 0,592458902 | CRY2 | FALSE | Downregulated |
| -0,824903551 | 0,592458902 | CXADR | FALSE | Downregulated |
| 0,486344017 | 0,592458902 | LRBA | FALSE | Upregulated |
| -0,530829129 | 0,592458902 | PRR3 | FALSE | Downregulated |
| 1,166960133 | 0,592458902 | RASA4 | FALSE | Upregulated |
| -0,603794878 | 0,592458902 | RPL39 | FALSE | Downregulated |
| 0,527922927 | 0,592458902 | VCL | FALSE | Upregulated |
| 0,650608034 | 0,592458902 | WSB1 | FALSE | Upregulated |
| -0,676157467 | 0,59246488 | GRB14 | FALSE | Downregulated |
| 0,700435793 | 0,592578572 | OSMR | FALSE | Upregulated |
| 0,501981874 | 0,593138287 | SLC17A9 | FALSE | Upregulated |
| 0,835840626 | 0,594565661 | ANKLE2 | FALSE | Upregulated |
| -0,493592447 | 0,594565661 | BEND3P3 | FALSE | Downregulated |
| 0,489091507 | 0,594565661 | CDK12 | FALSE | Upregulated |
| -0,781228666 | 0,594565661 | DBP | FALSE | Downregulated |
| -0,659642274 | 0,594565661 | ENTPD5 | FALSE | Downregulated |
| -0,665195363 | 0,594565661 | IGFBP5 | FALSE | Downregulated |
| -0,616934049 | 0,594565661 | KCTD21-AS1 | FALSE | Downregulated |
| 0,521527788 | 0,594565661 | PROSER2-AS1 | FALSE | Upregulated |
| 0,523087338 | 0,594954362 | ABCB11 | FALSE | Upregulated |
| -0,609695453 | 0,594954362 | BAZ1A | FALSE | Downregulated |
| -0,986339582 | 0,594954362 | C9orf116 | FALSE | Downregulated |
| 0,619648644 | 0,594954362 | IRAK1 | FALSE | Upregulated |
| -0,608580154 | 0,594954362 | LINC00242 | FALSE | Downregulated |
| -1,580428168 | 0,594954362 | LOC101927842 | FALSE | Downregulated |
| -0,527809208 | 0,594954362 | LOC148413 | FALSE | Downregulated |
| -0,542549214 | 0,594954362 | PAN3 | FALSE | Downregulated |
| -0,849470161 | 0,594954362 | RAI14 | FALSE | Downregulated |
| 0,57134522 | 0,594954362 | SPON1 | FALSE | Upregulated |
| 0,723333486 | 0,594954362 | TMEM204 | FALSE | Upregulated |
| -0,788086186 | 0,595078917 | DHRS1 | FALSE | Downregulated |
| -0,812383521 | 0,595078917 | RAD50 | FALSE | Downregulated |
| 0,774743699 | 0,595381497 | MAP3K14 | FALSE | Upregulated |
| -0,762321445 | 0,595381497 | NDFIP1 | FALSE | Downregulated |
| 0,714014241 | 0,595764858 | DOCK3 | FALSE | Upregulated |
| 0,589594687 | 0,595764858 | PPP1R9B | FALSE | Upregulated |
| 0,383100663 | 0,597326566 | HTATSF1P2 | FALSE | Upregulated |
| 0,885178621 | 0,597465264 | EEPD1 | FALSE | Upregulated |
| -0,913906881 | 0,597940857 | GTF3C3 | FALSE | Downregulated |
| -0,831000803 | 0,598978064 | AC016629.8 | FALSE | Downregulated |
| -0,934357609 | 0,598978064 | AKR1CL1 | FALSE | Downregulated |
| -0,592860346 | 0,598978064 | AKT3 | FALSE | Downregulated |
| -0,548692236 | 0,598978064 | APMAP | FALSE | Downregulated |
| 0,799466925 | 0,598978064 | ARPP19 | FALSE | Upregulated |
| 0,501617677 | 0,598978064 | ATP11A-AS1 | FALSE | Upregulated |
| -0,557268479 | 0,598978064 | CRYL1 | FALSE | Downregulated |
| 0,705979644 | 0,598978064 | CSNK1G2-AS1 | FALSE | Upregulated |
| -0,431119176 | 0,598978064 | CYP3A5 | FALSE | Downregulated |
| 0,84086029 | 0,598978064 | DMWD | FALSE | Upregulated |
| 0,612680058 | 0,598978064 | DNAJC14 | FALSE | Upregulated |
| -0,426050913 | 0,598978064 | EEF1G | FALSE | Downregulated |

| | | | | |
|---|---|---|---|---|
| 0,825885343 | 0,598978064 | ELAVL1 | FALSE | Upregulated |
| -0,618314565 | 0,598978064 | ENSA | FALSE | Downregulated |
| -0,900903414 | 0,598978064 | GABPA | FALSE | Downregulated |
| -0,53422207 | 0,598978064 | GRB10 | FALSE | Downregulated |
| -0,560792808 | 0,598978064 | LRRC8E | FALSE | Downregulated |
| -0,582499092 | 0,598978064 | MAN2A1 | FALSE | Downregulated |
| 0,620547664 | 0,598978064 | NBPF9 | FALSE | Upregulated |
| -0,63069805 | 0,598978064 | PAICS | FALSE | Downregulated |
| -0,740503503 | 0,598978064 | PBX2 | FALSE | Downregulated |
| 0,548142378 | 0,598978064 | PNPLA6 | FALSE | Upregulated |
| 0,614899543 | 0,598978064 | PPP1R18 | FALSE | Upregulated |
| 0,826095255 | 0,598978064 | SOGA1 | FALSE | Upregulated |
| -0,703485935 | 0,598978064 | SOX10 | FALSE | Downregulated |
| -0,498399273 | 0,598978064 | SRPK2 | FALSE | Downregulated |
| 0,888248528 | 0,598978064 | TAF5 | FALSE | Upregulated |
| 0,629449186 | 0,598978064 | VAC14 | FALSE | Upregulated |
| 0,540549249 | 0,598978064 | WDR1 | FALSE | Upregulated |
| 0,460560589 | 0,600425584 | CISD3 | FALSE | Upregulated |
| -0,402941206 | 0,60099964 | JMJD8 | FALSE | Downregulated |
| -0,821107377 | 0,60099964 | PSMD5 | FALSE | Downregulated |
| -0,504490846 | 0,60099964 | ZBTB12 | FALSE | Downregulated |
| 0,909648366 | 0,601380679 | FYB | FALSE | Upregulated |
| -0,655109474 | 0,601380679 | TST | FALSE | Downregulated |
| -0,577356391 | 0,601835527 | SPRYD4 | FALSE | Downregulated |
| 0,891072017 | 0,601835527 | TCHP | FALSE | Upregulated |
| -0,401476652 | 0,602068395 | C15orf62 | FALSE | Downregulated |
| -0,495111641 | 0,602068395 | GANAB | FALSE | Downregulated |
| -0,724302926 | 0,602183637 | CTPS1 | FALSE | Downregulated |
| -0,740732396 | 0,602183637 | GTPBP6 | FALSE | Downregulated |
| -0,762978181 | 0,602183637 | NQO2 | FALSE | Downregulated |
| -0,68714557 | 0,602183637 | SFXN1 | FALSE | Downregulated |
| -0,456297129 | 0,602391121 | ERBB3 | FALSE | Downregulated |
| 0,435969973 | 0,602391121 | LPIN2 | FALSE | Upregulated |
| -0,561053059 | 0,603160853 | RAPGEF2 | FALSE | Downregulated |
| -0,37493973 | 0,603160853 | RPL13A | FALSE | Downregulated |
| -0,724333411 | 0,60429119 | SMTN | FALSE | Downregulated |
| 0,735408893 | 0,604462672 | UNC5A | FALSE | Upregulated |
| -0,794584062 | 0,604793302 | RFC1 | FALSE | Downregulated |
| -0,533106376 | 0,605164656 | NCAPG | FALSE | Downregulated |
| -0,627801519 | 0,605164656 | OTC | FALSE | Downregulated |
| -0,704814462 | 0,605520586 | RNF11 | FALSE | Downregulated |
| 0,391753131 | 0,606258038 | ARHGEF10L | FALSE | Upregulated |
| -0,569432278 | 0,606258038 | SOX13 | FALSE | Downregulated |
| -0,861886797 | 0,606258038 | TRIB1 | FALSE | Downregulated |
| 0,609443089 | 0,606258038 | ZXDC | FALSE | Upregulated |
| -0,64407096 | 0,606484029 | WDR5B | FALSE | Downregulated |
| -0,625893305 | 0,606635315 | ACSS2 | FALSE | Downregulated |
| 0,778497083 | 0,606643613 | PTPN12 | FALSE | Upregulated |
| -0,568612479 | 0,606643613 | RPS15 | FALSE | Downregulated |
| 0,507726458 | 0,607847747 | AIDA | FALSE | Upregulated |
| -0,626917083 | 0,607847747 | LOC101928897 | FALSE | Downregulated |

| | | | | |
|---|---|---|---|---|
| 0,77761816 | 0,607847747 | TSPAN3 | FALSE | Upregulated |
| 0,651137964 | 0,60876048 | HIPK1 | FALSE | Upregulated |
| -0,369776694 | 0,60876048 | HIPK2 | FALSE | Downregulated |
| -0,831341526 | 0,609703905 | RING1 | FALSE | Downregulated |
| 0,519442001 | 0,609703905 | SMARCA5-AS1 | FALSE | Upregulated |
| 0,660948789 | 0,609976919 | TBC1D9 | FALSE | Upregulated |
| -0,790773709 | 0,610159943 | GABARAP | FALSE | Downregulated |
| -0,599850232 | 0,611357793 | PNPLA2 | FALSE | Downregulated |
| 0,576162018 | 0,61154654 | KANSL3 | FALSE | Upregulated |
| 0,438968886 | 0,61154654 | LOC101928524 | FALSE | Upregulated |
| 0,672437045 | 0,61154654 | TATDN2 | FALSE | Upregulated |
| -0,612420217 | 0,61154654 | ZNF227 | FALSE | Downregulated |
| 0,404028934 | 0,611710011 | DALRD3 | FALSE | Upregulated |
| -0,404043694 | 0,611710011 | TMPRSS6 | FALSE | Downregulated |
| 0,588768694 | 0,612192493 | LYST | FALSE | Upregulated |
| -0,516934273 | 0,612313992 | MOCS1 | FALSE | Downregulated |
| -0,64524467 | 0,612313992 | ZMYND11 | FALSE | Downregulated |
| 0,476348529 | 0,612361899 | SHISA5 | FALSE | Upregulated |
| -0,583065855 | 0,612361899 | TMEM51-AS1 | FALSE | Downregulated |
| -0,453594292 | 0,61248219 | A1BG | FALSE | Downregulated |
| 0,763008485 | 0,614141604 | BRAT1 | FALSE | Upregulated |
| 0,77587825 | 0,614141604 | TYW5 | FALSE | Upregulated |
| 0,558323491 | 0,614141604 | VAT1 | FALSE | Upregulated |
| -0,59150511 | 0,614485222 | FAM205A | FALSE | Downregulated |
| 0,447422757 | 0,614528947 | IL7 | FALSE | Upregulated |
| -0,469415749 | 0,614528947 | MORC2-AS1 | FALSE | Downregulated |
| -0,361787888 | 0,614528947 | SNED1 | FALSE | Downregulated |
| 0,661468927 | 0,614888657 | TNFRSF1B | FALSE | Upregulated |
| -0,448906566 | 0,615528775 | AKT2 | FALSE | Downregulated |
| 0,594893145 | 0,615528775 | ALPK2 | FALSE | Upregulated |
| -0,597831644 | 0,615528775 | BCKDK | FALSE | Downregulated |
| 0,751945561 | 0,615528775 | BCL2L12 | FALSE | Upregulated |
| 0,73644484 | 0,615528775 | CDK2AP2 | FALSE | Upregulated |
| 0,770242886 | 0,615528775 | DNM1P41 | FALSE | Upregulated |
| 0,702418668 | 0,615528775 | LATS2 | FALSE | Upregulated |
| 0,68458017 | 0,615528775 | LHFPL4 | FALSE | Upregulated |
| 0,478829751 | 0,615528775 | MSMP | FALSE | Upregulated |
| -0,955200238 | 0,615528775 | PAIP2 | FALSE | Downregulated |
| 0,756364238 | 0,615528775 | PHACTR2 | FALSE | Upregulated |
| -0,353643514 | 0,615528775 | RPL10 | FALSE | Downregulated |
| 0,742496665 | 0,615528775 | SLC39A13 | FALSE | Upregulated |
| -0,78605685 | 0,615528775 | SNHG16 | FALSE | Downregulated |
| -0,807430217 | 0,615528775 | YES1 | FALSE | Downregulated |
| -0,514026739 | 0,615553755 | DPYD | FALSE | Downregulated |
| -0,44812457 | 0,615553755 | RAD23B | FALSE | Downregulated |
| 0,535252112 | 0,615588131 | PRR14L | FALSE | Upregulated |
| -0,641445507 | 0,616636536 | PANK3 | FALSE | Downregulated |
| -0,604635546 | 0,617726353 | CTAGE7P | FALSE | Downregulated |
| 1,907458985 | 0,617835815 | CRP | FALSE | Upregulated |
| 0,615646806 | 0,617835815 | LOC101927178 | FALSE | Upregulated |
| 0,402687963 | 0,617835815 | LOC101929291 | FALSE | Upregulated |

| | | | | |
|---|---|---|---|---|
| 0,441644856 | 0,618767567 | C12orf76 | FALSE | Upregulated |
| -0,768085914 | 0,619323184 | DLG4 | FALSE | Downregulated |
| 0,492056317 | 0,620715882 | SGSM2 | FALSE | Upregulated |
| 0,598240839 | 0,620715882 | TAP1 | FALSE | Upregulated |
| -0,471870475 | 0,620925175 | NAPEPLD | FALSE | Downregulated |
| -0,69088108 | 0,621403798 | ADAMTSL3 | FALSE | Downregulated |
| 0,495410159 | 0,621983658 | TBC1D2B | FALSE | Upregulated |
| -0,478298178 | 0,621998809 | ALDH3A2 | FALSE | Downregulated |
| 0,77583446 | 0,622424026 | MTA2 | FALSE | Upregulated |
| -0,6990129 | 0,623332963 | HIST1H1C | FALSE | Downregulated |
| 0,72082637 | 0,623332963 | KDM4A-AS1 | FALSE | Upregulated |
| -0,364242094 | 0,623776881 | PPP6R2 | FALSE | Downregulated |
| -0,536158283 | 0,623778718 | EFNA1 | FALSE | Downregulated |
| -0,796424693 | 0,623895618 | EIF3H | FALSE | Downregulated |
| -0,856334401 | 0,624246116 | PSME1 | FALSE | Downregulated |
| -0,703539611 | 0,624260633 | GJA4 | FALSE | Downregulated |
| -0,654817498 | 0,624260633 | RABGEF1 | FALSE | Downregulated |
| -0,661916547 | 0,626067908 | RPLP2 | FALSE | Downregulated |
| 0,506806555 | 0,626122226 | FAM86A | FALSE | Upregulated |
| -0,653895779 | 0,626122226 | PRODH2 | FALSE | Downregulated |
| -0,395509848 | 0,626122226 | RPL32 | FALSE | Downregulated |
| 0,709358287 | 0,62669876 | ARHGEF5 | FALSE | Upregulated |
| -0,709097018 | 0,62669876 | FAM160B1 | FALSE | Downregulated |
| 0,727462715 | 0,62669876 | GBP1 | FALSE | Upregulated |
| 0,714957065 | 0,626740669 | KIAA1407 | FALSE | Upregulated |
| 0,903978884 | 0,626974263 | MIEN1 | FALSE | Upregulated |
| 0,817304445 | 0,626974263 | UNC5CL | FALSE | Upregulated |
| -0,592855855 | 0,627245706 | TMEM30B | FALSE | Downregulated |
| 0,878133113 | 0,62755868 | AP3M1 | FALSE | Upregulated |
| 0,697345557 | 0,62755868 | CAD | FALSE | Upregulated |
| -0,442017522 | 0,62755868 | LOC101927339 | FALSE | Downregulated |
| 0,459826271 | 0,62755868 | PDE4DIP | FALSE | Upregulated |
| -0,806405111 | 0,62755868 | ZNF770 | FALSE | Downregulated |
| 0,476834176 | 0,627684091 | PTPN14 | FALSE | Upregulated |
| 0,70555628 | 0,627684091 | RGP1 | FALSE | Upregulated |
| 0,722484994 | 0,628088767 | GNL3L | FALSE | Upregulated |
| 0,418585219 | 0,628588304 | VAMP1 | FALSE | Upregulated |
| -0,816628581 | 0,629444862 | PGAP3 | FALSE | Downregulated |
| -0,474951496 | 0,629772552 | CANX | FALSE | Downregulated |
| -0,636888468 | 0,63046465 | AKR7A2 | FALSE | Downregulated |
| -0,65298337 | 0,631364698 | PGAM1P5 | FALSE | Downregulated |
| 0,451435305 | 0,63137968 | NANOS1 | FALSE | Upregulated |
| -0,926120045 | 0,631608651 | AGPS | FALSE | Downregulated |
| 0,551779437 | 0,631608651 | SUPT7L | FALSE | Upregulated |
| 0,609154102 | 0,631608651 | ZFC3H1 | FALSE | Upregulated |
| 0,648252773 | 0,631615073 | KIAA0232 | FALSE | Upregulated |
| 0,450636156 | 0,631615073 | LOC100505767 | FALSE | Upregulated |
| -0,595744658 | 0,631615073 | NDUFV1 | FALSE | Downregulated |
| 0,361589198 | 0,631615073 | SPDYE5 | FALSE | Upregulated |
| 0,92768841 | 0,631915171 | ERAP2 | FALSE | Upregulated |
| 0,720543068 | 0,632363135 | EIF2S1 | FALSE | Upregulated |

| | | | | |
|---|---|---|---|---|
| -0,313471467 | 0,632731138 | COL18A1 | FALSE | Downregulated |
| 0,745820903 | 0,632731138 | ITPKB | FALSE | Upregulated |
| -0,738017229 | 0,632731138 | PAXBP1 | FALSE | Downregulated |
| -0,789371241 | 0,633759291 | TAF4 | FALSE | Downregulated |
| -0,628360215 | 0,633778312 | AKR7A3 | FALSE | Downregulated |
| -0,693516514 | 0,633778312 | ERCC1 | FALSE | Downregulated |
| -0,677978653 | 0,633778312 | INS | FALSE | Downregulated |
| 0,647125136 | 0,633778312 | PIKFYVE | FALSE | Upregulated |
| -0,490692343 | 0,633921785 | SLC3A1 | FALSE | Downregulated |
| -0,772209046 | 0,633921785 | TGFBR3 | FALSE | Downregulated |
| 0,858402072 | 0,63504548 | ETNK1 | FALSE | Upregulated |
| -0,615766204 | 0,63504548 | SCCPDH | FALSE | Downregulated |
| -0,525305456 | 0,635231546 | IFT140 | FALSE | Downregulated |
| -0,593237732 | 0,635231546 | SLC25A23 | FALSE | Downregulated |
| 0,652309619 | 0,635453244 | IARS | FALSE | Upregulated |
| 0,795589293 | 0,635583514 | SLC5A6 | FALSE | Upregulated |
| 0,684099108 | 0,6356054 | AC009948.5 | FALSE | Upregulated |
| 0,795828539 | 0,6356054 | ARMC7 | FALSE | Upregulated |
| 0,786829669 | 0,636314252 | CD93 | FALSE | Upregulated |
| 0,726668999 | 0,636314252 | PLA2G6 | FALSE | Upregulated |
| -0,805101306 | 0,636314252 | TMEM192 | FALSE | Downregulated |
| 0,449335971 | 0,636608236 | RNPEPL1 | FALSE | Upregulated |
| -0,533743701 | 0,637250359 | PDIA4 | FALSE | Downregulated |
| -0,418731784 | 0,637250359 | POLR2J4 | FALSE | Downregulated |
| 0,807551488 | 0,638307393 | FUT6 | FALSE | Upregulated |
| 0,629670789 | 0,638380158 | ITPRIPL2 | FALSE | Upregulated |
| -0,674726916 | 0,639469736 | MRC1 | FALSE | Downregulated |
| 0,467571923 | 0,640066645 | LRP2BP | FALSE | Upregulated |
| 0,516006156 | 0,640848927 | LOC101928552 | FALSE | Upregulated |
| 0,787237927 | 0,640848927 | MRPL4 | FALSE | Upregulated |
| 0,466046644 | 0,640848927 | PLIN2 | FALSE | Upregulated |
| -0,497692223 | 0,640848927 | PPP2R2D | FALSE | Downregulated |
| 0,697391602 | 0,640848927 | TNK2 | FALSE | Upregulated |
| 0,495318356 | 0,640921364 | FLJ32756 | FALSE | Upregulated |
| 0,629732992 | 0,641138515 | ALMS1 | FALSE | Upregulated |
| -0,520161913 | 0,641138515 | IRF2BPL | FALSE | Downregulated |
| -0,519683474 | 0,641485259 | SOWAHC | FALSE | Downregulated |
| 0,733960269 | 0,641661495 | GSS | FALSE | Upregulated |
| -0,585686827 | 0,641776778 | APH1A | FALSE | Downregulated |
| -0,416192093 | 0,64225423 | NADK2 | FALSE | Downregulated |
| -0,494834456 | 0,642400888 | STUB1 | FALSE | Downregulated |
| -0,658353527 | 0,642893195 | BAZ1B | FALSE | Downregulated |
| -0,422224235 | 0,642893195 | CHD2 | FALSE | Downregulated |
| -0,67790175 | 0,642893195 | CLVS1 | FALSE | Downregulated |
| 0,545852207 | 0,642893195 | FAM69A | FALSE | Upregulated |
| 0,626073742 | 0,642893195 | MRAP | FALSE | Upregulated |
| -0,764383963 | 0,642893195 | PPP3R1 | FALSE | Downregulated |
| 0,745790843 | 0,643314286 | CPEB3 | FALSE | Upregulated |
| 0,535210154 | 0,643314286 | DAZAP2 | FALSE | Upregulated |
| 0,685918978 | 0,643314286 | PRMT2 | FALSE | Upregulated |
| -0,48088937 | 0,643474151 | ABI2 | FALSE | Downregulated |

| | | | | |
|---|---|---|---|---|
| 0,660170716 | 0,643474151 | ANKRD40 | FALSE | Upregulated |
| -0,628854472 | 0,643474151 | ERF | FALSE | Downregulated |
| 0,610949371 | 0,643474151 | GRB2 | FALSE | Upregulated |
| -0,772489696 | 0,643551925 | FAM160A1 | FALSE | Downregulated |
| 0,436293063 | 0,64373122 | CUX1 | FALSE | Upregulated |
| 0,431930129 | 0,643931917 | BAZ2A | FALSE | Upregulated |
| 0,489436482 | 0,643931917 | DUOX1 | FALSE | Upregulated |
| -0,519014625 | 0,644252384 | CTDSPL2 | FALSE | Downregulated |
| -0,699309796 | 0,644252384 | RPL35A | FALSE | Downregulated |
| 0,482617704 | 0,644329087 | CTSA | FALSE | Upregulated |
| -0,576290492 | 0,644329087 | GLUD1P3 | FALSE | Downregulated |
| 0,570165614 | 0,644329087 | GOLPH3 | FALSE | Upregulated |
| 0,31643986 | 0,644329087 | H6PD | FALSE | Upregulated |
| -0,357866795 | 0,644329087 | IL6ST | FALSE | Downregulated |
| 0,844955277 | 0,644329087 | LOC101927420 | FALSE | Upregulated |
| -0,68119834 | 0,644329087 | SEC24B | FALSE | Downregulated |
| 0,46960944 | 0,644534 | ELK3 | FALSE | Upregulated |
| -0,514935762 | 0,644534 | LOC101928716 | FALSE | Downregulated |
| 0,538790039 | 0,644713449 | LOC441454 | FALSE | Upregulated |
| -0,437830551 | 0,645074594 | ARSD | FALSE | Downregulated |
| -0,892305199 | 0,645074594 | CBX1 | FALSE | Downregulated |
| -0,68972105 | 0,645074594 | FAM195A | FALSE | Downregulated |
| 0,554245197 | 0,645074594 | GNB4 | FALSE | Upregulated |
| -0,368773639 | 0,645074594 | MLXIPL | FALSE | Downregulated |
| -0,565050711 | 0,645074594 | RPL14 | FALSE | Downregulated |
| -0,434605373 | 0,645074594 | RPS7 | FALSE | Downregulated |
| -0,718412176 | 0,645074594 | UBE2L3 | FALSE | Downregulated |
| -0,592358789 | 0,645074594 | ZNF652 | FALSE | Downregulated |
| -0,516399736 | 0,645074594 | ZRANB1 | FALSE | Downregulated |
| 0,649869605 | 0,645331794 | FCHO2 | FALSE | Upregulated |
| -0,38410595 | 0,646525033 | SLC18A2 | FALSE | Downregulated |
| -0,745895472 | 0,646525033 | UTS2B | FALSE | Downregulated |
| -0,893991464 | 0,646942402 | AGXT2 | FALSE | Downregulated |
| -0,74397461 | 0,646942402 | AQP3 | FALSE | Downregulated |
| 0,51196486 | 0,646942402 | COL6A2 | FALSE | Upregulated |
| 0,820488768 | 0,646942402 | ENC1 | FALSE | Upregulated |
| -0,692016948 | 0,646942402 | FDFT1 | FALSE | Downregulated |
| 0,705925945 | 0,646942402 | LOC100499194 | FALSE | Upregulated |
| -0,814395295 | 0,646942402 | RSC1A1 | FALSE | Downregulated |
| 0,432341802 | 0,646942402 | SLC24A1 | FALSE | Upregulated |
| 0,693551285 | 0,646942402 | SMARCA5 | FALSE | Upregulated |
| 0,898095501 | 0,646942402 | VWF | FALSE | Upregulated |
| 0,542328924 | 0,647922159 | CFL1 | FALSE | Upregulated |
| -0,863069263 | 0,64898843 | DZIP1 | FALSE | Downregulated |
| -0,789156266 | 0,64905717 | MBTD1 | FALSE | Downregulated |
| 0,540162279 | 0,649216285 | NAV1 | FALSE | Upregulated |
| -0,644161617 | 0,649216285 | TMEM101 | FALSE | Downregulated |
| -0,486576306 | 0,650156481 | ATP5A1 | FALSE | Downregulated |
| 0,337060591 | 0,65027888 | ABHD2 | FALSE | Upregulated |
| 0,584875714 | 0,65027888 | EVI2A | FALSE | Upregulated |
| 0,663141362 | 0,65027888 | PRR15L | FALSE | Upregulated |

| | | | | |
|---|---|---|---|---|
| 0,613041432 | 0,650515984 | NAB2 | FALSE | Upregulated |
| 0,553332922 | 0,650904131 | ATAD5 | FALSE | Upregulated |
| 0,840400603 | 0,650904131 | C9orf66 | FALSE | Upregulated |
| 0,791281433 | 0,650904131 | FLOT2 | FALSE | Upregulated |
| -0,399677273 | 0,650904131 | KIAA1161 | FALSE | Downregulated |
| 0,511471208 | 0,650904131 | MFSD6 | FALSE | Upregulated |
| 0,658388298 | 0,650904131 | NF2 | FALSE | Upregulated |
| -0,381133418 | 0,650904131 | TAOK1 | FALSE | Downregulated |
| -0,657887194 | 0,65189885 | JADE2 | FALSE | Downregulated |
| -0,629353796 | 0,652077558 | AC004019.13 | FALSE | Downregulated |
| -0,539004245 | 0,652270719 | LOC101928660 | FALSE | Downregulated |
| -0,625542005 | 0,652270719 | NOP14-AS1 | FALSE | Downregulated |
| -0,719747156 | 0,652270719 | NUBP2 | FALSE | Downregulated |
| 0,781096125 | 0,652270719 | ZBTB37 | FALSE | Upregulated |
| 0,732150618 | 0,653231757 | STARD13-AS | FALSE | Upregulated |
| 0,323219022 | 0,653321897 | TTN | FALSE | Upregulated |
| -0,579283899 | 0,653480534 | ADH5 | FALSE | Downregulated |
| 0,423863717 | 0,654434037 | DIO1 | FALSE | Upregulated |
| 0,54723714 | 0,654434037 | TAF9B | FALSE | Upregulated |
| 0,743790299 | 0,654656283 | LOC101927028 | FALSE | Upregulated |
| -0,595374029 | 0,654967767 | ASPH | FALSE | Downregulated |
| 0,518246242 | 0,655244777 | NPLOC4 | FALSE | Upregulated |
| -0,407173795 | 0,655786134 | CALD1 | FALSE | Downregulated |
| -0,557332352 | 0,655792501 | SLFNL1-AS1 | FALSE | Downregulated |
| 0,509110867 | 0,655850157 | ATXN7L3 | FALSE | Upregulated |
| 0,394999789 | 0,655850157 | FANCA | FALSE | Upregulated |
| 0,664262434 | 0,656020891 | NOMO2 | FALSE | Upregulated |
| 0,590663941 | 0,656460207 | LRRFIP2 | FALSE | Upregulated |
| 0,545134371 | 0,656460207 | MTHFR | FALSE | Upregulated |
| 0,506697898 | 0,656460207 | PLXNC1 | FALSE | Upregulated |
| 0,612839969 | 0,656460207 | TRPV1 | FALSE | Upregulated |
| -0,653006619 | 0,657213946 | FLJ13197 | FALSE | Downregulated |
| 0,53286145 | 0,658150513 | ADCY6 | FALSE | Upregulated |
| 0,616236243 | 0,658460973 | ARHGAP31 | FALSE | Upregulated |
| -0,736554989 | 0,658475295 | ZNF551 | FALSE | Downregulated |
| -0,543161978 | 0,65958008 | KGFLP2 | FALSE | Downregulated |
| 0,448622438 | 0,661358172 | OGT | FALSE | Upregulated |
| 0,71498755 | 0,661876442 | RP5-914P20.5 | FALSE | Upregulated |
| 0,758106465 | 0,662067382 | ARSG | FALSE | Upregulated |
| -0,828957536 | 0,662067382 | ATP5J | FALSE | Downregulated |
| -0,42075705 | 0,662067382 | CES2 | FALSE | Downregulated |
| 0,683416068 | 0,662067382 | IPO9 | FALSE | Upregulated |
| -0,892415166 | 0,662067382 | LOC101926985 | FALSE | Downregulated |
| 0,538624062 | 0,662067382 | LOC166994 | FALSE | Upregulated |
| 0,51535361 | 0,662067382 | MINA | FALSE | Upregulated |
| 0,534153022 | 0,662067382 | NFRKB | FALSE | Upregulated |
| -0,662641758 | 0,662067382 | PKDREJ | FALSE | Downregulated |
| -0,587917078 | 0,662259588 | ADIPOR1 | FALSE | Downregulated |
| -0,531641012 | 0,662259588 | RNA45S5 | FALSE | Downregulated |
| -0,619707761 | 0,663163982 | BZW1 | FALSE | Downregulated |
| -0,605090394 | 0,663163982 | H3F3A | FALSE | Downregulated |

| | | | | |
|---|---|---|---|---|
| 0,828045968 | 0,663163982 | HIVEP2 | FALSE | Upregulated |
| -0,690209841 | 0,663163982 | LOC101928717 | FALSE | Downregulated |
| -0,420265506 | 0,663163982 | NCBP1 | FALSE | Downregulated |
| 0,483827965 | 0,663163982 | SDC3 | FALSE | Upregulated |
| 0,523924192 | 0,663163982 | SIDT2 | FALSE | Upregulated |
| 0,350508345 | 0,663334923 | CD163 | FALSE | Upregulated |
| 0,396126093 | 0,663334923 | DDX5 | FALSE | Upregulated |
| 0,609389985 | 0,663334923 | MICAL2 | FALSE | Upregulated |
| -0,371439544 | 0,663334923 | MUC6 | FALSE | Downregulated |
| -0,662657133 | 0,663334923 | NEU4 | FALSE | Downregulated |
| -0,565607594 | 0,663334923 | PBXIP1 | FALSE | Downregulated |
| 0,602376604 | 0,663334923 | ZNF587B | FALSE | Upregulated |
| 0,768442547 | 0,663377704 | CCAT1 | FALSE | Upregulated |
| 0,719806903 | 0,664183026 | DGCR9 | FALSE | Upregulated |
| -0,604765703 | 0,664442017 | SMARCA1 | FALSE | Downregulated |
| -0,715314886 | 0,664572946 | BTG3 | FALSE | Downregulated |
| -0,758472336 | 0,664572946 | C10orf95 | FALSE | Downregulated |
| 0,728338182 | 0,664629804 | MYBBP1A | FALSE | Upregulated |
| -0,343512899 | 0,664903629 | HEXIM2 | FALSE | Downregulated |
| -0,372316235 | 0,664903629 | ZSWIM8-AS1 | FALSE | Downregulated |
| 0,757931945 | 0,66512215 | SLC35D2 | FALSE | Upregulated |
| -0,71968951 | 0,665156372 | GRPEL1 | FALSE | Downregulated |
| -0,599738251 | 0,665403049 | STK19 | FALSE | Downregulated |
| 0,560309601 | 0,665403049 | TM7SF3 | FALSE | Upregulated |
| 0,731995081 | 0,665403049 | WDR13 | FALSE | Upregulated |
| 0,631067015 | 0,666472154 | UCHL5 | FALSE | Upregulated |
| -0,665116362 | 0,66906685 | CCDC47 | FALSE | Downregulated |
| -0,59725903 | 0,66906685 | CLDN14 | FALSE | Downregulated |
| -0,724328748 | 0,66906685 | LIN7C | FALSE | Downregulated |
| 0,534540014 | 0,66906685 | NSUN5 | FALSE | Upregulated |
| 0,558326403 | 0,669939825 | NUP98 | FALSE | Upregulated |
| 0,717658652 | 0,670948869 | CCM2 | FALSE | Upregulated |
| 0,413646587 | 0,670948869 | FKBP1AP1 | FALSE | Upregulated |
| 0,532576576 | 0,670948869 | HIP1R | FALSE | Upregulated |
| -0,631848904 | 0,670948869 | MED29 | FALSE | Downregulated |
| -0,68537886 | 0,670948869 | NPM1 | FALSE | Downregulated |
| 0,504543239 | 0,670948869 | SDC4 | FALSE | Upregulated |
| 0,598025259 | 0,670948869 | SPCS3 | FALSE | Upregulated |
| -0,63774556 | 0,670948869 | TBC1D10C | FALSE | Downregulated |
| -0,616756559 | 0,670948869 | THUMPD1 | FALSE | Downregulated |
| -0,479902061 | 0,670948869 | TUFM | FALSE | Downregulated |
| 0,61085467 | 0,670948869 | USP36 | FALSE | Upregulated |
| 0,4820114 | 0,672377777 | ADNP | FALSE | Upregulated |
| 0,567063906 | 0,672377777 | HYOU1 | FALSE | Upregulated |
| -0,453220333 | 0,672377777 | PTOV1 | FALSE | Downregulated |
| -0,791899478 | 0,672629509 | FGFR1OP | FALSE | Downregulated |
| -0,384421309 | 0,672629509 | TMEM63A | FALSE | Downregulated |
| 0,692698391 | 0,673237817 | TMEM180 | FALSE | Upregulated |
| 0,435806334 | 0,673585939 | RP11-133O22.6 | FALSE | Upregulated |
| -0,759708372 | 0,673669456 | MED31 | FALSE | Downregulated |
| 0,414086609 | 0,673689208 | MAB21L2 | FALSE | Upregulated |

| | | | | |
|---|---|---|---|---|
| -0,349061853 | 0,673843203 | LINC00657 | FALSE | Downregulated |
| 0,647902565 | 0,674305032 | C17orf70 | FALSE | Upregulated |
| -0,539772136 | 0,674476838 | ARGLU1 | FALSE | Downregulated |
| -0,589818593 | 0,674476838 | NPC1L1 | FALSE | Downregulated |
| 0,913689556 | 0,675044197 | SMA4 | FALSE | Upregulated |
| -0,673308297 | 0,675100831 | FIS1 | FALSE | Downregulated |
| -0,400558861 | 0,675100831 | NAIP | FALSE | Downregulated |
| 0,53471162 | 0,675100831 | SP2 | FALSE | Upregulated |
| -0,721441721 | 0,675100831 | TMUB1 | FALSE | Downregulated |
| -0,560056677 | 0,675414282 | CACNA1D | FALSE | Downregulated |
| 0,707724305 | 0,675816804 | MLK7-AS1 | FALSE | Upregulated |
| 0,514801957 | 0,675816804 | PTPN21 | FALSE | Upregulated |
| 0,783117404 | 0,676830209 | LOC101929714 | FALSE | Upregulated |
| 0,545014126 | 0,676830209 | NUP50 | FALSE | Upregulated |
| 0,450306898 | 0,676835095 | GNB1 | FALSE | Upregulated |
| -0,771132496 | 0,676835095 | HMGN3 | FALSE | Downregulated |
| 0,577383559 | 0,677225004 | PEAK1 | FALSE | Upregulated |
| 0,713375705 | 0,677379555 | ARAP2 | FALSE | Upregulated |
| 0,779134194 | 0,677379555 | HTRA1 | FALSE | Upregulated |
| 0,371473803 | 0,677379555 | LGALS8 | FALSE | Upregulated |
| 0,731307055 | 0,677379555 | LOC100128531 | FALSE | Upregulated |
| -0,757982764 | 0,677379555 | LOC101929173 | FALSE | Downregulated |
| 0,71839076 | 0,677379555 | MMP14 | FALSE | Upregulated |
| 0,637763085 | 0,677379555 | RHOU | FALSE | Upregulated |
| 0,651764413 | 0,677379555 | SECISBP2 | FALSE | Upregulated |
| 0,560703828 | 0,677379555 | ZNF862 | FALSE | Upregulated |
| 0,522965899 | 0,677450892 | CABIN1 | FALSE | Upregulated |
| -0,379440902 | 0,677450892 | COG5 | FALSE | Downregulated |
| -0,433072401 | 0,677450892 | EIF4H | FALSE | Downregulated |
| -0,53232499 | 0,677450892 | ERP29 | FALSE | Downregulated |
| 0,675213567 | 0,677450892 | ITPR1 | FALSE | Upregulated |
| -0,583054921 | 0,677450892 | LOC100506730 | FALSE | Downregulated |
| -0,49718601 | 0,677450892 | TRPM7 | FALSE | Downregulated |
| -0,336592199 | 0,677734485 | APOC2 | FALSE | Downregulated |
| 0,336782634 | 0,677734485 | TMEM140 | FALSE | Upregulated |
| 0,417226478 | 0,678068782 | TNS3 | FALSE | Upregulated |
| -0,775226767 | 0,67847575 | CHMP2A | FALSE | Downregulated |
| -0,53046006 | 0,67847575 | EFCAB14 | FALSE | Downregulated |
| 0,7572126 | 0,67847575 | NFATC2IP | FALSE | Upregulated |
| 0,552613038 | 0,67847575 | PRUNE2 | FALSE | Upregulated |
| -0,363622569 | 0,67847575 | RPLP0 | FALSE | Downregulated |
| 0,632114967 | 0,67847575 | TMEM8A | FALSE | Upregulated |
| -0,536006067 | 0,678663004 | CTSH | FALSE | Downregulated |
| -0,435705162 | 0,679016741 | ZNF154 | FALSE | Downregulated |
| -0,525547929 | 0,679147739 | ZNF395 | FALSE | Downregulated |
| -0,460091302 | 0,680236254 | RRAS2 | FALSE | Downregulated |
| 0,501364442 | 0,680815337 | TMEM127 | FALSE | Upregulated |
| -0,691047423 | 0,680815337 | USP3-AS1 | FALSE | Downregulated |
| 0,532208068 | 0,680965245 | MAMDC4 | FALSE | Upregulated |
| -0,77099244 | 0,680965245 | STOML2 | FALSE | Downregulated |
| -0,528238183 | 0,681203604 | RASSF5 | FALSE | Downregulated |

| | | | | |
|---|---|---|---|---|
| 0,747368226 | 0,682582174 | HLA-DPB1 | FALSE | Upregulated |
| 0,58399909 | 0,682582174 | NELFCD | FALSE | Upregulated |
| -0,643915824 | 0,68311162 | NTN4 | FALSE | Downregulated |
| 0,60477774 | 0,683406054 | IFNK | FALSE | Upregulated |
| -0,537857553 | 0,683754748 | NRD1 | FALSE | Downregulated |
| -0,446912899 | 0,684479115 | ACVR2A | FALSE | Downregulated |
| 0,711347187 | 0,684479115 | CPS1-IT1 | FALSE | Upregulated |
| 0,479192041 | 0,684479115 | DDX39B | FALSE | Upregulated |
| -0,471230624 | 0,684479115 | GPBP1L1 | FALSE | Downregulated |
| 0,487393142 | 0,684479115 | PAPPA | FALSE | Upregulated |
| 0,453038069 | 0,684479115 | SPATA13 | FALSE | Upregulated |
| 0,774582413 | 0,684479115 | TP53I3 | FALSE | Upregulated |
| 0,679005914 | 0,684479115 | ZNF780A | FALSE | Upregulated |
| -0,733734501 | 0,68547816 | AURKAIP1 | FALSE | Downregulated |
| 0,777275812 | 0,685550277 | LOC101927445 | FALSE | Upregulated |
| 0,64229851 | 0,685602595 | IDI2 | FALSE | Upregulated |
| -0,557440338 | 0,685817934 | CDH2 | FALSE | Downregulated |
| -0,395880208 | 0,685817934 | UGT2B15 | FALSE | Downregulated |
| -0,497556882 | 0,686018908 | H3F3B | FALSE | Downregulated |
| 0,458930338 | 0,686018908 | PITPNM2 | FALSE | Upregulated |
| 0,667212748 | 0,686018908 | REEP1 | FALSE | Upregulated |
| 0,470931168 | 0,686018908 | RP11-231I16.1 | FALSE | Upregulated |
| 0,367704687 | 0,686018908 | SRCAP | FALSE | Upregulated |
| 0,708238495 | 0,686018908 | TAX1BP1 | FALSE | Upregulated |
| -0,482536097 | 0,686018908 | TMEM220 | FALSE | Downregulated |
| -0,473663936 | 0,687679908 | TIMP3 | FALSE | Downregulated |
| 0,573729063 | 0,687689405 | AP2B1 | FALSE | Upregulated |
| -0,526985 | 0,688228675 | CCAR2 | FALSE | Downregulated |
| -0,551476024 | 0,688228675 | CCNG1 | FALSE | Downregulated |
| -0,60313111 | 0,688228675 | GTSE1-AS1 | FALSE | Downregulated |
| -0,596874057 | 0,688228675 | MYO9A | FALSE | Downregulated |
| -0,810905226 | 0,688228675 | ZBTB39 | FALSE | Downregulated |
| -0,510320777 | 0,68830206 | OSBP | FALSE | Downregulated |
| -0,668006414 | 0,688775203 | ASCC2 | FALSE | Downregulated |
| 0,331700974 | 0,688775203 | NDST1 | FALSE | Upregulated |
| 0,597799605 | 0,688907238 | SGSM3 | FALSE | Upregulated |
| 0,533354339 | 0,68914527 | EMILIN1 | FALSE | Upregulated |
| -0,427061487 | 0,68914527 | TMED10P1 | FALSE | Downregulated |
| -0,47146833 | 0,68914527 | ZNF584 | FALSE | Downregulated |
| -0,790712384 | 0,689294568 | DPP4 | FALSE | Downregulated |
| -0,592458116 | 0,689294568 | PORCN | FALSE | Downregulated |
| -0,538256029 | 0,689294568 | PROZ | FALSE | Downregulated |
| -0,621783987 | 0,689443197 | GOLGA8N | FALSE | Downregulated |
| 0,378297036 | 0,689443197 | HCFC1 | FALSE | Upregulated |
| -0,573438841 | 0,689443197 | SCARNA5 | FALSE | Downregulated |
| -0,605066329 | 0,689443197 | TYW1 | FALSE | Downregulated |
| 0,492849692 | 0,689655402 | LOC101929387 | FALSE | Upregulated |
| 0,678282337 | 0,690411474 | PQLC2 | FALSE | Upregulated |
| 0,62606534 | 0,690411474 | UBE2G2 | FALSE | Upregulated |
| 0,602922349 | 0,690634682 | GNA12 | FALSE | Upregulated |
| -0,675202097 | 0,69070862 | AC007038.7 | FALSE | Downregulated |

| | | | | |
|---|---|---|---|---|
| 0,549958939 | 0,691045532 | LYPD1 | FALSE | Upregulated |
| 0,477476252 | 0,691385719 | CLCN7 | FALSE | Upregulated |
| 0,526543496 | 0,691385719 | LTBP4 | FALSE | Upregulated |
| 0,594426818 | 0,692132141 | E4F1 | FALSE | Upregulated |
| -0,44270731 | 0,692132141 | IL13RA1 | FALSE | Downregulated |
| -0,749482265 | 0,692132141 | PSMB3 | FALSE | Downregulated |
| 0,36424616 | 0,692132141 | TPPP | FALSE | Upregulated |
| -0,606801293 | 0,692267673 | ATAD1 | FALSE | Downregulated |
| -0,462905794 | 0,692403197 | AP4S1 | FALSE | Downregulated |
| -0,530350326 | 0,69240339 | EEF1B2 | FALSE | Downregulated |
| 0,333144479 | 0,692747454 | GSN-AS1 | FALSE | Upregulated |
| 0,719534638 | 0,693074563 | TMCC1 | FALSE | Upregulated |
| -0,535301381 | 0,693540757 | AKTIP | FALSE | Downregulated |
| 0,40240162 | 0,693540757 | CNTD1 | FALSE | Upregulated |
| 0,427685949 | 0,693540757 | ITGB1 | FALSE | Upregulated |
| 0,582265358 | 0,693540757 | KLC1 | FALSE | Upregulated |
| 0,571107809 | 0,693540757 | LOC100506990 | FALSE | Upregulated |
| 0,680948263 | 0,693540757 | PARD3B | FALSE | Upregulated |
| 0,591034633 | 0,6940909 | PCNXL3 | FALSE | Upregulated |
| 0,550899857 | 0,694208268 | CSRP1 | FALSE | Upregulated |
| 0,377639441 | 0,69423809 | TUBGCP6 | FALSE | Upregulated |
| -0,736497515 | 0,694826464 | ABHD16A | FALSE | Downregulated |
| 0,70363791 | 0,694826464 | PLEKHG1 | FALSE | Upregulated |
| 0,413229121 | 0,695252388 | PRKDC | FALSE | Upregulated |
| 0,447171794 | 0,695252388 | ZNHIT3 | FALSE | Upregulated |
| -0,500930935 | 0,695385133 | EIF4A2 | FALSE | Downregulated |
| -0,40924504 | 0,695385133 | GSPT1 | FALSE | Downregulated |
| -0,759976524 | 0,69572473 | IKZF4 | FALSE | Downregulated |
| -0,334567563 | 0,69637025 | PLIN4 | FALSE | Downregulated |
| -1,195619285 | 0,696393265 | FKBP5 | FALSE | Downregulated |
| 0,411596683 | 0,696393265 | MAP4 | FALSE | Upregulated |
| 0,551701406 | 0,696401573 | GNPDA2 | FALSE | Upregulated |
| 0,709588223 | 0,696401573 | HN1L | FALSE | Upregulated |
| -0,714686969 | 0,69654322 | ZFYVE19 | FALSE | Downregulated |
| -0,585380544 | 0,696905022 | PNPLA3 | FALSE | Downregulated |
| -0,663906443 | 0,696919348 | ANAPC5 | FALSE | Downregulated |
| 0,721419446 | 0,697679139 | SEC23IP | FALSE | Upregulated |
| 0,428894003 | 0,697679139 | SHANK2 | FALSE | Upregulated |
| 0,547379471 | 0,698242959 | MDH1 | FALSE | Upregulated |
| 0,495527307 | 0,698242959 | NACC2 | FALSE | Upregulated |
| 0,368218293 | 0,698308846 | PVR | FALSE | Upregulated |
| 0,567439251 | 0,69849609 | CALB1 | FALSE | Upregulated |
| 0,459875206 | 0,69853217 | ALDOA | FALSE | Upregulated |
| -0,531127139 | 0,69853217 | APBA1 | FALSE | Downregulated |
| 0,541062134 | 0,69853217 | CABLES2 | FALSE | Upregulated |
| 0,480415539 | 0,69853217 | CFLAR-AS1 | FALSE | Upregulated |
| -0,45882213 | 0,69853217 | DIP2B | FALSE | Downregulated |
| -0,562761927 | 0,69853217 | DLC1 | FALSE | Downregulated |
| -0,553226983 | 0,69853217 | ELL2 | FALSE | Downregulated |
| -0,78934215 | 0,69853217 | EPRS | FALSE | Downregulated |
| -0,694327526 | 0,69853217 | LRRC45 | FALSE | Downregulated |

| | | | | |
|---|---|---|---|---|
| -0,619193677 | 0,69853217 | NR1H4 | FALSE | Downregulated |
| -0,432709937 | 0,69853217 | PRKCSH | FALSE | Downregulated |
| 0,366201168 | 0,69853217 | PSIP1 | FALSE | Upregulated |
| 0,58435427 | 0,69853217 | RASAL2 | FALSE | Upregulated |
| -0,496811413 | 0,69853217 | RPL36 | FALSE | Downregulated |
| -0,352371553 | 0,69853217 | SLC1A1 | FALSE | Downregulated |
| 0,661034269 | 0,69853217 | SNAP29 | FALSE | Upregulated |
| -0,584998797 | 0,69853217 | TIAM2 | FALSE | Downregulated |
| 0,731975392 | 0,69853217 | TMEM45B | FALSE | Upregulated |
| 0,457504163 | 0,69853217 | TSC22D2 | FALSE | Upregulated |
| -0,289949946 | 0,69853217 | UBR4 | FALSE | Downregulated |
| -0,652239734 | 0,69853217 | ZBTB48 | FALSE | Downregulated |
| 0,686066883 | 0,69853217 | ZFYVE27 | FALSE | Upregulated |
| 0,696623299 | 0,698705188 | ARPC1A | FALSE | Upregulated |
| 0,685244875 | 0,698705188 | CACNA1C | FALSE | Upregulated |
| -0,700077604 | 0,698705188 | DDI2 | FALSE | Downregulated |
| 0,52311083 | 0,698705188 | DYNC1LI2 | FALSE | Upregulated |
| 0,580437509 | 0,698705188 | FAM120B | FALSE | Upregulated |
| 0,559716595 | 0,698705188 | NSG1 | FALSE | Upregulated |
| -0,680375462 | 0,698705188 | PRDX4 | FALSE | Downregulated |
| -0,704263042 | 0,698705188 | PSMC5 | FALSE | Downregulated |
| 0,554851291 | 0,698705188 | PSPC1 | FALSE | Upregulated |
| -0,522276937 | 0,698705188 | RAB17 | FALSE | Downregulated |
| 0,712430073 | 0,698705188 | RP11-450K4.1 | FALSE | Upregulated |
| -0,439033023 | 0,698705188 | RTN4 | FALSE | Downregulated |
| -0,33150543 | 0,698705188 | SEC14L2 | FALSE | Downregulated |
| 0,454191422 | 0,698705188 | TRIM25 | FALSE | Upregulated |
| -0,395249613 | 0,698705188 | TUBB | FALSE | Downregulated |
| 0,433217848 | 0,699251769 | AFF4 | FALSE | Upregulated |
| -0,471766225 | 0,699251769 | APP | FALSE | Downregulated |
| 0,417426472 | 0,699302227 | BAP1 | FALSE | Upregulated |
| 0,486520508 | 0,699465121 | RP11-31F15.2 | FALSE | Upregulated |
| 0,5341296 | 0,700576319 | SLC1A4 | FALSE | Upregulated |
| 0,43677852 | 0,702171215 | CECR2 | FALSE | Upregulated |
| -0,386899785 | 0,702171215 | MKNK2 | FALSE | Downregulated |
| 0,385981182 | 0,702171215 | PDXK | FALSE | Upregulated |
| -0,385506281 | 0,702171215 | TMED10 | FALSE | Downregulated |
| 0,664862359 | 0,702198187 | FSCN1 | FALSE | Upregulated |
| -0,466725414 | 0,702789464 | MAN2C1 | FALSE | Downregulated |
| -0,438366379 | 0,702789464 | ZFYVE16 | FALSE | Downregulated |
| -0,363547676 | 0,703110109 | DIAPH1 | FALSE | Downregulated |
| 0,757281867 | 0,703440186 | C19orf54 | FALSE | Upregulated |
| 0,763159599 | 0,703454775 | AQP4-AS1 | FALSE | Upregulated |
| -0,459812018 | 0,703454775 | FGFR4 | FALSE | Downregulated |
| -0,581672771 | 0,703454775 | PREPL | FALSE | Downregulated |
| -0,496001185 | 0,703454775 | PTGES3 | FALSE | Downregulated |
| 0,798556404 | 0,703454775 | RARRES1 | FALSE | Upregulated |
| 0,613811231 | 0,704164793 | CAPNS1 | FALSE | Upregulated |
| 0,331596951 | 0,704164793 | TAPBP | FALSE | Upregulated |
| -0,42549491 | 0,704164793 | UBE2D3 | FALSE | Downregulated |
| -0,519296833 | 0,704527051 | DLEC1 | FALSE | Downregulated |

| | | | | |
|---|---|---|---|---|
| 0,762663028 | 0,704945552 | FAM163B | FALSE | Upregulated |
| 0,416951604 | 0,705143027 | C2orf68 | FALSE | Upregulated |
| 0,688677947 | 0,705220771 | EPN2 | FALSE | Upregulated |
| -0,440980073 | 0,705220771 | GSTK1 | FALSE | Downregulated |
| -0,580841664 | 0,705220771 | NFKBIZ | FALSE | Downregulated |
| 0,639370611 | 0,705220771 | PAXBP1-AS1 | FALSE | Upregulated |
| 0,445475365 | 0,705220771 | RNF113B | FALSE | Upregulated |
| 0,583562382 | 0,706571732 | ARRB1 | FALSE | Upregulated |
| 0,435796589 | 0,706740253 | PTPN3 | FALSE | Upregulated |
| 0,369901797 | 0,707471292 | AGBL5 | FALSE | Upregulated |
| 0,411629708 | 0,707471292 | DNM1L | FALSE | Upregulated |
| 0,474838992 | 0,707471292 | MEGF8 | FALSE | Upregulated |
| -0,404665996 | 0,707471292 | VPS54 | FALSE | Downregulated |
| 0,621639632 | 0,707691499 | CELSR1 | FALSE | Upregulated |
| -0,635803367 | 0,707691499 | HIBADH | FALSE | Downregulated |
| 0,650553257 | 0,707691499 | HPS5 | FALSE | Upregulated |
| -0,769033169 | 0,707691499 | LOC285593 | FALSE | Downregulated |
| 0,538995259 | 0,707691499 | RABL6 | FALSE | Upregulated |
| 0,495338948 | 0,707691499 | SF3B3 | FALSE | Upregulated |
| -0,620504006 | 0,707691499 | TCAIM | FALSE | Downregulated |
| -0,474379717 | 0,707691499 | UBE3A | FALSE | Downregulated |
| -0,416597026 | 0,707729163 | AKAP1 | FALSE | Downregulated |
| 0,694828308 | 0,707729163 | ARHGEF34P | FALSE | Upregulated |
| 0,621483267 | 0,707729163 | FNDC4 | FALSE | Upregulated |
| 0,50584722 | 0,707729163 | GATAD2A | FALSE | Upregulated |
| -0,61274902 | 0,707729163 | GSTO1 | FALSE | Downregulated |
| -0,349252041 | 0,707729163 | MYLK | FALSE | Downregulated |
| -0,565259123 | 0,707729163 | NET1 | FALSE | Downregulated |
| 0,67367297 | 0,707729163 | NEURL4 | FALSE | Upregulated |
| 0,512796699 | 0,707729163 | RAB23 | FALSE | Upregulated |
| -0,539241873 | 0,707729163 | RBM10 | FALSE | Downregulated |
| -0,453791709 | 0,707729163 | RNF123 | FALSE | Downregulated |
| -0,540395577 | 0,707796654 | SPHK2 | FALSE | Downregulated |
| 0,315110668 | 0,708146017 | SPATA21 | FALSE | Upregulated |
| -0,542949241 | 0,708478849 | GIGYF2 | FALSE | Downregulated |
| 0,417711494 | 0,708478849 | PRSS48 | FALSE | Upregulated |
| -0,518957339 | 0,708478849 | SNX33 | FALSE | Downregulated |
| 0,629971854 | 0,710056312 | NADSYN1 | FALSE | Upregulated |
| 0,696543191 | 0,710305602 | RP11-115J16.1 | FALSE | Upregulated |
| 0,74166932 | 0,710373722 | DENND5B-AS1 | FALSE | Upregulated |
| -0,458698312 | 0,710373722 | DMD | FALSE | Downregulated |
| -0,349501276 | 0,710564533 | HNRNPA2B1 | FALSE | Downregulated |
| 0,712179249 | 0,710990731 | PECAM1 | FALSE | Upregulated |
| -0,46634905 | 0,711579022 | RBM12B-AS1 | FALSE | Downregulated |
| 0,565871229 | 0,712307364 | GMPR2 | FALSE | Upregulated |
| 0,61669137 | 0,712307364 | TMEM9B | FALSE | Upregulated |
| 0,802606262 | 0,712866076 | EDNRB-AS1 | FALSE | Upregulated |
| -0,654605552 | 0,712866076 | LTBP1 | FALSE | Downregulated |
| 0,513499053 | 0,712866076 | MYO19 | FALSE | Upregulated |
| 0,54760387 | 0,712866076 | YPEL4 | FALSE | Upregulated |
| -0,483928708 | 0,713232149 | IL1R1 | FALSE | Downregulated |

| | | | | |
|---|---|---|---|---|
| -0,439809198 | 0,713232149 | MIB2 | FALSE | Downregulated |
| 0,338877139 | 0,713232149 | MRPS25 | FALSE | Upregulated |
| 0,456473319 | 0,713232149 | SAMD11 | FALSE | Upregulated |
| -0,502775406 | 0,713232149 | TOM1L1 | FALSE | Downregulated |
| 0,648451413 | 0,71331154 | CYP21A2 | FALSE | Upregulated |
| -0,39361442 | 0,714329727 | GNAS | FALSE | Downregulated |
| -0,392634579 | 0,714329727 | MED13 | FALSE | Downregulated |
| 0,579522124 | 0,714329727 | MPC1 | FALSE | Upregulated |
| 0,680785038 | 0,714366145 | UBFD1 | FALSE | Upregulated |
| -0,697310801 | 0,714580606 | SUMO3 | FALSE | Downregulated |
| -0,3601756 | 0,714972189 | CERS6-AS1 | FALSE | Downregulated |
| -0,466711007 | 0,714972189 | KDELR2 | FALSE | Downregulated |
| 0,596298221 | 0,71497983 | ITFG3 | FALSE | Upregulated |
| -0,593748179 | 0,715518155 | HSD3BP4 | FALSE | Downregulated |
| 0,69812649 | 0,715518155 | LOC102723992 | FALSE | Upregulated |
| -0,600904879 | 0,716203636 | LMLN | FALSE | Downregulated |
| 0,525332894 | 0,716225714 | DNMBP | FALSE | Upregulated |
| -0,508966705 | 0,716225714 | LINC00675 | FALSE | Downregulated |
| -0,333509797 | 0,716225714 | NCOA4 | FALSE | Downregulated |
| -0,564907923 | 0,716225714 | PDIA6 | FALSE | Downregulated |
| 0,605003993 | 0,716225714 | PIGQ | FALSE | Upregulated |
| 0,515711747 | 0,716225714 | TANC1 | FALSE | Upregulated |
| 0,680389113 | 0,716225714 | TRAPPC11 | FALSE | Upregulated |
| -0,457628062 | 0,716486545 | CAMSAP3 | FALSE | Downregulated |
| -0,548589455 | 0,716486545 | CHCHD10 | FALSE | Downregulated |
| -0,730274812 | 0,716486545 | CYP4F12 | FALSE | Downregulated |
| 0,584523863 | 0,716486545 | HEG1 | FALSE | Upregulated |
| -0,447919552 | 0,716486545 | ITM2B | FALSE | Downregulated |
| 0,567839536 | 0,716486545 | LOC102724466 | FALSE | Upregulated |
| -0,332603493 | 0,716486545 | SLC22A7 | FALSE | Downregulated |
| -0,513448653 | 0,716486545 | ST3GAL6 | FALSE | Downregulated |
| 0,646155735 | 0,716486545 | WARS | FALSE | Upregulated |
| -0,67767772 | 0,716486545 | WNK4 | FALSE | Downregulated |
| -0,592319942 | 0,717154503 | SPRED2 | FALSE | Downregulated |
| 0,45885225 | 0,71735217 | LOC102723520 | FALSE | Upregulated |
| 0,550844667 | 0,71735217 | LOC102724383 | FALSE | Upregulated |
| 0,605322986 | 0,71735217 | PLXDC2 | FALSE | Upregulated |
| -0,359226837 | 0,71735217 | RHOF | FALSE | Downregulated |
| 0,398851147 | 0,71735217 | SRR | FALSE | Upregulated |
| 0,689866557 | 0,71745297 | MTERFD3 | FALSE | Upregulated |
| 0,678336442 | 0,717674085 | ARHGAP17 | FALSE | Upregulated |
| 0,630579253 | 0,717674085 | CCDC130 | FALSE | Upregulated |
| -0,635443866 | 0,717674085 | FOXJ3 | FALSE | Downregulated |
| 0,684251055 | 0,717674085 | MVB12B | FALSE | Upregulated |
| -0,511508562 | 0,717674085 | RPS10 | FALSE | Downregulated |
| 0,631551939 | 0,717674085 | STARD9 | FALSE | Upregulated |
| -0,555539076 | 0,717674085 | UAP1 | FALSE | Downregulated |
| 0,610996418 | 0,717674085 | VPS36 | FALSE | Upregulated |
| -0,456043738 | 0,718012967 | ACAD11 | FALSE | Downregulated |
| 0,550906774 | 0,71843846 | NOD1 | FALSE | Upregulated |
| -0,643686679 | 0,71843846 | PAF1 | FALSE | Downregulated |

| | | | | |
|---|---|---|---|---|
| 0,371622771 | 0,718474736 | CST3 | FALSE | Upregulated |
| -0,434483293 | 0,718474736 | DCBLD2 | FALSE | Downregulated |
| 0,629421069 | 0,718474736 | DCP1A | FALSE | Upregulated |
| -0,571770044 | 0,718824501 | HS3ST3B1 | FALSE | Downregulated |
| 0,299927776 | 0,718824501 | MPRIP | FALSE | Upregulated |
| -0,422617341 | 0,718824501 | SLC40A1 | FALSE | Downregulated |
| -0,486602041 | 0,718824501 | SMEK1 | FALSE | Downregulated |
| -0,404282425 | 0,718848058 | C1orf192 | FALSE | Downregulated |
| 0,353341704 | 0,718888627 | AC005594.3 | FALSE | Upregulated |
| 0,655319719 | 0,718955987 | ARHGEF2 | FALSE | Upregulated |
| -0,484426272 | 0,718955987 | ARL6IP4 | FALSE | Downregulated |
| -0,451412767 | 0,718955987 | HMGB1 | FALSE | Downregulated |
| -0,311747523 | 0,71910248 | FAM208B | FALSE | Downregulated |
| 0,429523291 | 0,71910248 | SMAD3 | FALSE | Upregulated |
| -0,681188335 | 0,719420722 | CX3CL1 | FALSE | Downregulated |
| -0,343533781 | 0,719420722 | DDTL | FALSE | Downregulated |
| 0,519176254 | 0,719420722 | FNBP1 | FALSE | Upregulated |
| 0,604357257 | 0,719420722 | FNDC9 | FALSE | Upregulated |
| 0,28935816 | 0,719420722 | KCNQ1OT1 | FALSE | Upregulated |
| -0,526876375 | 0,719420722 | LOC101930137 | FALSE | Downregulated |
| -0,515615757 | 0,719420722 | LRPPRC | FALSE | Downregulated |
| 0,498416894 | 0,719420722 | RHOA | FALSE | Upregulated |
| -0,657055077 | 0,719420722 | SIRT3 | FALSE | Downregulated |
| -0,384556413 | 0,719430964 | RPS8 | FALSE | Downregulated |
| -0,515877132 | 0,719430964 | RUVBL1 | FALSE | Downregulated |
| -0,358943132 | 0,719531582 | B3GNT9 | FALSE | Downregulated |
| 0,62538818 | 0,719531582 | CLDN3 | FALSE | Upregulated |
| 0,561615307 | 0,719531582 | CUL9 | FALSE | Upregulated |
| -0,33670442 | 0,719531582 | HNRNPU-AS1 | FALSE | Downregulated |
| -0,31197687 | 0,719531582 | SPTB | FALSE | Downregulated |
| 0,430592305 | 0,719604199 | MBD6 | FALSE | Upregulated |
| 0,358254405 | 0,719604199 | TLN1 | FALSE | Upregulated |
| -0,256172645 | 0,720384893 | RN7SL1 | FALSE | Downregulated |
| -0,689378868 | 0,721034503 | TMEM135 | FALSE | Downregulated |
| 0,514383269 | 0,721312387 | GAPDHS | FALSE | Upregulated |
| -0,628644221 | 0,721575015 | JTB | FALSE | Downregulated |
| 0,613573878 | 0,721575015 | LOC101926967 | FALSE | Upregulated |
| -0,707680528 | 0,72181127 | DHCR7 | FALSE | Downregulated |
| 0,353496258 | 0,72181127 | MRS2 | FALSE | Upregulated |
| -0,392827387 | 0,72181127 | RPL23A | FALSE | Downregulated |
| -0,448537286 | 0,722229555 | C17orf103 | FALSE | Downregulated |
| -0,595808414 | 0,722642672 | ADHFE1 | FALSE | Downregulated |
| -0,396881363 | 0,722642672 | EDC3 | FALSE | Downregulated |
| 0,493171373 | 0,722642672 | FEM1B | FALSE | Upregulated |
| 0,669054755 | 0,722642672 | KIAA0753 | FALSE | Upregulated |
| -0,322020828 | 0,722642672 | LOC100505564 | FALSE | Downregulated |
| -0,387764537 | 0,722642672 | ZBTB7B | FALSE | Downregulated |
| -0,560575947 | 0,723062915 | AMACR | FALSE | Downregulated |
| -0,47651086 | 0,723561314 | ABCD3 | FALSE | Downregulated |
| 0,612801851 | 0,723561314 | C5orf24 | FALSE | Upregulated |
| -0,52964472 | 0,723561314 | FAM99A | FALSE | Downregulated |

| | | | | |
|---|---|---|---|---|
| -0,39717196 | 0,723561314 | FGFRL1 | FALSE | Downregulated |
| -0,567480882 | 0,723561314 | PURA | FALSE | Downregulated |
| -0,335271311 | 0,723561314 | SLC39A14 | FALSE | Downregulated |
| 0,313897306 | 0,723793478 | LPP | FALSE | Upregulated |
| 0,44818981 | 0,723793478 | MEGF9 | FALSE | Upregulated |
| -0,694970676 | 0,724286709 | STAP2 | FALSE | Downregulated |
| 0,415522452 | 0,725053957 | CARD14 | FALSE | Upregulated |
| 0,303672398 | 0,725053957 | IQSEC1 | FALSE | Upregulated |
| -0,443428212 | 0,725053957 | RP11-290F20.2 | FALSE | Downregulated |
| 0,377828681 | 0,725053957 | TNRC6A | FALSE | Upregulated |
| -0,475403747 | 0,725895428 | CTSL | FALSE | Downregulated |
| -0,573048862 | 0,726124543 | BRSK2 | FALSE | Downregulated |
| -0,62414242 | 0,726516829 | FLT1 | FALSE | Downregulated |
| -0,328279293 | 0,726516829 | RER1 | FALSE | Downregulated |
| -0,317526807 | 0,726577321 | CKAP5 | FALSE | Downregulated |
| -0,528780096 | 0,726577321 | LOC102723750 | FALSE | Downregulated |
| -0,50281848 | 0,726808589 | PRKAA1 | FALSE | Downregulated |
| -0,652773025 | 0,726877204 | TRIB3 | FALSE | Downregulated |
| 0,473702798 | 0,727328322 | ZBTB38 | FALSE | Upregulated |
| 0,62226353 | 0,727350219 | FUK | FALSE | Upregulated |
| -0,400649753 | 0,727350219 | R3HDM2 | FALSE | Downregulated |
| -0,621697463 | 0,727350219 | THOC2 | FALSE | Downregulated |
| 0,684228819 | 0,727772877 | TXLNG | FALSE | Upregulated |
| -0,453602216 | 0,727900179 | C1QC | FALSE | Downregulated |
| -0,662707812 | 0,727954903 | KIAA1143 | FALSE | Downregulated |
| -0,473157081 | 0,727954903 | LOC284837 | FALSE | Downregulated |
| -0,616033799 | 0,727954903 | ZNF83 | FALSE | Downregulated |
| 0,609007113 | 0,728049861 | FAM132B | FALSE | Upregulated |
| 0,673143811 | 0,728585173 | GALNT1 | FALSE | Upregulated |
| 0,489196794 | 0,728892857 | TACR1 | FALSE | Upregulated |
| 0,568411672 | 0,729338593 | AP2S1 | FALSE | Upregulated |
| -0,410259777 | 0,729338593 | OPLAH | FALSE | Downregulated |
| -0,579660071 | 0,729372123 | ADAM1A | FALSE | Downregulated |
| 0,65502382 | 0,729372123 | ATP6V0C | FALSE | Upregulated |
| -0,573267416 | 0,729372123 | TM9SF2 | FALSE | Downregulated |
| 0,615063253 | 0,72985572 | LUC7L | FALSE | Upregulated |
| 0,488152469 | 0,730338041 | ADCY3 | FALSE | Upregulated |
| -0,389224908 | 0,730338041 | LOC100130093 | FALSE | Downregulated |
| -0,343296153 | 0,730338041 | NFE2L1 | FALSE | Downregulated |
| 0,668660608 | 0,730338041 | STK10 | FALSE | Upregulated |
| -0,426440458 | 0,7308442 | C11orf49 | FALSE | Downregulated |
| -0,523473048 | 0,7308442 | CDRT15P1 | FALSE | Downregulated |
| -0,667812916 | 0,7308442 | DSG2 | FALSE | Downregulated |
| 0,494918038 | 0,7308442 | NUP214 | FALSE | Upregulated |
| -0,4429575 | 0,7308442 | OGDH | FALSE | Downregulated |
| -0,549633464 | 0,7308442 | PPP1R12B | FALSE | Downregulated |
| 0,409987042 | 0,7308442 | RC3H1 | FALSE | Upregulated |
| 0,559539374 | 0,7308442 | SH3GLB2 | FALSE | Upregulated |
| 0,575034644 | 0,731180544 | DHX16 | FALSE | Upregulated |
| 0,449963223 | 0,731389121 | IL17RB | FALSE | Upregulated |
| 0,422652425 | 0,731616947 | NMT1 | FALSE | Upregulated |

| | | | | |
|---|---|---|---|---|
| -0,581985889 | 0,731616947 | PRPF31 | FALSE | Downregulated |
| 0,392727707 | 0,731616947 | UTRN | FALSE | Upregulated |
| -0,446630346 | 0,731829135 | JUP | FALSE | Downregulated |
| 0,580856011 | 0,731829135 | NDUFA2 | FALSE | Upregulated |
| 0,627690394 | 0,731879508 | ELL3 | FALSE | Upregulated |
| 0,521219432 | 0,731891505 | ALOX12 | FALSE | Upregulated |
| -0,579143985 | 0,731891505 | ARSA | FALSE | Downregulated |
| -0,415509717 | 0,731891505 | BCL9 | FALSE | Downregulated |
| -0,406595834 | 0,731891505 | BID | FALSE | Downregulated |
| -0,426758235 | 0,731891505 | CGN | FALSE | Downregulated |
| -0,36731076 | 0,731891505 | CHRM5 | FALSE | Downregulated |
| 0,435865376 | 0,731891505 | CORO1B | FALSE | Upregulated |
| -0,597661405 | 0,731891505 | CRLS1 | FALSE | Downregulated |
| 0,449971196 | 0,731891505 | DYSF | FALSE | Upregulated |
| -0,428636876 | 0,731891505 | ELK4 | FALSE | Downregulated |
| 0,282707968 | 0,731891505 | FAM20A | FALSE | Upregulated |
| 0,490994644 | 0,731891505 | FGD5-AS1 | FALSE | Upregulated |
| -0,473691528 | 0,731891505 | FLCN | FALSE | Downregulated |
| 0,424213394 | 0,731891505 | FLJ31945 | FALSE | Upregulated |
| -0,669276861 | 0,731891505 | HLA-J | FALSE | Downregulated |
| 0,632665381 | 0,731891505 | LCP1 | FALSE | Upregulated |
| 0,650403788 | 0,731891505 | LOC102724811 | FALSE | Upregulated |
| 0,596068125 | 0,731891505 | LOC146880 | FALSE | Upregulated |
| -0,421584763 | 0,731891505 | LRRC14 | FALSE | Downregulated |
| -0,418393567 | 0,731891505 | MAGI1 | FALSE | Downregulated |
| -0,530523792 | 0,731891505 | MAPK14 | FALSE | Downregulated |
| 0,571675813 | 0,731891505 | MUC20 | FALSE | Upregulated |
| 0,500846901 | 0,731891505 | PRCD | FALSE | Upregulated |
| -0,454381824 | 0,731891505 | RAB14 | FALSE | Downregulated |
| 0,532414536 | 0,731891505 | SLMAP | FALSE | Upregulated |
| -0,382852059 | 0,731891505 | SREBF2 | FALSE | Downregulated |
| 0,524516831 | 0,731891505 | TUBA1B | FALSE | Upregulated |
| 0,438789478 | 0,731891505 | TUBB4B | FALSE | Upregulated |
| -0,447717578 | 0,731891505 | USP40 | FALSE | Downregulated |
| 0,619132475 | 0,73267047 | KXD1 | FALSE | Upregulated |
| 0,496262882 | 0,733022811 | USP32 | FALSE | Upregulated |
| -0,519258104 | 0,733026986 | C1orf111 | FALSE | Downregulated |
| 0,482991876 | 0,733026986 | DEF8 | FALSE | Upregulated |
| -0,485294725 | 0,733026986 | HDHD3 | FALSE | Downregulated |
| 0,457561305 | 0,733026986 | MUM1 | FALSE | Upregulated |
| -0,61666911 | 0,73366432 | GCLM | FALSE | Downregulated |
| 0,574725864 | 0,73366432 | LOC100506175 | FALSE | Upregulated |
| -0,577615499 | 0,73366432 | MLPH | FALSE | Downregulated |
| 0,432098855 | 0,73366432 | WDR26 | FALSE | Upregulated |
| -0,691144761 | 0,733797338 | ALDOC | FALSE | Downregulated |
| -0,423928628 | 0,733797338 | CLASP1 | FALSE | Downregulated |
| -0,454313645 | 0,733797338 | CMPK1 | FALSE | Downregulated |
| 0,516709427 | 0,733797338 | COG1 | FALSE | Upregulated |
| -0,493630752 | 0,733797338 | CYP2U1 | FALSE | Downregulated |
| -0,307930841 | 0,733797338 | DAPK1 | FALSE | Downregulated |
| -0,464132824 | 0,733797338 | ESPN | FALSE | Downregulated |

| | | | | |
|---|---|---|---|---|
| 0,399184611 | 0,733797338 | FOXK1 | FALSE | Upregulated |
| -0,333236221 | 0,733797338 | FTH1 | FALSE | Downregulated |
| 0,36339725 | 0,733797338 | GCN1L1 | FALSE | Upregulated |
| -0,694778375 | 0,733797338 | HOOK1 | FALSE | Downregulated |
| 0,673479247 | 0,733797338 | INPP5D | FALSE | Upregulated |
| -0,407295838 | 0,733797338 | LRIG1 | FALSE | Downregulated |
| -0,38841982 | 0,733797338 | LRRC37A | FALSE | Downregulated |
| -0,481893781 | 0,733797338 | LRRC3DN | FALSE | Downregulated |
| 0,498164385 | 0,733797338 | MROH1 | FALSE | Upregulated |
| -0,3735425 | 0,733797338 | MRPL19 | FALSE | Downregulated |
| -0,345569288 | 0,733797338 | MTMR10 | FALSE | Downregulated |
| 0,403267321 | 0,733797338 | PI4KA | FALSE | Upregulated |
| 0,585318869 | 0,733797338 | RP11-300J18.1 | FALSE | Upregulated |
| -0,39162075 | 0,733797338 | SNRNP70 | FALSE | Downregulated |
| -0,643586862 | 0,733797338 | TAC4 | FALSE | Downregulated |
| 0,4339296 | 0,733797338 | TUG1 | FALSE | Upregulated |
| -0,604047245 | 0,733797338 | USP7 | FALSE | Downregulated |
| 0,497852024 | 0,733797338 | YARS | FALSE | Upregulated |
| -0,576963983 | 0,733797338 | ZNF132 | FALSE | Downregulated |
| -0,510120374 | 0,733797338 | ZSCAN21 | FALSE | Downregulated |
| -0,405344075 | 0,734036697 | SLFNL1 | FALSE | Downregulated |
| 0,438123184 | 0,734780857 | SBF2 | FALSE | Upregulated |
| -0,422854919 | 0,73509427 | GAS8 | FALSE | Downregulated |
| -0,636148848 | 0,73509427 | GID8 | FALSE | Downregulated |
| 0,367710517 | 0,73509427 | PNN | FALSE | Upregulated |
| -0,486286053 | 0,73509427 | SF3B2 | FALSE | Downregulated |
| -0,425302087 | 0,73509427 | TUBGCP2 | FALSE | Downregulated |
| 0,378493855 | 0,735232639 | AMFR | FALSE | Upregulated |
| 0,540322485 | 0,736743439 | METTL7B | FALSE | Upregulated |
| 0,342865342 | 0,736835618 | OSGIN1 | FALSE | Upregulated |
| -0,360060149 | 0,736835618 | RPS5 | FALSE | Downregulated |
| -0,392755826 | 0,736835618 | ZNF592 | FALSE | Downregulated |
| -0,351667837 | 0,737195217 | LOC102724852 | FALSE | Downregulated |
| -0,403112886 | 0,737195217 | PRKD3 | FALSE | Downregulated |
| 0,52207993 | 0,737195217 | SEC24A | FALSE | Upregulated |
| -0,645412739 | 0,737821822 | BANP | FALSE | Downregulated |
| 0,47431081 | 0,737821822 | GPX2 | FALSE | Upregulated |
| 0,592366274 | 0,738023975 | FAM219A | FALSE | Upregulated |
| 0,520608981 | 0,738515553 | ASB14 | FALSE | Upregulated |
| 0,652812527 | 0,738515553 | TCF3 | FALSE | Upregulated |
| 0,553259039 | 0,738814112 | NOC2L | FALSE | Upregulated |
| -0,508882798 | 0,739043504 | FBXL17 | FALSE | Downregulated |
| 0,522357547 | 0,739043504 | GATAD2B | FALSE | Upregulated |
| -0,454478175 | 0,739043504 | SEC13 | FALSE | Downregulated |
| 0,383873789 | 0,73920483 | FAM65C | FALSE | Upregulated |
| 0,600644256 | 0,73920483 | FOXJ2 | FALSE | Upregulated |
| -0,562599047 | 0,73920483 | WRNIP1 | FALSE | Downregulated |
| -0,61952918 | 0,739307719 | ZDHHC17 | FALSE | Downregulated |
| -0,525963968 | 0,739352157 | NAT8 | FALSE | Downregulated |
| 0,336089138 | 0,739352157 | SLC38A10 | FALSE | Upregulated |
| -0,619352767 | 0,739673057 | CTCF | FALSE | Downregulated |

| | | | | |
|---|---|---|---|---|
| -0,613258146 | 0,739673057 | DAAM1 | FALSE | Downregulated |
| -0,530826351 | 0,739673057 | GOLGA2 | FALSE | Downregulated |
| -0,623979718 | 0,739711971 | AAMP | FALSE | Downregulated |
| 0,579805096 | 0,739711971 | AGFG1 | FALSE | Upregulated |
| -0,574281162 | 0,739711971 | BCCIP | FALSE | Downregulated |
| -0,55658647 | 0,739711971 | CAPN3 | FALSE | Downregulated |
| -0,465552717 | 0,739711971 | CTTN | FALSE | Downregulated |
| -0,392622603 | 0,739711971 | EPHB4 | FALSE | Downregulated |
| -0,494395417 | 0,739711971 | HNRNPDL | FALSE | Downregulated |
| -0,416425579 | 0,739711971 | IMMT | FALSE | Downregulated |
| -0,377055979 | 0,739711971 | LOC101928874 | FALSE | Downregulated |
| -0,433457481 | 0,739711971 | LOC102724345 | FALSE | Downregulated |
| -0,311468615 | 0,739711971 | LYL1 | FALSE | Downregulated |
| 0,54479947 | 0,739711971 | MEF2D | FALSE | Upregulated |
| 0,600968969 | 0,739711971 | PCGF5 | FALSE | Upregulated |
| -0,498484341 | 0,739711971 | PNRC1 | FALSE | Downregulated |
| -0,500789218 | 0,739711971 | RAB3GAP2 | FALSE | Downregulated |
| 0,730971958 | 0,739711971 | RNF138P1 | FALSE | Upregulated |
| 0,502903437 | 0,739711971 | ROCK1 | FALSE | Upregulated |
| 0,514673005 | 0,739711971 | SERHL | FALSE | Upregulated |
| -0,297472922 | 0,739711971 | SLC38A3 | FALSE | Downregulated |
| 0,260792568 | 0,739711971 | SON | FALSE | Upregulated |
| 0,368967784 | 0,739711971 | TPCN1 | FALSE | Upregulated |
| 0,64568838 | 0,739711971 | USP20 | FALSE | Upregulated |
| 0,63222527 | 0,739711971 | ZNF333 | FALSE | Upregulated |
| 0,466945556 | 0,739711971 | ZNF79 | FALSE | Upregulated |
| -0,484078636 | 0,739716024 | WDR59 | FALSE | Downregulated |
| 0,314966657 | 0,739966078 | TMOD1 | FALSE | Upregulated |
| -0,548225442 | 0,740753825 | SARS | FALSE | Downregulated |
| 0,50332706 | 0,741015527 | N4BP1 | FALSE | Upregulated |
| 0,560409789 | 0,741015527 | PEX5 | FALSE | Upregulated |
| -0,53999299 | 0,741015527 | TOMM20 | FALSE | Downregulated |
| 0,406976329 | 0,741136621 | DNHD1 | FALSE | Upregulated |
| -0,396686898 | 0,741136621 | ST7-AS1 | FALSE | Downregulated |
| 0,453973297 | 0,741313488 | ADCY9 | FALSE | Upregulated |
| 0,448009969 | 0,741879634 | EFR3A | FALSE | Upregulated |
| 0,438166565 | 0,742005853 | HOOK3 | FALSE | Upregulated |
| -0,501849127 | 0,742005853 | RPS13 | FALSE | Downregulated |
| 0,555044308 | 0,742511848 | CGGBP1 | FALSE | Upregulated |
| 0,308232797 | 0,742896338 | CHRD | FALSE | Upregulated |
| 0,534908374 | 0,743064075 | RABGAP1 | FALSE | Upregulated |
| -0,346020797 | 0,743502672 | PPAP2B | FALSE | Downregulated |
| -0,483406315 | 0,743755449 | SLC25A10 | FALSE | Downregulated |
| -0,403911773 | 0,74408704 | FUBP3 | FALSE | Downregulated |
| -0,368742755 | 0,744090399 | RAF1 | FALSE | Downregulated |
| -0,5547739 | 0,744114822 | ANAPC16 | FALSE | Downregulated |
| 0,464178358 | 0,744114822 | CC2D1A | FALSE | Upregulated |
| 0,591002645 | 0,744114822 | MOB3C | FALSE | Upregulated |
| 0,506906646 | 0,744114822 | ZNF407 | FALSE | Upregulated |
| 0,590623731 | 0,744597524 | LILRB5 | FALSE | Upregulated |
| -0,380538224 | 0,744868476 | LOC100289511 | FALSE | Downregulated |

| | | | | |
|---|---|---|---|---|
| -0,613869481 | 0,744966076 | SSRP1 | FALSE | Downregulated |
| -0,620212194 | 0,745044238 | DSCR3 | FALSE | Downregulated |
| -0,42142996 | 0,745044238 | SMCR8 | FALSE | Downregulated |
| -0,546040889 | 0,745610842 | MATN2 | FALSE | Downregulated |
| 0,517495996 | 0,745995396 | CD63 | FALSE | Upregulated |
| 0,3650004 | 0,745995396 | DNM2 | FALSE | Upregulated |
| -0,518369113 | 0,745995396 | SPATA6L | FALSE | Downregulated |
| 0,466529036 | 0,745995396 | WDR48 | FALSE | Upregulated |
| -0,269790767 | 0,746117115 | LOC101929264 | FALSE | Downregulated |
| -0,389631522 | 0,746117115 | PHKA2 | FALSE | Downregulated |
| 0,40192348 | 0,746759679 | LOC100506634 | FALSE | Upregulated |
| -0,405827633 | 0,746759679 | LOC102724144 | FALSE | Downregulated |
| 0,28569109 | 0,746759679 | LSR | FALSE | Upregulated |
| 0,388683048 | 0,746759679 | MFN2 | FALSE | Upregulated |
| -0,596123833 | 0,746759679 | PLCG2 | FALSE | Downregulated |
| 0,496590685 | 0,746759679 | PRC1 | FALSE | Upregulated |
| -0,536993859 | 0,746759679 | RPS27A | FALSE | Downregulated |
| -0,468982194 | 0,746759679 | SAMD14 | FALSE | Downregulated |
| -0,530913109 | 0,746759679 | SLC25A37 | FALSE | Downregulated |
| 0,534094271 | 0,746759679 | ZNF335 | FALSE | Upregulated |
| 0,605729637 | 0,747030853 | FBXO41 | FALSE | Upregulated |
| -0,406656283 | 0,747084033 | C10orf54 | FALSE | Downregulated |
| -0,649941508 | 0,747102323 | CHST13 | FALSE | Downregulated |
| -0,505522319 | 0,747102323 | LRCH4 | FALSE | Downregulated |
| -0,401232236 | 0,747398956 | CDKL1 | FALSE | Downregulated |
| -0,570348831 | 0,747398956 | HMGCR | FALSE | Downregulated |
| -0,680567122 | 0,748261519 | HEATR5B | FALSE | Downregulated |
| -0,624307771 | 0,748261519 | PGD | FALSE | Downregulated |
| 0,40990661 | 0,748507443 | MLANA | FALSE | Upregulated |
| -0,307337982 | 0,748701344 | ARID1A | FALSE | Downregulated |
| -0,503335288 | 0,748729543 | FAM84B | FALSE | Downregulated |
| -0,38398811 | 0,748729543 | GNPNAT1 | FALSE | Downregulated |
| -0,352606251 | 0,748729543 | JAK1 | FALSE | Downregulated |
| -0,604150178 | 0,748729543 | SYDE1 | FALSE | Downregulated |
| -0,358485821 | 0,748789734 | DEDD | FALSE | Downregulated |
| 0,332178027 | 0,748789734 | FAM120AOS | FALSE | Upregulated |
| 0,383543219 | 0,748789734 | KCNAB1 | FALSE | Upregulated |
| -0,297664593 | 0,748789734 | NFIC | FALSE | Downregulated |
| -0,375652893 | 0,748789734 | SELO | FALSE | Downregulated |
| -0,348833737 | 0,748789734 | SLC29A2 | FALSE | Downregulated |
| -0,643123948 | 0,748789734 | SMIM19 | FALSE | Downregulated |
| 0,33994632 | 0,748789734 | TMEM14E | FALSE | Upregulated |
| -0,520165705 | 0,748789734 | TPRG1L | FALSE | Downregulated |
| -0,57043226 | 0,749127495 | CNKSR3 | FALSE | Downregulated |
| 0,413416665 | 0,749127495 | TPP1 | FALSE | Upregulated |
| -0,54900815 | 0,749451065 | RNF13 | FALSE | Downregulated |
| 0,661311762 | 0,750333343 | GTF2H2B | FALSE | Upregulated |
| 0,540397637 | 0,750333343 | HIF1AN | FALSE | Upregulated |
| -0,569935549 | 0,750333343 | RANBP9 | FALSE | Downregulated |
| 0,422014527 | 0,750379436 | ANKRD61 | FALSE | Upregulated |
| -0,544257204 | 0,750704925 | NSUN2 | FALSE | Downregulated |

| | | | | |
|---|---|---|---|---|
| -0,29262274 | 0,750704925 | RPS3A | FALSE | Downregulated |
| -0,579433937 | 0,750704925 | SLC44A1 | FALSE | Downregulated |
| -0,325199549 | 0,751836871 | CDK13 | FALSE | Downregulated |
| 0,528125262 | 0,752374797 | TMEM33 | FALSE | Upregulated |
| -0,363023483 | 0,752820273 | ACKR4 | FALSE | Downregulated |
| -0,597343216 | 0,752820273 | LOC101060351 | FALSE | Downregulated |
| -0,324387829 | 0,752820273 | RP11-887P2.5 | FALSE | Downregulated |
| 0,543672025 | 0,752820273 | SCYL2 | FALSE | Upregulated |
| -0,334837007 | 0,752820273 | SETD6 | FALSE | Downregulated |
| 0,381467363 | 0,753325369 | LOC100506860 | FALSE | Upregulated |
| -0,662825406 | 0,753515325 | ARMC10 | FALSE | Downregulated |
| 0,424560741 | 0,753515325 | RALGAPA1 | FALSE | Upregulated |
| 0,333387087 | 0,754906264 | C1orf167 | FALSE | Upregulated |
| 0,459095848 | 0,755478963 | CD14 | FALSE | Upregulated |
| 0,44091431 | 0,755800821 | TMEM245 | FALSE | Upregulated |
| -0,566235834 | 0,756003165 | NDUFA13 | FALSE | Downregulated |
| -0,425851694 | 0,756252416 | ADAMTS13 | FALSE | Downregulated |
| -0,598026132 | 0,756252416 | STRN3 | FALSE | Downregulated |
| -0,340383921 | 0,75634616 | ERVW-1 | FALSE | Downregulated |
| 0,326593255 | 0,756487543 | CHD4 | FALSE | Upregulated |
| 0,533728095 | 0,756745385 | PDCD11 | FALSE | Upregulated |
| 0,43958894 | 0,756745385 | PPM1M | FALSE | Upregulated |
| 0,559086372 | 0,756745385 | TMX2 | FALSE | Upregulated |
| -0,456993318 | 0,757489693 | RPL17 | FALSE | Downregulated |
| 0,434949395 | 0,757582463 | ABCA9 | FALSE | Upregulated |
| 0,449153601 | 0,757582463 | CACFD1 | FALSE | Upregulated |
| 0,444740927 | 0,757582463 | RPTOR | FALSE | Upregulated |
| -0,460146451 | 0,757582463 | ZNF542 | FALSE | Downregulated |
| 0,539513364 | 0,757599175 | LOC102724187 | FALSE | Upregulated |
| 0,446765599 | 0,75774192 | ALCAM | FALSE | Upregulated |
| -0,398378368 | 0,758109904 | ALAS1 | FALSE | Downregulated |
| 0,450534418 | 0,758109904 | EML6 | FALSE | Upregulated |
| 0,569336965 | 0,758109904 | KMT2E-AS1 | FALSE | Upregulated |
| 0,310630259 | 0,758109904 | LOC101927469 | FALSE | Upregulated |
| 0,552884535 | 0,758109904 | LOC102723735 | FALSE | Upregulated |
| 0,389617287 | 0,758109904 | MAPKAPK2 | FALSE | Upregulated |
| 0,518865713 | 0,758109904 | OSGIN2 | FALSE | Upregulated |
| -0,506677072 | 0,758109904 | SRGAP1 | FALSE | Downregulated |
| 0,44267381 | 0,758109904 | SUN1 | FALSE | Upregulated |
| 0,548051942 | 0,758109904 | TTI2 | FALSE | Upregulated |
| 0,514500525 | 0,758109904 | ZC3H12B | FALSE | Upregulated |
| 0,585152217 | 0,759318144 | LOC101928081 | FALSE | Upregulated |
| 0,475015352 | 0,759672128 | SLC35E2 | FALSE | Upregulated |
| 0,533334447 | 0,759944803 | GSR | FALSE | Upregulated |
| -0,40420635 | 0,760887256 | AHR | FALSE | Downregulated |
| -0,301782357 | 0,760887256 | RMRP | FALSE | Downregulated |
| 0,602074328 | 0,761611732 | C15orf54 | FALSE | Upregulated |
| -0,426847823 | 0,761611732 | CLCN5 | FALSE | Downregulated |
| 0,339283898 | 0,76176281 | PDPK1 | FALSE | Upregulated |
| -0,587895864 | 0,762121861 | CFL2 | FALSE | Downregulated |
| -0,448496331 | 0,762121861 | MICA | FALSE | Downregulated |

| | | | | |
|---|---|---|---|---|
| 0,35093702 | 0,762121861 | PTOV1-AS1 | FALSE | Upregulated |
| -0,386553353 | 0,762302437 | HNRNPK | FALSE | Downregulated |
| -0,473165161 | 0,762302437 | PDK2 | FALSE | Downregulated |
| -0,409262345 | 0,762302437 | STAU1 | FALSE | Downregulated |
| -0,30329909 | 0,762328656 | ABCC6 | FALSE | Downregulated |
| -0,319834838 | 0,762514572 | C6orf62 | FALSE | Downregulated |
| 0,498483522 | 0,762514572 | LINC00476 | FALSE | Upregulated |
| -0,670739705 | 0,762679323 | LOC101929792 | FALSE | Downregulated |
| 0,291578188 | 0,762679323 | MAP3K13 | FALSE | Upregulated |
| -0,433043097 | 0,762756668 | EXOC3L4 | FALSE | Downregulated |
| 0,510550105 | 0,762849201 | TUB | FALSE | Upregulated |
| 0,36453232 | 0,763128238 | ATXN2L | FALSE | Upregulated |
| -0,452894409 | 0,763128238 | C10orf105 | FALSE | Downregulated |
| -0,62953458 | 0,763128238 | CARS2 | FALSE | Downregulated |
| 0,589655493 | 0,763128238 | CASP8 | FALSE | Upregulated |
| -0,316355267 | 0,763128238 | CFLAR | FALSE | Downregulated |
| 0,389934609 | 0,763128238 | DCLRE1C | FALSE | Upregulated |
| 0,651780273 | 0,763128238 | FAM21A | FALSE | Upregulated |
| 0,484939917 | 0,763128238 | FKBP9 | FALSE | Upregulated |
| 0,495388066 | 0,763128238 | HLA-F | FALSE | Upregulated |
| -0,332492509 | 0,763128238 | HNRNPUL1 | FALSE | Downregulated |
| -0,445234646 | 0,763128238 | LMAN2 | FALSE | Downregulated |
| 0,302405817 | 0,763128238 | LOC100128818 | FALSE | Upregulated |
| -0,534043588 | 0,763128238 | LOC102724552 | FALSE | Downregulated |
| -0,594665977 | 0,763128238 | MARVELD2 | FALSE | Downregulated |
| -0,353353483 | 0,763128238 | NEK6 | FALSE | Downregulated |
| 0,563939991 | 0,763128238 | OCLN | FALSE | Upregulated |
| 0,57685481 | 0,763128238 | PPM1F | FALSE | Upregulated |
| -0,517087175 | 0,763128238 | PTAR1 | FALSE | Downregulated |
| 0,486957668 | 0,763128238 | SDCBP | FALSE | Upregulated |
| 0,3928196 | 0,763128238 | TIMP2 | FALSE | Upregulated |
| -0,397013849 | 0,763128238 | WDR88 | FALSE | Downregulated |
| 0,500291315 | 0,763128238 | ZBTB40 | FALSE | Upregulated |
| 0,591734572 | 0,763128238 | ZNF266 | FALSE | Upregulated |
| -0,521456518 | 0,763243982 | FBXO38 | FALSE | Downregulated |
| 0,463216599 | 0,764066777 | CPSF6 | FALSE | Upregulated |
| 0,380959288 | 0,764362488 | GPR124 | FALSE | Upregulated |
| -0,521885921 | 0,764996231 | SIGIRR | FALSE | Downregulated |
| -0,354679044 | 0,765138756 | BAG6 | FALSE | Downregulated |
| -0,593455453 | 0,765856182 | SRP72 | FALSE | Downregulated |
| 0,507451478 | 0,765970798 | CD5 | FALSE | Upregulated |
| -0,428076115 | 0,765970798 | MBTPS1 | FALSE | Downregulated |
| 0,521085681 | 0,765970798 | TK2 | FALSE | Upregulated |
| -0,327082752 | 0,766202029 | FKBP8 | FALSE | Downregulated |
| 0,573680217 | 0,766330913 | ATP13A1 | FALSE | Upregulated |
| -0,465195423 | 0,766813171 | CEP164 | FALSE | Downregulated |
| -0,607023557 | 0,766813171 | ECSIT | FALSE | Downregulated |
| 0,343038205 | 0,766813171 | SPDYE2B | FALSE | Upregulated |
| -0,491031902 | 0,766813171 | TMEM66 | FALSE | Downregulated |
| 0,509182098 | 0,766813171 | TPM3 | FALSE | Upregulated |
| -0,347977514 | 0,767152496 | ABHD16B | FALSE | Downregulated |

| | | | | |
|---|---|---|---|---|
| -0,281962785 | 0,767152496 | CREBBP | FALSE | Downregulated |
| -0,462317834 | 0,76715908 | COLEC10 | FALSE | Downregulated |
| -0,572329964 | 0,76715908 | PYCRL | FALSE | Downregulated |
| -0,609088691 | 0,76715908 | SURF1 | FALSE | Downregulated |
| -0,387760283 | 0,76715908 | TSC2 | FALSE | Downregulated |
| 0,378159812 | 0,76715908 | TXLNA | FALSE | Upregulated |
| 0,458789163 | 0,76715908 | ZNF664 | FALSE | Upregulated |
| -0,49599968 | 0,768687139 | PRKAG2 | FALSE | Downregulated |
| -0,423406959 | 0,768892324 | LUC7L3 | FALSE | Downregulated |
| 0,377395962 | 0,768892324 | SLC2A12 | FALSE | Upregulated |
| -0,668252017 | 0,769130901 | YIPF3 | FALSE | Downregulated |
| 0,579594071 | 0,769130901 | ZNF787 | FALSE | Upregulated |
| 0,428194901 | 0,769218611 | EIF2AK4 | FALSE | Upregulated |
| -0,51790915 | 0,769218611 | TSHZ1 | FALSE | Downregulated |
| 0,630110966 | 0,769218611 | ZNF436 | FALSE | Upregulated |
| 0,494556365 | 0,769218611 | ZNF672 | FALSE | Upregulated |
| -0,400617827 | 0,769491416 | MAP3K15 | FALSE | Downregulated |
| 0,508325634 | 0,769491416 | NHP2 | FALSE | Upregulated |
| -0,436778806 | 0,769894065 | ELFN1 | FALSE | Downregulated |
| 0,467525139 | 0,769894065 | SH3BP4 | FALSE | Upregulated |
| 0,511524762 | 0,769894065 | SLC46A3 | FALSE | Upregulated |
| 0,606560364 | 0,769894065 | ZSWIM6 | FALSE | Upregulated |
| 0,551091658 | 0,770319864 | SPPL2A | FALSE | Upregulated |
| -0,360877281 | 0,770319864 | SRSF5 | FALSE | Downregulated |
| -0,490821599 | 0,770319864 | ZNF674 | FALSE | Downregulated |
| -0,460574086 | 0,770349822 | NOTCH4 | FALSE | Downregulated |
| 0,546123483 | 0,770547546 | ACTR3 | FALSE | Upregulated |
| -0,610121967 | 0,770547546 | CIPC | FALSE | Downregulated |
| 0,599071547 | 0,770821896 | IL10RA | FALSE | Upregulated |
| -0,367859146 | 0,770959104 | PDS5A | FALSE | Downregulated |
| 0,620489612 | 0,771017248 | AC010127.3 | FALSE | Upregulated |
| 0,598420044 | 0,771017248 | ALG13 | FALSE | Upregulated |
| -0,412934632 | 0,771017248 | DGCR8 | FALSE | Downregulated |
| 0,685424568 | 0,771017248 | GLTSCR1L | FALSE | Upregulated |
| -0,505469603 | 0,771017248 | SCARNA12 | FALSE | Downregulated |
| 0,505236176 | 0,771017248 | SOCS6 | FALSE | Upregulated |
| -0,515666359 | 0,771017248 | ZNF780B | FALSE | Downregulated |
| -0,443788911 | 0,771258674 | EIF2S3 | FALSE | Downregulated |
| 0,523162594 | 0,771258674 | EZR | FALSE | Upregulated |
| -0,51674961 | 0,771258674 | IGSF8 | FALSE | Downregulated |
| -0,621162986 | 0,771258674 | KIAA1958 | FALSE | Downregulated |
| -0,41761862 | 0,771258674 | LINC01061 | FALSE | Downregulated |
| 0,44099183 | 0,771258674 | LUZP1 | FALSE | Upregulated |
| 0,396274749 | 0,771258674 | LYRM2 | FALSE | Upregulated |
| -0,51851585 | 0,771258674 | POM121L10P | FALSE | Downregulated |
| -0,340195234 | 0,771258674 | PRKAR1A | FALSE | Downregulated |
| 0,292060387 | 0,771258674 | SLC7A6 | FALSE | Upregulated |
| -0,444218765 | 0,771258674 | STAR | FALSE | Downregulated |
| 0,5064801 | 0,771258674 | TRAK2 | FALSE | Upregulated |
| -0,449032442 | 0,771258674 | ZCWPW1 | FALSE | Downregulated |
| -0,365591502 | 0,771258674 | ZNF337 | FALSE | Downregulated |

| | | | | |
|---|---|---|---|---|
| 0,287019945 | 0,771515915 | CNP | FALSE | Upregulated |
| 0,464260232 | 0,771515915 | ZNF497 | FALSE | Upregulated |
| 0,541171273 | 0,771525954 | ARF4 | FALSE | Upregulated |
| -0,504157175 | 0,771525954 | PAK4 | FALSE | Downregulated |
| 0,4633097 | 0,771525954 | PKP2 | FALSE | Upregulated |
| 0,273023981 | 0,771525954 | SH2B3 | FALSE | Upregulated |
| -0,359973491 | 0,771525954 | SLC22A25 | FALSE | Downregulated |
| -0,428647102 | 0,771544432 | SHROOM2 | FALSE | Downregulated |
| 0,488527287 | 0,771690663 | ZNF318 | FALSE | Upregulated |
| -0,33543872 | 0,771759399 | GTPBP1 | FALSE | Downregulated |
| -0,338063528 | 0,771916957 | MSL1 | FALSE | Downregulated |
| -0,291110361 | 0,772506401 | RPL31 | FALSE | Downregulated |
| 0,407020835 | 0,772671157 | AKAP5 | FALSE | Upregulated |
| -0,340212578 | 0,772671157 | ALDH1A1 | FALSE | Downregulated |
| -0,497753029 | 0,772671157 | CBX7 | FALSE | Downregulated |
| -0,47351921 | 0,772671157 | FAM149A | FALSE | Downregulated |
| 0,351614133 | 0,772671157 | LOC100505736 | FALSE | Upregulated |
| -0,344182183 | 0,772671157 | PCGF2 | FALSE | Downregulated |
| 0,466356383 | 0,772671157 | RARB | FALSE | Upregulated |
| 0,266931878 | 0,772671157 | SQSTM1 | FALSE | Upregulated |
| 0,565644547 | 0,773022218 | CNN2 | FALSE | Upregulated |
| 0,401158155 | 0,773575206 | IMPAD1 | FALSE | Upregulated |
| 0,557228944 | 0,773575206 | JPH3 | FALSE | Upregulated |
| -0,523517204 | 0,773575206 | TMEM37 | FALSE | Downregulated |
| 0,468243573 | 0,773575206 | UBE3C | FALSE | Upregulated |
| -0,442490026 | 0,774018165 | SNAP47 | FALSE | Downregulated |
| -0,428540324 | 0,774223191 | NONO | FALSE | Downregulated |
| 0,539084315 | 0,774253146 | SNRNP35 | FALSE | Upregulated |
| -0,315513594 | 0,774253146 | SSFA2 | FALSE | Downregulated |
| 0,503816575 | 0,774404898 | MYO7B | FALSE | Upregulated |
| 0,503515597 | 0,774404898 | RAB40C | FALSE | Upregulated |
| 0,40258308 | 0,774404898 | RUSC1 | FALSE | Upregulated |
| 0,433996315 | 0,774404898 | TSPAN14 | FALSE | Upregulated |
| -0,552110581 | 0,774525784 | ARHGAP12 | FALSE | Downregulated |
| 0,51261718 | 0,774525784 | LEAP2 | FALSE | Upregulated |
| 0,47870448 | 0,774525784 | PEX26 | FALSE | Upregulated |
| 0,620287046 | 0,774525784 | PRKD2 | FALSE | Upregulated |
| -0,441120211 | 0,774525784 | THRB | FALSE | Downregulated |
| 0,297151201 | 0,774525784 | VPS13D | FALSE | Upregulated |
| 0,534103868 | 0,77497009 | FAM177A1 | FALSE | Upregulated |
| -0,427201446 | 0,77497009 | LOC729173 | FALSE | Downregulated |
| -0,486946905 | 0,775225032 | DDX54 | FALSE | Downregulated |
| 0,512849556 | 0,775225032 | MARK2 | FALSE | Upregulated |
| 0,548754426 | 0,775580939 | CCDC39 | FALSE | Upregulated |
| -0,409893608 | 0,775580939 | COQ6 | FALSE | Downregulated |
| -0,363832755 | 0,775580939 | DDAH1 | FALSE | Downregulated |
| -0,636277823 | 0,775580939 | GLUD1P7 | FALSE | Downregulated |
| 0,40288299 | 0,775580939 | IRF3 | FALSE | Upregulated |
| 0,465085613 | 0,775580939 | LOC102723769 | FALSE | Upregulated |
| 0,590010385 | 0,775580939 | LOC102723824 | FALSE | Upregulated |
| 0,639147355 | 0,775580939 | LOC154761 | FALSE | Upregulated |

| | | | | |
|---|---|---|---|---|
| -0,478974025 | 0,775580939 | LRRC3 | FALSE | Downregulated |
| 0,56693705 | 0,775580939 | RP11-4F22.2 | FALSE | Upregulated |
| 0,356531604 | 0,775580939 | SHC2 | FALSE | Upregulated |
| 0,232674401 | 0,775580939 | SPRN | FALSE | Upregulated |
| -0,434988638 | 0,775580939 | STAT5B | FALSE | Downregulated |
| -0,478565759 | 0,775580939 | UGT2B11 | FALSE | Downregulated |
| -0,468146236 | 0,775580939 | VIPR1 | FALSE | Downregulated |
| 0,477345423 | 0,775580939 | WWP2 | FALSE | Upregulated |
| -0,566841756 | 0,775580939 | ZNF197 | FALSE | Downregulated |
| 0,257349243 | 0,775606207 | KIF1C | FALSE | Upregulated |
| 0,374394812 | 0,775606207 | MTRF1L | FALSE | Upregulated |
| -0,457487253 | 0,775627669 | NHLRC2 | FALSE | Downregulated |
| 0,490714688 | 0,775865206 | LOC101928070 | FALSE | Upregulated |
| 0,413051864 | 0,775865206 | SEPT11 | FALSE | Upregulated |
| -0,445188213 | 0,775990638 | CYP2A7 | FALSE | Downregulated |
| 0,349492813 | 0,775990638 | RAB11FIP3 | FALSE | Upregulated |
| -0,353156498 | 0,776039327 | HDAC5 | FALSE | Downregulated |
| 0,51720995 | 0,776262935 | ARID2 | FALSE | Upregulated |
| 0,470941254 | 0,776262935 | AZIN1 | FALSE | Upregulated |
| -0,284705752 | 0,776262935 | CTDSP2 | FALSE | Downregulated |
| -0,361737987 | 0,776262935 | GABPB1-AS1 | FALSE | Downregulated |
| 0,365259552 | 0,776262935 | GPER1 | FALSE | Upregulated |
| -0,508967557 | 0,776262935 | KBTBD11 | FALSE | Downregulated |
| -0,59067026 | 0,776262935 | PDE2A | FALSE | Downregulated |
| 0,536412324 | 0,776262935 | STAG3L3 | FALSE | Upregulated |
| -0,404371544 | 0,776262935 | TEAD1 | FALSE | Downregulated |
| -0,463005188 | 0,776262935 | THSD4 | FALSE | Downregulated |
| 0,413262582 | 0,776262935 | UNKL | FALSE | Upregulated |
| 0,39627856 | 0,776262935 | YWHAG | FALSE | Upregulated |
| 0,59685718 | 0,776661613 | ACOT6 | FALSE | Upregulated |
| -0,459172656 | 0,777004422 | ATXN7L3B | FALSE | Downregulated |
| 0,569105321 | 0,777004422 | BOC | FALSE | Upregulated |
| -0,424619397 | 0,777004422 | KRI1 | FALSE | Downregulated |
| -0,40143799 | 0,777004422 | MIDN | FALSE | Downregulated |
| -0,304895444 | 0,777004422 | NUCB1 | FALSE | Downregulated |
| 0,485188849 | 0,777004422 | PML | FALSE | Upregulated |
| -0,397594154 | 0,777004422 | PROX2 | FALSE | Downregulated |
| 0,500342339 | 0,777004422 | PSMD2 | FALSE | Upregulated |
| -0,544927692 | 0,777004422 | RNF126 | FALSE | Downregulated |
| -0,402172642 | 0,777004422 | RPS24 | FALSE | Downregulated |
| -0,302156331 | 0,777004422 | TBC1D9B | FALSE | Downregulated |
| -0,320626736 | 0,777328405 | PGM3 | FALSE | Downregulated |
| -0,511685046 | 0,777475763 | ADAMTS17 | FALSE | Downregulated |
| 0,636358027 | 0,777475763 | AGAP2 | FALSE | Upregulated |
| 0,419620105 | 0,777475763 | CERKL | FALSE | Upregulated |
| 0,44778058 | 0,777475763 | EPHA1 | FALSE | Upregulated |
| 0,355752908 | 0,777679386 | DUSP3 | FALSE | Upregulated |
| 0,44350783 | 0,777679386 | LOC101929786 | FALSE | Upregulated |
| 0,354627636 | 0,777679386 | MSTO2P | FALSE | Upregulated |
| 0,582598502 | 0,77788182 | BRF1 | FALSE | Upregulated |
| -0,535315313 | 0,77788182 | COL4A3BP | FALSE | Downregulated |

| | | | | |
|---|---|---|---|---|
| -0,324836224 | 0,77788182 | FANCC | FALSE | Downregulated |
| -0,308729061 | 0,77788182 | HNRNPA1 | FALSE | Downregulated |
| -0,400696006 | 0,77788182 | PEX19 | FALSE | Downregulated |
| -0,493744809 | 0,77788182 | RSF1 | FALSE | Downregulated |
| -0,402317155 | 0,77788182 | TRABD | FALSE | Downregulated |
| 0,356125664 | 0,77788182 | WNT4 | FALSE | Upregulated |
| 0,4258045 | 0,777931052 | MUC4 | FALSE | Upregulated |
| 0,608734465 | 0,778072801 | PRKCE | FALSE | Upregulated |
| -0,443036132 | 0,77876789 | ARFGAP2 | FALSE | Downregulated |
| -0,561626468 | 0,77876789 | GYS1 | FALSE | Downregulated |
| 0,306863489 | 0,77915004 | LOC101928621 | FALSE | Upregulated |
| 0,419680921 | 0,779343939 | TMEM181 | FALSE | Upregulated |
| -0,489945376 | 0,780249746 | FAM211A-AS1 | FALSE | Downregulated |
| 0,570079176 | 0,780249746 | HMG20A | FALSE | Upregulated |
| -0,565710453 | 0,780249746 | MAMDC2 | FALSE | Downregulated |
| 0,485916099 | 0,780249746 | TESK2 | FALSE | Upregulated |
| 0,390988394 | 0,781327054 | SLC39A7 | FALSE | Upregulated |
| -0,543550466 | 0,781327054 | XAB2 | FALSE | Downregulated |
| -0,431478602 | 0,781525849 | CEP57 | FALSE | Downregulated |
| -0,386883444 | 0,781525849 | CHD6 | FALSE | Downregulated |
| -0,521342396 | 0,781525849 | SEC14L4 | FALSE | Downregulated |
| -0,266822776 | 0,781525849 | ZFP36L1 | FALSE | Downregulated |
| 0,321794783 | 0,78152944 | POM121C | FALSE | Upregulated |
| 0,340476561 | 0,782098199 | BTAF1 | FALSE | Upregulated |
| 0,44977428 | 0,782098199 | DENND2D | FALSE | Upregulated |
| 0,279670895 | 0,782098199 | EGLN1 | FALSE | Upregulated |
| -0,270822413 | 0,782098199 | LOC646652 | FALSE | Downregulated |
| 0,538963397 | 0,782098199 | MLX | FALSE | Upregulated |
| 0,587650758 | 0,782098199 | NCSTN | FALSE | Upregulated |
| -0,487168341 | 0,782098199 | OCRL | FALSE | Downregulated |
| 0,313122209 | 0,782098199 | PRICKLE2-AS1 | FALSE | Upregulated |
| 0,564487027 | 0,782098199 | PTPN4 | FALSE | Upregulated |
| 0,39447519 | 0,782098199 | RGS12 | FALSE | Upregulated |
| 0,344087387 | 0,782098199 | SYVN1 | FALSE | Upregulated |
| -0,547408295 | 0,782098199 | TP53I11 | FALSE | Downregulated |
| -0,520388957 | 0,782098199 | UACA | FALSE | Downregulated |
| 0,322921183 | 0,782098199 | UBA5 | FALSE | Upregulated |
| -0,415199056 | 0,782098199 | USP31 | FALSE | Downregulated |
| -0,333355581 | 0,782694091 | SAR1B | FALSE | Downregulated |
| -0,354605881 | 0,783116707 | AKR1C2 | FALSE | Downregulated |
| 0,421805883 | 0,783116707 | HDAC7 | FALSE | Upregulated |
| -0,577077763 | 0,783898538 | HNRNPA0 | FALSE | Downregulated |
| -0,244036795 | 0,783898538 | MYO18A | FALSE | Downregulated |
| 0,408686604 | 0,783910595 | KIAA1033 | FALSE | Upregulated |
| -0,552294493 | 0,783910595 | ZNF45 | FALSE | Downregulated |
| 0,455514069 | 0,783910618 | PLCE1 | FALSE | Upregulated |
| 0,426381477 | 0,784095844 | RBFOX2 | FALSE | Upregulated |
| 0,512737928 | 0,784461379 | STX3 | FALSE | Upregulated |
| -0,386048928 | 0,784461379 | THRAP3 | FALSE | Downregulated |
| -0,479931247 | 0,78473025 | GPRASP1 | FALSE | Downregulated |
| 0,57656514 | 0,78473025 | LINC00244 | FALSE | Upregulated |

| | | | | |
|---|---|---|---|---|
| 0,358677082 | 0,78473025 | ZNF513 | FALSE | Upregulated |
| 0,491853253 | 0,784890056 | CEP250 | FALSE | Upregulated |
| 0,462545581 | 0,784890056 | CYLD | FALSE | Upregulated |
| 0,347529872 | 0,784890056 | PCNT | FALSE | Upregulated |
| -0,434704243 | 0,785653507 | SATB1 | FALSE | Downregulated |
| -0,379332848 | 0,786112158 | STAG1 | FALSE | Downregulated |
| -0,312650987 | 0,786308235 | BRD2 | FALSE | Downregulated |
| 0,414164537 | 0,786308235 | EXT1 | FALSE | Upregulated |
| 0,450110854 | 0,786308235 | FDX1 | FALSE | Upregulated |
| 0,500353441 | 0,786308235 | MON1B | FALSE | Upregulated |
| -0,458559758 | 0,786308235 | NGFR | FALSE | Downregulated |
| 0,537082899 | 0,786308235 | PIM3 | FALSE | Upregulated |
| 0,487496543 | 0,786308235 | PROSER2 | FALSE | Upregulated |
| -0,476679436 | 0,786733141 | DMPK | FALSE | Downregulated |
| 0,301448292 | 0,786733141 | FBXO31 | FALSE | Upregulated |
| -0,308755666 | 0,786733141 | NDRG2 | FALSE | Downregulated |
| -0,376036034 | 0,786733141 | WAC | FALSE | Downregulated |
| 0,473189352 | 0,786733141 | ZNF692 | FALSE | Upregulated |
| 0,300864053 | 0,786895535 | LRFN4 | FALSE | Upregulated |
| 0,288605406 | 0,787689203 | PDZK1 | FALSE | Upregulated |
| -0,435782245 | 0,78776644 | ARHGAP29 | FALSE | Downregulated |
| -0,356393199 | 0,788118255 | CALM3 | FALSE | Downregulated |
| 0,341249948 | 0,788118255 | MGAT1 | FALSE | Upregulated |
| -0,390508071 | 0,788118255 | PNPLA7 | FALSE | Downregulated |
| 0,39076492 | 0,788264523 | ERO1LB | FALSE | Upregulated |
| -0,458595546 | 0,788264523 | HINT1 | FALSE | Downregulated |
| 0,45357995 | 0,788264523 | LOC101927898 | FALSE | Upregulated |
| -0,486252899 | 0,788264523 | RBM8A | FALSE | Downregulated |
| -0,40458997 | 0,788354696 | FBXW2 | FALSE | Downregulated |
| 0,341469602 | 0,788409644 | PNKD | FALSE | Upregulated |
| 0,532224517 | 0,788414721 | TMEM150A | FALSE | Upregulated |
| -0,453669876 | 0,788667932 | FAM222B | FALSE | Downregulated |
| -0,264057804 | 0,788667932 | GOLGB1 | FALSE | Downregulated |
| 0,401728162 | 0,788667932 | MLLT1 | FALSE | Upregulated |
| -0,357751461 | 0,788667932 | PAN2 | FALSE | Downregulated |
| 0,382065539 | 0,788667932 | RAPGEF1 | FALSE | Upregulated |
| -0,29911016 | 0,788667932 | SLC22A23 | FALSE | Downregulated |
| 0,445483638 | 0,788667932 | STAT5A | FALSE | Upregulated |
| -0,600718947 | 0,788714519 | ACOT1 | FALSE | Downregulated |
| -0,505612166 | 0,788752377 | FECH | FALSE | Downregulated |
| 0,38613102 | 0,788777733 | ITPR3 | FALSE | Upregulated |
| -0,50566694 | 0,789227092 | ATP5G3 | FALSE | Downregulated |
| 0,520437033 | 0,789227092 | C14orf1 | FALSE | Upregulated |
| 0,342736351 | 0,789227092 | CAPN12 | FALSE | Upregulated |
| 0,313248519 | 0,789227092 | NBPF12 | FALSE | Upregulated |
| 0,514844334 | 0,789227092 | NQO1 | FALSE | Upregulated |
| 0,47779637 | 0,789227092 | PEX5L | FALSE | Upregulated |
| -0,322935584 | 0,789227092 | PTPMT1 | FALSE | Downregulated |
| -0,50958597 | 0,789227092 | SRSF12 | FALSE | Downregulated |
| -0,378118405 | 0,789227092 | SRSF9 | FALSE | Downregulated |
| 0,389285236 | 0,789227092 | TNKS | FALSE | Upregulated |

| | | | | |
|---|---|---|---|---|
| -0,522728283 | 0,789227092 | TNPO1 | FALSE | Downregulated |
| 0,413482898 | 0,789612867 | ARMC6 | FALSE | Upregulated |
| 0,515316312 | 0,789612867 | EFEMP2 | FALSE | Upregulated |
| 0,394883728 | 0,789683044 | AGAP3 | FALSE | Upregulated |
| 0,27504831 | 0,790181063 | HEATR1 | FALSE | Upregulated |
| -0,447618909 | 0,790181063 | TXNDC11 | FALSE | Downregulated |
| -0,5009435 | 0,790288928 | FAM35A | FALSE | Downregulated |
| -0,649912063 | 0,790980649 | STARD4 | FALSE | Downregulated |
| -0,467478496 | 0,791074246 | CAND1 | FALSE | Downregulated |
| 0,346218944 | 0,79133241 | POLR2A | FALSE | Upregulated |
| -0,223839893 | 0,79142316 | MST1 | FALSE | Downregulated |
| 0,439614807 | 0,792396038 | ANKRD10 | FALSE | Upregulated |
| 0,36326112 | 0,792396038 | ARNT | FALSE | Upregulated |
| -0,494384187 | 0,792396038 | CYB561A3 | FALSE | Downregulated |
| -0,429030347 | 0,792396038 | FOXO1 | FALSE | Downregulated |
| 0,493538672 | 0,792396038 | HGSNAT | FALSE | Upregulated |
| 0,352773405 | 0,792396038 | HTT | FALSE | Upregulated |
| 0,337937795 | 0,792396038 | LOC101927385 | FALSE | Upregulated |
| -0,505144943 | 0,792396038 | LSM14A | FALSE | Downregulated |
| -0,538445702 | 0,792396038 | MKRN2 | FALSE | Downregulated |
| 0,384635687 | 0,792396038 | PMPCB | FALSE | Upregulated |
| -0,348866752 | 0,792396038 | SERF2 | FALSE | Downregulated |
| -0,539309016 | 0,792396038 | ZBED3 | FALSE | Downregulated |
| -0,501332216 | 0,792396038 | ZNF224 | FALSE | Downregulated |
| -0,404600717 | 0,792396038 | ZNF445 | FALSE | Downregulated |
| -0,426780237 | 0,793481914 | LGMN | FALSE | Downregulated |
| -0,325361297 | 0,794868055 | EIF5 | FALSE | Downregulated |
| -0,577414153 | 0,794868055 | FBXO32 | FALSE | Downregulated |
| -0,330926269 | 0,794868055 | FOXM1 | FALSE | Downregulated |
| -0,512784656 | 0,794868055 | FOXRED1 | FALSE | Downregulated |
| 0,526346023 | 0,794868055 | HS1BP3 | FALSE | Upregulated |
| -0,433400281 | 0,794868055 | IVNS1ABP | FALSE | Downregulated |
| -0,520019496 | 0,794868055 | KAT6B | FALSE | Downregulated |
| 0,368939493 | 0,794868055 | LIMD2 | FALSE | Upregulated |
| -0,50062298 | 0,794868055 | LTB4R2 | FALSE | Downregulated |
| -0,373897802 | 0,794868055 | NPEPPS | FALSE | Downregulated |
| -0,575641411 | 0,795436042 | HIRIP3 | FALSE | Downregulated |
| -0,500830544 | 0,795757761 | PPP2R5C | FALSE | Downregulated |
| 0,468888476 | 0,795757761 | UBOX5-AS1 | FALSE | Upregulated |
| -0,534783769 | 0,796304139 | AC091729.9 | FALSE | Downregulated |
| 0,411185189 | 0,796304139 | ANXA11 | FALSE | Upregulated |
| -0,283449933 | 0,796304139 | AQP9 | FALSE | Downregulated |
| 0,315689452 | 0,796304139 | ASB16 | FALSE | Upregulated |
| -0,411911922 | 0,796304139 | ASMTL | FALSE | Downregulated |
| 0,332157893 | 0,796304139 | AUTS2 | FALSE | Upregulated |
| 0,507109257 | 0,796304139 | BRD9 | FALSE | Upregulated |
| -0,330640367 | 0,796304139 | CDK4 | FALSE | Downregulated |
| 0,403128043 | 0,796304139 | CEACAM1 | FALSE | Upregulated |
| 0,501647884 | 0,796304139 | CSNK1E | FALSE | Upregulated |
| 0,507551838 | 0,796304139 | DHX8 | FALSE | Upregulated |
| 0,276050838 | 0,796304139 | DND1 | FALSE | Upregulated |

| | | | | |
|---|---|---|---|---|
| 0,520412829 | 0,796304139 | DYNC2LI1 | FALSE | Upregulated |
| 0,450681888 | 0,796304139 | FAM91A1 | FALSE | Upregulated |
| 0,488451088 | 0,796304139 | GDPD4 | FALSE | Upregulated |
| -0,322310704 | 0,796304139 | HLA-C | FALSE | Downregulated |
| -0,291915132 | 0,796304139 | INTS1 | FALSE | Downregulated |
| 0,381148573 | 0,796304139 | LAMC1 | FALSE | Upregulated |
| -0,297192523 | 0,796304139 | MAP3K11 | FALSE | Downregulated |
| -0,278148997 | 0,796304139 | MBL2 | FALSE | Downregulated |
| -0,429583276 | 0,796304139 | MGA | FALSE | Downregulated |
| 0,565532058 | 0,796304139 | PCBP4 | FALSE | Upregulated |
| -0,274619824 | 0,796304139 | PEX10 | FALSE | Downregulated |
| 0,409482548 | 0,796304139 | PMPCA | FALSE | Upregulated |
| -0,285679257 | 0,796304139 | PODXL | FALSE | Downregulated |
| -0,283494189 | 0,796304139 | PPP1R3B | FALSE | Downregulated |
| 0,549485664 | 0,796304139 | RNF141 | FALSE | Upregulated |
| -0,381232193 | 0,796304139 | RP1-60O19.1 | FALSE | Downregulated |
| -0,34715888 | 0,796304139 | RPS16 | FALSE | Downregulated |
| -0,553179907 | 0,796304139 | RTL1 | FALSE | Downregulated |
| -0,637038518 | 0,796304139 | SLC25A30 | FALSE | Downregulated |
| -0,418443969 | 0,796304139 | SSU72 | FALSE | Downregulated |
| 0,576682949 | 0,796304139 | TONSL | FALSE | Upregulated |
| 0,493565714 | 0,796304139 | TP73-AS1 | FALSE | Upregulated |
| 0,50952249 | 0,796304139 | ZBTB32 | FALSE | Upregulated |
| 0,36142231 | 0,796304139 | ZDHHC5 | FALSE | Upregulated |
| 0,408652323 | 0,796304139 | ZFP91 | FALSE | Upregulated |
| -0,337429592 | 0,796708318 | DHFR | FALSE | Downregulated |
| -0,31064418 | 0,796816027 | ANGPTL4 | FALSE | Downregulated |
| -0,508697671 | 0,796816027 | FADS1 | FALSE | Downregulated |
| 0,526118658 | 0,796816027 | FOXN3-AS1 | FALSE | Upregulated |
| 0,338629367 | 0,796816027 | LTBP2 | FALSE | Upregulated |
| 0,251064287 | 0,796816027 | NCOR2 | FALSE | Upregulated |
| -0,205131404 | 0,796816027 | THPO | FALSE | Downregulated |
| 0,335260556 | 0,796965657 | C22orf23 | FALSE | Upregulated |
| -0,361972059 | 0,796965657 | ERBB2IP | FALSE | Downregulated |
| 0,483007761 | 0,79853581 | ACVR1B | FALSE | Upregulated |
| 0,278806481 | 0,79853581 | GPR18 | FALSE | Upregulated |
| -0,514212539 | 0,799013022 | YPEL2 | FALSE | Downregulated |
| -0,492547462 | 0,799114555 | CCT4 | FALSE | Downregulated |
| -0,428416469 | 0,799114555 | LOC100287467 | FALSE | Downregulated |
| -0,352610465 | 0,799114555 | PUM2 | FALSE | Downregulated |
| 0,483222341 | 0,799450124 | ZNF696 | FALSE | Upregulated |
| -0,547372117 | 0,800240434 | LOC101927514 | FALSE | Downregulated |
| 0,543821631 | 0,800290727 | C2CD2 | FALSE | Upregulated |
| 0,52806116 | 0,800290727 | FMO2 | FALSE | Upregulated |
| -0,399724526 | 0,800290727 | TMEM205 | FALSE | Downregulated |
| 0,269726299 | 0,800533135 | BAIAP2L1 | FALSE | Upregulated |
| 0,281119516 | 0,800724946 | CYB5R3 | FALSE | Upregulated |
| 0,33474167 | 0,800724946 | IGFBP7 | FALSE | Upregulated |
| 0,432976454 | 0,800724946 | SYT9 | FALSE | Upregulated |
| -0,522972083 | 0,800987627 | IPO8 | FALSE | Downregulated |
| 0,456395752 | 0,800987627 | NAA30 | FALSE | Upregulated |

| | | | | |
|---|---|---|---|---|
| 0,349897469 | 0,80106697 | CD46 | FALSE | Upregulated |
| -0,560721923 | 0,801122464 | CBR4 | FALSE | Downregulated |
| -0,463833778 | 0,801122464 | LUC7L2 | FALSE | Downregulated |
| 0,362076658 | 0,801122464 | SNX17 | FALSE | Upregulated |
| -0,384177701 | 0,801122464 | USP19 | FALSE | Downregulated |
| 0,373788884 | 0,801122464 | ZNF529 | FALSE | Upregulated |
| 0,499720491 | 0,802032702 | HLCS | FALSE | Upregulated |
| 0,584245265 | 0,804101583 | LRRC46 | FALSE | Upregulated |
| 0,368988444 | 0,804469849 | YLPM1 | FALSE | Upregulated |
| -0,230558415 | 0,804715191 | OAF | FALSE | Downregulated |
| 0,305023912 | 0,804715191 | PFN1 | FALSE | Upregulated |
| 0,423976995 | 0,804715191 | PLEKHM1 | FALSE | Upregulated |
| 0,314807481 | 0,805046122 | ARFGEF2 | FALSE | Upregulated |
| -0,420393607 | 0,805070811 | PIK3C2G | FALSE | Downregulated |
| -0,383196645 | 0,805070811 | TTC17 | FALSE | Downregulated |
| -0,47491789 | 0,805397959 | CBY3 | FALSE | Downregulated |
| -0,343190429 | 0,805397959 | CCNT2-AS1 | FALSE | Downregulated |
| -0,342068613 | 0,805397959 | FAM63A | FALSE | Downregulated |
| 0,503100669 | 0,805397959 | FBXL20 | FALSE | Upregulated |
| 0,284116128 | 0,805397959 | GRINA | FALSE | Upregulated |
| -0,421488398 | 0,805397959 | SMARCD2 | FALSE | Downregulated |
| -0,416908553 | 0,805653032 | SLC5A9 | FALSE | Downregulated |
| -0,350938553 | 0,805771144 | PPM1A | FALSE | Downregulated |
| -0,543582067 | 0,805771144 | TFDP2 | FALSE | Downregulated |
| 0,255182801 | 0,805781933 | CTBP1 | FALSE | Upregulated |
| 0,336917311 | 0,805781933 | HIF1A-AS2 | FALSE | Upregulated |
| -0,446131025 | 0,805821494 | ISOC2 | FALSE | Downregulated |
| -0,413716732 | 0,806480408 | AKAP8L | FALSE | Downregulated |
| 0,283942836 | 0,806480408 | LOC102723342 | FALSE | Upregulated |
| 0,45368542 | 0,806760492 | ELF1 | FALSE | Upregulated |
| 0,519313182 | 0,806975202 | DOCK7 | FALSE | Upregulated |
| -0,382200607 | 0,806975202 | FAM92A1 | FALSE | Downregulated |
| -0,374728302 | 0,806975202 | G3BP1 | FALSE | Downregulated |
| -0,33201806 | 0,806975202 | KHSRP | FALSE | Downregulated |
| -0,275686895 | 0,807383129 | HSPA9 | FALSE | Downregulated |
| 0,440384482 | 0,807383129 | LOC102724748 | FALSE | Upregulated |
| -0,389141132 | 0,807383129 | MCCC2 | FALSE | Downregulated |
| 0,508679626 | 0,807383129 | MED24 | FALSE | Upregulated |
| -0,562785085 | 0,807383129 | NEURL1 | FALSE | Downregulated |
| 0,450108767 | 0,807383129 | SUMO1P3 | FALSE | Upregulated |
| -0,341296943 | 0,807383129 | TMEM44 | FALSE | Downregulated |
| 0,470363946 | 0,807671769 | CCNC | FALSE | Upregulated |
| 0,398897612 | 0,807671769 | SLC35A1 | FALSE | Upregulated |
| -0,451573946 | 0,807743641 | EDF1 | FALSE | Downregulated |
| -0,328394178 | 0,807828803 | ANO6 | FALSE | Downregulated |
| 0,44949731 | 0,807828803 | RASSF7 | FALSE | Upregulated |
| 0,259041637 | 0,807828803 | SDC1 | FALSE | Upregulated |
| 0,284696919 | 0,807947581 | BTBD9 | FALSE | Upregulated |
| 0,455243705 | 0,807947581 | CLCC1 | FALSE | Upregulated |
| 0,455730736 | 0,807947581 | FBN1 | FALSE | Upregulated |
| -0,332464067 | 0,807947581 | MRTO4 | FALSE | Downregulated |

| | | | | |
|---|---|---|---|---|
| 0,320130714 | 0,807947581 | POGZ | FALSE | Upregulated |
| -0,495432401 | 0,807947581 | PUS3 | FALSE | Downregulated |
| 0,475601552 | 0,807947581 | SEC14L1 | FALSE | Upregulated |
| 0,31308882 | 0,808315354 | PAN3-AS1 | FALSE | Upregulated |
| -0,284880798 | 0,808364246 | ATP13A3 | FALSE | Downregulated |
| 0,332453195 | 0,808364246 | C15orf27 | FALSE | Upregulated |
| -0,420037995 | 0,808364246 | CAMK1 | FALSE | Downregulated |
| -0,323626128 | 0,808364246 | CLIP1 | FALSE | Downregulated |
| 0,300173839 | 0,808364246 | IQCC | FALSE | Upregulated |
| 0,474950653 | 0,808364246 | MNT | FALSE | Upregulated |
| -0,238600073 | 0,808364246 | PRPF8 | FALSE | Downregulated |
| -0,314274324 | 0,808364246 | RBM47 | FALSE | Downregulated |
| -0,412157623 | 0,808364246 | SPATA2L | FALSE | Downregulated |
| -0,403668761 | 0,808364246 | UBR3 | FALSE | Downregulated |
| -0,283509485 | 0,808364246 | UBR5 | FALSE | Downregulated |
| -0,514780504 | 0,808364246 | USP5 | FALSE | Downregulated |
| -0,237212627 | 0,808592886 | AHSA2 | FALSE | Downregulated |
| -0,475128218 | 0,808592886 | B3GNT8 | FALSE | Downregulated |
| 0,424493793 | 0,808592886 | CLDN24 | FALSE | Upregulated |
| -0,272835014 | 0,808592886 | HAS3 | FALSE | Downregulated |
| 0,47250042 | 0,808592886 | RASGRF2 | FALSE | Upregulated |
| -0,344801435 | 0,808592886 | SF3A1 | FALSE | Downregulated |
| 0,461512344 | 0,808624162 | BNIP3L | FALSE | Upregulated |
| 0,465618332 | 0,808792669 | DBNL | FALSE | Upregulated |
| -0,292488482 | 0,808923687 | C6orf120 | FALSE | Downregulated |
| 0,401130054 | 0,808923687 | PCCA | FALSE | Upregulated |
| -0,436354463 | 0,808923687 | RP11-685M7.3 | FALSE | Downregulated |
| 0,438441367 | 0,808923687 | ZNF598 | FALSE | Upregulated |
| -0,492050172 | 0,809114783 | BLCAP | FALSE | Downregulated |
| 0,224836327 | 0,809114783 | CD302 | FALSE | Upregulated |
| 0,464224278 | 0,809114783 | RP11-760N9.1 | FALSE | Upregulated |
| -0,491406126 | 0,809377306 | PRPF19 | FALSE | Downregulated |
| -0,265186044 | 0,810323593 | RBM5-AS1 | FALSE | Downregulated |
| -0,333222063 | 0,810469083 | ABCG8 | FALSE | Downregulated |
| -0,479424482 | 0,810469083 | INHBE | FALSE | Downregulated |
| 0,237874018 | 0,811319577 | LOC100996746 | FALSE | Upregulated |
| -0,428186105 | 0,811319577 | NR1H3 | FALSE | Downregulated |
| -0,351297213 | 0,812244238 | AC012314.8 | FALSE | Downregulated |
| 0,466382586 | 0,812244238 | ANKDD1A | FALSE | Upregulated |
| 0,435281724 | 0,812244238 | CHPF | FALSE | Upregulated |
| -0,384716991 | 0,812244238 | FAM134A | FALSE | Downregulated |
| 0,385301751 | 0,812244238 | LARP4B | FALSE | Upregulated |
| 0,479478708 | 0,812244238 | MCTP1 | FALSE | Upregulated |
| 0,541784858 | 0,812244238 | REEP3 | FALSE | Upregulated |
| -0,330978829 | 0,812244238 | SCARNA10 | FALSE | Downregulated |
| 0,313658694 | 0,812244238 | SLC25A4 | FALSE | Upregulated |
| -0,345754487 | 0,812244238 | TFIP11 | FALSE | Downregulated |
| -0,346551003 | 0,812244238 | VPS51 | FALSE | Downregulated |
| 0,297347241 | 0,812244238 | ZNF7 | FALSE | Upregulated |
| 0,404674439 | 0,812374771 | TNPO2 | FALSE | Upregulated |
| 0,331517425 | 0,812550926 | CSF1R | FALSE | Upregulated |

| | | | | |
|---|---|---|---|---|
| 0,439285962 | 0,812550926 | GALNS | FALSE | Upregulated |
| 0,334286604 | 0,812550926 | IFITM3 | FALSE | Upregulated |
| -0,482439781 | 0,812550926 | MORC3 | FALSE | Downregulated |
| -0,343742882 | 0,812550926 | RPL21 | FALSE | Downregulated |
| -0,268536578 | 0,812550926 | TOMM70A | FALSE | Downregulated |
| 0,509653644 | 0,813242019 | AREL1 | FALSE | Upregulated |
| 0,440154707 | 0,813242019 | DCTN2 | FALSE | Upregulated |
| -0,397851539 | 0,813242019 | ERGIC3 | FALSE | Downregulated |
| 0,444929373 | 0,813242019 | SLC25A44 | FALSE | Upregulated |
| -0,500607518 | 0,813242019 | TRABD2B | FALSE | Downregulated |
| 0,331786125 | 0,813770412 | DDX51 | FALSE | Upregulated |
| -0,409372405 | 0,813772607 | C12orf43 | FALSE | Downregulated |
| 0,382601908 | 0,813772607 | CC2D2B | FALSE | Upregulated |
| -0,420965408 | 0,814057166 | LBX2-AS1 | FALSE | Downregulated |
| 0,382597916 | 0,814341481 | DNAH11 | FALSE | Upregulated |
| 0,307972675 | 0,814341481 | VAV2 | FALSE | Upregulated |
| -0,385251209 | 0,814341481 | ZYG11B | FALSE | Downregulated |
| -0,352737064 | 0,814548657 | FBXL15 | FALSE | Downregulated |
| -0,300999137 | 0,814548657 | LOC101928853 | FALSE | Downregulated |
| 0,31447988 | 0,814548657 | LOC101929330 | FALSE | Upregulated |
| 0,522497365 | 0,817788893 | PEX12 | FALSE | Upregulated |
| -0,352689327 | 0,818058626 | ANXA9 | FALSE | Downregulated |
| -0,378203015 | 0,818058626 | APOC4-APOC2 | FALSE | Downregulated |
| 0,383551072 | 0,818058626 | BCAM | FALSE | Upregulated |
| 0,27838729 | 0,818058626 | CCSAP | FALSE | Upregulated |
| 0,333507618 | 0,818058626 | ESYT1 | FALSE | Upregulated |
| 0,524053502 | 0,818058626 | GOLGA8M | FALSE | Upregulated |
| 0,382931647 | 0,818058626 | KCNJ9 | FALSE | Upregulated |
| 0,304479119 | 0,818058626 | KIF1B | FALSE | Upregulated |
| 0,355938378 | 0,818058626 | MAN2B1 | FALSE | Upregulated |
| 0,416426701 | 0,818058626 | MTRNR2L3 | FALSE | Upregulated |
| -0,299901427 | 0,818058626 | NCOA2 | FALSE | Downregulated |
| 0,327788026 | 0,818058626 | NOTCH2NL | FALSE | Upregulated |
| -0,396770087 | 0,818058626 | PCSK9 | FALSE | Downregulated |
| 0,451009088 | 0,818058626 | PLET1 | FALSE | Upregulated |
| 0,443761729 | 0,818058626 | PLXNA2 | FALSE | Upregulated |
| -0,476906233 | 0,818058626 | RAP2C-AS1 | FALSE | Downregulated |
| 0,414196529 | 0,818058626 | SATL1 | FALSE | Upregulated |
| 0,263259404 | 0,818058626 | TMEM123 | FALSE | Upregulated |
| 0,343538023 | 0,818058626 | ZNF562 | FALSE | Upregulated |
| 0,393631571 | 0,818102926 | DNAH17 | FALSE | Upregulated |
| -0,587524259 | 0,818102926 | EFHD1 | FALSE | Downregulated |
| 0,46968742 | 0,818102926 | NAA25 | FALSE | Upregulated |
| -0,262322959 | 0,818102926 | POLDIP2 | FALSE | Downregulated |
| 0,387664449 | 0,818102926 | RUFY2 | FALSE | Upregulated |
| 0,237487263 | 0,818102926 | SPDYE1 | FALSE | Upregulated |
| 0,495528823 | 0,818102926 | ST7L | FALSE | Upregulated |
| -0,341937065 | 0,818232512 | CLPTM1 | FALSE | Downregulated |
| -0,242460257 | 0,818232512 | KPNA1 | FALSE | Downregulated |
| -0,503772401 | 0,818232512 | NR4A3 | FALSE | Downregulated |
| 0,297796548 | 0,818232512 | OTUB2 | FALSE | Upregulated |

| | | | | |
|---|---|---|---|---|
| 0,508415951 | 0,818232512 | TMEM138 | FALSE | Upregulated |
| -0,289973091 | 0,818812792 | HDGF | FALSE | Downregulated |
| 0,337415061 | 0,819490308 | RBBP4 | FALSE | Upregulated |
| -0,400849406 | 0,819543401 | EIF3G | FALSE | Downregulated |
| -0,340664593 | 0,819543401 | SMPD4 | FALSE | Downregulated |
| 0,380810203 | 0,819700288 | ATG14 | FALSE | Upregulated |
| -0,487269731 | 0,819700288 | GATA4 | FALSE | Downregulated |
| 0,390979432 | 0,819700288 | LOC102723853 | FALSE | Upregulated |
| 0,449381566 | 0,819700288 | SNAPC4 | FALSE | Upregulated |
| -0,420532045 | 0,819700288 | TNRC6C-AS1 | FALSE | Downregulated |
| -0,401977049 | 0,819903526 | ENGASE | FALSE | Downregulated |
| -0,475619486 | 0,819908319 | BAG4 | FALSE | Downregulated |
| 0,498953613 | 0,819908319 | FAM32A | FALSE | Upregulated |
| -0,375961465 | 0,819964161 | GHITM | FALSE | Downregulated |
| 0,435279834 | 0,820231702 | DDOST | FALSE | Upregulated |
| 0,338226739 | 0,820991552 | C1orf63 | FALSE | Upregulated |
| 0,432515392 | 0,820991552 | PDLIM1 | FALSE | Upregulated |
| 0,407901471 | 0,821463286 | WDR82 | FALSE | Upregulated |
| -0,360781093 | 0,821637687 | LOC100288846 | FALSE | Downregulated |
| -0,381264535 | 0,822093655 | APOC1P1 | FALSE | Downregulated |
| 0,38243085 | 0,822234105 | TCEB3B | FALSE | Upregulated |
| 0,359512349 | 0,822673573 | HMCN1 | FALSE | Upregulated |
| 0,471427246 | 0,823214735 | EHD1 | FALSE | Upregulated |
| -0,446748672 | 0,823214735 | SDHAP1 | FALSE | Downregulated |
| -0,35409414 | 0,823398307 | WDTC1 | FALSE | Downregulated |
| 0,429455266 | 0,823409304 | CAPZA1 | FALSE | Upregulated |
| -0,37395824 | 0,82383205 | FLJ37453 | FALSE | Downregulated |
| 0,345097299 | 0,82383205 | PARD3 | FALSE | Upregulated |
| 0,352139178 | 0,82383205 | PRRT3 | FALSE | Upregulated |
| 0,51724779 | 0,824161162 | RIBC1 | FALSE | Upregulated |
| 0,391586862 | 0,82432807 | FAM193A | FALSE | Upregulated |
| 0,460750212 | 0,82432807 | LYRM4 | FALSE | Upregulated |
| 0,290190246 | 0,82432807 | MOV10 | FALSE | Upregulated |
| 0,533930413 | 0,82432807 | SSH2 | FALSE | Upregulated |
| 0,478744226 | 0,825285524 | AHNAK2 | FALSE | Upregulated |
| -0,359004935 | 0,825285524 | ATN1 | FALSE | Downregulated |
| -0,333509421 | 0,825285524 | COMT | FALSE | Downregulated |
| -0,467895672 | 0,825285524 | CREB3 | FALSE | Downregulated |
| -0,216407948 | 0,825285524 | LARP1 | FALSE | Downregulated |
| -0,401788142 | 0,825285524 | NDUFS1 | FALSE | Downregulated |
| -0,396145537 | 0,825285524 | PUM1 | FALSE | Downregulated |
| -0,345129125 | 0,825285524 | RIC8A | FALSE | Downregulated |
| -0,395101106 | 0,825285524 | RPS21 | FALSE | Downregulated |
| 0,409749917 | 0,825285524 | TTC37 | FALSE | Upregulated |
| 0,469967629 | 0,825494225 | C22orf29 | FALSE | Upregulated |
| 0,404760253 | 0,825909207 | ACTR1B | FALSE | Upregulated |
| -0,469549144 | 0,8265577 | FBXO8 | FALSE | Downregulated |
| -0,458989562 | 0,8265577 | GPR160 | FALSE | Downregulated |
| 0,409305419 | 0,8265577 | TRIM27 | FALSE | Upregulated |
| -0,3777156 | 0,826970401 | AUP1 | FALSE | Downregulated |
| 0,313874357 | 0,826970401 | WDR6 | FALSE | Upregulated |

| | | | | |
|---|---|---|---|---|
| -0,274172992 | 0,827480604 | PALLD | FALSE | Downregulated |
| 0,269048665 | 0,827480604 | ZNF512B | FALSE | Upregulated |
| -0,273857393 | 0,827487224 | ABTB2 | FALSE | Downregulated |
| -0,446954929 | 0,827487224 | CRHBP | FALSE | Downregulated |
| -0,379311833 | 0,827487224 | LOC100996724 | FALSE | Downregulated |
| -0,403528501 | 0,827487224 | LOC101929441 | FALSE | Downregulated |
| -0,378241182 | 0,827487224 | MST1L | FALSE | Downregulated |
| 0,52466354 | 0,827487224 | PRORSD1P | FALSE | Upregulated |
| -0,51068006 | 0,827487224 | SOS2 | FALSE | Downregulated |
| -0,442200481 | 0,827487224 | TCEA1 | FALSE | Downregulated |
| 0,271675351 | 0,827487224 | ZNF219 | FALSE | Upregulated |
| 0,381426301 | 0,827773934 | KLHL21 | FALSE | Upregulated |
| -0,276474519 | 0,828263797 | ACO1 | FALSE | Downregulated |
| -0,327208522 | 0,828666661 | UBN2 | FALSE | Downregulated |
| 0,331704846 | 0,82878179 | MAP3K3 | FALSE | Upregulated |
| -0,303647569 | 0,82878179 | TNFAIP8L2 | FALSE | Downregulated |
| 0,205134474 | 0,828933955 | ENTPD1 | FALSE | Upregulated |
| -0,276340621 | 0,828933955 | ITCH | FALSE | Downregulated |
| 0,369997088 | 0,828933955 | RP4-724E16.2 | FALSE | Upregulated |
| -0,473946221 | 0,828933955 | TIMM8B | FALSE | Downregulated |
| 0,401715157 | 0,82901094 | MMP25 | FALSE | Upregulated |
| -0,438688927 | 0,82901094 | ZNF586 | FALSE | Downregulated |
| -0,40672529 | 0,831594832 | ZNF275 | FALSE | Downregulated |
| 0,291879796 | 0,83179523 | PTPRJ | FALSE | Upregulated |
| 0,445619959 | 0,831988632 | MAPK13 | FALSE | Upregulated |
| -0,262866891 | 0,832215136 | RPL15 | FALSE | Downregulated |
| -0,334832519 | 0,832472529 | CREB3L3 | FALSE | Downregulated |
| 0,372460164 | 0,832472529 | DAZAP1 | FALSE | Upregulated |
| 0,350280104 | 0,832472529 | MAK | FALSE | Upregulated |
| -0,428158382 | 0,832472529 | NFYA | FALSE | Downregulated |
| -0,343756587 | 0,832472529 | PLD1 | FALSE | Downregulated |
| -0,402883846 | 0,832472529 | RPL13AP6 | FALSE | Downregulated |
| -0,254987613 | 0,832472529 | RPS11 | FALSE | Downregulated |
| -0,468417197 | 0,832472529 | SCAND1 | FALSE | Downregulated |
| -0,323460202 | 0,832472529 | SETD7 | FALSE | Downregulated |
| -0,36993219 | 0,832472529 | XRCC6 | FALSE | Downregulated |
| 0,229481389 | 0,832472529 | ZFHX3 | FALSE | Upregulated |
| 0,472362994 | 0,832472529 | ZNF641 | FALSE | Upregulated |
| -0,367759722 | 0,832647873 | AHCYL1 | FALSE | Downregulated |
| -0,525449724 | 0,832647873 | BCO2 | FALSE | Downregulated |
| -0,3138063 | 0,832647873 | COBL | FALSE | Downregulated |
| 0,39147089 | 0,832647873 | CROCCP3 | FALSE | Upregulated |
| 0,349114027 | 0,832647873 | DAGLB | FALSE | Upregulated |
| 0,315337204 | 0,832647873 | DGCR2 | FALSE | Upregulated |
| 0,335506276 | 0,832647873 | DUSP28 | FALSE | Upregulated |
| -0,336502058 | 0,832647873 | GPS1 | FALSE | Downregulated |
| -0,352718109 | 0,832647873 | HLA-L | FALSE | Downregulated |
| 0,379869718 | 0,832647873 | KIF13A | FALSE | Upregulated |
| -0,456739933 | 0,832647873 | LARS2-AS1 | FALSE | Downregulated |
| 0,284911616 | 0,832647873 | LOC101929533 | FALSE | Upregulated |
| -0,372632692 | 0,832647873 | LOC101929633 | FALSE | Downregulated |

| | | | | |
|---|---|---|---|---|
| -0,260087506 | 0,832647873 | OBSL1 | FALSE | Downregulated |
| -0,299162334 | 0,832647873 | PLGLB2 | FALSE | Downregulated |
| -0,261308775 | 0,832647873 | RBM33 | FALSE | Downregulated |
| 0,331463334 | 0,832647873 | RNF40 | FALSE | Upregulated |
| 0,434859803 | 0,832647873 | TMEM41A | FALSE | Upregulated |
| 0,321241527 | 0,832647873 | UBR2 | FALSE | Upregulated |
| -0,252261271 | 0,833029133 | C9orf131 | FALSE | Downregulated |
| -0,87053715 | 0,833029133 | CFHR1 | FALSE | Downregulated |
| 0,398772255 | 0,833029133 | FLOT1 | FALSE | Upregulated |
| 0,348961003 | 0,833029133 | KLHL5 | FALSE | Upregulated |
| -0,290057349 | 0,833029133 | PAPOLA | FALSE | Downregulated |
| -0,372685107 | 0,833029133 | RAP2C | FALSE | Downregulated |
| 0,322665806 | 0,833029133 | RIN3 | FALSE | Upregulated |
| 0,318949366 | 0,833029133 | SLC31A2 | FALSE | Upregulated |
| -0,237819529 | 0,833029133 | SNRNP200 | FALSE | Downregulated |
| 0,378845569 | 0,833029133 | TMSB4X | FALSE | Upregulated |
| -0,37467875 | 0,83314464 | TIAM1 | FALSE | Downregulated |
| 0,328196458 | 0,833732484 | GBF1 | FALSE | Upregulated |
| 0,422037183 | 0,833732484 | TYMS | FALSE | Upregulated |
| -0,505473966 | 0,833732484 | VPS28 | FALSE | Downregulated |
| -0,322056872 | 0,834176366 | SIN3A | FALSE | Downregulated |
| -0,405189414 | 0,834611214 | CDH5 | FALSE | Downregulated |
| 0,264731457 | 0,834611214 | CNDP2 | FALSE | Upregulated |
| 0,466059597 | 0,834611214 | CNTRL | FALSE | Upregulated |
| 0,432330065 | 0,834611214 | RAB22A | FALSE | Upregulated |
| -0,229064641 | 0,834611214 | RMDN1 | FALSE | Downregulated |
| 0,302011749 | 0,834611214 | SNX21 | FALSE | Upregulated |
| 0,385273642 | 0,834695169 | LOC100130075 | FALSE | Upregulated |
| -0,443211914 | 0,834695169 | TSPYL4 | FALSE | Downregulated |
| -0,502229066 | 0,834751834 | ATP5C1 | FALSE | Downregulated |
| 0,362737913 | 0,834751834 | BDP1 | FALSE | Upregulated |
| -0,372635639 | 0,834751834 | DENND4A | FALSE | Downregulated |
| 0,4183646 | 0,834751834 | DNAJB12 | FALSE | Upregulated |
| 0,488411819 | 0,834751834 | DNAJB14 | FALSE | Upregulated |
| 0,387441202 | 0,834751834 | EIF3J | FALSE | Upregulated |
| 0,270708016 | 0,834751834 | GOLM1 | FALSE | Upregulated |
| 0,379845292 | 0,834751834 | JOSD1 | FALSE | Upregulated |
| 0,414362517 | 0,834751834 | LINC00896 | FALSE | Upregulated |
| 0,462239427 | 0,834751834 | LOC101929384 | FALSE | Upregulated |
| 0,328935879 | 0,834751834 | LOC102724790 | FALSE | Upregulated |
| 0,344394864 | 0,834751834 | LRP10 | FALSE | Upregulated |
| 0,254060154 | 0,834751834 | MXRA8 | FALSE | Upregulated |
| 0,435987614 | 0,834751834 | PTBP3 | FALSE | Upregulated |
| 0,457174996 | 0,834751834 | RNF24 | FALSE | Upregulated |
| 0,23734096 | 0,834751834 | SNX22 | FALSE | Upregulated |
| -0,374981701 | 0,834751834 | ZHX2 | FALSE | Downregulated |
| 0,240521224 | 0,835660043 | CAST | FALSE | Upregulated |
| -0,382616137 | 0,835660043 | DNAJC11 | FALSE | Downregulated |
| 0,393092757 | 0,835693325 | BRICD5 | FALSE | Upregulated |
| -0,311538045 | 0,835693325 | CC2D2A | FALSE | Downregulated |
| 0,279740988 | 0,835981752 | SRSF6 | FALSE | Upregulated |

| | | | | |
|---|---|---|---|---|
| 0,352291771 | 0,835981752 | VAPA | FALSE | Upregulated |
| 0,435077703 | 0,836569918 | ZNF689 | FALSE | Upregulated |
| 0,286188002 | 0,836571799 | FARP2 | FALSE | Upregulated |
| 0,293974475 | 0,836571799 | LPAR6 | FALSE | Upregulated |
| -0,423989059 | 0,836571799 | UGT3A1 | FALSE | Downregulated |
| 0,420600699 | 0,836657959 | TSPAN11 | FALSE | Upregulated |
| -0,331898909 | 0,836749946 | APBB2 | FALSE | Downregulated |
| -0,325467238 | 0,836749946 | CYP2D6 | FALSE | Downregulated |
| -0,497257758 | 0,837073575 | GLRX | FALSE | Downregulated |
| -0,251224699 | 0,837400077 | ACO2 | FALSE | Downregulated |
| -0,405879926 | 0,837400077 | ADAM21 | FALSE | Downregulated |
| -0,425223148 | 0,837400077 | APOL4 | FALSE | Downregulated |
| -0,427142745 | 0,837400077 | ARID4A | FALSE | Downregulated |
| -0,397396376 | 0,837400077 | BBX | FALSE | Downregulated |
| 0,438763287 | 0,837400077 | CCDC125 | FALSE | Upregulated |
| 0,383316376 | 0,837400077 | CDC34 | FALSE | Upregulated |
| -0,479834966 | 0,837400077 | DEK | FALSE | Downregulated |
| -0,390534433 | 0,837400077 | DENND5B | FALSE | Downregulated |
| -0,254126168 | 0,837400077 | GAPDH | FALSE | Downregulated |
| -0,200472708 | 0,837400077 | HMHA1 | FALSE | Downregulated |
| 0,418706138 | 0,837400077 | ITGB3 | FALSE | Upregulated |
| -0,40022928 | 0,837400077 | IWS1 | FALSE | Downregulated |
| 0,36776176 | 0,837400077 | KDM4A | FALSE | Upregulated |
| 0,37996189 | 0,837400077 | LSM4 | FALSE | Upregulated |
| 0,302116678 | 0,837400077 | MED26 | FALSE | Upregulated |
| 0,216514754 | 0,837400077 | MLLT4 | FALSE | Upregulated |
| 0,308450046 | 0,837400077 | NF1 | FALSE | Upregulated |
| 0,371155285 | 0,837400077 | NFATC2 | FALSE | Upregulated |
| -0,283135515 | 0,837400077 | PPP1R10 | FALSE | Downregulated |
| -0,435274517 | 0,837400077 | RAC1 | FALSE | Downregulated |
| 0,473073462 | 0,837400077 | RIPK4 | FALSE | Upregulated |
| 0,274147419 | 0,837400077 | RPL23AP64 | FALSE | Upregulated |
| -0,364866684 | 0,837400077 | RPP38 | FALSE | Downregulated |
| 0,322591781 | 0,837400077 | SEC24C | FALSE | Upregulated |
| -0,294984904 | 0,837400077 | SEL1L | FALSE | Downregulated |
| 0,482195091 | 0,837400077 | SGPP2 | FALSE | Upregulated |
| 0,380996609 | 0,837400077 | SLC37A2 | FALSE | Upregulated |
| 0,511909144 | 0,837400077 | STRN | FALSE | Upregulated |
| 0,362624434 | 0,837400077 | TBC1D1 | FALSE | Upregulated |
| -0,345774203 | 0,837400077 | TPST2 | FALSE | Downregulated |
| -0,317404297 | 0,837400077 | TRAM2 | FALSE | Downregulated |
| 0,504668599 | 0,837400077 | UVRAG | FALSE | Upregulated |
| 0,413942928 | 0,837456964 | BICD2 | FALSE | Upregulated |
| 0,285203957 | 0,837456964 | XPO1 | FALSE | Upregulated |
| 0,464667473 | 0,837791367 | THNSL1 | FALSE | Upregulated |
| 0,272359941 | 0,837855001 | ASH1L | FALSE | Upregulated |
| -0,312267535 | 0,837855001 | LRG1 | FALSE | Downregulated |
| -0,410771413 | 0,838250135 | ZBED6CL | FALSE | Downregulated |
| 0,249937218 | 0,838548781 | BTBD18 | FALSE | Upregulated |
| -0,297370308 | 0,838548781 | FBXL22 | FALSE | Downregulated |
| -0,398702126 | 0,838548781 | GGT5 | FALSE | Downregulated |

| | | | | |
|---|---|---|---|---|
| -0,468610801 | 0,838548781 | SLC25A20 | FALSE | Downregulated |
| -0,468280049 | 0,838841802 | GAB1 | FALSE | Downregulated |
| 0,358465277 | 0,838841802 | LBP | FALSE | Upregulated |
| 0,35947007 | 0,838841802 | PDZD9 | FALSE | Upregulated |
| 0,347356633 | 0,839535294 | MAP2K3 | FALSE | Upregulated |
| 0,452344673 | 0,839806151 | PTPRA | FALSE | Upregulated |
| 0,330996117 | 0,839806151 | ZBTB4 | FALSE | Upregulated |
| -0,218264707 | 0,840178829 | CLTC | FALSE | Downregulated |
| -0,361562553 | 0,840178829 | CNPY4 | FALSE | Downregulated |
| 0,230702182 | 0,840178829 | COMMD5 | FALSE | Upregulated |
| 0,246278738 | 0,840178829 | FBXW5 | FALSE | Upregulated |
| 0,442889817 | 0,840489386 | EIF3D | FALSE | Upregulated |
| 0,224890807 | 0,840493467 | PARP14 | FALSE | Upregulated |
| -0,412584233 | 0,840659692 | HM13 | FALSE | Downregulated |
| -0,269284132 | 0,840659692 | MMP19 | FALSE | Downregulated |
| -0,275599393 | 0,840659692 | UPF1 | FALSE | Downregulated |
| -0,345172107 | 0,840957854 | MIB1 | FALSE | Downregulated |
| -0,467116941 | 0,840957854 | TIE1 | FALSE | Downregulated |
| 0,316483488 | 0,841002869 | RPS6KA2 | FALSE | Upregulated |
| 0,298532641 | 0,841326795 | AKNA | FALSE | Upregulated |
| 0,25321965 | 0,841568806 | SMARCC2 | FALSE | Upregulated |
| 0,382011393 | 0,841890984 | CLN8 | FALSE | Upregulated |
| -0,439755048 | 0,842017959 | ZNF76 | FALSE | Downregulated |
| 0,299329085 | 0,842371956 | ABHD12B | FALSE | Upregulated |
| 0,396283988 | 0,842371956 | ZC3HAV1 | FALSE | Upregulated |
| -0,39514314 | 0,842462285 | CROCCP2 | FALSE | Downregulated |
| -0,353401331 | 0,842462285 | LOC102723806 | FALSE | Downregulated |
| -0,351911269 | 0,842462285 | MIEF1 | FALSE | Downregulated |
| 0,427577655 | 0,842462285 | RAB10 | FALSE | Upregulated |
| -0,372856269 | 0,842462285 | RNF168 | FALSE | Downregulated |
| -0,220722215 | 0,842462285 | SETDB1 | FALSE | Downregulated |
| 0,278263282 | 0,842462285 | SPG11 | FALSE | Upregulated |
| 0,267726177 | 0,842462285 | UBTF | FALSE | Upregulated |
| -0,303978722 | 0,8425632 | NCKAP1 | FALSE | Downregulated |
| 0,302155709 | 0,8425632 | RARA-AS1 | FALSE | Upregulated |
| -0,373138554 | 0,8425632 | SEC24D | FALSE | Downregulated |
| -0,423584106 | 0,84280949 | ATP5SL | FALSE | Downregulated |
| -0,375738896 | 0,84280949 | PSMA7 | FALSE | Downregulated |
| -0,38703212 | 0,84387592 | ATP6V0E2 | FALSE | Downregulated |
| -0,393529703 | 0,84387592 | CREBL2 | FALSE | Downregulated |
| -0,216326274 | 0,84387592 | MAT2A | FALSE | Downregulated |
| 0,292883851 | 0,84387592 | MEF2A | FALSE | Upregulated |
| 0,158432542 | 0,84387592 | RN7SK | FALSE | Upregulated |
| 0,266475417 | 0,844499121 | PTPN23 | FALSE | Upregulated |
| 0,4355773 | 0,844499121 | RNF111 | FALSE | Upregulated |
| -0,442463318 | 0,844499121 | ZNF829 | FALSE | Downregulated |
| -0,357250334 | 0,845002225 | GIPC3 | FALSE | Downregulated |
| -0,293680591 | 0,845002225 | LOC100505658 | FALSE | Downregulated |
| 0,464375105 | 0,845461511 | HEATR2 | FALSE | Upregulated |
| 0,261047796 | 0,84556996 | RANBP10 | FALSE | Upregulated |
| -0,257696145 | 0,845626822 | C9orf114 | FALSE | Downregulated |

| | | | | |
|---|---|---|---|---|
| 0,206433159 | 0,845626822 | ZACN | FALSE | Upregulated |
| 0,414281333 | 0,846925934 | LOC101929347 | FALSE | Upregulated |
| -0,239800396 | 0,846925934 | SPEN | FALSE | Downregulated |
| 0,330612373 | 0,847021415 | PPFIA1 | FALSE | Upregulated |
| 0,241650665 | 0,847166147 | MYRF | FALSE | Upregulated |
| -0,201355062 | 0,848435643 | FAXDC2 | FALSE | Downregulated |
| -0,377951163 | 0,848435643 | HSPB1 | FALSE | Downregulated |
| 0,398954275 | 0,84856927 | BIN1 | FALSE | Upregulated |
| 0,364893869 | 0,84856927 | CCNK | FALSE | Upregulated |
| 0,309152533 | 0,84856927 | DOCK4 | FALSE | Upregulated |
| 0,227762582 | 0,84856927 | FARP1 | FALSE | Upregulated |
| 0,285306461 | 0,84856927 | GABARAPL1 | FALSE | Upregulated |
| -0,22440827 | 0,84856927 | IGFBP3 | FALSE | Downregulated |
| -0,322978968 | 0,84856927 | LINC00538 | FALSE | Downregulated |
| -0,492112705 | 0,84856927 | MOCOS | FALSE | Downregulated |
| 0,454269866 | 0,84856927 | ZNF69 | FALSE | Upregulated |
| 0,270788685 | 0,848767875 | GIGYF1 | FALSE | Upregulated |
| 0,325454189 | 0,848879175 | C7 | FALSE | Upregulated |
| 0,334705187 | 0,849697288 | KDM1A | FALSE | Upregulated |
| -0,251925716 | 0,849697288 | PSD4 | FALSE | Downregulated |
| 0,461455865 | 0,849697288 | SMIM12 | FALSE | Upregulated |
| 0,353532437 | 0,849697288 | TRAM1 | FALSE | Upregulated |
| 0,343273133 | 0,849848014 | ADPGK | FALSE | Upregulated |
| 0,364568675 | 0,849848014 | AKAP11 | FALSE | Upregulated |
| 0,273517215 | 0,849848014 | CDC14B | FALSE | Upregulated |
| 0,24092181 | 0,849848014 | USP22 | FALSE | Upregulated |
| 0,441984308 | 0,849979344 | TFE3 | FALSE | Upregulated |
| 0,391549068 | 0,85083094 | TAPBPL | FALSE | Upregulated |
| 0,311788787 | 0,85083094 | ZNF397 | FALSE | Upregulated |
| -0,439345309 | 0,851039495 | LEMD3 | FALSE | Downregulated |
| 0,387186127 | 0,851424033 | ABR | FALSE | Upregulated |
| -0,337513853 | 0,851424033 | ETFA | FALSE | Downregulated |
| 0,289023371 | 0,851424033 | HGS | FALSE | Upregulated |
| 0,341517814 | 0,851424033 | LOC102724499 | FALSE | Upregulated |
| 0,475534573 | 0,851424033 | PHYHIPL | FALSE | Upregulated |
| -0,394821189 | 0,851424033 | ZNF550 | FALSE | Downregulated |
| 0,365148285 | 0,851424033 | ZNF784 | FALSE | Upregulated |
| 0,409596595 | 0,851424033 | ZNF790 | FALSE | Upregulated |
| 0,317383953 | 0,851542209 | CREG1 | FALSE | Upregulated |
| -0,317025633 | 0,851965951 | IST1 | FALSE | Downregulated |
| -0,341385611 | 0,852087759 | SPP2 | FALSE | Downregulated |
| 0,231559591 | 0,852087759 | TMEM200B | FALSE | Upregulated |
| 0,217388963 | 0,852502895 | AKT1 | FALSE | Upregulated |
| -0,476676026 | 0,852502895 | DEPDC5 | FALSE | Downregulated |
| 0,385474643 | 0,852502895 | LCOR | FALSE | Upregulated |
| 0,454709677 | 0,852799483 | HYAL2 | FALSE | Upregulated |
| -0,283122074 | 0,853139959 | JAM2 | FALSE | Downregulated |
| -0,372242472 | 0,853679201 | ARID5A | FALSE | Downregulated |
| -0,460478486 | 0,853679201 | CPT1B | FALSE | Downregulated |
| 0,411307646 | 0,853679201 | CYP2D7P | FALSE | Upregulated |
| -0,266365498 | 0,853679201 | EIF1 | FALSE | Downregulated |

| | | | | |
|---|---|---|---|---|
| -0,395533385 | 0,853679201 | GTF3A | FALSE | Downregulated |
| 0,361861499 | 0,853679201 | JAG1 | FALSE | Upregulated |
| -0,391330147 | 0,854109888 | C14orf164 | FALSE | Downregulated |
| -0,270152825 | 0,854109888 | SPAG9 | FALSE | Downregulated |
| 0,353585593 | 0,854109888 | TNKS2 | FALSE | Upregulated |
| -0,243210141 | 0,854162372 | DHCR24 | FALSE | Downregulated |
| -0,286946306 | 0,854162372 | KLF3 | FALSE | Downregulated |
| -0,337917418 | 0,854316633 | SLC30A1 | FALSE | Downregulated |
| 0,353211683 | 0,854559347 | CMTR1 | FALSE | Upregulated |
| -0,296419952 | 0,854559347 | GPR68 | FALSE | Downregulated |
| 0,330899382 | 0,854559347 | TRAF4 | FALSE | Upregulated |
| 0,432584138 | 0,855469854 | GUSBP11 | FALSE | Upregulated |
| 0,359075087 | 0,856046095 | SCYL3 | FALSE | Upregulated |
| -0,324575744 | 0,856528399 | FOXP4 | FALSE | Downregulated |
| 0,341780603 | 0,856612212 | ZFP69 | FALSE | Upregulated |
| -0,375644123 | 0,85664942 | NRM | FALSE | Downregulated |
| 0,431814427 | 0,85664942 | STRADA | FALSE | Upregulated |
| -0,351144909 | 0,85687781 | ABHD14B | FALSE | Downregulated |
| -0,404569905 | 0,856960664 | FANCL | FALSE | Downregulated |
| -0,331392651 | 0,857186814 | FAM214A | FALSE | Downregulated |
| -0,378856823 | 0,857224698 | C1QTNF7 | FALSE | Downregulated |
| -0,351439978 | 0,857449899 | AC004893.11 | FALSE | Downregulated |
| -0,257844274 | 0,857449899 | ADORA2A | FALSE | Downregulated |
| 0,339986107 | 0,857449899 | MYO6 | FALSE | Upregulated |
| 0,4229934 | 0,857449899 | NAA60 | FALSE | Upregulated |
| -0,271134365 | 0,857449899 | NR3C1 | FALSE | Downregulated |
| -0,450577979 | 0,857449899 | PCTP | FALSE | Downregulated |
| 0,394777955 | 0,857449899 | PHF20L1 | FALSE | Upregulated |
| -0,314899375 | 0,857449899 | SHE | FALSE | Downregulated |
| 0,471287929 | 0,857449899 | STK4 | FALSE | Upregulated |
| 0,384582677 | 0,857449899 | TEX261 | FALSE | Upregulated |
| 0,232802955 | 0,857449899 | TRRAP | FALSE | Upregulated |
| -0,336665716 | 0,857449899 | USP2 | FALSE | Downregulated |
| -0,43726087 | 0,857612388 | CISD2 | FALSE | Downregulated |
| 0,34311418 | 0,857612388 | LEPROT | FALSE | Upregulated |
| -0,391512918 | 0,857612388 | LOC101928555 | FALSE | Downregulated |
| 0,393582902 | 0,857612388 | PELO | FALSE | Upregulated |
| -0,400725998 | 0,857612388 | SGTA | FALSE | Downregulated |
| -0,245779906 | 0,857612388 | STK25 | FALSE | Downregulated |
| -0,389111196 | 0,857612388 | ZFYVE9 | FALSE | Downregulated |
| -0,447253589 | 0,857922072 | ANAPC2 | FALSE | Downregulated |
| 0,329317222 | 0,858002421 | FAM134C | FALSE | Upregulated |
| 0,35630774 | 0,858002421 | TOM1 | FALSE | Upregulated |
| 0,35051989 | 0,858002421 | ZNF557 | FALSE | Upregulated |
| 0,241279744 | 0,858554784 | DOPEY1 | FALSE | Upregulated |
| -0,393701193 | 0,858554784 | LOC101929004 | FALSE | Downregulated |
| 0,363151909 | 0,858693325 | SNX18 | FALSE | Upregulated |
| -0,248757526 | 0,858695253 | AGPAT2 | FALSE | Downregulated |
| -0,449010723 | 0,858695253 | EXT2 | FALSE | Downregulated |
| -0,231761708 | 0,858695253 | LIFR | FALSE | Downregulated |
| 0,417158733 | 0,858695253 | LINC-PINT | FALSE | Upregulated |

| | | | | |
|---|---|---|---|---|
| -0,333447007 | 0,858695253 | THRB-AS1 | FALSE | Downregulated |
| -0,475724036 | 0,858695253 | UBE2J2 | FALSE | Downregulated |
| -0,312401547 | 0,858851039 | CDK5RAP3 | FALSE | Downregulated |
| 0,183358854 | 0,858851039 | HSPG2 | FALSE | Upregulated |
| 0,284157745 | 0,858851039 | SETBP1 | FALSE | Upregulated |
| 0,325736242 | 0,858851039 | SKIV2L | FALSE | Upregulated |
| -0,422959889 | 0,858977614 | ICAM3 | FALSE | Downregulated |
| -0,343927172 | 0,858977614 | LOC102723990 | FALSE | Downregulated |
| 0,381424853 | 0,858977614 | PLEKHM3 | FALSE | Upregulated |
| 0,249673936 | 0,858977614 | SLC16A4 | FALSE | Upregulated |
| 0,255513908 | 0,858977614 | TCF20 | FALSE | Upregulated |
| -0,279760616 | 0,858977614 | ZNF646 | FALSE | Downregulated |
| -0,370881224 | 0,858981441 | C10orf25 | FALSE | Downregulated |
| 0,364633508 | 0,858981441 | DESI1 | FALSE | Upregulated |
| 0,399350046 | 0,858981441 | LOC101928946 | FALSE | Upregulated |
| -0,35938666 | 0,858981441 | RAB7A | FALSE | Downregulated |
| -0,337583762 | 0,858981441 | TBC1D5 | FALSE | Downregulated |
| 0,202024273 | 0,859089685 | LOC100190986 | FALSE | Upregulated |
| 0,408432227 | 0,859187025 | ACOT4 | FALSE | Upregulated |
| 0,262014508 | 0,859187025 | ARVCF | FALSE | Upregulated |
| -0,330968116 | 0,859187025 | BHLHE40 | FALSE | Downregulated |
| -0,354213452 | 0,859187025 | PDXDC2P | FALSE | Downregulated |
| -0,480763762 | 0,859187025 | SLC25A38 | FALSE | Downregulated |
| -0,352599865 | 0,859187025 | SLC39A9 | FALSE | Downregulated |
| -0,27741939 | 0,859187025 | ZSCAN2 | FALSE | Downregulated |
| -0,293598126 | 0,860018458 | DLG1 | FALSE | Downregulated |
| 0,399380889 | 0,860018458 | DNASE1L1 | FALSE | Upregulated |
| 0,254118288 | 0,860018458 | INADL | FALSE | Upregulated |
| 0,245998678 | 0,860018458 | KIF13B | FALSE | Upregulated |
| 0,347555102 | 0,860018458 | LOC102723743 | FALSE | Upregulated |
| -0,30767625 | 0,860018458 | MRPL12 | FALSE | Downregulated |
| 0,302227579 | 0,860018458 | UHRF1BP1L | FALSE | Upregulated |
| 0,401478716 | 0,860018458 | ZNF532 | FALSE | Upregulated |
| -0,421773774 | 0,860112088 | IDH3B | FALSE | Downregulated |
| -0,304590463 | 0,860265759 | B9D2 | FALSE | Downregulated |
| -0,273880247 | 0,860461501 | FOXO3 | FALSE | Downregulated |
| 0,360939007 | 0,860461501 | TRAPPC9 | FALSE | Upregulated |
| 0,40056437 | 0,860569494 | PGAP2 | FALSE | Upregulated |
| -0,217712834 | 0,861013508 | PRRC2C | FALSE | Downregulated |
| -0,289719424 | 0,861320818 | XYLT2 | FALSE | Downregulated |
| 0,73566829 | 0,861636094 | CDR1 | FALSE | Upregulated |
| -0,381136374 | 0,861865952 | GPBP1 | FALSE | Downregulated |
| -0,229118635 | 0,862144517 | GPR161 | FALSE | Downregulated |
| 0,440423884 | 0,862589315 | LOC100506374 | FALSE | Upregulated |
| -0,264085503 | 0,862671729 | ASCC1 | FALSE | Downregulated |
| 0,352534495 | 0,862671729 | OGFOD3 | FALSE | Upregulated |
| 0,328102117 | 0,862875403 | GPR107 | FALSE | Upregulated |
| 0,30499517 | 0,862956219 | LOC101927903 | FALSE | Upregulated |
| -0,48782627 | 0,862956219 | SOX6 | FALSE | Downregulated |
| 0,358292228 | 0,863203849 | PRPH2 | FALSE | Upregulated |
| 0,374878191 | 0,863232633 | TMEM184C | FALSE | Upregulated |

| | | | | |
|---|---|---|---|---|
| -0,368333652 | 0,863296773 | ARID4B | FALSE | Downregulated |
| 0,290783155 | 0,863341744 | MAFB | FALSE | Upregulated |
| 0,400433584 | 0,864045418 | CLDN18 | FALSE | Upregulated |
| -0,215705695 | 0,864045418 | HNRNPA3 | FALSE | Downregulated |
| -0,26133804 | 0,864045418 | PEPD | FALSE | Downregulated |
| -0,272454842 | 0,86456184 | CACYBP | FALSE | Downregulated |
| -0,365515774 | 0,86456184 | CLCN3 | FALSE | Downregulated |
| 0,415349775 | 0,86456184 | CNOT6L | FALSE | Upregulated |
| -0,412130651 | 0,86456184 | SCAF4 | FALSE | Downregulated |
| 0,408726628 | 0,86456184 | SNX10 | FALSE | Upregulated |
| -0,218603865 | 0,865333139 | BOK | FALSE | Downregulated |
| -0,377038288 | 0,865333139 | SNF8 | FALSE | Downregulated |
| 0,427331657 | 0,865333139 | TNK1 | FALSE | Upregulated |
| 0,351782963 | 0,865663993 | NUP188 | FALSE | Upregulated |
| -0,326053028 | 0,865763412 | BZRAP1 | FALSE | Downregulated |
| 0,294782987 | 0,866050876 | ABL1 | FALSE | Upregulated |
| -0,302658547 | 0,866050876 | CYC1 | FALSE | Downregulated |
| 0,300037173 | 0,866050876 | RIPK1 | FALSE | Upregulated |
| -0,371443599 | 0,866535374 | LOC100506504 | FALSE | Downregulated |
| -0,277163972 | 0,866992654 | DNAJB7 | FALSE | Downregulated |
| 0,29029935 | 0,866992654 | TM4SF4 | FALSE | Upregulated |
| -0,390057825 | 0,867144295 | IP6K1 | FALSE | Downregulated |
| -0,291820938 | 0,867894159 | ABCA5 | FALSE | Downregulated |
| 0,383533835 | 0,867894159 | BAIAP3 | FALSE | Upregulated |
| -0,380930312 | 0,867894159 | NAALAD2 | FALSE | Downregulated |
| -0,403407969 | 0,867894159 | OXA1L | FALSE | Downregulated |
| -0,381835121 | 0,868204918 | C16orf45 | FALSE | Downregulated |
| -0,383077583 | 0,868204918 | PHF20 | FALSE | Downregulated |
| 0,383041754 | 0,868749678 | CALHM2 | FALSE | Upregulated |
| 0,275363986 | 0,868749678 | CAMKK2 | FALSE | Upregulated |
| 0,328470918 | 0,868749678 | CAPN1 | FALSE | Upregulated |
| 0,309701904 | 0,868749678 | LOC101928291 | FALSE | Upregulated |
| 0,259211805 | 0,868749678 | SDCBP2-AS1 | FALSE | Upregulated |
| -0,201868409 | 0,868749678 | SYT2 | FALSE | Downregulated |
| -0,186795073 | 0,868786764 | BACE1 | FALSE | Downregulated |
| -0,367918331 | 0,869100299 | EMC3 | FALSE | Downregulated |
| -0,291997525 | 0,869100299 | ICA1L | FALSE | Downregulated |
| -0,333185136 | 0,869100299 | SDHD | FALSE | Downregulated |
| -0,338335138 | 0,869289789 | PAPD7 | FALSE | Downregulated |
| -0,32101504 | 0,869680423 | CNST | FALSE | Downregulated |
| -0,23937083 | 0,870015076 | JMJD1C-AS1 | FALSE | Downregulated |
| 0,37412829 | 0,870482216 | ATXN10 | FALSE | Upregulated |
| 0,432021449 | 0,870482216 | CYBB | FALSE | Upregulated |
| -0,388351636 | 0,870482216 | DHX9 | FALSE | Downregulated |
| 0,273858558 | 0,870482216 | GORASP1 | FALSE | Upregulated |
| 0,253033155 | 0,870482216 | KDELR3 | FALSE | Upregulated |
| -0,177454236 | 0,870482216 | LOC102723521 | FALSE | Downregulated |
| -0,332287856 | 0,870482216 | POFUT1 | FALSE | Downregulated |
| -0,187947636 | 0,870482216 | PTP4A1 | FALSE | Downregulated |
| -0,241292411 | 0,870482216 | RBM39 | FALSE | Downregulated |
| -0,217702815 | 0,870482216 | UBN1 | FALSE | Downregulated |

| | | | | |
|---|---|---|---|---|
| 0,377559626 | 0,870482216 | VDAC2 | FALSE | Upregulated |
| -0,358297212 | 0,870653445 | LOC101928917 | FALSE | Downregulated |
| -0,407756557 | 0,870908408 | SERINC3 | FALSE | Downregulated |
| -0,369912066 | 0,872522568 | EXD3 | FALSE | Downregulated |
| -0,285993047 | 0,872533987 | LRRC37A3 | FALSE | Downregulated |
| 0,243147485 | 0,872859307 | KIAA2018 | FALSE | Upregulated |
| 0,444233027 | 0,872859307 | RP11-401P9.5 | FALSE | Upregulated |
| 0,41190928 | 0,872872096 | LOC100129917 | FALSE | Upregulated |
| -0,316143527 | 0,873533117 | MORF4L1 | FALSE | Downregulated |
| -0,30659097 | 0,873533117 | ORC4 | FALSE | Downregulated |
| 0,247123209 | 0,873533117 | PSMC3IP | FALSE | Upregulated |
| 0,343825182 | 0,873596932 | ALG12 | FALSE | Upregulated |
| 0,244536101 | 0,873596932 | IRF7 | FALSE | Upregulated |
| -0,304401729 | 0,873596932 | RAD23A | FALSE | Downregulated |
| 0,324519635 | 0,874515307 | NUDCD3 | FALSE | Upregulated |
| 0,379758036 | 0,874590101 | DOCK9 | FALSE | Upregulated |
| -0,405975406 | 0,874590101 | NLRC4 | FALSE | Downregulated |
| -0,279747914 | 0,874590101 | POU5F1P4 | FALSE | Downregulated |
| -0,178690615 | 0,874590101 | USP34 | FALSE | Downregulated |
| -0,353158514 | 0,874590101 | ZNF740 | FALSE | Downregulated |
| -0,233548103 | 0,874971561 | A2M-AS1 | FALSE | Downregulated |
| 0,353493537 | 0,874971561 | ARL5B | FALSE | Upregulated |
| 0,214415573 | 0,874971561 | BGN | FALSE | Upregulated |
| 0,309355877 | 0,874971561 | C16orf70 | FALSE | Upregulated |
| 0,361691782 | 0,874971561 | COTL1 | FALSE | Upregulated |
| 0,313281406 | 0,874971561 | EGLN2 | FALSE | Upregulated |
| 0,390622155 | 0,874971561 | GNA13 | FALSE | Upregulated |
| -0,701536417 | 0,874971561 | LOC101929309 | FALSE | Downregulated |
| 0,340299592 | 0,874971561 | LOC101929781 | FALSE | Upregulated |
| -0,296621005 | 0,874971561 | N4BP2 | FALSE | Downregulated |
| -0,208218679 | 0,874971561 | NRBP2 | FALSE | Downregulated |
| -0,310243549 | 0,874971561 | NSUN6 | FALSE | Downregulated |
| -0,331227619 | 0,874971561 | OPA1 | FALSE | Downregulated |
| 0,254668051 | 0,874971561 | PHC2 | FALSE | Upregulated |
| -0,251908993 | 0,874971561 | PJA2 | FALSE | Downregulated |
| -0,41095311 | 0,874971561 | PLN | FALSE | Downregulated |
| 0,331250822 | 0,874971561 | PPAP2A | FALSE | Upregulated |
| -0,424241745 | 0,874971561 | RNASE10 | FALSE | Downregulated |
| -0,321412951 | 0,874971561 | SHPRH | FALSE | Downregulated |
| 0,362541583 | 0,874971561 | TIRAP | FALSE | Upregulated |
| -0,19975439 | 0,874971561 | TRANK1 | FALSE | Downregulated |
| -0,330261071 | 0,874971561 | WBP2 | FALSE | Downregulated |
| -0,336229607 | 0,875408847 | BACH1 | FALSE | Downregulated |
| 0,222736006 | 0,875527149 | MTSS1L | FALSE | Upregulated |
| 0,315556011 | 0,87561645 | CAMK1D | FALSE | Upregulated |
| -0,39818719 | 0,87561645 | LOC389834 | FALSE | Downregulated |
| -0,221391017 | 0,87561645 | REPIN1 | FALSE | Downregulated |
| -0,278384825 | 0,875745402 | MYL6 | FALSE | Downregulated |
| -0,402869072 | 0,876143231 | ACTR8 | FALSE | Downregulated |
| -0,262785288 | 0,87619256 | ERCC6L | FALSE | Downregulated |
| 0,277723074 | 0,87619256 | PTPRG | FALSE | Upregulated |

| | | | | |
|---|---|---|---|---|
| -0,273838021 | 0,87619256 | RHOBTB3 | FALSE | Downregulated |
| -0,288813118 | 0,87619256 | UBXN4 | FALSE | Downregulated |
| 0,397386586 | 0,876203267 | ARPC4 | FALSE | Upregulated |
| -0,260416271 | 0,876203267 | GPC6 | FALSE | Downregulated |
| 0,250866446 | 0,876203267 | SHANK2-AS3 | FALSE | Upregulated |
| 0,224442393 | 0,876306288 | GDAP1L1 | FALSE | Upregulated |
| 0,310590427 | 0,876356339 | ACTR1A | FALSE | Upregulated |
| -0,310318329 | 0,876356339 | FAM210B | FALSE | Downregulated |
| 0,399631231 | 0,876356339 | HCP5 | FALSE | Upregulated |
| 0,375937495 | 0,876356339 | HPCAL1 | FALSE | Upregulated |
| 0,340951467 | 0,876356339 | IFITM2 | FALSE | Upregulated |
| 0,258540674 | 0,876356339 | POLRMT | FALSE | Upregulated |
| -0,237444601 | 0,876356339 | PTBP1 | FALSE | Downregulated |
| -0,220315374 | 0,876527992 | CLDN1 | FALSE | Downregulated |
| -0,28069549 | 0,876973095 | KLHDC10 | FALSE | Downregulated |
| -0,289385574 | 0,877583366 | SCYL1 | FALSE | Downregulated |
| -0,343322992 | 0,878366657 | MERTK | FALSE | Downregulated |
| 0,312911729 | 0,878366657 | NAGLU | FALSE | Upregulated |
| -0,447160433 | 0,878366657 | NDUFS7 | FALSE | Downregulated |
| 0,40408824 | 0,878366657 | RAB18 | FALSE | Upregulated |
| -0,397996383 | 0,878366657 | RPRD1B | FALSE | Downregulated |
| -0,352700804 | 0,878366657 | TXNRD2 | FALSE | Downregulated |
| -0,394825665 | 0,879365843 | DDT | FALSE | Downregulated |
| 0,251965471 | 0,879365843 | DHRS12 | FALSE | Upregulated |
| -0,16333201 | 0,879365843 | HUWE1 | FALSE | Downregulated |
| 0,434401179 | 0,879365843 | NRBP1 | FALSE | Upregulated |
| 0,19986585 | 0,879365843 | PLAC4 | FALSE | Upregulated |
| 0,244849136 | 0,879365843 | QKI | FALSE | Upregulated |
| -0,439219607 | 0,879365843 | SIPA1L2 | FALSE | Downregulated |
| 0,339658888 | 0,879365843 | THBS3 | FALSE | Upregulated |
| 0,396891965 | 0,879365843 | TSFM | FALSE | Upregulated |
| -0,346449491 | 0,879365843 | UBE2K | FALSE | Downregulated |
| 0,281772019 | 0,879557718 | GPM6B | FALSE | Upregulated |
| -0,319255832 | 0,879633647 | SMAD1-AS2 | FALSE | Downregulated |
| 0,405171052 | 0,880183818 | ATP6V1B2 | FALSE | Upregulated |
| 0,360763668 | 0,880183818 | FBXO21 | FALSE | Upregulated |
| 0,324691705 | 0,88030864 | TCTA | FALSE | Upregulated |
| 0,181977496 | 0,881126233 | MMP24 | FALSE | Upregulated |
| -0,255611025 | 0,881573291 | INSIG1 | FALSE | Downregulated |
| -0,317957598 | 0,881660557 | CPSF3L | FALSE | Downregulated |
| 0,274509655 | 0,881719923 | KAZN | FALSE | Upregulated |
| -0,385650644 | 0,881961981 | TXN2 | FALSE | Downregulated |
| 0,192877001 | 0,882036209 | PRR26 | FALSE | Upregulated |
| -0,382557322 | 0,882050679 | DDX20 | FALSE | Downregulated |
| -0,22836349 | 0,882050679 | ZBTB5 | FALSE | Downregulated |
| -0,336126843 | 0,882050679 | ZNF91 | FALSE | Downregulated |
| -0,287081078 | 0,882204259 | TMEM131 | FALSE | Downregulated |
| -0,472484887 | 0,882204259 | UPP2 | FALSE | Downregulated |
| -0,241774833 | 0,882378875 | GTF2IRD1P1 | FALSE | Downregulated |
| 0,276766326 | 0,882378875 | PIK3C2A | FALSE | Upregulated |
| 0,260448203 | 0,882378875 | TSC1 | FALSE | Upregulated |

| | | | | |
|---|---|---|---|---|
| 0,307906922 | 0,882414676 | C1QA | FALSE | Upregulated |
| 0,232128032 | 0,882414676 | C9orf84 | FALSE | Upregulated |
| 0,198640339 | 0,882414676 | ERAP1 | FALSE | Upregulated |
| 0,206756156 | 0,882414676 | KCNRG | FALSE | Upregulated |
| -0,311417544 | 0,882414676 | MED16 | FALSE | Downregulated |
| 0,260194216 | 0,882414676 | OGFR | FALSE | Upregulated |
| 0,349316613 | 0,882414676 | PLCH2 | FALSE | Upregulated |
| 0,169357651 | 0,882414676 | PLXNB2 | FALSE | Upregulated |
| 0,342006018 | 0,882414676 | PSMC4 | FALSE | Upregulated |
| 0,257573482 | 0,882414676 | RNF220 | FALSE | Upregulated |
| -0,269797636 | 0,882414676 | SLC43A1 | FALSE | Downregulated |
| 0,267796306 | 0,882414676 | TMEM248 | FALSE | Upregulated |
| 0,21323341 | 0,882414676 | USP35 | FALSE | Upregulated |
| -0,377228626 | 0,882523045 | FAM73B | FALSE | Downregulated |
| -0,254922199 | 0,882606673 | MGEA5 | FALSE | Downregulated |
| -0,27314951 | 0,883349813 | SH2B1 | FALSE | Downregulated |
| -0,291078415 | 0,883749074 | SAMD10 | FALSE | Downregulated |
| 0,332129266 | 0,883762933 | FAM129B | FALSE | Upregulated |
| -0,18826636 | 0,883775777 | LRRC37A4P | FALSE | Downregulated |
| -0,184951703 | 0,883780554 | COX20 | FALSE | Downregulated |
| 0,372899801 | 0,883780554 | MYL12B | FALSE | Upregulated |
| 0,250442748 | 0,883780554 | NRP1 | FALSE | Upregulated |
| 0,331110288 | 0,884051172 | ARHGAP26 | FALSE | Upregulated |
| 0,358283792 | 0,884171395 | ANKIB1 | FALSE | Upregulated |
| 0,335573479 | 0,884171395 | C17orf47 | FALSE | Upregulated |
| -0,32473664 | 0,884171395 | FAM219B | FALSE | Downregulated |
| 0,242393062 | 0,884171395 | MLEC | FALSE | Upregulated |
| 0,303658748 | 0,884171395 | NELFE | FALSE | Upregulated |
| 0,254274374 | 0,884171395 | NFX1 | FALSE | Upregulated |
| -0,333565496 | 0,884171395 | PABPC1L | FALSE | Downregulated |
| -0,170443463 | 0,884171395 | POM121 | FALSE | Downregulated |
| 0,322753216 | 0,884171395 | PP2D1 | FALSE | Upregulated |
| 0,39298865 | 0,884171395 | RFWD2 | FALSE | Upregulated |
| -0,675838538 | 0,884171395 | RNA28S5 | FALSE | Downregulated |
| -0,217624533 | 0,884171395 | RPL12 | FALSE | Downregulated |
| 0,345727561 | 0,884171395 | VPS53 | FALSE | Upregulated |
| -0,346564878 | 0,884171395 | ZNF33B | FALSE | Downregulated |
| 0,273467519 | 0,884171395 | ZNF655 | FALSE | Upregulated |
| 0,334279718 | 0,884171395 | ZSCAN16 | FALSE | Upregulated |
| -0,347990287 | 0,885821327 | BRD3 | FALSE | Downregulated |
| -0,297314641 | 0,885821327 | MLH3 | FALSE | Downregulated |
| -0,270718229 | 0,885821327 | PI4KB | FALSE | Downregulated |
| 0,313573851 | 0,885821327 | PPP1R14B | FALSE | Upregulated |
| -0,318647507 | 0,886006048 | CHCHD2 | FALSE | Downregulated |
| -0,376072975 | 0,88630385 | MEG3 | FALSE | Downregulated |
| -0,349028176 | 0,886420693 | DNAJB1 | FALSE | Downregulated |
| -0,282404215 | 0,886757105 | H2AFV | FALSE | Downregulated |
| -0,243103213 | 0,886757105 | KIAA1432 | FALSE | Downregulated |
| 0,353658626 | 0,886923512 | ARFRP1 | FALSE | Upregulated |
| -0,236737089 | 0,886923512 | C11orf57 | FALSE | Downregulated |
| 0,25614757 | 0,886923512 | CHD9 | FALSE | Upregulated |

| | | | | |
|---|---|---|---|---|
| 0,296689588 | 0,886923512 | COLGALT1 | FALSE | Upregulated |
| -0,217493332 | 0,886923512 | DENND1C | FALSE | Downregulated |
| 0,200545305 | 0,886923512 | GOLGA8B | FALSE | Upregulated |
| -0,322331036 | 0,886923512 | HEBP2 | FALSE | Downregulated |
| -0,300992668 | 0,886923512 | HNMT | FALSE | Downregulated |
| -0,28678123 | 0,886923512 | IFNAR1 | FALSE | Downregulated |
| 0,410307005 | 0,886923512 | JDP2 | FALSE | Upregulated |
| 0,33616455 | 0,886923512 | PERP | FALSE | Upregulated |
| -0,249571638 | 0,886923512 | RP11-61I13.3 | FALSE | Downregulated |
| -0,197913807 | 0,886923512 | RPS3 | FALSE | Downregulated |
| 0,384821806 | 0,886923512 | SRCIN1 | FALSE | Upregulated |
| -0,382080327 | 0,886923512 | TBC1D13 | FALSE | Downregulated |
| -0,299753036 | 0,886923512 | ZCCHC11 | FALSE | Downregulated |
| -0,318844656 | 0,886923512 | ZNF865 | FALSE | Downregulated |
| 0,321762628 | 0,887150369 | CEBPZ-AS1 | FALSE | Upregulated |
| 0,288050786 | 0,887419661 | DNAJC13 | FALSE | Upregulated |
| -0,215862253 | 0,887419661 | NTN1 | FALSE | Downregulated |
| -0,354844345 | 0,887805705 | LIX1L | FALSE | Downregulated |
| 0,252818955 | 0,887805705 | LOC101928862 | FALSE | Upregulated |
| -0,377461351 | 0,887805705 | MSANTD2 | FALSE | Downregulated |
| -0,347551733 | 0,887805705 | SEMA5A | FALSE | Downregulated |
| -0,372445972 | 0,887805705 | TMEM175 | FALSE | Downregulated |
| -0,390062664 | 0,888213716 | AHCTF1 | FALSE | Downregulated |
| 0,237632667 | 0,888213716 | CC2D1B | FALSE | Upregulated |
| -0,291696362 | 0,888213716 | EHD3 | FALSE | Downregulated |
| -0,372811723 | 0,888213716 | HBS1L | FALSE | Downregulated |
| -0,301893873 | 0,888213716 | HEMK1 | FALSE | Downregulated |
| 0,24426592 | 0,888213716 | JAZF1 | FALSE | Upregulated |
| 0,322027605 | 0,888213716 | LOC641367 | FALSE | Upregulated |
| 0,394897701 | 0,888213716 | NPIPA5 | FALSE | Upregulated |
| 0,231079752 | 0,888213716 | PRSS12 | FALSE | Upregulated |
| -0,350850625 | 0,888213716 | PWRN2 | FALSE | Downregulated |
| -0,29587782 | 0,888213716 | RLIM | FALSE | Downregulated |
| -0,336727441 | 0,888213716 | RP11-84C10.4 | FALSE | Downregulated |
| -0,345876818 | 0,888213716 | STARD13 | FALSE | Downregulated |
| 0,253941903 | 0,888213716 | TKT | FALSE | Upregulated |
| -0,22563757 | 0,888213716 | TTC38 | FALSE | Downregulated |
| 0,341580803 | 0,888213716 | YIPF2 | FALSE | Upregulated |
| -0,271018777 | 0,888213716 | YME1L1 | FALSE | Downregulated |
| 0,268120888 | 0,888213716 | ZBTB44 | FALSE | Upregulated |
| -0,32272316 | 0,888350499 | PKN2 | FALSE | Downregulated |
| 0,328144756 | 0,888368397 | NAP1L1 | FALSE | Upregulated |
| -0,250838586 | 0,888368397 | SERINC1 | FALSE | Downregulated |
| 0,259887756 | 0,888916196 | AIRN | FALSE | Upregulated |
| -0,37199969 | 0,889133357 | NID2 | FALSE | Downregulated |
| -0,208301042 | 0,889280092 | C20orf112 | FALSE | Downregulated |
| 0,397052052 | 0,889452102 | HNF1B | FALSE | Upregulated |
| -0,299065654 | 0,889452102 | PRKAR2A-AS1 | FALSE | Downregulated |
| 0,200954657 | 0,889452102 | SLC35E2B | FALSE | Upregulated |
| 0,261598947 | 0,889546773 | PKP4 | FALSE | Upregulated |
| 0,205021113 | 0,889553598 | LOC652276 | FALSE | Upregulated |

| | | | | |
|---|---|---|---|---|
| -0,352803164 | 0,889828668 | DIS3 | FALSE | Downregulated |
| -0,377990123 | 0,889828668 | SGK2 | FALSE | Downregulated |
| 0,35916931 | 0,890302425 | C17orf85 | FALSE | Upregulated |
| -0,317587628 | 0,890302425 | NAP1L5 | FALSE | Downregulated |
| 0,35053461 | 0,890302425 | ZNF704 | FALSE | Upregulated |
| 0,201453916 | 0,890454209 | C19orf44 | FALSE | Upregulated |
| -0,232731372 | 0,890454209 | CTC1 | FALSE | Downregulated |
| -0,254680629 | 0,890454209 | SIAH2 | FALSE | Downregulated |
| -0,395728437 | 0,890504396 | ANKRD26 | FALSE | Downregulated |
| 0,183624396 | 0,890504396 | APC2 | FALSE | Upregulated |
| 0,219313727 | 0,890504396 | ATP2A2 | FALSE | Upregulated |
| 0,324322783 | 0,890504396 | ATP9B | FALSE | Upregulated |
| -0,460273905 | 0,890504396 | BCL2L11 | FALSE | Downregulated |
| 0,259482849 | 0,890504396 | BSG | FALSE | Upregulated |
| -0,363567769 | 0,890504396 | CAPZB | FALSE | Downregulated |
| 0,18606076 | 0,890504396 | CASKIN1 | FALSE | Upregulated |
| 0,21086168 | 0,890504396 | CCNI | FALSE | Upregulated |
| 0,328778399 | 0,890504396 | CYB5D2 | FALSE | Upregulated |
| 0,222371971 | 0,890504396 | DLGAP4 | FALSE | Upregulated |
| -0,240759257 | 0,890504396 | DNMBP-AS1 | FALSE | Downregulated |
| 0,297156109 | 0,890504396 | ENPEP | FALSE | Upregulated |
| 0,282044081 | 0,890504396 | EPG5 | FALSE | Upregulated |
| 0,334809664 | 0,890504396 | EPM2AIP1 | FALSE | Upregulated |
| -0,313550976 | 0,890504396 | EVI5 | FALSE | Downregulated |
| 0,208639754 | 0,890504396 | FANCI | FALSE | Upregulated |
| 0,354128699 | 0,890504396 | FGD5 | FALSE | Upregulated |
| 0,354262413 | 0,890504396 | GMPPB | FALSE | Upregulated |
| 0,172457144 | 0,890504396 | GNAL | FALSE | Upregulated |
| 0,278077368 | 0,890504396 | GPR22 | FALSE | Upregulated |
| 0,310824535 | 0,890504396 | HERC4 | FALSE | Upregulated |
| 0,270406536 | 0,890504396 | HIATL1 | FALSE | Upregulated |
| -0,277679087 | 0,890504396 | IGFBP2 | FALSE | Downregulated |
| -0,238332785 | 0,890504396 | IL12RB2 | FALSE | Downregulated |
| 0,162409143 | 0,890504396 | INSR | FALSE | Upregulated |
| 0,349532156 | 0,890504396 | KIF2A | FALSE | Upregulated |
| -0,338485605 | 0,890504396 | LARS | FALSE | Downregulated |
| -0,323352703 | 0,890504396 | LINC00174 | FALSE | Downregulated |
| 0,251580554 | 0,890504396 | LOC102723600 | FALSE | Upregulated |
| 0,382155541 | 0,890504396 | LOC606724 | FALSE | Upregulated |
| -0,364198421 | 0,890504396 | LOC643072 | FALSE | Downregulated |
| 0,297186693 | 0,890504396 | LYPLA2 | FALSE | Upregulated |
| -0,287355032 | 0,890504396 | MARCH7 | FALSE | Downregulated |
| -0,326806949 | 0,890504396 | MRPL41 | FALSE | Downregulated |
| 0,320982076 | 0,890504396 | MRPS14 | FALSE | Upregulated |
| -0,363389087 | 0,890504396 | NAA50 | FALSE | Downregulated |
| -0,363869268 | 0,890504396 | NAPRT1 | FALSE | Downregulated |
| -0,225459029 | 0,890504396 | NUDT16 | FALSE | Downregulated |
| 0,229942448 | 0,890504396 | NUFIP2 | FALSE | Upregulated |
| -0,183803627 | 0,890504396 | P4HB | FALSE | Downregulated |
| 0,313050509 | 0,890504396 | PATZ1 | FALSE | Upregulated |
| 0,235506604 | 0,890504396 | PHACTR1 | FALSE | Upregulated |

| | | | | |
|---|---|---|---|---|
| 0,291202371 | 0,890504396 | PLEKHH1 | FALSE | Upregulated |
| 0,363739565 | 0,890504396 | PPM1E | FALSE | Upregulated |
| 0,237566603 | 0,890504396 | PTPRB | FALSE | Upregulated |
| 0,237305057 | 0,890504396 | RNF10 | FALSE | Upregulated |
| -0,231695343 | 0,890504396 | RP11-219B17.1 | FALSE | Downregulated |
| 0,231863801 | 0,890504396 | RUSC2 | FALSE | Upregulated |
| -0,343899002 | 0,890504396 | SIX5 | FALSE | Downregulated |
| -0,27908512 | 0,890504396 | SLC12A6 | FALSE | Downregulated |
| -0,298608498 | 0,890504396 | SLC43A3 | FALSE | Downregulated |
| -0,271740462 | 0,890504396 | SLC47A2 | FALSE | Downregulated |
| 0,243063218 | 0,890504396 | TP53INP1 | FALSE | Upregulated |
| -0,219263378 | 0,890504396 | TSSK3 | FALSE | Downregulated |
| 0,364538765 | 0,890504396 | WDR5 | FALSE | Upregulated |
| 0,198831084 | 0,890504396 | ZFP36L2 | FALSE | Upregulated |
| -0,373261135 | 0,890504396 | ZNF205 | FALSE | Downregulated |
| -0,374022756 | 0,890504396 | ZNF236 | FALSE | Downregulated |
| -0,226680409 | 0,890504396 | ZNF638 | FALSE | Downregulated |
| 0,244902739 | 0,890613969 | ATP8B1 | FALSE | Upregulated |
| 0,335189475 | 0,890613969 | GPATCH11 | FALSE | Upregulated |
| 0,283245598 | 0,890613969 | LOC101928188 | FALSE | Upregulated |
| 0,354018805 | 0,890613969 | LRRC41 | FALSE | Upregulated |
| -0,22070974 | 0,890613969 | ZHX3 | FALSE | Downregulated |
| 0,327541824 | 0,890643218 | ZNF417 | FALSE | Upregulated |
| 0,187078661 | 0,890974365 | DYNC1H1 | FALSE | Upregulated |
| -0,358176657 | 0,891229854 | CLPX | FALSE | Downregulated |
| -0,303184062 | 0,891229854 | SMC1A | FALSE | Downregulated |
| -0,332432552 | 0,891229854 | UBE2O | FALSE | Downregulated |
| -0,265853497 | 0,891385106 | RPS25 | FALSE | Downregulated |
| 0,301019108 | 0,89160006 | MYO1D | FALSE | Upregulated |
| -0,240699483 | 0,891774266 | ATP5B | FALSE | Downregulated |
| 0,328477157 | 0,891774266 | PDE8B | FALSE | Upregulated |
| 0,306001951 | 0,891774266 | USP10 | FALSE | Upregulated |
| -0,202023293 | 0,891774266 | ZC3H11A | FALSE | Downregulated |
| 0,337757294 | 0,89241365 | CCDC28B | FALSE | Upregulated |
| 0,304018289 | 0,892433353 | UBAP1 | FALSE | Upregulated |
| -0,389466017 | 0,892688237 | LOC101929986 | FALSE | Downregulated |
| -0,33265936 | 0,892688237 | UBAP2L | FALSE | Downregulated |
| -0,307491815 | 0,892779437 | AATK | FALSE | Downregulated |
| -0,187299765 | 0,892923098 | F10 | FALSE | Downregulated |
| -0,322910396 | 0,892923098 | FOXRED2 | FALSE | Downregulated |
| 0,261736679 | 0,893017998 | ETF1 | FALSE | Upregulated |
| 0,362334033 | 0,893195095 | CMTM3 | FALSE | Upregulated |
| -0,244662186 | 0,893231606 | RPL7 | FALSE | Downregulated |
| 0,333348725 | 0,893527652 | CHUK | FALSE | Upregulated |
| -0,317635694 | 0,893527652 | HHIP | FALSE | Downregulated |
| 0,286588728 | 0,893527652 | TRAF7 | FALSE | Upregulated |
| -0,279590603 | 0,8940421 | TNFRSF6B | FALSE | Downregulated |
| -0,216219581 | 0,894260834 | RAB15 | FALSE | Downregulated |
| 0,33265823 | 0,894260834 | SLK | FALSE | Upregulated |
| -0,230066237 | 0,89449414 | ABCA8 | FALSE | Downregulated |
| 0,335676701 | 0,89449414 | MTG2 | FALSE | Upregulated |

| | | | | |
|---|---|---|---|---|
| 0,250072936 | 0,89449414 | WASF2 | FALSE | Upregulated |
| 0,205356034 | 0,8945618 | ERGIC1 | FALSE | Upregulated |
| 0,319041899 | 0,8945618 | FAM110C | FALSE | Upregulated |
| -0,239190872 | 0,8945618 | TPI1 | FALSE | Downregulated |
| -0,327452972 | 0,894605042 | CERS6 | FALSE | Downregulated |
| 0,174931134 | 0,894605042 | LOC100288162 | FALSE | Upregulated |
| 0,329568287 | 0,894605042 | MKL2 | FALSE | Upregulated |
| 0,296023466 | 0,894605042 | PDDC1 | FALSE | Upregulated |
| 0,309595835 | 0,894605042 | RNF43 | FALSE | Upregulated |
| -0,305760522 | 0,894605042 | ZC3H14 | FALSE | Downregulated |
| -0,378352942 | 0,894618147 | TJP3 | FALSE | Downregulated |
| -0,376221687 | 0,894804257 | ALKBH7 | FALSE | Downregulated |
| -0,201400656 | 0,894804257 | MPHOSPH8 | FALSE | Downregulated |
| 0,313325608 | 0,894882078 | PRDM2 | FALSE | Upregulated |
| 0,245991966 | 0,894898371 | PRKCA | FALSE | Upregulated |
| -0,298318887 | 0,895093214 | LOC101928415 | FALSE | Downregulated |
| -0,376944308 | 0,895093214 | OSBPL11 | FALSE | Downregulated |
| -0,3248886 | 0,895284853 | MCCC1 | FALSE | Downregulated |
| -0,267885271 | 0,895365702 | SEC61A2 | FALSE | Downregulated |
| 0,229075703 | 0,895582831 | TRAPPC12 | FALSE | Upregulated |
| -0,222688902 | 0,895703782 | ATP7B | FALSE | Downregulated |
| 0,378033119 | 0,895703782 | CYP2B7P | FALSE | Upregulated |
| -0,24667606 | 0,895703782 | PTPLAD1 | FALSE | Downregulated |
| -0,25630653 | 0,897022726 | LMO7 | FALSE | Downregulated |
| -0,371667689 | 0,89722299 | A2M | FALSE | Downregulated |
| 0,327467574 | 0,89722299 | AMZ1 | FALSE | Upregulated |
| -0,306272381 | 0,89722299 | ANO9 | FALSE | Downregulated |
| -0,237864546 | 0,89722299 | ARRDC1 | FALSE | Downregulated |
| -0,304990716 | 0,89722299 | ASH2L | FALSE | Downregulated |
| 0,320920295 | 0,89722299 | ATP9A | FALSE | Upregulated |
| 0,248195629 | 0,89722299 | BRAF | FALSE | Upregulated |
| -0,207392488 | 0,89722299 | C1orf115 | FALSE | Downregulated |
| 0,345815606 | 0,89722299 | C5orf42 | FALSE | Upregulated |
| -0,319653229 | 0,89722299 | CARD8 | FALSE | Downregulated |
| -0,345750996 | 0,89722299 | CDK11B | FALSE | Downregulated |
| -0,351340032 | 0,89722299 | CELF3 | FALSE | Downregulated |
| 0,287825313 | 0,89722299 | CRYZ | FALSE | Upregulated |
| 0,30989615 | 0,89722299 | CYP2R1 | FALSE | Upregulated |
| -0,340675226 | 0,89722299 | EIF1AX | FALSE | Downregulated |
| -0,195036274 | 0,89722299 | FAM8A1 | FALSE | Downregulated |
| -0,353691621 | 0,89722299 | GTF3C2 | FALSE | Downregulated |
| 0,336232285 | 0,89722299 | HIF1A | FALSE | Upregulated |
| 0,284076621 | 0,89722299 | LOH12CR1 | FALSE | Upregulated |
| -0,207029378 | 0,89722299 | LTBP3 | FALSE | Downregulated |
| 0,26802503 | 0,89722299 | MBP | FALSE | Upregulated |
| 0,251534102 | 0,89722299 | METTL12 | FALSE | Upregulated |
| 0,335700358 | 0,89722299 | NUDT21 | FALSE | Upregulated |
| 0,350563036 | 0,89722299 | PSMB2 | FALSE | Upregulated |
| 0,291353688 | 0,89722299 | SDHAP2 | FALSE | Upregulated |
| -0,239726673 | 0,89722299 | SERPINA11 | FALSE | Downregulated |
| -0,332865081 | 0,89722299 | SNX2 | FALSE | Downregulated |

| | | | | |
|---|---|---|---|---|
| -0,133073288 | 0,89722299 | SRRM2 | FALSE | Downregulated |
| 0,234972442 | 0,89722299 | TBC1D16 | FALSE | Upregulated |
| 0,276073204 | 0,89722299 | TECPR1 | FALSE | Upregulated |
| -0,328258809 | 0,89722299 | TOP3A | FALSE | Downregulated |
| 0,329236636 | 0,89722299 | TSR3 | FALSE | Upregulated |
| -0,350474022 | 0,89722299 | UFM1 | FALSE | Downregulated |
| -0,174157281 | 0,89722299 | VASH1 | FALSE | Downregulated |
| -0,265415604 | 0,897735148 | TBC1D24 | FALSE | Downregulated |
| -0,233742233 | 0,897885005 | CEP350 | FALSE | Downregulated |
| -0,286842033 | 0,897885005 | LINC00671 | FALSE | Downregulated |
| 0,275720852 | 0,897885005 | MAN1B1 | FALSE | Upregulated |
| 0,217791609 | 0,897885005 | RP11-143J12.2 | FALSE | Upregulated |
| -0,249865258 | 0,897941579 | DICER1 | FALSE | Downregulated |
| 0,436616389 | 0,897941579 | RBMS3 | FALSE | Upregulated |
| 0,253882322 | 0,897941579 | TBC1D14 | FALSE | Upregulated |
| -0,336425976 | 0,898162377 | MSL2 | FALSE | Downregulated |
| -0,327330894 | 0,898211764 | TELO2 | FALSE | Downregulated |
| 0,320003826 | 0,89882178 | CCDC93 | FALSE | Upregulated |
| 0,19513794 | 0,89882178 | COL6A1 | FALSE | Upregulated |
| -0,27774412 | 0,89882178 | CTAGE8 | FALSE | Downregulated |
| -0,289507037 | 0,89882178 | GABPB1 | FALSE | Downregulated |
| 0,360668416 | 0,89882178 | NSUN5P1 | FALSE | Upregulated |
| 0,290391389 | 0,89882178 | ST3GAL2 | FALSE | Upregulated |
| 0,261455912 | 0,89882178 | TP53BP1 | FALSE | Upregulated |
| 0,260398072 | 0,898912843 | PDCD6IP | FALSE | Upregulated |
| -0,241146625 | 0,898912843 | PRPF6 | FALSE | Downregulated |
| -0,31244533 | 0,898912843 | SNAPC3 | FALSE | Downregulated |
| 0,237422163 | 0,898912843 | TLE1 | FALSE | Upregulated |
| 0,231543094 | 0,899185282 | CDS2 | FALSE | Upregulated |
| 0,18598494 | 0,899185282 | CTNND1 | FALSE | Upregulated |
| 0,238974594 | 0,899185282 | DTX3L | FALSE | Upregulated |
| -0,237432784 | 0,899185282 | FGD6 | FALSE | Downregulated |
| 0,245209591 | 0,899185282 | INTS6 | FALSE | Upregulated |
| -0,329019839 | 0,899185282 | MRPL38 | FALSE | Downregulated |
| 0,332469638 | 0,899185282 | NECAP2 | FALSE | Upregulated |
| -0,197346967 | 0,899185282 | PLEKHA6 | FALSE | Downregulated |
| -0,297104278 | 0,899185282 | PTP4A2 | FALSE | Downregulated |
| 0,344696048 | 0,899185282 | PWWP2A | FALSE | Upregulated |
| 0,275409697 | 0,899185282 | WTAP | FALSE | Upregulated |
| -0,220095399 | 0,899185282 | ZBTB20-AS1 | FALSE | Downregulated |
| 0,281396697 | 0,899185282 | ZNF654 | FALSE | Upregulated |
| 0,189769587 | 0,899521582 | HSP90AA1 | FALSE | Upregulated |
| 0,309753858 | 0,899521582 | LOC100293704 | FALSE | Upregulated |
| -0,239088192 | 0,899521582 | TMED8 | FALSE | Downregulated |
| -0,335953695 | 0,900785144 | F8 | FALSE | Downregulated |
| -0,288866298 | 0,900785144 | FKBP1A | FALSE | Downregulated |
| -0,337707587 | 0,900785144 | GTF2IRD2B | FALSE | Downregulated |
| -0,161437622 | 0,900785144 | ZSWIM8 | FALSE | Downregulated |
| 0,275091039 | 0,901537112 | RPRD2 | FALSE | Upregulated |
| -0,18911338 | 0,901537112 | SGPL1 | FALSE | Downregulated |
| -0,294649962 | 0,901537112 | TIA1 | FALSE | Downregulated |

| | | | | |
|---|---|---|---|---|
| -0,297454032 | 0,901610728 | AIFM1 | FALSE | Downregulated |
| -0,171199381 | 0,901610728 | AP1G2 | FALSE | Downregulated |
| -0,20311296 | 0,901610728 | RPL6 | FALSE | Downregulated |
| 0,144994945 | 0,90217011 | ANGPTL6 | FALSE | Upregulated |
| 0,204935071 | 0,90217011 | CRB2 | FALSE | Upregulated |
| -0,239692825 | 0,90217011 | NT5C2 | FALSE | Downregulated |
| -0,320106273 | 0,90217011 | ZKSCAN8 | FALSE | Downregulated |
| 0,221988074 | 0,90217011 | ZNF460 | FALSE | Upregulated |
| -0,306750534 | 0,902218964 | GUK1 | FALSE | Downregulated |
| -0,308178887 | 0,902228781 | LPIN1 | FALSE | Downregulated |
| 0,23013977 | 0,902522236 | ATF7IP | FALSE | Upregulated |
| -0,271141549 | 0,902522236 | CD37 | FALSE | Downregulated |
| -0,252985206 | 0,902522236 | CHTF8 | FALSE | Downregulated |
| -0,205160254 | 0,902522236 | EDEM1 | FALSE | Downregulated |
| 0,301264203 | 0,902522236 | FAM115C | FALSE | Upregulated |
| 0,241359065 | 0,902522236 | FLJ38668 | FALSE | Upregulated |
| 0,247865224 | 0,902522236 | ITPK1 | FALSE | Upregulated |
| -0,378031865 | 0,902522236 | LRRC39 | FALSE | Downregulated |
| -0,313110519 | 0,902522236 | MIF | FALSE | Downregulated |
| -0,308495768 | 0,902522236 | NDUFA6 | FALSE | Downregulated |
| -0,306983405 | 0,902522236 | NFYC-AS1 | FALSE | Downregulated |
| -0,172918396 | 0,902522236 | PHRF1 | FALSE | Downregulated |
| 0,304192949 | 0,902522236 | PIK3R3 | FALSE | Upregulated |
| 0,286192729 | 0,902522236 | RNF216 | FALSE | Upregulated |
| -0,308850591 | 0,902522236 | SS18 | FALSE | Downregulated |
| 0,180757 | 0,902522236 | STOM | FALSE | Upregulated |
| -0,241762036 | 0,902522236 | WNK3 | FALSE | Downregulated |
| 0,223301329 | 0,902522236 | ZBTB7A | FALSE | Upregulated |
| -0,268722461 | 0,902522236 | ZNF582 | FALSE | Downregulated |
| -0,294737991 | 0,902783267 | TOR3A | FALSE | Downregulated |
| -0,295032957 | 0,903558246 | ETS1 | FALSE | Downregulated |
| 0,283853868 | 0,903558246 | KCNIP4 | FALSE | Upregulated |
| -0,217589707 | 0,903558246 | MON2 | FALSE | Downregulated |
| 0,223582549 | 0,90357786 | TRAF3IP2 | FALSE | Upregulated |
| -0,230386829 | 0,903661529 | PPIB | FALSE | Downregulated |
| -0,248541624 | 0,903661529 | TCEB2 | FALSE | Downregulated |
| -0,310144309 | 0,903810609 | ANGEL1 | FALSE | Downregulated |
| 0,336262963 | 0,904041169 | NR0B2 | FALSE | Upregulated |
| 0,19870942 | 0,904162293 | C15orf60 | FALSE | Upregulated |
| -0,193470658 | 0,904162293 | MAN2A2 | FALSE | Downregulated |
| 0,314894454 | 0,904438102 | APOA1BP | FALSE | Upregulated |
| 0,235558906 | 0,904438102 | C2orf16 | FALSE | Upregulated |
| 0,239151156 | 0,904438102 | DLST | FALSE | Upregulated |
| -0,272984264 | 0,904438102 | FLJ10038 | FALSE | Downregulated |
| -0,258894291 | 0,904438102 | KLF12 | FALSE | Downregulated |
| 0,219809998 | 0,904438102 | LENG9 | FALSE | Upregulated |
| 0,304013472 | 0,904438102 | LOC100131270 | FALSE | Upregulated |
| 0,310230697 | 0,904438102 | LOC102724965 | FALSE | Upregulated |
| -0,302288026 | 0,904438102 | MAF1 | FALSE | Downregulated |
| -0,232438133 | 0,904438102 | PDE8A | FALSE | Downregulated |
| -0,24461946 | 0,904438102 | PXDC1 | FALSE | Downregulated |

| | | | | |
|---|---|---|---|---|
| -0,161944404 | 0,904438102 | TTPAL | FALSE | Downregulated |
| -0,315791553 | 0,904438102 | ZMYM2 | FALSE | Downregulated |
| 0,234975235 | 0,904589878 | LOC101929747 | FALSE | Upregulated |
| -0,232607653 | 0,904599348 | ATG2B | FALSE | Downregulated |
| -0,368161818 | 0,904599348 | CCDC92 | FALSE | Downregulated |
| -0,338690113 | 0,904599348 | CEP104 | FALSE | Downregulated |
| -0,324032097 | 0,904599348 | CPNE1 | FALSE | Downregulated |
| 0,263120356 | 0,904599348 | ITSN2 | FALSE | Upregulated |
| 0,251909994 | 0,904599348 | MYO1E | FALSE | Upregulated |
| -0,319209165 | 0,904599348 | NLRP9 | FALSE | Downregulated |
| 0,302968328 | 0,904599348 | PSMD5-AS1 | FALSE | Upregulated |
| -0,343455282 | 0,904599348 | PSME2 | FALSE | Downregulated |
| 0,284429482 | 0,904599348 | RAD9B | FALSE | Upregulated |
| 0,261898748 | 0,904599348 | RALY | FALSE | Upregulated |
| 0,23015285 | 0,904599348 | RPS6KL1 | FALSE | Upregulated |
| -0,28938841 | 0,904599348 | SAPCD2 | FALSE | Downregulated |
| -0,311798138 | 0,904599348 | SART1 | FALSE | Downregulated |
| 0,293069616 | 0,904599348 | UBE2Z | FALSE | Upregulated |
| -0,320319539 | 0,904963725 | SENP8 | FALSE | Downregulated |
| -0,323092919 | 0,9057938 | NUDC | FALSE | Downregulated |
| -0,26448401 | 0,9057938 | PLS3 | FALSE | Downregulated |
| -0,176175621 | 0,9057938 | ZNF3 | FALSE | Downregulated |
| -0,217695247 | 0,90581045 | TSPAN9 | FALSE | Downregulated |
| -0,230634152 | 0,90581301 | LDLRAD4 | FALSE | Downregulated |
| -0,211311348 | 0,905940329 | ACACA | FALSE | Downregulated |
| 0,24339595 | 0,905940329 | ACTR2 | FALSE | Upregulated |
| 0,286453523 | 0,905940329 | AGAP1 | FALSE | Upregulated |
| 0,327443875 | 0,905940329 | AGO4 | FALSE | Upregulated |
| 0,245301904 | 0,905940329 | AP4B1-AS1 | FALSE | Upregulated |
| 0,154539642 | 0,905940329 | CGNL1 | FALSE | Upregulated |
| -0,180747452 | 0,905940329 | CLSTN3 | FALSE | Downregulated |
| 0,257871909 | 0,905940329 | ELAC2 | FALSE | Upregulated |
| -0,234475059 | 0,905940329 | FAM126B | FALSE | Downregulated |
| -0,2210691 | 0,905940329 | LRPAP1 | FALSE | Downregulated |
| -0,343063213 | 0,905940329 | MTFR1 | FALSE | Downregulated |
| -0,177584673 | 0,905940329 | NFE2L3 | FALSE | Downregulated |
| -0,206101057 | 0,905940329 | PPP1R16A | FALSE | Downregulated |
| -0,176334752 | 0,905940329 | RPL27A | FALSE | Downregulated |
| 0,147955988 | 0,905940329 | SLC25A29 | FALSE | Upregulated |
| 0,317971657 | 0,905940329 | TBCEL | FALSE | Upregulated |
| 0,266625902 | 0,905940329 | ZNF644 | FALSE | Upregulated |
| 0,186531763 | 0,906264737 | ILF3 | FALSE | Upregulated |
| -0,290789828 | 0,906320861 | COCH | FALSE | Downregulated |
| 0,270906715 | 0,90643401 | IFIT2 | FALSE | Upregulated |
| -0,124294149 | 0,90643401 | PLEC | FALSE | Downregulated |
| -0,255927719 | 0,906464018 | MRPL49 | FALSE | Downregulated |
| 0,290135487 | 0,906704116 | BOLA3-AS1 | FALSE | Upregulated |
| 0,282966944 | 0,907192601 | LOC101927208 | FALSE | Upregulated |
| 0,181938565 | 0,907311551 | BRD4 | FALSE | Upregulated |
| -0,21189081 | 0,907598721 | CDH1 | FALSE | Downregulated |
| 0,256510759 | 0,907598721 | CEP170 | FALSE | Upregulated |

| | | | | |
|---|---|---|---|---|
| -0,20209457 | 0,907598721 | ENO1 | FALSE | Downregulated |
| -0,321743901 | 0,907598721 | LOC101927673 | FALSE | Downregulated |
| 0,236464542 | 0,907598721 | PXK | FALSE | Upregulated |
| -0,270623822 | 0,907598721 | USO1 | FALSE | Downregulated |
| -0,178089936 | 0,907667538 | ATRN | FALSE | Downregulated |
| 0,179800757 | 0,907667538 | SUPT6H | FALSE | Upregulated |
| -0,311343027 | 0,90802158 | NDUFA10 | FALSE | Downregulated |
| -0,2247502 | 0,908308039 | FAM87B | FALSE | Downregulated |
| -0,286654227 | 0,908559205 | RPL38 | FALSE | Downregulated |
| -0,194902605 | 0,908574724 | ATXN1 | FALSE | Downregulated |
| -0,297834765 | 0,908574724 | ELOVL5 | FALSE | Downregulated |
| -0,249457446 | 0,908574724 | ERN1 | FALSE | Downregulated |
| 0,208493393 | 0,908574724 | FYCO1 | FALSE | Upregulated |
| -0,211480417 | 0,908574724 | HEXDC | FALSE | Downregulated |
| -0,17710383 | 0,908574724 | KCNJ13 | FALSE | Downregulated |
| 0,179828552 | 0,908574724 | KCTD20 | FALSE | Upregulated |
| 0,276139387 | 0,908574724 | RNF219-AS1 | FALSE | Upregulated |
| -0,184101129 | 0,908574724 | ZNF106 | FALSE | Downregulated |
| -0,191526913 | 0,90903914 | GPT2 | FALSE | Downregulated |
| 0,258252297 | 0,910007657 | TCF7L2 | FALSE | Upregulated |
| 0,300780314 | 0,910175792 | PEG3 | FALSE | Upregulated |
| -0,24588875 | 0,910244344 | TM9SF3 | FALSE | Downregulated |
| 0,189695282 | 0,91035623 | CTNNB1 | FALSE | Upregulated |
| -0,308687474 | 0,91035623 | DNM3OS | FALSE | Downregulated |
| 0,200197563 | 0,91035623 | GGA3 | FALSE | Upregulated |
| -0,302531379 | 0,91035623 | GRSF1 | FALSE | Downregulated |
| -0,320391808 | 0,91035623 | HIST2H2AC | FALSE | Downregulated |
| 0,297997871 | 0,91035623 | INTS10 | FALSE | Upregulated |
| -0,289294279 | 0,91035623 | KEAP1 | FALSE | Downregulated |
| -0,24530667 | 0,91035623 | MTA1 | FALSE | Downregulated |
| -0,303655282 | 0,91035623 | MTFR1L | FALSE | Downregulated |
| 0,172751045 | 0,91035623 | NOTCH1 | FALSE | Upregulated |
| 0,252011664 | 0,91035623 | PDGFRB | FALSE | Upregulated |
| 0,28721146 | 0,91035623 | RRP1B | FALSE | Upregulated |
| -0,200204733 | 0,91035623 | SDF4 | FALSE | Downregulated |
| -0,340231722 | 0,91035623 | SMPD5 | FALSE | Downregulated |
| 0,244999501 | 0,91035623 | U2SURP | FALSE | Upregulated |
| -0,236163371 | 0,91035623 | VDAC1 | FALSE | Downregulated |
| -0,267543603 | 0,91035623 | ZADH2 | FALSE | Downregulated |
| 0,303650726 | 0,910591543 | PAX8 | FALSE | Upregulated |
| -0,199259134 | 0,911028779 | OAZ1 | FALSE | Downregulated |
| -0,212580872 | 0,911028779 | TMEM115 | FALSE | Downregulated |
| 0,252587415 | 0,91125387 | RDH14 | FALSE | Upregulated |
| -0,155445861 | 0,911605478 | MYH10 | FALSE | Downregulated |
| -0,207897179 | 0,911901796 | ARHGAP30 | FALSE | Downregulated |
| 0,208031816 | 0,911901796 | DENND4B | FALSE | Upregulated |
| 0,350828661 | 0,911901796 | ERCC3 | FALSE | Upregulated |
| 0,319033036 | 0,911901796 | MRPS18C | FALSE | Upregulated |
| 0,152637863 | 0,911901796 | SCAND2P | FALSE | Upregulated |
| -0,189435335 | 0,912013744 | SF1 | FALSE | Downregulated |
| -0,282865185 | 0,912393637 | CA14 | FALSE | Downregulated |

| | | | | |
|---|---|---|---|---|
| -0,336562661 | 0,912393637 | HSD17B12 | FALSE | Downregulated |
| 0,348458566 | 0,912988265 | ABCF1 | FALSE | Upregulated |
| 0,220560932 | 0,912988265 | C14orf93 | FALSE | Upregulated |
| 0,290125747 | 0,912988265 | LOC101928658 | FALSE | Upregulated |
| 0,256273784 | 0,912988265 | MCF2L | FALSE | Upregulated |
| -0,299153711 | 0,912988265 | PPIP5K2 | FALSE | Downregulated |
| 0,235360399 | 0,913187329 | CRIP3 | FALSE | Upregulated |
| -0,254974694 | 0,913512522 | AZI2 | FALSE | Downregulated |
| 0,22673667 | 0,913512522 | LOC101927057 | FALSE | Upregulated |
| 0,315825318 | 0,913512522 | LOC101927161 | FALSE | Upregulated |
| 0,229417584 | 0,913657071 | ADAMTSL2 | FALSE | Upregulated |
| -0,224548882 | 0,913657071 | DMTF1 | FALSE | Downregulated |
| 0,24323553 | 0,913657071 | HSF1 | FALSE | Upregulated |
| 0,283535827 | 0,913657071 | PLBD2 | FALSE | Upregulated |
| -0,189711643 | 0,913657071 | PON2 | FALSE | Downregulated |
| 0,172513337 | 0,913657071 | PXN | FALSE | Upregulated |
| 0,287795117 | 0,913657071 | SIKE1 | FALSE | Upregulated |
| -0,198234702 | 0,913657071 | SPSB3 | FALSE | Downregulated |
| -0,305323375 | 0,913657071 | TUBA8 | FALSE | Downregulated |
| 0,277284481 | 0,913657071 | UTP20 | FALSE | Upregulated |
| 0,301779378 | 0,914130213 | G3BP2 | FALSE | Upregulated |
| 0,173754933 | 0,914130213 | PCYT2 | FALSE | Upregulated |
| -0,174270038 | 0,914278452 | CLMN | FALSE | Downregulated |
| 0,275729179 | 0,914278452 | DLG5 | FALSE | Upregulated |
| -0,340864588 | 0,914278452 | FAM161B | FALSE | Downregulated |
| -0,137550424 | 0,914278452 | FRMD4A | FALSE | Downregulated |
| -0,190828372 | 0,914278452 | GCGR | FALSE | Downregulated |
| -0,222720908 | 0,914278452 | GNRHR | FALSE | Downregulated |
| -0,289943609 | 0,914278452 | RP11-242D8.1 | FALSE | Downregulated |
| -0,160480538 | 0,914278452 | TCF7 | FALSE | Downregulated |
| 0,305339445 | 0,915086164 | FBXO28 | FALSE | Upregulated |
| 0,312455995 | 0,915086164 | SLC39A11 | FALSE | Upregulated |
| -0,241135061 | 0,915514926 | BCL2L13 | FALSE | Downregulated |
| -0,288148722 | 0,915696095 | SLAIN2 | FALSE | Downregulated |
| -0,25102549 | 0,915820673 | SIM2 | FALSE | Downregulated |
| -0,286865961 | 0,916224593 | PALMD | FALSE | Downregulated |
| 0,314896662 | 0,916306165 | ACTA2 | FALSE | Upregulated |
| -0,293512356 | 0,916306165 | ZC3H18 | FALSE | Downregulated |
| -0,310337446 | 0,91631475 | CLASP2 | FALSE | Downregulated |
| 0,311795784 | 0,91631475 | LRRC37A6P | FALSE | Upregulated |
| -0,22936544 | 0,91631475 | PDIA3 | FALSE | Downregulated |
| -0,32718268 | 0,91631475 | SMIM15 | FALSE | Downregulated |
| 0,262202076 | 0,91631475 | YWHAB | FALSE | Upregulated |
| -0,372086027 | 0,916836554 | KCTD17 | FALSE | Downregulated |
| 0,175029739 | 0,916836554 | PIP4K2B | FALSE | Upregulated |
| 0,184732572 | 0,916836554 | PLEKHG6 | FALSE | Upregulated |
| -0,181133574 | 0,916836554 | RNF169 | FALSE | Downregulated |
| 0,284334102 | 0,916836554 | RRP7A | FALSE | Upregulated |
| 0,277978959 | 0,916836554 | USP4 | FALSE | Upregulated |
| 0,279168374 | 0,917345377 | HLA-F-AS1 | FALSE | Upregulated |
| -0,170846101 | 0,917345377 | NFE2L2 | FALSE | Downregulated |

| | | | | |
|---|---|---|---|---|
| -0,161621087 | 0,917530659 | KIAA1671 | FALSE | Downregulated |
| 0,245124965 | 0,917530659 | TCOF1 | FALSE | Upregulated |
| -0,226418183 | 0,9180018 | COX4I1 | FALSE | Downregulated |
| 0,266925248 | 0,9180018 | DIP2A | FALSE | Upregulated |
| 0,251992595 | 0,9180018 | FAM118B | FALSE | Upregulated |
| -0,222337822 | 0,9180018 | MAP2K2 | FALSE | Downregulated |
| -0,157364769 | 0,9180018 | RPL28 | FALSE | Downregulated |
| -0,200965991 | 0,9180018 | SRRT | FALSE | Downregulated |
| 0,199570756 | 0,9180018 | STIM1 | FALSE | Upregulated |
| -0,260448167 | 0,9180018 | TXNDC17 | FALSE | Downregulated |
| -0,274991504 | 0,918480378 | ZDHHC14 | FALSE | Downregulated |
| 0,156813761 | 0,918548776 | ARL17A | FALSE | Upregulated |
| 0,278965908 | 0,918548776 | MAP3K4 | FALSE | Upregulated |
| 0,308325124 | 0,918548776 | MARS | FALSE | Upregulated |
| 0,19838938 | 0,918548776 | MAST3 | FALSE | Upregulated |
| 0,193436653 | 0,918548776 | SYMPK | FALSE | Upregulated |
| 0,289537957 | 0,918548776 | TRMT5 | FALSE | Upregulated |
| 0,220384288 | 0,918548776 | XIAP | FALSE | Upregulated |
| -0,177372001 | 0,918548776 | ZBED1 | FALSE | Downregulated |
| 0,242825021 | 0,919039058 | GET4 | FALSE | Upregulated |
| 0,206824338 | 0,919039058 | HMGN2 | FALSE | Upregulated |
| 0,264888797 | 0,919039058 | PDE7B | FALSE | Upregulated |
| -0,291038711 | 0,919039058 | PIK3CA | FALSE | Downregulated |
| -0,21281551 | 0,919039058 | SEPT2 | FALSE | Downregulated |
| 0,321870421 | 0,919100552 | ABCC1 | FALSE | Upregulated |
| -0,187438358 | 0,919100552 | BBIP1 | FALSE | Downregulated |
| -0,311086323 | 0,919100552 | EARS2 | FALSE | Downregulated |
| -0,26659408 | 0,919100552 | ELMO2 | FALSE | Downregulated |
| -0,171554439 | 0,919100552 | MFSD11 | FALSE | Downregulated |
| 0,313031605 | 0,919100552 | PPP1R12A | FALSE | Upregulated |
| 0,283910424 | 0,919100552 | SLC39A1 | FALSE | Upregulated |
| -0,218315372 | 0,919100552 | SMLR1 | FALSE | Downregulated |
| -0,24481053 | 0,919199066 | AARS | FALSE | Downregulated |
| -0,181521088 | 0,919199066 | CYP2C18 | FALSE | Downregulated |
| 0,238739738 | 0,919199066 | LDB1 | FALSE | Upregulated |
| 0,307175212 | 0,919199066 | PTPRC | FALSE | Upregulated |
| 0,145877786 | 0,919199066 | SERINC4 | FALSE | Upregulated |
| 0,303898386 | 0,919199066 | TIPARP | FALSE | Upregulated |
| 0,190495254 | 0,919199066 | ZCCHC24 | FALSE | Upregulated |
| 0,301533049 | 0,920004459 | ANO8 | FALSE | Upregulated |
| -0,296799588 | 0,920004459 | TTC14 | FALSE | Downregulated |
| -0,255223272 | 0,920089118 | FNIP1 | FALSE | Downregulated |
| -0,225692263 | 0,920140996 | CD164 | FALSE | Downregulated |
| 0,286523545 | 0,920140996 | JRK | FALSE | Upregulated |
| -0,171589338 | 0,920140996 | TSKU | FALSE | Downregulated |
| -0,32061255 | 0,920205099 | ZMYND8 | FALSE | Downregulated |
| -0,296844359 | 0,92041201 | CLTA | FALSE | Downregulated |
| 0,210059526 | 0,920679511 | FAM102A | FALSE | Upregulated |
| 0,150601136 | 0,920743804 | GNPTAB | FALSE | Upregulated |
| -0,217281511 | 0,920888959 | BDNF | FALSE | Downregulated |
| 0,248550117 | 0,920888959 | GOSR1 | FALSE | Upregulated |

| | | | | |
|---|---|---|---|---|
| -0,285868198 | 0,920898377 | KIAA1731 | FALSE | Downregulated |
| 0,326278884 | 0,921077825 | CCSER2 | FALSE | Upregulated |
| -0,299127632 | 0,921077825 | CNOT2 | FALSE | Downregulated |
| -0,225413928 | 0,921077825 | EDEM3 | FALSE | Downregulated |
| -0,212040022 | 0,92122393 | RAPGEF3 | FALSE | Downregulated |
| 0,305963142 | 0,92142316 | AMMECR1L | FALSE | Upregulated |
| 0,216596619 | 0,92142316 | ODF3B | FALSE | Upregulated |
| -0,227983742 | 0,921453909 | GK3P | FALSE | Downregulated |
| -0,287825441 | 0,921453909 | KIAA1468 | FALSE | Downregulated |
| -0,286806756 | 0,921584732 | ADAM22 | FALSE | Downregulated |
| 0,230806072 | 0,921584732 | IPO7 | FALSE | Upregulated |
| -0,174916335 | 0,921587407 | TRIM62 | FALSE | Downregulated |
| 0,2308446 | 0,921587407 | ZNF507 | FALSE | Upregulated |
| -0,14007841 | 0,92185623 | ACOT13 | FALSE | Downregulated |
| -0,209506184 | 0,92185623 | C9orf173 | FALSE | Downregulated |
| -0,169966722 | 0,92185623 | GLDC | FALSE | Downregulated |
| 0,221341301 | 0,92185623 | LINC00674 | FALSE | Upregulated |
| 0,231747534 | 0,92185623 | LOC101929204 | FALSE | Upregulated |
| -0,3043742 | 0,92185623 | MMADHC | FALSE | Downregulated |
| -0,198997113 | 0,92185623 | MRFAP1 | FALSE | Downregulated |
| 0,369258182 | 0,92185623 | SLC8A1 | FALSE | Upregulated |
| -0,319692956 | 0,92185623 | TMEM170B | FALSE | Downregulated |
| 0,166912091 | 0,92185623 | USP45 | FALSE | Upregulated |
| 0,169846764 | 0,921946052 | HSPA8 | FALSE | Upregulated |
| -0,301276396 | 0,921946052 | SCAPER | FALSE | Downregulated |
| -0,278055126 | 0,921946052 | TMEM57 | FALSE | Downregulated |
| 0,192764792 | 0,92195526 | DUS1L | FALSE | Upregulated |
| 0,201626986 | 0,92195526 | LOC727978 | FALSE | Upregulated |
| -0,223246972 | 0,922086091 | FAM115A | FALSE | Downregulated |
| 0,235065449 | 0,922086091 | FUCA2 | FALSE | Upregulated |
| 0,254848632 | 0,922086091 | GNAQ | FALSE | Upregulated |
| 0,181777685 | 0,922086091 | GOLGA8H | FALSE | Upregulated |
| 0,202303982 | 0,922086091 | GUSB | FALSE | Upregulated |
| -0,181943936 | 0,922086091 | LOC101928042 | FALSE | Downregulated |
| 0,227422924 | 0,922086091 | NEDD4 | FALSE | Upregulated |
| 0,289294535 | 0,922086091 | RGL1 | FALSE | Upregulated |
| -0,193801835 | 0,922086091 | RNF217 | FALSE | Downregulated |
| 0,279202972 | 0,922086091 | RP11-864J10.4 | FALSE | Upregulated |
| -0,164833816 | 0,922086091 | SLCO1A2 | FALSE | Downregulated |
| -0,16675012 | 0,922086091 | TMEM80 | FALSE | Downregulated |
| 0,192453291 | 0,922086091 | USP54 | FALSE | Upregulated |
| -0,248407614 | 0,922260467 | GPR89A | FALSE | Downregulated |
| 0,242251665 | 0,922686389 | MLLT10 | FALSE | Upregulated |
| -0,259401157 | 0,922924403 | C22orf15 | FALSE | Downregulated |
| 0,242278789 | 0,922924403 | GAPVD1 | FALSE | Upregulated |
| -0,257687938 | 0,922924403 | LOC102723804 | FALSE | Downregulated |
| -0,252267541 | 0,922924403 | LOC102724262 | FALSE | Downregulated |
| 0,153449275 | 0,922924403 | NID1 | FALSE | Upregulated |
| 0,1307335 | 0,922924403 | PLXNB1 | FALSE | Upregulated |
| 0,233085293 | 0,922924403 | POLDIP3 | FALSE | Upregulated |
| 0,302416658 | 0,922924403 | RASSF1-AS1 | FALSE | Upregulated |

| | | | | |
|---|---|---|---|---|
| 0,223387511 | 0,923005057 | PTRH2 | FALSE | Upregulated |
| 0,234315659 | 0,923005057 | SPRYD3 | FALSE | Upregulated |
| 0,183463881 | 0,923035964 | AP2M1 | FALSE | Upregulated |
| 0,172403955 | 0,923035964 | ARFGEF1 | FALSE | Upregulated |
| -0,227515962 | 0,923035964 | DYRK1A | FALSE | Downregulated |
| 0,19885139 | 0,923035964 | STARD7 | FALSE | Upregulated |
| 0,178678044 | 0,923035964 | TRMT112 | FALSE | Upregulated |
| -0,264636047 | 0,923035964 | VPS52 | FALSE | Downregulated |
| 0,28425434 | 0,923035964 | ZDHHC3 | FALSE | Upregulated |
| 0,218572323 | 0,92358856 | ARIH2 | FALSE | Upregulated |
| -0,222717612 | 0,92358856 | ASB9P1 | FALSE | Downregulated |
| -0,315112626 | 0,92358856 | B3GNTL1 | FALSE | Downregulated |
| 0,264347917 | 0,92358856 | CAMK2N1 | FALSE | Upregulated |
| 0,262073331 | 0,92358856 | CDIPT | FALSE | Upregulated |
| 0,225122282 | 0,92358856 | FAM45A | FALSE | Upregulated |
| 0,286846139 | 0,92358856 | FGGY | FALSE | Upregulated |
| 0,26128786 | 0,92358856 | GFPT1 | FALSE | Upregulated |
| 0,208599044 | 0,92358856 | KAT2A | FALSE | Upregulated |
| 0,286347892 | 0,92358856 | LATS1 | FALSE | Upregulated |
| 0,257344615 | 0,92358856 | MYSM1 | FALSE | Upregulated |
| 0,250672979 | 0,92358856 | NOL8 | FALSE | Upregulated |
| 0,241326305 | 0,92358856 | PSMB8 | FALSE | Upregulated |
| -0,21735676 | 0,92358856 | TCF25 | FALSE | Downregulated |
| -0,297483688 | 0,92358856 | XPNPEP3 | FALSE | Downregulated |
| -0,269991809 | 0,92358856 | YTHDC1 | FALSE | Downregulated |
| -0,154306968 | 0,923880479 | TOR1AIP2 | FALSE | Downregulated |
| -0,119414054 | 0,923899602 | ANKH | FALSE | Downregulated |
| -0,219901015 | 0,923899602 | ANO1-AS2 | FALSE | Downregulated |
| -0,301631016 | 0,923899602 | BTBD7 | FALSE | Downregulated |
| -0,295459262 | 0,923899602 | CCT7 | FALSE | Downregulated |
| 0,28880259 | 0,923899602 | CKAP4 | FALSE | Upregulated |
| 0,184919182 | 0,923899602 | CRMP1 | FALSE | Upregulated |
| 0,172094144 | 0,923899602 | DGAT1 | FALSE | Upregulated |
| -0,255334323 | 0,923899602 | ESR2 | FALSE | Downregulated |
| -0,212729236 | 0,923899602 | GLIS2 | FALSE | Downregulated |
| 0,276859508 | 0,923899602 | GUF1 | FALSE | Upregulated |
| 0,228341568 | 0,923899602 | KIAA1656 | FALSE | Upregulated |
| 0,23092947 | 0,923899602 | MORF4L2 | FALSE | Upregulated |
| 0,287596158 | 0,923899602 | MYLK-AS1 | FALSE | Upregulated |
| 0,203290014 | 0,923899602 | NCOA3 | FALSE | Upregulated |
| -0,182492521 | 0,923899602 | OSBPL9 | FALSE | Downregulated |
| 0,257719611 | 0,923899602 | RALGAPB | FALSE | Upregulated |
| -0,187677161 | 0,923899602 | RBP5 | FALSE | Downregulated |
| -0,223534509 | 0,923899602 | RP11-28G8.1 | FALSE | Downregulated |
| 0,163528665 | 0,923899602 | SCAF11 | FALSE | Upregulated |
| -0,2426436 | 0,923899602 | TAB2 | FALSE | Downregulated |
| -0,223266253 | 0,923899602 | ZBTB45 | FALSE | Downregulated |
| 0,284627337 | 0,923899602 | ZXDB | FALSE | Upregulated |
| 0,215362343 | 0,924157613 | ARHGAP1 | FALSE | Upregulated |
| -0,23980811 | 0,924157613 | KIAA2026 | FALSE | Downregulated |
| -0,183767005 | 0,924157613 | LOC100996740 | FALSE | Downregulated |

| | | | | |
|---|---|---|---|---|
| -0,241200767 | 0,924157613 | PATL1 | FALSE | Downregulated |
| 0,258296992 | 0,924157613 | SLC35B4 | FALSE | Upregulated |
| -0,291600949 | 0,924157613 | ZNF217 | FALSE | Downregulated |
| -0,192184837 | 0,924417433 | ITGA9 | FALSE | Downregulated |
| -0,214243484 | 0,924819954 | GCH1 | FALSE | Downregulated |
| -0,186711783 | 0,925159784 | MTR | FALSE | Downregulated |
| 0,318454948 | 0,925569663 | ADAMTS5 | FALSE | Upregulated |
| 0,193269716 | 0,925569663 | AGPAT4 | FALSE | Upregulated |
| -0,171793798 | 0,925569663 | ARGFXP2 | FALSE | Downregulated |
| -0,243138676 | 0,925569663 | ATXN2 | FALSE | Downregulated |
| -0,278947723 | 0,925569663 | BBS1 | FALSE | Downregulated |
| 0,26239406 | 0,925569663 | C16orf62 | FALSE | Upregulated |
| -0,182743678 | 0,925569663 | C22orf26 | FALSE | Downregulated |
| 0,156017958 | 0,925569663 | CCDC85C | FALSE | Upregulated |
| 0,281939738 | 0,925569663 | DDX19A | FALSE | Upregulated |
| 0,306836031 | 0,925569663 | EVA1A | FALSE | Upregulated |
| 0,163844257 | 0,925569663 | EVI2B | FALSE | Upregulated |
| 0,275393276 | 0,925569663 | FLAD1 | FALSE | Upregulated |
| 0,209528022 | 0,925569663 | GALNT11 | FALSE | Upregulated |
| -0,312321828 | 0,925569663 | GRIN2B | FALSE | Downregulated |
| -0,27253482 | 0,925569663 | GTF3C5 | FALSE | Downregulated |
| -0,136962273 | 0,925569663 | IRS2 | FALSE | Downregulated |
| -0,192294934 | 0,925569663 | JPX | FALSE | Downregulated |
| -0,168743121 | 0,925569663 | KBTBD4 | FALSE | Downregulated |
| 0,150053094 | 0,925569663 | KIAA0100 | FALSE | Upregulated |
| -0,20884539 | 0,925569663 | KLHL24 | FALSE | Downregulated |
| -0,175768907 | 0,925569663 | LARP4 | FALSE | Downregulated |
| -0,137926894 | 0,925569663 | LOC100379224 | FALSE | Downregulated |
| 0,171447661 | 0,925569663 | LOC101927031 | FALSE | Upregulated |
| 0,241219099 | 0,925569663 | LOC101928604 | FALSE | Upregulated |
| -0,289422332 | 0,925569663 | LOC102723722 | FALSE | Downregulated |
| 0,302284124 | 0,925569663 | LOC102724364 | FALSE | Upregulated |
| 0,315296123 | 0,925569663 | LOC153684 | FALSE | Upregulated |
| -0,171961284 | 0,925569663 | MAP3K2 | FALSE | Downregulated |
| 0,167298696 | 0,925569663 | MARCH1 | FALSE | Upregulated |
| 0,270386494 | 0,925569663 | MEN1 | FALSE | Upregulated |
| 0,279315374 | 0,925569663 | NARS | FALSE | Upregulated |
| 0,189886151 | 0,925569663 | OTUD4 | FALSE | Upregulated |
| -0,194687948 | 0,925569663 | PTPN11 | FALSE | Downregulated |
| 0,176547604 | 0,925569663 | SAMD4B | FALSE | Upregulated |
| 0,233085984 | 0,925569663 | SLC13A4 | FALSE | Upregulated |
| 0,228121524 | 0,925569663 | SLC9A8 | FALSE | Upregulated |
| -0,151086646 | 0,925569663 | STARD7-AS1 | FALSE | Downregulated |
| 0,245383335 | 0,925569663 | SULF2 | FALSE | Upregulated |
| -0,182006076 | 0,925569663 | TGFBR2 | FALSE | Downregulated |
| -0,161338089 | 0,925569663 | TNKS1BP1 | FALSE | Downregulated |
| 0,229751232 | 0,925569663 | TP73 | FALSE | Upregulated |
| 0,283991827 | 0,925569663 | TRAF3 | FALSE | Upregulated |
| 0,223494898 | 0,925569663 | TRAPPC10 | FALSE | Upregulated |
| -0,25907873 | 0,925569663 | VHL | FALSE | Downregulated |
| -0,158211059 | 0,925569663 | ZCCHC14 | FALSE | Downregulated |

| | | | | |
|---|---|---|---|---|
| 0,268381208 | 0,925569663 | ZRANB3 | FALSE | Upregulated |
| 0,362542364 | 0,925570549 | IL21R-AS1 | FALSE | Upregulated |
| 0,24782146 | 0,925570549 | LYSMD1 | FALSE | Upregulated |
| 0,19306583 | 0,925570549 | MMRN2 | FALSE | Upregulated |
| 0,246377588 | 0,925570549 | PXDN | FALSE | Upregulated |
| -0,137637373 | 0,925570549 | TNFAIP1 | FALSE | Downregulated |
| -0,243245684 | 0,925570549 | ZDHHC23 | FALSE | Downregulated |
| 0,246576501 | 0,925648763 | ABCE1 | FALSE | Upregulated |
| -0,292264762 | 0,925648763 | ABCF3 | FALSE | Downregulated |
| 0,135552623 | 0,925648763 | ADAR | FALSE | Upregulated |
| -0,191774545 | 0,925648763 | AK4 | FALSE | Downregulated |
| 0,225933443 | 0,925648763 | AKR7L | FALSE | Upregulated |
| -0,250757155 | 0,925648763 | C4orf3 | FALSE | Downregulated |
| 0,181390132 | 0,925648763 | CHP1 | FALSE | Upregulated |
| 0,214252054 | 0,925648763 | CMTM6 | FALSE | Upregulated |
| -0,284687746 | 0,925648763 | EDNRB | FALSE | Downregulated |
| -0,283064585 | 0,925648763 | EIF1B-AS1 | FALSE | Downregulated |
| 0,271136631 | 0,925648763 | ELK1 | FALSE | Upregulated |
| 0,267934774 | 0,925648763 | ETV6 | FALSE | Upregulated |
| -0,242380525 | 0,925648763 | FANCF | FALSE | Downregulated |
| 0,178983769 | 0,925648763 | FBXO33 | FALSE | Upregulated |
| 0,274977669 | 0,925648763 | GDAP2 | FALSE | Upregulated |
| 0,174935899 | 0,925648763 | GGA1 | FALSE | Upregulated |
| 0,287367687 | 0,925648763 | IGSF23 | FALSE | Upregulated |
| -0,203374208 | 0,925648763 | ITGA1 | FALSE | Downregulated |
| 0,153750283 | 0,925648763 | KIAA0430 | FALSE | Upregulated |
| -0,170541082 | 0,925648763 | LOC100133091 | FALSE | Downregulated |
| -0,140830789 | 0,925648763 | PINK1 | FALSE | Downregulated |
| -0,193766386 | 0,925648763 | PMM2 | FALSE | Downregulated |
| 0,246304583 | 0,925648763 | POU2F1 | FALSE | Upregulated |
| 0,149207279 | 0,925648763 | PPAPDC1B | FALSE | Upregulated |
| -0,161764576 | 0,925648763 | PPP2R3A | FALSE | Downregulated |
| 0,240599954 | 0,925648763 | RNF146 | FALSE | Upregulated |
| 0,223102589 | 0,925648763 | SCAF8 | FALSE | Upregulated |
| -0,241678098 | 0,925648763 | SEPT4 | FALSE | Downregulated |
| -0,203428213 | 0,925648763 | SMAP2 | FALSE | Downregulated |
| 0,280274643 | 0,925648763 | SMIM6 | FALSE | Upregulated |
| 0,207550029 | 0,925648763 | TAGLN | FALSE | Upregulated |
| -0,192810868 | 0,925648763 | TFPI | FALSE | Downregulated |
| 0,249586834 | 0,925648763 | TMED9 | FALSE | Upregulated |
| 0,173588423 | 0,925648763 | TPI1P3 | FALSE | Upregulated |
| 0,232510486 | 0,925648763 | TRIM24 | FALSE | Upregulated |
| -0,346088707 | 0,925648763 | TSC22D3 | FALSE | Downregulated |
| 0,237126033 | 0,925648763 | TTC39C | FALSE | Upregulated |
| -0,237042366 | 0,925648763 | UCK1 | FALSE | Downregulated |
| -0,183328133 | 0,925965199 | AK2 | FALSE | Downregulated |
| -0,151923588 | 0,925965199 | CP | FALSE | Downregulated |
| 0,223665658 | 0,925965199 | FEM1A | FALSE | Upregulated |
| -0,227413319 | 0,925965199 | GIT1 | FALSE | Downregulated |
| 0,263079171 | 0,925965199 | MAP7D1 | FALSE | Upregulated |
| 0,234655384 | 0,925965199 | PKN1 | FALSE | Upregulated |

| | | | | |
|---|---|---|---|---|
| 0,190501592 | 0,925965199 | PPP6R1 | FALSE | Upregulated |
| -0,253530589 | 0,925965199 | SLC22A18 | FALSE | Downregulated |
| 0,195608529 | 0,925965199 | TRPC4AP | FALSE | Upregulated |
| 0,176954204 | 0,925965199 | ZNF516 | FALSE | Upregulated |
| 0,204303036 | 0,926442828 | API5 | FALSE | Upregulated |
| 0,165145865 | 0,926442828 | RBCK1 | FALSE | Upregulated |
| -0,154118277 | 0,926442828 | SFXN5 | FALSE | Downregulated |
| -0,175793629 | 0,926756679 | SMEK2 | FALSE | Downregulated |
| -0,244982783 | 0,927280601 | C14orf159 | FALSE | Downregulated |
| 0,118646892 | 0,927280601 | DDX17 | FALSE | Upregulated |
| -0,210226855 | 0,927485093 | LIMS2 | FALSE | Downregulated |
| -0,242603936 | 0,927879295 | PLGLB1 | FALSE | Downregulated |
| -0,250067603 | 0,927879295 | ZNF776 | FALSE | Downregulated |
| -0,19650185 | 0,927939733 | TUT1 | FALSE | Downregulated |
| -0,235645181 | 0,928182937 | EIF4ENIF1 | FALSE | Downregulated |
| 0,190674245 | 0,928322321 | UCKL1 | FALSE | Upregulated |
| 0,230651122 | 0,928322321 | UQCRQ | FALSE | Upregulated |
| 0,153573126 | 0,928814671 | MYH9 | FALSE | Upregulated |
| 0,240956891 | 0,92973497 | CCDC14 | FALSE | Upregulated |
| -0,182221776 | 0,92973497 | LOC101928892 | FALSE | Downregulated |
| 0,221261523 | 0,92973497 | RAD51B | FALSE | Upregulated |
| 0,150661026 | 0,92973497 | SUGP2 | FALSE | Upregulated |
| 0,245893016 | 0,929766182 | CRTAP | FALSE | Upregulated |
| 0,21600213 | 0,930164369 | FAM160A2 | FALSE | Upregulated |
| -0,214493152 | 0,930237825 | FOXK2 | FALSE | Downregulated |
| 0,285053763 | 0,930436071 | TSTA3 | FALSE | Upregulated |
| -0,210605196 | 0,93127894 | CRKL | FALSE | Downregulated |
| -0,191927432 | 0,93127894 | FZD5 | FALSE | Downregulated |
| 0,130217005 | 0,93127894 | HNRNPU | FALSE | Upregulated |
| 0,25952031 | 0,93127894 | NR1H2 | FALSE | Upregulated |
| 0,147264445 | 0,931569166 | LINC01043 | FALSE | Upregulated |
| -0,186014303 | 0,931569166 | SCTR | FALSE | Downregulated |
| 0,169444297 | 0,931660638 | CCDC42B | FALSE | Upregulated |
| -0,198247785 | 0,931660638 | CD99 | FALSE | Downregulated |
| -0,185149956 | 0,931660638 | FAM186B | FALSE | Downregulated |
| -0,234354502 | 0,931660638 | IGHMBP2 | FALSE | Downregulated |
| -0,147041015 | 0,931660638 | LOC101929079 | FALSE | Downregulated |
| 0,101423506 | 0,931660638 | LRP1 | FALSE | Upregulated |
| 0,151844004 | 0,931660638 | MXD3 | FALSE | Upregulated |
| 0,151719714 | 0,931660638 | REEP5 | FALSE | Upregulated |
| -0,183872838 | 0,931660638 | SRRM5 | FALSE | Downregulated |
| 0,254338712 | 0,931660638 | USP6NL | FALSE | Upregulated |
| 0,317596451 | 0,931724659 | WDR52 | FALSE | Upregulated |
| -0,132988591 | 0,931735415 | CARHSP1 | FALSE | Downregulated |
| -0,136982224 | 0,931775542 | DCAF8 | FALSE | Downregulated |
| -0,238627705 | 0,931775542 | IGF2BP3 | FALSE | Downregulated |
| -0,269322019 | 0,931775542 | MICU1 | FALSE | Downregulated |
| 0,125095812 | 0,931775542 | PKD1P1 | FALSE | Upregulated |
| 0,208967617 | 0,931775542 | PURB | FALSE | Upregulated |
| 0,153973039 | 0,931775542 | TNIP1 | FALSE | Upregulated |
| 0,136304209 | 0,931775542 | YPEL1 | FALSE | Upregulated |

| | | | | |
|---|---|---|---|---|
| 0,266250694 | 0,931775542 | ZNF700 | FALSE | Upregulated |
| 0,156138931 | 0,93183406 | ZNF687 | FALSE | Upregulated |
| 0,287828533 | 0,931959597 | ARHGAP44 | FALSE | Upregulated |
| -0,267648195 | 0,931959597 | PPP1R37 | FALSE | Downregulated |
| 0,22788141 | 0,931959597 | STT3A | FALSE | Upregulated |
| 0,123006969 | 0,931959597 | TNRC6B | FALSE | Upregulated |
| -0,270416163 | 0,931959597 | TOE1 | FALSE | Downregulated |
| -0,115418674 | 0,932573542 | MAP3K12 | FALSE | Downregulated |
| 0,280951253 | 0,932573542 | THEMIS2 | FALSE | Upregulated |
| -0,169645254 | 0,932949557 | RPLP1 | FALSE | Downregulated |
| -0,213225017 | 0,932949557 | USP30 | FALSE | Downregulated |
| 0,213761155 | 0,933123037 | CYB5D1 | FALSE | Upregulated |
| 0,16500672 | 0,933123037 | M6PR | FALSE | Upregulated |
| -0,259777215 | 0,933123037 | NUGGC | FALSE | Downregulated |
| 0,329319024 | 0,933123037 | PDK3 | FALSE | Upregulated |
| 0,164876153 | 0,933123037 | PITRM1-AS1 | FALSE | Upregulated |
| -0,195511849 | 0,933123037 | PUS10 | FALSE | Downregulated |
| 0,242160634 | 0,933123037 | RAD21 | FALSE | Upregulated |
| 0,270282443 | 0,933123037 | SLC22A9 | FALSE | Upregulated |
| -0,261612761 | 0,933153467 | LOC730668 | FALSE | Downregulated |
| -0,15447105 | 0,933215893 | ADD1 | FALSE | Downregulated |
| 0,23422197 | 0,933215893 | CABLES1 | FALSE | Upregulated |
| 0,27108767 | 0,933215893 | CTD-2201E9.1 | FALSE | Upregulated |
| 0,233042997 | 0,933215893 | RHD | FALSE | Upregulated |
| 0,236197863 | 0,933215893 | SYNE4 | FALSE | Upregulated |
| -0,215798564 | 0,933215893 | SYNRG | FALSE | Downregulated |
| -0,138903543 | 0,933324648 | GALNT10 | FALSE | Downregulated |
| -0,295293561 | 0,933324648 | HEIH | FALSE | Downregulated |
| -0,242844701 | 0,933324648 | RGS3 | FALSE | Downregulated |
| -0,159861684 | 0,933324648 | THEM5 | FALSE | Downregulated |
| -0,134883903 | 0,933563743 | FAM13A | FALSE | Downregulated |
| -0,238110329 | 0,933563743 | GNL1 | FALSE | Downregulated |
| -0,160633844 | 0,933638685 | LOC102723929 | FALSE | Downregulated |
| 0,196090576 | 0,933638685 | LRP11 | FALSE | Upregulated |
| 0,221736382 | 0,933638685 | PITPNB | FALSE | Upregulated |
| -0,162174408 | 0,933638685 | SAT1 | FALSE | Downregulated |
| -0,246762989 | 0,933638685 | ZNHIT2 | FALSE | Downregulated |
| 0,143210666 | 0,934011152 | APOL3 | FALSE | Upregulated |
| -0,175868142 | 0,934069778 | VCP | FALSE | Downregulated |
| 0,184147883 | 0,934228774 | BRD1 | FALSE | Upregulated |
| -0,247172988 | 0,934228774 | GPSM2 | FALSE | Downregulated |
| 0,200073025 | 0,934228774 | MTCH1 | FALSE | Upregulated |
| -0,183613551 | 0,934293995 | C16orf72 | FALSE | Downregulated |
| -0,214540466 | 0,934293995 | ENTPD3 | FALSE | Downregulated |
| 0,19198444 | 0,934293995 | ERVK13-1 | FALSE | Upregulated |
| 0,204831856 | 0,934293995 | FRYL | FALSE | Upregulated |
| 0,20276658 | 0,934293995 | GOPC | FALSE | Upregulated |
| 0,237112811 | 0,934293995 | ST7-AS2 | FALSE | Upregulated |
| -0,238045997 | 0,934293995 | UEVLD | FALSE | Downregulated |
| -0,194852542 | 0,934349374 | AADAC | FALSE | Downregulated |
| 0,170167109 | 0,934349374 | CPT2 | FALSE | Upregulated |

| | | | | |
|---|---|---|---|---|
| -0,117461515 | 0,934349374 | POLR3H | FALSE | Downregulated |
| 0,276161215 | 0,934349374 | PSAPL1 | FALSE | Upregulated |
| 0,260333032 | 0,934349374 | SH3GL1P2 | FALSE | Upregulated |
| 0,166223432 | 0,934349374 | USP8 | FALSE | Upregulated |
| -0,178194906 | 0,934349374 | WBP1L | FALSE | Downregulated |
| -0,231322597 | 0,934349374 | XRN2 | FALSE | Downregulated |
| 0,284265939 | 0,93470931 | CERS4 | FALSE | Upregulated |
| 0,15749452 | 0,93470931 | SEC31A | FALSE | Upregulated |
| -0,228605315 | 0,934750444 | CAB39 | FALSE | Downregulated |
| -0,237647687 | 0,935123701 | UBE3B | FALSE | Downregulated |
| 0,2443976 | 0,935207959 | ABCB1 | FALSE | Upregulated |
| -0,198129228 | 0,935207959 | ARCN1 | FALSE | Downregulated |
| -0,196383436 | 0,935207959 | ARHGEF37 | FALSE | Downregulated |
| -0,207702638 | 0,935213865 | CDC37 | FALSE | Downregulated |
| -0,25466344 | 0,935213865 | CEBPZ | FALSE | Downregulated |
| -0,162303335 | 0,935316113 | GPI | FALSE | Downregulated |
| 0,165849552 | 0,935316113 | KDM2B | FALSE | Upregulated |
| 0,155560259 | 0,935316113 | LOC102724391 | FALSE | Upregulated |
| 0,154856658 | 0,935316113 | TTC28-AS1 | FALSE | Upregulated |
| -0,171013187 | 0,935416869 | ZFHX4 | FALSE | Downregulated |
| -0,245485007 | 0,9356091 | LGALS3BP | FALSE | Downregulated |
| -0,146679555 | 0,93571828 | FGFR3 | FALSE | Downregulated |
| 0,201997729 | 0,935997742 | ATG4D | FALSE | Upregulated |
| 0,125956791 | 0,935997742 | CAPN15 | FALSE | Upregulated |
| 0,165258499 | 0,935997742 | CEACAM19 | FALSE | Upregulated |
| 0,126736814 | 0,935997742 | COL27A1 | FALSE | Upregulated |
| 0,148996369 | 0,935997742 | CREB1 | FALSE | Upregulated |
| -0,142102294 | 0,935997742 | GNRHR2 | FALSE | Downregulated |
| -0,227241176 | 0,935997742 | KIAA0922 | FALSE | Downregulated |
| 0,128820044 | 0,935997742 | LAMA5 | FALSE | Upregulated |
| 0,248833177 | 0,935997742 | LINC00612 | FALSE | Upregulated |
| -0,159960368 | 0,935997742 | LOC101928690 | FALSE | Downregulated |
| 0,186343223 | 0,935997742 | NME9 | FALSE | Upregulated |
| 0,149966274 | 0,935997742 | PAPLN | FALSE | Upregulated |
| -0,254907362 | 0,935997742 | PEX1 | FALSE | Downregulated |
| -0,223273596 | 0,935997742 | POLR2C | FALSE | Downregulated |
| 0,258687287 | 0,935997742 | RNF144B | FALSE | Upregulated |
| 0,188955235 | 0,935997742 | SCN8A | FALSE | Upregulated |
| 0,104158405 | 0,935997742 | SLCO2B1 | FALSE | Upregulated |
| 0,175314995 | 0,935997742 | STK24 | FALSE | Upregulated |
| -0,140403006 | 0,935997742 | UROC1 | FALSE | Downregulated |
| 0,217579709 | 0,936162478 | GOLGA1 | FALSE | Upregulated |
| -0,154585636 | 0,936216179 | FAM20C | FALSE | Downregulated |
| 0,239598407 | 0,936216179 | NEK4 | FALSE | Upregulated |
| -0,235204293 | 0,936216179 | PPP5C | FALSE | Downregulated |
| 0,191562459 | 0,936216179 | RGL2 | FALSE | Upregulated |
| 0,141801682 | 0,936362874 | SLC27A1 | FALSE | Upregulated |
| 0,148549165 | 0,936474388 | MAGI3 | FALSE | Upregulated |
| 0,215971006 | 0,936474388 | STK35 | FALSE | Upregulated |
| 0,11944247 | 0,936657099 | ASL | FALSE | Upregulated |
| 0,134187293 | 0,936657099 | BEST1 | FALSE | Upregulated |

| | | | | |
|---|---|---|---|---|
| 0,187495342 | 0,936657099 | CSNK1G2 | FALSE | Upregulated |
| -0,192892339 | 0,936657099 | LHX4 | FALSE | Downregulated |
| 0,160967628 | 0,936962679 | CASKIN2 | FALSE | Upregulated |
| 0,236040665 | 0,936962679 | POLR2L | FALSE | Upregulated |
| -0,189155535 | 0,937142713 | CARKD | FALSE | Downregulated |
| 0,140179584 | 0,937142713 | FAM120A | FALSE | Upregulated |
| -0,199806452 | 0,937142713 | PARP4 | FALSE | Downregulated |
| -0,192646203 | 0,937142713 | RP5-1033H22.2 | FALSE | Downregulated |
| 0,123061425 | 0,937142713 | RPL18A | FALSE | Upregulated |
| -0,148940741 | 0,937142713 | UBE2R2 | FALSE | Downregulated |
| -0,139874096 | 0,937251404 | STAG3L5P-PVRIG2P-PILRB | FALSE | Downregulated |
| -0,210727115 | 0,937386408 | ADRM1 | FALSE | Downregulated |
| 0,111782711 | 0,937386408 | LSMEM2 | FALSE | Upregulated |
| 0,188164529 | 0,937447454 | ATXN1L | FALSE | Upregulated |
| -0,233180122 | 0,937447454 | CNTNAP1 | FALSE | Downregulated |
| -0,254112044 | 0,937447454 | EXOC5 | FALSE | Downregulated |
| 0,14314291 | 0,937447454 | FAM211A | FALSE | Upregulated |
| 0,226795176 | 0,937447454 | FBXL13 | FALSE | Upregulated |
| 0,0993271 | 0,937447454 | FLNB | FALSE | Upregulated |
| 0,144093703 | 0,937447454 | KANK2 | FALSE | Upregulated |
| -0,171625916 | 0,937447454 | KIAA0355 | FALSE | Downregulated |
| -0,207259272 | 0,937447454 | MARCH8 | FALSE | Downregulated |
| -0,13391612 | 0,937447454 | TOLLIP | FALSE | Downregulated |
| -0,167115675 | 0,937447454 | UBA52 | FALSE | Downregulated |
| -0,184415867 | 0,937447454 | ZNF671 | FALSE | Downregulated |
| 0,18643853 | 0,937587361 | ATHL1 | FALSE | Upregulated |
| 0,158399043 | 0,937587361 | DGKD | FALSE | Upregulated |
| 0,149855675 | 0,937587361 | LOC101928958 | FALSE | Upregulated |
| -0,187929197 | 0,937587361 | NASP | FALSE | Downregulated |
| -0,17935696 | 0,937587361 | PPIA | FALSE | Downregulated |
| -0,243402814 | 0,937587361 | RBMS1 | FALSE | Downregulated |
| -0,230248272 | 0,937587361 | SART3 | FALSE | Downregulated |
| -0,224997696 | 0,937715533 | EYA3 | FALSE | Downregulated |
| -0,193924454 | 0,93784444 | ASMTL-AS1 | FALSE | Downregulated |
| 0,25752648 | 0,93784444 | NUB1 | FALSE | Upregulated |
| 0,142470425 | 0,93784444 | RAB11B | FALSE | Upregulated |
| 0,171202427 | 0,938085163 | CECR1 | FALSE | Upregulated |
| -0,231505513 | 0,938085163 | PIK3R2 | FALSE | Downregulated |
| -0,231267934 | 0,938329063 | LOC101929385 | FALSE | Downregulated |
| -0,231649852 | 0,938329063 | PANX1 | FALSE | Downregulated |
| -0,211969889 | 0,938375038 | CDC37L1 | FALSE | Downregulated |
| 0,33148199 | 0,938375038 | GUSBP1 | FALSE | Upregulated |
| 0,209861103 | 0,938375038 | SASH1 | FALSE | Upregulated |
| -0,189794356 | 0,938414114 | ALDH1L2 | FALSE | Downregulated |
| 0,179163963 | 0,938414114 | C21orf62 | FALSE | Upregulated |
| 0,138374648 | 0,938414114 | CIC | FALSE | Upregulated |
| -0,221826773 | 0,938414114 | DCUN1D4 | FALSE | Downregulated |
| 0,106067727 | 0,938414114 | EPAS1 | FALSE | Upregulated |
| 0,268400054 | 0,938414114 | FAM120C | FALSE | Upregulated |
| -0,149472658 | 0,938414114 | FBXO22-AS1 | FALSE | Downregulated |

| | | | | |
|---|---|---|---|---|
| 0,232233836 | 0,938414114 | KCNJ10 | FALSE | Upregulated |
| 0,148139965 | 0,938414114 | LOC101928445 | FALSE | Upregulated |
| 0,215108108 | 0,938414114 | LOC102723762 | FALSE | Upregulated |
| 0,255598584 | 0,938414114 | LOC441179 | FALSE | Upregulated |
| 0,197293942 | 0,938414114 | PRELID1 | FALSE | Upregulated |
| 0,207848747 | 0,938414114 | SPATA20 | FALSE | Upregulated |
| -0,198708098 | 0,939152559 | XPC | FALSE | Downregulated |
| 0,188791077 | 0,939152559 | ZCCHC2 | FALSE | Upregulated |
| 0,174045891 | 0,939590826 | WDR55 | FALSE | Upregulated |
| -0,207160446 | 0,939714868 | ELMO1 | FALSE | Downregulated |
| 0,150300997 | 0,939714868 | ITIH3 | FALSE | Upregulated |
| -0,235671229 | 0,939714868 | NUP43 | FALSE | Downregulated |
| 0,197611996 | 0,940476501 | LRRC24 | FALSE | Upregulated |
| 0,198605354 | 0,940476501 | RAPGEF5 | FALSE | Upregulated |
| -0,172146881 | 0,941226295 | HP1BP3 | FALSE | Downregulated |
| -0,17305948 | 0,941226295 | IDS | FALSE | Downregulated |
| -0,233146546 | 0,941226295 | SWT1 | FALSE | Downregulated |
| -0,187772725 | 0,941226295 | SYNJ2BP | FALSE | Downregulated |
| -0,197137248 | 0,941232072 | C1QL3 | FALSE | Downregulated |
| -0,167036269 | 0,941232072 | LARGE-AS1 | FALSE | Downregulated |
| 0,153723781 | 0,941232072 | MAST4 | FALSE | Upregulated |
| -0,141065229 | 0,941232072 | NBPF14 | FALSE | Downregulated |
| 0,237351871 | 0,941232072 | PSMD1 | FALSE | Upregulated |
| 0,15014749 | 0,941232072 | SCRIB | FALSE | Upregulated |
| -0,217153847 | 0,941232072 | SIL1 | FALSE | Downregulated |
| 0,193742594 | 0,941258364 | CRK | FALSE | Upregulated |
| -0,136439459 | 0,941258364 | LOC101930421 | FALSE | Downregulated |
| 0,174981688 | 0,941258364 | MKNK1-AS1 | FALSE | Upregulated |
| -0,189791017 | 0,941258364 | RNF167 | FALSE | Downregulated |
| -0,151095038 | 0,941638177 | ANKRD52 | FALSE | Downregulated |
| 0,185080336 | 0,941638177 | SNX9 | FALSE | Upregulated |
| -0,200813261 | 0,941638177 | TMEM214 | FALSE | Downregulated |
| 0,170802321 | 0,941828117 | C1QB | FALSE | Upregulated |
| 0,157540776 | 0,941828117 | DOC2A | FALSE | Upregulated |
| -0,298148078 | 0,941828117 | SLC25A34 | FALSE | Downregulated |
| 0,192174657 | 0,942242472 | APEH | FALSE | Upregulated |
| -0,141252554 | 0,942242472 | EHMT1 | FALSE | Downregulated |
| 0,145673429 | 0,942242472 | FAM149B1 | FALSE | Upregulated |
| 0,199255256 | 0,942242472 | MAGT1 | FALSE | Upregulated |
| -0,146909465 | 0,942242472 | ORAI2 | FALSE | Downregulated |
| 0,16067475 | 0,942305579 | DCAF5 | FALSE | Upregulated |
| 0,156888318 | 0,942360479 | BCL6 | FALSE | Upregulated |
| 0,184079246 | 0,942360479 | CCNY | FALSE | Upregulated |
| -0,23711682 | 0,942360479 | CENPN | FALSE | Downregulated |
| -0,204917058 | 0,942360479 | COG3 | FALSE | Downregulated |
| 0,151428497 | 0,942360479 | GOT1 | FALSE | Upregulated |
| 0,161504516 | 0,942360479 | KANSL1 | FALSE | Upregulated |
| 0,250920309 | 0,942360479 | LGALS4 | FALSE | Upregulated |
| -0,172643217 | 0,942360479 | LOC101928451 | FALSE | Downregulated |
| 0,202727972 | 0,942360479 | NRG4 | FALSE | Upregulated |
| -0,203870065 | 0,942360479 | OSBPL8 | FALSE | Downregulated |

| | | | | |
|---|---|---|---|---|
| -0,206032509 | 0,942360479 | PCID2 | FALSE | Downregulated |
| 0,178045973 | 0,942360479 | PDGFRA | FALSE | Upregulated |
| -0,140111603 | 0,942360479 | PNISR | FALSE | Downregulated |
| -0,228629928 | 0,942360479 | RBM42 | FALSE | Downregulated |
| -0,132243294 | 0,942360479 | SYT7 | FALSE | Downregulated |
| 0,172191051 | 0,942360479 | TMED1 | FALSE | Upregulated |
| 0,1448459 | 0,942360479 | TSPYL6 | FALSE | Upregulated |
| -0,23025064 | 0,942360479 | TTLL12 | FALSE | Downregulated |
| -0,161851526 | 0,942360479 | ZFAND5 | FALSE | Downregulated |
| 0,22569992 | 0,942360479 | ZIC4 | FALSE | Upregulated |
| -0,180251571 | 0,942409849 | RAB8A | FALSE | Downregulated |
| -0,192160598 | 0,942409849 | ZFP28 | FALSE | Downregulated |
| -0,135109402 | 0,942430304 | AGPAT3 | FALSE | Downregulated |
| 0,258219743 | 0,942430304 | ARHGEF35 | FALSE | Upregulated |
| -0,181340285 | 0,942430304 | BRCA2 | FALSE | Downregulated |
| -0,149070532 | 0,942430304 | EHBP1 | FALSE | Downregulated |
| 0,102989599 | 0,942430304 | KMT2D | FALSE | Upregulated |
| -0,216175084 | 0,942430304 | LOC101928429 | FALSE | Downregulated |
| 0,166736507 | 0,942520144 | EPB41L4A | FALSE | Upregulated |
| 0,175354892 | 0,942520144 | PHYKPL | FALSE | Upregulated |
| -0,156248644 | 0,942520144 | USP47 | FALSE | Downregulated |
| -0,237690017 | 0,942838706 | BCORL1 | FALSE | Downregulated |
| 0,183218566 | 0,942838706 | C3orf22 | FALSE | Upregulated |
| -0,231593838 | 0,942838706 | CLEC12A | FALSE | Downregulated |
| -0,163611895 | 0,942838706 | CRBN | FALSE | Downregulated |
| 0,094080934 | 0,942838706 | CTBP1-AS | FALSE | Upregulated |
| -0,160519197 | 0,942838706 | D2HGDH | FALSE | Downregulated |
| 0,183143367 | 0,942838706 | EIF3I | FALSE | Upregulated |
| -0,109578268 | 0,942838706 | EYS | FALSE | Downregulated |
| -0,138198853 | 0,942838706 | GOLGA4 | FALSE | Downregulated |
| 0,176981974 | 0,942838706 | GPR146 | FALSE | Upregulated |
| -0,192855946 | 0,942838706 | KANK3 | FALSE | Downregulated |
| -0,165718151 | 0,942838706 | LARGE | FALSE | Downregulated |
| -0,230297804 | 0,942838706 | LINC00861 | FALSE | Downregulated |
| 0,190686769 | 0,942838706 | LOC100132815 | FALSE | Upregulated |
| 0,155067898 | 0,942838706 | LOC101927190 | FALSE | Upregulated |
| 0,150285206 | 0,942838706 | MIR600HG | FALSE | Upregulated |
| 0,187219319 | 0,942838706 | NFYC | FALSE | Upregulated |
| -0,246421667 | 0,942838706 | NPY6R | FALSE | Downregulated |
| 0,117494748 | 0,942838706 | NUTM2D | FALSE | Upregulated |
| 0,209426192 | 0,942838706 | OSBP2 | FALSE | Upregulated |
| -0,163326391 | 0,942838706 | PABPC4 | FALSE | Downregulated |
| -0,159048563 | 0,942838706 | PANK2 | FALSE | Downregulated |
| -0,230223871 | 0,942838706 | RAB6B | FALSE | Downregulated |
| -0,234780123 | 0,942838706 | RBBP6 | FALSE | Downregulated |
| 0,170229441 | 0,942838706 | RCN3 | FALSE | Upregulated |
| 0,121631291 | 0,942838706 | RETSAT | FALSE | Upregulated |
| -0,227500053 | 0,942838706 | RSL1D1 | FALSE | Downregulated |
| -0,211452722 | 0,942838706 | SCARF1 | FALSE | Downregulated |
| -0,134691268 | 0,942838706 | SETD5 | FALSE | Downregulated |
| -0,196984105 | 0,942838706 | SGK494 | FALSE | Downregulated |

| | | | | |
|---|---|---|---|---|
| -0,11723564 | 0,942838706 | SHANK2-AS1 | FALSE | Downregulated |
| -0,232228985 | 0,942838706 | SLC19A3 | FALSE | Downregulated |
| -0,131916706 | 0,942838706 | SURF4 | FALSE | Downregulated |
| 0,187172604 | 0,942838706 | TSC22D1 | FALSE | Upregulated |
| 0,196533732 | 0,942838706 | UBA7 | FALSE | Upregulated |
| -0,149084375 | 0,942838706 | ZBTB46 | FALSE | Downregulated |
| -0,207911402 | 0,942838706 | ZFYVE20 | FALSE | Downregulated |
| 0,203568923 | 0,94288748 | CRTC1 | FALSE | Upregulated |
| -0,202203308 | 0,942952257 | AQP11 | FALSE | Downregulated |
| -0,177206234 | 0,942952257 | LOC100507520 | FALSE | Downregulated |
| 0,174833411 | 0,942952257 | LOC101927345 | FALSE | Upregulated |
| 0,193211915 | 0,942952257 | LOC102724281 | FALSE | Upregulated |
| -0,165914069 | 0,942952257 | PPP2R5A | FALSE | Downregulated |
| -0,224811469 | 0,942952257 | RP13-143G15.3 | FALSE | Downregulated |
| 0,131120674 | 0,942952257 | SKI | FALSE | Upregulated |
| -0,123152102 | 0,942952257 | WDR33 | FALSE | Downregulated |
| -0,234980044 | 0,942989169 | ANKS1B | FALSE | Downregulated |
| 0,118647337 | 0,942989169 | BCL9L | FALSE | Upregulated |
| -0,14016913 | 0,942989169 | CDC42BPA | FALSE | Downregulated |
| -0,166481847 | 0,942989169 | DDX42 | FALSE | Downregulated |
| -0,172960248 | 0,942989169 | INVS | FALSE | Downregulated |
| 0,245996774 | 0,942989169 | LOC102724254 | FALSE | Upregulated |
| -0,185969174 | 0,942989169 | NEMF | FALSE | Downregulated |
| 0,221777707 | 0,942989169 | PTPRG-AS1 | FALSE | Upregulated |
| 0,227929439 | 0,943208502 | LOC100507472 | FALSE | Upregulated |
| -0,214204003 | 0,943208502 | REPS2 | FALSE | Downregulated |
| 0,148834529 | 0,943368891 | APOL2 | FALSE | Upregulated |
| 0,180964425 | 0,943368891 | ARHGEF7 | FALSE | Upregulated |
| 0,152106148 | 0,943368891 | FZR1 | FALSE | Upregulated |
| -0,207026064 | 0,943368891 | LOC101060581 | FALSE | Downregulated |
| 0,186633441 | 0,943368891 | QRICH1 | FALSE | Upregulated |
| -0,183918571 | 0,943368891 | RPAIN | FALSE | Downregulated |
| -0,202719572 | 0,943368891 | SLC25A28 | FALSE | Downregulated |
| 0,11717993 | 0,943368891 | TMPRSS9 | FALSE | Upregulated |
| -0,22058374 | 0,943368891 | TOPORS-AS1 | FALSE | Downregulated |
| -0,095198652 | 0,943428311 | SZT2 | FALSE | Downregulated |
| 0,129180119 | 0,943548885 | RBMS2 | FALSE | Upregulated |
| -0,1502312 | 0,943666812 | ANGPT2 | FALSE | Downregulated |
| -0,200807518 | 0,943666812 | GPR113 | FALSE | Downregulated |
| 0,101318298 | 0,943666812 | KMT2C | FALSE | Upregulated |
| -0,135222367 | 0,943666812 | MSH6 | FALSE | Downregulated |
| -0,17777348 | 0,943666812 | PHKA2-AS1 | FALSE | Downregulated |
| 0,183843839 | 0,943666812 | SMPD1 | FALSE | Upregulated |
| 0,204382873 | 0,943731196 | DOK6 | FALSE | Upregulated |
| 0,194626855 | 0,943818985 | FAM57A | FALSE | Upregulated |
| 0,152542089 | 0,943908345 | PARP9 | FALSE | Upregulated |
| -0,110605032 | 0,944137262 | ABCC2 | FALSE | Downregulated |
| -0,230356622 | 0,944137262 | CLDN15 | FALSE | Downregulated |
| -0,176723251 | 0,944137262 | EPOR | FALSE | Downregulated |
| 0,106789492 | 0,944137262 | HECTD4 | FALSE | Upregulated |
| -0,221564963 | 0,944137262 | HIST1H2AC | FALSE | Downregulated |

| | | | | |
|---|---|---|---|---|
| -0,15977743 | 0,944137262 | IFIT1 | FALSE | Downregulated |
| 0,142483659 | 0,944137262 | PIK3AP1 | FALSE | Upregulated |
| -0,174169424 | 0,944137262 | SNRPD3 | FALSE | Downregulated |
| -0,142192734 | 0,944137262 | UFD1L | FALSE | Downregulated |
| -0,130229974 | 0,944534262 | RRS1 | FALSE | Downregulated |
| -0,118536944 | 0,944883536 | LRP6 | FALSE | Downregulated |
| -0,211345942 | 0,944883536 | PNPT1 | FALSE | Downregulated |
| 0,116940686 | 0,944883536 | STAB1 | FALSE | Upregulated |
| -0,137939974 | 0,944883536 | USP3 | FALSE | Downregulated |
| -0,154520848 | 0,944883536 | XRN1 | FALSE | Downregulated |
| -0,161891855 | 0,944916694 | BMP2K | FALSE | Downregulated |
| -0,200255424 | 0,945113036 | RP11-98I9.4 | FALSE | Downregulated |
| 0,163660548 | 0,945323203 | ACOT2 | FALSE | Upregulated |
| 0,195522994 | 0,945323203 | LETM1 | FALSE | Upregulated |
| 0,158996988 | 0,945323203 | PSKH1 | FALSE | Upregulated |
| -0,178357965 | 0,945323203 | WASL | FALSE | Downregulated |
| -0,187811009 | 0,945323203 | ZNF148 | FALSE | Downregulated |
| -0,169160605 | 0,945323203 | ZNF571-AS1 | FALSE | Downregulated |
| 0,206798838 | 0,945323203 | ZNRF3 | FALSE | Upregulated |
| 0,158287603 | 0,945852109 | C11orf24 | FALSE | Upregulated |
| 0,197483331 | 0,945852109 | NFYB | FALSE | Upregulated |
| -0,147575645 | 0,946056997 | AGO1 | FALSE | Downregulated |
| 0,164377586 | 0,946056997 | ASF1A | FALSE | Upregulated |
| -0,127695771 | 0,946056997 | BMP1 | FALSE | Downregulated |
| 0,229575066 | 0,946056997 | BTN3A2 | FALSE | Upregulated |
| 0,186829119 | 0,946056997 | C9orf172 | FALSE | Upregulated |
| 0,178272714 | 0,946056997 | C9orf37 | FALSE | Upregulated |
| -0,190830594 | 0,946056997 | CACTIN | FALSE | Downregulated |
| -0,150913234 | 0,946056997 | CACTIN-AS1 | FALSE | Downregulated |
| 0,192267008 | 0,946056997 | CEP95 | FALSE | Upregulated |
| -0,108095768 | 0,946056997 | EPN1 | FALSE | Downregulated |
| -0,118326778 | 0,946056997 | FNDC3A | FALSE | Downregulated |
| 0,135022699 | 0,946056997 | INTS3 | FALSE | Upregulated |
| -0,141815803 | 0,946056997 | KAT6A | FALSE | Downregulated |
| -0,164547018 | 0,946056997 | KIAA0141 | FALSE | Downregulated |
| 0,173480378 | 0,946056997 | LOXL3 | FALSE | Upregulated |
| -0,174487045 | 0,946056997 | LSM14B | FALSE | Downregulated |
| 0,144533438 | 0,946056997 | NAT9 | FALSE | Upregulated |
| 0,17524844 | 0,946056997 | NPHP3 | FALSE | Upregulated |
| -0,144070776 | 0,946056997 | PDPR | FALSE | Downregulated |
| -0,196327048 | 0,946056997 | PPP1R9A | FALSE | Downregulated |
| 0,151033145 | 0,946056997 | PPP6R3 | FALSE | Upregulated |
| -0,223454361 | 0,946056997 | PRSS8 | FALSE | Downregulated |
| -0,145081635 | 0,946056997 | PTMA | FALSE | Downregulated |
| 0,155433619 | 0,946056997 | RAD52 | FALSE | Upregulated |
| -0,095482041 | 0,946056997 | RPS2 | FALSE | Downregulated |
| 0,209571531 | 0,946056997 | SCRN3 | FALSE | Upregulated |
| 0,148756897 | 0,946056997 | SIPA1 | FALSE | Upregulated |
| -0,201206494 | 0,946056997 | THOP1 | FALSE | Downregulated |
| 0,170455089 | 0,946056997 | TMEM39A | FALSE | Upregulated |
| 0,173911054 | 0,946056997 | ZDHHC16 | FALSE | Upregulated |

| | | | | |
|---|---|---|---|---|
| 0,200355365 | 0,946056997 | ZDHHC20 | FALSE | Upregulated |
| -0,122151388 | 0,946056997 | ZNF750 | FALSE | Downregulated |
| -0,183945139 | 0,946361079 | RXRB | FALSE | Downregulated |
| 0,164972396 | 0,946361079 | SLC2A6 | FALSE | Upregulated |
| -0,156290866 | 0,946465986 | GPR64 | FALSE | Downregulated |
| -0,108169544 | 0,946465986 | RBM15B | FALSE | Downregulated |
| 0,142277008 | 0,946564797 | ASXL2 | FALSE | Upregulated |
| 0,173664264 | 0,946564797 | BST2 | FALSE | Upregulated |
| -0,162745809 | 0,946564797 | C4orf19 | FALSE | Downregulated |
| -0,139803512 | 0,946564797 | C6orf89 | FALSE | Downregulated |
| 0,225839659 | 0,946564797 | CCDC157 | FALSE | Upregulated |
| -0,155372594 | 0,946564797 | CDHR2 | FALSE | Downregulated |
| -0,207710325 | 0,946564797 | CHURC1 | FALSE | Downregulated |
| -0,183614324 | 0,946564797 | DCN | FALSE | Downregulated |
| 0,188585057 | 0,946564797 | EML3 | FALSE | Upregulated |
| 0,127627166 | 0,946564797 | FAM69B | FALSE | Upregulated |
| 0,084870146 | 0,946564797 | HPD | FALSE | Upregulated |
| 0,189141266 | 0,946564797 | IKBKAP | FALSE | Upregulated |
| -0,139242842 | 0,946564797 | LOC101929017 | FALSE | Downregulated |
| 0,12643844 | 0,946564797 | MPP5 | FALSE | Upregulated |
| 0,168802363 | 0,946564797 | NCLN | FALSE | Upregulated |
| -0,217477174 | 0,946564797 | P4HTM | FALSE | Downregulated |
| -0,206876477 | 0,946564797 | PHF1 | FALSE | Downregulated |
| 0,188130344 | 0,946564797 | RGAG4 | FALSE | Upregulated |
| 0,177722274 | 0,946564797 | VARS | FALSE | Upregulated |
| 0,091205234 | 0,946770455 | EP300 | FALSE | Upregulated |
| 0,124324462 | 0,946770455 | FZD4 | FALSE | Upregulated |
| 0,200203977 | 0,946911719 | GPAA1 | FALSE | Upregulated |
| -0,109093117 | 0,946911719 | TMEM259 | FALSE | Downregulated |
| 0,113756873 | 0,947022485 | CERS5 | FALSE | Upregulated |
| 0,16195711 | 0,947022485 | MGC16275 | FALSE | Upregulated |
| 0,13087078 | 0,947022485 | SMAD5-AS1 | FALSE | Upregulated |
| 0,219176977 | 0,947227288 | FBXW8 | FALSE | Upregulated |
| 0,092268346 | 0,947299873 | GATC | FALSE | Upregulated |
| -0,186938309 | 0,947299873 | MCMBP | FALSE | Downregulated |
| 0,204983006 | 0,947299873 | OLFM2 | FALSE | Upregulated |
| -0,110160341 | 0,947299873 | PDLIM5 | FALSE | Downregulated |
| -0,168869636 | 0,947299873 | SLC35E1 | FALSE | Downregulated |
| -0,169237528 | 0,947299873 | TSPAN4 | FALSE | Downregulated |
| 0,201606319 | 0,947299873 | USP33 | FALSE | Upregulated |
| -0,094470305 | 0,94741696 | FCGRT | FALSE | Downregulated |
| -0,138449321 | 0,94741696 | IL2RB | FALSE | Downregulated |
| 0,161440342 | 0,94741696 | SPATA1 | FALSE | Upregulated |
| 0,145580088 | 0,94764579 | ASPDH | FALSE | Upregulated |
| 0,19974865 | 0,94764579 | LOC101929040 | FALSE | Upregulated |
| -0,165571413 | 0,94764579 | MTX1 | FALSE | Downregulated |
| 0,196151314 | 0,94764579 | NPB | FALSE | Upregulated |
| 0,147011488 | 0,94764579 | PTPN1 | FALSE | Upregulated |
| -0,142109607 | 0,94764579 | SIK3 | FALSE | Downregulated |
| -0,208342892 | 0,94764579 | TOMM40 | FALSE | Downregulated |
| -0,159229182 | 0,947729071 | HECTD3 | FALSE | Downregulated |

| | | | | |
|---|---|---|---|---|
| -0,133242466 | 0,947729071 | RPS4X | FALSE | Downregulated |
| -0,166311043 | 0,947847147 | CAPS | FALSE | Downregulated |
| 0,142268286 | 0,947847147 | CSPP1 | FALSE | Upregulated |
| 0,132524357 | 0,947847147 | GTF3C1 | FALSE | Upregulated |
| -0,134293191 | 0,947847147 | MBD3 | FALSE | Downregulated |
| 0,141011864 | 0,947847147 | SLC12A4 | FALSE | Upregulated |
| -0,133975157 | 0,948167244 | SP3 | FALSE | Downregulated |
| 0,197517431 | 0,948167244 | ZC3H12C | FALSE | Upregulated |
| 0,121103322 | 0,948490227 | NNT | FALSE | Upregulated |
| 0,122768345 | 0,948667462 | MED1 | FALSE | Upregulated |
| -0,150554685 | 0,948667462 | MYL9 | FALSE | Downregulated |
| -0,182129696 | 0,948667462 | PRKAB1 | FALSE | Downregulated |
| -0,172380841 | 0,948737015 | EIF6 | FALSE | Downregulated |
| -0,168209779 | 0,948795913 | TRIM33 | FALSE | Downregulated |
| -0,139207003 | 0,949281192 | AP2A2 | FALSE | Downregulated |
| 0,083627678 | 0,949281192 | SPDYE2 | FALSE | Upregulated |
| 0,133232964 | 0,949309728 | LPHN2 | FALSE | Upregulated |
| 0,10331152 | 0,949309728 | PAQR3 | FALSE | Upregulated |
| 0,124745213 | 0,949309728 | STK11 | FALSE | Upregulated |
| -0,147032723 | 0,949309728 | USP49 | FALSE | Downregulated |
| -0,170652686 | 0,949320983 | PHF2 | FALSE | Downregulated |
| 0,12781268 | 0,949584521 | CLDN20 | FALSE | Upregulated |
| 0,151277784 | 0,949671561 | ARF6 | FALSE | Upregulated |
| -0,145962683 | 0,949736131 | CHID1 | FALSE | Downregulated |
| -0,144672967 | 0,949736131 | GPATCH8 | FALSE | Downregulated |
| -0,103106506 | 0,949736131 | KMO | FALSE | Downregulated |
| -0,090824089 | 0,949736131 | SBF1 | FALSE | Downregulated |
| -0,113912498 | 0,949844323 | BTBD2 | FALSE | Downregulated |
| -0,181589046 | 0,949844323 | TRIM52 | FALSE | Downregulated |
| 0,118912355 | 0,949922137 | PARP10 | FALSE | Upregulated |
| 0,149807495 | 0,949922137 | RNF103 | FALSE | Upregulated |
| -0,191909105 | 0,95017665 | RPS6KB1 | FALSE | Downregulated |
| -0,141988908 | 0,950185183 | DAP | FALSE | Downregulated |
| 0,092018581 | 0,950185183 | WNK1 | FALSE | Upregulated |
| -0,096972062 | 0,950268868 | FBRSL1 | FALSE | Downregulated |
| -0,124554066 | 0,950268868 | GLTPD2 | FALSE | Downregulated |
| 0,109263482 | 0,950268868 | NIT1 | FALSE | Upregulated |
| 0,165858931 | 0,950268868 | SPPL2B | FALSE | Upregulated |
| 0,097992523 | 0,950388011 | ANKRD36BP1 | FALSE | Upregulated |
| -0,163813407 | 0,950388011 | FBXL5 | FALSE | Downregulated |
| 0,125029159 | 0,950388011 | FSD2 | FALSE | Upregulated |
| 0,172888832 | 0,950388011 | HMG20B | FALSE | Upregulated |
| 0,122871703 | 0,950388011 | KIAA2013 | FALSE | Upregulated |
| 0,137724423 | 0,950388011 | LOC100996873 | FALSE | Upregulated |
| 0,208591016 | 0,950388011 | LOC101927124 | FALSE | Upregulated |
| -0,193093423 | 0,950388011 | METTL3 | FALSE | Downregulated |
| -0,110724692 | 0,950388011 | PDE7A | FALSE | Downregulated |
| -0,206431883 | 0,950388011 | RNASE7 | FALSE | Downregulated |
| -0,113021169 | 0,950388011 | SH3BP5-AS1 | FALSE | Downregulated |
| 0,165861305 | 0,950388011 | SSR3 | FALSE | Upregulated |
| 0,173992541 | 0,950388011 | TRIM35 | FALSE | Upregulated |

| | | | | |
|---|---|---|---|---|
| -0,177352823 | 0,950764538 | CD276 | FALSE | Downregulated |
| 0,118719794 | 0,950764538 | NEK9 | FALSE | Upregulated |
| 0,156839871 | 0,950848729 | KIF5B | FALSE | Upregulated |
| 0,185699349 | 0,950848729 | SMARCC1 | FALSE | Upregulated |
| -0,16108896 | 0,950892741 | AC104134.2 | FALSE | Downregulated |
| -0,135006952 | 0,951155145 | DCP2 | FALSE | Downregulated |
| -0,124069122 | 0,951155145 | LOC286190 | FALSE | Downregulated |
| 0,167165423 | 0,951155145 | PFKFB3 | FALSE | Upregulated |
| 0,18794575 | 0,951155145 | RBM26 | FALSE | Upregulated |
| 0,171445862 | 0,951329743 | MS4A6A | FALSE | Upregulated |
| -0,161583827 | 0,951329743 | RAET1E-AS1 | FALSE | Downregulated |
| 0,136441334 | 0,952253165 | EAF1 | FALSE | Upregulated |
| -0,14884668 | 0,952450066 | AMDHD1 | FALSE | Downregulated |
| 0,1739497 | 0,952450066 | ENDOU | FALSE | Upregulated |
| -0,117457929 | 0,952450066 | NBR1 | FALSE | Downregulated |
| -0,159046777 | 0,952450066 | NPRL3 | FALSE | Downregulated |
| -0,173721977 | 0,952450066 | PSMB4 | FALSE | Downregulated |
| 0,188887238 | 0,952450066 | XPO4 | FALSE | Upregulated |
| -0,143824414 | 0,952566177 | RBM14 | FALSE | Downregulated |
| -0,171503558 | 0,953001636 | SMCHD1 | FALSE | Downregulated |
| -0,13224557 | 0,953527711 | ZNF33A | FALSE | Downregulated |
| -0,132748372 | 0,953599899 | CDV3 | FALSE | Downregulated |
| -0,101624983 | 0,953599899 | ORMDL3 | FALSE | Downregulated |
| 0,176941186 | 0,954461589 | MFSD4 | FALSE | Upregulated |
| 0,198464697 | 0,954461589 | NS3BP | FALSE | Upregulated |
| 0,168947974 | 0,954461589 | PLCB3 | FALSE | Upregulated |
| 0,189550606 | 0,954775198 | LOC100506100 | FALSE | Upregulated |
| -0,14186772 | 0,95489734 | MXD4 | FALSE | Downregulated |
| 0,088644333 | 0,954932537 | TNS1 | FALSE | Upregulated |
| 0,117602765 | 0,955073394 | MRS2P2 | FALSE | Upregulated |
| -0,189192223 | 0,955365989 | CDPF1 | FALSE | Downregulated |
| -0,11642671 | 0,955365989 | HLF | FALSE | Downregulated |
| 0,137734667 | 0,95604062 | CNN3 | FALSE | Upregulated |
| -0,14552305 | 0,95604062 | COPB2 | FALSE | Downregulated |
| 0,127332688 | 0,95604062 | FNBP4 | FALSE | Upregulated |
| -0,111067688 | 0,956303858 | SCAF1 | FALSE | Downregulated |
| -0,156510903 | 0,956303858 | TAB3 | FALSE | Downregulated |
| -0,112975842 | 0,956329036 | CTIF | FALSE | Downregulated |
| 0,156307278 | 0,956329036 | LOC100287042 | FALSE | Upregulated |
| 0,118349426 | 0,956329036 | NR5A2 | FALSE | Upregulated |
| -0,174622177 | 0,956335853 | C16orf71 | FALSE | Downregulated |
| -0,094495683 | 0,956335853 | EIF2AK1 | FALSE | Downregulated |
| 0,180491063 | 0,956335853 | LOC101927066 | FALSE | Upregulated |
| 0,142365094 | 0,956335853 | MAP1LC3B | FALSE | Upregulated |
| -0,163958124 | 0,956335853 | PHB | FALSE | Downregulated |
| 0,148536674 | 0,956335853 | PSME4 | FALSE | Upregulated |
| 0,139245602 | 0,956335853 | TBCD | FALSE | Upregulated |
| -0,127584862 | 0,956335853 | WWC2 | FALSE | Downregulated |
| 0,142847229 | 0,956610691 | ATG9A | FALSE | Upregulated |
| -0,148717672 | 0,956610691 | INE2 | FALSE | Downregulated |
| 0,143574059 | 0,956610691 | SOS1 | FALSE | Upregulated |

| | | | | |
|---|---|---|---|---|
| -0,124872042 | 0,956715142 | SRGAP2-AS1 | FALSE | Downregulated |
| 0,12602958 | 0,956854484 | LOC101929211 | FALSE | Upregulated |
| -0,160444903 | 0,956854484 | PABPN1 | FALSE | Downregulated |
| -0,121433324 | 0,957292973 | ALPL | FALSE | Downregulated |
| -0,205218482 | 0,957292973 | APOOL | FALSE | Downregulated |
| 0,113802808 | 0,957292973 | CACNB2 | FALSE | Upregulated |
| -0,137413018 | 0,957292973 | CIAO1 | FALSE | Downregulated |
| -0,12314783 | 0,957292973 | HIPK3 | FALSE | Downregulated |
| 0,106689588 | 0,957292973 | KDM2A | FALSE | Upregulated |
| 0,100943154 | 0,957292973 | LOC102724696 | FALSE | Upregulated |
| 0,158125617 | 0,957292973 | MAK16 | FALSE | Upregulated |
| -0,175207979 | 0,957292973 | NDST3 | FALSE | Downregulated |
| 0,17874425 | 0,957292973 | PITPNC1 | FALSE | Upregulated |
| 0,198410107 | 0,957292973 | RTCB | FALSE | Upregulated |
| -0,095629434 | 0,957292973 | SCAP | FALSE | Downregulated |
| -0,166375429 | 0,957292973 | SIRT1 | FALSE | Downregulated |
| 0,122117954 | 0,957292973 | SLC25A3 | FALSE | Upregulated |
| -0,130448417 | 0,957292973 | SLC6A12 | FALSE | Downregulated |
| -0,098508847 | 0,957292973 | TXNIP | FALSE | Downregulated |
| 0,111772117 | 0,957387025 | LOC102724930 | FALSE | Upregulated |
| 0,090560158 | 0,958053082 | ANKRD11 | FALSE | Upregulated |
| 0,201883496 | 0,958053082 | ANKRD44 | FALSE | Upregulated |
| -0,098157806 | 0,958053082 | ARMC5 | FALSE | Downregulated |
| 0,1036152 | 0,958053082 | PBRM1 | FALSE | Upregulated |
| 0,121850012 | 0,958053082 | RCE1 | FALSE | Upregulated |
| -0,180176817 | 0,958053082 | SKP1 | FALSE | Downregulated |
| -0,161786334 | 0,958053082 | TANGO6 | FALSE | Downregulated |
| -0,125974268 | 0,958053082 | TNFRSF14 | FALSE | Downregulated |
| -0,132690071 | 0,958089052 | RBMX | FALSE | Downregulated |
| 0,094458725 | 0,958280227 | LIMD1-AS1 | FALSE | Upregulated |
| 0,132787328 | 0,958280227 | PTPN18 | FALSE | Upregulated |
| 0,13767632 | 0,959016731 | NBEAL1 | FALSE | Upregulated |
| 0,157335284 | 0,959171107 | ANK3 | FALSE | Upregulated |
| -0,156195849 | 0,959171107 | GOLGA8O | FALSE | Downregulated |
| -0,122957959 | 0,959171107 | SSR1 | FALSE | Downregulated |
| 0,140659763 | 0,959171107 | ZDHHC8 | FALSE | Upregulated |
| 0,140908605 | 0,959305853 | GIT2 | FALSE | Upregulated |
| 0,171710205 | 0,959305853 | PRPF40A | FALSE | Upregulated |
| -0,106531194 | 0,959442808 | CPT1A | FALSE | Downregulated |
| -0,215744851 | 0,959529421 | C14orf169 | FALSE | Downregulated |
| -0,134821617 | 0,959529421 | SPIDR | FALSE | Downregulated |
| 0,10316808 | 0,959931678 | PEA15 | FALSE | Upregulated |
| -0,160315243 | 0,960017053 | CABP7 | FALSE | Downregulated |
| -0,1173185 | 0,96007284 | LY6E | FALSE | Downregulated |
| 0,102776328 | 0,960226765 | ACIN1 | FALSE | Upregulated |
| -0,17298317 | 0,960226765 | ATP1A1OS | FALSE | Downregulated |
| -0,088152568 | 0,960226765 | ATP6V1C2 | FALSE | Downregulated |
| -0,135654376 | 0,960226765 | C11orf42 | FALSE | Downregulated |
| 0,108681945 | 0,960226765 | C3orf17 | FALSE | Upregulated |
| -0,109272746 | 0,960226765 | CTNNA1 | FALSE | Downregulated |
| 0,12455523 | 0,960226765 | DNAJC3 | FALSE | Upregulated |

| | | | | |
|---|---|---|---|---|
| -0,163841914 | 0,960226765 | EHMT2 | FALSE | Downregulated |
| 0,097449843 | 0,960226765 | FASN | FALSE | Upregulated |
| 0,122372172 | 0,960226765 | FUT2 | FALSE | Upregulated |
| 0,156425976 | 0,960226765 | LOC646127 | FALSE | Upregulated |
| -0,108441965 | 0,960226765 | ONECUT2 | FALSE | Downregulated |
| -0,126676155 | 0,960226765 | PCM1 | FALSE | Downregulated |
| 0,094414349 | 0,960226765 | SHANK3 | FALSE | Upregulated |
| 0,217006823 | 0,960226765 | SNX29 | FALSE | Upregulated |
| -0,172534717 | 0,960226765 | UQCRC2 | FALSE | Downregulated |
| -0,166533398 | 0,96030579 | FLT4 | FALSE | Downregulated |
| -0,090349719 | 0,96030579 | SDR39U1 | FALSE | Downregulated |
| 0,123115967 | 0,96030579 | ULK1 | FALSE | Upregulated |
| 0,113801584 | 0,96030579 | USF2 | FALSE | Upregulated |
| -0,22796347 | 0,960379234 | CCL23 | FALSE | Downregulated |
| 0,101164484 | 0,960509539 | NUTM2B | FALSE | Upregulated |
| -0,106379737 | 0,960552655 | SH3BP2 | FALSE | Downregulated |
| -0,16933462 | 0,960738501 | ATP2C1 | FALSE | Downregulated |
| -0,151990434 | 0,960738501 | KHDRBS1 | FALSE | Downregulated |
| 0,124956914 | 0,960738501 | PUF60 | FALSE | Upregulated |
| -0,166835631 | 0,960844273 | ATL2 | FALSE | Downregulated |
| -0,153318589 | 0,961445223 | DFNA5 | FALSE | Downregulated |
| 0,188783335 | 0,961445223 | GATS | FALSE | Upregulated |
| 0,153729207 | 0,961445223 | MMP24-AS1 | FALSE | Upregulated |
| 0,131646983 | 0,961445223 | NBPF26 | FALSE | Upregulated |
| -0,114310119 | 0,961445223 | OAZ2 | FALSE | Downregulated |
| 0,167265204 | 0,961445223 | SMYD4 | FALSE | Upregulated |
| 0,131206797 | 0,961445223 | SRRM1 | FALSE | Upregulated |
| -0,145866907 | 0,961445223 | TMPO | FALSE | Downregulated |
| 0,07331739 | 0,961445223 | VPS13B | FALSE | Upregulated |
| 0,113055684 | 0,961445223 | WDR24 | FALSE | Upregulated |
| 0,166591847 | 0,961494438 | AGPAT1 | FALSE | Upregulated |
| 0,098011652 | 0,961494438 | AP3D1 | FALSE | Upregulated |
| 0,130718246 | 0,961494438 | CTRL | FALSE | Upregulated |
| -0,085936373 | 0,961494438 | SH3BP5 | FALSE | Downregulated |
| -0,161213964 | 0,961504151 | KDM5D | FALSE | Downregulated |
| -0,118021787 | 0,961504151 | STT3B | FALSE | Downregulated |
| -0,153353145 | 0,961504151 | SUOX | FALSE | Downregulated |
| 0,084983838 | 0,961504151 | SYNC | FALSE | Upregulated |
| -0,188798459 | 0,961504151 | TTN-AS1 | FALSE | Downregulated |
| 0,126961444 | 0,961832428 | FKBP15 | FALSE | Upregulated |
| -0,091155175 | 0,962071941 | DHDDS | FALSE | Downregulated |
| -0,168137853 | 0,962486079 | C9orf69 | FALSE | Downregulated |
| 0,066490237 | 0,96258974 | MAVS | FALSE | Upregulated |
| 0,117478002 | 0,96258974 | VWA8 | FALSE | Upregulated |
| -0,167535747 | 0,962700262 | FTSJ3 | FALSE | Downregulated |
| -0,1270854 | 0,962700262 | HIP1 | FALSE | Downregulated |
| -0,166679287 | 0,962740922 | ACSF3 | FALSE | Downregulated |
| -0,143957644 | 0,962740922 | LOC101929648 | FALSE | Downregulated |
| 0,12854182 | 0,962740922 | PPP2R4 | FALSE | Upregulated |
| 0,12534283 | 0,962740922 | SREK1 | FALSE | Upregulated |
| -0,165213579 | 0,96280749 | KLF11 | FALSE | Downregulated |

| | | | | |
|---|---|---|---|---|
| 0,089899645 | 0,96280749 | KMT2A | FALSE | Upregulated |
| 0,0937671 | 0,96280749 | ZNF544 | FALSE | Upregulated |
| -0,117894313 | 0,963099993 | DNAAF2 | FALSE | Downregulated |
| -0,118169228 | 0,963165375 | ANKS1A | FALSE | Downregulated |
| -0,139851349 | 0,963764851 | ALX3 | FALSE | Downregulated |
| 0,146168021 | 0,963764851 | AMPD2 | FALSE | Upregulated |
| -0,080510334 | 0,963764851 | APLP2 | FALSE | Downregulated |
| 0,140428429 | 0,963764851 | CACUL1 | FALSE | Upregulated |
| 0,125247771 | 0,963764851 | CXCR6 | FALSE | Upregulated |
| -0,128466548 | 0,963764851 | GPR179 | FALSE | Downregulated |
| 0,206429715 | 0,963764851 | HGF | FALSE | Upregulated |
| 0,086656939 | 0,963764851 | LAMB2 | FALSE | Upregulated |
| -0,147073861 | 0,963764851 | LOC101928998 | FALSE | Downregulated |
| 0,106007243 | 0,963764851 | MCM3AP-AS1 | FALSE | Upregulated |
| 0,133591709 | 0,963764851 | NCBP2 | FALSE | Upregulated |
| 0,160711019 | 0,963764851 | PACSIN2 | FALSE | Upregulated |
| 0,116957864 | 0,963764851 | QARS | FALSE | Upregulated |
| -0,181950855 | 0,963764851 | RNU105A | FALSE | Downregulated |
| 0,151653005 | 0,963764851 | SCARNA13 | FALSE | Upregulated |
| 0,12196208 | 0,963764851 | SH2D3A | FALSE | Upregulated |
| 0,172130005 | 0,963764851 | STRN4 | FALSE | Upregulated |
| 0,167833968 | 0,963764851 | SYNGAP1 | FALSE | Upregulated |
| -0,126504767 | 0,963764851 | VRK2 | FALSE | Downregulated |
| -0,09968304 | 0,963764851 | ZNF506 | FALSE | Downregulated |
| -0,108853793 | 0,964455632 | RTEL1 | FALSE | Downregulated |
| 0,128576062 | 0,964510456 | RECQL5 | FALSE | Upregulated |
| 0,07240599 | 0,964510456 | SLC46A1 | FALSE | Upregulated |
| -0,153071567 | 0,964511491 | IDI1 | FALSE | Downregulated |
| -0,084559652 | 0,964565062 | ARID1B | FALSE | Downregulated |
| -0,123024788 | 0,964565062 | CBX5 | FALSE | Downregulated |
| -0,133684981 | 0,964565062 | CMIP | FALSE | Downregulated |
| 0,152450386 | 0,964565062 | CUL1 | FALSE | Upregulated |
| 0,145318471 | 0,964565062 | CYTH1 | FALSE | Upregulated |
| -0,120666798 | 0,964565062 | DNASE1 | FALSE | Downregulated |
| -0,152883357 | 0,964565062 | FBXW4 | FALSE | Downregulated |
| -0,098985463 | 0,964565062 | GPR17 | FALSE | Downregulated |
| -0,104648669 | 0,964565062 | GZF1 | FALSE | Downregulated |
| -0,172014767 | 0,964565062 | LOC100506621 | FALSE | Downregulated |
| -0,155788138 | 0,964565062 | MAPK6 | FALSE | Downregulated |
| -0,108507194 | 0,964565062 | NUTM2A | FALSE | Downregulated |
| 0,132570822 | 0,964565062 | RNF139-AS1 | FALSE | Upregulated |
| -0,084614395 | 0,964565062 | RREB1 | FALSE | Downregulated |
| 0,169172995 | 0,964565062 | SEC22B | FALSE | Upregulated |
| 0,103589999 | 0,964565062 | SMG5 | FALSE | Upregulated |
| -0,135806201 | 0,964565062 | TCF12 | FALSE | Downregulated |
| 0,153339117 | 0,964565062 | TRIM38 | FALSE | Upregulated |
| -0,139037913 | 0,964565062 | UBAP2 | FALSE | Downregulated |
| -0,12338314 | 0,964565062 | ZFAND3 | FALSE | Downregulated |
| 0,158295296 | 0,964984418 | DDX28 | FALSE | Upregulated |
| 0,131285603 | 0,965035637 | URGCP | FALSE | Upregulated |
| -0,118767487 | 0,965203916 | FAM175A | FALSE | Downregulated |

| | | | | |
|---|---|---|---|---|
| 0,12365808 | 0,965203916 | KIF3B | FALSE | Upregulated |
| -0,186782551 | 0,965203916 | LOC102723998 | FALSE | Downregulated |
| 0,113467182 | 0,965275878 | BAHCC1 | FALSE | Upregulated |
| 0,132886478 | 0,965275878 | LMO3 | FALSE | Upregulated |
| 0,126702129 | 0,965275878 | NCOA6 | FALSE | Upregulated |
| 0,155773314 | 0,965275878 | PRCP | FALSE | Upregulated |
| 0,149660464 | 0,965275878 | PRKAG3 | FALSE | Upregulated |
| 0,115874293 | 0,965275878 | SNN | FALSE | Upregulated |
| -0,112518839 | 0,965275878 | WDFY3 | FALSE | Downregulated |
| 0,109110519 | 0,965275878 | ZC2HC1C | FALSE | Upregulated |
| 0,081907638 | 0,965344649 | CCDC88A | FALSE | Upregulated |
| 0,094960843 | 0,965344649 | SRSF2 | FALSE | Upregulated |
| 0,162980823 | 0,965411123 | VPS9D1 | FALSE | Upregulated |
| -0,143647973 | 0,965514841 | ERLIN1 | FALSE | Downregulated |
| 0,110372711 | 0,965612786 | KDM6B | FALSE | Upregulated |
| -0,154011987 | 0,9659044 | PARVA | FALSE | Downregulated |
| 0,105719181 | 0,965970042 | C12orf61 | FALSE | Upregulated |
| -0,137185919 | 0,965970042 | CCDC57 | FALSE | Downregulated |
| -0,124739446 | 0,965970042 | CREBRF | FALSE | Downregulated |
| -0,160689171 | 0,965970042 | DIAPH2 | FALSE | Downregulated |
| -0,132731186 | 0,965970042 | EPS8 | FALSE | Downregulated |
| -0,146898305 | 0,965970042 | HDGFRP2 | FALSE | Downregulated |
| 0,166528088 | 0,965970042 | HPS4 | FALSE | Upregulated |
| 0,148784763 | 0,965970042 | KDM1B | FALSE | Upregulated |
| 0,108030025 | 0,965970042 | KIAA1522 | FALSE | Upregulated |
| 0,143077671 | 0,965970042 | KPNA4 | FALSE | Upregulated |
| 0,100172005 | 0,965970042 | LOC101927831 | FALSE | Upregulated |
| 0,13893547 | 0,965970042 | MCM7 | FALSE | Upregulated |
| -0,148564518 | 0,965970042 | MEX3D | FALSE | Downregulated |
| 0,084337393 | 0,965970042 | NNAT | FALSE | Upregulated |
| -0,112925724 | 0,965970042 | NRIP1 | FALSE | Downregulated |
| -0,102545111 | 0,965970042 | PRKAB2 | FALSE | Downregulated |
| 0,139656826 | 0,965970042 | PRKCZ | FALSE | Upregulated |
| -0,126858213 | 0,965970042 | RAB1B | FALSE | Downregulated |
| 0,103001658 | 0,965970042 | RAI1 | FALSE | Upregulated |
| -0,096614944 | 0,965970042 | REV3L | FALSE | Downregulated |
| -0,081840617 | 0,965970042 | SEPN1 | FALSE | Downregulated |
| 0,15712379 | 0,965970042 | SFMBT1 | FALSE | Upregulated |
| 0,115317652 | 0,965970042 | SLC25A18 | FALSE | Upregulated |
| 0,142037978 | 0,965970042 | SPAG5-AS1 | FALSE | Upregulated |
| 0,095569389 | 0,965970042 | TSHZ2 | FALSE | Upregulated |
| 0,101721077 | 0,965970042 | WHSC1 | FALSE | Upregulated |
| -0,120530108 | 0,966174131 | LOC102724611 | FALSE | Downregulated |
| -0,110490803 | 0,966437651 | BMPR2 | FALSE | Downregulated |
| 0,096555754 | 0,966437651 | EFCAB13 | FALSE | Upregulated |
| 0,13345638 | 0,966437651 | GALM | FALSE | Upregulated |
| -0,166330408 | 0,966437651 | GBP4 | FALSE | Downregulated |
| -0,168430819 | 0,966437651 | RFC5 | FALSE | Downregulated |
| -0,10767454 | 0,966437651 | TRIM56 | FALSE | Downregulated |
| 0,15009419 | 0,966603975 | C8G | FALSE | Upregulated |
| 0,116781035 | 0,966603975 | ZNF518A | FALSE | Upregulated |

| | | | | |
|---|---|---|---|---|
| -0,116978252 | 0,966662414 | DHX30 | FALSE | Downregulated |
| 0,115636776 | 0,966747234 | ADPGK-AS1 | FALSE | Upregulated |
| 0,129998446 | 0,966747234 | ANXA4 | FALSE | Upregulated |
| 0,131437732 | 0,966747234 | BCL2L14 | FALSE | Upregulated |
| -0,142944743 | 0,966747234 | CIZ1 | FALSE | Downregulated |
| 0,090497744 | 0,966747234 | IRF1 | FALSE | Upregulated |
| -0,099665396 | 0,966747234 | ZC3H4 | FALSE | Downregulated |
| 0,08677885 | 0,967334222 | ARAP1 | FALSE | Upregulated |
| -0,067901292 | 0,967632491 | ARHGEF12 | FALSE | Downregulated |
| -0,110554724 | 0,967722866 | FAM107B | FALSE | Downregulated |
| -0,123313807 | 0,96817987 | CALCOCO2 | FALSE | Downregulated |
| -0,088088776 | 0,96846372 | AKAP17A | FALSE | Downregulated |
| 0,147362238 | 0,96846372 | ANTXR2 | FALSE | Upregulated |
| 0,123810523 | 0,96846372 | BIRC2 | FALSE | Upregulated |
| -0,098472925 | 0,96846372 | CAMK2D | FALSE | Downregulated |
| 0,109502515 | 0,96846372 | CCPG1 | FALSE | Upregulated |
| 0,107684624 | 0,96846372 | DVL3 | FALSE | Upregulated |
| 0,067860026 | 0,96846372 | EIF4G1 | FALSE | Upregulated |
| 0,161225103 | 0,96846372 | FREM3 | FALSE | Upregulated |
| -0,088494962 | 0,96846372 | KIAA1109 | FALSE | Downregulated |
| -0,125599937 | 0,96846372 | LOC401296 | FALSE | Downregulated |
| 0,094209817 | 0,96846372 | MINK1 | FALSE | Upregulated |
| 0,095416888 | 0,96846372 | MMACHC | FALSE | Upregulated |
| -0,136561205 | 0,96846372 | NTN3 | FALSE | Downregulated |
| -0,151907762 | 0,96846372 | PFKFB1 | FALSE | Downregulated |
| -0,104861514 | 0,96846372 | PKD2L2 | FALSE | Downregulated |
| 0,097890442 | 0,96846372 | RANBP2 | FALSE | Upregulated |
| 0,088560735 | 0,96846372 | SCARB2 | FALSE | Upregulated |
| 0,138770715 | 0,96846372 | SNW1 | FALSE | Upregulated |
| 0,14765633 | 0,96846372 | SYNJ2 | FALSE | Upregulated |
| -0,103983512 | 0,96846372 | ZNF789 | FALSE | Downregulated |
| 0,117231823 | 0,96846372 | ZSCAN32 | FALSE | Upregulated |
| 0,088882509 | 0,968882603 | ANKRD17 | FALSE | Upregulated |
| -0,154256793 | 0,968882603 | EFNB1 | FALSE | Downregulated |
| 0,10131184 | 0,968882603 | SUMF2 | FALSE | Upregulated |
| 0,135548836 | 0,969097486 | WDFY1 | FALSE | Upregulated |
| -0,102810682 | 0,969166563 | AKAP8 | FALSE | Downregulated |
| 0,121903258 | 0,969166563 | EMC10 | FALSE | Upregulated |
| -0,146697955 | 0,969166563 | NPIPB9 | FALSE | Downregulated |
| -0,12315801 | 0,969166563 | TMX4 | FALSE | Downregulated |
| -0,070530805 | 0,969259379 | AMIGO3 | FALSE | Downregulated |
| 0,093554774 | 0,969259379 | AP1G1 | FALSE | Upregulated |
| 0,109735474 | 0,969259379 | BRD7P3 | FALSE | Upregulated |
| 0,072110693 | 0,969259379 | BRWD1 | FALSE | Upregulated |
| 0,138030502 | 0,969259379 | BTBD1 | FALSE | Upregulated |
| -0,132763286 | 0,969259379 | C19orf43 | FALSE | Downregulated |
| -0,088086081 | 0,969259379 | CALR | FALSE | Downregulated |
| -0,114312682 | 0,969259379 | CD99L2 | FALSE | Downregulated |
| 0,126113808 | 0,969259379 | CRCP | FALSE | Upregulated |
| 0,099675483 | 0,969259379 | CS | FALSE | Upregulated |
| 0,095196289 | 0,969259379 | CSRNP2 | FALSE | Upregulated |

| | | | | |
|---|---|---|---|---|
| 0,149820935 | 0,969259379 | DEPTOR | FALSE | Upregulated |
| 0,090254265 | 0,969259379 | DGKQ | FALSE | Upregulated |
| -0,083447655 | 0,969259379 | DHRS3 | FALSE | Downregulated |
| -0,137413269 | 0,969259379 | DNAJC4 | FALSE | Downregulated |
| -0,108731661 | 0,969259379 | EPN2-AS1 | FALSE | Downregulated |
| -0,108824805 | 0,969259379 | FBXL3 | FALSE | Downregulated |
| -0,080803048 | 0,969259379 | FSIP2 | FALSE | Downregulated |
| -0,105551559 | 0,969259379 | H1F0 | FALSE | Downregulated |
| -0,068153816 | 0,969259379 | HMBOX1 | FALSE | Downregulated |
| -0,155912536 | 0,969259379 | HSPA4 | FALSE | Downregulated |
| -0,139096743 | 0,969259379 | INPP5B | FALSE | Downregulated |
| 0,131291792 | 0,969259379 | KIF21A | FALSE | Upregulated |
| 0,079870891 | 0,969259379 | LIMD1 | FALSE | Upregulated |
| -0,096117308 | 0,969259379 | LOC100132356 | FALSE | Downregulated |
| -0,115316375 | 0,969259379 | LOC100506731 | FALSE | Downregulated |
| -0,145983066 | 0,969259379 | LOC101927513 | FALSE | Downregulated |
| -0,164525373 | 0,969259379 | LOC101928045 | FALSE | Downregulated |
| -0,105196116 | 0,969259379 | LOC101929751 | FALSE | Downregulated |
| 0,100561525 | 0,969259379 | LOC102724016 | FALSE | Upregulated |
| 0,059730353 | 0,969259379 | MACF1 | FALSE | Upregulated |
| 0,118188812 | 0,969259379 | MEPCE | FALSE | Upregulated |
| -0,139016809 | 0,969259379 | MPG | FALSE | Downregulated |
| -0,110177188 | 0,969259379 | NEIL1 | FALSE | Downregulated |
| 0,101016258 | 0,969259379 | NGEF | FALSE | Upregulated |
| -0,08095232 | 0,969259379 | PABPC1 | FALSE | Downregulated |
| -0,06782394 | 0,969259379 | PDXDC1 | FALSE | Downregulated |
| 0,144485981 | 0,969259379 | PIK3C2B | FALSE | Upregulated |
| -0,106010355 | 0,969259379 | PMCHL2 | FALSE | Downregulated |
| 0,101860473 | 0,969259379 | PPIL2 | FALSE | Upregulated |
| 0,075133214 | 0,969259379 | PSD3 | FALSE | Upregulated |
| 0,117034459 | 0,969259379 | PTK2 | FALSE | Upregulated |
| -0,059970763 | 0,969259379 | PTPRF | FALSE | Downregulated |
| -0,142514543 | 0,969259379 | PTRF | FALSE | Downregulated |
| -0,134338398 | 0,969259379 | RNF139 | FALSE | Downregulated |
| 0,101782748 | 0,969259379 | RPL7L1 | FALSE | Upregulated |
| 0,099931608 | 0,969259379 | SIPA1L1 | FALSE | Upregulated |
| -0,099919094 | 0,969259379 | SLC25A42 | FALSE | Downregulated |
| -0,115828311 | 0,969259379 | SPATC1 | FALSE | Downregulated |
| -0,128086222 | 0,969259379 | STARD8 | FALSE | Downregulated |
| -0,114323108 | 0,969259379 | SYCP3 | FALSE | Downregulated |
| -0,134021173 | 0,969259379 | TEN1 | FALSE | Downregulated |
| 0,098462357 | 0,969259379 | TMEM79 | FALSE | Upregulated |
| 0,137338091 | 0,969259379 | TPCN2 | FALSE | Upregulated |
| 0,14192642 | 0,969259379 | TPRN | FALSE | Upregulated |
| 0,10514869 | 0,969259379 | TRIM41 | FALSE | Upregulated |
| -0,135070224 | 0,969259379 | UBP1 | FALSE | Downregulated |
| 0,098860484 | 0,969259379 | UGGT1 | FALSE | Upregulated |
| 0,132913347 | 0,969259379 | USP14 | FALSE | Upregulated |
| -0,111011887 | 0,969259379 | ZFX | FALSE | Downregulated |
| 0,080450512 | 0,969259379 | ZNF609 | FALSE | Upregulated |
| -0,083630511 | 0,969315561 | AC009501.4 | FALSE | Downregulated |

| | | | | |
|---|---|---|---|---|
| -0,077009922 | 0,969315561 | AGK | FALSE | Downregulated |
| 0,096147684 | 0,969315561 | AMT | FALSE | Upregulated |
| 0,147829461 | 0,969315561 | AOC4P | FALSE | Upregulated |
| -0,105325949 | 0,969315561 | ARL8B | FALSE | Downregulated |
| 0,110704861 | 0,969315561 | ARSB | FALSE | Upregulated |
| 0,077882364 | 0,969315561 | ASXL1 | FALSE | Upregulated |
| 0,099834047 | 0,969315561 | ATF6 | FALSE | Upregulated |
| -0,119828792 | 0,969315561 | ATF6B | FALSE | Downregulated |
| 0,129218748 | 0,969315561 | BAG1 | FALSE | Upregulated |
| -0,10969162 | 0,969315561 | C17orf72 | FALSE | Downregulated |
| -0,134083404 | 0,969315561 | C22orf39 | FALSE | Downregulated |
| -0,074327942 | 0,969315561 | C6orf106 | FALSE | Downregulated |
| -0,080820812 | 0,969315561 | CARM1 | FALSE | Downregulated |
| 0,077386742 | 0,969315561 | CD3EAP | FALSE | Upregulated |
| -0,061350851 | 0,969315561 | CD81 | FALSE | Downregulated |
| -0,109448206 | 0,969315561 | CHD8 | FALSE | Downregulated |
| 0,066157794 | 0,969315561 | CHDH | FALSE | Upregulated |
| -0,109309893 | 0,969315561 | CLK1 | FALSE | Downregulated |
| -0,06314449 | 0,969315561 | COPA | FALSE | Downregulated |
| -0,115179216 | 0,969315561 | COPB1 | FALSE | Downregulated |
| -0,094027179 | 0,969315561 | CPD | FALSE | Downregulated |
| 0,09971409 | 0,969315561 | CSDC2 | FALSE | Upregulated |
| 0,108596751 | 0,969315561 | CUL4A | FALSE | Upregulated |
| 0,110084834 | 0,969315561 | CUL7 | FALSE | Upregulated |
| -0,124092403 | 0,969315561 | DCTD | FALSE | Downregulated |
| -0,100757404 | 0,969315561 | DENND1A | FALSE | Downregulated |
| -0,066848036 | 0,969315561 | DLEU1 | FALSE | Downregulated |
| 0,0955809 | 0,969315561 | DMXL1 | FALSE | Upregulated |
| -0,105002934 | 0,969315561 | DNAJA3 | FALSE | Downregulated |
| -0,083564784 | 0,969315561 | F2RL2 | FALSE | Downregulated |
| 0,093826744 | 0,969315561 | FAM105B | FALSE | Upregulated |
| 0,113456795 | 0,969315561 | FBXO17 | FALSE | Upregulated |
| -0,122687481 | 0,969315561 | FGD1 | FALSE | Downregulated |
| -0,120965736 | 0,969315561 | FRMD4B | FALSE | Downregulated |
| -0,092095413 | 0,969315561 | GLG1 | FALSE | Downregulated |
| 0,116589344 | 0,969315561 | GLT8D1 | FALSE | Upregulated |
| -0,120972437 | 0,969315561 | GTF2F1 | FALSE | Downregulated |
| -0,11753068 | 0,969315561 | HABP4 | FALSE | Downregulated |
| 0,084092248 | 0,969315561 | HLA-B | FALSE | Upregulated |
| -0,133105124 | 0,969315561 | HMGN3-AS1 | FALSE | Downregulated |
| -0,069247029 | 0,969315561 | HPS3 | FALSE | Downregulated |
| -0,123089309 | 0,969315561 | IFIT5 | FALSE | Downregulated |
| 0,174378785 | 0,969315561 | IGF2 | FALSE | Upregulated |
| -0,082476611 | 0,969315561 | INO80E | FALSE | Downregulated |
| 0,096225809 | 0,969315561 | ITGA5 | FALSE | Upregulated |
| 0,104344296 | 0,969315561 | KATNAL2 | FALSE | Upregulated |
| -0,101816012 | 0,969315561 | KDM5A | FALSE | Downregulated |
| 0,133930155 | 0,969315561 | KDM7A | FALSE | Upregulated |
| -0,102448514 | 0,969315561 | KIAA0947 | FALSE | Downregulated |
| -0,121543974 | 0,969315561 | LAP3 | FALSE | Downregulated |
| 0,126459845 | 0,969315561 | LHPP | FALSE | Upregulated |

| | | | | |
|---|---|---|---|---|
| 0,103623415 | 0,969315561 | LMTK2 | FALSE | Upregulated |
| 0,137794751 | 0,969315561 | LOC100129361 | FALSE | Upregulated |
| -0,109668422 | 0,969315561 | LOC100133331 | FALSE | Downregulated |
| -0,086509844 | 0,969315561 | LOC101927644 | FALSE | Downregulated |
| -0,126515789 | 0,969315561 | LOC101927894 | FALSE | Downregulated |
| 0,106040327 | 0,969315561 | LOC101928576 | FALSE | Upregulated |
| -0,088863452 | 0,969315561 | LOC101929340 | FALSE | Downregulated |
| -0,08570554 | 0,969315561 | LOC102723566 | FALSE | Downregulated |
| -0,070527731 | 0,969315561 | LOC102724484 | FALSE | Downregulated |
| -0,120981082 | 0,969315561 | LRRC58 | FALSE | Downregulated |
| 0,110102207 | 0,969315561 | MAU2 | FALSE | Upregulated |
| 0,06487682 | 0,969315561 | MCC | FALSE | Upregulated |
| -0,09210415 | 0,969315561 | MED13L | FALSE | Downregulated |
| 0,086442238 | 0,969315561 | MED23 | FALSE | Upregulated |
| -0,121561181 | 0,969315561 | MESDC1 | FALSE | Downregulated |
| -0,114920084 | 0,969315561 | MMS19 | FALSE | Downregulated |
| 0,106173692 | 0,969315561 | MOGS | FALSE | Upregulated |
| 0,107676452 | 0,969315561 | MSRB1 | FALSE | Upregulated |
| 0,123560102 | 0,969315561 | MTCH2 | FALSE | Upregulated |
| -0,091243786 | 0,969315561 | MUC5B | FALSE | Downregulated |
| -0,09247813 | 0,969315561 | NAMPT | FALSE | Downregulated |
| 0,077209705 | 0,969315561 | NBPF8 | FALSE | Upregulated |
| 0,077767114 | 0,969315561 | NFKBIA | FALSE | Upregulated |
| 0,092286187 | 0,969315561 | NLRC3 | FALSE | Upregulated |
| 0,114640348 | 0,969315561 | NME2 | FALSE | Upregulated |
| -0,091159707 | 0,969315561 | NT5E | FALSE | Downregulated |
| 0,136241355 | 0,969315561 | ORAI3 | FALSE | Upregulated |
| -0,091408786 | 0,969315561 | PCF11 | FALSE | Downregulated |
| -0,097396081 | 0,969315561 | PHACTR4 | FALSE | Downregulated |
| 0,136283897 | 0,969315561 | PHLPP2 | FALSE | Upregulated |
| 0,108161988 | 0,969315561 | PIGW | FALSE | Upregulated |
| 0,107650801 | 0,969315561 | PITPNA | FALSE | Upregulated |
| -0,112329787 | 0,969315561 | PLA2G12A | FALSE | Downregulated |
| 0,096123411 | 0,969315561 | PNRC2 | FALSE | Upregulated |
| 0,111714287 | 0,969315561 | POLR2B | FALSE | Upregulated |
| 0,140616415 | 0,969315561 | PPP1CA | FALSE | Upregulated |
| 0,131588629 | 0,969315561 | PRICKLE4 | FALSE | Upregulated |
| -0,108503977 | 0,969315561 | PRPF38A | FALSE | Downregulated |
| 0,105049097 | 0,969315561 | PRPF4B | FALSE | Upregulated |
| 0,054864362 | 0,969315561 | PRRC2B | FALSE | Upregulated |
| 0,097789071 | 0,969315561 | PSMF1 | FALSE | Upregulated |
| 0,088480869 | 0,969315561 | RABGAP1L | FALSE | Upregulated |
| 0,104196381 | 0,969315561 | RB1CC1 | FALSE | Upregulated |
| 0,09582482 | 0,969315561 | RELA | FALSE | Upregulated |
| -0,092306257 | 0,969315561 | RGPD8 | FALSE | Downregulated |
| 0,126963408 | 0,969315561 | RILP | FALSE | Upregulated |
| -0,112860576 | 0,969315561 | RILPL1 | FALSE | Downregulated |
| -0,072161035 | 0,969315561 | RNASEH2C | FALSE | Downregulated |
| -0,134084667 | 0,969315561 | RNF38 | FALSE | Downregulated |
| 0,104488872 | 0,969315561 | RNU4-2 | FALSE | Upregulated |
| -0,082008975 | 0,969315561 | RP11-545I5.3 | FALSE | Downregulated |

| | | | | |
|---|---|---|---|---|
| 0,121413579 | 0,969315561 | RPN2 | FALSE | Upregulated |
| 0,142311051 | 0,969315561 | RUVBL2 | FALSE | Upregulated |
| -0,122176918 | 0,969315561 | RYR3 | FALSE | Downregulated |
| -0,107552325 | 0,969315561 | SAR1A | FALSE | Downregulated |
| 0,112994561 | 0,969315561 | SCN9A | FALSE | Upregulated |
| -0,140570557 | 0,969315561 | SDSL | FALSE | Downregulated |
| -0,098594559 | 0,969315561 | SEMA4D | FALSE | Downregulated |
| 0,122223294 | 0,969315561 | SFT2D3 | FALSE | Upregulated |
| 0,106953736 | 0,969315561 | SIAE | FALSE | Upregulated |
| 0,127730077 | 0,969315561 | SLC25A46 | FALSE | Upregulated |
| -0,073242464 | 0,969315561 | SLC25A47 | FALSE | Downregulated |
| -0,100264743 | 0,969315561 | SLC28A1 | FALSE | Downregulated |
| 0,132383259 | 0,969315561 | SLC35G6 | FALSE | Upregulated |
| -0,11665113 | 0,969315561 | SLC5A3 | FALSE | Downregulated |
| -0,080229679 | 0,969315561 | SMAD4 | FALSE | Downregulated |
| -0,061038735 | 0,969315561 | SNRK-AS1 | FALSE | Downregulated |
| -0,085570464 | 0,969315561 | SP100 | FALSE | Downregulated |
| -0,084477642 | 0,969315561 | SPG7 | FALSE | Downregulated |
| 0,123312175 | 0,969315561 | SRP19 | FALSE | Upregulated |
| 0,132312854 | 0,969315561 | ST6GALNAC2 | FALSE | Upregulated |
| -0,108749461 | 0,969315561 | SUV39H1 | FALSE | Downregulated |
| 0,103443501 | 0,969315561 | SUV420H1 | FALSE | Upregulated |
| -0,095190597 | 0,969315561 | TADA3 | FALSE | Downregulated |
| 0,077769067 | 0,969315561 | TET2 | FALSE | Upregulated |
| -0,127943432 | 0,969315561 | THAP4 | FALSE | Downregulated |
| 0,132963767 | 0,969315561 | TMOD3 | FALSE | Upregulated |
| -0,107628929 | 0,969315561 | TOP2B | FALSE | Downregulated |
| -0,083208194 | 0,969315561 | TRIM28 | FALSE | Downregulated |
| -0,09514521 | 0,969315561 | TRIP11 | FALSE | Downregulated |
| -0,09503978 | 0,969315561 | TSR1 | FALSE | Downregulated |
| 0,094260711 | 0,969315561 | TVP23A | FALSE | Upregulated |
| 0,076258631 | 0,969315561 | UBA1 | FALSE | Upregulated |
| 0,103014213 | 0,969315561 | XPO5 | FALSE | Upregulated |
| 0,09161022 | 0,969315561 | ZBTB11 | FALSE | Upregulated |
| -0,123106358 | 0,969315561 | ZC3H6 | FALSE | Downregulated |
| 0,107945367 | 0,969315561 | ZNF546 | FALSE | Upregulated |
| -0,091341938 | 0,969315561 | ZNRF3-AS1 | FALSE | Downregulated |
| 0,078334344 | 0,969315561 | ZSCAN29 | FALSE | Upregulated |
| -0,071163554 | 0,969315561 | ZSCAN30 | FALSE | Downregulated |
| -0,108858913 | 0,969662707 | FAM20B | FALSE | Downregulated |
| 0,098082056 | 0,969662707 | LOC102724273 | FALSE | Upregulated |
| 0,12336068 | 0,969662707 | PHLDB1 | FALSE | Upregulated |
| 0,092867512 | 0,969662707 | SSH1 | FALSE | Upregulated |
| 0,058509669 | 0,969662707 | SUN2 | FALSE | Upregulated |
| -0,100500458 | 0,97033855 | SNAI3 | FALSE | Downregulated |
| -0,116404844 | 0,970473997 | EXOC3 | FALSE | Downregulated |
| -0,072065878 | 0,9705227 | FAM104A | FALSE | Downregulated |
| -0,1047103 | 0,9705227 | HNRNPUL2 | FALSE | Downregulated |
| -0,123232135 | 0,9705227 | KDELR1 | FALSE | Downregulated |
| 0,092880792 | 0,9705227 | MDM4 | FALSE | Upregulated |
| -0,07396601 | 0,9705227 | MYRIP | FALSE | Downregulated |

| | | | | |
|---|---|---|---|---|
| 0,108598625 | 0,9705227 | TAF1C | FALSE | Upregulated |
| -0,104230759 | 0,970645823 | ADAM10 | FALSE | Downregulated |
| 0,066744789 | 0,970645823 | BMP8A | FALSE | Upregulated |
| -0,111986377 | 0,970645823 | CDK6 | FALSE | Downregulated |
| -0,058041343 | 0,970645823 | IGF2R | FALSE | Downregulated |
| -0,087484664 | 0,970645823 | PRKACA | FALSE | Downregulated |
| 0,07914571 | 0,970645823 | RP11-142C4.6 | FALSE | Upregulated |
| 0,107848939 | 0,970645823 | SMAD9 | FALSE | Upregulated |
| -0,081640448 | 0,970957878 | NXF1 | FALSE | Downregulated |
| -0,074789316 | 0,971188493 | EIF3A | FALSE | Downregulated |
| -0,095423239 | 0,971188493 | SPTY2D1-AS1 | FALSE | Downregulated |
| -0,092339272 | 0,971257575 | CRAMP1L | FALSE | Downregulated |
| -0,089378219 | 0,971257575 | KPNA6 | FALSE | Downregulated |
| -0,1143686 | 0,971257575 | POLR3G | FALSE | Downregulated |
| 0,120812684 | 0,971257575 | UBAC2 | FALSE | Upregulated |
| 0,096821199 | 0,971257575 | VASH2 | FALSE | Upregulated |
| -0,066985115 | 0,971317577 | SETD1B | FALSE | Downregulated |
| -0,069868213 | 0,971634352 | AUNIP | FALSE | Downregulated |
| 0,093231698 | 0,971634352 | TBC1D10B | FALSE | Upregulated |
| -0,057327296 | 0,971665428 | MASP1 | FALSE | Downregulated |
| 0,081223142 | 0,971719544 | ROCK2 | FALSE | Upregulated |
| 0,105143494 | 0,971719544 | ZMYM3 | FALSE | Upregulated |
| 0,130194352 | 0,97197445 | LOC101928502 | FALSE | Upregulated |
| 0,072352455 | 0,97197445 | PP7080 | FALSE | Upregulated |
| -0,126916622 | 0,97197445 | ZDHHC9 | FALSE | Downregulated |
| -0,073055663 | 0,972070393 | C11orf95 | FALSE | Downregulated |
| 0,079634506 | 0,972070393 | HNRNPH1 | FALSE | Upregulated |
| 0,104886431 | 0,972670136 | CASZ1 | FALSE | Upregulated |
| 0,111003674 | 0,972670136 | CCDC18 | FALSE | Upregulated |
| -0,072771946 | 0,972670136 | EIF5A | FALSE | Downregulated |
| -0,081197832 | 0,972670136 | LIG3 | FALSE | Downregulated |
| -0,078311648 | 0,972670136 | LOC101927647 | FALSE | Downregulated |
| 0,068521964 | 0,972670136 | MEGF11 | FALSE | Upregulated |
| -0,069177202 | 0,972670136 | PQLC1 | FALSE | Downregulated |
| 0,095191389 | 0,972670136 | SBF2-AS1 | FALSE | Upregulated |
| -0,106110345 | 0,972937698 | TMEM43 | FALSE | Downregulated |
| -0,104748534 | 0,972954243 | GNPDA1 | FALSE | Downregulated |
| 0,059897611 | 0,97299636 | NME7 | FALSE | Upregulated |
| -0,070516742 | 0,97299636 | PARD6G | FALSE | Downregulated |
| 0,07742762 | 0,973498591 | ERC1 | FALSE | Upregulated |
| -0,145195311 | 0,973498591 | EVC | FALSE | Downregulated |
| 0,062140935 | 0,973498591 | GPRC5C | FALSE | Upregulated |
| 0,058362972 | 0,973498591 | LINC00094 | FALSE | Upregulated |
| 0,069481701 | 0,973498591 | LOC101927509 | FALSE | Upregulated |
| 0,090401948 | 0,973498591 | MDC1 | FALSE | Upregulated |
| 0,057515504 | 0,973498591 | MLLT6 | FALSE | Upregulated |
| 0,071383776 | 0,973498591 | PDE5A | FALSE | Upregulated |
| 0,13270874 | 0,973498591 | PDP2 | FALSE | Upregulated |
| -0,09064322 | 0,973498591 | RAB5B | FALSE | Downregulated |
| 0,088551956 | 0,973498591 | RUFY3 | FALSE | Upregulated |
| 0,077065835 | 0,973498591 | SIRT5 | FALSE | Upregulated |

| | | | | |
|---|---|---|---|---|
| -0,107713639 | 0,973498591 | TSC22D4 | FALSE | Downregulated |
| -0,072925123 | 0,973498591 | UHMK1 | FALSE | Downregulated |
| 0,093703712 | 0,973527998 | CBFA2T2 | FALSE | Upregulated |
| -0,071791252 | 0,973527998 | DENND4C | FALSE | Downregulated |
| 0,084037066 | 0,973527998 | GSN | FALSE | Upregulated |
| 0,109224741 | 0,973527998 | KBTBD2 | FALSE | Upregulated |
| -0,097392684 | 0,973527998 | LOC100996492 | FALSE | Downregulated |
| -0,072924234 | 0,973527998 | MZF1 | FALSE | Downregulated |
| 0,074838755 | 0,973527998 | NFIX | FALSE | Upregulated |
| 0,118824611 | 0,973527998 | RGS20 | FALSE | Upregulated |
| -0,066684759 | 0,973527998 | SNX1 | FALSE | Downregulated |
| 0,105639477 | 0,973527998 | XPO7 | FALSE | Upregulated |
| -0,10485485 | 0,973743532 | ATF2 | FALSE | Downregulated |
| 0,114452724 | 0,973913398 | FRY-AS1 | FALSE | Upregulated |
| 0,060173768 | 0,973913398 | GPLD1 | FALSE | Upregulated |
| 0,080279529 | 0,974024098 | GSDMD | FALSE | Upregulated |
| -0,061703127 | 0,974024098 | SLC25A15 | FALSE | Downregulated |
| 0,052152559 | 0,974024098 | TENC1 | FALSE | Upregulated |
| -0,07087669 | 0,974065459 | BSDC1 | FALSE | Downregulated |
| 0,098994221 | 0,974065459 | RHOT2 | FALSE | Upregulated |
| -0,090742649 | 0,974065459 | VCPIP1 | FALSE | Downregulated |
| 0,091079926 | 0,974109545 | MRPS26 | FALSE | Upregulated |
| 0,090032914 | 0,974157293 | ABL2 | FALSE | Upregulated |
| 0,076859626 | 0,974157293 | AFG3L2 | FALSE | Upregulated |
| 0,090580118 | 0,974157293 | AGPAT6 | FALSE | Upregulated |
| -0,098087441 | 0,974157293 | BCL3 | FALSE | Downregulated |
| -0,063924925 | 0,974157293 | BCR | FALSE | Downregulated |
| -0,078934219 | 0,974157293 | CALCOCO1 | FALSE | Downregulated |
| -0,088365152 | 0,974157293 | CLK3 | FALSE | Downregulated |
| 0,066894162 | 0,974157293 | DDX3X | FALSE | Upregulated |
| 0,096827439 | 0,974157293 | EIF3F | FALSE | Upregulated |
| -0,082798159 | 0,974157293 | EIF4E2 | FALSE | Downregulated |
| 0,053106519 | 0,974157293 | GGCX | FALSE | Upregulated |
| -0,057841976 | 0,974157293 | GNB2L1 | FALSE | Downregulated |
| -0,09277694 | 0,974157293 | IER2 | FALSE | Downregulated |
| 0,078207005 | 0,974157293 | KIDINS220 | FALSE | Upregulated |
| 0,071879177 | 0,974157293 | LAMB1 | FALSE | Upregulated |
| -0,086143101 | 0,974157293 | LOC100506922 | FALSE | Downregulated |
| 0,108915291 | 0,974157293 | LOC101928319 | FALSE | Upregulated |
| -0,07743819 | 0,974157293 | LOC440386 | FALSE | Downregulated |
| -0,086332513 | 0,974157293 | MPI | FALSE | Downregulated |
| 0,075592015 | 0,974157293 | PDCD2 | FALSE | Upregulated |
| 0,073941985 | 0,974157293 | RASGRP1 | FALSE | Upregulated |
| 0,098911674 | 0,974157293 | RNPEP | FALSE | Upregulated |
| -0,045095104 | 0,974157293 | RP11-367J11.2 | FALSE | Downregulated |
| -0,103276685 | 0,974157293 | SLC25A39 | FALSE | Downregulated |
| -0,098945511 | 0,974157293 | SOWAHB | FALSE | Downregulated |
| -0,077309047 | 0,974157293 | TMLHE | FALSE | Downregulated |
| 0,094757869 | 0,974157293 | TNPO3 | FALSE | Upregulated |
| 0,07355895 | 0,974157293 | TRAP1 | FALSE | Upregulated |
| 0,066960982 | 0,974157293 | TRMT10B | FALSE | Upregulated |

| | | | | |
|---|---|---|---|---|
| 0,085422046 | 0,974157293 | VKORC1L1 | FALSE | Upregulated |
| 0,104840256 | 0,974390636 | APC | FALSE | Upregulated |
| 0,048988555 | 0,974390636 | BIRC6 | FALSE | Upregulated |
| 0,116883362 | 0,974390636 | C11orf83 | FALSE | Upregulated |
| -0,120554245 | 0,974390636 | C1orf213 | FALSE | Downregulated |
| 0,075620251 | 0,974390636 | COPG1 | FALSE | Upregulated |
| 0,076852881 | 0,974390636 | DDX6 | FALSE | Upregulated |
| -0,075496707 | 0,974390636 | DECR2 | FALSE | Downregulated |
| -0,092017196 | 0,974390636 | DKFZP434H168 | FALSE | Downregulated |
| 0,072960089 | 0,974390636 | ESRRA | FALSE | Upregulated |
| -0,100459131 | 0,974390636 | FAM96A | FALSE | Downregulated |
| 0,093049607 | 0,974390636 | FAM99B | FALSE | Upregulated |
| -0,071307013 | 0,974390636 | GLTPD1 | FALSE | Downregulated |
| 0,113350885 | 0,974390636 | GNPTG | FALSE | Upregulated |
| 0,068589813 | 0,974390636 | GRTP1 | FALSE | Upregulated |
| 0,093472743 | 0,974390636 | HDAC4 | FALSE | Upregulated |
| -0,072391773 | 0,974390636 | HNRNPC | FALSE | Downregulated |
| -0,094313771 | 0,974390636 | ITGB7 | FALSE | Downregulated |
| -0,106074067 | 0,974390636 | LIN54 | FALSE | Downregulated |
| 0,071117645 | 0,974390636 | LMNA | FALSE | Upregulated |
| -0,046937923 | 0,974390636 | LOC100507577 | FALSE | Downregulated |
| -0,094751365 | 0,974390636 | LOC101928967 | FALSE | Downregulated |
| 0,052120417 | 0,974390636 | LRP5 | FALSE | Upregulated |
| -0,088909383 | 0,974390636 | NDST2 | FALSE | Downregulated |
| 0,055155429 | 0,974390636 | NEXN | FALSE | Upregulated |
| 0,047383743 | 0,974390636 | NUMA1 | FALSE | Upregulated |
| -0,076746364 | 0,974390636 | OTUD5 | FALSE | Downregulated |
| -0,069147676 | 0,974390636 | OXNAD1 | FALSE | Downregulated |
| 0,085330605 | 0,974390636 | PHF12 | FALSE | Upregulated |
| -0,066647931 | 0,974390636 | PHIP | FALSE | Downregulated |
| -0,048686392 | 0,974390636 | PKD1 | FALSE | Downregulated |
| -0,088557018 | 0,974390636 | PPP1R13B | FALSE | Downregulated |
| 0,080756954 | 0,974390636 | PPP1R15B | FALSE | Upregulated |
| -0,10569357 | 0,974390636 | PSMD4 | FALSE | Downregulated |
| 0,049427989 | 0,974390636 | RALGAPA2 | FALSE | Upregulated |
| -0,063299484 | 0,974390636 | RSBN1L-AS1 | FALSE | Downregulated |
| 0,089023994 | 0,974390636 | SCARNA7 | FALSE | Upregulated |
| -0,098527179 | 0,974390636 | SLC11A2 | FALSE | Downregulated |
| 0,101437852 | 0,974390636 | TARS | FALSE | Upregulated |
| 0,107126358 | 0,974390636 | TSPAN31 | FALSE | Upregulated |
| 0,094476402 | 0,974390636 | TUBB6 | FALSE | Upregulated |
| 0,104152875 | 0,974390636 | UTP14C | FALSE | Upregulated |
| -0,098321533 | 0,974390636 | YY1AP1 | FALSE | Downregulated |
| -0,070183517 | 0,974466972 | LOC100131496 | FALSE | Downregulated |
| -0,093075209 | 0,974587483 | DCAF13 | FALSE | Downregulated |
| -0,079214768 | 0,974587483 | EVL | FALSE | Downregulated |
| -0,072512666 | 0,974946704 | LOC102060414 | FALSE | Downregulated |
| 0,070336278 | 0,974946704 | MAP4K4 | FALSE | Upregulated |
| 0,092399935 | 0,974946704 | SERPINB6 | FALSE | Upregulated |
| 0,060724176 | 0,974946704 | SND1 | FALSE | Upregulated |
| 0,062632047 | 0,974946704 | TNRC6C | FALSE | Upregulated |

| | | | | |
|---|---|---|---|---|
| -0,082851472 | 0,975201245 | CNTLN | FALSE | Downregulated |
| 0,096540932 | 0,975201245 | INPP5E | FALSE | Upregulated |
| 0,111719252 | 0,975201245 | SDK1 | FALSE | Upregulated |
| 0,07894295 | 0,975563767 | RECQL | FALSE | Upregulated |
| -0,06671999 | 0,975563767 | TSTD2 | FALSE | Downregulated |
| 0,073417469 | 0,97601139 | KDM3B | FALSE | Upregulated |
| 0,069439601 | 0,976015373 | UBQLN1 | FALSE | Upregulated |
| -0,064613135 | 0,976364944 | IDH2 | FALSE | Downregulated |
| 0,066814993 | 0,976503047 | PLCG1 | FALSE | Upregulated |
| -0,07975763 | 0,976627091 | SIGLEC16 | FALSE | Downregulated |
| -0,055187044 | 0,976774435 | ACTN4 | FALSE | Downregulated |
| -0,056443351 | 0,976774435 | TEX2 | FALSE | Downregulated |
| -0,082192299 | 0,976774435 | WDR11 | FALSE | Downregulated |
| 0,081241114 | 0,976790399 | RAB5C | FALSE | Upregulated |
| -0,094834474 | 0,976954138 | XPA | FALSE | Downregulated |
| 0,05320338 | 0,977411908 | TINF2 | FALSE | Upregulated |
| -0,056259512 | 0,97745449 | CTBP2 | FALSE | Downregulated |
| -0,086712311 | 0,97745449 | PARP1 | FALSE | Downregulated |
| 0,098612752 | 0,97745449 | TAF6L | FALSE | Upregulated |
| -0,092384191 | 0,977499753 | DNM3 | FALSE | Downregulated |
| 0,085019622 | 0,97752943 | BCOR | FALSE | Upregulated |
| 0,080390085 | 0,97752943 | CBL | FALSE | Upregulated |
| -0,085540539 | 0,97752943 | CBX6 | FALSE | Downregulated |
| -0,082540268 | 0,97752943 | CHD1 | FALSE | Downregulated |
| -0,09770017 | 0,97752943 | CLCF1 | FALSE | Downregulated |
| 0,059522168 | 0,97752943 | CPSF7 | FALSE | Upregulated |
| -0,088916561 | 0,97752943 | DARS2 | FALSE | Downregulated |
| 0,078351388 | 0,97752943 | DHX38 | FALSE | Upregulated |
| -0,058980609 | 0,97752943 | ESYT3 | FALSE | Downregulated |
| 0,058080268 | 0,97752943 | FAM193B | FALSE | Upregulated |
| -0,065383889 | 0,97752943 | GAA | FALSE | Downregulated |
| 0,099810545 | 0,97752943 | GM2A | FALSE | Upregulated |
| 0,053163085 | 0,97752943 | HYAL1 | FALSE | Upregulated |
| -0,078888508 | 0,97752943 | KALRN | FALSE | Downregulated |
| 0,05831236 | 0,97752943 | KIAA1191 | FALSE | Upregulated |
| 0,065237988 | 0,97752943 | KIF24 | FALSE | Upregulated |
| -0,080058055 | 0,97752943 | LOC101926920 | FALSE | Downregulated |
| -0,096987997 | 0,97752943 | LOC102723863 | FALSE | Downregulated |
| -0,097733029 | 0,97752943 | LOC102724043 | FALSE | Downregulated |
| -0,090046023 | 0,97752943 | MASTL | FALSE | Downregulated |
| -0,052505001 | 0,97752943 | MCTS1 | FALSE | Downregulated |
| 0,057045124 | 0,97752943 | MYO7A | FALSE | Upregulated |
| 0,08873109 | 0,97752943 | NAA15 | FALSE | Upregulated |
| 0,048879615 | 0,97752943 | NRIP2 | FALSE | Upregulated |
| 0,075798943 | 0,97752943 | PELI3 | FALSE | Upregulated |
| 0,092760159 | 0,97752943 | POFUT2 | FALSE | Upregulated |
| -0,056746846 | 0,97752943 | POLR2E | FALSE | Downregulated |
| 0,07458956 | 0,97752943 | TM9SF4 | FALSE | Upregulated |
| 0,089417714 | 0,97752943 | USP12 | FALSE | Upregulated |
| -0,082296256 | 0,97752943 | ZNF496 | FALSE | Downregulated |
| -0,053545295 | 0,977586432 | ABCG5 | FALSE | Downregulated |

| | | | | |
|---|---|---|---|---|
| 0,070521017 | 0,977586432 | ATP1B1 | FALSE | Upregulated |
| 0,063203604 | 0,977586432 | BCLAF1 | FALSE | Upregulated |
| -0,064857621 | 0,977586432 | C6orf47 | FALSE | Downregulated |
| -0,044910985 | 0,977586432 | CDC42BPB | FALSE | Downregulated |
| -0,048020054 | 0,977586432 | CHERP | FALSE | Downregulated |
| 0,072933713 | 0,977586432 | COL14A1 | FALSE | Upregulated |
| -0,071810104 | 0,977586432 | DCTN1 | FALSE | Downregulated |
| -0,066154835 | 0,977586432 | DOCK6 | FALSE | Downregulated |
| 0,086860249 | 0,977586432 | DUSP6 | FALSE | Upregulated |
| 0,052330837 | 0,977586432 | EIF4G2 | FALSE | Upregulated |
| -0,050421234 | 0,977586432 | EML5 | FALSE | Downregulated |
| -0,086258736 | 0,977586432 | ERLEC1 | FALSE | Downregulated |
| 0,055877695 | 0,977586432 | GNAI2 | FALSE | Upregulated |
| -0,090802432 | 0,977586432 | IL3RA | FALSE | Downregulated |
| -0,099912267 | 0,977586432 | ING5 | FALSE | Downregulated |
| -0,088161298 | 0,977586432 | LDLR | FALSE | Downregulated |
| 0,036475062 | 0,977586432 | LDLRAD2 | FALSE | Upregulated |
| 0,090029483 | 0,977586432 | LEMD2 | FALSE | Upregulated |
| 0,05008738 | 0,977586432 | LOC101929844 | FALSE | Upregulated |
| -0,05623878 | 0,977586432 | LOC102724814 | FALSE | Downregulated |
| 0,085351546 | 0,977586432 | MARK4 | FALSE | Upregulated |
| -0,098264803 | 0,977586432 | MTF1 | FALSE | Downregulated |
| 0,052956843 | 0,977586432 | MYH14 | FALSE | Upregulated |
| 0,060476359 | 0,977586432 | NIPA2 | FALSE | Upregulated |
| -0,048540682 | 0,977586432 | NOTCH2 | FALSE | Downregulated |
| -0,056096678 | 0,977586432 | NRON | FALSE | Downregulated |
| 0,062415632 | 0,977586432 | NUP210 | FALSE | Upregulated |
| -0,054237048 | 0,977586432 | PNPO | FALSE | Downregulated |
| 0,080102193 | 0,977586432 | PPARGC1A | FALSE | Upregulated |
| -0,082199328 | 0,977586432 | PPFIBP2 | FALSE | Downregulated |
| 0,052772337 | 0,977586432 | PSAP | FALSE | Upregulated |
| 0,054566093 | 0,977586432 | PTCD3 | FALSE | Upregulated |
| -0,064001918 | 0,977586432 | SEC23A | FALSE | Downregulated |
| 0,070418321 | 0,977586432 | SETD1A | FALSE | Upregulated |
| -0,076704816 | 0,977586432 | SHF | FALSE | Downregulated |
| -0,087109649 | 0,977586432 | SLC27A3 | FALSE | Downregulated |
| -0,072854231 | 0,977586432 | SLC8B1 | FALSE | Downregulated |
| -0,088562916 | 0,977586432 | SRSF7 | FALSE | Downregulated |
| -0,051654444 | 0,977586432 | STK17A | FALSE | Downregulated |
| -0,051925447 | 0,977586432 | SYNE2 | FALSE | Downregulated |
| -0,083190182 | 0,977586432 | TAB1 | FALSE | Downregulated |
| 0,061925382 | 0,977586432 | TECR | FALSE | Upregulated |
| -0,057674609 | 0,977586432 | THUMPD3 | FALSE | Downregulated |
| 0,063741674 | 0,977586432 | TRAK1 | FALSE | Upregulated |
| -0,044678719 | 0,977586432 | TRIM13 | FALSE | Downregulated |
| -0,047242369 | 0,977586432 | USP24 | FALSE | Downregulated |
| 0,046217036 | 0,977586432 | WHSC1L1 | FALSE | Upregulated |
| 0,065635538 | 0,977586432 | WSB2 | FALSE | Upregulated |
| 0,065260026 | 0,977586432 | YTHDF3 | FALSE | Upregulated |
| 0,044743801 | 0,977586432 | ZNF552 | FALSE | Upregulated |
| 0,079571071 | 0,977693285 | CCDC142 | FALSE | Upregulated |

| | | | | |
|---|---|---|---|---|
| 0,067309534 | 0,977693285 | RPS6KA3 | FALSE | Upregulated |
| -0,067230859 | 0,977693285 | WIPI2 | FALSE | Downregulated |
| 0,056568427 | 0,977736921 | GNB2 | FALSE | Upregulated |
| -0,078214656 | 0,977736921 | PPP4R4 | FALSE | Downregulated |
| -0,056781681 | 0,9778815 | VPS26B | FALSE | Downregulated |
| 0,082768268 | 0,977901666 | PDXP | FALSE | Upregulated |
| 0,091535416 | 0,978013549 | GEN1 | FALSE | Upregulated |
| -0,049789997 | 0,978013549 | GJD3 | FALSE | Downregulated |
| -0,059166959 | 0,978162635 | CSPG4 | FALSE | Downregulated |
| 0,057076846 | 0,978374343 | LAT2 | FALSE | Upregulated |
| -0,045077624 | 0,978443497 | ATM | FALSE | Downregulated |
| 0,086007815 | 0,978467449 | RBPJ | FALSE | Upregulated |
| -0,045177953 | 0,978544665 | ALDH1B1 | FALSE | Downregulated |
| -0,057850259 | 0,978544665 | ANKRD6 | FALSE | Downregulated |
| -0,07837038 | 0,978544665 | EXOC4 | FALSE | Downregulated |
| -0,063634492 | 0,978544665 | HCG18 | FALSE | Downregulated |
| 0,047182186 | 0,978544665 | LAMP1 | FALSE | Upregulated |
| -0,062862811 | 0,978544665 | LOC100303749 | FALSE | Downregulated |
| 0,108197186 | 0,978544665 | LOC101927675 | FALSE | Upregulated |
| 0,048059676 | 0,978544665 | RELN | FALSE | Upregulated |
| -0,069692797 | 0,978544665 | RPL37 | FALSE | Downregulated |
| -0,057716614 | 0,978794479 | ATP1A1 | FALSE | Downregulated |
| 0,060401912 | 0,978794479 | FBXO11 | FALSE | Upregulated |
| 0,06183344 | 0,978794479 | INO80D | FALSE | Upregulated |
| 0,049388208 | 0,978794479 | LOC100288123 | FALSE | Upregulated |
| 0,038282048 | 0,978794479 | NCOR1 | FALSE | Upregulated |
| -0,075474077 | 0,978794479 | NOL9 | FALSE | Downregulated |
| -0,050749697 | 0,978794479 | SRPR | FALSE | Downregulated |
| -0,050348308 | 0,979266015 | PTEN | FALSE | Downregulated |
| 0,076828333 | 0,979509938 | ATOH8 | FALSE | Upregulated |
| 0,083286187 | 0,979509938 | CAND2 | FALSE | Upregulated |
| -0,050026528 | 0,979509938 | CPSF1 | FALSE | Downregulated |
| 0,048941874 | 0,979509938 | RASA1 | FALSE | Upregulated |
| -0,056820131 | 0,979754498 | ATXN7 | FALSE | Downregulated |
| -0,051376249 | 0,979983096 | CCNJ | FALSE | Downregulated |
| -0,058586602 | 0,979983096 | CRIM1 | FALSE | Downregulated |
| 0,051846694 | 0,979983096 | EMC1 | FALSE | Upregulated |
| 0,048039775 | 0,979983096 | LDHA | FALSE | Upregulated |
| 0,068354912 | 0,979983096 | LOC285074 | FALSE | Upregulated |
| 0,063905893 | 0,979983096 | TBL1XR1 | FALSE | Upregulated |
| -0,061079935 | 0,980070735 | LMF2 | FALSE | Downregulated |
| 0,074189998 | 0,980070735 | TPP2 | FALSE | Upregulated |
| 0,045144882 | 0,980070735 | ZNF710 | FALSE | Upregulated |
| 0,068785147 | 0,980226798 | AP1M1 | FALSE | Upregulated |
| -0,090346856 | 0,981011889 | CEBPG | FALSE | Downregulated |
| -0,045303995 | 0,981025278 | ENDOG | FALSE | Downregulated |
| 0,078306066 | 0,981025278 | MED14 | FALSE | Upregulated |
| -0,072501965 | 0,981025278 | RALBP1 | FALSE | Downregulated |
| -0,070053424 | 0,981025278 | SPSB2 | FALSE | Downregulated |
| 0,052625656 | 0,981118563 | AC093690.1 | FALSE | Upregulated |
| -0,080366284 | 0,981118563 | ARRB2 | FALSE | Downregulated |

| | | | | |
|---|---|---|---|---|
| -0,051381853 | 0,981118563 | SAMD4A | FALSE | Downregulated |
| 0,072691877 | 0,9811712 | ACOT12 | FALSE | Upregulated |
| 0,034760787 | 0,98128287 | NR2C2 | FALSE | Upregulated |
| -0,076721562 | 0,981588871 | RBM12B | FALSE | Downregulated |
| 0,052903291 | 0,981588871 | SEC16B | FALSE | Upregulated |
| -0,067999291 | 0,981588871 | SECISBP2L | FALSE | Downregulated |
| -0,047072863 | 0,981588871 | ZNF233 | FALSE | Downregulated |
| -0,077988291 | 0,981691236 | AGAP4 | FALSE | Downregulated |
| -0,067356873 | 0,981691236 | C9orf96 | FALSE | Downregulated |
| -0,065488661 | 0,981691236 | CMYA5 | FALSE | Downregulated |
| 0,039840705 | 0,981691236 | GDF11 | FALSE | Upregulated |
| -0,052662368 | 0,981691236 | MST1P2 | FALSE | Downregulated |
| -0,059874903 | 0,981691236 | NCOA1 | FALSE | Downregulated |
| -0,047000826 | 0,981691236 | PLEKHM2 | FALSE | Downregulated |
| 0,062622504 | 0,981691236 | RIF1 | FALSE | Upregulated |
| 0,06062343 | 0,981691236 | RTKN | FALSE | Upregulated |
| -0,047188579 | 0,981691236 | SETD2 | FALSE | Downregulated |
| -0,060247143 | 0,981691236 | UBE4B | FALSE | Downregulated |
| 0,079709618 | 0,981857107 | ACOT8 | FALSE | Upregulated |
| -0,05332324 | 0,981857107 | LYNX1 | FALSE | Downregulated |
| 0,057767301 | 0,981945644 | CCT6P3 | FALSE | Upregulated |
| -0,0595477 | 0,981945644 | SMARCD1 | FALSE | Downregulated |
| 0,04315088 | 0,981945644 | TRIP12 | FALSE | Upregulated |
| 0,046896293 | 0,982026469 | RHOB | FALSE | Upregulated |
| -0,075094914 | 0,982031393 | AKT1S1 | FALSE | Downregulated |
| -0,067716101 | 0,982031393 | ZBTB18 | FALSE | Downregulated |
| -0,048645638 | 0,982137643 | PGPEP1 | FALSE | Downregulated |
| 0,055845955 | 0,98224257 | HYLS1 | FALSE | Upregulated |
| -0,061925594 | 0,98224257 | MCMDC2 | FALSE | Downregulated |
| 0,052768105 | 0,982365277 | GPR155 | FALSE | Upregulated |
| -0,074310021 | 0,982365277 | LRRN1 | FALSE | Downregulated |
| 0,064071419 | 0,982365277 | SH3GLB1 | FALSE | Upregulated |
| -0,074411405 | 0,982403671 | LOC102724172 | FALSE | Downregulated |
| 0,069375504 | 0,98247617 | DDX46 | FALSE | Upregulated |
| 0,04602431 | 0,98247617 | ZC3H13 | FALSE | Upregulated |
| -0,06429693 | 0,98247617 | ZNF805 | FALSE | Downregulated |
| -0,046088416 | 0,982882376 | CREBZF | FALSE | Downregulated |
| -0,07245645 | 0,982882376 | DCUN1D1 | FALSE | Downregulated |
| 0,073690804 | 0,982882376 | DICER1-AS1 | FALSE | Upregulated |
| -0,056025097 | 0,982882376 | DIP2C | FALSE | Downregulated |
| -0,047789008 | 0,982882376 | FAM227B | FALSE | Downregulated |
| -0,04693929 | 0,982882376 | FAT4 | FALSE | Downregulated |
| 0,05103817 | 0,982882376 | FILIP1L | FALSE | Upregulated |
| -0,049457748 | 0,982882376 | FLJ31306 | FALSE | Downregulated |
| 0,063211159 | 0,982882376 | GCNT7 | FALSE | Upregulated |
| -0,046974315 | 0,982882376 | GGACT | FALSE | Downregulated |
| 0,058951785 | 0,982882376 | LOC101928061 | FALSE | Upregulated |
| -0,067607575 | 0,982882376 | MTMR12 | FALSE | Downregulated |
| -0,043067659 | 0,982882376 | NFIB | FALSE | Downregulated |
| -0,079130001 | 0,982882376 | PAFAH1B2 | FALSE | Downregulated |
| 0,057006962 | 0,982882376 | PMS2P1 | FALSE | Upregulated |

| | | | | |
|---|---|---|---|---|
| 0,040595681 | 0,982882376 | RBM4 | FALSE | Upregulated |
| 0,058973979 | 0,982882376 | RBM5 | FALSE | Upregulated |
| -0,071335762 | 0,982882376 | SGMS1 | FALSE | Downregulated |
| 0,076343321 | 0,982882376 | SLC16A8 | FALSE | Upregulated |
| -0,066140759 | 0,982882376 | TRAPPC8 | FALSE | Downregulated |
| -0,060972491 | 0,982882376 | USP53 | FALSE | Downregulated |
| -0,057426088 | 0,983027318 | SS18L1 | FALSE | Downregulated |
| -0,062261841 | 0,983193477 | KLHDC3 | FALSE | Downregulated |
| 0,057507979 | 0,983193477 | MYH7B | FALSE | Upregulated |
| -0,048462612 | 0,983193477 | TNFAIP8L1 | FALSE | Downregulated |
| 0,03781771 | 0,983219214 | IRF2BP2 | FALSE | Upregulated |
| 0,039734866 | 0,983219214 | WDR81 | FALSE | Upregulated |
| 0,05190934 | 0,983629719 | CCT3 | FALSE | Upregulated |
| 0,051653087 | 0,983629719 | ITSN1 | FALSE | Upregulated |
| 0,055200205 | 0,983629719 | MARK3 | FALSE | Upregulated |
| 0,064031016 | 0,983629719 | TBC1D4 | FALSE | Upregulated |
| -0,043995256 | 0,983993932 | SH3GL1 | FALSE | Downregulated |
| -0,048328183 | 0,984144704 | GFOD2 | FALSE | Downregulated |
| -0,074641737 | 0,984144704 | NEK11 | FALSE | Downregulated |
| 0,072190476 | 0,984223285 | COG4 | FALSE | Upregulated |
| -0,051457846 | 0,984223285 | CYP20A1 | FALSE | Downregulated |
| 0,058873871 | 0,984223285 | SMU1 | FALSE | Upregulated |
| 0,054613595 | 0,984223285 | TOM1L2 | FALSE | Upregulated |
| -0,052383856 | 0,984223285 | WAPAL | FALSE | Downregulated |
| -0,040788869 | 0,9842575 | ADIPOR2 | FALSE | Downregulated |
| -0,034503418 | 0,9842575 | AFF1 | FALSE | Downregulated |
| 0,063413075 | 0,9842575 | B3GAT2 | FALSE | Upregulated |
| 0,037673303 | 0,9842575 | EPS8L2 | FALSE | Upregulated |
| -0,063381989 | 0,9842575 | NUAK1 | FALSE | Downregulated |
| 0,072669277 | 0,9842575 | PHPT1 | FALSE | Upregulated |
| -0,067127741 | 0,9842575 | RUFY1 | FALSE | Downregulated |
| 0,068512582 | 0,984382653 | CCDC69 | FALSE | Upregulated |
| -0,059226914 | 0,984382653 | FSTL1 | FALSE | Downregulated |
| -0,048755766 | 0,984382653 | LOC100506710 | FALSE | Downregulated |
| 0,046167989 | 0,984382653 | LRRN4 | FALSE | Upregulated |
| -0,052281111 | 0,984382653 | MAPKAP1 | FALSE | Downregulated |
| 0,050725551 | 0,984382653 | MTMR2 | FALSE | Upregulated |
| -0,07144193 | 0,984382653 | PLEKHA1 | FALSE | Downregulated |
| 0,063466649 | 0,984382653 | RAVER2 | FALSE | Upregulated |
| -0,045230208 | 0,984436873 | BAZ2B | FALSE | Downregulated |
| -0,076376864 | 0,984540404 | IFIT3 | FALSE | Downregulated |
| -0,043290209 | 0,984540404 | PDE3B | FALSE | Downregulated |
| -0,051070951 | 0,984618311 | CASC4 | FALSE | Downregulated |
| 0,046913677 | 0,984618311 | HIVEP1 | FALSE | Upregulated |
| 0,05671568 | 0,985044572 | CNOT3 | FALSE | Upregulated |
| 0,069566279 | 0,985044572 | COL5A3 | FALSE | Upregulated |
| -0,063513379 | 0,985044572 | COX6B1 | FALSE | Downregulated |
| 0,05336699 | 0,985044572 | ELMSAN1 | FALSE | Upregulated |
| 0,066903701 | 0,985051484 | RP11-434D9.1 | FALSE | Upregulated |
| 0,035950755 | 0,985051484 | TENM1 | FALSE | Upregulated |
| -0,055938074 | 0,985080598 | CREB3L2 | FALSE | Downregulated |

| | | | | |
|---|---|---|---|---|
| 0,048572713 | 0,985080598 | CUL5 | FALSE | Upregulated |
| 0,056032875 | 0,985080598 | KDM4C | FALSE | Upregulated |
| 0,049899942 | 0,985080598 | KLHL36 | FALSE | Upregulated |
| -0,030395502 | 0,985080598 | LOC102723489 | FALSE | Downregulated |
| 0,055161161 | 0,985080598 | RNH1 | FALSE | Upregulated |
| 0,043475987 | 0,985080598 | SLC25A6 | FALSE | Upregulated |
| -0,046693545 | 0,985145505 | SRSF11 | FALSE | Downregulated |
| -0,039045667 | 0,985182674 | EWSR1 | FALSE | Downregulated |
| 0,034645693 | 0,985182674 | HERC2 | FALSE | Upregulated |
| 0,055942471 | 0,985182674 | INO80 | FALSE | Upregulated |
| -0,037980072 | 0,985182674 | LOC101930171 | FALSE | Downregulated |
| -0,031751766 | 0,985182674 | MTSS1 | FALSE | Downregulated |
| 0,049968753 | 0,985182674 | NTMT1 | FALSE | Upregulated |
| -0,050601136 | 0,985182674 | ST6GALNAC6 | FALSE | Downregulated |
| 0,059619693 | 0,985387057 | L3MBTL4 | FALSE | Upregulated |
| 0,045256295 | 0,985387057 | MAGI1-AS1 | FALSE | Upregulated |
| -0,033714348 | 0,985389146 | SMG1 | FALSE | Downregulated |
| 0,067032757 | 0,985389146 | ZFR | FALSE | Upregulated |
| 0,051263368 | 0,985408549 | FCHSD2 | FALSE | Upregulated |
| 0,041847969 | 0,985408549 | LRP3 | FALSE | Upregulated |
| 0,03893288 | 0,985408549 | LRRC61 | FALSE | Upregulated |
| -0,043395594 | 0,985408549 | MATR3 | FALSE | Downregulated |
| 0,058210743 | 0,985408549 | TLR4 | FALSE | Upregulated |
| -0,05101581 | 0,985609867 | SMAD2 | FALSE | Downregulated |
| -0,047644418 | 0,985721873 | UNK | FALSE | Downregulated |
| 0,055871354 | 0,985966565 | PAK2 | FALSE | Upregulated |
| 0,037717105 | 0,985966565 | PRR12 | FALSE | Upregulated |
| -0,038179918 | 0,986056487 | EXOC7 | FALSE | Downregulated |
| -0,032744025 | 0,986126488 | TMED5 | FALSE | Downregulated |
| 0,053233876 | 0,986126488 | TSEN54 | FALSE | Upregulated |
| 0,051344121 | 0,986126488 | WDR45B | FALSE | Upregulated |
| 0,063572375 | 0,98624207 | LOC101927940 | FALSE | Upregulated |
| -0,045566301 | 0,986258897 | DOK4 | FALSE | Downregulated |
| -0,029708188 | 0,986258897 | ZC3H7B | FALSE | Downregulated |
| -0,036275652 | 0,986383899 | ARF1 | FALSE | Downregulated |
| -0,034503272 | 0,986383899 | DSP | FALSE | Downregulated |
| 0,044184068 | 0,986383899 | FAM178A | FALSE | Upregulated |
| 0,050338434 | 0,98655222 | CSGALNACT1 | FALSE | Upregulated |
| 0,049302085 | 0,986589279 | GSDMB | FALSE | Upregulated |
| 0,051161408 | 0,986753392 | CPNE3 | FALSE | Upregulated |
| -0,043192698 | 0,986753392 | CSTF2T | FALSE | Downregulated |
| -0,048405116 | 0,986753392 | HEPN1 | FALSE | Downregulated |
| 0,045191549 | 0,986753392 | HNRNPL | FALSE | Upregulated |
| 0,035326627 | 0,986753392 | HSP90AB1 | FALSE | Upregulated |
| -0,03064065 | 0,986753392 | MIA3 | FALSE | Downregulated |
| 0,032441647 | 0,986753392 | OLFML1 | FALSE | Upregulated |
| -0,053211833 | 0,986753392 | PALM2 | FALSE | Downregulated |
| 0,052582365 | 0,986753392 | PRKAR2A | FALSE | Upregulated |
| 0,038615921 | 0,986753392 | RAD9A | FALSE | Upregulated |
| 0,02828941 | 0,986753392 | RPL4 | FALSE | Upregulated |
| -0,057909558 | 0,986753392 | S100PBP | FALSE | Downregulated |

| | | | | |
|---|---|---|---|---|
| -0,040785654 | 0,986753392 | SDHC | FALSE | Downregulated |
| 0,041343101 | 0,986753392 | UBE2D4 | FALSE | Upregulated |
| 0,050401514 | 0,986753392 | ZER1 | FALSE | Upregulated |
| 0,055159387 | 0,986862079 | PCED1A | FALSE | Upregulated |
| -0,041593007 | 0,986862079 | PPP2R1A | FALSE | Downregulated |
| 0,032715333 | 0,986862079 | RERE | FALSE | Upregulated |
| 0,046979983 | 0,986862079 | RHBDD1 | FALSE | Upregulated |
| -0,051241757 | 0,986862079 | RNF187 | FALSE | Downregulated |
| 0,04251793 | 0,986862079 | SYNCRIP | FALSE | Upregulated |
| 0,049351172 | 0,986862079 | ZNF292 | FALSE | Upregulated |
| -0,044359307 | 0,986889155 | TLK1 | FALSE | Downregulated |
| -0,050902108 | 0,987001089 | GNG12 | FALSE | Downregulated |
| -0,034176611 | 0,987041063 | TRIM44 | FALSE | Downregulated |
| -0,028300231 | 0,9874171 | P2RX5-TAX1BP3 | FALSE | Downregulated |
| 0,05929397 | 0,9874171 | ZNF510 | FALSE | Upregulated |
| 0,033453617 | 0,987507359 | SRSF1 | FALSE | Upregulated |
| -0,053696543 | 0,987507359 | TUBA4A | FALSE | Downregulated |
| 0,040635546 | 0,987759215 | C1orf43 | FALSE | Upregulated |
| 0,044809931 | 0,987759215 | TIMM13 | FALSE | Upregulated |
| 0,038404657 | 0,9879459 | LOC102723666 | FALSE | Upregulated |
| 0,029027178 | 0,988247942 | LOC102724559 | FALSE | Upregulated |
| -0,047805419 | 0,98830252 | TAOK3 | FALSE | Downregulated |
| -0,033457017 | 0,98839592 | HNRNPF | FALSE | Downregulated |
| -0,035922494 | 0,98839592 | KIAA1199 | FALSE | Downregulated |
| -0,044423252 | 0,988711439 | CAPN7 | FALSE | Downregulated |
| 0,049884769 | 0,989243108 | C15orf37 | FALSE | Upregulated |
| -0,034078156 | 0,989243108 | PYGL | FALSE | Downregulated |
| -0,034648191 | 0,989573419 | CSNK1A1 | FALSE | Downregulated |
| 0,04732118 | 0,989573419 | EEF2K | FALSE | Upregulated |
| 0,054857822 | 0,989573419 | MTMR3 | FALSE | Upregulated |
| -0,03661731 | 0,989573419 | RDX | FALSE | Downregulated |
| 0,045051917 | 0,989684449 | EIF2S2 | FALSE | Upregulated |
| 0,022268223 | 0,989686446 | ABCA1 | FALSE | Upregulated |
| 0,040083006 | 0,989686446 | ANKRD12 | FALSE | Upregulated |
| -0,044495823 | 0,989686446 | CNTN2 | FALSE | Downregulated |
| -0,04788446 | 0,989686446 | DNAJA4 | FALSE | Downregulated |
| -0,040944728 | 0,989686446 | PCDH1 | FALSE | Downregulated |
| 0,036926602 | 0,989686446 | PPP1CB | FALSE | Upregulated |
| -0,046722808 | 0,989686446 | RFX1 | FALSE | Downregulated |
| 0,024983807 | 0,989686446 | XRRA1 | FALSE | Upregulated |
| 0,039048701 | 0,989814508 | DNAJB5 | FALSE | Upregulated |
| -0,038879967 | 0,990042768 | AMBRA1 | FALSE | Downregulated |
| -0,039969171 | 0,990042768 | CANT1 | FALSE | Downregulated |
| -0,065000746 | 0,990042768 | CDC5L | FALSE | Downregulated |
| -0,046938248 | 0,990042768 | CLIC1 | FALSE | Downregulated |
| -0,045920403 | 0,990042768 | CXCR5 | FALSE | Downregulated |
| 0,047308911 | 0,990042768 | DPP7 | FALSE | Upregulated |
| -0,034099735 | 0,990042768 | ESYT2 | FALSE | Downregulated |
| 0,047920429 | 0,990042768 | FBXO9 | FALSE | Upregulated |
| -0,023307011 | 0,990042768 | HYI | FALSE | Downregulated |
| 0,045635097 | 0,990042768 | IKBKB | FALSE | Upregulated |

| | | | | |
|---|---|---|---|---|
| 0,045149368 | 0,990042768 | KDSR | FALSE | Upregulated |
| 0,018586282 | 0,990042768 | KHNYN | FALSE | Upregulated |
| -0,024695625 | 0,990042768 | KMT2E | FALSE | Downregulated |
| -0,048105802 | 0,990042768 | LARS2 | FALSE | Downregulated |
| -0,039318891 | 0,990042768 | LILRB3 | FALSE | Downregulated |
| -0,030673972 | 0,990042768 | LOC101927018 | FALSE | Downregulated |
| 0,040074488 | 0,990042768 | LOC101928185 | FALSE | Upregulated |
| -0,034771287 | 0,990042768 | LOC101929109 | FALSE | Downregulated |
| 0,033902833 | 0,990042768 | LOC642441 | FALSE | Upregulated |
| 0,042321059 | 0,990042768 | LRRK1 | FALSE | Upregulated |
| -0,030192812 | 0,990042768 | MKLN1 | FALSE | Downregulated |
| 0,031042907 | 0,990042768 | N4BP2L2 | FALSE | Upregulated |
| -0,038069274 | 0,990042768 | OGG1 | FALSE | Downregulated |
| 0,031742969 | 0,990042768 | PAFAH1B1 | FALSE | Upregulated |
| -0,044450538 | 0,990042768 | PGK1 | FALSE | Downregulated |
| -0,030900495 | 0,990042768 | PICALM | FALSE | Downregulated |
| -0,031075163 | 0,990042768 | PLOD1 | FALSE | Downregulated |
| 0,0307835 | 0,990042768 | POLD4 | FALSE | Upregulated |
| -0,033797664 | 0,990042768 | PTPRM | FALSE | Downregulated |
| 0,042991162 | 0,990042768 | RCAN1 | FALSE | Upregulated |
| 0,047963984 | 0,990042768 | RP11-7F17.5 | FALSE | Upregulated |
| 0,052590777 | 0,990042768 | SBSPON | FALSE | Upregulated |
| -0,022674357 | 0,990042768 | SCARNA2 | FALSE | Downregulated |
| -0,048514317 | 0,990042768 | SEPHS1 | FALSE | Downregulated |
| -0,035699015 | 0,990042768 | SESN3 | FALSE | Downregulated |
| 0,034353741 | 0,990042768 | ST5 | FALSE | Upregulated |
| -0,028311372 | 0,990042768 | STAT3 | FALSE | Downregulated |
| 0,031466628 | 0,990042768 | SUPT5H | FALSE | Upregulated |
| 0,043157565 | 0,990042768 | TIPARP-AS1 | FALSE | Upregulated |
| 0,046209808 | 0,990042768 | TSPAN6 | FALSE | Upregulated |
| -0,033520165 | 0,990042768 | TUBB1 | FALSE | Downregulated |
| 0,025563132 | 0,990042768 | WDR73 | FALSE | Upregulated |
| 0,028360289 | 0,990042768 | YBX1 | FALSE | Upregulated |
| 0,021702562 | 0,990105248 | AHNAK | FALSE | Upregulated |
| 0,02510841 | 0,990105248 | CTDSP1 | FALSE | Upregulated |
| 0,039330594 | 0,990105248 | HIRA | FALSE | Upregulated |
| -0,028825555 | 0,990105248 | NUDT13 | FALSE | Downregulated |
| 0,032314228 | 0,990105248 | SCO2 | FALSE | Upregulated |
| 0,024883837 | 0,990185013 | SVIL | FALSE | Upregulated |
| -0,021508514 | 0,990392809 | KIAA1147 | FALSE | Downregulated |
| -0,041713861 | 0,990392809 | LOC100652768 | FALSE | Downregulated |
| 0,038570294 | 0,990608358 | COASY | FALSE | Upregulated |
| 0,028129581 | 0,990608358 | GAK | FALSE | Upregulated |
| 0,032093542 | 0,990706091 | SBNO1 | FALSE | Upregulated |
| 0,020526404 | 0,990747126 | ASB6 | FALSE | Upregulated |
| -0,03623291 | 0,990747126 | ATMIN | FALSE | Downregulated |
| 0,035087899 | 0,990747126 | NRTN | FALSE | Upregulated |
| 0,024776516 | 0,990747126 | PRRC2A | FALSE | Upregulated |
| 0,020464264 | 0,990785092 | CNOT1 | FALSE | Upregulated |
| -0,03844496 | 0,990785092 | GOSR2 | FALSE | Downregulated |
| -0,037693438 | 0,991204542 | CRTC3 | FALSE | Downregulated |

| | | | | |
|---|---|---|---|---|
| 0,025793501 | 0,991204542 | PCDH12 | FALSE | Upregulated |
| 0,042677904 | 0,991204542 | RNF34 | FALSE | Upregulated |
| -0,037166817 | 0,991204542 | RP11-103C16.2 | FALSE | Downregulated |
| 0,021902446 | 0,991204542 | RPS19 | FALSE | Upregulated |
| -0,027571049 | 0,991204542 | TAOK2 | FALSE | Downregulated |
| -0,027795727 | 0,991258967 | ABHD1 | FALSE | Downregulated |
| 0,026994564 | 0,991258967 | AC137932.6 | FALSE | Upregulated |
| -0,022873939 | 0,991258967 | ADORA2A-AS1 | FALSE | Downregulated |
| -0,034404213 | 0,991258967 | ALG1 | FALSE | Downregulated |
| 0,032705883 | 0,991258967 | AMER1 | FALSE | Upregulated |
| -0,028821774 | 0,991258967 | ARHGEF17 | FALSE | Downregulated |
| -0,029391655 | 0,991258967 | BNIP2 | FALSE | Downregulated |
| -0,025376267 | 0,991258967 | BPTF | FALSE | Downregulated |
| 0,036374957 | 0,991258967 | CBX4 | FALSE | Upregulated |
| -0,030041695 | 0,991258967 | CYFIP2 | FALSE | Downregulated |
| 0,022870317 | 0,991258967 | DDX58 | FALSE | Upregulated |
| 0,021485861 | 0,991258967 | DVL1 | FALSE | Upregulated |
| -0,02978308 | 0,991258967 | ECHDC3 | FALSE | Downregulated |
| -0,034144079 | 0,991258967 | EEA1 | FALSE | Downregulated |
| -0,040634228 | 0,991258967 | EID1 | FALSE | Downregulated |
| 0,018608251 | 0,991258967 | EML4 | FALSE | Upregulated |
| -0,032391602 | 0,991258967 | EZH1 | FALSE | Downregulated |
| 0,026933837 | 0,991258967 | FAM168B | FALSE | Upregulated |
| 0,019768334 | 0,991258967 | FMNL3 | FALSE | Upregulated |
| 0,042411274 | 0,991258967 | GPR153 | FALSE | Upregulated |
| -0,040106022 | 0,991258967 | GUCY1A3 | FALSE | Downregulated |
| 0,02451719 | 0,991258967 | KIAA0319L | FALSE | Upregulated |
| -0,021398546 | 0,991258967 | LOC100505502 | FALSE | Downregulated |
| -0,016917706 | 0,991258967 | LOC101927556 | FALSE | Downregulated |
| 0,019359111 | 0,991258967 | LOC101928323 | FALSE | Upregulated |
| 0,027911155 | 0,991258967 | LRRN2 | FALSE | Upregulated |
| 0,030138758 | 0,991258967 | MAML1 | FALSE | Upregulated |
| 0,020212476 | 0,991258967 | MAN2B2 | FALSE | Upregulated |
| 0,040964243 | 0,991258967 | MAP3K5 | FALSE | Upregulated |
| -0,023707114 | 0,991258967 | MBNL1 | FALSE | Downregulated |
| -0,022547005 | 0,991258967 | MFSD12 | FALSE | Downregulated |
| -0,035499941 | 0,991258967 | NAT8B | FALSE | Downregulated |
| 0,024049356 | 0,991258967 | NMT2 | FALSE | Upregulated |
| -0,037675714 | 0,991258967 | NSFL1C | FALSE | Downregulated |
| -0,031813755 | 0,991258967 | NUP62 | FALSE | Downregulated |
| -0,02358739 | 0,991258967 | OGFRL1 | FALSE | Downregulated |
| 0,034112762 | 0,991258967 | OTUD7B | FALSE | Upregulated |
| -0,037918511 | 0,991258967 | PAX6 | FALSE | Downregulated |
| 0,021684526 | 0,991258967 | PCBP1 | FALSE | Upregulated |
| 0,031415889 | 0,991258967 | PCMTD1 | FALSE | Upregulated |
| -0,036493952 | 0,991258967 | PDHA1 | FALSE | Downregulated |
| 0,032499988 | 0,991258967 | PER1 | FALSE | Upregulated |
| 0,028101728 | 0,991258967 | PER3 | FALSE | Upregulated |
| -0,028483554 | 0,991258967 | PIAS1 | FALSE | Downregulated |
| 0,0275994 | 0,991258967 | PLEKHJ1 | FALSE | Upregulated |
| -0,042022642 | 0,991258967 | PREB | FALSE | Downregulated |

| | | | | |
|---|---|---|---|---|
| 0,033873452 | 0,991258967 | PSMC2 | FALSE | Upregulated |
| 0,028854372 | 0,991258967 | PSMD6-AS2 | FALSE | Upregulated |
| 0,031132769 | 0,991258967 | RAB13 | FALSE | Upregulated |
| -0,020928148 | 0,991258967 | RAB26 | FALSE | Downregulated |
| -0,033208027 | 0,991258967 | RBM25 | FALSE | Downregulated |
| 0,027312648 | 0,991258967 | REV1 | FALSE | Upregulated |
| -0,036840637 | 0,991258967 | RNF170 | FALSE | Downregulated |
| -0,030212249 | 0,991258967 | RPL27 | FALSE | Downregulated |
| -0,022270091 | 0,991258967 | RPSA | FALSE | Downregulated |
| -0,042439 | 0,991258967 | SCD | FALSE | Downregulated |
| 0,032365145 | 0,991258967 | SENP6 | FALSE | Upregulated |
| -0,022426528 | 0,991258967 | SKIV2L2 | FALSE | Downregulated |
| -0,016673707 | 0,991258967 | SMIM5 | FALSE | Downregulated |
| 0,024201222 | 0,991258967 | SRP9 | FALSE | Upregulated |
| -0,028522041 | 0,991258967 | SSBP3 | FALSE | Downregulated |
| -0,027871217 | 0,991258967 | STK40 | FALSE | Downregulated |
| -0,035178116 | 0,991258967 | STRADB | FALSE | Downregulated |
| 0,037497962 | 0,991258967 | TBC1D17 | FALSE | Upregulated |
| 0,031897073 | 0,991258967 | TCF7L1 | FALSE | Upregulated |
| -0,026358813 | 0,991258967 | THEM6 | FALSE | Downregulated |
| -0,030791058 | 0,991258967 | TMEM63B | FALSE | Downregulated |
| -0,040164087 | 0,991258967 | TOR1A | FALSE | Downregulated |
| -0,025423724 | 0,991258967 | UBC | FALSE | Downregulated |
| -0,023768986 | 0,991258967 | WIZ | FALSE | Downregulated |
| -0,034104266 | 0,991258967 | ZAK | FALSE | Downregulated |
| 0,032683872 | 0,991258967 | ZBED6 | FALSE | Upregulated |
| -0,040613701 | 0,991258967 | ZBTB42 | FALSE | Downregulated |
| -0,034849939 | 0,991258967 | ZBTB8A | FALSE | Downregulated |
| 0,039460432 | 0,991258967 | ZNF503-AS2 | FALSE | Upregulated |
| 0,033880046 | 0,991282301 | SEMA4B | FALSE | Upregulated |
| -0,03072991 | 0,99128255 | ANXA7 | FALSE | Downregulated |
| 0,029841419 | 0,99128255 | CHTOP | FALSE | Upregulated |
| -0,029608971 | 0,99128255 | CPSF4 | FALSE | Downregulated |
| -0,022049632 | 0,99128255 | KPNB1 | FALSE | Downregulated |
| 0,032589553 | 0,99128255 | MPHOSPH9 | FALSE | Upregulated |
| -0,029594176 | 0,991602738 | PI4K2B | FALSE | Downregulated |
| 0,021948614 | 0,991888232 | GNA11 | FALSE | Upregulated |
| 0,016097312 | 0,991888232 | USP9X | FALSE | Upregulated |
| -0,01909643 | 0,991926516 | MCM3AP | FALSE | Downregulated |
| 0,017218392 | 0,992012809 | AES | FALSE | Upregulated |
| 0,0239646 | 0,992064846 | LPPR2 | FALSE | Upregulated |
| 0,029165001 | 0,992179572 | FBRS | FALSE | Upregulated |
| 0,019962065 | 0,992179572 | RPL10A | FALSE | Upregulated |
| 0,0338864 | 0,992179572 | SCAMP2 | FALSE | Upregulated |
| -0,023827927 | 0,992179572 | SYNM | FALSE | Downregulated |
| -0,026198405 | 0,992582879 | NBAS | FALSE | Downregulated |
| 0,022580464 | 0,992582879 | TARBP2 | FALSE | Upregulated |
| 0,022782674 | 0,992582879 | UBE2H | FALSE | Upregulated |
| -0,025312726 | 0,992683518 | RANBP3 | FALSE | Downregulated |
| 0,016233748 | 0,992683518 | RFTN1 | FALSE | Upregulated |
| -0,028765604 | 0,992683518 | SEZ6 | FALSE | Downregulated |

| | | | | |
|---|---|---|---|---|
| 0,021265282 | 0,992683518 | TRIM66 | FALSE | Upregulated |
| -0,028077636 | 0,992683518 | YTHDF1 | FALSE | Downregulated |
| -0,029984039 | 0,992742339 | PRKAA2 | FALSE | Downregulated |
| -0,026082923 | 0,992832581 | MIER3 | FALSE | Downregulated |
| 0,018570897 | 0,992837198 | PFKL | FALSE | Upregulated |
| 0,026835799 | 0,992955535 | PHKB | FALSE | Upregulated |
| -0,022998017 | 0,993021765 | EGFL7 | FALSE | Downregulated |
| -0,021067814 | 0,993048602 | TAF1 | FALSE | Downregulated |
| 0,02779853 | 0,993260817 | WIPF2 | FALSE | Upregulated |
| 0,013289809 | 0,993344735 | CEP68 | FALSE | Upregulated |
| -0,023217838 | 0,993382267 | ATE1 | FALSE | Downregulated |
| 0,018777514 | 0,993382267 | GALNT2 | FALSE | Upregulated |
| 0,02113188 | 0,994367623 | ACP2 | FALSE | Upregulated |
| -0,021162095 | 0,994367623 | CLINT1 | FALSE | Downregulated |
| -0,029569685 | 0,994367623 | IL18BP | FALSE | Downregulated |
| 0,016197416 | 0,994367623 | LOC102724761 | FALSE | Upregulated |
| -0,02219955 | 0,994367623 | SLC6A1-AS1 | FALSE | Downregulated |
| 0,022532859 | 0,994367623 | SRSF10 | FALSE | Upregulated |
| 0,021706935 | 0,994427854 | EPS15 | FALSE | Upregulated |
| 0,015438234 | 0,994427854 | KDM5C | FALSE | Upregulated |
| 0,025327998 | 0,994569218 | ALS2 | FALSE | Upregulated |
| -0,016513041 | 0,994572552 | SPARC | FALSE | Downregulated |
| -0,01417161 | 0,994683241 | C1orf86 | FALSE | Downregulated |
| -0,010614748 | 0,994683241 | COL10A1 | FALSE | Downregulated |
| -0,025913474 | 0,994683241 | RP11-983P16.4 | FALSE | Downregulated |
| 0,013859666 | 0,99473218 | FOXN3-AS2 | FALSE | Upregulated |
| -0,020106764 | 0,99473218 | IRGQ | FALSE | Downregulated |
| 0,018587089 | 0,994743466 | ASH1L-AS1 | FALSE | Upregulated |
| -0,020041116 | 0,994783068 | ARHGAP32 | FALSE | Downregulated |
| 0,023945401 | 0,994783068 | BRPF1 | FALSE | Upregulated |
| -0,023493495 | 0,994783068 | EIF2B5 | FALSE | Downregulated |
| -0,013928547 | 0,994783068 | EIF3B | FALSE | Downregulated |
| -0,02488875 | 0,994783068 | LRP5L | FALSE | Downregulated |
| 0,02105505 | 0,994783068 | MBD1 | FALSE | Upregulated |
| -0,017762737 | 0,994783068 | MGAT2 | FALSE | Downregulated |
| 0,022281279 | 0,994783068 | OSGEPL1 | FALSE | Upregulated |
| 0,02309408 | 0,994783068 | RAB6A | FALSE | Upregulated |
| 0,015106221 | 0,994783068 | RBM4B | FALSE | Upregulated |
| 0,022618424 | 0,994783068 | SPIN1 | FALSE | Upregulated |
| 0,016445847 | 0,994783068 | TMEM30A | FALSE | Upregulated |
| 0,017153875 | 0,994783068 | ZNF316 | FALSE | Upregulated |
| -0,013319353 | 0,994783068 | ZNF587 | FALSE | Downregulated |
| -0,016173928 | 0,994835312 | BMP8B | FALSE | Downregulated |
| -0,016846203 | 0,995023389 | GLYR1 | FALSE | Downregulated |
| 0,013265596 | 0,995140932 | CAMTA2 | FALSE | Upregulated |
| -0,015409209 | 0,995140932 | TNFRSF1A | FALSE | Downregulated |
| -0,019700338 | 0,995194205 | NR1D2 | FALSE | Downregulated |
| 0,009829688 | 0,995194205 | ZBTB20 | FALSE | Upregulated |
| -0,016304481 | 0,995353663 | ABI3BP | FALSE | Downregulated |
| -0,013066514 | 0,995353663 | AIMP1 | FALSE | Downregulated |
| -0,014670762 | 0,995353663 | ARIH1 | FALSE | Downregulated |

| | | | | |
|---|---|---|---|---|
| 0,018720517 | 0,995353663 | DDX24 | FALSE | Upregulated |
| -0,015611568 | 0,995353663 | FAM211B | FALSE | Downregulated |
| -0,0171573 | 0,995353663 | GHRL | FALSE | Downregulated |
| -0,014090168 | 0,995353663 | GTF2I | FALSE | Downregulated |
| -0,017570413 | 0,995353663 | HARS | FALSE | Downregulated |
| 0,019379437 | 0,995353663 | HECA | FALSE | Upregulated |
| 0,011936521 | 0,995353663 | HLA-E | FALSE | Upregulated |
| 0,014911448 | 0,995353663 | IPO5 | FALSE | Upregulated |
| 0,014733531 | 0,995353663 | LTBR | FALSE | Upregulated |
| 0,009569114 | 0,995353663 | MASP2 | FALSE | Upregulated |
| -0,010194045 | 0,995353663 | MICAL3 | FALSE | Downregulated |
| 0,01613371 | 0,995353663 | MORC2 | FALSE | Upregulated |
| -0,014304061 | 0,995353663 | NAPA | FALSE | Downregulated |
| 0,019918566 | 0,995353663 | PTTG3P | FALSE | Upregulated |
| 0,020340847 | 0,995353663 | RGS14 | FALSE | Upregulated |
| -0,01812435 | 0,995353663 | RNPS1 | FALSE | Downregulated |
| 0,012492846 | 0,995353663 | RPS26 | FALSE | Upregulated |
| 0,014227809 | 0,995353663 | TRMT2A | FALSE | Upregulated |
| 0,014663534 | 0,995353663 | U2AF2 | FALSE | Upregulated |
| 0,013533628 | 0,995353663 | UBXN6 | FALSE | Upregulated |
| 0,018399827 | 0,995353663 | WDFY2 | FALSE | Upregulated |
| -0,016173719 | 0,995353663 | WDR20 | FALSE | Downregulated |
| -0,016405801 | 0,99536823 | CLEC16A | FALSE | Downregulated |
| 0,017645889 | 0,99536823 | RPF1 | FALSE | Upregulated |
| -0,015527345 | 0,99536823 | ZNF280D | FALSE | Downregulated |
| 0,010775967 | 0,995646126 | NIPBL | FALSE | Upregulated |
| 0,018008728 | 0,995680301 | RAB1A | FALSE | Upregulated |
| -0,018178731 | 0,995861052 | CCDC117 | FALSE | Downregulated |
| -0,013424865 | 0,995861052 | KIAA0195 | FALSE | Downregulated |
| -0,017266303 | 0,995861052 | SYNPO2 | FALSE | Downregulated |
| 0,016827115 | 0,995861052 | TPGS2 | FALSE | Upregulated |
| -0,016501086 | 0,99589662 | ANXA2P1 | FALSE | Downregulated |
| -0,011100742 | 0,99589662 | ARHGAP5 | FALSE | Downregulated |
| 0,01391838 | 0,99589662 | ATP2B1 | FALSE | Upregulated |
| 0,014211175 | 0,99589662 | ATRX | FALSE | Upregulated |
| -0,014348703 | 0,99589662 | DDX23 | FALSE | Downregulated |
| 0,007815528 | 0,99589662 | KANSL1L | FALSE | Upregulated |
| 0,017411302 | 0,99589662 | LOC100506497 | FALSE | Upregulated |
| 0,014370359 | 0,99589662 | NAB1 | FALSE | Upregulated |
| -0,013345224 | 0,99589662 | NUP153 | FALSE | Downregulated |
| -0,018419604 | 0,99589662 | PANX2 | FALSE | Downregulated |
| -0,012836229 | 0,99589662 | PSMD7 | FALSE | Downregulated |
| -0,014084887 | 0,99589662 | TECPR2 | FALSE | Downregulated |
| 0,008872252 | 0,996595275 | AIM1 | FALSE | Upregulated |
| 0,014315901 | 0,996595275 | BRWD1-IT2 | FALSE | Upregulated |
| -0,008966584 | 0,996595275 | CCDC50 | FALSE | Downregulated |
| 0,011956574 | 0,996595275 | FBXO7 | FALSE | Upregulated |
| 0,008847033 | 0,996595275 | GPC1 | FALSE | Upregulated |
| -0,007413467 | 0,996595275 | MGRN1 | FALSE | Downregulated |
| -0,00859091 | 0,996595275 | MYCBP2 | FALSE | Downregulated |
| 0,011116793 | 0,996595275 | NAGA | FALSE | Upregulated |

| | | | | |
|---|---|---|---|---|
| 0,013141421 | 0,996595275 | NAT10 | FALSE | Upregulated |
| -0,008234129 | 0,996595275 | ONECUT1 | FALSE | Downregulated |
| -0,011135945 | 0,996595275 | PAG1 | FALSE | Downregulated |
| 0,006323749 | 0,996595275 | PCGF3 | FALSE | Upregulated |
| -0,010558437 | 0,996595275 | RP11-266L9.2 | FALSE | Downregulated |
| 0,013026027 | 0,996595275 | RPL34 | FALSE | Upregulated |
| -0,012502103 | 0,996595275 | SMAD5 | FALSE | Downregulated |
| -0,012744818 | 0,996595275 | TBRG1 | FALSE | Downregulated |
| 0,015641801 | 0,996595275 | TUBA3D | FALSE | Upregulated |
| 0,01220031 | 0,996595275 | VAPB | FALSE | Upregulated |
| 0,009722855 | 0,996595275 | ZBTB11-AS1 | FALSE | Upregulated |
| 0,01037291 | 0,996755301 | ATAT1 | FALSE | Upregulated |
| -0,011173348 | 0,996755301 | BORA | FALSE | Downregulated |
| 0,015417255 | 0,996755301 | C19orf80 | FALSE | Upregulated |
| 0,011665566 | 0,996817581 | REST | FALSE | Upregulated |
| 0,006736022 | 0,996817581 | SYNPO | FALSE | Upregulated |
| 0,00601898 | 0,997040435 | ADAM17 | FALSE | Upregulated |
| -0,004899311 | 0,997040435 | ANKFY1 | FALSE | Downregulated |
| -0,007831706 | 0,997040435 | C3orf62 | FALSE | Downregulated |
| 0,008264984 | 0,997040435 | CCDC90B | FALSE | Upregulated |
| 0,006615297 | 0,997040435 | CDC42 | FALSE | Upregulated |
| 0,006712235 | 0,997040435 | CDK10 | FALSE | Upregulated |
| -0,007293946 | 0,997040435 | CKMT2 | FALSE | Downregulated |
| 0,005867358 | 0,997040435 | CLN5 | FALSE | Upregulated |
| -0,010681048 | 0,997040435 | COPE | FALSE | Downregulated |
| 0,009006149 | 0,997040435 | CXXC1 | FALSE | Upregulated |
| -0,007579844 | 0,997040435 | ETS2 | FALSE | Downregulated |
| -0,006893972 | 0,997040435 | FAM184B | FALSE | Downregulated |
| -0,009162739 | 0,997040435 | FUS | FALSE | Downregulated |
| -0,005573283 | 0,997040435 | HSPA5 | FALSE | Downregulated |
| -0,01731973 | 0,997040435 | LIFR-AS1 | FALSE | Downregulated |
| -0,006831574 | 0,997040435 | LOC101927377 | FALSE | Downregulated |
| 0,010957999 | 0,997040435 | LOC101927681 | FALSE | Upregulated |
| 0,005759056 | 0,997040435 | LOC101927838 | FALSE | Upregulated |
| 0,005845696 | 0,997040435 | LOC102724445 | FALSE | Upregulated |
| 0,007313748 | 0,997040435 | LONP1 | FALSE | Upregulated |
| 0,007104692 | 0,997040435 | LRRC19 | FALSE | Upregulated |
| 0,011077436 | 0,997040435 | MAP3K1 | FALSE | Upregulated |
| -0,005521018 | 0,997040435 | MTUS1 | FALSE | Downregulated |
| -0,011001743 | 0,997040435 | ORC2 | FALSE | Downregulated |
| 0,007996002 | 0,997040435 | PGAM1 | FALSE | Upregulated |
| 0,007118715 | 0,997040435 | PHF10 | FALSE | Upregulated |
| 0,007016193 | 0,997040435 | PLCD4 | FALSE | Upregulated |
| -0,005630929 | 0,997040435 | POR | FALSE | Downregulated |
| -0,005483495 | 0,997040435 | PPP1R12C | FALSE | Downregulated |
| -0,009796598 | 0,997040435 | PPP2R5D | FALSE | Downregulated |
| 0,007293824 | 0,997040435 | RCN1 | FALSE | Upregulated |
| -0,004930555 | 0,997040435 | REXO1 | FALSE | Downregulated |
| -0,005035686 | 0,997040435 | RHOQ | FALSE | Downregulated |
| -0,008079271 | 0,997040435 | RPS12 | FALSE | Downregulated |
| 0,005831072 | 0,997040435 | SERP1 | FALSE | Upregulated |

| | | | | |
|---|---|---|---|---|
| -0,009375459 | 0,997040435 | UBE4A | FALSE | Downregulated |
| 0,005761576 | 0,997040435 | UHRF1BP1 | FALSE | Upregulated |
| -0,007639431 | 0,997040435 | VEGFB | FALSE | Downregulated |
| -0,011561649 | 0,997040435 | ZFP41 | FALSE | Downregulated |
| 0,005912231 | 0,997077503 | ZNF207 | FALSE | Upregulated |
| -0,006983494 | 0,997232276 | ELF2 | FALSE | Downregulated |
| -0,005357165 | 0,997232276 | MAPKAPK5-AS1 | FALSE | Downregulated |
| 0,006468662 | 0,997232276 | SKIL | FALSE | Upregulated |
| -0,005496878 | 0,997488781 | ACAD10 | FALSE | Downregulated |
| 0,005073765 | 0,997488781 | B2M | FALSE | Upregulated |
| -0,007029065 | 0,997488781 | BCAP31 | FALSE | Downregulated |
| 0,004899405 | 0,997488781 | BEAN1 | FALSE | Upregulated |
| 0,003911672 | 0,997488781 | C12orf23 | FALSE | Upregulated |
| 0,003752819 | 0,997488781 | CAPRIN1 | FALSE | Upregulated |
| -0,004586642 | 0,997488781 | CENPB | FALSE | Downregulated |
| -0,004960017 | 0,997488781 | EDC4 | FALSE | Downregulated |
| -0,003781815 | 0,997488781 | LOC100506388 | FALSE | Downregulated |
| 0,004789936 | 0,997488781 | LOC101059949 | FALSE | Upregulated |
| 0,005271254 | 0,997488781 | MRPS16 | FALSE | Upregulated |
| 0,005410388 | 0,997488781 | MYO10 | FALSE | Upregulated |
| 0,005756895 | 0,997488781 | NT5C1B | FALSE | Upregulated |
| 0,005293669 | 0,997488781 | SLC23A1 | FALSE | Upregulated |
| 0,004730513 | 0,997488781 | SNRK | FALSE | Upregulated |
| 0,005171283 | 0,997488781 | SPIRE2 | FALSE | Upregulated |
| 0,006574653 | 0,997488781 | SYAP1 | FALSE | Upregulated |
| -0,004271266 | 0,997488781 | TP53INP2 | FALSE | Downregulated |
| 0,004468653 | 0,997488781 | ZFHX2 | FALSE | Upregulated |
| -0,004574407 | 0,997529532 | CPLX1 | FALSE | Downregulated |
| 0,00390344 | 0,997912974 | PITRM1 | FALSE | Upregulated |
| 0,003774275 | 0,997912974 | SAP30L | FALSE | Upregulated |
| -0,002727712 | 0,997912974 | SP1 | FALSE | Downregulated |
| 0,002753066 | 0,997960043 | PAWR | FALSE | Upregulated |
| 0,001907865 | 0,998400696 | CCRN4L | FALSE | Upregulated |
| -0,002155556 | 0,998400696 | DDB1 | FALSE | Downregulated |
| 0,002326122 | 0,998400696 | JADE1 | FALSE | Upregulated |
| 0,002319429 | 0,998400696 | LOC401286 | FALSE | Upregulated |
| 0,002569651 | 0,998400696 | NINL | FALSE | Upregulated |
| 0,002431681 | 0,998400696 | RIOK3 | FALSE | Upregulated |
| -0,002422713 | 0,998400696 | RNF32 | FALSE | Downregulated |
| -0,00221447 | 0,998400696 | SPECC1L | FALSE | Downregulated |
| 0,003647178 | 0,998400696 | ZNF394 | FALSE | Upregulated |
| -0,00181593 | 0,998793958 | KAT2B | FALSE | Downregulated |
| 0,000551363 | 0,999119452 | ALDH4A1 | FALSE | Upregulated |
| -0,001497413 | 0,999119452 | CDK9 | FALSE | Downregulated |
| 0,00134594 | 0,999119452 | CLDN12 | FALSE | Upregulated |
| 0,001229142 | 0,999119452 | DIS3L | FALSE | Upregulated |
| 0,001465125 | 0,999119452 | FMR1 | FALSE | Upregulated |
| 0,000864652 | 0,999119452 | TUBGCP4 | FALSE | Upregulated |
| -0,000785155 | 0,999119452 | ZNF814 | FALSE | Downregulated |
| 0,000968986 | 0,999309873 | IDNK | FALSE | Upregulated |
| -0,000613648 | 0,999484823 | ZMYM4 | FALSE | Downregulated |

| | | | | |
|---|---|---|---|---|
| -0,000311031 | 0,999688514 | ZNRF2 | FALSE | Downregulated |
| 0,000112463 | 0,999717719 | SF3B1 | FALSE | Upregulated |
| -0,000143273 | 0,999717719 | ZNF850 | FALSE | Downregulated |
| -0,007012033 | | AAAS | FALSE | Downregulated |
| -0,393340586 | | AADAT | FALSE | Downregulated |
| 0,404437617 | | AAGAB | FALSE | Upregulated |
| -0,337443285 | | AAR2 | FALSE | Downregulated |
| 0,176868373 | | AARS2 | FALSE | Upregulated |
| -0,236860288 | | ABCA10 | FALSE | Downregulated |
| 0,887284991 | | ABCA7 | FALSE | Upregulated |
| -0,89109055 | | ABCB10 | FALSE | Downregulated |
| -1,018981629 | | ABCB6 | | Downregulated |
| 1,028870088 | | ABCB8 | | Upregulated |
| 0,573940186 | | ABCC10 | FALSE | Upregulated |
| 0,052149295 | | ABCD1 | FALSE | Upregulated |
| -0,705405664 | | ABCD4 | FALSE | Downregulated |
| -0,273620959 | | ABCF2 | FALSE | Downregulated |
| -1,865977943 | | ABCG2 | | Downregulated |
| 0,929114605 | | ABHD11 | FALSE | Upregulated |
| 0,563823606 | | ABHD12 | FALSE | Upregulated |
| 1,27182413 | | ABHD13 | | Upregulated |
| -1,067229638 | | ABHD14A | | Downregulated |
| 0,904038959 | | ABHD4 | FALSE | Upregulated |
| -0,088478535 | | ABHD6 | FALSE | Downregulated |
| 0,319976413 | | ABI1 | FALSE | Upregulated |
| -0,900290057 | | ABT1 | FALSE | Downregulated |
| 0,296030028 | | ABTB1 | FALSE | Upregulated |
| 0,105797739 | | AC009264.1 | FALSE | Upregulated |
| -0,92836087 | | AC011747.7 | FALSE | Downregulated |
| 0,273078111 | | AC058791.1 | FALSE | Upregulated |
| -0,277454245 | | AC091878.1 | FALSE | Downregulated |
| 0,451245244 | | AC144449.1 | FALSE | Upregulated |
| -0,023807114 | | AC147651.4 | FALSE | Downregulated |
| 0,303709783 | | ACAD8 | FALSE | Upregulated |
| -0,40013554 | | ACAD9 | FALSE | Downregulated |
| 0,454684104 | | ACAP1 | FALSE | Upregulated |
| -0,159147543 | | ACBD3 | FALSE | Downregulated |
| 1,130461455 | | ACBD6 | | Upregulated |
| -0,87374675 | | ACCS | FALSE | Downregulated |
| -0,164900842 | | ACSL3 | FALSE | Downregulated |
| 0,349902822 | | ACSS1 | FALSE | Upregulated |
| 0,240786615 | | ACTL6A | FALSE | Upregulated |
| -1,026558651 | | ACTR10 | | Downregulated |
| 0,421122847 | | ACVR1 | FALSE | Upregulated |
| 1,087671994 | | ACVRL1 | | Upregulated |
| -1,044099181 | | ACYP2 | | Downregulated |
| -0,421321046 | | ADAL | FALSE | Downregulated |
| 0,015704157 | | ADAM15 | FALSE | Upregulated |
| 1,225099021 | | ADAM19 | | Upregulated |
| -0,503275174 | | ADAMTS16 | FALSE | Downregulated |
| -0,170860086 | | ADAMTS9 | FALSE | Downregulated |

| | | | |
|---|---|---|---|
| 0,736662954 | ADARB1 | FALSE | Upregulated |
| 1,097884784 | ADAT1 | | Upregulated |
| 0,366744179 | ADCK2 | FALSE | Upregulated |
| -0,678771698 | ADCK4 | FALSE | Downregulated |
| -1,563068293 | ADCY10 | | Downregulated |
| -0,317644303 | ADD3 | FALSE | Downregulated |
| -0,620089314 | ADIRF | FALSE | Downregulated |
| 1,201821451 | ADNP2 | | Upregulated |
| 0,475840755 | ADO | FALSE | Upregulated |
| -0,257383 | ADPRHL2 | FALSE | Downregulated |
| -0,137192702 | ADRA1B | FALSE | Downregulated |
| -0,055058416 | ADRB2 | FALSE | Downregulated |
| 0,450163496 | ADSSL1 | FALSE | Upregulated |
| -0,001031717 | AEBP2 | FALSE | Downregulated |
| -0,092813017 | AFAP1L1 | FALSE | Downregulated |
| 0,602015135 | AFF3 | FALSE | Upregulated |
| 0,722269546 | AFG3L1P | FALSE | Upregulated |
| 0,19496966 | AFTPH | FALSE | Upregulated |
| -0,012096736 | AGBL2 | FALSE | Downregulated |
| -0,080524644 | AGER | FALSE | Downregulated |
| -0,08219361 | AGGF1 | FALSE | Downregulated |
| -0,775334707 | AGMO | FALSE | Downregulated |
| 0,166648645 | AGO2 | FALSE | Upregulated |
| -1,445126546 | AGO3 | | Downregulated |
| 0,460849623 | AGTPBP1 | FALSE | Upregulated |
| 0,505569192 | AGTR1 | FALSE | Upregulated |
| -0,24896421 | AHCYL2 | FALSE | Downregulated |
| -0,120516198 | AHI1 | FALSE | Downregulated |
| 0,373940709 | AHSA1 | FALSE | Upregulated |
| 0,416292722 | AIF1L | FALSE | Upregulated |
| 0,247308291 | AIMP2 | FALSE | Upregulated |
| -0,658518096 | AIP | FALSE | Downregulated |
| -0,230566737 | AK1 | FALSE | Downregulated |
| 1,234549039 | AK5 | | Upregulated |
| 1,174622636 | AKIRIN1 | | Upregulated |
| 0,03766844 | AKIRIN2 | FALSE | Upregulated |
| -0,026733632 | ALDH16A1 | FALSE | Downregulated |
| 0,062308334 | ALDH18A1 | FALSE | Upregulated |
| 0,772217221 | ALDH1A3 | FALSE | Upregulated |
| 0,035501998 | ALDH3B1 | FALSE | Upregulated |
| 0,409318649 | ALG2 | FALSE | Upregulated |
| -0,212644144 | ALKBH3-AS1 | FALSE | Downregulated |
| 0,316856329 | ALOX12P2 | FALSE | Upregulated |
| 1,213167003 | ALX4 | | Upregulated |
| 0,83436276 | AMD1 | FALSE | Upregulated |
| 0,150276599 | AMDHD2 | FALSE | Upregulated |
| 0,725108545 | AMOT | FALSE | Upregulated |
| -1,41821884 | AMY2B | | Downregulated |
| -1,129357201 | AMZ2 | | Downregulated |
| 0,322166616 | ANAPC13 | FALSE | Upregulated |
| 0,576995644 | ANAPC7 | FALSE | Upregulated |

| | | | |
|---|---|---|---|
| -0,150253489 | ANKMY1 | FALSE | Downregulated |
| 0,235099756 | ANKRD13C | FALSE | Upregulated |
| 0,287605374 | ANKRD13D | FALSE | Upregulated |
| -1,022217646 | ANKRD24 | | Downregulated |
| 0,12875322 | ANKRD34A | FALSE | Upregulated |
| 1,002900385 | ANKRD36B | | Upregulated |
| 0,20657373 | ANKRD36C | FALSE | Upregulated |
| 0,24814338 | ANKRD37 | FALSE | Upregulated |
| -1,465300971 | ANKRD54 | | Downregulated |
| 0,754855789 | ANKRD9 | FALSE | Upregulated |
| -0,351566728 | ANKS3 | FALSE | Downregulated |
| -0,324067186 | ANKS4B | FALSE | Downregulated |
| 0,789640638 | ANKS6 | FALSE | Upregulated |
| -0,456838189 | ANKZF1 | FALSE | Downregulated |
| 0,854516341 | ANO10 | FALSE | Upregulated |
| 1,038457275 | AOAH | | Upregulated |
| 0,878417373 | AOC3 | FALSE | Upregulated |
| 0,020535871 | AP1AR | FALSE | Upregulated |
| 0,608122812 | AP1S1 | FALSE | Upregulated |
| 0,683662755 | AP1S2 | FALSE | Upregulated |
| -0,28464352 | AP3B1 | FALSE | Downregulated |
| 1,269458456 | AP5B1 | | Upregulated |
| 0,007838665 | AP5M1 | FALSE | Upregulated |
| 0,222939968 | AP5S1 | FALSE | Upregulated |
| 0,336442076 | AP5Z1 | FALSE | Upregulated |
| -0,25560742 | APBA3 | FALSE | Downregulated |
| 1,199620996 | APBB1 | | Upregulated |
| -0,159151204 | APBB1IP | FALSE | Downregulated |
| 1,513209888 | APBB3 | | Upregulated |
| 0,341688917 | APCDD1L | FALSE | Upregulated |
| -0,660738553 | APEX1 | FALSE | Downregulated |
| -0,482957742 | APLNR | FALSE | Downregulated |
| 0,439659676 | APPL1 | FALSE | Upregulated |
| 0,501131717 | APRT | FALSE | Upregulated |
| 0,406059125 | AQP1 | FALSE | Upregulated |
| -0,057463637 | AQP7P1 | FALSE | Downregulated |
| 0,239552753 | AQR | FALSE | Upregulated |
| -0,180919618 | ARAP3 | FALSE | Downregulated |
| -0,373803521 | ARF5 | FALSE | Downregulated |
| -0,693445182 | ARFGAP3 | FALSE | Downregulated |
| 0,646078667 | ARHGAP10 | FALSE | Upregulated |
| -0,404031337 | ARHGAP15 | FALSE | Downregulated |
| 0,04332819 | ARHGAP18 | FALSE | Upregulated |
| 0,367767451 | ARHGAP20 | FALSE | Upregulated |
| 1,020549286 | ARHGAP23 | | Upregulated |
| 0,771422296 | ARHGAP24 | FALSE | Upregulated |
| 1,461171051 | ARHGAP27 | | Upregulated |
| -0,187068639 | ARHGAP33 | FALSE | Downregulated |
| -0,022005756 | ARHGAP42 | FALSE | Downregulated |
| 1,083584137 | ARHGAP9 | | Upregulated |
| 0,190375454 | ARHGDIB | FALSE | Upregulated |

| | | | |
|---|---|---|---|
| -0,482802965 | ARHGEF10 | FALSE | Downregulated |
| 0,524555331 | ARHGEF15 | FALSE | Upregulated |
| 0,008378458 | ARHGEF33 | FALSE | Upregulated |
| -0,087757757 | ARHGEF4 | FALSE | Downregulated |
| -0,319686798 | ARHGEF6 | FALSE | Downregulated |
| 1,547449339 | ARID5B | | Upregulated |
| -0,21897345 | ARIH2OS | FALSE | Downregulated |
| -0,890063814 | ARL1 | FALSE | Downregulated |
| 0,292045617 | ARL13A | FALSE | Upregulated |
| -1,825481527 | ARL13B | | Downregulated |
| 0,312584137 | ARL16 | FALSE | Upregulated |
| -1,193766993 | ARL3 | | Downregulated |
| -0,595991501 | ARL5A | FALSE | Downregulated |
| 0,403693193 | ARL8A | FALSE | Upregulated |
| 0,482244709 | ARMC8 | FALSE | Upregulated |
| -0,119353972 | ARMCX3 | FALSE | Downregulated |
| 0,054115398 | ARMCX4 | FALSE | Upregulated |
| -0,407596949 | ARMCX5 | FALSE | Downregulated |
| -0,646310097 | ARNTL | FALSE | Downregulated |
| -0,104271368 | ARPC2 | FALSE | Downregulated |
| 1,06745611 | ARPC5 | | Upregulated |
| 0,401449019 | ARPC5L | FALSE | Upregulated |
| 0,789193762 | ARR3 | FALSE | Upregulated |
| -0,225284582 | ARSK | FALSE | Downregulated |
| -1,466168564 | ART4 | | Downregulated |
| -0,613217019 | ASAP2 | FALSE | Downregulated |
| 0,743170495 | ASB1 | FALSE | Upregulated |
| 0,501522608 | ASB7 | FALSE | Upregulated |
| 0,263725034 | ASB8 | FALSE | Upregulated |
| 0,40371125 | ASNA1 | FALSE | Upregulated |
| 0,011186745 | ASNSD1 | FALSE | Upregulated |
| 0,564469766 | ASPHD1 | FALSE | Upregulated |
| -0,131513408 | ASPHD2 | FALSE | Downregulated |
| 1,346984377 | ASPN | | Upregulated |
| -0,251227321 | ASPSCR1 | FALSE | Downregulated |
| 0,36410462 | ATAD3A | FALSE | Upregulated |
| 0,440458392 | ATE1-AS1 | FALSE | Upregulated |
| -0,366133867 | ATG12 | FALSE | Downregulated |
| 0,131206024 | ATG16L1 | FALSE | Upregulated |
| -1,235329978 | ATG3 | | Downregulated |
| 0,923342247 | ATG9B | FALSE | Upregulated |
| 0,402726558 | ATL1 | FALSE | Upregulated |
| 1,345028758 | ATP13A2 | | Upregulated |
| -0,020799631 | ATP5E | FALSE | Downregulated |
| -0,799579159 | ATP5F1 | FALSE | Downregulated |
| -0,165843361 | ATP5G2 | FALSE | Downregulated |
| -0,871705902 | ATP5H | FALSE | Downregulated |
| -0,899626418 | ATP5S | FALSE | Downregulated |
| 0,003709898 | ATP6AP2 | FALSE | Upregulated |
| 0,718403272 | ATP6V0A2 | FALSE | Upregulated |
| 0,96231511 | ATP6V0B | FALSE | Upregulated |

| | | | |
|---|---|---|---|
| 0,422827503 | ATP6V0D1 | FALSE | Upregulated |
| 0,157410836 | ATP6V0E1 | FALSE | Upregulated |
| -0,863830747 | ATP6V1A | FALSE | Downregulated |
| -0,007619081 | ATP6V1C1 | FALSE | Downregulated |
| -0,151510178 | ATP6V1D | FALSE | Downregulated |
| 0,170636486 | ATP6V1E1 | FALSE | Upregulated |
| -0,228112061 | ATP7A | FALSE | Downregulated |
| 1,475244492 | ATP8A1 | | Upregulated |
| 0,831523203 | ATP8B2 | FALSE | Upregulated |
| -0,915866739 | ATPAF1 | FALSE | Downregulated |
| 0,309426054 | ATRAID | FALSE | Upregulated |
| -0,088072399 | ATXN7L1 | FALSE | Downregulated |
| -1,188871791 | AVEN | | Downregulated |
| -0,171088049 | AVPI1 | FALSE | Downregulated |
| 1,026312003 | AVPR2 | | Upregulated |
| -1,031796159 | AZGP1P1 | | Downregulated |
| 0,019697028 | AZI1 | FALSE | Upregulated |
| 0,424016714 | B3GAT3 | FALSE | Upregulated |
| -0,820255225 | B3GNT1 | FALSE | Downregulated |
| 0,533631824 | B3GNT4 | FALSE | Upregulated |
| -0,225631072 | B4GALT2 | FALSE | Downregulated |
| -0,036537953 | B4GALT3 | FALSE | Downregulated |
| 0,794142734 | B4GALT5 | FALSE | Upregulated |
| -0,045803905 | BABAM1 | FALSE | Downregulated |
| 2,065071441 | BACH2 | | Upregulated |
| 0,249535011 | BAD | FALSE | Upregulated |
| -0,116760003 | BAG5 | FALSE | Downregulated |
| -0,625516903 | BAIAP2-AS1 | FALSE | Downregulated |
| 1,07831225 | BBS4 | | Upregulated |
| 0,715861936 | BBS5 | FALSE | Upregulated |
| -0,134763822 | BBS9 | FALSE | Downregulated |
| 0,513565773 | BCAP29 | FALSE | Upregulated |
| 0,593861998 | BCAR3 | FALSE | Upregulated |
| -0,654066086 | BCDIN3D | FALSE | Downregulated |
| 0,213409414 | BCL2L2 | FALSE | Upregulated |
| 0,57635755 | BCL6B | FALSE | Upregulated |
| -0,130433364 | BCL7C | FALSE | Downregulated |
| 0,097116619 | BCS1L | FALSE | Upregulated |
| -0,500074403 | BEND7 | FALSE | Downregulated |
| 0,025085533 | BET1L | FALSE | Upregulated |
| 0,185935398 | BFAR | FALSE | Upregulated |
| -1,709419765 | BIVM | | Downregulated |
| -0,242336094 | BLNK | FALSE | Downregulated |
| -0,24667711 | BLOC1S1 | FALSE | Downregulated |
| 0,198527701 | BLOC1S3 | FALSE | Upregulated |
| 0,692842773 | BLOC1S6 | FALSE | Upregulated |
| -0,473585081 | BLZF1 | FALSE | Downregulated |
| 0,466976537 | BMF | FALSE | Upregulated |
| 0,172686803 | BMPR1A | FALSE | Upregulated |
| 0,316977494 | BMS1 | FALSE | Upregulated |
| 0,254120999 | BMS1P4 | FALSE | Upregulated |

| | | | |
|---|---|---|---|
| 0,981710231 | BMS1P5 | FALSE | Upregulated |
| -1,095313736 | BOK-AS1 | | Downregulated |
| 0,260499044 | BOP1 | FALSE | Upregulated |
| -0,795409929 | BRAP | FALSE | Downregulated |
| -0,018390408 | BRCC3 | FALSE | Downregulated |
| 0,103122204 | BRD7 | FALSE | Upregulated |
| -0,571933906 | BRD8 | FALSE | Downregulated |
| -1,015522624 | BRF2 | | Downregulated |
| 0,317377131 | BRK1 | FALSE | Upregulated |
| -0,968863905 | BRWD3 | FALSE | Downregulated |
| 0,490883019 | BSPRY | FALSE | Upregulated |
| -1,17963817 | BTF3L4 | | Downregulated |
| -0,295330182 | BTN3A3 | FALSE | Downregulated |
| 0,850347915 | BTRC | FALSE | Upregulated |
| -0,341556716 | BUB3 | FALSE | Downregulated |
| 0,225091445 | C10orf118 | FALSE | Upregulated |
| 0,981379041 | C10orf12 | FALSE | Upregulated |
| -0,021588288 | C10orf76 | FALSE | Downregulated |
| 1,095685664 | C11orf16 | | Upregulated |
| 0,026858712 | C11orf30 | FALSE | Upregulated |
| -1,098012277 | C11orf35 | | Downregulated |
| 0,126099815 | C11orf68 | FALSE | Upregulated |
| -0,90479363 | C11orf71 | FALSE | Downregulated |
| 0,398902466 | C11orf96 | FALSE | Upregulated |
| -0,786873335 | C12orf29 | FALSE | Downregulated |
| 0,072968574 | C12orf65 | FALSE | Upregulated |
| -0,03914975 | C14orf132 | FALSE | Downregulated |
| -0,378382822 | C14orf166 | FALSE | Downregulated |
| 1,36498189 | C14orf79 | | Upregulated |
| -0,988290088 | C15orf40 | FALSE | Downregulated |
| 0,019328208 | C16orf52 | FALSE | Upregulated |
| -0,258085645 | C16orf86 | FALSE | Downregulated |
| -0,480767205 | C17orf104 | FALSE | Downregulated |
| 1,675833183 | C17orf78 | | Upregulated |
| -0,040731915 | C18orf25 | FALSE | Downregulated |
| 0,22150239 | C18orf8 | FALSE | Upregulated |
| -0,638210814 | C19orf10 | FALSE | Downregulated |
| 0,442909213 | C19orf25 | FALSE | Upregulated |
| -1,574016105 | C19orf35 | | Downregulated |
| 0,051148409 | C19orf53 | FALSE | Upregulated |
| -1,450777362 | C19orf60 | | Downregulated |
| -0,400986065 | C19orf68 | FALSE | Downregulated |
| -0,177527643 | C1GALT1 | FALSE | Downregulated |
| 0,210723855 | C1orf112 | FALSE | Upregulated |
| 1,048818692 | C1orf122 | | Upregulated |
| 0,01224677 | C1orf145 | FALSE | Upregulated |
| 0,447596037 | C1orf172 | FALSE | Upregulated |
| -0,72699724 | C1orf174 | FALSE | Downregulated |
| 0,163175196 | C1orf177 | FALSE | Upregulated |
| 1,546708293 | C1orf195 | | Upregulated |
| 1,684741341 | C1orf198 | | Upregulated |

| | | | |
|---|---|---|---|
| -0,555321615 | C1orf210 | FALSE | Downregulated |
| 0,410731851 | C1orf27 | FALSE | Upregulated |
| -0,003487421 | C1orf52 | FALSE | Downregulated |
| -0,314242631 | C1orf85 | FALSE | Downregulated |
| -0,022743538 | C1QBP | FALSE | Downregulated |
| 0,913546568 | C1QTNF6 | FALSE | Upregulated |
| 0,671455703 | C21orf58 | FALSE | Upregulated |
| 1,37454972 | C21orf67 | | Upregulated |
| 0,462845057 | C21orf91 | FALSE | Upregulated |
| -0,574045911 | C22orf46 | FALSE | Downregulated |
| 0,192110576 | C2CD3 | FALSE | Upregulated |
| 0,521682323 | C2CD5 | FALSE | Upregulated |
| 0,10726809 | C2orf43 | FALSE | Upregulated |
| 0,925651561 | C2orf49 | FALSE | Upregulated |
| 0,336346881 | C2orf69 | FALSE | Upregulated |
| 0,567564941 | C2orf82 | FALSE | Upregulated |
| -1,249663838 | C3orf58 | | Downregulated |
| 0,525442313 | C3orf83 | FALSE | Upregulated |
| -0,491388459 | C5orf15 | FALSE | Downregulated |
| 0,414673087 | C5orf51 | FALSE | Upregulated |
| 0,696742619 | C6orf132 | FALSE | Upregulated |
| -0,325635798 | C6orf136 | FALSE | Downregulated |
| 1,086271294 | C7orf43 | | Upregulated |
| 0,902589076 | C7orf50 | FALSE | Upregulated |
| -0,292225217 | C7orf71 | FALSE | Downregulated |
| -0,341900748 | C7orf73 | FALSE | Downregulated |
| -0,376144389 | C8orf33 | FALSE | Downregulated |
| 0,106420451 | C8orf46 | FALSE | Upregulated |
| -0,803878143 | C8orf58 | FALSE | Downregulated |
| -0,407939769 | C8orf59 | FALSE | Downregulated |
| 1,267012405 | C9orf16 | | Upregulated |
| -0,307516183 | C9orf3 | FALSE | Downregulated |
| -2,093804205 | C9orf41 | | Downregulated |
| 0,243141598 | C9orf64 | FALSE | Upregulated |
| -0,191424466 | C9orf78 | FALSE | Downregulated |
| -1,157735071 | CA5A | | Downregulated |
| 0,836496257 | CABYR | FALSE | Upregulated |
| 0,03722771 | CACNA1A | FALSE | Upregulated |
| 0,71964171 | CACNA2D4 | FALSE | Upregulated |
| -0,717071239 | CADPS2 | FALSE | Downregulated |
| -0,548882164 | CALCRL | FALSE | Downregulated |
| -0,829777583 | CALU | FALSE | Downregulated |
| -1,65449297 | CAMK2B | | Downregulated |
| 0,141916126 | CAMK2G | FALSE | Upregulated |
| 0,808246025 | CAMSAP1 | FALSE | Upregulated |
| 0,141548423 | CAMSAP2 | FALSE | Upregulated |
| 0,865035807 | CAPN10 | FALSE | Upregulated |
| 0,016744913 | CAPRIN2 | FALSE | Upregulated |
| 0,064375289 | CAPZA2 | FALSE | Upregulated |
| 0,043294022 | CARD6 | FALSE | Upregulated |
| -0,31833227 | CARS | FALSE | Downregulated |

| | | | |
|---|---|---|---|
| 0,511010095 | CASK | FALSE | Upregulated |
| 0,376730735 | CASP2 | FALSE | Upregulated |
| -0,015542985 | CASP3 | FALSE | Downregulated |
| 0,281697738 | CASP8AP2 | FALSE | Upregulated |
| 0,542227425 | CAV2 | FALSE | Upregulated |
| 1,133063858 | CBFB | | Upregulated |
| 0,292809233 | CBLB | FALSE | Upregulated |
| 0,194114698 | CBR3 | FALSE | Upregulated |
| 0,504903149 | CBWD5 | FALSE | Upregulated |
| -0,036706278 | CBX3 | FALSE | Downregulated |
| -0,313382256 | CCAR1 | FALSE | Downregulated |
| 0,125901977 | CCBE1 | FALSE | Upregulated |
| 0,06362417 | CCBL1 | FALSE | Upregulated |
| 0,290344591 | CCBL2 | FALSE | Upregulated |
| -0,733833764 | CCDC104 | FALSE | Downregulated |
| 0,074809587 | CCDC106 | FALSE | Upregulated |
| 1,085096893 | CCDC109B | | Upregulated |
| -0,352837603 | CCDC115 | FALSE | Downregulated |
| 0,653147788 | CCDC120 | FALSE | Upregulated |
| 0,523447503 | CCDC121 | FALSE | Upregulated |
| -0,054147975 | CCDC124 | FALSE | Downregulated |
| -0,037954256 | CCDC137 | FALSE | Downregulated |
| 1,225561226 | CCDC144B | | Upregulated |
| 0,276789251 | CCDC167 | FALSE | Upregulated |
| 0,799782384 | CCDC17 | FALSE | Upregulated |
| -0,541330916 | CCDC174 | FALSE | Downregulated |
| 0,207791945 | CCDC3 | FALSE | Upregulated |
| 0,474040341 | CCDC36 | FALSE | Upregulated |
| -0,68227629 | CCDC6 | FALSE | Downregulated |
| -1,598974052 | CCDC60 | | Downregulated |
| 1,027547672 | CCDC65 | | Upregulated |
| -0,408074578 | CCDC66 | FALSE | Downregulated |
| 2,043730699 | CCDC68 | | Upregulated |
| -0,627108117 | CCDC7 | FALSE | Downregulated |
| -0,939816887 | CCDC70 | FALSE | Downregulated |
| -0,920360931 | CCDC73 | FALSE | Downregulated |
| 1,40067502 | CCDC78 | | Upregulated |
| -0,705501726 | CCDC82 | FALSE | Downregulated |
| 0,661874734 | CCDC85B | FALSE | Upregulated |
| 0,53262306 | CCDC86 | FALSE | Upregulated |
| 0,617804299 | CCDC88C | FALSE | Upregulated |
| 0,02988699 | CCDC9 | FALSE | Upregulated |
| -0,177255935 | CCL16 | FALSE | Downregulated |
| 0,919849693 | CCL21 | FALSE | Upregulated |
| 0,265054699 | CCL22 | FALSE | Upregulated |
| 0,269243914 | CCND3 | FALSE | Upregulated |
| 0,38711186 | CCNE2 | FALSE | Upregulated |
| 0,072340389 | CCNH | FALSE | Upregulated |
| 0,611983723 | CCNI2 | FALSE | Upregulated |
| -0,130263064 | CCNT1 | FALSE | Downregulated |
| 0,612570424 | CCP110 | FALSE | Upregulated |

| | | | |
|---|---|---|---|
| 1,589488986 | CCR1 | | Upregulated |
| 0,484413209 | CCR9 | FALSE | Upregulated |
| -0,430720651 | CCS | FALSE | Downregulated |
| -0,215461178 | CCT2 | FALSE | Downregulated |
| -0,280191892 | CCT8 | FALSE | Downregulated |
| -0,138398974 | CD27 | FALSE | Downregulated |
| 0,273749492 | CD2AP | FALSE | Upregulated |
| 1,191284812 | CD300A | | Upregulated |
| 0,139813161 | CD36 | FALSE | Upregulated |
| 1,083084752 | CD44 | | Upregulated |
| -0,104159934 | CD47 | FALSE | Downregulated |
| 0,709854536 | CD6 | FALSE | Upregulated |
| 0,704713136 | CD80 | FALSE | Upregulated |
| -0,708661516 | CD82 | FALSE | Downregulated |
| 0,408556641 | CD97 | FALSE | Upregulated |
| -0,020355212 | CDC16 | FALSE | Downregulated |
| -0,445881986 | CDC26 | FALSE | Downregulated |
| 0,222119759 | CDC27 | FALSE | Upregulated |
| 0,184972929 | CDC40 | FALSE | Upregulated |
| -0,821201882 | CDC42P3 | FALSE | Downregulated |
| -0,640018028 | CDCA7L | FALSE | Downregulated |
| -1,082408875 | CDK14 | | Downregulated |
| -0,045169743 | CDK17 | FALSE | Downregulated |
| -0,424098347 | CDK19 | FALSE | Downregulated |
| -0,539098571 | CDK8 | FALSE | Downregulated |
| 0,561976802 | CDKAL1 | FALSE | Upregulated |
| -0,626511405 | CDKN1B | FALSE | Downregulated |
| 0,223230083 | CDKN2AIPNL | FALSE | Upregulated |
| 0,861485362 | CDR2 | FALSE | Upregulated |
| -0,950704236 | CDT1 | FALSE | Downregulated |
| -0,367201136 | CECR5 | FALSE | Downregulated |
| 0,504856993 | CELF4 | FALSE | Upregulated |
| 0,254921452 | CELF6 | FALSE | Upregulated |
| 0,645755796 | CELSR3 | FALSE | Upregulated |
| -0,050400439 | CENPBD1 | FALSE | Downregulated |
| -1,31691867 | CENPC | | Downregulated |
| -0,346314928 | CENPT | FALSE | Downregulated |
| -1,209914919 | CEP120 | | Downregulated |
| -0,13793783 | CEP290 | FALSE | Downregulated |
| -0,384722199 | CEP63 | FALSE | Downregulated |
| -0,329139733 | CEP70 | FALSE | Downregulated |
| 0,64457486 | CEP72 | FALSE | Upregulated |
| 1,130474595 | CEP85 | | Upregulated |
| 0,118063284 | CEPT1 | FALSE | Upregulated |
| -1,968540151 | CERS3 | | Downregulated |
| 0,787521913 | CFDP1 | FALSE | Upregulated |
| 0,878202832 | CHADL | FALSE | Upregulated |
| 0,331302687 | CHAF1A | FALSE | Upregulated |
| 0,254529594 | CHCHD3 | FALSE | Upregulated |
| -0,253929724 | CHD1L | FALSE | Downregulated |
| -1,526320436 | CHEK1 | | Downregulated |

| | | | |
|---|---|---|---|
| -1,973585986 | CHIC2 | | Downregulated |
| 0,450349184 | CHKB | FALSE | Upregulated |
| 0,292168027 | CHM | FALSE | Upregulated |
| 1,417472731 | CHML | | Upregulated |
| 0,343468925 | CHMP1A | FALSE | Upregulated |
| -0,898191701 | CHMP1B | FALSE | Downregulated |
| -0,124599088 | CHMP4A | FALSE | Downregulated |
| 0,168051656 | CHMP4B | FALSE | Upregulated |
| 0,332598894 | CHMP5 | FALSE | Upregulated |
| -0,554713546 | CHMP6 | FALSE | Downregulated |
| 0,443933116 | CHORDC1 | FALSE | Upregulated |
| 0,426335175 | CHPF2 | FALSE | Upregulated |
| 0,922897997 | CHRAC1 | FALSE | Upregulated |
| -0,322583551 | CHRNA2 | FALSE | Downregulated |
| 0,010481179 | CHRNB1 | FALSE | Upregulated |
| 0,103362689 | CHRNE | FALSE | Upregulated |
| 0,034585243 | CHRNG | FALSE | Upregulated |
| 1,156716851 | CHST12 | | Upregulated |
| 0,424812466 | CHST4 | FALSE | Upregulated |
| -1,606363406 | CHST7 | | Downregulated |
| 0,644403311 | CIB1 | FALSE | Upregulated |
| -0,548421341 | CKMT2-AS1 | FALSE | Downregulated |
| -0,507217872 | CKS1B | FALSE | Downregulated |
| 0,149346126 | CLCN6 | FALSE | Upregulated |
| -0,100706587 | CLDN5 | FALSE | Downregulated |
| -0,331299494 | CLEC14A | FALSE | Downregulated |
| -0,584969231 | CLEC1B | FALSE | Downregulated |
| 1,012447771 | CLEC2B | | Upregulated |
| -1,481808303 | CLEC4G | | Downregulated |
| 0,164974321 | CLIP2 | FALSE | Upregulated |
| 0,377023881 | CLIP3 | FALSE | Upregulated |
| 0,589463211 | CLK2 | FALSE | Upregulated |
| -0,539378252 | CLN6 | FALSE | Downregulated |
| 0,197480637 | CLNS1A | FALSE | Upregulated |
| 0,332409843 | CLPB | FALSE | Upregulated |
| -0,606081525 | CLPP | FALSE | Downregulated |
| -1,023290345 | CLRN1 | | Downregulated |
| 0,594301621 | CLSTN2 | FALSE | Upregulated |
| -0,275414909 | CLTB | FALSE | Downregulated |
| 0,278097236 | CLUAP1 | FALSE | Upregulated |
| -0,884863493 | CLYBL-AS2 | FALSE | Downregulated |
| 0,014398627 | CMC2 | FALSE | Upregulated |
| 1,022047011 | CMKLR1 | | Upregulated |
| -0,126916189 | CMTR2 | FALSE | Downregulated |
| -0,096521824 | CNGA1 | FALSE | Downregulated |
| 0,479889646 | CNNM2 | FALSE | Upregulated |
| -0,946738665 | CNOT4 | FALSE | Downregulated |
| -0,75824823 | CNOT8 | FALSE | Downregulated |
| -0,971746016 | CNPPD1 | FALSE | Downregulated |
| -0,82796692 | CNTNAP4 | FALSE | Downregulated |
| 0,271259133 | COA5 | FALSE | Upregulated |

| | | | |
|---|---|---|---|
| -0,327326881 | COA7 | FALSE | Downregulated |
| -0,18816293 | COG7 | FALSE | Downregulated |
| -1,219241132 | COL12A1 | | Downregulated |
| 1,425236163 | COL16A1 | | Upregulated |
| 0,668525738 | COL17A1 | FALSE | Upregulated |
| -1,126225007 | COL26A1 | | Downregulated |
| 0,508239935 | COL4A2-AS2 | FALSE | Upregulated |
| 0,053070735 | COL4A3 | FALSE | Upregulated |
| 0,608020571 | COL4A5 | FALSE | Upregulated |
| 1,118065911 | COL8A2 | | Upregulated |
| 1,47719361 | COMMD9 | | Upregulated |
| -1,053942679 | COPG2 | | Downregulated |
| 0,687455088 | COPG2IT1 | FALSE | Upregulated |
| -0,767830633 | COPRS | FALSE | Downregulated |
| 0,159516284 | COPS2 | FALSE | Upregulated |
| 0,586356962 | COPS5 | FALSE | Upregulated |
| 0,054658909 | COPS6 | FALSE | Upregulated |
| 0,372889073 | COPS7A | FALSE | Upregulated |
| 0,194610772 | COPZ1 | FALSE | Upregulated |
| 0,45695735 | COQ10A | FALSE | Upregulated |
| 1,027099843 | COQ4 | | Upregulated |
| -0,764755626 | COQ7 | FALSE | Downregulated |
| 0,378173888 | COQ9 | FALSE | Upregulated |
| 0,238581168 | CORIN | FALSE | Upregulated |
| 0,107626434 | CORO2A | FALSE | Upregulated |
| 0,292861516 | COX19 | FALSE | Upregulated |
| -1,393372369 | COX7A2L | | Downregulated |
| -0,858743498 | CPN1 | FALSE | Downregulated |
| -0,614102706 | CPNE8 | FALSE | Downregulated |
| -0,091251795 | CPOX | FALSE | Downregulated |
| 0,434643452 | CPQ | FALSE | Upregulated |
| -0,465582112 | CPSF2 | FALSE | Downregulated |
| 0,361926107 | CRB3 | FALSE | Upregulated |
| -0,614315348 | CRELD1 | FALSE | Downregulated |
| 0,437451298 | CRELD2 | FALSE | Upregulated |
| 0,493541284 | CRHR2 | FALSE | Upregulated |
| -0,524098239 | CROCC | FALSE | Downregulated |
| 0,093065463 | CRY1 | FALSE | Upregulated |
| -0,113010546 | CSE1L | FALSE | Downregulated |
| 0,162844059 | CSF3R | FALSE | Upregulated |
| 1,219275559 | CSGALNACT2 | | Upregulated |
| -0,942285787 | CSNK1G3 | FALSE | Downregulated |
| -1,169825292 | CSNK2A2 | | Downregulated |
| -0,038924754 | CSNK2B | FALSE | Downregulated |
| 0,967335883 | CSRNP1 | FALSE | Upregulated |
| -0,504527353 | CSRP2BP | FALSE | Downregulated |
| 0,398590456 | CST7 | FALSE | Upregulated |
| 0,35407419 | CSTF3 | FALSE | Upregulated |
| 0,124868356 | CTB-113P19.1 | FALSE | Upregulated |
| 0,043589845 | CTC-338M12.4 | FALSE | Upregulated |
| -0,558745818 | CTDNEP1 | FALSE | Downregulated |

| | | | |
|---|---|---|---|
| 0,038604341 | CTDP1 | FALSE | Upregulated |
| -0,099391162 | CTDSPL | FALSE | Downregulated |
| -0,852290809 | CTH | FALSE | Downregulated |
| -0,083943564 | CTHRC1 | FALSE | Downregulated |
| 0,090393626 | CTPS2 | FALSE | Upregulated |
| 0,618277894 | CTSC | FALSE | Upregulated |
| 0,54199493 | CTTNBP2NL | FALSE | Upregulated |
| 0,251329192 | CTU2 | FALSE | Upregulated |
| 1,513665137 | CUEDC1 | | Upregulated |
| -0,689611379 | CUEDC2 | FALSE | Downregulated |
| -0,122302549 | CUL2 | FALSE | Downregulated |
| -0,161005938 | CUL4B | FALSE | Downregulated |
| 0,141124948 | CUTA | FALSE | Upregulated |
| -0,64034922 | CWC25 | FALSE | Downregulated |
| -1,029002993 | CWF19L1 | | Downregulated |
| -0,445282651 | CXCL14 | FALSE | Downregulated |
| 0,724303346 | CXorf23 | FALSE | Upregulated |
| 0,614669059 | CXorf40B | FALSE | Upregulated |
| 0,156811337 | CXorf56 | FALSE | Upregulated |
| 0,245659851 | CYB561 | FALSE | Upregulated |
| 1,150714124 | CYB561D1 | | Upregulated |
| 0,892031965 | CYB561D2 | FALSE | Upregulated |
| -1,441749798 | CYB5B | | Downregulated |
| 0,695757867 | CYB5R2 | FALSE | Upregulated |
| 0,643866377 | CYBRD1 | FALSE | Upregulated |
| 0,220860534 | CYGB | FALSE | Upregulated |
| -0,027453641 | CYHR1 | FALSE | Downregulated |
| -0,489990082 | CYP1A1 | FALSE | Downregulated |
| 0,517032476 | CYP2W1 | FALSE | Upregulated |
| -2,112509701 | CYP39A1 | | Downregulated |
| 0,057566737 | CYP7B1 | FALSE | Upregulated |
| -0,097177579 | CYSTM1 | FALSE | Downregulated |
| 0,467059154 | CYTH2 | FALSE | Upregulated |
| 0,586302109 | CYTH3 | FALSE | Upregulated |
| 1,51412604 | CYTH4 | | Upregulated |
| -0,209420326 | DAP3 | FALSE | Downregulated |
| 0,471747158 | DAPK3 | FALSE | Upregulated |
| 0,339010415 | DBIL5P | FALSE | Upregulated |
| 1,245701974 | DCAF15 | | Upregulated |
| 0,534388928 | DCAF16 | FALSE | Upregulated |
| 0,421269385 | DCAF4L1 | FALSE | Upregulated |
| -0,863871612 | DCAKD | FALSE | Downregulated |
| 0,865673937 | DCBLD1 | FALSE | Upregulated |
| 1,096064008 | DCDC2B | | Upregulated |
| 0,828286161 | DCPS | FALSE | Upregulated |
| -1,31203139 | DCSTAMP | | Downregulated |
| -0,032552018 | DCTN4 | FALSE | Downregulated |
| -0,316008729 | DCUN1D3 | FALSE | Downregulated |
| 0,67593608 | DDAH2 | FALSE | Upregulated |
| 0,482306911 | DDHD2 | FALSE | Upregulated |
| 0,691760362 | DDIT3 | FALSE | Upregulated |

| | | | |
|---|---|---|---|
| -0,25753168 | DDIT4 | FALSE | Downregulated |
| 0,62875292 | DDO | FALSE | Upregulated |
| 0,736135915 | DDR2 | FALSE | Upregulated |
| -0,729458762 | DDX10 | FALSE | Downregulated |
| -0,093451205 | DDX11 | FALSE | Downregulated |
| -1,309351769 | DDX19B | | Downregulated |
| 0,33679337 | DDX21 | FALSE | Upregulated |
| 0,133230627 | DDX27 | FALSE | Upregulated |
| -0,471280598 | DDX31 | FALSE | Downregulated |
| -0,176706697 | DDX3Y | FALSE | Downregulated |
| -0,268528835 | DDX41 | FALSE | Downregulated |
| 0,008600285 | DDX55 | FALSE | Upregulated |
| 0,514983527 | DDX56 | FALSE | Upregulated |
| 0,60076431 | DDX59 | FALSE | Upregulated |
| 0,866177619 | DDX60 | FALSE | Upregulated |
| -0,087467523 | DEAF1 | FALSE | Downregulated |
| 0,475874747 | DEDD2 | FALSE | Upregulated |
| -0,404865095 | DEGS1 | FALSE | Downregulated |
| 0,538223904 | DENND1B | FALSE | Upregulated |
| 0,406524782 | DENND3 | FALSE | Upregulated |
| 0,130075212 | DENND6A | FALSE | Upregulated |
| -0,76164002 | DEPDC7 | FALSE | Downregulated |
| -0,01563387 | DERL1 | FALSE | Downregulated |
| -0,682103064 | DET1 | FALSE | Downregulated |
| 0,175633312 | DFFA | FALSE | Upregulated |
| -0,854471027 | DFFB | FALSE | Downregulated |
| 1,050163862 | DFNB31 | | Upregulated |
| -0,161368679 | DGCR14 | FALSE | Downregulated |
| 1,161487768 | DGCR5 | | Upregulated |
| -1,055810269 | DGKE | | Downregulated |
| -0,201495972 | DHFRL1 | FALSE | Downregulated |
| -0,106029903 | DHPS | FALSE | Downregulated |
| -1,515766509 | DHRS4L2 | | Downregulated |
| -0,230940316 | DHRSX | FALSE | Downregulated |
| 0,359071311 | DHX29 | FALSE | Upregulated |
| 0,47239218 | DHX32 | FALSE | Upregulated |
| 0,232397501 | DHX33 | FALSE | Upregulated |
| -0,203721863 | DHX36 | FALSE | Downregulated |
| 0,749483837 | DHX37 | FALSE | Upregulated |
| -0,687650067 | DHX40 | FALSE | Downregulated |
| 0,052017765 | DHX57 | FALSE | Upregulated |
| -1,009432889 | DHX58 | | Downregulated |
| -0,426890041 | DIEXF | FALSE | Downregulated |
| 0,017244028 | DIMT1 | FALSE | Upregulated |
| -0,314337534 | DIRAS3 | FALSE | Downregulated |
| -1,10676735 | DIS3L2 | | Downregulated |
| 0,572190799 | DISC2 | FALSE | Upregulated |
| 0,077850397 | DISP1 | FALSE | Upregulated |
| -0,492697962 | DISP2 | FALSE | Downregulated |
| -0,163655223 | DIXDC1 | FALSE | Downregulated |
| -0,466022299 | DKFZP434K028 | FALSE | Downregulated |

| | | | |
|---|---|---|---|
| -1,599439176 | DKFZP586I1420 | | Downregulated |
| -1,107464889 | DKK3 | | Downregulated |
| 0,872036748 | DLAT | FALSE | Upregulated |
| -1,07352982 | DLD | | Downregulated |
| -0,396593603 | DLEU2 | FALSE | Downregulated |
| 0,17387038 | DLGAP2 | FALSE | Upregulated |
| -0,159145304 | DMAP1 | FALSE | Downregulated |
| 0,996555768 | DMRTA1 | FALSE | Upregulated |
| -0,71833328 | DNAH14 | FALSE | Downregulated |
| -0,596853356 | DNAH5 | FALSE | Downregulated |
| -1,070418696 | DNAH6 | | Downregulated |
| -0,122430743 | DNAJB2 | FALSE | Downregulated |
| -0,37137809 | DNAJB9 | FALSE | Downregulated |
| -0,34505053 | DNAJC19 | FALSE | Downregulated |
| -0,792911777 | DNAJC24 | FALSE | Downregulated |
| -1,391533028 | DNAJC8 | | Downregulated |
| -0,443637341 | DNAJC9 | FALSE | Downregulated |
| -0,156757787 | DNAJC9-AS1 | FALSE | Downregulated |
| 0,939578249 | DNASE2 | FALSE | Upregulated |
| -0,0802728 | DNM1 | FALSE | Downregulated |
| 1,006540692 | DNMT1 | | Upregulated |
| -0,047130816 | DNPEP | FALSE | Downregulated |
| -0,880549379 | DNPH1 | FALSE | Downregulated |
| 1,062853228 | DOK1 | | Upregulated |
| 0,814373508 | DOK7 | FALSE | Upregulated |
| 0,078759273 | DOLPP1 | FALSE | Upregulated |
| 0,142320055 | DONSON | FALSE | Upregulated |
| 0,099345024 | DPCD | FALSE | Upregulated |
| 1,88177765 | DPEP2 | | Upregulated |
| -0,588700781 | DPF2 | FALSE | Downregulated |
| -0,604085274 | DPH1 | FALSE | Downregulated |
| -0,44551454 | DPH3P1 | FALSE | Downregulated |
| -0,134672112 | DPH7 | FALSE | Downregulated |
| 0,727679594 | DPP8 | FALSE | Upregulated |
| 0,527252153 | DPY19L1P1 | FALSE | Upregulated |
| -1,091670337 | DPY19L2P2 | | Downregulated |
| -0,738686545 | DPY19L2P4 | FALSE | Downregulated |
| -0,168439523 | DPY19L3 | FALSE | Downregulated |
| -0,570339671 | DPY19L4 | FALSE | Downregulated |
| -0,261238938 | DPYD-AS2 | FALSE | Downregulated |
| 0,315424994 | DPYSL3 | FALSE | Upregulated |
| -0,459797043 | DR1 | FALSE | Downregulated |
| 0,002162874 | DRAP1 | FALSE | Upregulated |
| -0,792399944 | DRD5 | FALSE | Downregulated |
| -1,778850946 | DRG1 | | Downregulated |
| -0,031071846 | DROSHA | FALSE | Downregulated |
| 0,97052485 | DSERG1 | FALSE | Upregulated |
| -2,03713999 | DSG1 | | Downregulated |
| -0,107956654 | DSTN | FALSE | Downregulated |
| 0,04586729 | DSTYK | FALSE | Upregulated |
| 0,519655589 | DTX1 | FALSE | Upregulated |

| | | | |
|---|---|---|---|
| 0,913350228 | DTX2 | FALSE | Upregulated |
| 0,230800099 | DUS3L | FALSE | Upregulated |
| -0,230919992 | DUSP14 | FALSE | Downregulated |
| 0,421962584 | DUSP22 | FALSE | Upregulated |
| 0,351969529 | DVL2 | FALSE | Upregulated |
| 0,007886001 | DYNC1I2 | FALSE | Upregulated |
| 0,120093395 | DYNC1LI1 | FALSE | Upregulated |
| 0,470517358 | DYNC2H1 | FALSE | Upregulated |
| 0,628641925 | DYNLL1 | FALSE | Upregulated |
| 0,163213668 | DYNLL1-AS1 | FALSE | Upregulated |
| 0,326316062 | DYNLL2 | FALSE | Upregulated |
| -0,955840825 | DYNLT3 | FALSE | Downregulated |
| 0,65809654 | DYRK2 | FALSE | Upregulated |
| 1,178201487 | E2F4 | | Upregulated |
| -0,519831581 | E2F5 | FALSE | Downregulated |
| -0,678487412 | EBF4 | FALSE | Downregulated |
| -0,14364354 | ECD | FALSE | Downregulated |
| -0,094673575 | EDAR | FALSE | Downregulated |
| -0,401281122 | EDEM2 | FALSE | Downregulated |
| -0,246178083 | EDRF1 | FALSE | Downregulated |
| -0,180042631 | EED | FALSE | Downregulated |
| -0,012234042 | EEFSEC | FALSE | Downregulated |
| -0,526605012 | EFCAB2 | FALSE | Downregulated |
| 0,452273814 | EFCAB4A | FALSE | Upregulated |
| -0,069724342 | EFHC1 | FALSE | Downregulated |
| 0,553352389 | EFHD2 | FALSE | Upregulated |
| -0,179006448 | EFTUD2 | FALSE | Downregulated |
| 0,565295582 | EGOT | FALSE | Upregulated |
| 0,961496575 | EGR2 | FALSE | Upregulated |
| 0,179674612 | EHD2 | FALSE | Upregulated |
| -0,112127062 | EIF2AK2 | FALSE | Downregulated |
| 0,484274811 | EIF2B1 | FALSE | Upregulated |
| -0,677227396 | EIF2B3 | FALSE | Downregulated |
| 0,85371007 | EIF2B4 | FALSE | Upregulated |
| -0,157586737 | EIF2D | FALSE | Downregulated |
| -0,03006467 | EIF3K | FALSE | Downregulated |
| -0,181490347 | EIF3M | FALSE | Downregulated |
| -0,212311574 | EIF4A3 | FALSE | Downregulated |
| -0,449339821 | EIF4E3 | FALSE | Downregulated |
| -0,039084935 | ELAC1 | FALSE | Downregulated |
| -0,037528022 | ELL | FALSE | Downregulated |
| -0,579309894 | ELMO3 | FALSE | Downregulated |
| -0,361783695 | ELOF1 | FALSE | Downregulated |
| -0,256125615 | ELOVL6 | FALSE | Downregulated |
| 1,200503502 | ELP6 | | Upregulated |
| -2,245918574 | EMBP1 | | Downregulated |
| -0,016489885 | EMC8 | FALSE | Downregulated |
| -0,538481515 | EMC9 | FALSE | Downregulated |
| 0,243557718 | EME1 | FALSE | Upregulated |
| 0,297248248 | EMG1 | FALSE | Upregulated |
| -0,083142937 | EMID1 | FALSE | Downregulated |

| | | | |
|---|---|---|---|
| -0,131763353 | EMR2 | FALSE | Downregulated |
| 0,820175232 | ENDOD1 | FALSE | Upregulated |
| 0,603175533 | ENTHD2 | FALSE | Upregulated |
| 1,32541145 | ENTPD1-AS1 | | Upregulated |
| 1,305556418 | EPB41L3 | | Upregulated |
| 0,070216733 | EPB41L4A-AS2 | FALSE | Upregulated |
| -0,485340325 | EPC1 | FALSE | Downregulated |
| 0,038514097 | EPC2 | FALSE | Upregulated |
| 0,777611829 | EPM2A | FALSE | Upregulated |
| 1,864202597 | EPPK1 | | Upregulated |
| 0,409624615 | EPS15L1 | FALSE | Upregulated |
| 0,463216393 | EPX | FALSE | Upregulated |
| -0,564024511 | ERAL1 | FALSE | Downregulated |
| 0,657892646 | ERCC2 | FALSE | Upregulated |
| 0,069083946 | ERCC6 | FALSE | Upregulated |
| 0,072099157 | ERCC6L2 | FALSE | Upregulated |
| -0,795717591 | ERH | FALSE | Downregulated |
| 0,756431229 | ERI3 | FALSE | Upregulated |
| -0,002940097 | ESCO1 | FALSE | Downregulated |
| -0,198039683 | ETAA1 | FALSE | Downregulated |
| 0,85414522 | ETV3 | FALSE | Upregulated |
| 0,109837572 | ETV4 | FALSE | Upregulated |
| 0,500927127 | EVA1C | FALSE | Upregulated |
| -0,379680021 | EXD2 | FALSE | Downregulated |
| -0,170287449 | EXO5 | FALSE | Downregulated |
| 0,313111087 | EXOC6B | FALSE | Upregulated |
| 0,084499683 | EXOC8 | FALSE | Upregulated |
| -0,74044078 | EXOSC10 | FALSE | Downregulated |
| -0,620749472 | EXOSC2 | FALSE | Downregulated |
| -0,860698289 | EXOSC3 | FALSE | Downregulated |
| 0,045875804 | EXOSC6 | FALSE | Upregulated |
| -0,157398136 | EXOSC8 | FALSE | Downregulated |
| -0,344447488 | EXTL3-AS1 | FALSE | Downregulated |
| -0,082993239 | F2R | FALSE | Downregulated |
| -0,41098286 | FAAH | FALSE | Downregulated |
| -1,0207543 | FABP2 | | Downregulated |
| 0,311948193 | FABP3 | FALSE | Upregulated |
| 0,512895319 | FABP5 | FALSE | Upregulated |
| 0,935786964 | FADS3 | FALSE | Upregulated |
| 0,05291225 | FAF1 | FALSE | Upregulated |
| -0,340527391 | FAF2 | FALSE | Downregulated |
| -1,240744496 | FAM114A2 | | Downregulated |
| 0,582360767 | FAM118A | FALSE | Upregulated |
| 1,637230465 | FAM122B | | Upregulated |
| 1,014958449 | FAM122C | | Upregulated |
| -0,000511867 | FAM126A | FALSE | Downregulated |
| 1,11518825 | FAM129A | | Upregulated |
| -0,505632306 | FAM131A | FALSE | Downregulated |
| 1,782182655 | FAM131B | | Upregulated |
| -0,127726944 | FAM134B | FALSE | Downregulated |
| 0,737657098 | FAM135A | FALSE | Upregulated |

| | | | |
|---|---|---|---|
| -0,555869401 | FAM136A | FALSE | Downregulated |
| 0,381098297 | FAM13B | FALSE | Upregulated |
| -0,370299115 | FAM13C | FALSE | Downregulated |
| -1,530690267 | FAM162A | | Downregulated |
| -0,704382097 | FAM173A | FALSE | Downregulated |
| -1,206722832 | FAM174B | | Downregulated |
| -0,578819559 | FAM178B | FALSE | Downregulated |
| 0,14095966 | FAM179B | FALSE | Upregulated |
| -1,00209984 | FAM184A | | Downregulated |
| 0,053752847 | FAM188A | FALSE | Upregulated |
| 0,510935654 | FAM189B | FALSE | Upregulated |
| -0,648302264 | FAM192A | FALSE | Downregulated |
| -0,259055097 | FAM195B | FALSE | Downregulated |
| 1,152676847 | FAM199X | | Upregulated |
| 0,00222386 | FAM205B | FALSE | Upregulated |
| 0,869094116 | FAM206A | FALSE | Upregulated |
| -1,400880807 | FAM210A | | Downregulated |
| -0,524401707 | FAM212A | FALSE | Downregulated |
| 0,613412093 | FAM214B | FALSE | Upregulated |
| 0,012841011 | FAM221A | FALSE | Upregulated |
| 0,667669129 | FAM222A | FALSE | Upregulated |
| -0,456193407 | FAM227A | FALSE | Downregulated |
| 0,003951365 | FAM3C | FALSE | Upregulated |
| 0,253855342 | FAM46A | FALSE | Upregulated |
| 1,203946047 | FAM53B-AS1 | | Upregulated |
| -0,2573689 | FAM64A | FALSE | Downregulated |
| 0,006543994 | FAM73A | FALSE | Upregulated |
| 0,120823816 | FAM86B1 | FALSE | Upregulated |
| -0,710007392 | FAM86C1 | FALSE | Downregulated |
| -0,099118717 | FAM86FP | FALSE | Downregulated |
| -0,445973299 | FAM96B | FALSE | Downregulated |
| 0,37853509 | FAM98B | FALSE | Upregulated |
| 0,398978462 | FANCM | FALSE | Upregulated |
| -0,02491568 | FAR1 | FALSE | Downregulated |
| -0,276577551 | FARS2 | FALSE | Downregulated |
| 0,411577845 | FASTK | FALSE | Upregulated |
| -1,148486369 | FBL | | Downregulated |
| 0,801507524 | FBLN2 | FALSE | Upregulated |
| -0,689788082 | FBXL12 | FALSE | Downregulated |
| 1,153495408 | FBXL18 | | Upregulated |
| 0,420535095 | FBXL21 | FALSE | Upregulated |
| -0,700861489 | FBXL4 | FALSE | Downregulated |
| 0,057356553 | FBXO22 | FALSE | Upregulated |
| -0,036108546 | FBXO24 | FALSE | Downregulated |
| 0,378808497 | FBXO30 | FALSE | Upregulated |
| 0,232245039 | FBXO34 | FALSE | Upregulated |
| -0,360637497 | FBXO42 | FALSE | Downregulated |
| 1,093653301 | FBXO44 | | Upregulated |
| 0,279888588 | FBXO46 | FALSE | Upregulated |
| -0,308618007 | FBXO6 | FALSE | Downregulated |
| -1,489817183 | FBXW7 | | Downregulated |

| | | | |
|---|---|---|---|
| 1,119086031 | FCGR1A | | Upregulated |
| 0,543732256 | FCGR2A | FALSE | Upregulated |
| -0,948625046 | FCGR2B | FALSE | Downregulated |
| 0,197752481 | FDXR | FALSE | Upregulated |
| 0,545206525 | FEM1C | FALSE | Upregulated |
| -0,45896063 | FER | FALSE | Downregulated |
| 0,130611873 | FER1L5 | FALSE | Upregulated |
| 0,336277356 | FES | FALSE | Upregulated |
| 0,318313352 | FEZ2 | FALSE | Upregulated |
| -0,288206979 | FFAR4 | FALSE | Downregulated |
| -1,739398149 | FGF11 | | Downregulated |
| 1,517657274 | FGF14 | | Upregulated |
| -0,027525843 | FGF14-AS2 | FALSE | Downregulated |
| -0,002942411 | FGFR1 | FALSE | Downregulated |
| 0,042776459 | FGFR1OP2 | FALSE | Upregulated |
| -0,253085772 | FIGN | FALSE | Downregulated |
| 0,493086676 | FIGNL2 | FALSE | Upregulated |
| -0,335233662 | FILIP1 | FALSE | Downregulated |
| 0,487256973 | FKBP10 | FALSE | Upregulated |
| -0,210937925 | FKBP14 | FALSE | Downregulated |
| -0,45046209 | FKBP3 | FALSE | Downregulated |
| -0,426811145 | FKBP7 | FALSE | Downregulated |
| -0,242403715 | FKRP | FALSE | Downregulated |
| 0,446695985 | FLJ20464 | FALSE | Upregulated |
| 0,090193383 | FLJ30403 | FALSE | Upregulated |
| 0,056825904 | FLJ38717 | FALSE | Upregulated |
| 1,333886957 | FLJ39095 | | Upregulated |
| -0,763885643 | FLJ41481 | FALSE | Downregulated |
| -0,094411801 | FLJ44342 | FALSE | Downregulated |
| 0,511438378 | FLJ46906 | FALSE | Upregulated |
| 1,019226115 | FLRT2 | | Upregulated |
| -0,843754955 | FLVCR1 | FALSE | Downregulated |
| 1,91846062 | FMN1 | | Upregulated |
| 0,350384681 | FN3K | FALSE | Upregulated |
| -0,214748088 | FN3KRP | FALSE | Downregulated |
| -0,16928201 | FNBP1L | FALSE | Downregulated |
| 0,074137314 | FNDC8 | FALSE | Upregulated |
| 0,498083769 | FOCAD | FALSE | Upregulated |
| -0,791625445 | FOLH1 | FALSE | Downregulated |
| 1,305597994 | FOLR1 | | Upregulated |
| -1,126011644 | FOXN4 | | Downregulated |
| -0,017014354 | FOXO4 | FALSE | Downregulated |
| 2,001642277 | FPR3 | | Upregulated |
| -0,883471699 | FRAT1 | FALSE | Downregulated |
| 0,881192413 | FRAT2 | FALSE | Upregulated |
| 0,032457083 | FRK | FALSE | Upregulated |
| -1,791631162 | FRRS1 | | Downregulated |
| 0,327111658 | FRS2 | FALSE | Upregulated |
| 0,302590899 | FST | FALSE | Upregulated |
| -0,890460055 | FTO | FALSE | Downregulated |
| 0,257701298 | FTSJ1 | FALSE | Upregulated |

| | | | |
|---|---|---|---|
| 0,64991426 | FTX | FALSE | Upregulated |
| -0,095851644 | FUCA1 | FALSE | Downregulated |
| -0,744474262 | FUNDC2 | FALSE | Downregulated |
| 0,013865034 | FUT11 | FALSE | Upregulated |
| 0,460388598 | FUZ | FALSE | Upregulated |
| -0,004007019 | FXN | FALSE | Downregulated |
| -0,575190906 | FXR2 | FALSE | Downregulated |
| 1,508976856 | FXYD5 | | Upregulated |
| -0,558063404 | FXYD6 | FALSE | Downregulated |
| -0,515626245 | FYN | FALSE | Downregulated |
| 0,624158678 | FYTTD1 | FALSE | Upregulated |
| 0,005985518 | FZD1 | FALSE | Upregulated |
| 0,657447359 | FZD6 | FALSE | Upregulated |
| -0,117854052 | G6PD | FALSE | Downregulated |
| -0,116645042 | GAB3 | FALSE | Downregulated |
| 0,126152046 | GABARAPL2 | FALSE | Upregulated |
| -0,743792359 | GABBR1 | FALSE | Downregulated |
| -0,087817155 | GABPB2 | FALSE | Downregulated |
| -0,666946293 | GALE | FALSE | Downregulated |
| 0,355373064 | GALNT18 | FALSE | Upregulated |
| -1,503228843 | GAREM | | Downregulated |
| 0,596634073 | GAS2L1 | FALSE | Upregulated |
| 1,060158106 | GATA6 | | Upregulated |
| 0,237445059 | GBP2 | FALSE | Upregulated |
| -0,251096624 | GBP3 | FALSE | Downregulated |
| -2,33924864 | GCAT | | Downregulated |
| -0,169573952 | GCC1 | FALSE | Downregulated |
| -0,975156716 | GDE1 | FALSE | Downregulated |
| -0,141426435 | GDF2 | FALSE | Downregulated |
| -0,35816866 | GEMIN2 | FALSE | Downregulated |
| -0,299181323 | GEMIN4 | FALSE | Downregulated |
| -0,300195622 | GEMIN5 | FALSE | Downregulated |
| -0,508970904 | GEMIN7 | FALSE | Downregulated |
| 1,229245619 | GEMIN8 | | Upregulated |
| -0,603174727 | GEMIN8P4 | FALSE | Downregulated |
| -0,564206997 | GFER | FALSE | Downregulated |
| -0,200925616 | GFM1 | FALSE | Downregulated |
| 0,898361298 | GFM2 | FALSE | Upregulated |
| -0,403247876 | GGT7 | FALSE | Downregulated |
| -0,445733654 | GHDC | FALSE | Downregulated |
| -1,183240687 | GHRLOS | | Downregulated |
| -0,16298347 | GID4 | FALSE | Downregulated |
| -0,770276537 | GIMAP6 | FALSE | Downregulated |
| -0,359435094 | GIMAP8 | FALSE | Downregulated |
| -0,229192105 | GINS2 | FALSE | Downregulated |
| 0,897323908 | GIPC1 | FALSE | Upregulated |
| 0,958743112 | GK5 | FALSE | Upregulated |
| -0,252047725 | GLB1 | FALSE | Downregulated |
| 0,603401394 | GLCCI1 | FALSE | Upregulated |
| -0,358534371 | GLCE | FALSE | Downregulated |
| 0,292090491 | GLE1 | FALSE | Upregulated |

| | | | |
|---|---|---|---|
| 1,49750504 | GLI1 | | Upregulated |
| 0,466163114 | GLIPR1L1 | FALSE | Upregulated |
| -0,533544764 | GLO1 | FALSE | Downregulated |
| -0,17130252 | GLOD4 | FALSE | Downregulated |
| -0,343438542 | GLT1D1 | FALSE | Downregulated |
| 1,250309863 | GLTP | | Upregulated |
| -0,576494055 | GLUD2 | FALSE | Downregulated |
| 0,365904546 | GMEB2 | FALSE | Upregulated |
| -0,651745324 | GMPPA | FALSE | Downregulated |
| 0,939811249 | GNA14 | FALSE | Upregulated |
| -1,995146321 | GNAI1 | | Downregulated |
| 0,489383537 | GNB5 | FALSE | Upregulated |
| 0,186641598 | GNG5 | FALSE | Upregulated |
| -1,239255819 | GNG7 | | Downregulated |
| -0,078622039 | GNL2 | FALSE | Downregulated |
| -0,138261886 | GNPAT | FALSE | Downregulated |
| -0,899992027 | GOLGA5 | FALSE | Downregulated |
| -0,59995845 | GOLGA6L5P | FALSE | Downregulated |
| -0,40235235 | GOLGA7 | FALSE | Downregulated |
| 0,205298596 | GOLT1A | FALSE | Upregulated |
| 1,107460076 | GOLT1B | | Upregulated |
| -0,331797862 | GORASP2 | FALSE | Downregulated |
| 0,136699264 | GPANK1 | FALSE | Upregulated |
| 0,614132039 | GPAT2 | FALSE | Upregulated |
| 0,528159451 | GPATCH2 | FALSE | Upregulated |
| -0,127586608 | GPATCH4 | FALSE | Downregulated |
| -0,230332869 | GPBAR1 | FALSE | Downregulated |
| 1,110231658 | GPCPD1 | | Upregulated |
| 0,423549908 | GPD2 | FALSE | Upregulated |
| 0,29469474 | GPN3 | FALSE | Upregulated |
| 0,778784468 | GPR108 | FALSE | Upregulated |
| -1,38009857 | GPR128 | | Downregulated |
| 0,7167037 | GPR137B | FALSE | Upregulated |
| -0,651282868 | GPR180 | FALSE | Downregulated |
| 1,057837469 | GPR39 | | Upregulated |
| -0,035513456 | GPR61 | FALSE | Downregulated |
| -0,663616098 | GPR75 | FALSE | Downregulated |
| -0,250604375 | GPRASP2 | FALSE | Downregulated |
| -0,148646553 | GPRC5D | FALSE | Downregulated |
| 0,582129917 | GPS2 | FALSE | Upregulated |
| 0,38503322 | GPSM3 | FALSE | Upregulated |
| 0,996120588 | GRAMD1A | FALSE | Upregulated |
| -0,925649018 | GRAMD3 | FALSE | Downregulated |
| -0,387619594 | GRASP | FALSE | Downregulated |
| 0,058607452 | GRB7 | FALSE | Upregulated |
| 0,493969441 | GREB1 | FALSE | Upregulated |
| 1,290760386 | GRIK1 | | Upregulated |
| 1,087480985 | GRIP1 | | Upregulated |
| 0,050370157 | GRIP2 | FALSE | Upregulated |
| -0,945281234 | GRIPAP1 | FALSE | Downregulated |
| 0,513200906 | GRK5 | FALSE | Upregulated |

| | | | |
|---|---|---|---|
| 1,055452058 | GRK6 | | Upregulated |
| 0,5489478 | GRWD1 | FALSE | Upregulated |
| 0,473883611 | GSAP | FALSE | Upregulated |
| -0,059278804 | GSK3A | FALSE | Downregulated |
| -0,124649992 | GSTM3 | FALSE | Downregulated |
| -0,700383355 | GSTM4 | FALSE | Downregulated |
| -0,839371004 | GSTO2 | FALSE | Downregulated |
| 0,555321654 | GSTP1 | FALSE | Upregulated |
| 0,159197338 | GTF2A1 | FALSE | Upregulated |
| -0,356944202 | GTF2H1 | FALSE | Downregulated |
| 0,153623464 | GTF3C4 | FALSE | Upregulated |
| -0,901514053 | GTPBP10 | FALSE | Downregulated |
| 0,235171558 | GTPBP3 | FALSE | Upregulated |
| 0,629033063 | GUCY1A2 | FALSE | Upregulated |
| -0,181325291 | GVINP1 | FALSE | Downregulated |
| -0,059755156 | GYG2 | FALSE | Downregulated |
| 0,010259418 | H2AFJ | FALSE | Upregulated |
| 0,720874704 | HAND2 | FALSE | Upregulated |
| 0,464154249 | HAPLN4 | FALSE | Upregulated |
| 0,621023842 | HAUS6 | FALSE | Upregulated |
| 2,36724293 | HCFC1R1 | | Upregulated |
| -0,472548226 | HCFC2 | FALSE | Downregulated |
| 0,22060579 | HCG4B | FALSE | Upregulated |
| 0,315907445 | HCG9 | FALSE | Upregulated |
| 0,92479566 | HCLS1 | FALSE | Upregulated |
| 0,692334221 | HCRTR1 | FALSE | Upregulated |
| -0,767771752 | HDAC1 | FALSE | Downregulated |
| -0,258007174 | HDAC2 | FALSE | Downregulated |
| -1,165317822 | HDAC3 | | Downregulated |
| -0,796737946 | HDAC8 | FALSE | Downregulated |
| -0,028696454 | HDDC3 | FALSE | Downregulated |
| -0,356298866 | HDHD2 | FALSE | Downregulated |
| 0,254736491 | HEATR5A | FALSE | Upregulated |
| 0,121706981 | HECTD2 | FALSE | Upregulated |
| -0,089379145 | HECW2 | FALSE | Downregulated |
| 0,241404729 | HEPHL1 | FALSE | Upregulated |
| -0,043930095 | HERC3 | FALSE | Downregulated |
| -1,832973093 | HERC5 | | Downregulated |
| 0,584329806 | HERPUD2 | FALSE | Upregulated |
| 0,593395161 | HES1 | FALSE | Upregulated |
| 0,150767264 | HES4 | FALSE | Upregulated |
| 0,537202774 | HFE | FALSE | Upregulated |
| -0,432427527 | HHEX | FALSE | Downregulated |
| -1,677317151 | HIBCH | | Downregulated |
| 0,872340872 | HIC1 | FALSE | Upregulated |
| 0,687629529 | HIC2 | FALSE | Upregulated |
| 0,462553323 | HIGD1B | FALSE | Upregulated |
| 0,178789095 | HIGD1C | FALSE | Upregulated |
| -0,44047962 | HIGD2A | FALSE | Downregulated |
| 0,733864489 | HIPK4 | FALSE | Upregulated |
| 1,311421329 | HIST1H2APS1 | | Upregulated |

| | | | |
|---|---|---|---|
| 0,825480645 | HIST1H2BA | FALSE | Upregulated |
| 0,515350157 | HIST1H2BK | FALSE | Upregulated |
| -0,562712332 | HIST1H4C | FALSE | Downregulated |
| -1,011867524 | HIST2H2BE | | Downregulated |
| 1,391713416 | HK1 | | Upregulated |
| 0,277331555 | HK2 | FALSE | Upregulated |
| 0,983291229 | HK3 | FALSE | Upregulated |
| -0,336964767 | HKR1 | FALSE | Downregulated |
| 1,344836727 | HLA-DMB | | Upregulated |
| 0,837541162 | HLA-DQA1 | FALSE | Upregulated |
| -0,181167948 | HLTF | FALSE | Downregulated |
| 0,176661463 | HMCES | FALSE | Upregulated |
| 1,602252667 | HMGA1 | | Upregulated |
| 0,909956073 | HMGB3 | FALSE | Upregulated |
| -0,667770616 | HMGN1 | FALSE | Downregulated |
| 0,645262983 | HMOX1 | FALSE | Upregulated |
| 1,709351469 | HN1 | | Upregulated |
| 0,841336908 | HNRNPA1P10 | FALSE | Upregulated |
| -0,388343037 | HNRNPD | FALSE | Downregulated |
| 0,419128493 | HNRNPR | FALSE | Upregulated |
| 0,875301471 | HOMER3 | FALSE | Upregulated |
| 0,018796769 | HOOK2 | FALSE | Upregulated |
| -0,109807741 | HOXB3 | FALSE | Downregulated |
| 0,170036984 | HPS1 | FALSE | Upregulated |
| 0,735291807 | HPS6 | FALSE | Upregulated |
| 0,040643688 | HRAS | FALSE | Upregulated |
| -0,591439536 | HS3ST3A1 | FALSE | Downregulated |
| -0,004980217 | HSBP1 | FALSE | Downregulated |
| 0,964430907 | HSD11B2 | FALSE | Upregulated |
| -1,708812078 | HSD17B14 | | Downregulated |
| -0,219994495 | HSD17B2 | FALSE | Downregulated |
| 0,590456432 | HSF4 | FALSE | Upregulated |
| 0,122245205 | HSPA14 | FALSE | Upregulated |
| 0,374459723 | HSPA4L | FALSE | Upregulated |
| 0,33055141 | HSPB6 | FALSE | Upregulated |
| 0,387142673 | HSPBAP1 | FALSE | Upregulated |
| -0,228620819 | HSPH1 | FALSE | Downregulated |
| 1,157103544 | HTR6 | | Upregulated |
| 0,794265879 | HTRA2 | FALSE | Upregulated |
| 1,113857962 | HULC | | Upregulated |
| 0,0562565 | HYPK | FALSE | Upregulated |
| -0,642966156 | IBA57 | FALSE | Downregulated |
| -1,000614996 | ICK | | Downregulated |
| 1,973752974 | ID1 | | Upregulated |
| -0,267156214 | IDE | FALSE | Downregulated |
| -1,560731095 | IDI2-AS1 | | Downregulated |
| 0,217629576 | IFFO1 | FALSE | Upregulated |
| 0,052098224 | IFI27 | FALSE | Upregulated |
| 0,455997116 | IFI35 | FALSE | Upregulated |
| -0,41353578 | IFIH1 | FALSE | Downregulated |
| -0,844426768 | IFIT1B | FALSE | Downregulated |

| | | | |
|---|---|---|---|
| 0,361143439 | IFITM1 | FALSE | Upregulated |
| -0,209093507 | IFITM10 | FALSE | Downregulated |
| -0,110597247 | IFNGR1 | FALSE | Downregulated |
| 0,251738453 | IFNLR1 | FALSE | Upregulated |
| -0,074822857 | IFRD1 | FALSE | Downregulated |
| 0,206260617 | IFRD2 | FALSE | Upregulated |
| -0,083699828 | IFT122 | FALSE | Downregulated |
| -1,070700511 | IFT20 | | Downregulated |
| -0,266283795 | IFT46 | FALSE | Downregulated |
| -0,177957431 | IFT88 | FALSE | Downregulated |
| -0,731269795 | IGBP1P1 | FALSE | Downregulated |
| 0,471839671 | IGF1R | FALSE | Upregulated |
| -0,19885566 | IGIP | FALSE | Downregulated |
| 1,383160754 | IGLON5 | | Upregulated |
| 0,631184438 | IGSF9 | FALSE | Upregulated |
| 0,165623215 | IK | FALSE | Upregulated |
| 1,297489273 | IKBKG | | Upregulated |
| 0,165784947 | IKZF1 | FALSE | Upregulated |
| -0,319596495 | IL10RB | FALSE | Downregulated |
| -0,135703846 | IL11RA | FALSE | Downregulated |
| 1,527875877 | IL15RA | | Upregulated |
| -1,184553658 | IL17D | | Downregulated |
| -0,435228177 | ILF2 | FALSE | Downregulated |
| -0,390409035 | ILF3-AS1 | FALSE | Downregulated |
| 0,300376746 | IMMP2L | FALSE | Upregulated |
| 0,103845063 | IMP4 | FALSE | Upregulated |
| -0,359772993 | IMPA1 | FALSE | Downregulated |
| 1,580558819 | IMPDH1 | | Upregulated |
| 0,330743363 | IMPG1 | FALSE | Upregulated |
| 0,021332379 | INCA1 | FALSE | Upregulated |
| -0,280915932 | ING1 | FALSE | Downregulated |
| -0,142147472 | ING4 | FALSE | Downregulated |
| 0,463522813 | INIP | FALSE | Upregulated |
| 1,315349666 | INMT | | Upregulated |
| -0,60977797 | INPP5A | FALSE | Downregulated |
| -0,722132487 | INTS4 | FALSE | Downregulated |
| 0,59861382 | INTS5 | FALSE | Upregulated |
| -0,503561549 | INTS9 | FALSE | Downregulated |
| -0,367721705 | IP6K2 | FALSE | Downregulated |
| 0,137333871 | IPO13 | FALSE | Upregulated |
| -0,111040267 | IPP | FALSE | Downregulated |
| 0,059323419 | IQCG | FALSE | Upregulated |
| 0,076474894 | IQCH | FALSE | Upregulated |
| 0,332037791 | IQSEC2 | FALSE | Upregulated |
| -0,904562125 | IQSEC3 | FALSE | Downregulated |
| 0,546871382 | IRAK4 | FALSE | Upregulated |
| -0,089790757 | IRF2 | FALSE | Downregulated |
| 0,455190596 | IRF5 | FALSE | Upregulated |
| 1,154841284 | IRF8 | | Upregulated |
| 0,387862856 | IRF9 | FALSE | Upregulated |
| 0,127004873 | ISCU | FALSE | Upregulated |

| | | | |
|---|---|---|---|
| -0,071636039 | ISG20L2 | FALSE | Downregulated |
| -1,623296043 | ISM1 | | Downregulated |
| -0,60976867 | ISM1-AS1 | FALSE | Downregulated |
| -1,527527378 | ISPD | | Downregulated |
| -1,048781903 | ISPD-AS1 | | Downregulated |
| 0,787834681 | ISYNA1 | FALSE | Upregulated |
| 0,621199445 | ITFG2 | FALSE | Upregulated |
| 0,410714896 | ITGA4 | FALSE | Upregulated |
| 2,403697302 | ITGAX | | Upregulated |
| 0,712101779 | ITGB1BP1 | FALSE | Upregulated |
| 0,803055274 | ITGB2-AS1 | FALSE | Upregulated |
| 0,451442406 | ITGBL1 | FALSE | Upregulated |
| -0,265731783 | ITPA | FALSE | Downregulated |
| 0,636442139 | JAK2 | FALSE | Upregulated |
| 0,553124035 | JAK3 | FALSE | Upregulated |
| 0,487283279 | JARID2 | FALSE | Upregulated |
| -0,63160802 | JARID2-AS1 | FALSE | Downregulated |
| 0,112105083 | JKAMP | FALSE | Upregulated |
| -0,947832074 | JMJD6 | FALSE | Downregulated |
| -0,340210337 | KAT8 | FALSE | Downregulated |
| -0,006745938 | KATNA1 | FALSE | Downregulated |
| -0,916321308 | KBTBD7 | FALSE | Downregulated |
| -0,167817303 | KCMF1 | FALSE | Downregulated |
| 0,654772907 | KCNC3 | FALSE | Upregulated |
| -2,439088813 | KCNC4 | | Downregulated |
| 0,306907108 | KCNH3 | FALSE | Upregulated |
| -0,496159005 | KCNIP2 | FALSE | Downregulated |
| 0,040854975 | KCNK1 | FALSE | Upregulated |
| 0,326149788 | KCNT2 | FALSE | Upregulated |
| -0,910071257 | KCTD1 | FALSE | Downregulated |
| 0,88844426 | KCTD15 | FALSE | Upregulated |
| -0,065250272 | KCTD18 | FALSE | Downregulated |
| 0,190931449 | KCTD2 | FALSE | Upregulated |
| -0,095814392 | KCTD7 | FALSE | Downregulated |
| 0,579132058 | KDM4E | FALSE | Upregulated |
| -0,336943079 | KDM5B | FALSE | Downregulated |
| 0,172459949 | KDM6A | FALSE | Upregulated |
| 0,749678111 | KHDRBS3 | FALSE | Upregulated |
| 0,262024679 | KIAA0196 | FALSE | Upregulated |
| -0,908955574 | KIAA0319 | FALSE | Downregulated |
| -0,014168456 | KIAA0391 | FALSE | Downregulated |
| 1,655071946 | KIAA0513 | | Upregulated |
| 0,483977511 | KIAA0586 | FALSE | Upregulated |
| 0,164969581 | KIAA0895 | FALSE | Upregulated |
| 0,062371559 | KIAA0895L | FALSE | Upregulated |
| 0,257419275 | KIAA0907 | FALSE | Upregulated |
| -0,446679747 | KIAA1244 | FALSE | Downregulated |
| -0,088463591 | KIAA1430 | FALSE | Downregulated |
| -0,610096734 | KIAA1456 | FALSE | Downregulated |
| -0,744651803 | KIAA1652 | FALSE | Downregulated |
| -0,34192171 | KIAA1683 | FALSE | Downregulated |

| | | | |
|---|---|---|---|
| -0,136067769 | KIAA1715 | FALSE | Downregulated |
| -1,511007855 | KIF12 | | Downregulated |
| 0,334332358 | KIF16B | FALSE | Upregulated |
| 2,050566568 | KIF21B | | Upregulated |
| 0,579973766 | KIF26B | FALSE | Upregulated |
| 0,223617877 | KIF9 | FALSE | Upregulated |
| 0,043441559 | KIRREL3 | FALSE | Upregulated |
| 0,105337677 | KLC2 | FALSE | Upregulated |
| 0,061176449 | KLC3 | FALSE | Upregulated |
| 0,064461785 | KLF10 | FALSE | Upregulated |
| 1,101608456 | KLF7 | | Upregulated |
| 0,719738144 | KLHDC7A | FALSE | Upregulated |
| -0,167001294 | KLHL12 | FALSE | Downregulated |
| 0,189531874 | KLHL15 | FALSE | Upregulated |
| -0,358959279 | KLHL17 | FALSE | Downregulated |
| -0,221931254 | KLHL2 | FALSE | Downregulated |
| -1,302787293 | KLHL20 | | Downregulated |
| 1,698859513 | KLHL25 | | Upregulated |
| 1,639998221 | KLHL42 | | Upregulated |
| 0,81545785 | KLHL6 | FALSE | Upregulated |
| -1,467600723 | KLHL7 | | Downregulated |
| -0,652311 | KLHL8 | FALSE | Downregulated |
| -0,271672644 | KLLN | FALSE | Downregulated |
| 1,955066609 | KNTC1 | | Upregulated |
| 0,766731792 | KPNA3 | FALSE | Upregulated |
| -0,022998559 | KRBOX4 | FALSE | Downregulated |
| 0,503616971 | KREMEN1 | FALSE | Upregulated |
| 0,11255813 | KRIT1 | FALSE | Upregulated |
| 1,08066297 | KSR1 | | Upregulated |
| 0,051886555 | KY | FALSE | Upregulated |
| -0,252822572 | KYNU | FALSE | Downregulated |
| -1,055076915 | L2HGDH | | Downregulated |
| 0,816123307 | LACC1 | FALSE | Upregulated |
| -0,140365854 | LACTB | FALSE | Downregulated |
| -1,037259385 | LAD1 | | Downregulated |
| -0,532977695 | LAMA4 | FALSE | Downregulated |
| 0,51473783 | LAMTOR4 | FALSE | Upregulated |
| 0,381637231 | LANCL1 | FALSE | Upregulated |
| -0,644800594 | LAPTM4B | FALSE | Downregulated |
| -0,317151185 | LAS1L | FALSE | Downregulated |
| -0,763662721 | LBR | FALSE | Downregulated |
| 0,399708154 | LCA5L | FALSE | Upregulated |
| 0,857295626 | LCN12 | FALSE | Upregulated |
| -1,044201822 | LCT | | Downregulated |
| -0,850113804 | LDHAL6A | FALSE | Downregulated |
| 0,469367113 | LDLRAP1 | FALSE | Upregulated |
| 0,905528236 | LDOC1L | FALSE | Upregulated |
| -0,292036902 | LECT1 | FALSE | Downregulated |
| 0,344273795 | LEKR1 | FALSE | Upregulated |
| -0,705941704 | LENEP | FALSE | Downregulated |
| -0,529489151 | LETM2 | FALSE | Downregulated |

| | | | |
|---|---|---|---|
| 0,311587842 | LETMD1 | FALSE | Upregulated |
| 0,182091779 | LGALS1 | FALSE | Upregulated |
| 0,845322998 | LGALS8-AS1 | FALSE | Upregulated |
| -0,734839987 | LHFP | FALSE | Downregulated |
| -0,148805997 | LIG1 | FALSE | Downregulated |
| -0,502678804 | LIG4 | FALSE | Downregulated |
| -0,031478123 | LILRA6 | FALSE | Downregulated |
| 0,775703854 | LILRB1 | FALSE | Upregulated |
| 0,648649548 | LIMA1 | FALSE | Upregulated |
| -0,90962014 | LIMCH1 | FALSE | Downregulated |
| -0,228212073 | LIMS1 | FALSE | Downregulated |
| -0,759026839 | LIMS3L | FALSE | Downregulated |
| -0,783837799 | LIN7A | FALSE | Downregulated |
| -0,708584836 | LINC00092 | FALSE | Downregulated |
| -0,171024365 | LINC00102 | FALSE | Downregulated |
| 0,083098396 | LINC00161 | FALSE | Upregulated |
| -1,20864145 | LINC00282 | | Downregulated |
| 0,02258873 | LINC00294 | FALSE | Upregulated |
| 0,225139285 | LINC00342 | FALSE | Upregulated |
| 0,296292314 | LINC00478 | FALSE | Upregulated |
| 1,103919872 | LINC00482 | | Upregulated |
| 0,106023347 | LINC00574 | FALSE | Upregulated |
| 0,235699666 | LINC00641 | FALSE | Upregulated |
| -0,469990392 | LINC00847 | FALSE | Downregulated |
| 0,169048213 | LINC00883 | FALSE | Upregulated |
| 0,059507877 | LINC00886 | FALSE | Upregulated |
| 0,315311593 | LINC00921 | FALSE | Upregulated |
| 0,244747839 | LINC01126 | FALSE | Upregulated |
| 0,325969423 | LINC01128 | FALSE | Upregulated |
| 0,340277996 | LINGO3 | FALSE | Upregulated |
| 0,724987715 | LIPE | FALSE | Upregulated |
| 0,284457075 | LIPT1 | FALSE | Upregulated |
| 1,61824552 | LLGL1 | | Upregulated |
| -0,438346362 | LMBRD1 | FALSE | Downregulated |
| -0,029763289 | LMBRD2 | FALSE | Downregulated |
| 0,342501646 | LMF1 | FALSE | Upregulated |
| -0,301331573 | LMO4 | FALSE | Downregulated |
| -0,667747319 | LMOD1 | FALSE | Downregulated |
| 0,998906106 | LNPEP | FALSE | Upregulated |
| -0,454121475 | LNX1 | FALSE | Downregulated |
| -0,008749468 | LNX2 | FALSE | Downregulated |
| -0,101646196 | LOC100126582 | FALSE | Downregulated |
| 0,409184266 | LOC100128573 | FALSE | Upregulated |
| -1,342263588 | LOC100129148 | | Downregulated |
| 0,628130192 | LOC100129518 | FALSE | Upregulated |
| -0,34777141 | LOC100129697 | FALSE | Downregulated |
| -0,442501532 | LOC100129722 | FALSE | Downregulated |
| -0,250731514 | LOC100130872 | FALSE | Downregulated |
| -1,50226548 | LOC100130876 | | Downregulated |
| 0,56628674 | LOC100132352 | FALSE | Upregulated |
| -0,315427124 | LOC100132731 | FALSE | Downregulated |

| | | | |
|---|---|---|---|
| 0,560224172 | LOC100132832 | FALSE | Upregulated |
| 0,607834992 | LOC100134822 | FALSE | Upregulated |
| -0,692474162 | LOC100287166 | FALSE | Downregulated |
| 0,15988919 | LOC100287808 | FALSE | Upregulated |
| 0,48816293 | LOC100287934 | FALSE | Upregulated |
| -0,43561258 | LOC100288152 | FALSE | Downregulated |
| 0,837739983 | LOC100289097 | FALSE | Upregulated |
| 1,038386324 | LOC100289455 | | Upregulated |
| 0,415969281 | LOC100499489 | FALSE | Upregulated |
| -0,30335674 | LOC100505570 | FALSE | Downregulated |
| -0,850725681 | LOC100506083 | FALSE | Downregulated |
| 0,20369614 | LOC100506114 | FALSE | Upregulated |
| -0,530390788 | LOC100506548 | FALSE | Downregulated |
| 0,367087502 | LOC100506734 | FALSE | Upregulated |
| 0,613236996 | LOC100507071 | FALSE | Upregulated |
| 0,781235199 | LOC100507156 | FALSE | Upregulated |
| -0,664292136 | LOC100507191 | FALSE | Downregulated |
| 0,213001037 | LOC100507217 | FALSE | Upregulated |
| 0,032682393 | LOC100507346 | FALSE | Upregulated |
| -0,226440537 | LOC100507373 | FALSE | Downregulated |
| 0,346933178 | LOC100996255 | FALSE | Upregulated |
| -0,717490919 | LOC100996273 | FALSE | Downregulated |
| 0,06566237 | LOC100996573 | FALSE | Upregulated |
| -0,764931906 | LOC100996761 | FALSE | Downregulated |
| 0,219497625 | LOC101059906 | FALSE | Upregulated |
| 1,412482706 | LOC101060553 | | Upregulated |
| -0,354988374 | LOC101593348 | FALSE | Downregulated |
| -0,051257808 | LOC101926934 | FALSE | Downregulated |
| 0,617065885 | LOC101927000 | FALSE | Upregulated |
| 0,530992581 | LOC101927002 | FALSE | Upregulated |
| 1,079256126 | LOC101927044 | | Upregulated |
| 1,167662732 | LOC101927051 | | Upregulated |
| -1,053219332 | LOC101927163 | | Downregulated |
| -0,144057225 | LOC101927185 | FALSE | Downregulated |
| -1,482124555 | LOC101927187 | | Downregulated |
| -0,492155695 | LOC101927371 | FALSE | Downregulated |
| -0,125091517 | LOC101927437 | FALSE | Downregulated |
| -0,371791445 | LOC101927451 | FALSE | Downregulated |
| 0,830524671 | LOC101927479 | FALSE | Upregulated |
| -0,24215087 | LOC101927559 | FALSE | Downregulated |
| 0,606379683 | LOC101927578 | FALSE | Upregulated |
| 0,390206118 | LOC101927719 | FALSE | Upregulated |
| 1,256220761 | LOC101927727 | | Upregulated |
| -0,616850275 | LOC101927741 | FALSE | Downregulated |
| -0,112048836 | LOC101927761 | FALSE | Downregulated |
| -1,152291603 | LOC101927790 | | Downregulated |
| -0,386089377 | LOC101927793 | FALSE | Downregulated |
| -1,174680033 | LOC101927806 | | Downregulated |
| -0,666517281 | LOC101927858 | FALSE | Downregulated |
| 0,917465806 | LOC101927864 | FALSE | Upregulated |
| 1,528556502 | LOC101927955 | | Upregulated |

| | | | |
|---|---|---|---|
| 1,215974725 | LOC101928038 | | Upregulated |
| -0,914237954 | LOC101928059 | FALSE | Downregulated |
| 0,786516832 | LOC101928068 | FALSE | Upregulated |
| -0,202497833 | LOC101928069 | FALSE | Downregulated |
| 0,639629739 | LOC101928089 | FALSE | Upregulated |
| 0,338782116 | LOC101928098 | FALSE | Upregulated |
| -1,185816034 | LOC101928143 | | Downregulated |
| -0,066996327 | LOC101928170 | FALSE | Downregulated |
| 1,932225839 | LOC101928173 | | Upregulated |
| 0,552852197 | LOC101928206 | FALSE | Upregulated |
| -0,683271037 | LOC101928355 | FALSE | Downregulated |
| -2,728167404 | LOC101928384 | | Downregulated |
| 0,02359112 | LOC101928408 | FALSE | Upregulated |
| -0,255115199 | LOC101928431 | FALSE | Downregulated |
| 0,051058123 | LOC101928439 | FALSE | Upregulated |
| 0,584823983 | LOC101928466 | FALSE | Upregulated |
| 2,687755139 | LOC101928491 | | Upregulated |
| 0,558728636 | LOC101928499 | FALSE | Upregulated |
| 0,113957114 | LOC101928512 | FALSE | Upregulated |
| -0,03049978 | LOC101928517 | FALSE | Downregulated |
| 0,408786768 | LOC101928548 | FALSE | Upregulated |
| -0,001434599 | LOC101928574 | FALSE | Downregulated |
| 0,678167649 | LOC101928588 | FALSE | Upregulated |
| 0,408276617 | LOC101928678 | FALSE | Upregulated |
| 0,574479058 | LOC101928707 | FALSE | Upregulated |
| -0,339227396 | LOC101928714 | FALSE | Downregulated |
| 1,769801117 | LOC101928745 | | Upregulated |
| -0,078385984 | LOC101928762 | FALSE | Downregulated |
| 0,390810539 | LOC101928789 | FALSE | Upregulated |
| -1,281350774 | LOC101928926 | | Downregulated |
| -0,306316544 | LOC101928927 | FALSE | Downregulated |
| 0,77597816 | LOC101929005 | FALSE | Upregulated |
| 0,564516978 | LOC101929018 | FALSE | Upregulated |
| 0,302544687 | LOC101929046 | FALSE | Upregulated |
| -0,175975987 | LOC101929051 | FALSE | Downregulated |
| -0,565801259 | LOC101929072 | FALSE | Downregulated |
| 0,005480409 | LOC101929112 | FALSE | Upregulated |
| 0,148322566 | LOC101929130 | FALSE | Upregulated |
| 0,127618536 | LOC101929140 | FALSE | Upregulated |
| -0,639476553 | LOC101929188 | FALSE | Downregulated |
| -1,949802779 | LOC101929195 | | Downregulated |
| -1,170555856 | LOC101929263 | | Downregulated |
| 0,322713996 | LOC101929270 | FALSE | Upregulated |
| 0,998100212 | LOC101929272 | FALSE | Upregulated |
| -0,682109518 | LOC101929354 | FALSE | Downregulated |
| 0,416622358 | LOC101929425 | FALSE | Upregulated |
| 0,678240006 | LOC101929433 | FALSE | Upregulated |
| 0,502116304 | LOC101929519 | FALSE | Upregulated |
| 0,671003207 | LOC101929636 | FALSE | Upregulated |
| -1,292111181 | LOC101929657 | | Downregulated |
| -0,524173349 | LOC101929691 | FALSE | Downregulated |

| | | | |
|---|---|---|---|
| 0,632803516 | LOC101929837 | FALSE | Upregulated |
| 0,530910996 | LOC102723309 | FALSE | Upregulated |
| 1,039280788 | LOC102723331 | | Upregulated |
| 0,023505065 | LOC102723382 | FALSE | Upregulated |
| -0,001457034 | LOC102723539 | FALSE | Downregulated |
| 0,174175985 | LOC102723694 | FALSE | Upregulated |
| 0,202862152 | LOC102723708 | FALSE | Upregulated |
| 0,578467489 | LOC102723758 | FALSE | Upregulated |
| -0,808683604 | LOC102723772 | FALSE | Downregulated |
| 0,201307223 | LOC102723773 | FALSE | Upregulated |
| 0,583441458 | LOC102723809 | FALSE | Upregulated |
| -0,913378502 | LOC102723820 | FALSE | Downregulated |
| -1,726773826 | LOC102723886 | | Downregulated |
| 0,03480352 | LOC102723954 | FALSE | Upregulated |
| -0,754091668 | LOC102723972 | FALSE | Downregulated |
| -0,665244285 | LOC102723997 | FALSE | Downregulated |
| 0,311513254 | LOC102724009 | FALSE | Upregulated |
| 0,524797743 | LOC102724103 | FALSE | Upregulated |
| 0,799575353 | LOC102724133 | FALSE | Upregulated |
| 0,55379365 | LOC102724141 | FALSE | Upregulated |
| -0,529939522 | LOC102724157 | FALSE | Downregulated |
| -0,117062891 | LOC102724217 | FALSE | Downregulated |
| 0,819905839 | LOC102724314 | FALSE | Upregulated |
| -0,398409781 | LOC102724316 | FALSE | Downregulated |
| -0,822698729 | LOC102724322 | FALSE | Downregulated |
| 0,53264455 | LOC102724350 | FALSE | Upregulated |
| 0,860829369 | LOC102724455 | FALSE | Upregulated |
| -0,45051938 | LOC102724474 | FALSE | Downregulated |
| 0,381922114 | LOC102724490 | FALSE | Upregulated |
| 0,357361756 | LOC102724549 | FALSE | Upregulated |
| -0,258216927 | LOC102724634 | FALSE | Downregulated |
| -0,796071034 | LOC102724672 | FALSE | Downregulated |
| 1,197710841 | LOC102724709 | | Upregulated |
| 1,634097081 | LOC102724733 | | Upregulated |
| 1,102831608 | LOC102724782 | | Upregulated |
| -1,219403328 | LOC102724807 | | Downregulated |
| 1,170671391 | LOC115110 | | Upregulated |
| 0,136022504 | LOC155060 | FALSE | Upregulated |
| 1,099898539 | LOC202181 | | Upregulated |
| 0,821275016 | LOC283788 | FALSE | Upregulated |
| 0,538097152 | LOC339803 | FALSE | Upregulated |
| 1,408981343 | LOC388906 | | Upregulated |
| 2,333746755 | LOC389332 | | Upregulated |
| -0,133146335 | LOC389906 | FALSE | Downregulated |
| 0,263642863 | LOC390937 | FALSE | Upregulated |
| -0,541181136 | LOC400043 | FALSE | Downregulated |
| 0,004544869 | LOC401320 | FALSE | Upregulated |
| -0,188772988 | LOC642361 | FALSE | Downregulated |
| 1,381677287 | LOC642648 | | Upregulated |
| -0,354078425 | LOC644662 | FALSE | Downregulated |
| 0,142829818 | LOC645967 | FALSE | Upregulated |

| | | | |
|---|---|---|---|
| 0,867949496 | LOC728613 | FALSE | Upregulated |
| -0,552580804 | LOC728763 | FALSE | Downregulated |
| 0,165115544 | LOC729652 | FALSE | Upregulated |
| -0,27056338 | LOC729658 | FALSE | Downregulated |
| 1,114738723 | LOC729683 | | Upregulated |
| 0,085230839 | LOC729887 | FALSE | Upregulated |
| 0,198059261 | LOC730102 | FALSE | Upregulated |
| -1,03436739 | LOC730183 | | Downregulated |
| 0,862666411 | LOC731656 | FALSE | Upregulated |
| 0,483246428 | LOC90784 | FALSE | Upregulated |
| 0,191657352 | LONRF1 | FALSE | Upregulated |
| -0,577245102 | LONRF3 | FALSE | Downregulated |
| 0,170441663 | LOX | FALSE | Upregulated |
| -0,285701239 | LOXL2 | FALSE | Downregulated |
| 1,301633022 | LPCAT1 | | Upregulated |
| 0,273596429 | LPP-AS2 | FALSE | Upregulated |
| 0,229124054 | LPPR3 | FALSE | Upregulated |
| 0,577078835 | LRCH1 | FALSE | Upregulated |
| 0,149798176 | LRCH3 | FALSE | Upregulated |
| -0,586782692 | LRRC15 | FALSE | Downregulated |
| 0,656921317 | LRRC16A | FALSE | Upregulated |
| 1,29015242 | LRRC18 | | Upregulated |
| -0,401476278 | LRRC2 | FALSE | Downregulated |
| -0,089172608 | LRRC2-AS1 | FALSE | Downregulated |
| 0,132666876 | LRRC32 | FALSE | Upregulated |
| -0,093616191 | LRRC47 | FALSE | Downregulated |
| 1,092406075 | LRRC4B | | Upregulated |
| 0,148540768 | LRRC56 | FALSE | Upregulated |
| -0,779312715 | LRRC57 | FALSE | Downregulated |
| 0,373099802 | LRRC8B | FALSE | Upregulated |
| 0,953510017 | LRRC8C | FALSE | Upregulated |
| -0,52883738 | LRRC8D | FALSE | Downregulated |
| -1,558664682 | LRRK2 | | Downregulated |
| 0,380737874 | LRSAM1 | FALSE | Upregulated |
| 0,118629715 | LRWD1 | FALSE | Upregulated |
| 0,522667588 | LSG1 | FALSE | Upregulated |
| 1,216256067 | LSM8 | | Upregulated |
| 0,589143212 | LTA4H | FALSE | Upregulated |
| -0,431948941 | LTB4R | FALSE | Downregulated |
| 0,163315133 | LTN1 | FALSE | Upregulated |
| 0,423111858 | LURAP1L | FALSE | Upregulated |
| 0,714949577 | LY9 | FALSE | Upregulated |
| -0,585611977 | LYG2 | FALSE | Downregulated |
| -0,648366777 | LYPLA1 | FALSE | Downregulated |
| -0,094194492 | LYPLAL1 | FALSE | Downregulated |
| 0,044538364 | LYSMD3 | FALSE | Upregulated |
| -0,384502212 | LZTFL1 | FALSE | Downregulated |
| 1,389858228 | LZTS2 | | Upregulated |
| -0,210208323 | LZTS3 | FALSE | Downregulated |
| -0,357756939 | MACROD1 | FALSE | Downregulated |
| -0,242573432 | MAD1L1 | FALSE | Downregulated |

| | | | |
|---|---|---|---|
| 0,933203546 | MAEA | FALSE | Upregulated |
| 0,981642052 | MAFF | FALSE | Upregulated |
| -0,246551098 | MAGED2 | FALSE | Downregulated |
| -0,10382275 | MAGEF1 | FALSE | Downregulated |
| -0,871343465 | MAGI2 | FALSE | Downregulated |
| 0,074217114 | MALT1 | FALSE | Upregulated |
| 0,28041587 | MAML2 | FALSE | Upregulated |
| 0,860760735 | MANEAL | FALSE | Upregulated |
| 0,852759635 | MANF | FALSE | Upregulated |
| -0,911097565 | MAP10 | FALSE | Downregulated |
| 1,376668188 | MAP1A | | Upregulated |
| 0,464096137 | MAP1B | FALSE | Upregulated |
| 0,291746007 | MAP1LC3A | FALSE | Upregulated |
| -0,089279025 | MAP2 | FALSE | Downregulated |
| 0,262909938 | MAP2K5 | FALSE | Upregulated |
| 0,23528987 | MAP3K10 | FALSE | Upregulated |
| 0,999544295 | MAP3K14-AS1 | FALSE | Upregulated |
| -0,668849305 | MAP3K19 | FALSE | Downregulated |
| 1,028450605 | MAP3K6 | | Upregulated |
| -0,376152646 | MAP3K7 | FALSE | Downregulated |
| 0,144870153 | MAP3K9 | FALSE | Upregulated |
| -0,308107494 | MAP4K3 | FALSE | Downregulated |
| 0,731415339 | MAP4K5 | FALSE | Upregulated |
| -0,467914971 | MAP6 | FALSE | Downregulated |
| 1,133353972 | MAPK10 | | Upregulated |
| -0,002725136 | MAPK11 | FALSE | Downregulated |
| 0,289395255 | MAPK8 | FALSE | Upregulated |
| -0,484379903 | MAPK8IP1 | FALSE | Downregulated |
| 0,802213838 | MAPK9 | FALSE | Upregulated |
| -0,016718882 | MAPKAPK5 | FALSE | Downregulated |
| 0,45202961 | MAPKBP1 | FALSE | Upregulated |
| 0,882080687 | MAPRE1 | FALSE | Upregulated |
| -0,536139439 | MAPRE2 | FALSE | Downregulated |
| -1,185799523 | MAPRE3 | | Downregulated |
| -0,01415854 | MARCH2 | FALSE | Downregulated |
| -0,542647181 | MARCH5 | FALSE | Downregulated |
| -0,121715842 | MARVELD3 | FALSE | Downregulated |
| -0,433676169 | MAT2B | FALSE | Downregulated |
| -0,478671896 | MAX | FALSE | Downregulated |
| 1,123863976 | MBD2 | | Upregulated |
| 0,301207049 | MBD4 | FALSE | Upregulated |
| -0,556424564 | MBLAC1 | FALSE | Downregulated |
| -0,518232437 | MBLAC2 | FALSE | Downregulated |
| 0,564045039 | MBOAT7 | FALSE | Upregulated |
| -0,941878169 | MBTPS2 | FALSE | Downregulated |
| 1,027922145 | MCAM | | Upregulated |
| 0,420767349 | MCCD1 | FALSE | Upregulated |
| 1,168922905 | MCM3 | | Upregulated |
| 0,881434715 | MCM5 | FALSE | Upregulated |
| 0,189920079 | MCM9 | FALSE | Upregulated |
| 0,680689541 | MCPH1 | FALSE | Upregulated |

| | | | |
|---|---|---|---|
| 0,44636692 | MCRS1 | FALSE | Upregulated |
| 0,194283673 | MDFIC | FALSE | Upregulated |
| -0,555678809 | ME2 | FALSE | Downregulated |
| -0,271611426 | ME3 | FALSE | Downregulated |
| -0,937190428 | MEA1 | FALSE | Downregulated |
| 0,226764422 | MECR | FALSE | Upregulated |
| 0,10704091 | MED11 | FALSE | Upregulated |
| -0,57302378 | MED12 | FALSE | Downregulated |
| 1,529270903 | MED12L | | Upregulated |
| 1,326190006 | MED19 | | Upregulated |
| 0,285289971 | MED20 | FALSE | Upregulated |
| -0,83938155 | MED21 | FALSE | Downregulated |
| 0,321561482 | MED25 | FALSE | Upregulated |
| -0,560777098 | MED28 | FALSE | Downregulated |
| -0,259862764 | MEIS1-AS2 | FALSE | Downregulated |
| -0,14095155 | MEIS2 | FALSE | Downregulated |
| 0,157931347 | MEIS3P1 | FALSE | Upregulated |
| 0,514512886 | MEMO1 | FALSE | Upregulated |
| 0,800757933 | MESDC2 | FALSE | Upregulated |
| 0,186221235 | METAP1 | FALSE | Upregulated |
| -1,170781247 | METAP2 | | Downregulated |
| -0,370824117 | METRN | FALSE | Downregulated |
| 0,126253286 | METTL13 | FALSE | Upregulated |
| 0,464823992 | METTL14 | FALSE | Upregulated |
| 0,434810232 | METTL20 | FALSE | Upregulated |
| 0,008591188 | METTL22 | FALSE | Upregulated |
| 0,084286371 | METTL23 | FALSE | Upregulated |
| -0,789249087 | METTL2B | FALSE | Downregulated |
| -0,136707297 | METTL8 | FALSE | Downregulated |
| 0,826076169 | MEX3C | FALSE | Upregulated |
| -0,670600872 | MFAP1 | FALSE | Downregulated |
| -0,715175342 | MFAP3 | FALSE | Downregulated |
| -0,201561365 | MFN1 | FALSE | Downregulated |
| 0,525978875 | MFSD10 | FALSE | Upregulated |
| 0,089146846 | MFSD9 | FALSE | Upregulated |
| 0,838812043 | MGAT4A | FALSE | Upregulated |
| -0,887269088 | MGC12916 | FALSE | Downregulated |
| -0,451251105 | MGC27345 | FALSE | Downregulated |
| -0,628891272 | MID1IP1 | FALSE | Downregulated |
| 1,123242261 | MIF4GD | | Upregulated |
| 0,611906127 | MIR181A2HG | FALSE | Upregulated |
| -0,562907103 | MIR210HG | FALSE | Downregulated |
| 1,491976274 | MIR22HG | | Upregulated |
| -0,724847331 | MIS12 | FALSE | Downregulated |
| 0,133889311 | MIS18BP1 | FALSE | Upregulated |
| -0,277652358 | MKKS | FALSE | Downregulated |
| 0,430812539 | MKNK1 | FALSE | Upregulated |
| 0,387050261 | MLH1 | FALSE | Upregulated |
| 1,108193181 | MLLT11 | | Upregulated |
| -1,827366612 | MLLT4-AS1 | | Downregulated |
| -0,487983856 | MLST8 | FALSE | Downregulated |

| | | | |
|---|---|---|---|
| -0,251097902 | MLYCD | FALSE | Downregulated |
| -0,897340219 | MMAA | FALSE | Downregulated |
| 1,181733563 | MMEL1 | | Upregulated |
| 0,572963566 | MMP2 | FALSE | Upregulated |
| 0,999595352 | MMP23B | FALSE | Upregulated |
| 0,371868368 | MOB1B | FALSE | Upregulated |
| 1,082936015 | MOB3A | | Upregulated |
| -0,325133964 | MOCS2 | FALSE | Downregulated |
| -1,301618587 | MOGAT2 | | Downregulated |
| 0,099831859 | MOK | FALSE | Upregulated |
| 0,567627886 | MPDU1 | FALSE | Upregulated |
| -0,436958136 | MPL | FALSE | Downregulated |
| -0,456742873 | MPLKIP | FALSE | Downregulated |
| -0,327080724 | MPND | FALSE | Downregulated |
| 1,073679264 | MPP1 | | Upregulated |
| -0,760214905 | MPP6 | FALSE | Downregulated |
| 0,861861835 | MPPE1 | FALSE | Upregulated |
| 0,628934069 | MR1 | FALSE | Upregulated |
| -0,236155888 | MRE11A | FALSE | Downregulated |
| 0,302638274 | MREG | FALSE | Upregulated |
| -0,102449192 | MRFAP1L1 | FALSE | Downregulated |
| -0,441452062 | MRGBP | FALSE | Downregulated |
| -1,212216309 | MRI1 | | Downregulated |
| -0,434916498 | MRO | FALSE | Downregulated |
| -0,473722498 | MRP63 | FALSE | Downregulated |
| -0,574940209 | MRPL14 | FALSE | Downregulated |
| -0,075163345 | MRPL16 | FALSE | Downregulated |
| 0,147847566 | MRPL17 | FALSE | Upregulated |
| -1,034083958 | MRPL3 | | Downregulated |
| -0,798307324 | MRPL34 | FALSE | Downregulated |
| -0,803583366 | MRPL35 | FALSE | Downregulated |
| 0,044788022 | MRPL40 | FALSE | Upregulated |
| -1,114467899 | MRPL42 | | Downregulated |
| 0,760674357 | MRPL44 | FALSE | Upregulated |
| -0,896708187 | MRPL45P2 | FALSE | Downregulated |
| -0,661805449 | MRPS10 | FALSE | Downregulated |
| -0,43596583 | MRPS11 | FALSE | Downregulated |
| 0,102226074 | MRPS12 | FALSE | Upregulated |
| 0,204217282 | MRPS2 | FALSE | Upregulated |
| 0,486080834 | MRPS22 | FALSE | Upregulated |
| -0,710423824 | MRPS34 | FALSE | Downregulated |
| 0,311135249 | MRPS35 | FALSE | Upregulated |
| -0,770038755 | MRPS5 | FALSE | Downregulated |
| -0,607355016 | MRPS7 | FALSE | Downregulated |
| -0,700606035 | MRRF | FALSE | Downregulated |
| 0,874702588 | MS4A7 | FALSE | Upregulated |
| -0,811338521 | MSH3 | FALSE | Downregulated |
| 0,663317922 | MSI2 | FALSE | Upregulated |
| -1,470232104 | MSRA | | Downregulated |
| 0,288046247 | MSRB2 | FALSE | Upregulated |
| 0,095006427 | MSTO1 | FALSE | Upregulated |

| | | | |
|---|---|---|---|
| -0,831627666 | MTAP | FALSE | Downregulated |
| 0,253961032 | MTCP1 | FALSE | Upregulated |
| -0,908056242 | MTF2 | FALSE | Downregulated |
| -0,405687192 | MTFMT | FALSE | Downregulated |
| 0,194426565 | MTG1 | FALSE | Upregulated |
| -0,723223093 | MTHFSD | FALSE | Downregulated |
| -1,197525691 | MTIF2 | | Downregulated |
| 0,758816334 | MTIF3 | FALSE | Upregulated |
| 0,00813858 | MTMR1 | FALSE | Upregulated |
| 0,109352091 | MTMR14 | FALSE | Upregulated |
| -0,288821319 | MTO1 | FALSE | Downregulated |
| 1,276833047 | MTOR-AS1 | | Upregulated |
| -1,040193466 | MTRF1 | | Downregulated |
| 0,751169607 | MTRNR2L4 | FALSE | Upregulated |
| 0,767285293 | MTRNR2L8 | FALSE | Upregulated |
| 0,320935216 | MTRR | FALSE | Upregulated |
| 0,146557957 | MTX3 | FALSE | Upregulated |
| 1,562206529 | MUC12 | | Upregulated |
| 0,364848006 | MUS81 | FALSE | Upregulated |
| -0,555016242 | MUSTN1 | FALSE | Downregulated |
| 0,417772272 | MXRA5 | FALSE | Upregulated |
| 0,3510011 | MYADM | FALSE | Upregulated |
| 1,291039003 | MYC | | Upregulated |
| -1,317688465 | MYCL | | Downregulated |
| -0,089135231 | MYH3 | FALSE | Downregulated |
| -0,930487133 | MYL6B | FALSE | Downregulated |
| -0,35000771 | MYNN | FALSE | Downregulated |
| -0,217916587 | MYO15A | FALSE | Downregulated |
| 0,950283281 | MYO1G | FALSE | Upregulated |
| 0,01550872 | MYO1H | FALSE | Upregulated |
| 0,185696372 | MYO5C | FALSE | Upregulated |
| -0,902803596 | MYOZ3 | FALSE | Downregulated |
| -0,024005347 | MZT2A | FALSE | Downregulated |
| -0,090818045 | N4BP2L2-IT2 | FALSE | Downregulated |
| 0,596882754 | NAA16 | FALSE | Upregulated |
| 0,489414098 | NAA20 | FALSE | Upregulated |
| 1,425527353 | NAA40 | | Upregulated |
| 2,048993084 | NABP1 | | Upregulated |
| 1,054056779 | NAGK | | Upregulated |
| 0,728889126 | NAGPA | FALSE | Upregulated |
| -0,29423051 | NAP1L4 | FALSE | Downregulated |
| 0,225389144 | NARF | FALSE | Upregulated |
| 0,812279064 | NARFL | FALSE | Upregulated |
| 0,742229866 | NARG2 | FALSE | Upregulated |
| 1,908448289 | NBEA | | Upregulated |
| -0,25232675 | NBPF10 | FALSE | Downregulated |
| -0,785079536 | NBPF19 | FALSE | Downregulated |
| 0,787160988 | NBPF20 | FALSE | Upregulated |
| 0,335940751 | NBPF3 | FALSE | Upregulated |
| -0,282278784 | NCALD | FALSE | Downregulated |
| 0,540032905 | NCAPD3 | FALSE | Upregulated |

| | | | |
|---|---|---|---|
| -0,177931221 | NCDN | FALSE | Downregulated |
| -0,753854308 | NCKAP5 | FALSE | Downregulated |
| -0,368237083 | NCKIPSD | FALSE | Downregulated |
| -0,162974663 | NDC1 | FALSE | Downregulated |
| -1,280044173 | NDUFA11 | | Downregulated |
| -0,991474432 | NDUFA4 | FALSE | Downregulated |
| -0,522293274 | NDUFA4L2 | FALSE | Downregulated |
| -0,974912464 | NDUFA5 | FALSE | Downregulated |
| -0,043080093 | NDUFAF3 | FALSE | Downregulated |
| -1,230696157 | NDUFB3 | | Downregulated |
| -0,21732251 | NDUFB9 | FALSE | Downregulated |
| -0,180617013 | NECAB3 | FALSE | Downregulated |
| 0,031564031 | NECAP1 | FALSE | Upregulated |
| -0,331775599 | NEDD8 | FALSE | Downregulated |
| 0,221904794 | NEFH | FALSE | Upregulated |
| -0,263443561 | NEIL2 | FALSE | Downregulated |
| -0,201014236 | NEK1 | FALSE | Downregulated |
| -0,15698161 | NEK7 | FALSE | Downregulated |
| -0,20949445 | NELFA | FALSE | Downregulated |
| 0,557154102 | NELFB | FALSE | Upregulated |
| -1,2097806 | NENF | | Downregulated |
| -0,659361523 | NEO1 | FALSE | Downregulated |
| 0,106117396 | NES | FALSE | Upregulated |
| -0,174907173 | NEU3 | FALSE | Downregulated |
| -0,343306118 | NEURL2 | FALSE | Downregulated |
| 1,100757213 | NEXN-AS1 | | Upregulated |
| 0,241680921 | NFKB1 | FALSE | Upregulated |
| -0,411593453 | NFKBIB | FALSE | Downregulated |
| -1,028393997 | NFXL1 | | Downregulated |
| -0,430349699 | NGLY1 | FALSE | Downregulated |
| 0,128541409 | NGRN | FALSE | Upregulated |
| -0,162765764 | NHLRC3 | FALSE | Downregulated |
| 0,250242887 | NHP2L1 | FALSE | Upregulated |
| -0,88605989 | NIF3L1 | FALSE | Downregulated |
| 0,653155927 | NIP7 | FALSE | Upregulated |
| 0,484322592 | NIPA1 | FALSE | Upregulated |
| 1,453609905 | NIPAL2 | | Upregulated |
| -0,401469184 | NIPAL3 | FALSE | Downregulated |
| 2,039426699 | NIPAL4 | | Upregulated |
| 0,458933248 | NIPSNAP3A | FALSE | Upregulated |
| 1,15276744 | NKD1 | | Upregulated |
| 0,074256746 | NKIRAS1 | FALSE | Upregulated |
| 0,229779959 | NKIRAS2 | FALSE | Upregulated |
| 0,804121397 | NKPD1 | FALSE | Upregulated |
| 0,207696002 | NLGN2 | FALSE | Upregulated |
| -0,227781926 | NLK | FALSE | Downregulated |
| -0,105745656 | NLN | FALSE | Downregulated |
| 0,478443824 | NLRP1 | FALSE | Upregulated |
| 0,316391658 | NLRX1 | FALSE | Upregulated |
| -0,216944719 | NME6 | FALSE | Downregulated |
| 2,015568243 | NMNAT1 | | Upregulated |

| | | | |
|---|---|---|---|
| 0,488047393 | NMNAT3 | FALSE | Upregulated |
| -0,562202341 | NMRK1 | FALSE | Downregulated |
| -1,040523191 | NOA1 | | Downregulated |
| -0,086013933 | NOB1 | FALSE | Downregulated |
| 0,061509703 | NOL12 | FALSE | Upregulated |
| 0,645137622 | NOL3 | FALSE | Upregulated |
| -0,101677176 | NOL6 | FALSE | Downregulated |
| -0,725166618 | NOLC1 | FALSE | Downregulated |
| -0,075456003 | NOM1 | FALSE | Downregulated |
| 1,577413147 | NOP2 | | Upregulated |
| 0,02460699 | NOP56 | FALSE | Upregulated |
| -0,011674791 | NOS3 | FALSE | Downregulated |
| 1,106409276 | NOTCH3 | | Upregulated |
| 0,441375252 | NOXO1 | FALSE | Upregulated |
| 0,915369414 | NPC2 | FALSE | Upregulated |
| 0,249264533 | NPDC1 | FALSE | Upregulated |
| -0,473393611 | NPHP4 | FALSE | Downregulated |
| -0,928002671 | NPR1 | FALSE | Downregulated |
| 1,442429766 | NPR2 | | Upregulated |
| 0,447145552 | NR1D1 | FALSE | Upregulated |
| -1,179053997 | NR2C1 | | Downregulated |
| -0,010090014 | NR2C2AP | FALSE | Downregulated |
| -0,935044456 | NR6A1 | FALSE | Downregulated |
| -0,678063635 | NRADDP | FALSE | Downregulated |
| -1,120009012 | NRARP | | Downregulated |
| 0,606907813 | NRAS | FALSE | Upregulated |
| -0,287360631 | NRDE2 | FALSE | Downregulated |
| 0,545823816 | NRIP3 | FALSE | Upregulated |
| -0,50427615 | NRL | FALSE | Downregulated |
| 0,20497053 | NRP2 | FALSE | Upregulated |
| 0,702190432 | NRSN2 | FALSE | Upregulated |
| -1,033363196 | NSA2 | | Downregulated |
| -0,417581635 | NSF | FALSE | Downregulated |
| 0,341956342 | NSL1 | FALSE | Upregulated |
| 0,323343042 | NSUN4 | FALSE | Upregulated |
| -0,740079451 | NSUN5P2 | FALSE | Downregulated |
| 0,52152754 | NSUN7 | FALSE | Upregulated |
| 0,427599235 | NT5C | FALSE | Upregulated |
| 0,296675755 | NT5DC2 | FALSE | Upregulated |
| -1,75199077 | NT5DC3 | | Downregulated |
| 0,38527686 | NTAN1 | FALSE | Upregulated |
| -1,109045517 | NTHL1 | | Downregulated |
| 0,119983955 | NUBP1 | FALSE | Upregulated |
| -2,140432396 | NUCB2 | | Downregulated |
| 0,506818264 | NUDT15 | FALSE | Upregulated |
| -0,054196438 | NUDT16L1 | FALSE | Downregulated |
| -1,174939634 | NUDT16P1 | | Downregulated |
| -1,078428532 | NUDT19 | | Downregulated |
| 0,407026416 | NUDT22 | FALSE | Upregulated |
| 0,653002844 | NUP133 | FALSE | Upregulated |
| -0,276279329 | NUP160 | FALSE | Downregulated |

| | | | |
|---|---|---|---|
| -0,023194468 | NUP205 | FALSE | Downregulated |
| -0,060959396 | NUP85 | FALSE | Downregulated |
| -0,687007692 | NUP88 | FALSE | Downregulated |
| 0,078671661 | NUSAP1 | FALSE | Upregulated |
| -0,498127384 | NUTM2F | FALSE | Downregulated |
| -1,496010755 | NXN | | Downregulated |
| 0,54634355 | NXPH3 | FALSE | Upregulated |
| 0,310009253 | OAS1 | FALSE | Upregulated |
| 0,997436956 | OAS2 | FALSE | Upregulated |
| -0,864114844 | OCEL1 | FALSE | Downregulated |
| 0,110491274 | ODF2 | FALSE | Upregulated |
| 0,508988474 | ODF2L | FALSE | Upregulated |
| 0,494942403 | OFD1 | FALSE | Upregulated |
| 0,57483147 | OGFOD2 | FALSE | Upregulated |
| -1,376779675 | OIP5 | | Downregulated |
| -1,598632865 | OR10J5 | | Downregulated |
| -0,327826016 | OR2A9P | FALSE | Downregulated |
| 0,118063586 | OR52W1 | FALSE | Upregulated |
| 0,02614327 | ORMDL1 | FALSE | Upregulated |
| 0,315411604 | ORMDL2 | FALSE | Upregulated |
| 0,672967271 | OSBPL10 | FALSE | Upregulated |
| 0,701272854 | OSBPL1A | FALSE | Upregulated |
| -0,503372061 | OSGEP | FALSE | Downregulated |
| -0,785865315 | OST4 | FALSE | Downregulated |
| -0,778681909 | OTUD1 | FALSE | Downregulated |
| -0,416337929 | OXR1 | FALSE | Downregulated |
| 0,341614072 | OXSR1 | FALSE | Upregulated |
| 0,075396441 | P2RX2 | FALSE | Upregulated |
| 1,27735998 | P2RX4 | | Upregulated |
| 0,821376862 | P4HA2-AS1 | FALSE | Upregulated |
| -0,290752336 | PA2G4 | FALSE | Downregulated |
| -0,203904457 | PAFAH2 | FALSE | Downregulated |
| 0,360170007 | PAIP1 | FALSE | Upregulated |
| 0,046398868 | PAK1 | FALSE | Upregulated |
| -0,202752895 | PALM | FALSE | Downregulated |
| 0,346253761 | PAM | FALSE | Upregulated |
| 0,275807952 | PANK4 | FALSE | Upregulated |
| 0,442671943 | PAPD4 | FALSE | Upregulated |
| -0,597799985 | PAPD5 | FALSE | Downregulated |
| 2,147256039 | PAPPA-AS1 | | Upregulated |
| -0,552997364 | PAQR7 | FALSE | Downregulated |
| 2,011588408 | PAQR8 | | Upregulated |
| -0,126963742 | PAQR9 | FALSE | Downregulated |
| 0,034233157 | PARD6B | FALSE | Upregulated |
| -0,112855851 | PARD6G-AS1 | FALSE | Downregulated |
| 0,138573126 | PARP3 | FALSE | Upregulated |
| 0,159434074 | PARP6 | FALSE | Upregulated |
| -0,124859456 | PASK | FALSE | Downregulated |
| 0,915445386 | PAXIP1-AS2 | FALSE | Upregulated |
| -0,553877978 | PCDH18 | FALSE | Downregulated |
| -0,308829389 | PCGF1 | FALSE | Downregulated |

| | | | |
|---|---|---|---|
| -0,277140767 | PCMTD2 | FALSE | Downregulated |
| -0,234286156 | PCNXL4 | FALSE | Downregulated |
| -1,024402852 | PCOLCE-AS1 | | Downregulated |
| -0,368256126 | PCP4L1 | FALSE | Downregulated |
| -0,089446422 | PCSK4 | FALSE | Downregulated |
| 0,13080022 | PDC | FALSE | Upregulated |
| 0,598634268 | PDCD1LG2 | FALSE | Upregulated |
| -1,063552007 | PDCD5 | | Downregulated |
| -0,07262988 | PDCD7 | FALSE | Downregulated |
| 0,4889284 | PDE12 | FALSE | Upregulated |
| -0,59081007 | PDE3A | FALSE | Downregulated |
| 0,076669027 | PDE4A | FALSE | Upregulated |
| -1,243861055 | PDE4C | | Downregulated |
| 1,327563592 | PDE4D | | Upregulated |
| 0,720901933 | PDE6A | FALSE | Upregulated |
| -0,08564188 | PDGFC | FALSE | Downregulated |
| 0,094276627 | PDHB | FALSE | Upregulated |
| -0,322512002 | PDHX | FALSE | Downregulated |
| -0,077366469 | PDS5B | FALSE | Downregulated |
| 0,300963112 | PEAR1 | FALSE | Upregulated |
| 0,281385648 | PELP1 | FALSE | Upregulated |
| -0,538681661 | PEMT | FALSE | Downregulated |
| -0,153208479 | PET112 | FALSE | Downregulated |
| -2,026708985 | PEX11A | | Downregulated |
| 0,127122006 | PEX11G | FALSE | Upregulated |
| -0,433937033 | PEX13 | FALSE | Downregulated |
| -1,133393768 | PEX14 | | Downregulated |
| 0,741220369 | PFAS | FALSE | Upregulated |
| 0,829108747 | PFDN1 | FALSE | Upregulated |
| 1,06105586 | PFKM | | Upregulated |
| 0,099002585 | PFN2 | FALSE | Upregulated |
| 0,554488135 | PGAM4 | FALSE | Upregulated |
| 1,328876316 | PGBD2 | | Upregulated |
| -0,120620032 | PGBD5 | FALSE | Downregulated |
| -0,340074895 | PGGT1B | FALSE | Downregulated |
| 0,143632025 | PGLS | FALSE | Upregulated |
| 0,673140412 | PGM5P2 | FALSE | Upregulated |
| -0,222555503 | PGP | FALSE | Downregulated |
| -0,302581547 | PGR | FALSE | Downregulated |
| 1,308315507 | PHF11 | | Upregulated |
| 0,27905422 | PHF14 | FALSE | Upregulated |
| -0,232621517 | PHF19 | FALSE | Downregulated |
| -0,668665398 | PHF21A | FALSE | Downregulated |
| -1,013118916 | PHF23 | | Downregulated |
| 0,007493655 | PHKA1 | FALSE | Upregulated |
| -0,94131534 | PHKG2 | FALSE | Downregulated |
| 2,009084225 | PHLDA1 | | Upregulated |
| -0,330577712 | PHYHD1 | FALSE | Downregulated |
| 1,094666019 | PI4KAP1 | | Upregulated |
| 0,014723936 | PIANP | FALSE | Upregulated |
| 0,557226038 | PIAS3 | FALSE | Upregulated |

| | | | |
|---|---|---|---|
| 0,304883939 | PIAS4 | FALSE | Upregulated |
| 1,482332768 | PIGG | | Upregulated |
| -0,532270742 | PIGH | FALSE | Downregulated |
| -0,201623113 | PIGL | FALSE | Downregulated |
| 0,037187343 | PIGM | FALSE | Upregulated |
| -0,406261817 | PIGO | FALSE | Downregulated |
| 0,994713762 | PIGZ | FALSE | Upregulated |
| 0,330318598 | PIK3C3 | FALSE | Upregulated |
| 0,099245432 | PIK3CD | FALSE | Upregulated |
| 1,026216089 | PIK3R5 | | Upregulated |
| 0,086704647 | PIN1 | FALSE | Upregulated |
| 0,325535327 | PIN4 | FALSE | Upregulated |
| 0,304089735 | PIP4K2A | FALSE | Upregulated |
| 0,132856325 | PIP5K1A | FALSE | Upregulated |
| -0,397869653 | PIP5K1P1 | FALSE | Downregulated |
| 0,483544388 | PIPSL | FALSE | Upregulated |
| -1,032092458 | PITPNA-AS1 | | Downregulated |
| -0,838035732 | PKD2 | FALSE | Downregulated |
| 1,195227945 | PKDCC | | Upregulated |
| -0,11042254 | PKIG | FALSE | Downregulated |
| 1,533961364 | PKNOX1 | | Upregulated |
| 1,207113572 | PLAA | | Upregulated |
| -0,146042209 | PLAGL2 | FALSE | Downregulated |
| 0,391674166 | PLAT | FALSE | Upregulated |
| 0,658236912 | PLBD1 | FALSE | Upregulated |
| 0,030440774 | PLCB1 | FALSE | Upregulated |
| -0,65081225 | PLD6 | FALSE | Downregulated |
| 0,960809266 | PLEK2 | FALSE | Upregulated |
| 0,6400087 | PLEKHA2 | FALSE | Upregulated |
| 0,727439317 | PLEKHA4 | FALSE | Upregulated |
| 0,557180243 | PLEKHB1 | FALSE | Upregulated |
| 0,971234751 | PLEKHB2 | FALSE | Upregulated |
| 0,092472085 | PLEKHF1 | FALSE | Upregulated |
| 0,920214942 | PLEKHG2 | FALSE | Upregulated |
| -0,820439294 | PLEKHH2 | FALSE | Downregulated |
| -0,817388394 | PLEKHH3 | FALSE | Downregulated |
| -0,190673506 | PLK1S1 | FALSE | Downregulated |
| -0,863535667 | PLOD2 | FALSE | Downregulated |
| -0,311520548 | PLSCR4 | FALSE | Downregulated |
| 1,910172534 | PLXNA3 | | Upregulated |
| -0,987904705 | PMF1 | FALSE | Downregulated |
| -0,364408174 | PMM1 | FALSE | Downregulated |
| -0,175753675 | PMS1 | FALSE | Downregulated |
| 0,013740304 | PMS2CL | FALSE | Upregulated |
| 0,649708389 | PMS2P5 | FALSE | Upregulated |
| 0,201288228 | PNKP | FALSE | Upregulated |
| 0,658369912 | PNMA6A | FALSE | Upregulated |
| 0,744753398 | PNMAL2 | FALSE | Upregulated |
| 0,206511053 | PNO1 | FALSE | Upregulated |
| -0,011050372 | PODN | FALSE | Downregulated |
| 0,071520222 | PODNL1 | FALSE | Upregulated |

| | | | |
|---|---|---|---|
| 1,078924111 | POGLUT1 | | Upregulated |
| -0,151199856 | POLA2 | FALSE | Downregulated |
| -0,465519063 | POLI | FALSE | Downregulated |
| 0,033111601 | POLK | FALSE | Upregulated |
| -0,464226196 | POLL | FALSE | Downregulated |
| -0,265772323 | POLM | FALSE | Downregulated |
| 0,017408172 | POLR1B | FALSE | Upregulated |
| -0,12931632 | POLR1D | FALSE | Downregulated |
| 0,38323343 | POLR1E | FALSE | Upregulated |
| -1,470249557 | POLR2F | | Downregulated |
| 0,64550603 | POLR2J | FALSE | Upregulated |
| 0,169684879 | POLR3A | FALSE | Upregulated |
| -0,225335732 | POLR3B | FALSE | Downregulated |
| -0,303345923 | POM121L9P | FALSE | Downregulated |
| -0,00616957 | POMGNT2 | FALSE | Downregulated |
| 0,619403574 | POMK | FALSE | Upregulated |
| 0,368061057 | POMT2 | FALSE | Upregulated |
| 0,235376579 | POMZP3 | FALSE | Upregulated |
| -0,474735212 | POT1 | FALSE | Downregulated |
| -0,24380645 | POU2F2 | FALSE | Downregulated |
| 0,761023469 | POU5F1 | FALSE | Upregulated |
| 0,649118675 | PP14571 | FALSE | Upregulated |
| -0,203596411 | PPA1 | FALSE | Downregulated |
| -0,023451866 | PPARGC1B | FALSE | Downregulated |
| -0,793131519 | PPAT | FALSE | Downregulated |
| 0,061091874 | PPCDC | FALSE | Upregulated |
| 1,127542053 | PPHLN1 | | Upregulated |
| 0,672173224 | PPIC | FALSE | Upregulated |
| 1,023014686 | PPIE | | Upregulated |
| -2,027615553 | PPM1K | | Downregulated |
| 0,496786418 | PPM1L | FALSE | Upregulated |
| -0,823083279 | PPME1 | FALSE | Downregulated |
| 0,226701913 | PPP1CC | FALSE | Upregulated |
| -0,629707128 | PPP1R11 | FALSE | Downregulated |
| 0,010224576 | PPP1R13L | FALSE | Upregulated |
| 0,492471757 | PPP1R15A | FALSE | Upregulated |
| 0,015692931 | PPP1R16B | FALSE | Upregulated |
| 0,824870827 | PPP1R2 | FALSE | Upregulated |
| 1,210297675 | PPP1R21 | | Upregulated |
| 0,675708037 | PPP1R35 | FALSE | Upregulated |
| 0,902897948 | PPP1R3F | FALSE | Upregulated |
| -0,063900671 | PPP1R7 | FALSE | Downregulated |
| -0,417998296 | PPP2CA | FALSE | Downregulated |
| -0,340061386 | PPP2R2A | FALSE | Downregulated |
| -1,166932005 | PPP2R3B | | Downregulated |
| 0,138940034 | PPP3CA | FALSE | Upregulated |
| 0,386173034 | PPP4C | FALSE | Upregulated |
| 1,336743273 | PPP4R1 | | Upregulated |
| 0,993146958 | PPTC7 | FALSE | Upregulated |
| -0,511859494 | PPWD1 | FALSE | Downregulated |
| 0,471876203 | PQLC3 | FALSE | Upregulated |

| | | | |
|---|---|---|---|
| 0,474858224 | PRAM1 | FALSE | Upregulated |
| 0,273749935 | PRCC | FALSE | Upregulated |
| -0,768419293 | PRDM10 | FALSE | Downregulated |
| 1,485772966 | PRDM15 | | Upregulated |
| 0,645729138 | PRDM4 | FALSE | Upregulated |
| -0,176657905 | PRDX5 | FALSE | Downregulated |
| -0,201957191 | PRELP | FALSE | Downregulated |
| 0,09902288 | PREP | FALSE | Upregulated |
| -0,362531896 | PRH1-PRR4 | FALSE | Downregulated |
| 0,227258157 | PRICKLE2 | FALSE | Upregulated |
| 1,206345373 | PRICKLE2-AS3 | | Upregulated |
| 0,301167075 | PRIMPOL | FALSE | Upregulated |
| -0,27109684 | PRKAR1B | FALSE | Downregulated |
| 0,887924252 | PRKCB | FALSE | Upregulated |
| -0,227848459 | PRKCH | FALSE | Downregulated |
| 0,379893308 | PRKCI | FALSE | Upregulated |
| 0,553833139 | PRKCQ | FALSE | Upregulated |
| -0,007191993 | PRKG1-AS1 | FALSE | Downregulated |
| -0,752177468 | PRKRIR | FALSE | Downregulated |
| 0,130634086 | PRKXP1 | FALSE | Upregulated |
| -0,00302851 | PRMT1 | FALSE | Downregulated |
| 0,049084443 | PRMT3 | FALSE | Upregulated |
| 0,39404598 | PRMT5 | FALSE | Upregulated |
| 0,151733731 | PRMT7 | FALSE | Upregulated |
| 0,191850526 | PRMT9 | FALSE | Upregulated |
| 0,187683904 | PRNP | FALSE | Upregulated |
| 0,260483518 | PROCA1 | FALSE | Upregulated |
| 1,196620641 | PROSER1 | | Upregulated |
| -1,318317154 | PROX1-AS1 | | Downregulated |
| -0,07461755 | PRPF38B | FALSE | Downregulated |
| -0,150486556 | PRPF39 | FALSE | Downregulated |
| -0,058365687 | PRPF4 | FALSE | Downregulated |
| -0,715492355 | PRPS1 | FALSE | Downregulated |
| -0,315695948 | PRR13 | FALSE | Downregulated |
| 0,222720161 | PRR14 | FALSE | Upregulated |
| 0,454420057 | PRR18 | FALSE | Upregulated |
| -0,584241637 | PRR5 | FALSE | Downregulated |
| 1,593010812 | PRR5L | | Upregulated |
| 1,08659545 | PRR7-AS1 | | Upregulated |
| -0,409612057 | PRRG1 | FALSE | Downregulated |
| 0,844751572 | PRSS23 | FALSE | Upregulated |
| -0,017132616 | PRSS37 | FALSE | Downregulated |
| -0,974245101 | PRSS53 | FALSE | Downregulated |
| 1,822722005 | PSD | | Upregulated |
| 0,07091658 | PSMA1 | FALSE | Upregulated |
| 0,177533858 | PSMA4 | FALSE | Upregulated |
| 0,197741902 | PSMA5 | FALSE | Upregulated |
| -0,971886685 | PSMA6 | FALSE | Downregulated |
| -0,177605394 | PSMC6 | FALSE | Downregulated |
| 0,588476882 | PSMD6 | FALSE | Upregulated |
| 0,168481141 | PSMD9 | FALSE | Upregulated |

| | | | |
|---|---|---|---|
| 0,528659686 | PSME3 | FALSE | Upregulated |
| -0,489506048 | PTBP2 | FALSE | Downregulated |
| -0,022440322 | PTCHD3P1 | FALSE | Downregulated |
| -0,309285194 | PTDSS1 | FALSE | Downregulated |
| -0,16197331 | PTDSS2 | FALSE | Downregulated |
| -0,190754043 | PTGDR2 | FALSE | Downregulated |
| 1,226562099 | PTGER2 | | Upregulated |
| -0,638616573 | PTGES2 | FALSE | Downregulated |
| 0,111647038 | PTK6 | FALSE | Upregulated |
| 0,964813124 | PTPLAD2 | FALSE | Upregulated |
| -0,195882096 | PTPN2 | FALSE | Downregulated |
| -0,152982569 | PTPN6 | FALSE | Downregulated |
| 0,050598341 | PTPN9 | FALSE | Upregulated |
| -0,436837903 | PTPRCAP | FALSE | Downregulated |
| -0,612855517 | PUS1 | FALSE | Downregulated |
| -0,171633834 | PVRL1 | FALSE | Downregulated |
| 0,902752204 | PWWP2B | FALSE | Upregulated |
| 0,49736148 | PYCR2 | FALSE | Upregulated |
| -0,411804049 | PYGO2 | FALSE | Downregulated |
| 0,372733352 | PYURF | FALSE | Upregulated |
| -0,936537896 | QPCTL | FALSE | Downregulated |
| -0,699057064 | QRICH2 | FALSE | Downregulated |
| 0,604101433 | QSOX2 | FALSE | Upregulated |
| -0,456136886 | QTRT1 | FALSE | Downregulated |
| 0,499161081 | R3HCC1L | FALSE | Upregulated |
| 0,876466613 | R3HDM4 | FALSE | Upregulated |
| 0,068338201 | RAB11A | FALSE | Upregulated |
| -0,974573101 | RAB11B-AS1 | FALSE | Downregulated |
| 0,285177111 | RAB11FIP4 | FALSE | Upregulated |
| 0,118015851 | RAB11FIP5 | FALSE | Upregulated |
| 1,068112959 | RAB12 | | Upregulated |
| 0,413990154 | RAB20 | FALSE | Upregulated |
| 0,326487227 | RAB21 | FALSE | Upregulated |
| 0,035569593 | RAB24 | FALSE | Upregulated |
| -0,738697059 | RAB27A | FALSE | Downregulated |
| -0,300148534 | RAB2A | FALSE | Downregulated |
| -0,191654378 | RAB30 | FALSE | Downregulated |
| -0,598065572 | RAB37 | FALSE | Downregulated |
| -0,279608248 | RAB3GAP1 | FALSE | Downregulated |
| 0,918917449 | RAB3IP | FALSE | Upregulated |
| 0,356182545 | RAB4A | FALSE | Upregulated |
| 0,473135121 | RABEP2 | FALSE | Upregulated |
| -0,491762538 | RABEPK | FALSE | Downregulated |
| 0,177808525 | RABGGTA | FALSE | Upregulated |
| 0,027291613 | RABIF | FALSE | Upregulated |
| 0,432144495 | RABL5 | FALSE | Upregulated |
| -0,218658245 | RAD1 | FALSE | Downregulated |
| -0,212685771 | RAD17 | FALSE | Downregulated |
| 1,168971358 | RAD51-AS1 | | Upregulated |
| 0,064976989 | RAET1E | FALSE | Upregulated |
| 0,706778556 | RALB | FALSE | Upregulated |

| | | | |
|---|---|---|---|
| 1,339177575 | RAMP1 | | Upregulated |
| -2,660665309 | RANBP3L | | Downregulated |
| 1,022633325 | RANGRF | | Upregulated |
| 0,635178645 | RAP1A | FALSE | Upregulated |
| 1,207664286 | RAP1GAP | | Upregulated |
| -0,961458038 | RAPGEF4 | FALSE | Downregulated |
| 1,186057971 | RAPGEF6 | | Upregulated |
| -0,601273173 | RAPGEFL1 | FALSE | Downregulated |
| -0,027620028 | RARG | FALSE | Downregulated |
| 0,150602827 | RARS2 | FALSE | Upregulated |
| 1,85587416 | RASAL3 | | Upregulated |
| -0,548316505 | RASGEF1B | FALSE | Downregulated |
| 0,544356978 | RASGRP4 | FALSE | Upregulated |
| -0,730994405 | RAVER1 | FALSE | Downregulated |
| 0,934840935 | RB1 | FALSE | Upregulated |
| -0,414736975 | RBAK | FALSE | Downregulated |
| 0,804803876 | RBFA | FALSE | Upregulated |
| 0,032344077 | RBM23 | FALSE | Upregulated |
| 0,540729021 | RBM27 | FALSE | Upregulated |
| -0,192975485 | RBM38 | FALSE | Downregulated |
| -0,990182097 | RBM43 | FALSE | Downregulated |
| 0,542612442 | RBM7 | FALSE | Upregulated |
| -0,114297892 | RBMXL1 | FALSE | Downregulated |
| -0,33988441 | RBP1 | FALSE | Downregulated |
| -0,100393123 | RC3H2 | FALSE | Downregulated |
| 1,487570813 | RCBTB1 | | Upregulated |
| 0,745209442 | RCBTB2 | FALSE | Upregulated |
| -0,182192497 | RCC1 | FALSE | Downregulated |
| 0,306095401 | RCHY1 | FALSE | Upregulated |
| -0,077320867 | RCOR1 | FALSE | Downregulated |
| 0,778304422 | RCSD1 | FALSE | Upregulated |
| -0,259509069 | RDH13 | FALSE | Downregulated |
| -0,394920504 | REL | FALSE | Downregulated |
| 0,220833842 | RELT | FALSE | Upregulated |
| -0,029543537 | REXO4 | FALSE | Downregulated |
| -0,43057404 | RFK | FALSE | Downregulated |
| -0,269753689 | RFTN2 | FALSE | Downregulated |
| 1,333196242 | RFX5 | | Upregulated |
| -0,593643585 | RFX7 | FALSE | Downregulated |
| -0,123766239 | RFXANK | FALSE | Downregulated |
| 0,708965226 | RGL3 | FALSE | Upregulated |
| 0,785916379 | RGMB | FALSE | Upregulated |
| -0,775790647 | RGS17 | FALSE | Downregulated |
| -1,182091385 | RGS5 | | Downregulated |
| -0,080612368 | RHBDD3 | FALSE | Downregulated |
| 1,875069849 | RHBDF2 | | Upregulated |
| 0,267069828 | RHCG | FALSE | Upregulated |
| 0,315733239 | RHOG | FALSE | Upregulated |
| 0,109978995 | RHPN2 | FALSE | Upregulated |
| 0,594486624 | RIN2 | FALSE | Upregulated |
| 1,090972838 | RIT1 | | Upregulated |

| | | | |
|---|---|---|---|
| -0,450962387 | RITA1 | FALSE | Downregulated |
| 0,277801584 | RLF | FALSE | Upregulated |
| -0,440215845 | RMDN3 | FALSE | Downregulated |
| -0,142395392 | RMI1 | FALSE | Downregulated |
| 0,708619032 | RMND1 | FALSE | Upregulated |
| 0,078788206 | RMND5B | FALSE | Upregulated |
| -0,571251013 | RNASET2 | FALSE | Downregulated |
| 0,475114013 | RND3 | FALSE | Upregulated |
| -0,038349871 | RNF114 | FALSE | Downregulated |
| 0,25485633 | RNF121 | FALSE | Upregulated |
| -0,566545886 | RNF125 | FALSE | Downregulated |
| 1,270031457 | RNF135 | | Upregulated |
| 0,058300452 | RNF138 | FALSE | Upregulated |
| -0,251277214 | RNF14 | FALSE | Downregulated |
| 0,205361029 | RNF149 | FALSE | Upregulated |
| 0,383365046 | RNF165 | FALSE | Upregulated |
| -0,514377513 | RNF19B | FALSE | Downregulated |
| 0,156472598 | RNF20 | FALSE | Upregulated |
| 0,725489287 | RNF207 | FALSE | Upregulated |
| -0,148309825 | RNF214 | FALSE | Downregulated |
| 0,222009684 | RNF222 | FALSE | Upregulated |
| 0,136367197 | RNF25 | FALSE | Upregulated |
| 0,628208971 | RNF31 | FALSE | Upregulated |
| 0,027667513 | RNF41 | FALSE | Upregulated |
| 0,088282696 | RNF6 | FALSE | Upregulated |
| 0,388418546 | RNF7 | FALSE | Upregulated |
| -0,631685825 | RNFT1 | FALSE | Downregulated |
| -0,007030414 | RNMTL1 | FALSE | Downregulated |
| -0,393178025 | RNU2-2P | FALSE | Downregulated |
| 0,332402185 | RNU4-1 | FALSE | Upregulated |
| 0,310640763 | ROBO4 | FALSE | Upregulated |
| -0,351546848 | ROGDI | FALSE | Downregulated |
| -1,102678933 | ROM1 | | Downregulated |
| 0,510908078 | RP1-1J6.2 | FALSE | Upregulated |
| 0,48488004 | RP1-256G22.2 | FALSE | Upregulated |
| -0,050822774 | RP11-1007G5.2 | FALSE | Downregulated |
| -0,103286292 | RP11-1072A3.2 | FALSE | Downregulated |
| -1,562356272 | RP11-1096D5.1 | | Downregulated |
| -0,013837892 | RP11-118E18.4 | FALSE | Downregulated |
| 0,536979301 | RP11-1H15.2 | FALSE | Upregulated |
| -0,467278978 | RP11-203B9.4 | FALSE | Downregulated |
| -0,39487454 | RP11-234K24.3 | FALSE | Downregulated |
| 0,124775606 | RP11-266K4.9 | FALSE | Upregulated |
| 0,597182677 | RP11-279D17.2 | FALSE | Upregulated |
| 0,060225655 | RP11-365O16.6 | FALSE | Upregulated |
| 0,664221415 | RP11-379J5.5 | FALSE | Upregulated |
| 0,367481632 | RP11-387D10.3 | FALSE | Upregulated |
| 0,731865291 | RP11-41O4.2 | FALSE | Upregulated |
| 0,573016875 | RP11-43N5.1 | FALSE | Upregulated |
| -0,697358186 | RP11-473M20.7 | FALSE | Downregulated |
| 0,915805055 | RP11-480I12.7 | FALSE | Upregulated |

| | | | |
|---|---|---|---|
| 0,123385768 | RP11-508N12.2 | FALSE | Upregulated |
| 0,107478187 | RP11-521B24.3 | FALSE | Upregulated |
| 0,652550936 | RP11-538P18.2 | FALSE | Upregulated |
| -0,79040154 | RP11-622A1.2 | FALSE | Downregulated |
| -1,082220207 | RP11-665G4.1 | | Downregulated |
| 0,047564642 | RP11-890B15.2 | FALSE | Upregulated |
| 1,899230175 | RP11-996F15.2 | | Upregulated |
| -0,659293701 | RP2 | FALSE | Downregulated |
| -0,28047497 | RP3-395M20.9 | FALSE | Downregulated |
| 1,06006234 | RP3-400N23.6 | | Upregulated |
| 0,558406152 | RP4-777D9.2 | FALSE | Upregulated |
| 0,177923228 | RP5-1029K10.2 | FALSE | Upregulated |
| 0,228673347 | RP5-1180C10.2 | FALSE | Upregulated |
| 0,792872945 | RP6-109B7.3 | FALSE | Upregulated |
| 0,807931583 | RPA1 | FALSE | Upregulated |
| 0,232929551 | RPAP1 | FALSE | Upregulated |
| 1,097183944 | RPAP2 | | Upregulated |
| -0,265634561 | RPH3AL | FALSE | Downregulated |
| -0,072874863 | RPL31P11 | FALSE | Downregulated |
| -0,239595563 | RPP14 | FALSE | Downregulated |
| -0,126018259 | RPS18P9 | FALSE | Downregulated |
| -0,157291803 | RPS19BP1 | FALSE | Downregulated |
| -0,0085391 | RPS2P32 | FALSE | Downregulated |
| 2,255769398 | RPS6KA1 | | Upregulated |
| 0,935303436 | RPS6KA4 | FALSE | Upregulated |
| -0,146794317 | RPS6KB2 | FALSE | Downregulated |
| 0,613396975 | RPS6KC1 | FALSE | Upregulated |
| 0,284182423 | RRAGC | FALSE | Upregulated |
| 0,001383923 | RRN3 | FALSE | Upregulated |
| 0,054842991 | RRNAD1 | FALSE | Upregulated |
| 0,880046631 | RRP7B | FALSE | Upregulated |
| -1,031099789 | RRP8 | | Downregulated |
| -0,891217928 | RSAD1 | FALSE | Downregulated |
| 0,694121821 | RSBN1 | FALSE | Upregulated |
| 0,195769863 | RSBN1L | FALSE | Upregulated |
| -0,216290612 | RSL24D1 | FALSE | Downregulated |
| -0,061704597 | RSPH3 | FALSE | Downregulated |
| 0,816558172 | RSPH9 | FALSE | Upregulated |
| -0,331958577 | RSPRY1 | FALSE | Downregulated |
| 0,204421027 | RSRC2 | FALSE | Upregulated |
| 0,166982894 | RTF1 | FALSE | Upregulated |
| -0,1713116 | RTP3 | FALSE | Downregulated |
| 0,085103573 | RTTN | FALSE | Upregulated |
| -1,152073506 | RWDD2B | | Downregulated |
| 0,128697574 | RYBP | FALSE | Upregulated |
| 1,612445106 | S100A10 | | Upregulated |
| 0,483796372 | SACM1L | FALSE | Upregulated |
| -0,438142865 | SAMD5 | FALSE | Downregulated |
| 0,993522827 | SAMD8 | FALSE | Upregulated |
| 0,031414565 | SAP130 | FALSE | Upregulated |
| -0,899134699 | SAP18 | FALSE | Downregulated |

| | | | |
|---|---|---|---|
| -0,574949607 | SARNP | FALSE | Downregulated |
| 0,171262086 | SASS6 | FALSE | Upregulated |
| -0,482355114 | SAV1 | FALSE | Downregulated |
| -0,123857749 | SBDS | FALSE | Downregulated |
| 0,511807363 | SCAMP1 | FALSE | Upregulated |
| -1,963801483 | SCAMP3 | | Downregulated |
| 0,011035587 | SCAMP4 | FALSE | Upregulated |
| 0,949487053 | SCARF2 | FALSE | Upregulated |
| -0,17239492 | SCARNA6 | FALSE | Downregulated |
| 0,800400359 | SCFD1 | FALSE | Upregulated |
| 0,195733757 | SCFD2 | FALSE | Upregulated |
| 0,694416892 | SCG3 | FALSE | Upregulated |
| 0,436798788 | SCML1 | FALSE | Upregulated |
| -0,303521357 | SCNN1A | FALSE | Downregulated |
| -0,150953721 | SCNN1D | FALSE | Downregulated |
| -0,150924584 | SCOC | FALSE | Downregulated |
| 0,043506715 | SCRN2 | FALSE | Upregulated |
| 0,216468693 | SCT | FALSE | Upregulated |
| 0,756832281 | SCXB | FALSE | Upregulated |
| 0,116560758 | SDAD1 | FALSE | Upregulated |
| 0,081359519 | SDCCAG3 | FALSE | Upregulated |
| -0,12591835 | SDCCAG8 | FALSE | Downregulated |
| 0,172017691 | SDHAF1 | FALSE | Upregulated |
| -0,861875989 | SDPR | FALSE | Downregulated |
| -0,460290733 | SEC11A | FALSE | Downregulated |
| -0,349213 | SEC1P | FALSE | Downregulated |
| 1,12957862 | SEC22C | | Upregulated |
| 0,755368 | SEC23B | FALSE | Upregulated |
| 0,134026102 | SEC61B | FALSE | Upregulated |
| 0,817423221 | SEL1L3 | FALSE | Upregulated |
| 0,926716514 | SELT | FALSE | Upregulated |
| -0,04784048 | SEP15 | FALSE | Downregulated |
| -1,758868353 | SEPT10 | | Downregulated |
| -0,038672325 | SEPT6 | FALSE | Downregulated |
| 0,875650817 | SEPT7 | FALSE | Upregulated |
| 0,887700664 | SEPT8 | FALSE | Upregulated |
| -0,143185582 | SEPW1 | FALSE | Downregulated |
| 0,548761046 | SERAC1 | FALSE | Upregulated |
| 0,967403451 | SERPINB1 | FALSE | Upregulated |
| 0,394380747 | SERPINH1 | FALSE | Upregulated |
| -0,579386079 | SERTAD2 | FALSE | Downregulated |
| 0,284225079 | SESN1 | FALSE | Upregulated |
| -0,079343723 | SESN2 | FALSE | Downregulated |
| 0,671965745 | SESTD1 | FALSE | Upregulated |
| -0,500814517 | SETD3 | FALSE | Downregulated |
| 0,764281254 | SETD8 | FALSE | Upregulated |
| -0,240402776 | SETDB2 | FALSE | Downregulated |
| -0,310872445 | SF3A2 | FALSE | Downregulated |
| 0,802457215 | SF3A3 | FALSE | Upregulated |
| 1,55012981 | SF3B4 | | Upregulated |
| 0,360428184 | SF3B5 | FALSE | Upregulated |

| | | | |
|---|---|---|---|
| 0,175133835 | SFI1 | FALSE | Upregulated |
| 1,429010255 | SFMBT2 | | Upregulated |
| -0,7378619 | SFSWAP | FALSE | Downregulated |
| -0,839944071 | SFTPD | FALSE | Downregulated |
| -0,448843475 | SFXN2 | FALSE | Downregulated |
| 0,159126288 | SFXN4 | FALSE | Upregulated |
| -0,570969212 | SGCE | FALSE | Downregulated |
| 2,160671223 | SGK223 | | Upregulated |
| 0,781656595 | SGOL1 | FALSE | Upregulated |
| 0,429761948 | SGPP1 | FALSE | Upregulated |
| 0,682703727 | SGTB | FALSE | Upregulated |
| 0,141445555 | SH2D4A | FALSE | Upregulated |
| 0,957379135 | SH3BGRL3 | FALSE | Upregulated |
| 1,062741632 | SH3BP5L | | Upregulated |
| -0,249227073 | SH3KBP1 | FALSE | Downregulated |
| -0,069916258 | SH3RF1 | FALSE | Downregulated |
| 1,813956914 | SH3RF3 | | Upregulated |
| 0,361475858 | SHBG | FALSE | Upregulated |
| -0,369885749 | SHH | FALSE | Downregulated |
| -1,193404531 | SHISA6 | | Downregulated |
| -0,646305313 | SHISA9 | FALSE | Downregulated |
| 1,662641287 | SHKBP1 | | Upregulated |
| 0,042477134 | SHOC2 | FALSE | Upregulated |
| -1,290165428 | SIDT1 | | Downregulated |
| -0,883449221 | SIMC1 | FALSE | Downregulated |
| -0,204166683 | SIRT2 | FALSE | Downregulated |
| -1,192322573 | SIRT7 | | Downregulated |
| 0,140334261 | SIVA1 | FALSE | Upregulated |
| -0,051408604 | SKP2 | FALSE | Downregulated |
| -1,138141552 | SLC10A5 | | Downregulated |
| 0,43602037 | SLC11A1 | FALSE | Upregulated |
| 1,048139525 | SLC12A2 | | Upregulated |
| -0,461858896 | SLC12A5 | FALSE | Downregulated |
| 0,139558118 | SLC12A9 | FALSE | Upregulated |
| -0,644658 | SLC15A1 | FALSE | Downregulated |
| -1,032216687 | SLC16A11 | | Downregulated |
| 1,215884697 | SLC16A13 | | Upregulated |
| 0,615782497 | SLC16A6 | FALSE | Upregulated |
| 1,487674559 | SLC16A7 | | Upregulated |
| -0,280115341 | SLC17A1 | FALSE | Downregulated |
| 0,328125091 | SLC17A5 | FALSE | Upregulated |
| -1,25141071 | SLC19A1 | | Downregulated |
| -1,548830725 | SLC19A2 | | Downregulated |
| 1,391667205 | SLC22A18AS | | Upregulated |
| 0,489406639 | SLC22A5 | FALSE | Upregulated |
| 0,003884766 | SLC25A27 | FALSE | Upregulated |
| -0,964762705 | SLC25A32 | FALSE | Downregulated |
| 1,694285597 | SLC25A33 | | Upregulated |
| 0,730381534 | SLC25A35 | FALSE | Upregulated |
| -0,072273151 | SLC25A40 | FALSE | Downregulated |
| 1,280635196 | SLC25A45 | | Upregulated |

| | | | |
|---|---|---|---|
| -0,605934271 | SLC25A51 | FALSE | Downregulated |
| 1,118418107 | SLC26A11 | | Upregulated |
| -0,214660694 | SLC26A8 | FALSE | Downregulated |
| -0,729593277 | SLC28A2 | FALSE | Downregulated |
| -0,834424096 | SLC29A1 | FALSE | Downregulated |
| -0,648764938 | SLC2A11 | FALSE | Downregulated |
| -0,117440598 | SLC2A8 | FALSE | Downregulated |
| 0,121808771 | SLC30A5 | FALSE | Upregulated |
| 0,347301514 | SLC30A9 | FALSE | Upregulated |
| 0,120759078 | SLC33A1 | FALSE | Upregulated |
| 1,165381183 | SLC35A4 | | Upregulated |
| -0,264727393 | SLC35B3 | FALSE | Downregulated |
| 0,581620814 | SLC35C2 | FALSE | Upregulated |
| 0,575655413 | SLC35E3 | FALSE | Upregulated |
| -0,161462115 | SLC35F5 | FALSE | Downregulated |
| 0,752165805 | SLC35F6 | FALSE | Upregulated |
| 0,987235266 | SLC37A1 | FALSE | Upregulated |
| 0,933844472 | SLC38A1 | FALSE | Upregulated |
| -0,108280268 | SLC38A11 | FALSE | Downregulated |
| -0,270755469 | SLC3A2 | FALSE | Downregulated |
| 1,356951253 | SLC43A2 | | Upregulated |
| 0,389706778 | SLC44A2 | FALSE | Upregulated |
| 1,433955312 | SLC44A3 | | Upregulated |
| -1,455932056 | SLC45A3 | | Downregulated |
| -0,070721732 | SLC4A1AP | FALSE | Downregulated |
| 1,826882814 | SLC4A5 | | Upregulated |
| -0,148725961 | SLC4A7 | FALSE | Downregulated |
| 0,294775887 | SLC50A1 | FALSE | Upregulated |
| -1,346212531 | SLC6A13 | | Downregulated |
| 2,094265641 | SLC6A6 | | Upregulated |
| 0,273915301 | SLC7A10 | FALSE | Upregulated |
| 1,702141964 | SLC7A5 | | Upregulated |
| 1,827866726 | SLC7A5P2 | | Upregulated |
| 0,423483154 | SLC7A8 | FALSE | Upregulated |
| 0,765283714 | SLC8A1-AS1 | FALSE | Upregulated |
| 0,867186 | SLC8A3 | FALSE | Upregulated |
| -0,993141102 | SLC9A9 | FALSE | Downregulated |
| -1,011815336 | SLC9B2 | | Downregulated |
| 0,84331211 | SLFN14 | FALSE | Upregulated |
| 0,714325109 | SLFN5 | FALSE | Upregulated |
| 1,215721091 | SLIT1 | | Upregulated |
| -0,019931466 | SLIT2 | FALSE | Downregulated |
| 0,574234951 | SLPI | FALSE | Upregulated |
| -0,115732463 | SLU7 | FALSE | Downregulated |
| 0,206940044 | SLX1B | FALSE | Upregulated |
| 0,075799316 | SLX4 | FALSE | Upregulated |
| 0,185601575 | SMAD6 | FALSE | Upregulated |
| 0,678556487 | SMAGP | FALSE | Upregulated |
| 0,507713355 | SMAP1 | FALSE | Upregulated |
| 0,405417756 | SMARCAL1 | FALSE | Upregulated |
| 0,349444762 | SMC1B | FALSE | Upregulated |

| | | | |
|---|---|---|---|
| -1,10953113 | SMC3 | | Downregulated |
| 0,844245154 | SMC4 | FALSE | Upregulated |
| -0,729250226 | SMC5 | FALSE | Downregulated |
| 0,108894059 | SMC6 | FALSE | Upregulated |
| 0,094080768 | SMDT1 | FALSE | Upregulated |
| -0,496084319 | SMIM13 | FALSE | Downregulated |
| -0,73133055 | SMIM18 | FALSE | Downregulated |
| 1,062267173 | SMTNL1 | | Upregulated |
| -0,08789326 | SMURF2 | FALSE | Downregulated |
| 0,432702552 | SMYD2 | FALSE | Upregulated |
| 0,199974658 | SMYD5 | FALSE | Upregulated |
| 0,588607622 | SNAP23 | FALSE | Upregulated |
| 0,512743194 | SNAP25 | FALSE | Upregulated |
| -0,568903453 | SNAPIN | FALSE | Downregulated |
| -0,66582535 | SND1-IT1 | FALSE | Downregulated |
| -0,754669268 | SNHG7 | FALSE | Downregulated |
| 0,942773296 | SNORA48 | FALSE | Upregulated |
| -1,050107551 | SNORD17 | | Downregulated |
| 0,645505227 | SNPH | FALSE | Upregulated |
| -0,606348196 | SNRNP25 | FALSE | Downregulated |
| -0,286323862 | SNRPC | FALSE | Downregulated |
| -0,024057321 | SNTA1 | FALSE | Downregulated |
| 0,084575753 | SNUPN | FALSE | Upregulated |
| -0,001194868 | SNX11 | FALSE | Downregulated |
| -0,014005711 | SNX12 | FALSE | Downregulated |
| -0,200151094 | SNX13 | FALSE | Downregulated |
| -0,254719939 | SNX14 | FALSE | Downregulated |
| 0,234213492 | SNX20 | FALSE | Upregulated |
| -0,146995755 | SNX25 | FALSE | Downregulated |
| -0,603578785 | SNX27 | FALSE | Downregulated |
| -0,734372009 | SNX3 | FALSE | Downregulated |
| 0,98646005 | SNX30 | FALSE | Upregulated |
| 0,857905999 | SNX32 | FALSE | Upregulated |
| -0,377387354 | SNX4 | FALSE | Downregulated |
| -0,070205838 | SNX5 | FALSE | Downregulated |
| -0,076305434 | SNX6 | FALSE | Downregulated |
| 0,533290964 | SNX8 | FALSE | Upregulated |
| -0,525752197 | SOAT1 | FALSE | Downregulated |
| -2,124827434 | SOCS2 | | Downregulated |
| 0,927609039 | SOCS3 | FALSE | Upregulated |
| 0,634006556 | SOCS4 | FALSE | Upregulated |
| 1,308552213 | SOCS7 | | Upregulated |
| 2,211509082 | SOX9 | | Upregulated |
| -0,944485723 | SP110 | FALSE | Downregulated |
| 0,776329889 | SP140L | FALSE | Upregulated |
| -0,562046759 | SP7 | FALSE | Downregulated |
| 0,865508994 | SPACA6P-AS | FALSE | Upregulated |
| 0,788339449 | SPAG1 | FALSE | Upregulated |
| 0,802738965 | SPAG5 | FALSE | Upregulated |
| 0,854101319 | SPAG8 | FALSE | Upregulated |
| -0,002654319 | SPAST | FALSE | Downregulated |

| | | | |
|---|---|---|---|
| 0,651870229 | SPATA25 | FALSE | Upregulated |
| 0,030235637 | SPATA6 | FALSE | Upregulated |
| 0,044721789 | SPATA7 | FALSE | Upregulated |
| 0,465508489 | SPCS1 | FALSE | Upregulated |
| 0,066142863 | SPCS2 | FALSE | Upregulated |
| 0,090289547 | SPDYA | FALSE | Upregulated |
| -0,025289496 | SPDYE4 | FALSE | Downregulated |
| -0,25019913 | SPDYE7P | FALSE | Downregulated |
| 1,047778957 | SPECC1 | | Upregulated |
| 0,705440046 | SPEG | FALSE | Upregulated |
| 0,333261896 | SPG21 | FALSE | Upregulated |
| 1,3587017 | SPI1 | | Upregulated |
| -0,498644323 | SPICE1 | FALSE | Downregulated |
| 0,3749502 | SPIN3 | FALSE | Upregulated |
| 1,053443818 | SPIRE1 | | Upregulated |
| 0,094482369 | SPOP | FALSE | Upregulated |
| 0,316941287 | SPOPL | FALSE | Upregulated |
| 0,014573421 | SPPL3 | FALSE | Upregulated |
| -0,047134651 | SPR | FALSE | Downregulated |
| 0,731476053 | SPRED1 | FALSE | Upregulated |
| 0,761312869 | SPRY1 | FALSE | Upregulated |
| -0,495640955 | SPRY2 | FALSE | Downregulated |
| -0,542260297 | SPRY3 | FALSE | Downregulated |
| 1,139284592 | SPRY4 | | Upregulated |
| 2,148615714 | SPSB1 | | Upregulated |
| -0,284953607 | SPTBN5 | FALSE | Downregulated |
| -0,581280484 | SPTLC1 | FALSE | Downregulated |
| 0,234961949 | SPTLC2 | FALSE | Upregulated |
| 0,928726467 | SPTLC3 | FALSE | Upregulated |
| -0,53416001 | SPTY2D1 | FALSE | Downregulated |
| -0,497507856 | SREK1IP1 | FALSE | Downregulated |
| -0,392591886 | SRF | FALSE | Downregulated |
| 0,219327461 | SRM | FALSE | Upregulated |
| 0,395813223 | SRP14 | FALSE | Upregulated |
| -0,502430311 | SRP68 | FALSE | Downregulated |
| -0,716237689 | SRRD | FALSE | Downregulated |
| 0,318534926 | SRSF3 | FALSE | Upregulated |
| -0,160638467 | SRSF8 | FALSE | Downregulated |
| 1,617489208 | SSBP2 | | Upregulated |
| 0,587594422 | SSBP4 | FALSE | Upregulated |
| 0,809965478 | SSC5D | FALSE | Upregulated |
| 1,039441468 | SSH3 | | Upregulated |
| -0,507570781 | SSPO | FALSE | Downregulated |
| -0,298160224 | SSR2 | FALSE | Downregulated |
| -0,630777074 | SSR4 | FALSE | Downregulated |
| 2,699579482 | ST14 | | Upregulated |
| -0,399073331 | ST3GAL3 | FALSE | Downregulated |
| 0,489296323 | ST3GAL4 | FALSE | Upregulated |
| -0,926148727 | ST7 | FALSE | Downregulated |
| -0,247885676 | STAM2 | FALSE | Downregulated |
| -0,599597087 | STAMBP | FALSE | Downregulated |

| | | | |
|---|---|---|---|
| 0,198174034 | STAU2 | FALSE | Upregulated |
| -1,120710008 | STAU2-AS1 | | Downregulated |
| -0,370675769 | STEAP2 | FALSE | Downregulated |
| 0,099914821 | STEAP4 | FALSE | Upregulated |
| 0,080194788 | STK16 | FALSE | Upregulated |
| 0,070895181 | STK36 | FALSE | Upregulated |
| 0,209350041 | STK38 | FALSE | Upregulated |
| -0,231606418 | STK38L | FALSE | Downregulated |
| 0,689531341 | STON2 | FALSE | Upregulated |
| 0,384811979 | STRA13 | FALSE | Upregulated |
| -0,179072664 | STRAP | FALSE | Downregulated |
| -0,095225871 | STRBP | FALSE | Downregulated |
| 0,139116402 | STRIP1 | FALSE | Upregulated |
| -0,919026028 | STS | FALSE | Downregulated |
| -0,008541852 | STX12 | FALSE | Downregulated |
| 0,068559857 | STX18 | FALSE | Upregulated |
| 0,410891321 | STX1A | FALSE | Upregulated |
| -0,647739285 | STX1B | FALSE | Downregulated |
| 0,519058081 | STX2 | FALSE | Upregulated |
| 0,802333069 | STX5 | FALSE | Upregulated |
| 0,259669138 | STX7 | FALSE | Upregulated |
| -0,139735714 | STXBP2 | FALSE | Downregulated |
| -0,234811149 | STXBP3 | FALSE | Downregulated |
| 0,768726734 | SUB1 | FALSE | Upregulated |
| -0,704523828 | SUCLG1 | FALSE | Downregulated |
| 0,63533305 | SUCO | FALSE | Upregulated |
| -0,581979892 | SUDS3 | FALSE | Downregulated |
| -0,043141703 | SUFU | FALSE | Downregulated |
| -0,789131471 | SUGP1 | FALSE | Downregulated |
| -0,07347687 | SUMF1 | FALSE | Downregulated |
| -1,628463497 | SUMO1 | | Downregulated |
| -0,428240759 | SUMO2 | FALSE | Downregulated |
| -0,534980219 | SUPT16H | FALSE | Downregulated |
| 1,516215388 | SUPT20H | | Upregulated |
| 0,140414797 | SURF6 | FALSE | Upregulated |
| 0,440280899 | SUSD3 | FALSE | Upregulated |
| 1,204233762 | SUV39H2 | | Upregulated |
| -0,759285329 | SVIP | FALSE | Downregulated |
| 0,904667884 | SWAP70 | FALSE | Upregulated |
| 1,015512629 | SYK | | Upregulated |
| -0,131649819 | SYNGR1 | FALSE | Downregulated |
| 0,844963975 | SYNGR2 | FALSE | Upregulated |
| -0,344527053 | SYNJ1 | FALSE | Downregulated |
| 0,319020476 | SYPL1 | FALSE | Upregulated |
| -0,480463039 | SYPL2 | FALSE | Downregulated |
| -0,299334919 | SYT1 | FALSE | Downregulated |
| 1,481871571 | SYT12 | | Upregulated |
| -0,805959302 | SYT17 | FALSE | Downregulated |
| 1,235367855 | SYTL2 | | Upregulated |
| -0,829626368 | TADA1 | FALSE | Downregulated |
| 0,70831178 | TAF10 | FALSE | Upregulated |

| | | | |
|---|---|---|---|
| -0,541771764 | TAF15 | FALSE | Downregulated |
| 0,043168855 | TAF1D | FALSE | Upregulated |
| -0,698234101 | TAF3 | FALSE | Downregulated |
| -0,173005637 | TAF6 | FALSE | Downregulated |
| 0,369494668 | TAF9 | FALSE | Upregulated |
| 0,957385768 | TANC2 | FALSE | Upregulated |
| 0,847531716 | TANGO2 | FALSE | Upregulated |
| -0,331019299 | TANK | FALSE | Downregulated |
| 0,798353276 | TAP2 | FALSE | Upregulated |
| -0,811826811 | TAPT1 | FALSE | Downregulated |
| -0,701848663 | TAPT1-AS1 | FALSE | Downregulated |
| -0,222946295 | TARS2 | FALSE | Downregulated |
| 0,362144107 | TAS1R3 | FALSE | Upregulated |
| 0,457733247 | TAZ | FALSE | Upregulated |
| 0,322775018 | TBC1D2 | FALSE | Upregulated |
| 0,759817625 | TBC1D20 | FALSE | Upregulated |
| 0,960635413 | TBC1D22B | FALSE | Upregulated |
| -0,998221196 | TBC1D23 | FALSE | Downregulated |
| 0,255395131 | TBC1D25 | FALSE | Upregulated |
| -0,153428961 | TBC1D8B | FALSE | Downregulated |
| 0,101986893 | TBCB | FALSE | Upregulated |
| 0,572234489 | TBCCD1 | FALSE | Upregulated |
| 0,857420036 | TBKBP1 | FALSE | Upregulated |
| 0,583286874 | TBL2 | FALSE | Upregulated |
| -0,376325158 | TBRG4 | FALSE | Downregulated |
| 0,619956011 | TBX2 | FALSE | Upregulated |
| -0,444527302 | TCEAL3 | FALSE | Downregulated |
| 0,544746143 | TCERG1 | FALSE | Upregulated |
| -0,133273122 | TCF4 | FALSE | Downregulated |
| 0,255597375 | TCN2 | FALSE | Upregulated |
| 0,222550465 | TCP11L1 | FALSE | Upregulated |
| 0,442346193 | TEAD3 | FALSE | Upregulated |
| 0,370446728 | TEFM | FALSE | Upregulated |
| 0,704036053 | TENM2 | FALSE | Upregulated |
| -0,594801548 | TERF2 | FALSE | Downregulated |
| -0,425310586 | TET1 | FALSE | Downregulated |
| -1,07559261 | TEX10 | | Downregulated |
| -1,471636369 | TEX264 | | Downregulated |
| -0,265192874 | TFAM | FALSE | Downregulated |
| 0,362132573 | TFAP2E | FALSE | Upregulated |
| 1,201006508 | TFB1M | | Upregulated |
| -0,173606096 | TFCP2 | FALSE | Downregulated |
| 0,01725247 | TFDP1 | FALSE | Upregulated |
| -0,003688532 | TFEB | FALSE | Downregulated |
| -0,725051053 | TFG | FALSE | Downregulated |
| 0,940513639 | TGFB3 | FALSE | Upregulated |
| -0,013697681 | TGFBRAP1 | FALSE | Downregulated |
| 0,247283116 | TGIF1 | FALSE | Upregulated |
| -0,436912089 | TGIF2 | FALSE | Downregulated |
| -0,967157519 | TGS1 | FALSE | Downregulated |
| 0,395263487 | THAP11 | FALSE | Upregulated |

| | | | |
|---|---|---|---|
| 0,910444215 | THAP2 | FALSE | Upregulated |
| -1,094680644 | THAP7-AS1 | | Downregulated |
| 1,158668745 | THAP9 | | Upregulated |
| -0,590591464 | THEM4 | FALSE | Downregulated |
| -0,17019737 | THNSL2 | FALSE | Downregulated |
| 0,331237707 | THOC1 | FALSE | Upregulated |
| -0,925736221 | THOC5 | FALSE | Downregulated |
| -0,269346658 | THOC7-AS1 | FALSE | Downregulated |
| -0,773211845 | THTPA | FALSE | Downregulated |
| 0,206820887 | THYN1 | FALSE | Upregulated |
| 0,256928837 | TICAM1 | FALSE | Upregulated |
| 0,942402784 | TICRR | FALSE | Upregulated |
| -0,767078939 | TIGD2 | FALSE | Downregulated |
| 0,496842755 | TIGD5 | FALSE | Upregulated |
| 0,245072243 | TIMELESS | FALSE | Upregulated |
| 0,722201154 | TIMM10B | FALSE | Upregulated |
| 0,313344284 | TIMM44 | FALSE | Upregulated |
| -0,441375676 | TIMM50 | FALSE | Downregulated |
| 0,302469739 | TIMMDC1 | FALSE | Upregulated |
| -0,771897229 | TINAGL1 | FALSE | Downregulated |
| -0,400756077 | TIPRL | FALSE | Downregulated |
| 0,323218191 | TJAP1 | FALSE | Upregulated |
| 0,84989838 | TLCD1 | FALSE | Upregulated |
| 0,404641116 | TLDC1 | FALSE | Upregulated |
| -1,496807648 | TLE2 | | Downregulated |
| 0,492591888 | TLE3 | FALSE | Upregulated |
| -0,332104816 | TLE4 | FALSE | Downregulated |
| 1,105078331 | TLR1 | | Upregulated |
| -0,344743971 | TLR9 | FALSE | Downregulated |
| 1,191449225 | TM4SF20 | | Upregulated |
| -0,469975115 | TM9SF1 | FALSE | Downregulated |
| 0,356641654 | TMBIM4 | FALSE | Upregulated |
| 0,245725588 | TMCO1 | FALSE | Upregulated |
| -0,119811774 | TMCO4 | FALSE | Downregulated |
| 0,66596122 | TMCO6 | FALSE | Upregulated |
| -0,677668696 | TMED4 | FALSE | Downregulated |
| 0,468733076 | TMEM106A | FALSE | Upregulated |
| -0,791580936 | TMEM109 | FALSE | Downregulated |
| -0,55049053 | TMEM120A | FALSE | Downregulated |
| -0,206298361 | TMEM125 | FALSE | Downregulated |
| 0,900960968 | TMEM126A | FALSE | Upregulated |
| -0,474133793 | TMEM132D | FALSE | Downregulated |
| 0,436747571 | TMEM134 | FALSE | Upregulated |
| -0,262641515 | TMEM139 | FALSE | Downregulated |
| -0,311491915 | TMEM161A | FALSE | Downregulated |
| -0,277206646 | TMEM168 | FALSE | Downregulated |
| 1,247538387 | TMEM173 | | Upregulated |
| 0,354265539 | TMEM183A | FALSE | Upregulated |
| 1,359614484 | TMEM184B | | Upregulated |
| 0,862705726 | TMEM185B | FALSE | Upregulated |
| -0,483171815 | TMEM186 | FALSE | Downregulated |

| | | | |
|---|---|---|---|
| 0,044741635 | TMEM194A | FALSE | Upregulated |
| 0,228974999 | TMEM194B | FALSE | Upregulated |
| -1,733332202 | TMEM203 | | Downregulated |
| 0,443152011 | TMEM208 | FALSE | Upregulated |
| -0,36399381 | TMEM209 | FALSE | Downregulated |
| 0,303873809 | TMEM219 | FALSE | Upregulated |
| 1,301917091 | TMEM222 | | Upregulated |
| 0,084991303 | TMEM223 | FALSE | Upregulated |
| -0,377999822 | TMEM230 | FALSE | Downregulated |
| 0,006254748 | TMEM242 | FALSE | Upregulated |
| 0,210158046 | TMEM243 | FALSE | Upregulated |
| -0,528497934 | TMEM25 | FALSE | Downregulated |
| -1,227181653 | TMEM253 | | Downregulated |
| 0,610035292 | TMEM255A | FALSE | Upregulated |
| 0,084428933 | TMEM260 | FALSE | Upregulated |
| 0,916335519 | TMEM44-AS1 | FALSE | Upregulated |
| -0,298816068 | TMEM47 | FALSE | Downregulated |
| 0,920823844 | TMEM50A | FALSE | Upregulated |
| 0,815124114 | TMEM50B | FALSE | Upregulated |
| 0,636728756 | TMEM53 | FALSE | Upregulated |
| -0,187470732 | TMEM55B | FALSE | Downregulated |
| -0,787950207 | TMEM60 | FALSE | Downregulated |
| 1,633782401 | TMEM62 | | Upregulated |
| -0,039878301 | TMEM68 | FALSE | Downregulated |
| 0,291950157 | TMEM69 | FALSE | Upregulated |
| -1,173501885 | TMEM82 | | Downregulated |
| 1,501244453 | TMEM86A | | Upregulated |
| -1,065310941 | TMEM86B | | Downregulated |
| -0,338740692 | TMEM87A | FALSE | Downregulated |
| 0,432175936 | TMEM87B | FALSE | Upregulated |
| 0,20735292 | TMEM9 | FALSE | Upregulated |
| -0,033558416 | TMF1 | FALSE | Downregulated |
| 0,670496481 | TMLHE-AS1 | FALSE | Upregulated |
| 0,854779018 | TMPPE | FALSE | Upregulated |
| -0,119489789 | TMPRSS11D | FALSE | Downregulated |
| -0,00858201 | TMTC3 | FALSE | Downregulated |
| -0,036503587 | TMUB2 | FALSE | Downregulated |
| 0,083563979 | TMX1 | FALSE | Upregulated |
| 0,398725813 | TNIP2 | FALSE | Upregulated |
| -0,603565063 | TNN | FALSE | Downregulated |
| 0,049516439 | TOMM22 | FALSE | Upregulated |
| 0,857835266 | TOMM34 | FALSE | Upregulated |
| -0,587791855 | TOMM7 | FALSE | Downregulated |
| -0,167026024 | TOP1 | FALSE | Downregulated |
| 0,055294446 | TOP3B | FALSE | Upregulated |
| 0,04944707 | TOPBP1 | FALSE | Upregulated |
| -0,155315649 | TOPORS | FALSE | Downregulated |
| 1,109327244 | TOR4A | | Upregulated |
| 0,960574519 | TP53 | FALSE | Upregulated |
| 0,822072277 | TP53BP2 | FALSE | Upregulated |
| 0,23800157 | TP53TG5 | FALSE | Upregulated |

| | | | |
|---|---|---|---|
| 0,160319409 | TPD52 | FALSE | Upregulated |
| 0,838734309 | TPRA1 | FALSE | Upregulated |
| 0,043723199 | TPRG1 | FALSE | Upregulated |
| -0,159722585 | TPT1-AS1 | FALSE | Downregulated |
| 0,44228674 | TRA2A | FALSE | Upregulated |
| 1,625703812 | TRAF1 | | Upregulated |
| -0,571863635 | TRAF6 | FALSE | Downregulated |
| 0,385114275 | TRAPPC2 | FALSE | Upregulated |
| -0,32425794 | TRAPPC3 | FALSE | Downregulated |
| -0,095386848 | TRAPPC4 | FALSE | Downregulated |
| 0,358483148 | TRAPPC6B | FALSE | Upregulated |
| -1,130798471 | TRHDE-AS1 | | Downregulated |
| 0,47120724 | TRIAP1 | FALSE | Upregulated |
| 1,132523742 | TRIB2 | | Upregulated |
| 1,69652058 | TRIM11 | | Upregulated |
| 0,447403176 | TRIM15 | FALSE | Upregulated |
| 0,106349518 | TRIM16L | FALSE | Upregulated |
| -0,200802696 | TRIM23 | FALSE | Downregulated |
| -0,117147703 | TRIM32 | FALSE | Downregulated |
| 1,096971793 | TRIM37 | | Upregulated |
| -0,226153334 | TRIM4 | FALSE | Downregulated |
| 0,473438261 | TRIM45 | FALSE | Upregulated |
| 0,122590693 | TRIM5 | FALSE | Upregulated |
| 1,498632511 | TRIM55 | | Upregulated |
| 0,609579492 | TRIM59 | FALSE | Upregulated |
| 0,534435284 | TRIM65 | FALSE | Upregulated |
| 0,748084313 | TRIM67 | FALSE | Upregulated |
| -0,061431204 | TRIM7 | FALSE | Downregulated |
| 0,468975662 | TRIP10 | FALSE | Upregulated |
| -0,122350746 | TRMT1 | FALSE | Downregulated |
| 0,532468841 | TRMT12 | FALSE | Upregulated |
| -0,588301808 | TRMT1L | FALSE | Downregulated |
| -0,151920232 | TRMT2B | FALSE | Downregulated |
| 0,205492588 | TRMT44 | FALSE | Upregulated |
| 0,266400068 | TRMT61A | FALSE | Upregulated |
| 1,714632701 | TRNAU1AP | | Upregulated |
| 0,208641151 | TROVE2 | FALSE | Upregulated |
| 0,546935671 | TRPA1 | FALSE | Upregulated |
| 0,508106071 | TRPC5 | FALSE | Upregulated |
| 1,799335913 | TRPM2 | | Upregulated |
| 0,914767352 | TRPM4 | FALSE | Upregulated |
| 1,630755253 | TRPS1 | | Upregulated |
| 0,079222 | TSC22D1-AS1 | FALSE | Upregulated |
| 0,493607464 | TSG101 | FALSE | Upregulated |
| 0,278463727 | TSN | FALSE | Upregulated |
| 0,124051831 | TSNAX | FALSE | Upregulated |
| -1,308043227 | TSPAN12 | | Downregulated |
| -0,60921212 | TSPAN18 | FALSE | Downregulated |
| -0,423545602 | TSPAN33 | FALSE | Downregulated |
| -0,704262798 | TSPYL2 | FALSE | Downregulated |
| -0,022687963 | TSR2 | FALSE | Downregulated |

| | | | |
|---|---|---|---|
| 0,847832066 | TSSC1 | FALSE | Upregulated |
| 0,064780302 | TSSC4 | FALSE | Upregulated |
| 0,201655365 | TSSK2 | FALSE | Upregulated |
| 0,569788569 | TSTD3 | FALSE | Upregulated |
| -0,794710189 | TTBK1 | FALSE | Downregulated |
| -0,422262473 | TTC1 | FALSE | Downregulated |
| 0,222717915 | TTC18 | FALSE | Upregulated |
| 0,257813234 | TTC21B | FALSE | Upregulated |
| 1,507370062 | TTC21B-AS1 | | Upregulated |
| 0,00723298 | TTC3-AS1 | FALSE | Upregulated |
| 0,357361136 | TTC33 | FALSE | Upregulated |
| 0,464021726 | TTC7B | FALSE | Upregulated |
| 1,965433358 | TTL | | Upregulated |
| -0,218435702 | TTLL9 | FALSE | Downregulated |
| 0,456428558 | TTYH1 | FALSE | Upregulated |
| -0,064156516 | TUBA1A | FALSE | Downregulated |
| 1,219606083 | TUBA1C | | Upregulated |
| -0,495384967 | TUBE1 | FALSE | Downregulated |
| -0,75913621 | TUBG2 | FALSE | Downregulated |
| -0,960545758 | TULP3 | FALSE | Downregulated |
| 0,074969752 | TVP23C | FALSE | Upregulated |
| 0,677363978 | TWF1 | FALSE | Upregulated |
| 0,665727003 | TWF2 | FALSE | Upregulated |
| -0,087746199 | TXNDC12 | FALSE | Downregulated |
| -0,355996502 | TXNDC9 | FALSE | Downregulated |
| -0,388781794 | TXNL1 | FALSE | Downregulated |
| -0,524926896 | TXNL4A | FALSE | Downregulated |
| -0,477816482 | TXNL4B | FALSE | Downregulated |
| 0,657677096 | TXNRD1 | FALSE | Upregulated |
| -0,467504409 | TXNRD3 | FALSE | Downregulated |
| 0,204259149 | TYSND1 | FALSE | Upregulated |
| -0,085354249 | UBA2 | FALSE | Downregulated |
| 0,273193668 | UBAC1 | FALSE | Upregulated |
| 0,614295271 | UBALD1 | FALSE | Upregulated |
| 0,001385905 | UBALD2 | FALSE | Upregulated |
| -0,629473357 | UBE2A | FALSE | Downregulated |
| 0,036630461 | UBE2B | FALSE | Upregulated |
| -0,128002226 | UBE2D2 | FALSE | Downregulated |
| -0,21527361 | UBE2E3 | FALSE | Downregulated |
| 0,02153939 | UBE2F | FALSE | Upregulated |
| 0,146726005 | UBE2G1 | FALSE | Upregulated |
| -0,097287534 | UBE2I | FALSE | Downregulated |
| 0,195430709 | UBE2J1 | FALSE | Upregulated |
| -1,41264488 | UBE2M | | Downregulated |
| -0,608466464 | UBE2Q1 | FALSE | Downregulated |
| -0,235150667 | UBE2Q2 | FALSE | Downregulated |
| 0,742044512 | UBIAD1 | FALSE | Upregulated |
| 0,506534764 | UBL4A | FALSE | Upregulated |
| 0,524474081 | UBL7 | FALSE | Upregulated |
| -0,305947502 | UBQLN2 | FALSE | Downregulated |
| 0,206097135 | UBQLN4 | FALSE | Upregulated |

| | | | |
|---|---|---|---|
| -0,178345612 | UBR7 | FALSE | Downregulated |
| -0,233551147 | UBXN1 | FALSE | Downregulated |
| 0,318192322 | UBXN2B | FALSE | Upregulated |
| -0,25586772 | UBXN7 | FALSE | Downregulated |
| -0,838781584 | UFSP2 | FALSE | Downregulated |
| 0,571089723 | UGCG | FALSE | Upregulated |
| 1,115672518 | UHRF1 | | Upregulated |
| 0,334910499 | UHRF2 | FALSE | Upregulated |
| -0,913733097 | UIMC1 | FALSE | Downregulated |
| 1,62619592 | ULK2 | | Upregulated |
| 0,360468209 | ULK3 | FALSE | Upregulated |
| -0,06692815 | ULK4 | FALSE | Downregulated |
| -0,85964626 | UMPS | FALSE | Downregulated |
| -0,020100833 | UNC119B | FALSE | Downregulated |
| 0,061126993 | UNC13D | FALSE | Upregulated |
| 0,602054798 | UNC45A | FALSE | Upregulated |
| 0,293814309 | UNC5C | FALSE | Upregulated |
| 0,792439974 | UNC93B1 | FALSE | Upregulated |
| -0,596924675 | UPF2 | FALSE | Downregulated |
| 0,580521755 | UPF3A | FALSE | Upregulated |
| -0,893154215 | UPF3B | FALSE | Downregulated |
| 1,493281684 | UPP1 | | Upregulated |
| -0,25131172 | UQCC1 | FALSE | Downregulated |
| -1,290012533 | UQCC2 | | Downregulated |
| -0,334343591 | UQCRBP1 | FALSE | Downregulated |
| 0,118559496 | URI1 | FALSE | Upregulated |
| -1,052094286 | UROD | | Downregulated |
| -0,427804177 | UROS | FALSE | Downregulated |
| 0,673068561 | USB1 | FALSE | Upregulated |
| 0,103607203 | USF1 | FALSE | Upregulated |
| -0,43850002 | USP1 | FALSE | Downregulated |
| 1,012764093 | USP11 | | Upregulated |
| 0,834752188 | USP13 | FALSE | Upregulated |
| 0,116920387 | USP16 | FALSE | Upregulated |
| 0,164214946 | USP25 | FALSE | Upregulated |
| -0,51105989 | USP30-AS1 | FALSE | Downregulated |
| 0,714049327 | USP38 | FALSE | Upregulated |
| 0,553439194 | USP42 | FALSE | Upregulated |
| -0,915401961 | USP43 | FALSE | Downregulated |
| 0,046141663 | USP44 | FALSE | Upregulated |
| -0,314444626 | USP48 | FALSE | Downregulated |
| -0,151880271 | USP50 | FALSE | Downregulated |
| -0,775549876 | USP6 | FALSE | Downregulated |
| -1,566739844 | USPL1 | | Downregulated |
| -0,372681683 | UTP14A | FALSE | Downregulated |
| -0,602241245 | UTP18 | FALSE | Downregulated |
| -0,065024791 | UVSSA | FALSE | Downregulated |
| 0,477530371 | VAC14-AS1 | FALSE | Upregulated |
| -0,03251216 | VAMP2 | FALSE | Downregulated |
| -0,261311503 | VAMP3 | FALSE | Downregulated |
| -0,820057907 | VCAM1 | FALSE | Downregulated |

| | | | |
|---|---|---|---|
| 0,892692578 | VEZF1 | FALSE | Upregulated |
| -0,564576904 | VEZT | FALSE | Downregulated |
| -0,06507794 | VKORC1 | FALSE | Downregulated |
| -0,295294118 | VMAC | FALSE | Downregulated |
| 1,432628141 | VOPP1 | | Upregulated |
| 1,082274742 | VPS18 | | Upregulated |
| 0,625851958 | VPS29 | FALSE | Upregulated |
| -0,136237931 | VPS33B | FALSE | Downregulated |
| 0,78700343 | VPS37A | FALSE | Upregulated |
| -0,116652709 | VPS41 | FALSE | Downregulated |
| 0,456986646 | VPS45 | FALSE | Upregulated |
| -0,57468763 | VPS4B | FALSE | Downregulated |
| 0,205885406 | VRK3 | FALSE | Upregulated |
| 2,143510474 | VSIG10L | | Upregulated |
| -0,510644571 | VSNL1 | FALSE | Downregulated |
| -0,566023383 | VTI1B | FALSE | Downregulated |
| -0,363493633 | VWA9 | FALSE | Downregulated |
| 0,329022753 | VWCE | FALSE | Upregulated |
| 0,089538605 | WAC-AS1 | FALSE | Upregulated |
| -0,884786503 | WARS2 | FALSE | Downregulated |
| 0,123282021 | WASH2P | FALSE | Upregulated |
| -0,527044871 | WBP11 | FALSE | Downregulated |
| -0,669081045 | WDR19 | FALSE | Downregulated |
| 0,118881159 | WDR27 | FALSE | Upregulated |
| -1,878537008 | WDR31 | | Downregulated |
| -0,47732236 | WDR34 | FALSE | Downregulated |
| 0,756326437 | WDR36 | FALSE | Upregulated |
| 0,195272173 | WDR37 | FALSE | Upregulated |
| 0,729868672 | WDR41 | FALSE | Upregulated |
| 0,076637908 | WDR46 | FALSE | Upregulated |
| 0,257219449 | WDR54 | FALSE | Upregulated |
| -0,106977646 | WDR83 | FALSE | Downregulated |
| 0,64805517 | WDR83OS | FALSE | Upregulated |
| 0,242846333 | WEE1 | FALSE | Upregulated |
| 0,661628366 | WEE2 | FALSE | Upregulated |
| -0,778784673 | WFIKKN1 | FALSE | Downregulated |
| -0,189002077 | WFS1 | FALSE | Downregulated |
| 0,227841929 | WHAMMP3 | FALSE | Upregulated |
| 0,662067354 | WIPF1 | FALSE | Upregulated |
| 0,720018113 | WIPI1 | FALSE | Upregulated |
| -0,14586881 | WNT2B | FALSE | Downregulated |
| -0,072185068 | WRN | FALSE | Downregulated |
| -0,306790625 | WWC2-AS2 | FALSE | Downregulated |
| 0,488738615 | WWC3 | FALSE | Upregulated |
| -0,525423757 | WWTR1 | FALSE | Downregulated |
| -1,689772829 | XKR9 | | Downregulated |
| -0,066642688 | XPNPEP1 | FALSE | Downregulated |
| -0,646316195 | XPOT | FALSE | Downregulated |
| 0,947664027 | XPR1 | FALSE | Upregulated |
| 0,09358301 | XRCC1 | FALSE | Upregulated |
| 0,926366561 | XYLT1 | FALSE | Upregulated |

| | | | |
|---|---|---|---|
| 0,072569333 | YAF2 | FALSE | Upregulated |
| -1,237440867 | YBX3 | | Downregulated |
| 0,218113884 | YIF1A | FALSE | Upregulated |
| -0,146665869 | YIF1B | FALSE | Downregulated |
| 0,010264327 | YIPF5 | FALSE | Upregulated |
| 0,925021744 | YKT6 | FALSE | Upregulated |
| -1,402050447 | YPEL3 | | Downregulated |
| -0,20736379 | YPEL5 | FALSE | Downregulated |
| 0,337033282 | YTHDF2 | FALSE | Upregulated |
| 0,056021036 | ZBTB1 | FALSE | Upregulated |
| -0,466497427 | ZBTB14 | FALSE | Downregulated |
| -0,599323457 | ZBTB17 | FALSE | Downregulated |
| -0,194818984 | ZBTB21 | FALSE | Downregulated |
| -0,056432194 | ZBTB3 | FALSE | Downregulated |
| -0,37841213 | ZBTB34 | FALSE | Downregulated |
| 0,89835722 | ZBTB43 | FALSE | Upregulated |
| 0,532244973 | ZBTB47 | FALSE | Upregulated |
| -0,693058154 | ZC3H7A | FALSE | Downregulated |
| -0,089388384 | ZCCHC3 | FALSE | Downregulated |
| -0,655530156 | ZCCHC8 | FALSE | Downregulated |
| 0,453578182 | ZDBF2 | FALSE | Upregulated |
| 0,269634454 | ZDHHC2 | FALSE | Upregulated |
| 0,175889266 | ZDHHC21 | FALSE | Upregulated |
| 0,460656855 | ZDHHC6 | FALSE | Upregulated |
| 0,324519834 | ZDHHC7 | FALSE | Upregulated |
| -0,099147567 | ZEB1 | FALSE | Downregulated |
| 0,17265204 | ZFAND2B | FALSE | Upregulated |
| 0,041111044 | ZFAND4 | FALSE | Upregulated |
| 0,929695498 | ZFAT-AS1 | FALSE | Upregulated |
| -0,575344235 | ZFP14 | FALSE | Downregulated |
| 0,799554207 | ZFP30 | FALSE | Upregulated |
| -0,06968628 | ZFP62 | FALSE | Downregulated |
| 0,357542223 | ZFP64 | FALSE | Upregulated |
| -0,669881232 | ZFP90 | FALSE | Downregulated |
| -0,484503494 | ZFPL1 | FALSE | Downregulated |
| 0,28773445 | ZFPM2 | FALSE | Upregulated |
| 0,131322582 | ZFYVE1 | FALSE | Upregulated |
| -0,262686371 | ZFYVE21 | FALSE | Downregulated |
| 0,755789909 | ZFYVE28 | FALSE | Upregulated |
| -0,1401935 | ZKSCAN3 | FALSE | Downregulated |
| 0,851102568 | ZKSCAN4 | FALSE | Upregulated |
| -0,279380308 | ZMAT1 | FALSE | Downregulated |
| 0,300772736 | ZMAT2 | FALSE | Upregulated |
| -0,0108207 | ZMYM6 | FALSE | Downregulated |
| 0,006577686 | ZMYND19 | FALSE | Upregulated |
| 1,198831897 | ZNF117 | | Upregulated |
| -0,006257093 | ZNF12 | FALSE | Downregulated |
| -1,465530051 | ZNF141 | | Downregulated |
| -0,641764963 | ZNF146 | FALSE | Downregulated |
| -0,053583686 | ZNF175 | FALSE | Downregulated |
| -0,843952851 | ZNF177 | FALSE | Downregulated |

| | | | |
|---|---|---|---|
| -0,407605148 | ZNF182 | FALSE | Downregulated |
| 0,41041868 | ZNF189 | FALSE | Upregulated |
| 0,343538371 | ZNF195 | FALSE | Upregulated |
| 1,027444598 | ZNF213 | | Upregulated |
| -0,836003127 | ZNF221 | FALSE | Downregulated |
| -0,18805813 | ZNF225 | FALSE | Downregulated |
| 1,091568174 | ZNF234 | | Upregulated |
| 0,073232692 | ZNF235 | FALSE | Upregulated |
| 0,647237446 | ZNF248 | FALSE | Upregulated |
| -0,249136082 | ZNF251 | FALSE | Downregulated |
| -0,306684423 | ZNF252P | FALSE | Downregulated |
| -0,629161826 | ZNF259 | FALSE | Downregulated |
| -0,474678298 | ZNF268 | FALSE | Downregulated |
| -1,314205311 | ZNF277 | | Downregulated |
| 0,474437742 | ZNF282 | FALSE | Upregulated |
| -0,54539279 | ZNF287 | FALSE | Downregulated |
| -0,192509316 | ZNF296 | FALSE | Downregulated |
| -0,012183094 | ZNF30 | FALSE | Downregulated |
| 0,353844606 | ZNF302 | FALSE | Upregulated |
| -0,110415289 | ZNF319 | FALSE | Downregulated |
| -0,118908068 | ZNF32 | FALSE | Downregulated |
| 1,174118488 | ZNF320 | | Upregulated |
| 0,975580655 | ZNF322 | FALSE | Upregulated |
| 0,331475835 | ZNF324B | FALSE | Upregulated |
| 0,006457269 | ZNF343 | FALSE | Upregulated |
| -0,064021403 | ZNF345 | FALSE | Downregulated |
| -0,112674261 | ZNF35 | FALSE | Downregulated |
| -1,278633408 | ZNF350 | | Downregulated |
| -0,038171234 | ZNF362 | FALSE | Downregulated |
| 0,223586778 | ZNF367 | FALSE | Upregulated |
| 0,075909743 | ZNF37A | FALSE | Upregulated |
| -0,317273595 | ZNF37BP | FALSE | Downregulated |
| -0,772698895 | ZNF384 | FALSE | Downregulated |
| -0,473474509 | ZNF385C | FALSE | Downregulated |
| -0,151646396 | ZNF398 | FALSE | Downregulated |
| -1,054982528 | ZNF404 | | Downregulated |
| -0,34579602 | ZNF408 | FALSE | Downregulated |
| 0,415972812 | ZNF41 | FALSE | Upregulated |
| 0,641312839 | ZNF410 | FALSE | Upregulated |
| -0,276117438 | ZNF428 | FALSE | Downregulated |
| -0,063596176 | ZNF438 | FALSE | Downregulated |
| 0,210309683 | ZNF44 | FALSE | Upregulated |
| 0,08046142 | ZNF440 | FALSE | Upregulated |
| 0,716487706 | ZNF441 | FALSE | Upregulated |
| 0,271894981 | ZNF449 | FALSE | Upregulated |
| -0,33211554 | ZNF462 | FALSE | Downregulated |
| -0,009221755 | ZNF467 | FALSE | Downregulated |
| 0,027161252 | ZNF48 | FALSE | Upregulated |
| -0,192728666 | ZNF490 | FALSE | Downregulated |
| 0,588760854 | ZNF493 | FALSE | Upregulated |
| 0,488167232 | ZNF500 | FALSE | Upregulated |

| | | | |
|---|---|---|---|
| -0,103986754 | ZNF512 | FALSE | Downregulated |
| 0,011219892 | ZNF514 | FALSE | Upregulated |
| -0,324089228 | ZNF524 | FALSE | Downregulated |
| 0,366025775 | ZNF526 | FALSE | Upregulated |
| -0,2816895 | ZNF540 | FALSE | Downregulated |
| -0,462456332 | ZNF548 | FALSE | Downregulated |
| 0,99907075 | ZNF555 | FALSE | Upregulated |
| 0,026889861 | ZNF558 | FALSE | Upregulated |
| 0,40854762 | ZNF561 | FALSE | Upregulated |
| 0,571426425 | ZNF564 | FALSE | Upregulated |
| -1,34787338 | ZNF568 | | Downregulated |
| -0,348191727 | ZNF569 | FALSE | Downregulated |
| 0,367390416 | ZNF574 | FALSE | Upregulated |
| -0,154306152 | ZNF575 | FALSE | Downregulated |
| 0,451971636 | ZNF576 | FALSE | Upregulated |
| 0,461455265 | ZNF580 | FALSE | Upregulated |
| -0,323766209 | ZNF585B | FALSE | Downregulated |
| 0,727469802 | ZNF589 | FALSE | Upregulated |
| -0,180675252 | ZNF593 | FALSE | Downregulated |
| -0,475487094 | ZNF594 | FALSE | Downregulated |
| 0,469892247 | ZNF605 | FALSE | Upregulated |
| -0,053367708 | ZNF606 | FALSE | Downregulated |
| -0,759135863 | ZNF608 | FALSE | Downregulated |
| -0,797856993 | ZNF615 | FALSE | Downregulated |
| -1,028980658 | ZNF616 | | Downregulated |
| 0,163456511 | ZNF620 | FALSE | Upregulated |
| 0,660342322 | ZNF623 | FALSE | Upregulated |
| 0,076774678 | ZNF627 | FALSE | Upregulated |
| -0,08256278 | ZNF628 | FALSE | Downregulated |
| 1,235904909 | ZNF638-IT1 | | Upregulated |
| -0,483527928 | ZNF660 | FALSE | Downregulated |
| 0,896607954 | ZNF662 | FALSE | Upregulated |
| 0,459280881 | ZNF668 | FALSE | Upregulated |
| -0,178757183 | ZNF680 | FALSE | Downregulated |
| -0,637330441 | ZNF688 | FALSE | Downregulated |
| 0,20061206 | ZNF717 | FALSE | Upregulated |
| 1,021892838 | ZNF720 | | Upregulated |
| -1,006341788 | ZNF721 | | Downregulated |
| -0,333793528 | ZNF737 | FALSE | Downregulated |
| -0,306229598 | ZNF74 | FALSE | Downregulated |
| -0,110542365 | ZNF747 | FALSE | Downregulated |
| 0,957415577 | ZNF75A | FALSE | Upregulated |
| -0,282668384 | ZNF75D | FALSE | Downregulated |
| -0,639795822 | ZNF761 | FALSE | Downregulated |
| -0,236677239 | ZNF764 | FALSE | Downregulated |
| 0,904446886 | ZNF766 | FALSE | Upregulated |
| 0,517673891 | ZNF767 | FALSE | Upregulated |
| -0,646629404 | ZNF768 | FALSE | Downregulated |
| -0,179643894 | ZNF772 | FALSE | Downregulated |
| 0,165826651 | ZNF782 | FALSE | Upregulated |
| 1,029451993 | ZNF785 | | Upregulated |

-0,466878045 ZNF786 FALSE Downregulated
-0,100177399 ZNF791 FALSE Downregulated
-0,006252561 ZNF8 FALSE Downregulated
-0,043431678 ZNF800 FALSE Downregulated
-0,120533249 ZNF808 FALSE Downregulated
0,16853403 ZNF81 FALSE Upregulated
0,013676346 ZNF821 FALSE Upregulated
0,075550993 ZNF823 FALSE Upregulated
-0,750034044 ZNF841 FALSE Downregulated
-1,060115077 ZNF843 Downregulated
-1,305673825 ZNF891 Downregulated
0,035310163 ZNRF2P1 FALSE Upregulated
-0,120967915 ZRANB2 FALSE Downregulated
0,096742923 ZRANB2-AS1 FALSE Upregulated
-0,301539466 ZSCAN12 FALSE Downregulated
-0,327770854 ZSCAN18 FALSE Downregulated
0,932154889 ZSWIM4 FALSE Upregulated
0,047341046 ZSWIM7 FALSE Upregulated
-1,160602074 ZYG11A Downregulated

#### Suppl. Table 16: Pathway enrichment of differentially expressed genes of chronic ductopenic rejection as compared to controls

##### Upregulated

| q-value | Pathway name | Pathway ID | | q-value | pathway | external_id |
|---|---|---|---|---|---|---|
| 0.018941504 | morphogenesis of an epithelial sheet | GO:0002011 | | 0.000230204 | Collagen formation | R-HSA-14742 |
| 0.018941504 | epithelial cell apoptotic process | GO:1904019 | | 0.000284607 | Collagen biosynthesis and modifying enzymes | R-HSA-16508 |
| 0.018941504 | sphingolipid translocation | GO:0099039 | | 0.000533981 | Extracellular matrix organization | R-HSA-14742 |
| 0.018941504 | cellular response to amino acid stimulus | GO:0071230 | | 0.001161245 | Focal adhesion - Homo sapiens (human) | path:hsa0451 |
| 0.019357076 | protein localization to cell surface | GO:0034394 | | 0.001161245 | AGE-RAGE signaling pathway in diabetic complications - Homo sapiens (human) | path:hsa0493 |
| 0.076601095 | collagen-activated tyrosine kinase receptor signaling pathway | GO:0038063 | | 0.004435135 | Collagen chain trimerization | R-HSA-89482 |
| 0.076601095 | acylglycerol biosynthetic process | GO:0046463 | | 0.005235178 | Scavenging by Class A Receptors | R-HSA-30004 |
| 0.076601095 | organic acid transport | GO:0015849 | | 0.011685460 | RUNX1 regulates transcription of genes involved in WNT signaling | R-HSA-89392 |
| 0.086716938 | positive regulation of cell-cell adhesion | GO:0022409 | | 0.011685460 | MicroRNAs in cancer - Homo sapiens (human) | path:hsa0520 |
| 0.095913713 | regulation of integrin activation | GO:0033623 | | 0.016718064 | p53 signaling pathway - Homo sapiens (human) | path:hsa0411 |
| 0.098128137 | positive regulation of actin filament bundle assembly | GO:0032233 | | 0.016718064 | RUNX3 regulates p14-ARF | R-HSA-89519 |
| 0.098128137 | positive regulation of cellular component organization | GO:0051130 | | 0.016718064 | Chronic myeloid leukemia - Homo sapiens (human) | path:hsa0522 |
| 0.098128137 | pyrimidine nucleobase metabolic process | GO:0006206 | | 0.016718064 | Pancreatic cancer - Homo sapiens (human) | path:hsa0521 |
| 0.098128137 | alpha-beta T cell lineage commitment | GO:0002363 | | 0.017441512 | PI3K-Akt signaling pathway - Homo sapiens (human) | path:hsa0415 |
| 0.098128137 | CD4-positive or CD8-positive, alpha-beta T cell lineage commitment | GO:0043369 | | 0.018791509 | Proteoglycans in cancer - Homo sapiens (human) | path:hsa0520 |
| 0.098128137 | striated muscle cell development | GO:0055002 | | 0.018791509 | RUNX1 and FOXP3 control the development of regulatory T lymphocytes (Tregs) | R-HSA-88773 |
| 0.098128137 | anion transport | GO:0006820 | | 0.019646960 | Colorectal cancer - Homo sapiens (human) | path:hsa0521 |
| 0.098128137 | establishment of skin barrier | GO:0061436 | | 0.019646960 | Bladder cancer - Homo sapiens (human) | path:hsa0521 |
| 0.098128137 | fatty acid beta-oxidation | GO:0006635 | | 0.019646960 | ECM-receptor interaction - Homo sapiens (human) | path:hsa0451 |
| 0.098128137 | positive regulation of response to wounding | GO:1903036 | | 0.019646960 | Binding and Uptake of Ligands by Scavenger Receptors | R-HSA-21737 |
| 0.098128137 | integrin activation | GO:0033622 | | 0.029134782 | Transcriptional regulation by RUNX3 | R-HSA-88781 |
| 0.099799982 | water homeostasis | GO:0030104 | | 0.029410855 | Protein digestion and absorption - Homo sapiens (human) | path:hsa0497 |
| 0.099799982 | actin filament bundle organization | GO:0061572 | | 0.029410855 | Tight junction - Homo sapiens (human) | path:hsa0453 |
| 0.099799982 | positive regulation of collagen metabolic process | GO:0010714 | | 0.030122224 | Human papillomavirus infection - Homo sapiens (human) | path:hsa0516 |
| 0.101022595 | eye development | GO:0001654 | | 0.030122224 | Th17 cell differentiation - Homo sapiens (human) | path:hsa0465 |
| 0.101022595 | positive regulation of cell differentiation | GO:0045597 | | 0.030122224 | Cell-extracellular matrix interactions | R-HSA-44635 |
| | | | | 0.032746025 | Regulation of RUNX1 Expression and Activity | R-HSA-89345 |
| | | | | 0.050764219 | Relaxin signaling pathway - Homo sapiens (human) | path:hsa0492 |
| | | | | 0.050764219 | Acute myeloid leukemia - Homo sapiens (human) | path:hsa0522 |

##### Downregulated

| q-value | Pathway name | Pathway ID |
|---|---|---|
| 2,26E-49 | carboxylic acid metabolic process | GO:0019752 |
| 1,22E-21 | carboxylic acid catabolic process | GO:0046395 |
| 3,17E-13 | carboxylic acid biosynthetic process | GO:0046394 |
| 4,43E-09 | fatty acid catabolic process | GO:0009062 |
| 3,13E-07 | long-chain fatty acid metabolic process | GO:0001676 |
| 6,37E-06 | acute inflammatory response | GO:0002526 |
| 1,01E-05 | terpenoid metabolic process | GO:0006721 |
| 1,33E-05 | fatty acid beta-oxidation | GO:0006635 |
| 2,07E-04 | cholesterol metabolic process | GO:0008203 |
| 6,23E-01 | steroid biosynthetic process | GO:0006694 |
| 1,03E+00 | unsaturated fatty acid metabolic process | GO:0033559 |
| 6,35E+00 | regulation of lipid metabolic process | GO:0019216 |
| 1,31E+01 | protein processing | GO:0016485 |
| 1,82E+01 | arachidonic acid metabolic process | GO:0019369 |
| 2,00E+01 | fatty acid biosynthetic process | GO:0006633 |
| 8,03E+01 | acylglycerol metabolic process | GO:0006639 |
| 1,87E+02 | alpha-amino acid catabolic process | GO:1901606 |
| 1,07E+03 | aspartate family amino acid metabolic process | GO:0009066 |
| 2,32E+03 | alpha-amino acid biosynthetic process | GO:1901607 |
| 2,80E+03 | exogenous drug catabolic process | GO:0042738 |
| 2,80E+03 | regulation of lipid biosynthetic process | GO:0046890 |
| 3,57E+03 | protein-lipid complex remodeling | GO:0034368 |
| 5,22E+03 | negative regulation of blood coagulation | GO:0030195 |
| 7,04E+03 | nucleoside bisphosphate metabolic process | GO:0033865 |
| 7,04E+03 | purine nucleoside bisphosphate metabolic process | GO:0034032 |
| 8,66E+03 | negative regulation of coagulation | GO:0050819 |
| 8,87E+03 | regulation of complement activation | GO:0030449 |
| 1,03E+04 | regulation of humoral immune response | GO:0002920 |
| 1,63E+04 | lipid transport | GO:0006869 |
| 2,34E+04 | regulation of cholesterol metabolic process | GO:0090181 |
| 5,72E+04 | regulation of fibrinolysis | GO:0051917 |
| 1,28E+04 | secondary alcohol biosynthetic process | GO:1902653 |
| 1,73E+05 | negative regulation of response to wounding | GO:1903035 |
| 2,74E+05 | sterol biosynthetic process | GO:0016126 |
| 3,06E+05 | nucleotide metabolic process | GO:0009117 |
| 5,83E+05 | response to cadmium ion | GO:0046686 |
| 6,30E+05 | coenzyme biosynthetic process | GO:0009108 |
| 7,76E+05 | cellular modified amino acid catabolic process | GO:0042219 |
| 8,22E+05 | very-low-density lipoprotein particle remodeling | GO:0034372 |
| 9,57E+04 | glucose metabolic process | GO:0006006 |
| 9,71E+04 | ethanol metabolic process | GO:0006067 |
| 1,04E+06 | response to oleic acid | GO:0034201 |
| 1,63E+06 | negative regulation of hydrolase activity | GO:0051346 |
| 2,43E+06 | glycerolipid catabolic process | GO:0046503 |
| 2,43E+06 | regulation of lipase activity | GO:0060191 |
| 2,54E+06 | steroid catabolic process | GO:0006706 |
| 2,68E+06 | organic hydroxy compound transport | GO:0015850 |
| 2,68E+06 | ribonucleotide metabolic process | GO:0009259 |
| 2,68E+06 | lipid homeostasis | GO:0055088 |
| 3,37E+06 | steroid esterification | GO:0034433 |
| 3,44E+05 | regulation of cellular ketone metabolic process | GO:0010565 |
| 3,69E+06 | regulation of inflammatory response | GO:0050727 |
| 3,69E+06 | lipid hydroxylation | GO:0002933 |
| 3,69E+06 | amino-acid betaine catabolic process | GO:0006579 |
| 3,90E+06 | response to corticosteroid | GO:0031960 |
| 4,14E+06 | cellular response to dexamethasone stimulus | GO:0071549 |
| 4,16E+06 | regulation of lipid catabolic process | GO:0050994 |
| 7,22E+06 | neutral lipid catabolic process | GO:0046461 |
| 7,32E+06 | methionine metabolic process | GO:0006555 |
| 8,94E+06 | organic anion transport | GO:0015711 |
| 9,93E+06 | cellular response to ketone | GO:1901655 |
| 9,94E+06 | protein-lipid complex assembly | GO:0065005 |
| 0.000104343 | purine nucleotide metabolic process | GO:0006163 |
| 0.000115165 | amine catabolic process | GO:0009310 |
| 0.000115361 | response to purine-containing compound | GO:0014074 |
| 0.000119427 | farnesyl diphosphate biosynthetic process | GO:0045337 |
| 0.000131675 | regulation of lipid transport | GO:0032368 |
| 0.000138548 | branched-chain amino acid catabolic process | GO:0009083 |
| 0.000148707 | bile acid biosynthetic process | GO:0006699 |
| 0.000154028 | regulation of tube diameter | GO:0035296 |
| 0.000154028 | regulation of blood vessel size | GO:0050880 |
| 0.000165135 | regulation of response to wounding | GO:1903034 |
| 0.000207441 | negative regulation of small molecule metabolic process | GO:0062014 |
| 0.000259804 | response to aluminum ion | GO:0010044 |
| 0.000259804 | farnesyl diphosphate metabolic process | GO:0045338 |
| 0.000280318 | pyridine nucleotide metabolic process | GO:0019362 |
| 0.000342853 | medium-chain fatty acid metabolic process | GO:0051791 |
| 0.000456071 | terpenoid biosynthetic process | GO:0016114 |
| 0.000466511 | anion transport | GO:0006820 |
| 0.000472059 | hexose biosynthetic process | GO:0019319 |
| 0.000481119 | positive regulation of blood coagulation | GO:0030194 |
| 0.000541110 | positive regulation of lipid metabolic process | GO:0045834 |
| 0.000556531 | positive regulation of coagulation | GO:0050820 |
| 0.000603209 | oxidoreduction coenzyme metabolic process | GO:0006733 |
| 0.000605771 | negative regulation of lipid metabolic process | GO:0045833 |
| 0.000616801 | cellular modified amino acid biosynthetic process | GO:0042398 |
| 0.000696177 | sulfur amino acid biosynthetic process | GO:0000097 |
| 0.000769239 | regulation of protein metabolic process | GO:0051246 |
| 0.000964723 | very long-chain fatty acid metabolic process | GO:0000038 |
| 0.001322007 | coenzyme catabolic process | GO:0009109 |
| 0.001359113 | regulation of triglyceride metabolic process | GO:0090207 |
| 0.001359113 | response to glucagon | GO:0033762 |
| 0.001359113 | nucleoside phosphate catabolic process | GO:1901292 |
| 0.001359113 | coumarin catabolic process | GO:0046226 |
| 0.001359113 | lauric acid metabolic process | GO:0048252 |
| 0.001359113 | creatine biosynthetic process | GO:0006601 |
| 0.001359113 | isoprenoid biosynthetic process via mevalonate | GO:1902767 |
| 0.001359113 | propionate catabolic process | GO:0019543 |
| 0.001370897 | regulation of defense response | GO:0031347 |
| 0.001370897 | response to zinc ion | GO:0010043 |
| 0.001438037 | pyrimidine nucleobase metabolic process | GO:0006206 |
| 0.001477478 | cellular biogenic amine metabolic process | GO:0006576 |
| 0.001519366 | cytolysis by host of symbiont cells | GO:0051838 |
| 0.002566458 | cellular response to organic cyclic compound | GO:0071407 |
| 0.002788265 | glucuronate metabolic process | GO:0019585 |
| 0.002848506 | negative regulation of response to external stimulus | GO:0032102 |
| 0.003195566 | regulation of heterotypic cell-cell adhesion | GO:0034114 |
| 0.003379895 | regulation of systemic arterial blood pressure mediated by a chemical signal | GO:0003044 |
| 0.003570599 | carnitine metabolic process, CoA-linked | GO:0019254 |
| 0.003570599 | regulation of exo-alpha-sialidase activity | GO:1903015 |
| 0.003570599 | regulation of cholesterol esterification | GO:0010872 |
| 0.003707147 | ribose phosphate biosynthetic process | GO:0046390 |
| 0.003937614 | acylglycerol biosynthetic process | GO:0046463 |
| 0.004069579 | nucleoside phosphate biosynthetic process | GO:1901293 |
| 0.004430544 | midgut development | GO:0007494 |
| 0.004876505 | organic acid transport | GO:0015849 |
| 0.005220907 | purine-containing compound biosynthetic process | GO:0072522 |
| 0.005362481 | positive regulation of response to wounding | GO:1903036 |
| 0.005408731 | regulation of carbohydrate metabolic process | GO:0006109 |
| 0.005793699 | liver development | GO:0001889 |
| 0.006291229 | cellular response to fatty acid | GO:0071398 |
| 0.006291229 | negative regulation of very-low-density lipoprotein particle remodeling | GO:0010903 |
| 0.006291229 | regulation of very-low-density lipoprotein particle clearance | GO:0010915 |
| 0.006291229 | propionate metabolic process | GO:0019541 |
| 0.006291229 | cellular response to peptide hormone stimulus | GO:0071375 |
| 0.006291229 | fructose metabolic process | GO:0006000 |
| 0.006574162 | regulation of cellular carbohydrate metabolic process | GO:0010675 |
| 0.007421602 | primary alcohol catabolic process | GO:0034310 |
| 0.007421602 | sulfur amino acid catabolic process | GO:0000098 |
| 0.007421602 | hepatocyte differentiation | GO:0070365 |
| 0.007440530 | icosanoid biosynthetic process | GO:0046456 |
| 0.007470652 | regulation of reactive oxygen species metabolic process | GO:2000377 |
| 0.007568456 | glycolytic process | GO:0006096 |
| 0.007760416 | positive regulation of small molecule metabolic process | GO:0062013 |
| 0.008386921 | pyridine-containing compound biosynthetic process | GO:0072525 |
| 0.008518927 | short-chain fatty acid metabolic process | GO:0046459 |
| 0.008525303 | regulation of glucose metabolic process | GO:0010906 |
| 0.009102242 | fructose catabolic process to hydroxyacetone phosphate and glyceraldehyde-3-phosphate | GO:0061624 |
| 0.009102242 | response to phenylpropanoid | GO:0080184 |
| 0.009102242 | leukotriene B4 catabolic process | GO:0036101 |
| 0.009102242 | glucuronate catabolic process | GO:0006064 |
| 0.009102242 | xylulose 5-phosphate biosynthetic process | GO:1901159 |
| 0.009614152 | cellular response to metal ion | GO:0071248 |
| 0.010059278 | organic acid transmembrane transport | GO:1903825 |
| 0.010488659 | polyol biosynthetic process | GO:0046173 |
| 0.010977386 | tissue homeostasis | GO:0001894 |
| 0.010977386 | adipose tissue development | GO:0060612 |
| 0.011700576 | ERK1 and ERK2 cascade | GO:0070371 |
| 0.011892731 | response to growth hormone | GO:0060416 |
| 0.012523800 | male genitalia development | GO:0030539 |
| 0.012711986 | isoleucine metabolic process | GO:0006549 |
| 0.012886481 | response to insulin | GO:0032868 |
| 0.014074403 | cellular response to sterol | GO:0036315 |
| 0.014074403 | glycerol metabolic process | GO:0006071 |
| 0.014270658 | cellular response to steroid hormone stimulus | GO:0071383 |
| 0.014660130 | mammary gland development | GO:0030879 |
| 0.014660130 | positive regulation of immune response | GO:0050778 |
| 0.015031497 | B cell mediated immunity | GO:0019724 |
| 0.015482751 | response to lead ion | GO:0010288 |
| 0.015482751 | regulation of systemic arterial blood pressure by renin-angiotensin | GO:0003081 |
| 0.015482751 | male gonad development | GO:0008584 |
| 0.016223415 | amino-acid betaine biosynthetic process | GO:0006578 |
| 0.016223415 | valine metabolic process | GO:0006573 |
| 0.016223415 | alditol biosynthetic process | GO:0019401 |
| 0.016345983 | cellular response to lipid | GO:0071396 |
| 0.019329355 | regulation of alcohol biosynthetic process | GO:1902930 |
| 0.019329355 | glutamine family amino acid metabolic process | GO:0009064 |
| 0.020540068 | citrulline metabolic process | GO:0000052 |
| 0.020540068 | leucine metabolic process | GO:0006551 |
| 0.020540068 | icosanoid catabolic process | GO:1901523 |
| 0.020540068 | trans-synaptic signaling by trans-synaptic complex | GO:0099545 |
| 0.020844244 | cellular response to peptide | GO:1901653 |
| 0.021910989 | epithelial cell apoptotic process | GO:1904019 |
| 0.022824552 | serine family amino acid metabolic process | GO:0009069 |
| 0.024201089 | phospholipid catabolic process | GO:0009395 |
| 0.024201089 | regulation of bone remodeling | GO:0046850 |
| 0.024341352 | organophosphate ester transport | GO:0015748 |
| 0.024432611 | phosphatidylinositol-mediated signaling | GO:0048015 |
| 0.024432611 | disruption of cells of other organism involved in symbiotic interaction | GO:0051818 |
| 0.024432611 | response to cobalt ion | GO:0032025 |
| 0.024432611 | modulation of molecular function in other organism involved in symbiotic interaction | GO:0052205 |
| 0.024432611 | chylomicron remodeling | GO:0034371 |
| 0.024432611 | negative regulation of lipoprotein particle clearance | GO:0010985 |
| 0.025966317 | protein complex oligomerization | GO:0051259 |
| 0.026562510 | response to iron ion | GO:0010039 |
| 0.026562510 | aromatic amino acid family catabolic process | GO:0009074 |
| 0.028572871 | response to monosaccharide | GO:0034284 |
| 0.029199451 | nucleobase catabolic process | GO:0046113 |
| 0.029199451 | thyroid hormone transport | GO:0070327 |
| 0.029199451 | hexose catabolic process | GO:0019320 |
| 0.029376444 | negative regulation of catabolic process | GO:0009895 |
| 0.029752801 | regulation of cofactor metabolic process | GO:0051193 |

| q-value | pathway | external_id |
|---|---|---|
| 4,07E-28 | Metabolism | R-HSA-14307 |
| 1,38E-12 | Complement and coagulation cascades - Homo sapiens (human) | path:hsa0461 |
| 1,23E-08 | Fatty acid degradation - Homo sapiens (human) | path:hsa0007 |
| 3,08E-06 | Biological oxidations | R-HSA-21185 |
| 1,20E-05 | Phase I - Functionalization of compounds | R-HSA-21194 |
| 1,42E-05 | Drug metabolism - cytochrome P450 - Homo sapiens (human) | path:hsa0098 |
| 7,55E-05 | Valine, leucine and isoleucine degradation - Homo sapiens (human) | path:hsa0028 |
| 5,34E-04 | Metabolism of lipids | R-HSA-55683 |
| 2,86E-03 | Fatty acid metabolism | R-HSA-89788 |
| 1,07E-02 | PPAR signaling pathway - Homo sapiens (human) | path:hsa0332 |
| 2,52E-02 | Pyruvate metabolism - Homo sapiens (human) | path:hsa0062 |
| 7,82E-02 | Chemical carcinogenesis - Homo sapiens (human) | path:hsa0520 |
| 2,26E-01 | Metabolism of xenobiotics by cytochrome P450 - Homo sapiens (human) | path:hsa0098 |
| 5,00E-01 | Retinol metabolism - Homo sapiens (human) | path:hsa0083 |
| 1,02E+01 | Metabolism of amino acids and derivatives | R-HSA-71291 |
| 4,24E+01 | Tryptophan metabolism - Homo sapiens (human) | path:hsa0038 |
| 1,68E+02 | Butanoate metabolism - Homo sapiens (human) | path:hsa0065 |
| 1,68E+02 | Formation of Fibrin Clot (Clotting Cascade) | R-HSA-14087 |
| 1,71E+02 | Platelet degranulation | R-HSA-11460 |
| 3,07E+02 | Response to elevated platelet cytosolic Ca2+ | R-HSA-76005 |
| 1,60E+03 | Cytochrome P450 - arranged by substrate type | R-HSA-21189 |
| 2,80E+03 | Arachidonic acid metabolism | R-HSA-21427 |
| 2,80E+03 | Biosynthesis of maresins | R-HSA-90186 |
| 3,64E+03 | Common Pathway of Fibrin Clot Formation | R-HSA-14087 |
| 5,54E+03 | Xenobiotics | R-HSA-21198 |
| 5,81E+03 | Glycine, serine and threonine metabolism - Homo sapiens (human) | path:hsa0026 |
| 7,04E+03 | Synthesis of (16-20)-hydroxyeicosatetraenoic acids (HETE) | R-HSA-21428 |
| 8,55E+02 | Glycolysis / Gluconeogenesis - Homo sapiens (human) | path:hsa0001 |
| 8,90E+02 | Peroxisome - Homo sapiens (human) | path:hsa0414 |
| 2,43E+04 | Biosynthesis of maresin-like SPMs | R-HSA-90273 |
| 2,43E+04 | Regulation of Insulin-like Growth Factor (IGF) transport and uptake by Insulin-like Growth Factor Binding Proteins (IGFBPs) | R-HSA-38142 |
| 2,43E+04 | Metabolism of steroids | R-HSA-89573 |
| 2,87E+03 | Tyrosine metabolism - Homo sapiens (human) | path:hsa0035 |
| 2,98E+04 | CYP2E1 reactions | R-HSA-21199 |
| 4,75E+04 | Peroxisomal protein import | R-HSA-90332 |
| 5,16E+04 | Biosynthesis of specialized proresolving mediators (SPMs) | R-HSA-90186 |
| 5,24E+04 | Scavenging of heme from plasma | R-HSA-21688 |
| 5,24E+04 | Ethanol oxidation | R-HSA-71384 |
| 8,79E+04 | beta-Alanine metabolism - Homo sapiens (human) | path:hsa0041 |
| 1,67E+05 | Synthesis of epoxy (EET) and dihydroxyeicosatrienoic acids (DHET) | R-HSA-21426 |
| 1,88E+04 | Regulation of Complement cascade | R-HSA-97760 |
| 2,21E+05 | Platelet activation, signaling and aggregation | R-HSA-76002 |
| 2,62E+05 | Steroid hormone biosynthesis - Homo sapiens (human) | path:hsa0014 |
| 3,64E+04 | Mitochondrial Fatty Acid Beta-Oxidation | R-HSA-77289 |
| 4,41E+05 | Complement cascade | R-HSA-16665 |
| 5,35E+05 | Biosynthesis of DHA-derived SPMs | R-HSA-90186 |
| 1,06E+06 | Post-translational protein phosphorylation | R-HSA-89572 |
| 1,10E+06 | mitochondrial fatty acid beta-oxidation of saturated fatty acids | R-HSA-77286 |
| 1,39E+06 | Bile acid and bile salt metabolism | R-HSA-19406 |
| 1,47E+06 | Synthesis of bile acids and bile salts via 7alpha-hydroxycholesterol | R-HSA-19336 |
| 1,87E+06 | Choline catabolism | R-HSA-67981 |
| 1,94E+06 | Branched-chain amino acid catabolism | R-HSA-70895 |
| 2,92E+06 | Propanoate metabolism - Homo sapiens (human) | path:hsa0064 |
| 3,46E+06 | Protein localization | R-HSA-96095 |
| 3,99E+06 | Terminal pathway of complement | R-HSA-16666 |
| 7,72E+05 | Conjugation of salicylate with glycine | R-HSA-17712 |
| 0.000112313 | Primary bile acid biosynthesis - Homo sapiens (human) | path:hsa0012 |
| 0.000117168 | Binding and Uptake of Ligands by Scavenger Receptors | R-HSA-21737 |
| 0.000125243 | HDL remodeling | R-HSA-89640 |
| 0.000125243 | Conjugation of carboxylic acids | R-HSA-15942 |
| 0.000125243 | Amino Acid conjugation | R-HSA-15658 |
| 0.000125243 | Linoleic acid metabolism - Homo sapiens (human) | path:hsa0059 |
| 0.000144009 | Plasma lipoprotein assembly, remodeling, and clearance | R-HSA-17482 |
| 0.000149219 | Synthesis of bile acids and bile salts | R-HSA-19210 |
| 0.000157989 | Arachidonic acid metabolism - Homo sapiens (human) | path:hsa0059 |
| 0.000168371 | Drug metabolism - other enzymes - Homo sapiens (human) | path:hsa0098 |
| 0.000194965 | Lysine degradation - Homo sapiens (human) | path:hsa0031 |
| 0.000310593 | Coronavirus disease - COVID-19 - Homo sapiens (human) | path:hsa0517 |
| 0.000315028 | Cysteine and methionine metabolism - Homo sapiens (human) | path:hsa0027 |
| 0.000315028 | Cholesterol metabolism - Homo sapiens (human) | path:hsa0497 |
| 0.000349545 | Terpenoid backbone biosynthesis - Homo sapiens (human) | path:hsa0090 |
| 0.000349545 | RA biosynthesis pathway | R-HSA-53658 |
| 0.000377210 | Phase II - Conjugation of compounds | R-HSA-15658 |
| 0.000438880 | Beta oxidation of hexanoyl-CoA to butanoyl-CoA | R-HSA-77350 |
| 0.000438880 | Beta oxidation of lauroyl-CoA to decanoyl-CoA-CoA | R-HSA-77310 |
| 0.000438880 | Beta oxidation of octanoyl-CoA to hexanoyl-CoA | R-HSA-77348 |
| 0.000732686 | Synthesis of bile acids and bile salts via 24-hydroxycholesterol | R-HSA-19377 |
| 0.000752602 | Plasma lipoprotein remodeling | R-HSA-89638 |
| 0.000752602 | Hemostasis | R-HSA-10958 |
| 0.000821927 | Beta oxidation of decanoyl-CoA to octanoyl-CoA-CoA | R-HSA-77346 |
| 0.000880045 | Sulfur amino acid metabolism | R-HSA-16146 |
| 0.000901974 | Fatty acids | R-HSA-21193 |
| 0.000901974 | p130Cas linkage to MAPK signaling for integrins | R-HSA-37270 |
| 0.000901974 | GRB2:SOS provides linkage to MAPK signaling for Integrins | R-HSA-35419 |
| 0.000934497 | Fat digestion and absorption - Homo sapiens (human) | path:hsa0497 |
| 0.001052320 | Signaling by Retinoic Acid | R-HSA-53625 |
| 0.001102994 | Metabolism of vitamins and cofactors | R-HSA-19685 |
| 0.001154516 | Metabolism of porphyrins | R-HSA-18944 |
| 0.001274415 | Fructose metabolism | R-HSA-56520 |
| 0.001318757 | Peroxisomal lipid metabolism | R-HSA-39091 |
| 0.001318757 | Glyoxylate and dicarboxylate metabolism - Homo sapiens (human) | path:hsa0063 |
| 0.001363122 | Intrinsic Pathway of Fibrin Clot Formation | R-HSA-14083 |
| 0.001363122 | Heme degradation | R-HSA-18948 |
| 0.001363122 | Plasma lipoprotein assembly | R-HSA-89638 |
| 0.001729620 | Glyoxylate metabolism and glycine degradation | R-HSA-38966 |
| 0.001738233 | Bile secretion - Homo sapiens (human) | path:hsa0497 |
| 0.001906348 | Arginine and proline metabolism - Homo sapiens (human) | path:hsa0033 |
| 0.002698874 | Chylomicron assembly | R-HSA-89638 |
| 0.003062339 | Synthesis of Leukotrienes (LT) and Eoxins (EX) | R-HSA-21426 |
| 0.003287296 | Platelet Aggregation (Plug Formation) | R-HSA-76009 |
| 0.003613370 | Arginine biosynthesis - Homo sapiens (human) | path:hsa0022 |
| 0.003616018 | Synthesis and degradation of ketone bodies - Homo sapiens (human) | path:hsa0007 |
| 0.003616018 | Abacavir transport and metabolism | R-HSA-21615 |
| 0.004183829 | Proximal tubule bicarbonate reclamation - Homo sapiens (human) | path:hsa0496 |
| 0.004807777 | Synthesis of bile acids and bile salts via 27-hydroxycholesterol | R-HSA-19380 |
| 0.004993999 | Beta oxidation of myristoyl-CoA to lauroyl-CoA | R-HSA-77285 |
| 0.005058657 | Signaling by BRAF and RAF fusions | R-HSA-68029 |
| 0.005731343 | Metabolism of carbohydrates | R-HSA-71387 |
| 0.005924112 | Transport of organic anions | R-HSA-87951 |
| 0.005924112 | Lysine catabolism | R-HSA-71064 |
| 0.005924112 | Eicosanoids | R-HSA-21197 |
| 0.005924112 | Miscellaneous substrates | R-HSA-21195 |
| 0.006211617 | Activation of gene expression by SREBF (SREBP) | R-HSA-24261 |
| 0.006440032 | Transport of small molecules | R-HSA-38255 |
| 0.006986356 | Integrin signaling | R-HSA-35419 |
| 0.006986356 | Retinoid metabolism and transport | R-HSA-97563 |
| 0.006986356 | Integrin cell surface interactions | R-HSA-21608 |
| 0.007145169 | alpha-linolenic acid (ALA) metabolism | R-HSA-20461 |
| 0.007145169 | alpha-linolenic (omega3) and linoleic (omega6) acid metabolism | R-HSA-20461 |
| 0.007771794 | Signaling by high-kinase activity BRAF mutants | R-HSA-68029 |
| 0.008033662 | Glucose metabolism | R-HSA-70326 |
| 0.008424867 | Oncogenic MAPK signaling | R-HSA-68029 |
| 0.009684738 | Metabolism of fat-soluble vitamins | R-HSA-68066 |
| 0.009766537 | Ascorbate and aldarate metabolism - Homo sapiens (human) | path:hsa0005 |
| 0.009857410 | Innate Immune System | R-HSA-16824 |
| 0.010869839 | Regulation of cholesterol biosynthesis by SREBP (SREBF) | R-HSA-16558 |
| 0.012460399 | Recycling of bile acids and salts | R-HSA-15941 |
| 0.012460399 | Regulation of TLR by endogenous ligand | R-HSA-56869 |
| 0.012738887 | Abacavir metabolism | R-HSA-21615 |
| 0.012738887 | Beta oxidation of butanoyl-CoA to acetyl-CoA | R-HSA-77352 |
| 0.012738887 | Defects of contact activation system (CAS) and kallikrein/kinin system (KKS) | R-HSA-96514 |
| 0.012738887 | Diseases of hemostasis | R-HSA-96717 |
| 0.012738887 | Fructose catabolism | R-HSA-70350 |
| 0.012738887 | VLDL assembly | R-HSA-88664 |
| 0.012738887 | Formation of xylulose-5-phosphate | R-HSA-56612 |
| 0.013517979 | Visual phototransduction | R-HSA-21873 |
| 0.013780264 | Nitrogen metabolism - Homo sapiens (human) | path:hsa0091 |
| 0.013780264 | Phenylalanine metabolism - Homo sapiens (human) | path:hsa0036 |
| 0.013780264 | Gluconeogenesis | R-HSA-70263 |
| 0.013780264 | Pentose and glucuronate interconversions - Homo sapiens (human) | path:hsa0004 |
| 0.017934842 | Caffeine metabolism - Homo sapiens (human) | path:hsa0023 |
| 0.017934842 | Conjugation of benzoate with glycine | R-HSA-17713 |
| 0.018363780 | Alanine, aspartate and glutamate metabolism - Homo sapiens (human) | path:hsa0025 |
| 0.019222520 | Metabolism of water-soluble vitamins and cofactors | R-HSA-19684 |
| 0.019938797 | MAP2K and MAPK activation | R-HSA-56741 |
| 0.023834090 | TYSND1 cleaves peroxisomal proteins | R-HSA-90335 |
| 0.023834090 | Pantothenate and CoA biosynthesis - Homo sapiens (human) | path:hsa0077 |
| 0.023834090 | The canonical retinoid cycle in rods (twilight vision) | R-HSA-24539 |
| 0.026807077 | Histidine metabolism - Homo sapiens (human) | path:hsa0034 |
| 0.026807077 | Signaling downstream of RAS mutants | R-HSA-96499 |
| 0.026807077 | Signaling by RAS mutants | R-HSA-68029 |
| 0.026807077 | Paradoxical activation of RAF signaling by kinase inactive BRAF | R-HSA-68029 |
| 0.026807077 | Signaling by moderate kinase activity BRAF mutants | R-HSA-68029 |
| 0.026807077 | Signaling by RAF1 mutants | R-HSA-96562 |
| 0.029895545 | Synthesis of Ketone Bodies | R-HSA-77111 |
| 0.030038939 | Staphylococcus aureus infection - Homo sapiens (human) | path:hsa0515 |

**Suppl. Table 17: Validated differentially expressed genes per phenotype in the Barcelona cohort**

**cABMR upregulated**

ACHE
ADRA2A
AEBP1
AIM1L
AMICA1
ANO9
ATP10A
ATP2A3
BDKRB2
C16orf54
C7
CACNA1C
CALHM2
CCL19
CCL21
CCR7
CD19
CD200
CD22
CD37
CD6
CH25H
CILP2
CLDN11
CLMP
CLSTN2
COL8A2
CRISPLD2
CTHRC1
CTSE
CXCL6
CXCR4
DARC
DCDC2
DGKA
DKK3
DPYSL3
DTNA
DUSP8
EFEMP1
EPCAM
F13A1
F3
FAT1
FBLN5
FER1L4

FNDC1
FOXL1
GEM
ID4
IL8
ITGA2
ITGB2-AS1
KCNN4
KIAA1199
KIAA1644
KLHL29
KRT19
KRT23
LAMC3
LBH
LIF
LINC00313
LINC00319
LOXL1
LOXL4
LRRC1
LTBP2
LUM
MAP1A
MDFI
MEX3B
MICAL1
MMP14
MMP7
MOXD1
MS4A1
MTMR11
MUC13
NALCN
NFASC
OTOGL
P2RY10
PAPLN
PCDP1
PDCD1
PDGFA
PLCXD3
PTGDS
PTPRCAP
RAB3B
RGS4
RHOF
RLTPR
SCN7A
SHISA3
SIT1

SLC7A6
SMOC2
SPINT1
SPOCK2
ST8SIA1
STMN2
STMN3
SUSD2
SYTL1
TBC1D10C
THBS2
TIAM2
TMEM132A
TMEM132C
TMEM200A
TNFRSF12A
TTC9
VIM
VWF
WISP2
WNT10A
ZNF683

**cABMR downregulated**
AC004862.6
AC074289.1
AC097468.4
ACOT13
ACSM3
ANKRD23
AQP7P1
AQP7P3
ART5
ASPDH
ATM
ATP5L2
BBOX1
BRCA1
C14orf37
C1orf173
C5orf27
CHCHD10
CNGA1
COL28A1
COX6C
CTC-558O2.1
CYP2C19
DCDC5
DHRS4L2
DKFZP434I0714
DNM1

EFCAB1
EIF5AL1
EMBP1
FBXO40
GLYATL3
GNMT
HAO2
HAP1
HMGCS1
HSD17B14
IGF1
INTS6-AS1
KCNE1
KCNMA1
KCNN2
KLHL41
LINC00086
LINC01093
LOC100505985
LOC100509620
LOC100996723
LOC101927331
LOC101928230
LOC101928816
LOC101929633
LOC102723583
LOC102723845
LOC102724645
LOC643327
LPA
LRRC39
MEIOB
MME
MPPED2
MT1G
MURC
MYH3
NCAM2
NEURL1
NPIPB6
NRAP
PGAM2
PTH1R
RP11-10L7.1
RP11-12A2.3
RP11-242D8.1
RP11-458D21.1
RP11-75N4.2
SCG5
SLC9A3R2
SLCO4C1

SMIM6
SNHG9
SNTG1
SPOCK3
SQLE
SULT1B1
TCP10L
TENM3
TRHDE-AS1
USH2A
UXT-AS1
ZSCAN16-AS1

**clinTCMR upregulated**
AKR1B10
ALDH3A1
ALOX15B
AMICA1
ANLN
ANXA2
APOBEC3B
ASF1B
AURKA
BCAT1
BUB1
BUB1B
C12orf5
CAPG
CCAT1
CCL18
CCL19
CCL20
CCL4
CCNB1
CCNE1
CD19
CD27
CD300LF
CD37
CD3D
CD52
CD53
CD84
CD8A
CDC20
CDC25A
CDC6
CDCA2
CDCA5
CDCA8
CDKN1A

CDT1
CECR1
CENPA
CENPF
CENPI
CENPK
CENPM
CENPU
CHEK1
CHIT1
CLEC4E
CLSPN
CRIP1
CSF2RA
CTSE
CXCL10
CXCL11
CXCL9
CXCR4
CYTIP
DBNDD1
DEPDC1
DIAPH3
DLGAP5
DNAJC5B
DTL
DUSP2
DUSP8
E2F1
E2F2
E2F7
E2F8
EMILIN2
FABP5
FAM111B
FAM169A
FAM26F
FANCD2
FCGR1A
FOXM1
GAS7
GCNT3
GDF15
GINS1
GINS2
GLDN
GPNMB
GPRIN1
GTSE1
GZMK
HAPLN3

HAVCR2
HCST
HJURP
HKDC1
HLA-DMB
HLA-DQA1
HLA-DQB1
HLA-DRA
HMMR
HS1BP3-IT1
HS3ST2
ICOS
IFI30
IL1RL1
IL32
IL8
IQGAP3
ITGAX
ITM2A
KCNJ5
KIF18B
KIF20A
KIF2C
KIF4A
KLHL6
KPNA2
KRT18
KRT23
KRT8
LAIR1
LCK
LGALS3
LINC00152
LINC00313
LOC101059954
LOC102724385
LOC102724895
LOXL4
LRRC25
LYZ
MCM2
MCM3
MCM6
MELK
MICAL1
MKI67
MMP7
MMP9
MTHFD2
MUC13
MYBL2

NCAPG
NDC80
NEK2
NRIP3
ORC1
ORC6
OTOA
PAQR4
PBK
PCDP1
PCNA
PDGFA
PKMYT1
PLA2G7
PLXNC1
POLQ
PRC1
PTTG1
RAB3B
RAB42
RACGAP1
RARRES1
RGS1
RMI2
RNASE6
RPS6KL1
RRM2
SCIN
SERPINB8
SLA
SLAMF8
SLC1A3
SLC29A4
SLC51B
SOCS1
SPATA21
SPC24
SPOCK2
SPP1
SPSB1
SRGN
STMN1
STMN2
THBD
TIMP1
TK1
TNFRSF12A
TNFRSF9
TOP2A
TP73
TPX2

TREM2
TYMS
UBD
UBE2C
UBE2S
UBE2T
UBXN10
UHRF1
VAMP5
WDR76
ZBED2
ZNF367

**clinTCMR downregulated**
AC074138.3
AC097468.4
ACSM3
ADAM12
ADCY10P1
ADRB1
ASB4
ASXL3
AVPR1A
BCRP3
C19orf71
C1orf101
C2orf16
C2orf54
CELF6
CFHR4
CLEC4GP1
COL11A2
COPG2IT1
CRABP2
CTD-3080P12.3
CTNNA3
CXXC4
CYP2C19
CYS1
DCDC5
DGKB
DIO3OS
DKFZp434J0226
DLGAP2
DLK2
DNAH12
DNM1
DOK6
DPP10
DRD1
DSCAML1

EFCAB1
ENHO
FAM19A2
FAM9B
FLJ13197
FLJ34503
FRAS1
GLUD1P3
GOLGA8A
GSTA7P
HAO2-IT1
HERC2P7
HSD17B14
IDO2
IQUB
KCNIP3
LGI1
LGSN
LINC01093
LINC01118
LINC01144
LOC100133669
LOC100134822
LOC100287175
LOC100288974
LOC100996419
LOC100996813
LOC101059949
LOC101060212
LOC101926960
LOC101926980
LOC101927032
LOC101927121
LOC101927170
LOC101927595
LOC101927610
LOC101927635
LOC101928669
LOC101929055
LOC101929177
LOC101929443
LOC101929552
LOC101929633
LOC102546294
LOC102723356
LOC102723580
LOC102723845
LOC102723953
LOC102724329
LOC102724483
LOC102724603

LOC102724645
LOC102724660
LOC102724921
LOC339240
LOC646903
LOC730101
LRIT3
LRRC16B
MAST1
MBL1P
MCM3AP-AS1
MDN1
MEIOB
MST1L
MUSTN1
MYH3
NCAM2
NDRG1
NFKBIZ
NIPSNAP3B
NLRP14
NPIPB6
NPIPB9
PCDH19
PCK1
PCOLCE-AS1
PCOLCE2
PDE8B
PITPNM3
PLIN1
PP13
PRIMA1
RAD51AP2
RAMP2-AS1
RANBP3L
RGPD8
RP1-234P15.4
RP11-10L7.1
RP11-12A2.3
RP11-15A1.7
RP11-205M3.3
RP11-242D8.1
RP11-31F15.2
RP11-439L18.3
RP11-458D21.1
RP11-75N4.2
RP11-996F15.2
SCG5
SCN11A
SCN2A
SEC14L3

SEMA5B
SGK494
SLC18A2
SLC28A2
SLC30A10
SLC45A2
SLCO1A2
SNX29P2
TRHDE
TRHDE-AS1
TUBA3FP
TUBB1
UXT-AS1
YPEL2
ZFP2

#### Suppl. Table 18: Pathway enrichment of differentially expressed genes in the Barcelona validation cohort

##### cABMR upregulated

| q-value | name | ID | | q-value | name | ID |
|---|---|---|---|---|---|---|
| 0.00264074520482 | positive regulation of antigen processing and presentation | GO:0002579 | | 2,12E+05 | Extracellular matrix organization | R-HSA-1474244 |
| 0.00264074520482 | regulation of dendritic cell antigen processing and presentation | GO:0002604 | | 0.00249097869256 | Chemokine receptors bind chemokines | R-HSA-380108 |
| 0.00264074520482 | establishment of T cell polarity | GO:0001768 | | 0.00635697185409 | Complement and coagulation cascades - Homo sapiens (human) | path:hsa04610 |
| 0.00264074520482 | positive regulation of cellular component movement | GO:0051272 | | 0.00635697185409 | Assembly of collagen fibrils and other multimeric structures | R-HSA-2022090 |
| 0.0043251915703 | regulation of lymphocyte activation | GO:0051249 | | 0.00806184078958 | Hematopoietic cell lineage - Homo sapiens (human) | path:hsa04640 |
| 0.0043251915703 | chemokine-mediated signaling pathway | GO:0070098 | | 0.00806184078958 | Viral protein interaction with cytokine and cytokine receptor - Homo sapiens (human) | path:hsa04061 |
| 0.0043251915703 | cation transport | GO:0006812 | | 0.0130125157113 | G alpha (i) signalling events | R-HSA-418594 |
| 0.00584274220822 | response to chemokine | GO:1990868 | | 0.0133339356504 | Molecules associated with elastic fibres | R-HSA-2129379 |
| 0.00858531178963 | positive regulation of response to external stimulus | GO:0032103 | | 0.0148329045794 | Signaling by GPCR | R-HSA-372790 |
| 0.0092276724318 | regulation of antigen receptor-mediated signaling pathway | GO:0050854 | | 0.0148329045794 | Class A/1 (Rhodopsin-like receptors) | R-HSA-373076 |
| 0.0098402721712 | regulation of response to wounding | GO:1903034 | | 0.0148329045794 | Elastic fibre formation | R-HSA-1566948 |
| 0.0098402721712 | ion transmembrane transport | GO:0034220 | | 0.0148329045794 | Formation of Fibrin Clot (Clotting Cascade) | R-HSA-140877 |
| 0.0131942179334 | calcium ion transmembrane import into cytosol | GO:0097553 | | 0.0148329045794 | GPCR downstream signalling | R-HSA-388396 |
| 0.0131942179334 | cellular cation homeostasis | GO:0030003 | | 0.0148329045794 | Crosslinking of collagen fibrils | R-HSA-2243919 |
| 0.0131942179334 | positive regulation of response to wounding | GO:1903036 | | 0.0148329045794 | Peptide ligand-binding receptors | R-HSA-375276 |
| 0.0141191458909 | positive regulation of leukocyte migration | GO:0002687 | | 0.0151851496229 | ECM-receptor interaction - Homo sapiens (human) | path:hsa04512 |
| 0.0141191458909 | regulation of plasma membrane bounded cell projection assembly | GO:0120032 | | 0.0151851496229 | Cell adhesion molecules - Homo sapiens (human) | path:hsa04514 |
| 0.0181720972629 | ion homeostasis | GO:0050801 | | 0.0151851496229 | Non-integrin membrane-ECM interactions | R-HSA-3000171 |
| 0.0181720972629 | regulation of cell projection organization | GO:0031344 | | 0.0151851496229 | Cytokine-cytokine receptor interaction - Homo sapiens (human) | path:hsa04060 |
| 0.0186393568017 | regulation of neutrophil migration | GO:1902622 | | 0.0155075896893 | Collagen formation | R-HSA-1474290 |
| 0.0215209788652 | regulation of lipid metabolic process | GO:0019216 | | 0.0155075896893 | GPCR ligand binding | R-HSA-500792 |
| 0.0220601131921 | regulation of glycoprotein metabolic process | GO:1903018 | | 0.0168640602863 | Platelet Adhesion to exposed collagen | R-HSA-75892 |
| 0.0242175793829 | leukocyte apoptotic process | GO:0071887 | | 0.0254058190987 | Signal Transduction | R-HSA-162582 |
| 0.0242175793829 | neutrophil chemotaxis | GO:0030593 | | 0.0357382701758 | Chemokine signaling pathway - Homo sapiens (human) | path:hsa04062 |
| 0.0258035168092 | positive regulation of plasma membrane bounded cell projection assembly | GO:0120034 | | 0.0399164117813 | Focal adhesion - Homo sapiens (human) | path:hsa04510 |
| 0.0332564420551 | pseudopodium organization | GO:0031268 | | 0.0399164117813 | Phase 0 - rapid depolarisation | R-HSA-5576892 |
| 0.0332564420551 | vascular wound healing | GO:0061042 | | 0.0399164117813 | Hemostasis | R-HSA-109582 |
| 0.0332564420551 | gliogenesis | GO:0042063 | | 0.0413136940279 | Laminin interactions | R-HSA-3000157 |
| 0.0347554790487 | regulation of ion transport | GO:0043269 | | | | |
| 0.0377290131159 | neutrophil migration | GO:1990266 | | | | |
| 0.0412838605753 | regulation of intracellular signal transduction | GO:1902531 | | | | |
| 0.0425693030709 | positive regulation of cell adhesion mediated by integrin | GO:0033630 | | | | |
| 0.0425693030709 | peptide secretion | GO:0002790 | | | | |
| 0.0425693030709 | calcium-mediated signaling | GO:0019722 | | | | |
| 0.0437748510244 | astrocyte differentiation | GO:0048708 | | | | |
| 0.0448859886649 | positive regulation of cell communication | GO:0010647 | | | | |
| 0.0468103866801 | ERK1 and ERK2 cascade | GO:0070371 | | | | |
| 0.0478193945711 | acute inflammatory response | GO:0002526 | | | | |
| 0.0513781390994 | positive regulation of lipid metabolic process | GO:0045834 | | | | |
| 0.0562024834064 | positive regulation of hydrolase activity | GO:0051345 | | | | |
| 0.0562024834064 | positive regulation of collagen metabolic process | GO:0010714 | | | | |
| 0.0573897041421 | glycoprotein biosynthetic process | GO:0009101 | | | | |

##### cABMR downregulated

| q-value | name | ID | | q-value | name | ID |
|---|---|---|---|---|---|---|
| 0.00185762461822 | ribonucleotide metabolic process | GO:0009259 | | 0.00635464305046 | Acetylcholine inhibits contraction of outer hair cells | R-HSA-9667769 |
| 0.00185762461822 | nucleotide metabolic process | GO:0009117 | | 0.0113568866713 | Ca2+ activated K+ channels | R-HSA-1296052 |
| 0.00185762461822 | purine nucleotide metabolic process | GO:0006163 | | 0.0256111783015 | Fatty acid metabolism | R-HSA-8978868 |
| 0.00484170390762 | striated muscle contraction | GO:0006941 | | 0.0256111783015 | Metabolism | R-HSA-1430728 |
| 0.0142498438951 | amyloid-beta metabolic process | GO:0050435 | | 0.0256111783015 | Metabolism of lipids | R-HSA-556833 |
| 0.0142498438951 | nucleoside bisphosphate metabolic process | GO:0033865 | | 0.0256111783015 | Cholesterol biosynthesis | R-HSA-191273 |
| 0.0142498438951 | purine nucleoside bisphosphate metabolic process | GO:0034032 | | 0.0256111783015 | Activation of gene expression by SREBF (SREBP) | R-HSA-2426168 |
| 0.0142498438951 | striated muscle cell development | GO:0055002 | | 0.0256111783015 | Resolution of D-loop Structures through Synthesis-Dependent Strand Annealing (SDSA) | R-HSA-5693554 |
| 0.0232234828373 | regulation of alcohol biosynthetic process | GO:1902930 | | 0.0256111783015 | Butanoate metabolism - Homo sapiens (human) | path:hsa00650 |
| 0.037461403488 | DNA double-strand break processing | GO:0000729 | | 0.0256111783015 | Regulation of cholesterol biosynthesis by SREBP (SREBF) | R-HSA-1655829 |
| 0.037461403488 | regulation of carbohydrate biosynthetic process | GO:0043255 | | 0.0256111783015 | Resolution of D-loop Structures through Holliday Junction Intermediates | R-HSA-5693568 |
| 0.0498414339407 | nucleoside triphosphate metabolic process | GO:0009141 | | 0.0256111783015 | Resolution of D-Loop Structures | R-HSA-5693537 |
| 0.0536074828525 | nucleoside monophosphate metabolic process | GO:0009123 | | 0.0256111783015 | Mitochondrial Fatty Acid Beta-Oxidation | R-HSA-77289 |
| 0.0536074828525 | G2 DNA damage checkpoint | GO:0031572 | | 0.0256111783015 | HDR through Single Strand Annealing (SSA) | R-HSA-5685938 |
| 0.0558065042052 | sarcomere organization | GO:0045214 | | 0.0256111783015 | Aldosterone-regulated sodium reabsorption - Homo sapiens (human) | path:hsa04960 |
| 0.0600165964907 | striated muscle cell differentiation | GO:0051146 | | 0.0256111783015 | Metabolism of steroids | R-HSA-8957322 |
| 0.0746823847997 | regulation of cellular carbohydrate metabolic process | GO:0010675 | | 0.0256111783015 | Presynaptic phase of homologous DNA pairing and strand exchange | R-HSA-5693616 |
| 0.080471315162 | pyridine-containing compound biosynthetic process | GO:0072525 | | 0.0256111783015 | Glycine, serine and threonine metabolism - Homo sapiens (human) | path:hsa00260 |
| 0.0884519239509 | amyloid precursor protein catabolic process | GO:0042987 | | 0.0256111783015 | Homologous recombination - Homo sapiens (human) | path:hsa03440 |
| | | | | 0.0256111783015 | Homologous DNA Pairing and Strand Exchange | R-HSA-5693579 |
| | | | | 0.0342141651074 | Sensory processing of sound by outer hair cells of the cochlea | R-HSA-9662361 |
| | | | | 0.036543084496 | Endocrine and other factor-regulated calcium reabsorption - Homo sapiens (human) | path:hsa04961 |

#### **clinTCMR upregulated**

| q-value | name | ID |
|---|---|---|
| 5,52E-06 | nuclear division | GO:0000280 |
| 1,10E-04 | regulation of cell cycle process | GO:0010564 |
| 2,57E-04 | regulation of mitotic cell cycle | GO:0007346 |
| 2,74E-02 | G1/S transition of mitotic cell cycle | GO:0000082 |
| 2,68E+01 | DNA replication | GO:0006260 |
| 2,86E+01 | negative regulation of cell cycle | GO:0045786 |
| 8,42E+03 | signal transduction involved in DNA integrity checkpoint | GO:0072401 |
| 8,42E+03 | signal transduction involved in DNA damage checkpoint | GO:0072422 |
| 1,62E+04 | DNA replication initiation | GO:0006270 |
| 1,64E+04 | G2/M transition of mitotic cell cycle | GO:0000086 |
| 1,91E+04 | positive regulation of cell cycle | GO:0045787 |
| 4,59E+04 | cellular response to cytokine stimulus | GO:0071345 |
| 6,09E+04 | negative regulation of chromosome segregation | GO:0051985 |
| 1,35E+05 | mitotic DNA damage checkpoint | GO:0044773 |
| 1,42E+05 | mitotic spindle assembly checkpoint | GO:0007094 |
| 1,70E+05 | centromere complex assembly | GO:0034508 |
| 3,56E+05 | G1 DNA damage checkpoint | GO:0044783 |
| 4,15E+05 | chemokine-mediated signaling pathway | GO:0070098 |
| 7,65E+05 | regulation of organelle organization | GO:0033043 |
| 8,58E+05 | response to chemokine | GO:1990868 |
| 8,91E+04 | antigen processing and presentation of exogenous peptide antigen via MHC class II | GO:0019886 |
| 1,19E+06 | DNA conformation change | GO:0071103 |
| 1,46E+06 | DNA damage response, signal transduction by p53 class mediator | GO:0030330 |
| 1,51E+06 | DNA replication-independent nucleosome organization | GO:0034724 |
| 1,66E+06 | protein-DNA complex assembly | GO:0065004 |
| 4,80E+06 | neutrophil migration | GO:1990266 |
| 5,17E+06 | response to interferon-gamma | GO:0034341 |
| 5,95E+06 | regulation of syncytium formation by plasma membrane fusion | GO:0060142 |
| 8,50E+06 | chromatin remodeling at centromere | GO:0031055 |
| 8,50E+06 | regulation of protein metabolic process | GO:0051246 |
| 8,63E+06 | neutrophil chemotaxis | GO:0030593 |
| 8,64E+06 | phosphorylation | GO:0016310 |
| 8,64E+06 | regulation of cellular protein metabolic process | GO:0032268 |
| 0.000115705212636 | positive regulation of transferase activity | GO:0051347 |
| 0.000142746201899 | response to UV | GO:0009411 |
| 0.000159560329614 | negative regulation of cellular component organization | GO:0051129 |
| 0.000174948120454 | regulation of T cell activation via T cell receptor contact with antigen bound to MHC molecule on antigen presenting cell | GO:2001188 |
| 0.000218127345154 | histone exchange | GO:0043486 |
| 0.000412160356908 | DNA repair | GO:0006281 |
| 0.000490756111528 | regulation of phosphorus metabolic process | GO:0051174 |
| 0.000518660519696 | cellular response to light stimulus | GO:0071482 |
| 0.0008577719409 | nucleosome assembly | GO:0006334 |
| 0.0008577719409 | positive regulation of cellular component movement | GO:0051272 |
| 0.0008577719409 | mitotic DNA replication checkpoint | GO:0033314 |
| 0.000901875060233 | regulation of fibroblast proliferation | GO:0048145 |
| 0.00116933222393 | exit from mitosis | GO:0010458 |
| 0.00121629744275 | DNA biosynthetic process | GO:0071897 |
| 0.00123028882369 | cellular protein-containing complex assembly | GO:0034622 |
| 0.00126378674604 | DNA strand elongation | GO:0022616 |
| 0.00178774251865 | ATP-dependent chromatin remodeling | GO:0043044 |
| 0.00179718042944 | regulation of cell cycle checkpoint | GO:1901976 |
| 0.00182569859157 | interferon-gamma-mediated signaling pathway | GO:0060333 |
| 0.00194104341485 | negative regulation of transcription involved in G1/S transition of mitotic cell cycle | GO:0071930 |
| 0.00232978958737 | regulation of DNA metabolic process | GO:0051052 |
| 0.00232978958737 | positive regulation of chromosome segregation | GO:0051984 |
| 0.00232978958737 | G2 DNA damage checkpoint | GO:0031572 |
| 0.00260099597164 | hemopoiesis | GO:0030097 |
| 0.00265293395635 | positive regulation of leukocyte migration | GO:0002687 |
| 0.00281232271194 | hepatocyte differentiation | GO:0070365 |
| 0.00281232271194 | negative regulation of cell division | GO:0051782 |
| 0.002880068373 | regulation of cyclin-dependent protein kinase activity | GO:1904029 |
| 0.00323211258744 | regulation of lymphocyte activation | GO:0051249 |
| 0.00333176364916 | positive regulation of cellular component organization | GO:0051130 |
| 0.00358414336196 | negative regulation of cell activation | GO:0050866 |
| 0.00372916354493 | morphogenesis of an endothelium | GO:0003159 |
| 0.00441068210267 | DNA recombination | GO:0006310 |
| 0.00529312024009 | response to tumor necrosis factor | GO:0034612 |
| 0.00602075650543 | regulation of leukocyte cell-cell adhesion | GO:1903037 |
| 0.00628029370833 | cellular protein modification process | GO:0006464 |
| 0.00628029370833 | dendritic cell differentiation | GO:0097028 |
| 0.00669173888962 | regulation of mononuclear cell migration | GO:0071675 |
| 0.00694376002288 | regulation of CD40 signaling pathway | GO:2000348 |
| 0.00694376002288 | negative regulation of hydrolase activity | GO:0051346 |
| 0.00739053206871 | cellular protein complex disassembly | GO:0043624 |
| 0.0089741139563 | calcium ion transmembrane import into cytosol | GO:0097553 |
| 0.00923601047808 | columnar/cuboidal epithelial cell maturation | GO:0002069 |
| 0.0105742896418 | positive regulation of leukocyte activation | GO:0002696 |
| 0.0107982380527 | liver development | GO:0001889 |
| 0.0131097831602 | positive regulation of cell activation | GO:0050867 |
| 0.0136378660481 | positive regulation of cell proliferation | GO:0008284 |
| 0.0146674125449 | positive regulation of antigen processing and presentation | GO:0002579 |
| 0.014846688703 | liver regeneration | GO:0097421 |
| 0.0151809624172 | positive regulation of hemopoiesis | GO:1903708 |
| 0.0155780697647 | ERK1 and ERK2 cascade | GO:0070371 |
| 0.016386176523 | positive regulation of response to external stimulus | GO:0032103 |
| 0.0184416012974 | regulation of intracellular signal transduction | GO:1902531 |
| 0.0190806640019 | positive regulation of macromolecule metabolic process | GO:0010604 |
| 0.0214865070361 | positive regulation of nitrogen compound metabolic process | GO:0051173 |
| 0.0217540693128 | positive regulation of cell-cell adhesion | GO:0022409 |
| 0.0237756809142 | intrinsic apoptotic signaling pathway by p53 class mediator | GO:0072332 |
| 0.0241712107607 | regulation of leukocyte differentiation | GO:1902105 |
| 0.0249716396665 | positive regulation of cell differentiation | GO:0045597 |
| 0.0249716396665 | regulation of leukocyte chemotaxis | GO:0002688 |
| 0.0266958219706 | regulation of peroxisome proliferator activated receptor signaling pathway | GO:0035358 |
| 0.0275325629852 | positive regulation of cell communication | GO:0010647 |
| 0.0275918103957 | T-helper 1 type immune response | GO:0042088 |
| 0.0275918103957 | positive regulation of signal transduction | GO:0009967 |
| 0.0275918103957 | positive regulation of cellular metabolic process | GO:0031325 |
| 0.0285353349604 | negative regulation of leukocyte activation | GO:0002695 |
| 0.0290909889982 | deoxyribonucleotide biosynthetic process | GO:0009263 |
| 0.0310530916864 | regulation of response to wounding | GO:1903034 |
| 0.032655714633 | regulation of chemokine production | GO:0032642 |
| 0.0359053357367 | regulation of glycoprotein metabolic process | GO:1903018 |

| q-value | name | ID |
|---|---|---|
| 2,03E-14 | Cell Cycle | R-HSA-1640170 |
| 4,92E-14 | Cell Cycle, Mitotic | R-HSA-69278 |
| 1,13E-08 | Cell Cycle Checkpoints | R-HSA-69620 |
| 2,93E-03 | Mitotic G1 phase and G1/S transition | R-HSA-453279 |
| 3,60E-03 | Cell cycle - Homo sapiens (human) | path:hsa04110 |
| 8,17E-02 | G1/S Transition | R-HSA-69206 |
| 4,78E-02 | G1/S-Specific Transcription | R-HSA-69205 |
| 4,78E-02 | S Phase | R-HSA-69242 |
| 1,11E+00 | Separation of Sister Chromatids | R-HSA-2467813 |
| 1,24E+01 | Resolution of Sister Chromatid Cohesion | R-HSA-2500257 |
| 1,89E+01 | Synthesis of DNA | R-HSA-69239 |
| 2,02E+01 | Mitotic Spindle Checkpoint | R-HSA-69618 |
| 3,72E+01 | DNA Replication | R-HSA-69306 |
| 5,68E+01 | Mitotic Anaphase | R-HSA-68882 |
| 5,77E+01 | Mitotic Metaphase and Anaphase | R-HSA-2555396 |
| 2,62E+02 | Amplification of signal from unattached kinetochores via a MAD2 inhibitory signal | R-HSA-141444 |
| 2,62E+02 | Amplification of signal from the kinetochores | R-HSA-141424 |
| 3,16E+02 | Mitotic Prometaphase | R-HSA-68877 |
| 3,35E+02 | Switching of origins to a post-replicative state | R-HSA-69052 |
| 3,76E+01 | EML4 and NUDC in mitotic spindle formation | R-HSA-9648025 |
| 4,32E+02 | RHO GTPases Activate Formins | R-HSA-5663220 |
| 6,66E+02 | Activation of ATR in response to replication stress | R-HSA-176187 |
| 2,95E+03 | G2/M Checkpoints | R-HSA-69481 |
| 3,32E+03 | Assembly of the pre-replicative complex | R-HSA-68867 |
| 9,80E+03 | M Phase | R-HSA-68886 |
| 1,81E+04 | G0 and Early G1 | R-HSA-1538133 |
| 1,85E+04 | Mitotic G2-G2/M phases | R-HSA-453274 |
| 2,27E+04 | Orc1 removal from chromatin | R-HSA-68949 |
| 2,51E+04 | Polo-like kinase mediated events | R-HSA-156711 |
| 5,38E+04 | APC/C-mediated degradation of cell cycle proteins | R-HSA-174143 |
| 5,38E+04 | Regulation of mitotic cell cycle | R-HSA-453276 |
| 7,27E+04 | Human T-cell leukemia virus 1 infection - Homo sapiens (human) | path:hsa05166 |
| 9,28E+04 | APC/C:Cdc20 mediated degradation of mitotic proteins | R-HSA-176409 |
| 1,07E+05 | G2/M Transition | R-HSA-69275 |
| 1,07E+05 | Activation of APC/C and APC/C:Cdc20 mediated degradation of mitotic proteins | R-HSA-176814 |
| 1,07E+05 | Activation of the pre-replicative complex | R-HSA-68962 |
| 1,51E+05 | TP53 Regulates Transcription of Cell Cycle Genes | R-HSA-6791312 |
| 1,66E+05 | Unwinding of DNA | R-HSA-176974 |
| 2,28E+05 | DNA Replication Pre-Initiation | R-HSA-69002 |
| 2,53E+05 | Hematopoietic cell lineage - Homo sapiens (human) | path:hsa04640 |
| 3,81E+05 | TP53 Regulates Transcription of Genes Involved in G1 Cell Cycle Arrest | R-HSA-6804116 |
| 7,96E+05 | Transcription of E2F targets under negative control by DREAM complex | R-HSA-1362277 |
| 1,04E+06 | RHO GTPase Effectors | R-HSA-195258 |
| 1,59E+06 | DNA strand elongation | R-HSA-69190 |
| 2,32E+05 | Cytokine-cytokine receptor interaction - Homo sapiens (human) | path:hsa04060 |
| 3,14E+06 | Signaling by Rho GTPases, Miro GTPases and RHOBTB3 | R-HSA-9716542 |
| 6,85E+06 | Signaling by Rho GTPases | R-HSA-194315 |
| 6,87E+06 | Chemokine receptors bind chemokines | R-HSA-380108 |
| 8,44E+06 | Deposition of new CENPA-containing nucleosomes at the centromere | R-HSA-606279 |
| 8,44E+06 | Nucleosome assembly | R-HSA-774815 |
| 0.000108819701133 | APC-Cdc20 mediated degradation of Nek2A | R-HSA-179409 |
| 0.000126184135098 | CDT1 association with the CDC6:ORC:origin complex | R-HSA-68827 |
| 0.000126233405762 | Translocation of ZAP-70 to Immunological synapse | R-HSA-202430 |
| 0.000132277946223 | Intestinal immune network for IgA production - Homo sapiens (human) | path:hsa04672 |
| 0.000169870396328 | APC:Cdc20 mediated degradation of cell cycle proteins prior to satisfation of the cell cycle checkpoint | R-HSA-179419 |
| 0.000169870396328 | APC/C:Cdh1 mediated degradation of Cdc20 and other APC/C:Cdh1 targeted proteins in late mitosis/early G1 | R-HSA-174178 |
| 0.000169938376661 | p53 signaling pathway - Homo sapiens (human) | path:hsa04115 |
| 0.000169938376661 | Viral protein interaction with cytokine and cytokine receptor - Homo sapiens (human) | path:hsa04061 |
| 0.00019036510984 | Phosphorylation of CD3 and TCR zeta chains | R-HSA-202427 |
| 0.000194330616616 | Epstein-Barr virus infection - Homo sapiens (human) | path:hsa05169 |
| 0.000216093672648 | PD-1 signaling | R-HSA-389948 |
| 0.000341998363913 | Signal Transduction | R-HSA-162582 |
| 0.000424536386603 | Primary immunodeficiency - Homo sapiens (human) | path:hsa05340 |
| 0.000424536386603 | Regulation of APC/C activators between G1/S and early anaphase | R-HSA-176408 |
| 0.000449925989046 | MHC class II antigen presentation | R-HSA-2132295 |
| 0.000490914172612 | CDC6 association with the ORC:origin complex | R-HSA-68689 |
| 0.000547976660609 | G1/S DNA Damage Checkpoints | R-HSA-69615 |
| 0.000585556935176 | Immune System | R-HSA-168256 |
| 0.000594853204197 | Cellular senescence - Homo sapiens (human) | path:hsa04218 |
| 0.000617370507502 | E2F mediated regulation of DNA replication | R-HSA-113510 |
| 0.000617370507502 | Inactivation of APC/C via direct inhibition of the APC/C complex | R-HSA-141430 |
| 0.000617370507502 | Inhibition of the proteolytic activity of APC/C required for the onset of anaphase by mitotic spindle checkpoint components | R-HSA-141405 |
| 0.000659828234506 | E2F-enabled inhibition of pre-replication complex formation | R-HSA-113507 |
| 0.000670289446418 | Staphylococcus aureus infection - Homo sapiens (human) | path:hsa05150 |
| 0.000673259458685 | Generation of second messenger molecules | R-HSA-202433 |
| 0.000734844605618 | Adaptive Immune System | R-HSA-1280218 |
| 0.000734844605618 | Kinesins | R-HSA-983189 |
| 0.000802035944237 | Cyclin A/B1/B2 associated events during G2/M transition | R-HSA-69273 |
| 0.000802035944237 | APC/C:Cdc20 mediated degradation of Securin | R-HSA-174154 |
| 0.000934001104706 | APC/C:Cdc20 mediated degradation of Cyclin B | R-HSA-174048 |
| 0.00114737616371 | The role of GTSE1 in G2/M progression after G2 checkpoint | R-HSA-8852276 |
| 0.0014256630665 | Costimulation by the CD28 family | R-HSA-388841 |
| 0.0015107452374 | Antigen processing and presentation - Homo sapiens (human) | path:hsa04612 |
| 0.00158930621951 | Cdc20:Phospho-APC/C mediated degradation of Cyclin A | R-HSA-174184 |
| 0.00158930621951 | CDK-mediated phosphorylation and removal of Cdc6 | R-HSA-69017 |
| 0.0016270797262 | Downstream TCR signaling | R-HSA-202424 |
| 0.00177483637974 | Asthma - Homo sapiens (human) | path:hsa05310 |
| 0.0017970089733 | Transcriptional Regulation by TP53 | R-HSA-3700989 |
| 0.00238044163944 | Cytokine Signaling in Immune system | R-HSA-1280215 |
| 0.003365354994 | Th1 and Th2 cell differentiation - Homo sapiens (human) | path:hsa04658 |
| 0.00344674416176 | DNA replication - Homo sapiens (human) | path:hsa03030 |
| 0.00369762717729 | Oocyte meiosis - Homo sapiens (human) | path:hsa04114 |
| 0.00374601051733 | Allograft rejection - Homo sapiens (human) | path:hsa05330 |
| 0.00402602376769 | Interferon gamma signaling | R-HSA-877300 |
| 0.0042043892177 | Prostate cancer - Homo sapiens (human) | path:hsa05215 |
| 0.00423628377316 | Chromosome Maintenance | R-HSA-73886 |
| 0.00445150724322 | TP53 Regulates Transcription of Genes Involved in G2 Cell Cycle Arrest | R-HSA-6804114 |
| 0.00502891483338 | G2/M DNA replication checkpoint | R-HSA-69478 |
| 0.00519670133611 | Bladder cancer - Homo sapiens (human) | path:hsa05219 |
| 0.00563313324156 | Graft-versus-host disease - Homo sapiens (human) | path:hsa05332 |
| 0.00603221236807 | Type I diabetes mellitus - Homo sapiens (human) | path:hsa04940 |
| 0.00603221236807 | Signaling by SCF-KIT | R-HSA-1433557 |
| 0.00636127526772 | TCR signaling | R-HSA-202403 |
| 0.00636127526772 | Th17 cell differentiation - Homo sapiens (human) | path:hsa04659 |
| 0.00646465216203 | Phosphorylation of the APC/C | R-HSA-176412 |
| 0.00646465216203 | Conversion from APC/C:Cdc20 to APC/C:Cdh1 in late anaphase | R-HSA-176407 |
| 0.00679037147615 | Phosphorylation of Emi1 | R-HSA-176417 |
| 0.00679037147615 | Assembly of the ORC complex at the origin of replication | R-HSA-68616 |
| 0.00722777207015 | Leishmaniasis - Homo sapiens (human) | path:hsa05140 |
| 0.00744535211438 | Chemokine signaling pathway - Homo sapiens (human) | path:hsa04062 |
| 0.00901355062396 | Ubiquitin Mediated Degradation of Phosphorylated Cdc25A | R-HSA-69601 |
| 0.00901355062396 | p53-Independent DNA Damage Response | R-HSA-69610 |
| 0.00901355062396 | p53-Independent G1/S DNA damage checkpoint | R-HSA-69613 |
| 0.00963079597295 | COPI-dependent Golgi-to-ER retrograde traffic | R-HSA-6811434 |
| 0.0107878138924 | Autoimmune thyroid disease - Homo sapiens (human) | path:hsa05320 |
| 0.0135817895376 | MicroRNAs in cancer - Homo sapiens (human) | path:hsa05206 |
| 0.013929584497 | Immunoregulatory interactions between a Lymphoid and a non-Lymphoid cell | R-HSA-198933 |
| 0.0144935759684 | Activation of PUMA and translocation to mitochondria | R-HSA-139915 |
| 0.0144935759684 | Cyclin E associated events during G1/S transition | R-HSA-69202 |
| 0.0144935759684 | Cyclin A:Cdk2-associated events at S phase entry | R-HSA-69656 |
| 0.0144935759684 | Activation of Matrix Metalloproteinases | R-HSA-1592389 |
| 0.0147269916476 | Rheumatoid arthritis - Homo sapiens (human) | path:hsa05323 |
| 0.0158830222322 | Viral myocarditis - Homo sapiens (human) | path:hsa05416 |
| 0.0170931097448 | Synthesis of active ubiquitin: roles of E1 and E2 enzymes | R-HSA-8866652 |
| 0.0171066186758 | Tuberculosis - Homo sapiens (human) | path:hsa05152 |
| 0.0202119341114 | Progesterone-mediated oocyte maturation - Homo sapiens (human) | path:hsa04914 |
| 0.0203452816738 | Transcription of E2F targets under negative control by p107 (RBL1) and p130 (RBL2) in complex with HDAC1 | R-HSA-1362300 |
| 0.0214701060735 | Inflammatory bowel disease - Homo sapiens (human) | path:hsa05321 |
| 0.023026325876 | Transcriptional Regulation by E2F6 | R-HSA-8953750 |
| 0.023026325876 | Toll-like receptor signaling pathway - Homo sapiens (human) | path:hsa04620 |
| 0.0234813592517 | Condensation of Prometaphase Chromosomes | R-HSA-2514853 |
| 0.0243303542096 | Cell adhesion molecules - Homo sapiens (human) | path:hsa04514 |
| 0.0243303542096 | PI3K/AKT Signaling in Cancer | R-HSA-2219528 |

#### **clinTCMR downregulated**

| q-value | name | ID | | q-value | pathway | external_id |
|---|---|---|---|---|---|---|
| 0.148942366467 | adenylate cyclase-activating adrenergic receptor signaling pathway | GO:0071880 | | 0.0560918060293 | Phase 0 - rapid depolarisation | R-HSA-5576892 |
| 0.148942366467 | amine transport | GO:0015837 | | 0.0560918060293 | Muscle contraction | R-HSA-397014 |
| 0.148942366467 | regulation of lipid biosynthetic process | GO:0046890 | | 0.0560918060293 | Gap junction - Homo sapiens (human) | path:hsa04540 |
| 0.148942366467 | neuronal action potential | GO:0019228 | | 0.0560918060293 | Interaction between L1 and Ankyrins | R-HSA-445095 |
| 0.148942366467 | membrane depolarization during action potential | GO:0086010 | | 0.0560918060293 | L1CAM interactions | R-HSA-373760 |
| 0.148942366467 | embryonic limb morphogenesis | GO:0030326 | | 0.0560918060293 | Cardiac conduction | R-HSA-5576891 |
| | | | | 0.0560918060293 | Axon guidance | R-HSA-422475 |
| | | | | 0.0560918060293 | Nervous system development | R-HSA-9675108 |
| | | | | 0.0560918060293 | SLC-mediated transmembrane transport | R-HSA-425407 |
| | | | | 0.0560918060293 | Amine ligand-binding receptors | R-HSA-375280 |
| | | | | 0.0560918060293 | Transport of vitamins, nucleosides, and related molecules | R-HSA-425397 |

[13] Barascuk N, Veidal SS, Larsen L, et al. A novel assay for extracellular matrix remodeling associated with liver fibrosis: An enzyme-linked immunosorbent

assay (ELISA) for a MMP-9 proteolytically revealed neo-epitope of type III collagen. Clin Biochem 2010;43:899–904. https://doi.org/10.1016/j.clinbiochem.2010.03.012.
[14] Sand JM, Larsen L, Hogaboam C, et al. MMP Mediated Degradation of Type IV Collagen Alpha 1 and Alpha 3 Chains Reflects Basement Membrane Remodeling in Experimental and Clinical Fibrosis – Validation of Two Novel Biomarker Assays. PLoS One 2013;8:e84934. https://doi.org/10.1371/journal.pone.0084934.
[15] Vassiliadis E, Oliveira CP, Alvares-da-Silva MR, et al. Circulating levels of citrullinated and MMP-degraded vimentin (VICM) in liver fibrosis related pathology. Am J Transl Res 2012;4:403–14.
[16] Jensen C, Sinkeviciute D, Madsen DH, et al. Granzyme B Degraded Type IV Collagen Products in Serum Identify Melanoma Patients Responding to Immune Checkpoint Blockade. Cancers (Basel) 2020;12:2786. https://doi.org/10.3390/cancers12102786.
